# Hundreds of cardiac MRI traits derived using 3D diffusion autoencoders share a common genetic architecture

**DOI:** 10.1101/2024.11.04.24316700

**Authors:** Sara Ometto, Soumick Chatterjee, Andrea Mario Vergani, Arianna Landini, Sodbo Sharapov, Edoardo Giacopuzzi, Alessia Visconti, Emanuele Bianchi, Federica Santonastaso, Emanuel M. Soda, Francesco Cisternino, Carlo Andrea Pivato, Francesca Ieva, Emanuele Di Angelantonio, Nicola Pirastu, Craig A. Glastonbury

## Abstract

Biobank-scale imaging provides an unprecedented opportunity to characterise thousands of organ phenotypes, how they vary in populations and how they relate to disease outcomes. However, deriving specific phenotypes from imaging data, such as Magnetic Resonance Imaging (MRI), requires time-consuming expert annotation, limiting scalability, and does not exploit how information-dense such image acquisitions are. In this study, we developed a 3D diffusion autoencoder to derive latent phenotypes from temporally resolved cardiac MRI data of 71,021 UK Biobank participants. These phenotypes were reproducible, heritable (*h*^2^ = [4 - 18%]), and significantly associated with cardiometabolic traits and outcomes, including atrial fibrillation (*P* = 8.5 × 10^−29^) and myocardial infarction (*P* = 3.7 × 10^−12^). By using latent space manipulation techniques, we were able to learn, directly interpret and visualise what specific latent phenotypes are capturing in a given MRI. To establish the genetic basis of such traits, we performed a genome-wide association study, identifying 89 significant common variants (*P <* 2.3 × 10^−9^) across 42 loci, including seven novel loci. Extensive multi-trait colocalisation analyses (PP.H_4_ *>* 0.8) linked variants across phenotypic scales, from intermediate cardiac traits to cardiac disease endpoints. For example, rs142556838 that falls in *CCDC141* colocalises with a latent imaging phenotype and a diastolic blood pressure locus. Using single-cell RNA-sequencing data we map *CCDC141* expression specifically to a population of ventricular cardiomyocytes. Finally, Polygenic Risk Scores (PRS) derived from latent phenotypes demonstrated predictive power for a range of cardiometabolic diseases and enabled us to successfully stratify the individuals into different risk groups. In conclusion, this study showcases the use of diffusion autoencoding methods as powerful tools for unsupervised phenotyping, genetic discovery and disease risk prediction using cardiac MRI data.

## Main

Biobank scale non-invasive medical imaging coupled with advances in unsupervised machine learning provides an unprecedented opportunity for unbiased phenotyping at scale. For example, UK Biobank (UKB) Magnetic Resonance Imaging (MRI) acquisitions contain a plethora of quantifiable structural and functional phenotypes that remain untouched. Previous research efforts, using cardiac MRIs, have been used to extract single quantitative phenotypes (e.g. Left Ventricle Ejection Fraction (LVEF)) [1, 2]. These traits, termed “*image-derived phenotypes*” (IDPs) and their variability within a population, can be linked with electronic healthcare records to understand coincident diseases, as well as common and rare genetic variation through Genome Wide Association Studies (GWAS). However, each derived trait requires time-consuming expert annotation and specific model development, limiting both the scalability of phenotypic quantification and their downstream genetic characterisation. Moreover, 3D or temporal MRI acquisitions are extremely information rich and therefore deriving single phenotypes of organ function is reductive for such complex systems. Recent breakthroughs in machine learning (e.g. diffusion) allow interpretable phenotypes to be learnt from images without any specific annotations. These models learn to compress and represent high-dimensional imaging data into a low-dimensional representation (*z*) [3], where each dimension of *z* can be thought of as capturing a specific imaging feature or “latent phenotype”. Whilst such approaches have been applied to 2D natural images (e.g. faces, scenic photography, etc) [4], they have not been extended to work with time resolved medical imaging data. Importantly, approaches using non-diffusion methods, suffer from the inability to explain or interpret what each latent phenotype is capturing in the image. Here, we make a step change, to enable both high throughput and interpretable phenotyping by extending diffusion autoencoders to work with 3D data, enabling us to learn phenotypes from cardiac MRIs captured over 50 time points (**1A**).

We demonstrate that these learnt latent phenotypes recapitulate both known and novel cardiac phenotype and disease associations, both epidemiologically and genetically (**1B-C**). By utilising extensive multi-trait colocalisation analyses, we provide for the first time, a high resolution map of shared common causal variants that influence traits across phenotypic scales, from low-level cardiac endophenotypes (e.g. heart rate), risk factors (LDL cholesterol levels) and cardiac disease outcomes (myocardial infarction) (**1D**). Finally, as imaging provides a means of unbiased prognostic modelling, we show that polygenic risk scores built from image derived latent factors can pinpoint individuals with up to a 26x increased cumulative hazard for several common cardiac-relevant diseases (**1F**). In summary, we have conducted the largest systematic characterisation of cardiac imaging, intermediate cardiac traits and cardiac diseases to date and demonstrate that unsupervised phenotyping of cardiac MRIs using diffusion processes provides a tractable means for novel genetic discovery, risk prediction and interpretation.

## Results

### Unsupervised phenotyping by 3D diffusion autoencoders of cardiac MRIs

To derive unbiased phenotypes in a high-throughput manner, we exploited 4 chamber view Electrocardiogram (ECG) synchronised temporal cardiac MRI acquisitions in UK Biobank, a modality that provides a detailed characterisation of overall cardiac function. Whilst multiple studies have focused on deriving single phenotypes from such images [5, 6, 7], unsupervised and self-supervised representation learning methods have increasingly been shown to be capable of representing complex images as a compressed vector of latent factors, in which each dimension of the latent vector corresponds to a specific phenotype or feature present in the image (here referred to as *latent phenotype*) [8, 9, 10, 11]. Therefore, we set out to extract latent phenotypes from all 71,021 UK Biobank individuals with available cardiac MRIs, which we can use for diagnostic and prognostic risk modelling as well as genetically characterising the basis of hundreds of cardiometabolically relevant traits.

After quality control steps to remove noisy or failed acquisitions along with breathing artefacts, images were cropped, centering on the heart (see methods). As the data are dynamic 2D MRIs captured over 50 time points, we extended the diffusion autoencoder framework to 3D (3DDiffAE) by stacking them across time points as a single 3D volume, allowing us to capture time as the 3rd dimension. This allowed us to model the heart through one cardiac cycle. We extracted 128 latent phenotypes for each subject, and to ensure their replicability and stability, we took forward latent phenotypes that were present over multiple independent model runs (R ≥ 0.8), resulting in 182 latent phenotypes that can be robustly derived from cardiac MRIs. We denote latent phenotypes by their index and the model run they were discovered in. For example, Z10_S1 refers to the 10th latent phenotype (Z10), discovered in the first (S1) of five model runs. As DiffAE is based on a diffusion process, we observed excellent image reconstruction quality (**Supplementary Figure 1**), with a median structural similarity index measure (SSIM) of 0.96 ± 0.02, a metric that quantifies how similar the reconstructed image is to that of the original input image (**Supplementary Figure 2**) (see methods). Together, our ability to extract unbiased phenotypes at scale and reconstruct cardiac MRIs with high fidelity enables us to characterise cardiac structure and function in whole populations without the need for time-consuming annotations.

**Figure 1:**
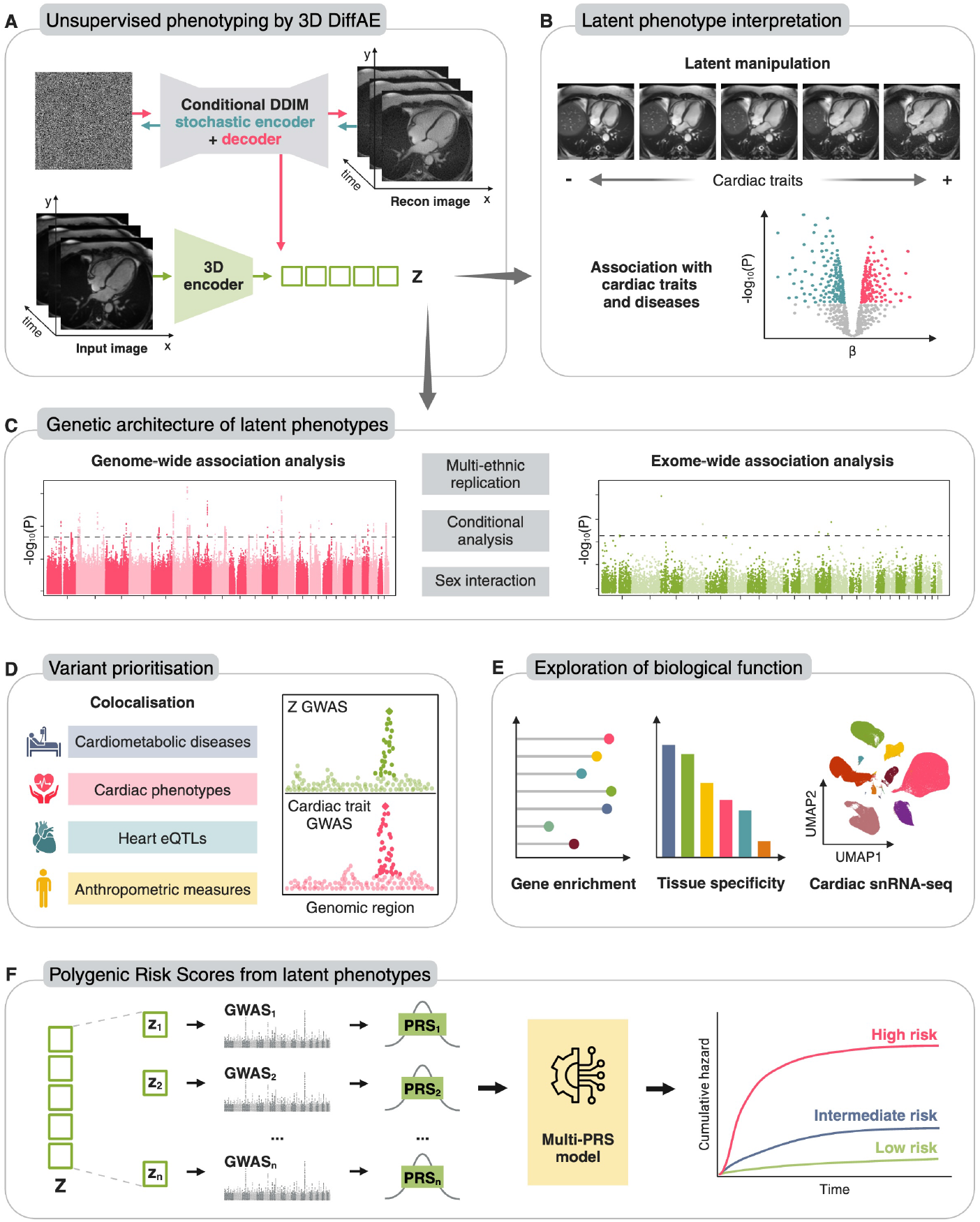
Schematic overview of our study. **A**. We fit a 3D diffusion autoencoding model to learn a vector of latent phenotypes (*z*) from temporal cardiac MRIs in UKB. **B**. These learnt latent phenotypes are associated with specific cardiac outcomes and can be interpreted via latent manipulation and visualisation of image reconstructions. **C**. The genetic architecture of latent phenotypes was investigated through both common variant GWAS, rare-variant and burden-based exome-wide analysis. **D**. Fine-mapping, conditional analysis and colocalisation was employed to understand causal variant sharing across latent phenotypes, cardiac traits and diseases. **E**. Gene, tissue and single-cell enrichment analyses were carried out to understand underlying target genes. **F**. Polygenic risk scores were built to demonstrate the prognostic predictive value of latent phenotypes.

**Figure 2:**
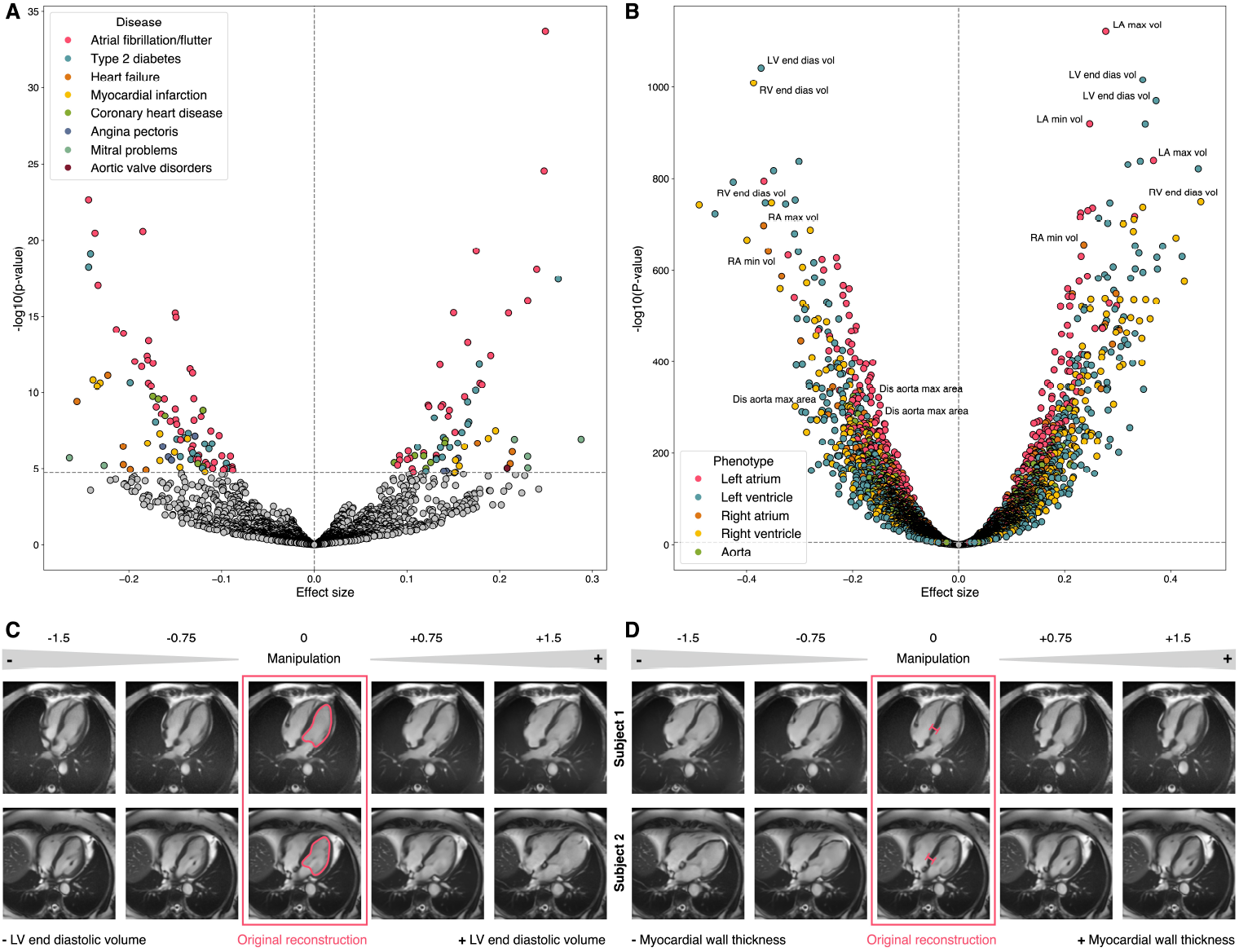
**A-B** Associations between latent phenotypes and diseases (**A**) and continuous traits (**B**): volcano plot of effect sizes and significance. Bonferroni adjusted P-value threshold: *p* < 1.96 × 10^−05^ for diseases, *p* < 9.8 × 10^−06^ for continuous traits. The top 3 continuous traits per each category having the most significant associations with latent phenotypes are labelled. LV: left ventricle, RV: right ventricle, LA: left atrium, RA: right atrium. **C-D** Latent manipulation of left ventricle end diastolic volume (**C**) and myocardial wall thickness (**D**) for two subjects, with increasing manipulation magnitude from left to right.

### Latent phenotypes associate with multiple cardiometabolic traits and outcomes

To explore the diagnostic and prognostic potential of our cardiac MRI latent phenotypes, we leveraged the UK Biobank’s rich and concurrently collected electronic healthcare records. In particular, we considered diseases present in at least 200 donors from GP clinical event records (UKB data-field ID: 42040), hospital inpatient (HI) admissions (UKB data-field ID: 41270) and self reported (SR) medical conditions (UKB data-field ID: 20002). We performed multiple linear regression to test for association between the selected latent phenotypes and 14 relevant diseases, adjusting for several covariates (see methods). This comprehensive approach revealed 166 significant associations (P-value ≤ 1.96 × 10^−5^) between latent phenotypes and diseases, with 57% of all latent phenotypes demonstrating at least one significant association (**Figure 2A**).

The most significant latent phenotype - diagnosis association was with atrial fibrillation, a widespread cardiac arrhythmia characterised by irregular and often rapid heartbeats originating from the atria. Unsurprisingly, Type 2 diabetes, a common cardiometabolic condition known to increase the risk of cardiovascular disease (CVD), was the second most frequently associated disease, with 33 significant latent phenotype associations. 17 latent phenotypes were associated with myocardial infarction. Importantly, several latent phenotypes demonstrated specificity for particular cardiac diseases. For example, latent phenotype Z49_S1 was exclusively associated with: myocardial infarction (*β* = − 0.23, P-value = 2.45 × 10^−11^), coronary artery disease (CAD) (*β* = − 0.17, P-value = 2.6 × 10^−10^) and angina pectoris (*β* = − 0.16, P-value = 3.6 × 10^−7^) (See **Supplementary Table 1** for the full list of associations), highlighting that these traits manifest similarly in a cardiac MRI and can be captured by a single latent phenotype.

To bridge the gap between imaging and endpoint diseases, we further assessed how well our latent phenotypes simultaneously captured specific heart and aortic endophenotypes [12], such as left ventricular ejection fraction and aorta diameter. Remarkably, despite using no labels or annotations, all 182 latent phenotypes demonstrated robust associations with at least one aortic or cardiac trait (**Figure 2B, Supplementary Table 2**). In particular, the most associated trait category was left ventricle traits, collectively accounting for up to 34% of all significant associations, reinforcing the ability of our latent phenotypes to reflect detailed structural and functional cardiac features.

Interestingly, we see latent phenotype associations spanning both cardiac diseases and MRI-derived traits (Z82_S1). This latent phenotype was associated with several MRI-derived measurements of the ventricles and atria, including Left Atrium (LA) maximum volume (*β* = *−*0.37, P-value = 1.86 × 10^−354^) and Left Ventricle (LV) stroke volume (*β* = *−*0.43, P-value = 1.47 × 10^−339^) along with 17 other cardiac phenotypes (P-values ≤ × 10^−100^). Additionally, Z82_S1 was significantly associated with type 2 diabetes (*β* = 0.26, P-value = 3.74 × 10^−18^), myocardial infarction (*β* = *−*0.23, P-value = 3.93 × 10^−11^), coronary artery disease (CAD) (*β* = *−*0.17, P-value=1.75 × 10^−10^), atrial fibrillation (*β* = *−*0.17, P-value = 8.47 × 10^−10^), angina pectoris (*β* = *−*0.16, P-value = 1.74 × 10^−6^) and mitral problems (*β* = *−*0.26, P-value = 2 × 10^−6^).

In summary, these latent phenotypes robustly capture novel links between low-level heart endophenotypes (e.g. myocardial wall thickness), intermediate traits (pulse pressure) but also more complex end stage disease outcomes (e.g. MI). This demonstrates their potential as powerful tools for integrating information across imaging biomarkers and clinical cardiometabolic conditions.

### Latent phenotypes encode semantic concepts relevant to disease that can be interpreted using latent manipulation

As our 3DDiffAE model is both generative and has high fidelity image reconstructions, we can modify the magnitude of the latent phenotypes to understand what they are specifically capturing in a cardiac MRI. To do this, we trained linear and logistic Lasso regression models, predicting multiple cardiac traits and diseases. We see that latent phenotypes were broadly predictive for several time-dynamic traits such as left ventricle end diastolic volume (mean cross-validation *R*^2^ = 0.58), demonstrating the model captures temporal aspects of cardiac function (**Supplementary Figure 5, Supplementary Table 3**). For cardiac diseases, latent phenotypes achieved a predictive performance comparable to traditional derived cardiac measures (mean cross-validation *AUC* = [0.63 − 0.77], **Supplementary Tables 4**,**5**). Together, these results confirm that learnt latent phenotypes are predictive of both dynamic cardiac traits and clinical outcomes.

To further investigate the biological relevance of these latent phenotypes, we used the trained linear models to identify those predictive of a given trait or disease. By manipulating these latent phenotypes and decoding the modified latent vector to obtain image reconstructions, we can directly visualise what is changing in the MRI (see methods). For example, **Figure 2C** shows the manipulation results of latent phenotypes selected by the Lasso model to predict LV end diastolic volume in two subjects. Strikingly, increasing the manipulation magnitude *m* [-1.5, 1.5] resulted in a progressive increase in LV end diastolic volume, with a visibly larger heart in the reconstructed MRIs (additional example subjects in **Supplementary Figure 4** and **Supplementary Videos 1-5**). Specifically, the mean of LV end diastolic volume (measured in millilitres with the area-length method) across the 5 analysed subjects reported in **Supplementary Figure 4** decreased from 138.95 ±29.65 to 127.82 ±23.87 with manipulation magnitude *m* = 0.75, and got to 118.00 ± 23.65 with *m* = −1.5; accordingly, LV end diastolic volume mean increased to 145.53± 31.96 when *m* = 0.75, and to 167.13 ± 23.96 when *m* = 1.5. We obtained similar findings with other cardiac-related traits. For example, **Figure 2D** illustrates manipulation of latent phenotypes predictive of LV mean myocardial wall thickness in two subjects, where increasing manipulation magnitude produced a clear increase in myocardial thickness (additional subjects in **Supplementary Figure 4** and **Supplementary Videos 6-10**). Again, we quantified the trait changes in **Supplementary Figure 4**, observing mean myocardial wall thickness decreasing by *−*4.09%*±*7.84% with manipulation magnitude *m* = *−*1.5, and increasing by 15.59% *±* 9.16% when *m* = 1.5; mean changes with *m* = *±*0.75 were modest in this case, but still in the expected direction (*−*2.30% *±* 6.19% with *m* = *−*0.75, and 0.30% *±*10.96% with *m* = 0.75). To our knowledge, this is the first clear demonstration of interpretation of latent phenotypes in large-scale MRI datasets, overcoming limitations posed by variational autoencoding methods [9]. Our framework paves the way for high-throughput phenotyping with a means of direct interpretation and visualisation of the captured features, something that has been lacking from previous unsupervised phenotyping approaches.

### Latent phenotypes are heritable and are associated with novel genome-wide significant variants

We have robustly demonstrated that latent phenotypes capture multivariate aspects of the structure and function of the heart. Here, we leverage concurrently collected genotyping data to map the genetic architecture of latent phenotypes and how they share common causal variants with traits that span biological scales, from eQTLs, endophenotypes, intermediate traits and disease outcomes. First, we saw that the heritability of inferred latent phenotypes ranged from 4% to 18%, with a mean *h*^2^ of 9.6% (SD = 0.9). Notably, the 182 prioritised latent phenotypes (Pearson’s R ≥ 0.8) chosen for their robustness and replicability across model runs, on average exhibited a significantly higher heritability compared to the remaining 458 latent phenotypes (One-sided Mann-Whitney U test P-value = 6.5 × 10^−3^, **Supplementary Figure 8**). Next, to assess whether any of our latent phenotypes are associated with common genetic variants, we sought to carry out a large-scale Genome-Wide Association Study (GWAS). Following image quality control (**Supplementary Figure 7**), the discovery cohort included 47,854 Caucasian individuals with baseline clinical information, with covariate characteristics presented in **Supplementary Table 23**. We conducted 182 latent phenotype GWAS on 47,740 subjects passing genotype QC, testing 42.8 million imputed common genetic variants (MAF >1%) using the REGENIE pipeline (see methods for full details). All GWAS analyses showed no evidence of inflation driven by either population stratification or cryptic relatedness (mean *λ* = 1.023).

Across the 182 latent phenotypes tested, we detected a total of 95 significant associations at a study-wide significance threshold of 2.27 × 10^−9^, accounting for the number of independent traits tested [13]. Additionally, 366 SNPs reached significance at the standard genome-wide threshold of 5 × 10^−8^. Through conditional analysis, we further refined these associations, identifying 89 conditionally independent trait-wise associations (**Supplementary Table 6**) mapping to 44 lead SNPs within 42 unique genomic loci across all latent phenotypes (**Supplementary Table 7**, see methods). The Manhattan plot in **Figure 3A** provides an overview of the 182 GWAS results, highlighting all significant loci annotated with their nearest or overlapping genes. Notably, 7 of the 44 lead SNPs were previously unreported in the GWAS Catalog [14]. To categorise significant loci, we assigned a label based on the most prevalent trait association category reported in the GWAS Catalog. The dotplot in **Figure 3B** reveals that the majority of lead variants have been previously associated with cardiac traits and diseases, followed by anthropometric measures, biomarkers and other cardiometabolic risk factors. Some variants are also associated with lung or bone phenotypes, which is not unexpected given that both lungs, spinal cord and skeletal structures are visible in the cropped version of the cardiac MRIs. Importantly, four of our lead variants were recently described in a smaller study that performed GWAS on latent embeddings learnt from a cross-modal autoencoder trained on ECG and MRI data [15] (labelled as “MRI latents” in **Figure 3B**). This overlap supports the robustness of our unsupervised approach, while the additional variants discovered here highlight the power of our large-scale dataset and analytical framework to identify new genetic links to cardiac structure and function.

**Figure 3:**
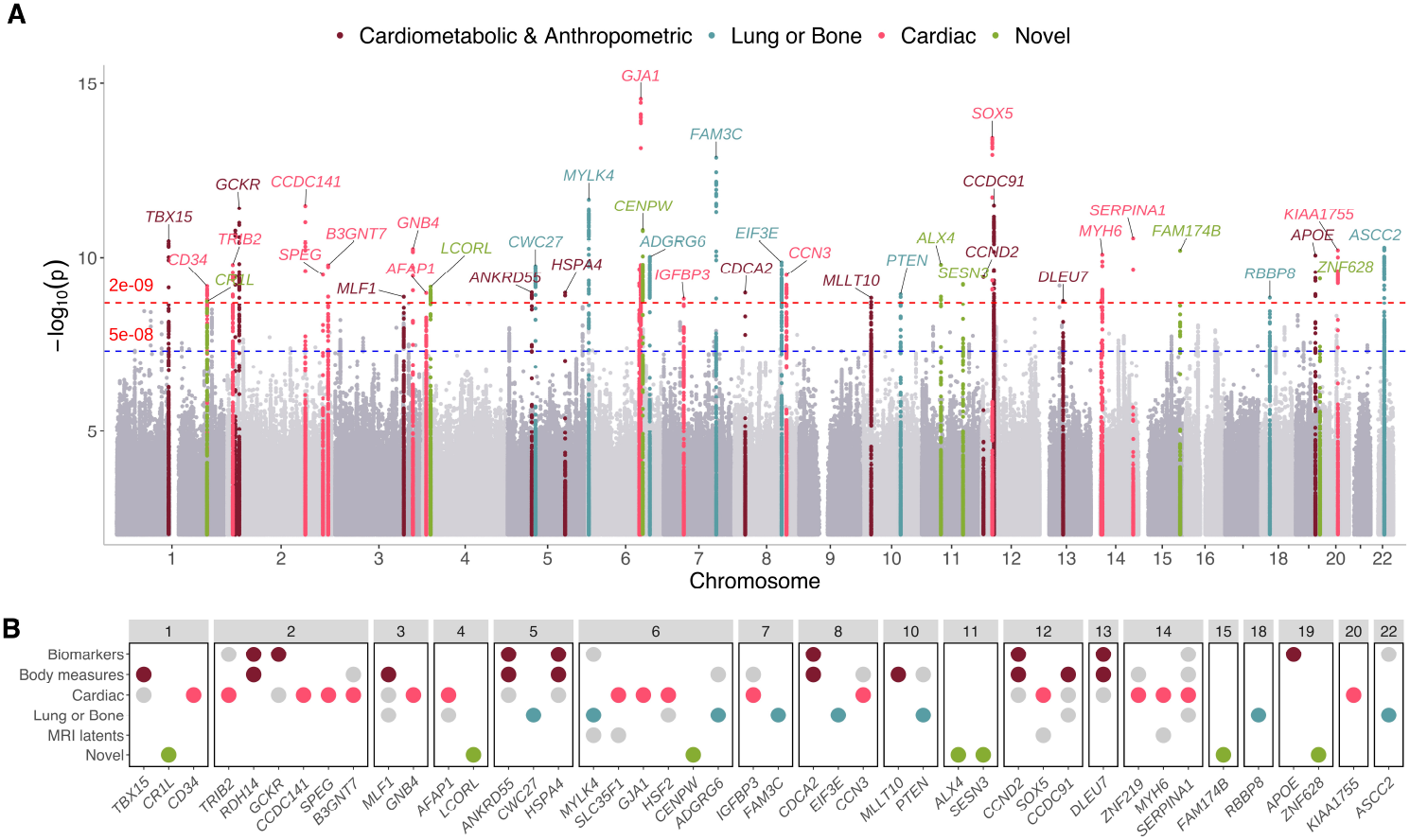
GWAS for 182 DiffAE latent phenotypes. **A**. Manhattan plot summarising genome-wide associations across 182 latent phenotypes. Blue line: genome-wide significance (5 × 10^−8^). Red line: study-wide significance (2.27 × 10^−9^). The 44 lead SNPs are annotated with the nearest or overlapping gene and highlighted according to the most frequent category of previously reported associations. **B**. Dotplot showing the categories of traits previously associated with our lead variants, according to the GWAS Catalog. “MRI latents” refers to the unsupervised GWAS of cross-modal representations from the study by Radhakrishnan et al. [15] Previously unreported loci are labelled as novel.

Interestingly, the two previously mentioned latent phenotypes Z49_S1 and Z82_S1, having robust associations with multiple cardiac diseases and various MRI-derived cardiac traits, also exhibited significant associations within the *SOX5* locus. The lead variant, rs4963772 (minimum P value = 3.55 × 10^−14^), has also been identified as significant in other GWAS for blood pressure, ECG traits and atrial fibrillation. Beyond the SOX5 locus, Z82_S1 showed associations in several other loci, including the novel *CRL1* locus, as well as *CCDC141, SLC35F1, HSF2, MYH6* and *KIAA1755*, The lead variants within these loci have significant associations with cardiac-related phenotypes in prior GWAS, demonstrating that latent phenotypes robustly identify both novel and known genetic loci relevant to cardiac traits and diseases.

Considering the closest gene to each significant variant, several well characterised cardiac genes are present. For example, Myosin heavy chain 6 (*MYH6*), responsible for forming thick filaments responsible for heart contraction, has been previously associated with atrial fibrillation (rs422068, P-value = 3.5 × 10^−12^ [16]). Gap Junction Alpha 1 (*GJA1*), also known as Connexin 43 (Cx43), has been previously associated with pulse rate and resting heart rate (rs9388001, P-value = 4.4 × 10^−110^ [16]) and is responsible for electrical coupling between adjacent cardiomyocytes. Interestingly, we also see novel associations for variants near the gene Ligand dependent nuclear receptor corepressor-like (*LCORL*). Whilst the intronic deletion TG to T at 4:17956213 in *LCORL* has not previously been associated with cardiac traits in humans, Knockout mice for *LCORL* display a decreased heart rate and a prolonged RR interval [17], warranting further investigation into *LCORL’s* precise role.

Epidemiological evidence suggests that many cardiovascular diseases are sexually dimorphic [18], motivating us to investigate sex-specific genetic effects of cardiac latent phenotypes. We performed a genome-wide SNP-by-sex interaction analysis in the full discovery cohort (see **Supplementary Table 23**) across the 182 selected latent phenotypes. At the study-wide significance threshold of 2.27 × 10^−9^, we identified one significant locus on chromosome 11, with the lead variant, rs72925197 (P-value = 1.34 × 10^−11^), being an intronic SNP in *NRXN2* (**Supplementary Figure 9**). Although not previously linked to cardiac or sex-specific traits, we see that this SNP is an eQTL for *VEGFB* in heart and adipose tissues [19] and has been implicated in the early development of the cardiovascular system [20]. Additionally, nine other loci demonstrate suggestive associations at 5 × 10^−8^ (see **Supplementary Table 8**).

We replicated our GWAS findings in a held out cohort in UKB of *N* = 12,396 participants (**Supplementary Table 24**), with 84/89 conditionally independent significant trait-SNP associations had concordant effect directions (**Supplementary Figure 11, Supplementary Table 9**), and 51 variants replicating (P < 0.05).

Additionally, we wanted to investigate whether latent phenotypes can capture the same genetic effects as specific cardiac derived measures, cardiac diseases and risk factors. To do this, we used multivariable LD-Score regression to assess genetic correlation [21] (**Supplementary Figure 12**). Notably, latent phenotype Z82_S1 exhibited strong positive genetic correlation with resting heart rate (*r* = 0.78 ±0.08,*p* = 8.7 × 10^−25^) and pulse rate (*r* = 0.83 ±0.09,*p* = 8 × 10^−21^), while showing negative correlations with several MRI-derived cardiac measurements. This latent phenotype has been previously mentioned for having strong linear associations with multiple cardiac diseases, including atrial fibrillation (P-value = 1.6 × 10^−11^) and myocardial infarction (P-value = 5.8 × 10^−10^). As expected, no significant genetic correlation was observed with our negative control phenotype, hair colour (Mean absolute correlation = 0.054 and minimum P-value = 0.135, see **Supplementary Figure 13**). Overall, we identify replicable genetic associations with derived latent phenotypes and demonstrate their ability to robustly capture biologically meaningful features such as pulse rate, with extremely high genetic correlation. Combining high-throughput unsupervised phenotyping, which does not require time-consuming expert annotation and redefines traditional trait boundaries, with the power to identify novel and existing phenotypes and genetic associations, allows us to capture the full gambit of variation effecting the cardiac structure and function.

### Genes linked to latent phenotypes are enriched for heart-related functions and are expressed in cardiac specific cell types

To infer the biological implications of the significant variants and prioritise the most likely target genes, several techniques were utilised. First, to prioritise genes at our identified genomic loci, we combined positional mapping with eQTL mapping in cardiac tissues using the FUMA platform [22]. This approach linked a total of 82 protein-coding genes to the 44 lead SNPs. **Figure 4A** presents the significant results of an over-representation analysis conducted on these genes using *Enrichr* [23], highlighting a clear and significant enrichment for traits such as heart rate, cardiac contraction cardiomyopathies and directly to cardiomyocytes (full results in **Supplementary Table 10**). Using Multi-marker Analysis of GenoMic Annotation (MAGMA) [24] to perform a gene-set enrichment analysis, we are able to demonstrate variants are significantly enriched for heart morphogenesis and atrial septation defects (**Supplementary Figure 15** and **Supplementary Table 11**). To assess tissue specificity of the latent phenotypes, we also applied MAGMA’s gene-property analysis on GTEx v8 gene expression data (**Figure 4B** and **Supplementary Table 12**), revealing significant enrichment in both left ventricle and right atrial appendage tissues, as well as colon sigmoid, a tissue which contains a significant proportion of muscularis propria. Finally, we applied the Chromatin Element Enrichment Ranking by Specificity (CHEERS) analysis [25], showing enrichment of latent phenotype-associated variants in ATAC-seq peaks active in fibroblasts and atrial cardiomyocytes (**Supplementary Figure 14**), suggesting regulatory activity in these cardiac cell types.

**Figure 4:**
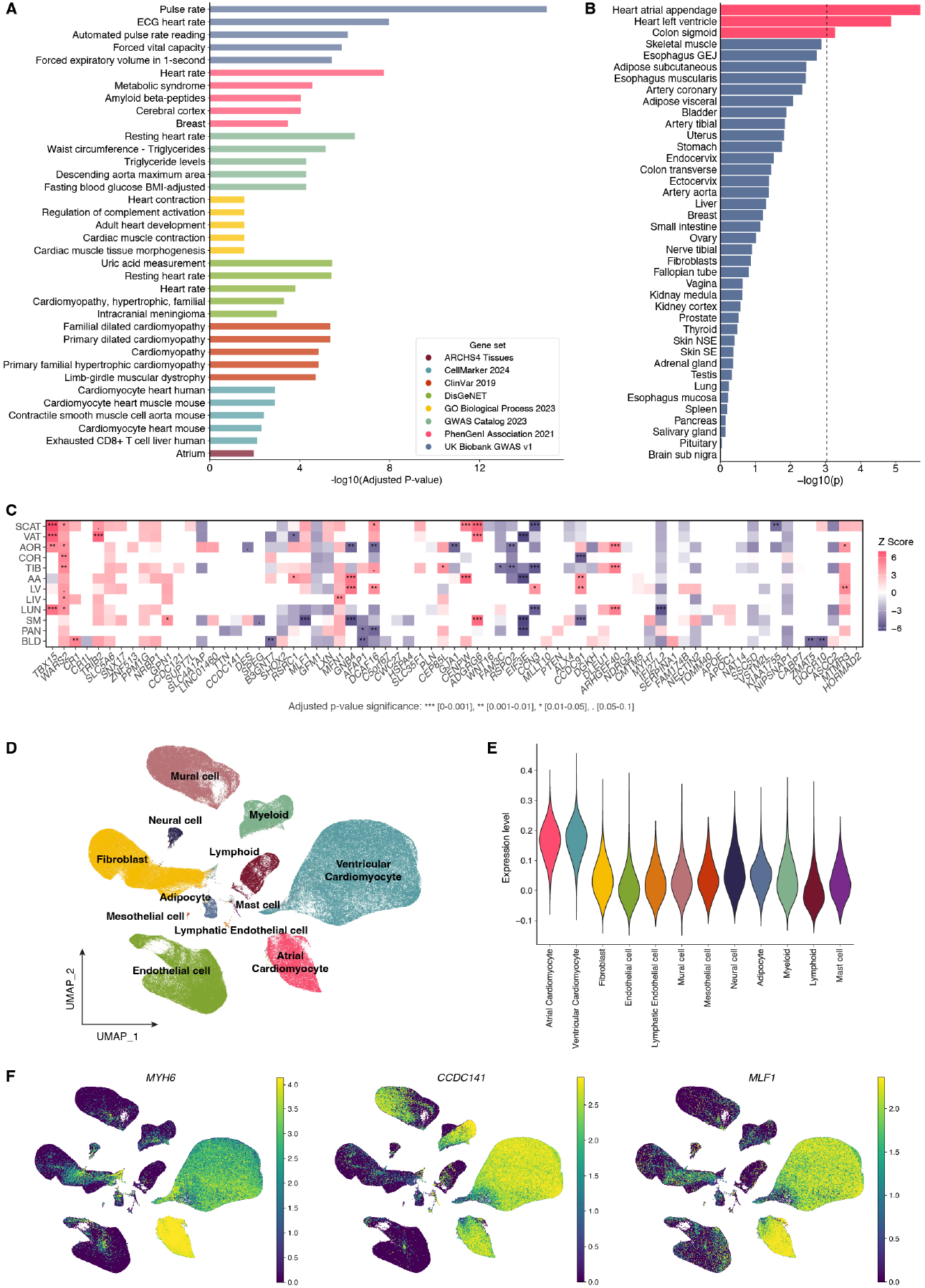
**A**. EnrichR gene set enrichment barplot, showing only terms with significant adjusted P value in log scale. **B**. MAGMA tissue enrichment analysis, using GTEx data for 53 tissue types. Only tissues with − *log*10(*p*) > 0.01 are shown. **C**. TWAS heatmap of 82 genes prioritised by FUMA. The colour indicate direction of effects. The asterisks represent significant gene-tissue pairs. SCAT adipose subcutaneous, VAT adipose visceral, AOR aorta, COR coronary artery, TIB tibial artery, AA heart atrial appendage, LV heart left ventricle, LIV liver, LUN lung, SM skeletal muscle, PAN pancreas, BLD blood. **D**. Uniform manifold approximation and projection (UMAP) visualisation of human heart single-nucleus sequencing data. **E**. Expression levels of MAGMA-prioritised latent phenotypes-related genes in human heart single-nucleus sequencing data. **F**. UMAP of human heart snRNA-seq data for *MYH6, MLF1, CCDC141*.

To tie variants to targets and resolve precisely which specific cell subtypes they are expressed in, we leveraged a transcriptome-wide association study (TWAS) using FUSION (see Methods). TWAS analyses identified significant associations between the expression of 82 protein-coding genes in the right atrial appendage and 81 genes in the left ventricle with latent phenotypes, of which 14 genes demonstrated concordant effects across both tissues. Among the top TWAS hits was *BCKDHA*, expressed specifically in heart atrial appendage (*Z* = 7.28, *P* = 1.86 × 10^−9^). *BCKDHA* encodes an enzyme involved in branched-chain aminoacid (BCAA) degradation, a process previously linked to heart failure [26, 27]. Other highly prioritised genes in atrial appendage included *WWP2* (*Z* = −7.16, *P* = 4.79 × 10^−9^), a E3 ubiquitin protein ligase known to regulate cardiac fibrosis [28, 29], and *SCN5A* (*Z* = −6.12, *P* = 5.39 × 10^−6^), which encodes the main cardiac sodium channel, with mutations extensively linked to cardiomyopathies [30]. Notably, 27 of the protein-coding genes identified as significant by TWAS overlapped with those prioritised by FUMA, as illustrated in **Figure 4C**. By computing π_1_, we show that many TWAS genes have significant overlap between tissues (**Supplementary Figure 16B**). Full TWAS results can be found in **Supplementary Table 13**.

To determine whether the genes linked to the latent phenotypes were expressed in specific cardiac cell types, we leveraged publicly available single-nucleus RNA sequencing (snRNA-seq) data from the human heart [31]. This analysis demonstrated that the 82 protein-coding genes prioritised by MAGMA showed significantly higher expression in both atrial and ventricular cardiomyocytes (**Figure 4E**). Notably, key marker genes for these cell types were identified, with *MYH6* and *MYH7* serving as dominant markers for atrial (ACMs) and ventricular (VCMs) cardiomyocytes, respectively, while *TTN* was strongly expressed in both (**Supplementary Table 14**). Suprisingly, we were able to map several prioritised genes linked to our latent phenotypes to specific cardiomyocyte populations (**Figure 4F**). For instance, *MLF1* showed a log fold change (LFC) of 2.68 in ACMs and 3.0 in VCMs. Similarly, *CCDC141* demonstrated a significant fold enrichment for VCMs (LFC = 4.08) compared to ACMs (LFC = 1.38). In summary, we demonstrate that target genes linked to variants associated with our latent phenotypes are significantly enriched in cardiac relevant processes and diseases, but importantly, we are also able to resolve target gene expression to specific atrial and ventricular cardiomyocyte populations, enabling the design of functional validation studies in the correct cell types that are more likely to succeed.

### Latent phenotypes associated loci colocalise with cardiac endophenotypes, cardiometabolic traits, eQTLs and disease outcomes

Given many of our lead variants have been previously linked to cardiometabolic traits and diseases including heart rate, ECG measures, and atrial fibrillation, we sought to determine whether the same causal genetic variants underlie both latent phenotypes and these complex traits. For instance, our lead SNP rs142556838 (MAF = 0.09), an intron variant of *CCDC141* (with *TTN* as nearest gene), is also a lead variant in a diastolic blood pressure GWAS [32] and has been associated with pulse rate, heart rate and ECG measures. Interestingly, we establish here that rs142556838 falls within a intron that overlaps a DNAse hypersentitivity peak in cardiac right atrium tissue as well falling into H3K4me1 and H3K27ac CHIP-seq peaks in heart left ventricle, ascending aorta and right atrium auricular tissue [33]. Indeed, it seems rs142556838 is part of an active enhancer, specific to right atrium auricular tissue, implicating it in controlling the regulatory activity of neighbouring gene *TTN* [33]. Additionally, we see that rs72801474 (MAF = 0.09) is a highly pleiotropic variant near *HSPA4*, and is associated with several cardiometabolic risk factors, including triglyceride levels and hypertension and sits in a candidate *cis*-regulatory element (cCRE) [33]. Motivated by these example discoveries, we decided to comprehensively characterise and interpret the common variants associated with each latent phenotype by performing multi-trait colocalisation across 54 existing GWAS datasets and eQTL data spanning 12 tissues (**Supplementary Table 15**). These traits include structural and functional cardiac endophenotypes (MRI-derived), cardiac diseases, blood pressure traits, ECG measures, and other non-cardiac phenotypes.

We performed colocalisation analysis for the 42 genomic loci containing 44 unique lead SNPs associated with the latent phenotypes. As illustrated in **Figure 5A** and **B**, 34 genomic regions exhibited significant colocalisation (*PP*.*H*_4_ > 0.8), which are detailed in **Supplementary Table 16**. Of these 34, 19 loci colocalised with at least one MRI-derived cardiac phenotype, ECG trait or blood pressure measure, while 10 loci colocalised with cardiac eQTLs (from heart and/or artery), and 7 with cardiac diseases. Of particular interest, 9 loci were classified as cardiac-specific, meaning the latent phenotypes colocalised exclusively with cardiac eQTLs or cardiac GWAS traits. For example, the locus near *KIAA1755* colocalised with left and right ventricular stroke volume, pulse pressure, PR interval and resting heart rate (**Figure 5B**). Similarly, at the *MYH6* locus, the causal variant is shared between three latent phenotypes, descending aorta maximum and minimum area, stroke volume of both left and right ventricles (LV/RV), right ventricle ejection fraction, pulse pressure, and atrial fibrillation (**Supplementary Figure 17**). Among these colocalising latent phenotypes was Z82_S1, which not only showed linear associations with cardiac diseases and MRI-derived cardiac measures, but - as discussed in the GWAS results - exhibited strong positive genetic correlations with pulse rate and heart rate, along with multiple common variant associations identified in the GWAS. This is effectively the first large-scale effort to integrate genetic information across scales, linking various categories of cardiac traits through unsupervised phenotyping and therefore uncovering multivariate relationships in cardiovascular genetics.

**Figure 5:**
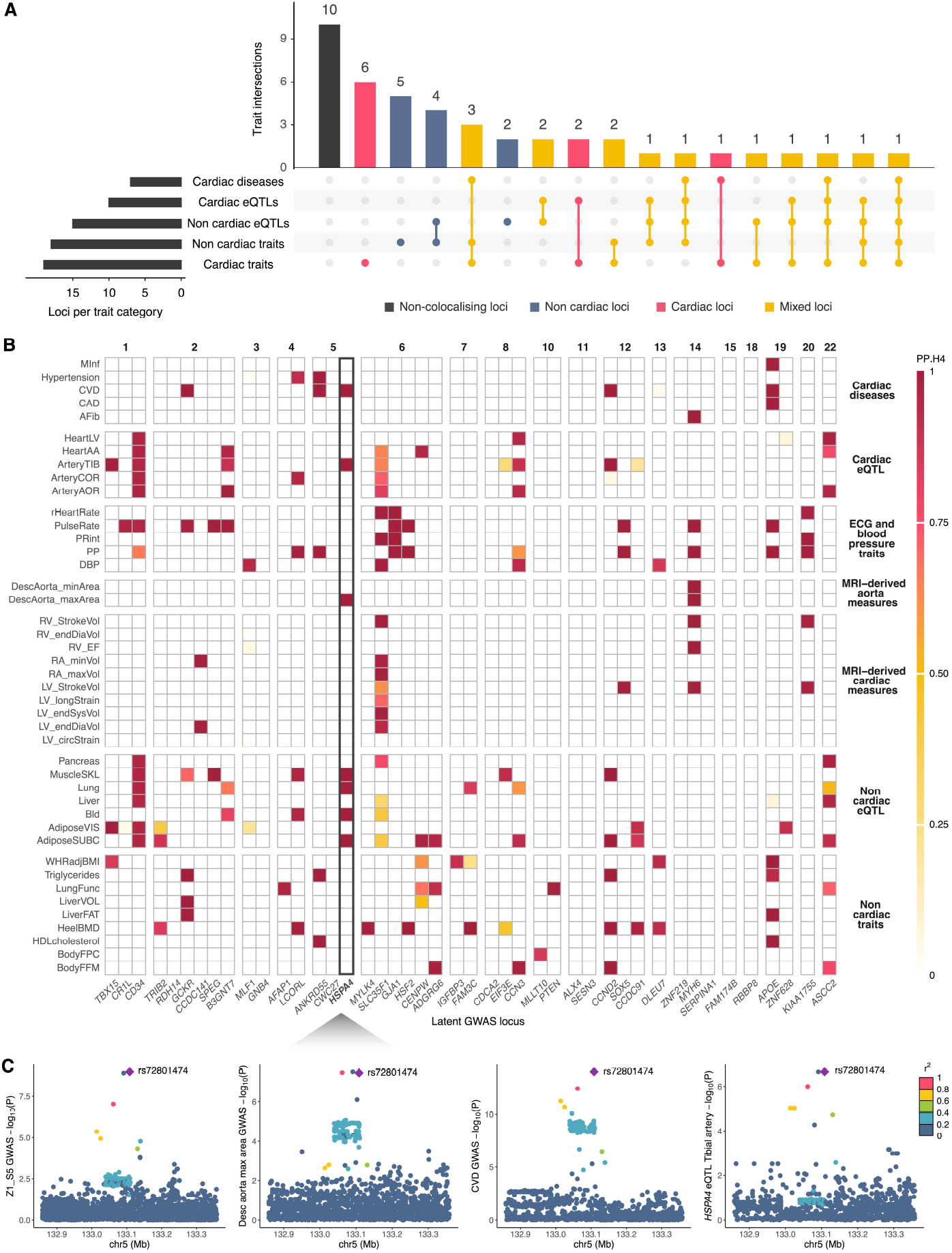
Colocalisation analysis for the 44 lead SNPs from latent phenotypes GWAS in 54 GWAS of relevant traits and 12 GTEx tissues. **A**. Upset plot of the 44 latent phenotypes lead variants mapped to an association in a cardiac eQTL, non cardiac eQTL, cardiac disease GWAS, cardiac trait GWAS (ECG and blood pressure traits, MRI-derived heart and aorta phenotypes) or non cardiac trait GWAS (body, liver and lung measures, biomarkers) with a colocalisation PP.H_4_ > 0.8 (Methods). **B**. Heatmap summarising the colocalisation results, coloured by posterior probabilities (H4). On the x axis, latent phenotypes lead SNPs are indicated using their closest gene, grouped by chromosome. On the y axis, the GWAS traits and eQTL tissues with at least one significant colocalisation, grouped by category. **C** Colocalising traits at the *HSPA4* locus.

**Figure 6:**
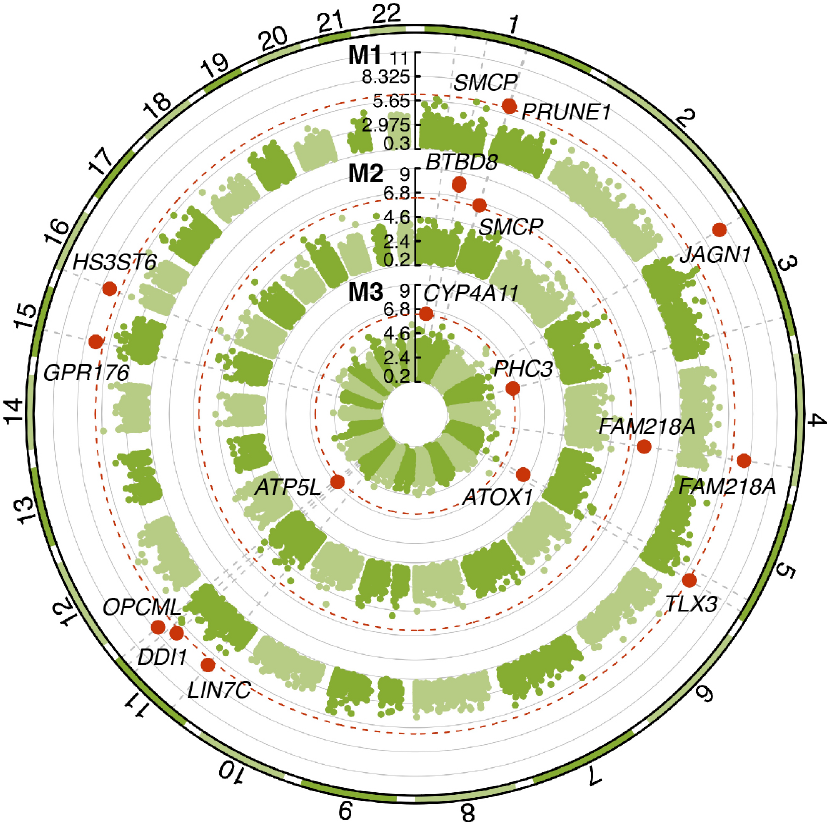
SKATO circular Manhattan plot representing three different masks: M1 LoF, M2 LoF and missense (5/5), M3 LoF and missense (>=1/5). On the y axis -log10(P-value), with significant genes annotated and marked in red according to the P-value threshold of 4.7 × 10^−7^ (dashed red line).

Beyond known loci, our analysis identified three novel genomic regions from our latent phenotype GWAS that colocalised with various traits, enabling us to characterise them by linking them to known cardiac phenotypes (**Supplementary Figure 18**). For instance, rs11118288 on chromosome 1 colocalised with the lead variant of a pulse rate locus. Another novel intronic deletion (TG to T at 4:17956213) in *LCORL* colocalised with hypertension, pulse pressure and bone mineral density, and the same variant is an eQTL for *DCAF16* expression in coronary artery, blood and skeletal muscle tissues. Lastly, the locus on chromosome 6 colocalised with eQTLs for *CENPW* in heart atrial appendage and subcutaneous adipose tissue.

A key advantage of our approach is its ability to reveal pleiotropic genetic effects that span multiple cardiovascular traits and risk factors. Approximately one-third of discovered lead variants (14/44) exhibited colocalisation across multiple trait categories, including cardiac diseases (e.g coronary artery disease), known cardiac risk factors (e.g Triglycerides) and cardiovascular traits such as heart rate, and pulse pressure (**Supplementary Figure 19**). For instance, at the *HSPA4* locus, we were able to demonstrate that the latent phenotype Z1_S5 shares the same causal variant (rs72801474) with cardiovascular disease (CVD), hypertension, pulse pressure, triglycerides and HDL cholesterol, as shown in **Figure 5C**.

These results demonstrate the power of MRI-derived latent phenotypes to capture multivariate aspects of cardiac function and to uncover relevant genetic signals that bridge different biological scales. Our approach offers a more comprehensive understanding of the shared genetic architecture of cardiometabolic phenotypes.

### Exome-wide gene based burden tests of latent phenotypes implicates known cardiac genes

We wanted to investigate whether any latent phenotype was explained by large effect rare variation in the population. To do this, we conducted a single-variant exome-wide association study (EWAS) on 46,034 individuals from the discovery cohort, conditioning on the independent significant common variants identified in the corresponding GWAS. Of the five associations that reached exome-wide significance, four variants had MAF ≤ 0.01 (**Supplementary Table 26**). However, due to the low minor allele count, additional evidence is warranted to support our findings.

To gain power and assess whether aggregated rare variants (MAF ≤ 0.01) were associated with any latent phenotype, we conducted gene based burden tests in 46,134 individuals from the discovery cohort. Using the SKATO-ACAT test implemented in REGENIE, three different masks were incorporated, to include both LoF and missense rare variants. In total, 17 significant genes were identified at the exome-wide threshold of 4.7 × 10^−07^ (**Figure 6** and **Supplementary Table 17**), four of which have not been previously described in Genebass [34] (*SMCP, PRUNE1, FAM218A, LINC7*). Specifically, pLoF mutations in *JAGN1* have been associated with stroke as well as vascular or heart problems (Genebass). Mice knockouts of *ATP5L, PHC3* and *BTBD8* showed an enlarged heart and / or abnormal heart morphology [17]. Another study reported that knockout mice of *ATP5L* exhibited increased cardiomyocyte necrosis and markedly larger infarcts compared to controls [35]. This gene, encoding subunit G of the mitochondrial ATP synthase, is also specifically expressed in cardiac derived tissues (GTEx [19]). Of the four novel genes, *PRUNE1* - encoding a member of the DHH protein superfamily of phosphoesterases - has been linked to hypertrophic cardiomyopathy by the Human Phenotype Ontology (HPO) project [36]. Taken together, these results suggest that rare variants may contribute to variation in cardiac phenotypes captured by cardiac MRI imaging, but larger cohorts and replication are warranted.

### Polygenic risk scores of latent phenotypes are predictive for a range of cardiometabolic diseases

As our cardiac imaging latent phenotypes capture a variety of cardiometabolic risk factors, we sought to assess whether latent phenotype Polygenic Risk Scores (PRS) could be used to stratify donors into distinct risk groups for multiple cardiac relevant diseases (**Supplementary Table 27**). After creating single PRS for each latent phenotype, we firstly assessed whether those in the top risk percentiles in the most predictive PRS for that particular disease had a higher incidence of cardiac relevant diseases. Across all diseases considered, we observed that the mean cohort disease prevalence was 14.7%, with a higher prevalence in males (18.6%) compared to females (11.5%). Strikingly, the average increase in prevalence when comparing the *bottom 5%* to the *top 5%* PRS percentiles for atherosclerotic conditions was substantial, at 26.5% in the combined population, 25.1% in males, and 26.6% in females (**Fig. 7A**). Similarly **Fig. 7D** shows it for the metabolic syndrome, whilst all other diseases are presented in the **Supplementary Figures 22, 23**. Overall, we see an average increase in prevalence across all diseases considered of 17.8% from the *bottom 5%* to the *top 5%* PRS percentiles, with males showing an increase of 17.5% and females 18.2%. These findings highlight the significant role that disease agnostic imaging derived PRS can play in identifying individuals at elevated genetic risk for multiple cardiac diseases, thereby informing targeted prevention and intervention strategies.

**Figure 7:**
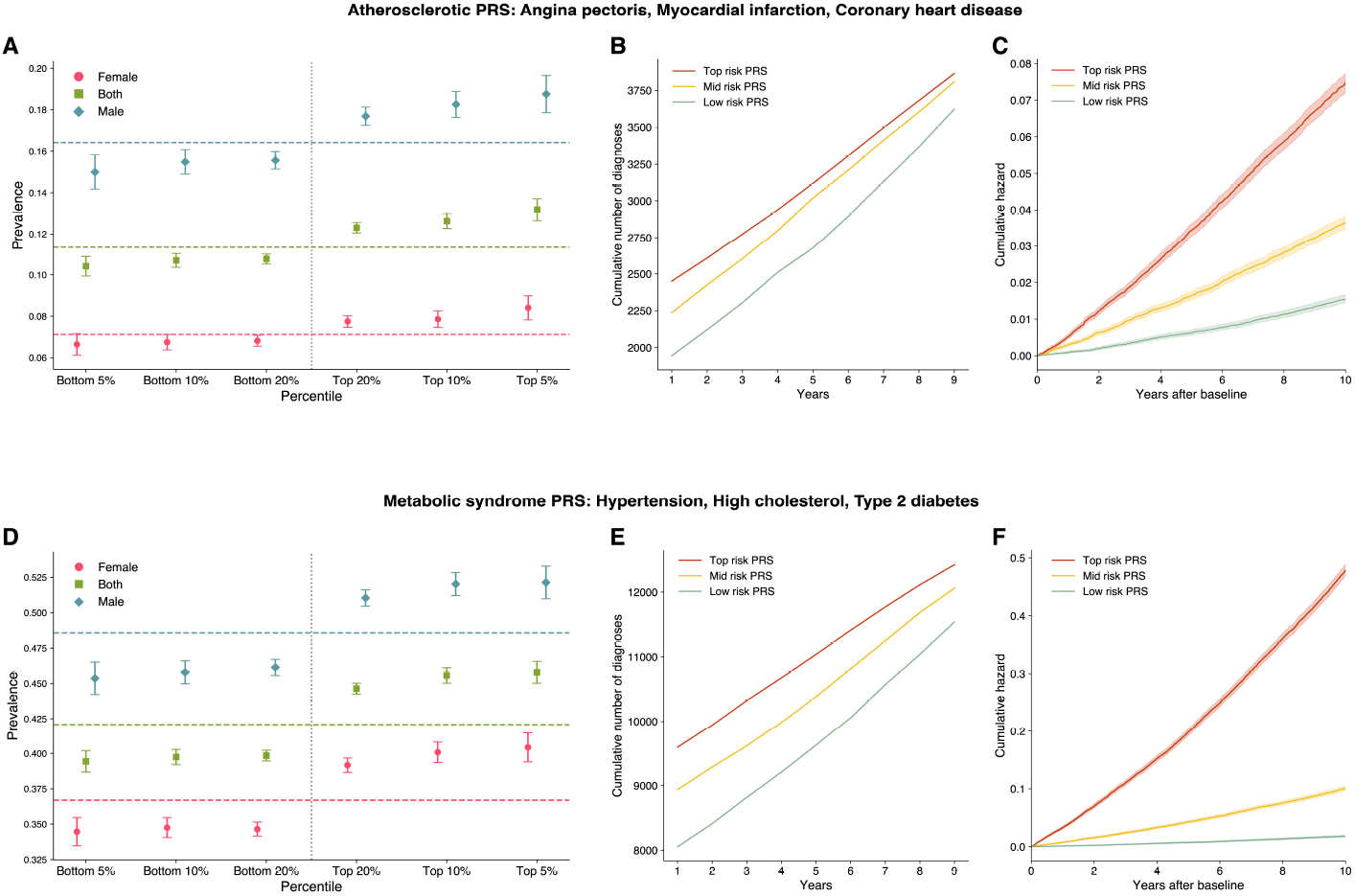
**A**,**D**. PRS prevalence plot stratified by sex. **B-C, E-F**. Cumulative disease burden and cumulative hazard by PRS risk quantiles. Many diseases show a significant increase of cumulative disease burden as their PRS risk increases.

Given there are significant differences in disease prevalence across the PRS distribution, we investigated whether there’s a difference in both the cumulative diagnoses and hazard rates up to ten years after the collection of baseline characteristics. To do so, we fit disease-specific Generalised Linear Models (GLMs) using each latent PRS as a predictor (one model for each latent PRS). **Fig. 7B** and **C** respectively present the cumulative number of new diagnoses and the cumulative hazard rate for atherosclerotic diseasesa, and **Fig. 7E** and **F** respectively for metabolic syndrome, whilst the results for all other diseases are presented in **Supplementary Figures 22, 23**. Across all diseases, myocardial infarction showed the largest increase (20.6%) in cumulative diagnoses when comparing the high risk group to the low risk group. Focusing on atherosclerotic diseases, the average increase across time points was 16.1%, with a maximum increase of 26%. These results indicate that individuals in the *Top risk* group experienced a significantly higher cumulative incidence of new diagnoses compared to those in the *Low risk* group. In terms of cumulative hazard rates, we also see that individuals in the top-risk group demonstrate a 28.2-fold increase for metabolic syndrome at two-years post baseline relative to those in the low risk group category. This substantial increase remained relatively consistent over time, with a 26.5-fold increase at five years and a 26.2-fold increase at ten years. For atherosclerotic diseases, the fold increases were 6.32 at two years, 5.5 at five years and 4.8 at ten years. These results indicate that individuals in the higher risk groups, as determined by the PRS-based GLMs, exhibit substantially higher cumulative hazard rates over time compared to those in the lower risk groups. These findings indicate that unbiased latent phenotypes learnt directly from cardiac MRIs can be used to construct powerful risk scores for prognostication of several relevant cardiac diseases.

Finally, to assess the combined predictive power of all the latent PRS, we combined the individual PRS obtained for each latent phenotype using a LASSO model to create a multi-latent PRS, and compared its performance against each of the single PRS and against a baseline of age, sex, BMI and smoking status. For predicting several diseases, we saw modest, but statistically significant improvements (P < 0.05) in the AUC for both disease diagnosis and prognosis over a strong baseline model (See box plots for AUC in **Supplementary Figure 21A** and **B**, and Supplementary Tables 19-21 for full diagnostic, prognostic, and whole cohort results).

## Discussion

In this study, we leveraged a 3D diffusion autoencoder (DiffAE) to analyse cardiac MRI data from 71,021 participants in the UK Biobank, in order to derive latent phenotypes representing cardiac structure and function. To our knowledge, this is the first implementation of a 3D DiffAE model applied to temporally resolved MRI data. Our approach enabled the extraction of latent cardiac phenotypes at the population level without the need for manual annotation or segmentation, thereby capturing a comprehensive representation of cardiac morphology and dynamics. By integrating these latent phenotypes with clinical and genetic data, we identified significant associations with various cardiometabolic traits and diseases, including atrial fibrillation and myocardial infarction. Furthermore, we demonstrated that the latent phenotypes are predictive of cardiac traits such as left ventricular volume and myocardial mass, as well as disease outcomes. Through genome-wide association studies (GWAS), we uncovered 89 significant common variant associations across 42 loci, including seven novel loci not previously associated with cardiac phenotypes. We thoroughly characterised the genomic variants and demonstrated their biological relevance at different levels through enrichment analyses and TWAS. Our findings highlight the potential of unsupervised deep learning models, specifically diffusion autoencoders, in extracting meaningful and clinically relevant features from large-scale imaging data. The significant associations with cardiometabolic diseases and the ability to predict cardiac traits underscore the cardiac relevance of the latent phenotypes. For instance, latent phenotype Z49_S1 was significantly associated with myocardial infarction and coronary artery disease, as well as multiple MRI-derived cardiac traits such as left ventricular cardiac output. Additionally, our genetic analyses, including GWAS and our multi-trait colocalisation integration, revealed a pervasive shared genetic architecture between the latent phenotypes and known cardiac traits and diseases across scales.

One of the key strengths of our approach is the ability to interpret the latent phenotypes through latent space manipulation and visualisation of image reconstructions. This allows for direct interpretation of how specific latent phenotypes influence cardiac structure and function. For example, by manipulating latent factors associated with left ventricular end diastolic volume, we could demonstrate an increase or decrease in the size of the left ventricle in the reconstructed images. Similarly, manipulation of latent factors related to myocardial wall thickness allowed us to change wall thickness, which is clinically relevant for conditions such as hypertrophy. Moreover, the latent polygenic risk scores demonstrated predictive power for a range of cardiometabolic diseases and showed that individuals in the top risk quantiles had a significantly higher cumulative hazard for diseases such as metabolic syndrome and atherosclerotic diseases, with up to a 26-fold increase compared to those in the low-risk quantiles at the 10-year mark, suggesting utility in risk stratification and personalised medicine.

Whilst the framework we propose here was applied to cardiac MRI, it is completely generalisable to any other imaging modality. Utilising other organ MRI acquisitions, such as brain or abdominal imaging, would enable the characterisation of multi-organ phenotypes, their genetic underpinnings, and how they are co-regulated, a line of work we are actively pursuing. Additionally, applying our methodology to other large-scale imaging cohorts, as they become available, would allow for external validation and assessment of generalisability across different populations and imaging protocols.

Finally, integrating longitudinal imaging data, when available, could enable the study of temporal changes in cardiac structure and function, potentially identifying early imaging biomarkers of disease progression. Longitudinal analysis might reveal latent phenotypes that predict the development of heart failure or other cardiomyopathies before clinical symptoms arise. In conclusion, our study demonstrates the utility of diffusion based autoencoding models for phenotyping and genetic discovery. By capturing complex imaging features without manual annotation, our approach provides a scalable and efficient means to deepen our understanding of the genetic architecture and pervasive pleiotropy of cardiac-specific genes, traits and diseases. The identification of novel genetic loci, coupled with the ability to visualise and interpret latent phenotypes, underscores the potential of this methodology in advancing our understanding of the heart’s structure and function in relation to disease.

## Materials and Methods

### Study participants

The UK Biobank is a prospective cohort study including over 500,000 individuals aged between 40 and 70 living across the United Kingdom at the time of recruitment (2006-2010) [37, 38]. The UK Biobank received ethical approval from the UK Biobank Research Ethics Committee (reference number: 11/NW/0382).

### Cardiac MRI

Cardiac magnetic resonance imaging (CMR) was performed at 1.5 Tesla using a MAGNETOM Aera scanner by Siemens Healthcare, Erlangen, Germany. Long axis CMRs (UK Biobank data-field ID: 20208) were available for 71,017 subjects with ongoing consent (release: October 2023). These images were acquired in sagittal views using the TRUFI (True Fast Imaging with Steady-State Free Precession) sequence. Full details regarding the CMR protocol and acquisition can be found elsewhere [39, 40]. The dataset includes 2D dynamic images capturing 50 distinct cardiac phases throughout one cardiac cycle per subject, obtained via retrospective electrocardiogram (ECG) gating. The Long-axis CMRs were measured in three directions: the two-chamber (2Ch), three-chamber (3Ch), and four-chamber (4Ch) views, further categorised into three planes (transverse, coronal, and sagittal). This study focuses specifically on the four-chamber view, as its angle providing visibility to both the atria and ventricles. For consistency across subjects, only the transverse plane, representing the most common orientation, was used in the analysis. The images were cropped at the heart using a segmentation mask as guide, with a final dimension of 128 x 128 x 50 time steps. MRIs were normalised dividing the intensity by maximum value for that subject. 8,455 subjects were discarded for shape mismatch during cropping, 4Ch transversal plane not present and less than 50 time steps. The majority of participants were imaged only once, either at instance 2 (UKB encoding of the first imaging visit) or at instance 3 (repeated imaging visit). For the subjects having two repeated acquisitions, only the MRIs from the first visit were kept.

To detect and remove low-quality images (in terms of image acquisition), we used a pre-trained ResNet-18 model from Torchvision (trained on ImageNet) to extract features from each slice. We then applied principal component analysis (PCA) and uniform manifold approximation and projection (UMAP) to generate a final two-dimensional embedding for each image, with outliers identified by applying k-nearest neighbours (KNN) to remove the top 5% with the largest distances to their fifth nearest neighbour, resulting in the removal of 71 images. The remaining 62,491 subjects were then divided into 48,427 and 14,064 discovery and replication latent-phenotyped cohorts, respectively.

Regarding UK Biobank’s phenotypic data, subjects with missing values were removed. Then, those exhibiting outlier values for a certain phenotype were excluded utilising the Elliptic Envelope algorithm [41], which operates under the assumption that the outlier-free data follows a Gaussian distribution and ascertains the deviation of a data point from the central tendency using the Mahalanobis distance. The algorithm was applied with a contamination factor of 0.01 − i.e., it assumes that 1% of the data points are anomalies and adjusts the decision function accordingly to accurately identify outlier boundaries, and with a support fraction of 0.7, indicating that a randomly selected 70% of the data points would be used for the estimation of the covariance matrix.

Considering only the subjects from the latent-phenotyped cohort that also had phenotypic data available in the UKB, the final discovery and replication cohorts resulted in *N*_*disc*_ = 47854 (**Supplementary Table 23**) and *N*_*replic*_ = 12396 (**Supplementary Table 24**) subjects, respectively.

The discovery cohort includes only Caucasian subjects determined by genetic ancestry based on a PCA of the genotypes (UK Biobank data-field ID: 22006, as described in [42]), which were released before June 2023. The replication cohort includes subjects of all ancestries, regardless of their release date (N=8319) and Caucasian subjects released between June 2023 and October 2023 (N=4077). Full ethnicity breakup is presented in **Supplementary Table 25**.

### Inferring latent phenotypes

Given the CMR data acquired at different time points, let **X**_*t*_ ∈ ℝ^*m*× *n*^ represent the MRI slice at time point *t*, where *m n* is the dimensionality of the image, and a set of such MRI slices over time with *T* time points can be represented as **X** = {**X**_1_, **X**_2_,. .., **X**_*T*_}. The objective is to learn a latent representation **Z ∈** ℝ^*d*^ (with *d* being the dimensionality of the latent space) such that **Z** captures the underlying structural information and temporal dynamics across all time points. This can be formulated as **Z** = *f*(**X**_1_, **X**_2_,. .., **X**_*T*_ ; *θ*_2D_), where the function *f*(·), with parameters *θ*_2D_, maps the set of MRI slices over time to the latent space **Z**. We could represent the MRI slices over time as a 3D volume by stacking the slices along with temporal axis as **V** = [**X**_1_, **X**_2_,. .., **X**_T_ ] *∈* ℝ^*m*× *n*× *T*^ , and re-formulate the task as **Z** = *f*(**V**; *θ*_3*D*_). The latent representation can be expressed as **Z** = [*z*_1_, *z*_2_,. .., *z*_*d*_]^>^, where each *z*_*i*_ ∈ ℝ is a latent factor.

**Z**, or any of its subsets, may be used for downstream tasks such as disease diagnosis prediction. More importantly, each *z*_i_ can be used to perform GWAS by considering it as a latent phenotype, without the need for any hand-crafted features or annotation. Multiple methods can be employed to learn this function *f*(·), such as Autoencoders (AEs) and Variational Autoencoders (VAEs). Diffusion Autoencoders (DiffAE) have demonstrated superior performance in both latent representation and image generation tasks [4], utilising a diffusion-based approach to generate high-quality images while learning a meaningful latent representation, outperforming traditional methods like AE and VAE in reconstructing fine details and preserving global image structures. Hence, in this research, we have implemented a 3D diffusion autoencoder to learn a 3D feature set *θ*_3D_ for *f*(·), in order to obtain the latent representation **Z** from a given **V**.

Diffusion probabilistic models (DPMs) or simply diffusion models belong to the generative model family, which model the target distribution by learning a process of denoising noise at different levels, starting from an arbitrary Gaussian noise map and reconstructing it into an image after *T* timesteps of denoising. A typical DPM [43] model learns a denoising process that reverses the diffusion process of adding Gaussian noise *T* times to an input image *x*_0_ by learning a function *ε*_*θ*_ (*x*_t_, *t*) and by optimising the loss term:

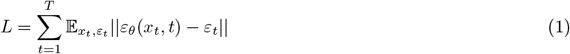

where *x*_t_ is the noisy image at timestep *t* (*t* ∈ *T*), obtained by adding *ε*_*t*_ noise to *x*_*t*−1_. The latents generated by such a model are stochastic and only represent the sequence of adding Gaussian noise to the input image. Denoising Diffusion Implicit Model (DDIM) [44] extends this idea to include a deterministic generative process and a novel inference distribution, while maintaining the marginal distribution of the original DPM, presented in Eqs. 2, 3, and 4, respectively. The DDIM can be considered as an image decoder, as the generative process can be deterministically reversed to get the noise map *x*_*T*_ , representing a latent representation of the input image that can be decoded to obtain the reconstruction of the input image *x*_0_. Even though the reconstruction quality of such a model is high, *x*_*T*_ does not contain high-level semantics of the image [4].

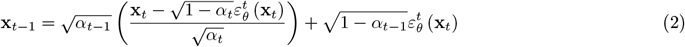

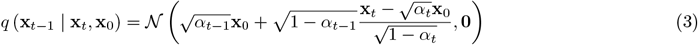

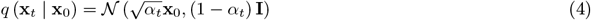

To address the problem of DDIM and being able to encode meaningful high-level semantic features *z*_sem_, while also encoding low-level stochastic latents *x*_*T*_ , the approach of DDIM was further extended as the Diffusion Autoencoder (DiffAE) [4]. DiffAE uses a convolutional semantic encoder to learn the semantic features (semantic latents) from the input image *x*_0_ (*z*_*sem*_ = *Enc*_*ϕ*_ (*x*_0_), where *ϕ* is the set of trainable parameters of the encoder network), a DDIM-based image decoder *p* (**x**_*t*−1_ | **x**_t_, **z**_*sem*_) conditioned on the semantic latent *z*_*sem*_ that decodes the latent variable *z*, that comprises of a semantic latent *z*_*sem*_ and a stochastic latent *x*_*T*_ . The decoder models *p*_*θ*_(*x*_t−1_|*x*_t_, *z*_sem_) matching the inference distribution *q*(*x*_*t*−1_|*x*_*t*_, *x*_0_) as a reverse generative process, following the equation:

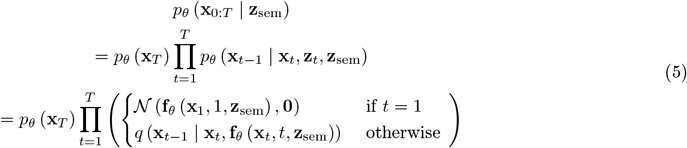

where the noise prediction network *f*_*θ*_ is given by the equation:

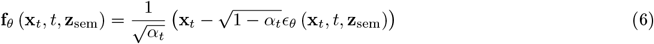

The final objective of the DiffAE is given by:

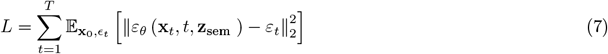

### Implementation, training and evaluation of 3D DiffAE

We used 62,562 subjects with 4-chamber view CMRs along the long axis of the heart, having 50 time points, to train the models, and then infer the embeddings using the trained models. The dataset was divided into training (*N*_*train*_ = 23, 776), validation (*N*_*validation*_ = 3, 840)), and held-out test sets *N*_*test*_ = 35, 714). Before providing the images as input to the model, the data was pre-processed using a two-step normalisation process: first, min-max normalisation is applied based on the input volume’s own range, followed by z-score normalisation with a mean of 0.5 and a standard deviation of 0.5.

In our implementation of the 3D DiffAE, we set the network’s base channel size to 32 and the dimension of the latent space to 128. The implementation was carried out using PyTorch, and the complete code is publicly available (see Code Availability). We trained the 3D DiffAE model, which comprises approximately 20.8 million trainable parameters, by optimising the objective function (see Eq. 7) using the Adam optimiser, with a batch size of two, for 200 epochs on an Nvidia Tesla V100-SXM2-32GB GPU, utilising 16-bit automatic mixed precision (AMP). We performed gradient clipping during training, which involves clipping the gradient norm of the model parameters when the norm of the gradients across all parameters exceeds one, to ensure that the gradients remain within a controlled range, preventing excessively large updates during backpropagation and promoting more stable training. Rather than using the entire dataset for each epoch, we randomly sampled 25% of the training and 2.5% validation samples during each epoch, thereby introducing additional randomness into the gradient estimation, which acts as a form of implicit regularisation. Furthermore, we applied minimalist data augmentation by randomly flipping images horizontally with a probability of 0.5, in order to enhance the model’s generalisation capability.

Each epoch of training and validation took an average of 42 minutes, resulting in a total training time of approximately 140 hours (around six days). Upon completion of training, we selected the model state that achieved the best validation loss as the final model to avoid any possible overfitting, which was subsequently used for inference on the entire dataset. As the inference step (and the validation step during training) is feedforward-only and does not require gradient calculations, it demands significantly less memory. This allowed us to increase the batch size to 16 during validation and inference on the same GPU. The full inference process, involving the acquisition of both embeddings and reconstructions using `t_step` = 20, took approximately 12.5 hours for 10,000 subjects. To obtain only the latent embeddings from the encoder, without the reconstruction steps, the process took around half an hour for the complete set of 62,562 subjects.

Owing to random initialisations and other stochastic processes, each training run may result in different latent representations, even when the same model and data are used. It is well-established in the literature that factors models, including autoencoders are non-identifiable, thereby causing the latent factors to capture different features from the image during a given model run [45, 46]. This variability highlights the importance of conducting multiple runs, as reliance on a single run may introduce bias and lead to suboptimal representations that do not generalise effectively. By performing several runs, we can capture a broader range of latent features, enabling us to determine which aspects of the representations are robust across runs and which are more sensitive to initial conditions. This approach facilitates a more comprehensive exploration of the latent representation of the given input. Consequently, in our research, we initialised the model’s weights using five different seeds: (1701, 1993, 2023, 42, and 1994), thereby obtaining five sets of latent factors.

We evaluated the perceptual quality of the reconstructions from the trained models across five seeds using the Structural Similarity Index (SSIM) [47], which accounts for changes in structural information, luminance, and contrast. In a given image patch from the images *x* and *y*, with local means *µ*_*x*_ and *µ*_*y*_, standard deviations *σ*_*x*_ and *σ*_*y*_, the cross-covariance for images *σ*_*xy*_, and the dynamic range of the pixel-values *L*, SSIM can be calculated using the following formula:

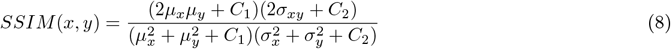

where *C*_1_ = (*k*_1_*L*)^2^ and *C*_2_ = (*k*_2_*L*)^2^ (with *k*_1_ = 0.01 and *k*_2_ = 0.03) are constants used to stabilise the division when the denominators are close to zero.

Additionally, we quantified the pixel-wise difference between the original and reconstructed images by measuring the distortion (or noise) introduced during reconstruction, employing the Peak Signal-to-Noise Ratio (PSNR), which is calculated as follows:

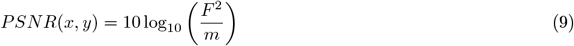

where *m* represents the mean-squared error between the images *x* and *y*, and *F* denotes the maximum fluctuation in the pixel values of the input images.

### Selection of Latent phenotypes across model runs

We obtained five sets of 128-dimensional latent factors from the entire dataset by training the models with five different seeds, resulting in a total of 640 latent factors. Across these sets of representations, while some features may be captured more frequently than others, certain factors might be identified uniquely in a single model run. To maximise reproducibility and select the most robust factors, we focused on latent factors consistently identified across multiple model runs for downstream analyses. The correlation coefficients between pairs of latent factors across the model runs were computed, and we subsequently selected those latent factors that exhibited a paired correlation of 0.8 or higher (see Supplementary Algorithm 2). This process resulted in 182 latent factors, which we considered the final set of latent phenotypes used for downstream analyses. We refer to them using the following nomenclature: Z*n*_S*i* where *n* is the latent factor number from the original training (1 to 128), and *i* is a number from 1 to 5 to distinguish the model run.

### Association of latent phenotypes with diseases, MRI-derived cardiac traits and body measures

Curated diseases were defined using three different data sources available in UK Biobank: GP clinical event records (UKB data-field ID: 42040), hospital inpatient (HI) admissions (UKB data-field ID: 41270) and self reported (SR) medical conditions (UKB data-field ID: 20002), converting ICD-9 codes to ICD-10 and discarding the mismatches. Tables were manually filtered, selecting records related to diseases of the cardiovascular system (including hypertension), Type 2 Diabetes and hypercholesterolaemia. Records of heart failure, left ventricular failure and cardiomegaly were grouped into “Heart failure” category, while “Conduction block” included Tawara branches, atrioventricular block and fascicular block. Individuals without any record of these diagnoses were considered healthy. For the continuous traits, 28 MRI-derived structural and functional cardiac and aortic endophenotypes were downloaded from UK Biobank (category ID: 157) [12]. Linear regression with ordinary least squares (OLS) was employed to test for associations between each latent phenotype and incident diseases or continuous traits in the discovery cohort (*N* = 48482), including genetic sex (UKB data-field ID: 22006), age at acquisition (computed from the MRI scan date and the birth year/month, UKB data-field IDs: 34 and 52 respectively), standing height, waist circumference (data-field ID: 48), body surface area (computed from weight and standing height, UKB data-field IDs: 21002, 50 respectively), MRI visit (Instance 2 or 3, as explained above) and MRI centre (taken from the DICOM header) as covariates. An additional analysis was performed removing weight and standing height from the covariates, results are shown in **Supplementary Figure 3**. Only diseases with a minimum of 200 diagnosed subjects within 5 years before and one year after the MRI acquisition were considered. Associations with very small magnitude effect sizes (< |1 × 10^−6^|) were discarded. The P-value threshold for significance was 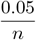, with *n* equal to the number of latent-trait pairs.

Linear and logistic regression models with L1 penalty were trained to predict the traits and diseases from the latent phenotypes and from single seeded DiffAE embeddings (see **Supplementary Table 22**). We trained the models on the discovery cohort with 5 fold cross-validation, and tested on the replication cohort. To include only non-colinear latent phenotypes as covariates, we iteratively removed all latent phenotypes with a variance inflation factor > 10. Moreover, features and continuous outcomes were standard scaled. Additional models were trained conditioning on LV end diastolic volume.

We performed latent manipulation for a certain target phenotype and latent factor **z** = (**z**_**sem**_, *x*_T_) following the formula:

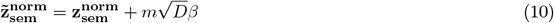

where *m* denotes the manipulation magnitude, *D* the dimension of **z**_**sem**_ (128 in our case), *β* the normalised regression coefficients for the considered target (possibly conditioned); the superscript *norm* is used to indicate standard scaled vectors [4]. The manipulated latent 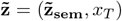 was decoded, and the reconstruction was visually inspected. We repeated the latent manipulation process for different *m*, **z**, and targets.

### UK Biobank genotyping and quality control

Full details of UK Biobank genotyping and quality control have been described elsewhere [42]. Briefly, the samples were genotyped using a combination of two arrays: the UK Biobank Axiom array from Affymetrix and the UK BiLEVE array, then imputed with the Haplotype Reference Consortium and the UK10K +1000 Genomes haplotype panels. Quality control involved discarding genotyped variants with call rate < 95%, imputed variants with INFO score < 0.3, and variants with minor allele count (MAC) < 100 - corresponding to a minor allele frequency (MAF) of about 0.001. Variants with severe Hardy-Weinberg equilibrium (HWE) violations were removed (test P-value < 10^−30^). After variant-level QC, 42,795,932 autosomal SNPs remained for the analysis, with positions reported in GRCh37 coordinates. Subjects without imputed genetic data, high missing rate (≥10%) or having discordant sex annotations (self reported and inferred) were excluded from the analysis.

### GWAS of inferred latent phenotypes

Genome-wide Association study (GWAS) on the discovery cohort was performed using an automated Nextflow pipeline developed in house based on REGENIE [48] (v3.3.0). Autosomes were tested assuming an additive genetic model and including as covariates: genetic sex (UKB data-field ID: 22006), age at acquisition (computed from the MRI scan date and the birth year/month, having UKB data-field IDs: 34 and 52 respectively) , genotyping batch (UKB data-field ID: 22000), body surface area (BSA, computed from weight and standing height, with the formula: BSA m^2^ = 0.007184 × Weight(kg)^0.425^ × Height(cm)^0.725^ [49]), year in which the imaging was performed (2014-2023), assessment centre of the imaging visit, and visit number. All the information regarding the MRI acquisitions were extracted from the DICOM images. REGENIE performs genome-wide association studies in two steps. In the first step, a whole genome regression model which captures part of the phenotypic variance due to genetic effects is fit using a subset of variants. This subset, which included 417,400 SNPs, was generated as suggested in [50], *i*.*e*., by using only genotyped data, with minor allele frequency (MAF) ≥ 1%, call rate ≥ 99%, HWE test P-value > 10^−15^, followed by linkage-disequilibrium (LD) pruning (1000 variant windows, 100 sliding windows and *R*^2^ > 0.8, performed with PLINK [51], version 1.9). In the second step, REGENIE tests genetic variants for association conditioning on the prediction generated by the regression model fit in the first step, using a leave one chromosome out (LOCO) scheme, thus avoiding proximal contamination. The autosomal panel of 42,795,932 common SNPs that passed QC having minor allele frequency ≥ 1% was used in our genome-wide association study. Variants in the major histocompatibility complex (MHC) region (GRCh37 chr6:28,477,797-33,448,354) were excluded due to its long-range LD structure.

The GWAS discovery cohort included 47,740 Caucasian individuals that passed both phenotypic and genotype QC (**Supplementary Figure 7**). 182 latent phenotypes were tested after quantile-normalisation. The associations were considered significant if the discovery P-value in Step 2 was lower than 5× 10^−8^/22 = 2.27× 10^−9^ , where 22 is the effective number of independent tests computed using the approach proposed by Li & Ji [13], taking into account the strong correlation among latent phenotypes. Genome-wide Manhattan plots were generated with *topr* R package [52] (v2.0.1). We annotate the nearest gene or the overlapping gene using the ‘closest’ function from *bedtools* (v2.31.0). Positions are reported in GRCh37 coordinates.

### Heritability and genetic correlation

SNP-based heritability of the latent phenotypes and genetic correlations against cardiac traits and diseases were computed using the R package *GenomicSEM* [53] (v0.0.5c) on the latent phenotypes GWAS summary statistics. *GenomicSEM* relies on a two-stage structural equation modelling approach based on a multivariable extension of *LDSC* [21] that accounts for varying and potentially unknown degrees of sample overlap. The set of relevant cardiometabolic traits and diseases is the same as the one used for colocalisation analysis, explained below. The Bonferroni-adjusted P-value threshold to define significance was computed as 0.05/(22× 54) = 4.21× 10^−05^, where 22 is the effective number of independent tests for the latent phenotypes and 54 is the number of traits included in the analysis.

### Statistical fine mapping

The reported number of 95 genome-wide significant associations refers to the last steps of the GWAS pipeline, which performs clumping with PLINK [51] (version 1.9). The genome-wide significance P-value was used as index variant threshold, including in the clump all the SNPs within 250 kb having *r*^2^ larger than 0.5. Significant variants were annotated with the closest gene using bedtools [54] (version 2.30). To dissect all the independent signals associated to each latent phenotype, conditional analysis was performed on raw summary statistics with GCTA’s Conditional & joint (COJO) analysis [55] (v1.94.0 beta), considering a window of 10 Mb, collinearity = 0.9 and variants with a MAF > 1%. Genotypes from 30,000 randomly selected UKB unrelated white British participants were used as reference panel for LD computations. The 89 variants having an association Pj ≤2.27 × 10^−9^ in the COJO joint model were considered significant and independent (**Supplementary Table 6**). To define a number of globally significant loci across traits, genome-wide significant genomic regions overlapping among different latent phenotypes were merged considering linkage disequilibrium. LD was computed with PLINK for each variant within a window of 500kb from an independent SNP. If two significant and independent SNPs inside the same genomic window were in LD (*r*^2^ > 0.1), only one signal was considered and the variant with strongest P-value was selected as lead. On the other hand, if inside the same region two SNPs were not in high LD, they were both kept as independent signals. This led to the definition of 44 lead independent SNPs in 42 genomic loci (**Supplementary Table 7**).

### GWAS Replication

Association study on the replication cohort was performed using REGENIE with the same configuration and set of covariates used in the discovery GWAS. The cohort included a total of 12,396 subjects of all ancestries, specifically 4,077 Caucasian and 8,319 non-Caucasian (**Supplementary Table 25** details the ethnicity breakdown, while (**Supplementary Figure 7**) illustrates the filtering process). The allele frequency comparison with the discovery cohort is shown in **Supplementary Figure 10**. Plots were created with the Python package *GwasLab* (v3.4.24) [56]. The 11,907 individuals passing genotype QC were included in the genetic analysis. Given the limitations of our replication cohort, characterised by both underpowered sample size and increased ethnic heterogeneity compared to the discovery cohort, we adopted a comprehensive approach to assess variant replication: lenient - how many variants have a concordant effect direction in the replication cohort, nominal - how many variants replicate at P-value ≤0.05 with consistent direction of effect and conservative - how many variants replicate with replication P-value ≤ 0.05/44 = 1.1 × 10^−3^, where 44 is the number of independent loci identified in the previous step. Not only sentinal lead SNP replication was considered, but also SNPs in high LD (*r*^2^ > 0.6) with the significant variants (**Supplementary Table 9**).

### Curation of variants

We interrogated the NHGRI-EBI GWAS Catalog database through the *LDtrait* tool (inside *LDlink* [57] R package, v1.1.2) to identify any overlap between significantly associated SNPs discovered in our study (or in high linkage disequilibrium with them; *r*^2^ ≥0.6) and previously reported associations in published summary statistics. Associations were considered novel if previously unreported in GWAS Catalog database. Additionally, Open Targets Genetics [16, 58] database (https://genetics.opentargets.org release October 2022, v22.10) was used to manually explore the lead variants.

### Gene-by-sex interaction GWAS

To examine sex differences in the genetic architecture of latent phenotypes, a gene-by sex interaction analysis was performed separately with REGENIE, testing for the interaction effect between being male and SNPs in the discovery cohort (**Table ??**). We corrected for the same covariates as the discovery GWAS, excluding genetic variants that had a minor allele frequency < 0.01. Also the same Bonferroni-corrected threshold as discovery GWAS was used to determine significance. Conditional analysis was performed with the same configuration of the discovery GWAS, considering the suggestive significance of 5 × 10^−8^ as threshold (**Supplementary Table 8**).

### Use of additional published GWAS summary statistics for colocalisation

A total of 27 summary statistics from published studies were downloaded from GWAS Catalog [14]. The downloaded traits span multiple biological scales and phenotypic categories, including: cardiac, ECG and blood pressure traits, cardiac diseases and non-cardiac traits related to body measures and other organs. The summary statistics of “Dark brown hair colour” from UKB was included as a negative control. In addition, 27 summary statistics were produced with REGENIE for the MRI-derived cardiac and aortic endophenotypes from UK Biobank (Field ID: 157) [12], using the same process as the one previously described for GWAS on latent phenotypes. **Supplementary Table 15** lists all the relevant phenotypes used in downstream analyses with the respective number of significant loci, cohort size, proportion of cases and controls, PMID and URL for the download. All the study cohorts include European UKB individuals, however some of the summary statistics derive from meta-analysis including also other cohorts. For 24 traits more than 80% of the genetic variants are shared with the UKB genotype data, whilst 3 GWAS summary statistics have an overlap of approximately 70%. Summary statistics were formatted and lifted to the hg38 reference genome build.

### Conditional analysis and colocalisation

In addition to the 54 GWAS summary statistics described above, we included GTEx eQTLs v8 from 12 cardiovascular-related tissues and other organs (Heart - atrial appendage, Heart - left ventricle, Artery - aorta, Artery - coronary, Artery - tibial, Adipose - subcutaneous, Adipose - visceral omentum, Muscle - skeletal, Whole blood, Lung, Liver, Pancreas) in the colocalisation analysis. GWAS and eQTL summary statistics were harmonised and variants were filtered by minor allele frequency (MAF > 0.01).

Multi-trait colocalisation test was based on the Bayesian framework of *coloc* described by Giambartolomei et al. (2014) [59].

We defined trait-specific genomic regions by selecting all SNPs with p-value threshold of 1 × 10^−5^ and setting loci boundaries between SNPs more than 250kb apart. We enlarged each genomic region by 100kb on both sides. Among such identified loci, only those having at least one SNP with p-value < 2 × 10^−9^ for latent phenotypes, < 2.5 × 10^−7^ for GTEx (as suggested in their original paper [19]) and < 5 × 10^−8^ for the downloaded disease/trait GWAS, were considered significant and carried out to the following analyses. The loci which encompassed the MHC II region (GRCh38 chr6:28,510,120-33,480,577) were excluded from the analysis due to the difficulties in properly fine-mapping them.

For each locus, we tested if secondary conditionally independent associations existed and which were shared with other traits. To this end, we developed a statistical approach that divides each GWAS-significant genomic region into its component signals and then uses colocalisation across different traits to group association signals likely caused by the same SNP into single modulatory modules. We performed conditional analysis of each significant locus using GCTA-COJO software [55] in two steps. First, a stepwise forward conditional regression procedure was applied to select the independently associated SNPs at each locus identified as described above. The stopping criterion employed was that all conditional and joint p-values (pJ) were larger than 1 × 10^−4^. This resulted in the identification of independent SNPs within the genomic region boundary. Then, for each of the identified variants, association analysis was performed conditional on all the other independent signals except for the target one (leave one out approach), generating a conditional dataset having only one independent association signal. This procedure is similar to that used by Robinson et al. but instead of using the step-wise conditioned datasets, it uses an ‘all but one’ approach.

True independent signals were defined as those being genome-wide significant (according to the thresholds listed above) in the original GWAS and with a joint p-value < 1 × 10^−6^ or having a genome-wide significant joint p-value. When possible, we used in-sample LD for the conditional analysis. For the downloaded GWAS tables, the UKB LD reference containing our latent phenotyped individuals was used and DENTIST [60] was applied prior to COJO to detect and remove variants showing heterogeneity between the GWAS and LD reference samples. We thus generated for each trait and locus a set of conditional datasets where the association signal is driven by a single causal variant. We then fine-mapped each conditional dataset obtained by computing the 99% credible set, using the ‘finemap.abf’ function from the R package *coloc* (v5.2.3) [59].

To understand which loci were pleiotropic between traits, we ran pairwise colocalisation using *coloc* [59] for each combination of conditional datasets having at least one SNP in their 99% credible set in common. The colocalisation tests resulting in PP.H_4_ > 0.8 (posterior probability for the hypothesis of same underlying causal variant contributing to both traits variation) were considered significant. Finally, since the *coloc* package allows only for pairwise testing, we followed-up colocalisation analysis by network analysis (using the R package *igraph* v2.0.2 [61]) to identify, among multiple pairs of colocalising conditional datasets (PP.H_4_ > 0.8), larger regulatory modules of traits all sharing the same causal variant. Locuszoom plots were created adapting the functions from *LocusCompareR* package (v1.0.0)

### Functional enrichments

To facilitate coherent and computationally feasible post-GWAS functional analyses, we consolidated multiple sets of GWAS summary statistics into a single dataset. This was achieved by selecting the minimum P-value for each SNP across the 182 GWAS summary statistics, referred to here as ’minP GWAS’, which represents the strongest association for each SNP across all phenotypes. This approach prioritises the most significant genetic signals, irrespective of the phenotype in which they appear, thereby maximising the likelihood of identifying SNPs with true associations. Gene, gene-set and tissue expression analysis were performed with the MAGMA tool (v1.08) [24] as implemented inside FUMA software (https://fuma.ctglab.nl/v1.5.2) [22], using the SNP2GENE process on the aggregated minP GWAS summary statistic. A total of 17005 gene sets were tested, obtained from MSigDB (v2023.1Hs) for “Curated gene sets” and “GO terms”, with window size set to ± 1kb and using the full distribution of SNP p-values. Gene sets with Bonferroni-adjusted P value ≤0.05 were considered significant. To assess tissue specificity, we used MAGMA’s gene-property analysis to test the relationships between latent-gene associations and tissue-specific expression profiles (RPKM) from 53 tissue types in GTEx v8. This one-sided test evaluated average gene expression per tissue, conditioned on the average expression across all tissues. Tissue types with Bonferroni-adjusted P value ≤ 0.05 were considered significant.

FUMA was also employed to map genes through position (maximum distance 10 kb) and eQTL (tissues: heart atrial appendage and left ventricle). The resulting list of 85 mapped protein coding genes was used as input for an over-representation analysis with Enrichr [23], performed through the Python package *GSEApy* (v1.1.3) [63].

The Chromatin Element Enrichment Ranking by Specificity (CHEERS) method [25] was employed to assess whether latent-associated variants were enriched in chromatin accessible regions of nine cardiac cell types. We used the read counts per peak matrix from a snATAC-seq cardiac atlas comprising atrial and ventricular cardiomyocyte, smooth muscle, endothelial, adipocyte, macrophage, fibroblast, lymphocyte and nervous cells [64]; together with the 42 genomic risk loci (LD *r*^2^ > 0.1) of 500 kb from the GWAS summary statistics of latent phenotypes. Before computing the enrichment, CHEERS removed the bottom decile of peaks with the lowest read counts and obtained a cell type specificity score through Euclidean normalisation. The resulting enrichment one-sided P-value was corrected for the number of cell types tested (0.05/9).

### TWAS

Transciptome-wide association study was performed to identify associations between our latent phenotypes and the imputed tissue-specific gene expression from GTEx v8 [19]. The FUSION tool [65] was used on the aggregated minP GWAS summary statistic with its default settings, downloading the precomputed transcript expression reference weights of 12 tissues (Heart - atrial appendage, Heart - left ventricle, Artery - aorta, Artery - coronary, Artery - tibial, Adipose - subcutaneous, Adipose - visceral omentum, Muscle - skeletal, Whole blood, Lung, Liver, Pancreas) for European samples from FUSION authors’ website http://gusevlab.org/projects/fusion/. To account for multiple hypotheses, a Bonferroni-corrected P value threshold (0.05/*n* of genes tested) was used to define significant TWAS associations. The π_1_ statistic was computed to assess the degree of overlap in significant genes between tissue types. For each tissue pair, we filtered the significant genes in the target tissue to those overlapping with the reference tissue. We then computed the q-value object for this subset of P-values and derived the π_1_ as 1 − π_0_, where π_0_ represents the proportion of null hypotheses that are true.

### Heart single-nucleus RNA sequencing

Single-nucleus RNA sequencing (snRNA-seq) data of six anatomical adult heart regions (left and right atrium, left and right ventricles, apex and interventricular septum) was obtained from a study conducted by Litviňuková et al. [31]. The clustering and annotation of the cell types were available inside the log-normalised H5AD file provided by the authors. The primary analysis and visualisation were conducted using the Python package *Scanpy* (v1.10.1). We computed the gene set score of the 82 MAGMA prioritised protein-coding genes using the ’score_genes’ function, then we tested whether these 82 genes were differentially expressed in the two cardiomyocytes populations compared to the other cell types with the ’rank_genes_groups’ function using the T-test with overestimated variance.

### UK Biobank exome sequencing

Single-variant exome-wide association study (EWAS) and rare variant association tests, utilising the whole exome sequencing (WES) data (UKB data-field ID: 23159) on the discovery cohort, were performed on the UKB Research Analysis Platform (RAP, cf. https://ukbiobank.dnanexus.com) using REGENIE [48] (version 3.4.1). WES data available on the RAP are the raw calls after merging sample-level data with GLNexus [66], and QC [67, 68] with bcftools [69] was performed before using them. Low-quality genotypes were set to missing, respectively SNVs with a depth (DP) less than 7 and indels with a DP less than 10. Selected variants met the following criteria: at least 75% of non-missing genotypes with quality ≥20, ratio of balanced heterozygous calls to total heterozygous calls ≥0.5 missing call rate < 10%, and HWE test P-value > 10^−15^. Subjects with sex discordance were also removed. A total of 26,388,600 variants aligned to GRCh38 passed QC and were utilised for EWAS and the rare variant tests.

### Rare-variant association tests

Regarding the exome wide-association study, the first step of REGENIE was identical to the GWAS (as described in the previous sub-section). For the second step, the WES release tranche (UKB data-field ID: 32050) was used as an additional covariate to correct for batch effects, along with the covariates used for GWAS. EWAS was performed on variants with *MAC* > 5, integrating the conditionally independent genome-wide significant variants obtained after performing GWAS as covariates for each latent-chromosome pair (for the latent-chromosome pairs without any significant hit, no conditioning was performed). A total of 46,034 individuals from the discovery cohort passing genotype QC were included in the EWAS analysis, while for the burden tests 46,134 subjects were included (see **Supplementary Figure 7**). For the burden tests, helper files (available on the UKB RAP) containing variant annotations and gene sets collapsing rare variants (*MAC* > 1) were utilised. Specifically, variants were annotated using SnpEff [70] based on the Ensembl 85 gene model, selecting the most severe consequence for each variant across all protein-coding transcripts [67, 68]. For each gene, three categories of rare variant masks were considered: M1 with Loss-of-Function (LoF) variants only [LoF], M2 with LoF variants and likely deleterious missense variants - predicted to be deleterious by all five scoring algorithms - [LoF, missense (5/5)], and finally, M3 with LoF variants and possibly deleterious missense variants (predicted to be deleterious by at least one of the five scoring algorithms) [M3 LoF, missense (>=1/5)]. For each of these categories, two different burden masks per gene were built using the maximum number of alternative alleles across sites based on the frequency of the alternative allele of the variants: *AAF* < 0.01 and singletons only. Furthermore, the SKATO-ACAT test, which combines the strengths of burden tests and sequence kernel association tests (SKAT), allowing for robust detection of genetic associations regardless of variant effect directions, was performed for those three mask categories using variants with *AAF* < 0.01. When performing gene burden tests on whole exome sequencing (WES) data, the conventional genome-wide significance threshold of *p* < 5 × 10^−08^ may be overly stringent, as the analysis is focused on a subset of the genome−the exome−and the tests are conducted on gene sets rather than individual variants. Therefore, we recalculated the significance threshold using a method based on the distribution of the minimum p-values across different phenotypes [34]. The number of independent tests was estimated by taking the median of the minimum inverted p-values across all phenotypes, and then dividing the nominal significance level of 0.05 by this value. This resulted in an adjusted P-value threshold of *p* < 4.7 × 10^−07^ for SKATO-ACAT. **Supplementary Figure 20** illustrates the results for the burden test analysis, with an adjusted significance threshold of *p* < 1.5 × 10^−07^. Significant variants from the burden test are listed in (**Supplementary Table 18**).

### Polygenic risk score calculation and disease prediction

Polygenic risk scores (PRS) for each latent were computed for 314,774 Caucasian subjects (the entire UKB Caucasian cohort with available baseline data and genotypes, excluding subjects with a kinship coefficient > 0.0625 (computed using PLINK version 2.0 and subjects used for the discovery GWAS) using LDPred2-auto [71, 72, 73]. Similar to previous approaches [72], variants with a missing call rate < 0.1, *INFO* > 0.4 and *MAF* > 0.01 were pruned using PLINK [51] (version 2.0) using a window size of 250kb, a step size of 5, and an unphased hardcall *R*^2^ threshold of 0.5, resulting in 1,281,751 variants. Subsequently, all conditionally independent genome-wide significant variants were included, leading to a final total of 1,281,790 variants that were used for computing the PRS from the summary statistics of the discovery GWAS.

Furthermore, multi-PRS [74] for ten cardiac (and related) diseases were computed by applying Lasso (stratified five-fold cross-validation using *LogisticRegressionCV* function of *scikit-learn* [75] with the L1 penalty and saga solver). Multi-PRS models were trained by supplying the individual PRS computed for each latent factor, along with baseline characteristics (age, sex, BMI, and smoking status) as input and whether the subject was a positive or negative control case with respect to the particular disease as the target.

To evaluate the predictive ability of individual latent-specific PRS and multi-PRS, binary disease cohorts were created for angina pectoris, atherosclerotic conditions, conduction block, coronary heart disease, heart failure, high cholesterol, hypertension, metabolic syndrome, myocardial infraction, and type 2 diabetes, by taking positive cases from the entire UKB Caucasian cohort (excluding the cohort used for the discovery GWAS) and an equal number of negative super controls (i.e., subjects without any cardiac or metabolic disease marked as positive) matched for sex and age. Two types of predictive task were evaluated: diagnostic and prognostic, with respect to the date of collection of the baseline characteristics. If the disease was reported on or before the date of baseline data collection, the subject was included in the diagnostic cohort, while if the disease was reported between one and ten years after the collection of the baseline data, the subject was included in the prognostic cohort. The number of subjects for each disease is reported in the **Supplementary Table 27**. Five-fold cross-validation was performed for each model to evaluate them without data selection bias. For the latent-specific PRS models, separate predictors were trained for each by applying linear regression on the latent-specific PRS and the set of baseline characteristics, and then the predictor demonstrating the best Area Under the Receiver Operating Characteristic Curve (ROC-AUC, or simply AUC) was selected, referred to here as max(Single PRS + Baseline). The performances of the max(Single PRS + Baseline) and Multi PRS + Baseline for those ten diseases were benchmarked against a linear regression predictor trained solely using the baseline characteristics.

## Resource availability

Requests for further information and resources should be directed to and will be fulfilled by the lead contact, Craig A. Glastonbury.

## Data availability

All summary statistics will be deposited with GWAS Catalog. All data used to conduct the study is available to any UK biobank approved researcher. Information and URLs of the publicly available GWAS summary statistics used for colocalisation analysis are detailed in Supplementary Table 15. The full cis-eQTL summary statistics from GTEx v8 were obtained from on 11/06/2024 under dbGaP application phs000424.v2.p1. Single nucleus RNA sequencing data objects were downloaded via the webportal.

## Code availability

For detailed information, including video presentations, see our project page at https://glastonburygroup.github.io/CardiacDiffAE_GWAS. The codebase of this project (except for the phenotyping using 3D DiffAE) is available at https://github.com/GlastonburyGroup/CardiacDiffAE_GWAS, while the pipeline for phenotyping using 3D DiffAE is available at https://github.com/GlastonburyGroup/ImLatent. For the original source code of DiffAE (2D, for RGB images), see https://github.com/phizaz/DiffAE. The in-house GWAS pipeline based on REGENIE is available at yses using WES were conducted on the UKB Research Analysis Platform (https://ukbiobank.dnanexus.com).

## Acknowledgements

This research has been conducted using the UK Biobank Resource under application number 82779. We would like to thank all UKB participants for their invaluable data that make studies like these, and many more, possible.

## Authors Contributions

C.A.G. conceived the study design. S.C. implemented 3DDiffAE, the image processing and result analysis pipelines, ran the GWAS, EWAS, and burden test pipelines, and performed polygenic risk score modelling and analysis. S.O carried out all genetic analyses, including GWAS, finemapping, colocalisation, and heritability, and performed the post-GWAS enrichment analyses. A.M.V performed the latent-phenotype association and latent manipulation experiments. E.G implemented the Regenie GWAS pipeline. A.L, S.S and N.P implemented the multi-trait colocalisation pipeline. F.C. provided advice on image segmentation and latent selection and helped with the enrichment analysis. E.S helped with figure design. F.S provided advice on polygenic risk scores. E.B helped to QC the clinical data.

C.A.P. quantified the changes in the manipulation analyses and assisted in the disease grouping criteria. C.A.G, S.O, S.C, A.M.V and A.V wrote and edited the manuscript. E.A and F.I read and reviewed the manuscript.

## References

[1] B. Gomes, A. Singh, J. W. O’Sullivan, T. M. Schnurr, P. C. Goddard, S. Loong, D. Amar, J. W. Hughes, M. Kostur, F. Haddad, et al., “Genetic architecture of cardiac dynamic flow volumes,” Nature Genetics, vol. 56, no. 2, pp. 245–257, 2024.

[2] J. P. Pirruccello, M. D. Chaffin, E. L. Chou, S. J. Fleming, H. Lin, M. Nekoui, S. Khurshid, S. F. Friedman, A. G. Bick, A. Arduini, et al., “Deep learning enables genetic analysis of the human thoracic aorta,” Nature genetics, vol. 54, no. 1, pp. 40–51, 2022.

[3] D. E. Rumelhart, G. E. Hinton, and R. J. Williams, “Learning internal representations by error propagation, parallel distributed processing, explorations in the microstructure of cognition, ed. de rumelhart and j. mcclelland. vol. 1. 1986,” Biometrika, vol. 71, no. 599-607, p. 6, 1986.

[4] K. Preechakul, N. Chatthee, S. Wizadwongsa, and S. Suwajanakorn, “Diffusion autoencoders: Toward a meaningful and decodable representation,” in Proceedings of the IEEE/CVF Conference on Computer Vision and Pattern Recognition, pp. 10619–10629, 2022.

[5] C. A. Glastonbury, S. L. Pulit, J. Honecker, J. C. Censin, S. Laber, H. Yaghootkar, N. Rahmioglu, E. Pastel, K. Kos, A. Pitt, et al., “Machine learning based histology phenotyping to investigate the epidemiologic and genetic basis of adipocyte morphology and cardiometabolic traits,” PLoS computational biology, vol. 16, no. 8, p. e1008044, 2020.

[6] Y. Liu, N. Basty, B. Whitcher, J. D. Bell, E. P. Sorokin, N. van Bruggen, E. L. Thomas, and M. Cule, “Genetic architecture of 11 organ traits derived from abdominal mri using deep learning,” Elife, vol. 10, p. e65554, 2021.

[7] B. Alipanahi, F. Hormozdiari, B. Behsaz, J. Cosentino, Z. R. McCaw, E. Schorsch, D. Sculley, E. H. Dorfman, P. J. Foster, L. H. Peng, et al., “Large-scale machine-learning-based phenotyping significantly improves genomic discovery for optic nerve head morphology,” The American Journal of Human Genetics, vol. 108, no. 7, pp. 1217–1230, 2021.

[8] F. Cisternino, S. Ometto, S. Chatterjee, E. Giacopuzzi, A. P. Levine, and C. A. Glastonbury, “Self-supervised learning for characterising histomorphological diversity and spatial rna expression prediction across 23 human tissue types,” Nature Communications, vol. 15, no. 1, p. 5906, 2024.

[9] M. Kirchler, S. Konigorski, M. Norden, C. Meltendorf, M. Kloft, C. Schurmann, and C. Lippert, “transfergwas: Gwas of images using deep transfer learning,” Bioinformatics, vol. 38, no. 14, pp. 3621–3628, 2022.

[10] R. Bonazzola, E. Ferrante, N. Ravikumar, Y. Xia, B. Keavney, S. Plein, T. Syeda-Mahmood, and A. F. Frangi, “Unsupervised ensemble-based phenotyping enhances discoverability of genes related to left-ventricular morphology,” Nature Machine Intelligence, vol. 6, no. 3, pp. 291–306, 2024.

[11] K. Patel, Z. Xie, H. Yuan, S. M. S. Islam, Y. Xie, W. He, W. Zhang, A. Gottlieb, H. Chen, L. Giancardo, et al., “Unsupervised deep representation learning enables phenotype discovery for genetic association studies of brain imaging,” Communications Biology, vol. 7, no. 1, p. 414, 2024.

[12] W. Bai, H. Suzuki, J. Huang, C. Francis, S. Wang, G. Tarroni, F. Guitton, N. Aung, K. Fung, S. E. Petersen, et al., “A population-based phenome-wide association study of cardiac and aortic structure and function,” Nature medicine, vol. 26, no. 10, pp. 1654–1662, 2020.

[13] J. Li and L. Ji, “Adjusting multiple testing in multilocus analyses using the eigenvalues of a correlation matrix,” Heredity, vol. 95, no. 3, pp. 221–227, 2005.

[14] J. MacArthur, E. Bowler, M. Cerezo, L. Gil, P. Hall, E. Hastings, H. Junkins, A. McMahon, A. Milano, J. Morales, et al., “The new nhgri-ebi catalog of published genome-wide association studies (gwas catalog),” Nucleic acids research, vol. 45, no. D1, pp. D896–D901, 2017.

[15] A. Radhakrishnan, S. F. Friedman, S. Khurshid, K. Ng, P. Batra, S. A. Lubitz, A. A. Philippakis, and C. Uhler, “Cross-modal autoencoder framework learns holistic representations of cardiovascular state,” Nature Communications, vol. 14, no. 1, p. 2436, 2023.

[16] M. Ghoussaini, E. Mountjoy, M. Carmona, G. Peat, E. M. Schmidt, A. Hercules, L. Fumis, A. Miranda, D. Carvalho-Silva, A. Buniello, et al., “Open targets genetics: systematic identification of trait-associated genes using large-scale genetics and functional genomics,” Nucleic acids research, vol. 49, no. D1, pp. D1311–D1320, 2021.

[17] T. Groza, F. L. Gomez, H. H. Mashhadi, V. Muñoz-Fuentes, O. Gunes, R. Wilson, P. Cacheiro, A. Frost, P. Keskivali-Bond, B. Vardal, et al., “The international mouse phenotyping consortium: comprehensive knockout phenotyping underpinning the study of human disease,” Nucleic acids research, vol. 51, no. D1, pp. D1038–D1045, 2023.

[18] E. Gerdts and V. Regitz-Zagrosek, “Sex differences in cardiometabolic disorders,” Nature medicine, vol. 25, no. 11, pp. 1657–1666, 2019.

[19] J. Lonsdale, J. Thomas, M. Salvatore, R. Phillips, E. Lo, S. Shad, R. Hasz, G. Walters, F. Garcia, N. Young, et al., “The genotype-tissue expression (gtex) project,” Nature genetics, vol. 45, no. 6, pp. 580–585, 2013.

[20] Y. Zhou, X. Zhu, H. Cui, J. Shi, G. Yuan, S. Shi, and Y. Hu, “The role of the vegf family in coronary heart disease,” Frontiers in cardiovascular medicine, vol. 8, p. 738325, 2021.

[21] B. K. Bulik-Sullivan, P.-R. Loh, H. K. Finucane, S. Ripke, J. Yang, S. W. G. of the Psychiatric Genomics Consortium, N. Patterson, M. J. Daly, A. L. Price, and B. M. Neale, “Ld score regression distinguishes confounding from polygenicity in genome-wide association studies,” Nature genetics, vol. 47, no. 3, pp. 291–295, 2015.

[22] K. Watanabe, E. Taskesen, A. Van Bochoven, and D. Posthuma, “Functional mapping and annotation of genetic associations with fuma,” Nature communications, vol. 8, no. 1, p. 1826, 2017.

[23] E. Y. Chen, C. M. Tan, Y. Kou, Q. Duan, Z. Wang, G. V. Meirelles, N. R. Clark, and A. Ma’ayan, “Enrichr: interactive and collaborative html5 gene list enrichment analysis tool,” BMC bioinformatics, vol. 14, pp. 1–14, 2013.

[24] C. A. de Leeuw, J. M. Mooij, T. Heskes, and D. Posthuma, “Magma: generalized gene-set analysis of gwas data,” PLoS computational biology, vol. 11, no. 4, p. e1004219, 2015.

[25] B. Soskic, E. Cano-Gamez, D. J. Smyth, W. C. Rowan, N. Nakic, J. Esparza-Gordillo, L. Bossini-Castillo, D. F. Tough, C. G. Larminie, P. G. Bronson, et al., “Chromatin activity at gwas loci identifies t cell states driving complex immune diseases,” Nature genetics, vol. 51, no. 10, pp. 1486–1493, 2019.

[26] H. Sun, K. C. Olson, C. Gao, D. A. Prosdocimo, M. Zhou, Z. Wang, D. Jeyaraj, J.-Y. Youn, S. Ren, Y. Liu, et al., “Catabolic defect of branched-chain amino acids promotes heart failure,” Circulation, vol. 133, no. 21, pp. 2038–2049, 2016.

[27] J.-y. Yu, N. Cao, C. D. Rau, R.-P. Lee, J. Yang, R. J. R. Flach, L. Petersen, C. Zhu, Y.-L. Pak, R. A. Miller, et al., “Cell-autonomous effect of cardiomyocyte branched-chain amino acid catabolism in heart failure in mice,” Acta Pharmacologica Sinica, vol. 44, no. 7, pp. 1380–1390, 2023.

[28] H. Chen, A. Moreno-Moral, F. Pesce, N. Devapragash, M. Mancini, E. L. Heng, M. Rotival, P. K. Srivastava, N. Harmston, K. Shkura, et al., “Wwp2 regulates pathological cardiac fibrosis by modulating smad2 signaling,” Nature communications, vol. 10, no. 1, p. 3616, 2019.

[29] H. Chen, G. Chew, N. Devapragash, J. Z. Loh, K. Y. Huang, J. Guo, S. Liu, E. L. S. Tan, S. Chen, N. G. Z. Tee, et al., “The e3 ubiquitin ligase wwp2 regulates pro-fibrogenic monocyte infiltration and activity in heart fibrosis,” Nature Communications, vol. 13, no. 1, p. 7375, 2022.

[30] T. Zimmer and R. Surber, “Scn5a channelopathies–an update on mutations and mechanisms,” Progress in biophysics and molecular biology, vol. 98, no. 2-3, pp. 120–136, 2008.

[31] M. Litviňuková, C. Talavera-López, H. Maatz, D. Reichart, C. L. Worth, E. L. Lindberg, M. Kanda, K. Polanski, M. Heinig, M. Lee, et al., “Cells of the adult human heart,” Nature, vol. 588, no. 7838, pp. 466–472, 2020.

[32] S. Sakaue, M. Kanai, Y. Tanigawa, J. Karjalainen, M. Kurki, S. Koshiba, A. Narita, T. Konuma, K. Yamamoto, M. Akiyama, et al., “A cross-population atlas of genetic associations for 220 human phenotypes,” Nature genetics, vol. 53, no. 10, pp. 1415–1424, 2021.

[33] S. Dong, N. Zhao, E. Spragins, M. S. Kagda, M. Li, P. Assis, O. Jolanki, Y. Luo, J. M. Cherry, A. P. Boyle, et al., “Annotating and prioritizing human non-coding variants with regulomedb v. 2,” Nature genetics, vol. 55, no. 5, pp. 724–726, 2023.

[34] K. J. Karczewski, M. Solomonson, K. R. Chao, J. K. Goodrich, G. Tiao, W. Lu, B. M. Riley-Gillis, E. A. Tsai, H. I. Kim, X. Zheng, et al., “Systematic single-variant and gene-based association testing of thousands of phenotypes in 394,841 uk biobank exomes,” Cell Genomics, vol. 2, no. 9, 2022.

[35] R. Pekson, F. G. Liang, J. L. Axelrod, J. Lee, D. Qin, A. J. Wittig, V. M. Paulino, M. Zheng, P. M. Peixoto, and R. N. Kitsis, “The mitochondrial atp synthase is a negative regulator of the mitochondrial permeability transition pore,” Proceedings of the National Academy of Sciences, vol. 120, no. 51, p. e2303713120, 2023.

[36] M. A. Gargano, N. Matentzoglu, B. Coleman, E. B. Addo-Lartey, A. V. Anagnostopoulos, J. Anderton, P. Avillach, A. M. Bagley, E. Bakštein, J. P. Balhoff, G. Baynam, S. M. Bello, M. Berk, H. Bertram, S. Bishop, H. Blau, D. F. Bodenstein, P. Botas, K. Boztug, J. Čady, T. J. Callahan, R. Cameron, S. J. Carbon, F. Castellanos, J. H. Caufield, L. E. Chan, C. G. Chute, J. Cruz-Rojo, N. Dahan-Oliel, J. R. Davids, M. de Dieuleveult, V. de Souza, B. B. A. de Vries, E. de Vries, J. R. DePaulo, B. Derfalvi, F. Dhombres, C. Diaz-Byrd, A. J. M. Dingemans, B. Donadille, M. Duyzend, R. Elfeky, S. Essaid, C. Fabrizzi, G. Fico, H. V. Firth, Y. Freudenberg-Hua, J. M. Fullerton, D. L. Gabriel, K. Gilmour, J. Giordano, F. S. Goes, R. G. Moses, I. Green, M. Griese, T. Groza, W. Gu, J. Guthrie, B. Gyori, A. Hamosh, M. Hanauer, K. Hanušová, Y. O. He, H. Hegde, I. Helbig, K. Holasová, C. T. Hoyt, S. Huang, E. Hurwitz, J. O. B. Jacobsen, X. Jiang, L. Joseph, K. Keramatian, B. King, K. Knoflach, D. A. Koolen, M. L. Kraus, C. Kroll, M. Kusters, M. S. Ladewig, D. Lagorce, M.-C. Lai, P. Lapunzina, B. Laraway, D. Lewis-Smith, X. Li, C. Lucano, M. Majd, M. L. Marazita, V. Martinez-Glez, T. H. McHenry, M. G. McInnis, J. A. McMurry, M. Mihulová, C. E. Millett, P. B. Mitchell, V. Moslerová, K. Narutomi, S. Nematollahi, J. Nevado, A. A. Nierenberg, N. N. Čajbiková, J. Nurnberger, John I, S. Ogishima, D. Olson, A. Ortiz, H. Pachajoa, G. Perez de Nanclares, A. Peters, T. Putman, C. K. Rapp, A. Rath, J. Reese, L. Rekerle, A. M. Roberts, S. Roy, S. J. Sanders, C. Schuetz, E. C. Schulte, T. G. Schulze, M. Schwarz, K. Scott, D. Seelow, B. Seitz, Y. Shen, M. N. Similuk, E. S. Simon, B. Singh, D. Smedley, C. L. Smith, J. T. Smolinsky, S. Sperry, E. Stafford, R. Stefancsik, R. Steinhaus, R. Strawbridge, J. C. Sundaramurthi, P. Talapova, J. A. Tenorio Castano, P. Tesner, R. H. Thomas, A. Thurm, M. Turnovec, M. E. van Gijn, N. A. Vasilevsky, M. Vlčková, A. Walden, K. Wang, R. Wapner, J. S. Ware, A. A. Wiafe, S. A. Wiafe, L. D. Wiggins, A. E. Williams, C. Wu, M. J. Wyrwoll, H. Xiong, N. Yalin, Y. Yamamoto, L. N. Yatham, A. K. Yocum, A. H. Young, Z. Yüksel, P. P. Zandi, A. Zankl, I. Zarante, M. Zvolský, S. Toro, L. C. Carmody, N. L. Harris, M. C. Munoz-Torres, D. Danis, C. J. Mungall, S. Köhler, M. A. Haendel, and P. N. Robinson, “The human phenotype ontology in 2024: phenotypes around the world,” Nucleic Acids Research, vol. 52, pp. D1333–D1346, 11 2023.

[37] C. Sudlow, J. Gallacher, N. Allen, V. Beral, P. Burton, J. Danesh, P. Downey, P. Elliott, J. Green, M. Landray, et al., “Uk biobank: an open access resource for identifying the causes of a wide range of complex diseases of middle and old age,” PLoS medicine, vol. 12, no. 3, p. e1001779, 2015.

[38] T. J. Littlejohns, C. Sudlow, N. E. Allen, and R. Collins, “Uk biobank: opportunities for cardiovascular research,” European heart journal, vol. 40, no. 14, pp. 1158–1166, 2019.

[39] S. E. Petersen, P. M. Matthews, F. Bamberg, D. A. Bluemke, J. M. Francis, M. G. Friedrich, P. Leeson, E. Nagel, S. Plein, F. E. Rademakers, et al., “Imaging in population science: cardiovascular magnetic resonance in 100,000 participants of uk biobank-rationale, challenges and approaches,” Journal of Cardiovascular Magnetic Resonance, vol. 15, no. 1, pp. 1–10, 2013.

[40] S. E. Petersen, P. M. Matthews, J. M. Francis, M. D. Robson, F. Zemrak, R. Boubertakh, A. A. Young, S. Hudson, P. Weale, S. Garratt, et al., “Uk biobank’s cardiovascular magnetic resonance protocol,” Journal of cardiovascular magnetic resonance, vol. 18, no. 1, pp. 1–7, 2015.

[41] P. J. Rousseeuw and K. V. Driessen, “A fast algorithm for the minimum covariance determinant estimator,” Technometrics, vol. 41, no. 3, pp. 212–223, 1999.

[42] C. Bycroft, C. Freeman, D. Petkova, G. Band, L. T. Elliott, K. Sharp, A. Motyer, D. Vukcevic, O. Delaneau, J. O’Connell, et al., “The uk biobank resource with deep phenotyping and genomic data,” Nature, vol. 562, no. 7726, pp. 203–209, 2018.

[43] J. Ho, A. Jain, and P. Abbeel, “Denoising diffusion probabilistic models,” Advances in neural information processing systems, vol. 33, pp. 6840–6851, 2020.

[44] J. Song, C. Meng, and S. Ermon, “Denoising diffusion implicit models,” in International Conference on Learning Representations, 2020.

[45] K. Godfrey and J. DiStefano III, “Identifiability of model parameter,” IFAC Proceedings Volumes, vol. 18, no. 5, pp. 89–114, 1985.

[46] Y. Wang, D. Blei, and J. P. Cunningham, “Posterior collapse and latent variable non-identifiability,” Advances in Neural Information Processing Systems, vol. 34, pp. 5443–5455, 2021.

[47] Z. Wang, A. C. Bovik, H. R. Sheikh, and E. P. Simoncelli, “Image quality assessment: from error visibility to structural similarity,” IEEE transactions on image processing, vol. 13, no. 4, pp. 600–612, 2004.

[48] J. Mbatchou, L. Barnard, J. Backman, A. Marcketta, J. A. Kosmicki, A. Ziyatdinov, C. Benner, C. O’Dushlaine, M. Barber, B. Boutkov, et al., “Computationally efficient whole-genome regression for quantitative and binary traits,” Nature genetics, vol. 53, no. 7, pp. 1097–1103, 2021.

[49] E. H. Livingston and S. Lee, “Body surface area prediction in normal-weight and obese patients,” American Journal of Physiology-Endocrinology and Metabolism, vol. 281, no. 3, pp. E586–E591, 2001.

[50] B. B. Sun, J. Chiou, M. Traylor, C. Benner, Y.-H. Hsu, T. G. Richardson, P. Surendran, A. Mahajan, C. Robins, S. G. Vasquez-Grinnell, et al., “Genetic regulation of the human plasma proteome in 54,306 uk biobank participants,” BioRxiv, pp. 2022–06, 2022.

[51] C. C. Chang, C. C. Chow, L. C. Tellier, S. Vattikuti, S. M. Purcell, and J. J. Lee, “Second-generation plink: rising to the challenge of larger and richer datasets,” Gigascience, vol. 4, no. 1, pp. s13742–015, 2015.

[52] T. Juliusdottir, “topr: an r package for viewing and annotating genetic association results,” BMC bioinformatics, vol. 24, no. 1, p. 268, 2023.

[53] A. D. Grotzinger, M. Rhemtulla, R. de Vlaming, S. J. Ritchie, T. T. Mallard, W. D. Hill, H. F. Ip, R. E. Marioni, A. M. McIntosh, I. J. Deary, et al., “Genomic structural equation modelling provides insights into the multivariate genetic architecture of complex traits,” Nature human behaviour, vol. 3, no. 5, pp. 513–525, 2019.

[54] A. R. Quinlan and I. M. Hall, “Bedtools: a flexible suite of utilities for comparing genomic features,” Bioinformatics, vol. 26, no. 6, pp. 841–842, 2010.

[55] J. Yang, T. Ferreira, A. P. Morris, S. E. Medland, G. I. of ANthropometric Traits (GIANT) Consortium, D. G. Replication, M. analysis (DIAGRAM) Consortium, P. A. Madden, A. C. Heath, N. G. Martin, G. W. Montgomery, et al., “Conditional and joint multiple-snp analysis of gwas summary statistics identifies additional variants influencing complex traits,” Nature genetics, vol. 44, no. 4, pp. 369–375, 2012.

[56] Y. He, M. Koido, Y. Shimmori, and Y. Kamatani, “Gwaslab: a python package for processing and visualizing gwas summary statistics,” Jxiv, 05 2023.

[57] M. J. Machiela and S. J. Chanock, “Ldlink: a web-based application for exploring population-specific haplotype structure and linking correlated alleles of possible functional variants,” Bioinformatics, vol. 31, no. 21, pp. 3555–3557, 2015.

[58] E. Mountjoy, E. M. Schmidt, M. Carmona, J. Schwartzentruber, G. Peat, A. Miranda, L. Fumis, J. Hayhurst, A. Buniello, M. A. Karim, et al., “An open approach to systematically prioritize causal variants and genes at all published human gwas trait-associated loci,” Nature genetics, vol. 53, no. 11, pp. 1527–1533, 2021.

[59] C. Giambartolomei, D. Vukcevic, E. E. Schadt, L. Franke, A. D. Hingorani, C. Wallace, and V. Plagnol, “Bayesian test for colocalisation between pairs of genetic association studies using summary statistics,” PLoS genetics, vol. 10, no. 5, p. e1004383, 2014.

[60] W. Chen, Y. Wu, Z. Zheng, T. Qi, P. M. Visscher, Z. Zhu, and J. Yang, “Improved analyses of gwas summary statistics by reducing data heterogeneity and errors,” Nature Communications, vol. 12, no. 1, p. 7117, 2021.

[61] G. Csardi and T. Nepusz, “The igraph software,” Complex syst, vol. 1695, pp. 1–9, 2006.

[62] B. Liu, M. J. Gloudemans, A. S. Rao, E. Ingelsson, and S. B. Montgomery, “Abundant associations with gene expression complicate gwas follow-up,” Nature genetics, vol. 51, no. 5, pp. 768–769, 2019.

[63] Z. Fang, X. Liu, and G. Peltz, “Gseapy: a comprehensive package for performing gene set enrichment analysis in python,” Bioinformatics, vol. 39, no. 1, p. btac757, 2023.

[64] J. D. Hocker, O. B. Poirion, F. Zhu, J. Buchanan, K. Zhang, J. Chiou, T.-M. Wang, Q. Zhang, X. Hou, Y. E. Li, et al., “Cardiac cell type–specific gene regulatory programs and disease risk association,” Science advances, vol. 7, no. 20, p. eabf1444, 2021.

[65] A. Gusev, A. Ko, H. Shi, G. Bhatia, W. Chung, B. W. Penninx, R. Jansen, E. J. De Geus, D. I. Boomsma, F. A. Wright, et al., “Integrative approaches for large-scale transcriptome-wide association studies,” Nature genetics, vol. 48, no. 3, pp. 245–252, 2016.

[66] M. F. Lin, O. Rodeh, J. Penn, X. Bai, J. G. Reid, O. Krasheninina, and W. J. Salerno, “Glnexus: joint variant calling for large cohort sequencing,” BioRxiv, p. 343970, 2018.

[67] J. D. Szustakowski, S. Balasubramanian, E. Kvikstad, S. Khalid, P. G. Bronson, A. Sasson, E. Wong, D. Liu, J. Wade Davis, C. Haefliger, et al., “Advancing human genetics research and drug discovery through exome sequencing of the uk biobank,” Nature genetics, vol. 53, no. 7, pp. 942–948, 2021.

[68] J. D. Backman, A. H. Li, A. Marcketta, D. Sun, J. Mbatchou, M. D. Kessler, C. Benner, D. Liu, A. E. Locke, S. Balasubramanian, et al., “Exome sequencing and analysis of 454,787 uk biobank participants,” Nature, vol. 599, no. 7886, pp. 628–634, 2021.

[69] P. Danecek, J. K. Bonfield, J. Liddle, J. Marshall, V. Ohan, M. O. Pollard, A. Whitwham, T. Keane, S. A. McCarthy, R. M. Davies, et al., “Twelve years of samtools and bcftools,” Gigascience, vol. 10, no. 2, p. giab008, 2021.

[70] P. Cingolani, A. Platts, L. L. Wang, M. Coon, T. Nguyen, L. Wang, S. J. Land, X. Lu, and D. M. Ruden, “A program for annotating and predicting the effects of single nucleotide polymorphisms, snpeff: Snps in the genome of drosophila melanogaster strain w1118; iso-2; iso-3,” fly, vol. 6, no. 2, pp. 80–92, 2012.

[71] F. Privé, J. Arbel, and B. J. Vilhjálmsson, “Ldpred2: better, faster, stronger,” Bioinformatics, vol. 36, no. 22-23, pp. 5424–5431, 2020.

[72] Y. Xu, D. Vuckovic, S. C. Ritchie, P. Akbari, T. Jiang, J. Grealey, A. S. Butterworth, W. H. Ouwehand, D. J. Roberts, E. Di Angelantonio, et al., “Machine learning optimized polygenic scores for blood cell traits identify sex-specific trajectories and genetic correlations with disease,” Cell Genomics, vol. 2, no. 1, 2022.

[73] S. Zhang, H. Shu, J. Zhou, J. Rubin-Sigler, X. Yang, Y. Liu, J. Cooper-Knock, E. Monte, C. Zhu, S. Tu, et al., “Deconvolution of polygenic risk score in single cells unravels cellular and molecular heterogeneity of complex human diseases,” bioRxiv, 2024.

[74] C. Albiñana, Z. Zhu, A. J. Schork, A. Ingason, H. Aschard, I. Brikell, C. M. Bulik, L. V. Petersen, E. Agerbo, J. Grove, et al., “Multi-pgs enhances polygenic prediction by combining 937 polygenic scores,” Nature communications, vol. 14, no. 1, p. 4702, 2023.

[75] F. Pedregosa, G. Varoquaux, A. Gramfort, V. Michel, B. Thirion, O. Grisel, M. Blondel, P. Prettenhofer, R. Weiss, V. Dubourg, J. Vanderplas, A. Passos, D. Cournapeau, M. Brucher, M. Perrot, and E. Duchesnay, “Scikit-learn: Machine learning in Python,” Journal of Machine Learning Research, vol. 12, pp. 2825–2830, 2011.

