## Supplementary Table 27 for "Hundreds of cardiac MRI traits derived using 3D diffusion autoencoders share a common genetic architecture"

SNP CHR BP EA EAFREQ BETA SE P START END LOCUS NearestGENE fullSNPIDs
Latents
rs10802069 1 119517357 C 0.61501 -0.0385983 0.00582362 3,41E-06 119267357
119767357 1 TBX15 rs10802069_T_C S1993_Z97
rs11118288 1 207835812 A 0.132778 0.0566408 0.0094198 1,82E-04 207585812
208085812 2.1 CR1L rs11118288_G_A S1701_Z82
rs11579568 1 208130627 G 0.33753 -0.0365552 0.00592219 6,72E-05 207880627
208380627 2.2 CD34 rs11579568_T_G S42_Z10
rs35330522 2 12877060 A 0.47981 0.0393238 0.00615015 1,62E-05 12627060
13127060 3 TRIB2 rs35330522_G_A S2023_Z86
rs7558413 2 18721662 A 0.583498 0.0405233 0.00602172 1,70E-06 18471662
18971662 4 RDH14 rs7558413_G_A S1701_Z29, S1994_Z5
rs1260326 2 27730940 C 0.606543 0.0417682 0.00601653 3,86E-07 27480940
27980940 5 GCKR rs1260326_T_C S1701_Z53
rs142556838 2 179747068 T 0.0909455 0.0765574 0.01099 3,26E-07 179497068
179997068 6 CCDC141 rs142556838_C_T S1701_Z82
rs13386459 2 220299018 C 0.390501 -0.0381575 0.00606589 3,16E-05 220049018
220549018 7 SPEG rs13386459_T_C S1701_Z29
rs6742228 2 232267116 G 0.325574 0.0292572 0.00457815 1,65E-05 232017116
232517116 8 B3GNT7 rs6742228_A_G S1994_Z111, S1994_Z31
rs1656376 3 158284681 C 0.556862 0.033081 0.00545708 1,34E-04 158034681
158534681 9 MLF1 rs1656376_G_C S2023_Z77
rs2339798 3 179173620 A 0.139424 -0.0427439 0.00652487 5,72E-06 178923620
179423620 10 GNB4 rs2339798_C_A S1994_Z127, S1994_Z35, S1994_Z94, S42_Z105,
S42_Z106, S42_Z117, S42_Z12, S42_Z4, S42_Z51, S42_Z89
rs28568794 4 7852093 C 0.442003 0.0256081 0.00419636 1,04E-04 7602093
8102093 11 AFAP1 rs28568794_G_C S2023_Z95
4:17956213_TG_T 4 17956213 T 0.738469 0.042988 0.00697231 7,02E-05 17706213
18206213 12 LCORL 4:17956213_TG_T_TG_T S1993_Z14, S2023_Z86
rs464605 5 55807370 T 0.745499 0.0415688 0.00680494 1,00E-04 55557370
56057370 13 ANKRD55 rs464605_C_T S1701_Z111
rs1309546 5 64290004 C 0.446979 0.0313195 0.00490859 1,76E-05 64040004
64540004 14 CWC27 rs1309546_T_C S1701_Z78
rs72801474 5 132444128 A 0.0915467 0.0610096 0.00999342 1,03E-04 132194128
132694128 15 HSPA4 rs72801474_G_A S1994_Z1
rs9503212 6 2501535 G 0.519455 0.0368997 0.00525301 2,15E-07 2251535
2751535 16 MYLK4 rs9503212_A_G S1701_Z118, S1993_Z22, S1993_Z28, S1993_Z6,
S1993_Z97, S1994_Z108, S2023_Z44, S2023_Z95
rs11153730 6 118667522 C 0.492803 -0.0399759 0.00629537 2,15E-05 118417522
118917522 17 SLC35F1 rs11153730_T_C S1701_Z82, S1994_Z31
rs7759673 6 121771621 T 0.540029 0.0272367 0.00450771 1,52E-04 121521621
122021621 18.1 GJA1 rs7759673_A_T S42_Z12
rs9388001 6 122092897 A 0.099312 0.0830179 0.0105093 2,80E-10 121842897
122342897 18.2 GJA1 rs9388001_G_A S1701_Z82, S1994_Z92, S42_Z10
rs9388487 6 126678268 T 0.469791 0.0399413 0.00593037 1,64E-06 126428268
126928268 19 CENPW rs9388487_G_T S1701_Z29, S2023_Z87, S42_Z125
rs263182 6 142862612 C 0.289586 0.0390443 0.00603027 9,50E-06 142612612
143112612 20 ADGRG6 rs263182_T_C S2023_Z100
rs35164779 7 46560250 C 0.0952039 -0.0627024 0.0103758 1,51E-04 46310250
46810250 21 IGFBP3 rs35164779_G_C S42_Z127
rs12531355 7 121027434 T 0.265773 0.0509644 0.00688814 1,37E-08 120777434
121277434 22 FAM3C rs12531355_C_T S2023_Z86, S2023_Z86
rs73221948 8 25464670 T 0.292074 0.0418033 0.00684593 1,02E-04 25214670
25714670 23 CDCA2 rs73221948_G_T S1701_Z111
rs610891 8 109161003 G 0.460489 -0.0393593 0.00613103 1,37E-05 108911003

109411003 24 EIF3E rs610891_A_G S2023_Z86
rs2100837 8 120392313 C 0.250743 0.0400673 0.00637153 3,21E-06 120142313 120642313 25 CCN3 rs2100837_A_C S1701_Z80
rs10828265 10 22019212 T 0.592916 -0.0331427 0.00551699 1,89E-04 21769212 22269212 26 MLLT10 rs10828265_C_T S1993_Z14
rs806676 10 89787009 T 0.168264 0.0477705 0.00784666 1,14E-04 89537009 90037009 27 PTEN rs806676_C_T S1994_Z1
rs4755800 11 44287681 A 0.653418 -0.0397696 0.0062171 1,59E-05 44037681 44537681 28 ALX4 rs4755800_G_A S2023_Z26
rs77282531 11 95023373 A 0.0210414 -0.127922 0.0206515 5,85E-05 94773373 95273373 29 SESN3 rs77282531_G_A S1701_Z29, S1994_Z5, S42_Z125
rs76895963 12 4384844 G 0.0196553 0.139134 0.0222016 3,68E-05 4134844 4634844 30 CCND2 rs76895963_T_G S1994_Z5
rs4963772 12 24758480 A 0.151465 -0.0672216 0.00887219 3,55E-09 24508480 25008480 31 SOX5 rs4963772_G_A S1701_Z122, S1701_Z49, S1701_Z82, S1994_Z85, S42_Z10
rs3741760 12 28544464 A 0.757005 0.0492741 0.00706832 3,14E-08 28294464 28794464 32 CCDC91 rs3741760_G_A S2023_Z77, S2023_Z86
rs9535455 13 51088547 A 0.213153 -0.043368 0.00720741 1,78E-04 50838547 51338547 33 DLEU7 rs9535455_G_A S1701_Z53
rs2319625 14 21565146 T 0.174677 0.0354542 0.00588682 1,72E-04 21315146 21815146 34 ZNF219 rs2319625_C_T S42_Z12
rs422068 14 23864804 C 0.358255 -0.0380976 0.00586422 8,21E-06 23614804 24114804 35 MYH6 rs422068_T_C S1701_Z122, S1701_Z82, S1994_Z111
rs28929474 14 94844947 T 0.0205905 -0.145459 0.0218588 2,84E-06 94594947 95094947 36 SERPINA1 rs28929474_C_T S1701_Z111, S1994_Z38, S2023_Z77, S2023_Z86, S42_Z125
rs139583754 15 93331251 G 0.0107545 -0.177189 0.0271119 6,34E-06 93081251 93581251 37 FAM174B rs139583754_A_G S1701_Z32, S1701_Z80
rs11664030 18 20261639 T 0.542083 0.0354844 0.00586224 1,42E-04 20011639 20511639 38 RBBP8 rs11664030_C_T S1701_Z20
rs429358 19 45411941 C 0.15751 -0.0490037 0.0075527 8,68E-06 45161941 45661941 39 APOE rs429358_T_C S2023_Z87, S42_Z28
rs147110934 19 55993436 T 0.0246819 0.116306 0.0186035 4,06E-06 55743436 56243436 40 ZNF628 rs147110934_G_T S42_Z125
rs3746471 20 36841914 A 0.463428 -0.0413851 0.00632912 6,20E-07 36591914 37091914 41 KIAA1755 rs3746471_G_A S1701_Z82
rs1076135 22 30238317 T 0.478591 -0.0387768 0.00590672 5,21E-06 29988317 30488317 42 ASCC2 rs1076135_A_T S1701_Z93, S1993_Z27, S1993_Z28, S1993_Z41, S1993_Z66, S1994_Z
