## Supplementary Table 26 for "Hundreds of cardiac MRI traits derived using 3D diffusion autoencoders share a common genetic architecture"

| Latent | Chr | SNP | BP | EA | EAFreq | BETA | SE | P | N | NearestGENE | fullSNPIDs | LOCUS |
|---|---|---|---|---|---|---|---|---|---|---|---|---|
| S1993_Z97 | 1 | rs10802069 | 119517357 | C | 0.61501 | -0.0385983 | 0.00582362 | 3,41E-06 | 48368.4 | TBX15 | rs10802069_T_C | 1 |
| S1701_Z82 | 1 | rs11118288 | 207835812 | A | 0.132778 | 0.0566408 | 0.0094198 | 1,82E-04 | 46881.5 | CR1L | rs11118288_G_A | 2.1 |
| S42_Z10 | 1 | rs11579568 | 208130627 | G | 0.33753 | -0.0365552 | 0.00592219 | 6,72E-05 | 48197.7 | CD34 | rs11579568_T_G | 2.2 |
| S2023_Z86 | 2 | rs35330522 | 12877060 | A | 0.47981 | 0.0393238 | 0.00615015 | 1,62E-05 | 47592.2 | TRIB2 | rs35330522_G_A | 3 |
| S1701_Z29 | 2 | rs7558413 | 18721662 | A | 0.583498 | 0.0405233 | 0.00602172 | 1,70E-06 | 48319.6 | RDH14 | rs7558413_G_A | 4 |
| S1994_Z5 | 2 | rs4832605 | 18722424 | C | 0.583698 | 0.03699 | 0.0057724 | 1,47E-05 | 48279.6 | RDH14 | rs4832605_T_C | 4 |
| S1701_Z53 | 2 | rs1260326 | 27730940 | C | 0.606543 | 0.0417682 | 0.00601653 | 3,86E-07 | 48385.1 | GCKR | rs1260326_T_C | 5 |
| S1701_Z82 | 2 | rs142556838 | 179747068 | T | 0.0909455 | 0.0765574 | 0.01099 | 3,26E-07 | 47958.9 | CCDC141 | rs142556838_C_T | 6 |
| S1701_Z29 | 2 | rs13386459 | 220299018 | C | 0.390501 | -0.0381575 | 0.00606589 | 3,16E-05 | 48628.4 | SPEG | rs13386459_T_C | 7 |
| S1994_Z111 | 2 | rs763302497 | 232252196 | A | 0.71681 | -0.0352841 | 0.0058624 | 1,76E-05 | 47362.7 | B3GNT7 | rs763302497_ATTGACCCAGCTCGGG_A | 8 |
| S1994_Z31 | 2 | rs6742228 | 232267116 | G | 0.325574 | 0.0292572 | 0.00457815 | 1,65E-05 | 48301.3 | B3GNT7 | rs6742228_A_G | 8 |
| S2023_Z77 | 3 | rs1656376 | 158284681 | C | 0.556862 | 0.033081 | 0.00545708 | 1,34E-04 | 48824 | MLF1 | rs1656376_G_C | 9 |
| S1994_Z127 | 3 | rs2339798 | 179173620 | A | 0.139424 | 0.0387883 | 0.00644027 | 1,71E-04 | 47672.4 | GNB4 | rs2339798_C_A | 10 |
| S1994_Z35 | 3 | rs2339798 | 179173620 | A | 0.139424 | -0.0395575 | 0.00655498 | 1,59E-04 | 47645.9 | GNB4 | rs2339798_C_A | 10 |
| S1994_Z94 | 3 | rs2339798 | 179173620 | A | 0.139424 | 0.0397286 | 0.00653979 | 1,24E-04 | 47677.1 | GNB4 | rs2339798_C_A | 10 |
| S42_Z105 | 3 | rs2339798 | 179173620 | A | 0.139424 | -0.0425265 | 0.00693605 | 8,72E-05 | 47643.2 | GNB4 | rs2339798_C_A | 10 |
| S42_Z106 | 3 | rs2339798 | 179173620 | A | 0.139424 | 0.0389536 | 0.00646758 | 1,71E-04 | 47632.2 | GNB4 | rs2339798_C_A | 10 |
| S42_Z117 | 3 | rs2339798 | 179173620 | A | 0.139424 | -0.0405872 | 0.00621921 | 6,75E-06 | 47639.7 | GNB4 | rs2339798_C_A | 10 |
| S42_Z12 | 3 | rs2339798 | 179173620 | A | 0.139424 | -0.0427439 | 0.00652487 | 5,72E-06 | 47634.4 | GNB4 | rs2339798_C_A | 10 |
| S42_Z4 | 3 | rs2339798 | 179173620 | A | 0.139424 | -0.0424266 | 0.0066587 | 1,87E-05 | 47652 | GNB4 | rs2339798_C_A | 10 |
| S42_Z51 | 3 | rs2339798 | 179173620 | A | 0.139424 | -0.0399834 | 0.00649709 | 7,55E-05 | 47638.5 | GNB4 | rs2339798_C_A | 10 |
| S42_Z89 | 3 | rs2339798 | 179173620 | A | 0.139424 | -0.043684 | 0.00718714 | 1,22E-04 | 47637.7 | GNB4 | rs2339798_C_A | 10 |
| S2023_Z95 | 4 | rs28568794 | 7852093 | C | 0.442003 | 0.0256081 | 0.00419636 | 1,04E-04 | 48428.8 | AFAP1 | rs28568794_G_C | 11 |
| S1993_Z14 | 4 | 4:17956213_TG_T | 17956213 | T | 0.738469 | 0.0371831 | 0.00618021 | 1,78E-04 | 47842.1 | LCORL | 4:17956213_TG_T_TG_T | 12 |
| S2023_Z86 | 4 | 4:17956213_TG_T | 17956213 | T | 0.738469 | 0.042988 | 0.00697231 | 7,02E-05 | 47858.4 | LCORL | 4:17956213_TG_T_TG_T | 12 |
| S1701_Z111 | 5 | rs464605 | 55807370 | T | 0.745499 | 0.0415688 | 0.00680494 | 1,00E-04 | 48877.5 | ANKRD55 | rs464605_C_T | 13 |
| S1701_Z78 | 5 | rs1309546 | 64290004 | C | 0.446979 | 0.0313195 | 0.00490859 | 1,76E-05 | 48090.5 | CWC27 | rs1309546_T_C | 14 |

S1994_Z1 5 rs72801474 132444128 A 0.0915467 0.0610096 0.00999342 1,03E-04 49378.7 HSPA4 rs72801474_G_A 15
S1993_Z97 6 rs6920875 2478975 A 0.483184 0.0389913 0.00567112 6,18E-07 48357.4 MYLK4 rs6920875_G_A 16
S1701_Z118 6 rs10458143 2479086 A 0.4828 0.0365618 0.00592435 6,77E-05 48392.3 MYLK4 rs10458143_C_A 16
S1993_Z22 6 rs9503212 2501535 G 0.519455 0.0368997 0.00525301 2,15E-07 48143.5 MYLK4 rs9503212_A_G 16
S1993_Z28 6 rs4959678 2503173 A 0.484052 0.0356157 0.0058948 1,52E-04 48279.1 MYLK4 rs4959678_C_A 16
S1994_Z108 6 rs4959678 2503173 A 0.484052 0.0358017 0.00542328 4,07E-07 48259.4 MYLK4 rs4959678_C_A 16
S2023_Z44 6 rs4959678 2503173 A 0.484052 0.0364636 0.00580553 3,37E-06 48264.3 MYLK4 rs4959678_C_A 16
S1993_Z6 6 rs11242779 2507901 C 0.489813 0.0289993 0.00442518 5,63E-06 48107 MYLK4 rs11242779_T_C 16
S2023_Z95 6 rs11242779 2507901 C 0.489813 0.0280391 0.0041838 2,06E-06 48076.6 MYLK4 rs11242779_T_C 16
S1994_Z31 6 rs3951016 118559658 A 0.469273 -0.0265563 0.00431029 7,22E-05 48043.8 SLC35F1 rs3951016_T_A 17
S1701_Z82 6 rs11153730 118667522 C 0.492803 -0.0399759 0.00629537 2,15E-05 48352.7 SLC35F1 rs11153730_T_C 17
S42_Z12 6 rs7759673 121771621 T 0.540029 0.0272367 0.00450771 1,52E-04 48216.3 GJA1 rs7759673_A_T 18.1
S1994_Z92 6 rs58730006 122089704 AT 0.0986941 -0.0745815 0.0102927 4,29E-08 48440.4 GJA1 rs58730006_A_AT 18.2
S1701_Z82 6 rs9388001 122092897 A 0.099312 0.0830179 0.0105093 2,80E-10 48461 GJA1 rs9388001_G_A 18.2
S42_Z10 6 rs9388001 122092897 A 0.099312 0.0619263 0.00933304 3,24E-07 48506.1 GJA1 rs9388001_G_A 18.2
S1701_Z29 6 rs9388487 126678268 T 0.469791 0.0399413 0.00593037 1,64E-06 48608 CENPW rs9388487_G_T 19
S42_Z125 6 6:126707845_CT_C 126707845 C 0.457082 -0.0364688 0.00584112 4,28E-05 48589.7 CENPW 6:126707845_CT_C_CT_C 19
S2023_Z87 6 rs2184968 126760994 C 0.453074 -0.0338303 0.00540086 3,76E-05 48702 CENPW rs2184968_T_C 19
S2023_Z100 6 rs263182 142862612 C 0.289586 0.0390443 0.00603027 9,50E-06 48314.1 ADGRG6 rs263182_T_C 20
S42_Z127 7 rs35164779 46560250 C 0.0952039 -0.0627024 0.0103758 1,51E-04 47428.3 IGFBP3 rs35164779_G_C 21
S2023_Z86 7 rs35348547 120785124 CTG 0.615928 0.0353766 0.00625661 1,57E-03 48529.4 CPED1 rs35348547_C_CTG 22
S2023_Z86 7 rs12531355 121027434 T 0.265773 0.0509644 0.00688814 1,37E-08 48515.1 FAM3C rs12531355_C_T 22
S1701_Z111 8 rs73221948 25464670 T 0.292074 0.0418033 0.00684593 1,02E-04 44311.3 CDCA2 rs73221948_G_T 23
S2023_Z86 8 rs610891 109161003 G 0.460489 -0.0393593 0.00613103 1,37E-05 48112 EIF3E rs610891_A_G 24
S1701_Z80 8 rs2100837 120392313 C 0.250743 0.0400673 0.00637153 3,21E-06 48350.1 CCN3 rs2100837_A_C 25
S1993_Z14 10 rs10828265 22019212 T 0.592916 -0.0331427 0.00551699 1,89E-04 48038.9 MLLT10 rs10828265_C_T 26
S1994_Z1 10 rs806676 89787009 T 0.168264 0.0477705 0.00784666 1,14E-04 47594.5 PTEN rs806676_C_T 27
S2023_Z26 11 rs4755800 44287681 A 0.653418 -0.0397696 0.0062171 1,59E-05 48295.8 ALX4 rs4755800_G_A 28

S1994_Z5 11 rs77282531 95023373 A 0.0210414 0.123793 0.020129 7,75E-05
46839.1 SESN3 rs77282531_G_A 29
S42_Z125 11 rs77282531 95023373 A 0.0210414 -0.127922 0.0206515 5,85E-05
46829.1 SESN3 rs77282531_G_A 29
S1701_Z29 11 rs145910347 95035065 T 0.0210915 0.126837 0.0209358 1,38E-04
47061 SESN3 rs145910347_G_T 29
S1994_Z5 12 rs76895963 4384844 G 0.0196553 0.139134 0.0222016 3,68E-05
41153 CCND2 rs76895963_T_G 30
S1994_Z5 12 rs76895963 4384844 G 0.0196553 0.139134 0.0222016 3,68E-05
41153 ENSG00000285901 rs76895963_T_G 30
S1701_Z82 12 rs4963772 24758480 A 0.151465 -0.0672216 0.00887219 3,55E-09
47326.7 SOX5 rs4963772_G_A 31
S42_Z10 12 rs11047527 24762501 C 0.151465 -0.0474349 0.0078774 1,73E-04
47395.2 SOX5 rs11047527_A_C 31
S1994_Z85 12 rs11610461 24774691 G 0.177265 0.0412598 0.00636129 8,81E-06
47714.1 SOX5 rs11610461_A_G 31
S1701_Z122 12 rs11047539 24781446 G 0.150412 0.0523152 0.00793713 4,36E-07
47523.5 SOX5 rs11047539_A_G 31
S1701_Z49 12 rs11047543 24788339 A 0.150913 -0.0570262 0.00868876 5,27E-06
47493.1 SOX5 rs11047543_G_A 31
S2023_Z77 12 rs2881860 28516201 T 0.22905 -0.0387257 0.00645341 1,96E-04
48787.8 CCDC91 rs2881860_A_T 32
S2023_Z86 12 rs3741760 28544464 A 0.757005 0.0492741 0.00706832 3,14E-08
48882.3 CCDC91 rs3741760_G_A 32
S1701_Z53 13 rs9535455 51088547 A 0.213153 -0.043368 0.00720741 1,78E-04
47987.5 DLEU7 rs9535455_G_A 33
S42_Z12 14 rs2319625 21565146 T 0.174677 0.0354542 0.00588682 1,72E-04
48712.2 ZNF219 rs2319625_C_T 34
S1701_Z122 14 rs422068 23864804 C 0.358255 -0.0380976 0.00586422 8,21E-06
48391.6 MYH6 rs422068_T_C 35
S1994_Z111 14 rs422068 23864804 C 0.358255 -0.0329689 0.0054491 1,45E-04
48402.7 MYH6 rs422068_T_C 35
S1701_Z82 14 rs2284651 23882144 C 0.378645 0.0397303 0.00647605 8,52E-05
48545.2 MYH7 rs2284651_T_C 35
S1701_Z111 14 rs28929474 94844947 T 0.0205905 0.136465 0.0213698 1,70E-05
46624.4 SERPINA1 rs28929474_C_T 36
S1994_Z38 14 rs28929474 94844947 T 0.0205905 -0.125626 0.0208489 1,69E-05
46631 SERPINA1 rs28929474_C_T 36
S2023_Z77 14 rs28929474 94844947 T 0.0205905 -0.117298 0.0195331 1,91E-04
46629.3 SERPINA1 rs28929474_C_T 36
S2023_Z86 14 rs28929474 94844947 T 0.0205905 -0.145459 0.0218588 2,84E-06
46624.7 SERPINA1 rs28929474_C_T 36
S42_Z125 14 rs28929474 94844947 T 0.0205905 -0.137261 0.0209143 5,27E-06
46633.1 SERPINA1 rs28929474_C_T 36
S1701_Z32 15 rs139583754 93331251 G 0.0107545 -0.164908 0.0265246 5,06E-05
47128.7 FAM174B rs139583754_A_G 37
S1701_Z80 15 rs139583754 93331251 G 0.0107545 -0.177189 0.0271119 6,34E-06
47151.3 FAM174B rs139583754_A_G 37
S1701_Z20 18 rs11664030 20261639 T 0.542083 0.0354844 0.00586224 1,42E-04
48610.6 RBBP8 rs11664030_C_T 38
S2023_Z87 19 rs429358 45411941 C 0.15751 -0.0490037 0.0075527 8,68E-06
46499.8 APOE rs429358_T_C 39
S42_Z28 19 rs429358 45411941 C 0.15751 -0.0531432 0.00820887 9,55E-06
46511.4 APOE rs429358_T_C 39
S42_Z125 19 rs147110934 55993436 T 0.0246819 0.116306 0.0186035 4,06E-06
49380.5 ZNF628 rs147110934_G_T 40

S1701_Z82 20 rs3746471 36841914 A 0.463428 -0.0413851 0.00632912 6,20E-07
48083.1 KIAA1755 rs3746471_G_A 41
S1993_Z27 22 rs140120 30138548 C 0.513393 0.0394055 0.00623701 2,65E-05
48227.9 ZMAT5 rs140120_T_C 42
S1993_Z41 22 rs140120 30138548 C 0.513393 0.0387455 0.00624356 5,45E-05
48241.9 ZMAT5 rs140120_T_C 42
S1994_Z1 22 rs131272 30153655 G 0.504743 -0.035868 0.00581765 7,03E-05
48473.5 ZMAT5 rs131272_A_G 42
S1701_Z93 22 rs131299 30180153 AT 0.648191 0.0398552 0.00651926 9,75E-05
46534.7 ASCC2 rs131299_A_AT 42
S1993_Z66 22 rs5752961 30188084 T 0.492402 -0.0368714 0.00589093 3,87E-05
48344.2 ASCC2 rs5752961_C_T 42
S1993_Z28 22 rs1076135 30238317 T 0.478591 -0.0387768 0.00590672 5,21E-06
48117.1 ASCC2 rs1076135_A_T 4
