## Supplementary Table 25 for "Hundreds of cardiac MRI traits derived using 3D diffusion autoencoders share a common genetic architecture"

Number of subjects in each type of cohort used in the PRS analyses;;;
Disease;Full cohort;Prognosis (1-10 years);Diagnosis
Hypertension;111853;26663;75251
High cholesterol;66734;16409;43997
Coronary heart disease;27939;12832;10135
Type 2 diabetes;22631;10653;8220
Angina pectoris;22203;6879;12866
Myocardial infarction;16433;5984;7838
Heart failure;14172;8142;2553
Conduction block;9936;5781;1227
Metabolic syndrome [Hypertension;  High cholesterol; Type 2 diabetes];131187;45681;91964
Atherosclerotic [Angina pectoris; Myocardial infarction; Coronary heart disease];36178;16845;17299
