## Supplementary Table 24 for "Hundreds of cardiac MRI traits derived using 3D diffusion autoencoders share a common genetic architecture"

| Gene | SNP | Chr | BP | EA | MAC | Beta | SE | P value |
| --- | --- | --- | --- | --- | --- | --- | --- | --- |
| TRIM43B | rs1292677 | 2 | 95481764 | G | 7 | -1.361 | 0.222 | 8.73E-10 |
| GPX1 | rs5644507 | 3 | 49357945 | ACGGCACC | 18 | -1.393 | 0.229 | 1.10E-09 |
| PROM1 | rs7456316 | 4 | 16025406 | C | 6 | -2.146 | 0.352 | 1.12E-09 |
| NOTCH1 | rs7751289 | 9 | 1.37E+08 | C | 7 | -1.538 | 0.251 | 9.08E-10 |
| MLLT10 | rs1802669 | 10 | 21538867 | A | 31109 | -0.038 | 0.006 | 8.56E-10 |
