## Supplementary Table 23 for "Hundreds of cardiac MRI traits derived using 3D diffusion autoencoders share a common genetic architecture"

| Ethnicity | Count | Percentage |
|---|---|---|
| British | 6739 | 54.36 |
| Any other w | 1889 | 15.24 |
| Irish | 1551 | 12.51 |
| Indian | 501 | 4.04 |
| Other ethn | 341 | 2.75 |
| Caribbean | 256 | 2.07 |
| African | 184 | 1.48 |
| Chinese | 169 | 1.36 |
| Prefer not t | 159 | 1.28 |
| Pakistani | 110 | 0.89 |
| Any other A | 109 | 0.88 |
| Any other m | 106 | 0.86 |
| White and | 89 | 0.72 |
| White and | 67 | 0.54 |
| White | 42 | 0.34 |
| White and | 33 | 0.27 |
| Do not kno | 31 | 0.25 |
| Bangladesh | 9 | 0.07 |
| Any other B | 6 | 0.05 |
| Mixed | 4 | 0.03 |
| Asian or As | 1 | 0.01 |
| Total | 12396 | 100 |
