## Supplementary Table 22 for "Hundreds of cardiac MRI traits derived using 3D diffusion autoencoders share a common genetic architecture"

| Characteris | Women (N | Men (N = 6233) |
|---|---|---|
| Age (years) | 66.49 ± 8.0 | 67.06 ± 8.30 |
| BSA (m^2) | 1.74 ± 0.16 | 2.00 ± 0.16 |
| First imagir | 6154 | 6224 |
| Repeated i | 9 | 9 |
| BiLEVE arra | 442 | 502 |
| Axiom arra | 5721 | 5731 |
