## Supplementary Table 21 for "Hundreds of cardiac MRI traits derived using 3D diffusion autoencoders share a common genetic architecture"

| Characteris | Women (N | Men (N = 25164) |
|---|---|---|
| Age (years) | 65.47 ± 7.5 | 66.54 ± 7.73 |
| BSA (m^2) | 1.75 ± 0.15 | 2.01 ± 0.16 |
| First imagir | 22646 | 25091 |
| Repeated i | 44 | 73 |
| BiLEVE arra | 2051 | 2715 |
| Axiom arra | 20639 | 22449 |
