## Supplementary Table 20 for "Hundreds of cardiac MRI traits derived using 3D diffusion autoencoders share a common genetic architecture"

| Trait | Model | Conditione | predefined | L1_penaliz | CV_OR_He | Fold_num | NumCoeffi | NumSignifi |
|---|---|---|---|---|---|---|---|---|
| Ascending_ | LinRegrs | FALSE | FALSE | | HeldoutTest | | 24 | 19 |
| Ascending_ | Lasso | FALSE | FALSE | 0.001047 | HeldoutTest | | 24 | 22 |
| Ascending_ | Lasso | FALSE | TRUE | 0.05 | HeldoutTest | | 24 | 12 |
| Ascending_ | LinRegrs | FALSE | FALSE | | CV | 0 | 24 | 17 |
| Ascending_ | LinRegrs | FALSE | FALSE | | CV | 1 | 24 | 19 |
| Ascending_ | LinRegrs | FALSE | FALSE | | CV | 2 | 24 | 24 |
| Ascending_ | LinRegrs | FALSE | FALSE | | CV | 3 | 24 | 18 |
| Ascending_ | LinRegrs | FALSE | FALSE | | CV | 4 | 24 | 18 |
| Ascending_ | Lasso | FALSE | FALSE | 0.001 | CV | 0 | 24 | 20 |
| Ascending_ | Lasso | FALSE | FALSE | 0.001196 | CV | 1 | 24 | 22 |
| Ascending_ | Lasso | FALSE | FALSE | 0.00021 | CV | 2 | 24 | 24 |
| Ascending_ | Lasso | FALSE | FALSE | 0.001667 | CV | 3 | 24 | 21 |
| Ascending_ | Lasso | FALSE | FALSE | 0.000685 | CV | 4 | 24 | 23 |
| Ascending_ | Lasso | FALSE | TRUE | 0.05 | CV | 0 | 24 | 11 |
| Ascending_ | Lasso | FALSE | TRUE | 0.05 | CV | 1 | 24 | 12 |
| Ascending_ | Lasso | FALSE | TRUE | 0.05 | CV | 2 | 24 | 12 |
| Ascending_ | Lasso | FALSE | TRUE | 0.05 | CV | 3 | 24 | 12 |
| Ascending_ | Lasso | FALSE | TRUE | 0.05 | CV | 4 | 24 | 12 |
| Ascending_ | LinRegrs | FALSE | FALSE | | HeldoutTest | | 24 | 19 |
| Ascending_ | Lasso | FALSE | FALSE | 0.001433 | HeldoutTest | | 24 | 21 |
| Ascending_ | Lasso | FALSE | TRUE | 0.05 | HeldoutTest | | 24 | 12 |
| Ascending_ | LinRegrs | FALSE | FALSE | | CV | 0 | 24 | 17 |
| Ascending_ | LinRegrs | FALSE | FALSE | | CV | 1 | 24 | 19 |
| Ascending_ | LinRegrs | FALSE | FALSE | | CV | 2 | 24 | 24 |
| Ascending_ | LinRegrs | FALSE | FALSE | | CV | 3 | 24 | 17 |
| Ascending_ | LinRegrs | FALSE | FALSE | | CV | 4 | 24 | 19 |
| Ascending_ | Lasso | FALSE | FALSE | 0.000901 | CV | 0 | 24 | 22 |
| Ascending_ | Lasso | FALSE | FALSE | 0.00163 | CV | 1 | 24 | 20 |
| Ascending_ | Lasso | FALSE | FALSE | 0.000211 | CV | 2 | 24 | 24 |
| Ascending_ | Lasso | FALSE | FALSE | 0.001605 | CV | 3 | 24 | 20 |
| Ascending_ | Lasso | FALSE | FALSE | 0.001081 | CV | 4 | 24 | 21 |
| Ascending_ | Lasso | FALSE | TRUE | 0.05 | CV | 0 | 24 | 12 |
| Ascending_ | Lasso | FALSE | TRUE | 0.05 | CV | 1 | 24 | 12 |
| Ascending_ | Lasso | FALSE | TRUE | 0.05 | CV | 2 | 24 | 13 |
| Ascending_ | Lasso | FALSE | TRUE | 0.05 | CV | 3 | 24 | 12 |
| Ascending_ | Lasso | FALSE | TRUE | 0.05 | CV | 4 | 24 | 11 |
| Ascending_ | LinRegrs | FALSE | FALSE | | HeldoutTest | | 24 | 22 |
| Ascending_ | Lasso | FALSE | FALSE | 0.000403 | HeldoutTest | | 24 | 24 |
| Ascending_ | Lasso | FALSE | TRUE | 0.05 | HeldoutTest | | 24 | 9 |
| Ascending_ | LinRegrs | FALSE | FALSE | | CV | 0 | 24 | 22 |
| Ascending_ | LinRegrs | FALSE | FALSE | | CV | 1 | 24 | 19 |
| Ascending_ | LinRegrs | FALSE | FALSE | | CV | 2 | 24 | 22 |
| Ascending_ | LinRegrs | FALSE | FALSE | | CV | 3 | 24 | 22 |
| Ascending_ | LinRegrs | FALSE | FALSE | | CV | 4 | 24 | 21 |

| | | | | | | | | |
|---|---|---|---|---|---|---|---|---|
| Ascending_ | Lasso | FALSE | FALSE | 0.000771 | CV | 0 | 24 | 24 |
| Ascending_ | Lasso | FALSE | FALSE | 0.000522 | CV | 1 | 24 | 22 |
| Ascending_ | Lasso | FALSE | FALSE | 0.000218 | CV | 2 | 24 | 24 |
| Ascending_ | Lasso | FALSE | FALSE | 0.000343 | CV | 3 | 24 | 23 |
| Ascending_ | Lasso | FALSE | FALSE | 0.00035 | CV | 4 | 24 | 22 |
| Ascending_ | Lasso | FALSE | TRUE | 0.05 | CV | 0 | 24 | 9 |
| Ascending_ | Lasso | FALSE | TRUE | 0.05 | CV | 1 | 24 | 9 |
| Ascending_ | Lasso | FALSE | TRUE | 0.05 | CV | 2 | 24 | 9 |
| Ascending_ | Lasso | FALSE | TRUE | 0.05 | CV | 3 | 24 | 9 |
| Ascending_ | Lasso | FALSE | TRUE | 0.05 | CV | 4 | 24 | 8 |
| Descending | LinRegrs | FALSE | FALSE | | HeldoutTest | | 24 | 24 |
| Descending | Lasso | FALSE | FALSE | 0.000392 | HeldoutTest | | 24 | 24 |
| Descending | Lasso | FALSE | TRUE | 0.05 | HeldoutTest | | 24 | 13 |
| Descending | LinRegrs | FALSE | FALSE | | CV | 0 | 24 | 24 |
| Descending | LinRegrs | FALSE | FALSE | | CV | 1 | 24 | 24 |
| Descending | LinRegrs | FALSE | FALSE | | CV | 2 | 24 | 24 |
| Descending | LinRegrs | FALSE | FALSE | | CV | 3 | 24 | 23 |
| Descending | LinRegrs | FALSE | FALSE | | CV | 4 | 24 | 24 |
| Descending | Lasso | FALSE | FALSE | 0.000397 | CV | 0 | 24 | 24 |
| Descending | Lasso | FALSE | FALSE | 0.000391 | CV | 1 | 24 | 24 |
| Descending | Lasso | FALSE | FALSE | 0.000244 | CV | 2 | 24 | 24 |
| Descending | Lasso | FALSE | FALSE | 0.000386 | CV | 3 | 24 | 24 |
| Descending | Lasso | FALSE | FALSE | 0.000392 | CV | 4 | 24 | 24 |
| Descending | Lasso | FALSE | TRUE | 0.05 | CV | 0 | 24 | 14 |
| Descending | Lasso | FALSE | TRUE | 0.05 | CV | 1 | 24 | 13 |
| Descending | Lasso | FALSE | TRUE | 0.05 | CV | 2 | 24 | 15 |
| Descending | Lasso | FALSE | TRUE | 0.05 | CV | 3 | 24 | 13 |
| Descending | Lasso | FALSE | TRUE | 0.05 | CV | 4 | 24 | 12 |
| Descending | LinRegrs | FALSE | FALSE | | HeldoutTest | | 24 | 23 |
| Descending | Lasso | FALSE | FALSE | 0.000369 | HeldoutTest | | 24 | 24 |
| Descending | Lasso | FALSE | TRUE | 0.05 | HeldoutTest | | 24 | 11 |
| Descending | LinRegrs | FALSE | FALSE | | CV | 0 | 24 | 23 |
| Descending | LinRegrs | FALSE | FALSE | | CV | 1 | 24 | 23 |
| Descending | LinRegrs | FALSE | FALSE | | CV | 2 | 24 | 24 |
| Descending | LinRegrs | FALSE | FALSE | | CV | 3 | 24 | 23 |
| Descending | LinRegrs | FALSE | FALSE | | CV | 4 | 24 | 24 |
| Descending | Lasso | FALSE | FALSE | 0.000374 | CV | 0 | 24 | 23 |
| Descending | Lasso | FALSE | FALSE | 0.000366 | CV | 1 | 24 | 24 |
| Descending | Lasso | FALSE | FALSE | 0.000244 | CV | 2 | 24 | 24 |
| Descending | Lasso | FALSE | FALSE | 0.000363 | CV | 3 | 24 | 24 |
| Descending | Lasso | FALSE | FALSE | 0.000369 | CV | 4 | 24 | 24 |
| Descending | Lasso | FALSE | TRUE | 0.05 | CV | 0 | 24 | 13 |
| Descending | Lasso | FALSE | TRUE | 0.05 | CV | 1 | 24 | 11 |
| Descending | Lasso | FALSE | TRUE | 0.05 | CV | 2 | 24 | 14 |
| Descending | Lasso | FALSE | TRUE | 0.05 | CV | 3 | 24 | 12 |

| | | | | | | | | |
|---|---|---|---|---|---|---|---|---|
| Descending | Lasso | FALSE | TRUE | 0.05 | CV | 4 | 24 | 11 |
| Descending | LinRegrs | FALSE | FALSE | | HeldoutTest | | 24 | 22 |
| Descending | Lasso | FALSE | FALSE | 0.000307 | HeldoutTest | | 24 | 24 |
| Descending | Lasso | FALSE | TRUE | 0.05 | HeldoutTest | | 24 | 7 |
| Descending | LinRegrs | FALSE | FALSE | | CV | 0 | 24 | 22 |
| Descending | LinRegrs | FALSE | FALSE | | CV | 1 | 24 | 22 |
| Descending | LinRegrs | FALSE | FALSE | | CV | 2 | 24 | 21 |
| Descending | LinRegrs | FALSE | FALSE | | CV | 3 | 24 | 22 |
| Descending | LinRegrs | FALSE | FALSE | | CV | 4 | 24 | 22 |
| Descending | Lasso | FALSE | FALSE | 0.000355 | CV | 0 | 24 | 24 |
| Descending | Lasso | FALSE | FALSE | 0.000462 | CV | 1 | 24 | 24 |
| Descending | Lasso | FALSE | FALSE | 0.000539 | CV | 2 | 24 | 23 |
| Descending | Lasso | FALSE | FALSE | 0.00037 | CV | 3 | 24 | 23 |
| Descending | Lasso | FALSE | FALSE | 0.000236 | CV | 4 | 24 | 24 |
| Descending | Lasso | FALSE | TRUE | 0.05 | CV | 0 | 24 | 7 |
| Descending | Lasso | FALSE | TRUE | 0.05 | CV | 1 | 24 | 7 |
| Descending | Lasso | FALSE | TRUE | 0.05 | CV | 2 | 24 | 8 |
| Descending | Lasso | FALSE | TRUE | 0.05 | CV | 3 | 24 | 8 |
| Descending | Lasso | FALSE | TRUE | 0.05 | CV | 4 | 24 | 7 |
| Ascending_ | LinRegrs | TRUE | FALSE | | HeldoutTest | | 24 | 21 |
| Ascending_ | Lasso | TRUE | FALSE | 0.00026 | HeldoutTest | | 24 | 23 |
| Ascending_ | Lasso | TRUE | TRUE | 0.05 | HeldoutTest | | 24 | 9 |
| Ascending_ | LinRegrs | TRUE | FALSE | | CV | 0 | 24 | 22 |
| Ascending_ | LinRegrs | TRUE | FALSE | | CV | 1 | 24 | 21 |
| Ascending_ | LinRegrs | TRUE | FALSE | | CV | 2 | 24 | 23 |
| Ascending_ | LinRegrs | TRUE | FALSE | | CV | 3 | 24 | 21 |
| Ascending_ | LinRegrs | TRUE | FALSE | | CV | 4 | 24 | 23 |
| Ascending_ | Lasso | TRUE | FALSE | 0.000245 | CV | 0 | 24 | 24 |
| Ascending_ | Lasso | TRUE | FALSE | 0.000264 | CV | 1 | 24 | 24 |
| Ascending_ | Lasso | TRUE | FALSE | 0.000247 | CV | 2 | 24 | 24 |
| Ascending_ | Lasso | TRUE | FALSE | 0.000444 | CV | 3 | 24 | 23 |
| Ascending_ | Lasso | TRUE | FALSE | 0.000238 | CV | 4 | 24 | 24 |
| Ascending_ | Lasso | TRUE | TRUE | 0.05 | CV | 0 | 24 | 9 |
| Ascending_ | Lasso | TRUE | TRUE | 0.05 | CV | 1 | 24 | 9 |
| Ascending_ | Lasso | TRUE | TRUE | 0.05 | CV | 2 | 24 | 7 |
| Ascending_ | Lasso | TRUE | TRUE | 0.05 | CV | 3 | 24 | 9 |
| Ascending_ | Lasso | TRUE | TRUE | 0.05 | CV | 4 | 24 | 9 |
| Ascending_ | LinRegrs | TRUE | FALSE | | HeldoutTest | | 24 | 23 |
| Ascending_ | Lasso | TRUE | FALSE | 0.000242 | HeldoutTest | | 24 | 24 |
| Ascending_ | Lasso | TRUE | TRUE | 0.05 | HeldoutTest | | 24 | 8 |
| Ascending_ | LinRegrs | TRUE | FALSE | | CV | 0 | 24 | 22 |
| Ascending_ | LinRegrs | TRUE | FALSE | | CV | 1 | 24 | 23 |
| Ascending_ | LinRegrs | TRUE | FALSE | | CV | 2 | 24 | 24 |
| Ascending_ | LinRegrs | TRUE | FALSE | | CV | 3 | 24 | 22 |
| Ascending_ | LinRegrs | TRUE | FALSE | | CV | 4 | 24 | 22 |

| | | | | | | | | |
|---|---|---|---|---|---|---|---|---|
| Ascending_ | Lasso | TRUE | FALSE | 0.000245 | CV | 0 | 24 | 24 |
| Ascending_ | Lasso | TRUE | FALSE | 0.000246 | CV | 1 | 24 | 24 |
| Ascending_ | Lasso | TRUE | FALSE | 0.000247 | CV | 2 | 24 | 24 |
| Ascending_ | Lasso | TRUE | FALSE | 0.000341 | CV | 3 | 24 | 24 |
| Ascending_ | Lasso | TRUE | FALSE | 0.000242 | CV | 4 | 24 | 24 |
| Ascending_ | Lasso | TRUE | TRUE | 0.05 | CV | 0 | 24 | 8 |
| Ascending_ | Lasso | TRUE | TRUE | 0.05 | CV | 1 | 24 | 8 |
| Ascending_ | Lasso | TRUE | TRUE | 0.05 | CV | 2 | 24 | 7 |
| Ascending_ | Lasso | TRUE | TRUE | 0.05 | CV | 3 | 24 | 8 |
| Ascending_ | Lasso | TRUE | TRUE | 0.05 | CV | 4 | 24 | 8 |
| Ascending_ | LinRegrs | TRUE | FALSE | | HeldoutTest | | 24 | 22 |
| Ascending_ | Lasso | TRUE | FALSE | 0.000286 | HeldoutTest | | 24 | 22 |
| Ascending_ | Lasso | TRUE | TRUE | 0.05 | HeldoutTest | | 24 | 9 |
| Ascending_ | LinRegrs | TRUE | FALSE | | CV | 0 | 24 | 20 |
| Ascending_ | LinRegrs | TRUE | FALSE | | CV | 1 | 24 | 21 |
| Ascending_ | LinRegrs | TRUE | FALSE | | CV | 2 | 24 | 21 |
| Ascending_ | LinRegrs | TRUE | FALSE | | CV | 3 | 24 | 21 |
| Ascending_ | LinRegrs | TRUE | FALSE | | CV | 4 | 24 | 21 |
| Ascending_ | Lasso | TRUE | FALSE | 0.000336 | CV | 0 | 24 | 24 |
| Ascending_ | Lasso | TRUE | FALSE | 0.000246 | CV | 1 | 24 | 23 |
| Ascending_ | Lasso | TRUE | FALSE | 0.000219 | CV | 2 | 24 | 24 |
| Ascending_ | Lasso | TRUE | FALSE | 0.000343 | CV | 3 | 24 | 22 |
| Ascending_ | Lasso | TRUE | FALSE | 0.000287 | CV | 4 | 24 | 24 |
| Ascending_ | Lasso | TRUE | TRUE | 0.05 | CV | 0 | 24 | 10 |
| Ascending_ | Lasso | TRUE | TRUE | 0.05 | CV | 1 | 24 | 9 |
| Ascending_ | Lasso | TRUE | TRUE | 0.05 | CV | 2 | 24 | 10 |
| Ascending_ | Lasso | TRUE | TRUE | 0.05 | CV | 3 | 24 | 10 |
| Ascending_ | Lasso | TRUE | TRUE | 0.05 | CV | 4 | 24 | 9 |
| Descending | LinRegrs | TRUE | FALSE | | HeldoutTest | | 24 | 23 |
| Descending | Lasso | TRUE | FALSE | 0.000485 | HeldoutTest | | 24 | 24 |
| Descending | Lasso | TRUE | TRUE | 0.05 | HeldoutTest | | 24 | 12 |
| Descending | LinRegrs | TRUE | FALSE | | CV | 0 | 24 | 20 |
| Descending | LinRegrs | TRUE | FALSE | | CV | 1 | 24 | 24 |
| Descending | LinRegrs | TRUE | FALSE | | CV | 2 | 24 | 22 |
| Descending | LinRegrs | TRUE | FALSE | | CV | 3 | 24 | 24 |
| Descending | LinRegrs | TRUE | FALSE | | CV | 4 | 24 | 22 |
| Descending | Lasso | TRUE | FALSE | 0.000673 | CV | 0 | 24 | 23 |
| Descending | Lasso | TRUE | FALSE | 0.000243 | CV | 1 | 24 | 24 |
| Descending | Lasso | TRUE | FALSE | 0.00056 | CV | 2 | 24 | 23 |
| Descending | Lasso | TRUE | FALSE | 0.000242 | CV | 3 | 24 | 24 |
| Descending | Lasso | TRUE | FALSE | 0.000605 | CV | 4 | 24 | 23 |
| Descending | Lasso | TRUE | TRUE | 0.05 | CV | 0 | 24 | 12 |
| Descending | Lasso | TRUE | TRUE | 0.05 | CV | 1 | 24 | 12 |
| Descending | Lasso | TRUE | TRUE | 0.05 | CV | 2 | 24 | 7 |
| Descending | Lasso | TRUE | TRUE | 0.05 | CV | 3 | 24 | 12 |

| | | | | | | | | |
|---|---|---|---|---|---|---|---|---|
| Descending | Lasso | TRUE | TRUE | 0.05 | CV | 4 | 24 | 11 |
| Descending | LinRegrs | TRUE | FALSE | | HeldoutTest | | 24 | 23 |
| Descending | Lasso | TRUE | FALSE | 0.000252 | HeldoutTest | | 24 | 24 |
| Descending | Lasso | TRUE | TRUE | 0.05 | HeldoutTest | | 24 | 10 |
| Descending | LinRegrs | TRUE | FALSE | | CV | 0 | 24 | 22 |
| Descending | LinRegrs | TRUE | FALSE | | CV | 1 | 24 | 23 |
| Descending | LinRegrs | TRUE | FALSE | | CV | 2 | 24 | 22 |
| Descending | LinRegrs | TRUE | FALSE | | CV | 3 | 24 | 23 |
| Descending | LinRegrs | TRUE | FALSE | | CV | 4 | 24 | 23 |
| Descending | Lasso | TRUE | FALSE | 0.00035 | CV | 0 | 24 | 23 |
| Descending | Lasso | TRUE | FALSE | 0.000253 | CV | 1 | 24 | 24 |
| Descending | Lasso | TRUE | FALSE | 0.000252 | CV | 2 | 24 | 23 |
| Descending | Lasso | TRUE | FALSE | 0.00041 | CV | 3 | 24 | 24 |
| Descending | Lasso | TRUE | FALSE | 0.000512 | CV | 4 | 24 | 24 |
| Descending | Lasso | TRUE | TRUE | 0.05 | CV | 0 | 24 | 10 |
| Descending | Lasso | TRUE | TRUE | 0.05 | CV | 1 | 24 | 10 |
| Descending | Lasso | TRUE | TRUE | 0.05 | CV | 2 | 24 | 9 |
| Descending | Lasso | TRUE | TRUE | 0.05 | CV | 3 | 24 | 10 |
| Descending | Lasso | TRUE | TRUE | 0.05 | CV | 4 | 24 | 10 |
| Descending | LinRegrs | TRUE | FALSE | | HeldoutTest | | 24 | 20 |
| Descending | Lasso | TRUE | FALSE | 0.000657 | HeldoutTest | | 24 | 24 |
| Descending | Lasso | TRUE | TRUE | 0.05 | HeldoutTest | | 24 | 11 |
| Descending | LinRegrs | TRUE | FALSE | | CV | 0 | 24 | 20 |
| Descending | LinRegrs | TRUE | FALSE | | CV | 1 | 24 | 21 |
| Descending | LinRegrs | TRUE | FALSE | | CV | 2 | 24 | 21 |
| Descending | LinRegrs | TRUE | FALSE | | CV | 3 | 24 | 21 |
| Descending | LinRegrs | TRUE | FALSE | | CV | 4 | 24 | 21 |
| Descending | Lasso | TRUE | FALSE | 0.000757 | CV | 0 | 24 | 23 |
| Descending | Lasso | TRUE | FALSE | 0.000615 | CV | 1 | 24 | 23 |
| Descending | Lasso | TRUE | FALSE | 0.000231 | CV | 2 | 24 | 23 |
| Descending | Lasso | TRUE | FALSE | 0.00045 | CV | 3 | 24 | 23 |
| Descending | Lasso | TRUE | FALSE | 0.000412 | CV | 4 | 24 | 23 |
| Descending | Lasso | TRUE | TRUE | 0.05 | CV | 0 | 24 | 12 |
| Descending | Lasso | TRUE | TRUE | 0.05 | CV | 1 | 24 | 12 |
| Descending | Lasso | TRUE | TRUE | 0.05 | CV | 2 | 24 | 10 |
| Descending | Lasso | TRUE | TRUE | 0.05 | CV | 3 | 24 | 11 |
| Descending | Lasso | TRUE | TRUE | 0.05 | CV | 4 | 24 | 11 |
| RA_maxim | LinRegrs | FALSE | FALSE | | HeldoutTest | | 24 | 23 |
| RA_maxim | Lasso | FALSE | FALSE | 0.000323 | HeldoutTest | | 24 | 24 |
| RA_maxim | Lasso | FALSE | TRUE | 0.05 | HeldoutTest | | 24 | 15 |
| RA_maxim | LinRegrs | FALSE | FALSE | | CV | 0 | 24 | 23 |
| RA_maxim | LinRegrs | FALSE | FALSE | | CV | 1 | 24 | 23 |
| RA_maxim | LinRegrs | FALSE | FALSE | | CV | 2 | 24 | 23 |
| RA_maxim | LinRegrs | FALSE | FALSE | | CV | 3 | 24 | 23 |
| RA_maxim | LinRegrs | FALSE | FALSE | | CV | 4 | 24 | 24 |

| | | | | | | | | |
|---|---|---|---|---|---|---|---|---|
| RA_maxim | Lasso | FALSE | FALSE | 0.000322 | CV | 0 | 24 | 24 |
| RA_maxim | Lasso | FALSE | FALSE | 0.000321 | CV | 1 | 24 | 24 |
| RA_maxim | Lasso | FALSE | FALSE | 0.000324 | CV | 2 | 24 | 24 |
| RA_maxim | Lasso | FALSE | FALSE | 0.000326 | CV | 3 | 24 | 23 |
| RA_maxim | Lasso | FALSE | FALSE | 0.000322 | CV | 4 | 24 | 24 |
| RA_maxim | Lasso | FALSE | TRUE | 0.05 | CV | 0 | 24 | 15 |
| RA_maxim | Lasso | FALSE | TRUE | 0.05 | CV | 1 | 24 | 15 |
| RA_maxim | Lasso | FALSE | TRUE | 0.05 | CV | 2 | 24 | 15 |
| RA_maxim | Lasso | FALSE | TRUE | 0.05 | CV | 3 | 24 | 15 |
| RA_maxim | Lasso | FALSE | TRUE | 0.05 | CV | 4 | 24 | 13 |
| RA_minimu | LinRegrs | FALSE | FALSE | | HeldoutTest | | 24 | 24 |
| RA_minimu | Lasso | FALSE | FALSE | 0.000337 | HeldoutTest | | 24 | 24 |
| RA_minimu | Lasso | FALSE | TRUE | 0.05 | HeldoutTest | | 24 | 16 |
| RA_minimu | LinRegrs | FALSE | FALSE | | CV | 0 | 24 | 24 |
| RA_minimu | LinRegrs | FALSE | FALSE | | CV | 1 | 24 | 23 |
| RA_minimu | LinRegrs | FALSE | FALSE | | CV | 2 | 24 | 23 |
| RA_minimu | LinRegrs | FALSE | FALSE | | CV | 3 | 24 | 24 |
| RA_minimu | LinRegrs | FALSE | FALSE | | CV | 4 | 24 | 23 |
| RA_minimu | Lasso | FALSE | FALSE | 0.000341 | CV | 0 | 24 | 24 |
| RA_minimu | Lasso | FALSE | FALSE | 0.000338 | CV | 1 | 24 | 24 |
| RA_minimu | Lasso | FALSE | FALSE | 0.00034 | CV | 2 | 24 | 24 |
| RA_minimu | Lasso | FALSE | FALSE | 0.000332 | CV | 3 | 24 | 24 |
| RA_minimu | Lasso | FALSE | FALSE | 0.000333 | CV | 4 | 24 | 24 |
| RA_minimu | Lasso | FALSE | TRUE | 0.05 | CV | 0 | 24 | 16 |
| RA_minimu | Lasso | FALSE | TRUE | 0.05 | CV | 1 | 24 | 16 |
| RA_minimu | Lasso | FALSE | TRUE | 0.05 | CV | 2 | 24 | 16 |
| RA_minimu | Lasso | FALSE | TRUE | 0.05 | CV | 3 | 24 | 16 |
| RA_minimu | Lasso | FALSE | TRUE | 0.05 | CV | 4 | 24 | 15 |
| RA_stroke_ | LinRegrs | FALSE | FALSE | | HeldoutTest | | 24 | 24 |
| RA_stroke_ | Lasso | FALSE | FALSE | 0.000283 | HeldoutTest | | 24 | 24 |
| RA_stroke_ | Lasso | FALSE | TRUE | 0.05 | HeldoutTest | | 24 | 11 |
| RA_stroke_ | LinRegrs | FALSE | FALSE | | CV | 0 | 24 | 24 |
| RA_stroke_ | LinRegrs | FALSE | FALSE | | CV | 1 | 24 | 24 |
| RA_stroke_ | LinRegrs | FALSE | FALSE | | CV | 2 | 24 | 24 |
| RA_stroke_ | LinRegrs | FALSE | FALSE | | CV | 3 | 24 | 24 |
| RA_stroke_ | LinRegrs | FALSE | FALSE | | CV | 4 | 24 | 24 |
| RA_stroke_ | Lasso | FALSE | FALSE | 0.000282 | CV | 0 | 24 | 24 |
| RA_stroke_ | Lasso | FALSE | FALSE | 0.000283 | CV | 1 | 24 | 24 |
| RA_stroke_ | Lasso | FALSE | FALSE | 0.000284 | CV | 2 | 24 | 24 |
| RA_stroke_ | Lasso | FALSE | FALSE | 0.000286 | CV | 3 | 24 | 24 |
| RA_stroke_ | Lasso | FALSE | FALSE | 0.00028 | CV | 4 | 24 | 24 |
| RA_stroke_ | Lasso | FALSE | TRUE | 0.05 | CV | 0 | 24 | 11 |
| RA_stroke_ | Lasso | FALSE | TRUE | 0.05 | CV | 1 | 24 | 10 |
| RA_stroke_ | Lasso | FALSE | TRUE | 0.05 | CV | 2 | 24 | 11 |
| RA_stroke_ | Lasso | FALSE | TRUE | 0.05 | CV | 3 | 24 | 10 |

| | | | | | | | | |
|---|---|---|---|---|---|---|---|---|
| RA_stroke_ | Lasso | FALSE | TRUE | 0.05 | CV | 4 | 24 | 9 |
| RA_ejectio | LinRegrs | FALSE | FALSE | | HeldoutTest | | 24 | 24 |
| RA_ejectio | Lasso | FALSE | FALSE | 0.000232 | HeldoutTest | | 24 | 24 |
| RA_ejectio | Lasso | FALSE | TRUE | 0.05 | HeldoutTest | | 24 | 10 |
| RA_ejectio | LinRegrs | FALSE | FALSE | | CV | 0 | 24 | 24 |
| RA_ejectio | LinRegrs | FALSE | FALSE | | CV | 1 | 24 | 24 |
| RA_ejectio | LinRegrs | FALSE | FALSE | | CV | 2 | 24 | 24 |
| RA_ejectio | LinRegrs | FALSE | FALSE | | CV | 3 | 24 | 24 |
| RA_ejectio | LinRegrs | FALSE | FALSE | | CV | 4 | 24 | 24 |
| RA_ejectio | Lasso | FALSE | FALSE | 0.000237 | CV | 0 | 24 | 24 |
| RA_ejectio | Lasso | FALSE | FALSE | 0.000237 | CV | 1 | 24 | 24 |
| RA_ejectio | Lasso | FALSE | FALSE | 0.000232 | CV | 2 | 24 | 24 |
| RA_ejectio | Lasso | FALSE | FALSE | 0.000226 | CV | 3 | 24 | 24 |
| RA_ejectio | Lasso | FALSE | FALSE | 0.000226 | CV | 4 | 24 | 24 |
| RA_ejectio | Lasso | FALSE | TRUE | 0.05 | CV | 0 | 24 | 10 |
| RA_ejectio | Lasso | FALSE | TRUE | 0.05 | CV | 1 | 24 | 9 |
| RA_ejectio | Lasso | FALSE | TRUE | 0.05 | CV | 2 | 24 | 10 |
| RA_ejectio | Lasso | FALSE | TRUE | 0.05 | CV | 3 | 24 | 10 |
| RA_ejectio | Lasso | FALSE | TRUE | 0.05 | CV | 4 | 24 | 7 |
| LV_myocar | LinRegrs | FALSE | FALSE | | HeldoutTest | | 24 | 24 |
| LV_myocar | Lasso | FALSE | FALSE | 0.000474 | HeldoutTest | | 24 | 24 |
| LV_myocar | Lasso | FALSE | TRUE | 0.05 | HeldoutTest | | 24 | 16 |
| LV_myocar | LinRegrs | FALSE | FALSE | | CV | 0 | 24 | 24 |
| LV_myocar | LinRegrs | FALSE | FALSE | | CV | 1 | 24 | 24 |
| LV_myocar | LinRegrs | FALSE | FALSE | | CV | 2 | 24 | 24 |
| LV_myocar | LinRegrs | FALSE | FALSE | | CV | 3 | 24 | 24 |
| LV_myocar | LinRegrs | FALSE | FALSE | | CV | 4 | 24 | 23 |
| LV_myocar | Lasso | FALSE | FALSE | 0.000478 | CV | 0 | 24 | 24 |
| LV_myocar | Lasso | FALSE | FALSE | 0.000474 | CV | 1 | 24 | 24 |
| LV_myocar | Lasso | FALSE | FALSE | 0.000472 | CV | 2 | 24 | 24 |
| LV_myocar | Lasso | FALSE | FALSE | 0.000473 | CV | 3 | 24 | 24 |
| LV_myocar | Lasso | FALSE | FALSE | 0.000475 | CV | 4 | 24 | 23 |
| LV_myocar | Lasso | FALSE | TRUE | 0.05 | CV | 0 | 24 | 16 |
| LV_myocar | Lasso | FALSE | TRUE | 0.05 | CV | 1 | 24 | 15 |
| LV_myocar | Lasso | FALSE | TRUE | 0.05 | CV | 2 | 24 | 16 |
| LV_myocar | Lasso | FALSE | TRUE | 0.05 | CV | 3 | 24 | 16 |
| LV_myocar | Lasso | FALSE | TRUE | 0.05 | CV | 4 | 24 | 15 |
| LV_mean_ | LinRegrs | FALSE | FALSE | | HeldoutTest | | 24 | 22 |
| LV_mean_ | Lasso | FALSE | FALSE | 0.000343 | HeldoutTest | | 24 | 24 |
| LV_mean_ | Lasso | FALSE | TRUE | 0.05 | HeldoutTest | | 24 | 11 |
| LV_mean_ | LinRegrs | FALSE | FALSE | | CV | 0 | 24 | 23 |
| LV_mean_ | LinRegrs | FALSE | FALSE | | CV | 1 | 24 | 24 |
| LV_mean_ | LinRegrs | FALSE | FALSE | | CV | 2 | 24 | 22 |
| LV_mean_ | LinRegrs | FALSE | FALSE | | CV | 3 | 24 | 22 |
| LV_mean_ | LinRegrs | FALSE | FALSE | | CV | 4 | 24 | 24 |

| | | | | | | | | |
|---|---|---|---|---|---|---|---|---|
| LV_mean_ı | Lasso | FALSE | FALSE | 0.000347 | CV | 0 | 24 | 24 |
| LV_mean_ı | Lasso | FALSE | FALSE | 0.000342 | CV | 1 | 24 | 24 |
| LV_mean_ı | Lasso | FALSE | FALSE | 0.000342 | CV | 2 | 24 | 23 |
| LV_mean_ı | Lasso | FALSE | FALSE | 0.000364 | CV | 3 | 24 | 23 |
| LV_mean_ı | Lasso | FALSE | FALSE | 0.000344 | CV | 4 | 24 | 24 |
| LV_mean_ı | Lasso | FALSE | TRUE | 0.05 | CV | 0 | 24 | 11 |
| LV_mean_ı | Lasso | FALSE | TRUE | 0.05 | CV | 1 | 24 | 11 |
| LV_mean_ı | Lasso | FALSE | TRUE | 0.05 | CV | 2 | 24 | 12 |
| LV_mean_ı | Lasso | FALSE | TRUE | 0.05 | CV | 3 | 24 | 11 |
| LV_mean_ı | Lasso | FALSE | TRUE | 0.05 | CV | 4 | 24 | 11 |
| LV_mean_ı | LinRegrs | FALSE | FALSE | | HeldoutTest | | 24 | 22 |
| LV_mean_ı | Lasso | FALSE | FALSE | 0.000327 | HeldoutTest | | 24 | 24 |
| LV_mean_ı | Lasso | FALSE | TRUE | 0.05 | HeldoutTest | | 24 | 11 |
| LV_mean_ı | LinRegrs | FALSE | FALSE | | CV | 0 | 24 | 21 |
| LV_mean_ı | LinRegrs | FALSE | FALSE | | CV | 1 | 24 | 22 |
| LV_mean_ı | LinRegrs | FALSE | FALSE | | CV | 2 | 24 | 23 |
| LV_mean_ı | LinRegrs | FALSE | FALSE | | CV | 3 | 24 | 22 |
| LV_mean_ı | LinRegrs | FALSE | FALSE | | CV | 4 | 24 | 23 |
| LV_mean_ı | Lasso | FALSE | FALSE | 0.000472 | CV | 0 | 24 | 24 |
| LV_mean_ı | Lasso | FALSE | FALSE | 0.000248 | CV | 1 | 24 | 24 |
| LV_mean_ı | Lasso | FALSE | FALSE | 0.000242 | CV | 2 | 24 | 24 |
| LV_mean_ı | Lasso | FALSE | FALSE | 0.000246 | CV | 3 | 24 | 24 |
| LV_mean_ı | Lasso | FALSE | FALSE | 0.00025 | CV | 4 | 24 | 24 |
| LV_mean_ı | Lasso | FALSE | TRUE | 0.05 | CV | 0 | 24 | 10 |
| LV_mean_ı | Lasso | FALSE | TRUE | 0.05 | CV | 1 | 24 | 11 |
| LV_mean_ı | Lasso | FALSE | TRUE | 0.05 | CV | 2 | 24 | 11 |
| LV_mean_ı | Lasso | FALSE | TRUE | 0.05 | CV | 3 | 24 | 11 |
| LV_mean_ı | Lasso | FALSE | TRUE | 0.05 | CV | 4 | 24 | 11 |
| LV_mean_ı | LinRegrs | FALSE | FALSE | | HeldoutTest | | 24 | 24 |
| LV_mean_ı | Lasso | FALSE | FALSE | 0.000207 | HeldoutTest | | 24 | 24 |
| LV_mean_ı | Lasso | FALSE | TRUE | 0.05 | HeldoutTest | | 24 | 10 |
| LV_mean_ı | LinRegrs | FALSE | FALSE | | CV | 0 | 24 | 23 |
| LV_mean_ı | LinRegrs | FALSE | FALSE | | CV | 1 | 24 | 23 |
| LV_mean_ı | LinRegrs | FALSE | FALSE | | CV | 2 | 24 | 23 |
| LV_mean_ı | LinRegrs | FALSE | FALSE | | CV | 3 | 24 | 21 |
| LV_mean_ı | LinRegrs | FALSE | FALSE | | CV | 4 | 24 | 21 |
| LV_mean_ı | Lasso | FALSE | FALSE | 0.00021 | CV | 0 | 24 | 24 |
| LV_mean_ı | Lasso | FALSE | FALSE | 0.000241 | CV | 1 | 24 | 24 |
| LV_mean_ı | Lasso | FALSE | FALSE | 0.000356 | CV | 2 | 24 | 23 |
| LV_mean_ı | Lasso | FALSE | FALSE | 0.00031 | CV | 3 | 24 | 24 |
| LV_mean_ı | Lasso | FALSE | FALSE | 0.000422 | CV | 4 | 24 | 23 |
| LV_mean_ı | Lasso | FALSE | TRUE | 0.05 | CV | 0 | 24 | 11 |
| LV_mean_ı | Lasso | FALSE | TRUE | 0.05 | CV | 1 | 24 | 10 |
| LV_mean_ı | Lasso | FALSE | TRUE | 0.05 | CV | 2 | 24 | 9 |
| LV_mean_ı | Lasso | FALSE | TRUE | 0.05 | CV | 3 | 24 | 9 |

| | | | | | | | | |
|---|---|---|---|---|---|---|---|---|
| LV_mean_ı | Lasso | FALSE | TRUE | 0.05 | CV | 4 | 24 | 11 |
| LV_mean_ı | LinRegrs | FALSE | FALSE | | HeldoutTest | | 24 | 23 |
| LV_mean_ı | Lasso | FALSE | FALSE | 0.000322 | HeldoutTest | | 24 | 23 |
| LV_mean_ı | Lasso | FALSE | TRUE | 0.05 | HeldoutTest | | 24 | 12 |
| LV_mean_ı | LinRegrs | FALSE | FALSE | | CV | 0 | 24 | 22 |
| LV_mean_ı | LinRegrs | FALSE | FALSE | | CV | 1 | 24 | 23 |
| LV_mean_ı | LinRegrs | FALSE | FALSE | | CV | 2 | 24 | 22 |
| LV_mean_ı | LinRegrs | FALSE | FALSE | | CV | 3 | 24 | 23 |
| LV_mean_ı | LinRegrs | FALSE | FALSE | | CV | 4 | 24 | 24 |
| LV_mean_ı | Lasso | FALSE | FALSE | 0.000326 | CV | 0 | 24 | 24 |
| LV_mean_ı | Lasso | FALSE | FALSE | 0.000322 | CV | 1 | 24 | 24 |
| LV_mean_ı | Lasso | FALSE | FALSE | 0.000318 | CV | 2 | 24 | 24 |
| LV_mean_ı | Lasso | FALSE | FALSE | 0.000318 | CV | 3 | 24 | 24 |
| LV_mean_ı | Lasso | FALSE | FALSE | 0.000323 | CV | 4 | 24 | 24 |
| LV_mean_ı | Lasso | FALSE | TRUE | 0.05 | CV | 0 | 24 | 12 |
| LV_mean_ı | Lasso | FALSE | TRUE | 0.05 | CV | 1 | 24 | 13 |
| LV_mean_ı | Lasso | FALSE | TRUE | 0.05 | CV | 2 | 24 | 12 |
| LV_mean_ı | Lasso | FALSE | TRUE | 0.05 | CV | 3 | 24 | 12 |
| LV_mean_ı | Lasso | FALSE | TRUE | 0.05 | CV | 4 | 24 | 13 |
| LV_mean_ı | LinRegrs | FALSE | FALSE | | HeldoutTest | | 24 | 22 |
| LV_mean_ı | Lasso | FALSE | FALSE | 0.000364 | HeldoutTest | | 24 | 24 |
| LV_mean_ı | Lasso | FALSE | TRUE | 0.05 | HeldoutTest | | 24 | 15 |
| LV_mean_ı | LinRegrs | FALSE | FALSE | | CV | 0 | 24 | 22 |
| LV_mean_ı | LinRegrs | FALSE | FALSE | | CV | 1 | 24 | 22 |
| LV_mean_ı | LinRegrs | FALSE | FALSE | | CV | 2 | 24 | 23 |
| LV_mean_ı | LinRegrs | FALSE | FALSE | | CV | 3 | 24 | 22 |
| LV_mean_ı | LinRegrs | FALSE | FALSE | | CV | 4 | 24 | 24 |
| LV_mean_ı | Lasso | FALSE | FALSE | 0.000366 | CV | 0 | 24 | 24 |
| LV_mean_ı | Lasso | FALSE | FALSE | 0.000364 | CV | 1 | 24 | 24 |
| LV_mean_ı | Lasso | FALSE | FALSE | 0.000361 | CV | 2 | 24 | 24 |
| LV_mean_ı | Lasso | FALSE | FALSE | 0.000364 | CV | 3 | 24 | 24 |
| LV_mean_ı | Lasso | FALSE | FALSE | 0.000366 | CV | 4 | 24 | 24 |
| LV_mean_ı | Lasso | FALSE | TRUE | 0.05 | CV | 0 | 24 | 13 |
| LV_mean_ı | Lasso | FALSE | TRUE | 0.05 | CV | 1 | 24 | 15 |
| LV_mean_ı | Lasso | FALSE | TRUE | 0.05 | CV | 2 | 24 | 15 |
| LV_mean_ı | Lasso | FALSE | TRUE | 0.05 | CV | 3 | 24 | 15 |
| LV_mean_ı | Lasso | FALSE | TRUE | 0.05 | CV | 4 | 24 | 15 |
| LV_mean_ı | LinRegrs | FALSE | FALSE | | HeldoutTest | | 24 | 22 |
| LV_mean_ı | Lasso | FALSE | FALSE | 0.000373 | HeldoutTest | | 24 | 24 |
| LV_mean_ı | Lasso | FALSE | TRUE | 0.05 | HeldoutTest | | 24 | 12 |
| LV_mean_ı | LinRegrs | FALSE | FALSE | | CV | 0 | 24 | 23 |
| LV_mean_ı | LinRegrs | FALSE | FALSE | | CV | 1 | 24 | 22 |
| LV_mean_ı | LinRegrs | FALSE | FALSE | | CV | 2 | 24 | 23 |
| LV_mean_ı | LinRegrs | FALSE | FALSE | | CV | 3 | 24 | 24 |
| LV_mean_ı | LinRegrs | FALSE | FALSE | | CV | 4 | 24 | 23 |

| | | | | | | | | |
|---|---|---|---|---|---|---|---|---|
| LV_mean_ı | Lasso | FALSE | FALSE | 0.000373 | CV | 0 | 24 | 24 |
| LV_mean_ı | Lasso | FALSE | FALSE | 0.000371 | CV | 1 | 24 | 24 |
| LV_mean_ı | Lasso | FALSE | FALSE | 0.000372 | CV | 2 | 24 | 24 |
| LV_mean_ı | Lasso | FALSE | FALSE | 0.000374 | CV | 3 | 24 | 24 |
| LV_mean_ı | Lasso | FALSE | FALSE | 0.000373 | CV | 4 | 24 | 24 |
| LV_mean_ı | Lasso | FALSE | TRUE | 0.05 | CV | 0 | 24 | 12 |
| LV_mean_ı | Lasso | FALSE | TRUE | 0.05 | CV | 1 | 24 | 12 |
| LV_mean_ı | Lasso | FALSE | TRUE | 0.05 | CV | 2 | 24 | 13 |
| LV_mean_ı | Lasso | FALSE | TRUE | 0.05 | CV | 3 | 24 | 13 |
| LV_mean_ı | Lasso | FALSE | TRUE | 0.05 | CV | 4 | 24 | 13 |
| LV_mean_ı | LinRegrs | FALSE | FALSE | | HeldoutTest | | 24 | 24 |
| LV_mean_ı | Lasso | FALSE | FALSE | 0.000415 | HeldoutTest | | 24 | 24 |
| LV_mean_ı | Lasso | FALSE | TRUE | 0.05 | HeldoutTest | | 24 | 14 |
| LV_mean_ı | LinRegrs | FALSE | FALSE | | CV | 0 | 24 | 23 |
| LV_mean_ı | LinRegrs | FALSE | FALSE | | CV | 1 | 24 | 23 |
| LV_mean_ı | LinRegrs | FALSE | FALSE | | CV | 2 | 24 | 24 |
| LV_mean_ı | LinRegrs | FALSE | FALSE | | CV | 3 | 24 | 24 |
| LV_mean_ı | LinRegrs | FALSE | FALSE | | CV | 4 | 24 | 24 |
| LV_mean_ı | Lasso | FALSE | FALSE | 0.000418 | CV | 0 | 24 | 24 |
| LV_mean_ı | Lasso | FALSE | FALSE | 0.000584 | CV | 1 | 24 | 24 |
| LV_mean_ı | Lasso | FALSE | FALSE | 0.000677 | CV | 2 | 24 | 24 |
| LV_mean_ı | Lasso | FALSE | FALSE | 0.000416 | CV | 3 | 24 | 24 |
| LV_mean_ı | Lasso | FALSE | FALSE | 0.000413 | CV | 4 | 24 | 24 |
| LV_mean_ı | Lasso | FALSE | TRUE | 0.05 | CV | 0 | 24 | 14 |
| LV_mean_ı | Lasso | FALSE | TRUE | 0.05 | CV | 1 | 24 | 14 |
| LV_mean_ı | Lasso | FALSE | TRUE | 0.05 | CV | 2 | 24 | 14 |
| LV_mean_ı | Lasso | FALSE | TRUE | 0.05 | CV | 3 | 24 | 13 |
| LV_mean_ı | Lasso | FALSE | TRUE | 0.05 | CV | 4 | 24 | 15 |
| LV_mean_ı | LinRegrs | FALSE | FALSE | | HeldoutTest | | 24 | 23 |
| LV_mean_ı | Lasso | FALSE | FALSE | 0.000612 | HeldoutTest | | 24 | 24 |
| LV_mean_ı | Lasso | FALSE | TRUE | 0.05 | HeldoutTest | | 24 | 13 |
| LV_mean_ı | LinRegrs | FALSE | FALSE | | CV | 0 | 24 | 22 |
| LV_mean_ı | LinRegrs | FALSE | FALSE | | CV | 1 | 24 | 21 |
| LV_mean_ı | LinRegrs | FALSE | FALSE | | CV | 2 | 24 | 22 |
| LV_mean_ı | LinRegrs | FALSE | FALSE | | CV | 3 | 24 | 22 |
| LV_mean_ı | LinRegrs | FALSE | FALSE | | CV | 4 | 24 | 23 |
| LV_mean_ı | Lasso | FALSE | FALSE | 0.000377 | CV | 0 | 24 | 24 |
| LV_mean_ı | Lasso | FALSE | FALSE | 0.000605 | CV | 1 | 24 | 23 |
| LV_mean_ı | Lasso | FALSE | FALSE | 0.00057 | CV | 2 | 24 | 23 |
| LV_mean_ı | Lasso | FALSE | FALSE | 0.000379 | CV | 3 | 24 | 23 |
| LV_mean_ı | Lasso | FALSE | FALSE | 0.000375 | CV | 4 | 24 | 24 |
| LV_mean_ı | Lasso | FALSE | TRUE | 0.05 | CV | 0 | 24 | 12 |
| LV_mean_ı | Lasso | FALSE | TRUE | 0.05 | CV | 1 | 24 | 13 |
| LV_mean_ı | Lasso | FALSE | TRUE | 0.05 | CV | 2 | 24 | 13 |
| LV_mean_ı | Lasso | FALSE | TRUE | 0.05 | CV | 3 | 24 | 12 |

| | | | | | | | | |
|---|---|---|---|---|---|---|---|---|
| LV_mean_ | Lasso | FALSE | TRUE | 0.05 | CV | 4 | 24 | 13 |
| LV_mean_ | LinRegrs | FALSE | FALSE | | HeldoutTest | | 24 | 24 |
| LV_mean_ | Lasso | FALSE | FALSE | 0.000454 | HeldoutTest | | 24 | 24 |
| LV_mean_ | Lasso | FALSE | TRUE | 0.05 | HeldoutTest | | 24 | 13 |
| LV_mean_ | LinRegrs | FALSE | FALSE | | CV | 0 | 24 | 24 |
| LV_mean_ | LinRegrs | FALSE | FALSE | | CV | 1 | 24 | 24 |
| LV_mean_ | LinRegrs | FALSE | FALSE | | CV | 2 | 24 | 24 |
| LV_mean_ | LinRegrs | FALSE | FALSE | | CV | 3 | 24 | 24 |
| LV_mean_ | LinRegrs | FALSE | FALSE | | CV | 4 | 24 | 23 |
| LV_mean_ | Lasso | FALSE | FALSE | 0.000457 | CV | 0 | 24 | 24 |
| LV_mean_ | Lasso | FALSE | FALSE | 0.000452 | CV | 1 | 24 | 24 |
| LV_mean_ | Lasso | FALSE | FALSE | 0.000454 | CV | 2 | 24 | 24 |
| LV_mean_ | Lasso | FALSE | FALSE | 0.000456 | CV | 3 | 24 | 24 |
| LV_mean_ | Lasso | FALSE | FALSE | 0.000453 | CV | 4 | 24 | 24 |
| LV_mean_ | Lasso | FALSE | TRUE | 0.05 | CV | 0 | 24 | 13 |
| LV_mean_ | Lasso | FALSE | TRUE | 0.05 | CV | 1 | 24 | 13 |
| LV_mean_ | Lasso | FALSE | TRUE | 0.05 | CV | 2 | 24 | 13 |
| LV_mean_ | Lasso | FALSE | TRUE | 0.05 | CV | 3 | 24 | 13 |
| LV_mean_ | Lasso | FALSE | TRUE | 0.05 | CV | 4 | 24 | 14 |
| LV_mean_ | LinRegrs | FALSE | FALSE | | HeldoutTest | | 24 | 24 |
| LV_mean_ | Lasso | FALSE | FALSE | 0.00047 | HeldoutTest | | 24 | 24 |
| LV_mean_ | Lasso | FALSE | TRUE | 0.05 | HeldoutTest | | 24 | 13 |
| LV_mean_ | LinRegrs | FALSE | FALSE | | CV | 0 | 24 | 24 |
| LV_mean_ | LinRegrs | FALSE | FALSE | | CV | 1 | 24 | 24 |
| LV_mean_ | LinRegrs | FALSE | FALSE | | CV | 2 | 24 | 24 |
| LV_mean_ | LinRegrs | FALSE | FALSE | | CV | 3 | 24 | 24 |
| LV_mean_ | LinRegrs | FALSE | FALSE | | CV | 4 | 24 | 23 |
| LV_mean_ | Lasso | FALSE | FALSE | 0.000472 | CV | 0 | 24 | 24 |
| LV_mean_ | Lasso | FALSE | FALSE | 0.000468 | CV | 1 | 24 | 24 |
| LV_mean_ | Lasso | FALSE | FALSE | 0.000469 | CV | 2 | 24 | 24 |
| LV_mean_ | Lasso | FALSE | FALSE | 0.000471 | CV | 3 | 24 | 24 |
| LV_mean_ | Lasso | FALSE | FALSE | 0.000471 | CV | 4 | 24 | 23 |
| LV_mean_ | Lasso | FALSE | TRUE | 0.05 | CV | 0 | 24 | 13 |
| LV_mean_ | Lasso | FALSE | TRUE | 0.05 | CV | 1 | 24 | 13 |
| LV_mean_ | Lasso | FALSE | TRUE | 0.05 | CV | 2 | 24 | 13 |
| LV_mean_ | Lasso | FALSE | TRUE | 0.05 | CV | 3 | 24 | 13 |
| LV_mean_ | Lasso | FALSE | TRUE | 0.05 | CV | 4 | 24 | 13 |
| LV_mean_ | LinRegrs | FALSE | FALSE | | HeldoutTest | | 24 | 24 |
| LV_mean_ | Lasso | FALSE | FALSE | 0.000473 | HeldoutTest | | 24 | 24 |
| LV_mean_ | Lasso | FALSE | TRUE | 0.05 | HeldoutTest | | 24 | 13 |
| LV_mean_ | LinRegrs | FALSE | FALSE | | CV | 0 | 24 | 24 |
| LV_mean_ | LinRegrs | FALSE | FALSE | | CV | 1 | 24 | 24 |
| LV_mean_ | LinRegrs | FALSE | FALSE | | CV | 2 | 24 | 24 |
| LV_mean_ | LinRegrs | FALSE | FALSE | | CV | 3 | 24 | 24 |
| LV_mean_ | LinRegrs | FALSE | FALSE | | CV | 4 | 24 | 24 |

| | | | | | | | | |
|---|---|---|---|---|---|---|---|---|
| LV_mean_ı | Lasso | FALSE | FALSE | 0.000478 | CV | 0 | 24 | 24 |
| LV_mean_ı | Lasso | FALSE | FALSE | 0.000471 | CV | 1 | 24 | 24 |
| LV_mean_ı | Lasso | FALSE | FALSE | 0.00047 | CV | 2 | 24 | 24 |
| LV_mean_ı | Lasso | FALSE | FALSE | 0.000474 | CV | 3 | 24 | 24 |
| LV_mean_ı | Lasso | FALSE | FALSE | 0.000472 | CV | 4 | 24 | 24 |
| LV_mean_ı | Lasso | FALSE | TRUE | 0.05 | CV | 0 | 24 | 13 |
| LV_mean_ı | Lasso | FALSE | TRUE | 0.05 | CV | 1 | 24 | 12 |
| LV_mean_ı | Lasso | FALSE | TRUE | 0.05 | CV | 2 | 24 | 13 |
| LV_mean_ı | Lasso | FALSE | TRUE | 0.05 | CV | 3 | 24 | 13 |
| LV_mean_ı | Lasso | FALSE | TRUE | 0.05 | CV | 4 | 24 | 13 |
| LV_mean_ı | LinRegrs | FALSE | FALSE | | HeldoutTest | | 24 | 24 |
| LV_mean_ı | Lasso | FALSE | FALSE | 0.000445 | HeldoutTest | | 24 | 24 |
| LV_mean_ı | Lasso | FALSE | TRUE | 0.05 | HeldoutTest | | 24 | 14 |
| LV_mean_ı | LinRegrs | FALSE | FALSE | | CV | 0 | 24 | 24 |
| LV_mean_ı | LinRegrs | FALSE | FALSE | | CV | 1 | 24 | 24 |
| LV_mean_ı | LinRegrs | FALSE | FALSE | | CV | 2 | 24 | 24 |
| LV_mean_ı | LinRegrs | FALSE | FALSE | | CV | 3 | 24 | 23 |
| LV_mean_ı | LinRegrs | FALSE | FALSE | | CV | 4 | 24 | 23 |
| LV_mean_ı | Lasso | FALSE | FALSE | 0.000449 | CV | 0 | 24 | 24 |
| LV_mean_ı | Lasso | FALSE | FALSE | 0.000444 | CV | 1 | 24 | 24 |
| LV_mean_ı | Lasso | FALSE | FALSE | 0.000442 | CV | 2 | 24 | 24 |
| LV_mean_ı | Lasso | FALSE | FALSE | 0.000447 | CV | 3 | 24 | 24 |
| LV_mean_ı | Lasso | FALSE | FALSE | 0.000444 | CV | 4 | 24 | 24 |
| LV_mean_ı | Lasso | FALSE | TRUE | 0.05 | CV | 0 | 24 | 14 |
| LV_mean_ı | Lasso | FALSE | TRUE | 0.05 | CV | 1 | 24 | 13 |
| LV_mean_ı | Lasso | FALSE | TRUE | 0.05 | CV | 2 | 24 | 14 |
| LV_mean_ı | Lasso | FALSE | TRUE | 0.05 | CV | 3 | 24 | 14 |
| LV_mean_ı | Lasso | FALSE | TRUE | 0.05 | CV | 4 | 24 | 14 |
| LV_mean_ı | LinRegrs | FALSE | FALSE | | HeldoutTest | | 24 | 23 |
| LV_mean_ı | Lasso | FALSE | FALSE | 0.000413 | HeldoutTest | | 24 | 24 |
| LV_mean_ı | Lasso | FALSE | TRUE | 0.05 | HeldoutTest | | 24 | 14 |
| LV_mean_ı | LinRegrs | FALSE | FALSE | | CV | 0 | 24 | 23 |
| LV_mean_ı | LinRegrs | FALSE | FALSE | | CV | 1 | 24 | 22 |
| LV_mean_ı | LinRegrs | FALSE | FALSE | | CV | 2 | 24 | 23 |
| LV_mean_ı | LinRegrs | FALSE | FALSE | | CV | 3 | 24 | 23 |
| LV_mean_ı | LinRegrs | FALSE | FALSE | | CV | 4 | 24 | 23 |
| LV_mean_ı | Lasso | FALSE | FALSE | 0.000413 | CV | 0 | 24 | 24 |
| LV_mean_ı | Lasso | FALSE | FALSE | 0.000441 | CV | 1 | 24 | 24 |
| LV_mean_ı | Lasso | FALSE | FALSE | 0.000413 | CV | 2 | 24 | 24 |
| LV_mean_ı | Lasso | FALSE | FALSE | 0.000413 | CV | 3 | 24 | 23 |
| LV_mean_ı | Lasso | FALSE | FALSE | 0.000416 | CV | 4 | 24 | 24 |
| LV_mean_ı | Lasso | FALSE | TRUE | 0.05 | CV | 0 | 24 | 14 |
| LV_mean_ı | Lasso | FALSE | TRUE | 0.05 | CV | 1 | 24 | 13 |
| LV_mean_ı | Lasso | FALSE | TRUE | 0.05 | CV | 2 | 24 | 13 |
| LV_mean_ı | Lasso | FALSE | TRUE | 0.05 | CV | 3 | 24 | 13 |

| | | | | | | | | |
|---|---|---|---|---|---|---|---|---|
| LV_mean_ | Lasso | FALSE | TRUE | 0.05 | CV | 4 | 24 | 14 |
| LV_mean_ | LinRegrs | FALSE | FALSE | | HeldoutTest | | 24 | 21 |
| LV_mean_ | Lasso | FALSE | FALSE | 0.000445 | HeldoutTest | | 24 | 22 |
| LV_mean_ | Lasso | FALSE | TRUE | 0.05 | HeldoutTest | | 24 | 13 |
| LV_mean_ | LinRegrs | FALSE | FALSE | | CV | 0 | 24 | 22 |
| LV_mean_ | LinRegrs | FALSE | FALSE | | CV | 1 | 24 | 22 |
| LV_mean_ | LinRegrs | FALSE | FALSE | | CV | 2 | 24 | 21 |
| LV_mean_ | LinRegrs | FALSE | FALSE | | CV | 3 | 24 | 21 |
| LV_mean_ | LinRegrs | FALSE | FALSE | | CV | 4 | 24 | 22 |
| LV_mean_ | Lasso | FALSE | FALSE | 0.000482 | CV | 0 | 24 | 24 |
| LV_mean_ | Lasso | FALSE | FALSE | 0.000442 | CV | 1 | 24 | 23 |
| LV_mean_ | Lasso | FALSE | FALSE | 0.000676 | CV | 2 | 24 | 22 |
| LV_mean_ | Lasso | FALSE | FALSE | 0.000511 | CV | 3 | 24 | 22 |
| LV_mean_ | Lasso | FALSE | FALSE | 0.000446 | CV | 4 | 24 | 23 |
| LV_mean_ | Lasso | FALSE | TRUE | 0.05 | CV | 0 | 24 | 13 |
| LV_mean_ | Lasso | FALSE | TRUE | 0.05 | CV | 1 | 24 | 13 |
| LV_mean_ | Lasso | FALSE | TRUE | 0.05 | CV | 2 | 24 | 13 |
| LV_mean_ | Lasso | FALSE | TRUE | 0.05 | CV | 3 | 24 | 12 |
| LV_mean_ | Lasso | FALSE | TRUE | 0.05 | CV | 4 | 24 | 14 |
| LV_mean_ | LinRegrs | FALSE | FALSE | | HeldoutTest | | 24 | 24 |
| LV_mean_ | Lasso | FALSE | FALSE | 0.000448 | HeldoutTest | | 24 | 24 |
| LV_mean_ | Lasso | FALSE | TRUE | 0.05 | HeldoutTest | | 24 | 11 |
| LV_mean_ | LinRegrs | FALSE | FALSE | | CV | 0 | 24 | 24 |
| LV_mean_ | LinRegrs | FALSE | FALSE | | CV | 1 | 24 | 24 |
| LV_mean_ | LinRegrs | FALSE | FALSE | | CV | 2 | 24 | 24 |
| LV_mean_ | LinRegrs | FALSE | FALSE | | CV | 3 | 24 | 24 |
| LV_mean_ | LinRegrs | FALSE | FALSE | | CV | 4 | 24 | 24 |
| LV_mean_ | Lasso | FALSE | FALSE | 0.000447 | CV | 0 | 24 | 24 |
| LV_mean_ | Lasso | FALSE | FALSE | 0.000443 | CV | 1 | 24 | 24 |
| LV_mean_ | Lasso | FALSE | FALSE | 0.000448 | CV | 2 | 24 | 24 |
| LV_mean_ | Lasso | FALSE | FALSE | 0.000451 | CV | 3 | 24 | 24 |
| LV_mean_ | Lasso | FALSE | FALSE | 0.000449 | CV | 4 | 24 | 24 |
| LV_mean_ | Lasso | FALSE | TRUE | 0.05 | CV | 0 | 24 | 11 |
| LV_mean_ | Lasso | FALSE | TRUE | 0.05 | CV | 1 | 24 | 11 |
| LV_mean_ | Lasso | FALSE | TRUE | 0.05 | CV | 2 | 24 | 11 |
| LV_mean_ | Lasso | FALSE | TRUE | 0.05 | CV | 3 | 24 | 11 |
| LV_mean_ | Lasso | FALSE | TRUE | 0.05 | CV | 4 | 24 | 12 |
| LV_mean_ | LinRegrs | FALSE | FALSE | | HeldoutTest | | 24 | 23 |
| LV_mean_ | Lasso | FALSE | FALSE | 0.000442 | HeldoutTest | | 24 | 24 |
| LV_mean_ | Lasso | FALSE | TRUE | 0.05 | HeldoutTest | | 24 | 14 |
| LV_mean_ | LinRegrs | FALSE | FALSE | | CV | 0 | 24 | 22 |
| LV_mean_ | LinRegrs | FALSE | FALSE | | CV | 1 | 24 | 22 |
| LV_mean_ | LinRegrs | FALSE | FALSE | | CV | 2 | 24 | 24 |
| LV_mean_ | LinRegrs | FALSE | FALSE | | CV | 3 | 24 | 24 |
| LV_mean_ | LinRegrs | FALSE | FALSE | | CV | 4 | 24 | 23 |

| | | | | | | | | |
|---|---|---|---|---|---|---|---|---|
| LV_mean_ | Lasso | FALSE | FALSE | 0.000443 | CV | 0 | 24 | 24 |
| LV_mean_ | Lasso | FALSE | FALSE | 0.00044 | CV | 1 | 24 | 23 |
| LV_mean_ | Lasso | FALSE | FALSE | 0.000441 | CV | 2 | 24 | 24 |
| LV_mean_ | Lasso | FALSE | FALSE | 0.000444 | CV | 3 | 24 | 24 |
| LV_mean_ | Lasso | FALSE | FALSE | 0.000442 | CV | 4 | 24 | 24 |
| LV_mean_ | Lasso | FALSE | TRUE | 0.05 | CV | 0 | 24 | 14 |
| LV_mean_ | Lasso | FALSE | TRUE | 0.05 | CV | 1 | 24 | 14 |
| LV_mean_ | Lasso | FALSE | TRUE | 0.05 | CV | 2 | 24 | 14 |
| LV_mean_ | Lasso | FALSE | TRUE | 0.05 | CV | 3 | 24 | 14 |
| LV_mean_ | Lasso | FALSE | TRUE | 0.05 | CV | 4 | 24 | 14 |
| LV_mean_ | LinRegrs | FALSE | FALSE | | HeldoutTest | | 24 | 24 |
| LV_mean_ | Lasso | FALSE | FALSE | 0.000466 | HeldoutTest | | 24 | 24 |
| LV_mean_ | Lasso | FALSE | TRUE | 0.05 | HeldoutTest | | 24 | 14 |
| LV_mean_ | LinRegrs | FALSE | FALSE | | CV | 0 | 24 | 24 |
| LV_mean_ | LinRegrs | FALSE | FALSE | | CV | 1 | 24 | 24 |
| LV_mean_ | LinRegrs | FALSE | FALSE | | CV | 2 | 24 | 24 |
| LV_mean_ | LinRegrs | FALSE | FALSE | | CV | 3 | 24 | 24 |
| LV_mean_ | LinRegrs | FALSE | FALSE | | CV | 4 | 24 | 24 |
| LV_mean_ | Lasso | FALSE | FALSE | 0.000468 | CV | 0 | 24 | 24 |
| LV_mean_ | Lasso | FALSE | FALSE | 0.000464 | CV | 1 | 24 | 24 |
| LV_mean_ | Lasso | FALSE | FALSE | 0.000463 | CV | 2 | 24 | 24 |
| LV_mean_ | Lasso | FALSE | FALSE | 0.000465 | CV | 3 | 24 | 24 |
| LV_mean_ | Lasso | FALSE | FALSE | 0.000467 | CV | 4 | 24 | 24 |
| LV_mean_ | Lasso | FALSE | TRUE | 0.05 | CV | 0 | 24 | 14 |
| LV_mean_ | Lasso | FALSE | TRUE | 0.05 | CV | 1 | 24 | 14 |
| LV_mean_ | Lasso | FALSE | TRUE | 0.05 | CV | 2 | 24 | 15 |
| LV_mean_ | Lasso | FALSE | TRUE | 0.05 | CV | 3 | 24 | 13 |
| LV_mean_ | Lasso | FALSE | TRUE | 0.05 | CV | 4 | 24 | 14 |
| LV_circumf | LinRegrs | FALSE | FALSE | | HeldoutTest | | 24 | 21 |
| LV_circumf | Lasso | FALSE | FALSE | 0.000333 | HeldoutTest | | 24 | 21 |
| LV_circumf | Lasso | FALSE | TRUE | 0.05 | HeldoutTest | | 24 | 6 |
| LV_circumf | LinRegrs | FALSE | FALSE | | CV | 0 | 24 | 20 |
| LV_circumf | LinRegrs | FALSE | FALSE | | CV | 1 | 24 | 21 |
| LV_circumf | LinRegrs | FALSE | FALSE | | CV | 2 | 24 | 22 |
| LV_circumf | LinRegrs | FALSE | FALSE | | CV | 3 | 24 | 20 |
| LV_circumf | LinRegrs | FALSE | FALSE | | CV | 4 | 24 | 20 |
| LV_circumf | Lasso | FALSE | FALSE | 0.000287 | CV | 0 | 24 | 24 |
| LV_circumf | Lasso | FALSE | FALSE | 0.000273 | CV | 1 | 24 | 24 |
| LV_circumf | Lasso | FALSE | FALSE | 0.000505 | CV | 2 | 24 | 23 |
| LV_circumf | Lasso | FALSE | FALSE | 0.000358 | CV | 3 | 24 | 24 |
| LV_circumf | Lasso | FALSE | FALSE | 0.000276 | CV | 4 | 24 | 22 |
| LV_circumf | Lasso | FALSE | TRUE | 0.05 | CV | 0 | 24 | 5 |
| LV_circumf | Lasso | FALSE | TRUE | 0.05 | CV | 1 | 24 | 5 |
| LV_circumf | Lasso | FALSE | TRUE | 0.05 | CV | 2 | 24 | 4 |
| LV_circumf | Lasso | FALSE | TRUE | 0.05 | CV | 3 | 24 | 6 |

| | | | | | | | | |
|---|---|---|---|---|---|---|---|---|
| LV_circumf | Lasso | FALSE | TRUE | 0.05 | CV | 4 | 24 | 6 |
| LV_circumf | LinRegrs | FALSE | FALSE | | HeldoutTest | | 24 | 24 |
| LV_circumf | Lasso | FALSE | FALSE | 0.00023 | HeldoutTest | | 24 | 24 |
| LV_circumf | Lasso | FALSE | TRUE | 0.05 | HeldoutTest | | 24 | 12 |
| LV_circumf | LinRegrs | FALSE | FALSE | | CV | 0 | 24 | 23 |
| LV_circumf | LinRegrs | FALSE | FALSE | | CV | 1 | 24 | 23 |
| LV_circumf | LinRegrs | FALSE | FALSE | | CV | 2 | 24 | 24 |
| LV_circumf | LinRegrs | FALSE | FALSE | | CV | 3 | 24 | 23 |
| LV_circumf | LinRegrs | FALSE | FALSE | | CV | 4 | 24 | 23 |
| LV_circumf | Lasso | FALSE | FALSE | 0.000236 | CV | 0 | 24 | 24 |
| LV_circumf | Lasso | FALSE | FALSE | 0.00023 | CV | 1 | 24 | 24 |
| LV_circumf | Lasso | FALSE | FALSE | 0.00023 | CV | 2 | 24 | 24 |
| LV_circumf | Lasso | FALSE | FALSE | 0.000227 | CV | 3 | 24 | 24 |
| LV_circumf | Lasso | FALSE | FALSE | 0.000228 | CV | 4 | 24 | 24 |
| LV_circumf | Lasso | FALSE | TRUE | 0.05 | CV | 0 | 24 | 12 |
| LV_circumf | Lasso | FALSE | TRUE | 0.05 | CV | 1 | 24 | 12 |
| LV_circumf | Lasso | FALSE | TRUE | 0.05 | CV | 2 | 24 | 12 |
| LV_circumf | Lasso | FALSE | TRUE | 0.05 | CV | 3 | 24 | 13 |
| LV_circumf | Lasso | FALSE | TRUE | 0.05 | CV | 4 | 24 | 12 |
| LV_circumf | LinRegrs | FALSE | FALSE | | HeldoutTest | | 24 | 21 |
| LV_circumf | Lasso | FALSE | FALSE | 0.000174 | HeldoutTest | | 24 | 23 |
| LV_circumf | Lasso | FALSE | TRUE | 0.05 | HeldoutTest | | 24 | 6 |
| LV_circumf | LinRegrs | FALSE | FALSE | | CV | 0 | 24 | 21 |
| LV_circumf | LinRegrs | FALSE | FALSE | | CV | 1 | 24 | 20 |
| LV_circumf | LinRegrs | FALSE | FALSE | | CV | 2 | 24 | 19 |
| LV_circumf | LinRegrs | FALSE | FALSE | | CV | 3 | 24 | 20 |
| LV_circumf | LinRegrs | FALSE | FALSE | | CV | 4 | 24 | 22 |
| LV_circumf | Lasso | FALSE | FALSE | 0.000231 | CV | 0 | 24 | 21 |
| LV_circumf | Lasso | FALSE | FALSE | 0.000691 | CV | 1 | 24 | 23 |
| LV_circumf | Lasso | FALSE | FALSE | 0.000209 | CV | 2 | 24 | 24 |
| LV_circumf | Lasso | FALSE | FALSE | 0.000715 | CV | 3 | 24 | 24 |
| LV_circumf | Lasso | FALSE | FALSE | 0.000108 | CV | 4 | 24 | 24 |
| LV_circumf | Lasso | FALSE | TRUE | 0.05 | CV | 0 | 24 | 7 |
| LV_circumf | Lasso | FALSE | TRUE | 0.05 | CV | 1 | 24 | 6 |
| LV_circumf | Lasso | FALSE | TRUE | 0.05 | CV | 2 | 24 | 6 |
| LV_circumf | Lasso | FALSE | TRUE | 0.05 | CV | 3 | 24 | 6 |
| LV_circumf | Lasso | FALSE | TRUE | 0.05 | CV | 4 | 24 | 6 |
| LV_circumf | LinRegrs | FALSE | FALSE | | HeldoutTest | | 24 | 18 |
| LV_circumf | Lasso | FALSE | FALSE | 0.000867 | HeldoutTest | | 24 | 21 |
| LV_circumf | Lasso | FALSE | TRUE | 0.05 | HeldoutTest | | 24 | 4 |
| LV_circumf | LinRegrs | FALSE | FALSE | | CV | 0 | 24 | 16 |
| LV_circumf | LinRegrs | FALSE | FALSE | | CV | 1 | 24 | 17 |
| LV_circumf | LinRegrs | FALSE | FALSE | | CV | 2 | 24 | 16 |
| LV_circumf | LinRegrs | FALSE | FALSE | | CV | 3 | 24 | 20 |
| LV_circumf | LinRegrs | FALSE | FALSE | | CV | 4 | 24 | 23 |

| | | | | | | | | |
|---|---|---|---|---|---|---|---|---|
| LV_circumf | Lasso | FALSE | FALSE | 0.001601 | CV | 0 | 24 | 20 |
| LV_circumf | Lasso | FALSE | FALSE | 0.000737 | CV | 1 | 24 | 24 |
| LV_circumf | Lasso | FALSE | FALSE | 0.001593 | CV | 2 | 24 | 21 |
| LV_circumf | Lasso | FALSE | FALSE | 0.000966 | CV | 3 | 24 | 21 |
| LV_circumf | Lasso | FALSE | FALSE | 0.000109 | CV | 4 | 24 | 24 |
| LV_circumf | Lasso | FALSE | TRUE | 0.05 | CV | 0 | 24 | 4 |
| LV_circumf | Lasso | FALSE | TRUE | 0.05 | CV | 1 | 24 | 4 |
| LV_circumf | Lasso | FALSE | TRUE | 0.05 | CV | 2 | 24 | 4 |
| LV_circumf | Lasso | FALSE | TRUE | 0.05 | CV | 3 | 24 | 4 |
| LV_circumf | Lasso | FALSE | TRUE | 0.05 | CV | 4 | 24 | 4 |
| LV_circumf | LinRegrs | FALSE | FALSE | | HeldoutTest | | 24 | 21 |
| LV_circumf | Lasso | FALSE | FALSE | 0.000349 | HeldoutTest | | 24 | 22 |
| LV_circumf | Lasso | FALSE | TRUE | 0.05 | HeldoutTest | | 24 | 9 |
| LV_circumf | LinRegrs | FALSE | FALSE | | CV | 0 | 24 | 22 |
| LV_circumf | LinRegrs | FALSE | FALSE | | CV | 1 | 24 | 21 |
| LV_circumf | LinRegrs | FALSE | FALSE | | CV | 2 | 24 | 21 |
| LV_circumf | LinRegrs | FALSE | FALSE | | CV | 3 | 24 | 21 |
| LV_circumf | LinRegrs | FALSE | FALSE | | CV | 4 | 24 | 24 |
| LV_circumf | Lasso | FALSE | FALSE | 0.000255 | CV | 0 | 24 | 23 |
| LV_circumf | Lasso | FALSE | FALSE | 0.000381 | CV | 1 | 24 | 23 |
| LV_circumf | Lasso | FALSE | FALSE | 0.000241 | CV | 2 | 24 | 23 |
| LV_circumf | Lasso | FALSE | FALSE | 0.000175 | CV | 3 | 24 | 24 |
| LV_circumf | Lasso | FALSE | FALSE | 0.000177 | CV | 4 | 24 | 24 |
| LV_circumf | Lasso | FALSE | TRUE | 0.05 | CV | 0 | 24 | 11 |
| LV_circumf | Lasso | FALSE | TRUE | 0.05 | CV | 1 | 24 | 9 |
| LV_circumf | Lasso | FALSE | TRUE | 0.05 | CV | 2 | 24 | 9 |
| LV_circumf | Lasso | FALSE | TRUE | 0.05 | CV | 3 | 24 | 9 |
| LV_circumf | Lasso | FALSE | TRUE | 0.05 | CV | 4 | 24 | 9 |
| LV_circumf | LinRegrs | FALSE | FALSE | | HeldoutTest | | 24 | 22 |
| LV_circumf | Lasso | FALSE | FALSE | 0.000105 | HeldoutTest | | 24 | 24 |
| LV_circumf | Lasso | FALSE | TRUE | 0.05 | HeldoutTest | | 24 | 4 |
| LV_circumf | LinRegrs | FALSE | FALSE | | CV | 0 | 24 | 23 |
| LV_circumf | LinRegrs | FALSE | FALSE | | CV | 1 | 24 | 22 |
| LV_circumf | LinRegrs | FALSE | FALSE | | CV | 2 | 24 | 23 |
| LV_circumf | LinRegrs | FALSE | FALSE | | CV | 3 | 24 | 22 |
| LV_circumf | LinRegrs | FALSE | FALSE | | CV | 4 | 24 | 23 |
| LV_circumf | Lasso | FALSE | FALSE | 9.98E-05 | CV | 0 | 24 | 24 |
| LV_circumf | Lasso | FALSE | FALSE | 0.000203 | CV | 1 | 24 | 24 |
| LV_circumf | Lasso | FALSE | FALSE | 0.000119 | CV | 2 | 24 | 24 |
| LV_circumf | Lasso | FALSE | FALSE | 0.000309 | CV | 3 | 24 | 24 |
| LV_circumf | Lasso | FALSE | FALSE | 0.000103 | CV | 4 | 24 | 24 |
| LV_circumf | Lasso | FALSE | TRUE | 0.05 | CV | 0 | 24 | 4 |
| LV_circumf | Lasso | FALSE | TRUE | 0.05 | CV | 1 | 24 | 5 |
| LV_circumf | Lasso | FALSE | TRUE | 0.05 | CV | 2 | 24 | 3 |
| LV_circumf | Lasso | FALSE | TRUE | 0.05 | CV | 3 | 24 | 4 |

| | | | | | | | | |
|---|---|---|---|---|---|---|---|---|
| LV_circumf | Lasso | FALSE | TRUE | 0.05 | CV | 4 | 24 | 5 |
| LV_circumf | LinRegrs | FALSE | FALSE | | HeldoutTest | | 24 | 22 |
| LV_circumf | Lasso | FALSE | FALSE | 0.00015 | HeldoutTest | | 24 | 24 |
| LV_circumf | Lasso | FALSE | TRUE | 0.05 | HeldoutTest | | 24 | 12 |
| LV_circumf | LinRegrs | FALSE | FALSE | | CV | 0 | 24 | 23 |
| LV_circumf | LinRegrs | FALSE | FALSE | | CV | 1 | 24 | 21 |
| LV_circumf | LinRegrs | FALSE | FALSE | | CV | 2 | 24 | 22 |
| LV_circumf | LinRegrs | FALSE | FALSE | | CV | 3 | 24 | 22 |
| LV_circumf | LinRegrs | FALSE | FALSE | | CV | 4 | 24 | 21 |
| LV_circumf | Lasso | FALSE | FALSE | 0.000154 | CV | 0 | 24 | 24 |
| LV_circumf | Lasso | FALSE | FALSE | 0.000187 | CV | 1 | 24 | 24 |
| LV_circumf | Lasso | FALSE | FALSE | 0.000149 | CV | 2 | 24 | 24 |
| LV_circumf | Lasso | FALSE | FALSE | 0.000147 | CV | 3 | 24 | 24 |
| LV_circumf | Lasso | FALSE | FALSE | 0.000224 | CV | 4 | 24 | 24 |
| LV_circumf | Lasso | FALSE | TRUE | 0.05 | CV | 0 | 24 | 12 |
| LV_circumf | Lasso | FALSE | TRUE | 0.05 | CV | 1 | 24 | 12 |
| LV_circumf | Lasso | FALSE | TRUE | 0.05 | CV | 2 | 24 | 12 |
| LV_circumf | Lasso | FALSE | TRUE | 0.05 | CV | 3 | 24 | 12 |
| LV_circumf | Lasso | FALSE | TRUE | 0.05 | CV | 4 | 24 | 8 |
| LV_circumf | LinRegrs | FALSE | FALSE | | HeldoutTest | | 24 | 24 |
| LV_circumf | Lasso | FALSE | FALSE | 0.000189 | HeldoutTest | | 24 | 24 |
| LV_circumf | Lasso | FALSE | TRUE | 0.05 | HeldoutTest | | 24 | 10 |
| LV_circumf | LinRegrs | FALSE | FALSE | | CV | 0 | 24 | 23 |
| LV_circumf | LinRegrs | FALSE | FALSE | | CV | 1 | 24 | 24 |
| LV_circumf | LinRegrs | FALSE | FALSE | | CV | 2 | 24 | 23 |
| LV_circumf | LinRegrs | FALSE | FALSE | | CV | 3 | 24 | 24 |
| LV_circumf | LinRegrs | FALSE | FALSE | | CV | 4 | 24 | 21 |
| LV_circumf | Lasso | FALSE | FALSE | 0.000192 | CV | 0 | 24 | 24 |
| LV_circumf | Lasso | FALSE | FALSE | 0.000189 | CV | 1 | 24 | 24 |
| LV_circumf | Lasso | FALSE | FALSE | 0.000183 | CV | 2 | 24 | 24 |
| LV_circumf | Lasso | FALSE | FALSE | 0.000419 | CV | 3 | 24 | 24 |
| LV_circumf | Lasso | FALSE | FALSE | 0.000652 | CV | 4 | 24 | 22 |
| LV_circumf | Lasso | FALSE | TRUE | 0.05 | CV | 0 | 24 | 10 |
| LV_circumf | Lasso | FALSE | TRUE | 0.05 | CV | 1 | 24 | 10 |
| LV_circumf | Lasso | FALSE | TRUE | 0.05 | CV | 2 | 24 | 10 |
| LV_circumf | Lasso | FALSE | TRUE | 0.05 | CV | 3 | 24 | 10 |
| LV_circumf | Lasso | FALSE | TRUE | 0.05 | CV | 4 | 24 | 8 |
| LV_circumf | LinRegrs | FALSE | FALSE | | HeldoutTest | | 24 | 21 |
| LV_circumf | Lasso | FALSE | FALSE | 0.000133 | HeldoutTest | | 24 | 24 |
| LV_circumf | Lasso | FALSE | TRUE | 0.05 | HeldoutTest | | 24 | 5 |
| LV_circumf | LinRegrs | FALSE | FALSE | | CV | 0 | 24 | 21 |
| LV_circumf | LinRegrs | FALSE | FALSE | | CV | 1 | 24 | 21 |
| LV_circumf | LinRegrs | FALSE | FALSE | | CV | 2 | 24 | 19 |
| LV_circumf | LinRegrs | FALSE | FALSE | | CV | 3 | 24 | 21 |
| LV_circumf | LinRegrs | FALSE | FALSE | | CV | 4 | 24 | 22 |

| | | | | | | | | |
|---|---|---|---|---|---|---|---|---|
| LV_circumf | Lasso | FALSE | FALSE | 0.000134 | CV | 0 | 24 | 23 |
| LV_circumf | Lasso | FALSE | FALSE | 0.00022 | CV | 1 | 24 | 24 |
| LV_circumf | Lasso | FALSE | FALSE | 0.000174 | CV | 2 | 24 | 23 |
| LV_circumf | Lasso | FALSE | FALSE | 0.000238 | CV | 3 | 24 | 24 |
| LV_circumf | Lasso | FALSE | FALSE | 0.000136 | CV | 4 | 24 | 24 |
| LV_circumf | Lasso | FALSE | TRUE | 0.05 | CV | 0 | 24 | 6 |
| LV_circumf | Lasso | FALSE | TRUE | 0.05 | CV | 1 | 24 | 5 |
| LV_circumf | Lasso | FALSE | TRUE | 0.05 | CV | 2 | 24 | 5 |
| LV_circumf | Lasso | FALSE | TRUE | 0.05 | CV | 3 | 24 | 5 |
| LV_circumf | Lasso | FALSE | TRUE | 0.05 | CV | 4 | 24 | 6 |
| LV_circumf | LinRegrs | FALSE | FALSE | | HeldoutTest | | 24 | 22 |
| LV_circumf | Lasso | FALSE | FALSE | 0.000741 | HeldoutTest | | 24 | 23 |
| LV_circumf | Lasso | FALSE | TRUE | 0.05 | HeldoutTest | | 24 | 3 |
| LV_circumf | LinRegrs | FALSE | FALSE | | CV | 0 | 24 | 18 |
| LV_circumf | LinRegrs | FALSE | FALSE | | CV | 1 | 24 | 21 |
| LV_circumf | LinRegrs | FALSE | FALSE | | CV | 2 | 24 | 20 |
| LV_circumf | LinRegrs | FALSE | FALSE | | CV | 3 | 24 | 19 |
| LV_circumf | LinRegrs | FALSE | FALSE | | CV | 4 | 24 | 21 |
| LV_circumf | Lasso | FALSE | FALSE | 0.001013 | CV | 0 | 24 | 23 |
| LV_circumf | Lasso | FALSE | FALSE | 0.000427 | CV | 1 | 24 | 24 |
| LV_circumf | Lasso | FALSE | FALSE | 0.000715 | CV | 2 | 24 | 23 |
| LV_circumf | Lasso | FALSE | FALSE | 0.002103 | CV | 3 | 24 | 17 |
| LV_circumf | Lasso | FALSE | FALSE | 0.000599 | CV | 4 | 24 | 24 |
| LV_circumf | Lasso | FALSE | TRUE | 0.05 | CV | 0 | 24 | 3 |
| LV_circumf | Lasso | FALSE | TRUE | 0.05 | CV | 1 | 24 | 3 |
| LV_circumf | Lasso | FALSE | TRUE | 0.05 | CV | 2 | 24 | 3 |
| LV_circumf | Lasso | FALSE | TRUE | 0.05 | CV | 3 | 24 | 3 |
| LV_circumf | Lasso | FALSE | TRUE | 0.05 | CV | 4 | 24 | 3 |
| LV_circumf | LinRegrs | FALSE | FALSE | | HeldoutTest | | 24 | 24 |
| LV_circumf | Lasso | FALSE | FALSE | 0.000125 | HeldoutTest | | 24 | 24 |
| LV_circumf | Lasso | FALSE | TRUE | 0.05 | HeldoutTest | | 24 | 8 |
| LV_circumf | LinRegrs | FALSE | FALSE | | CV | 0 | 24 | 23 |
| LV_circumf | LinRegrs | FALSE | FALSE | | CV | 1 | 24 | 24 |
| LV_circumf | LinRegrs | FALSE | FALSE | | CV | 2 | 24 | 24 |
| LV_circumf | LinRegrs | FALSE | FALSE | | CV | 3 | 24 | 24 |
| LV_circumf | LinRegrs | FALSE | FALSE | | CV | 4 | 24 | 24 |
| LV_circumf | Lasso | FALSE | FALSE | 0.000131 | CV | 0 | 24 | 24 |
| LV_circumf | Lasso | FALSE | FALSE | 0.000126 | CV | 1 | 24 | 24 |
| LV_circumf | Lasso | FALSE | FALSE | 0.000123 | CV | 2 | 24 | 24 |
| LV_circumf | Lasso | FALSE | FALSE | 0.000122 | CV | 3 | 24 | 24 |
| LV_circumf | Lasso | FALSE | FALSE | 0.000123 | CV | 4 | 24 | 24 |
| LV_circumf | Lasso | FALSE | TRUE | 0.05 | CV | 0 | 24 | 7 |
| LV_circumf | Lasso | FALSE | TRUE | 0.05 | CV | 1 | 24 | 8 |
| LV_circumf | Lasso | FALSE | TRUE | 0.05 | CV | 2 | 24 | 8 |
| LV_circumf | Lasso | FALSE | TRUE | 0.05 | CV | 3 | 24 | 8 |

| | | | | | | | | |
|---|---|---|---|---|---|---|---|---|
| LV_circumf | Lasso | FALSE | TRUE | 0.05 | CV | 4 | 24 | 8 |
| LV_circumf | LinRegrs | FALSE | FALSE | | HeldoutTest | | 24 | 24 |
| LV_circumf | Lasso | FALSE | FALSE | 0.000113 | HeldoutTest | | 24 | 24 |
| LV_circumf | Lasso | FALSE | TRUE | 0.05 | HeldoutTest | | 24 | 9 |
| LV_circumf | LinRegrs | FALSE | FALSE | | CV | 0 | 24 | 23 |
| LV_circumf | LinRegrs | FALSE | FALSE | | CV | 1 | 24 | 23 |
| LV_circumf | LinRegrs | FALSE | FALSE | | CV | 2 | 24 | 22 |
| LV_circumf | LinRegrs | FALSE | FALSE | | CV | 3 | 24 | 23 |
| LV_circumf | LinRegrs | FALSE | FALSE | | CV | 4 | 24 | 22 |
| LV_circumf | Lasso | FALSE | FALSE | 0.000113 | CV | 0 | 24 | 24 |
| LV_circumf | Lasso | FALSE | FALSE | 0.000113 | CV | 1 | 24 | 24 |
| LV_circumf | Lasso | FALSE | FALSE | 0.000114 | CV | 2 | 24 | 24 |
| LV_circumf | Lasso | FALSE | FALSE | 0.000113 | CV | 3 | 24 | 24 |
| LV_circumf | Lasso | FALSE | FALSE | 0.000115 | CV | 4 | 24 | 24 |
| LV_circumf | Lasso | FALSE | TRUE | 0.05 | CV | 0 | 24 | 8 |
| LV_circumf | Lasso | FALSE | TRUE | 0.05 | CV | 1 | 24 | 10 |
| LV_circumf | Lasso | FALSE | TRUE | 0.05 | CV | 2 | 24 | 9 |
| LV_circumf | Lasso | FALSE | TRUE | 0.05 | CV | 3 | 24 | 9 |
| LV_circumf | Lasso | FALSE | TRUE | 0.05 | CV | 4 | 24 | 8 |
| LV_circumf | LinRegrs | FALSE | FALSE | | HeldoutTest | | 24 | 23 |
| LV_circumf | Lasso | FALSE | FALSE | 0.000149 | HeldoutTest | | 24 | 24 |
| LV_circumf | Lasso | FALSE | TRUE | 0.05 | HeldoutTest | | 24 | 12 |
| LV_circumf | LinRegrs | FALSE | FALSE | | CV | 0 | 24 | 23 |
| LV_circumf | LinRegrs | FALSE | FALSE | | CV | 1 | 24 | 23 |
| LV_circumf | LinRegrs | FALSE | FALSE | | CV | 2 | 24 | 24 |
| LV_circumf | LinRegrs | FALSE | FALSE | | CV | 3 | 24 | 23 |
| LV_circumf | LinRegrs | FALSE | FALSE | | CV | 4 | 24 | 23 |
| LV_circumf | Lasso | FALSE | FALSE | 0.000154 | CV | 0 | 24 | 24 |
| LV_circumf | Lasso | FALSE | FALSE | 0.000154 | CV | 1 | 24 | 24 |
| LV_circumf | Lasso | FALSE | FALSE | 0.00015 | CV | 2 | 24 | 24 |
| LV_circumf | Lasso | FALSE | FALSE | 0.000146 | CV | 3 | 24 | 24 |
| LV_circumf | Lasso | FALSE | FALSE | 0.000149 | CV | 4 | 24 | 24 |
| LV_circumf | Lasso | FALSE | TRUE | 0.05 | CV | 0 | 24 | 12 |
| LV_circumf | Lasso | FALSE | TRUE | 0.05 | CV | 1 | 24 | 12 |
| LV_circumf | Lasso | FALSE | TRUE | 0.05 | CV | 2 | 24 | 12 |
| LV_circumf | Lasso | FALSE | TRUE | 0.05 | CV | 3 | 24 | 12 |
| LV_circumf | Lasso | FALSE | TRUE | 0.05 | CV | 4 | 24 | 12 |
| LV_circumf | LinRegrs | FALSE | FALSE | | HeldoutTest | | 24 | 22 |
| LV_circumf | Lasso | FALSE | FALSE | 0.000181 | HeldoutTest | | 24 | 24 |
| LV_circumf | Lasso | FALSE | TRUE | 0.05 | HeldoutTest | | 24 | 9 |
| LV_circumf | LinRegrs | FALSE | FALSE | | CV | 0 | 24 | 22 |
| LV_circumf | LinRegrs | FALSE | FALSE | | CV | 1 | 24 | 23 |
| LV_circumf | LinRegrs | FALSE | FALSE | | CV | 2 | 24 | 22 |
| LV_circumf | LinRegrs | FALSE | FALSE | | CV | 3 | 24 | 23 |
| LV_circumf | LinRegrs | FALSE | FALSE | | CV | 4 | 24 | 22 |

| | | | | | | | | |
|---|---|---|---|---|---|---|---|---|
| LV_circumf | Lasso | FALSE | FALSE | 0.000183 | CV | 0 | 24 | 24 |
| LV_circumf | Lasso | FALSE | FALSE | 0.00018 | CV | 1 | 24 | 24 |
| LV_circumf | Lasso | FALSE | FALSE | 0.000204 | CV | 2 | 24 | 24 |
| LV_circumf | Lasso | FALSE | FALSE | 0.000177 | CV | 3 | 24 | 24 |
| LV_circumf | Lasso | FALSE | FALSE | 0.000185 | CV | 4 | 24 | 24 |
| LV_circumf | Lasso | FALSE | TRUE | 0.05 | CV | 0 | 24 | 9 |
| LV_circumf | Lasso | FALSE | TRUE | 0.05 | CV | 1 | 24 | 10 |
| LV_circumf | Lasso | FALSE | TRUE | 0.05 | CV | 2 | 24 | 9 |
| LV_circumf | Lasso | FALSE | TRUE | 0.05 | CV | 3 | 24 | 9 |
| LV_circumf | Lasso | FALSE | TRUE | 0.05 | CV | 4 | 24 | 9 |
| LV_circumf | LinRegrs | FALSE | FALSE | | HeldoutTest | | 24 | 23 |
| LV_circumf | Lasso | FALSE | FALSE | 0.000102 | HeldoutTest | | 24 | 24 |
| LV_circumf | Lasso | FALSE | TRUE | 0.05 | HeldoutTest | | 24 | 9 |
| LV_circumf | LinRegrs | FALSE | FALSE | | CV | 0 | 24 | 22 |
| LV_circumf | LinRegrs | FALSE | FALSE | | CV | 1 | 24 | 22 |
| LV_circumf | LinRegrs | FALSE | FALSE | | CV | 2 | 24 | 22 |
| LV_circumf | LinRegrs | FALSE | FALSE | | CV | 3 | 24 | 22 |
| LV_circumf | LinRegrs | FALSE | FALSE | | CV | 4 | 24 | 20 |
| LV_circumf | Lasso | FALSE | FALSE | 9.70E-05 | CV | 0 | 24 | 24 |
| LV_circumf | Lasso | FALSE | FALSE | 0.000172 | CV | 1 | 24 | 24 |
| LV_circumf | Lasso | FALSE | FALSE | 0.000101 | CV | 2 | 24 | 24 |
| LV_circumf | Lasso | FALSE | FALSE | 0.000275 | CV | 3 | 24 | 24 |
| LV_circumf | Lasso | FALSE | FALSE | 0.00012 | CV | 4 | 24 | 23 |
| LV_circumf | Lasso | FALSE | TRUE | 0.05 | CV | 0 | 24 | 11 |
| LV_circumf | Lasso | FALSE | TRUE | 0.05 | CV | 1 | 24 | 9 |
| LV_circumf | Lasso | FALSE | TRUE | 0.05 | CV | 2 | 24 | 9 |
| LV_circumf | Lasso | FALSE | TRUE | 0.05 | CV | 3 | 24 | 9 |
| LV_circumf | Lasso | FALSE | TRUE | 0.05 | CV | 4 | 24 | 8 |
| LV_circumf | LinRegrs | FALSE | FALSE | | HeldoutTest | | 24 | 23 |
| LV_circumf | Lasso | FALSE | FALSE | 0.000143 | HeldoutTest | | 24 | 24 |
| LV_circumf | Lasso | FALSE | TRUE | 0.05 | HeldoutTest | | 24 | 10 |
| LV_circumf | LinRegrs | FALSE | FALSE | | CV | 0 | 24 | 23 |
| LV_circumf | LinRegrs | FALSE | FALSE | | CV | 1 | 24 | 23 |
| LV_circumf | LinRegrs | FALSE | FALSE | | CV | 2 | 24 | 24 |
| LV_circumf | LinRegrs | FALSE | FALSE | | CV | 3 | 24 | 23 |
| LV_circumf | LinRegrs | FALSE | FALSE | | CV | 4 | 24 | 23 |
| LV_circumf | Lasso | FALSE | FALSE | 0.000147 | CV | 0 | 24 | 24 |
| LV_circumf | Lasso | FALSE | FALSE | 0.000141 | CV | 1 | 24 | 24 |
| LV_circumf | Lasso | FALSE | FALSE | 0.000141 | CV | 2 | 24 | 24 |
| LV_circumf | Lasso | FALSE | FALSE | 0.000142 | CV | 3 | 24 | 24 |
| LV_circumf | Lasso | FALSE | FALSE | 0.000143 | CV | 4 | 24 | 24 |
| LV_circumf | Lasso | FALSE | TRUE | 0.05 | CV | 0 | 24 | 10 |
| LV_circumf | Lasso | FALSE | TRUE | 0.05 | CV | 1 | 24 | 9 |
| LV_circumf | Lasso | FALSE | TRUE | 0.05 | CV | 2 | 24 | 9 |
| LV_circumf | Lasso | FALSE | TRUE | 0.05 | CV | 3 | 24 | 10 |

| | | | | | | | | |
|---|---|---|---|---|---|---|---|---|
| LV_circumf | Lasso | FALSE | TRUE | 0.05 | CV | 4 | 24 | 9 |
| LV_circumf | LinRegrs | FALSE | FALSE | | HeldoutTest | | 24 | 24 |
| LV_circumf | Lasso | FALSE | FALSE | 0.000195 | HeldoutTest | | 24 | 24 |
| LV_circumf | Lasso | FALSE | TRUE | 0.05 | HeldoutTest | | 24 | 13 |
| LV_circumf | LinRegrs | FALSE | FALSE | | CV | 0 | 24 | 24 |
| LV_circumf | LinRegrs | FALSE | FALSE | | CV | 1 | 24 | 24 |
| LV_circumf | LinRegrs | FALSE | FALSE | | CV | 2 | 24 | 24 |
| LV_circumf | LinRegrs | FALSE | FALSE | | CV | 3 | 24 | 23 |
| LV_circumf | LinRegrs | FALSE | FALSE | | CV | 4 | 24 | 24 |
| LV_circumf | Lasso | FALSE | FALSE | 0.000198 | CV | 0 | 24 | 24 |
| LV_circumf | Lasso | FALSE | FALSE | 0.000194 | CV | 1 | 24 | 24 |
| LV_circumf | Lasso | FALSE | FALSE | 0.000194 | CV | 2 | 24 | 24 |
| LV_circumf | Lasso | FALSE | FALSE | 0.000193 | CV | 3 | 24 | 24 |
| LV_circumf | Lasso | FALSE | FALSE | 0.000199 | CV | 4 | 24 | 24 |
| LV_circumf | Lasso | FALSE | TRUE | 0.05 | CV | 0 | 24 | 12 |
| LV_circumf | Lasso | FALSE | TRUE | 0.05 | CV | 1 | 24 | 13 |
| LV_circumf | Lasso | FALSE | TRUE | 0.05 | CV | 2 | 24 | 12 |
| LV_circumf | Lasso | FALSE | TRUE | 0.05 | CV | 3 | 24 | 15 |
| LV_circumf | Lasso | FALSE | TRUE | 0.05 | CV | 4 | 24 | 12 |
| RV_end_di | LinRegrs | FALSE | FALSE | | HeldoutTest | | 24 | 24 |
| RV_end_di | Lasso | FALSE | FALSE | 0.000414 | HeldoutTest | | 24 | 23 |
| RV_end_di | Lasso | FALSE | TRUE | 0.05 | HeldoutTest | | 24 | 15 |
| RV_end_di | LinRegrs | FALSE | FALSE | | CV | 0 | 24 | 24 |
| RV_end_di | LinRegrs | FALSE | FALSE | | CV | 1 | 24 | 24 |
| RV_end_di | LinRegrs | FALSE | FALSE | | CV | 2 | 24 | 24 |
| RV_end_di | LinRegrs | FALSE | FALSE | | CV | 3 | 24 | 23 |
| RV_end_di | LinRegrs | FALSE | FALSE | | CV | 4 | 24 | 24 |
| RV_end_di | Lasso | FALSE | FALSE | 0.000417 | CV | 0 | 24 | 23 |
| RV_end_di | Lasso | FALSE | FALSE | 0.000412 | CV | 1 | 24 | 24 |
| RV_end_di | Lasso | FALSE | FALSE | 0.000414 | CV | 2 | 24 | 23 |
| RV_end_di | Lasso | FALSE | FALSE | 0.000414 | CV | 3 | 24 | 24 |
| RV_end_di | Lasso | FALSE | FALSE | 0.000414 | CV | 4 | 24 | 24 |
| RV_end_di | Lasso | FALSE | TRUE | 0.05 | CV | 0 | 24 | 15 |
| RV_end_di | Lasso | FALSE | TRUE | 0.05 | CV | 1 | 24 | 15 |
| RV_end_di | Lasso | FALSE | TRUE | 0.05 | CV | 2 | 24 | 15 |
| RV_end_di | Lasso | FALSE | TRUE | 0.05 | CV | 3 | 24 | 15 |
| RV_end_di | Lasso | FALSE | TRUE | 0.05 | CV | 4 | 24 | 13 |
| RV_end_sy | LinRegrs | FALSE | FALSE | | HeldoutTest | | 24 | 21 |
| RV_end_sy | Lasso | FALSE | FALSE | 0.000387 | HeldoutTest | | 24 | 24 |
| RV_end_sy | Lasso | FALSE | TRUE | 0.05 | HeldoutTest | | 24 | 16 |
| RV_end_sy | LinRegrs | FALSE | FALSE | | CV | 0 | 24 | 20 |
| RV_end_sy | LinRegrs | FALSE | FALSE | | CV | 1 | 24 | 21 |
| RV_end_sy | LinRegrs | FALSE | FALSE | | CV | 2 | 24 | 22 |
| RV_end_sy | LinRegrs | FALSE | FALSE | | CV | 3 | 24 | 21 |
| RV_end_sy | LinRegrs | FALSE | FALSE | | CV | 4 | 24 | 23 |

| | | | | | | | | |
|---|---|---|---|---|---|---|---|---|
| RV_end_sy | Lasso | FALSE | FALSE | 0.000387 | CV | 0 | 24 | 23 |
| RV_end_sy | Lasso | FALSE | FALSE | 0.000386 | CV | 1 | 24 | 23 |
| RV_end_sy | Lasso | FALSE | FALSE | 0.000386 | CV | 2 | 24 | 23 |
| RV_end_sy | Lasso | FALSE | FALSE | 0.000387 | CV | 3 | 24 | 24 |
| RV_end_sy | Lasso | FALSE | FALSE | 0.000388 | CV | 4 | 24 | 24 |
| RV_end_sy | Lasso | FALSE | TRUE | 0.05 | CV | 0 | 24 | 16 |
| RV_end_sy | Lasso | FALSE | TRUE | 0.05 | CV | 1 | 24 | 16 |
| RV_end_sy | Lasso | FALSE | TRUE | 0.05 | CV | 2 | 24 | 16 |
| RV_end_sy | Lasso | FALSE | TRUE | 0.05 | CV | 3 | 24 | 16 |
| RV_end_sy | Lasso | FALSE | TRUE | 0.05 | CV | 4 | 24 | 15 |
| RV_stroke_ | LinRegrs | FALSE | FALSE | | HeldoutTest | | 24 | 24 |
| RV_stroke_ | Lasso | FALSE | FALSE | 0.000358 | HeldoutTest | | 24 | 24 |
| RV_stroke_ | Lasso | FALSE | TRUE | 0.05 | HeldoutTest | | 24 | 13 |
| RV_stroke_ | LinRegrs | FALSE | FALSE | | CV | 0 | 24 | 24 |
| RV_stroke_ | LinRegrs | FALSE | FALSE | | CV | 1 | 24 | 24 |
| RV_stroke_ | LinRegrs | FALSE | FALSE | | CV | 2 | 24 | 24 |
| RV_stroke_ | LinRegrs | FALSE | FALSE | | CV | 3 | 24 | 24 |
| RV_stroke_ | LinRegrs | FALSE | FALSE | | CV | 4 | 24 | 24 |
| RV_stroke_ | Lasso | FALSE | FALSE | 0.000364 | CV | 0 | 24 | 24 |
| RV_stroke_ | Lasso | FALSE | FALSE | 0.000358 | CV | 1 | 24 | 24 |
| RV_stroke_ | Lasso | FALSE | FALSE | 0.000358 | CV | 2 | 24 | 24 |
| RV_stroke_ | Lasso | FALSE | FALSE | 0.000356 | CV | 3 | 24 | 24 |
| RV_stroke_ | Lasso | FALSE | FALSE | 0.000355 | CV | 4 | 24 | 24 |
| RV_stroke_ | Lasso | FALSE | TRUE | 0.05 | CV | 0 | 24 | 13 |
| RV_stroke_ | Lasso | FALSE | TRUE | 0.05 | CV | 1 | 24 | 13 |
| RV_stroke_ | Lasso | FALSE | TRUE | 0.05 | CV | 2 | 24 | 12 |
| RV_stroke_ | Lasso | FALSE | TRUE | 0.05 | CV | 3 | 24 | 13 |
| RV_stroke_ | Lasso | FALSE | TRUE | 0.05 | CV | 4 | 24 | 11 |
| RV_ejectio | LinRegrs | FALSE | FALSE | | HeldoutTest | | 24 | 23 |
| RV_ejectio | Lasso | FALSE | FALSE | 0.000329 | HeldoutTest | | 24 | 24 |
| RV_ejectio | Lasso | FALSE | TRUE | 0.05 | HeldoutTest | | 24 | 11 |
| RV_ejectio | LinRegrs | FALSE | FALSE | | CV | 0 | 24 | 24 |
| RV_ejectio | LinRegrs | FALSE | FALSE | | CV | 1 | 24 | 23 |
| RV_ejectio | LinRegrs | FALSE | FALSE | | CV | 2 | 24 | 22 |
| RV_ejectio | LinRegrs | FALSE | FALSE | | CV | 3 | 24 | 22 |
| RV_ejectio | LinRegrs | FALSE | FALSE | | CV | 4 | 24 | 21 |
| RV_ejectio | Lasso | FALSE | FALSE | 0.000191 | CV | 0 | 24 | 24 |
| RV_ejectio | Lasso | FALSE | FALSE | 0.000247 | CV | 1 | 24 | 24 |
| RV_ejectio | Lasso | FALSE | FALSE | 0.000381 | CV | 2 | 24 | 24 |
| RV_ejectio | Lasso | FALSE | FALSE | 0.000398 | CV | 3 | 24 | 24 |
| RV_ejectio | Lasso | FALSE | FALSE | 0.000217 | CV | 4 | 24 | 24 |
| RV_ejectio | Lasso | FALSE | TRUE | 0.05 | CV | 0 | 24 | 11 |
| RV_ejectio | Lasso | FALSE | TRUE | 0.05 | CV | 1 | 24 | 11 |
| RV_ejectio | Lasso | FALSE | TRUE | 0.05 | CV | 2 | 24 | 11 |
| RV_ejectio | Lasso | FALSE | TRUE | 0.05 | CV | 3 | 24 | 10 |

| | | | | | | | | |
|---|---|---|---|---|---|---|---|---|
| RV_ejectio | Lasso | FALSE | TRUE | 0.05 | CV | 4 | 24 | 11 |
| LV_longitu | LinRegrs | FALSE | FALSE | | HeldoutTest | | 24 | 21 |
| LV_longitu | Lasso | FALSE | FALSE | 0.000648 | HeldoutTest | | 24 | 23 |
| LV_longitu | Lasso | FALSE | TRUE | 0.05 | HeldoutTest | | 24 | 9 |
| LV_longitu | LinRegrs | FALSE | FALSE | | CV | 0 | 24 | 21 |
| LV_longitu | LinRegrs | FALSE | FALSE | | CV | 1 | 24 | 21 |
| LV_longitu | LinRegrs | FALSE | FALSE | | CV | 2 | 24 | 22 |
| LV_longitu | LinRegrs | FALSE | FALSE | | CV | 3 | 24 | 21 |
| LV_longitu | LinRegrs | FALSE | FALSE | | CV | 4 | 24 | 21 |
| LV_longitu | Lasso | FALSE | FALSE | 0.00044 | CV | 0 | 24 | 23 |
| LV_longitu | Lasso | FALSE | FALSE | 0.000586 | CV | 1 | 24 | 24 |
| LV_longitu | Lasso | FALSE | FALSE | 0.00079 | CV | 2 | 24 | 23 |
| LV_longitu | Lasso | FALSE | FALSE | 0.00019 | CV | 3 | 24 | 24 |
| LV_longitu | Lasso | FALSE | FALSE | 0.000517 | CV | 4 | 24 | 24 |
| LV_longitu | Lasso | FALSE | TRUE | 0.05 | CV | 0 | 24 | 8 |
| LV_longitu | Lasso | FALSE | TRUE | 0.05 | CV | 1 | 24 | 8 |
| LV_longitu | Lasso | FALSE | TRUE | 0.05 | CV | 2 | 24 | 8 |
| LV_longitu | Lasso | FALSE | TRUE | 0.05 | CV | 3 | 24 | 8 |
| LV_longitu | Lasso | FALSE | TRUE | 0.05 | CV | 4 | 24 | 7 |
| LV_longitu | LinRegrs | FALSE | FALSE | | HeldoutTest | | 24 | 24 |
| LV_longitu | Lasso | FALSE | FALSE | 0.000227 | HeldoutTest | | 24 | 24 |
| LV_longitu | Lasso | FALSE | TRUE | 0.05 | HeldoutTest | | 24 | 7 |
| LV_longitu | LinRegrs | FALSE | FALSE | | CV | 0 | 24 | 24 |
| LV_longitu | LinRegrs | FALSE | FALSE | | CV | 1 | 24 | 19 |
| LV_longitu | LinRegrs | FALSE | FALSE | | CV | 2 | 24 | 24 |
| LV_longitu | LinRegrs | FALSE | FALSE | | CV | 3 | 24 | 21 |
| LV_longitu | LinRegrs | FALSE | FALSE | | CV | 4 | 24 | 22 |
| LV_longitu | Lasso | FALSE | FALSE | 0.000227 | CV | 0 | 24 | 24 |
| LV_longitu | Lasso | FALSE | FALSE | 0.000322 | CV | 1 | 24 | 23 |
| LV_longitu | Lasso | FALSE | FALSE | 0.000225 | CV | 2 | 24 | 24 |
| LV_longitu | Lasso | FALSE | FALSE | 0.000529 | CV | 3 | 24 | 23 |
| LV_longitu | Lasso | FALSE | FALSE | 0.000305 | CV | 4 | 24 | 24 |
| LV_longitu | Lasso | FALSE | TRUE | 0.05 | CV | 0 | 24 | 7 |
| LV_longitu | Lasso | FALSE | TRUE | 0.05 | CV | 1 | 24 | 7 |
| LV_longitu | Lasso | FALSE | TRUE | 0.05 | CV | 2 | 24 | 7 |
| LV_longitu | Lasso | FALSE | TRUE | 0.05 | CV | 3 | 24 | 7 |
| LV_longitu | Lasso | FALSE | TRUE | 0.05 | CV | 4 | 24 | 10 |
| LV_longitu | LinRegrs | FALSE | FALSE | | HeldoutTest | | 24 | 22 |
| LV_longitu | Lasso | FALSE | FALSE | 0.000254 | HeldoutTest | | 24 | 24 |
| LV_longitu | Lasso | FALSE | TRUE | 0.05 | HeldoutTest | | 24 | 5 |
| LV_longitu | LinRegrs | FALSE | FALSE | | CV | 0 | 24 | 21 |
| LV_longitu | LinRegrs | FALSE | FALSE | | CV | 1 | 24 | 22 |
| LV_longitu | LinRegrs | FALSE | FALSE | | CV | 2 | 24 | 22 |
| LV_longitu | LinRegrs | FALSE | FALSE | | CV | 3 | 24 | 21 |
| LV_longitu | LinRegrs | FALSE | FALSE | | CV | 4 | 24 | 22 |

| | | | | | | | | |
|---|---|---|---|---|---|---|---|---|
| LV_longitu | Lasso | FALSE | FALSE | 0.000359 | CV | 0 | 24 | 24 |
| LV_longitu | Lasso | FALSE | FALSE | 0.000238 | CV | 1 | 24 | 23 |
| LV_longitu | Lasso | FALSE | FALSE | 0.000238 | CV | 2 | 24 | 23 |
| LV_longitu | Lasso | FALSE | FALSE | 0.00024 | CV | 3 | 24 | 23 |
| LV_longitu | Lasso | FALSE | FALSE | 0.000241 | CV | 4 | 24 | 23 |
| LV_longitu | Lasso | FALSE | TRUE | 0.05 | CV | 0 | 24 | 5 |
| LV_longitu | Lasso | FALSE | TRUE | 0.05 | CV | 1 | 24 | 5 |
| LV_longitu | Lasso | FALSE | TRUE | 0.05 | CV | 2 | 24 | 6 |
| LV_longitu | Lasso | FALSE | TRUE | 0.05 | CV | 3 | 24 | 6 |
| LV_longitu | Lasso | FALSE | TRUE | 0.05 | CV | 4 | 24 | 6 |
| LV_longitu | LinRegrs | FALSE | FALSE | | HeldoutTest | | 24 | 23 |
| LV_longitu | Lasso | FALSE | FALSE | 0.000191 | HeldoutTest | | 24 | 24 |
| LV_longitu | Lasso | FALSE | TRUE | 0.05 | HeldoutTest | | 24 | 8 |
| LV_longitu | LinRegrs | FALSE | FALSE | | CV | 0 | 24 | 23 |
| LV_longitu | LinRegrs | FALSE | FALSE | | CV | 1 | 24 | 24 |
| LV_longitu | LinRegrs | FALSE | FALSE | | CV | 2 | 24 | 23 |
| LV_longitu | LinRegrs | FALSE | FALSE | | CV | 3 | 24 | 23 |
| LV_longitu | LinRegrs | FALSE | FALSE | | CV | 4 | 24 | 23 |
| LV_longitu | Lasso | FALSE | FALSE | 0.000191 | CV | 0 | 24 | 24 |
| LV_longitu | Lasso | FALSE | FALSE | 0.000194 | CV | 1 | 24 | 24 |
| LV_longitu | Lasso | FALSE | FALSE | 0.000188 | CV | 2 | 24 | 24 |
| LV_longitu | Lasso | FALSE | FALSE | 0.000191 | CV | 3 | 24 | 24 |
| LV_longitu | Lasso | FALSE | FALSE | 0.000191 | CV | 4 | 24 | 24 |
| LV_longitu | Lasso | FALSE | TRUE | 0.05 | CV | 0 | 24 | 7 |
| LV_longitu | Lasso | FALSE | TRUE | 0.05 | CV | 1 | 24 | 9 |
| LV_longitu | Lasso | FALSE | TRUE | 0.05 | CV | 2 | 24 | 8 |
| LV_longitu | Lasso | FALSE | TRUE | 0.05 | CV | 3 | 24 | 8 |
| LV_longitu | Lasso | FALSE | TRUE | 0.05 | CV | 4 | 24 | 7 |
| LV_longitu | LinRegrs | FALSE | FALSE | | HeldoutTest | | 24 | 21 |
| LV_longitu | Lasso | FALSE | FALSE | 0.000221 | HeldoutTest | | 24 | 23 |
| LV_longitu | Lasso | FALSE | TRUE | 0.05 | HeldoutTest | | 24 | 4 |
| LV_longitu | LinRegrs | FALSE | FALSE | | CV | 0 | 24 | 19 |
| LV_longitu | LinRegrs | FALSE | FALSE | | CV | 1 | 24 | 20 |
| LV_longitu | LinRegrs | FALSE | FALSE | | CV | 2 | 24 | 20 |
| LV_longitu | LinRegrs | FALSE | FALSE | | CV | 3 | 24 | 21 |
| LV_longitu | LinRegrs | FALSE | FALSE | | CV | 4 | 24 | 19 |
| LV_longitu | Lasso | FALSE | FALSE | 0.000285 | CV | 0 | 24 | 24 |
| LV_longitu | Lasso | FALSE | FALSE | 0.000316 | CV | 1 | 24 | 24 |
| LV_longitu | Lasso | FALSE | FALSE | 0.000366 | CV | 2 | 24 | 23 |
| LV_longitu | Lasso | FALSE | FALSE | 0.000224 | CV | 3 | 24 | 23 |
| LV_longitu | Lasso | FALSE | FALSE | 0.000468 | CV | 4 | 24 | 24 |
| LV_longitu | Lasso | FALSE | TRUE | 0.05 | CV | 0 | 24 | 4 |
| LV_longitu | Lasso | FALSE | TRUE | 0.05 | CV | 1 | 24 | 4 |
| LV_longitu | Lasso | FALSE | TRUE | 0.05 | CV | 2 | 24 | 5 |
| LV_longitu | Lasso | FALSE | TRUE | 0.05 | CV | 3 | 24 | 4 |

| | | | | | | | | |
|---|---|---|---|---|---|---|---|---|
| LV_longitu | Lasso | FALSE | TRUE | 0.05 | CV | 4 | 24 | 5 |
| LV_longitu | LinRegrs | FALSE | FALSE | | HeldoutTest | | 24 | 22 |
| LV_longitu | Lasso | FALSE | FALSE | 0.00015 | HeldoutTest | | 24 | 24 |
| LV_longitu | Lasso | FALSE | TRUE | 0.05 | HeldoutTest | | 24 | 11 |
| LV_longitu | LinRegrs | FALSE | FALSE | | CV | 0 | 24 | 23 |
| LV_longitu | LinRegrs | FALSE | FALSE | | CV | 1 | 24 | 22 |
| LV_longitu | LinRegrs | FALSE | FALSE | | CV | 2 | 24 | 21 |
| LV_longitu | LinRegrs | FALSE | FALSE | | CV | 3 | 24 | 21 |
| LV_longitu | LinRegrs | FALSE | FALSE | | CV | 4 | 24 | 23 |
| LV_longitu | Lasso | FALSE | FALSE | 0.000146 | CV | 0 | 24 | 23 |
| LV_longitu | Lasso | FALSE | FALSE | 0.000178 | CV | 1 | 24 | 24 |
| LV_longitu | Lasso | FALSE | FALSE | 0.000151 | CV | 2 | 24 | 24 |
| LV_longitu | Lasso | FALSE | FALSE | 0.00015 | CV | 3 | 24 | 23 |
| LV_longitu | Lasso | FALSE | FALSE | 0.000149 | CV | 4 | 24 | 24 |
| LV_longitu | Lasso | FALSE | TRUE | 0.05 | CV | 0 | 24 | 11 |
| LV_longitu | Lasso | FALSE | TRUE | 0.05 | CV | 1 | 24 | 11 |
| LV_longitu | Lasso | FALSE | TRUE | 0.05 | CV | 2 | 24 | 11 |
| LV_longitu | Lasso | FALSE | TRUE | 0.05 | CV | 3 | 24 | 11 |
| LV_longitu | Lasso | FALSE | TRUE | 0.05 | CV | 4 | 24 | 11 |
| LV_longitu | LinRegrs | FALSE | FALSE | | HeldoutTest | | 24 | 22 |
| LV_longitu | Lasso | FALSE | FALSE | 0.000216 | HeldoutTest | | 24 | 24 |
| LV_longitu | Lasso | FALSE | TRUE | 0.05 | HeldoutTest | | 24 | 10 |
| LV_longitu | LinRegrs | FALSE | FALSE | | CV | 0 | 24 | 22 |
| LV_longitu | LinRegrs | FALSE | FALSE | | CV | 1 | 24 | 22 |
| LV_longitu | LinRegrs | FALSE | FALSE | | CV | 2 | 24 | 22 |
| LV_longitu | LinRegrs | FALSE | FALSE | | CV | 3 | 24 | 23 |
| LV_longitu | LinRegrs | FALSE | FALSE | | CV | 4 | 24 | 24 |
| LV_longitu | Lasso | FALSE | FALSE | 0.000217 | CV | 0 | 24 | 24 |
| LV_longitu | Lasso | FALSE | FALSE | 0.000286 | CV | 1 | 24 | 23 |
| LV_longitu | Lasso | FALSE | FALSE | 0.00023 | CV | 2 | 24 | 24 |
| LV_longitu | Lasso | FALSE | FALSE | 0.000219 | CV | 3 | 24 | 24 |
| LV_longitu | Lasso | FALSE | FALSE | 0.00022 | CV | 4 | 24 | 24 |
| LV_longitu | Lasso | FALSE | TRUE | 0.05 | CV | 0 | 24 | 9 |
| LV_longitu | Lasso | FALSE | TRUE | 0.05 | CV | 1 | 24 | 10 |
| LV_longitu | Lasso | FALSE | TRUE | 0.05 | CV | 2 | 24 | 10 |
| LV_longitu | Lasso | FALSE | TRUE | 0.05 | CV | 3 | 24 | 10 |
| LV_longitu | Lasso | FALSE | TRUE | 0.05 | CV | 4 | 24 | 11 |
| LV_radial_s | LinRegrs | FALSE | FALSE | | HeldoutTest | | 24 | 20 |
| LV_radial_s | Lasso | FALSE | FALSE | 0.000462 | HeldoutTest | | 24 | 23 |
| LV_radial_s | Lasso | FALSE | TRUE | 0.05 | HeldoutTest | | 24 | 4 |
| LV_radial_s | LinRegrs | FALSE | FALSE | | CV | 0 | 24 | 24 |
| LV_radial_s | LinRegrs | FALSE | FALSE | | CV | 1 | 24 | 18 |
| LV_radial_s | LinRegrs | FALSE | FALSE | | CV | 2 | 24 | 22 |
| LV_radial_s | LinRegrs | FALSE | FALSE | | CV | 3 | 24 | 19 |
| LV_radial_s | LinRegrs | FALSE | FALSE | | CV | 4 | 24 | 21 |

| | | | | | | | | |
|---|---|---|---|---|---|---|---|---|
| LV_radial_s | Lasso | FALSE | FALSE | 0.00013 | CV | 0 | 24 | 24 |
| LV_radial_s | Lasso | FALSE | FALSE | 0.000852 | CV | 1 | 24 | 20 |
| LV_radial_s | Lasso | FALSE | FALSE | 0.000482 | CV | 2 | 24 | 23 |
| LV_radial_s | Lasso | FALSE | FALSE | 0.000914 | CV | 3 | 24 | 23 |
| LV_radial_s | Lasso | FALSE | FALSE | 0.000139 | CV | 4 | 24 | 24 |
| LV_radial_s | Lasso | FALSE | TRUE | 0.05 | CV | 0 | 24 | 4 |
| LV_radial_s | Lasso | FALSE | TRUE | 0.05 | CV | 1 | 24 | 4 |
| LV_radial_s | Lasso | FALSE | TRUE | 0.05 | CV | 2 | 24 | 5 |
| LV_radial_s | Lasso | FALSE | TRUE | 0.05 | CV | 3 | 24 | 5 |
| LV_radial_s | Lasso | FALSE | TRUE | 0.05 | CV | 4 | 24 | 5 |
| LV_radial_s | LinRegrs | FALSE | FALSE | | HeldoutTest | | 24 | 23 |
| LV_radial_s | Lasso | FALSE | FALSE | 0.000178 | HeldoutTest | | 24 | 24 |
| LV_radial_s | Lasso | FALSE | TRUE | 0.05 | HeldoutTest | | 24 | 9 |
| LV_radial_s | LinRegrs | FALSE | FALSE | | CV | 0 | 24 | 21 |
| LV_radial_s | LinRegrs | FALSE | FALSE | | CV | 1 | 24 | 21 |
| LV_radial_s | LinRegrs | FALSE | FALSE | | CV | 2 | 24 | 23 |
| LV_radial_s | LinRegrs | FALSE | FALSE | | CV | 3 | 24 | 21 |
| LV_radial_s | LinRegrs | FALSE | FALSE | | CV | 4 | 24 | 23 |
| LV_radial_s | Lasso | FALSE | FALSE | 0.000222 | CV | 0 | 24 | 24 |
| LV_radial_s | Lasso | FALSE | FALSE | 0.000188 | CV | 1 | 24 | 24 |
| LV_radial_s | Lasso | FALSE | FALSE | 0.000181 | CV | 2 | 24 | 24 |
| LV_radial_s | Lasso | FALSE | FALSE | 0.000179 | CV | 3 | 24 | 24 |
| LV_radial_s | Lasso | FALSE | FALSE | 0.000174 | CV | 4 | 24 | 24 |
| LV_radial_s | Lasso | FALSE | TRUE | 0.05 | CV | 0 | 24 | 9 |
| LV_radial_s | Lasso | FALSE | TRUE | 0.05 | CV | 1 | 24 | 8 |
| LV_radial_s | Lasso | FALSE | TRUE | 0.05 | CV | 2 | 24 | 8 |
| LV_radial_s | Lasso | FALSE | TRUE | 0.05 | CV | 3 | 24 | 9 |
| LV_radial_s | Lasso | FALSE | TRUE | 0.05 | CV | 4 | 24 | 9 |
| LV_radial_s | LinRegrs | FALSE | FALSE | | HeldoutTest | | 24 | 22 |
| LV_radial_s | Lasso | FALSE | FALSE | 0.000256 | HeldoutTest | | 24 | 24 |
| LV_radial_s | Lasso | FALSE | TRUE | 0.05 | HeldoutTest | | 24 | 12 |
| LV_radial_s | LinRegrs | FALSE | FALSE | | CV | 0 | 24 | 21 |
| LV_radial_s | LinRegrs | FALSE | FALSE | | CV | 1 | 24 | 22 |
| LV_radial_s | LinRegrs | FALSE | FALSE | | CV | 2 | 24 | 23 |
| LV_radial_s | LinRegrs | FALSE | FALSE | | CV | 3 | 24 | 22 |
| LV_radial_s | LinRegrs | FALSE | FALSE | | CV | 4 | 24 | 21 |
| LV_radial_s | Lasso | FALSE | FALSE | 0.000255 | CV | 0 | 24 | 24 |
| LV_radial_s | Lasso | FALSE | FALSE | 0.000258 | CV | 1 | 24 | 24 |
| LV_radial_s | Lasso | FALSE | FALSE | 0.000258 | CV | 2 | 24 | 24 |
| LV_radial_s | Lasso | FALSE | FALSE | 0.000255 | CV | 3 | 24 | 24 |
| LV_radial_s | Lasso | FALSE | FALSE | 0.000291 | CV | 4 | 24 | 24 |
| LV_radial_s | Lasso | FALSE | TRUE | 0.05 | CV | 0 | 24 | 12 |
| LV_radial_s | Lasso | FALSE | TRUE | 0.05 | CV | 1 | 24 | 12 |
| LV_radial_s | Lasso | FALSE | TRUE | 0.05 | CV | 2 | 24 | 12 |
| LV_radial_s | Lasso | FALSE | TRUE | 0.05 | CV | 3 | 24 | 12 |

| | | | | | | | | |
|---|---|---|---|---|---|---|---|---|
| LV_radial_s | Lasso | FALSE | TRUE | 0.05 | CV | 4 | 24 | 11 |
| LV_radial_s | LinRegrs | FALSE | FALSE | | HeldoutTest | | 24 | 21 |
| LV_radial_s | Lasso | FALSE | FALSE | 0.000552 | HeldoutTest | | 24 | 24 |
| LV_radial_s | Lasso | FALSE | TRUE | 0.05 | HeldoutTest | | 24 | 9 |
| LV_radial_s | LinRegrs | FALSE | FALSE | | CV | 0 | 24 | 21 |
| LV_radial_s | LinRegrs | FALSE | FALSE | | CV | 1 | 24 | 20 |
| LV_radial_s | LinRegrs | FALSE | FALSE | | CV | 2 | 24 | 21 |
| LV_radial_s | LinRegrs | FALSE | FALSE | | CV | 3 | 24 | 21 |
| LV_radial_s | LinRegrs | FALSE | FALSE | | CV | 4 | 24 | 19 |
| LV_radial_s | Lasso | FALSE | FALSE | 0.000779 | CV | 0 | 24 | 22 |
| LV_radial_s | Lasso | FALSE | FALSE | 0.000559 | CV | 1 | 24 | 24 |
| LV_radial_s | Lasso | FALSE | FALSE | 0.000544 | CV | 2 | 24 | 23 |
| LV_radial_s | Lasso | FALSE | FALSE | 0.000341 | CV | 3 | 24 | 24 |
| LV_radial_s | Lasso | FALSE | FALSE | 0.00073 | CV | 4 | 24 | 23 |
| LV_radial_s | Lasso | FALSE | TRUE | 0.05 | CV | 0 | 24 | 9 |
| LV_radial_s | Lasso | FALSE | TRUE | 0.05 | CV | 1 | 24 | 9 |
| LV_radial_s | Lasso | FALSE | TRUE | 0.05 | CV | 2 | 24 | 9 |
| LV_radial_s | Lasso | FALSE | TRUE | 0.05 | CV | 3 | 24 | 9 |
| LV_radial_s | Lasso | FALSE | TRUE | 0.05 | CV | 4 | 24 | 9 |
| LV_radial_s | LinRegrs | FALSE | FALSE | | HeldoutTest | | 24 | 22 |
| LV_radial_s | Lasso | FALSE | FALSE | 0.000309 | HeldoutTest | | 24 | 24 |
| LV_radial_s | Lasso | FALSE | TRUE | 0.05 | HeldoutTest | | 24 | 8 |
| LV_radial_s | LinRegrs | FALSE | FALSE | | CV | 0 | 24 | 21 |
| LV_radial_s | LinRegrs | FALSE | FALSE | | CV | 1 | 24 | 19 |
| LV_radial_s | LinRegrs | FALSE | FALSE | | CV | 2 | 24 | 21 |
| LV_radial_s | LinRegrs | FALSE | FALSE | | CV | 3 | 24 | 21 |
| LV_radial_s | LinRegrs | FALSE | FALSE | | CV | 4 | 24 | 20 |
| LV_radial_s | Lasso | FALSE | FALSE | 0.000713 | CV | 0 | 24 | 22 |
| LV_radial_s | Lasso | FALSE | FALSE | 0.000508 | CV | 1 | 24 | 24 |
| LV_radial_s | Lasso | FALSE | FALSE | 0.000493 | CV | 2 | 24 | 24 |
| LV_radial_s | Lasso | FALSE | FALSE | 0.000253 | CV | 3 | 24 | 24 |
| LV_radial_s | Lasso | FALSE | FALSE | 0.000829 | CV | 4 | 24 | 22 |
| LV_radial_s | Lasso | FALSE | TRUE | 0.05 | CV | 0 | 24 | 9 |
| LV_radial_s | Lasso | FALSE | TRUE | 0.05 | CV | 1 | 24 | 7 |
| LV_radial_s | Lasso | FALSE | TRUE | 0.05 | CV | 2 | 24 | 8 |
| LV_radial_s | Lasso | FALSE | TRUE | 0.05 | CV | 3 | 24 | 8 |
| LV_radial_s | Lasso | FALSE | TRUE | 0.05 | CV | 4 | 24 | 7 |
| LV_radial_s | LinRegrs | FALSE | FALSE | | HeldoutTest | | 24 | 21 |
| LV_radial_s | Lasso | FALSE | FALSE | 0.000509 | HeldoutTest | | 24 | 23 |
| LV_radial_s | Lasso | FALSE | TRUE | 0.05 | HeldoutTest | | 24 | 8 |
| LV_radial_s | LinRegrs | FALSE | FALSE | | CV | 0 | 24 | 22 |
| LV_radial_s | LinRegrs | FALSE | FALSE | | CV | 1 | 24 | 21 |
| LV_radial_s | LinRegrs | FALSE | FALSE | | CV | 2 | 24 | 20 |
| LV_radial_s | LinRegrs | FALSE | FALSE | | CV | 3 | 24 | 20 |
| LV_radial_s | LinRegrs | FALSE | FALSE | | CV | 4 | 24 | 23 |

| | | | | | | | | |
|---|---|---|---|---|---|---|---|---|
| LV_radial_ | Lasso | FALSE | FALSE | 0.000285 | CV | 0 | 24 | 24 |
| LV_radial_ | Lasso | FALSE | FALSE | 0.000576 | CV | 1 | 24 | 23 |
| LV_radial_ | Lasso | FALSE | FALSE | 0.000332 | CV | 2 | 24 | 24 |
| LV_radial_ | Lasso | FALSE | FALSE | 0.000599 | CV | 3 | 24 | 23 |
| LV_radial_ | Lasso | FALSE | FALSE | 0.001833 | CV | 4 | 24 | 20 |
| LV_radial_ | Lasso | FALSE | TRUE | 0.05 | CV | 0 | 24 | 8 |
| LV_radial_ | Lasso | FALSE | TRUE | 0.05 | CV | 1 | 24 | 8 |
| LV_radial_ | Lasso | FALSE | TRUE | 0.05 | CV | 2 | 24 | 8 |
| LV_radial_ | Lasso | FALSE | TRUE | 0.05 | CV | 3 | 24 | 7 |
| LV_radial_ | Lasso | FALSE | TRUE | 0.05 | CV | 4 | 24 | 8 |
| LV_radial_ | LinRegrs | FALSE | FALSE | | HeldoutTest | | 24 | 19 |
| LV_radial_ | Lasso | FALSE | FALSE | 0.000442 | HeldoutTest | | 24 | 23 |
| LV_radial_ | Lasso | FALSE | TRUE | 0.05 | HeldoutTest | | 24 | 9 |
| LV_radial_ | LinRegrs | FALSE | FALSE | | CV | 0 | 24 | 19 |
| LV_radial_ | LinRegrs | FALSE | FALSE | | CV | 1 | 24 | 19 |
| LV_radial_ | LinRegrs | FALSE | FALSE | | CV | 2 | 24 | 21 |
| LV_radial_ | LinRegrs | FALSE | FALSE | | CV | 3 | 24 | 20 |
| LV_radial_ | LinRegrs | FALSE | FALSE | | CV | 4 | 24 | 21 |
| LV_radial_ | Lasso | FALSE | FALSE | 0.0005 | CV | 0 | 24 | 23 |
| LV_radial_ | Lasso | FALSE | FALSE | 0.00031 | CV | 1 | 24 | 24 |
| LV_radial_ | Lasso | FALSE | FALSE | 0.000356 | CV | 2 | 24 | 24 |
| LV_radial_ | Lasso | FALSE | FALSE | 0.000472 | CV | 3 | 24 | 23 |
| LV_radial_ | Lasso | FALSE | FALSE | 0.00037 | CV | 4 | 24 | 24 |
| LV_radial_ | Lasso | FALSE | TRUE | 0.05 | CV | 0 | 24 | 8 |
| LV_radial_ | Lasso | FALSE | TRUE | 0.05 | CV | 1 | 24 | 10 |
| LV_radial_ | Lasso | FALSE | TRUE | 0.05 | CV | 2 | 24 | 8 |
| LV_radial_ | Lasso | FALSE | TRUE | 0.05 | CV | 3 | 24 | 9 |
| LV_radial_ | Lasso | FALSE | TRUE | 0.05 | CV | 4 | 24 | 8 |
| LV_radial_ | LinRegrs | FALSE | FALSE | | HeldoutTest | | 24 | 22 |
| LV_radial_ | Lasso | FALSE | FALSE | 0.000248 | HeldoutTest | | 24 | 24 |
| LV_radial_ | Lasso | FALSE | TRUE | 0.05 | HeldoutTest | | 24 | 5 |
| LV_radial_ | LinRegrs | FALSE | FALSE | | CV | 0 | 24 | 24 |
| LV_radial_ | LinRegrs | FALSE | FALSE | | CV | 1 | 24 | 22 |
| LV_radial_ | LinRegrs | FALSE | FALSE | | CV | 2 | 24 | 22 |
| LV_radial_ | LinRegrs | FALSE | FALSE | | CV | 3 | 24 | 22 |
| LV_radial_ | LinRegrs | FALSE | FALSE | | CV | 4 | 24 | 22 |
| LV_radial_ | Lasso | FALSE | FALSE | 0.000135 | CV | 0 | 24 | 24 |
| LV_radial_ | Lasso | FALSE | FALSE | 0.000699 | CV | 1 | 24 | 24 |
| LV_radial_ | Lasso | FALSE | FALSE | 0.000143 | CV | 2 | 24 | 24 |
| LV_radial_ | Lasso | FALSE | FALSE | 0.000451 | CV | 3 | 24 | 23 |
| LV_radial_ | Lasso | FALSE | FALSE | 0.000144 | CV | 4 | 24 | 24 |
| LV_radial_ | Lasso | FALSE | TRUE | 0.05 | CV | 0 | 24 | 6 |
| LV_radial_ | Lasso | FALSE | TRUE | 0.05 | CV | 1 | 24 | 5 |
| LV_radial_ | Lasso | FALSE | TRUE | 0.05 | CV | 2 | 24 | 5 |
| LV_radial_ | Lasso | FALSE | TRUE | 0.05 | CV | 3 | 24 | 4 |

| | | | | | | | | |
|---|---|---|---|---|---|---|---|---|
| LV_radial_s | Lasso | FALSE | TRUE | 0.05 | CV | 4 | 24 | 5 |
| LV_radial_s | LinRegrs | FALSE | FALSE | | HeldoutTest | | 24 | 22 |
| LV_radial_s | Lasso | FALSE | FALSE | 0.000236 | HeldoutTest | | 24 | 23 |
| LV_radial_s | Lasso | FALSE | TRUE | 0.05 | HeldoutTest | | 24 | 7 |
| LV_radial_s | LinRegrs | FALSE | FALSE | | CV | 0 | 24 | 22 |
| LV_radial_s | LinRegrs | FALSE | FALSE | | CV | 1 | 24 | 23 |
| LV_radial_s | LinRegrs | FALSE | FALSE | | CV | 2 | 24 | 22 |
| LV_radial_s | LinRegrs | FALSE | FALSE | | CV | 3 | 24 | 22 |
| LV_radial_s | LinRegrs | FALSE | FALSE | | CV | 4 | 24 | 23 |
| LV_radial_s | Lasso | FALSE | FALSE | 0.000162 | CV | 0 | 24 | 24 |
| LV_radial_s | Lasso | FALSE | FALSE | 0.000147 | CV | 1 | 24 | 24 |
| LV_radial_s | Lasso | FALSE | FALSE | 0.000331 | CV | 2 | 24 | 23 |
| LV_radial_s | Lasso | FALSE | FALSE | 0.000482 | CV | 3 | 24 | 22 |
| LV_radial_s | Lasso | FALSE | FALSE | 0.000145 | CV | 4 | 24 | 24 |
| LV_radial_s | Lasso | FALSE | TRUE | 0.05 | CV | 0 | 24 | 7 |
| LV_radial_s | Lasso | FALSE | TRUE | 0.05 | CV | 1 | 24 | 7 |
| LV_radial_s | Lasso | FALSE | TRUE | 0.05 | CV | 2 | 24 | 7 |
| LV_radial_s | Lasso | FALSE | TRUE | 0.05 | CV | 3 | 24 | 8 |
| LV_radial_s | Lasso | FALSE | TRUE | 0.05 | CV | 4 | 24 | 8 |
| LV_radial_s | LinRegrs | FALSE | FALSE | | HeldoutTest | | 24 | 22 |
| LV_radial_s | Lasso | FALSE | FALSE | 0.000235 | HeldoutTest | | 24 | 23 |
| LV_radial_s | Lasso | FALSE | TRUE | 0.05 | HeldoutTest | | 24 | 10 |
| LV_radial_s | LinRegrs | FALSE | FALSE | | CV | 0 | 24 | 21 |
| LV_radial_s | LinRegrs | FALSE | FALSE | | CV | 1 | 24 | 20 |
| LV_radial_s | LinRegrs | FALSE | FALSE | | CV | 2 | 24 | 22 |
| LV_radial_s | LinRegrs | FALSE | FALSE | | CV | 3 | 24 | 19 |
| LV_radial_s | LinRegrs | FALSE | FALSE | | CV | 4 | 24 | 21 |
| LV_radial_s | Lasso | FALSE | FALSE | 0.000464 | CV | 0 | 24 | 23 |
| LV_radial_s | Lasso | FALSE | FALSE | 0.00023 | CV | 1 | 24 | 23 |
| LV_radial_s | Lasso | FALSE | FALSE | 0.00038 | CV | 2 | 24 | 23 |
| LV_radial_s | Lasso | FALSE | FALSE | 0.000706 | CV | 3 | 24 | 23 |
| LV_radial_s | Lasso | FALSE | FALSE | 0.000248 | CV | 4 | 24 | 24 |
| LV_radial_s | Lasso | FALSE | TRUE | 0.05 | CV | 0 | 24 | 10 |
| LV_radial_s | Lasso | FALSE | TRUE | 0.05 | CV | 1 | 24 | 11 |
| LV_radial_s | Lasso | FALSE | TRUE | 0.05 | CV | 2 | 24 | 10 |
| LV_radial_s | Lasso | FALSE | TRUE | 0.05 | CV | 3 | 24 | 10 |
| LV_radial_s | Lasso | FALSE | TRUE | 0.05 | CV | 4 | 24 | 13 |
| LV_radial_s | LinRegrs | FALSE | FALSE | | HeldoutTest | | 24 | 23 |
| LV_radial_s | Lasso | FALSE | FALSE | 0.000147 | HeldoutTest | | 24 | 24 |
| LV_radial_s | Lasso | FALSE | TRUE | 0.05 | HeldoutTest | | 24 | 13 |
| LV_radial_s | LinRegrs | FALSE | FALSE | | CV | 0 | 24 | 23 |
| LV_radial_s | LinRegrs | FALSE | FALSE | | CV | 1 | 24 | 23 |
| LV_radial_s | LinRegrs | FALSE | FALSE | | CV | 2 | 24 | 21 |
| LV_radial_s | LinRegrs | FALSE | FALSE | | CV | 3 | 24 | 24 |
| LV_radial_s | LinRegrs | FALSE | FALSE | | CV | 4 | 24 | 23 |

| | | | | | | | | |
|---|---|---|---|---|---|---|---|---|
| LV_radial_s | Lasso | FALSE | FALSE | 0.000145 | CV | 0 | 24 | 24 |
| LV_radial_s | Lasso | FALSE | FALSE | 0.000149 | CV | 1 | 24 | 24 |
| LV_radial_s | Lasso | FALSE | FALSE | 0.0004 | CV | 2 | 24 | 24 |
| LV_radial_s | Lasso | FALSE | FALSE | 0.000149 | CV | 3 | 24 | 24 |
| LV_radial_s | Lasso | FALSE | FALSE | 0.000148 | CV | 4 | 24 | 24 |
| LV_radial_s | Lasso | FALSE | TRUE | 0.05 | CV | 0 | 24 | 13 |
| LV_radial_s | Lasso | FALSE | TRUE | 0.05 | CV | 1 | 24 | 14 |
| LV_radial_s | Lasso | FALSE | TRUE | 0.05 | CV | 2 | 24 | 13 |
| LV_radial_s | Lasso | FALSE | TRUE | 0.05 | CV | 3 | 24 | 13 |
| LV_radial_s | Lasso | FALSE | TRUE | 0.05 | CV | 4 | 24 | 11 |
| LV_radial_s | LinRegrs | FALSE | FALSE | | HeldoutTest | | 24 | 23 |
| LV_radial_s | Lasso | FALSE | FALSE | 0.00021 | HeldoutTest | | 24 | 23 |
| LV_radial_s | Lasso | FALSE | TRUE | 0.05 | HeldoutTest | | 24 | 13 |
| LV_radial_s | LinRegrs | FALSE | FALSE | | CV | 0 | 24 | 21 |
| LV_radial_s | LinRegrs | FALSE | FALSE | | CV | 1 | 24 | 23 |
| LV_radial_s | LinRegrs | FALSE | FALSE | | CV | 2 | 24 | 22 |
| LV_radial_s | LinRegrs | FALSE | FALSE | | CV | 3 | 24 | 23 |
| LV_radial_s | LinRegrs | FALSE | FALSE | | CV | 4 | 24 | 24 |
| LV_radial_s | Lasso | FALSE | FALSE | 0.000744 | CV | 0 | 24 | 24 |
| LV_radial_s | Lasso | FALSE | FALSE | 0.000299 | CV | 1 | 24 | 24 |
| LV_radial_s | Lasso | FALSE | FALSE | 0.001367 | CV | 2 | 24 | 21 |
| LV_radial_s | Lasso | FALSE | FALSE | 0.000146 | CV | 3 | 24 | 24 |
| LV_radial_s | Lasso | FALSE | FALSE | 0.000336 | CV | 4 | 24 | 24 |
| LV_radial_s | Lasso | FALSE | TRUE | 0.05 | CV | 0 | 24 | 13 |
| LV_radial_s | Lasso | FALSE | TRUE | 0.05 | CV | 1 | 24 | 13 |
| LV_radial_s | Lasso | FALSE | TRUE | 0.05 | CV | 2 | 24 | 13 |
| LV_radial_s | Lasso | FALSE | TRUE | 0.05 | CV | 3 | 24 | 13 |
| LV_radial_s | Lasso | FALSE | TRUE | 0.05 | CV | 4 | 24 | 11 |
| LV_radial_s | LinRegrs | FALSE | FALSE | | HeldoutTest | | 24 | 21 |
| LV_radial_s | Lasso | FALSE | FALSE | 0.000277 | HeldoutTest | | 24 | 24 |
| LV_radial_s | Lasso | FALSE | TRUE | 0.05 | HeldoutTest | | 24 | 4 |
| LV_radial_s | LinRegrs | FALSE | FALSE | | CV | 0 | 24 | 19 |
| LV_radial_s | LinRegrs | FALSE | FALSE | | CV | 1 | 24 | 21 |
| LV_radial_s | LinRegrs | FALSE | FALSE | | CV | 2 | 24 | 21 |
| LV_radial_s | LinRegrs | FALSE | FALSE | | CV | 3 | 24 | 21 |
| LV_radial_s | LinRegrs | FALSE | FALSE | | CV | 4 | 24 | 21 |
| LV_radial_s | Lasso | FALSE | FALSE | 0.000172 | CV | 0 | 24 | 23 |
| LV_radial_s | Lasso | FALSE | FALSE | 0.000123 | CV | 1 | 24 | 24 |
| LV_radial_s | Lasso | FALSE | FALSE | 0.000278 | CV | 2 | 24 | 24 |
| LV_radial_s | Lasso | FALSE | FALSE | 0.000344 | CV | 3 | 24 | 23 |
| LV_radial_s | Lasso | FALSE | FALSE | 0.000116 | CV | 4 | 24 | 24 |
| LV_radial_s | Lasso | FALSE | TRUE | 0.05 | CV | 0 | 24 | 3 |
| LV_radial_s | Lasso | FALSE | TRUE | 0.05 | CV | 1 | 24 | 4 |
| LV_radial_s | Lasso | FALSE | TRUE | 0.05 | CV | 2 | 24 | 3 |
| LV_radial_s | Lasso | FALSE | TRUE | 0.05 | CV | 3 | 24 | 4 |

| | | | | | | | | |
|---|---|---|---|---|---|---|---|---|
| LV_radial_s | Lasso | FALSE | TRUE | 0.05 | CV | 4 | 24 | 6 |
| LV_radial_s | LinRegrs | FALSE | FALSE | | HeldoutTest | | 24 | 18 |
| LV_radial_s | Lasso | FALSE | FALSE | 0.000687 | HeldoutTest | | 24 | 23 |
| LV_radial_s | Lasso | FALSE | TRUE | 0.05 | HeldoutTest | | 24 | 7 |
| LV_radial_s | LinRegrs | FALSE | FALSE | | CV | 0 | 24 | 20 |
| LV_radial_s | LinRegrs | FALSE | FALSE | | CV | 1 | 24 | 19 |
| LV_radial_s | LinRegrs | FALSE | FALSE | | CV | 2 | 24 | 20 |
| LV_radial_s | LinRegrs | FALSE | FALSE | | CV | 3 | 24 | 20 |
| LV_radial_s | LinRegrs | FALSE | FALSE | | CV | 4 | 24 | 21 |
| LV_radial_s | Lasso | FALSE | FALSE | 0.00053 | CV | 0 | 24 | 23 |
| LV_radial_s | Lasso | FALSE | FALSE | 0.001039 | CV | 1 | 24 | 22 |
| LV_radial_s | Lasso | FALSE | FALSE | 0.001124 | CV | 2 | 24 | 23 |
| LV_radial_s | Lasso | FALSE | FALSE | 0.000615 | CV | 3 | 24 | 22 |
| LV_radial_s | Lasso | FALSE | FALSE | 0.000496 | CV | 4 | 24 | 23 |
| LV_radial_s | Lasso | FALSE | TRUE | 0.05 | CV | 0 | 24 | 7 |
| LV_radial_s | Lasso | FALSE | TRUE | 0.05 | CV | 1 | 24 | 6 |
| LV_radial_s | Lasso | FALSE | TRUE | 0.05 | CV | 2 | 24 | 7 |
| LV_radial_s | Lasso | FALSE | TRUE | 0.05 | CV | 3 | 24 | 6 |
| LV_radial_s | Lasso | FALSE | TRUE | 0.05 | CV | 4 | 24 | 8 |
| LV_radial_s | LinRegrs | FALSE | FALSE | | HeldoutTest | | 24 | 22 |
| LV_radial_s | Lasso | FALSE | FALSE | 0.000477 | HeldoutTest | | 24 | 23 |
| LV_radial_s | Lasso | FALSE | TRUE | 0.05 | HeldoutTest | | 24 | 11 |
| LV_radial_s | LinRegrs | FALSE | FALSE | | CV | 0 | 24 | 22 |
| LV_radial_s | LinRegrs | FALSE | FALSE | | CV | 1 | 24 | 22 |
| LV_radial_s | LinRegrs | FALSE | FALSE | | CV | 2 | 24 | 22 |
| LV_radial_s | LinRegrs | FALSE | FALSE | | CV | 3 | 24 | 22 |
| LV_radial_s | LinRegrs | FALSE | FALSE | | CV | 4 | 24 | 21 |
| LV_radial_s | Lasso | FALSE | FALSE | 0.000109 | CV | 0 | 24 | 24 |
| LV_radial_s | Lasso | FALSE | FALSE | 0.000629 | CV | 1 | 24 | 23 |
| LV_radial_s | Lasso | FALSE | FALSE | 0.000138 | CV | 2 | 24 | 24 |
| LV_radial_s | Lasso | FALSE | FALSE | 0.000507 | CV | 3 | 24 | 24 |
| LV_radial_s | Lasso | FALSE | FALSE | 0.00043 | CV | 4 | 24 | 24 |
| LV_radial_s | Lasso | FALSE | TRUE | 0.05 | CV | 0 | 24 | 9 |
| LV_radial_s | Lasso | FALSE | TRUE | 0.05 | CV | 1 | 24 | 10 |
| LV_radial_s | Lasso | FALSE | TRUE | 0.05 | CV | 2 | 24 | 11 |
| LV_radial_s | Lasso | FALSE | TRUE | 0.05 | CV | 3 | 24 | 9 |
| LV_radial_s | Lasso | FALSE | TRUE | 0.05 | CV | 4 | 24 | 9 |
| LV_radial_s | LinRegrs | FALSE | FALSE | | HeldoutTest | | 24 | 21 |
| LV_radial_s | Lasso | FALSE | FALSE | 0.000239 | HeldoutTest | | 24 | 24 |
| LV_radial_s | Lasso | FALSE | TRUE | 0.05 | HeldoutTest | | 24 | 6 |
| LV_radial_s | LinRegrs | FALSE | FALSE | | CV | 0 | 24 | 22 |
| LV_radial_s | LinRegrs | FALSE | FALSE | | CV | 1 | 24 | 21 |
| LV_radial_s | LinRegrs | FALSE | FALSE | | CV | 2 | 24 | 19 |
| LV_radial_s | LinRegrs | FALSE | FALSE | | CV | 3 | 24 | 20 |
| LV_radial_s | LinRegrs | FALSE | FALSE | | CV | 4 | 24 | 22 |

| | | | | | | | | |
|---|---|---|---|---|---|---|---|---|
| LV_radial_s | Lasso | FALSE | FALSE | 0.000181 | CV | 0 | 24 | 24 |
| LV_radial_s | Lasso | FALSE | FALSE | 0.000613 | CV | 1 | 24 | 23 |
| LV_radial_s | Lasso | FALSE | FALSE | 0.000964 | CV | 2 | 24 | 22 |
| LV_radial_s | Lasso | FALSE | FALSE | 0.000265 | CV | 3 | 24 | 24 |
| LV_radial_s | Lasso | FALSE | FALSE | 0.000193 | CV | 4 | 24 | 24 |
| LV_radial_s | Lasso | FALSE | TRUE | 0.05 | CV | 0 | 24 | 6 |
| LV_radial_s | Lasso | FALSE | TRUE | 0.05 | CV | 1 | 24 | 6 |
| LV_radial_s | Lasso | FALSE | TRUE | 0.05 | CV | 2 | 24 | 6 |
| LV_radial_s | Lasso | FALSE | TRUE | 0.05 | CV | 3 | 24 | 8 |
| LV_radial_s | Lasso | FALSE | TRUE | 0.05 | CV | 4 | 24 | 5 |
| LV_radial_s | LinRegrs | FALSE | FALSE | | HeldoutTest | | 24 | 23 |
| LV_radial_s | Lasso | FALSE | FALSE | 0.000198 | HeldoutTest | | 24 | 24 |
| LV_radial_s | Lasso | FALSE | TRUE | 0.05 | HeldoutTest | | 24 | 12 |
| LV_radial_s | LinRegrs | FALSE | FALSE | | CV | 0 | 24 | 22 |
| LV_radial_s | LinRegrs | FALSE | FALSE | | CV | 1 | 24 | 23 |
| LV_radial_s | LinRegrs | FALSE | FALSE | | CV | 2 | 24 | 23 |
| LV_radial_s | LinRegrs | FALSE | FALSE | | CV | 3 | 24 | 22 |
| LV_radial_s | LinRegrs | FALSE | FALSE | | CV | 4 | 24 | 22 |
| LV_radial_s | Lasso | FALSE | FALSE | 0.000191 | CV | 0 | 24 | 24 |
| LV_radial_s | Lasso | FALSE | FALSE | 0.000199 | CV | 1 | 24 | 24 |
| LV_radial_s | Lasso | FALSE | FALSE | 0.000197 | CV | 2 | 24 | 24 |
| LV_radial_s | Lasso | FALSE | FALSE | 0.000433 | CV | 3 | 24 | 23 |
| LV_radial_s | Lasso | FALSE | FALSE | 0.000203 | CV | 4 | 24 | 24 |
| LV_radial_s | Lasso | FALSE | TRUE | 0.05 | CV | 0 | 24 | 12 |
| LV_radial_s | Lasso | FALSE | TRUE | 0.05 | CV | 1 | 24 | 13 |
| LV_radial_s | Lasso | FALSE | TRUE | 0.05 | CV | 2 | 24 | 12 |
| LV_radial_s | Lasso | FALSE | TRUE | 0.05 | CV | 3 | 24 | 12 |
| LV_radial_s | Lasso | FALSE | TRUE | 0.05 | CV | 4 | 24 | 14 |
| LA_maximu | LinRegrs | FALSE | FALSE | | HeldoutTest | | 24 | 23 |
| LA_maximu | Lasso | FALSE | FALSE | 0.000387 | HeldoutTest | | 24 | 24 |
| LA_maximu | Lasso | FALSE | TRUE | 0.05 | HeldoutTest | | 24 | 11 |
| LA_maximu | LinRegrs | FALSE | FALSE | | CV | 0 | 24 | 23 |
| LA_maximu | LinRegrs | FALSE | FALSE | | CV | 1 | 24 | 23 |
| LA_maximu | LinRegrs | FALSE | FALSE | | CV | 2 | 24 | 23 |
| LA_maximu | LinRegrs | FALSE | FALSE | | CV | 3 | 24 | 22 |
| LA_maximu | LinRegrs | FALSE | FALSE | | CV | 4 | 24 | 22 |
| LA_maximu | Lasso | FALSE | FALSE | 0.000334 | CV | 0 | 24 | 24 |
| LA_maximu | Lasso | FALSE | FALSE | 0.000394 | CV | 1 | 24 | 24 |
| LA_maximu | Lasso | FALSE | FALSE | 0.000335 | CV | 2 | 24 | 24 |
| LA_maximu | Lasso | FALSE | FALSE | 0.000587 | CV | 3 | 24 | 22 |
| LA_maximu | Lasso | FALSE | FALSE | 0.000338 | CV | 4 | 24 | 24 |
| LA_maximu | Lasso | FALSE | TRUE | 0.05 | CV | 0 | 24 | 11 |
| LA_maximu | Lasso | FALSE | TRUE | 0.05 | CV | 1 | 24 | 11 |
| LA_maximu | Lasso | FALSE | TRUE | 0.05 | CV | 2 | 24 | 11 |
| LA_maximu | Lasso | FALSE | TRUE | 0.05 | CV | 3 | 24 | 11 |

| | | | | | | | | |
|---|---|---|---|---|---|---|---|---|
| LA_maximu | Lasso | FALSE | TRUE | 0.05 | CV | 4 | 24 | 11 |
| LA_minimu | LinRegrs | FALSE | FALSE | | HeldoutTest | | 24 | 19 |
| LA_minimu | Lasso | FALSE | FALSE | 0.000755 | HeldoutTest | | 24 | 22 |
| LA_minimu | Lasso | FALSE | TRUE | 0.05 | HeldoutTest | | 24 | 13 |
| LA_minimu | LinRegrs | FALSE | FALSE | | CV | 0 | 24 | 20 |
| LA_minimu | LinRegrs | FALSE | FALSE | | CV | 1 | 24 | 19 |
| LA_minimu | LinRegrs | FALSE | FALSE | | CV | 2 | 24 | 21 |
| LA_minimu | LinRegrs | FALSE | FALSE | | CV | 3 | 24 | 20 |
| LA_minimu | LinRegrs | FALSE | FALSE | | CV | 4 | 24 | 22 |
| LA_minimu | Lasso | FALSE | FALSE | 0.000752 | CV | 0 | 24 | 21 |
| LA_minimu | Lasso | FALSE | FALSE | 0.000872 | CV | 1 | 24 | 22 |
| LA_minimu | Lasso | FALSE | FALSE | 0.00081 | CV | 2 | 24 | 24 |
| LA_minimu | Lasso | FALSE | FALSE | 0.000816 | CV | 3 | 24 | 23 |
| LA_minimu | Lasso | FALSE | FALSE | 0.000302 | CV | 4 | 24 | 24 |
| LA_minimu | Lasso | FALSE | TRUE | 0.05 | CV | 0 | 24 | 13 |
| LA_minimu | Lasso | FALSE | TRUE | 0.05 | CV | 1 | 24 | 14 |
| LA_minimu | Lasso | FALSE | TRUE | 0.05 | CV | 2 | 24 | 13 |
| LA_minimu | Lasso | FALSE | TRUE | 0.05 | CV | 3 | 24 | 12 |
| LA_minimu | Lasso | FALSE | TRUE | 0.05 | CV | 4 | 24 | 11 |
| LA_stroke_ | LinRegrs | FALSE | FALSE | | HeldoutTest | | 24 | 21 |
| LA_stroke_ | Lasso | FALSE | FALSE | 0.000346 | HeldoutTest | | 24 | 23 |
| LA_stroke_ | Lasso | FALSE | TRUE | 0.05 | HeldoutTest | | 24 | 10 |
| LA_stroke_ | LinRegrs | FALSE | FALSE | | CV | 0 | 24 | 22 |
| LA_stroke_ | LinRegrs | FALSE | FALSE | | CV | 1 | 24 | 21 |
| LA_stroke_ | LinRegrs | FALSE | FALSE | | CV | 2 | 24 | 22 |
| LA_stroke_ | LinRegrs | FALSE | FALSE | | CV | 3 | 24 | 22 |
| LA_stroke_ | LinRegrs | FALSE | FALSE | | CV | 4 | 24 | 23 |
| LA_stroke_ | Lasso | FALSE | FALSE | 0.000346 | CV | 0 | 24 | 24 |
| LA_stroke_ | Lasso | FALSE | FALSE | 0.000352 | CV | 1 | 24 | 24 |
| LA_stroke_ | Lasso | FALSE | FALSE | 0.000369 | CV | 2 | 24 | 24 |
| LA_stroke_ | Lasso | FALSE | FALSE | 0.00045 | CV | 3 | 24 | 24 |
| LA_stroke_ | Lasso | FALSE | FALSE | 0.000349 | CV | 4 | 24 | 23 |
| LA_stroke_ | Lasso | FALSE | TRUE | 0.05 | CV | 0 | 24 | 10 |
| LA_stroke_ | Lasso | FALSE | TRUE | 0.05 | CV | 1 | 24 | 10 |
| LA_stroke_ | Lasso | FALSE | TRUE | 0.05 | CV | 2 | 24 | 10 |
| LA_stroke_ | Lasso | FALSE | TRUE | 0.05 | CV | 3 | 24 | 10 |
| LA_stroke_ | Lasso | FALSE | TRUE | 0.05 | CV | 4 | 24 | 10 |
| LA_ejectior | LinRegrs | FALSE | FALSE | | HeldoutTest | | 24 | 21 |
| LA_ejectior | Lasso | FALSE | FALSE | 0.000194 | HeldoutTest | | 24 | 24 |
| LA_ejectior | Lasso | FALSE | TRUE | 0.05 | HeldoutTest | | 24 | 10 |
| LA_ejectior | LinRegrs | FALSE | FALSE | | CV | 0 | 24 | 22 |
| LA_ejectior | LinRegrs | FALSE | FALSE | | CV | 1 | 24 | 22 |
| LA_ejectior | LinRegrs | FALSE | FALSE | | CV | 2 | 24 | 21 |
| LA_ejectior | LinRegrs | FALSE | FALSE | | CV | 3 | 24 | 23 |
| LA_ejectior | LinRegrs | FALSE | FALSE | | CV | 4 | 24 | 21 |

| | | | | | | | | |
|---|---|---|---|---|---|---|---|---|
| LA_ejectior | Lasso | FALSE | FALSE | 0.000182 | CV | 0 | 24 | 24 |
| LA_ejectior | Lasso | FALSE | FALSE | 0.000337 | CV | 1 | 24 | 23 |
| LA_ejectior | Lasso | FALSE | FALSE | 0.000481 | CV | 2 | 24 | 24 |
| LA_ejectior | Lasso | FALSE | FALSE | 0.00069 | CV | 3 | 24 | 23 |
| LA_ejectior | Lasso | FALSE | FALSE | 0.000726 | CV | 4 | 24 | 22 |
| LA_ejectior | Lasso | FALSE | TRUE | 0.05 | CV | 0 | 24 | 11 |
| LA_ejectior | Lasso | FALSE | TRUE | 0.05 | CV | 1 | 24 | 10 |
| LA_ejectior | Lasso | FALSE | TRUE | 0.05 | CV | 2 | 24 | 10 |
| LA_ejectior | Lasso | FALSE | TRUE | 0.05 | CV | 3 | 24 | 10 |
| LA_ejectior | Lasso | FALSE | TRUE | 0.05 | CV | 4 | 24 | 10 |
| LV_end_di | LinRegrs | FALSE | FALSE | | HeldoutTest | | 24 | 23 |
| LV_end_di | Lasso | FALSE | FALSE | 0.000419 | HeldoutTest | | 24 | 24 |
| LV_end_di | Lasso | FALSE | TRUE | 0.05 | HeldoutTest | | 24 | 16 |
| LV_end_di | LinRegrs | FALSE | FALSE | | CV | 0 | 24 | 23 |
| LV_end_di | LinRegrs | FALSE | FALSE | | CV | 1 | 24 | 23 |
| LV_end_di | LinRegrs | FALSE | FALSE | | CV | 2 | 24 | 23 |
| LV_end_di | LinRegrs | FALSE | FALSE | | CV | 3 | 24 | 23 |
| LV_end_di | LinRegrs | FALSE | FALSE | | CV | 4 | 24 | 24 |
| LV_end_di | Lasso | FALSE | FALSE | 0.000454 | CV | 0 | 24 | 24 |
| LV_end_di | Lasso | FALSE | FALSE | 0.00042 | CV | 1 | 24 | 23 |
| LV_end_di | Lasso | FALSE | FALSE | 0.000417 | CV | 2 | 24 | 24 |
| LV_end_di | Lasso | FALSE | FALSE | 0.000418 | CV | 3 | 24 | 24 |
| LV_end_di | Lasso | FALSE | FALSE | 0.000418 | CV | 4 | 24 | 24 |
| LV_end_di | Lasso | FALSE | TRUE | 0.05 | CV | 0 | 24 | 16 |
| LV_end_di | Lasso | FALSE | TRUE | 0.05 | CV | 1 | 24 | 16 |
| LV_end_di | Lasso | FALSE | TRUE | 0.05 | CV | 2 | 24 | 15 |
| LV_end_di | Lasso | FALSE | TRUE | 0.05 | CV | 3 | 24 | 16 |
| LV_end_di | Lasso | FALSE | TRUE | 0.05 | CV | 4 | 24 | 13 |
| LV_end_sy | LinRegrs | FALSE | FALSE | | HeldoutTest | | 24 | 24 |
| LV_end_sy | Lasso | FALSE | FALSE | 0.000371 | HeldoutTest | | 24 | 24 |
| LV_end_sy | Lasso | FALSE | TRUE | 0.05 | HeldoutTest | | 24 | 15 |
| LV_end_sy | LinRegrs | FALSE | FALSE | | CV | 0 | 24 | 23 |
| LV_end_sy | LinRegrs | FALSE | FALSE | | CV | 1 | 24 | 24 |
| LV_end_sy | LinRegrs | FALSE | FALSE | | CV | 2 | 24 | 24 |
| LV_end_sy | LinRegrs | FALSE | FALSE | | CV | 3 | 24 | 24 |
| LV_end_sy | LinRegrs | FALSE | FALSE | | CV | 4 | 24 | 24 |
| LV_end_sy | Lasso | FALSE | FALSE | 0.000377 | CV | 0 | 24 | 24 |
| LV_end_sy | Lasso | FALSE | FALSE | 0.000372 | CV | 1 | 24 | 24 |
| LV_end_sy | Lasso | FALSE | FALSE | 0.000368 | CV | 2 | 24 | 24 |
| LV_end_sy | Lasso | FALSE | FALSE | 0.000369 | CV | 3 | 24 | 24 |
| LV_end_sy | Lasso | FALSE | FALSE | 0.000371 | CV | 4 | 24 | 24 |
| LV_end_sy | Lasso | FALSE | TRUE | 0.05 | CV | 0 | 24 | 16 |
| LV_end_sy | Lasso | FALSE | TRUE | 0.05 | CV | 1 | 24 | 15 |
| LV_end_sy | Lasso | FALSE | TRUE | 0.05 | CV | 2 | 24 | 14 |
| LV_end_sy | Lasso | FALSE | TRUE | 0.05 | CV | 3 | 24 | 16 |

| | | | | | | | | |
|---|---|---|---|---|---|---|---|---|
| LV_end_sy | Lasso | FALSE | TRUE | 0.05 | CV | 4 | 24 | 15 |
| LV_stroke_ | LinRegrs | FALSE | FALSE | | HeldoutTest | | 24 | 24 |
| LV_stroke_ | Lasso | FALSE | FALSE | 0.000369 | HeldoutTest | | 24 | 24 |
| LV_stroke_ | Lasso | FALSE | TRUE | 0.05 | HeldoutTest | | 24 | 12 |
| LV_stroke_ | LinRegrs | FALSE | FALSE | | CV | 0 | 24 | 22 |
| LV_stroke_ | LinRegrs | FALSE | FALSE | | CV | 1 | 24 | 23 |
| LV_stroke_ | LinRegrs | FALSE | FALSE | | CV | 2 | 24 | 23 |
| LV_stroke_ | LinRegrs | FALSE | FALSE | | CV | 3 | 24 | 23 |
| LV_stroke_ | LinRegrs | FALSE | FALSE | | CV | 4 | 24 | 24 |
| LV_stroke_ | Lasso | FALSE | FALSE | 0.000372 | CV | 0 | 24 | 23 |
| LV_stroke_ | Lasso | FALSE | FALSE | 0.00037 | CV | 1 | 24 | 24 |
| LV_stroke_ | Lasso | FALSE | FALSE | 0.000368 | CV | 2 | 24 | 24 |
| LV_stroke_ | Lasso | FALSE | FALSE | 0.000368 | CV | 3 | 24 | 24 |
| LV_stroke_ | Lasso | FALSE | FALSE | 0.000367 | CV | 4 | 24 | 24 |
| LV_stroke_ | Lasso | FALSE | TRUE | 0.05 | CV | 0 | 24 | 12 |
| LV_stroke_ | Lasso | FALSE | TRUE | 0.05 | CV | 1 | 24 | 12 |
| LV_stroke_ | Lasso | FALSE | TRUE | 0.05 | CV | 2 | 24 | 12 |
| LV_stroke_ | Lasso | FALSE | TRUE | 0.05 | CV | 3 | 24 | 12 |
| LV_stroke_ | Lasso | FALSE | TRUE | 0.05 | CV | 4 | 24 | 11 |
| LV_ejectior | LinRegrs | FALSE | FALSE | | HeldoutTest | | 24 | 23 |
| LV_ejectior | Lasso | FALSE | FALSE | 0.000153 | HeldoutTest | | 24 | 24 |
| LV_ejectior | Lasso | FALSE | TRUE | 0.05 | HeldoutTest | | 24 | 12 |
| LV_ejectior | LinRegrs | FALSE | FALSE | | CV | 0 | 24 | 23 |
| LV_ejectior | LinRegrs | FALSE | FALSE | | CV | 1 | 24 | 23 |
| LV_ejectior | LinRegrs | FALSE | FALSE | | CV | 2 | 24 | 23 |
| LV_ejectior | LinRegrs | FALSE | FALSE | | CV | 3 | 24 | 23 |
| LV_ejectior | LinRegrs | FALSE | FALSE | | CV | 4 | 24 | 23 |
| LV_ejectior | Lasso | FALSE | FALSE | 0.000146 | CV | 0 | 24 | 24 |
| LV_ejectior | Lasso | FALSE | FALSE | 0.000144 | CV | 1 | 24 | 24 |
| LV_ejectior | Lasso | FALSE | FALSE | 0.000142 | CV | 2 | 24 | 24 |
| LV_ejectior | Lasso | FALSE | FALSE | 0.000204 | CV | 3 | 24 | 24 |
| LV_ejectior | Lasso | FALSE | FALSE | 0.000146 | CV | 4 | 24 | 24 |
| LV_ejectior | Lasso | FALSE | TRUE | 0.05 | CV | 0 | 24 | 12 |
| LV_ejectior | Lasso | FALSE | TRUE | 0.05 | CV | 1 | 24 | 12 |
| LV_ejectior | Lasso | FALSE | TRUE | 0.05 | CV | 2 | 24 | 11 |
| LV_ejectior | Lasso | FALSE | TRUE | 0.05 | CV | 3 | 24 | 11 |
| LV_ejectior | Lasso | FALSE | TRUE | 0.05 | CV | 4 | 24 | 13 |
| LV_cardiac | LinRegrs | FALSE | FALSE | | HeldoutTest | | 24 | 24 |
| LV_cardiac | Lasso | FALSE | FALSE | 0.000215 | HeldoutTest | | 24 | 24 |
| LV_cardiac | Lasso | FALSE | TRUE | 0.05 | HeldoutTest | | 24 | 15 |
| LV_cardiac | LinRegrs | FALSE | FALSE | | CV | 0 | 24 | 24 |
| LV_cardiac | LinRegrs | FALSE | FALSE | | CV | 1 | 24 | 24 |
| LV_cardiac | LinRegrs | FALSE | FALSE | | CV | 2 | 24 | 24 |
| LV_cardiac | LinRegrs | FALSE | FALSE | | CV | 3 | 24 | 23 |
| LV_cardiac | LinRegrs | FALSE | FALSE | | CV | 4 | 24 | 23 |

| | | | | | | | | |
|---|---|---|---|---|---|---|---|---|
| LV_cardiac | Lasso | FALSE | FALSE | 0.000218 | CV | 0 | 24 | 24 |
| LV_cardiac | Lasso | FALSE | FALSE | 0.000214 | CV | 1 | 24 | 24 |
| LV_cardiac | Lasso | FALSE | FALSE | 0.000214 | CV | 2 | 24 | 24 |
| LV_cardiac | Lasso | FALSE | FALSE | 0.000326 | CV | 3 | 24 | 24 |
| LV_cardiac | Lasso | FALSE | FALSE | 0.000214 | CV | 4 | 24 | 24 |
| LV_cardiac | Lasso | FALSE | TRUE | 0.05 | CV | 0 | 24 | 15 |
| LV_cardiac | Lasso | FALSE | TRUE | 0.05 | CV | 1 | 24 | 15 |
| LV_cardiac | Lasso | FALSE | TRUE | 0.05 | CV | 2 | 24 | 13 |
| LV_cardiac | Lasso | FALSE | TRUE | 0.05 | CV | 3 | 24 | 15 |
| LV_cardiac | Lasso | FALSE | TRUE | 0.05 | CV | 4 | 24 | 13 |
| RA_maxim | LinRegrs | TRUE | FALSE | | HeldoutTest | | 24 | 22 |
| RA_maxim | Lasso | TRUE | FALSE | 0.000474 | HeldoutTest | | 24 | 24 |
| RA_maxim | Lasso | TRUE | TRUE | 0.05 | HeldoutTest | | 24 | 12 |
| RA_maxim | LinRegrs | TRUE | FALSE | | CV | 0 | 24 | 19 |
| RA_maxim | LinRegrs | TRUE | FALSE | | CV | 1 | 24 | 20 |
| RA_maxim | LinRegrs | TRUE | FALSE | | CV | 2 | 24 | 21 |
| RA_maxim | LinRegrs | TRUE | FALSE | | CV | 3 | 24 | 22 |
| RA_maxim | LinRegrs | TRUE | FALSE | | CV | 4 | 24 | 21 |
| RA_maxim | Lasso | TRUE | FALSE | 0.000444 | CV | 0 | 24 | 23 |
| RA_maxim | Lasso | TRUE | FALSE | 0.000469 | CV | 1 | 24 | 24 |
| RA_maxim | Lasso | TRUE | FALSE | 0.000623 | CV | 2 | 24 | 23 |
| RA_maxim | Lasso | TRUE | FALSE | 0.000551 | CV | 3 | 24 | 23 |
| RA_maxim | Lasso | TRUE | FALSE | 0.000411 | CV | 4 | 24 | 24 |
| RA_maxim | Lasso | TRUE | TRUE | 0.05 | CV | 0 | 24 | 12 |
| RA_maxim | Lasso | TRUE | TRUE | 0.05 | CV | 1 | 24 | 12 |
| RA_maxim | Lasso | TRUE | TRUE | 0.05 | CV | 2 | 24 | 12 |
| RA_maxim | Lasso | TRUE | TRUE | 0.05 | CV | 3 | 24 | 12 |
| RA_maxim | Lasso | TRUE | TRUE | 0.05 | CV | 4 | 24 | 11 |
| RA_minimu | LinRegrs | TRUE | FALSE | | HeldoutTest | | 24 | 24 |
| RA_minimu | Lasso | TRUE | FALSE | 0.000288 | HeldoutTest | | 24 | 24 |
| RA_minimu | Lasso | TRUE | TRUE | 0.05 | HeldoutTest | | 24 | 13 |
| RA_minimu | LinRegrs | TRUE | FALSE | | CV | 0 | 24 | 23 |
| RA_minimu | LinRegrs | TRUE | FALSE | | CV | 1 | 24 | 23 |
| RA_minimu | LinRegrs | TRUE | FALSE | | CV | 2 | 24 | 24 |
| RA_minimu | LinRegrs | TRUE | FALSE | | CV | 3 | 24 | 23 |
| RA_minimu | LinRegrs | TRUE | FALSE | | CV | 4 | 24 | 24 |
| RA_minimu | Lasso | TRUE | FALSE | 0.00031 | CV | 0 | 24 | 24 |
| RA_minimu | Lasso | TRUE | FALSE | 0.00023 | CV | 1 | 24 | 24 |
| RA_minimu | Lasso | TRUE | FALSE | 0.000818 | CV | 2 | 24 | 24 |
| RA_minimu | Lasso | TRUE | FALSE | 0.000251 | CV | 3 | 24 | 24 |
| RA_minimu | Lasso | TRUE | FALSE | 0.000236 | CV | 4 | 24 | 24 |
| RA_minimu | Lasso | TRUE | TRUE | 0.05 | CV | 0 | 24 | 13 |
| RA_minimu | Lasso | TRUE | TRUE | 0.05 | CV | 1 | 24 | 13 |
| RA_minimu | Lasso | TRUE | TRUE | 0.05 | CV | 2 | 24 | 13 |
| RA_minimu | Lasso | TRUE | TRUE | 0.05 | CV | 3 | 24 | 13 |

| | | | | | | | | |
|---|---|---|---|---|---|---|---|---|
| RA_minimu | Lasso | TRUE | TRUE | 0.05 | CV | 4 | 24 | 12 |
| RA_stroke_ | LinRegrs | TRUE | FALSE | | HeldoutTest | | 24 | 23 |
| RA_stroke_ | Lasso | TRUE | FALSE | 0.000213 | HeldoutTest | | 24 | 23 |
| RA_stroke_ | Lasso | TRUE | TRUE | 0.05 | HeldoutTest | | 24 | 8 |
| RA_stroke_ | LinRegrs | TRUE | FALSE | | CV | 0 | 24 | 23 |
| RA_stroke_ | LinRegrs | TRUE | FALSE | | CV | 1 | 24 | 23 |
| RA_stroke_ | LinRegrs | TRUE | FALSE | | CV | 2 | 24 | 23 |
| RA_stroke_ | LinRegrs | TRUE | FALSE | | CV | 3 | 24 | 22 |
| RA_stroke_ | LinRegrs | TRUE | FALSE | | CV | 4 | 24 | 23 |
| RA_stroke_ | Lasso | TRUE | FALSE | 0.00024 | CV | 0 | 24 | 24 |
| RA_stroke_ | Lasso | TRUE | FALSE | 0.000215 | CV | 1 | 24 | 24 |
| RA_stroke_ | Lasso | TRUE | FALSE | 0.000212 | CV | 2 | 24 | 24 |
| RA_stroke_ | Lasso | TRUE | FALSE | 0.000218 | CV | 3 | 24 | 24 |
| RA_stroke_ | Lasso | TRUE | FALSE | 0.00021 | CV | 4 | 24 | 24 |
| RA_stroke_ | Lasso | TRUE | TRUE | 0.05 | CV | 0 | 24 | 9 |
| RA_stroke_ | Lasso | TRUE | TRUE | 0.05 | CV | 1 | 24 | 8 |
| RA_stroke_ | Lasso | TRUE | TRUE | 0.05 | CV | 2 | 24 | 8 |
| RA_stroke_ | Lasso | TRUE | TRUE | 0.05 | CV | 3 | 24 | 8 |
| RA_stroke_ | Lasso | TRUE | TRUE | 0.05 | CV | 4 | 24 | 8 |
| RA_ejectio | LinRegrs | TRUE | FALSE | | HeldoutTest | | 24 | 24 |
| RA_ejectio | Lasso | TRUE | FALSE | 0.000403 | HeldoutTest | | 24 | 24 |
| RA_ejectio | Lasso | TRUE | TRUE | 0.05 | HeldoutTest | | 24 | 16 |
| RA_ejectio | LinRegrs | TRUE | FALSE | | CV | 0 | 24 | 24 |
| RA_ejectio | LinRegrs | TRUE | FALSE | | CV | 1 | 24 | 24 |
| RA_ejectio | LinRegrs | TRUE | FALSE | | CV | 2 | 24 | 24 |
| RA_ejectio | LinRegrs | TRUE | FALSE | | CV | 3 | 24 | 24 |
| RA_ejectio | LinRegrs | TRUE | FALSE | | CV | 4 | 24 | 22 |
| RA_ejectio | Lasso | TRUE | FALSE | 0.000407 | CV | 0 | 24 | 24 |
| RA_ejectio | Lasso | TRUE | FALSE | 0.000405 | CV | 1 | 24 | 24 |
| RA_ejectio | Lasso | TRUE | FALSE | 0.000404 | CV | 2 | 24 | 24 |
| RA_ejectio | Lasso | TRUE | FALSE | 0.000398 | CV | 3 | 24 | 24 |
| RA_ejectio | Lasso | TRUE | FALSE | 0.000701 | CV | 4 | 24 | 22 |
| RA_ejectio | Lasso | TRUE | TRUE | 0.05 | CV | 0 | 24 | 16 |
| RA_ejectio | Lasso | TRUE | TRUE | 0.05 | CV | 1 | 24 | 16 |
| RA_ejectio | Lasso | TRUE | TRUE | 0.05 | CV | 2 | 24 | 16 |
| RA_ejectio | Lasso | TRUE | TRUE | 0.05 | CV | 3 | 24 | 16 |
| RA_ejectio | Lasso | TRUE | TRUE | 0.05 | CV | 4 | 24 | 13 |
| LV_myocar | LinRegrs | TRUE | FALSE | | HeldoutTest | | 24 | 23 |
| LV_myocar | Lasso | TRUE | FALSE | 0.000319 | HeldoutTest | | 24 | 24 |
| LV_myocar | Lasso | TRUE | TRUE | 0.05 | HeldoutTest | | 24 | 15 |
| LV_myocar | LinRegrs | TRUE | FALSE | | CV | 0 | 24 | 22 |
| LV_myocar | LinRegrs | TRUE | FALSE | | CV | 1 | 24 | 24 |
| LV_myocar | LinRegrs | TRUE | FALSE | | CV | 2 | 24 | 23 |
| LV_myocar | LinRegrs | TRUE | FALSE | | CV | 3 | 24 | 22 |
| LV_myocar | LinRegrs | TRUE | FALSE | | CV | 4 | 24 | 24 |

| | | | | | | | | |
|---|---|---|---|---|---|---|---|---|
| LV_myocar | Lasso | TRUE | FALSE | 0.000344 | CV | 0 | 24 | 24 |
| LV_myocar | Lasso | TRUE | FALSE | 0.000315 | CV | 1 | 24 | 24 |
| LV_myocar | Lasso | TRUE | FALSE | 0.000518 | CV | 2 | 24 | 24 |
| LV_myocar | Lasso | TRUE | FALSE | 0.00032 | CV | 3 | 24 | 24 |
| LV_myocar | Lasso | TRUE | FALSE | 0.00032 | CV | 4 | 24 | 24 |
| LV_myocar | Lasso | TRUE | TRUE | 0.05 | CV | 0 | 24 | 14 |
| LV_myocar | Lasso | TRUE | TRUE | 0.05 | CV | 1 | 24 | 14 |
| LV_myocar | Lasso | TRUE | TRUE | 0.05 | CV | 2 | 24 | 15 |
| LV_myocar | Lasso | TRUE | TRUE | 0.05 | CV | 3 | 24 | 14 |
| LV_myocar | Lasso | TRUE | TRUE | 0.05 | CV | 4 | 24 | 13 |
| LV_mean_ı | LinRegrs | TRUE | FALSE | | HeldoutTest | | 24 | 23 |
| LV_mean_ı | Lasso | TRUE | FALSE | 0.000324 | HeldoutTest | | 24 | 24 |
| LV_mean_ı | Lasso | TRUE | TRUE | 0.05 | HeldoutTest | | 24 | 14 |
| LV_mean_ı | LinRegrs | TRUE | FALSE | | CV | 0 | 24 | 23 |
| LV_mean_ı | LinRegrs | TRUE | FALSE | | CV | 1 | 24 | 23 |
| LV_mean_ı | LinRegrs | TRUE | FALSE | | CV | 2 | 24 | 22 |
| LV_mean_ı | LinRegrs | TRUE | FALSE | | CV | 3 | 24 | 23 |
| LV_mean_ı | LinRegrs | TRUE | FALSE | | CV | 4 | 24 | 20 |
| LV_mean_ı | Lasso | TRUE | FALSE | 0.000328 | CV | 0 | 24 | 24 |
| LV_mean_ı | Lasso | TRUE | FALSE | 0.000324 | CV | 1 | 24 | 24 |
| LV_mean_ı | Lasso | TRUE | FALSE | 0.000325 | CV | 2 | 24 | 24 |
| LV_mean_ı | Lasso | TRUE | FALSE | 0.000319 | CV | 3 | 24 | 24 |
| LV_mean_ı | Lasso | TRUE | FALSE | 0.000802 | CV | 4 | 24 | 21 |
| LV_mean_ı | Lasso | TRUE | TRUE | 0.05 | CV | 0 | 24 | 13 |
| LV_mean_ı | Lasso | TRUE | TRUE | 0.05 | CV | 1 | 24 | 14 |
| LV_mean_ı | Lasso | TRUE | TRUE | 0.05 | CV | 2 | 24 | 13 |
| LV_mean_ı | Lasso | TRUE | TRUE | 0.05 | CV | 3 | 24 | 13 |
| LV_mean_ı | Lasso | TRUE | TRUE | 0.05 | CV | 4 | 24 | 13 |
| LV_mean_ı | LinRegrs | TRUE | FALSE | | HeldoutTest | | 24 | 21 |
| LV_mean_ı | Lasso | TRUE | FALSE | 0.00028 | HeldoutTest | | 24 | 24 |
| LV_mean_ı | Lasso | TRUE | TRUE | 0.05 | HeldoutTest | | 24 | 11 |
| LV_mean_ı | LinRegrs | TRUE | FALSE | | CV | 0 | 24 | 23 |
| LV_mean_ı | LinRegrs | TRUE | FALSE | | CV | 1 | 24 | 20 |
| LV_mean_ı | LinRegrs | TRUE | FALSE | | CV | 2 | 24 | 21 |
| LV_mean_ı | LinRegrs | TRUE | FALSE | | CV | 3 | 24 | 21 |
| LV_mean_ı | LinRegrs | TRUE | FALSE | | CV | 4 | 24 | 20 |
| LV_mean_ı | Lasso | TRUE | FALSE | 0.000283 | CV | 0 | 24 | 24 |
| LV_mean_ı | Lasso | TRUE | FALSE | 0.000351 | CV | 1 | 24 | 24 |
| LV_mean_ı | Lasso | TRUE | FALSE | 0.000372 | CV | 2 | 24 | 23 |
| LV_mean_ı | Lasso | TRUE | FALSE | 0.000272 | CV | 3 | 24 | 24 |
| LV_mean_ı | Lasso | TRUE | FALSE | 0.000561 | CV | 4 | 24 | 23 |
| LV_mean_ı | Lasso | TRUE | TRUE | 0.05 | CV | 0 | 24 | 11 |
| LV_mean_ı | Lasso | TRUE | TRUE | 0.05 | CV | 1 | 24 | 11 |
| LV_mean_ı | Lasso | TRUE | TRUE | 0.05 | CV | 2 | 24 | 11 |
| LV_mean_ı | Lasso | TRUE | TRUE | 0.05 | CV | 3 | 24 | 11 |

| | | | | | | | | |
|---|---|---|---|---|---|---|---|---|
| LV_mean_ı | Lasso | TRUE | TRUE | 0.05 | CV | 4 | 24 | 12 |
| LV_mean_ı | LinRegrs | TRUE | FALSE | | HeldoutTest | | 24 | 24 |
| LV_mean_ı | Lasso | TRUE | FALSE | 0.000291 | HeldoutTest | | 24 | 24 |
| LV_mean_ı | Lasso | TRUE | TRUE | 0.05 | HeldoutTest | | 24 | 10 |
| LV_mean_ı | LinRegrs | TRUE | FALSE | | CV | 0 | 24 | 23 |
| LV_mean_ı | LinRegrs | TRUE | FALSE | | CV | 1 | 24 | 22 |
| LV_mean_ı | LinRegrs | TRUE | FALSE | | CV | 2 | 24 | 22 |
| LV_mean_ı | LinRegrs | TRUE | FALSE | | CV | 3 | 24 | 23 |
| LV_mean_ı | LinRegrs | TRUE | FALSE | | CV | 4 | 24 | 21 |
| LV_mean_ı | Lasso | TRUE | FALSE | 0.000838 | CV | 0 | 24 | 23 |
| LV_mean_ı | Lasso | TRUE | FALSE | 0.000415 | CV | 1 | 24 | 24 |
| LV_mean_ı | Lasso | TRUE | FALSE | 0.000444 | CV | 2 | 24 | 24 |
| LV_mean_ı | Lasso | TRUE | FALSE | 0.000288 | CV | 3 | 24 | 24 |
| LV_mean_ı | Lasso | TRUE | FALSE | 0.000772 | CV | 4 | 24 | 23 |
| LV_mean_ı | Lasso | TRUE | TRUE | 0.05 | CV | 0 | 24 | 11 |
| LV_mean_ı | Lasso | TRUE | TRUE | 0.05 | CV | 1 | 24 | 10 |
| LV_mean_ı | Lasso | TRUE | TRUE | 0.05 | CV | 2 | 24 | 11 |
| LV_mean_ı | Lasso | TRUE | TRUE | 0.05 | CV | 3 | 24 | 10 |
| LV_mean_ı | Lasso | TRUE | TRUE | 0.05 | CV | 4 | 24 | 9 |
| LV_mean_ı | LinRegrs | TRUE | FALSE | | HeldoutTest | | 24 | 23 |
| LV_mean_ı | Lasso | TRUE | FALSE | 0.000421 | HeldoutTest | | 24 | 24 |
| LV_mean_ı | Lasso | TRUE | TRUE | 0.05 | HeldoutTest | | 24 | 11 |
| LV_mean_ı | LinRegrs | TRUE | FALSE | | CV | 0 | 24 | 23 |
| LV_mean_ı | LinRegrs | TRUE | FALSE | | CV | 1 | 24 | 21 |
| LV_mean_ı | LinRegrs | TRUE | FALSE | | CV | 2 | 24 | 23 |
| LV_mean_ı | LinRegrs | TRUE | FALSE | | CV | 3 | 24 | 22 |
| LV_mean_ı | LinRegrs | TRUE | FALSE | | CV | 4 | 24 | 24 |
| LV_mean_ı | Lasso | TRUE | FALSE | 0.000987 | CV | 0 | 24 | 21 |
| LV_mean_ı | Lasso | TRUE | FALSE | 0.000483 | CV | 1 | 24 | 23 |
| LV_mean_ı | Lasso | TRUE | FALSE | 0.000319 | CV | 2 | 24 | 24 |
| LV_mean_ı | Lasso | TRUE | FALSE | 0.000417 | CV | 3 | 24 | 24 |
| LV_mean_ı | Lasso | TRUE | FALSE | 0.000317 | CV | 4 | 24 | 24 |
| LV_mean_ı | Lasso | TRUE | TRUE | 0.05 | CV | 0 | 24 | 11 |
| LV_mean_ı | Lasso | TRUE | TRUE | 0.05 | CV | 1 | 24 | 11 |
| LV_mean_ı | Lasso | TRUE | TRUE | 0.05 | CV | 2 | 24 | 11 |
| LV_mean_ı | Lasso | TRUE | TRUE | 0.05 | CV | 3 | 24 | 11 |
| LV_mean_ı | Lasso | TRUE | TRUE | 0.05 | CV | 4 | 24 | 9 |
| LV_mean_ı | LinRegrs | TRUE | FALSE | | HeldoutTest | | 24 | 23 |
| LV_mean_ı | Lasso | TRUE | FALSE | 0.000382 | HeldoutTest | | 24 | 23 |
| LV_mean_ı | Lasso | TRUE | TRUE | 0.05 | HeldoutTest | | 24 | 13 |
| LV_mean_ı | LinRegrs | TRUE | FALSE | | CV | 0 | 24 | 21 |
| LV_mean_ı | LinRegrs | TRUE | FALSE | | CV | 1 | 24 | 21 |
| LV_mean_ı | LinRegrs | TRUE | FALSE | | CV | 2 | 24 | 22 |
| LV_mean_ı | LinRegrs | TRUE | FALSE | | CV | 3 | 24 | 22 |
| LV_mean_ı | LinRegrs | TRUE | FALSE | | CV | 4 | 24 | 23 |

| | | | | | | | | |
|---|---|---|---|---|---|---|---|---|
| LV_mean_ı | Lasso | TRUE | FALSE | 0.000416 | CV | 0 | 24 | 22 |
| LV_mean_ı | Lasso | TRUE | FALSE | 0.000355 | CV | 1 | 24 | 23 |
| LV_mean_ı | Lasso | TRUE | FALSE | 0.000356 | CV | 2 | 24 | 22 |
| LV_mean_ı | Lasso | TRUE | FALSE | 0.0005 | CV | 3 | 24 | 23 |
| LV_mean_ı | Lasso | TRUE | FALSE | 0.000355 | CV | 4 | 24 | 23 |
| LV_mean_ı | Lasso | TRUE | TRUE | 0.05 | CV | 0 | 24 | 13 |
| LV_mean_ı | Lasso | TRUE | TRUE | 0.05 | CV | 1 | 24 | 13 |
| LV_mean_ı | Lasso | TRUE | TRUE | 0.05 | CV | 2 | 24 | 13 |
| LV_mean_ı | Lasso | TRUE | TRUE | 0.05 | CV | 3 | 24 | 13 |
| LV_mean_ı | Lasso | TRUE | TRUE | 0.05 | CV | 4 | 24 | 12 |
| LV_mean_ı | LinRegrs | TRUE | FALSE | | HeldoutTest | | 24 | 24 |
| LV_mean_ı | Lasso | TRUE | FALSE | 0.000363 | HeldoutTest | | 24 | 24 |
| LV_mean_ı | Lasso | TRUE | TRUE | 0.05 | HeldoutTest | | 24 | 13 |
| LV_mean_ı | LinRegrs | TRUE | FALSE | | CV | 0 | 24 | 24 |
| LV_mean_ı | LinRegrs | TRUE | FALSE | | CV | 1 | 24 | 23 |
| LV_mean_ı | LinRegrs | TRUE | FALSE | | CV | 2 | 24 | 24 |
| LV_mean_ı | LinRegrs | TRUE | FALSE | | CV | 3 | 24 | 24 |
| LV_mean_ı | LinRegrs | TRUE | FALSE | | CV | 4 | 24 | 24 |
| LV_mean_ı | Lasso | TRUE | FALSE | 0.000367 | CV | 0 | 24 | 24 |
| LV_mean_ı | Lasso | TRUE | FALSE | 0.000362 | CV | 1 | 24 | 24 |
| LV_mean_ı | Lasso | TRUE | FALSE | 0.000364 | CV | 2 | 24 | 24 |
| LV_mean_ı | Lasso | TRUE | FALSE | 0.000359 | CV | 3 | 24 | 24 |
| LV_mean_ı | Lasso | TRUE | FALSE | 0.000363 | CV | 4 | 24 | 24 |
| LV_mean_ı | Lasso | TRUE | TRUE | 0.05 | CV | 0 | 24 | 13 |
| LV_mean_ı | Lasso | TRUE | TRUE | 0.05 | CV | 1 | 24 | 13 |
| LV_mean_ı | Lasso | TRUE | TRUE | 0.05 | CV | 2 | 24 | 13 |
| LV_mean_ı | Lasso | TRUE | TRUE | 0.05 | CV | 3 | 24 | 13 |
| LV_mean_ı | Lasso | TRUE | TRUE | 0.05 | CV | 4 | 24 | 12 |
| LV_mean_ı | LinRegrs | TRUE | FALSE | | HeldoutTest | | 24 | 23 |
| LV_mean_ı | Lasso | TRUE | FALSE | 0.000349 | HeldoutTest | | 24 | 24 |
| LV_mean_ı | Lasso | TRUE | TRUE | 0.05 | HeldoutTest | | 24 | 11 |
| LV_mean_ı | LinRegrs | TRUE | FALSE | | CV | 0 | 24 | 24 |
| LV_mean_ı | LinRegrs | TRUE | FALSE | | CV | 1 | 24 | 24 |
| LV_mean_ı | LinRegrs | TRUE | FALSE | | CV | 2 | 24 | 24 |
| LV_mean_ı | LinRegrs | TRUE | FALSE | | CV | 3 | 24 | 23 |
| LV_mean_ı | LinRegrs | TRUE | FALSE | | CV | 4 | 24 | 22 |
| LV_mean_ı | Lasso | TRUE | FALSE | 0.000351 | CV | 0 | 24 | 24 |
| LV_mean_ı | Lasso | TRUE | FALSE | 0.000347 | CV | 1 | 24 | 24 |
| LV_mean_ı | Lasso | TRUE | FALSE | 0.000351 | CV | 2 | 24 | 24 |
| LV_mean_ı | Lasso | TRUE | FALSE | 0.000347 | CV | 3 | 24 | 24 |
| LV_mean_ı | Lasso | TRUE | FALSE | 0.000351 | CV | 4 | 24 | 24 |
| LV_mean_ı | Lasso | TRUE | TRUE | 0.05 | CV | 0 | 24 | 11 |
| LV_mean_ı | Lasso | TRUE | TRUE | 0.05 | CV | 1 | 24 | 11 |
| LV_mean_ı | Lasso | TRUE | TRUE | 0.05 | CV | 2 | 24 | 11 |
| LV_mean_ı | Lasso | TRUE | TRUE | 0.05 | CV | 3 | 24 | 11 |

| | | | | | | | | |
|---|---|---|---|---|---|---|---|---|
| LV_mean_ı | Lasso | TRUE | TRUE | 0.05 | CV | 4 | 24 | 9 |
| LV_mean_ı | LinRegrs | TRUE | FALSE | | HeldoutTest | | 24 | 24 |
| LV_mean_ı | Lasso | TRUE | FALSE | 0.000337 | HeldoutTest | | 24 | 24 |
| LV_mean_ı | Lasso | TRUE | TRUE | 0.05 | HeldoutTest | | 24 | 12 |
| LV_mean_ı | LinRegrs | TRUE | FALSE | | CV | 0 | 24 | 24 |
| LV_mean_ı | LinRegrs | TRUE | FALSE | | CV | 1 | 24 | 24 |
| LV_mean_ı | LinRegrs | TRUE | FALSE | | CV | 2 | 24 | 24 |
| LV_mean_ı | LinRegrs | TRUE | FALSE | | CV | 3 | 24 | 23 |
| LV_mean_ı | LinRegrs | TRUE | FALSE | | CV | 4 | 24 | 22 |
| LV_mean_ı | Lasso | TRUE | FALSE | 0.000338 | CV | 0 | 24 | 24 |
| LV_mean_ı | Lasso | TRUE | FALSE | 0.000335 | CV | 1 | 24 | 24 |
| LV_mean_ı | Lasso | TRUE | FALSE | 0.000339 | CV | 2 | 24 | 24 |
| LV_mean_ı | Lasso | TRUE | FALSE | 0.000334 | CV | 3 | 24 | 24 |
| LV_mean_ı | Lasso | TRUE | FALSE | 0.000337 | CV | 4 | 24 | 24 |
| LV_mean_ı | Lasso | TRUE | TRUE | 0.05 | CV | 0 | 24 | 12 |
| LV_mean_ı | Lasso | TRUE | TRUE | 0.05 | CV | 1 | 24 | 12 |
| LV_mean_ı | Lasso | TRUE | TRUE | 0.05 | CV | 2 | 24 | 12 |
| LV_mean_ı | Lasso | TRUE | TRUE | 0.05 | CV | 3 | 24 | 12 |
| LV_mean_ı | Lasso | TRUE | TRUE | 0.05 | CV | 4 | 24 | 9 |
| LV_mean_ı | LinRegrs | TRUE | FALSE | | HeldoutTest | | 24 | 23 |
| LV_mean_ı | Lasso | TRUE | FALSE | 0.000309 | HeldoutTest | | 24 | 24 |
| LV_mean_ı | Lasso | TRUE | TRUE | 0.05 | HeldoutTest | | 24 | 12 |
| LV_mean_ı | LinRegrs | TRUE | FALSE | | CV | 0 | 24 | 23 |
| LV_mean_ı | LinRegrs | TRUE | FALSE | | CV | 1 | 24 | 23 |
| LV_mean_ı | LinRegrs | TRUE | FALSE | | CV | 2 | 24 | 23 |
| LV_mean_ı | LinRegrs | TRUE | FALSE | | CV | 3 | 24 | 23 |
| LV_mean_ı | LinRegrs | TRUE | FALSE | | CV | 4 | 24 | 22 |
| LV_mean_ı | Lasso | TRUE | FALSE | 0.000311 | CV | 0 | 24 | 24 |
| LV_mean_ı | Lasso | TRUE | FALSE | 0.000307 | CV | 1 | 24 | 24 |
| LV_mean_ı | Lasso | TRUE | FALSE | 0.000312 | CV | 2 | 24 | 24 |
| LV_mean_ı | Lasso | TRUE | FALSE | 0.000306 | CV | 3 | 24 | 24 |
| LV_mean_ı | Lasso | TRUE | FALSE | 0.000309 | CV | 4 | 24 | 24 |
| LV_mean_ı | Lasso | TRUE | TRUE | 0.05 | CV | 0 | 24 | 12 |
| LV_mean_ı | Lasso | TRUE | TRUE | 0.05 | CV | 1 | 24 | 12 |
| LV_mean_ı | Lasso | TRUE | TRUE | 0.05 | CV | 2 | 24 | 12 |
| LV_mean_ı | Lasso | TRUE | TRUE | 0.05 | CV | 3 | 24 | 12 |
| LV_mean_ı | Lasso | TRUE | TRUE | 0.05 | CV | 4 | 24 | 11 |
| LV_mean_ı | LinRegrs | TRUE | FALSE | | HeldoutTest | | 24 | 24 |
| LV_mean_ı | Lasso | TRUE | FALSE | 0.000314 | HeldoutTest | | 24 | 24 |
| LV_mean_ı | Lasso | TRUE | TRUE | 0.05 | HeldoutTest | | 24 | 12 |
| LV_mean_ı | LinRegrs | TRUE | FALSE | | CV | 0 | 24 | 24 |
| LV_mean_ı | LinRegrs | TRUE | FALSE | | CV | 1 | 24 | 24 |
| LV_mean_ı | LinRegrs | TRUE | FALSE | | CV | 2 | 24 | 24 |
| LV_mean_ı | LinRegrs | TRUE | FALSE | | CV | 3 | 24 | 24 |
| LV_mean_ı | LinRegrs | TRUE | FALSE | | CV | 4 | 24 | 22 |

| | | | | | | | | |
|---|---|---|---|---|---|---|---|---|
| LV_mean_ | Lasso | TRUE | FALSE | 0.000316 | CV | 0 | 24 | 24 |
| LV_mean_ | Lasso | TRUE | FALSE | 0.000314 | CV | 1 | 24 | 24 |
| LV_mean_ | Lasso | TRUE | FALSE | 0.000315 | CV | 2 | 24 | 24 |
| LV_mean_ | Lasso | TRUE | FALSE | 0.000312 | CV | 3 | 24 | 24 |
| LV_mean_ | Lasso | TRUE | FALSE | 0.000314 | CV | 4 | 24 | 23 |
| LV_mean_ | Lasso | TRUE | TRUE | 0.05 | CV | 0 | 24 | 12 |
| LV_mean_ | Lasso | TRUE | TRUE | 0.05 | CV | 1 | 24 | 12 |
| LV_mean_ | Lasso | TRUE | TRUE | 0.05 | CV | 2 | 24 | 12 |
| LV_mean_ | Lasso | TRUE | TRUE | 0.05 | CV | 3 | 24 | 12 |
| LV_mean_ | Lasso | TRUE | TRUE | 0.05 | CV | 4 | 24 | 11 |
| LV_mean_ | LinRegrs | TRUE | FALSE | | HeldoutTest | | 24 | 21 |
| LV_mean_ | Lasso | TRUE | FALSE | 0.000326 | HeldoutTest | | 24 | 22 |
| LV_mean_ | Lasso | TRUE | TRUE | 0.05 | HeldoutTest | | 24 | 13 |
| LV_mean_ | LinRegrs | TRUE | FALSE | | CV | 0 | 24 | 21 |
| LV_mean_ | LinRegrs | TRUE | FALSE | | CV | 1 | 24 | 21 |
| LV_mean_ | LinRegrs | TRUE | FALSE | | CV | 2 | 24 | 21 |
| LV_mean_ | LinRegrs | TRUE | FALSE | | CV | 3 | 24 | 21 |
| LV_mean_ | LinRegrs | TRUE | FALSE | | CV | 4 | 24 | 22 |
| LV_mean_ | Lasso | TRUE | FALSE | 0.000353 | CV | 0 | 24 | 23 |
| LV_mean_ | Lasso | TRUE | FALSE | 0.000325 | CV | 1 | 24 | 24 |
| LV_mean_ | Lasso | TRUE | FALSE | 0.000327 | CV | 2 | 24 | 23 |
| LV_mean_ | Lasso | TRUE | FALSE | 0.000322 | CV | 3 | 24 | 24 |
| LV_mean_ | Lasso | TRUE | FALSE | 0.000327 | CV | 4 | 24 | 24 |
| LV_mean_ | Lasso | TRUE | TRUE | 0.05 | CV | 0 | 24 | 13 |
| LV_mean_ | Lasso | TRUE | TRUE | 0.05 | CV | 1 | 24 | 13 |
| LV_mean_ | Lasso | TRUE | TRUE | 0.05 | CV | 2 | 24 | 13 |
| LV_mean_ | Lasso | TRUE | TRUE | 0.05 | CV | 3 | 24 | 13 |
| LV_mean_ | Lasso | TRUE | TRUE | 0.05 | CV | 4 | 24 | 13 |
| LV_mean_ | LinRegrs | TRUE | FALSE | | HeldoutTest | | 24 | 23 |
| LV_mean_ | Lasso | TRUE | FALSE | 0.000345 | HeldoutTest | | 24 | 24 |
| LV_mean_ | Lasso | TRUE | TRUE | 0.05 | HeldoutTest | | 24 | 12 |
| LV_mean_ | LinRegrs | TRUE | FALSE | | CV | 0 | 24 | 23 |
| LV_mean_ | LinRegrs | TRUE | FALSE | | CV | 1 | 24 | 23 |
| LV_mean_ | LinRegrs | TRUE | FALSE | | CV | 2 | 24 | 23 |
| LV_mean_ | LinRegrs | TRUE | FALSE | | CV | 3 | 24 | 23 |
| LV_mean_ | LinRegrs | TRUE | FALSE | | CV | 4 | 24 | 22 |
| LV_mean_ | Lasso | TRUE | FALSE | 0.000348 | CV | 0 | 24 | 24 |
| LV_mean_ | Lasso | TRUE | FALSE | 0.000343 | CV | 1 | 24 | 23 |
| LV_mean_ | Lasso | TRUE | FALSE | 0.000347 | CV | 2 | 24 | 24 |
| LV_mean_ | Lasso | TRUE | FALSE | 0.00034 | CV | 3 | 24 | 24 |
| LV_mean_ | Lasso | TRUE | FALSE | 0.000345 | CV | 4 | 24 | 23 |
| LV_mean_ | Lasso | TRUE | TRUE | 0.05 | CV | 0 | 24 | 12 |
| LV_mean_ | Lasso | TRUE | TRUE | 0.05 | CV | 1 | 24 | 12 |
| LV_mean_ | Lasso | TRUE | TRUE | 0.05 | CV | 2 | 24 | 12 |
| LV_mean_ | Lasso | TRUE | TRUE | 0.05 | CV | 3 | 24 | 12 |

| | | | | | | | | |
|---|---|---|---|---|---|---|---|---|
| LV_mean_ı | Lasso | TRUE | TRUE | 0.05 | CV | 4 | 24 | 11 |
| LV_mean_ı | LinRegrs | TRUE | FALSE | | HeldoutTest | | 24 | 23 |
| LV_mean_ı | Lasso | TRUE | FALSE | 0.000342 | HeldoutTest | | 24 | 23 |
| LV_mean_ı | Lasso | TRUE | TRUE | 0.05 | HeldoutTest | | 24 | 11 |
| LV_mean_ı | LinRegrs | TRUE | FALSE | | CV | 0 | 24 | 23 |
| LV_mean_ı | LinRegrs | TRUE | FALSE | | CV | 1 | 24 | 23 |
| LV_mean_ı | LinRegrs | TRUE | FALSE | | CV | 2 | 24 | 23 |
| LV_mean_ı | LinRegrs | TRUE | FALSE | | CV | 3 | 24 | 23 |
| LV_mean_ı | LinRegrs | TRUE | FALSE | | CV | 4 | 24 | 23 |
| LV_mean_ı | Lasso | TRUE | FALSE | 0.000343 | CV | 0 | 24 | 24 |
| LV_mean_ı | Lasso | TRUE | FALSE | 0.000343 | CV | 1 | 24 | 24 |
| LV_mean_ı | Lasso | TRUE | FALSE | 0.000339 | CV | 2 | 24 | 24 |
| LV_mean_ı | Lasso | TRUE | FALSE | 0.000342 | CV | 3 | 24 | 23 |
| LV_mean_ı | Lasso | TRUE | FALSE | 0.000344 | CV | 4 | 24 | 24 |
| LV_mean_ı | Lasso | TRUE | TRUE | 0.05 | CV | 0 | 24 | 11 |
| LV_mean_ı | Lasso | TRUE | TRUE | 0.05 | CV | 1 | 24 | 11 |
| LV_mean_ı | Lasso | TRUE | TRUE | 0.05 | CV | 2 | 24 | 11 |
| LV_mean_ı | Lasso | TRUE | TRUE | 0.05 | CV | 3 | 24 | 11 |
| LV_mean_ı | Lasso | TRUE | TRUE | 0.05 | CV | 4 | 24 | 10 |
| LV_mean_ı | LinRegrs | TRUE | FALSE | | HeldoutTest | | 24 | 24 |
| LV_mean_ı | Lasso | TRUE | FALSE | 0.000304 | HeldoutTest | | 24 | 24 |
| LV_mean_ı | Lasso | TRUE | TRUE | 0.05 | HeldoutTest | | 24 | 11 |
| LV_mean_ı | LinRegrs | TRUE | FALSE | | CV | 0 | 24 | 24 |
| LV_mean_ı | LinRegrs | TRUE | FALSE | | CV | 1 | 24 | 24 |
| LV_mean_ı | LinRegrs | TRUE | FALSE | | CV | 2 | 24 | 24 |
| LV_mean_ı | LinRegrs | TRUE | FALSE | | CV | 3 | 24 | 24 |
| LV_mean_ı | LinRegrs | TRUE | FALSE | | CV | 4 | 24 | 23 |
| LV_mean_ı | Lasso | TRUE | FALSE | 0.000306 | CV | 0 | 24 | 24 |
| LV_mean_ı | Lasso | TRUE | FALSE | 0.000303 | CV | 1 | 24 | 24 |
| LV_mean_ı | Lasso | TRUE | FALSE | 0.000304 | CV | 2 | 24 | 24 |
| LV_mean_ı | Lasso | TRUE | FALSE | 0.0003 | CV | 3 | 24 | 24 |
| LV_mean_ı | Lasso | TRUE | FALSE | 0.000327 | CV | 4 | 24 | 24 |
| LV_mean_ı | Lasso | TRUE | TRUE | 0.05 | CV | 0 | 24 | 11 |
| LV_mean_ı | Lasso | TRUE | TRUE | 0.05 | CV | 1 | 24 | 11 |
| LV_mean_ı | Lasso | TRUE | TRUE | 0.05 | CV | 2 | 24 | 11 |
| LV_mean_ı | Lasso | TRUE | TRUE | 0.05 | CV | 3 | 24 | 11 |
| LV_mean_ı | Lasso | TRUE | TRUE | 0.05 | CV | 4 | 24 | 9 |
| LV_mean_ı | LinRegrs | TRUE | FALSE | | HeldoutTest | | 24 | 21 |
| LV_mean_ı | Lasso | TRUE | FALSE | 0.000285 | HeldoutTest | | 24 | 24 |
| LV_mean_ı | Lasso | TRUE | TRUE | 0.05 | HeldoutTest | | 24 | 12 |
| LV_mean_ı | LinRegrs | TRUE | FALSE | | CV | 0 | 24 | 21 |
| LV_mean_ı | LinRegrs | TRUE | FALSE | | CV | 1 | 24 | 22 |
| LV_mean_ı | LinRegrs | TRUE | FALSE | | CV | 2 | 24 | 22 |
| LV_mean_ı | LinRegrs | TRUE | FALSE | | CV | 3 | 24 | 22 |
| LV_mean_ı | LinRegrs | TRUE | FALSE | | CV | 4 | 24 | 22 |

| | | | | | | | | |
|---|---|---|---|---|---|---|---|---|
| LV_mean_ | Lasso | TRUE | FALSE | 0.000306 | CV | 0 | 24 | 23 |
| LV_mean_ | Lasso | TRUE | FALSE | 0.000325 | CV | 1 | 24 | 22 |
| LV_mean_ | Lasso | TRUE | FALSE | 0.00029 | CV | 2 | 24 | 24 |
| LV_mean_ | Lasso | TRUE | FALSE | 0.000287 | CV | 3 | 24 | 24 |
| LV_mean_ | Lasso | TRUE | FALSE | 0.000286 | CV | 4 | 24 | 24 |
| LV_mean_ | Lasso | TRUE | TRUE | 0.05 | CV | 0 | 24 | 12 |
| LV_mean_ | Lasso | TRUE | TRUE | 0.05 | CV | 1 | 24 | 12 |
| LV_mean_ | Lasso | TRUE | TRUE | 0.05 | CV | 2 | 24 | 12 |
| LV_mean_ | Lasso | TRUE | TRUE | 0.05 | CV | 3 | 24 | 12 |
| LV_mean_ | Lasso | TRUE | TRUE | 0.05 | CV | 4 | 24 | 10 |
| LV_mean_ | LinRegrs | TRUE | FALSE | | HeldoutTest | | 24 | 24 |
| LV_mean_ | Lasso | TRUE | FALSE | 0.000315 | HeldoutTest | | 24 | 24 |
| LV_mean_ | Lasso | TRUE | TRUE | 0.05 | HeldoutTest | | 24 | 11 |
| LV_mean_ | LinRegrs | TRUE | FALSE | | CV | 0 | 24 | 24 |
| LV_mean_ | LinRegrs | TRUE | FALSE | | CV | 1 | 24 | 23 |
| LV_mean_ | LinRegrs | TRUE | FALSE | | CV | 2 | 24 | 23 |
| LV_mean_ | LinRegrs | TRUE | FALSE | | CV | 3 | 24 | 24 |
| LV_mean_ | LinRegrs | TRUE | FALSE | | CV | 4 | 24 | 23 |
| LV_mean_ | Lasso | TRUE | FALSE | 0.000316 | CV | 0 | 24 | 24 |
| LV_mean_ | Lasso | TRUE | FALSE | 0.000316 | CV | 1 | 24 | 24 |
| LV_mean_ | Lasso | TRUE | FALSE | 0.000315 | CV | 2 | 24 | 23 |
| LV_mean_ | Lasso | TRUE | FALSE | 0.000315 | CV | 3 | 24 | 24 |
| LV_mean_ | Lasso | TRUE | FALSE | 0.000313 | CV | 4 | 24 | 24 |
| LV_mean_ | Lasso | TRUE | TRUE | 0.05 | CV | 0 | 24 | 11 |
| LV_mean_ | Lasso | TRUE | TRUE | 0.05 | CV | 1 | 24 | 11 |
| LV_mean_ | Lasso | TRUE | TRUE | 0.05 | CV | 2 | 24 | 11 |
| LV_mean_ | Lasso | TRUE | TRUE | 0.05 | CV | 3 | 24 | 11 |
| LV_mean_ | Lasso | TRUE | TRUE | 0.05 | CV | 4 | 24 | 10 |
| LV_mean_ | LinRegrs | TRUE | FALSE | | HeldoutTest | | 24 | 23 |
| LV_mean_ | Lasso | TRUE | FALSE | 0.000363 | HeldoutTest | | 24 | 24 |
| LV_mean_ | Lasso | TRUE | TRUE | 0.05 | HeldoutTest | | 24 | 12 |
| LV_mean_ | LinRegrs | TRUE | FALSE | | CV | 0 | 24 | 23 |
| LV_mean_ | LinRegrs | TRUE | FALSE | | CV | 1 | 24 | 22 |
| LV_mean_ | LinRegrs | TRUE | FALSE | | CV | 2 | 24 | 23 |
| LV_mean_ | LinRegrs | TRUE | FALSE | | CV | 3 | 24 | 23 |
| LV_mean_ | LinRegrs | TRUE | FALSE | | CV | 4 | 24 | 22 |
| LV_mean_ | Lasso | TRUE | FALSE | 0.000367 | CV | 0 | 24 | 24 |
| LV_mean_ | Lasso | TRUE | FALSE | 0.000363 | CV | 1 | 24 | 23 |
| LV_mean_ | Lasso | TRUE | FALSE | 0.000364 | CV | 2 | 24 | 24 |
| LV_mean_ | Lasso | TRUE | FALSE | 0.000359 | CV | 3 | 24 | 23 |
| LV_mean_ | Lasso | TRUE | FALSE | 0.000363 | CV | 4 | 24 | 24 |
| LV_mean_ | Lasso | TRUE | TRUE | 0.05 | CV | 0 | 24 | 12 |
| LV_mean_ | Lasso | TRUE | TRUE | 0.05 | CV | 1 | 24 | 13 |
| LV_mean_ | Lasso | TRUE | TRUE | 0.05 | CV | 2 | 24 | 13 |
| LV_mean_ | Lasso | TRUE | TRUE | 0.05 | CV | 3 | 24 | 12 |

| | | | | | | | | |
|---|---|---|---|---|---|---|---|---|
| LV_mean_ | Lasso | TRUE | TRUE | 0.05 | CV | 4 | 24 | 11 |
| LV_circumf | LinRegrs | TRUE | FALSE | | HeldoutTest | | 24 | 21 |
| LV_circumf | Lasso | TRUE | FALSE | 0.00041 | HeldoutTest | | 24 | 23 |
| LV_circumf | Lasso | TRUE | TRUE | 0.05 | HeldoutTest | | 24 | 13 |
| LV_circumf | LinRegrs | TRUE | FALSE | | CV | 0 | 24 | 21 |
| LV_circumf | LinRegrs | TRUE | FALSE | | CV | 1 | 24 | 22 |
| LV_circumf | LinRegrs | TRUE | FALSE | | CV | 2 | 24 | 21 |
| LV_circumf | LinRegrs | TRUE | FALSE | | CV | 3 | 24 | 20 |
| LV_circumf | LinRegrs | TRUE | FALSE | | CV | 4 | 24 | 24 |
| LV_circumf | Lasso | TRUE | FALSE | 0.000417 | CV | 0 | 24 | 23 |
| LV_circumf | Lasso | TRUE | FALSE | 0.000831 | CV | 1 | 24 | 23 |
| LV_circumf | Lasso | TRUE | FALSE | 0.000437 | CV | 2 | 24 | 23 |
| LV_circumf | Lasso | TRUE | FALSE | 0.000434 | CV | 3 | 24 | 22 |
| LV_circumf | Lasso | TRUE | FALSE | 0.000288 | CV | 4 | 24 | 24 |
| LV_circumf | Lasso | TRUE | TRUE | 0.05 | CV | 0 | 24 | 14 |
| LV_circumf | Lasso | TRUE | TRUE | 0.05 | CV | 1 | 24 | 13 |
| LV_circumf | Lasso | TRUE | TRUE | 0.05 | CV | 2 | 24 | 13 |
| LV_circumf | Lasso | TRUE | TRUE | 0.05 | CV | 3 | 24 | 13 |
| LV_circumf | Lasso | TRUE | TRUE | 0.05 | CV | 4 | 24 | 12 |
| LV_circumf | LinRegrs | TRUE | FALSE | | HeldoutTest | | 24 | 23 |
| LV_circumf | Lasso | TRUE | FALSE | 0.000327 | HeldoutTest | | 24 | 24 |
| LV_circumf | Lasso | TRUE | TRUE | 0.05 | HeldoutTest | | 24 | 13 |
| LV_circumf | LinRegrs | TRUE | FALSE | | CV | 0 | 24 | 23 |
| LV_circumf | LinRegrs | TRUE | FALSE | | CV | 1 | 24 | 23 |
| LV_circumf | LinRegrs | TRUE | FALSE | | CV | 2 | 24 | 23 |
| LV_circumf | LinRegrs | TRUE | FALSE | | CV | 3 | 24 | 23 |
| LV_circumf | LinRegrs | TRUE | FALSE | | CV | 4 | 24 | 24 |
| LV_circumf | Lasso | TRUE | FALSE | 0.00033 | CV | 0 | 24 | 23 |
| LV_circumf | Lasso | TRUE | FALSE | 0.000327 | CV | 1 | 24 | 24 |
| LV_circumf | Lasso | TRUE | FALSE | 0.000327 | CV | 2 | 24 | 24 |
| LV_circumf | Lasso | TRUE | FALSE | 0.000325 | CV | 3 | 24 | 24 |
| LV_circumf | Lasso | TRUE | FALSE | 0.000325 | CV | 4 | 24 | 24 |
| LV_circumf | Lasso | TRUE | TRUE | 0.05 | CV | 0 | 24 | 13 |
| LV_circumf | Lasso | TRUE | TRUE | 0.05 | CV | 1 | 24 | 13 |
| LV_circumf | Lasso | TRUE | TRUE | 0.05 | CV | 2 | 24 | 12 |
| LV_circumf | Lasso | TRUE | TRUE | 0.05 | CV | 3 | 24 | 13 |
| LV_circumf | Lasso | TRUE | TRUE | 0.05 | CV | 4 | 24 | 14 |
| LV_circumf | LinRegrs | TRUE | FALSE | | HeldoutTest | | 24 | 22 |
| LV_circumf | Lasso | TRUE | FALSE | 0.000349 | HeldoutTest | | 24 | 23 |
| LV_circumf | Lasso | TRUE | TRUE | 0.05 | HeldoutTest | | 24 | 12 |
| LV_circumf | LinRegrs | TRUE | FALSE | | CV | 0 | 24 | 23 |
| LV_circumf | LinRegrs | TRUE | FALSE | | CV | 1 | 24 | 22 |
| LV_circumf | LinRegrs | TRUE | FALSE | | CV | 2 | 24 | 22 |
| LV_circumf | LinRegrs | TRUE | FALSE | | CV | 3 | 24 | 22 |
| LV_circumf | LinRegrs | TRUE | FALSE | | CV | 4 | 24 | 22 |

| | | | | | | | | |
|---|---|---|---|---|---|---|---|---|
| LV_circumf | Lasso | TRUE | FALSE | 0.000283 | CV | 0 | 24 | 24 |
| LV_circumf | Lasso | TRUE | FALSE | 0.000325 | CV | 1 | 24 | 24 |
| LV_circumf | Lasso | TRUE | FALSE | 0.000303 | CV | 2 | 24 | 24 |
| LV_circumf | Lasso | TRUE | FALSE | 0.000464 | CV | 3 | 24 | 24 |
| LV_circumf | Lasso | TRUE | FALSE | 0.000264 | CV | 4 | 24 | 23 |
| LV_circumf | Lasso | TRUE | TRUE | 0.05 | CV | 0 | 24 | 12 |
| LV_circumf | Lasso | TRUE | TRUE | 0.05 | CV | 1 | 24 | 12 |
| LV_circumf | Lasso | TRUE | TRUE | 0.05 | CV | 2 | 24 | 12 |
| LV_circumf | Lasso | TRUE | TRUE | 0.05 | CV | 3 | 24 | 12 |
| LV_circumf | Lasso | TRUE | TRUE | 0.05 | CV | 4 | 24 | 12 |
| LV_circumf | LinRegrs | TRUE | FALSE | | HeldoutTest | | 24 | 23 |
| LV_circumf | Lasso | TRUE | FALSE | 0.000272 | HeldoutTest | | 24 | 24 |
| LV_circumf | Lasso | TRUE | TRUE | 0.05 | HeldoutTest | | 24 | 14 |
| LV_circumf | LinRegrs | TRUE | FALSE | | CV | 0 | 24 | 23 |
| LV_circumf | LinRegrs | TRUE | FALSE | | CV | 1 | 24 | 23 |
| LV_circumf | LinRegrs | TRUE | FALSE | | CV | 2 | 24 | 24 |
| LV_circumf | LinRegrs | TRUE | FALSE | | CV | 3 | 24 | 23 |
| LV_circumf | LinRegrs | TRUE | FALSE | | CV | 4 | 24 | 22 |
| LV_circumf | Lasso | TRUE | FALSE | 0.000281 | CV | 0 | 24 | 24 |
| LV_circumf | Lasso | TRUE | FALSE | 0.000353 | CV | 1 | 24 | 24 |
| LV_circumf | Lasso | TRUE | FALSE | 0.000269 | CV | 2 | 24 | 24 |
| LV_circumf | Lasso | TRUE | FALSE | 0.000332 | CV | 3 | 24 | 24 |
| LV_circumf | Lasso | TRUE | FALSE | 0.000508 | CV | 4 | 24 | 23 |
| LV_circumf | Lasso | TRUE | TRUE | 0.05 | CV | 0 | 24 | 13 |
| LV_circumf | Lasso | TRUE | TRUE | 0.05 | CV | 1 | 24 | 14 |
| LV_circumf | Lasso | TRUE | TRUE | 0.05 | CV | 2 | 24 | 14 |
| LV_circumf | Lasso | TRUE | TRUE | 0.05 | CV | 3 | 24 | 14 |
| LV_circumf | Lasso | TRUE | TRUE | 0.05 | CV | 4 | 24 | 10 |
| LV_circumf | LinRegrs | TRUE | FALSE | | HeldoutTest | | 24 | 24 |
| LV_circumf | Lasso | TRUE | FALSE | 0.000356 | HeldoutTest | | 24 | 24 |
| LV_circumf | Lasso | TRUE | TRUE | 0.05 | HeldoutTest | | 24 | 15 |
| LV_circumf | LinRegrs | TRUE | FALSE | | CV | 0 | 24 | 24 |
| LV_circumf | LinRegrs | TRUE | FALSE | | CV | 1 | 24 | 24 |
| LV_circumf | LinRegrs | TRUE | FALSE | | CV | 2 | 24 | 24 |
| LV_circumf | LinRegrs | TRUE | FALSE | | CV | 3 | 24 | 24 |
| LV_circumf | LinRegrs | TRUE | FALSE | | CV | 4 | 24 | 23 |
| LV_circumf | Lasso | TRUE | FALSE | 0.00036 | CV | 0 | 24 | 24 |
| LV_circumf | Lasso | TRUE | FALSE | 0.000356 | CV | 1 | 24 | 24 |
| LV_circumf | Lasso | TRUE | FALSE | 0.000359 | CV | 2 | 24 | 24 |
| LV_circumf | Lasso | TRUE | FALSE | 0.000353 | CV | 3 | 24 | 24 |
| LV_circumf | Lasso | TRUE | FALSE | 0.000436 | CV | 4 | 24 | 23 |
| LV_circumf | Lasso | TRUE | TRUE | 0.05 | CV | 0 | 24 | 15 |
| LV_circumf | Lasso | TRUE | TRUE | 0.05 | CV | 1 | 24 | 15 |
| LV_circumf | Lasso | TRUE | TRUE | 0.05 | CV | 2 | 24 | 15 |
| LV_circumf | Lasso | TRUE | TRUE | 0.05 | CV | 3 | 24 | 16 |

| | | | | | | | | |
|---|---|---|---|---|---|---|---|---|
| LV_circumf | Lasso | TRUE | TRUE | 0.05 | CV | 4 | 24 | 14 |
| LV_circumf | LinRegrs | TRUE | FALSE | | HeldoutTest | | 24 | 23 |
| LV_circumf | Lasso | TRUE | FALSE | 0.000289 | HeldoutTest | | 24 | 23 |
| LV_circumf | Lasso | TRUE | TRUE | 0.05 | HeldoutTest | | 24 | 14 |
| LV_circumf | LinRegrs | TRUE | FALSE | | CV | 0 | 24 | 22 |
| LV_circumf | LinRegrs | TRUE | FALSE | | CV | 1 | 24 | 22 |
| LV_circumf | LinRegrs | TRUE | FALSE | | CV | 2 | 24 | 23 |
| LV_circumf | LinRegrs | TRUE | FALSE | | CV | 3 | 24 | 22 |
| LV_circumf | LinRegrs | TRUE | FALSE | | CV | 4 | 24 | 23 |
| LV_circumf | Lasso | TRUE | FALSE | 0.000361 | CV | 0 | 24 | 24 |
| LV_circumf | Lasso | TRUE | FALSE | 0.000333 | CV | 1 | 24 | 24 |
| LV_circumf | Lasso | TRUE | FALSE | 0.000289 | CV | 2 | 24 | 24 |
| LV_circumf | Lasso | TRUE | FALSE | 0.00047 | CV | 3 | 24 | 24 |
| LV_circumf | Lasso | TRUE | FALSE | 0.000292 | CV | 4 | 24 | 24 |
| LV_circumf | Lasso | TRUE | TRUE | 0.05 | CV | 0 | 24 | 14 |
| LV_circumf | Lasso | TRUE | TRUE | 0.05 | CV | 1 | 24 | 14 |
| LV_circumf | Lasso | TRUE | TRUE | 0.05 | CV | 2 | 24 | 13 |
| LV_circumf | Lasso | TRUE | TRUE | 0.05 | CV | 3 | 24 | 14 |
| LV_circumf | Lasso | TRUE | TRUE | 0.05 | CV | 4 | 24 | 11 |
| LV_circumf | LinRegrs | TRUE | FALSE | | HeldoutTest | | 24 | 24 |
| LV_circumf | Lasso | TRUE | FALSE | 0.000339 | HeldoutTest | | 24 | 24 |
| LV_circumf | Lasso | TRUE | TRUE | 0.05 | HeldoutTest | | 24 | 15 |
| LV_circumf | LinRegrs | TRUE | FALSE | | CV | 0 | 24 | 24 |
| LV_circumf | LinRegrs | TRUE | FALSE | | CV | 1 | 24 | 24 |
| LV_circumf | LinRegrs | TRUE | FALSE | | CV | 2 | 24 | 24 |
| LV_circumf | LinRegrs | TRUE | FALSE | | CV | 3 | 24 | 24 |
| LV_circumf | LinRegrs | TRUE | FALSE | | CV | 4 | 24 | 21 |
| LV_circumf | Lasso | TRUE | FALSE | 0.000346 | CV | 0 | 24 | 24 |
| LV_circumf | Lasso | TRUE | FALSE | 0.000338 | CV | 1 | 24 | 24 |
| LV_circumf | Lasso | TRUE | FALSE | 0.000338 | CV | 2 | 24 | 24 |
| LV_circumf | Lasso | TRUE | FALSE | 0.000336 | CV | 3 | 24 | 24 |
| LV_circumf | Lasso | TRUE | FALSE | 0.000334 | CV | 4 | 24 | 23 |
| LV_circumf | Lasso | TRUE | TRUE | 0.05 | CV | 0 | 24 | 15 |
| LV_circumf | Lasso | TRUE | TRUE | 0.05 | CV | 1 | 24 | 15 |
| LV_circumf | Lasso | TRUE | TRUE | 0.05 | CV | 2 | 24 | 14 |
| LV_circumf | Lasso | TRUE | TRUE | 0.05 | CV | 3 | 24 | 15 |
| LV_circumf | Lasso | TRUE | TRUE | 0.05 | CV | 4 | 24 | 16 |
| LV_circumf | LinRegrs | TRUE | FALSE | | HeldoutTest | | 24 | 24 |
| LV_circumf | Lasso | TRUE | FALSE | 0.000331 | HeldoutTest | | 24 | 24 |
| LV_circumf | Lasso | TRUE | TRUE | 0.05 | HeldoutTest | | 24 | 13 |
| LV_circumf | LinRegrs | TRUE | FALSE | | CV | 0 | 24 | 24 |
| LV_circumf | LinRegrs | TRUE | FALSE | | CV | 1 | 24 | 24 |
| LV_circumf | LinRegrs | TRUE | FALSE | | CV | 2 | 24 | 24 |
| LV_circumf | LinRegrs | TRUE | FALSE | | CV | 3 | 24 | 24 |
| LV_circumf | LinRegrs | TRUE | FALSE | | CV | 4 | 24 | 24 |

| | | | | | | | | |
|---|---|---|---|---|---|---|---|---|
| LV_circumf | Lasso | TRUE | FALSE | 0.000335 | CV | 0 | 24 | 24 |
| LV_circumf | Lasso | TRUE | FALSE | 0.000328 | CV | 1 | 24 | 24 |
| LV_circumf | Lasso | TRUE | FALSE | 0.00033 | CV | 2 | 24 | 24 |
| LV_circumf | Lasso | TRUE | FALSE | 0.00033 | CV | 3 | 24 | 24 |
| LV_circumf | Lasso | TRUE | FALSE | 0.000331 | CV | 4 | 24 | 24 |
| LV_circumf | Lasso | TRUE | TRUE | 0.05 | CV | 0 | 24 | 14 |
| LV_circumf | Lasso | TRUE | TRUE | 0.05 | CV | 1 | 24 | 13 |
| LV_circumf | Lasso | TRUE | TRUE | 0.05 | CV | 2 | 24 | 14 |
| LV_circumf | Lasso | TRUE | TRUE | 0.05 | CV | 3 | 24 | 13 |
| LV_circumf | Lasso | TRUE | TRUE | 0.05 | CV | 4 | 24 | 16 |
| LV_circumf | LinRegrs | TRUE | FALSE | | HeldoutTest | | 24 | 22 |
| LV_circumf | Lasso | TRUE | FALSE | 0.000263 | HeldoutTest | | 24 | 23 |
| LV_circumf | Lasso | TRUE | TRUE | 0.05 | HeldoutTest | | 24 | 11 |
| LV_circumf | LinRegrs | TRUE | FALSE | | CV | 0 | 24 | 22 |
| LV_circumf | LinRegrs | TRUE | FALSE | | CV | 1 | 24 | 22 |
| LV_circumf | LinRegrs | TRUE | FALSE | | CV | 2 | 24 | 22 |
| LV_circumf | LinRegrs | TRUE | FALSE | | CV | 3 | 24 | 23 |
| LV_circumf | LinRegrs | TRUE | FALSE | | CV | 4 | 24 | 24 |
| LV_circumf | Lasso | TRUE | FALSE | 0.000266 | CV | 0 | 24 | 23 |
| LV_circumf | Lasso | TRUE | FALSE | 0.000263 | CV | 1 | 24 | 24 |
| LV_circumf | Lasso | TRUE | FALSE | 0.000302 | CV | 2 | 24 | 22 |
| LV_circumf | Lasso | TRUE | FALSE | 0.000259 | CV | 3 | 24 | 24 |
| LV_circumf | Lasso | TRUE | FALSE | 0.000264 | CV | 4 | 24 | 24 |
| LV_circumf | Lasso | TRUE | TRUE | 0.05 | CV | 0 | 24 | 11 |
| LV_circumf | Lasso | TRUE | TRUE | 0.05 | CV | 1 | 24 | 11 |
| LV_circumf | Lasso | TRUE | TRUE | 0.05 | CV | 2 | 24 | 12 |
| LV_circumf | Lasso | TRUE | TRUE | 0.05 | CV | 3 | 24 | 11 |
| LV_circumf | Lasso | TRUE | TRUE | 0.05 | CV | 4 | 24 | 11 |
| LV_circumf | LinRegrs | TRUE | FALSE | | HeldoutTest | | 24 | 23 |
| LV_circumf | Lasso | TRUE | FALSE | 0.000303 | HeldoutTest | | 24 | 24 |
| LV_circumf | Lasso | TRUE | TRUE | 0.05 | HeldoutTest | | 24 | 12 |
| LV_circumf | LinRegrs | TRUE | FALSE | | CV | 0 | 24 | 22 |
| LV_circumf | LinRegrs | TRUE | FALSE | | CV | 1 | 24 | 22 |
| LV_circumf | LinRegrs | TRUE | FALSE | | CV | 2 | 24 | 23 |
| LV_circumf | LinRegrs | TRUE | FALSE | | CV | 3 | 24 | 21 |
| LV_circumf | LinRegrs | TRUE | FALSE | | CV | 4 | 24 | 22 |
| LV_circumf | Lasso | TRUE | FALSE | 0.000714 | CV | 0 | 24 | 22 |
| LV_circumf | Lasso | TRUE | FALSE | 0.000644 | CV | 1 | 24 | 24 |
| LV_circumf | Lasso | TRUE | FALSE | 0.000282 | CV | 2 | 24 | 23 |
| LV_circumf | Lasso | TRUE | FALSE | 0.000327 | CV | 3 | 24 | 23 |
| LV_circumf | Lasso | TRUE | FALSE | 0.000425 | CV | 4 | 24 | 24 |
| LV_circumf | Lasso | TRUE | TRUE | 0.05 | CV | 0 | 24 | 12 |
| LV_circumf | Lasso | TRUE | TRUE | 0.05 | CV | 1 | 24 | 12 |
| LV_circumf | Lasso | TRUE | TRUE | 0.05 | CV | 2 | 24 | 12 |
| LV_circumf | Lasso | TRUE | TRUE | 0.05 | CV | 3 | 24 | 13 |

| | | | | | | | | |
|---|---|---|---|---|---|---|---|---|
| LV_circumf | Lasso | TRUE | TRUE | 0.05 | CV | 4 | 24 | 9 |
| LV_circumf | LinRegrs | TRUE | FALSE | | HeldoutTest | | 24 | 23 |
| LV_circumf | Lasso | TRUE | FALSE | 0.000335 | HeldoutTest | | 24 | 24 |
| LV_circumf | Lasso | TRUE | TRUE | 0.05 | HeldoutTest | | 24 | 14 |
| LV_circumf | LinRegrs | TRUE | FALSE | | CV | 0 | 24 | 23 |
| LV_circumf | LinRegrs | TRUE | FALSE | | CV | 1 | 24 | 23 |
| LV_circumf | LinRegrs | TRUE | FALSE | | CV | 2 | 24 | 23 |
| LV_circumf | LinRegrs | TRUE | FALSE | | CV | 3 | 24 | 23 |
| LV_circumf | LinRegrs | TRUE | FALSE | | CV | 4 | 24 | 22 |
| LV_circumf | Lasso | TRUE | FALSE | 0.000341 | CV | 0 | 24 | 23 |
| LV_circumf | Lasso | TRUE | FALSE | 0.000336 | CV | 1 | 24 | 23 |
| LV_circumf | Lasso | TRUE | FALSE | 0.000333 | CV | 2 | 24 | 24 |
| LV_circumf | Lasso | TRUE | FALSE | 0.000333 | CV | 3 | 24 | 24 |
| LV_circumf | Lasso | TRUE | FALSE | 0.00041 | CV | 4 | 24 | 24 |
| LV_circumf | Lasso | TRUE | TRUE | 0.05 | CV | 0 | 24 | 13 |
| LV_circumf | Lasso | TRUE | TRUE | 0.05 | CV | 1 | 24 | 14 |
| LV_circumf | Lasso | TRUE | TRUE | 0.05 | CV | 2 | 24 | 14 |
| LV_circumf | Lasso | TRUE | TRUE | 0.05 | CV | 3 | 24 | 13 |
| LV_circumf | Lasso | TRUE | TRUE | 0.05 | CV | 4 | 24 | 12 |
| LV_circumf | LinRegrs | TRUE | FALSE | | HeldoutTest | | 24 | 24 |
| LV_circumf | Lasso | TRUE | FALSE | 0.000313 | HeldoutTest | | 24 | 24 |
| LV_circumf | Lasso | TRUE | TRUE | 0.05 | HeldoutTest | | 24 | 12 |
| LV_circumf | LinRegrs | TRUE | FALSE | | CV | 0 | 24 | 23 |
| LV_circumf | LinRegrs | TRUE | FALSE | | CV | 1 | 24 | 24 |
| LV_circumf | LinRegrs | TRUE | FALSE | | CV | 2 | 24 | 24 |
| LV_circumf | LinRegrs | TRUE | FALSE | | CV | 3 | 24 | 24 |
| LV_circumf | LinRegrs | TRUE | FALSE | | CV | 4 | 24 | 22 |
| LV_circumf | Lasso | TRUE | FALSE | 0.000314 | CV | 0 | 24 | 23 |
| LV_circumf | Lasso | TRUE | FALSE | 0.000312 | CV | 1 | 24 | 24 |
| LV_circumf | Lasso | TRUE | FALSE | 0.000313 | CV | 2 | 24 | 24 |
| LV_circumf | Lasso | TRUE | FALSE | 0.000312 | CV | 3 | 24 | 24 |
| LV_circumf | Lasso | TRUE | FALSE | 0.000312 | CV | 4 | 24 | 23 |
| LV_circumf | Lasso | TRUE | TRUE | 0.05 | CV | 0 | 24 | 12 |
| LV_circumf | Lasso | TRUE | TRUE | 0.05 | CV | 1 | 24 | 12 |
| LV_circumf | Lasso | TRUE | TRUE | 0.05 | CV | 2 | 24 | 12 |
| LV_circumf | Lasso | TRUE | TRUE | 0.05 | CV | 3 | 24 | 12 |
| LV_circumf | Lasso | TRUE | TRUE | 0.05 | CV | 4 | 24 | 13 |
| LV_circumf | LinRegrs | TRUE | FALSE | | HeldoutTest | | 24 | 24 |
| LV_circumf | Lasso | TRUE | FALSE | 0.00036 | HeldoutTest | | 24 | 24 |
| LV_circumf | Lasso | TRUE | TRUE | 0.05 | HeldoutTest | | 24 | 15 |
| LV_circumf | LinRegrs | TRUE | FALSE | | CV | 0 | 24 | 23 |
| LV_circumf | LinRegrs | TRUE | FALSE | | CV | 1 | 24 | 23 |
| LV_circumf | LinRegrs | TRUE | FALSE | | CV | 2 | 24 | 23 |
| LV_circumf | LinRegrs | TRUE | FALSE | | CV | 3 | 24 | 23 |
| LV_circumf | LinRegrs | TRUE | FALSE | | CV | 4 | 24 | 20 |

| | | | | | | | | |
|---|---|---|---|---|---|---|---|---|
| LV_circumf | Lasso | TRUE | FALSE | 0.000365 | CV | 0 | 24 | 24 |
| LV_circumf | Lasso | TRUE | FALSE | 0.000356 | CV | 1 | 24 | 24 |
| LV_circumf | Lasso | TRUE | FALSE | 0.000361 | CV | 2 | 24 | 24 |
| LV_circumf | Lasso | TRUE | FALSE | 0.000358 | CV | 3 | 24 | 24 |
| LV_circumf | Lasso | TRUE | FALSE | 0.00036 | CV | 4 | 24 | 23 |
| LV_circumf | Lasso | TRUE | TRUE | 0.05 | CV | 0 | 24 | 16 |
| LV_circumf | Lasso | TRUE | TRUE | 0.05 | CV | 1 | 24 | 15 |
| LV_circumf | Lasso | TRUE | TRUE | 0.05 | CV | 2 | 24 | 15 |
| LV_circumf | Lasso | TRUE | TRUE | 0.05 | CV | 3 | 24 | 14 |
| LV_circumf | Lasso | TRUE | TRUE | 0.05 | CV | 4 | 24 | 15 |
| LV_circumf | LinRegrs | TRUE | FALSE | | HeldoutTest | | 24 | 22 |
| LV_circumf | Lasso | TRUE | FALSE | 0.000341 | HeldoutTest | | 24 | 24 |
| LV_circumf | Lasso | TRUE | TRUE | 0.05 | HeldoutTest | | 24 | 17 |
| LV_circumf | LinRegrs | TRUE | FALSE | | CV | 0 | 24 | 22 |
| LV_circumf | LinRegrs | TRUE | FALSE | | CV | 1 | 24 | 22 |
| LV_circumf | LinRegrs | TRUE | FALSE | | CV | 2 | 24 | 23 |
| LV_circumf | LinRegrs | TRUE | FALSE | | CV | 3 | 24 | 22 |
| LV_circumf | LinRegrs | TRUE | FALSE | | CV | 4 | 24 | 22 |
| LV_circumf | Lasso | TRUE | FALSE | 0.000345 | CV | 0 | 24 | 23 |
| LV_circumf | Lasso | TRUE | FALSE | 0.000339 | CV | 1 | 24 | 24 |
| LV_circumf | Lasso | TRUE | FALSE | 0.000341 | CV | 2 | 24 | 24 |
| LV_circumf | Lasso | TRUE | FALSE | 0.000339 | CV | 3 | 24 | 24 |
| LV_circumf | Lasso | TRUE | FALSE | 0.000342 | CV | 4 | 24 | 23 |
| LV_circumf | Lasso | TRUE | TRUE | 0.05 | CV | 0 | 24 | 16 |
| LV_circumf | Lasso | TRUE | TRUE | 0.05 | CV | 1 | 24 | 17 |
| LV_circumf | Lasso | TRUE | TRUE | 0.05 | CV | 2 | 24 | 17 |
| LV_circumf | Lasso | TRUE | TRUE | 0.05 | CV | 3 | 24 | 16 |
| LV_circumf | Lasso | TRUE | TRUE | 0.05 | CV | 4 | 24 | 16 |
| LV_circumf | LinRegrs | TRUE | FALSE | | HeldoutTest | | 24 | 23 |
| LV_circumf | Lasso | TRUE | FALSE | 0.000266 | HeldoutTest | | 24 | 24 |
| LV_circumf | Lasso | TRUE | TRUE | 0.05 | HeldoutTest | | 24 | 12 |
| LV_circumf | LinRegrs | TRUE | FALSE | | CV | 0 | 24 | 22 |
| LV_circumf | LinRegrs | TRUE | FALSE | | CV | 1 | 24 | 22 |
| LV_circumf | LinRegrs | TRUE | FALSE | | CV | 2 | 24 | 23 |
| LV_circumf | LinRegrs | TRUE | FALSE | | CV | 3 | 24 | 23 |
| LV_circumf | LinRegrs | TRUE | FALSE | | CV | 4 | 24 | 23 |
| LV_circumf | Lasso | TRUE | FALSE | 0.000271 | CV | 0 | 24 | 24 |
| LV_circumf | Lasso | TRUE | FALSE | 0.000429 | CV | 1 | 24 | 23 |
| LV_circumf | Lasso | TRUE | FALSE | 0.000265 | CV | 2 | 24 | 23 |
| LV_circumf | Lasso | TRUE | FALSE | 0.000266 | CV | 3 | 24 | 23 |
| LV_circumf | Lasso | TRUE | FALSE | 0.000265 | CV | 4 | 24 | 23 |
| LV_circumf | Lasso | TRUE | TRUE | 0.05 | CV | 0 | 24 | 12 |
| LV_circumf | Lasso | TRUE | TRUE | 0.05 | CV | 1 | 24 | 12 |
| LV_circumf | Lasso | TRUE | TRUE | 0.05 | CV | 2 | 24 | 12 |
| LV_circumf | Lasso | TRUE | TRUE | 0.05 | CV | 3 | 24 | 12 |

| | | | | | | | | |
|---|---|---|---|---|---|---|---|---|
| LV_circumf | Lasso | TRUE | TRUE | 0.05 | CV | 4 | 24 | 12 |
| LV_circumf | LinRegrs | TRUE | FALSE | | HeldoutTest | | 24 | 22 |
| LV_circumf | Lasso | TRUE | FALSE | 0.000322 | HeldoutTest | | 24 | 24 |
| LV_circumf | Lasso | TRUE | TRUE | 0.05 | HeldoutTest | | 24 | 13 |
| LV_circumf | LinRegrs | TRUE | FALSE | | CV | 0 | 24 | 22 |
| LV_circumf | LinRegrs | TRUE | FALSE | | CV | 1 | 24 | 23 |
| LV_circumf | LinRegrs | TRUE | FALSE | | CV | 2 | 24 | 21 |
| LV_circumf | LinRegrs | TRUE | FALSE | | CV | 3 | 24 | 21 |
| LV_circumf | LinRegrs | TRUE | FALSE | | CV | 4 | 24 | 22 |
| LV_circumf | Lasso | TRUE | FALSE | 0.000403 | CV | 0 | 24 | 24 |
| LV_circumf | Lasso | TRUE | FALSE | 0.00032 | CV | 1 | 24 | 24 |
| LV_circumf | Lasso | TRUE | FALSE | 0.000323 | CV | 2 | 24 | 24 |
| LV_circumf | Lasso | TRUE | FALSE | 0.000318 | CV | 3 | 24 | 24 |
| LV_circumf | Lasso | TRUE | FALSE | 0.000321 | CV | 4 | 24 | 23 |
| LV_circumf | Lasso | TRUE | TRUE | 0.05 | CV | 0 | 24 | 13 |
| LV_circumf | Lasso | TRUE | TRUE | 0.05 | CV | 1 | 24 | 13 |
| LV_circumf | Lasso | TRUE | TRUE | 0.05 | CV | 2 | 24 | 13 |
| LV_circumf | Lasso | TRUE | TRUE | 0.05 | CV | 3 | 24 | 13 |
| LV_circumf | Lasso | TRUE | TRUE | 0.05 | CV | 4 | 24 | 13 |
| LV_circumf | LinRegrs | TRUE | FALSE | | HeldoutTest | | 24 | 23 |
| LV_circumf | Lasso | TRUE | FALSE | 0.000381 | HeldoutTest | | 24 | 24 |
| LV_circumf | Lasso | TRUE | TRUE | 0.05 | HeldoutTest | | 24 | 17 |
| LV_circumf | LinRegrs | TRUE | FALSE | | CV | 0 | 24 | 22 |
| LV_circumf | LinRegrs | TRUE | FALSE | | CV | 1 | 24 | 23 |
| LV_circumf | LinRegrs | TRUE | FALSE | | CV | 2 | 24 | 23 |
| LV_circumf | LinRegrs | TRUE | FALSE | | CV | 3 | 24 | 23 |
| LV_circumf | LinRegrs | TRUE | FALSE | | CV | 4 | 24 | 20 |
| LV_circumf | Lasso | TRUE | FALSE | 0.000387 | CV | 0 | 24 | 24 |
| LV_circumf | Lasso | TRUE | FALSE | 0.000379 | CV | 1 | 24 | 24 |
| LV_circumf | Lasso | TRUE | FALSE | 0.00038 | CV | 2 | 24 | 24 |
| LV_circumf | Lasso | TRUE | FALSE | 0.000379 | CV | 3 | 24 | 24 |
| LV_circumf | Lasso | TRUE | FALSE | 0.000379 | CV | 4 | 24 | 21 |
| LV_circumf | Lasso | TRUE | TRUE | 0.05 | CV | 0 | 24 | 16 |
| LV_circumf | Lasso | TRUE | TRUE | 0.05 | CV | 1 | 24 | 18 |
| LV_circumf | Lasso | TRUE | TRUE | 0.05 | CV | 2 | 24 | 15 |
| LV_circumf | Lasso | TRUE | TRUE | 0.05 | CV | 3 | 24 | 17 |
| LV_circumf | Lasso | TRUE | TRUE | 0.05 | CV | 4 | 24 | 16 |
| RV_end_di | LinRegrs | TRUE | FALSE | | HeldoutTest | | 24 | 22 |
| RV_end_di | Lasso | TRUE | FALSE | 0.000145 | HeldoutTest | | 24 | 24 |
| RV_end_di | Lasso | TRUE | TRUE | 0.05 | HeldoutTest | | 24 | 5 |
| RV_end_di | LinRegrs | TRUE | FALSE | | CV | 0 | 24 | 20 |
| RV_end_di | LinRegrs | TRUE | FALSE | | CV | 1 | 24 | 22 |
| RV_end_di | LinRegrs | TRUE | FALSE | | CV | 2 | 24 | 21 |
| RV_end_di | LinRegrs | TRUE | FALSE | | CV | 3 | 24 | 20 |
| RV_end_di | LinRegrs | TRUE | FALSE | | CV | 4 | 24 | 22 |

| | | | | | | | | |
|---|---|---|---|---|---|---|---|---|
| RV_end_di | Lasso | TRUE | FALSE | 0.000589 | CV | 0 | 24 | 24 |
| RV_end_di | Lasso | TRUE | FALSE | 0.000144 | CV | 1 | 24 | 24 |
| RV_end_di | Lasso | TRUE | FALSE | 0.000382 | CV | 2 | 24 | 23 |
| RV_end_di | Lasso | TRUE | FALSE | 0.000364 | CV | 3 | 24 | 23 |
| RV_end_di | Lasso | TRUE | FALSE | 0.000442 | CV | 4 | 24 | 24 |
| RV_end_di | Lasso | TRUE | TRUE | 0.05 | CV | 0 | 24 | 5 |
| RV_end_di | Lasso | TRUE | TRUE | 0.05 | CV | 1 | 24 | 5 |
| RV_end_di | Lasso | TRUE | TRUE | 0.05 | CV | 2 | 24 | 5 |
| RV_end_di | Lasso | TRUE | TRUE | 0.05 | CV | 3 | 24 | 5 |
| RV_end_di | Lasso | TRUE | TRUE | 0.05 | CV | 4 | 24 | 5 |
| RV_end_sy | LinRegrs | TRUE | FALSE | | HeldoutTest | | 24 | 20 |
| RV_end_sy | Lasso | TRUE | FALSE | 0.000256 | HeldoutTest | | 24 | 24 |
| RV_end_sy | Lasso | TRUE | TRUE | 0.05 | HeldoutTest | | 24 | 9 |
| RV_end_sy | LinRegrs | TRUE | FALSE | | CV | 0 | 24 | 19 |
| RV_end_sy | LinRegrs | TRUE | FALSE | | CV | 1 | 24 | 20 |
| RV_end_sy | LinRegrs | TRUE | FALSE | | CV | 2 | 24 | 20 |
| RV_end_sy | LinRegrs | TRUE | FALSE | | CV | 3 | 24 | 19 |
| RV_end_sy | LinRegrs | TRUE | FALSE | | CV | 4 | 24 | 22 |
| RV_end_sy | Lasso | TRUE | FALSE | 0.000641 | CV | 0 | 24 | 23 |
| RV_end_sy | Lasso | TRUE | FALSE | 0.000221 | CV | 1 | 24 | 24 |
| RV_end_sy | Lasso | TRUE | FALSE | 0.000223 | CV | 2 | 24 | 24 |
| RV_end_sy | Lasso | TRUE | FALSE | 0.000509 | CV | 3 | 24 | 23 |
| RV_end_sy | Lasso | TRUE | FALSE | 0.000222 | CV | 4 | 24 | 24 |
| RV_end_sy | Lasso | TRUE | TRUE | 0.05 | CV | 0 | 24 | 9 |
| RV_end_sy | Lasso | TRUE | TRUE | 0.05 | CV | 1 | 24 | 10 |
| RV_end_sy | Lasso | TRUE | TRUE | 0.05 | CV | 2 | 24 | 10 |
| RV_end_sy | Lasso | TRUE | TRUE | 0.05 | CV | 3 | 24 | 9 |
| RV_end_sy | Lasso | TRUE | TRUE | 0.05 | CV | 4 | 24 | 10 |
| RV_stroke_ | LinRegrs | TRUE | FALSE | | HeldoutTest | | 24 | 23 |
| RV_stroke_ | Lasso | TRUE | FALSE | 0.000277 | HeldoutTest | | 24 | 24 |
| RV_stroke_ | Lasso | TRUE | TRUE | 0.05 | HeldoutTest | | 24 | 7 |
| RV_stroke_ | LinRegrs | TRUE | FALSE | | CV | 0 | 24 | 23 |
| RV_stroke_ | LinRegrs | TRUE | FALSE | | CV | 1 | 24 | 22 |
| RV_stroke_ | LinRegrs | TRUE | FALSE | | CV | 2 | 24 | 23 |
| RV_stroke_ | LinRegrs | TRUE | FALSE | | CV | 3 | 24 | 22 |
| RV_stroke_ | LinRegrs | TRUE | FALSE | | CV | 4 | 24 | 21 |
| RV_stroke_ | Lasso | TRUE | FALSE | 0.000226 | CV | 0 | 24 | 24 |
| RV_stroke_ | Lasso | TRUE | FALSE | 0.00046 | CV | 1 | 24 | 24 |
| RV_stroke_ | Lasso | TRUE | FALSE | 0.000191 | CV | 2 | 24 | 24 |
| RV_stroke_ | Lasso | TRUE | FALSE | 0.000275 | CV | 3 | 24 | 24 |
| RV_stroke_ | Lasso | TRUE | FALSE | 0.000446 | CV | 4 | 24 | 23 |
| RV_stroke_ | Lasso | TRUE | TRUE | 0.05 | CV | 0 | 24 | 7 |
| RV_stroke_ | Lasso | TRUE | TRUE | 0.05 | CV | 1 | 24 | 7 |
| RV_stroke_ | Lasso | TRUE | TRUE | 0.05 | CV | 2 | 24 | 6 |
| RV_stroke_ | Lasso | TRUE | TRUE | 0.05 | CV | 3 | 24 | 6 |

| | | | | | | | | |
|---|---|---|---|---|---|---|---|---|
| RV_stroke_ | Lasso | TRUE | TRUE | 0.05 | CV | 4 | 24 | 7 |
| RV_ejectio | LinRegrs | TRUE | FALSE | | HeldoutTest | | 24 | 22 |
| RV_ejectio | Lasso | TRUE | FALSE | 0.000403 | HeldoutTest | | 24 | 24 |
| RV_ejectio | Lasso | TRUE | TRUE | 0.05 | HeldoutTest | | 24 | 15 |
| RV_ejectio | LinRegrs | TRUE | FALSE | | CV | 0 | 24 | 22 |
| RV_ejectio | LinRegrs | TRUE | FALSE | | CV | 1 | 24 | 22 |
| RV_ejectio | LinRegrs | TRUE | FALSE | | CV | 2 | 24 | 22 |
| RV_ejectio | LinRegrs | TRUE | FALSE | | CV | 3 | 24 | 21 |
| RV_ejectio | LinRegrs | TRUE | FALSE | | CV | 4 | 24 | 23 |
| RV_ejectio | Lasso | TRUE | FALSE | 0.000407 | CV | 0 | 24 | 24 |
| RV_ejectio | Lasso | TRUE | FALSE | 0.000402 | CV | 1 | 24 | 24 |
| RV_ejectio | Lasso | TRUE | FALSE | 0.000402 | CV | 2 | 24 | 23 |
| RV_ejectio | Lasso | TRUE | FALSE | 0.000402 | CV | 3 | 24 | 24 |
| RV_ejectio | Lasso | TRUE | FALSE | 0.000404 | CV | 4 | 24 | 24 |
| RV_ejectio | Lasso | TRUE | TRUE | 0.05 | CV | 0 | 24 | 16 |
| RV_ejectio | Lasso | TRUE | TRUE | 0.05 | CV | 1 | 24 | 15 |
| RV_ejectio | Lasso | TRUE | TRUE | 0.05 | CV | 2 | 24 | 15 |
| RV_ejectio | Lasso | TRUE | TRUE | 0.05 | CV | 3 | 24 | 16 |
| RV_ejectio | Lasso | TRUE | TRUE | 0.05 | CV | 4 | 24 | 13 |
| LV_longitu | LinRegrs | TRUE | FALSE | | HeldoutTest | | 24 | 20 |
| LV_longitu | Lasso | TRUE | FALSE | 0.000228 | HeldoutTest | | 24 | 24 |
| LV_longitu | Lasso | TRUE | TRUE | 0.05 | HeldoutTest | | 24 | 12 |
| LV_longitu | LinRegrs | TRUE | FALSE | | CV | 0 | 24 | 21 |
| LV_longitu | LinRegrs | TRUE | FALSE | | CV | 1 | 24 | 21 |
| LV_longitu | LinRegrs | TRUE | FALSE | | CV | 2 | 24 | 22 |
| LV_longitu | LinRegrs | TRUE | FALSE | | CV | 3 | 24 | 20 |
| LV_longitu | LinRegrs | TRUE | FALSE | | CV | 4 | 24 | 24 |
| LV_longitu | Lasso | TRUE | FALSE | 0.000231 | CV | 0 | 24 | 24 |
| LV_longitu | Lasso | TRUE | FALSE | 0.000266 | CV | 1 | 24 | 24 |
| LV_longitu | Lasso | TRUE | FALSE | 0.000453 | CV | 2 | 24 | 23 |
| LV_longitu | Lasso | TRUE | FALSE | 0.00028 | CV | 3 | 24 | 23 |
| LV_longitu | Lasso | TRUE | FALSE | 0.000227 | CV | 4 | 24 | 24 |
| LV_longitu | Lasso | TRUE | TRUE | 0.05 | CV | 0 | 24 | 12 |
| LV_longitu | Lasso | TRUE | TRUE | 0.05 | CV | 1 | 24 | 11 |
| LV_longitu | Lasso | TRUE | TRUE | 0.05 | CV | 2 | 24 | 11 |
| LV_longitu | Lasso | TRUE | TRUE | 0.05 | CV | 3 | 24 | 12 |
| LV_longitu | Lasso | TRUE | TRUE | 0.05 | CV | 4 | 24 | 11 |
| LV_longitu | LinRegrs | TRUE | FALSE | | HeldoutTest | | 24 | 22 |
| LV_longitu | Lasso | TRUE | FALSE | 0.00034 | HeldoutTest | | 24 | 23 |
| LV_longitu | Lasso | TRUE | TRUE | 0.05 | HeldoutTest | | 24 | 14 |
| LV_longitu | LinRegrs | TRUE | FALSE | | CV | 0 | 24 | 22 |
| LV_longitu | LinRegrs | TRUE | FALSE | | CV | 1 | 24 | 22 |
| LV_longitu | LinRegrs | TRUE | FALSE | | CV | 2 | 24 | 21 |
| LV_longitu | LinRegrs | TRUE | FALSE | | CV | 3 | 24 | 21 |
| LV_longitu | LinRegrs | TRUE | FALSE | | CV | 4 | 24 | 24 |

| | | | | | | | | |
|---|---|---|---|---|---|---|---|---|
| LV_longitu | Lasso | TRUE | FALSE | 0.000341 | CV | 0 | 24 | 23 |
| LV_longitu | Lasso | TRUE | FALSE | 0.000338 | CV | 1 | 24 | 24 |
| LV_longitu | Lasso | TRUE | FALSE | 0.000338 | CV | 2 | 24 | 24 |
| LV_longitu | Lasso | TRUE | FALSE | 0.000342 | CV | 3 | 24 | 24 |
| LV_longitu | Lasso | TRUE | FALSE | 0.000338 | CV | 4 | 24 | 24 |
| LV_longitu | Lasso | TRUE | TRUE | 0.05 | CV | 0 | 24 | 13 |
| LV_longitu | Lasso | TRUE | TRUE | 0.05 | CV | 1 | 24 | 13 |
| LV_longitu | Lasso | TRUE | TRUE | 0.05 | CV | 2 | 24 | 13 |
| LV_longitu | Lasso | TRUE | TRUE | 0.05 | CV | 3 | 24 | 14 |
| LV_longitu | Lasso | TRUE | TRUE | 0.05 | CV | 4 | 24 | 13 |
| LV_longitu | LinRegrs | TRUE | FALSE | | HeldoutTest | | 24 | 23 |
| LV_longitu | Lasso | TRUE | FALSE | 0.000275 | HeldoutTest | | 24 | 24 |
| LV_longitu | Lasso | TRUE | TRUE | 0.05 | HeldoutTest | | 24 | 15 |
| LV_longitu | LinRegrs | TRUE | FALSE | | CV | 0 | 24 | 23 |
| LV_longitu | LinRegrs | TRUE | FALSE | | CV | 1 | 24 | 22 |
| LV_longitu | LinRegrs | TRUE | FALSE | | CV | 2 | 24 | 23 |
| LV_longitu | LinRegrs | TRUE | FALSE | | CV | 3 | 24 | 23 |
| LV_longitu | LinRegrs | TRUE | FALSE | | CV | 4 | 24 | 20 |
| LV_longitu | Lasso | TRUE | FALSE | 0.000279 | CV | 0 | 24 | 24 |
| LV_longitu | Lasso | TRUE | FALSE | 0.000272 | CV | 1 | 24 | 24 |
| LV_longitu | Lasso | TRUE | FALSE | 0.000272 | CV | 2 | 24 | 24 |
| LV_longitu | Lasso | TRUE | FALSE | 0.000276 | CV | 3 | 24 | 24 |
| LV_longitu | Lasso | TRUE | FALSE | 0.000479 | CV | 4 | 24 | 24 |
| LV_longitu | Lasso | TRUE | TRUE | 0.05 | CV | 0 | 24 | 15 |
| LV_longitu | Lasso | TRUE | TRUE | 0.05 | CV | 1 | 24 | 15 |
| LV_longitu | Lasso | TRUE | TRUE | 0.05 | CV | 2 | 24 | 16 |
| LV_longitu | Lasso | TRUE | TRUE | 0.05 | CV | 3 | 24 | 15 |
| LV_longitu | Lasso | TRUE | TRUE | 0.05 | CV | 4 | 24 | 14 |
| LV_longitu | LinRegrs | TRUE | FALSE | | HeldoutTest | | 24 | 23 |
| LV_longitu | Lasso | TRUE | FALSE | 0.000271 | HeldoutTest | | 24 | 24 |
| LV_longitu | Lasso | TRUE | TRUE | 0.05 | HeldoutTest | | 24 | 13 |
| LV_longitu | LinRegrs | TRUE | FALSE | | CV | 0 | 24 | 22 |
| LV_longitu | LinRegrs | TRUE | FALSE | | CV | 1 | 24 | 23 |
| LV_longitu | LinRegrs | TRUE | FALSE | | CV | 2 | 24 | 23 |
| LV_longitu | LinRegrs | TRUE | FALSE | | CV | 3 | 24 | 22 |
| LV_longitu | LinRegrs | TRUE | FALSE | | CV | 4 | 24 | 23 |
| LV_longitu | Lasso | TRUE | FALSE | 0.000271 | CV | 0 | 24 | 23 |
| LV_longitu | Lasso | TRUE | FALSE | 0.000271 | CV | 1 | 24 | 24 |
| LV_longitu | Lasso | TRUE | FALSE | 0.00027 | CV | 2 | 24 | 24 |
| LV_longitu | Lasso | TRUE | FALSE | 0.000269 | CV | 3 | 24 | 24 |
| LV_longitu | Lasso | TRUE | FALSE | 0.000272 | CV | 4 | 24 | 24 |
| LV_longitu | Lasso | TRUE | TRUE | 0.05 | CV | 0 | 24 | 13 |
| LV_longitu | Lasso | TRUE | TRUE | 0.05 | CV | 1 | 24 | 13 |
| LV_longitu | Lasso | TRUE | TRUE | 0.05 | CV | 2 | 24 | 13 |
| LV_longitu | Lasso | TRUE | TRUE | 0.05 | CV | 3 | 24 | 13 |

| | | | | | | | | |
|---|---|---|---|---|---|---|---|---|
| LV_longitu | Lasso | TRUE | TRUE | 0.05 | CV | 4 | 24 | 12 |
| LV_longitu | LinRegrs | TRUE | FALSE | | HeldoutTest | | 24 | 24 |
| LV_longitu | Lasso | TRUE | FALSE | 0.000256 | HeldoutTest | | 24 | 24 |
| LV_longitu | Lasso | TRUE | TRUE | 0.05 | HeldoutTest | | 24 | 12 |
| LV_longitu | LinRegrs | TRUE | FALSE | | CV | 0 | 24 | 22 |
| LV_longitu | LinRegrs | TRUE | FALSE | | CV | 1 | 24 | 23 |
| LV_longitu | LinRegrs | TRUE | FALSE | | CV | 2 | 24 | 24 |
| LV_longitu | LinRegrs | TRUE | FALSE | | CV | 3 | 24 | 24 |
| LV_longitu | LinRegrs | TRUE | FALSE | | CV | 4 | 24 | 21 |
| LV_longitu | Lasso | TRUE | FALSE | 0.000259 | CV | 0 | 24 | 24 |
| LV_longitu | Lasso | TRUE | FALSE | 0.000253 | CV | 1 | 24 | 24 |
| LV_longitu | Lasso | TRUE | FALSE | 0.000255 | CV | 2 | 24 | 24 |
| LV_longitu | Lasso | TRUE | FALSE | 0.000275 | CV | 3 | 24 | 24 |
| LV_longitu | Lasso | TRUE | FALSE | 0.000548 | CV | 4 | 24 | 23 |
| LV_longitu | Lasso | TRUE | TRUE | 0.05 | CV | 0 | 24 | 13 |
| LV_longitu | Lasso | TRUE | TRUE | 0.05 | CV | 1 | 24 | 12 |
| LV_longitu | Lasso | TRUE | TRUE | 0.05 | CV | 2 | 24 | 13 |
| LV_longitu | Lasso | TRUE | TRUE | 0.05 | CV | 3 | 24 | 12 |
| LV_longitu | Lasso | TRUE | TRUE | 0.05 | CV | 4 | 24 | 13 |
| LV_longitu | LinRegrs | TRUE | FALSE | | HeldoutTest | | 24 | 23 |
| LV_longitu | Lasso | TRUE | FALSE | 0.000322 | HeldoutTest | | 24 | 24 |
| LV_longitu | Lasso | TRUE | TRUE | 0.05 | HeldoutTest | | 24 | 15 |
| LV_longitu | LinRegrs | TRUE | FALSE | | CV | 0 | 24 | 23 |
| LV_longitu | LinRegrs | TRUE | FALSE | | CV | 1 | 24 | 23 |
| LV_longitu | LinRegrs | TRUE | FALSE | | CV | 2 | 24 | 23 |
| LV_longitu | LinRegrs | TRUE | FALSE | | CV | 3 | 24 | 23 |
| LV_longitu | LinRegrs | TRUE | FALSE | | CV | 4 | 24 | 22 |
| LV_longitu | Lasso | TRUE | FALSE | 0.000323 | CV | 0 | 24 | 24 |
| LV_longitu | Lasso | TRUE | FALSE | 0.000318 | CV | 1 | 24 | 24 |
| LV_longitu | Lasso | TRUE | FALSE | 0.000325 | CV | 2 | 24 | 24 |
| LV_longitu | Lasso | TRUE | FALSE | 0.000322 | CV | 3 | 24 | 24 |
| LV_longitu | Lasso | TRUE | FALSE | 0.000393 | CV | 4 | 24 | 23 |
| LV_longitu | Lasso | TRUE | TRUE | 0.05 | CV | 0 | 24 | 15 |
| LV_longitu | Lasso | TRUE | TRUE | 0.05 | CV | 1 | 24 | 15 |
| LV_longitu | Lasso | TRUE | TRUE | 0.05 | CV | 2 | 24 | 16 |
| LV_longitu | Lasso | TRUE | TRUE | 0.05 | CV | 3 | 24 | 15 |
| LV_longitu | Lasso | TRUE | TRUE | 0.05 | CV | 4 | 24 | 15 |
| LV_longitu | LinRegrs | TRUE | FALSE | | HeldoutTest | | 24 | 22 |
| LV_longitu | Lasso | TRUE | FALSE | 0.000397 | HeldoutTest | | 24 | 24 |
| LV_longitu | Lasso | TRUE | TRUE | 0.05 | HeldoutTest | | 24 | 15 |
| LV_longitu | LinRegrs | TRUE | FALSE | | CV | 0 | 24 | 22 |
| LV_longitu | LinRegrs | TRUE | FALSE | | CV | 1 | 24 | 23 |
| LV_longitu | LinRegrs | TRUE | FALSE | | CV | 2 | 24 | 23 |
| LV_longitu | LinRegrs | TRUE | FALSE | | CV | 3 | 24 | 22 |
| LV_longitu | LinRegrs | TRUE | FALSE | | CV | 4 | 24 | 22 |

| | | | | | | | | |
|---|---|---|---|---|---|---|---|---|
| LV_longitu | Lasso | TRUE | FALSE | 0.000399 | CV | 0 | 24 | 23 |
| LV_longitu | Lasso | TRUE | FALSE | 0.000395 | CV | 1 | 24 | 24 |
| LV_longitu | Lasso | TRUE | FALSE | 0.000396 | CV | 2 | 24 | 23 |
| LV_longitu | Lasso | TRUE | FALSE | 0.000398 | CV | 3 | 24 | 24 |
| LV_longitu | Lasso | TRUE | FALSE | 0.000396 | CV | 4 | 24 | 24 |
| LV_longitu | Lasso | TRUE | TRUE | 0.05 | CV | 0 | 24 | 15 |
| LV_longitu | Lasso | TRUE | TRUE | 0.05 | CV | 1 | 24 | 17 |
| LV_longitu | Lasso | TRUE | TRUE | 0.05 | CV | 2 | 24 | 16 |
| LV_longitu | Lasso | TRUE | TRUE | 0.05 | CV | 3 | 24 | 16 |
| LV_longitu | Lasso | TRUE | TRUE | 0.05 | CV | 4 | 24 | 14 |
| LV_radial_s | LinRegrs | TRUE | FALSE | | HeldoutTest | | 24 | 20 |
| LV_radial_s | Lasso | TRUE | FALSE | 0.000712 | HeldoutTest | | 24 | 21 |
| LV_radial_s | Lasso | TRUE | TRUE | 0.05 | HeldoutTest | | 24 | 12 |
| LV_radial_s | LinRegrs | TRUE | FALSE | | CV | 0 | 24 | 20 |
| LV_radial_s | LinRegrs | TRUE | FALSE | | CV | 1 | 24 | 21 |
| LV_radial_s | LinRegrs | TRUE | FALSE | | CV | 2 | 24 | 21 |
| LV_radial_s | LinRegrs | TRUE | FALSE | | CV | 3 | 24 | 19 |
| LV_radial_s | LinRegrs | TRUE | FALSE | | CV | 4 | 24 | 23 |
| LV_radial_s | Lasso | TRUE | FALSE | 0.000896 | CV | 0 | 24 | 21 |
| LV_radial_s | Lasso | TRUE | FALSE | 0.000623 | CV | 1 | 24 | 22 |
| LV_radial_s | Lasso | TRUE | FALSE | 0.000891 | CV | 2 | 24 | 22 |
| LV_radial_s | Lasso | TRUE | FALSE | 0.000698 | CV | 3 | 24 | 21 |
| LV_radial_s | Lasso | TRUE | FALSE | 0.000261 | CV | 4 | 24 | 24 |
| LV_radial_s | Lasso | TRUE | TRUE | 0.05 | CV | 0 | 24 | 12 |
| LV_radial_s | Lasso | TRUE | TRUE | 0.05 | CV | 1 | 24 | 11 |
| LV_radial_s | Lasso | TRUE | TRUE | 0.05 | CV | 2 | 24 | 12 |
| LV_radial_s | Lasso | TRUE | TRUE | 0.05 | CV | 3 | 24 | 11 |
| LV_radial_s | Lasso | TRUE | TRUE | 0.05 | CV | 4 | 24 | 10 |
| LV_radial_s | LinRegrs | TRUE | FALSE | | HeldoutTest | | 24 | 21 |
| LV_radial_s | Lasso | TRUE | FALSE | 0.000279 | HeldoutTest | | 24 | 24 |
| LV_radial_s | Lasso | TRUE | TRUE | 0.05 | HeldoutTest | | 24 | 11 |
| LV_radial_s | LinRegrs | TRUE | FALSE | | CV | 0 | 24 | 22 |
| LV_radial_s | LinRegrs | TRUE | FALSE | | CV | 1 | 24 | 20 |
| LV_radial_s | LinRegrs | TRUE | FALSE | | CV | 2 | 24 | 21 |
| LV_radial_s | LinRegrs | TRUE | FALSE | | CV | 3 | 24 | 22 |
| LV_radial_s | LinRegrs | TRUE | FALSE | | CV | 4 | 24 | 21 |
| LV_radial_s | Lasso | TRUE | FALSE | 0.000281 | CV | 0 | 24 | 24 |
| LV_radial_s | Lasso | TRUE | FALSE | 0.000348 | CV | 1 | 24 | 24 |
| LV_radial_s | Lasso | TRUE | FALSE | 0.000372 | CV | 2 | 24 | 24 |
| LV_radial_s | Lasso | TRUE | FALSE | 0.000276 | CV | 3 | 24 | 24 |
| LV_radial_s | Lasso | TRUE | FALSE | 0.00039 | CV | 4 | 24 | 24 |
| LV_radial_s | Lasso | TRUE | TRUE | 0.05 | CV | 0 | 24 | 12 |
| LV_radial_s | Lasso | TRUE | TRUE | 0.05 | CV | 1 | 24 | 11 |
| LV_radial_s | Lasso | TRUE | TRUE | 0.05 | CV | 2 | 24 | 11 |
| LV_radial_s | Lasso | TRUE | TRUE | 0.05 | CV | 3 | 24 | 12 |

| | | | | | | | | |
|---|---|---|---|---|---|---|---|---|
| LV_radial_s | Lasso | TRUE | TRUE | 0.05 | CV | 4 | 24 | 10 |
| LV_radial_s | LinRegrs | TRUE | FALSE | | HeldoutTest | | 24 | 23 |
| LV_radial_s | Lasso | TRUE | FALSE | 0.000312 | HeldoutTest | | 24 | 23 |
| LV_radial_s | Lasso | TRUE | TRUE | 0.05 | HeldoutTest | | 24 | 14 |
| LV_radial_s | LinRegrs | TRUE | FALSE | | CV | 0 | 24 | 23 |
| LV_radial_s | LinRegrs | TRUE | FALSE | | CV | 1 | 24 | 23 |
| LV_radial_s | LinRegrs | TRUE | FALSE | | CV | 2 | 24 | 23 |
| LV_radial_s | LinRegrs | TRUE | FALSE | | CV | 3 | 24 | 23 |
| LV_radial_s | LinRegrs | TRUE | FALSE | | CV | 4 | 24 | 23 |
| LV_radial_s | Lasso | TRUE | FALSE | 0.000313 | CV | 0 | 24 | 23 |
| LV_radial_s | Lasso | TRUE | FALSE | 0.000315 | CV | 1 | 24 | 24 |
| LV_radial_s | Lasso | TRUE | FALSE | 0.000312 | CV | 2 | 24 | 24 |
| LV_radial_s | Lasso | TRUE | FALSE | 0.000311 | CV | 3 | 24 | 23 |
| LV_radial_s | Lasso | TRUE | FALSE | 0.00031 | CV | 4 | 24 | 23 |
| LV_radial_s | Lasso | TRUE | TRUE | 0.05 | CV | 0 | 24 | 14 |
| LV_radial_s | Lasso | TRUE | TRUE | 0.05 | CV | 1 | 24 | 14 |
| LV_radial_s | Lasso | TRUE | TRUE | 0.05 | CV | 2 | 24 | 14 |
| LV_radial_s | Lasso | TRUE | TRUE | 0.05 | CV | 3 | 24 | 14 |
| LV_radial_s | Lasso | TRUE | TRUE | 0.05 | CV | 4 | 24 | 12 |
| LV_radial_s | LinRegrs | TRUE | FALSE | | HeldoutTest | | 24 | 24 |
| LV_radial_s | Lasso | TRUE | FALSE | 0.000335 | HeldoutTest | | 24 | 24 |
| LV_radial_s | Lasso | TRUE | TRUE | 0.05 | HeldoutTest | | 24 | 16 |
| LV_radial_s | LinRegrs | TRUE | FALSE | | CV | 0 | 24 | 24 |
| LV_radial_s | LinRegrs | TRUE | FALSE | | CV | 1 | 24 | 24 |
| LV_radial_s | LinRegrs | TRUE | FALSE | | CV | 2 | 24 | 24 |
| LV_radial_s | LinRegrs | TRUE | FALSE | | CV | 3 | 24 | 24 |
| LV_radial_s | LinRegrs | TRUE | FALSE | | CV | 4 | 24 | 24 |
| LV_radial_s | Lasso | TRUE | FALSE | 0.000334 | CV | 0 | 24 | 24 |
| LV_radial_s | Lasso | TRUE | FALSE | 0.000337 | CV | 1 | 24 | 24 |
| LV_radial_s | Lasso | TRUE | FALSE | 0.000331 | CV | 2 | 24 | 24 |
| LV_radial_s | Lasso | TRUE | FALSE | 0.000335 | CV | 3 | 24 | 24 |
| LV_radial_s | Lasso | TRUE | FALSE | 0.000338 | CV | 4 | 24 | 24 |
| LV_radial_s | Lasso | TRUE | TRUE | 0.05 | CV | 0 | 24 | 17 |
| LV_radial_s | Lasso | TRUE | TRUE | 0.05 | CV | 1 | 24 | 16 |
| LV_radial_s | Lasso | TRUE | TRUE | 0.05 | CV | 2 | 24 | 16 |
| LV_radial_s | Lasso | TRUE | TRUE | 0.05 | CV | 3 | 24 | 16 |
| LV_radial_s | Lasso | TRUE | TRUE | 0.05 | CV | 4 | 24 | 15 |
| LV_radial_s | LinRegrs | TRUE | FALSE | | HeldoutTest | | 24 | 23 |
| LV_radial_s | Lasso | TRUE | FALSE | 0.00035 | HeldoutTest | | 24 | 24 |
| LV_radial_s | Lasso | TRUE | TRUE | 0.05 | HeldoutTest | | 24 | 14 |
| LV_radial_s | LinRegrs | TRUE | FALSE | | CV | 0 | 24 | 23 |
| LV_radial_s | LinRegrs | TRUE | FALSE | | CV | 1 | 24 | 23 |
| LV_radial_s | LinRegrs | TRUE | FALSE | | CV | 2 | 24 | 23 |
| LV_radial_s | LinRegrs | TRUE | FALSE | | CV | 3 | 24 | 24 |
| LV_radial_s | LinRegrs | TRUE | FALSE | | CV | 4 | 24 | 23 |

| | | | | | | | | |
|---|---|---|---|---|---|---|---|---|
| LV_radial_s | Lasso | TRUE | FALSE | 0.000349 | CV | 0 | 24 | 24 |
| LV_radial_s | Lasso | TRUE | FALSE | 0.000351 | CV | 1 | 24 | 24 |
| LV_radial_s | Lasso | TRUE | FALSE | 0.00035 | CV | 2 | 24 | 24 |
| LV_radial_s | Lasso | TRUE | FALSE | 0.000352 | CV | 3 | 24 | 24 |
| LV_radial_s | Lasso | TRUE | FALSE | 0.000376 | CV | 4 | 24 | 24 |
| LV_radial_s | Lasso | TRUE | TRUE | 0.05 | CV | 0 | 24 | 15 |
| LV_radial_s | Lasso | TRUE | TRUE | 0.05 | CV | 1 | 24 | 15 |
| LV_radial_s | Lasso | TRUE | TRUE | 0.05 | CV | 2 | 24 | 13 |
| LV_radial_s | Lasso | TRUE | TRUE | 0.05 | CV | 3 | 24 | 14 |
| LV_radial_s | Lasso | TRUE | TRUE | 0.05 | CV | 4 | 24 | 12 |
| LV_radial_s | LinRegrs | TRUE | FALSE | | HeldoutTest | | 24 | 24 |
| LV_radial_s | Lasso | TRUE | FALSE | 0.000263 | HeldoutTest | | 24 | 24 |
| LV_radial_s | Lasso | TRUE | TRUE | 0.05 | HeldoutTest | | 24 | 10 |
| LV_radial_s | LinRegrs | TRUE | FALSE | | CV | 0 | 24 | 24 |
| LV_radial_s | LinRegrs | TRUE | FALSE | | CV | 1 | 24 | 24 |
| LV_radial_s | LinRegrs | TRUE | FALSE | | CV | 2 | 24 | 24 |
| LV_radial_s | LinRegrs | TRUE | FALSE | | CV | 3 | 24 | 24 |
| LV_radial_s | LinRegrs | TRUE | FALSE | | CV | 4 | 24 | 23 |
| LV_radial_s | Lasso | TRUE | FALSE | 0.000264 | CV | 0 | 24 | 24 |
| LV_radial_s | Lasso | TRUE | FALSE | 0.000264 | CV | 1 | 24 | 24 |
| LV_radial_s | Lasso | TRUE | FALSE | 0.000261 | CV | 2 | 24 | 24 |
| LV_radial_s | Lasso | TRUE | FALSE | 0.000263 | CV | 3 | 24 | 24 |
| LV_radial_s | Lasso | TRUE | FALSE | 0.000262 | CV | 4 | 24 | 24 |
| LV_radial_s | Lasso | TRUE | TRUE | 0.05 | CV | 0 | 24 | 9 |
| LV_radial_s | Lasso | TRUE | TRUE | 0.05 | CV | 1 | 24 | 9 |
| LV_radial_s | Lasso | TRUE | TRUE | 0.05 | CV | 2 | 24 | 10 |
| LV_radial_s | Lasso | TRUE | TRUE | 0.05 | CV | 3 | 24 | 9 |
| LV_radial_s | Lasso | TRUE | TRUE | 0.05 | CV | 4 | 24 | 10 |
| LV_radial_s | LinRegrs | TRUE | FALSE | | HeldoutTest | | 24 | 23 |
| LV_radial_s | Lasso | TRUE | FALSE | 0.000336 | HeldoutTest | | 24 | 24 |
| LV_radial_s | Lasso | TRUE | TRUE | 0.05 | HeldoutTest | | 24 | 15 |
| LV_radial_s | LinRegrs | TRUE | FALSE | | CV | 0 | 24 | 23 |
| LV_radial_s | LinRegrs | TRUE | FALSE | | CV | 1 | 24 | 23 |
| LV_radial_s | LinRegrs | TRUE | FALSE | | CV | 2 | 24 | 21 |
| LV_radial_s | LinRegrs | TRUE | FALSE | | CV | 3 | 24 | 23 |
| LV_radial_s | LinRegrs | TRUE | FALSE | | CV | 4 | 24 | 21 |
| LV_radial_s | Lasso | TRUE | FALSE | 0.000341 | CV | 0 | 24 | 24 |
| LV_radial_s | Lasso | TRUE | FALSE | 0.000337 | CV | 1 | 24 | 24 |
| LV_radial_s | Lasso | TRUE | FALSE | 0.000891 | CV | 2 | 24 | 22 |
| LV_radial_s | Lasso | TRUE | FALSE | 0.000333 | CV | 3 | 24 | 24 |
| LV_radial_s | Lasso | TRUE | FALSE | 0.000335 | CV | 4 | 24 | 24 |
| LV_radial_s | Lasso | TRUE | TRUE | 0.05 | CV | 0 | 24 | 16 |
| LV_radial_s | Lasso | TRUE | TRUE | 0.05 | CV | 1 | 24 | 16 |
| LV_radial_s | Lasso | TRUE | TRUE | 0.05 | CV | 2 | 24 | 15 |
| LV_radial_s | Lasso | TRUE | TRUE | 0.05 | CV | 3 | 24 | 14 |

| | | | | | | | | |
|---|---|---|---|---|---|---|---|---|
| LV_radial_s | Lasso | TRUE | TRUE | 0.05 | CV | 4 | 24 | 11 |
| LV_radial_s | LinRegrs | TRUE | FALSE | | HeldoutTest | | 24 | 24 |
| LV_radial_s | Lasso | TRUE | FALSE | 0.000349 | HeldoutTest | | 24 | 24 |
| LV_radial_s | Lasso | TRUE | TRUE | 0.05 | HeldoutTest | | 24 | 14 |
| LV_radial_s | LinRegrs | TRUE | FALSE | | CV | 0 | 24 | 24 |
| LV_radial_s | LinRegrs | TRUE | FALSE | | CV | 1 | 24 | 24 |
| LV_radial_s | LinRegrs | TRUE | FALSE | | CV | 2 | 24 | 22 |
| LV_radial_s | LinRegrs | TRUE | FALSE | | CV | 3 | 24 | 24 |
| LV_radial_s | LinRegrs | TRUE | FALSE | | CV | 4 | 24 | 23 |
| LV_radial_s | Lasso | TRUE | FALSE | 0.000352 | CV | 0 | 24 | 24 |
| LV_radial_s | Lasso | TRUE | FALSE | 0.000375 | CV | 1 | 24 | 24 |
| LV_radial_s | Lasso | TRUE | FALSE | 0.000699 | CV | 2 | 24 | 22 |
| LV_radial_s | Lasso | TRUE | FALSE | 0.000398 | CV | 3 | 24 | 24 |
| LV_radial_s | Lasso | TRUE | FALSE | 0.000348 | CV | 4 | 24 | 24 |
| LV_radial_s | Lasso | TRUE | TRUE | 0.05 | CV | 0 | 24 | 14 |
| LV_radial_s | Lasso | TRUE | TRUE | 0.05 | CV | 1 | 24 | 14 |
| LV_radial_s | Lasso | TRUE | TRUE | 0.05 | CV | 2 | 24 | 14 |
| LV_radial_s | Lasso | TRUE | TRUE | 0.05 | CV | 3 | 24 | 14 |
| LV_radial_s | Lasso | TRUE | TRUE | 0.05 | CV | 4 | 24 | 9 |
| LV_radial_s | LinRegrs | TRUE | FALSE | | HeldoutTest | | 24 | 23 |
| LV_radial_s | Lasso | TRUE | FALSE | 0.000329 | HeldoutTest | | 24 | 24 |
| LV_radial_s | Lasso | TRUE | TRUE | 0.05 | HeldoutTest | | 24 | 16 |
| LV_radial_s | LinRegrs | TRUE | FALSE | | CV | 0 | 24 | 22 |
| LV_radial_s | LinRegrs | TRUE | FALSE | | CV | 1 | 24 | 24 |
| LV_radial_s | LinRegrs | TRUE | FALSE | | CV | 2 | 24 | 22 |
| LV_radial_s | LinRegrs | TRUE | FALSE | | CV | 3 | 24 | 23 |
| LV_radial_s | LinRegrs | TRUE | FALSE | | CV | 4 | 24 | 22 |
| LV_radial_s | Lasso | TRUE | FALSE | 0.000335 | CV | 0 | 24 | 24 |
| LV_radial_s | Lasso | TRUE | FALSE | 0.000326 | CV | 1 | 24 | 24 |
| LV_radial_s | Lasso | TRUE | FALSE | 0.000706 | CV | 2 | 24 | 23 |
| LV_radial_s | Lasso | TRUE | FALSE | 0.000376 | CV | 3 | 24 | 24 |
| LV_radial_s | Lasso | TRUE | FALSE | 0.000328 | CV | 4 | 24 | 23 |
| LV_radial_s | Lasso | TRUE | TRUE | 0.05 | CV | 0 | 24 | 16 |
| LV_radial_s | Lasso | TRUE | TRUE | 0.05 | CV | 1 | 24 | 16 |
| LV_radial_s | Lasso | TRUE | TRUE | 0.05 | CV | 2 | 24 | 15 |
| LV_radial_s | Lasso | TRUE | TRUE | 0.05 | CV | 3 | 24 | 16 |
| LV_radial_s | Lasso | TRUE | TRUE | 0.05 | CV | 4 | 24 | 11 |
| LV_radial_s | LinRegrs | TRUE | FALSE | | HeldoutTest | | 24 | 23 |
| LV_radial_s | Lasso | TRUE | FALSE | 0.000325 | HeldoutTest | | 24 | 24 |
| LV_radial_s | Lasso | TRUE | TRUE | 0.05 | HeldoutTest | | 24 | 13 |
| LV_radial_s | LinRegrs | TRUE | FALSE | | CV | 0 | 24 | 23 |
| LV_radial_s | LinRegrs | TRUE | FALSE | | CV | 1 | 24 | 23 |
| LV_radial_s | LinRegrs | TRUE | FALSE | | CV | 2 | 24 | 23 |
| LV_radial_s | LinRegrs | TRUE | FALSE | | CV | 3 | 24 | 22 |
| LV_radial_s | LinRegrs | TRUE | FALSE | | CV | 4 | 24 | 23 |

| | | | | | | | | |
|---|---|---|---|---|---|---|---|---|
| LV_radial_s | Lasso | TRUE | FALSE | 0.000333 | CV | 0 | 24 | 24 |
| LV_radial_s | Lasso | TRUE | FALSE | 0.000324 | CV | 1 | 24 | 24 |
| LV_radial_s | Lasso | TRUE | FALSE | 0.000325 | CV | 2 | 24 | 24 |
| LV_radial_s | Lasso | TRUE | FALSE | 0.000346 | CV | 3 | 24 | 24 |
| LV_radial_s | Lasso | TRUE | FALSE | 0.000324 | CV | 4 | 24 | 24 |
| LV_radial_s | Lasso | TRUE | TRUE | 0.05 | CV | 0 | 24 | 13 |
| LV_radial_s | Lasso | TRUE | TRUE | 0.05 | CV | 1 | 24 | 13 |
| LV_radial_s | Lasso | TRUE | TRUE | 0.05 | CV | 2 | 24 | 13 |
| LV_radial_s | Lasso | TRUE | TRUE | 0.05 | CV | 3 | 24 | 13 |
| LV_radial_s | Lasso | TRUE | TRUE | 0.05 | CV | 4 | 24 | 15 |
| LV_radial_s | LinRegrs | TRUE | FALSE | | HeldoutTest | | 24 | 23 |
| LV_radial_s | Lasso | TRUE | FALSE | 0.000339 | HeldoutTest | | 24 | 24 |
| LV_radial_s | Lasso | TRUE | TRUE | 0.05 | HeldoutTest | | 24 | 15 |
| LV_radial_s | LinRegrs | TRUE | FALSE | | CV | 0 | 24 | 24 |
| LV_radial_s | LinRegrs | TRUE | FALSE | | CV | 1 | 24 | 23 |
| LV_radial_s | LinRegrs | TRUE | FALSE | | CV | 2 | 24 | 24 |
| LV_radial_s | LinRegrs | TRUE | FALSE | | CV | 3 | 24 | 24 |
| LV_radial_s | LinRegrs | TRUE | FALSE | | CV | 4 | 24 | 22 |
| LV_radial_s | Lasso | TRUE | FALSE | 0.000348 | CV | 0 | 24 | 24 |
| LV_radial_s | Lasso | TRUE | FALSE | 0.000337 | CV | 1 | 24 | 24 |
| LV_radial_s | Lasso | TRUE | FALSE | 0.000339 | CV | 2 | 24 | 24 |
| LV_radial_s | Lasso | TRUE | FALSE | 0.000336 | CV | 3 | 24 | 24 |
| LV_radial_s | Lasso | TRUE | FALSE | 0.000337 | CV | 4 | 24 | 24 |
| LV_radial_s | Lasso | TRUE | TRUE | 0.05 | CV | 0 | 24 | 15 |
| LV_radial_s | Lasso | TRUE | TRUE | 0.05 | CV | 1 | 24 | 16 |
| LV_radial_s | Lasso | TRUE | TRUE | 0.05 | CV | 2 | 24 | 15 |
| LV_radial_s | Lasso | TRUE | TRUE | 0.05 | CV | 3 | 24 | 15 |
| LV_radial_s | Lasso | TRUE | TRUE | 0.05 | CV | 4 | 24 | 14 |
| LV_radial_s | LinRegrs | TRUE | FALSE | | HeldoutTest | | 24 | 23 |
| LV_radial_s | Lasso | TRUE | FALSE | 0.000337 | HeldoutTest | | 24 | 23 |
| LV_radial_s | Lasso | TRUE | TRUE | 0.05 | HeldoutTest | | 24 | 16 |
| LV_radial_s | LinRegrs | TRUE | FALSE | | CV | 0 | 24 | 23 |
| LV_radial_s | LinRegrs | TRUE | FALSE | | CV | 1 | 24 | 22 |
| LV_radial_s | LinRegrs | TRUE | FALSE | | CV | 2 | 24 | 22 |
| LV_radial_s | LinRegrs | TRUE | FALSE | | CV | 3 | 24 | 22 |
| LV_radial_s | LinRegrs | TRUE | FALSE | | CV | 4 | 24 | 21 |
| LV_radial_s | Lasso | TRUE | FALSE | 0.000345 | CV | 0 | 24 | 24 |
| LV_radial_s | Lasso | TRUE | FALSE | 0.000777 | CV | 1 | 24 | 23 |
| LV_radial_s | Lasso | TRUE | FALSE | 0.000337 | CV | 2 | 24 | 24 |
| LV_radial_s | Lasso | TRUE | FALSE | 0.000333 | CV | 3 | 24 | 24 |
| LV_radial_s | Lasso | TRUE | FALSE | 0.000359 | CV | 4 | 24 | 24 |
| LV_radial_s | Lasso | TRUE | TRUE | 0.05 | CV | 0 | 24 | 16 |
| LV_radial_s | Lasso | TRUE | TRUE | 0.05 | CV | 1 | 24 | 16 |
| LV_radial_s | Lasso | TRUE | TRUE | 0.05 | CV | 2 | 24 | 16 |
| LV_radial_s | Lasso | TRUE | TRUE | 0.05 | CV | 3 | 24 | 15 |

| | | | | | | | | |
|---|---|---|---|---|---|---|---|---|
| LV_radial_s | Lasso | TRUE | TRUE | 0.05 | CV | 4 | 24 | 15 |
| LV_radial_s | LinRegrs | TRUE | FALSE | | HeldoutTest | | 24 | 22 |
| LV_radial_s | Lasso | TRUE | FALSE | 0.0003 | HeldoutTest | | 24 | 24 |
| LV_radial_s | Lasso | TRUE | TRUE | 0.05 | HeldoutTest | | 24 | 13 |
| LV_radial_s | LinRegrs | TRUE | FALSE | | CV | 0 | 24 | 22 |
| LV_radial_s | LinRegrs | TRUE | FALSE | | CV | 1 | 24 | 24 |
| LV_radial_s | LinRegrs | TRUE | FALSE | | CV | 2 | 24 | 23 |
| LV_radial_s | LinRegrs | TRUE | FALSE | | CV | 3 | 24 | 22 |
| LV_radial_s | LinRegrs | TRUE | FALSE | | CV | 4 | 24 | 24 |
| LV_radial_s | Lasso | TRUE | FALSE | 0.000495 | CV | 0 | 24 | 23 |
| LV_radial_s | Lasso | TRUE | FALSE | 0.000738 | CV | 1 | 24 | 23 |
| LV_radial_s | Lasso | TRUE | FALSE | 0.000524 | CV | 2 | 24 | 23 |
| LV_radial_s | Lasso | TRUE | FALSE | 0.000363 | CV | 3 | 24 | 24 |
| LV_radial_s | Lasso | TRUE | FALSE | 0.000302 | CV | 4 | 24 | 24 |
| LV_radial_s | Lasso | TRUE | TRUE | 0.05 | CV | 0 | 24 | 13 |
| LV_radial_s | Lasso | TRUE | TRUE | 0.05 | CV | 1 | 24 | 13 |
| LV_radial_s | Lasso | TRUE | TRUE | 0.05 | CV | 2 | 24 | 13 |
| LV_radial_s | Lasso | TRUE | TRUE | 0.05 | CV | 3 | 24 | 13 |
| LV_radial_s | Lasso | TRUE | TRUE | 0.05 | CV | 4 | 24 | 9 |
| LV_radial_s | LinRegrs | TRUE | FALSE | | HeldoutTest | | 24 | 24 |
| LV_radial_s | Lasso | TRUE | FALSE | 0.000309 | HeldoutTest | | 24 | 24 |
| LV_radial_s | Lasso | TRUE | TRUE | 0.05 | HeldoutTest | | 24 | 12 |
| LV_radial_s | LinRegrs | TRUE | FALSE | | CV | 0 | 24 | 24 |
| LV_radial_s | LinRegrs | TRUE | FALSE | | CV | 1 | 24 | 24 |
| LV_radial_s | LinRegrs | TRUE | FALSE | | CV | 2 | 24 | 24 |
| LV_radial_s | LinRegrs | TRUE | FALSE | | CV | 3 | 24 | 24 |
| LV_radial_s | LinRegrs | TRUE | FALSE | | CV | 4 | 24 | 23 |
| LV_radial_s | Lasso | TRUE | FALSE | 0.000314 | CV | 0 | 24 | 24 |
| LV_radial_s | Lasso | TRUE | FALSE | 0.000307 | CV | 1 | 24 | 24 |
| LV_radial_s | Lasso | TRUE | FALSE | 0.000308 | CV | 2 | 24 | 24 |
| LV_radial_s | Lasso | TRUE | FALSE | 0.000307 | CV | 3 | 24 | 24 |
| LV_radial_s | Lasso | TRUE | FALSE | 0.000309 | CV | 4 | 24 | 24 |
| LV_radial_s | Lasso | TRUE | TRUE | 0.05 | CV | 0 | 24 | 13 |
| LV_radial_s | Lasso | TRUE | TRUE | 0.05 | CV | 1 | 24 | 12 |
| LV_radial_s | Lasso | TRUE | TRUE | 0.05 | CV | 2 | 24 | 12 |
| LV_radial_s | Lasso | TRUE | TRUE | 0.05 | CV | 3 | 24 | 12 |
| LV_radial_s | Lasso | TRUE | TRUE | 0.05 | CV | 4 | 24 | 12 |
| LV_radial_s | LinRegrs | TRUE | FALSE | | HeldoutTest | | 24 | 22 |
| LV_radial_s | Lasso | TRUE | FALSE | 0.000427 | HeldoutTest | | 24 | 23 |
| LV_radial_s | Lasso | TRUE | TRUE | 0.05 | HeldoutTest | | 24 | 15 |
| LV_radial_s | LinRegrs | TRUE | FALSE | | CV | 0 | 24 | 22 |
| LV_radial_s | LinRegrs | TRUE | FALSE | | CV | 1 | 24 | 22 |
| LV_radial_s | LinRegrs | TRUE | FALSE | | CV | 2 | 24 | 23 |
| LV_radial_s | LinRegrs | TRUE | FALSE | | CV | 3 | 24 | 22 |
| LV_radial_s | LinRegrs | TRUE | FALSE | | CV | 4 | 24 | 22 |

| | | | | | | | | |
|---|---|---|---|---|---|---|---|---|
| LV_radial_s | Lasso | TRUE | FALSE | 0.000439 | CV | 0 | 24 | 23 |
| LV_radial_s | Lasso | TRUE | FALSE | 0.000321 | CV | 1 | 24 | 24 |
| LV_radial_s | Lasso | TRUE | FALSE | 0.000323 | CV | 2 | 24 | 24 |
| LV_radial_s | Lasso | TRUE | FALSE | 0.000557 | CV | 3 | 24 | 22 |
| LV_radial_s | Lasso | TRUE | FALSE | 0.000425 | CV | 4 | 24 | 23 |
| LV_radial_s | Lasso | TRUE | TRUE | 0.05 | CV | 0 | 24 | 15 |
| LV_radial_s | Lasso | TRUE | TRUE | 0.05 | CV | 1 | 24 | 15 |
| LV_radial_s | Lasso | TRUE | TRUE | 0.05 | CV | 2 | 24 | 15 |
| LV_radial_s | Lasso | TRUE | TRUE | 0.05 | CV | 3 | 24 | 15 |
| LV_radial_s | Lasso | TRUE | TRUE | 0.05 | CV | 4 | 24 | 15 |
| LV_radial_s | LinRegrs | TRUE | FALSE | | HeldoutTest | | 24 | 23 |
| LV_radial_s | Lasso | TRUE | FALSE | 0.000321 | HeldoutTest | | 24 | 23 |
| LV_radial_s | Lasso | TRUE | TRUE | 0.05 | HeldoutTest | | 24 | 15 |
| LV_radial_s | LinRegrs | TRUE | FALSE | | CV | 0 | 24 | 23 |
| LV_radial_s | LinRegrs | TRUE | FALSE | | CV | 1 | 24 | 23 |
| LV_radial_s | LinRegrs | TRUE | FALSE | | CV | 2 | 24 | 23 |
| LV_radial_s | LinRegrs | TRUE | FALSE | | CV | 3 | 24 | 23 |
| LV_radial_s | LinRegrs | TRUE | FALSE | | CV | 4 | 24 | 24 |
| LV_radial_s | Lasso | TRUE | FALSE | 0.000325 | CV | 0 | 24 | 23 |
| LV_radial_s | Lasso | TRUE | FALSE | 0.000321 | CV | 1 | 24 | 23 |
| LV_radial_s | Lasso | TRUE | FALSE | 0.000321 | CV | 2 | 24 | 24 |
| LV_radial_s | Lasso | TRUE | FALSE | 0.000319 | CV | 3 | 24 | 23 |
| LV_radial_s | Lasso | TRUE | FALSE | 0.000322 | CV | 4 | 24 | 24 |
| LV_radial_s | Lasso | TRUE | TRUE | 0.05 | CV | 0 | 24 | 16 |
| LV_radial_s | Lasso | TRUE | TRUE | 0.05 | CV | 1 | 24 | 15 |
| LV_radial_s | Lasso | TRUE | TRUE | 0.05 | CV | 2 | 24 | 15 |
| LV_radial_s | Lasso | TRUE | TRUE | 0.05 | CV | 3 | 24 | 15 |
| LV_radial_s | Lasso | TRUE | TRUE | 0.05 | CV | 4 | 24 | 12 |
| LV_radial_s | LinRegrs | TRUE | FALSE | | HeldoutTest | | 24 | 24 |
| LV_radial_s | Lasso | TRUE | FALSE | 0.00038 | HeldoutTest | | 24 | 24 |
| LV_radial_s | Lasso | TRUE | TRUE | 0.05 | HeldoutTest | | 24 | 18 |
| LV_radial_s | LinRegrs | TRUE | FALSE | | CV | 0 | 24 | 24 |
| LV_radial_s | LinRegrs | TRUE | FALSE | | CV | 1 | 24 | 24 |
| LV_radial_s | LinRegrs | TRUE | FALSE | | CV | 2 | 24 | 24 |
| LV_radial_s | LinRegrs | TRUE | FALSE | | CV | 3 | 24 | 23 |
| LV_radial_s | LinRegrs | TRUE | FALSE | | CV | 4 | 24 | 22 |
| LV_radial_s | Lasso | TRUE | FALSE | 0.000386 | CV | 0 | 24 | 24 |
| LV_radial_s | Lasso | TRUE | FALSE | 0.00038 | CV | 1 | 24 | 24 |
| LV_radial_s | Lasso | TRUE | FALSE | 0.00038 | CV | 2 | 24 | 24 |
| LV_radial_s | Lasso | TRUE | FALSE | 0.000377 | CV | 3 | 24 | 24 |
| LV_radial_s | Lasso | TRUE | FALSE | 0.000379 | CV | 4 | 24 | 24 |
| LV_radial_s | Lasso | TRUE | TRUE | 0.05 | CV | 0 | 24 | 18 |
| LV_radial_s | Lasso | TRUE | TRUE | 0.05 | CV | 1 | 24 | 16 |
| LV_radial_s | Lasso | TRUE | TRUE | 0.05 | CV | 2 | 24 | 16 |
| LV_radial_s | Lasso | TRUE | TRUE | 0.05 | CV | 3 | 24 | 17 |

| | | | | | | | | |
|---|---|---|---|---|---|---|---|---|
| LV_radial_s | Lasso | TRUE | TRUE | 0.05 | CV | 4 | 24 | 17 |
| LA_maximu | LinRegrs | TRUE | FALSE | | HeldoutTest | | 24 | 18 |
| LA_maximu | Lasso | TRUE | FALSE | 0.000926 | HeldoutTest | | 24 | 21 |
| LA_maximu | Lasso | TRUE | TRUE | 0.05 | HeldoutTest | | 24 | 6 |
| LA_maximu | LinRegrs | TRUE | FALSE | | CV | 0 | 24 | 19 |
| LA_maximu | LinRegrs | TRUE | FALSE | | CV | 1 | 24 | 18 |
| LA_maximu | LinRegrs | TRUE | FALSE | | CV | 2 | 24 | 18 |
| LA_maximu | LinRegrs | TRUE | FALSE | | CV | 3 | 24 | 20 |
| LA_maximu | LinRegrs | TRUE | FALSE | | CV | 4 | 24 | 22 |
| LA_maximu | Lasso | TRUE | FALSE | 0.00052 | CV | 0 | 24 | 22 |
| LA_maximu | Lasso | TRUE | FALSE | 0.000996 | CV | 1 | 24 | 20 |
| LA_maximu | Lasso | TRUE | FALSE | 0.001063 | CV | 2 | 24 | 22 |
| LA_maximu | Lasso | TRUE | FALSE | 0.000868 | CV | 3 | 24 | 23 |
| LA_maximu | Lasso | TRUE | FALSE | 0.000287 | CV | 4 | 24 | 24 |
| LA_maximu | Lasso | TRUE | TRUE | 0.05 | CV | 0 | 24 | 5 |
| LA_maximu | Lasso | TRUE | TRUE | 0.05 | CV | 1 | 24 | 6 |
| LA_maximu | Lasso | TRUE | TRUE | 0.05 | CV | 2 | 24 | 6 |
| LA_maximu | Lasso | TRUE | TRUE | 0.05 | CV | 3 | 24 | 6 |
| LA_maximu | Lasso | TRUE | TRUE | 0.05 | CV | 4 | 24 | 6 |
| LA_minimu | LinRegrs | TRUE | FALSE | | HeldoutTest | | 24 | 20 |
| LA_minimu | Lasso | TRUE | FALSE | 0.000382 | HeldoutTest | | 24 | 23 |
| LA_minimu | Lasso | TRUE | TRUE | 0.05 | HeldoutTest | | 24 | 10 |
| LA_minimu | LinRegrs | TRUE | FALSE | | CV | 0 | 24 | 20 |
| LA_minimu | LinRegrs | TRUE | FALSE | | CV | 1 | 24 | 19 |
| LA_minimu | LinRegrs | TRUE | FALSE | | CV | 2 | 24 | 18 |
| LA_minimu | LinRegrs | TRUE | FALSE | | CV | 3 | 24 | 20 |
| LA_minimu | LinRegrs | TRUE | FALSE | | CV | 4 | 24 | 21 |
| LA_minimu | Lasso | TRUE | FALSE | 0.000235 | CV | 0 | 24 | 24 |
| LA_minimu | Lasso | TRUE | FALSE | 0.000268 | CV | 1 | 24 | 23 |
| LA_minimu | Lasso | TRUE | FALSE | 0.001252 | CV | 2 | 24 | 21 |
| LA_minimu | Lasso | TRUE | FALSE | 0.000588 | CV | 3 | 24 | 24 |
| LA_minimu | Lasso | TRUE | FALSE | 0.000431 | CV | 4 | 24 | 24 |
| LA_minimu | Lasso | TRUE | TRUE | 0.05 | CV | 0 | 24 | 10 |
| LA_minimu | Lasso | TRUE | TRUE | 0.05 | CV | 1 | 24 | 10 |
| LA_minimu | Lasso | TRUE | TRUE | 0.05 | CV | 2 | 24 | 10 |
| LA_minimu | Lasso | TRUE | TRUE | 0.05 | CV | 3 | 24 | 10 |
| LA_minimu | Lasso | TRUE | TRUE | 0.05 | CV | 4 | 24 | 9 |
| LA_stroke_ | LinRegrs | TRUE | FALSE | | HeldoutTest | | 24 | 22 |
| LA_stroke_ | Lasso | TRUE | FALSE | 0.000274 | HeldoutTest | | 24 | 23 |
| LA_stroke_ | Lasso | TRUE | TRUE | 0.05 | HeldoutTest | | 24 | 10 |
| LA_stroke_ | LinRegrs | TRUE | FALSE | | CV | 0 | 24 | 21 |
| LA_stroke_ | LinRegrs | TRUE | FALSE | | CV | 1 | 24 | 21 |
| LA_stroke_ | LinRegrs | TRUE | FALSE | | CV | 2 | 24 | 20 |
| LA_stroke_ | LinRegrs | TRUE | FALSE | | CV | 3 | 24 | 20 |
| LA_stroke_ | LinRegrs | TRUE | FALSE | | CV | 4 | 24 | 22 |

| | | | | | | | | |
|---|---|---|---|---|---|---|---|---|
| LA_stroke_ | Lasso | TRUE | FALSE | 0.000335 | CV | 0 | 24 | 23 |
| LA_stroke_ | Lasso | TRUE | FALSE | 0.00045 | CV | 1 | 24 | 23 |
| LA_stroke_ | Lasso | TRUE | FALSE | 0.000418 | CV | 2 | 24 | 24 |
| LA_stroke_ | Lasso | TRUE | FALSE | 0.000413 | CV | 3 | 24 | 24 |
| LA_stroke_ | Lasso | TRUE | FALSE | 0.000278 | CV | 4 | 24 | 24 |
| LA_stroke_ | Lasso | TRUE | TRUE | 0.05 | CV | 0 | 24 | 10 |
| LA_stroke_ | Lasso | TRUE | TRUE | 0.05 | CV | 1 | 24 | 10 |
| LA_stroke_ | Lasso | TRUE | TRUE | 0.05 | CV | 2 | 24 | 10 |
| LA_stroke_ | Lasso | TRUE | TRUE | 0.05 | CV | 3 | 24 | 10 |
| LA_stroke_ | Lasso | TRUE | TRUE | 0.05 | CV | 4 | 24 | 9 |
| LA_ejectio | LinRegrs | TRUE | FALSE | | HeldoutTest | | 24 | 24 |
| LA_ejectio | Lasso | TRUE | FALSE | 0.00038 | HeldoutTest | | 24 | 24 |
| LA_ejectio | Lasso | TRUE | TRUE | 0.05 | HeldoutTest | | 24 | 15 |
| LA_ejectio | LinRegrs | TRUE | FALSE | | CV | 0 | 24 | 24 |
| LA_ejectio | LinRegrs | TRUE | FALSE | | CV | 1 | 24 | 24 |
| LA_ejectio | LinRegrs | TRUE | FALSE | | CV | 2 | 24 | 23 |
| LA_ejectio | LinRegrs | TRUE | FALSE | | CV | 3 | 24 | 24 |
| LA_ejectio | LinRegrs | TRUE | FALSE | | CV | 4 | 24 | 24 |
| LA_ejectio | Lasso | TRUE | FALSE | 0.000384 | CV | 0 | 24 | 24 |
| LA_ejectio | Lasso | TRUE | FALSE | 0.00054 | CV | 1 | 24 | 24 |
| LA_ejectio | Lasso | TRUE | FALSE | 0.000377 | CV | 2 | 24 | 24 |
| LA_ejectio | Lasso | TRUE | FALSE | 0.000382 | CV | 3 | 24 | 24 |
| LA_ejectio | Lasso | TRUE | FALSE | 0.000379 | CV | 4 | 24 | 24 |
| LA_ejectio | Lasso | TRUE | TRUE | 0.05 | CV | 0 | 24 | 15 |
| LA_ejectio | Lasso | TRUE | TRUE | 0.05 | CV | 1 | 24 | 15 |
| LA_ejectio | Lasso | TRUE | TRUE | 0.05 | CV | 2 | 24 | 15 |
| LA_ejectio | Lasso | TRUE | TRUE | 0.05 | CV | 3 | 24 | 15 |
| LA_ejectio | Lasso | TRUE | TRUE | 0.05 | CV | 4 | 24 | 10 |
| LV_end_di | LinRegrs | TRUE | FALSE | | HeldoutTest | | 24 | 0 |
| LV_end_di | Lasso | TRUE | FALSE | 1.00E-15 | HeldoutTest | | 24 | 0 |
| LV_end_di | Lasso | TRUE | TRUE | 0.05 | HeldoutTest | | 24 | 0 |
| LV_end_di | LinRegrs | TRUE | FALSE | | CV | 0 | 24 | 0 |
| LV_end_di | LinRegrs | TRUE | FALSE | | CV | 1 | 24 | 0 |
| LV_end_di | LinRegrs | TRUE | FALSE | | CV | 2 | 24 | 0 |
| LV_end_di | LinRegrs | TRUE | FALSE | | CV | 3 | 24 | 0 |
| LV_end_di | LinRegrs | TRUE | FALSE | | CV | 4 | 24 | 0 |
| LV_end_di | Lasso | TRUE | FALSE | 1.00E-15 | CV | 0 | 24 | 0 |
| LV_end_di | Lasso | TRUE | FALSE | 1.00E-15 | CV | 1 | 24 | 0 |
| LV_end_di | Lasso | TRUE | FALSE | 1.00E-15 | CV | 2 | 24 | 0 |
| LV_end_di | Lasso | TRUE | FALSE | 1.00E-15 | CV | 3 | 24 | 0 |
| LV_end_di | Lasso | TRUE | FALSE | 1.00E-15 | CV | 4 | 24 | 0 |
| LV_end_di | Lasso | TRUE | TRUE | 0.05 | CV | 0 | 24 | 0 |
| LV_end_di | Lasso | TRUE | TRUE | 0.05 | CV | 1 | 24 | 0 |
| LV_end_di | Lasso | TRUE | TRUE | 0.05 | CV | 2 | 24 | 0 |
| LV_end_di | Lasso | TRUE | TRUE | 0.05 | CV | 3 | 24 | 0 |

| | | | | | | | | |
|---|---|---|---|---|---|---|---|---|
| LV_end_di | Lasso | TRUE | TRUE | 0.05 | CV | 4 | 24 | 0 |
| LV_end_sy | LinRegrs | TRUE | FALSE | | HeldoutTest | | 24 | 23 |
| LV_end_sy | Lasso | TRUE | FALSE | 0.000153 | HeldoutTest | | 24 | 24 |
| LV_end_sy | Lasso | TRUE | TRUE | 0.05 | HeldoutTest | | 24 | 12 |
| LV_end_sy | LinRegrs | TRUE | FALSE | | CV | 0 | 24 | 23 |
| LV_end_sy | LinRegrs | TRUE | FALSE | | CV | 1 | 24 | 23 |
| LV_end_sy | LinRegrs | TRUE | FALSE | | CV | 2 | 24 | 23 |
| LV_end_sy | LinRegrs | TRUE | FALSE | | CV | 3 | 24 | 23 |
| LV_end_sy | LinRegrs | TRUE | FALSE | | CV | 4 | 24 | 23 |
| LV_end_sy | Lasso | TRUE | FALSE | 0.000146 | CV | 0 | 24 | 24 |
| LV_end_sy | Lasso | TRUE | FALSE | 0.000144 | CV | 1 | 24 | 24 |
| LV_end_sy | Lasso | TRUE | FALSE | 0.000142 | CV | 2 | 24 | 24 |
| LV_end_sy | Lasso | TRUE | FALSE | 0.000204 | CV | 3 | 24 | 24 |
| LV_end_sy | Lasso | TRUE | FALSE | 0.000146 | CV | 4 | 24 | 24 |
| LV_end_sy | Lasso | TRUE | TRUE | 0.05 | CV | 0 | 24 | 12 |
| LV_end_sy | Lasso | TRUE | TRUE | 0.05 | CV | 1 | 24 | 12 |
| LV_end_sy | Lasso | TRUE | TRUE | 0.05 | CV | 2 | 24 | 11 |
| LV_end_sy | Lasso | TRUE | TRUE | 0.05 | CV | 3 | 24 | 11 |
| LV_end_sy | Lasso | TRUE | TRUE | 0.05 | CV | 4 | 24 | 13 |
| LV_stroke_ | LinRegrs | TRUE | FALSE | | HeldoutTest | | 24 | 23 |
| LV_stroke_ | Lasso | TRUE | FALSE | 0.000153 | HeldoutTest | | 24 | 24 |
| LV_stroke_ | Lasso | TRUE | TRUE | 0.05 | HeldoutTest | | 24 | 12 |
| LV_stroke_ | LinRegrs | TRUE | FALSE | | CV | 0 | 24 | 23 |
| LV_stroke_ | LinRegrs | TRUE | FALSE | | CV | 1 | 24 | 23 |
| LV_stroke_ | LinRegrs | TRUE | FALSE | | CV | 2 | 24 | 23 |
| LV_stroke_ | LinRegrs | TRUE | FALSE | | CV | 3 | 24 | 23 |
| LV_stroke_ | LinRegrs | TRUE | FALSE | | CV | 4 | 24 | 23 |
| LV_stroke_ | Lasso | TRUE | FALSE | 0.000146 | CV | 0 | 24 | 24 |
| LV_stroke_ | Lasso | TRUE | FALSE | 0.000144 | CV | 1 | 24 | 24 |
| LV_stroke_ | Lasso | TRUE | FALSE | 0.000142 | CV | 2 | 24 | 24 |
| LV_stroke_ | Lasso | TRUE | FALSE | 0.000204 | CV | 3 | 24 | 24 |
| LV_stroke_ | Lasso | TRUE | FALSE | 0.000146 | CV | 4 | 24 | 24 |
| LV_stroke_ | Lasso | TRUE | TRUE | 0.05 | CV | 0 | 24 | 12 |
| LV_stroke_ | Lasso | TRUE | TRUE | 0.05 | CV | 1 | 24 | 12 |
| LV_stroke_ | Lasso | TRUE | TRUE | 0.05 | CV | 2 | 24 | 11 |
| LV_stroke_ | Lasso | TRUE | TRUE | 0.05 | CV | 3 | 24 | 11 |
| LV_stroke_ | Lasso | TRUE | TRUE | 0.05 | CV | 4 | 24 | 13 |
| LV_ejectior | LinRegrs | TRUE | FALSE | | HeldoutTest | | 24 | 24 |
| LV_ejectior | Lasso | TRUE | FALSE | 0.000406 | HeldoutTest | | 24 | 24 |
| LV_ejectior | Lasso | TRUE | TRUE | 0.05 | HeldoutTest | | 24 | 14 |
| LV_ejectior | LinRegrs | TRUE | FALSE | | CV | 0 | 24 | 23 |
| LV_ejectior | LinRegrs | TRUE | FALSE | | CV | 1 | 24 | 24 |
| LV_ejectior | LinRegrs | TRUE | FALSE | | CV | 2 | 24 | 24 |
| LV_ejectior | LinRegrs | TRUE | FALSE | | CV | 3 | 24 | 23 |
| LV_ejectior | LinRegrs | TRUE | FALSE | | CV | 4 | 24 | 23 |

| | | | | | | | | |
|---|---|---|---|---|---|---|---|---|
| LV_ejectior | Lasso | TRUE | FALSE | 0.000411 | CV | 0 | 24 | 23 |
| LV_ejectior | Lasso | TRUE | FALSE | 0.000405 | CV | 1 | 24 | 24 |
| LV_ejectior | Lasso | TRUE | FALSE | 0.000404 | CV | 2 | 24 | 24 |
| LV_ejectior | Lasso | TRUE | FALSE | 0.000404 | CV | 3 | 24 | 24 |
| LV_ejectior | Lasso | TRUE | FALSE | 0.000405 | CV | 4 | 24 | 24 |
| LV_ejectior | Lasso | TRUE | TRUE | 0.05 | CV | 0 | 24 | 15 |
| LV_ejectior | Lasso | TRUE | TRUE | 0.05 | CV | 1 | 24 | 13 |
| LV_ejectior | Lasso | TRUE | TRUE | 0.05 | CV | 2 | 24 | 15 |
| LV_ejectior | Lasso | TRUE | TRUE | 0.05 | CV | 3 | 24 | 15 |
| LV_ejectior | Lasso | TRUE | TRUE | 0.05 | CV | 4 | 24 | 15 |
| LV_cardiac | LinRegrs | TRUE | FALSE | | HeldoutTest | | 24 | 23 |
| LV_cardiac | Lasso | TRUE | FALSE | 0.000405 | HeldoutTest | | 24 | 24 |
| LV_cardiac | Lasso | TRUE | TRUE | 0.05 | HeldoutTest | | 24 | 10 |
| LV_cardiac | LinRegrs | TRUE | FALSE | | CV | 0 | 24 | 23 |
| LV_cardiac | LinRegrs | TRUE | FALSE | | CV | 1 | 24 | 23 |
| LV_cardiac | LinRegrs | TRUE | FALSE | | CV | 2 | 24 | 23 |
| LV_cardiac | LinRegrs | TRUE | FALSE | | CV | 3 | 24 | 24 |
| LV_cardiac | LinRegrs | TRUE | FALSE | | CV | 4 | 24 | 23 |
| LV_cardiac | Lasso | TRUE | FALSE | 0.0004 | CV | 0 | 24 | 23 |
| LV_cardiac | Lasso | TRUE | FALSE | 0.000408 | CV | 1 | 24 | 23 |
| LV_cardiac | Lasso | TRUE | FALSE | 0.000404 | CV | 2 | 24 | 24 |
| LV_cardiac | Lasso | TRUE | FALSE | 0.000409 | CV | 3 | 24 | 24 |
| LV_cardiac | Lasso | TRUE | FALSE | 0.000406 | CV | 4 | 24 | 24 |
| LV_cardiac | Lasso | TRUE | TRUE | 0.05 | CV | 0 | 24 | 11 |
| LV_cardiac | Lasso | TRUE | TRUE | 0.05 | CV | 1 | 24 | 10 |
| LV_cardiac | Lasso | TRUE | TRUE | 0.05 | CV | 2 | 24 | 11 |
| LV_cardiac | Lasso | TRUE | TRUE | 0.05 | CV | 3 | 24 | 12 |
| LV_cardiac | Lasso | TRUE | TRUE | 0.05 | CV | 4 | 24 | 10 |

| MSE | R-squared | TrainSize | TestSize |
|---|---|---|---|
| 0.697039 | 0.256471 | 23462 | 3401 |
| 0.696934 | 0.256582 | 23462 | 3401 |
| 0.735617 | 0.21532 | 23462 | 3401 |
| 0.750789 | 0.232389 | 18769 | 4693 |
| 0.73374 | 0.240808 | 18769 | 4693 |
| 0.761371 | 0.262509 | 18770 | 4692 |
| 0.746911 | 0.265059 | 18770 | 4692 |
| 0.756222 | 0.249148 | 18770 | 4692 |
| 0.750423 | 0.232763 | 18769 | 4693 |
| 0.733526 | 0.24103 | 18769 | 4693 |
| 0.761433 | 0.262449 | 18770 | 4692 |
| 0.747834 | 0.26415 | 18770 | 4692 |
| 0.756157 | 0.249213 | 18770 | 4692 |
| 0.782789 | 0.199672 | 18769 | 4693 |
| 0.768536 | 0.204806 | 18769 | 4693 |
| 0.83883 | 0.18748 | 18770 | 4692 |
| 0.796979 | 0.215793 | 18770 | 4692 |
| 0.803577 | 0.20213 | 18770 | 4692 |
| 0.722887 | 0.239565 | 23462 | 3401 |
| 0.723267 | 0.239166 | 23462 | 3401 |
| 0.765002 | 0.195263 | 23462 | 3401 |
| 0.765146 | 0.2146 | 18769 | 4693 |
| 0.746047 | 0.224792 | 18769 | 4693 |
| 0.78296 | 0.245518 | 18770 | 4692 |
| 0.76216 | 0.248045 | 18770 | 4692 |
| 0.775068 | 0.234575 | 18770 | 4692 |
| 0.764758 | 0.214998 | 18769 | 4693 |
| 0.745864 | 0.224982 | 18769 | 4693 |
| 0.783028 | 0.245453 | 18770 | 4692 |
| 0.763001 | 0.247216 | 18770 | 4692 |
| 0.775135 | 0.234508 | 18770 | 4692 |
| 0.798573 | 0.180288 | 18769 | 4693 |
| 0.779831 | 0.189687 | 18769 | 4693 |
| 0.858013 | 0.173195 | 18770 | 4692 |
| 0.810331 | 0.200519 | 18770 | 4692 |
| 0.823094 | 0.187146 | 18770 | 4692 |
| 1.07236 | 0.082969 | 23462 | 3401 |
| 1.072716 | 0.082664 | 23462 | 3401 |
| 1.105978 | 0.05422 | 23462 | 3401 |
| 0.929714 | 0.099123 | 18769 | 4693 |
| 0.838525 | 0.110988 | 18769 | 4693 |
| 0.898804 | 0.109581 | 18770 | 4692 |
| 0.855544 | 0.115561 | 18770 | 4692 |
| 0.927186 | 0.115719 | 18770 | 4692 |

| | | | |
|---|---|---|---|
| 0.929767 | 0.099072 | 18769 | 4693 |
| 0.838684 | 0.110819 | 18769 | 4693 |
| 0.898665 | 0.109719 | 18770 | 4692 |
| 0.855506 | 0.1156 | 18770 | 4692 |
| 0.927121 | 0.11578 | 18770 | 4692 |
| 0.955295 | 0.074336 | 18769 | 4693 |
| 0.862973 | 0.085068 | 18769 | 4693 |
| 0.930555 | 0.078126 | 18770 | 4692 |
| 0.880031 | 0.090247 | 18770 | 4692 |
| 0.960717 | 0.083739 | 18770 | 4692 |
| 0.568363 | 0.408922 | 23462 | 3401 |
| 0.568213 | 0.409078 | 23462 | 3401 |
| 0.613769 | 0.361702 | 23462 | 3401 |
| 0.576603 | 0.415979 | 18769 | 4693 |
| 0.567532 | 0.430006 | 18769 | 4693 |
| 0.552671 | 0.452129 | 18770 | 4692 |
| 0.571051 | 0.429891 | 18770 | 4692 |
| 0.585085 | 0.418619 | 18770 | 4692 |
| 0.576451 | 0.416134 | 18769 | 4693 |
| 0.567615 | 0.429923 | 18769 | 4693 |
| 0.552739 | 0.452062 | 18770 | 4692 |
| 0.571072 | 0.42987 | 18770 | 4692 |
| 0.585058 | 0.418646 | 18770 | 4692 |
| 0.623715 | 0.368262 | 18769 | 4693 |
| 0.622159 | 0.375141 | 18769 | 4693 |
| 0.651014 | 0.354641 | 18770 | 4692 |
| 0.622791 | 0.378236 | 18770 | 4692 |
| 0.646411 | 0.357681 | 18770 | 4692 |
| 0.584145 | 0.380722 | 23462 | 3401 |
| 0.583966 | 0.380912 | 23462 | 3401 |
| 0.624295 | 0.338158 | 23462 | 3401 |
| 0.600686 | 0.390781 | 18769 | 4693 |
| 0.587239 | 0.407358 | 18769 | 4693 |
| 0.582128 | 0.425553 | 18770 | 4692 |
| 0.590398 | 0.406276 | 18770 | 4692 |
| 0.618215 | 0.391065 | 18770 | 4692 |
| 0.600485 | 0.390985 | 18769 | 4693 |
| 0.587322 | 0.407275 | 18769 | 4693 |
| 0.582188 | 0.425495 | 18770 | 4692 |
| 0.590395 | 0.406278 | 18770 | 4692 |
| 0.618198 | 0.391082 | 18770 | 4692 |
| 0.645255 | 0.345579 | 18769 | 4693 |
| 0.639156 | 0.354964 | 18769 | 4693 |
| 0.676506 | 0.33242 | 18770 | 4692 |
| 0.637328 | 0.359081 | 18770 | 4692 |

| | | | |
|---|---|---|---|
| 0.674301 | 0.335821 | 18770 | 4692 |
| 0.978359 | 0.095403 | 23462 | 3401 |
| 0.978353 | 0.095408 | 23462 | 3401 |
| 1.002307 | 0.07326 | 23462 | 3401 |
| 0.910737 | 0.119493 | 18769 | 4693 |
| 0.879524 | 0.119923 | 18769 | 4693 |
| 0.858916 | 0.121816 | 18770 | 4692 |
| 0.870525 | 0.125974 | 18770 | 4692 |
| 0.875771 | 0.116912 | 18770 | 4692 |
| 0.910877 | 0.119358 | 18769 | 4693 |
| 0.879516 | 0.119931 | 18769 | 4693 |
| 0.858584 | 0.122155 | 18770 | 4692 |
| 0.870266 | 0.126234 | 18770 | 4692 |
| 0.875751 | 0.116932 | 18770 | 4692 |
| 0.937562 | 0.093559 | 18769 | 4693 |
| 0.905168 | 0.094262 | 18769 | 4693 |
| 0.882232 | 0.097977 | 18770 | 4692 |
| 0.894069 | 0.102336 | 18770 | 4692 |
| 0.903654 | 0.088796 | 18770 | 4692 |
| 0.80958 | 0.236423 | 23462 | 3401 |
| 0.809793 | 0.236222 | 23462 | 3401 |
| 0.868854 | 0.180517 | 23462 | 3401 |
| 0.795316 | 0.178288 | 18769 | 4693 |
| 0.798753 | 0.196165 | 18769 | 4693 |
| 0.854875 | 0.169054 | 18770 | 4692 |
| 0.760758 | 0.202793 | 18770 | 4692 |
| 0.841684 | 0.202523 | 18770 | 4692 |
| 0.795334 | 0.178268 | 18769 | 4693 |
| 0.798725 | 0.196193 | 18769 | 4693 |
| 0.85472 | 0.169205 | 18770 | 4692 |
| 0.760819 | 0.202729 | 18770 | 4692 |
| 0.841763 | 0.202448 | 18770 | 4692 |
| 0.832285 | 0.140091 | 18769 | 4693 |
| 0.845496 | 0.149125 | 18769 | 4693 |
| 0.888754 | 0.136124 | 18770 | 4692 |
| 0.799291 | 0.162414 | 18770 | 4692 |
| 0.893476 | 0.15345 | 18770 | 4692 |
| 0.813264 | 0.238346 | 23462 | 3401 |
| 0.813487 | 0.238137 | 23462 | 3401 |
| 0.870901 | 0.184367 | 23462 | 3401 |
| 0.785897 | 0.179903 | 18769 | 4693 |
| 0.790443 | 0.200984 | 18769 | 4693 |
| 0.855219 | 0.174781 | 18770 | 4692 |
| 0.758949 | 0.208411 | 18770 | 4692 |
| 0.835073 | 0.21042 | 18770 | 4692 |

| | | | |
|---|---|---|---|
| 0.785934 | 0.179865 | 18769 | 4693 |
| 0.79039 | 0.201038 | 18769 | 4693 |
| 0.855071 | 0.174924 | 18770 | 4692 |
| 0.758975 | 0.208384 | 18770 | 4692 |
| 0.835183 | 0.210316 | 18770 | 4692 |
| 0.819077 | 0.145279 | 18769 | 4693 |
| 0.835359 | 0.155581 | 18769 | 4693 |
| 0.889797 | 0.141416 | 18770 | 4692 |
| 0.795603 | 0.170181 | 18770 | 4692 |
| 0.886942 | 0.161377 | 18770 | 4692 |
| 1.196328 | 0.074833 | 23462 | 3401 |
| 1.19627 | 0.074877 | 23462 | 3401 |
| 1.250611 | 0.032853 | 23462 | 3401 |
| 0.945057 | 0.097991 | 18769 | 4693 |
| 0.841071 | 0.100971 | 18769 | 4693 |
| 0.916424 | 0.1149 | 18770 | 4692 |
| 0.846924 | 0.108738 | 18770 | 4692 |
| 0.930322 | 0.099077 | 18770 | 4692 |
| 0.945085 | 0.097965 | 18769 | 4693 |
| 0.841061 | 0.100982 | 18769 | 4693 |
| 0.916381 | 0.114941 | 18770 | 4692 |
| 0.846913 | 0.10875 | 18770 | 4692 |
| 0.930203 | 0.099193 | 18770 | 4692 |
| 0.982663 | 0.062099 | 18769 | 4693 |
| 0.872157 | 0.067744 | 18769 | 4693 |
| 0.96701 | 0.066043 | 18770 | 4692 |
| 0.878875 | 0.075114 | 18770 | 4692 |
| 0.963556 | 0.066894 | 18770 | 4692 |
| 0.802413 | 0.203722 | 23462 | 3401 |
| 0.80252 | 0.203615 | 23462 | 3401 |
| 0.834867 | 0.171516 | 23462 | 3401 |
| 0.824718 | 0.193646 | 18769 | 4693 |
| 0.789811 | 0.19749 | 18769 | 4693 |
| 0.790031 | 0.186689 | 18770 | 4692 |
| 0.794891 | 0.197741 | 18770 | 4692 |
| 0.828967 | 0.195965 | 18770 | 4692 |
| 0.824975 | 0.193396 | 18769 | 4693 |
| 0.789741 | 0.197561 | 18769 | 4693 |
| 0.789942 | 0.18678 | 18770 | 4692 |
| 0.794835 | 0.197798 | 18770 | 4692 |
| 0.829025 | 0.195908 | 18770 | 4692 |
| 0.859508 | 0.159631 | 18769 | 4693 |
| 0.830534 | 0.156111 | 18769 | 4693 |
| 0.810396 | 0.165724 | 18770 | 4692 |
| 0.827717 | 0.164611 | 18770 | 4692 |

| | | | |
|---|---|---|---|
| 0.870966 | 0.155229 | 18770 | 4692 |
| 0.789232 | 0.215962 | 23462 | 3401 |
| 0.789279 | 0.215915 | 23462 | 3401 |
| 0.818218 | 0.187166 | 23462 | 3401 |
| 0.80816 | 0.20616 | 18769 | 4693 |
| 0.772452 | 0.215142 | 18769 | 4693 |
| 0.77951 | 0.202507 | 18770 | 4692 |
| 0.776017 | 0.213178 | 18770 | 4692 |
| 0.816574 | 0.210367 | 18770 | 4692 |
| 0.808239 | 0.206082 | 18769 | 4693 |
| 0.772371 | 0.215225 | 18769 | 4693 |
| 0.779471 | 0.202547 | 18770 | 4692 |
| 0.775907 | 0.213289 | 18770 | 4692 |
| 0.816729 | 0.210217 | 18770 | 4692 |
| 0.838598 | 0.176261 | 18769 | 4693 |
| 0.81061 | 0.176372 | 18769 | 4693 |
| 0.799429 | 0.182129 | 18770 | 4692 |
| 0.803639 | 0.185171 | 18770 | 4692 |
| 0.854782 | 0.17342 | 18770 | 4692 |
| 1.059525 | 0.108608 | 23462 | 3401 |
| 1.059434 | 0.108685 | 23462 | 3401 |
| 1.108707 | 0.067231 | 23462 | 3401 |
| 0.885379 | 0.128417 | 18769 | 4693 |
| 0.876652 | 0.125599 | 18769 | 4693 |
| 0.832412 | 0.146448 | 18770 | 4692 |
| 0.846031 | 0.142717 | 18770 | 4692 |
| 0.892315 | 0.124529 | 18770 | 4692 |
| 0.885035 | 0.128756 | 18769 | 4693 |
| 0.876629 | 0.125622 | 18769 | 4693 |
| 0.832386 | 0.146474 | 18770 | 4692 |
| 0.845887 | 0.142863 | 18770 | 4692 |
| 0.892317 | 0.124526 | 18770 | 4692 |
| 0.922307 | 0.092065 | 18769 | 4693 |
| 0.910842 | 0.091497 | 18769 | 4693 |
| 0.888432 | 0.089005 | 18770 | 4692 |
| 0.884888 | 0.103343 | 18770 | 4692 |
| 0.930294 | 0.087267 | 18770 | 4692 |
| 0.454022 | 0.555959 | 27542 | 4025 |
| 0.454046 | 0.555935 | 27542 | 4025 |
| 0.543796 | 0.468159 | 27542 | 4025 |
| 0.446878 | 0.532651 | 22033 | 5509 |
| 0.452276 | 0.540022 | 22033 | 5509 |
| 0.465914 | 0.534042 | 22034 | 5508 |
| 0.457821 | 0.561366 | 22034 | 5508 |
| 0.462724 | 0.544889 | 22034 | 5508 |

| | | | |
|---|---|---|---|
| 0.446754 | 0.532781 | 22033 | 5509 |
| 0.452008 | 0.540295 | 22033 | 5509 |
| 0.465942 | 0.534014 | 22034 | 5508 |
| 0.457942 | 0.56125 | 22034 | 5508 |
| 0.462809 | 0.544805 | 22034 | 5508 |
| 0.535236 | 0.440246 | 22033 | 5509 |
| 0.528862 | 0.462132 | 22033 | 5509 |
| 0.550554 | 0.449393 | 22034 | 5508 |
| 0.554622 | 0.468623 | 22034 | 5508 |
| 0.565417 | 0.443886 | 22034 | 5508 |
| 0.451854 | 0.541142 | 27542 | 4025 |
| 0.451817 | 0.54118 | 27542 | 4025 |
| 0.534213 | 0.457507 | 27542 | 4025 |
| 0.461124 | 0.516825 | 22033 | 5509 |
| 0.453936 | 0.531494 | 22033 | 5509 |
| 0.484034 | 0.522833 | 22034 | 5508 |
| 0.467867 | 0.550219 | 22034 | 5508 |
| 0.478016 | 0.532281 | 22034 | 5508 |
| 0.461044 | 0.516908 | 22033 | 5509 |
| 0.453847 | 0.531586 | 22033 | 5509 |
| 0.484146 | 0.522723 | 22034 | 5508 |
| 0.46787 | 0.550217 | 22034 | 5508 |
| 0.478059 | 0.532238 | 22034 | 5508 |
| 0.54392 | 0.430068 | 22033 | 5509 |
| 0.528635 | 0.454397 | 22033 | 5509 |
| 0.571087 | 0.437015 | 22034 | 5508 |
| 0.563457 | 0.458325 | 22034 | 5508 |
| 0.593838 | 0.418953 | 22034 | 5508 |
| 0.68343 | 0.333901 | 27542 | 4025 |
| 0.683473 | 0.333859 | 27542 | 4025 |
| 0.748315 | 0.270661 | 27542 | 4025 |
| 0.677268 | 0.304391 | 22033 | 5509 |
| 0.694743 | 0.3011 | 22033 | 5509 |
| 0.707147 | 0.298769 | 22034 | 5508 |
| 0.682521 | 0.322378 | 22034 | 5508 |
| 0.693672 | 0.317528 | 22034 | 5508 |
| 0.677196 | 0.304465 | 22033 | 5509 |
| 0.694585 | 0.301259 | 22033 | 5509 |
| 0.707185 | 0.29873 | 22034 | 5508 |
| 0.682626 | 0.322275 | 22034 | 5508 |
| 0.693772 | 0.317429 | 22034 | 5508 |
| 0.733649 | 0.246483 | 22033 | 5509 |
| 0.744397 | 0.251148 | 22033 | 5509 |
| 0.763738 | 0.242651 | 22034 | 5508 |
| 0.745118 | 0.260231 | 22034 | 5508 |

| | | | |
|---|---|---|---|
| 0.754801 | 0.257386 | 22034 | 5508 |
| 0.823097 | 0.159738 | 27542 | 4025 |
| 0.822996 | 0.159841 | 27542 | 4025 |
| 0.869038 | 0.112839 | 27542 | 4025 |
| 0.828881 | 0.158701 | 22033 | 5509 |
| 0.820842 | 0.156527 | 22033 | 5509 |
| 0.850294 | 0.175557 | 22034 | 5508 |
| 0.814011 | 0.175088 | 22034 | 5508 |
| 0.837151 | 0.182192 | 22034 | 5508 |
| 0.828821 | 0.158761 | 22033 | 5509 |
| 0.820887 | 0.156481 | 22033 | 5509 |
| 0.850485 | 0.175371 | 22034 | 5508 |
| 0.81396 | 0.17514 | 22034 | 5508 |
| 0.837092 | 0.182249 | 22034 | 5508 |
| 0.87842 | 0.10842 | 22033 | 5509 |
| 0.86954 | 0.106487 | 22033 | 5509 |
| 0.913564 | 0.11421 | 22034 | 5508 |
| 0.864777 | 0.123642 | 22034 | 5508 |
| 0.904788 | 0.116117 | 22034 | 5508 |
| 0.387493 | 0.612012 | 27542 | 4025 |
| 0.387622 | 0.611883 | 27542 | 4025 |
| 0.455557 | 0.543861 | 27542 | 4025 |
| 0.377185 | 0.613311 | 22033 | 5509 |
| 0.364898 | 0.627357 | 22033 | 5509 |
| 0.368927 | 0.630177 | 22034 | 5508 |
| 0.372249 | 0.630885 | 22034 | 5508 |
| 0.386647 | 0.62784 | 22034 | 5508 |
| 0.377135 | 0.613362 | 22033 | 5509 |
| 0.364868 | 0.627388 | 22033 | 5509 |
| 0.369004 | 0.6301 | 22034 | 5508 |
| 0.372273 | 0.630862 | 22034 | 5508 |
| 0.386803 | 0.62769 | 22034 | 5508 |
| 0.437183 | 0.551802 | 22033 | 5509 |
| 0.431136 | 0.559713 | 22033 | 5509 |
| 0.435801 | 0.563141 | 22034 | 5508 |
| 0.440191 | 0.563516 | 22034 | 5508 |
| 0.476178 | 0.541664 | 22034 | 5508 |
| 0.713555 | 0.279012 | 27542 | 4025 |
| 0.713449 | 0.279119 | 27542 | 4025 |
| 0.756038 | 0.236087 | 27542 | 4025 |
| 0.711741 | 0.278313 | 22033 | 5509 |
| 0.707548 | 0.286897 | 22033 | 5509 |
| 0.716465 | 0.291404 | 22034 | 5508 |
| 0.690269 | 0.29729 | 22034 | 5508 |
| 0.72402 | 0.295779 | 22034 | 5508 |

| | | | |
|---|---|---|---|
| 0.711593 | 0.278463 | 22033 | 5509 |
| 0.707316 | 0.28713 | 22033 | 5509 |
| 0.716532 | 0.291337 | 22034 | 5508 |
| 0.690404 | 0.297153 | 22034 | 5508 |
| 0.724141 | 0.295661 | 22034 | 5508 |
| 0.75605 | 0.233385 | 22033 | 5509 |
| 0.753101 | 0.240986 | 22033 | 5509 |
| 0.76426 | 0.244134 | 22034 | 5508 |
| 0.741829 | 0.244801 | 22034 | 5508 |
| 0.785816 | 0.235672 | 22034 | 5508 |
| 0.848077 | 0.156 | 27542 | 4025 |
| 0.84808 | 0.155997 | 27542 | 4025 |
| 0.886251 | 0.118009 | 27542 | 4025 |
| 0.816123 | 0.172917 | 22033 | 5509 |
| 0.835821 | 0.160514 | 22033 | 5509 |
| 0.822471 | 0.191232 | 22034 | 5508 |
| 0.786585 | 0.163808 | 22034 | 5508 |
| 0.877487 | 0.173007 | 22034 | 5508 |
| 0.816164 | 0.172876 | 22033 | 5509 |
| 0.83562 | 0.160717 | 22033 | 5509 |
| 0.82257 | 0.191135 | 22034 | 5508 |
| 0.786509 | 0.163889 | 22034 | 5508 |
| 0.877575 | 0.172923 | 22034 | 5508 |
| 0.859375 | 0.129084 | 22033 | 5509 |
| 0.87019 | 0.125995 | 22033 | 5509 |
| 0.871136 | 0.143378 | 22034 | 5508 |
| 0.818663 | 0.129707 | 22034 | 5508 |
| 0.93628 | 0.117597 | 22034 | 5508 |
| 0.934846 | 0.100444 | 27542 | 4025 |
| 0.934853 | 0.100438 | 27542 | 4025 |
| 0.967156 | 0.069354 | 27542 | 4025 |
| 0.878592 | 0.116158 | 22033 | 5509 |
| 0.926815 | 0.09651 | 22033 | 5509 |
| 0.865164 | 0.121572 | 22034 | 5508 |
| 0.859591 | 0.121524 | 22034 | 5508 |
| 0.90013 | 0.113711 | 22034 | 5508 |
| 0.878539 | 0.116212 | 22033 | 5509 |
| 0.926807 | 0.096518 | 22033 | 5509 |
| 0.865183 | 0.121552 | 22034 | 5508 |
| 0.859592 | 0.121523 | 22034 | 5508 |
| 0.900201 | 0.113641 | 22034 | 5508 |
| 0.920939 | 0.073558 | 22033 | 5509 |
| 0.956729 | 0.067349 | 22033 | 5509 |
| 0.905364 | 0.080755 | 22034 | 5508 |
| 0.903079 | 0.077081 | 22034 | 5508 |

| | | | |
|---|---|---|---|
| 0.944894 | 0.069635 | 22034 | 5508 |
| 0.740341 | 0.265384 | 27542 | 4025 |
| 0.740356 | 0.265369 | 27542 | 4025 |
| 0.787186 | 0.218901 | 27542 | 4025 |
| 0.716824 | 0.278176 | 22033 | 5509 |
| 0.72418 | 0.259582 | 22033 | 5509 |
| 0.702684 | 0.283181 | 22034 | 5508 |
| 0.716028 | 0.291678 | 22034 | 5508 |
| 0.746928 | 0.280051 | 22034 | 5508 |
| 0.716639 | 0.278363 | 22033 | 5509 |
| 0.724098 | 0.259665 | 22033 | 5509 |
| 0.702701 | 0.283165 | 22034 | 5508 |
| 0.716088 | 0.291619 | 22034 | 5508 |
| 0.747104 | 0.279882 | 22034 | 5508 |
| 0.768839 | 0.225799 | 22033 | 5509 |
| 0.76562 | 0.217212 | 22033 | 5509 |
| 0.750701 | 0.234198 | 22034 | 5508 |
| 0.775421 | 0.232924 | 22034 | 5508 |
| 0.808361 | 0.220837 | 22034 | 5508 |
| 0.724493 | 0.278418 | 27542 | 4025 |
| 0.72456 | 0.278351 | 27542 | 4025 |
| 0.772676 | 0.230428 | 27542 | 4025 |
| 0.690361 | 0.316574 | 22033 | 5509 |
| 0.666614 | 0.313553 | 22033 | 5509 |
| 0.669247 | 0.319624 | 22034 | 5508 |
| 0.671989 | 0.329096 | 22034 | 5508 |
| 0.715668 | 0.307513 | 22034 | 5508 |
| 0.690136 | 0.316797 | 22033 | 5509 |
| 0.66674 | 0.313424 | 22033 | 5509 |
| 0.669222 | 0.319649 | 22034 | 5508 |
| 0.672118 | 0.328967 | 22034 | 5508 |
| 0.715648 | 0.307532 | 22034 | 5508 |
| 0.746061 | 0.261434 | 22033 | 5509 |
| 0.718424 | 0.260202 | 22033 | 5509 |
| 0.71827 | 0.269786 | 22034 | 5508 |
| 0.734993 | 0.266195 | 22034 | 5508 |
| 0.778203 | 0.247004 | 22034 | 5508 |
| 0.753763 | 0.264379 | 27542 | 4025 |
| 0.753822 | 0.264321 | 27542 | 4025 |
| 0.792643 | 0.226434 | 27542 | 4025 |
| 0.675139 | 0.308213 | 22033 | 5509 |
| 0.673744 | 0.309001 | 22033 | 5509 |
| 0.702523 | 0.306826 | 22034 | 5508 |
| 0.706712 | 0.30583 | 22034 | 5508 |
| 0.714294 | 0.297994 | 22034 | 5508 |

| | | | |
|---|---|---|---|
| 0.674939 | 0.308418 | 22033 | 5509 |
| 0.673724 | 0.309022 | 22033 | 5509 |
| 0.702636 | 0.306715 | 22034 | 5508 |
| 0.706705 | 0.305837 | 22034 | 5508 |
| 0.714351 | 0.297938 | 22034 | 5508 |
| 0.724749 | 0.25738 | 22033 | 5509 |
| 0.722589 | 0.258906 | 22033 | 5509 |
| 0.753598 | 0.25643 | 22034 | 5508 |
| 0.762925 | 0.250615 | 22034 | 5508 |
| 0.768595 | 0.244627 | 22034 | 5508 |
| 0.702511 | 0.33971 | 27542 | 4025 |
| 0.702932 | 0.339314 | 27542 | 4025 |
| 0.755339 | 0.290057 | 27542 | 4025 |
| 0.637135 | 0.35047 | 22033 | 5509 |
| 0.628131 | 0.36919 | 22033 | 5509 |
| 0.637672 | 0.360801 | 22034 | 5508 |
| 0.644321 | 0.36607 | 22034 | 5508 |
| 0.65697 | 0.348942 | 22034 | 5508 |
| 0.636816 | 0.350796 | 22033 | 5509 |
| 0.628262 | 0.369058 | 22033 | 5509 |
| 0.637876 | 0.360597 | 22034 | 5508 |
| 0.644431 | 0.365962 | 22034 | 5508 |
| 0.656998 | 0.348915 | 22034 | 5508 |
| 0.680851 | 0.305903 | 22033 | 5509 |
| 0.68046 | 0.316638 | 22033 | 5509 |
| 0.683421 | 0.314943 | 22034 | 5508 |
| 0.702411 | 0.308917 | 22034 | 5508 |
| 0.70281 | 0.303515 | 22034 | 5508 |
| 0.729922 | 0.303303 | 27542 | 4025 |
| 0.73054 | 0.302714 | 27542 | 4025 |
| 0.783479 | 0.252184 | 27542 | 4025 |
| 0.699577 | 0.308775 | 22033 | 5509 |
| 0.648799 | 0.333102 | 22033 | 5509 |
| 0.654654 | 0.335041 | 22034 | 5508 |
| 0.693411 | 0.329031 | 22034 | 5508 |
| 0.679459 | 0.318556 | 22034 | 5508 |
| 0.699236 | 0.309112 | 22033 | 5509 |
| 0.649069 | 0.332825 | 22033 | 5509 |
| 0.654882 | 0.334809 | 22034 | 5508 |
| 0.693386 | 0.329056 | 22034 | 5508 |
| 0.67946 | 0.318555 | 22034 | 5508 |
| 0.738588 | 0.270229 | 22033 | 5509 |
| 0.697754 | 0.282782 | 22033 | 5509 |
| 0.703957 | 0.284962 | 22034 | 5508 |
| 0.750859 | 0.273443 | 22034 | 5508 |

| | | | |
|---|---|---|---|
| 0.725236 | 0.272645 | 22034 | 5508 |
| 0.603335 | 0.410049 | 27542 | 4025 |
| 0.603591 | 0.409798 | 27542 | 4025 |
| 0.656728 | 0.35784 | 27542 | 4025 |
| 0.571067 | 0.418008 | 22033 | 5509 |
| 0.541715 | 0.450323 | 22033 | 5509 |
| 0.565851 | 0.442107 | 22034 | 5508 |
| 0.57209 | 0.436499 | 22034 | 5508 |
| 0.558587 | 0.443428 | 22034 | 5508 |
| 0.570672 | 0.418411 | 22033 | 5509 |
| 0.541871 | 0.450165 | 22033 | 5509 |
| 0.565918 | 0.44204 | 22034 | 5508 |
| 0.572243 | 0.436347 | 22034 | 5508 |
| 0.558708 | 0.443307 | 22034 | 5508 |
| 0.609879 | 0.378454 | 22033 | 5509 |
| 0.597933 | 0.393279 | 22033 | 5509 |
| 0.618825 | 0.389877 | 22034 | 5508 |
| 0.62984 | 0.379615 | 22034 | 5508 |
| 0.618556 | 0.383674 | 22034 | 5508 |
| 0.545031 | 0.452512 | 27542 | 4025 |
| 0.545206 | 0.452336 | 27542 | 4025 |
| 0.598674 | 0.398627 | 27542 | 4025 |
| 0.524573 | 0.459108 | 22033 | 5509 |
| 0.513671 | 0.484051 | 22033 | 5509 |
| 0.534439 | 0.473064 | 22034 | 5508 |
| 0.544919 | 0.465958 | 22034 | 5508 |
| 0.530934 | 0.468912 | 22034 | 5508 |
| 0.524348 | 0.45934 | 22033 | 5509 |
| 0.513693 | 0.484029 | 22033 | 5509 |
| 0.534552 | 0.472953 | 22034 | 5508 |
| 0.544985 | 0.465894 | 22034 | 5508 |
| 0.531079 | 0.468766 | 22034 | 5508 |
| 0.569387 | 0.412899 | 22033 | 5509 |
| 0.570886 | 0.426583 | 22033 | 5509 |
| 0.588725 | 0.419541 | 22034 | 5508 |
| 0.601476 | 0.410531 | 22034 | 5508 |
| 0.598302 | 0.401524 | 22034 | 5508 |
| 0.580318 | 0.420357 | 27542 | 4025 |
| 0.580493 | 0.420183 | 27542 | 4025 |
| 0.623157 | 0.377568 | 27542 | 4025 |
| 0.566129 | 0.430375 | 22033 | 5509 |
| 0.527302 | 0.456333 | 22033 | 5509 |
| 0.555332 | 0.456665 | 22034 | 5508 |
| 0.539191 | 0.448053 | 22034 | 5508 |
| 0.571771 | 0.448849 | 22034 | 5508 |

| | | | |
|---|---|---|---|
| 0.565934 | 0.430571 | 22033 | 5509 |
| 0.527212 | 0.456426 | 22033 | 5509 |
| 0.55547 | 0.45653 | 22034 | 5508 |
| 0.539223 | 0.448019 | 22034 | 5508 |
| 0.57204 | 0.44859 | 22034 | 5508 |
| 0.609032 | 0.387207 | 22033 | 5509 |
| 0.575454 | 0.406686 | 22033 | 5509 |
| 0.602302 | 0.41071 | 22034 | 5508 |
| 0.587908 | 0.398183 | 22034 | 5508 |
| 0.635959 | 0.386975 | 22034 | 5508 |
| 0.66209 | 0.358613 | 27542 | 4025 |
| 0.662303 | 0.358407 | 27542 | 4025 |
| 0.707688 | 0.314441 | 27542 | 4025 |
| 0.613702 | 0.381889 | 22033 | 5509 |
| 0.599223 | 0.393585 | 22033 | 5509 |
| 0.595692 | 0.40412 | 22034 | 5508 |
| 0.592863 | 0.395892 | 22034 | 5508 |
| 0.636514 | 0.38678 | 22034 | 5508 |
| 0.613521 | 0.382071 | 22033 | 5509 |
| 0.59919 | 0.393618 | 22033 | 5509 |
| 0.595741 | 0.40407 | 22034 | 5508 |
| 0.593017 | 0.395736 | 22034 | 5508 |
| 0.636636 | 0.386662 | 22034 | 5508 |
| 0.660606 | 0.334647 | 22033 | 5509 |
| 0.648168 | 0.344052 | 22033 | 5509 |
| 0.639971 | 0.359826 | 22034 | 5508 |
| 0.647072 | 0.340656 | 22034 | 5508 |
| 0.693728 | 0.331659 | 22034 | 5508 |
| 0.709539 | 0.314787 | 27542 | 4025 |
| 0.709833 | 0.314502 | 27542 | 4025 |
| 0.763757 | 0.262427 | 27542 | 4025 |
| 0.676948 | 0.322519 | 22033 | 5509 |
| 0.639549 | 0.350928 | 22033 | 5509 |
| 0.669251 | 0.331853 | 22034 | 5508 |
| 0.670536 | 0.33321 | 22034 | 5508 |
| 0.68907 | 0.315772 | 22034 | 5508 |
| 0.676956 | 0.322511 | 22033 | 5509 |
| 0.639559 | 0.350918 | 22033 | 5509 |
| 0.669331 | 0.331774 | 22034 | 5508 |
| 0.67053 | 0.333216 | 22034 | 5508 |
| 0.689158 | 0.315685 | 22034 | 5508 |
| 0.719178 | 0.280257 | 22033 | 5509 |
| 0.695741 | 0.2939 | 22033 | 5509 |
| 0.716971 | 0.284212 | 22034 | 5508 |
| 0.725252 | 0.278799 | 22034 | 5508 |

| | | | |
|---|---|---|---|
| 0.735515 | 0.269653 | 22034 | 5508 |
| 0.627231 | 0.393059 | 27542 | 4025 |
| 0.62742 | 0.392877 | 27542 | 4025 |
| 0.678352 | 0.343592 | 27542 | 4025 |
| 0.598563 | 0.394725 | 22033 | 5509 |
| 0.575985 | 0.415902 | 22033 | 5509 |
| 0.588594 | 0.407831 | 22034 | 5508 |
| 0.595499 | 0.405829 | 22034 | 5508 |
| 0.614256 | 0.402812 | 22034 | 5508 |
| 0.598546 | 0.394743 | 22033 | 5509 |
| 0.575878 | 0.41601 | 22033 | 5509 |
| 0.588679 | 0.407745 | 22034 | 5508 |
| 0.595525 | 0.405804 | 22034 | 5508 |
| 0.614325 | 0.402744 | 22034 | 5508 |
| 0.641571 | 0.351235 | 22033 | 5509 |
| 0.622151 | 0.369085 | 22033 | 5509 |
| 0.634286 | 0.361861 | 22034 | 5508 |
| 0.647388 | 0.354056 | 22034 | 5508 |
| 0.668443 | 0.35013 | 22034 | 5508 |
| 0.644111 | 0.372019 | 27542 | 4025 |
| 0.644393 | 0.371745 | 27542 | 4025 |
| 0.69215 | 0.325183 | 27542 | 4025 |
| 0.613994 | 0.378404 | 22033 | 5509 |
| 0.569763 | 0.418864 | 22033 | 5509 |
| 0.623333 | 0.387508 | 22034 | 5508 |
| 0.619023 | 0.381936 | 22034 | 5508 |
| 0.623909 | 0.383531 | 22034 | 5508 |
| 0.613952 | 0.378447 | 22033 | 5509 |
| 0.569787 | 0.41884 | 22033 | 5509 |
| 0.623387 | 0.387455 | 22034 | 5508 |
| 0.619013 | 0.381946 | 22034 | 5508 |
| 0.624011 | 0.38343 | 22034 | 5508 |
| 0.650709 | 0.341235 | 22033 | 5509 |
| 0.618702 | 0.368948 | 22033 | 5509 |
| 0.667713 | 0.343901 | 22034 | 5508 |
| 0.66652 | 0.334512 | 22034 | 5508 |
| 0.679391 | 0.32871 | 22034 | 5508 |
| 0.679192 | 0.350239 | 27542 | 4025 |
| 0.679482 | 0.349962 | 27542 | 4025 |
| 0.731173 | 0.30051 | 27542 | 4025 |
| 0.641734 | 0.352392 | 22033 | 5509 |
| 0.62848 | 0.379871 | 22033 | 5509 |
| 0.643212 | 0.362038 | 22034 | 5508 |
| 0.630658 | 0.358648 | 22034 | 5508 |
| 0.643559 | 0.358876 | 22034 | 5508 |

| | | | |
|---|---|---|---|
| 0.641617 | 0.35251 | 22033 | 5509 |
| 0.628433 | 0.379916 | 22033 | 5509 |
| 0.643331 | 0.36192 | 22034 | 5508 |
| 0.630609 | 0.358698 | 22034 | 5508 |
| 0.643689 | 0.358746 | 22034 | 5508 |
| 0.6787 | 0.315088 | 22033 | 5509 |
| 0.68311 | 0.325966 | 22033 | 5509 |
| 0.687582 | 0.31803 | 22034 | 5508 |
| 0.678388 | 0.310109 | 22034 | 5508 |
| 0.69198 | 0.310637 | 22034 | 5508 |
| 0.592618 | 0.422154 | 27542 | 4025 |
| 0.59274 | 0.422036 | 27542 | 4025 |
| 0.645439 | 0.37065 | 27542 | 4025 |
| 0.537925 | 0.451394 | 22033 | 5509 |
| 0.535999 | 0.454763 | 22033 | 5509 |
| 0.541229 | 0.464867 | 22034 | 5508 |
| 0.540456 | 0.458078 | 22034 | 5508 |
| 0.567027 | 0.448191 | 22034 | 5508 |
| 0.537683 | 0.451642 | 22033 | 5509 |
| 0.535955 | 0.454807 | 22033 | 5509 |
| 0.541365 | 0.464733 | 22034 | 5508 |
| 0.540568 | 0.457965 | 22034 | 5508 |
| 0.567233 | 0.44799 | 22034 | 5508 |
| 0.588756 | 0.399554 | 22033 | 5509 |
| 0.587893 | 0.401974 | 22033 | 5509 |
| 0.595141 | 0.411562 | 22034 | 5508 |
| 0.600727 | 0.397643 | 22034 | 5508 |
| 0.635007 | 0.382035 | 22034 | 5508 |
| 0.958621 | 0.032971 | 27542 | 4025 |
| 0.958847 | 0.032743 | 27542 | 4025 |
| 0.975381 | 0.016063 | 27542 | 4025 |
| 0.938231 | 0.037745 | 22033 | 5509 |
| 0.970806 | 0.033261 | 22033 | 5509 |
| 0.946681 | 0.034736 | 22034 | 5508 |
| 0.985172 | 0.0349 | 22034 | 5508 |
| 0.98416 | 0.033934 | 22034 | 5508 |
| 0.938288 | 0.037686 | 22033 | 5509 |
| 0.97076 | 0.033308 | 22033 | 5509 |
| 0.946683 | 0.034734 | 22034 | 5508 |
| 0.985008 | 0.035061 | 22034 | 5508 |
| 0.984226 | 0.033869 | 22034 | 5508 |
| 0.960595 | 0.014809 | 22033 | 5509 |
| 0.990641 | 0.01351 | 22033 | 5509 |
| 0.967034 | 0.013983 | 22034 | 5508 |
| 1.005618 | 0.014871 | 22034 | 5508 |

| | | | |
|---|---|---|---|
| 1.004883 | 0.013592 | 22034 | 5508 |
| 0.793674 | 0.209549 | 27542 | 4025 |
| 0.793769 | 0.209455 | 27542 | 4025 |
| 0.830327 | 0.173044 | 27542 | 4025 |
| 0.831607 | 0.20696 | 22033 | 5509 |
| 0.777794 | 0.220918 | 22033 | 5509 |
| 0.775535 | 0.195441 | 22034 | 5508 |
| 0.771662 | 0.220107 | 22034 | 5508 |
| 0.798528 | 0.201462 | 22034 | 5508 |
| 0.831585 | 0.206981 | 22033 | 5509 |
| 0.777946 | 0.220766 | 22033 | 5509 |
| 0.775497 | 0.195481 | 22034 | 5508 |
| 0.77168 | 0.22009 | 22034 | 5508 |
| 0.798564 | 0.201425 | 22034 | 5508 |
| 0.87597 | 0.164655 | 22033 | 5509 |
| 0.825725 | 0.172908 | 22033 | 5509 |
| 0.803995 | 0.165915 | 22034 | 5508 |
| 0.816365 | 0.174928 | 22034 | 5508 |
| 0.837478 | 0.162511 | 22034 | 5508 |
| 0.933861 | 0.047048 | 27542 | 4025 |
| 0.933848 | 0.047061 | 27542 | 4025 |
| 0.967319 | 0.012906 | 27542 | 4025 |
| 0.935792 | 0.047952 | 22033 | 5509 |
| 0.953963 | 0.06228 | 22033 | 5509 |
| 0.935335 | 0.064366 | 22034 | 5508 |
| 0.925945 | 0.071566 | 22034 | 5508 |
| 0.953626 | 0.048551 | 22034 | 5508 |
| 0.935685 | 0.048061 | 22033 | 5509 |
| 0.954333 | 0.061916 | 22033 | 5509 |
| 0.935377 | 0.064325 | 22034 | 5508 |
| 0.926212 | 0.071299 | 22034 | 5508 |
| 0.953522 | 0.048656 | 22034 | 5508 |
| 0.967597 | 0.015595 | 22033 | 5509 |
| 0.997251 | 0.019729 | 22033 | 5509 |
| 0.980981 | 0.018706 | 22034 | 5508 |
| 0.981687 | 0.015675 | 22034 | 5508 |
| 0.985155 | 0.017095 | 22034 | 5508 |
| 0.944949 | 0.016785 | 27542 | 4025 |
| 0.944322 | 0.017436 | 27542 | 4025 |
| 0.951106 | 0.010378 | 27542 | 4025 |
| 0.982997 | 0.01521 | 22033 | 5509 |
| 0.981381 | 0.02764 | 22033 | 5509 |
| 0.945004 | 0.031714 | 22034 | 5508 |
| 0.963427 | 0.028159 | 22034 | 5508 |
| 0.996314 | 0.027947 | 22034 | 5508 |

| | | | |
|---|---|---|---|
| 0.982888 | 0.015319 | 22033 | 5509 |
| 0.981261 | 0.027758 | 22033 | 5509 |
| 0.945456 | 0.031251 | 22034 | 5508 |
| 0.963076 | 0.028514 | 22034 | 5508 |
| 0.996302 | 0.027959 | 22034 | 5508 |
| 0.987317 | 0.010881 | 22033 | 5509 |
| 0.994926 | 0.014219 | 22033 | 5509 |
| 0.961327 | 0.014989 | 22034 | 5508 |
| 0.976494 | 0.014978 | 22034 | 5508 |
| 1.011108 | 0.013513 | 22034 | 5508 |
| 0.895571 | 0.088377 | 27542 | 4025 |
| 0.895543 | 0.088405 | 27542 | 4025 |
| 0.924637 | 0.05879 | 27542 | 4025 |
| 0.912015 | 0.103441 | 22033 | 5509 |
| 0.881778 | 0.099497 | 22033 | 5509 |
| 0.853432 | 0.102875 | 22034 | 5508 |
| 0.942944 | 0.112874 | 22034 | 5508 |
| 0.885775 | 0.105809 | 22034 | 5508 |
| 0.911852 | 0.103601 | 22033 | 5509 |
| 0.881748 | 0.099527 | 22033 | 5509 |
| 0.853369 | 0.102942 | 22034 | 5508 |
| 0.943014 | 0.112809 | 22034 | 5508 |
| 0.885744 | 0.10584 | 22034 | 5508 |
| 0.946003 | 0.070029 | 22033 | 5509 |
| 0.914311 | 0.066273 | 22033 | 5509 |
| 0.887278 | 0.067297 | 22034 | 5508 |
| 0.992413 | 0.066333 | 22034 | 5508 |
| 0.925394 | 0.065813 | 22034 | 5508 |
| 0.927777 | 0.044472 | 27542 | 4025 |
| 0.927739 | 0.044511 | 27542 | 4025 |
| 0.947815 | 0.023835 | 27542 | 4025 |
| 0.932199 | 0.056098 | 22033 | 5509 |
| 0.961426 | 0.028349 | 22033 | 5509 |
| 0.925061 | 0.046155 | 22034 | 5508 |
| 0.979559 | 0.044278 | 22034 | 5508 |
| 0.988238 | 0.038929 | 22034 | 5508 |
| 0.932235 | 0.056062 | 22033 | 5509 |
| 0.961288 | 0.028489 | 22033 | 5509 |
| 0.925093 | 0.046122 | 22034 | 5508 |
| 0.979607 | 0.044231 | 22034 | 5508 |
| 0.988156 | 0.039008 | 22034 | 5508 |
| 0.963775 | 0.024126 | 22033 | 5509 |
| 0.971132 | 0.018539 | 22033 | 5509 |
| 0.94779 | 0.022718 | 22034 | 5508 |
| 1.003815 | 0.020612 | 22034 | 5508 |

| | | | |
|---|---|---|---|
| 1.01161 | 0.016199 | 22034 | 5508 |
| 0.846099 | 0.121169 | 27542 | 4025 |
| 0.846034 | 0.121237 | 27542 | 4025 |
| 0.881298 | 0.084608 | 27542 | 4025 |
| 0.890102 | 0.124891 | 22033 | 5509 |
| 0.846662 | 0.121897 | 22033 | 5509 |
| 0.885231 | 0.115966 | 22034 | 5508 |
| 0.882421 | 0.123268 | 22034 | 5508 |
| 0.882714 | 0.126358 | 22034 | 5508 |
| 0.890053 | 0.124938 | 22033 | 5509 |
| 0.846727 | 0.12183 | 22033 | 5509 |
| 0.885111 | 0.116086 | 22034 | 5508 |
| 0.882524 | 0.123166 | 22034 | 5508 |
| 0.882628 | 0.126443 | 22034 | 5508 |
| 0.937676 | 0.078118 | 22033 | 5509 |
| 0.885566 | 0.081549 | 22033 | 5509 |
| 0.92181 | 0.079437 | 22034 | 5508 |
| 0.92729 | 0.078689 | 22034 | 5508 |
| 0.932649 | 0.076937 | 22034 | 5508 |
| 0.868085 | 0.129418 | 27542 | 4025 |
| 0.868174 | 0.12933 | 27542 | 4025 |
| 0.923482 | 0.073862 | 27542 | 4025 |
| 0.925476 | 0.112882 | 22033 | 5509 |
| 0.912403 | 0.120706 | 22033 | 5509 |
| 0.847966 | 0.1214 | 22034 | 5508 |
| 0.863548 | 0.117562 | 22034 | 5508 |
| 0.855696 | 0.123234 | 22034 | 5508 |
| 0.925408 | 0.112946 | 22033 | 5509 |
| 0.912533 | 0.120581 | 22033 | 5509 |
| 0.847888 | 0.121481 | 22034 | 5508 |
| 0.863618 | 0.117491 | 22034 | 5508 |
| 0.855893 | 0.123032 | 22034 | 5508 |
| 0.972536 | 0.067772 | 22033 | 5509 |
| 0.966376 | 0.068692 | 22033 | 5509 |
| 0.891099 | 0.076708 | 22034 | 5508 |
| 0.911568 | 0.068492 | 22034 | 5508 |
| 0.915483 | 0.061975 | 22034 | 5508 |
| 0.929799 | 0.073988 | 27542 | 4025 |
| 0.929867 | 0.073921 | 27542 | 4025 |
| 0.966043 | 0.037891 | 27542 | 4025 |
| 0.921939 | 0.065719 | 22033 | 5509 |
| 0.933878 | 0.072142 | 22033 | 5509 |
| 0.915601 | 0.081666 | 22034 | 5508 |
| 0.938878 | 0.078608 | 22034 | 5508 |
| 0.92477 | 0.065587 | 22034 | 5508 |

| | | | |
|---|---|---|---|
| 0.921906 | 0.065752 | 22033 | 5509 |
| 0.933905 | 0.072115 | 22033 | 5509 |
| 0.915738 | 0.081529 | 22034 | 5508 |
| 0.938766 | 0.078718 | 22034 | 5508 |
| 0.92465 | 0.065708 | 22034 | 5508 |
| 0.950099 | 0.037182 | 22033 | 5509 |
| 0.967751 | 0.038487 | 22033 | 5509 |
| 0.958538 | 0.038601 | 22034 | 5508 |
| 0.978077 | 0.040139 | 22034 | 5508 |
| 0.95354 | 0.036518 | 22034 | 5508 |
| 0.93716 | 0.030417 | 27542 | 4025 |
| 0.937547 | 0.030017 | 27542 | 4025 |
| 0.959087 | 0.007732 | 27542 | 4025 |
| 0.971954 | 0.027504 | 22033 | 5509 |
| 0.979325 | 0.025976 | 22033 | 5509 |
| 0.968121 | 0.016163 | 22034 | 5508 |
| 0.982284 | 0.02141 | 22034 | 5508 |
| 0.982769 | 0.023534 | 22034 | 5508 |
| 0.972205 | 0.027254 | 22033 | 5509 |
| 0.979354 | 0.025947 | 22033 | 5509 |
| 0.967697 | 0.016594 | 22034 | 5508 |
| 0.982065 | 0.021629 | 22034 | 5508 |
| 0.98285 | 0.023453 | 22034 | 5508 |
| 0.990798 | 0.008649 | 22033 | 5509 |
| 0.997665 | 0.007736 | 22033 | 5509 |
| 0.977246 | 0.00689 | 22034 | 5508 |
| 0.996695 | 0.007054 | 22034 | 5508 |
| 0.998606 | 0.007798 | 22034 | 5508 |
| 0.92125 | 0.083164 | 27542 | 4025 |
| 0.921338 | 0.083076 | 27542 | 4025 |
| 0.962909 | 0.041704 | 27542 | 4025 |
| 0.946939 | 0.065556 | 22033 | 5509 |
| 0.911482 | 0.085793 | 22033 | 5509 |
| 0.923871 | 0.078078 | 22034 | 5508 |
| 0.932827 | 0.08188 | 22034 | 5508 |
| 0.902068 | 0.071088 | 22034 | 5508 |
| 0.94678 | 0.065713 | 22033 | 5509 |
| 0.911481 | 0.085794 | 22033 | 5509 |
| 0.923942 | 0.078007 | 22034 | 5508 |
| 0.932856 | 0.081852 | 22034 | 5508 |
| 0.902077 | 0.071078 | 22034 | 5508 |
| 0.977458 | 0.03544 | 22033 | 5509 |
| 0.958399 | 0.038736 | 22033 | 5509 |
| 0.963893 | 0.03814 | 22034 | 5508 |
| 0.974924 | 0.040447 | 22034 | 5508 |

| | | | |
|---|---|---|---|
| 0.93489 | 0.037289 | 22034 | 5508 |
| 0.933838 | 0.072122 | 27542 | 4025 |
| 0.933811 | 0.07215 | 27542 | 4025 |
| 0.972246 | 0.03396 | 27542 | 4025 |
| 0.902209 | 0.084216 | 22033 | 5509 |
| 0.929591 | 0.082692 | 22033 | 5509 |
| 0.919846 | 0.08884 | 22034 | 5508 |
| 0.919668 | 0.087089 | 22034 | 5508 |
| 0.900839 | 0.08472 | 22034 | 5508 |
| 0.902112 | 0.084314 | 22033 | 5509 |
| 0.929593 | 0.08269 | 22033 | 5509 |
| 0.919933 | 0.088753 | 22034 | 5508 |
| 0.919638 | 0.087118 | 22034 | 5508 |
| 0.900874 | 0.084684 | 22034 | 5508 |
| 0.946963 | 0.038788 | 22033 | 5509 |
| 0.976374 | 0.036527 | 22033 | 5509 |
| 0.971476 | 0.037697 | 22034 | 5508 |
| 0.970245 | 0.036884 | 22034 | 5508 |
| 0.95002 | 0.034751 | 22034 | 5508 |
| 0.892207 | 0.110541 | 27542 | 4025 |
| 0.892261 | 0.110487 | 27542 | 4025 |
| 0.941151 | 0.061747 | 27542 | 4025 |
| 0.905886 | 0.093646 | 22033 | 5509 |
| 0.881615 | 0.101273 | 22033 | 5509 |
| 0.915396 | 0.111873 | 22034 | 5508 |
| 0.926054 | 0.115427 | 22034 | 5508 |
| 0.828332 | 0.120417 | 22034 | 5508 |
| 0.905863 | 0.093668 | 22033 | 5509 |
| 0.881666 | 0.101221 | 22033 | 5509 |
| 0.915306 | 0.11196 | 22034 | 5508 |
| 0.926058 | 0.115423 | 22034 | 5508 |
| 0.828378 | 0.120369 | 22034 | 5508 |
| 0.943341 | 0.056171 | 22033 | 5509 |
| 0.923522 | 0.058552 | 22033 | 5509 |
| 0.963561 | 0.065143 | 22034 | 5508 |
| 0.979218 | 0.064644 | 22034 | 5508 |
| 0.883124 | 0.062235 | 22034 | 5508 |
| 0.92517 | 0.088734 | 27542 | 4025 |
| 0.925248 | 0.088657 | 27542 | 4025 |
| 0.953024 | 0.061299 | 27542 | 4025 |
| 0.909264 | 0.086009 | 22033 | 5509 |
| 0.900573 | 0.088618 | 22033 | 5509 |
| 0.93304 | 0.106259 | 22034 | 5508 |
| 0.860229 | 0.102521 | 22034 | 5508 |
| 0.920998 | 0.092441 | 22034 | 5508 |

| | | | |
|---|---|---|---|
| 0.909324 | 0.085949 | 22033 | 5509 |
| 0.900612 | 0.088579 | 22033 | 5509 |
| 0.933058 | 0.106242 | 22034 | 5508 |
| 0.86024 | 0.102509 | 22034 | 5508 |
| 0.920932 | 0.092506 | 22034 | 5508 |
| 0.939479 | 0.055637 | 22033 | 5509 |
| 0.928075 | 0.060786 | 22033 | 5509 |
| 0.97774 | 0.063441 | 22034 | 5508 |
| 0.896033 | 0.065167 | 22034 | 5508 |
| 0.959994 | 0.054015 | 22034 | 5508 |
| 0.986325 | 0.040729 | 27542 | 4025 |
| 0.986393 | 0.040662 | 27542 | 4025 |
| 1.009213 | 0.018469 | 27542 | 4025 |
| 0.984113 | 0.038017 | 22033 | 5509 |
| 0.926816 | 0.042749 | 22033 | 5509 |
| 0.954405 | 0.047255 | 22034 | 5508 |
| 0.971378 | 0.050193 | 22034 | 5508 |
| 0.942416 | 0.042113 | 22034 | 5508 |
| 0.984028 | 0.0381 | 22033 | 5509 |
| 0.926832 | 0.042733 | 22033 | 5509 |
| 0.954411 | 0.04725 | 22034 | 5508 |
| 0.971402 | 0.05017 | 22034 | 5508 |
| 0.942432 | 0.042097 | 22034 | 5508 |
| 0.999611 | 0.022867 | 22033 | 5509 |
| 0.945482 | 0.023471 | 22033 | 5509 |
| 0.976784 | 0.024916 | 22034 | 5508 |
| 0.998689 | 0.023489 | 22034 | 5508 |
| 0.962769 | 0.021425 | 22034 | 5508 |
| 0.956141 | 0.064206 | 27542 | 4025 |
| 0.956177 | 0.06417 | 27542 | 4025 |
| 0.985831 | 0.035147 | 27542 | 4025 |
| 0.959608 | 0.062291 | 22033 | 5509 |
| 0.894035 | 0.069992 | 22033 | 5509 |
| 0.943756 | 0.092844 | 22034 | 5508 |
| 0.923132 | 0.080065 | 22034 | 5508 |
| 0.897442 | 0.076451 | 22034 | 5508 |
| 0.959531 | 0.062367 | 22033 | 5509 |
| 0.894033 | 0.069994 | 22033 | 5509 |
| 0.943907 | 0.092699 | 22034 | 5508 |
| 0.923085 | 0.080112 | 22034 | 5508 |
| 0.897437 | 0.076456 | 22034 | 5508 |
| 0.985065 | 0.037415 | 22033 | 5509 |
| 0.923121 | 0.039735 | 22033 | 5509 |
| 0.995369 | 0.043232 | 22034 | 5508 |
| 0.963545 | 0.039792 | 22034 | 5508 |

| | | | |
|---|---|---|---|
| 0.935105 | 0.037692 | 22034 | 5508 |
| 0.80504 | 0.184159 | 27542 | 4025 |
| 0.805108 | 0.18409 | 27542 | 4025 |
| 0.861817 | 0.12662 | 27542 | 4025 |
| 0.829844 | 0.188789 | 22033 | 5509 |
| 0.810618 | 0.192618 | 22033 | 5509 |
| 0.79196 | 0.199338 | 22034 | 5508 |
| 0.815775 | 0.199878 | 22034 | 5508 |
| 0.776553 | 0.194496 | 22034 | 5508 |
| 0.829785 | 0.188847 | 22033 | 5509 |
| 0.810681 | 0.192555 | 22033 | 5509 |
| 0.791934 | 0.199364 | 22034 | 5508 |
| 0.81582 | 0.199833 | 22034 | 5508 |
| 0.776549 | 0.194499 | 22034 | 5508 |
| 0.891892 | 0.128135 | 22033 | 5509 |
| 0.873567 | 0.12992 | 22033 | 5509 |
| 0.851918 | 0.138721 | 22034 | 5508 |
| 0.88221 | 0.134718 | 22034 | 5508 |
| 0.844959 | 0.123539 | 22034 | 5508 |
| 0.360627 | 0.642716 | 27542 | 4025 |
| 0.360891 | 0.642454 | 27542 | 4025 |
| 0.432027 | 0.571977 | 27542 | 4025 |
| 0.377364 | 0.624674 | 22033 | 5509 |
| 0.346896 | 0.643047 | 22033 | 5509 |
| 0.354922 | 0.637026 | 22034 | 5508 |
| 0.362111 | 0.649496 | 22034 | 5508 |
| 0.362826 | 0.641191 | 22034 | 5508 |
| 0.377539 | 0.6245 | 22033 | 5509 |
| 0.346762 | 0.643185 | 22033 | 5509 |
| 0.35499 | 0.636956 | 22034 | 5508 |
| 0.362186 | 0.649423 | 22034 | 5508 |
| 0.362784 | 0.641233 | 22034 | 5508 |
| 0.458177 | 0.544297 | 22033 | 5509 |
| 0.422655 | 0.565091 | 22033 | 5509 |
| 0.429542 | 0.560712 | 22034 | 5508 |
| 0.441873 | 0.572291 | 22034 | 5508 |
| 0.449491 | 0.555485 | 22034 | 5508 |
| 0.439534 | 0.555426 | 27542 | 4025 |
| 0.439582 | 0.555377 | 27542 | 4025 |
| 0.505583 | 0.488619 | 27542 | 4025 |
| 0.44567 | 0.55957 | 22033 | 5509 |
| 0.425606 | 0.565433 | 22033 | 5509 |
| 0.430363 | 0.56113 | 22034 | 5508 |
| 0.428796 | 0.583575 | 22034 | 5508 |
| 0.429003 | 0.57016 | 22034 | 5508 |

| | | | |
|---|---|---|---|
| 0.445834 | 0.559408 | 22033 | 5509 |
| 0.425503 | 0.565538 | 22033 | 5509 |
| 0.430439 | 0.561052 | 22034 | 5508 |
| 0.428727 | 0.583642 | 22034 | 5508 |
| 0.428878 | 0.570285 | 22034 | 5508 |
| 0.530756 | 0.475484 | 22033 | 5509 |
| 0.495635 | 0.493929 | 22033 | 5509 |
| 0.503292 | 0.486759 | 22034 | 5508 |
| 0.510757 | 0.503979 | 22034 | 5508 |
| 0.516541 | 0.482451 | 22034 | 5508 |
| 0.495136 | 0.513465 | 27542 | 4025 |
| 0.49524 | 0.513363 | 27542 | 4025 |
| 0.551388 | 0.458191 | 27542 | 4025 |
| 0.53709 | 0.467174 | 22033 | 5509 |
| 0.481811 | 0.497507 | 22033 | 5509 |
| 0.505754 | 0.487526 | 22034 | 5508 |
| 0.519373 | 0.495473 | 22034 | 5508 |
| 0.51475 | 0.493454 | 22034 | 5508 |
| 0.537221 | 0.467044 | 22033 | 5509 |
| 0.481761 | 0.49756 | 22033 | 5509 |
| 0.505811 | 0.487469 | 22034 | 5508 |
| 0.519556 | 0.495295 | 22034 | 5508 |
| 0.514764 | 0.493439 | 22034 | 5508 |
| 0.589429 | 0.415251 | 22033 | 5509 |
| 0.534603 | 0.44245 | 22033 | 5509 |
| 0.560026 | 0.432533 | 22034 | 5508 |
| 0.576397 | 0.440079 | 22034 | 5508 |
| 0.573864 | 0.435281 | 22034 | 5508 |
| 0.852349 | 0.125816 | 27542 | 4025 |
| 0.852298 | 0.125868 | 27542 | 4025 |
| 0.892863 | 0.084264 | 27542 | 4025 |
| 0.871016 | 0.14841 | 22033 | 5509 |
| 0.827789 | 0.146675 | 22033 | 5509 |
| 0.836831 | 0.146612 | 22034 | 5508 |
| 0.865618 | 0.164691 | 22034 | 5508 |
| 0.842902 | 0.149062 | 22034 | 5508 |
| 0.870989 | 0.148436 | 22033 | 5509 |
| 0.827856 | 0.146606 | 22033 | 5509 |
| 0.836737 | 0.146708 | 22034 | 5508 |
| 0.865827 | 0.16449 | 22034 | 5508 |
| 0.842782 | 0.149183 | 22034 | 5508 |
| 0.929741 | 0.090995 | 22033 | 5509 |
| 0.872264 | 0.100828 | 22033 | 5509 |
| 0.885223 | 0.097263 | 22034 | 5508 |
| 0.931806 | 0.100821 | 22034 | 5508 |

| | | | |
|---|---|---|---|
| 0.901529 | 0.089876 | 22034 | 5508 |
| 0.928904 | 0.056066 | 27542 | 4025 |
| 0.928793 | 0.056179 | 27542 | 4025 |
| 0.960064 | 0.024402 | 27542 | 4025 |
| 0.970903 | 0.041966 | 22033 | 5509 |
| 0.950131 | 0.063763 | 22033 | 5509 |
| 0.915927 | 0.050489 | 22034 | 5508 |
| 0.935924 | 0.055173 | 22034 | 5508 |
| 0.958039 | 0.057791 | 22034 | 5508 |
| 0.970854 | 0.042015 | 22033 | 5509 |
| 0.950255 | 0.063641 | 22033 | 5509 |
| 0.915965 | 0.050449 | 22034 | 5508 |
| 0.935921 | 0.055176 | 22034 | 5508 |
| 0.958063 | 0.057767 | 22034 | 5508 |
| 0.993435 | 0.019733 | 22033 | 5509 |
| 0.990051 | 0.024426 | 22033 | 5509 |
| 0.942354 | 0.023093 | 22034 | 5508 |
| 0.968412 | 0.022375 | 22034 | 5508 |
| 0.99314 | 0.02327 | 22034 | 5508 |
| 0.843156 | 0.142995 | 27542 | 4025 |
| 0.843308 | 0.142841 | 27542 | 4025 |
| 0.881098 | 0.10443 | 27542 | 4025 |
| 0.864271 | 0.152175 | 22033 | 5509 |
| 0.861415 | 0.144453 | 22033 | 5509 |
| 0.842588 | 0.145127 | 22034 | 5508 |
| 0.86606 | 0.145547 | 22034 | 5508 |
| 0.834374 | 0.143519 | 22034 | 5508 |
| 0.864322 | 0.152125 | 22033 | 5509 |
| 0.861523 | 0.144345 | 22033 | 5509 |
| 0.842554 | 0.145161 | 22034 | 5508 |
| 0.865982 | 0.145624 | 22034 | 5508 |
| 0.834311 | 0.143584 | 22034 | 5508 |
| 0.906772 | 0.110483 | 22033 | 5509 |
| 0.899645 | 0.106483 | 22033 | 5509 |
| 0.880337 | 0.106827 | 22034 | 5508 |
| 0.905278 | 0.106855 | 22034 | 5508 |
| 0.891204 | 0.085183 | 22034 | 5508 |
| 0.880737 | 0.141733 | 27542 | 4025 |
| 0.880814 | 0.141658 | 27542 | 4025 |
| 0.913379 | 0.109924 | 27542 | 4025 |
| 0.915132 | 0.133546 | 22033 | 5509 |
| 0.854202 | 0.134888 | 22033 | 5509 |
| 0.822271 | 0.149196 | 22034 | 5508 |
| 0.862019 | 0.13027 | 22034 | 5508 |
| 0.866863 | 0.13245 | 22034 | 5508 |

| | | | |
|---|---|---|---|
| 0.914916 | 0.133751 | 22033 | 5509 |
| 0.85422 | 0.13487 | 22033 | 5509 |
| 0.822351 | 0.149113 | 22034 | 5508 |
| 0.86214 | 0.130148 | 22034 | 5508 |
| 0.866809 | 0.132505 | 22034 | 5508 |
| 0.95028 | 0.100268 | 22033 | 5509 |
| 0.886419 | 0.10226 | 22033 | 5509 |
| 0.858932 | 0.111262 | 22034 | 5508 |
| 0.897483 | 0.094489 | 22034 | 5508 |
| 0.900313 | 0.098974 | 22034 | 5508 |
| 0.898125 | 0.116079 | 27542 | 4025 |
| 0.898142 | 0.116062 | 27542 | 4025 |
| 0.941281 | 0.073606 | 27542 | 4025 |
| 0.878701 | 0.118499 | 22033 | 5509 |
| 0.869379 | 0.112721 | 22033 | 5509 |
| 0.874227 | 0.120961 | 22034 | 5508 |
| 0.915256 | 0.118511 | 22034 | 5508 |
| 0.873923 | 0.117186 | 22034 | 5508 |
| 0.878575 | 0.118625 | 22033 | 5509 |
| 0.869516 | 0.112582 | 22033 | 5509 |
| 0.874307 | 0.120881 | 22034 | 5508 |
| 0.915124 | 0.118639 | 22034 | 5508 |
| 0.874094 | 0.117013 | 22034 | 5508 |
| 0.920519 | 0.076547 | 22033 | 5509 |
| 0.911505 | 0.069728 | 22033 | 5509 |
| 0.917364 | 0.077587 | 22034 | 5508 |
| 0.960725 | 0.07472 | 22034 | 5508 |
| 0.919024 | 0.071626 | 22034 | 5508 |
| 0.896533 | 0.0679 | 27542 | 4025 |
| 0.896406 | 0.068032 | 27542 | 4025 |
| 0.930103 | 0.032998 | 27542 | 4025 |
| 0.934598 | 0.059826 | 22033 | 5509 |
| 0.918637 | 0.062738 | 22033 | 5509 |
| 0.920404 | 0.049564 | 22034 | 5508 |
| 0.981299 | 0.062023 | 22034 | 5508 |
| 0.937862 | 0.071565 | 22034 | 5508 |
| 0.93444 | 0.059984 | 22033 | 5509 |
| 0.918535 | 0.062843 | 22033 | 5509 |
| 0.920168 | 0.049808 | 22034 | 5508 |
| 0.981509 | 0.061823 | 22034 | 5508 |
| 0.938131 | 0.071298 | 22034 | 5508 |
| 0.961929 | 0.032332 | 22033 | 5509 |
| 0.953813 | 0.026849 | 22033 | 5509 |
| 0.942382 | 0.026869 | 22034 | 5508 |
| 1.017445 | 0.027473 | 22034 | 5508 |

| | | | |
|---|---|---|---|
| 0.977199 | 0.032623 | 22034 | 5508 |
| 0.925963 | 0.095663 | 27542 | 4025 |
| 0.926052 | 0.095576 | 27542 | 4025 |
| 0.968791 | 0.053836 | 27542 | 4025 |
| 0.885125 | 0.097543 | 22033 | 5509 |
| 0.892605 | 0.08178 | 22033 | 5509 |
| 0.931488 | 0.082807 | 22034 | 5508 |
| 0.900986 | 0.08734 | 22034 | 5508 |
| 0.936743 | 0.102673 | 22034 | 5508 |
| 0.88525 | 0.097416 | 22033 | 5509 |
| 0.892581 | 0.081805 | 22033 | 5509 |
| 0.931408 | 0.082885 | 22034 | 5508 |
| 0.900993 | 0.087333 | 22034 | 5508 |
| 0.936803 | 0.102615 | 22034 | 5508 |
| 0.926934 | 0.054916 | 22033 | 5509 |
| 0.924349 | 0.049125 | 22033 | 5509 |
| 0.966402 | 0.048429 | 22034 | 5508 |
| 0.937644 | 0.050207 | 22034 | 5508 |
| 0.987888 | 0.053679 | 22034 | 5508 |
| 0.789217 | 0.176828 | 27542 | 4025 |
| 0.789182 | 0.176864 | 27542 | 4025 |
| 0.83657 | 0.127437 | 27542 | 4025 |
| 0.826092 | 0.179539 | 22033 | 5509 |
| 0.814706 | 0.183537 | 22033 | 5509 |
| 0.806969 | 0.171953 | 22034 | 5508 |
| 0.818246 | 0.178788 | 22034 | 5508 |
| 0.850558 | 0.169492 | 22034 | 5508 |
| 0.825968 | 0.179662 | 22033 | 5509 |
| 0.814998 | 0.183244 | 22033 | 5509 |
| 0.806875 | 0.172049 | 22034 | 5508 |
| 0.818213 | 0.178822 | 22034 | 5508 |
| 0.850628 | 0.169424 | 22034 | 5508 |
| 0.875249 | 0.130717 | 22033 | 5509 |
| 0.872238 | 0.125881 | 22033 | 5509 |
| 0.849507 | 0.128303 | 22034 | 5508 |
| 0.881162 | 0.115645 | 22034 | 5508 |
| 0.923715 | 0.09806 | 22034 | 5508 |
| 0.92197 | 0.050208 | 27542 | 4025 |
| 0.922454 | 0.04971 | 27542 | 4025 |
| 0.940712 | 0.030901 | 27542 | 4025 |
| 0.97786 | 0.047255 | 22033 | 5509 |
| 0.953861 | 0.046381 | 22033 | 5509 |
| 0.948208 | 0.035557 | 22034 | 5508 |
| 0.931454 | 0.037974 | 22034 | 5508 |
| 0.970068 | 0.051 | 22034 | 5508 |

| | | | |
|---|---|---|---|
| 0.977771 | 0.047342 | 22033 | 5509 |
| 0.954096 | 0.046146 | 22033 | 5509 |
| 0.948138 | 0.035628 | 22034 | 5508 |
| 0.930607 | 0.038849 | 22034 | 5508 |
| 0.970219 | 0.050852 | 22034 | 5508 |
| 0.998983 | 0.026674 | 22033 | 5509 |
| 0.974324 | 0.025923 | 22033 | 5509 |
| 0.961929 | 0.021601 | 22034 | 5508 |
| 0.943604 | 0.025425 | 22034 | 5508 |
| 0.992751 | 0.02881 | 22034 | 5508 |
| 0.870523 | 0.095054 | 27542 | 4025 |
| 0.870621 | 0.094953 | 27542 | 4025 |
| 0.908189 | 0.055899 | 27542 | 4025 |
| 0.908281 | 0.094468 | 22033 | 5509 |
| 0.899722 | 0.100591 | 22033 | 5509 |
| 0.886543 | 0.092117 | 22034 | 5508 |
| 0.910117 | 0.113173 | 22034 | 5508 |
| 0.895054 | 0.098969 | 22034 | 5508 |
| 0.908111 | 0.094637 | 22033 | 5509 |
| 0.899867 | 0.100446 | 22033 | 5509 |
| 0.886528 | 0.092133 | 22034 | 5508 |
| 0.910191 | 0.113101 | 22034 | 5508 |
| 0.894975 | 0.099049 | 22034 | 5508 |
| 0.941952 | 0.060899 | 22033 | 5509 |
| 0.940525 | 0.059803 | 22033 | 5509 |
| 0.922838 | 0.054948 | 22034 | 5508 |
| 0.961427 | 0.063177 | 22034 | 5508 |
| 0.932088 | 0.061687 | 22034 | 5508 |
| 0.800149 | 0.172744 | 27542 | 4025 |
| 0.800212 | 0.172679 | 27542 | 4025 |
| 0.84021 | 0.131325 | 27542 | 4025 |
| 0.840062 | 0.171589 | 22033 | 5509 |
| 0.83949 | 0.184936 | 22033 | 5509 |
| 0.800114 | 0.173169 | 22034 | 5508 |
| 0.82923 | 0.187967 | 22034 | 5508 |
| 0.795676 | 0.176474 | 22034 | 5508 |
| 0.839914 | 0.171736 | 22033 | 5509 |
| 0.839689 | 0.184742 | 22033 | 5509 |
| 0.800033 | 0.173253 | 22034 | 5508 |
| 0.829285 | 0.187913 | 22034 | 5508 |
| 0.79551 | 0.176646 | 22034 | 5508 |
| 0.877378 | 0.134791 | 22033 | 5509 |
| 0.897612 | 0.128505 | 22033 | 5509 |
| 0.841396 | 0.130509 | 22034 | 5508 |
| 0.88164 | 0.136644 | 22034 | 5508 |

| | | | |
|---|---|---|---|
| 0.839911 | 0.13069 | 22034 | 5508 |
| 0.833403 | 0.178512 | 27542 | 4025 |
| 0.833507 | 0.17841 | 27542 | 4025 |
| 0.873296 | 0.139189 | 27542 | 4025 |
| 0.819234 | 0.180344 | 22033 | 5509 |
| 0.832049 | 0.176672 | 22033 | 5509 |
| 0.799217 | 0.187422 | 22034 | 5508 |
| 0.825703 | 0.187596 | 22034 | 5508 |
| 0.815727 | 0.175816 | 22034 | 5508 |
| 0.818822 | 0.180756 | 22033 | 5509 |
| 0.83257 | 0.176156 | 22033 | 5509 |
| 0.799186 | 0.187453 | 22034 | 5508 |
| 0.82579 | 0.187511 | 22034 | 5508 |
| 0.815737 | 0.175806 | 22034 | 5508 |
| 0.859764 | 0.139793 | 22033 | 5509 |
| 0.880854 | 0.128377 | 22033 | 5509 |
| 0.83959 | 0.146374 | 22034 | 5508 |
| 0.880085 | 0.13409 | 22034 | 5508 |
| 0.860625 | 0.130453 | 22034 | 5508 |
| 0.897553 | 0.118829 | 27542 | 4025 |
| 0.897621 | 0.118762 | 27542 | 4025 |
| 0.932468 | 0.084552 | 27542 | 4025 |
| 0.891222 | 0.122948 | 22033 | 5509 |
| 0.885126 | 0.107472 | 22033 | 5509 |
| 0.864252 | 0.123625 | 22034 | 5508 |
| 0.900079 | 0.119504 | 22034 | 5508 |
| 0.866532 | 0.119123 | 22034 | 5508 |
| 0.891288 | 0.122883 | 22033 | 5509 |
| 0.885011 | 0.107589 | 22033 | 5509 |
| 0.86421 | 0.123668 | 22034 | 5508 |
| 0.900047 | 0.119536 | 22034 | 5508 |
| 0.866677 | 0.118975 | 22034 | 5508 |
| 0.926646 | 0.088086 | 22033 | 5509 |
| 0.911382 | 0.080996 | 22033 | 5509 |
| 0.893218 | 0.094252 | 22034 | 5508 |
| 0.938123 | 0.082288 | 22034 | 5508 |
| 0.908657 | 0.076301 | 22034 | 5508 |
| 0.916966 | 0.090678 | 27542 | 4025 |
| 0.916959 | 0.090685 | 27542 | 4025 |
| 0.960459 | 0.047548 | 27542 | 4025 |
| 0.902237 | 0.086766 | 22033 | 5509 |
| 0.923009 | 0.076151 | 22033 | 5509 |
| 0.912683 | 0.091191 | 22034 | 5508 |
| 0.932982 | 0.070943 | 22034 | 5508 |
| 0.927018 | 0.076567 | 22034 | 5508 |

| | | | |
|---|---|---|---|
| 0.902186 | 0.086817 | 22033 | 5509 |
| 0.922971 | 0.076189 | 22033 | 5509 |
| 0.912895 | 0.09098 | 22034 | 5508 |
| 0.933033 | 0.070893 | 22034 | 5508 |
| 0.927089 | 0.076495 | 22034 | 5508 |
| 0.939513 | 0.049035 | 22033 | 5509 |
| 0.954115 | 0.045017 | 22033 | 5509 |
| 0.956073 | 0.047986 | 22034 | 5508 |
| 0.96169 | 0.042356 | 22034 | 5508 |
| 0.95387 | 0.049818 | 22034 | 5508 |
| 0.959332 | 0.073709 | 27542 | 4025 |
| 0.95967 | 0.073383 | 27542 | 4025 |
| 0.989212 | 0.044859 | 27542 | 4025 |
| 0.912158 | 0.076723 | 22033 | 5509 |
| 0.964581 | 0.074602 | 22033 | 5509 |
| 0.880407 | 0.078678 | 22034 | 5508 |
| 0.941222 | 0.069492 | 22034 | 5508 |
| 0.928054 | 0.074353 | 22034 | 5508 |
| 0.912345 | 0.076535 | 22033 | 5509 |
| 0.964491 | 0.074689 | 22033 | 5509 |
| 0.880541 | 0.078537 | 22034 | 5508 |
| 0.940703 | 0.070005 | 22034 | 5508 |
| 0.928028 | 0.07438 | 22034 | 5508 |
| 0.943124 | 0.04538 | 22033 | 5509 |
| 0.994066 | 0.046315 | 22033 | 5509 |
| 0.910452 | 0.047236 | 22034 | 5508 |
| 0.963076 | 0.047886 | 22034 | 5508 |
| 0.960742 | 0.041751 | 22034 | 5508 |
| 0.977808 | 0.055095 | 27542 | 4025 |
| 0.977608 | 0.055289 | 27542 | 4025 |
| 0.994812 | 0.038664 | 27542 | 4025 |
| 0.949282 | 0.065366 | 22033 | 5509 |
| 0.963096 | 0.056939 | 22033 | 5509 |
| 0.919986 | 0.055206 | 22034 | 5508 |
| 0.962041 | 0.051823 | 22034 | 5508 |
| 0.923079 | 0.052945 | 22034 | 5508 |
| 0.949281 | 0.065367 | 22033 | 5509 |
| 0.963025 | 0.057009 | 22033 | 5509 |
| 0.919972 | 0.055221 | 22034 | 5508 |
| 0.962123 | 0.051742 | 22034 | 5508 |
| 0.923083 | 0.052941 | 22034 | 5508 |
| 0.980058 | 0.035066 | 22033 | 5509 |
| 0.987753 | 0.032794 | 22033 | 5509 |
| 0.942292 | 0.032298 | 22034 | 5508 |
| 0.984578 | 0.02961 | 22034 | 5508 |

| | | | |
|---|---|---|---|
| 0.94536 | 0.030085 | 22034 | 5508 |
| 0.954836 | 0.058423 | 27542 | 4025 |
| 0.954872 | 0.058387 | 27542 | 4025 |
| 0.978551 | 0.035037 | 27542 | 4025 |
| 0.920739 | 0.075061 | 22033 | 5509 |
| 0.909898 | 0.077859 | 22033 | 5509 |
| 0.922138 | 0.076304 | 22034 | 5508 |
| 0.944925 | 0.072141 | 22034 | 5508 |
| 0.923563 | 0.077106 | 22034 | 5508 |
| 0.920704 | 0.075096 | 22033 | 5509 |
| 0.909827 | 0.07793 | 22033 | 5509 |
| 0.922127 | 0.076315 | 22034 | 5508 |
| 0.944841 | 0.072223 | 22034 | 5508 |
| 0.923568 | 0.077101 | 22034 | 5508 |
| 0.957602 | 0.038029 | 22033 | 5509 |
| 0.94637 | 0.040897 | 22033 | 5509 |
| 0.957911 | 0.04047 | 22034 | 5508 |
| 0.979897 | 0.0378 | 22034 | 5508 |
| 0.965654 | 0.035046 | 22034 | 5508 |
| 0.917561 | 0.090166 | 27542 | 4025 |
| 0.917683 | 0.090045 | 27542 | 4025 |
| 0.958035 | 0.050033 | 27542 | 4025 |
| 0.886235 | 0.091792 | 22033 | 5509 |
| 0.90859 | 0.115748 | 22033 | 5509 |
| 0.921269 | 0.096425 | 22034 | 5508 |
| 0.879933 | 0.116028 | 22034 | 5508 |
| 0.874711 | 0.108876 | 22034 | 5508 |
| 0.886059 | 0.091972 | 22033 | 5509 |
| 0.908631 | 0.115709 | 22033 | 5509 |
| 0.921208 | 0.096485 | 22034 | 5508 |
| 0.880135 | 0.115825 | 22034 | 5508 |
| 0.874696 | 0.108891 | 22034 | 5508 |
| 0.924764 | 0.052308 | 22033 | 5509 |
| 0.966365 | 0.059521 | 22033 | 5509 |
| 0.963347 | 0.055155 | 22034 | 5508 |
| 0.935781 | 0.059924 | 22034 | 5508 |
| 0.9339 | 0.048576 | 22034 | 5508 |
| 0.927446 | 0.096714 | 27542 | 4025 |
| 0.927503 | 0.096658 | 27542 | 4025 |
| 0.96352 | 0.06158 | 27542 | 4025 |
| 0.87922 | 0.108971 | 22033 | 5509 |
| 0.906852 | 0.120937 | 22033 | 5509 |
| 0.881264 | 0.11311 | 22034 | 5508 |
| 0.894215 | 0.111199 | 22034 | 5508 |
| 0.865198 | 0.118979 | 22034 | 5508 |

| | | | |
|---|---|---|---|
| 0.879204 | 0.108987 | 22033 | 5509 |
| 0.90684 | 0.120948 | 22033 | 5509 |
| 0.881436 | 0.112937 | 22034 | 5508 |
| 0.894245 | 0.111169 | 22034 | 5508 |
| 0.865135 | 0.119044 | 22034 | 5508 |
| 0.916006 | 0.071691 | 22033 | 5509 |
| 0.955843 | 0.073447 | 22033 | 5509 |
| 0.922935 | 0.071173 | 22034 | 5508 |
| 0.931999 | 0.073643 | 22034 | 5508 |
| 0.911733 | 0.071593 | 22034 | 5508 |
| 0.926756 | 0.113501 | 27542 | 4025 |
| 0.926771 | 0.113486 | 27542 | 4025 |
| 0.960979 | 0.080764 | 27542 | 4025 |
| 0.866931 | 0.113896 | 22033 | 5509 |
| 0.907014 | 0.126419 | 22033 | 5509 |
| 0.847125 | 0.131997 | 22034 | 5508 |
| 0.852931 | 0.126719 | 22034 | 5508 |
| 0.906545 | 0.12095 | 22034 | 5508 |
| 0.866994 | 0.113831 | 22033 | 5509 |
| 0.90696 | 0.126471 | 22033 | 5509 |
| 0.847469 | 0.131644 | 22034 | 5508 |
| 0.852892 | 0.126759 | 22034 | 5508 |
| 0.906487 | 0.121006 | 22034 | 5508 |
| 0.899229 | 0.080884 | 22033 | 5509 |
| 0.946436 | 0.088449 | 22033 | 5509 |
| 0.887869 | 0.090249 | 22034 | 5508 |
| 0.89007 | 0.088695 | 22034 | 5508 |
| 0.95526 | 0.073712 | 22034 | 5508 |
| 0.97444 | 0.048544 | 27542 | 4025 |
| 0.974458 | 0.048526 | 27542 | 4025 |
| 1.002177 | 0.021461 | 27542 | 4025 |
| 0.965592 | 0.047835 | 22033 | 5509 |
| 0.945586 | 0.053404 | 22033 | 5509 |
| 0.937764 | 0.042308 | 22034 | 5508 |
| 0.959047 | 0.048358 | 22034 | 5508 |
| 0.956603 | 0.042986 | 22034 | 5508 |
| 0.965518 | 0.047907 | 22033 | 5509 |
| 0.945601 | 0.053388 | 22033 | 5509 |
| 0.937687 | 0.042386 | 22034 | 5508 |
| 0.958993 | 0.048411 | 22034 | 5508 |
| 0.956577 | 0.043012 | 22034 | 5508 |
| 0.992226 | 0.021572 | 22033 | 5509 |
| 0.97463 | 0.024328 | 22033 | 5509 |
| 0.959037 | 0.020582 | 22034 | 5508 |
| 0.986242 | 0.021372 | 22034 | 5508 |

| | | | |
|---|---|---|---|
| 0.979655 | 0.019924 | 22034 | 5508 |
| 0.955423 | 0.060288 | 27542 | 4025 |
| 0.955611 | 0.060103 | 27542 | 4025 |
| 0.987496 | 0.028743 | 27542 | 4025 |
| 0.97267 | 0.05385 | 22033 | 5509 |
| 0.95846 | 0.055587 | 22033 | 5509 |
| 0.924499 | 0.049207 | 22034 | 5508 |
| 0.938788 | 0.043042 | 22034 | 5508 |
| 0.958435 | 0.044317 | 22034 | 5508 |
| 0.972789 | 0.053734 | 22033 | 5509 |
| 0.958482 | 0.055565 | 22033 | 5509 |
| 0.924477 | 0.04923 | 22034 | 5508 |
| 0.938443 | 0.043394 | 22034 | 5508 |
| 0.958479 | 0.044274 | 22034 | 5508 |
| 1.00142 | 0.025883 | 22033 | 5509 |
| 0.988702 | 0.025788 | 22033 | 5509 |
| 0.946906 | 0.026163 | 22034 | 5508 |
| 0.95812 | 0.023336 | 22034 | 5508 |
| 0.9788 | 0.024011 | 22034 | 5508 |
| 0.908387 | 0.059725 | 27542 | 4025 |
| 0.908187 | 0.059932 | 27542 | 4025 |
| 0.934374 | 0.032826 | 27542 | 4025 |
| 1.000964 | 0.066134 | 22033 | 5509 |
| 0.915865 | 0.072115 | 22033 | 5509 |
| 0.885442 | 0.077337 | 22034 | 5508 |
| 0.928814 | 0.076241 | 22034 | 5508 |
| 0.901725 | 0.07706 | 22034 | 5508 |
| 1.000944 | 0.066153 | 22033 | 5509 |
| 0.915859 | 0.072121 | 22033 | 5509 |
| 0.885478 | 0.0773 | 22034 | 5508 |
| 0.928904 | 0.076152 | 22034 | 5508 |
| 0.901662 | 0.077125 | 22034 | 5508 |
| 1.03801 | 0.031571 | 22033 | 5509 |
| 0.951524 | 0.035988 | 22033 | 5509 |
| 0.924954 | 0.036164 | 22034 | 5508 |
| 0.967987 | 0.037282 | 22034 | 5508 |
| 0.945478 | 0.032278 | 22034 | 5508 |
| 0.969412 | 0.035183 | 27542 | 4025 |
| 0.969204 | 0.035389 | 27542 | 4025 |
| 0.987105 | 0.017573 | 27542 | 4025 |
| 0.965242 | 0.047432 | 22033 | 5509 |
| 0.930936 | 0.053952 | 22033 | 5509 |
| 0.920418 | 0.056792 | 22034 | 5508 |
| 0.974005 | 0.049999 | 22034 | 5508 |
| 0.949467 | 0.052048 | 22034 | 5508 |

| | | | |
|---|---|---|---|
| 0.965123 | 0.04755 | 22033 | 5509 |
| 0.931165 | 0.053721 | 22033 | 5509 |
| 0.920569 | 0.056637 | 22034 | 5508 |
| 0.974104 | 0.049902 | 22034 | 5508 |
| 0.949402 | 0.052113 | 22034 | 5508 |
| 0.990725 | 0.022283 | 22033 | 5509 |
| 0.959098 | 0.025334 | 22033 | 5509 |
| 0.952395 | 0.024023 | 22034 | 5508 |
| 1.00267 | 0.02204 | 22034 | 5508 |
| 0.979106 | 0.022456 | 22034 | 5508 |
| 0.844013 | 0.147997 | 27542 | 4025 |
| 0.843923 | 0.148088 | 27542 | 4025 |
| 0.879067 | 0.112612 | 27542 | 4025 |
| 0.841883 | 0.168242 | 22033 | 5509 |
| 0.847031 | 0.172976 | 22033 | 5509 |
| 0.800907 | 0.166019 | 22034 | 5508 |
| 0.852261 | 0.168784 | 22034 | 5508 |
| 0.815018 | 0.166939 | 22034 | 5508 |
| 0.841831 | 0.168293 | 22033 | 5509 |
| 0.847112 | 0.172897 | 22033 | 5509 |
| 0.800845 | 0.166083 | 22034 | 5508 |
| 0.852498 | 0.168553 | 22034 | 5508 |
| 0.814948 | 0.16701 | 22034 | 5508 |
| 0.889474 | 0.121223 | 22033 | 5509 |
| 0.898687 | 0.12254 | 22033 | 5509 |
| 0.845865 | 0.119204 | 22034 | 5508 |
| 0.904347 | 0.117985 | 22034 | 5508 |
| 0.879694 | 0.100831 | 22034 | 5508 |
| 0.569891 | 0.401782 | 27542 | 4025 |
| 0.569923 | 0.401748 | 27542 | 4025 |
| 0.615669 | 0.353729 | 27542 | 4025 |
| 0.551066 | 0.426394 | 22033 | 5509 |
| 0.57118 | 0.42373 | 22033 | 5509 |
| 0.572083 | 0.422626 | 22034 | 5508 |
| 0.582883 | 0.431478 | 22034 | 5508 |
| 0.568857 | 0.448824 | 22034 | 5508 |
| 0.550889 | 0.426578 | 22033 | 5509 |
| 0.571241 | 0.423668 | 22033 | 5509 |
| 0.571968 | 0.422743 | 22034 | 5508 |
| 0.583176 | 0.431193 | 22034 | 5508 |
| 0.568965 | 0.44872 | 22034 | 5508 |
| 0.597002 | 0.378579 | 22033 | 5509 |
| 0.637425 | 0.356895 | 22033 | 5509 |
| 0.622948 | 0.371291 | 22034 | 5508 |
| 0.647829 | 0.368133 | 22034 | 5508 |

| | | | |
|---|---|---|---|
| 0.659121 | 0.361366 | 22034 | 5508 |
| 0.644924 | 0.297077 | 27542 | 4025 |
| 0.644591 | 0.297441 | 27542 | 4025 |
| 0.679115 | 0.259812 | 27542 | 4025 |
| 0.653855 | 0.326444 | 22033 | 5509 |
| 0.665453 | 0.341056 | 22033 | 5509 |
| 0.641822 | 0.340368 | 22034 | 5508 |
| 0.692026 | 0.332371 | 22034 | 5508 |
| 0.630352 | 0.37596 | 22034 | 5508 |
| 0.653307 | 0.327008 | 22033 | 5509 |
| 0.665532 | 0.340978 | 22033 | 5509 |
| 0.641529 | 0.340669 | 22034 | 5508 |
| 0.692411 | 0.331999 | 22034 | 5508 |
| 0.630472 | 0.375842 | 22034 | 5508 |
| 0.695021 | 0.284038 | 22033 | 5509 |
| 0.730275 | 0.276869 | 22033 | 5509 |
| 0.692972 | 0.287799 | 22034 | 5508 |
| 0.755636 | 0.271003 | 22034 | 5508 |
| 0.723215 | 0.284027 | 22034 | 5508 |
| 0.628608 | 0.360161 | 27542 | 4025 |
| 0.628695 | 0.360072 | 27542 | 4025 |
| 0.681539 | 0.306284 | 27542 | 4025 |
| 0.631661 | 0.355261 | 22033 | 5509 |
| 0.613916 | 0.349352 | 22033 | 5509 |
| 0.663707 | 0.345872 | 22034 | 5508 |
| 0.648206 | 0.365601 | 22034 | 5508 |
| 0.671287 | 0.354898 | 22034 | 5508 |
| 0.631606 | 0.355317 | 22033 | 5509 |
| 0.613968 | 0.349296 | 22033 | 5509 |
| 0.663684 | 0.345894 | 22034 | 5508 |
| 0.648209 | 0.365599 | 22034 | 5508 |
| 0.671379 | 0.354809 | 22034 | 5508 |
| 0.678205 | 0.307754 | 22033 | 5509 |
| 0.667552 | 0.292506 | 22033 | 5509 |
| 0.710344 | 0.299907 | 22034 | 5508 |
| 0.70343 | 0.311554 | 22034 | 5508 |
| 0.741522 | 0.287403 | 22034 | 5508 |
| 0.844922 | 0.081755 | 27542 | 4025 |
| 0.844761 | 0.08193 | 27542 | 4025 |
| 0.85458 | 0.071258 | 27542 | 4025 |
| 0.90463 | 0.112066 | 22033 | 5509 |
| 0.843689 | 0.135455 | 22033 | 5509 |
| 0.865939 | 0.13226 | 22034 | 5508 |
| 0.900861 | 0.118986 | 22034 | 5508 |
| 0.834019 | 0.153251 | 22034 | 5508 |

| | | | |
|---|---|---|---|
| 0.904454 | 0.11224 | 22033 | 5509 |
| 0.843626 | 0.135519 | 22033 | 5509 |
| 0.866201 | 0.131998 | 22034 | 5508 |
| 0.900572 | 0.119268 | 22034 | 5508 |
| 0.834309 | 0.152957 | 22034 | 5508 |
| 0.930689 | 0.086489 | 22033 | 5509 |
| 0.882385 | 0.095802 | 22033 | 5509 |
| 0.905939 | 0.092177 | 22034 | 5508 |
| 0.93316 | 0.087398 | 22034 | 5508 |
| 0.903026 | 0.083191 | 22034 | 5508 |
| 0.360904 | 0.641097 | 27542 | 4025 |
| 0.361059 | 0.640943 | 27542 | 4025 |
| 0.425107 | 0.577251 | 27542 | 4025 |
| 0.378764 | 0.61924 | 22033 | 5509 |
| 0.35679 | 0.634454 | 22033 | 5509 |
| 0.370039 | 0.627227 | 22034 | 5508 |
| 0.35407 | 0.648491 | 22034 | 5508 |
| 0.380126 | 0.630388 | 22034 | 5508 |
| 0.378798 | 0.619205 | 22033 | 5509 |
| 0.356922 | 0.634318 | 22033 | 5509 |
| 0.370003 | 0.627263 | 22034 | 5508 |
| 0.354076 | 0.648485 | 22034 | 5508 |
| 0.380166 | 0.63035 | 22034 | 5508 |
| 0.443341 | 0.554322 | 22033 | 5509 |
| 0.429637 | 0.559819 | 22033 | 5509 |
| 0.435287 | 0.561497 | 22034 | 5508 |
| 0.424498 | 0.578572 | 22034 | 5508 |
| 0.469952 | 0.543047 | 22034 | 5508 |
| 0.468306 | 0.516571 | 27542 | 4025 |
| 0.468291 | 0.516586 | 27542 | 4025 |
| 0.533226 | 0.449555 | 27542 | 4025 |
| 0.49045 | 0.510369 | 22033 | 5509 |
| 0.501566 | 0.507354 | 22033 | 5509 |
| 0.474795 | 0.517672 | 22034 | 5508 |
| 0.458106 | 0.534343 | 22034 | 5508 |
| 0.487412 | 0.518011 | 22034 | 5508 |
| 0.490369 | 0.51045 | 22033 | 5509 |
| 0.501691 | 0.507231 | 22033 | 5509 |
| 0.47487 | 0.517595 | 22034 | 5508 |
| 0.458139 | 0.53431 | 22034 | 5508 |
| 0.487491 | 0.517932 | 22034 | 5508 |
| 0.563553 | 0.437388 | 22033 | 5509 |
| 0.580777 | 0.429552 | 22033 | 5509 |
| 0.546296 | 0.445037 | 22034 | 5508 |
| 0.537051 | 0.454097 | 22034 | 5508 |

| | | | |
|---|---|---|---|
| 0.593395 | 0.413206 | 22034 | 5508 |
| 0.488639 | 0.525189 | 27542 | 4025 |
| 0.488944 | 0.524893 | 27542 | 4025 |
| 0.542293 | 0.473053 | 27542 | 4025 |
| 0.508528 | 0.4871 | 22033 | 5509 |
| 0.464956 | 0.51037 | 22033 | 5509 |
| 0.495322 | 0.501006 | 22034 | 5508 |
| 0.495175 | 0.51903 | 22034 | 5508 |
| 0.51263 | 0.505526 | 22034 | 5508 |
| 0.508534 | 0.487094 | 22033 | 5509 |
| 0.465089 | 0.51023 | 22033 | 5509 |
| 0.495204 | 0.501125 | 22034 | 5508 |
| 0.495309 | 0.5189 | 22034 | 5508 |
| 0.512655 | 0.505502 | 22034 | 5508 |
| 0.553192 | 0.442052 | 22033 | 5509 |
| 0.513249 | 0.459515 | 22033 | 5509 |
| 0.544777 | 0.451184 | 22034 | 5508 |
| 0.548751 | 0.46699 | 22034 | 5508 |
| 0.570783 | 0.449433 | 22034 | 5508 |
| 0.857045 | 0.133716 | 27542 | 4025 |
| 0.856993 | 0.133769 | 27542 | 4025 |
| 0.902309 | 0.087964 | 27542 | 4025 |
| 0.867778 | 0.146372 | 22033 | 5509 |
| 0.873428 | 0.138491 | 22033 | 5509 |
| 0.832719 | 0.145275 | 22034 | 5508 |
| 0.847965 | 0.151631 | 22034 | 5508 |
| 0.854135 | 0.141683 | 22034 | 5508 |
| 0.867778 | 0.146372 | 22033 | 5509 |
| 0.873443 | 0.138476 | 22033 | 5509 |
| 0.832639 | 0.145357 | 22034 | 5508 |
| 0.848075 | 0.151521 | 22034 | 5508 |
| 0.854119 | 0.1417 | 22034 | 5508 |
| 0.926099 | 0.089002 | 22033 | 5509 |
| 0.923751 | 0.088854 | 22033 | 5509 |
| 0.88369 | 0.092957 | 22034 | 5508 |
| 0.910878 | 0.088688 | 22034 | 5508 |
| 0.921744 | 0.073744 | 22034 | 5508 |
| 0.694997 | 0.299279 | 27542 | 4025 |
| 0.695054 | 0.299222 | 27542 | 4025 |
| 0.76932 | 0.224344 | 27542 | 4025 |
| 0.720313 | 0.285188 | 22033 | 5509 |
| 0.685833 | 0.278248 | 22033 | 5509 |
| 0.733354 | 0.276204 | 22034 | 5508 |
| 0.722213 | 0.27074 | 22034 | 5508 |
| 0.746079 | 0.281323 | 22034 | 5508 |

| | | | |
|---|---|---|---|
| 0.72033 | 0.285171 | 22033 | 5509 |
| 0.685874 | 0.278205 | 22033 | 5509 |
| 0.733362 | 0.276196 | 22034 | 5508 |
| 0.722138 | 0.270815 | 22034 | 5508 |
| 0.746094 | 0.281309 | 22034 | 5508 |
| 0.796833 | 0.209252 | 22033 | 5509 |
| 0.75557 | 0.204859 | 22033 | 5509 |
| 0.810006 | 0.200551 | 22034 | 5508 |
| 0.786911 | 0.205411 | 22034 | 5508 |
| 0.839785 | 0.191059 | 22034 | 5508 |
| 0.80867 | 0.225069 | 27542 | 4025 |
| 0.808801 | 0.224943 | 27542 | 4025 |
| 0.865309 | 0.170793 | 27542 | 4025 |
| 0.808898 | 0.205962 | 22033 | 5509 |
| 0.763008 | 0.216501 | 22033 | 5509 |
| 0.825958 | 0.204788 | 22034 | 5508 |
| 0.783331 | 0.211059 | 22034 | 5508 |
| 0.760887 | 0.220671 | 22034 | 5508 |
| 0.808963 | 0.205898 | 22033 | 5509 |
| 0.762775 | 0.216741 | 22033 | 5509 |
| 0.825939 | 0.204806 | 22034 | 5508 |
| 0.783384 | 0.211005 | 22034 | 5508 |
| 0.760874 | 0.220684 | 22034 | 5508 |
| 0.859154 | 0.15663 | 22033 | 5509 |
| 0.805344 | 0.173028 | 22033 | 5509 |
| 0.869218 | 0.163138 | 22034 | 5508 |
| 0.834181 | 0.159845 | 22034 | 5508 |
| 0.814455 | 0.165805 | 22034 | 5508 |
| 0.774383 | 0.224176 | 27542 | 4025 |
| 0.774293 | 0.224266 | 27542 | 4025 |
| 0.83854 | 0.1599 | 27542 | 4025 |
| 0.790542 | 0.214539 | 22033 | 5509 |
| 0.746672 | 0.227191 | 22033 | 5509 |
| 0.795221 | 0.217041 | 22034 | 5508 |
| 0.779129 | 0.230964 | 22034 | 5508 |
| 0.773232 | 0.225613 | 22034 | 5508 |
| 0.790602 | 0.21448 | 22033 | 5509 |
| 0.746679 | 0.227184 | 22033 | 5509 |
| 0.795418 | 0.216847 | 22034 | 5508 |
| 0.779055 | 0.231037 | 22034 | 5508 |
| 0.773224 | 0.225622 | 22034 | 5508 |
| 0.850389 | 0.155077 | 22033 | 5509 |
| 0.804175 | 0.167676 | 22033 | 5509 |
| 0.850232 | 0.162878 | 22034 | 5508 |
| 0.845545 | 0.165408 | 22034 | 5508 |

| | | | |
|---|---|---|---|
| 0.839901 | 0.158845 | 22034 | 5508 |
| 0.858737 | 0.175905 | 27542 | 4025 |
| 0.858668 | 0.175971 | 27542 | 4025 |
| 0.903631 | 0.132822 | 27542 | 4025 |
| 0.851925 | 0.15657 | 22033 | 5509 |
| 0.818328 | 0.157659 | 22033 | 5509 |
| 0.886348 | 0.161815 | 22034 | 5508 |
| 0.825258 | 0.15445 | 22034 | 5508 |
| 0.807154 | 0.181301 | 22034 | 5508 |
| 0.851863 | 0.156631 | 22033 | 5509 |
| 0.818254 | 0.157735 | 22033 | 5509 |
| 0.886418 | 0.161749 | 22034 | 5508 |
| 0.825261 | 0.154448 | 22034 | 5508 |
| 0.807245 | 0.181209 | 22034 | 5508 |
| 0.889213 | 0.119653 | 22033 | 5509 |
| 0.856712 | 0.118149 | 22033 | 5509 |
| 0.931436 | 0.119178 | 22034 | 5508 |
| 0.86554 | 0.113178 | 22034 | 5508 |
| 0.857696 | 0.130037 | 22034 | 5508 |
| 0.48715 | 0.532272 | 27542 | 4025 |
| 0.487169 | 0.532254 | 27542 | 4025 |
| 0.568372 | 0.454288 | 27542 | 4025 |
| 0.474617 | 0.512887 | 22033 | 5509 |
| 0.447429 | 0.535597 | 22033 | 5509 |
| 0.461451 | 0.524742 | 22034 | 5508 |
| 0.486617 | 0.537939 | 22034 | 5508 |
| 0.486945 | 0.531172 | 22034 | 5508 |
| 0.474437 | 0.513071 | 22033 | 5509 |
| 0.447565 | 0.535456 | 22033 | 5509 |
| 0.461548 | 0.524642 | 22034 | 5508 |
| 0.486629 | 0.537927 | 22034 | 5508 |
| 0.487183 | 0.530944 | 22034 | 5508 |
| 0.543702 | 0.441982 | 22033 | 5509 |
| 0.527348 | 0.452647 | 22033 | 5509 |
| 0.537311 | 0.446612 | 22034 | 5508 |
| 0.569466 | 0.459271 | 22034 | 5508 |
| 0.586174 | 0.435636 | 22034 | 5508 |
| 0.751437 | 0.274886 | 27542 | 4025 |
| 0.751402 | 0.274919 | 27542 | 4025 |
| 0.786949 | 0.240618 | 27542 | 4025 |
| 0.685848 | 0.31439 | 22033 | 5509 |
| 0.678274 | 0.311521 | 22033 | 5509 |
| 0.698168 | 0.305026 | 22034 | 5508 |
| 0.681358 | 0.309748 | 22034 | 5508 |
| 0.714344 | 0.301532 | 22034 | 5508 |

| | | | |
|---|---|---|---|
| 0.685865 | 0.314374 | 22033 | 5509 |
| 0.678195 | 0.311601 | 22033 | 5509 |
| 0.698059 | 0.305135 | 22034 | 5508 |
| 0.681438 | 0.309667 | 22034 | 5508 |
| 0.714412 | 0.301465 | 22034 | 5508 |
| 0.72701 | 0.273243 | 22033 | 5509 |
| 0.722056 | 0.26708 | 22033 | 5509 |
| 0.733943 | 0.269415 | 22034 | 5508 |
| 0.730186 | 0.260282 | 22034 | 5508 |
| 0.75111 | 0.265583 | 22034 | 5508 |
| 0.758598 | 0.283653 | 27542 | 4025 |
| 0.758631 | 0.283623 | 27542 | 4025 |
| 0.798262 | 0.246199 | 27542 | 4025 |
| 0.718391 | 0.292899 | 22033 | 5509 |
| 0.706706 | 0.291976 | 22033 | 5509 |
| 0.698253 | 0.28156 | 22034 | 5508 |
| 0.689925 | 0.304801 | 22034 | 5508 |
| 0.725596 | 0.289177 | 22034 | 5508 |
| 0.7183 | 0.292987 | 22033 | 5509 |
| 0.706667 | 0.292015 | 22033 | 5509 |
| 0.698114 | 0.281703 | 22034 | 5508 |
| 0.690075 | 0.30465 | 22034 | 5508 |
| 0.725838 | 0.288941 | 22034 | 5508 |
| 0.76894 | 0.243143 | 22033 | 5509 |
| 0.75345 | 0.245145 | 22033 | 5509 |
| 0.739772 | 0.238841 | 22034 | 5508 |
| 0.739654 | 0.254692 | 22034 | 5508 |
| 0.77943 | 0.23644 | 22034 | 5508 |
| 0.872933 | 0.187141 | 27542 | 4025 |
| 0.872959 | 0.187117 | 27542 | 4025 |
| 0.909145 | 0.153421 | 27542 | 4025 |
| 0.806922 | 0.186342 | 22033 | 5509 |
| 0.812191 | 0.193065 | 22033 | 5509 |
| 0.782216 | 0.183057 | 22034 | 5508 |
| 0.773154 | 0.211653 | 22034 | 5508 |
| 0.86232 | 0.189674 | 22034 | 5508 |
| 0.806828 | 0.186437 | 22033 | 5509 |
| 0.812175 | 0.193081 | 22033 | 5509 |
| 0.78226 | 0.183011 | 22034 | 5508 |
| 0.773153 | 0.211654 | 22034 | 5508 |
| 0.862505 | 0.1895 | 22034 | 5508 |
| 0.844586 | 0.148364 | 22033 | 5509 |
| 0.856407 | 0.149135 | 22033 | 5509 |
| 0.817601 | 0.146101 | 22034 | 5508 |
| 0.816262 | 0.167698 | 22034 | 5508 |

| | | | |
|---|---|---|---|
| 0.908246 | 0.146517 | 22034 | 5508 |
| 0.820922 | 0.249823 | 27542 | 4025 |
| 0.82097 | 0.24978 | 27542 | 4025 |
| 0.866916 | 0.207793 | 27542 | 4025 |
| 0.751127 | 0.243089 | 22033 | 5509 |
| 0.758042 | 0.259162 | 22033 | 5509 |
| 0.739849 | 0.232285 | 22034 | 5508 |
| 0.717229 | 0.259611 | 22034 | 5508 |
| 0.793231 | 0.246347 | 22034 | 5508 |
| 0.751026 | 0.24319 | 22033 | 5509 |
| 0.758217 | 0.258991 | 22033 | 5509 |
| 0.739685 | 0.232456 | 22034 | 5508 |
| 0.717247 | 0.259592 | 22034 | 5508 |
| 0.793552 | 0.246042 | 22034 | 5508 |
| 0.790991 | 0.202917 | 22033 | 5509 |
| 0.813229 | 0.205228 | 22033 | 5509 |
| 0.769118 | 0.201914 | 22034 | 5508 |
| 0.76402 | 0.211308 | 22034 | 5508 |
| 0.854959 | 0.187699 | 22034 | 5508 |
| 0.711166 | 0.338796 | 27542 | 4025 |
| 0.71122 | 0.338746 | 27542 | 4025 |
| 0.757308 | 0.295896 | 27542 | 4025 |
| 0.689119 | 0.314127 | 22033 | 5509 |
| 0.671519 | 0.329838 | 22033 | 5509 |
| 0.680865 | 0.304536 | 22034 | 5508 |
| 0.660705 | 0.327044 | 22034 | 5508 |
| 0.706838 | 0.315402 | 22034 | 5508 |
| 0.689261 | 0.313985 | 22033 | 5509 |
| 0.67171 | 0.329648 | 22033 | 5509 |
| 0.680575 | 0.304833 | 22034 | 5508 |
| 0.660833 | 0.326915 | 22034 | 5508 |
| 0.706958 | 0.315286 | 22034 | 5508 |
| 0.734915 | 0.268547 | 22033 | 5509 |
| 0.72724 | 0.27423 | 22033 | 5509 |
| 0.712298 | 0.27243 | 22034 | 5508 |
| 0.709607 | 0.277236 | 22034 | 5508 |
| 0.762804 | 0.261197 | 22034 | 5508 |
| 0.673565 | 0.379888 | 27542 | 4025 |
| 0.673721 | 0.379744 | 27542 | 4025 |
| 0.723029 | 0.334349 | 27542 | 4025 |
| 0.655836 | 0.357032 | 22033 | 5509 |
| 0.624532 | 0.375288 | 22033 | 5509 |
| 0.62072 | 0.352403 | 22034 | 5508 |
| 0.633829 | 0.368389 | 22034 | 5508 |
| 0.647965 | 0.363614 | 22034 | 5508 |

| | | | |
|---|---|---|---|
| 0.655907 | 0.356963 | 22033 | 5509 |
| 0.624549 | 0.375271 | 22033 | 5509 |
| 0.620379 | 0.352758 | 22034 | 5508 |
| 0.633875 | 0.368342 | 22034 | 5508 |
| 0.648168 | 0.363415 | 22034 | 5508 |
| 0.705365 | 0.308476 | 22033 | 5509 |
| 0.677086 | 0.322718 | 22033 | 5509 |
| 0.654873 | 0.316771 | 22034 | 5508 |
| 0.682995 | 0.319394 | 22034 | 5508 |
| 0.706133 | 0.306485 | 22034 | 5508 |
| 0.668419 | 0.385473 | 27542 | 4025 |
| 0.668505 | 0.385394 | 27542 | 4025 |
| 0.712359 | 0.345075 | 27542 | 4025 |
| 0.625942 | 0.376545 | 22033 | 5509 |
| 0.60682 | 0.39277 | 22033 | 5509 |
| 0.616793 | 0.361094 | 22034 | 5508 |
| 0.614846 | 0.386561 | 22034 | 5508 |
| 0.642441 | 0.375608 | 22034 | 5508 |
| 0.625953 | 0.376535 | 22033 | 5509 |
| 0.606926 | 0.392664 | 22033 | 5509 |
| 0.616482 | 0.361416 | 22034 | 5508 |
| 0.615019 | 0.386389 | 22034 | 5508 |
| 0.642612 | 0.375442 | 22034 | 5508 |
| 0.674124 | 0.328555 | 22033 | 5509 |
| 0.660232 | 0.339322 | 22033 | 5509 |
| 0.650854 | 0.325812 | 22034 | 5508 |
| 0.664489 | 0.337032 | 22034 | 5508 |
| 0.701292 | 0.31841 | 22034 | 5508 |
| 0.701944 | 0.387916 | 27542 | 4025 |
| 0.701966 | 0.387897 | 27542 | 4025 |
| 0.746819 | 0.348787 | 27542 | 4025 |
| 0.610193 | 0.397273 | 22033 | 5509 |
| 0.588723 | 0.410955 | 22033 | 5509 |
| 0.611223 | 0.368743 | 22034 | 5508 |
| 0.587408 | 0.412326 | 22034 | 5508 |
| 0.631668 | 0.380464 | 22034 | 5508 |
| 0.610283 | 0.397184 | 22033 | 5509 |
| 0.588799 | 0.410879 | 22033 | 5509 |
| 0.610915 | 0.369061 | 22034 | 5508 |
| 0.587425 | 0.412308 | 22034 | 5508 |
| 0.631762 | 0.380372 | 22034 | 5508 |
| 0.656009 | 0.352018 | 22033 | 5509 |
| 0.640314 | 0.359335 | 22033 | 5509 |
| 0.642319 | 0.336628 | 22034 | 5508 |
| 0.636477 | 0.363234 | 22034 | 5508 |

| | | | |
|---|---|---|---|
| 0.682069 | 0.331032 | 22034 | 5508 |
| 0.689269 | 0.388973 | 27542 | 4025 |
| 0.689333 | 0.388916 | 27542 | 4025 |
| 0.742878 | 0.341449 | 27542 | 4025 |
| 0.635745 | 0.381313 | 22033 | 5509 |
| 0.598177 | 0.397226 | 22033 | 5509 |
| 0.610135 | 0.364574 | 22034 | 5508 |
| 0.617059 | 0.389225 | 22034 | 5508 |
| 0.636691 | 0.369171 | 22034 | 5508 |
| 0.635828 | 0.381232 | 22033 | 5509 |
| 0.598259 | 0.397143 | 22033 | 5509 |
| 0.609838 | 0.364883 | 22034 | 5508 |
| 0.617155 | 0.38913 | 22034 | 5508 |
| 0.636791 | 0.369072 | 22034 | 5508 |
| 0.682869 | 0.335453 | 22033 | 5509 |
| 0.652595 | 0.34239 | 22033 | 5509 |
| 0.644844 | 0.328426 | 22034 | 5508 |
| 0.666748 | 0.340041 | 22034 | 5508 |
| 0.690934 | 0.315427 | 22034 | 5508 |
| 0.734492 | 0.350253 | 27542 | 4025 |
| 0.734597 | 0.35016 | 27542 | 4025 |
| 0.776531 | 0.313065 | 27542 | 4025 |
| 0.652051 | 0.367118 | 22033 | 5509 |
| 0.617365 | 0.364703 | 22033 | 5509 |
| 0.651568 | 0.334004 | 22034 | 5508 |
| 0.648864 | 0.36106 | 22034 | 5508 |
| 0.6496 | 0.352692 | 22034 | 5508 |
| 0.652144 | 0.367027 | 22033 | 5509 |
| 0.617462 | 0.364604 | 22033 | 5509 |
| 0.651268 | 0.33431 | 22034 | 5508 |
| 0.648891 | 0.361033 | 22034 | 5508 |
| 0.649758 | 0.352534 | 22034 | 5508 |
| 0.695648 | 0.324802 | 22033 | 5509 |
| 0.658438 | 0.322437 | 22033 | 5509 |
| 0.680484 | 0.304447 | 22034 | 5508 |
| 0.688885 | 0.321652 | 22034 | 5508 |
| 0.690944 | 0.311493 | 22034 | 5508 |
| 0.7095 | 0.366596 | 27542 | 4025 |
| 0.70959 | 0.366516 | 27542 | 4025 |
| 0.752212 | 0.328465 | 27542 | 4025 |
| 0.64043 | 0.370618 | 22033 | 5509 |
| 0.624426 | 0.370151 | 22033 | 5509 |
| 0.632434 | 0.348721 | 22034 | 5508 |
| 0.632591 | 0.371106 | 22034 | 5508 |
| 0.647735 | 0.361051 | 22034 | 5508 |

| | | | |
|---|---|---|---|
| 0.640494 | 0.370556 | 22033 | 5509 |
| 0.624568 | 0.370008 | 22033 | 5509 |
| 0.632145 | 0.349018 | 22034 | 5508 |
| 0.632668 | 0.371029 | 22034 | 5508 |
| 0.647896 | 0.360892 | 22034 | 5508 |
| 0.684924 | 0.326893 | 22033 | 5509 |
| 0.670607 | 0.323569 | 22033 | 5509 |
| 0.664626 | 0.315569 | 22034 | 5508 |
| 0.676703 | 0.327251 | 22034 | 5508 |
| 0.692426 | 0.316966 | 22034 | 5508 |
| 0.669973 | 0.379968 | 27542 | 4025 |
| 0.669963 | 0.379977 | 27542 | 4025 |
| 0.713959 | 0.339261 | 27542 | 4025 |
| 0.657967 | 0.368795 | 22033 | 5509 |
| 0.60941 | 0.386715 | 22033 | 5509 |
| 0.621117 | 0.352074 | 22034 | 5508 |
| 0.60832 | 0.391425 | 22034 | 5508 |
| 0.631288 | 0.372155 | 22034 | 5508 |
| 0.658038 | 0.368728 | 22033 | 5509 |
| 0.609448 | 0.386677 | 22033 | 5509 |
| 0.620871 | 0.35233 | 22034 | 5508 |
| 0.608328 | 0.391416 | 22034 | 5508 |
| 0.631436 | 0.372009 | 22034 | 5508 |
| 0.705882 | 0.322829 | 22033 | 5509 |
| 0.659455 | 0.336352 | 22033 | 5509 |
| 0.654881 | 0.316852 | 22034 | 5508 |
| 0.657739 | 0.341985 | 22034 | 5508 |
| 0.68349 | 0.320238 | 22034 | 5508 |
| 0.69382 | 0.372496 | 27542 | 4025 |
| 0.69384 | 0.372478 | 27542 | 4025 |
| 0.731886 | 0.338068 | 27542 | 4025 |
| 0.62537 | 0.379975 | 22033 | 5509 |
| 0.618943 | 0.398104 | 22033 | 5509 |
| 0.616851 | 0.357127 | 22034 | 5508 |
| 0.605518 | 0.396656 | 22034 | 5508 |
| 0.624776 | 0.374839 | 22034 | 5508 |
| 0.625348 | 0.379997 | 22033 | 5509 |
| 0.619131 | 0.397921 | 22033 | 5509 |
| 0.61662 | 0.357367 | 22034 | 5508 |
| 0.605528 | 0.396646 | 22034 | 5508 |
| 0.624964 | 0.374652 | 22034 | 5508 |
| 0.66913 | 0.336589 | 22033 | 5509 |
| 0.673791 | 0.344766 | 22033 | 5509 |
| 0.648407 | 0.324239 | 22034 | 5508 |
| 0.652756 | 0.349588 | 22034 | 5508 |

| | | | |
|---|---|---|---|
| 0.67567 | 0.323915 | 22034 | 5508 |
| 0.688452 | 0.352694 | 27542 | 4025 |
| 0.688548 | 0.352603 | 27542 | 4025 |
| 0.734904 | 0.309017 | 27542 | 4025 |
| 0.662151 | 0.355413 | 22033 | 5509 |
| 0.635673 | 0.353719 | 22033 | 5509 |
| 0.643837 | 0.331408 | 22034 | 5508 |
| 0.652937 | 0.359883 | 22034 | 5508 |
| 0.681906 | 0.322107 | 22034 | 5508 |
| 0.662304 | 0.355265 | 22033 | 5509 |
| 0.635765 | 0.353626 | 22033 | 5509 |
| 0.643525 | 0.331732 | 22034 | 5508 |
| 0.653055 | 0.359767 | 22034 | 5508 |
| 0.682073 | 0.32194 | 22034 | 5508 |
| 0.712263 | 0.30663 | 22033 | 5509 |
| 0.685653 | 0.302905 | 22033 | 5509 |
| 0.676548 | 0.29744 | 22034 | 5508 |
| 0.703255 | 0.310553 | 22034 | 5508 |
| 0.72695 | 0.277328 | 22034 | 5508 |
| 0.745405 | 0.320077 | 27542 | 4025 |
| 0.745552 | 0.319943 | 27542 | 4025 |
| 0.792357 | 0.277249 | 27542 | 4025 |
| 0.675184 | 0.332274 | 22033 | 5509 |
| 0.698851 | 0.322403 | 22033 | 5509 |
| 0.668988 | 0.302224 | 22034 | 5508 |
| 0.677368 | 0.327118 | 22034 | 5508 |
| 0.691272 | 0.303023 | 22034 | 5508 |
| 0.675225 | 0.332234 | 22033 | 5509 |
| 0.698933 | 0.322324 | 22033 | 5509 |
| 0.668833 | 0.302386 | 22034 | 5508 |
| 0.677416 | 0.32707 | 22034 | 5508 |
| 0.691414 | 0.302879 | 22034 | 5508 |
| 0.722385 | 0.285595 | 22033 | 5509 |
| 0.748304 | 0.274454 | 22033 | 5509 |
| 0.700649 | 0.2692 | 22034 | 5508 |
| 0.721925 | 0.282855 | 22034 | 5508 |
| 0.730865 | 0.263103 | 22034 | 5508 |
| 0.792054 | 0.284203 | 27542 | 4025 |
| 0.792143 | 0.284123 | 27542 | 4025 |
| 0.82949 | 0.250371 | 27542 | 4025 |
| 0.727734 | 0.296028 | 22033 | 5509 |
| 0.701239 | 0.293891 | 22033 | 5509 |
| 0.726818 | 0.261044 | 22034 | 5508 |
| 0.701009 | 0.291756 | 22034 | 5508 |
| 0.724569 | 0.275081 | 22034 | 5508 |

| | | | |
|---|---|---|---|
| 0.727691 | 0.29607 | 22033 | 5509 |
| 0.701381 | 0.293748 | 22033 | 5509 |
| 0.726617 | 0.261249 | 22034 | 5508 |
| 0.701031 | 0.291734 | 22034 | 5508 |
| 0.724649 | 0.275 | 22034 | 5508 |
| 0.768601 | 0.256495 | 22033 | 5509 |
| 0.743257 | 0.251581 | 22033 | 5509 |
| 0.755385 | 0.232 | 22034 | 5508 |
| 0.740674 | 0.251681 | 22034 | 5508 |
| 0.7609 | 0.238732 | 22034 | 5508 |
| 0.731414 | 0.315542 | 27542 | 4025 |
| 0.731526 | 0.315436 | 27542 | 4025 |
| 0.771271 | 0.278243 | 27542 | 4025 |
| 0.695739 | 0.318474 | 22033 | 5509 |
| 0.683121 | 0.322648 | 22033 | 5509 |
| 0.686569 | 0.293265 | 22034 | 5508 |
| 0.662553 | 0.337198 | 22034 | 5508 |
| 0.691332 | 0.308179 | 22034 | 5508 |
| 0.69568 | 0.318532 | 22033 | 5509 |
| 0.683336 | 0.322435 | 22033 | 5509 |
| 0.686402 | 0.293437 | 22034 | 5508 |
| 0.662617 | 0.337135 | 22034 | 5508 |
| 0.691426 | 0.308084 | 22034 | 5508 |
| 0.736905 | 0.27815 | 22033 | 5509 |
| 0.733492 | 0.272703 | 22033 | 5509 |
| 0.716215 | 0.262748 | 22034 | 5508 |
| 0.711965 | 0.287768 | 22034 | 5508 |
| 0.732897 | 0.266585 | 22034 | 5508 |
| 0.66211 | 0.396183 | 27542 | 4025 |
| 0.662231 | 0.396072 | 27542 | 4025 |
| 0.707162 | 0.355097 | 27542 | 4025 |
| 0.614192 | 0.395437 | 22033 | 5509 |
| 0.594423 | 0.406601 | 22033 | 5509 |
| 0.599678 | 0.375827 | 22034 | 5508 |
| 0.588519 | 0.41001 | 22034 | 5508 |
| 0.627186 | 0.387398 | 22034 | 5508 |
| 0.614199 | 0.39543 | 22033 | 5509 |
| 0.594528 | 0.406497 | 22033 | 5509 |
| 0.599345 | 0.376174 | 22034 | 5508 |
| 0.588623 | 0.409905 | 22034 | 5508 |
| 0.627476 | 0.387114 | 22034 | 5508 |
| 0.660928 | 0.349433 | 22033 | 5509 |
| 0.647581 | 0.353535 | 22033 | 5509 |
| 0.633143 | 0.340995 | 22034 | 5508 |
| 0.638572 | 0.359831 | 22034 | 5508 |

| | | | |
|---|---|---|---|
| 0.682486 | 0.333383 | 22034 | 5508 |
| 0.723407 | 0.302022 | 27542 | 4025 |
| 0.723485 | 0.301947 | 27542 | 4025 |
| 0.787198 | 0.240473 | 27542 | 4025 |
| 0.668441 | 0.330271 | 22033 | 5509 |
| 0.694283 | 0.307934 | 22033 | 5509 |
| 0.657864 | 0.306085 | 22034 | 5508 |
| 0.70497 | 0.320987 | 22034 | 5508 |
| 0.704849 | 0.303167 | 22034 | 5508 |
| 0.668407 | 0.330304 | 22033 | 5509 |
| 0.694313 | 0.307904 | 22033 | 5509 |
| 0.657571 | 0.306394 | 22034 | 5508 |
| 0.705112 | 0.32085 | 22034 | 5508 |
| 0.705005 | 0.303012 | 22034 | 5508 |
| 0.744797 | 0.253767 | 22033 | 5509 |
| 0.758167 | 0.244253 | 22033 | 5509 |
| 0.709434 | 0.251689 | 22034 | 5508 |
| 0.779578 | 0.249126 | 22034 | 5508 |
| 0.780197 | 0.228675 | 22034 | 5508 |
| 0.626552 | 0.409771 | 27542 | 4025 |
| 0.626714 | 0.409618 | 27542 | 4025 |
| 0.695248 | 0.345057 | 27542 | 4025 |
| 0.62839 | 0.395146 | 22033 | 5509 |
| 0.588419 | 0.419951 | 22033 | 5509 |
| 0.586213 | 0.389361 | 22034 | 5508 |
| 0.582494 | 0.422602 | 22034 | 5508 |
| 0.58897 | 0.397619 | 22034 | 5508 |
| 0.628354 | 0.395181 | 22033 | 5509 |
| 0.588522 | 0.41985 | 22033 | 5509 |
| 0.586074 | 0.389506 | 22034 | 5508 |
| 0.58262 | 0.422477 | 22034 | 5508 |
| 0.588969 | 0.39762 | 22034 | 5508 |
| 0.691552 | 0.33435 | 22033 | 5509 |
| 0.66253 | 0.346895 | 22033 | 5509 |
| 0.635723 | 0.337789 | 22034 | 5508 |
| 0.651787 | 0.353916 | 22034 | 5508 |
| 0.666493 | 0.318331 | 22034 | 5508 |
| 0.7731 | 0.252225 | 27542 | 4025 |
| 0.773143 | 0.252184 | 27542 | 4025 |
| 0.830498 | 0.196708 | 27542 | 4025 |
| 0.727703 | 0.278536 | 22033 | 5509 |
| 0.736 | 0.271988 | 22033 | 5509 |
| 0.699648 | 0.274315 | 22034 | 5508 |
| 0.724585 | 0.292194 | 22034 | 5508 |
| 0.739846 | 0.254089 | 22034 | 5508 |

| | | | |
|---|---|---|---|
| 0.727543 | 0.278695 | 22033 | 5509 |
| 0.736152 | 0.271837 | 22033 | 5509 |
| 0.699613 | 0.274351 | 22034 | 5508 |
| 0.724765 | 0.292018 | 22034 | 5508 |
| 0.739756 | 0.254179 | 22034 | 5508 |
| 0.794089 | 0.212719 | 22033 | 5509 |
| 0.799425 | 0.209251 | 22033 | 5509 |
| 0.756156 | 0.215704 | 22034 | 5508 |
| 0.80389 | 0.214726 | 22034 | 5508 |
| 0.798184 | 0.195272 | 22034 | 5508 |
| 0.78642 | 0.22905 | 27542 | 4025 |
| 0.786333 | 0.229135 | 27542 | 4025 |
| 0.84192 | 0.174641 | 27542 | 4025 |
| 0.799426 | 0.218512 | 22033 | 5509 |
| 0.776415 | 0.241015 | 22033 | 5509 |
| 0.741086 | 0.23687 | 22034 | 5508 |
| 0.767508 | 0.227239 | 22034 | 5508 |
| 0.767548 | 0.224207 | 22034 | 5508 |
| 0.799466 | 0.218473 | 22033 | 5509 |
| 0.776418 | 0.241011 | 22033 | 5509 |
| 0.741104 | 0.236851 | 22034 | 5508 |
| 0.767415 | 0.227333 | 22034 | 5508 |
| 0.767603 | 0.224151 | 22034 | 5508 |
| 0.860327 | 0.158977 | 22033 | 5509 |
| 0.842025 | 0.176877 | 22033 | 5509 |
| 0.796018 | 0.180303 | 22034 | 5508 |
| 0.819431 | 0.17496 | 22034 | 5508 |
| 0.830867 | 0.160208 | 22034 | 5508 |
| 0.569889 | 0.440924 | 27542 | 4025 |
| 0.56983 | 0.440982 | 27542 | 4025 |
| 0.63122 | 0.380756 | 27542 | 4025 |
| 0.561671 | 0.445385 | 22033 | 5509 |
| 0.539261 | 0.456973 | 22033 | 5509 |
| 0.523874 | 0.445018 | 22034 | 5508 |
| 0.564148 | 0.467438 | 22034 | 5508 |
| 0.534077 | 0.461311 | 22034 | 5508 |
| 0.56161 | 0.445446 | 22033 | 5509 |
| 0.539268 | 0.456965 | 22033 | 5509 |
| 0.523775 | 0.445124 | 22034 | 5508 |
| 0.564269 | 0.467324 | 22034 | 5508 |
| 0.534125 | 0.461262 | 22034 | 5508 |
| 0.628135 | 0.379757 | 22033 | 5509 |
| 0.608029 | 0.387724 | 22033 | 5509 |
| 0.581783 | 0.383671 | 22034 | 5508 |
| 0.643192 | 0.392819 | 22034 | 5508 |

| | | | |
|---|---|---|---|
| 0.621293 | 0.373341 | 22034 | 5508 |
| 0.634239 | 0.381704 | 27542 | 4025 |
| 0.634177 | 0.381765 | 27542 | 4025 |
| 0.70353 | 0.314155 | 27542 | 4025 |
| 0.625885 | 0.394922 | 22033 | 5509 |
| 0.604882 | 0.380597 | 22033 | 5509 |
| 0.598317 | 0.371438 | 22034 | 5508 |
| 0.642487 | 0.387156 | 22034 | 5508 |
| 0.628987 | 0.363947 | 22034 | 5508 |
| 0.626031 | 0.394781 | 22033 | 5509 |
| 0.604758 | 0.380725 | 22033 | 5509 |
| 0.598249 | 0.371509 | 22034 | 5508 |
| 0.642422 | 0.387219 | 22034 | 5508 |
| 0.628967 | 0.363967 | 22034 | 5508 |
| 0.716658 | 0.307167 | 22033 | 5509 |
| 0.67219 | 0.311673 | 22033 | 5509 |
| 0.657791 | 0.308957 | 22034 | 5508 |
| 0.71752 | 0.315586 | 22034 | 5508 |
| 0.71609 | 0.275866 | 22034 | 5508 |
| 0.592439 | 0.440897 | 27542 | 4025 |
| 0.592457 | 0.440879 | 27542 | 4025 |
| 0.661881 | 0.375361 | 27542 | 4025 |
| 0.58388 | 0.427575 | 22033 | 5509 |
| 0.560841 | 0.438275 | 22033 | 5509 |
| 0.542711 | 0.428744 | 22034 | 5508 |
| 0.575885 | 0.44331 | 22034 | 5508 |
| 0.55371 | 0.444473 | 22034 | 5508 |
| 0.583922 | 0.427534 | 22033 | 5509 |
| 0.56085 | 0.438266 | 22033 | 5509 |
| 0.54253 | 0.428934 | 22034 | 5508 |
| 0.576014 | 0.443186 | 22034 | 5508 |
| 0.553659 | 0.444524 | 22034 | 5508 |
| 0.65958 | 0.35336 | 22033 | 5509 |
| 0.631699 | 0.367306 | 22033 | 5509 |
| 0.599882 | 0.368566 | 22034 | 5508 |
| 0.651095 | 0.370607 | 22034 | 5508 |
| 0.646658 | 0.35122 | 22034 | 5508 |
| 0.594744 | 0.447714 | 27542 | 4025 |
| 0.594908 | 0.447561 | 27542 | 4025 |
| 0.678434 | 0.369998 | 27542 | 4025 |
| 0.595421 | 0.422498 | 22033 | 5509 |
| 0.590017 | 0.434404 | 22033 | 5509 |
| 0.54601 | 0.430178 | 22034 | 5508 |
| 0.564006 | 0.439909 | 22034 | 5508 |
| 0.543147 | 0.434646 | 22034 | 5508 |

| | | | |
|---|---|---|---|
| 0.595384 | 0.422535 | 22033 | 5509 |
| 0.590107 | 0.434318 | 22033 | 5509 |
| 0.545916 | 0.430277 | 22034 | 5508 |
| 0.563978 | 0.439936 | 22034 | 5508 |
| 0.543196 | 0.434595 | 22034 | 5508 |
| 0.668869 | 0.351261 | 22033 | 5509 |
| 0.671803 | 0.356003 | 22033 | 5509 |
| 0.610024 | 0.363373 | 22034 | 5508 |
| 0.638248 | 0.366182 | 22034 | 5508 |
| 0.633501 | 0.340599 | 22034 | 5508 |
| 0.800078 | 0.250547 | 27542 | 4025 |
| 0.800151 | 0.250478 | 27542 | 4025 |
| 0.857137 | 0.197098 | 27542 | 4025 |
| 0.739774 | 0.269532 | 22033 | 5509 |
| 0.745515 | 0.25752 | 22033 | 5509 |
| 0.706148 | 0.268533 | 22034 | 5508 |
| 0.745099 | 0.28083 | 22034 | 5508 |
| 0.729546 | 0.256118 | 22034 | 5508 |
| 0.739686 | 0.26962 | 22033 | 5509 |
| 0.745585 | 0.25745 | 22033 | 5509 |
| 0.706145 | 0.268536 | 22034 | 5508 |
| 0.745047 | 0.28088 | 22034 | 5508 |
| 0.729573 | 0.25609 | 22034 | 5508 |
| 0.803374 | 0.206733 | 22033 | 5509 |
| 0.8 | 0.203257 | 22033 | 5509 |
| 0.763721 | 0.208895 | 22034 | 5508 |
| 0.81132 | 0.216913 | 22034 | 5508 |
| 0.78727 | 0.197259 | 22034 | 5508 |
| 0.81493 | 0.245891 | 27542 | 4025 |
| 0.815103 | 0.24573 | 27542 | 4025 |
| 0.880237 | 0.185458 | 27542 | 4025 |
| 0.792856 | 0.22982 | 22033 | 5509 |
| 0.780253 | 0.243689 | 22033 | 5509 |
| 0.765347 | 0.205257 | 22034 | 5508 |
| 0.783409 | 0.220161 | 22034 | 5508 |
| 0.751447 | 0.22618 | 22034 | 5508 |
| 0.792951 | 0.229727 | 22033 | 5509 |
| 0.780414 | 0.243532 | 22033 | 5509 |
| 0.765096 | 0.205518 | 22034 | 5508 |
| 0.783443 | 0.220127 | 22034 | 5508 |
| 0.751514 | 0.226111 | 22034 | 5508 |
| 0.854802 | 0.169645 | 22033 | 5509 |
| 0.848892 | 0.177156 | 22033 | 5509 |
| 0.798764 | 0.170556 | 22034 | 5508 |
| 0.834863 | 0.168941 | 22034 | 5508 |

| | | | |
|---|---|---|---|
| 0.811511 | 0.164329 | 22034 | 5508 |
| 0.645093 | 0.386248 | 27542 | 4025 |
| 0.645214 | 0.386132 | 27542 | 4025 |
| 0.713742 | 0.320933 | 27542 | 4025 |
| 0.64297 | 0.363022 | 22033 | 5509 |
| 0.628274 | 0.392534 | 22033 | 5509 |
| 0.610407 | 0.365882 | 22034 | 5508 |
| 0.628981 | 0.386934 | 22034 | 5508 |
| 0.597015 | 0.382814 | 22034 | 5508 |
| 0.642747 | 0.363243 | 22033 | 5509 |
| 0.628378 | 0.392434 | 22033 | 5509 |
| 0.6104 | 0.365889 | 22034 | 5508 |
| 0.628937 | 0.386976 | 22034 | 5508 |
| 0.597168 | 0.382655 | 22034 | 5508 |
| 0.702685 | 0.303863 | 22033 | 5509 |
| 0.707546 | 0.315888 | 22033 | 5509 |
| 0.665102 | 0.309062 | 22034 | 5508 |
| 0.693419 | 0.324126 | 22034 | 5508 |
| 0.665347 | 0.312173 | 22034 | 5508 |
| 0.68238 | 0.346145 | 27542 | 4025 |
| 0.682363 | 0.346161 | 27542 | 4025 |
| 0.744192 | 0.286917 | 27542 | 4025 |
| 0.637035 | 0.355117 | 22033 | 5509 |
| 0.647161 | 0.350882 | 22033 | 5509 |
| 0.641973 | 0.34736 | 22034 | 5508 |
| 0.667062 | 0.363647 | 22034 | 5508 |
| 0.632389 | 0.35668 | 22034 | 5508 |
| 0.636925 | 0.355229 | 22033 | 5509 |
| 0.647166 | 0.350878 | 22033 | 5509 |
| 0.641977 | 0.347356 | 22034 | 5508 |
| 0.667108 | 0.363603 | 22034 | 5508 |
| 0.632379 | 0.35669 | 22034 | 5508 |
| 0.697898 | 0.293505 | 22033 | 5509 |
| 0.712867 | 0.284978 | 22033 | 5509 |
| 0.705534 | 0.282743 | 22034 | 5508 |
| 0.737609 | 0.296347 | 22034 | 5508 |
| 0.714083 | 0.273574 | 22034 | 5508 |
| 0.592211 | 0.42579 | 27542 | 4025 |
| 0.59226 | 0.425741 | 27542 | 4025 |
| 0.6639 | 0.356279 | 27542 | 4025 |
| 0.592811 | 0.415913 | 22033 | 5509 |
| 0.572382 | 0.4265 | 22033 | 5509 |
| 0.568134 | 0.413658 | 22034 | 5508 |
| 0.593374 | 0.430507 | 22034 | 5508 |
| 0.55001 | 0.436221 | 22034 | 5508 |

| | | | |
|---|---|---|---|
| 0.592741 | 0.415981 | 22033 | 5509 |
| 0.572428 | 0.426454 | 22033 | 5509 |
| 0.567975 | 0.413822 | 22034 | 5508 |
| 0.59341 | 0.430472 | 22034 | 5508 |
| 0.550193 | 0.436033 | 22034 | 5508 |
| 0.657968 | 0.351714 | 22033 | 5509 |
| 0.642278 | 0.356468 | 22033 | 5509 |
| 0.628027 | 0.351845 | 22034 | 5508 |
| 0.669825 | 0.357133 | 22034 | 5508 |
| 0.643311 | 0.340584 | 22034 | 5508 |
| 0.643776 | 0.384083 | 27542 | 4025 |
| 0.643754 | 0.384105 | 27542 | 4025 |
| 0.704911 | 0.325594 | 27542 | 4025 |
| 0.616022 | 0.396661 | 22033 | 5509 |
| 0.603452 | 0.400715 | 22033 | 5509 |
| 0.595354 | 0.392839 | 22034 | 5508 |
| 0.582722 | 0.410025 | 22034 | 5508 |
| 0.611013 | 0.390875 | 22034 | 5508 |
| 0.616049 | 0.396634 | 22033 | 5509 |
| 0.603474 | 0.400693 | 22033 | 5509 |
| 0.595251 | 0.392943 | 22034 | 5508 |
| 0.58267 | 0.410078 | 22034 | 5508 |
| 0.611055 | 0.390832 | 22034 | 5508 |
| 0.686738 | 0.327401 | 22033 | 5509 |
| 0.673055 | 0.331592 | 22033 | 5509 |
| 0.660693 | 0.326203 | 22034 | 5508 |
| 0.650597 | 0.341306 | 22034 | 5508 |
| 0.692865 | 0.309275 | 22034 | 5508 |
| 0.854009 | 0.211187 | 27542 | 4025 |
| 0.854011 | 0.211185 | 27542 | 4025 |
| 0.909521 | 0.159912 | 27542 | 4025 |
| 0.838943 | 0.21382 | 22033 | 5509 |
| 0.776084 | 0.225875 | 22033 | 5509 |
| 0.747316 | 0.216463 | 22034 | 5508 |
| 0.782185 | 0.222476 | 22034 | 5508 |
| 0.750673 | 0.227087 | 22034 | 5508 |
| 0.838826 | 0.213929 | 22033 | 5509 |
| 0.776156 | 0.225804 | 22033 | 5509 |
| 0.747246 | 0.216536 | 22034 | 5508 |
| 0.782109 | 0.222552 | 22034 | 5508 |
| 0.750934 | 0.226819 | 22034 | 5508 |
| 0.89534 | 0.16097 | 22033 | 5509 |
| 0.8301 | 0.171996 | 22033 | 5509 |
| 0.789044 | 0.172712 | 22034 | 5508 |
| 0.835897 | 0.169084 | 22034 | 5508 |

| | | | |
|---|---|---|---|
| 0.81183 | 0.164119 | 22034 | 5508 |
| 0.684733 | 0.335293 | 27542 | 4025 |
| 0.684755 | 0.335272 | 27542 | 4025 |
| 0.740847 | 0.28082 | 27542 | 4025 |
| 0.675136 | 0.33567 | 22033 | 5509 |
| 0.651683 | 0.35319 | 22033 | 5509 |
| 0.643614 | 0.340753 | 22034 | 5508 |
| 0.654496 | 0.362017 | 22034 | 5508 |
| 0.623193 | 0.359799 | 22034 | 5508 |
| 0.675024 | 0.33578 | 22033 | 5509 |
| 0.651702 | 0.353171 | 22033 | 5509 |
| 0.643733 | 0.340632 | 22034 | 5508 |
| 0.654431 | 0.362081 | 22034 | 5508 |
| 0.623399 | 0.359588 | 22034 | 5508 |
| 0.732721 | 0.279006 | 22033 | 5509 |
| 0.711315 | 0.294004 | 22033 | 5509 |
| 0.698781 | 0.284246 | 22034 | 5508 |
| 0.716372 | 0.301703 | 22034 | 5508 |
| 0.691014 | 0.290127 | 22034 | 5508 |
| 0.528465 | 0.496309 | 27542 | 4025 |
| 0.528498 | 0.496277 | 27542 | 4025 |
| 0.601903 | 0.426314 | 27542 | 4025 |
| 0.519345 | 0.49522 | 22033 | 5509 |
| 0.501791 | 0.507242 | 22033 | 5509 |
| 0.481034 | 0.49576 | 22034 | 5508 |
| 0.499582 | 0.512351 | 22034 | 5508 |
| 0.482384 | 0.504732 | 22034 | 5508 |
| 0.519319 | 0.495245 | 22033 | 5509 |
| 0.501848 | 0.507186 | 22033 | 5509 |
| 0.480869 | 0.495932 | 22034 | 5508 |
| 0.499648 | 0.512287 | 22034 | 5508 |
| 0.482545 | 0.504567 | 22034 | 5508 |
| 0.595226 | 0.421467 | 22033 | 5509 |
| 0.580789 | 0.429666 | 22033 | 5509 |
| 0.543331 | 0.430456 | 22034 | 5508 |
| 0.577062 | 0.436722 | 22034 | 5508 |
| 0.574971 | 0.409672 | 22034 | 5508 |
| 0.976809 | 0.061621 | 27542 | 4025 |
| 0.976891 | 0.061542 | 27542 | 4025 |
| 1.007215 | 0.032411 | 27542 | 4025 |
| 0.985565 | 0.05312 | 22033 | 5509 |
| 0.915664 | 0.048684 | 22033 | 5509 |
| 0.938766 | 0.053604 | 22034 | 5508 |
| 0.957481 | 0.048832 | 22034 | 5508 |
| 0.938376 | 0.059736 | 22034 | 5508 |

| | | | |
|---|---|---|---|
| 0.985697 | 0.052994 | 22033 | 5509 |
| 0.915465 | 0.048891 | 22033 | 5509 |
| 0.938908 | 0.053461 | 22034 | 5508 |
| 0.957478 | 0.048836 | 22034 | 5508 |
| 0.93847 | 0.059641 | 22034 | 5508 |
| 1.011009 | 0.028676 | 22033 | 5509 |
| 0.933036 | 0.030636 | 22033 | 5509 |
| 0.961226 | 0.030962 | 22034 | 5508 |
| 0.979849 | 0.026612 | 22034 | 5508 |
| 0.969399 | 0.028651 | 22034 | 5508 |
| 0.884479 | 0.134924 | 27542 | 4025 |
| 0.884471 | 0.134933 | 27542 | 4025 |
| 0.925548 | 0.094756 | 27542 | 4025 |
| 0.87719 | 0.151083 | 22033 | 5509 |
| 0.818923 | 0.141156 | 22033 | 5509 |
| 0.859819 | 0.143549 | 22034 | 5508 |
| 0.854511 | 0.156852 | 22034 | 5508 |
| 0.851204 | 0.145601 | 22034 | 5508 |
| 0.877423 | 0.150857 | 22033 | 5509 |
| 0.818831 | 0.141252 | 22033 | 5509 |
| 0.859744 | 0.143623 | 22034 | 5508 |
| 0.8546 | 0.156765 | 22034 | 5508 |
| 0.851054 | 0.145752 | 22034 | 5508 |
| 0.933342 | 0.09674 | 22033 | 5509 |
| 0.852751 | 0.105679 | 22033 | 5509 |
| 0.899991 | 0.103534 | 22034 | 5508 |
| 0.909674 | 0.102422 | 22034 | 5508 |
| 0.895191 | 0.101449 | 22034 | 5508 |
| 0.947242 | 0.042483 | 27542 | 4025 |
| 0.947174 | 0.042551 | 27542 | 4025 |
| 0.971293 | 0.018171 | 27542 | 4025 |
| 0.977843 | 0.045898 | 22033 | 5509 |
| 0.943509 | 0.05508 | 22033 | 5509 |
| 0.924341 | 0.052226 | 22034 | 5508 |
| 0.959144 | 0.058705 | 22034 | 5508 |
| 0.923549 | 0.059367 | 22034 | 5508 |
| 0.97779 | 0.04595 | 22033 | 5509 |
| 0.943512 | 0.055077 | 22033 | 5509 |
| 0.924296 | 0.052272 | 22034 | 5508 |
| 0.959275 | 0.058578 | 22034 | 5508 |
| 0.923685 | 0.059229 | 22034 | 5508 |
| 1.001813 | 0.02251 | 22033 | 5509 |
| 0.974437 | 0.024106 | 22033 | 5509 |
| 0.953565 | 0.022261 | 22034 | 5508 |
| 0.99586 | 0.022673 | 22034 | 5508 |

| | | | |
|---|---|---|---|
| 0.965979 | 0.016153 | 22034 | 5508 |
| 0.429708 | 0.579361 | 27542 | 4025 |
| 0.429599 | 0.579468 | 27542 | 4025 |
| 0.495658 | 0.514804 | 27542 | 4025 |
| 0.422426 | 0.579079 | 22033 | 5509 |
| 0.412098 | 0.59031 | 22033 | 5509 |
| 0.384862 | 0.589084 | 22034 | 5508 |
| 0.414607 | 0.604316 | 22034 | 5508 |
| 0.41542 | 0.587148 | 22034 | 5508 |
| 0.422401 | 0.579104 | 22033 | 5509 |
| 0.412129 | 0.590279 | 22033 | 5509 |
| 0.384724 | 0.589232 | 22034 | 5508 |
| 0.4147 | 0.604228 | 22034 | 5508 |
| 0.415463 | 0.587105 | 22034 | 5508 |
| 0.494733 | 0.50703 | 22033 | 5509 |
| 0.486255 | 0.516586 | 22033 | 5509 |
| 0.445052 | 0.52482 | 22034 | 5508 |
| 0.494022 | 0.528526 | 22034 | 5508 |
| 0.509528 | 0.493621 | 22034 | 5508 |
| 0.827567 | 0.216229 | 27542 | 4025 |
| 0.82748 | 0.216311 | 27542 | 4025 |
| 0.86829 | 0.177662 | 27542 | 4025 |
| 0.790519 | 0.222894 | 22033 | 5509 |
| 0.769278 | 0.220032 | 22033 | 5509 |
| 0.735554 | 0.229739 | 22034 | 5508 |
| 0.771197 | 0.240321 | 22034 | 5508 |
| 0.79292 | 0.227811 | 22034 | 5508 |
| 0.790453 | 0.222958 | 22033 | 5509 |
| 0.76926 | 0.22005 | 22033 | 5509 |
| 0.735483 | 0.229812 | 22034 | 5508 |
| 0.771226 | 0.240293 | 22034 | 5508 |
| 0.793031 | 0.227703 | 22034 | 5508 |
| 0.830357 | 0.183731 | 22033 | 5509 |
| 0.812098 | 0.176617 | 22033 | 5509 |
| 0.765171 | 0.198723 | 22034 | 5508 |
| 0.823261 | 0.189035 | 22034 | 5508 |
| 0.846804 | 0.175336 | 22034 | 5508 |
| 0.597801 | 0.446566 | 27542 | 4025 |
| 0.597843 | 0.446527 | 27542 | 4025 |
| 0.656706 | 0.392033 | 27542 | 4025 |
| 0.559861 | 0.437443 | 22033 | 5509 |
| 0.566015 | 0.442632 | 22033 | 5509 |
| 0.54304 | 0.438731 | 22034 | 5508 |
| 0.563542 | 0.442375 | 22034 | 5508 |
| 0.578192 | 0.427862 | 22034 | 5508 |

| | | | |
|---|---|---|---|
| 0.559771 | 0.437534 | 22033 | 5509 |
| 0.566148 | 0.442501 | 22033 | 5509 |
| 0.542842 | 0.438934 | 22034 | 5508 |
| 0.563592 | 0.442325 | 22034 | 5508 |
| 0.578095 | 0.427958 | 22034 | 5508 |
| 0.605089 | 0.391997 | 22033 | 5509 |
| 0.622558 | 0.386953 | 22033 | 5509 |
| 0.583436 | 0.396978 | 22034 | 5508 |
| 0.619404 | 0.387099 | 22034 | 5508 |
| 0.654859 | 0.351997 | 22034 | 5508 |
| 0.752357 | 0.287253 | 27542 | 4025 |
| 0.752513 | 0.287105 | 27542 | 4025 |
| 0.812746 | 0.230043 | 27542 | 4025 |
| 0.774058 | 0.276857 | 22033 | 5509 |
| 0.731204 | 0.278609 | 22033 | 5509 |
| 0.673778 | 0.278787 | 22034 | 5508 |
| 0.713941 | 0.285749 | 22034 | 5508 |
| 0.700998 | 0.286698 | 22034 | 5508 |
| 0.773972 | 0.276938 | 22033 | 5509 |
| 0.731228 | 0.278584 | 22033 | 5509 |
| 0.673721 | 0.278848 | 22034 | 5508 |
| 0.714017 | 0.285674 | 22034 | 5508 |
| 0.70111 | 0.286583 | 22034 | 5508 |
| 0.846089 | 0.209563 | 22033 | 5509 |
| 0.791392 | 0.219228 | 22033 | 5509 |
| 0.729979 | 0.218629 | 22034 | 5508 |
| 0.785953 | 0.213706 | 22034 | 5508 |
| 0.776886 | 0.209477 | 22034 | 5508 |
| 0.786353 | 0.266068 | 27542 | 4025 |
| 0.786446 | 0.265982 | 27542 | 4025 |
| 0.852317 | 0.204502 | 27542 | 4025 |
| 0.721887 | 0.285273 | 22033 | 5509 |
| 0.702634 | 0.280425 | 22033 | 5509 |
| 0.692698 | 0.276952 | 22034 | 5508 |
| 0.743083 | 0.292335 | 22034 | 5508 |
| 0.716572 | 0.286944 | 22034 | 5508 |
| 0.721754 | 0.285404 | 22033 | 5509 |
| 0.702786 | 0.280269 | 22033 | 5509 |
| 0.692665 | 0.276987 | 22034 | 5508 |
| 0.742987 | 0.292427 | 22034 | 5508 |
| 0.716731 | 0.286786 | 22034 | 5508 |
| 0.784485 | 0.223296 | 22033 | 5509 |
| 0.760276 | 0.221393 | 22033 | 5509 |
| 0.747052 | 0.220217 | 22034 | 5508 |
| 0.809792 | 0.228806 | 22034 | 5508 |

| | | | |
|---|---|---|---|
| 0.797627 | 0.206286 | 22034 | 5508 |
| 0.809141 | 0.219841 | 27542 | 4025 |
| 0.809037 | 0.219941 | 27542 | 4025 |
| 0.871951 | 0.15928 | 27542 | 4025 |
| 0.823603 | 0.199972 | 22033 | 5509 |
| 0.792444 | 0.206815 | 22033 | 5509 |
| 0.782676 | 0.192483 | 22034 | 5508 |
| 0.809579 | 0.19765 | 22034 | 5508 |
| 0.779804 | 0.214499 | 22034 | 5508 |
| 0.823531 | 0.200041 | 22033 | 5509 |
| 0.792332 | 0.206927 | 22033 | 5509 |
| 0.782663 | 0.192497 | 22034 | 5508 |
| 0.80969 | 0.19754 | 22034 | 5508 |
| 0.780214 | 0.214086 | 22034 | 5508 |
| 0.877447 | 0.147668 | 22033 | 5509 |
| 0.849241 | 0.149965 | 22033 | 5509 |
| 0.826423 | 0.147348 | 22034 | 5508 |
| 0.865579 | 0.142151 | 22034 | 5508 |
| 0.852579 | 0.141192 | 22034 | 5508 |
| 0.705115 | 0.318293 | 27542 | 4025 |
| 0.705171 | 0.318239 | 27542 | 4025 |
| 0.776404 | 0.24937 | 27542 | 4025 |
| 0.689034 | 0.32542 | 22033 | 5509 |
| 0.672204 | 0.328872 | 22033 | 5509 |
| 0.669431 | 0.308598 | 22034 | 5508 |
| 0.67093 | 0.329361 | 22034 | 5508 |
| 0.671438 | 0.333611 | 22034 | 5508 |
| 0.689047 | 0.325407 | 22033 | 5509 |
| 0.672226 | 0.32885 | 22033 | 5509 |
| 0.669339 | 0.308693 | 22034 | 5508 |
| 0.670961 | 0.32933 | 22034 | 5508 |
| 0.671608 | 0.333442 | 22034 | 5508 |
| 0.761504 | 0.25447 | 22033 | 5509 |
| 0.739109 | 0.262075 | 22033 | 5509 |
| 0.727741 | 0.248375 | 22034 | 5508 |
| 0.742836 | 0.257486 | 22034 | 5508 |
| 0.761026 | 0.244696 | 22034 | 5508 |
| 0.497523 | 0.527651 | 27542 | 4025 |
| 0.497523 | 0.52765 | 27542 | 4025 |
| 0.574444 | 0.454621 | 27542 | 4025 |
| 0.4903 | 0.524004 | 22033 | 5509 |
| 0.467038 | 0.53328 | 22033 | 5509 |
| 0.452415 | 0.52135 | 22034 | 5508 |
| 0.460885 | 0.541736 | 22034 | 5508 |
| 0.481362 | 0.527304 | 22034 | 5508 |

| | | | |
|---|---|---|---|
| 0.490096 | 0.524201 | 22033 | 5509 |
| 0.467159 | 0.533159 | 22033 | 5509 |
| 0.452207 | 0.521571 | 22034 | 5508 |
| 0.460936 | 0.541686 | 22034 | 5508 |
| 0.481636 | 0.527035 | 22034 | 5508 |
| 0.562635 | 0.453778 | 22033 | 5509 |
| 0.542132 | 0.458237 | 22033 | 5509 |
| 0.509476 | 0.460981 | 22034 | 5508 |
| 0.540967 | 0.46211 | 22034 | 5508 |
| 0.579128 | 0.431299 | 22034 | 5508 |
| 0.754495 | 0.237527 | 27542 | 4025 |
| 0.754291 | 0.237733 | 27542 | 4025 |
| 0.792412 | 0.199208 | 27542 | 4025 |
| 0.779452 | 0.248006 | 22033 | 5509 |
| 0.745628 | 0.246349 | 22033 | 5509 |
| 0.737204 | 0.22311 | 22034 | 5508 |
| 0.738031 | 0.254945 | 22034 | 5508 |
| 0.779939 | 0.246542 | 22034 | 5508 |
| 0.779741 | 0.247727 | 22033 | 5509 |
| 0.745484 | 0.246495 | 22033 | 5509 |
| 0.736682 | 0.22366 | 22034 | 5508 |
| 0.738131 | 0.254844 | 22034 | 5508 |
| 0.779955 | 0.246526 | 22034 | 5508 |
| 0.834246 | 0.195143 | 22033 | 5509 |
| 0.79203 | 0.199448 | 22033 | 5509 |
| 0.77098 | 0.187516 | 22034 | 5508 |
| 0.788989 | 0.203502 | 22034 | 5508 |
| 0.840092 | 0.188431 | 22034 | 5508 |
| 0.731888 | 0.269603 | 27542 | 4025 |
| 0.731923 | 0.269568 | 27542 | 4025 |
| 0.783583 | 0.218014 | 27542 | 4025 |
| 0.735998 | 0.27078 | 22033 | 5509 |
| 0.728888 | 0.278548 | 22033 | 5509 |
| 0.703162 | 0.265317 | 22034 | 5508 |
| 0.722492 | 0.298994 | 22034 | 5508 |
| 0.721607 | 0.272675 | 22034 | 5508 |
| 0.735877 | 0.2709 | 22033 | 5509 |
| 0.729073 | 0.278365 | 22033 | 5509 |
| 0.703007 | 0.265478 | 22034 | 5508 |
| 0.722615 | 0.298875 | 22034 | 5508 |
| 0.721401 | 0.272883 | 22034 | 5508 |
| 0.783815 | 0.223404 | 22033 | 5509 |
| 0.785474 | 0.222539 | 22033 | 5509 |
| 0.746074 | 0.220481 | 22034 | 5508 |
| 0.786827 | 0.236572 | 22034 | 5508 |

| | | | |
|---|---|---|---|
| 0.777359 | 0.216481 | 22034 | 5508 |
| 0.658072 | 0.359107 | 27542 | 4025 |
| 0.658208 | 0.358974 | 27542 | 4025 |
| 0.720846 | 0.297972 | 27542 | 4025 |
| 0.658666 | 0.348226 | 22033 | 5509 |
| 0.66779 | 0.36962 | 22033 | 5509 |
| 0.611426 | 0.355259 | 22034 | 5508 |
| 0.650775 | 0.378091 | 22034 | 5508 |
| 0.597707 | 0.361367 | 22034 | 5508 |
| 0.658539 | 0.348352 | 22033 | 5509 |
| 0.66803 | 0.369394 | 22033 | 5509 |
| 0.611312 | 0.355379 | 22034 | 5508 |
| 0.650935 | 0.377939 | 22034 | 5508 |
| 0.597597 | 0.361485 | 22034 | 5508 |
| 0.708974 | 0.298445 | 22033 | 5509 |
| 0.747191 | 0.294667 | 22033 | 5509 |
| 0.66148 | 0.302478 | 22034 | 5508 |
| 0.724097 | 0.308022 | 22034 | 5508 |
| 0.663605 | 0.290957 | 22034 | 5508 |
| 0.612352 | 0.403466 | 27542 | 4025 |
| 0.612366 | 0.403452 | 27542 | 4025 |
| 0.68335 | 0.334301 | 27542 | 4025 |
| 0.574104 | 0.413551 | 22033 | 5509 |
| 0.605781 | 0.415711 | 22033 | 5509 |
| 0.56447 | 0.420889 | 22034 | 5508 |
| 0.597656 | 0.425404 | 22034 | 5508 |
| 0.560557 | 0.421931 | 22034 | 5508 |
| 0.573992 | 0.413666 | 22033 | 5509 |
| 0.605951 | 0.415547 | 22033 | 5509 |
| 0.564435 | 0.420926 | 22034 | 5508 |
| 0.597703 | 0.425359 | 22034 | 5508 |
| 0.560531 | 0.421957 | 22034 | 5508 |
| 0.638605 | 0.347663 | 22033 | 5509 |
| 0.689387 | 0.335071 | 22033 | 5509 |
| 0.63322 | 0.350357 | 22034 | 5508 |
| 0.68063 | 0.345632 | 22034 | 5508 |
| 0.651275 | 0.328379 | 22034 | 5508 |
| 0.653284 | 0.370491 | 27542 | 4025 |
| 0.653253 | 0.37052 | 27542 | 4025 |
| 0.714942 | 0.311077 | 27542 | 4025 |
| 0.603469 | 0.397313 | 22033 | 5509 |
| 0.611498 | 0.392318 | 22033 | 5509 |
| 0.574346 | 0.393277 | 22034 | 5508 |
| 0.635277 | 0.396538 | 22034 | 5508 |
| 0.587883 | 0.408149 | 22034 | 5508 |

| | | | |
|---|---|---|---|
| 0.603463 | 0.39732 | 22033 | 5509 |
| 0.611476 | 0.392339 | 22033 | 5509 |
| 0.574239 | 0.39339 | 22034 | 5508 |
| 0.635264 | 0.39655 | 22034 | 5508 |
| 0.587976 | 0.408056 | 22034 | 5508 |
| 0.669213 | 0.331655 | 22033 | 5509 |
| 0.673492 | 0.33071 | 22033 | 5509 |
| 0.629012 | 0.335529 | 22034 | 5508 |
| 0.707939 | 0.327516 | 22034 | 5508 |
| 0.676131 | 0.319306 | 22034 | 5508 |
| 0.798755 | 0.225778 | 27542 | 4025 |
| 0.798683 | 0.225847 | 27542 | 4025 |
| 0.849212 | 0.176871 | 27542 | 4025 |
| 0.757183 | 0.241904 | 22033 | 5509 |
| 0.764743 | 0.235076 | 22033 | 5509 |
| 0.751923 | 0.238984 | 22034 | 5508 |
| 0.792117 | 0.223014 | 22034 | 5508 |
| 0.759444 | 0.235316 | 22034 | 5508 |
| 0.757151 | 0.241935 | 22033 | 5509 |
| 0.764783 | 0.235036 | 22033 | 5509 |
| 0.751979 | 0.238928 | 22034 | 5508 |
| 0.792142 | 0.222988 | 22034 | 5508 |
| 0.759391 | 0.23537 | 22034 | 5508 |
| 0.818931 | 0.18008 | 22033 | 5509 |
| 0.818269 | 0.181538 | 22033 | 5509 |
| 0.809089 | 0.181127 | 22034 | 5508 |
| 0.843354 | 0.172755 | 22034 | 5508 |
| 0.822662 | 0.171662 | 22034 | 5508 |
| 0.708386 | 0.338789 | 27542 | 4025 |
| 0.70849 | 0.338692 | 27542 | 4025 |
| 0.767538 | 0.283576 | 27542 | 4025 |
| 0.640211 | 0.354904 | 22033 | 5509 |
| 0.687395 | 0.339058 | 22033 | 5509 |
| 0.637647 | 0.342845 | 22034 | 5508 |
| 0.647772 | 0.350545 | 22034 | 5508 |
| 0.653042 | 0.346536 | 22034 | 5508 |
| 0.640236 | 0.354879 | 22033 | 5509 |
| 0.687358 | 0.339093 | 22033 | 5509 |
| 0.637779 | 0.342709 | 22034 | 5508 |
| 0.647685 | 0.350632 | 22034 | 5508 |
| 0.6531 | 0.346479 | 22034 | 5508 |
| 0.703725 | 0.290906 | 22033 | 5509 |
| 0.749509 | 0.279334 | 22033 | 5509 |
| 0.692482 | 0.286332 | 22034 | 5508 |
| 0.709495 | 0.288661 | 22034 | 5508 |

| | | | |
|---|---|---|---|
| 0.726413 | 0.273118 | 22034 | 5508 |
| 0.67612 | 0.370169 | 27542 | 4025 |
| 0.675961 | 0.370317 | 27542 | 4025 |
| 0.727428 | 0.322374 | 27542 | 4025 |
| 0.629894 | 0.386573 | 22033 | 5509 |
| 0.65336 | 0.373534 | 22033 | 5509 |
| 0.59857 | 0.368923 | 22034 | 5508 |
| 0.625181 | 0.375851 | 22034 | 5508 |
| 0.608033 | 0.379694 | 22034 | 5508 |
| 0.629881 | 0.386586 | 22033 | 5509 |
| 0.653316 | 0.373576 | 22033 | 5509 |
| 0.598342 | 0.369163 | 22034 | 5508 |
| 0.625149 | 0.375883 | 22034 | 5508 |
| 0.608052 | 0.379674 | 22034 | 5508 |
| 0.697357 | 0.320874 | 22033 | 5509 |
| 0.722259 | 0.307472 | 22033 | 5509 |
| 0.652123 | 0.312462 | 22034 | 5508 |
| 0.686263 | 0.31487 | 22034 | 5508 |
| 0.678226 | 0.308084 | 22034 | 5508 |
| 0.648264 | 0.36741 | 27542 | 4025 |
| 0.64829 | 0.367385 | 27542 | 4025 |
| 0.703999 | 0.313023 | 27542 | 4025 |
| 0.62981 | 0.383321 | 22033 | 5509 |
| 0.614855 | 0.389484 | 22033 | 5509 |
| 0.604955 | 0.378288 | 22034 | 5508 |
| 0.600844 | 0.395121 | 22034 | 5508 |
| 0.607074 | 0.395682 | 22034 | 5508 |
| 0.629768 | 0.383362 | 22033 | 5509 |
| 0.614811 | 0.389527 | 22033 | 5509 |
| 0.604932 | 0.378312 | 22034 | 5508 |
| 0.600799 | 0.395167 | 22034 | 5508 |
| 0.60719 | 0.395566 | 22034 | 5508 |
| 0.697294 | 0.317244 | 22033 | 5509 |
| 0.686872 | 0.317975 | 22033 | 5509 |
| 0.665481 | 0.316086 | 22034 | 5508 |
| 0.667201 | 0.328319 | 22034 | 5508 |
| 0.692258 | 0.310885 | 22034 | 5508 |
| 0.650206 | 0.352856 | 27542 | 4025 |
| 0.65026 | 0.352803 | 27542 | 4025 |
| 0.708095 | 0.29524 | 27542 | 4025 |
| 0.630915 | 0.356201 | 22033 | 5509 |
| 0.638411 | 0.378173 | 22033 | 5509 |
| 0.636484 | 0.347741 | 22034 | 5508 |
| 0.620251 | 0.388773 | 22034 | 5508 |
| 0.62382 | 0.377461 | 22034 | 5508 |

| | | | |
|---|---|---|---|
| 0.630794 | 0.356324 | 22033 | 5509 |
| 0.638425 | 0.37816 | 22033 | 5509 |
| 0.636351 | 0.347877 | 22034 | 5508 |
| 0.62033 | 0.388695 | 22034 | 5508 |
| 0.623908 | 0.377372 | 22034 | 5508 |
| 0.689705 | 0.29621 | 22033 | 5509 |
| 0.714452 | 0.304108 | 22033 | 5509 |
| 0.689473 | 0.293439 | 22034 | 5508 |
| 0.695523 | 0.314596 | 22034 | 5508 |
| 0.720405 | 0.281073 | 22034 | 5508 |
| 0.591572 | 0.412504 | 27542 | 4025 |
| 0.591568 | 0.412508 | 27542 | 4025 |
| 0.653277 | 0.351224 | 27542 | 4025 |
| 0.577829 | 0.410622 | 22033 | 5509 |
| 0.595364 | 0.432095 | 22033 | 5509 |
| 0.554502 | 0.414055 | 22034 | 5508 |
| 0.591678 | 0.431541 | 22034 | 5508 |
| 0.553794 | 0.437487 | 22034 | 5508 |
| 0.577715 | 0.410739 | 22033 | 5509 |
| 0.595429 | 0.432032 | 22033 | 5509 |
| 0.554453 | 0.414107 | 22034 | 5508 |
| 0.59173 | 0.431491 | 22034 | 5508 |
| 0.553744 | 0.437537 | 22034 | 5508 |
| 0.638691 | 0.348544 | 22033 | 5509 |
| 0.675164 | 0.355975 | 22033 | 5509 |
| 0.613048 | 0.35219 | 22034 | 5508 |
| 0.666459 | 0.359695 | 22034 | 5508 |
| 0.646671 | 0.343148 | 22034 | 5508 |
| 0.662919 | 0.383102 | 27542 | 4025 |
| 0.662896 | 0.383123 | 27542 | 4025 |
| 0.727623 | 0.32289 | 27542 | 4025 |
| 0.607662 | 0.382944 | 22033 | 5509 |
| 0.632158 | 0.396441 | 22033 | 5509 |
| 0.583093 | 0.395923 | 22034 | 5508 |
| 0.593332 | 0.406304 | 22034 | 5508 |
| 0.608596 | 0.393507 | 22034 | 5508 |
| 0.607604 | 0.383003 | 22033 | 5509 |
| 0.632357 | 0.396251 | 22033 | 5509 |
| 0.582982 | 0.396039 | 22034 | 5508 |
| 0.593352 | 0.406284 | 22034 | 5508 |
| 0.608513 | 0.393589 | 22034 | 5508 |
| 0.673231 | 0.316362 | 22033 | 5509 |
| 0.702896 | 0.328903 | 22033 | 5509 |
| 0.645299 | 0.33148 | 22034 | 5508 |
| 0.66545 | 0.334142 | 22034 | 5508 |

| | | | |
|---|---|---|---|
| 0.691637 | 0.310752 | 22034 | 5508 |
| 0.735462 | 0.311088 | 27542 | 4025 |
| 0.735633 | 0.310927 | 27542 | 4025 |
| 0.793645 | 0.256587 | 27542 | 4025 |
| 0.706678 | 0.311646 | 22033 | 5509 |
| 0.69852 | 0.307503 | 22033 | 5509 |
| 0.663053 | 0.304049 | 22034 | 5508 |
| 0.692882 | 0.320486 | 22034 | 5508 |
| 0.695867 | 0.298577 | 22034 | 5508 |
| 0.706693 | 0.311631 | 22033 | 5509 |
| 0.698496 | 0.307527 | 22033 | 5509 |
| 0.662948 | 0.30416 | 22034 | 5508 |
| 0.692802 | 0.320565 | 22034 | 5508 |
| 0.695809 | 0.298635 | 22034 | 5508 |
| 0.770045 | 0.249922 | 22033 | 5509 |
| 0.758376 | 0.248164 | 22033 | 5509 |
| 0.715706 | 0.248784 | 22034 | 5508 |
| 0.759541 | 0.255113 | 22034 | 5508 |
| 0.763109 | 0.230797 | 22034 | 5508 |
| 0.74351 | 0.313083 | 27542 | 4025 |
| 0.743613 | 0.312988 | 27542 | 4025 |
| 0.790449 | 0.269717 | 27542 | 4025 |
| 0.709297 | 0.322562 | 22033 | 5509 |
| 0.709847 | 0.318314 | 22033 | 5509 |
| 0.654317 | 0.314719 | 22034 | 5508 |
| 0.657509 | 0.322091 | 22034 | 5508 |
| 0.671277 | 0.319915 | 22034 | 5508 |
| 0.709386 | 0.322477 | 22033 | 5509 |
| 0.709887 | 0.318275 | 22033 | 5509 |
| 0.654245 | 0.314794 | 22034 | 5508 |
| 0.657402 | 0.322201 | 22034 | 5508 |
| 0.671442 | 0.319748 | 22034 | 5508 |
| 0.770182 | 0.264411 | 22033 | 5509 |
| 0.764921 | 0.265425 | 22033 | 5509 |
| 0.701101 | 0.265721 | 22034 | 5508 |
| 0.706705 | 0.271369 | 22034 | 5508 |
| 0.740648 | 0.249633 | 22034 | 5508 |
| 0.622203 | 0.36781 | 27542 | 4025 |
| 0.622076 | 0.367939 | 27542 | 4025 |
| 0.677714 | 0.311409 | 27542 | 4025 |
| 0.654527 | 0.36272 | 22033 | 5509 |
| 0.622049 | 0.385199 | 22033 | 5509 |
| 0.59404 | 0.373565 | 22034 | 5508 |
| 0.62073 | 0.390563 | 22034 | 5508 |
| 0.615169 | 0.381438 | 22034 | 5508 |

| | | | |
|---|---|---|---|
| 0.654462 | 0.362783 | 22033 | 5509 |
| 0.622036 | 0.385212 | 22033 | 5509 |
| 0.593998 | 0.37361 | 22034 | 5508 |
| 0.620684 | 0.390608 | 22034 | 5508 |
| 0.615349 | 0.381257 | 22034 | 5508 |
| 0.721993 | 0.297032 | 22033 | 5509 |
| 0.693215 | 0.314863 | 22033 | 5509 |
| 0.655144 | 0.309129 | 22034 | 5508 |
| 0.692829 | 0.319775 | 22034 | 5508 |
| 0.708333 | 0.28776 | 22034 | 5508 |
| 0.652754 | 0.350751 | 27542 | 4025 |
| 0.652647 | 0.350857 | 27542 | 4025 |
| 0.706778 | 0.297017 | 27542 | 4025 |
| 0.640782 | 0.365073 | 22033 | 5509 |
| 0.631996 | 0.368885 | 22033 | 5509 |
| 0.602287 | 0.366756 | 22034 | 5508 |
| 0.652433 | 0.368382 | 22034 | 5508 |
| 0.646705 | 0.356478 | 22034 | 5508 |
| 0.640749 | 0.365105 | 22033 | 5509 |
| 0.632064 | 0.368817 | 22033 | 5509 |
| 0.602182 | 0.366866 | 22034 | 5508 |
| 0.652476 | 0.36834 | 22034 | 5508 |
| 0.646676 | 0.356507 | 22034 | 5508 |
| 0.708422 | 0.298051 | 22033 | 5509 |
| 0.702953 | 0.298027 | 22033 | 5509 |
| 0.663688 | 0.302199 | 22034 | 5508 |
| 0.727771 | 0.295448 | 22034 | 5508 |
| 0.723602 | 0.27996 | 22034 | 5508 |
| 0.511011 | 0.495216 | 27542 | 4025 |
| 0.510962 | 0.495264 | 27542 | 4025 |
| 0.572896 | 0.434084 | 27542 | 4025 |
| 0.502967 | 0.501528 | 22033 | 5509 |
| 0.50617 | 0.508701 | 22033 | 5509 |
| 0.476397 | 0.497807 | 22034 | 5508 |
| 0.495733 | 0.514627 | 22034 | 5508 |
| 0.488839 | 0.506491 | 22034 | 5508 |
| 0.502913 | 0.501581 | 22033 | 5509 |
| 0.506238 | 0.508634 | 22033 | 5509 |
| 0.476277 | 0.497933 | 22034 | 5508 |
| 0.495798 | 0.514563 | 22034 | 5508 |
| 0.488917 | 0.506412 | 22034 | 5508 |
| 0.573638 | 0.431488 | 22033 | 5509 |
| 0.584735 | 0.432444 | 22033 | 5509 |
| 0.537783 | 0.433097 | 22034 | 5508 |
| 0.572815 | 0.439156 | 22034 | 5508 |

| | | | |
|---|---|---|---|
| 0.584185 | 0.410234 | 22034 | 5508 |
| 0.894012 | 0.090368 | 27542 | 4025 |
| 0.893687 | 0.090699 | 27542 | 4025 |
| 0.906349 | 0.077816 | 27542 | 4025 |
| 0.870664 | 0.136389 | 22033 | 5509 |
| 0.874715 | 0.13402 | 22033 | 5509 |
| 0.848969 | 0.130772 | 22034 | 5508 |
| 0.90793 | 0.121087 | 22034 | 5508 |
| 0.833235 | 0.142539 | 22034 | 5508 |
| 0.870512 | 0.13654 | 22033 | 5509 |
| 0.874719 | 0.134016 | 22033 | 5509 |
| 0.848882 | 0.130861 | 22034 | 5508 |
| 0.907461 | 0.121541 | 22034 | 5508 |
| 0.833369 | 0.142401 | 22034 | 5508 |
| 0.903338 | 0.10398 | 22033 | 5509 |
| 0.911806 | 0.097299 | 22033 | 5509 |
| 0.881654 | 0.097308 | 22034 | 5508 |
| 0.934099 | 0.095755 | 22034 | 5508 |
| 0.880064 | 0.094348 | 22034 | 5508 |
| 0.897124 | 0.076107 | 27542 | 4025 |
| 0.896885 | 0.076354 | 27542 | 4025 |
| 0.904702 | 0.068304 | 27542 | 4025 |
| 0.904829 | 0.112814 | 22033 | 5509 |
| 0.867255 | 0.13141 | 22033 | 5509 |
| 0.824369 | 0.122842 | 22034 | 5508 |
| 0.97671 | 0.107942 | 22034 | 5508 |
| 0.812008 | 0.144893 | 22034 | 5508 |
| 0.904713 | 0.112928 | 22033 | 5509 |
| 0.867242 | 0.131423 | 22033 | 5509 |
| 0.824721 | 0.122468 | 22034 | 5508 |
| 0.976425 | 0.108202 | 22034 | 5508 |
| 0.812101 | 0.144795 | 22034 | 5508 |
| 0.929049 | 0.089067 | 22033 | 5509 |
| 0.907566 | 0.091038 | 22033 | 5509 |
| 0.854828 | 0.090433 | 22034 | 5508 |
| 1.004468 | 0.082591 | 22034 | 5508 |
| 0.868102 | 0.085822 | 22034 | 5508 |
| 0.862018 | 0.108707 | 27542 | 4025 |
| 0.862087 | 0.108636 | 27542 | 4025 |
| 0.882072 | 0.087972 | 27542 | 4025 |
| 0.872263 | 0.143977 | 22033 | 5509 |
| 0.860314 | 0.124984 | 22033 | 5509 |
| 0.875563 | 0.133175 | 22034 | 5508 |
| 0.851574 | 0.131682 | 22034 | 5508 |
| 0.873846 | 0.131576 | 22034 | 5508 |

| | | | |
|---|---|---|---|
| 0.872349 | 0.143893 | 22033 | 5509 |
| 0.860218 | 0.12508 | 22033 | 5509 |
| 0.875606 | 0.133133 | 22034 | 5508 |
| 0.851491 | 0.131767 | 22034 | 5508 |
| 0.873938 | 0.131485 | 22034 | 5508 |
| 0.914132 | 0.102888 | 22033 | 5509 |
| 0.888702 | 0.09611 | 22033 | 5509 |
| 0.911856 | 0.097244 | 22034 | 5508 |
| 0.881101 | 0.101574 | 22034 | 5508 |
| 0.913139 | 0.092527 | 22034 | 5508 |
| 0.442489 | 0.556275 | 27542 | 4025 |
| 0.442458 | 0.556306 | 27542 | 4025 |
| 0.503938 | 0.494654 | 27542 | 4025 |
| 0.438403 | 0.556725 | 22033 | 5509 |
| 0.439655 | 0.57347 | 22033 | 5509 |
| 0.407236 | 0.57178 | 22034 | 5508 |
| 0.419847 | 0.583431 | 22034 | 5508 |
| 0.423221 | 0.58559 | 22034 | 5508 |
| 0.438193 | 0.556937 | 22033 | 5509 |
| 0.439785 | 0.573344 | 22033 | 5509 |
| 0.40716 | 0.57186 | 22034 | 5508 |
| 0.419878 | 0.583401 | 22034 | 5508 |
| 0.423469 | 0.585347 | 22034 | 5508 |
| 0.501865 | 0.492557 | 22033 | 5509 |
| 0.519906 | 0.495615 | 22033 | 5509 |
| 0.469547 | 0.506258 | 22034 | 5508 |
| 0.497412 | 0.506472 | 22034 | 5508 |
| 0.524494 | 0.486427 | 22034 | 5508 |
| 0 | 1 | 27542 | 4025 |
| 0 | 1 | 27542 | 4025 |
| 0 | 1 | 27542 | 4025 |
| 0 | 1 | 22033 | 5509 |
| 0 | 1 | 22033 | 5509 |
| 0 | 1 | 22034 | 5508 |
| 0 | 1 | 22034 | 5508 |
| 0 | 1 | 22034 | 5508 |
| 0 | 1 | 22033 | 5509 |
| 0 | 1 | 22033 | 5509 |
| 0 | 1 | 22034 | 5508 |
| 0 | 1 | 22034 | 5508 |
| 0 | 1 | 22034 | 5508 |
| 0 | 1 | 22033 | 5509 |
| 0 | 1 | 22033 | 5509 |
| 0 | 1 | 22034 | 5508 |
| 0 | 1 | 22034 | 5508 |

| | | | |
|---|---|---|---|
| 0 | 1 | 22034 | 5508 |
| 0.857046 | 0.133716 | 27542 | 4025 |
| 0.856993 | 0.133769 | 27542 | 4025 |
| 0.90231 | 0.087964 | 27542 | 4025 |
| 0.867778 | 0.146372 | 22033 | 5509 |
| 0.873429 | 0.138491 | 22033 | 5509 |
| 0.832718 | 0.145275 | 22034 | 5508 |
| 0.847965 | 0.151631 | 22034 | 5508 |
| 0.854135 | 0.141683 | 22034 | 5508 |
| 0.867779 | 0.146372 | 22033 | 5509 |
| 0.873443 | 0.138476 | 22033 | 5509 |
| 0.832638 | 0.145357 | 22034 | 5508 |
| 0.848075 | 0.151521 | 22034 | 5508 |
| 0.854118 | 0.1417 | 22034 | 5508 |
| 0.926099 | 0.089002 | 22033 | 5509 |
| 0.923752 | 0.088854 | 22033 | 5509 |
| 0.883689 | 0.092958 | 22034 | 5508 |
| 0.910878 | 0.088688 | 22034 | 5508 |
| 0.921743 | 0.073744 | 22034 | 5508 |
| 0.857046 | 0.133716 | 27542 | 4025 |
| 0.856993 | 0.133769 | 27542 | 4025 |
| 0.90231 | 0.087964 | 27542 | 4025 |
| 0.867778 | 0.146372 | 22033 | 5509 |
| 0.873426 | 0.138491 | 22033 | 5509 |
| 0.83272 | 0.145275 | 22034 | 5508 |
| 0.847965 | 0.151631 | 22034 | 5508 |
| 0.854136 | 0.141683 | 22034 | 5508 |
| 0.867779 | 0.146371 | 22033 | 5509 |
| 0.873441 | 0.138476 | 22033 | 5509 |
| 0.83264 | 0.145357 | 22034 | 5508 |
| 0.848075 | 0.151521 | 22034 | 5508 |
| 0.85412 | 0.141699 | 22034 | 5508 |
| 0.926099 | 0.089002 | 22033 | 5509 |
| 0.92375 | 0.088854 | 22033 | 5509 |
| 0.883691 | 0.092957 | 22034 | 5508 |
| 0.910878 | 0.088688 | 22034 | 5508 |
| 0.921745 | 0.073743 | 22034 | 5508 |
| 0.457855 | 0.566254 | 27542 | 4025 |
| 0.457858 | 0.566252 | 27542 | 4025 |
| 0.531738 | 0.496262 | 27542 | 4025 |
| 0.447591 | 0.56104 | 22033 | 5509 |
| 0.435178 | 0.571054 | 22033 | 5509 |
| 0.408865 | 0.566986 | 22034 | 5508 |
| 0.426458 | 0.582671 | 22034 | 5508 |
| 0.431496 | 0.568119 | 22034 | 5508 |

| | | | |
|---|---|---|---|
| 0.447515 | 0.561114 | 22033 | 5509 |
| 0.435217 | 0.571015 | 22033 | 5509 |
| 0.408708 | 0.567152 | 22034 | 5508 |
| 0.426531 | 0.582599 | 22034 | 5508 |
| 0.431547 | 0.568068 | 22034 | 5508 |
| 0.524121 | 0.485986 | 22033 | 5509 |
| 0.514425 | 0.492942 | 22033 | 5509 |
| 0.47222 | 0.499889 | 22034 | 5508 |
| 0.504928 | 0.50588 | 22034 | 5508 |
| 0.533798 | 0.465725 | 22034 | 5508 |
| 0.599013 | 0.376062 | 27542 | 4025 |
| 0.598768 | 0.376318 | 27542 | 4025 |
| 0.628448 | 0.345402 | 27542 | 4025 |
| 0.610241 | 0.38259 | 22033 | 5509 |
| 0.62524 | 0.370689 | 22033 | 5509 |
| 0.610577 | 0.376661 | 22034 | 5508 |
| 0.634822 | 0.368999 | 22034 | 5508 |
| 0.639091 | 0.380955 | 22034 | 5508 |
| 0.61015 | 0.382682 | 22033 | 5509 |
| 0.62527 | 0.37066 | 22033 | 5509 |
| 0.610629 | 0.376608 | 22034 | 5508 |
| 0.634805 | 0.369016 | 22034 | 5508 |
| 0.639168 | 0.38088 | 22034 | 5508 |
| 0.649421 | 0.34295 | 22033 | 5509 |
| 0.665921 | 0.329744 | 22033 | 5509 |
| 0.655785 | 0.330507 | 22034 | 5508 |
| 0.676416 | 0.327655 | 22034 | 5508 |
| 0.685681 | 0.335826 | 22034 | 5508 |
