## Supplementary Table 19 for "Hundreds of cardiac MRI traits derived using 3D diffusion autoencoders share a common genetic architecture"

| Disease | Model | Sex | AUC | logOR | F1 | logOR_pan | nPatients | AUC_impro |
|---|---|---|---|---|---|---|---|---|
| angina pec | Multi PRS + | Both | 0.683656 | 1.091635 | 0.629017 | 1.111893 | 22203 | 0.342718 |
| angina pec | Multi PRS + | Female | 0.680272 | 1.203681 | 0.618051 | 1.198096 | 22203 | -0.11825 |
| angina pec | Multi PRS + | Male | 0.688198 | 1.031176 | 0.632911 | 1.00778 | 22203 | 0.364215 |
| angina pec | max(Single | Both | 0.683279 | 1.09474 | 0.627958 | 1.114817 | 22203 | 0.287265 |
| angina pec | max(Single | Female | 0.681537 | 1.206223 | 0.619522 | 1.200541 | 22203 | 0.067471 |
| angina pec | max(Single | Male | 0.687935 | 1.033736 | 0.630963 | 1.010413 | 22203 | 0.325886 |
| atheroscle | Multi PRS + | Both | 0.667748 | 0.987911 | 0.609907 | 1.017556 | 36178 | 0.213052 |
| atheroscle | Multi PRS + | Female | 0.673259 | 1.091514 | 0.605586 | 1.091503 | 36178 | 0.462193 |
| atheroscle | Multi PRS + | Male | 0.667584 | 0.932525 | 0.612617 | 0.919224 | 36178 | 0.172332 |
| atheroscle | max(Single | Both | 0.667695 | 0.984683 | 0.610891 | 1.017901 | 36178 | 0.205114 |
| atheroscle | max(Single | Female | 0.67247 | 1.085551 | 0.607279 | 1.09169 | 36178 | 0.344363 |
| atheroscle | max(Single | Male | 0.667076 | 0.922395 | 0.611087 | 0.919131 | 36178 | 0.096021 |
| conduction | Multi PRS + | Both | 0.641058 | 0.841348 | 0.593782 | 0.863028 | 9936 | 0.085811 |
| conduction | Multi PRS + | Female | 0.615715 | 0.912848 | 0.567692 | 0.93875 | 9936 | 0.343166 |
| conduction | Multi PRS + | Male | 0.658257 | 0.805046 | 0.602228 | 0.775852 | 9936 | -0.35467 |
| conduction | max(Single | Both | 0.641335 | 0.84814 | 0.593349 | 0.868009 | 9936 | 0.129162 |
| conduction | max(Single | Female | 0.615062 | 0.912211 | 0.562784 | 0.941505 | 9936 | 0.236683 |
| conduction | max(Single | Male | 0.661063 | 0.80929 | 0.602168 | 0.780765 | 9936 | 0.070163 |
| coronary h | Multi PRS + | Both | 0.666923 | 0.978838 | 0.611587 | 1.02557 | 27939 | 0.061327 |
| coronary h | Multi PRS + | Female | 0.671983 | 1.100236 | 0.609375 | 1.100517 | 27939 | -0.23228 |
| coronary h | Multi PRS + | Male | 0.664419 | 0.933187 | 0.614944 | 0.929101 | 27939 | 0.182339 |
| coronary h | max(Single | Both | 0.667308 | 0.978058 | 0.611726 | 1.024667 | 27939 | 0.118995 |
| coronary h | max(Single | Female | 0.67372 | 1.100074 | 0.613038 | 1.099931 | 27939 | 0.025676 |
| coronary h | max(Single | Male | 0.66496 | 0.927117 | 0.612458 | 0.927619 | 27939 | 0.263878 |
| heart failur | Multi PRS + | Both | 0.683615 | 1.102006 | 0.623442 | 1.063715 | 14172 | 0.084045 |
| heart failur | Multi PRS + | Female | 0.677412 | 1.227723 | 0.6144 | 1.153747 | 14172 | 0.089133 |
| heart failur | Multi PRS + | Male | 0.684585 | 1.033989 | 0.626573 | 0.960238 | 14172 | 0.030137 |
| heart failur | max(Single | Both | 0.683986 | 1.110047 | 0.62379 | 1.070789 | 14172 | 0.138258 |
| heart failur | max(Single | Female | 0.677985 | 1.230796 | 0.620479 | 1.158857 | 14172 | 0.173853 |
| heart failur | max(Single | Male | 0.68542 | 1.039793 | 0.62679 | 0.967932 | 14172 | 0.15213 |
| high choles | Multi PRS + | Both | 0.670871 | 0.965101 | 0.614889 | 1.059456 | 66734 | 0.395179 |
| high choles | Multi PRS + | Female | 0.659031 | 1.047558 | 0.604577 | 1.127432 | 66734 | 0.378951 |
| high choles | Multi PRS + | Male | 0.680752 | 0.908269 | 0.62489 | 0.964639 | 66734 | 0.446114 |
| high choles | max(Single | Both | 0.670424 | 0.96579 | 0.61456 | 1.055515 | 66734 | 0.328208 |
| high choles | max(Single | Female | 0.658578 | 1.046089 | 0.603415 | 1.122511 | 66734 | 0.309951 |
| high choles | max(Single | Male | 0.680288 | 0.902758 | 0.624088 | 0.961438 | 66734 | 0.377735 |
| hypertensi | Multi PRS + | Both | 0.705296 | 1.093214 | 0.650422 | 1.22692 | 111853 | 0.138184 |
| hypertensi | Multi PRS + | Female | 0.660502 | 1.132735 | 0.615948 | 1.295763 | 111853 | 0.154914 |
| hypertensi | Multi PRS + | Male | 0.744061 | 1.058328 | 0.680279 | 1.058928 | 111853 | 0.126793 |
| hypertensi | max(Single | Both | 0.704903 | 1.091304 | 0.650207 | 1.227404 | 111853 | 0.082403 |
| hypertensi | max(Single | Female | 0.660027 | 1.12892 | 0.614432 | 1.293389 | 111853 | 0.082848 |
| hypertensi | max(Single | Male | 0.74388 | 1.061538 | 0.681496 | 1.062636 | 111853 | 0.102515 |
| metabolic s | Multi PRS + | Both | 0.713742 | 1.104246 | 0.672851 | 1.318155 | 131187 | 0.095006 |
| metabolic s | Multi PRS + | Female | 0.667454 | 1.103737 | 0.622496 | 1.395981 | 131187 | 0.098852 |

| | | | | | | | | |
|---|---|---|---|---|---|---|---|---|
| metabolic s | Multi PRS + | Male | 0.754554 | 1.081335 | 0.718035 | 1.053237 | 131187 | 0.106237 |
| metabolic s | max(Single | Both | 0.713516 | 1.102649 | 0.672352 | 1.315676 | 131187 | 0.06325 |
| metabolic s | max(Single | Female | 0.667208 | 1.102189 | 0.623224 | 1.393753 | 131187 | 0.061949 |
| metabolic s | max(Single | Male | 0.754292 | 1.076094 | 0.718483 | 1.051337 | 131187 | 0.071578 |
| myocardial | Multi PRS + | Both | 0.677801 | 1.01908 | 0.626184 | 1.067945 | 16433 | 0.082361 |
| myocardial | Multi PRS + | Female | 0.673912 | 1.15095 | 0.615676 | 1.13831 | 16433 | 0.114416 |
| myocardial | Multi PRS + | Male | 0.679617 | 0.979751 | 0.626436 | 0.979197 | 16433 | 0.179885 |
| myocardial | max(Single | Both | 0.678715 | 1.025945 | 0.623602 | 1.066466 | 16433 | 0.217371 |
| myocardial | max(Single | Female | 0.675038 | 1.156064 | 0.612667 | 1.140336 | 16433 | 0.281632 |
| myocardial | max(Single | Male | 0.680077 | 0.979 | 0.626516 | 0.974934 | 16433 | 0.247658 |
| type 2 diab | Multi PRS + | Both | 0.784483 | 1.82675 | 0.708909 | 1.715002 | 22631 | 0.218981 |
| type 2 diab | Multi PRS + | Female | 0.791649 | 2.033735 | 0.715889 | 1.87625 | 22631 | 0.14228 |
| type 2 diab | Multi PRS + | Male | 0.775509 | 1.717789 | 0.704924 | 1.528279 | 22631 | 0.146086 |
| type 2 diab | max(Single | Both | 0.783667 | 1.825677 | 0.710577 | 1.702044 | 22631 | 0.114819 |
| type 2 diab | max(Single | Female | 0.791455 | 2.030405 | 0.72 | 1.862706 | 22631 | 0.117774 |
| type 2 diab | max(Single | Male | 0.775423 | 1.708816 | 0.704041 | 1.519782 | 22631 | 0.134949 |

| p_value_DeLong_AUC |
|---|
| 0.00025 |
| 0.00025 |
| 0.00025 |
| 0.000168 |
| 2.69E-05 |
| 0.000831 |
| 0.041117 |
| 0.041117 |
| 0.041117 |
| 0.003556 |
| 0.000196 |
| 0.017482 |
| 0.579666 |
| 0.579666 |
| 0.579666 |
| 0.099129 |
| 0.001391 |
| 0.588237 |
| 0.032875 |
| 0.032875 |
| 0.032875 |
| 0.052369 |
| 0.000115 |
| 0.052369 |
| 0.792013 |
| 0.792013 |
| 0.792013 |
| 0.010202 |
| 0.001572 |
| 0.10278 |
| 2.46E-12 |
| 2.46E-12 |
| 2.46E-12 |
| 1.19E-09 |
| 1.63E-09 |
| 1.19E-09 |
| 2.28E-13 |
| 2.28E-13 |
| 2.28E-13 |
| 3.98E-09 |
| 1.77E-10 |
| 1.81E-05 |
| 1.47E-14 |
| 1.47E-14 |

| |
|---|
| 1.47E-14 |
| 2.87E-10 |
| 1.56E-10 |
| 5.26E-09 |
| 0.96106 |
| 0.96106 |
| 0.96106 |
| 0.031595 |
| 0.000296 |
| 0.52114 |
| 7.10E-05 |
| 7.10E-05 |
| 7.10E-05 |
| 1.03E-05 |
| 4.07E-06 |
| 0.003168 |
