## Supplementary Table 18 for "Hundreds of cardiac MRI traits derived using 3D diffusion autoencoders share a common genetic architecture"

| Disease | Model | Sex | AUC | logOR | F1 | logOR_pan | nPatients | AUC_impro |
|---|---|---|---|---|---|---|---|---|
| angina pec | Multi PRS + | Both | 0.653188 | 0.877925 | 0.594635 | 0.932443 | 6879 | 0.078645 |
| angina pec | Multi PRS + | Female | 0.658933 | 0.978246 | 0.6037 | 1.012744 | 6879 | 0.084318 |
| angina pec | Multi PRS + | Male | 0.648611 | 0.829256 | 0.594334 | 0.844357 | 6879 | -0.28974 |
| angina pec | max(Single | Both | 0.654002 | 0.898529 | 0.597907 | 0.950018 | 6879 | 0.203295 |
| angina pec | max(Single | Female | 0.661309 | 0.98327 | 0.609195 | 1.02775 | 6879 | 0.445073 |
| angina pec | max(Single | Male | 0.651008 | 0.844817 | 0.590769 | 0.862808 | 6879 | 0.078653 |
| atheroscle | Multi PRS + | Both | 0.650155 | 0.903865 | 0.596737 | 0.933227 | 16845 | 0.121358 |
| atheroscle | Multi PRS + | Female | 0.653456 | 0.990748 | 0.592969 | 1.00776 | 16845 | 0.189323 |
| atheroscle | Multi PRS + | Male | 0.649153 | 0.855692 | 0.601126 | 0.847635 | 16845 | 0.125905 |
| atheroscle | max(Single | Both | 0.650394 | 0.895736 | 0.598689 | 0.940334 | 16845 | 0.158167 |
| atheroscle | max(Single | Female | 0.653889 | 0.998339 | 0.599149 | 1.013928 | 16845 | 0.255696 |
| atheroscle | max(Single | Male | 0.64918 | 0.86401 | 0.602351 | 0.855161 | 16845 | 0.130059 |
| conduction | Multi PRS + | Both | 0.649271 | 0.855187 | 0.593258 | 0.856885 | 5781 | -0.09152 |
| conduction | Multi PRS + | Female | 0.627402 | 0.940819 | 0.552 | 0.932404 | 5781 | 0.108942 |
| conduction | Multi PRS + | Male | 0.660111 | 0.79765 | 0.606019 | 0.772819 | 5781 | 0.18598 |
| conduction | max(Single | Both | 0.650293 | 0.860682 | 0.595249 | 0.866243 | 5781 | 0.065638 |
| conduction | max(Single | Female | 0.629195 | 0.940499 | 0.574386 | 0.93761 | 5781 | 0.395044 |
| conduction | max(Single | Male | 0.660145 | 0.807703 | 0.611342 | 0.782392 | 5781 | 0.191169 |
| coronary h | Multi PRS + | Both | 0.656775 | 0.914808 | 0.596314 | 0.935092 | 12832 | -0.2045 |
| coronary h | Multi PRS + | Female | 0.666427 | 0.981728 | 0.608019 | 1.006501 | 12832 | -0.07366 |
| coronary h | Multi PRS + | Male | 0.648819 | 0.875326 | 0.589751 | 0.852878 | 12832 | -0.23141 |
| coronary h | max(Single | Both | 0.658718 | 0.912781 | 0.598249 | 0.931078 | 12832 | 0.090762 |
| coronary h | max(Single | Female | 0.668311 | 0.991085 | 0.61194 | 1.00504 | 12832 | 0.20876 |
| coronary h | max(Single | Male | 0.651498 | 0.86611 | 0.591549 | 0.846263 | 12832 | 0.180456 |
| heart failur | Multi PRS + | Both | 0.69315 | 1.150783 | 0.632008 | 1.121235 | 8142 | 0.06706 |
| heart failur | Multi PRS + | Female | 0.672777 | 1.27727 | 0.617261 | 1.210131 | 8142 | 0.166435 |
| heart failur | Multi PRS + | Male | 0.696468 | 1.082493 | 0.629666 | 1.019373 | 8142 | 0.098833 |
| heart failur | max(Single | Both | 0.694657 | 1.159358 | 0.633525 | 1.129382 | 8142 | 0.28454 |
| heart failur | max(Single | Female | 0.673545 | 1.282376 | 0.621849 | 1.219411 | 8142 | 0.280914 |
| heart failur | max(Single | Male | 0.697042 | 1.093986 | 0.625984 | 1.026728 | 8142 | 0.181357 |
| high choles | Multi PRS + | Both | 0.635976 | 0.790501 | 0.581554 | 0.820462 | 16409 | 0.119617 |
| high choles | Multi PRS + | Female | 0.638859 | 0.858719 | 0.575664 | 0.887204 | 16409 | 0.165487 |
| high choles | Multi PRS + | Male | 0.632954 | 0.744783 | 0.585338 | 0.744197 | 16409 | 0.217978 |
| high choles | max(Single | Both | 0.636505 | 0.799163 | 0.583162 | 0.825473 | 16409 | 0.203001 |
| high choles | max(Single | Female | 0.639504 | 0.850368 | 0.578965 | 0.894039 | 16409 | 0.266567 |
| high choles | max(Single | Male | 0.633509 | 0.739549 | 0.582659 | 0.749361 | 16409 | 0.305992 |
| hypertensi | Multi PRS + | Both | 0.633648 | 0.752895 | 0.578715 | 0.792792 | 26663 | 0.10278 |
| hypertensi | Multi PRS + | Female | 0.63356 | 0.819843 | 0.579505 | 0.860096 | 26663 | 0.195264 |
| hypertensi | Multi PRS + | Male | 0.624109 | 0.687725 | 0.573607 | 0.716577 | 26663 | 0.079445 |
| hypertensi | max(Single | Both | 0.633637 | 0.755933 | 0.579035 | 0.791678 | 26663 | 0.101036 |
| hypertensi | max(Single | Female | 0.633641 | 0.822066 | 0.578958 | 0.859733 | 26663 | 0.208139 |
| hypertensi | max(Single | Male | 0.624778 | 0.691214 | 0.574658 | 0.714031 | 26663 | 0.186796 |
| metabolic s | Multi PRS + | Both | 0.653662 | 0.925569 | 0.595579 | 0.934545 | 45681 | 0.178768 |
| metabolic s | Multi PRS + | Female | 0.65096 | 1.008875 | 0.592467 | 1.013163 | 45681 | 0.198777 |

| | | | | | | | | | |
|---|---|---|---|---|---|---|---|---|---|
| metabolic s | Multi PRS + | Male | 0.656628 | 0.845695 | 0.600646 | 0.839428 | | 45681 | 0.163264 |
| metabolic s | max(Single | Both | 0.653578 | 0.922141 | 0.597288 | 0.939912 | | 45681 | 0.165886 |
| metabolic s | max(Single | Female | 0.650898 | 1.009457 | 0.5941 | 1.018775 | | 45681 | 0.1892 |
| metabolic s | max(Single | Male | 0.656575 | 0.847853 | 0.599029 | 0.844224 | | 45681 | 0.155267 |
| myocardial | Multi PRS + | Both | 0.657146 | 0.899038 | 0.6 | 0.935294 | | 5984 | -0.02841 |
| myocardial | Multi PRS + | Female | 0.64946 | 0.969803 | 0.598886 | 1.004504 | | 5984 | 0.010541 |
| myocardial | Multi PRS + | Male | 0.65634 | 0.86041 | 0.603214 | 0.859223 | | 5984 | 0.012039 |
| myocardial | max(Single | Both | 0.658881 | 0.940088 | 0.603478 | 0.959772 | | 5984 | 0.235548 |
| myocardial | max(Single | Female | 0.65162 | 1.0078 | 0.60989 | 1.020458 | | 5984 | 0.343164 |
| myocardial | max(Single | Male | 0.657289 | 0.8918 | 0.602757 | 0.892551 | | 5984 | 0.15673 |
| type 2 diab | Multi PRS + | Both | 0.782791 | 1.824318 | 0.713208 | 1.738506 | | 10653 | 0.320782 |
| type 2 diab | Multi PRS + | Female | 0.796825 | 2.003828 | 0.714738 | 1.89773 | | 10653 | 0.003577 |
| type 2 diab | Multi PRS + | Male | 0.777253 | 1.715181 | 0.704646 | 1.563053 | | 10653 | 0.312622 |
| type 2 diab | max(Single | Both | 0.781494 | 1.827702 | 0.713072 | 1.732996 | | 10653 | 0.154544 |
| type 2 diab | max(Single | Female | 0.797928 | 2.013697 | 0.721311 | 1.894749 | | 10653 | 0.141998 |
| type 2 diab | max(Single | Male | 0.776964 | 1.697514 | 0.704247 | 1.557491 | | 10653 | 0.275306 |

| p_value_DeLong_AUC |
|---|
| 0.722255 |
| 0.722255 |
| 0.722255 |
| 0.038777 |
| 0.002412 |
| 0.22023 |
| 0.439758 |
| 0.439758 |
| 0.439758 |
| 0.005446 |
| 0.001191 |
| 0.89906 |
| 0.672234 |
| 0.672234 |
| 0.672234 |
| 0.002137 |
| 4.29E-05 |
| 0.261268 |
| 0.395332 |
| 0.395332 |
| 0.395332 |
| 0.018025 |
| 1.18E-05 |
| 0.634325 |
| 0.635357 |
| 0.635357 |
| 0.635357 |
| 8.66E-05 |
| 8.66E-05 |
| 0.195004 |
| 0.155464 |
| 0.155464 |
| 0.155464 |
| 0.003744 |
| 0.000117 |
| 0.01928 |
| 0.49627 |
| 0.49627 |
| 0.49627 |
| 0.00746 |
| 0.000245 |
| 0.149233 |
| 0.128087 |
| 0.128087 |

| |
|---|
| 0.128087 |
| 0.001813 |
| 0.000274 |
| 0.090194 |
| 0.936144 |
| 0.936144 |
| 0.936144 |
| 0.033692 |
| 0.001329 |
| 0.779906 |
| 0.010701 |
| 0.010701 |
| 0.010701 |
| 0.001624 |
| 9.72E-05 |
| 0.210106 |
