## Supplementary Table 17 for "Hundreds of cardiac MRI traits derived using 3D diffusion autoencoders share a common genetic architecture"

| Disease | Model | Sex | AUC | logOR | F1 | logOR_pan | nPatients | AUC_impro |
|---|---|---|---|---|---|---|---|---|
| angina pec | Multi PRS + | Both | 0.712285 | 1.227109 | 0.65393 | 1.241715 | 12866 | 0.220871 |
| angina pec | Multi PRS + | Female | 0.712365 | 1.36755 | 0.629534 | 1.338712 | 12866 | 0.917568 |
| angina pec | Multi PRS + | Male | 0.713226 | 1.159588 | 0.661871 | 1.129332 | 12866 | 0.367831 |
| angina pec | max(Single | Both | 0.712373 | 1.238255 | 0.651431 | 1.256136 | 12866 | 0.23321 |
| angina pec | max(Single | Female | 0.710234 | 1.37854 | 0.646986 | 1.350688 | 12866 | 0.615713 |
| angina pec | max(Single | Male | 0.71313 | 1.163649 | 0.655284 | 1.145308 | 12866 | 0.35437 |
| atheroscler | Multi PRS + | Both | 0.698334 | 1.134687 | 0.636323 | 1.145408 | 17299 | 0.304961 |
| atheroscler | Multi PRS + | Female | 0.698224 | 1.295929 | 0.62268 | 1.241186 | 17299 | -0.03987 |
| atheroscler | Multi PRS + | Male | 0.699808 | 1.06919 | 0.641882 | 1.032761 | 17299 | 0.258976 |
| atheroscler | max(Single | Both | 0.699252 | 1.14399 | 0.637451 | 1.182802 | 17299 | 0.436826 |
| atheroscler | max(Single | Female | 0.700425 | 1.294361 | 0.635827 | 1.272737 | 17299 | 0.275275 |
| atheroscler | max(Single | Male | 0.700116 | 1.082168 | 0.637161 | 1.074078 | 17299 | 0.30307 |
| conduction | Multi PRS + | Both | 0.620354 | 0.577845 | 0.577495 | 0.600042 | 1227 | -0.33856 |
| conduction | Multi PRS + | Female | 0.563291 | 0.675824 | 0.484848 | 0.665597 | 1227 | -0.30338 |
| conduction | Multi PRS + | Male | 0.622974 | 0.527665 | 0.598802 | 0.533761 | 1227 | -1.77225 |
| conduction | max(Single | Both | 0.625879 | 0.708863 | 0.568966 | 0.696507 | 1227 | 0.549158 |
| conduction | max(Single | Female | 0.573376 | 0.770321 | 0.5375 | 0.758203 | 1227 | 1.481529 |
| conduction | max(Single | Male | 0.635892 | 0.674184 | 0.60119 | 0.631003 | 1227 | 0.26459 |
| coronary h | Multi PRS + | Both | 0.699574 | 1.144131 | 0.641787 | 1.211295 | 10135 | -0.41992 |
| coronary h | Multi PRS + | Female | 0.701868 | 1.344873 | 0.631808 | 1.298561 | 10135 | 0.009391 |
| coronary h | Multi PRS + | Male | 0.698779 | 1.089254 | 0.642088 | 1.107519 | 10135 | -0.02114 |
| coronary h | max(Single | Both | 0.704196 | 1.184713 | 0.642132 | 1.213516 | 10135 | 0.237931 |
| coronary h | max(Single | Female | 0.704204 | 1.352372 | 0.644031 | 1.30311 | 10135 | 0.342285 |
| coronary h | max(Single | Male | 0.7003 | 1.128522 | 0.644645 | 1.106976 | 10135 | 0.196434 |
| heart failur | Multi PRS + | Both | 0.691283 | 1.132814 | 0.635015 | 1.152329 | 2553 | 0.198256 |
| heart failur | Multi PRS + | Female | 0.659134 | 1.312166 | 0.611321 | 1.2607 | 2553 | 0.31264 |
| heart failur | Multi PRS + | Male | 0.706223 | 1.05938 | 0.64099 | 1.037868 | 2553 | -0.4137 |
| heart failur | max(Single | Both | 0.691842 | 1.179999 | 0.640919 | 1.187151 | 2553 | 0.279236 |
| heart failur | max(Single | Female | 0.660895 | 1.362148 | 0.635294 | 1.289857 | 2553 | 0.580616 |
| heart failur | max(Single | Male | 0.710701 | 1.09924 | 0.6498 | 1.074997 | 2553 | 0.217757 |
| high choles | Multi PRS + | Both | 0.68199 | 1.065882 | 0.621093 | 1.120644 | 43997 | 0.296227 |
| high choles | Multi PRS + | Female | 0.678944 | 1.173998 | 0.614954 | 1.210533 | 43997 | 0.510763 |
| high choles | Multi PRS + | Male | 0.684658 | 0.99185 | 0.624085 | 1.009056 | 43997 | 0.123007 |
| high choles | max(Single | Both | 0.681602 | 1.064064 | 0.620258 | 1.114815 | 43997 | 0.239156 |
| high choles | max(Single | Female | 0.677829 | 1.166465 | 0.616034 | 1.204649 | 43997 | 0.345648 |
| high choles | max(Single | Male | 0.68499 | 0.986228 | 0.624219 | 1.001549 | 43997 | 0.171534 |
| hypertensi | Multi PRS + | Both | 0.69397 | 1.14285 | 0.634406 | 1.15581 | 75251 | 0.315895 |
| hypertensi | Multi PRS + | Female | 0.6838 | 1.242317 | 0.625046 | 1.243924 | 75251 | 0.418036 |
| hypertensi | Multi PRS + | Male | 0.70396 | 1.046158 | 0.642277 | 1.040864 | 75251 | 0.223156 |
| hypertensi | max(Single | Both | 0.692932 | 1.13741 | 0.631601 | 1.151434 | 75251 | 0.165768 |
| hypertensi | max(Single | Female | 0.682401 | 1.235914 | 0.621498 | 1.240554 | 75251 | 0.212462 |
| hypertensi | max(Single | Male | 0.703641 | 1.041918 | 0.640893 | 1.034552 | 75251 | 0.177725 |
| metabolic s | Multi PRS + | Both | 0.694729 | 1.094869 | 0.64002 | 1.184883 | 91964 | 0.292914 |
| metabolic s | Multi PRS + | Female | 0.66618 | 1.177819 | 0.614536 | 1.251647 | 91964 | 0.301557 |

| | | | | | | | | |
|---|---|---|---|---|---|---|---|---|
| metabolic s | Multi PRS + | Male | 0.719154 | 1.033578 | 0.659113 | 1.07536 | 91964 | 0.106392 |
| metabolic s | max(Single | Both | 0.694138 | 1.090592 | 0.639861 | 1.179345 | 91964 | 0.207662 |
| metabolic s | max(Single | Female | 0.66506 | 1.171481 | 0.612405 | 1.246611 | 91964 | 0.132913 |
| metabolic s | max(Single | Male | 0.719536 | 1.024449 | 0.65886 | 1.06846 | 91964 | 0.159548 |
| myocardial | Multi PRS + | Both | 0.713209 | 1.21678 | 0.651862 | 1.255147 | 7838 | -0.38998 |
| myocardial | Multi PRS + | Female | 0.699891 | 1.360811 | 0.634711 | 1.340365 | 7838 | -0.40481 |
| myocardial | Multi PRS + | Male | 0.716316 | 1.184744 | 0.657724 | 1.153038 | 7838 | 0.412108 |
| myocardial | max(Single | Both | 0.718095 | 1.220882 | 0.652542 | 1.245744 | 7838 | 0.292396 |
| myocardial | max(Single | Female | 0.705668 | 1.381006 | 0.641196 | 1.332282 | 7838 | 0.417169 |
| myocardial | max(Single | Male | 0.716687 | 1.168118 | 0.658758 | 1.142294 | 7838 | 0.464032 |
| type 2 diab | Multi PRS + | Both | 0.791156 | 1.927482 | 0.714822 | 1.814177 | 8220 | 0.092882 |
| type 2 diab | Multi PRS + | Female | 0.801294 | 2.165311 | 0.725792 | 1.984158 | 8220 | 0.56661 |
| type 2 diab | Multi PRS + | Male | 0.788265 | 1.798723 | 0.70898 | 1.628797 | 8220 | 0.564605 |
| type 2 diab | max(Single | Both | 0.792128 | 1.911684 | 0.717643 | 1.791748 | 8220 | 0.215913 |
| type 2 diab | max(Single | Female | 0.798777 | 2.138391 | 0.723404 | 1.962308 | 8220 | 0.250782 |
| type 2 diab | max(Single | Male | 0.787461 | 1.785268 | 0.712621 | 1.608593 | 8220 | 0.461986 |

| p_value_DeLong_AUC |
| --- |
| 0.317073 |
| 0.317073 |
| 0.317073 |
| 0.0046 |
| 3.57E-07 |
| 0.079824 |
| 0.800627 |
| 0.800627 |
| 0.800627 |
| 0.000341 |
| 1.65E-05 |
| 0.003463 |
| 0.310946 |
| 0.310946 |
| 0.310946 |
| 0.0229 |
| 0.009158 |
| 0.119148 |
| 0.452586 |
| 0.452586 |
| 0.452586 |
| 0.000452 |
| 2.10E-06 |
| 0.027658 |
| 0.18387 |
| 0.18387 |
| 0.18387 |
| 0.021267 |
| 0.002617 |
| 0.189784 |
| 7.97E-07 |
| 7.97E-07 |
| 7.97E-07 |
| 4.00E-07 |
| 1.41E-07 |
| 4.00E-07 |
| 1.78E-14 |
| 1.78E-14 |
| 1.78E-14 |
| 6.11E-08 |
| 1.57E-09 |
| 4.51E-05 |
| 1.74E-18 |
| 1.74E-18 |

| |
|---|
| 1.74E-18 |
| 7.70E-12 |
| 5.05E-13 |
| 6.47E-09 |
| 0.982224 |
| 0.982224 |
| 0.982224 |
| 0.068681 |
| 0.00016 |
| 0.206881 |
| 0.009041 |
| 0.009041 |
| 0.009041 |
| 0.000252 |
| 0.000375 |
| 0.035106 |
