## Supplementary Table 16 for "Hundreds of cardiac MRI traits derived using 3D diffusion autoencoders share a common genetic architecture"

| CHROM | GENPOS | A1FREQ | N | TEST | BETA | SE | CHISQ | P |
|---|---|---|---|---|---|---|---|---|
| 1 | 46930031 | 0.002418 | 46113 | ADD | 0.313392 | 0.057832 | 29.3656 | 5.99E-08 |
| 3 | 9890723 | 1.08E-05 | 46113 | ADD | 4.91382 | 0.903584 | 29.5734 | 5.38E-08 |
| 3 | 9890723 | 1.08E-05 | 46113 | ADD | -4.77162 | 0.726198 | 43.1739 | 5.01E-11 |
| 3 | 9890723 | 1.08E-05 | 46113 | ADD | 4.76643 | 0.73157 | 42.4497 | 7.25E-11 |
| 3 | 9890723 | 1.08E-05 | 46113 | ADD | -4.81218 | 0.788664 | 37.2306 | 1.05E-09 |
| 3 | 9890723 | 1.08E-05 | 46113 | ADD | -4.87921 | 0.786243 | 38.5111 | 5.44E-10 |
| 3 | 9890723 | 1.08E-05 | 46113 | ADD | 4.91465 | 0.804892 | 37.2829 | 1.02E-09 |
| 3 | 9890723 | 1.08E-05 | 46113 | ADD | -4.81038 | 0.738163 | 42.4674 | 7.19E-11 |
| 4 | 1.65E+08 | 1.08E-05 | 46113 | ADD | 3.92658 | 0.700049 | 31.461 | 2.03E-08 |
| 4 | 1.65E+08 | 1.08E-05 | 46113 | ADD | 3.92658 | 0.700049 | 31.461 | 2.03E-08 |
| 5 | 1.52E+08 | 3.25E-05 | 46113 | ADD | 2.6096 | 0.440342 | 35.1212 | 3.10E-09 |
| 11 | 27498652 | 1.08E-05 | 46109 | ADD | -4.13414 | 0.779273 | 28.1444 | 1.13E-07 |
| 11 | 1.32E+08 | 3.25E-05 | 46113 | ADD | 2.44091 | 0.429315 | 32.3259 | 1.30E-08 |
| 15 | 39801133 | 1.08E-05 | 46111 | ADD | -3.56959 | 0.660786 | 29.182 | 6.59E-08 |
| 15 | 39801133 | 1.08E-05 | 46111 | ADD | -3.56959 | 0.660786 | 29.182 | 6.59E-08 |
| 16 | 1911590 | 1.08E-05 | 46113 | ADD | -3.67061 | 0.664316 | 30.53 | 3.29E-08 |

| LOG10P | mask_nam | AAF_cutoff | gene_nam( | gene_id | Latent |
|---|---|---|---|---|---|
| 7.22234 | M3 | 0.01 | CYP4A11 | ENSG0000( | Z74_S4 |
| 7.26892 | M1 | 0.01 | JAGN1 | ENSG0000( | Z53_S2 |
| 10.3003 | M1 | 0.01 | JAGN1 | ENSG0000( | Z56_S5 |
| 10.1395 | M1 | 0.01 | JAGN1 | ENSG0000( | Z60_S2 |
| 8.979 | M1 | 0.01 | JAGN1 | ENSG0000( | Z70_S5 |
| 9.26405 | M1 | 0.01 | JAGN1 | ENSG0000( | Z99_S5 |
| 8.99066 | M1 | 0.01 | JAGN1 | ENSG0000( | Z92_S1 |
| 10.1434 | M1 | 0.01 | JAGN1 | ENSG0000( | Z122_S5 |
| 7.69146 | M1 | 0.01 | FAM218A | ENSG0000( | Z44_S4 |
| 7.69146 | M2 | 0.01 | FAM218A | ENSG0000( | Z44_S4 |
| 8.5089 | M3 | 0.01 | ATOX1 | ENSG0000( | Z89_S4 |
| 6.94848 | M1 | 0.01 | LIN7C | ENSG0000( | Z3_S2 |
| 7.88483 | M1 | 0.01 | OPCML | ENSG0000( | Z57_S1 |
| 7.18118 | M1 | 0.01 | AC023908. | ENSG0000( | Z31_S5 |
| 7.18118 | M1 | 0.01 | GPR176 | ENSG0000( | Z31_S5 |
| 7.48315 | M1 | 0.01 | HS3ST6 | ENSG0000( | Z1_S2 |
