## Supplementary Table 15 for "Hundreds of cardiac MRI traits derived using 3D diffusion autoencoders share a common genetic architecture"

| CHROM | GENPOS | N | TEST | CHISQ | P | LOG10P | mask_nam | AAF_cutoff |
|---|---|---|---|---|---|---|---|---|
| 1 | 46930031 | 46134 | ADD-SKAT( | 25.7677 | 3.85E-07 | 6.41444 | M3 | 0.01 |
| 1 | 92080572 | 46134 | ADD-SKAT( | 26.8252 | 2.23E-07 | 6.65224 | M2 | 0.01 |
| 1 | 92080572 | 46134 | ADD-SKAT( | 32.1853 | 1.40E-08 | 7.85341 | M2 | 0.01 |
| 1 | 92080572 | 46134 | ADD-SKAT( | 27.4589 | 1.60E-07 | 6.79461 | M2 | 0.01 |
| 1 | 92080572 | 46134 | ADD-SKAT( | 30.9339 | 2.67E-08 | 7.57355 | M2 | 0.01 |
| 1 | 92167859 | 46134 | ADD-SKAT( | 28.0321 | 1.19E-07 | 6.92328 | M2 | 0.01 |
| 1 | 92167859 | 46134 | ADD-SKAT( | 32.973 | 9.34E-09 | 8.02944 | M2 | 0.01 |
| 1 | 92167859 | 46134 | ADD-SKAT( | 27.6807 | 1.43E-07 | 6.8444 | M2 | 0.01 |
| 1 | 92167859 | 46134 | ADD-SKAT( | 31.3075 | 2.20E-08 | 7.65713 | M2 | 0.01 |
| 1 | 1.51E+08 | 46134 | ADD-SKAT( | 26.8284 | 2.22E-07 | 6.65298 | M1 | 0.01 |
| 1 | 1.53E+08 | 46134 | ADD-SKAT( | 26.2858 | 2.94E-07 | 6.531 | M1 | 0.01 |
| 1 | 1.53E+08 | 46134 | ADD-SKAT( | 26.2858 | 2.94E-07 | 6.531 | M2 | 0.01 |
| 3 | 9890723 | 46134 | ADD-SKAT( | 29.5734 | 5.38E-08 | 7.26892 | M1 | 0.01 |
| 3 | 9890723 | 46134 | ADD-SKAT( | 43.1739 | 5.01E-11 | 10.3003 | M1 | 0.01 |
| 3 | 9890723 | 46134 | ADD-SKAT( | 42.4497 | 7.25E-11 | 10.1395 | M1 | 0.01 |
| 3 | 9890723 | 46134 | ADD-SKAT( | 37.2306 | 1.05E-09 | 8.979 | M1 | 0.01 |
| 3 | 9890723 | 46134 | ADD-SKAT( | 38.5111 | 5.44E-10 | 9.26405 | M1 | 0.01 |
| 3 | 9890723 | 46134 | ADD-SKAT( | 37.2829 | 1.02E-09 | 8.99066 | M1 | 0.01 |
| 3 | 9890723 | 46134 | ADD-SKAT( | 42.4674 | 7.19E-11 | 10.1434 | M1 | 0.01 |
| 3 | 1.7E+08 | 46134 | ADD-SKAT( | 25.4337 | 4.58E-07 | 6.33928 | M3 | 0.01 |
| 3 | 1.7E+08 | 46134 | ADD-SKAT( | 25.8758 | 3.64E-07 | 6.43877 | M3 | 0.01 |
| 4 | 1.65E+08 | 46134 | ADD-SKAT( | 31.461 | 2.03E-08 | 7.69146 | M1 | 0.01 |
| 4 | 1.65E+08 | 46134 | ADD-SKAT( | 31.461 | 2.03E-08 | 7.69146 | M2 | 0.01 |
| 4 | 1.65E+08 | 46134 | ADD-SKAT( | 26.1071 | 3.23E-07 | 6.49081 | M1 | 0.01 |
| 4 | 1.65E+08 | 46134 | ADD-SKAT( | 26.1071 | 3.23E-07 | 6.49081 | M2 | 0.01 |
| 4 | 1.65E+08 | 46134 | ADD-SKAT( | 26.2732 | 2.96E-07 | 6.52817 | M1 | 0.01 |
| 4 | 1.65E+08 | 46134 | ADD-SKAT( | 26.2732 | 2.96E-07 | 6.52817 | M2 | 0.01 |
| 5 | 1.52E+08 | 46134 | ADD-SKAT( | 35.1212 | 3.10E-09 | 8.5089 | M3 | 0.01 |
| 5 | 1.71E+08 | 46134 | ADD-SKAT( | 25.8634 | 3.66E-07 | 6.43598 | M1 | 0.01 |
| 11 | 27498652 | 46134 | ADD-SKAT( | 28.1444 | 1.13E-07 | 6.94848 | M1 | 0.01 |
| 11 | 1.04E+08 | 46134 | ADD-SKAT( | 27.3797 | 1.67E-07 | 6.77683 | M1 | 0.01 |
| 11 | 1.18E+08 | 46134 | ADD-SKAT( | 26.6351 | 2.46E-07 | 6.60952 | M3 | 0.01 |
| 11 | 1.32E+08 | 46134 | ADD-SKAT( | 26.5923 | 2.51E-07 | 6.59992 | M1 | 0.01 |
| 11 | 1.32E+08 | 46134 | ADD-SKAT( | 32.3259 | 1.30E-08 | 7.88483 | M1 | 0.01 |
| 15 | 39801133 | 46134 | ADD-SKAT( | 29.182 | 6.59E-08 | 7.18118 | M1 | 0.01 |
| 15 | 39801133 | 46134 | ADD-SKAT( | 29.182 | 6.59E-08 | 7.18118 | M1 | 0.01 |
| 16 | 1911590 | 46134 | ADD-SKAT( | 26.5572 | 2.56E-07 | 6.59201 | M1 | 0.01 |
| 16 | 1911590 | 46134 | ADD-SKAT( | 30.53 | 3.29E-08 | 7.48315 | M1 | 0.01 |

| gene_nam( | gene_id | Latent |
|---|---|---|
| CYP4A11 | ENSG0000( | Z74_S4 |
| BTBD8 | ENSG0000( | Z122_S2 |
| BTBD8 | ENSG0000( | Z73_S1 |
| BTBD8 | ENSG0000( | Z78_S1 |
| BTBD8 | ENSG0000( | Z13_S1 |
| KIAA1107 | ENSG0000( | Z122_S2 |
| KIAA1107 | ENSG0000( | Z73_S1 |
| KIAA1107 | ENSG0000( | Z78_S1 |
| KIAA1107 | ENSG0000( | Z13_S1 |
| PRUNE1 | ENSG0000( | Z62_S4 |
| SMCP | ENSG0000( | Z37_S2 |
| SMCP | ENSG0000( | Z37_S2 |
| JAGN1 | ENSG0000( | Z53_S2 |
| JAGN1 | ENSG0000( | Z56_S5 |
| JAGN1 | ENSG0000( | Z60_S2 |
| JAGN1 | ENSG0000( | Z70_S5 |
| JAGN1 | ENSG0000( | Z99_S5 |
| JAGN1 | ENSG0000( | Z92_S1 |
| JAGN1 | ENSG0000( | Z122_S5 |
| PHC3 | ENSG0000( | Z24_S5 |
| PHC3 | ENSG0000( | Z111_S1 |
| FAM218A | ENSG0000( | Z44_S4 |
| FAM218A | ENSG0000( | Z44_S4 |
| FAM218A | ENSG0000( | Z80_S4 |
| FAM218A | ENSG0000( | Z80_S4 |
| FAM218A | ENSG0000( | Z57_S1 |
| FAM218A | ENSG0000( | Z57_S1 |
| ATOX1 | ENSG0000( | Z89_S4 |
| TLX3 | ENSG0000( | Z118_S1 |
| LIN7C | ENSG0000( | Z3_S2 |
| DDI1 | ENSG0000( | Z1_S4 |
| ATP5L | ENSG0000( | Z53_S1 |
| OPCML | ENSG0000( | Z73_S1 |
| OPCML | ENSG0000( | Z57_S1 |
| AC023908. | ENSG0000( | Z31_S5 |
| GPR176 | ENSG0000( | Z31_S5 |
| HS3ST6 | ENSG0000( | Z20_S2 |
| HS3ST6 | ENSG0000( | Z1_S2 |
