## Supplementary Table 14 for "Hundreds of cardiac MRI traits derived using 3D diffusion autoencoders share a common genetic architecture"

| t1_study_i | hit1 | t2_study_i | hit2 | nsnps | PP.H0.abf | PP.H1.abf | PP.H2.abf | PP.H3.abf |
|---|---|---|---|---|---|---|---|---|
| S1993_Z97 | chr1:11897 | AdiposeVIS | chr1:11900 | 503 | 1.18E-12 | 1.18E-07 | 3.79E-08 | 0.002796 |
| S1993_Z97 | chr1:11897 | ArteryTIB | chr1:11900 | 495 | 4.12E-16 | 7.81E-06 | 2.27E-13 | 0.003306 |
| S1993_Z97 | chr1:11897 | WHRadjBN | chr1:11898 | 449 | 1.31E-35 | 1.77E-31 | 1.16E-05 | 0.155001 |
| S1701_Z82 | chr1:20766 | PulseRate | chr1:20781 | 574 | 1.30E-34 | 1.50E-31 | 3.16E-05 | 0.035417 |
| S42_Z10 | chr1:20795 | AdiposeSU | chr1:20796 | 387 | 5.33E-55 | 3.63E-06 | 5.17E-51 | 0.034318 |
| S42_Z10 | chr1:20795 | AdiposeVIS | chr1:20795 | 386 | 7.34E-34 | 3.57E-06 | 7.13E-30 | 0.03373 |
| S42_Z10 | chr1:20795 | ArteryAOR | chr1:20796 | 388 | 8.68E-26 | 2.32E-06 | 8.43E-22 | 0.021514 |
| S42_Z10 | chr1:20795 | ArteryCOR | chr1:20795 | 351 | 1.04E-12 | 3.56E-06 | 1.01E-08 | 0.033651 |
| S42_Z10 | chr1:20795 | ArteryTIB | chr1:20796 | 389 | 1.29E-24 | 3.93E-06 | 1.25E-20 | 0.037219 |
| S42_Z10 | chr1:20795 | HeartAA | chr1:20795 | 392 | 5.31E-45 | 3.85E-06 | 5.15E-41 | 0.036467 |
| S42_Z10 | chr1:20795 | HeartLV | chr1:20795 | 387 | 5.69E-29 | 3.72E-06 | 5.53E-25 | 0.035201 |
| S42_Z10 | chr1:20795 | Liver | chr1:20792 | 327 | 1.39E-07 | 4.73E-06 | 0.001261 | 0.042022 |
| S42_Z10 | chr1:20795 | Lung | chr1:20795 | 384 | 1.71E-23 | 3.58E-06 | 1.63E-19 | 0.03297 |
| S42_Z10 | chr1:20795 | MuscleSKL | chr1:20795 | 386 | 5.28E-75 | 3.53E-06 | 5.12E-71 | 0.033362 |
| S42_Z10 | chr1:20795 | Pancreas | chr1:20795 | 387 | 1.20E-18 | 3.56E-06 | 1.16E-14 | 0.033599 |
| S42_Z10 | chr1:20795 | PulseRate | chr1:20795 | 438 | 6.91E-29 | 7.27E-25 | 3.74E-06 | 0.038319 |
| S2023_Z86 | chr2:12736 | AdiposeSU | chr2:12717 | 417 | 1.55E-09 | 1.07E-05 | 1.78E-05 | 0.121925 |
| S2023_Z86 | chr2:12736 | HeelBMD | chr2:12769 | 451 | 4.23E-09 | 4.87E-05 | 1.36E-05 | 0.156103 |
| S1701_Z29 | chr2:18540 | S1994_Z5 | chr2:18541 | 371 | 4.31E-12 | 3.84E-07 | 5.11E-08 | 0.003558 |
| S1701_Z53 | chr2:27508 | CVD | chr2:27508 | 495 | 1.34E-14 | 3.06E-09 | 1.05E-08 | 0.001405 |
| S1701_Z53 | chr2:27508 | LiverFAT | chr2:27531 | 330 | 1.10E-09 | 0.000251 | 2.18E-08 | 0.003967 |
| S1701_Z53 | chr2:27508 | LiverVOL | chr2:27508 | 495 | 9.11E-21 | 2.08E-15 | 9.16E-09 | 0.001093 |
| S1701_Z53 | chr2:27508 | PulseRate | chr2:27543 | 493 | 1.05E-19 | 2.62E-15 | 8.17E-07 | 0.019371 |
| S1701_Z53 | chr2:27508 | Triglyceride | chr2:27508 | 493 | 1.97E-106 | 4.50E-101 | 8.87E-09 | 0.001027 |
| S1701_Z82 | chr2:17885 | LV_endDia | chr2:17892 | 330 | 0.000234 | 0.001692 | 0.000691 | 0.00401 |
| S1701_Z82 | chr2:17885 | RA_minVol | chr2:17890 | 373 | 0.000157 | 0.00114 | 0.000917 | 0.005656 |
| S1701_Z29 | chr2:21943 | MuscleSKL | chr2:21943 | 159 | 2.73E-14 | 4.19E-11 | 6.55E-07 | 2.80E-06 |
| S1701_Z29 | chr2:21943 | PulseRate | chr2:21943 | 235 | 3.81E-25 | 5.84E-22 | 6.53E-07 | 8.54E-07 |
| S1994_Z11 | chr2:23138 | ArteryAOR | chr2:23139 | 300 | 3.14E-33 | 2.02E-05 | 9.00E-31 | 0.004812 |
| S1994_Z11 | chr2:23138 | ArteryTIB | chr2:23142 | 302 | 1.77E-10 | 0.000367 | 5.07E-08 | 0.10434 |
| S1994_Z11 | chr2:23138 | HeartAA | chr2:23140 | 299 | 6.73E-08 | 1.93E-05 | 0.000163 | 0.04589 |
| S1994_Z11 | chr2:23138 | PulseRate | chr2:23140 | 392 | 1.23E-21 | 3.83E-19 | 2.43E-05 | 0.006561 |
| S1994_Z11 | chr2:23138 | S1994_Z31 | chr2:23140 | 391 | 8.16E-06 | 0.004814 | 1.24E-05 | 0.00632 |
| S1994_Z31 | chr2:23140 | HeartAA | chr2:23140 | 266 | 2.22E-05 | 2.93E-05 | 0.053897 | 0.070271 |
| S2023_Z77 | chr3:15856 | DBP | chr3:15874 | 1139 | 1.01E-18 | 2.96E-15 | 2.13E-05 | 0.061257 |
| S2023_Z77 | chr3:15856 | DBP | chr3:15822 | 1094 | 2.02E-28 | 5.91E-25 | 1.94E-05 | 0.05576 |
| S1994_Z12 | chr3:17945 | S1994_Z35 | chr3:17945 | 507 | 6.46E-09 | 3.53E-06 | 3.82E-06 | 0.001089 |
| S1994_Z12 | chr3:17945 | S1994_Z94 | chr3:17945 | 538 | 4.98E-09 | 2.73E-06 | 3.77E-06 | 0.001066 |
| S1994_Z35 | chr3:17945 | S1994_Z94 | chr3:17945 | 507 | 4.67E-09 | 2.76E-06 | 3.54E-06 | 0.001089 |
| S1994_Z35 | chr3:17945 | S42_Z105 | chr3:17945 | 487 | 2.86E-09 | 1.69E-06 | 3.90E-06 | 0.001304 |
| S1994_Z94 | chr3:17945 | S42_Z105 | chr3:17945 | 487 | 2.20E-09 | 1.67E-06 | 3.00E-06 | 0.001278 |
| S1994_Z12 | chr3:17945 | S42_Z105 | chr3:17945 | 487 | 3.05E-09 | 1.67E-06 | 4.16E-06 | 0.001278 |
| S42_Z105 | chr3:17945 | S42_Z106 | chr3:17945 | 487 | 3.04E-09 | 4.15E-06 | 1.69E-06 | 0.001307 |
| S1994_Z12 | chr3:17945 | S42_Z106 | chr3:17945 | 526 | 6.87E-09 | 3.76E-06 | 3.82E-06 | 0.001091 |

| | | | | | | | | |
|---|---|---|---|---|---|---|---|---|
| S1994_Z35 | chr3:17945 | S42_Z106 | chr3:17945 | 491 | 6.44E-09 | 3.80E-06 | 3.58E-06 | 0.001115 |
| S1994_Z94 | chr3:17945 | S42_Z106 | chr3:17945 | 522 | 4.97E-09 | 3.76E-06 | 2.76E-06 | 0.001093 |
| S1994_Z35 | chr3:17945 | S42_Z117 | chr3:17945 | 489 | 2.73E-10 | 1.61E-07 | 3.62E-06 | 0.001142 |
| S1994_Z94 | chr3:17945 | S42_Z117 | chr3:17945 | 520 | 2.10E-10 | 1.59E-07 | 2.79E-06 | 0.001117 |
| S1994_Z12 | chr3:17945 | S42_Z117 | chr3:17945 | 524 | 2.91E-10 | 1.59E-07 | 3.86E-06 | 0.001115 |
| S42_Z106 | chr3:17945 | S42_Z117 | chr3:17945 | 524 | 2.90E-10 | 1.61E-07 | 3.85E-06 | 0.001142 |
| S42_Z105 | chr3:17945 | S42_Z117 | chr3:17945 | 487 | 1.27E-10 | 1.74E-07 | 1.69E-06 | 0.001306 |
| S1994_Z94 | chr3:17945 | S42_Z12 | chr3:17945 | 520 | 1.68E-10 | 1.28E-07 | 2.74E-06 | 0.001079 |
| S1994_Z35 | chr3:17945 | S42_Z12 | chr3:17945 | 489 | 2.19E-10 | 1.29E-07 | 3.56E-06 | 0.001107 |
| S1994_Z12 | chr3:17945 | S42_Z12 | chr3:17945 | 524 | 2.33E-10 | 1.27E-07 | 3.79E-06 | 0.001076 |
| S42_Z106 | chr3:17945 | S42_Z12 | chr3:17945 | 524 | 2.32E-10 | 1.29E-07 | 3.78E-06 | 0.001103 |
| S42_Z117 | chr3:17945 | S42_Z12 | chr3:17945 | 524 | 9.78E-12 | 1.30E-07 | 1.59E-07 | 0.001115 |
| S42_Z105 | chr3:17945 | S42_Z12 | chr3:17945 | 487 | 1.02E-10 | 1.39E-07 | 1.66E-06 | 0.001268 |
| S42_Z117 | chr3:17945 | S42_Z4 | chr3:17945 | 524 | 3.04E-11 | 4.04E-07 | 1.67E-07 | 0.001219 |
| S42_Z12 | chr3:17945 | S42_Z4 | chr3:17945 | 524 | 2.44E-11 | 3.97E-07 | 1.34E-07 | 0.001179 |
| S1994_Z35 | chr3:17945 | S42_Z4 | chr3:17945 | 489 | 6.81E-10 | 4.02E-07 | 3.74E-06 | 0.001209 |
| S1994_Z94 | chr3:17945 | S42_Z4 | chr3:17945 | 520 | 5.25E-10 | 3.98E-07 | 2.88E-06 | 0.001184 |
| S1994_Z12 | chr3:17945 | S42_Z4 | chr3:17945 | 524 | 7.27E-10 | 3.98E-07 | 3.99E-06 | 0.001183 |
| S42_Z106 | chr3:17945 | S42_Z4 | chr3:17945 | 524 | 7.24E-10 | 4.03E-07 | 3.98E-06 | 0.001211 |
| S42_Z105 | chr3:17945 | S42_Z4 | chr3:17945 | 487 | 3.15E-10 | 4.30E-07 | 1.73E-06 | 0.00136 |
| S42_Z12 | chr3:17945 | S42_Z51 | chr3:17945 | 524 | 9.86E-11 | 1.60E-06 | 1.29E-07 | 0.001096 |
| S1994_Z94 | chr3:17945 | S42_Z51 | chr3:17945 | 538 | 2.12E-09 | 1.61E-06 | 2.77E-06 | 0.001099 |
| S42_Z106 | chr3:17945 | S42_Z51 | chr3:17945 | 526 | 2.93E-09 | 1.63E-06 | 3.82E-06 | 0.001124 |
| S1994_Z12 | chr3:17945 | S42_Z51 | chr3:17945 | 542 | 2.93E-09 | 1.60E-06 | 3.83E-06 | 0.001097 |
| S42_Z4 | chr3:17945 | S42_Z51 | chr3:17945 | 524 | 3.07E-10 | 1.69E-06 | 4.01E-07 | 0.001202 |
| S1994_Z35 | chr3:17945 | S42_Z51 | chr3:17945 | 507 | 2.75E-09 | 1.63E-06 | 3.60E-06 | 0.001126 |
| S42_Z117 | chr3:17945 | S42_Z51 | chr3:17945 | 524 | 1.23E-10 | 1.64E-06 | 1.61E-07 | 0.001138 |
| S42_Z105 | chr3:17945 | S42_Z51 | chr3:17945 | 487 | 1.28E-09 | 1.75E-06 | 1.68E-06 | 0.001291 |
| S42_Z4 | chr3:17945 | S42_Z89 | chr3:17945 | 524 | 4.49E-10 | 2.46E-06 | 4.03E-07 | 0.001213 |
| S1994_Z94 | chr3:17945 | S42_Z89 | chr3:17945 | 538 | 3.10E-09 | 2.35E-06 | 2.78E-06 | 0.001108 |
| S42_Z105 | chr3:17945 | S42_Z89 | chr3:17945 | 487 | 1.88E-09 | 2.56E-06 | 1.69E-06 | 0.001301 |
| S42_Z12 | chr3:17945 | S42_Z89 | chr3:17945 | 524 | 1.44E-10 | 2.35E-06 | 1.29E-07 | 0.001108 |
| S42_Z106 | chr3:17945 | S42_Z89 | chr3:17945 | 526 | 4.27E-09 | 2.37E-06 | 3.84E-06 | 0.001134 |
| S1994_Z12 | chr3:17945 | S42_Z89 | chr3:17945 | 542 | 4.29E-09 | 2.34E-06 | 3.85E-06 | 0.001106 |
| S42_Z117 | chr3:17945 | S42_Z89 | chr3:17945 | 524 | 1.80E-10 | 2.39E-06 | 1.62E-07 | 0.001148 |
| S42_Z51 | chr3:17945 | S42_Z89 | chr3:17945 | 542 | 1.82E-09 | 2.37E-06 | 1.63E-06 | 0.00113 |
| S1994_Z35 | chr3:17945 | S42_Z89 | chr3:17945 | 507 | 4.02E-09 | 2.38E-06 | 3.61E-06 | 0.001134 |
| S2023_Z95 | chr4:78503 | LungFunc | chr4:78771 | 742 | 9.01E-08 | 8.77E-05 | 3.79E-05 | 0.035885 |
| S2023_Z86 | chr4:17954 | ArteryCOR | chr4:17858 | 303 | 2.48E-07 | 5.93E-05 | 0.00019 | 0.044537 |
| S1993_Z14 | chr4:17954 | ArteryCOR | chr4:17858 | 303 | 5.52E-07 | 0.000132 | 0.00019 | 0.044421 |
| S1993_Z14 | chr4:17954 | Bld | chr4:17858 | 338 | 1.02E-05 | 0.000104 | 0.003501 | 0.034786 |
| S2023_Z86 | chr4:17954 | Bld | chr4:17858 | 338 | 4.57E-06 | 4.67E-05 | 0.003509 | 0.034875 |
| S1993_Z14 | chr4:17954 | HeelBMD | chr4:17916 | 376 | 3.63E-09 | 5.93E-06 | 8.75E-06 | 0.013305 |
| S2023_Z86 | chr4:17954 | HeelBMD | chr4:17916 | 376 | 1.48E-09 | 5.59E-06 | 3.57E-06 | 0.012483 |
| S1993_Z14 | chr4:17954 | Hypertensi | chr4:17852 | 43 | 7.51E-06 | 0.009671 | 6.37E-05 | 0.081212 |

| | | | | | | | | |
|---|---|---|---|---|---|---|---|---|
| S2023_Z86 | chr4:17954 | Hypertensi | chr4:17852 | 43 | 2.90E-06 | 0.008711 | 2.46E-05 | 0.073051 |
| S1993_Z14 | chr4:17954 | MuscleSKL | chr4:17858 | 353 | 1.32E-05 | 0.004539 | 8.91E-05 | 0.029688 |
| S2023_Z86 | chr4:17954 | MuscleSKL | chr4:17858 | 353 | 5.90E-06 | 0.004528 | 3.98E-05 | 0.029615 |
| S1993_Z14 | chr4:17954 | PP | chr4:18006 | 340 | 9.61E-11 | 1.57E-07 | 5.93E-06 | 0.008692 |
| S2023_Z86 | chr4:17954 | PP | chr4:18006 | 340 | 4.33E-11 | 1.63E-07 | 2.67E-06 | 0.00909 |
| S1993_Z14 | chr4:17954 | S2023_Z86 | chr4:17954 | 547 | 1.12E-09 | 2.27E-06 | 5.40E-06 | 0.00999 |
| S1701_Z11 | chr5:56513 | CVD | chr5:56512 | 441 | 4.09E-10 | 3.43E-07 | 2.54E-06 | 0.001131 |
| S1701_Z11 | chr5:56513 | HDLcholest | chr5:56512 | 419 | 1.37E-08 | 1.15E-05 | 2.45E-06 | 0.001062 |
| S1701_Z11 | chr5:56513 | Hypertensi | chr5:56521 | 40 | 5.95E-05 | 0.001105 | 5.38E-05 | 5.64E-07 |
| S1701_Z11 | chr5:56513 | PP | chr5:56511 | 404 | 6.94E-10 | 5.83E-07 | 2.45E-06 | 0.00106 |
| S1701_Z11 | chr5:56513 | Triglyceride | chr5:56512 | 441 | 1.51E-19 | 1.27E-16 | 2.45E-06 | 0.001056 |
| S1994_Z1 | chr5:13310 | AdiposeSU | chr5:13310 | 298 | 2.64E-17 | 1.33E-06 | 2.03E-14 | 2.03E-05 |
| S1994_Z1 | chr5:13310 | ArteryTIB | chr5:13310 | 314 | 2.54E-09 | 1.48E-06 | 1.95E-06 | 0.000137 |
| S1994_Z1 | chr5:13310 | Bld | chr5:13310 | 325 | 6.82E-16 | 1.32E-06 | 5.25E-13 | 1.75E-05 |
| S1994_Z1 | chr5:13310 | CVD | chr5:13310 | 416 | 3.91E-13 | 3.01E-10 | 1.35E-06 | 3.85E-05 |
| S1994_Z1 | chr5:13310 | DescAorta_ | chr5:13309 | 489 | 2.55E-08 | 3.59E-05 | 2.20E-06 | 0.002091 |
| S1994_Z1 | chr5:13310 | Lung | chr5:13306 | 324 | 7.33E-08 | 6.73E-06 | 5.64E-05 | 0.004184 |
| S1994_Z1 | chr5:13310 | MuscleSKL | chr5:13310 | 314 | 4.03E-08 | 3.10E-05 | 2.53E-06 | 0.000951 |
| S1701_Z11 | chr6:24788 | HeelBMD | chr6:24928 | 388 | 2.42E-33 | 1.35E-29 | 3.98E-06 | 0.021109 |
| S1701_Z11 | chr6:24788 | S1993_Z28 | chr6:25029 | 513 | 1.97E-09 | 1.31E-05 | 1.98E-06 | 0.012147 |
| S1701_Z11 | chr6:24788 | S1993_Z6 | chr6:25076 | 513 | 1.31E-10 | 8.66E-07 | 2.69E-06 | 0.016851 |
| S1701_Z11 | chr6:24788 | S1993_Z97 | chr6:24913 | 513 | 3.46E-12 | 2.29E-08 | 2.00E-06 | 0.012252 |
| S1701_Z11 | chr6:24788 | S1994_Z10 | chr6:25029 | 513 | 4.97E-11 | 3.29E-07 | 2.00E-06 | 0.012242 |
| S1701_Z11 | chr6:24788 | S2023_Z44 | chr6:25029 | 513 | 2.09E-10 | 1.38E-06 | 1.98E-06 | 0.012104 |
| S1701_Z11 | chr6:24788 | S2023_Z95 | chr6:25076 | 513 | 7.53E-11 | 4.99E-07 | 4.04E-06 | 0.025764 |
| S1993_Z97 | chr6:24913 | HeelBMD | chr6:24928 | 399 | 2.25E-35 | 1.06E-29 | 3.69E-08 | 0.016392 |
| S1993_Z97 | chr6:24913 | S1994_Z10 | chr6:25029 | 515 | 5.39E-13 | 3.11E-07 | 2.17E-08 | 0.011512 |
| S1993_Z97 | chr6:24913 | S2023_Z44 | chr6:25029 | 513 | 2.15E-12 | 1.24E-06 | 2.04E-08 | 0.010765 |
| S1993_Z97 | chr6:24913 | S2023_Z95 | chr6:25076 | 515 | 1.75E-12 | 1.01E-06 | 9.40E-08 | 0.053318 |
| S2023_Z44 | chr6:25029 | HeelBMD | chr6:24928 | 388 | 1.34E-33 | 1.12E-29 | 2.19E-06 | 0.017391 |
| S1994_Z10 | chr6:25029 | HeelBMD | chr6:24928 | 390 | 2.06E-34 | 7.56E-30 | 3.38E-07 | 0.011427 |
| S1993_Z28 | chr6:25029 | HeelBMD | chr6:24928 | 390 | 8.77E-33 | 7.58E-30 | 1.44E-05 | 0.011449 |
| S1993_Z28 | chr6:25029 | S1993_Z6 | chr6:25076 | 515 | 5.11E-10 | 5.14E-07 | 1.05E-05 | 0.009595 |
| S1993_Z28 | chr6:25029 | S1993_Z97 | chr6:24913 | 515 | 1.92E-11 | 1.93E-08 | 1.11E-05 | 0.010158 |
| S1993_Z28 | chr6:25029 | S1994_Z10 | chr6:25029 | 515 | 2.02E-10 | 2.03E-07 | 8.11E-06 | 0.007168 |
| S1993_Z28 | chr6:25029 | S2023_Z44 | chr6:25029 | 513 | 1.10E-09 | 1.10E-06 | 1.04E-05 | 0.009447 |
| S1994_Z10 | chr6:25029 | S2023_Z44 | chr6:25029 | 513 | 2.91E-11 | 1.17E-06 | 2.75E-07 | 0.010078 |
| S2023_Z44 | chr6:25029 | S2023_Z95 | chr6:25076 | 513 | 6.17E-11 | 5.83E-07 | 3.31E-06 | 0.030298 |
| S1994_Z10 | chr6:25029 | S2023_Z95 | chr6:25076 | 515 | 7.37E-12 | 2.96E-07 | 3.95E-07 | 0.014888 |
| S1993_Z28 | chr6:25029 | S2023_Z95 | chr6:25076 | 515 | 4.00E-10 | 4.03E-07 | 2.15E-05 | 0.020617 |
| S1993_Z22 | chr6:25038 | S1993_Z6 | chr6:25076 | 523 | 2.16E-12 | 7.67E-07 | 4.46E-08 | 0.01481 |
| S1993_Z22 | chr6:25038 | S2023_Z95 | chr6:25076 | 515 | 4.99E-13 | 1.77E-07 | 2.68E-08 | 0.008501 |
| S2023_Z95 | chr6:25076 | HeelBMD | chr6:24928 | 390 | 7.74E-34 | 4.13E-29 | 1.27E-06 | 0.066936 |
| S1993_Z6 | chr6:25076 | HeelBMD | chr6:24928 | 397 | 7.96E-34 | 1.61E-29 | 1.31E-06 | 0.025535 |
| S1993_Z6 | chr6:25076 | S1993_Z97 | chr6:24913 | 523 | 2.04E-12 | 4.20E-08 | 1.18E-06 | 0.023239 |

| | | | | | | | | |
|---|---|---|---|---|---|---|---|---|
| S1993_Z6 | chr6:25076 | S1994_Z10 | chr6:25029 | 515 | 1.17E-11 | 2.41E-07 | 4.71E-07 | 0.008696 |
| S1993_Z6 | chr6:25076 | S2023_Z44 | chr6:25029 | 513 | 8.67E-11 | 1.78E-06 | 8.20E-07 | 0.015891 |
| S1993_Z6 | chr6:25076 | S2023_Z95 | chr6:25076 | 515 | 6.07E-12 | 1.25E-07 | 3.25E-07 | 0.005701 |
| S1994_Z31 | chr6:11823 | RA_maxVo | chr6:11823 | 1017 | 9.73E-08 | 4.23E-06 | 0.000124 | 0.004376 |
| S1994_Z31 | chr6:11823 | RA_minVol | chr6:11823 | 1009 | 6.67E-05 | 0.002898 | 0.000442 | 0.01821 |
| S1994_Z31 | chr6:11823 | RV_Stroke\ | chr6:11833 | 1088 | 1.27E-05 | 0.000551 | 0.003142 | 0.1356 |
| S1701_Z82 | chr6:11834 | ArteryAOR | chr6:11865 | 706 | 1.24E-09 | 4.66E-05 | 4.52E-06 | 0.168658 |
| S1701_Z82 | chr6:11834 | DBP | chr6:11836 | 783 | 1.03E-08 | 4.71E-05 | 1.83E-06 | 0.007401 |
| S1701_Z82 | chr6:11834 | LV_endDia | chr6:11822 | 1063 | 3.85E-06 | 0.017899 | 9.46E-06 | 0.043048 |
| S1701_Z82 | chr6:11834 | LV_endSys | chr6:11835 | 1065 | 8.51E-08 | 0.000396 | 2.82E-06 | 0.012104 |
| S1701_Z82 | chr6:11834 | PRint | chr6:11834 | 147 | 3.07E-51 | 1.40E-47 | 5.18E-07 | 0.001356 |
| S1701_Z82 | chr6:11834 | RA_maxVo | chr6:11823 | 1026 | 1.23E-09 | 5.71E-06 | 1.57E-06 | 0.006318 |
| S1701_Z82 | chr6:11834 | RA_minVol | chr6:11823 | 1009 | 5.22E-06 | 0.02425 | 3.45E-05 | 0.159737 |
| S1701_Z82 | chr6:11834 | rHeartRate | chr6:11834 | 207 | 4.04E-21 | 1.82E-17 | 1.05E-06 | 0.003737 |
| S1701_Z82 | chr6:11834 | RV_Stroke\ | chr6:11833 | 1117 | 5.67E-09 | 2.64E-05 | 1.41E-06 | 0.005541 |
| S1701_Z82 | chr6:11834 | S1994_Z31 | chr6:11823 | 1088 | 6.29E-08 | 0.000292 | 2.73E-06 | 0.011713 |
| S1994_Z92 | chr6:12142 | PulseRate | chr6:12139 | 430 | 2.16E-05 | 0.0002 | 0.000238 | 0.0012 |
| S1994_Z92 | chr6:12142 | rHeartRate | chr6:12140 | 41 | 4.63E-09 | 1.19E-07 | 9.07E-05 | 0.001341 |
| S42_Z12 | chr6:12145 | PP | chr6:12145 | 431 | 6.04E-08 | 4.22E-06 | 6.71E-05 | 0.00369 |
| S42_Z12 | chr6:12145 | PRint | chr6:12145 | 47 | 4.52E-08 | 2.18E-06 | 5.92E-05 | 0.001858 |
| S42_Z12 | chr6:12145 | PulseRate | chr6:12143 | 342 | 0.000408 | 0.001678 | 0.010787 | 0.04341 |
| S42_Z12 | chr6:12145 | PulseRate | chr6:12146 | 488 | 3.67E-06 | 1.87E-05 | 0.005344 | 0.026173 |
| S42_Z12 | chr6:12145 | rHeartRate | chr6:12145 | 57 | 1.71E-19 | 7.64E-18 | 4.90E-05 | 0.001192 |
| S1701_Z82 | chr6:12177 | HeelBMD | chr6:12177 | 608 | 4.59E-19 | 4.60E-11 | 2.65E-11 | 0.001662 |
| S1994_Z92 | chr6:12177 | HeelBMD | chr6:12177 | 571 | 1.51E-16 | 4.05E-11 | 3.85E-09 | 3.00E-05 |
| S42_Z10 | chr6:12177 | HeelBMD | chr6:12177 | 596 | 1.05E-15 | 5.16E-11 | 6.04E-08 | 0.001986 |
| S1994_Z92 | chr6:12177 | PP | chr6:12186 | 539 | 1.91E-26 | 5.12E-21 | 3.84E-09 | 2.69E-05 |
| S1701_Z82 | chr6:12177 | PP | chr6:12186 | 576 | 5.10E-29 | 5.11E-21 | 2.87E-11 | 0.001884 |
| S42_Z10 | chr6:12177 | PP | chr6:12186 | 564 | 1.09E-25 | 5.40E-21 | 6.17E-08 | 0.002047 |
| S42_Z10 | chr6:12177 | PulseRate | chr6:12186 | 654 | 2.49E-66 | 1.24E-61 | 7.14E-08 | 0.002547 |
| S1701_Z82 | chr6:12177 | PulseRate | chr6:12186 | 659 | 1.32E-69 | 1.32E-61 | 3.78E-11 | 0.00279 |
| S1701_Z82 | chr6:12177 | S1994_Z92 | chr6:12177 | 741 | 6.51E-17 | 3.85E-09 | 1.75E-11 | 3.51E-05 |
| S1701_Z82 | chr6:12177 | S42_Z10 | chr6:12177 | 794 | 5.68E-16 | 5.70E-08 | 2.83E-11 | 0.001841 |
| S1994_Z92 | chr6:12177 | S42_Z10 | chr6:12177 | 726 | 1.85E-13 | 4.99E-08 | 4.33E-09 | 0.000166 |
| S1701_Z29 | chr6:12638 | AdiposeSU | chr6:12638 | 330 | 1.51E-12 | 1.37E-06 | 3.20E-08 | 0.028051 |
| S42_Z125 | chr6:12638 | AdiposeSU | chr6:12638 | 330 | 6.60E-12 | 5.99E-06 | 4.12E-08 | 0.036393 |
| S42_Z125 | chr6:12638 | HeartAA | chr6:12644 | 260 | 2.73E-08 | 6.75E-06 | 0.00017 | 0.041117 |
| S1701_Z29 | chr6:12638 | HeartAA | chr6:12644 | 260 | 7.69E-09 | 1.90E-06 | 0.000163 | 0.039246 |
| S1701_Z29 | chr6:12638 | S2023_Z87 | chr6:12643 | 478 | 5.88E-11 | 2.18E-06 | 9.96E-07 | 0.035962 |
| S1701_Z29 | chr6:12638 | S42_Z125 | chr6:12638 | 478 | 8.54E-11 | 3.16E-06 | 8.39E-07 | 0.030124 |
| S2023_Z87 | chr6:12643 | AdiposeSU | chr6:12638 | 330 | 3.34E-12 | 3.03E-06 | 4.11E-08 | 0.036366 |
| S2023_Z87 | chr6:12643 | HeartAA | chr6:12644 | 260 | 1.33E-08 | 3.28E-06 | 0.000164 | 0.039485 |
| S2023_Z87 | chr6:12643 | S42_Z125 | chr6:12638 | 478 | 2.09E-10 | 3.54E-06 | 2.05E-06 | 0.033771 |
| S2023_Z10 | chr6:14254 | AdiposeSU | chr6:14233 | 365 | 9.33E-12 | 7.83E-07 | 5.77E-07 | 0.047531 |
| S2023_Z10 | chr6:14254 | BodyFFM | chr6:14232 | 159 | 1.68E-35 | 1.43E-31 | 1.45E-06 | 0.01143 |

| | | | | | | | | |
|---|---|---|---|---|---|---|---|---|
| S2023_Z10 | chr6:14254 | LungFunc | chr6:14234 | 423 | 3.85E-73 | 2.39E-68 | 1.47E-06 | 0.090072 |
| S42_Z127 | chr7:46520 | WHRadjBM | chr7:46533 | 762 | 2.58E-06 | 0.006294 | 3.85E-05 | 0.092755 |
| S2023_Z86 | chr7:12114 | Lung | chr7:12108 | 326 | 1.06E-08 | 8.72E-07 | 0.001897 | 0.155498 |
| S2023_Z86 | chr7:12137 | HeelBMD | chr7:12139 | 244 | 1.23E-164 | 2.25E-156 | 5.46E-12 | 1.17E-11 |
| S2023_Z86 | chr8:10814 | MuscleSKL | chr8:10812 | 885 | 1.64E-10 | 8.51E-06 | 1.53E-06 | 0.078228 |
| S1701_Z80 | chr8:11943 | AdiposeSU | chr8:11938 | 294 | 3.07E-11 | 9.09E-06 | 2.52E-07 | 0.073484 |
| S1701_Z80 | chr8:11943 | ArteryAOR | chr8:11940 | 309 | 3.41E-13 | 7.05E-06 | 2.79E-09 | 0.056768 |
| S1701_Z80 | chr8:11943 | ArteryTIB | chr8:11945 | 296 | 1.28E-08 | 1.23E-05 | 0.000105 | 0.099722 |
| S1701_Z80 | chr8:11943 | BodyFFM | chr8:11940 | 318 | 3.29E-45 | 6.39E-43 | 7.41E-05 | 0.01341 |
| S1701_Z80 | chr8:11943 | DBP | chr8:11938 | 411 | 2.00E-18 | 1.70E-14 | 8.13E-06 | 0.068102 |
| S1701_Z80 | chr8:11943 | HeartLV | chr8:11945 | 307 | 6.91E-08 | 0.000566 | 6.22E-06 | 0.050001 |
| S1993_Z14 | chr10:2174 | BodyFPC | chr10:2153 | 451 | 1.14E-24 | 4.40E-22 | 0.000381 | 0.145875 |
| S1994_Z1 | chr10:8805 | LungFunc | chr10:8806 | 285 | 4.62E-08 | 3.94E-05 | 1.93E-05 | 0.015501 |
| S1701_Z29 | chr11:9529 | S1994_Z5 | chr11:9529 | 333 | 7.10E-10 | 6.72E-07 | 1.52E-06 | 0.000444 |
| S1701_Z29 | chr11:9529 | S42_Z125 | chr11:9529 | 333 | 5.40E-10 | 5.12E-07 | 1.43E-06 | 0.000357 |
| S1994_Z5 | chr11:9529 | S42_Z125 | chr11:9529 | 333 | 2.85E-10 | 6.11E-07 | 7.54E-07 | 0.00062 |
| S1994_Z5 | chr12:4275 | AdiposeSU | chr12:4275 | 199 | 8.18E-07 | 3.18E-07 | 0.002652 | 3.34E-05 |
| S1994_Z5 | chr12:4275 | ArteryTIB | chr12:4275 | 200 | 7.26E-10 | 3.08E-07 | 2.36E-06 | 2.15E-07 |
| S1994_Z5 | chr12:4275 | BodyFFM | chr12:4275 | 227 | 2.53E-134 | 8.19E-131 | 3.08E-07 | 1.54E-09 |
| S1994_Z5 | chr12:4275 | CVD | chr12:4275 | 262 | 3.93E-09 | 1.28E-05 | 3.08E-07 | 1.38E-07 |
| S1994_Z5 | chr12:4275 | HeelBMD | chr12:4275 | 273 | 2.44E-26 | 7.92E-23 | 3.08E-07 | 8.74E-08 |
| S1994_Z5 | chr12:4275 | MuscleSKL | chr12:4275 | 200 | 1.70E-11 | 5.51E-08 | 3.08E-07 | 8.80E-08 |
| S1994_Z5 | chr12:4275 | Triglyceride | chr12:4275 | 326 | 6.06E-10 | 1.96E-06 | 3.09E-07 | 4.51E-07 |
| S1701_Z82 | chr12:2460 | LV_Stroke\ | chr12:2463 | 346 | 1.05E-12 | 6.32E-05 | 9.86E-11 | 0.004962 |
| S42_Z10 | chr12:2460 | LV_Stroke\ | chr12:2463 | 346 | 6.34E-08 | 6.70E-05 | 5.98E-06 | 0.005319 |
| S42_Z10 | chr12:2460 | PP | chr12:2461 | 303 | 6.32E-11 | 5.70E-08 | 6.64E-06 | 0.004995 |
| S1701_Z82 | chr12:2460 | PP | chr12:2461 | 303 | 1.10E-15 | 5.35E-08 | 1.16E-10 | 0.00463 |
| S42_Z10 | chr12:2460 | PulseRate | chr12:2460 | 437 | 3.59E-88 | 3.78E-85 | 3.85E-06 | 0.003068 |
| S1701_Z82 | chr12:2460 | PulseRate | chr12:2460 | 423 | 7.22E-93 | 4.36E-85 | 7.76E-11 | 0.003689 |
| S1701_Z82 | chr12:2460 | S1994_Z85 | chr12:2463 | 375 | 5.52E-15 | 3.34E-07 | 1.07E-10 | 0.005451 |
| S1701_Z82 | chr12:2460 | S42_Z10 | chr12:2460 | 493 | 8.32E-14 | 5.03E-06 | 8.79E-11 | 0.004315 |
| S1701_Z12 | chr12:2462 | LV_Stroke\ | chr12:2463 | 346 | 1.07E-09 | 6.05E-05 | 1.01E-07 | 0.004709 |
| S1701_Z12 | chr12:2462 | PP | chr12:2461 | 306 | 9.49E-13 | 4.35E-08 | 9.97E-08 | 0.003579 |
| S1701_Z12 | chr12:2462 | PulseRate | chr12:2460 | 362 | 1.27E-89 | 7.19E-85 | 1.36E-07 | 0.006736 |
| S1701_Z12 | chr12:2462 | S1701_Z49 | chr12:2463 | 381 | 1.79E-12 | 1.01E-07 | 1.05E-07 | 0.004959 |
| S1701_Z12 | chr12:2462 | S1701_Z82 | chr12:2460 | 420 | 1.83E-15 | 1.04E-10 | 1.10E-07 | 0.005264 |
| S1701_Z12 | chr12:2462 | S1994_Z85 | chr12:2463 | 379 | 4.96E-12 | 2.82E-07 | 9.59E-08 | 0.004444 |
| S1701_Z12 | chr12:2462 | S42_Z10 | chr12:2460 | 420 | 1.16E-10 | 6.59E-06 | 1.23E-07 | 0.005966 |
| S1994_Z85 | chr12:2463 | LV_Stroke\ | chr12:2463 | 346 | 3.16E-09 | 6.10E-05 | 2.98E-07 | 0.004753 |
| S1994_Z85 | chr12:2463 | PP | chr12:2461 | 306 | 2.75E-12 | 4.33E-08 | 2.89E-07 | 0.003558 |
| S1994_Z85 | chr12:2463 | PulseRate | chr12:2460 | 321 | 4.06E-89 | 7.85E-85 | 4.36E-07 | 0.00744 |
| S1994_Z85 | chr12:2463 | S42_Z10 | chr12:2460 | 375 | 3.55E-10 | 6.86E-06 | 3.75E-07 | 0.006257 |
| S1701_Z49 | chr12:2463 | LV_Stroke\ | chr12:2463 | 346 | 1.09E-09 | 6.40E-05 | 1.03E-07 | 0.005038 |
| S1701_Z49 | chr12:2463 | PP | chr12:2461 | 303 | 9.63E-13 | 4.60E-08 | 1.01E-07 | 0.003842 |
| S1701_Z49 | chr12:2463 | PulseRate | chr12:2460 | 323 | 1.07E-89 | 6.28E-85 | 1.15E-07 | 0.005755 |

| | | | | | | | | |
|---|---|---|---|---|---|---|---|---|
| S1701_Z49 | chr12:2463 | S1701_Z82 | chr12:2460 | 381 | 1.76E-15 | 1.03E-10 | 1.06E-07 | 0.005242 |
| S1701_Z49 | chr12:2463 | S1994_Z85 | chr12:2463 | 375 | 5.27E-12 | 3.09E-07 | 1.02E-07 | 0.004974 |
| S1701_Z49 | chr12:2463 | S42_Z10 | chr12:2460 | 381 | 1.02E-10 | 5.99E-06 | 1.08E-07 | 0.005329 |
| S2023_Z77 | chr12:2825 | AdiposeSU | chr12:2812 | 717 | 1.38E-07 | 7.41E-05 | 0.000238 | 0.126765 |
| S2023_Z77 | chr12:2825 | AdiposeVIS | chr12:2818 | 704 | 3.15E-07 | 5.16E-05 | 0.000543 | 0.088057 |
| S2023_Z77 | chr12:2825 | HeelBMD | chr12:2846 | 879 | 4.73E-22 | 1.32E-18 | 1.50E-05 | 0.040824 |
| S2023_Z77 | chr12:2825 | S2023_Z86 | chr12:2839 | 1105 | 4.02E-12 | 2.31E-08 | 1.52E-06 | 0.007752 |
| S2023_Z86 | chr12:2839 | AdiposeSU | chr12:2812 | 717 | 1.70E-09 | 9.12E-07 | 0.000261 | 0.138617 |
| S2023_Z86 | chr12:2839 | AdiposeVIS | chr12:2818 | 704 | 4.20E-09 | 6.88E-07 | 0.000642 | 0.104312 |
| S2023_Z86 | chr12:2839 | HeelBMD | chr12:2846 | 879 | 3.58E-24 | 9.68E-19 | 1.13E-07 | 0.029669 |
| S1994_Z38 | chr13:4216 | BodyFFM | chr13:4218 | 431 | 1.62E-39 | 4.14E-38 | 0.00025 | 0.005389 |
| S1701_Z53 | chr13:5051 | DBP | chr13:5052 | 352 | 1.68E-06 | 0.001061 | 0.000223 | 0.139649 |
| S1701_Z53 | chr13:5051 | HeelBMD | chr13:5053 | 367 | 4.48E-09 | 2.43E-06 | 0.000134 | 0.07199 |
| S1701_Z53 | chr13:5051 | WHRadjBM | chr13:5055 | 306 | 5.98E-05 | 0.028773 | 8.34E-05 | 0.039227 |
| S1701_Z12 | chr14:2339 | AFib | chr14:2339 | 254 | 1.00E-09 | 9.12E-06 | 2.41E-07 | 0.001196 |
| S1994_Z11 | chr14:2339 | AFib | chr14:2339 | 235 | 1.87E-08 | 8.81E-06 | 4.50E-06 | 0.001121 |
| S1994_Z11 | chr14:2339 | DescAorta_ | chr14:2339 | 297 | 3.97E-11 | 1.89E-08 | 4.30E-06 | 0.001041 |
| S1701_Z12 | chr14:2339 | DescAorta_ | chr14:2339 | 318 | 2.09E-12 | 1.91E-08 | 2.26E-07 | 0.001066 |
| S1994_Z11 | chr14:2339 | DescAorta_ | chr14:2339 | 297 | 9.32E-09 | 4.42E-06 | 4.70E-06 | 0.001231 |
| S1701_Z12 | chr14:2339 | DescAorta_ | chr14:2339 | 318 | 4.89E-10 | 4.46E-06 | 2.47E-07 | 0.001252 |
| S1701_Z12 | chr14:2339 | LV_StrokeV | chr14:2339 | 318 | 6.05E-10 | 5.52E-06 | 5.94E-07 | 0.004421 |
| S1994_Z11 | chr14:2339 | LV_StrokeV | chr14:2339 | 297 | 1.14E-08 | 5.43E-06 | 1.12E-05 | 0.004331 |
| S1994_Z11 | chr14:2339 | PP | chr14:2339 | 217 | 1.06E-34 | 4.99E-32 | 3.59E-06 | 0.000692 |
| S1701_Z12 | chr14:2339 | PP | chr14:2339 | 232 | 5.80E-36 | 5.28E-32 | 1.97E-07 | 0.000793 |
| S1994_Z11 | chr14:2339 | PulseRate | chr14:2339 | 204 | 3.46E-46 | 1.64E-43 | 3.49E-06 | 0.000655 |
| S1701_Z12 | chr14:2339 | PulseRate | chr14:2339 | 220 | 1.91E-47 | 1.75E-43 | 1.93E-07 | 0.000761 |
| S1994_Z11 | chr14:2339 | RV_EF | chr14:2339 | 297 | 1.30E-07 | 6.17E-05 | 3.99E-06 | 0.000893 |
| S1701_Z12 | chr14:2339 | RV_EF | chr14:2339 | 297 | 6.96E-09 | 6.35E-05 | 2.14E-07 | 0.00095 |
| S1994_Z11 | chr14:2339 | RV_StrokeV | chr14:2339 | 297 | 1.10E-09 | 5.21E-07 | 4.49E-06 | 0.00113 |
| S1701_Z12 | chr14:2339 | RV_StrokeV | chr14:2339 | 318 | 5.88E-11 | 5.36E-07 | 2.40E-07 | 0.001193 |
| S1701_Z12 | chr14:2339 | S1701_Z82 | chr14:2341 | 318 | 1.12E-09 | 1.02E-05 | 4.37E-06 | 0.038893 |
| S1701_Z12 | chr14:2339 | S1994_Z11 | chr14:2339 | 297 | 4.35E-10 | 3.97E-06 | 2.07E-07 | 0.000885 |
| S1701_Z82 | chr14:2341 | AFib | chr14:2339 | 257 | 4.77E-08 | 0.000184 | 1.15E-05 | 0.043454 |
| S1701_Z82 | chr14:2341 | DescAorta_ | chr14:2339 | 322 | 8.00E-11 | 3.11E-07 | 8.65E-06 | 0.032724 |
| S1701_Z82 | chr14:2341 | DescAorta_ | chr14:2339 | 321 | 1.56E-08 | 6.07E-05 | 7.86E-06 | 0.029644 |
| S1701_Z82 | chr14:2341 | LV_StrokeV | chr14:2339 | 322 | 2.00E-08 | 7.78E-05 | 1.96E-05 | 0.075335 |
| S1701_Z82 | chr14:2341 | PP | chr14:2339 | 235 | 2.93E-34 | 1.13E-30 | 9.93E-06 | 0.037456 |
| S1701_Z82 | chr14:2341 | PulseRate | chr14:2339 | 223 | 9.76E-46 | 3.80E-42 | 9.84E-06 | 0.037364 |
| S1701_Z82 | chr14:2341 | RV_EF | chr14:2339 | 297 | 3.22E-07 | 0.001254 | 9.88E-06 | 0.037524 |
| S1701_Z82 | chr14:2341 | RV_StrokeV | chr14:2339 | 322 | 1.93E-09 | 7.53E-06 | 7.90E-06 | 0.029808 |
| S1701_Z82 | chr14:2341 | S1994_Z11 | chr14:2339 | 297 | 2.15E-08 | 8.36E-05 | 1.02E-05 | 0.0387 |
| S1701_Z32 | chr15:9280 | S1701_Z80 | chr15:9280 | 471 | 2.22E-08 | 4.06E-06 | 2.54E-05 | 0.003639 |
| S42_Z28 | chr19:4490 | CAD | chr19:4488 | 387 | 1.91E-12 | 1.36E-08 | 3.47E-07 | 0.001471 |
| S2023_Z87 | chr19:4490 | CAD | chr19:4488 | 387 | 1.70E-12 | 1.75E-08 | 3.08E-07 | 0.002175 |
| S42_Z28 | chr19:4490 | CVD | chr19:4490 | 427 | 1.47E-35 | 1.05E-31 | 1.54E-07 | 0.000101 |

| | | | | | | | | |
|---|---|---|---|---|---|---|---|---|
| S2023_Z87 | chr19:4490 | CVD | chr19:4490 | 427 | 1.44E-35 | 1.49E-31 | 1.51E-07 | 0.000558 |
| S42_Z28 | chr19:4490 | HDLcholest | chr19:4491 | 426 | 2.70E-13 | 1.92E-09 | 6.08E-07 | 0.00334 |
| S2023_Z87 | chr19:4490 | HDLcholest | chr19:4491 | 426 | 2.61E-13 | 2.69E-09 | 5.89E-07 | 0.005068 |
| S42_Z28 | chr19:4490 | LiverFAT | chr19:4490 | 427 | 1.66E-30 | 1.18E-26 | 1.54E-07 | 0.000101 |
| S2023_Z87 | chr19:4490 | LiverFAT | chr19:4490 | 427 | 1.63E-30 | 1.67E-26 | 1.51E-07 | 0.000558 |
| S2023_Z87 | chr19:4490 | MInf | chr19:4492 | 384 | 1.66E-11 | 1.71E-07 | 9.29E-07 | 0.008561 |
| S2023_Z87 | chr19:4490 | PP | chr19:4490 | 311 | 1.14E-07 | 0.001169 | 1.62E-07 | 0.000667 |
| S42_Z28 | chr19:4490 | PP | chr19:4490 | 311 | 1.16E-07 | 0.00083 | 1.66E-07 | 0.000183 |
| S42_Z28 | chr19:4490 | PulseRate | chr19:4491 | 397 | 1.59E-08 | 0.000116 | 8.70E-07 | 0.005379 |
| S2023_Z87 | chr19:4490 | PulseRate | chr19:4491 | 397 | 8.80E-09 | 0.000112 | 4.82E-07 | 0.005165 |
| S2023_Z87 | chr19:4490 | S42_Z28 | chr19:4490 | 480 | 2.26E-11 | 2.89E-07 | 1.66E-07 | 0.001117 |
| S2023_Z87 | chr19:4490 | Triglyceride | chr19:4491 | 422 | 1.31E-09 | 1.35E-05 | 6.72E-06 | 0.06818 |
| S42_Z28 | chr19:4490 | WHRadjBM | chr19:4490 | 363 | 6.37E-22 | 4.54E-18 | 1.69E-07 | 0.000207 |
| S2023_Z87 | chr19:4490 | WHRadjBM | chr19:4490 | 363 | 6.24E-22 | 6.42E-18 | 1.66E-07 | 0.000707 |
| S42_Z125 | chr19:5553 | AdiposeVIS | chr19:5547 | 326 | 0.000999 | 0.00017 | 0.084379 | 0.013463 |
| S1701_Z82 | chr20:3821 | LV_Stroke\ | chr20:3821 | 395 | 9.15E-11 | 0.000144 | 8.97E-09 | 0.013134 |
| S1701_Z82 | chr20:3821 | PP | chr20:3821 | 319 | 4.33E-12 | 6.58E-06 | 5.26E-09 | 0.006998 |
| S1701_Z82 | chr20:3821 | PRint | chr20:3821 | 44 | 2.49E-14 | 3.78E-08 | 5.01E-09 | 0.006625 |
| S1701_Z82 | chr20:3821 | rHeartRate | chr20:3821 | 63 | 5.03E-171 | 7.62E-165 | 1.09E-08 | 0.015604 |
| S1701_Z82 | chr20:3821 | RV_Stroke\ | chr20:3821 | 398 | 4.12E-11 | 6.50E-05 | 9.82E-09 | 0.014477 |
| S1701_Z93 | chr22:2972 | AdiposeSU | chr22:2973 | 386 | 2.79E-06 | 0.000237 | 0.001043 | 0.087908 |
| S1701_Z93 | chr22:2972 | ArteryAOR | chr22:2981 | 524 | 1.38E-07 | 0.000322 | 5.19E-05 | 0.120163 |
| S1701_Z93 | chr22:2972 | HeartLV | chr22:3000 | 385 | 1.89E-07 | 0.000153 | 7.06E-05 | 0.056473 |
| S1701_Z93 | chr22:2972 | Liver | chr22:2951 | 139 | 1.73E-05 | 0.00574 | 0.000243 | 0.079717 |
| S1701_Z93 | chr22:2972 | Pancreas | chr22:2973 | 331 | 3.22E-06 | 0.000111 | 0.001203 | 0.040577 |
| S1701_Z93 | chr22:2972 | S1993_Z27 | chr22:2974 | 828 | 2.53E-09 | 1.10E-06 | 4.24E-05 | 0.017359 |
| S1701_Z93 | chr22:2972 | S1993_Z28 | chr22:2976 | 832 | 9.81E-10 | 4.25E-07 | 4.90E-05 | 0.020248 |
| S1701_Z93 | chr22:2972 | S1993_Z41 | chr22:2974 | 829 | 1.23E-08 | 5.34E-06 | 5.97E-05 | 0.024876 |
| S1701_Z93 | chr22:2972 | S1993_Z66 | chr22:2976 | 829 | 8.59E-09 | 3.72E-06 | 6.65E-05 | 0.027832 |
| S1701_Z93 | chr22:2972 | S1994_Z1 | chr22:2974 | 832 | 9.61E-09 | 4.16E-06 | 5.78E-05 | 0.024057 |
| S1994_Z1 | chr22:2974 | ArteryAOR | chr22:2981 | 524 | 1.12E-08 | 2.62E-05 | 5.70E-05 | 0.132108 |
| S1993_Z41 | chr22:2974 | ArteryAOR | chr22:2981 | 524 | 8.75E-09 | 2.04E-05 | 3.63E-05 | 0.083854 |
| S1993_Z27 | chr22:2974 | ArteryAOR | chr22:2981 | 524 | 1.72E-09 | 4.01E-06 | 2.48E-05 | 0.056939 |
| S1994_Z1 | chr22:2974 | HeartLV | chr22:3000 | 385 | 7.15E-09 | 5.82E-06 | 3.62E-05 | 0.028455 |
| S1993_Z41 | chr22:2974 | HeartLV | chr22:3000 | 385 | 8.50E-09 | 6.91E-06 | 3.53E-05 | 0.027722 |
| S1993_Z27 | chr22:2974 | HeartLV | chr22:3000 | 385 | 2.65E-09 | 2.16E-06 | 3.83E-05 | 0.030175 |
| S1993_Z41 | chr22:2974 | Liver | chr22:2951 | 137 | 7.79E-07 | 0.002927 | 1.09E-05 | 0.040162 |
| S1994_Z1 | chr22:2974 | Liver | chr22:2951 | 201 | 5.98E-07 | 0.002816 | 1.83E-05 | 0.085049 |
| S1993_Z27 | chr22:2974 | Liver | chr22:2951 | 136 | 2.61E-07 | 0.003095 | 3.67E-06 | 0.042521 |
| S1994_Z1 | chr22:2974 | Pancreas | chr22:2973 | 331 | 1.18E-07 | 4.07E-06 | 0.000596 | 0.019588 |
| S1993_Z41 | chr22:2974 | Pancreas | chr22:2973 | 331 | 1.44E-07 | 4.96E-06 | 0.000596 | 0.019604 |
| S1993_Z27 | chr22:2974 | Pancreas | chr22:2973 | 331 | 4.63E-08 | 1.60E-06 | 0.000669 | 0.022102 |
| S1993_Z27 | chr22:2974 | S1993_Z28 | chr22:2976 | 828 | 1.77E-11 | 2.96E-07 | 8.84E-07 | 0.013819 |
| S1993_Z27 | chr22:2974 | S1993_Z41 | chr22:2974 | 828 | 1.69E-10 | 2.82E-06 | 8.17E-07 | 0.012692 |
| S1993_Z41 | chr22:2974 | S1993_Z66 | chr22:2976 | 829 | 4.23E-10 | 2.05E-06 | 3.28E-06 | 0.014895 |

| | | | | | | | | | |
|---|---|---|---|---|---|---|---|---|---|
| S1993_Z27 | chr22:2974 | S1993_Z66 | chr22:2976 | | 828 | 1.16E-10 | 1.94E-06 | 8.99E-07 | 0.014065 |
| S1993_Z41 | chr22:2974 | S1994_Z1 | chr22:2974 | | 829 | 4.66E-10 | 2.26E-06 | 2.81E-06 | 0.012609 |
| S1993_Z27 | chr22:2974 | S1994_Z1 | chr22:2974 | | 828 | 1.45E-10 | 2.43E-06 | 8.75E-07 | 0.013669 |
| S1993_Z28 | chr22:2976 | ArteryAOR | chr22:2981 | | 528 | 6.55E-10 | 1.53E-06 | 2.79E-05 | 0.064243 |
| S1993_Z66 | chr22:2976 | ArteryAOR | chr22:2981 | | 524 | 2.83E-09 | 6.61E-06 | 1.90E-05 | 0.043468 |
| S1993_Z28 | chr22:2976 | HeartLV | chr22:3000 | | 385 | 7.86E-10 | 6.39E-07 | 3.34E-05 | 0.0262 |
| S1993_Z66 | chr22:2976 | HeartLV | chr22:3000 | | 385 | 4.15E-09 | 3.38E-06 | 2.79E-05 | 0.021739 |
| S1993_Z66 | chr22:2976 | Liver | chr22:2951 | | 137 | 6.17E-07 | 0.002904 | 8.67E-06 | 0.039838 |
| S1993_Z28 | chr22:2976 | Liver | chr22:2951 | | 139 | 8.91E-08 | 0.003046 | 1.25E-06 | 0.041827 |
| S1993_Z28 | chr22:2976 | Pancreas | chr22:2973 | | 331 | 1.60E-08 | 5.52E-07 | 0.00068 | 0.022495 |
| S1993_Z66 | chr22:2976 | Pancreas | chr22:2973 | | 331 | 1.07E-07 | 3.70E-06 | 0.000721 | 0.023926 |
| S1993_Z28 | chr22:2976 | S1993_Z41 | chr22:2974 | | 829 | 6.07E-11 | 3.04E-06 | 2.94E-07 | 0.013724 |
| S1993_Z28 | chr22:2976 | S1993_Z66 | chr22:2976 | | 829 | 4.04E-11 | 2.02E-06 | 3.13E-07 | 0.014676 |
| S1993_Z28 | chr22:2976 | S1994_Z1 | chr22:2974 | | 832 | 5.12E-11 | 2.56E-06 | 3.08E-07 | 0.014435 |
| S1993_Z66 | chr22:2976 | S1994_Z1 | chr22:2974 | | 829 | 3.43E-10 | 2.66E-06 | 2.07E-06 | 0.015016 |

| PP.H4.abf | sub_locus | chr1 | pos1 | t1 | t2 | category | hit1_snp | locus |
|---|---|---|---|---|---|---|---|---|
| 0.997204 | 390 | 1 | 1.19E+08 | S1993_Z97 | AdiposeVIS | Non cardia | rs1080206 | 1 |
| 0.996687 | 390 | 1 | 1.19E+08 | S1993_Z97 | ArteryTIB_ | Cardiac eQ | rs1080206 | 1 |
| 0.844987 | 390 | 1 | 1.19E+08 | S1993_Z97 | WHRadjBM | Non cardia | rs1080206 | 1 |
| 0.964551 | 7624 | 1 | 2.08E+08 | S1701_Z82 | PulseRate | ECG and bl | rs1111828 | 2 |
| 0.965678 | 275 | 1 | 2.08E+08 | S42_Z10 | AdiposeSU | Non cardia | rs1157956 | 3 |
| 0.966267 | 275 | 1 | 2.08E+08 | S42_Z10 | AdiposeVIS | Non cardia | rs1157956 | 3 |
| 0.978483 | 275 | 1 | 2.08E+08 | S42_Z10 | ArteryAOR_ | Cardiac eQ | rs1157956 | 3 |
| 0.966345 | 275 | 1 | 2.08E+08 | S42_Z10 | ArteryCOR_ | Cardiac eQ | rs1157956 | 3 |
| 0.962777 | 275 | 1 | 2.08E+08 | S42_Z10 | ArteryTIB_ | Cardiac eQ | rs1157956 | 3 |
| 0.963529 | 275 | 1 | 2.08E+08 | S42_Z10 | HeartAA_c | Cardiac eQ | rs1157956 | 3 |
| 0.964795 | 275 | 1 | 2.08E+08 | S42_Z10 | HeartLV_ch | Cardiac eQ | rs1157956 | 3 |
| 0.956712 | 275 | 1 | 2.08E+08 | S42_Z10 | Liver_chr1_ | Non cardia | rs1157956 | 3 |
| 0.967026 | 275 | 1 | 2.08E+08 | S42_Z10 | Lung_chr1_ | Non cardia | rs1157956 | 3 |
| 0.966635 | 275 | 1 | 2.08E+08 | S42_Z10 | MuscleSKL_ | Non cardia | rs1157956 | 3 |
| 0.966397 | 275 | 1 | 2.08E+08 | S42_Z10 | Pancreas_c | Non cardia | rs1157956 | 3 |
| 0.961677 | 275 | 1 | 2.08E+08 | S42_Z10 | PulseRate | ECG and bl | rs1157956 | 3 |
| 0.878047 | 3418 | 2 | 12736934 | S2023_Z86 | AdiposeSU | Non cardia | rs3533052 | 4 |
| 0.843835 | 3418 | 2 | 12736934 | S2023_Z86 | HeelBMD | Non cardia | rs3533052 | 4 |
| 0.996442 | 7977 | 2 | 18540396 | S1701_Z29 | S1994_Z5 | Latents | rs7558413_ | 5 |
| 0.998595 | 1807 | 2 | 27508073 | S1701_Z53 | CVD | Cardiac dis | rs1260326_ | 6 |
| 0.995782 | 1807 | 2 | 27508073 | S1701_Z53 | LiverFAT | Non cardia | rs1260326_ | 6 |
| 0.998907 | 1807 | 2 | 27508073 | S1701_Z53 | LiverVOL | Non cardia | rs1260326_ | 6 |
| 0.980628 | 1807 | 2 | 27508073 | S1701_Z53 | PulseRate | ECG and bl | rs1260326_ | 6 |
| 0.998973 | 1807 | 2 | 27508073 | S1701_Z53 | Triglyceride | Non cardia | rs1260326_ | 6 |
| 0.993373 | 8018 | 2 | 1.79E+08 | S1701_Z82 | LV_endDia | MRI-derive | rs1736258 | 7 |
| 0.992129 | 8018 | 2 | 1.79E+08 | S1701_Z82 | RA_minVol | MRI-derive | rs1736258 | 7 |
| 0.999997 | 8035 | 2 | 2.19E+08 | S1701_Z29 | MuscleSKL_ | Non cardia | rs1338645 | 8 |
| 0.999998 | 8035 | 2 | 2.19E+08 | S1701_Z29 | PulseRate | ECG and bl | rs1338645 | 8 |
| 0.995168 | 6543 | 2 | 2.31E+08 | S1994_Z11 | ArteryAOR_ | Cardiac eQ | rs7633024 | 9 |
| 0.895293 | 6543 | 2 | 2.31E+08 | S1994_Z11 | ArteryTIB_ | Cardiac eQ | rs7633024 | 9 |
| 0.953927 | 6543 | 2 | 2.31E+08 | S1994_Z11 | HeartAA_c | Cardiac eQ | rs7633024 | 9 |
| 0.993415 | 6543 | 2 | 2.31E+08 | S1994_Z11 | PulseRate | ECG and bl | rs7633024 | 9 |
| 0.988845 | 6543 | 2 | 2.31E+08 | S1994_Z11 | S1994_Z31 | Latents | rs7633024 | 9 |
| 0.875781 | 6543 | 2 | 2.31E+08 | S1994_Z31 | HeartAA_c | Cardiac eQ | rs6733349_ | 9 |
| 0.938722 | 7183 | 3 | 1.59E+08 | S2023_Z77 | DBP | ECG and bl | rs1656376_ | 10 |
| 0.94422 | 7183 | 3 | 1.59E+08 | S2023_Z77 | DBP | ECG and bl | rs1656376_ | 10 |
| 0.998904 | 7169 | 3 | 1.79E+08 | S1994_Z12 | S1994_Z35 | Latents | rs2339798_ | 11 |
| 0.998927 | 7169 | 3 | 1.79E+08 | S1994_Z12 | S1994_Z94 | Latents | rs2339798_ | 11 |
| 0.998904 | 7169 | 3 | 1.79E+08 | S1994_Z35 | S1994_Z94 | Latents | rs2339798_ | 11 |
| 0.99869 | 7169 | 3 | 1.79E+08 | S1994_Z35 | S42_Z105 | Latents | rs2339798_ | 11 |
| 0.998718 | 7169 | 3 | 1.79E+08 | S1994_Z94 | S42_Z105 | Latents | rs2339798_ | 11 |
| 0.998716 | 7169 | 3 | 1.79E+08 | S1994_Z12 | S42_Z105 | Latents | rs2339798_ | 11 |
| 0.998688 | 7169 | 3 | 1.79E+08 | S42_Z105 | S42_Z106 | Latents | rs2339798_ | 11 |
| 0.998901 | 7169 | 3 | 1.79E+08 | S1994_Z12 | S42_Z106 | Latents | rs2339798_ | 11 |

| | | | | | | | | |
|---|---|---|---|---|---|---|---|---|
| 0.998877 | 7169 | 3 | 1.79E+08 | S1994_Z35 | S42_Z106 | Latents | rs2339798_ | 11 |
| 0.998901 | 7169 | 3 | 1.79E+08 | S1994_Z94 | S42_Z106 | Latents | rs2339798_ | 11 |
| 0.998854 | 7169 | 3 | 1.79E+08 | S1994_Z35 | S42_Z117 | Latents | rs2339798_ | 11 |
| 0.99888 | 7169 | 3 | 1.79E+08 | S1994_Z94 | S42_Z117 | Latents | rs2339798_ | 11 |
| 0.998881 | 7169 | 3 | 1.79E+08 | S1994_Z12 | S42_Z117 | Latents | rs2339798_ | 11 |
| 0.998854 | 7169 | 3 | 1.79E+08 | S42_Z106 | S42_Z117 | Latents | rs2339798_ | 11 |
| 0.998692 | 7169 | 3 | 1.79E+08 | S42_Z105 | S42_Z117 | Latents | rs2339798_ | 11 |
| 0.998918 | 7169 | 3 | 1.79E+08 | S1994_Z94 | S42_Z12 | Latents | rs2339798_ | 11 |
| 0.998889 | 7169 | 3 | 1.79E+08 | S1994_Z35 | S42_Z12 | Latents | rs2339798_ | 11 |
| 0.99892 | 7169 | 3 | 1.79E+08 | S1994_Z12 | S42_Z12 | Latents | rs2339798_ | 11 |
| 0.998893 | 7169 | 3 | 1.79E+08 | S42_Z106 | S42_Z12 | Latents | rs2339798_ | 11 |
| 0.998884 | 7169 | 3 | 1.79E+08 | S42_Z117 | S42_Z12 | Latents | rs2339798_ | 11 |
| 0.99873 | 7169 | 3 | 1.79E+08 | S42_Z105 | S42_Z12 | Latents | rs2339798_ | 11 |
| 0.998781 | 7169 | 3 | 1.79E+08 | S42_Z117 | S42_Z4 | Latents | rs2339798_ | 11 |
| 0.99882 | 7169 | 3 | 1.79E+08 | S42_Z12 | S42_Z4 | Latents | rs2339798_ | 11 |
| 0.998787 | 7169 | 3 | 1.79E+08 | S1994_Z35 | S42_Z4 | Latents | rs2339798_ | 11 |
| 0.998813 | 7169 | 3 | 1.79E+08 | S1994_Z94 | S42_Z4 | Latents | rs2339798_ | 11 |
| 0.998813 | 7169 | 3 | 1.79E+08 | S1994_Z12 | S42_Z4 | Latents | rs2339798_ | 11 |
| 0.998785 | 7169 | 3 | 1.79E+08 | S42_Z106 | S42_Z4 | Latents | rs2339798_ | 11 |
| 0.998638 | 7169 | 3 | 1.79E+08 | S42_Z105 | S42_Z4 | Latents | rs2339798_ | 11 |
| 0.998902 | 7169 | 3 | 1.79E+08 | S42_Z12 | S42_Z51 | Latents | rs2339798_ | 11 |
| 0.998897 | 7169 | 3 | 1.79E+08 | S1994_Z94 | S42_Z51 | Latents | rs2339798_ | 11 |
| 0.99887 | 7169 | 3 | 1.79E+08 | S42_Z106 | S42_Z51 | Latents | rs2339798_ | 11 |
| 0.998898 | 7169 | 3 | 1.79E+08 | S1994_Z12 | S42_Z51 | Latents | rs2339798_ | 11 |
| 0.998796 | 7169 | 3 | 1.79E+08 | S42_Z4 | S42_Z51 | Latents | rs2339798_ | 11 |
| 0.998869 | 7169 | 3 | 1.79E+08 | S1994_Z35 | S42_Z51 | Latents | rs2339798_ | 11 |
| 0.99886 | 7169 | 3 | 1.79E+08 | S42_Z117 | S42_Z51 | Latents | rs2339798_ | 11 |
| 0.998706 | 7169 | 3 | 1.79E+08 | S42_Z105 | S42_Z51 | Latents | rs2339798_ | 11 |
| 0.998785 | 7169 | 3 | 1.79E+08 | S42_Z4 | S42_Z89 | Latents | rs2339798_ | 11 |
| 0.998887 | 7169 | 3 | 1.79E+08 | S1994_Z94 | S42_Z89 | Latents | rs2339798_ | 11 |
| 0.998694 | 7169 | 3 | 1.79E+08 | S42_Z105 | S42_Z89 | Latents | rs2339798_ | 11 |
| 0.99889 | 7169 | 3 | 1.79E+08 | S42_Z12 | S42_Z89 | Latents | rs2339798_ | 11 |
| 0.99886 | 7169 | 3 | 1.79E+08 | S42_Z106 | S42_Z89 | Latents | rs2339798_ | 11 |
| 0.998888 | 7169 | 3 | 1.79E+08 | S1994_Z12 | S42_Z89 | Latents | rs2339798_ | 11 |
| 0.998849 | 7169 | 3 | 1.79E+08 | S42_Z117 | S42_Z89 | Latents | rs2339798_ | 11 |
| 0.998866 | 7169 | 3 | 1.79E+08 | S42_Z51 | S42_Z89 | Latents | rs2339798_ | 11 |
| 0.99886 | 7169 | 3 | 1.79E+08 | S1994_Z35 | S42_Z89 | Latents | rs2339798_ | 11 |
| 0.96399 | 7465 | 4 | 7850366 | S2023_Z95 | LungFunc | Non cardia | rs2856879 | 12 |
| 0.955213 | 2087 | 4 | 17954590 | S2023_Z86 | ArteryCOR_ | Cardiac eQ | 4:1795621 | 13 |
| 0.955257 | 2087 | 4 | 17954590 | S1993_Z14 | ArteryCOR_ | Cardiac eQ | 4:1795621 | 13 |
| 0.961599 | 2087 | 4 | 17954590 | S1993_Z14 | Bld_chr4_e | Non cardia | 4:1795621 | 13 |
| 0.961565 | 2087 | 4 | 17954590 | S2023_Z86 | Bld_chr4_e | Non cardia | 4:1795621 | 13 |
| 0.98668 | 2087 | 4 | 17954590 | S1993_Z14 | HeelBMD | Non cardia | 4:1795621 | 13 |
| 0.987508 | 2087 | 4 | 17954590 | S2023_Z86 | HeelBMD | Non cardia | 4:1795621 | 13 |
| 0.909046 | 2087 | 4 | 17954590 | S1993_Z14 | Hypertensi | Cardiac dis | 4:1795621 | 13 |

| | | | | | | | | |
|---|---|---|---|---|---|---|---|---|
| 0.91821 | 2087 | 4 | 17954590 | S2023_Z86 | Hypertensi | Cardiac dis | 4:1795621: | 13 |
| 0.96567 | 2087 | 4 | 17954590 | S1993_Z14 | MuscleSKL_ | Non cardia | 4:1795621: | 13 |
| 0.965811 | 2087 | 4 | 17954590 | S2023_Z86 | MuscleSKL_ | Non cardia | 4:1795621: | 13 |
| 0.991302 | 2087 | 4 | 17954590 | S1993_Z14 | PP | ECG and bl | 4:1795621: | 13 |
| 0.990908 | 2087 | 4 | 17954590 | S2023_Z86 | PP | ECG and bl | 4:1795621: | 13 |
| 0.990002 | 2087 | 4 | 17954590 | S1993_Z14 | S2023_Z86 | Latents | 4:1795621: | 13 |
| 0.998866 | 3462 | 5 | 56513311 | S1701_Z11 | CVD | Cardiac dis | rs256903_( | 14 |
| 0.998924 | 3462 | 5 | 56513311 | S1701_Z11 | HDLcholest | Non cardia | rs256903_( | 14 |
| 0.998781 | 3462 | 5 | 56513311 | S1701_Z11 | Hypertensi | Cardiac dis | rs256903_( | 14 |
| 0.998937 | 3462 | 5 | 56513311 | S1701_Z11 | PP | ECG and bl | rs256903_( | 14 |
| 0.998941 | 3462 | 5 | 56513311 | S1701_Z11 | Triglyceride | Non cardia | rs256903_( | 14 |
| 0.999978 | 3464 | 5 | 1.33E+08 | S1994_Z1 | AdiposeSU | Non cardia | rs7280147- | 16 |
| 0.99986 | 3464 | 5 | 1.33E+08 | S1994_Z1 | ArteryTIB_( | Cardiac eQ | rs7280147- | 16 |
| 0.999981 | 3464 | 5 | 1.33E+08 | S1994_Z1 | Bld_chr5_e | Non cardia | rs7280147- | 16 |
| 0.99996 | 3464 | 5 | 1.33E+08 | S1994_Z1 | CVD | Cardiac dis | rs7280147- | 16 |
| 0.997871 | 3464 | 5 | 1.33E+08 | S1994_Z1 | DescAorta_ | MRI-derive | rs7280147- | 16 |
| 0.995753 | 3464 | 5 | 1.33E+08 | S1994_Z1 | Lung_chr5_ | Non cardia | rs7280147- | 16 |
| 0.999016 | 3464 | 5 | 1.33E+08 | S1994_Z1 | MuscleSKL_ | Non cardia | rs7280147- | 16 |
| 0.978887 | 5549 | 6 | 2478852 | S1701_Z11 | HeelBMD | Non cardia | rs1045814: | 17 |
| 0.987838 | 5549 | 6 | 2478852 | S1701_Z11 | S1993_Z28 | Latents | rs1045814: | 17 |
| 0.983145 | 5549 | 6 | 2478852 | S1701_Z11 | S1993_Z6 | Latents | rs1045814: | 17 |
| 0.987746 | 5549 | 6 | 2478852 | S1701_Z11 | S1993_Z97 | Latents | rs1045814: | 17 |
| 0.987756 | 5549 | 6 | 2478852 | S1701_Z11 | S1994_Z10 | Latents | rs1045814: | 17 |
| 0.987893 | 5549 | 6 | 2478852 | S1701_Z11 | S2023_Z44 | Latents | rs1045814: | 17 |
| 0.974232 | 5549 | 6 | 2478852 | S1701_Z11 | S2023_Z95 | Latents | rs1045814: | 17 |
| 0.983607 | 5549 | 6 | 2491373 | S1993_Z97 | HeelBMD | Non cardia | 6:2491607_ | 17 |
| 0.988488 | 5549 | 6 | 2491373 | S1993_Z97 | S1994_Z10 | Latents | 6:2491607_ | 17 |
| 0.989234 | 5549 | 6 | 2491373 | S1993_Z97 | S2023_Z44 | Latents | 6:2491607_ | 17 |
| 0.946681 | 5549 | 6 | 2491373 | S1993_Z97 | S2023_Z95 | Latents | 6:2491607_ | 17 |
| 0.982607 | 5549 | 6 | 2502939 | S2023_Z44 | HeelBMD | Non cardia | rs4959678_ | 17 |
| 0.988573 | 5549 | 6 | 2502939 | S1994_Z10 | HeelBMD | Non cardia | rs4959678_ | 17 |
| 0.988537 | 5549 | 6 | 2502939 | S1993_Z28 | HeelBMD | Non cardia | rs4959678_ | 17 |
| 0.990394 | 5549 | 6 | 2502939 | S1993_Z28 | S1993_Z6 | Latents | rs4959678_ | 17 |
| 0.989831 | 5549 | 6 | 2502939 | S1993_Z28 | S1993_Z97 | Latents | rs4959678_ | 17 |
| 0.992824 | 5549 | 6 | 2502939 | S1993_Z28 | S1994_Z10 | Latents | rs4959678_ | 17 |
| 0.990541 | 5549 | 6 | 2502939 | S1993_Z28 | S2023_Z44 | Latents | rs4959678_ | 17 |
| 0.989921 | 5549 | 6 | 2502939 | S1994_Z10 | S2023_Z44 | Latents | rs4959678_ | 17 |
| 0.969698 | 5549 | 6 | 2502939 | S2023_Z44 | S2023_Z95 | Latents | rs4959678_ | 17 |
| 0.985111 | 5549 | 6 | 2502939 | S1994_Z10 | S2023_Z95 | Latents | rs4959678_ | 17 |
| 0.979362 | 5549 | 6 | 2502939 | S1993_Z28 | S2023_Z95 | Latents | rs4959678_ | 17 |
| 0.985189 | 5549 | 6 | 2503817 | S1993_Z22 | S1993_Z6 | Latents | rs1680832_ | 17 |
| 0.991498 | 5549 | 6 | 2503817 | S1993_Z22 | S2023_Z95 | Latents | rs1680832_ | 17 |
| 0.933062 | 5549 | 6 | 2507667 | S2023_Z95 | HeelBMD | Non cardia | rs1124277( | 17 |
| 0.974464 | 5549 | 6 | 2507667 | S1993_Z6 | HeelBMD | Non cardia | rs1124277( | 17 |
| 0.97676 | 5549 | 6 | 2507667 | S1993_Z6 | S1993_Z97 | Latents | rs1124277( | 17 |

| | | | | | | | | |
|---|---|---|---|---|---|---|---|---|
| 0.991303 | 5549 | 6 | 2507667 | S1993_Z6 | S1994_Z10 | Latents | rs1124277! | 17 |
| 0.984107 | 5549 | 6 | 2507667 | S1993_Z6 | S2023_Z44 | Latents | rs1124277! | 17 |
| 0.994298 | 5549 | 6 | 2507667 | S1993_Z6 | S2023_Z95 | Latents | rs1124277! | 17 |
| 0.995496 | 5367 | 6 | 1.18E+08 | S1994_Z31 | RA_maxVo | MRI-derive | rs3951016_ | 18 |
| 0.978383 | 5367 | 6 | 1.18E+08 | S1994_Z31 | RA_minVol | MRI-derive | rs3951016_ | 18 |
| 0.860695 | 5367 | 6 | 1.18E+08 | S1994_Z31 | RV_Stroke\ | MRI-derive | rs3951016_ | 18 |
| 0.831291 | 5367 | 6 | 1.18E+08 | S1701_Z82 | ArteryAOR_ | Cardiac eQ | rs1115373( | 18 |
| 0.99255 | 5367 | 6 | 1.18E+08 | S1701_Z82 | DBP | ECG and bl | rs1115373( | 18 |
| 0.93904 | 5367 | 6 | 1.18E+08 | S1701_Z82 | LV_endDia | MRI-derive | rs1115373( | 18 |
| 0.987497 | 5367 | 6 | 1.18E+08 | S1701_Z82 | LV_endSys | MRI-derive | rs1115373( | 18 |
| 0.998644 | 5367 | 6 | 1.18E+08 | S1701_Z82 | PRint | ECG and bl | rs1115373( | 18 |
| 0.993675 | 5367 | 6 | 1.18E+08 | S1701_Z82 | RA_maxVo | MRI-derive | rs1115373( | 18 |
| 0.815973 | 5367 | 6 | 1.18E+08 | S1701_Z82 | RA_minVol | MRI-derive | rs1115373( | 18 |
| 0.996262 | 5367 | 6 | 1.18E+08 | S1701_Z82 | rHeartRate | ECG and bl | rs1115373( | 18 |
| 0.994432 | 5367 | 6 | 1.18E+08 | S1701_Z82 | RV_Stroke\ | MRI-derive | rs1115373( | 18 |
| 0.987992 | 5367 | 6 | 1.18E+08 | S1701_Z82 | S1994_Z31 | Latents | rs1115373( | 18 |
| 0.998341 | 7774 | 6 | 1.21E+08 | S1994_Z92 | PulseRate | ECG and bl | rs6900506_ | 19 |
| 0.998568 | 7774 | 6 | 1.21E+08 | S1994_Z92 | rHeartRate | ECG and bl | rs6900506_ | 19 |
| 0.996238 | 7761 | 6 | 1.21E+08 | S42_Z12 | PP | ECG and bl | 6:1217771 | 19 |
| 0.998081 | 7761 | 6 | 1.21E+08 | S42_Z12 | PRint | ECG and bl | 6:1217771 | 19 |
| 0.943716 | 7761 | 6 | 1.21E+08 | S42_Z12 | PulseRate | ECG and bl | 6:1217771 | 19 |
| 0.968461 | 7761 | 6 | 1.21E+08 | S42_Z12 | PulseRate | ECG and bl | 6:1217771 | 19 |
| 0.998759 | 7761 | 6 | 1.21E+08 | S42_Z12 | rHeartRate | ECG and bl | 6:1217771 | 19 |
| 0.998338 | 5551 | 6 | 1.22E+08 | S1701_Z82 | HeelBMD | Non cardia | rs9388001_ | 20 |
| 0.99997 | 5551 | 6 | 1.22E+08 | S1994_Z92 | HeelBMD | Non cardia | rs9388001_ | 20 |
| 0.998014 | 5551 | 6 | 1.22E+08 | S42_Z10 | HeelBMD | Non cardia | rs9388001_ | 20 |
| 0.999973 | 5551 | 6 | 1.22E+08 | S1994_Z92 | PP | ECG and bl | rs9388001_ | 20 |
| 0.998116 | 5551 | 6 | 1.22E+08 | S1701_Z82 | PP | ECG and bl | rs9388001_ | 20 |
| 0.997953 | 5551 | 6 | 1.22E+08 | S42_Z10 | PP | ECG and bl | rs9388001_ | 20 |
| 0.997453 | 5551 | 6 | 1.22E+08 | S42_Z10 | PulseRate | ECG and bl | rs9388001_ | 20 |
| 0.99721 | 5551 | 6 | 1.22E+08 | S1701_Z82 | PulseRate | ECG and bl | rs9388001_ | 20 |
| 0.999965 | 5551 | 6 | 1.22E+08 | S1701_Z82 | S1994_Z92 | Latents | rs9388001_ | 20 |
| 0.998159 | 5551 | 6 | 1.22E+08 | S1701_Z82 | S42_Z10 | Latents | rs9388001_ | 20 |
| 0.999834 | 5551 | 6 | 1.22E+08 | S1994_Z92 | S42_Z10 | Latents | rs9388001_ | 20 |
| 0.971948 | 5547 | 6 | 1.26E+08 | S1701_Z29 | AdiposeSU | Non cardia | 6:1267078 | 21 |
| 0.963601 | 5547 | 6 | 1.26E+08 | S42_Z125 | AdiposeSU | Non cardia | 6:1267078 | 21 |
| 0.958706 | 5547 | 6 | 1.26E+08 | S42_Z125 | HeartAA_c | Cardiac eQ | 6:1267078 | 21 |
| 0.96059 | 5547 | 6 | 1.26E+08 | S1701_Z29 | HeartAA_c | Cardiac eQ | 6:1267078 | 21 |
| 0.964035 | 5547 | 6 | 1.26E+08 | S1701_Z29 | S2023_Z87 | Latents | 6:1267078 | 21 |
| 0.969872 | 5547 | 6 | 1.26E+08 | S1701_Z29 | S42_Z125 | Latents | 6:1267078 | 21 |
| 0.963631 | 5547 | 6 | 1.26E+08 | S2023_Z87 | AdiposeSU | Non cardia | rs2184968_ | 21 |
| 0.960348 | 5547 | 6 | 1.26E+08 | S2023_Z87 | HeartAA_c | Cardiac eQ | rs2184968_ | 21 |
| 0.966223 | 5547 | 6 | 1.26E+08 | S2023_Z87 | S42_Z125 | Latents | rs2184968_ | 21 |
| 0.952467 | 5552 | 6 | 1.43E+08 | S2023_Z10 | AdiposeSU | Non cardia | rs263182_ | 22 |
| 0.988568 | 5552 | 6 | 1.43E+08 | S2023_Z10 | BodyFFM | Non cardia | rs263182_ | 22 |

| | | | | | | | | |
|---|---|---|---|---|---|---|---|---|
| 0.909926 | 5552 | 6 | 1.43E+08 | S2023_Z10 | LungFunc | Non cardia | rs263182_ | 22 |
| 0.900911 | 9271 | 7 | 46520652 | S42_Z127 | WHRadjBM | Non cardia | rs3516477 | 23 |
| 0.842604 | 9192 | 7 | 1.21E+08 | S2023_Z86 | Lung_chr7_ | Non cardia | rs1026167 | 24 |
| 1 | 9246 | 7 | 1.21E+08 | S2023_Z86 | HeelBMD | Non cardia | rs5636461 | 24 |
| 0.921762 | 6053 | 8 | 1.08E+08 | S2023_Z86 | MuscleSKL_ | Non cardia | rs640068_ | 26 |
| 0.926507 | 5938 | 8 | 1.19E+08 | S1701_Z80 | AdiposeSU | Non cardia | rs7012790_ | 27 |
| 0.943225 | 5938 | 8 | 1.19E+08 | S1701_Z80 | ArteryAOR_ | Cardiac eQ | rs7012790_ | 27 |
| 0.900161 | 5938 | 8 | 1.19E+08 | S1701_Z80 | ArteryTIB_ | Cardiac eQ | rs7012790_ | 27 |
| 0.986516 | 5938 | 8 | 1.19E+08 | S1701_Z80 | BodyFFM | Non cardia | rs7012790_ | 27 |
| 0.93189 | 5938 | 8 | 1.19E+08 | S1701_Z80 | DBP | ECG and bl | rs7012790_ | 27 |
| 0.949427 | 5938 | 8 | 1.19E+08 | S1701_Z80 | HeartLV_ch | Cardiac eQ | rs7012790_ | 27 |
| 0.853744 | 10094 | 10 | 21741274 | S1993_Z14 | BodyFPC | Non cardia | 10:220302 | 28 |
| 0.98444 | 10108 | 10 | 88059229 | S1994_Z1 | LungFunc | Non cardia | rs2975112_ | 29 |
| 0.999554 | 3059 | 11 | 95290209 | S1701_Z29 | S1994_Z5 | Latents | rs7728253 | 31 |
| 0.999641 | 3059 | 11 | 95290209 | S1701_Z29 | S42_Z125 | Latents | rs7728253 | 31 |
| 0.999379 | 3059 | 11 | 95290209 | S1994_Z5 | S42_Z125 | Latents | rs7728253 | 31 |
| 0.997314 | 5704 | 12 | 4275678 | S1994_Z5 | AdiposeSU | Non cardia | rs7689596 | 32 |
| 0.999997 | 5704 | 12 | 4275678 | S1994_Z5 | ArteryTIB_ | Cardiac eQ | rs7689596 | 32 |
| 1 | 5704 | 12 | 4275678 | S1994_Z5 | BodyFFM | Non cardia | rs7689596 | 32 |
| 0.999987 | 5704 | 12 | 4275678 | S1994_Z5 | CVD | Cardiac dis | rs7689596 | 32 |
| 1 | 5704 | 12 | 4275678 | S1994_Z5 | HeelBMD | Non cardia | rs7689596 | 32 |
| 1 | 5704 | 12 | 4275678 | S1994_Z5 | MuscleSKL_ | Non cardia | rs7689596 | 32 |
| 0.999997 | 5704 | 12 | 4275678 | S1994_Z5 | Triglycerid | Non cardia | rs7689596 | 32 |
| 0.994975 | 9782 | 12 | 24605546 | S1701_Z82 | LV_Stroke\ | MRI-derive | rs4963772_ | 33 |
| 0.994608 | 9782 | 12 | 24605546 | S42_Z10 | LV_Stroke\ | MRI-derive | rs4963772_ | 33 |
| 0.994998 | 9782 | 12 | 24605546 | S42_Z10 | PP | ECG and bl | rs4963772_ | 33 |
| 0.99537 | 9782 | 12 | 24605546 | S1701_Z82 | PP | ECG and bl | rs4963772_ | 33 |
| 0.996928 | 9782 | 12 | 24605546 | S42_Z10 | PulseRate | ECG and bl | rs4963772_ | 33 |
| 0.996311 | 9782 | 12 | 24605546 | S1701_Z82 | PulseRate | ECG and bl | rs4963772_ | 33 |
| 0.994549 | 9782 | 12 | 24605546 | S1701_Z82 | S1994_Z85 | Latents | rs4963772_ | 33 |
| 0.99568 | 9782 | 12 | 24605546 | S1701_Z82 | S42_Z10 | Latents | rs4963772_ | 33 |
| 0.995231 | 9782 | 12 | 24620985 | S1701_Z12 | LV_Stroke\ | MRI-derive | rs1126307 | 33 |
| 0.996421 | 9782 | 12 | 24620985 | S1701_Z12 | PP | ECG and bl | rs1126307 | 33 |
| 0.993264 | 9782 | 12 | 24620985 | S1701_Z12 | PulseRate | ECG and bl | rs1126307 | 33 |
| 0.99504 | 9782 | 12 | 24620985 | S1701_Z12 | S1701_Z49 | Latents | rs1126307 | 33 |
| 0.994736 | 9782 | 12 | 24620985 | S1701_Z12 | S1701_Z82 | Latents | rs1126307 | 33 |
| 0.995555 | 9782 | 12 | 24620985 | S1701_Z12 | S1994_Z85 | Latents | rs1126307 | 33 |
| 0.994028 | 9782 | 12 | 24620985 | S1701_Z12 | S42_Z10 | Latents | rs1126307 | 33 |
| 0.995185 | 9782 | 12 | 24631205 | S1994_Z85 | LV_Stroke\ | MRI-derive | rs4246224_ | 33 |
| 0.996441 | 9782 | 12 | 24631205 | S1994_Z85 | PP | ECG and bl | rs4246224_ | 33 |
| 0.99256 | 9782 | 12 | 24631205 | S1994_Z85 | PulseRate | ECG and bl | rs4246224_ | 33 |
| 0.993736 | 9782 | 12 | 24631205 | S1994_Z85 | S42_Z10 | Latents | rs4246224_ | 33 |
| 0.994898 | 9782 | 12 | 24635405 | S1701_Z49 | LV_Stroke\ | MRI-derive | rs1104754 | 33 |
| 0.996158 | 9782 | 12 | 24635405 | S1701_Z49 | PP | ECG and bl | rs1104754 | 33 |
| 0.994245 | 9782 | 12 | 24635405 | S1701_Z49 | PulseRate | ECG and bl | rs1104754 | 33 |

| | | | | | | | | | |
|---|---|---|---|---|---|---|---|---|---|
| 0.994758 | 9782 | 12 | 24635405 | S1701_Z49 | S1701_Z82 | Latents | rs1104754: | | 33 |
| 0.995025 | 9782 | 12 | 24635405 | S1701_Z49 | S1994_Z85 | Latents | rs1104754: | | 33 |
| 0.994665 | 9782 | 12 | 24635405 | S1701_Z49 | S42_Z10 | Latents | rs1104754: | | 33 |
| 0.872922 | 4095 | 12 | 28259482 | S2023_Z77 | AdiposeSU | Non cardia | rs1104948! | | 34 |
| 0.911349 | 4095 | 12 | 28259482 | S2023_Z77 | AdiposeVIS | Non cardia | rs1104948! | | 34 |
| 0.959161 | 4095 | 12 | 28259482 | S2023_Z77 | HeelBMD | Non cardia | rs1104948! | | 34 |
| 0.992247 | 4095 | 12 | 28259482 | S2023_Z77 | S2023_Z86 | Latents | rs1104948! | | 34 |
| 0.861121 | 4095 | 12 | 28391531 | S2023_Z86 | AdiposeSU | Non cardia | rs3741760_ | | 34 |
| 0.895045 | 4095 | 12 | 28391531 | S2023_Z86 | AdiposeVIS | Non cardia | rs3741760_ | | 34 |
| 0.970331 | 4095 | 12 | 28391531 | S2023_Z86 | HeelBMD | Non cardia | rs3741760_ | | 34 |
| 0.994362 | 4307 | 13 | 42163132 | S1994_Z38 | BodyFFM | Non cardia | rs9594689_ | NA | |
| 0.859066 | 4310 | 13 | 50514411 | S1701_Z53 | DBP | ECG and bl | rs9535455_ | | 35 |
| 0.927873 | 4310 | 13 | 50514411 | S1701_Z53 | HeelBMD | Non cardia | rs9535455_ | | 35 |
| 0.931857 | 4310 | 13 | 50514411 | S1701_Z53 | WHRadjBM | Non cardia | rs9535455_ | | 35 |
| 0.998795 | 3888 | 14 | 23395595 | S1701_Z12 | AFib | Cardiac dis | rs422068_T | | 37 |
| 0.998866 | 3888 | 14 | 23395595 | S1994_Z11 | AFib | Cardiac dis | rs422068_T | | 37 |
| 0.998955 | 3888 | 14 | 23395595 | S1994_Z11 | DescAorta_ | MRI-derive | rs422068_T | | 37 |
| 0.998934 | 3888 | 14 | 23395595 | S1701_Z12 | DescAorta_ | MRI-derive | rs422068_T | | 37 |
| 0.99876 | 3888 | 14 | 23395595 | S1994_Z11 | DescAorta_ | MRI-derive | rs422068_T | | 37 |
| 0.998743 | 3888 | 14 | 23395595 | S1701_Z12 | DescAorta_ | MRI-derive | rs422068_T | | 37 |
| 0.995573 | 3888 | 14 | 23395595 | S1701_Z12 | LV_Stroke\ | MRI-derive | rs422068_T | | 37 |
| 0.995652 | 3888 | 14 | 23395595 | S1994_Z11 | LV_Stroke\ | MRI-derive | rs422068_T | | 37 |
| 0.999305 | 3888 | 14 | 23395595 | S1994_Z11 | PP | ECG and bl | rs422068_T | | 37 |
| 0.999207 | 3888 | 14 | 23395595 | S1701_Z12 | PP | ECG and bl | rs422068_T | | 37 |
| 0.999342 | 3888 | 14 | 23395595 | S1994_Z11 | PulseRate | ECG and bl | rs422068_T | | 37 |
| 0.999239 | 3888 | 14 | 23395595 | S1701_Z12 | PulseRate | ECG and bl | rs422068_T | | 37 |
| 0.999041 | 3888 | 14 | 23395595 | S1994_Z11 | RV_EF | MRI-derive | rs422068_T | | 37 |
| 0.998986 | 3888 | 14 | 23395595 | S1701_Z12 | RV_EF | MRI-derive | rs422068_T | | 37 |
| 0.998865 | 3888 | 14 | 23395595 | S1994_Z11 | RV_Stroke\ | MRI-derive | rs422068_T | | 37 |
| 0.998806 | 3888 | 14 | 23395595 | S1701_Z12 | RV_Stroke\ | MRI-derive | rs422068_T | | 37 |
| 0.961092 | 3888 | 14 | 23395595 | S1701_Z12 | S1701_Z82 | Latents | rs422068_T | | 37 |
| 0.999111 | 3888 | 14 | 23395595 | S1701_Z12 | S1994_Z11 | Latents | rs422068_T | | 37 |
| 0.95635 | 3888 | 14 | 23412935 | S1701_Z82 | AFib | Cardiac dis | rs2284651_ | | 37 |
| 0.967267 | 3888 | 14 | 23412935 | S1701_Z82 | DescAorta_ | MRI-derive | rs2284651_ | | 37 |
| 0.970288 | 3888 | 14 | 23412935 | S1701_Z82 | DescAorta_ | MRI-derive | rs2284651_ | | 37 |
| 0.924568 | 3888 | 14 | 23412935 | S1701_Z82 | LV_Stroke\ | MRI-derive | rs2284651_ | | 37 |
| 0.962534 | 3888 | 14 | 23412935 | S1701_Z82 | PP | ECG and bl | rs2284651_ | | 37 |
| 0.962626 | 3888 | 14 | 23412935 | S1701_Z82 | PulseRate | ECG and bl | rs2284651_ | | 37 |
| 0.961211 | 3888 | 14 | 23412935 | S1701_Z82 | RV_EF | MRI-derive | rs2284651_ | | 37 |
| 0.970177 | 3888 | 14 | 23412935 | S1701_Z82 | RV_Stroke\ | MRI-derive | rs2284651_ | | 37 |
| 0.961206 | 3888 | 14 | 23412935 | S1701_Z82 | S1994_Z11 | Latents | rs2284651_ | | 37 |
| 0.996331 | 5688 | 15 | 92801441 | S1701_Z32 | S1701_Z80 | Latents | rs4404018_ | | 39 |
| 0.998528 | 708 | 19 | 44908684 | S42_Z28 | CAD | Cardiac dis | rs429358_T | | 41 |
| 0.997825 | 708 | 19 | 44908684 | S2023_Z87 | CAD | Cardiac dis | rs429358_T | | 41 |
| 0.999899 | 708 | 19 | 44908684 | S42_Z28 | CVD | Cardiac dis | rs429358_T | | 41 |

| | | | | | | | | |
|---|---|---|---|---|---|---|---|---|
| 0.999442 | 708 | 19 | 44908684 | S2023_Z87 | CVD | Cardiac dis | rs429358_ | 41 |
| 0.996659 | 708 | 19 | 44908684 | S42_Z28 | HDLcholest | Non cardia | rs429358_ | 41 |
| 0.994932 | 708 | 19 | 44908684 | S2023_Z87 | HDLcholest | Non cardia | rs429358_ | 41 |
| 0.999899 | 708 | 19 | 44908684 | S42_Z28 | LiverFAT | Non cardia | rs429358_ | 41 |
| 0.999442 | 708 | 19 | 44908684 | S2023_Z87 | LiverFAT | Non cardia | rs429358_ | 41 |
| 0.991438 | 708 | 19 | 44908684 | S2023_Z87 | MInf | Cardiac dis | rs429358_ | 41 |
| 0.998163 | 708 | 19 | 44908684 | S2023_Z87 | PP | ECG and bl | rs429358_ | 41 |
| 0.998987 | 708 | 19 | 44908684 | S42_Z28 | PP | ECG and bl | rs429358_ | 41 |
| 0.994504 | 708 | 19 | 44908684 | S42_Z28 | PulseRate | ECG and bl | rs429358_ | 41 |
| 0.994722 | 708 | 19 | 44908684 | S2023_Z87 | PulseRate | ECG and bl | rs429358_ | 41 |
| 0.998882 | 708 | 19 | 44908684 | S2023_Z87 | S42_Z28 | Latents | rs429358_ | 41 |
| 0.9318 | 708 | 19 | 44908684 | S2023_Z87 | Triglyceride | Non cardia | rs429358_ | 41 |
| 0.999793 | 708 | 19 | 44908684 | S42_Z28 | WHRadjBM | Non cardia | rs429358_ | 41 |
| 0.999293 | 708 | 19 | 44908684 | S2023_Z87 | WHRadjBM | Non cardia | rs429358_ | 41 |
| 0.900988 | 691 | 19 | 55534345 | S42_Z125 | AdiposeVIS | Non cardia | rs7306110 | 42 |
| 0.986722 | 8665 | 20 | 38213512 | S1701_Z82 | LV_Stroke\ | MRI-derive | rs3746471_ | 43 |
| 0.992996 | 8665 | 20 | 38213512 | S1701_Z82 | PP | ECG and bl | rs3746471_ | 43 |
| 0.993375 | 8665 | 20 | 38213512 | S1701_Z82 | PRint | ECG and bl | rs3746471_ | 43 |
| 0.984396 | 8665 | 20 | 38213512 | S1701_Z82 | rHeartRate | ECG and bl | rs3746471_ | 43 |
| 0.985458 | 8665 | 20 | 38213512 | S1701_Z82 | RV_Stroke\ | MRI-derive | rs3746471_ | 43 |
| 0.910809 | 4674 | 22 | 29728844 | S1701_Z93 | AdiposeSU | Non cardia | rs140097_ | 44 |
| 0.879463 | 4674 | 22 | 29728844 | S1701_Z93 | ArteryAOR_ | Cardiac eQ | rs140097_ | 44 |
| 0.943302 | 4674 | 22 | 29728844 | S1701_Z93 | HeartLV_ch | Cardiac eQ | rs140097_ | 44 |
| 0.914283 | 4674 | 22 | 29728844 | S1701_Z93 | Liver_chr2 | Non cardia | rs140097_ | 44 |
| 0.958106 | 4674 | 22 | 29728844 | S1701_Z93 | Pancreas_c | Non cardia | rs140097_ | 44 |
| 0.982597 | 4674 | 22 | 29728844 | S1701_Z93 | S1993_Z27 | Latents | rs140097_ | 44 |
| 0.979702 | 4674 | 22 | 29728844 | S1701_Z93 | S1993_Z28 | Latents | rs140097_ | 44 |
| 0.975059 | 4674 | 22 | 29728844 | S1701_Z93 | S1993_Z41 | Latents | rs140097_ | 44 |
| 0.972098 | 4674 | 22 | 29728844 | S1701_Z93 | S1993_Z66 | Latents | rs140097_ | 44 |
| 0.975881 | 4674 | 22 | 29728844 | S1701_Z93 | S1994_Z1 | Latents | rs140097_ | 44 |
| 0.867808 | 4674 | 22 | 29742559 | S1994_Z1 | ArteryAOR_ | Cardiac eQ | rs140120_ | 44 |
| 0.91609 | 4674 | 22 | 29742559 | S1993_Z41 | ArteryAOR_ | Cardiac eQ | rs140120_ | 44 |
| 0.943033 | 4674 | 22 | 29742559 | S1993_Z27 | ArteryAOR_ | Cardiac eQ | rs140120_ | 44 |
| 0.971503 | 4674 | 22 | 29742559 | S1994_Z1 | HeartLV_ch | Cardiac eQ | rs140120_ | 44 |
| 0.972236 | 4674 | 22 | 29742559 | S1993_Z41 | HeartLV_ch | Cardiac eQ | rs140120_ | 44 |
| 0.969785 | 4674 | 22 | 29742559 | S1993_Z27 | HeartLV_ch | Cardiac eQ | rs140120_ | 44 |
| 0.956899 | 4674 | 22 | 29742559 | S1993_Z41 | Liver_chr2 | Non cardia | rs140120_ | 44 |
| 0.912116 | 4674 | 22 | 29742559 | S1994_Z1 | Liver_chr2 | Non cardia | rs140120_ | 44 |
| 0.95438 | 4674 | 22 | 29742559 | S1993_Z27 | Liver_chr2 | Non cardia | rs140120_ | 44 |
| 0.979812 | 4674 | 22 | 29742559 | S1994_Z1 | Pancreas_c | Non cardia | rs140120_ | 44 |
| 0.979795 | 4674 | 22 | 29742559 | S1993_Z41 | Pancreas_c | Non cardia | rs140120_ | 44 |
| 0.977227 | 4674 | 22 | 29742559 | S1993_Z27 | Pancreas_c | Non cardia | rs140120_ | 44 |
| 0.98618 | 4674 | 22 | 29742559 | S1993_Z27 | S1993_Z28 | Latents | rs140120_ | 44 |
| 0.987304 | 4674 | 22 | 29742559 | S1993_Z27 | S1993_Z41 | Latents | rs140120_ | 44 |
| 0.9851 | 4674 | 22 | 29742559 | S1993_Z41 | S1993_Z66 | Latents | rs140120_ | 44 |

| | | | | | | | | |
|---|---|---|---|---|---|---|---|---|
| 0.985933 | 4674 | 22 | 29742559 | S1993_Z27 | S1993_Z66 | Latents | rs140120_ | 44 |
| 0.987386 | 4674 | 22 | 29742559 | S1993_Z41 | S1994_Z1 | Latents | rs140120_ | 44 |
| 0.986327 | 4674 | 22 | 29742559 | S1993_Z27 | S1994_Z1 | Latents | rs140120_ | 44 |
| 0.935727 | 4674 | 22 | 29761951 | S1993_Z28 | ArteryAOR_ | Cardiac eQ | rs131285_ | 44 |
| 0.956506 | 4674 | 22 | 29761951 | S1993_Z66 | ArteryAOR_ | Cardiac eQ | rs131285_ | 44 |
| 0.973766 | 4674 | 22 | 29761951 | S1993_Z28 | HeartLV_cł | Cardiac eQ | rs131285_ | 44 |
| 0.97823 | 4674 | 22 | 29761951 | S1993_Z66 | HeartLV_cł | Cardiac eQ | rs131285_ | 44 |
| 0.957248 | 4674 | 22 | 29761951 | S1993_Z66 | Liver_chr2 | Non cardia | rs131285_ | 44 |
| 0.955126 | 4674 | 22 | 29761951 | S1993_Z28 | Liver_chr2 | Non cardia | rs131285_ | 44 |
| 0.976825 | 4674 | 22 | 29761951 | S1993_Z28 | Pancreas_c | Non cardia | rs131285_ | 44 |
| 0.975349 | 4674 | 22 | 29761951 | S1993_Z66 | Pancreas_c | Non cardia | rs131285_ | 44 |
| 0.986273 | 4674 | 22 | 29761951 | S1993_Z28 | S1993_Z41 | Latents | rs131285_ | 44 |
| 0.985321 | 4674 | 22 | 29761951 | S1993_Z28 | S1993_Z66 | Latents | rs131285_ | 44 |
| 0.985562 | 4674 | 22 | 29761951 | S1993_Z28 | S1994_Z1 | Latents | rs131285_ | 44 |
| 0.98498 | 4674 | 22 | 29761951 | S1993_Z66 | S1994_Z1 | Latents | rs131285_ | 44 |
