## Supplementary Table 13 for "Hundreds of cardiac MRI traits derived using 3D diffusion autoencoders share a common genetic architecture"

Trait;Significant loci;Cases;Controls;Total N;PMID;Download link;
Non ischaemic cardiomiopathy - NICM;1;1816;388326;390142;30586722;http://ftp.ebi.ac.uk/pub/data
Heart failure - HF;1;6504;387652;394156;30586722;http://ftp.ebi.ac.uk/pub/databases/gwas/summar
Atrial fibrillation - Afib;28;8404;447944;456348;34737426;http://ftp.ebi.ac.uk/pub/databases/gwas/su
Myocardial infarction - Minf;12;8528;447820;456348;34737426;http://ftp.ebi.ac.uk/pub/databases/gw
Pulmonary heart disease - PulmHD;10;2971;453377;456348;34737426;http://ftp.ebi.ac.uk/pub/databa
Coronary atherosclerosis - CAD;39;16041;440307;456348;34737426;http://ftp.ebi.ac.uk/pub/database
Cardiovascular disease - CVD;224;177923 ;306675;306675;33959723;http://ftp.ebi.ac.uk/pub/databas
Cardiac arrythmia;9;7207;477391;484598;33959723;http://ftp.ebi.ac.uk/pub/databases/gwas/summa
LV myocardial fractal dimension;4;;;18096;32814899;http://ftp.ebi.ac.uk/pub/databases/gwas/summa
hypertension;285;144793;313761;458554;30940143;http://ftp.ebi.ac.uk/pub/databases/gwas/summa
PR interval;199;;;271570;32439900;https://personal.broadinstitute.org/ryank/PR_1000g_GWAS_EUR.
Resting heart rate - rHR;339;;;458969;30940143;http://ftp.ebi.ac.uk/pub/databases/gwas/summary_s
Pulse pressure;514;;;757601;30224653;http://ftp.ebi.ac.uk/pub/databases/gwas/summary_statistics/
Pulse rate;250;;;340162;;https://broad-ukb-sumstats-us-east-1.s3.amazonaws.com/round2/additive-ts
Diastolic blood pressure - DBP;215;;;340162;;https://broad-ukb-sumstats-us-east-1.s3.amazonaws.com
Systolic blood pressure - SBP;85;;;340159;;https://broad-ukb-sumstats-us-east-1.s3.amazonaws.com/r
Lung function (FEV1/FVC ratio);349;;;321047;30804560;http://ftp.ebi.ac.uk/pub/databases/gwas/sum
Heel bone mineral density - BMD;571;;;426824;30598549;http://ftp.ebi.ac.uk/pub/databases/gwas/su
Waist to hip ratio (BMI adjusted) - WHRadjBMI;439;;;694649;30239722;Index of /pub/databases/gwas
Body fat free mass - FFM;579;;;155961;30593698;http://ftp.ebi.ac.uk/pub/databases/gwas/summary_
Body fat percentage - FPC;72;;;155961;30593698;http://ftp.ebi.ac.uk/pub/databases/gwas/summary_
HDL cholesterol;76;;;115082;35213538;http://ftp.ebi.ac.uk/pub/databases/gwas/summary_statistics/
Triglyceride levels;62;;;115082;35213538;http://ftp.ebi.ac.uk/pub/databases/gwas/summary_statistics
Liver fat percentage;10;;;32858;34128465;http://ftp.ebi.ac.uk/pub/databases/gwas/summary_statistic
Liver iron content;3;;;32858;34128465;http://ftp.ebi.ac.uk/pub/databases/gwas/summary_statistics/G
Liver volume;12;;;32860;34128465;http://ftp.ebi.ac.uk/pub/databases/gwas/summary_statistics/GCST
Hair colour - Dark brown hair;119;134627;360270;494897;;https://broad-ukb-sumstats-us-east-1.s3.an
Ascending aorta distensibility;6;;;28587;;REGENIE in-house on UKB Data-field 157;
Ascending aorta max area;45;;;28587;;REGENIE in-house on UKB Data-field 157;
Ascending aorta min area;48;;;28587;;REGENIE in-house on UKB Data-field 157;
Descending aorta distensibility;5;;;28587;;REGENIE in-house on UKB Data-field 157;
Descending aorta max area;28;;;28587;;REGENIE in-house on UKB Data-field 157;
Descending aorta min area;26;;;28587;;REGENIE in-house on UKB Data-field 157;
LA ejection fraction;3;;;31921;;REGENIE in-house on UKB Data-field 157;
LA max volume;2;;;31921;;REGENIE in-house on UKB Data-field 157;
LA min volume;3;;;31921;;REGENIE in-house on UKB Data-field 157;
LA stroke volume;2;;;31921;;REGENIE in-house on UKB Data-field 157;
LV circumferential strain;24;;;31921;;REGENIE in-house on UKB Data-field 157;
LV ejection fraction;10;;;31921;;REGENIE in-house on UKB Data-field 157;
LV end diastolic volume;12;;;31921;;REGENIE in-house on UKB Data-field 157;
LV end systolic volume;24;;;31921;;REGENIE in-house on UKB Data-field 157;
LV longitudinal strain;4;;;31921;;REGENIE in-house on UKB Data-field 157;
LV mean myocardial wall thickness;15;;;31921;;REGENIE in-house on UKB Data-field 157;
LV myocardial mass;8;;;31921;;REGENIE in-house on UKB Data-field 157;

LV radial strain;13;;;31921;;REGENIE in-house on UKB Data-field 157;
LV cardiac output;0;;;31921;;REGENIE in-house on UKB Data-field 157;
LV stroke volume;9;;;31921;;REGENIE in-house on UKB Data-field 157;
RA ejection fraction;3;;;31921;;REGENIE in-house on UKB Data-field 157;
RA max volume;4;;;31921;;REGENIE in-house on UKB Data-field 157;
RA min volume;7;;;31921;;REGENIE in-house on UKB Data-field 157;
RA stroke volume;2;;;31921;;REGENIE in-house on UKB Data-field 157;
RV ejection fraction;13;;;31921;;REGENIE in-house on UKB Data-field 157;
RV end diastolic volume;9;;;31921;;REGENIE in-house on UKB Data-field 157;
RV end systolic volume;17;;;31921;;REGENIE in-house on UKB Data-field 157;
RV stroke volume;6;;;31921;;REGENIE in-house on UKB Data-field 157;

abases/gwas/summary_statistics/GCST007001-GCST008000/GCST007714/NICM_HRC_GWAS_UKBB_EU
y_statistics/GCST007001-GCST008000/GCST007715/HF_HRC_GWAS_UKBB_EUR.txt.gz;
immary_statistics/GCST90043001-GCST90044000/GCST90043977/GCST90043977_buildGRCh37.tsv.gz;
vas/summary_statistics/GCST90043001-GCST90044000/GCST90043954/GCST90043954_buildGRCh37.t
ases/gwas/summary_statistics/GCST90043001-GCST90044000/GCST90043962/GCST90043962_buildGF
s/gwas/summary_statistics/GCST90043001-GCST90044000/GCST90043957/GCST90043957_buildGRCh
es/gwas/summary_statistics/GCST90038001-GCST90039000/GCST90038595;
ry_statistics/GCST90038001-GCST90039000/GCST90038611;
iry_statistics/GCST90000001-GCST90001000/GCST90000296/;
iry_statistics/GCST007001-GCST008000/GCST007610;
zip;
tatistics/GCST007001-GCST008000/GCST007609;
GCST006001-GCST007000/GCST006629;
svs/102_irnt.gwas.imputed_v3.both_sexes.tsv.bgz;
n/round2/additive-tsvs/4079_irnt.gwas.imputed_v3.both_sexes.tsv.bgz;
ound2/additive-tsvs/4080_irnt.gwas.imputed_v3.both_sexes.tsv.bgz;
mary_statistics/GCST007001-GCST008000/GCST007431/harmonised/30804560-GCST007431-EFO_000
mmary_statistics/GCST006001-GCST007000/GCST006979/Biobank2-British-Bmd-As-C-Gwas-SumStats.t
/summary_statistics/GCST008001-GCST009000/GCST008999 (ebi.ac.uk);
statistics/GCST007001-GCST008000/GCST007063/FFM_c_maf0.01_meta_pos.txt;
statistics/GCST007001-GCST008000/GCST007064/BFPC_h_maf0.01_meta_pos.txt;
GCST90092001-GCST90093000/GCST90092822;
s/GCST90092001-GCST90093000/GCST90092992;
cs/GCST90016001-GCST90017000/GCST90016673/GCST90016673_buildGRCh37.tsv.gz;
GCST90016001-GCST90017000/GCST90016674;
T90016001-GCST90017000/GCST90016666/GCST90016666_buildGRCh37.tsv.gz;
nazonaws.com/round2/additive-tsvs/1747_4.gwas.imputed_v3.both_sexes.tsv.bgz;

IR.txt.gz;

:sv.gz;
RCh37.tsv.gz;
h37.tsv.gz;

4713-build37.f.tsv.gz;
txt.gz;
