## Supplementary Table 12 for "Hundreds of cardiac MRI traits derived using 3D diffusion autoencoders share a common genetic architecture"

| | group | names | scores | logfoldchar | pvals | pvals_adj |
|---|---|---|---|---|---|---|
| 0 | Atrial Cardi | MYH6 | 571.1518 | 6.050993 | 0 | 0 |
| 1 | Atrial Cardi | TTN | 565.713 | 4.842293 | 0 | 0 |
| 2 | Atrial Cardi | PLN | 178.6273 | 2.272183 | 0 | 0 |
| 3 | Atrial Cardi | DES | 175.944 | 2.153101 | 0 | 0 |
| 4 | Atrial Cardi | MLF1 | 171.2207 | 2.678606 | 0 | 0 |
| 5 | Atrial Cardi | VSTM2L | 107.4507 | 4.603875 | 0 | 0 |
| 6 | Atrial Cardi | CCDC141 | 106.6184 | 1.376574 | 0 | 0 |
| 7 | Atrial Cardi | MLLT10 | 88.48335 | 0.983779 | 0 | 0 |
| 8 | Atrial Cardi | SPEG | 88.36505 | 1.967986 | 0 | 0 |
| 9 | Atrial Cardi | MFN1 | 66.6959 | 1.091875 | 0 | 0 |
| 10 | Atrial Cardi | CCDC91 | 48.74859 | 0.577819 | 0 | 0 |
| 11 | Atrial Cardi | RSRC1 | 35.68112 | 0.483832 | 1.32E-275 | 3.49E-275 |
| 12 | Atrial Cardi | MTMR3 | 33.12246 | 0.461664 | 3.67E-238 | 9.40E-238 |
| 13 | Atrial Cardi | CEP85L | 27.42109 | 0.356268 | 2.08E-164 | 4.87E-164 |
| 14 | Atrial Cardi | WARS2 | 23.78091 | 0.527931 | 2.41E-124 | 5.50E-124 |
| 15 | Atrial Cardi | CWC27 | 22.48053 | 0.353335 | 2.11E-111 | 4.56E-111 |
| 16 | Atrial Cardi | SUPT7L | 22.45193 | 0.615563 | 4.16E-111 | 8.74E-111 |
| 17 | Atrial Cardi | GJA1 | 21.72331 | 0.383725 | 3.53E-104 | 7.06E-104 |
| 18 | Atrial Cardi | NIPSNAP1 | 21.69355 | 0.983818 | 7.09E-104 | 1.39E-103 |
| 19 | Atrial Cardi | DCAF16 | 21.60987 | 0.52917 | 4.08E-103 | 7.78E-103 |
| 20 | Atrial Cardi | AFAP1 | 21.58222 | 0.451955 | 7.17E-103 | 1.34E-102 |
| 21 | Atrial Cardi | GFM1 | 17.92543 | 0.325851 | 1.21E-71 | 2.11E-71 |
| 22 | Atrial Cardi | ASCC2 | 17.38231 | 0.351465 | 1.72E-67 | 2.88E-67 |
| 23 | Atrial Cardi | CMTM5 | 17.00704 | 2.224686 | 1.12E-64 | 1.84E-64 |
| 24 | Atrial Cardi | CR1L | 16.10498 | 1.076128 | 3.31E-58 | 5.22E-58 |
| 25 | Atrial Cardi | RSPO2 | 15.61293 | 1.116393 | 8.05E-55 | 1.25E-54 |
| 26 | Atrial Cardi | DNAJC1 | 14.76932 | 0.183006 | 2.87E-49 | 4.21E-49 |
| 27 | Atrial Cardi | SKIDA1 | 12.19775 | 1.020907 | 3.58E-34 | 5.06E-34 |
| 28 | Atrial Cardi | C2orf16 | 7.800368 | 1.084508 | 6.29E-15 | 8.32E-15 |
| 29 | Atrial Cardi | HSPA4 | 7.026895 | 0.134842 | 2.14E-12 | 2.78E-12 |
| 30 | Atrial Cardi | FAM174B | 6.990696 | 0.173626 | 2.77E-12 | 3.55E-12 |
| 31 | Atrial Cardi | LCORL | 6.150843 | 0.101816 | 7.76E-10 | 9.50E-10 |
| 32 | Atrial Cardi | SHOX2 | 6.125534 | 0.372885 | 9.10E-10 | 1.10E-09 |
| 33 | Atrial Cardi | ZNF219 | 5.680629 | 0.23175 | 1.35E-08 | 1.57E-08 |
| 34 | Atrial Cardi | HORMAD2 | 3.097201 | 0.452964 | 0.001955 | 0.002166 |
| 35 | Atrial Cardi | ZNF512 | -2.19516 | -0.04806 | 0.028156 | 0.030379 |
| 36 | Atrial Cardi | UQCR10 | -2.38906 | -0.04635 | 0.016895 | 0.018472 |
| 37 | Atrial Cardi | WNT16 | -4.70235 | -1.46655 | 2.58E-06 | 2.89E-06 |
| 38 | Atrial Cardi | ZMAT5 | -5.49146 | -0.2115 | 4.00E-08 | 4.56E-08 |
| 39 | Atrial Cardi | ZNF628 | -5.67974 | -0.45745 | 1.36E-08 | 1.57E-08 |
| 40 | Atrial Cardi | TOMM40 | -5.90796 | -0.14971 | 3.48E-09 | 4.14E-09 |
| 41 | Atrial Cardi | SLC5A6 | -6.17625 | -0.27595 | 6.61E-10 | 8.21E-10 |
| 42 | Atrial Cardi | CCDC121 | -6.83252 | -0.50649 | 8.42E-12 | 1.06E-11 |
| 43 | Atrial Cardi | NAT14 | -8.71232 | -0.57902 | 3.05E-18 | 4.10E-18 |

| | | | | | | |
|---|---|---|---|---|---|---|
| 44 | Atrial Cardi | B3GNT7 | -10.7219 | -0.9912 | 8.47E-27 | 1.16E-26 |
| 45 | Atrial Cardi | EIF2B4 | -10.7642 | -0.30552 | 5.39E-27 | 7.50E-27 |
| 46 | Atrial Cardi | CENPW | -13.3152 | -1.42509 | 2.12E-40 | 3.04E-40 |
| 47 | Atrial Cardi | GNB4 | -15.1289 | -0.29079 | 1.31E-51 | 1.96E-51 |
| 48 | Atrial Cardi | ZCCHC10 | -15.3203 | -0.3274 | 7.14E-53 | 1.08E-52 |
| 49 | Atrial Cardi | ZNF513 | -16.8643 | -0.82498 | 1.13E-63 | 1.82E-63 |
| 50 | Atrial Cardi | NCAPG | -17.7425 | -1.75861 | 2.63E-70 | 4.50E-70 |
| 51 | Atrial Cardi | SLC35F1 | -18.4769 | -0.49411 | 5.23E-76 | 9.33E-76 |
| 52 | Atrial Cardi | LXN | -21.577 | -0.6049 | 7.38E-103 | 1.34E-102 |
| 53 | Atrial Cardi | GPN1 | -22.0029 | -0.83422 | 6.88E-107 | 1.41E-106 |
| 54 | Atrial Cardi | DGKH | -22.8968 | -0.33093 | 1.60E-115 | 3.55E-115 |
| 55 | Atrial Cardi | NRBP1 | -27.798 | -0.61047 | 5.56E-169 | 1.34E-168 |
| 56 | Atrial Cardi | NDRG2 | -33.0326 | -0.57934 | 3.60E-237 | 8.96E-237 |
| 57 | Atrial Cardi | APOC1 | -38.7016 | -3.18763 | 0 | 0 |
| 58 | Atrial Cardi | FAM3C | -40.9689 | -0.76451 | 0 | 0 |
| 59 | Atrial Cardi | ARHGEF40 | -43.0737 | -1.109 | 0 | 0 |
| 60 | Atrial Cardi | PPM1G | -43.744 | -0.89735 | 0 | 0 |
| 61 | Atrial Cardi | EIF3E | -45.4445 | -0.63076 | 0 | 0 |
| 62 | Atrial Cardi | MYH7 | -47.4052 | -0.67454 | 0 | 0 |
| 63 | Atrial Cardi | SNX17 | -57.7379 | -1.55281 | 0 | 0 |
| 64 | Atrial Cardi | ADGRG6 | -58.4507 | -2.29528 | 0 | 0 |
| 65 | Atrial Cardi | PTEN | -58.5725 | -0.63063 | 0 | 0 |
| 66 | Atrial Cardi | NECTIN2 | -72.5625 | -1.24011 | 0 | 0 |
| 67 | Atrial Cardi | IFI27L2 | -76.3215 | -1.91073 | 0 | 0 |
| 68 | Atrial Cardi | TBX15 | -76.9035 | -3.71147 | 0 | 0 |
| 69 | Atrial Cardi | CCN3 | -77.3716 | -5.03774 | 0 | 0 |
| 70 | Atrial Cardi | CR1 | -83.1074 | -3.64751 | 0 | 0 |
| 71 | Atrial Cardi | SERPINA1 | -85.5823 | -5.50601 | 0 | 0 |
| 72 | Atrial Cardi | TRIB2 | -101.236 | -2.30015 | 0 | 0 |
| 73 | Atrial Cardi | KIAA1755 | -112.584 | -4.24868 | 0 | 0 |
| 74 | Atrial Cardi | APOE | -138.543 | -4.06906 | 0 | 0 |
| 75 | Atrial Cardi | CD34 | -225.051 | -4.20456 | 0 | 0 |
| 76 | Ventricular | TTN | 1476.945 | 7.145225 | 0 | 0 |
| 77 | Ventricular | MYH7 | 893.6967 | 6.36412 | 0 | 0 |
| 78 | Ventricular | DES | 661.6328 | 4.342802 | 0 | 0 |
| 79 | Ventricular | PLN | 559.9172 | 4.301789 | 0 | 0 |
| 80 | Ventricular | CCDC141 | 532.696 | 4.082849 | 0 | 0 |
| 81 | Ventricular | MLF1 | 298.2622 | 3.027637 | 0 | 0 |
| 82 | Ventricular | CEP85L | 216.3748 | 1.650192 | 0 | 0 |
| 83 | Ventricular | MYH6 | 211.1994 | 1.975667 | 0 | 0 |
| 84 | Ventricular | SPEG | 187.7031 | 2.76389 | 0 | 0 |
| 85 | Ventricular | MLLT10 | 175.3368 | 1.129616 | 0 | 0 |
| 86 | Ventricular | GJA1 | 144.967 | 1.527995 | 0 | 0 |
| 87 | Ventricular | CCDC91 | 118.9838 | 0.814718 | 0 | 0 |
| 88 | Ventricular | SLC35F1 | 113.3278 | 1.768625 | 0 | 0 |

| | | | | | |
|---|---|---|---|---|---|
| 89 | Ventricular | MTMR3 | 89.70687 | 0.732406 | 0 | 0 |
| 90 | Ventricular | FAM174B | 60.36589 | 0.876731 | 0 | 0 |
| 91 | Ventricular | MFN1 | 59.86271 | 0.576349 | 0 | 0 |
| 92 | Ventricular | CWC27 | 49.99966 | 0.460192 | 0 | 0 |
| 93 | Ventricular | RSPO2 | 35.37435 | 1.437949 | 1.81E-273 | 3.23E-273 |
| 94 | Ventricular | SKIDA1 | 34.78425 | 1.808701 | 1.88E-264 | 3.27E-264 |
| 95 | Ventricular | NIPSNAP1 | 27.69562 | 0.733753 | 1.26E-168 | 2.02E-168 |
| 96 | Ventricular | DCAF16 | 26.08455 | 0.379988 | 7.52E-150 | 1.19E-149 |
| 97 | Ventricular | SUPT7L | 24.50313 | 0.391384 | 1.76E-132 | 2.72E-132 |
| 98 | Ventricular | ASCC2 | 22.39585 | 0.267012 | 5.10E-111 | 7.75E-111 |
| 99 | Ventricular | VSTM2L | 17.50838 | 0.426707 | 1.33E-68 | 1.88E-68 |
| 100 | Ventricular | WARS2 | 17.18095 | 0.220122 | 3.91E-66 | 5.43E-66 |
| 101 | Ventricular | CR1L | 15.03372 | 0.542503 | 4.58E-51 | 5.96E-51 |
| 102 | Ventricular | GFM1 | 14.43932 | 0.153421 | 3.01E-47 | 3.80E-47 |
| 103 | Ventricular | C2orf16 | 7.592148 | 0.608809 | 3.15E-14 | 3.80E-14 |
| 104 | Ventricular | LCORL | 6.45466 | 0.062789 | 1.09E-10 | 1.25E-10 |
| 105 | Ventricular | TMEM253 | 4.193552 | 0.782494 | 2.75E-05 | 3.04E-05 |
| 106 | Ventricular | ALX4 | 4.081035 | 0.301698 | 4.48E-05 | 4.90E-05 |
| 107 | Ventricular | ZNF219 | 3.795154 | 0.091266 | 0.000148 | 0.000157 |
| 108 | Ventricular | HORMAD2 | 2.356149 | 0.191399 | 0.018466 | 0.019167 |
| 109 | Ventricular | CABP7 | 2.150072 | 0.52464 | 0.03155 | 0.032339 |
| 110 | Ventricular | B3GNT7 | -3.46309 | -0.18397 | 0.000534 | 0.000561 |
| 111 | Ventricular | SLC4A1AP | -3.81381 | -0.0624 | 0.000137 | 0.000148 |
| 112 | Ventricular | CCDC121 | -4.98963 | -0.21712 | 6.05E-07 | 6.80E-07 |
| 113 | Ventricular | SSC5D | -5.99282 | -0.14254 | 2.06E-09 | 2.35E-09 |
| 114 | Ventricular | WNT16 | -7.16858 | -1.5646 | 7.59E-13 | 8.89E-13 |
| 115 | Ventricular | TOMM40 | -7.29414 | -0.10552 | 3.01E-13 | 3.58E-13 |
| 116 | Ventricular | ZNF628 | -10.3457 | -0.48561 | 4.40E-25 | 5.39E-25 |
| 117 | Ventricular | CMTM5 | -13.4405 | -1.2738 | 3.55E-41 | 4.41E-41 |
| 118 | Ventricular | UQCR10 | -14.8246 | -0.1536 | 1.05E-49 | 1.34E-49 |
| 119 | Ventricular | DNAJC1 | -15.1965 | -0.11054 | 3.85E-52 | 5.09E-52 |
| 120 | Ventricular | HSPA4 | -16.3324 | -0.18592 | 6.05E-60 | 8.13E-60 |
| 121 | Ventricular | EIF2B4 | -17.1639 | -0.28395 | 5.20E-66 | 7.10E-66 |
| 122 | Ventricular | ZCCHC10 | -17.6512 | -0.21865 | 1.05E-69 | 1.52E-69 |
| 123 | Ventricular | NCAPG | -20.4358 | -1.33895 | 8.63E-93 | 1.26E-92 |
| 124 | Ventricular | SLC5A6 | -21.1077 | -0.55305 | 7.48E-99 | 1.12E-98 |
| 125 | Ventricular | NAT14 | -28.1026 | -1.20031 | 1.18E-173 | 1.94E-173 |
| 126 | Ventricular | ZNF512 | -29.4407 | -0.38932 | 2.46E-190 | 4.11E-190 |
| 127 | Ventricular | CENPW | -32.6014 | -2.39013 | 5.86E-233 | 1.00E-232 |
| 128 | Ventricular | LXN | -37.045 | -0.61115 | 5.67E-300 | 1.03E-299 |
| 129 | Ventricular | ZNF513 | -43.6148 | -1.37324 | 0 | 0 |
| 130 | Ventricular | GPN1 | -46.5813 | -1.089 | 0 | 0 |
| 131 | Ventricular | GNB4 | -51.391 | -0.59708 | 0 | 0 |
| 132 | Ventricular | APOC1 | -53.8753 | -3.97531 | 0 | 0 |
| 133 | Ventricular | NRBP1 | -54.0059 | -0.70482 | 0 | 0 |

| | | | | | | |
|---|---|---|---|---|---|---|
| 134 | Ventricular | ARHGEF40 | -63.8443 | -1.03049 | 0 | 0 |
| 135 | Ventricular | NDRG2 | -73.2622 | -0.77945 | 0 | 0 |
| 136 | Ventricular | RSRC1 | -77.1554 | -0.61934 | 0 | 0 |
| 137 | Ventricular | SHOX2 | -77.2486 | -6.10511 | 0 | 0 |
| 138 | Ventricular | PPM1G | -79.1886 | -0.97462 | 0 | 0 |
| 139 | Ventricular | TRIB2 | -88.927 | -1.30065 | 0 | 0 |
| 140 | Ventricular | CCN3 | -92.1552 | -7.58316 | 0 | 0 |
| 141 | Ventricular | SERPINA1 | -92.1943 | -5.77907 | 0 | 0 |
| 142 | Ventricular | FAM3C | -103.931 | -1.21402 | 0 | 0 |
| 143 | Ventricular | TBX15 | -103.989 | -4.55175 | 0 | 0 |
| 144 | Ventricular | ADGRG6 | -111.11 | -4.12294 | 0 | 0 |
| 145 | Ventricular | SNX17 | -111.156 | -1.93001 | 0 | 0 |
| 146 | Ventricular | CR1 | -111.468 | -4.56642 | 0 | 0 |
| 147 | Ventricular | DGKH | -122.698 | -1.10101 | 0 | 0 |
| 148 | Ventricular | KIAA1755 | -138.089 | -4.79974 | 0 | 0 |
| 149 | Ventricular | EIF3E | -138.8 | -1.16812 | 0 | 0 |
| 150 | Ventricular | APOE | -160.161 | -4.04487 | 0 | 0 |
| 151 | Ventricular | NECTIN2 | -164.992 | -1.83321 | 0 | 0 |
| 152 | Ventricular | IFI27L2 | -175.629 | -3.32474 | 0 | 0 |
| 153 | Ventricular | AFAP1 | -185.592 | -3.09076 | 0 | 0 |
| 154 | Ventricular | PTEN | -190.647 | -1.28546 | 0 | 0 |
| 155 | Ventricular | CD34 | -303.24 | -7.1804 | 0 | 0 |
