## Supplementary Table 11 for "Hundreds of cardiac MRI traits derived using 3D diffusion autoencoders share a common genetic architecture"

| TISSUE | GENE.NAM | GENE.ID | CHR | HSQ | BEST.GWA | BEST.GWA | EQTL.ID | EQTL.R2 |
|---|---|---|---|---|---|---|---|---|
| Heart_Atri | GNPDA2 | ENSG00000 | 4 | 0.412 | rs6812769 | -5.66 | rs1249996 | 0.273772 |
| Heart_Atri | PRDM5 | ENSG00000 | 4 | 0.1998 | rs1250723 | -7.12 | rs3789141 | 0.086393 |
| Heart_Atri | GUCY1A1 | ENSG00000 | 4 | 0.1804 | rs1173525 | -4.53 | rs1155422 | 0.021731 |
| Heart_Atri | SERPINB6 | ENSG00000 | 6 | 0.2325 | rs1994124 | 6.93 | rs1054132 | 0.0888 |
| Heart_Atri | CRIP3 | ENSG00000 | 6 | 0.1665 | rs1221276 | 6.53 | rs6919440 | 0.133 |
| Heart_Atri | CLIC5 | ENSG00000 | 6 | 0.1972 | rs6458479 | -4.37 | rs6458479 | 0.0614 |
| Heart_Atri | CENPW | ENSG00000 | 6 | 0.0986 | rs1591805 | -6.48 | rs1361108 | 0.0741 |
| Heart_Atri | SCN5A | ENSG00000 | 3 | 0.1207 | rs194706 | 9.27 | rs3922843 | 0.0228 |
| Heart_Atri | NCKIPSD | ENSG00000 | 3 | 0.2468 | rs1249039 | -4.93 | rs4858798 | 0.145 |
| Heart_Atri | P4HTM | ENSG00000 | 3 | 0.072 | rs1249039 | -4.93 | rs885592 | 0.00957 |
| Heart_Atri | QRICH1 | ENSG00000 | 3 | 0.0896 | rs1249039 | -4.93 | rs4858828 | 0.0616 |
| Heart_Atri | IHO1 | ENSG00000 | 3 | 0.1532 | rs1249039 | -4.93 | rs1171558 | 0.136 |
| Heart_Atri | USP4 | ENSG00000 | 3 | 0.0349 | rs1249039 | -4.93 | rs9860055 | 0.015 |
| Heart_Atri | NICN1 | ENSG00000 | 3 | 0.0706 | rs1249039 | -4.93 | rs6809851 | 0.0474 |
| Heart_Atri | RSRC1 | ENSG00000 | 3 | 0.0856 | rs1210359 | 5.84 | rs1768485 | 0.0296 |
| Heart_Atri | GNB4 | ENSG00000 | 3 | 0.6677 | rs1191837 | 8.08 | rs7612445 | 0.51 |
| Heart_Atri | BICD2 | ENSG00000 | 9 | 0.2828 | rs1099263 | -5.66 | rs1255462 | 0.049735 |
| Heart_Atri | HMCN2 | ENSG00000 | 9 | 0.2147 | rs1178952 | 7.49 | rs1124387 | 0.01663 |
| Heart_Atri | HAPLN1 | ENSG00000 | 5 | 0.1335 | rs2441010 | -9.16 | rs6898404 | 0.004979 |
| Heart_Atri | PTGR2 | ENSG00000 | 14 | 0.3177 | rs2109750 | -5.12 | rs8019765 | 0.112 |
| Heart_Atri | NCOR1 | ENSG00000 | 17 | 0.0938 | rs1186922 | -9.78 | rs1005380 | -0.00193 |
| Heart_Atri | RASL10B | ENSG00000 | 17 | 0.1439 | rs4500794 | 8.95 | rs321618 | 0.033221 |
| Heart_Atri | ACBD4 | ENSG00000 | 17 | 0.1477 | rs3744759 | -6.31 | rs4986169 | 0.043886 |
| Heart_Atri | BACH1 | ENSG00000 | 21 | 0.0666 | rs2832279 | 4.87 | rs1153284 | 0.059065 |
| Heart_Atri | PRDM15 | ENSG00000 | 21 | 0.3931 | rs420737 | 10.67 | rs915832 | 0.033708 |
| Heart_Atri | GEN1 | ENSG00000 | 2 | 0.4753 | rs368430 | -5.3 | rs6761104 | 0.193213 |
| Heart_Atri | STRN | ENSG00000 | 2 | 0.1265 | rs888083 | -5.58 | rs3845780 | 0.090318 |
| Heart_Atri | ANKRD36B | ENSG00000 | 2 | 0.3074 | rs1299119 | 7.11 | rs3906948 | 0.193741 |
| Heart_Atri | ACTR1B | ENSG00000 | 2 | 0.079 | rs1168085 | 6.92 | rs1470625 | 0.037415 |
| Heart_Atri | LIPT1 | ENSG00000 | 2 | 0.1787 | rs958778 | -5.56 | rs1168318 | 0.254502 |
| Heart_Atri | MITD1 | ENSG00000 | 2 | 0.1485 | rs2048748 | -6.76 | rs1168318 | 0.137776 |
| Heart_Atri | NHEJ1 | ENSG00000 | 2 | 0.0717 | rs1155634 | -7.87 | rs1996992 | 0.010473 |
| Heart_Atri | VSIR | ENSG00000 | 10 | 0.1961 | rs1099995 | 6.33 | rs7918082 | 0.0274 |
| Heart_Atri | SFTPD | ENSG00000 | 10 | 0.3889 | rs6413520 | -5.09 | rs1120101 | 0.293 |
| Heart_Atri | NRAP | ENSG00000 | 10 | 0.1243 | rs4918833 | 5.02 | rs3121491 | -0.00198 |
| Heart_Atri | FAAP20 | ENSG00000 | 1 | 0.4874 | rs7548829 | 4.04 | rs2503701 | 0.203 |
| Heart_Atri | SLC2A5 | ENSG00000 | 1 | 0.0937 | rs1702718 | -6.81 | rs2274327 | -0.00317 |
| Heart_Atri | MFAP2 | ENSG00000 | 1 | 0.1737 | rs9435734 | -5.04 | rs4920608 | 0.0944 |
| Heart_Atri | WNT4 | ENSG00000 | 1 | 0.1176 | rs1079972 | 5.92 | rs1204208 | -0.00085 |
| Heart_Atri | EFNA3 | ENSG00000 | 1 | 0.0839 | rs4845663 | -5.25 | rs2066981 | -0.00195 |
| Heart_Atri | NMNAT2 | ENSG00000 | 1 | 0.1887 | rs789191 | -7.64 | rs588492 | 0.0181 |
| Heart_Atri | EIF3E | ENSG00000 | 8 | 0.0384 | rs610891 | 6.42 | rs7815105 | 0.002742 |
| Heart_Atri | CDK8 | ENSG00000 | 13 | 0.2111 | rs1359310 | 11.42 | rs1286413 | 0.14842 |
| Heart_Atri | CCZ1 | ENSG00000 | 7 | 0.4979 | rs1025541 | 5.01 | rs6975026 | 0.166689 |

| | | | | | | | | |
|---|---|---|---|---|---|---|---|---|
| Heart_Atri | ANKMY2 | ENSG00000 | 7 | 0.3227 | rs7783093 | 6.38 | rs7156 | 0.061145 |
| Heart_Atri | HIBADH | ENSG00000 | 7 | 0.197 | rs864745 | -4.07 | rs1270083 | 0.116108 |
| Heart_Atri | H2AZ2 | ENSG00000 | 7 | 0.1892 | rs1253118 | -9.49 | rs7016 | 0.004279 |
| Heart_Atri | TBL2 | ENSG00000 | 7 | 0.3334 | rs875342 | 8.91 | rs1714572 | 0.09375 |
| Heart_Atri | SPDYE12P | ENSG00000 | 7 | 0.3323 | rs1028076 | -15.06 | rs3500543 | 0.096162 |
| Heart_Atri | STYXL1 | ENSG00000 | 7 | 0.5323 | rs1253245 | -7.37 | rs7788926 | 0.408378 |
| Heart_Atri | GIGYF1 | ENSG00000 | 7 | 0.1802 | rs1330696 | -4.89 | rs221786 | 0.102858 |
| Heart_Atri | TMEM139 | ENSG00000 | 7 | 0.1053 | rs8175963 | 6.43 | rs8175963 | -0.00221 |
| Heart_Atri | SPX | ENSG00000 | 12 | 0.1928 | rs768314 | 15.68 | rs7976688 | 0.012286 |
| Heart_Atri | CCDC91 | ENSG00000 | 12 | 0.1516 | rs1133028 | -13.23 | rs1104940 | 0.086009 |
| Heart_Atri | MCRS1 | ENSG00000 | 12 | 0.0705 | rs1183106 | 7.47 | rs7965658 | 0.03696 |
| Heart_Atri | CERS5 | ENSG00000 | 12 | 0.1734 | rs1183106 | 9.13 | rs3184122 | 0.162595 |
| Heart_Atri | CDK2AP1 | ENSG00000 | 12 | 0.0565 | rs3531484 | 10.28 | rs1879380 | 0.057881 |
| Heart_Atri | KMT5A | ENSG00000 | 12 | 0.0402 | rs3531484 | 11 | rs2856988 | 0.023944 |
| Heart_Atri | CCDC92 | ENSG00000 | 12 | 0.1858 | rs1257967 | -6.19 | rs9863 | 0.125163 |
| Heart_Atri | MGRN1 | ENSG00000 | 16 | 0.7024 | rs1164579 | -11.05 | rs841225 | 0.468 |
| Heart_Atri | NPIPA2 | ENSG00000 | 16 | 0.1821 | rs3572250 | 5.72 | rs3198697 | 0.0632 |
| Heart_Atri | INO80E | ENSG00000 | 16 | 0.176 | rs4541091 | 7.08 | rs1292199 | 0.192 |
| Heart_Atri | DOC2A | ENSG00000 | 16 | 0.1008 | rs4541091 | 6.02 | rs4787491 | 0.0416 |
| Heart_Atri | YPEL3 | ENSG00000 | 16 | 0.2023 | rs1164274 | 5.72 | rs7205802 | 0.201 |
| Heart_Atri | ZNF768 | ENSG00000 | 16 | 0.0375 | rs3597408 | -12.18 | rs8050463 | 0.0705 |
| Heart_Atri | NOB1 | ENSG00000 | 16 | 0.0768 | rs2291959 | -5.54 | rs1469908 | 0.0723 |
| Heart_Atri | WWP2 | ENSG00000 | 16 | 0.1661 | rs2291959 | -5.54 | rs1729708 | 0.0047 |
| Heart_Atri | CDH15 | ENSG00000 | 16 | 0.1636 | rs555812 | 9 | rs1164613 | 0.0806 |
| Heart_Atri | VEGFB | ENSG00000 | 11 | 0.4279 | rs1089747 | 6.03 | rs3516979 | 0.206 |
| Heart_Atri | ACAT1 | ENSG00000 | 11 | 0.1888 | rs1257779 | 3.92 | rs1257779 | 0.0535 |
| Heart_Atri | FXYD6 | ENSG00000 | 11 | 0.1441 | rs2925768 | 6.26 | rs4938445 | 0.00394 |
| Heart_Atri | ZNF100 | ENSG00000 | 19 | 0.7482 | rs1041066 | 4.53 | rs6511286 | 0.427 |
| Heart_Atri | PDCD5 | ENSG00000 | 19 | 0.2884 | rs1775465 | 10.93 | rs1215097 | 0.142 |
| Heart_Atri | PEPD | ENSG00000 | 19 | 0.2551 | rs1246250 | -5.43 | rs1423062 | 0.0555 |
| Heart_Atri | TMEM91 | ENSG00000 | 19 | 0.3418 | rs3848568 | 6.21 | rs12602 | 0.0235 |
| Heart_Atri | BCKDHA | ENSG00000 | 19 | 0.2128 | rs3848568 | 6.67 | rs1187909 | 0.117 |
| Heart_Atri | CARD8 | ENSG00000 | 19 | 0.2104 | rs1698213 | -5.84 | rs1698183 | 0.00478 |
| Heart_Atri | NTN5 | ENSG00000 | 19 | 0.1715 | rs2270941 | 7.47 | rs1041803 | 0.0527 |
| Heart_Atri | ATF5 | ENSG00000 | 19 | 0.1666 | rs1042568 | 6.68 | rs1152237 | 0.00298 |
| Heart_Atri | MMP24OS | ENSG00000 | 20 | 0.2083 | rs7280 | -5.85 | rs2425049 | 0.129588 |
| Heart_Atri | EIF6 | ENSG00000 | 20 | 0.3705 | rs7280 | -5.85 | rs2425044 | 0.073007 |
| Heart_Atri | UQCC1 | ENSG00000 | 20 | 0.0993 | rs7280 | -5.85 | rs224331 | 0.063954 |
| Heart_Left | CAPS | ENSG00000 | 19 | 0.0908 | rs1040213 | 6.27 | rs420458 | 0.0699 |
| Heart_Left | CD320 | ENSG00000 | 19 | 0.2016 | rs7247382 | 9.45 | rs4147644 | 0.0216 |
| Heart_Left | IQCN | ENSG00000 | 19 | 0.2927 | rs7254275 | 7.06 | rs1246170 | 0.182 |
| Heart_Left | ZNF93 | ENSG00000 | 19 | 0.1723 | rs1599797 | 6.3 | rs2859006 | 0.0499 |
| Heart_Left | PPP1R15A | ENSG00000 | 19 | 0.1286 | rs1698213 | -6.52 | rs609985 | 0.0853 |
| Heart_Left | CCDC91 | ENSG00000 | 12 | 0.0673 | rs1133028 | -13.23 | rs1084309 | 0.053053 |
| Heart_Left | ALG10 | ENSG00000 | 12 | 0.0357 | rs7311406 | 9.59 | rs4001713 | 0.004987 |

| | | | | | | | | |
|---|---|---|---|---|---|---|---|---|
| Heart_Left | CPNE8 | ENSG00000( | 12 | 0.0856 | rs1516556 | -8.42 | rs2630779 | 0.040043 |
| Heart_Left | RPS26 | ENSG00000( | 12 | 0.6925 | rs14483 | -8.38 | rs1087686 | 0.633264 |
| Heart_Left | POC1B | ENSG00000( | 12 | 0.1946 | rs796036 | -4.93 | rs6538188 | 0.126575 |
| Heart_Left | MYO1H | ENSG00000( | 12 | 0.137 | rs1183122( | -6.08 | rs2075434 | 0.009478 |
| Heart_Left | CDK2AP1 | ENSG00000( | 12 | 0.1244 | rs3531484 | 10.37 | rs1084650 | 0.043131 |
| Heart_Left | CCDC92 | ENSG00000( | 12 | 0.3144 | rs1257967( | -6.13 | rs1231111 | 0.193554 |
| Heart_Left | JPT2 | ENSG00000( | 16 | 0.1207 | rs7191794 | 8.8 | rs2235642 | 0.0529 |
| Heart_Left | MGRN1 | ENSG00000( | 16 | 0.4984 | rs1164579 | -10.97 | rs841225 | 0.37 |
| Heart_Left | INO80E | ENSG00000( | 16 | 0.1266 | rs4541091 | 7.08 | rs9932702 | 0.191 |
| Heart_Left | DOC2A | ENSG00000( | 16 | 0.0873 | rs4541091 | 6.02 | rs1293357 | 0.0439 |
| Heart_Left | TBX6 | ENSG00000( | 16 | 0.283 | rs1164274( | 5.72 | rs3809624 | 0.105 |
| Heart_Left | YPEL3 | ENSG00000( | 16 | 0.1583 | rs1164274( | 5.72 | rs8060511 | 0.178 |
| Heart_Left | FTO | ENSG00000( | 16 | 0.0987 | rs1421090 | 5.22 | rs9930506 | -0.00185 |
| Heart_Left | DDX19A | ENSG00000( | 16 | 0.101 | rs775212 | 10.66 | rs2270844 | 0.00154 |
| Heart_Left | ANGPT2 | ENSG00000( | 8 | 0.1217 | rs1707810 | 9.25 | rs1707800 | -0.00188 |
| Heart_Left | PTK2B | ENSG00000( | 8 | 0.2425 | rs9314347 | 4.74 | rs7813625 | 0.0545 |
| Heart_Left | FGFR1 | ENSG00000( | 8 | 0.1546 | rs1773643 | 8.94 | rs1802267 | -0.00206 |
| Heart_Left | RPS20 | ENSG00000( | 8 | 0.1569 | rs3403795 | 5.17 | rs1781408 | 0.141 |
| Heart_Left | CCN3 | ENSG00000( | 8 | 0.1334 | rs7012790 | -6.15 | rs2071519 | 0.0339 |
| Heart_Left | PLEC | ENSG00000( | 8 | 0.1245 | rs1178689( | 5.04 | rs1178689( | 0.151 |
| Heart_Left | HEATR4 | ENSG00000( | 14 | 0.1658 | rs8018725 | -5.57 | rs8018967 | 0.027375 |
| Heart_Left | DCAF16 | ENSG00000( | 4 | 0.0898 | rs1049601 | 6.68 | rs7667864 | 0.0182 |
| Heart_Left | PRDM5 | ENSG00000( | 4 | 0.1154 | rs1250723 | -6.55 | rs3789141 | 0.0636 |
| Heart_Left | TMEM144 | ENSG00000( | 4 | 0.3494 | rs1110013( | 5.5 | rs2881373 | 0.217 |
| Heart_Left | TCTN3 | ENSG00000( | 10 | 0.155 | rs7907476 | -8.39 | rs7074866 | 0.075116 |
| Heart_Left | CACUL1 | ENSG00000( | 10 | 0.2684 | rs1709796 | 4.39 | rs1247142 | 0.087128 |
| Heart_Left | CPXM2 | ENSG00000( | 10 | 0.3602 | rs7092456 | 6.82 | rs7909120 | 0.147844 |
| Heart_Left | SNTG2 | ENSG00000( | 2 | 0.1407 | rs2724853 | 11.88 | rs4971437 | 0.0507 |
| Heart_Left | PXDN | ENSG00000( | 2 | 0.1109 | rs1341432 | 7.46 | rs1342422 | 0.00561 |
| Heart_Left | SLC35E2B | ENSG00000( | 1 | 0.8264 | rs2887286 | 7.44 | rs4648786 | 0.121 |
| Heart_Left | FAAP20 | ENSG00000( | 1 | 0.3777 | rs7548829 | 4.04 | rs2460002 | 0.188 |
| Heart_Left | KIAA2013 | ENSG00000( | 1 | 0.134 | rs6683331 | -6.02 | rs2038028 | -0.00294 |
| Heart_Left | CROCC | ENSG00000( | 1 | 0.3718 | rs9435734 | -5.04 | rs6691985 | 0.29 |
| Heart_Left | CEP85 | ENSG00000( | 1 | 0.2871 | rs9438620 | -6.44 | rs1090272 | 0.143 |
| Heart_Left | SCMH1 | ENSG00000( | 1 | 0.078 | rs1735872 | 8.72 | rs4453027 | 0.00121 |
| Heart_Left | MTMR11 | ENSG00000( | 1 | 0.1746 | rs7534365 | -6.45 | rs1868992 | 0.12 |
| Heart_Left | ARNT | ENSG00000( | 1 | 0.1101 | rs1088838 | 6.35 | rs1256875 | 0.0984 |
| Heart_Left | METTL25B | ENSG00000( | 1 | 0.2291 | rs983227 | 4.66 | rs1214043 | 0.131 |
| Heart_Left | NUAK2 | ENSG00000( | 1 | 0.1345 | rs2275869 | -6.74 | rs4951243 | 0.0119 |
| Heart_Left | EIF6 | ENSG00000( | 20 | 0.107 | rs7280 | -5.85 | rs2425044 | 0.0749 |
| Heart_Left | GSTT2B | ENSG00000( | 22 | 0.6694 | rs9620291 | -6.9 | rs9624364 | 0.344786 |
| Heart_Left | MTMR3 | ENSG00000( | 22 | 0.116 | rs131272 | 6.48 | rs140104 | 0.070271 |
| Heart_Left | MAPK12 | ENSG00000( | 22 | 0.4569 | rs2867267 | 4.13 | rs1129880 | 0.2138 |
| Heart_Left | DUOX1 | ENSG00000( | 15 | 0.2289 | rs1695266 | 6.9 | rs1693975 | 0.108 |
| Heart_Left | ZNF609 | ENSG00000( | 15 | 0.1422 | rs677561 | -11.07 | rs1185860 | 0.0377 |

| | | | | | | | | |
|---|---|---|---|---|---|---|---|---|
| Heart_Left | EFL1 | ENSG00000 | 15 | 0.0911 | rs1185316 | 6.81 | rs4725 | 0.00272 |
| Heart_Left | SAXO2 | ENSG00000 | 15 | 0.5643 | rs1185316 | 6.81 | rs9972386 | 0.298 |
| Heart_Left | LYSMD4 | ENSG00000 | 15 | 0.5344 | rs7170513 | -5.8 | rs6598265 | 0.166 |
| Heart_Left | CTDNEP1 | ENSG00000 | 17 | 0.1358 | rs2269459 | 8.75 | rs222852 | 0.102 |
| Heart_Left | TTC19 | ENSG00000 | 17 | 0.1201 | rs178791 | 4.29 | rs2015353 | 0.101 |
| Heart_Left | TMEM199 | ENSG00000 | 17 | 0.0624 | rs2125844 | -6.54 | rs1780273 | 0.037 |
| Heart_Left | HHATL | ENSG00000 | 3 | 0.2629 | rs9311319 | 6.28 | rs1723879 | 0.04499 |
| Heart_Left | ARIH2 | ENSG00000 | 3 | 0.0685 | rs1249039 | -4.93 | rs6796790 | 0.041767 |
| Heart_Left | NDUFAF3 | ENSG00000 | 3 | 0.0889 | rs1249039 | -4.93 | rs7100 | 0.00129 |
| Heart_Left | QRICH1 | ENSG00000 | 3 | 0.0638 | rs1249039 | -4.93 | rs4974088 | 0.076023 |
| Heart_Left | IHO1 | ENSG00000 | 3 | 0.195 | rs1249039 | -4.93 | rs1263198 | 0.134276 |
| Heart_Left | RAD54L2 | ENSG00000 | 3 | 0.0954 | rs323887 | 8.34 | rs3749316 | 0.010652 |
| Heart_Left | GK5 | ENSG00000 | 3 | 0.1186 | rs4484207 | -4.91 | rs1171561 | -0.00308 |
| Heart_Left | GNB4 | ENSG00000 | 3 | 0.5516 | rs2339798 | 6.55 | rs7612445 | 0.466945 |
| Heart_Left | ILRUN | ENSG00000 | 6 | 0.087 | rs2820239 | -6.01 | rs1689495 | 0.0593 |
| Heart_Left | TCF21 | ENSG00000 | 6 | 0.0922 | rs4424102 | 5.79 | rs1029212 | 0.03 |
| Heart_Left | OLIG1 | ENSG00000 | 21 | 0.1442 | rs9982759 | -11.33 | rs2834132 | 0.01219 |
| Heart_Left | CBR1 | ENSG00000 | 21 | 0.486 | rs2835109 | 5.02 | rs2835269 | 0.13771 |
| Heart_Left | MRPL23 | ENSG00000 | 11 | 0.5807 | rs2234283 | 9.51 | rs7395920 | 0.106846 |
| Heart_Left | MADD | ENSG00000 | 11 | 0.1414 | rs7939069 | 4.47 | rs1076925 | 0.057026 |
| Heart_Left | FAM89B | ENSG00000 | 11 | 0.2846 | rs551523 | 8.41 | rs749112 | 0.044216 |
| Heart_Left | RADIL | ENSG00000 | 7 | 0.2917 | rs1198345 | 4.36 | rs4724137 | 0.037453 |
| Heart_Left | GTF2IRD2B | ENSG00000 | 7 | 0.0852 | rs1253222 | -12.78 | rs1167794 | -0.00306 |
| Heart_Left | STYXL1 | ENSG00000 | 7 | 0.4285 | rs7799638 | -5.92 | rs1155309 | 0.337271 |
| Heart_Left | ADCK2 | ENSG00000 | 7 | 0.1098 | rs1044121 | 6.8 | rs7802537 | 0.017576 |
| Heart_Left | SLC9A3 | ENSG00000 | 5 | 0.6782 | rs6869655 | -5.52 | rs2672744 | 0.070163 |
| Heart_Left | GCNT4 | ENSG00000 | 5 | 0.1964 | rs6883614 | 5.88 | rs1195117 | 0.027591 |
| Heart_Left | TMEM232 | ENSG00000 | 5 | 0.1144 | rs1304889 | 7.32 | rs151784 | 0.020856 |
| Heart_Left | RNF14 | ENSG00000 | 5 | 0.0954 | rs6580243 | -9.01 | rs108593 | -0.00157 |
| Artery_Cor | GIGYF1 | ENSG00000 | 7 | 0.0811 | rs1330696 | -4.89 | rs314353 | 0.002982 |
| Artery_Cor | ACHE | ENSG00000 | 7 | 0.4283 | rs1025451 | -6.19 | rs6706 | 0.303004 |
| Artery_Cor | ACADVL | ENSG00000 | 17 | 0.2852 | rs2269459 | 8.13 | rs390200 | 0.08244 |
| Artery_Cor | CHRNB1 | ENSG00000 | 17 | 0.2895 | rs2269459 | 8.16 | rs2302764 | 0.08858 |
| Artery_Cor | TBC1D26 | ENSG00000 | 17 | 0.2334 | rs7215851 | -4.29 | rs3865264 | 0.07095 |
| Artery_Cor | SHMT1 | ENSG00000 | 17 | 0.3235 | rs4925174 | 6.82 | rs7207306 | 0.14563 |
| Artery_Cor | SKA2 | ENSG00000 | 17 | 0.1491 | rs1694332 | 7.6 | rs1294913 | 0.06813 |
| Artery_Cor | DAP | ENSG00000 | 5 | 0.2794 | rs1779037 | 6.23 | rs1531842 | 0.149659 |
| Artery_Cor | CDO1 | ENSG00000 | 5 | 0.4285 | rs1713925 | 4.49 | rs1422290 | 0.011352 |
| Artery_Cor | ADCY4 | ENSG00000 | 14 | 0.1796 | rs1157450 | -4.22 | rs751877 | 0.027134 |
| Artery_Cor | DCAF4 | ENSG00000 | 14 | 0.3513 | rs1013993 | 5.98 | rs6574106 | 0.087525 |
| Artery_Cor | COQ6 | ENSG00000 | 14 | 0.1424 | rs730384 | -5.19 | rs2058391 | 0.039811 |
| Artery_Cor | EIF2B2 | ENSG00000 | 14 | 0.2245 | rs7148230 | -4.73 | rs175016 | 0.129943 |
| Artery_Cor | TARS3 | ENSG00000 | 15 | 0.1682 | rs3784491 | -4.32 | rs507903 | 0.082358 |
| Artery_Cor | TPGS2 | ENSG00000 | 18 | 0.334 | rs903733 | 6.82 | rs1045781 | 0.216 |
| Artery_Cor | PTK7 | ENSG00000 | 6 | 0.1853 | rs1221276 | 6.33 | rs1537638 | 0.0475 |

| | | | | | | | | |
|---|---|---|---|---|---|---|---|---|
| Artery_Cor | TCF21 | ENSG0000( | 6 | 0.2136 | rs7767123 | 7.06 | rs1219397: | 0.063 |
| Artery_Cor | HMOX2 | ENSG0000( | 16 | 0.154 | rs1164579: | -8.6 | rs4786501 | -0.00511 |
| Artery_Cor | INO80E | ENSG0000( | 16 | 0.1597 | rs4541091 | 7.08 | rs4787491 | 0.180937 |
| Artery_Cor | TBX6 | ENSG0000( | 16 | 0.3324 | rs1164274( | 5.72 | rs1115058‹ | 0.124875 |
| Artery_Cor | YPEL3 | ENSG0000( | 16 | 0.218 | rs1164274( | 5.72 | rs7205802 | 0.210383 |
| Artery_Cor | MBOAT2 | ENSG0000( | 2 | 0.26 | rs2666205 | -6.26 | rs4459703 | 0.005711 |
| Artery_Cor | GEN1 | ENSG0000( | 2 | 0.3926 | rs368430 | -5.3 | rs6761104 | 0.199287 |
| Artery_Cor | DYNC2LI1 | ENSG0000( | 2 | 0.3534 | rs7585346 | 6.23 | rs1189185! | 0.10719 |
| Artery_Cor | ANKRD36B | ENSG0000( | 2 | 0.5039 | rs1299119 | 7.11 | rs6718109 | 0.286748 |
| Artery_Cor | UPF3A | ENSG0000( | 13 | 0.4454 | rs6602905 | 3.93 | rs7320104 | 0.178 |
| Artery_Cor | SYN2 | ENSG0000( | 3 | 0.286 | rs4135285 | 7.21 | rs373827 | 0.09907 |
| Artery_Cor | NCKIPSD | ENSG0000( | 3 | 0.0924 | rs1249039: | -4.93 | rs1249357! | 0.005738 |
| Artery_Cor | DALRD3 | ENSG0000( | 3 | 0.1249 | rs1249039: | -4.93 | rs7653408 | 0.11578 |
| Artery_Cor | QRICH1 | ENSG0000( | 3 | 0.0724 | rs1759541( | 7.38 | rs990211 | 0.043414 |
| Artery_Cor | IHO1 | ENSG0000( | 3 | 0.4194 | rs1249039: | -4.93 | rs1171558: | 0.143009 |
| Artery_Cor | AMT | ENSG0000( | 3 | 0.3173 | rs1249039: | -4.93 | rs6769821 | 0.388918 |
| Artery_Cor | NICN1 | ENSG0000( | 3 | 0.1746 | rs1249039: | -4.93 | rs6769821 | 0.164761 |
| Artery_Cor | ZMYND10 | ENSG0000( | 3 | 0.0805 | rs2239751 | 25.31 | rs762898 | -0.00578 |
| Artery_Cor | RAD54L2 | ENSG0000( | 3 | 0.3835 | rs323887 | 8.34 | rs4441646 | 0.111657 |
| Artery_Cor | TOPBP1 | ENSG0000( | 3 | 0.2265 | rs6772354 | 7.55 | rs6802154 | 0.045643 |
| Artery_Cor | NAALAD2 | ENSG0000( | 11 | 0.1808 | rs1076527( | 4.02 | rs1691762: | 0.0232 |
| Artery_Cor | SLC35E2B | ENSG0000( | 1 | 0.8416 | rs2887286 | 7.44 | rs4648786 | 0.102813 |
| Artery_Cor | FAAP20 | ENSG0000( | 1 | 0.2997 | rs4648640 | 3.86 | rs2460002 | 0.130892 |
| Artery_Cor | MFAP2 | ENSG0000( | 1 | 0.2722 | rs9435734 | -5.04 | rs6691985 | 0.049719 |
| Artery_Cor | WARS2 | ENSG0000( | 1 | 0.7058 | rs984225 | -6.58 | rs2645303 | 0.405881 |
| Artery_Cor | HAO2 | ENSG0000( | 1 | 0.1508 | rs984225 | -6.58 | rs2885226 | 0.001882 |
| Artery_Cor | CTSK | ENSG0000( | 1 | 0.2384 | rs9733 | 4.49 | rs3738483 | 0.146683 |
| Artery_Cor | ARNT | ENSG0000( | 1 | 0.1318 | rs1088838: | 6.48 | rs9733 | 0.117434 |
| Artery_Cor | ASB13 | ENSG0000( | 10 | 0.2322 | rs942200 | 6.07 | rs7915861 | 0.137016 |
| Artery_Cor | MALRD1 | ENSG0000( | 10 | 0.5385 | rs1225405: | -5.15 | rs2151234 | 0.118973 |
| Artery_Cor | CASP7 | ENSG0000( | 10 | 0.5751 | rs1709099: | -5.32 | rs4353229 | 0.218014 |
| Artery_Cor | CCDC91 | ENSG0000( | 12 | 0.1112 | rs1133028 | -13.23 | rs1104951: | 0.0299 |
| Artery_Cor | ASIC1 | ENSG0000( | 12 | 0.0886 | rs1183106: | 9.39 | rs3812825 | 0.0719 |
| Artery_Cor | ATF1 | ENSG0000( | 12 | 0.1211 | rs1116974! | 4.64 | rs1078338 | 0.1 |
| Artery_Cor | C12orf73 | ENSG0000( | 12 | 0.2701 | rs7301863 | 11.23 | rs2722195 | 0.161 |
| Artery_Cor | VPS37B | ENSG0000( | 12 | 0.1372 | rs1260317 | -7.52 | rs967281 | 0.0654 |
| Artery_Cor | H3C6 | ENSG0000( | 6 | 0.14 | rs6910993 | 8.65 | rs7773163 | 0.0137 |
| Artery_Cor | ADGRL1 | ENSG0000( | 19 | 0.156 | rs8107052 | -12.62 | rs3745462 | -0.00491 |
| Artery_Cor | ZNF100 | ENSG0000( | 19 | 0.76 | rs1041066( | 4.53 | rs6511286 | 0.453 |
| Artery_Cor | PEPD | ENSG0000( | 19 | 0.213 | rs1246250! | -5.43 | rs8111294 | 0.0189 |
| Artery_Cor | HRC | ENSG0000( | 19 | 0.204 | rs2270941 | 5.79 | rs3745299 | 0.0541 |
| Artery_Cor | MMP24OS | ENSG0000( | 20 | 0.2348 | rs7280 | -5.85 | rs6060341 | 0.13355 |
| Artery_Cor | ADNP | ENSG0000( | 20 | 0.1617 | rs756433 | -7.86 | rs2426159 | 0.016075 |
| Artery_Aor | SCAMP4 | ENSG0000( | 19 | 0.135 | rs1166721: | -6.85 | rs8730 | 0.094922 |
| Artery_Aor | ZNF100 | ENSG0000( | 19 | 0.8363 | rs1041066( | 4.53 | rs6511286 | 0.385584 |

| | | | | | | | | |
|---|---|---|---|---|---|---|---|---|
| Artery_Aor | ZNF208 | ENSG0000( | 19 | 0.2819 | rs1041066( | 6.32 | rs7255049 | 0.087348 |
| Artery_Aor | GEMIN7 | ENSG0000( | 19 | 0.2778 | rs4420638 | 6.07 | rs1040585! | 0.107734 |
| Artery_Aor | PLXNB1 | ENSG0000( | 3 | 0.0417 | rs1191861! | -4.81 | rs7434077 | -0.00112 |
| Artery_Aor | P4HTM | ENSG0000( | 3 | 0.1277 | rs1249039: | -4.93 | rs4955411 | 0.141 |
| Artery_Aor | NDUFAF3 | ENSG0000( | 3 | 0.0474 | rs1249039: | -4.93 | rs7653408 | 0.0313 |
| Artery_Aor | DALRD3 | ENSG0000( | 3 | 0.0537 | rs1249039: | -4.93 | rs4955411 | 0.0498 |
| Artery_Aor | QRICH1 | ENSG0000( | 3 | 0.0477 | rs1759541( | 6.08 | rs4974088 | 0.0275 |
| Artery_Aor | C3orf62 | ENSG0000( | 3 | 0.0804 | rs1249039: | -4.93 | rs4279134 | 0.0528 |
| Artery_Aor | GPX1 | ENSG0000( | 3 | 0.0593 | rs1249039: | -4.93 | rs1309647 | 0.0631 |
| Artery_Aor | AMT | ENSG0000( | 3 | 0.4807 | rs1249039: | -4.93 | rs3448 | 0.481 |
| Artery_Aor | NICN1 | ENSG0000( | 3 | 0.1951 | rs1249039: | -4.93 | rs1309647 | 0.211 |
| Artery_Aor | RNF123 | ENSG0000( | 3 | 0.3553 | rs1759541( | -5.75 | rs7648987 | 0.144 |
| Artery_Aor | DOCK3 | ENSG0000( | 3 | 0.208 | rs2239751 | 20.48 | rs1192044: | 0.104 |
| Artery_Aor | RAD54L2 | ENSG0000( | 3 | 0.1407 | rs323887 | 8.34 | rs4441646 | 0.0566 |
| Artery_Aor | ADAMTS9 | ENSG0000( | 3 | 0.1691 | rs9311910 | -5.47 | rs1772706 | 0.0533 |
| Artery_Aor | TOMM70 | ENSG0000( | 3 | 0.1003 | rs1263645: | -4.67 | rs1486308 | 0.015 |
| Artery_Aor | RAB7A | ENSG0000( | 3 | 0.1592 | rs6795608 | -4.51 | rs9847178 | 0.00773 |
| Artery_Aor | DNAJC13 | ENSG0000( | 3 | 0.0846 | rs6777846 | -4.91 | rs1263925 | 0.0507 |
| Artery_Aor | GNB4 | ENSG0000( | 3 | 0.0746 | rs1191837: | 8.08 | rs4855074 | 0.0295 |
| Artery_Aor | ZDHHC11 | ENSG0000( | 5 | 0.2449 | rs1007266 | 4.24 | rs2671898 | -0.002 |
| Artery_Aor | ARHGEF28 | ENSG0000( | 5 | 0.0935 | rs2973927 | 6.09 | rs255601 | 0.00219 |
| Artery_Aor | ACADVL | ENSG0000( | 17 | 0.2013 | rs2269459 | 8.13 | rs446994 | 0.222 |
| Artery_Aor | DCAF16 | ENSG0000( | 4 | 0.1213 | rs1049601 | 6.68 | rs6842303 | 0.10926 |
| Artery_Aor | SRD5A3 | ENSG0000( | 4 | 0.4636 | rs6837735 | 7.99 | rs1250083 | 0.099236 |
| Artery_Aor | METAP1 | ENSG0000( | 4 | 0.3251 | rs1700894 | 5.87 | rs7681427 | 0.08665 |
| Artery_Aor | BTN3A3 | ENSG0000( | 6 | 0.1886 | rs2754715 | 12.15 | rs6929846 | -0.00016 |
| Artery_Aor | TBL2 | ENSG0000( | 7 | 0.7534 | rs875342 | 8.91 | rs1714572 | 0.293944 |
| Artery_Aor | RHBDD2 | ENSG0000( | 7 | 0.2749 | rs1253222 | -7.26 | rs7807392 | 0.057411 |
| Artery_Aor | TMEM120/ | ENSG0000( | 7 | 0.131 | rs1253222 | -6.94 | rs7788926 | 0.077224 |
| Artery_Aor | STYXL1 | ENSG0000( | 7 | 0.5648 | rs1253245 | -7.37 | rs1155309( | 0.296766 |
| Artery_Aor | MDH2 | ENSG0000( | 7 | 0.0691 | rs1253245 | -7.37 | rs1253245 | -0.00277 |
| Artery_Aor | PEX1 | ENSG0000( | 7 | 0.0735 | rs1005346 | -5.02 | rs424 | 0.013354 |
| Artery_Aor | RASA4 | ENSG0000( | 7 | 0.2415 | rs1129271 | 8.27 | rs4729790 | 0.083061 |
| Artery_Aor | GABRB3 | ENSG0000( | 15 | 0.3813 | rs2045151 | -8.78 | rs768899 | 0.132498 |
| Artery_Aor | NDUFAF1 | ENSG0000( | 15 | 0.3767 | rs1206842 | 4.36 | rs3759794 | 0.022305 |
| Artery_Aor | TPM1 | ENSG0000( | 15 | 0.0848 | rs6494367 | 5.8 | rs4775613 | 0.02825 |
| Artery_Aor | CSNK1G1 | ENSG0000( | 15 | 0.0549 | rs677561 | -10.14 | rs7403071 | 0.013024 |
| Artery_Aor | TRIP4 | ENSG0000( | 15 | 0.2505 | rs677561 | -10.15 | rs1163521: | 0.281292 |
| Artery_Aor | CYP2C8 | ENSG0000( | 10 | 0.3314 | rs7918235 | -5.16 | rs1049814 | 0.129497 |
| Artery_Aor | PDCD11 | ENSG0000( | 10 | 0.3149 | rs1225980: | -6.36 | rs1225980: | 0.063452 |
| Artery_Aor | CASP7 | ENSG0000( | 10 | 0.4836 | rs1709099: | -5.32 | rs4353229 | 0.274648 |
| Artery_Aor | TM7SF3 | ENSG0000( | 12 | 0.6169 | rs2306852 | -9.69 | rs6487582 | 0.171045 |
| Artery_Aor | CPNE8 | ENSG0000( | 12 | 0.1668 | rs1516556 | -8.42 | rs2630779 | 0.096785 |
| Artery_Aor | FKBP11 | ENSG0000( | 12 | 0.2189 | rs4760645 | -6.69 | rs1242463 | 0.169343 |
| Artery_Aor | PRPH | ENSG0000( | 12 | 0.1241 | rs1116908 | 11.44 | rs1116898: | -0.00172 |

| | | | | | | | | |
|---|---|---|---|---|---|---|---|---|
| Artery_Aor | RXYLT1 | ENSG00000( | 12 | 0.4173 | rs1710009: | 5.47 | rs1232051( | 0.192511 |
| Artery_Aor | TRIAP1 | ENSG00000( | 12 | 0.121 | rs1084977 | 5.79 | rs2235217 | 0.032632 |
| Artery_Aor | OGFOD2 | ENSG00000( | 12 | 0.0343 | rs3531484 | 10.15 | rs2856988 | 0.026096 |
| Artery_Aor | CDK2AP1 | ENSG00000( | 12 | 0.2076 | rs3531484 | 10.37 | rs1231613 | 0.195741 |
| Artery_Aor | MTMR3 | ENSG00000( | 22 | 0.1362 | rs131272 | 6.48 | rs713718 | 0.063948 |
| Artery_Aor | ARHGEF40 | ENSG00000( | 14 | 0.4282 | rs2319627 | -6.01 | rs8019890 | 0.396042 |
| Artery_Aor | GZMB | ENSG00000( | 14 | 0.0821 | rs854344 | 7.61 | rs7154849 | 0.012299 |
| Artery_Aor | ACOT1 | ENSG00000( | 14 | 0.6588 | rs8018725 | -5.77 | rs2041073 | 0.143153 |
| Artery_Aor | PTGR2 | ENSG00000( | 14 | 0.5193 | rs2109750 | -5.12 | rs6574158 | 0.195394 |
| Artery_Aor | COQ6 | ENSG00000( | 14 | 0.1636 | rs730384 | -5.19 | rs2159177 | 0.129343 |
| Artery_Aor | FLRT2 | ENSG00000( | 14 | 0.1521 | rs6574859 | -6.71 | rs7144022 | 0.007431 |
| Artery_Aor | CEP170B | ENSG00000( | 14 | 0.1872 | rs2498806 | 6.45 | rs2028416 | 0.033609 |
| Artery_Aor | DKK3 | ENSG00000( | 11 | 0.6143 | rs2641942 | 6.38 | rs1102211 | 0.294 |
| Artery_Aor | VPS11 | ENSG00000( | 11 | 0.2584 | rs589925 | -4.92 | rs589925 | 0.208 |
| Artery_Aor | ADAMTS8 | ENSG00000( | 11 | 0.1668 | rs4937554 | -8.71 | rs747249 | 0.0619 |
| Artery_Aor | KCTD12 | ENSG00000( | 13 | 0.108 | rs8001997 | -5.67 | rs7319157 | 0.01124 |
| Artery_Aor | SLC35E2B | ENSG00000( | 1 | 0.8307 | rs2887286 | 7.44 | rs4648786 | 0.188 |
| Artery_Aor | FAAP20 | ENSG00000( | 1 | 0.2918 | rs4648640 | 3.86 | rs2460002 | 0.108 |
| Artery_Aor | AGTRAP | ENSG00000( | 1 | 0.2302 | rs1208985 | -6.71 | rs1209551 | 0.0794 |
| Artery_Aor | CEP85 | ENSG00000( | 1 | 0.2509 | rs9438620 | -6.44 | rs4585968 | 0.154 |
| Artery_Aor | CTBS | ENSG00000( | 1 | 0.3701 | rs1205763 | 5.85 | rs7551329 | 0.152 |
| Artery_Aor | TSHB | ENSG00000( | 1 | 0.0985 | rs2268793 | 8.25 | rs4478805 | 0.0118 |
| Artery_Aor | TBX15 | ENSG00000( | 1 | 0.097 | rs984225 | -6.58 | rs6701188 | 0.00419 |
| Artery_Aor | WARS2 | ENSG00000( | 1 | 0.4684 | rs984225 | -6.58 | rs2645303 | 0.346 |
| Artery_Aor | LAMC1 | ENSG00000( | 1 | 0.071 | rs9943111 | -6.19 | rs1091119 | 0.0104 |
| Artery_Aor | LYPLAL1 | ENSG00000( | 1 | 0.1325 | rs6675807 | 13.03 | rs6660443 | 0.0715 |
| Artery_Aor | PCNX2 | ENSG00000( | 1 | 0.2066 | rs6667845 | 7.9 | rs6667845 | 0.0678 |
| Artery_Aor | SLX4 | ENSG00000( | 16 | 0.0594 | rs2531995 | 4.13 | rs2741907 | -0.0016 |
| Artery_Aor | NPIPB12 | ENSG00000( | 16 | 0.0549 | rs1115057( | 4.88 | rs7201384 | 0.00921 |
| Artery_Aor | INO80E | ENSG00000( | 16 | 0.2329 | rs4541091 | 7.08 | rs4787491 | 0.243286 |
| Artery_Aor | DOC2A | ENSG00000( | 16 | 0.1328 | rs4541091 | 6.02 | rs8043883 | 0.061359 |
| Artery_Aor | YPEL3 | ENSG00000( | 16 | 0.2846 | rs1164274( | 5.72 | rs7205802 | 0.312011 |
| Artery_Aor | BCAR1 | ENSG00000( | 16 | 0.1828 | rs1078197( | 5.41 | rs1293328 | 0.19019 |
| Artery_Aor | MYLK4 | ENSG00000( | 6 | 0.5061 | rs1994124 | 6.93 | rs2038760 | 0.299 |
| Artery_Aor | TBC1D7 | ENSG00000( | 6 | 0.3194 | rs6923878 | -5.8 | rs2496132 | 0.11 |
| Artery_Aor | MAN1A1 | ENSG00000( | 6 | 0.1354 | rs6929403 | -4.06 | rs6935126 | 0.0696 |
| Artery_Aor | GJA1 | ENSG00000( | 6 | 0.1936 | rs1012020 | -7.77 | rs1252664( | 0.0186 |
| Artery_Aor | TCF21 | ENSG00000( | 6 | 0.1252 | rs7767123 | 7.06 | rs2327429 | 0.0959 |
| Artery_Aor | TNFRSF10A | ENSG00000( | 8 | 0.56 | rs1708935 | 4.56 | rs4242394 | 0.208219 |
| Artery_Aor | PTK2B | ENSG00000( | 8 | 0.1041 | rs9314347 | 4.74 | rs4733058 | 0.002519 |
| Artery_Aor | SDCBP | ENSG00000( | 8 | 0.0828 | rs4737534 | 9.13 | rs1267456 | 0.025042 |
| Artery_Aor | RSPO2 | ENSG00000( | 8 | 0.0804 | rs610891 | 6.42 | rs399085 | 0.031876 |
| Artery_Aor | GAREM2 | ENSG00000( | 2 | 0.2395 | rs1112649 | 8.23 | rs1465720 | 0.0125 |
| Artery_Aor | MAT2A | ENSG00000( | 2 | 0.0901 | rs6720075 | 4.98 | rs6733550 | 0.0418 |
| Artery_Aor | ASTL | ENSG00000( | 2 | 0.1004 | rs3580943 | 6.26 | rs997547 | 0.00062 |

| | | | | | | | | |
|---|---|---|---|---|---|---|---|---|
| Artery_Aor | TMEM127 | ENSG0000( | 2 | 0.0701 | rs3580943: | 6.8 | rs2301707 | 0.0203 |
| Artery_Aor | ANKRD36B | ENSG0000( | 2 | 0.394 | rs1299119 | 7.11 | rs6718109 | 0.385 |
| Artery_Aor | ZAP70 | ENSG0000( | 2 | 0.0492 | rs1168085! | 6.96 | rs1189360: | 0.0426 |
| Artery_Aor | ORC4 | ENSG0000( | 2 | 0.2087 | rs1682871! | -3.99 | rs1721879! | 0.0128 |
| Artery_Aor | PLA2R1 | ENSG0000( | 2 | 0.2323 | rs4665145 | -4.44 | rs1457240 | 0.0783 |
| Artery_Aor | FIGN | ENSG0000( | 2 | 0.2783 | rs988990 | -6.3 | rs1303123( | 0.121 |
| Artery_Aor | URB1 | ENSG0000( | 21 | 0.1731 | rs7276508 | 4.39 | rs2282097 | 0.059527 |
| Artery_Aor | PAK5 | ENSG0000( | 20 | 0.19 | rs926475 | -4.56 | rs6133737 | 0.023302 |
| Artery_Aor | MYH7B | ENSG0000( | 20 | 0.316 | rs7280 | -5.85 | rs6088662 | 0.156706 |
| Artery_Aor | MMP24OS | ENSG0000( | 20 | 0.2802 | rs7280 | -5.85 | rs6060341 | 0.173647 |
| Artery_Aor | SLC2A10 | ENSG0000( | 20 | 0.2341 | rs2073171 | 9.7 | rs998422 | 0.092634 |
| Artery_Aor | LPAR1 | ENSG0000( | 9 | 0.1329 | rs1098045: | 6.07 | rs1234861 | 0.0107 |
| Artery_Tibi | TCP11L1 | ENSG0000( | 11 | 0.3557 | rs2447524 | 4.72 | rs3802790 | 0.381919 |
| Artery_Tibi | NR1H3 | ENSG0000( | 11 | 0.0587 | rs7120737 | 4 | rs830085 | 0.002222 |
| Artery_Tibi | C1QTNF4 | ENSG0000( | 11 | 0.1947 | rs1681630 | 4.24 | rs1083875 | 0.133407 |
| Artery_Tibi | VPS11 | ENSG0000( | 11 | 0.2757 | rs1157466! | 6.7 | rs17075 | 0.238805 |
| Artery_Tibi | FAM47E | ENSG0000( | 4 | 0.3894 | rs7664889 | -4.61 | rs1876539 | 0.202712 |
| Artery_Tibi | HPSE | ENSG0000( | 4 | 0.122 | rs6851716 | -7.55 | rs6535451 | 0.018965 |
| Artery_Tibi | PRDM5 | ENSG0000( | 4 | 0.1527 | rs1705115: | -5.66 | rs4833682 | 0.041043 |
| Artery_Tibi | GUCY1A1 | ENSG0000( | 4 | 0.1077 | rs1001106( | -6.78 | rs3796592 | 0.048053 |
| Artery_Tibi | GABRB3 | ENSG0000( | 15 | 0.3419 | rs2045151 | -8.78 | rs1290711( | 0.193923 |
| Artery_Tibi | ARHGAP11 | ENSG0000( | 15 | 0.2097 | rs543354 | 12.99 | rs1210164: | 0.049088 |
| Artery_Tibi | TRIP4 | ENSG0000( | 15 | 0.2077 | rs677561 | -10.15 | rs673931 | 0.246997 |
| Artery_Tibi | PDCD7 | ENSG0000( | 15 | 0.0619 | rs1163711 | -13.92 | rs936860 | 0.001813 |
| Artery_Tibi | LYSMD4 | ENSG0000( | 15 | 0.4863 | rs7170513 | -5.8 | rs6598265 | 0.295905 |
| Artery_Tibi | NSUN5 | ENSG0000( | 7 | 0.093 | rs875342 | 8.04 | rs6971390 | 0.00554 |
| Artery_Tibi | TBL2 | ENSG0000( | 7 | 0.5535 | rs875342 | 8.91 | rs1714572: | 0.237 |
| Artery_Tibi | RHBDD2 | ENSG0000( | 7 | 0.1742 | rs1253222 | -7.26 | rs7807392 | 0.000117 |
| Artery_Tibi | GIGYF1 | ENSG0000( | 7 | 0.2049 | rs1330696! | -4.89 | rs1734907 | 0.1 |
| Artery_Tibi | TRIP6 | ENSG0000( | 7 | 0.1216 | rs1155912: | 6.72 | rs314330 | 0.0959 |
| Artery_Tibi | CPED1 | ENSG0000( | 7 | 0.103 | rs7776725 | -7.19 | rs7776725 | 0.00608 |
| Artery_Tibi | FAM3C | ENSG0000( | 7 | 0.1065 | rs7776725 | -7.19 | rs4727922 | 0.0133 |
| Artery_Tibi | STRIP2 | ENSG0000( | 7 | 0.0815 | rs9607 | -4.99 | rs676947 | 0.00699 |
| Artery_Tibi | LRGUK | ENSG0000( | 7 | 0.5461 | rs1646650 | -9.17 | rs7457999 | 0.0675 |
| Artery_Tibi | SCAMP4 | ENSG0000( | 19 | 0.0924 | rs1166721 | -6.85 | rs8109669 | 0.00384 |
| Artery_Tibi | ZNF100 | ENSG0000( | 19 | 0.8631 | rs1041066( | 4.53 | rs6511286 | 0.464 |
| Artery_Tibi | ZNF208 | ENSG0000( | 19 | 0.2397 | rs1041066( | 6.32 | rs1987570 | 0.13 |
| Artery_Tibi | PEPD | ENSG0000( | 19 | 0.241 | rs1246250! | -5.43 | rs3556 | 0.148 |
| Artery_Tibi | C19orf47 | ENSG0000( | 19 | 0.1479 | rs399405 | -4.67 | rs1991823 | 0.0559 |
| Artery_Tibi | TMEM91 | ENSG0000( | 19 | 0.2468 | rs3848568 | 6.21 | rs4674 | 0.0224 |
| Artery_Tibi | BCKDHA | ENSG0000( | 19 | 0.3156 | rs3848568 | 6.67 | rs1187909( | 0.0952 |
| Artery_Tibi | PAFAH1B3 | ENSG0000( | 19 | 0.101 | rs3582285: | 6.11 | rs3610483! | -0.00189 |
| Artery_Tibi | ZNF155 | ENSG0000( | 19 | 0.5276 | rs239940 | 5.18 | rs453932 | 0.121 |
| Artery_Tibi | GEMIN7 | ENSG0000( | 19 | 0.2012 | rs4420638 | 6.07 | rs1040585! | 0.0849 |
| Artery_Tibi | TMEM160 | ENSG0000( | 19 | 0.052 | rs2694568 | 9.11 | rs1167338( | 0.0041 |

| | | | | | | | | |
|---|---|---|---|---|---|---|---|---|
| Artery_Tibi | PLEKHA4 | ENSG0000( | 19 | 0.1066 | rs1698213: | -6.77 | rs638050 | -0.00039 |
| Artery_Tibi | ITM2B | ENSG0000( | 13 | 0.0796 | rs6561461 | -6.66 | rs1555722 | -0.00106 |
| Artery_Tibi | PHF11 | ENSG0000( | 13 | 0.0552 | rs9568353 | -5.39 | rs1925742 | 0.00747 |
| Artery_Tibi | ALKAL2 | ENSG0000( | 2 | 0.09 | rs1317824 | 5.14 | rs2685230 | 0.0179 |
| Artery_Tibi | CENPO | ENSG0000( | 2 | 0.186 | rs1018970: | -7.26 | rs6545800 | 0.107 |
| Artery_Tibi | ADCY3 | ENSG0000( | 2 | 0.3466 | rs1018970: | -6.95 | rs7576788 | 0.212 |
| Artery_Tibi | PKDCC | ENSG0000( | 2 | 0.1306 | rs1703868! | 7.04 | rs1341659! | 0.02 |
| Artery_Tibi | ANKRD36B | ENSG0000( | 2 | 0.4583 | rs1299119 | 7.11 | rs6718109 | 0.317 |
| Artery_Tibi | C2orf92 | ENSG0000( | 2 | 0.2292 | rs1168085! | 6.92 | rs1742640: | -0.00097 |
| Artery_Tibi | ZAP70 | ENSG0000( | 2 | 0.0815 | rs1168085! | 6.96 | rs1189465: | 0.0544 |
| Artery_Tibi | TSGA10 | ENSG0000( | 2 | 0.0719 | rs958778 | -5.56 | rs1261362 | 0.0522 |
| Artery_Tibi | LYG1 | ENSG0000( | 2 | 0.1265 | rs1261551! | 4.85 | rs1189652( | 0.0733 |
| Artery_Tibi | TANC1 | ENSG0000( | 2 | 0.1779 | rs6745896 | -4.8 | rs264655 | 0.00662 |
| Artery_Tibi | PHOSPHO2 | ENSG0000( | 2 | 0.1988 | rs7591026 | -4.72 | rs7575494 | 0.217 |
| Artery_Tibi | SPATS2L | ENSG0000( | 2 | 0.1188 | rs3754800 | 6.94 | rs3754800 | 0.0272 |
| Artery_Tibi | NYAP2 | ENSG0000( | 2 | 0.1517 | rs6436519 | -7.18 | rs7580086 | 0.0423 |
| Artery_Tibi | SLC9A3 | ENSG0000( | 5 | 0.6852 | rs6869655 | -5.52 | rs1113405 | 0.169 |
| Artery_Tibi | IRX1 | ENSG0000( | 5 | 0.1658 | rs2398646 | -5.86 | rs1703290 | 0.0852 |
| Artery_Tibi | GHR | ENSG0000( | 5 | 0.0412 | rs4146624 | -4.77 | rs1251841 | 0.00286 |
| Artery_Tibi | PCYOX1L | ENSG0000( | 5 | 0.2118 | rs6887404 | -4.64 | rs783777 | 0.0902 |
| Artery_Tibi | SLC35E2B | ENSG0000( | 1 | 0.7508 | rs2887286 | 7.44 | rs4648786 | 0.189 |
| Artery_Tibi | FAAP20 | ENSG0000( | 1 | 0.475 | rs4648640 | 3.86 | rs2503706 | 0.129 |
| Artery_Tibi | CROCC | ENSG0000( | 1 | 0.3916 | rs978528 | -5.08 | rs6691985 | 0.225 |
| Artery_Tibi | MFAP2 | ENSG0000( | 1 | 0.2414 | rs9435734 | -5.04 | rs761422 | 0.127 |
| Artery_Tibi | CEP85 | ENSG0000( | 1 | 0.3426 | rs9438620 | -6.44 | rs4585968 | 0.192 |
| Artery_Tibi | SDC3 | ENSG0000( | 1 | 0.6349 | rs3124326 | 6.14 | rs1272686: | 0.326 |
| Artery_Tibi | ATG4C | ENSG0000( | 1 | 0.0684 | rs6657139 | 5.23 | rs7540030 | 0.00222 |
| Artery_Tibi | MIER1 | ENSG0000( | 1 | 0.3201 | rs1209095! | 8.48 | rs2755246 | 0.218 |
| Artery_Tibi | FUBP1 | ENSG0000( | 1 | 0.1893 | rs1739169 | -5.73 | rs1739169 | 0.116 |
| Artery_Tibi | MAN1A2 | ENSG0000( | 1 | 0.0936 | rs1703768( | 11.59 | rs1256200: | 0.0381 |
| Artery_Tibi | WARS2 | ENSG0000( | 1 | 0.578 | rs984225 | -6.58 | rs2645303 | 0.485 |
| Artery_Tibi | ARNT | ENSG0000( | 1 | 0.1481 | rs1088838: | 6.44 | rs1256875 | 0.169 |
| Artery_Tibi | METTL25B | ENSG0000( | 1 | 0.1085 | rs1683807 | 4.66 | rs1214043 | 0.0664 |
| Artery_Tibi | OLFML2B | ENSG0000( | 1 | 0.1123 | rs1207379: | 8.27 | rs2499835 | 0.0181 |
| Artery_Tibi | MR1 | ENSG0000( | 1 | 0.0901 | rs1207969( | -5.49 | rs1075321 | 0.00181 |
| Artery_Tibi | SYT2 | ENSG0000( | 1 | 0.1135 | rs1092054: | 8 | rs4950858 | 0.0055 |
| Artery_Tibi | PCNX2 | ENSG0000( | 1 | 0.1551 | rs6667845 | 7.9 | rs6667845 | 0.0711 |
| Artery_Tibi | CBR1 | ENSG0000( | 21 | 0.1089 | rs2835109 | 5.02 | rs762360 | 0.00981 |
| Artery_Tibi | PRDM15 | ENSG0000( | 21 | 0.581 | rs420737 | 10.7 | rs9983923 | 0.0843 |
| Artery_Tibi | C2CD2 | ENSG0000( | 21 | 0.1353 | rs420737 | 7.22 | rs7280126 | 0.0103 |
| Artery_Tibi | TPGS2 | ENSG0000( | 18 | 0.363 | rs903733 | 6.82 | rs2303508 | 0.283 |
| Artery_Tibi | KDSR | ENSG0000( | 18 | 0.5019 | rs4987797 | -5.17 | rs9946122 | 0.0927 |
| Artery_Tibi | HSBP1L1 | ENSG0000( | 18 | 0.2577 | rs732771 | -7.65 | rs4799113 | 0.229 |
| Artery_Tibi | TPSB2 | ENSG0000( | 16 | 0.2073 | rs7191794 | 5.19 | rs4984637 | 0.129 |
| Artery_Tibi | TMEM219 | ENSG0000( | 16 | 0.0432 | rs4541091 | 7.76 | rs8060511 | 0.0219 |

| | | | | | | | | |
|---|---|---|---|---|---|---|---|---|
| Artery_Tibi | INO80E | ENSG00000( | 16 | 0.2114 | rs4541091 | 7.08 | rs4787491 | 0.253 |
| Artery_Tibi | DOC2A | ENSG00000( | 16 | 0.0443 | rs4541091 | 6.02 | rs1293357! | 0.0239 |
| Artery_Tibi | TBX6 | ENSG00000( | 16 | 0.1672 | rs1164274( | 5.72 | rs9928448 | 0.108 |
| Artery_Tibi | YPEL3 | ENSG00000( | 16 | 0.2475 | rs1164274( | 5.72 | rs7205802 | 0.362 |
| Artery_Tibi | GDPD3 | ENSG00000( | 16 | 0.2558 | rs1164274( | 5.72 | rs3809624 | 0.0255 |
| Artery_Tibi | ZNF720 | ENSG00000( | 16 | 0.1041 | rs1034614 | -5.87 | rs1783951! | 0.101 |
| Artery_Tibi | UTP4 | ENSG00000( | 16 | 0.2989 | rs1333017( | -5.82 | rs8061222 | 0.0625 |
| Artery_Tibi | WWP2 | ENSG00000( | 16 | 0.1368 | rs2291959 | -5.54 | rs1566452 | 0.0611 |
| Artery_Tibi | DBNDD1 | ENSG00000( | 16 | 0.5555 | rs1333267: | 7.62 | rs1805007 | 0.155 |
| Artery_Tibi | ARHGEF40 | ENSG00000( | 14 | 0.3326 | rs2319627 | -6.01 | rs8019890 | 0.275906 |
| Artery_Tibi | TRMT5 | ENSG00000( | 14 | 0.0701 | rs4899015 | 6.53 | rs1289345( | -0.00017 |
| Artery_Tibi | DCAF4 | ENSG00000( | 14 | 0.5212 | rs1013993 | 5.98 | rs1076458 | 0.102565 |
| Artery_Tibi | ACOT2 | ENSG00000( | 14 | 0.6589 | rs1709110( | -5.71 | rs6574128 | 0.22818 |
| Artery_Tibi | PTGR2 | ENSG00000( | 14 | 0.5665 | rs2109750 | -5.12 | rs1162881: | 0.200282 |
| Artery_Tibi | SYNDIG1L | ENSG00000( | 14 | 0.0688 | rs730384 | -5.19 | rs4903240 | -0.00207 |
| Artery_Tibi | FLRT2 | ENSG00000( | 14 | 0.142 | rs6574859 | -6.71 | rs3519542: | 0.032102 |
| Artery_Tibi | ACADVL | ENSG00000( | 17 | 0.1941 | rs2269459 | 8.13 | rs446994 | 0.192 |
| Artery_Tibi | CLDN7 | ENSG00000( | 17 | 0.0943 | rs2269459 | 8.28 | rs222857 | 0.0575 |
| Artery_Tibi | MPDU1 | ENSG00000( | 17 | 0.1114 | rs2269459 | 8.15 | rs1155270! | 0.027 |
| Artery_Tibi | TTLL6 | ENSG00000( | 17 | 0.0763 | rs1054072 | -4.41 | rs8067245 | 0.0112 |
| Artery_Tibi | GORASP1 | ENSG00000( | 3 | 0.1127 | rs1703875: | 5.99 | rs1170710: | 0.0121 |
| Artery_Tibi | PRKAR2A | ENSG00000( | 3 | 0.0896 | rs1249039: | -4.93 | rs4955426 | 0.00846 |
| Artery_Tibi | P4HTM | ENSG00000( | 3 | 0.0625 | rs1249039: | -4.93 | rs9284885 | 0.0416 |
| Artery_Tibi | DALRD3 | ENSG00000( | 3 | 0.0317 | rs1249039: | -4.93 | rs4974081 | 0.0245 |
| Artery_Tibi | GPX1 | ENSG00000( | 3 | 0.0199 | rs1249039: | -4.93 | rs1309647 | 0.00726 |
| Artery_Tibi | TCTA | ENSG00000( | 3 | 0.0377 | rs1249039: | -4.93 | rs6809851 | 0.0409 |
| Artery_Tibi | AMT | ENSG00000( | 3 | 0.4324 | rs1249039: | -4.93 | rs3448 | 0.514 |
| Artery_Tibi | NICN1 | ENSG00000( | 3 | 0.179 | rs1249039: | -4.93 | rs3448 | 0.194 |
| Artery_Tibi | DOCK3 | ENSG00000( | 3 | 0.452 | rs2239751 | 20.48 | rs1192838! | 0.221 |
| Artery_Tibi | RBM15B | ENSG00000( | 3 | 0.3226 | rs1263950: | 6.85 | rs1263799 | 0.159 |
| Artery_Tibi | FAM107A | ENSG00000( | 3 | 0.3188 | rs7640235 | -13.26 | rs2306671 | 0.212 |
| Artery_Tibi | ALCAM | ENSG00000( | 3 | 0.1064 | rs1685090: | -4.12 | rs1685068 | 0.00807 |
| Artery_Tibi | ADPRH | ENSG00000( | 3 | 0.3891 | rs3814056 | 3.88 | rs1485331 | 0.0776 |
| Artery_Tibi | DNAJC13 | ENSG00000( | 3 | 0.0789 | rs1734129: | 6.58 | rs6800166 | 0.0286 |
| Artery_Tibi | SIRT5 | ENSG00000( | 6 | 0.0695 | rs9474418 | -4.28 | rs2328676 | 0.0302 |
| Artery_Tibi | ILRUN | ENSG00000( | 6 | 0.1197 | rs2820239 | -6.01 | rs1175363 | 0.0329 |
| Artery_Tibi | C6orf226 | ENSG00000( | 6 | 0.0822 | rs2016128 | -5.74 | rs9471938 | 0.0892 |
| Artery_Tibi | SRSF12 | ENSG00000( | 6 | 0.1685 | rs6942204 | 5.31 | rs2150820 | 0.103 |
| Artery_Tibi | CEP85L | ENSG00000( | 6 | 0.0528 | rs4374854 | -7.9 | rs9481842 | 0.0512 |
| Artery_Tibi | L3MBTL3 | ENSG00000( | 6 | 0.1268 | rs7755865 | -7.18 | rs7769599 | 0.0496 |
| Artery_Tibi | TCF21 | ENSG00000( | 6 | 0.0857 | rs7767123 | 7.06 | rs1029212 | 0.0328 |
| Artery_Tibi | SLC5A4 | ENSG00000( | 22 | 0.2675 | rs5998188 | -6.53 | rs5754035 | 0.0576 |
| Artery_Tibi | COL13A1 | ENSG00000( | 10 | 0.1294 | rs1241283( | -5.15 | rs2683560 | 0.00285 |
| Artery_Tibi | SEC24C | ENSG00000( | 10 | 0.0442 | rs7098158 | 6.6 | rs7069592 | 0.000231 |
| Artery_Tibi | ALDH18A1 | ENSG00000( | 10 | 0.2291 | rs7907476 | -8.39 | rs1053905 | 0.143 |

| | | | | | | | | |
|---|---|---|---|---|---|---|---|---|
| Artery_Tibi | WBP1L | ENSG00000( | 10 | 0.1539 | rs1119145 | -4.8 | rs486955 | 0.019 |
| Artery_Tibi | CASP7 | ENSG00000( | 10 | 0.6131 | rs1709099 | -5.32 | rs3814231 | 0.358 |
| Artery_Tibi | PHF19 | ENSG00000( | 9 | 0.1479 | rs1017119 | -7.01 | rs7853645 | 0.0177 |
| Artery_Tibi | RBM18 | ENSG00000( | 9 | 0.1077 | rs7035536 | -6.14 | rs7035313 | 0.0856 |
| Artery_Tibi | NIBAN2 | ENSG00000( | 9 | 0.1584 | rs4836593 | 3.89 | rs4837165 | 0.0123 |
| Artery_Tibi | TM7SF3 | ENSG00000( | 12 | 0.5925 | rs2306852 | -9.69 | rs6487582 | 0.131 |
| Artery_Tibi | FKBP11 | ENSG00000( | 12 | 0.1347 | rs4760645 | -6.69 | rs1087590 | 0.0714 |
| Artery_Tibi | CERS5 | ENSG00000( | 12 | 0.0826 | rs1183106 | 9.13 | rs3184122 | 0.075 |
| Artery_Tibi | ALDH2 | ENSG00000( | 12 | 0.1361 | rs4767293 | -5.31 | rs4646777 | 0.0853 |
| Artery_Tibi | MTRFR | ENSG00000( | 12 | 0.0692 | rs3531484 | 8.77 | rs940904 | 0.0615 |
| Artery_Tibi | KMT5A | ENSG00000( | 12 | 0.0641 | rs3531484 | 11.07 | rs1236820 | 0.0216 |
| Artery_Tibi | DNAH10 | ENSG00000( | 12 | 0.2043 | rs9668827 | -5.35 | rs7973683 | 0.143 |
| Artery_Tibi | UQCC1 | ENSG00000( | 20 | 0.0628 | rs7280 | -5.85 | rs224331 | 0.0332 |
| Artery_Tibi | PTK2B | ENSG00000( | 8 | 0.1918 | rs9314347 | 4.74 | rs7813625 | 0.0712 |
| Artery_Tibi | NECAB1 | ENSG00000( | 8 | 0.116 | rs7819987 | 9.35 | rs1011243 | 0.00342 |
| Artery_Tibi | RSPO2 | ENSG00000( | 8 | 0.1061 | rs610891 | 6.42 | rs7357448 | 0.0529 |
| Artery_Tibi | CCN3 | ENSG00000( | 8 | 0.0774 | rs7012790 | -6.15 | rs7816205 | 0.0365 |
| Whole_Blo | COX14 | ENSG00000( | 12 | 0.0839 | rs1183106 | 8.51 | rs7972465 | 0.0487 |
| Whole_Blo | SUOX | ENSG00000( | 12 | 0.2161 | rs14483 | -6.95 | rs705700 | 0.179 |
| Whole_Blo | SMARCC2 | ENSG00000( | 12 | 0.079 | rs14483 | -7.2 | rs14483 | 0.0141 |
| Whole_Blo | HSP90B1 | ENSG00000( | 12 | 0.0801 | rs7301863 | 12.31 | rs2888810 | 0.00367 |
| Whole_Blo | MTRFR | ENSG00000( | 12 | 0.0842 | rs3531484 | 8.77 | rs2853203 | 0.0595 |
| Whole_Blo | CDK2AP1 | ENSG00000( | 12 | 0.2701 | rs3531484 | 10.37 | rs7972811 | 0.234 |
| Whole_Blo | CCDC92 | ENSG00000( | 12 | 0.1694 | rs1257967 | -6.13 | rs825457 | 0.0332 |
| Whole_Blo | ZNF664 | ENSG00000( | 12 | 0.0665 | rs1257967 | -6.08 | rs6488910 | 0.0203 |
| Whole_Blo | RBM23 | ENSG00000( | 14 | 0.2574 | rs8022177 | -8.33 | rs8022177 | 0.128289 |
| Whole_Blo | PSEN1 | ENSG00000( | 14 | 0.2244 | rs1712019 | 8.39 | rs7523 | 0.162185 |
| Whole_Blo | PTGR2 | ENSG00000( | 14 | 0.2447 | rs2109750 | -5.12 | rs4243645 | 0.118552 |
| Whole_Blo | POLI | ENSG00000( | 18 | 0.4918 | rs1259807 | -4.52 | rs3730783 | 0.23258 |
| Whole_Blo | P4HTM | ENSG00000( | 3 | 0.0372 | rs1249039 | -4.93 | rs6766238 | 0.0344 |
| Whole_Blo | QRICH1 | ENSG00000( | 3 | 0.058 | rs1249039 | -4.93 | rs4955426 | 0.0819 |
| Whole_Blo | LAMB2 | ENSG00000( | 3 | 0.0191 | rs1249039 | -4.93 | rs1263198 | 0.0225 |
| Whole_Blo | AMT | ENSG00000( | 3 | 0.2171 | rs1249039 | -4.93 | rs3905330 | 0.303 |
| Whole_Blo | RBM15B | ENSG00000( | 3 | 0.1332 | rs1263950 | 6.85 | rs7634991 | 0.0246 |
| Whole_Blo | NT5DC2 | ENSG00000( | 3 | 0.2952 | rs1139106 | -4.43 | rs1541495 | 0.0811 |
| Whole_Blo | CFAP44 | ENSG00000( | 3 | 0.1251 | rs1686066 | 5.97 | rs6438140 | 0.039 |
| Whole_Blo | CD86 | ENSG00000( | 3 | 0.0584 | rs1683240 | -9.62 | rs2681408 | 0.000934 |
| Whole_Blo | EFCAB12 | ENSG00000( | 3 | 0.1834 | rs6787488 | 6.19 | rs3138345 | 0.0148 |
| Whole_Blo | MAP3K13 | ENSG00000( | 3 | 0.2092 | rs6786934 | -9.15 | rs1191846 | 0.243 |
| Whole_Blo | MELTF | ENSG00000( | 3 | 0.2208 | rs9843585 | -5.27 | rs9861658 | 0.15 |
| Whole_Blo | BRWD1 | ENSG00000( | 21 | 0.1494 | rs2836866 | 5.52 | rs2836939 | 0.0133 |
| Whole_Blo | TRAPPC4 | ENSG00000( | 11 | 0.3634 | rs1157466 | 6.72 | rs4938621 | 0.259441 |
| Whole_Blo | AFAP1 | ENSG00000( | 4 | 0.6163 | rs1193378 | -7.14 | rs1173317 | 0.578 |
| Whole_Blo | C4orf36 | ENSG00000( | 4 | 0.0725 | rs1173679 | -5.11 | rs7657530 | 0.0202 |
| Whole_Blo | HSD17B11 | ENSG00000( | 4 | 0.2273 | rs7657530 | -4.57 | rs6811010 | 0.024 |

| | | | | | | | | |
|---|---|---|---|---|---|---|---|---|
| Whole_Blo | PCYOX1L | ENSG0000( | 5 | 0.3927 | rs6887404 | -4.64 | rs2242376 | 0.263 |
| Whole_Blo | PTCH1 | ENSG0000( | 9 | 0.1134 | rs2282040 | 5.19 | rs2853553( | 0.0335 |
| Whole_Blo | SNAPC4 | ENSG0000( | 9 | 0.4022 | rs1725077{ | 6.72 | rs1012164( | 0.0389 |
| Whole_Blo | ICA1 | ENSG0000( | 7 | 0.1725 | rs7800813 | -6.9 | rs1197332( | 0.0792 |
| Whole_Blo | POM121C | ENSG0000( | 7 | 0.1819 | rs1253245 | -11.26 | rs1253590 | 0.0125 |
| Whole_Blo | STYXL1 | ENSG0000( | 7 | 0.4665 | rs1253245 | -7.37 | rs1155309( | 0.458 |
| Whole_Blo | POLR2J2 | ENSG0000( | 7 | 0.3974 | rs1129271 | 7.89 | rs1198121! | 0.149 |
| Whole_Blo | DLD | ENSG0000( | 7 | 0.0412 | rs4730271 | 5.65 | rs3735602 | 0.0104 |
| Whole_Blo | PLXNA4 | ENSG0000( | 7 | 0.0532 | rs1323572! | 4.23 | rs1023368! | -0.0018 |
| Whole_Blo | GIMAP4 | ENSG0000( | 7 | 0.1656 | rs6950421 | -4.65 | rs1851434 | 0.106 |
| Whole_Blo | MSRA | ENSG0000( | 8 | 0.1653 | rs7009513 | -7.32 | rs4448276 | 0.0313 |
| Whole_Blo | RPS20 | ENSG0000( | 8 | 0.0813 | rs3403795 | 5.17 | rs6988900 | 0.0151 |
| Whole_Blo | TOP1MT | ENSG0000( | 8 | 0.1556 | rs2450763 | -6.26 | rs3814772 | 0.00537 |
| Whole_Blo | SLC35E2B | ENSG0000( | 1 | 0.683 | rs2887286 | 7.44 | rs4648786 | 0.139 |
| Whole_Blo | AKR7A2 | ENSG0000( | 1 | 0.1453 | rs2027508 | 3.97 | rs859218 | 0.0453 |
| Whole_Blo | ID3 | ENSG0000( | 1 | 0.0605 | rs4514282 | 4.45 | rs909536 | 0.00134 |
| Whole_Blo | XKR8 | ENSG0000( | 1 | 0.0682 | rs1890462 | 7.67 | rs1090268 | 0.0147 |
| Whole_Blo | SHISAL2A | ENSG0000( | 1 | 0.1028 | rs1736460( | -9.28 | rs6686035 | -0.00178 |
| Whole_Blo | CD84 | ENSG0000( | 1 | 0.0794 | rs1126543 | -5.66 | rs1213439 | 0.013 |
| Whole_Blo | CREG1 | ENSG0000( | 1 | 0.1087 | rs909935 | 7.72 | rs909935 | 0.0106 |
| Whole_Blo | CR1 | ENSG0000( | 1 | 0.0602 | rs1157916 | 6.09 | rs2796249 | 0.00144 |
| Whole_Blo | AHCTF1 | ENSG0000( | 1 | 0.3532 | rs2799179 | 5.92 | rs1092487 | 0.0833 |
| Whole_Blo | KBTBD6 | ENSG0000( | 13 | 0.0719 | rs1706151 | -8.69 | rs1572018 | 0.002161 |
| Whole_Blo | MYH7B | ENSG0000( | 20 | 0.0559 | rs7280 | -5.85 | rs6088764 | 0.0256 |
| Whole_Blo | MMP24OS | ENSG0000( | 20 | 0.1782 | rs7280 | -5.85 | rs6060341 | 0.102 |
| Whole_Blo | UQCC1 | ENSG0000( | 20 | 0.1009 | rs7280 | -5.85 | rs1540927 | 0.113 |
| Whole_Blo | CAPN15 | ENSG0000( | 16 | 0.1419 | rs1015319 | -10.5 | rs1186465 | 0.0664 |
| Whole_Blo | ZNF500 | ENSG0000( | 16 | 0.2418 | rs6500627 | -5.02 | rs1259976 | 0.0619 |
| Whole_Blo | NAGPA | ENSG0000( | 16 | 0.214 | rs4786590 | 5.2 | rs887854 | 0.107 |
| Whole_Blo | NFATC2IP | ENSG0000( | 16 | 0.0603 | rs34837 | -4.65 | rs252297 | -0.00056 |
| Whole_Blo | BOLA2 | ENSG0000( | 16 | 0.0882 | rs1115057( | 4.88 | rs1164611 | 0.0527 |
| Whole_Blo | INO80E | ENSG0000( | 16 | 0.0665 | rs4541091 | 7.08 | rs9932702 | 0.091 |
| Whole_Blo | TBX6 | ENSG0000( | 16 | 0.2422 | rs1164274( | 5.72 | rs3809624 | 0.143 |
| Whole_Blo | YPEL3 | ENSG0000( | 16 | 0.0997 | rs1164274( | 5.72 | rs7205802 | 0.116 |
| Whole_Blo | ZNRF1 | ENSG0000( | 16 | 0.0834 | rs1078197( | 5.41 | rs8053898 | 0.0144 |
| Whole_Blo | SFMBT2 | ENSG0000( | 10 | 0.1987 | rs1244454 | -6.58 | rs2762612 | 0.115 |
| Whole_Blo | TMEM273 | ENSG0000( | 10 | 0.2764 | rs2377877 | -6.16 | rs1224937( | 0.243 |
| Whole_Blo | MRPS16 | ENSG0000( | 10 | 0.1234 | rs4746139 | 4.16 | rs3740293 | 0.0129 |
| Whole_Blo | NDST2 | ENSG0000( | 10 | 0.116 | rs4746160 | -6.59 | rs2242258 | 0.0735 |
| Whole_Blo | XKR3 | ENSG0000( | 22 | 0.1964 | rs5749011 | 5.07 | rs1807512 | 0.0127 |
| Whole_Blo | AIFM3 | ENSG0000( | 22 | 0.4934 | rs362089 | 6.73 | rs178255 | 0.193 |
| Whole_Blo | ZMAT5 | ENSG0000( | 22 | 0.0387 | rs131272 | 6.48 | rs140104 | 0.0283 |
| Whole_Blo | UQCR10 | ENSG0000( | 22 | 0.0438 | rs131272 | 6.48 | rs737787 | 0.005 |
| Whole_Blo | PRKD3 | ENSG0000( | 2 | 0.1625 | rs2373001 | -6.55 | rs2041840 | 0.114 |
| Whole_Blo | CD8A | ENSG0000( | 2 | 0.0705 | rs1340401( | 4.73 | rs938487 | 0.0178 |

| | | | | | | | | |
|---|---|---|---|---|---|---|---|---|
| Whole_Blo | ARID5A | ENSG00000( | 2 | 0.0647 | rs3580943: | 5.49 | rs2118836 | 0.0169 |
| Whole_Blo | ANKRD36B | ENSG00000( | 2 | 0.0801 | rs1299119 | 7.11 | rs3906948 | 0.099 |
| Whole_Blo | TSGA10 | ENSG00000( | 2 | 0.1407 | rs958778 | -5.56 | rs1168318 | 0.228 |
| Whole_Blo | MITD1 | ENSG00000( | 2 | 0.0539 | rs2048748 | -6.76 | rs13798 | 0.0425 |
| Whole_Blo | REV1 | ENSG00000( | 2 | 0.0785 | rs1261551 | 4.85 | rs1011633 | 0.0679 |
| Whole_Blo | SH3RF3 | ENSG00000( | 2 | 0.256 | rs1169110 | -5.11 | rs4676266 | 0.0708 |
| Whole_Blo | OSBPL6 | ENSG00000( | 2 | 0.0435 | rs1735499 | -10.72 | rs1246705 | 0.0175 |
| Whole_Blo | SESTD1 | ENSG00000( | 2 | 0.2193 | rs1736258 | -6.61 | rs1020987 | 0.0746 |
| Whole_Blo | PNKD | ENSG00000( | 2 | 0.1715 | rs3092968 | 4.77 | rs4672884 | 0.135 |
| Whole_Blo | B3GNT7 | ENSG00000( | 2 | 0.0566 | rs6705872 | 13.95 | rs4973397 | 0.0459 |
| Whole_Blo | TUBGCP5 | ENSG00000( | 15 | 0.0636 | rs1259472 | -6.05 | rs8031642 | -0.00149 |
| Whole_Blo | NDUFAF1 | ENSG00000( | 15 | 0.252 | rs4924533 | -4.87 | rs1107032 | 0.0466 |
| Whole_Blo | SNX1 | ENSG00000( | 15 | 0.0976 | rs2253557 | -4.7 | rs1210220 | 0.047 |
| Whole_Blo | PPIB | ENSG00000( | 15 | 0.0317 | rs677561 | -6.73 | rs7165405 | 0.000114 |
| Whole_Blo | RAB11A | ENSG00000( | 15 | 0.0535 | rs2292114 | -6.46 | rs4514623 | 0.0377 |
| Whole_Blo | UBE2Q2 | ENSG00000( | 15 | 0.0502 | rs4886489 | 3.81 | rs1742754 | 0.0435 |
| Whole_Blo | TSPAN3 | ENSG00000( | 15 | 0.5263 | rs1696862 | -4.13 | rs1163919 | 0.138 |
| Whole_Blo | ZNF592 | ENSG00000( | 15 | 0.0724 | rs2242047 | 5.72 | rs2059927 | 0.0395 |
| Whole_Blo | LYSMD4 | ENSG00000( | 15 | 0.3072 | rs7170513 | -5.8 | rs8041078 | 0.132 |
| Whole_Blo | ACADVL | ENSG00000( | 17 | 0.1451 | rs2269459 | 8.13 | rs446994 | 0.149 |
| Whole_Blo | TNFSF12 | ENSG00000( | 17 | 0.2324 | rs2269459 | 9.72 | rs9899183 | 0.194 |
| Whole_Blo | NUFIP2 | ENSG00000( | 17 | 0.0573 | rs8070400 | 5.58 | rs565977 | 0.0101 |
| Whole_Blo | PLEKHM1 | ENSG00000( | 17 | 0.0619 | rs3744759 | -6.28 | rs1694066 | 0.00401 |
| Whole_Blo | POLRMT | ENSG00000( | 19 | 0.1429 | rs9304925 | -9.04 | rs2863108 | 0.0281 |
| Whole_Blo | GNG7 | ENSG00000( | 19 | 0.0748 | rs1041591 | -6.94 | rs2317323 | 0.0545 |
| Whole_Blo | AP1M2 | ENSG00000( | 19 | 0.4074 | rs1246200 | 3.86 | rs7252007 | 0.117 |
| Whole_Blo | SLC44A2 | ENSG00000( | 19 | 0.2957 | rs2228671 | -4.03 | rs8106664 | 0.128 |
| Whole_Blo | WDR83OS | ENSG00000( | 19 | 0.0679 | rs1041161 | -10.04 | rs1188195 | 0.0107 |
| Whole_Blo | ANKRD27 | ENSG00000( | 19 | 0.445 | rs1775465 | 8.49 | rs7250780 | 0.169 |
| Whole_Blo | ZNF529 | ENSG00000( | 19 | 0.0949 | rs2967436 | 4.63 | rs2967465 | 0.0256 |
| Whole_Blo | TMEM91 | ENSG00000( | 19 | 0.5163 | rs3848568 | 6.21 | rs1187909 | 0.0823 |
| Whole_Blo | LYPD3 | ENSG00000( | 19 | 0.0785 | rs7256718 | 8.32 | rs2599437 | 0.0058 |
| Whole_Blo | PHACTR1 | ENSG00000( | 6 | 0.1879 | rs6905435 | -7.16 | rs1219827 | 0.103213 |
| Whole_Blo | ILRUN | ENSG00000( | 6 | 0.1488 | rs2820239 | -6.01 | rs1689495 | 0.091791 |
| Whole_Blo | CALHM6 | ENSG00000( | 6 | 0.6569 | rs4946203 | -13.87 | rs1219212 | 0.105417 |
| Whole_Blo | TBPL1 | ENSG00000( | 6 | 0.0789 | rs4424102 | 5.97 | rs1155366 | 0.00703 |
| Whole_Blo | NHSL1 | ENSG00000( | 6 | 0.0587 | rs410543 | -5.25 | rs4397246 | 0.011144 |
| Lung | GNB1L | ENSG00000( | 22 | 0.1824 | rs759575 | 6.97 | rs1053001 | 0.048349 |
| Lung | MORC2 | ENSG00000( | 22 | 0.0935 | rs8135379 | 9.62 | rs9621153 | 0.041873 |
| Lung | APOL4 | ENSG00000( | 22 | 0.4199 | rs1699691 | -12.95 | rs132734 | 0.1459 |
| Lung | NDUFA6 | ENSG00000( | 22 | 0.6416 | rs1700251 | -8.19 | rs2142695 | 0.22739 |
| Lung | CCZ1 | ENSG00000( | 7 | 0.591 | rs852488 | 6.44 | rs6975026 | 0.161352 |
| Lung | NDUFA4 | ENSG00000( | 7 | 0.0696 | rs2189546 | 4.98 | rs218979 | -0.00228 |
| Lung | STYXL1 | ENSG00000( | 7 | 0.4069 | rs1253245 | -7.37 | rs1155309 | 0.3863 |
| Lung | ASB13 | ENSG00000( | 10 | 0.1417 | rs942200 | 6.07 | rs6602236 | 0.045769 |

| | | | | | | | | |
|---|---|---|---|---|---|---|---|---|
| Lung | ATP5F1C | ENSG00000( | 10 | 0.1161 | rs1079554! | 5.75 | rs1244418 | 0.068073 |
| Lung | TRIM2 | ENSG00000( | 4 | 0.1666 | rs1312436! | 7.11 | rs7686229 | 0.010102 |
| Lung | CDH10 | ENSG00000( | 5 | 0.3503 | rs1316281! | 6.71 | rs6888249 | 0.212543 |
| Lung | C6 | ENSG00000( | 5 | 0.0589 | rs404223 | 6.9 | rs751138 | -0.00149 |
| Lung | HSPB3 | ENSG00000( | 5 | 0.1522 | rs1007425: | 6.5 | rs4865832 | 0.002096 |
| Lung | KIAA1191 | ENSG00000( | 5 | 0.0588 | rs1252027: | -5.48 | rs4868630 | 0.015971 |
| Lung | MRPS26 | ENSG00000( | 20 | 0.1245 | rs6051348 | 8.22 | rs6084203 | 0.035048 |
| Lung | MYH7B | ENSG00000( | 20 | 0.2591 | rs7280 | -5.85 | rs6120777 | 0.103177 |
| Lung | MMP24OS | ENSG00000( | 20 | 0.1645 | rs7280 | -5.85 | rs6060341 | 0.130501 |
| Lung | GDF5 | ENSG00000( | 20 | 0.1026 | rs7280 | -5.85 | rs6060402 | 0.060693 |
| Lung | KCNB1 | ENSG00000( | 20 | 0.1814 | rs4810985 | -4.69 | rs6090978 | 0.000612 |
| Lung | SLC38A4 | ENSG00000( | 12 | 0.0897 | rs1709750( | -4.95 | rs1709746 | -0.00166 |
| Lung | CERS5 | ENSG00000( | 12 | 0.0741 | rs1183106: | 9.13 | rs3184122 | 0.070401 |
| Lung | HOXC8 | ENSG00000( | 12 | 0.1823 | rs2706251 | -6.74 | rs746423 | 0.070888 |
| Lung | AGAP2 | ENSG00000( | 12 | 0.0599 | rs3556937: | -7.86 | rs2269720 | -0.00159 |
| Lung | KCTD10 | ENSG00000( | 12 | 0.1399 | rs1183122( | -6.32 | rs1183237: | 0.003644 |
| Lung | SUDS3 | ENSG00000( | 12 | 0.1194 | rs1761956! | 7.25 | rs1761956! | 0.047868 |
| Lung | MPHOSPH! | ENSG00000( | 12 | 0.0399 | rs3531484: | 9.25 | rs7132277 | 0.037065 |
| Lung | CDK2AP1 | ENSG00000( | 12 | 0.1168 | rs3531484: | 10.37 | rs1879380 | 0.121762 |
| Lung | ZNF140 | ENSG00000( | 12 | 0.0402 | rs1078165: | -4.52 | rs1114724! | 0.035763 |
| Lung | GABRB3 | ENSG00000( | 15 | 0.3053 | rs2045151 | -8.78 | rs768899 | 0.0806 |
| Lung | PCLAF | ENSG00000( | 15 | 0.0527 | rs677561 | -10.15 | rs673931 | 0.004687 |
| Lung | TRIP4 | ENSG00000( | 15 | 0.1281 | rs677561 | -10.15 | rs673931 | 0.13861 |
| Lung | RCCD1 | ENSG00000( | 15 | 0.2445 | rs7601 | -5.1 | rs2290202 | 0.10873 |
| Lung | TNFRSF10A | ENSG00000( | 8 | 0.2489 | rs1708935: | 4.56 | rs4242394 | 0.091987 |
| Lung | EBF2 | ENSG00000( | 8 | 0.1348 | rs6985478 | 4.68 | rs1477548 | 0.036169 |
| Lung | PLEKHF2 | ENSG00000( | 8 | 0.1205 | rs7011044 | 6.55 | rs4735307 | -0.0021 |
| Lung | CCN3 | ENSG00000( | 8 | 0.1874 | rs7012790 | -6.15 | rs7816205 | 0.180148 |
| Lung | PLK5 | ENSG00000( | 19 | 0.1001 | rs7258839 | -5.21 | rs1108491 | 0.022 |
| Lung | IQCN | ENSG00000( | 19 | 0.2622 | rs3212700 | -6.54 | rs1075403 | 0.126 |
| Lung | ZNF100 | ENSG00000( | 19 | 0.8543 | rs1041066( | 4.53 | rs6511286 | 0.456 |
| Lung | ZNF208 | ENSG00000( | 19 | 0.1841 | rs1041066( | 6.32 | rs1745924( | 0.0539 |
| Lung | ZNF529 | ENSG00000( | 19 | 0.1779 | rs2967436 | 4.63 | rs2967465 | 0.0651 |
| Lung | TMEM91 | ENSG00000( | 19 | 0.3879 | rs3848568 | 6.21 | rs1187909( | 0.0194 |
| Lung | GEMIN7 | ENSG00000( | 19 | 0.2306 | rs4420638 | 6.07 | rs1040585! | 0.0771 |
| Lung | DHX34 | ENSG00000( | 19 | 0.1873 | rs3544588: | -6.89 | rs2694557 | 0.066 |
| Lung | PLEKHA4 | ENSG00000( | 19 | 0.1426 | rs1698213: | -6.77 | rs609985 | 0.0797 |
| Lung | ZNF468 | ENSG00000( | 19 | 0.3212 | rs1108423! | -5.41 | rs7258677 | 0.144 |
| Lung | PRPF31 | ENSG00000( | 19 | 0.1752 | rs1698547! | -7.85 | rs4806711 | 0.00585 |
| Lung | SYN2 | ENSG00000( | 3 | 0.2161 | rs9875338 | -5.68 | rs307560 | 0.119 |
| Lung | TIMP4 | ENSG00000( | 3 | 0.0435 | rs9875338 | -5.68 | rs310749 | 0.0394 |
| Lung | DALRD3 | ENSG00000( | 3 | 0.035 | rs1249039: | -4.93 | rs7100 | 0.0139 |
| Lung | QRICH1 | ENSG00000( | 3 | 0.0889 | rs1249039: | -4.93 | rs4955426 | 0.109 |
| Lung | KLHDC8B | ENSG00000( | 3 | 0.0418 | rs1249039: | -4.93 | rs1171043 | 0.0377 |
| Lung | C3orf62 | ENSG00000( | 3 | 0.028 | rs1249039: | -4.93 | rs1307894! | 0.0203 |

| | | | | | | | | |
|---|---|---|---|---|---|---|---|---|
| Lung | AMT | ENSG00000( | 3 | 0.3557 | rs1249039: | -4.93 | rs974495 | 0.511 |
| Lung | HEMK1 | ENSG00000( | 3 | 0.0901 | rs743753 | 23.7 | rs4438692 | 0.0554 |
| Lung | DOCK3 | ENSG00000( | 3 | 0.1928 | rs743753 | 20.48 | rs1192044: | 0.0668 |
| Lung | STIMATE | ENSG00000( | 3 | 0.2523 | rs1705220( | -6.13 | rs1191585: | 0.0922 |
| Lung | CPNE4 | ENSG00000( | 3 | 0.1848 | rs3610159: | -7.12 | rs3905934 | 0.0699 |
| Lung | ILRUN | ENSG00000( | 6 | 0.0786 | rs2820239 | -6.01 | rs1175526( | 0.0796 |
| Lung | TCF21 | ENSG00000( | 6 | 0.1406 | rs4424102 | 5.79 | rs969282 | 0.0701 |
| Lung | SCUBE2 | ENSG00000( | 11 | 0.2923 | rs1883099 | -4.45 | rs1228771: | 0.037613 |
| Lung | CRYAB | ENSG00000( | 11 | 0.0689 | rs360726 | -12.07 | rs1182308! | 0.006302 |
| Lung | THY1 | ENSG00000( | 11 | 0.2417 | rs589925 | -4.92 | rs7130716 | 0.117034 |
| Lung | TMEM219 | ENSG00000( | 16 | 0.0468 | rs4541091 | 7.76 | rs4788204 | 0.044355 |
| Lung | INO80E | ENSG00000( | 16 | 0.2639 | rs4541091 | 7.08 | rs4787491 | 0.253466 |
| Lung | DOC2A | ENSG00000( | 16 | 0.0673 | rs4541091 | 6.02 | rs7205802 | 0.032172 |
| Lung | YPEL3 | ENSG00000( | 16 | 0.2345 | rs1164274( | 5.72 | rs7205802 | 0.308287 |
| Lung | GDPD3 | ENSG00000( | 16 | 0.2241 | rs1164274( | 5.72 | rs6565174 | 0.045743 |
| Lung | ZNF720 | ENSG00000( | 16 | 0.0909 | rs1034614 | -5.87 | rs1783951! | 0.030161 |
| Lung | FTO | ENSG00000( | 16 | 0.0839 | rs1421090 | 5.22 | rs1125338 | -0.0002 |
| Lung | ADAD2 | ENSG00000( | 16 | 0.1291 | rs907037 | 7.92 | rs2278043 | 0.022617 |
| Lung | DEF8 | ENSG00000( | 16 | 0.1099 | rs1333267: | 6.96 | rs7192165 | 0.005011 |
| Lung | ARHGEF40 | ENSG00000( | 14 | 0.1638 | rs2319627 | -6.01 | rs1952151 | 0.115 |
| Lung | RBM23 | ENSG00000( | 14 | 0.2215 | rs8022177 | -8.33 | rs8022177 | 0.106 |
| Lung | KHNYN | ENSG00000( | 14 | 0.4631 | rs1015127: | -7.39 | rs1014675! | 0.289 |
| Lung | MIS18BP1 | ENSG00000( | 14 | 0.1077 | rs1711623( | 7.93 | rs1014957! | 0.0385 |
| Lung | ACOT2 | ENSG00000( | 14 | 0.4972 | rs1709110( | -5.71 | rs6574128 | 0.181 |
| Lung | PTGR2 | ENSG00000( | 14 | 0.4662 | rs2109750 | -5.12 | rs7148485 | 0.242 |
| Lung | IFI27L2 | ENSG00000( | 14 | 0.4348 | rs1950969 | 6.08 | rs4905164 | 0.133 |
| Lung | DEGS2 | ENSG00000( | 14 | 0.1031 | rs8016700 | -4.21 | rs1243388! | -0.00099 |
| Lung | CAMTA2 | ENSG00000( | 17 | 0.2081 | rs1156812! | 7.17 | rs418305 | 0.0734 |
| Lung | USP6 | ENSG00000( | 17 | 0.5508 | rs1156812! | 11.45 | rs2585266 | 0.24 |
| Lung | ZNF232 | ENSG00000( | 17 | 0.4386 | rs1156812! | 10.62 | rs2585266 | 0.212 |
| Lung | ACADVL | ENSG00000( | 17 | 0.1212 | rs2269459 | 8.13 | rs446994 | 0.0589 |
| Lung | HEXIM2 | ENSG00000( | 17 | 0.0689 | rs3744759 | -6.28 | rs2129732 | -0.00165 |
| Lung | TMEM235 | ENSG00000( | 17 | 0.103 | rs1051261( | -8.32 | rs1764127! | 0.0105 |
| Lung | OGFOD3 | ENSG00000( | 17 | 0.2858 | rs1785547( | -7.73 | rs4074069 | 0.0754 |
| Lung | B3GNTL1 | ENSG00000( | 17 | 0.1954 | rs1051855 | -3.84 | rs9911217 | 0.032 |
| Lung | SLC35E2B | ENSG00000( | 1 | 0.7591 | rs2887286 | 7.44 | rs4648786 | 0.168 |
| Lung | FAAP20 | ENSG00000( | 1 | 0.3559 | rs7548829 | 4.04 | rs2460002 | 0.0683 |
| Lung | H6PD | ENSG00000( | 1 | 0.212 | rs1256663' | 5.99 | rs2268175 | 0.0303 |
| Lung | CROCC | ENSG00000( | 1 | 0.3097 | rs978528 | -5.08 | rs6691985 | 0.205 |
| Lung | MFAP2 | ENSG00000( | 1 | 0.316 | rs9435734 | -5.04 | rs7513616 | 0.239 |
| Lung | BMP8A | ENSG00000( | 1 | 0.0888 | rs2182022 | -8.08 | rs1212011: | 0.0553 |
| Lung | SLC2A1 | ENSG00000( | 1 | 0.0971 | rs1121084: | 10.85 | rs710222 | 0.0504 |
| Lung | FUBP1 | ENSG00000( | 1 | 0.1142 | rs1739169⁄ | -5.73 | rs1739169⁄ | 0.0755 |
| Lung | GFI1 | ENSG00000( | 1 | 0.1919 | rs1158687( | 7.58 | rs6662618 | 0.0354 |
| Lung | TBX15 | ENSG00000( | 1 | 0.1512 | rs984225 | -6.58 | rs1092371⁄ | 0.0451 |

| | | | | | | | | |
|---|---|---|---|---|---|---|---|---|
| Lung | WARS2 | ENSG00000( | 1 | 0.6898 | rs984225 | -6.58 | rs2645303 | 0.345 |
| Lung | METTL25B | ENSG00000( | 1 | 0.1142 | rs1683807‹ | 4.66 | rs1214043‹ | 0.0545 |
| Lung | DPT | ENSG00000( | 1 | 0.0936 | rs7549700 | 4.43 | rs607484 | -0.00224 |
| Lung | C1orf21 | ENSG00000( | 1 | 0.1238 | rs1205749‹ | -10.47 | rs6696816 | 0.0188 |
| Lung | LEFTY2 | ENSG00000( | 1 | 0.1315 | rs2840956 | 4.21 | rs1204665( | 0.0403 |
| Lung | ADCY3 | ENSG00000( | 2 | 0.083 | rs1018970‹ | -6.95 | rs1541984 | 0.026297 |
| Lung | RPIA | ENSG00000( | 2 | 0.1596 | rs2306676 | 4.56 | rs1017952‹ | 0.012218 |
| Lung | ANKRD36B | ENSG00000( | 2 | 0.4763 | rs1299119‹ | 7.11 | rs6718109 | 0.387692 |
| Lung | C2orf92 | ENSG00000( | 2 | 0.1783 | rs1168085‹ | 6.92 | rs298913 | 0.031142 |
| Lung | TSGA10 | ENSG00000( | 2 | 0.2044 | rs958778 | -5.56 | rs1168318‹ | 0.20448 |
| Lung | LIPT1 | ENSG00000( | 2 | 0.2267 | rs958778 | -5.56 | rs1168318‹ | 0.249721 |
| Lung | GPD2 | ENSG00000( | 2 | 0.1038 | rs295797 | 4.77 | rs298233 | 0.02236 |
| Lung | PHF24 | ENSG00000( | 9 | 0.0669 | rs1097223‹ | -6.5 | rs3802426 | 0.009705 |
| Lung | ECM2 | ENSG00000( | 9 | 0.0969 | rs9886781 | -4.82 | rs1738065‹ | 0.049494 |
| Lung | PTCH1 | ENSG00000( | 9 | 0.0632 | rs2282040 | 5.19 | rs2852036( | 0.02328 |
| Lung | SVEP1 | ENSG00000( | 9 | 0.1393 | rs7875689 | 4.85 | rs1998974 | 0.00199 |
| Lung | SNAPC4 | ENSG00000( | 9 | 0.3665 | rs1085819‹ | -6.12 | rs1268427‹ | 0.070058 |
| Adipose_Su | SMCO3 | ENSG00000( | 12 | 0.0908 | rs1105598‹ | 7 | rs4763397 | 0.0246 |
| Adipose_Su | PTHLH | ENSG00000( | 12 | 0.0714 | rs1133028 | -16.29 | rs7316831 | 0.0489 |
| Adipose_Su | ALG10 | ENSG00000( | 12 | 0.028 | rs7311406 | 9.59 | rs2218650 | 0.0306 |
| Adipose_Su | CPNE8 | ENSG00000( | 12 | 0.0701 | rs1516556 | -8.42 | rs2653741 | 0.0245 |
| Adipose_Su | METAP2 | ENSG00000( | 12 | 0.1058 | rs1436127 | -7.93 | rs1110809‹ | 0.00148 |
| Adipose_Su | KCTD10 | ENSG00000( | 12 | 0.2639 | rs1183122( | -6.32 | rs1183237‹ | 0.127 |
| Adipose_Su | NAA25 | ENSG00000( | 12 | 0.0275 | rs1106598‹ | 5.24 | rs632650 | -2.94E-05 |
| Adipose_Su | SUDS3 | ENSG00000( | 12 | 0.0643 | rs1761956‹ | 7.25 | rs1106892‹ | 0.0336 |
| Adipose_Su | CDK2AP1 | ENSG00000( | 12 | 0.2721 | rs1260317 | -8.34 | rs1879380 | 0.298 |
| Adipose_Su | DNAH10 | ENSG00000( | 12 | 0.2307 | rs9668827 | -5.35 | rs7133378 | 0.0315 |
| Adipose_Su | CCDC92 | ENSG00000( | 12 | 0.2736 | rs1257967( | -6.13 | rs863750 | 0.119 |
| Adipose_Su | NOC4L | ENSG00000( | 12 | 0.3616 | rs2852882‹ | -5.44 | rs1124696‹ | 0.0197 |
| Adipose_Su | ZNF268 | ENSG00000( | 12 | 0.1214 | rs1078165‹ | -4.39 | rs1161465‹ | 0.0119 |
| Adipose_Su | MMP24OS | ENSG00000( | 20 | 0.1659 | rs7280 | -5.85 | rs6060341 | 0.117 |
| Adipose_Su | UQCC1 | ENSG00000( | 20 | 0.1787 | rs7280 | -5.85 | rs4911494 | 0.147 |
| Adipose_Su | KIAA1755 | ENSG00000( | 20 | 0.0829 | rs5743539 | 7.02 | rs1934915 | 0.0386 |
| Adipose_Su | SCD | ENSG00000( | 10 | 0.0885 | rs3894173 | -5.21 | rs603424 | 0.0437 |
| Adipose_Su | BTN3A3 | ENSG00000( | 6 | 0.384 | rs2754715 | 12.15 | rs7751645 | 0.020887 |
| Adipose_Su | TRAPPC13 | ENSG00000( | 5 | 0.185 | rs1689340‹ | -10.71 | rs149514 | 0.035093 |
| Adipose_Su | ARHGEF28 | ENSG00000( | 5 | 0.0577 | rs2973927 | 6.09 | rs1687097‹ | 0.002705 |
| Adipose_Su | SPATA9 | ENSG00000( | 5 | 0.255 | rs1047664‹ | 4.21 | rs150548 | 0.066244 |
| Adipose_Su | CDC42SE2 | ENSG00000( | 5 | 0.1525 | rs1007266‹ | 12.52 | rs1291602 | 0.061288 |
| Adipose_Su | GPRIN1 | ENSG00000( | 5 | 0.187 | rs1252027‹ | -5.05 | rs4868660 | 0.075147 |
| Adipose_Su | TBC1D9B | ENSG00000( | 5 | 0.3517 | rs1707980‹ | -4.14 | rs42414 | 0.090772 |
| Adipose_Su | RACK1 | ENSG00000( | 5 | 0.061 | rs2545098 | -5.04 | rs888709 | 0.012682 |
| Adipose_Su | CAPN15 | ENSG00000( | 16 | 0.1587 | rs1015319‹ | -10.5 | rs1186465‹ | 0.0963 |
| Adipose_Su | ACSM5 | ENSG00000( | 16 | 0.2335 | rs2285834 | -13.88 | rs1186002‹ | 0.0443 |
| Adipose_Su | KCTD13 | ENSG00000( | 16 | 0.0404 | rs4541091 | 7.83 | rs1115057( | 0.0179 |

| | | | | | | | | |
|---|---|---|---|---|---|---|---|---|
| Adipose_Su | INO80E | ENSG00000 | 16 | 0.2061 | rs4541091 | 7.08 | rs4787491 | 0.275 |
| Adipose_Su | TBX6 | ENSG00000 | 16 | 0.2395 | rs1164274 | 5.72 | rs1186508 | 0.0818 |
| Adipose_Su | YPEL3 | ENSG00000 | 16 | 0.3898 | rs1164274 | 5.72 | rs7205802 | 0.519 |
| Adipose_Su | WWP2 | ENSG00000 | 16 | 0.0598 | rs2291959 | -5.54 | rs4985377 | 0.00992 |
| Adipose_Su | TXNL4B | ENSG00000 | 16 | 0.0808 | rs1946768 | -5.03 | rs1820248 | 0.0232 |
| Adipose_Su | TMEM170A | ENSG00000 | 16 | 0.0503 | rs1010632 | 6.21 | rs4888380 | -0.00067 |
| Adipose_Su | CHMP1A | ENSG00000 | 16 | 0.4214 | rs8056585 | 5.59 | rs258322 | 0.0268 |
| Adipose_Su | CDK10 | ENSG00000 | 16 | 0.5654 | rs1333267 | 5.41 | rs1292457 | 0.26 |
| Adipose_Su | EVA1C | ENSG00000 | 21 | 0.0952 | rs679317 | 7.1 | rs2011344 | 0.02734 |
| Adipose_Su | CBR1 | ENSG00000 | 21 | 0.3579 | rs2835109 | 5.02 | rs2835269 | 0.11149 |
| Adipose_Su | PRDM15 | ENSG00000 | 21 | 0.4895 | rs420737 | 10.7 | rs9983923 | 0.24352 |
| Adipose_Su | RADIL | ENSG00000 | 7 | 0.2142 | rs1198345 | 4.36 | rs7800598 | 0.0182 |
| Adipose_Su | ZNF92 | ENSG00000 | 7 | 0.0657 | rs3846972 | 4.73 | rs9638226 | -7.00E-04 |
| Adipose_Su | FKBP6 | ENSG00000 | 7 | 0.1982 | rs875342 | 8.43 | rs1733909 | 0.0459 |
| Adipose_Su | TBL2 | ENSG00000 | 7 | 0.4655 | rs875342 | 8.91 | rs1714572 | 0.142 |
| Adipose_Su | BUD23 | ENSG00000 | 7 | 0.0756 | rs875342 | 9.74 | rs1215477 | 0.0316 |
| Adipose_Su | RCC1L | ENSG00000 | 7 | 0.1363 | rs1253222 | -13.47 | rs1720719 | 0.0239 |
| Adipose_Su | MDH2 | ENSG00000 | 7 | 0.0268 | rs1253245 | -7.37 | rs1025559 | 0.00567 |
| Adipose_Su | EPHB4 | ENSG00000 | 7 | 0.0659 | rs1155912 | 6.61 | rs6465776 | 0.0104 |
| Adipose_Su | CRYL1 | ENSG00000 | 13 | 0.1379 | rs5023264 | -7.55 | rs4381447 | 0.0274 |
| Adipose_Su | RASL11A | ENSG00000 | 13 | 0.1603 | rs1218879 | -4.25 | rs9512603 | 0.0492 |
| Adipose_Su | UGGT2 | ENSG00000 | 13 | 0.3018 | rs9590320 | 3.91 | rs816142 | 0.167 |
| Adipose_Su | TPGS2 | ENSG00000 | 18 | 0.2661 | rs903733 | 6.82 | rs1050267 | 0.130378 |
| Adipose_Su | TIMP4 | ENSG00000 | 3 | 0.1165 | rs9875338 | -5.68 | rs3773364 | 0.031 |
| Adipose_Su | P4HTM | ENSG00000 | 3 | 0.1543 | rs1249039 | -4.93 | rs7616815 | 0.0647 |
| Adipose_Su | NDUFAF3 | ENSG00000 | 3 | 0.0783 | rs1249039 | -4.93 | rs4974083 | 0.0278 |
| Adipose_Su | IHO1 | ENSG00000 | 3 | 0.1458 | rs1249039 | -4.93 | rs1263757 | 0.125 |
| Adipose_Su | TCTA | ENSG00000 | 3 | 0.0386 | rs1249039 | -4.93 | rs6769821 | 0.0348 |
| Adipose_Su | AMT | ENSG00000 | 3 | 0.3407 | rs1249039 | -4.93 | rs3448 | 0.435 |
| Adipose_Su | NICN1 | ENSG00000 | 3 | 0.1654 | rs1249039 | -4.93 | rs3448 | 0.183 |
| Adipose_Su | RBM15B | ENSG00000 | 3 | 0.1548 | rs1263950 | 6.85 | rs1263799 | 0.0538 |
| Adipose_Su | RAD54L2 | ENSG00000 | 3 | 0.3806 | rs323887 | 8.34 | rs1192827 | 0.107 |
| Adipose_Su | RPL29 | ENSG00000 | 3 | 0.1058 | rs7641255 | -9.45 | rs323893 | 0.0945 |
| Adipose_Su | ISY1 | ENSG00000 | 3 | 0.0345 | rs7427129 | 6.22 | rs6787488 | 0.0121 |
| Adipose_Su | ZBTB38 | ENSG00000 | 3 | 0.075 | rs6440031 | -6.3 | rs6785073 | 0.0804 |
| Adipose_Su | CEP19 | ENSG00000 | 3 | 0.2201 | rs1562 | 7.62 | rs1463628 | 0.0906 |
| Adipose_Su | SH3YL1 | ENSG00000 | 2 | 0.4034 | rs1317824 | 5.14 | rs2290911 | 0.134345 |
| Adipose_Su | CENPO | ENSG00000 | 2 | 0.1003 | rs1018970 | -7.26 | rs1298661 | 0.068198 |
| Adipose_Su | ADCY3 | ENSG00000 | 2 | 0.1773 | rs1018970 | -6.95 | rs1865689 | 0.102483 |
| Adipose_Su | ZNF638 | ENSG00000 | 2 | 0.1583 | rs1338754 | -5.12 | rs7604412 | 0.045858 |
| Adipose_Su | ANKRD36B | ENSG00000 | 2 | 0.484 | rs1299119 | 7.11 | rs6718109 | 0.377401 |
| Adipose_Su | ZAP70 | ENSG00000 | 2 | 0.1786 | rs1168085 | 6.96 | rs1189465 | 0.026513 |
| Adipose_Su | LYG1 | ENSG00000 | 2 | 0.1443 | rs1261551 | 4.85 | rs1955393 | 0.118905 |
| Adipose_Su | SH3RF3 | ENSG00000 | 2 | 0.05 | rs1169110 | -5.11 | rs4676283 | 0.00163 |
| Adipose_Su | RFTN2 | ENSG00000 | 2 | 0.0602 | rs700639 | -5.02 | rs4850808 | 0.07388 |

| | | | | | | | | |
|---|---|---|---|---|---|---|---|---|
| Adipose_Su | EHD4 | ENSG00000 | 15 | 0.1548 | rs1704401 | -5.25 | rs1614170 | 0.0314 |
| Adipose_Su | CSNK1G1 | ENSG00000 | 15 | 0.0555 | rs677561 | -10.14 | rs8034610 | 0.0301 |
| Adipose_Su | PIAS1 | ENSG00000 | 15 | 0.5469 | rs4408487 | -4.49 | rs1695152 | 0.223 |
| Adipose_Su | CLN6 | ENSG00000 | 15 | 0.0763 | rs1695235 | 7.62 | rs450466 | 0.0307 |
| Adipose_Su | LYSMD4 | ENSG00000 | 15 | 0.5098 | rs7170513 | -5.8 | rs6598265 | 0.288 |
| Adipose_Su | TBC1D7 | ENSG00000 | 6 | 0.2893 | rs6923878 | -5.8 | rs2496132 | -0.00205 |
| Adipose_Su | ILRUN | ENSG00000 | 6 | 0.1139 | rs2820239 | -6.01 | rs2744972 | 0.0706 |
| Adipose_Su | CEP57L1 | ENSG00000 | 6 | 0.0572 | rs1322814 | -9.49 | rs3734649 | 0.00678 |
| Adipose_Su | CENPW | ENSG00000 | 6 | 0.056 | rs7768545 | -9.51 | rs9375435 | 0.0758 |
| Adipose_Su | TCF21 | ENSG00000 | 6 | 0.2038 | rs4424102 | 5.79 | rs2327429 | 0.0868 |
| Adipose_Su | ADGRG6 | ENSG00000 | 6 | 0.0878 | rs263182 | -6.47 | rs2050157 | 0.0651 |
| Adipose_Su | NDUFA11 | ENSG00000 | 19 | 0.1839 | rs1040213 | 6.41 | rs6510861 | 0.109 |
| Adipose_Su | ZNF100 | ENSG00000 | 19 | 0.8527 | rs1041066 | 4.53 | rs6511286 | 0.529 |
| Adipose_Su | ANKRD27 | ENSG00000 | 19 | 0.2543 | rs1775465 | 8.49 | rs7250780 | 0.108 |
| Adipose_Su | ZNF529 | ENSG00000 | 19 | 0.179 | rs2967436 | 4.63 | rs2967465 | 0.103 |
| Adipose_Su | FBXO17 | ENSG00000 | 19 | 0.2723 | rs674440 | -11.91 | rs8108375 | 0.133 |
| Adipose_Su | CYP2A6 | ENSG00000 | 19 | 0.0689 | rs338600 | 5.44 | rs3852872 | 0.000951 |
| Adipose_Su | TMEM91 | ENSG00000 | 19 | 0.3545 | rs3848568 | 6.21 | rs1187909 | 0.0201 |
| Adipose_Su | ZNF155 | ENSG00000 | 19 | 0.5345 | rs239940 | 5.18 | rs1042367 | 0.0481 |
| Adipose_Su | GEMIN7 | ENSG00000 | 19 | 0.21 | rs4420638 | 6.07 | rs1040585 | 0.126 |
| Adipose_Su | PLEKHA4 | ENSG00000 | 19 | 0.0874 | rs1698213 | -6.77 | rs610308 | 0.0527 |
| Adipose_Su | ZNF761 | ENSG00000 | 19 | 0.3285 | rs4803026 | 12.85 | rs4803113 | 0.09 |
| Adipose_Su | PEG3 | ENSG00000 | 19 | 0.1161 | rs1468773 | 7.14 | rs7260649 | 0.00301 |
| Adipose_Su | ESS2 | ENSG00000 | 22 | 0.5191 | rs5992372 | -4.41 | rs715544 | 0.119 |
| Adipose_Su | PMM1 | ENSG00000 | 22 | 0.0875 | rs7291704 | -7.09 | rs203320 | 0.00379 |
| Adipose_Su | SLC35E2B | ENSG00000 | 1 | 0.7661 | rs2887286 | 7.44 | rs4648786 | 0.202 |
| Adipose_Su | FAAP20 | ENSG00000 | 1 | 0.3986 | rs4648640 | 3.86 | rs2503706 | 0.113 |
| Adipose_Su | CROCC | ENSG00000 | 1 | 0.325 | rs978528 | -5.08 | rs6691985 | 0.306 |
| Adipose_Su | MFAP2 | ENSG00000 | 1 | 0.1551 | rs9435734 | -5.04 | rs3738814 | 0.113 |
| Adipose_Su | ZBTB40 | ENSG00000 | 1 | 0.0683 | rs9187 | -5.34 | rs2007200 | 5.67E-05 |
| Adipose_Su | PNRC2 | ENSG00000 | 1 | 0.1228 | rs1124902 | 5.14 | rs6689056 | -4.42E-05 |
| Adipose_Su | AKIRIN1 | ENSG00000 | 1 | 0.1057 | rs7553434 | -6.35 | rs1203659 | 0.0188 |
| Adipose_Su | MROH7 | ENSG00000 | 1 | 0.1659 | rs505151 | -4.96 | rs1368883 | 0.0345 |
| Adipose_Su | FUBP1 | ENSG00000 | 1 | 0.1228 | rs1739169 | -5.73 | rs1739169 | 0.0975 |
| Adipose_Su | TBX15 | ENSG00000 | 1 | 0.0532 | rs984225 | -6.58 | rs984225 | 0.0516 |
| Adipose_Su | WARS2 | ENSG00000 | 1 | 0.573 | rs984225 | -6.58 | rs2645303 | 0.527 |
| Adipose_Su | THBS3 | ENSG00000 | 1 | 0.1062 | rs4845663 | -5.25 | rs4971079 | 0.0301 |
| Adipose_Su | METTL25B | ENSG00000 | 1 | 0.2371 | rs1683807 | 4.66 | rs1214574 | 0.121 |
| Adipose_Su | ITPKB | ENSG00000 | 1 | 0.0452 | rs603609 | 7.95 | rs6662583 | -0.00016 |
| Adipose_Su | AK3 | ENSG00000 | 9 | 0.3347 | rs7020509 | -5.55 | rs3808857 | 0.0632 |
| Adipose_Su | SAXO1 | ENSG00000 | 9 | 0.1345 | rs1049155 | -4.92 | rs1012340 | 0.0655 |
| Adipose_Su | BARX1 | ENSG00000 | 9 | 0.0812 | rs1234335 | -6.59 | rs1933680 | -0.0004 |
| Adipose_Su | NIBAN2 | ENSG00000 | 9 | 0.2334 | rs4836593 | 3.89 | rs2417053 | 0.0371 |
| Adipose_Su | ACADVL | ENSG00000 | 17 | 0.1485 | rs2269459 | 8.13 | rs446994 | 0.098898 |
| Adipose_Su | SLC2A4 | ENSG00000 | 17 | 0.0822 | rs2269459 | 8.02 | rs222836 | 0.011934 |

| | | | | | | | | |
|---|---|---|---|---|---|---|---|---|
| Adipose_Su | HS3ST3A1 | ENSG00000 | 17 | 0.1907 | rs1107814 | 5.32 | rs9904647 | 0.017749 |
| Adipose_Su | NT5M | ENSG00000 | 17 | 0.2617 | rs4646364 | 6.51 | rs1696151 | 0.052913 |
| Adipose_Su | ABHD15 | ENSG00000 | 17 | 0.0776 | rs8070400 | 6.1 | rs3110496 | 0.032266 |
| Adipose_Su | PLCD3 | ENSG00000 | 17 | 0.4021 | rs3744759 | -6.31 | rs2129732 | 0.155053 |
| Adipose_Su | LRRC37A2 | ENSG00000 | 17 | 0.5412 | rs2074405 | 6.03 | rs169201 | 0.562407 |
| Adipose_Su | CDC27 | ENSG00000 | 17 | 0.1792 | rs2074405 | 6.69 | rs1157057 | 0.0257 |
| Adipose_Su | COPZ2 | ENSG00000 | 17 | 0.1452 | rs1695708 | 4.34 | rs757352 | 0.034761 |
| Adipose_Su | MYCBPAP | ENSG00000 | 17 | 0.3157 | rs1694876 | 5.4 | rs1186971 | 0.120465 |
| Adipose_Su | NUCB2 | ENSG00000 | 11 | 0.0939 | rs1102410 | 4.44 | rs214091 | 0.039 |
| Adipose_Su | ANO3 | ENSG00000 | 11 | 0.1066 | rs1381187 | 5.73 | rs1724309 | 0.0321 |
| Adipose_Su | MS4A2 | ENSG00000 | 11 | 0.0671 | rs1229235 | 10.93 | rs502581 | 0.00365 |
| Adipose_Su | VEGFB | ENSG00000 | 11 | 0.3233 | rs1089747 | 6.03 | rs2845885 | 0.111 |
| Adipose_Su | DCAF16 | ENSG00000 | 4 | 0.2812 | rs1049601 | 6.68 | rs7667864 | 0.214 |
| Adipose_Su | OCIAD1 | ENSG00000 | 4 | 0.0513 | rs1765664 | 5.11 | rs7665209 | 0.0432 |
| Adipose_Su | ADH1B | ENSG00000 | 4 | 0.2241 | rs1426736 | 7.3 | rs1826907 | 0.106 |
| Adipose_Su | FAM149A | ENSG00000 | 4 | 0.3112 | rs2306989 | 5.69 | rs6816903 | 0.156 |
| Adipose_Su | FLRT2 | ENSG00000 | 14 | 0.0842 | rs6574859 | -6.71 | rs1289482 | 0.013678 |
| Adipose_Su | XRCC3 | ENSG00000 | 14 | 0.1881 | rs8007670 | 19.98 | rs1288480 | 0.150786 |
| Adipose_Su | CLDN23 | ENSG00000 | 8 | 0.2762 | rs1199354 | -10.27 | rs559534 | 0.050571 |
| Adipose_Su | MTUS1 | ENSG00000 | 8 | 0.1458 | rs2027922 | 9.22 | rs454100 | 0.049943 |
| Adipose_Su | TNFRSF10A | ENSG00000 | 8 | 0.2184 | rs1708935 | 4.56 | rs4242394 | 0.069265 |
| Adipose_Su | RDH10 | ENSG00000 | 8 | 0.0736 | rs1693838 | 5.11 | rs6472767 | 0.001382 |
| Adipose_Su | CCN3 | ENSG00000 | 8 | 0.0693 | rs7012790 | -6.15 | rs3411216 | 0.0619 |
| Adipose_Su | MROH6 | ENSG00000 | 8 | 0.5093 | rs2450763 | -7.05 | rs1154978 | 0.08676 |
| Adipose_Vi | WRN | ENSG00000 | 8 | 0.3192 | rs6997014 | 4.37 | rs3213202 | 0.072 |
| Adipose_Vi | EIF3E | ENSG00000 | 8 | 0.0454 | rs610891 | 6.42 | rs610891 | 0.0236 |
| Adipose_Vi | SBF2 | ENSG00000 | 11 | 0.0828 | rs1241818 | 7.12 | rs7111987 | 0.020531 |
| Adipose_Vi | TRIB2 | ENSG00000 | 2 | 0.1232 | rs2380426 | -7.21 | rs2059428 | 0.0319 |
| Adipose_Vi | CENPO | ENSG00000 | 2 | 0.1466 | rs1018970 | -7.26 | rs7576788 | 0.0977 |
| Adipose_Vi | ANKRD36B | ENSG00000 | 2 | 0.406 | rs1299119 | 7.11 | rs3906948 | 0.413 |
| Adipose_Vi | TMEM131 | ENSG00000 | 2 | 0.1049 | rs1168085 | 7.05 | rs1189360 | 0.0504 |
| Adipose_Vi | TSGA10 | ENSG00000 | 2 | 0.1217 | rs958778 | -5.56 | rs1168318 | 0.117 |
| Adipose_Vi | ZEB2 | ENSG00000 | 2 | 0.131 | rs6710774 | 10.84 | rs6740731 | 0.0257 |
| Adipose_Vi | SLC38A11 | ENSG00000 | 2 | 0.2958 | rs1190117 | -7.25 | rs1018412 | 0.226 |
| Adipose_Vi | SLC25A12 | ENSG00000 | 2 | 0.1101 | rs4972760 | 4.33 | rs1269297 | 0.0274 |
| Adipose_Vi | CHN1 | ENSG00000 | 2 | 0.0751 | rs6712279 | 4.28 | rs1728821 | 0.0383 |
| Adipose_Vi | RFTN2 | ENSG00000 | 2 | 0.075 | rs700639 | -5.02 | rs1682314 | 0.0314 |
| Adipose_Vi | MMP24OS | ENSG00000 | 20 | 0.092 | rs7280 | -5.85 | rs2425049 | 0.061112 |
| Adipose_Vi | BACE2 | ENSG00000 | 21 | 0.2804 | rs420737 | 8.96 | rs2837996 | 0.020481 |
| Adipose_Vi | PRDM15 | ENSG00000 | 21 | 0.4256 | rs420737 | 10.67 | rs9983923 | 0.176859 |
| Adipose_Vi | RAP2A | ENSG00000 | 13 | 0.0797 | rs7334826 | -6.57 | rs1258474 | -0.00192 |
| Adipose_Vi | PSMG4 | ENSG00000 | 6 | 0.1492 | rs9501920 | 8.82 | rs4421240 | 0.091862 |
| Adipose_Vi | SENP6 | ENSG00000 | 6 | 0.0679 | rs1323069 | 5.89 | rs1321371 | -0.00178 |
| Adipose_Vi | MANEA | ENSG00000 | 6 | 0.3391 | rs1175694 | -6.67 | rs1115325 | 0.176058 |
| Adipose_Vi | TCF21 | ENSG00000 | 6 | 0.1096 | rs7767123 | 7.06 | rs9375992 | 0.032834 |

| | | | | | | | | |
|---|---|---|---|---|---|---|---|---|
| Adipose_V | ADGRG6 | ENSG00000 | 6 | 0.1112 | rs263182 | -6.47 | rs972982 | 0.012818 |
| Adipose_V | AHRR | ENSG00000 | 5 | 0.171 | rs1574220 | -5.73 | rs6555242 | 0.012063 |
| Adipose_V | RAPGEF6 | ENSG00000 | 5 | 0.0515 | rs1006069 | 13.69 | rs6872533 | 0.016469 |
| Adipose_V | PCDHA10 | ENSG00000 | 5 | 0.2708 | rs1709694 | 4.3 | rs1174715 | 0.092943 |
| Adipose_V | PCDHB6 | ENSG00000 | 5 | 0.0912 | rs1709694 | 4.3 | rs2338530 | 0.008171 |
| Adipose_V | APOL4 | ENSG00000 | 22 | 0.524 | rs1699691 | -13.09 | rs132717 | 0.302905 |
| Adipose_V | TUBGCP6 | ENSG00000 | 22 | 0.075 | rs6010129 | -4.8 | rs5771104 | 0.004168 |
| Adipose_V | DCAF10 | ENSG00000 | 9 | 0.0852 | rs1156779 | -4.83 | rs7040503 | 0.0107 |
| Adipose_V | HSD17B3 | ENSG00000 | 9 | 0.2243 | rs1691148 | 9.32 | rs4743709 | 0.0367 |
| Adipose_V | COL15A1 | ENSG00000 | 9 | 0.1209 | rs2808529 | 5.7 | rs927468 | 0.0584 |
| Adipose_V | ALAD | ENSG00000 | 9 | 0.2837 | rs1179466 | -5.28 | rs818706 | 0.15 |
| Adipose_V | ALG10 | ENSG00000 | 12 | 0.106 | rs7311406 | 9.59 | rs1105286 | 0.055 |
| Adipose_V | DRAM1 | ENSG00000 | 12 | 0.1432 | rs1111112 | 6.34 | rs4764814 | 0.0634 |
| Adipose_V | ABCB9 | ENSG00000 | 12 | 0.0775 | rs1260317 | -10.24 | rs7953894 | 0.0129 |
| Adipose_V | H3C1 | ENSG00000 | 6 | 0.055 | rs6910993 | 5.64 | rs1436306 | 0.00117 |
| Adipose_V | TBL2 | ENSG00000 | 7 | 0.2162 | rs875342 | 8.91 | rs1714572 | 0.0115 |
| Adipose_V | CLDN4 | ENSG00000 | 7 | 0.0835 | rs875342 | 8.99 | rs9647713 | 0.0442 |
| Adipose_V | RCC1L | ENSG00000 | 7 | 0.1244 | rs1253222 | -13.47 | rs1746657 | 0.0161 |
| Adipose_V | STYXL1 | ENSG00000 | 7 | 0.5376 | rs1253245 | -7.37 | rs7788926 | 0.376 |
| Adipose_V | MDH2 | ENSG00000 | 7 | 0.09 | rs1253245 | -7.37 | rs1023508 | 0.00488 |
| Adipose_V | ADCK2 | ENSG00000 | 7 | 0.2629 | rs1044121 | 6.8 | rs2930320 | 0.0737 |
| Adipose_V | GABRB3 | ENSG00000 | 15 | 0.2752 | rs2045151 | -8.78 | rs768899 | 0.193528 |
| Adipose_V | TPM1 | ENSG00000 | 15 | 0.0915 | rs6494367 | 5.8 | rs7170462 | 0.033492 |
| Adipose_V | TRIP4 | ENSG00000 | 15 | 0.1219 | rs677561 | -10.15 | rs1244285 | 0.175836 |
| Adipose_V | HAPLN3 | ENSG00000 | 15 | 0.1855 | rs7176941 | 4.47 | rs1694244 | 0.01947 |
| Adipose_V | SLC35E2B | ENSG00000 | 1 | 0.7879 | rs2887286 | 7.44 | rs4648786 | 0.11 |
| Adipose_V | CENPS | ENSG00000 | 1 | 0.1681 | rs6660223 | -4.78 | rs1307497 | 0.0889 |
| Adipose_V | MFAP2 | ENSG00000 | 1 | 0.2627 | rs9435734 | -5.04 | rs9435734 | 0.189 |
| Adipose_V | RHD | ENSG00000 | 1 | 0.6871 | rs311437 | -3.84 | rs9689 | 0.249 |
| Adipose_V | PHACTR4 | ENSG00000 | 1 | 0.3012 | rs6679755 | 8.74 | rs1079451 | 0.136 |
| Adipose_V | TBX15 | ENSG00000 | 1 | 0.1233 | rs984225 | -6.58 | rs1092372 | 0.0892 |
| Adipose_V | SETDB1 | ENSG00000 | 1 | 0.0424 | rs1088838 | 5.59 | rs1241086 | 0.00452 |
| Adipose_V | LYSMD1 | ENSG00000 | 1 | 0.2198 | rs1120484 | 4.64 | rs6691701 | 0.0109 |
| Adipose_V | NUCKS1 | ENSG00000 | 1 | 0.1407 | rs2153904 | 4.69 | rs823121 | 0.0943 |
| Adipose_V | ANGPTL6 | ENSG00000 | 19 | 0.0824 | rs4239546 | -4.89 | rs1246084 | -0.00092 |
| Adipose_V | ZNF100 | ENSG00000 | 19 | 0.8577 | rs1041066 | 4.53 | rs6511286 | 0.472 |
| Adipose_V | ZNF208 | ENSG00000 | 19 | 0.2012 | rs1041066 | 6.32 | rs7255049 | 0.0752 |
| Adipose_V | RHPN2 | ENSG00000 | 19 | 0.1717 | rs1775465 | 8.06 | rs7256470 | 0.0901 |
| Adipose_V | PEPD | ENSG00000 | 19 | 0.2415 | rs1246250 | -5.43 | rs1040446 | 0.183 |
| Adipose_V | ZNF529 | ENSG00000 | 19 | 0.1474 | rs2967436 | 4.63 | rs2967465 | 0.093 |
| Adipose_V | TMEM91 | ENSG00000 | 19 | 0.2261 | rs3848568 | 6.21 | rs1046909 | 0.0199 |
| Adipose_V | PLEKHA4 | ENSG00000 | 19 | 0.1521 | rs1698213 | -6.77 | rs609985 | 0.0846 |
| Adipose_V | TULP2 | ENSG00000 | 19 | 0.1234 | rs1698213 | -5.96 | rs500079 | 0.0701 |
| Adipose_V | ZNF415 | ENSG00000 | 19 | 0.2308 | rs7258967 | -5.39 | rs6509724 | 0.00252 |
| Adipose_V | CAPN15 | ENSG00000 | 16 | 0.1658 | rs1015319 | -10.5 | rs1186465 | 0.101817 |

| | | | | | | | | |
|---|---|---|---|---|---|---|---|---|
| Adipose_V | INO80E | ENSG00000 | 16 | 0.1814 | rs4541091 | 7.08 | rs1115057 | 0.28653 |
| Adipose_V | DOC2A | ENSG00000 | 16 | 0.0357 | rs4541091 | 6.02 | rs1293357 | 0.039309 |
| Adipose_V | PPP4C | ENSG00000 | 16 | 0.1095 | rs1164274 | 5.72 | rs7205802 | 0.01148 |
| Adipose_V | TBX6 | ENSG00000 | 16 | 0.2437 | rs1164274 | 5.72 | rs1186508 | 0.0515 |
| Adipose_V | YPEL3 | ENSG00000 | 16 | 0.2051 | rs1164274 | 5.72 | rs8060511 | 0.288974 |
| Adipose_V | AKTIP | ENSG00000 | 16 | 0.0555 | rs7185783 | -4.76 | rs3095635 | 0.012641 |
| Adipose_V | CLEC18A | ENSG00000 | 16 | 0.5979 | rs2291959 | -5.54 | rs1808417 | 0.319727 |
| Adipose_V | DBNDD1 | ENSG00000 | 16 | 0.4909 | rs1333267 | 7.62 | rs1805007 | 0.197136 |
| Adipose_V | MED28 | ENSG00000 | 4 | 0.2228 | rs2191612 | -7.19 | rs3815414 | 0.232119 |
| Adipose_V | ADH1C | ENSG00000 | 4 | 0.2195 | rs1002108 | -5.91 | rs1789924 | 0.085925 |
| Adipose_V | ANKRD50 | ENSG00000 | 4 | 0.076 | rs6825123 | -7.78 | rs2036203 | 0.027027 |
| Adipose_V | TIMP4 | ENSG00000 | 3 | 0.1658 | rs9875338 | -5.68 | rs4488811 | 0.0416 |
| Adipose_V | PLCL2 | ENSG00000 | 3 | 0.1203 | rs1704226 | -7.08 | rs6442644 | -8.28E-05 |
| Adipose_V | P4HTM | ENSG00000 | 3 | 0.0851 | rs1249039 | -4.93 | rs1192025 | 0.0549 |
| Adipose_V | NDUFAF3 | ENSG00000 | 3 | 0.0233 | rs1249039 | -4.93 | rs3774628 | 0.00336 |
| Adipose_V | IHO1 | ENSG00000 | 3 | 0.1283 | rs1249039 | -4.93 | rs4955418 | 0.167 |
| Adipose_V | AMT | ENSG00000 | 3 | 0.3461 | rs1249039 | -4.93 | rs6769821 | 0.457 |
| Adipose_V | NICN1 | ENSG00000 | 3 | 0.1804 | rs1249039 | -4.93 | rs1171043 | 0.202 |
| Adipose_V | RBM15B | ENSG00000 | 3 | 0.1581 | rs1263950 | 6.85 | rs1263799 | 0.0286 |
| Adipose_V | RAD54L2 | ENSG00000 | 3 | 0.2464 | rs323887 | 8.34 | rs3749316 | 0.103 |
| Adipose_V | MBNL1 | ENSG00000 | 3 | 0.0765 | rs1715648 | -4.54 | rs1400612 | 0.00332 |
| Adipose_V | RSRC1 | ENSG00000 | 3 | 0.1537 | rs1210359 | 5.84 | rs9827781 | 0.129 |
| Adipose_V | DVL3 | ENSG00000 | 3 | 0.1425 | rs7630982 | 5.41 | rs1709642 | 0.0141 |
| Adipose_V | PTGR2 | ENSG00000 | 14 | 0.556 | rs2109750 | -5.12 | rs8021337 | 0.196714 |
| Adipose_V | PGF | ENSG00000 | 14 | 0.139 | rs8010077 | 11.08 | rs8013023 | 0.007934 |
| Adipose_V | CEP170B | ENSG00000 | 14 | 0.1116 | rs2498806 | 6.45 | rs2028416 | 0.047309 |
| Adipose_V | ZNF37A | ENSG00000 | 10 | 0.0976 | rs7100316 | -14.09 | rs2472177 | 0.0129 |
| Adipose_V | FUT11 | ENSG00000 | 10 | 0.1026 | rs7098158 | 6.48 | rs2894040 | 0.0759 |
| Adipose_V | SCD | ENSG00000 | 10 | 0.1535 | rs3894173 | -5.21 | rs603424 | 0.0733 |
| Adipose_V | TRPV3 | ENSG00000 | 17 | 0.2491 | rs3744683 | 6.42 | rs401643 | 0.028781 |
| Adipose_V | MPDU1 | ENSG00000 | 17 | 0.1432 | rs2269459 | 8.15 | rs3809824 | -0.00192 |
| Adipose_V | PSMB3 | ENSG00000 | 17 | 0.1084 | rs16529 | -4.98 | rs1043515 | 0.032265 |
| Adipose_V | KRT27 | ENSG00000 | 17 | 0.1902 | rs1107895 | -7.34 | rs719866 | 0.07782 |
| Muscle_Sk | MSRA | ENSG00000 | 8 | 0.0451 | rs7009513 | -7.32 | rs3750314 | 0.0209 |
| Muscle_Sk | RPS20 | ENSG00000 | 8 | 0.1048 | rs3403795 | 5.17 | rs2668007 | 0.145 |
| Muscle_Sk | EIF3E | ENSG00000 | 8 | 0.0457 | rs610891 | 6.42 | rs640068 | 0.0352 |
| Muscle_Sk | EMC2 | ENSG00000 | 8 | 0.0515 | rs610891 | 6.42 | rs7817783 | 0.0261 |
| Muscle_Sk | RAD21 | ENSG00000 | 8 | 0.1209 | rs885205 | 11.58 | rs1008157 | 0.0645 |
| Muscle_Sk | TATDN1 | ENSG00000 | 8 | 0.1942 | rs6990136 | -5.86 | rs3829038 | 0.0792 |
| Muscle_Sk | TBL2 | ENSG00000 | 7 | 0.1983 | rs875342 | 8.91 | rs1714572 | 0.048624 |
| Muscle_Sk | STYXL1 | ENSG00000 | 7 | 0.2967 | rs1253245 | -7.37 | rs7788926 | 0.217484 |
| Muscle_Sk | PTPN12 | ENSG00000 | 7 | 0.1123 | rs3093277 | -11.12 | rs6976860 | 0.052752 |
| Muscle_Sk | ABCB4 | ENSG00000 | 7 | 0.1729 | rs1725275 | -6.12 | rs6946119 | 0.034019 |
| Muscle_Sk | CPA4 | ENSG00000 | 7 | 0.1163 | rs901799 | 6.56 | rs1253425 | 0.000964 |
| Muscle_Sk | GRIN3B | ENSG00000 | 19 | 0.1448 | rs9304925 | -8.7 | rs1108523 | 0.0627 |

| | | | | | | | | |
|---|---|---|---|---|---|---|---|---|
| Muscle_Sk | HDGFL2 | ENSG00000 | 19 | 0.0446 | rs9807813 | 7.76 | rs8887 | 0.00503 |
| Muscle_Sk | ZNF100 | ENSG00000 | 19 | 0.542 | rs1041066 | 4.53 | rs6511286 | 0.238 |
| Muscle_Sk | RHPN2 | ENSG00000 | 19 | 0.1045 | rs1775465 | 8.06 | rs7256470 | 0.0855 |
| Muscle_Sk | ZNF229 | ENSG00000 | 19 | 0.1041 | rs769451 | -6.09 | rs2722683 | 0.0258 |
| Muscle_Sk | GEMIN7 | ENSG00000 | 19 | 0.2618 | rs4420638 | 6.07 | rs1040585 | 0.143 |
| Muscle_Sk | SMIM17 | ENSG00000 | 19 | 0.0595 | rs1042511 | -6.23 | rs1041221 | 0.0154 |
| Muscle_Sk | TANGO2 | ENSG00000 | 22 | 0.0788 | rs759575 | 7.14 | rs696885 | 0.00409 |
| Muscle_Sk | LIF | ENSG00000 | 22 | 0.0506 | rs8135379 | 6.62 | rs2412970 | 0.0333 |
| Muscle_Sk | FOXRED2 | ENSG00000 | 22 | 0.1983 | rs1699691 | -9.76 | rs1170471 | 0.0569 |
| Muscle_Sk | UPK3A | ENSG00000 | 22 | 0.498 | rs1699336 | 9.4 | rs1135360 | 0.243 |
| Muscle_Sk | IFITM2 | ENSG00000 | 11 | 0.1341 | rs1715606 | -6.98 | rs909098 | 0.018 |
| Muscle_Sk | APBB1 | ENSG00000 | 11 | 0.247 | rs1160178 | 4.69 | rs7129298 | 0.135 |
| Muscle_Sk | ATG13 | ENSG00000 | 11 | 0.0544 | rs1076918 | -4.03 | rs7951870 | 0.0268 |
| Muscle_Sk | FEN1 | ENSG00000 | 11 | 0.0436 | rs1123094 | -7.55 | rs4246215 | 0.00417 |
| Muscle_Sk | VEGFB | ENSG00000 | 11 | 0.1013 | rs1089747 | 6.03 | rs3516979 | 0.0443 |
| Muscle_Sk | RPP30 | ENSG00000 | 10 | 0.0596 | rs1710643 | 5.76 | rs2000322 | 0.027016 |
| Muscle_Sk | FNDC4 | ENSG00000 | 2 | 0.0472 | rs1260326 | 6.94 | rs1275539 | -0.00095 |
| Muscle_Sk | GPN1 | ENSG00000 | 2 | 0.1524 | rs1260326 | 6.94 | rs2068834 | 0.11 |
| Muscle_Sk | LTBP1 | ENSG00000 | 2 | 0.0961 | rs150721 | 7.74 | rs1979151 | 0.00654 |
| Muscle_Sk | VRK2 | ENSG00000 | 2 | 0.0811 | rs6712575 | 8.15 | rs1439269 | 0.0453 |
| Muscle_Sk | C2orf42 | ENSG00000 | 2 | 0.0969 | rs3771492 | -7.35 | rs14234 | 0.0307 |
| Muscle_Sk | ANKRD36B | ENSG00000 | 2 | 0.0868 | rs1299119 | 7.11 | rs6718109 | 0.12 |
| Muscle_Sk | C2orf92 | ENSG00000 | 2 | 0.1598 | rs1168085 | 6.92 | rs1168085 | 0.162 |
| Muscle_Sk | LIPT1 | ENSG00000 | 2 | 0.1209 | rs958778 | -5.56 | rs1168318 | 0.191 |
| Muscle_Sk | SLC38A11 | ENSG00000 | 2 | 0.7049 | rs1190117 | -7.25 | rs1018412 | 0.487 |
| Muscle_Sk | RAPGEF4 | ENSG00000 | 2 | 0.0976 | rs2113807 | -9.71 | rs4151454 | 0.00089 |
| Muscle_Sk | SGO2 | ENSG00000 | 2 | 0.0489 | rs3754800 | 6.58 | rs7597713 | 0.0119 |
| Muscle_Sk | FARSB | ENSG00000 | 2 | 0.3215 | rs7592038 | -4.76 | rs1168432 | 0.152 |
| Muscle_Sk | MLPH | ENSG00000 | 2 | 0.1909 | rs7588511 | -6.3 | rs6705903 | 0.0983 |
| Muscle_Sk | CCDC91 | ENSG00000 | 12 | 0.2136 | rs1133028 | -13.23 | rs4931086 | 0.0284 |
| Muscle_Sk | CERS5 | ENSG00000 | 12 | 0.1525 | rs1183106 | 9.13 | rs7397849 | 0.134 |
| Muscle_Sk | LIMA1 | ENSG00000 | 12 | 0.0699 | rs1183106 | 9.67 | rs1712455 | 0.0535 |
| Muscle_Sk | SNRPF | ENSG00000 | 12 | 0.0895 | rs7970079 | -4.39 | rs1050707 | 0.0121 |
| Muscle_Sk | GIT2 | ENSG00000 | 12 | 0.0461 | rs7314708 | 7.53 | rs1045802 | 0.0179 |
| Muscle_Sk | ANAPC7 | ENSG00000 | 12 | 0.2293 | rs7961976 | -7.88 | rs1693976 | 0.0509 |
| Muscle_Sk | CAMKK2 | ENSG00000 | 12 | 0.3938 | rs2840577 | -6.47 | rs4980993 | 0.194 |
| Muscle_Sk | NOC4L | ENSG00000 | 12 | 0.5501 | rs2852882 | -5.44 | rs7397057 | 0.0105 |
| Muscle_Sk | PLA2G4E | ENSG00000 | 15 | 0.1681 | rs1704405 | 4.8 | rs1704370 | 0.081109 |
| Muscle_Sk | GNB5 | ENSG00000 | 15 | 0.2198 | rs7167590 | 8.08 | rs4776017 | 0.065136 |
| Muscle_Sk | HACD3 | ENSG00000 | 15 | 0.0558 | rs4514623 | 4.52 | rs1435126 | 0.000324 |
| Muscle_Sk | CYP11A1 | ENSG00000 | 15 | 0.3272 | rs9806234 | 4.7 | rs2277603 | 0.058233 |
| Muscle_Sk | DOP1B | ENSG00000 | 21 | 0.2833 | rs2835109 | 4.5 | rs1573306 | 0.100947 |
| Muscle_Sk | DLG4 | ENSG00000 | 17 | 0.0966 | rs2269459 | 8.06 | rs390200 | 0.083871 |
| Muscle_Sk | CHRNB1 | ENSG00000 | 17 | 0.4551 | rs2269459 | 8.16 | rs4151121 | 0.390108 |
| Muscle_Sk | TNFSF13 | ENSG00000 | 17 | 0.0453 | rs2269459 | 9.6 | rs6259 | 0.035076 |

| | | | | | | | | |
|---|---|---|---|---|---|---|---|---|
| Muscle_Sk | SEPTIN4 | ENSG00000 | 17 | 0.0372 | rs1694332 | 5.55 | rs2429369 | 0.009261 |
| Muscle_Sk | PRKAR2A | ENSG00000 | 3 | 0.037 | rs1249039 | -4.93 | rs4955411 | 0.029971 |
| Muscle_Sk | NDUFAF3 | ENSG00000 | 3 | 0.0371 | rs1249039 | -4.93 | rs4974080 | 0.029229 |
| Muscle_Sk | DALRD3 | ENSG00000 | 3 | 0.0212 | rs1249039 | -4.93 | rs1262975 | 0.032592 |
| Muscle_Sk | USP4 | ENSG00000 | 3 | 0.0369 | rs1249039 | -4.93 | rs974495 | 0.00948 |
| Muscle_Sk | AMT | ENSG00000 | 3 | 0.2012 | rs1249039 | -4.93 | rs3905330 | 0.307878 |
| Muscle_Sk | RAD54L2 | ENSG00000 | 3 | 0.1123 | rs323887 | 8.34 | rs3749316 | 0.060074 |
| Muscle_Sk | THOC7 | ENSG00000 | 3 | 0.0916 | rs853250 | 4.3 | rs7615475 | 0.006071 |
| Muscle_Sk | DHFR2 | ENSG00000 | 3 | 0.0571 | rs9837294 | -4.24 | rs9860972 | -0.00169 |
| Muscle_Sk | WDR5B | ENSG00000 | 3 | 0.1983 | rs1248759 | 4.49 | rs3749213 | 0.125746 |
| Muscle_Sk | PLCH1 | ENSG00000 | 3 | 0.0932 | rs6802192 | -7.82 | rs1682511 | 0.013307 |
| Muscle_Sk | MLF1 | ENSG00000 | 3 | 0.2726 | rs1210359 | 5.84 | rs7628293 | 0.039164 |
| Muscle_Sk | NAALADL2 | ENSG00000 | 3 | 0.1426 | rs6765689 | 7.52 | rs9290515 | 0.094185 |
| Muscle_Sk | GNB4 | ENSG00000 | 3 | 0.0522 | rs1191837 | 8.08 | rs1004928 | 0.031235 |
| Muscle_Sk | SLC35E2B | ENSG00000 | 1 | 0.7798 | rs2887286 | 7.44 | rs6699975 | 0.186 |
| Muscle_Sk | CCDC27 | ENSG00000 | 1 | 0.1241 | rs4292923 | 5.65 | rs2298222 | 0.048 |
| Muscle_Sk | CROCC | ENSG00000 | 1 | 0.2389 | rs978528 | -5.08 | rs6691985 | 0.236 |
| Muscle_Sk | MFAP2 | ENSG00000 | 1 | 0.1197 | rs9435734 | -5.04 | rs761422 | 0.0888 |
| Muscle_Sk | SDHB | ENSG00000 | 1 | 0.0401 | rs9435734 | -5.04 | rs1158904 | -0.00128 |
| Muscle_Sk | RHD | ENSG00000 | 1 | 0.6099 | rs3093647 | -7.15 | rs3091242 | 0.244 |
| Muscle_Sk | PDZK1IP1 | ENSG00000 | 1 | 0.0724 | rs1105456 | -9.39 | rs2248907 | 0.00303 |
| Muscle_Sk | FUBP1 | ENSG00000 | 1 | 0.0557 | rs1739169 | -5.73 | rs1739169 | 0.0459 |
| Muscle_Sk | AMPD1 | ENSG00000 | 1 | 0.4757 | rs7541732 | -8.3 | rs1760272 | 0.391 |
| Muscle_Sk | METTL25B | ENSG00000 | 1 | 0.2874 | rs1683807 | 4.66 | rs1216142 | 0.169 |
| Muscle_Sk | C1orf112 | ENSG00000 | 1 | 0.1231 | rs1204014 | -5.58 | rs1048917 | 0.00445 |
| Muscle_Sk | ANGPTL1 | ENSG00000 | 1 | 0.1539 | rs871631 | 4.25 | rs871631 | 0.0803 |
| Muscle_Sk | SMG7 | ENSG00000 | 1 | 0.1379 | rs789191 | -7.6 | rs1275366 | 0.0104 |
| Muscle_Sk | PIK3C2B | ENSG00000 | 1 | 0.048 | rs4951095 | -5.01 | rs3747636 | 0.00297 |
| Muscle_Sk | ETNPPL | ENSG00000 | 4 | 0.0805 | rs1000632 | 6.46 | rs1703959 | 0.007132 |
| Muscle_Sk | DDX60L | ENSG00000 | 4 | 0.0552 | rs1761411 | -4.05 | rs660223 | 0.003816 |
| Muscle_Sk | TTC39C | ENSG00000 | 18 | 0.076 | rs2282558 | 7.05 | rs8094549 | 0.0144 |
| Muscle_Sk | ACO1 | ENSG00000 | 9 | 0.2257 | rs1097094 | 7.33 | rs4879586 | 0.075014 |
| Muscle_Sk | GNA14 | ENSG00000 | 9 | 0.0378 | rs1114535 | 4.34 | rs6560613 | 0.005235 |
| Muscle_Sk | ECM2 | ENSG00000 | 9 | 0.0947 | rs9886781 | -4.82 | rs7043114 | 0.069762 |
| Muscle_Sk | FPGS | ENSG00000 | 9 | 0.0739 | rs7026544 | 4.54 | rs3780652 | -0.00053 |
| Muscle_Sk | TBC1D7 | ENSG00000 | 6 | 0.3598 | rs6923878 | -5.8 | rs2496132 | 0.0332 |
| Muscle_Sk | UHRF1BP1 | ENSG00000 | 6 | 0.365 | rs2820239 | -5.83 | rs9469917 | 0.346 |
| Muscle_Sk | FRS3 | ENSG00000 | 6 | 0.0761 | rs7762770 | 6.18 | rs1220221 | 0.0266 |
| Muscle_Sk | C6orf226 | ENSG00000 | 6 | 0.1884 | rs2016128 | -5.74 | rs9471938 | 0.135 |
| Muscle_Sk | TCF21 | ENSG00000 | 6 | 0.1636 | rs7767123 | 7.06 | rs1967917 | 0.0806 |
| Muscle_Sk | ADGRG6 | ENSG00000 | 6 | 0.0377 | rs263182 | -6.47 | rs9385994 | 0.00516 |
| Muscle_Sk | ULBP2 | ENSG00000 | 6 | 0.0858 | rs9766101 | 7.24 | rs237008 | -0.00108 |
| Muscle_Sk | RBM23 | ENSG00000 | 14 | 0.2352 | rs8022177 | -8.33 | rs8022177 | 0.058112 |
| Muscle_Sk | C14orf39 | ENSG00000 | 14 | 0.2124 | rs1254324 | -5.74 | rs761557 | 0.205349 |
| Muscle_Sk | COQ6 | ENSG00000 | 14 | 0.0991 | rs730384 | -5.19 | rs2159177 | 0.094911 |

| | | | | | | | | |
|---|---|---|---|---|---|---|---|---|
| Muscle_Sk | C14orf180 | ENSG00000 | 14 | 0.3064 | rs2498806 | 8.11 | rs7155652 | 0.049778 |
| Muscle_Sk | IRX1 | ENSG00000 | 5 | 0.1699 | rs2398646 | -5.86 | rs1113403 | 0.132 |
| Muscle_Sk | LIFR | ENSG00000 | 5 | 0.1504 | rs890909 | 9.78 | rs1046224 | 0.0262 |
| Muscle_Sk | MIER3 | ENSG00000 | 5 | 0.1255 | rs40271 | 6.87 | rs2662027 | -0.0008 |
| Muscle_Sk | GCNT4 | ENSG00000 | 5 | 0.259 | rs300259 | 4.25 | rs1195117 | 0.0218 |
| Muscle_Sk | LNPEP | ENSG00000 | 5 | 0.0891 | rs316201 | -4.53 | rs2549796 | 0.0393 |
| Muscle_Sk | PCDHB6 | ENSG00000 | 5 | 0.0806 | rs1784430 | -5.68 | rs7701616 | 0.00333 |
| Muscle_Sk | SIMC1 | ENSG00000 | 5 | 0.0587 | rs1251898 | 4.42 | rs3915381 | -0.00106 |
| Muscle_Sk | MMP24OS | ENSG00000 | 20 | 0.1131 | rs7280 | -5.85 | rs6060341 | 0.09709 |
| Muscle_Sk | UQCC1 | ENSG00000 | 20 | 0.2266 | rs7280 | -5.85 | rs224331 | 0.209891 |
| Muscle_Sk | ZBTB46 | ENSG00000 | 20 | 0.0829 | rs8126295 | -5.57 | rs6062317 | -0.00011 |
| Muscle_Sk | RPS2 | ENSG00000 | 16 | 0.361 | rs1759677 | -9.19 | rs1133099 | 0.021295 |
| Muscle_Sk | MGRN1 | ENSG00000 | 16 | 0.3954 | rs1164579 | -10.97 | rs1659493 | 0.134425 |
| Muscle_Sk | INO80E | ENSG00000 | 16 | 0.0546 | rs4541091 | 7.08 | rs7190185 | 0.04875 |
| Muscle_Sk | TLCD3B | ENSG00000 | 16 | 0.1175 | rs1164274 | 5.72 | rs1115058 | 0.136465 |
| Muscle_Sk | TBX6 | ENSG00000 | 16 | 0.2802 | rs1164274 | 5.72 | rs1186508 | 0.100472 |
| Muscle_Sk | YPEL3 | ENSG00000 | 16 | 0.0393 | rs1164274 | 5.72 | rs7205802 | 0.018225 |
| Muscle_Sk | GDPD3 | ENSG00000 | 16 | 0.0693 | rs1164274 | 5.72 | rs1186093 | 0.00635 |
| Muscle_Sk | ZNF720 | ENSG00000 | 16 | 0.0601 | rs1034614 | -5.87 | rs1783951 | 0.029378 |
| Muscle_Sk | CFDP1 | ENSG00000 | 16 | 0.0665 | rs1010632 | 6.04 | rs1291765 | 0.074475 |
| Liver | COPS7A | ENSG00000 | 12 | 0.2559 | rs1106409 | -10.46 | rs1257857 | 0.118152 |
| Liver | TM7SF3 | ENSG00000 | 12 | 0.5093 | rs2306852 | -9.69 | rs6487582 | 0.207218 |
| Liver | RESF1 | ENSG00000 | 12 | 0.2502 | rs2651364 | -5.35 | rs1050607 | -0.0052 |
| Liver | FKBP11 | ENSG00000 | 12 | 0.2873 | rs4760645 | -6.69 | rs1242463 | 0.087874 |
| Liver | COX14 | ENSG00000 | 12 | 0.152 | rs1183106 | 8.51 | rs7138945 | 0.09258 |
| Liver | CDK2AP1 | ENSG00000 | 12 | 0.1722 | rs3531484 | 10.37 | rs1727309 | 0.13817 |
| Liver | CCDC92 | ENSG00000 | 12 | 0.2288 | rs1257967 | -6.13 | rs1054852 | 0.176841 |
| Liver | VSNL1 | ENSG00000 | 2 | 0.2276 | rs368430 | -5.3 | rs650275 | 0.112051 |
| Liver | ANKRD36B | ENSG00000 | 2 | 0.3645 | rs1299119 | 7.11 | rs6718109 | 0.179066 |
| Liver | TSGA10 | ENSG00000 | 2 | 0.2702 | rs958778 | -5.42 | rs1168318 | 0.270285 |
| Liver | LIPT1 | ENSG00000 | 2 | 0.1595 | rs958778 | -5.42 | rs1168318 | 0.168972 |
| Liver | ACTR3 | ENSG00000 | 2 | 0.2017 | rs1704813 | 5.19 | rs1168628 | 0.026418 |
| Liver | SLC29A4 | ENSG00000 | 7 | 0.4514 | rs852405 | -4.57 | rs1027879 | 0.205255 |
| Liver | CCZ1 | ENSG00000 | 7 | 0.3464 | rs852488 | 6.44 | rs6975026 | 0.121626 |
| Liver | STYXL1 | ENSG00000 | 7 | 0.295 | rs1253245 | -7.37 | rs3801472 | 0.184194 |
| Liver | SRRT | ENSG00000 | 7 | 0.1771 | rs1155912 | 6.69 | rs1788458 | 0.129571 |
| Liver | PWP2 | ENSG00000 | 21 | 0.8083 | rs1131999 | -4.42 | rs2277806 | 0.496 |
| Liver | KDM4B | ENSG00000 | 19 | 0.2058 | rs1034863 | 6.33 | rs2249152 | 0.147 |
| Liver | ZNF763 | ENSG00000 | 19 | 0.1863 | rs1040469 | -5.46 | rs279237 | 0.0292 |
| Liver | ZNF100 | ENSG00000 | 19 | 0.6703 | rs1041066 | 4.53 | rs6511286 | 0.234 |
| Liver | FBXO17 | ENSG00000 | 19 | 0.3497 | rs9676843 | -4.51 | rs498586 | 0.00996 |
| Liver | PRKD2 | ENSG00000 | 19 | 0.2216 | rs312181 | 9.15 | rs432157 | 0.0935 |
| Liver | ZNF584 | ENSG00000 | 19 | 0.3836 | rs3425166 | -8.58 | rs1167087 | 0.277 |
| Liver | MAEA | ENSG00000 | 4 | 0.41 | rs1716468 | 5.14 | rs1193385 | 0.172091 |
| Liver | SRD5A3 | ENSG00000 | 4 | 0.669 | rs6837735 | 7.99 | rs1264188 | 0.129447 |

| | | | | | | | | |
|---|---|---|---|---|---|---|---|---|
| Liver | PROCR | ENSG00000( | 20 | 0.2122 | rs7280 | -5.85 | rs6120849 | 0.125558 |
| Liver | MMP24OS | ENSG00000( | 20 | 0.4618 | rs7280 | -5.85 | rs6060341 | 0.191518 |
| Liver | CCDC13 | ENSG00000( | 3 | 0.1852 | rs7650470 | -7.4 | rs3846063 | 0.042293 |
| Liver | P4HTM | ENSG00000( | 3 | 0.0713 | rs1249039: | -4.93 | rs4974078 | -0.00516 |
| Liver | DALRD3 | ENSG00000( | 3 | 0.0885 | rs1249039: | -4.93 | rs9850134 | 0.043111 |
| Liver | CSTA | ENSG00000( | 3 | 0.2922 | rs1248759: | 4.49 | rs1512048 | -0.00531 |
| Liver | NPHP3 | ENSG00000( | 3 | 0.5588 | rs3439194: | -10.79 | rs2369832 | 0.231247 |
| Liver | MFN1 | ENSG00000( | 3 | 0.1826 | rs2339798 | 6.55 | rs1046088: | 0.038238 |
| Liver | CDH23 | ENSG00000( | 10 | 0.196 | rs1099995: | 6.72 | rs1771252: | 0.081512 |
| Liver | TSPAN14 | ENSG00000( | 10 | 0.0911 | rs7915814 | 4.38 | rs4934167 | 0.011082 |
| Liver | BBIP1 | ENSG00000( | 10 | 0.2288 | rs1225752: | -4.66 | rs4918538 | -0.00498 |
| Liver | CACNA1H | ENSG00000( | 16 | 0.2264 | rs3809663 | 6.56 | rs4984636 | 0.015239 |
| Liver | CLCN7 | ENSG00000( | 16 | 0.2871 | rs7191794 | 6.48 | rs1292608: | 0.02297 |
| Liver | INO80E | ENSG00000( | 16 | 0.1607 | rs4541091 | 7.19 | rs4787491 | 0.188829 |
| Liver | YPEL3 | ENSG00000( | 16 | 0.1751 | rs1164274( | 5.72 | rs9928448 | 0.103799 |
| Liver | SHBG | ENSG00000( | 17 | 0.18 | rs2269459 | 7.62 | rs4602096 | -0.00485 |
| Liver | ZSWIM7 | ENSG00000( | 17 | 0.4309 | rs178791 | 4.29 | rs3826360 | 0.163198 |
| Liver | SYNRG | ENSG00000( | 17 | 0.3334 | rs9906814 | -4.37 | rs1294945: | 0.108707 |
| Liver | ARL17A | ENSG00000( | 17 | 0.3712 | rs2074405 | 6.23 | rs1769212: | 0.179158 |
| Liver | OGFOD3 | ENSG00000( | 17 | 0.4641 | rs1785547( | -7.73 | rs4074069 | 0.111949 |
| Liver | HEATR4 | ENSG00000( | 14 | 0.6201 | rs8018725 | -5.54 | rs8018967 | 0.1626 |
| Liver | MTMR6 | ENSG00000( | 13 | 0.191 | rs9507413 | -8.35 | rs7330216 | 0.05816 |
| Liver | EBPL | ENSG00000( | 13 | 0.33 | rs9568353 | -5.61 | rs2273816 | 0.01824 |
| Liver | PROZ | ENSG00000( | 13 | 0.699 | rs2297800 | -3.94 | rs513479 | 0.16439 |
| Liver | GOLGA8A | ENSG00000( | 15 | 0.4645 | rs661968 | 7.21 | rs4299123 | 0.112352 |
| Liver | MESP2 | ENSG00000( | 15 | 0.2015 | rs7165704 | -4.01 | rs7172694 | -0.00463 |
| Liver | MED19 | ENSG00000( | 11 | 0.1996 | rs3741089 | 3.97 | rs1613592 | 0.133438 |
| Liver | PPP6R3 | ENSG00000( | 11 | 0.164 | rs1089637: | -4.96 | rs901824 | -0.00543 |
| Liver | SLC35E2B | ENSG00000( | 1 | 0.6945 | rs2887286 | 7.44 | rs4648786 | 0.064279 |
| Liver | IGSF21 | ENSG00000( | 1 | 0.5232 | rs7549273 | 7.14 | rs930851 | 0.00407 |
| Liver | TRIM63 | ENSG00000( | 1 | 0.1093 | rs9438621 | -7.21 | rs3008226 | 0.079842 |
| Liver | RPA2 | ENSG00000( | 1 | 0.3323 | rs1890462 | 7.57 | rs1725725: | 0.181913 |
| Liver | IQCC | ENSG00000( | 1 | 0.3087 | rs7539490 | 10.83 | rs1683498( | -0.00553 |
| Liver | ZMYM1 | ENSG00000( | 1 | 0.1191 | rs3814303 | -6.88 | rs561905 | 0.005235 |
| Liver | WARS2 | ENSG00000( | 1 | 0.2643 | rs984225 | -6.58 | rs2645303 | 0.137209 |
| Liver | MTMR11 | ENSG00000( | 1 | 0.1376 | rs7534365 | -6.45 | rs3767627 | 0.079425 |
| Liver | CTSK | ENSG00000( | 1 | 0.1771 | rs9733 | 4.49 | rs1256875: | 0.18263 |
| Liver | ATF6 | ENSG00000( | 1 | 0.1597 | rs1207379: | 8.69 | rs3554152: | 0.018715 |
| Liver | SUCO | ENSG00000( | 1 | 0.2524 | rs4916266 | 4.53 | rs2516064 | 0.088197 |
| Liver | PSMG4 | ENSG00000( | 6 | 0.2426 | rs9501920 | 8.82 | rs4602754 | 0.0987 |
| Liver | ILRUN | ENSG00000( | 6 | 0.1742 | rs2820239 | -6.01 | rs2814983 | 0.0843 |
| Liver | CRIP3 | ENSG00000( | 6 | 0.1941 | rs2396004 | 4.98 | rs1574430 | 0.112 |
| Liver | SLC25A27 | ENSG00000( | 6 | 0.3215 | rs613870 | 7.11 | rs2270450 | 0.141 |
| Liver | FAM162B | ENSG00000( | 6 | 0.4595 | rs4946203 | -9.3 | rs548101 | 0.317 |
| Liver | L3MBTL3 | ENSG00000( | 6 | 0.4442 | rs7755865 | -7.18 | rs6569648 | 0.313 |

| | | | | | | | | |
|---|---|---|---|---|---|---|---|---|
| Liver | WDR97 | ENSG0000( | 8 | 0.4891 | rs1178689( | 5.04 | rs4977165 | 0.277726 |
| Pancreas | AKR1C1 | ENSG0000( | 10 | 0.3313 | rs7903936 | -7.32 | rs7358080 | 0.146569 |
| Pancreas | AS3MT | ENSG0000( | 10 | 0.4917 | rs4919632 | 6.02 | rs1119138! | 0.124957 |
| Pancreas | PLEKHA1 | ENSG0000( | 10 | 0.166 | rs2837672( | -7.99 | rs1224600! | 0.042707 |
| Pancreas | CLDN23 | ENSG0000( | 8 | 0.2904 | rs1199354: | -10.27 | rs1268129! | 0.0319 |
| Pancreas | EIF3E | ENSG0000( | 8 | 0.0727 | rs610891 | 6.42 | rs2514844 | 0.0139 |
| Pancreas | MAF1 | ENSG0000( | 8 | 0.1371 | rs1178689( | 5.04 | rs1255072! | 0.0174 |
| Pancreas | GPR63 | ENSG0000( | 6 | 0.2222 | rs1413524 | 4.38 | rs7765182 | 0.015172 |
| Pancreas | MMS22L | ENSG0000( | 6 | 0.1129 | rs9320388 | -4.29 | rs1780658! | 0.022878 |
| Pancreas | SAMD5 | ENSG0000( | 6 | 0.1431 | rs508902 | -4.12 | rs7763189 | 0.048451 |
| Pancreas | MYRIP | ENSG0000( | 3 | 0.1903 | rs1918028 | 4.5 | rs6783267 | 0.085459 |
| Pancreas | CTNNB1 | ENSG0000( | 3 | 0.1238 | rs6802052 | -6.24 | rs1705727( | -0.00201 |
| Pancreas | SHISA5 | ENSG0000( | 3 | 0.1417 | rs1191861! | -4.75 | rs6442120 | 0.016164 |
| Pancreas | P4HTM | ENSG0000( | 3 | 0.1943 | rs1249039: | -4.93 | rs4279134 | 0.117196 |
| Pancreas | QRICH1 | ENSG0000( | 3 | 0.1221 | rs1759541( | 6.08 | rs4974083 | 0.131456 |
| Pancreas | AMT | ENSG0000( | 3 | 0.1937 | rs1249039: | -4.93 | rs4955426 | 0.16339 |
| Pancreas | RBM15B | ENSG0000( | 3 | 0.151 | rs1263950: | 6.85 | rs1263799: | 0.078007 |
| Pancreas | ALDH1L1 | ENSG0000( | 3 | 0.283 | rs3772410 | 10.18 | rs2077523 | 0.028957 |
| Pancreas | MSL2 | ENSG0000( | 3 | 0.1623 | rs3480724( | 4.79 | rs9845762 | -0.00093 |
| Pancreas | MMP24OS | ENSG0000( | 20 | 0.3525 | rs7280 | -5.85 | rs6060341 | 0.278026 |
| Pancreas | UQCC1 | ENSG0000( | 20 | 0.097 | rs7280 | -5.85 | rs1540927 | 0.098681 |
| Pancreas | CEP250 | ENSG0000( | 20 | 0.0617 | rs7280 | -5.85 | rs1570841 | 0.010752 |
| Pancreas | CCDC152 | ENSG0000( | 5 | 0.1463 | rs1252075! | 4.52 | rs1423651 | 0.129053 |
| Pancreas | SPATA24 | ENSG0000( | 5 | 0.0702 | rs889021 | -7.84 | rs2351905 | 0.004683 |
| Pancreas | PCDHAC1 | ENSG0000( | 5 | 0.1613 | rs1709694! | 4.3 | rs31746 | 0.007658 |
| Pancreas | PTGR2 | ENSG0000( | 14 | 0.3405 | rs2109750 | -5.12 | rs7148485 | 0.275426 |
| Pancreas | HMCN2 | ENSG0000( | 9 | 0.419 | rs1178952! | 7.5 | rs1233538: | 0.155785 |
| Pancreas | CCZ1 | ENSG0000( | 7 | 0.3501 | rs852488 | 6.44 | rs6975026 | 0.094136 |
| Pancreas | STYXL1 | ENSG0000( | 7 | 0.3545 | rs1253245: | -7.37 | rs1155309( | 0.229017 |
| Pancreas | ASB4 | ENSG0000( | 7 | 0.3282 | rs1527719 | 4.52 | rs1176407! | 0.186648 |
| Pancreas | TRIP6 | ENSG0000( | 7 | 0.1466 | rs1155912: | 6.72 | rs314330 | 0.079011 |
| Pancreas | PDIA4 | ENSG0000( | 7 | 0.1682 | rs6958740 | 8.94 | rs1198188! | -0.00134 |
| Pancreas | AFAP1 | ENSG0000( | 4 | 0.1624 | rs1193378: | -7.14 | rs6818156 | 0.000984 |
| Pancreas | LDB2 | ENSG0000( | 4 | 0.2835 | rs2191612 | -9.01 | rs1093969: | 0.185529 |
| Pancreas | DCAF16 | ENSG0000( | 4 | 0.1961 | rs1049601 | 6.68 | rs7667864 | 0.090274 |
| Pancreas | PRDM5 | ENSG0000( | 4 | 0.2342 | rs1250723! | -6.53 | rs343192 | 0.124494 |
| Pancreas | TPSB2 | ENSG0000( | 16 | 0.3742 | rs7191794 | 5.19 | rs4984637 | 0.316708 |
| Pancreas | INO80E | ENSG0000( | 16 | 0.1303 | rs1143695 | -9.36 | rs4787491 | 0.159591 |
| Pancreas | DOC2A | ENSG0000( | 16 | 0.0792 | rs1143695 | -8.79 | rs1293357! | 0.052736 |
| Pancreas | YPEL3 | ENSG0000( | 16 | 0.2097 | rs1143695 | -9.33 | rs7205802 | 0.263893 |
| Pancreas | WWP2 | ENSG0000( | 16 | 0.0865 | rs2291959 | -5.54 | rs3748388 | 0.06237 |
| Pancreas | CLEC18A | ENSG0000( | 16 | 0.5204 | rs2291959 | -5.54 | rs1808417 | 0.383187 |
| Pancreas | TXNL4B | ENSG0000( | 16 | 0.1176 | rs1946768 | -5.03 | rs1820248 | 0.012271 |
| Pancreas | TXNL4A | ENSG0000( | 18 | 0.1789 | rs732771 | -6.61 | rs8095413 | 0.156731 |
| Pancreas | LAPTM4A | ENSG0000( | 2 | 0.4929 | rs6750471 | 6.73 | rs7578149 | 0.264 |

| | | | | | | | | |
|---|---|---|---|---|---|---|---|---|
| Pancreas | RPIA | ENSG00000( | 2 | 0.0854 | rs2306676 | 4.56 | rs1299577: | 0.085 |
| Pancreas | ANKRD36B | ENSG00000( | 2 | 0.482 | rs1299119 | 7.11 | rs6718109 | 0.378 |
| Pancreas | ZAP70 | ENSG00000( | 2 | 0.107 | rs1168085 | 6.96 | rs1189360 | 0.125 |
| Pancreas | TSGA10 | ENSG00000( | 2 | 0.3862 | rs958778 | -5.56 | rs1168318 | 0.501 |
| Pancreas | MITD1 | ENSG00000( | 2 | 0.0789 | rs2048748 | -6.76 | rs13798 | 0.0616 |
| Pancreas | AFF3 | ENSG00000( | 2 | 0.2696 | rs4851250 | -4.56 | rs1169578 | 0.069 |
| Pancreas | RFTN2 | ENSG00000( | 2 | 0.0807 | rs700639 | -5.02 | rs1064213 | 0.0192 |
| Pancreas | TRIP4 | ENSG00000( | 15 | 0.0824 | rs677561 | -10.15 | rs4777499 | 0.093958 |
| Pancreas | LYSMD4 | ENSG00000( | 15 | 0.6739 | rs899947 | -4.36 | rs6598265 | 0.313873 |
| Pancreas | LRP4 | ENSG00000( | 11 | 0.3143 | rs3561959 | 5.55 | rs747650 | 0.341 |
| Pancreas | PCNX3 | ENSG00000( | 11 | 0.1129 | rs551523 | 8.61 | rs1182006 | -0.00239 |
| Pancreas | RAD9A | ENSG00000( | 11 | 0.1436 | rs1228804 | 4.88 | rs7952436 | 0.126 |
| Pancreas | VPS11 | ENSG00000( | 11 | 0.2164 | rs1157466 | 6.7 | rs549893 | 0.105 |
| Pancreas | ADAMTSL5 | ENSG00000( | 19 | 0.2243 | rs7258839 | -5.3 | rs7258839 | 0.263 |
| Pancreas | ZNF100 | ENSG00000( | 19 | 0.7867 | rs1041066 | 4.53 | rs6511286 | 0.52 |
| Pancreas | FAAP24 | ENSG00000( | 19 | 0.1573 | rs1775465 | 8.61 | rs3764633 | 0.0799 |
| Pancreas | CYTH2 | ENSG00000( | 19 | 0.3202 | rs2307278 | 5.85 | rs1799270 | 0.141 |
| Pancreas | ATAD3A | ENSG00000( | 1 | 0.2416 | rs1158618 | -7.45 | rs819980 | 0.055473 |
| Pancreas | SLC35E2B | ENSG00000( | 1 | 0.7307 | rs2887286 | 7.44 | rs4648786 | 0.0684 |
| Pancreas | RPA2 | ENSG00000( | 1 | 0.1774 | rs1890462 | 7.49 | rs1725725 | 0.09274 |
| Pancreas | XKR8 | ENSG00000( | 1 | 0.1356 | rs1890462 | 7.67 | rs1090268 | 0.053072 |
| Pancreas | PCSK9 | ENSG00000( | 1 | 0.2308 | rs2864123 | 6.49 | rs499718 | -0.00319 |
| Pancreas | LYPLAL1 | ENSG00000( | 1 | 0.0829 | rs6675807 | 13.03 | rs642836 | 0.005502 |
| Pancreas | PTHLH | ENSG00000( | 12 | 0.1307 | rs1133028 | -16.29 | rs4931081 | 0.031544 |
| Pancreas | CERS5 | ENSG00000( | 12 | 0.0806 | rs1183106 | 9.13 | rs3184122 | 0.041689 |
| Pancreas | ACSS3 | ENSG00000( | 12 | 0.4406 | rs4489813 | 10.85 | rs6539557 | 0.163913 |
| Pancreas | CDK2AP1 | ENSG00000( | 12 | 0.2525 | rs3531484 | 10.37 | rs1060105 | 0.319053 |
| Pancreas | CCDC92 | ENSG00000( | 12 | 0.0892 | rs1257967 | -6.23 | rs1054852 | 0.114256 |
| Pancreas | GGACT | ENSG00000( | 13 | 0.1351 | rs7997419 | -5.75 | rs1695748 | 0.031969 |

| EQTL.Z | EQTL.GWA | NSNP | NWGT | MODEL | MODELCV. | MODELCV. | TWAS.Z | TWAS.P |
|---|---|---|---|---|---|---|---|---|
| 9.36 | 2.81 | 375 | 375 | susie | 0.3 | 1.70E-26 | 4.59242 | 4.38E-06 |
| 6.37 | -4.49 | 451 | 10 | lasso | 0.096 | 1.00E-08 | -4.52355 | 6.08E-06 |
| 4.45 | 4.294 | 434 | 434 | susie | 0.022 | 0.0044 | 4.73436 | 2.20E-06 |
| 6 | -2.772 | 674 | 23 | enet | 0.14 | 2.10E-12 | -4.5044 | 6.66E-06 |
| 6.61 | 4.934 | 379 | 3 | lasso | 0.14 | 4.00E-12 | 4.9452 | 7.61E-07 |
| -6.11 | -4.373 | 462 | 13 | enet | 0.16 | 2.10E-13 | 4.617 | 3.89E-06 |
| -5.16 | -6.18 | 182 | 1 | top1 | 0.074 | 5.40E-07 | 6.18 | 6.41E-10 |
| 4.27 | -4.995 | 453 | 24 | enet | 0.057 | 1.00E-05 | -6.12069 | 9.32E-10 |
| -6.91 | -4.032 | 262 | 262 | susie | 0.15 | 1.40E-12 | 4.54484 | 5.50E-06 |
| 4.01 | 3.687 | 260 | 260 | susie | 0.027 | 0.0021 | 4.69861 | 2.62E-06 |
| 4.76 | -4.03 | 253 | 15 | enet | 0.064 | 2.90E-06 | -5.19534 | 2.04E-07 |
| -7.18 | -4.915 | 259 | 259 | susie | 0.14 | 1.80E-12 | 4.84547 | 1.26E-06 |
| 3.55 | -4.675 | 278 | 278 | susie | 0.02 | 0.0067 | -4.81306 | 1.49E-06 |
| -4.11 | -4.573 | 288 | 1 | top1 | 0.047 | 5.70E-05 | 4.573 | 4.81E-06 |
| -4.68 | -2.962 | 319 | 20 | enet | 0.044 | 0.00011 | 4.46689 | 7.94E-06 |
| 12.74 | 5.783 | 368 | 368 | susie | 0.51 | 7.60E-51 | 5.78332 | 7.32E-09 |
| -4.98 | -3.986 | 407 | 10 | lasso | 0.1 | 2.40E-09 | 4.5912 | 4.41E-06 |
| -4.48 | 2.211 | 475 | 33 | enet | 0.079 | 2.10E-07 | 5.0852 | 3.67E-07 |
| 4.58 | -2.578 | 373 | 35 | enet | 0.044 | 1.00E-04 | -4.9268 | 8.36E-07 |
| 7.46 | -3.885 | 384 | 384 | susie | 0.32 | 8.40E-29 | -4.68013 | 2.87E-06 |
| 4.01 | 2.855 | 299 | 25 | enet | 0.016 | 0.015 | 5.6531 | 1.58E-08 |
| -4.66 | 2.682 | 390 | 33 | enet | 0.083 | 1.10E-07 | -4.6489 | 3.34E-06 |
| 4.96 | 4.345 | 280 | 16 | enet | 0.12 | 2.30E-10 | 4.6255 | 3.74E-06 |
| -4.5 | 4.5 | 448 | 7 | enet | 0.063 | 3.90E-06 | -4.606 | 4.10E-06 |
| -5.32 | -2.86 | 807 | 28 | enet | 0.18 | 1.00E-15 | 4.7891 | 1.67E-06 |
| 8.66 | -3.641 | 472 | 472 | susie | 0.32 | 5.80E-28 | -4.80187 | 1.57E-06 |
| 6.07 | -5.474 | 529 | 529 | susie | 0.097 | 9.90E-09 | -5.53603 | 3.09E-08 |
| 8.01 | 5.7572 | 118 | 1 | top1 | 0.19 | 1.30E-16 | 5.7572 | 8.55E-09 |
| -4.62 | 5.79557 | 143 | 12 | enet | 0.051 | 3.20E-05 | -4.7557 | 1.98E-06 |
| -9.15 | -4.524 | 297 | 297 | susie | 0.26 | 2.30E-22 | 4.51815 | 6.24E-06 |
| 7.5 | -4.524 | 302 | 302 | susie | 0.15 | 3.20E-13 | -4.52803 | 5.95E-06 |
| 3.72 | -4.238 | 377 | 6 | lasso | 0.023 | 0.0038 | -4.50944 | 6.50E-06 |
| 5.55 | 4.3266 | 614 | 19 | enet | 0.077 | 3.40E-07 | 4.6801 | 2.87E-06 |
| -10.25 | 3.479 | 305 | 305 | susie | 0.39 | 1.90E-35 | -4.7148 | 2.42E-06 |
| 3.73 | -2.658 | 596 | 5 | lasso | 0.00087 | 0.26 | -4.5393 | 5.64E-06 |
| 8.1 | -2.687 | 376 | 376 | susie | 0.31 | 6.50E-27 | -5.4269 | 5.74E-08 |
| -3.49 | -2.524 | 410 | 17 | enet | 0.016 | 0.013 | 6.5392 | 6.19E-11 |
| -6.63 | -4.228 | 344 | 344 | susie | 0.1 | 4.30E-09 | 4.515 | 6.33E-06 |
| -3.77 | -3.734 | 477 | 32 | enet | 0.034 | 0.00055 | 4.8534 | 1.21E-06 |
| 3.75 | 3.277 | 373 | 7 | lasso | 0.019 | 0.0081 | 4.9112 | 9.05E-07 |
| 3.88 | 3.205 | 488 | 22 | enet | 0.029 | 0.0014 | 4.9371 | 7.93E-07 |
| 3.22 | -5.158 | 343 | 343 | susie | 0.011 | 0.037 | -6.2704 | 3.60E-10 |
| -6.96 | -4.455 | 499 | 1 | top1 | 0.15 | 7.70E-13 | 4.455 | 8.39E-06 |
| 7.43 | 3.796 | 341 | 341 | susie | 0.22 | 7.20E-19 | 4.57231 | 4.82E-06 |

| | | | | | | | | |
|---|---|---|---|---|---|---|---|---|
| -5.29 | 3.267 | 622 | 48 | enet | 0.16 | 1.80E-13 | -5.90871 | 3.45E-09 |
| -6.23 | 3.849 | 486 | 486 | susie | 0.15 | 5.50E-13 | -4.47384 | 7.68E-06 |
| 3.85 | -1.542 | 429 | 48 | enet | 0.033 | 0.00074 | -5.73335 | 9.85E-09 |
| 5.84 | 3.213 | 232 | 34 | enet | 0.18 | 4.10E-15 | 6.33222 | 2.42E-10 |
| 5.87 | 3.044 | 87 | 87 | susie | 0.16 | 3.60E-14 | 5.6286 | 1.82E-08 |
| 11.36 | 4.263 | 311 | 22 | enet | 0.52 | 7.40E-52 | 4.66657 | 3.06E-06 |
| 6.89 | -2.927 | 320 | 320 | susie | 0.19 | 1.10E-16 | -4.44993 | 8.59E-06 |
| 3.42 | 6.427 | 339 | 5 | lasso | 0.0078 | 0.064 | 6.23744 | 4.45E-10 |
| 4.99 | 2.919 | 539 | 539 | susie | 0.11 | 1.30E-09 | 4.8568 | 1.19E-06 |
| 5.9 | 4.726 | 458 | 458 | susie | 0.091 | 2.90E-08 | 4.7893 | 1.67E-06 |
| -4.45 | -4.477 | 317 | 1 | top1 | 0.037 | 0.00035 | 4.477 | 7.57E-06 |
| -7.52 | -4.815 | 305 | 1 | top1 | 0.16 | 5.30E-14 | 4.815 | 1.47E-06 |
| -4.65 | -4.898 | 341 | 1 | top1 | 0.058 | 9.10E-06 | 4.898 | 9.68E-07 |
| 4 | -4.692 | 340 | 1 | top1 | 0.024 | 0.0034 | -4.692 | 2.71E-06 |
| 6.84 | -3.88 | 432 | 20 | enet | 0.15 | 9.70E-13 | -5.194 | 2.06E-07 |
| -12.57 | 3.117 | 417 | 21 | enet | 0.62 | 1.30E-67 | -4.5967 | 4.29E-06 |
| 5.77 | 2.05 | 146 | 20 | enet | 0.098 | 7.80E-09 | 5.6982 | 1.21E-08 |
| -8.33 | 4.804 | 203 | 203 | susie | 0.21 | 1.10E-17 | -4.8484 | 1.24E-06 |
| 4.54 | -4.78 | 215 | 215 | susie | 0.064 | 3.20E-06 | -4.9551 | 7.23E-07 |
| 8.13 | 4.954 | 225 | 2 | lasso | 0.2 | 2.20E-17 | 4.9557 | 7.21E-07 |
| 4.91 | -5.6784 | 185 | 1 | top1 | 0.071 | 1.00E-06 | -5.6784 | 1.36E-08 |
| -5.24 | 4.461 | 254 | 1 | top1 | 0.072 | 7.40E-07 | -4.461 | 8.16E-06 |
| -3.71 | 3.329 | 252 | 13 | enet | 0.052 | 2.50E-05 | -7.1565 | 8.28E-13 |
| 5.65 | 2.77 | 421 | 13 | enet | 0.1 | 3.60E-09 | 4.4679 | 7.90E-06 |
| -8.58 | 4.945 | 342 | 342 | susie | 0.22 | 1.80E-18 | -4.95 | 7.60E-07 |
| -4.32 | 3.92 | 357 | 29 | enet | 0.069 | 1.40E-06 | -4.52 | 6.15E-06 |
| -3.86 | 2.737 | 672 | 34 | enet | 0.074 | 5.50E-07 | -5.47 | 4.45E-08 |
| 11.62 | -2.604 | 298 | 298 | susie | 0.53 | 2.60E-53 | -4.7132 | 2.44E-06 |
| 6.84 | 2.621 | 403 | 33 | enet | 0.18 | 2.00E-15 | 4.5507 | 5.35E-06 |
| 5.94 | -2.484 | 466 | 466 | susie | 0.11 | 1.60E-09 | -4.7708 | 1.83E-06 |
| 4.58 | 3.922 | 376 | 54 | enet | 0.18 | 1.40E-15 | 5.7139 | 1.10E-08 |
| 6.29 | 3.152 | 375 | 375 | susie | 0.12 | 1.30E-10 | 7.2847 | 3.22E-13 |
| -3.97 | 4.015 | 516 | 6 | lasso | 0.015 | 0.017 | -5.5546 | 2.78E-08 |
| 4.58 | -1.984 | 459 | 459 | susie | 0.094 | 1.50E-08 | -4.8044 | 1.55E-06 |
| 4.19 | 3.35 | 336 | 22 | enet | 0.029 | 0.0015 | 4.7928 | 1.64E-06 |
| -6.51 | -4.697 | 345 | 1 | top1 | 0.13 | 2.50E-11 | 4.697 | 2.64E-06 |
| 5.34 | -5.001 | 344 | 1 | top1 | 0.073 | 6.50E-07 | -5.001 | 5.70E-07 |
| -4.88 | -5.06 | 331 | 1 | top1 | 0.064 | 3.20E-06 | 5.06 | 4.19E-07 |
| -5 | -2.933 | 384 | 19 | enet | 0.077 | 1.90E-07 | 4.5694 | 4.89E-06 |
| -4.72 | -3.671 | 371 | 55 | enet | 0.046 | 5.70E-05 | 5.3528 | 8.66E-08 |
| -8.59 | -2.495 | 411 | 411 | susie | 0.28 | 3.30E-25 | 4.82 | 1.44E-06 |
| 5.09 | 3.16 | 299 | 10 | lasso | 0.087 | 3.30E-08 | 4.555 | 5.24E-06 |
| 5.76 | -4.696 | 415 | 415 | susie | 0.088 | 2.90E-08 | -4.7668 | 1.87E-06 |
| 5.45 | 4.667 | 458 | 458 | susie | 0.065 | 1.80E-06 | 4.7832 | 1.72E-06 |
| -3.74 | -4.375 | 225 | 225 | susie | 0.017 | 0.011 | 4.4466 | 8.72E-06 |

| | | | | | | | | |
|---|---|---|---|---|---|---|---|---|
| 4.19 | -4.844 | 325 | 1 | top1 | 0.04 | 0.00016 | -4.8436 | 1.28E-06 |
| 14.43 | -3.516 | 290 | 24 | enet | 0.65 | 4.70E-77 | -4.6531 | 3.27E-06 |
| 6.63 | 3.256 | 356 | 356 | susie | 0.17 | 4.00E-15 | 4.651 | 3.30E-06 |
| -4.18 | 2.648 | 433 | 21 | enet | 0.03 | 0.00095 | -4.5548 | 5.24E-06 |
| 5.09 | -4.415 | 347 | 24 | enet | 0.063 | 2.60E-06 | -5.7866 | 7.18E-09 |
| 8.73 | -4.43 | 433 | 24 | enet | 0.28 | 7.40E-25 | -4.6767 | 2.91E-06 |
| -5.61 | -3.556 | 476 | 14 | enet | 0.083 | 6.50E-08 | 4.8231 | 1.41E-06 |
| -11.15 | 3.117 | 417 | 417 | susie | 0.53 | 8.80E-56 | -4.8522 | 1.22E-06 |
| -8.03 | 4.862 | 203 | 1 | top1 | 0.19 | 7.00E-17 | -4.862 | 1.16E-06 |
| -4.28 | 5.093 | 215 | 1 | top1 | 0.044 | 8.00E-05 | -5.093 | 3.52E-07 |
| -6.72 | -4.353 | 225 | 44 | enet | 0.17 | 3.90E-15 | 5.0738 | 3.90E-07 |
| 7.79 | 5.024 | 225 | 1 | top1 | 0.18 | 9.70E-16 | 5.024 | 5.06E-07 |
| 3.42 | 4.176 | 455 | 455 | susie | 0.0012 | 0.24 | 4.7397 | 2.14E-06 |
| -3.33 | 2.081 | 191 | 191 | susie | 0.0024 | 0.18 | -4.6788 | 2.89E-06 |
| -4.36 | -5.46 | 934 | 934 | susie | 0.0051 | 0.1 | 5.56102 | 2.68E-08 |
| -5.71 | 4.08 | 626 | 21 | enet | 0.12 | 9.60E-11 | -5.29329 | 1.20E-07 |
| 3.27 | -2.29 | 282 | 6 | lasso | -0.00036 | 0.35 | -4.58782 | 4.48E-06 |
| 7.04 | -4.68 | 367 | 1 | top1 | 0.14 | 1.30E-12 | -4.68 | 2.87E-06 |
| -4.75 | -6.14 | 475 | 9 | lasso | 0.041 | 0.00014 | 4.54098 | 5.60E-06 |
| -7.2 | 5.04 | 292 | 3 | lasso | 0.17 | 1.20E-14 | -4.99481 | 5.89E-07 |
| -5.16 | -3.932 | 361 | 17 | enet | 0.039 | 2.00E-04 | 4.439 | 9.02E-06 |
| 4.16 | 4.78 | 375 | 1 | top1 | 0.018 | 0.0084 | 4.775 | 1.80E-06 |
| 5.66 | -4.49 | 453 | 7 | lasso | 0.095 | 8.00E-09 | -4.6519 | 3.29E-06 |
| -9.1 | 3.34 | 296 | 32 | enet | 0.32 | 2.80E-29 | -4.5419 | 5.57E-06 |
| 6.69 | -2.641 | 506 | 26 | enet | 0.13 | 1.10E-11 | -4.4255 | 9.62E-06 |
| 6.05 | -2.996 | 509 | 509 | susie | 0.2 | 1.60E-17 | -5.6713 | 1.42E-08 |
| 7.09 | 2.78 | 546 | 546 | susie | 0.17 | 6.00E-15 | 4.4728 | 7.72E-06 |
| -5.05 | 3 | 498 | 18 | enet | 0.1 | 2.40E-09 | -5.5351 | 3.11E-08 |
| -3.64 | -1.98 | 490 | 31 | enet | 0.021 | 0.0052 | 4.6535 | 3.26E-06 |
| -7.34 | -0.9 | 306 | 306 | susie | 0.44 | 1.90E-42 | 5.9193 | 3.23E-09 |
| 8.24 | -3.255 | 374 | 374 | susie | 0.21 | 1.30E-18 | -4.76969 | 1.85E-06 |
| 3.22 | 4.195 | 463 | 66 | enet | 0.015 | 0.014 | 5.38327 | 7.31E-08 |
| 9.83 | -4.258 | 351 | 23 | enet | 0.3 | 3.10E-27 | -5.03188 | 4.86E-07 |
| 6.97 | 3.869 | 425 | 425 | susie | 0.18 | 1.20E-15 | 5.14442 | 2.68E-07 |
| 3.29 | -2.727 | 427 | 3 | lasso | 0.017 | 0.011 | -5.08189 | 3.74E-07 |
| 6.39 | 4.319 | 137 | 3 | lasso | 0.12 | 2.90E-11 | 5.05395 | 4.33E-07 |
| 6.44 | 4.449 | 302 | 302 | susie | 0.11 | 7.00E-10 | 4.43411 | 9.25E-06 |
| 7.05 | -4.58 | 448 | 448 | susie | 0.14 | 1.50E-12 | -4.46373 | 8.05E-06 |
| 4.36 | 3.044 | 576 | 28 | enet | 0.052 | 1.80E-05 | 4.90172 | 9.50E-07 |
| 5.42 | -5.001 | 344 | 1 | top1 | 0.075 | 3.00E-07 | -5.001 | 5.70E-07 |
| 10.64 | -2.52 | 426 | 426 | susie | 0.54 | 2.30E-57 | -5.0476 | 4.47E-07 |
| -5.5 | -5.01 | 391 | 1 | top1 | 0.07 | 6.90E-07 | 5.006 | 5.56E-07 |
| 8.57 | 3.25 | 408 | 408 | susie | 0.32 | 5.00E-29 | 5.1001 | 3.39E-07 |
| -6.21 | -1.97 | 293 | 19 | enet | 0.12 | 9.10E-11 | 5.2084 | 1.90E-07 |
| -4.03 | 2.123 | 275 | 4 | lasso | 0.051 | 2.20E-05 | 5.5111 | 3.56E-08 |

| | | | | | | | | |
|---|---|---|---|---|---|---|---|---|
| 4.56 | -2.169 | 322 | 28 | enet | 0.028 | 0.0014 | -5.2753 | 1.33E-07 |
| 9.91 | 2.674 | 322 | 322 | susie | 0.39 | 2.00E-37 | 4.6526 | 3.28E-06 |
| 8.64 | 2.58 | 506 | 46 | enet | 0.34 | 1.40E-31 | 5.4898 | 4.02E-08 |
| -5.88 | 4.626 | 469 | 1 | top1 | 0.1 | 2.10E-09 | -4.626 | 3.73E-06 |
| -6.27 | 4.174 | 279 | 279 | susie | 0.11 | 8.10E-10 | -4.5056 | 6.62E-06 |
| 4.69 | 2.843 | 268 | 19 | enet | 0.051 | 2.30E-05 | 4.431 | 9.38E-06 |
| -5.26 | 3.228 | 465 | 36 | enet | 0.079 | 1.50E-07 | -5.981 | 2.22E-09 |
| -4.27 | -4.608 | 263 | 263 | susie | 0.047 | 4.70E-05 | 4.6638 | 3.10E-06 |
| -3.87 | -4.457 | 262 | 262 | susie | 0.014 | 0.017 | 4.495 | 6.96E-06 |
| 5.18 | -4.598 | 253 | 1 | top1 | 0.076 | 2.40E-07 | -4.598 | 4.27E-06 |
| -6.77 | -4.887 | 259 | 1 | top1 | 0.13 | 4.80E-12 | 4.887 | 1.02E-06 |
| 3.72 | -4.528 | 237 | 1 | top1 | 0.011 | 0.034 | -4.528 | 5.95E-06 |
| 3.11 | 2.013 | 325 | 31 | enet | 0.032 | 0.00063 | -4.7487 | 2.05E-06 |
| 12.36 | 5.783 | 367 | 1 | top1 | 0.47 | 1.60E-46 | 5.783 | 7.34E-09 |
| 5.03 | 4.494 | 335 | 1 | top1 | 0.059 | 5.00E-06 | 4.494 | 6.99E-06 |
| -4.25 | 4.855 | 429 | 1 | top1 | 0.03 | 0.00097 | -4.855 | 1.20E-06 |
| -3.56 | 3.16 | 478 | 39 | enet | 0.048 | 4.00E-05 | -7.0422 | 1.89E-12 |
| -7.19 | -2.41 | 510 | 510 | susie | 0.28 | 9.10E-25 | 4.8346 | 1.33E-06 |
| -6.4 | -2.074 | 422 | 422 | susie | 0.38 | 8.90E-36 | 4.4871 | 7.22E-06 |
| -4.68 | 2.647 | 302 | 20 | enet | 0.064 | 2.00E-06 | -4.51 | 6.48E-06 |
| 5.06 | 1.934 | 356 | 53 | enet | 0.074 | 3.50E-07 | -6.4402 | 1.19E-10 |
| -4.71 | 1.658 | 377 | 55 | enet | 0.13 | 6.10E-12 | -5.5189 | 3.41E-08 |
| -3.04 | 2.76 | 76 | 11 | enet | -0.0012 | 0.43 | 4.7446 | 2.09E-06 |
| 10.53 | 4.49 | 318 | 318 | susie | 0.38 | 7.00E-36 | 6.0527 | 1.42E-09 |
| -3.65 | -4.0343 | 359 | 4 | lasso | 0.027 | 0.0016 | 4.4487 | 8.64E-06 |
| 7.07 | 3.177 | 342 | 342 | susie | 0.48 | 2.40E-48 | 4.8631 | 1.16E-06 |
| -4.92 | -3.776 | 375 | 35 | enet | 0.054 | 1.30E-05 | 4.7774 | 1.78E-06 |
| 5.01 | 3.191 | 332 | 26 | enet | 0.061 | 3.40E-06 | 5.8261 | 5.67E-09 |
| 3.99 | -2.547 | 540 | 11 | enet | 0.0056 | 0.094 | -5.0053 | 5.58E-07 |
| -4.01 | 3.746 | 319 | 15 | enet | 0.015 | 0.057 | -4.33796 | 1.44E-05 |
| 7.35 | 4.4 | 356 | 1 | top1 | 0.3 | 1.80E-15 | 4.4 | 1.08E-05 |
| 5.17 | 4.66 | 465 | 465 | susie | 0.094 | 2.20E-05 | 4.7804 | 1.75E-06 |
| -4.93 | -2.451 | 480 | 17 | enet | 0.18 | 3.90E-09 | 5.1315 | 2.87E-07 |
| -4.74 | 3.007 | 344 | 20 | enet | 0.11 | 2.90E-06 | -4.75 | 2.03E-06 |
| 5.36 | -2.194 | 255 | 30 | enet | 0.18 | 3.40E-09 | -4.3945 | 1.11E-05 |
| -5.2 | -3.593 | 293 | 18 | enet | 0.073 | 0.00017 | 4.4914 | 7.08E-06 |
| -5.43 | 4.15 | 628 | 628 | susie | 0.17 | 1.40E-08 | -4.3669 | 1.26E-05 |
| 4.8 | 1.73 | 534 | 20 | enet | 0.12 | 2.30E-06 | 5.5914 | 2.25E-08 |
| 4.11 | -4.024 | 518 | 23 | enet | 0.099 | 1.30E-05 | -5.5059 | 3.67E-08 |
| 5.23 | 4.528 | 464 | 464 | susie | 0.11 | 6.20E-06 | 4.5007 | 6.77E-06 |
| -4.33 | -4.534 | 408 | 6 | lasso | 0.043 | 0.0035 | 4.6848 | 2.80E-06 |
| -5.41 | 4.118 | 414 | 414 | susie | 0.16 | 2.70E-08 | -4.6303 | 3.65E-06 |
| 4.82 | -3.782 | 383 | 7 | lasso | 0.1 | 1.10E-05 | -4.3555 | 1.33E-05 |
| -6.66 | 4.331 | 431 | 16 | enet | 0.25 | 6.20E-13 | -4.926 | 8.39E-07 |
| 4.26 | 3.478 | 371 | 16 | enet | 0.073 | 0.00018 | 4.7424 | 2.11E-06 |

| | | | | | | | | |
|---|---|---|---|---|---|---|---|---|
| 5.14 | -4.494 | 430 | 430 | susie | 0.087 | 4.20E-05 | -5.1487 | 2.62E-07 |
| 4.34 | -3.629 | 433 | 15 | enet | 0.019 | 0.038 | -4.5434 | 5.53E-06 |
| 5.88 | -4.78 | 203 | 1 | top1 | 0.18 | 2.60E-09 | -4.78 | 1.75E-06 |
| 5.92 | 5.151 | 224 | 224 | susie | 0.28 | 3.30E-14 | 7.157 | 8.24E-13 |
| 6.4 | 4.954 | 224 | 224 | susie | 0.21 | 7.80E-11 | 5.1348 | 2.83E-07 |
| 3.36 | 3.656 | 386 | 386 | susie | 0.064 | 0.00041 | 5.7984 | 6.69E-09 |
| 6.54 | -3.641 | 471 | 471 | susie | 0.25 | 6.80E-13 | -4.68157 | 2.85E-06 |
| -5.48 | -4.594 | 472 | 472 | susie | 0.11 | 3.10E-06 | 4.41796 | 9.96E-06 |
| 7.16 | 5.576 | 118 | 1 | top1 | 0.29 | 1.40E-14 | 5.57628 | 2.46E-08 |
| 5.87 | -3.06 | 217 | 217 | susie | 0.24 | 2.00E-12 | 4.5852 | 4.53E-06 |
| 5.29 | 4.589 | 552 | 552 | susie | 0.11 | 6.40E-06 | 4.4109 | 1.03E-05 |
| -3.79 | -4.072 | 261 | 261 | susie | 0.023 | 0.024 | 4.6718 | 2.99E-06 |
| -4.82 | -4.652 | 262 | 1 | top1 | 0.12 | 2.40E-06 | 4.652 | 3.29E-06 |
| 3.64 | -3.596 | 253 | 253 | susie | 0.046 | 0.0025 | -4.744 | 2.10E-06 |
| -5.38 | -4.915 | 259 | 1 | top1 | 0.14 | 1.50E-07 | 4.915 | 8.88E-07 |
| -8.38 | -4.384 | 288 | 1 | top1 | 0.39 | 1.80E-20 | 4.384 | 1.17E-05 |
| -5.68 | -4.384 | 288 | 1 | top1 | 0.16 | 1.50E-08 | 4.384 | 1.17E-05 |
| 3.44 | 3.426 | 336 | 51 | enet | 0.021 | 0.032 | 4.9816 | 6.31E-07 |
| 4.94 | -4.568 | 236 | 1 | top1 | 0.11 | 3.70E-06 | -4.568 | 4.92E-06 |
| 4.26 | 1.89 | 498 | 38 | enet | 0.074 | 0.00015 | 4.9017 | 9.50E-07 |
| 3.6 | 2.11 | 259 | 31 | enet | 0.033 | 0.0094 | 5.78832 | 7.11E-09 |
| -5.36 | -0.9 | 306 | 30 | enet | 0.41 | 5.70E-22 | 4.3567 | 1.32E-05 |
| 5.08 | -3.255 | 375 | 375 | susie | 0.2 | 2.90E-10 | -5.8423 | 5.15E-09 |
| -4.55 | -4.258 | 344 | 6 | lasso | 0.19 | 1.20E-09 | 4.6923 | 2.70E-06 |
| -8.51 | -4.929 | 417 | 417 | susie | 0.42 | 3.00E-22 | 5.1071 | 3.27E-07 |
| -3.5 | 3.693 | 440 | 25 | enet | 0.02 | 0.033 | -5.1335 | 2.84E-07 |
| -5.67 | -4.357 | 294 | 294 | susie | 0.17 | 1.40E-08 | 4.3345 | 1.46E-05 |
| 5.06 | 4.493 | 303 | 1 | top1 | 0.12 | 2.10E-06 | 4.493 | 7.02E-06 |
| -5.09 | -4.444 | 669 | 1 | top1 | 0.14 | 2.80E-07 | 4.444 | 8.83E-06 |
| 5.85 | -3.914 | 552 | 28 | enet | 0.27 | 1.20E-13 | -5.0871 | 3.64E-07 |
| -7.36 | 3.839 | 597 | 597 | susie | 0.4 | 2.70E-21 | -5.2418 | 1.59E-07 |
| -4.2 | 5.641 | 458 | 1 | top1 | 0.03 | 0.013 | -5.641 | 1.69E-08 |
| -3.99 | -4.634 | 316 | 1 | top1 | 0.072 | 2.00E-04 | 4.634 | 3.59E-06 |
| 4.88 | -3.621 | 309 | 309 | susie | 0.12 | 2.10E-06 | -4.3994 | 1.09E-05 |
| 6.19 | 4.212 | 637 | 18 | enet | 0.17 | 5.60E-09 | 4.3767 | 1.20E-05 |
| 4.49 | -4.64 | 309 | 1 | top1 | 0.065 | 0.00037 | -4.6397 | 3.49E-06 |
| 4.12 | 2.86 | 642 | 18 | enet | 0.036 | 0.0071 | 4.5373 | 5.70E-06 |
| 3.66 | -3.38 | 314 | 7 | lasso | 0.0059 | 0.16 | -4.5168 | 6.28E-06 |
| 8.94 | -2.6 | 298 | 298 | susie | 0.59 | 1.40E-35 | -4.705 | 2.54E-06 |
| 4.39 | -3.23 | 465 | 9 | lasso | 0.047 | 0.0022 | -5.2072 | 1.92E-07 |
| 4.21 | 4.3 | 392 | 4 | lasso | 0.07 | 0.00025 | 5.8076 | 6.34E-09 |
| -5.47 | -4.841 | 345 | 345 | susie | 0.15 | 7.20E-08 | 4.5686 | 4.91E-06 |
| -4.02 | 4.609 | 475 | 12 | enet | 0.025 | 0.02 | -4.9537 | 7.28E-07 |
| 5.7 | 3.308 | 364 | 9 | enet | 0.12 | 4.10E-11 | 4.79935 | 1.59E-06 |
| 11.28 | -2.604 | 298 | 33 | enet | 0.63 | 2.10E-73 | -4.79604 | 1.62E-06 |

| | | | | | | | | |
|---|---|---|---|---|---|---|---|---|
| -6.54 | 2.888 | 307 | 43 | enet | 0.18 | 4.50E-16 | -5.31957 | 1.04E-07 |
| 6.8 | -3.148 | 364 | 364 | susie | 0.21 | 2.60E-18 | -4.52957 | 5.91E-06 |
| -3.15 | -1.153 | 254 | 254 | susie | 0.026 | 0.0019 | 4.7955 | 1.62E-06 |
| -7.19 | -4.595 | 260 | 1 | top1 | 0.14 | 1.00E-12 | 4.595 | 4.33E-06 |
| -4.31 | -4.652 | 263 | 263 | susie | 0.041 | 0.00013 | 4.6135 | 3.96E-06 |
| -4.8 | -4.595 | 263 | 1 | top1 | 0.05 | 2.60E-05 | 4.595 | 4.33E-06 |
| 4.3 | -4.598 | 254 | 3 | lasso | 0.037 | 0.00028 | -4.6315 | 3.63E-06 |
| -5.41 | -4.345 | 278 | 9 | lasso | 0.058 | 5.50E-06 | 4.586 | 4.52E-06 |
| 5.06 | -4.656 | 278 | 1 | top1 | 0.063 | 2.40E-06 | -4.656 | 3.22E-06 |
| -12.66 | -4.576 | 289 | 56 | enet | 0.5 | 8.40E-51 | 4.9077 | 9.22E-07 |
| -8.66 | -4.656 | 289 | 289 | susie | 0.23 | 5.60E-21 | 4.5353 | 5.75E-06 |
| 7.33 | -1.713 | 318 | 79 | enet | 0.15 | 9.20E-14 | 5.2807 | 1.29E-07 |
| 6.02 | -4.984 | 311 | 1 | top1 | 0.1 | 1.40E-09 | -4.984 | 6.23E-07 |
| 5.54 | -4.568 | 237 | 4 | enet | 0.083 | 6.10E-08 | -4.7612 | 1.92E-06 |
| -5.54 | -5.19 | 675 | 675 | susie | 0.067 | 1.20E-06 | 5.0568 | 4.26E-07 |
| 4.32 | -2.55 | 383 | 28 | enet | 0.028 | 0.0013 | -4.5523 | 5.31E-06 |
| -3.66 | 1.363 | 345 | 51 | enet | 0.063 | 2.30E-06 | -4.9144 | 8.90E-07 |
| -4.94 | -3.453 | 464 | 464 | susie | 0.06 | 4.00E-06 | 4.6513 | 3.30E-06 |
| -4.73 | 6.28 | 366 | 8 | enet | 0.059 | 5.30E-06 | -5.1869 | 2.14E-07 |
| -4.09 | 3.517 | 357 | 10 | lasso | 0.011 | 0.03 | -4.8141 | 1.48E-06 |
| 3.9 | -3.823 | 545 | 545 | susie | 0.015 | 0.016 | -4.602 | 4.19E-06 |
| 8.59 | 4.753 | 467 | 1 | top1 | 0.22 | 8.80E-20 | 4.753 | 2.00E-06 |
| 6.36 | -4.949 | 375 | 1 | top1 | 0.11 | 4.80E-10 | -4.949 | 7.46E-07 |
| 6.7 | -2.277 | 401 | 101 | enet | 0.27 | 1.00E-24 | -5.3399 | 9.30E-08 |
| 6.67 | -2.896 | 505 | 505 | susie | 0.21 | 5.30E-19 | -4.6618 | 3.13E-06 |
| -4.23 | 3.056 | 489 | 58 | enet | 0.068 | 1.00E-06 | 8.329 | 8.15E-17 |
| 9.9 | 3.213 | 232 | 232 | susie | 0.4 | 3.90E-38 | 5.73512 | 9.74E-09 |
| -5.23 | 4.035 | 295 | 295 | susie | 0.093 | 9.10E-09 | -4.72445 | 2.31E-06 |
| -5.63 | 4.263 | 327 | 327 | susie | 0.087 | 2.90E-08 | -4.7194 | 2.37E-06 |
| 10.79 | 4.49 | 317 | 317 | susie | 0.49 | 2.90E-50 | 4.53528 | 5.75E-06 |
| -4.5 | -7.366 | 317 | 317 | susie | 0.013 | 0.023 | 7.23903 | 4.52E-13 |
| 3.96 | 2.94 | 313 | 46 | enet | 0.018 | 0.0082 | 4.86056 | 1.17E-06 |
| 5.44 | 7.594 | 211 | 7 | lasso | 0.11 | 2.00E-10 | 7.56198 | 3.97E-14 |
| 7.29 | -2.651 | 461 | 461 | susie | 0.25 | 1.60E-22 | -5.165 | 2.40E-07 |
| 4.58 | -2.434 | 298 | 298 | susie | 0.17 | 3.40E-15 | 5.914 | 3.35E-09 |
| 4.74 | 4.615 | 514 | 514 | susie | 0.045 | 5.90E-05 | 4.669 | 3.03E-06 |
| 3.86 | -4.7165 | 291 | 1 | top1 | 0.013 | 0.022 | -4.716 | 2.40E-06 |
| 9.7 | -4.6449 | 286 | 1 | top1 | 0.28 | 1.80E-25 | -4.645 | 3.40E-06 |
| -7.19 | -2.745 | 486 | 486 | susie | 0.18 | 6.00E-16 | 4.6766 | 2.92E-06 |
| 6.06 | -6.358 | 352 | 4 | lasso | 0.11 | 2.30E-10 | -4.5747 | 4.77E-06 |
| -9.68 | 3.839 | 598 | 49 | enet | 0.38 | 1.10E-35 | -4.533 | 5.81E-06 |
| 7.55 | -3.585 | 526 | 526 | susie | 0.32 | 2.50E-29 | -7.1351 | 9.67E-13 |
| 5.83 | -4.844 | 338 | 1 | top1 | 0.097 | 4.90E-09 | -4.8436 | 1.28E-06 |
| -7.71 | 4.679 | 333 | 333 | susie | 0.2 | 6.00E-18 | -4.9548 | 7.24E-07 |
| -3.82 | -3.343 | 296 | 296 | susie | 0.014 | 0.018 | 5.0775 | 3.82E-07 |

| | | | | | | | | |
|---|---|---|---|---|---|---|---|---|
| 8.17 | 3.13 | 310 | 34 | enet | 0.2 | 3.40E-18 | 4.5706 | 4.86E-06 |
| 4.7 | -3.37 | 390 | 4 | lasso | 0.079 | 1.30E-07 | -4.7848 | 1.71E-06 |
| 3.87 | -4.692 | 320 | 1 | top1 | 0.026 | 0.0019 | -4.692 | 2.71E-06 |
| -8.29 | -5.088 | 347 | 7 | lasso | 0.23 | 9.50E-21 | 5.8957 | 3.73E-09 |
| -5.67 | -4.25 | 390 | 27 | enet | 0.077 | 1.90E-07 | 4.5102 | 6.48E-06 |
| 11.68 | 5.454 | 531 | 6 | enet | 0.41 | 1.70E-39 | 5.4444 | 5.20E-08 |
| 4.07 | 2.75 | 604 | 28 | enet | 0.026 | 0.0021 | 5.1431 | 2.70E-07 |
| -7.82 | 4.014 | 355 | 355 | susie | 0.36 | 2.40E-33 | 4.5188 | 6.22E-06 |
| -9.15 | 4.736 | 387 | 45 | enet | 0.47 | 1.10E-46 | -4.5355 | 5.75E-06 |
| -6.94 | -4.423 | 408 | 22 | enet | 0.15 | 2.00E-13 | 4.6931 | 2.69E-06 |
| 4.35 | -3.033 | 508 | 16 | enet | 0.051 | 2.20E-05 | -4.8755 | 1.09E-06 |
| 5.26 | 2.547 | 340 | 340 | susie | 0.18 | 3.30E-16 | 4.9413 | 7.76E-07 |
| 9.97 | 4.158 | 769 | 37 | enet | 0.35 | 1.00E-32 | 5.7027 | 1.18E-08 |
| 8.9 | -4.923 | 376 | 376 | susie | 0.22 | 2.10E-19 | -4.9127 | 8.98E-07 |
| -5.01 | 4.487 | 518 | 1 | top1 | 0.062 | 2.90E-06 | -4.487 | 7.22E-06 |
| -4.05 | -2.862 | 449 | 8 | enet | 0.013 | 0.022 | 4.7445 | 2.09E-06 |
| -7.98 | -0.9002 | 305 | 26 | enet | 0.44 | 1.30E-43 | 5.27761 | 1.31E-07 |
| 6.11 | -3.255 | 373 | 373 | susie | 0.18 | 3.20E-16 | -6.16599 | 7.00E-10 |
| -5.3 | -3.071 | 482 | 14 | enet | 0.12 | 1.50E-10 | 4.89025 | 1.01E-06 |
| 7.19 | 3.8 | 426 | 21 | enet | 0.19 | 1.50E-16 | 5.22599 | 1.73E-07 |
| 7.87 | -3.837 | 535 | 535 | susie | 0.22 | 2.20E-19 | -4.9694 | 6.72E-07 |
| -3.81 | 2.472 | 480 | 8 | lasso | 0.021 | 0.0052 | -4.64647 | 3.38E-06 |
| -3.64 | -5.732 | 428 | 428 | susie | 0.011 | 0.03 | 4.95578 | 7.20E-07 |
| -10.71 | -4.929 | 415 | 5 | lasso | 0.38 | 2.00E-36 | 4.66301 | 3.12E-06 |
| -3.95 | -3.216 | 434 | 18 | enet | 0.027 | 0.0016 | 4.67044 | 3.01E-06 |
| -5.17 | 4.701 | 482 | 1 | top1 | 0.072 | 5.10E-07 | -4.701 | 2.59E-06 |
| -5.64 | 7.8994 | 591 | 4 | lasso | 0.092 | 1.10E-08 | -6.30835 | 2.82E-10 |
| 2.93 | -3.689 | 364 | 364 | susie | 0.01 | 0.038 | -4.68685 | 2.77E-06 |
| -3.68 | 4.151 | 170 | 5 | lasso | 0.016 | 0.012 | -4.79627 | 1.62E-06 |
| 9.06 | -4.78 | 203 | 29 | enet | 0.26 | 4.90E-23 | -5.41594 | 6.10E-08 |
| -5.12 | 5.2 | 215 | 21 | enet | 0.064 | 2.20E-06 | -5.87862 | 4.14E-09 |
| 10.15 | 4.954 | 225 | 1 | top1 | 0.31 | 1.30E-28 | 4.954 | 7.27E-07 |
| -8.03 | 5.072 | 378 | 1 | top1 | 0.19 | 6.30E-17 | -5.072 | 3.94E-07 |
| 9.96 | 3.17 | 581 | 581 | susie | 0.37 | 2.20E-34 | 4.8574 | 1.19E-06 |
| 6.41 | 2.59 | 512 | 512 | susie | 0.13 | 1.40E-11 | 5.105 | 3.31E-07 |
| 5.33 | 3.69 | 400 | 28 | enet | 0.087 | 2.90E-08 | 5.0796 | 3.78E-07 |
| -4.7 | 3.19 | 343 | 32 | enet | 0.053 | 1.40E-05 | -5.248 | 1.54E-07 |
| 5.87 | -4.8 | 431 | 1 | top1 | 0.096 | 5.80E-09 | -4.798 | 1.60E-06 |
| 8.33 | 2.109 | 622 | 622 | susie | 0.36 | 4.90E-34 | 5.1349 | 2.82E-07 |
| 4.21 | -2.899 | 625 | 8 | enet | 0.0042 | 0.12 | -5.0373 | 4.72E-07 |
| 4.7 | 2.601 | 524 | 10 | enet | 0.029 | 0.0011 | 5.199 | 2.00E-07 |
| -4.23 | 3.539 | 381 | 381 | susie | 0.037 | 0.00026 | -4.8705 | 1.11E-06 |
| -4.11 | 2.601 | 342 | 45 | enet | 0.042 | 0.00011 | -6.57041 | 5.02E-11 |
| 4.82 | -4.117 | 480 | 4 | lasso | 0.052 | 1.70E-05 | -4.85264 | 1.22E-06 |
| 4.15 | -2.255 | 141 | 21 | enet | 0.021 | 0.0048 | -4.91622 | 8.82E-07 |

| | | | | | | | | |
|---|---|---|---|---|---|---|---|---|
| 4.08 | 3.998 | 154 | 22 | enet | 0.036 | 0.00029 | 5.56531 | 2.62E-08 |
| 11.32 | 5.5763 | 118 | 4 | lasso | 0.41 | 2.80E-39 | 5.33517 | 9.55E-08 |
| 4.18 | 5.7095 | 166 | 1 | top1 | 0.043 | 9.60E-05 | 5.70951 | 1.13E-08 |
| -4.74 | -2.194 | 328 | 28 | enet | 0.06 | 4.20E-06 | 4.63951 | 3.49E-06 |
| -5.95 | -3.342 | 426 | 24 | enet | 0.1 | 1.10E-09 | 4.70144 | 2.58E-06 |
| -6.55 | -3.48 | 330 | 330 | susie | 0.18 | 3.20E-16 | 5.29188 | 1.21E-07 |
| -5.49 | -1.81 | 484 | 34 | enet | 0.12 | 1.10E-10 | 5.155 | 2.54E-07 |
| 5.07 | -2.452 | 682 | 22 | enet | 0.05 | 2.50E-05 | -5.77452 | 7.72E-09 |
| 7.67 | -3.321 | 352 | 8 | lasso | 0.21 | 2.40E-18 | -4.66204 | 3.13E-06 |
| -7.74 | -4.841 | 345 | 345 | susie | 0.17 | 1.70E-15 | 4.74414 | 2.09E-06 |
| -5.8 | 3.067 | 570 | 35 | enet | 0.13 | 1.40E-11 | -4.58567 | 4.53E-06 |
| -4.13 | -1.607 | 521 | 28 | enet | 0.025 | 0.0024 | 4.5925 | 4.38E-06 |
| 13.72 | 3.545 | 307 | 307 | susie | 0.42 | 6.20E-58 | 4.9496 | 7.44E-07 |
| 3.41 | 3.284 | 296 | 296 | susie | 0.0046 | 0.075 | 4.8464 | 1.26E-06 |
| -8.69 | -3.331 | 286 | 17 | enet | 0.17 | 1.30E-21 | 4.6271 | 3.71E-06 |
| 11.05 | -4.69 | 379 | 379 | susie | 0.25 | 5.70E-32 | -4.7423 | 2.11E-06 |
| -9.94 | 3.351 | 482 | 23 | enet | 0.24 | 1.10E-30 | -4.7206 | 2.35E-06 |
| -4.45 | 2.361 | 386 | 21 | enet | 0.04 | 6.70E-06 | -6.0052 | 1.91E-09 |
| 6 | -3.884 | 451 | 22 | enet | 0.065 | 1.10E-08 | -5.3727 | 7.76E-08 |
| 5.41 | 1.678 | 438 | 41 | enet | 0.063 | 1.90E-08 | 4.7712 | 1.83E-06 |
| 10.13 | -2.498 | 460 | 460 | susie | 0.31 | 4.60E-40 | -4.87 | 1.14E-06 |
| -6.2 | -2.799 | 403 | 24 | enet | 0.079 | 2.50E-10 | 5.47 | 4.58E-08 |
| 10.91 | -4.0119 | 286 | 31 | enet | 0.26 | 9.80E-33 | -4.85 | 1.25E-06 |
| 4.38 | -13.9217 | 311 | 311 | susie | 0.0036 | 0.1 | -13.8 | 4.42E-43 |
| 11.94 | 2.58 | 505 | 505 | susie | 0.42 | 8.20E-59 | 5.13 | 2.95E-07 |
| 3.33 | -2.901 | 147 | 21 | enet | 0.025 | 0.00028 | 4.540109 | 5.62E-06 |
| 10.67 | 3.213 | 232 | 232 | susie | 0.32 | 4.40E-41 | 5.592735 | 2.24E-08 |
| -4.11 | 4.035 | 294 | 9 | lasso | 0.012 | 0.0088 | -5.13675 | 2.80E-07 |
| 8.79 | -3.905 | 320 | 320 | susie | 0.19 | 5.50E-24 | -5.02464 | 5.04E-07 |
| -7.35 | 3.709 | 360 | 19 | enet | 0.11 | 1.90E-13 | -4.6388 | 3.50E-06 |
| 4.6 | -7.188 | 352 | 14 | enet | 0.034 | 2.80E-05 | -4.97563 | 6.50E-07 |
| -5.28 | 3.047 | 365 | 22 | enet | 0.055 | 1.50E-07 | -4.65549 | 3.23E-06 |
| -4.25 | -2.692 | 330 | 33 | enet | 0.013 | 0.0081 | 5.830231 | 5.54E-09 |
| -6.42 | 2.847 | 437 | 61 | enet | 0.13 | 7.60E-17 | -6.62296 | 3.52E-11 |
| 4.66 | 2.035 | 365 | 15 | enet | 0.072 | 1.50E-09 | 5.3273 | 9.97E-08 |
| 14.91 | -2.604 | 298 | 298 | susie | 0.62 | 5.80E-103 | -4.6193 | 3.85E-06 |
| -8.25 | 2.456 | 307 | 307 | susie | 0.22 | 1.50E-27 | -4.6362 | 3.55E-06 |
| 8.81 | -4.175 | 467 | 467 | susie | 0.19 | 2.30E-23 | -5.1047 | 3.31E-07 |
| -6.35 | -4.023 | 380 | 380 | susie | 0.075 | 7.90E-10 | 4.9019 | 9.49E-07 |
| 4.59 | 3.819 | 376 | 376 | susie | 0.2 | 1.20E-24 | 9.4516 | 3.34E-21 |
| 6.8 | 3.152 | 377 | 377 | susie | 0.15 | 6.00E-19 | 7.4956 | 6.60E-14 |
| -3.61 | 2.305 | 170 | 17 | enet | 0.038 | 1.20E-05 | -5.0029 | 5.65E-07 |
| -8.45 | 1.726 | 470 | 470 | susie | 0.3 | 1.70E-39 | -5.241 | 1.60E-07 |
| 7.34 | -3.148 | 364 | 364 | susie | 0.16 | 3.50E-20 | -4.5446 | 5.50E-06 |
| -3.95 | 2.871 | 326 | 4 | lasso | 0.037 | 1.40E-05 | -4.5336 | 5.80E-06 |

| | | | | | | | | |
|---|---|---|---|---|---|---|---|---|
| 4.1 | -4.854 | 417 | 417 | susie | 0.0095 | 0.019 | -4.7161 | 2.40E-06 |
| 3.97 | -2.214 | 348 | 5 | lasso | 0.0086 | 0.024 | -5.0162 | 5.27E-07 |
| -3.9 | -4.081 | 398 | 4 | lasso | 0.015 | 0.0039 | 5.0046 | 5.60E-07 |
| -4.37 | -4.034 | 388 | 388 | susie | 0.027 | 0.00017 | 5.2178 | 1.81E-07 |
| 8.14 | -4.403 | 311 | 311 | susie | 0.17 | 5.20E-21 | -4.9597 | 7.06E-07 |
| 10.35 | -4.571 | 319 | 23 | enet | 0.27 | 3.00E-34 | -5.5973 | 2.18E-08 |
| -4.83 | -4.034 | 555 | 8 | lasso | 0.028 | 0.00014 | 5.5381 | 3.06E-08 |
| 12.35 | 5.576 | 118 | 118 | susie | 0.33 | 8.60E-43 | 5.2313 | 1.68E-07 |
| 3.74 | -2.595 | 143 | 34 | enet | 0.041 | 4.70E-06 | -7.8562 | 3.96E-15 |
| 6.26 | 5.71 | 166 | 166 | susie | 0.071 | 2.00E-09 | 5.8451 | 5.06E-09 |
| -5.53 | -4.454 | 298 | 3 | lasso | 0.058 | 5.50E-08 | 4.5397 | 5.63E-06 |
| 6.51 | 4.788 | 312 | 17 | enet | 0.086 | 4.10E-11 | 4.8714 | 1.11E-06 |
| 4.86 | -4.611 | 512 | 27 | enet | 0.062 | 2.20E-08 | -6.1951 | 5.82E-10 |
| -10.27 | 3.609 | 466 | 43 | enet | 0.23 | 1.70E-28 | -4.6368 | 3.54E-06 |
| -5.25 | 6.94 | 506 | 506 | susie | 0.041 | 5.50E-06 | -6.5107 | 7.48E-11 |
| 5.06 | 2.154 | 402 | 50 | enet | 0.078 | 3.00E-10 | 6.0306 | 1.63E-09 |
| -9.04 | -2.178 | 343 | 343 | susie | 0.45 | 2.00E-64 | 4.78503 | 1.71E-06 |
| 6.69 | -5.14 | 688 | 1 | top1 | 0.085 | 5.10E-11 | -5.14 | 2.75E-07 |
| 4.05 | 3.912 | 276 | 17 | enet | 0.021 | 0.00082 | 6.22455 | 4.83E-10 |
| -7.57 | -2.801 | 576 | 576 | susie | 0.17 | 1.20E-21 | 5.30651 | 1.12E-07 |
| -9.68 | -0.90025 | 306 | 28 | enet | 0.42 | 1.20E-57 | 5.0538 | 4.33E-07 |
| 8.45 | -3.276 | 374 | 34 | enet | 0.22 | 2.30E-27 | -4.93902 | 7.85E-07 |
| 10.46 | -4.258 | 353 | 40 | enet | 0.3 | 3.60E-38 | -6.16358 | 7.11E-10 |
| 8.03 | 3.905 | 344 | 16 | enet | 0.13 | 9.30E-17 | 4.83738 | 1.32E-06 |
| 9.7 | 3.8 | 425 | 425 | susie | 0.26 | 7.50E-33 | 5.49961 | 3.81E-08 |
| 12.56 | -3.666 | 501 | 25 | enet | 0.36 | 1.90E-48 | -4.66211 | 3.13E-06 |
| -4.04 | 2.652 | 506 | 14 | enet | 0.015 | 0.0044 | -5.4173 | 6.05E-08 |
| 10.45 | -2.338 | 547 | 547 | susie | 0.27 | 3.80E-34 | -5.62105 | 1.90E-08 |
| -7.48 | -5.731 | 332 | 1 | top1 | 0.12 | 1.40E-14 | 5.731 | 9.98E-09 |
| 5.4 | 2.651 | 484 | 25 | enet | 0.052 | 3.10E-07 | 4.66559 | 3.08E-06 |
| -15.25 | -4.929 | 415 | 13 | lasso | 0.51 | 5.70E-76 | 4.89717 | 9.72E-07 |
| 9.12 | 4.449 | 303 | 303 | susie | 0.18 | 2.00E-22 | 4.78324 | 1.72E-06 |
| 6.14 | -4.58 | 449 | 449 | susie | 0.073 | 1.40E-09 | -4.59358 | 4.36E-06 |
| 5.19 | -3.638 | 566 | 566 | susie | 0.068 | 4.60E-09 | -4.56867 | 4.91E-06 |
| 4.04 | -2.285 | 468 | 28 | enet | 0.016 | 0.0035 | -4.55042 | 5.35E-06 |
| -4.05 | -3.165 | 448 | 40 | enet | 0.013 | 0.0066 | 7.2534 | 4.06E-13 |
| -5.99 | 7.89939 | 591 | 33 | enet | 0.1 | 1.00E-12 | -4.84138 | 1.29E-06 |
| 4.35 | 2.246 | 510 | 27 | enet | 0.049 | 5.90E-07 | 5.4294 | 5.65E-08 |
| 8.59 | 2.702 | 806 | 28 | enet | 0.39 | 3.20E-53 | 5.1127 | 3.18E-07 |
| -4.57 | 2.228 | 822 | 18 | enet | 0.023 | 0.00054 | -4.7533 | 2.00E-06 |
| -11.65 | 3.377 | 434 | 434 | susie | 0.36 | 7.80E-49 | -4.9123 | 9.00E-07 |
| 7.96 | -2.782 | 507 | 507 | susie | 0.34 | 1.70E-45 | -5.8853 | 3.97E-09 |
| 10.64 | -3.513 | 315 | 315 | susie | 0.25 | 8.60E-32 | -4.8014 | 1.58E-06 |
| -8.37 | 4.582 | 446 | 446 | susie | 0.13 | 1.00E-16 | -4.5512 | 5.33E-06 |
| -4.23 | 5.024 | 183 | 183 | susie | 0.022 | 0.00071 | -5.3479 | 8.90E-08 |

| | | | | | | | | |
|---|---|---|---|---|---|---|---|---|
| 11.31 | -4.78 | 203 | 21 | enet | 0.26 | 2.20E-33 | -4.565 | 4.99E-06 |
| -4.37 | 5.093 | 215 | 215 | susie | 0.028 | 0.00013 | -5.1723 | 2.31E-07 |
| 7.74 | 5.069 | 225 | 225 | susie | 0.12 | 1.70E-15 | 6.4528 | 1.10E-10 |
| 13.4 | 4.954 | 225 | 4 | lasso | 0.37 | 3.90E-50 | 5.1045 | 3.32E-07 |
| 5.45 | -4.353 | 226 | 59 | enet | 0.13 | 4.80E-16 | 5.8855 | 3.97E-09 |
| -7.07 | -5.6689 | 209 | 209 | susie | 0.1 | 6.90E-13 | 5.5225 | 3.34E-08 |
| -6.63 | -2.3619 | 323 | 31 | enet | 0.12 | 1.90E-15 | 5.9057 | 3.51E-09 |
| -5.55 | -5.03 | 252 | 1 | top1 | 0.061 | 2.80E-08 | 5.03 | 4.90E-07 |
| 9.58 | -3.509 | 276 | 40 | enet | 0.33 | 1.10E-42 | -5.4061 | 6.44E-08 |
| 11.53 | 5.454 | 539 | 2 | lasso | 0.28 | 2.00E-35 | 5.4725 | 4.44E-08 |
| -3.25 | -2.779 | 313 | 22 | enet | 0.019 | 0.0014 | 4.9403 | 7.80E-07 |
| 8.53 | 2.88 | 464 | 464 | susie | 0.35 | 4.40E-46 | 5.0122 | 5.38E-07 |
| 10.54 | 3.161 | 360 | 85 | enet | 0.43 | 1.60E-60 | 6.218 | 5.03E-10 |
| 10.78 | -3.992 | 387 | 387 | susie | 0.49 | 1.30E-70 | -4.864 | 1.15E-06 |
| 3.36 | 3.582 | 384 | 10 | enet | 0.0072 | 0.036 | 5.5093 | 3.60E-08 |
| -6.19 | 5.145 | 508 | 5 | lasso | 0.058 | 5.80E-08 | -5.3483 | 8.88E-08 |
| 9.6 | 4.753 | 465 | 1 | top1 | 0.19 | 5.70E-24 | 4.753 | 2.00E-06 |
| 6.4 | 4.824 | 463 | 463 | susie | 0.059 | 5.30E-08 | 4.5643 | 5.01E-06 |
| 4.9 | 3.734 | 476 | 5 | lasso | 0.05 | 4.50E-07 | 6.1024 | 1.05E-09 |
| -4.39 | -3.424 | 394 | 19 | enet | 0.015 | 0.0041 | 5.4121 | 6.23E-08 |
| -4.54 | 3.513 | 472 | 25 | enet | 0.039 | 7.90E-06 | -4.9363 | 7.96E-07 |
| 4.56 | -4.468 | 271 | 11 | enet | 0.024 | 0.00045 | -5.4741 | 4.40E-08 |
| -5.64 | -4.673 | 259 | 14 | enet | 0.054 | 1.70E-07 | 4.9077 | 9.22E-07 |
| -4.28 | -4.58 | 262 | 262 | susie | 0.027 | 2.00E-04 | 4.7644 | 1.89E-06 |
| 3.91 | -4.656 | 277 | 277 | susie | 0.019 | 0.0017 | -4.7814 | 1.74E-06 |
| -4.74 | -4.573 | 288 | 1 | top1 | 0.041 | 5.20E-06 | 4.573 | 4.81E-06 |
| -15.92 | -4.576 | 288 | 288 | susie | 0.54 | 2.60E-81 | 4.6892 | 2.74E-06 |
| -9.96 | -4.576 | 288 | 288 | susie | 0.24 | 6.50E-30 | 4.9669 | 6.80E-07 |
| 10.29 | -5.131 | 314 | 314 | susie | 0.22 | 6.00E-28 | -5.5955 | 2.20E-08 |
| 9.08 | -4.81 | 247 | 247 | susie | 0.16 | 2.10E-20 | -4.6721 | 2.98E-06 |
| -10.21 | -3.7 | 385 | 21 | enet | 0.21 | 7.10E-27 | 5.0834 | 3.71E-07 |
| 4.28 | 2.235 | 435 | 24 | enet | 0.016 | 0.0029 | 4.758 | 1.96E-06 |
| -6.24 | -2.496 | 514 | 73 | enet | 0.13 | 8.30E-16 | -4.6311 | 3.64E-06 |
| -4.69 | -3.669 | 465 | 20 | enet | 0.032 | 5.70E-05 | 6.3556 | 2.08E-10 |
| -4.99 | 1.744 | 508 | 508 | susie | 0.038 | 9.70E-06 | -6.35685 | 2.06E-10 |
| 5.78 | -3.437 | 338 | 8 | lasso | 0.066 | 8.10E-09 | -5.50937 | 3.60E-08 |
| -6.68 | 5.334 | 357 | 357 | susie | 0.089 | 1.70E-11 | -5.3342 | 9.60E-08 |
| -7.29 | -3.533 | 404 | 404 | susie | 0.11 | 8.20E-14 | 4.610755 | 4.01E-06 |
| -5.09 | -4.692 | 392 | 1 | top1 | 0.051 | 3.60E-07 | 4.692 | 2.71E-06 |
| 5.65 | 4.448 | 561 | 561 | susie | 0.058 | 6.90E-08 | 5.242407 | 1.58E-07 |
| -5.95 | 4.855 | 435 | 16 | enet | 0.061 | 3.20E-08 | -5.34614 | 8.98E-08 |
| 6.19 | 2.121 | 576 | 36 | enet | 0.14 | 8.40E-18 | 4.8077 | 1.53E-06 |
| -3.79 | 2.328 | 753 | 45 | enet | 0.0077 | 0.031 | -5.3982 | 6.73E-08 |
| 3.32 | 3.185 | 243 | 243 | susie | 0.015 | 0.0049 | 4.63344 | 3.60E-06 |
| -9.3 | 3.662 | 500 | 500 | susie | 0.19 | 2.80E-23 | -5.07051 | 3.97E-07 |

| | | | | | | | | |
|---|---|---|---|---|---|---|---|---|
| -4.85 | -2.982 | 358 | 44 | enet | 0.038 | 9.70E-06 | 6.28171 | 3.35E-10 |
| -13.17 | 3.84 | 598 | 598 | susie | 0.56 | 6.70E-86 | -5.49966 | 3.81E-08 |
| -4.31 | -2.082 | 357 | 39 | enet | 0.054 | 1.60E-07 | 4.6295 | 3.67E-06 |
| -6.66 | -3.308 | 531 | 26 | enet | 0.13 | 1.10E-16 | 5.3809 | 7.41E-08 |
| -4.13 | 2.336 | 356 | 42 | enet | 0.045 | 1.90E-06 | -5.5866 | 2.32E-08 |
| 9.13 | -3.585 | 526 | 526 | susie | 0.36 | 8.90E-49 | -7.2192 | 5.23E-13 |
| -7.23 | 3.293 | 333 | 7 | lasso | 0.12 | 2.60E-15 | -4.8249 | 1.40E-06 |
| -6.13 | -4.815 | 308 | 1 | top1 | 0.075 | 7.40E-10 | 4.815 | 1.47E-06 |
| -7.06 | -3.807 | 220 | 6 | lasso | 0.098 | 1.60E-12 | 4.7922 | 1.65E-06 |
| -5.66 | 4.889 | 346 | 1 | top1 | 0.061 | 2.50E-08 | -4.889 | 1.01E-06 |
| -4.22 | -4.075 | 346 | 4 | lasso | 0.04 | 7.00E-06 | 5.5158 | 3.47E-08 |
| 8.41 | -4.858 | 410 | 1 | top1 | 0.14 | 7.00E-18 | -4.858 | 1.19E-06 |
| -4.89 | -5.06 | 331 | 1 | top1 | 0.033 | 3.80E-05 | 5.06 | 4.19E-07 |
| -6.45 | 4.083 | 626 | 27 | enet | 0.12 | 3.00E-15 | -5.0849 | 3.68E-07 |
| -3.6 | -2.622 | 303 | 25 | enet | 0.024 | 0.00039 | 6.4283 | 1.29E-10 |
| 6.61 | -4.395 | 382 | 382 | susie | 0.093 | 6.40E-12 | -4.9635 | 6.92E-07 |
| 5.27 | -5.452 | 475 | 475 | susie | 0.041 | 5.30E-06 | -5.7449 | 9.20E-09 |
| 6.09 | -4.357 | 308 | 13 | enet | 0.071 | 8.70E-11 | -5.2102 | 1.89E-07 |
| -10.23 | -3.425 | 293 | 45 | enet | 0.19 | 6.20E-27 | 5.5657 | 2.61E-08 |
| 4.16 | -7.196 | 269 | 20 | enet | 0.039 | 1.60E-06 | -6.8179 | 9.24E-12 |
| 4.1 | -3.392 | 614 | 45 | enet | 0.03 | 2.60E-05 | 5.3314 | 9.74E-08 |
| 6.39 | 5.02 | 346 | 1 | top1 | 0.06 | 3.20E-09 | 5.02 | 5.17E-07 |
| 11.73 | 5.118 | 347 | 347 | susie | 0.26 | 1.70E-38 | 5.3002 | 1.16E-07 |
| 6.04 | -4.691 | 433 | 27 | enet | 0.08 | 6.60E-12 | -4.6048 | 4.13E-06 |
| -4.94 | -3.515 | 432 | 23 | enet | 0.033 | 1.00E-05 | 4.8359 | 1.33E-06 |
| -8.55 | -8.335 | 572 | 7 | lasso | 0.13 | 4.70E-19 | 6.6154 | 3.71E-11 |
| 9.91 | 3.073 | 406 | 7 | lasso | 0.27 | 6.10E-40 | 5.0187 | 5.20E-07 |
| -8.22 | 4.688 | 387 | 387 | susie | 0.24 | 8.30E-35 | -4.5313 | 5.86E-06 |
| -11.45 | -2.797 | 383 | 383 | susie | 0.27 | 6.30E-40 | 4.5737 | 4.79E-06 |
| -5.36 | -4.607 | 259 | 3 | lasso | 0.04 | 1.00E-06 | 4.59519 | 4.32E-06 |
| 7.17 | -4.468 | 253 | 253 | susie | 0.086 | 9.80E-13 | -4.71892 | 2.37E-06 |
| 3.93 | -4.887 | 253 | 1 | top1 | 0.023 | 0.00022 | -4.887 | 1.02E-06 |
| -13.2 | -4.702 | 288 | 38 | enet | 0.31 | 9.70E-47 | 4.68111 | 2.85E-06 |
| 5.28 | -4.439 | 247 | 247 | susie | 0.036 | 4.30E-06 | -4.74356 | 2.10E-06 |
| 8.19 | -2.444 | 369 | 369 | susie | 0.17 | 1.30E-24 | -4.53241 | 5.83E-06 |
| -6.6 | -2.711 | 437 | 31 | enet | 0.076 | 2.10E-11 | 4.79123 | 1.66E-06 |
| 4.23 | -3.732 | 436 | 18 | enet | 0.039 | 1.50E-06 | -6.57743 | 4.79E-11 |
| 5.27 | 3.356 | 269 | 45 | enet | 0.093 | 1.20E-13 | 5.06067 | 4.18E-07 |
| -11.89 | -4.518 | 339 | 1 | top1 | 0.24 | 9.70E-36 | 4.518 | 6.24E-06 |
| -9.65 | -4.42 | 478 | 478 | susie | 0.17 | 4.20E-24 | 4.52275 | 6.10E-06 |
| 5.8 | 3.625 | 673 | 36 | enet | 0.063 | 1.10E-09 | 5.397 | 6.78E-08 |
| 12.94 | -2.941 | 382 | 382 | susie | 0.34 | 8.50E-52 | -4.6467 | 3.37E-06 |
| 17.96 | -4.429 | 733 | 8 | lasso | 0.61 | 5.30E-116 | -5.05631 | 4.27E-07 |
| -5 | -4.573 | 321 | 11 | enet | 0.022 | 0.00026 | 5.12935 | 2.91E-07 |
| 4.99 | -2.408 | 456 | 93 | enet | 0.092 | 1.70E-13 | -4.77206 | 1.82E-06 |

| | | | | | | | | |
|---|---|---|---|---|---|---|---|---|
| -12.98 | -3.555 | 576 | 576 | susie | 0.34 | 6.80E-52 | 4.8806 | 1.06E-06 |
| 5.73 | 4.716 | 417 | 417 | susie | 0.048 | 8.90E-08 | 5.05331 | 4.34E-07 |
| 5.83 | -2.162 | 365 | 365 | susie | 0.22 | 7.30E-32 | 9.88261 | 4.95E-23 |
| -6.78 | -6.465 | 725 | 725 | susie | 0.11 | 3.20E-16 | 7.444668 | 9.72E-14 |
| 4.5 | -3.242 | 198 | 198 | susie | 0.093 | 9.30E-14 | 8.175985 | 2.93E-16 |
| 15.99 | 4.49 | 317 | 317 | susie | 0.5 | 5.00E-86 | 5.780845 | 7.43E-09 |
| 9.39 | -2.313 | 207 | 36 | enet | 0.22 | 1.40E-31 | -5.08791 | 3.62E-07 |
| -4.33 | 2.692 | 499 | 15 | enet | 0.019 | 0.00073 | -4.79301 | 1.64E-06 |
| -3.62 | -2.951 | 463 | 11 | enet | 0.003 | 0.1 | 5.027857 | 4.96E-07 |
| -7.87 | 2.692 | 431 | 431 | susie | 0.15 | 3.80E-22 | -4.5864 | 4.51E-06 |
| 4.66 | 3.227 | 620 | 13 | enet | 0.037 | 3.10E-06 | 4.53454 | 5.77E-06 |
| -5.15 | -3.741 | 372 | 13 | lasso | 0.065 | 6.30E-10 | 4.57422 | 4.78E-06 |
| -5.21 | 2.315 | 369 | 23 | enet | 0.053 | 2.40E-08 | -4.78978 | 1.67E-06 |
| -8.86 | -0.9002 | 306 | 306 | susie | 0.35 | 2.50E-54 | 5.80958 | 6.26E-09 |
| -5.49 | 2.234 | 487 | 487 | susie | 0.086 | 8.70E-13 | -4.6473 | 3.36E-06 |
| -3.58 | -2.79 | 291 | 31 | enet | 0.017 | 0.0012 | 5.4792 | 4.27E-08 |
| -4.46 | 3.932 | 249 | 12 | enet | 0.036 | 3.90E-06 | -5.01772 | 5.23E-07 |
| 3.79 | -1.705 | 325 | 35 | enet | 0.0037 | 0.081 | -5.02489 | 5.04E-07 |
| 4.9 | -5.2719 | 457 | 3 | lasso | 0.038 | 2.20E-06 | -5.0064 | 5.55E-07 |
| 4.75 | 7.7205 | 584 | 13 | enet | 0.012 | 0.0055 | 5.97308 | 2.33E-09 |
| 3.57 | 4.331 | 387 | 40 | enet | 0.022 | 0.00026 | 5.16931 | 2.35E-07 |
| 7 | 2.562 | 446 | 446 | susie | 0.21 | 6.60E-30 | 6.08574 | 1.16E-09 |
| -4.24 | 2.54 | 309 | 28 | enet | 0.01 | 0.01 | -6.23207 | 4.60E-10 |
| 4.99 | -4.468 | 351 | 3 | lasso | 0.031 | 1.70E-05 | -5.1471 | 2.65E-07 |
| -7.86 | -4.841 | 345 | 1 | top1 | 0.1 | 6.80E-15 | 4.841 | 1.29E-06 |
| -8.16 | -5.448 | 331 | 1 | top1 | 0.11 | 2.30E-16 | 5.448 | 5.09E-08 |
| -7.12 | -5.7838 | 455 | 15 | enet | 0.098 | 2.50E-14 | 4.61 | 3.94E-06 |
| 7.24 | 3.692 | 387 | 36 | enet | 0.14 | 1.20E-20 | 5.75 | 9.17E-09 |
| -8.29 | -3.163 | 439 | 439 | susie | 0.16 | 9.30E-24 | -5.05 | 4.32E-07 |
| 3.16 | 2.399 | 184 | 11 | enet | 0.0039 | 0.076 | 4.71 | 2.45E-06 |
| -5.59 | 3.456 | 165 | 14 | enet | 0.074 | 3.30E-11 | -4.58 | 4.70E-06 |
| -7.24 | 4.862 | 203 | 1 | top1 | 0.091 | 2.00E-13 | -4.86 | 1.16E-06 |
| -9.13 | -4.353 | 225 | 38 | enet | 0.18 | 1.80E-25 | 5.27 | 1.39E-07 |
| 8.4 | 4.954 | 225 | 225 | susie | 0.12 | 2.20E-17 | 5.16 | 2.47E-07 |
| 4.68 | 1.704 | 397 | 22 | enet | 0.041 | 9.20E-07 | 4.77 | 1.83E-06 |
| 8.43 | 3.329 | 645 | 645 | susie | 0.19 | 8.80E-27 | 4.5629 | 5.05E-06 |
| 11.69 | -3.425 | 636 | 636 | susie | 0.31 | 1.40E-47 | -5.3124 | 1.08E-07 |
| 4.31 | 4.098 | 217 | 6 | lasso | 0.031 | 1.50E-05 | 4.4967 | 6.90E-06 |
| -7.1 | -2.818 | 238 | 19 | enet | 0.12 | 1.00E-16 | 4.6933 | 2.69E-06 |
| -5.32 | -2.176 | 356 | 356 | susie | 0.16 | 3.80E-23 | 4.91383 | 8.93E-07 |
| -10.44 | 3.05 | 380 | 380 | susie | 0.34 | 2.90E-52 | -4.81242 | 1.49E-06 |
| 4.68 | -5.006 | 371 | 1 | top1 | 0.028 | 3.80E-05 | -5.006 | 5.56E-07 |
| -4.56 | 4.48 | 371 | 5 | enet | 0.023 | 0.00018 | -5.24395 | 1.57E-07 |
| 8.89 | 3.999 | 471 | 19 | enet | 0.15 | 1.20E-21 | 4.64402 | 3.42E-06 |
| -4.11 | -2.29 | 208 | 32 | enet | 0.039 | 1.50E-06 | 5.67513 | 1.39E-08 |

| | | | | | | | | |
|---|---|---|---|---|---|---|---|---|
| -4.94 | -0.9381 | 213 | 36 | enet | 0.033 | 9.30E-06 | 4.86506 | 1.14E-06 |
| 7.76 | 5.7572 | 118 | 118 | susie | 0.1 | 6.10E-15 | 5.69764 | 1.21E-08 |
| -11.34 | -4.524 | 297 | 1 | top1 | 0.23 | 2.60E-33 | 4.524 | 6.07E-06 |
| 5.36 | -4.421 | 302 | 3 | lasso | 0.046 | 1.80E-07 | -4.5268 | 5.99E-06 |
| -6.75 | 4.658 | 324 | 1 | top1 | 0.068 | 2.50E-10 | -4.658 | 3.19E-06 |
| -8.28 | -3.6423 | 384 | 384 | susie | 0.18 | 2.00E-26 | 4.756 | 1.97E-06 |
| -4.16 | 3.68 | 426 | 5 | lasso | 0.018 | 0.00075 | -4.8974 | 9.71E-07 |
| 7.6 | -1.354 | 510 | 27 | enet | 0.15 | 3.10E-22 | -4.58967 | 4.44E-06 |
| -9.33 | 3.582 | 433 | 19 | enet | 0.14 | 5.50E-20 | -4.49821 | 6.85E-06 |
| -5.57 | 4.914 | 384 | 1 | top1 | 0.046 | 2.00E-07 | -4.914 | 8.92E-07 |
| -3.38 | -2.316 | 227 | 18 | enet | -0.0011 | 0.53 | 5.3531 | 8.64E-08 |
| 6.01 | 1.878 | 300 | 300 | susie | 0.097 | 3.60E-14 | 5.3983 | 6.73E-08 |
| 6.12 | -3.185 | 289 | 12 | enet | 0.071 | 1.10E-10 | -4.7084 | 2.50E-06 |
| 3.5 | -4.8 | 285 | 285 | susie | 0.006 | 0.037 | -4.6489 | 3.34E-06 |
| 4.81 | 4.521 | 386 | 1 | top1 | 0.038 | 2.30E-06 | 4.521 | 6.15E-06 |
| -5.09 | 3.611 | 282 | 282 | susie | 0.054 | 1.90E-08 | -4.7648 | 1.89E-06 |
| 9.7 | 2.131 | 255 | 39 | enet | 0.28 | 1.90E-41 | -4.5437 | 5.53E-06 |
| 5.48 | 2.022 | 359 | 11 | enet | 0.062 | 1.70E-09 | 4.8719 | 1.11E-06 |
| 9.54 | 3.637 | 505 | 505 | susie | 0.3 | 8.90E-45 | 4.9511 | 7.38E-07 |
| 9.7 | 4.753 | 464 | 1 | top1 | 0.15 | 1.70E-21 | 4.753 | 2.00E-06 |
| 10.45 | -4.623 | 477 | 7 | lasso | 0.22 | 1.90E-31 | -5.08431 | 3.69E-07 |
| 4.25 | 2.074 | 334 | 27 | enet | 0.041 | 9.20E-07 | 4.50853 | 6.53E-06 |
| -4.34 | 3.531 | 158 | 158 | susie | 0.012 | 0.0059 | -4.49293 | 7.02E-06 |
| 5.61 | -3.26 | 458 | 6 | lasso | 0.041 | 9.00E-07 | -5.1374 | 2.79E-07 |
| -6.01 | 2.921 | 437 | 437 | susie | 0.067 | 3.40E-10 | -6.0104 | 1.85E-09 |
| -9.09 | -2.227 | 356 | 356 | susie | 0.32 | 3.00E-49 | 4.7355 | 2.18E-06 |
| -8.64 | -2.109 | 356 | 356 | susie | 0.24 | 7.70E-36 | 5.3173 | 1.05E-07 |
| -4.07 | 3.966 | 298 | 28 | enet | 0.023 | 2.00E-04 | -4.502 | 6.73E-06 |
| 9.79 | -3.75 | 397 | 45 | enet | 0.26 | 3.20E-38 | -4.6026 | 4.17E-06 |
| -4.57 | 4.348 | 303 | 23 | enet | 0.047 | 1.20E-07 | -6.4079 | 1.48E-10 |
| 6.96 | 3.152 | 377 | 377 | susie | 0.28 | 9.00E-42 | 9.1707 | 4.70E-20 |
| 3.96 | 2.481 | 337 | 21 | enet | 0.024 | 0.00012 | 4.6483 | 3.35E-06 |
| -7.72 | 2.712 | 504 | 33 | enet | 0.12 | 3.90E-17 | -5.2777 | 1.31E-07 |
| 7.51 | 4.494 | 338 | 1 | top1 | 0.092 | 1.60E-13 | 4.494 | 6.99E-06 |
| 8.95 | -1.482 | 378 | 53 | enet | 0.37 | 1.60E-57 | -4.97449 | 6.54E-07 |
| -3.86 | -3.569 | 433 | 433 | susie | 0.018 | 0.00089 | 4.63712 | 3.53E-06 |
| -4.24 | -2.897 | 443 | 11 | enet | 0.016 | 0.0018 | 4.88857 | 1.02E-06 |
| 5.52 | -3.687 | 565 | 565 | susie | 0.16 | 6.20E-18 | -6.3964 | 1.59E-10 |
| 4.84 | -3.059 | 382 | 37 | enet | 0.061 | 1.10E-07 | -5.0349 | 4.78E-07 |
| -8.13 | 2.407 | 492 | 492 | susie | 0.29 | 6.40E-35 | -5.264 | 1.41E-07 |
| 10.11 | 2.837 | 341 | 48 | enet | 0.32 | 6.50E-38 | 4.5 | 6.80E-06 |
| 8.53 | 3.796 | 343 | 343 | susie | 0.22 | 5.40E-25 | 4.6067 | 4.09E-06 |
| -3.54 | -3.165 | 466 | 4 | lasso | -0.0019 | 0.66 | 4.503 | 6.70E-06 |
| 13.01 | 4.49 | 319 | 319 | susie | 0.43 | 2.20E-55 | 5.9103 | 3.42E-09 |
| -5.19 | -3.957 | 671 | 23 | enet | 0.06 | 1.30E-07 | 5.32235 | 1.02E-07 |

| | | | | | | | | |
|---|---|---|---|---|---|---|---|---|
| -5.64 | 3.13 | 613 | 613 | susie | 0.075 | 3.80E-09 | -5.51051 | 3.58E-08 |
| -4.27 | 3.206 | 441 | 14 | enet | 0.039 | 1.90E-05 | -5.4262 | 5.76E-08 |
| 9.7 | 3.282 | 342 | 342 | susie | 0.29 | 5.30E-35 | 4.5761 | 4.74E-06 |
| -3.59 | 2.977 | 465 | 23 | enet | 0.012 | 0.012 | -4.5211 | 6.15E-06 |
| 4.22 | -2.792 | 580 | 21 | enet | 0.05 | 1.50E-06 | -4.7383 | 2.16E-06 |
| 4.63 | -3.246 | 314 | 12 | enet | 0.035 | 5.10E-05 | -4.5021 | 6.73E-06 |
| 4.66 | -2.011 | 435 | 32 | enet | 0.07 | 1.20E-08 | -5.5469 | 2.91E-08 |
| 6.96 | -3.406 | 351 | 11 | lasso | 0.12 | 2.60E-14 | -4.5475 | 5.43E-06 |
| -7.83 | -4.841 | 345 | 1 | top1 | 0.13 | 4.20E-15 | 4.841 | 1.29E-06 |
| -5.64 | -5.329 | 328 | 1 | top1 | 0.061 | 1.10E-07 | 5.329 | 9.88E-08 |
| 4.3 | 3.254 | 427 | 18 | enet | 0.11 | 7.00E-13 | 6.3863 | 1.70E-10 |
| -3.35 | -1.466 | 374 | 26 | enet | 0.039 | 1.70E-05 | 4.59583 | 4.31E-06 |
| -6.1 | -4.815 | 310 | 1 | top1 | 0.07 | 1.10E-08 | 4.815 | 1.47E-06 |
| 6.47 | 3.01 | 411 | 37 | enet | 0.093 | 4.40E-11 | 5.04898 | 4.44E-07 |
| 3.87 | 3.239 | 340 | 20 | enet | 0.0059 | 0.058 | 5.35087 | 8.75E-08 |
| -4.26 | -3.352 | 427 | 8 | lasso | 0.049 | 1.70E-06 | 5.29386 | 1.20E-07 |
| 5.44 | 7.25 | 446 | 446 | susie | 0.058 | 2.00E-07 | 6.3751 | 1.83E-10 |
| 4.6 | 4.532 | 350 | 1 | top1 | 0.037 | 3.10E-05 | 4.532 | 5.84E-06 |
| -7.8 | -4.898 | 347 | 3 | lasso | 0.13 | 1.00E-14 | 5.63713 | 1.73E-08 |
| -4.18 | -3.18 | 179 | 14 | enet | 0.036 | 4.10E-05 | 5.11099 | 3.20E-07 |
| 7.58 | -2.651 | 461 | 461 | susie | 0.18 | 9.80E-21 | -5.2861 | 1.25E-07 |
| 3.73 | -4.0119 | 286 | 286 | susie | 0.012 | 0.012 | -5.4234 | 5.85E-08 |
| 8.32 | -4.0119 | 286 | 286 | susie | 0.14 | 2.90E-16 | -4.5157 | 6.31E-06 |
| 7.57 | -3.119 | 476 | 5 | lasso | 0.17 | 1.10E-19 | -4.5552 | 5.23E-06 |
| 6.84 | 2.109 | 622 | 622 | susie | 0.13 | 3.30E-15 | 4.5251 | 6.04E-06 |
| 6.05 | -3.531 | 560 | 16 | enet | 0.067 | 2.60E-08 | -4.6142 | 3.95E-06 |
| -3.77 | -2.475 | 561 | 26 | enet | 0.019 | 0.0022 | 5.108 | 3.26E-07 |
| 9.16 | -5.452 | 475 | 475 | susie | 0.18 | 4.60E-21 | -5.4612 | 4.73E-08 |
| -3.93 | -2.654 | 383 | 5 | lasso | 0.039 | 1.80E-05 | 4.9366 | 7.95E-07 |
| 9.03 | 5.498 | 410 | 410 | susie | 0.21 | 2.60E-24 | 4.8857 | 1.03E-06 |
| 14.11 | -2.604 | 298 | 298 | susie | 0.64 | 3.00E-99 | -5.18807 | 2.12E-07 |
| -6.16 | 3.056 | 307 | 307 | susie | 0.19 | 2.40E-21 | -5.4602 | 4.76E-08 |
| -5.48 | 4.348 | 303 | 303 | susie | 0.075 | 3.60E-09 | -5.26186 | 1.43E-07 |
| 4.55 | 3.152 | 376 | 376 | susie | 0.26 | 7.90E-30 | 9.41609 | 4.68E-21 |
| 6.81 | -3.148 | 365 | 365 | susie | 0.15 | 8.00E-18 | -4.54646 | 5.46E-06 |
| 5.55 | 2.2959 | 349 | 19 | enet | 0.08 | 1.20E-09 | 4.61088 | 4.01E-06 |
| 6.41 | -4.696 | 418 | 1 | top1 | 0.08 | 1.20E-09 | -4.696 | 2.65E-06 |
| -8.15 | 2.323 | 467 | 467 | susie | 0.2 | 3.10E-23 | -4.79445 | 1.63E-06 |
| -4.16 | 2.669 | 442 | 442 | susie | 0.015 | 0.0054 | -4.57551 | 4.75E-06 |
| 7.85 | 4.603 | 553 | 553 | susie | 0.12 | 6.60E-14 | 4.54983 | 5.37E-06 |
| -4.35 | 5.072 | 512 | 2 | lasso | 0.04 | 1.60E-05 | -5.02983 | 4.91E-07 |
| -4.3 | -4.457 | 262 | 262 | susie | 0.025 | 0.00054 | 4.83237 | 1.35E-06 |
| 7.63 | -4.468 | 253 | 5 | lasso | 0.12 | 2.90E-14 | -4.90239 | 9.47E-07 |
| 5 | -4.673 | 262 | 10 | lasso | 0.043 | 7.50E-06 | -4.7744 | 1.80E-06 |
| 4.25 | 3.22 | 277 | 277 | susie | 0.026 | 0.00044 | 4.54119 | 5.59E-06 |

| | | | | | | | | |
|---|---|---|---|---|---|---|---|---|
| -14.98 | -4.658 | 288 | 288 | susie | 0.51 | 3.70E-70 | 4.66721 | 3.05E-06 |
| -6.11 | -3.966 | 287 | 287 | susie | 0.078 | 1.80E-09 | 5.71123 | 1.12E-08 |
| 5.7 | -4.984 | 289 | 1 | top1 | 0.067 | 2.60E-08 | -4.984 | 6.23E-07 |
| 7.73 | 2.656 | 378 | 34 | enet | 0.12 | 6.10E-14 | 4.52347 | 6.08E-06 |
| 5.74 | 4.82 | 485 | 1 | top1 | 0.07 | 1.20E-08 | 4.82 | 1.44E-06 |
| 6.11 | -4.046 | 335 | 6 | lasso | 0.089 | 1.30E-10 | -4.6427 | 3.44E-06 |
| -6.51 | 4.52 | 429 | 429 | susie | 0.088 | 1.80E-10 | -4.9924 | 5.96E-07 |
| 5.2 | 2.518 | 364 | 36 | enet | 0.11 | 5.30E-13 | 5.2401 | 1.61E-07 |
| 3.7 | 3.624 | 346 | 17 | enet | 0.027 | 0.00032 | 5.0423 | 4.60E-07 |
| 7.61 | -3.653 | 430 | 430 | susie | 0.13 | 1.60E-15 | -4.5381 | 5.68E-06 |
| -5.36 | 4.852 | 183 | 183 | susie | 0.046 | 3.70E-06 | -4.8947 | 9.85E-07 |
| 10.59 | -4.78 | 203 | 1 | top1 | 0.25 | 1.30E-29 | -4.78 | 1.75E-06 |
| -4.65 | 4.954 | 215 | 215 | susie | 0.037 | 3.40E-05 | -5.597 | 2.18E-08 |
| 11.63 | 4.954 | 225 | 1 | top1 | 0.31 | 7.60E-37 | 4.954 | 7.27E-07 |
| -5.76 | 3.355 | 226 | 226 | susie | 0.091 | 8.50E-11 | -6.5834 | 4.60E-11 |
| -4.73 | -5.6689 | 209 | 209 | susie | 0.045 | 4.70E-06 | 5.5862 | 2.32E-08 |
| -3.23 | -3.223 | 456 | 456 | susie | 0.015 | 0.006 | 4.6993 | 2.61E-06 |
| -4.74 | 1.575 | 966 | 54 | enet | 0.031 | 0.00014 | -7.562 | 3.97E-14 |
| 3.66 | -3.67 | 294 | 23 | enet | 0.013 | 0.0091 | -5.22 | 1.79E-07 |
| 7.17 | 5.373 | 539 | 3 | lasso | 0.12 | 9.90E-14 | 5.413 | 6.20E-08 |
| -7.87 | -8.335 | 571 | 571 | susie | 0.19 | 2.40E-21 | 7.2178 | 5.28E-13 |
| 11.41 | 4.199 | 555 | 555 | susie | 0.34 | 1.90E-41 | 5.4764 | 4.34E-08 |
| -5.2 | 2.96 | 232 | 25 | enet | 0.085 | 3.60E-10 | -4.7689 | 1.85E-06 |
| 8.99 | 3.161 | 360 | 71 | enet | 0.33 | 3.10E-39 | 6.5133 | 7.35E-11 |
| 11.37 | -3.918 | 387 | 387 | susie | 0.5 | 2.80E-67 | -4.8728 | 1.10E-06 |
| -7.91 | 1.887 | 750 | 750 | susie | 0.28 | 1.40E-32 | -5.9354 | 2.93E-09 |
| 3.08 | -1.951 | 404 | 39 | enet | 0.034 | 6.30E-05 | -5.8208 | 5.86E-09 |
| 7 | 2.157 | 426 | 34 | enet | 0.16 | 2.20E-18 | 4.69898 | 2.61E-06 |
| 10.35 | -2.719 | 431 | 73 | enet | 0.31 | 4.70E-37 | -4.56713 | 4.94E-06 |
| 9.8 | -2.719 | 435 | 54 | enet | 0.3 | 2.30E-35 | -5.27541 | 1.32E-07 |
| 5.73 | 4.753 | 463 | 463 | susie | 0.064 | 5.50E-08 | 5.37493 | 7.66E-08 |
| 3.75 | -3.811 | 277 | 277 | susie | -0.0011 | 0.47 | -4.72782 | 2.27E-06 |
| 4.38 | 1.561 | 505 | 20 | enet | 0.03 | 0.00016 | 4.81883 | 1.44E-06 |
| -7.43 | -2.23 | 364 | 364 | susie | 0.22 | 6.20E-25 | 5.35215 | 8.69E-08 |
| -4.47 | -2.306 | 252 | 46 | enet | 0.066 | 3.40E-08 | 6.87656 | 6.13E-12 |
| -8.62 | -0.9002 | 303 | 303 | susie | 0.36 | 7.30E-44 | 5.419365 | 5.98E-08 |
| 6.16 | -3.255 | 374 | 374 | susie | 0.17 | 1.80E-19 | -6.47721 | 9.34E-11 |
| -5.9 | 3.44 | 427 | 45 | enet | 0.094 | 3.40E-11 | -4.73557 | 2.18E-06 |
| 9.58 | -4.258 | 351 | 21 | enet | 0.25 | 1.40E-28 | -4.61021 | 4.02E-06 |
| 10.62 | 3.459 | 344 | 12 | enet | 0.27 | 3.40E-32 | 4.718033 | 2.38E-06 |
| -5.45 | -3.454 | 368 | 368 | susie | 0.061 | 1.20E-07 | 5.558044 | 2.73E-08 |
| 4.91 | 4.553 | 434 | 1 | top1 | 0.05 | 1.30E-06 | 4.553 | 5.29E-06 |
| -6.33 | -5.731 | 332 | 332 | susie | 0.076 | 3.00E-09 | 5.73101 | 9.98E-09 |
| -4.6 | -3.048 | 270 | 22 | enet | 0.067 | 2.70E-08 | 4.688224 | 2.76E-06 |
| 5.05 | 4.204 | 421 | 10 | lasso | 0.054 | 5.30E-07 | 5.847544 | 4.99E-09 |

| | | | | | | | | |
|---|---|---|---|---|---|---|---|---|
| -12.98 | -4.929 | 408 | 33 | enet | 0.48 | 1.40E-64 | 4.561552 | 5.08E-06 |
| 5.02 | -4.58 | 449 | 1 | top1 | 0.054 | 5.00E-07 | -4.58 | 4.65E-06 |
| 3.56 | -3.502 | 581 | 581 | susie | 0.03 | 0.00016 | -6.13494 | 8.52E-10 |
| 4.95 | 2.019 | 488 | 16 | enet | 0.041 | 1.10E-05 | 4.541152 | 5.59E-06 |
| 5.25 | 2.572 | 378 | 6 | lasso | 0.11 | 2.20E-12 | 4.820261 | 1.43E-06 |
| 4.7 | -4.506 | 320 | 7 | lasso | 0.04 | 1.60E-05 | -5.36989 | 7.88E-08 |
| -4.88 | 2.942 | 196 | 36 | enet | 0.072 | 6.90E-09 | -5.03358 | 4.81E-07 |
| 13.09 | 5.5763 | 118 | 10 | lasso | 0.39 | 1.00E-48 | 5.68157 | 1.33E-08 |
| 5.09 | 2.992 | 143 | 21 | enet | 0.043 | 7.10E-06 | 4.90641 | 9.28E-07 |
| -9.57 | -4.524 | 298 | 1 | top1 | 0.2 | 1.40E-23 | 4.524 | 6.07E-06 |
| -10.83 | -4.524 | 298 | 298 | susie | 0.26 | 5.00E-31 | 4.53121 | 5.86E-06 |
| 4.71 | -2.961 | 305 | 40 | enet | 0.033 | 8.70E-05 | -4.65126 | 3.30E-06 |
| -3.94 | 1.733 | 348 | 21 | enet | 0.02 | 0.0016 | -4.87254 | 1.10E-06 |
| -5.46 | -4.042 | 382 | 382 | susie | 0.06 | 1.40E-07 | 4.6919 | 2.71E-06 |
| 4.1 | 4.713 | 417 | 1 | top1 | 0.023 | 0.00081 | 4.713 | 2.44E-06 |
| -3.36 | -2.347 | 655 | 10 | lasso | 0.0036 | 0.11 | 4.91666 | 8.80E-07 |
| 6.41 | 2.583 | 364 | 364 | susie | 0.12 | 1.20E-13 | -7.56971 | 3.74E-14 |
| 4.54 | 2.851 | 366 | 17 | enet | 0.027 | 0.00019 | 5.01103 | 5.41E-07 |
| -5.5 | 5.364 | 486 | 1 | top1 | 0.049 | 6.10E-07 | -5.364 | 8.14E-08 |
| -4.85 | -4.148 | 226 | 226 | susie | 0.038 | 1.20E-05 | 4.55495 | 5.24E-06 |
| 4.39 | -5.039 | 339 | 1 | top1 | 0.025 | 0.00034 | -5.03851 | 4.69E-07 |
| -3.86 | 2.503 | 584 | 31 | enet | 0.0093 | 0.02 | -5.37066 | 7.84E-08 |
| -7.95 | -3.352 | 427 | 427 | susie | 0.16 | 1.20E-20 | 4.53286 | 5.82E-06 |
| 3.8 | -3.809 | 203 | 8 | enet | 0.0037 | 0.095 | -4.81581 | 1.47E-06 |
| 4.16 | 6.129 | 446 | 11 | enet | 0.036 | 1.70E-05 | 6.01168 | 1.84E-09 |
| -12 | -4.898 | 346 | 1 | top1 | 0.3 | 1.10E-38 | 4.898 | 9.68E-07 |
| 5.81 | -5.126 | 409 | 15 | enet | 0.09 | 1.40E-11 | -5.60036 | 2.14E-08 |
| 8.11 | -3.417 | 433 | 37 | enet | 0.23 | 1.80E-29 | -5.75767 | 8.53E-09 |
| 4.28 | 2.589 | 317 | 62 | enet | 0.1 | 4.90E-13 | 7.5818 | 3.41E-14 |
| 4.34 | 2.75 | 129 | 6 | enet | 0.016 | 0.0034 | 4.6969 | 2.64E-06 |
| -7.62 | -4.841 | 345 | 1 | top1 | 0.12 | 7.30E-15 | 4.841 | 1.29E-06 |
| -8.43 | 5.465 | 331 | 1 | top1 | 0.15 | 2.00E-18 | -5.465 | 4.63E-08 |
| 6.06 | -3.71 | 497 | 12 | enet | 0.051 | 3.30E-07 | -4.91393 | 8.93E-07 |
| -4.88 | -4.527 | 406 | 1 | top1 | 0.044 | 2.30E-06 | 4.52684 | 5.99E-06 |
| -5.73 | 0.451 | 495 | 47 | enet | 0.12 | 9.60E-15 | 7.831 | 4.84E-15 |
| 5.46 | -2.178 | 389 | 389 | susie | 0.05 | 4.50E-07 | 4.67871 | 2.89E-06 |
| -3.91 | -2.648 | 545 | 34 | enet | 0.0099 | 0.017 | 4.94831 | 7.49E-07 |
| -6.38 | -3.414 | 398 | 398 | susie | 0.13 | 2.60E-16 | 4.53364 | 5.80E-06 |
| -5.62 | -2.124 | 272 | 54 | enet | 0.072 | 1.40E-09 | -6.35422 | 2.09E-10 |
| 6.68 | -3.203 | 405 | 23 | enet | 0.081 | 1.20E-10 | -4.69232 | 2.70E-06 |
| 7.04 | -2.492 | 438 | 438 | susie | 0.2 | 1.60E-24 | -5.02232 | 5.11E-07 |
| -3.78 | -4.906 | 192 | 4 | lasso | 0.016 | 0.0035 | 5.362 | 8.23E-08 |
| -7.24 | -5.7838 | 454 | 454 | susie | 0.11 | 1.10E-13 | 5.3289 | 9.88E-08 |
| -5.74 | 2.928 | 398 | 58 | enet | 0.13 | 2.60E-16 | -5.7551 | 8.66E-09 |
| 4.04 | 4.878 | 181 | 181 | susie | 0.024 | 0.00037 | 4.7261 | 2.29E-06 |

| | | | | | | | | |
|---|---|---|---|---|---|---|---|---|
| 11.54 | -4.78 | 203 | 27 | enet | 0.28 | 6.60E-36 | -5.0559 | 4.28E-07 |
| -7.06 | -0.2388 | 225 | 34 | enet | 0.14 | 5.50E-18 | 5.2576 | 1.46E-07 |
| 15.81 | 4.954 | 225 | 225 | susie | 0.52 | 9.60E-79 | 5.0017 | 5.68E-07 |
| 3.65 | -4.855 | 252 | 16 | enet | 0.01 | 0.015 | -6.1069 | 1.02E-09 |
| 5.04 | 3.965 | 349 | 16 | enet | 0.028 | 0.00016 | 4.5303 | 5.89E-06 |
| 3.82 | 5.047 | 349 | 349 | susie | 0.0052 | 0.063 | 5.165 | 2.40E-07 |
| -5.35 | 2.707 | 428 | 57 | enet | 0.12 | 7.80E-15 | 4.8217 | 1.42E-06 |
| -12.16 | -1.811 | 418 | 45 | enet | 0.47 | 2.50E-67 | 4.771 | 1.83E-06 |
| -4.92 | 2.13 | 492 | 19 | enet | 0.039 | 8.30E-06 | -5.0469 | 4.49E-07 |
| -7.56 | -2.41 | 510 | 510 | susie | 0.14 | 1.10E-17 | 4.696 | 2.65E-06 |
| 10.87 | 2.7 | 807 | 12 | enet | 0.31 | 3.10E-40 | 4.5579 | 5.17E-06 |
| 5.27 | -0.2708 | 376 | 376 | susie | 0.083 | 7.80E-11 | -6.19976 | 5.65E-10 |
| 3.19 | -2.081 | 160 | 20 | enet | 0.008 | 0.028 | -4.62447 | 3.76E-06 |
| 4.94 | -2.562 | 151 | 36 | enet | 0.062 | 2.00E-08 | -6.386 | 1.70E-10 |
| 8.33 | 3.213 | 232 | 232 | susie | 0.22 | 3.20E-27 | 5.95604 | 2.58E-09 |
| 4.95 | 2.995 | 268 | 33 | enet | 0.046 | 1.20E-06 | 5.48458 | 4.14E-08 |
| -4.51 | 3.344 | 72 | 31 | enet | 0.063 | 1.40E-08 | 4.67729 | 2.91E-06 |
| -4.21 | -2.312 | 314 | 314 | susie | 0.017 | 0.0024 | 6.96381 | 3.31E-12 |
| -4.33 | 3.875 | 355 | 23 | enet | 0.035 | 2.20E-05 | -5.30003 | 1.16E-07 |
| 6.16 | -2.558 | 500 | 26 | enet | 0.064 | 1.30E-08 | -6.1654 | 7.03E-10 |
| -6.31 | 3.986 | 597 | 597 | susie | 0.069 | 3.40E-09 | -4.5327 | 5.82E-06 |
| -10.14 | 3.052 | 327 | 327 | susie | 0.3 | 1.80E-39 | -4.7758 | 1.79E-06 |
| -8.67 | 4.14 | 434 | 434 | susie | 0.2 | 8.40E-26 | -5.9787 | 2.25E-09 |
| 5.55 | 3.505 | 511 | 31 | enet | 0.054 | 1.60E-07 | 5.7999 | 6.63E-09 |
| -6.44 | -4.073 | 261 | 21 | enet | 0.078 | 3.40E-10 | 5.1627 | 2.43E-07 |
| -4.57 | -4.568 | 264 | 16 | enet | 0.04 | 5.90E-06 | 4.9814 | 6.31E-07 |
| -7.94 | -4.862 | 260 | 1 | top1 | 0.12 | 9.20E-16 | 4.862 | 1.16E-06 |
| -5.11 | -4.384 | 289 | 289 | susie | 0.042 | 3.50E-06 | 4.692 | 2.71E-06 |
| -14.73 | -4.576 | 289 | 289 | susie | 0.44 | 8.90E-63 | 4.6701 | 3.01E-06 |
| -9.7 | -4.576 | 289 | 289 | susie | 0.2 | 3.60E-25 | 4.9655 | 6.85E-07 |
| 5.68 | -4.81 | 247 | 1 | top1 | 0.054 | 1.70E-07 | -4.81 | 1.51E-06 |
| 7.42 | -4.498 | 237 | 237 | susie | 0.11 | 9.70E-14 | -4.8559 | 1.20E-06 |
| -6.87 | -5.28 | 252 | 252 | susie | 0.095 | 3.70E-12 | 5.2797 | 1.29E-07 |
| -4.1 | 5.968 | 307 | 307 | susie | 0.017 | 0.0024 | -4.5675 | 4.94E-06 |
| 6.4 | 4.671 | 405 | 1 | top1 | 0.08 | 1.60E-10 | 4.671 | 3.00E-06 |
| 6.79 | -2.729 | 403 | 31 | enet | 0.095 | 3.50E-12 | -4.535 | 5.76E-06 |
| 8.27 | -1.264 | 371 | 60 | enet | 0.27 | 4.70E-34 | 5.7217 | 1.05E-08 |
| 6.72 | -4.584 | 311 | 7 | lasso | 0.072 | 1.60E-09 | -4.9235 | 8.50E-07 |
| 8.4 | -4.452 | 319 | 10 | lasso | 0.12 | 1.80E-15 | -4.995 | 5.88E-07 |
| 5.62 | 3.12 | 482 | 482 | susie | 0.085 | 5.10E-11 | 5.4018 | 6.60E-08 |
| 13.49 | 5.5763 | 118 | 15 | enet | 0.39 | 5.50E-53 | 4.9554 | 7.22E-07 |
| 4.55 | 5.7095 | 166 | 1 | top1 | 0.027 | 2.00E-04 | 5.7095 | 1.13E-08 |
| 8.18 | 3.827 | 312 | 21 | enet | 0.14 | 6.60E-18 | 4.9344 | 8.04E-07 |
| 3.65 | 3.801 | 384 | 5 | lasso | 0.0055 | 0.057 | 4.779 | 1.76E-06 |
| 6.31 | -4.054 | 317 | 15 | enet | 0.075 | 5.90E-10 | -4.6361 | 3.55E-06 |

| | | | | | | | | |
|---|---|---|---|---|---|---|---|---|
| -5.7 | 4.344 | 430 | 8 | enet | 0.045 | 1.50E-06 | -5.40681 | 6.42E-08 |
| 4.63 | -5.2903 | 291 | 291 | susie | 0.034 | 2.70E-05 | -4.90675 | 9.26E-07 |
| 10.54 | 2.138 | 450 | 31 | enet | 0.28 | 1.40E-35 | 4.58607 | 4.52E-06 |
| -4.12 | -2.673 | 483 | 21 | enet | 0.038 | 1.00E-05 | 5.29609 | 1.18E-07 |
| 11.82 | 2.58 | 506 | 506 | susie | 0.43 | 2.10E-60 | 4.65398 | 3.26E-06 |
| 4.93 | 2.59 | 504 | 504 | susie | 0.12 | 3.30E-15 | 5.82095 | 5.85E-09 |
| 6.11 | -4.376 | 339 | 339 | susie | 0.072 | 1.40E-09 | -5.03931 | 4.67E-07 |
| 3.77 | -2.908 | 411 | 14 | enet | 0.014 | 0.0057 | -4.98918 | 6.06E-07 |
| -6.63 | -6.31 | 185 | 185 | susie | 0.081 | 1.40E-10 | 5.36085 | 8.28E-08 |
| 6.71 | -4.798 | 433 | 433 | susie | 0.12 | 4.50E-15 | -6.10296 | 1.04E-09 |
| -6.34 | -6.055 | 383 | 383 | susie | 0.07 | 2.50E-09 | 6.25948 | 3.86E-10 |
| -8.27 | -2.684 | 381 | 381 | susie | 0.16 | 1.60E-19 | 5.16355 | 2.42E-07 |
| 15.92 | -2.604 | 298 | 298 | susie | 0.7 | 1.30E-127 | -4.82912 | 1.37E-06 |
| 7.43 | -3.75 | 404 | 32 | enet | 0.15 | 3.40E-18 | -4.78995 | 1.67E-06 |
| -7.1 | 4.348 | 303 | 303 | susie | 0.13 | 2.50E-16 | -5.78288 | 7.34E-09 |
| 8.02 | -2.295 | 393 | 19 | enet | 0.15 | 3.00E-18 | -4.98785 | 6.11E-07 |
| -3.76 | 3.423 | 426 | 6 | lasso | 0.0082 | 0.026 | -4.5409 | 5.60E-06 |
| 4.63 | 3.152 | 376 | 376 | susie | 0.19 | 1.00E-23 | 9.39035 | 5.98E-21 |
| -7.2 | 1.738 | 474 | 474 | susie | 0.31 | 1.90E-40 | -5.38943 | 7.07E-08 |
| 8.01 | -3.148 | 365 | 365 | susie | 0.22 | 1.00E-27 | -4.55931 | 5.13E-06 |
| 5.19 | -4.597 | 417 | 1 | top1 | 0.053 | 2.30E-07 | -4.597 | 4.29E-06 |
| 6.7 | 2.668 | 488 | 83 | enet | 0.18 | 3.30E-22 | 5.20058 | 1.99E-07 |
| 4.19 | -3.676 | 498 | 4 | lasso | 0.019 | 0.0016 | -4.64978 | 3.32E-06 |
| -7.62 | -2.302 | 377 | 50 | enet | 0.16 | 1.20E-19 | 4.9704 | 6.68E-07 |
| -4.29 | -3.922 | 309 | 10 | lasso | 0.02 | 0.00098 | 4.6187 | 3.86E-06 |
| -9.87 | -0.9002 | 306 | 306 | susie | 0.48 | 2.30E-69 | 5.35435 | 8.59E-08 |
| 7.48 | -3.276 | 374 | 374 | susie | 0.17 | 6.80E-22 | -6.31437 | 2.71E-10 |
| 12.14 | -4.258 | 353 | 20 | enet | 0.31 | 7.50E-41 | -4.89766 | 9.70E-07 |
| -7.54 | -4.848 | 344 | 1 | top1 | 0.11 | 2.20E-14 | 4.848 | 1.25E-06 |
| -3.29 | -3.768 | 498 | 4 | lasso | 0.0017 | 0.18 | 5.09157 | 3.55E-07 |
| -4.2 | 2.278 | 400 | 40 | enet | 0.015 | 0.0042 | -5.57343 | 2.50E-08 |
| -4.78 | -6.2881 | 394 | 1 | top1 | 0.019 | 0.0015 | 6.28808 | 3.21E-10 |
| -5 | 3.082 | 555 | 555 | susie | 0.044 | 2.20E-06 | -4.78203 | 1.74E-06 |
| -6.95 | -5.731 | 332 | 332 | susie | 0.098 | 1.30E-12 | 5.731 | 9.98E-09 |
| -5.14 | -6.58 | 428 | 4 | lasso | 0.054 | 1.40E-07 | 7.02112 | 2.20E-12 |
| -15.93 | -4.929 | 417 | 4 | lasso | 0.58 | 7.30E-92 | 4.67011 | 3.01E-06 |
| 5.07 | -4.114 | 335 | 28 | enet | 0.038 | 1.10E-05 | -6.00908 | 1.87E-09 |
| 7.7 | -4.553 | 449 | 1 | top1 | 0.12 | 2.90E-15 | -4.553 | 5.29E-06 |
| -3.86 | 2.164 | 469 | 18 | enet | 0.012 | 0.0083 | -4.91519 | 8.87E-07 |
| -6.8 | -2.174 | 628 | 628 | susie | 0.16 | 5.10E-20 | 4.83245 | 1.35E-06 |
| -6.88 | 3.18 | 627 | 21 | enet | 0.091 | 9.00E-12 | -4.88286 | 1.05E-06 |
| -3.44 | 3.931 | 438 | 438 | susie | 0.0082 | 0.027 | -4.52919 | 5.92E-06 |
| 4.59 | -3.342 | 356 | 356 | susie | 0.059 | 4.30E-08 | -5.62444 | 1.86E-08 |
| 7.1 | 4.753 | 468 | 1 | top1 | 0.099 | 1.10E-12 | 4.753 | 2.00E-06 |
| -4.31 | 3.762 | 464 | 3 | lasso | 0.012 | 0.0085 | -4.96442 | 6.89E-07 |

| | | | | | | | | |
|---|---|---|---|---|---|---|---|---|
| 4.88 | -3.507 | 610 | 34 | enet | 0.059 | 4.90E-08 | -4.94759 | 7.51E-07 |
| 7.32 | 2.986 | 408 | 11 | enet | 0.12 | 9.70E-15 | 5.26349 | 1.41E-07 |
| 4.75 | 1.662 | 315 | 71 | enet | 0.034 | 2.90E-05 | 5.04283 | 4.59E-07 |
| 9.9 | -3.811 | 284 | 284 | susie | 0.3 | 2.40E-39 | -4.81938 | 1.44E-06 |
| 16.45 | 3.671 | 135 | 40 | enet | 0.57 | 2.30E-90 | 4.63259 | 3.61E-06 |
| 4.1 | 2.83 | 331 | 45 | enet | 0.14 | 4.10E-17 | 4.94115 | 7.77E-07 |
| 4.91 | -3.092 | 413 | 17 | enet | 0.036 | 1.50E-05 | -5.14649 | 2.65E-07 |
| -8.57 | 1.664 | 473 | 40 | enet | 0.19 | 3.10E-24 | -4.55167 | 5.32E-06 |
| -5.3 | 2.764 | 536 | 21 | enet | 0.046 | 1.40E-06 | -5.07659 | 3.84E-07 |
| 4.92 | 4.505 | 548 | 5 | lasso | 0.054 | 1.60E-07 | 5.09968 | 3.40E-07 |
| 4.33 | -3.471 | 408 | 15 | enet | 0.019 | 0.0013 | -4.80342 | 1.56E-06 |
| -8.67 | 3.745 | 342 | 21 | enet | 0.15 | 5.00E-19 | -5.03191 | 4.86E-07 |
| 10.22 | 4.775 | 378 | 5 | lasso | 0.22 | 3.40E-27 | 4.67269 | 2.97E-06 |
| -5.23 | 2.819 | 221 | 14 | enet | 0.059 | 4.00E-08 | -4.5913 | 4.40E-06 |
| -7.26 | 3.496 | 494 | 494 | susie | 0.14 | 4.70E-18 | -4.83153 | 1.35E-06 |
| -8.83 | 3.201 | 594 | 594 | susie | 0.22 | 3.30E-27 | -5.1538 | 2.55E-07 |
| -4.94 | 4.871 | 508 | 508 | susie | 0.022 | 0.00075 | -5.14562 | 2.67E-07 |
| 8.99 | -3.987 | 345 | 49 | enet | 0.17 | 3.70E-21 | -5.88393 | 4.01E-09 |
| -6.08 | 2.813 | 627 | 45 | enet | 0.15 | 2.70E-18 | -6.2989 | 3.00E-10 |
| 5.64 | 3.442 | 877 | 28 | enet | 0.063 | 1.40E-08 | 4.6133 | 3.96E-06 |
| 5.97 | 2.109 | 622 | 622 | susie | 0.11 | 4.10E-14 | 5.6124 | 2.00E-08 |
| -3.77 | 2.096 | 543 | 543 | susie | 0.0053 | 0.059 | -5.5434 | 2.97E-08 |
| 6.13 | -5.39 | 474 | 474 | susie | 0.068 | 4.20E-09 | -5.6678 | 1.45E-08 |
| 7.36 | -2.092 | 341 | 59 | enet | 0.36 | 4.20E-48 | -4.7561 | 1.97E-06 |
| 7.09 | 2.353 | 372 | 372 | susie | 0.19 | 2.80E-20 | 4.4952 | 6.95E-06 |
| -4.32 | 6.42 | 344 | 1 | top1 | 0.024 | 0.0013 | -6.42 | 1.36E-10 |
| 4.74 | 3.506 | 488 | 22 | enet | 0.036 | 8.70E-05 | 5.6339 | 1.76E-08 |
| 5.43 | 4.308 | 465 | 23 | enet | 0.064 | 2.00E-07 | 6.2691 | 3.63E-10 |
| 6.32 | -4.571 | 312 | 1 | top1 | 0.098 | 1.40E-10 | -4.571 | 4.85E-06 |
| 12.82 | 5.757 | 118 | 4 | lasso | 0.44 | 5.00E-51 | 5.5185 | 3.42E-08 |
| -4.63 | 5.813 | 251 | 1 | top1 | 0.05 | 4.20E-06 | -5.813 | 6.13E-09 |
| -6.96 | -4.524 | 298 | 1 | top1 | 0.12 | 1.80E-12 | 4.524 | 6.07E-06 |
| -4.39 | 4.977 | 263 | 1 | top1 | 0.026 | 0.00084 | -4.977 | 6.46E-07 |
| 9.55 | 2.972 | 402 | 402 | susie | 0.24 | 6.70E-25 | 4.7845 | 1.71E-06 |
| -5.05 | 3.166 | 474 | 36 | enet | 0.054 | 2.00E-06 | -4.9285 | 8.29E-07 |
| -4.65 | -2.945 | 338 | 17 | enet | 0.046 | 9.90E-06 | 5.0648 | 4.09E-07 |
| 4.95 | -2.936 | 317 | 25 | enet | 0.038 | 5.50E-05 | -5.5747 | 2.48E-08 |
| -5.06 | -4.697 | 345 | 1 | top1 | 0.061 | 4.20E-07 | 4.697 | 2.64E-06 |
| -5.53 | -2.124 | 628 | 628 | susie | 0.2 | 1.70E-20 | 4.8879 | 1.02E-06 |
| 8.47 | 2.702 | 808 | 36 | enet | 0.21 | 1.80E-22 | 5.2396 | 1.61E-07 |
| 3.36 | 1.44 | 410 | 410 | susie | 0.02 | 0.0028 | -5.03301 | 4.83E-07 |
| -6.76 | -3.936 | 611 | 5 | lasso | 0.098 | 1.40E-10 | 4.6903 | 2.73E-06 |
| 3.74 | 3.678 | 334 | 27 | enet | 0.02 | 0.0032 | 5.134 | 2.84E-07 |
| 8.47 | -2.013 | 293 | 60 | enet | 0.23 | 2.30E-24 | -5.1714 | 2.32E-07 |
| -4.6 | 4.096 | 435 | 30 | enet | 0.088 | 1.20E-09 | -6.4622 | 1.03E-10 |

| | | | | | | | | |
|---|---|---|---|---|---|---|---|---|
| -4.35 | -6.131 | 382 | 382 | susie | 0.031 | 0.00028 | 5.5671 | 2.59E-08 |
| 4.07 | 3.16 | 290 | 38 | enet | 0.061 | 3.90E-07 | 4.60695 | 4.09E-06 |
| 4.3 | -1.837 | 286 | 24 | enet | 0.019 | 0.0038 | -4.59041 | 4.42E-06 |
| -6.59 | 2.298 | 377 | 377 | susie | 0.14 | 2.00E-14 | -4.83982 | 1.30E-06 |
| 4.36 | -1.987 | 448 | 17 | enet | 0.033 | 0.00017 | -4.49038 | 7.11E-06 |
| -10.98 | 2.339 | 495 | 495 | susie | 0.41 | 2.30E-47 | -4.4945 | 6.97E-06 |
| 3.81 | 2.922 | 413 | 413 | susie | 0.026 | 0.00077 | 4.5068 | 6.58E-06 |
| 4.12 | -2.57 | 494 | 8 | lasso | 0.019 | 0.0033 | -4.7736 | 1.81E-06 |
| -5.23 | -1.659 | 384 | 42 | enet | 0.07 | 5.60E-08 | -5.0237 | 5.07E-07 |
| -5.03 | 2.584 | 554 | 19 | enet | 0.06 | 5.20E-07 | -5.2035 | 1.96E-07 |
| -7.9 | 3.02 | 570 | 69 | enet | 0.16 | 3.50E-17 | -4.5168 | 6.28E-06 |
| -5.38 | -4.629 | 229 | 1 | top1 | 0.055 | 1.60E-06 | 4.629 | 3.67E-06 |
| 5.74 | -2.803 | 445 | 20 | enet | 0.077 | 1.40E-08 | -4.7676 | 1.86E-06 |
| 4.46 | -2.457 | 318 | 318 | susie | 0.039 | 4.60E-05 | -4.6135 | 3.96E-06 |
| -3.6 | 3.833 | 676 | 7 | lasso | 0.026 | 0.00073 | -5.05 | 4.43E-07 |
| 4.27 | 3.213 | 232 | 25 | enet | 0.049 | 5.60E-06 | 6.853 | 7.23E-12 |
| 4.99 | -2.203 | 290 | 4 | enet | 0.067 | 1.10E-07 | -4.6062 | 4.10E-06 |
| 3.84 | 3.03 | 72 | 19 | enet | 0.036 | 9.30E-05 | 7.1944 | 6.27E-13 |
| 12.56 | 4.263 | 319 | 319 | susie | 0.54 | 5.40E-68 | 6.3 | 2.98E-10 |
| -5.01 | -2.297 | 319 | 14 | enet | 0.046 | 1.20E-05 | 6.4677 | 9.95E-11 |
| -5.85 | -2.873 | 358 | 358 | susie | 0.1 | 4.00E-11 | 4.6585 | 3.19E-06 |
| 8.99 | -2.651 | 460 | 460 | susie | 0.25 | 8.30E-27 | -4.7113 | 2.46E-06 |
| 4.75 | -4.642 | 516 | 1 | top1 | 0.033 | 0.00016 | -4.642 | 3.45E-06 |
| 8.52 | -4.629 | 286 | 3 | lasso | 0.18 | 8.40E-19 | -4.5996 | 4.23E-06 |
| -4.79 | -2.347 | 443 | 19 | enet | 0.036 | 9.80E-05 | 4.7345 | 2.20E-06 |
| -7.67 | -0.90025 | 306 | 39 | enet | 0.47 | 1.30E-55 | 4.73802 | 2.16E-06 |
| 6.4 | -3.177 | 382 | 16 | enet | 0.09 | 8.40E-10 | -4.78305 | 1.73E-06 |
| -9.69 | -5.036 | 344 | 20 | enet | 0.23 | 4.70E-24 | 4.87203 | 1.10E-06 |
| 10.12 | 2.197 | 350 | 350 | susie | 0.51 | 2.00E-63 | 4.49877 | 6.83E-06 |
| -7.49 | 2.46 | 277 | 46 | enet | 0.16 | 3.60E-17 | 5.23025 | 1.69E-07 |
| -6.61 | -6.441 | 428 | 428 | susie | 0.094 | 3.60E-10 | 6.47118 | 9.72E-11 |
| -3.39 | 3.264 | 303 | 303 | susie | 0.0082 | 0.04 | -4.94232 | 7.72E-07 |
| 4.76 | 1.479 | 307 | 40 | enet | 0.079 | 8.00E-09 | -5.34912 | 8.84E-08 |
| -6.56 | -4.094 | 441 | 441 | susie | 0.12 | 1.30E-12 | 5.86212 | 4.57E-09 |
| -3.72 | 1.851 | 359 | 28 | enet | -0.00075 | 0.4 | -4.7967 | 1.61E-06 |
| 13.67 | -2.604 | 298 | 298 | susie | 0.61 | 1.90E-81 | -4.6523 | 3.28E-06 |
| -6.03 | 2.888 | 307 | 307 | susie | 0.14 | 3.20E-15 | -5.1002 | 3.39E-07 |
| -6.38 | -4.712 | 441 | 1 | top1 | 0.09 | 7.60E-10 | 4.7117 | 2.46E-06 |
| 8.52 | -1.389 | 466 | 466 | susie | 0.22 | 6.00E-23 | -5.6277 | 1.83E-08 |
| -6.23 | 4.348 | 303 | 7 | lasso | 0.099 | 1.10E-10 | -4.9606 | 7.03E-07 |
| 4.73 | 2.745 | 376 | 376 | susie | 0.17 | 4.90E-18 | 9.3867 | 6.19E-21 |
| 6.25 | -4.696 | 417 | 27 | enet | 0.11 | 5.10E-12 | -6.2188 | 5.01E-10 |
| 5.81 | -4.558 | 420 | 420 | susie | 0.075 | 2.00E-08 | -4.5572 | 5.19E-06 |
| -5.42 | 1.781 | 491 | 34 | enet | 0.11 | 1.10E-11 | -4.6837 | 2.82E-06 |
| -6.47 | -5.7838 | 454 | 5 | lasso | 0.1 | 3.20E-11 | 4.9745 | 6.54E-07 |

| | | | | | | | | |
|---|---|---|---|---|---|---|---|---|
| -10.88 | 4.878 | 203 | 27 | enet | 0.3 | 5.40E-32 | -4.5269 | 5.98E-06 |
| -4.31 | 5.093 | 215 | 1 | top1 | 0.039 | 4.50E-05 | -5.093 | 3.52E-07 |
| -4.06 | 4.954 | 223 | 223 | susie | 0.014 | 0.011 | -5.3166 | 1.06E-07 |
| -5.78 | -0.2388 | 225 | 22 | enet | 0.15 | 1.70E-15 | 6.2082 | 5.36E-10 |
| 10.76 | 5.024 | 225 | 3 | lasso | 0.29 | 1.00E-31 | 5.0719 | 3.94E-07 |
| 4.09 | 2.426 | 399 | 12 | enet | 0.013 | 0.013 | 4.6336 | 3.59E-06 |
| -11.34 | 3.084 | 209 | 31 | enet | 0.43 | 3.30E-50 | -4.6964 | 2.65E-06 |
| 8.82 | -3.509 | 276 | 24 | enet | 0.23 | 1.80E-24 | -5.2184 | 1.80E-07 |
| -9.88 | -3.288 | 417 | 32 | enet | 0.24 | 1.50E-25 | 4.6035 | 4.16E-06 |
| -6.54 | 3.157 | 505 | 43 | enet | 0.12 | 7.70E-13 | -5.1523 | 2.57E-07 |
| -4.6 | -1.92 | 385 | 22 | enet | 0.042 | 2.40E-05 | 4.535 | 5.76E-06 |
| -5.53 | -4.298 | 513 | 6 | lasso | 0.11 | 5.20E-12 | 5.8347 | 5.39E-09 |
| 3.5 | 2.294 | 481 | 39 | enet | 0.0063 | 0.062 | -5.0565 | 4.27E-07 |
| -5.49 | -3.975 | 261 | 261 | susie | 0.061 | 4.30E-07 | 4.5206 | 6.17E-06 |
| -3.58 | -4.2 | 264 | 264 | susie | 0.017 | 0.0062 | 4.7242 | 2.31E-06 |
| -8.16 | -4.91 | 260 | 1 | top1 | 0.17 | 1.90E-17 | 4.91 | 9.11E-07 |
| -13.6 | -4.384 | 289 | 14 | lasso | 0.46 | 7.60E-55 | 4.6661 | 3.07E-06 |
| -9.19 | -4.673 | 289 | 289 | susie | 0.22 | 4.70E-23 | 4.9502 | 7.41E-07 |
| 4.69 | -4.81 | 247 | 25 | enet | 0.034 | 0.00015 | -5.056 | 4.28E-07 |
| 7.04 | -4.528 | 237 | 1 | top1 | 0.1 | 4.40E-11 | -4.528 | 5.95E-06 |
| -3.86 | 2.186 | 364 | 24 | enet | 0.025 | 0.001 | -5.4332 | 5.53E-08 |
| -7.41 | 5.068 | 319 | 319 | susie | 0.14 | 7.00E-15 | -4.7422 | 2.11E-06 |
| 4.31 | 3.728 | 489 | 27 | enet | 0.032 | 0.00021 | 5.5948 | 2.21E-08 |
| -9.81 | 3.994 | 387 | 387 | susie | 0.49 | 5.90E-60 | -6.8439 | 7.71E-12 |
| 3.59 | -1.0282 | 400 | 25 | enet | 0.034 | 0.00014 | -5.8165 | 6.01E-09 |
| 5.09 | 2.547 | 341 | 26 | enet | 0.05 | 5.00E-06 | 5.7326 | 9.89E-09 |
| -3.87 | 1.889 | 240 | 7 | lasso | 0.015 | 0.0081 | 4.7028 | 2.57E-06 |
| -6.11 | 3.533 | 240 | 42 | enet | 0.11 | 5.20E-12 | -5.0495 | 4.43E-07 |
| -5.65 | -4.527 | 411 | 1 | top1 | 0.073 | 2.90E-08 | 4.5268 | 5.99E-06 |
| -5.85 | 2.288 | 519 | 28 | enet | 0.14 | 1.20E-14 | -4.6537 | 3.26E-06 |
| 3.36 | 2.373 | 476 | 476 | susie | 0.027 | 0.00067 | 4.6053 | 4.12E-06 |
| 4.78 | -3.149 | 347 | 30 | enet | 0.064 | 2.20E-07 | -4.811 | 1.50E-06 |
| -6.02 | 3.045 | 406 | 19 | enet | 0.099 | 1.20E-10 | -5.1227 | 3.01E-07 |
| 4.73 | -6.93 | 620 | 4 | lasso | 0.049 | 3.80E-08 | -4.96494 | 6.87E-07 |
| 9.43 | -4.592 | 372 | 372 | susie | 0.15 | 2.40E-22 | -4.64912 | 3.33E-06 |
| -5.69 | 6.392 | 344 | 344 | susie | 0.037 | 1.40E-06 | -6.35544 | 2.08E-10 |
| 4.9 | -4.645 | 345 | 345 | susie | 0.033 | 5.50E-06 | -4.57383 | 4.79E-06 |
| -6.56 | -2.299 | 464 | 14 | enet | 0.086 | 2.50E-13 | 4.78005 | 1.75E-06 |
| -6.89 | -2.549 | 525 | 13 | enet | 0.098 | 5.50E-15 | 4.93195 | 8.14E-07 |
| 5.54 | 3.213 | 232 | 232 | susie | 0.12 | 9.50E-18 | 6.50216 | 7.92E-11 |
| 11.48 | 4.263 | 317 | 317 | susie | 0.31 | 7.70E-50 | 6.29261 | 3.12E-10 |
| -5.72 | -2.766 | 372 | 23 | enet | 0.053 | 9.50E-09 | 4.64262 | 3.44E-06 |
| 5.21 | -2.144 | 440 | 29 | enet | 0.074 | 1.20E-11 | 4.94649 | 7.56E-07 |
| 3.74 | 2.48 | 377 | 42 | enet | 0.037 | 1.40E-06 | 5.1243 | 2.99E-07 |
| 6.48 | -2.606 | 464 | 26 | enet | 0.097 | 5.70E-15 | -5.04914 | 4.44E-07 |

| | | | | | | | | |
|---|---|---|---|---|---|---|---|---|
| -4.37 | 3.226 | 393 | 15 | enet | 0.0091 | 0.012 | -4.96359 | 6.92E-07 |
| 11.86 | -2.604 | 298 | 298 | susie | 0.45 | 5.60E-78 | -5.29411 | 1.20E-07 |
| -7.51 | -4.7117 | 439 | 1 | top1 | 0.086 | 2.80E-13 | 4.71168 | 2.46E-06 |
| 4.56 | -2.253 | 446 | 446 | susie | 0.034 | 3.60E-06 | -5.22384 | 1.75E-07 |
| 10.08 | -3.148 | 365 | 365 | susie | 0.27 | 2.30E-41 | -4.50854 | 6.53E-06 |
| -4.02 | 2.247 | 500 | 27 | enet | 0.038 | 1.30E-06 | -5.77885 | 7.52E-09 |
| 3.72 | -2.208 | 548 | 5 | lasso | 0.013 | 0.003 | -4.54244 | 5.56E-06 |
| 4.56 | 4.869 | 430 | 1 | top1 | 0.033 | 5.00E-06 | 4.869 | 1.12E-06 |
| 6.91 | -2.959 | 637 | 637 | susie | 0.14 | 7.80E-22 | -4.83531 | 1.33E-06 |
| 12.02 | 2.838 | 627 | 627 | susie | 0.35 | 1.60E-56 | 4.70772 | 2.51E-06 |
| -4.83 | -2.102 | 324 | 29 | enet | 0.061 | 8.50E-10 | 4.55565 | 5.22E-06 |
| 9.21 | 3.141 | 638 | 39 | enet | 0.15 | 3.00E-23 | 4.62358 | 3.77E-06 |
| -5.06 | -2.606 | 235 | 21 | enet | 0.03 | 1.50E-05 | 5.49276 | 3.96E-08 |
| -4.26 | -4.58 | 403 | 403 | susie | 0.012 | 0.0048 | 4.57676 | 4.72E-06 |
| -5.68 | 4.945 | 344 | 1 | top1 | 0.044 | 1.50E-07 | -4.945 | 7.61E-07 |
| -5.21 | 1.506 | 452 | 29 | enet | 0.039 | 8.80E-07 | -4.5263 | 6.00E-06 |
| -3.21 | -4.222 | 285 | 285 | susie | 0.0045 | 0.055 | 4.9998 | 5.74E-07 |
| 8.11 | 5.725 | 284 | 15 | enet | 0.11 | 2.40E-17 | 4.7028 | 2.57E-06 |
| 4.34 | 3.857 | 487 | 25 | enet | 0.019 | 0.00054 | 4.7161 | 2.40E-06 |
| -5.97 | 2.756 | 381 | 23 | enet | 0.057 | 2.80E-09 | -6.6617 | 2.71E-11 |
| -4.8 | -3.06 | 424 | 47 | enet | 0.031 | 1.10E-05 | 4.5094 | 6.50E-06 |
| 8.49 | 5.576 | 118 | 1 | top1 | 0.12 | 2.90E-18 | 5.5763 | 2.46E-08 |
| 9.93 | 6.921 | 143 | 1 | top1 | 0.16 | 1.70E-24 | 6.9209 | 4.49E-12 |
| -10.93 | -4.524 | 298 | 1 | top1 | 0.19 | 5.00E-29 | 4.524 | 6.07E-06 |
| 16.98 | 2.972 | 397 | 397 | susie | 0.53 | 2.20E-98 | 4.5345 | 5.77E-06 |
| 4.26 | -2.031 | 493 | 28 | enet | 0.048 | 4.60E-08 | -4.8753 | 1.09E-06 |
| 4.62 | 2.26 | 482 | 3 | enet | 0.019 | 0.00046 | 4.6062 | 4.10E-06 |
| -9.8 | 3.552 | 444 | 28 | enet | 0.16 | 3.50E-24 | -4.7016 | 2.58E-06 |
| 8.18 | -4.765 | 511 | 511 | susie | 0.099 | 3.70E-15 | -5.3565 | 8.48E-08 |
| -4.42 | 4.804 | 458 | 41 | enet | 0.035 | 2.90E-06 | -5.30585 | 1.12E-07 |
| 9.05 | 4.234 | 310 | 310 | susie | 0.17 | 2.00E-25 | 4.89504 | 9.83E-07 |
| -5.74 | -3.399 | 313 | 13 | enet | 0.055 | 5.70E-09 | 4.52195 | 6.13E-06 |
| 5.13 | -3.295 | 583 | 583 | susie | 0.039 | 8.90E-07 | 4.76155 | 1.92E-06 |
| -4.27 | 3.507 | 306 | 6 | lasso | 0.018 | 0.00059 | -4.98054 | 6.34E-07 |
| -7.53 | -3.296 | 234 | 234 | susie | 0.13 | 1.10E-19 | 4.68872 | 2.75E-06 |
| 11.15 | -4.577 | 395 | 395 | susie | 0.22 | 1.00E-33 | -5.42507 | 5.79E-08 |
| -5.44 | -1.84 | 317 | 317 | susie | 0.13 | 1.00E-19 | 6.61175 | 3.80E-11 |
| 7.13 | 3.114 | 402 | 16 | enet | 0.087 | 1.60E-13 | 4.97202 | 6.63E-07 |
| -6.68 | 3.3889 | 385 | 29 | enet | 0.074 | 1.30E-11 | -5.11503 | 3.14E-07 |
| 4.23 | 3.44 | 312 | 11 | enet | 0.013 | 0.0038 | 4.50731 | 6.57E-06 |
| 6.7 | 1.923 | 378 | 378 | susie | 0.15 | 2.50E-22 | 4.81115 | 1.50E-06 |
| -7.85 | -3.73 | 505 | 505 | susie | 0.14 | 7.80E-22 | 4.659 | 3.18E-06 |
| -7.56 | 4.66 | 471 | 12 | enet | 0.086 | 2.70E-13 | -4.71 | 2.42E-06 |
| -15.19 | -3.455 | 485 | 22 | enet | 0.49 | 6.60E-87 | 4.58 | 4.61E-06 |
| -4.97 | 4.338 | 479 | 479 | susie | 0.041 | 3.90E-07 | -4.58 | 4.55E-06 |

| | | | | | | | | |
|---|---|---|---|---|---|---|---|---|
| 3.67 | 4.589 | 330 | 1 | top1 | 0.0093 | 0.011 | 4.59 | 4.45E-06 |
| 4.55 | -4.595 | 271 | 1 | top1 | 0.03 | 1.40E-05 | -4.595 | 4.33E-06 |
| -5.5 | -4.16 | 262 | 2 | lasso | 0.033 | 4.80E-06 | 4.64851 | 3.34E-06 |
| -4.53 | -4.849 | 262 | 1 | top1 | 0.033 | 6.30E-06 | 4.849 | 1.24E-06 |
| 4.02 | -4.658 | 278 | 1 | top1 | 0.0095 | 0.01 | -4.658 | 3.19E-06 |
| -13.64 | -4.702 | 288 | 288 | susie | 0.31 | 5.60E-50 | 4.80943 | 1.51E-06 |
| 6.11 | -4.528 | 237 | 1 | top1 | 0.06 | 1.00E-09 | -4.528 | 5.95E-06 |
| -4.51 | 2.897 | 599 | 41 | enet | 0.037 | 1.70E-06 | -4.66859 | 3.03E-06 |
| -3.48 | -2.385 | 195 | 17 | enet | 0.0022 | 0.13 | 4.56431 | 5.01E-06 |
| 10.12 | 3.965 | 461 | 461 | susie | 0.21 | 5.50E-32 | 5.33497 | 9.56E-08 |
| -4.61 | -3.177 | 352 | 8 | enet | 0.028 | 2.30E-05 | 4.86953 | 1.12E-06 |
| 6.9 | -2.671 | 389 | 69 | enet | 0.15 | 2.80E-22 | -5.82493 | 5.71E-09 |
| 7.66 | -2.197 | 434 | 26 | enet | 0.098 | 4.50E-15 | -4.55711 | 5.19E-06 |
| -4.93 | 5.191 | 368 | 368 | susie | 0.035 | 3.00E-06 | -6.052 | 1.43E-09 |
| 10.55 | 3.852 | 306 | 40 | enet | 0.43 | 2.40E-73 | 5.1564 | 2.52E-07 |
| -6.27 | -2.001 | 544 | 17 | enet | 0.061 | 8.00E-10 | 4.8237 | 1.41E-06 |
| 11.84 | -4.258 | 353 | 353 | susie | 0.25 | 3.80E-38 | -4.5607 | 5.10E-06 |
| 7.96 | 3.905 | 344 | 9 | enet | 0.1 | 2.40E-15 | 4.5501 | 5.36E-06 |
| 3.38 | 3.68 | 360 | 360 | susie | 0.00058 | 0.25 | 5.2047 | 1.94E-07 |
| 12.33 | 2.051 | 353 | 33 | enet | 0.51 | 6.80E-94 | 4.6622 | 3.13E-06 |
| -4.18 | -2.104 | 413 | 22 | enet | 0.037 | 1.60E-06 | 5.6184 | 1.93E-08 |
| -5.5 | -5.731 | 332 | 1 | top1 | 0.046 | 9.20E-08 | 5.731 | 9.98E-09 |
| 15.17 | -5.123 | 412 | 412 | susie | 0.39 | 8.10E-66 | -4.8572 | 1.19E-06 |
| 10.19 | -4.547 | 449 | 69 | enet | 0.19 | 2.70E-28 | -5.8779 | 4.15E-09 |
| -4.48 | -3.012 | 529 | 36 | enet | 0.025 | 7.90E-05 | 4.7946 | 1.63E-06 |
| 6.97 | 4.249 | 426 | 10 | lasso | 0.11 | 3.70E-17 | 4.6316 | 3.63E-06 |
| -4.04 | -2.63 | 514 | 22 | enet | 0.03 | 1.30E-05 | 4.8979 | 9.69E-07 |
| 4.12 | -3.129 | 614 | 24 | enet | 0.017 | 0.00099 | -6.1198 | 9.37E-10 |
| -4.68 | -3.287 | 434 | 25 | enet | 0.013 | 0.0031 | 5.6751 | 1.39E-08 |
| -4.21 | -1.61 | 524 | 17 | enet | 0.013 | 0.0032 | 4.6042 | 4.14E-06 |
| 4.61 | -3.375 | 414 | 7 | lasso | 0.016 | 0.0013 | -4.8622 | 1.16E-06 |
| 7.05 | 1.927 | 463 | 40 | enet | 0.086 | 2.60E-13 | -5.17351 | 2.30E-07 |
| -3.74 | -2.062 | 383 | 383 | susie | 0.0055 | 0.039 | 4.53553 | 5.75E-06 |
| -6.72 | -4.795 | 383 | 1 | top1 | 0.07 | 4.60E-11 | 4.795 | 1.63E-06 |
| -3.81 | -2.964 | 377 | 28 | enet | 0.02 | 0.00036 | 5.34928 | 8.83E-08 |
| 5.77 | 2.59 | 512 | 512 | susie | 0.16 | 8.90E-25 | 4.6261 | 3.73E-06 |
| 14.35 | 3.179 | 354 | 354 | susie | 0.4 | 1.60E-66 | 5.8271 | 5.64E-09 |
| 5.17 | -3.193 | 503 | 503 | susie | 0.082 | 1.00E-12 | -4.5898 | 4.44E-06 |
| -9.11 | 5.334 | 357 | 357 | susie | 0.13 | 2.00E-20 | -5.3343 | 9.59E-08 |
| 7.32 | -5.204 | 435 | 30 | enet | 0.092 | 3.30E-14 | -5.779 | 7.51E-09 |
| -3.88 | -4.046 | 383 | 383 | susie | 0.018 | 0.00075 | 6.1443 | 8.03E-10 |
| 3.43 | -2.343 | 526 | 30 | enet | 0.024 | 0.00011 | -5.0205 | 5.15E-07 |
| -7.09 | -8.33465 | 571 | 50 | enet | 0.13 | 2.50E-19 | 4.5427 | 5.55E-06 |
| -11.08 | -5.118 | 272 | 24 | enet | 0.23 | 1.80E-35 | 4.927 | 8.35E-07 |
| -7.94 | -4.423 | 408 | 408 | susie | 0.096 | 8.80E-15 | 4.6191 | 3.85E-06 |

| | | | | | | | | | |
|---|---|---|---|---|---|---|---|---|---|
| -5.65 | -1.544 | 355 | 355 | susie | | 0.15 | 5.70E-22 | 5.1547 | 2.54E-07 |
| 9.13 | -5.079 | 687 | 687 | susie | | 0.13 | 2.80E-20 | -5.10904 | 3.24E-07 |
| 5.89 | -2.768 | 453 | 27 | enet | | 0.077 | 4.00E-12 | -5.25425 | 1.49E-07 |
| -3.25 | -3.106 | 478 | 42 | enet | | 0.012 | 0.0051 | 5.58123 | 2.39E-08 |
| -4.75 | -3.776 | 374 | 43 | enet | | 0.11 | 2.90E-16 | 5.65663 | 1.54E-08 |
| -5.34 | -3.227 | 495 | 495 | susie | | 0.053 | 9.00E-09 | 4.65791 | 3.19E-06 |
| 3.8 | 2.346 | 448 | 34 | enet | | 0.016 | 0.0011 | 4.76232 | 1.91E-06 |
| -3.1 | 3.212 | 353 | 353 | susie | | 0.013 | 0.0037 | -4.5079 | 6.55E-06 |
| -7.8 | -4.841 | 345 | 1 | top1 | | 0.097 | 6.30E-15 | 4.841 | 1.29E-06 |
| -11.17 | -5.06 | 331 | 1 | top1 | | 0.21 | 4.60E-32 | 5.06 | 4.19E-07 |
| 3.99 | -2.445 | 433 | 39 | enet | | 0.013 | 0.0037 | -4.6909 | 2.72E-06 |
| -5.84 | 2.226 | 440 | 30 | enet | | 0.17 | 6.20E-26 | -4.575 | 4.76E-06 |
| -10.58 | 3.281 | 420 | 18 | enet | | 0.39 | 3.30E-65 | -5.7455 | 9.17E-09 |
| -5.73 | 4.845 | 203 | 1 | top1 | | 0.049 | 3.80E-08 | -4.845 | 1.27E-06 |
| -9.23 | 5.151 | 220 | 220 | susie | | 0.14 | 8.90E-21 | -5.1548 | 2.54E-07 |
| -9.25 | -0.2388 | 225 | 225 | susie | | 0.21 | 4.40E-32 | 7.1997 | 6.04E-13 |
| 4.74 | 4.954 | 225 | 225 | susie | | 0.027 | 4.30E-05 | 5.2378 | 1.62E-07 |
| -3.58 | -2.496 | 226 | 25 | enet | | 0.019 | 0.00049 | -4.7763 | 1.79E-06 |
| -5.22 | -5.6689 | 209 | 33 | enet | | 0.043 | 2.20E-07 | 5.1111 | 3.20E-07 |
| 6.98 | 5.109 | 348 | 4 | lasso | | 0.081 | 1.30E-12 | 4.8403 | 1.30E-06 |
| 4.94 | 4.671 | 263 | 1 | top1 | | 0.12 | 1.60E-06 | 4.671 | 3.00E-06 |
| 6.51 | -3.585 | 526 | 526 | susie | | 0.34 | 1.40E-17 | -6.3715 | 1.87E-10 |
| 3.5 | -2.643 | 525 | 20 | enet | | 0.023 | 0.024 | -4.7205 | 2.35E-06 |
| -5.17 | 4.679 | 333 | 13 | lasso | | 0.18 | 1.60E-09 | -4.9995 | 5.75E-07 |
| 4.62 | 4.455 | 310 | 1 | top1 | | 0.093 | 2.20E-05 | 4.455 | 8.39E-06 |
| -5.45 | 4.98 | 347 | 1 | top1 | | 0.14 | 1.90E-07 | -4.98 | 6.36E-07 |
| 5.82 | -4.922 | 433 | 1 | top1 | | 0.18 | 3.10E-09 | -4.922 | 8.57E-07 |
| 5.56 | -5.037 | 416 | 416 | susie | | 0.17 | 7.80E-09 | -4.86919 | 1.12E-06 |
| 6.43 | 5.576 | 118 | 3 | lasso | | 0.24 | 2.00E-12 | 5.2202 | 1.79E-07 |
| -7.03 | -4.524 | 295 | 1 | top1 | | 0.27 | 6.20E-14 | 4.524 | 6.07E-06 |
| -5.69 | -4.524 | 295 | 295 | susie | | 0.18 | 3.30E-09 | 4.51562 | 6.31E-06 |
| -4.05 | -2.975 | 288 | 25 | enet | | 0.058 | 0.00071 | 4.3581 | 1.31E-05 |
| 6.29 | 2.598 | 361 | 22 | enet | | 0.24 | 2.10E-12 | 4.6827 | 2.83E-06 |
| 4.77 | 3.796 | 343 | 343 | susie | | 0.16 | 1.80E-08 | 4.8485 | 1.24E-06 |
| 6 | 4.374 | 321 | 321 | susie | | 0.2 | 2.60E-10 | 5.9271 | 3.08E-09 |
| 5.19 | -4.409 | 358 | 1 | top1 | | 0.13 | 4.80E-07 | -4.409 | 1.04E-05 |
| 9.45 | -4.11 | 340 | 340 | susie | | 0.58 | 3.20E-35 | -4.628 | 3.69E-06 |
| 5.71 | 2.456 | 415 | 11 | enet | | 0.2 | 2.10E-10 | 4.35296 | 1.34E-05 |
| -4.38 | 3.2429 | 309 | 40 | enet | | 0.077 | 1.00E-04 | -4.32768 | 1.51E-05 |
| 6.56 | -2.604 | 298 | 298 | susie | | 0.42 | 1.80E-22 | -5.19546 | 2.04E-07 |
| 3.79 | -2.554 | 397 | 45 | enet | | 0.036 | 0.0063 | -5.33917 | 9.34E-08 |
| 5.09 | -2.951 | 351 | 10 | enet | | 0.16 | 2.80E-08 | -4.39114 | 1.13E-05 |
| -7.1 | -2.367 | 311 | 311 | susie | | 0.29 | 8.60E-15 | 6.03789 | 1.56E-09 |
| -6.19 | -2.798 | 388 | 388 | susie | | 0.3 | 1.10E-15 | 4.4133 | 1.02E-05 |
| 5.79 | -3.069 | 400 | 20 | lasso | | 0.4 | 2.00E-21 | -4.4788 | 7.51E-06 |

| | | | | | | | | |
|---|---|---|---|---|---|---|---|---|
| 5.18 | 4.054 | 352 | 2 | lasso | 0.13 | 3.60E-07 | 4.737 | 2.17E-06 |
| -6.85 | -4.841 | 344 | 344 | susie | 0.25 | 4.40E-13 | 4.75636 | 1.97E-06 |
| -4.17 | 2.464 | 447 | 21 | enet | 0.093 | 2.00E-05 | -4.6538 | 3.26E-06 |
| -3.31 | -4.014 | 258 | 258 | susie | 0.02 | 0.034 | 4.9235 | 8.50E-07 |
| -4.24 | -4.144 | 261 | 261 | susie | 0.068 | 0.00027 | 4.4646 | 8.02E-06 |
| 3.72 | -3.424 | 472 | 8 | lasso | -0.0029 | 0.48 | -5.2658 | 1.40E-07 |
| -7.35 | -3.913 | 401 | 401 | susie | 0.28 | 1.30E-14 | 4.3726 | 1.23E-05 |
| 4.4 | 4.743 | 348 | 348 | susie | 0.058 | 0.00073 | 4.9596 | 7.07E-07 |
| 4.55 | -2.285 | 691 | 22 | enet | 0.14 | 2.70E-07 | -5.20822 | 1.91E-07 |
| 3.79 | 1.879 | 546 | 18 | enet | 0.027 | 0.017 | 4.32646 | 1.52E-05 |
| -3.44 | 2.683 | 504 | 13 | enet | 0.014 | 0.06 | -5.18662 | 2.14E-07 |
| 4.28 | 3.454 | 457 | 18 | enet | 0.036 | 0.0064 | 4.90619 | 9.29E-07 |
| -4.06 | -3.966 | 478 | 11 | enet | 0.069 | 0.00024 | 5.82342 | 5.77E-09 |
| 6.19 | -4.78 | 201 | 1 | top1 | 0.19 | 8.10E-10 | -4.78 | 1.75E-06 |
| 4.87 | 5.069 | 223 | 223 | susie | 0.12 | 2.20E-06 | 5.37269 | 7.76E-08 |
| -3.69 | 3.192 | 497 | 497 | susie | 0.04 | 0.0042 | -5.7552 | 8.66E-09 |
| -5.89 | 4.071 | 276 | 9 | lasso | 0.2 | 1.70E-10 | -4.3602 | 1.30E-05 |
| -4.64 | -3.979 | 304 | 304 | susie | 0.12 | 9.00E-07 | 5.0824 | 3.73E-07 |
| -6.47 | -2.864 | 139 | 24 | enet | 0.33 | 2.30E-17 | 5.2553 | 1.48E-07 |
| -5.31 | -2.23 | 363 | 11 | lasso | 0.2 | 1.40E-10 | 5.3258 | 1.01E-07 |
| -6.29 | -3.932 | 360 | 27 | enet | 0.21 | 1.10E-10 | 5.1 | 3.40E-07 |
| 4.27 | -2.69 | 482 | 6 | lasso | 0.061 | 0.00051 | -4.8788 | 1.07E-06 |
| -4.06 | -2.37 | 398 | 15 | enet | 0.054 | 0.0011 | 5.3348 | 9.57E-08 |
| -6.25 | -3.25 | 433 | 433 | susie | 0.47 | 4.70E-26 | 4.6635 | 3.11E-06 |
| 4.59 | 1.87 | 415 | 60 | enet | 0.15 | 4.50E-08 | 5.06144 | 4.16E-07 |
| 3.25 | 3.544 | 392 | 15 | lasso | -0.003 | 0.5 | 4.55884 | 5.14E-06 |
| 5.69 | 3.378 | 386 | 30 | enet | 0.16 | 1.80E-08 | 4.4672 | 7.92E-06 |
| -3.75 | -4.244 | 358 | 2 | lasso | 0.03 | 0.012 | 5.1051 | 3.31E-07 |
| -4.47 | -0.9002 | 306 | 41 | enet | 0.25 | 4.30E-13 | 4.9173 | 8.77E-07 |
| -3.22 | -1.789 | 650 | 82 | enet | 0.012 | 0.077 | -4.5213 | 6.14E-06 |
| 4.44 | -4.407 | 452 | 1 | top1 | 0.08 | 7.80E-05 | -4.407 | 1.05E-05 |
| 5.75 | 4.49 | 238 | 4 | lasso | 0.22 | 3.10E-11 | 4.3964 | 1.10E-05 |
| 3.21 | 1.629 | 212 | 43 | enet | 0.081 | 7.20E-05 | 4.7075 | 2.51E-06 |
| 3.52 | -5.1092 | 303 | 303 | susie | 0.021 | 0.029 | -5.3423 | 9.18E-08 |
| -5.56 | -4.929 | 415 | 415 | susie | 0.15 | 4.60E-08 | 4.5053 | 6.63E-06 |
| 4.94 | 4.076 | 137 | 3 | enet | 0.095 | 1.70E-05 | 4.5921 | 4.39E-06 |
| 6.3 | 4.449 | 293 | 35 | enet | 0.19 | 8.00E-10 | 4.7782 | 1.77E-06 |
| 4.3 | -4.8225 | 487 | 487 | susie | 0.039 | 0.0048 | -4.5016 | 6.75E-06 |
| -4.54 | -2.123 | 392 | 392 | susie | 0.11 | 2.90E-06 | 4.4156 | 1.01E-05 |
| -4.75 | -3.976 | 610 | 610 | susie | 0.1 | 9.40E-06 | 4.31288 | 1.61E-05 |
| 4.48 | -4.333 | 339 | 1 | top1 | 0.084 | 5.00E-05 | -4.333 | 1.47E-05 |
| 4.86 | 4.345 | 381 | 381 | susie | 0.12 | 2.10E-06 | 4.47493 | 7.64E-06 |
| 5.8 | -3.164 | 497 | 27 | enet | 0.23 | 1.00E-11 | -4.56839 | 4.91E-06 |
| 7.62 | 4.344 | 375 | 1 | top1 | 0.32 | 1.70E-16 | 4.344 | 1.40E-05 |
| 7.71 | 4.367 | 561 | 561 | susie | 0.32 | 2.00E-16 | 4.37749 | 1.20E-05 |

| | | | | | | | | |
|---|---|---|---|---|---|---|---|---|
| -7.38 | -4.016 | 264 | 17 | enet | 0.32 | 1.80E-16 | 4.3414 | 1.42E-05 |
| 6.21 | -3.793 | 654 | 38 | enet | 0.2 | 1.70E-13 | -5.7733 | 7.77E-09 |
| 6.79 | -2.423 | 344 | 21 | enet | 0.27 | 1.20E-18 | -5.2837 | 1.27E-07 |
| 4.92 | -3.424 | 613 | 31 | enet | 0.092 | 9.00E-07 | -4.8211 | 1.43E-06 |
| 4.62 | -3.787 | 627 | 627 | susie | 0.05 | 0.00027 | -5.00296 | 5.65E-07 |
| 4.02 | -6.091 | 343 | 1 | top1 | 0.014 | 0.037 | -6.091 | 1.12E-09 |
| -4.1 | -3.753 | 258 | 11 | enet | 0.043 | 7.00E-04 | 4.718925 | 2.37E-06 |
| -4.34 | -3.009 | 407 | 38 | enet | 0.049 | 3.00E-04 | 4.87519 | 1.09E-06 |
| -4.26 | 3.276 | 332 | 28 | enet | 0.042 | 0.00074 | -4.81804 | 1.45E-06 |
| -4.21 | 2.958 | 482 | 33 | enet | 0.063 | 4.30E-05 | -4.50824 | 6.54E-06 |
| 5.18 | 2.3 | 407 | 29 | enet | 0.1 | 1.40E-07 | -4.6234 | 3.78E-06 |
| 3.66 | 2.354 | 378 | 26 | enet | 0.024 | 0.0089 | 5.855 | 4.77E-09 |
| -3.66 | 3.763 | 251 | 251 | susie | 0.038 | 0.0014 | -4.5502 | 5.36E-06 |
| -6.99 | -4.345 | 260 | 11 | lasso | 0.15 | 1.50E-10 | 4.9763 | 6.48E-07 |
| 6.01 | -4.568 | 254 | 1 | top1 | 0.13 | 3.50E-09 | -4.568 | 4.92E-06 |
| -6.8 | -4.468 | 289 | 11 | lasso | 0.17 | 9.30E-12 | 4.844 | 1.27E-06 |
| 4.7 | -4.81 | 247 | 1 | top1 | 0.078 | 5.90E-06 | -4.81 | 1.51E-06 |
| 4.74 | -2.538 | 466 | 74 | enet | 0.07 | 1.70E-05 | 4.4592 | 8.23E-06 |
| 3.31 | -4.075 | 284 | 284 | susie | 0.021 | 0.013 | -4.9156 | 8.85E-07 |
| -8.37 | -4.841 | 345 | 1 | top1 | 0.28 | 5.20E-19 | 4.841 | 1.29E-06 |
| -5.14 | -5.448 | 331 | 1 | top1 | 0.099 | 3.40E-07 | 5.448 | 5.09E-08 |
| 3.8 | 2.984 | 326 | 21 | enet | 0.027 | 0.0058 | 4.84287 | 1.28E-06 |
| 6.34 | 4.455 | 315 | 315 | susie | 0.14 | 1.20E-09 | 4.428157 | 9.50E-06 |
| 3.27 | 2.013 | 272 | 6 | enet | 0.014 | 0.038 | 4.536315 | 5.72E-06 |
| 4.11 | 1.779 | 401 | 30 | enet | 0.067 | 2.70E-05 | 5.940332 | 2.84E-09 |
| 8.58 | -3.918 | 386 | 386 | susie | 0.34 | 7.60E-24 | -4.8074 | 1.53E-06 |
| -6.63 | -2.563 | 474 | 15 | enet | 0.23 | 1.60E-15 | 4.7271 | 2.28E-06 |
| 5.13 | 3.796 | 342 | 342 | susie | 0.17 | 6.70E-12 | 5.0864 | 3.65E-07 |
| 7.57 | 4.49 | 314 | 314 | susie | 0.25 | 6.70E-17 | 6.1246 | 9.09E-10 |
| 7.7 | -2.743 | 473 | 35 | enet | 0.23 | 1.70E-15 | -4.4267 | 9.57E-06 |
| -5.9 | 3.709 | 360 | 25 | enet | 0.13 | 4.80E-09 | -5.3064 | 1.12E-07 |
| 3.32 | 4.383 | 386 | 386 | susie | 0.025 | 0.0079 | 4.8333 | 1.34E-06 |
| -3.92 | -1.957 | 732 | 20 | enet | 0.047 | 4.00E-04 | -4.63583 | 3.56E-06 |
| -6.79 | 4.159 | 535 | 13 | enet | 0.23 | 1.90E-15 | -5.02876 | 4.94E-07 |
| 5.06 | 4.775 | 376 | 34 | enet | 0.098 | 3.90E-07 | -5.18517 | 2.16E-07 |
| 5.67 | -4.477 | 452 | 1 | top1 | 0.12 | 9.30E-09 | -4.477 | 7.57E-06 |
| -8.92 | 4.582 | 446 | 446 | susie | 0.33 | 1.10E-22 | -4.47208 | 7.75E-06 |
| 6.31 | -4.78 | 203 | 1 | top1 | 0.16 | 6.00E-11 | -4.78 | 1.75E-06 |
| -4.58 | 5.093 | 215 | 215 | susie | 0.054 | 0.00016 | -5.46479 | 4.63E-08 |
| 8.06 | 4.954 | 225 | 1 | top1 | 0.26 | 5.50E-18 | 4.954 | 7.27E-07 |
| -4.19 | -4.553 | 252 | 1 | top1 | 0.062 | 4.90E-05 | 4.553 | 5.29E-06 |
| -9.71 | 3.084 | 209 | 31 | enet | 0.49 | 2.80E-37 | -5.54038 | 3.02E-08 |
| 3.91 | 3.965 | 349 | 18 | enet | 0.043 | 0.00071 | 5.13734 | 2.79E-07 |
| 6.96 | -3.513 | 281 | 19 | enet | 0.19 | 8.60E-13 | -4.44923 | 8.62E-06 |
| -8.13 | 2.475 | 606 | 43 | enet | 0.29 | 1.50E-19 | -4.6514 | 3.30E-06 |

| | | | | | | | | |
|---|---|---|---|---|---|---|---|---|
| -5.02 | 4.419 | 191 | 3 | lasso | 0.097 | 4.40E-07 | -4.7258 | 2.29E-06 |
| 9.62 | 5.576 | 118 | 1 | top1 | 0.38 | 7.20E-27 | 5.5763 | 2.46E-08 |
| 5.83 | 5.71 | 166 | 1 | top1 | 0.12 | 9.10E-09 | 5.7095 | 1.13E-08 |
| -11.03 | -4.524 | 297 | 1 | top1 | 0.5 | 1.80E-38 | 4.524 | 6.07E-06 |
| 4.44 | -4.421 | 302 | 1 | top1 | 0.062 | 5.40E-05 | -4.421 | 9.82E-06 |
| -5.13 | 3.103 | 452 | 4 | lasso | 0.096 | 5.10E-07 | -4.4621 | 8.12E-06 |
| 4.65 | -4.237 | 317 | 317 | susie | 0.037 | 0.0015 | -4.4652 | 8.00E-06 |
| 5.58 | -5.23 | 286 | 3 | lasso | 0.099 | 3.40E-07 | -5.1187 | 3.08E-07 |
| 8.82 | 2.58 | 504 | 504 | susie | 0.5 | 2.90E-38 | 4.7653 | 1.89E-06 |
| -9.15 | 3.725 | 279 | 279 | susie | 0.34 | 4.50E-24 | -4.8916 | 1.00E-06 |
| 3.93 | -2.294 | 346 | 12 | enet | 0.032 | 0.0031 | -7.6175 | 2.59E-14 |
| 5.77 | -4.724 | 263 | 1 | top1 | 0.13 | 7.20E-09 | -4.724 | 2.31E-06 |
| 5.92 | -4.32 | 378 | 378 | susie | 0.11 | 1.20E-07 | -4.5183 | 6.23E-06 |
| 8.05 | -5.304 | 389 | 389 | susie | 0.27 | 3.90E-18 | -5.25113 | 1.51E-07 |
| 11.26 | -2.604 | 296 | 296 | susie | 0.66 | 3.40E-58 | -4.58616 | 4.51E-06 |
| -5.04 | -2.043 | 442 | 26 | enet | 0.084 | 2.70E-06 | 5.13326 | 2.85E-07 |
| -6.11 | -3.534 | 529 | 529 | susie | 0.23 | 2.20E-15 | 6.24451 | 4.25E-10 |
| -5.08 | 5.473 | 278 | 278 | susie | 0.058 | 8.90E-05 | -5.3448 | 9.05E-08 |
| -6.05 | -0.9 | 305 | 305 | susie | 0.35 | 2.10E-24 | 5.03454 | 4.79E-07 |
| 5.67 | 4.49 | 255 | 255 | susie | 0.096 | 5.20E-07 | 4.77044 | 1.84E-06 |
| -5.49 | 3.932 | 248 | 6 | enet | 0.091 | 1.00E-06 | -5.04222 | 4.60E-07 |
| -3.86 | 2.658 | 615 | 11 | lasso | 0.01 | 0.064 | -4.42837 | 9.49E-06 |
| -4.04 | 5.752 | 482 | 482 | susie | 0.04 | 0.001 | -4.80383 | 1.56E-06 |
| 4.4 | 5.168 | 486 | 486 | susie | 0.038 | 0.0013 | 4.60282 | 4.17E-06 |
| -4.28 | -4.815 | 306 | 306 | susie | 0.047 | 0.00037 | 4.66548 | 3.08E-06 |
| 7.47 | 4.165 | 411 | 411 | susie | 0.22 | 7.70E-15 | 4.55453 | 5.25E-06 |
| -8.88 | 5.075 | 346 | 1 | top1 | 0.32 | 4.20E-22 | -5.075 | 3.87E-07 |
| 5.36 | -4.922 | 431 | 2 | lasso | 0.13 | 8.10E-09 | -4.99485 | 5.89E-07 |
| -4.57 | 2.67 | 506 | 34 | enet | 0.055 | 0.00013 | -4.6536 | 3.26E-06 |

| TWAS.P.ADJ |
| --- |
| 0.025321 |
| 0.035148 |
| 0.012718 |
| 0.038501 |
| 0.004399 |
| 0.022488 |
| 3.71E-06 |
| 5.39E-06 |
| 0.031796 |
| 0.015146 |
| 0.001179 |
| 0.007284 |
| 0.008614 |
| 0.027807 |
| 0.045901 |
| 4.23E-05 |
| 0.025494 |
| 0.002122 |
| 0.004833 |
| 0.016591 |
| 9.13E-05 |
| 0.019309 |
| 0.021621 |
| 0.023702 |
| 0.009654 |
| 0.009076 |
| 0.000179 |
| 4.94E-05 |
| 0.011446 |
| 0.036073 |
| 0.034397 |
| 0.037577 |
| 0.016591 |
| 0.01399 |
| 0.032605 |
| 0.000332 |
| 3.58E-07 |
| 0.036594 |
| 0.006995 |
| 0.005232 |
| 0.004584 |
| 2.08E-06 |
| 0.048503 |
| 0.027864 |

| |
|---|
| 1.99E-05 |
| 0.044398 |
| 5.69E-05 |
| 1.40E-06 |
| 0.000105 |
| 0.01769 |
| 0.049659 |
| 2.57E-06 |
| 0.006879 |
| 0.009654 |
| 0.043762 |
| 0.008498 |
| 0.005596 |
| 0.015667 |
| 0.001191 |
| 0.0248 |
| 7.00E-05 |
| 0.007168 |
| 0.00418 |
| 0.004168 |
| 7.86E-05 |
| 0.047173 |
| 4.79E-09 |
| 0.04567 |
| 0.004394 |
| 0.035553 |
| 0.000257 |
| 0.014106 |
| 0.030928 |
| 0.010579 |
| 6.36E-05 |
| 1.86E-09 |
| 0.000161 |
| 0.008961 |
| 0.009481 |
| 0.015262 |
| 0.003295 |
| 0.002422 |
| 0.025144 |
| 0.000445 |
| 0.007404 |
| 0.026944 |
| 0.009616 |
| 0.008844 |
| 0.044838 |

| |
|---|
| 0.006582 |
| 0.016814 |
| 0.016969 |
| 0.026944 |
| 3.69E-05 |
| 0.014963 |
| 0.00725 |
| 0.006273 |
| 0.005965 |
| 0.00181 |
| 0.002005 |
| 0.002602 |
| 0.011004 |
| 0.01486 |
| 0.000138 |
| 0.000617 |
| 0.023036 |
| 0.014758 |
| 0.028795 |
| 0.003029 |
| 0.046381 |
| 0.009256 |
| 0.016917 |
| 0.028641 |
| 0.049466 |
| 7.30E-05 |
| 0.039696 |
| 0.00016 |
| 0.016763 |
| 1.66E-05 |
| 0.009513 |
| 0.000376 |
| 0.002499 |
| 0.001378 |
| 0.001923 |
| 0.002226 |
| 0.047564 |
| 0.041393 |
| 0.004885 |
| 0.002931 |
| 0.002298 |
| 0.002859 |
| 0.001743 |
| 0.000977 |
| 0.000183 |

| |
|---|
| 0.000684 |
| 0.016866 |
| 0.000207 |
| 0.01918 |
| 0.03404 |
| 0.048232 |
| 1.14E-05 |
| 0.01594 |
| 0.035788 |
| 0.021956 |
| 0.005245 |
| 0.030595 |
| 0.010541 |
| 3.77E-05 |
| 0.035943 |
| 0.00617 |
| 9.72E-09 |
| 0.006839 |
| 0.037125 |
| 0.03332 |
| 6.12E-07 |
| 0.000175 |
| 0.010747 |
| 7.30E-06 |
| 0.044427 |
| 0.005965 |
| 0.009153 |
| 2.92E-05 |
| 0.002869 |
| 0.046915 |
| 0.035186 |
| 0.005702 |
| 0.000935 |
| 0.006614 |
| 0.036164 |
| 0.023067 |
| 0.041051 |
| 7.33E-05 |
| 0.00012 |
| 0.022057 |
| 0.009122 |
| 0.011892 |
| 0.043331 |
| 0.002733 |
| 0.006874 |

| |
|---|
| 0.000854 |
| 0.018017 |
| 0.005702 |
| 2.68E-09 |
| 0.000922 |
| 2.18E-05 |
| 0.009285 |
| 0.03245 |
| 8.01E-05 |
| 0.014759 |
| 0.033557 |
| 0.009741 |
| 0.010719 |
| 0.006842 |
| 0.002893 |
| 0.038119 |
| 0.038119 |
| 0.002056 |
| 0.016029 |
| 0.003095 |
| 2.32E-05 |
| 0.043006 |
| 1.68E-05 |
| 0.008797 |
| 0.001065 |
| 0.000925 |
| 0.047567 |
| 0.022871 |
| 0.028768 |
| 0.001186 |
| 0.000518 |
| 5.51E-05 |
| 0.011696 |
| 0.035512 |
| 0.039096 |
| 0.01137 |
| 0.018571 |
| 0.02046 |
| 0.008275 |
| 0.000626 |
| 2.07E-05 |
| 0.015997 |
| 0.002372 |
| 0.010659 |
| 0.01086 |

| |
|---|
| 0.000697 |
| 0.039621 |
| 0.01086 |
| 0.029028 |
| 0.026548 |
| 0.029028 |
| 0.024336 |
| 0.030302 |
| 0.021587 |
| 0.006181 |
| 0.038548 |
| 0.000865 |
| 0.004177 |
| 0.012872 |
| 0.002856 |
| 0.035598 |
| 0.005967 |
| 0.022123 |
| 0.001435 |
| 0.009922 |
| 0.02809 |
| 0.013408 |
| 0.005001 |
| 0.000623 |
| 0.020984 |
| 5.46E-13 |
| 6.53E-05 |
| 0.015486 |
| 0.015888 |
| 0.038548 |
| 3.03E-09 |
| 0.007844 |
| 2.66E-10 |
| 0.001609 |
| 2.25E-05 |
| 0.020313 |
| 0.01609 |
| 0.022794 |
| 0.019576 |
| 0.031978 |
| 0.03895 |
| 6.48E-09 |
| 0.008581 |
| 0.004854 |
| 0.002561 |

| |
|---|
| 0.032581 |
| 0.011464 |
| 0.018168 |
| 2.50E-05 |
| 0.043442 |
| 0.000349 |
| 0.00181 |
| 0.041699 |
| 0.038548 |
| 0.018034 |
| 0.007307 |
| 0.005202 |
| 7.91E-05 |
| 0.00602 |
| 0.048403 |
| 0.014011 |
| 0.000878 |
| 4.69E-06 |
| 0.006771 |
| 0.00116 |
| 0.004505 |
| 0.02266 |
| 0.004827 |
| 0.020916 |
| 0.020179 |
| 0.017363 |
| 1.89E-06 |
| 0.01857 |
| 0.01086 |
| 0.000409 |
| 2.78E-05 |
| 0.004874 |
| 0.002641 |
| 0.007978 |
| 0.002219 |
| 0.002534 |
| 0.001032 |
| 0.010726 |
| 0.001891 |
| 0.003164 |
| 0.001341 |
| 0.007441 |
| 3.37E-07 |
| 0.008179 |
| 0.005913 |

| |
|---|
| 0.000176 |
| 0.00064 |
| 7.58E-05 |
| 0.023397 |
| 0.017296 |
| 0.000811 |
| 0.001703 |
| 5.18E-05 |
| 0.020984 |
| 0.014011 |
| 0.030369 |
| 0.029364 |
| 0.006122 |
| 0.010369 |
| 0.03053 |
| 0.017363 |
| 0.019338 |
| 1.57E-05 |
| 0.000639 |
| 0.015059 |
| 0.009381 |
| 0.000377 |
| 0.010286 |
| 3.64E-39 |
| 0.002428 |
| 0.046247 |
| 0.000184 |
| 0.002304 |
| 0.004147 |
| 0.028802 |
| 0.005349 |
| 0.02658 |
| 4.56E-05 |
| 2.90E-07 |
| 0.00082 |
| 0.031682 |
| 0.029213 |
| 0.002724 |
| 0.007809 |
| 2.75E-17 |
| 5.43E-10 |
| 0.004649 |
| 0.001317 |
| 0.04526 |
| 0.047728 |

| |
|---|
| 0.01975 |
| 0.004337 |
| 0.004608 |
| 0.001489 |
| 0.00581 |
| 0.000179 |
| 0.000252 |
| 0.001382 |
| 3.26E-11 |
| 4.16E-05 |
| 0.046329 |
| 0.009134 |
| 4.79E-06 |
| 0.029131 |
| 6.16E-07 |
| 1.34E-05 |
| 0.014072 |
| 0.002263 |
| 3.97E-06 |
| 0.000922 |
| 0.003563 |
| 0.00646 |
| 5.85E-06 |
| 0.010862 |
| 0.000314 |
| 0.025757 |
| 0.000498 |
| 0.000156 |
| 8.21E-05 |
| 0.025345 |
| 0.007999 |
| 0.014154 |
| 0.035878 |
| 0.040404 |
| 0.044025 |
| 3.34E-09 |
| 0.010615 |
| 0.000465 |
| 0.002617 |
| 0.016458 |
| 0.007406 |
| 3.27E-05 |
| 0.013002 |
| 0.043861 |
| 0.000732 |

| |
|---|
| 0.041063 |
| 0.001901 |
| 9.05E-07 |
| 0.002732 |
| 3.27E-05 |
| 0.000275 |
| 2.89E-05 |
| 0.004032 |
| 0.00053 |
| 0.000365 |
| 0.006419 |
| 0.004427 |
| 4.14E-06 |
| 0.009463 |
| 0.000296 |
| 0.000731 |
| 0.016458 |
| 0.041227 |
| 8.64E-06 |
| 0.000513 |
| 0.00655 |
| 0.000362 |
| 0.007587 |
| 0.015553 |
| 0.014318 |
| 0.039581 |
| 0.022547 |
| 0.005596 |
| 0.000181 |
| 0.024522 |
| 0.003053 |
| 0.016129 |
| 0.029954 |
| 1.71E-06 |
| 1.70E-06 |
| 0.000296 |
| 0.00079 |
| 0.032998 |
| 0.022301 |
| 0.0013 |
| 0.000739 |
| 0.01259 |
| 0.000554 |
| 0.029624 |
| 0.003267 |

| |
|---|
| 2.76E-06 |
| 0.000314 |
| 0.0302 |
| 0.00061 |
| 0.000191 |
| 4.30E-09 |
| 0.011521 |
| 0.012097 |
| 0.013578 |
| 0.008311 |
| 0.000286 |
| 0.009793 |
| 0.003448 |
| 0.003028 |
| 1.06E-06 |
| 0.005694 |
| 7.57E-05 |
| 0.001334 |
| 0.000184 |
| 6.52E-08 |
| 0.000687 |
| 0.003648 |
| 0.000818 |
| 0.029141 |
| 0.009384 |
| 2.62E-07 |
| 0.003669 |
| 0.041348 |
| 0.033798 |
| 0.030482 |
| 0.016723 |
| 0.007197 |
| 0.02011 |
| 0.014818 |
| 0.041136 |
| 0.011713 |
| 3.38E-07 |
| 0.002949 |
| 0.044029 |
| 0.043042 |
| 0.000478 |
| 0.023779 |
| 0.003013 |
| 0.002053 |
| 0.012842 |

| |
|---|
| 0.007479 |
| 0.003062 |
| 3.49E-19 |
| 6.86E-10 |
| 2.07E-12 |
| 5.24E-05 |
| 0.002554 |
| 0.011572 |
| 0.0035 |
| 0.031823 |
| 0.040713 |
| 0.033728 |
| 0.011784 |
| 4.42E-05 |
| 0.023708 |
| 0.000301 |
| 0.00369 |
| 0.003556 |
| 0.003916 |
| 1.64E-05 |
| 0.001658 |
| 8.18E-06 |
| 3.25E-06 |
| 0.00187 |
| 0.009102 |
| 0.000359 |
| 0.027801 |
| 6.47E-05 |
| 0.003048 |
| 0.017287 |
| 0.033163 |
| 0.008185 |
| 0.000981 |
| 0.001743 |
| 0.012912 |
| 0.035633 |
| 0.000762 |
| 0.048686 |
| 0.018981 |
| 0.006301 |
| 0.010513 |
| 0.003923 |
| 0.001108 |
| 0.024132 |
| 9.81E-05 |

| |
|---|
| 0.008044 |
| 8.54E-05 |
| 0.04283 |
| 0.042265 |
| 0.022509 |
| 0.0139 |
| 0.006851 |
| 0.031329 |
| 0.048334 |
| 0.006294 |
| 0.00061 |
| 0.000475 |
| 0.01764 |
| 0.023567 |
| 0.043394 |
| 0.013336 |
| 0.03902 |
| 0.007832 |
| 0.005207 |
| 0.014112 |
| 0.002604 |
| 0.046076 |
| 0.049533 |
| 0.001969 |
| 1.31E-05 |
| 0.015382 |
| 0.000741 |
| 0.047487 |
| 0.029424 |
| 1.04E-06 |
| 3.32E-16 |
| 0.023638 |
| 0.000924 |
| 0.049321 |
| 0.004615 |
| 0.024908 |
| 0.007197 |
| 1.17E-06 |
| 0.003509 |
| 0.001035 |
| 0.049926 |
| 0.030029 |
| 0.049191 |
| 2.51E-05 |
| 0.000749 |

| |
|---|
| 0.000263 |
| 0.000423 |
| 0.034801 |
| 0.045153 |
| 0.015859 |
| 0.049412 |
| 0.000214 |
| 0.039867 |
| 0.009471 |
| 0.000725 |
| 1.25E-06 |
| 0.031644 |
| 0.010793 |
| 0.00326 |
| 0.000642 |
| 0.000881 |
| 1.34E-06 |
| 0.042877 |
| 0.000127 |
| 0.002349 |
| 0.000918 |
| 0.00043 |
| 0.046328 |
| 0.038399 |
| 0.044346 |
| 0.029001 |
| 0.002393 |
| 0.000347 |
| 0.005837 |
| 0.007562 |
| 0.001557 |
| 0.000349 |
| 0.00105 |
| 3.44E-17 |
| 0.040087 |
| 0.029441 |
| 0.019456 |
| 0.011967 |
| 0.034875 |
| 0.039427 |
| 0.003605 |
| 0.009912 |
| 0.006953 |
| 0.013216 |
| 0.041042 |

| |
|---|
| 0.022393 |
| 8.22E-05 |
| 0.004574 |
| 0.044639 |
| 0.010572 |
| 0.025256 |
| 0.004376 |
| 0.001182 |
| 0.003377 |
| 0.041703 |
| 0.007232 |
| 0.012849 |
| 0.00016 |
| 0.005338 |
| 3.38E-07 |
| 0.00017 |
| 0.019163 |
| 2.91E-10 |
| 0.001314 |
| 0.000455 |
| 3.88E-09 |
| 0.000319 |
| 0.013583 |
| 5.40E-07 |
| 0.008076 |
| 2.15E-05 |
| 4.30E-05 |
| 0.019163 |
| 0.036269 |
| 0.000969 |
| 0.000562 |
| 0.016666 |
| 0.010572 |
| 0.000638 |
| 4.50E-08 |
| 0.000439 |
| 6.86E-07 |
| 0.016006 |
| 0.029515 |
| 0.017474 |
| 0.0002 |
| 0.038839 |
| 7.33E-05 |
| 0.020264 |
| 3.66E-05 |

| |
|---|
| 0.037297 |
| 0.03414 |
| 6.26E-06 |
| 0.041042 |
| 0.010499 |
| 0.000579 |
| 0.003532 |
| 9.76E-05 |
| 0.006813 |
| 0.044566 |
| 0.043024 |
| 0.024229 |
| 0.008076 |
| 0.019897 |
| 0.017914 |
| 0.006461 |
| 2.75E-10 |
| 0.004471 |
| 0.000673 |
| 0.043303 |
| 0.003876 |
| 0.000648 |
| 0.048096 |
| 0.012148 |
| 1.52E-05 |
| 0.008 |
| 0.000177 |
| 7.05E-05 |
| 2.82E-10 |
| 0.021817 |
| 0.010661 |
| 0.000383 |
| 0.00738 |
| 0.049501 |
| 4.00E-11 |
| 0.023883 |
| 0.00619 |
| 0.047931 |
| 1.73E-06 |
| 0.022313 |
| 0.004223 |
| 0.00068 |
| 0.000816 |
| 7.16E-05 |
| 0.018925 |

| |
|---|
| 0.003537 |
| 0.001207 |
| 0.004694 |
| 8.43E-06 |
| 0.048675 |
| 0.001983 |
| 0.011735 |
| 0.015123 |
| 0.003711 |
| 0.0219 |
| 0.042725 |
| 4.67E-06 |
| 0.031073 |
| 1.40E-06 |
| 2.13E-05 |
| 0.000342 |
| 0.024048 |
| 2.74E-08 |
| 0.000959 |
| 5.81E-06 |
| 0.048096 |
| 0.014793 |
| 1.86E-05 |
| 5.48E-05 |
| 0.002008 |
| 0.005215 |
| 0.009586 |
| 0.022395 |
| 0.024875 |
| 0.005661 |
| 0.012479 |
| 0.009917 |
| 0.001066 |
| 0.040824 |
| 0.024792 |
| 0.047601 |
| 8.68E-05 |
| 0.007024 |
| 0.004859 |
| 0.000545 |
| 0.005967 |
| 9.34E-05 |
| 0.006644 |
| 0.014545 |
| 0.029337 |

| |
|---|
| 0.000531 |
| 0.007652 |
| 0.037353 |
| 0.000975 |
| 0.026941 |
| 4.83E-05 |
| 0.003859 |
| 0.005008 |
| 0.000684 |
| 8.59E-06 |
| 3.19E-06 |
| 0.002 |
| 0.011322 |
| 0.013801 |
| 6.07E-05 |
| 0.005049 |
| 0.046278 |
| 4.94E-17 |
| 0.000584 |
| 0.042394 |
| 0.035453 |
| 0.001645 |
| 0.027436 |
| 0.00552 |
| 0.031899 |
| 0.00071 |
| 2.24E-06 |
| 0.008016 |
| 0.01033 |
| 0.002934 |
| 0.000207 |
| 2.65E-06 |
| 0.014379 |
| 8.25E-05 |
| 1.82E-08 |
| 0.024875 |
| 1.55E-05 |
| 0.043717 |
| 0.00733 |
| 0.011156 |
| 0.008677 |
| 0.048923 |
| 0.000154 |
| 0.016528 |
| 0.005694 |

| |
|---|
| 0.006206 |
| 0.001165 |
| 0.003793 |
| 0.0119 |
| 0.029833 |
| 0.006421 |
| 0.00219 |
| 0.043964 |
| 0.003173 |
| 0.00281 |
| 0.012892 |
| 0.004016 |
| 0.024544 |
| 0.036362 |
| 0.011156 |
| 0.002107 |
| 0.002206 |
| 3.31E-05 |
| 2.48E-06 |
| 0.032725 |
| 0.000165 |
| 0.000245 |
| 0.00012 |
| 0.01628 |
| 0.046092 |
| 9.02E-07 |
| 0.000117 |
| 2.41E-06 |
| 0.032165 |
| 0.000227 |
| 4.07E-05 |
| 0.040256 |
| 0.004284 |
| 0.011341 |
| 0.005498 |
| 0.002712 |
| 0.000164 |
| 0.017508 |
| 0.006765 |
| 0.001068 |
| 0.003203 |
| 0.018105 |
| 0.001883 |
| 0.001539 |
| 6.83E-07 |

| |
|---|
| 0.000172 |
| 0.027125 |
| 0.029313 |
| 0.008622 |
| 0.047154 |
| 0.046225 |
| 0.043639 |
| 0.012004 |
| 0.003362 |
| 0.0013 |
| 0.041649 |
| 0.024339 |
| 0.012336 |
| 0.026263 |
| 0.002938 |
| 4.79E-08 |
| 0.027191 |
| 4.16E-09 |
| 1.98E-06 |
| 6.60E-07 |
| 0.021156 |
| 0.016315 |
| 0.02288 |
| 0.028053 |
| 0.01459 |
| 0.014325 |
| 0.011473 |
| 0.007295 |
| 0.045297 |
| 0.001121 |
| 6.45E-07 |
| 0.00512 |
| 0.000586 |
| 3.03E-05 |
| 0.010678 |
| 0.021753 |
| 0.002248 |
| 0.016315 |
| 0.000121 |
| 0.004662 |
| 4.11E-17 |
| 3.32E-06 |
| 0.03442 |
| 0.018702 |
| 0.004337 |

| |
|---|
| 0.039659 |
| 0.002334 |
| 0.000703 |
| 3.55E-06 |
| 0.002613 |
| 0.023809 |
| 0.017575 |
| 0.001194 |
| 0.027589 |
| 0.001704 |
| 0.0382 |
| 3.57E-05 |
| 0.002832 |
| 0.040919 |
| 0.01532 |
| 0.006042 |
| 0.02036 |
| 0.004914 |
| 0.002838 |
| 0.03946 |
| 0.000367 |
| 0.013994 |
| 0.000147 |
| 5.11E-08 |
| 3.99E-05 |
| 6.56E-05 |
| 0.017044 |
| 0.002938 |
| 0.039726 |
| 0.02162 |
| 0.027324 |
| 0.009948 |
| 0.001996 |
| 0.005123 |
| 0.024832 |
| 1.55E-06 |
| 0.035719 |
| 0.01305 |
| 0.00607 |
| 5.91E-07 |
| 2.33E-06 |
| 0.025652 |
| 0.005637 |
| 0.00223 |
| 0.003311 |

| |
|---|
| 0.00516 |
| 0.000895 |
| 0.018344 |
| 0.001305 |
| 0.048694 |
| 5.61E-05 |
| 0.041461 |
| 0.008352 |
| 0.009918 |
| 0.018717 |
| 0.038926 |
| 0.028113 |
| 0.000295 |
| 0.035197 |
| 0.005675 |
| 0.044742 |
| 0.00428 |
| 0.019164 |
| 0.017897 |
| 2.02E-07 |
| 0.048471 |
| 0.000183 |
| 3.35E-08 |
| 0.045264 |
| 0.043027 |
| 0.008128 |
| 0.030574 |
| 0.019239 |
| 0.000632 |
| 0.000835 |
| 0.00733 |
| 0.045711 |
| 0.014317 |
| 0.004728 |
| 0.020507 |
| 0.000432 |
| 2.83E-07 |
| 0.004944 |
| 0.002341 |
| 0.048992 |
| 0.011186 |
| 0.023713 |
| 0.018046 |
| 0.034377 |
| 0.033929 |

| |
|---|
| 0.033184 |
| 0.032289 |
| 0.024906 |
| 0.009247 |
| 0.023788 |
| 0.01126 |
| 0.044369 |
| 0.022595 |
| 0.03736 |
| 0.000713 |
| 0.008352 |
| 4.26E-05 |
| 0.038702 |
| 1.07E-05 |
| 0.001879 |
| 0.010514 |
| 0.038031 |
| 0.03997 |
| 0.001447 |
| 0.02334 |
| 0.000144 |
| 7.44E-05 |
| 0.008874 |
| 3.09E-05 |
| 0.012155 |
| 0.027069 |
| 0.007226 |
| 6.99E-06 |
| 0.000104 |
| 0.030872 |
| 0.00865 |
| 0.001715 |
| 0.042878 |
| 0.012155 |
| 0.000658 |
| 0.027815 |
| 4.21E-05 |
| 0.033109 |
| 0.000715 |
| 5.60E-05 |
| 5.99E-06 |
| 0.00384 |
| 0.041386 |
| 0.006227 |
| 0.028709 |

| |
|---|
| 0.001894 |
| 0.002416 |
| 0.001111 |
| 0.000178 |
| 0.000115 |
| 0.023788 |
| 0.014243 |
| 0.048843 |
| 0.00962 |
| 0.003124 |
| 0.020283 |
| 0.035495 |
| 6.84E-05 |
| 0.00947 |
| 0.001894 |
| 4.50E-09 |
| 0.001208 |
| 0.013348 |
| 0.002386 |
| 0.009694 |
| 0.009234 |
| 5.76E-07 |
| 0.007233 |
| 0.00177 |
| 0.025824 |
| 0.001958 |
| 0.002638 |
| 0.003447 |
| 0.000551 |
| 0.018683 |
| 0.019422 |
| 0.040322 |
| 0.008711 |
| 0.003817 |
| 9.48E-06 |
| 0.032011 |
| 0.011358 |
| 0.041245 |
| 0.046478 |
| 0.000628 |
| 0.000287 |
| 0.034781 |
| 4.80E-06 |
| 0.031396 |
| 0.023116 |

| |
|---|
| 0.006679 |
| 0.006064 |
| 0.010034 |
| 0.002616 |
| 0.024686 |
| 0.000431 |
| 0.037859 |
| 0.002176 |
| 0.000588 |
| 0.046786 |
| 0.000659 |
| 0.002859 |
| 1.78E-05 |
| 0.005387 |
| 0.000239 |
| 2.67E-05 |
| 0.040014 |
| 0.001148 |
| 0.000456 |
| 0.000311 |
| 0.001047 |
| 0.003293 |
| 0.000295 |
| 0.009573 |
| 0.00128 |
| 0.015821 |
| 0.024378 |
| 0.001019 |
| 0.002699 |
| 0.018899 |
| 0.032319 |
| 0.033858 |
| 0.007726 |
| 0.000283 |
| 0.020407 |
| 0.013512 |
| 0.005448 |
| 0.020777 |
| 0.031088 |
| 0.049556 |
| 0.045247 |
| 0.023516 |
| 0.015113 |
| 0.043092 |
| 0.036936 |

| |
|---|
| 0.043708 |
| 3.85E-05 |
| 0.000629 |
| 0.007086 |
| 0.0028 |
| 5.55E-06 |
| 0.011743 |
| 0.005401 |
| 0.007185 |
| 0.032406 |
| 0.01873 |
| 2.36E-05 |
| 0.026559 |
| 0.003211 |
| 0.024379 |
| 0.006293 |
| 0.007482 |
| 0.04078 |
| 0.004385 |
| 0.006392 |
| 0.000252 |
| 0.006342 |
| 0.047073 |
| 0.028343 |
| 1.41E-05 |
| 0.007581 |
| 0.011297 |
| 0.001809 |
| 4.50E-06 |
| 0.047419 |
| 0.000555 |
| 0.00664 |
| 0.01764 |
| 0.002448 |
| 0.00107 |
| 0.037509 |
| 0.038401 |
| 0.008671 |
| 0.000229 |
| 0.003602 |
| 0.026212 |
| 0.00015 |
| 0.001382 |
| 0.042712 |
| 0.016352 |

| |
|---|
| 0.011347 |
| 0.000122 |
| 5.60E-05 |
| 0.030077 |
| 0.048658 |
| 0.040235 |
| 0.03964 |
| 0.001526 |
| 0.009365 |
| 0.004955 |
| 1.28E-10 |
| 0.011446 |
| 0.03087 |
| 0.000748 |
| 0.022347 |
| 0.001412 |
| 2.11E-06 |
| 0.000448 |
| 0.002373 |
| 0.009117 |
| 0.002279 |
| 0.047023 |
| 0.00773 |
| 0.020662 |
| 0.015261 |
| 0.026014 |
| 0.001918 |
| 0.002918 |
| 0.016153 |
