## Supplementary Table 10 for "Hundreds of cardiac MRI traits derived using 3D diffusion autoencoders share a common genetic architecture"

| TISSUE | NGENES | BETA | BETA_STD | SE | P | ADJP |
|---|---|---|---|---|---|---|
| Heart_Atri | 17127 | 0.026077 | 0.046754 | 0.005656 | 2.02E-06 | 0.000109 |
| Heart_Left_ | 17127 | 0.022556 | 0.037603 | 0.005375 | 1.36E-05 | 0.000736 |
| Colon_Sign | 17127 | 0.024362 | 0.047598 | 0.007449 | 0.000538 | 0.02906 |
| Muscle_Sk | 17127 | 0.012407 | 0.023459 | 0.004125 | 0.001317 | 0.07114 |
| Esophagus_ | 17127 | 0.022086 | 0.043418 | 0.007589 | 0.001808 | 0.097616 |
| Adipose_Su | 17127 | 0.017259 | 0.034393 | 0.006411 | 0.003552 | 0.191813 |
| Esophagus_ | 17127 | 0.019699 | 0.038805 | 0.007356 | 0.003709 | 0.200308 |
| Artery_Cor | 17127 | 0.018385 | 0.036544 | 0.00706 | 0.004609 | 0.248875 |
| Adipose_Vi | 17127 | 0.015889 | 0.03081 | 0.006651 | 0.008455 | 0.456586 |
| Bladder | 17127 | 0.017295 | 0.033657 | 0.007771 | 0.013028 | 0.703512 |
| Artery_Tibi | 17127 | 0.012927 | 0.026588 | 0.005912 | 0.014393 | 0.777222 |
| Uterus | 17127 | 0.013946 | 0.02838 | 0.006442 | 0.015205 | 0.82107 |
| Stomach | 17127 | 0.014948 | 0.02711 | 0.007094 | 0.017558 | 0.948132 |
| Cervix_End | 17127 | 0.01279 | 0.025283 | 0.006815 | 0.030275 | 1.63485 |
| Colon_Tran | 17127 | 0.012377 | 0.022718 | 0.006878 | 0.035977 | 1.942758 |
| Cervix_Ecto | 17127 | 0.012419 | 0.024305 | 0.00715 | 0.041207 | 2.225178 |
| Artery_Aor | 17127 | 0.010509 | 0.02142 | 0.006076 | 0.041876 | 2.261304 |
| Liver | 17127 | 0.006284 | 0.011302 | 0.003807 | 0.04942 | 2.66868 |
| Breast_Ma | 17127 | 0.011637 | 0.022342 | 0.007528 | 0.061066 | 3.297564 |
| Small_Intes | 17127 | 0.008292 | 0.015219 | 0.005687 | 0.072414 | 3.910356 |
| Ovary | 17127 | 0.007473 | 0.015132 | 0.005736 | 0.096319 | 5.201226 |
| Nerve_Tibi | 17127 | 0.006851 | 0.013769 | 0.005942 | 0.12448 | 6.72192 |
| Cells_Cultu | 17127 | 0.004296 | 0.009199 | 0.003912 | 0.13609 | 7.34886 |
| Fallopian_T | 17127 | 0.007113 | 0.013776 | 0.007023 | 0.15559 | 8.40186 |
| Vagina | 17127 | 0.00478 | 0.009127 | 0.006518 | 0.23167 | 12.51018 |
| Kidney_Me | 17127 | 0.003988 | 0.007359 | 0.005549 | 0.23619 | 12.75426 |
| Kidney_Co | 17127 | 0.003398 | 0.005958 | 0.005406 | 0.26483 | 14.30082 |
| Prostate | 17127 | 0.003808 | 0.007212 | 0.007175 | 0.29782 | 16.08228 |
| Thyroid | 17127 | 0.00256 | 0.005063 | 0.005763 | 0.32847 | 17.73738 |
| Skin_Not_S | 17127 | 0.001196 | 0.002303 | 0.004568 | 0.39669 | 21.42126 |
| Skin_Sun_E | 17127 | 0.000869 | 0.001684 | 0.004585 | 0.42484 | 22.94136 |
| Adrenal_Gl | 17127 | 0.001024 | 0.001972 | 0.006083 | 0.43319 | 23.39226 |
| Testis | 17127 | 0.000238 | 0.00041 | 0.003326 | 0.4715 | 25.461 |
| Lung | 17127 | -0.00093 | -0.0018 | 0.005445 | 0.56808 | 30.67632 |
| Esophagus_ | 17127 | -0.00118 | -0.00229 | 0.004524 | 0.6033 | 32.5782 |
| Spleen | 17127 | -0.00144 | -0.00284 | 0.004336 | 0.62974 | 34.00596 |
| Pancreas | 17127 | -0.00286 | -0.00478 | 0.004994 | 0.7166 | 38.6964 |
| Minor_Sali | 17127 | -0.00331 | -0.00615 | 0.00571 | 0.71883 | 38.81682 |
| Pituitary | 17127 | -0.00704 | -0.01309 | 0.005339 | 0.90628 | 48.93912 |
| Brain_Subs | 17127 | -0.00957 | -0.01646 | 0.005063 | 0.97067 | 52.41618 |
| Cells_EBV-t | 17127 | -0.00607 | -0.01322 | 0.002908 | 0.98152 | 53.00208 |
| Brain_Puta | 17127 | -0.00988 | -0.01679 | 0.004682 | 0.98256 | 53.05824 |
| Brain_Amy | 17127 | -0.00983 | -0.01672 | 0.004632 | 0.98307 | 53.08578 |
| Whole_Blo | 17127 | -0.00733 | -0.0132 | 0.003316 | 0.98641 | 53.26614 |

| | | | | | | |
|---|---|---|---|---|---|---|
| Brain_Hipp | 17127 | -0.01078 | -0.01823 | 0.00471 | 0.98894 | 53.40276 |
| Brain_Spin | 17127 | -0.01168 | -0.02096 | 0.005098 | 0.98901 | 53.40654 |
| Brain_Cauc | 17127 | -0.01092 | -0.01883 | 0.004658 | 0.99044 | 53.48376 |
| Brain_Ante | 17127 | -0.01006 | -0.01774 | 0.004277 | 0.99063 | 53.49402 |
| Brain_Hypo | 17127 | -0.01153 | -0.0198 | 0.004783 | 0.99201 | 53.56854 |
| Brain_Cort | 17127 | -0.01111 | -0.02023 | 0.004233 | 0.99565 | 53.7651 |
| Brain_Nucl | 17127 | -0.01211 | -0.02098 | 0.00449 | 0.99649 | 53.81046 |
| Brain_Fron | 17127 | -0.01106 | -0.02035 | 0.004082 | 0.99662 | 53.81748 |
| Brain_Cere | 17127 | -0.01179 | -0.02339 | 0.003739 | 0.99919 | 53.95626 |
| Brain_Cere | 17127 | -0.0118 | -0.02366 | 0.003624 | 0.99943 | 53.96922 |
