## Supplementary Table 9 for "Hundreds of cardiac MRI traits derived using 3D diffusion autoencoders share a common genetic architecture"

| GENE_SET | NGENES | BETA | BETA_STD | SE | P | ADJP |
|---|---|---|---|---|---|---|
| BRUNEAU_ | 5 | 1.1953 | 0.01945 | 0.24874 | 7.79E-07 | 0.013255 |
| GOBP_HEA | 251 | 0.16225 | 0.018584 | 0.033953 | 8.90E-07 | 0.015132 |
| GOBP_CHR | 581 | 0.095053 | 0.016417 | 0.02018 | 1.25E-06 | 0.0212 |
| GOBP_HEA | 568 | 0.097487 | 0.016654 | 0.021813 | 3.95E-06 | 0.067205 |
| GOBP_CAR | 35 | 0.42007 | 0.018071 | 0.094602 | 4.52E-06 | 0.076834 |
| GOBP_CAR | 58 | 0.32055 | 0.01774 | 0.072565 | 5.03E-06 | 0.085482 |
| GOBP_CAR | 122 | 0.21452 | 0.01719 | 0.048671 | 5.26E-06 | 0.089451 |
| GOBP_CIR( | 1058 | 0.071731 | 0.016499 | 0.016377 | 5.97E-06 | 0.101596 |
| GOBP_REG | 17 | 0.55666 | 0.016697 | 0.13297 | 1.42E-05 | 0.242219 |
| GOBP_MU: | 393 | 0.10617 | 0.015159 | 0.025442 | 1.51E-05 | 0.256776 |
| WP_DIFFEI | 25 | 0.509 | 0.018511 | 0.12231 | 1.59E-05 | 0.270209 |
| GOCC_SPLI | 172 | 0.15032 | 0.014283 | 0.036229 | 1.68E-05 | 0.285072 |
| GOBP_MIT | 276 | 0.12005 | 0.01441 | 0.028958 | 1.70E-05 | 0.289391 |
| GOBP_MEI | 57 | 0.26208 | 0.014379 | 0.063239 | 1.71E-05 | 0.291347 |
| GOMF_HEI | 160 | 0.17568 | 0.016105 | 0.042405 | 1.72E-05 | 0.292911 |
| GOBP_NEG | 1379 | 0.057647 | 0.015001 | 0.013938 | 1.78E-05 | 0.302179 |
| GOBP_ATR | 22 | 0.48501 | 0.016547 | 0.11751 | 1.84E-05 | 0.313436 |
| KEGG_SPLI | 113 | 0.18114 | 0.013973 | 0.044255 | 2.14E-05 | 0.363533 |
| GOBP_REG | 8 | 0.88416 | 0.018197 | 0.21609 | 2.15E-05 | 0.365982 |
| GOBP_MU: | 332 | 0.11408 | 0.014995 | 0.028016 | 2.34E-05 | 0.398053 |
| GOBP_MIT | 58 | 0.25658 | 0.0142 | 0.063092 | 2.39E-05 | 0.407066 |
| GOBP_NEG | 1454 | 0.055085 | 0.014687 | 0.013637 | 2.69E-05 | 0.457571 |
| GOBP_CAR | 70 | 0.26644 | 0.016195 | 0.066222 | 2.88E-05 | 0.489744 |
| GOCC_U2_ | 84 | 0.21592 | 0.014371 | 0.053846 | 3.05E-05 | 0.518635 |
| GOBP_PRC | 4 | 1.1435 | 0.016643 | 0.28534 | 3.08E-05 | 0.524383 |
| GOBP_SKEI | 505 | 0.097637 | 0.015754 | 0.024528 | 3.45E-05 | 0.58686 |
| GOBP_REG | 120 | 0.19332 | 0.015364 | 0.048691 | 3.60E-05 | 0.612758 |
| GOBP_SAR | 43 | 0.30192 | 0.014393 | 0.076405 | 3.90E-05 | 0.662889 |
| REACTOMI | 653 | 0.071346 | 0.013038 | 0.018286 | 4.80E-05 | 0.815713 |
| GOBP_CAR | 123 | 0.19452 | 0.01565 | 0.04986 | 4.80E-05 | 0.816716 |
| GOBP_CAR | 162 | 0.16433 | 0.015157 | 0.04241 | 5.36E-05 | 0.91089 |
| GOBP_MU: | 77 | 0.23942 | 0.01526 | 0.061848 | 5.44E-05 | 0.924494 |
| GOMF_PAT | 8 | 0.88534 | 0.018222 | 0.22889 | 5.51E-05 | 0.936431 |
| GOBP_NEG | 5 | 0.79573 | 0.012948 | 0.20623 | 5.73E-05 | 0.973876 |
| GOBP_CHC | 111 | 0.20014 | 0.015302 | 0.051901 | 5.78E-05 | 0.98277 |
| GOBP_ANII | 987 | 0.067489 | 0.015023 | 0.017531 | 5.94E-05 | 1.009672 |
| GOCC_AMI | 13 | 0.51339 | 0.013468 | 0.13347 | 6.01E-05 | 1.022834 |
| GOMF_MII | 9 | 0.59845 | 0.013064 | 0.15613 | 6.35E-05 | 1.079988 |
| GOBP_ACT | 16 | 0.47989 | 0.013965 | 0.1267 | 7.64E-05 | 1.29857 |
| IIZUKA_LIV | 14 | 0.4415 | 0.012019 | 0.11661 | 7.68E-05 | 1.305865 |
| GOBP_MIC | 17 | 0.45891 | 0.013765 | 0.12132 | 7.79E-05 | 1.324332 |
| REACTOMI | 192 | 0.12411 | 0.012453 | 0.032839 | 7.89E-05 | 1.341388 |
| GOBP_MES | 117 | 0.17695 | 0.013887 | 0.046922 | 8.15E-05 | 1.386554 |
| GOBP_STR | 236 | 0.12721 | 0.014134 | 0.033847 | 8.58E-05 | 1.458485 |

| | | | | | | |
|---|---|---|---|---|---|---|
| REACTOME | 154 | 0.15314 | 0.013775 | 0.040915 | 9.13E-05 | 1.552199 |
| GOBP_THY | 29 | 0.35786 | 0.014015 | 0.096041 | 9.76E-05 | 1.658957 |
| IVANOVA_ | 527 | 0.080707 | 0.013295 | 0.021768 | 0.000105 | 1.784845 |
| GOCC_CON | 231 | 0.12626 | 0.013881 | 0.03411 | 0.000108 | 1.828548 |
| GOBP_ESC | 10 | 0.6167 | 0.01419 | 0.16758 | 0.000117 | 1.989585 |
| GOBP_PRE | 6 | 0.76619 | 0.013657 | 0.20927 | 0.000126 | 2.140249 |
| GOBP_POS | 5 | 1.0109 | 0.01645 | 0.27627 | 0.000127 | 2.157764 |
| GAUSSMAI | 174 | 0.14473 | 0.013831 | 0.039562 | 0.000127 | 2.165417 |
| GOBP_STR | 70 | 0.21864 | 0.013289 | 0.05988 | 0.000131 | 2.224934 |
| GOMF_RNA | 328 | 0.10261 | 0.013408 | 0.028133 | 0.000133 | 2.259114 |
| LIU_VAV3_ | 81 | 0.20578 | 0.01345 | 0.056564 | 0.000138 | 2.344139 |
| GOBP_REG | 57 | 0.23392 | 0.012834 | 0.064369 | 0.00014 | 2.37985 |
| GOBP_GAS | 176 | 0.14265 | 0.013709 | 0.039258 | 0.00014 | 2.382911 |
| KEGG_DILA | 85 | 0.20845 | 0.013956 | 0.057422 | 0.000142 | 2.41522 |
| GOCC_CHR | 1738 | 0.045369 | 0.013117 | 0.012536 | 0.000148 | 2.520481 |
| GOBP_HEN | 21 | 0.37427 | 0.012476 | 0.10345 | 0.000149 | 2.533915 |
| GOBP_POS | 52 | 0.24788 | 0.012992 | 0.0686 | 0.000152 | 2.576428 |
| GOBP_CAR | 28 | 0.38718 | 0.0149 | 0.10726 | 0.000154 | 2.612988 |
| GOBP_LYS( | 15 | 0.45798 | 0.012905 | 0.12688 | 0.000154 | 2.614689 |
| JEPSEN_SM | 30 | 0.29179 | 0.011623 | 0.080921 | 0.000156 | 2.65329 |
| GOBP_NEG | 5 | 0.92886 | 0.015115 | 0.25896 | 0.000168 | 2.852929 |
| GOBP_TEL( | 161 | 0.13147 | 0.012089 | 0.036789 | 0.000177 | 3.002233 |
| GOBP_RNA | 264 | 0.10439 | 0.012258 | 0.029241 | 0.000179 | 3.043555 |
| GOBP_END | 133 | 0.16687 | 0.013957 | 0.046909 | 0.000188 | 3.193879 |
| GOBP_TEL( | 76 | 0.19744 | 0.012502 | 0.055523 | 0.000189 | 3.210374 |
| GOBP_CON | 264 | 0.11692 | 0.013729 | 0.032885 | 0.000189 | 3.218196 |
| GOBP_CHR | 42 | 0.24796 | 0.011683 | 0.069746 | 0.000189 | 3.221087 |
| GOMF_TRA | 1587 | 0.048479 | 0.013452 | 0.013717 | 0.000205 | 3.488236 |
| GOBP_REG | 8 | 0.72277 | 0.014876 | 0.20462 | 0.000207 | 3.512893 |
| GOBP_ANT | 25 | 0.34034 | 0.012377 | 0.096885 | 0.000222 | 3.780382 |
| GOBP_PER | 18 | 0.46994 | 0.014504 | 0.13387 | 0.000224 | 3.813881 |
| GOBP_NEG | 1163 | 0.053348 | 0.012827 | 0.015251 | 0.000235 | 3.996005 |
| GOMF_REC | 75 | 0.19596 | 0.012327 | 0.05616 | 0.000243 | 4.127114 |
| GOBP_REG | 6 | 0.81999 | 0.014616 | 0.23508 | 0.000244 | 4.147349 |
| GOBP_NEG | 11 | 0.58203 | 0.014045 | 0.16699 | 0.000246 | 4.188672 |
| GOMF_SEC | 1449 | 0.050698 | 0.013496 | 0.014568 | 0.000251 | 4.273867 |
| GOMF_SM | 74 | 0.21346 | 0.013339 | 0.061349 | 0.000252 | 4.282199 |
| GOBP_NEG | 23 | 0.37252 | 0.012995 | 0.10714 | 0.000254 | 4.323861 |
| GOBP_POS | 5 | 0.9456 | 0.015387 | 0.27228 | 0.000258 | 4.38848 |
| DELACROIX | 355 | 0.094276 | 0.012806 | 0.027216 | 0.000267 | 4.537444 |
| GOCC_KINI | 20 | 0.36538 | 0.011886 | 0.10549 | 0.000267 | 4.543736 |
| GOCC_AMI | 11 | 0.48528 | 0.011711 | 0.14019 | 0.000269 | 4.578086 |
| GOBP_REG | 10 | 0.62983 | 0.014492 | 0.18224 | 0.000275 | 4.672634 |
| GOBP_ORG | 154 | 0.15153 | 0.01363 | 0.04393 | 0.000282 | 4.791329 |
| FEVR_CTNI | 520 | 0.075267 | 0.012319 | 0.021821 | 0.000282 | 4.791499 |

| | | | | | | |
|---|---|---|---|---|---|---|
| GOMF_PR( | 36 | 0.2937 | 0.012813 | 0.085237 | 0.000286 | 4.856118 |
| GOBP_MET | 73 | 0.19632 | 0.012184 | 0.056987 | 0.000286 | 4.868021 |
| GOBP_VEN | 45 | 0.28242 | 0.013772 | 0.082016 | 0.000288 | 4.895059 |
| GOBP_VEN | 70 | 0.22456 | 0.013649 | 0.065271 | 0.000291 | 4.951006 |
| GOBP_REN | 306 | 0.10867 | 0.013722 | 0.031585 | 0.000291 | 4.951176 |
| GOBP_REG | 10 | 0.56346 | 0.012965 | 0.1641 | 0.000299 | 5.076843 |
| GOBP_NON | 14 | 0.45217 | 0.012309 | 0.13169 | 0.000299 | 5.079394 |
| GOBP_VEN | 54 | 0.25634 | 0.013691 | 0.074718 | 0.000302 | 5.128538 |
| REACTOME | 256 | 0.097722 | 0.011302 | 0.028497 | 0.000303 | 5.160167 |
| GOBP_CER | 4 | 0.92954 | 0.013529 | 0.27137 | 0.000308 | 5.231588 |
| GOBP_MUS | 193 | 0.12879 | 0.012956 | 0.037624 | 0.00031 | 5.273931 |
| GOMF_RN/ | 1169 | 0.055979 | 0.013492 | 0.016363 | 0.000313 | 5.317464 |
| REACTOME | 15 | 0.49245 | 0.013876 | 0.14409 | 0.000317 | 5.383273 |
| GOBP_REG | 27 | 0.33184 | 0.012541 | 0.097429 | 0.00033 | 5.617772 |
| GOMF_DN/ | 443 | 0.083268 | 0.012605 | 0.024452 | 0.000331 | 5.632396 |
| BOUDOUKI | 97 | 0.17252 | 0.012335 | 0.050738 | 0.000338 | 5.739698 |
| GOBP_NEG | 9 | 0.59235 | 0.012931 | 0.17426 | 0.000339 | 5.758573 |
| IBRAHIM_N | 132 | 0.14063 | 0.011718 | 0.041419 | 0.000344 | 5.845129 |
| GOBP_END | 17 | 0.41007 | 0.0123 | 0.12086 | 0.000346 | 5.890532 |
| GOBP_SEQ | 4 | 0.94234 | 0.013716 | 0.27805 | 0.000351 | 5.975217 |
| GOBP_POS | 1698 | 0.043274 | 0.012381 | 0.012769 | 0.000352 | 5.980829 |
| GOBP_CAR | 48 | 0.2709 | 0.013643 | 0.079999 | 0.000355 | 6.037625 |
| GOMF_FRU | 4 | 0.67224 | 0.009784 | 0.1989 | 0.000363 | 6.181147 |
| GOBP_GLA | 421 | 0.086749 | 0.012809 | 0.025705 | 0.00037 | 6.295081 |
| GOBP_REG | 4 | 1.0423 | 0.01517 | 0.30889 | 0.000371 | 6.306304 |
| GOBP_NEG | 12 | 0.62066 | 0.015643 | 0.18414 | 0.000376 | 6.39439 |
| GOBP_TISS | 598 | 0.073227 | 0.012825 | 0.021761 | 0.000383 | 6.521247 |
| GOBP_CELI | 24 | 0.38847 | 0.013842 | 0.11544 | 0.000384 | 6.521588 |
| REACTOME | 156 | 0.1278 | 0.011569 | 0.038052 | 0.000393 | 6.677013 |
| GOBP_SIGI | 161 | 0.12784 | 0.011755 | 0.038096 | 0.000397 | 6.747414 |
| GOBP_NEP | 143 | 0.15852 | 0.013744 | 0.047274 | 0.0004 | 6.808462 |
| GOBP_URC | 63 | 0.23406 | 0.013499 | 0.069812 | 0.000401 | 6.817815 |
| GOBP_MIT | 191 | 0.11857 | 0.011866 | 0.035379 | 0.000403 | 6.854205 |
| GOMF_GLY | 222 | 0.11909 | 0.012838 | 0.035538 | 0.000403 | 6.857776 |
| GOBP_CELI | 12 | 0.5129 | 0.012927 | 0.15334 | 0.000413 | 7.015073 |
| GOBP_REG | 27 | 0.30312 | 0.011455 | 0.090665 | 0.000415 | 7.055034 |
| GOBP_EMI | 5 | 0.98764 | 0.016071 | 0.29561 | 0.000418 | 7.112681 |
| GOBP_REG | 5 | 0.80691 | 0.01313 | 0.24177 | 0.000423 | 7.201447 |
| GOBP_REG | 99 | 0.1571 | 0.011347 | 0.047082 | 0.000425 | 7.224234 |
| GOBP_NEG | 13 | 0.44561 | 0.011689 | 0.13355 | 0.000425 | 7.225254 |
| CAIRO_LIVI | 151 | 0.14011 | 0.012481 | 0.042055 | 0.000433 | 7.358234 |
| GOBP_POS | 62 | 0.20875 | 0.011943 | 0.062777 | 0.000443 | 7.527093 |
| GOBP_CYT( | 144 | 0.12497 | 0.010873 | 0.037631 | 0.000449 | 7.642047 |
| GOBP_TEL( | 148 | 0.1292 | 0.011395 | 0.038928 | 0.000453 | 7.697483 |
| REACTOME | 38 | 0.26751 | 0.01199 | 0.080641 | 0.000455 | 7.743227 |

| | | | | | | |
|---|---|---|---|---|---|---|
| FLOTHO_PI | 52 | 0.21717 | 0.011382 | 0.06578 | 0.000482 | 8.19573 |
| BURTON_A | 38 | 0.25333 | 0.011354 | 0.077007 | 0.000503 | 8.545013 |
| GOBP_REG | 350 | 0.089079 | 0.012016 | 0.027101 | 0.000508 | 8.630548 |
| REACTOME | 31 | 0.30123 | 0.012197 | 0.091804 | 0.000518 | 8.805019 |
| GOBP_FOR | 13 | 0.50685 | 0.013296 | 0.15487 | 0.000534 | 9.076589 |
| GOBP_RES | 531 | 0.074386 | 0.012299 | 0.022741 | 0.000537 | 9.130835 |
| GOBP_POS | 6 | 0.81358 | 0.014502 | 0.24921 | 0.000549 | 9.338976 |
| GOBP_PAR | 5 | 0.87571 | 0.01425 | 0.26873 | 0.000561 | 9.537424 |
| GOBP_GAS | 24 | 0.35415 | 0.012619 | 0.10883 | 0.00057 | 9.69115 |
| REACTOME | 362 | 0.083256 | 0.011418 | 0.025642 | 0.000585 | 9.941803 |
| GOMF_SUL | 4 | 0.64839 | 0.009437 | 0.19999 | 0.000594 | 10.10845 |
| BLALOCK_A | 1552 | 0.041393 | 0.01137 | 0.012774 | 0.000598 | 10.1678 |
| WANG_TU | 372 | 0.080031 | 0.011123 | 0.024791 | 0.000624 | 10.61367 |
| GOCC_AXC | 13 | 0.57446 | 0.01507 | 0.17813 | 0.000631 | 10.72947 |
| GOBP_GRA | 36 | 0.27291 | 0.011906 | 0.084634 | 0.000632 | 10.74546 |
| GOBP_CAR | 192 | 0.12193 | 0.012234 | 0.037872 | 0.000643 | 10.93728 |
| GOBP_REG | 11 | 0.53665 | 0.01295 | 0.16683 | 0.00065 | 11.0456 |
| GOBP_POS | 33 | 0.30613 | 0.012788 | 0.095555 | 0.00068 | 11.55609 |
| GOBP_VAC | 12 | 0.42494 | 0.01071 | 0.13281 | 0.000689 | 11.72104 |
| GUO_HEX_ | 72 | 0.19322 | 0.01191 | 0.060417 | 0.000693 | 11.7831 |
| GOBP_REG | 5 | 0.79134 | 0.012877 | 0.2475 | 0.000695 | 11.8166 |
| GOBP_CELI | 54 | 0.24522 | 0.013096 | 0.0769 | 0.000716 | 12.17048 |
| GOMF_SEL | 9 | 0.51293 | 0.011197 | 0.16087 | 0.000717 | 12.18578 |
| GOBP_REG | 155 | 0.13095 | 0.011817 | 0.041154 | 0.000733 | 12.45667 |
| GOBP_REG | 31 | 0.28973 | 0.011731 | 0.091233 | 0.000749 | 12.73113 |
| GOBP_SIST | 241 | 0.099081 | 0.011123 | 0.031201 | 0.000749 | 12.73589 |
| GOCC_SPIN | 76 | 0.17325 | 0.01097 | 0.054609 | 0.000757 | 12.87449 |
| GOBP_NEG | 5 | 0.60936 | 0.009916 | 0.19227 | 0.000766 | 13.01886 |
| GOBP_EXP | 27 | 0.28969 | 0.010948 | 0.091461 | 0.00077 | 13.10116 |
| LEIN_LOCA | 17 | 0.37314 | 0.011192 | 0.11784 | 0.000773 | 13.1418 |
| GRYDER_PA | 981 | 0.055592 | 0.012339 | 0.017608 | 0.000798 | 13.56778 |
| GOBP_MET | 87 | 0.18612 | 0.012606 | 0.058956 | 0.000799 | 13.57866 |
| GOCC_CHR | 1207 | 0.047793 | 0.011692 | 0.015154 | 0.000807 | 13.72729 |
| SARTIPY_N | 18 | 0.38195 | 0.011789 | 0.12113 | 0.000809 | 13.75807 |
| HOSHIDA_I | 54 | 0.20351 | 0.010869 | 0.06456 | 0.000811 | 13.79905 |
| GOBP_NUC | 24 | 0.28742 | 0.010242 | 0.091208 | 0.000814 | 13.84785 |
| GOBP_NEP | 107 | 0.17091 | 0.012831 | 0.054304 | 0.000825 | 14.03219 |
| GOBP_REG | 21 | 0.35319 | 0.011773 | 0.11227 | 0.00083 | 14.10837 |
| GOBP_REG | 36 | 0.29998 | 0.013087 | 0.095373 | 0.000831 | 14.13201 |
| GOBP_PRC | 28 | 0.28837 | 0.011098 | 0.091757 | 0.000838 | 14.25325 |
| KONDO_EZ | 214 | 0.11241 | 0.011901 | 0.035787 | 0.000843 | 14.33045 |
| GOBP_BRC | 52 | 0.21669 | 0.011357 | 0.069168 | 0.000867 | 14.74793 |
| GOBP_MET | 15 | 0.46832 | 0.013196 | 0.15013 | 0.000907 | 15.42694 |
| NUMATA_( | 21 | 0.33807 | 0.011269 | 0.10842 | 0.000911 | 15.49615 |
| GOBP_REG | 6 | 0.69354 | 0.012362 | 0.22265 | 0.000922 | 15.67215 |

| | | | | | | |
|---|---|---|---|---|---|---|
| GOBP_VIRA | 27 | 0.28864 | 0.010908 | 0.092725 | 0.000928 | 15.78353 |
| GOBP_SMC | 24 | 0.35665 | 0.012708 | 0.11462 | 0.000932 | 15.8439 |
| GOBP_EMI | 585 | 0.069639 | 0.012068 | 0.022407 | 0.000944 | 16.04456 |
| REACTOME | 73 | 0.17468 | 0.010841 | 0.056221 | 0.000947 | 16.09931 |
| WP_MAMI | 31 | 0.28713 | 0.011626 | 0.0926 | 0.000967 | 16.44213 |
| GOBP_REG | 15 | 0.47575 | 0.013405 | 0.1537 | 0.000984 | 16.73938 |
| ICHIBA_GR | 96 | 0.15638 | 0.011124 | 0.050524 | 0.000985 | 16.74601 |
| GOMF_MC | 7 | 0.63855 | 0.012294 | 0.20653 | 0.000996 | 16.94514 |
| BOYERINAS | 10 | 0.48515 | 0.011163 | 0.15734 | 0.001025 | 17.42502 |
| GOMF_RNA | 1503 | 0.039386 | 0.010662 | 0.012785 | 0.001034 | 17.58997 |
| REACTOME | 224 | 0.099732 | 0.010799 | 0.032432 | 0.001054 | 17.92327 |
| REACTOME | 9 | 0.61803 | 0.013491 | 0.20108 | 0.00106 | 18.0168 |
| GOBP_LOC | 15 | 0.376 | 0.010595 | 0.12237 | 0.001062 | 18.06101 |
| GOBP_MUS | 637 | 0.062447 | 0.011276 | 0.020339 | 0.001071 | 18.21065 |
| GOBP_RIBC | 187 | 0.10495 | 0.010393 | 0.034234 | 0.001088 | 18.49974 |
| GOBP_VEN | 40 | 0.28117 | 0.012929 | 0.091809 | 0.001099 | 18.6902 |
| GOBP_NEG | 12 | 0.56316 | 0.014194 | 0.18426 | 0.001122 | 19.08301 |
| GOCC_I_BA | 132 | 0.13984 | 0.011653 | 0.045846 | 0.001145 | 19.47073 |
| GOBP_TRA | 10 | 0.4189 | 0.009639 | 0.1375 | 0.00116 | 19.7173 |
| GOBP_RNA | 67 | 0.18053 | 0.010736 | 0.059276 | 0.001163 | 19.77001 |
| DING_LUN | 19 | 0.37661 | 0.011942 | 0.1239 | 0.001186 | 20.16623 |
| GOMF_TRC | 5 | 0.81126 | 0.013201 | 0.26691 | 0.001187 | 20.18153 |
| GOBP_CELI | 102 | 0.16676 | 0.012224 | 0.05488 | 0.00119 | 20.24275 |
| GOBP_NEU | 5 | 0.58952 | 0.009593 | 0.19402 | 0.001191 | 20.24785 |
| GOBP_NEG | 7 | 0.48207 | 0.009281 | 0.15882 | 0.001203 | 20.45702 |
| GOBP_NEG | 26 | 0.29983 | 0.01112 | 0.098891 | 0.001217 | 20.69168 |
| MARTORIA | 489 | 0.068896 | 0.010944 | 0.022725 | 0.001218 | 20.70869 |
| GOCC_RIBC | 639 | 0.055996 | 0.010126 | 0.018478 | 0.001223 | 20.79541 |
| WP_MEVA | 13 | 0.34711 | 0.009106 | 0.11463 | 0.001233 | 20.96376 |
| RIZ_ERYTH | 37 | 0.24054 | 0.010638 | 0.079465 | 0.001237 | 21.03689 |
| GOMF_ATF | 44 | 0.22362 | 0.010783 | 0.0739 | 0.001241 | 21.10831 |
| GOBP_POS | 36 | 0.24626 | 0.010744 | 0.081426 | 0.001248 | 21.21714 |
| GOBP_NUC | 35 | 0.22189 | 0.009545 | 0.073648 | 0.001296 | 22.04018 |
| GOBP_POS | 1770 | 0.037705 | 0.010991 | 0.012517 | 0.001299 | 22.08609 |
| GOBP_ATR | 15 | 0.43532 | 0.012266 | 0.14493 | 0.001336 | 22.71018 |
| GOBP_MES | 31 | 0.26836 | 0.010866 | 0.089398 | 0.001344 | 22.84622 |
| GOCC_SPEI | 9 | 0.44901 | 0.009802 | 0.14972 | 0.001356 | 23.06388 |
| GOBP_ESTA | 9 | 0.60798 | 0.013272 | 0.20286 | 0.001366 | 23.22033 |
| LIN_MELAI | 41 | 0.24082 | 0.011211 | 0.080397 | 0.001373 | 23.34446 |
| GOMF_PHC | 6 | 0.62089 | 0.011067 | 0.20741 | 0.001381 | 23.4805 |
| GOBP_NEG | 13 | 0.41477 | 0.010881 | 0.13877 | 0.001402 | 23.83931 |
| GOBP_EMI | 6 | 0.69911 | 0.012462 | 0.23485 | 0.001458 | 24.79839 |
| GOBP_CHR | 412 | 0.071414 | 0.010434 | 0.024013 | 0.001472 | 25.02626 |
| GOBP_END | 76 | 0.18311 | 0.011595 | 0.061628 | 0.001485 | 25.25583 |
| BUCKANO\ | 19 | 0.37807 | 0.011988 | 0.1273 | 0.001491 | 25.36126 |

| | | | | | | |
|---|---|---|---|---|---|---|
| GOBP_STR | 162 | 0.12062 | 0.011126 | 0.040659 | 0.001507 | 25.62994 |
| GOBP_CELI | 261 | 0.10124 | 0.011822 | 0.034126 | 0.001507 | 25.63334 |
| GOBP_CER | 33 | 0.26156 | 0.010926 | 0.088167 | 0.001508 | 25.63844 |
| GOCC_U1_ | 18 | 0.41404 | 0.012779 | 0.13973 | 0.001524 | 25.92242 |
| GOBP_ART | 98 | 0.16455 | 0.011825 | 0.055584 | 0.001538 | 26.15369 |
| KEGG_HYP | 79 | 0.16988 | 0.010966 | 0.05742 | 0.001548 | 26.32204 |
| BIOCARTA_ | 77 | 0.15827 | 0.010087 | 0.053581 | 0.001572 | 26.72676 |
| WANG_ML | 277 | 0.095083 | 0.011433 | 0.032207 | 0.00158 | 26.8594 |
| GOBP_DIEI | 70 | 0.19929 | 0.012113 | 0.067531 | 0.001586 | 26.96313 |
| GOBP_RES | 33 | 0.26661 | 0.011137 | 0.090376 | 0.001591 | 27.06006 |
| GOBP_POS | 483 | 0.06904 | 0.010901 | 0.023448 | 0.00162 | 27.5549 |
| WP_MESO | 137 | 0.13357 | 0.011337 | 0.045369 | 0.001622 | 27.58041 |
| GOBP_INTI | 41 | 0.23086 | 0.010747 | 0.078503 | 0.001639 | 27.8746 |
| GOBP_DEF | 19 | 0.35229 | 0.011171 | 0.12022 | 0.001695 | 28.82518 |
| GOBP_CELI | 26 | 0.31224 | 0.01158 | 0.10658 | 0.001699 | 28.8915 |
| GOCC_CHR | 357 | 0.073492 | 0.01001 | 0.025105 | 0.001712 | 29.10576 |
| GOMF_BET | 4 | 0.70288 | 0.01023 | 0.24018 | 0.001717 | 29.19078 |
| GOBP_REG | 163 | 0.11145 | 0.010311 | 0.038086 | 0.001718 | 29.20609 |
| GOBP_EMI | 124 | 0.15754 | 0.012726 | 0.053837 | 0.001718 | 29.21629 |
| GOBP_ORG | 477 | 0.06444 | 0.010113 | 0.022098 | 0.001775 | 30.17707 |
| GOBP_REG | 82 | 0.17679 | 0.011627 | 0.06072 | 0.0018 | 30.6124 |
| GOMF_MC | 15 | 0.35735 | 0.010069 | 0.12304 | 0.001842 | 31.32661 |
| GOBP_CD4 | 15 | 0.40149 | 0.011313 | 0.1383 | 0.00185 | 31.46095 |
| GOBP_MIT | 23 | 0.28347 | 0.009888 | 0.097646 | 0.001851 | 31.46775 |
| GOBP_REG | 7 | 0.55463 | 0.010678 | 0.19109 | 0.001854 | 31.51877 |
| GOBP_RES | 8 | 0.54747 | 0.011268 | 0.18877 | 0.001867 | 31.75174 |
| GOBP_DIG | 48 | 0.23099 | 0.011633 | 0.079708 | 0.001881 | 31.9847 |
| GOBP_NEG | 137 | 0.13141 | 0.011154 | 0.045353 | 0.001883 | 32.02212 |
| GOMF_DEA | 11 | 0.45256 | 0.010921 | 0.15641 | 0.001908 | 32.44044 |
| KIM_HYPO | 17 | 0.36654 | 0.010994 | 0.12677 | 0.001921 | 32.6581 |
| GOBP_EMI | 91 | 0.18021 | 0.012481 | 0.062354 | 0.001928 | 32.79244 |
| GOBP_GLC | 5 | 0.57021 | 0.009279 | 0.19734 | 0.001932 | 32.85706 |
| GOBP_ATR | 20 | 0.37216 | 0.012107 | 0.12885 | 0.00194 | 32.9829 |
| GOBP_REG | 53 | 0.22237 | 0.011766 | 0.077215 | 0.001992 | 33.87566 |
| GOMF_TRA | 23 | 0.29396 | 0.010254 | 0.10209 | 0.001995 | 33.92838 |
| GOBP_REG | 97 | 0.14183 | 0.01014 | 0.049259 | 0.001996 | 33.93518 |
| GOBP_LATI | 21 | 0.29421 | 0.009807 | 0.10227 | 0.002011 | 34.18855 |
| GOMF_UBI | 23 | 0.28052 | 0.009786 | 0.097766 | 0.002059 | 35.0184 |
| GOBP_NUC | 324 | 0.077303 | 0.01004 | 0.026957 | 0.002071 | 35.21055 |
| WILCOX_RI | 144 | 0.12098 | 0.010525 | 0.042196 | 0.002075 | 35.28027 |
| GOBP_REG | 45 | 0.20382 | 0.009939 | 0.071105 | 0.002078 | 35.33639 |
| GOCC_CILI | 45 | 0.19861 | 0.009685 | 0.069321 | 0.002087 | 35.49454 |
| AFFAR_YY1 | 237 | 0.093834 | 0.010448 | 0.032758 | 0.002091 | 35.55746 |
| GOCC_SMA | 9 | 0.55338 | 0.01208 | 0.19319 | 0.002091 | 35.56256 |
| GOBP_TRA | 13 | 0.43742 | 0.011475 | 0.1528 | 0.002104 | 35.77002 |

| | | | | | | |
|---|---|---|---|---|---|---|
| GOBP_MU: | 320 | 0.084293 | 0.010881 | 0.029452 | 0.002107 | 35.82954 |
| BOQUEST_ | 399 | 0.078379 | 0.011274 | 0.027393 | 0.002112 | 35.91626 |
| GOMF_PR( | 628 | 0.056562 | 0.010143 | 0.019779 | 0.002123 | 36.09991 |
| GOBP_POS | 7 | 0.64468 | 0.012412 | 0.2256 | 0.002137 | 36.33458 |
| REACTOME | 20 | 0.29163 | 0.009487 | 0.10217 | 0.002159 | 36.7189 |
| GOBP_KIDI | 137 | 0.13719 | 0.011645 | 0.04808 | 0.002165 | 36.81412 |
| GOBP_NEG | 36 | 0.25992 | 0.01134 | 0.091141 | 0.002176 | 37.00288 |
| GOBP_GTP | 10 | 0.38178 | 0.008785 | 0.13389 | 0.002178 | 37.04369 |
| GOBP_REG | 16 | 0.36801 | 0.010709 | 0.12913 | 0.00219 | 37.23415 |
| GOBP_CELI | 9 | 0.56692 | 0.012375 | 0.19895 | 0.002191 | 37.26476 |
| MISIAK_AN | 18 | 0.37643 | 0.011618 | 0.13219 | 0.002205 | 37.48922 |
| GOBP_REG | 9 | 0.38163 | 0.008331 | 0.13405 | 0.00221 | 37.58445 |
| GOBP_REG | 24 | 0.30147 | 0.010742 | 0.10602 | 0.002235 | 37.99767 |
| REACTOME | 30 | 0.23573 | 0.00939 | 0.082914 | 0.002236 | 38.02998 |
| GOBP_MIT | 866 | 0.048491 | 0.010145 | 0.01706 | 0.002242 | 38.13031 |
| GOBP_HIST | 8 | 0.48281 | 0.009937 | 0.16988 | 0.002243 | 38.14902 |
| REACTOME | 8 | 0.48281 | 0.009937 | 0.16988 | 0.002243 | 38.14902 |
| GOBP_REG | 12 | 0.48448 | 0.012211 | 0.1709 | 0.002295 | 39.03158 |
| WP_MECP: | 68 | 0.18267 | 0.010943 | 0.064498 | 0.002314 | 39.35637 |
| GOBP_FOR | 115 | 0.13664 | 0.010632 | 0.04826 | 0.00232 | 39.4584 |
| BHATTACH | 78 | 0.15336 | 0.009838 | 0.054231 | 0.002345 | 39.87502 |
| MYLLYKAN | 5 | 0.95183 | 0.015488 | 0.33661 | 0.002347 | 39.90903 |
| GOBP_REG | 11 | 0.50135 | 0.012098 | 0.17732 | 0.002349 | 39.94985 |
| GOMF_WD | 4 | 0.67112 | 0.009768 | 0.23772 | 0.002381 | 40.4838 |
| GOBP_REG | 11 | 0.46952 | 0.01133 | 0.16634 | 0.002385 | 40.54842 |
| STARK_PRE | 493 | 0.063467 | 0.010122 | 0.022511 | 0.002409 | 40.96164 |
| GOBP_NEG | 17 | 0.30429 | 0.009127 | 0.10808 | 0.002438 | 41.46499 |
| GOBP_NEG | 8 | 0.57193 | 0.011771 | 0.20326 | 0.00245 | 41.66905 |
| BONOME_( | 463 | 0.063087 | 0.009758 | 0.022421 | 0.002451 | 41.68096 |
| REACTOME | 56 | 0.18545 | 0.010086 | 0.065931 | 0.002458 | 41.79829 |
| GOBP_REG | 11 | 0.37939 | 0.009155 | 0.1349 | 0.002462 | 41.86631 |
| GOBP_REG | 8 | 0.49326 | 0.010152 | 0.17548 | 0.002473 | 42.04656 |
| GOBP_PLA: | 27 | 0.2692 | 0.010174 | 0.095845 | 0.00249 | 42.33565 |
| CHEN_LVA | 97 | 0.14939 | 0.010681 | 0.053213 | 0.0025 | 42.5142 |
| GOBP_CHO | 32 | 0.28093 | 0.011557 | 0.10014 | 0.002515 | 42.76077 |
| GRYDER_P. | 438 | 0.07168 | 0.010791 | 0.025613 | 0.002569 | 43.68414 |
| STEARMAN | 35 | 0.21884 | 0.009414 | 0.078212 | 0.002573 | 43.75557 |
| GOMF_SUI | 254 | 0.090753 | 0.010456 | 0.032452 | 0.002586 | 43.97153 |
| KHETCHOU | 44 | 0.23375 | 0.011272 | 0.083588 | 0.002586 | 43.97833 |
| GOBP_RNA | 860 | 0.045184 | 0.009422 | 0.016176 | 0.002612 | 44.41536 |
| GOBP_LOV | 6 | 0.49912 | 0.008897 | 0.17884 | 0.002632 | 44.75716 |
| GOBP_PRO | 46 | 0.23364 | 0.011519 | 0.083811 | 0.002657 | 45.18229 |
| GOBP_RNA | 389 | 0.06654 | 0.009453 | 0.023905 | 0.002692 | 45.78256 |
| WP_MARK | 55 | 0.21179 | 0.011415 | 0.076122 | 0.002702 | 45.95431 |
| GOMF_UBI | 9 | 0.37768 | 0.008245 | 0.13601 | 0.002747 | 46.71274 |

| | | | | | | |
|---|---|---|---|---|---|---|
| GOBP_REG | 85 | 0.1517 | 0.010156 | 0.054703 | 0.002779 | 47.25349 |
| KEGG_END | 50 | 0.20659 | 0.010618 | 0.074508 | 0.002783 | 47.31641 |
| GOBP_POS | 7 | 0.47578 | 0.00916 | 0.17181 | 0.002813 | 47.82996 |
| REACTOME | 10 | 0.50918 | 0.011716 | 0.18409 | 0.002841 | 48.31801 |
| GOBP_CAR | 103 | 0.148 | 0.010902 | 0.053513 | 0.002843 | 48.34522 |
| GOBP_INTI | 7 | 0.53038 | 0.010211 | 0.19179 | 0.002846 | 48.39623 |
| GOBP_THA | 11 | 0.45076 | 0.010878 | 0.16314 | 0.002867 | 48.75504 |
| GOBP_SKEI | 65 | 0.16212 | 0.009496 | 0.058682 | 0.00287 | 48.80605 |
| GOBP_POS | 3 | 1.2746 | 0.016067 | 0.46165 | 0.002884 | 49.03732 |
| GOMF_TEL | 22 | 0.28412 | 0.009694 | 0.10304 | 0.002917 | 49.60359 |
| BURTON_A | 56 | 0.18874 | 0.010265 | 0.068597 | 0.00297 | 50.49805 |
| GOBP_REG | 64 | 0.18407 | 0.010699 | 0.067016 | 0.003014 | 51.24457 |
| GOMF_HIS | 13 | 0.39353 | 0.010323 | 0.14328 | 0.003015 | 51.27008 |
| LI_WILMS_ | 25 | 0.30428 | 0.011066 | 0.1108 | 0.003018 | 51.31939 |
| GOBP_VIRI | 36 | 0.21091 | 0.009201 | 0.076898 | 0.00305 | 51.86695 |
| BIOCARTA_ | 16 | 0.37138 | 0.010807 | 0.13555 | 0.003076 | 52.31418 |
| GOBP_POS | 28 | 0.25479 | 0.009805 | 0.093038 | 0.003089 | 52.52674 |
| GOBP_REG | 56 | 0.17888 | 0.009728 | 0.065352 | 0.003102 | 52.74271 |
| REACTOME | 14 | 0.32712 | 0.008905 | 0.11951 | 0.003102 | 52.75291 |
| GOBP_REG | 37 | 0.24038 | 0.010632 | 0.087904 | 0.003126 | 53.16273 |
| WP_REGUI | 14 | 0.35121 | 0.009561 | 0.12853 | 0.003146 | 53.49433 |
| WP_TGFBE | 17 | 0.33476 | 0.010041 | 0.12256 | 0.003155 | 53.65588 |
| GOBP_NEG | 7 | 0.57124 | 0.010998 | 0.20916 | 0.003159 | 53.7239 |
| GOBP_NEG | 12 | 0.33666 | 0.008485 | 0.12332 | 0.00317 | 53.91265 |
| GOBP_MES | 69 | 0.16809 | 0.010143 | 0.061601 | 0.003183 | 54.13202 |
| REICHERT_ | 27 | 0.22693 | 0.008576 | 0.083261 | 0.003213 | 54.64047 |
| GOBP_HEA | 80 | 0.16413 | 0.010662 | 0.060223 | 0.003215 | 54.66427 |
| GOBP_CELI | 7 | 0.55168 | 0.010621 | 0.2027 | 0.003251 | 55.28836 |
| GOBP_NEG | 58 | 0.19123 | 0.010583 | 0.070309 | 0.00327 | 55.60125 |
| GOBP_CELI | 1185 | 0.039753 | 0.009642 | 0.01462 | 0.003276 | 55.71008 |
| WOO_LIVE | 98 | 0.14737 | 0.010591 | 0.054252 | 0.003303 | 56.16581 |
| GOMF_EXT | 54 | 0.20129 | 0.01075 | 0.074101 | 0.003304 | 56.17602 |
| GOMF_CAI | 18 | 0.31025 | 0.009575 | 0.11429 | 0.003322 | 56.48551 |
| GOBP_CAR | 123 | 0.12682 | 0.010203 | 0.046722 | 0.003324 | 56.51952 |
| GOCC_SPLI | 60 | 0.16925 | 0.009527 | 0.062411 | 0.003348 | 56.93784 |
| GOBP_NEG | 8 | 0.53828 | 0.011079 | 0.19854 | 0.003356 | 57.06538 |
| GOMF_GAI | 5 | 0.60521 | 0.009848 | 0.22326 | 0.00336 | 57.1419 |
| LEE_CALOF | 66 | 0.15998 | 0.009443 | 0.059101 | 0.003399 | 57.79489 |
| GOBP_NEG | 858 | 0.047584 | 0.009911 | 0.017595 | 0.003424 | 58.23022 |
| GOBP_OSS | 379 | 0.072678 | 0.010194 | 0.02689 | 0.003442 | 58.52271 |
| REACTOME | 16 | 0.38146 | 0.011101 | 0.14117 | 0.003449 | 58.64684 |
| GOBP_REG | 16 | 0.37842 | 0.011012 | 0.14013 | 0.003466 | 58.94103 |
| GOBP_REG | 10 | 0.41958 | 0.009654 | 0.15542 | 0.003473 | 59.06347 |
| GOMF_ADI | 12 | 0.35814 | 0.009027 | 0.13266 | 0.003475 | 59.08727 |
| GOBP_VAS | 59 | 0.18196 | 0.010157 | 0.067418 | 0.00348 | 59.1842 |

| | | | | | | |
|---|---|---|---|---|---|---|
| GOCC_SUP | 955 | 0.047374 | 0.010382 | 0.017571 | 0.00351 | 59.69265 |
| GOBP_NEG | 6 | 0.63435 | 0.011307 | 0.23531 | 0.003515 | 59.76407 |
| GRYDER_P/ | 430 | 0.072615 | 0.010834 | 0.026952 | 0.003531 | 60.04125 |
| GOBP_REG | 10 | 0.39637 | 0.00912 | 0.1472 | 0.003547 | 60.32184 |
| GOBP_CAR | 69 | 0.18518 | 0.011175 | 0.068776 | 0.00355 | 60.36605 |
| BOWIE_RE: | 15 | 0.35216 | 0.009923 | 0.1308 | 0.003551 | 60.38135 |
| MARSON_I | 653 | 0.053707 | 0.009814 | 0.019949 | 0.003553 | 60.41536 |
| GOMF_CHI | 519 | 0.059197 | 0.00968 | 0.022004 | 0.003573 | 60.76057 |
| GOMF_LAN | 28 | 0.26165 | 0.010069 | 0.097345 | 0.003599 | 61.19419 |
| GOBP_NEG | 841 | 0.049014 | 0.010112 | 0.018244 | 0.003613 | 61.43566 |
| GOBP_HEA | 770 | 0.051956 | 0.010277 | 0.019345 | 0.003622 | 61.59891 |
| GOMF_DIA | 10 | 0.50454 | 0.011609 | 0.18793 | 0.003632 | 61.76556 |
| GOBP_NEG | 8 | 0.45545 | 0.009374 | 0.16967 | 0.003637 | 61.84378 |
| GOBP_NEG | 17 | 0.28929 | 0.008677 | 0.10782 | 0.003651 | 62.09036 |
| MARKEY_R | 104 | 0.12913 | 0.009558 | 0.048131 | 0.003654 | 62.13457 |
| REACTOME | 104 | 0.11725 | 0.008678 | 0.04371 | 0.003659 | 62.2213 |
| GOBP_SON | 81 | 0.15903 | 0.010394 | 0.059312 | 0.003672 | 62.43556 |
| GOBP_LATI | 15 | 0.37448 | 0.010552 | 0.13973 | 0.003685 | 62.66343 |
| TSAI_DNAJ | 6 | 0.51416 | 0.009165 | 0.19192 | 0.003695 | 62.83518 |
| BURTON_A | 31 | 0.28035 | 0.011351 | 0.10469 | 0.003709 | 63.06984 |
| GOBP_ECT | 17 | 0.32993 | 0.009896 | 0.12336 | 0.003745 | 63.67522 |
| GOBP_NEG | 6 | 0.56174 | 0.010013 | 0.21016 | 0.003763 | 63.98641 |
| GOBP_REG | 68 | 0.16801 | 0.010066 | 0.062861 | 0.003765 | 64.01872 |
| REACTOME | 20 | 0.26378 | 0.008581 | 0.098844 | 0.003812 | 64.82136 |
| GOBP_NUC | 89 | 0.14017 | 0.009602 | 0.052536 | 0.003818 | 64.92849 |
| WP_MEVA | 7 | 0.46816 | 0.009013 | 0.17574 | 0.003866 | 65.73963 |
| GOBP_MES | 60 | 0.19789 | 0.011139 | 0.074322 | 0.00388 | 65.9811 |
| KEGG_BAS/ | 55 | 0.179 | 0.009648 | 0.067288 | 0.003908 | 66.45894 |
| GOBP_TRIC | 5 | 0.68704 | 0.01118 | 0.2583 | 0.003912 | 66.52356 |
| GOBP_REG | 6 | 0.55725 | 0.009933 | 0.2098 | 0.003956 | 67.26328 |
| GOBP_APP | 141 | 0.12465 | 0.010733 | 0.046934 | 0.003958 | 67.30579 |
| GOBP_WO | 406 | 0.069565 | 0.010091 | 0.026196 | 0.003963 | 67.38231 |
| GOBP_MRI | 422 | 0.061109 | 0.009034 | 0.023073 | 0.004046 | 68.80733 |
| GOCC_CHR | 225 | 0.083801 | 0.009094 | 0.03166 | 0.004065 | 69.13043 |
| GOBP_VIR/ | 27 | 0.24493 | 0.009256 | 0.092564 | 0.004075 | 69.29367 |
| GOBP_DEV | 596 | 0.056905 | 0.00995 | 0.021545 | 0.004134 | 70.30547 |
| GOBP_ROC | 89 | 0.15349 | 0.010514 | 0.058195 | 0.004179 | 71.0673 |
| GOBP_RES | 1614 | 0.034627 | 0.009682 | 0.013137 | 0.004202 | 71.44821 |
| GOBP_REC | 67 | 0.15597 | 0.009275 | 0.05918 | 0.004205 | 71.50773 |
| GOBP_MES | 290 | 0.080376 | 0.009885 | 0.030565 | 0.004277 | 72.73209 |
| KIM_MYC_ | 94 | 0.15332 | 0.010792 | 0.05838 | 0.00432 | 73.4565 |
| GOBP_CHR | 92 | 0.13197 | 0.00919 | 0.05027 | 0.004334 | 73.69967 |
| WP_ESTRA | 5 | 0.7104 | 0.01156 | 0.27084 | 0.004362 | 74.17751 |
| GOBP_REG | 790 | 0.047626 | 0.009537 | 0.01818 | 0.004405 | 74.90362 |
| GOMF_HEI | 18 | 0.30559 | 0.009432 | 0.11671 | 0.004421 | 75.17911 |

| | | | | | | |
|---|---|---|---|---|---|---|
| GOCC_NUC | 1105 | 0.039127 | 0.009185 | 0.014946 | 0.004429 | 75.30664 |
| GOBP_REG | 12 | 0.364 | 0.009175 | 0.13925 | 0.004478 | 76.15349 |
| GOMF_NU | 38 | 0.20312 | 0.009104 | 0.077723 | 0.004487 | 76.29463 |
| TIEN_INTES | 536 | 0.055077 | 0.009148 | 0.021075 | 0.004487 | 76.29463 |
| GOCC_NUC | 748 | 0.04727 | 0.009221 | 0.018134 | 0.004575 | 77.79277 |
| ACEVEDO_ | 97 | 0.13083 | 0.009354 | 0.050207 | 0.004588 | 78.01044 |
| GOBP_NEG | 57 | 0.17418 | 0.009556 | 0.066854 | 0.004594 | 78.11417 |
| GOBP_POS | 105 | 0.1221 | 0.009081 | 0.046889 | 0.004612 | 78.43046 |
| GOBP_REG | 167 | 0.11045 | 0.010343 | 0.042443 | 0.004633 | 78.78927 |
| REACTOME | 22 | 0.30586 | 0.010435 | 0.11757 | 0.004643 | 78.95932 |
| GOBP_CEN | 994 | 0.044606 | 0.009962 | 0.017145 | 0.004643 | 78.96102 |
| GOMF_PRC | 89 | 0.14012 | 0.009598 | 0.053881 | 0.004658 | 79.20589 |
| GOBP_DIG | 137 | 0.11278 | 0.009572 | 0.043385 | 0.004673 | 79.46777 |
| GOBP_STA | 26 | 0.26723 | 0.009911 | 0.10281 | 0.004674 | 79.47287 |
| GOBP_PER | 19 | 0.34604 | 0.010973 | 0.13317 | 0.004685 | 79.67013 |
| GOBP_NEG | 53 | 0.17956 | 0.009501 | 0.06911 | 0.00469 | 79.75175 |
| REACTOME | 110 | 0.11155 | 0.00849 | 0.042965 | 0.004717 | 80.20918 |
| GOBP_DET | 8 | 0.45727 | 0.009411 | 0.17614 | 0.004719 | 80.24319 |
| GOBP_HIN | 8 | 0.47939 | 0.009867 | 0.18469 | 0.004725 | 80.34863 |
| GOBP_NEP | 73 | 0.17073 | 0.010596 | 0.065804 | 0.00474 | 80.6003 |
| GOBP_CELI | 15 | 0.32875 | 0.009263 | 0.12675 | 0.004751 | 80.78735 |
| GALIE_TUN | 6 | 0.51236 | 0.009133 | 0.19758 | 0.00476 | 80.9353 |
| GOBP_CELI | 21 | 0.32227 | 0.010743 | 0.12435 | 0.004782 | 81.31451 |
| WHITESIDE | 11 | 0.35899 | 0.008663 | 0.1387 | 0.004826 | 82.06953 |
| GOBP_REG | 6 | 0.61639 | 0.010987 | 0.23816 | 0.004829 | 82.12225 |
| GOBP_MEI | 5 | 0.71211 | 0.011588 | 0.27537 | 0.004859 | 82.6307 |
| GOBP_RES | 46 | 0.19121 | 0.009427 | 0.073952 | 0.004865 | 82.72082 |
| ONGUSAHA | 11 | 0.38464 | 0.009282 | 0.14878 | 0.00487 | 82.80585 |
| GOBP_LIPI | 15 | 0.38183 | 0.010759 | 0.14774 | 0.00488 | 82.9912 |
| REN_ALVEC | 96 | 0.13509 | 0.009609 | 0.052272 | 0.004881 | 83.00821 |
| GOMF_UBI | 20 | 0.27475 | 0.008938 | 0.10631 | 0.004883 | 83.03882 |
| GOBP_REG | 259 | 0.084921 | 0.009878 | 0.032882 | 0.004907 | 83.44354 |
| GOBP_REG | 44 | 0.20887 | 0.010072 | 0.080902 | 0.00492 | 83.6578 |
| GOBP_REG | 27 | 0.26302 | 0.00994 | 0.10194 | 0.004942 | 84.04211 |
| GOMF_PHC | 10 | 0.38358 | 0.008826 | 0.14886 | 0.004989 | 84.84475 |
| GOMF_COI | 63 | 0.16798 | 0.009688 | 0.065221 | 0.005008 | 85.15424 |
| KEGG_TERI | 15 | 0.29774 | 0.008389 | 0.11562 | 0.005013 | 85.23756 |
| GOBP_EXIT | 28 | 0.22485 | 0.008653 | 0.087339 | 0.005024 | 85.42972 |
| MCMURRA | 24 | 0.25254 | 0.008999 | 0.09819 | 0.005061 | 86.0589 |
| WP_SARSC | 8 | 0.4742 | 0.00976 | 0.18444 | 0.005075 | 86.30378 |
| GAZDA_DIA | 23 | 0.26392 | 0.009206 | 0.10266 | 0.005077 | 86.33098 |
| GOMF_TRA | 66 | 0.16897 | 0.009974 | 0.065762 | 0.005097 | 86.67278 |
| GOBP_NOD | 16 | 0.34566 | 0.010059 | 0.13461 | 0.005121 | 87.0792 |
| WP_HEDGI | 43 | 0.21214 | 0.010113 | 0.082753 | 0.005186 | 88.18453 |
| GOBP_LON | 26 | 0.26681 | 0.009895 | 0.10425 | 0.005249 | 89.26605 |

| | | | | | | |
|---|---|---|---|---|---|---|
| GOBP_ANT | 5 | 0.53284 | 0.008671 | 0.2083 | 0.005268 | 89.57894 |
| TCGA_GLIC | 29 | 0.28028 | 0.010977 | 0.1096 | 0.005277 | 89.73709 |
| GOBP_STR | 68 | 0.16437 | 0.009847 | 0.064341 | 0.005319 | 90.44619 |
| GOBP_SPLI | 5 | 0.49184 | 0.008003 | 0.19267 | 0.005349 | 90.95975 |
| GOBP_NEG | 16 | 0.33701 | 0.009807 | 0.13204 | 0.005356 | 91.07028 |
| GOBP_CELI | 16 | 0.35525 | 0.010338 | 0.13924 | 0.00537 | 91.31175 |
| FARMER_B | 32 | 0.22016 | 0.009057 | 0.086315 | 0.00538 | 91.4886 |
| GOBP_CELI | 33 | 0.24318 | 0.010158 | 0.095374 | 0.005394 | 91.73007 |
| REACTOME | 20 | 0.25234 | 0.008209 | 0.099074 | 0.005438 | 92.46469 |
| GOMF_ADI | 18 | 0.28097 | 0.008672 | 0.11037 | 0.005457 | 92.79969 |
| BROWNE_I | 217 | 0.08264 | 0.008809 | 0.032484 | 0.005484 | 93.24862 |
| GOMF_TRA | 558 | 0.055775 | 0.009446 | 0.021928 | 0.005491 | 93.37956 |
| GOBP_GEN | 5 | 0.54664 | 0.008895 | 0.21505 | 0.005517 | 93.81148 |
| REACTOME | 8 | 0.35398 | 0.007285 | 0.13945 | 0.005572 | 94.75526 |
| GUTIERREZ | 34 | 0.20406 | 0.008652 | 0.080413 | 0.005584 | 94.96272 |
| GOBP_EMI | 436 | 0.065848 | 0.009891 | 0.025954 | 0.005593 | 95.11067 |
| SILIGAN_TA | 17 | 0.37113 | 0.011132 | 0.1464 | 0.005628 | 95.70074 |
| GOBP_REG | 11 | 0.42516 | 0.01026 | 0.16781 | 0.00565 | 96.07315 |
| GOBP_REG | 26 | 0.22579 | 0.008374 | 0.089359 | 0.00576 | 97.9437 |
| GOCC_U5_ | 19 | 0.30428 | 0.009648 | 0.12042 | 0.00576 | 97.9437 |
| GOBP_POS | 8 | 0.36411 | 0.007494 | 0.14413 | 0.005768 | 98.08314 |
| GOBP_PUR | 41 | 0.1881 | 0.008757 | 0.074503 | 0.005793 | 98.51677 |
| GOBP_URA | 12 | 0.36732 | 0.009258 | 0.14561 | 0.005828 | 99.10854 |
| GOBP_ORG | 25 | 0.21663 | 0.007878 | 0.085912 | 0.005846 | 99.41293 |
| SENESE_HI | 214 | 0.087842 | 0.009299 | 0.034843 | 0.005854 | 99.55067 |
| CALVET_IR | 5 | 0.4373 | 0.007116 | 0.17348 | 0.005861 | 99.6612 |
| GOBP_NEG | 1036 | 0.040944 | 0.009325 | 0.016255 | 0.00589 | 100.1629 |
| GOCC_CON | 257 | 0.076155 | 0.008825 | 0.030235 | 0.005893 | 100.202 |
| REACTOME | 24 | 0.23196 | 0.008265 | 0.09214 | 0.005915 | 100.5795 |
| GOBP_PUR | 20 | 0.24746 | 0.00805 | 0.098325 | 0.005927 | 100.7818 |
| GOBP_MAI | 7 | 0.54634 | 0.010519 | 0.21712 | 0.005935 | 100.9213 |
| GOBP_REG | 4 | 0.55674 | 0.008103 | 0.22131 | 0.005945 | 101.0862 |
| GOBP_LOC | 15 | 0.33626 | 0.009475 | 0.13367 | 0.005946 | 101.1066 |
| GOBP_HIN | 43 | 0.19977 | 0.009523 | 0.079432 | 0.005957 | 101.2903 |
| GOBP_REG | 134 | 0.11483 | 0.00964 | 0.045675 | 0.00597 | 101.5267 |
| GOBP_CELI | 19 | 0.30899 | 0.009798 | 0.12298 | 0.005996 | 101.9688 |
| WP_COHES | 31 | 0.21248 | 0.008603 | 0.08458 | 0.006005 | 102.1133 |
| GOMF_DN. | 1137 | 0.041273 | 0.009819 | 0.016444 | 0.006044 | 102.7799 |
| KRISHNAN_ | 9 | 0.38579 | 0.008422 | 0.15372 | 0.006047 | 102.8309 |
| GOBP_POS | 5 | 0.5668 | 0.009223 | 0.22627 | 0.006129 | 104.2151 |
| GOCC_CON | 162 | 0.094208 | 0.00869 | 0.037612 | 0.006132 | 104.2747 |
| GOBP_PRO | 43 | 0.18098 | 0.008628 | 0.072285 | 0.006149 | 104.5586 |
| GOBP_REG | 5 | 0.56144 | 0.009136 | 0.22424 | 0.006149 | 104.5603 |
| BLANCO_M | 37 | 0.21663 | 0.009581 | 0.086546 | 0.006162 | 104.7814 |
| GOMF_CO_ | 13 | 0.32454 | 0.008514 | 0.12976 | 0.006195 | 105.3528 |

| | | | | | | |
|---|---|---|---|---|---|---|
| VERHAAK_ | 158 | 0.10227 | 0.009317 | 0.040903 | 0.006207 | 105.5551 |
| GOBP_REG | 22 | 0.31253 | 0.010663 | 0.12506 | 0.006229 | 105.9309 |
| AGUIRRE_F | 290 | 0.075959 | 0.009342 | 0.030398 | 0.006236 | 106.0415 |
| GOBP_MET | 9 | 0.52294 | 0.011415 | 0.20932 | 0.006245 | 106.1945 |
| GOBP_REG | 15 | 0.33761 | 0.009513 | 0.13514 | 0.006247 | 106.2234 |
| SENESE_HD | 453 | 0.06164 | 0.009433 | 0.024677 | 0.006252 | 106.3204 |
| GOBP_REG | 6 | 0.633 | 0.011283 | 0.2536 | 0.006285 | 106.873 |
| SMID_BREA | 37 | 0.21776 | 0.009631 | 0.087271 | 0.006299 | 107.1145 |
| GOCC_CAT | 83 | 0.12717 | 0.008414 | 0.050991 | 0.006322 | 107.509 |
| SCHAEFFEF | 53 | 0.17361 | 0.009186 | 0.069653 | 0.006348 | 107.9477 |
| GOBP_NEG | 5 | 0.57004 | 0.009276 | 0.22881 | 0.006368 | 108.2929 |
| GOBP_MEI | 8 | 0.51448 | 0.010589 | 0.20655 | 0.006378 | 108.4545 |
| GOBP_NEG | 9 | 0.40571 | 0.008856 | 0.16298 | 0.006405 | 108.917 |
| GOBP_HIN | 34 | 0.23592 | 0.010003 | 0.094805 | 0.006419 | 109.1568 |
| GOBP_SKEI | 167 | 0.092909 | 0.0087 | 0.037344 | 0.006429 | 109.3251 |
| GOBP_REG | 136 | 0.11048 | 0.009344 | 0.044431 | 0.006454 | 109.7537 |
| BIOCARTA_ | 24 | 0.27417 | 0.009769 | 0.11027 | 0.006457 | 109.8064 |
| GOMF_JUN | 11 | 0.36241 | 0.008746 | 0.14576 | 0.006458 | 109.8132 |
| GOBP_NEG | 918 | 0.042227 | 0.009083 | 0.01699 | 0.006477 | 110.1346 |
| GOBP_REG | 96 | 0.13347 | 0.009494 | 0.053846 | 0.006597 | 112.182 |
| DURAND_S | 268 | 0.081545 | 0.009647 | 0.032921 | 0.00663 | 112.7397 |
| GOCC_NUC | 40 | 0.17499 | 0.008047 | 0.070674 | 0.006647 | 113.0339 |
| GOBP_NEG | 19 | 0.29078 | 0.00922 | 0.11751 | 0.006676 | 113.5254 |
| REACTOME | 4 | 0.64454 | 0.009381 | 0.2605 | 0.006681 | 113.6053 |
| GOCC_PRE | 16 | 0.31286 | 0.009104 | 0.12646 | 0.006684 | 113.6648 |
| GOBP_REG | 31 | 0.22581 | 0.009143 | 0.091337 | 0.006718 | 114.2379 |
| GOCC_MYE | 45 | 0.19058 | 0.009294 | 0.077134 | 0.006747 | 114.731 |
| WP_ENDOC | 63 | 0.1716 | 0.009897 | 0.069466 | 0.006755 | 114.8705 |
| WP_ENDOC | 63 | 0.1716 | 0.009897 | 0.069466 | 0.006755 | 114.8705 |
| GOBP_NEG | 9 | 0.33046 | 0.007214 | 0.13378 | 0.006756 | 114.8892 |
| GOBP_ACT | 194 | 0.089157 | 0.008992 | 0.036105 | 0.006773 | 115.1664 |
| GOBP_DIRI | 7 | 0.49292 | 0.00949 | 0.19979 | 0.006814 | 115.8687 |
| GOCC_SM_ | 77 | 0.13786 | 0.008786 | 0.055909 | 0.006842 | 116.3482 |
| GOBP_REG | 25 | 0.27912 | 0.010151 | 0.11322 | 0.006849 | 116.474 |
| GOBP_MU | 143 | 0.10567 | 0.009162 | 0.042919 | 0.006913 | 117.5624 |
| GOBP_TRA | 199 | 0.090785 | 0.009272 | 0.036904 | 0.006951 | 118.2069 |
| GOBP_RIBO | 402 | 0.056687 | 0.008184 | 0.023051 | 0.006969 | 118.4993 |
| GOMF_DN/ | 16 | 0.27126 | 0.007894 | 0.11046 | 0.007032 | 119.5809 |
| CHIARADO | 106 | 0.12771 | 0.009543 | 0.052028 | 0.007057 | 120.0043 |
| CREIGHTOI | 275 | 0.075887 | 0.009092 | 0.03093 | 0.007079 | 120.3733 |
| NOUZOVA_ | 127 | 0.10475 | 0.008563 | 0.042713 | 0.007099 | 120.7219 |
| GOBP_REG | 5 | 0.6857 | 0.011158 | 0.28004 | 0.007176 | 122.0228 |
| WANG_BA | 22 | 0.28714 | 0.009797 | 0.11746 | 0.007255 | 123.3781 |
| GOBP_AXIS | 85 | 0.13687 | 0.009163 | 0.056029 | 0.007292 | 124.0039 |
| GOBP_POS | 5 | 0.46286 | 0.007532 | 0.18954 | 0.007309 | 124.2861 |

| | | | | | | |
|---|---|---|---|---|---|---|
| GOBP_NEG | 49 | 0.18488 | 0.009407 | 0.07572 | 0.007315 | 124.3933 |
| GOBP_MIT | 716 | 0.045523 | 0.008696 | 0.018665 | 0.00737 | 125.32 |
| GOBP_OLF | 7 | 0.45192 | 0.008701 | 0.18539 | 0.007397 | 125.7928 |
| GOBP_STR | 6 | 0.59435 | 0.010594 | 0.24384 | 0.0074 | 125.8387 |
| VETTER_TA | 16 | 0.35952 | 0.010462 | 0.14766 | 0.007456 | 126.7944 |
| GOCC_MEI | 307 | 0.072712 | 0.009197 | 0.029871 | 0.007467 | 126.9831 |
| HSIAO_HO | 354 | 0.06076 | 0.008242 | 0.024968 | 0.007482 | 127.2314 |
| GOBP_REG | 42 | 0.18448 | 0.008692 | 0.075834 | 0.007499 | 127.5222 |
| CAIRO_HEI | 579 | 0.049653 | 0.008561 | 0.020419 | 0.007519 | 127.8657 |
| REACTOMI | 8 | 0.52379 | 0.01078 | 0.21556 | 0.007555 | 128.4796 |
| MANALO_I | 278 | 0.069211 | 0.008337 | 0.028485 | 0.00756 | 128.5578 |
| GOMF_PRO | 27 | 0.2207 | 0.008341 | 0.090969 | 0.007637 | 129.8621 |
| PLASARI_T | 100 | 0.12155 | 0.008824 | 0.050152 | 0.007687 | 130.7123 |
| GOBP_REG | 28 | 0.23265 | 0.008953 | 0.095997 | 0.007691 | 130.7787 |
| MARTINEZ_ | 263 | 0.075281 | 0.008824 | 0.031097 | 0.007748 | 131.7496 |
| PHONG_TN | 161 | 0.10275 | 0.009448 | 0.042479 | 0.007792 | 132.5047 |
| GOBP_TRA | 350 | 0.067628 | 0.009123 | 0.027968 | 0.007808 | 132.7716 |
| PID_IL2_1F | 54 | 0.1721 | 0.009191 | 0.071189 | 0.00782 | 132.9706 |
| GOBP_VAS | 684 | 0.049629 | 0.009274 | 0.020532 | 0.007828 | 133.1083 |
| GOBP_RES | 7 | 0.38457 | 0.007404 | 0.15916 | 0.007846 | 133.4212 |
| GOBP_PEP | 30 | 0.22853 | 0.009103 | 0.094717 | 0.007922 | 134.7102 |
| GOBP_EMI | 1070 | 0.038869 | 0.008988 | 0.01612 | 0.007955 | 135.2765 |
| REACTOMI | 8 | 0.49937 | 0.010278 | 0.20725 | 0.007993 | 135.9142 |
| MOROSETT | 20 | 0.29259 | 0.009518 | 0.12144 | 0.007995 | 135.9533 |
| GOBP_AMI | 9 | 0.39469 | 0.008616 | 0.16409 | 0.008086 | 137.4956 |
| GOMF_TYF | 5 | 0.66572 | 0.010833 | 0.27681 | 0.008092 | 137.6062 |
| GOMF_GRO | 127 | 0.11478 | 0.009382 | 0.047747 | 0.008117 | 138.0228 |
| GOBP_REG | 10 | 0.40147 | 0.009238 | 0.16706 | 0.008134 | 138.3187 |
| GOBP_GLC | 64 | 0.17394 | 0.01011 | 0.072448 | 0.008185 | 139.1791 |
| GOBP_NEG | 4 | 0.67169 | 0.009776 | 0.27981 | 0.008191 | 139.2931 |
| GOMF_SH2 | 40 | 0.19437 | 0.008937 | 0.080974 | 0.008195 | 139.3509 |
| GOBP_REG | 69 | 0.13678 | 0.008254 | 0.057013 | 0.008225 | 139.8695 |
| GOMF_P53 | 62 | 0.1484 | 0.008491 | 0.061877 | 0.00824 | 140.1263 |
| GOBP_SIST | 51 | 0.1724 | 0.008948 | 0.071952 | 0.008293 | 141.0276 |
| NADELLA_I | 7 | 0.4948 | 0.009526 | 0.20671 | 0.008345 | 141.905 |
| REACTOMI | 10 | 0.40674 | 0.009359 | 0.16992 | 0.008346 | 141.9271 |
| PASQUALU | 161 | 0.092791 | 0.008533 | 0.038776 | 0.008361 | 142.1771 |
| FOROUTAN | 189 | 0.091959 | 0.009155 | 0.038488 | 0.008446 | 143.6191 |
| GOBP_REG | 68 | 0.15928 | 0.009542 | 0.066673 | 0.008454 | 143.7552 |
| GOMF_HIS | 9 | 0.40779 | 0.008902 | 0.17081 | 0.008491 | 144.3929 |
| GOCC_ANA | 21 | 0.2158 | 0.007194 | 0.090577 | 0.008602 | 146.2821 |
| REACTOMI | 77 | 0.13011 | 0.008293 | 0.054675 | 0.008667 | 147.3823 |
| GOCC_PIN | 7 | 0.46112 | 0.008878 | 0.19381 | 0.008681 | 147.6272 |
| GOBP_NEG | 21 | 0.2408 | 0.008027 | 0.10131 | 0.008736 | 148.5625 |
| GOBP_REG | 28 | 0.25252 | 0.009718 | 0.10627 | 0.00875 | 148.7869 |

| | | | | | | |
|---|---|---|---|---|---|---|
| GOMF_PRO | 32 | 0.23758 | 0.009773 | 0.10005 | 0.008786 | 149.4093 |
| GOBP_NEG | 616 | 0.050637 | 0.008997 | 0.021338 | 0.008826 | 150.0878 |
| GOBP_REG | 290 | 0.073743 | 0.009069 | 0.031112 | 0.008894 | 151.2442 |
| GOBP_POS | 98 | 0.11568 | 0.008313 | 0.048811 | 0.008899 | 151.3292 |
| GOBP_POS | 21 | 0.28624 | 0.009542 | 0.12082 | 0.008919 | 151.6727 |
| WEST_ADR | 22 | 0.26588 | 0.009071 | 0.11223 | 0.008924 | 151.7577 |
| GOBP_POS | 898 | 0.040444 | 0.008609 | 0.017078 | 0.008942 | 152.0587 |
| GOBP_POS | 22 | 0.29913 | 0.010206 | 0.12657 | 0.009063 | 154.1129 |
| GOBP_NEG | 7 | 0.45322 | 0.008726 | 0.19183 | 0.009078 | 154.3748 |
| GOBP_NEG | 88 | 0.13087 | 0.008915 | 0.055427 | 0.009114 | 154.9819 |
| KRIEG_KDN | 184 | 0.084879 | 0.008339 | 0.035948 | 0.009115 | 154.9921 |
| GOBP_ANI | 65 | 0.16091 | 0.009426 | 0.068156 | 0.009121 | 155.106 |
| WHITFIELD | 116 | 0.11012 | 0.008606 | 0.046688 | 0.009176 | 156.0379 |
| SHETH_LIV | 241 | 0.07487 | 0.008405 | 0.031764 | 0.009216 | 156.7215 |
| GOBP_MIT | 19 | 0.24363 | 0.007725 | 0.10336 | 0.009217 | 156.7402 |
| GOCC_NUC | 937 | 0.038532 | 0.008369 | 0.016357 | 0.009251 | 157.3065 |
| HELLEBREK | 73 | 0.14232 | 0.008833 | 0.060488 | 0.009321 | 158.4968 |
| KOINUMA_ | 8 | 0.42896 | 0.008829 | 0.18234 | 0.009329 | 158.6379 |
| GOBP_MAI | 155 | 0.099642 | 0.008992 | 0.042362 | 0.009338 | 158.7995 |
| GOBP_NEG | 16 | 0.34005 | 0.009896 | 0.14462 | 0.009358 | 159.1328 |
| GOBP_MET | 15 | 0.34194 | 0.009635 | 0.14544 | 0.009367 | 159.2909 |
| GOBP_EMI | 35 | 0.20915 | 0.008997 | 0.089013 | 0.0094 | 159.8538 |
| MOOTHA_ | 19 | 0.23646 | 0.007498 | 0.10065 | 0.00941 | 160.0171 |
| WP_BREAS | 150 | 0.098846 | 0.008776 | 0.042088 | 0.009429 | 160.3469 |
| GOBP_REG | 9 | 0.45014 | 0.009826 | 0.19194 | 0.009514 | 161.7771 |
| GOBP_NEG | 644 | 0.047837 | 0.008684 | 0.02042 | 0.009578 | 162.8739 |
| GOBP_REG | 25 | 0.23513 | 0.008551 | 0.10042 | 0.00961 | 163.4232 |
| GOBP_POS | 21 | 0.28421 | 0.009474 | 0.12139 | 0.009612 | 163.4453 |
| MARTINEZ_ | 67 | 0.14437 | 0.008585 | 0.061777 | 0.009728 | 165.4297 |
| REACTOME | 21 | 0.23011 | 0.007671 | 0.098553 | 0.00978 | 166.3055 |
| GOBP_POS | 170 | 0.089944 | 0.008497 | 0.038552 | 0.009829 | 167.1455 |
| YOSHIMUR | 358 | 0.061947 | 0.008449 | 0.026569 | 0.009869 | 167.8223 |
| GOBP_DOF | 40 | 0.20072 | 0.00923 | 0.086169 | 0.009925 | 168.7763 |
| GOBP_MET | 39 | 0.20465 | 0.009292 | 0.087902 | 0.00996 | 169.3613 |
| GOBP_ALD | 16 | 0.2888 | 0.008404 | 0.12438 | 0.010126 | 172.1926 |
| GOBP_SYN | 14 | 0.35326 | 0.009617 | 0.1522 | 0.01015 | 172.6008 |
| AMIT_DEL | 18 | 0.28157 | 0.008691 | 0.12133 | 0.010158 | 172.7368 |
| GALLUZZI_ | 13 | 0.28981 | 0.007603 | 0.1249 | 0.010165 | 172.8558 |
| CAFFAREL_ | 5 | 0.61348 | 0.009983 | 0.26453 | 0.0102 | 173.451 |
| GOBP_DOF | 23 | 0.26329 | 0.009185 | 0.11354 | 0.010204 | 173.519 |
| GOCC_SPLI | 35 | 0.19897 | 0.008559 | 0.085809 | 0.01021 | 173.6211 |
| YAO_TEMP | 31 | 0.18525 | 0.007501 | 0.079891 | 0.010211 | 173.6381 |
| GOBP_HOF | 226 | 0.083131 | 0.009041 | 0.035876 | 0.010254 | 174.3693 |
| GOBP_FIBF | 52 | 0.17486 | 0.009165 | 0.075481 | 0.010268 | 174.6073 |
| GOBP_REG | 242 | 0.072424 | 0.008147 | 0.031269 | 0.01028 | 174.8114 |

| | | | | | | |
|---|---|---|---|---|---|---|
| GUO_TARG | 91 | 0.11575 | 0.008017 | 0.049975 | 0.010283 | 174.8624 |
| GOMF_TRA | 17 | 0.2686 | 0.008057 | 0.11599 | 0.01029 | 174.9815 |
| GOBP_POS | 36 | 0.21282 | 0.009285 | 0.091924 | 0.010308 | 175.2875 |
| GOBP_POS | 5 | 0.64283 | 0.01046 | 0.27776 | 0.010331 | 175.6787 |
| GOCC_TRA | 4 | 0.63447 | 0.009235 | 0.27416 | 0.010334 | 175.7297 |
| GOBP_KIDI | 90 | 0.13574 | 0.00935 | 0.058664 | 0.010343 | 175.8827 |
| REACTOME | 22 | 0.20872 | 0.007121 | 0.090228 | 0.010359 | 176.1548 |
| GOBP_REG | 44 | 0.19351 | 0.009332 | 0.083666 | 0.01037 | 176.3419 |
| GOBP_NEG | 30 | 0.19075 | 0.007598 | 0.082484 | 0.010378 | 176.4779 |
| FOSTER_TC | 397 | 0.061611 | 0.00884 | 0.02665 | 0.010399 | 176.835 |
| GOBP_NEG | 10 | 0.40666 | 0.009357 | 0.17594 | 0.010412 | 177.0561 |
| GOBP_CELI | 7 | 0.42573 | 0.008197 | 0.1842 | 0.010416 | 177.1241 |
| REACTOME | 10 | 0.42137 | 0.009695 | 0.18233 | 0.010421 | 177.2091 |
| ZWANG_D( | 223 | 0.074869 | 0.008089 | 0.032397 | 0.010424 | 177.2601 |
| GOMF_GAI | 5 | 0.59839 | 0.009737 | 0.25905 | 0.01045 | 177.7023 |
| GOBP_CYT( | 58 | 0.14254 | 0.007889 | 0.061734 | 0.010477 | 178.1614 |
| GOMF_U1_ | 8 | 0.46611 | 0.009593 | 0.2019 | 0.010489 | 178.3654 |
| IBRAHIM_N | 144 | 0.090277 | 0.007855 | 0.039123 | 0.010519 | 178.8756 |
| GOBP_NEG | 34 | 0.18994 | 0.008054 | 0.082339 | 0.010538 | 179.1987 |
| GOBP_POS | 17 | 0.26943 | 0.008082 | 0.11691 | 0.010603 | 180.304 |
| GOBP_ELAS | 6 | 0.47221 | 0.008417 | 0.20499 | 0.01063 | 180.7632 |
| WP_IL6_SI( | 42 | 0.19058 | 0.008979 | 0.082738 | 0.010635 | 180.8482 |
| GOBP_SEQ | 10 | 0.42168 | 0.009703 | 0.18309 | 0.010642 | 180.9672 |
| GOBP_GLA | 118 | 0.11565 | 0.009115 | 0.050256 | 0.010695 | 181.8685 |
| GOBP_ADR | 27 | 0.24766 | 0.009359 | 0.10771 | 0.010748 | 182.7697 |
| GOBP_LATI | 35 | 0.18738 | 0.008061 | 0.081518 | 0.010768 | 183.1098 |
| GOBP_NUC | 38 | 0.16842 | 0.007548 | 0.073289 | 0.010788 | 183.4499 |
| MILI_PSEU | 43 | 0.16723 | 0.007972 | 0.072788 | 0.010802 | 183.688 |
| PID_ERBB_ | 15 | 0.33259 | 0.009371 | 0.1448 | 0.010817 | 183.9431 |
| GOBP_NOI | 70 | 0.13753 | 0.008359 | 0.059912 | 0.010857 | 184.6233 |
| GOBP_DIAI | 5 | 0.64157 | 0.01044 | 0.2799 | 0.010955 | 186.2898 |
| GOBP_MES | 95 | 0.13115 | 0.009281 | 0.057241 | 0.01098 | 186.7149 |
| MIKKELSEN | 10 | 0.36911 | 0.008493 | 0.16112 | 0.010989 | 186.8679 |
| GOBP_TRA | 7 | 0.51427 | 0.009901 | 0.22456 | 0.011012 | 187.2591 |
| GOBP_INTI | 8 | 0.39958 | 0.008224 | 0.17454 | 0.011035 | 187.6502 |
| GOBP_MES | 13 | 0.29797 | 0.007817 | 0.13019 | 0.011053 | 187.9563 |
| GOBP_END | 30 | 0.24364 | 0.009705 | 0.10646 | 0.011059 | 188.0583 |
| GOMF_G_I | 35 | 0.22024 | 0.009474 | 0.096271 | 0.011085 | 188.5004 |
| GOBP_VAS | 78 | 0.13673 | 0.008771 | 0.059781 | 0.011098 | 188.7215 |
| GOBP_EPIT | 157 | 0.092637 | 0.008413 | 0.040514 | 0.011118 | 189.0616 |
| REACTOME | 28 | 0.19652 | 0.007563 | 0.085952 | 0.011121 | 189.1126 |
| GOBP_CELI | 15 | 0.2749 | 0.007746 | 0.12031 | 0.011165 | 189.8608 |
| MARSON_I | 9 | 0.39866 | 0.008703 | 0.17462 | 0.011222 | 190.8301 |
| GOBP_REG | 28 | 0.24754 | 0.009526 | 0.10857 | 0.011312 | 192.3606 |
| GOBP_MET | 5 | 0.54076 | 0.0088 | 0.23722 | 0.011321 | 192.5136 |

| | | | | | | |
|---|---|---|---|---|---|---|
| VANDESLU | 14 | 0.35108 | 0.009557 | 0.15404 | 0.011335 | 192.7517 |
| GOBP_NEG | 20 | 0.25515 | 0.008301 | 0.112 | 0.011367 | 193.2958 |
| GOBP_REP | 283 | 0.070912 | 0.008617 | 0.031139 | 0.011391 | 193.704 |
| GOBP_REG | 23 | 0.26728 | 0.009324 | 0.1174 | 0.011412 | 194.0611 |
| GOCC_NUC | 379 | 0.055474 | 0.007781 | 0.024407 | 0.011522 | 195.9316 |
| GOBP_ATR | 5 | 0.55333 | 0.009004 | 0.24345 | 0.011524 | 195.9656 |
| GOBP_POS | 13 | 0.32418 | 0.008504 | 0.14264 | 0.01153 | 196.0677 |
| WP_HIPPO | 116 | 0.11118 | 0.008689 | 0.048954 | 0.011574 | 196.8159 |
| WARTERS_ | 72 | 0.13388 | 0.008252 | 0.058971 | 0.011601 | 197.275 |
| GOBP_MAI | 33 | 0.18731 | 0.007825 | 0.082595 | 0.011676 | 198.5504 |
| GOMF_TR/ | 447 | 0.052856 | 0.008036 | 0.023308 | 0.011681 | 198.6354 |
| GOBP_NUC | 17 | 0.25608 | 0.007681 | 0.11293 | 0.011684 | 198.6864 |
| GOBP_POS | 393 | 0.058985 | 0.008422 | 0.026012 | 0.011684 | 198.6864 |
| LOPEZ_MB | 3 | 0.6455 | 0.008137 | 0.28493 | 0.01175 | 199.8088 |
| GOBP_NEG | 11 | 0.36293 | 0.008758 | 0.16027 | 0.011777 | 200.2679 |
| REACTOME | 17 | 0.22637 | 0.00679 | 0.10007 | 0.011856 | 201.6113 |
| GRESHOCK | 318 | 0.064695 | 0.008326 | 0.028625 | 0.011914 | 202.5976 |
| GOBP_MIC | 152 | 0.086593 | 0.007739 | 0.038334 | 0.011951 | 203.2268 |
| DOUGLAS_ | 528 | 0.049664 | 0.008189 | 0.021995 | 0.011981 | 203.7369 |
| GOCC_PRE | 5 | 0.43718 | 0.007114 | 0.19369 | 0.012006 | 204.162 |
| REACTOME | 11 | 0.37976 | 0.009164 | 0.16828 | 0.01202 | 204.4001 |
| GOBP_REG | 57 | 0.16195 | 0.008886 | 0.071817 | 0.012072 | 205.2844 |
| MAYBURD_ | 54 | 0.1506 | 0.008043 | 0.066795 | 0.012084 | 205.4884 |
| GOBP_ANT | 48 | 0.16339 | 0.008228 | 0.072515 | 0.012132 | 206.3047 |
| REACTOME | 46 | 0.16318 | 0.008045 | 0.072498 | 0.012206 | 207.563 |
| GOBP_FEM | 12 | 0.27561 | 0.006947 | 0.12247 | 0.012216 | 207.7331 |
| GOMF_GAI | 4 | 0.57879 | 0.008424 | 0.2577 | 0.012359 | 210.1648 |
| GOCC_FC_ | 5 | 0.4224 | 0.006873 | 0.18815 | 0.012389 | 210.6749 |
| MORI_LAR | 83 | 0.11886 | 0.007864 | 0.053007 | 0.012478 | 212.1884 |
| REACTOME | 27 | 0.18744 | 0.007084 | 0.083626 | 0.012505 | 212.6475 |
| PID_BARD1 | 29 | 0.20857 | 0.008168 | 0.09311 | 0.012552 | 213.4468 |
| HOUNKPE_ | 1083 | 0.03266 | 0.007595 | 0.014605 | 0.012676 | 215.5554 |
| GOBP_HEP | 8 | 0.47009 | 0.009675 | 0.21022 | 0.012677 | 215.5724 |
| WAESCH_A | 13 | 0.24906 | 0.006534 | 0.11138 | 0.01268 | 215.6234 |
| GOBP_NEU | 71 | 0.13437 | 0.008225 | 0.060106 | 0.012695 | 215.8785 |
| LI_INDUCE | 128 | 0.098651 | 0.008096 | 0.044146 | 0.012727 | 216.4226 |
| GOBP_GTP | 24 | 0.19199 | 0.006841 | 0.086003 | 0.012802 | 217.698 |
| GOBP_MRI | 643 | 0.042265 | 0.007666 | 0.018937 | 0.012818 | 217.9701 |
| IVANOVA_ | 480 | 0.051126 | 0.008048 | 0.022914 | 0.012838 | 218.3102 |
| GOBP_CAR | 6 | 0.51524 | 0.009184 | 0.23095 | 0.012848 | 218.4802 |
| GOMF_MIT | 9 | 0.42497 | 0.009277 | 0.19049 | 0.01285 | 218.5143 |
| GOBP_SKE | 221 | 0.083494 | 0.008981 | 0.037441 | 0.012879 | 219.0074 |
| GOBP_REG | 8 | 0.47258 | 0.009726 | 0.21196 | 0.012895 | 219.2795 |
| GOBP_ALP | 9 | 0.39117 | 0.008539 | 0.17556 | 0.012941 | 220.0617 |
| GOMF_PRO | 136 | 0.092771 | 0.007846 | 0.041663 | 0.012991 | 220.912 |

| | | | | | | |
|---|---|---|---|---|---|---|
| GOCC_AXC | 21 | 0.24122 | 0.008041 | 0.10838 | 0.013023 | 221.4561 |
| BENNETT_S | 27 | 0.21999 | 0.008314 | 0.098939 | 0.013096 | 222.6975 |
| GOBP_NEU | 15 | 0.29987 | 0.008449 | 0.13486 | 0.013098 | 222.7315 |
| GOBP_RES | 257 | 0.073093 | 0.00847 | 0.032895 | 0.013147 | 223.5647 |
| GOBP_ANT | 6 | 0.4398 | 0.007839 | 0.19805 | 0.013195 | 224.381 |
| KORKOLA_ | 42 | 0.19519 | 0.009197 | 0.087942 | 0.01323 | 224.9762 |
| CHICAS_RE | 530 | 0.049798 | 0.008226 | 0.022449 | 0.013274 | 225.7244 |
| GOBP_RES | 670 | 0.04545 | 0.008409 | 0.02049 | 0.013278 | 225.7924 |
| SESTO_RES | 51 | 0.1621 | 0.008414 | 0.073082 | 0.013282 | 225.8604 |
| GOBP_SON | 9 | 0.34045 | 0.007432 | 0.15351 | 0.013293 | 226.0475 |
| GOBP_REN | 92 | 0.13021 | 0.009068 | 0.058726 | 0.013309 | 226.3195 |
| WP_PATHV | 16 | 0.24804 | 0.007218 | 0.11191 | 0.01334 | 226.8467 |
| GOBP_EST/ | 248 | 0.065726 | 0.007484 | 0.029675 | 0.013393 | 227.748 |
| ZHU_CMV_ | 47 | 0.17288 | 0.008616 | 0.078071 | 0.013407 | 227.986 |
| DODD_NAS | 1299 | 0.031862 | 0.008065 | 0.014391 | 0.01342 | 228.2071 |
| FOURNIER_ | 11 | 0.36783 | 0.008876 | 0.16618 | 0.013442 | 228.5812 |
| GOBP_SPIN | 121 | 0.096563 | 0.007706 | 0.043632 | 0.013451 | 228.7343 |
| GOBP_MIT | 9 | 0.32444 | 0.007082 | 0.14666 | 0.013487 | 229.3464 |
| GOBP_MIT | 65 | 0.12916 | 0.007566 | 0.058396 | 0.013499 | 229.5505 |
| GOMF_BH_ | 9 | 0.36736 | 0.008019 | 0.16611 | 0.013508 | 229.7035 |
| KORKOLA_ | 60 | 0.14945 | 0.008412 | 0.06761 | 0.013545 | 230.3327 |
| KAUFFMAN | 6 | 0.39897 | 0.007112 | 0.18057 | 0.013577 | 230.8769 |
| GOMF_SUL | 11 | 0.30248 | 0.0073 | 0.13692 | 0.013588 | 231.0639 |
| RIZ_ERYTH | 34 | 0.20067 | 0.008508 | 0.090854 | 0.013604 | 231.336 |
| GOBP_REC | 43 | 0.16598 | 0.007913 | 0.075186 | 0.013644 | 232.0162 |
| KRASNOSE | 36 | 0.18153 | 0.00792 | 0.082231 | 0.013644 | 232.0162 |
| GAUSSMAI | 90 | 0.12561 | 0.008653 | 0.05693 | 0.013684 | 232.6964 |
| GOBP_PLA | 6 | 0.50649 | 0.009028 | 0.22963 | 0.01371 | 233.1386 |
| GOBP_CER | 23 | 0.23568 | 0.008221 | 0.10697 | 0.013797 | 234.618 |
| GOBP_REG | 25 | 0.24123 | 0.008773 | 0.10951 | 0.013814 | 234.9071 |
| BOYLAN_M | 27 | 0.22485 | 0.008498 | 0.10211 | 0.013833 | 235.2302 |
| SMID_BRE/ | 276 | 0.070757 | 0.008493 | 0.032146 | 0.01387 | 235.8594 |
| GOBP_MET | 11 | 0.33055 | 0.007977 | 0.15041 | 0.013991 | 237.917 |
| SCHUHMA( | 7 | 0.4297 | 0.008273 | 0.19554 | 0.013997 | 238.019 |
| GOBP_RES | 21 | 0.24775 | 0.008259 | 0.11278 | 0.014027 | 238.5291 |
| TOMIDA_M | 17 | 0.24906 | 0.007471 | 0.11339 | 0.014035 | 238.6652 |
| FONTAINE_ | 61 | 0.14433 | 0.008191 | 0.065714 | 0.014042 | 238.7842 |
| FLORIO_NE | 180 | 0.08372 | 0.008136 | 0.038134 | 0.014074 | 239.3284 |
| GOBP_POS | 6 | 0.54256 | 0.009671 | 0.24715 | 0.014079 | 239.4134 |
| REACTOME | 25 | 0.1974 | 0.007179 | 0.089943 | 0.014097 | 239.7195 |
| BENPORAT | 912 | 0.041591 | 0.008918 | 0.018958 | 0.014132 | 240.3147 |
| SCHMIDT_I | 26 | 0.21227 | 0.007872 | 0.096808 | 0.014172 | 240.9949 |
| GOBP_MO | 187 | 0.089337 | 0.008847 | 0.040748 | 0.014181 | 241.1479 |
| BIOCARTA_ | 26 | 0.22762 | 0.008442 | 0.10383 | 0.014191 | 241.318 |
| KORKOLA_ | 43 | 0.1865 | 0.008891 | 0.085083 | 0.014196 | 241.403 |

| | | | | | | |
|---|---|---|---|---|---|---|
| BERNARD_ | 36 | 0.1783 | 0.007779 | 0.08136 | 0.014216 | 241.7431 |
| WP_MAPK_ | 31 | 0.19041 | 0.00771 | 0.086894 | 0.014224 | 241.8791 |
| GOBP_CAR | 35 | 0.22315 | 0.0096 | 0.10187 | 0.014253 | 242.3723 |
| REACTOME | 13 | 0.34914 | 0.009159 | 0.15941 | 0.014264 | 242.5593 |
| REACTOME | 22 | 0.26822 | 0.009151 | 0.12247 | 0.014266 | 242.5933 |
| GOBP_RIB( | 31 | 0.17902 | 0.007249 | 0.081759 | 0.014282 | 242.8654 |
| REACTOME | 10 | 0.31131 | 0.007163 | 0.14219 | 0.014293 | 243.0525 |
| GOBP_CELI | 13 | 0.28601 | 0.007503 | 0.13064 | 0.014294 | 243.0695 |
| GOMF_CHI | 4 | 0.44428 | 0.006466 | 0.20297 | 0.014308 | 243.3075 |
| GOBP_MUS | 179 | 0.083144 | 0.008058 | 0.037987 | 0.014315 | 243.4266 |
| GOMF_TRA | 206 | 0.07181 | 0.00746 | 0.032815 | 0.014329 | 243.6646 |
| GOBP_STR | 8 | 0.36605 | 0.007534 | 0.1673 | 0.01434 | 243.8517 |
| GOBP_HIST | 10 | 0.32851 | 0.007559 | 0.15027 | 0.01441 | 245.0421 |
| GOBP_NEG | 29 | 0.21354 | 0.008363 | 0.097787 | 0.014497 | 246.5215 |
| GOBP_GAS | 12 | 0.28752 | 0.007247 | 0.13171 | 0.014522 | 246.9466 |
| RIZKI_TUM | 60 | 0.14839 | 0.008353 | 0.068002 | 0.014554 | 247.4908 |
| GOMF_MA | 17 | 0.26656 | 0.007996 | 0.12216 | 0.014557 | 247.5418 |
| WP_OSTEC | 16 | 0.3184 | 0.009266 | 0.14597 | 0.014585 | 248.0179 |
| GOCC_DEN | 7 | 0.45082 | 0.00868 | 0.20673 | 0.014608 | 248.409 |
| GOBP_REC | 10 | 0.38991 | 0.008972 | 0.17902 | 0.01471 | 250.1436 |
| GOBP_GEN | 6 | 0.44122 | 0.007865 | 0.20276 | 0.014781 | 251.3509 |
| GOMF_HYI | 8 | 0.41441 | 0.008529 | 0.19044 | 0.014781 | 251.3509 |
| GOMF_PR( | 6 | 0.45523 | 0.008114 | 0.20934 | 0.014836 | 252.2862 |
| GOBP_ACT | 41 | 0.19035 | 0.008861 | 0.087588 | 0.014889 | 253.1874 |
| GOBP_AOF | 59 | 0.15594 | 0.008704 | 0.071799 | 0.01494 | 254.0547 |
| ROZANOV_ | 33 | 0.20952 | 0.008752 | 0.096488 | 0.014955 | 254.3098 |
| REACTOME | 21 | 0.25088 | 0.008363 | 0.11554 | 0.014957 | 254.3438 |
| GOBP_CER | 49 | 0.14249 | 0.00725 | 0.065676 | 0.015026 | 255.5171 |
| GOCC_SUP | 1294 | 0.032176 | 0.00813 | 0.014836 | 0.015057 | 256.0443 |
| GOBP_REG | 99 | 0.11059 | 0.007987 | 0.050994 | 0.015062 | 256.1293 |
| REACTOME | 16 | 0.32372 | 0.00942 | 0.14938 | 0.015119 | 257.0986 |
| GOBP_ADE | 132 | 0.10161 | 0.008467 | 0.046922 | 0.015183 | 258.1869 |
| GOMF_PR( | 118 | 0.099882 | 0.007872 | 0.046155 | 0.015237 | 259.1052 |
| REACTOME | 544 | 0.047142 | 0.007886 | 0.021791 | 0.015264 | 259.5643 |
| GOBP_NU( | 16 | 0.2502 | 0.007281 | 0.11573 | 0.015317 | 260.4656 |
| GOBP_REG | 190 | 0.081952 | 0.00818 | 0.037944 | 0.0154 | 261.877 |
| GOBP_PRC | 7 | 0.43277 | 0.008332 | 0.20046 | 0.015439 | 262.5402 |
| WP_MALIG | 417 | 0.0544 | 0.007995 | 0.025204 | 0.015455 | 262.8123 |
| FLECHNER_ | 137 | 0.090212 | 0.007657 | 0.041796 | 0.015456 | 262.8293 |
| ZHU_CMV_ | 114 | 0.10811 | 0.008376 | 0.050092 | 0.015464 | 262.9653 |
| GOMF_LIG | 11 | 0.29692 | 0.007165 | 0.13763 | 0.015497 | 263.5265 |
| BIOCARTA_ | 21 | 0.23475 | 0.007825 | 0.10882 | 0.015501 | 263.5945 |
| GOBP_KIDI | 18 | 0.28987 | 0.008947 | 0.13441 | 0.015524 | 263.9856 |
| DING_LUN( | 6 | 0.42771 | 0.007624 | 0.19849 | 0.015594 | 265.176 |
| REACTOME | 26 | 0.19556 | 0.007252 | 0.090761 | 0.015602 | 265.312 |

| | | | | | | |
|---|---|---|---|---|---|---|
| GOBP_POS | 32 | 0.1979 | 0.008141 | 0.091944 | 0.015689 | 266.7914 |
| JINESH_BLI | 251 | 0.07 | 0.008018 | 0.032538 | 0.015732 | 267.5227 |
| APPEL_IMA | 31 | 0.18133 | 0.007342 | 0.084299 | 0.015746 | 267.7607 |
| WP_TRANS | 17 | 0.28278 | 0.008482 | 0.13147 | 0.015747 | 267.7777 |
| GOBP_NEG | 1499 | 0.028985 | 0.007837 | 0.013481 | 0.015783 | 268.3899 |
| GOBP_POS | 18 | 0.2569 | 0.007929 | 0.11951 | 0.0158 | 268.679 |
| GOBP_CELI | 125 | 0.096744 | 0.007846 | 0.045015 | 0.015818 | 268.9851 |
| GOBP_AND | 27 | 0.19981 | 0.007551 | 0.092992 | 0.015836 | 269.2912 |
| WHITEFOR | 114 | 0.097061 | 0.00752 | 0.045176 | 0.015845 | 269.4442 |
| GOBP_DIAI | 3 | 0.81466 | 0.010269 | 0.37925 | 0.01586 | 269.6993 |
| GOBP_EQU | 7 | 0.44125 | 0.008495 | 0.20541 | 0.015861 | 269.7163 |
| GOBP_NEG | 133 | 0.089045 | 0.007448 | 0.041498 | 0.015952 | 271.2638 |
| REACTOME | 7 | 0.41983 | 0.008083 | 0.19565 | 0.015952 | 271.2638 |
| GOBP_CIRC | 66 | 0.12615 | 0.007446 | 0.058851 | 0.01604 | 272.7602 |
| GOCC_END | 26 | 0.20307 | 0.007531 | 0.094798 | 0.016099 | 273.7635 |
| VILIMAS_N | 24 | 0.21845 | 0.007784 | 0.10198 | 0.016102 | 273.8145 |
| BLANCO_M | 102 | 0.10148 | 0.007439 | 0.047384 | 0.016117 | 274.0696 |
| GOMF_5_D | 5 | 0.39411 | 0.006413 | 0.18402 | 0.016118 | 274.0866 |
| PID_CMYB_ | 83 | 0.12288 | 0.00813 | 0.057438 | 0.016213 | 275.7021 |
| CUI_TCF21_ | 29 | 0.23371 | 0.009153 | 0.1093 | 0.016253 | 276.3823 |
| PID_HIV_N | 32 | 0.1737 | 0.007146 | 0.081272 | 0.016295 | 277.0965 |
| GOBP_RET | 21 | 0.26605 | 0.008869 | 0.12448 | 0.016296 | 277.1135 |
| CORRE_MU | 69 | 0.1353 | 0.008165 | 0.063313 | 0.016309 | 277.3345 |
| WP_OSTEO | 116 | 0.10766 | 0.008413 | 0.050383 | 0.016312 | 277.3856 |
| GOBP_TELO | 39 | 0.15775 | 0.007163 | 0.073825 | 0.016316 | 277.4536 |
| GOCC_MIT | 1546 | 0.027623 | 0.007574 | 0.012937 | 0.016381 | 278.5589 |
| SHARMA_F | 26 | 0.2363 | 0.008763 | 0.11069 | 0.016396 | 278.814 |
| GOBP_CELI | 20 | 0.27313 | 0.008886 | 0.12795 | 0.016398 | 278.848 |
| GOMF_ER_ | 5 | 0.41746 | 0.006793 | 0.19556 | 0.016401 | 278.899 |
| GOCC_MYO | 25 | 0.20837 | 0.007578 | 0.097626 | 0.016415 | 279.1371 |
| GOBP_ANA | 24 | 0.18672 | 0.006653 | 0.087487 | 0.01642 | 279.2221 |
| REACTOME | 27 | 0.20064 | 0.007583 | 0.094072 | 0.016476 | 280.1744 |
| GOCC_PLA | 526 | 0.048988 | 0.008063 | 0.022986 | 0.016544 | 281.3307 |
| GOMF_MC | 81 | 0.12307 | 0.008044 | 0.057766 | 0.016575 | 281.8579 |
| GOBP_ATR | 8 | 0.44576 | 0.009175 | 0.20928 | 0.016592 | 282.147 |
| GOCC_JUXT | 10 | 0.35339 | 0.008131 | 0.16597 | 0.016623 | 282.6741 |
| KEGG_CAR | 68 | 0.12364 | 0.007407 | 0.058103 | 0.016679 | 283.6264 |
| SMID_BREA | 595 | 0.04553 | 0.007955 | 0.021397 | 0.016681 | 283.6604 |
| GOBP_MES | 235 | 0.07194 | 0.007976 | 0.033811 | 0.016687 | 283.7624 |
| GOBP_ABS | 6 | 0.53032 | 0.009453 | 0.24933 | 0.016719 | 284.3066 |
| GOBP_DEN | 13 | 0.28775 | 0.007549 | 0.13542 | 0.016804 | 285.752 |
| GOBP_SMO | 135 | 0.094889 | 0.007996 | 0.044658 | 0.016809 | 285.837 |
| KEGG_RIBO | 80 | 0.10553 | 0.006855 | 0.049685 | 0.016844 | 286.4322 |
| GOBP_POS | 6 | 0.4519 | 0.008055 | 0.21278 | 0.01685 | 286.5343 |
| GOBP_REG | 220 | 0.07515 | 0.008065 | 0.035396 | 0.01688 | 287.0444 |

| | | | | | | |
|---|---|---|---|---|---|---|
| GOBP_MIT | 121 | 0.089613 | 0.007151 | 0.042218 | 0.0169 | 287.3845 |
| GOBP_GOL | 39 | 0.15688 | 0.007123 | 0.073927 | 0.016921 | 287.7416 |
| GOBP_BON | 219 | 0.075743 | 0.008111 | 0.035696 | 0.01693 | 287.8947 |
| GOBP_MRI | 6 | 0.51709 | 0.009217 | 0.2437 | 0.016933 | 287.9457 |
| PIEPOLI_LG | 12 | 0.3408 | 0.00859 | 0.16068 | 0.016969 | 288.5578 |
| GOBP_ISOI | 5 | 0.44319 | 0.007212 | 0.20908 | 0.017024 | 289.4931 |
| GOBP_POS | 43 | 0.1528 | 0.007285 | 0.072116 | 0.017058 | 290.0713 |
| JEON_SMA | 19 | 0.21305 | 0.006756 | 0.10055 | 0.017061 | 290.1223 |
| GOBP_MES | 6 | 0.44137 | 0.007867 | 0.20846 | 0.017125 | 291.2106 |
| GOBP_NEG | 7 | 0.3667 | 0.00706 | 0.17331 | 0.017185 | 292.2309 |
| GOBP_MUS | 401 | 0.056062 | 0.008084 | 0.026503 | 0.017208 | 292.622 |
| GOBP_PYR | 7 | 0.50541 | 0.009731 | 0.23896 | 0.01722 | 292.8261 |
| GAJATE_RE | 20 | 0.23871 | 0.007766 | 0.11294 | 0.017284 | 293.9144 |
| MILI_PSEU | 477 | 0.048917 | 0.007677 | 0.023168 | 0.017378 | 295.5129 |
| GOBP_NEG | 12 | 0.33731 | 0.008502 | 0.15985 | 0.017428 | 296.3631 |
| GOBP_RIBO | 55 | 0.1376 | 0.007416 | 0.065225 | 0.017454 | 296.8053 |
| GOBP_NEG | 73 | 0.11783 | 0.007313 | 0.05586 | 0.017461 | 296.9243 |
| GOBP_NEG | 30 | 0.18802 | 0.007489 | 0.08915 | 0.017479 | 297.2304 |
| GOBP_POS | 44 | 0.1617 | 0.007797 | 0.076679 | 0.017491 | 297.4345 |
| GERHOLD_ | 47 | 0.15358 | 0.007654 | 0.072831 | 0.017493 | 297.4685 |
| WP_MIRNA | 29 | 0.2136 | 0.008365 | 0.1013 | 0.017498 | 297.5535 |
| REACTOME | 52 | 0.15588 | 0.00817 | 0.073941 | 0.017514 | 297.8256 |
| GOBP_NEG | 22 | 0.20164 | 0.006879 | 0.095687 | 0.017555 | 298.5228 |
| GOBP_GLC | 13 | 0.31215 | 0.008189 | 0.14813 | 0.017558 | 298.5738 |
| PICCALUGA | 200 | 0.078226 | 0.008009 | 0.037127 | 0.017567 | 298.7268 |
| YAO_TEMP | 166 | 0.076218 | 0.007116 | 0.03618 | 0.017582 | 298.9819 |
| GOBP_NEG | 6 | 0.44081 | 0.007857 | 0.20933 | 0.017619 | 299.6111 |
| KONG_E2F | 10 | 0.32324 | 0.007438 | 0.15353 | 0.017635 | 299.8832 |
| GOBP_POS | 727 | 0.040283 | 0.007752 | 0.019141 | 0.017672 | 300.5124 |
| NUTT_GBM | 45 | 0.14827 | 0.00723 | 0.070454 | 0.017677 | 300.5974 |
| SEAVEY_EP | 90 | 0.11226 | 0.007733 | 0.053363 | 0.01771 | 301.1586 |
| MISSIAGLIA | 110 | 0.090577 | 0.006894 | 0.043069 | 0.017736 | 301.6007 |
| LEE_NEURA | 139 | 0.092824 | 0.007936 | 0.044141 | 0.017745 | 301.7537 |
| GOMF_5_3 | 8 | 0.3017 | 0.006209 | 0.14372 | 0.017906 | 304.4915 |
| GOBP_RES | 6 | 0.36511 | 0.006508 | 0.17401 | 0.017951 | 305.2568 |
| GOBP_REG | 5 | 0.51143 | 0.008322 | 0.24375 | 0.017954 | 305.3078 |
| KEGG_NON | 13 | 0.29278 | 0.00768 | 0.13959 | 0.017988 | 305.8859 |
| GOBP_POS | 7 | 0.43216 | 0.00832 | 0.20608 | 0.018004 | 306.158 |
| WP_HEART | 44 | 0.17558 | 0.008467 | 0.083736 | 0.018012 | 306.2941 |
| GOMF_STR | 705 | 0.04099 | 0.007772 | 0.019551 | 0.018027 | 306.5491 |
| GOBP_CAR | 10 | 0.36197 | 0.008329 | 0.17277 | 0.018086 | 307.5524 |
| GOBP_INTI | 59 | 0.14993 | 0.008369 | 0.071572 | 0.018099 | 307.7735 |
| REACTOME | 15 | 0.30536 | 0.008604 | 0.14578 | 0.01811 | 307.9606 |
| DELACROIX | 45 | 0.16872 | 0.008228 | 0.080584 | 0.018151 | 308.6578 |
| GOBP_BLO | 582 | 0.04622 | 0.00799 | 0.02208 | 0.018169 | 308.9638 |

| | | | | | | |
|---|---|---|---|---|---|---|
| GOBP_NEG | 55 | 0.13899 | 0.007491 | 0.066416 | 0.018193 | 309.372 |
| GOMF_I_SI | 14 | 0.3133 | 0.008529 | 0.14971 | 0.018194 | 309.389 |
| GOBP_POS | 31 | 0.20864 | 0.008448 | 0.099747 | 0.018241 | 310.1882 |
| ZWANG_CI | 472 | 0.049592 | 0.007743 | 0.023735 | 0.018344 | 311.9397 |
| PECE_MAN | 132 | 0.086689 | 0.007224 | 0.041518 | 0.018406 | 312.994 |
| GOMF_MIS | 13 | 0.26758 | 0.00702 | 0.12821 | 0.018446 | 313.6742 |
| GOBP_REG | 16 | 0.2754 | 0.008014 | 0.13198 | 0.018464 | 313.9803 |
| GOBP_CIRC | 554 | 0.04761 | 0.008035 | 0.022817 | 0.018469 | 314.0653 |
| HOLLEMAN | 20 | 0.22431 | 0.007297 | 0.10752 | 0.018484 | 314.3204 |
| GOBP_REG | 39 | 0.15352 | 0.006971 | 0.073614 | 0.01852 | 314.9326 |
| SIMBULAN_ | 31 | 0.18291 | 0.007406 | 0.087738 | 0.018554 | 315.5108 |
| GOBP_VIRA | 395 | 0.053353 | 0.007636 | 0.02563 | 0.018694 | 317.8915 |
| GOBP_REG | 159 | 0.086752 | 0.007928 | 0.041692 | 0.018735 | 318.5887 |
| GOCC_DYS | 7 | 0.54519 | 0.010496 | 0.26209 | 0.018762 | 319.0478 |
| GOBP_REG | 6 | 0.43579 | 0.007768 | 0.20954 | 0.018781 | 319.3709 |
| GOBP_INSU | 47 | 0.15494 | 0.007722 | 0.074511 | 0.018796 | 319.626 |
| VARELA_ZN | 41 | 0.16839 | 0.007839 | 0.080981 | 0.018797 | 319.643 |
| GOBP_APP | 176 | 0.084997 | 0.008169 | 0.04091 | 0.018878 | 321.0204 |
| KEGG_END | 168 | 0.078898 | 0.00741 | 0.038011 | 0.018969 | 322.5678 |
| GOBP_EMB | 653 | 0.042018 | 0.007678 | 0.020263 | 0.019064 | 324.1833 |
| GOCC_GAP | 30 | 0.20323 | 0.008095 | 0.098033 | 0.019089 | 324.6084 |
| REACTOME | 186 | 0.068953 | 0.006811 | 0.033263 | 0.019095 | 324.7105 |
| PUJANA_CI | 713 | 0.038189 | 0.00728 | 0.018434 | 0.019153 | 325.6968 |
| GOBP_PRO | 22 | 0.19647 | 0.006703 | 0.094849 | 0.01917 | 325.9859 |
| GOBP_ACY | 35 | 0.16262 | 0.006996 | 0.07857 | 0.019248 | 327.3122 |
| GOMF_CHI | 108 | 0.0962 | 0.007255 | 0.046494 | 0.019276 | 327.7884 |
| GOBP_HIG | 15 | 0.29883 | 0.00842 | 0.14443 | 0.019277 | 327.8054 |
| WP_HEMA | 19 | 0.23014 | 0.007298 | 0.11124 | 0.019286 | 327.9584 |
| GOBP_POS | 7 | 0.38707 | 0.007452 | 0.18713 | 0.019306 | 328.2985 |
| GOBP_LINC | 20 | 0.21631 | 0.007037 | 0.10458 | 0.019317 | 328.4856 |
| REACTOME | 57 | 0.12933 | 0.007096 | 0.062536 | 0.019321 | 328.5536 |
| GOBP_REG | 18 | 0.23937 | 0.007388 | 0.11576 | 0.01934 | 328.8767 |
| BENPORAT | 939 | 0.037816 | 0.008222 | 0.018295 | 0.01937 | 329.3869 |
| GOBP_AMI | 408 | 0.053131 | 0.007726 | 0.025705 | 0.019378 | 329.5229 |
| YAGI_AML_ | 121 | 0.095711 | 0.007638 | 0.046328 | 0.019424 | 330.3051 |
| GOMF_IGE | 5 | 0.41525 | 0.006757 | 0.20106 | 0.019456 | 330.8493 |
| GOBP_OLF | 12 | 0.31243 | 0.007875 | 0.1513 | 0.019471 | 331.1044 |
| GOBP_POS | 18 | 0.25196 | 0.007777 | 0.12206 | 0.019508 | 331.7335 |
| PID_HNF3A | 43 | 0.16611 | 0.007919 | 0.080492 | 0.019531 | 332.1247 |
| ODONNELL | 117 | 0.091313 | 0.007166 | 0.04425 | 0.019537 | 332.2267 |
| GOBP_REC | 47 | 0.14364 | 0.007158 | 0.069609 | 0.01954 | 332.2777 |
| BURTON_A | 30 | 0.19714 | 0.007853 | 0.09554 | 0.019544 | 332.3457 |
| REACTOME | 86 | 0.098352 | 0.006623 | 0.047675 | 0.019566 | 332.7198 |
| WP_ETHAN | 9 | 0.3462 | 0.007557 | 0.16784 | 0.019581 | 332.9749 |
| ZHAN_V2_ | 39 | 0.1582 | 0.007183 | 0.076703 | 0.019588 | 333.0939 |

| | | | | | | |
|---|---|---|---|---|---|---|
| MILI_PSEU | 77 | 0.10936 | 0.00697 | 0.053048 | 0.019633 | 333.8592 |
| SASAI_RESI | 48 | 0.15682 | 0.007898 | 0.076078 | 0.019642 | 334.0122 |
| GOBP_POS | 1145 | 0.031809 | 0.007593 | 0.015434 | 0.019657 | 334.2673 |
| GOBP_POS | 5 | 0.59241 | 0.00964 | 0.2875 | 0.019682 | 334.6924 |
| GOCC_CAR | 6 | 0.34222 | 0.0061 | 0.16613 | 0.019713 | 335.2196 |
| GOBP_POS | 24 | 0.25108 | 0.008947 | 0.1219 | 0.019721 | 335.3556 |
| DELPUECH_ | 38 | 0.1537 | 0.006889 | 0.07465 | 0.01976 | 336.0188 |
| GOBP_REG | 44 | 0.17542 | 0.008459 | 0.085213 | 0.019776 | 336.2909 |
| GOBP_LYSO | 122 | 0.091745 | 0.007352 | 0.044578 | 0.019799 | 336.682 |
| GOCC_EXO | 9 | 0.36872 | 0.008049 | 0.1792 | 0.019824 | 337.1071 |
| FOROUTAN | 182 | 0.080104 | 0.007827 | 0.039007 | 0.020016 | 340.3721 |
| GOMF_GD | 14 | 0.24256 | 0.006603 | 0.11824 | 0.020121 | 342.1576 |
| GOBP_MU | 32 | 0.16486 | 0.006782 | 0.080382 | 0.020142 | 342.5147 |
| REACTOME | 16 | 0.28304 | 0.008237 | 0.13804 | 0.020172 | 343.0249 |
| MYLLYKAN | 6 | 0.37608 | 0.006704 | 0.1835 | 0.020215 | 343.7561 |
| TONKS_TAI | 16 | 0.24171 | 0.007034 | 0.11796 | 0.020235 | 344.0962 |
| ZHAN_V1_ | 14 | 0.27938 | 0.007605 | 0.1364 | 0.020279 | 344.8444 |
| REACTOME | 77 | 0.12749 | 0.008126 | 0.062272 | 0.020322 | 345.5756 |
| GOBP_CELI | 605 | 0.04038 | 0.007112 | 0.019724 | 0.020324 | 345.6096 |
| SESTO_RES | 96 | 0.099137 | 0.007052 | 0.048486 | 0.020452 | 347.7863 |
| GOBP_ANT | 9 | 0.33383 | 0.007287 | 0.16332 | 0.020484 | 348.3304 |
| GOBP_RET | 20 | 0.21641 | 0.00704 | 0.10589 | 0.020502 | 348.6365 |
| PEDERSEN_ | 46 | 0.1531 | 0.007548 | 0.074936 | 0.020531 | 349.1297 |
| GOBP_DOU | 6 | 0.45313 | 0.008077 | 0.2218 | 0.020537 | 349.2317 |
| WP_HEDGI | 16 | 0.26197 | 0.007623 | 0.12824 | 0.020541 | 349.2997 |
| GOBP_CAR | 229 | 0.066705 | 0.007302 | 0.032656 | 0.020551 | 349.4698 |
| GOBP_REG | 8 | 0.42653 | 0.008779 | 0.20882 | 0.020555 | 349.5378 |
| GOCC_GLY | 3 | 0.5926 | 0.00747 | 0.29013 | 0.020557 | 349.5718 |
| GOBP_TRA | 7 | 0.44192 | 0.008508 | 0.21643 | 0.020591 | 350.15 |
| GOBP_NEU | 33 | 0.20148 | 0.008417 | 0.098717 | 0.020632 | 350.8472 |
| GOMF_TYF | 9 | 0.32146 | 0.007017 | 0.1576 | 0.020695 | 351.9185 |
| GOBP_FIBF | 24 | 0.21725 | 0.007741 | 0.10656 | 0.020748 | 352.8197 |
| GOBP_ELA: | 3 | 0.54028 | 0.00681 | 0.26521 | 0.020825 | 354.1291 |
| GOBP_NEG | 42 | 0.14793 | 0.00697 | 0.072618 | 0.020827 | 354.1631 |
| REACTOME | 33 | 0.16784 | 0.007011 | 0.082401 | 0.020838 | 354.3502 |
| GOBP_NEG | 4 | 0.59633 | 0.008679 | 0.29277 | 0.020842 | 354.4182 |
| REACTOME | 195 | 0.078328 | 0.00792 | 0.038478 | 0.020902 | 355.4385 |
| GOCC_FEM | 15 | 0.30575 | 0.008615 | 0.15022 | 0.020919 | 355.7276 |
| REACTOME | 5 | 0.46207 | 0.007519 | 0.22707 | 0.020937 | 356.0337 |
| GOBP_REG | 8 | 0.41877 | 0.008619 | 0.2058 | 0.02094 | 356.0847 |
| GOMF_CIS_ | 38 | 0.14488 | 0.006494 | 0.071205 | 0.020952 | 356.2888 |
| GOBP_POS | 795 | 0.037104 | 0.007452 | 0.018243 | 0.020992 | 356.969 |
| GOBP_POS | 64 | 0.12246 | 0.007118 | 0.060278 | 0.021109 | 358.9585 |
| GOBP_NEG | 19 | 0.22879 | 0.007255 | 0.11275 | 0.021226 | 360.9481 |
| GOBP_FRU | 11 | 0.3214 | 0.007756 | 0.1584 | 0.021237 | 361.1352 |

| | | | | | | |
|---|---|---|---|---|---|---|
| GOMF_PRO | 130 | 0.088903 | 0.007352 | 0.043827 | 0.021262 | 361.5603 |
| GOBP_NEG | 28 | 0.18378 | 0.007073 | 0.090639 | 0.021305 | 362.2915 |
| PID_HDAC_ | 59 | 0.13112 | 0.007319 | 0.064674 | 0.021316 | 362.4786 |
| MIKKELSEN | 11 | 0.28076 | 0.006775 | 0.13859 | 0.021395 | 363.822 |
| REACTOME | 276 | 0.05559 | 0.006672 | 0.027447 | 0.021423 | 364.2981 |
| GOBP_MES | 7 | 0.42291 | 0.008142 | 0.20882 | 0.021428 | 364.3831 |
| GOBP_CELI | 12 | 0.30224 | 0.007618 | 0.14924 | 0.021433 | 364.4682 |
| GOBP_RES | 159 | 0.087415 | 0.007989 | 0.043169 | 0.021445 | 364.6722 |
| WP_ENDOI | 61 | 0.13599 | 0.007718 | 0.067171 | 0.021466 | 365.0293 |
| REACTOME | 9 | 0.3797 | 0.008289 | 0.18792 | 0.021673 | 368.5494 |
| REACTOME | 104 | 0.087675 | 0.00649 | 0.043413 | 0.021722 | 369.3826 |
| GOBP_ERB | 18 | 0.25434 | 0.00785 | 0.12594 | 0.021726 | 369.4506 |
| GOBP_PRO | 6 | 0.42187 | 0.00752 | 0.20916 | 0.021855 | 371.6443 |
| GOBP_CELI | 8 | 0.43037 | 0.008858 | 0.21346 | 0.021899 | 372.3925 |
| BUYTAERT_ | 605 | 0.040619 | 0.007154 | 0.020159 | 0.021964 | 373.4978 |
| WP_PROST | 30 | 0.19365 | 0.007714 | 0.096114 | 0.021968 | 373.5658 |
| GOBP_MO | 492 | 0.048376 | 0.007707 | 0.02401 | 0.02197 | 373.5999 |
| GOBP_SPLI | 66 | 0.11774 | 0.00695 | 0.058439 | 0.021972 | 373.6339 |
| WP_CYTOF | 79 | 0.10212 | 0.006592 | 0.05069 | 0.021984 | 373.8379 |
| REACTOME | 156 | 0.075901 | 0.006871 | 0.037688 | 0.022016 | 374.3821 |
| GOBP_HEA | 71 | 0.12747 | 0.007803 | 0.063322 | 0.022061 | 375.1473 |
| TANG_SEN | 52 | 0.12849 | 0.006735 | 0.063839 | 0.022077 | 375.4194 |
| PID_ERA_G | 63 | 0.12138 | 0.007 | 0.060332 | 0.022126 | 376.2526 |
| GOMF_SNO | 32 | 0.16422 | 0.006755 | 0.081674 | 0.022189 | 377.3239 |
| GOCC_STR | 21 | 0.2201 | 0.007337 | 0.10959 | 0.022311 | 379.3986 |
| GOBP_ART | 69 | 0.13215 | 0.007975 | 0.06582 | 0.022342 | 379.9257 |
| VERRECCHI | 35 | 0.1687 | 0.007257 | 0.084037 | 0.022359 | 380.2148 |
| REACTOME | 16 | 0.22277 | 0.006483 | 0.11099 | 0.022374 | 380.4699 |
| GOCC_END | 957 | 0.032275 | 0.00708 | 0.016101 | 0.022517 | 382.9016 |
| GOBP_NEG | 225 | 0.068851 | 0.007472 | 0.034356 | 0.022542 | 383.3267 |
| GOBP_SPE | 36 | 0.18535 | 0.008086 | 0.092524 | 0.022582 | 384.0069 |
| REACTOME | 20 | 0.19621 | 0.006383 | 0.09797 | 0.022611 | 384.5001 |
| REACTOME | 91 | 0.10959 | 0.007591 | 0.054793 | 0.02275 | 386.8638 |
| GOCC_LOW | 11 | 0.25208 | 0.006083 | 0.12605 | 0.022764 | 387.1018 |
| REACTOME | 580 | 0.043255 | 0.007464 | 0.02164 | 0.022822 | 388.0881 |
| REACTOME | 26 | 0.19167 | 0.007108 | 0.095893 | 0.022823 | 388.1051 |
| GOMF_VIT | 54 | 0.13402 | 0.007158 | 0.067061 | 0.022837 | 388.3432 |
| GOBP_NEG | 7 | 0.36631 | 0.007052 | 0.18329 | 0.02284 | 388.3942 |
| RIGGINS_T | 55 | 0.14115 | 0.007608 | 0.070633 | 0.022845 | 388.4792 |
| GOBP_REG | 13 | 0.26624 | 0.006984 | 0.13324 | 0.022851 | 388.5813 |
| GOBP_INTI | 77 | 0.1108 | 0.007062 | 0.05547 | 0.022893 | 389.2955 |
| IGLESIAS_E | 135 | 0.091291 | 0.007692 | 0.045705 | 0.022899 | 389.3975 |
| GOBP_VAS | 25 | 0.2451 | 0.008913 | 0.12275 | 0.02294 | 390.0947 |
| WP_PHOTO | 48 | 0.14189 | 0.007146 | 0.071072 | 0.022949 | 390.2477 |
| SHETH_LIV | 266 | 0.061337 | 0.00723 | 0.030725 | 0.022958 | 390.4008 |

| | | | | | | |
|---|---|---|---|---|---|---|
| GOBP_DEN | 16 | 0.28688 | 0.008348 | 0.14375 | 0.022991 | 390.962 |
| DELACROIX | 428 | 0.048486 | 0.007217 | 0.024303 | 0.023025 | 391.5401 |
| GOBP_REG | 20 | 0.22829 | 0.007427 | 0.11447 | 0.023067 | 392.2543 |
| GOBP_END | 8 | 0.41402 | 0.008521 | 0.20762 | 0.023077 | 392.4244 |
| REACTOME | 13 | 0.24466 | 0.006418 | 0.12276 | 0.023143 | 393.5467 |
| WP_PURIN | 11 | 0.30721 | 0.007414 | 0.15422 | 0.023192 | 394.38 |
| GOBP_FAR | 6 | 0.33296 | 0.005935 | 0.16725 | 0.023257 | 395.4853 |
| GOBP_CELI | 39 | 0.15879 | 0.00721 | 0.079761 | 0.02326 | 395.5363 |
| GOBP_HYP | 27 | 0.19715 | 0.007451 | 0.099036 | 0.023263 | 395.5873 |
| GOCC_U2_ | 30 | 0.19171 | 0.007636 | 0.096304 | 0.023266 | 395.6383 |
| GOMF_TEL | 37 | 0.15674 | 0.006932 | 0.078739 | 0.023268 | 395.6723 |
| GOBP_PER | 8 | 0.44593 | 0.009178 | 0.22405 | 0.023285 | 395.9614 |
| PLASARI_N | 21 | 0.24676 | 0.008226 | 0.12398 | 0.023289 | 396.0294 |
| GOMF_KIN | 6 | 0.4199 | 0.007485 | 0.21104 | 0.023324 | 396.6246 |
| GOBP_MUS | 371 | 0.053064 | 0.007366 | 0.026676 | 0.023347 | 397.0157 |
| GOBP_MRI | 31 | 0.172 | 0.006964 | 0.086474 | 0.023358 | 397.2028 |
| GOBP_REG | 62 | 0.1394 | 0.007976 | 0.070093 | 0.023371 | 397.4239 |
| REACTOME | 5 | 0.42284 | 0.006881 | 0.21274 | 0.023436 | 398.5292 |
| GOBP_EME | 43 | 0.17125 | 0.008164 | 0.086172 | 0.023453 | 398.8183 |
| REACTOME | 95 | 0.096924 | 0.006858 | 0.048776 | 0.023462 | 398.9713 |
| SENGUPTA | 291 | 0.060934 | 0.007507 | 0.030671 | 0.023486 | 399.3794 |
| GOBP_POS | 14 | 0.28887 | 0.007864 | 0.14542 | 0.023503 | 399.6685 |
| REACTOME | 732 | 0.03561 | 0.006875 | 0.017944 | 0.023611 | 401.5051 |
| GOCC_M_E | 19 | 0.24104 | 0.007643 | 0.12157 | 0.023701 | 403.0355 |
| GOMF_ALP | 27 | 0.20736 | 0.007836 | 0.10459 | 0.023717 | 403.3076 |
| GOBP_PYR | 13 | 0.2467 | 0.006472 | 0.12449 | 0.023765 | 404.1238 |
| PAPASPYRI | 42 | 0.16161 | 0.007614 | 0.081562 | 0.02378 | 404.3789 |
| REACTOME | 180 | 0.072364 | 0.007032 | 0.036524 | 0.023788 | 404.5149 |
| GOBP_SEPT | 5 | 0.49237 | 0.008012 | 0.24852 | 0.023794 | 404.617 |
| GOMF_U1_ | 6 | 0.42341 | 0.007547 | 0.21388 | 0.023876 | 406.0114 |
| GOMF_SIG | 1330 | 0.029421 | 0.007529 | 0.014864 | 0.023896 | 406.3515 |
| GOBP_SEX_ | 18 | 0.24029 | 0.007416 | 0.12142 | 0.023917 | 406.7086 |
| GOMF_PHO | 60 | 0.13128 | 0.00739 | 0.066407 | 0.024031 | 408.6472 |
| GOBP_VAS | 8 | 0.37687 | 0.007757 | 0.19064 | 0.024037 | 408.7492 |
| WP_HAIR_ | 30 | 0.2011 | 0.00801 | 0.1018 | 0.024122 | 410.1946 |
| GOMF_STE | 6 | 0.45512 | 0.008112 | 0.23047 | 0.024155 | 410.7558 |
| GOBP_CELI | 1076 | 0.029589 | 0.00686 | 0.014986 | 0.024177 | 411.1299 |
| GOBP_LIM | 5 | 0.54412 | 0.008854 | 0.27563 | 0.024193 | 411.402 |
| GU_PDEF_T | 39 | 0.17326 | 0.007867 | 0.087817 | 0.024257 | 412.4903 |
| GOCC_RIBO | 169 | 0.066523 | 0.006266 | 0.033721 | 0.024269 | 412.6943 |
| GOBP_VOL | 4 | 0.54138 | 0.00788 | 0.27445 | 0.024279 | 412.8644 |
| GOBP_CHA | 15 | 0.2713 | 0.007645 | 0.13756 | 0.024295 | 413.1365 |
| GOCC_ACT | 72 | 0.12413 | 0.007652 | 0.062953 | 0.024322 | 413.5956 |
| WP_NEPHR | 18 | 0.24785 | 0.00765 | 0.12574 | 0.024362 | 414.2758 |
| GOBP_REG | 11 | 0.29883 | 0.007211 | 0.15164 | 0.024391 | 414.769 |

| | | | | | | |
|---|---|---|---|---|---|---|
| GOBP_DEV | 6 | 0.41995 | 0.007486 | 0.21323 | 0.024457 | 415.8913 |
| GOBP_CAR | 14 | 0.29669 | 0.008077 | 0.15066 | 0.024471 | 416.1294 |
| GOBP_POS | 15 | 0.2444 | 0.006886 | 0.12414 | 0.024499 | 416.6055 |
| GOBP_SER | 14 | 0.23857 | 0.006494 | 0.12118 | 0.024506 | 416.7245 |
| ZHU_CMV_ | 81 | 0.1165 | 0.007615 | 0.059179 | 0.024508 | 416.7585 |
| GOBP_HIN | 144 | 0.081954 | 0.00713 | 0.041646 | 0.024549 | 417.4557 |
| KORKOLA_ | 6 | 0.37914 | 0.006758 | 0.1928 | 0.024632 | 418.8672 |
| BUYTAERT_ | 782 | 0.035562 | 0.007087 | 0.018093 | 0.024685 | 419.7684 |
| GOBP_NEG | 52 | 0.14824 | 0.007769 | 0.075435 | 0.02471 | 420.1936 |
| GOBP_PEP | 663 | 0.036161 | 0.006657 | 0.018414 | 0.02479 | 421.554 |
| BOQUEST_ | 240 | 0.06969 | 0.007808 | 0.0355 | 0.024825 | 422.1491 |
| REACTOME | 89 | 0.098053 | 0.006717 | 0.04995 | 0.02483 | 422.2342 |
| GOMF_TYP | 5 | 0.49706 | 0.008088 | 0.25331 | 0.024873 | 422.9654 |
| MA_RAT_A | 147 | 0.082598 | 0.00726 | 0.042114 | 0.02493 | 423.9347 |
| GOBP_PRO | 24 | 0.20423 | 0.007277 | 0.10413 | 0.024931 | 423.9517 |
| GOBP_DOU | 6 | 0.34813 | 0.006205 | 0.17752 | 0.024944 | 424.1727 |
| BENPORAT | 595 | 0.046082 | 0.008051 | 0.023502 | 0.02496 | 424.4448 |
| GOBP_REG | 12 | 0.25454 | 0.006416 | 0.12989 | 0.025024 | 425.5331 |
| SMID_BREA | 80 | 0.11565 | 0.007512 | 0.059028 | 0.025054 | 426.0433 |
| REACTOME | 58 | 0.12987 | 0.007187 | 0.066294 | 0.025068 | 426.2813 |
| GOBP_POS | 4 | 0.5636 | 0.008203 | 0.28774 | 0.025083 | 426.5364 |
| REACTOME | 57 | 0.13298 | 0.007296 | 0.067912 | 0.025117 | 427.1146 |
| GOBP_MET | 27 | 0.19172 | 0.007245 | 0.097964 | 0.025181 | 428.2029 |
| HUMMERIC | 95 | 0.096839 | 0.006852 | 0.04949 | 0.025199 | 428.509 |
| GOCC_END | 17 | 0.23281 | 0.006983 | 0.11908 | 0.025299 | 430.2095 |
| ROSS_AML | 66 | 0.11458 | 0.006763 | 0.058612 | 0.025306 | 430.3285 |
| BERENJENC | 377 | 0.050639 | 0.007084 | 0.025907 | 0.025321 | 430.5836 |
| BIOCARTA_ | 5 | 0.36936 | 0.00601 | 0.18897 | 0.025325 | 430.6516 |
| GOBP_ACT | 5 | 0.42051 | 0.006843 | 0.21515 | 0.025331 | 430.7537 |
| GOMF_ATP | 4 | 0.47835 | 0.006962 | 0.2448 | 0.025357 | 431.1958 |
| GOBP_POS | 14 | 0.26278 | 0.007154 | 0.13455 | 0.025418 | 432.2331 |
| GOBP_REG | 13 | 0.27766 | 0.007284 | 0.14222 | 0.025459 | 432.9303 |
| GOBP_EMB | 117 | 0.10049 | 0.007887 | 0.051477 | 0.025465 | 433.0323 |
| GOBP_NEG | 6 | 0.49041 | 0.008742 | 0.25121 | 0.025467 | 433.0663 |
| GOCC_EUC | 55 | 0.12607 | 0.006795 | 0.06462 | 0.025544 | 434.3757 |
| GRAESSMA | 717 | 0.035992 | 0.00688 | 0.018451 | 0.025555 | 434.5628 |
| GOBP_ORG | 23 | 0.23007 | 0.008026 | 0.11795 | 0.025563 | 434.6988 |
| MARKEY_R | 205 | 0.066926 | 0.006936 | 0.034314 | 0.025573 | 434.8689 |
| SANA_RESI | 63 | 0.12087 | 0.006971 | 0.061977 | 0.025583 | 435.0389 |
| WP_MAPK_ | 230 | 0.066124 | 0.007254 | 0.033908 | 0.025591 | 435.175 |
| MAHAJAN_ | 71 | 0.11972 | 0.007328 | 0.061409 | 0.025626 | 435.7701 |
| GOBP_ER_ | 45 | 0.15115 | 0.007371 | 0.077559 | 0.025664 | 436.4163 |
| GOBP_POS | 32 | 0.17351 | 0.007138 | 0.08904 | 0.025674 | 436.5864 |
| GOBP_CAR | 15 | 0.27715 | 0.007809 | 0.14232 | 0.025757 | 437.9978 |
| GOBP_POS | 1347 | 0.027724 | 0.007137 | 0.014245 | 0.025819 | 439.0521 |

| | | | | | | |
|---|---|---|---|---|---|---|
| GOBP_NEG | 5 | 0.50413 | 0.008203 | 0.25909 | 0.025848 | 439.5452 |
| GOBP_REG | 15 | 0.23664 | 0.006668 | 0.12166 | 0.025888 | 440.2254 |
| KIM_WT1_ | 207 | 0.067152 | 0.006993 | 0.034525 | 0.025894 | 440.3275 |
| YAGUE_PR | 13 | 0.2788 | 0.007314 | 0.1435 | 0.026021 | 442.4871 |
| GOMF_DIS | 29 | 0.17957 | 0.007033 | 0.092424 | 0.026026 | 442.5721 |
| GOBP_NEG | 12 | 0.31059 | 0.007828 | 0.15989 | 0.026041 | 442.8272 |
| GOBP_REG | 803 | 0.035949 | 0.007255 | 0.018507 | 0.026046 | 442.9122 |
| WP_MICRO | 4 | 0.43965 | 0.006399 | 0.22634 | 0.026047 | 442.9292 |
| JOSEPH_RE | 27 | 0.20568 | 0.007773 | 0.1059 | 0.026061 | 443.1673 |
| REACTOME | 37 | 0.17546 | 0.00776 | 0.090342 | 0.026067 | 443.2693 |
| GOMF_CUI | 23 | 0.23408 | 0.008166 | 0.12055 | 0.026091 | 443.6775 |
| GOBP_REG | 18 | 0.23859 | 0.007364 | 0.12288 | 0.026095 | 443.7455 |
| NIKOLSKY_ | 31 | 0.35166 | 0.014239 | 0.18117 | 0.026138 | 444.4767 |
| VANDESLU | 5 | 0.41931 | 0.006823 | 0.21618 | 0.026222 | 445.9051 |
| DUAN_PRD | 73 | 0.11281 | 0.007001 | 0.058174 | 0.026252 | 446.4153 |
| GOBP_REG | 39 | 0.16256 | 0.007381 | 0.083841 | 0.026263 | 446.6023 |
| REACTOME | 6 | 0.43993 | 0.007842 | 0.2269 | 0.026269 | 446.7043 |
| GOCC_GAN | 15 | 0.22057 | 0.006215 | 0.11384 | 0.02635 | 448.0818 |
| PID_IL2_ST | 27 | 0.19611 | 0.007411 | 0.10122 | 0.026358 | 448.2178 |
| ANASTASSI | 62 | 0.14106 | 0.008071 | 0.072824 | 0.026382 | 448.6259 |
| BIOCARTA_ | 11 | 0.31119 | 0.00751 | 0.16077 | 0.026463 | 450.0033 |
| REACTOME | 8 | 0.39423 | 0.008114 | 0.20368 | 0.026472 | 450.1564 |
| WEBER_MI | 18 | 0.23236 | 0.007172 | 0.12008 | 0.026503 | 450.6835 |
| GOBP_PRO | 8 | 0.37079 | 0.007631 | 0.19167 | 0.026533 | 451.1937 |
| GOBP_AST | 10 | 0.29867 | 0.006872 | 0.15441 | 0.026547 | 451.4317 |
| GOBP_CELI | 20 | 0.23875 | 0.007767 | 0.12345 | 0.026572 | 451.8569 |
| GOBP_CELI | 1672 | 0.024231 | 0.006884 | 0.012533 | 0.026605 | 452.418 |
| GOBP_EPO | 5 | 0.41354 | 0.006729 | 0.2139 | 0.026605 | 452.418 |
| GOBP_NEG | 95 | 0.099037 | 0.007008 | 0.051258 | 0.02668 | 453.6934 |
| REACTOME | 14 | 0.22673 | 0.006172 | 0.11736 | 0.026696 | 453.9655 |
| GOMF_CAI | 18 | 0.23654 | 0.007301 | 0.12245 | 0.026704 | 454.1015 |
| GOCC_PRE | 48 | 0.1387 | 0.006985 | 0.071802 | 0.02671 | 454.2036 |
| GOBP_EMI | 64 | 0.12739 | 0.007405 | 0.065962 | 0.026734 | 454.6117 |
| GOBP_REG | 88 | 0.10472 | 0.007133 | 0.054227 | 0.026738 | 454.6797 |
| KEGG_PAN | 66 | 0.11943 | 0.007049 | 0.061871 | 0.026792 | 455.598 |
| GOMF_ICO | 13 | 0.28325 | 0.00743 | 0.14679 | 0.026836 | 456.3462 |
| GOBP_REG | 73 | 0.11459 | 0.007112 | 0.059392 | 0.026854 | 456.6523 |
| SAMOLS_T | 8 | 0.33319 | 0.006858 | 0.17275 | 0.026887 | 457.2134 |
| GOBP_AOF | 4 | 0.49957 | 0.007271 | 0.25919 | 0.02697 | 458.6249 |
| GOMF_NU | 66 | 0.10948 | 0.006462 | 0.056887 | 0.027158 | 461.8218 |
| GOBP_CYT | 33 | 0.14875 | 0.006214 | 0.077301 | 0.027164 | 461.9238 |
| KREPPEL_C | 7 | 0.36891 | 0.007102 | 0.19171 | 0.02717 | 462.0259 |
| REACTOME | 1268 | 0.02873 | 0.007192 | 0.014941 | 0.02725 | 463.3863 |
| GOCC_END | 5 | 0.47452 | 0.007722 | 0.24677 | 0.027253 | 463.4373 |
| FIGUEROA_ | 12 | 0.2927 | 0.007377 | 0.15222 | 0.027256 | 463.4883 |

| | | | | | | |
|---|---|---|---|---|---|---|
| GARGALOV | 26 | 0.17798 | 0.006601 | 0.092617 | 0.027329 | 464.7296 |
| GOBP_DOU | 65 | 0.11535 | 0.006757 | 0.060029 | 0.027341 | 464.9337 |
| WP_P38_M | 32 | 0.15718 | 0.006466 | 0.08194 | 0.027546 | 468.4197 |
| GOBP_NEG | 7 | 0.37553 | 0.00723 | 0.19587 | 0.027613 | 469.5591 |
| GOBP_EMB | 285 | 0.062038 | 0.007565 | 0.03236 | 0.027622 | 469.7121 |
| COLINA_TA | 331 | 0.052346 | 0.00687 | 0.027309 | 0.027639 | 470.0012 |
| GOBP_ANT | 198 | 0.076785 | 0.007823 | 0.040063 | 0.027652 | 470.2223 |
| GOBP_EMB | 32 | 0.18851 | 0.007755 | 0.098357 | 0.027652 | 470.2223 |
| GOBP_NEG | 31 | 0.17328 | 0.007016 | 0.090428 | 0.027676 | 470.6304 |
| WILCOX_RE | 55 | 0.13911 | 0.007497 | 0.072606 | 0.027697 | 470.9875 |
| GOCC_MU | 65 | 0.1115 | 0.006531 | 0.058205 | 0.027715 | 471.2936 |
| GOCC_GAT | 5 | 0.39132 | 0.006368 | 0.20428 | 0.027719 | 471.3616 |
| GOCC_BAS | 6 | 0.45269 | 0.008069 | 0.23643 | 0.027775 | 472.3139 |
| GOBP_BOX | 8 | 0.29669 | 0.006106 | 0.15501 | 0.027815 | 472.9941 |
| GOBP_COL | 16 | 0.25195 | 0.007332 | 0.13169 | 0.027867 | 473.8783 |
| GOBP_NON | 369 | 0.047017 | 0.006509 | 0.024577 | 0.027879 | 474.0824 |
| GOBP_CD8 | 26 | 0.1788 | 0.006631 | 0.093468 | 0.027888 | 474.2354 |
| GOBP_REG | 14 | 0.22571 | 0.006144 | 0.11799 | 0.027891 | 474.2865 |
| REACTOME | 53 | 0.11595 | 0.006135 | 0.060614 | 0.027891 | 474.2865 |
| MYLLYKAN | 5 | 0.50986 | 0.008297 | 0.26656 | 0.027901 | 474.4565 |
| GOBP_BAS | 14 | 0.19642 | 0.005347 | 0.10271 | 0.027925 | 474.8646 |
| GOMF_PHO | 341 | 0.051348 | 0.006839 | 0.026863 | 0.02798 | 475.7999 |
| GOBP_CELL | 13 | 0.26854 | 0.007044 | 0.14049 | 0.027988 | 475.9359 |
| REACTOME | 5 | 0.40209 | 0.006543 | 0.2104 | 0.028006 | 476.242 |
| GOBP_HEA | 235 | 0.065297 | 0.00724 | 0.034178 | 0.028045 | 476.9052 |
| MARZEC_IL | 108 | 0.097173 | 0.007329 | 0.05087 | 0.028062 | 477.1943 |
| WANG_RES | 9 | 0.27027 | 0.0059 | 0.14157 | 0.028133 | 478.4017 |
| DING_LUN | 24 | 0.19908 | 0.007094 | 0.1043 | 0.028155 | 478.7758 |
| BIOCARTA_ | 15 | 0.25402 | 0.007158 | 0.13309 | 0.028158 | 478.8268 |
| GOBP_EPIT | 417 | 0.04843 | 0.007118 | 0.025381 | 0.028199 | 479.524 |
| GOBP_AMY | 33 | 0.16087 | 0.00672 | 0.084369 | 0.028286 | 481.0034 |
| GOBP_NUC | 140 | 0.075748 | 0.006499 | 0.039732 | 0.028303 | 481.2925 |
| CAFFAREL_ | 28 | 0.18872 | 0.007263 | 0.099026 | 0.028349 | 482.0747 |
| CLIMENT_B | 8 | 0.3646 | 0.007504 | 0.19134 | 0.028363 | 482.3128 |
| GOBP_ATR | 13 | 0.32016 | 0.008399 | 0.16804 | 0.028383 | 482.6529 |
| WP_AMINO | 6 | 0.37057 | 0.006605 | 0.19454 | 0.028408 | 483.078 |
| SENESE_HD | 110 | 0.093824 | 0.007141 | 0.049301 | 0.028522 | 485.0166 |
| YU_MYC_T | 44 | 0.13669 | 0.006592 | 0.071914 | 0.028675 | 487.6184 |
| GOBP_PRO | 6 | 0.28838 | 0.00514 | 0.15176 | 0.028707 | 488.1625 |
| GOBP_END | 69 | 0.11321 | 0.006832 | 0.059618 | 0.028799 | 489.727 |
| GOCC_L_TY | 12 | 0.31557 | 0.007954 | 0.16619 | 0.0288 | 489.744 |
| BIOCARTA_ | 17 | 0.25322 | 0.007595 | 0.13337 | 0.028811 | 489.9311 |
| KIM_PTEN_ | 5 | 0.46022 | 0.007489 | 0.24241 | 0.028822 | 490.1181 |
| WINNEPEN | 37 | 0.15915 | 0.007039 | 0.08383 | 0.028824 | 490.1521 |
| GOMF_ATP | 5 | 0.41596 | 0.006769 | 0.21911 | 0.028832 | 490.2882 |

| | | | | | | |
|---|---|---|---|---|---|---|
| REACTOME | 8 | 0.38824 | 0.00799 | 0.20466 | 0.028924 | 491.8526 |
| GOMF_MU | 14 | 0.25422 | 0.006921 | 0.13403 | 0.028933 | 492.0057 |
| GOBP_COF | 5 | 0.37654 | 0.006127 | 0.19865 | 0.029022 | 493.5191 |
| REACTOME | 85 | 0.10592 | 0.007091 | 0.055883 | 0.029028 | 493.6211 |
| GOBP_ARG | 4 | 0.48454 | 0.007052 | 0.2557 | 0.029055 | 494.0803 |
| YAUCH_HE | 241 | 0.065079 | 0.007306 | 0.034355 | 0.029102 | 494.8795 |
| ZAMORA_N | 53 | 0.12103 | 0.006404 | 0.063915 | 0.02915 | 495.6958 |
| WP_ALZHE | 246 | 0.059275 | 0.006722 | 0.031316 | 0.029201 | 496.563 |
| WP_ALZHE | 246 | 0.059275 | 0.006722 | 0.031316 | 0.029201 | 496.563 |
| GOCC_EXT | 9 | 0.32503 | 0.007095 | 0.17173 | 0.029209 | 496.699 |
| GOBP_PEP | 11 | 0.28677 | 0.00692 | 0.15155 | 0.02924 | 497.2262 |
| GOBP_REL | 30 | 0.17684 | 0.007044 | 0.093458 | 0.029243 | 497.2772 |
| GOCC_TRA | 474 | 0.044333 | 0.006936 | 0.023431 | 0.029248 | 497.3622 |
| GOBP_NEG | 18 | 0.21451 | 0.006621 | 0.1135 | 0.029392 | 499.811 |
| WP_NEOLA | 11 | 0.31384 | 0.007574 | 0.16606 | 0.029392 | 499.811 |
| GOBP_DEV | 211 | 0.067479 | 0.007094 | 0.035707 | 0.029402 | 499.981 |
| BOYLAN_M | 131 | 0.082753 | 0.00687 | 0.043806 | 0.029448 | 500.7632 |
| GOBP_SON | 63 | 0.12398 | 0.00715 | 0.065641 | 0.029473 | 501.1884 |
| REACTOME | 118 | 0.093885 | 0.007399 | 0.049725 | 0.029516 | 501.9196 |
| GOBP_EPIT | 1151 | 0.030478 | 0.007293 | 0.016145 | 0.029541 | 502.3447 |
| IWANAGA_ | 112 | 0.086291 | 0.006627 | 0.045737 | 0.029612 | 503.5521 |
| GOBP_NUC | 41 | 0.13645 | 0.006352 | 0.072344 | 0.029648 | 504.1642 |
| GOBP_REG | 1437 | 0.026383 | 0.006997 | 0.013989 | 0.029655 | 504.2833 |
| GOBP_POS | 5 | 0.445 | 0.007241 | 0.23599 | 0.029682 | 504.7424 |
| TSENG_AD | 40 | 0.15438 | 0.007099 | 0.081889 | 0.029706 | 505.1505 |
| GOBP_NUC | 56 | 0.11307 | 0.006149 | 0.059996 | 0.029748 | 505.8647 |
| GOBP_MAI | 128 | 0.086003 | 0.007058 | 0.045663 | 0.02983 | 507.2592 |
| GOBP_REG | 8 | 0.30729 | 0.006325 | 0.16323 | 0.029892 | 508.3135 |
| TIAN_TNF_ | 19 | 0.22916 | 0.007267 | 0.12173 | 0.029892 | 508.3135 |
| KINSEY_TA | 309 | 0.0545 | 0.006915 | 0.028959 | 0.029929 | 508.9426 |
| PUJANA_AT | 1261 | 0.026262 | 0.006557 | 0.013956 | 0.029943 | 509.1807 |
| GOCC_CAL | 75 | 0.11295 | 0.007106 | 0.06003 | 0.029951 | 509.3168 |
| JOHANSSO | 7 | 0.38699 | 0.007451 | 0.20576 | 0.030007 | 510.269 |
| GOBP_EPIT | 6 | 0.45869 | 0.008176 | 0.24424 | 0.030195 | 513.466 |
| SA_PTEN_F | 17 | 0.22617 | 0.006784 | 0.12043 | 0.030201 | 513.568 |
| GOBP_PRC | 194 | 0.065564 | 0.006612 | 0.034913 | 0.030204 | 513.619 |
| GOBP_VITA | 18 | 0.20962 | 0.00647 | 0.1117 | 0.030294 | 515.1495 |
| DOANE_BR | 103 | 0.098921 | 0.007287 | 0.052724 | 0.030323 | 515.6426 |
| REACTOME | 35 | 0.16183 | 0.006962 | 0.086281 | 0.030359 | 516.2548 |
| GOBP_SMA | 10 | 0.27381 | 0.0063 | 0.14601 | 0.030386 | 516.7139 |
| GOBP_POS | 28 | 0.16147 | 0.006214 | 0.086114 | 0.030399 | 516.935 |
| GOBP_MID | 19 | 0.23746 | 0.00753 | 0.12669 | 0.030448 | 517.7682 |
| WP_ONECA | 29 | 0.16692 | 0.006537 | 0.089079 | 0.030483 | 518.3634 |
| NIKOLSKY_ | 108 | 0.17536 | 0.013226 | 0.09364 | 0.030564 | 519.7408 |
| PID_IL3_PA | 25 | 0.18609 | 0.006768 | 0.099386 | 0.030583 | 520.0639 |

| | | | | | | |
|---|---|---|---|---|---|---|
| GOBP_CEN | 8 | 0.28915 | 0.005951 | 0.15447 | 0.030623 | 520.7441 |
| SMID_BREA | 6 | 0.46957 | 0.00837 | 0.25092 | 0.030653 | 521.2543 |
| GOBP_REG | 85 | 0.10688 | 0.007156 | 0.057128 | 0.030691 | 521.9005 |
| FERRANDO | 14 | 0.2603 | 0.007086 | 0.13918 | 0.030738 | 522.6997 |
| GOBP_REC | 26 | 0.1731 | 0.006419 | 0.092561 | 0.030745 | 522.8187 |
| GOBP_POS | 53 | 0.14096 | 0.007458 | 0.075397 | 0.030781 | 523.4309 |
| BOYLAN_M | 46 | 0.15078 | 0.007434 | 0.080686 | 0.030837 | 524.3832 |
| GOMF_BET | 4 | 0.41398 | 0.006025 | 0.22166 | 0.030913 | 525.6756 |
| GOMF_ALP | 6 | 0.46265 | 0.008247 | 0.24781 | 0.030961 | 526.4918 |
| REACTOME | 8 | 0.38066 | 0.007834 | 0.20394 | 0.030993 | 527.036 |
| REACTOME | 31 | 0.1655 | 0.006701 | 0.088714 | 0.03106 | 528.1753 |
| GOBP_RES | 42 | 0.13431 | 0.006328 | 0.072029 | 0.031122 | 529.2296 |
| GOBP_NEG | 34 | 0.16459 | 0.006979 | 0.088297 | 0.031164 | 529.9438 |
| GOBP_MUS | 80 | 0.11534 | 0.007493 | 0.061883 | 0.031178 | 530.1819 |
| PASTURAL_ | 7 | 0.28826 | 0.00555 | 0.15469 | 0.031207 | 530.675 |
| GOBP_PYR | 19 | 0.24642 | 0.007814 | 0.13228 | 0.031251 | 531.4233 |
| BENPORAT | 613 | 0.03764 | 0.006672 | 0.020215 | 0.031316 | 532.5286 |
| GOBP_CHR | 68 | 0.10586 | 0.006342 | 0.056872 | 0.03135 | 533.1068 |
| GOBP_PHA | 28 | 0.20397 | 0.00785 | 0.10962 | 0.031402 | 533.991 |
| WP_RETT_ | 40 | 0.15104 | 0.006945 | 0.081209 | 0.031463 | 535.0283 |
| GOBP_SPO | 8 | 0.4209 | 0.008663 | 0.22635 | 0.031487 | 535.4364 |
| CHICAS_RB | 88 | 0.097548 | 0.006645 | 0.052471 | 0.031516 | 535.9296 |
| GOBP_PYR | 21 | 0.1912 | 0.006374 | 0.10286 | 0.031531 | 536.1847 |
| FOROUTAN | 118 | 0.094498 | 0.007448 | 0.050869 | 0.031615 | 537.6131 |
| REACTOME | 14 | 0.28444 | 0.007743 | 0.1532 | 0.031685 | 538.8034 |
| GOMF_PHO | 84 | 0.09796 | 0.00652 | 0.052778 | 0.031731 | 539.5857 |
| GOBP_SIN( | 12 | 0.32654 | 0.00823 | 0.17596 | 0.031752 | 539.9428 |
| GOBP_GMI | 13 | 0.21853 | 0.005733 | 0.11781 | 0.031813 | 540.9801 |
| GOMF_FIB | 31 | 0.18064 | 0.007314 | 0.097414 | 0.031851 | 541.6263 |
| GOBP_NEG | 24 | 0.18728 | 0.006673 | 0.10102 | 0.031886 | 542.2214 |
| GOBP_MAT | 34 | 0.17248 | 0.007313 | 0.093043 | 0.031893 | 542.3405 |
| GOBP_URE | 9 | 0.28656 | 0.006255 | 0.15458 | 0.031894 | 542.3575 |
| HAN_JNK_S | 34 | 0.16995 | 0.007206 | 0.091684 | 0.031904 | 542.5275 |
| REACTOME | 87 | 0.094439 | 0.006396 | 0.050948 | 0.031906 | 542.5615 |
| REACTOME | 10 | 0.30299 | 0.006972 | 0.16346 | 0.031908 | 542.5955 |
| GOMF_TRA | 20 | 0.19395 | 0.00631 | 0.10464 | 0.031921 | 542.8166 |
| GOBP_NUC | 9 | 0.33487 | 0.00731 | 0.18074 | 0.031969 | 543.6328 |
| GRAESSMA | 585 | 0.036921 | 0.006398 | 0.019928 | 0.031973 | 543.7009 |
| GOBP_OLF | 10 | 0.35368 | 0.008138 | 0.191 | 0.032039 | 544.8232 |
| GOBP_MET | 104 | 0.086224 | 0.006382 | 0.046571 | 0.032061 | 545.1973 |
| GOBP_CAR | 36 | 0.17365 | 0.007576 | 0.093931 | 0.032262 | 548.6153 |
| REACTOME | 62 | 0.11144 | 0.006376 | 0.06029 | 0.032279 | 548.9044 |
| GOBP_PLA | 124 | 0.086166 | 0.006961 | 0.046641 | 0.032351 | 550.1288 |
| GOBP_MES | 8 | 0.36183 | 0.007447 | 0.19588 | 0.032369 | 550.4348 |
| REACTOME | 36 | 0.1464 | 0.006387 | 0.079258 | 0.032373 | 550.5029 |

| | | | | | | |
|---|---|---|---|---|---|---|
| MCCLUNG_ | 53 | 0.13729 | 0.007264 | 0.074329 | 0.032381 | 550.6389 |
| NUTT_GBM | 39 | 0.14915 | 0.006772 | 0.080754 | 0.032389 | 550.7749 |
| GOBP_SA_ | 13 | 0.30247 | 0.007935 | 0.16377 | 0.03239 | 550.792 |
| GOBP_TRA | 6 | 0.40453 | 0.007211 | 0.21905 | 0.032403 | 551.013 |
| MIKKELSEN | 131 | 0.082756 | 0.00687 | 0.04482 | 0.032427 | 551.4211 |
| GOBP_REG | 8 | 0.37177 | 0.007652 | 0.20138 | 0.032448 | 551.7782 |
| GOBP_MES | 59 | 0.13957 | 0.007791 | 0.075615 | 0.032466 | 552.0843 |
| GOBP_INTI | 15 | 0.27971 | 0.007882 | 0.15155 | 0.032475 | 552.2374 |
| GOBP_THY | 6 | 0.42201 | 0.007522 | 0.22868 | 0.032499 | 552.6455 |
| GOMF_ACI | 7 | 0.30828 | 0.005935 | 0.16707 | 0.032515 | 552.9176 |
| GOMF_MI( | 252 | 0.056563 | 0.006491 | 0.030662 | 0.03255 | 553.5128 |
| GOBP_DNA | 8 | 0.32204 | 0.006628 | 0.17476 | 0.032688 | 555.8594 |
| KEGG_ARR | 70 | 0.12176 | 0.007401 | 0.066084 | 0.032707 | 556.1825 |
| WP_ARRHY | 70 | 0.12176 | 0.007401 | 0.066084 | 0.032707 | 556.1825 |
| LI_WILMS_ | 27 | 0.18101 | 0.006841 | 0.098241 | 0.032712 | 556.2676 |
| GOBP_CAR | 10 | 0.32485 | 0.007475 | 0.17634 | 0.032731 | 556.5907 |
| REACTOME | 31 | 0.15021 | 0.006082 | 0.08157 | 0.032786 | 557.5259 |
| GOBP_MIT | 45 | 0.14127 | 0.006889 | 0.076732 | 0.032813 | 557.9851 |
| KANG_AR_ | 18 | 0.23743 | 0.007328 | 0.12896 | 0.032815 | 558.0191 |
| GOBP_MES | 19 | 0.24546 | 0.007783 | 0.13333 | 0.032824 | 558.1721 |
| GOBP_POS | 33 | 0.15117 | 0.006315 | 0.082289 | 0.03311 | 563.0356 |
| REACTOME | 117 | 0.089146 | 0.006996 | 0.048557 | 0.033194 | 564.464 |
| WP_CYTOS | 7 | 0.37844 | 0.007286 | 0.20614 | 0.0332 | 564.566 |
| GOBP_G1_ | 20 | 0.18579 | 0.006044 | 0.10124 | 0.03325 | 565.4163 |
| GOBP_REG | 280 | 0.053112 | 0.00642 | 0.028944 | 0.033263 | 565.6373 |
| GOBP_SON | 5 | 0.43407 | 0.007063 | 0.23671 | 0.033356 | 567.2188 |
| REACTOME | 49 | 0.13085 | 0.006658 | 0.07136 | 0.033362 | 567.3208 |
| REACTOME | 11 | 0.29391 | 0.007093 | 0.16031 | 0.033385 | 567.7119 |
| BIOCARTA_ | 28 | 0.1844 | 0.007097 | 0.10063 | 0.033449 | 568.8002 |
| HU_GENOT | 33 | 0.1427 | 0.005961 | 0.07787 | 0.033449 | 568.8002 |
| GOMF_PRC | 27 | 0.20157 | 0.007618 | 0.11005 | 0.033506 | 569.7695 |
| GOBP_OST | 213 | 0.065437 | 0.006912 | 0.035733 | 0.033537 | 570.2967 |
| REACTOME | 9 | 0.3372 | 0.007361 | 0.18413 | 0.033538 | 570.3137 |
| GOBP_POS | 40 | 0.1636 | 0.007523 | 0.089391 | 0.033625 | 571.7931 |
| GOBP_GOL | 19 | 0.20096 | 0.006372 | 0.10982 | 0.03364 | 572.0482 |
| GOBP_NEU | 1556 | 0.02484 | 0.006831 | 0.013578 | 0.033681 | 572.7454 |
| GOCC_SMA | 36 | 0.14604 | 0.006371 | 0.079832 | 0.033683 | 572.7794 |
| WANG_IM | 29 | 0.17978 | 0.007041 | 0.09829 | 0.033706 | 573.1705 |
| STEARMAN | 56 | 0.12191 | 0.00663 | 0.066663 | 0.033731 | 573.5957 |
| GOMF_PH( | 20 | 0.1997 | 0.006497 | 0.10921 | 0.033744 | 573.8167 |
| REACTOME | 16 | 0.20992 | 0.006109 | 0.11483 | 0.033772 | 574.2929 |
| OUILLETTE_ | 73 | 0.10016 | 0.006217 | 0.054801 | 0.033801 | 574.786 |
| GOCC_GAN | 6 | 0.35894 | 0.006398 | 0.19643 | 0.033837 | 575.3982 |
| GOBP_POS | 13 | 0.26152 | 0.006861 | 0.14314 | 0.033855 | 575.7043 |
| GOBP_CGN | 33 | 0.16165 | 0.006753 | 0.088478 | 0.033857 | 575.7383 |

| | | | | | | |
|---|---|---|---|---|---|---|
| GOBP_REG | 7 | 0.37356 | 0.007192 | 0.20455 | 0.033916 | 576.7416 |
| GOCC_MUS | 15 | 0.261 | 0.007354 | 0.14293 | 0.033927 | 576.9286 |
| DAZARD_U | 140 | 0.077044 | 0.00661 | 0.0422 | 0.033956 | 577.4218 |
| REACTOME | 14 | 0.27133 | 0.007386 | 0.14864 | 0.033976 | 577.7619 |
| GOBP_CER | 17 | 0.21358 | 0.006406 | 0.11704 | 0.034017 | 578.4591 |
| REACTOME | 29 | 0.16642 | 0.006518 | 0.091263 | 0.034119 | 580.1936 |
| GOCC_CYT | 87 | 0.086897 | 0.005886 | 0.047662 | 0.034145 | 580.6357 |
| GOMF_TRA | 171 | 0.069692 | 0.006603 | 0.038235 | 0.034181 | 581.2479 |
| GOBP_POS | 31 | 0.18479 | 0.007482 | 0.10138 | 0.034187 | 581.3499 |
| GOBP_EMB | 16 | 0.25506 | 0.007422 | 0.13995 | 0.0342 | 581.571 |
| GOBP_LON | 102 | 0.08794 | 0.006447 | 0.048268 | 0.034245 | 582.3362 |
| GOBP_NUC | 71 | 0.10281 | 0.006293 | 0.056441 | 0.034271 | 582.7784 |
| GOBP_FOR | 10 | 0.3466 | 0.007975 | 0.19044 | 0.034394 | 584.87 |
| REACTOME | 33 | 0.17207 | 0.007188 | 0.094556 | 0.034408 | 585.108 |
| GOBP_SEX_ | 258 | 0.059043 | 0.006855 | 0.032455 | 0.034449 | 585.8052 |
| GOBP_REG | 68 | 0.11283 | 0.006759 | 0.062025 | 0.034461 | 586.0093 |
| BRUINS_UV | 93 | 0.092824 | 0.006499 | 0.05105 | 0.034519 | 586.9956 |
| GOMF_K63 | 21 | 0.19491 | 0.006497 | 0.10721 | 0.03453 | 587.1827 |
| EGUCHI_CE | 22 | 0.17652 | 0.006022 | 0.097087 | 0.034531 | 587.1997 |
| GOBP_LAB | 17 | 0.21623 | 0.006486 | 0.11894 | 0.034541 | 587.3697 |
| GOBP_NEG | 21 | 0.20188 | 0.00673 | 0.11105 | 0.034544 | 587.4207 |
| REACTOME | 6 | 0.35579 | 0.006342 | 0.19571 | 0.034546 | 587.4547 |
| REACTOME | 8 | 0.3517 | 0.007239 | 0.19349 | 0.034565 | 587.7778 |
| GOCC_CAT | 184 | 0.072215 | 0.007095 | 0.039735 | 0.034585 | 588.1179 |
| GOBP_CELL | 72 | 0.10929 | 0.006737 | 0.060168 | 0.034658 | 589.3593 |
| GOMF_NEU | 10 | 0.27335 | 0.00629 | 0.15049 | 0.034665 | 589.4783 |
| GOBP_MES | 42 | 0.16521 | 0.007784 | 0.090966 | 0.03468 | 589.7334 |
| GOMF_STR | 150 | 0.067011 | 0.00595 | 0.036936 | 0.034828 | 592.2501 |
| WP_GPR40 | 15 | 0.20221 | 0.005698 | 0.11146 | 0.03484 | 592.4542 |
| REACTOME | 18 | 0.22242 | 0.006865 | 0.12264 | 0.034882 | 593.1684 |
| MARTENS_ | 738 | 0.031677 | 0.00614 | 0.017473 | 0.034931 | 594.0017 |
| REACTOME | 1305 | 0.025647 | 0.006506 | 0.01415 | 0.034965 | 594.5798 |
| GOBP_REG | 9 | 0.32056 | 0.006998 | 0.17686 | 0.034966 | 594.5968 |
| GOCC_LAT | 274 | 0.053891 | 0.006445 | 0.029734 | 0.03497 | 594.6649 |
| XIE_ST_HSC | 181 | 0.072057 | 0.007022 | 0.039763 | 0.034992 | 595.039 |
| GOBP_PRO | 197 | 0.061932 | 0.006294 | 0.034177 | 0.034994 | 595.073 |
| CASORELLI_ | 616 | 0.035615 | 0.006328 | 0.019658 | 0.035022 | 595.5491 |
| GOBP_MAI | 28 | 0.1904 | 0.007327 | 0.1051 | 0.03504 | 595.8552 |
| GOBP_POS | 19 | 0.21269 | 0.006744 | 0.11746 | 0.035103 | 596.9265 |
| MATZUK_E | 18 | 0.22695 | 0.007005 | 0.12535 | 0.035108 | 597.0115 |
| GOMF_3_5 | 17 | 0.22895 | 0.006867 | 0.12649 | 0.035154 | 597.7938 |
| GOBP_OTIC | 15 | 0.26693 | 0.007521 | 0.14749 | 0.035176 | 598.1679 |
| GOBP_ASY | 5 | 0.32019 | 0.00521 | 0.17696 | 0.035201 | 598.593 |
| GOBP_REG | 7 | 0.39919 | 0.007685 | 0.22063 | 0.035209 | 598.729 |
| GOBP_REG | 57 | 0.1274 | 0.00699 | 0.070432 | 0.035249 | 599.4092 |

| | | | | | | |
|---|---|---|---|---|---|---|
| LIU_OVARI | 1433 | 0.026756 | 0.007086 | 0.014798 | 0.035309 | 600.4295 |
| WP_EICOS/ | 28 | 0.16096 | 0.006194 | 0.089026 | 0.035311 | 600.4636 |
| JAZAERI_BI | 40 | 0.14799 | 0.006805 | 0.081866 | 0.035338 | 600.9227 |
| GOBP_PITU | 39 | 0.16706 | 0.007586 | 0.092443 | 0.035372 | 601.5009 |
| GOBP_END | 12 | 0.21319 | 0.005373 | 0.11807 | 0.035499 | 603.6605 |
| GOBP_CELI | 29 | 0.16276 | 0.006375 | 0.090151 | 0.035511 | 603.8646 |
| GOBP_REG | 36 | 0.17252 | 0.007527 | 0.095566 | 0.035529 | 604.1706 |
| PID_HDAC_ | 31 | 0.15388 | 0.00623 | 0.085259 | 0.035561 | 604.7148 |
| LOPEZ_TRA | 35 | 0.14892 | 0.006407 | 0.082593 | 0.035694 | 606.9765 |
| WP_FAMIL | 21 | 0.18448 | 0.006149 | 0.10231 | 0.035697 | 607.0275 |
| GOBP_REG | 38 | 0.1386 | 0.006212 | 0.076895 | 0.035744 | 607.8267 |
| GOBP_EMI | 57 | 0.13317 | 0.007307 | 0.073895 | 0.035767 | 608.2178 |
| GOBP_FOR | 7 | 0.44251 | 0.008519 | 0.24575 | 0.035887 | 610.2584 |
| GOBP_END | 52 | 0.13023 | 0.006825 | 0.072327 | 0.035899 | 610.4625 |
| GOMF_STR | 47 | 0.13172 | 0.006564 | 0.073173 | 0.035928 | 610.9556 |
| GOBP_NEG | 7 | 0.37437 | 0.007208 | 0.2082 | 0.036087 | 613.6594 |
| GOBP_CD8 | 15 | 0.22092 | 0.006225 | 0.12287 | 0.036101 | 613.8975 |
| GOBP_POS | 5 | 0.44483 | 0.007238 | 0.24745 | 0.036128 | 614.3566 |
| GOBP_REG | 139 | 0.07758 | 0.006632 | 0.043159 | 0.036134 | 614.4587 |
| GOBP_REG | 191 | 0.064376 | 0.006443 | 0.035822 | 0.036169 | 615.0538 |
| GOBP_END | 53 | 0.13621 | 0.007207 | 0.0758 | 0.036176 | 615.1729 |
| REACTOME | 34 | 0.14295 | 0.006061 | 0.079569 | 0.036211 | 615.7681 |
| KARLSSON_ | 118 | 0.082288 | 0.006485 | 0.045812 | 0.03624 | 616.2612 |
| GOBP_ANA | 1106 | 0.028529 | 0.0067 | 0.015885 | 0.036257 | 616.5503 |
| GOBP_MES | 17 | 0.21438 | 0.00643 | 0.11937 | 0.03627 | 616.7714 |
| MATZUK_II | 19 | 0.21053 | 0.006676 | 0.11724 | 0.036279 | 616.9244 |
| REACTOME | 70 | 0.10282 | 0.006249 | 0.057274 | 0.036321 | 617.6386 |
| GOBP_TISS | 298 | 0.05377 | 0.006702 | 0.029958 | 0.036351 | 618.1488 |
| GOBP_NEG | 7 | 0.34849 | 0.006709 | 0.19426 | 0.036423 | 619.3731 |
| GOMF_HIS | 6 | 0.4391 | 0.007827 | 0.24498 | 0.036544 | 621.4307 |
| WP_GENES | 12 | 0.2988 | 0.007531 | 0.16678 | 0.036608 | 622.519 |
| GOBP_SPO | 13 | 0.31071 | 0.008151 | 0.17343 | 0.036609 | 622.536 |
| REACTOME | 415 | 0.04389 | 0.006436 | 0.024502 | 0.036631 | 622.9102 |
| GOBP_SM/ | 1708 | 0.02263 | 0.006492 | 0.012636 | 0.036657 | 623.3523 |
| REACTOME | 18 | 0.18797 | 0.005801 | 0.10496 | 0.036662 | 623.4373 |
| GOBP_NEG | 153 | 0.072002 | 0.006456 | 0.040211 | 0.036687 | 623.8624 |
| GOBP_NOT | 166 | 0.0711 | 0.006638 | 0.03972 | 0.036735 | 624.6787 |
| GOCC_NUC | 11 | 0.25034 | 0.006041 | 0.13987 | 0.036752 | 624.9678 |
| REACTOME | 6 | 0.36803 | 0.00656 | 0.20563 | 0.036756 | 625.0358 |
| GOMF_5_3 | 17 | 0.18018 | 0.005405 | 0.1007 | 0.036794 | 625.682 |
| REACTOME | 53 | 0.13064 | 0.006912 | 0.073011 | 0.036795 | 625.699 |
| GOMF_S_A | 152 | 0.067609 | 0.006042 | 0.037791 | 0.036815 | 626.0391 |
| GOBP_FOR | 374 | 0.050395 | 0.007023 | 0.02819 | 0.036921 | 627.8416 |
| GOMF_G_I | 7 | 0.3647 | 0.007021 | 0.20405 | 0.036953 | 628.3858 |
| GRAESSMA | 1097 | 0.026669 | 0.006239 | 0.014924 | 0.036984 | 628.9129 |

| | | | | | | |
|---|---|---|---|---|---|---|
| GOCC_REC | 86 | 0.088491 | 0.005959 | 0.049541 | 0.037041 | 629.8822 |
| PID_MYC_I | 62 | 0.11645 | 0.006663 | 0.065269 | 0.037208 | 632.722 |
| BIOCARTA_ | 16 | 0.2151 | 0.006259 | 0.12056 | 0.037213 | 632.8071 |
| RAHMAN_ | 20 | 0.19728 | 0.006418 | 0.11061 | 0.037257 | 633.5553 |
| GOBP_NEG | 26 | 0.18689 | 0.006931 | 0.10482 | 0.037306 | 634.3885 |
| BACOLOD_ | 23 | 0.19376 | 0.006759 | 0.10868 | 0.037307 | 634.4055 |
| BROWNE_I | 94 | 0.10131 | 0.007131 | 0.056833 | 0.03733 | 634.7967 |
| GOCC_CYT | 5 | 0.46662 | 0.007593 | 0.26181 | 0.03736 | 635.3068 |
| GOBP_PRIN | 9 | 0.33829 | 0.007385 | 0.18989 | 0.037427 | 636.4461 |
| BENPORAT | 714 | 0.031651 | 0.006038 | 0.01777 | 0.037451 | 636.8543 |
| GOBP_GOL | 12 | 0.26766 | 0.006746 | 0.15032 | 0.037493 | 637.5685 |
| LIU_PROST | 452 | 0.043695 | 0.00668 | 0.024547 | 0.037541 | 638.3847 |
| KONG_E2F | 93 | 0.092983 | 0.00651 | 0.052261 | 0.037614 | 639.6261 |
| DARWICHE | 185 | 0.060666 | 0.005976 | 0.034106 | 0.037651 | 640.2553 |
| GOBP_NEG | 25 | 0.18672 | 0.006791 | 0.10498 | 0.037659 | 640.3913 |
| GOBP_FAT | 7 | 0.33545 | 0.006458 | 0.18863 | 0.037685 | 640.8334 |
| GOMF_VEF | 7 | 0.33545 | 0.006458 | 0.18863 | 0.037685 | 640.8334 |
| BYSTRYKH_ | 149 | 0.072783 | 0.006441 | 0.040939 | 0.037725 | 641.5136 |
| GOBP_NUC | 9 | 0.34986 | 0.007637 | 0.1968 | 0.03773 | 641.5987 |
| GOBP_PRO | 16 | 0.20424 | 0.005944 | 0.11489 | 0.037732 | 641.6327 |
| REACTOME | 32 | 0.13133 | 0.005403 | 0.073896 | 0.037775 | 642.3639 |
| GOBP_ER_ | 12 | 0.30686 | 0.007734 | 0.1727 | 0.037809 | 642.942 |
| GOBP_PRIN | 14 | 0.26569 | 0.007233 | 0.1496 | 0.037873 | 644.0304 |
| GOBP_DEN | 19 | 0.21496 | 0.006816 | 0.12105 | 0.037889 | 644.3024 |
| MODY_HIP | 43 | 0.12606 | 0.006009 | 0.070986 | 0.037892 | 644.3535 |
| MORI_EMU | 108 | 0.078476 | 0.005919 | 0.04421 | 0.037951 | 645.3568 |
| GOBP_REG | 115 | 0.084394 | 0.006567 | 0.047556 | 0.03799 | 646.02 |
| GOCC_CHR | 7 | 0.26163 | 0.005037 | 0.14743 | 0.037993 | 646.071 |
| ZHAN_MUI | 40 | 0.15148 | 0.006966 | 0.085433 | 0.038115 | 648.1456 |
| GOBP_POS | 83 | 0.10095 | 0.006679 | 0.056935 | 0.03812 | 648.2306 |
| GOBP_NEG | 26 | 0.15918 | 0.005903 | 0.089786 | 0.038133 | 648.4517 |
| GOBP_EPIT | 19 | 0.23816 | 0.007552 | 0.13438 | 0.038181 | 649.2679 |
| GOBP_AMI | 792 | 0.030297 | 0.006074 | 0.017095 | 0.038182 | 649.2849 |
| GOBP_NEG | 10 | 0.25233 | 0.005806 | 0.14244 | 0.038251 | 650.4583 |
| GOBP_FEM | 32 | 0.13695 | 0.005634 | 0.077316 | 0.038269 | 650.7643 |
| GOBP_STR | 103 | 0.089731 | 0.00661 | 0.050664 | 0.03828 | 650.9514 |
| GOBP_NEG | 8 | 0.3949 | 0.008128 | 0.22297 | 0.038284 | 651.0194 |
| GOBP_CHR | 8 | 0.36445 | 0.007501 | 0.20599 | 0.038437 | 653.6212 |
| WANG_RES | 28 | 0.17774 | 0.00684 | 0.10048 | 0.03846 | 654.0123 |
| GOBP_EXO | 44 | 0.14187 | 0.006841 | 0.08023 | 0.038518 | 654.9986 |
| REACTOME | 118 | 0.080059 | 0.00631 | 0.045277 | 0.038523 | 655.0836 |
| GOBP_POS | 8 | 0.33413 | 0.006877 | 0.18897 | 0.03853 | 655.2027 |
| GOBP_NUC | 13 | 0.23619 | 0.006196 | 0.1336 | 0.038551 | 655.5598 |
| GOBP_RAP | 12 | 0.27631 | 0.006964 | 0.15632 | 0.038573 | 655.9339 |
| GOCC_SAR | 40 | 0.13406 | 0.006164 | 0.075874 | 0.038638 | 657.0392 |

| | | | | | | |
|---|---|---|---|---|---|---|
| GOBP_NEG | 1261 | 0.025941 | 0.006477 | 0.014687 | 0.038687 | 657.8724 |
| GOBP_NEU | 145 | 0.074184 | 0.006477 | 0.042003 | 0.038693 | 657.9745 |
| GAUSSMAI | 161 | 0.070493 | 0.006482 | 0.039936 | 0.038778 | 659.4199 |
| GOBP_REG | 95 | 0.091356 | 0.006464 | 0.051757 | 0.038782 | 659.4879 |
| REACTOME | 78 | 0.095789 | 0.006145 | 0.054286 | 0.038832 | 660.3382 |
| GOBP_CELI | 58 | 0.13066 | 0.007231 | 0.074069 | 0.038874 | 661.0524 |
| LIN_APC_T | 66 | 0.10197 | 0.006019 | 0.057838 | 0.038959 | 662.4978 |
| GOMF_MA | 22 | 0.17333 | 0.005914 | 0.098342 | 0.038995 | 663.11 |
| GOBP_PRC | 117 | 0.076893 | 0.006035 | 0.043654 | 0.039092 | 664.7595 |
| GOBP_REG | 90 | 0.09439 | 0.006502 | 0.053589 | 0.039097 | 664.8445 |
| WP_WNT_ | 51 | 0.12211 | 0.006338 | 0.069352 | 0.039158 | 665.8818 |
| GOBP_GAN | 17 | 0.2291 | 0.006872 | 0.13012 | 0.039161 | 665.9328 |
| JOHANSSO | 43 | 0.12662 | 0.006036 | 0.071938 | 0.039197 | 666.545 |
| GOBP_RES | 54 | 0.1263 | 0.006745 | 0.071803 | 0.039305 | 668.3815 |
| KEGG_ERB | 84 | 0.10073 | 0.006704 | 0.057269 | 0.039314 | 668.5346 |
| GOBP_PRC | 1600 | 0.022444 | 0.006251 | 0.012761 | 0.039321 | 668.6536 |
| GOMF_NO | 12 | 0.28745 | 0.007245 | 0.16348 | 0.039359 | 669.2998 |
| REACTOME | 21 | 0.21154 | 0.007052 | 0.12036 | 0.039419 | 670.3201 |
| GOBP_REG | 22 | 0.19621 | 0.006694 | 0.11167 | 0.039465 | 671.1023 |
| IIZUKA_LIV | 9 | 0.26999 | 0.005894 | 0.15372 | 0.039523 | 672.0886 |
| REACTOME | 9 | 0.2914 | 0.006361 | 0.16592 | 0.039529 | 672.1906 |
| GOBP_POS | 10 | 0.24793 | 0.005705 | 0.14119 | 0.039555 | 672.6328 |
| GOMF_HIS | 22 | 0.18083 | 0.006169 | 0.10299 | 0.03957 | 672.8879 |
| GOMF_INT | 11 | 0.25878 | 0.006245 | 0.14745 | 0.03964 | 674.0782 |
| GOBP_MU | 15 | 0.23591 | 0.006647 | 0.13446 | 0.039679 | 674.7414 |
| GOBP_NEG | 11 | 0.27796 | 0.006708 | 0.15851 | 0.039758 | 676.0848 |
| GOMF_BH3 | 6 | 0.35803 | 0.006382 | 0.20418 | 0.039769 | 676.2718 |
| GOMF_STE | 58 | 0.12148 | 0.006723 | 0.069279 | 0.03977 | 676.2889 |
| WP_CCL18 | 32 | 0.16502 | 0.006788 | 0.094124 | 0.039795 | 676.714 |
| MYLLYKAN | 6 | 0.31182 | 0.005558 | 0.17787 | 0.0398 | 676.799 |
| GOBP_BLO | 11 | 0.28098 | 0.006781 | 0.16037 | 0.039892 | 678.3635 |
| GOBP_RET | 6 | 0.49776 | 0.008873 | 0.28411 | 0.039895 | 678.4145 |
| REACTOME | 44 | 0.13578 | 0.006547 | 0.077545 | 0.039985 | 679.9449 |
| PID_WNT_ | 27 | 0.15749 | 0.005952 | 0.089956 | 0.040009 | 680.353 |
| BILANGES_ | 65 | 0.096894 | 0.005676 | 0.055356 | 0.040035 | 680.7952 |
| GOBP_REG | 32 | 0.1444 | 0.00594 | 0.082508 | 0.040059 | 681.2033 |
| GOBP_REG | 16 | 0.21347 | 0.006212 | 0.12202 | 0.040115 | 682.1556 |
| REACTOME | 200 | 0.059687 | 0.006111 | 0.034131 | 0.040173 | 683.1419 |
| GOBP_EOS | 4 | 0.49764 | 0.007243 | 0.28478 | 0.040287 | 685.0804 |
| GOCC_GLY | 20 | 0.24099 | 0.00784 | 0.13791 | 0.040289 | 685.1144 |
| PUJANA_BI | 1493 | 0.022887 | 0.006177 | 0.013101 | 0.040335 | 685.8967 |
| GOMF_3_H | 6 | 0.36184 | 0.00645 | 0.20722 | 0.040403 | 687.053 |
| MYLLYKAN | 7 | 0.3201 | 0.006163 | 0.18337 | 0.040445 | 687.7672 |
| BIOCARTA_ | 6 | 0.33496 | 0.005971 | 0.19197 | 0.040514 | 688.9406 |
| REACTOME | 93 | 0.083426 | 0.005841 | 0.047847 | 0.040626 | 690.8451 |

| | | | | | | |
|---|---|---|---|---|---|---|
| JIANG_AGI | 33 | 0.14507 | 0.00606 | 0.083239 | 0.040688 | 691.8994 |
| GOCC_VOL | 53 | 0.12997 | 0.006877 | 0.074578 | 0.040693 | 691.9845 |
| GOCC_LEA | 174 | 0.068255 | 0.006523 | 0.039169 | 0.040712 | 692.3076 |
| REACTOME | 17 | 0.22433 | 0.006729 | 0.12875 | 0.040733 | 692.6647 |
| GOBP_END | 149 | 0.075337 | 0.006667 | 0.043244 | 0.040751 | 692.9708 |
| GOCC_COH | 8 | 0.30802 | 0.00634 | 0.17688 | 0.040817 | 694.0931 |
| BYSTRYKH_ | 8 | 0.26418 | 0.005437 | 0.15173 | 0.040843 | 694.5352 |
| PID_NOTCH | 57 | 0.1157 | 0.006348 | 0.066492 | 0.040935 | 696.0997 |
| GOBP_POS | 26 | 0.17762 | 0.006587 | 0.10209 | 0.040953 | 696.4058 |
| REACTOME | 15 | 0.21228 | 0.005981 | 0.12203 | 0.040972 | 696.7289 |
| GOBP_NEG | 9 | 0.24316 | 0.005308 | 0.13979 | 0.040987 | 696.9839 |
| CHIANG_LI | 158 | 0.071108 | 0.006478 | 0.040891 | 0.04103 | 697.7152 |
| GOBP_SPLI | 37 | 0.14646 | 0.006478 | 0.084255 | 0.041085 | 698.6504 |
| GOBP_PRO | 8 | 0.27358 | 0.005631 | 0.1575 | 0.041199 | 700.589 |
| BLANCO_M | 12 | 0.24985 | 0.006297 | 0.14386 | 0.041226 | 701.0481 |
| GOCC_PAL | 10 | 0.28368 | 0.006527 | 0.16337 | 0.041255 | 701.5413 |
| GOCC_PRE | 70 | 0.096572 | 0.00587 | 0.055644 | 0.041333 | 702.8677 |
| GOCC_KAT | 5 | 0.48515 | 0.007895 | 0.27957 | 0.041348 | 703.1227 |
| LIM_MAMI | 93 | 0.10526 | 0.00737 | 0.060663 | 0.041361 | 703.3438 |
| BIOCARTA_ | 11 | 0.26001 | 0.006275 | 0.14984 | 0.041364 | 703.3948 |
| GOCC_MET | 13 | 0.30379 | 0.007969 | 0.17509 | 0.041377 | 703.6159 |
| GOMF_MIC | 21 | 0.1993 | 0.006644 | 0.11489 | 0.041401 | 704.024 |
| CHEOK_RES | 21 | 0.18312 | 0.006104 | 0.10559 | 0.041441 | 704.7042 |
| KEGG_MAP | 250 | 0.055725 | 0.00637 | 0.032134 | 0.041454 | 704.9253 |
| GOBP_NEU | 7 | 0.30572 | 0.005886 | 0.17633 | 0.041491 | 705.5545 |
| GOBP_REG | 1531 | 0.023507 | 0.006417 | 0.013568 | 0.041603 | 707.459 |
| GOBP_VIRA | 298 | 0.050988 | 0.006356 | 0.029437 | 0.041637 | 708.0372 |
| GOBP_DEV | 34 | 0.15881 | 0.006733 | 0.091694 | 0.041655 | 708.3433 |
| MOLENAAR | 62 | 0.10643 | 0.006089 | 0.061457 | 0.041667 | 708.5473 |
| NAKAYAMA | 42 | 0.14149 | 0.006667 | 0.081716 | 0.041689 | 708.9214 |
| GOBP_GLU | 23 | 0.18998 | 0.006627 | 0.10977 | 0.041768 | 710.2648 |
| YANG_BRE | 28 | 0.14729 | 0.005668 | 0.085111 | 0.041776 | 710.4009 |
| GOBP_POS | 489 | 0.038518 | 0.006119 | 0.022277 | 0.041906 | 712.6115 |
| GOBP_REG | 125 | 0.075139 | 0.006094 | 0.043456 | 0.041906 | 712.6115 |
| WP_ATM_S | 41 | 0.12061 | 0.005615 | 0.069768 | 0.041938 | 713.1557 |
| GOBP_AXIS | 26 | 0.17781 | 0.006594 | 0.10286 | 0.041945 | 713.2747 |
| REACTOME | 5 | 0.43736 | 0.007117 | 0.25304 | 0.041962 | 713.5638 |
| GOBP_MET | 23 | 0.18342 | 0.006398 | 0.10619 | 0.042069 | 715.3833 |
| MMS_MOU | 29 | 0.1506 | 0.005898 | 0.087196 | 0.042081 | 715.5874 |
| REACTOME | 20 | 0.1959 | 0.006373 | 0.11348 | 0.042155 | 716.8458 |
| REACTOME | 5 | 0.34907 | 0.00568 | 0.20224 | 0.042183 | 717.3219 |
| GOBP_CAR | 15 | 0.26595 | 0.007494 | 0.1541 | 0.042194 | 717.509 |
| LAIHO_COL | 109 | 0.084553 | 0.006406 | 0.048995 | 0.042204 | 717.679 |
| DIRMEIER_ | 24 | 0.17401 | 0.006201 | 0.10084 | 0.042211 | 717.7981 |
| GOMF_FLA | 6 | 0.26573 | 0.004737 | 0.15401 | 0.042239 | 718.2742 |

| | | | | | | |
|---|---|---|---|---|---|---|
| GOBP_STR | 274 | 0.053017 | 0.006341 | 0.030728 | 0.04224 | 718.2912 |
| WP_HYPOT | 24 | 0.20402 | 0.00727 | 0.11826 | 0.042251 | 718.4783 |
| GOBP_CELI | 426 | 0.043652 | 0.006483 | 0.02531 | 0.0423 | 719.3115 |
| SCHLOSSEF | 113 | 0.076863 | 0.005929 | 0.044568 | 0.042308 | 719.4475 |
| REACTOME | 47 | 0.12627 | 0.006293 | 0.073224 | 0.042325 | 719.7366 |
| GOBP_OLE | 149 | 0.073185 | 0.006476 | 0.04244 | 0.042326 | 719.7536 |
| GOMF_NEU | 8 | 0.28906 | 0.005949 | 0.16764 | 0.042338 | 719.9577 |
| GOBP_REG | 12 | 0.22031 | 0.005553 | 0.12779 | 0.042357 | 720.2808 |
| GOMF_SIG | 7 | 0.28441 | 0.005476 | 0.1651 | 0.042485 | 722.4574 |
| GOMF_DN/ | 7 | 0.35891 | 0.00691 | 0.20843 | 0.04255 | 723.5628 |
| GOBP_ACT | 95 | 0.096159 | 0.006804 | 0.055847 | 0.04256 | 723.7328 |
| REACTOME | 38 | 0.12992 | 0.005823 | 0.075474 | 0.042594 | 724.311 |
| GOBP_NEG | 24 | 0.16999 | 0.006057 | 0.098773 | 0.04263 | 724.9232 |
| GOBP_CYT | 39 | 0.12141 | 0.005513 | 0.070573 | 0.042692 | 725.9775 |
| CORRE_MU | 54 | 0.12296 | 0.006567 | 0.071505 | 0.042758 | 727.0998 |
| TIMOFEEV/ | 16 | 0.21818 | 0.006349 | 0.12692 | 0.042816 | 728.0861 |
| PYEON_CA | 160 | 0.070089 | 0.006425 | 0.04079 | 0.042881 | 729.1914 |
| KEGG_HED | 55 | 0.1221 | 0.006581 | 0.071081 | 0.042933 | 730.0757 |
| ZHU_SKIL_ | 20 | 0.21156 | 0.006883 | 0.12319 | 0.04296 | 730.5348 |
| GOBP_NEG | 40 | 0.14295 | 0.006573 | 0.083242 | 0.042978 | 730.8409 |
| GOCC_PRC | 7 | 0.37484 | 0.007217 | 0.21833 | 0.043011 | 731.4021 |
| GOBP_POS | 19 | 0.18539 | 0.005879 | 0.10798 | 0.043013 | 731.4361 |
| WP_PILOC | 6 | 0.38834 | 0.006922 | 0.2263 | 0.043085 | 732.6604 |
| GOBP_TER | 22 | 0.20392 | 0.006957 | 0.11883 | 0.043089 | 732.7284 |
| KAAB_FAIL | 39 | 0.12947 | 0.005879 | 0.075486 | 0.043167 | 734.0548 |
| GOBP_CHR | 7 | 0.30601 | 0.005892 | 0.17844 | 0.043185 | 734.3609 |
| GOBP_CER | 13 | 0.23338 | 0.006122 | 0.13609 | 0.04319 | 734.446 |
| GOBP_NEG | 10 | 0.27307 | 0.006283 | 0.15927 | 0.043229 | 735.1091 |
| GOBP_NEG | 34 | 0.14351 | 0.006085 | 0.083738 | 0.043297 | 736.2655 |
| LOPES_ME | 28 | 0.17737 | 0.006826 | 0.10352 | 0.043335 | 736.9117 |
| BOSCO_EP | 59 | 0.11271 | 0.006291 | 0.065813 | 0.043406 | 738.119 |
| GOBP_REG | 74 | 0.1009 | 0.006305 | 0.058922 | 0.043418 | 738.3231 |
| REACTOME | 11 | 0.22483 | 0.005426 | 0.13134 | 0.043471 | 739.2244 |
| BORCZUK_ | 283 | 0.05098 | 0.006195 | 0.029799 | 0.043567 | 740.8568 |
| GOCC_SAR | 132 | 0.081597 | 0.006799 | 0.047703 | 0.043594 | 741.316 |
| ALFANO_M | 228 | 0.058812 | 0.006424 | 0.034408 | 0.043711 | 743.3056 |
| GOBP_POS | 128 | 0.076468 | 0.006275 | 0.044739 | 0.043716 | 743.3906 |
| GOBP_REG | 8 | 0.3008 | 0.006191 | 0.17612 | 0.043836 | 745.4312 |
| SANA_RESI | 83 | 0.09136 | 0.006045 | 0.053505 | 0.043873 | 746.0604 |
| GOCC_NUC | 209 | 0.056952 | 0.005959 | 0.033354 | 0.043875 | 746.0944 |
| SARRIO_EP | 134 | 0.072995 | 0.006128 | 0.04275 | 0.043875 | 746.0944 |
| GOMF_UBI | 5 | 0.30313 | 0.004933 | 0.17756 | 0.043902 | 746.5535 |
| WONG_EN | 71 | 0.11516 | 0.007049 | 0.067479 | 0.043957 | 747.4888 |
| GOBP_MYE | 8 | 0.31936 | 0.006573 | 0.18715 | 0.043971 | 747.7269 |
| GOBP_HIST | 15 | 0.2114 | 0.005957 | 0.12394 | 0.044043 | 748.9512 |

| | | | | | | |
|---|---|---|---|---|---|---|
| REACTOME | 4 | 0.58361 | 0.008494 | 0.34233 | 0.044124 | 750.3286 |
| BIOCARTA_ | 12 | 0.26515 | 0.006683 | 0.15554 | 0.044137 | 750.5497 |
| BREUHAHN | 15 | 0.25817 | 0.007274 | 0.15152 | 0.044212 | 751.8251 |
| WP_WNT_ | 36 | 0.13626 | 0.005945 | 0.079992 | 0.044253 | 752.5223 |
| GOBP_END | 71 | 0.099308 | 0.006079 | 0.058302 | 0.044261 | 752.6583 |
| REACTOME | 47 | 0.13341 | 0.006648 | 0.078331 | 0.044279 | 752.9644 |
| GOBERT_O | 544 | 0.035917 | 0.006009 | 0.021098 | 0.044354 | 754.2398 |
| STANELLE_ | 27 | 0.16611 | 0.006277 | 0.097578 | 0.04436 | 754.3418 |
| GOMF_BET | 6 | 0.34123 | 0.006083 | 0.20052 | 0.044414 | 755.2601 |
| ROESSLER_ | 101 | 0.080956 | 0.005906 | 0.047577 | 0.044429 | 755.5151 |
| REACTOME | 27 | 0.15429 | 0.005831 | 0.0907 | 0.044469 | 756.1953 |
| GOMF_HYI | 10 | 0.22777 | 0.005241 | 0.1339 | 0.044484 | 756.4504 |
| REACTOME | 29 | 0.16231 | 0.006357 | 0.09546 | 0.044548 | 757.5387 |
| PID_TGFBR | 52 | 0.12587 | 0.006597 | 0.074064 | 0.044626 | 758.8651 |
| PICCALUGA | 122 | 0.076439 | 0.006125 | 0.044994 | 0.044681 | 759.8004 |
| GOBP_NEG | 18 | 0.17987 | 0.005551 | 0.1059 | 0.044729 | 760.6166 |
| GOBP_REG | 6 | 0.39167 | 0.006981 | 0.23068 | 0.044773 | 761.3649 |
| REACTOME | 17 | 0.23317 | 0.006994 | 0.13737 | 0.044821 | 762.1811 |
| GOBP_VIRA | 45 | 0.12376 | 0.006035 | 0.072924 | 0.044854 | 762.7423 |
| REACTOME | 20 | 0.22212 | 0.007226 | 0.13092 | 0.044892 | 763.3885 |
| GOBP_VAC | 155 | 0.067072 | 0.006053 | 0.039536 | 0.044908 | 763.6605 |
| GOBP_MIT | 20 | 0.20405 | 0.006638 | 0.12032 | 0.044956 | 764.4768 |
| GOBP_POS | 8 | 0.32007 | 0.006588 | 0.18873 | 0.04496 | 764.5448 |
| GOBP_REG | 39 | 0.13057 | 0.005929 | 0.077018 | 0.045016 | 765.4971 |
| GOBP_REG | 314 | 0.046145 | 0.005902 | 0.027222 | 0.045034 | 765.8032 |
| BRUNEAU_ | 10 | 0.2748 | 0.006323 | 0.16222 | 0.045143 | 767.6567 |
| GOBP_PRII | 4 | 0.47199 | 0.00687 | 0.27863 | 0.045144 | 767.6737 |
| GOBP_POS | 5 | 0.43065 | 0.007008 | 0.25428 | 0.04518 | 768.2859 |
| REACTOME | 5 | 0.3638 | 0.00592 | 0.21481 | 0.045182 | 768.3199 |
| WP_15Q11 | 6 | 0.33669 | 0.006002 | 0.1988 | 0.045182 | 768.3199 |
| GOBP_IMP | 4 | 0.37756 | 0.005495 | 0.22299 | 0.045219 | 768.9491 |
| GOBP_REG | 17 | 0.23004 | 0.0069 | 0.13589 | 0.045251 | 769.4933 |
| GOBP_HEA | 58 | 0.11885 | 0.006578 | 0.070223 | 0.045289 | 770.1394 |
| BIOCARTA_ | 11 | 0.25606 | 0.006179 | 0.15131 | 0.045304 | 770.3945 |
| BIOCARTA_ | 11 | 0.2301 | 0.005553 | 0.13598 | 0.045315 | 770.5816 |
| GOBP_REG | 15 | 0.22564 | 0.006358 | 0.13335 | 0.045319 | 770.6496 |
| GOCC_REC | 182 | 0.058106 | 0.005678 | 0.03434 | 0.045325 | 770.7516 |
| GOBP_POS | 37 | 0.15083 | 0.006671 | 0.089173 | 0.045384 | 771.7549 |
| GOBP_RES | 12 | 0.31247 | 0.007876 | 0.18476 | 0.045407 | 772.146 |
| REACTOME | 16 | 0.2417 | 0.007034 | 0.14292 | 0.045409 | 772.18 |
| GOBP_REG | 23 | 0.17655 | 0.006159 | 0.10447 | 0.045531 | 774.2547 |
| WP_GLIOB | 80 | 0.095331 | 0.006193 | 0.05642 | 0.045555 | 774.6628 |
| GOBP_PRE | 12 | 0.24481 | 0.00617 | 0.14489 | 0.045559 | 774.7308 |
| MATTHEW | 17 | 0.17689 | 0.005306 | 0.1047 | 0.045581 | 775.1049 |
| GOBP_REG | 18 | 0.22239 | 0.006864 | 0.13165 | 0.045601 | 775.445 |

| | | | | | | |
|---|---|---|---|---|---|---|
| WP_MICRC | 46 | 0.11598 | 0.005718 | 0.068672 | 0.045634 | 776.0062 |
| GOBP_REG | 5 | 0.31829 | 0.005179 | 0.18848 | 0.045643 | 776.1592 |
| GOBP_TUB | 809 | 0.031876 | 0.006456 | 0.018884 | 0.045717 | 777.4176 |
| BRUINS_U\ | 292 | 0.049521 | 0.006111 | 0.029339 | 0.045728 | 777.6046 |
| GOMF_STE | 94 | 0.089387 | 0.006292 | 0.052959 | 0.045728 | 777.6046 |
| MURATA_\ | 20 | 0.19046 | 0.006196 | 0.11285 | 0.045743 | 777.8597 |
| REACTOME | 18 | 0.18975 | 0.005856 | 0.11243 | 0.045748 | 777.9447 |
| WP_UNFOI | 22 | 0.19837 | 0.006768 | 0.11755 | 0.045764 | 778.2168 |
| GOCC_HIG | 25 | 0.18301 | 0.006655 | 0.10846 | 0.045771 | 778.3359 |
| GOMF_KIN | 726 | 0.03132 | 0.006023 | 0.018565 | 0.045808 | 778.965 |
| GOBP_NEG | 16 | 0.2342 | 0.006815 | 0.13888 | 0.045876 | 780.1214 |
| GOBP_FOR | 7 | 0.36291 | 0.006987 | 0.21529 | 0.045936 | 781.1417 |
| RORIE_TAF | 23 | 0.19439 | 0.006781 | 0.11535 | 0.04599 | 782.06 |
| GAZDA_DI/ | 34 | 0.1596 | 0.006767 | 0.094725 | 0.046012 | 782.4341 |
| BOYAULT_I | 179 | 0.060248 | 0.005839 | 0.035759 | 0.04602 | 782.5701 |
| GOBP_REG | 73 | 0.095092 | 0.005902 | 0.05644 | 0.046021 | 782.5871 |
| GOBP_REG | 30 | 0.14991 | 0.005971 | 0.088998 | 0.046065 | 783.3353 |
| GOBP_REG | 26 | 0.15842 | 0.005875 | 0.094098 | 0.04614 | 784.6107 |
| HERNANDE | 26 | 0.17309 | 0.006419 | 0.10283 | 0.04618 | 785.2909 |
| GOBP_REG | 130 | 0.076997 | 0.006368 | 0.045751 | 0.046201 | 785.648 |
| BASAKI_YB | 272 | 0.05019 | 0.005981 | 0.029835 | 0.04627 | 786.8214 |
| GOBP_CEN | 170 | 0.068867 | 0.006506 | 0.040951 | 0.046324 | 787.7396 |
| TURASHVIL | 67 | 0.10746 | 0.006391 | 0.063905 | 0.046337 | 787.9607 |
| GOBP_NEG | 32 | 0.13819 | 0.005685 | 0.082208 | 0.046395 | 788.947 |
| BOQUEST_ | 31 | 0.16042 | 0.006496 | 0.095459 | 0.046433 | 789.5932 |
| GOBP_GRC | 12 | 0.24549 | 0.006188 | 0.14609 | 0.046451 | 789.8993 |
| SMIRNOV_ | 108 | 0.080319 | 0.006058 | 0.047799 | 0.046453 | 789.9333 |
| GOMF_INT | 5 | 0.44498 | 0.007241 | 0.26486 | 0.046484 | 790.4604 |
| GOBP_CHR | 330 | 0.045864 | 0.006011 | 0.027314 | 0.046573 | 791.9739 |
| REACTOME | 57 | 0.10333 | 0.005669 | 0.061554 | 0.046622 | 792.8071 |
| GOBP_VIR/ | 92 | 0.086576 | 0.006029 | 0.051581 | 0.046639 | 793.0962 |
| GOBP_NOT | 18 | 0.20854 | 0.006436 | 0.12429 | 0.046702 | 794.1675 |
| GOBP_NEG | 45 | 0.13304 | 0.006488 | 0.079345 | 0.046801 | 795.851 |
| GOBP_REG | 17 | 0.19613 | 0.005883 | 0.11699 | 0.046834 | 796.4122 |
| GOBP_SKEI | 9 | 0.29076 | 0.006347 | 0.17344 | 0.046841 | 796.5312 |
| GOBP_POS | 15 | 0.1977 | 0.005571 | 0.11794 | 0.046855 | 796.7693 |
| GOMF_SEF | 26 | 0.17761 | 0.006587 | 0.10599 | 0.046913 | 797.7556 |
| REACTOME | 14 | 0.20386 | 0.00555 | 0.12171 | 0.04697 | 798.7249 |
| GESERICK_ | 20 | 0.21989 | 0.007154 | 0.13132 | 0.04703 | 799.7452 |
| REACTOME | 19 | 0.21276 | 0.006747 | 0.12718 | 0.047176 | 802.2279 |
| GOBP_PRC | 7 | 0.31119 | 0.005991 | 0.18611 | 0.047263 | 803.7073 |
| MORI_PRE_ | 74 | 0.092912 | 0.005806 | 0.055571 | 0.047277 | 803.9454 |
| MARSON_I | 62 | 0.10652 | 0.006095 | 0.063723 | 0.047303 | 804.3875 |
| ZHANG_AN | 41 | 0.13547 | 0.006307 | 0.081054 | 0.047334 | 804.9147 |
| KEGG_CHR | 71 | 0.099702 | 0.006103 | 0.059688 | 0.047432 | 806.5812 |

| | | | | | | |
|---|---|---|---|---|---|---|
| GOBP_NEG | 29 | 0.16117 | 0.006312 | 0.096498 | 0.04745 | 806.8873 |
| WP_HIPPO | 97 | 0.089898 | 0.006427 | 0.053855 | 0.047542 | 808.4517 |
| GOBP_NEU | 149 | 0.067291 | 0.005955 | 0.040328 | 0.047608 | 809.574 |
| MEISSNER_ | 259 | 0.055869 | 0.006499 | 0.033483 | 0.047611 | 809.6251 |
| REACTOME | 25 | 0.16585 | 0.006032 | 0.099425 | 0.047655 | 810.3733 |
| PEART_HD/ | 67 | 0.098924 | 0.005883 | 0.059313 | 0.047686 | 810.9004 |
| REACTOME | 65 | 0.11691 | 0.006849 | 0.070102 | 0.047691 | 810.9855 |
| NAKAMUR/ | 8 | 0.2758 | 0.005676 | 0.16538 | 0.047703 | 811.1895 |
| ODONNELL | 42 | 0.12473 | 0.005877 | 0.074909 | 0.047951 | 815.4068 |
| GOBP_MAI | 18 | 0.21804 | 0.00673 | 0.13096 | 0.047974 | 815.7979 |
| GOBP_STEI | 106 | 0.082638 | 0.006175 | 0.049658 | 0.04805 | 817.0903 |
| CHIANG_LI | 84 | 0.10023 | 0.006671 | 0.060235 | 0.048069 | 817.4133 |
| NIKOLSKY_ | 7 | 0.51617 | 0.009938 | 0.31025 | 0.048092 | 817.8045 |
| GOBP_PUR | 6 | 0.33424 | 0.005958 | 0.20093 | 0.048121 | 818.2976 |
| FOSTER_KE | 232 | 0.057457 | 0.00633 | 0.034551 | 0.048169 | 819.1138 |
| REACTOME | 39 | 0.1334 | 0.006057 | 0.080237 | 0.048207 | 819.76 |
| GOBP_PRC | 10 | 0.31509 | 0.00725 | 0.18952 | 0.04821 | 819.8111 |
| GOBP_REG | 160 | 0.066947 | 0.006137 | 0.040272 | 0.048228 | 820.1171 |
| GRAESSMA | 527 | 0.03652 | 0.006016 | 0.021971 | 0.048242 | 820.3552 |
| GOBP_VAS | 34 | 0.1563 | 0.006627 | 0.094041 | 0.048261 | 820.6783 |
| GOBP_NEG | 912 | 0.028522 | 0.006116 | 0.017162 | 0.048274 | 820.8994 |
| GOBP_POS | 86 | 0.091024 | 0.00613 | 0.054804 | 0.048378 | 822.6679 |
| KEGG_ONE | 17 | 0.1876 | 0.005627 | 0.11297 | 0.048405 | 823.127 |
| GOBP_GLU | 15 | 0.23765 | 0.006696 | 0.14315 | 0.048449 | 823.8752 |
| GOBP_SEG | 96 | 0.087368 | 0.006215 | 0.052676 | 0.048608 | 826.579 |
| MEISSNER_ | 472 | 0.039567 | 0.006178 | 0.023866 | 0.04868 | 827.8034 |
| WP_APOPT | 80 | 0.092648 | 0.006018 | 0.05589 | 0.0487 | 828.1435 |
| GOBP_CELI | 30 | 0.15381 | 0.006127 | 0.092802 | 0.048729 | 828.6366 |
| ZHOU_INFI | 392 | 0.043079 | 0.006143 | 0.025992 | 0.048734 | 828.7217 |
| REACTOME | 38 | 0.13419 | 0.006014 | 0.08101 | 0.048825 | 830.2691 |
| GOMF_OXI | 7 | 0.2745 | 0.005285 | 0.16584 | 0.048951 | 832.4118 |
| GOBP_PRC | 26 | 0.18524 | 0.00687 | 0.11205 | 0.049155 | 835.8808 |
| KEGG_GAP | 85 | 0.098462 | 0.006592 | 0.059564 | 0.049171 | 836.1529 |
| GOBP_REG | 65 | 0.099401 | 0.005823 | 0.06014 | 0.049191 | 836.493 |
| GOMF_C_/ | 20 | 0.18256 | 0.005939 | 0.11045 | 0.049196 | 836.578 |
| GOBP_REG | 11 | 0.22045 | 0.00532 | 0.13344 | 0.049273 | 837.8874 |
| BROWNE_I | 94 | 0.081893 | 0.005764 | 0.049573 | 0.049281 | 838.0234 |
| GOBP_NEG | 8 | 0.26331 | 0.005419 | 0.15949 | 0.049384 | 839.7749 |
| REACTOME | 12 | 0.24988 | 0.006298 | 0.15144 | 0.049476 | 841.3394 |
| GOBP_POS | 6 | 0.26187 | 0.004668 | 0.15872 | 0.049488 | 841.5434 |
| GOBP_REG | 6 | 0.32406 | 0.005776 | 0.19647 | 0.049545 | 842.5127 |
| GOMF_KIN | 133 | 0.073455 | 0.006144 | 0.044537 | 0.049554 | 842.6658 |
| KEGG_COL | 61 | 0.1136 | 0.006447 | 0.068889 | 0.049578 | 843.0739 |
| GOMF_KIN | 199 | 0.058097 | 0.005933 | 0.035232 | 0.049583 | 843.1589 |
| GOBP_RET | 10 | 0.30909 | 0.007112 | 0.18745 | 0.049594 | 843.346 |

| | | | | | | |
|---|---|---|---|---|---|---|
| REACTOME | 291 | 0.049851 | 0.006142 | 0.030245 | 0.049663 | 844.5193 |
| GOMF_G_I | 10 | 0.28887 | 0.006647 | 0.17529 | 0.049686 | 844.9104 |
| MEISSNER_ | 1048 | 0.02851 | 0.006528 | 0.017301 | 0.049692 | 845.0125 |
| REACTOME | 502 | 0.035597 | 0.005727 | 0.021602 | 0.049696 | 845.0805 |
| GOBP_NEG | 17 | 0.18401 | 0.005519 | 0.11167 | 0.049705 | 845.2335 |
| GOBP_LOC | 630 | 0.032294 | 0.0058 | 0.019599 | 0.049713 | 845.3696 |
| GOBP_REG | 5 | 0.36885 | 0.006002 | 0.22386 | 0.04972 | 845.4886 |
| KIM_WT1_ | 161 | 0.064693 | 0.005949 | 0.039278 | 0.049782 | 846.5429 |
| GOMF_C_A | 7 | 0.23852 | 0.004592 | 0.14484 | 0.049811 | 847.0361 |
| GOCC_NEU | 56 | 0.12695 | 0.006904 | 0.077089 | 0.049816 | 847.1211 |
| MARSON_I | 1164 | 0.025671 | 0.006175 | 0.01559 | 0.049827 | 847.3081 |
| GOMF_EPC | 6 | 0.3567 | 0.006358 | 0.21666 | 0.049849 | 847.6822 |
| GOBP_PAR | 17 | 0.23487 | 0.007045 | 0.14267 | 0.049867 | 847.9883 |
| GOBP_NEG | 7 | 0.30061 | 0.005788 | 0.18263 | 0.049892 | 848.4135 |
| GOBP_REG | 68 | 0.10301 | 0.006171 | 0.062582 | 0.049896 | 848.4815 |
| REACTOME | 17 | 0.22302 | 0.00669 | 0.13551 | 0.049919 | 848.8726 |
| GOBP_PLA | 70 | 0.098903 | 0.006011 | 0.060104 | 0.049939 | 849.2127 |
| GOMF_TRA | 78 | 0.10766 | 0.006906 | 0.065443 | 0.049985 | 849.9949 |
| GOBP_MET | 331 | 0.04432 | 0.005817 | 0.026947 | 0.050024 | 850.6581 |
| BIOCARTA_ | 11 | 0.26296 | 0.006346 | 0.15991 | 0.050052 | 851.1343 |
| PID_VEGF_ | 9 | 0.29394 | 0.006417 | 0.17877 | 0.050072 | 851.4744 |
| GENTILE_U | 60 | 0.11643 | 0.006554 | 0.070828 | 0.050107 | 852.0695 |
| BIOCARTA_ | 10 | 0.29038 | 0.006682 | 0.17664 | 0.050108 | 852.0865 |
| GOMF_ERE | 5 | 0.32341 | 0.005263 | 0.19679 | 0.050153 | 852.8518 |
| GOCC_CLA | 59 | 0.10841 | 0.006051 | 0.065968 | 0.050165 | 853.0558 |
| SERVITJA_I | 158 | 0.070504 | 0.006423 | 0.042903 | 0.050168 | 853.1068 |
| GOBP_PRC | 66 | 0.092452 | 0.005457 | 0.056276 | 0.050217 | 853.9401 |
| REACTOME | 30 | 0.14226 | 0.005666 | 0.086635 | 0.050304 | 855.4195 |
| GOCC_FIBF | 135 | 0.067722 | 0.005706 | 0.041255 | 0.050351 | 856.2188 |
| GOBP_REG | 53 | 0.11618 | 0.006147 | 0.070832 | 0.050484 | 858.4804 |
| GOCC_FAC | 6 | 0.33365 | 0.005947 | 0.20355 | 0.050602 | 860.487 |
| GOBP_RET | 5 | 0.40994 | 0.006671 | 0.25012 | 0.050616 | 860.7251 |
| GOCC_MIC | 432 | 0.039119 | 0.00585 | 0.023886 | 0.050745 | 862.9187 |
| GOMF_ACT | 14 | 0.26541 | 0.007225 | 0.16209 | 0.050784 | 863.5819 |
| GOBP_REG | 7 | 0.3081 | 0.005932 | 0.18819 | 0.050805 | 863.939 |
| GOBP_NEG | 13 | 0.24911 | 0.006535 | 0.15216 | 0.050813 | 864.0751 |
| KEGG_PRO | 85 | 0.088751 | 0.005942 | 0.054214 | 0.050821 | 864.2111 |
| GOCC_END | 8 | 0.29873 | 0.006148 | 0.18252 | 0.050861 | 864.8913 |
| GOBP_SON | 60 | 0.1089 | 0.006129 | 0.066541 | 0.050876 | 865.1464 |
| GOCC_BRC | 6 | 0.39693 | 0.007075 | 0.24259 | 0.050904 | 865.6225 |
| GOMF_7SK | 9 | 0.34546 | 0.007541 | 0.21118 | 0.050944 | 866.3027 |
| GOBP_OUT | 78 | 0.10324 | 0.006622 | 0.063118 | 0.050966 | 866.6768 |
| BIOCARTA_ | 34 | 0.12659 | 0.005367 | 0.077424 | 0.051034 | 867.8332 |
| REACTOME | 32 | 0.13578 | 0.005586 | 0.08308 | 0.051106 | 869.0575 |
| GOBP_NEG | 7 | 0.35627 | 0.006859 | 0.21807 | 0.051165 | 870.0608 |

| | | | | | | |
|---|---|---|---|---|---|---|
| TERAMOTO | 26 | 0.16081 | 0.005964 | 0.098436 | 0.05117 | 870.1459 |
| BIOCARTA_ | 12 | 0.22836 | 0.005756 | 0.13978 | 0.051174 | 870.2139 |
| GOCC_EQU | 4 | 0.36577 | 0.005324 | 0.22396 | 0.051223 | 871.0471 |
| GOMF_PH( | 163 | 0.063865 | 0.005909 | 0.039115 | 0.051268 | 871.8123 |
| GOBP_PUR | 14 | 0.21752 | 0.005922 | 0.13323 | 0.051277 | 871.9654 |
| GOBP_END | 43 | 0.14298 | 0.006816 | 0.087606 | 0.051341 | 873.0537 |
| GOMF_TRA | 28 | 0.14053 | 0.005408 | 0.08611 | 0.051345 | 873.1217 |
| REACTOME | 9 | 0.27817 | 0.006072 | 0.17052 | 0.051421 | 874.4141 |
| GARGALOV | 16 | 0.21369 | 0.006219 | 0.13102 | 0.051457 | 875.0263 |
| GOBP_REG | 35 | 0.14893 | 0.006407 | 0.091338 | 0.051504 | 875.8255 |
| LEE_AGING | 73 | 0.095257 | 0.005912 | 0.058442 | 0.051566 | 876.8798 |
| GOMF_TR( | 9 | 0.32381 | 0.007069 | 0.19867 | 0.051568 | 876.9138 |
| WP_NANO | 28 | 0.17185 | 0.006613 | 0.10551 | 0.051694 | 879.0565 |
| GOBP_PUR | 10 | 0.25292 | 0.00582 | 0.15544 | 0.051866 | 881.9813 |
| GOMF_PR( | 11 | 0.26282 | 0.006342 | 0.16167 | 0.052022 | 884.6341 |
| KOYAMA_S | 336 | 0.04663 | 0.006165 | 0.028685 | 0.052029 | 884.7531 |
| GOCC_CHA | 10 | 0.26694 | 0.006142 | 0.16423 | 0.052053 | 885.1613 |
| GOBP_RRN | 238 | 0.048359 | 0.005396 | 0.029755 | 0.052065 | 885.3653 |
| ACEVEDO_ | 518 | 0.035494 | 0.005798 | 0.021843 | 0.052098 | 885.9265 |
| ZHAN_MUI | 36 | 0.13775 | 0.00601 | 0.084787 | 0.052132 | 886.5047 |
| SEITZ_NEO | 70 | 0.10824 | 0.006579 | 0.066627 | 0.052138 | 886.6067 |
| GOMF_SPH | 27 | 0.13656 | 0.005161 | 0.084095 | 0.052206 | 887.763 |
| GOMF_VOI | 51 | 0.12723 | 0.006604 | 0.078378 | 0.052279 | 889.0044 |
| GOCC_CHA | 39 | 0.12416 | 0.005637 | 0.07651 | 0.052328 | 889.8376 |
| REACTOME | 12 | 0.24484 | 0.006171 | 0.15088 | 0.052333 | 889.9227 |
| GOBP_PRC | 182 | 0.059815 | 0.005845 | 0.036873 | 0.052388 | 890.8579 |
| GOMF_PR( | 89 | 0.086186 | 0.005904 | 0.053137 | 0.052418 | 891.3681 |
| REACTOME | 21 | 0.17792 | 0.005931 | 0.10971 | 0.052441 | 891.7592 |
| GOBP_CELI | 2 | 0.66288 | 0.006823 | 0.40876 | 0.052445 | 891.8272 |
| GOBP_REG | 7 | 0.29398 | 0.00566 | 0.18129 | 0.052453 | 891.9633 |
| GOMF_DIA | 5 | 0.38871 | 0.006325 | 0.23975 | 0.052484 | 892.4904 |
| GOBP_GLC | 24 | 0.18599 | 0.006627 | 0.11473 | 0.052506 | 892.8645 |
| GOBP_FOR | 39 | 0.13798 | 0.006265 | 0.085117 | 0.05251 | 892.9326 |
| GOBP_RES | 29 | 0.15881 | 0.00622 | 0.098 | 0.05257 | 893.9529 |
| REACTOME | 10 | 0.28417 | 0.006539 | 0.1754 | 0.052614 | 894.7011 |
| GOBP_REG | 47 | 0.12597 | 0.006278 | 0.077771 | 0.052647 | 895.2622 |
| GOBP_NEC | 58 | 0.10518 | 0.005821 | 0.064938 | 0.052656 | 895.4153 |
| HEDENFALI | 19 | 0.17062 | 0.00541 | 0.10534 | 0.052657 | 895.4323 |
| GOBP_REG | 5 | 0.39061 | 0.006356 | 0.24117 | 0.052662 | 895.5173 |
| REACTOME | 45 | 0.1059 | 0.005164 | 0.06542 | 0.052754 | 897.0818 |
| GOCC_INTI | 87 | 0.081835 | 0.005543 | 0.050559 | 0.052776 | 897.4559 |
| GOMF_FAT | 13 | 0.23511 | 0.006168 | 0.14529 | 0.052827 | 898.3231 |
| GOBP_OD( | 130 | 0.077296 | 0.006392 | 0.047809 | 0.052974 | 900.8229 |
| MARKEY_R | 221 | 0.054044 | 0.005813 | 0.033429 | 0.052986 | 901.0269 |
| GOBP_RES | 47 | 0.12093 | 0.006027 | 0.074812 | 0.053008 | 901.401 |

| | | | | | | |
|---|---|---|---|---|---|---|
| GOCC_TRA | 17 | 0.19134 | 0.005739 | 0.11841 | 0.053067 | 902.4043 |
| WP_SARSC | 4 | 0.42756 | 0.006223 | 0.26461 | 0.05308 | 902.6254 |
| GU_PDEF_ | 69 | 0.10681 | 0.006445 | 0.066105 | 0.053091 | 902.8125 |
| REACTOME | 7 | 0.24462 | 0.00471 | 0.1514 | 0.053091 | 902.8125 |
| REACTOME | 21 | 0.18919 | 0.006307 | 0.1171 | 0.053097 | 902.9145 |
| GOBP_SER | 36 | 0.15109 | 0.006592 | 0.093524 | 0.053108 | 903.1015 |
| REACTOME | 103 | 0.085597 | 0.006305 | 0.05302 | 0.053226 | 905.1081 |
| GOBP_NAD | 4 | 0.42839 | 0.006235 | 0.26542 | 0.053276 | 905.9584 |
| WU_APOP | 51 | 0.1168 | 0.006063 | 0.072388 | 0.053318 | 906.6726 |
| RHEIN_ALL | 334 | 0.043875 | 0.005784 | 0.027202 | 0.053391 | 907.914 |
| GOBP_CEL | 61 | 0.10805 | 0.006132 | 0.067 | 0.053413 | 908.2881 |
| COULOUAF | 133 | 0.067655 | 0.005659 | 0.041988 | 0.053567 | 910.9068 |
| WP_INFLAI | 28 | 0.16209 | 0.006238 | 0.10061 | 0.053587 | 911.2469 |
| GOMF_EXC | 56 | 0.090532 | 0.004924 | 0.056201 | 0.053614 | 911.7061 |
| GOBP_REG | 1251 | 0.02354 | 0.005856 | 0.014618 | 0.053677 | 912.7774 |
| GOBP_REG | 117 | 0.072159 | 0.005663 | 0.044823 | 0.053723 | 913.5596 |
| GOBP_REG | 45 | 0.11603 | 0.005658 | 0.072075 | 0.053723 | 913.5596 |
| MASRI_RES | 17 | 0.22045 | 0.006613 | 0.13695 | 0.053742 | 913.8827 |
| FERRARI_R | 5 | 0.38547 | 0.006273 | 0.23975 | 0.053947 | 917.3687 |
| BIOCARTA_ | 16 | 0.21268 | 0.006189 | 0.13228 | 0.053953 | 917.4708 |
| REACTOME | 5 | 0.46 | 0.007485 | 0.2862 | 0.054006 | 918.372 |
| ICHIBA_GR | 117 | 0.072083 | 0.005657 | 0.044859 | 0.054049 | 919.1032 |
| GOBP_GON | 17 | 0.2089 | 0.006266 | 0.13002 | 0.054064 | 919.3583 |
| GOBP_REG | 77 | 0.096152 | 0.006128 | 0.059853 | 0.054096 | 919.9025 |
| BYSTRYKH_ | 121 | 0.072095 | 0.005753 | 0.04489 | 0.054142 | 920.6847 |
| GOBP_DET | 16 | 0.19714 | 0.005737 | 0.12278 | 0.054188 | 921.4669 |
| GOBP_NEG | 6 | 0.25471 | 0.00454 | 0.15868 | 0.054233 | 922.2322 |
| GOBP_POS | 92 | 0.082699 | 0.005759 | 0.051548 | 0.054331 | 923.8987 |
| WINNEPEN | 157 | 0.060197 | 0.005467 | 0.037542 | 0.054429 | 925.5651 |
| GOMF_RNA | 8 | 0.26636 | 0.005482 | 0.16614 | 0.054455 | 926.0073 |
| GOBP_POS | 15 | 0.21793 | 0.006141 | 0.13594 | 0.054459 | 926.0753 |
| REACTOME | 12 | 0.27545 | 0.006943 | 0.17182 | 0.054462 | 926.1263 |
| GOBP_UNS | 105 | 0.077958 | 0.005798 | 0.048635 | 0.054485 | 926.5174 |
| GOBP_REG | 94 | 0.079271 | 0.00558 | 0.049459 | 0.054504 | 926.8405 |
| GOBP_POL | 7 | 0.33076 | 0.006368 | 0.20637 | 0.054507 | 926.8915 |
| GOBP_POS | 22 | 0.17727 | 0.006048 | 0.11062 | 0.054523 | 927.1636 |
| REACTOME | 24 | 0.18458 | 0.006577 | 0.11519 | 0.05455 | 927.6228 |
| MIKKELSEN | 9 | 0.30259 | 0.006605 | 0.1889 | 0.054602 | 928.507 |
| GOBP_POS | 106 | 0.078033 | 0.005831 | 0.048723 | 0.054637 | 929.1022 |
| REACTOME | 18 | 0.19736 | 0.006091 | 0.12327 | 0.054693 | 930.0545 |
| REACTOME | 30 | 0.14607 | 0.005818 | 0.091259 | 0.054744 | 930.9217 |
| GOMF_PRO | 6 | 0.51143 | 0.009116 | 0.31954 | 0.054749 | 931.0067 |
| GOMF_O_I | 19 | 0.18115 | 0.005744 | 0.11324 | 0.05484 | 932.5542 |
| GOBP_CAV | 5 | 0.42674 | 0.006944 | 0.26679 | 0.05486 | 932.8943 |
| GOBP_PLA | 5 | 0.42674 | 0.006944 | 0.26679 | 0.05486 | 932.8943 |

| | | | | | | |
|---|---|---|---|---|---|---|
| GOBP_PEP | 782 | 0.027479 | 0.005476 | 0.01718 | 0.054867 | 933.0133 |
| CAMPS_CC | 85 | 0.094331 | 0.006316 | 0.058993 | 0.054917 | 933.8636 |
| KORKOLA_ | 17 | 0.1939 | 0.005816 | 0.12127 | 0.054929 | 934.0676 |
| REACTOME | 5 | 0.35318 | 0.005747 | 0.22094 | 0.054974 | 934.8329 |
| GOBP_DE_ | 37 | 0.12065 | 0.005336 | 0.07548 | 0.054975 | 934.8499 |
| GOBP_NEG | 12 | 0.20534 | 0.005175 | 0.12848 | 0.055009 | 935.428 |
| KEGG_MISI | 22 | 0.16212 | 0.005531 | 0.10146 | 0.055044 | 936.0232 |
| WP_STRIAT | 36 | 0.13135 | 0.005731 | 0.082239 | 0.055116 | 937.2476 |
| GOMF_DO | 16 | 0.17382 | 0.005058 | 0.10887 | 0.055188 | 938.4719 |
| GOBP_REG | 98 | 0.089997 | 0.006468 | 0.056378 | 0.055217 | 938.9651 |
| GOBP_NEG | 70 | 0.094775 | 0.005761 | 0.059391 | 0.055279 | 940.0194 |
| REACTOME | 124 | 0.078855 | 0.00637 | 0.049428 | 0.055325 | 940.8016 |
| GOBP_PYR | 17 | 0.17195 | 0.005158 | 0.10778 | 0.055331 | 940.9037 |
| HAMAI_AP | 181 | 0.057591 | 0.005612 | 0.036101 | 0.055339 | 941.0397 |
| WP_MITOC | 30 | 0.12524 | 0.004989 | 0.078526 | 0.055382 | 941.7709 |
| GOBP_VEN | 6 | 0.30805 | 0.005491 | 0.19335 | 0.05556 | 944.7978 |
| GOBP_EPIT | 45 | 0.13722 | 0.006692 | 0.086132 | 0.055572 | 945.0019 |
| GOCC_PER | 5 | 0.37544 | 0.006109 | 0.23574 | 0.055633 | 946.0392 |
| GOBP_COF | 10 | 0.2917 | 0.006712 | 0.18327 | 0.055742 | 947.8927 |
| REACTOME | 7 | 0.38109 | 0.007337 | 0.23944 | 0.055745 | 947.9437 |
| GOBP_EXC | 8 | 0.25481 | 0.005244 | 0.16011 | 0.05576 | 948.1988 |
| TARTE_PLA | 289 | 0.045538 | 0.005591 | 0.028619 | 0.05579 | 948.709 |
| MASRI_RES | 18 | 0.19786 | 0.006107 | 0.12439 | 0.055857 | 949.8483 |
| GOMF_CHO | 49 | 0.11802 | 0.006005 | 0.074218 | 0.055914 | 950.8176 |
| REACTOME | 69 | 0.090039 | 0.005434 | 0.056629 | 0.055928 | 951.0556 |
| GOBP_END | 56 | 0.10972 | 0.005967 | 0.069017 | 0.055959 | 951.5828 |
| BANG_VER | 14 | 0.20768 | 0.005654 | 0.13068 | 0.056006 | 952.382 |
| GOBP_REG | 7 | 0.37467 | 0.007213 | 0.23593 | 0.056147 | 954.7797 |
| TIEN_INTES | 162 | 0.063419 | 0.00585 | 0.039949 | 0.056209 | 955.834 |
| GOBP_POS | 8 | 0.22491 | 0.004629 | 0.14168 | 0.056214 | 955.9191 |
| GOCC_CON | 8 | 0.22491 | 0.004629 | 0.14168 | 0.056214 | 955.9191 |
| GOCC_MAI | 62 | 0.1092 | 0.006248 | 0.068794 | 0.056223 | 956.0721 |
| GOCC_MU | 6 | 0.36152 | 0.006444 | 0.22783 | 0.056293 | 957.2625 |
| MURAKAM | 10 | 0.25543 | 0.005877 | 0.16098 | 0.056303 | 957.4325 |
| GOBP_FAT | 12 | 0.21837 | 0.005504 | 0.13763 | 0.056308 | 957.5175 |
| GOCC_MIS | 7 | 0.36487 | 0.007025 | 0.23002 | 0.056352 | 958.2658 |
| GOCC_PER | 687 | 0.029653 | 0.005553 | 0.018697 | 0.056376 | 958.6739 |
| LEIN_MEDI | 79 | 0.086068 | 0.005556 | 0.054285 | 0.056437 | 959.7112 |
| GOBP_POS | 48 | 0.12611 | 0.006351 | 0.079583 | 0.056533 | 961.3437 |
| LASTOWSK | 780 | 0.033612 | 0.00669 | 0.021216 | 0.056576 | 962.0749 |
| GOBP_REG | 14 | 0.25668 | 0.006987 | 0.16206 | 0.056624 | 962.8911 |
| OSWALD_H | 209 | 0.056783 | 0.005942 | 0.035866 | 0.056698 | 964.1495 |
| GOBP_RES | 117 | 0.077003 | 0.006043 | 0.048642 | 0.056714 | 964.4216 |
| BIOCARTA_ | 12 | 0.23462 | 0.005913 | 0.14824 | 0.056754 | 965.1018 |
| GOMF_TUI | 9 | 0.30564 | 0.006672 | 0.19315 | 0.05679 | 965.714 |

| | | | | | | |
|---|---|---|---|---|---|---|
| ABBUD_LIF | 13 | 0.23499 | 0.006165 | 0.14851 | 0.056795 | 965.799 |
| GOMF_SUL | 48 | 0.12747 | 0.006419 | 0.080557 | 0.056799 | 965.867 |
| GOCC_SMA | 65 | 0.08779 | 0.005143 | 0.055491 | 0.056827 | 966.3431 |
| GOMF_TAT | 10 | 0.31786 | 0.007314 | 0.20095 | 0.056862 | 966.9383 |
| BIOCARTA_ | 10 | 0.24412 | 0.005617 | 0.15438 | 0.056917 | 967.8736 |
| GOMF_PHO | 19 | 0.17866 | 0.005665 | 0.11301 | 0.05695 | 968.4348 |
| GOBP_POS | 5 | 0.36925 | 0.006009 | 0.23361 | 0.056988 | 969.0809 |
| GOBP_REG | 660 | 0.031385 | 0.005765 | 0.019859 | 0.05702 | 969.6251 |
| GOBP_MAI | 6 | 0.37863 | 0.006749 | 0.2396 | 0.057033 | 969.8462 |
| WP_ENVEL | 44 | 0.12725 | 0.006136 | 0.080523 | 0.057034 | 969.8632 |
| MOLENAAI | 54 | 0.10828 | 0.005783 | 0.068524 | 0.057046 | 970.0672 |
| PLASARI_N | 26 | 0.1765 | 0.006546 | 0.11172 | 0.05708 | 970.6454 |
| REACTOME | 6 | 0.35388 | 0.006308 | 0.22411 | 0.057171 | 972.1929 |
| BROWNE_I | 136 | 0.070037 | 0.005923 | 0.044371 | 0.057242 | 973.4002 |
| GOBP_GLY | 5 | 0.34959 | 0.005689 | 0.22152 | 0.057278 | 974.0124 |
| GOBP_POS | 11 | 0.25253 | 0.006094 | 0.16003 | 0.057288 | 974.1824 |
| GOBP_PAR | 6 | 0.38032 | 0.006779 | 0.24101 | 0.05729 | 974.2165 |
| YAMAZAKI_ | 187 | 0.060702 | 0.006012 | 0.038469 | 0.057296 | 974.3185 |
| GOBP_NEU | 49 | 0.10883 | 0.005537 | 0.068971 | 0.057304 | 974.4545 |
| REACTOME | 8 | 0.3459 | 0.007119 | 0.21925 | 0.057333 | 974.9477 |
| RADAEVA_ | 10 | 0.2522 | 0.005803 | 0.1599 | 0.057384 | 975.8149 |
| GOBP_ESTA | 5 | 0.44455 | 0.007234 | 0.28186 | 0.057385 | 975.8319 |
| GOBP_ANT | 11 | 0.2995 | 0.007228 | 0.1899 | 0.057392 | 975.951 |
| GOBP_REG | 234 | 0.054682 | 0.00605 | 0.034674 | 0.057406 | 976.189 |
| GOMF_EXO | 80 | 0.07607 | 0.004942 | 0.04824 | 0.057419 | 976.4101 |
| REACTOME | 11 | 0.22741 | 0.005488 | 0.14427 | 0.057489 | 977.6004 |
| GOMF_ME | 24 | 0.17415 | 0.006205 | 0.1105 | 0.057532 | 978.3317 |
| GOBP_POS | 31 | 0.14391 | 0.005827 | 0.091349 | 0.057599 | 979.471 |
| CROMER_M | 57 | 0.10802 | 0.005927 | 0.068576 | 0.057614 | 979.7261 |
| BIOCARTA_ | 19 | 0.16656 | 0.005281 | 0.10584 | 0.05779 | 982.719 |
| GOBP_ART | 6 | 0.38438 | 0.006852 | 0.24427 | 0.057801 | 982.906 |
| GOBP_TRA | 6 | 0.26609 | 0.004743 | 0.16914 | 0.057845 | 983.6542 |
| WP_NSP1_ | 14 | 0.21553 | 0.005867 | 0.13703 | 0.057885 | 984.3344 |
| NADLER_H | 57 | 0.10775 | 0.005912 | 0.068524 | 0.057943 | 985.3207 |
| GOBP_REG | 6 | 0.28469 | 0.005075 | 0.18106 | 0.057948 | 985.4057 |
| ISHIDA_E2I | 50 | 0.10337 | 0.005313 | 0.065749 | 0.057955 | 985.5248 |
| BIOCARTA_ | 21 | 0.16786 | 0.005595 | 0.10682 | 0.058062 | 987.3443 |
| ROSS_AML | 71 | 0.093377 | 0.005716 | 0.059452 | 0.058145 | 988.7557 |
| VERRECCHI | 11 | 0.22323 | 0.005387 | 0.14217 | 0.058188 | 989.4869 |
| BROWNE_I | 131 | 0.06626 | 0.0055 | 0.042198 | 0.058195 | 989.606 |
| MEISSNER_ | 22 | 0.15932 | 0.005436 | 0.10149 | 0.058243 | 990.4222 |
| LAIHO_COI | 66 | 0.089678 | 0.005293 | 0.057133 | 0.058261 | 990.7283 |
| CHEMNITZ_ | 136 | 0.067522 | 0.00571 | 0.043049 | 0.058391 | 992.939 |
| GOBP_POS | 83 | 0.083092 | 0.005498 | 0.052983 | 0.058417 | 993.3811 |
| GOMF_INT | 5 | 0.33084 | 0.005384 | 0.21096 | 0.058424 | 993.5001 |

| | | | | | | |
|---|---|---|---|---|---|---|
| GOBP_NEU | 47 | 0.112 | 0.005581 | 0.071422 | 0.05844 | 993.7722 |
| GOBP_POS | 40 | 0.11299 | 0.005196 | 0.072064 | 0.058454 | 994.0103 |
| GOMF_AR/ | 18 | 0.21498 | 0.006635 | 0.13716 | 0.058524 | 995.2006 |
| TSAI_RESP | 31 | 0.14509 | 0.005875 | 0.09257 | 0.058525 | 995.2176 |
| GOBP_HEP | 11 | 0.27914 | 0.006736 | 0.17815 | 0.058585 | 996.2379 |
| KEGG_VIR/ | 48 | 0.12016 | 0.006051 | 0.07669 | 0.058592 | 996.357 |
| KIM_GASTI | 85 | 0.087563 | 0.005862 | 0.055899 | 0.05863 | 997.0032 |
| CHIANG_LI | 166 | 0.060255 | 0.005625 | 0.038476 | 0.058678 | 997.8194 |
| WP_LEPTIN | 9 | 0.24669 | 0.005385 | 0.15753 | 0.058682 | 997.8874 |
| WP_FAMIL | 13 | 0.205 | 0.005378 | 0.13091 | 0.058685 | 997.9384 |
| GOBP_REG | 17 | 0.18559 | 0.005567 | 0.11854 | 0.058725 | 998.6186 |
| GOBP_ENZ | 922 | 0.027014 | 0.005823 | 0.017257 | 0.058748 | 999.0097 |
| GOMF_SNI | 9 | 0.25331 | 0.00553 | 0.16182 | 0.058753 | 999.0948 |
| GOBP_REG | 4 | 0.40568 | 0.005905 | 0.25924 | 0.058811 | 1000.081 |
| GOBP_REG | 6 | 0.4048 | 0.007216 | 0.25872 | 0.058845 | 1000.659 |
| BREDEMEY | 53 | 0.10549 | 0.005582 | 0.067443 | 0.058897 | 1001.543 |
| DUNNE_TA | 18 | 0.17717 | 0.005468 | 0.11331 | 0.058969 | 1002.768 |
| GOBP_PAT | 60 | 0.10186 | 0.005733 | 0.065196 | 0.059119 | 1005.319 |
| GOBP_BLO | 5 | 0.37209 | 0.006055 | 0.23818 | 0.059129 | 1005.489 |
| GOBP_NEG | 17 | 0.18684 | 0.005604 | 0.11961 | 0.059149 | 1005.829 |
| GOMF_VOI | 135 | 0.076745 | 0.006467 | 0.049133 | 0.059154 | 1005.914 |
| REACTOME | 16 | 0.2096 | 0.006099 | 0.13422 | 0.0592 | 1006.696 |
| LIU_TARGE | 15 | 0.21224 | 0.00598 | 0.13592 | 0.059204 | 1006.764 |
| GOMF_CEF | 18 | 0.15975 | 0.004931 | 0.1023 | 0.059207 | 1006.815 |
| GOMF_RET | 10 | 0.29045 | 0.006683 | 0.18604 | 0.05924 | 1007.376 |
| GOBP_SON | 13 | 0.18885 | 0.004954 | 0.12096 | 0.059242 | 1007.41 |
| GOBP_REG | 74 | 0.086573 | 0.00541 | 0.055455 | 0.059255 | 1007.631 |
| GOBP_COF | 6 | 0.31212 | 0.005563 | 0.20001 | 0.059332 | 1008.941 |
| XU_HGF_T/ | 8 | 0.28283 | 0.005821 | 0.18125 | 0.059338 | 1009.043 |
| GOBP_CAL | 187 | 0.060648 | 0.006006 | 0.038866 | 0.05934 | 1009.077 |
| LEE_CALOF | 45 | 0.11292 | 0.005507 | 0.072365 | 0.059342 | 1009.111 |
| GOMF_G_I | 38 | 0.11722 | 0.005254 | 0.075165 | 0.059449 | 1010.93 |
| GOMF_NEU | 6 | 0.26032 | 0.00464 | 0.16696 | 0.059488 | 1011.593 |
| GOBP_POS | 29 | 0.17276 | 0.006766 | 0.11083 | 0.05954 | 1012.478 |
| PEDRIOLI_I | 193 | 0.055551 | 0.005588 | 0.035639 | 0.059542 | 1012.512 |
| GOBP_ADE | 23 | 0.18928 | 0.006603 | 0.12146 | 0.059574 | 1013.056 |
| GOBP_REG | 17 | 0.19012 | 0.005703 | 0.122 | 0.059589 | 1013.311 |
| GOBP_REG | 77 | 0.092894 | 0.005921 | 0.059646 | 0.059693 | 1015.079 |
| GOMF_POI | 22 | 0.1742 | 0.005943 | 0.11187 | 0.059728 | 1015.675 |
| GOBP_POS | 9 | 0.28029 | 0.006119 | 0.18001 | 0.059732 | 1015.743 |
| REACTOME | 43 | 0.10966 | 0.005228 | 0.070429 | 0.059739 | 1015.862 |
| INAMURA_ | 12 | 0.25325 | 0.006383 | 0.16265 | 0.059746 | 1015.981 |
| GOBP_CAR | 10 | 0.30326 | 0.006978 | 0.19483 | 0.059798 | 1016.865 |
| GOMF_FRU | 6 | 0.35208 | 0.006276 | 0.22624 | 0.059835 | 1017.494 |
| DURCHDEV | 82 | 0.080363 | 0.005285 | 0.051646 | 0.059861 | 1017.936 |

| | | | | | | |
|---|---|---|---|---|---|---|
| GOBP_TEN | 5 | 0.41565 | 0.006764 | 0.26717 | 0.059893 | 1018.48 |
| GOBP_ISOI | 30 | 0.13389 | 0.005333 | 0.086071 | 0.059918 | 1018.906 |
| WP_HISTO | 46 | 0.10196 | 0.005027 | 0.065564 | 0.059959 | 1019.603 |
| TERAO_AO | 6 | 0.32982 | 0.005879 | 0.21208 | 0.059962 | 1019.654 |
| GOBP_DE_ | 6 | 0.37887 | 0.006753 | 0.24371 | 0.060033 | 1020.861 |
| REACTOME | 22 | 0.18536 | 0.006324 | 0.11924 | 0.060035 | 1020.895 |
| KUNINGER_ | 78 | 0.084812 | 0.00544 | 0.05458 | 0.060114 | 1022.239 |
| KEGG_BAS | 32 | 0.11949 | 0.004915 | 0.076909 | 0.060145 | 1022.766 |
| GOBP_NEG | 35 | 0.12916 | 0.005556 | 0.083135 | 0.060149 | 1022.834 |
| GOBP_NEG | 19 | 0.16989 | 0.005387 | 0.10939 | 0.060207 | 1023.82 |
| WP_EXTRA | 30 | 0.14892 | 0.005932 | 0.095912 | 0.060255 | 1024.636 |
| GOBP_HOI | 28 | 0.16963 | 0.006528 | 0.10927 | 0.060291 | 1025.248 |
| GOBP_POS | 30 | 0.13776 | 0.005487 | 0.088772 | 0.060363 | 1026.473 |
| REACTOME | 42 | 0.12082 | 0.005692 | 0.077864 | 0.060383 | 1026.813 |
| GOBP_POS | 12 | 0.20535 | 0.005176 | 0.13238 | 0.060436 | 1027.714 |
| GOCC_DNA | 4 | 0.39832 | 0.005797 | 0.2569 | 0.060522 | 1029.177 |
| BOWIE_RE | 17 | 0.19388 | 0.005815 | 0.12511 | 0.060621 | 1030.86 |
| BIOCARTA_ | 5 | 0.42458 | 0.006909 | 0.27416 | 0.060738 | 1032.85 |
| GOBP_FAT | 145 | 0.062999 | 0.0055 | 0.040693 | 0.060801 | 1033.921 |
| SNIJDERS_ | 37 | 0.12473 | 0.005517 | 0.080626 | 0.060936 | 1036.217 |
| GOMF_GLY | 5 | 0.36629 | 0.00596 | 0.23679 | 0.060951 | 1036.472 |
| GOCC_MEI | 1082 | 0.023737 | 0.005518 | 0.015349 | 0.060998 | 1037.271 |
| REACTOME | 38 | 0.14169 | 0.006351 | 0.091713 | 0.061195 | 1040.621 |
| GOBP_CER | 26 | 0.12968 | 0.004809 | 0.083947 | 0.06121 | 1040.876 |
| GOMF_ALD | 10 | 0.22027 | 0.005068 | 0.1426 | 0.061232 | 1041.25 |
| REACTOME | 18 | 0.1653 | 0.005102 | 0.10703 | 0.061256 | 1041.658 |
| GOBP_MIC | 32 | 0.13707 | 0.005638 | 0.088762 | 0.06128 | 1042.066 |
| GOMF_TRA | 46 | 0.10637 | 0.005244 | 0.068886 | 0.061294 | 1042.304 |
| GOBP_ISOI | 10 | 0.26542 | 0.006107 | 0.17189 | 0.061295 | 1042.321 |
| GOBP_NEG | 10 | 0.20825 | 0.004792 | 0.1349 | 0.061337 | 1043.036 |
| REACTOME | 84 | 0.082612 | 0.005498 | 0.053522 | 0.061365 | 1043.512 |
| XU_AKT1_ | 26 | 0.13269 | 0.004921 | 0.086022 | 0.061483 | 1045.518 |
| GOBP_REG | 22 | 0.16742 | 0.005712 | 0.10855 | 0.061499 | 1045.79 |
| AMIT_EGF_ | 43 | 0.13099 | 0.006245 | 0.084936 | 0.061516 | 1046.08 |
| KEGG_HOI | 27 | 0.14263 | 0.00539 | 0.09251 | 0.061567 | 1046.947 |
| GOBP_NEU | 12 | 0.23725 | 0.00598 | 0.15393 | 0.061636 | 1048.12 |
| BIOCARTA_ | 19 | 0.17374 | 0.005509 | 0.11278 | 0.061732 | 1049.753 |
| REACTOME | 109 | 0.070846 | 0.005368 | 0.045993 | 0.061744 | 1049.957 |
| GOBP_AMI | 10 | 0.27449 | 0.006316 | 0.17822 | 0.061765 | 1050.314 |
| MARTINELI | 9 | 0.25826 | 0.005638 | 0.16771 | 0.061793 | 1050.79 |
| SHIN_B_CE | 26 | 0.14529 | 0.005388 | 0.094386 | 0.061878 | 1052.235 |
| GOCC_COF | 58 | 0.11778 | 0.006518 | 0.076533 | 0.061918 | 1052.916 |
| WP_DEVEL | 56 | 0.11801 | 0.006418 | 0.076689 | 0.061938 | 1053.256 |
| GOMF_ANA | 9 | 0.22911 | 0.005001 | 0.14902 | 0.062102 | 1056.045 |
| GOCC_HET | 74 | 0.084835 | 0.005301 | 0.055181 | 0.062107 | 1056.13 |

| | | | | | | |
|---|---|---|---|---|---|---|
| REACTOME | 34 | 0.1226 | 0.005199 | 0.079751 | 0.062114 | 1056.249 |
| GOBP_REG | 1494 | 0.02086 | 0.005631 | 0.013572 | 0.06216 | 1057.031 |
| BONCI_TAI | 91 | 0.080015 | 0.005542 | 0.052061 | 0.062164 | 1057.099 |
| GOBP_ACT | 121 | 0.07462 | 0.005955 | 0.04856 | 0.062199 | 1057.694 |
| REACTOME | 7 | 0.31045 | 0.005977 | 0.20208 | 0.06225 | 1058.561 |
| GOBP_REG | 290 | 0.047415 | 0.005832 | 0.030869 | 0.062278 | 1059.037 |
| WP_CANN/ | 29 | 0.14495 | 0.005677 | 0.094383 | 0.062302 | 1059.446 |
| GOCC_MIT | 724 | 0.027318 | 0.005246 | 0.017789 | 0.062325 | 1059.837 |
| GOBP_MEI | 13 | 0.19547 | 0.005128 | 0.12732 | 0.06236 | 1060.432 |
| GOBP_ALTI | 66 | 0.089908 | 0.005307 | 0.058566 | 0.062381 | 1060.789 |
| GOBP_DIA( | 26 | 0.16487 | 0.006114 | 0.10741 | 0.062401 | 1061.129 |
| GOBP_NEG | 126 | 0.068799 | 0.005602 | 0.044821 | 0.062407 | 1061.231 |
| CROONQU | 17 | 0.17831 | 0.005348 | 0.11619 | 0.062448 | 1061.928 |
| GOBP_AST | 37 | 0.14198 | 0.006279 | 0.092549 | 0.062515 | 1063.068 |
| GOCC_MIC | 143 | 0.057844 | 0.005015 | 0.037707 | 0.062521 | 1063.17 |
| GOBP_COF | 19 | 0.17342 | 0.005499 | 0.11305 | 0.062527 | 1063.272 |
| REACTOME | 18 | 0.18417 | 0.005684 | 0.12007 | 0.062545 | 1063.578 |
| GOBP_VER | 6 | 0.32232 | 0.005745 | 0.21015 | 0.062552 | 1063.697 |
| REACTOME | 7 | 0.30445 | 0.005862 | 0.19851 | 0.062567 | 1063.952 |
| GOBP_REG | 109 | 0.077274 | 0.005855 | 0.050435 | 0.062751 | 1067.081 |
| GOMF_STA | 10 | 0.24588 | 0.005658 | 0.16054 | 0.06282 | 1068.254 |
| REACTOME | 17 | 0.20304 | 0.00609 | 0.13258 | 0.062838 | 1068.56 |
| GOBP_DEV | 874 | 0.026583 | 0.005586 | 0.017371 | 0.062975 | 1070.89 |
| LINDGREN_ | 356 | 0.039925 | 0.005431 | 0.026091 | 0.062988 | 1071.111 |
| MIKKELSEN | 37 | 0.13063 | 0.005777 | 0.085369 | 0.063 | 1071.315 |
| NIKOLSKY_ | 37 | 0.2011 | 0.008894 | 0.13148 | 0.063073 | 1072.556 |
| GOCC_FIBF | 11 | 0.27042 | 0.006526 | 0.17681 | 0.063083 | 1072.726 |
| GOBP_AUT | 10 | 0.22388 | 0.005151 | 0.14647 | 0.063209 | 1074.869 |
| GOMF_HIS | 2 | 0.36038 | 0.003709 | 0.23586 | 0.063273 | 1075.957 |
| GOBP_VAS | 25 | 0.15768 | 0.005734 | 0.1032 | 0.063274 | 1075.974 |
| FAELT_B_C | 44 | 0.11063 | 0.005335 | 0.072456 | 0.063409 | 1078.27 |
| GOBP_REG | 90 | 0.080389 | 0.005537 | 0.052671 | 0.063483 | 1079.528 |
| MULLIGHA | 119 | 0.069348 | 0.005489 | 0.045439 | 0.063493 | 1079.698 |
| HAN_SATB | 399 | 0.040339 | 0.005802 | 0.026435 | 0.063518 | 1080.124 |
| CHAUHAN_ | 89 | 0.076024 | 0.005208 | 0.049822 | 0.063526 | 1080.26 |
| GOMF_CYT | 5 | 0.37375 | 0.006082 | 0.24495 | 0.063534 | 1080.396 |
| GOBP_POS | 145 | 0.068133 | 0.005948 | 0.044667 | 0.063597 | 1081.467 |
| GOBP_SIGI | 37 | 0.13033 | 0.005764 | 0.085462 | 0.063644 | 1082.266 |
| WP_TRANS | 45 | 0.11535 | 0.005625 | 0.075674 | 0.063726 | 1083.661 |
| GOBP_MES | 6 | 0.3549 | 0.006326 | 0.23291 | 0.063795 | 1084.834 |
| GOCC_GLY | 7 | 0.31308 | 0.006028 | 0.20547 | 0.063798 | 1084.885 |
| GOBP_OST | 11 | 0.22323 | 0.005387 | 0.14652 | 0.06382 | 1085.259 |
| GOBP_POS | 8 | 0.26396 | 0.005433 | 0.17326 | 0.063831 | 1085.446 |
| GOBP_TEL( | 9 | 0.21398 | 0.004671 | 0.14047 | 0.06385 | 1085.769 |
| REACTOME | 19 | 0.18789 | 0.005958 | 0.12335 | 0.063856 | 1085.871 |

| | | | | | | |
|---|---|---|---|---|---|---|
| GOBP_REG | 12 | 0.24347 | 0.006137 | 0.15992 | 0.063961 | 1087.657 |
| GOBP_URE | 6 | 0.3337 | 0.005948 | 0.21928 | 0.064041 | 1089.017 |
| GOCC_END | 488 | 0.033995 | 0.005395 | 0.022346 | 0.064099 | 1090.003 |
| GOBP_EPID | 35 | 0.1359 | 0.005846 | 0.089346 | 0.064138 | 1090.667 |
| GOBP_BRA | 142 | 0.070258 | 0.00607 | 0.046199 | 0.064169 | 1091.194 |
| REACTOME | 353 | 0.039207 | 0.005311 | 0.025782 | 0.064172 | 1091.245 |
| SENESE_HD | 113 | 0.072768 | 0.005613 | 0.047869 | 0.064248 | 1092.537 |
| REACTOME | 34 | 0.12745 | 0.005404 | 0.083862 | 0.064302 | 1093.456 |
| PID_DNA_I | 15 | 0.1983 | 0.005588 | 0.1305 | 0.064319 | 1093.745 |
| GOBP_STEI | 300 | 0.046078 | 0.005762 | 0.030324 | 0.064328 | 1093.898 |
| GOBP_PER | 7 | 0.32855 | 0.006326 | 0.21629 | 0.064385 | 1094.867 |
| GOBP_ATR | 27 | 0.16182 | 0.006115 | 0.10657 | 0.064473 | 1096.363 |
| CHIANG_LI | 169 | 0.057799 | 0.005444 | 0.038078 | 0.064528 | 1097.299 |
| TSENG_IRS | 118 | 0.073166 | 0.005767 | 0.04823 | 0.064638 | 1099.169 |
| WP_SOMA | 13 | 0.20972 | 0.005502 | 0.13827 | 0.06467 | 1099.713 |
| VANDESLU | 35 | 0.1441 | 0.006199 | 0.09504 | 0.064738 | 1100.87 |
| GOBP_PAT | 442 | 0.039837 | 0.006024 | 0.026276 | 0.064755 | 1101.159 |
| GOMF_PR( | 10 | 0.26082 | 0.006001 | 0.17208 | 0.064807 | 1102.043 |
| WP_CELLT | 13 | 0.24782 | 0.006501 | 0.1636 | 0.064917 | 1103.914 |
| GOMF_MY | 71 | 0.089616 | 0.005486 | 0.059166 | 0.06494 | 1104.305 |
| SENESE_HD | 504 | 0.03467 | 0.005589 | 0.022893 | 0.064969 | 1104.798 |
| GOBP_NEG | 5 | 0.44915 | 0.007309 | 0.29667 | 0.065026 | 1105.767 |
| HANN_RES | 45 | 0.11094 | 0.00541 | 0.073277 | 0.065028 | 1105.801 |
| GOBP_POS | 13 | 0.24405 | 0.006402 | 0.16121 | 0.06504 | 1106.005 |
| WP_PANCI | 84 | 0.084933 | 0.005653 | 0.056109 | 0.065058 | 1106.311 |
| GOBP_NOT | 7 | 0.26554 | 0.005112 | 0.17544 | 0.065082 | 1106.719 |
| BIOCARTA_ | 23 | 0.15929 | 0.005557 | 0.10534 | 0.06526 | 1109.746 |
| GOBP_REG | 676 | 0.028981 | 0.005385 | 0.019172 | 0.065328 | 1110.903 |
| GOBP_C_5_ | 5 | 0.41141 | 0.006695 | 0.27219 | 0.065347 | 1111.226 |
| GOBP_DNA | 5 | 0.41141 | 0.006695 | 0.27219 | 0.065347 | 1111.226 |
| REACTOME | 29 | 0.14448 | 0.005659 | 0.095623 | 0.065407 | 1112.246 |
| AMIT_EGF_ | 36 | 0.13453 | 0.005869 | 0.089043 | 0.065425 | 1112.552 |
| HOLLEMAN | 12 | 0.18975 | 0.004783 | 0.1256 | 0.065442 | 1112.841 |
| GOBP_CELI | 52 | 0.10789 | 0.005655 | 0.071418 | 0.065447 | 1112.926 |
| TONKS_TAI | 196 | 0.053414 | 0.005414 | 0.035363 | 0.065471 | 1113.334 |
| GOBP_NEG | 7 | 0.22659 | 0.004362 | 0.15006 | 0.065531 | 1114.355 |
| MULLIGHA | 243 | 0.048554 | 0.005473 | 0.032156 | 0.065536 | 1114.44 |
| GOBP_REG | 40 | 0.13591 | 0.006249 | 0.090015 | 0.065553 | 1114.729 |
| GOMF_CR( | 7 | 0.23565 | 0.004537 | 0.15611 | 0.06559 | 1115.358 |
| GOMF_HEI | 14 | 0.24097 | 0.00656 | 0.15965 | 0.065616 | 1115.8 |
| GOMF_PR( | 8 | 0.29627 | 0.006098 | 0.1963 | 0.065631 | 1116.055 |
| REACTOME | 20 | 0.17291 | 0.005625 | 0.1146 | 0.065688 | 1117.024 |
| GOCC_HIST | 6 | 0.50369 | 0.008978 | 0.33394 | 0.065746 | 1118.011 |
| GOMF_MA | 15 | 0.19011 | 0.005357 | 0.12606 | 0.065773 | 1118.47 |
| GROSS_HY | 149 | 0.064835 | 0.005737 | 0.042996 | 0.065794 | 1118.827 |

| | | | | | | |
|---|---|---|---|---|---|---|
| GOBP_HEN | 215 | 0.054442 | 0.005777 | 0.036105 | 0.065804 | 1118.997 |
| REACTOME | 11 | 0.21199 | 0.005116 | 0.14059 | 0.06581 | 1119.099 |
| GOCC_MPI | 5 | 0.40972 | 0.006667 | 0.27174 | 0.065818 | 1119.235 |
| REACTOME | 110 | 0.06632 | 0.005048 | 0.043993 | 0.06585 | 1119.779 |
| GOBP_POS | 32 | 0.14107 | 0.005803 | 0.093583 | 0.065856 | 1119.881 |
| GROSS_HY | 132 | 0.068859 | 0.005738 | 0.045692 | 0.065914 | 1120.868 |
| VANASSE_I | 57 | 0.091588 | 0.005025 | 0.060787 | 0.065954 | 1121.548 |
| GOBP_LIPC | 5 | 0.30634 | 0.004985 | 0.20338 | 0.066007 | 1122.449 |
| WP_AEROE | 11 | 0.24765 | 0.005976 | 0.16442 | 0.066011 | 1122.517 |
| GOBP_POS | 27 | 0.15211 | 0.005748 | 0.10101 | 0.06607 | 1123.52 |
| GOMF_TYF | 11 | 0.24424 | 0.005894 | 0.16223 | 0.066108 | 1124.167 |
| GOBP_MO | 19 | 0.18051 | 0.005724 | 0.1199 | 0.066115 | 1124.286 |
| GARGALOV | 26 | 0.15749 | 0.005841 | 0.10464 | 0.066157 | 1125 |
| GOBP_CAR | 143 | 0.064362 | 0.00558 | 0.042763 | 0.066163 | 1125.102 |
| RICKMAN_ | 36 | 0.14394 | 0.00628 | 0.095666 | 0.066225 | 1126.156 |
| MARSHALL | 28 | 0.13443 | 0.005173 | 0.089353 | 0.066237 | 1126.36 |
| GOBP_RET | 90 | 0.073767 | 0.005081 | 0.049035 | 0.066253 | 1126.632 |
| PID_TELON | 65 | 0.095233 | 0.005579 | 0.063329 | 0.066329 | 1127.925 |
| GOCC_TRIC | 18 | 0.19801 | 0.006111 | 0.13169 | 0.066352 | 1128.316 |
| GOBP_REG | 504 | 0.034674 | 0.005589 | 0.023061 | 0.066358 | 1128.418 |
| GOCC_MU | 28 | 0.13397 | 0.005156 | 0.089115 | 0.066388 | 1128.928 |
| GOBP_SEN | 553 | 0.034519 | 0.005821 | 0.022977 | 0.066511 | 1131.02 |
| GOBP_SPEI | 5 | 0.33627 | 0.005472 | 0.22383 | 0.066511 | 1131.02 |
| GOBP_BON | 12 | 0.23046 | 0.005809 | 0.15349 | 0.066629 | 1133.026 |
| GOBP_NEG | 6 | 0.28472 | 0.005075 | 0.18967 | 0.066664 | 1133.621 |
| YAO_TEMF | 14 | 0.17376 | 0.00473 | 0.11578 | 0.066716 | 1134.506 |
| GOMF_DEI | 8 | 0.30296 | 0.006235 | 0.20189 | 0.066736 | 1134.846 |
| GOCC_PLA | 110 | 0.076148 | 0.005796 | 0.050747 | 0.066746 | 1135.016 |
| GOBP_SEC | 25 | 0.16599 | 0.006036 | 0.11065 | 0.066798 | 1135.9 |
| WP_FACTC | 36 | 0.1381 | 0.006025 | 0.092061 | 0.066809 | 1136.087 |
| GOBP_RES | 53 | 0.11393 | 0.006028 | 0.075956 | 0.066817 | 1136.223 |
| GOBP_FAT | 12 | 0.21444 | 0.005405 | 0.14298 | 0.066837 | 1136.563 |
| ACEVEDO_ | 893 | 0.025094 | 0.005327 | 0.016732 | 0.066847 | 1136.733 |
| LI_WILMS_ | 46 | 0.12058 | 0.005945 | 0.08044 | 0.06695 | 1138.485 |
| REACTOME | 25 | 0.1388 | 0.005048 | 0.092615 | 0.066982 | 1139.029 |
| GOCC_RDN | 4 | 0.31802 | 0.004629 | 0.21221 | 0.067002 | 1139.369 |
| REACTOME | 26 | 0.21328 | 0.00791 | 0.14236 | 0.067062 | 1140.389 |
| MIKKELSEN | 119 | 0.072194 | 0.005714 | 0.048191 | 0.067064 | 1140.423 |
| GOBP_HEP | 14 | 0.23899 | 0.006506 | 0.15954 | 0.067085 | 1140.78 |
| REACTOME | 150 | 0.058702 | 0.005212 | 0.03922 | 0.067242 | 1143.45 |
| GOBP_REG | 27 | 0.14186 | 0.005361 | 0.094794 | 0.067272 | 1143.96 |
| DAVICIONI_ | 170 | 0.060965 | 0.005759 | 0.040744 | 0.067297 | 1144.385 |
| GOBP_REG | 37 | 0.14114 | 0.006243 | 0.094367 | 0.067376 | 1145.729 |
| GOBP_REG | 130 | 0.068778 | 0.005688 | 0.045997 | 0.067432 | 1146.681 |
| REACTOME | 32 | 0.1398 | 0.005751 | 0.093574 | 0.067601 | 1149.555 |

| | | | | | | |
|---|---|---|---|---|---|---|
| GOCC_JUN | 6 | 0.32125 | 0.005726 | 0.2151 | 0.067665 | 1150.643 |
| GOCC_RUF | 99 | 0.073109 | 0.00528 | 0.048958 | 0.06769 | 1151.068 |
| GOCC_DEN | 7 | 0.2895 | 0.005574 | 0.1939 | 0.067721 | 1151.596 |
| GOBP_REG | 36 | 0.11614 | 0.005067 | 0.077789 | 0.067726 | 1151.681 |
| GOBP_POS | 36 | 0.14341 | 0.006257 | 0.096081 | 0.067776 | 1152.531 |
| GOCC_CHR | 150 | 0.056802 | 0.005043 | 0.038064 | 0.067819 | 1153.262 |
| CAFFAREL_ | 10 | 0.23826 | 0.005482 | 0.1597 | 0.067875 | 1154.214 |
| GOBP_MIT | 34 | 0.1073 | 0.00455 | 0.071934 | 0.067898 | 1154.605 |
| GOBP_REG | 41 | 0.13174 | 0.006133 | 0.088339 | 0.067954 | 1155.558 |
| GOCC_PRP | 13 | 0.20857 | 0.005471 | 0.13988 | 0.067979 | 1155.983 |
| GOCC_SPEI | 5 | 0.31375 | 0.005106 | 0.21045 | 0.068007 | 1156.459 |
| GOBP_IN_U | 380 | 0.037625 | 0.005284 | 0.025239 | 0.068027 | 1156.799 |
| GOMF_MA | 13 | 0.20811 | 0.005459 | 0.13961 | 0.068041 | 1157.037 |
| REACTOME | 25 | 0.12866 | 0.004679 | 0.086329 | 0.068078 | 1157.666 |
| GOBP_MEI | 69 | 0.10321 | 0.006229 | 0.069261 | 0.068095 | 1157.955 |
| GOBP_IMM | 5 | 0.31799 | 0.005174 | 0.21339 | 0.068097 | 1157.989 |
| GOBP_COF | 12 | 0.23528 | 0.00593 | 0.15795 | 0.068172 | 1159.265 |
| TCGA_GLIC | 8 | 0.27537 | 0.005668 | 0.1849 | 0.068212 | 1159.945 |
| GOBP_CAR | 20 | 0.17959 | 0.005842 | 0.12059 | 0.068225 | 1160.166 |
| GOBP_ADU | 12 | 0.23092 | 0.00582 | 0.15506 | 0.068227 | 1160.2 |
| GOMF_PRO | 5 | 0.36741 | 0.005979 | 0.24675 | 0.068255 | 1160.676 |
| KOKKINAKI | 68 | 0.087593 | 0.005248 | 0.05885 | 0.06833 | 1161.952 |
| GOBP_POS | 1050 | 0.023765 | 0.005447 | 0.015976 | 0.068435 | 1163.737 |
| APPIERTO_ | 49 | 0.10568 | 0.005377 | 0.071053 | 0.068465 | 1164.247 |
| GOBP_DEV | 9 | 0.26926 | 0.005878 | 0.18108 | 0.068525 | 1165.268 |
| GOBP_LIM | 107 | 0.078065 | 0.005861 | 0.052527 | 0.068624 | 1166.951 |
| GOBP_STEI | 119 | 0.074444 | 0.005892 | 0.050094 | 0.068637 | 1167.172 |
| MCBRYAN_ | 35 | 0.13183 | 0.005671 | 0.088721 | 0.068662 | 1167.597 |
| GOBP_CAR | 18 | 0.19129 | 0.005904 | 0.12875 | 0.068682 | 1167.937 |
| GOBP_BAS | 41 | 0.10099 | 0.004701 | 0.067987 | 0.068728 | 1168.72 |
| FISCHER_G | 185 | 0.05449 | 0.005368 | 0.036692 | 0.068773 | 1169.485 |
| GOMF_AM | 5 | 0.34858 | 0.005672 | 0.23472 | 0.068773 | 1169.485 |
| GOMF_STF | 5 | 0.3127 | 0.005088 | 0.21061 | 0.068816 | 1170.216 |
| ACEVEDO_ | 261 | 0.042849 | 0.005003 | 0.028903 | 0.069111 | 1175.233 |
| GOBP_NEG | 23 | 0.14006 | 0.004886 | 0.09453 | 0.069234 | 1177.324 |
| GOBP_NEG | 45 | 0.10561 | 0.00515 | 0.071292 | 0.069267 | 1177.885 |
| GOBP_REG | 19 | 0.14698 | 0.004661 | 0.099226 | 0.069272 | 1177.97 |
| GOBP_REG | 30 | 0.15102 | 0.006016 | 0.10197 | 0.069301 | 1178.464 |
| PETRETTO_ | 26 | 0.15405 | 0.005713 | 0.1041 | 0.069467 | 1181.286 |
| GOCC_SPIN | 394 | 0.036754 | 0.005254 | 0.024841 | 0.069502 | 1181.882 |
| GOMF_MR | 15 | 0.17549 | 0.004945 | 0.11861 | 0.069508 | 1181.984 |
| GOBP_REN | 30 | 0.16298 | 0.006492 | 0.11018 | 0.069543 | 1182.579 |
| WP_OVARI | 35 | 0.13201 | 0.005679 | 0.089246 | 0.06955 | 1182.698 |
| GOBP_REG | 193 | 0.053732 | 0.005405 | 0.036348 | 0.069678 | 1184.874 |
| REACTOME | 100 | 0.07231 | 0.005249 | 0.048926 | 0.069719 | 1185.572 |

| | | | | | | |
|---|---|---|---|---|---|---|
| GOMF_OX | 11 | 0.2321 | 0.005601 | 0.15711 | 0.069812 | 1187.153 |
| GOMF_PR | 40 | 0.11222 | 0.00516 | 0.076052 | 0.070039 | 1191.013 |
| AMIT_SER | 63 | 0.09497 | 0.005477 | 0.064366 | 0.070053 | 1191.251 |
| GOBP_LIM | 11 | 0.26124 | 0.006304 | 0.17707 | 0.070068 | 1191.506 |
| GOBP_TRA | 57 | 0.094983 | 0.005211 | 0.064382 | 0.070075 | 1191.625 |
| WP_LEUK | 13 | 0.1926 | 0.005052 | 0.13055 | 0.070081 | 1191.727 |
| UROSEVIC_ | 18 | 0.15862 | 0.004896 | 0.10758 | 0.070196 | 1193.683 |
| GOMF_AC | 34 | 0.13155 | 0.005578 | 0.089233 | 0.070222 | 1194.125 |
| GOBP_B_C | 6 | 0.3064 | 0.005462 | 0.20793 | 0.070308 | 1195.588 |
| GOMF_AX | 10 | 0.23463 | 0.005399 | 0.15925 | 0.070331 | 1195.979 |
| WP_PHYSI | 63 | 0.094079 | 0.005426 | 0.063852 | 0.070333 | 1196.013 |
| KAAB_FAIL | 136 | 0.061179 | 0.005174 | 0.041531 | 0.070374 | 1196.71 |
| GOBP_POS | 5 | 0.32262 | 0.00525 | 0.21904 | 0.070402 | 1197.186 |
| BIOCARTA_ | 9 | 0.24294 | 0.005303 | 0.16499 | 0.070453 | 1198.053 |
| GOBP_PRO | 302 | 0.040254 | 0.005051 | 0.02735 | 0.070545 | 1199.618 |
| GOBP_GM | 25 | 0.14104 | 0.005129 | 0.09584 | 0.07057 | 1200.043 |
| WP_VASO | 43 | 0.11396 | 0.005433 | 0.077476 | 0.070666 | 1201.675 |
| GOBP_REG | 89 | 0.079853 | 0.00547 | 0.0543 | 0.07071 | 1202.424 |
| BIOCARTA_ | 23 | 0.15514 | 0.005412 | 0.10549 | 0.070712 | 1202.458 |
| GOBP_NEG | 5 | 0.30895 | 0.005027 | 0.21018 | 0.070801 | 1203.971 |
| GOBP_SAR | 34 | 0.12606 | 0.005345 | 0.085766 | 0.070816 | 1204.226 |
| REACTOME | 14 | 0.20354 | 0.005541 | 0.13849 | 0.070827 | 1204.413 |
| GOBP_CEL | 27 | 0.13665 | 0.005164 | 0.092988 | 0.070857 | 1204.923 |
| GOBP_NEG | 24 | 0.15456 | 0.005508 | 0.10518 | 0.070861 | 1204.991 |
| XU_HGF_S | 19 | 0.15743 | 0.004992 | 0.10716 | 0.070916 | 1205.927 |
| GOBP_SM | 67 | 0.10269 | 0.006107 | 0.069911 | 0.070945 | 1206.42 |
| GOBP_L_S | 5 | 0.28618 | 0.004657 | 0.19485 | 0.070968 | 1206.811 |
| GOBP_ADE | 8 | 0.28064 | 0.005776 | 0.1911 | 0.070981 | 1207.032 |
| BURTON_A | 42 | 0.10901 | 0.005136 | 0.074249 | 0.07103 | 1207.865 |
| FERREIRA_ | 160 | 0.057087 | 0.005233 | 0.038882 | 0.071032 | 1207.899 |
| BOYLAN_M | 77 | 0.092481 | 0.005894 | 0.062999 | 0.071064 | 1208.443 |
| GOBP_REG | 8 | 0.25436 | 0.005235 | 0.17333 | 0.071133 | 1209.617 |
| GOBP_THI | 6 | 0.29791 | 0.00531 | 0.20303 | 0.071158 | 1210.042 |
| GOMF_MA | 21 | 0.15591 | 0.005197 | 0.10626 | 0.071159 | 1210.059 |
| GOMF_HIS | 6 | 0.37374 | 0.006662 | 0.25486 | 0.071269 | 1211.929 |
| GOBP_EM | 27 | 0.16867 | 0.006374 | 0.11508 | 0.071371 | 1213.664 |
| GOMF_NU | 15 | 0.20264 | 0.00571 | 0.13825 | 0.071379 | 1213.8 |
| GOBP_MIT | 15 | 0.20366 | 0.005738 | 0.13898 | 0.071414 | 1214.395 |
| GOMF_PH | 28 | 0.12023 | 0.004627 | 0.082045 | 0.071418 | 1214.463 |
| VERRECCH | 5 | 0.34682 | 0.005644 | 0.23684 | 0.071552 | 1216.742 |
| GOBP_MIT | 31 | 0.12289 | 0.004976 | 0.083928 | 0.071571 | 1217.065 |
| BLANCO_M | 584 | 0.031026 | 0.005372 | 0.021195 | 0.071625 | 1217.983 |
| BIOCARTA_ | 10 | 0.26106 | 0.006007 | 0.17843 | 0.071729 | 1219.752 |
| FIGUEROA_ | 112 | 0.071841 | 0.005517 | 0.049108 | 0.071755 | 1220.194 |
| AMIT_EGF_ | 10 | 0.26617 | 0.006124 | 0.18206 | 0.071886 | 1222.421 |

| | | | | | | |
|---|---|---|---|---|---|---|
| GOBP_POS | 5 | 0.32966 | 0.005364 | 0.2255 | 0.07189 | 1222.489 |
| GOBP_ADE | 7 | 0.30254 | 0.005825 | 0.20697 | 0.071908 | 1222.796 |
| GOMF_DEA | 12 | 0.24598 | 0.0062 | 0.1683 | 0.071941 | 1223.357 |
| GOBP_REG | 80 | 0.085261 | 0.005539 | 0.05835 | 0.071989 | 1224.173 |
| GOBP_SOD | 10 | 0.22591 | 0.005198 | 0.15463 | 0.072027 | 1224.819 |
| REACTOME | 7 | 0.25292 | 0.004869 | 0.17315 | 0.072063 | 1225.431 |
| GOBP_PER | 14 | 0.20772 | 0.005655 | 0.14224 | 0.072104 | 1226.129 |
| REACTOME | 33 | 0.12845 | 0.005366 | 0.087978 | 0.07215 | 1226.911 |
| GOMF_ME | 10 | 0.26572 | 0.006114 | 0.18207 | 0.072228 | 1228.237 |
| GOBP_RET | 6 | 0.33079 | 0.005896 | 0.22668 | 0.072254 | 1228.679 |
| REACTOME | 15 | 0.20616 | 0.005809 | 0.14128 | 0.072265 | 1228.866 |
| GOBP_RES | 7 | 0.29051 | 0.005593 | 0.19911 | 0.072287 | 1229.24 |
| FIGUEROA_ | 101 | 0.072585 | 0.005295 | 0.049763 | 0.072346 | 1230.244 |
| REACTOME | 20 | 0.16316 | 0.005308 | 0.11186 | 0.072347 | 1230.261 |
| YANAGIHA | 24 | 0.15834 | 0.005642 | 0.10858 | 0.072394 | 1231.06 |
| DOANE_BR | 64 | 0.095759 | 0.005566 | 0.065685 | 0.072448 | 1231.978 |
| GOBP_NEG | 38 | 0.12578 | 0.005637 | 0.086282 | 0.072466 | 1232.284 |
| GOBP_POS | 14 | 0.18417 | 0.005014 | 0.12639 | 0.072547 | 1233.662 |
| GOBP_MEI | 7 | 0.32296 | 0.006218 | 0.22168 | 0.072585 | 1234.308 |
| REACTOME | 47 | 0.10811 | 0.005388 | 0.074252 | 0.072712 | 1236.468 |
| GOBP_REG | 57 | 0.090791 | 0.004981 | 0.06238 | 0.072782 | 1237.658 |
| KYNG_WEI | 19 | 0.17483 | 0.005544 | 0.12013 | 0.072801 | 1237.981 |
| GOMF_FIB | 23 | 0.16674 | 0.005817 | 0.11464 | 0.072915 | 1239.92 |
| PID_RET_P | 39 | 0.12519 | 0.005684 | 0.086088 | 0.072953 | 1240.566 |
| GOBP_RES | 117 | 0.066841 | 0.005246 | 0.045986 | 0.07305 | 1242.215 |
| GARGALOV | 60 | 0.093434 | 0.005259 | 0.064336 | 0.073222 | 1245.14 |
| GOCC_CILI | 156 | 0.057934 | 0.005245 | 0.039892 | 0.073224 | 1245.174 |
| GOMF_DN | 20 | 0.14243 | 0.004634 | 0.098159 | 0.073392 | 1248.031 |
| GOBP_EMI | 37 | 0.12481 | 0.00552 | 0.086062 | 0.073506 | 1249.97 |
| LI_AMPLIFI | 172 | 0.053594 | 0.005092 | 0.036957 | 0.073515 | 1250.123 |
| RODRIGUE | 616 | 0.029724 | 0.005281 | 0.020497 | 0.073524 | 1250.276 |
| GOBP_THY | 20 | 0.18279 | 0.005946 | 0.12613 | 0.073652 | 1252.452 |
| GOBP_NUC | 37 | 0.12212 | 0.005401 | 0.084279 | 0.073679 | 1252.911 |
| WP_MRNA | 118 | 0.064138 | 0.005055 | 0.0443 | 0.073844 | 1255.717 |
| WP_DNA_I | 77 | 0.078344 | 0.004993 | 0.054113 | 0.07385 | 1255.819 |
| GOBP_REG | 95 | 0.070895 | 0.005017 | 0.048973 | 0.07387 | 1256.159 |
| GOBP_DET | 5 | 0.33246 | 0.00541 | 0.22971 | 0.073921 | 1257.027 |
| GOBP_MIT | 10 | 0.19594 | 0.004508 | 0.13539 | 0.073931 | 1257.197 |
| GRAESSMA | 201 | 0.051531 | 0.005289 | 0.035627 | 0.074041 | 1259.067 |
| GOBP_NEG | 22 | 0.16256 | 0.005546 | 0.1124 | 0.074052 | 1259.254 |
| REACTOME | 5 | 0.34883 | 0.005676 | 0.24126 | 0.074119 | 1260.394 |
| GOMF_DEA | 8 | 0.28902 | 0.005948 | 0.19992 | 0.074146 | 1260.853 |
| REACTOME | 49 | 0.11223 | 0.00571 | 0.077633 | 0.074151 | 1260.938 |
| GOBP_REG | 42 | 0.12226 | 0.005761 | 0.0846 | 0.07421 | 1261.941 |
| GOMF_TRI | 8 | 0.24041 | 0.004948 | 0.16635 | 0.074217 | 1262.06 |

| | | | | | | |
|---|---|---|---|---|---|---|
| GOMF_QU | 49 | 0.0956 | 0.004864 | 0.066152 | 0.074218 | 1262.077 |
| SWEET_KR. | 83 | 0.076711 | 0.005075 | 0.053092 | 0.074255 | 1262.706 |
| GOBP_SON | 11 | 0.22774 | 0.005496 | 0.15764 | 0.074279 | 1263.114 |
| GOBP_MIC | 7 | 0.29336 | 0.005648 | 0.20315 | 0.074373 | 1264.713 |
| GOBP_REG | 23 | 0.16218 | 0.005658 | 0.11231 | 0.074377 | 1264.781 |
| GOCC_PRC | 17 | 0.16431 | 0.004929 | 0.1138 | 0.0744 | 1265.172 |
| GOBP_PRC | 22 | 0.14334 | 0.004891 | 0.099291 | 0.074426 | 1265.614 |
| PID_ERBB1 | 35 | 0.11515 | 0.004954 | 0.079776 | 0.074455 | 1266.107 |
| LEE_METAS | 66 | 0.091853 | 0.005422 | 0.063668 | 0.074565 | 1267.978 |
| PID_P53_R | 57 | 0.092577 | 0.005079 | 0.064195 | 0.074644 | 1269.321 |
| GOBP_CELI | 17 | 0.19389 | 0.005816 | 0.13446 | 0.074663 | 1269.644 |
| GOCC_MIC | 5 | 0.34501 | 0.005614 | 0.23941 | 0.074789 | 1271.787 |
| SHIPP_DLB | 44 | 0.10214 | 0.004926 | 0.070911 | 0.074882 | 1273.368 |
| GOBP_NEG | 105 | 0.075357 | 0.005604 | 0.052321 | 0.074902 | 1273.709 |
| MIYAGAW/ | 218 | 0.053397 | 0.005705 | 0.037074 | 0.074904 | 1273.743 |
| GOBP_FIBF | 75 | 0.088485 | 0.005566 | 0.061477 | 0.075039 | 1276.038 |
| KEGG_PAT | 313 | 0.042237 | 0.005393 | 0.029346 | 0.075042 | 1276.089 |
| AMIT_EGF_ | 41 | 0.11979 | 0.005577 | 0.083259 | 0.075114 | 1277.314 |
| WP_VITAN | 7 | 0.26604 | 0.005122 | 0.18491 | 0.075119 | 1277.399 |
| WP_IL2_SI | 41 | 0.12461 | 0.005801 | 0.086631 | 0.075169 | 1278.249 |
| WP_ETHER | 20 | 0.14188 | 0.004616 | 0.098666 | 0.075234 | 1279.354 |
| NIKOLSKY_ | 5 | 0.56269 | 0.009156 | 0.39132 | 0.075237 | 1279.405 |
| GOBP_CHC | 5 | 0.26946 | 0.004385 | 0.1874 | 0.075241 | 1279.473 |
| GOCC_CYT | 34 | 0.10744 | 0.004555 | 0.074723 | 0.075257 | 1279.745 |
| BIOCARTA_ | 22 | 0.15676 | 0.005348 | 0.10905 | 0.075303 | 1280.528 |
| GOBP_ADF | 28 | 0.15906 | 0.006121 | 0.11066 | 0.075319 | 1280.8 |
| GOBP_DET | 13 | 0.18821 | 0.004937 | 0.13094 | 0.07532 | 1280.817 |
| GOBP_NEG | 5 | 0.28105 | 0.004573 | 0.19557 | 0.075357 | 1281.446 |
| GOBP_PRC | 26 | 0.16992 | 0.006302 | 0.11825 | 0.075369 | 1281.65 |
| WP_TYROE | 56 | 0.094001 | 0.005112 | 0.065419 | 0.075382 | 1281.871 |
| GOBP_FUS | 6 | 0.37202 | 0.006631 | 0.25892 | 0.075393 | 1282.058 |
| WP_ERK_P | 14 | 0.20185 | 0.005495 | 0.1405 | 0.075421 | 1282.534 |
| GOBP_PEX | 6 | 0.25137 | 0.004481 | 0.17499 | 0.075446 | 1282.959 |
| MARIADAS | 37 | 0.11585 | 0.005124 | 0.080659 | 0.075463 | 1283.248 |
| GOCC_SER | 5 | 0.37864 | 0.006161 | 0.26372 | 0.075545 | 1284.643 |
| GOMF_SEF | 5 | 0.37864 | 0.006161 | 0.26372 | 0.075545 | 1284.643 |
| GOBP_NEG | 32 | 0.12261 | 0.005044 | 0.085442 | 0.075655 | 1286.513 |
| GOMF_MR | 24 | 0.13135 | 0.00468 | 0.091553 | 0.075702 | 1287.313 |
| WP_TGIF_I | 9 | 0.27046 | 0.005904 | 0.18859 | 0.075775 | 1288.554 |
| GOBP_POS | 18 | 0.19313 | 0.005961 | 0.13468 | 0.07579 | 1288.809 |
| GOCC_FIBF | 8 | 0.28105 | 0.005784 | 0.19599 | 0.075791 | 1288.826 |
| GOBP_REG | 39 | 0.12483 | 0.005668 | 0.087058 | 0.075818 | 1289.285 |
| GAUTSCHI_ | 8 | 0.27038 | 0.005565 | 0.1886 | 0.075855 | 1289.914 |
| REACTOME | 1075 | 0.024048 | 0.005573 | 0.016777 | 0.075889 | 1290.492 |
| GOBP_MET | 6 | 0.33213 | 0.00592 | 0.23175 | 0.075913 | 1290.901 |

| | | | | | | |
|---|---|---|---|---|---|---|
| GOBP_SEN | 382 | 0.039374 | 0.005544 | 0.027492 | 0.076055 | 1293.315 |
| GOMF_INS | 5 | 0.35955 | 0.005851 | 0.25121 | 0.076184 | 1295.509 |
| PID_KIT_PA | 52 | 0.10322 | 0.00541 | 0.072132 | 0.076218 | 1296.087 |
| MATZUK_F | 13 | 0.23255 | 0.0061 | 0.16269 | 0.076454 | 1300.1 |
| GOCC_POS | 5 | 0.27765 | 0.004518 | 0.19427 | 0.076479 | 1300.525 |
| REACTOME | 31 | 0.11715 | 0.004743 | 0.081972 | 0.076495 | 1300.797 |
| GOBP_CELI | 13 | 0.22918 | 0.006012 | 0.16038 | 0.076505 | 1300.968 |
| TAKADA_G | 29 | 0.15093 | 0.005911 | 0.10563 | 0.076525 | 1301.308 |
| BIOCARTA_ | 13 | 0.18827 | 0.004939 | 0.13177 | 0.076552 | 1301.767 |
| GOBP_VIRA | 120 | 0.067065 | 0.00533 | 0.046952 | 0.076604 | 1302.651 |
| GOBP_ACT | 23 | 0.17138 | 0.005978 | 0.12001 | 0.076648 | 1303.399 |
| SIMBULAN_ | 16 | 0.16057 | 0.004673 | 0.11244 | 0.076656 | 1303.535 |
| GOBP_ORG | 1605 | 0.018022 | 0.005027 | 0.012621 | 0.076657 | 1303.552 |
| GOMF_NEU | 5 | 0.31016 | 0.005047 | 0.21721 | 0.076667 | 1303.722 |
| GOCC_SER | 8 | 0.31337 | 0.00645 | 0.2195 | 0.076704 | 1304.352 |
| GOBP_POS | 8 | 0.27737 | 0.005709 | 0.19431 | 0.076736 | 1304.896 |
| GOBP_OVI | 4 | 0.43597 | 0.006346 | 0.30545 | 0.076755 | 1305.219 |
| GOBP_NEG | 56 | 0.086835 | 0.004723 | 0.060865 | 0.076846 | 1306.766 |
| GOBP_FOR | 6 | 0.32673 | 0.005824 | 0.22929 | 0.077093 | 1310.966 |
| DURCHDEV | 253 | 0.04605 | 0.005295 | 0.032319 | 0.07711 | 1311.256 |
| GOBP_LEPT | 10 | 0.25645 | 0.005901 | 0.17999 | 0.077122 | 1311.46 |
| GOBP_REG | 51 | 0.097169 | 0.005044 | 0.0682 | 0.077123 | 1311.477 |
| GOMF_MC | 14 | 0.18377 | 0.005003 | 0.12903 | 0.077207 | 1312.905 |
| MORI_IMM | 86 | 0.070854 | 0.004771 | 0.049761 | 0.07725 | 1313.636 |
| BERENJENG | 514 | 0.03039 | 0.004946 | 0.021343 | 0.077251 | 1313.653 |
| GOBP_NEG | 5 | 0.25937 | 0.004221 | 0.18222 | 0.077323 | 1314.878 |
| GOBP_POS | 5 | 0.2737 | 0.004454 | 0.1923 | 0.077333 | 1315.048 |
| GOBP_NEG | 29 | 0.13644 | 0.005344 | 0.095865 | 0.077339 | 1315.15 |
| GOBP_CELI | 515 | 0.031251 | 0.005091 | 0.021959 | 0.077356 | 1315.439 |
| GOBP_POS | 8 | 0.26836 | 0.005523 | 0.18863 | 0.077431 | 1316.714 |
| REACTOME | 14 | 0.15865 | 0.004319 | 0.11152 | 0.077436 | 1316.799 |
| GOBP_NEG | 220 | 0.047168 | 0.005062 | 0.033159 | 0.077454 | 1317.105 |
| GOBP_RES | 150 | 0.057596 | 0.005114 | 0.040497 | 0.07749 | 1317.717 |
| HASLINGEF | 23 | 0.14518 | 0.005064 | 0.10209 | 0.07752 | 1318.228 |
| GOBP_MO | 3 | 0.4196 | 0.005289 | 0.29508 | 0.077521 | 1318.245 |
| GOBP_END | 45 | 0.10836 | 0.005284 | 0.076217 | 0.077558 | 1318.874 |
| REACTOME | 110 | 0.066541 | 0.005065 | 0.046812 | 0.077601 | 1319.605 |
| CAIRO_HEF | 195 | 0.052263 | 0.005284 | 0.036784 | 0.077695 | 1321.203 |
| PID_AUROI | 38 | 0.10818 | 0.004849 | 0.076166 | 0.077766 | 1322.411 |
| GOBP_REG | 7 | 0.27718 | 0.005336 | 0.19519 | 0.077809 | 1323.142 |
| GOBP_RIG_ | 25 | 0.12962 | 0.004714 | 0.091327 | 0.07791 | 1324.86 |
| GOBP_KER | 19 | 0.15516 | 0.00492 | 0.10938 | 0.078021 | 1326.747 |
| VILIMAS_N | 49 | 0.1101 | 0.005602 | 0.077619 | 0.078029 | 1326.883 |
| GOBP_NEG | 22 | 0.16303 | 0.005562 | 0.11494 | 0.078056 | 1327.342 |
| REACTOME | 26 | 0.13362 | 0.004956 | 0.094234 | 0.078108 | 1328.227 |

| | | | | | | |
|---|---|---|---|---|---|---|
| REACTOME | 72 | 0.093368 | 0.005755 | 0.065847 | 0.078113 | 1328.312 |
| GOBP_NEU | 7 | 0.27896 | 0.005371 | 0.19681 | 0.078198 | 1329.757 |
| THUM_MIF | 17 | 0.1922 | 0.005765 | 0.13562 | 0.078228 | 1330.267 |
| REACTOME | 16 | 0.16982 | 0.004942 | 0.1199 | 0.078346 | 1332.274 |
| REACTOME | 8 | 0.22305 | 0.004591 | 0.15751 | 0.078386 | 1332.954 |
| GOMF_NU | 40 | 0.10723 | 0.004931 | 0.075725 | 0.078389 | 1333.005 |
| WP_TRANS | 21 | 0.16561 | 0.00552 | 0.11695 | 0.078392 | 1333.056 |
| GOCC_MLL | 9 | 0.25195 | 0.0055 | 0.178 | 0.078481 | 1334.569 |
| GOBP_NEG | 5 | 0.39555 | 0.006437 | 0.27953 | 0.078538 | 1335.539 |
| GOBP_POS | 8 | 0.26899 | 0.005536 | 0.19012 | 0.078564 | 1335.981 |
| GOBP_MIC | 14 | 0.18016 | 0.004904 | 0.12734 | 0.078574 | 1336.151 |
| GOBP_MO | 90 | 0.076071 | 0.00524 | 0.05378 | 0.078621 | 1336.95 |
| GOBP_CELL | 19 | 0.1621 | 0.00514 | 0.1146 | 0.078631 | 1337.12 |
| SCHRAETS_ | 31 | 0.14235 | 0.005764 | 0.10065 | 0.078658 | 1337.579 |
| GOMF_PRO | 70 | 0.081374 | 0.004946 | 0.057552 | 0.0787 | 1338.294 |
| GOBP_REG | 3 | 0.46762 | 0.005894 | 0.33081 | 0.078755 | 1339.229 |
| KANG_DOX | 18 | 0.17188 | 0.005305 | 0.12161 | 0.078784 | 1339.722 |
| GOBP_HEA | 33 | 0.12747 | 0.005325 | 0.090278 | 0.078991 | 1343.242 |
| SCIAN_INV | 10 | 0.2111 | 0.004857 | 0.14951 | 0.078992 | 1343.259 |
| TARTE_PLA | 348 | 0.037308 | 0.005019 | 0.026431 | 0.079055 | 1344.33 |
| GOBP_OUT | 26 | 0.16637 | 0.00617 | 0.11788 | 0.079083 | 1344.806 |
| WATANABE | 102 | 0.068872 | 0.005049 | 0.048813 | 0.079142 | 1345.81 |
| GOBP_NEG | 36 | 0.1142 | 0.004982 | 0.080965 | 0.079204 | 1346.864 |
| GOBP_TYP | 6 | 0.28886 | 0.005149 | 0.20488 | 0.079295 | 1348.411 |
| GOBP_REG | 47 | 0.11056 | 0.00551 | 0.078431 | 0.079336 | 1349.109 |
| LINDVALL_ | 61 | 0.094662 | 0.005372 | 0.067156 | 0.079341 | 1349.194 |
| GOBP_CHO | 22 | 0.1641 | 0.005599 | 0.11643 | 0.079356 | 1349.449 |
| GOBP_STEI | 27 | 0.13994 | 0.005289 | 0.099304 | 0.079394 | 1350.095 |
| GOBP_NEG | 259 | 0.046762 | 0.00544 | 0.033187 | 0.079417 | 1350.486 |
| GOCC_CELL | 32 | 0.14068 | 0.005787 | 0.099844 | 0.079426 | 1350.639 |
| GOBP_POS | 53 | 0.092979 | 0.00492 | 0.065995 | 0.079444 | 1350.945 |
| GOBP_3_U | 17 | 0.16176 | 0.004852 | 0.11486 | 0.079522 | 1352.272 |
| RASHI_RES | 121 | 0.064232 | 0.005126 | 0.045613 | 0.079546 | 1352.68 |
| GOBP_PUR | 5 | 0.31684 | 0.005156 | 0.22503 | 0.079583 | 1353.309 |
| KEGG_PRIN | 29 | 0.13355 | 0.005231 | 0.094901 | 0.07968 | 1354.958 |
| WP_MITOC | 15 | 0.15299 | 0.004311 | 0.10875 | 0.079754 | 1356.217 |
| GOBP_NEG | 5 | 0.27728 | 0.004512 | 0.19713 | 0.079789 | 1356.812 |
| GOCC_MIC | 11 | 0.18826 | 0.004543 | 0.13389 | 0.079866 | 1358.121 |
| GOBP_AXO | 440 | 0.035082 | 0.005293 | 0.024955 | 0.079898 | 1358.665 |
| BENPORAT | 35 | 0.10485 | 0.004511 | 0.074586 | 0.079902 | 1358.734 |
| REACTOME | 28 | 0.13246 | 0.005097 | 0.094291 | 0.080056 | 1361.352 |
| GOBP_EXT | 25 | 0.13477 | 0.004901 | 0.095942 | 0.080067 | 1361.539 |
| GOMF_ATF | 341 | 0.038161 | 0.005082 | 0.027167 | 0.080067 | 1361.539 |
| GOCC_MIT | 14 | 0.17622 | 0.004797 | 0.12549 | 0.080128 | 1362.577 |
| LU_IL4_SIG | 86 | 0.078754 | 0.005303 | 0.056091 | 0.080161 | 1363.138 |

| | | | | | | |
|---|---|---|---|---|---|---|
| GOBP_MET | 5 | 0.37847 | 0.006159 | 0.26959 | 0.08019 | 1363.631 |
| WEST_ADR | 491 | 0.031569 | 0.005025 | 0.022489 | 0.080203 | 1363.852 |
| GOMF_PRC | 18 | 0.18343 | 0.005661 | 0.13068 | 0.080226 | 1364.243 |
| GOBP_LIPC | 16 | 0.1602 | 0.004662 | 0.11414 | 0.080236 | 1364.413 |
| REACTOME | 30 | 0.13567 | 0.005404 | 0.09667 | 0.080253 | 1364.702 |
| KALMA_E2 | 11 | 0.15464 | 0.003732 | 0.11019 | 0.080262 | 1364.855 |
| GOMF_ROU | 8 | 0.25673 | 0.005284 | 0.18297 | 0.080297 | 1365.45 |
| GOBP_PYR | 16 | 0.16321 | 0.004749 | 0.11634 | 0.080339 | 1366.165 |
| GRUETZMA | 182 | 0.052019 | 0.005083 | 0.037087 | 0.080373 | 1366.743 |
| GOBP_HYP | 12 | 0.2194 | 0.00553 | 0.15646 | 0.080424 | 1367.61 |
| GOBP_NEG | 16 | 0.18115 | 0.005272 | 0.12921 | 0.080472 | 1368.426 |
| GOMF_OXI | 14 | 0.20727 | 0.005643 | 0.14786 | 0.080498 | 1368.868 |
| GOBP_REG | 81 | 0.079936 | 0.005225 | 0.057025 | 0.080502 | 1368.937 |
| ACEVEDO_ | 795 | 0.024309 | 0.004882 | 0.017347 | 0.080566 | 1370.025 |
| KENNY_CTI | 50 | 0.097744 | 0.005024 | 0.069758 | 0.080589 | 1370.416 |
| GOBP_SMC | 12 | 0.2221 | 0.005598 | 0.15853 | 0.080614 | 1370.841 |
| GOBP_WO | 20 | 0.15877 | 0.005165 | 0.11334 | 0.080638 | 1371.249 |
| GOBP_PRC | 33 | 0.14128 | 0.005902 | 0.1009 | 0.080742 | 1373.018 |
| REACTOME | 94 | 0.07231 | 0.00509 | 0.051659 | 0.080802 | 1374.038 |
| GOBP_L_SI | 12 | 0.1898 | 0.004784 | 0.13567 | 0.080918 | 1376.011 |
| REACTOME | 11 | 0.22979 | 0.005545 | 0.16426 | 0.080923 | 1376.096 |
| GOBP_NEG | 312 | 0.040278 | 0.005135 | 0.028792 | 0.080928 | 1376.181 |
| GOBP_PRC | 862 | 0.023825 | 0.004973 | 0.017036 | 0.080994 | 1377.303 |
| KEGG_BIOS | 21 | 0.14439 | 0.004813 | 0.10325 | 0.080996 | 1377.337 |
| GOBP_END | 13 | 0.16958 | 0.004449 | 0.12131 | 0.081073 | 1378.646 |
| GOBP_TUB | 1006 | 0.023595 | 0.0053 | 0.016881 | 0.0811 | 1379.106 |
| GOBP_MID | 17 | 0.18063 | 0.005418 | 0.12925 | 0.081139 | 1379.769 |
| GOMF_LOV | 17 | 0.18915 | 0.005674 | 0.13535 | 0.08115 | 1379.956 |
| GOBP_NEG | 9 | 0.22974 | 0.005015 | 0.16443 | 0.081192 | 1380.67 |
| REACTOME | 25 | 0.12243 | 0.004452 | 0.087635 | 0.081213 | 1381.027 |
| GOBP_CYC | 82 | 0.078421 | 0.005157 | 0.056143 | 0.081246 | 1381.588 |
| GOBP_HIS_ | 7 | 0.32693 | 0.006294 | 0.23417 | 0.081351 | 1383.374 |
| ZHOU_INFI | 438 | 0.03537 | 0.005325 | 0.025349 | 0.081465 | 1385.312 |
| GOBP_POS | 58 | 0.096493 | 0.00534 | 0.069165 | 0.0815 | 1385.908 |
| KAMMINGA | 41 | 0.095911 | 0.004465 | 0.06875 | 0.081507 | 1386.027 |
| GOMF_WN | 31 | 0.12446 | 0.005039 | 0.089215 | 0.081514 | 1386.146 |
| NIKOLSKY_ | 45 | 0.16062 | 0.007833 | 0.11517 | 0.081565 | 1387.013 |
| GOBP_REG | 39 | 0.12739 | 0.005784 | 0.091357 | 0.081603 | 1387.659 |
| GROSS_HY | 106 | 0.07151 | 0.005343 | 0.05129 | 0.081632 | 1388.152 |
| GOBP_FAT | 5 | 0.25002 | 0.004068 | 0.17935 | 0.081668 | 1388.764 |
| GOBP_MIC | 5 | 0.33851 | 0.005508 | 0.24287 | 0.081698 | 1389.274 |
| GOBP_MUS | 20 | 0.16397 | 0.005334 | 0.11768 | 0.081756 | 1390.261 |
| BIOCARTA_ | 8 | 0.19879 | 0.004092 | 0.14268 | 0.081769 | 1390.482 |
| REACTOME | 56 | 0.085939 | 0.004674 | 0.061697 | 0.08183 | 1391.519 |
| WP_PROGI | 21 | 0.16654 | 0.005551 | 0.11965 | 0.08199 | 1394.24 |

| | | | | | | |
|---|---|---|---|---|---|---|
| REACTOME | 119 | 0.062556 | 0.004951 | 0.044953 | 0.08203 | 1394.92 |
| GOMF_DN. | 292 | 0.042325 | 0.005223 | 0.030415 | 0.082035 | 1395.005 |
| REACTOME | 306 | 0.040527 | 0.005118 | 0.029156 | 0.082273 | 1399.052 |
| GOBP_EPIT | 10 | 0.2238 | 0.00515 | 0.16103 | 0.082301 | 1399.529 |
| GOBP_NEG | 5 | 0.28708 | 0.004672 | 0.20657 | 0.082309 | 1399.665 |
| GOBP_CELI | 720 | 0.027028 | 0.005177 | 0.019449 | 0.082313 | 1399.733 |
| PID_IL2_PI: | 33 | 0.11821 | 0.004938 | 0.085088 | 0.082382 | 1400.906 |
| VANTVEER_ | 211 | 0.04931 | 0.005184 | 0.035504 | 0.082446 | 1401.994 |
| REACTOME | 4 | 0.31087 | 0.004525 | 0.22384 | 0.082449 | 1402.045 |
| KIM_BIPOL | 78 | 0.075419 | 0.004838 | 0.054311 | 0.082477 | 1402.521 |
| PLASARI_T( | 31 | 0.13429 | 0.005437 | 0.096705 | 0.082485 | 1402.657 |
| GOBP_SPIN | 184 | 0.04845 | 0.00476 | 0.034892 | 0.082488 | 1402.708 |
| REACTOME | 21 | 0.1327 | 0.004424 | 0.095597 | 0.082553 | 1403.814 |
| GOBP_REG | 1025 | 0.021837 | 0.004948 | 0.015736 | 0.082625 | 1405.038 |
| GOBP_NEG | 43 | 0.10625 | 0.005065 | 0.076575 | 0.082644 | 1405.361 |
| GOBP_REG | 22 | 0.13815 | 0.004713 | 0.099624 | 0.082771 | 1407.521 |
| GOBP_NEU | 31 | 0.13426 | 0.005436 | 0.096834 | 0.082814 | 1408.252 |
| GOBP_VEN | 45 | 0.11409 | 0.005564 | 0.08231 | 0.082868 | 1409.17 |
| REACTOME | 129 | 0.06191 | 0.0051 | 0.044673 | 0.082905 | 1409.8 |
| GOBP_REG | 26 | 0.14256 | 0.005287 | 0.10288 | 0.08292 | 1410.055 |
| IBRAHIM_I | 13 | 0.16638 | 0.004365 | 0.12007 | 0.08293 | 1410.225 |
| GOBP_REG | 86 | 0.076542 | 0.005154 | 0.05524 | 0.082941 | 1410.412 |
| GOERING_ | 13 | 0.19722 | 0.005174 | 0.14235 | 0.082966 | 1410.837 |
| REACTOME | 37 | 0.11092 | 0.004906 | 0.080075 | 0.083003 | 1411.466 |
| KEGG_GLIC | 64 | 0.087151 | 0.005066 | 0.062928 | 0.083046 | 1412.197 |
| AFFAR_YY1 | 186 | 0.050295 | 0.004968 | 0.036318 | 0.08306 | 1412.435 |
| REACTOME | 28 | 0.13974 | 0.005378 | 0.10092 | 0.083091 | 1412.962 |
| GOBP_RES | 1054 | 0.022063 | 0.005066 | 0.015943 | 0.083206 | 1414.918 |
| GOBP_POS | 12 | 0.20791 | 0.00524 | 0.15026 | 0.083238 | 1415.462 |
| VISALA_RE | 14 | 0.1663 | 0.004527 | 0.1202 | 0.083251 | 1415.683 |
| REACTOME | 26 | 0.15338 | 0.005688 | 0.11087 | 0.083275 | 1416.091 |
| GOMF_INT | 10 | 0.2254 | 0.005186 | 0.16294 | 0.083282 | 1416.21 |
| JECHLINGE | 64 | 0.080791 | 0.004696 | 0.058407 | 0.083303 | 1416.568 |
| LEIN_MIDE | 72 | 0.078585 | 0.004844 | 0.056833 | 0.083381 | 1417.894 |
| EBAUER_M | 45 | 0.10431 | 0.005087 | 0.075442 | 0.083391 | 1418.064 |
| KYNG_DNA | 33 | 0.12247 | 0.005116 | 0.088576 | 0.083402 | 1418.251 |
| BROWNE_I | 431 | 0.033672 | 0.00503 | 0.024357 | 0.083429 | 1418.71 |
| PARK_HSC_ | 14 | 0.17456 | 0.004752 | 0.12628 | 0.083455 | 1419.152 |
| WP_OVERL | 52 | 0.096219 | 0.005043 | 0.069637 | 0.083537 | 1420.547 |
| GOMF_LIPI | 6 | 0.30474 | 0.005432 | 0.22055 | 0.08354 | 1420.598 |
| GOBP_MID | 10 | 0.25917 | 0.005963 | 0.18759 | 0.083565 | 1421.023 |
| GOBP_REG | 77 | 0.081446 | 0.005191 | 0.058953 | 0.083565 | 1421.023 |
| WILLIAMS_ | 6 | 0.30437 | 0.005425 | 0.22034 | 0.083599 | 1421.601 |
| WORSCHE( | 14 | 0.1755 | 0.004778 | 0.12706 | 0.083611 | 1421.805 |
| REACTOME | 38 | 0.0986 | 0.004419 | 0.071389 | 0.083624 | 1422.026 |

| | | | | | | |
|---|---|---|---|---|---|---|
| LI_CISPLAT | 29 | 0.13767 | 0.005392 | 0.099691 | 0.083653 | 1422.519 |
| GOBP_NEG | 34 | 0.11964 | 0.005073 | 0.086635 | 0.083661 | 1422.655 |
| REACTOME | 7 | 0.28882 | 0.005561 | 0.20924 | 0.083755 | 1424.254 |
| GOBP_MIT | 31 | 0.11672 | 0.004726 | 0.084583 | 0.083808 | 1425.155 |
| CAFFAREL_ | 29 | 0.12929 | 0.005064 | 0.093699 | 0.083823 | 1425.41 |
| GOBP_REG | 183 | 0.053435 | 0.005236 | 0.038725 | 0.083825 | 1425.444 |
| GOBP_FAT | 368 | 0.035696 | 0.004935 | 0.025871 | 0.083832 | 1425.563 |
| SCHLESING | 10 | 0.2304 | 0.005301 | 0.167 | 0.083857 | 1425.988 |
| GOBP_CELI | 931 | 0.022188 | 0.004804 | 0.016084 | 0.083878 | 1426.345 |
| LEIN_LOCA | 34 | 0.11498 | 0.004875 | 0.083368 | 0.08393 | 1427.23 |
| GOBP_CRA | 68 | 0.091957 | 0.005509 | 0.066723 | 0.084082 | 1429.814 |
| HERNANDE | 35 | 0.11586 | 0.004984 | 0.084079 | 0.084108 | 1430.257 |
| GOBP_NEG | 5 | 0.30701 | 0.004996 | 0.22287 | 0.084182 | 1431.515 |
| GOCC_FILT | 8 | 0.2604 | 0.00536 | 0.18906 | 0.084208 | 1431.957 |
| GOBP_NON | 63 | 0.081734 | 0.004714 | 0.059363 | 0.084289 | 1433.334 |
| GOMF_NEU | 7 | 0.2869 | 0.005524 | 0.2084 | 0.084317 | 1433.811 |
| GOCC_OUT | 21 | 0.13378 | 0.00446 | 0.09729 | 0.084559 | 1437.926 |
| GOBP_CAR | 92 | 0.081315 | 0.005663 | 0.059176 | 0.08471 | 1440.494 |
| BECKER_TA | 51 | 0.10651 | 0.005529 | 0.077517 | 0.084729 | 1440.817 |
| GOMF_PRO | 88 | 0.070474 | 0.0048 | 0.051364 | 0.085035 | 1446.02 |
| AMIT_EGF_ | 44 | 0.11659 | 0.005622 | 0.084981 | 0.085055 | 1446.36 |
| REACTOME | 7 | 0.26868 | 0.005173 | 0.19585 | 0.085061 | 1446.462 |
| GOBP_RNA | 175 | 0.048996 | 0.004696 | 0.035715 | 0.085065 | 1446.53 |
| TRAYNOR_ | 13 | 0.19116 | 0.005015 | 0.13938 | 0.085125 | 1447.551 |
| GOBP_NEG | 13 | 0.19708 | 0.00517 | 0.14374 | 0.08519 | 1448.656 |
| GOMF_MIS | 23 | 0.14018 | 0.00489 | 0.10224 | 0.085194 | 1448.724 |
| GOBP_POS | 5 | 0.27224 | 0.00443 | 0.19862 | 0.085251 | 1449.693 |
| BENPORAT | 1024 | 0.024332 | 0.005511 | 0.017768 | 0.085436 | 1452.839 |
| GERHOLD_ | 13 | 0.20754 | 0.005444 | 0.15156 | 0.085456 | 1453.179 |
| GOCC_CAJ | 56 | 0.083165 | 0.004523 | 0.060733 | 0.085456 | 1453.179 |
| GOBP_REG | 174 | 0.054089 | 0.005169 | 0.039508 | 0.085498 | 1453.893 |
| GOCC_DNA | 109 | 0.063614 | 0.00482 | 0.046467 | 0.085503 | 1453.979 |
| GOBP_NEG | 12 | 0.20607 | 0.005194 | 0.15054 | 0.085523 | 1454.319 |
| REACTOME | 13 | 0.1676 | 0.004397 | 0.12246 | 0.085563 | 1454.999 |
| GOCC_MIT | 7 | 0.29035 | 0.00559 | 0.21217 | 0.085587 | 1455.407 |
| BENPORAT | 933 | 0.022858 | 0.004955 | 0.016707 | 0.085631 | 1456.155 |
| GOBP_REG | 6 | 0.28841 | 0.005141 | 0.21082 | 0.085657 | 1456.597 |
| FORTSCHE | 719 | 0.025283 | 0.004839 | 0.018487 | 0.085724 | 1457.737 |
| REACTOME | 18 | 0.17177 | 0.005302 | 0.1256 | 0.085727 | 1457.788 |
| GOBP_REG | 5 | 0.35304 | 0.005745 | 0.25815 | 0.085732 | 1457.873 |
| GOCC_MEI | 35 | 0.11056 | 0.004756 | 0.080851 | 0.085755 | 1458.264 |
| GOBP_REG | 12 | 0.21657 | 0.005458 | 0.15843 | 0.085824 | 1459.437 |
| GOBP_NEG | 259 | 0.042051 | 0.004892 | 0.030768 | 0.085865 | 1460.134 |
| REACTOME | 24 | 0.14478 | 0.005159 | 0.10595 | 0.085902 | 1460.764 |
| GOBP_REG | 11 | 0.20941 | 0.005054 | 0.15326 | 0.085918 | 1461.036 |

| | | | | | | |
|---|---|---|---|---|---|---|
| GOBP_NEG | 8 | 0.18821 | 0.003874 | 0.13775 | 0.085933 | 1461.291 |
| SCHLOSSER | 46 | 0.09593 | 0.00473 | 0.070232 | 0.085996 | 1462.362 |
| REACTOME | 9 | 0.21574 | 0.00471 | 0.15799 | 0.08605 | 1463.28 |
| GOBP_SUL | 9 | 0.23507 | 0.005131 | 0.17221 | 0.086137 | 1464.76 |
| GOBP_HIST | 24 | 0.11955 | 0.00426 | 0.087599 | 0.086184 | 1465.559 |
| GOBP_NEG | 6 | 0.27952 | 0.004982 | 0.20483 | 0.086195 | 1465.746 |
| WP_NEURA | 95 | 0.074486 | 0.005271 | 0.054584 | 0.086198 | 1465.797 |
| GOBP_MIC | 20 | 0.16606 | 0.005402 | 0.12176 | 0.086319 | 1467.855 |
| GOBP_AOR | 33 | 0.12614 | 0.005269 | 0.092512 | 0.086367 | 1468.671 |
| GOBP_REG | 127 | 0.064802 | 0.005297 | 0.047525 | 0.086369 | 1468.705 |
| GOBP_REG | 20 | 0.1442 | 0.004691 | 0.10576 | 0.086379 | 1468.875 |
| GOBP_ANT | 11 | 0.20894 | 0.005042 | 0.15326 | 0.086406 | 1469.334 |
| LIU_TOPBP | 13 | 0.22138 | 0.005807 | 0.16242 | 0.086445 | 1469.997 |
| GOBP_BRA | 13 | 0.20245 | 0.005311 | 0.14854 | 0.086462 | 1470.286 |
| REACTOME | 15 | 0.21719 | 0.00612 | 0.15936 | 0.086477 | 1470.541 |
| HOQUE_M | 56 | 0.095437 | 0.00519 | 0.070042 | 0.086518 | 1471.239 |
| GOBP_POS | 182 | 0.049139 | 0.004802 | 0.036071 | 0.086562 | 1471.987 |
| REACTOME | 5 | 0.30416 | 0.004949 | 0.22342 | 0.086705 | 1474.419 |
| GOBP_MEI | 6 | 0.20056 | 0.003575 | 0.14732 | 0.086707 | 1474.453 |
| GOBP_REG | 271 | 0.043268 | 0.005147 | 0.031783 | 0.086714 | 1474.572 |
| REACTOME | 105 | 0.06386 | 0.004749 | 0.046916 | 0.086741 | 1475.031 |
| GOBP_POS | 13 | 0.17021 | 0.004465 | 0.1251 | 0.086828 | 1476.51 |
| GOBP_POS | 6 | 0.36373 | 0.006484 | 0.26733 | 0.086828 | 1476.51 |
| GOBP_REG | 5 | 0.37762 | 0.006145 | 0.2776 | 0.086876 | 1477.326 |
| GOBP_AXO | 15 | 0.18058 | 0.005088 | 0.13286 | 0.087065 | 1480.54 |
| GOBP_REG | 11 | 0.1859 | 0.004486 | 0.1368 | 0.087096 | 1481.067 |
| SCHUETZ_E | 82 | 0.080376 | 0.005286 | 0.059149 | 0.087103 | 1481.187 |
| GOBP_BIOI | 106 | 0.064646 | 0.004831 | 0.047575 | 0.087113 | 1481.357 |
| GOBP_PRO | 231 | 0.043592 | 0.004793 | 0.032083 | 0.087125 | 1481.561 |
| REACTOME | 17 | 0.18154 | 0.005445 | 0.13362 | 0.087142 | 1481.85 |
| GOMF_SEC | 14 | 0.18748 | 0.005104 | 0.138 | 0.087152 | 1482.02 |
| GOBP_POS | 107 | 0.066919 | 0.005024 | 0.049265 | 0.087185 | 1482.581 |
| GOMF_MR | 285 | 0.038134 | 0.00465 | 0.028085 | 0.087272 | 1484.06 |
| WP_INSULI | 8 | 0.25453 | 0.005239 | 0.18747 | 0.087283 | 1484.247 |
| WP_INSULI | 8 | 0.25453 | 0.005239 | 0.18747 | 0.087283 | 1484.247 |
| GOBP_OST | 33 | 0.11891 | 0.004967 | 0.087605 | 0.087352 | 1485.421 |
| GOBP_NUC | 28 | 0.12895 | 0.004963 | 0.095018 | 0.087375 | 1485.812 |
| CAFFAREL_ | 8 | 0.22108 | 0.00455 | 0.16294 | 0.087427 | 1486.696 |
| CHIANG_LI | 165 | 0.053309 | 0.004962 | 0.039292 | 0.087445 | 1487.002 |
| GOBP_SPIN | 29 | 0.14087 | 0.005517 | 0.10385 | 0.087483 | 1487.648 |
| GOBP_REG | 127 | 0.062323 | 0.005095 | 0.045948 | 0.087498 | 1487.903 |
| WP_ENDOI | 129 | 0.065061 | 0.00536 | 0.04797 | 0.087511 | 1488.125 |
| GOBP_REG | 15 | 0.1795 | 0.005058 | 0.13236 | 0.087533 | 1488.499 |
| ELVIDGE_H | 128 | 0.064385 | 0.005284 | 0.047484 | 0.087571 | 1489.145 |
| WP_THIAM | 8 | 0.25136 | 0.005173 | 0.18541 | 0.087604 | 1489.706 |

| | | | | | | |
|---|---|---|---|---|---|---|
| OHASHI_AU | 7 | 0.24895 | 0.004793 | 0.18364 | 0.08762 | 1489.978 |
| REACTOME | 59 | 0.079522 | 0.004439 | 0.058663 | 0.087628 | 1490.114 |
| BURTON_A | 94 | 0.069707 | 0.004907 | 0.051436 | 0.087683 | 1491.049 |
| GOBP_LUN | 8 | 0.26327 | 0.005419 | 0.19442 | 0.087859 | 1494.042 |
| GOBP_HIG | 10 | 0.24027 | 0.005528 | 0.1775 | 0.087935 | 1495.335 |
| MITSIADES | 404 | 0.033983 | 0.004918 | 0.025107 | 0.087954 | 1495.658 |
| GOBP_C_T | 13 | 0.21424 | 0.00562 | 0.15831 | 0.087991 | 1496.287 |
| GOCC_REP | 14 | 0.16399 | 0.004464 | 0.1212 | 0.088033 | 1497.001 |
| NUYTTEN_ | 798 | 0.023986 | 0.004826 | 0.017727 | 0.088033 | 1497.001 |
| GOBP_INTI | 24 | 0.13479 | 0.004803 | 0.099631 | 0.088056 | 1497.392 |
| GOBP_PRC | 477 | 0.030359 | 0.004765 | 0.022441 | 0.088065 | 1497.545 |
| WP_H19_A | 14 | 0.16598 | 0.004519 | 0.1227 | 0.088077 | 1497.749 |
| GOBP_DNA | 5 | 0.28244 | 0.004596 | 0.20881 | 0.088101 | 1498.158 |
| REACTOME | 43 | 0.098993 | 0.004719 | 0.073198 | 0.088132 | 1498.685 |
| GOBP_NUC | 16 | 0.15377 | 0.004475 | 0.1137 | 0.088133 | 1498.702 |
| MOOTHA_ | 398 | 0.032663 | 0.004692 | 0.024154 | 0.088148 | 1498.957 |
| GOBP_RES | 426 | 0.033702 | 0.005005 | 0.024923 | 0.088156 | 1499.093 |
| LEIN_CERE | 77 | 0.08042 | 0.005126 | 0.059486 | 0.088211 | 1500.028 |
| GOBP_REG | 9 | 0.2119 | 0.004626 | 0.15677 | 0.088249 | 1500.674 |
| BLANCO_M | 166 | 0.048663 | 0.004543 | 0.036011 | 0.088304 | 1501.61 |
| GOMF_CYC | 33 | 0.11194 | 0.004676 | 0.082843 | 0.088324 | 1501.95 |
| GOBP_NCR | 401 | 0.0311 | 0.004484 | 0.023023 | 0.088392 | 1503.106 |
| GOBP_WN | 11 | 0.21615 | 0.005216 | 0.16005 | 0.088434 | 1503.82 |
| GOCC_MIC | 23 | 0.13115 | 0.004575 | 0.09712 | 0.088447 | 1504.041 |
| GOMF_WII | 22 | 0.13947 | 0.004758 | 0.10331 | 0.088515 | 1505.198 |
| GOBP_CILI | 5 | 0.30271 | 0.004926 | 0.22435 | 0.088633 | 1507.204 |
| WP_BENZE | 6 | 0.24529 | 0.004372 | 0.18184 | 0.088694 | 1508.241 |
| GOBP_POS | 6 | 0.30926 | 0.005513 | 0.22937 | 0.088785 | 1509.789 |
| GOBP_RES | 21 | 0.16407 | 0.005469 | 0.1217 | 0.088815 | 1510.299 |
| GOMF_DN | 33 | 0.1174 | 0.004904 | 0.087084 | 0.088825 | 1510.469 |
| GOBP_PRC | 5 | 0.60207 | 0.009797 | 0.44681 | 0.088921 | 1512.102 |
| GOBP_PYR | 14 | 0.16774 | 0.004566 | 0.1245 | 0.08895 | 1512.595 |
| REACTOME | 28 | 0.12639 | 0.004864 | 0.09382 | 0.088977 | 1513.054 |
| GOBP_REG | 7 | 0.2131 | 0.004103 | 0.1582 | 0.088993 | 1513.326 |
| GOBP_POS | 20 | 0.16329 | 0.005312 | 0.12129 | 0.089107 | 1515.265 |
| GOBP_NUC | 5 | 0.27519 | 0.004478 | 0.20444 | 0.089151 | 1516.013 |
| GOBP_MES | 4 | 0.3222 | 0.00469 | 0.2394 | 0.089178 | 1516.472 |
| GOBP_STEI | 159 | 0.05474 | 0.005003 | 0.040676 | 0.0892 | 1516.846 |
| OHM_MET | 27 | 0.1276 | 0.004822 | 0.094831 | 0.089231 | 1517.373 |
| GOBP_REG | 1122 | 0.020676 | 0.004889 | 0.015367 | 0.089249 | 1517.679 |
| GOBP_POS | 9 | 0.21529 | 0.0047 | 0.16001 | 0.089253 | 1517.747 |
| GOBP_REG | 3 | 0.33072 | 0.004169 | 0.24584 | 0.089281 | 1518.223 |
| GOBP_REG | 11 | 0.2416 | 0.00583 | 0.17962 | 0.089318 | 1518.853 |
| REACTOME | 49 | 0.10575 | 0.005381 | 0.078641 | 0.08936 | 1519.567 |
| GOBP_POS | 12 | 0.20028 | 0.005048 | 0.14898 | 0.089432 | 1520.791 |

| | | | | | | |
|---|---|---|---|---|---|---|
| HASEGAWA | 8 | 0.2201 | 0.00453 | 0.16376 | 0.089481 | 1521.624 |
| KEGG_VAL | 43 | 0.10027 | 0.00478 | 0.074619 | 0.089515 | 1522.203 |
| WP_CHOLE | 70 | 0.082069 | 0.004988 | 0.061077 | 0.089532 | 1522.492 |
| AGARWAL_ | 10 | 0.22306 | 0.005132 | 0.16601 | 0.089538 | 1522.594 |
| GOBP_POS | 5 | 0.3209 | 0.005222 | 0.23883 | 0.089548 | 1522.764 |
| GRAESSMA | 80 | 0.072525 | 0.004711 | 0.053991 | 0.089598 | 1523.614 |
| GOBP_REG | 13 | 0.18081 | 0.004743 | 0.13461 | 0.089614 | 1523.886 |
| GOBP_BRA | 12 | 0.224 | 0.005646 | 0.1668 | 0.089659 | 1524.651 |
| HAMAI_AP | 619 | 0.028295 | 0.005039 | 0.021073 | 0.089691 | 1525.195 |
| REACTOME | 6 | 0.25323 | 0.004514 | 0.18861 | 0.089709 | 1525.502 |
| GOBP_NEG | 25 | 0.12715 | 0.004624 | 0.09472 | 0.089739 | 1526.012 |
| GOBP_REC | 56 | 0.090764 | 0.004936 | 0.067627 | 0.089789 | 1526.862 |
| GOBP_DEH | 9 | 0.23761 | 0.005187 | 0.17706 | 0.089808 | 1527.185 |
| WP_HOSTF | 20 | 0.15565 | 0.005063 | 0.11602 | 0.089873 | 1528.29 |
| WP_DNA_I | 109 | 0.06528 | 0.004946 | 0.048662 | 0.08989 | 1528.579 |
| REACTOME | 36 | 0.11888 | 0.005186 | 0.088649 | 0.089973 | 1529.991 |
| VANDESLU | 76 | 0.076946 | 0.004872 | 0.057381 | 0.089974 | 1530.008 |
| GOBP_POS | 483 | 0.029249 | 0.004618 | 0.021819 | 0.090049 | 1531.283 |
| GOMF_OU | 13 | 0.21547 | 0.005652 | 0.16076 | 0.09009 | 1531.98 |
| HUMMEL_ | 41 | 0.12878 | 0.005995 | 0.096085 | 0.090091 | 1531.997 |
| GOMF_RDI | 10 | 0.23118 | 0.005319 | 0.17252 | 0.090134 | 1532.729 |
| GOMF_TRA | 61 | 0.097814 | 0.005551 | 0.073013 | 0.090184 | 1533.579 |
| GOBP_VER | 33 | 0.10728 | 0.004481 | 0.080118 | 0.090292 | 1535.415 |
| GOBP_EXT | 79 | 0.078522 | 0.005069 | 0.058651 | 0.090325 | 1535.977 |
| REACTOME | 9 | 0.27309 | 0.005961 | 0.20398 | 0.090326 | 1535.994 |
| WP_HOMC | 13 | 0.18986 | 0.004981 | 0.14185 | 0.090381 | 1536.929 |
| GOCC_INSU | 5 | 0.25072 | 0.00408 | 0.18736 | 0.09043 | 1537.762 |
| GOMF_NEU | 5 | 0.25738 | 0.004188 | 0.19235 | 0.090442 | 1537.966 |
| GOBP_REG | 97 | 0.065719 | 0.004699 | 0.049116 | 0.090453 | 1538.153 |
| RAFFEL_VE | 5 | 0.3911 | 0.006364 | 0.29233 | 0.090475 | 1538.527 |
| YIH_RESPO | 15 | 0.18115 | 0.005104 | 0.13541 | 0.090493 | 1538.833 |
| GOBP_LATI | 26 | 0.12226 | 0.004534 | 0.091393 | 0.090495 | 1538.867 |
| WOTTON_I | 20 | 0.15687 | 0.005103 | 0.11728 | 0.090533 | 1539.514 |
| INGRAM_S | 119 | 0.064553 | 0.005109 | 0.048268 | 0.090556 | 1539.905 |
| GOBP_INTI | 10 | 0.2124 | 0.004887 | 0.15882 | 0.090567 | 1540.092 |
| JISON_SICK | 165 | 0.049557 | 0.004613 | 0.037062 | 0.090596 | 1540.585 |
| FRIDMAN_ | 11 | 0.19863 | 0.004793 | 0.14868 | 0.0908 | 1544.054 |
| STEGER_AI | 21 | 0.15874 | 0.005292 | 0.119 | 0.09111 | 1549.326 |
| GOBP_LUN | 50 | 0.1037 | 0.00533 | 0.077751 | 0.091155 | 1550.091 |
| DIRMEIER_ | 58 | 0.084933 | 0.004701 | 0.063704 | 0.091237 | 1551.485 |
| GOBP_BRA | 17 | 0.17752 | 0.005325 | 0.13323 | 0.091363 | 1553.628 |
| GOBP_BRA | 20 | 0.16008 | 0.005208 | 0.12018 | 0.091444 | 1555.005 |
| KRIGE_RES | 836 | 0.022934 | 0.004718 | 0.017226 | 0.091553 | 1556.859 |
| REACTOME | 56 | 0.085782 | 0.004665 | 0.064457 | 0.091628 | 1558.134 |
| GOBP_INTI | 10 | 0.20935 | 0.004817 | 0.15732 | 0.091647 | 1558.457 |

| | | | | | | |
|---|---|---|---|---|---|---|
| GOBP_CELI | 15 | 0.18778 | 0.005291 | 0.14115 | 0.091713 | 1559.58 |
| JEON_SMA | 22 | 0.17072 | 0.005825 | 0.12837 | 0.091773 | 1560.6 |
| GOBP_RES | 104 | 0.069712 | 0.00516 | 0.052418 | 0.091781 | 1560.736 |
| REACTOME | 164 | 0.049498 | 0.004594 | 0.037226 | 0.09182 | 1561.399 |
| GOBP_CELI | 218 | 0.046339 | 0.004951 | 0.034865 | 0.091915 | 1563.015 |
| GOBP_NEG | 69 | 0.078379 | 0.00473 | 0.058972 | 0.09192 | 1563.1 |
| REACTOME | 39 | 0.10643 | 0.004832 | 0.080123 | 0.092055 | 1565.395 |
| BARIS_THY | 53 | 0.094944 | 0.005024 | 0.071538 | 0.092233 | 1568.422 |
| BIOCARTA_ | 5 | 0.31077 | 0.005057 | 0.23417 | 0.092241 | 1568.558 |
| REACTOME | 48 | 0.088729 | 0.004468 | 0.066868 | 0.092274 | 1569.119 |
| REACTOME | 12 | 0.22574 | 0.00569 | 0.17013 | 0.092285 | 1569.306 |
| REACTOME | 70 | 0.077957 | 0.004738 | 0.058763 | 0.092323 | 1569.953 |
| MEBARKI_I | 39 | 0.11545 | 0.005242 | 0.08704 | 0.092365 | 1570.667 |
| HUPER_BR | 51 | 0.098169 | 0.005096 | 0.074013 | 0.092368 | 1570.718 |
| GOBP_NEG | 21 | 0.14104 | 0.004701 | 0.10634 | 0.092371 | 1570.769 |
| ACEVEDO_ | 159 | 0.04994 | 0.004564 | 0.037661 | 0.092424 | 1571.67 |
| GOBP_NEG | 4 | 0.45013 | 0.006552 | 0.33946 | 0.09243 | 1571.772 |
| GOBP_RNA | 167 | 0.048009 | 0.004496 | 0.036219 | 0.092506 | 1573.065 |
| GOMF_MY | 15 | 0.15353 | 0.004326 | 0.11584 | 0.092538 | 1573.609 |
| GOBP_REG | 9 | 0.24897 | 0.005435 | 0.18785 | 0.092539 | 1573.626 |
| WP_BASE_ | 30 | 0.10688 | 0.004257 | 0.080655 | 0.092572 | 1574.187 |
| REACTOME | 5 | 0.36005 | 0.005859 | 0.27188 | 0.092712 | 1576.568 |
| WP_MELAI | 65 | 0.083469 | 0.004889 | 0.063054 | 0.092799 | 1578.047 |
| BILANGES_ | 31 | 0.11248 | 0.004554 | 0.084999 | 0.092879 | 1579.407 |
| GOBP_NEG | 18 | 0.16775 | 0.005177 | 0.12678 | 0.092894 | 1579.662 |
| SPIELMAN_ | 446 | 0.030312 | 0.004604 | 0.022919 | 0.092999 | 1581.448 |
| GOBP_NEG | 12 | 0.1916 | 0.004829 | 0.14487 | 0.093004 | 1581.533 |
| GOBP_L_C' | 4 | 0.31976 | 0.004654 | 0.24181 | 0.093037 | 1582.094 |
| GOBP_POS | 18 | 0.15625 | 0.004823 | 0.11818 | 0.093061 | 1582.502 |
| GOLUB_AL | 22 | 0.13613 | 0.004645 | 0.103 | 0.093147 | 1583.965 |
| GOBP_NEG | 4 | 0.29819 | 0.00434 | 0.22567 | 0.093196 | 1584.798 |
| CEBALLOS_ | 18 | 0.14866 | 0.004588 | 0.11253 | 0.093242 | 1585.58 |
| GOBP_MU | 46 | 0.096121 | 0.004739 | 0.072789 | 0.093336 | 1587.179 |
| GOBP_REG | 128 | 0.058944 | 0.004837 | 0.044648 | 0.093397 | 1588.216 |
| GOBP_HYD | 7 | 0.25688 | 0.004946 | 0.19465 | 0.093479 | 1589.61 |
| MONTERO_ | 10 | 0.23451 | 0.005396 | 0.17772 | 0.0935 | 1589.968 |
| GOBP_RES | 50 | 0.092906 | 0.004775 | 0.070409 | 0.093508 | 1590.104 |
| GOBP_EMI | 23 | 0.14662 | 0.005115 | 0.11114 | 0.093541 | 1590.665 |
| GOBP_URE | 11 | 0.18796 | 0.004536 | 0.14256 | 0.093675 | 1592.943 |
| GOBP_CELI | 52 | 0.095155 | 0.004987 | 0.072227 | 0.093854 | 1595.987 |
| GOBP_SMA | 543 | 0.027736 | 0.004636 | 0.021057 | 0.093898 | 1596.735 |
| GOCC_ENV | 1155 | 0.019042 | 0.004564 | 0.014467 | 0.094059 | 1599.473 |
| GOBP_MAI | 43 | 0.10656 | 0.00508 | 0.080965 | 0.094076 | 1599.762 |
| REACTOME | 70 | 0.082046 | 0.004987 | 0.062348 | 0.094106 | 1600.273 |
| GOBP_POS | 6 | 0.30295 | 0.0054 | 0.2303 | 0.094184 | 1601.599 |

| | | | | | | |
|---|---|---|---|---|---|---|
| GOBP_APC | 39 | 0.10708 | 0.004862 | 0.081433 | 0.094271 | 1603.078 |
| COLLER_M | 7 | 0.3042 | 0.005857 | 0.23138 | 0.09431 | 1603.742 |
| GOBP_ADI | 51 | 0.09797 | 0.005085 | 0.074534 | 0.094358 | 1604.558 |
| GOBP_NEG | 14 | 0.16777 | 0.004567 | 0.12766 | 0.094407 | 1605.391 |
| GOBP_MIN | 11 | 0.21 | 0.005068 | 0.15981 | 0.094415 | 1605.527 |
| GOBP_AMI | 23 | 0.1246 | 0.004347 | 0.094839 | 0.094458 | 1606.258 |
| GOMF_5AL | 6 | 0.23914 | 0.004263 | 0.18202 | 0.094463 | 1606.343 |
| GOBP_REG | 13 | 0.20771 | 0.005449 | 0.15817 | 0.094574 | 1608.231 |
| CONRAD_S | 36 | 0.11514 | 0.005024 | 0.087693 | 0.094593 | 1608.554 |
| CHANDRAN | 182 | 0.049071 | 0.004795 | 0.037385 | 0.094669 | 1609.846 |
| GOBP_NEG | 6 | 0.24783 | 0.004418 | 0.18883 | 0.094689 | 1610.186 |
| GOBP_CHR | 556 | 0.027719 | 0.004687 | 0.021123 | 0.094723 | 1610.765 |
| GOBP_NEG | 6 | 0.29798 | 0.005311 | 0.22709 | 0.094745 | 1611.139 |
| GOBP_POS | 5 | 0.23109 | 0.00376 | 0.17612 | 0.094747 | 1611.173 |
| GOBP_NEG | 81 | 0.076096 | 0.004974 | 0.058 | 0.094767 | 1611.513 |
| REACTOME | 195 | 0.046926 | 0.004745 | 0.035772 | 0.094803 | 1612.125 |
| GOBP_NEU | 22 | 0.15142 | 0.005166 | 0.11545 | 0.094834 | 1612.652 |
| REACTOME | 25 | 0.12207 | 0.004439 | 0.093076 | 0.094844 | 1612.822 |
| GOBP_MYE | 383 | 0.033822 | 0.004768 | 0.025789 | 0.094855 | 1613.009 |
| GOBP_SPIN | 13 | 0.1797 | 0.004714 | 0.13704 | 0.09489 | 1613.604 |
| BLANCO_M | 247 | 0.041806 | 0.004751 | 0.031884 | 0.094906 | 1613.877 |
| GOBP_REG | 81 | 0.078287 | 0.005117 | 0.059707 | 0.094909 | 1613.928 |
| GOBP_CELI | 28 | 0.13258 | 0.005102 | 0.10114 | 0.094945 | 1614.54 |
| VILLANUEV | 59 | 0.079083 | 0.004414 | 0.060336 | 0.094987 | 1615.254 |
| KIM_WT1_ | 213 | 0.047364 | 0.005003 | 0.036147 | 0.095053 | 1616.376 |
| GOBP_HEA | 16 | 0.19376 | 0.005638 | 0.14797 | 0.095209 | 1619.029 |
| GOBP_ANT | 9 | 0.21588 | 0.004713 | 0.16498 | 0.09535 | 1621.427 |
| GOBP_POS | 28 | 0.10633 | 0.004092 | 0.081267 | 0.095366 | 1621.699 |
| GOMF_TRA | 5 | 0.40612 | 0.006609 | 0.31039 | 0.095376 | 1621.869 |
| RODRIGUE | 727 | 0.024784 | 0.004769 | 0.018943 | 0.095385 | 1622.022 |
| GOBP_EPIE | 35 | 0.11806 | 0.005079 | 0.090246 | 0.09542 | 1622.617 |
| GOBP_REG | 26 | 0.13151 | 0.004877 | 0.10055 | 0.09546 | 1623.297 |
| KAAB_FAIL | 31 | 0.12335 | 0.004995 | 0.094353 | 0.095552 | 1624.862 |
| ONKEN_UV | 493 | 0.029695 | 0.004736 | 0.022715 | 0.09557 | 1625.168 |
| REACTOME | 28 | 0.12295 | 0.004731 | 0.094074 | 0.095633 | 1626.239 |
| BRUINS_UV | 1070 | 0.020472 | 0.004734 | 0.015667 | 0.095677 | 1626.987 |
| GOBP_RRN | 10 | 0.18561 | 0.004271 | 0.14211 | 0.095764 | 1628.467 |
| WP_NCRNA | 83 | 0.070639 | 0.004674 | 0.054087 | 0.095783 | 1628.79 |
| WP_VALPR | 11 | 0.19822 | 0.004783 | 0.15179 | 0.095801 | 1629.096 |
| GOMF_C_F | 5 | 0.32339 | 0.005262 | 0.24765 | 0.095811 | 1629.266 |
| GOBP_DET | 20 | 0.14564 | 0.004738 | 0.11159 | 0.095943 | 1631.511 |
| GOBP_REG | 225 | 0.044742 | 0.004855 | 0.034293 | 0.096008 | 1632.616 |
| GOBP_GLC | 9 | 0.2613 | 0.005704 | 0.2003 | 0.09603 | 1632.99 |
| WP_GLUCC | 8 | 0.27015 | 0.00556 | 0.2071 | 0.096045 | 1633.245 |
| THUM_MIF | 7 | 0.26415 | 0.005086 | 0.20252 | 0.096069 | 1633.653 |

| | | | | | | |
|---|---|---|---|---|---|---|
| GOBP_POS | 38 | 0.10339 | 0.004634 | 0.079269 | 0.09608 | 1633.84 |
| KRIGE_RES | 962 | 0.020448 | 0.004497 | 0.015679 | 0.096095 | 1634.095 |
| WP_TYPE_ | 19 | 0.14525 | 0.004606 | 0.11139 | 0.096129 | 1634.674 |
| AMIT_SERU | 16 | 0.16968 | 0.004938 | 0.13014 | 0.096148 | 1634.997 |
| REACTOME | 14 | 0.18419 | 0.005014 | 0.14129 | 0.096186 | 1635.643 |
| DOANE_RE | 228 | 0.043808 | 0.004785 | 0.033628 | 0.096342 | 1638.296 |
| GOBP_REG | 294 | 0.036547 | 0.004525 | 0.028055 | 0.096352 | 1638.466 |
| GOBP_POS | 10 | 0.23382 | 0.00538 | 0.17949 | 0.096353 | 1638.483 |
| SMID_BRE/ | 6 | 0.25682 | 0.004578 | 0.19717 | 0.096382 | 1638.976 |
| TURASHVIL | 43 | 0.098675 | 0.004704 | 0.075762 | 0.096392 | 1639.146 |
| WP_PHOT( | 27 | 0.12569 | 0.00475 | 0.096521 | 0.09644 | 1639.962 |
| WP_CHOLE | 14 | 0.16175 | 0.004403 | 0.12424 | 0.09647 | 1640.472 |
| GOBP_POS | 35 | 0.11694 | 0.00503 | 0.089814 | 0.096472 | 1640.506 |
| LE_EGR2_T | 103 | 0.068004 | 0.005009 | 0.052242 | 0.096518 | 1641.289 |
| RIZKI_TUM | 244 | 0.039643 | 0.004478 | 0.030472 | 0.096641 | 1643.38 |
| GOBP_NU( | 7 | 0.29829 | 0.005743 | 0.22929 | 0.096654 | 1643.601 |
| TCGA_GLIC | 72 | 0.10159 | 0.006262 | 0.078103 | 0.096695 | 1644.298 |
| GOBP_PUR | 9 | 0.17199 | 0.003755 | 0.13228 | 0.096777 | 1645.693 |
| FUJII_YBX1 | 37 | 0.10138 | 0.004484 | 0.077976 | 0.09678 | 1645.744 |
| GOBP_NEG | 931 | 0.021203 | 0.004591 | 0.016309 | 0.096797 | 1646.033 |
| GOCC_MIC | 13 | 0.19504 | 0.005116 | 0.15005 | 0.096833 | 1646.645 |
| GOBP_REG | 31 | 0.14022 | 0.005678 | 0.10788 | 0.096845 | 1646.849 |
| GOBP_PEP | 8 | 0.24796 | 0.005103 | 0.19087 | 0.096967 | 1648.924 |
| GOBP_PLU | 7 | 0.21697 | 0.004177 | 0.16703 | 0.09698 | 1649.145 |
| BIOCARTA_ | 18 | 0.14304 | 0.004415 | 0.11016 | 0.09706 | 1650.505 |
| GOBP_POS | 31 | 0.1276 | 0.005166 | 0.098334 | 0.097222 | 1653.26 |
| WP_NANO | 19 | 0.1494 | 0.004737 | 0.11515 | 0.097252 | 1653.77 |
| COLLIS_PRI | 16 | 0.17428 | 0.005072 | 0.1344 | 0.097379 | 1655.93 |
| GOBP_POS | 87 | 0.071506 | 0.004843 | 0.055148 | 0.097391 | 1656.134 |
| GOCC_INTI | 11 | 0.21616 | 0.005216 | 0.16673 | 0.097414 | 1656.525 |
| BOYAULT_l | 10 | 0.17972 | 0.004135 | 0.13868 | 0.097511 | 1658.175 |
| GOBP_APC | 26 | 0.13534 | 0.005019 | 0.10444 | 0.097514 | 1658.226 |
| GOBP_IRE1 | 20 | 0.1507 | 0.004903 | 0.11629 | 0.097514 | 1658.226 |
| GOBP_MO | 74 | 0.074882 | 0.004679 | 0.057787 | 0.097526 | 1658.43 |
| GOMF_G_( | 11 | 0.21375 | 0.005158 | 0.16499 | 0.097575 | 1659.263 |
| GOBP_EST/ | 54 | 0.092864 | 0.00496 | 0.071695 | 0.097624 | 1660.096 |
| BROWNE_l | 150 | 0.055205 | 0.004901 | 0.042623 | 0.097639 | 1660.351 |
| REACTOME | 295 | 0.036929 | 0.00458 | 0.028532 | 0.097792 | 1662.953 |
| GOBP_CRD | 11 | 0.22467 | 0.005422 | 0.1736 | 0.09781 | 1663.259 |
| SCHLINGEN | 38 | 0.11379 | 0.0051 | 0.08793 | 0.097826 | 1663.531 |
| GOBP_MAI | 6 | 0.30998 | 0.005526 | 0.23955 | 0.097838 | 1663.735 |
| WANG_ME | 22 | 0.13005 | 0.004437 | 0.10051 | 0.097865 | 1664.194 |
| GOBP_GAF | 15 | 0.20218 | 0.005697 | 0.15627 | 0.097882 | 1664.483 |
| SASAKI_AD | 156 | 0.051075 | 0.004624 | 0.039489 | 0.097943 | 1665.521 |
| GOBP_REG | 9 | 0.2629 | 0.005739 | 0.2033 | 0.097986 | 1666.252 |

| | | | | | | |
|---|---|---|---|---|---|---|
| BHATI_G2I | 116 | 0.058886 | 0.004602 | 0.04556 | 0.098101 | 1668.208 |
| BIOCARTA_ | 5 | 0.28939 | 0.004709 | 0.22393 | 0.098137 | 1668.82 |
| GOCC_CAV | 82 | 0.075497 | 0.004965 | 0.058508 | 0.098469 | 1674.465 |
| GOBP_REG | 23 | 0.11346 | 0.003958 | 0.087928 | 0.098479 | 1674.635 |
| WP_ERBB_ | 87 | 0.07209 | 0.004883 | 0.055872 | 0.098487 | 1674.771 |
| GOMF_PH( | 181 | 0.047972 | 0.004675 | 0.037184 | 0.098511 | 1675.18 |
| GOBP_REG | 12 | 0.19974 | 0.005034 | 0.15486 | 0.098559 | 1675.996 |
| GOBP_AUT | 58 | 0.082245 | 0.004552 | 0.063763 | 0.098562 | 1676.047 |
| GOBP_MAI | 22 | 0.15637 | 0.005335 | 0.12123 | 0.098563 | 1676.064 |
| GOBP_INTI | 23 | 0.15266 | 0.005325 | 0.11837 | 0.098582 | 1676.387 |
| WP_DEREG | 14 | 0.16611 | 0.004522 | 0.12881 | 0.098604 | 1676.761 |
| GOBP_HEA | 18 | 0.16219 | 0.005006 | 0.1258 | 0.098664 | 1677.781 |
| MAHADEV/ | 15 | 0.18676 | 0.005263 | 0.14489 | 0.098712 | 1678.598 |
| GOMF_CHI | 27 | 0.124 | 0.004686 | 0.096209 | 0.098727 | 1678.853 |
| GOBP_POS | 43 | 0.10349 | 0.004934 | 0.080315 | 0.098787 | 1679.873 |
| SANCHEZ_I | 11 | 0.19212 | 0.004636 | 0.14911 | 0.098808 | 1680.23 |
| GOMF_ENI | 8 | 0.20595 | 0.004239 | 0.15989 | 0.09888 | 1681.454 |
| GOBP_SCH | 36 | 0.11026 | 0.004811 | 0.085609 | 0.098885 | 1681.539 |
| GOBP_RHY | 46 | 0.098752 | 0.004869 | 0.076677 | 0.098899 | 1681.777 |
| SCHAEFFEF | 44 | 0.10562 | 0.005093 | 0.082029 | 0.098944 | 1682.543 |
| SAGIV_CD2 | 45 | 0.089077 | 0.004344 | 0.069187 | 0.098971 | 1683.002 |
| GOBP_RES | 26 | 0.13105 | 0.00486 | 0.1018 | 0.098993 | 1683.376 |
| GOMF_HIG | 23 | 0.15058 | 0.005253 | 0.11706 | 0.099162 | 1686.25 |
| WP_HFE_E | 7 | 0.18823 | 0.003624 | 0.14634 | 0.099191 | 1686.743 |
| DANG_MY( | 29 | 0.12794 | 0.005011 | 0.099498 | 0.099249 | 1687.729 |
| GOBP_REG | 9 | 0.22063 | 0.004816 | 0.17162 | 0.099309 | 1688.75 |
| GOMF_GU/ | 7 | 0.2768 | 0.005329 | 0.21536 | 0.099354 | 1689.515 |
| GOCC_FILA | 26 | 0.12328 | 0.004572 | 0.095919 | 0.099361 | 1689.634 |
| GOBP_VES | 8 | 0.22776 | 0.004688 | 0.17727 | 0.099429 | 1690.79 |
| WP_LINOLI | 5 | 0.32028 | 0.005212 | 0.24934 | 0.099488 | 1691.793 |
| GOBP_POS | 5 | 0.39883 | 0.00649 | 0.3105 | 0.099492 | 1691.861 |
| REACTOME | 91 | 0.07015 | 0.004859 | 0.054622 | 0.099531 | 1692.525 |
| DORSAM_I | 31 | 0.11253 | 0.004557 | 0.087627 | 0.099539 | 1692.661 |
| GOBP_PIGI | 36 | 0.10135 | 0.004422 | 0.078927 | 0.099558 | 1692.984 |
| LEI_HOXC8 | 17 | 0.1511 | 0.004532 | 0.11768 | 0.099586 | 1693.46 |
| GOBP_REG | 9 | 0.21565 | 0.004707 | 0.16797 | 0.099603 | 1693.749 |
| GOCC_CAT | 10 | 0.21583 | 0.004966 | 0.16817 | 0.09968 | 1695.058 |
| GOMF_C2I | 11 | 0.18098 | 0.004367 | 0.14103 | 0.099703 | 1695.45 |
| WP_PATH\ | 57 | 0.085418 | 0.004687 | 0.066563 | 0.099708 | 1695.535 |
| CHARAFE_I | 360 | 0.033184 | 0.004539 | 0.025861 | 0.099728 | 1695.875 |
| ZHENG_GL | 233 | 0.042573 | 0.004701 | 0.033181 | 0.099746 | 1696.181 |
| GOBP_REG | 14 | 0.16089 | 0.00438 | 0.12548 | 0.099894 | 1698.697 |
| MARTIN_N | 42 | 0.10496 | 0.004945 | 0.081882 | 0.099955 | 1699.735 |
| KATSANOU | 162 | 0.052577 | 0.00485 | 0.041023 | 0.099993 | 1700.381 |
| GOBP_UBI( | 5 | 0.35177 | 0.005724 | 0.27479 | 0.10026 | 1704.921 |

| | | | | | | |
|---|---|---|---|---|---|---|
| GARY_CD5 | 405 | 0.032376 | 0.004691 | 0.025296 | 0.1003 | 1705.602 |
| GOBP_FOR | 7 | 0.28244 | 0.005438 | 0.22068 | 0.10031 | 1705.772 |
| NAKAMUR. | 572 | 0.027305 | 0.00468 | 0.021338 | 0.10034 | 1706.282 |
| BIDUS_ME | 197 | 0.045341 | 0.004608 | 0.035444 | 0.10042 | 1707.642 |
| GOBP_MO | 21 | 0.13828 | 0.00461 | 0.10812 | 0.10045 | 1708.152 |
| GOBP_CEL | 12 | 0.20313 | 0.00512 | 0.15887 | 0.10053 | 1709.513 |
| REACTOME | 19 | 0.14792 | 0.00469 | 0.1157 | 0.10054 | 1709.683 |
| GOBP_GEN | 1344 | 0.018592 | 0.004781 | 0.014544 | 0.10058 | 1710.363 |
| GOBP_HIST | 6 | 0.30668 | 0.005467 | 0.23993 | 0.1006 | 1710.703 |
| GOBP_NEU | 7 | 0.24674 | 0.00475 | 0.19308 | 0.10065 | 1711.553 |
| GOMF_MU | 7 | 0.19169 | 0.003691 | 0.15 | 0.10065 | 1711.553 |
| REACTOME | 26 | 0.12474 | 0.004626 | 0.097639 | 0.10071 | 1712.574 |
| GOBP_NEU | 211 | 0.047489 | 0.004993 | 0.037179 | 0.10076 | 1713.424 |
| ONGUSAH/ | 33 | 0.11328 | 0.004732 | 0.088685 | 0.10076 | 1713.424 |
| REACTOME | 5 | 0.29054 | 0.004728 | 0.22752 | 0.10081 | 1714.274 |
| GOBP_POS | 17 | 0.14628 | 0.004388 | 0.11459 | 0.10088 | 1715.464 |
| GOBP_REG | 8 | 0.25007 | 0.005147 | 0.1959 | 0.10089 | 1715.634 |
| GOBP_END | 92 | 0.064127 | 0.004466 | 0.050251 | 0.10096 | 1716.825 |
| GOMF_ARI | 9 | 0.17512 | 0.003823 | 0.13724 | 0.10098 | 1717.165 |
| MIKKELSEN | 5 | 0.24956 | 0.004061 | 0.1956 | 0.10101 | 1717.675 |
| GOBP_POS | 129 | 0.057831 | 0.004764 | 0.04534 | 0.10108 | 1718.865 |
| YU_MYC_T | 43 | 0.088668 | 0.004227 | 0.06953 | 0.10112 | 1719.546 |
| JI_CARCIN( | 15 | 0.19106 | 0.005384 | 0.14984 | 0.10114 | 1719.886 |
| GOBP_POS | 14 | 0.18 | 0.0049 | 0.14118 | 0.10117 | 1720.396 |
| BURTON_A | 44 | 0.098793 | 0.004764 | 0.077491 | 0.10118 | 1720.566 |
| KORKOLA_ | 20 | 0.1446 | 0.004704 | 0.11346 | 0.10126 | 1721.926 |
| REACTOME | 6 | 0.24496 | 0.004367 | 0.19221 | 0.10126 | 1721.926 |
| GOBP_POS | 25 | 0.14022 | 0.005099 | 0.11004 | 0.1013 | 1722.607 |
| KYNG_RESI | 40 | 0.094597 | 0.00435 | 0.074257 | 0.10136 | 1723.627 |
| WP_CALOF | 8 | 0.21035 | 0.004329 | 0.16513 | 0.10136 | 1723.627 |
| GOBP_POS | 14 | 0.18472 | 0.005029 | 0.14501 | 0.10137 | 1723.797 |
| GOBP_REG | 121 | 0.056758 | 0.00453 | 0.044561 | 0.10139 | 1724.137 |
| GOBP_WN | 5 | 0.35086 | 0.005709 | 0.27547 | 0.1014 | 1724.307 |
| REACTOME | 9 | 0.21944 | 0.00479 | 0.17232 | 0.10144 | 1724.987 |
| GOBP_ERB | 5 | 0.34239 | 0.005572 | 0.26894 | 0.1015 | 1726.008 |
| PID_BMP_I | 39 | 0.11672 | 0.0053 | 0.091698 | 0.10154 | 1726.688 |
| GOBP_REG | 22 | 0.13821 | 0.004716 | 0.10859 | 0.10155 | 1726.858 |
| GOBP_PRC | 122 | 0.055727 | 0.004465 | 0.043788 | 0.10158 | 1727.368 |
| GOCC_MIT | 167 | 0.049379 | 0.004624 | 0.038803 | 0.1016 | 1727.708 |
| GOBP_REG | 12 | 0.15086 | 0.003802 | 0.11857 | 0.10163 | 1728.218 |
| CARRILLOR | 157 | 0.054924 | 0.004988 | 0.043171 | 0.10166 | 1728.728 |
| WANG_RE( | 16 | 0.16415 | 0.004777 | 0.12903 | 0.10167 | 1728.898 |
| KEGG_GLY( | 23 | 0.12805 | 0.004467 | 0.10067 | 0.1017 | 1729.409 |
| GOBP_NCR | 567 | 0.025119 | 0.004288 | 0.019749 | 0.10171 | 1729.579 |
| SESTO_RES | 38 | 0.094871 | 0.004252 | 0.07461 | 0.10177 | 1730.599 |

| | | | | | | |
|---|---|---|---|---|---|---|
| GOMF_OXI | 59 | 0.076626 | 0.004277 | 0.060277 | 0.10183 | 1731.619 |
| KRIEG_HYP | 50 | 0.09488 | 0.004877 | 0.074644 | 0.10186 | 1732.129 |
| GOBP_HAR | 8 | 0.26583 | 0.005471 | 0.20916 | 0.10189 | 1732.639 |
| REACTOME | 142 | 0.051084 | 0.004414 | 0.040201 | 0.10193 | 1733.32 |
| GOBP_NEG | 15 | 0.16254 | 0.00458 | 0.12793 | 0.10195 | 1733.66 |
| SAKAI_CHR | 74 | 0.070092 | 0.00438 | 0.055165 | 0.10195 | 1733.66 |
| ZWANG_CI | 214 | 0.045832 | 0.004852 | 0.036072 | 0.10195 | 1733.66 |
| REACTOME | 34 | 0.11941 | 0.005063 | 0.093991 | 0.10196 | 1733.83 |
| GOBP_MEI | 23 | 0.15462 | 0.005394 | 0.1217 | 0.10197 | 1734 |
| AMIT_SERU | 34 | 0.10622 | 0.004504 | 0.08367 | 0.10213 | 1736.721 |
| LEE_NEURA | 104 | 0.070626 | 0.005228 | 0.055651 | 0.10221 | 1738.081 |
| GOMF_GLU | 10 | 0.17729 | 0.004079 | 0.13971 | 0.10223 | 1738.421 |
| GOBP_REG | 465 | 0.028313 | 0.004389 | 0.022321 | 0.10233 | 1740.122 |
| GOBP_SYN | 8 | 0.2265 | 0.004662 | 0.17865 | 0.10245 | 1742.162 |
| UEDA_PER | 167 | 0.049716 | 0.004655 | 0.039219 | 0.10247 | 1742.502 |
| GOBP_CEN | 6 | 0.19867 | 0.003541 | 0.15675 | 0.1025 | 1743.013 |
| GOBP_REG | 10 | 0.20671 | 0.004756 | 0.16314 | 0.10258 | 1744.373 |
| GOBP_SIGI | 60 | 0.089346 | 0.005029 | 0.070529 | 0.10262 | 1745.053 |
| GOBP_NEG | 8 | 0.23284 | 0.004792 | 0.18389 | 0.10273 | 1746.924 |
| GOCC_DEN | 11 | 0.18727 | 0.004519 | 0.14795 | 0.10281 | 1748.284 |
| DING_LUN | 11 | 0.18272 | 0.00441 | 0.14444 | 0.10293 | 1750.325 |
| GOBP_RES | 18 | 0.15124 | 0.004668 | 0.11966 | 0.10313 | 1753.726 |
| WONG_AD | 674 | 0.025962 | 0.004817 | 0.020542 | 0.10315 | 1754.066 |
| WP_EPITHI | 156 | 0.052681 | 0.004769 | 0.04169 | 0.10319 | 1754.746 |
| GOBP_GRC | 850 | 0.022624 | 0.004691 | 0.017906 | 0.10322 | 1755.256 |
| GOCC_KINI | 45 | 0.087522 | 0.004268 | 0.069291 | 0.10328 | 1756.276 |
| GOCC_MIC | 198 | 0.0443 | 0.004513 | 0.035074 | 0.1033 | 1756.617 |
| GOCC_SEH | 8 | 0.207 | 0.00426 | 0.16392 | 0.10333 | 1757.127 |
| GOMF_NO | 23 | 0.13688 | 0.004775 | 0.10843 | 0.10342 | 1758.657 |
| GOMF_ATF | 29 | 0.11094 | 0.004345 | 0.08794 | 0.10357 | 1761.208 |
| GOBP_POS | 190 | 0.045871 | 0.004579 | 0.036367 | 0.1036 | 1761.718 |
| KEGG_MEL | 100 | 0.063255 | 0.004592 | 0.050153 | 0.10362 | 1762.058 |
| TIEN_INTES | 202 | 0.043605 | 0.004486 | 0.034581 | 0.10367 | 1762.908 |
| YANG_BRE | 35 | 0.12714 | 0.00547 | 0.10084 | 0.10368 | 1763.078 |
| GOBP_CELI | 38 | 0.11584 | 0.005192 | 0.091922 | 0.10382 | 1765.459 |
| GOBP_REG | 6 | 0.32029 | 0.005709 | 0.25422 | 0.10386 | 1766.139 |
| GOBP_NEG | 197 | 0.042459 | 0.004315 | 0.033708 | 0.10391 | 1766.99 |
| GOBP_VEN | 7 | 0.27847 | 0.005361 | 0.22135 | 0.10419 | 1771.751 |
| GOBP_POS | 5 | 0.23729 | 0.003861 | 0.18869 | 0.10428 | 1773.281 |
| GOBP_CELI | 14 | 0.16145 | 0.004395 | 0.12839 | 0.10429 | 1773.451 |
| GOBP_SKEI | 5 | 0.35957 | 0.005851 | 0.28593 | 0.10429 | 1773.451 |
| GOBP_SUC | 6 | 0.26267 | 0.004682 | 0.20889 | 0.10431 | 1773.792 |
| GOMF_SUC | 6 | 0.26267 | 0.004682 | 0.20889 | 0.10431 | 1773.792 |
| GOBP_DEC | 14 | 0.1572 | 0.004279 | 0.12506 | 0.10439 | 1775.152 |
| LE_EGR2_T | 103 | 0.06332 | 0.004664 | 0.050382 | 0.10442 | 1775.662 |

| | | | | | | |
|---|---|---|---|---|---|---|
| REACTOME | 7 | 0.24968 | 0.004807 | 0.19872 | 0.10449 | 1776.852 |
| GOCC_CLA | 77 | 0.071205 | 0.004538 | 0.056744 | 0.10478 | 1781.784 |
| REACTOME | 32 | 0.11321 | 0.004657 | 0.090226 | 0.1048 | 1782.124 |
| REACTOME | 10 | 0.18636 | 0.004288 | 0.14855 | 0.10484 | 1782.804 |
| GOBP_POS | 25 | 0.14026 | 0.005101 | 0.11183 | 0.10488 | 1783.484 |
| WP_MAMI | 24 | 0.13875 | 0.004944 | 0.11065 | 0.10492 | 1784.165 |
| BOYAULT_ | 15 | 0.1703 | 0.004799 | 0.13582 | 0.10494 | 1784.505 |
| GOMF_SCA | 45 | 0.097254 | 0.004743 | 0.077576 | 0.10499 | 1785.355 |
| REACTOME | 24 | 0.1279 | 0.004558 | 0.10203 | 0.10501 | 1785.695 |
| GOBP_REG | 10 | 0.20088 | 0.004622 | 0.16029 | 0.10508 | 1786.885 |
| GOBP_POS | 31 | 0.12525 | 0.005071 | 0.099949 | 0.10509 | 1787.055 |
| GOBP_PRC | 166 | 0.048054 | 0.004486 | 0.03835 | 0.10511 | 1787.396 |
| PID_RANBI | 11 | 0.18306 | 0.004418 | 0.14609 | 0.10511 | 1787.396 |
| GOMF_OXY | 7 | 0.2612 | 0.005029 | 0.20855 | 0.10521 | 1789.096 |
| GOBP_SIGI | 22 | 0.13481 | 0.0046 | 0.10771 | 0.10536 | 1791.647 |
| REACTOME | 34 | 0.098583 | 0.00418 | 0.078766 | 0.10537 | 1791.817 |
| GOBP_GLU | 12 | 0.18753 | 0.004726 | 0.14986 | 0.10542 | 1792.667 |
| REACTOME | 103 | 0.06136 | 0.00452 | 0.049037 | 0.10543 | 1792.837 |
| WP_MAMI | 14 | 0.18128 | 0.004935 | 0.14489 | 0.10545 | 1793.177 |
| OHM_EMB | 6 | 0.23064 | 0.004111 | 0.18437 | 0.10548 | 1793.687 |
| GOBP_REG | 10 | 0.21938 | 0.005048 | 0.17539 | 0.10552 | 1794.368 |
| REACTOME | 10 | 0.23397 | 0.005384 | 0.18713 | 0.1056 | 1795.728 |
| GOBP_MYE | 36 | 0.099309 | 0.004333 | 0.079431 | 0.10561 | 1795.898 |
| GOBP_RES | 28 | 0.13212 | 0.005085 | 0.10568 | 0.10562 | 1796.068 |
| GOBP_REN | 18 | 0.16715 | 0.005159 | 0.13373 | 0.10567 | 1796.918 |
| SENGUPTA | 369 | 0.034223 | 0.004738 | 0.027392 | 0.10577 | 1798.619 |
| GOBP_MA( | 7 | 0.27617 | 0.005317 | 0.22121 | 0.10594 | 1801.51 |
| GOBP_EMI | 86 | 0.071447 | 0.004811 | 0.057235 | 0.10597 | 1802.02 |
| GOBP_VIT/ | 5 | 0.29958 | 0.004875 | 0.24001 | 0.10598 | 1802.19 |
| REACTOME | 50 | 0.088698 | 0.004559 | 0.071061 | 0.10599 | 1802.36 |
| GOCC_TRA | 74 | 0.071722 | 0.004482 | 0.057464 | 0.106 | 1802.53 |
| YAO_HOXA | 75 | 0.07372 | 0.004637 | 0.059089 | 0.1061 | 1804.231 |
| GOBP_NEG | 40 | 0.10167 | 0.004675 | 0.081522 | 0.10618 | 1805.591 |
| REACTOME | 134 | 0.051978 | 0.004364 | 0.041678 | 0.10618 | 1805.591 |
| GOMF_CAI | 7 | 0.26637 | 0.005128 | 0.21359 | 0.10619 | 1805.761 |
| YAMAZAKI_ | 158 | 0.051621 | 0.004703 | 0.041399 | 0.10622 | 1806.271 |
| WALLACE_ | 5 | 0.28929 | 0.004708 | 0.23208 | 0.10629 | 1807.461 |
| WEST_ADR | 20 | 0.12731 | 0.004142 | 0.10218 | 0.1064 | 1809.332 |
| GOBP_NEG | 10 | 0.21403 | 0.004925 | 0.17179 | 0.10641 | 1809.502 |
| REACTOME | 15 | 0.14315 | 0.004033 | 0.11492 | 0.10646 | 1810.352 |
| GOMF_CHI | 8 | 0.18326 | 0.003772 | 0.14716 | 0.10652 | 1811.373 |
| GOBP_PEP | 18 | 0.16588 | 0.00512 | 0.13323 | 0.10655 | 1811.883 |
| SMITH_TEI | 130 | 0.05614 | 0.004643 | 0.045092 | 0.10657 | 1812.223 |
| GOMF_PH( | 18 | 0.1482 | 0.004574 | 0.11907 | 0.10664 | 1813.413 |
| WP_MECH | 44 | 0.095311 | 0.004596 | 0.076584 | 0.10666 | 1813.753 |

| | | | | | | |
|---|---|---|---|---|---|---|
| GOBP_POS | 9 | 0.1945 | 0.004246 | 0.1563 | 0.10669 | 1814.263 |
| PID_EPO_F | 32 | 0.10709 | 0.004405 | 0.08611 | 0.10683 | 1816.644 |
| PURBEY_TA | 70 | 0.069765 | 0.00424 | 0.056159 | 0.10708 | 1820.895 |
| GOBP_NEG | 28 | 0.12031 | 0.00463 | 0.096872 | 0.10714 | 1821.916 |
| GOMF_1_4 | 5 | 0.22773 | 0.003706 | 0.18346 | 0.10726 | 1823.956 |
| REACTOME | 19 | 0.13059 | 0.004141 | 0.10524 | 0.10733 | 1825.147 |
| GOCC_NUC | 89 | 0.062846 | 0.004305 | 0.050664 | 0.10741 | 1826.507 |
| GOMF_FRI | 36 | 0.096767 | 0.004222 | 0.07801 | 0.10742 | 1826.677 |
| GOBP_RES | 348 | 0.034167 | 0.004596 | 0.02756 | 0.10754 | 1828.718 |
| GOBP_NEG | 47 | 0.099586 | 0.004963 | 0.080377 | 0.10768 | 1831.098 |
| NAKAJIMA_ | 42 | 0.091228 | 0.004298 | 0.073642 | 0.10772 | 1831.779 |
| IKEDA_MIR | 53 | 0.09044 | 0.004785 | 0.073012 | 0.10774 | 1832.119 |
| GOBP_POS | 15 | 0.17159 | 0.004835 | 0.13862 | 0.10788 | 1834.499 |
| GOBP_NEG | 101 | 0.064426 | 0.0047 | 0.052066 | 0.10798 | 1836.2 |
| REACTOME | 72 | 0.072326 | 0.004458 | 0.058461 | 0.10802 | 1836.88 |
| GOMF_LAF | 5 | 0.21618 | 0.003518 | 0.17475 | 0.10803 | 1837.05 |
| GOBP_MET | 14 | 0.19712 | 0.005366 | 0.15937 | 0.10808 | 1837.9 |
| GOBP_NEG | 172 | 0.047485 | 0.004512 | 0.038392 | 0.10808 | 1837.9 |
| GOMF_ARG | 23 | 0.15554 | 0.005426 | 0.12577 | 0.10811 | 1838.411 |
| GOBP_SMC | 140 | 0.052423 | 0.004498 | 0.042391 | 0.10812 | 1838.581 |
| GOCC_END | 291 | 0.0372 | 0.004583 | 0.030082 | 0.10812 | 1838.581 |
| GOBP_GOL | 58 | 0.078436 | 0.004341 | 0.063449 | 0.1082 | 1839.941 |
| BIOCARTA_ | 6 | 0.25426 | 0.004532 | 0.20572 | 0.10824 | 1840.621 |
| GOCC_LAR | 107 | 0.051375 | 0.003857 | 0.041568 | 0.10825 | 1840.791 |
| GOBP_SPIN | 13 | 0.16346 | 0.004288 | 0.13229 | 0.1083 | 1841.642 |
| GOMF_STE | 32 | 0.11455 | 0.004712 | 0.092742 | 0.1084 | 1843.342 |
| FIGUEROA_ | 8 | 0.24925 | 0.00513 | 0.20185 | 0.10845 | 1844.192 |
| GOBP_MYE | 213 | 0.044563 | 0.004707 | 0.036091 | 0.10847 | 1844.532 |
| MACAEVA_ | 95 | 0.061116 | 0.004325 | 0.049498 | 0.10848 | 1844.702 |
| GOCC_CYT | 52 | 0.076391 | 0.004004 | 0.061873 | 0.10849 | 1844.872 |
| GOBP_CHO | 9 | 0.2206 | 0.004815 | 0.1787 | 0.10853 | 1845.553 |
| GOBP_APO | 43 | 0.096312 | 0.004591 | 0.078056 | 0.10863 | 1847.253 |
| GOMF_SIG | 491 | 0.029842 | 0.00475 | 0.024205 | 0.10882 | 1850.484 |
| GOBP_REG | 14 | 0.16962 | 0.004618 | 0.13763 | 0.10889 | 1851.674 |
| REACTOME | 57 | 0.08423 | 0.004621 | 0.06834 | 0.10889 | 1851.674 |
| PETRETTO_ | 7 | 0.26973 | 0.005193 | 0.21888 | 0.10893 | 1852.355 |
| WP_GLYOX | 11 | 0.18594 | 0.004487 | 0.1509 | 0.10893 | 1852.355 |
| TESAR_ALK | 5 | 0.2836 | 0.004615 | 0.23033 | 0.10912 | 1855.586 |
| GOBP_END | 87 | 0.070154 | 0.004752 | 0.056994 | 0.10919 | 1856.776 |
| GOBP_HET | 51 | 0.098811 | 0.005129 | 0.080287 | 0.10922 | 1857.286 |
| GOBP_PRO | 5 | 0.30715 | 0.004998 | 0.24957 | 0.10922 | 1857.286 |
| BYSTRYKH_ | 65 | 0.078406 | 0.004593 | 0.063718 | 0.10926 | 1857.966 |
| GOBP_SMC | 21 | 0.13664 | 0.004555 | 0.11104 | 0.10926 | 1857.966 |
| AUJLA_IL22 | 9 | 0.18127 | 0.003957 | 0.14736 | 0.10934 | 1859.327 |
| MATZUK_M | 8 | 0.22157 | 0.00456 | 0.18012 | 0.10934 | 1859.327 |

| | | | | | | |
|---|---|---|---|---|---|---|
| GOBP_POS | 5 | 0.26694 | 0.004344 | 0.21704 | 0.10937 | 1859.837 |
| SESTO_RES | 20 | 0.14444 | 0.004699 | 0.11746 | 0.10942 | 1860.687 |
| GOBP_GLU | 3 | 0.39878 | 0.005027 | 0.32435 | 0.10945 | 1861.197 |
| GOBP_POS | 5 | 0.32864 | 0.005348 | 0.26732 | 0.10947 | 1861.537 |
| GOBP_MAI | 13 | 0.14575 | 0.003823 | 0.11866 | 0.10968 | 1865.108 |
| VANOEVEL | 210 | 0.040504 | 0.004248 | 0.032993 | 0.1098 | 1867.149 |
| GOBP_REG | 7 | 0.22559 | 0.004343 | 0.18376 | 0.10981 | 1867.319 |
| GOBP_URE | 16 | 0.17302 | 0.005035 | 0.14097 | 0.10985 | 1867.999 |
| DACOSTA_ | 286 | 0.033617 | 0.004106 | 0.027393 | 0.10988 | 1868.509 |
| GOMF_N_/ | 5 | 0.27487 | 0.004473 | 0.22399 | 0.10989 | 1868.679 |
| URS_ADIPC | 31 | 0.11778 | 0.004769 | 0.095995 | 0.10993 | 1869.36 |
| GOBP_MRI | 6 | 0.19593 | 0.003493 | 0.15982 | 0.11011 | 1872.421 |
| DACOSTA_ | 64 | 0.067882 | 0.003946 | 0.055375 | 0.11013 | 1872.761 |
| GOMF_RN/ | 6 | 0.29441 | 0.005248 | 0.24019 | 0.11016 | 1873.271 |
| REACTOME | 19 | 0.12731 | 0.004037 | 0.1039 | 0.11024 | 1874.631 |
| KEGG_MEL | 68 | 0.076822 | 0.004602 | 0.062707 | 0.11028 | 1875.311 |
| GOMF_ME | 24 | 0.14062 | 0.005011 | 0.11484 | 0.11039 | 1877.182 |
| GOCC_CELI | 407 | 0.03024 | 0.004392 | 0.024702 | 0.11044 | 1878.032 |
| GOBP_HIST | 15 | 0.16231 | 0.004573 | 0.1326 | 0.11048 | 1878.712 |
| GOBP_NEP | 12 | 0.20887 | 0.005264 | 0.17068 | 0.11053 | 1879.563 |
| WESTON_\ | 59 | 0.081943 | 0.004574 | 0.066984 | 0.11061 | 1880.923 |
| GOBP_POS | 15 | 0.15405 | 0.004341 | 0.12597 | 0.11068 | 1882.113 |
| GOBP_NEG | 44 | 0.089749 | 0.004328 | 0.073397 | 0.11072 | 1882.794 |
| GOBP_REG | 37 | 0.10587 | 0.004683 | 0.086596 | 0.11075 | 1883.304 |
| GOCC_CHY | 12 | 0.17858 | 0.004501 | 0.14607 | 0.11076 | 1883.474 |
| DANG_BOU | 974 | 0.018905 | 0.004182 | 0.015466 | 0.1108 | 1884.154 |
| GOBP_CELI | 8 | 0.2037 | 0.004193 | 0.16669 | 0.11085 | 1885.004 |
| GOBP_CAL | 15 | 0.1779 | 0.005013 | 0.1456 | 0.11089 | 1885.684 |
| GOBP_POS | 7 | 0.22112 | 0.004257 | 0.181 | 0.11093 | 1886.365 |
| GOBP_POS | 11 | 0.17108 | 0.004129 | 0.14016 | 0.11112 | 1889.596 |
| MARTIN_N | 11 | 0.16691 | 0.004028 | 0.13675 | 0.11115 | 1890.106 |
| GOBP_POS | 15 | 0.14924 | 0.004205 | 0.1223 | 0.11119 | 1890.786 |
| HERNANDE | 20 | 0.14623 | 0.004757 | 0.11985 | 0.11121 | 1891.126 |
| BIOCARTA_ | 18 | 0.14735 | 0.004548 | 0.12081 | 0.1113 | 1892.657 |
| GOBP_MAS | 14 | 0.16815 | 0.004578 | 0.13788 | 0.11132 | 1892.997 |
| FORTSCHE( | 251 | 0.037976 | 0.00435 | 0.031151 | 0.11141 | 1894.527 |
| GOCC_SPEI | 16 | 0.15292 | 0.00445 | 0.12544 | 0.11141 | 1894.527 |
| REACTOME | 19 | 0.16812 | 0.005331 | 0.13795 | 0.11148 | 1895.717 |
| LI_WILMS_ | 157 | 0.047992 | 0.004358 | 0.039383 | 0.11151 | 1896.228 |
| SYED_ESTR | 15 | 0.1721 | 0.004849 | 0.14123 | 0.11151 | 1896.228 |
| GOBP_SKEI | 10 | 0.19744 | 0.004543 | 0.16211 | 0.11163 | 1898.268 |
| GOBP_PRC | 13 | 0.16525 | 0.004335 | 0.1357 | 0.11167 | 1898.948 |
| GOBP_PRC | 17 | 0.13343 | 0.004002 | 0.10967 | 0.11188 | 1902.519 |
| GOCC_PRC | 37 | 0.10059 | 0.004449 | 0.08272 | 0.11199 | 1904.39 |
| KEGG_FOC | 190 | 0.045584 | 0.00455 | 0.03749 | 0.11202 | 1904.9 |

| | | | | | | |
|---|---|---|---|---|---|---|
| GOMF_HM | 14 | 0.18681 | 0.005085 | 0.15365 | 0.11204 | 1905.24 |
| REACTOME | 12 | 0.17258 | 0.00435 | 0.14195 | 0.11206 | 1905.58 |
| WINTER_H | 220 | 0.041475 | 0.004451 | 0.034127 | 0.11213 | 1906.771 |
| VANDESLU | 16 | 0.14381 | 0.004185 | 0.11834 | 0.11215 | 1907.111 |
| GOMF_SOI | 13 | 0.17785 | 0.004666 | 0.14637 | 0.11218 | 1907.621 |
| HORIUCHI_ | 297 | 0.033947 | 0.004224 | 0.027942 | 0.11221 | 1908.131 |
| NOJIMA_SI | 29 | 0.11728 | 0.004593 | 0.096542 | 0.11223 | 1908.471 |
| KANG_FLU | 21 | 0.12795 | 0.004265 | 0.10535 | 0.11227 | 1909.151 |
| REACTOME | 12 | 0.21094 | 0.005317 | 0.17369 | 0.1123 | 1909.662 |
| REACTOME | 6 | 0.27752 | 0.004947 | 0.22855 | 0.11233 | 1910.172 |
| NAGASHIM | 55 | 0.081292 | 0.004381 | 0.066958 | 0.11237 | 1910.852 |
| REACTOME | 4 | 0.30158 | 0.00439 | 0.24841 | 0.11237 | 1910.852 |
| GOMF_PHO | 101 | 0.061691 | 0.0045 | 0.050818 | 0.11239 | 1911.192 |
| GRANDVAU | 13 | 0.1734 | 0.004549 | 0.14286 | 0.11243 | 1911.872 |
| GOMF_PHO | 26 | 0.11872 | 0.004403 | 0.097822 | 0.11245 | 1912.212 |
| GOCC_DEN | 14 | 0.16935 | 0.00461 | 0.13956 | 0.11248 | 1912.722 |
| CASTELLAN | 6 | 0.24292 | 0.00433 | 0.2002 | 0.1125 | 1913.063 |
| GOBP_BIOI | 287 | 0.035947 | 0.004398 | 0.029625 | 0.1125 | 1913.063 |
| GOBP_NEG | 4 | 0.32724 | 0.004763 | 0.2698 | 0.11259 | 1914.593 |
| TAKAYAMA | 10 | 0.20396 | 0.004693 | 0.1682 | 0.11265 | 1915.613 |
| MEBARKI_I | 22 | 0.15942 | 0.005439 | 0.13148 | 0.11267 | 1915.953 |
| PARK_HSC_ | 43 | 0.083657 | 0.003988 | 0.068997 | 0.11267 | 1915.953 |
| GOBP_AXO | 6 | 0.19291 | 0.003439 | 0.15917 | 0.11277 | 1917.654 |
| GOBP_REG | 25 | 0.12844 | 0.004671 | 0.10598 | 0.11277 | 1917.654 |
| REACTOME | 190 | 0.043456 | 0.004338 | 0.035857 | 0.11278 | 1917.824 |
| BIOCARTA_ | 12 | 0.18982 | 0.004784 | 0.15665 | 0.11281 | 1918.334 |
| BASSO_CD | 67 | 0.073454 | 0.004368 | 0.060638 | 0.11289 | 1919.694 |
| STARK_PRE | 186 | 0.045562 | 0.0045 | 0.037619 | 0.11293 | 1920.375 |
| GOBP_CELI | 89 | 0.063848 | 0.004374 | 0.052719 | 0.11294 | 1920.545 |
| GOCC_HRD | 6 | 0.23031 | 0.004105 | 0.19021 | 0.113 | 1921.565 |
| GOMF_GD | 4 | 0.27435 | 0.003993 | 0.2266 | 0.11301 | 1921.735 |
| HUANG_GA | 71 | 0.068537 | 0.004195 | 0.056609 | 0.11301 | 1921.735 |
| REACTOME | 29 | 0.13131 | 0.005143 | 0.10845 | 0.11301 | 1921.735 |
| GOMF_PRO | 5 | 0.29322 | 0.004771 | 0.24219 | 0.11302 | 1921.905 |
| GOBP_REG | 16 | 0.16124 | 0.004692 | 0.13322 | 0.11307 | 1922.755 |
| REACTOME | 26 | 0.11903 | 0.004414 | 0.098358 | 0.11312 | 1923.606 |
| GOBP_MYO | 13 | 0.16071 | 0.004216 | 0.13285 | 0.1132 | 1924.966 |
| GOMF_MR | 93 | 0.061185 | 0.004284 | 0.050588 | 0.11325 | 1925.816 |
| GOBP_BRA | 10 | 0.22488 | 0.005174 | 0.18598 | 0.11331 | 1926.837 |
| GOBP_MAI | 7 | 0.23362 | 0.004498 | 0.19321 | 0.11331 | 1926.837 |
| GENTILE_U | 34 | 0.11104 | 0.004708 | 0.091849 | 0.11335 | 1927.517 |
| MARTINEZ_ | 574 | 0.025705 | 0.004414 | 0.021262 | 0.11335 | 1927.517 |
| WP_GDNFI | 21 | 0.1543 | 0.005144 | 0.12766 | 0.1134 | 1928.367 |
| KIM_WT1_ | 158 | 0.050159 | 0.00457 | 0.0415 | 0.11341 | 1928.537 |
| GOBP_HOM | 49 | 0.083177 | 0.004232 | 0.06882 | 0.11342 | 1928.707 |

| | | | | | | |
|---|---|---|---|---|---|---|
| GOMF_MR | 26 | 0.13259 | 0.004917 | 0.10975 | 0.11352 | 1930.408 |
| BROWNE_I | 47 | 0.090661 | 0.004518 | 0.075125 | 0.11376 | 1934.489 |
| GOBP_VEN | 16 | 0.16437 | 0.004783 | 0.13623 | 0.11381 | 1935.339 |
| GOBP_PAR | 7 | 0.25894 | 0.004985 | 0.21464 | 0.11385 | 1936.019 |
| GOBP_POS | 43 | 0.095653 | 0.00456 | 0.079292 | 0.11385 | 1936.019 |
| GOBP_PUR | 20 | 0.15056 | 0.004898 | 0.12483 | 0.11388 | 1936.529 |
| GOBP_NEG | 13 | 0.16657 | 0.00437 | 0.13812 | 0.11393 | 1937.38 |
| REACTOME | 23 | 0.13679 | 0.004772 | 0.11343 | 0.11394 | 1937.55 |
| OKAWA_N | 22 | 0.23843 | 0.008135 | 0.1978 | 0.11403 | 1939.08 |
| TONKS_TA | 174 | 0.045337 | 0.004333 | 0.037619 | 0.11408 | 1939.93 |
| GOBP_POS | 45 | 0.090764 | 0.004426 | 0.075349 | 0.11419 | 1941.801 |
| GOCC_CEN | 148 | 0.048031 | 0.004236 | 0.039889 | 0.11428 | 1943.331 |
| REACTOME | 7 | 0.2112 | 0.004066 | 0.1754 | 0.11428 | 1943.331 |
| LINDSTEDT | 58 | 0.075317 | 0.004168 | 0.062565 | 0.11434 | 1944.352 |
| GOMF_PH( | 24 | 0.10664 | 0.0038 | 0.088623 | 0.11445 | 1946.222 |
| BRUINS_U\ | 86 | 0.067906 | 0.004573 | 0.056454 | 0.11452 | 1947.413 |
| WP_METAI | 6 | 0.24239 | 0.004321 | 0.20152 | 0.11454 | 1947.753 |
| GOBP_POS | 7 | 0.22697 | 0.00437 | 0.18875 | 0.1146 | 1948.773 |
| MEISSNER_ | 78 | 0.075361 | 0.004834 | 0.062671 | 0.1146 | 1948.773 |
| GOBP_END | 208 | 0.042769 | 0.004465 | 0.035571 | 0.11462 | 1949.113 |
| GRAESSMA | 1658 | 0.015374 | 0.004352 | 0.012792 | 0.11473 | 1950.984 |
| OUELLET_( | 11 | 0.20965 | 0.005059 | 0.1746 | 0.11493 | 1954.385 |
| GOBP_NEG | 8 | 0.19472 | 0.004008 | 0.16217 | 0.11494 | 1954.555 |
| REACTOME | 24 | 0.11671 | 0.004159 | 0.097221 | 0.11499 | 1955.405 |
| GOBP_PAL | 171 | 0.049107 | 0.004653 | 0.04091 | 0.11501 | 1955.745 |
| GOMF_G_I | 21 | 0.13884 | 0.004628 | 0.11574 | 0.11517 | 1958.466 |
| GOBP_EXT | 12 | 0.18729 | 0.004721 | 0.15614 | 0.11518 | 1958.636 |
| GOMF_FOI | 5 | 0.2997 | 0.004877 | 0.24988 | 0.1152 | 1958.976 |
| GOBP_CAN | 9 | 0.20162 | 0.004401 | 0.1682 | 0.11534 | 1961.357 |
| PID_AVB3_ | 71 | 0.074255 | 0.004545 | 0.06195 | 0.11534 | 1961.357 |
| REACTOME | 10 | 0.19553 | 0.004499 | 0.16312 | 0.11534 | 1961.357 |
| GOBP_PRC | 6 | 0.21708 | 0.003869 | 0.18113 | 0.11537 | 1961.867 |
| CHARAFE_I | 427 | 0.02867 | 0.004263 | 0.02393 | 0.11545 | 1963.227 |
| GOBP_CAR | 1017 | 0.019053 | 0.004302 | 0.015905 | 0.11548 | 1963.737 |
| GOBP_RES | 304 | 0.034448 | 0.004336 | 0.028757 | 0.11549 | 1963.907 |
| GOBP_ERB | 10 | 0.21178 | 0.004873 | 0.17686 | 0.11557 | 1965.268 |
| ZHOU_INFI | 337 | 0.033899 | 0.004489 | 0.028317 | 0.11564 | 1966.458 |
| DARWICHE | 160 | 0.045465 | 0.004168 | 0.037988 | 0.11569 | 1967.308 |
| GOBP_DN/ | 971 | 0.018954 | 0.004187 | 0.01584 | 0.11574 | 1968.159 |
| WACKER_H | 13 | 0.181 | 0.004748 | 0.15126 | 0.11574 | 1968.159 |
| GOBP_POS | 109 | 0.057695 | 0.004371 | 0.048233 | 0.11582 | 1969.519 |
| REACTOME | 19 | 0.13308 | 0.00422 | 0.1113 | 0.11591 | 1971.05 |
| GOMF_HYI | 60 | 0.080367 | 0.004524 | 0.06722 | 0.11594 | 1971.56 |
| GOCC_ATP | 23 | 0.13 | 0.004535 | 0.10877 | 0.11603 | 1973.09 |
| GOBP_MEI | 6 | 0.22655 | 0.004038 | 0.18962 | 0.11611 | 1974.451 |

| | | | | | | |
|---|---|---|---|---|---|---|
| REACTOME | 5 | 0.19753 | 0.003214 | 0.16536 | 0.11614 | 1974.961 |
| GOBP_FAT | 12 | 0.14417 | 0.003634 | 0.12072 | 0.11618 | 1975.641 |
| BYSTROEM | 38 | 0.097546 | 0.004372 | 0.081679 | 0.1162 | 1975.981 |
| REACTOME | 12 | 0.18049 | 0.004549 | 0.15125 | 0.11638 | 1979.042 |
| PRAMOON | 48 | 0.083057 | 0.004183 | 0.069621 | 0.11645 | 1980.232 |
| REACTOME | 25 | 0.14389 | 0.005233 | 0.12063 | 0.11648 | 1980.742 |
| REN_ALVE( | 372 | 0.031915 | 0.004436 | 0.026756 | 0.11648 | 1980.742 |
| GOBP_POS | 8 | 0.22779 | 0.004688 | 0.191 | 0.11651 | 1981.253 |
| CROONQU | 37 | 0.098077 | 0.004338 | 0.082291 | 0.11667 | 1983.973 |
| GOCC_MIC | 786 | 0.021302 | 0.004255 | 0.017873 | 0.11667 | 1983.973 |
| GOBP_REG | 14 | 0.16476 | 0.004485 | 0.13832 | 0.1168 | 1986.184 |
| CHO_NR4A | 29 | 0.10316 | 0.00404 | 0.086609 | 0.11681 | 1986.354 |
| REACTOME | 5 | 0.24931 | 0.004057 | 0.20934 | 0.11685 | 1987.034 |
| REACTOME | 101 | 0.065838 | 0.004803 | 0.055295 | 0.1169 | 1987.885 |
| GOBP_NEG | 97 | 0.061322 | 0.004384 | 0.051513 | 0.11695 | 1988.735 |
| HOFMANN | 19 | 0.15106 | 0.00479 | 0.1269 | 0.11695 | 1988.735 |
| BROWNE_I | 48 | 0.087287 | 0.004396 | 0.073345 | 0.11702 | 1989.925 |
| GOCC_CON | 73 | 0.065148 | 0.004043 | 0.05475 | 0.11705 | 1990.435 |
| GOBP_CELI | 48 | 0.084401 | 0.004251 | 0.070961 | 0.11715 | 1992.136 |
| REACTOME | 11 | 0.20122 | 0.004856 | 0.16924 | 0.11724 | 1993.666 |
| GOMF_DO | 9 | 0.20969 | 0.004578 | 0.1764 | 0.11727 | 1994.176 |
| GOBP_DIAI | 8 | 0.22095 | 0.004547 | 0.18589 | 0.11731 | 1994.857 |
| GOMF_D1_ | 8 | 0.1633 | 0.003361 | 0.13739 | 0.11731 | 1994.857 |
| GOBP_SMA | 13 | 0.1975 | 0.005181 | 0.16622 | 0.11739 | 1996.217 |
| GOBP_POS | 6 | 0.20708 | 0.003691 | 0.17429 | 0.11741 | 1996.557 |
| GOMF_ACE | 20 | 0.14415 | 0.004689 | 0.12134 | 0.11743 | 1996.897 |
| GOBP_POS | 9 | 0.17181 | 0.00375 | 0.14463 | 0.11744 | 1997.067 |
| REACTOME | 40 | 0.093452 | 0.004297 | 0.078683 | 0.11749 | 1997.917 |
| GOBP_INTI | 50 | 0.079188 | 0.00407 | 0.066687 | 0.11753 | 1998.598 |
| GOMF_LIPI | 50 | 0.079188 | 0.00407 | 0.066687 | 0.11753 | 1998.598 |
| GOBP_PAR | 14 | 0.15727 | 0.004281 | 0.1325 | 0.11762 | 2000.128 |
| GOMF_STR | 23 | 0.1306 | 0.004556 | 0.11003 | 0.11764 | 2000.468 |
| GOBP_RIB( | 9 | 0.21939 | 0.004789 | 0.18499 | 0.11782 | 2003.529 |
| GOBP_CHC | 6 | 0.23954 | 0.00427 | 0.20203 | 0.11788 | 2004.549 |
| GOMF_OXI | 8 | 0.22341 | 0.004598 | 0.18843 | 0.1179 | 2004.89 |
| FALVELLA_ | 74 | 0.068115 | 0.004256 | 0.057492 | 0.11806 | 2007.61 |
| GOBP_CELI | 58 | 0.080214 | 0.004439 | 0.067717 | 0.11811 | 2008.461 |
| MCBRYAN_ | 251 | 0.038728 | 0.004436 | 0.032699 | 0.11813 | 2008.801 |
| GOBP_REG | 49 | 0.085118 | 0.004331 | 0.071883 | 0.11819 | 2009.821 |
| GOMF_PR( | 9 | 0.20386 | 0.00445 | 0.1722 | 0.11825 | 2010.841 |
| GOBP_CELI | 18 | 0.13523 | 0.004174 | 0.11433 | 0.11845 | 2014.242 |
| REACTOME | 14 | 0.15873 | 0.004321 | 0.13426 | 0.11857 | 2016.283 |
| WP_ARACH | 7 | 0.19151 | 0.003687 | 0.162 | 0.11858 | 2016.453 |
| GOBP_DNA | 18 | 0.1366 | 0.004216 | 0.11559 | 0.11865 | 2017.643 |
| GOBP_WN | 12 | 0.17926 | 0.004518 | 0.15169 | 0.11867 | 2017.983 |

| | | | | | | |
|---|---|---|---|---|---|---|
| GOMF_ADI | 8 | 0.25079 | 0.005162 | 0.21223 | 0.11867 | 2017.983 |
| REACTOME | 8 | 0.25079 | 0.005162 | 0.21223 | 0.11867 | 2017.983 |
| GAZDA_DI/ | 57 | 0.074365 | 0.00408 | 0.062933 | 0.11868 | 2018.153 |
| WANG_HC | 103 | 0.058239 | 0.00429 | 0.049299 | 0.11874 | 2019.174 |
| GOBP_MAI | 11 | 0.18739 | 0.004522 | 0.15876 | 0.11894 | 2022.575 |
| GOBP_VOL | 7 | 0.21038 | 0.00405 | 0.17824 | 0.11895 | 2022.745 |
| GOBP_MIT | 25 | 0.12027 | 0.004374 | 0.10191 | 0.11897 | 2023.085 |
| SHEN_SMA | 267 | 0.035688 | 0.004214 | 0.030246 | 0.11902 | 2023.935 |
| REACTOME | 79 | 0.065689 | 0.004241 | 0.055683 | 0.11907 | 2024.785 |
| REACTOME | 69 | 0.074513 | 0.004497 | 0.063175 | 0.11911 | 2025.466 |
| GOBP_DOF | 92 | 0.066489 | 0.00463 | 0.056381 | 0.11915 | 2026.146 |
| GOBP_VAS | 4 | 0.36482 | 0.00531 | 0.30941 | 0.11919 | 2026.826 |
| GOBP_5S_( | 6 | 0.19577 | 0.00349 | 0.16604 | 0.11921 | 2027.166 |
| GOCC_TRA | 6 | 0.19577 | 0.00349 | 0.16604 | 0.11921 | 2027.166 |
| REACTOME | 41 | 0.10478 | 0.004878 | 0.088887 | 0.11924 | 2027.676 |
| BIOCARTA_ | 14 | 0.18702 | 0.005091 | 0.15866 | 0.11926 | 2028.016 |
| GOBP_NEG | 7 | 0.22317 | 0.004297 | 0.18933 | 0.11926 | 2028.016 |
| GOMF_PR( | 13 | 0.17885 | 0.004692 | 0.1518 | 0.11937 | 2029.887 |
| GOCC_MAI | 5 | 0.21069 | 0.003429 | 0.17894 | 0.11951 | 2032.268 |
| GOMF_2_/ | 6 | 0.26133 | 0.004658 | 0.22198 | 0.11955 | 2032.948 |
| LIU_SOX4_ | 293 | 0.034457 | 0.004259 | 0.02927 | 0.11956 | 2033.118 |
| GOBP_CIR( | 197 | 0.041473 | 0.004214 | 0.035234 | 0.1196 | 2033.798 |
| WANG_CIS | 208 | 0.038724 | 0.004042 | 0.032921 | 0.11975 | 2036.349 |
| GOBP_CYT | 175 | 0.043064 | 0.004127 | 0.036638 | 0.11992 | 2039.24 |
| GOBP_RRN | 199 | 0.038071 | 0.003888 | 0.032396 | 0.11996 | 2039.92 |
| GOBP_REG | 8 | 0.19478 | 0.004009 | 0.16578 | 0.12001 | 2040.77 |
| GOBP_REG | 16 | 0.14109 | 0.004106 | 0.1201 | 0.12005 | 2041.45 |
| LENAOUR_ | 109 | 0.057474 | 0.004355 | 0.048934 | 0.1201 | 2042.301 |
| XU_GH1_A | 124 | 0.055976 | 0.004522 | 0.047663 | 0.12012 | 2042.641 |
| GOBP_FAT | 36 | 0.09469 | 0.004131 | 0.08063 | 0.12013 | 2042.811 |
| GOBP_SOF | 4 | 0.34706 | 0.005051 | 0.29556 | 0.12016 | 2043.321 |
| REACTOME | 12 | 0.17791 | 0.004484 | 0.15152 | 0.12017 | 2043.491 |
| GOBP_ELA: | 8 | 0.19043 | 0.003919 | 0.16218 | 0.12018 | 2043.661 |
| HOLLMANI | 190 | 0.043979 | 0.00439 | 0.037466 | 0.12024 | 2044.681 |
| MATZUK_S | 69 | 0.067816 | 0.004093 | 0.057777 | 0.12025 | 2044.851 |
| GOZGIT_ES | 132 | 0.054522 | 0.004543 | 0.046462 | 0.12031 | 2045.872 |
| GOBP_POS | 88 | 0.062286 | 0.004243 | 0.053098 | 0.1204 | 2047.402 |
| WP_MIR51 | 5 | 0.21152 | 0.003442 | 0.18045 | 0.12057 | 2050.293 |
| GOBP_ACII | 5 | 0.28043 | 0.004563 | 0.23926 | 0.12059 | 2050.633 |
| GOBP_COF | 43 | 0.10729 | 0.005115 | 0.091539 | 0.12059 | 2050.633 |
| GOBP_REG | 7 | 0.23028 | 0.004433 | 0.19649 | 0.12062 | 2051.143 |
| PID_SYNDE | 41 | 0.097624 | 0.004545 | 0.083314 | 0.12065 | 2051.653 |
| GOBP_THY | 26 | 0.12958 | 0.004806 | 0.11067 | 0.12082 | 2054.544 |
| SMITH_LIV | 42 | 0.089365 | 0.004211 | 0.076342 | 0.12089 | 2055.734 |
| GOBP_PRE | 6 | 0.2098 | 0.00374 | 0.17924 | 0.12091 | 2056.075 |

| | | | | | | |
|---|---|---|---|---|---|---|
| GOBP_POS | 29 | 0.099539 | 0.003898 | 0.085068 | 0.12099 | 2057.435 |
| GOBP_EXT | 7 | 0.24821 | 0.004779 | 0.21223 | 0.12111 | 2059.476 |
| BIOCARTA_ | 43 | 0.094001 | 0.004481 | 0.080398 | 0.12117 | 2060.496 |
| GOMF_STF | 18 | 0.14069 | 0.004342 | 0.12033 | 0.12117 | 2060.496 |
| MARTIN_IN | 38 | 0.090653 | 0.004063 | 0.077536 | 0.12118 | 2060.666 |
| OZANNE_A | 14 | 0.14029 | 0.003819 | 0.12 | 0.12119 | 2060.836 |
| LUI_THYRC | 43 | 0.083631 | 0.003987 | 0.071542 | 0.12121 | 2061.176 |
| GOBP_NEG | 361 | 0.031109 | 0.004261 | 0.026615 | 0.12124 | 2061.686 |
| GOBP_REG | 18 | 0.13316 | 0.00411 | 0.11392 | 0.12124 | 2061.686 |
| WP_PHOSF | 44 | 0.087055 | 0.004198 | 0.074482 | 0.12125 | 2061.856 |
| GOBP_TOL | 60 | 0.079806 | 0.004492 | 0.068297 | 0.12131 | 2062.877 |
| SCHLESING | 82 | 0.071255 | 0.004686 | 0.060987 | 0.12134 | 2063.387 |
| GOBP_TRA | 107 | 0.054213 | 0.00407 | 0.046418 | 0.12143 | 2064.917 |
| THEILGAAF | 211 | 0.039849 | 0.004189 | 0.034122 | 0.12144 | 2065.087 |
| FRIDMAN_ | 33 | 0.098227 | 0.004103 | 0.084115 | 0.12146 | 2065.427 |
| GOBP_MO | 88 | 0.059711 | 0.004067 | 0.051136 | 0.12147 | 2065.597 |
| GOBP_AMI | 275 | 0.035191 | 0.004216 | 0.030141 | 0.1215 | 2066.108 |
| GOBP_OXY | 5 | 0.27681 | 0.004504 | 0.23717 | 0.12159 | 2067.638 |
| WP_BONE_ | 11 | 0.19786 | 0.004775 | 0.16953 | 0.12159 | 2067.638 |
| WP_LNCRN | 91 | 0.059847 | 0.004145 | 0.051284 | 0.12162 | 2068.148 |
| WP_EDA_S | 11 | 0.17438 | 0.004208 | 0.14943 | 0.12163 | 2068.318 |
| GOBP_CON | 5 | 0.31448 | 0.005117 | 0.26952 | 0.12165 | 2068.658 |
| WP_PLURII | 47 | 0.08926 | 0.004448 | 0.076514 | 0.1217 | 2069.509 |
| GOMF_EXT | 19 | 0.15435 | 0.004894 | 0.13237 | 0.12179 | 2071.039 |
| GOBP_ERB | 127 | 0.052961 | 0.004329 | 0.045428 | 0.12185 | 2072.059 |
| GOBP_MAI | 65 | 0.077271 | 0.004526 | 0.066279 | 0.12185 | 2072.059 |
| GOCC_POL | 28 | 0.10572 | 0.004069 | 0.090687 | 0.12186 | 2072.229 |
| GOBP_G_P | 53 | 0.086987 | 0.004603 | 0.074646 | 0.12195 | 2073.76 |
| GOBP_RES | 20 | 0.12075 | 0.003928 | 0.10363 | 0.12198 | 2074.27 |
| GOBP_ANI | 29 | 0.11502 | 0.004505 | 0.098727 | 0.12202 | 2074.95 |
| GOBP_MIN | 13 | 0.1743 | 0.004572 | 0.14963 | 0.12204 | 2075.29 |
| LUCAS_HN | 53 | 0.080799 | 0.004275 | 0.069383 | 0.12211 | 2076.481 |
| REACTOME | 145 | 0.046903 | 0.004095 | 0.040278 | 0.12212 | 2076.651 |
| GOBP_GLU | 172 | 0.042055 | 0.003996 | 0.036129 | 0.12221 | 2078.181 |
| GOBP_PRC | 237 | 0.038401 | 0.004276 | 0.03299 | 0.12222 | 2078.351 |
| REACTOME | 145 | 0.044875 | 0.003918 | 0.038555 | 0.12224 | 2078.691 |
| GOBP_POS | 14 | 0.16949 | 0.004614 | 0.14563 | 0.12226 | 2079.031 |
| GOBP_RES | 5 | 0.28488 | 0.004636 | 0.24482 | 0.12229 | 2079.541 |
| GOMF_DN/ | 14 | 0.13211 | 0.003596 | 0.11356 | 0.12237 | 2080.902 |
| SMID_BRE/ | 76 | 0.071438 | 0.004524 | 0.061419 | 0.1224 | 2081.412 |
| GOCC_TRN | 6 | 0.17056 | 0.00304 | 0.14666 | 0.12243 | 2081.922 |
| GOBP_GLU | 21 | 0.12877 | 0.004292 | 0.11078 | 0.12254 | 2083.793 |
| HWANG_P | 26 | 0.11547 | 0.004282 | 0.099351 | 0.12257 | 2084.303 |
| GOBP_CAR | 4 | 0.37121 | 0.005403 | 0.31944 | 0.12261 | 2084.983 |
| STOSSI_RES | 41 | 0.10085 | 0.004695 | 0.086799 | 0.12265 | 2085.663 |

| | | | | | | |
|---|---|---|---|---|---|---|
| BIOCARTA_ | 32 | 0.10582 | 0.004353 | 0.091091 | 0.12268 | 2086.173 |
| MYLLYKAN | 14 | 0.1492 | 0.004061 | 0.12846 | 0.12274 | 2087.194 |
| GOBP_PRC | 6 | 0.27265 | 0.00486 | 0.23495 | 0.12293 | 2090.425 |
| GOBP_POS | 33 | 0.097632 | 0.004078 | 0.084132 | 0.12294 | 2090.595 |
| GOBP_POS | 21 | 0.14409 | 0.004803 | 0.12418 | 0.12297 | 2091.105 |
| FIGUEROA_ | 15 | 0.16279 | 0.004587 | 0.14032 | 0.12301 | 2091.785 |
| FRASOR_T/ | 10 | 0.18767 | 0.004318 | 0.16177 | 0.12302 | 2091.955 |
| CUI_TCF21 | 403 | 0.029272 | 0.004231 | 0.02524 | 0.12309 | 2093.145 |
| BURTON_A | 43 | 0.097256 | 0.004636 | 0.083873 | 0.12312 | 2093.656 |
| GOCC_CELI | 332 | 0.032032 | 0.00421 | 0.02765 | 0.12334 | 2097.397 |
| BORLAK_LI | 47 | 0.089815 | 0.004476 | 0.07754 | 0.12338 | 2098.077 |
| GOMF_CYC | 17 | 0.1342 | 0.004025 | 0.11588 | 0.12342 | 2098.757 |
| GOBP_RES | 50 | 0.083055 | 0.004269 | 0.071729 | 0.12346 | 2099.437 |
| GOBP_REG | 47 | 0.081314 | 0.004052 | 0.07023 | 0.12348 | 2099.777 |
| GOMF_HYI | 37 | 0.092834 | 0.004106 | 0.080207 | 0.12356 | 2101.138 |
| GOBP_TRN | 19 | 0.11169 | 0.003542 | 0.096546 | 0.12367 | 2103.008 |
| CHIANG_LI | 23 | 0.12912 | 0.004504 | 0.11165 | 0.12376 | 2104.539 |
| JAZAG_TGI | 33 | 0.088866 | 0.003712 | 0.07686 | 0.12381 | 2105.389 |
| WAKABAY/ | 144 | 0.044463 | 0.003869 | 0.038457 | 0.12381 | 2105.389 |
| AZARE_ST/ | 23 | 0.12718 | 0.004437 | 0.11005 | 0.12391 | 2107.09 |
| REACTOME | 94 | 0.05873 | 0.004134 | 0.05082 | 0.12392 | 2107.26 |
| WP_ENDOI | 42 | 0.086516 | 0.004076 | 0.074874 | 0.12395 | 2107.77 |
| GOBP_MES | 7 | 0.24954 | 0.004804 | 0.21601 | 0.124 | 2108.62 |
| REACTOME | 23 | 0.12143 | 0.004236 | 0.10515 | 0.1241 | 2110.321 |
| REACTOME | 84 | 0.065246 | 0.004343 | 0.056504 | 0.12411 | 2110.491 |
| WP_HEMA | 51 | 0.08283 | 0.004299 | 0.071731 | 0.12411 | 2110.491 |
| WP_METAI | 17 | 0.15465 | 0.004639 | 0.13394 | 0.12412 | 2110.661 |
| KEGG_ADH | 70 | 0.074997 | 0.004558 | 0.064953 | 0.12413 | 2110.831 |
| GOBP_REG | 6 | 0.27758 | 0.004948 | 0.24042 | 0.12414 | 2111.001 |
| GOBP_RNA | 76 | 0.063055 | 0.003993 | 0.054616 | 0.12415 | 2111.171 |
| GOBP_RIB( | 10 | 0.15414 | 0.003547 | 0.13352 | 0.12416 | 2111.341 |
| ALTEMEIEF | 115 | 0.05684 | 0.004423 | 0.049236 | 0.12417 | 2111.511 |
| FUKUSHIM | 15 | 0.17508 | 0.004933 | 0.15174 | 0.12429 | 2113.551 |
| GOBP_SM/ | 80 | 0.066718 | 0.004334 | 0.057828 | 0.12431 | 2113.892 |
| GOBP_NEU | 38 | 0.10747 | 0.004817 | 0.093165 | 0.12435 | 2114.572 |
| GOBP_POS | 6 | 0.2405 | 0.004287 | 0.20855 | 0.12442 | 2115.762 |
| GOBP_RES | 9 | 0.2095 | 0.004573 | 0.18167 | 0.12443 | 2115.932 |
| GOBP_ALC | 129 | 0.048859 | 0.004025 | 0.042373 | 0.12445 | 2116.272 |
| GOBP_POS | 7 | 0.24784 | 0.004772 | 0.21496 | 0.12448 | 2116.782 |
| REACTOME | 106 | 0.054921 | 0.004104 | 0.047636 | 0.12448 | 2116.782 |
| GOBP_NEU | 6 | 0.28402 | 0.005063 | 0.2464 | 0.12453 | 2117.633 |
| GOMF_CD4 | 7 | 0.25093 | 0.004831 | 0.21771 | 0.12455 | 2117.973 |
| GOBP_NEG | 13 | 0.15457 | 0.004055 | 0.13417 | 0.12466 | 2119.843 |
| GOMF_FM | 14 | 0.14519 | 0.003953 | 0.12608 | 0.12475 | 2121.374 |
| QI_PLASM/ | 238 | 0.039552 | 0.004413 | 0.034351 | 0.12479 | 2122.054 |

| | | | | | | |
|---|---|---|---|---|---|---|
| GOBP_ERB | 8 | 0.24585 | 0.00506 | 0.21374 | 0.12503 | 2126.135 |
| GOBP_MIT | 5 | 0.31546 | 0.005133 | 0.27431 | 0.12507 | 2126.815 |
| GOBP_MID | 86 | 0.061545 | 0.004145 | 0.053517 | 0.12508 | 2126.985 |
| GOBP_RES | 78 | 0.072142 | 0.004628 | 0.062732 | 0.12508 | 2126.985 |
| GOBP_ADE | 7 | 0.20064 | 0.003863 | 0.17457 | 0.12521 | 2129.196 |
| SANSOM_\ | 61 | 0.073428 | 0.004167 | 0.063903 | 0.12527 | 2130.216 |
| NADLER_O | 56 | 0.071372 | 0.003882 | 0.062119 | 0.1253 | 2130.727 |
| GOMF_LYS | 57 | 0.072932 | 0.004002 | 0.063485 | 0.12533 | 2131.237 |
| GOBP_POS | 6 | 0.22267 | 0.003969 | 0.19392 | 0.12544 | 2133.107 |
| GOBP_REG | 7 | 0.24124 | 0.004645 | 0.21023 | 0.12559 | 2135.658 |
| KUMAR_T/ | 375 | 0.030873 | 0.004308 | 0.026905 | 0.1256 | 2135.828 |
| REACTOME | 23 | 0.14903 | 0.005199 | 0.12988 | 0.12562 | 2136.168 |
| LE_SKI_TAI | 6 | 0.2908 | 0.005184 | 0.25359 | 0.12575 | 2138.379 |
| GOMF_AM | 38 | 0.10207 | 0.004575 | 0.089041 | 0.12585 | 2140.079 |
| GOBP_EAR | 215 | 0.042193 | 0.004477 | 0.036827 | 0.12596 | 2141.95 |
| GOBP_PRC | 32 | 0.10773 | 0.004432 | 0.09403 | 0.12596 | 2141.95 |
| GYORFFY_I | 35 | 0.10696 | 0.004601 | 0.093385 | 0.12604 | 2143.31 |
| KUWANO_ | 7 | 0.22518 | 0.004335 | 0.19663 | 0.12607 | 2143.82 |
| GOMF_PRE | 2 | 0.62861 | 0.00647 | 0.54894 | 0.12609 | 2144.16 |
| GOBP_PYR | 33 | 0.092117 | 0.003848 | 0.080458 | 0.12613 | 2144.841 |
| REACTOME | 37 | 0.096176 | 0.004254 | 0.084007 | 0.12614 | 2145.011 |
| GOBP_OST | 17 | 0.14521 | 0.004356 | 0.12687 | 0.12619 | 2145.861 |
| REACTOME | 41 | 0.085351 | 0.003973 | 0.074567 | 0.12619 | 2145.861 |
| GOBP_OTC | 4 | 0.35269 | 0.005133 | 0.3082 | 0.12625 | 2146.881 |
| GOBP_PRC | 176 | 0.043547 | 0.004185 | 0.038057 | 0.12627 | 2147.221 |
| REACTOME | 10 | 0.17168 | 0.00395 | 0.15004 | 0.12627 | 2147.221 |
| ZHANG_G/ | 63 | 0.073994 | 0.004267 | 0.064666 | 0.12627 | 2147.221 |
| GOBP_POS | 6 | 0.22913 | 0.004084 | 0.20027 | 0.12629 | 2147.561 |
| GOBP_EST/ | 70 | 0.069291 | 0.004212 | 0.060573 | 0.12633 | 2148.242 |
| GOBP_REG | 89 | 0.061029 | 0.004181 | 0.053375 | 0.12644 | 2150.112 |
| BIOCARTA_ | 24 | 0.1185 | 0.004223 | 0.10365 | 0.12646 | 2150.452 |
| GOBP_NEP | 4 | 0.35371 | 0.005148 | 0.30945 | 0.12652 | 2151.473 |
| GOBP_POS | 97 | 0.059835 | 0.004278 | 0.05236 | 0.12657 | 2152.323 |
| ISHIKAWA_ | 9 | 0.17422 | 0.003803 | 0.15255 | 0.12673 | 2155.044 |
| GOBP_HIN | 5 | 0.186 | 0.003027 | 0.16287 | 0.12674 | 2155.214 |
| OISHI_CHC | 328 | 0.032294 | 0.00422 | 0.028284 | 0.12679 | 2156.064 |
| TURJANSKI | 11 | 0.20911 | 0.005046 | 0.18315 | 0.12679 | 2156.064 |
| REACTOME | 54 | 0.078221 | 0.004178 | 0.06853 | 0.12686 | 2157.254 |
| REACTOME | 109 | 0.05699 | 0.004318 | 0.049945 | 0.12693 | 2158.445 |
| SHEDDEN_ | 255 | 0.03522 | 0.004066 | 0.030867 | 0.12694 | 2158.615 |
| GOBP_CHC | 54 | 0.086704 | 0.004631 | 0.075993 | 0.12695 | 2158.785 |
| BEGUM_T/ | 45 | 0.090968 | 0.004436 | 0.079735 | 0.12697 | 2159.125 |
| WP_SEROT | 18 | 0.14242 | 0.004396 | 0.12487 | 0.12704 | 2160.315 |
| GOBP_SUL | 34 | 0.084561 | 0.003585 | 0.074151 | 0.12707 | 2160.825 |
| VERNELL_F | 67 | 0.06757 | 0.004018 | 0.059276 | 0.12717 | 2162.526 |

| | | | | | | |
|---|---|---|---|---|---|---|
| MELLMAN_ | 47 | 0.076637 | 0.003819 | 0.06724 | 0.1272 | 2163.036 |
| KARLSSON_ | 184 | 0.040547 | 0.003984 | 0.035578 | 0.12722 | 2163.376 |
| HERNANDE | 19 | 0.13615 | 0.004317 | 0.1195 | 0.1273 | 2164.737 |
| REACTOME | 14 | 0.15087 | 0.004107 | 0.13242 | 0.1273 | 2164.737 |
| BAELDE_DI | 408 | 0.030307 | 0.004407 | 0.026611 | 0.12738 | 2166.097 |
| MURAKAM | 19 | 0.12361 | 0.00392 | 0.10854 | 0.12739 | 2166.267 |
| REACTOME | 626 | 0.022133 | 0.003963 | 0.019438 | 0.12743 | 2166.947 |
| WESTON_V | 22 | 0.1416 | 0.004831 | 0.12437 | 0.12746 | 2167.457 |
| GOBP_EPIT | 683 | 0.023883 | 0.00446 | 0.020978 | 0.12748 | 2167.797 |
| GOCC_A_B | 32 | 0.10112 | 0.00416 | 0.088833 | 0.12751 | 2168.308 |
| GOMF_PUI | 20 | 0.12376 | 0.004026 | 0.10872 | 0.12751 | 2168.308 |
| DEMAGALH | 14 | 0.14673 | 0.003994 | 0.1289 | 0.12752 | 2168.478 |
| KAN_RESP | 109 | 0.057153 | 0.00433 | 0.050212 | 0.12752 | 2168.478 |
| COLLIS_PRI | 12 | 0.19163 | 0.00483 | 0.16839 | 0.12757 | 2169.328 |
| AZARE_NE | 116 | 0.057465 | 0.004491 | 0.050509 | 0.12763 | 2170.348 |
| GOCC_RIB | 198 | 0.036339 | 0.003702 | 0.03195 | 0.1277 | 2171.539 |
| MCGARVEY | 41 | 0.092858 | 0.004323 | 0.081648 | 0.12772 | 2171.879 |
| GOMF_THI | 7 | 0.22199 | 0.004274 | 0.19527 | 0.12781 | 2173.409 |
| REACTOME | 8 | 0.19091 | 0.003929 | 0.16795 | 0.12784 | 2173.919 |
| GOBP_VOC | 15 | 0.16045 | 0.004521 | 0.14118 | 0.12788 | 2174.599 |
| GOBP_NEG | 21 | 0.11942 | 0.003981 | 0.1051 | 0.12793 | 2175.45 |
| WP_PROLA | 74 | 0.069254 | 0.004327 | 0.060956 | 0.12796 | 2175.96 |
| GOBP_EXE | 69 | 0.070726 | 0.004268 | 0.062256 | 0.12798 | 2176.3 |
| GOBP_ORG | 226 | 0.03814 | 0.004148 | 0.033573 | 0.12798 | 2176.3 |
| GOBP_RES | 47 | 0.084536 | 0.004213 | 0.074423 | 0.12801 | 2176.81 |
| GOMF_HEI | 7 | 0.25809 | 0.004969 | 0.22728 | 0.12808 | 2178 |
| GOBP_CELI | 14 | 0.16307 | 0.004439 | 0.14366 | 0.12819 | 2179.871 |
| MIKKELSEN | 7 | 0.2523 | 0.004857 | 0.22236 | 0.12827 | 2181.231 |
| WANG_RES | 22 | 0.14037 | 0.004789 | 0.12372 | 0.12827 | 2181.231 |
| GOBP_UTP | 9 | 0.16697 | 0.003645 | 0.14718 | 0.1283 | 2181.742 |
| REACTOME | 383 | 0.02938 | 0.004142 | 0.025898 | 0.12831 | 2181.912 |
| GOBP_PEP | 38 | 0.085031 | 0.003811 | 0.074969 | 0.12836 | 2182.762 |
| GOBP_POS | 58 | 0.077403 | 0.004284 | 0.068242 | 0.12836 | 2182.762 |
| GOBP_POS | 7 | 0.32835 | 0.006322 | 0.2895 | 0.12836 | 2182.762 |
| GOBP_EST | 60 | 0.070698 | 0.003979 | 0.062356 | 0.12845 | 2184.292 |
| BLANCO_M | 135 | 0.048778 | 0.00411 | 0.043031 | 0.1285 | 2185.143 |
| LANDIS_BR | 69 | 0.073147 | 0.004414 | 0.064535 | 0.12852 | 2185.483 |
| EBAUER_T | 180 | 0.040829 | 0.003968 | 0.036023 | 0.12853 | 2185.653 |
| GOBP_REG | 6 | 0.22127 | 0.003944 | 0.19524 | 0.12855 | 2185.993 |
| GOCC_TRO | 8 | 0.18545 | 0.003817 | 0.16363 | 0.12855 | 2185.993 |
| GOBP_REG | 6 | 0.2443 | 0.004355 | 0.21558 | 0.12858 | 2186.503 |
| ZHAN_MUI | 29 | 0.098631 | 0.003863 | 0.087082 | 0.12869 | 2188.373 |
| GOBP_REG | 58 | 0.079855 | 0.00442 | 0.070536 | 0.1288 | 2190.244 |
| GOBP_PRIM | 8 | 0.22957 | 0.004725 | 0.20285 | 0.12889 | 2191.774 |
| RODRIGUE | 662 | 0.02215 | 0.004074 | 0.019576 | 0.12894 | 2192.625 |

| | | | | | | |
|---|---|---|---|---|---|---|
| GOBP_MAS | 6 | 0.23786 | 0.00424 | 0.21024 | 0.12896 | 2192.965 |
| GOBP_EMI | 14 | 0.16252 | 0.004424 | 0.1437 | 0.12904 | 2194.325 |
| GOBP_INTI | 24 | 0.11721 | 0.004177 | 0.10368 | 0.12914 | 2196.026 |
| GOCC_CAN | 5 | 0.35025 | 0.005699 | 0.30987 | 0.12918 | 2196.706 |
| REACTOME | 29 | 0.10613 | 0.004157 | 0.093904 | 0.1292 | 2197.046 |
| GOBP_REG | 8 | 0.23245 | 0.004784 | 0.20585 | 0.12941 | 2200.617 |
| GOBP_MES | 13 | 0.16155 | 0.004238 | 0.14308 | 0.12943 | 2200.957 |
| GOBP_NEU | 29 | 0.11694 | 0.00458 | 0.1036 | 0.1295 | 2202.148 |
| REACTOME | 50 | 0.076674 | 0.003941 | 0.067937 | 0.12954 | 2202.828 |
| KEGG_THY | 27 | 0.11093 | 0.004192 | 0.098335 | 0.12964 | 2204.528 |
| GOBP_POS | 3 | 0.28249 | 0.003561 | 0.25045 | 0.12968 | 2205.208 |
| GOBP_RES | 143 | 0.0487 | 0.004223 | 0.04318 | 0.1297 | 2205.549 |
| GOCC_GLY | 6 | 0.23116 | 0.00412 | 0.20496 | 0.12971 | 2205.719 |
| GOCC_MIT | 10 | 0.14281 | 0.003286 | 0.12666 | 0.12977 | 2206.739 |
| GOBP_LAB | 43 | 0.09174 | 0.004373 | 0.081386 | 0.12983 | 2207.759 |
| GOBP_POS | 1003 | 0.017924 | 0.00402 | 0.015902 | 0.12985 | 2208.099 |
| GOCC_MCI | 5 | 0.31218 | 0.00508 | 0.27697 | 0.12985 | 2208.099 |
| GOBP_POS | 100 | 0.056504 | 0.004102 | 0.05015 | 0.12994 | 2209.63 |
| KORKOLA_ | 18 | 0.14195 | 0.004381 | 0.12599 | 0.12995 | 2209.8 |
| GOBP_LUT | 10 | 0.18962 | 0.004363 | 0.16831 | 0.12997 | 2210.14 |
| REACTOME | 15 | 0.16342 | 0.004605 | 0.14508 | 0.13001 | 2210.82 |
| GOBP_APC | 7 | 0.25215 | 0.004855 | 0.22398 | 0.13013 | 2212.861 |
| GOBP_EPIT | 14 | 0.17476 | 0.004758 | 0.15524 | 0.13014 | 2213.031 |
| GOBP_REG | 7 | 0.2218 | 0.00427 | 0.19707 | 0.1302 | 2214.051 |
| GOBP_T_C | 7 | 0.2218 | 0.00427 | 0.19707 | 0.1302 | 2214.051 |
| WP_CONG | 18 | 0.13994 | 0.004319 | 0.12434 | 0.13021 | 2214.221 |
| MARTINEZ_ | 48 | 0.089579 | 0.004511 | 0.079709 | 0.13055 | 2220.003 |
| REACTOME | 12 | 0.13884 | 0.003499 | 0.12355 | 0.13056 | 2220.173 |
| GOBP_REG | 10 | 0.17331 | 0.003988 | 0.15435 | 0.13075 | 2223.404 |
| BIOCARTA_ | 52 | 0.074002 | 0.003879 | 0.065911 | 0.13078 | 2223.914 |
| PID_BETA_ | 17 | 0.12341 | 0.003702 | 0.10993 | 0.1308 | 2224.254 |
| GOCC_SUP | 5 | 0.22462 | 0.003655 | 0.20016 | 0.1309 | 2225.955 |
| GOBP_END | 12 | 0.18012 | 0.00454 | 0.16052 | 0.13092 | 2226.295 |
| GOMF_PLA | 11 | 0.197 | 0.004754 | 0.17558 | 0.13094 | 2226.635 |
| REACTOME | 23 | 0.10688 | 0.003728 | 0.095258 | 0.13095 | 2226.805 |
| GOBP_POS | 9 | 0.22368 | 0.004883 | 0.19941 | 0.131 | 2227.655 |
| GOBP_REP | 1369 | 0.015711 | 0.004075 | 0.014008 | 0.13103 | 2228.165 |
| GOCC_SEPT | 15 | 0.14417 | 0.004062 | 0.12857 | 0.13107 | 2228.845 |
| GOBP_COR | 5 | 0.22321 | 0.003632 | 0.19916 | 0.1312 | 2231.056 |
| GOBP_REG | 119 | 0.050105 | 0.003966 | 0.044722 | 0.13129 | 2232.586 |
| PLASARI_T | 33 | 0.11196 | 0.004677 | 0.099939 | 0.1313 | 2232.757 |
| ZWANG_E( | 56 | 0.068188 | 0.003708 | 0.060901 | 0.13144 | 2235.137 |
| HOFMANN | 8 | 0.17892 | 0.003683 | 0.15984 | 0.1315 | 2236.158 |
| GOBP_REG | 5 | 0.30449 | 0.004955 | 0.2721 | 0.13157 | 2237.348 |
| GOBP_MIT | 423 | 0.026868 | 0.003977 | 0.024011 | 0.13159 | 2237.688 |

| | | | | | | |
|---|---|---|---|---|---|---|
| GOBP_POS | 27 | 0.12187 | 0.004606 | 0.10892 | 0.13161 | 2238.028 |
| GOBP_REG | 16 | 0.14611 | 0.004252 | 0.13059 | 0.13161 | 2238.028 |
| GOBP_UBI( | 34 | 0.087095 | 0.003693 | 0.077894 | 0.13176 | 2240.579 |
| WP_HIF1A_ | 8 | 0.20537 | 0.004227 | 0.18371 | 0.13181 | 2241.429 |
| GOCC_OUT | 219 | 0.036549 | 0.003914 | 0.032704 | 0.13189 | 2242.789 |
| PID_ALK1_ | 24 | 0.12047 | 0.004293 | 0.10781 | 0.13192 | 2243.3 |
| GOBP_POS | 17 | 0.13357 | 0.004007 | 0.11955 | 0.13195 | 2243.81 |
| GOBP_REG | 32 | 0.10009 | 0.004117 | 0.089615 | 0.13204 | 2245.34 |
| GOMF_FOI | 13 | 0.16287 | 0.004273 | 0.14585 | 0.13207 | 2245.85 |
| GOBP_EXT | 58 | 0.078906 | 0.004367 | 0.07067 | 0.1321 | 2246.361 |
| MOREAUX_ | 171 | 0.041564 | 0.003938 | 0.037241 | 0.1322 | 2248.061 |
| GOMF_MY | 7 | 0.22026 | 0.004241 | 0.19737 | 0.13222 | 2248.401 |
| REACTOME | 53 | 0.075747 | 0.004008 | 0.067881 | 0.13225 | 2248.911 |
| GOBP_ATR | 8 | 0.23627 | 0.004863 | 0.21181 | 0.13233 | 2250.272 |
| KASLER_HI | 16 | 0.14416 | 0.004195 | 0.12928 | 0.13241 | 2251.632 |
| GOBP_NEG | 8 | 0.29161 | 0.006002 | 0.26156 | 0.13246 | 2252.482 |
| GOBP_VEN | 9 | 0.19244 | 0.004201 | 0.17263 | 0.13248 | 2252.822 |
| BIOCARTA_ | 20 | 0.12753 | 0.004149 | 0.11442 | 0.13254 | 2253.843 |
| GOBP_LEA | 144 | 0.048379 | 0.004209 | 0.043411 | 0.13255 | 2254.013 |
| GOBP_NEG | 5 | 0.26213 | 0.004266 | 0.23521 | 0.13255 | 2254.013 |
| GOBP_NEG | 11 | 0.17532 | 0.004231 | 0.15731 | 0.13255 | 2254.013 |
| REACTOME | 11 | 0.23599 | 0.005695 | 0.21189 | 0.1327 | 2256.564 |
| GOBP_NEG | 20 | 0.11359 | 0.003695 | 0.10199 | 0.13271 | 2256.734 |
| GOBP_RES | 8 | 0.21703 | 0.004467 | 0.19488 | 0.13271 | 2256.734 |
| REACTOME | 79 | 0.063813 | 0.004119 | 0.057297 | 0.13271 | 2256.734 |
| GOBP_POS | 8 | 0.17402 | 0.003582 | 0.1563 | 0.13279 | 2258.094 |
| GOBP_GPI_ | 31 | 0.095605 | 0.003871 | 0.085894 | 0.13285 | 2259.114 |
| GOMF_P_T | 8 | 0.22634 | 0.004658 | 0.20345 | 0.13297 | 2261.155 |
| GOBP_REG | 3 | 0.38057 | 0.004797 | 0.34214 | 0.13301 | 2261.835 |
| GOBP_POS | 12 | 0.16762 | 0.004225 | 0.15072 | 0.13305 | 2262.515 |
| GOBP_N_A | 6 | 0.23977 | 0.004274 | 0.21563 | 0.13309 | 2263.195 |
| GOBP_CELI | 7 | 0.20729 | 0.003991 | 0.18643 | 0.1331 | 2263.366 |
| GOBP_POS | 6 | 0.22345 | 0.003983 | 0.20103 | 0.13318 | 2264.726 |
| KEGG_NON | 51 | 0.077673 | 0.004032 | 0.069897 | 0.13324 | 2265.746 |
| GOMF_ADI | 13 | 0.14789 | 0.00388 | 0.1331 | 0.13327 | 2266.256 |
| GOBP_SUP | 755 | 0.020929 | 0.004101 | 0.018838 | 0.1333 | 2266.767 |
| GOBP_MIT | 11 | 0.1682 | 0.004059 | 0.15147 | 0.1334 | 2268.467 |
| GOBP_POS | 109 | 0.054794 | 0.004152 | 0.049342 | 0.1334 | 2268.467 |
| REACTOME | 20 | 0.12765 | 0.004153 | 0.11496 | 0.13342 | 2268.807 |
| LIU_SOX4_ | 129 | 0.048059 | 0.003959 | 0.043282 | 0.13343 | 2268.977 |
| GOBP_PER | 74 | 0.068654 | 0.00429 | 0.061836 | 0.13345 | 2269.317 |
| GOBP_GER | 8 | 0.23985 | 0.004936 | 0.21605 | 0.13348 | 2269.827 |
| GOBP_GAS | 7 | 0.23111 | 0.004449 | 0.20822 | 0.13352 | 2270.508 |
| GOBP_NEG | 11 | 0.15989 | 0.003859 | 0.14407 | 0.13354 | 2270.848 |
| WP_ANGIC | 27 | 0.12162 | 0.004596 | 0.10967 | 0.13372 | 2273.909 |

| | | | | | | |
|---|---|---|---|---|---|---|
| GARGALOV | 79 | 0.067506 | 0.004358 | 0.060884 | 0.13378 | 2274.929 |
| REACTOME | 9 | 0.16475 | 0.003597 | 0.14859 | 0.13378 | 2274.929 |
| GOMF_DIP | 16 | 0.13543 | 0.003941 | 0.12218 | 0.13383 | 2275.779 |
| CARRILLOR | 151 | 0.051427 | 0.004581 | 0.046397 | 0.13385 | 2276.119 |
| WP_NONH | 10 | 0.17149 | 0.003946 | 0.15484 | 0.13404 | 2279.35 |
| GOBP_POS | 237 | 0.035607 | 0.003965 | 0.032165 | 0.13415 | 2281.221 |
| GOBP_NUC | 121 | 0.046794 | 0.003734 | 0.042271 | 0.13416 | 2281.391 |
| GOMF_POI | 27 | 0.10842 | 0.004097 | 0.098019 | 0.13436 | 2284.792 |
| GOBP_POS | 155 | 0.046423 | 0.004189 | 0.041976 | 0.13438 | 2285.132 |
| SAUL_SEN_ | 116 | 0.055402 | 0.00433 | 0.050097 | 0.13439 | 2285.302 |
| GOMF_VOI | 20 | 0.13528 | 0.004401 | 0.12235 | 0.13444 | 2286.152 |
| WHITE_NE | 19 | 0.19047 | 0.006039 | 0.17226 | 0.13444 | 2286.152 |
| NIKOLSKY_ | 52 | 0.13827 | 0.007247 | 0.12507 | 0.13447 | 2286.662 |
| GOBP_POS | 37 | 0.10735 | 0.004748 | 0.097104 | 0.13448 | 2286.832 |
| GOBP_PLA | 9 | 0.21564 | 0.004707 | 0.19507 | 0.13449 | 2287.002 |
| GOBP_NEG | 32 | 0.10312 | 0.004242 | 0.093296 | 0.13451 | 2287.343 |
| REACTOME | 75 | 0.061496 | 0.003869 | 0.055639 | 0.13453 | 2287.683 |
| GOBP_ALD | 5 | 0.23306 | 0.003793 | 0.21102 | 0.1347 | 2290.574 |
| NICK_RESP | 4 | 0.302 | 0.004396 | 0.27356 | 0.13482 | 2292.614 |
| REACTOME | 17 | 0.12995 | 0.003898 | 0.11776 | 0.13491 | 2294.145 |
| WP_EPO_F | 26 | 0.10852 | 0.004025 | 0.098341 | 0.13491 | 2294.145 |
| YAO_HOXA | 18 | 0.13539 | 0.004179 | 0.1227 | 0.13493 | 2294.485 |
| GOBP_MA( | 35 | 0.086876 | 0.003737 | 0.078817 | 0.13518 | 2298.736 |
| GOBP_NUC | 14 | 0.14026 | 0.003818 | 0.1273 | 0.13527 | 2300.266 |
| GOBP_REP | 44 | 0.082127 | 0.00396 | 0.074536 | 0.13527 | 2300.266 |
| GOBP_ECT | 17 | 0.13265 | 0.003979 | 0.12049 | 0.13547 | 2303.667 |
| GOMF_N_/ | 5 | 0.23626 | 0.003845 | 0.21464 | 0.13551 | 2304.348 |
| GOBP_CIR( | 5 | 0.22513 | 0.003663 | 0.20453 | 0.13553 | 2304.688 |
| GOCC_PRC | 8 | 0.18093 | 0.003724 | 0.16439 | 0.13554 | 2304.858 |
| BIOCARTA_ | 20 | 0.13159 | 0.004281 | 0.11958 | 0.13559 | 2305.708 |
| LI_ADIPOG | 18 | 0.1329 | 0.004102 | 0.12082 | 0.13568 | 2307.238 |
| REACTOME | 19 | 0.11108 | 0.003522 | 0.10099 | 0.13571 | 2307.749 |
| GOBP_POS | 12 | 0.13203 | 0.003328 | 0.12005 | 0.13572 | 2307.919 |
| GOBP_MO | 12 | 0.18989 | 0.004786 | 0.1727 | 0.13577 | 2308.769 |
| GOBP_TRN | 184 | 0.037574 | 0.003691 | 0.034174 | 0.13579 | 2309.109 |
| GOBP_CER | 117 | 0.054473 | 0.004275 | 0.049548 | 0.1358 | 2309.279 |
| REACTOME | 22 | 0.11887 | 0.004056 | 0.10817 | 0.1359 | 2310.98 |
| GOMF_TAU | 41 | 0.081895 | 0.003812 | 0.07453 | 0.13593 | 2311.49 |
| GOCC_POL | 716 | 0.022311 | 0.004262 | 0.020305 | 0.13594 | 2311.66 |
| BERTUCCI_ | 189 | 0.040378 | 0.00402 | 0.036749 | 0.13595 | 2311.83 |
| MYLLYKAN | 5 | 0.25466 | 0.004144 | 0.23177 | 0.13595 | 2311.83 |
| GOBP_FAT_ | 225 | 0.037467 | 0.004066 | 0.034116 | 0.13606 | 2313.7 |
| GOBP_EMI | 8 | 0.20983 | 0.004319 | 0.1911 | 0.13611 | 2314.551 |
| GOMF_INT | 28 | 0.10207 | 0.003928 | 0.092978 | 0.13616 | 2315.401 |
| BIOCARTA_ | 12 | 0.16712 | 0.004212 | 0.15231 | 0.13629 | 2317.611 |

| | | | | | | |
|---|---|---|---|---|---|---|
| LY_AGING_ | 13 | 0.16615 | 0.004359 | 0.15148 | 0.13636 | 2318.802 |
| GOMF_PR( | 7 | 0.24605 | 0.004737 | 0.22433 | 0.13637 | 2318.972 |
| BRUNEAU_ | 8 | 0.22997 | 0.004733 | 0.20969 | 0.13639 | 2319.312 |
| GOCC_BLEI | 9 | 0.19023 | 0.004153 | 0.17352 | 0.13647 | 2320.672 |
| LE_SKI_TAI | 17 | 0.13467 | 0.004039 | 0.12287 | 0.13654 | 2321.863 |
| GOBP_SPH | 11 | 0.19482 | 0.004701 | 0.17776 | 0.13655 | 2322.033 |
| GOBP_IMP | 17 | 0.11837 | 0.003551 | 0.10801 | 0.13657 | 2322.373 |
| GOMF_RN/ | 7 | 0.20268 | 0.003902 | 0.18496 | 0.13659 | 2322.713 |
| GOBP_PRC | 216 | 0.037572 | 0.003996 | 0.034288 | 0.1366 | 2322.883 |
| BAE_BRCA: | 29 | 0.10977 | 0.004299 | 0.10019 | 0.13661 | 2323.053 |
| GOBP_NEU | 60 | 0.067704 | 0.003811 | 0.061791 | 0.13661 | 2323.053 |
| GOMF_ANI | 4 | 0.25527 | 0.003715 | 0.23297 | 0.13661 | 2323.053 |
| GRADE_CO | 35 | 0.10489 | 0.004512 | 0.095786 | 0.13676 | 2325.604 |
| GOBP_POS | 9 | 0.18317 | 0.003999 | 0.16732 | 0.13682 | 2326.624 |
| PID_EPHA2 | 19 | 0.11669 | 0.0037 | 0.10661 | 0.13685 | 2327.134 |
| GOBP_REG | 4 | 0.28005 | 0.004076 | 0.25595 | 0.13695 | 2328.835 |
| GOBP_PRC | 6 | 0.2525 | 0.004501 | 0.23086 | 0.13704 | 2330.365 |
| REACTOMI | 6 | 0.2525 | 0.004501 | 0.23086 | 0.13704 | 2330.365 |
| BRUINS_U\ | 259 | 0.033347 | 0.003879 | 0.030493 | 0.13708 | 2331.045 |
| REACTOMI | 21 | 0.13023 | 0.004341 | 0.11908 | 0.13708 | 2331.045 |
| MODY_HIP | 30 | 0.10949 | 0.004361 | 0.10012 | 0.13709 | 2331.215 |
| GOBP_RES | 24 | 0.11608 | 0.004136 | 0.10618 | 0.13714 | 2332.066 |
| GOBP_SER | 10 | 0.17597 | 0.004049 | 0.16099 | 0.1372 | 2333.086 |
| GOMF_SEF | 10 | 0.17597 | 0.004049 | 0.16099 | 0.1372 | 2333.086 |
| GOBP_CELI | 68 | 0.069495 | 0.004163 | 0.063589 | 0.13723 | 2333.596 |
| GOMF_VOI | 179 | 0.045019 | 0.004363 | 0.041195 | 0.13724 | 2333.766 |
| REACTOMI | 26 | 0.10986 | 0.004074 | 0.10061 | 0.13742 | 2336.827 |
| GOBP_REG | 5 | 0.29339 | 0.004774 | 0.26874 | 0.13749 | 2338.017 |
| GOCC_NU( | 12 | 0.15839 | 0.003992 | 0.1451 | 0.1375 | 2338.188 |
| PANGAS_T | 139 | 0.048276 | 0.004127 | 0.044223 | 0.1375 | 2338.188 |
| REACTOMI | 57 | 0.072502 | 0.003978 | 0.066427 | 0.13754 | 2338.868 |
| BIOCARTA_ | 18 | 0.11745 | 0.003625 | 0.10761 | 0.13755 | 2339.038 |
| KOKKINAKI | 57 | 0.068934 | 0.003782 | 0.06316 | 0.13755 | 2339.038 |
| GOBP_RUF | 53 | 0.076496 | 0.004048 | 0.0701 | 0.13759 | 2339.718 |
| PID_VEGFF | 26 | 0.10502 | 0.003895 | 0.096246 | 0.13761 | 2340.058 |
| CHANGOLK | 40 | 0.078116 | 0.003592 | 0.071597 | 0.13763 | 2340.398 |
| GOBP_REG | 43 | 0.084497 | 0.004028 | 0.077463 | 0.13769 | 2341.418 |
| GOBP_REG | 8 | 0.19141 | 0.00394 | 0.1755 | 0.13772 | 2341.929 |
| GOBP_MA! | 11 | 0.16936 | 0.004087 | 0.15529 | 0.13773 | 2342.099 |
| GOBP_REG | 6 | 0.30189 | 0.005381 | 0.27686 | 0.13777 | 2342.779 |
| PID_P38_A | 36 | 0.092052 | 0.004016 | 0.084461 | 0.13789 | 2344.819 |
| GOBP_RES | 198 | 0.0408 | 0.004157 | 0.037436 | 0.1379 | 2344.99 |
| GOBP_FAT | 15 | 0.12135 | 0.003419 | 0.11136 | 0.13792 | 2345.33 |
| GOBP_NAE | 15 | 0.1493 | 0.004207 | 0.13709 | 0.13807 | 2347.88 |
| GOMF_CCI | 4 | 0.33344 | 0.004853 | 0.30618 | 0.13807 | 2347.88 |

| | | | | | | |
|---|---|---|---|---|---|---|
| GOBP_MU: | 67 | 0.065293 | 0.003883 | 0.05996 | 0.1381 | 2348.391 |
| GOBP_G_P | 15 | 0.15747 | 0.004437 | 0.14464 | 0.13816 | 2349.411 |
| GOBP_RES | 167 | 0.041774 | 0.003912 | 0.038371 | 0.13816 | 2349.411 |
| GOBP_REG | 12 | 0.15764 | 0.003973 | 0.14484 | 0.13822 | 2350.431 |
| BIOCARTA_ | 24 | 0.10237 | 0.003648 | 0.094071 | 0.13826 | 2351.111 |
| GOBP_REG | 22 | 0.11541 | 0.003938 | 0.10606 | 0.13827 | 2351.281 |
| CASTELLAN | 7 | 0.20079 | 0.003866 | 0.18455 | 0.1383 | 2351.792 |
| BENPORAT | 95 | 0.056816 | 0.00402 | 0.052224 | 0.13832 | 2352.132 |
| BALLIF_DE\ | 13 | 0.17025 | 0.004466 | 0.15657 | 0.13844 | 2354.172 |
| WP_EMBR\ | 110 | 0.055347 | 0.004213 | 0.050907 | 0.13848 | 2354.852 |
| ICHIBA_GR | 41 | 0.091205 | 0.004246 | 0.083907 | 0.13853 | 2355.703 |
| REACTOME | 44 | 0.076885 | 0.003708 | 0.070739 | 0.13855 | 2356.043 |
| WP_HOSTF | 35 | 0.095928 | 0.004127 | 0.088289 | 0.13863 | 2357.403 |
| GOBP_REG | 97 | 0.061075 | 0.004367 | 0.056226 | 0.1387 | 2358.594 |
| GOBP_REG | 16 | 0.1329 | 0.003867 | 0.12236 | 0.13871 | 2358.764 |
| REACTOME | 10 | 0.16733 | 0.00385 | 0.15411 | 0.1388 | 2360.294 |
| TONKS_TAI | 9 | 0.17528 | 0.003826 | 0.16145 | 0.13881 | 2360.464 |
| TAKEDA_T/ | 179 | 0.043106 | 0.004178 | 0.039711 | 0.13886 | 2361.314 |
| PID_ERBB2 | 44 | 0.081484 | 0.003929 | 0.07509 | 0.13894 | 2362.675 |
| RICKMAN_ | 119 | 0.050734 | 0.004015 | 0.046759 | 0.13897 | 2363.185 |
| KEGG_LON | 66 | 0.066666 | 0.003935 | 0.061446 | 0.13898 | 2363.355 |
| REACTOME | 31 | 0.10158 | 0.004113 | 0.093655 | 0.13906 | 2364.715 |
| GOBP_REG | 115 | 0.052049 | 0.00405 | 0.048019 | 0.13921 | 2367.266 |
| MCCLUNG_ | 45 | 0.085167 | 0.004153 | 0.078574 | 0.13921 | 2367.266 |
| GOBP_MO | 202 | 0.038253 | 0.003936 | 0.035299 | 0.13926 | 2368.116 |
| GARGALOV | 11 | 0.18552 | 0.004477 | 0.17122 | 0.1393 | 2368.797 |
| GOBP_ATT | 21 | 0.10246 | 0.003415 | 0.094605 | 0.13941 | 2370.667 |
| SCHLOSSEF | 43 | 0.082836 | 0.003949 | 0.076489 | 0.13942 | 2370.837 |
| REACTOME | 16 | 0.11143 | 0.003243 | 0.10292 | 0.13948 | 2371.857 |
| GOBP_ALD | 14 | 0.14723 | 0.004008 | 0.136 | 0.13951 | 2372.368 |
| CHUANG_( | 11 | 0.14164 | 0.003418 | 0.13086 | 0.13955 | 2373.048 |
| REACTOME | 21 | 0.13435 | 0.004478 | 0.12419 | 0.13968 | 2375.258 |
| GOBP_REG | 12 | 0.14445 | 0.003641 | 0.13356 | 0.13973 | 2376.109 |
| GOMF_G_( | 5 | 0.23535 | 0.00383 | 0.21766 | 0.1398 | 2377.299 |
| XU_HGF_T/ | 82 | 0.05673 | 0.003731 | 0.052524 | 0.14006 | 2381.72 |
| GOBP_REG | 8 | 0.20542 | 0.004228 | 0.19021 | 0.14008 | 2382.06 |
| REACTOME | 10 | 0.18313 | 0.004214 | 0.1696 | 0.14013 | 2382.911 |
| REACTOME | 8 | 0.16162 | 0.003326 | 0.14968 | 0.14014 | 2383.081 |
| GOBP_POS | 29 | 0.096636 | 0.003785 | 0.089504 | 0.14015 | 2383.251 |
| HOFMANN | 20 | 0.11952 | 0.003888 | 0.11072 | 0.1402 | 2384.101 |
| GOBP_PYR | 83 | 0.059267 | 0.003921 | 0.054924 | 0.14029 | 2385.631 |
| GOMF_PH( | 24 | 0.1036 | 0.003692 | 0.096017 | 0.1403 | 2385.802 |
| GOMF_PH( | 7 | 0.19064 | 0.00367 | 0.17669 | 0.14032 | 2386.142 |
| GOCC_COF | 7 | 0.17241 | 0.003319 | 0.15988 | 0.14044 | 2388.182 |
| GOMF_NEU | 11 | 0.15055 | 0.003633 | 0.13962 | 0.14045 | 2388.352 |

| | | | | | | |
|---|---|---|---|---|---|---|
| GOBP_POS | 8 | 0.18223 | 0.003751 | 0.16904 | 0.14052 | 2389.543 |
| SIMBULAN_ | 29 | 0.1159 | 0.004539 | 0.10755 | 0.14062 | 2391.243 |
| GOBP_RES | 360 | 0.029383 | 0.004019 | 0.02727 | 0.14064 | 2391.583 |
| REACTOME | 19 | 0.11257 | 0.003569 | 0.10449 | 0.14068 | 2392.263 |
| GOBP_L_SI | 8 | 0.20864 | 0.004294 | 0.1937 | 0.14073 | 2393.114 |
| GOMF_L_S | 8 | 0.20864 | 0.004294 | 0.1937 | 0.14073 | 2393.114 |
| GOBP_NEG | 48 | 0.081407 | 0.0041 | 0.075594 | 0.14077 | 2393.794 |
| GOBP_POS | 7 | 0.22725 | 0.004375 | 0.2111 | 0.14086 | 2395.324 |
| GOCC_PRO | 19 | 0.12311 | 0.003904 | 0.11439 | 0.14091 | 2396.175 |
| BERTUCCI_ | 24 | 0.10157 | 0.003619 | 0.094427 | 0.14104 | 2398.385 |
| GOMF_INT | 8 | 0.18497 | 0.003807 | 0.17206 | 0.14119 | 2400.936 |
| GOBP_REG | 47 | 0.090151 | 0.004493 | 0.083885 | 0.14126 | 2402.126 |
| SWEET_LU | 433 | 0.025512 | 0.003819 | 0.023751 | 0.14139 | 2404.337 |
| LOCKWOO | 213 | 0.042138 | 0.004451 | 0.039234 | 0.14142 | 2404.847 |
| MALONEY_ | 73 | 0.05995 | 0.003721 | 0.055837 | 0.14149 | 2406.037 |
| GAL_LEUKE | 117 | 0.048417 | 0.0038 | 0.045108 | 0.14156 | 2407.228 |
| GOBP_PUR | 16 | 0.12897 | 0.003753 | 0.12021 | 0.14168 | 2409.268 |
| GOBP_DIPE | 7 | 0.17987 | 0.003463 | 0.1677 | 0.14174 | 2410.289 |
| GOBP_EPIT | 10 | 0.19797 | 0.004555 | 0.18459 | 0.14175 | 2410.459 |
| GOBP_NEG | 12 | 0.17521 | 0.004416 | 0.16337 | 0.14177 | 2410.799 |
| GOMF_PHO | 78 | 0.062053 | 0.00398 | 0.057895 | 0.14191 | 2413.18 |
| GOBP_NEG | 131 | 0.048115 | 0.003994 | 0.044898 | 0.14194 | 2413.69 |
| ZHENG_FO | 36 | 0.092251 | 0.004025 | 0.086107 | 0.14202 | 2415.05 |
| GOBP_NEG | 22 | 0.11006 | 0.003755 | 0.10274 | 0.14205 | 2415.56 |
| REACTOME | 38 | 0.083983 | 0.003764 | 0.078403 | 0.14205 | 2415.56 |
| BURTON_A | 69 | 0.062156 | 0.003751 | 0.058032 | 0.14208 | 2416.07 |
| BENPORAT | 169 | 0.040874 | 0.00385 | 0.038169 | 0.14212 | 2416.751 |
| DASU_IL6_ | 54 | 0.067602 | 0.00361 | 0.063136 | 0.14215 | 2417.261 |
| CHIARADO | 48 | 0.086254 | 0.004344 | 0.080561 | 0.14217 | 2417.601 |
| PAL_PRMT | 186 | 0.037855 | 0.003739 | 0.035356 | 0.14217 | 2417.601 |
| GOBP_REG | 16 | 0.11951 | 0.003478 | 0.11164 | 0.1422 | 2418.111 |
| GOBP_NEG | 7 | 0.151 | 0.002907 | 0.14107 | 0.14223 | 2418.621 |
| KEGG_NEU | 120 | 0.048629 | 0.003865 | 0.045433 | 0.14224 | 2418.791 |
| WP_PTDIN | 10 | 0.16257 | 0.003741 | 0.1519 | 0.14227 | 2419.301 |
| GOMF_UBI | 110 | 0.04988 | 0.003796 | 0.046624 | 0.14236 | 2420.832 |
| GOBP_NEG | 85 | 0.058444 | 0.003913 | 0.054641 | 0.14241 | 2421.682 |
| ROZANOV_ | 250 | 0.033538 | 0.003834 | 0.031357 | 0.14241 | 2421.682 |
| BRUNO_HE | 63 | 0.065787 | 0.003794 | 0.06152 | 0.14246 | 2422.532 |
| KORKOLA_ | 6 | 0.26667 | 0.004753 | 0.2494 | 0.14249 | 2423.042 |
| GOBP_MIT | 39 | 0.071812 | 0.003261 | 0.067167 | 0.14251 | 2423.383 |
| REACTOME | 273 | 0.032464 | 0.003876 | 0.030377 | 0.14261 | 2425.083 |
| GOBP_THY | 22 | 0.12282 | 0.00419 | 0.11494 | 0.14264 | 2425.593 |
| WP_TUMO | 31 | 0.095447 | 0.003865 | 0.089377 | 0.14279 | 2428.144 |
| REACTOME | 8 | 0.19573 | 0.004028 | 0.18329 | 0.1428 | 2428.314 |
| GOBP_REG | 566 | 0.023183 | 0.003954 | 0.021711 | 0.14282 | 2428.654 |

| | | | | | | |
|---|---|---|---|---|---|---|
| CHIBA_RES | 48 | 0.080755 | 0.004067 | 0.075634 | 0.14283 | 2428.824 |
| REACTOME | 19 | 0.13594 | 0.00431 | 0.12733 | 0.14285 | 2429.164 |
| GOMF_GU. | 365 | 0.028016 | 0.003858 | 0.026242 | 0.14286 | 2429.334 |
| GOBP_NEG | 161 | 0.044984 | 0.004137 | 0.042144 | 0.14291 | 2430.185 |
| GOBP_REG | 82 | 0.055502 | 0.00365 | 0.052011 | 0.14296 | 2431.035 |
| GOBP_CHA | 64 | 0.063958 | 0.003718 | 0.05994 | 0.14299 | 2431.545 |
| REACTOME | 28 | 0.10041 | 0.003864 | 0.094141 | 0.14309 | 2433.245 |
| GOCC_POT | 87 | 0.064069 | 0.004339 | 0.060107 | 0.14324 | 2435.796 |
| GOMF_G_I | 24 | 0.10692 | 0.00381 | 0.1003 | 0.14324 | 2435.796 |
| GOMF_N_I | 94 | 0.054046 | 0.003804 | 0.050733 | 0.14338 | 2438.177 |
| GOMF_ION | 21 | 0.11422 | 0.003808 | 0.10723 | 0.14339 | 2438.347 |
| GOBP_APC | 5 | 0.19849 | 0.00323 | 0.18634 | 0.1434 | 2438.517 |
| KAYO_CAL( | 88 | 0.055885 | 0.003807 | 0.052479 | 0.14347 | 2439.707 |
| GOCC_HIST | 73 | 0.06279 | 0.003897 | 0.058982 | 0.14355 | 2441.068 |
| GOCC_SPEI | 6 | 0.18907 | 0.00337 | 0.17763 | 0.14358 | 2441.578 |
| WP_SOMA | 6 | 0.2092 | 0.003729 | 0.19659 | 0.14364 | 2442.598 |
| GOBP_CELI | 105 | 0.051878 | 0.003858 | 0.048759 | 0.14368 | 2443.278 |
| GOBP_REG | 445 | 0.024084 | 0.003654 | 0.022637 | 0.14369 | 2443.448 |
| GOBP_RNA | 15 | 0.16288 | 0.00459 | 0.15314 | 0.14376 | 2444.639 |
| GOBP_MET | 6 | 0.21274 | 0.003792 | 0.20009 | 0.14384 | 2445.999 |
| WP_ANDR( | 85 | 0.058552 | 0.00392 | 0.055075 | 0.14387 | 2446.509 |
| GOMF_DE/ | 32 | 0.099374 | 0.004088 | 0.093491 | 0.14391 | 2447.19 |
| RIGGI_EWI | 179 | 0.04238 | 0.004107 | 0.039871 | 0.14392 | 2447.36 |
| PID_ARF_3 | 19 | 0.11097 | 0.003519 | 0.1044 | 0.14393 | 2447.53 |
| WP_MITO( | 5 | 0.30681 | 0.004993 | 0.28868 | 0.14394 | 2447.7 |
| GOBP_REG | 7 | 0.21879 | 0.004212 | 0.20591 | 0.144 | 2448.72 |
| GOBP_SEN | 8 | 0.19199 | 0.003952 | 0.18078 | 0.14411 | 2450.591 |
| GOBP_FAT | 8 | 0.19207 | 0.003953 | 0.18086 | 0.14412 | 2450.761 |
| GOBP_CELI | 10 | 0.16337 | 0.003759 | 0.15395 | 0.1443 | 2453.822 |
| MANNE_C( | 170 | 0.03787 | 0.003578 | 0.03569 | 0.14433 | 2454.332 |
| WILSON_P | 5 | 0.22177 | 0.003609 | 0.20905 | 0.14438 | 2455.182 |
| TESAR_ALK | 6 | 0.22902 | 0.004082 | 0.21601 | 0.14453 | 2457.733 |
| TESAR_ALK | 6 | 0.22902 | 0.004082 | 0.21601 | 0.14453 | 2457.733 |
| GOBP_REG | 2 | 0.30992 | 0.00319 | 0.29233 | 0.14454 | 2457.903 |
| GOMF_DN. | 9 | 0.14375 | 0.003138 | 0.13561 | 0.14457 | 2458.413 |
| GOBP_ANC | 34 | 0.093823 | 0.003978 | 0.088512 | 0.14458 | 2458.583 |
| GOBP_COF | 5 | 0.22445 | 0.003652 | 0.21179 | 0.14463 | 2459.433 |
| NAM_FXYD | 17 | 0.12984 | 0.003895 | 0.12254 | 0.14469 | 2460.453 |
| GOBP_GLI/ | 109 | 0.053651 | 0.004065 | 0.050642 | 0.14471 | 2460.794 |
| GOMF_CAI | 13 | 0.13087 | 0.003433 | 0.12353 | 0.14471 | 2460.794 |
| FRIDMAN_ | 76 | 0.063051 | 0.003993 | 0.059539 | 0.14481 | 2462.494 |
| GOMF_SUI | 11 | 0.14828 | 0.003578 | 0.14012 | 0.14498 | 2465.385 |
| WP_MET_I | 55 | 0.071861 | 0.003873 | 0.067905 | 0.14498 | 2465.385 |
| GOBP_PRC | 9 | 0.13969 | 0.003049 | 0.13206 | 0.14507 | 2466.915 |
| GOCC_EM( | 9 | 0.13969 | 0.003049 | 0.13206 | 0.14507 | 2466.915 |

| | | | | | | |
|---|---|---|---|---|---|---|
| GOBP_PYR | 20 | 0.11286 | 0.003672 | 0.10671 | 0.14512 | 2467.766 |
| HOFFMANI | 61 | 0.065588 | 0.003722 | 0.062014 | 0.14512 | 2467.766 |
| REACTOME | 21 | 0.11834 | 0.003945 | 0.11191 | 0.14516 | 2468.446 |
| GOBP_ACT | 6 | 0.26211 | 0.004672 | 0.24795 | 0.14524 | 2469.806 |
| GALIE_TUN | 8 | 0.21045 | 0.004331 | 0.1991 | 0.14525 | 2469.976 |
| GOMF_DIP | 12 | 0.16822 | 0.00424 | 0.15914 | 0.14526 | 2470.146 |
| BROWNE_I | 170 | 0.04209 | 0.003976 | 0.039824 | 0.14528 | 2470.486 |
| DING_LUN | 100 | 0.05217 | 0.003787 | 0.049376 | 0.14536 | 2471.847 |
| WP_BURN_ | 94 | 0.055896 | 0.003934 | 0.052933 | 0.1455 | 2474.228 |
| WP_MONC | 30 | 0.098636 | 0.003929 | 0.093408 | 0.1455 | 2474.228 |
| GOMF_PH( | 7 | 0.17852 | 0.003437 | 0.16907 | 0.14552 | 2474.568 |
| WP_NEUR( | 9 | 0.17981 | 0.003925 | 0.17033 | 0.14556 | 2475.248 |
| GOBP_CELI | 11 | 0.16105 | 0.003886 | 0.15256 | 0.14558 | 2475.588 |
| GOCC_DEN | 27 | 0.099923 | 0.003776 | 0.094718 | 0.14573 | 2478.139 |
| GOCC_NUC | 12 | 0.13013 | 0.00328 | 0.12339 | 0.1458 | 2479.329 |
| REACTOME | 21 | 0.11358 | 0.003786 | 0.1077 | 0.14582 | 2479.669 |
| GOBP_HIST | 12 | 0.14541 | 0.003665 | 0.13789 | 0.14583 | 2479.839 |
| GOBP_T_C | 24 | 0.10223 | 0.003643 | 0.096941 | 0.14583 | 2479.839 |
| GOBP_PRC | 715 | 0.019479 | 0.003718 | 0.018482 | 0.14596 | 2482.05 |
| GOBP_BON | 25 | 0.11031 | 0.004012 | 0.10469 | 0.14603 | 2483.24 |
| GOBP_BER | 11 | 0.19127 | 0.004616 | 0.18154 | 0.14604 | 2483.41 |
| GOMF_OXI | 17 | 0.12961 | 0.003888 | 0.12307 | 0.14614 | 2485.111 |
| BLANCO_M | 196 | 0.035758 | 0.003625 | 0.033963 | 0.14622 | 2486.471 |
| SCHAEFFEF | 452 | 0.02471 | 0.003778 | 0.023471 | 0.14623 | 2486.641 |
| GOBP_PRC | 9 | 0.1691 | 0.003691 | 0.16066 | 0.14628 | 2487.491 |
| GOBP_HEA | 14 | 0.15633 | 0.004256 | 0.14856 | 0.14634 | 2488.512 |
| GOCC_LAT | 12 | 0.16097 | 0.004057 | 0.15299 | 0.14637 | 2489.022 |
| REACTOME | 25 | 0.099858 | 0.003632 | 0.094912 | 0.14638 | 2489.192 |
| WP_BENZC | 9 | 0.17993 | 0.003928 | 0.17105 | 0.14642 | 2489.872 |
| REACTOME | 14 | 0.1417 | 0.003857 | 0.13471 | 0.14644 | 2490.212 |
| GOBP_RIB( | 22 | 0.10423 | 0.003556 | 0.099104 | 0.14648 | 2490.892 |
| GOBP_LIPII | 6 | 0.20677 | 0.003686 | 0.19663 | 0.1465 | 2491.233 |
| GOBP_HON | 91 | 0.056802 | 0.003934 | 0.054029 | 0.14656 | 2492.253 |
| GOBP_RES | 17 | 0.11337 | 0.003401 | 0.10784 | 0.14657 | 2492.423 |
| REACTOME | 53 | 0.068974 | 0.00365 | 0.065616 | 0.1466 | 2492.933 |
| WP_IL9_SI( | 15 | 0.16729 | 0.004714 | 0.1592 | 0.14668 | 2494.293 |
| SHETH_LIV | 67 | 0.067806 | 0.004032 | 0.064537 | 0.14672 | 2494.974 |
| GOBP_MAI | 18 | 0.13886 | 0.004286 | 0.13217 | 0.14673 | 2495.144 |
| PANGAS_T | 127 | 0.047945 | 0.003919 | 0.04565 | 0.1468 | 2496.334 |
| GOBP_MIC | 8 | 0.18705 | 0.00385 | 0.17816 | 0.14689 | 2497.864 |
| REACTOME | 20 | 0.12147 | 0.003952 | 0.1157 | 0.1469 | 2498.035 |
| ENGELMAN | 62 | 0.066375 | 0.003798 | 0.063234 | 0.14694 | 2498.715 |
| GOMF_CAI | 76 | 0.060306 | 0.003819 | 0.057465 | 0.147 | 2499.735 |
| GOCC_MIT | 36 | 0.094536 | 0.004124 | 0.090106 | 0.14706 | 2500.755 |
| GOBP_BRA | 20 | 0.13784 | 0.004484 | 0.1314 | 0.1471 | 2501.436 |

| | | | | | | |
|---|---|---|---|---|---|---|
| KYNG_WEF | 76 | 0.05578 | 0.003532 | 0.053195 | 0.14719 | 2502.966 |
| GOBP_PRC | 63 | 0.058868 | 0.003395 | 0.056145 | 0.14721 | 2503.306 |
| GOBP_ANA | 15 | 0.14392 | 0.004055 | 0.13728 | 0.14725 | 2503.986 |
| GOMF_REC | 15 | 0.12376 | 0.003487 | 0.11807 | 0.14728 | 2504.496 |
| ZHONG_SE | 62 | 0.06724 | 0.003847 | 0.064147 | 0.14728 | 2504.496 |
| GOBP_GLIA | 10 | 0.18884 | 0.004345 | 0.18019 | 0.14733 | 2505.347 |
| LEI_HOXC8 | 10 | 0.1919 | 0.004416 | 0.18319 | 0.14743 | 2507.047 |
| GOBP_STEI | 40 | 0.090242 | 0.00415 | 0.086152 | 0.14745 | 2507.387 |
| NADELLA_I | 9 | 0.18147 | 0.003961 | 0.17325 | 0.14745 | 2507.387 |
| REACTOME | 63 | 0.069148 | 0.003988 | 0.06604 | 0.14754 | 2508.918 |
| GOBP_POS | 14 | 0.14721 | 0.004008 | 0.14061 | 0.14757 | 2509.428 |
| KIM_MYCL | 13 | 0.14765 | 0.003873 | 0.14105 | 0.14761 | 2510.108 |
| TERAMOTC | 18 | 0.11349 | 0.003503 | 0.10842 | 0.14762 | 2510.278 |
| SAKAI_TUN | 76 | 0.055636 | 0.003523 | 0.053185 | 0.14777 | 2512.829 |
| GOBP_EPO | 18 | 0.14518 | 0.004481 | 0.1388 | 0.1478 | 2513.339 |
| GOBP_FAT | 12 | 0.16055 | 0.004047 | 0.15356 | 0.14789 | 2514.869 |
| WP_REGUL | 17 | 0.125 | 0.003749 | 0.11956 | 0.14791 | 2515.21 |
| GOBP_END | 551 | 0.022359 | 0.003764 | 0.021388 | 0.14793 | 2515.55 |
| GOBP_NEG | 59 | 0.067488 | 0.003767 | 0.064573 | 0.14798 | 2516.4 |
| CARRILLOR | 1023 | 0.01718 | 0.00389 | 0.016441 | 0.14802 | 2517.08 |
| GOBP_REG | 17 | 0.12879 | 0.003863 | 0.12326 | 0.14804 | 2517.42 |
| GOMF_ATF | 82 | 0.059731 | 0.003928 | 0.057193 | 0.14817 | 2519.631 |
| GOBP_POS | 55 | 0.075153 | 0.004051 | 0.071979 | 0.14823 | 2520.651 |
| MAINA_VH | 16 | 0.13643 | 0.00397 | 0.1307 | 0.14828 | 2521.501 |
| WP_FOCAL | 190 | 0.039035 | 0.003896 | 0.037394 | 0.14828 | 2521.501 |
| GOCC_AUT | 47 | 0.072213 | 0.003599 | 0.069196 | 0.14834 | 2522.522 |
| GOBP_MES | 12 | 0.1784 | 0.004497 | 0.17096 | 0.14835 | 2522.692 |
| WP_NETRI | 50 | 0.07548 | 0.003879 | 0.072328 | 0.14835 | 2522.692 |
| REACTOME | 13 | 0.16758 | 0.004396 | 0.16068 | 0.1485 | 2525.243 |
| WP_RALA_ | 12 | 0.14579 | 0.003675 | 0.13981 | 0.14853 | 2525.753 |
| GOBP_RES | 6 | 0.17497 | 0.003119 | 0.1678 | 0.14855 | 2526.093 |
| REACTOME | 6 | 0.17127 | 0.003053 | 0.16427 | 0.14858 | 2526.603 |
| ZHU_CMV_ | 99 | 0.050665 | 0.003659 | 0.048605 | 0.14862 | 2527.283 |
| GOMF_CYT | 919 | 0.017432 | 0.003751 | 0.016729 | 0.14871 | 2528.814 |
| DARWICHE | 176 | 0.037058 | 0.003561 | 0.035573 | 0.14877 | 2529.834 |
| GOBP_GLY | 14 | 0.15994 | 0.004354 | 0.15358 | 0.14885 | 2531.194 |
| GOBP_MO | 20 | 0.10444 | 0.003398 | 0.10029 | 0.14886 | 2531.364 |
| GOBP_REG | 26 | 0.094284 | 0.003497 | 0.090542 | 0.14887 | 2531.534 |
| GOBP_RIB | 17 | 0.11653 | 0.003495 | 0.11201 | 0.1491 | 2535.446 |
| WHITFIELD | 158 | 0.039815 | 0.003627 | 0.038285 | 0.14918 | 2536.806 |
| WP_RELAT | 25 | 0.10128 | 0.003683 | 0.097413 | 0.14926 | 2538.166 |
| GOBP_HEN | 6 | 0.20879 | 0.003722 | 0.20084 | 0.14927 | 2538.336 |
| GOBP_STEI | 236 | 0.034754 | 0.003862 | 0.033432 | 0.14928 | 2538.506 |
| GOBP_CELI | 7 | 0.22267 | 0.004287 | 0.2142 | 0.14929 | 2538.676 |
| GOBP_NEG | 11 | 0.15476 | 0.003735 | 0.14889 | 0.14932 | 2539.187 |

| | | | | | | |
|---|---|---|---|---|---|---|
| LINDGREN_ | 352 | 0.029456 | 0.003985 | 0.028341 | 0.14933 | 2539.357 |
| GOBP_MIS | 31 | 0.08997 | 0.003643 | 0.086571 | 0.14935 | 2539.697 |
| REACTOME | 6 | 0.19195 | 0.003422 | 0.18471 | 0.14935 | 2539.697 |
| GOBP_GAN | 5 | 0.25365 | 0.004128 | 0.2441 | 0.14938 | 2540.207 |
| GOBP_REG | 500 | 0.023174 | 0.003721 | 0.022316 | 0.14954 | 2542.928 |
| BIOCARTA_ | 36 | 0.088873 | 0.003877 | 0.085588 | 0.14955 | 2543.098 |
| WP_16P11 | 69 | 0.076127 | 0.004594 | 0.073316 | 0.14956 | 2543.268 |
| GOBP_REG | 17 | 0.12613 | 0.003783 | 0.12152 | 0.14967 | 2545.138 |
| GOBP_TAIL | 14 | 0.11619 | 0.003163 | 0.11196 | 0.14969 | 2545.478 |
| MUELLER_ | 29 | 0.097467 | 0.003817 | 0.093925 | 0.14971 | 2545.819 |
| GOMF_OXI | 53 | 0.076616 | 0.004054 | 0.073878 | 0.14986 | 2548.369 |
| GOBP_POS | 27 | 0.089907 | 0.003398 | 0.08673 | 0.14996 | 2550.07 |
| GOBP_STE | 29 | 0.10136 | 0.00397 | 0.097816 | 0.15005 | 2551.6 |
| GOMF_G_I | 10 | 0.1561 | 0.003592 | 0.15066 | 0.15009 | 2552.28 |
| BERNARD_ | 56 | 0.073132 | 0.003977 | 0.070592 | 0.15011 | 2552.621 |
| SCHOEN_N | 34 | 0.097178 | 0.00412 | 0.093801 | 0.15011 | 2552.621 |
| GOBP_REG | 21 | 0.10951 | 0.00365 | 0.10574 | 0.15021 | 2554.321 |
| GOCC_END | 117 | 0.044514 | 0.003494 | 0.042985 | 0.15021 | 2554.321 |
| REACTOME | 6 | 0.20799 | 0.003708 | 0.20085 | 0.15021 | 2554.321 |
| GOBP_REG | 6 | 0.20535 | 0.00366 | 0.19837 | 0.15031 | 2556.022 |
| FARMER_B | 18 | 0.12956 | 0.003999 | 0.12517 | 0.15033 | 2556.362 |
| GOBP_REG | 18 | 0.10989 | 0.003392 | 0.10618 | 0.15035 | 2556.702 |
| GOMF_LOV | 24 | 0.10647 | 0.003794 | 0.10288 | 0.15038 | 2557.212 |
| GOBP_ENG | 14 | 0.13947 | 0.003797 | 0.13478 | 0.15039 | 2557.382 |
| GOBP_SPLI | 35 | 0.09593 | 0.004127 | 0.092724 | 0.15044 | 2558.232 |
| GOBP_GLU | 8 | 0.18037 | 0.003712 | 0.17436 | 0.15047 | 2558.742 |
| GOBP_PUR | 7 | 0.19985 | 0.003848 | 0.19323 | 0.15051 | 2559.423 |
| GOBP_EST | 25 | 0.10857 | 0.003949 | 0.105 | 0.15056 | 2560.273 |
| GOCC_MIC | 1267 | 0.014678 | 0.003673 | 0.014195 | 0.15058 | 2560.613 |
| BIOCARTA_ | 11 | 0.15576 | 0.003759 | 0.15071 | 0.15069 | 2562.483 |
| GOBP_NEU | 46 | 0.087477 | 0.004313 | 0.084674 | 0.15078 | 2564.014 |
| LI_PROSTA | 30 | 0.094297 | 0.003756 | 0.091277 | 0.15079 | 2564.184 |
| GOBP_POS | 39 | 0.090679 | 0.004117 | 0.087778 | 0.1508 | 2564.354 |
| ALONSO_M | 33 | 0.088402 | 0.003693 | 0.085594 | 0.15086 | 2565.374 |
| GOBP_CELI | 1194 | 0.015678 | 0.003816 | 0.015189 | 0.15099 | 2567.585 |
| GOBP_RES | 11 | 0.19612 | 0.004733 | 0.19001 | 0.15101 | 2567.925 |
| GOBP_STA | 17 | 0.13689 | 0.004106 | 0.13266 | 0.15106 | 2568.775 |
| PARENT_M | 521 | 0.022145 | 0.003628 | 0.021463 | 0.1511 | 2569.456 |
| GOBP_CELI | 651 | 0.020466 | 0.003734 | 0.019837 | 0.15111 | 2569.626 |
| GOBP_REG | 7 | 0.2112 | 0.004066 | 0.20476 | 0.15117 | 2570.646 |
| WP_FGFR3 | 27 | 0.11131 | 0.004207 | 0.10795 | 0.15124 | 2571.836 |
| REACTOME | 21 | 0.10803 | 0.003601 | 0.1048 | 0.15133 | 2573.367 |
| GOBP_MIC | 8 | 0.19475 | 0.004008 | 0.18903 | 0.15145 | 2575.407 |
| GOBP_NEU | 13 | 0.13141 | 0.003447 | 0.12755 | 0.15145 | 2575.407 |
| REACTOME | 58 | 0.072694 | 0.004023 | 0.07058 | 0.15152 | 2576.598 |

| | | | | | | |
|---|---|---|---|---|---|---|
| GOBP_MO | 57 | 0.071293 | 0.003912 | 0.069244 | 0.15161 | 2578.128 |
| GOBP_FRU | 10 | 0.18293 | 0.004209 | 0.17769 | 0.15164 | 2578.638 |
| VALK_AML | 27 | 0.11113 | 0.0042 | 0.10795 | 0.15164 | 2578.638 |
| GOBP_POS | 6 | 0.19135 | 0.003411 | 0.1859 | 0.15167 | 2579.148 |
| GOBP_COL | 113 | 0.051763 | 0.003993 | 0.050304 | 0.15175 | 2580.509 |
| GOBP_REG | 5 | 0.27579 | 0.004488 | 0.26802 | 0.15175 | 2580.509 |
| GOBP_NEG | 21 | 0.11807 | 0.003936 | 0.11475 | 0.15176 | 2580.679 |
| KEGG_FRU | 32 | 0.082632 | 0.003399 | 0.080332 | 0.15184 | 2582.039 |
| WP_PLATE | 14 | 0.14096 | 0.003837 | 0.13706 | 0.15187 | 2582.549 |
| GOBP_REG | 15 | 0.13739 | 0.003871 | 0.13368 | 0.15204 | 2585.44 |
| GAUSSMAI | 173 | 0.041429 | 0.003948 | 0.040321 | 0.15211 | 2586.631 |
| GOCC_CON | 14 | 0.14374 | 0.003913 | 0.13994 | 0.15218 | 2587.821 |
| GOBP_REG | 27 | 0.096897 | 0.003662 | 0.094358 | 0.15224 | 2588.841 |
| REACTOME | 12 | 0.15387 | 0.003878 | 0.14985 | 0.15227 | 2589.351 |
| GOBP_POS | 6 | 0.25667 | 0.004575 | 0.25003 | 0.15232 | 2590.202 |
| GOBP_OUT | 13 | 0.13082 | 0.003432 | 0.12747 | 0.15239 | 2591.392 |
| GOBP_MAI | 51 | 0.064645 | 0.003356 | 0.063003 | 0.15244 | 2592.242 |
| CAFFAREL_ | 14 | 0.13092 | 0.003564 | 0.1276 | 0.15246 | 2592.582 |
| MISHRA_C/ | 23 | 0.11091 | 0.003869 | 0.1081 | 0.15247 | 2592.752 |
| GOMF_TR/ | 5 | 0.28033 | 0.004562 | 0.27325 | 0.15248 | 2592.922 |
| REACTOME | 16 | 0.11681 | 0.003399 | 0.11386 | 0.15248 | 2592.922 |
| BILANGES_ | 20 | 0.10959 | 0.003565 | 0.10688 | 0.15262 | 2595.303 |
| TURASHVIL | 64 | 0.07004 | 0.004071 | 0.068326 | 0.15267 | 2596.153 |
| GOMF_LIM | 6 | 0.18789 | 0.003349 | 0.18334 | 0.15272 | 2597.004 |
| GOBP_REG | 6 | 0.21148 | 0.00377 | 0.20638 | 0.15276 | 2597.684 |
| GOBP_CAN | 287 | 0.031818 | 0.003893 | 0.031053 | 0.15278 | 2598.024 |
| GOMF_RS_ | 5 | 0.23872 | 0.003885 | 0.23317 | 0.15297 | 2601.255 |
| GOCC_RN/ | 15 | 0.15127 | 0.004263 | 0.1478 | 0.15304 | 2602.445 |
| HANSON_H | 24 | 0.10278 | 0.003662 | 0.10043 | 0.15307 | 2602.955 |
| GOCC_SPIN | 161 | 0.040168 | 0.003694 | 0.039254 | 0.15309 | 2603.295 |
| GOBP_DOU | 289 | 0.02882 | 0.003538 | 0.028182 | 0.15325 | 2606.016 |
| GOMF_PH( | 256 | 0.031826 | 0.003681 | 0.031129 | 0.15331 | 2607.037 |
| GOBP_VES | 32 | 0.092773 | 0.003816 | 0.09077 | 0.15338 | 2608.227 |
| REACTOME | 108 | 0.051878 | 0.003913 | 0.050761 | 0.1534 | 2608.567 |
| WP_DNA_I | 23 | 0.10216 | 0.003564 | 0.099977 | 0.15344 | 2609.247 |
| CHIANG_LI | 162 | 0.04224 | 0.003896 | 0.041347 | 0.1535 | 2610.268 |
| ZHENG_GL | 57 | 0.072247 | 0.003964 | 0.070728 | 0.15352 | 2610.608 |
| GOBP_POS | 9 | 0.20291 | 0.004429 | 0.19872 | 0.15362 | 2612.308 |
| REACTOME | 94 | 0.05233 | 0.003683 | 0.051277 | 0.15374 | 2614.349 |
| GOBP_R_L( | 6 | 0.2095 | 0.003734 | 0.20529 | 0.15375 | 2614.519 |
| GOBP_SM( | 7 | 0.20871 | 0.004018 | 0.20451 | 0.15375 | 2614.519 |
| GOBP_REG | 55 | 0.067847 | 0.003657 | 0.066489 | 0.15377 | 2614.859 |
| GOCC_ACT | 6 | 0.29394 | 0.005239 | 0.28805 | 0.15377 | 2614.859 |
| GOBP_STR/ | 6 | 0.18841 | 0.003358 | 0.18464 | 0.15378 | 2615.029 |
| GOBP_ADE | 16 | 0.1515 | 0.004409 | 0.14854 | 0.15389 | 2616.899 |

| | | | | | | |
|---|---|---|---|---|---|---|
| GOBP_REG | 6 | 0.20895 | 0.003725 | 0.20494 | 0.15398 | 2618.43 |
| KIM_MYCL | 19 | 0.11471 | 0.003637 | 0.11253 | 0.15401 | 2618.94 |
| GOBP_FAT | 66 | 0.059392 | 0.003506 | 0.058288 | 0.15412 | 2620.811 |
| GOBP_PYR | 11 | 0.15015 | 0.003624 | 0.14737 | 0.15413 | 2620.981 |
| WP_EXTRA | 19 | 0.13065 | 0.004143 | 0.12823 | 0.15414 | 2621.151 |
| GOBP_BLO | 12 | 0.18 | 0.004537 | 0.17669 | 0.15417 | 2621.661 |
| WAKABAYA | 616 | 0.020745 | 0.003686 | 0.020363 | 0.15417 | 2621.661 |
| GOBP_INTI | 90 | 0.066181 | 0.004559 | 0.064972 | 0.1542 | 2622.171 |
| GOCC_ACT | 74 | 0.062429 | 0.003901 | 0.0613 | 0.15425 | 2623.021 |
| MORI_SMA | 73 | 0.058752 | 0.003646 | 0.057695 | 0.15427 | 2623.361 |
| WP_DISRU | 28 | 0.097289 | 0.003744 | 0.095557 | 0.15432 | 2624.212 |
| GOBP_PER | 34 | 0.077049 | 0.003267 | 0.075701 | 0.15439 | 2625.402 |
| REACTOME | 42 | 0.080592 | 0.003797 | 0.079206 | 0.15446 | 2626.592 |
| GOBP_UBI | 6 | 0.21126 | 0.003766 | 0.20767 | 0.15451 | 2627.443 |
| ZHAN_MUI | 42 | 0.082371 | 0.003881 | 0.080987 | 0.15456 | 2628.293 |
| REACTOME | 200 | 0.03693 | 0.003781 | 0.036321 | 0.15464 | 2629.653 |
| REACTOME | 7 | 0.20064 | 0.003863 | 0.19737 | 0.15469 | 2630.503 |
| GOCC_XY_ | 14 | 0.14563 | 0.003964 | 0.14327 | 0.1547 | 2630.674 |
| GOBP_REG | 7 | 0.17733 | 0.003414 | 0.17447 | 0.15473 | 2631.184 |
| GOBP_REG | 13 | 0.16407 | 0.004304 | 0.16145 | 0.15477 | 2631.864 |
| GOMF_SOI | 26 | 0.10045 | 0.003725 | 0.098855 | 0.15478 | 2632.034 |
| WONG_EM | 317 | 0.027436 | 0.003525 | 0.027004 | 0.15482 | 2632.714 |
| REACTOME | 10 | 0.18549 | 0.004268 | 0.18258 | 0.15483 | 2632.884 |
| KAYO_AGI | 116 | 0.043435 | 0.003394 | 0.042758 | 0.15486 | 2633.394 |
| KAUFFMAN | 210 | 0.033051 | 0.003467 | 0.032537 | 0.15487 | 2633.564 |
| BIOCARTA_ | 20 | 0.11792 | 0.003836 | 0.11615 | 0.155 | 2635.775 |
| GOMF_HIS | 8 | 0.15209 | 0.00313 | 0.14983 | 0.15505 | 2636.625 |
| GOBP_REG | 75 | 0.057063 | 0.00359 | 0.056242 | 0.15515 | 2638.326 |
| GOBP_LAT | 9 | 0.21003 | 0.004585 | 0.20702 | 0.15517 | 2638.666 |
| GOMF_HYI | 129 | 0.044888 | 0.003698 | 0.044244 | 0.15517 | 2638.666 |
| GOBP_ADP | 5 | 0.23389 | 0.003806 | 0.23056 | 0.1552 | 2639.176 |
| GOBP_REG | 202 | 0.03507 | 0.003608 | 0.034572 | 0.1552 | 2639.176 |
| GOBP_POS | 44 | 0.076802 | 0.003704 | 0.075714 | 0.15521 | 2639.346 |
| SAENZ_DE | 14 | 0.13577 | 0.003696 | 0.13385 | 0.15522 | 2639.516 |
| GOCC_CAT | 17 | 0.1371 | 0.004112 | 0.13527 | 0.15541 | 2642.747 |
| SIMBULAN_ | 31 | 0.084227 | 0.00341 | 0.083101 | 0.15541 | 2642.747 |
| KEGG_OOC | 107 | 0.048165 | 0.003616 | 0.047523 | 0.15542 | 2642.917 |
| QI_HYPOXI | 128 | 0.045895 | 0.003766 | 0.045302 | 0.15552 | 2644.618 |
| REACTOME | 13 | 0.14666 | 0.003847 | 0.14477 | 0.15552 | 2644.618 |
| GOBP_NEG | 6 | 0.2498 | 0.004453 | 0.2466 | 0.15555 | 2645.128 |
| GOBP_PYR | 29 | 0.089741 | 0.003515 | 0.08861 | 0.15559 | 2645.808 |
| GOBP_PSEI | 12 | 0.12112 | 0.003053 | 0.1196 | 0.15561 | 2646.148 |
| REACTOME | 77 | 0.055285 | 0.003524 | 0.054599 | 0.15564 | 2646.658 |
| GOBP_PEN | 9 | 0.15264 | 0.003332 | 0.15075 | 0.15565 | 2646.828 |
| GOBP_REG | 163 | 0.0396 | 0.003664 | 0.039117 | 0.15569 | 2647.508 |

| | | | | | | |
|---|---|---|---|---|---|---|
| WESTON_\ | 87 | 0.056889 | 0.003853 | 0.056199 | 0.15571 | 2647.849 |
| HOEGERKC | 11 | 0.14218 | 0.003431 | 0.14048 | 0.15574 | 2648.359 |
| GOBP_POS | 34 | 0.0831 | 0.003524 | 0.082112 | 0.15577 | 2648.869 |
| GOBP_RIB( | 53 | 0.06401 | 0.003387 | 0.063254 | 0.15579 | 2649.209 |
| WP_MITO( | 28 | 0.088395 | 0.003402 | 0.087364 | 0.15582 | 2649.719 |
| GOBP_NEG | 89 | 0.051217 | 0.003508 | 0.050621 | 0.15583 | 2649.889 |
| GOBP_POS | 23 | 0.12307 | 0.004293 | 0.1217 | 0.15595 | 2651.93 |
| REACTOME | 33 | 0.084355 | 0.003524 | 0.083417 | 0.15596 | 2652.1 |
| KEGG_GLY( | 15 | 0.12458 | 0.00351 | 0.12322 | 0.15601 | 2652.95 |
| GOBP_HEN | 94 | 0.05395 | 0.003798 | 0.053376 | 0.15607 | 2653.97 |
| GOBP_POS | 10 | 0.20788 | 0.004783 | 0.20579 | 0.15621 | 2656.351 |
| GOBP_POL | 24 | 0.10307 | 0.003673 | 0.10204 | 0.15623 | 2656.691 |
| GOBP_TEL( | 18 | 0.11826 | 0.00365 | 0.11708 | 0.15623 | 2656.691 |
| GOBP_RES | 20 | 0.12604 | 0.0041 | 0.1249 | 0.15646 | 2660.602 |
| GOCC_EAR | 397 | 0.024648 | 0.003537 | 0.024435 | 0.15656 | 2662.303 |
| GOCC_NU( | 9 | 0.14796 | 0.00323 | 0.14668 | 0.15657 | 2662.473 |
| GOMF_SEN | 22 | 0.10431 | 0.003559 | 0.10344 | 0.15664 | 2663.663 |
| ZERBINI_RI | 6 | 0.19927 | 0.003552 | 0.19764 | 0.15667 | 2664.173 |
| GOBP_REG | 14 | 0.13256 | 0.003609 | 0.13149 | 0.15669 | 2664.513 |
| WEST_ADR | 289 | 0.029576 | 0.003631 | 0.029338 | 0.1567 | 2664.684 |
| REACTOME | 33 | 0.083838 | 0.003502 | 0.083176 | 0.15674 | 2665.364 |
| GOBP_VAL | 7 | 0.19834 | 0.003819 | 0.1968 | 0.15678 | 2666.044 |
| GOMF_AB( | 8 | 0.19228 | 0.003957 | 0.1908 | 0.15679 | 2666.214 |
| GOBP_WA | 6 | 0.19267 | 0.003434 | 0.1912 | 0.15681 | 2666.554 |
| GOBP_RES | 1669 | 0.01302 | 0.003696 | 0.012923 | 0.15685 | 2667.234 |
| NIKOLSKY_ | 37 | 0.15737 | 0.00696 | 0.15624 | 0.15692 | 2668.425 |
| GOMF_CX( | 5 | 0.27509 | 0.004476 | 0.27316 | 0.15696 | 2669.105 |
| HOLLERN_ | 30 | 0.10305 | 0.004105 | 0.10233 | 0.15697 | 2669.275 |
| GOMF_TR/ | 16 | 0.15109 | 0.004397 | 0.1501 | 0.15707 | 2670.975 |
| GOMF_INT | 8 | 0.17193 | 0.003539 | 0.17081 | 0.15709 | 2671.315 |
| GOBP_POS | 10 | 0.18795 | 0.004325 | 0.18675 | 0.15711 | 2671.656 |
| TERAO_AO | 28 | 0.080342 | 0.003092 | 0.079845 | 0.15716 | 2672.506 |
| GOBP_REG | 1351 | 0.01411 | 0.003637 | 0.014037 | 0.15741 | 2676.757 |
| GOMF_OXI | 40 | 0.085494 | 0.003931 | 0.085093 | 0.15753 | 2678.798 |
| WP_PRIMA | 71 | 0.063169 | 0.003867 | 0.062889 | 0.15759 | 2679.818 |
| GOBP_MRI | 23 | 0.10296 | 0.003592 | 0.10251 | 0.15762 | 2680.328 |
| GOBP_REG | 7 | 0.18171 | 0.003498 | 0.18093 | 0.15763 | 2680.498 |
| THUM_SYS | 377 | 0.026284 | 0.003677 | 0.026173 | 0.15763 | 2680.498 |
| HELLER_HI | 272 | 0.029852 | 0.003557 | 0.029732 | 0.15769 | 2681.518 |
| GOBP_POS | 26 | 0.10796 | 0.004004 | 0.10754 | 0.15771 | 2681.859 |
| GOBP_REG | 145 | 0.041956 | 0.003663 | 0.041798 | 0.15775 | 2682.539 |
| GOMF_INC | 5 | 0.25048 | 0.004076 | 0.24956 | 0.15777 | 2682.879 |
| GOBP_NEG | 41 | 0.075313 | 0.003506 | 0.075045 | 0.1578 | 2683.389 |
| GOBP_RES | 68 | 0.065552 | 0.003927 | 0.065367 | 0.15798 | 2686.45 |
| REACTOME | 11 | 0.20699 | 0.004995 | 0.20641 | 0.15798 | 2686.45 |

| | | | | | | |
|---|---|---|---|---|---|---|
| LIM_MAMI | 458 | 0.024768 | 0.003811 | 0.02471 | 0.15809 | 2688.32 |
| REACTOME | 9 | 0.1611 | 0.003517 | 0.16078 | 0.15817 | 2689.681 |
| RICKMAN_ | 99 | 0.051778 | 0.00374 | 0.05168 | 0.15821 | 2690.361 |
| GOBP_NEG | 11 | 0.14739 | 0.003557 | 0.14715 | 0.15827 | 2691.381 |
| NIKOLSKY_ | 17 | 0.2359 | 0.007076 | 0.23568 | 0.15844 | 2694.272 |
| GOMF_VOI | 9 | 0.17963 | 0.003921 | 0.17955 | 0.15855 | 2696.143 |
| GOBP_MA( | 53 | 0.071925 | 0.003806 | 0.071908 | 0.1586 | 2696.993 |
| GOBP_CAR | 8 | 0.17225 | 0.003545 | 0.17225 | 0.15866 | 2698.013 |
| GOBP_SKEI | 5 | 0.23916 | 0.003892 | 0.23917 | 0.15868 | 2698.353 |
| GOBP_INTI | 14 | 0.13729 | 0.003737 | 0.13733 | 0.15874 | 2699.374 |
| GOBP_POS | 36 | 0.082345 | 0.003593 | 0.082382 | 0.15877 | 2699.884 |
| MIKKELSEN | 7 | 0.16038 | 0.003088 | 0.16049 | 0.15883 | 2700.904 |
| DALESSIO_ | 17 | 0.13567 | 0.004069 | 0.13577 | 0.15885 | 2701.244 |
| GOBP_REG | 12 | 0.14946 | 0.003767 | 0.14962 | 0.15892 | 2702.435 |
| GOCC_BCL_ | 10 | 0.15838 | 0.003644 | 0.15865 | 0.15907 | 2704.985 |
| GOBP_NEG | 56 | 0.072931 | 0.003966 | 0.073086 | 0.15918 | 2706.856 |
| WP_METAS | 6 | 0.21595 | 0.003849 | 0.21643 | 0.15919 | 2707.026 |
| GOMF_TRA | 17 | 0.14661 | 0.004398 | 0.14697 | 0.15925 | 2708.046 |
| GOBP_MRI | 110 | 0.043977 | 0.003347 | 0.044092 | 0.15929 | 2708.726 |
| REACTOME | 11 | 0.14644 | 0.003534 | 0.14682 | 0.15929 | 2708.726 |
| FU_INTERA | 13 | 0.12297 | 0.003226 | 0.12331 | 0.15933 | 2709.407 |
| GOCC_EAR | 167 | 0.037843 | 0.003544 | 0.037947 | 0.15933 | 2709.407 |
| GOBP_REG | 18 | 0.1359 | 0.004194 | 0.13633 | 0.15942 | 2710.937 |
| GOBP_BLA | 21 | 0.11051 | 0.003684 | 0.11087 | 0.15946 | 2711.617 |
| GOBP_DEC | 5 | 0.19466 | 0.003168 | 0.19531 | 0.15947 | 2711.787 |
| BIOCARTA_ | 17 | 0.12032 | 0.003609 | 0.12076 | 0.15956 | 2713.318 |
| GOBP_SYN | 28 | 0.094114 | 0.003622 | 0.094508 | 0.15967 | 2715.188 |
| KYNG_DNA | 171 | 0.037608 | 0.003563 | 0.037767 | 0.15968 | 2715.358 |
| GOBP_RES | 12 | 0.16361 | 0.004124 | 0.16431 | 0.15969 | 2715.528 |
| TONKS_TAI | 21 | 0.096146 | 0.003205 | 0.096574 | 0.15974 | 2716.379 |
| LEI_MYB_T | 301 | 0.028874 | 0.003617 | 0.029015 | 0.15984 | 2718.079 |
| GOMF_CYT | 83 | 0.059695 | 0.00395 | 0.059989 | 0.15985 | 2718.249 |
| CHEN_HOX | 9 | 0.1891 | 0.004128 | 0.19006 | 0.15989 | 2718.929 |
| GOBP_SUP | 12 | 0.14299 | 0.003604 | 0.14372 | 0.15989 | 2718.929 |
| SUH_COEX | 18 | 0.12734 | 0.00393 | 0.128 | 0.15991 | 2719.27 |
| REACTOME | 181 | 0.036933 | 0.003599 | 0.037128 | 0.15994 | 2719.78 |
| JIANG_AGI | 31 | 0.086381 | 0.003498 | 0.086852 | 0.15998 | 2720.46 |
| GOBP_CELI | 200 | 0.034395 | 0.003522 | 0.034588 | 0.16001 | 2720.97 |
| GOBP_DET | 13 | 0.14387 | 0.003774 | 0.14471 | 0.16007 | 2721.99 |
| SCHAEFFEF | 101 | 0.052018 | 0.003795 | 0.052329 | 0.1601 | 2722.501 |
| GOBP_REG | 210 | 0.033863 | 0.003552 | 0.034077 | 0.16019 | 2724.031 |
| GOMF_GLU | 6 | 0.20572 | 0.003667 | 0.2071 | 0.16027 | 2725.391 |
| GOBP_POS | 9 | 0.15573 | 0.0034 | 0.15684 | 0.16038 | 2727.262 |
| REACTOME | 8 | 0.13881 | 0.002857 | 0.13982 | 0.16042 | 2727.942 |
| WP_INTER/ | 15 | 0.1358 | 0.003827 | 0.13691 | 0.16063 | 2731.513 |

| | | | | | | |
|---|---|---|---|---|---|---|
| GOCC_CHR | 6 | 0.18331 | 0.003268 | 0.18481 | 0.16064 | 2731.683 |
| GOBP_NUC | 10 | 0.13577 | 0.003124 | 0.13696 | 0.16076 | 2733.724 |
| GOBP_ANT | 5 | 0.18539 | 0.003017 | 0.18702 | 0.16078 | 2734.064 |
| OISHI_CHC | 263 | 0.030249 | 0.003545 | 0.030517 | 0.1608 | 2734.404 |
| REACTOME | 14 | 0.14014 | 0.003815 | 0.14139 | 0.1608 | 2734.404 |
| REACTOME | 5 | 0.25575 | 0.004162 | 0.25804 | 0.16082 | 2734.744 |
| MEDINA_S | 35 | 0.07801 | 0.003356 | 0.07871 | 0.16083 | 2734.914 |
| MIKKELSEN | 132 | 0.040647 | 0.003387 | 0.041021 | 0.16088 | 2735.764 |
| REACTOME | 29 | 0.095514 | 0.003741 | 0.096421 | 0.16095 | 2736.955 |
| GOCC_NUF | 16 | 0.14757 | 0.004294 | 0.14899 | 0.16099 | 2737.635 |
| REACTOME | 73 | 0.058569 | 0.003635 | 0.059136 | 0.16099 | 2737.635 |
| REACTOME | 8 | 0.18487 | 0.003805 | 0.18672 | 0.16107 | 2738.995 |
| MARIADAS | 45 | 0.074473 | 0.003632 | 0.075264 | 0.16122 | 2741.546 |
| GOMF_ENC | 7 | 0.18428 | 0.003548 | 0.18626 | 0.16125 | 2742.056 |
| GOBP_HIPI | 80 | 0.059047 | 0.003836 | 0.059695 | 0.1613 | 2742.907 |
| GOBP_POS | 105 | 0.048543 | 0.00361 | 0.049085 | 0.16135 | 2743.757 |
| WP_GENET | 32 | 0.08552 | 0.003518 | 0.086482 | 0.16137 | 2744.097 |
| GOBP_NEG | 18 | 0.12645 | 0.003903 | 0.12788 | 0.16139 | 2744.437 |
| GOBP_NEG | 215 | 0.033824 | 0.003589 | 0.034221 | 0.16148 | 2745.967 |
| GOBP_REG | 1390 | 0.013746 | 0.00359 | 0.013907 | 0.16148 | 2745.967 |
| GOBP_TER | 17 | 0.11132 | 0.003339 | 0.11265 | 0.16152 | 2746.648 |
| GOBP_REG | 85 | 0.056233 | 0.003765 | 0.056916 | 0.16158 | 2747.668 |
| GOMF_NU | 24 | 0.098473 | 0.003509 | 0.099681 | 0.16161 | 2748.178 |
| WP_MED_ | 6 | 0.21029 | 0.003748 | 0.21287 | 0.16162 | 2748.348 |
| GOBP_ASS | 82 | 0.058897 | 0.003873 | 0.059628 | 0.16165 | 2748.858 |
| REACTOME | 44 | 0.076074 | 0.003668 | 0.077026 | 0.16167 | 2749.198 |
| GOBP_OLE | 23 | 0.11814 | 0.004121 | 0.11965 | 0.16175 | 2750.559 |
| GOBP_POS | 232 | 0.033391 | 0.003679 | 0.033823 | 0.16177 | 2750.899 |
| GOBP_REG | 66 | 0.060523 | 0.003572 | 0.061324 | 0.16184 | 2752.089 |
| REACTOME | 145 | 0.041944 | 0.003662 | 0.042509 | 0.1619 | 2753.11 |
| REACTOME | 5 | 0.24656 | 0.004012 | 0.24997 | 0.16199 | 2754.64 |
| REACTOME | 62 | 0.057037 | 0.003263 | 0.057834 | 0.16202 | 2755.15 |
| REACTOME | 88 | 0.052122 | 0.00355 | 0.052872 | 0.16212 | 2756.851 |
| GOBP_ERB | 6 | 0.27026 | 0.004817 | 0.27427 | 0.16223 | 2758.721 |
| GOMF_HYI | 700 | 0.018581 | 0.003511 | 0.01886 | 0.16228 | 2759.571 |
| GOBP_CELI | 8 | 0.19016 | 0.003914 | 0.19305 | 0.16232 | 2760.252 |
| GOMF_KEF | 5 | 0.18744 | 0.00305 | 0.19033 | 0.16237 | 2761.102 |
| WAMUNYC | 177 | 0.037784 | 0.003641 | 0.038368 | 0.16237 | 2761.102 |
| GOMF_INS | 19 | 0.12491 | 0.003961 | 0.12684 | 0.16238 | 2761.272 |
| MIKKELSEN | 8 | 0.14935 | 0.003074 | 0.15168 | 0.1624 | 2761.612 |
| AMIT_SERU | 56 | 0.071248 | 0.003875 | 0.072378 | 0.16247 | 2762.802 |
| MARCHINI_ | 45 | 0.073599 | 0.003589 | 0.074767 | 0.16247 | 2762.802 |
| SMID_BRE/ | 506 | 0.023633 | 0.003817 | 0.024014 | 0.16253 | 2763.823 |
| SHEPARD_( | 182 | 0.036265 | 0.003544 | 0.036851 | 0.16254 | 2763.993 |
| GOBP_IRES | 10 | 0.14769 | 0.003398 | 0.15012 | 0.16262 | 2765.353 |

| | | | | | | |
|---|---|---|---|---|---|---|
| GOBP_REG | 5 | 0.20481 | 0.003333 | 0.2082 | 0.16263 | 2765.523 |
| GOBP_PUR | 5 | 0.24677 | 0.004016 | 0.2509 | 0.16267 | 2766.203 |
| WP_FRAGI | 115 | 0.046419 | 0.003612 | 0.047193 | 0.16267 | 2766.203 |
| WANG_SM | 335 | 0.028956 | 0.003823 | 0.029444 | 0.16271 | 2766.884 |
| REACTOME | 30 | 0.098878 | 0.003939 | 0.1006 | 0.16284 | 2769.094 |
| GOBP_PRC | 215 | 0.032884 | 0.003489 | 0.033492 | 0.16309 | 2773.345 |
| KEGG_TGF_ | 83 | 0.054822 | 0.003627 | 0.055847 | 0.16315 | 2774.366 |
| GOCC_CEN | 9 | 0.17988 | 0.003927 | 0.18325 | 0.16316 | 2774.536 |
| GOCC_STEI | 6 | 0.22534 | 0.004017 | 0.22957 | 0.16316 | 2774.536 |
| GOBP_PRC | 27 | 0.10066 | 0.003804 | 0.10257 | 0.1632 | 2775.216 |
| REACTOME | 10 | 0.14648 | 0.003371 | 0.14929 | 0.16325 | 2776.066 |
| BIOCARTA_ | 6 | 0.15529 | 0.002768 | 0.15827 | 0.16326 | 2776.236 |
| GOLDRATH | 62 | 0.067395 | 0.003856 | 0.068692 | 0.16327 | 2776.406 |
| REACTOME | 41 | 0.083983 | 0.00391 | 0.085619 | 0.16333 | 2777.427 |
| GOBP_PML | 6 | 0.1858 | 0.003312 | 0.18948 | 0.1634 | 2778.617 |
| ZEILSTRA_( | 8 | 0.19527 | 0.004019 | 0.19919 | 0.16348 | 2779.977 |
| GOBP_MRI | 56 | 0.059517 | 0.003237 | 0.06072 | 0.1635 | 2780.318 |
| GOBP_NEG | 34 | 0.085392 | 0.003621 | 0.087186 | 0.16369 | 2783.548 |
| REACTOME | 51 | 0.070518 | 0.00366 | 0.072001 | 0.1637 | 2783.719 |
| GOBP_MA( | 12 | 0.13664 | 0.003444 | 0.13957 | 0.16378 | 2785.079 |
| REACTOME | 183 | 0.036295 | 0.003556 | 0.037079 | 0.16383 | 2785.929 |
| PID_ERBB4 | 38 | 0.083024 | 0.003721 | 0.084828 | 0.16386 | 2786.439 |
| REACTOME | 42 | 0.075099 | 0.003538 | 0.07677 | 0.16398 | 2788.48 |
| WP_MONC | 31 | 0.096962 | 0.003926 | 0.099152 | 0.16407 | 2790.01 |
| KYNG_RESI | 63 | 0.056631 | 0.003266 | 0.057919 | 0.1641 | 2790.521 |
| REACTOME | 36 | 0.077944 | 0.003401 | 0.079751 | 0.16421 | 2792.391 |
| GOBP_POI | 7 | 0.21335 | 0.004108 | 0.21833 | 0.16424 | 2792.901 |
| GOBP_MAI | 10 | 0.17906 | 0.00412 | 0.18328 | 0.1643 | 2793.922 |
| YAO_TEMP | 63 | 0.065219 | 0.003761 | 0.066837 | 0.16459 | 2798.853 |
| GOBP_NEG | 70 | 0.057446 | 0.003492 | 0.058883 | 0.16464 | 2799.703 |
| BROWNE_I | 102 | 0.049465 | 0.003626 | 0.050708 | 0.16467 | 2800.213 |
| YORDY_RE( | 67 | 0.065368 | 0.003887 | 0.067012 | 0.16467 | 2800.213 |
| GOBP_BILE | 47 | 0.077763 | 0.003875 | 0.079733 | 0.16471 | 2800.894 |
| GOBP_REG | 12 | 0.15189 | 0.003828 | 0.15574 | 0.16472 | 2801.064 |
| GOBP_RIB( | 271 | 0.027111 | 0.003225 | 0.027799 | 0.16473 | 2801.234 |
| GOBP_MO | 13 | 0.14753 | 0.00387 | 0.15129 | 0.16476 | 2801.744 |
| PHONG_TN | 214 | 0.033682 | 0.003566 | 0.034578 | 0.16501 | 2805.995 |
| GOMF_SYN | 5 | 0.21429 | 0.003487 | 0.22 | 0.16502 | 2806.165 |
| GOMF_LIP( | 29 | 0.090513 | 0.003545 | 0.092941 | 0.16507 | 2807.015 |
| GOBP_CAL | 28 | 0.099879 | 0.003844 | 0.10257 | 0.16509 | 2807.355 |
| GOBP_CHA | 5 | 0.24301 | 0.003954 | 0.24961 | 0.16514 | 2808.206 |
| BHAT_ESR1 | 87 | 0.055547 | 0.003762 | 0.057064 | 0.16518 | 2808.886 |
| DELASERN/ | 11 | 0.12762 | 0.00308 | 0.13111 | 0.16518 | 2808.886 |
| REACTOME | 9 | 0.16722 | 0.00365 | 0.17183 | 0.16523 | 2809.736 |
| GOBP_CON | 11 | 0.18303 | 0.004417 | 0.18816 | 0.16535 | 2811.777 |

| | | | | | | |
|---|---|---|---|---|---|---|
| GOBP_NEP | 18 | 0.12928 | 0.00399 | 0.13295 | 0.16542 | 2812.967 |
| REACTOME | 17 | 0.11477 | 0.003443 | 0.11802 | 0.16542 | 2812.967 |
| REACTOME | 11 | 0.14572 | 0.003517 | 0.14985 | 0.16543 | 2813.137 |
| GOBP_BLA | 111 | 0.04308 | 0.003294 | 0.04433 | 0.16558 | 2815.688 |
| REACTOME | 16 | 0.10386 | 0.003022 | 0.10687 | 0.16558 | 2815.688 |
| GOBP_CRA | 59 | 0.068036 | 0.003798 | 0.070041 | 0.16569 | 2817.558 |
| GOBP_RET | 6 | 0.1959 | 0.003492 | 0.20168 | 0.16569 | 2817.558 |
| GOBP_MU | 53 | 0.066855 | 0.003537 | 0.068846 | 0.16576 | 2818.749 |
| GOBP_FOR | 15 | 0.125 | 0.003522 | 0.12875 | 0.16581 | 2819.599 |
| LEE_LIVER_ | 63 | 0.063354 | 0.003654 | 0.065256 | 0.16582 | 2819.769 |
| GOBP_END | 231 | 0.031104 | 0.00342 | 0.032043 | 0.16585 | 2820.279 |
| BIOCARTA_ | 13 | 0.12979 | 0.003405 | 0.13373 | 0.16588 | 2820.789 |
| GOBP_NUC | 19 | 0.12101 | 0.003837 | 0.12469 | 0.16591 | 2821.3 |
| GOBP_REG | 13 | 0.12286 | 0.003223 | 0.12662 | 0.16597 | 2822.32 |
| GOBP_MO | 111 | 0.047356 | 0.003621 | 0.048811 | 0.16598 | 2822.49 |
| GOBP_CELI | 19 | 0.12049 | 0.003821 | 0.1242 | 0.16599 | 2822.66 |
| GOCC_SEX_ | 3 | 0.26679 | 0.003363 | 0.27505 | 0.16604 | 2823.51 |
| MATZUK_S | 25 | 0.10433 | 0.003794 | 0.10756 | 0.16604 | 2823.51 |
| GOBP_REG | 23 | 0.092745 | 0.003235 | 0.095639 | 0.16609 | 2824.36 |
| CHNG_MU | 52 | 0.058945 | 0.003089 | 0.060809 | 0.16619 | 2826.061 |
| REACTOME | 6 | 0.2051 | 0.003656 | 0.21171 | 0.16634 | 2828.612 |
| IIZUKA_LIV | 13 | 0.12297 | 0.003226 | 0.12695 | 0.16636 | 2828.952 |
| GOBP_LIPII | 147 | 0.040369 | 0.003548 | 0.041678 | 0.16638 | 2829.292 |
| REACTOME | 6 | 0.20294 | 0.003617 | 0.20955 | 0.16641 | 2829.802 |
| BONOME_( | 29 | 0.094908 | 0.003717 | 0.098031 | 0.16649 | 2831.162 |
| HELLER_HD | 263 | 0.030254 | 0.003546 | 0.031255 | 0.16654 | 2832.013 |
| GOMF_HIS | 5 | 0.23191 | 0.003774 | 0.23967 | 0.16662 | 2833.373 |
| GOMF_HEI | 8 | 0.1587 | 0.003266 | 0.16403 | 0.16665 | 2833.883 |
| GOBP_DNA | 6 | 0.22957 | 0.004092 | 0.23743 | 0.16681 | 2836.604 |
| GOMF_CAI | 12 | 0.13889 | 0.003501 | 0.14365 | 0.16681 | 2836.604 |
| KEGG_VAS | 42 | 0.077965 | 0.003673 | 0.080637 | 0.16681 | 2836.604 |
| GOBP_THR | 13 | 0.14354 | 0.003765 | 0.14848 | 0.16685 | 2837.284 |
| GOBP_HIST | 12 | 0.13726 | 0.00346 | 0.14201 | 0.16689 | 2837.964 |
| GOBP_POS | 52 | 0.071123 | 0.003728 | 0.073597 | 0.16693 | 2838.645 |
| GOMF_WN | 17 | 0.10864 | 0.003259 | 0.11245 | 0.16699 | 2839.665 |
| GOMF_GAI | 7 | 0.19449 | 0.003745 | 0.20134 | 0.16703 | 2840.345 |
| LIANG_HEI | 38 | 0.072116 | 0.003232 | 0.074656 | 0.16704 | 2840.515 |
| WU_HBX_T | 21 | 0.11419 | 0.003807 | 0.11825 | 0.1671 | 2841.536 |
| BERENJENC | 27 | 0.096547 | 0.003649 | 0.10003 | 0.16722 | 2843.576 |
| GOBP_REG | 41 | 0.078243 | 0.003642 | 0.081075 | 0.16726 | 2844.256 |
| GOBP_NEG | 52 | 0.070383 | 0.003689 | 0.07297 | 0.16739 | 2846.467 |
| GOBP_NEG | 7 | 0.17361 | 0.003342 | 0.18 | 0.1674 | 2846.637 |
| GOBP_MYE | 50 | 0.065389 | 0.003361 | 0.067799 | 0.16742 | 2846.977 |
| GOCC_MEI | 5 | 0.24569 | 0.003998 | 0.25478 | 0.16745 | 2847.487 |
| HOFFMAN_ | 9 | 0.18137 | 0.003959 | 0.1882 | 0.16761 | 2850.208 |

| | | | | | | |
|---|---|---|---|---|---|---|
| ADDYA_ER | 69 | 0.058488 | 0.00353 | 0.060768 | 0.16791 | 2855.31 |
| GOBP_AMY | 7 | 0.16002 | 0.003081 | 0.16632 | 0.16799 | 2856.67 |
| MIKKELSEN | 12 | 0.1517 | 0.003824 | 0.15778 | 0.16816 | 2859.561 |
| GOMF_TRA | 26 | 0.098429 | 0.00365 | 0.10238 | 0.16817 | 2859.731 |
| WANG_BA | 47 | 0.069049 | 0.003441 | 0.071819 | 0.16817 | 2859.731 |
| GAUSSMAN | 29 | 0.081202 | 0.00318 | 0.084487 | 0.16825 | 2861.091 |
| SENESE_HD | 223 | 0.032543 | 0.003516 | 0.033863 | 0.16828 | 2861.601 |
| GOBP_MO | 41 | 0.078522 | 0.003655 | 0.081753 | 0.16841 | 2863.812 |
| GOBP_QUI | 5 | 0.20781 | 0.003382 | 0.21637 | 0.16842 | 2863.982 |
| GOBP_NEG | 4 | 0.23847 | 0.003471 | 0.24832 | 0.16845 | 2864.492 |
| REACTOME | 36 | 0.086338 | 0.003767 | 0.089907 | 0.16846 | 2864.662 |
| KRIEG_HYP | 687 | 0.018743 | 0.00351 | 0.019522 | 0.1685 | 2865.343 |
| GRAHAM_( | 11 | 0.12312 | 0.002971 | 0.12831 | 0.16865 | 2867.893 |
| REACTOME | 5 | 0.22806 | 0.003711 | 0.23778 | 0.16876 | 2869.764 |
| GOBP_REG | 4 | 0.23904 | 0.003479 | 0.24927 | 0.16879 | 2870.274 |
| GOBP_TELI | 256 | 0.032182 | 0.003722 | 0.033558 | 0.16879 | 2870.274 |
| OLSSON_E | 25 | 0.11262 | 0.004096 | 0.11744 | 0.16881 | 2870.614 |
| GOBP_MEI | 756 | 0.017117 | 0.003356 | 0.017854 | 0.16885 | 2871.294 |
| WP_SEROT | 18 | 0.11266 | 0.003477 | 0.11752 | 0.16887 | 2871.634 |
| BENPORAT | 697 | 0.018241 | 0.00344 | 0.019036 | 0.16898 | 2873.505 |
| BIOCARTA_ | 14 | 0.15559 | 0.004236 | 0.16242 | 0.16905 | 2874.695 |
| KEGG_AMI | 39 | 0.07868 | 0.003573 | 0.082144 | 0.16908 | 2875.205 |
| LOPEZ_MB | 884 | 0.016014 | 0.003383 | 0.016725 | 0.16916 | 2876.566 |
| GOMF_CAI | 25 | 0.10403 | 0.003783 | 0.10866 | 0.16919 | 2877.076 |
| MULLIGHA | 151 | 0.041357 | 0.003684 | 0.0432 | 0.1692 | 2877.246 |
| TSAI_RESP | 138 | 0.042135 | 0.003589 | 0.044014 | 0.16921 | 2877.416 |
| GHANDHI_ | 33 | 0.086186 | 0.0036 | 0.09005 | 0.16927 | 2878.436 |
| GOBP_REG | 3 | 0.27442 | 0.003459 | 0.28682 | 0.16935 | 2879.797 |
| GOBP_PHC | 7 | 0.19802 | 0.003812 | 0.20703 | 0.16942 | 2880.987 |
| GOCC_GOL | 9 | 0.15893 | 0.003469 | 0.16621 | 0.16949 | 2882.177 |
| WP_APOPT | 82 | 0.054716 | 0.003598 | 0.057231 | 0.16953 | 2882.858 |
| SANA_TNF_ | 76 | 0.056316 | 0.003566 | 0.058907 | 0.16954 | 2883.028 |
| HARRIS_BR | 40 | 0.078561 | 0.003612 | 0.082268 | 0.16981 | 2887.619 |
| GOBP_CELI | 1006 | 0.015822 | 0.003554 | 0.01657 | 0.16983 | 2887.959 |
| GOBP_PRC | 325 | 0.026074 | 0.003392 | 0.027313 | 0.16989 | 2888.979 |
| REACTOME | 9 | 0.16716 | 0.003649 | 0.17514 | 0.16994 | 2889.83 |
| GOBP_NEU | 45 | 0.076491 | 0.00373 | 0.080166 | 0.17001 | 2891.02 |
| GOBP_REG | 235 | 0.031323 | 0.003473 | 0.032831 | 0.17003 | 2891.36 |
| GOCC_FICC | 118 | 0.041032 | 0.003234 | 0.043014 | 0.17007 | 2892.04 |
| GOBP_RES | 25 | 0.096503 | 0.00351 | 0.10118 | 0.17011 | 2892.721 |
| WENG_POI | 19 | 0.10462 | 0.003317 | 0.10973 | 0.1702 | 2894.251 |
| REACTOME | 14 | 0.12553 | 0.003417 | 0.13172 | 0.1703 | 2895.952 |
| GOBP_POS | 6 | 0.21909 | 0.003905 | 0.22991 | 0.17032 | 2896.292 |
| GOBP_POS | 29 | 0.089561 | 0.003508 | 0.094014 | 0.1704 | 2897.652 |
| GOBP_RES | 7 | 0.18156 | 0.003496 | 0.1906 | 0.1704 | 2897.652 |

| | | | | | | |
|---|---|---|---|---|---|---|
| GOBP_POS | 23 | 0.097279 | 0.003393 | 0.10214 | 0.17045 | 2898.502 |
| GOBP_MYI | 13 | 0.14156 | 0.003714 | 0.14864 | 0.17046 | 2898.672 |
| WP_GASTF | 30 | 0.086486 | 0.003445 | 0.090814 | 0.17047 | 2898.842 |
| MCCOLLUN | 7 | 0.17772 | 0.003422 | 0.18672 | 0.1706 | 2901.053 |
| GOBP_BMI | 6 | 0.24162 | 0.004307 | 0.25402 | 0.17076 | 2903.774 |
| GOBP_CELI | 6 | 0.15825 | 0.002821 | 0.16638 | 0.17077 | 2903.944 |
| GOBP_IOD | 6 | 0.22091 | 0.003938 | 0.23226 | 0.17078 | 2904.114 |
| REACTOME | 11 | 0.13564 | 0.003273 | 0.14263 | 0.17081 | 2904.624 |
| WP_HEPAT | 33 | 0.085591 | 0.003575 | 0.090006 | 0.17082 | 2904.794 |
| SILIGAN_B | 44 | 0.081099 | 0.003911 | 0.085291 | 0.17085 | 2905.304 |
| ROZANOV_ | 10 | 0.18713 | 0.004306 | 0.19687 | 0.17093 | 2906.665 |
| REACTOME | 10 | 0.14781 | 0.003401 | 0.15559 | 0.17107 | 2909.045 |
| MIKKELSEN | 575 | 0.021249 | 0.003652 | 0.02237 | 0.17109 | 2909.385 |
| GOCC_PRC | 19 | 0.088075 | 0.002793 | 0.092747 | 0.17116 | 2910.576 |
| GOBP_REG | 54 | 0.065154 | 0.00348 | 0.068642 | 0.17127 | 2912.446 |
| GOBP_REG | 118 | 0.046161 | 0.003638 | 0.048637 | 0.17129 | 2912.786 |
| GOBP_HIST | 12 | 0.11505 | 0.0029 | 0.12127 | 0.17139 | 2914.487 |
| GOBP_NEG | 48 | 0.067098 | 0.003379 | 0.070725 | 0.17139 | 2914.487 |
| JACKSON_I | 78 | 0.051966 | 0.003333 | 0.05478 | 0.17141 | 2914.827 |
| GOBP_CYT | 168 | 0.036109 | 0.003391 | 0.038067 | 0.17143 | 2915.167 |
| GOBP_NEG | 6 | 0.24371 | 0.004344 | 0.25695 | 0.17145 | 2915.507 |
| AGUIRRE_I | 230 | 0.033345 | 0.003658 | 0.035167 | 0.17152 | 2916.698 |
| GOCC_WEI | 5 | 0.19756 | 0.003215 | 0.20842 | 0.1716 | 2918.058 |
| GOBP_MET | 24 | 0.1023 | 0.003645 | 0.10793 | 0.17161 | 2918.228 |
| GOBP_REG | 4 | 0.30399 | 0.004424 | 0.32071 | 0.17161 | 2918.228 |
| GOBP_POS | 71 | 0.053438 | 0.003271 | 0.056405 | 0.17173 | 2920.269 |
| GOBP_POS | 6 | 0.16657 | 0.002969 | 0.17586 | 0.17177 | 2920.949 |
| GOBP_NEG | 10 | 0.14128 | 0.003251 | 0.14916 | 0.17178 | 2921.119 |
| GOCC_ESCI | 27 | 0.079812 | 0.003016 | 0.084263 | 0.17178 | 2921.119 |
| GOCC_DNA | 7 | 0.1496 | 0.00288 | 0.15794 | 0.17179 | 2921.289 |
| WOO_LIVE | 77 | 0.052839 | 0.003368 | 0.055803 | 0.17186 | 2922.479 |
| GOMF_HYI | 125 | 0.04025 | 0.003264 | 0.04251 | 0.17187 | 2922.649 |
| JIANG_HYP | 57 | 0.065993 | 0.003621 | 0.069728 | 0.17197 | 2924.35 |
| REACTOME | 6 | 0.18277 | 0.003258 | 0.19318 | 0.17204 | 2925.54 |
| REACTOME | 9 | 0.16647 | 0.003634 | 0.17598 | 0.1721 | 2926.561 |
| PEDERSEN_ | 365 | 0.025378 | 0.003495 | 0.026842 | 0.17222 | 2928.601 |
| GOCC_PRC | 9 | 0.2166 | 0.004728 | 0.22916 | 0.1723 | 2929.962 |
| WP_GLYCC | 43 | 0.076039 | 0.003625 | 0.080541 | 0.17257 | 2934.553 |
| GOMF_PR | 267 | 0.029166 | 0.003444 | 0.030897 | 0.1726 | 2935.063 |
| WP_PENTC | 6 | 0.18199 | 0.003244 | 0.19287 | 0.1727 | 2936.764 |
| GOBP_TRA | 25 | 0.090336 | 0.003285 | 0.095741 | 0.17271 | 2936.934 |
| GOMF_AR | 8 | 0.16047 | 0.003303 | 0.17013 | 0.17279 | 2938.294 |
| GOBP_SEQ | 127 | 0.042942 | 0.00351 | 0.045531 | 0.17282 | 2938.804 |
| BIOCARTA_ | 9 | 0.12958 | 0.002829 | 0.1374 | 0.17283 | 2938.974 |
| GOBP_NEG | 7 | 0.1758 | 0.003385 | 0.18654 | 0.17299 | 2941.695 |

| | | | | | | |
|---|---|---|---|---|---|---|
| GOMF_DEI | 27 | 0.10276 | 0.003883 | 0.10906 | 0.17305 | 2942.715 |
| REACTOME | 8 | 0.15723 | 0.003236 | 0.16689 | 0.17308 | 2943.225 |
| GOBP_EST/ | 36 | 0.069228 | 0.00302 | 0.07352 | 0.1732 | 2945.266 |
| CARRILLOR | 780 | 0.017615 | 0.003506 | 0.018708 | 0.17321 | 2945.436 |
| KAUFFMAN | 134 | 0.039813 | 0.003342 | 0.042285 | 0.17322 | 2945.606 |
| GARCIA_T/ | 158 | 0.034898 | 0.003179 | 0.037068 | 0.17324 | 2945.946 |
| NIKOLSKY_ | 15 | 0.20508 | 0.005779 | 0.21784 | 0.17326 | 2946.286 |
| GOMF_TR/ | 131 | 0.037226 | 0.00309 | 0.039551 | 0.17331 | 2947.137 |
| REACTOME | 19 | 0.10995 | 0.003487 | 0.11691 | 0.17349 | 2950.197 |
| GOBP_VIT/ | 5 | 0.18495 | 0.00301 | 0.19674 | 0.1736 | 2952.068 |
| NAKAMUR. | 95 | 0.049228 | 0.003483 | 0.052376 | 0.17364 | 2952.748 |
| GOBP_FAT | 15 | 0.11814 | 0.003329 | 0.1257 | 0.17365 | 2952.918 |
| REACTOME | 40 | 0.062711 | 0.002884 | 0.066726 | 0.17366 | 2953.088 |
| TAKEDA_T/ | 130 | 0.04083 | 0.003377 | 0.043444 | 0.17366 | 2953.088 |
| BYSTRYKH_ | 7 | 0.20191 | 0.003887 | 0.21499 | 0.17382 | 2955.809 |
| GOBP_SEM | 46 | 0.072908 | 0.003595 | 0.077634 | 0.17384 | 2956.149 |
| GOBP_NEG | 13 | 0.13011 | 0.003413 | 0.13857 | 0.17388 | 2956.829 |
| KIM_BIPOL | 629 | 0.018138 | 0.003255 | 0.01932 | 0.17392 | 2957.51 |
| GOBP_LEU | 112 | 0.04331 | 0.003326 | 0.046149 | 0.17401 | 2959.04 |
| GOBP_NEG | 6 | 0.20311 | 0.00362 | 0.21643 | 0.17402 | 2959.21 |
| GOBP_ORG | 513 | 0.021474 | 0.003492 | 0.022894 | 0.17413 | 2961.081 |
| CLASPER_L | 20 | 0.10745 | 0.003496 | 0.11459 | 0.1742 | 2962.271 |
| GOBP_POS | 4 | 0.23574 | 0.003431 | 0.25141 | 0.17421 | 2962.441 |
| REACTOME | 61 | 0.059343 | 0.003368 | 0.063293 | 0.17423 | 2962.781 |
| GOBP_EMI | 10 | 0.15319 | 0.003525 | 0.16339 | 0.17424 | 2962.951 |
| GOBP_DEN | 12 | 0.12504 | 0.003152 | 0.13341 | 0.17433 | 2964.482 |
| GOMF_ME | 13 | 0.1051 | 0.002757 | 0.11216 | 0.17437 | 2965.162 |
| GOBP_NEG | 8 | 0.16097 | 0.003313 | 0.17181 | 0.17442 | 2966.012 |
| WP_CHOLE | 17 | 0.10901 | 0.00327 | 0.11636 | 0.17442 | 2966.012 |
| GOBP_NEG | 7 | 0.20389 | 0.003925 | 0.21769 | 0.17449 | 2967.202 |
| GOBP_DOS | 25 | 0.10437 | 0.003796 | 0.1115 | 0.17463 | 2969.583 |
| GOMF_MA | 7 | 0.21634 | 0.004165 | 0.23114 | 0.17465 | 2969.923 |
| LIM_MAMI | 14 | 0.12603 | 0.003431 | 0.13466 | 0.17466 | 2970.093 |
| GOCC_PRC | 6 | 0.15235 | 0.002716 | 0.1628 | 0.17468 | 2970.433 |
| DORSAM_I | 35 | 0.07375 | 0.003173 | 0.078819 | 0.17472 | 2971.114 |
| GOCC_PCG | 32 | 0.082492 | 0.003393 | 0.088167 | 0.17474 | 2971.454 |
| GOMF_MD | 11 | 0.12668 | 0.003057 | 0.13544 | 0.17482 | 2972.814 |
| GOBP_NEG | 87 | 0.050689 | 0.003433 | 0.054199 | 0.17484 | 2973.154 |
| GOBP_COF | 5 | 0.24033 | 0.003911 | 0.25706 | 0.17492 | 2974.515 |
| GOMF_ALF | 3 | 0.26295 | 0.003315 | 0.2813 | 0.17497 | 2975.365 |
| GOBP_REG | 30 | 0.095234 | 0.003793 | 0.1019 | 0.17501 | 2976.045 |
| REACTOME | 7 | 0.18762 | 0.003612 | 0.20076 | 0.17502 | 2976.215 |
| GOBP_POS | 43 | 0.073502 | 0.003504 | 0.078671 | 0.17508 | 2977.235 |
| GOCC_GLY | 6 | 0.19012 | 0.003389 | 0.20351 | 0.17511 | 2977.746 |
| GOBP_PHC | 28 | 0.078713 | 0.003029 | 0.084306 | 0.17525 | 2980.126 |

| | | | | | | |
|---|---|---|---|---|---|---|
| KEGG_CAL( | 167 | 0.038484 | 0.003604 | 0.041225 | 0.17529 | 2980.806 |
| GOBP_ERA | 103 | 0.045359 | 0.003341 | 0.048611 | 0.17539 | 2982.507 |
| REACTOME | 146 | 0.034519 | 0.003024 | 0.037041 | 0.1757 | 2987.779 |
| GOBP_CELI | 49 | 0.076132 | 0.003874 | 0.081713 | 0.17575 | 2988.629 |
| GOBP_NEG | 20 | 0.11268 | 0.003666 | 0.12095 | 0.17578 | 2989.139 |
| GOCC_RNA | 8 | 0.17378 | 0.003577 | 0.18659 | 0.17585 | 2990.329 |
| BIOCARTA_ | 6 | 0.18441 | 0.003287 | 0.19803 | 0.17587 | 2990.669 |
| GOBP_PRC | 25 | 0.095471 | 0.003472 | 0.10253 | 0.1759 | 2991.18 |
| ASTON_MA | 147 | 0.038613 | 0.003394 | 0.041473 | 0.17592 | 2991.52 |
| GOBP_POS | 193 | 0.03597 | 0.003618 | 0.03864 | 0.17596 | 2992.2 |
| GOMF_SIG | 24 | 0.091618 | 0.003265 | 0.098422 | 0.17597 | 2992.37 |
| GOBP_IMN | 8 | 0.14668 | 0.003019 | 0.1576 | 0.17602 | 2993.22 |
| KERLEY_RE | 5 | 0.23951 | 0.003897 | 0.25738 | 0.17605 | 2993.73 |
| WANG_LSD | 33 | 0.087595 | 0.003659 | 0.094229 | 0.1763 | 2997.982 |
| GOBP_POS | 5 | 0.22476 | 0.003657 | 0.2418 | 0.17631 | 2998.152 |
| GOBP_EGG | 11 | 0.13306 | 0.003211 | 0.14316 | 0.17634 | 2998.662 |
| GOBP_VITA | 9 | 0.14566 | 0.00318 | 0.15677 | 0.17642 | 3000.022 |
| HERNANDE | 16 | 0.11268 | 0.003279 | 0.1213 | 0.17646 | 3000.702 |
| GOBP_TETI | 9 | 0.13005 | 0.002839 | 0.14001 | 0.17649 | 3001.212 |
| GOMF_UN | 8 | 0.13169 | 0.00271 | 0.14179 | 0.17651 | 3001.553 |
| GOBP_NEG | 3 | 0.27758 | 0.003499 | 0.29903 | 0.17664 | 3003.763 |
| GOBP_DET | 6 | 0.22646 | 0.004037 | 0.24397 | 0.17665 | 3003.933 |
| MEISSNER_ | 17 | 0.10518 | 0.003155 | 0.11331 | 0.17665 | 3003.933 |
| FUNG_IL2_ | 5 | 0.27318 | 0.004445 | 0.29436 | 0.17669 | 3004.613 |
| GOCC_COF | 25 | 0.085614 | 0.003114 | 0.092311 | 0.17685 | 3007.334 |
| GOBP_REG | 6 | 0.21265 | 0.003791 | 0.22937 | 0.17694 | 3008.865 |
| GOBP_NU( | 8 | 0.15359 | 0.003161 | 0.1657 | 0.17699 | 3009.715 |
| WP_FAMIL | 14 | 0.12322 | 0.003354 | 0.13299 | 0.17708 | 3011.245 |
| GOMF_SIN | 77 | 0.051012 | 0.003251 | 0.055077 | 0.17718 | 3012.946 |
| REACTOME | 33 | 0.076156 | 0.003181 | 0.082246 | 0.17724 | 3013.966 |
| GOBP_PAN | 77 | 0.056489 | 0.0036 | 0.06106 | 0.17745 | 3017.537 |
| REACTOME | 116 | 0.04149 | 0.003242 | 0.044856 | 0.1775 | 3018.388 |
| BRACHAT_ | 24 | 0.093473 | 0.003331 | 0.10109 | 0.17757 | 3019.578 |
| GOBP_NEG | 8 | 0.15896 | 0.003272 | 0.17198 | 0.17767 | 3021.278 |
| REACTOME | 47 | 0.065617 | 0.00327 | 0.071 | 0.1777 | 3021.789 |
| GOBP_CYT( | 1398 | 0.012871 | 0.003371 | 0.013929 | 0.17774 | 3022.469 |
| GOBP_NEG | 11 | 0.12574 | 0.003034 | 0.13608 | 0.17774 | 3022.469 |
| REACTOME | 39 | 0.077582 | 0.003523 | 0.083978 | 0.17779 | 3023.319 |
| GOBP_REC | 118 | 0.04232 | 0.003335 | 0.04585 | 0.17801 | 3027.06 |
| GOBP_ME( | 50 | 0.061768 | 0.003175 | 0.066925 | 0.17802 | 3027.23 |
| LIANG_HEI | 33 | 0.076595 | 0.0032 | 0.083016 | 0.1781 | 3028.591 |
| FINETTI_BF | 16 | 0.1239 | 0.003606 | 0.13431 | 0.17814 | 3029.271 |
| REACTOME | 45 | 0.063234 | 0.003084 | 0.068549 | 0.17815 | 3029.441 |
| REACTOME | 8 | 0.21134 | 0.00435 | 0.22918 | 0.17823 | 3030.801 |
| BILD_E2F3_ | 237 | 0.030794 | 0.003429 | 0.033395 | 0.17824 | 3030.971 |

| | | | | | | |
|---|---|---|---|---|---|---|
| REACTOME | 5 | 0.22226 | 0.003617 | 0.24113 | 0.17834 | 3032.672 |
| SIG_PIP3_S | 33 | 0.083873 | 0.003504 | 0.090994 | 0.17834 | 3032.672 |
| GOBP_CELI | 535 | 0.020687 | 0.003433 | 0.022446 | 0.17836 | 3033.012 |
| GOBP_REG | 54 | 0.062511 | 0.003339 | 0.067839 | 0.17841 | 3033.862 |
| WP_CAMK | 32 | 0.083048 | 0.003416 | 0.090136 | 0.17844 | 3034.372 |
| NIKOLSKY_ | 11 | 0.28942 | 0.006984 | 0.31415 | 0.17845 | 3034.542 |
| REACTOME | 40 | 0.075142 | 0.003455 | 0.081572 | 0.17849 | 3035.222 |
| GOBP_NEG | 184 | 0.033156 | 0.003257 | 0.035997 | 0.17851 | 3035.563 |
| LIANG_HEI | 38 | 0.076352 | 0.003422 | 0.082908 | 0.17855 | 3036.243 |
| NATSUME_ | 51 | 0.063925 | 0.003318 | 0.069415 | 0.17856 | 3036.413 |
| GOBP_PIGI | 52 | 0.061664 | 0.003232 | 0.066964 | 0.17857 | 3036.583 |
| REACTOME | 100 | 0.047824 | 0.003472 | 0.051936 | 0.17858 | 3036.753 |
| GOBP_XEN | 13 | 0.14905 | 0.00391 | 0.16187 | 0.17859 | 3036.923 |
| GOCC_MIC | 33 | 0.078693 | 0.003287 | 0.0855 | 0.17869 | 3038.623 |
| WP_PI3KAI | 326 | 0.026413 | 0.003441 | 0.0287 | 0.17871 | 3038.964 |
| GOCC_PMI | 94 | 0.048401 | 0.003407 | 0.052612 | 0.1788 | 3040.494 |
| GOBP_TRN | 8 | 0.13331 | 0.002744 | 0.14492 | 0.17883 | 3041.004 |
| MILI_PSEU | 393 | 0.022451 | 0.003205 | 0.024409 | 0.17885 | 3041.344 |
| GOBP_TETI | 31 | 0.074544 | 0.003018 | 0.081059 | 0.17889 | 3042.024 |
| PID_WNT_ | 31 | 0.077296 | 0.00313 | 0.084055 | 0.1789 | 3042.195 |
| GOBP_OLF | 35 | 0.082758 | 0.00356 | 0.090055 | 0.17906 | 3044.915 |
| GOMF_DIS | 38 | 0.07775 | 0.003485 | 0.084604 | 0.17906 | 3044.915 |
| GOBP_REG | 5 | 0.23985 | 0.003903 | 0.26105 | 0.1791 | 3045.596 |
| GOBP_PRC | 11 | 0.14197 | 0.003426 | 0.15464 | 0.17931 | 3049.167 |
| GOBP_NEU | 13 | 0.1291 | 0.003387 | 0.14075 | 0.17952 | 3052.738 |
| GOMF_ALK | 17 | 0.12452 | 0.003735 | 0.13584 | 0.17967 | 3055.288 |
| GOBP_NEG | 16 | 0.14333 | 0.004171 | 0.15637 | 0.17968 | 3055.458 |
| JIANG_AGI | 46 | 0.065038 | 0.003207 | 0.071041 | 0.17997 | 3060.39 |
| GOBP_ADE | 222 | 0.033628 | 0.003625 | 0.036737 | 0.18 | 3060.9 |
| MULLIGHA | 72 | 0.055876 | 0.003444 | 0.061055 | 0.18006 | 3061.92 |
| GOBP_RHY | 291 | 0.026915 | 0.003316 | 0.029411 | 0.18007 | 3062.09 |
| GOBP_POS | 25 | 0.09807 | 0.003567 | 0.10718 | 0.18011 | 3062.771 |
| GOCC_SEX_ | 27 | 0.089555 | 0.003384 | 0.097878 | 0.18011 | 3062.771 |
| GOBP_HEP | 8 | 0.17004 | 0.0035 | 0.18595 | 0.18026 | 3065.321 |
| GOBP_FEA | 42 | 0.07511 | 0.003539 | 0.082162 | 0.18032 | 3066.342 |
| MEBARKI_I | 386 | 0.024104 | 0.003411 | 0.026371 | 0.18035 | 3066.852 |
| BREDEMEY | 47 | 0.070127 | 0.003495 | 0.076726 | 0.18037 | 3067.192 |
| GOCC_STEI | 6 | 0.19274 | 0.003436 | 0.2109 | 0.18039 | 3067.532 |
| GOBP_ORG | 903 | 0.015535 | 0.003315 | 0.017001 | 0.18043 | 3068.212 |
| DELASERN/ | 86 | 0.049927 | 0.003362 | 0.054655 | 0.1805 | 3069.403 |
| GOBP_NEG | 4 | 0.19889 | 0.002895 | 0.21774 | 0.18052 | 3069.743 |
| WENG_POI | 23 | 0.090715 | 0.003165 | 0.099319 | 0.18053 | 3069.913 |
| LEIN_CHOF | 100 | 0.047377 | 0.003439 | 0.051879 | 0.18057 | 3070.593 |
| GAVIN_IL2_ | 5 | 0.2375 | 0.003865 | 0.26027 | 0.18076 | 3073.824 |
| REACTOME | 16 | 0.11478 | 0.00334 | 0.12579 | 0.18076 | 3073.824 |

| | | | | | | |
|---|---|---|---|---|---|---|
| REACTOME | 245 | 0.029402 | 0.003328 | 0.032223 | 0.18077 | 3073.994 |
| KEGG_PHC | 74 | 0.053379 | 0.003336 | 0.058605 | 0.1812 | 3081.306 |
| GOBP_REG | 905 | 0.015232 | 0.003254 | 0.01673 | 0.1813 | 3083.007 |
| REACTOME | 81 | 0.050582 | 0.003306 | 0.055559 | 0.18131 | 3083.177 |
| GOCC_GER | 65 | 0.055995 | 0.00328 | 0.061509 | 0.18132 | 3083.347 |
| YAO_TEMF | 67 | 0.050592 | 0.003009 | 0.055576 | 0.18134 | 3083.687 |
| REACTOME | 54 | 0.060095 | 0.00321 | 0.066026 | 0.18137 | 3084.197 |
| GOCC_PAR | 8 | 0.13875 | 0.002856 | 0.15248 | 0.18143 | 3085.217 |
| SHEDDEN_ | 437 | 0.021361 | 0.003212 | 0.02349 | 0.18159 | 3087.938 |
| GOBP_MRI | 30 | 0.074338 | 0.002961 | 0.081806 | 0.18176 | 3090.829 |
| GOBP_POS | 169 | 0.035894 | 0.003381 | 0.039501 | 0.18176 | 3090.829 |
| REACTOME | 9 | 0.1434 | 0.00313 | 0.15784 | 0.18181 | 3091.679 |
| SABATES_( | 261 | 0.030692 | 0.003584 | 0.033801 | 0.18194 | 3093.89 |
| GOMF_AM | 26 | 0.10055 | 0.003729 | 0.11074 | 0.18196 | 3094.23 |
| BREDEMEY | 62 | 0.055505 | 0.003176 | 0.061139 | 0.18199 | 3094.74 |
| GOBP_FAC | 48 | 0.068881 | 0.003469 | 0.075878 | 0.182 | 3094.91 |
| BLANCO_M | 105 | 0.044529 | 0.003312 | 0.049056 | 0.18202 | 3095.25 |
| MATZUK_E | 12 | 0.16331 | 0.004116 | 0.18003 | 0.18218 | 3097.971 |
| GOBP_POS | 88 | 0.051649 | 0.003518 | 0.056981 | 0.18236 | 3101.032 |
| GOBP_NEG | 41 | 0.064349 | 0.002996 | 0.071018 | 0.18245 | 3102.562 |
| GOBP_EST/ | 39 | 0.072719 | 0.003302 | 0.080272 | 0.1825 | 3103.413 |
| CAIRO_HEF | 254 | 0.02924 | 0.003369 | 0.03228 | 0.18251 | 3103.583 |
| CURSONS_ | 20 | 0.10853 | 0.003531 | 0.11983 | 0.18255 | 3104.263 |
| GOBP_REG | 5 | 0.25956 | 0.004224 | 0.28664 | 0.18259 | 3104.943 |
| IGLESIAS_E | 13 | 0.1332 | 0.003494 | 0.14711 | 0.18261 | 3105.283 |
| GOBP_POS | 2 | 0.32824 | 0.003378 | 0.36256 | 0.18264 | 3105.793 |
| LIM_MAMI | 400 | 0.022237 | 0.003202 | 0.024574 | 0.18276 | 3107.834 |
| GOCC_HFE | 8 | 0.13005 | 0.002677 | 0.14373 | 0.18279 | 3108.344 |
| DANG_MY( | 122 | 0.037723 | 0.003023 | 0.041704 | 0.18286 | 3109.534 |
| GOBP_DET | 10 | 0.13395 | 0.003082 | 0.14814 | 0.18295 | 3111.065 |
| REACTOME | 13 | 0.14094 | 0.003697 | 0.15588 | 0.18296 | 3111.235 |
| GOBP_REG | 82 | 0.052249 | 0.003436 | 0.057794 | 0.18299 | 3111.745 |
| WP_BMP_S | 20 | 0.12106 | 0.003938 | 0.13397 | 0.18309 | 3113.445 |
| GOBP_POS | 46 | 0.057123 | 0.002816 | 0.063215 | 0.1831 | 3113.616 |
| GOBP_POL | 10 | 0.16292 | 0.003749 | 0.18036 | 0.18319 | 3115.146 |
| EPPERT_LS | 35 | 0.079606 | 0.003425 | 0.088153 | 0.18326 | 3116.336 |
| GOBP_POR | 20 | 0.097812 | 0.003182 | 0.10835 | 0.18333 | 3117.527 |
| HOFMANN | 18 | 0.10167 | 0.003138 | 0.11264 | 0.18337 | 3118.207 |
| GOMF_LIPI | 793 | 0.016276 | 0.003265 | 0.018037 | 0.18343 | 3119.227 |
| GOCC_MIT | 266 | 0.02532 | 0.002984 | 0.028065 | 0.18349 | 3120.247 |
| CHENG_IM | 104 | 0.047479 | 0.003514 | 0.05263 | 0.1835 | 3120.418 |
| GOBP_POS | 6 | 0.22331 | 0.003981 | 0.24755 | 0.18352 | 3120.758 |
| GOBP_CELI | 8 | 0.16526 | 0.003401 | 0.18323 | 0.18355 | 3121.268 |
| GOBP_POL | 20 | 0.099213 | 0.003228 | 0.11003 | 0.18362 | 3122.458 |
| REACTOME | 5 | 0.17005 | 0.002767 | 0.18859 | 0.18362 | 3122.458 |

| | | | | | | |
|---|---|---|---|---|---|---|
| GOBP_REG | 138 | 0.037141 | 0.003164 | 0.041197 | 0.18365 | 3122.968 |
| GOBP_LEFT | 14 | 0.11574 | 0.003151 | 0.1284 | 0.18369 | 3123.648 |
| GOBP_ORG | 240 | 0.02739 | 0.003069 | 0.030386 | 0.18369 | 3123.648 |
| GOBP_CAN | 123 | 0.043518 | 0.003501 | 0.048299 | 0.1838 | 3125.519 |
| GOBP_GER | 10 | 0.14447 | 0.003324 | 0.16035 | 0.18381 | 3125.689 |
| CHUNG_BL | 126 | 0.036622 | 0.002982 | 0.040649 | 0.18382 | 3125.859 |
| GOMF_DE( | 54 | 0.056583 | 0.003022 | 0.062814 | 0.18385 | 3126.369 |
| REACTOME | 9 | 0.14263 | 0.003114 | 0.15842 | 0.18397 | 3128.41 |
| GOBP_NEG | 8 | 0.16106 | 0.003315 | 0.17889 | 0.18398 | 3128.58 |
| REACTOME | 19 | 0.11776 | 0.003734 | 0.13083 | 0.18404 | 3129.6 |
| PIONTEK_F | 40 | 0.06533 | 0.003004 | 0.07259 | 0.18407 | 3130.11 |
| GOBP_N_A | 11 | 0.12119 | 0.002925 | 0.13469 | 0.18413 | 3131.131 |
| WP_FAS_LI | 40 | 0.069043 | 0.003175 | 0.076742 | 0.18415 | 3131.471 |
| GOMF_MI( | 12 | 0.13169 | 0.003319 | 0.14645 | 0.18427 | 3133.511 |
| WP_VEGFA | 411 | 0.022397 | 0.003269 | 0.024917 | 0.18437 | 3135.212 |
| GOBP_MIN | 7 | 0.15473 | 0.002979 | 0.17219 | 0.18444 | 3136.402 |
| GOBP_REG | 22 | 0.10233 | 0.003491 | 0.11389 | 0.18447 | 3136.912 |
| BOYLAN_M | 34 | 0.071741 | 0.003042 | 0.079873 | 0.18455 | 3138.273 |
| WP_CHROI | 72 | 0.05794 | 0.003571 | 0.064525 | 0.18461 | 3139.293 |
| FISCHER_G | 225 | 0.029964 | 0.003252 | 0.033374 | 0.18465 | 3139.973 |
| GOBP_CAR | 112 | 0.042445 | 0.00326 | 0.047277 | 0.18465 | 3139.973 |
| GOBP_POS | 22 | 0.10271 | 0.003504 | 0.11441 | 0.18468 | 3140.483 |
| GOBP_POT | 45 | 0.072957 | 0.003558 | 0.081279 | 0.1847 | 3140.824 |
| BARRIER_C | 31 | 0.087946 | 0.003561 | 0.09805 | 0.18488 | 3143.884 |
| GOMF_FOU | 16 | 0.11119 | 0.003236 | 0.1241 | 0.18515 | 3148.476 |
| RUAN_RES | 23 | 0.10023 | 0.003496 | 0.11188 | 0.18517 | 3148.816 |
| BHAT_ESR1 | 82 | 0.053167 | 0.003497 | 0.059351 | 0.18518 | 3148.986 |
| REACTOME | 6 | 0.18656 | 0.003325 | 0.20836 | 0.1853 | 3151.027 |
| GOBP_POS | 13 | 0.11638 | 0.003053 | 0.12998 | 0.18531 | 3151.197 |
| AMIT_EGF_ | 17 | 0.10469 | 0.00314 | 0.11696 | 0.18536 | 3152.047 |
| GOMF_PR( | 30 | 0.069672 | 0.002775 | 0.077841 | 0.18538 | 3152.387 |
| GOBP_REG | 5 | 0.1712 | 0.002786 | 0.19128 | 0.18539 | 3152.557 |
| GOBP_ERK | 306 | 0.027257 | 0.003442 | 0.030461 | 0.18544 | 3153.407 |
| REACTOME | 88 | 0.046624 | 0.003176 | 0.052117 | 0.18551 | 3154.598 |
| SMID_BRE/ | 34 | 0.075176 | 0.003188 | 0.084049 | 0.18555 | 3155.278 |
| GOBP_CELI | 8 | 0.1475 | 0.003036 | 0.165 | 0.18568 | 3157.488 |
| REACTOME | 25 | 0.077772 | 0.002828 | 0.087035 | 0.18578 | 3159.189 |
| GOBP_REG | 18 | 0.099111 | 0.003059 | 0.11092 | 0.18579 | 3159.359 |
| GOBP_TRA | 38 | 0.074381 | 0.003334 | 0.083246 | 0.1858 | 3159.529 |
| GOMF_UBI | 291 | 0.025484 | 0.00314 | 0.028525 | 0.18583 | 3160.039 |
| GOMF_STR | 40 | 0.069424 | 0.003192 | 0.077717 | 0.18586 | 3160.549 |
| GOBP_TAN | 7 | 0.16872 | 0.003248 | 0.18889 | 0.18588 | 3160.889 |
| GOBP_NEG | 6 | 0.18658 | 0.003326 | 0.20891 | 0.1859 | 3161.23 |
| VECCHI_GA | 164 | 0.037424 | 0.003473 | 0.041912 | 0.18596 | 3162.25 |
| TONKS_TA | 41 | 0.067756 | 0.003154 | 0.075889 | 0.18598 | 3162.59 |

| | | | | | | |
|---|---|---|---|---|---|---|
| DIRMEIER_ | 54 | 0.06232 | 0.003328 | 0.06981 | 0.18601 | 3163.1 |
| GOBP_TRA | 18 | 0.090535 | 0.002794 | 0.10143 | 0.18606 | 3163.95 |
| GOMF_DN. | 6 | 0.17923 | 0.003195 | 0.20087 | 0.18613 | 3165.141 |
| MATZUK_( | 15 | 0.13781 | 0.003883 | 0.15445 | 0.18614 | 3165.311 |
| SCHAEFFEF | 417 | 0.022453 | 0.0033 | 0.025182 | 0.18631 | 3168.202 |
| GOBP_SALI | 32 | 0.086052 | 0.00354 | 0.09656 | 0.18643 | 3170.242 |
| GOBP_NOI | 7 | 0.17173 | 0.003306 | 0.19273 | 0.18647 | 3170.922 |
| ZIRN_TRET | 6 | 0.19886 | 0.003545 | 0.22324 | 0.18653 | 3171.943 |
| GOCC_EAR | 5 | 0.15297 | 0.002489 | 0.17178 | 0.1866 | 3173.133 |
| GOBP_NEU | 16 | 0.13355 | 0.003886 | 0.15 | 0.18665 | 3173.983 |
| GOBP_REG | 5 | 0.2428 | 0.003951 | 0.27271 | 0.18665 | 3173.983 |
| GOMF_GAI | 15 | 0.12371 | 0.003486 | 0.13895 | 0.18665 | 3173.983 |
| GOBP_NEG | 175 | 0.034804 | 0.003335 | 0.039102 | 0.18672 | 3175.174 |
| GOBP_HYP | 22 | 0.10587 | 0.003612 | 0.11896 | 0.18674 | 3175.514 |
| LASTOWSK | 177 | 0.033747 | 0.003252 | 0.037922 | 0.18676 | 3175.854 |
| WP_COMN | 38 | 0.079876 | 0.00358 | 0.089785 | 0.18684 | 3177.214 |
| GOBP_POS | 4 | 0.16627 | 0.00242 | 0.18702 | 0.18699 | 3179.765 |
| BROWN_M | 157 | 0.036224 | 0.00329 | 0.040747 | 0.18701 | 3180.105 |
| WP_RETIN | 82 | 0.047158 | 0.003101 | 0.053056 | 0.18705 | 3180.785 |
| GOBP_REG | 5 | 0.24082 | 0.003919 | 0.27096 | 0.18707 | 3181.125 |
| NAISHIRO_ | 8 | 0.1724 | 0.003548 | 0.19399 | 0.18708 | 3181.295 |
| GOBP_TRA | 20 | 0.09839 | 0.003201 | 0.11076 | 0.1872 | 3183.336 |
| HEIDENBL/ | 39 | 0.075608 | 0.003433 | 0.085127 | 0.18723 | 3183.846 |
| DER_IFN_A | 5 | 0.17381 | 0.002828 | 0.19571 | 0.18726 | 3184.356 |
| GOBP_POS | 22 | 0.1026 | 0.0035 | 0.11554 | 0.18728 | 3184.696 |
| GOBP_REG | 473 | 0.0202 | 0.003157 | 0.02276 | 0.1874 | 3186.737 |
| LIU_IL13_N | 17 | 0.096051 | 0.002881 | 0.10822 | 0.18741 | 3186.907 |
| GOCC_PRC | 168 | 0.033589 | 0.003155 | 0.037854 | 0.18746 | 3187.757 |
| GOMF_VOI | 14 | 0.13439 | 0.003658 | 0.15146 | 0.18746 | 3187.757 |
| GOCC_GRC | 8 | 0.14732 | 0.003032 | 0.16604 | 0.18748 | 3188.097 |
| GOBP_POS | 18 | 0.11963 | 0.003692 | 0.13485 | 0.18751 | 3188.608 |
| REACTOME | 19 | 0.11517 | 0.003652 | 0.12984 | 0.18754 | 3189.118 |
| GOBP_POS | 15 | 0.121 | 0.003409 | 0.13645 | 0.18761 | 3190.308 |
| PID_HEDGI | 22 | 0.093348 | 0.003185 | 0.1053 | 0.18769 | 3191.668 |
| GOBP_NEG | 6 | 0.18271 | 0.003257 | 0.20613 | 0.18771 | 3192.009 |
| GOBP_BRA | 26 | 0.085328 | 0.003165 | 0.096317 | 0.18784 | 3194.219 |
| HOFFMANI | 145 | 0.034289 | 0.002994 | 0.038733 | 0.18801 | 3197.11 |
| REACTOME | 700 | 0.017208 | 0.003252 | 0.019438 | 0.18801 | 3197.11 |
| DANG_REG | 235 | 0.030028 | 0.003329 | 0.033929 | 0.18808 | 3198.3 |
| GOBP_NEG | 54 | 0.060883 | 0.003252 | 0.068845 | 0.18826 | 3201.361 |
| REACTOME | 88 | 0.044313 | 0.003018 | 0.050117 | 0.18831 | 3202.212 |
| GOBP_POS | 15 | 0.13593 | 0.00383 | 0.15377 | 0.18836 | 3203.062 |
| GOMF_THI | 6 | 0.20025 | 0.00357 | 0.22659 | 0.18841 | 3203.912 |
| GOCC_ESC | 8 | 0.16455 | 0.003387 | 0.18621 | 0.18845 | 3204.592 |
| BIOCARTA_ | 18 | 0.098536 | 0.003041 | 0.11153 | 0.18848 | 3205.102 |

| | | | | | | |
|---|---|---|---|---|---|---|
| PECE_MAN | 124 | 0.040581 | 0.003278 | 0.045934 | 0.1885 | 3205.443 |
| GOBP_PRC | 14 | 0.1096 | 0.002984 | 0.12407 | 0.18852 | 3205.783 |
| WP_TROP2 | 48 | 0.060301 | 0.003037 | 0.068265 | 0.18853 | 3205.953 |
| ZHAN_MUI | 42 | 0.066545 | 0.003135 | 0.075333 | 0.18853 | 3205.953 |
| GOBP_POS | 5 | 0.16276 | 0.002649 | 0.18441 | 0.18873 | 3209.354 |
| GOCC_DYS | 17 | 0.13834 | 0.004149 | 0.15675 | 0.18875 | 3209.694 |
| GOBP_GRA | 5 | 0.18821 | 0.003063 | 0.21333 | 0.18884 | 3211.224 |
| GOBP_KER | 7 | 0.14292 | 0.002752 | 0.16201 | 0.18884 | 3211.224 |
| GOBP_RES | 870 | 0.015551 | 0.003261 | 0.017635 | 0.18895 | 3213.095 |
| REACTOME | 93 | 0.044117 | 0.003089 | 0.050041 | 0.189 | 3213.945 |
| GOBP_REG | 189 | 0.032809 | 0.003266 | 0.037225 | 0.18906 | 3214.965 |
| GOMF_CAI | 13 | 0.12643 | 0.003317 | 0.1435 | 0.18915 | 3216.496 |
| GOBP_POS | 32 | 0.074697 | 0.003073 | 0.084788 | 0.18917 | 3216.836 |
| GOBP_PRC | 11 | 0.13468 | 0.00325 | 0.15288 | 0.18919 | 3217.176 |
| GOBP_REG | 196 | 0.032378 | 0.003282 | 0.036755 | 0.18919 | 3217.176 |
| GOMF_ALC | 85 | 0.047951 | 0.00321 | 0.054439 | 0.18922 | 3217.686 |
| GOBP_PRC | 11 | 0.13569 | 0.003274 | 0.1541 | 0.1893 | 3219.047 |
| GOBP_DET | 30 | 0.077169 | 0.003074 | 0.087677 | 0.18939 | 3220.577 |
| GOCC_MIC | 63 | 0.049888 | 0.002877 | 0.05669 | 0.18943 | 3221.257 |
| HOLLEMAN | 28 | 0.081118 | 0.003122 | 0.092186 | 0.18945 | 3221.597 |
| GOMF_OXI | 10 | 0.13382 | 0.003079 | 0.15211 | 0.18949 | 3222.277 |
| GOBP_REC | 6 | 0.15708 | 0.0028 | 0.17858 | 0.18955 | 3223.298 |
| GOMF_LIG | 39 | 0.070401 | 0.003197 | 0.080039 | 0.18955 | 3223.298 |
| GOBP_AND | 5 | 0.17592 | 0.002863 | 0.2001 | 0.18966 | 3225.168 |
| GOBP_POS | 5 | 0.23715 | 0.003859 | 0.26977 | 0.18969 | 3225.678 |
| VECCHI_GA | 133 | 0.037256 | 0.003116 | 0.042383 | 0.1897 | 3225.849 |
| HEIDENBLA | 53 | 0.070332 | 0.003721 | 0.080015 | 0.18971 | 3226.019 |
| HONMA_D | 34 | 0.075838 | 0.003216 | 0.0863 | 0.18977 | 3227.039 |
| GOBP_POS | 5 | 0.22667 | 0.003688 | 0.25794 | 0.18978 | 3227.209 |
| GOCC_INN | 5 | 0.1988 | 0.003235 | 0.22629 | 0.18984 | 3228.229 |
| NIKOLSKY_ | 326 | 0.041762 | 0.00544 | 0.047544 | 0.18987 | 3228.739 |
| GOBP_REG | 48 | 0.062775 | 0.003161 | 0.071469 | 0.18988 | 3228.909 |
| GOBP_MES | 5 | 0.23573 | 0.003836 | 0.26844 | 0.18994 | 3229.93 |
| SENESE_HD | 235 | 0.028415 | 0.003151 | 0.032365 | 0.18999 | 3230.78 |
| GOBP_NEG | 7 | 0.15351 | 0.002956 | 0.17487 | 0.19002 | 3231.29 |
| GOBP_IMN | 104 | 0.04391 | 0.00325 | 0.050022 | 0.19003 | 3231.46 |
| REACTOME | 27 | 0.081151 | 0.003067 | 0.092452 | 0.19004 | 3231.63 |
| GOBP_REC | 245 | 0.027597 | 0.003123 | 0.031441 | 0.19005 | 3231.8 |
| GOBP_SEN | 260 | 0.029862 | 0.00348 | 0.034022 | 0.19005 | 3231.8 |
| RIGGINS_T | 210 | 0.031934 | 0.003349 | 0.036395 | 0.19013 | 3233.161 |
| ONDER_CD | 79 | 0.053932 | 0.003482 | 0.06149 | 0.19023 | 3234.861 |
| ZHAN_MUI | 62 | 0.055163 | 0.003156 | 0.062896 | 0.19024 | 3235.031 |
| GOBP_CELI | 12 | 0.13588 | 0.003425 | 0.15496 | 0.19028 | 3235.711 |
| GOBP_REN | 7 | 0.1859 | 0.003579 | 0.212 | 0.19028 | 3235.711 |
| GOBP_REG | 10 | 0.15402 | 0.003544 | 0.17566 | 0.1903 | 3236.052 |

| | | | | | | |
|---|---|---|---|---|---|---|
| GOBP_CHA | 28 | 0.075151 | 0.002892 | 0.08572 | 0.19033 | 3236.562 |
| GOBP_POS | 8 | 0.12606 | 0.002595 | 0.14381 | 0.19036 | 3237.072 |
| GOBP_NEU | 341 | 0.023438 | 0.003122 | 0.026739 | 0.19038 | 3237.412 |
| GOBP_O_G | 41 | 0.067159 | 0.003126 | 0.076629 | 0.19041 | 3237.922 |
| DORSEY_G | 28 | 0.093807 | 0.00361 | 0.10704 | 0.19043 | 3238.262 |
| GOBP_NEG | 4 | 0.23469 | 0.003416 | 0.26784 | 0.19046 | 3238.772 |
| GOBP_PAR | 10 | 0.15301 | 0.003521 | 0.17465 | 0.19049 | 3239.282 |
| GOBP_PRO | 42 | 0.06551 | 0.003087 | 0.074791 | 0.19055 | 3240.303 |
| GOBP_POS | 18 | 0.11556 | 0.003567 | 0.13196 | 0.1906 | 3241.153 |
| GOBP_MAI | 26 | 0.091149 | 0.00338 | 0.10409 | 0.19061 | 3241.323 |
| REACTOME | 151 | 0.033232 | 0.00296 | 0.03796 | 0.19068 | 3242.513 |
| REACTOME | 19 | 0.098082 | 0.00311 | 0.11206 | 0.19071 | 3243.024 |
| GOBP_EXC | 31 | 0.081614 | 0.003305 | 0.093305 | 0.19088 | 3245.914 |
| GOMF_BM | 18 | 0.10215 | 0.003153 | 0.11678 | 0.19088 | 3245.914 |
| GOBP_GM | 4 | 0.19969 | 0.002906 | 0.22838 | 0.19097 | 3247.445 |
| ZHENG_RE | 11 | 0.12774 | 0.003083 | 0.14611 | 0.191 | 3247.955 |
| GOCC_MET | 82 | 0.046208 | 0.003039 | 0.052867 | 0.19105 | 3248.805 |
| NUNODA_I | 29 | 0.073572 | 0.002881 | 0.084198 | 0.19112 | 3249.996 |
| GOBP_CYT | 433 | 0.022816 | 0.003416 | 0.026117 | 0.19117 | 3250.846 |
| PID_FCER1 | 57 | 0.055697 | 0.003056 | 0.063769 | 0.19122 | 3251.696 |
| GOBP_TER | 8 | 0.16874 | 0.003473 | 0.19323 | 0.19126 | 3252.376 |
| GOCC_EUK | 12 | 0.10796 | 0.002721 | 0.12362 | 0.19126 | 3252.376 |
| CARRILLOR | 2 | 0.38471 | 0.00396 | 0.44063 | 0.19132 | 3253.397 |
| GOBP_POS | 6 | 0.16717 | 0.00298 | 0.19148 | 0.19133 | 3253.567 |
| TSUDA_AL | 10 | 0.12991 | 0.002989 | 0.1488 | 0.19133 | 3253.567 |
| GOCC_SYN | 21 | 0.1021 | 0.003403 | 0.11697 | 0.19136 | 3254.077 |
| GOBP_T_C | 50 | 0.067409 | 0.003465 | 0.077232 | 0.19139 | 3254.587 |
| GOBP_BRA | 37 | 0.080014 | 0.003539 | 0.091678 | 0.1914 | 3254.757 |
| GOBP_PRO | 96 | 0.042296 | 0.003009 | 0.048465 | 0.19141 | 3254.927 |
| GAUSSMAI | 21 | 0.098981 | 0.003299 | 0.11346 | 0.1915 | 3256.458 |
| REACTOME | 43 | 0.069055 | 0.003292 | 0.079173 | 0.19155 | 3257.308 |
| GOBP_TER | 6 | 0.17626 | 0.003142 | 0.20226 | 0.19176 | 3260.879 |
| PID_A6B1_ | 45 | 0.067004 | 0.003268 | 0.076909 | 0.19183 | 3262.069 |
| GOMF_CAI | 79 | 0.048411 | 0.003125 | 0.05557 | 0.19184 | 3262.239 |
| NUYTTEN_ | 947 | 0.014265 | 0.003114 | 0.016375 | 0.19185 | 3262.409 |
| GOBP_NEU | 8 | 0.15637 | 0.003218 | 0.17953 | 0.19188 | 3262.919 |
| GOBP_REG | 13 | 0.11249 | 0.002951 | 0.12917 | 0.19191 | 3263.43 |
| REACTOME | 90 | 0.048526 | 0.003343 | 0.055719 | 0.19191 | 3263.43 |
| GOBP_IMN | 1482 | 0.012045 | 0.00324 | 0.013843 | 0.19213 | 3267.171 |
| WANG_RE | 16 | 0.1142 | 0.003323 | 0.13125 | 0.19213 | 3267.171 |
| GOBP_AMI | 42 | 0.067131 | 0.003163 | 0.077181 | 0.19221 | 3268.531 |
| NAKAMUR | 259 | 0.027309 | 0.003177 | 0.031406 | 0.19228 | 3269.721 |
| GOBP_REG | 125 | 0.037956 | 0.003078 | 0.043659 | 0.19232 | 3270.402 |
| REACTOME | 42 | 0.063173 | 0.002976 | 0.072673 | 0.19235 | 3270.912 |
| GOMF_NU | 15 | 0.11609 | 0.003271 | 0.1336 | 0.19244 | 3272.442 |

| | | | | | | |
|---|---|---|---|---|---|---|
| REACTOME | 74 | 0.044671 | 0.002791 | 0.051408 | 0.19244 | 3272.442 |
| NEMETH_I | 75 | 0.052165 | 0.003282 | 0.060035 | 0.19245 | 3272.612 |
| WP_2Q13_ | 59 | 0.06831 | 0.003813 | 0.078635 | 0.19251 | 3273.633 |
| GOBP_DET | 20 | 0.10158 | 0.003305 | 0.11698 | 0.19261 | 3275.333 |
| GOBP_LIVE | 23 | 0.097568 | 0.003404 | 0.1124 | 0.19269 | 3276.693 |
| GOBP_MAI | 170 | 0.032847 | 0.003103 | 0.037842 | 0.19271 | 3277.034 |
| BIOCARTA_ | 9 | 0.1343 | 0.002932 | 0.15478 | 0.19279 | 3278.394 |
| GOMF_PEF | 10 | 0.14782 | 0.003401 | 0.1704 | 0.19285 | 3279.414 |
| GOBP_NEG | 2 | 0.39465 | 0.004062 | 0.455 | 0.19287 | 3279.754 |
| REACTOME | 19 | 0.094644 | 0.003001 | 0.10912 | 0.19288 | 3279.924 |
| GOBP_GLC | 12 | 0.14724 | 0.003711 | 0.16976 | 0.19289 | 3280.094 |
| GOMF_WV | 23 | 0.090987 | 0.003174 | 0.10493 | 0.19293 | 3280.775 |
| HUMMEL_ | 15 | 0.13114 | 0.003695 | 0.15129 | 0.19303 | 3282.475 |
| HORTON_S | 23 | 0.08705 | 0.003037 | 0.10043 | 0.19304 | 3282.645 |
| REACTOME | 5 | 0.22403 | 0.003645 | 0.25851 | 0.19308 | 3283.325 |
| PID_HNF3E | 44 | 0.070716 | 0.00341 | 0.081618 | 0.19313 | 3284.176 |
| GOBP_REG | 50 | 0.061427 | 0.003157 | 0.070925 | 0.19323 | 3285.876 |
| GOBP_RES | 99 | 0.043499 | 0.003142 | 0.050227 | 0.19324 | 3286.046 |
| PROVENZA | 137 | 0.035278 | 0.002994 | 0.040758 | 0.19338 | 3288.427 |
| WP_UREA_ | 19 | 0.08844 | 0.002804 | 0.10229 | 0.19365 | 3293.018 |
| GOBP_REG | 10 | 0.13548 | 0.003117 | 0.15673 | 0.19367 | 3293.358 |
| GOBP_REG | 5 | 0.19478 | 0.00317 | 0.22535 | 0.1937 | 3293.869 |
| REACTOME | 49 | 0.062247 | 0.003167 | 0.072023 | 0.19373 | 3294.379 |
| REACTOME | 10 | 0.14291 | 0.003288 | 0.16538 | 0.19375 | 3294.719 |
| GOBP_REG | 5 | 0.19212 | 0.003126 | 0.22242 | 0.19386 | 3296.589 |
| BRUECKNE | 73 | 0.052319 | 0.003247 | 0.060583 | 0.19391 | 3297.44 |
| WP_PARKII | 62 | 0.054207 | 0.003101 | 0.062772 | 0.19392 | 3297.61 |
| GOBP_COI | 6 | 0.18044 | 0.003216 | 0.20909 | 0.19409 | 3300.5 |
| BRACHAT_ | 39 | 0.068802 | 0.003124 | 0.079766 | 0.1942 | 3302.371 |
| GOBP_PYR | 31 | 0.075026 | 0.003038 | 0.086988 | 0.19422 | 3302.711 |
| GOCC_SOD | 11 | 0.13795 | 0.003329 | 0.15995 | 0.19423 | 3302.881 |
| CHYLA_CBI | 367 | 0.022696 | 0.003134 | 0.02633 | 0.19436 | 3305.092 |
| RAY_ALZHE | 9 | 0.14064 | 0.00307 | 0.16316 | 0.19436 | 3305.092 |
| GOBP_DEV | 103 | 0.044327 | 0.003265 | 0.051428 | 0.19437 | 3305.262 |
| REACTOME | 40 | 0.068435 | 0.003147 | 0.079406 | 0.1944 | 3305.772 |
| SCHLOSSEF | 102 | 0.038892 | 0.002851 | 0.04514 | 0.19446 | 3306.792 |
| GOBP_REG | 14 | 0.11719 | 0.00319 | 0.13609 | 0.19458 | 3308.833 |
| GOBP_NEU | 32 | 0.087958 | 0.003618 | 0.1023 | 0.19495 | 3315.125 |
| GOCC_RNA | 230 | 0.028336 | 0.003109 | 0.032955 | 0.19495 | 3315.125 |
| DER_IFN_B | 6 | 0.15588 | 0.002779 | 0.1813 | 0.19496 | 3315.295 |
| GOBP_POS | 37 | 0.075863 | 0.003355 | 0.088253 | 0.19501 | 3316.145 |
| BIOCARTA_ | 17 | 0.11726 | 0.003517 | 0.13643 | 0.19505 | 3316.825 |
| GOMF_GAI | 5 | 0.18618 | 0.00303 | 0.21664 | 0.19507 | 3317.165 |
| GOBP_ANA | 236 | 0.028532 | 0.00317 | 0.033202 | 0.19509 | 3317.505 |
| GOBP_ESTA | 40 | 0.061639 | 0.002834 | 0.071773 | 0.19523 | 3319.886 |

| | | | | | | |
|---|---|---|---|---|---|---|
| GOBP_PLA | 11 | 0.1171 | 0.002826 | 0.13636 | 0.19526 | 3320.396 |
| DAZARD_U | 23 | 0.088979 | 0.003104 | 0.10364 | 0.19529 | 3320.906 |
| REACTOME | 7 | 0.14772 | 0.002844 | 0.17211 | 0.19537 | 3322.267 |
| WP_RAS_S | 174 | 0.033414 | 0.003193 | 0.038934 | 0.1954 | 3322.777 |
| GOBP_POS | 8 | 0.13932 | 0.002867 | 0.16236 | 0.19544 | 3323.457 |
| WP_ESTRO | 13 | 0.12781 | 0.003353 | 0.149 | 0.19551 | 3324.648 |
| FARMER_B | 14 | 0.12741 | 0.003468 | 0.14855 | 0.19553 | 3324.988 |
| GOBP_DOF | 10 | 0.15479 | 0.003562 | 0.18048 | 0.19555 | 3325.328 |
| REACTOME | 129 | 0.036671 | 0.003021 | 0.042758 | 0.19555 | 3325.328 |
| WANG_IM | 24 | 0.096022 | 0.003422 | 0.11196 | 0.19555 | 3325.328 |
| GOBP_DNA | 269 | 0.02588 | 0.003067 | 0.030185 | 0.19562 | 3326.518 |
| MOOTHA_ | 84 | 0.041744 | 0.002778 | 0.048704 | 0.1957 | 3327.879 |
| GOBP_RRN | 30 | 0.076147 | 0.003033 | 0.088861 | 0.19575 | 3328.729 |
| GOBP_POS | 7 | 0.1595 | 0.003071 | 0.18615 | 0.19578 | 3329.239 |
| REACTOME | 32 | 0.079783 | 0.003282 | 0.093115 | 0.19578 | 3329.239 |
| GOBP_SNC | 12 | 0.13294 | 0.003351 | 0.15517 | 0.1958 | 3329.579 |
| PID_P38_M | 26 | 0.077234 | 0.002864 | 0.090147 | 0.1958 | 3329.579 |
| GOBP_PRC | 10 | 0.14629 | 0.003366 | 0.17082 | 0.1959 | 3331.28 |
| GOMF_DN/ | 19 | 0.093106 | 0.002952 | 0.10873 | 0.19591 | 3331.45 |
| GOBP_OXA | 9 | 0.1301 | 0.00284 | 0.15202 | 0.19606 | 3334 |
| GOBP_ERR | 11 | 0.13422 | 0.003239 | 0.15685 | 0.19608 | 3334.34 |
| KEGG_PAT | 52 | 0.058816 | 0.003083 | 0.068733 | 0.19608 | 3334.34 |
| WP_PATHC | 52 | 0.058816 | 0.003083 | 0.068733 | 0.19608 | 3334.34 |
| REACTOME | 25 | 0.077226 | 0.002809 | 0.090274 | 0.19616 | 3335.701 |
| GOBP_INH | 5 | 0.21591 | 0.003513 | 0.25249 | 0.19624 | 3337.061 |
| GOBP_MEI | 267 | 0.024998 | 0.002952 | 0.029234 | 0.19625 | 3337.231 |
| GOBP_TYR | 6 | 0.16924 | 0.003017 | 0.19795 | 0.19629 | 3337.911 |
| STEGER_AI | 23 | 0.090367 | 0.003152 | 0.10572 | 0.19634 | 3338.762 |
| TAKEDA_T/ | 81 | 0.05294 | 0.00346 | 0.061937 | 0.19635 | 3338.932 |
| GENTILE_U | 53 | 0.05552 | 0.002938 | 0.064957 | 0.19636 | 3339.102 |
| CHESLER_E | 8 | 0.18343 | 0.003775 | 0.21469 | 0.19646 | 3340.802 |
| PID_LYMPH | 24 | 0.090713 | 0.003232 | 0.10621 | 0.19652 | 3341.823 |
| SIG_CD40P | 32 | 0.070929 | 0.002918 | 0.083057 | 0.19656 | 3342.503 |
| GOBP_REG | 6 | 0.2161 | 0.003852 | 0.25311 | 0.19662 | 3343.523 |
| GOBP_NEG | 113 | 0.041287 | 0.003185 | 0.04837 | 0.19668 | 3344.543 |
| GOMF_GLU | 26 | 0.092868 | 0.003444 | 0.10881 | 0.1967 | 3344.884 |
| DAUER_ST/ | 46 | 0.063217 | 0.003117 | 0.074077 | 0.19673 | 3345.394 |
| GOBP_ERB | 5 | 0.20182 | 0.003284 | 0.23659 | 0.19683 | 3347.094 |
| GOCC_ER_ | 9 | 0.12389 | 0.002704 | 0.14529 | 0.19693 | 3348.795 |
| GOBP_TRN | 91 | 0.038584 | 0.002672 | 0.045257 | 0.19696 | 3349.305 |
| FIGUEROA_ | 38 | 0.068588 | 0.003074 | 0.080454 | 0.19697 | 3349.475 |
| COLDREN_ | 214 | 0.02972 | 0.003146 | 0.034867 | 0.19701 | 3350.155 |
| WP_MODU | 14 | 0.1236 | 0.003365 | 0.14501 | 0.19702 | 3350.325 |
| GOMF_INS | 16 | 0.10831 | 0.003152 | 0.12717 | 0.19719 | 3353.216 |
| GOBP_KER | 18 | 0.090275 | 0.002786 | 0.10603 | 0.19727 | 3354.576 |

| | | | | | | |
|---|---|---|---|---|---|---|
| GOBP_PH_ | 5 | 0.19095 | 0.003107 | 0.22435 | 0.19736 | 3356.107 |
| REACTOME | 11 | 0.14848 | 0.003583 | 0.17452 | 0.19745 | 3357.637 |
| CROONQU | 47 | 0.066597 | 0.003319 | 0.078289 | 0.19748 | 3358.147 |
| GOBP_REG | 6 | 0.21162 | 0.003772 | 0.24879 | 0.1975 | 3358.488 |
| WP_GENES | 42 | 0.075182 | 0.003542 | 0.088395 | 0.19752 | 3358.828 |
| TONKS_TA | 179 | 0.032907 | 0.003189 | 0.038691 | 0.19753 | 3358.998 |
| GOMF_PRO | 55 | 0.054248 | 0.002924 | 0.063785 | 0.19754 | 3359.168 |
| GOBP_RNA | 14 | 0.09902 | 0.002696 | 0.11644 | 0.19756 | 3359.508 |
| GOBP_CHO | 10 | 0.14149 | 0.003256 | 0.16642 | 0.19761 | 3360.358 |
| GOMF_PHO | 33 | 0.068025 | 0.002842 | 0.080051 | 0.19773 | 3362.399 |
| GOBP_REG | 207 | 0.031221 | 0.003251 | 0.036744 | 0.19775 | 3362.739 |
| GOBP_PEP | 19 | 0.10659 | 0.00338 | 0.12545 | 0.19777 | 3363.079 |
| REACTOME | 14 | 0.10279 | 0.002798 | 0.12099 | 0.19778 | 3363.249 |
| GOCC_LAT | 140 | 0.036098 | 0.003097 | 0.04249 | 0.19779 | 3363.419 |
| GOBP_REG | 21 | 0.089299 | 0.002977 | 0.10514 | 0.19785 | 3364.439 |
| GOBP_RNA | 6 | 0.14534 | 0.002591 | 0.17116 | 0.19789 | 3365.119 |
| GOBP_POS | 8 | 0.12566 | 0.002586 | 0.148 | 0.19792 | 3365.63 |
| GOBP_GOL | 6 | 0.21993 | 0.00392 | 0.25906 | 0.19796 | 3366.31 |
| GOBP_INTE | 38 | 0.065587 | 0.00294 | 0.077272 | 0.19801 | 3367.16 |
| BLANCO_M | 310 | 0.025054 | 0.003184 | 0.029523 | 0.19805 | 3367.84 |
| BIOCARTA_ | 26 | 0.082172 | 0.003047 | 0.096845 | 0.19809 | 3368.52 |
| BIOCARTA_ | 25 | 0.082996 | 0.003018 | 0.097832 | 0.19813 | 3369.201 |
| GOBP_ARG | 17 | 0.10759 | 0.003227 | 0.12684 | 0.19815 | 3369.541 |
| GOBP_REG | 1121 | 0.012742 | 0.003012 | 0.015022 | 0.19816 | 3369.711 |
| GOMF_MO | 72 | 0.052245 | 0.00322 | 0.061622 | 0.19827 | 3371.581 |
| GARGALOV | 31 | 0.076105 | 0.003082 | 0.08982 | 0.19842 | 3374.132 |
| GOBP_PLA | 7 | 0.17335 | 0.003338 | 0.20464 | 0.19847 | 3374.982 |
| FONTAINE_ | 64 | 0.054099 | 0.003145 | 0.063878 | 0.19853 | 3376.003 |
| BIOCARTA_ | 17 | 0.096346 | 0.00289 | 0.11377 | 0.19855 | 3376.343 |
| GOBP_POS | 28 | 0.074782 | 0.002878 | 0.088315 | 0.19857 | 3376.683 |
| REACTOME | 27 | 0.080969 | 0.00306 | 0.095643 | 0.19862 | 3377.533 |
| PID_ERB_G | 14 | 0.11664 | 0.003175 | 0.13779 | 0.19864 | 3377.873 |
| GOCC_NUC | 10 | 0.14232 | 0.003275 | 0.16816 | 0.19869 | 3378.723 |
| GOBP_END | 58 | 0.058402 | 0.003232 | 0.069029 | 0.19877 | 3380.084 |
| GOBP_EXO | 7 | 0.17104 | 0.003293 | 0.20217 | 0.19879 | 3380.424 |
| LIEN_BREA | 32 | 0.077136 | 0.003173 | 0.091221 | 0.1989 | 3382.295 |
| GOBP_POS | 9 | 0.15235 | 0.003326 | 0.18019 | 0.19893 | 3382.805 |
| KOHN_EMT | 14 | 0.10798 | 0.002939 | 0.12774 | 0.19897 | 3383.485 |
| GOBP_AMI | 5 | 0.21174 | 0.003446 | 0.25052 | 0.19901 | 3384.165 |
| BIOCARTA_ | 15 | 0.1242 | 0.0035 | 0.14697 | 0.19903 | 3384.505 |
| ZHAN_MUI | 52 | 0.062571 | 0.003279 | 0.07406 | 0.1991 | 3385.696 |
| CHEN_LIVE | 92 | 0.046263 | 0.003222 | 0.054763 | 0.19912 | 3386.036 |
| WP_RESOL | 12 | 0.11189 | 0.00282 | 0.13247 | 0.19917 | 3386.886 |
| GOBP_GLC | 6 | 0.20394 | 0.003635 | 0.24155 | 0.19926 | 3388.416 |
| GOMF_CAI | 12 | 0.11909 | 0.003002 | 0.14106 | 0.19926 | 3388.416 |

| | | | | | | |
|---|---|---|---|---|---|---|
| KASLER_HD | 33 | 0.074669 | 0.003119 | 0.088442 | 0.19926 | 3388.416 |
| GOBP_CELI | 142 | 0.034949 | 0.00302 | 0.041451 | 0.19959 | 3394.028 |
| GOBP_REG | 35 | 0.072018 | 0.003098 | 0.085424 | 0.1996 | 3394.198 |
| GOBP_TRIF | 8 | 0.15292 | 0.003147 | 0.18153 | 0.19979 | 3397.429 |
| TURASHVIL | 64 | 0.055278 | 0.003213 | 0.065629 | 0.19982 | 3397.939 |
| GOBP_EPIT | 4 | 0.26185 | 0.003811 | 0.31096 | 0.19988 | 3398.959 |
| GOBP_DIPI | 2 | 0.40442 | 0.004162 | 0.48043 | 0.19996 | 3400.32 |
| GOMF_ENI | 2 | 0.40442 | 0.004162 | 0.48043 | 0.19996 | 3400.32 |
| GOBP_RRN | 22 | 0.082746 | 0.002823 | 0.098304 | 0.19997 | 3400.49 |
| GOBP_NEG | 13 | 0.12868 | 0.003376 | 0.15291 | 0.20002 | 3401.34 |
| GOBP_SOD | 21 | 0.094505 | 0.00315 | 0.1123 | 0.20003 | 3401.51 |
| GOBP_ANT | 11 | 0.11939 | 0.002881 | 0.14192 | 0.20011 | 3402.871 |
| GOCC_SPIN | 36 | 0.069427 | 0.003029 | 0.082568 | 0.20022 | 3404.741 |
| GOMF_MC | 1803 | 0.010561 | 0.003104 | 0.012562 | 0.20026 | 3405.421 |
| GOCC_CELI | 72 | 0.048281 | 0.002976 | 0.057434 | 0.20028 | 3405.761 |
| ZHAN_MUI | 56 | 0.052145 | 0.002836 | 0.06203 | 0.20028 | 3405.761 |
| GOBP_CELI | 32 | 0.085742 | 0.003527 | 0.102 | 0.2003 | 3406.102 |
| PYEON_HP | 85 | 0.04497 | 0.003011 | 0.053502 | 0.20031 | 3406.272 |
| GOBP_SUG | 6 | 0.15778 | 0.002813 | 0.18777 | 0.20038 | 3407.462 |
| GOBP_4FE_ | 5 | 0.25663 | 0.004176 | 0.30549 | 0.20044 | 3408.482 |
| GOBP_TISS | 67 | 0.053181 | 0.003163 | 0.063305 | 0.20044 | 3408.482 |
| CHOI_ATL_ | 18 | 0.093529 | 0.002887 | 0.11137 | 0.20052 | 3409.843 |
| GOBP_REG | 31 | 0.081882 | 0.003315 | 0.097511 | 0.20054 | 3410.183 |
| MCBRYAN_ | 73 | 0.046055 | 0.002858 | 0.054845 | 0.20054 | 3410.183 |
| GOBP_REG | 10 | 0.13935 | 0.003206 | 0.16604 | 0.20067 | 3412.393 |
| GOBP_POS | 6 | 0.20167 | 0.003595 | 0.24037 | 0.20074 | 3413.584 |
| REACTOME | 107 | 0.039647 | 0.002976 | 0.047259 | 0.20076 | 3413.924 |
| WP_RANKI | 54 | 0.05467 | 0.00292 | 0.065187 | 0.20083 | 3415.114 |
| GOBP_POS | 17 | 0.10255 | 0.003076 | 0.12239 | 0.20106 | 3419.025 |
| GOBP_DEN | 14 | 0.12913 | 0.003515 | 0.15414 | 0.20109 | 3419.535 |
| GOBP_NEG | 10 | 0.14075 | 0.003239 | 0.16801 | 0.20109 | 3419.535 |
| WOTTON_I | 29 | 0.087567 | 0.00343 | 0.10457 | 0.2012 | 3421.406 |
| WP_HIPPO | 27 | 0.087351 | 0.003301 | 0.10432 | 0.20122 | 3421.746 |
| GOBP_LIPII | 47 | 0.061096 | 0.003045 | 0.072973 | 0.20124 | 3422.086 |
| GOMF_LOV | 15 | 0.10334 | 0.002912 | 0.12349 | 0.20136 | 3424.127 |
| BOCHKIS_F | 414 | 0.019847 | 0.002907 | 0.023721 | 0.20139 | 3424.637 |
| GOCC_MRI | 14 | 0.11523 | 0.003137 | 0.13777 | 0.20147 | 3425.997 |
| HONRADO_ | 17 | 0.10248 | 0.003074 | 0.12254 | 0.20148 | 3426.167 |
| WP_FBXL1 | 21 | 0.085102 | 0.002837 | 0.10176 | 0.20149 | 3426.337 |
| GOBP_NEG | 209 | 0.028735 | 0.003007 | 0.034363 | 0.20152 | 3426.848 |
| GOBP_POS | 17 | 0.11654 | 0.003496 | 0.13938 | 0.20155 | 3427.358 |
| REACTOME | 20 | 0.10294 | 0.003349 | 0.12312 | 0.20156 | 3427.528 |
| GOBP_REG | 71 | 0.052408 | 0.003208 | 0.062698 | 0.20162 | 3428.548 |
| TIEN_INTES | 46 | 0.057901 | 0.002855 | 0.069332 | 0.20183 | 3432.119 |
| GOBP_MA( | 1260 | 0.011684 | 0.002916 | 0.013995 | 0.20192 | 3433.65 |

| | | | | | | |
|---|---|---|---|---|---|---|
| PID_IGF1_I | 29 | 0.082647 | 0.003237 | 0.099024 | 0.20197 | 3434.5 |
| GOBP_BLA | 44 | 0.058508 | 0.002821 | 0.070146 | 0.20212 | 3437.051 |
| GOMF_PRC | 339 | 0.022361 | 0.00297 | 0.026809 | 0.20212 | 3437.051 |
| HASLINGEF | 21 | 0.093958 | 0.003132 | 0.11265 | 0.20214 | 3437.391 |
| ENK_UV_R | 276 | 0.025213 | 0.003026 | 0.030233 | 0.20215 | 3437.561 |
| MAHAJAN_ | 65 | 0.049621 | 0.002907 | 0.059501 | 0.20216 | 3437.731 |
| GOBP_RES | 45 | 0.05817 | 0.002837 | 0.069757 | 0.20217 | 3437.901 |
| REACTOME | 7 | 0.17032 | 0.003279 | 0.20424 | 0.20217 | 3437.901 |
| YANG_BRE | 22 | 0.092473 | 0.003155 | 0.11099 | 0.20239 | 3441.642 |
| GOBP_TOR | 55 | 0.055384 | 0.002985 | 0.066479 | 0.2024 | 3441.812 |
| OSADA_AS | 23 | 0.0943 | 0.00329 | 0.11319 | 0.2024 | 3441.812 |
| CHEN_MET | 1159 | 0.012559 | 0.003015 | 0.015086 | 0.20256 | 3444.533 |
| GOBP_POS | 5 | 0.24324 | 0.003958 | 0.29221 | 0.20259 | 3445.043 |
| GOBP_SPIN | 97 | 0.045062 | 0.003222 | 0.054133 | 0.20259 | 3445.043 |
| GOBP_LON | 24 | 0.084975 | 0.003028 | 0.10213 | 0.20269 | 3446.743 |
| GOBP_REG | 11 | 0.12386 | 0.002989 | 0.14886 | 0.2027 | 3446.914 |
| GOBP_PRC | 6 | 0.22003 | 0.003922 | 0.26446 | 0.20271 | 3447.084 |
| GOBP_ATR | 15 | 0.12148 | 0.003423 | 0.14604 | 0.20277 | 3448.104 |
| RIZKI_TUM | 63 | 0.050422 | 0.002908 | 0.060632 | 0.20282 | 3448.954 |
| GOBP_AMI | 1064 | 0.012543 | 0.002893 | 0.01509 | 0.20292 | 3450.655 |
| GOBP_NEG | 5 | 0.21995 | 0.003579 | 0.26466 | 0.20297 | 3451.505 |
| GOBP_NEG | 68 | 0.049571 | 0.00297 | 0.059646 | 0.20297 | 3451.505 |
| GOBP_CAN | 52 | 0.058518 | 0.003067 | 0.070456 | 0.20312 | 3454.056 |
| GOCC_ADH | 174 | 0.033808 | 0.003231 | 0.040713 | 0.20316 | 3454.736 |
| GOBP_POS | 34 | 0.069345 | 0.00294 | 0.083537 | 0.20324 | 3456.096 |
| GOBP_PRC | 7 | 0.17514 | 0.003372 | 0.21106 | 0.20334 | 3457.797 |
| REACTOME | 7 | 0.15468 | 0.002978 | 0.1865 | 0.20345 | 3459.667 |
| GOBP_NEG | 4 | 0.24233 | 0.003527 | 0.29221 | 0.20347 | 3460.007 |
| GOBP_PRC | 291 | 0.023737 | 0.002924 | 0.02864 | 0.20361 | 3462.388 |
| REACTOME | 9 | 0.14405 | 0.003144 | 0.17385 | 0.20368 | 3463.578 |
| LIEN_BREA | 76 | 0.051304 | 0.003249 | 0.061935 | 0.20374 | 3464.599 |
| DAVICIONI_ | 19 | 0.099485 | 0.003155 | 0.12016 | 0.20386 | 3466.639 |
| BIOCARTA_ | 27 | 0.079142 | 0.002991 | 0.095621 | 0.20394 | 3468 |
| NAKAMURA | 40 | 0.061378 | 0.002822 | 0.074184 | 0.20402 | 3469.36 |
| ENK_UV_R | 485 | 0.017919 | 0.002835 | 0.021664 | 0.20409 | 3470.55 |
| GOMF_CHA | 98 | 0.038993 | 0.002802 | 0.047145 | 0.2041 | 3470.721 |
| GOBP_NEG | 29 | 0.081396 | 0.003188 | 0.098449 | 0.20419 | 3472.251 |
| GOBP_CELI | 262 | 0.030865 | 0.003611 | 0.037337 | 0.20423 | 3472.931 |
| GOBP_POS | 9 | 0.13337 | 0.002911 | 0.16135 | 0.20425 | 3473.271 |
| GOBP_NEG | 4 | 0.17242 | 0.00251 | 0.2086 | 0.20426 | 3473.441 |
| GOBP_REG | 5 | 0.1743 | 0.002836 | 0.21098 | 0.20437 | 3475.312 |
| GOMF_ATF | 69 | 0.045988 | 0.002775 | 0.055669 | 0.20438 | 3475.482 |
| ROZANOV_ | 32 | 0.078047 | 0.003211 | 0.094499 | 0.20444 | 3476.502 |
| GOBP_SPIN | 5 | 0.22113 | 0.003598 | 0.26777 | 0.20446 | 3476.842 |
| MELLMAN_ | 18 | 0.086571 | 0.002672 | 0.10485 | 0.20451 | 3477.693 |

| | | | | | | |
|---|---|---|---|---|---|---|
| FIGUEROA_ | 130 | 0.036836 | 0.003046 | 0.044619 | 0.20453 | 3478.033 |
| REACTOME | 27 | 0.093699 | 0.003541 | 0.11352 | 0.20457 | 3478.713 |
| GOBP_PRC | 19 | 0.084027 | 0.002664 | 0.10181 | 0.20461 | 3479.393 |
| WP_SYNTH | 22 | 0.097547 | 0.003328 | 0.11821 | 0.20464 | 3479.903 |
| GOBP_ASP | 7 | 0.1996 | 0.003843 | 0.24198 | 0.20472 | 3481.264 |
| GOBP_NEG | 431 | 0.020236 | 0.003023 | 0.02455 | 0.2049 | 3484.325 |
| GOBP_MIT | 107 | 0.036343 | 0.002728 | 0.044105 | 0.20497 | 3485.515 |
| GOBP_DEF | 1568 | 0.01107 | 0.003055 | 0.013438 | 0.20505 | 3486.875 |
| GOBP_ISO | 7 | 0.16998 | 0.003273 | 0.20637 | 0.20507 | 3487.215 |
| GOMF_STF | 101 | 0.043432 | 0.003168 | 0.052731 | 0.20507 | 3487.215 |
| GOBP_MIT | 6 | 0.192 | 0.003422 | 0.23313 | 0.2051 | 3487.726 |
| GOMF_PRC | 12 | 0.10514 | 0.00265 | 0.12769 | 0.20514 | 3488.406 |
| GOMF_BET | 12 | 0.11248 | 0.002835 | 0.13662 | 0.20518 | 3489.086 |
| GOBP_REG | 60 | 0.055443 | 0.003121 | 0.067354 | 0.20522 | 3489.766 |
| GOBP_NEG | 23 | 0.084369 | 0.002943 | 0.10254 | 0.20533 | 3491.637 |
| GOBP_SKEI | 7 | 0.16749 | 0.003225 | 0.20357 | 0.20533 | 3491.637 |
| GOCC_VES | 309 | 0.022974 | 0.002915 | 0.027925 | 0.20534 | 3491.807 |
| GOMF_KIN | 44 | 0.059474 | 0.002868 | 0.072294 | 0.20535 | 3491.977 |
| REACTOME | 40 | 0.060831 | 0.002797 | 0.073941 | 0.20535 | 3491.977 |
| OUYANG_F | 20 | 0.092826 | 0.00302 | 0.11284 | 0.20536 | 3492.147 |
| GOBP_MIR | 44 | 0.05826 | 0.002809 | 0.070824 | 0.20537 | 3492.317 |
| GOBP_EST/ | 26 | 0.087067 | 0.003229 | 0.10586 | 0.20541 | 3492.997 |
| GOBP_PRC | 9 | 0.11158 | 0.002436 | 0.13567 | 0.20542 | 3493.167 |
| GOBP_HOF | 62 | 0.055054 | 0.00315 | 0.066967 | 0.20551 | 3494.698 |
| GOBP_REG | 5 | 0.25713 | 0.004184 | 0.31282 | 0.20555 | 3495.378 |
| WP_DISOR | 19 | 0.08504 | 0.002697 | 0.10352 | 0.2057 | 3497.929 |
| SHEPARD_ | 193 | 0.029081 | 0.002925 | 0.035421 | 0.20583 | 3500.139 |
| GOCC_WAS | 9 | 0.19911 | 0.004347 | 0.24263 | 0.20593 | 3501.84 |
| GOBP_REG | 433 | 0.021269 | 0.003184 | 0.025921 | 0.20597 | 3502.52 |
| DACOSTA_ | 8 | 0.12288 | 0.002529 | 0.14993 | 0.20622 | 3506.771 |
| MIZUKAMI | 6 | 0.17296 | 0.003083 | 0.2111 | 0.2063 | 3508.132 |
| MORI_SMA | 75 | 0.048743 | 0.003066 | 0.0595 | 0.20634 | 3508.812 |
| GOMF_FIB | 5 | 0.17728 | 0.002885 | 0.21642 | 0.20635 | 3508.982 |
| GOBP_EMI | 82 | 0.047294 | 0.00311 | 0.057739 | 0.20637 | 3509.322 |
| GOBP_POS | 68 | 0.051495 | 0.003085 | 0.062872 | 0.20639 | 3509.662 |
| GOBP_NEG | 31 | 0.065811 | 0.002665 | 0.080368 | 0.20644 | 3510.512 |
| GOBP_HEN | 869 | 0.014448 | 0.003028 | 0.017653 | 0.20656 | 3512.553 |
| GOBP_LEU | 7 | 0.16535 | 0.003184 | 0.20206 | 0.20659 | 3513.063 |
| MIKKELSEN | 146 | 0.033131 | 0.002902 | 0.040505 | 0.2067 | 3514.934 |
| GOBP_REG | 15 | 0.1183 | 0.003333 | 0.14467 | 0.20678 | 3516.294 |
| GOMF_FUC | 8 | 0.17722 | 0.003647 | 0.21675 | 0.20679 | 3516.464 |
| LOPEZ_ME | 8 | 0.13293 | 0.002736 | 0.16258 | 0.20679 | 3516.464 |
| GOBP_REG | 4 | 0.25128 | 0.003657 | 0.30737 | 0.20682 | 3516.974 |
| REACTOME | 15 | 0.1113 | 0.003136 | 0.13615 | 0.20682 | 3516.974 |
| GOMF_POI | 47 | 0.058644 | 0.002923 | 0.071737 | 0.20683 | 3517.144 |

| | | | | | | |
|---|---|---|---|---|---|---|
| GOBP_NEG | 12 | 0.11508 | 0.002901 | 0.14084 | 0.20694 | 3519.015 |
| TURASHVIL | 72 | 0.047539 | 0.00293 | 0.058183 | 0.20695 | 3519.185 |
| WP_CARDI | 51 | 0.061411 | 0.003188 | 0.07516 | 0.20695 | 3519.185 |
| RAMJAUN_ | 10 | 0.13528 | 0.003113 | 0.1656 | 0.207 | 3520.035 |
| GOBP_GDF | 8 | 0.16604 | 0.003417 | 0.2034 | 0.20716 | 3522.756 |
| GOBP_LUN | 43 | 0.066461 | 0.003168 | 0.081434 | 0.20722 | 3523.776 |
| GOMF_ALF | 5 | 0.15411 | 0.002508 | 0.18885 | 0.20725 | 3524.286 |
| GOBP_EST/ | 17 | 0.097041 | 0.002911 | 0.11893 | 0.20728 | 3524.796 |
| GOBP_ENU | 10 | 0.10819 | 0.002489 | 0.13267 | 0.2074 | 3526.837 |
| BOYAULT_I | 43 | 0.062098 | 0.00296 | 0.076162 | 0.20745 | 3527.687 |
| GOMF_NF_ | 3 | 0.25407 | 0.003203 | 0.31165 | 0.20748 | 3528.197 |
| GOBP_PEP | 5 | 0.14555 | 0.002368 | 0.17855 | 0.20749 | 3528.367 |
| WP_NRP1T | 52 | 0.061647 | 0.003231 | 0.075625 | 0.20749 | 3528.367 |
| GOBP_POS | 8 | 0.17143 | 0.003528 | 0.21036 | 0.20755 | 3529.388 |
| REACTOME | 11 | 0.12397 | 0.002992 | 0.15213 | 0.20757 | 3529.728 |
| GOBP_LEU | 4 | 0.20888 | 0.00304 | 0.25634 | 0.20759 | 3530.068 |
| GOBP_BIOI | 49 | 0.062131 | 0.003161 | 0.076293 | 0.20772 | 3532.279 |
| MARTINEZ_ | 647 | 0.016027 | 0.002916 | 0.019685 | 0.20777 | 3533.129 |
| GOBP_GLY | 8 | 0.12866 | 0.002648 | 0.15804 | 0.20779 | 3533.469 |
| GOBP_PYR | 49 | 0.05465 | 0.002781 | 0.067133 | 0.20781 | 3533.809 |
| REACTOME | 5 | 0.16649 | 0.002709 | 0.20453 | 0.20782 | 3533.979 |
| GOMF_NU | 38 | 0.068782 | 0.003083 | 0.084532 | 0.20792 | 3535.68 |
| ANDERSEN | 72 | 0.046571 | 0.002871 | 0.057239 | 0.20793 | 3535.85 |
| GOBP_INFL | 15 | 0.1076 | 0.003032 | 0.13226 | 0.20797 | 3536.53 |
| GOBP_HIST | 6 | 0.16336 | 0.002912 | 0.20082 | 0.20799 | 3536.87 |
| KEGG_GLY( | 28 | 0.0754 | 0.002902 | 0.09272 | 0.20806 | 3538.06 |
| REACTOME | 21 | 0.085418 | 0.002847 | 0.10505 | 0.20807 | 3538.23 |
| MORI_MAT | 72 | 0.049111 | 0.003027 | 0.060404 | 0.2081 | 3538.741 |
| WP_NUCLE | 61 | 0.045415 | 0.002577 | 0.055868 | 0.20815 | 3539.591 |
| YAMASHIT/ | 50 | 0.05749 | 0.002955 | 0.070811 | 0.20843 | 3544.352 |
| REACTOME | 35 | 0.066214 | 0.002848 | 0.08158 | 0.2085 | 3545.543 |
| CASORELLI_ | 38 | 0.078079 | 0.0035 | 0.096213 | 0.20854 | 3546.223 |
| GOBP_REL/ | 17 | 0.10673 | 0.003201 | 0.13156 | 0.20862 | 3547.583 |
| REACTOME | 14 | 0.12642 | 0.003441 | 0.15584 | 0.20864 | 3547.923 |
| GOBP_REG | 4 | 0.18076 | 0.002631 | 0.22302 | 0.20883 | 3551.154 |
| GOBP_REG | 341 | 0.023314 | 0.003105 | 0.028784 | 0.20899 | 3553.875 |
| WP_EICOS/ | 9 | 0.1349 | 0.002945 | 0.16657 | 0.209 | 3554.045 |
| GOBP_CAL | 330 | 0.023274 | 0.00305 | 0.028738 | 0.20901 | 3554.215 |
| SASAI_TAR | 12 | 0.10878 | 0.002742 | 0.13435 | 0.20907 | 3555.235 |
| GOBP_DAN | 7 | 0.167 | 0.003215 | 0.20626 | 0.20908 | 3555.405 |
| GOBP_MRI | 17 | 0.086407 | 0.002592 | 0.10676 | 0.20916 | 3556.766 |
| KONDO_CC | 25 | 0.084876 | 0.003087 | 0.10489 | 0.20922 | 3557.786 |
| GOBP_SYN | 420 | 0.020605 | 0.003039 | 0.025466 | 0.20923 | 3557.956 |
| GOBP_LYN | 24 | 0.092423 | 0.003293 | 0.11424 | 0.20925 | 3558.296 |
| GOBP_REG | 179 | 0.030602 | 0.002966 | 0.037839 | 0.20934 | 3559.827 |

| | | | | | | |
|---|---|---|---|---|---|---|
| REACTOME | 17 | 0.10153 | 0.003045 | 0.12555 | 0.20936 | 3560.167 |
| PASINI_SU: | 292 | 0.024919 | 0.003075 | 0.030824 | 0.20942 | 3561.187 |
| GOCC_COF | 5 | 0.19552 | 0.003182 | 0.24192 | 0.20949 | 3562.377 |
| GOMF_VAS | 13 | 0.12523 | 0.003285 | 0.15498 | 0.20953 | 3563.058 |
| REACTOME | 11 | 0.14582 | 0.003519 | 0.1805 | 0.20958 | 3563.908 |
| ZHOU_CELI | 80 | 0.043662 | 0.002836 | 0.054044 | 0.20958 | 3563.908 |
| GOBP_COL | 59 | 0.056165 | 0.003135 | 0.069536 | 0.20963 | 3564.758 |
| BIOCARTA_ | 26 | 0.078106 | 0.002897 | 0.096738 | 0.20972 | 3566.289 |
| REACTOME | 35 | 0.07098 | 0.003053 | 0.087917 | 0.20974 | 3566.629 |
| GOCC_CELI | 400 | 0.020763 | 0.00299 | 0.025723 | 0.20979 | 3567.479 |
| GOMF_UBI | 31 | 0.075974 | 0.003076 | 0.09419 | 0.20995 | 3570.2 |
| GOBP_CAL | 8 | 0.14589 | 0.003003 | 0.18091 | 0.21 | 3571.05 |
| WP_PURIN | 20 | 0.081747 | 0.002659 | 0.10138 | 0.21003 | 3571.56 |
| KASLER_HI | 187 | 0.029838 | 0.002955 | 0.037026 | 0.21016 | 3573.771 |
| REACTOME | 51 | 0.057984 | 0.00301 | 0.071962 | 0.2102 | 3574.451 |
| PID_ARF6_ | 15 | 0.10129 | 0.002854 | 0.12584 | 0.21043 | 3578.362 |
| GOBP_XMI | 7 | 0.12158 | 0.002341 | 0.15112 | 0.21056 | 3580.573 |
| GOBP_PYR | 27 | 0.076306 | 0.002884 | 0.094938 | 0.21078 | 3584.314 |
| ZHU_CMV_ | 77 | 0.043095 | 0.002747 | 0.053619 | 0.21079 | 3584.484 |
| GOBP_ACT | 154 | 0.033287 | 0.002994 | 0.041428 | 0.21085 | 3585.504 |
| GOBP_NEG | 475 | 0.018264 | 0.00286 | 0.022747 | 0.21101 | 3588.225 |
| GOMF_OXI | 12 | 0.096245 | 0.002426 | 0.11988 | 0.21104 | 3588.735 |
| GOBP_PRC | 11 | 0.097603 | 0.002355 | 0.12158 | 0.21105 | 3588.905 |
| CUI_TCF21 | 795 | 0.01521 | 0.003055 | 0.018952 | 0.21111 | 3589.926 |
| KYNG_NOF | 18 | 0.096315 | 0.002973 | 0.12003 | 0.21117 | 3590.946 |
| NAKAMUR | 34 | 0.075847 | 0.003216 | 0.094567 | 0.21127 | 3592.646 |
| GOMF_ACY | 7 | 0.13999 | 0.002695 | 0.17461 | 0.21137 | 3594.347 |
| GOBP_PHC | 100 | 0.041451 | 0.003009 | 0.051711 | 0.2114 | 3594.857 |
| GOBP_GLU | 6 | 0.19978 | 0.003561 | 0.2493 | 0.21146 | 3595.877 |
| PID_REELII | 28 | 0.072848 | 0.002804 | 0.090914 | 0.21149 | 3596.387 |
| GOBP_SKEI | 10 | 0.11246 | 0.002588 | 0.14038 | 0.21153 | 3597.068 |
| GOMF_GLY | 4 | 0.22512 | 0.003277 | 0.281 | 0.21153 | 3597.068 |
| WP_GLUC( | 12 | 0.12597 | 0.003175 | 0.15728 | 0.21158 | 3597.918 |
| GOBP_POS | 145 | 0.034838 | 0.003042 | 0.043496 | 0.21159 | 3598.088 |
| GOBP_PIE( | 10 | 0.13076 | 0.003009 | 0.1633 | 0.21165 | 3599.108 |
| GOMF_DN | 20 | 0.08504 | 0.002767 | 0.10624 | 0.21173 | 3600.469 |
| GOBP_EST | 13 | 0.097138 | 0.002548 | 0.12136 | 0.21175 | 3600.809 |
| LIU_SMAR( | 46 | 0.063148 | 0.003113 | 0.078905 | 0.21177 | 3601.149 |
| GERHOLD_ | 6 | 0.1708 | 0.003045 | 0.21344 | 0.21179 | 3601.489 |
| DAIRKEE_C | 51 | 0.053793 | 0.002792 | 0.067227 | 0.21181 | 3601.829 |
| GOBP_PLA | 25 | 0.087231 | 0.003172 | 0.10902 | 0.21183 | 3602.169 |
| GOMF_POI | 13 | 0.086262 | 0.002263 | 0.10781 | 0.21183 | 3602.169 |
| GOBP_PRC | 53 | 0.05304 | 0.002806 | 0.06633 | 0.21197 | 3604.55 |
| WP_WNT_ | 111 | 0.038184 | 0.002919 | 0.047767 | 0.21204 | 3605.74 |
| SCHAEFFEF | 5 | 0.15275 | 0.002486 | 0.19117 | 0.21213 | 3607.271 |

| | | | | | | |
|---|---|---|---|---|---|---|
| GOCC_PAN | 5 | 0.16502 | 0.002685 | 0.20664 | 0.21227 | 3609.651 |
| GOMF_OXI | 13 | 0.090929 | 0.002385 | 0.11388 | 0.21231 | 3610.332 |
| TSUTSUMI_ | 8 | 0.15246 | 0.003138 | 0.19098 | 0.21235 | 3611.012 |
| GOBP_REG | 5 | 0.20023 | 0.003258 | 0.2509 | 0.21242 | 3612.202 |
| REACTOME | 22 | 0.087074 | 0.002971 | 0.10911 | 0.21242 | 3612.202 |
| GOBP_UTP | 11 | 0.10969 | 0.002647 | 0.13747 | 0.21246 | 3612.882 |
| GOBP_ENE | 80 | 0.044498 | 0.002891 | 0.05577 | 0.21248 | 3613.222 |
| GOBP_INTI | 70 | 0.068255 | 0.004149 | 0.085546 | 0.21248 | 3613.222 |
| LENAOUR_ | 114 | 0.03816 | 0.002956 | 0.047838 | 0.21253 | 3614.073 |
| GAZDA_DI | 37 | 0.062542 | 0.002766 | 0.078497 | 0.21281 | 3618.834 |
| REACTOME | 25 | 0.080931 | 0.002943 | 0.10159 | 0.21283 | 3619.174 |
| GOMF_CDI | 8 | 0.1165 | 0.002398 | 0.14625 | 0.21285 | 3619.514 |
| BIOCARTA_ | 18 | 0.10107 | 0.003119 | 0.127 | 0.21307 | 3623.255 |
| MARCINIAI | 21 | 0.099542 | 0.003318 | 0.12509 | 0.21309 | 3623.595 |
| HUANG_FC | 47 | 0.060483 | 0.003014 | 0.076037 | 0.21318 | 3625.126 |
| WEIGEL_O | 31 | 0.065713 | 0.002661 | 0.08261 | 0.21318 | 3625.126 |
| WP_NUCLE | 8 | 0.1212 | 0.002495 | 0.1524 | 0.21323 | 3625.976 |
| GOBP_TRA | 12 | 0.12617 | 0.00318 | 0.15865 | 0.21324 | 3626.146 |
| GOMF_SIN | 114 | 0.036295 | 0.002812 | 0.045646 | 0.21327 | 3626.656 |
| PUJANA_XI | 158 | 0.030416 | 0.002771 | 0.038257 | 0.2133 | 3627.167 |
| GOMF_INT | 9 | 0.12296 | 0.002684 | 0.15478 | 0.21348 | 3630.227 |
| WP_NUCLE | 35 | 0.069098 | 0.002973 | 0.086987 | 0.2135 | 3630.568 |
| REACTOME | 6 | 0.12484 | 0.002225 | 0.15725 | 0.21364 | 3632.948 |
| GOBP_NEG | 260 | 0.024561 | 0.002863 | 0.030947 | 0.21371 | 3634.139 |
| REACTOME | 11 | 0.11071 | 0.002672 | 0.13949 | 0.21371 | 3634.139 |
| GOMF_TPF | 6 | 0.1237 | 0.002205 | 0.1559 | 0.21375 | 3634.819 |
| WEBER_MI | 7 | 0.14085 | 0.002712 | 0.17757 | 0.21383 | 3636.179 |
| AIGNER_ZE | 33 | 0.070972 | 0.002965 | 0.089491 | 0.21388 | 3637.029 |
| GOBP_CELI | 1808 | 0.00961 | 0.002828 | 0.01212 | 0.21393 | 3637.88 |
| WP_EGFR_ | 83 | 0.046548 | 0.00308 | 0.058709 | 0.21394 | 3638.05 |
| REACTOME | 10 | 0.12857 | 0.002958 | 0.16221 | 0.214 | 3639.07 |
| GOBP_POS | 16 | 0.098481 | 0.002866 | 0.1243 | 0.2141 | 3640.771 |
| BENPORAT | 207 | 0.026167 | 0.002725 | 0.033043 | 0.21421 | 3642.641 |
| GOBP_POS | 5 | 0.17927 | 0.002917 | 0.22642 | 0.21426 | 3643.491 |
| GOBP_NEG | 5 | 0.19868 | 0.003233 | 0.25103 | 0.21434 | 3644.852 |
| GOBP_REN | 8 | 0.15127 | 0.003113 | 0.19135 | 0.21461 | 3649.443 |
| VALK_AML | 24 | 0.085518 | 0.003047 | 0.10825 | 0.21476 | 3651.994 |
| GOBP_NEG | 28 | 0.075327 | 0.002899 | 0.09537 | 0.21481 | 3652.844 |
| GOBP_TRN | 129 | 0.030823 | 0.002539 | 0.039047 | 0.21494 | 3655.055 |
| GOBP_PLA | 14 | 0.10626 | 0.002893 | 0.13467 | 0.21506 | 3657.095 |
| MARTINEZ_ | 577 | 0.01668 | 0.002871 | 0.021152 | 0.21519 | 3659.306 |
| REACTOME | 7 | 0.1191 | 0.002293 | 0.15107 | 0.21524 | 3660.156 |
| RODRIGUE | 14 | 0.11684 | 0.003181 | 0.1483 | 0.21538 | 3662.537 |
| MYLLYKAN | 8 | 0.14616 | 0.003008 | 0.18551 | 0.2154 | 3662.877 |
| GOBP_NEG | 10 | 0.1237 | 0.002846 | 0.15712 | 0.21555 | 3665.428 |

| | | | | | | |
|---|---|---|---|---|---|---|
| GOCC_UBI( | 287 | 0.022427 | 0.002744 | 0.028489 | 0.21558 | 3665.938 |
| GOCC_SOR | 6 | 0.1585 | 0.002825 | 0.20137 | 0.21562 | 3666.618 |
| GOBP_POS | 77 | 0.048691 | 0.003103 | 0.061865 | 0.21563 | 3666.788 |
| TESAR_ALK | 5 | 0.24271 | 0.00395 | 0.30839 | 0.21564 | 3666.958 |
| REACTOME | 13 | 0.12208 | 0.003202 | 0.15513 | 0.21567 | 3667.468 |
| BIOCARTA_ | 14 | 0.095294 | 0.002594 | 0.12111 | 0.21569 | 3667.808 |
| GOBP_REG | 16 | 0.10179 | 0.002962 | 0.12939 | 0.21574 | 3668.659 |
| GOBP_REG | 10 | 0.11037 | 0.00254 | 0.14037 | 0.21586 | 3670.699 |
| GOCC_SAR | 71 | 0.045469 | 0.002783 | 0.057839 | 0.2159 | 3671.38 |
| KAPOSI_LIV | 18 | 0.092183 | 0.002845 | 0.11729 | 0.21595 | 3672.23 |
| REACTOME | 28 | 0.084419 | 0.003249 | 0.10743 | 0.216 | 3673.08 |
| GOBP_MAI | 5 | 0.17429 | 0.002836 | 0.22195 | 0.21615 | 3675.631 |
| GOBP_PEP | 5 | 0.19193 | 0.003123 | 0.24442 | 0.21616 | 3675.801 |
| GOBP_AMI | 11 | 0.11637 | 0.002808 | 0.14821 | 0.21619 | 3676.311 |
| HAHTOLA_ | 15 | 0.096863 | 0.002729 | 0.12343 | 0.21631 | 3678.352 |
| MIKKELSEN | 65 | 0.045098 | 0.002642 | 0.057478 | 0.21635 | 3679.032 |
| MEISSNER_ | 55 | 0.05517 | 0.002974 | 0.070321 | 0.21637 | 3679.372 |
| SAKAI_TUM | 26 | 0.07516 | 0.002787 | 0.095836 | 0.21645 | 3680.732 |
| LI_LUNG_C | 39 | 0.058887 | 0.002674 | 0.075119 | 0.21655 | 3682.433 |
| GOBP_PHC | 19 | 0.090651 | 0.002874 | 0.11566 | 0.2166 | 3683.283 |
| GOBP_7_M | 8 | 0.20295 | 0.004177 | 0.25896 | 0.21661 | 3683.453 |
| GOBP_NEU | 4 | 0.16489 | 0.0024 | 0.2104 | 0.21661 | 3683.453 |
| GARGALOV | 14 | 0.11714 | 0.003189 | 0.14951 | 0.21669 | 3684.813 |
| GOMF_ME | 63 | 0.05278 | 0.003044 | 0.067375 | 0.21671 | 3685.154 |
| GOMF_GLU | 19 | 0.0828 | 0.002626 | 0.10573 | 0.21677 | 3686.174 |
| APRELIKOV | 45 | 0.056379 | 0.002749 | 0.07201 | 0.21684 | 3687.364 |
| GOBP_MRI | 222 | 0.02486 | 0.00268 | 0.031752 | 0.21684 | 3687.364 |
| NABA_COR | 256 | 0.026909 | 0.003112 | 0.03438 | 0.21691 | 3688.555 |
| GOBP_PRC | 8 | 0.15178 | 0.003124 | 0.19402 | 0.21703 | 3690.595 |
| GOBP_REG | 555 | 0.017793 | 0.003006 | 0.022746 | 0.21704 | 3690.765 |
| BIOCARTA_ | 28 | 0.071317 | 0.002745 | 0.091262 | 0.21727 | 3694.676 |
| GOBP_PRC | 72 | 0.046592 | 0.002872 | 0.059631 | 0.21731 | 3695.357 |
| MIKKELSEN | 24 | 0.076798 | 0.002737 | 0.098363 | 0.21748 | 3698.247 |
| GOMF_TITI | 13 | 0.12737 | 0.003341 | 0.16318 | 0.21754 | 3699.268 |
| GOCC_SCH | 86 | 0.043292 | 0.002915 | 0.055498 | 0.21768 | 3701.648 |
| GOMF_M7 | 9 | 0.12424 | 0.002712 | 0.15928 | 0.21769 | 3701.818 |
| GOBP_BRA | 4 | 0.1998 | 0.002908 | 0.25623 | 0.21777 | 3703.179 |
| REACTOME | 9 | 0.12584 | 0.002747 | 0.16146 | 0.21789 | 3705.219 |
| GOMF_FIB | 24 | 0.091449 | 0.003259 | 0.11739 | 0.21798 | 3706.75 |
| WP_LEUCII | 23 | 0.079422 | 0.002771 | 0.10206 | 0.21822 | 3710.831 |
| GOBP_GAS | 5 | 0.18007 | 0.00293 | 0.23141 | 0.21825 | 3711.341 |
| GOBP_ISOI | 6 | 0.15049 | 0.002682 | 0.19342 | 0.21828 | 3711.851 |
| HELLER_HD | 275 | 0.023611 | 0.002829 | 0.03036 | 0.21838 | 3713.552 |
| REACTOME | 5 | 0.17326 | 0.002819 | 0.22279 | 0.21839 | 3713.722 |
| GOCC_LAM | 16 | 0.10428 | 0.003035 | 0.13414 | 0.21847 | 3715.082 |

| | | | | | | |
|---|---|---|---|---|---|---|
| GOMF_C_X | 7 | 0.13753 | 0.002648 | 0.17704 | 0.21863 | 3717.803 |
| KAAB_HEA | 235 | 0.026333 | 0.00292 | 0.033902 | 0.21867 | 3718.483 |
| WP_TGFBE | 127 | 0.035303 | 0.002886 | 0.045458 | 0.2187 | 3718.994 |
| GOBP_POS | 7 | 0.12387 | 0.002385 | 0.15951 | 0.21871 | 3719.164 |
| GOBP_BRA | 31 | 0.078587 | 0.003182 | 0.10122 | 0.21876 | 3720.014 |
| GOBP_PRC | 2 | 0.21283 | 0.002191 | 0.27412 | 0.21876 | 3720.014 |
| GOBP_COF | 7 | 0.16045 | 0.003089 | 0.20678 | 0.2189 | 3722.395 |
| KUUSELO_I | 32 | 0.14343 | 0.0059 | 0.18486 | 0.21892 | 3722.735 |
| GOBP_POS | 8 | 0.13801 | 0.002841 | 0.17796 | 0.21902 | 3724.435 |
| KANG_CISF | 8 | 0.13033 | 0.002682 | 0.16806 | 0.21903 | 3724.605 |
| GOBP_AUT | 82 | 0.041808 | 0.002749 | 0.053917 | 0.21906 | 3725.115 |
| GOMF_U3_ | 11 | 0.13749 | 0.003318 | 0.17736 | 0.21911 | 3725.966 |
| CHEN_NEU | 50 | 0.070091 | 0.003602 | 0.090424 | 0.21914 | 3726.476 |
| REACTOME | 35 | 0.065457 | 0.002816 | 0.084448 | 0.21914 | 3726.476 |
| KOKKINAKI | 127 | 0.035308 | 0.002886 | 0.04558 | 0.21928 | 3728.856 |
| LIU_CMYB_ | 6 | 0.19023 | 0.003391 | 0.24557 | 0.21928 | 3728.856 |
| GOBP_C21_ | 20 | 0.095709 | 0.003114 | 0.12356 | 0.21929 | 3729.026 |
| GOBP_ORG | 141 | 0.033099 | 0.00285 | 0.042733 | 0.21931 | 3729.367 |
| GOBP_ODC | 88 | 0.045253 | 0.003082 | 0.058428 | 0.21932 | 3729.537 |
| REACTOME | 119 | 0.034465 | 0.002728 | 0.044499 | 0.21932 | 3729.537 |
| GOBP_PLA | 27 | 0.069879 | 0.002641 | 0.090262 | 0.21942 | 3731.237 |
| GOBP_REG | 22 | 0.09889 | 0.003374 | 0.12778 | 0.21949 | 3732.427 |
| MARTORIA | 220 | 0.0263 | 0.002823 | 0.033993 | 0.21956 | 3733.618 |
| GOBP_NEG | 8 | 0.13578 | 0.002795 | 0.17554 | 0.21961 | 3734.468 |
| BLANCO_M | 106 | 0.034253 | 0.00256 | 0.044321 | 0.21982 | 3738.039 |
| REACTOME | 5 | 0.23038 | 0.003749 | 0.29809 | 0.21982 | 3738.039 |
| GOBP_RRN | 37 | 0.059309 | 0.002623 | 0.076771 | 0.2199 | 3739.4 |
| GOBP_NEG | 3 | 0.26321 | 0.003318 | 0.34073 | 0.21992 | 3739.74 |
| GOBP_PUR | 514 | 0.016702 | 0.002718 | 0.021621 | 0.21992 | 3739.74 |
| REACTOME | 26 | 0.092916 | 0.003446 | 0.12031 | 0.21997 | 3740.59 |
| GOBP_AXC | 74 | 0.042468 | 0.002654 | 0.054991 | 0.21998 | 3740.76 |
| VERRECCHI | 19 | 0.089568 | 0.00284 | 0.116 | 0.22002 | 3741.44 |
| WP_COMP | 98 | 0.042087 | 0.003025 | 0.054517 | 0.22006 | 3742.12 |
| GOBP_HEP | 14 | 0.10761 | 0.002929 | 0.1394 | 0.22008 | 3742.46 |
| GOBP_PRC | 34 | 0.063627 | 0.002698 | 0.082433 | 0.2201 | 3742.801 |
| GOBP_TRIF | 16 | 0.10177 | 0.002962 | 0.13188 | 0.22015 | 3743.651 |
| REACTOME | 5 | 0.15047 | 0.002449 | 0.195 | 0.22016 | 3743.821 |
| YEMELYAN | 10 | 0.1181 | 0.002717 | 0.15304 | 0.22016 | 3743.821 |
| WP_SPLICI | 41 | 0.064478 | 0.003002 | 0.083576 | 0.22022 | 3744.841 |
| REACTOME | 82 | 0.045414 | 0.002987 | 0.05889 | 0.22031 | 3746.372 |
| GOBP_SUB | 40 | 0.055268 | 0.002541 | 0.071692 | 0.22038 | 3747.562 |
| BONOME_ | 18 | 0.067479 | 0.002083 | 0.087539 | 0.22041 | 3748.072 |
| GOBP_REN | 7 | 0.15203 | 0.002927 | 0.1973 | 0.22049 | 3749.432 |
| GOBP_BRA | 5 | 0.17348 | 0.002823 | 0.22545 | 0.2208 | 3754.704 |
| PARK_APL_ | 13 | 0.10927 | 0.002867 | 0.14201 | 0.22081 | 3754.874 |

| | | | | | | |
|---|---|---|---|---|---|---|
| WP_IMMU | 20 | 0.089544 | 0.002913 | 0.11637 | 0.22082 | 3755.044 |
| ABRAHAM_ | 26 | 0.077356 | 0.002869 | 0.10055 | 0.22085 | 3755.554 |
| GOBP_SEM | 13 | 0.12887 | 0.003381 | 0.16755 | 0.22091 | 3756.575 |
| REACTOME | 165 | 0.030053 | 0.002797 | 0.039088 | 0.22099 | 3757.935 |
| GOBP_REP | 6 | 0.22835 | 0.00407 | 0.29703 | 0.22102 | 3758.445 |
| GOCC_DES | 24 | 0.082171 | 0.002928 | 0.10688 | 0.22102 | 3758.445 |
| GOCC_EUK | 6 | 0.14322 | 0.002553 | 0.18638 | 0.22112 | 3760.146 |
| REACTOME | 114 | 0.037946 | 0.00294 | 0.049388 | 0.22115 | 3760.656 |
| JIANG_HYP | 193 | 0.026474 | 0.002663 | 0.034465 | 0.2212 | 3761.506 |
| GOCC_U2A | 4 | 0.18777 | 0.002733 | 0.24453 | 0.22129 | 3763.036 |
| WP_RAC1F | 66 | 0.048741 | 0.002877 | 0.063497 | 0.22136 | 3764.227 |
| REACTOME | 34 | 0.059857 | 0.002538 | 0.078001 | 0.22143 | 3765.417 |
| GOBP_LIPI | 7 | 0.16462 | 0.00317 | 0.21455 | 0.22146 | 3765.927 |
| WILLERT_V | 21 | 0.09801 | 0.003267 | 0.12776 | 0.22151 | 3766.778 |
| GOBP_CELI | 13 | 0.099138 | 0.002601 | 0.12926 | 0.22155 | 3767.458 |
| WIEMANN_ | 4 | 0.1833 | 0.002668 | 0.23902 | 0.22158 | 3767.968 |
| RIZ_ERYTH | 37 | 0.068609 | 0.003034 | 0.089483 | 0.22163 | 3768.818 |
| GOBP_MAI | 17 | 0.10657 | 0.003197 | 0.13914 | 0.22187 | 3772.899 |
| BROWNE_I | 79 | 0.045729 | 0.002952 | 0.059733 | 0.22198 | 3774.77 |
| GOCC_NOI | 16 | 0.10791 | 0.00314 | 0.141 | 0.22204 | 3775.79 |
| GOBP_POS | 9 | 0.12617 | 0.002754 | 0.16489 | 0.22209 | 3776.64 |
| REACTOME | 5 | 0.15966 | 0.002598 | 0.20867 | 0.2221 | 3776.811 |
| WP_TYROS | 5 | 0.15966 | 0.002598 | 0.20867 | 0.2221 | 3776.811 |
| GOBP_CIR( | 7 | 0.15607 | 0.003005 | 0.20407 | 0.2222 | 3778.511 |
| GOBP_DOU | 8 | 0.10877 | 0.002239 | 0.14224 | 0.22224 | 3779.191 |
| GOBP_ADE | 7 | 0.1399 | 0.002694 | 0.18304 | 0.22234 | 3780.892 |
| BIOCARTA_ | 24 | 0.087346 | 0.003112 | 0.1143 | 0.22238 | 3781.572 |
| GOBP_POS | 51 | 0.05254 | 0.002727 | 0.068784 | 0.22249 | 3783.442 |
| REACTOME | 110 | 0.030958 | 0.002356 | 0.04054 | 0.22255 | 3784.463 |
| BOYAULT_I | 18 | 0.089284 | 0.002756 | 0.11693 | 0.22256 | 3784.633 |
| GOMF_SIG | 46 | 0.050547 | 0.002492 | 0.066208 | 0.2226 | 3785.313 |
| GOBP_ENT | 29 | 0.061643 | 0.002414 | 0.080749 | 0.22262 | 3785.653 |
| GOBP_POS | 34 | 0.068159 | 0.00289 | 0.089321 | 0.22271 | 3787.184 |
| GOBP_GLY | 367 | 0.019834 | 0.002738 | 0.025993 | 0.22272 | 3787.354 |
| GOBP_PYR | 22 | 0.074376 | 0.002538 | 0.097504 | 0.2228 | 3788.714 |
| GOBP_SYN | 177 | 0.030185 | 0.002909 | 0.039576 | 0.22283 | 3789.224 |
| GOCC_EUK | 14 | 0.099657 | 0.002713 | 0.13068 | 0.22286 | 3789.734 |
| GOBP_AOF | 38 | 0.065758 | 0.002947 | 0.086235 | 0.22287 | 3789.904 |
| GOCC_PHC | 49 | 0.052963 | 0.002695 | 0.069474 | 0.22293 | 3790.925 |
| GOBP_POS | 21 | 0.079453 | 0.002649 | 0.10427 | 0.22303 | 3792.625 |
| REACTOME | 51 | 0.053659 | 0.002785 | 0.070431 | 0.22307 | 3793.305 |
| GOBP_ANT | 8 | 0.13379 | 0.002754 | 0.17564 | 0.22312 | 3794.156 |
| GOBP_DNA | 69 | 0.04589 | 0.002769 | 0.060269 | 0.22321 | 3795.686 |
| GOBP_DOL | 6 | 0.14023 | 0.0025 | 0.18419 | 0.22323 | 3796.026 |
| GOBP_REG | 14 | 0.089667 | 0.002441 | 0.11781 | 0.2233 | 3797.217 |

| | | | | | | |
|---|---|---|---|---|---|---|
| GOBP_REG | 8 | 0.14635 | 0.003012 | 0.19235 | 0.22338 | 3798.577 |
| SA_MMP_( | 14 | 0.10135 | 0.002759 | 0.13322 | 0.2234 | 3798.917 |
| GOBP_POS | 130 | 0.032365 | 0.002677 | 0.042556 | 0.22348 | 3800.277 |
| GOBP_IMP | 10 | 0.098528 | 0.002267 | 0.1296 | 0.22355 | 3801.468 |
| MIKKELSEN | 53 | 0.050739 | 0.002685 | 0.06674 | 0.22356 | 3801.638 |
| BIOCARTA_ | 19 | 0.083254 | 0.00264 | 0.10954 | 0.22362 | 3802.658 |
| SCHAEFFER | 383 | 0.020939 | 0.002952 | 0.027552 | 0.22363 | 3802.828 |
| WP_ADIPO | 127 | 0.034711 | 0.002837 | 0.045684 | 0.22369 | 3803.848 |
| KEGG_INSU | 129 | 0.032347 | 0.002665 | 0.042587 | 0.22377 | 3805.209 |
| GOBP_ANT | 15 | 0.10518 | 0.002964 | 0.1385 | 0.22381 | 3805.889 |
| GOMF_SIG | 82 | 0.040267 | 0.002648 | 0.053048 | 0.22391 | 3807.59 |
| WANG_CIS | 168 | 0.030294 | 0.002845 | 0.039911 | 0.22392 | 3807.76 |
| GOMF_NA/ | 5 | 0.14563 | 0.00237 | 0.19188 | 0.22393 | 3807.93 |
| REACTOME | 15 | 0.10171 | 0.002866 | 0.13403 | 0.22398 | 3808.78 |
| GOMF_OR( | 29 | 0.068869 | 0.002697 | 0.090782 | 0.22405 | 3809.97 |
| WP_CORTI | 88 | 0.038347 | 0.002612 | 0.050572 | 0.22415 | 3811.671 |
| GOBP_CD4 | 23 | 0.085534 | 0.002984 | 0.11283 | 0.2242 | 3812.521 |
| GOMF_INT | 14 | 0.09176 | 0.002498 | 0.12106 | 0.22424 | 3813.201 |
| GOBP_POS | 15 | 0.10134 | 0.002856 | 0.13378 | 0.22437 | 3815.412 |
| GOBP_CAL | 8 | 0.15868 | 0.003266 | 0.20961 | 0.22452 | 3817.963 |
| GOBP_POS | 40 | 0.065642 | 0.003018 | 0.086715 | 0.22453 | 3818.133 |
| HOWLIN_C | 16 | 0.11045 | 0.003214 | 0.14593 | 0.22458 | 3818.983 |
| REACTOME | 28 | 0.071048 | 0.002734 | 0.09397 | 0.22481 | 3822.894 |
| GOBP_GDF | 7 | 0.12614 | 0.002429 | 0.16688 | 0.22487 | 3823.914 |
| GOMF_FRU | 9 | 0.13569 | 0.002962 | 0.17954 | 0.22491 | 3824.595 |
| RAMPON_I | 14 | 0.094509 | 0.002573 | 0.12507 | 0.22493 | 3824.935 |
| RODRIGUE | 5 | 0.18155 | 0.002954 | 0.24038 | 0.22505 | 3826.975 |
| WP_STATIN | 29 | 0.071856 | 0.002814 | 0.095216 | 0.22523 | 3830.036 |
| GOBP_POS | 5 | 0.15358 | 0.002499 | 0.20354 | 0.22527 | 3830.716 |
| GOBP_REG | 24 | 0.08193 | 0.002919 | 0.10859 | 0.22529 | 3831.056 |
| GOBP_PRIN | 101 | 0.039595 | 0.002888 | 0.052498 | 0.22536 | 3832.247 |
| GOCC_PRE | 15 | 0.080762 | 0.002276 | 0.10708 | 0.22537 | 3832.417 |
| REACTOME | 286 | 0.022318 | 0.002726 | 0.029602 | 0.22545 | 3833.777 |
| REACTOME | 7 | 0.15838 | 0.003049 | 0.21013 | 0.22551 | 3834.798 |
| REACTOME | 4 | 0.18104 | 0.002635 | 0.24019 | 0.22551 | 3834.798 |
| GOBP_TRN | 8 | 0.12004 | 0.002471 | 0.15927 | 0.22552 | 3834.968 |
| GOCC_NUC | 1100 | 0.011446 | 0.002681 | 0.015197 | 0.22567 | 3837.518 |
| GOBP_PRC | 4 | 0.18058 | 0.002628 | 0.23996 | 0.22587 | 3840.919 |
| GOBP_PRC | 10 | 0.14068 | 0.003237 | 0.18697 | 0.22592 | 3841.77 |
| TERAMOTC | 7 | 0.1601 | 0.003082 | 0.21282 | 0.22594 | 3842.11 |
| BANDRES_ | 148 | 0.030026 | 0.002648 | 0.039929 | 0.22603 | 3843.64 |
| GRASEMAN | 10 | 0.13201 | 0.003038 | 0.17555 | 0.22603 | 3843.64 |
| RODRIGUE | 72 | 0.049707 | 0.003064 | 0.066151 | 0.22621 | 3846.701 |
| GOBP_POS | 57 | 0.050779 | 0.002786 | 0.067588 | 0.22624 | 3847.211 |
| BIDUS_ME | 140 | 0.031967 | 0.002743 | 0.042563 | 0.22631 | 3848.402 |

| | | | | | | |
|---|---|---|---|---|---|---|
| REACTOME | 147 | 0.029997 | 0.002637 | 0.039946 | 0.22635 | 3849.082 |
| GOBP_PRC | 3 | 0.24881 | 0.003136 | 0.33139 | 0.22639 | 3849.762 |
| BIOCARTA_ | 8 | 0.1172 | 0.002412 | 0.15615 | 0.22645 | 3850.782 |
| REACTOME | 194 | 0.026198 | 0.002642 | 0.034907 | 0.22648 | 3851.292 |
| GOCC_FILC | 16 | 0.093365 | 0.002717 | 0.12444 | 0.22654 | 3852.313 |
| WP_LEUKC | 30 | 0.06955 | 0.00277 | 0.092757 | 0.22669 | 3854.863 |
| GOMF_UBI | 93 | 0.037792 | 0.002646 | 0.050406 | 0.22671 | 3855.204 |
| REACTOME | 8 | 0.14932 | 0.003073 | 0.19925 | 0.22681 | 3856.904 |
| GOBP_GAL | 5 | 0.18808 | 0.003061 | 0.25105 | 0.22688 | 3858.094 |
| GOBP_LEYI | 10 | 0.10353 | 0.002382 | 0.1382 | 0.22689 | 3858.264 |
| GOBP_RED | 6 | 0.15293 | 0.002726 | 0.20418 | 0.22693 | 3858.945 |
| SA_PROGR | 11 | 0.11744 | 0.002834 | 0.15683 | 0.227 | 3860.135 |
| WUNDER_I | 9 | 0.11712 | 0.002557 | 0.15645 | 0.22705 | 3860.985 |
| GOBP_ESTA | 416 | 0.017981 | 0.00264 | 0.024023 | 0.22708 | 3861.495 |
| GOBP_INN | 7 | 0.17184 | 0.003308 | 0.22958 | 0.22709 | 3861.665 |
| KAUFFMAN | 60 | 0.048241 | 0.002715 | 0.064492 | 0.22723 | 3864.046 |
| GOBP_NEG | 34 | 0.061739 | 0.002618 | 0.08255 | 0.22727 | 3864.726 |
| GOBP_NEG | 65 | 0.046837 | 0.002744 | 0.062657 | 0.22738 | 3866.597 |
| REACTOME | 19 | 0.072175 | 0.002289 | 0.096619 | 0.22753 | 3869.148 |
| FERRANDO | 83 | 0.037139 | 0.002457 | 0.049719 | 0.22754 | 3869.318 |
| GOMF_MA | 62 | 0.045409 | 0.002598 | 0.060791 | 0.22755 | 3869.488 |
| GOBP_POS | 5 | 0.16382 | 0.002666 | 0.21932 | 0.22756 | 3869.658 |
| JU_AGING_ | 6 | 0.17756 | 0.003165 | 0.23773 | 0.22757 | 3869.828 |
| REACTOME | 15 | 0.099599 | 0.002806 | 0.13336 | 0.22758 | 3869.998 |
| GOBP_EXT | 12 | 0.12199 | 0.003075 | 0.16336 | 0.2276 | 3870.338 |
| GOCC_BET | 13 | 0.10891 | 0.002857 | 0.14594 | 0.22776 | 3873.059 |
| REACTOME | 7 | 0.12114 | 0.002332 | 0.16242 | 0.22789 | 3875.269 |
| FERNANDE | 172 | 0.027275 | 0.002592 | 0.036572 | 0.22791 | 3875.61 |
| OHM_EMB | 8 | 0.13431 | 0.002764 | 0.18016 | 0.22798 | 3876.8 |
| GOBP_CELI | 463 | 0.01758 | 0.002719 | 0.02359 | 0.22807 | 3878.33 |
| GOBP_REG | 213 | 0.024469 | 0.002584 | 0.032844 | 0.22814 | 3879.521 |
| GOCC_ORG | 86 | 0.035059 | 0.002361 | 0.04706 | 0.22815 | 3879.691 |
| LIU_BREAS | 26 | 0.068629 | 0.002545 | 0.092128 | 0.22816 | 3879.861 |
| REACTOME | 51 | 0.050796 | 0.002637 | 0.068191 | 0.22817 | 3880.031 |
| PID_SYNDE | 31 | 0.071804 | 0.002907 | 0.096462 | 0.22833 | 3882.752 |
| GOBP_FOL | 8 | 0.15736 | 0.003239 | 0.21152 | 0.22845 | 3884.792 |
| PID_LPA4_ | 14 | 0.12218 | 0.003326 | 0.16428 | 0.22851 | 3885.813 |
| REACTOME | 30 | 0.071197 | 0.002836 | 0.09575 | 0.22857 | 3886.833 |
| WP_LACTC | 6 | 0.17466 | 0.003113 | 0.2349 | 0.22858 | 3887.003 |
| HELLER_SIL | 235 | 0.024853 | 0.002756 | 0.033427 | 0.2286 | 3887.343 |
| REACTOME | 90 | 0.040319 | 0.002777 | 0.054231 | 0.2286 | 3887.343 |
| GOBP_B_C | 6 | 0.15064 | 0.002685 | 0.20267 | 0.22866 | 3888.363 |
| PID_SMAD | 16 | 0.097418 | 0.002835 | 0.13111 | 0.22873 | 3889.554 |
| GOMF_TUI | 347 | 0.019589 | 0.002631 | 0.026373 | 0.22882 | 3891.084 |
| HOFMANN | 37 | 0.059224 | 0.002619 | 0.079738 | 0.22883 | 3891.254 |

| | | | | | | |
|---|---|---|---|---|---|---|
| BIOCARTA_ | 14 | 0.087908 | 0.002393 | 0.11847 | 0.22904 | 3894.825 |
| PALOMERC | 4 | 0.20477 | 0.00298 | 0.27614 | 0.22919 | 3897.376 |
| GOBP_G1_ | 6 | 0.1372 | 0.002446 | 0.18504 | 0.22921 | 3897.716 |
| WP_MITOC | 8 | 0.16615 | 0.00342 | 0.22426 | 0.2294 | 3900.947 |
| GOCC_PLA | 175 | 0.028686 | 0.002749 | 0.038738 | 0.2295 | 3902.648 |
| GOBP_T_C | 29 | 0.074745 | 0.002927 | 0.10094 | 0.22952 | 3902.988 |
| WP_BIOGE | 13 | 0.097699 | 0.002563 | 0.13194 | 0.22952 | 3902.988 |
| GOBP_RES | 7 | 0.12016 | 0.002313 | 0.16231 | 0.22957 | 3903.838 |
| GOMF_AM | 50 | 0.048306 | 0.002483 | 0.065278 | 0.22965 | 3905.198 |
| GOCC_NEU | 81 | 0.039877 | 0.002607 | 0.053901 | 0.22971 | 3906.219 |
| KEGG_GNF | 98 | 0.039243 | 0.00282 | 0.053052 | 0.22974 | 3906.729 |
| PID_P53_D | 134 | 0.031673 | 0.002659 | 0.042823 | 0.22977 | 3907.239 |
| GOBP_POS | 199 | 0.027986 | 0.002858 | 0.037844 | 0.2298 | 3907.749 |
| GOBP_CELL | 806 | 0.012765 | 0.002581 | 0.017281 | 0.23006 | 3912.17 |
| REACTOME | 13 | 0.089692 | 0.002353 | 0.12143 | 0.23006 | 3912.17 |
| GOBP_SPH | 15 | 0.099826 | 0.002813 | 0.13515 | 0.23007 | 3912.34 |
| BIOCARTA_ | 6 | 0.17218 | 0.003069 | 0.23323 | 0.2302 | 3914.551 |
| GOBP_CELL | 40 | 0.053335 | 0.002453 | 0.072265 | 0.23025 | 3915.401 |
| GOBP_CELL | 193 | 0.027103 | 0.002726 | 0.036725 | 0.23026 | 3915.571 |
| GOBP_DIST | 5 | 0.1717 | 0.002794 | 0.2327 | 0.2303 | 3916.252 |
| GOBP_REG | 132 | 0.033821 | 0.002818 | 0.045841 | 0.23033 | 3916.762 |
| GOBP_GLY | 27 | 0.062587 | 0.002365 | 0.084844 | 0.23036 | 3917.272 |
| PID_INTEG | 26 | 0.074175 | 0.002751 | 0.10055 | 0.23036 | 3917.272 |
| GOBP_REG | 24 | 0.065731 | 0.002342 | 0.089111 | 0.23038 | 3917.612 |
| WESTON_V | 46 | 0.060705 | 0.002993 | 0.082326 | 0.23045 | 3918.802 |
| GOBP_HET | 92 | 0.038049 | 0.00265 | 0.051634 | 0.2306 | 3921.353 |
| GOCC_CAS | 10 | 0.105 | 0.002416 | 0.1425 | 0.23062 | 3921.693 |
| GOBP_END | 12 | 0.11794 | 0.002973 | 0.16009 | 0.23064 | 3922.033 |
| GOBP_UTE | 22 | 0.083117 | 0.002836 | 0.11284 | 0.23069 | 3922.883 |
| OSAWA_TN | 9 | 0.1073 | 0.002342 | 0.14571 | 0.23075 | 3923.904 |
| REACTOME | 7 | 0.13518 | 0.002603 | 0.18359 | 0.23077 | 3924.244 |
| GOBP_NEG | 11 | 0.097162 | 0.002345 | 0.13196 | 0.23078 | 3924.414 |
| YAGI_AML_ | 375 | 0.018455 | 0.002575 | 0.025068 | 0.23082 | 3925.094 |
| WANG_ESC | 110 | 0.036499 | 0.002778 | 0.049587 | 0.23085 | 3925.604 |
| GOCC_PRC | 14 | 0.094304 | 0.002567 | 0.12813 | 0.23086 | 3925.774 |
| GOBP_NEG | 6 | 0.24032 | 0.004284 | 0.32667 | 0.23097 | 3927.645 |
| GOCC_MHC | 3 | 0.1445 | 0.001821 | 0.19641 | 0.23097 | 3927.645 |
| GOBP_DET | 10 | 0.12798 | 0.002945 | 0.174 | 0.23102 | 3928.495 |
| KIM_LRRC3 | 20 | 0.073439 | 0.002389 | 0.099877 | 0.23109 | 3929.685 |
| REACTOME | 9 | 0.12583 | 0.002747 | 0.17116 | 0.23112 | 3930.196 |
| GOBP_REG | 765 | 0.014009 | 0.002762 | 0.019056 | 0.23114 | 3930.536 |
| GOCC_SCH | 11 | 0.11296 | 0.002726 | 0.15366 | 0.23114 | 3930.536 |
| GOBP_SUL | 31 | 0.069951 | 0.002832 | 0.095171 | 0.23117 | 3931.046 |
| GOBP_RES | 5 | 0.17808 | 0.002898 | 0.2423 | 0.23119 | 3931.386 |
| REACTOME | 79 | 0.036944 | 0.002385 | 0.050268 | 0.23119 | 3931.386 |

| | | | | | | |
|---|---|---|---|---|---|---|
| REACTOME | 82 | 0.041924 | 0.002757 | 0.057044 | 0.23119 | 3931.386 |
| GOBP_REG | 5 | 0.17437 | 0.002838 | 0.23727 | 0.2312 | 3931.556 |
| WP_OLIGO | 27 | 0.081139 | 0.003066 | 0.11043 | 0.23125 | 3932.406 |
| REACTOME | 57 | 0.046159 | 0.002533 | 0.06286 | 0.23139 | 3934.787 |
| GOCC_EXT | 517 | 0.017285 | 0.002821 | 0.023542 | 0.23141 | 3935.127 |
| JI_RESPON | 63 | 0.045964 | 0.002651 | 0.062611 | 0.23144 | 3935.637 |
| WP_ROBO | 6 | 0.14051 | 0.002505 | 0.19143 | 0.23147 | 3936.147 |
| REACTOME | 20 | 0.082062 | 0.00267 | 0.11188 | 0.23164 | 3939.038 |
| GOBP_PRE | 15 | 0.091768 | 0.002586 | 0.12517 | 0.23173 | 3940.569 |
| GOBP_NEG | 14 | 0.095651 | 0.002604 | 0.13048 | 0.23176 | 3941.079 |
| GOBP_CER | 12 | 0.1161 | 0.002926 | 0.15842 | 0.23183 | 3942.269 |
| GOBP_CELL | 20 | 0.092883 | 0.003022 | 0.12675 | 0.23185 | 3942.609 |
| GOBP_ORG | 267 | 0.023187 | 0.002738 | 0.031643 | 0.23186 | 3942.779 |
| SCHLINGEN | 24 | 0.065925 | 0.002349 | 0.089987 | 0.2319 | 3943.46 |
| GOBP_MEL | 5 | 0.17785 | 0.002894 | 0.24276 | 0.23191 | 3943.63 |
| GOBP_NEG | 11 | 0.11424 | 0.002757 | 0.15594 | 0.23191 | 3943.63 |
| WELCSH_B | 128 | 0.030872 | 0.002533 | 0.04216 | 0.23201 | 3945.33 |
| GOBP_POS | 9 | 0.13532 | 0.002954 | 0.18483 | 0.23205 | 3946.01 |
| REACTOME | 14 | 0.096455 | 0.002626 | 0.13175 | 0.23206 | 3946.18 |
| GOBP_RNA | 82 | 0.037179 | 0.002445 | 0.050788 | 0.23208 | 3946.52 |
| REACTOME | 20 | 0.088627 | 0.002883 | 0.12117 | 0.23227 | 3949.751 |
| GOBP_RES | 15 | 0.096923 | 0.002731 | 0.13254 | 0.23232 | 3950.602 |
| GOBP_RES | 13 | 0.086974 | 0.002282 | 0.11898 | 0.23239 | 3951.792 |
| GOBP_CELL | 29 | 0.073607 | 0.002883 | 0.10071 | 0.23243 | 3952.472 |
| DAVICIONI_ | 14 | 0.106 | 0.002886 | 0.14507 | 0.23249 | 3953.492 |
| GOBP_NEG | 14 | 0.10391 | 0.002829 | 0.14228 | 0.2326 | 3955.363 |
| GOBP_NEU | 66 | 0.052419 | 0.003094 | 0.071794 | 0.23266 | 3956.383 |
| BOSCO_AL | 140 | 0.033059 | 0.002836 | 0.045299 | 0.23276 | 3958.084 |
| WP_BDNFT | 33 | 0.06087 | 0.002543 | 0.083419 | 0.23279 | 3958.594 |
| GOBP_POS | 166 | 0.028424 | 0.002654 | 0.038971 | 0.23289 | 3960.294 |
| REACTOME | 31 | 0.061786 | 0.002502 | 0.084711 | 0.23289 | 3960.294 |
| WP_CONVE | 5 | 0.22227 | 0.003617 | 0.30478 | 0.23292 | 3960.805 |
| GOBP_PRO | 24 | 0.068349 | 0.002436 | 0.093752 | 0.23299 | 3961.995 |
| GOBP_POS | 20 | 0.084123 | 0.002737 | 0.11542 | 0.23306 | 3963.185 |
| WP_LIPID_ | 10 | 0.12621 | 0.002904 | 0.17316 | 0.23306 | 3963.185 |
| MATZUK_M | 25 | 0.070998 | 0.002582 | 0.097451 | 0.23314 | 3964.546 |
| LAU_APOP | 55 | 0.051421 | 0.002772 | 0.070583 | 0.23315 | 3964.716 |
| POS_RESPO | 12 | 0.10127 | 0.002553 | 0.13902 | 0.23316 | 3964.886 |
| GOBP_GLY | 7 | 0.11828 | 0.002277 | 0.16238 | 0.23318 | 3965.226 |
| BIOCARTA_ | 27 | 0.06732 | 0.002544 | 0.092433 | 0.23322 | 3965.906 |
| GOCC_COL | 386 | 0.019431 | 0.00275 | 0.026681 | 0.23323 | 3966.076 |
| GOMF_CHI | 5 | 0.19292 | 0.003139 | 0.2649 | 0.23323 | 3966.076 |
| GOBP_OBS | 7 | 0.13135 | 0.002529 | 0.18045 | 0.23333 | 3967.777 |
| GOBP_BRA | 7 | 0.13093 | 0.002521 | 0.17993 | 0.23342 | 3969.307 |
| GOBP_CELL | 28 | 0.076089 | 0.002928 | 0.10463 | 0.23356 | 3971.688 |

| | | | | | | |
|---|---|---|---|---|---|---|
| GOBP_HIST | 23 | 0.078832 | 0.00275 | 0.10841 | 0.23357 | 3971.858 |
| GOBP_POS | 307 | 0.020439 | 0.002585 | 0.028112 | 0.23361 | 3972.538 |
| GOCC_PWI | 5 | 0.12292 | 0.002 | 0.16914 | 0.23371 | 3974.239 |
| GOBP_CAN | 7 | 0.15507 | 0.002986 | 0.2134 | 0.23372 | 3974.409 |
| GOBP_EOS | 6 | 0.14731 | 0.002626 | 0.20272 | 0.23372 | 3974.409 |
| IKEDA_MIF | 6 | 0.19367 | 0.003452 | 0.26661 | 0.2338 | 3975.769 |
| GOMF_CAL | 5 | 0.21488 | 0.003497 | 0.29583 | 0.23381 | 3975.939 |
| BIOCARTA_ | 7 | 0.15676 | 0.003018 | 0.21582 | 0.23382 | 3976.109 |
| REACTOME | 22 | 0.086858 | 0.002963 | 0.11962 | 0.23389 | 3977.299 |
| GOBP_RES | 14 | 0.09903 | 0.002696 | 0.13656 | 0.23418 | 3982.231 |
| GOBP_REG | 27 | 0.071895 | 0.002717 | 0.099151 | 0.2342 | 3982.571 |
| HOLLEMAN | 7 | 0.1303 | 0.002509 | 0.17971 | 0.23421 | 3982.741 |
| CHARAFE_I | 440 | 0.018616 | 0.002809 | 0.025676 | 0.23422 | 3982.911 |
| GOMF_CHI | 14 | 0.089937 | 0.002448 | 0.12406 | 0.23424 | 3983.251 |
| GOBP_VES | 38 | 0.057064 | 0.002558 | 0.078742 | 0.23432 | 3984.612 |
| GOMF_NIT | 12 | 0.10784 | 0.002718 | 0.14891 | 0.23447 | 3987.162 |
| POMEROY_ | 43 | 0.052042 | 0.002481 | 0.071888 | 0.23456 | 3988.693 |
| REACTOME | 49 | 0.055454 | 0.002822 | 0.076602 | 0.23456 | 3988.693 |
| GOBP_NEG | 3 | 0.20338 | 0.002564 | 0.28103 | 0.23463 | 3989.883 |
| GOBP_RRN | 21 | 0.078767 | 0.002626 | 0.10886 | 0.23467 | 3990.563 |
| GOMF_SOI | 7 | 0.18005 | 0.003466 | 0.24885 | 0.23468 | 3990.733 |
| GOBP_REG | 10 | 0.14709 | 0.003384 | 0.20335 | 0.23475 | 3991.924 |
| GOBP_DEF | 283 | 0.021732 | 0.002641 | 0.030062 | 0.23487 | 3993.964 |
| GOBP_NEG | 8 | 0.12678 | 0.002609 | 0.17543 | 0.23495 | 3995.325 |
| REACTOME | 9 | 0.13121 | 0.002864 | 0.18157 | 0.23496 | 3995.495 |
| STAMBOLS | 45 | 0.055409 | 0.002702 | 0.076676 | 0.23496 | 3995.495 |
| REACTOME | 106 | 0.03621 | 0.002706 | 0.050111 | 0.23497 | 3995.665 |
| REACTOME | 121 | 0.032979 | 0.002632 | 0.04564 | 0.23497 | 3995.665 |
| REACTOME | 8 | 0.11996 | 0.002469 | 0.16601 | 0.23497 | 3995.665 |
| REACTOME | 8 | 0.13662 | 0.002812 | 0.18918 | 0.2351 | 3997.876 |
| GOBP_NEG | 91 | 0.037205 | 0.002577 | 0.051527 | 0.23514 | 3998.556 |
| REACTOME | 80 | 0.039023 | 0.002535 | 0.05407 | 0.23524 | 4000.256 |
| GOBP_REG | 11 | 0.091607 | 0.002211 | 0.12701 | 0.23538 | 4002.637 |
| GOBP_CELI | 32 | 0.065866 | 0.00271 | 0.091338 | 0.23542 | 4003.317 |
| GOBP_POS | 9 | 0.13233 | 0.002889 | 0.18363 | 0.23556 | 4005.698 |
| GOBP_REG | 418 | 0.018146 | 0.00267 | 0.025188 | 0.23564 | 4007.058 |
| GOMF_CYT | 242 | 0.025373 | 0.002854 | 0.035224 | 0.23566 | 4007.398 |
| GOBP_REG | 300 | 0.02033 | 0.002542 | 0.028228 | 0.2357 | 4008.079 |
| ELVIDGE_H | 162 | 0.030536 | 0.002817 | 0.042406 | 0.23574 | 4008.759 |
| GOMF_PRO | 15 | 0.082327 | 0.00232 | 0.11434 | 0.23577 | 4009.269 |
| WP_KIT_RE | 57 | 0.04974 | 0.002729 | 0.069101 | 0.23582 | 4010.119 |
| GOBP_PRO | 35 | 0.064081 | 0.002757 | 0.089118 | 0.23606 | 4014.2 |
| LINDGREN_ | 213 | 0.024668 | 0.002606 | 0.034306 | 0.23606 | 4014.2 |
| GOBP_REN | 8 | 0.14951 | 0.003077 | 0.20796 | 0.2361 | 4014.881 |
| GOBP_REG | 15 | 0.095105 | 0.00268 | 0.13229 | 0.23611 | 4015.051 |

| | | | | | | |
|---|---|---|---|---|---|---|
| GOBP_REG | 174 | 0.026 | 0.002485 | 0.03617 | 0.23613 | 4015.391 |
| GOBP_HIN | 8 | 0.15182 | 0.003125 | 0.21121 | 0.23614 | 4015.561 |
| GOBP_REG | 21 | 0.089817 | 0.002994 | 0.12496 | 0.23614 | 4015.561 |
| REACTOME | 11 | 0.096081 | 0.002319 | 0.13367 | 0.23614 | 4015.561 |
| REACTOME | 6 | 0.12666 | 0.002258 | 0.17625 | 0.23619 | 4016.411 |
| GOBP_REG | 28 | 0.072211 | 0.002779 | 0.1005 | 0.23622 | 4016.921 |
| GOMF_NU | 45 | 0.053425 | 0.002605 | 0.074353 | 0.23622 | 4016.921 |
| GOBP_POS | 861 | 0.012853 | 0.002682 | 0.017891 | 0.23625 | 4017.431 |
| GOBP_RES | 7 | 0.13406 | 0.002581 | 0.1866 | 0.23626 | 4017.601 |
| GOCC_CON | 5 | 0.20267 | 0.003298 | 0.2821 | 0.23626 | 4017.601 |
| REACTOME | 48 | 0.046459 | 0.00234 | 0.064681 | 0.2363 | 4018.282 |
| KAMIKUBC | 27 | 0.079078 | 0.002989 | 0.11011 | 0.23633 | 4018.792 |
| WP_TRANS | 10 | 0.11488 | 0.002643 | 0.1601 | 0.23652 | 4022.023 |
| REACTOME | 43 | 0.056474 | 0.002692 | 0.078733 | 0.2366 | 4023.383 |
| GOBP_LIPI | 20 | 0.090237 | 0.002936 | 0.12583 | 0.23665 | 4024.233 |
| GOBP_HON | 158 | 0.036683 | 0.003342 | 0.051199 | 0.23686 | 4027.804 |
| GOMF_DN | 441 | 0.017745 | 0.00268 | 0.02477 | 0.23688 | 4028.144 |
| WP_CLOCK | 78 | 0.040375 | 0.00259 | 0.056364 | 0.2369 | 4028.485 |
| GOMF_PRC | 124 | 0.029659 | 0.002396 | 0.041423 | 0.237 | 4030.185 |
| GOBP_DIST | 12 | 0.09932 | 0.002503 | 0.13872 | 0.23701 | 4030.355 |
| GOBP_KET | 43 | 0.05339 | 0.002545 | 0.074593 | 0.23708 | 4031.545 |
| GOBP_REG | 85 | 0.038453 | 0.002575 | 0.053726 | 0.23708 | 4031.545 |
| GOBP_TOC | 7 | 0.14128 | 0.00272 | 0.1974 | 0.23708 | 4031.545 |
| GOBP_PEP | 308 | 0.02096 | 0.002655 | 0.029293 | 0.23715 | 4032.736 |
| GOMF_NA | 5 | 0.21746 | 0.003539 | 0.30412 | 0.2373 | 4035.287 |
| GROSS_HY | 42 | 0.0497 | 0.002342 | 0.069525 | 0.23735 | 4036.137 |
| MASSARW | 534 | 0.015786 | 0.002617 | 0.022082 | 0.23735 | 4036.137 |
| GOMF_CAT | 577 | 0.014084 | 0.002424 | 0.019703 | 0.23736 | 4036.307 |
| WP_PHASE | 8 | 0.14871 | 0.003061 | 0.20803 | 0.23736 | 4036.307 |
| GOMF_CYT | 118 | 0.035731 | 0.002816 | 0.050011 | 0.23748 | 4038.347 |
| MULLIGHA | 263 | 0.022576 | 0.002646 | 0.031601 | 0.23749 | 4038.517 |
| GOCC_AUT | 10 | 0.14282 | 0.003286 | 0.19994 | 0.23752 | 4039.028 |
| GOMF_ME | 8 | 0.14106 | 0.002903 | 0.19753 | 0.23758 | 4040.048 |
| GOMF_LYA | 193 | 0.025801 | 0.002595 | 0.036139 | 0.23764 | 4041.068 |
| LINDVALL_ | 68 | 0.046394 | 0.002779 | 0.064987 | 0.23765 | 4041.238 |
| GOCC_SAR | 88 | 0.037042 | 0.002523 | 0.051893 | 0.23768 | 4041.748 |
| GOBP_MIR | 15 | 0.093539 | 0.002636 | 0.13109 | 0.23776 | 4043.109 |
| GOBP_VES | 79 | 0.039028 | 0.002519 | 0.054697 | 0.23777 | 4043.279 |
| GOBP_POS | 30 | 0.062257 | 0.00248 | 0.087283 | 0.23784 | 4044.469 |
| KEGG_WN | 145 | 0.028731 | 0.002508 | 0.040283 | 0.23785 | 4044.639 |
| BIOCARTA_ | 16 | 0.091182 | 0.002653 | 0.12785 | 0.23787 | 4044.979 |
| OSMAN_BI | 376 | 0.018162 | 0.002538 | 0.025467 | 0.23788 | 4045.149 |
| GOBP_SHC | 7 | 0.12066 | 0.002323 | 0.16926 | 0.23796 | 4046.51 |
| BASSO_HA | 76 | 0.041311 | 0.002616 | 0.057967 | 0.23803 | 4047.7 |
| GOBP_NEG | 74 | 0.040434 | 0.002527 | 0.056735 | 0.23803 | 4047.7 |

| | | | | | | |
|---|---|---|---|---|---|---|
| GENTLES_L | 28 | 0.080426 | 0.003095 | 0.11286 | 0.23804 | 4047.87 |
| REACTOME | 19 | 0.098322 | 0.003118 | 0.13799 | 0.23807 | 4048.38 |
| GOBP_OPE | 5 | 0.18849 | 0.003067 | 0.26457 | 0.2381 | 4048.891 |
| GOBP_POS | 6 | 0.15614 | 0.002783 | 0.21921 | 0.23815 | 4049.741 |
| DAVICIONI_ | 62 | 0.049253 | 0.002818 | 0.069163 | 0.23819 | 4050.421 |
| GOMF_NU | 24 | 0.077682 | 0.002768 | 0.10913 | 0.23829 | 4052.121 |
| GOBP_REG | 17 | 0.07976 | 0.002392 | 0.11206 | 0.23832 | 4052.632 |
| SHIPP_DLB | 40 | 0.051464 | 0.002367 | 0.072345 | 0.23843 | 4054.502 |
| GOMF_UBI | 307 | 0.020513 | 0.002595 | 0.028842 | 0.23848 | 4055.352 |
| GOMF_NU | 9 | 0.11351 | 0.002478 | 0.15961 | 0.2385 | 4055.693 |
| GOBP_INTI | 1471 | 0.009325 | 0.0025 | 0.013116 | 0.23856 | 4056.713 |
| GOBP_REG | 151 | 0.03042 | 0.00271 | 0.042794 | 0.23859 | 4057.223 |
| GOBP_POS | 9 | 0.12194 | 0.002662 | 0.17155 | 0.2386 | 4057.393 |
| GOBP_REG | 116 | 0.031631 | 0.002472 | 0.044503 | 0.23862 | 4057.733 |
| GOBP_EPIT | 28 | 0.076741 | 0.002953 | 0.10799 | 0.23866 | 4058.413 |
| MASSARW | 61 | 0.050461 | 0.002864 | 0.071015 | 0.23868 | 4058.753 |
| DORN_ADE | 11 | 0.10206 | 0.002463 | 0.14367 | 0.23874 | 4059.774 |
| GOBP_POS | 5 | 0.15015 | 0.002443 | 0.2115 | 0.23888 | 4062.154 |
| GOMF_AM | 20 | 0.089932 | 0.002926 | 0.12671 | 0.23893 | 4063.005 |
| GOBP_VIRA | 17 | 0.079922 | 0.002397 | 0.11261 | 0.23894 | 4063.175 |
| BIOCARTA_ | 13 | 0.10021 | 0.002629 | 0.14123 | 0.23899 | 4064.025 |
| WP_MRNA | 8 | 0.12179 | 0.002507 | 0.17168 | 0.23905 | 4065.045 |
| GOBP_LEN | 84 | 0.043606 | 0.002902 | 0.061479 | 0.23908 | 4065.555 |
| GOBP_STR | 40 | 0.054887 | 0.002524 | 0.077407 | 0.23915 | 4066.746 |
| GOBP_NEG | 54 | 0.04635 | 0.002475 | 0.06537 | 0.23916 | 4066.916 |
| VART_KSHV | 129 | 0.033184 | 0.002734 | 0.046807 | 0.23918 | 4067.256 |
| GOBP_NEG | 9 | 0.13137 | 0.002868 | 0.18532 | 0.2392 | 4067.596 |
| GOBP_NEG | 4 | 0.202 | 0.00294 | 0.28503 | 0.23926 | 4068.616 |
| CHIANG_LI | 47 | 0.051525 | 0.002568 | 0.072714 | 0.23929 | 4069.126 |
| GOBP_RES | 101 | 0.036041 | 0.002629 | 0.050891 | 0.23942 | 4071.337 |
| ZHOU_INFI | 263 | 0.021539 | 0.002525 | 0.03043 | 0.23953 | 4073.208 |
| GOBP_CELI | 13 | 0.10545 | 0.002766 | 0.14899 | 0.23957 | 4073.888 |
| BIOCARTA_ | 27 | 0.064913 | 0.002453 | 0.091759 | 0.23965 | 4075.248 |
| REACTOME | 10 | 0.10711 | 0.002465 | 0.15141 | 0.23965 | 4075.248 |
| GOBP_NAT | 68 | 0.044811 | 0.002685 | 0.063357 | 0.23971 | 4076.269 |
| REACTOME | 15 | 0.084339 | 0.002376 | 0.11927 | 0.23975 | 4076.949 |
| GOBP_PUR | 25 | 0.073252 | 0.002664 | 0.10361 | 0.23978 | 4077.459 |
| PID_GLYPIC | 27 | 0.0766 | 0.002895 | 0.10837 | 0.23983 | 4078.309 |
| GOBP_STEI | 6 | 0.15042 | 0.002681 | 0.21288 | 0.23992 | 4079.84 |
| GOBP_LIPII | 113 | 0.031449 | 0.002426 | 0.044513 | 0.23994 | 4080.18 |
| GOBP_ESTI | 35 | 0.06586 | 0.002833 | 0.093239 | 0.23999 | 4081.03 |
| REACTOME | 9 | 0.10671 | 0.002329 | 0.15108 | 0.24001 | 4081.37 |
| GOBP_KILL | 25 | 0.070377 | 0.002559 | 0.099659 | 0.24004 | 4081.88 |
| GOBP_DNA | 187 | 0.02496 | 0.002472 | 0.035367 | 0.24018 | 4084.261 |
| REACTOME | 6 | 0.14039 | 0.002503 | 0.19899 | 0.24025 | 4085.451 |

| | | | | | | |
|---|---|---|---|---|---|---|
| REACTOME | 177 | 0.025781 | 0.002485 | 0.036579 | 0.24047 | 4089.192 |
| PLASARI_T | 237 | 0.024219 | 0.002697 | 0.034386 | 0.24062 | 4091.743 |
| MIKKELSEN | 425 | 0.018476 | 0.002741 | 0.02624 | 0.24069 | 4092.933 |
| GOCC_PRC | 12 | 0.086534 | 0.002181 | 0.12299 | 0.24085 | 4095.654 |
| REACTOME | 21 | 0.071413 | 0.002381 | 0.10151 | 0.24088 | 4096.164 |
| YAO_TEMP | 70 | 0.040406 | 0.002456 | 0.057449 | 0.24093 | 4097.015 |
| SHIN_B_CE | 17 | 0.088178 | 0.002645 | 0.12544 | 0.24104 | 4098.885 |
| GOBP_REG | 217 | 0.024515 | 0.002613 | 0.034898 | 0.2412 | 4101.606 |
| GOBP_PRC | 7 | 0.16483 | 0.003173 | 0.23466 | 0.24122 | 4101.946 |
| SABATES_C | 19 | 0.080046 | 0.002538 | 0.11398 | 0.24125 | 4102.456 |
| GOMF_STR | 36 | 0.074625 | 0.003256 | 0.10627 | 0.24128 | 4102.966 |
| REACTOME | 35 | 0.062854 | 0.002704 | 0.08953 | 0.24133 | 4103.817 |
| GOBP_POS | 48 | 0.046801 | 0.002357 | 0.066671 | 0.24136 | 4104.327 |
| GOBP_AMI | 106 | 0.032693 | 0.002443 | 0.046612 | 0.24154 | 4107.388 |
| GOBP_POS | 37 | 0.06171 | 0.002729 | 0.087989 | 0.24155 | 4107.558 |
| GOBP_NEG | 116 | 0.033293 | 0.002602 | 0.047506 | 0.24171 | 4110.279 |
| GOMF_VOI | 84 | 0.043875 | 0.00292 | 0.062624 | 0.24178 | 4111.469 |
| DUNNE_TA | 50 | 0.050671 | 0.002604 | 0.072338 | 0.24182 | 4112.149 |
| GOMF_UBI | 11 | 0.10708 | 0.002584 | 0.15288 | 0.24184 | 4112.489 |
| GOBP_LIPC | 132 | 0.029985 | 0.002499 | 0.042814 | 0.24185 | 4112.659 |
| BOYLAN_M | 89 | 0.038004 | 0.002603 | 0.054273 | 0.24189 | 4113.339 |
| ACEVEDO_ | 339 | 0.017953 | 0.002384 | 0.025641 | 0.24192 | 4113.85 |
| GOBP_POS | 5 | 0.18245 | 0.002969 | 0.2606 | 0.24193 | 4114.02 |
| BOYAULT_ | 16 | 0.088839 | 0.002585 | 0.12692 | 0.24197 | 4114.7 |
| GOMF_K63 | 10 | 0.11679 | 0.002687 | 0.16689 | 0.24203 | 4115.72 |
| GOCC_POS | 617 | 0.014814 | 0.002634 | 0.021177 | 0.24211 | 4117.081 |
| GOBP_POS | 28 | 0.069905 | 0.00269 | 0.099943 | 0.24214 | 4117.591 |
| GOBP_POT | 221 | 0.025589 | 0.002752 | 0.036605 | 0.24227 | 4119.801 |
| REACTOME | 10 | 0.11609 | 0.002671 | 0.16613 | 0.24235 | 4121.162 |
| GOBP_REG | 41 | 0.060491 | 0.002816 | 0.086581 | 0.24239 | 4121.842 |
| IVANOVA_ | 180 | 0.026904 | 0.002615 | 0.038528 | 0.24249 | 4123.542 |
| GOBP_REG | 13 | 0.10512 | 0.002758 | 0.15054 | 0.24251 | 4123.883 |
| BIOCARTA_ | 20 | 0.085568 | 0.002784 | 0.12263 | 0.24266 | 4126.433 |
| GOBP_GOI | 2 | 0.21819 | 0.002246 | 0.31277 | 0.24271 | 4127.284 |
| GOBP_BON | 96 | 0.037681 | 0.00268 | 0.054021 | 0.24274 | 4127.794 |
| GOBP_REG | 67 | 0.042685 | 0.002538 | 0.061217 | 0.24282 | 4129.154 |
| JACKSON_I | 20 | 0.081209 | 0.002642 | 0.11649 | 0.24287 | 4130.004 |
| REACTOME | 13 | 0.12206 | 0.003202 | 0.1751 | 0.24288 | 4130.174 |
| GOMF_TRA | 252 | 0.02142 | 0.002458 | 0.03073 | 0.24289 | 4130.344 |
| GOCC_GPI_ | 5 | 0.15376 | 0.002502 | 0.22061 | 0.2429 | 4130.515 |
| GOBP_REG | 18 | 0.085667 | 0.002644 | 0.12293 | 0.24295 | 4131.365 |
| MIKKELSEN | 160 | 0.026494 | 0.002429 | 0.038021 | 0.24296 | 4131.535 |
| FIRESTEIN_ | 26 | 0.075424 | 0.002797 | 0.10826 | 0.24301 | 4132.385 |
| REACTOME | 9 | 0.12372 | 0.002701 | 0.1776 | 0.24304 | 4132.895 |
| HASINA_N | 11 | 0.10697 | 0.002581 | 0.15358 | 0.24307 | 4133.405 |

| | | | | | | |
|---|---|---|---|---|---|---|
| BURTON_A | 108 | 0.030107 | 0.002271 | 0.043261 | 0.24324 | 4136.296 |
| GOBP_GEN | 44 | 0.056505 | 0.002725 | 0.081206 | 0.24327 | 4136.806 |
| GOBP_NOF | 4 | 0.215 | 0.003129 | 0.30909 | 0.24335 | 4138.167 |
| GOBP_CAR | 201 | 0.024126 | 0.002476 | 0.034697 | 0.24343 | 4139.527 |
| GOBP_REG | 30 | 0.067252 | 0.002679 | 0.096733 | 0.24346 | 4140.037 |
| GOBP_CELI | 4 | 0.17194 | 0.002503 | 0.24743 | 0.24357 | 4141.908 |
| GOBP_RES | 4 | 0.17194 | 0.002503 | 0.24743 | 0.24357 | 4141.908 |
| WP_ARSEN | 9 | 0.11349 | 0.002478 | 0.16338 | 0.24363 | 4142.928 |
| SASSON_RI | 80 | 0.03913 | 0.002542 | 0.056335 | 0.24366 | 4143.438 |
| REACTOME | 9 | 0.12668 | 0.002765 | 0.18249 | 0.24379 | 4145.649 |
| GOBP_RES | 241 | 0.022486 | 0.002524 | 0.03241 | 0.2439 | 4147.52 |
| GOBP_RES | 14 | 0.1013 | 0.002758 | 0.14607 | 0.24401 | 4149.39 |
| BIOCARTA_ | 6 | 0.12462 | 0.002221 | 0.17973 | 0.24404 | 4149.9 |
| GOMF_OXI | 37 | 0.067689 | 0.002994 | 0.09763 | 0.24406 | 4150.24 |
| GOMF_IMI | 6 | 0.11981 | 0.002136 | 0.17284 | 0.24409 | 4150.75 |
| GOBP_PRC | 270 | 0.020473 | 0.002431 | 0.029534 | 0.2441 | 4150.921 |
| GOCC_PTW | 6 | 0.12021 | 0.002143 | 0.17349 | 0.24419 | 4152.451 |
| GOBP_POS | 41 | 0.053394 | 0.002486 | 0.077077 | 0.24424 | 4153.301 |
| GOBP_POS | 163 | 0.026234 | 0.002427 | 0.037913 | 0.24449 | 4157.552 |
| WP_MICRC | 79 | 0.03876 | 0.002502 | 0.056021 | 0.24451 | 4157.893 |
| GOBP_SKEI | 7 | 0.14614 | 0.002814 | 0.21123 | 0.24452 | 4158.063 |
| GOBP_SYN | 22 | 0.065798 | 0.002245 | 0.095103 | 0.24452 | 4158.063 |
| GOBP_REG | 19 | 0.068527 | 0.002173 | 0.099072 | 0.24457 | 4158.913 |
| GOCC_SMI | 14 | 0.11272 | 0.003069 | 0.16299 | 0.2446 | 4159.423 |
| GOBP_POS | 15 | 0.079761 | 0.002247 | 0.11542 | 0.24478 | 4162.484 |
| GOBP_MET | 30 | 0.066933 | 0.002666 | 0.096867 | 0.2448 | 4162.824 |
| GOBP_PRC | 6 | 0.1271 | 0.002266 | 0.18395 | 0.2448 | 4162.824 |
| GOBP_POS | 10 | 0.11854 | 0.002728 | 0.17169 | 0.24497 | 4165.715 |
| GOBP_POS | 25 | 0.067021 | 0.002437 | 0.097116 | 0.24507 | 4167.415 |
| REACTOME | 9 | 0.13044 | 0.002847 | 0.18913 | 0.24521 | 4169.796 |
| GOBP_ECT | 4 | 0.19639 | 0.002858 | 0.28482 | 0.24525 | 4170.476 |
| WP_TRANS | 13 | 0.10361 | 0.002718 | 0.15031 | 0.24531 | 4171.497 |
| SHEPARD_ | 202 | 0.023393 | 0.002407 | 0.033938 | 0.24533 | 4171.837 |
| PID_INTEG | 63 | 0.047766 | 0.002755 | 0.069315 | 0.24538 | 4172.687 |
| GOBP_INTI | 10 | 0.10543 | 0.002426 | 0.15307 | 0.24548 | 4174.387 |
| GOBP_EYE_ | 152 | 0.030355 | 0.002713 | 0.044073 | 0.2455 | 4174.728 |
| BIOCARTA_ | 10 | 0.10294 | 0.002369 | 0.14951 | 0.24557 | 4175.918 |
| GOBP_IMP | 5 | 0.14995 | 0.00244 | 0.21781 | 0.2456 | 4176.428 |
| GOMF_MA | 10 | 0.10033 | 0.002309 | 0.14575 | 0.24561 | 4176.598 |
| GOBP_HEX | 38 | 0.054671 | 0.00245 | 0.079425 | 0.24562 | 4176.768 |
| REACTOME | 76 | 0.041365 | 0.002619 | 0.060114 | 0.2457 | 4178.129 |
| MATTHEW | 17 | 0.086643 | 0.002599 | 0.12592 | 0.24571 | 4178.299 |
| GOBP_PRC | 23 | 0.070233 | 0.00245 | 0.1021 | 0.24577 | 4179.319 |
| VALK_AML | 35 | 0.064169 | 0.002761 | 0.093289 | 0.24578 | 4179.489 |
| NABA_ECM | 183 | 0.027486 | 0.002693 | 0.039975 | 0.24586 | 4180.849 |

| | | | | | | |
|---|---|---|---|---|---|---|
| LI_CYTIDIN | 17 | 0.087513 | 0.002625 | 0.12737 | 0.24602 | 4183.57 |
| GOBP_GLC | 17 | 0.099369 | 0.002981 | 0.14463 | 0.24603 | 4183.74 |
| GOBP_POS | 111 | 0.032784 | 0.002507 | 0.047734 | 0.24611 | 4185.101 |
| YANAGISAV | 6 | 0.15452 | 0.002754 | 0.22502 | 0.24614 | 4185.611 |
| CHANG_IM | 58 | 0.044349 | 0.002455 | 0.064606 | 0.24622 | 4186.971 |
| GOBP_RES | 5 | 0.15878 | 0.002584 | 0.23154 | 0.24644 | 4190.712 |
| GOBP_L_PI | 8 | 0.12325 | 0.002537 | 0.17975 | 0.24646 | 4191.052 |
| WP_KALLM | 23 | 0.072863 | 0.002542 | 0.10626 | 0.24646 | 4191.052 |
| KEGG_N_G | 45 | 0.049012 | 0.00239 | 0.071531 | 0.24662 | 4193.773 |
| GOCC_JUN | 9 | 0.13876 | 0.003029 | 0.20259 | 0.24669 | 4194.963 |
| GOCC_BAR | 5 | 0.16438 | 0.002675 | 0.24013 | 0.24683 | 4197.344 |
| GOBP_REG | 19 | 0.076111 | 0.002413 | 0.1112 | 0.24686 | 4197.854 |
| REACTOME | 7 | 0.11167 | 0.00215 | 0.16316 | 0.24686 | 4197.854 |
| GOBP_IROI | 27 | 0.067331 | 0.002545 | 0.098409 | 0.24693 | 4199.045 |
| GOBP_REG | 7 | 0.12759 | 0.002457 | 0.18649 | 0.24694 | 4199.215 |
| MYLLYKAN | 19 | 0.076324 | 0.00242 | 0.11158 | 0.24698 | 4199.895 |
| WILLIAMS_ | 28 | 0.057992 | 0.002232 | 0.08478 | 0.24698 | 4199.895 |
| GOBP_HIPI | 7 | 0.13571 | 0.002613 | 0.19842 | 0.24702 | 4200.575 |
| REACTOME | 72 | 0.040398 | 0.00249 | 0.05907 | 0.24702 | 4200.575 |
| WP_INSULI | 154 | 0.027859 | 0.002506 | 0.040775 | 0.24723 | 4204.146 |
| GOBP_DNA | 6 | 0.11158 | 0.001989 | 0.16332 | 0.24724 | 4204.316 |
| GOBP_PUR | 30 | 0.062136 | 0.002475 | 0.090962 | 0.24728 | 4204.996 |
| PID_NEPHI | 31 | 0.059241 | 0.002399 | 0.086749 | 0.24734 | 4206.017 |
| WP_APOPT | 50 | 0.049665 | 0.002553 | 0.072752 | 0.24742 | 4207.377 |
| GOBP_REG | 70 | 0.043181 | 0.002625 | 0.063281 | 0.24751 | 4208.908 |
| KEGG_CYT | 239 | 0.02531 | 0.00283 | 0.037096 | 0.24753 | 4209.248 |
| WP_VITAM | 34 | 0.058053 | 0.002462 | 0.085083 | 0.24753 | 4209.248 |
| GOBP_SIN | 20 | 0.085886 | 0.002794 | 0.12589 | 0.24756 | 4209.758 |
| GOBP_RES | 85 | 0.0373 | 0.002497 | 0.054697 | 0.24765 | 4211.288 |
| GOBP_NUC | 14 | 0.086752 | 0.002362 | 0.12727 | 0.24773 | 4212.649 |
| REACTOME | 11 | 0.11018 | 0.002659 | 0.16166 | 0.24777 | 4213.329 |
| WP_PHOT | 35 | 0.058474 | 0.002516 | 0.085795 | 0.24777 | 4213.329 |
| GOBP_PIN | 23 | 0.06677 | 0.002329 | 0.098041 | 0.24793 | 4216.05 |
| GOBP_CELI | 90 | 0.035642 | 0.002455 | 0.05234 | 0.24795 | 4216.39 |
| DOANE_BR | 33 | 0.062541 | 0.002613 | 0.091876 | 0.24803 | 4217.75 |
| GOBP_CRA | 29 | 0.066803 | 0.002616 | 0.098145 | 0.24805 | 4218.09 |
| MCDOWEL | 41 | 0.051787 | 0.002411 | 0.07609 | 0.24806 | 4218.26 |
| KEGG_VEG | 76 | 0.039086 | 0.002475 | 0.057454 | 0.24816 | 4219.961 |
| GOBP_EST | 142 | 0.026944 | 0.002328 | 0.039624 | 0.24826 | 4221.661 |
| DAWSON_ | 57 | 0.054646 | 0.002998 | 0.080379 | 0.2483 | 4222.342 |
| GOBP_NEG | 57 | 0.045534 | 0.002498 | 0.066987 | 0.24834 | 4223.022 |
| REACTOME | 5 | 0.20253 | 0.003296 | 0.29796 | 0.24834 | 4223.022 |
| GOMF_PH | 4 | 0.15888 | 0.002313 | 0.23381 | 0.2484 | 4224.042 |
| REACTOME | 62 | 0.046615 | 0.002667 | 0.06864 | 0.24853 | 4226.253 |
| GOMF_PR | 5 | 0.18373 | 0.00299 | 0.27055 | 0.24854 | 4226.423 |

| | | | | | | |
|---|---|---|---|---|---|---|
| GOBP_ACT | 7 | 0.1322 | 0.002545 | 0.19469 | 0.24856 | 4226.763 |
| LAMB_CCN | 17 | 0.077137 | 0.002314 | 0.11367 | 0.24869 | 4228.973 |
| GOBP_POS | 13 | 0.089984 | 0.002361 | 0.13265 | 0.24877 | 4230.334 |
| GOBP_NEP | 9 | 0.14023 | 0.003061 | 0.20682 | 0.24887 | 4232.034 |
| WP_TGFBE | 51 | 0.051195 | 0.002657 | 0.07551 | 0.24889 | 4232.374 |
| GOMF_LYS | 8 | 0.11597 | 0.002387 | 0.17107 | 0.24892 | 4232.885 |
| GOBP_NEG | 43 | 0.049313 | 0.002351 | 0.07276 | 0.24897 | 4233.735 |
| ZHENG_FO | 194 | 0.02646 | 0.002669 | 0.039058 | 0.24906 | 4235.265 |
| DARWICHE | 174 | 0.024669 | 0.002358 | 0.036421 | 0.2491 | 4235.946 |
| PODAR_RE | 18 | 0.078409 | 0.00242 | 0.11578 | 0.24914 | 4236.626 |
| WU_HBX_T | 21 | 0.078742 | 0.002625 | 0.11629 | 0.24917 | 4237.136 |
| GOBP_ICOS | 7 | 0.10941 | 0.002107 | 0.1616 | 0.24919 | 4237.476 |
| GOMF_SIN | 10 | 0.13218 | 0.003041 | 0.19522 | 0.24919 | 4237.476 |
| HERNANDE | 58 | 0.042668 | 0.002361 | 0.063065 | 0.24934 | 4240.027 |
| GOBP_NEG | 62 | 0.043408 | 0.002484 | 0.064205 | 0.2495 | 4242.748 |
| GOBP_MO | 90 | 0.036414 | 0.002508 | 0.053875 | 0.24955 | 4243.598 |
| MEISSNER_ | 14 | 0.080479 | 0.002191 | 0.11911 | 0.24963 | 4244.958 |
| GOBP_CELI | 12 | 0.10542 | 0.002657 | 0.15603 | 0.24965 | 4245.298 |
| REACTOME | 6 | 0.11964 | 0.002133 | 0.17713 | 0.24971 | 4246.319 |
| GOBP_REG | 38 | 0.056506 | 0.002533 | 0.083723 | 0.24987 | 4249.039 |
| GOBP_POS | 14 | 0.087957 | 0.002394 | 0.13033 | 0.24988 | 4249.209 |
| GOCC_MU | 7 | 0.13556 | 0.00261 | 0.20096 | 0.24998 | 4250.91 |
| GOBP_VITA | 11 | 0.11651 | 0.002812 | 0.17273 | 0.25 | 4251.25 |
| GOBP_TEST | 6 | 0.15935 | 0.00284 | 0.23626 | 0.25001 | 4251.42 |
| GOBP_REG | 24 | 0.077808 | 0.002773 | 0.11541 | 0.2501 | 4252.951 |
| PID_IL5_PA | 14 | 0.092998 | 0.002532 | 0.13795 | 0.25011 | 4253.121 |
| GOBP_POS | 33 | 0.053029 | 0.002215 | 0.078697 | 0.25021 | 4254.821 |
| GOBP_FEM | 117 | 0.032567 | 0.002556 | 0.048334 | 0.25023 | 4255.161 |
| GOBP_LEU | 5 | 0.13559 | 0.002206 | 0.20127 | 0.25026 | 4255.671 |
| GOBP_MO | 19 | 0.079571 | 0.002523 | 0.11834 | 0.25066 | 4262.473 |
| GHANDHI_ | 11 | 0.12731 | 0.003072 | 0.18935 | 0.25067 | 4262.643 |
| GOBP_PRO | 5 | 0.1479 | 0.002407 | 0.22005 | 0.25076 | 4264.174 |
| GOBP_REG | 540 | 0.015401 | 0.002567 | 0.022913 | 0.25076 | 4264.174 |
| GOBP_OOC | 29 | 0.058824 | 0.002304 | 0.087553 | 0.25084 | 4265.534 |
| CADWELL_ | 100 | 0.03464 | 0.002515 | 0.051606 | 0.25104 | 4268.935 |
| HUANG_DA | 68 | 0.041921 | 0.002511 | 0.062461 | 0.25107 | 4269.445 |
| GOBP_POS | 19 | 0.090643 | 0.002874 | 0.13507 | 0.25108 | 4269.615 |
| GOBP_RES | 156 | 0.027808 | 0.002517 | 0.041467 | 0.25124 | 4272.336 |
| GOBP_BON | 37 | 0.0558 | 0.002468 | 0.083222 | 0.25128 | 4273.016 |
| GOMF_CHI | 26 | 0.064848 | 0.002405 | 0.096761 | 0.25137 | 4274.547 |
| GOBP_REG | 41 | 0.055112 | 0.002566 | 0.082236 | 0.25138 | 4274.717 |
| GOBP_NEG | 20 | 0.080583 | 0.002622 | 0.12025 | 0.25139 | 4274.887 |
| REACTOME | 36 | 0.052241 | 0.002279 | 0.077963 | 0.25141 | 4275.227 |
| GOBP_REG | 18 | 0.075525 | 0.002331 | 0.11275 | 0.25148 | 4276.417 |
| WP_ALPHA | 33 | 0.06311 | 0.002636 | 0.094241 | 0.25154 | 4277.438 |

| | | | | | | |
|---|---|---|---|---|---|---|
| REACTOME | 103 | 0.03073 | 0.002264 | 0.045899 | 0.25159 | 4278.288 |
| GOBP_NEG | 13 | 0.084349 | 0.002213 | 0.126 | 0.25162 | 4278.798 |
| REACTOME | 112 | 0.03165 | 0.002431 | 0.047288 | 0.25166 | 4279.478 |
| GOBP_POS | 11 | 0.10113 | 0.002441 | 0.15117 | 0.25175 | 4281.009 |
| GOMF_HEX | 5 | 0.1479 | 0.002407 | 0.22112 | 0.25179 | 4281.689 |
| BLANCO_M | 272 | 0.021415 | 0.002552 | 0.032017 | 0.2518 | 4281.859 |
| GOCC_VOL | 8 | 0.13114 | 0.002699 | 0.19615 | 0.25189 | 4283.389 |
| REACTOME | 17 | 0.08031 | 0.002409 | 0.12015 | 0.25194 | 4284.24 |
| GOBP_REG | 8 | 0.10976 | 0.002259 | 0.16429 | 0.25205 | 4286.11 |
| PRAMOON | 48 | 0.049222 | 0.002479 | 0.073706 | 0.25213 | 4287.471 |
| GOBP_ACT | 20 | 0.077354 | 0.002517 | 0.11586 | 0.25218 | 4288.321 |
| WP_TGFBE | 55 | 0.047924 | 0.002583 | 0.071782 | 0.25219 | 4288.491 |
| GOBP_PRC | 182 | 0.025024 | 0.002445 | 0.037522 | 0.25242 | 4292.402 |
| GOBP_NEG | 8 | 0.1409 | 0.0029 | 0.21128 | 0.25243 | 4292.572 |
| WONG_MI | 202 | 0.021841 | 0.002247 | 0.032753 | 0.25244 | 4292.742 |
| GOBP_CUT | 5 | 0.13743 | 0.002236 | 0.20611 | 0.25245 | 4292.912 |
| GOBP_NEG | 80 | 0.040241 | 0.002614 | 0.060387 | 0.25259 | 4295.293 |
| GOMF_NU | 130 | 0.028943 | 0.002394 | 0.043442 | 0.25263 | 4295.973 |
| REACTOME | 132 | 0.028511 | 0.002376 | 0.042803 | 0.25268 | 4296.823 |
| KEGG_CELL | 121 | 0.028709 | 0.002291 | 0.043124 | 0.25279 | 4298.694 |
| REACTOME | 34 | 0.05798 | 0.002458 | 0.087146 | 0.25293 | 4301.075 |
| GOBP_POS | 1495 | 0.009247 | 0.002497 | 0.013905 | 0.25303 | 4302.775 |
| GOMF_PUI | 15 | 0.082869 | 0.002335 | 0.12464 | 0.25308 | 4303.625 |
| REACTOME | 188 | 0.025368 | 0.002519 | 0.038159 | 0.2531 | 4303.966 |
| GOBP_LEU | 30 | 0.059931 | 0.002387 | 0.090195 | 0.2532 | 4305.666 |
| GOBP_NEG | 93 | 0.033338 | 0.002334 | 0.050186 | 0.25326 | 4306.686 |
| GOBP_REG | 27 | 0.070524 | 0.002665 | 0.10617 | 0.25327 | 4306.856 |
| GOBP_CAR | 82 | 0.038505 | 0.002532 | 0.057971 | 0.25328 | 4307.026 |
| GOBP_ECT | 12 | 0.097802 | 0.002465 | 0.14724 | 0.25328 | 4307.026 |
| GOCC_UBI | 7 | 0.097032 | 0.001868 | 0.14627 | 0.25355 | 4311.618 |
| GOBP_RES | 65 | 0.040624 | 0.00238 | 0.061252 | 0.2536 | 4312.468 |
| GOBP_INTE | 4 | 0.19876 | 0.002893 | 0.29983 | 0.25369 | 4313.998 |
| GOBP_NEG | 591 | 0.014069 | 0.00245 | 0.021223 | 0.25369 | 4313.998 |
| GOBP_SCH | 9 | 0.13743 | 0.003 | 0.20731 | 0.25369 | 4313.998 |
| WP_FOXP3 | 13 | 0.10627 | 0.002788 | 0.16031 | 0.2537 | 4314.169 |
| GOBP_REG | 6 | 0.13021 | 0.002321 | 0.19644 | 0.25372 | 4314.509 |
| GOCC_EXO | 19 | 0.074649 | 0.002367 | 0.11263 | 0.25373 | 4314.679 |
| GARY_CD5 | 437 | 0.015238 | 0.002292 | 0.022992 | 0.25374 | 4314.849 |
| BIOCARTA_ | 22 | 0.069851 | 0.002383 | 0.10541 | 0.25377 | 4315.359 |
| GOBP_DEP | 5 | 0.11988 | 0.001951 | 0.18095 | 0.25382 | 4316.209 |
| YANG_BCL | 8 | 0.10604 | 0.002183 | 0.16007 | 0.25383 | 4316.379 |
| GOCC_MYC | 24 | 0.073369 | 0.002614 | 0.11078 | 0.2539 | 4317.57 |
| MCMURRA | 65 | 0.04348 | 0.002547 | 0.065689 | 0.25402 | 4319.61 |
| HATADA_M | 32 | 0.064697 | 0.002661 | 0.097747 | 0.25403 | 4319.78 |
| GUTIERREZ | 9 | 0.11085 | 0.00242 | 0.16749 | 0.25405 | 4320.12 |

| | | | | | | |
|---|---|---|---|---|---|---|
| GOBP_T_H | 18 | 0.084385 | 0.002604 | 0.12751 | 0.25406 | 4320.29 |
| GOBP_NEU | 5 | 0.16947 | 0.002758 | 0.25619 | 0.25414 | 4321.651 |
| HIRSCH_CE | 96 | 0.033075 | 0.002353 | 0.050005 | 0.25417 | 4322.161 |
| GOMF_BM | 7 | 0.14245 | 0.002743 | 0.21543 | 0.25424 | 4323.351 |
| BAKER_HEI | 7 | 0.14532 | 0.002798 | 0.21989 | 0.25435 | 4325.222 |
| GOCC_NUC | 10 | 0.092272 | 0.002123 | 0.13963 | 0.25436 | 4325.392 |
| PUJANA_BI | 110 | 0.030359 | 0.002311 | 0.045938 | 0.25436 | 4325.392 |
| BIOCARTA_ | 21 | 0.073324 | 0.002444 | 0.11099 | 0.25443 | 4326.582 |
| WIERENGA | 190 | 0.024126 | 0.002408 | 0.036522 | 0.25444 | 4326.752 |
| MOREAUX_ | 75 | 0.034979 | 0.0022 | 0.052954 | 0.25445 | 4326.922 |
| GOBP_NEG | 53 | 0.052167 | 0.00276 | 0.07899 | 0.2545 | 4327.773 |
| REACTOME | 40 | 0.054174 | 0.002491 | 0.082041 | 0.25453 | 4328.283 |
| REACTOME | 11 | 0.10125 | 0.002444 | 0.15335 | 0.25454 | 4328.453 |
| GOBP_MO | 127 | 0.026716 | 0.002184 | 0.04047 | 0.25459 | 4329.303 |
| WP_OXIDA | 37 | 0.054882 | 0.002427 | 0.083169 | 0.25467 | 4330.663 |
| JINESH_BLI | 159 | 0.028143 | 0.002572 | 0.042655 | 0.2547 | 4331.174 |
| WP_LEPTIN | 72 | 0.038165 | 0.002353 | 0.057873 | 0.2548 | 4332.874 |
| DAIRKEE_C | 28 | 0.062977 | 0.002424 | 0.095501 | 0.25481 | 4333.044 |
| REACTOME | 50 | 0.047634 | 0.002448 | 0.072255 | 0.25487 | 4334.064 |
| REACTOME | 6 | 0.15219 | 0.002713 | 0.23088 | 0.25489 | 4334.404 |
| GOBP_AST | 8 | 0.12756 | 0.002625 | 0.19373 | 0.25513 | 4338.486 |
| GOBP_PRC | 825 | 0.0115 | 0.002351 | 0.01748 | 0.2553 | 4341.377 |
| GOBP_BAS | 33 | 0.063881 | 0.002669 | 0.097099 | 0.25531 | 4341.547 |
| BIOCARTA_ | 8 | 0.12463 | 0.002565 | 0.18949 | 0.25536 | 4342.397 |
| GOBP_IRIS | 9 | 0.11728 | 0.00256 | 0.17834 | 0.25539 | 4342.907 |
| YAO_TEMP | 13 | 0.083035 | 0.002178 | 0.12634 | 0.25551 | 4344.948 |
| KOBAYASH | 7 | 0.12154 | 0.00234 | 0.18498 | 0.25558 | 4346.138 |
| GOBP_WH | 17 | 0.080418 | 0.002412 | 0.12243 | 0.25564 | 4347.158 |
| GOBP_CELI | 268 | 0.019119 | 0.002262 | 0.029108 | 0.25565 | 4347.328 |
| KEGG_PEN | 23 | 0.070887 | 0.002473 | 0.10792 | 0.25565 | 4347.328 |
| PID_FGF_P | 55 | 0.044359 | 0.002391 | 0.067546 | 0.25568 | 4347.838 |
| PID_GMCS | 35 | 0.058013 | 0.002496 | 0.088342 | 0.25569 | 4348.008 |
| REACTOME | 122 | 0.028831 | 0.00231 | 0.043905 | 0.2557 | 4348.179 |
| BIOCARTA_ | 20 | 0.070877 | 0.002306 | 0.10796 | 0.25574 | 4348.859 |
| GOBP_EST/ | 16 | 0.075381 | 0.002194 | 0.11482 | 0.25574 | 4348.859 |
| GOBP_REG | 8 | 0.12023 | 0.002475 | 0.18321 | 0.25584 | 4350.559 |
| GOBP_PUR | 6 | 0.13127 | 0.00234 | 0.20014 | 0.25595 | 4352.43 |
| GOBP_POS | 6 | 0.15283 | 0.002724 | 0.23323 | 0.25616 | 4356.001 |
| GOBP_OVA | 56 | 0.043773 | 0.002381 | 0.066828 | 0.25623 | 4357.191 |
| GOBP_ACT | 5 | 0.18533 | 0.003016 | 0.28297 | 0.25626 | 4357.701 |
| BIOCARTA_ | 21 | 0.065314 | 0.002177 | 0.099768 | 0.25635 | 4359.232 |
| REACTOME | 46 | 0.045807 | 0.002258 | 0.069997 | 0.25643 | 4360.592 |
| GOBP_NEG | 7 | 0.14212 | 0.002736 | 0.21723 | 0.25648 | 4361.442 |
| GOBP_N_A | 16 | 0.076553 | 0.002228 | 0.11701 | 0.25649 | 4361.612 |
| GOBP_RES | 8 | 0.11691 | 0.002406 | 0.17879 | 0.25659 | 4363.313 |

| | | | | | | |
|---|---|---|---|---|---|---|
| PID_RHOA_ | 42 | 0.048147 | 0.002268 | 0.073653 | 0.25666 | 4364.503 |
| GOBP_SCH | 30 | 0.061874 | 0.002465 | 0.094681 | 0.25672 | 4365.524 |
| GOBP_RET | 59 | 0.046392 | 0.002589 | 0.071023 | 0.25682 | 4367.224 |
| GOMF_GLU | 5 | 0.12466 | 0.002029 | 0.19088 | 0.25685 | 4367.734 |
| GOBP_REG | 11 | 0.11336 | 0.002736 | 0.17361 | 0.25689 | 4368.414 |
| WP_MIRNA | 21 | 0.083757 | 0.002792 | 0.12831 | 0.25696 | 4369.605 |
| WP_ONCO | 65 | 0.043365 | 0.00254 | 0.066459 | 0.25704 | 4370.965 |
| GOBP_SEC | 11 | 0.10141 | 0.002447 | 0.15545 | 0.25709 | 4371.815 |
| GOBP_POS | 60 | 0.041946 | 0.002361 | 0.064316 | 0.25715 | 4372.836 |
| GOBP_PUR | 10 | 0.10301 | 0.00237 | 0.15795 | 0.25715 | 4372.836 |
| GOBP_TRN | 6 | 0.19973 | 0.00356 | 0.3063 | 0.25718 | 4373.346 |
| GOBP_INO | 164 | 0.026945 | 0.002501 | 0.041324 | 0.25719 | 4373.516 |
| GOBP_NEG | 11 | 0.092495 | 0.002232 | 0.14187 | 0.25722 | 4374.026 |
| GOBP_REG | 15 | 0.083772 | 0.00236 | 0.12854 | 0.25729 | 4375.216 |
| CHIARADO | 117 | 0.031326 | 0.002459 | 0.04809 | 0.2574 | 4377.087 |
| GOBP_POS | 5 | 0.17121 | 0.002786 | 0.26286 | 0.25742 | 4377.427 |
| GOBP_VITA | 18 | 0.077589 | 0.002395 | 0.11919 | 0.25753 | 4379.298 |
| GOBP_NEG | 8 | 0.13537 | 0.002786 | 0.20796 | 0.25756 | 4379.808 |
| BIOCARTA_ | 14 | 0.092212 | 0.00251 | 0.1417 | 0.25761 | 4380.658 |
| MIKKELSEN | 77 | 0.036734 | 0.002341 | 0.056453 | 0.25762 | 4380.828 |
| BIOCARTA_ | 17 | 0.084282 | 0.002528 | 0.12963 | 0.2578 | 4383.889 |
| GOBP_HOM | 58 | 0.040882 | 0.002263 | 0.062893 | 0.25784 | 4384.569 |
| REACTOME | 74 | 0.037862 | 0.002366 | 0.05825 | 0.25785 | 4384.739 |
| GOBP_END | 10 | 0.1179 | 0.002713 | 0.18139 | 0.25786 | 4384.909 |
| PID_INTEG | 22 | 0.069978 | 0.002388 | 0.1077 | 0.25793 | 4386.1 |
| GOBP_REG | 129 | 0.027642 | 0.002277 | 0.042545 | 0.25795 | 4386.44 |
| GOBP_GLIA | 223 | 0.023744 | 0.002565 | 0.036575 | 0.25811 | 4389.161 |
| VANHARAN | 40 | 0.056541 | 0.0026 | 0.087115 | 0.25816 | 4390.011 |
| LY_AGING_ | 6 | 0.14299 | 0.002549 | 0.22038 | 0.25822 | 4391.031 |
| GOBP_END | 120 | 0.02988 | 0.002375 | 0.046055 | 0.25824 | 4391.371 |
| RICKMAN_ | 15 | 0.074035 | 0.002086 | 0.11416 | 0.25834 | 4393.072 |
| GOBP_INN | 5 | 0.15735 | 0.002561 | 0.24271 | 0.2584 | 4394.092 |
| REACTOME | 60 | 0.042215 | 0.002376 | 0.065121 | 0.25841 | 4394.262 |
| WP_EICOSA | 23 | 0.067213 | 0.002345 | 0.10369 | 0.25844 | 4394.772 |
| GOBP_POS | 24 | 0.079394 | 0.002829 | 0.12251 | 0.25847 | 4395.282 |
| GOBP_NEG | 13 | 0.10328 | 0.002709 | 0.15937 | 0.25848 | 4395.452 |
| GOBP_REG | 13 | 0.087089 | 0.002285 | 0.13441 | 0.25853 | 4396.303 |
| GOBP_MET | 5 | 0.16893 | 0.002749 | 0.26095 | 0.25871 | 4399.364 |
| CHEBOTAE | 113 | 0.031712 | 0.002446 | 0.048998 | 0.25875 | 4400.044 |
| PLASARI_T | 192 | 0.025364 | 0.002545 | 0.039191 | 0.25876 | 4400.214 |
| GOBP_SMO | 57 | 0.047648 | 0.002614 | 0.073625 | 0.25877 | 4400.384 |
| GOBP_ORG | 151 | 0.025264 | 0.002251 | 0.039058 | 0.25887 | 4402.084 |
| PID_PLK1_ | 42 | 0.044662 | 0.002104 | 0.06907 | 0.25894 | 4403.275 |
| GOCC_CON | 19 | 0.085435 | 0.002709 | 0.13215 | 0.25898 | 4403.955 |
| GOBP_NEG | 14 | 0.089137 | 0.002427 | 0.13788 | 0.25899 | 4404.125 |

| | | | | | | |
|---|---|---|---|---|---|---|
| GOBP_URE | 7 | 0.13483 | 0.002596 | 0.20856 | 0.25899 | 4404.125 |
| GOBP_TOX | 17 | 0.078007 | 0.00234 | 0.12067 | 0.25901 | 4404.465 |
| GOBP_CELI | 5 | 0.1528 | 0.002486 | 0.23639 | 0.25902 | 4404.635 |
| GOMF_PRC | 14 | 0.072704 | 0.001979 | 0.11251 | 0.25908 | 4405.655 |
| PID_TCR_R | 13 | 0.095379 | 0.002502 | 0.1476 | 0.25908 | 4405.655 |
| REACTOME | 11 | 0.099378 | 0.002398 | 0.1538 | 0.25909 | 4405.825 |
| WAKABAYA | 145 | 0.025116 | 0.002193 | 0.038875 | 0.25912 | 4406.336 |
| JIANG_AGI | 47 | 0.045875 | 0.002286 | 0.071025 | 0.25918 | 4407.356 |
| RIZ_ERYTH | 25 | 0.073524 | 0.002674 | 0.11386 | 0.25923 | 4408.206 |
| GOCC_CYT | 33 | 0.048952 | 0.002045 | 0.075831 | 0.25929 | 4409.226 |
| MORI_LAR | 51 | 0.041975 | 0.002179 | 0.065038 | 0.25934 | 4410.077 |
| HOLLEMAN | 21 | 0.069224 | 0.002308 | 0.10726 | 0.25935 | 4410.247 |
| YAO_TEMP | 178 | 0.022765 | 0.0022 | 0.035294 | 0.25946 | 4412.117 |
| GOBP_GLU | 26 | 0.059391 | 0.002203 | 0.092103 | 0.25952 | 4413.138 |
| SHIN_B_CE | 11 | 0.10229 | 0.002468 | 0.15866 | 0.25957 | 4413.988 |
| KEGG_APO | 79 | 0.036198 | 0.002337 | 0.056158 | 0.25961 | 4414.668 |
| GOCC_ACT | 93 | 0.032286 | 0.002261 | 0.050109 | 0.25969 | 4416.028 |
| VALK_AML | 31 | 0.060902 | 0.002466 | 0.09456 | 0.25977 | 4417.389 |
| GOBP_EXT | 297 | 0.019987 | 0.002487 | 0.03104 | 0.25982 | 4418.239 |
| GOBP_OTI | 10 | 0.12282 | 0.002826 | 0.1909 | 0.26 | 4421.3 |
| GOBP_NEG | 375 | 0.016292 | 0.002273 | 0.025324 | 0.26001 | 4421.47 |
| GOBP_SRP | 17 | 0.072705 | 0.002181 | 0.11301 | 0.26001 | 4421.47 |
| HADDAD_T | 72 | 0.037927 | 0.002338 | 0.058963 | 0.26004 | 4421.98 |
| WP_SILDEN | 22 | 0.072762 | 0.002483 | 0.11312 | 0.26004 | 4421.98 |
| WP_SOMIT | 9 | 0.094494 | 0.002063 | 0.14696 | 0.26012 | 4423.341 |
| GOBP_NUC | 303 | 0.017724 | 0.002227 | 0.027569 | 0.26015 | 4423.851 |
| QI_HYPOXI | 33 | 0.055401 | 0.002314 | 0.086268 | 0.26038 | 4427.762 |
| REACTOME | 292 | 0.019007 | 0.002346 | 0.029599 | 0.26039 | 4427.932 |
| GOBP_TRU | 5 | 0.13137 | 0.002138 | 0.20461 | 0.26042 | 4428.442 |
| GOBP_RES | 16 | 0.072668 | 0.002115 | 0.1132 | 0.26047 | 4429.292 |
| GOBP_POS | 5 | 0.13452 | 0.002189 | 0.20981 | 0.26071 | 4433.374 |
| GOBP_D_A | 6 | 0.16631 | 0.002965 | 0.25939 | 0.26072 | 4433.544 |
| GOBP_NEG | 5 | 0.12837 | 0.002089 | 0.20023 | 0.26072 | 4433.544 |
| KEGG_ALZI | 151 | 0.024054 | 0.002143 | 0.03752 | 0.26074 | 4433.884 |
| POS_HISTA | 30 | 0.063346 | 0.002523 | 0.098816 | 0.26075 | 4434.054 |
| GOBP_REG | 11 | 0.095535 | 0.002306 | 0.14904 | 0.26076 | 4434.224 |
| GOMF_HIS | 4 | 0.14753 | 0.002147 | 0.23017 | 0.26077 | 4434.394 |
| KIM_ALL_D | 517 | 0.014014 | 0.002287 | 0.021876 | 0.2609 | 4436.605 |
| REACTOME | 16 | 0.098095 | 0.002855 | 0.15328 | 0.2611 | 4440.006 |
| MANN_RES | 9 | 0.10915 | 0.002383 | 0.1706 | 0.26116 | 4441.026 |
| HASLINGEF | 22 | 0.06972 | 0.002379 | 0.109 | 0.26121 | 4441.876 |
| TRAYNOR_ | 37 | 0.05775 | 0.002554 | 0.090291 | 0.26122 | 4442.046 |
| GOBP_CELI | 223 | 0.022063 | 0.002384 | 0.034497 | 0.26123 | 4442.216 |
| GOCC_90S_ | 26 | 0.05535 | 0.002053 | 0.086548 | 0.26125 | 4442.556 |
| GOBP_MET | 6 | 0.13432 | 0.002394 | 0.21005 | 0.26127 | 4442.896 |

| | | | | | | |
|---|---|---|---|---|---|---|
| GOCC_ATP | 6 | 0.11695 | 0.002085 | 0.18293 | 0.26132 | 4443.747 |
| GOMF_CAT | 375 | 0.015483 | 0.00216 | 0.024234 | 0.26145 | 4445.957 |
| GOBP_REG | 4 | 0.18275 | 0.00266 | 0.28609 | 0.26148 | 4446.467 |
| GOBP_POS | 11 | 0.08964 | 0.002163 | 0.14041 | 0.2616 | 4448.508 |
| BERENJENC | 28 | 0.060254 | 0.002319 | 0.094397 | 0.26164 | 4449.188 |
| GOBP_TRA | 5 | 0.1033 | 0.001681 | 0.16186 | 0.26168 | 4449.868 |
| GOBP_TRN | 5 | 0.11611 | 0.001889 | 0.18207 | 0.26183 | 4452.419 |
| GOMF_OXI | 125 | 0.029806 | 0.002417 | 0.046747 | 0.26187 | 4453.099 |
| GOCC_COL | 4 | 0.17751 | 0.002584 | 0.27846 | 0.26191 | 4453.78 |
| PID_WNT_ | 20 | 0.06475 | 0.002106 | 0.10163 | 0.26203 | 4455.82 |
| GOBP_PRC | 44 | 0.051801 | 0.002498 | 0.081313 | 0.26205 | 4456.16 |
| MARTINEZ_ | 584 | 0.013211 | 0.002287 | 0.020743 | 0.2621 | 4457.011 |
| GOMF_NU | 17 | 0.069972 | 0.002099 | 0.10987 | 0.26211 | 4457.181 |
| GOBP_HIST | 15 | 0.087226 | 0.002458 | 0.13698 | 0.26214 | 4457.691 |
| PID_TCPTP | 42 | 0.053538 | 0.002522 | 0.084102 | 0.2622 | 4458.711 |
| GOBP_TOL | 17 | 0.077239 | 0.002317 | 0.12134 | 0.26221 | 4458.881 |
| GOMF_DAI | 71 | 0.035582 | 0.002178 | 0.055901 | 0.26222 | 4459.051 |
| GOMF_RNA | 39 | 0.041774 | 0.001897 | 0.06563 | 0.26223 | 4459.221 |
| GOMF_L_G | 7 | 0.11562 | 0.002226 | 0.18168 | 0.26226 | 4459.731 |
| GOBP_FOL | 10 | 0.10674 | 0.002456 | 0.16774 | 0.26228 | 4460.071 |
| TAKEDA_TA | 115 | 0.029357 | 0.002284 | 0.046156 | 0.26238 | 4461.772 |
| GOCC_MYE | 6 | 0.15306 | 0.002728 | 0.24073 | 0.26244 | 4462.792 |
| GOBP_NEG | 6 | 0.12321 | 0.002196 | 0.19377 | 0.26245 | 4462.962 |
| GOBP_REC | 10 | 0.12054 | 0.002774 | 0.18962 | 0.26248 | 4463.472 |
| GOBP_ISOI | 113 | 0.029991 | 0.002313 | 0.047193 | 0.26256 | 4464.833 |
| GOBP_PRC | 16 | 0.089626 | 0.002608 | 0.14105 | 0.26259 | 4465.343 |
| WP_NONG | 69 | 0.041049 | 0.002477 | 0.064611 | 0.26262 | 4465.853 |
| GOCC_POR | 24 | 0.061949 | 0.002207 | 0.097531 | 0.26266 | 4466.533 |
| REACTOME | 54 | 0.044885 | 0.002397 | 0.070733 | 0.26286 | 4469.934 |
| REACTOME | 11 | 0.092124 | 0.002223 | 0.14518 | 0.26286 | 4469.934 |
| MALIK_REF | 13 | 0.086195 | 0.002261 | 0.13586 | 0.2629 | 4470.615 |
| GOBP_MIC | 28 | 0.062369 | 0.0024 | 0.098352 | 0.263 | 4472.315 |
| CHICAS_RB | 522 | 0.014105 | 0.002313 | 0.022246 | 0.26302 | 4472.655 |
| GOCC_NAS | 4 | 0.14067 | 0.002047 | 0.22185 | 0.26303 | 4472.825 |
| PID_LIS1_P | 27 | 0.054744 | 0.002069 | 0.086369 | 0.2631 | 4474.016 |
| GOMF_ZIN | 742 | 0.011748 | 0.002283 | 0.018539 | 0.26314 | 4474.696 |
| GOBP_POS | 56 | 0.042422 | 0.002307 | 0.06696 | 0.26319 | 4475.546 |
| GOBP_POS | 5 | 0.15794 | 0.00257 | 0.24929 | 0.26319 | 4475.546 |
| GOBP_POS | 27 | 0.065498 | 0.002475 | 0.10352 | 0.26348 | 4480.477 |
| REACTOME | 41 | 0.053609 | 0.002496 | 0.084737 | 0.26349 | 4480.647 |
| BANDRES_ | 9 | 0.12207 | 0.002665 | 0.19298 | 0.26351 | 4480.988 |
| GOMF_CYS | 12 | 0.088732 | 0.002236 | 0.14036 | 0.26364 | 4483.198 |
| GOBP_ACT | 750 | 0.011881 | 0.002321 | 0.018798 | 0.26368 | 4483.878 |
| GOMF_REC | 4 | 0.18864 | 0.002746 | 0.29848 | 0.2637 | 4484.219 |
| GOBP_NOI | 10 | 0.1176 | 0.002706 | 0.18642 | 0.26407 | 4490.51 |

| | | | | | | |
|---|---|---|---|---|---|---|
| BIOCARTA_ | 28 | 0.055982 | 0.002154 | 0.088752 | 0.2641 | 4491.021 |
| GOBP_NEG | 58 | 0.041555 | 0.0023 | 0.06588 | 0.2641 | 4491.021 |
| REACTOME | 7 | 0.1232 | 0.002372 | 0.19546 | 0.26425 | 4493.571 |
| GOCC_MH | 4 | 0.11253 | 0.001638 | 0.17854 | 0.26426 | 4493.741 |
| GOMF_C_C | 5 | 0.13045 | 0.002123 | 0.20706 | 0.26434 | 4495.102 |
| WP_DISOR | 5 | 0.14888 | 0.002423 | 0.23653 | 0.26455 | 4498.673 |
| GOCC_CHR | 12 | 0.092855 | 0.00234 | 0.14754 | 0.26457 | 4499.013 |
| GOBP_LAT | 3 | 0.1448 | 0.001825 | 0.23015 | 0.26464 | 4500.203 |
| GOBP_REG | 49 | 0.046024 | 0.002342 | 0.073179 | 0.2647 | 4501.224 |
| GOBP_HEA | 34 | 0.055284 | 0.002344 | 0.087956 | 0.26483 | 4503.434 |
| GOBP_DOF | 8 | 0.1244 | 0.00256 | 0.1981 | 0.26501 | 4506.495 |
| GOBP_FIBF | 27 | 0.060573 | 0.002289 | 0.096485 | 0.26507 | 4507.515 |
| GOBP_EME | 35 | 0.055804 | 0.002401 | 0.088892 | 0.26508 | 4507.685 |
| GOBP_MIT | 11 | 0.076133 | 0.001837 | 0.12129 | 0.26511 | 4508.196 |
| PID_ANTHI | 16 | 0.089143 | 0.002594 | 0.14202 | 0.26511 | 4508.196 |
| GOBP_MIT | 18 | 0.067853 | 0.002094 | 0.10812 | 0.26515 | 4508.876 |
| GOBP_REL | 6 | 0.13453 | 0.002398 | 0.21441 | 0.26519 | 4509.556 |
| GOERING_ | 10 | 0.08495 | 0.001955 | 0.13547 | 0.2653 | 4511.427 |
| HOLLERN_ | 14 | 0.096964 | 0.00264 | 0.15465 | 0.26533 | 4511.937 |
| HOEGERKC | 24 | 0.070322 | 0.002506 | 0.1122 | 0.2654 | 4513.127 |
| JINESH_BL | 353 | 0.017282 | 0.002341 | 0.027575 | 0.26542 | 4513.467 |
| GOBP_NEG | 18 | 0.072168 | 0.002227 | 0.11524 | 0.26558 | 4516.188 |
| GOBP_SM | 372 | 0.015772 | 0.002192 | 0.025187 | 0.2656 | 4516.528 |
| GOBP_BOD | 45 | 0.047534 | 0.002318 | 0.075948 | 0.2657 | 4518.229 |
| REACTOME | 56 | 0.04253 | 0.002313 | 0.067952 | 0.2657 | 4518.229 |
| ENGELMAN | 47 | 0.046618 | 0.002323 | 0.074488 | 0.26571 | 4518.399 |
| GOCC_RNA | 10 | 0.083634 | 0.001924 | 0.13365 | 0.26574 | 4518.909 |
| GOBP_NEG | 85 | 0.03358 | 0.002248 | 0.053667 | 0.26575 | 4519.079 |
| REACTOME | 5 | 0.1735 | 0.002823 | 0.27767 | 0.26604 | 4524.01 |
| GOBP_CAV | 11 | 0.09792 | 0.002363 | 0.15673 | 0.26606 | 4524.35 |
| REACTOME | 50 | 0.043465 | 0.002234 | 0.069577 | 0.26608 | 4524.69 |
| VALK_AML | 29 | 0.058417 | 0.002288 | 0.093517 | 0.2661 | 4525.031 |
| KEGG_PER | 76 | 0.031773 | 0.002012 | 0.050866 | 0.26611 | 4525.201 |
| GOBP_CHC | 5 | 0.12358 | 0.002011 | 0.19785 | 0.26613 | 4525.541 |
| GOMF_CHI | 11 | 0.10858 | 0.00262 | 0.17388 | 0.26617 | 4526.221 |
| GOBP_REG | 8 | 0.10022 | 0.002063 | 0.16052 | 0.26621 | 4526.901 |
| GOBP_CAN | 23 | 0.07063 | 0.002464 | 0.11315 | 0.26624 | 4527.411 |
| WU_ALZHE | 17 | 0.076665 | 0.0023 | 0.12288 | 0.26634 | 4529.112 |
| GOBP_HYP | 13 | 0.093858 | 0.002462 | 0.1505 | 0.26643 | 4530.642 |
| GOBP_NEG | 126 | 0.029475 | 0.0024 | 0.047269 | 0.26646 | 4531.152 |
| GOBP_REG | 38 | 0.048289 | 0.002164 | 0.077441 | 0.26646 | 4531.152 |
| GOBP_POS | 5 | 0.12535 | 0.00204 | 0.20108 | 0.26652 | 4532.173 |
| GOBP_CEL | 6 | 0.12529 | 0.002233 | 0.20099 | 0.26653 | 4532.343 |
| TAKAO_RE | 94 | 0.029748 | 0.002094 | 0.047747 | 0.26664 | 4534.213 |
| WP_NANO | 11 | 0.095914 | 0.002315 | 0.15399 | 0.2667 | 4535.234 |

| | | | | | | |
|---|---|---|---|---|---|---|
| REACTOME | 142 | 0.025985 | 0.002245 | 0.041734 | 0.26677 | 4536.424 |
| GOBP_NEG | 51 | 0.046345 | 0.002406 | 0.074437 | 0.26678 | 4536.594 |
| PID_FAK_P | 57 | 0.041216 | 0.002261 | 0.066199 | 0.26678 | 4536.594 |
| REACTOME | 14 | 0.080554 | 0.002193 | 0.12941 | 0.26682 | 4537.274 |
| GOMF_HYI | 67 | 0.038297 | 0.002277 | 0.061567 | 0.26696 | 4539.655 |
| WP_GENE_ | 11 | 0.083982 | 0.002027 | 0.13503 | 0.26699 | 4540.165 |
| WP_APOPT | 17 | 0.072504 | 0.002175 | 0.11663 | 0.26709 | 4541.865 |
| GOBP_SELI | 89 | 0.032986 | 0.00226 | 0.053075 | 0.26714 | 4542.716 |
| GOMF_PR( | 6 | 0.14708 | 0.002622 | 0.2367 | 0.26718 | 4543.396 |
| GOBP_POI | 11 | 0.088664 | 0.00214 | 0.14275 | 0.26726 | 4544.756 |
| GOBP_NEG | 74 | 0.036796 | 0.002299 | 0.05925 | 0.26729 | 4545.266 |
| GOBP_NEG | 9 | 0.094978 | 0.002073 | 0.15295 | 0.26731 | 4545.607 |
| RIZ_ERYTH | 74 | 0.033441 | 0.00209 | 0.053858 | 0.26733 | 4545.947 |
| CHIARADO | 56 | 0.042455 | 0.002309 | 0.068391 | 0.26738 | 4546.797 |
| GOBP_POS | 8 | 0.13252 | 0.002727 | 0.21349 | 0.26739 | 4546.967 |
| FARMER_B | 6 | 0.19309 | 0.003442 | 0.3111 | 0.26741 | 4547.307 |
| GOBP_POS | 10 | 0.11246 | 0.002588 | 0.18121 | 0.26743 | 4547.647 |
| GOMF_GLU | 5 | 0.15468 | 0.002517 | 0.24931 | 0.26749 | 4548.667 |
| REACTOME | 23 | 0.065878 | 0.002298 | 0.10623 | 0.26759 | 4550.368 |
| GOCC_COF | 17 | 0.06626 | 0.001988 | 0.10688 | 0.26766 | 4551.558 |
| LUI_THYRC | 45 | 0.048315 | 0.002356 | 0.077957 | 0.26771 | 4552.409 |
| REACTOME | 36 | 0.051746 | 0.002258 | 0.083494 | 0.26771 | 4552.409 |
| GOBP_PRC | 101 | 0.031045 | 0.002265 | 0.050095 | 0.26772 | 4552.579 |
| GOCC_AXC | 36 | 0.048862 | 0.002132 | 0.07888 | 0.26782 | 4554.279 |
| GOMF_C4_ | 16 | 0.07765 | 0.00226 | 0.12536 | 0.26782 | 4554.279 |
| REACTOME | 28 | 0.059663 | 0.002296 | 0.096388 | 0.26797 | 4556.83 |
| GOMF_UBI | 7 | 0.10612 | 0.002043 | 0.17147 | 0.26801 | 4557.51 |
| FLECHNER_ | 517 | 0.013758 | 0.002245 | 0.022235 | 0.26805 | 4558.19 |
| GOBP_POS | 7 | 0.13653 | 0.002629 | 0.2208 | 0.26818 | 4560.401 |
| NIKOLSKY_ | 54 | 0.07213 | 0.003852 | 0.11667 | 0.26822 | 4561.081 |
| GOBP_PYR | 15 | 0.076658 | 0.00216 | 0.12402 | 0.26825 | 4561.591 |
| BROCKE_A | 141 | 0.026445 | 0.002277 | 0.042794 | 0.26831 | 4562.612 |
| GOMF_HYI | 699 | 0.011778 | 0.002224 | 0.019069 | 0.2684 | 4564.142 |
| GOBP_EXO | 13 | 0.095077 | 0.002494 | 0.15402 | 0.26852 | 4566.183 |
| GOBP_PRC | 15 | 0.089218 | 0.002514 | 0.1446 | 0.26863 | 4568.053 |
| TAKEDA_T/ | 180 | 0.023692 | 0.002302 | 0.038422 | 0.26875 | 4570.094 |
| PETROVA_ | 25 | 0.065256 | 0.002373 | 0.10595 | 0.26899 | 4574.175 |
| GOBP_GLY | 8 | 0.096385 | 0.001984 | 0.15657 | 0.26908 | 4575.705 |
| REACTOME | 6 | 0.13182 | 0.00235 | 0.21424 | 0.26919 | 4577.576 |
| GOBP_POS | 9 | 0.10499 | 0.002292 | 0.17076 | 0.26933 | 4579.957 |
| GOBP_VAS | 43 | 0.049518 | 0.002361 | 0.080547 | 0.26936 | 4580.467 |
| REACTOME | 6 | 0.1189 | 0.002119 | 0.19341 | 0.26936 | 4580.467 |
| REACTOME | 14 | 0.073844 | 0.00201 | 0.12012 | 0.26937 | 4580.637 |
| GENTILE_U | 37 | 0.053151 | 0.002351 | 0.086528 | 0.26953 | 4583.358 |
| GOMF_DE/ | 36 | 0.049289 | 0.00215 | 0.080241 | 0.26953 | 4583.358 |

| | | | | | | |
|---|---|---|---|---|---|---|
| SHANK_TA | 10 | 0.10401 | 0.002393 | 0.16933 | 0.26953 | 4583.358 |
| GOBP_INTI | 94 | 0.034808 | 0.00245 | 0.056684 | 0.26959 | 4584.378 |
| GOCC_POS | 10 | 0.082036 | 0.001888 | 0.13362 | 0.26962 | 4584.888 |
| GOMF_RRI | 61 | 0.035256 | 0.002001 | 0.057455 | 0.26973 | 4586.759 |
| GOBP_REG | 378 | 0.016353 | 0.002291 | 0.026653 | 0.26975 | 4587.099 |
| GOMF_FIL/ | 14 | 0.087742 | 0.002389 | 0.14303 | 0.26979 | 4587.779 |
| REACTOME | 11 | 0.090485 | 0.002184 | 0.1475 | 0.26979 | 4587.779 |
| HU_ANGIO | 36 | 0.054044 | 0.002358 | 0.088103 | 0.2698 | 4587.949 |
| KRIGE_RES | 871 | 0.010006 | 0.002099 | 0.016315 | 0.26984 | 4588.629 |
| GOBP_NEG | 5 | 0.16048 | 0.002611 | 0.26176 | 0.26992 | 4589.99 |
| GOBP_DEP | 319 | 0.016802 | 0.002166 | 0.027409 | 0.26994 | 4590.33 |
| GOMF_THI | 7 | 0.11092 | 0.002136 | 0.18094 | 0.26994 | 4590.33 |
| GOBP_REG | 89 | 0.032761 | 0.002244 | 0.053445 | 0.26995 | 4590.5 |
| GOBP_SPH | 22 | 0.069563 | 0.002373 | 0.1135 | 0.26997 | 4590.84 |
| GOBP_PRC | 6 | 0.13341 | 0.002378 | 0.21772 | 0.27003 | 4591.86 |
| GOBP_NOS | 15 | 0.078521 | 0.002213 | 0.12816 | 0.27005 | 4592.2 |
| GOBP_PRC | 39 | 0.049291 | 0.002238 | 0.080495 | 0.27016 | 4594.071 |
| MIKKELSEN | 32 | 0.058363 | 0.002401 | 0.095365 | 0.27027 | 4595.941 |
| GRAESSMA | 229 | 0.019604 | 0.002146 | 0.032038 | 0.2703 | 4596.452 |
| GOBP_REG | 115 | 0.026373 | 0.002052 | 0.043121 | 0.27041 | 4598.322 |
| GOBP_SEN | 99 | 0.032605 | 0.002355 | 0.053316 | 0.27042 | 4598.492 |
| REACTOME | 24 | 0.067407 | 0.002402 | 0.11027 | 0.27052 | 4600.193 |
| BURTON_A | 46 | 0.051151 | 0.002522 | 0.083777 | 0.27075 | 4604.104 |
| GOBP_PRC | 8 | 0.086572 | 0.001782 | 0.1418 | 0.27076 | 4604.274 |
| ABE_VEGF/ | 31 | 0.053367 | 0.002161 | 0.087427 | 0.2708 | 4604.954 |
| GOBP_HEA | 15 | 0.088237 | 0.002486 | 0.14457 | 0.27082 | 4605.294 |
| GOCC_MIT | 10 | 0.079669 | 0.001833 | 0.13057 | 0.27087 | 4606.144 |
| GOBP_ERY | 127 | 0.026325 | 0.002152 | 0.043149 | 0.2709 | 4606.655 |
| GOMF_MIT | 6 | 0.13118 | 0.002338 | 0.21501 | 0.2709 | 4606.655 |
| GOBP_POS | 4 | 0.14531 | 0.002115 | 0.23823 | 0.27096 | 4607.675 |
| REACTOME | 5 | 0.17983 | 0.002926 | 0.2949 | 0.271 | 4608.355 |
| MARKS_HD | 13 | 0.089572 | 0.00235 | 0.1469 | 0.27101 | 4608.525 |
| GOCC_T_TI | 52 | 0.04543 | 0.002381 | 0.074514 | 0.27104 | 4609.035 |
| KYNG_DNA | 56 | 0.03942 | 0.002144 | 0.064678 | 0.27111 | 4610.226 |
| GOCC_END | 23 | 0.055443 | 0.001934 | 0.090984 | 0.27114 | 4610.736 |
| REACTOME | 45 | 0.045231 | 0.002206 | 0.074232 | 0.27116 | 4611.076 |
| REACTOME | 66 | 0.035098 | 0.002072 | 0.057608 | 0.27118 | 4611.416 |
| BOYAULT_I | 46 | 0.043394 | 0.002139 | 0.071242 | 0.27123 | 4612.266 |
| PID_IL6_7_ | 46 | 0.046782 | 0.002307 | 0.076803 | 0.27123 | 4612.266 |
| REACTOME | 22 | 0.066786 | 0.002279 | 0.10964 | 0.27123 | 4612.266 |
| ABRAMSOI | 42 | 0.04089 | 0.001927 | 0.067166 | 0.27134 | 4614.137 |
| GOBP_INTI | 96 | 0.030363 | 0.00216 | 0.049884 | 0.27138 | 4614.817 |
| WP_MIR5C | 17 | 0.085979 | 0.002579 | 0.14126 | 0.27138 | 4614.817 |
| CHICAS_RE | 229 | 0.020042 | 0.002194 | 0.032942 | 0.27147 | 4616.347 |
| GOBP_BIOI | 160 | 0.025286 | 0.002318 | 0.041601 | 0.27166 | 4619.578 |

| | | | | | | |
|---|---|---|---|---|---|---|
| REACTOME | 6 | 0.13087 | 0.002333 | 0.21533 | 0.27168 | 4619.918 |
| SIG_IL4REC | 27 | 0.060682 | 0.002293 | 0.099851 | 0.27169 | 4620.088 |
| GOBP_SUB | 97 | 0.031864 | 0.002278 | 0.052444 | 0.27174 | 4620.939 |
| GOCC_EUK | 7 | 0.11858 | 0.002283 | 0.19527 | 0.27184 | 4622.639 |
| GOBP_NEG | 937 | 0.010312 | 0.00224 | 0.016988 | 0.27192 | 4624 |
| GOBP_FER | 4 | 0.17452 | 0.00254 | 0.28758 | 0.27197 | 4624.85 |
| GOBP_L_G | 3 | 0.1835 | 0.002313 | 0.30244 | 0.27202 | 4625.7 |
| GOMF_G_I | 258 | 0.019362 | 0.002248 | 0.031916 | 0.27204 | 4626.04 |
| WIKMAN_A | 14 | 0.092213 | 0.00251 | 0.15214 | 0.27223 | 4629.271 |
| GOBP_NEG | 7 | 0.11179 | 0.002152 | 0.18449 | 0.27228 | 4630.121 |
| REACTOME | 34 | 0.04782 | 0.002028 | 0.078923 | 0.27229 | 4630.291 |
| GENTILE_U | 26 | 0.059762 | 0.002216 | 0.098682 | 0.27239 | 4631.992 |
| GOBP_ESTA | 10 | 0.1054 | 0.002425 | 0.17416 | 0.27254 | 4634.543 |
| SHEPARD_I | 71 | 0.034589 | 0.002117 | 0.057158 | 0.27255 | 4634.713 |
| WANG_CLI | 168 | 0.022499 | 0.002113 | 0.037189 | 0.2726 | 4635.563 |
| GOBP_CAR | 16 | 0.085509 | 0.002488 | 0.14139 | 0.27267 | 4636.753 |
| GOBP_REG | 111 | 0.028065 | 0.002146 | 0.046407 | 0.27268 | 4636.923 |
| WANG_TU | 199 | 0.019928 | 0.002035 | 0.032973 | 0.2728 | 4638.964 |
| PLASARI_T | 31 | 0.05951 | 0.00241 | 0.098535 | 0.27294 | 4641.345 |
| GOBP_NEG | 8 | 0.10818 | 0.002226 | 0.17916 | 0.27299 | 4642.195 |
| GOBP_NEG | 43 | 0.047974 | 0.002287 | 0.079474 | 0.27304 | 4643.045 |
| GOBP_POS | 27 | 0.065701 | 0.002483 | 0.10886 | 0.27307 | 4643.555 |
| GOBP_POS | 6 | 0.10576 | 0.001885 | 0.17531 | 0.27317 | 4645.256 |
| GOBP_REG | 126 | 0.028016 | 0.002281 | 0.046451 | 0.27322 | 4646.106 |
| STONER_ES | 34 | 0.050739 | 0.002151 | 0.084129 | 0.27322 | 4646.106 |
| ELVIDGE_H | 142 | 0.024005 | 0.002074 | 0.039804 | 0.27324 | 4646.446 |
| GOBP_RES | 92 | 0.031313 | 0.002181 | 0.051935 | 0.27328 | 4647.126 |
| GOBP_REG | 115 | 0.029952 | 0.002331 | 0.049723 | 0.27347 | 4650.357 |
| GOBP_NEG | 5 | 0.14036 | 0.002284 | 0.23311 | 0.27355 | 4651.718 |
| REACTOME | 6 | 0.13692 | 0.002441 | 0.22744 | 0.27359 | 4652.398 |
| GOBP_PEPT | 11 | 0.08258 | 0.001993 | 0.13731 | 0.27378 | 4655.629 |
| WANG_LM | 330 | 0.016524 | 0.002166 | 0.027473 | 0.27378 | 4655.629 |
| GOBP_CELI | 94 | 0.029109 | 0.002049 | 0.048419 | 0.27386 | 4656.989 |
| GOBP_PRC | 55 | 0.038175 | 0.002058 | 0.063504 | 0.27387 | 4657.159 |
| GOBP_POS | 84 | 0.031746 | 0.002113 | 0.052809 | 0.27388 | 4657.329 |
| GOCC_APIC | 15 | 0.085842 | 0.002419 | 0.14286 | 0.27397 | 4658.86 |
| GOBP_REG | 53 | 0.036551 | 0.001934 | 0.060854 | 0.27405 | 4660.22 |
| NUYTTEN_ | 975 | 0.009934 | 0.002198 | 0.016549 | 0.27417 | 4662.261 |
| GOBP_PAT | 9 | 0.12002 | 0.00262 | 0.20007 | 0.27429 | 4664.301 |
| REACTOME | 11 | 0.08159 | 0.001969 | 0.13603 | 0.27432 | 4664.812 |
| WP_CARDI | 50 | 0.043091 | 0.002215 | 0.071863 | 0.27438 | 4665.832 |
| GOMF_MA | 8 | 0.12098 | 0.00249 | 0.20182 | 0.27443 | 4666.682 |
| GOBP_REG | 1650 | 0.007792 | 0.002201 | 0.013001 | 0.27448 | 4667.532 |
| GOMF_CEL | 50 | 0.045508 | 0.002339 | 0.075944 | 0.27451 | 4668.043 |
| GOBP_SNR | 6 | 0.12684 | 0.002261 | 0.21168 | 0.27452 | 4668.213 |

| | | | | | | |
|---|---|---|---|---|---|---|
| GOBP_REG | 17 | 0.071611 | 0.002148 | 0.11957 | 0.27463 | 4670.083 |
| GOBP_REG | 6 | 0.14298 | 0.002549 | 0.23878 | 0.27466 | 4670.593 |
| GOBP_INFL | 22 | 0.063011 | 0.00215 | 0.10526 | 0.27472 | 4671.614 |
| GOBP_STEI | 23 | 0.060968 | 0.002127 | 0.10188 | 0.27478 | 4672.634 |
| REACTOME | 6 | 0.11639 | 0.002075 | 0.19452 | 0.27482 | 4673.314 |
| GOBP_GLIA | 54 | 0.043353 | 0.002315 | 0.072469 | 0.27485 | 4673.824 |
| REACTOME | 11 | 0.081101 | 0.001957 | 0.13557 | 0.27486 | 4673.994 |
| WATANABI | 19 | 0.075926 | 0.002408 | 0.12696 | 0.27491 | 4674.845 |
| GOBP_ORG | 14 | 0.085724 | 0.002334 | 0.14337 | 0.27495 | 4675.525 |
| GOBP_MAT | 24 | 0.060987 | 0.002173 | 0.10204 | 0.27503 | 4676.885 |
| YAGI_AML_ | 317 | 0.016848 | 0.002165 | 0.028201 | 0.27512 | 4678.416 |
| GOBP_HIST | 8 | 0.1138 | 0.002342 | 0.19054 | 0.27518 | 4679.436 |
| GOMF_PHO | 11 | 0.082431 | 0.001989 | 0.13803 | 0.27519 | 4679.606 |
| IKEDA_MIR | 111 | 0.029962 | 0.002291 | 0.050176 | 0.27521 | 4679.946 |
| REACTOME | 5 | 0.11925 | 0.001941 | 0.1997 | 0.27521 | 4679.946 |
| GOBP_PEP | 108 | 0.028117 | 0.002121 | 0.047094 | 0.27525 | 4680.626 |
| GOCC_EXT | 317 | 0.018185 | 0.002337 | 0.03046 | 0.27526 | 4680.796 |
| MYLLYKAN | 11 | 0.076945 | 0.001857 | 0.12894 | 0.27535 | 4682.327 |
| GOBP_NEG | 54 | 0.045006 | 0.002404 | 0.075426 | 0.27536 | 4682.497 |
| RAFFEL_VE | 9 | 0.12169 | 0.002657 | 0.20406 | 0.27547 | 4684.367 |
| GOMF_PEF | 12 | 0.089137 | 0.002247 | 0.14948 | 0.27549 | 4684.707 |
| GOMF_RIB | 3 | 0.13754 | 0.001734 | 0.23069 | 0.27552 | 4685.218 |
| GOBP_POS | 128 | 0.028298 | 0.002322 | 0.047469 | 0.27555 | 4685.728 |
| GOBP_REG | 15 | 0.094152 | 0.002653 | 0.15795 | 0.27556 | 4685.898 |
| TIEN_INTES | 85 | 0.033928 | 0.002272 | 0.056945 | 0.27566 | 4687.598 |
| GOBP_BAS | 19 | 0.071504 | 0.002267 | 0.12002 | 0.27568 | 4687.938 |
| GOBP_EPIC | 157 | 0.023305 | 0.002117 | 0.039136 | 0.27576 | 4689.299 |
| GOBP_CAL | 306 | 0.017844 | 0.002253 | 0.029982 | 0.27587 | 4691.169 |
| GOBP_REG | 19 | 0.064293 | 0.002039 | 0.10811 | 0.27603 | 4693.89 |
| GOBP_ALP | 6 | 0.127 | 0.002264 | 0.21357 | 0.27605 | 4694.23 |
| LEE_TARGE | 54 | 0.041097 | 0.002195 | 0.069179 | 0.27624 | 4697.461 |
| GOBP_REG | 15 | 0.074587 | 0.002102 | 0.12556 | 0.27625 | 4697.631 |
| WP_HEPAT | 49 | 0.047808 | 0.002433 | 0.080489 | 0.27627 | 4697.971 |
| GOMF_CYC | 7 | 0.11947 | 0.0023 | 0.20116 | 0.27628 | 4698.141 |
| KIM_PTEN_ | 17 | 0.082465 | 0.002474 | 0.13887 | 0.27632 | 4698.822 |
| GOMF_LBD | 5 | 0.17788 | 0.002895 | 0.29963 | 0.27637 | 4699.672 |
| GOBP_AMI | 10 | 0.096991 | 0.002232 | 0.16341 | 0.27641 | 4700.352 |
| GOMF_AM | 10 | 0.096991 | 0.002232 | 0.16341 | 0.27641 | 4700.352 |
| GOBP_MET | 5 | 0.13608 | 0.002214 | 0.22937 | 0.2765 | 4701.883 |
| GOMF_MA | 24 | 0.068498 | 0.002441 | 0.11546 | 0.27651 | 4702.053 |
| GOBP_B_C | 144 | 0.027037 | 0.002352 | 0.045593 | 0.27659 | 4703.413 |
| GOBP_CELI | 11 | 0.10051 | 0.002426 | 0.16963 | 0.27675 | 4706.134 |
| WP_PROST | 45 | 0.048451 | 0.002363 | 0.081797 | 0.27682 | 4707.324 |
| AMIT_EGF_ | 40 | 0.043196 | 0.001986 | 0.072939 | 0.27685 | 4707.834 |
| WP_CORI_ | 15 | 0.070404 | 0.001984 | 0.11895 | 0.27696 | 4709.705 |

| | | | | | | |
|---|---|---|---|---|---|---|
| GOBP_POS | 13 | 0.081342 | 0.002134 | 0.13744 | 0.27698 | 4710.045 |
| GOCC_CON | 8 | 0.099746 | 0.002053 | 0.16856 | 0.27701 | 4710.555 |
| GOMF_AM | 6 | 0.13479 | 0.002403 | 0.22783 | 0.27704 | 4711.065 |
| GOBP_VIR/ | 29 | 0.058958 | 0.002309 | 0.099681 | 0.27711 | 4712.256 |
| REACTOME | 16 | 0.06531 | 0.001901 | 0.11042 | 0.27711 | 4712.256 |
| GOBP_POS | 17 | 0.061805 | 0.001854 | 0.10455 | 0.27721 | 4713.956 |
| LEE_AGING | 70 | 0.035692 | 0.002169 | 0.060379 | 0.27722 | 4714.126 |
| GOBP_IP_1 | 4 | 0.14667 | 0.002135 | 0.24826 | 0.27734 | 4716.167 |
| GOBP_CGN | 15 | 0.08778 | 0.002473 | 0.14865 | 0.27743 | 4717.697 |
| WP_CELL_( | 117 | 0.025933 | 0.002035 | 0.043937 | 0.27753 | 4719.398 |
| GOMF_PH( | 447 | 0.014276 | 0.002171 | 0.024212 | 0.27772 | 4722.629 |
| GOBP_NUC | 240 | 0.017765 | 0.00199 | 0.030129 | 0.27773 | 4722.799 |
| REACTOME | 12 | 0.10199 | 0.002571 | 0.17306 | 0.27783 | 4724.499 |
| MCLACHLA | 71 | 0.03643 | 0.00223 | 0.061843 | 0.27791 | 4725.86 |
| FERRANDO | 22 | 0.060583 | 0.002067 | 0.1029 | 0.27801 | 4727.56 |
| GOBP_REG | 668 | 0.01137 | 0.002101 | 0.01933 | 0.2782 | 4730.791 |
| GOBP_INTI | 18 | 0.075492 | 0.00233 | 0.12839 | 0.27827 | 4731.981 |
| GOBP_NUC | 36 | 0.049946 | 0.002179 | 0.084982 | 0.27836 | 4733.512 |
| REACTOME | 29 | 0.05656 | 0.002215 | 0.09625 | 0.27839 | 4734.022 |
| WP_ULTRA | 2 | 0.2676 | 0.002754 | 0.45588 | 0.2786 | 4737.593 |
| BIOCARTA_ | 14 | 0.083166 | 0.002264 | 0.1417 | 0.27863 | 4738.103 |
| HOOI_ST7_ | 103 | 0.030289 | 0.002231 | 0.051618 | 0.27867 | 4738.783 |
| HAHTOLA_ | 163 | 0.022578 | 0.002089 | 0.038487 | 0.27873 | 4739.804 |
| GOBP_MO | 4 | 0.16263 | 0.002367 | 0.27729 | 0.27878 | 4740.654 |
| GOBP_RNA | 7 | 0.10168 | 0.001958 | 0.1734 | 0.2788 | 4740.994 |
| BENPORAT | 8 | 0.098122 | 0.00202 | 0.16742 | 0.27891 | 4742.865 |
| GOCC_FEN | 10 | 0.087411 | 0.002011 | 0.14914 | 0.27891 | 4742.865 |
| GOBP_RES | 10 | 0.088981 | 0.002047 | 0.1519 | 0.27901 | 4744.565 |
| GOBP_CELI | 41 | 0.052111 | 0.002426 | 0.088969 | 0.27904 | 4745.075 |
| GOBP_NEG | 15 | 0.076663 | 0.00216 | 0.1309 | 0.27905 | 4745.245 |
| GOBP_NEG | 12 | 0.094841 | 0.00239 | 0.16199 | 0.27912 | 4746.436 |
| GOMF_PEF | 5 | 0.12507 | 0.002035 | 0.21364 | 0.27914 | 4746.776 |
| GOBP_POS | 26 | 0.053778 | 0.001994 | 0.091978 | 0.27938 | 4750.857 |
| MONNIER_ | 382 | 0.014184 | 0.001997 | 0.024272 | 0.27949 | 4752.727 |
| MORI_IMN | 45 | 0.039873 | 0.001944 | 0.068272 | 0.27961 | 4754.768 |
| GOBP_RES | 5 | 0.13475 | 0.002193 | 0.23076 | 0.27963 | 4755.108 |
| GOBP_MIT | 500 | 0.012546 | 0.002015 | 0.021492 | 0.27969 | 4756.128 |
| GOCC_GOL | 10 | 0.089585 | 0.002061 | 0.15348 | 0.27971 | 4756.469 |
| GOBP_ORG | 1250 | 0.008343 | 0.002075 | 0.014294 | 0.27972 | 4756.639 |
| HASLINGEF | 15 | 0.065523 | 0.001846 | 0.11232 | 0.27983 | 4758.509 |
| KANNAN_T | 57 | 0.035491 | 0.001947 | 0.06084 | 0.27983 | 4758.509 |
| NELSON_R | 17 | 0.074363 | 0.002231 | 0.12751 | 0.27989 | 4759.529 |
| GOBP_POS | 30 | 0.05544 | 0.002208 | 0.095068 | 0.2799 | 4759.7 |
| GOBP_RES | 483 | 0.013401 | 0.002116 | 0.022981 | 0.27991 | 4759.87 |
| YIH_RESPO | 14 | 0.079868 | 0.002174 | 0.13702 | 0.27998 | 4761.06 |

| | | | | | | |
|---|---|---|---|---|---|---|
| GOBP_MIT | 9 | 0.078657 | 0.001717 | 0.13508 | 0.28018 | 4764.461 |
| GOBP_NEG | 5 | 0.15698 | 0.002554 | 0.26961 | 0.2802 | 4764.801 |
| GOBP_REG | 12 | 0.071915 | 0.001813 | 0.12353 | 0.28023 | 4765.311 |
| WP_CANO | 17 | 0.081785 | 0.002453 | 0.14061 | 0.2804 | 4768.202 |
| GOBP_CELI | 335 | 0.01647 | 0.002175 | 0.028352 | 0.28065 | 4772.453 |
| PID_PTP1B | 52 | 0.043447 | 0.002277 | 0.074803 | 0.28068 | 4772.963 |
| GOBP_EAR | 119 | 0.029409 | 0.002328 | 0.050636 | 0.28069 | 4773.133 |
| GOBP_REG | 5 | 0.12478 | 0.00203 | 0.21488 | 0.28073 | 4773.814 |
| HOSHIDA_ | 108 | 0.026533 | 0.002001 | 0.045708 | 0.28079 | 4774.834 |
| GOBP_NEG | 4 | 0.15794 | 0.002299 | 0.27226 | 0.28093 | 4777.215 |
| KUMAR_PA | 244 | 0.017616 | 0.00199 | 0.030376 | 0.28098 | 4778.065 |
| GOBP_CELI | 178 | 0.022057 | 0.002132 | 0.038041 | 0.28102 | 4778.745 |
| GOCC_PLA | 5 | 0.15226 | 0.002478 | 0.2626 | 0.28102 | 4778.745 |
| GOBP_LYS( | 12 | 0.079162 | 0.001995 | 0.13657 | 0.28108 | 4779.765 |
| ZHONG_SE | 70 | 0.035655 | 0.002167 | 0.06151 | 0.28108 | 4779.765 |
| GOBP_MA( | 6 | 0.11691 | 0.002084 | 0.20178 | 0.28116 | 4781.126 |
| REACTOME | 17 | 0.068249 | 0.002047 | 0.11781 | 0.2812 | 4781.806 |
| GOBP_RET | 17 | 0.076721 | 0.002301 | 0.13246 | 0.28122 | 4782.146 |
| GOCC_PRE | 17 | 0.080136 | 0.002404 | 0.13839 | 0.28128 | 4783.166 |
| REACTOME | 28 | 0.060778 | 0.002339 | 0.10496 | 0.28128 | 4783.166 |
| DURAND_S | 166 | 0.023069 | 0.002154 | 0.039843 | 0.2813 | 4783.507 |
| GALLUZZI_ | 39 | 0.045542 | 0.002068 | 0.078667 | 0.28132 | 4783.847 |
| GOBP_PRC | 31 | 0.049371 | 0.001999 | 0.085362 | 0.28151 | 4787.078 |
| KOBAYASH | 245 | 0.017743 | 0.002008 | 0.030678 | 0.28152 | 4787.248 |
| WP_TRANS | 46 | 0.038911 | 0.001918 | 0.067298 | 0.28157 | 4788.098 |
| GOBP_FRU | 10 | 0.097199 | 0.002237 | 0.16814 | 0.28161 | 4788.778 |
| BIOCARTA_ | 10 | 0.09335 | 0.002148 | 0.16154 | 0.28167 | 4789.798 |
| GOCC_PRC | 14 | 0.069567 | 0.001894 | 0.12039 | 0.28169 | 4790.138 |
| PID_TCR_P | 57 | 0.038913 | 0.002135 | 0.06736 | 0.28174 | 4790.989 |
| GOBP_PRC | 7 | 0.10116 | 0.001948 | 0.17517 | 0.28181 | 4792.179 |
| PID_INTEG | 39 | 0.052636 | 0.00239 | 0.091149 | 0.28182 | 4792.349 |
| ZHONG_SE | 121 | 0.026414 | 0.002108 | 0.045755 | 0.28187 | 4793.199 |
| GOBP_REG | 738 | 0.010548 | 0.002044 | 0.018281 | 0.28198 | 4795.07 |
| GOBP_B_1 | 5 | 0.11935 | 0.001942 | 0.2069 | 0.28203 | 4795.92 |
| GOBP_NEU | 8 | 0.12717 | 0.002617 | 0.22048 | 0.28204 | 4796.09 |
| REACTOME | 41 | 0.04618 | 0.00215 | 0.080061 | 0.28204 | 4796.09 |
| GOBP_REG | 8 | 0.10097 | 0.002078 | 0.17508 | 0.28207 | 4796.6 |
| GENTILE_U | 19 | 0.067686 | 0.002146 | 0.1174 | 0.28212 | 4797.451 |
| GOCC_9PLI | 121 | 0.025935 | 0.00207 | 0.044982 | 0.28212 | 4797.451 |
| LEE_EARLY | 99 | 0.028481 | 0.002057 | 0.049426 | 0.28223 | 4799.321 |
| GOBP_RNA | 43 | 0.036824 | 0.001756 | 0.063938 | 0.28233 | 4801.022 |
| GOBP_RES | 32 | 0.047303 | 0.001946 | 0.082183 | 0.28245 | 4803.062 |
| GOBP_POS | 26 | 0.052681 | 0.001954 | 0.091596 | 0.2826 | 4805.613 |
| GOBP_SIGI | 173 | 0.022687 | 0.002162 | 0.039446 | 0.2826 | 4805.613 |
| GOBP_POS | 784 | 0.010721 | 0.002139 | 0.018641 | 0.28261 | 4805.783 |

| | | | | | | |
|---|---|---|---|---|---|---|
| GOBP_EMI | 27 | 0.05946 | 0.002247 | 0.10341 | 0.28265 | 4806.463 |
| GOBP_REG | 6 | 0.11216 | 0.001999 | 0.19506 | 0.28265 | 4806.463 |
| HOLLEMAN | 22 | 0.054184 | 0.001849 | 0.094244 | 0.28267 | 4806.803 |
| GOBP_POS | 5 | 0.14833 | 0.002414 | 0.25814 | 0.28278 | 4808.674 |
| LOPEZ_ME | 13 | 0.07389 | 0.001938 | 0.12862 | 0.28283 | 4809.524 |
| REACTOME | 7 | 0.10096 | 0.001944 | 0.17576 | 0.28285 | 4809.864 |
| GOBP_DTN | 6 | 0.09898 | 0.001764 | 0.17235 | 0.28289 | 4810.544 |
| LEE_DOUB | 24 | 0.063622 | 0.002267 | 0.11079 | 0.2829 | 4810.715 |
| DORN_ADE | 30 | 0.047064 | 0.001875 | 0.08198 | 0.28296 | 4811.735 |
| REACTOME | 28 | 0.059859 | 0.002304 | 0.10436 | 0.28312 | 4814.456 |
| GOBP_NEG | 2 | 0.25192 | 0.002593 | 0.43924 | 0.28314 | 4814.796 |
| GOBP_MIC | 34 | 0.042591 | 0.001806 | 0.074263 | 0.28315 | 4814.966 |
| GOMF_INS | 21 | 0.070575 | 0.002353 | 0.12306 | 0.28316 | 4815.136 |
| LI_INDUCE | 296 | 0.017227 | 0.00214 | 0.03005 | 0.28323 | 4816.326 |
| GOBP_REG | 169 | 0.02301 | 0.002167 | 0.040147 | 0.28328 | 4817.176 |
| ODONNELL | 337 | 0.015766 | 0.002088 | 0.027511 | 0.2833 | 4817.517 |
| GOBP_BRA | 6 | 0.12043 | 0.002147 | 0.21021 | 0.28336 | 4818.537 |
| LUI_THYRC | 7 | 0.088398 | 0.001702 | 0.15442 | 0.28351 | 4821.088 |
| GOBP_INTI | 7 | 0.11419 | 0.002198 | 0.19958 | 0.28362 | 4822.958 |
| GOMF_INT | 10 | 0.098678 | 0.002271 | 0.17256 | 0.28371 | 4824.489 |
| GOBP_RAS | 304 | 0.016368 | 0.00206 | 0.028627 | 0.28374 | 4824.999 |
| GOCC_NON | 153 | 0.022771 | 0.002042 | 0.039841 | 0.28382 | 4826.359 |
| GOBERT_C | 40 | 0.044946 | 0.002067 | 0.078665 | 0.28388 | 4827.379 |
| KEGG_BLAI | 40 | 0.04491 | 0.002065 | 0.078602 | 0.28388 | 4827.379 |
| GOBP_REG | 360 | 0.015308 | 0.002094 | 0.026802 | 0.28395 | 4828.57 |
| GOMF_HYI | 35 | 0.048552 | 0.002089 | 0.085035 | 0.28402 | 4829.76 |
| GOMF_VIT | 144 | 0.023588 | 0.002052 | 0.041313 | 0.28402 | 4829.76 |
| REACTOME | 18 | 0.071448 | 0.002205 | 0.12515 | 0.28403 | 4829.93 |
| GOBP_POS | 13 | 0.087498 | 0.002295 | 0.15334 | 0.28413 | 4831.631 |
| GLINSKY_C | 6 | 0.11777 | 0.002099 | 0.20641 | 0.28415 | 4831.971 |
| GOMF_PRC | 48 | 0.046201 | 0.002327 | 0.08098 | 0.28416 | 4832.141 |
| GOBP_CELI | 5 | 0.1573 | 0.00256 | 0.27584 | 0.28426 | 4833.841 |
| GOBP_NEG | 97 | 0.029165 | 0.002085 | 0.051145 | 0.28426 | 4833.841 |
| REACTOME | 9 | 0.09395 | 0.002051 | 0.16476 | 0.28426 | 4833.841 |
| GOBP_GOL | 10 | 0.081625 | 0.001878 | 0.14317 | 0.2843 | 4834.522 |
| GOBP_REG | 59 | 0.037847 | 0.002113 | 0.066482 | 0.28458 | 4839.283 |
| GOBP_REG | 327 | 0.015639 | 0.00204 | 0.027475 | 0.28462 | 4839.963 |
| GOBP_PRC | 9 | 0.083167 | 0.001816 | 0.14617 | 0.28469 | 4841.153 |
| GOBP_RES | 6 | 0.097731 | 0.001742 | 0.17186 | 0.2848 | 4843.024 |
| GOBP_HEP | 11 | 0.090088 | 0.002174 | 0.15849 | 0.28489 | 4844.554 |
| GOBP_REG | 9 | 0.10759 | 0.002349 | 0.18931 | 0.28491 | 4844.895 |
| THUM_SYS | 193 | 0.020873 | 0.0021 | 0.036726 | 0.28491 | 4844.895 |
| GOBP_REG | 12 | 0.087176 | 0.002197 | 0.15342 | 0.28495 | 4845.575 |
| SARRIO_EP | 160 | 0.02185 | 0.002003 | 0.038458 | 0.28497 | 4845.915 |
| WP_MACR | 9 | 0.082875 | 0.001809 | 0.14594 | 0.28507 | 4847.615 |

| | | | | | | |
|---|---|---|---|---|---|---|
| WP_MIR22 | 4 | 0.141 | 0.002052 | 0.24832 | 0.28509 | 4847.955 |
| GOBP_NEG | 59 | 0.035554 | 0.001985 | 0.06265 | 0.28519 | 4849.656 |
| GOBP_CAR | 11 | 0.096966 | 0.00234 | 0.17092 | 0.28525 | 4850.676 |
| NAKAMUR | 24 | 0.060743 | 0.002165 | 0.10707 | 0.28526 | 4850.846 |
| REACTOME | 7 | 0.092021 | 0.001772 | 0.16222 | 0.28527 | 4851.016 |
| GOBP_ENE | 44 | 0.041675 | 0.00201 | 0.073492 | 0.28534 | 4852.207 |
| GOBP_SUC | 11 | 0.081212 | 0.00196 | 0.14331 | 0.28546 | 4854.247 |
| BIOCARTA_ | 14 | 0.073414 | 0.001999 | 0.12955 | 0.28547 | 4854.417 |
| PID_CD8_T | 61 | 0.041979 | 0.002382 | 0.074094 | 0.28551 | 4855.098 |
| GOBP_POL | 9 | 0.082045 | 0.001791 | 0.14491 | 0.28563 | 4857.138 |
| GAUSSMAI | 7 | 0.1247 | 0.002401 | 0.22027 | 0.28566 | 4857.648 |
| GOBP_REG | 7 | 0.089522 | 0.001724 | 0.15823 | 0.28578 | 4859.689 |
| REACTOME | 61 | 0.035404 | 0.002009 | 0.062596 | 0.28584 | 4860.709 |
| GOBP_POS | 34 | 0.048546 | 0.002058 | 0.085852 | 0.28589 | 4861.559 |
| GOBP_RES | 838 | 0.010271 | 0.002115 | 0.018197 | 0.28624 | 4867.511 |
| GOBP_AST | 79 | 0.035561 | 0.002296 | 0.063036 | 0.28633 | 4869.042 |
| REACTOME | 10 | 0.09106 | 0.002095 | 0.16147 | 0.2864 | 4870.232 |
| GOBP_NEG | 53 | 0.038155 | 0.002019 | 0.067661 | 0.28641 | 4870.402 |
| GOBP_FAC | 29 | 0.05344 | 0.002093 | 0.094786 | 0.28645 | 4871.082 |
| GOMF_RNA | 8 | 0.10185 | 0.002096 | 0.18066 | 0.28645 | 4871.082 |
| GOMF_GLY | 9 | 0.10686 | 0.002333 | 0.18956 | 0.28647 | 4871.422 |
| KEGG_PRO | 80 | 0.030238 | 0.001964 | 0.05365 | 0.28651 | 4872.103 |
| LEE_RECEN | 212 | 0.019774 | 0.002084 | 0.035086 | 0.28652 | 4872.273 |
| GOBP_NEG | 9 | 0.097563 | 0.00213 | 0.17333 | 0.28676 | 4876.354 |
| GOBP_REG | 21 | 0.057914 | 0.001931 | 0.1029 | 0.28678 | 4876.694 |
| GOBP_NEG | 5 | 0.14082 | 0.002292 | 0.25024 | 0.28681 | 4877.204 |
| GOBP_REG | 9 | 0.092871 | 0.002027 | 0.16513 | 0.28692 | 4879.075 |
| REACTOME | 33 | 0.0466 | 0.001947 | 0.082875 | 0.28696 | 4879.755 |
| GOBP_REG | 5 | 0.15153 | 0.002466 | 0.26966 | 0.28708 | 4881.795 |
| YAO_TEMP | 74 | 0.033405 | 0.002087 | 0.059455 | 0.28711 | 4882.306 |
| GOBP_TON | 10 | 0.094161 | 0.002167 | 0.16775 | 0.2873 | 4885.537 |
| GOBP_AMY | 36 | 0.046742 | 0.002039 | 0.083297 | 0.28735 | 4886.387 |
| WP_ALANI | 12 | 0.081211 | 0.002047 | 0.14474 | 0.28738 | 4886.897 |
| REACTOME | 231 | 0.018647 | 0.00205 | 0.033238 | 0.2874 | 4887.237 |
| WP_SPHIN | 30 | 0.05951 | 0.00237 | 0.10608 | 0.28741 | 4887.407 |
| GOBP_INN | 185 | 0.021304 | 0.002099 | 0.037993 | 0.28749 | 4888.767 |
| GOBP_SKE | 22 | 0.060912 | 0.002078 | 0.10865 | 0.28754 | 4889.618 |
| GOBP_REG | 11 | 0.085332 | 0.002059 | 0.15231 | 0.28766 | 4891.658 |
| REACTOME | 190 | 0.020479 | 0.002044 | 0.036556 | 0.28767 | 4891.828 |
| BIOCARTA_ | 7 | 0.095111 | 0.001831 | 0.16987 | 0.28778 | 4893.699 |
| YANG_BCL | 309 | 0.015675 | 0.001989 | 0.028014 | 0.2879 | 4895.74 |
| MIKKELSEN | 71 | 0.032636 | 0.001998 | 0.058357 | 0.288 | 4897.44 |
| GARGALOV | 15 | 0.070894 | 0.001998 | 0.12683 | 0.2881 | 4899.141 |
| REACTOME | 16 | 0.071309 | 0.002075 | 0.12759 | 0.28812 | 4899.481 |
| GOBP_INTI | 57 | 0.039573 | 0.002171 | 0.070847 | 0.28823 | 4901.351 |

| | | | | | | |
|---|---|---|---|---|---|---|
| GOMF_WA | 13 | 0.090066 | 0.002363 | 0.16135 | 0.28835 | 4903.392 |
| GOBP_TRA | 11 | 0.081169 | 0.001959 | 0.1455 | 0.28847 | 4905.432 |
| GOBP_PUR | 5 | 0.10702 | 0.001742 | 0.19195 | 0.28858 | 4907.303 |
| GOMF_EPH | 28 | 0.053245 | 0.002049 | 0.095514 | 0.28861 | 4907.813 |
| REACTOME | 41 | 0.041996 | 0.001955 | 0.075336 | 0.28861 | 4907.813 |
| REACTOME | 20 | 0.063818 | 0.002076 | 0.1145 | 0.28864 | 4908.323 |
| GOBP_SEM | 40 | 0.044938 | 0.002066 | 0.080627 | 0.28865 | 4908.493 |
| GOBP_CELI | 22 | 0.060347 | 0.002059 | 0.1083 | 0.28869 | 4909.173 |
| GOBP_AMI | 12 | 0.067223 | 0.001694 | 0.12067 | 0.28873 | 4909.854 |
| GOBP_MES | 5 | 0.12159 | 0.001979 | 0.21826 | 0.28874 | 4910.024 |
| GOBP_DEN | 82 | 0.0311 | 0.002045 | 0.055828 | 0.28875 | 4910.194 |
| GOMF_PHO | 15 | 0.070008 | 0.001973 | 0.12572 | 0.28882 | 4911.384 |
| GOBP_POS | 39 | 0.046542 | 0.002113 | 0.083633 | 0.28894 | 4913.425 |
| WP_G_PRO | 90 | 0.031222 | 0.002151 | 0.056112 | 0.28897 | 4913.935 |
| GOBP_NEG | 17 | 0.072874 | 0.002186 | 0.131 | 0.28901 | 4914.615 |
| GOCC_PLA | 14 | 0.082403 | 0.002243 | 0.14815 | 0.28904 | 4915.125 |
| GOMF_UBI | 36 | 0.046156 | 0.002014 | 0.082989 | 0.28905 | 4915.295 |
| GOBP_PRO | 6 | 0.10026 | 0.001787 | 0.1804 | 0.28919 | 4917.676 |
| GOBP_OLIG | 5 | 0.1239 | 0.002016 | 0.22294 | 0.2892 | 4917.846 |
| GOBP_END | 12 | 0.093716 | 0.002362 | 0.1687 | 0.28927 | 4919.036 |
| MUELLER_ | 286 | 0.015585 | 0.001904 | 0.028061 | 0.28931 | 4919.717 |
| GOCC_PER | 22 | 0.060477 | 0.002063 | 0.10891 | 0.28934 | 4920.227 |
| GOBP_POT | 17 | 0.077905 | 0.002337 | 0.1403 | 0.28935 | 4920.397 |
| REACTOME | 3 | 0.18981 | 0.002393 | 0.34203 | 0.28947 | 4922.437 |
| MYLLYKAN | 6 | 0.095598 | 0.001704 | 0.17227 | 0.28948 | 4922.607 |
| KEGG_TYPI | 45 | 0.042673 | 0.002081 | 0.076929 | 0.28955 | 4923.798 |
| GOBP_GLY | 8 | 0.10505 | 0.002162 | 0.18955 | 0.28972 | 4926.689 |
| GOMF_PDZ | 83 | 0.032128 | 0.002126 | 0.057971 | 0.28972 | 4926.689 |
| GOBP_TRIG | 91 | 0.029282 | 0.002028 | 0.052845 | 0.28975 | 4927.199 |
| GOBP_POS | 82 | 0.030163 | 0.001984 | 0.054468 | 0.28987 | 4929.239 |
| GOMF_PRE | 8 | 0.10664 | 0.002195 | 0.19258 | 0.28987 | 4929.239 |
| GOBP_RES | 29 | 0.048075 | 0.001883 | 0.086842 | 0.28993 | 4930.26 |
| REACTOME | 76 | 0.032792 | 0.002077 | 0.059246 | 0.28997 | 4930.94 |
| GOMF_POT | 16 | 0.078573 | 0.002287 | 0.14205 | 0.29009 | 4932.98 |
| REACTOME | 37 | 0.043203 | 0.001911 | 0.078123 | 0.29013 | 4933.661 |
| GOBP_GLIG | 299 | 0.017308 | 0.002161 | 0.031309 | 0.2902 | 4934.851 |
| GOBP_VEN | 10 | 0.10484 | 0.002412 | 0.18969 | 0.29024 | 4935.531 |
| BLALOCK_A | 359 | 0.014045 | 0.001918 | 0.025435 | 0.29042 | 4938.592 |
| GOMF_TES | 9 | 0.086834 | 0.001896 | 0.15727 | 0.29043 | 4938.762 |
| GOBP_REG | 8 | 0.10043 | 0.002067 | 0.18191 | 0.29045 | 4939.102 |
| VERHAAK_ | 231 | 0.01895 | 0.002083 | 0.03433 | 0.29048 | 4939.612 |
| GOBP_NEG | 16 | 0.067105 | 0.001953 | 0.12157 | 0.29049 | 4939.782 |
| SASSON_RI | 78 | 0.031736 | 0.002036 | 0.057502 | 0.29051 | 4940.123 |
| MIKKELSEN | 339 | 0.016668 | 0.002213 | 0.030213 | 0.29059 | 4941.483 |
| BIOCARTA_ | 5 | 0.16023 | 0.002607 | 0.29061 | 0.2907 | 4943.354 |

| | | | | | | |
|---|---|---|---|---|---|---|
| WP_NOCG | 42 | 0.042993 | 0.002026 | 0.078001 | 0.29075 | 4944.204 |
| OSMAN_BI | 379 | 0.013545 | 0.0019 | 0.02459 | 0.29087 | 4946.244 |
| GOBP_CELI | 1536 | 0.007458 | 0.002039 | 0.013542 | 0.29091 | 4946.925 |
| GOBP_T_C | 9 | 0.096562 | 0.002108 | 0.17538 | 0.29096 | 4947.775 |
| GOCC_SECI | 828 | 0.009644 | 0.001975 | 0.017523 | 0.29104 | 4949.135 |
| GOBP_SHC | 15 | 0.071201 | 0.002006 | 0.12944 | 0.29114 | 4950.836 |
| GOBP_RIB( | 34 | 0.05325 | 0.002258 | 0.096882 | 0.29129 | 4953.386 |
| GOBP_FRE | 9 | 0.083038 | 0.001813 | 0.1511 | 0.29131 | 4953.727 |
| GOBP_MET | 11 | 0.077191 | 0.001863 | 0.14048 | 0.29134 | 4954.237 |
| GOBP_CYT( | 110 | 0.024337 | 0.001852 | 0.044313 | 0.29144 | 4955.937 |
| LAU_APOP | 5 | 0.10998 | 0.00179 | 0.20034 | 0.29152 | 4957.298 |
| HU_GENOT | 37 | 0.042523 | 0.001881 | 0.077493 | 0.2916 | 4958.658 |
| GOBP_VEN | 14 | 0.081459 | 0.002218 | 0.14848 | 0.29164 | 4959.338 |
| SA_TRKA_I | 16 | 0.071375 | 0.002077 | 0.13014 | 0.29169 | 4960.188 |
| GOBP_POS | 17 | 0.068971 | 0.002069 | 0.12576 | 0.2917 | 4960.359 |
| WP_EICOS/ | 31 | 0.048828 | 0.001977 | 0.089049 | 0.29174 | 4961.039 |
| GOBP_MIT | 29 | 0.047351 | 0.001855 | 0.086368 | 0.29176 | 4961.379 |
| GOBP_PEP | 5 | 0.13956 | 0.002271 | 0.25457 | 0.29177 | 4961.549 |
| GOMF_PR( | 5 | 0.13956 | 0.002271 | 0.25457 | 0.29177 | 4961.549 |
| GOBP_COF | 6 | 0.12004 | 0.00214 | 0.21898 | 0.29178 | 4961.719 |
| GOMF_VAS | 7 | 0.12808 | 0.002466 | 0.23363 | 0.29178 | 4961.719 |
| GOCC_MIT | 55 | 0.030717 | 0.001656 | 0.056049 | 0.29184 | 4962.739 |
| GOBP_LYM | 76 | 0.030115 | 0.001907 | 0.054954 | 0.29185 | 4962.909 |
| IZADPANAI | 102 | 0.029167 | 0.002138 | 0.053229 | 0.29187 | 4963.249 |
| GOBP_NEG | 5 | 0.14925 | 0.002429 | 0.27247 | 0.29192 | 4964.1 |
| WP_17Q12 | 46 | 0.070061 | 0.003454 | 0.12794 | 0.29198 | 4965.12 |
| WP_PREIM | 56 | 0.037519 | 0.00204 | 0.068513 | 0.29198 | 4965.12 |
| GOBP_NEG | 8 | 0.090312 | 0.001859 | 0.16494 | 0.29201 | 4965.63 |
| GOBP_SYM | 22 | 0.062835 | 0.002144 | 0.11476 | 0.29201 | 4965.63 |
| GOMF_THI | 6 | 0.10654 | 0.001899 | 0.19459 | 0.29203 | 4965.97 |
| GOCC_CELI | 6 | 0.13324 | 0.002375 | 0.24342 | 0.29207 | 4966.65 |
| SHEN_SMA | 408 | 0.013908 | 0.002022 | 0.025408 | 0.29207 | 4966.65 |
| GOBP_SYM | 9 | 0.094823 | 0.00207 | 0.17325 | 0.29208 | 4966.82 |
| GOMF_DYS | 10 | 0.08204 | 0.001888 | 0.14994 | 0.29214 | 4967.841 |
| GOMF_CYC | 13 | 0.070835 | 0.001858 | 0.12949 | 0.29218 | 4968.521 |
| GINESTIER_ | 300 | 0.01468 | 0.001836 | 0.026854 | 0.29232 | 4970.902 |
| REACTOME | 87 | 0.029446 | 0.001994 | 0.053889 | 0.29239 | 4972.092 |
| GOMF_MI( | 71 | 0.032595 | 0.001995 | 0.059698 | 0.29254 | 4974.643 |
| GOCC_CUL | 4 | 0.19196 | 0.002794 | 0.35165 | 0.29258 | 4975.323 |
| NIKOLSKY_ | 150 | 0.037182 | 0.003301 | 0.068116 | 0.29258 | 4975.323 |
| GOMF_ARY | 6 | 0.12897 | 0.002299 | 0.23641 | 0.29269 | 4977.193 |
| GOMF_PH( | 5 | 0.11031 | 0.001795 | 0.20221 | 0.2927 | 4977.364 |
| WP_CANCE | 21 | 0.067112 | 0.002237 | 0.1231 | 0.29281 | 4979.234 |
| GOBP_HOS | 8 | 0.084382 | 0.001737 | 0.15478 | 0.29282 | 4979.404 |
| GOBP_DEC | 9 | 0.088753 | 0.001937 | 0.16282 | 0.29285 | 4979.914 |

| | | | | | | |
|---|---|---|---|---|---|---|
| GOBP_NEG | 30 | 0.05785 | 0.002304 | 0.10616 | 0.2929 | 4980.765 |
| GOBP_HEA | 3 | 0.17108 | 0.002157 | 0.31412 | 0.29301 | 4982.635 |
| GOBP_T_C | 3 | 0.16041 | 0.002022 | 0.29456 | 0.29303 | 4982.975 |
| GOMF_ATI | 10 | 0.08831 | 0.002032 | 0.16219 | 0.29305 | 4983.315 |
| GOBP_POS | 9 | 0.10806 | 0.002359 | 0.19847 | 0.29306 | 4983.485 |
| GOBP_GLC | 11 | 0.086836 | 0.002096 | 0.1595 | 0.29308 | 4983.825 |
| GOBP_TAU | 9 | 0.091205 | 0.001991 | 0.16758 | 0.29314 | 4984.846 |
| GOBP_REG | 7 | 0.12654 | 0.002436 | 0.23256 | 0.29319 | 4985.696 |
| GOBP_FUN | 5 | 0.1251 | 0.002036 | 0.22999 | 0.29324 | 4986.546 |
| BIOCARTA_ | 11 | 0.090472 | 0.002183 | 0.16633 | 0.29325 | 4986.716 |
| NAGASHIM | 52 | 0.042531 | 0.002229 | 0.078221 | 0.29332 | 4987.907 |
| GOMF_TUI | 25 | 0.055688 | 0.002025 | 0.10245 | 0.29338 | 4988.927 |
| RORIE_TAF | 25 | 0.060675 | 0.002207 | 0.11164 | 0.2934 | 4989.267 |
| TORCHIA_T | 18 | 0.061072 | 0.001885 | 0.11237 | 0.2934 | 4989.267 |
| BECKER_TA | 53 | 0.036255 | 0.001918 | 0.066752 | 0.29352 | 4991.308 |
| GOBP_GAN | 686 | 0.010399 | 0.001946 | 0.019145 | 0.29352 | 4991.308 |
| CUI_TCF21 | 35 | 0.045716 | 0.001967 | 0.084211 | 0.29361 | 4992.838 |
| GOBP_BEH | 5 | 0.10886 | 0.001771 | 0.20053 | 0.29361 | 4992.838 |
| WHITFIELD | 44 | 0.040596 | 0.001958 | 0.074788 | 0.29363 | 4993.178 |
| GOBP_NEG | 10 | 0.099518 | 0.00229 | 0.18337 | 0.29366 | 4993.688 |
| REACTOME | 12 | 0.075063 | 0.001892 | 0.13835 | 0.29372 | 4994.709 |
| GOBP_NEG | 8 | 0.098474 | 0.002027 | 0.18167 | 0.2939 | 4997.77 |
| GOMF_PHO | 6 | 0.089711 | 0.001599 | 0.16555 | 0.29394 | 4998.45 |
| IGARASHI_ | 4 | 0.16142 | 0.002349 | 0.29789 | 0.29396 | 4998.79 |
| GOMF_DN/ | 35 | 0.044817 | 0.001928 | 0.082721 | 0.29399 | 4999.3 |
| GOBP_MRI | 5 | 0.11884 | 0.001934 | 0.21942 | 0.29404 | 5000.15 |
| NAKAYAM/ | 82 | 0.028242 | 0.001857 | 0.052143 | 0.29404 | 5000.15 |
| GOMF_LRF | 15 | 0.069006 | 0.001944 | 0.12742 | 0.29406 | 5000.49 |
| GOBP_POS | 62 | 0.035191 | 0.002013 | 0.064998 | 0.29412 | 5001.511 |
| REACTOME | 36 | 0.044338 | 0.001934 | 0.0819 | 0.29413 | 5001.681 |
| GOCC_SPEI | 48 | 0.039205 | 0.001974 | 0.072433 | 0.29417 | 5002.361 |
| GOMF_CAI | 8 | 0.097213 | 0.002001 | 0.17973 | 0.29429 | 5004.401 |
| REACTOME | 62 | 0.04046 | 0.002315 | 0.074813 | 0.29432 | 5004.912 |
| GOBP_POS | 11 | 0.079792 | 0.001926 | 0.14756 | 0.29434 | 5005.252 |
| GOBP_NUC | 195 | 0.018795 | 0.0019 | 0.034762 | 0.29436 | 5005.592 |
| GOBP_REG | 13 | 0.071081 | 0.001865 | 0.13153 | 0.29446 | 5007.292 |
| KEGG_ARA | 57 | 0.036244 | 0.001989 | 0.067074 | 0.29448 | 5007.632 |
| GOBP_2FE_ | 9 | 0.10621 | 0.002318 | 0.1966 | 0.29453 | 5008.483 |
| GOMF_EST | 8 | 0.10706 | 0.002204 | 0.19829 | 0.29463 | 5010.183 |
| WAMUNYO | 132 | 0.024036 | 0.002003 | 0.044541 | 0.29473 | 5011.884 |
| WP_OXIDA | 60 | 0.038766 | 0.002182 | 0.071836 | 0.29473 | 5011.884 |
| GOMF_FK5 | 10 | 0.078225 | 0.0018 | 0.14499 | 0.29477 | 5012.564 |
| GOBP_PEP | 7 | 0.086499 | 0.001665 | 0.16036 | 0.29481 | 5013.244 |
| WP_22Q11 | 116 | 0.028154 | 0.0022 | 0.052204 | 0.29484 | 5013.754 |
| ZHANG_BR | 134 | 0.022691 | 0.001905 | 0.042084 | 0.29488 | 5014.434 |

| | | | | | | |
|---|---|---|---|---|---|---|
| XU_HGF_T/ | 13 | 0.078409 | 0.002057 | 0.14547 | 0.29494 | 5015.455 |
| PHESSE_TA | 17 | 0.063615 | 0.001908 | 0.11808 | 0.29503 | 5016.985 |
| GOBP_BIOl | 193 | 0.019463 | 0.001958 | 0.036187 | 0.29534 | 5022.257 |
| GOCC_CLO | 4 | 0.14416 | 0.002098 | 0.26812 | 0.29541 | 5023.447 |
| CLIMENT_E | 23 | 0.059762 | 0.002085 | 0.11139 | 0.2958 | 5030.079 |
| JOHANSSO | 21 | 0.05303 | 0.001768 | 0.098848 | 0.29582 | 5030.419 |
| YAO_TEMP | 49 | 0.040019 | 0.002036 | 0.074601 | 0.29583 | 5030.589 |
| REACTOME | 39 | 0.045352 | 0.002059 | 0.08461 | 0.29598 | 5033.14 |
| FARMER_B | 303 | 0.015694 | 0.001972 | 0.029288 | 0.29603 | 5033.99 |
| CHANG_PC | 8 | 0.095473 | 0.001965 | 0.17826 | 0.29613 | 5035.691 |
| KEGG_TYPI | 21 | 0.066479 | 0.002216 | 0.12414 | 0.29614 | 5035.861 |
| ODONNELL | 71 | 0.03224 | 0.001973 | 0.06021 | 0.29617 | 5036.371 |
| GARGALOV | 22 | 0.053881 | 0.001838 | 0.10064 | 0.2962 | 5036.881 |
| GOBP_TRIC | 12 | 0.092795 | 0.002339 | 0.17333 | 0.2962 | 5036.881 |
| LEE_LIVER_ | 169 | 0.020018 | 0.001886 | 0.03739 | 0.2962 | 5036.881 |
| GOCC_CRD | 6 | 0.1178 | 0.0021 | 0.22017 | 0.29631 | 5038.752 |
| GOBP_MU | 87 | 0.029985 | 0.002031 | 0.056056 | 0.29636 | 5039.602 |
| REACTOME | 60 | 0.035093 | 0.001975 | 0.065618 | 0.2964 | 5040.282 |
| GOBP_REG | 9 | 0.093786 | 0.002047 | 0.1754 | 0.29643 | 5040.792 |
| GOBP_ROL | 5 | 0.14878 | 0.002421 | 0.27828 | 0.29645 | 5041.132 |
| REACTOME | 23 | 0.057596 | 0.002009 | 0.10777 | 0.29652 | 5042.323 |
| GOBP_ICOS | 114 | 0.025019 | 0.001938 | 0.046847 | 0.29665 | 5044.533 |
| GOCC_GOL | 95 | 0.029466 | 0.002085 | 0.055187 | 0.2967 | 5045.384 |
| GOBP_RNA | 5 | 0.11822 | 0.001924 | 0.22156 | 0.29683 | 5047.594 |
| GOBP_REG | 7 | 0.1044 | 0.00201 | 0.19571 | 0.29687 | 5048.274 |
| MYLLYKAN | 12 | 0.082762 | 0.002086 | 0.15516 | 0.29689 | 5048.614 |
| WP_PDGF_ | 38 | 0.043605 | 0.001954 | 0.081754 | 0.29689 | 5048.614 |
| PID_S1P_S | 14 | 0.066308 | 0.001805 | 0.12441 | 0.29702 | 5050.825 |
| GOMF_NEI | 6 | 0.10638 | 0.001896 | 0.19979 | 0.2972 | 5053.886 |
| GOBP_POS | 7 | 0.1184 | 0.00228 | 0.22238 | 0.29723 | 5054.396 |
| KEGG_AUT | 31 | 0.063938 | 0.002589 | 0.12015 | 0.29731 | 5055.757 |
| GOBP_BIOI | 11 | 0.073209 | 0.001767 | 0.13757 | 0.29732 | 5055.927 |
| WP_INTERI | 54 | 0.037357 | 0.001995 | 0.070208 | 0.29734 | 5056.267 |
| GOBP_CELI | 18 | 0.070432 | 0.002174 | 0.13241 | 0.29739 | 5057.117 |
| SEITZ_NEO | 29 | 0.049496 | 0.001939 | 0.093057 | 0.2974 | 5057.287 |
| GOBP_DNA | 36 | 0.040106 | 0.00175 | 0.075421 | 0.29745 | 5058.137 |
| GOBP_CELI | 24 | 0.068938 | 0.002456 | 0.12965 | 0.29746 | 5058.307 |
| GOBP_PEN | 6 | 0.10072 | 0.001795 | 0.18945 | 0.2975 | 5058.988 |
| GOBP_RIBC | 424 | 0.012694 | 0.001881 | 0.02388 | 0.29751 | 5059.158 |
| GOBP_REG | 7 | 0.091244 | 0.001757 | 0.17167 | 0.29754 | 5059.668 |
| GOBP_NEG | 10 | 0.096062 | 0.00221 | 0.18079 | 0.29759 | 5060.518 |
| GOBP_PRC | 4 | 0.1434 | 0.002087 | 0.26991 | 0.29761 | 5060.858 |
| GOBP_POS | 5 | 0.1406 | 0.002288 | 0.26494 | 0.29782 | 5064.429 |
| GOBP_POS | 17 | 0.061939 | 0.001858 | 0.11674 | 0.29786 | 5065.109 |
| GOMF_UD | 11 | 0.074301 | 0.001793 | 0.14004 | 0.29786 | 5065.109 |

| | | | | | | |
|---|---|---|---|---|---|---|
| VANTVEER_ | 32 | 0.040997 | 0.001687 | 0.077282 | 0.29789 | 5065.619 |
| HERNANDE | 65 | 0.034161 | 0.002001 | 0.064398 | 0.2979 | 5065.79 |
| CREIGHTOI | 468 | 0.012695 | 0.001974 | 0.023934 | 0.29792 | 5066.13 |
| REACTOME | 35 | 0.045428 | 0.001954 | 0.085663 | 0.29795 | 5066.64 |
| GARGALOV | 49 | 0.037443 | 0.001905 | 0.070645 | 0.29805 | 5068.34 |
| GOBP_AV_ | 12 | 0.086333 | 0.002176 | 0.16296 | 0.29813 | 5069.701 |
| KIM_ALL_D | 8 | 0.10064 | 0.002071 | 0.19002 | 0.29819 | 5070.721 |
| GOBP_NEU | 6 | 0.1043 | 0.001859 | 0.19698 | 0.29824 | 5071.571 |
| GOBP_BRA | 4 | 0.11761 | 0.001712 | 0.22219 | 0.29829 | 5072.421 |
| GOBP_REG | 23 | 0.052649 | 0.001837 | 0.099473 | 0.29831 | 5072.762 |
| GOBP_POS | 8 | 0.10161 | 0.002091 | 0.19203 | 0.29837 | 5073.782 |
| GOBP_NEG | 7 | 0.09421 | 0.001814 | 0.17808 | 0.29839 | 5074.122 |
| GOBP_REG | 59 | 0.038709 | 0.002161 | 0.073168 | 0.29839 | 5074.122 |
| FARMER_B | 307 | 0.015868 | 0.002007 | 0.029999 | 0.29843 | 5074.802 |
| GOBP_REG | 15 | 0.066599 | 0.001877 | 0.12599 | 0.29854 | 5076.673 |
| GOBP_REG | 38 | 0.04228 | 0.001895 | 0.080013 | 0.29861 | 5077.863 |
| GOBP_LIPC | 96 | 0.026063 | 0.001854 | 0.049332 | 0.29864 | 5078.373 |
| GOBP_HIST | 26 | 0.048553 | 0.001801 | 0.091933 | 0.29871 | 5079.564 |
| GOBP_PHC | 27 | 0.050536 | 0.00191 | 0.095741 | 0.29881 | 5081.264 |
| REACTOME | 75 | 0.029993 | 0.001887 | 0.056834 | 0.29884 | 5081.774 |
| BROWNE_I | 138 | 0.022689 | 0.001933 | 0.043017 | 0.29894 | 5083.475 |
| GOBP_CEN | 4 | 0.14378 | 0.002093 | 0.27259 | 0.29894 | 5083.475 |
| PLASARI_T | 4 | 0.16169 | 0.002353 | 0.30658 | 0.29896 | 5083.815 |
| GOBP_EME | 26 | 0.053417 | 0.001981 | 0.10133 | 0.29904 | 5085.175 |
| GOBP_GAF | 6 | 0.11 | 0.001961 | 0.20866 | 0.29904 | 5085.175 |
| GOCC_SPIN | 13 | 0.07467 | 0.001959 | 0.14167 | 0.29908 | 5085.855 |
| LEE_LIVER_ | 53 | 0.034847 | 0.001844 | 0.06612 | 0.29909 | 5086.025 |
| REACTOME | 10 | 0.089774 | 0.002066 | 0.17037 | 0.29912 | 5086.536 |
| GOBP_FUC | 9 | 0.081001 | 0.001768 | 0.1538 | 0.29922 | 5088.236 |
| OLSSON_E: | 36 | 0.04388 | 0.001914 | 0.083362 | 0.29932 | 5089.937 |
| GOCC_SYN | 33 | 0.045144 | 0.001886 | 0.085775 | 0.29934 | 5090.277 |
| OUELLET_C | 31 | 0.047806 | 0.001936 | 0.090865 | 0.29941 | 5091.467 |
| WANG_LSD | 23 | 0.062223 | 0.002171 | 0.11834 | 0.29951 | 5093.168 |
| KEGG_NIC( | 24 | 0.051052 | 0.001819 | 0.097097 | 0.29952 | 5093.338 |
| GOBP_NEG | 11 | 0.087516 | 0.002112 | 0.16647 | 0.29955 | 5093.848 |
| GOBP_C21_ | 36 | 0.048596 | 0.00212 | 0.092461 | 0.29959 | 5094.528 |
| GOBP_REG | 175 | 0.01943 | 0.001862 | 0.036975 | 0.29962 | 5095.038 |
| CARRILLOR | 19 | 0.060821 | 0.001929 | 0.11576 | 0.29966 | 5095.718 |
| GOBP_POS | 8 | 0.083046 | 0.001709 | 0.15808 | 0.29968 | 5096.058 |
| GOBP_INTI | 6 | 0.13065 | 0.002329 | 0.2488 | 0.29975 | 5097.249 |
| GOBP_REG | 239 | 0.0176 | 0.001968 | 0.033518 | 0.29977 | 5097.589 |
| GOCC_NUC | 33 | 0.043174 | 0.001804 | 0.082237 | 0.2998 | 5098.099 |
| GOBP_DET | 15 | 0.074113 | 0.002088 | 0.14118 | 0.29981 | 5098.269 |
| GOBP_PER | 22 | 0.047214 | 0.001611 | 0.090017 | 0.29997 | 5100.99 |
| GOBP_MIR | 11 | 0.078774 | 0.001901 | 0.15025 | 0.30004 | 5102.18 |

| | | | | | | |
|---|---|---|---|---|---|---|
| GOBP_CELI | 42 | 0.037839 | 0.001783 | 0.072191 | 0.30009 | 5103.03 |
| REACTOME | 8 | 0.10054 | 0.002069 | 0.19182 | 0.30009 | 5103.03 |
| GOBP_POS | 5 | 0.17423 | 0.002835 | 0.33245 | 0.30011 | 5103.371 |
| GOMF_GLY | 10 | 0.079988 | 0.001841 | 0.15265 | 0.30015 | 5104.051 |
| GOBP_NEG | 14 | 0.067491 | 0.001837 | 0.12881 | 0.30016 | 5104.221 |
| REACTOME | 381 | 0.014453 | 0.002032 | 0.027603 | 0.30028 | 5106.261 |
| GOBP_HEN | 9 | 0.078282 | 0.001709 | 0.14956 | 0.30035 | 5107.452 |
| BROWNE_I | 97 | 0.026415 | 0.001889 | 0.050528 | 0.30057 | 5111.193 |
| GOMF_PR( | 665 | 0.010276 | 0.001894 | 0.019677 | 0.30076 | 5114.424 |
| WP_GPCRS | 22 | 0.062198 | 0.002122 | 0.11913 | 0.30081 | 5115.274 |
| WP_MICRC | 7 | 0.090725 | 0.001747 | 0.17379 | 0.30083 | 5115.614 |
| DUTERTRE_ | 89 | 0.029225 | 0.002002 | 0.056094 | 0.30118 | 5121.566 |
| GOBP_NEG | 153 | 0.021534 | 0.001931 | 0.041339 | 0.30121 | 5122.076 |
| WP_THYRC | 23 | 0.054545 | 0.001903 | 0.10471 | 0.30122 | 5122.246 |
| REACTOME | 56 | 0.034916 | 0.001899 | 0.067034 | 0.30123 | 5122.416 |
| GOBP_MEI | 25 | 0.059352 | 0.002158 | 0.11402 | 0.30135 | 5124.457 |
| GOMF_PH( | 169 | 0.020104 | 0.001894 | 0.038624 | 0.30136 | 5124.627 |
| GOBP_REG | 6 | 0.11261 | 0.002007 | 0.21637 | 0.30139 | 5125.137 |
| REACTOME | 77 | 0.031805 | 0.002027 | 0.06112 | 0.30141 | 5125.477 |
| LOPEZ_ME | 12 | 0.072217 | 0.00182 | 0.1388 | 0.30143 | 5125.817 |
| GOBP_NEG | 276 | 0.016088 | 0.001931 | 0.030949 | 0.3016 | 5128.708 |
| WP_MAMI | 12 | 0.076133 | 0.001919 | 0.14651 | 0.30166 | 5129.728 |
| GOBP_POS | 6 | 0.092613 | 0.001651 | 0.17825 | 0.30169 | 5130.238 |
| GOBP_REG | 7 | 0.10776 | 0.002075 | 0.20749 | 0.30176 | 5131.429 |
| SARTIPY_B | 17 | 0.069163 | 0.002075 | 0.13328 | 0.30191 | 5133.98 |
| ABDELMOH | 16 | 0.059729 | 0.001738 | 0.11512 | 0.30194 | 5134.49 |
| INGRAM_S | 58 | 0.036501 | 0.00202 | 0.070352 | 0.30194 | 5134.49 |
| GOBP_GLY | 15 | 0.062143 | 0.001751 | 0.11981 | 0.30199 | 5135.34 |
| GOMF_G_I | 25 | 0.056007 | 0.002037 | 0.10799 | 0.30202 | 5135.85 |
| GOBP_RES | 13 | 0.085059 | 0.002231 | 0.16408 | 0.30209 | 5137.04 |
| GOBP_REN | 117 | 0.025265 | 0.001983 | 0.048787 | 0.30228 | 5140.271 |
| GOBP_POS | 9 | 0.092136 | 0.002011 | 0.17801 | 0.30237 | 5141.802 |
| REACTOME | 32 | 0.041406 | 0.001703 | 0.080008 | 0.3024 | 5142.312 |
| GOBP_NEG | 12 | 0.080841 | 0.002038 | 0.15628 | 0.30248 | 5143.672 |
| JIANG_COF | 3 | 0.14424 | 0.001818 | 0.27909 | 0.30264 | 5146.393 |
| GOBP_CELI | 14 | 0.072368 | 0.00197 | 0.14004 | 0.30267 | 5146.903 |
| KEGG_UBI( | 126 | 0.022707 | 0.001849 | 0.043949 | 0.30269 | 5147.243 |
| GYORFFY_I | 32 | 0.047792 | 0.001966 | 0.092549 | 0.30279 | 5148.944 |
| REACTOME | 8 | 0.099454 | 0.002047 | 0.1927 | 0.3029 | 5150.815 |
| GOBP_OVU | 9 | 0.10679 | 0.002331 | 0.20701 | 0.30297 | 5152.005 |
| BIOCARTA_ | 13 | 0.072781 | 0.001909 | 0.14113 | 0.30304 | 5153.195 |
| GOBP_REG | 160 | 0.018742 | 0.001718 | 0.036344 | 0.30304 | 5153.195 |
| GOCC_AUT | 106 | 0.0232 | 0.001734 | 0.045005 | 0.30311 | 5154.386 |
| HUI_MAPK | 18 | 0.06145 | 0.001897 | 0.11922 | 0.30312 | 5154.556 |
| YUAN_ZNF | 19 | 0.050057 | 0.001587 | 0.097173 | 0.30323 | 5156.426 |

| | | | | | | |
|---|---|---|---|---|---|---|
| GOBP_POS | 9 | 0.079009 | 0.001725 | 0.15342 | 0.30328 | 5157.276 |
| GOBP_PRC | 14 | 0.062785 | 0.001709 | 0.12202 | 0.30344 | 5159.997 |
| GOBP_POS | 6 | 0.12268 | 0.002187 | 0.2386 | 0.30356 | 5162.038 |
| MITSIADES | 239 | 0.016556 | 0.001851 | 0.032225 | 0.30372 | 5164.759 |
| REACTOME | 5 | 0.13775 | 0.002241 | 0.26817 | 0.30375 | 5165.269 |
| WP_SYNAF | 49 | 0.036802 | 0.001873 | 0.071651 | 0.30376 | 5165.439 |
| GOBP_POS | 102 | 0.026634 | 0.001952 | 0.05186 | 0.30378 | 5165.779 |
| GOBP_REG | 7 | 0.10343 | 0.001991 | 0.20143 | 0.30382 | 5166.459 |
| KEGG_VAS | 112 | 0.02548 | 0.001957 | 0.049628 | 0.30383 | 5166.629 |
| GOBP_REG | 21 | 0.049977 | 0.001666 | 0.097345 | 0.30384 | 5166.799 |
| GOBP_POS | 19 | 0.057706 | 0.00183 | 0.11243 | 0.30388 | 5167.479 |
| GOBP_CELI | 245 | 0.015715 | 0.001779 | 0.030639 | 0.30401 | 5169.69 |
| WP_G13_S | 35 | 0.040279 | 0.001733 | 0.078531 | 0.30401 | 5169.69 |
| WU_HBX_T | 13 | 0.07829 | 0.002054 | 0.15266 | 0.30404 | 5170.2 |
| GOBP_SUB | 25 | 0.05084 | 0.001849 | 0.099145 | 0.30405 | 5170.37 |
| GOBP_DET | 47 | 0.03776 | 0.001882 | 0.073649 | 0.30408 | 5170.88 |
| HINATA_NI | 80 | 0.028086 | 0.001825 | 0.054824 | 0.30422 | 5173.261 |
| GOMF_AM | 12 | 0.061637 | 0.001554 | 0.12034 | 0.30426 | 5173.941 |
| REACTOME | 7 | 0.11416 | 0.002198 | 0.22296 | 0.30432 | 5174.962 |
| REACTOME | 5 | 0.13686 | 0.002227 | 0.26729 | 0.30432 | 5174.962 |
| GOBP_PRC | 125 | 0.021444 | 0.001739 | 0.041886 | 0.30434 | 5175.302 |
| VARELA_ZN | 38 | 0.040228 | 0.001803 | 0.078628 | 0.30446 | 5177.342 |
| MIKKELSEN | 115 | 0.024476 | 0.001905 | 0.04788 | 0.30461 | 5179.893 |
| GOMF_HSF | 41 | 0.039918 | 0.001858 | 0.078108 | 0.30466 | 5180.743 |
| REACTOME | 25 | 0.048896 | 0.001778 | 0.095688 | 0.30468 | 5181.083 |
| GOMF_HYI | 4 | 0.11525 | 0.001678 | 0.22563 | 0.30475 | 5182.274 |
| GOBP_ESTI | 17 | 0.060758 | 0.001823 | 0.11897 | 0.30478 | 5182.784 |
| REACTOME | 5 | 0.14472 | 0.002355 | 0.28338 | 0.30479 | 5182.954 |
| GOMF_CYT | 214 | 0.020207 | 0.002139 | 0.039581 | 0.30485 | 5183.974 |
| GOBP_UDF | 9 | 0.11511 | 0.002513 | 0.22552 | 0.30488 | 5184.484 |
| GOBP_PHC | 8 | 0.10953 | 0.002254 | 0.21474 | 0.305 | 5186.525 |
| GOBP_REG | 7 | 0.099231 | 0.001911 | 0.19456 | 0.30502 | 5186.865 |
| GOBP_MET | 7 | 0.084616 | 0.001629 | 0.16594 | 0.30506 | 5187.545 |
| FIRESTEIN_ | 6 | 0.13554 | 0.002416 | 0.26584 | 0.30508 | 5187.885 |
| GOBP_REG | 4 | 0.16942 | 0.002466 | 0.3323 | 0.30509 | 5188.055 |
| GOBP_POS | 1377 | 0.007228 | 0.00188 | 0.014184 | 0.30517 | 5189.416 |
| GOCC_CAT | 13 | 0.068438 | 0.001795 | 0.13435 | 0.30523 | 5190.436 |
| ZHANG_RE | 100 | 0.026673 | 0.001936 | 0.052405 | 0.30538 | 5192.987 |
| GOMF_OX( | 6 | 0.088996 | 0.001586 | 0.17491 | 0.30544 | 5194.007 |
| GOBP_POS | 4 | 0.11858 | 0.001726 | 0.23307 | 0.30545 | 5194.177 |
| GOBP_RES | 11 | 0.07655 | 0.001847 | 0.15048 | 0.30548 | 5194.687 |
| GOBP_BEH | 37 | 0.042451 | 0.001878 | 0.083451 | 0.30549 | 5194.857 |
| GOBP_NIK_ | 115 | 0.023932 | 0.001862 | 0.047047 | 0.30549 | 5194.857 |
| KONDO_PF | 92 | 0.028834 | 0.002008 | 0.056717 | 0.3056 | 5196.728 |
| REACTOME | 16 | 0.058033 | 0.001689 | 0.11417 | 0.30563 | 5197.238 |

| | | | | | | |
|---|---|---|---|---|---|---|
| NOUSHME | 7 | 0.11203 | 0.002157 | 0.22053 | 0.30573 | 5198.939 |
| GOBP_POS | 10 | 0.070157 | 0.001614 | 0.13813 | 0.30576 | 5199.449 |
| GROSS_HY | 77 | 0.027361 | 0.001744 | 0.053868 | 0.30576 | 5199.449 |
| GOBP_POS | 122 | 0.023788 | 0.001906 | 0.046853 | 0.30583 | 5200.639 |
| GOMF_NF/ | 5 | 0.13165 | 0.002142 | 0.25935 | 0.30586 | 5201.149 |
| GOMF_PYF | 5 | 0.10911 | 0.001776 | 0.21515 | 0.30603 | 5204.04 |
| GOBP_PRC | 926 | 0.008333 | 0.0018 | 0.016439 | 0.30612 | 5205.571 |
| REACTOME | 4 | 0.15466 | 0.002251 | 0.30529 | 0.30623 | 5207.441 |
| GOBP_RES | 28 | 0.047194 | 0.001816 | 0.093247 | 0.30639 | 5210.162 |
| GOMF_MC | 149 | 0.020607 | 0.001824 | 0.040755 | 0.30656 | 5213.053 |
| REACTOME | 182 | 0.018756 | 0.001833 | 0.037099 | 0.30659 | 5213.563 |
| GOBP_FOA | 38 | 0.043435 | 0.001947 | 0.085929 | 0.30662 | 5214.073 |
| GOMF_CY( | 26 | 0.049608 | 0.00184 | 0.098168 | 0.30666 | 5214.753 |
| GOBP_RIB( | 72 | 0.028029 | 0.001728 | 0.055476 | 0.3067 | 5215.434 |
| WP_SERINI | 8 | 0.082053 | 0.001689 | 0.16243 | 0.30672 | 5215.774 |
| GOBP_AMI | 12 | 0.082206 | 0.002072 | 0.16278 | 0.30678 | 5216.794 |
| GOBP_VES | 347 | 0.013349 | 0.001793 | 0.02645 | 0.30689 | 5218.664 |
| GOBP_NAT | 25 | 0.049421 | 0.001797 | 0.097956 | 0.30695 | 5219.685 |
| GOBP_PLA: | 155 | 0.020362 | 0.001837 | 0.040369 | 0.307 | 5220.535 |
| GOBP_LEN | 37 | 0.045883 | 0.002029 | 0.090991 | 0.30704 | 5221.215 |
| KEGG_DNA | 34 | 0.041123 | 0.001744 | 0.081553 | 0.30705 | 5221.385 |
| GOCC_MIT | 6 | 0.10174 | 0.001813 | 0.20179 | 0.30708 | 5221.895 |
| GOMF_INV | 26 | 0.056117 | 0.002081 | 0.11131 | 0.30708 | 5221.895 |
| GOBP_PEP | 7 | 0.11133 | 0.002144 | 0.22088 | 0.30712 | 5222.576 |
| GOBP_NU( | 225 | 0.015643 | 0.001698 | 0.031053 | 0.30722 | 5224.276 |
| GOBP_NEG | 23 | 0.052995 | 0.001849 | 0.10522 | 0.30726 | 5224.956 |
| KRIGE_AM | 29 | 0.050895 | 0.001993 | 0.10113 | 0.3074 | 5227.337 |
| BIOCARTA_ | 19 | 0.064903 | 0.002058 | 0.12899 | 0.30743 | 5227.847 |
| ROPERO_H | 97 | 0.02604 | 0.001862 | 0.051756 | 0.30744 | 5228.017 |
| GOBP_POS | 10 | 0.090366 | 0.002079 | 0.17963 | 0.30746 | 5228.357 |
| GOBP_REG | 5 | 0.11629 | 0.001892 | 0.23118 | 0.30747 | 5228.527 |
| GROSS_HY | 8 | 0.087704 | 0.001805 | 0.17436 | 0.30748 | 5228.697 |
| GOBP_CYC | 39 | 0.044121 | 0.002003 | 0.087728 | 0.30751 | 5229.208 |
| GOBP_POS | 7 | 0.090675 | 0.001746 | 0.18031 | 0.30752 | 5229.378 |
| REACTOME | 14 | 0.072054 | 0.001962 | 0.14333 | 0.30758 | 5230.398 |
| GOMF_CAT | 226 | 0.015551 | 0.001691 | 0.030954 | 0.3077 | 5232.439 |
| GOBP_REG | 168 | 0.019426 | 0.001824 | 0.038674 | 0.30773 | 5232.949 |
| GOBP_SEC | 272 | 0.015655 | 0.001866 | 0.031198 | 0.30791 | 5236.01 |
| GOCC_ELA: | 5 | 0.11934 | 0.001942 | 0.23797 | 0.30801 | 5237.71 |
| KOKKINAKI | 115 | 0.024184 | 0.001882 | 0.048247 | 0.3081 | 5239.241 |
| GOBP_POS | 6 | 0.13683 | 0.002439 | 0.273 | 0.30811 | 5239.411 |
| GOBP_PRC | 19 | 0.050819 | 0.001611 | 0.1014 | 0.30813 | 5239.751 |
| GOBP_MO | 42 | 0.038556 | 0.001817 | 0.076957 | 0.30819 | 5240.771 |
| GOBP_TEL( | 32 | 0.040145 | 0.001651 | 0.08021 | 0.30836 | 5243.662 |
| GOBP_PEP | 33 | 0.047154 | 0.00197 | 0.094224 | 0.30838 | 5244.002 |

| | | | | | | |
|---|---|---|---|---|---|---|
| GOBP_REG | 154 | 0.020969 | 0.001886 | 0.041929 | 0.3085 | 5246.043 |
| WP_EXERC | 46 | 0.036086 | 0.001779 | 0.07218 | 0.30856 | 5247.063 |
| GOMF_MIT | 29 | 0.045235 | 0.001772 | 0.09057 | 0.30874 | 5250.124 |
| GOBP_NEU | 116 | 0.023371 | 0.001826 | 0.046811 | 0.3088 | 5251.144 |
| GOBP_HEP | 136 | 0.021412 | 0.001811 | 0.04289 | 0.30881 | 5251.314 |
| CHEMELLO | 17 | 0.058093 | 0.001743 | 0.11645 | 0.30895 | 5253.695 |
| GOBP_POS | 5 | 0.1271 | 0.002068 | 0.2548 | 0.30895 | 5253.695 |
| BIOCARTA_ | 9 | 0.099829 | 0.002179 | 0.20016 | 0.30898 | 5254.205 |
| GOBP_POS | 1208 | 0.007645 | 0.001871 | 0.015331 | 0.30902 | 5254.885 |
| BIOCARTA_ | 21 | 0.048142 | 0.001605 | 0.096553 | 0.30903 | 5255.055 |
| FERRANDO | 78 | 0.028672 | 0.001839 | 0.057503 | 0.30903 | 5255.055 |
| GOBP_POS | 18 | 0.069313 | 0.002139 | 0.13901 | 0.30903 | 5255.055 |
| GOBP_NEG | 15 | 0.059561 | 0.001678 | 0.11952 | 0.30913 | 5256.756 |
| KEGG_PHE | 16 | 0.065222 | 0.001898 | 0.13091 | 0.30918 | 5257.606 |
| WP_OSTEC | 14 | 0.070262 | 0.001913 | 0.14108 | 0.30923 | 5258.456 |
| GOBP_PLA | 38 | 0.04396 | 0.00197 | 0.088284 | 0.30927 | 5259.136 |
| PLASARI_T | 54 | 0.037268 | 0.00199 | 0.074848 | 0.30927 | 5259.136 |
| GOMF_ACT | 5 | 0.1368 | 0.002226 | 0.27521 | 0.30957 | 5264.238 |
| NOUZOVA_ | 58 | 0.035391 | 0.001959 | 0.071202 | 0.30958 | 5264.408 |
| GAUSSMAN | 24 | 0.054213 | 0.001932 | 0.10918 | 0.30975 | 5267.299 |
| PID_NCADI | 33 | 0.039609 | 0.001655 | 0.079765 | 0.30975 | 5267.299 |
| MOOTHA_ | 19 | 0.057045 | 0.001809 | 0.11491 | 0.3098 | 5268.149 |
| REACTOME | 21 | 0.05541 | 0.001847 | 0.11162 | 0.3098 | 5268.149 |
| BARRIER_C | 27 | 0.046669 | 0.001764 | 0.09402 | 0.30982 | 5268.489 |
| GOCC_RNA | 9 | 0.070641 | 0.001542 | 0.14235 | 0.30986 | 5269.169 |
| GOCC_AZU | 55 | 0.032444 | 0.001749 | 0.065407 | 0.30994 | 5270.53 |
| REACTOME | 42 | 0.035776 | 0.001686 | 0.072128 | 0.30995 | 5270.7 |
| GOBP_POS | 25 | 0.051351 | 0.001868 | 0.10356 | 0.30999 | 5271.38 |
| GOBP_REG | 10 | 0.093211 | 0.002145 | 0.18801 | 0.31003 | 5272.06 |
| GOBP_EAT | 40 | 0.044993 | 0.002069 | 0.09077 | 0.31006 | 5272.57 |
| REACTOME | 150 | 0.020069 | 0.001782 | 0.040496 | 0.3101 | 5273.251 |
| GOBP_POS | 5 | 0.12524 | 0.002038 | 0.25283 | 0.31017 | 5274.441 |
| GOBP_INTI | 138 | 0.020969 | 0.001786 | 0.042362 | 0.31031 | 5276.822 |
| GOBP_AXC | 9 | 0.082347 | 0.001798 | 0.16642 | 0.31037 | 5277.842 |
| GOBP_SNC | 16 | 0.062848 | 0.001829 | 0.12705 | 0.31042 | 5278.692 |
| TURASHVIL | 92 | 0.025767 | 0.001794 | 0.05209 | 0.31042 | 5278.692 |
| GOBP_NEG | 203 | 0.017207 | 0.001775 | 0.034786 | 0.31043 | 5278.862 |
| GOMF_UBI | 6 | 0.077659 | 0.001384 | 0.157 | 0.31043 | 5278.862 |
| GOBP_POS | 2 | 0.1968 | 0.002026 | 0.398 | 0.31049 | 5279.882 |
| ULE_SPLICI | 42 | 0.038267 | 0.001803 | 0.077414 | 0.31055 | 5280.903 |
| GOBP_NUC | 617 | 0.009951 | 0.001769 | 0.020147 | 0.31069 | 5283.283 |
| GOBP_OXA | 10 | 0.072853 | 0.001676 | 0.14755 | 0.31075 | 5284.304 |
| GOBP_POS | 7 | 0.10504 | 0.002022 | 0.21281 | 0.3108 | 5285.154 |
| GOMF_AM | 16 | 0.062723 | 0.001825 | 0.12711 | 0.31085 | 5286.004 |
| HOLLEMAN | 19 | 0.055943 | 0.001774 | 0.11337 | 0.31085 | 5286.004 |

| | | | | | | |
|---|---|---|---|---|---|---|
| GOBP_AMI | 5 | 0.12037 | 0.001959 | 0.24397 | 0.31087 | 5286.344 |
| REACTOME | 57 | 0.028713 | 0.001575 | 0.058206 | 0.3109 | 5286.855 |
| REACTOME | 8 | 0.077751 | 0.0016 | 0.15765 | 0.31094 | 5287.535 |
| GOBP_REG | 6 | 0.12266 | 0.002186 | 0.24876 | 0.31098 | 5288.215 |
| WP_NEOV/ | 36 | 0.04372 | 0.001907 | 0.088669 | 0.31099 | 5288.385 |
| GOBP_THR | 5 | 0.098025 | 0.001595 | 0.19886 | 0.31103 | 5289.065 |
| GOBP_NEG | 76 | 0.028059 | 0.001777 | 0.056927 | 0.31104 | 5289.235 |
| GOBP_CAR | 552 | 0.010128 | 0.001706 | 0.02056 | 0.31116 | 5291.276 |
| GOBP_CLE/ | 28 | 0.044351 | 0.001707 | 0.090092 | 0.31126 | 5292.976 |
| GOMF_TEL | 6 | 0.10169 | 0.001813 | 0.20659 | 0.31128 | 5293.316 |
| GOMF_CYT | 112 | 0.024196 | 0.001858 | 0.04916 | 0.3113 | 5293.657 |
| GOBP_FAT | 102 | 0.022205 | 0.001628 | 0.045131 | 0.31136 | 5294.677 |
| GOBP_SEXI | 812 | 0.00873 | 0.001771 | 0.017752 | 0.31144 | 5296.037 |
| GOBP_POS | 31 | 0.042961 | 0.00174 | 0.087426 | 0.31157 | 5298.248 |
| GOBP_NEG | 11 | 0.080413 | 0.001941 | 0.16372 | 0.31165 | 5299.608 |
| GOBP_PRC | 21 | 0.051289 | 0.00171 | 0.10442 | 0.31166 | 5299.778 |
| WP_OMEG | 13 | 0.072712 | 0.001907 | 0.14814 | 0.31177 | 5301.649 |
| GOBP_NEG | 6 | 0.1102 | 0.001964 | 0.22456 | 0.31181 | 5302.329 |
| REACTOME | 18 | 0.058265 | 0.001798 | 0.11875 | 0.31183 | 5302.669 |
| GOMF_OXI | 9 | 0.093306 | 0.002037 | 0.19023 | 0.3119 | 5303.86 |
| GOBP_CHR | 16 | 0.064871 | 0.001888 | 0.13232 | 0.31198 | 5305.22 |
| REACTOME | 2 | 0.17338 | 0.001785 | 0.35367 | 0.31199 | 5305.39 |
| BAFNA_MU | 3 | 0.15332 | 0.001933 | 0.31287 | 0.31206 | 5306.58 |
| WIERENGA | 51 | 0.037149 | 0.001928 | 0.075819 | 0.31208 | 5306.92 |
| GOCC_THC | 4 | 0.085193 | 0.00124 | 0.17393 | 0.31214 | 5307.941 |
| GOBP_U4_ | 8 | 0.11941 | 0.002458 | 0.24381 | 0.31215 | 5308.111 |
| NAGASHIM | 167 | 0.019152 | 0.001793 | 0.039115 | 0.3122 | 5308.961 |
| GOBP_POS | 8 | 0.07818 | 0.001609 | 0.15974 | 0.31227 | 5310.151 |
| CHEN_HOX | 218 | 0.017334 | 0.001852 | 0.035424 | 0.31231 | 5310.832 |
| GOMF_EXT | 154 | 0.021146 | 0.001902 | 0.043217 | 0.31232 | 5311.002 |
| LANDEMAI | 21 | 0.054337 | 0.001811 | 0.11106 | 0.31233 | 5311.172 |
| TAKEDA_T/ | 173 | 0.020352 | 0.001939 | 0.041603 | 0.31235 | 5311.512 |
| WELCSH_B | 184 | 0.017686 | 0.001738 | 0.036195 | 0.31255 | 5314.913 |
| GOBP_BIOI | 1477 | 0.006693 | 0.001797 | 0.013702 | 0.31262 | 5316.103 |
| GOMF_OLI | 6 | 0.10136 | 0.001807 | 0.20773 | 0.3128 | 5319.164 |
| WP_STERO | 67 | 0.028042 | 0.001668 | 0.057469 | 0.3128 | 5319.164 |
| GOBP_REG | 8 | 0.074351 | 0.00153 | 0.15239 | 0.31281 | 5319.334 |
| GOBP_RES | 6 | 0.12267 | 0.002187 | 0.25177 | 0.31305 | 5323.415 |
| GREENBAU | 30 | 0.04555 | 0.001814 | 0.093525 | 0.31312 | 5324.606 |
| PEDERSEN_ | 11 | 0.076059 | 0.001835 | 0.15618 | 0.31314 | 5324.946 |
| PID_PI3KCI | 46 | 0.034587 | 0.001705 | 0.071075 | 0.31327 | 5327.156 |
| GARGALOV | 73 | 0.030984 | 0.001923 | 0.063679 | 0.31329 | 5327.496 |
| GOBP_GLY | 19 | 0.053068 | 0.001683 | 0.10922 | 0.31353 | 5331.578 |
| GOBP_REC | 4 | 0.13999 | 0.002038 | 0.28824 | 0.31361 | 5332.938 |
| GOBP_NEG | 5 | 0.11749 | 0.001912 | 0.24197 | 0.31363 | 5333.278 |

| | | | | | | |
|---|---|---|---|---|---|---|
| GOCC_CYT | 99 | 0.023569 | 0.001702 | 0.048536 | 0.31363 | 5333.278 |
| GOBP_UBI | 77 | 0.02646 | 0.001686 | 0.05452 | 0.31373 | 5334.979 |
| REACTOME | 5 | 0.090713 | 0.001476 | 0.18707 | 0.31387 | 5337.359 |
| GOBP_REG | 234 | 0.016444 | 0.001819 | 0.033916 | 0.3139 | 5337.87 |
| GOBP_DEF | 1026 | 0.007863 | 0.001783 | 0.016243 | 0.31416 | 5342.291 |
| GOBP_NEG | 400 | 0.012245 | 0.001763 | 0.025312 | 0.31428 | 5344.331 |
| ZHAN_MU | 36 | 0.040511 | 0.001767 | 0.083745 | 0.31429 | 5344.501 |
| GOBP_PRC | 27 | 0.044883 | 0.001696 | 0.092804 | 0.31433 | 5345.182 |
| SCHLESING | 26 | 0.053304 | 0.001977 | 0.11026 | 0.31439 | 5346.202 |
| GOBP_CEL | 118 | 0.021818 | 0.00172 | 0.045167 | 0.31453 | 5348.583 |
| WP_IL4_SI | 51 | 0.03376 | 0.001752 | 0.069917 | 0.3146 | 5349.773 |
| WNT_SIGN | 87 | 0.025983 | 0.00176 | 0.053825 | 0.31464 | 5350.453 |
| GOMF_DIH | 6 | 0.10433 | 0.00186 | 0.21615 | 0.31468 | 5351.133 |
| NOUSHME | 9 | 0.084296 | 0.00184 | 0.17482 | 0.31484 | 5353.854 |
| GOBP_MYI | 17 | 0.061489 | 0.001844 | 0.12754 | 0.31487 | 5354.364 |
| GOCC_PLA | 81 | 0.02821 | 0.001844 | 0.05856 | 0.315 | 5356.575 |
| GOBP_RNA | 62 | 0.030677 | 0.001755 | 0.063709 | 0.31508 | 5357.935 |
| RUIZ_TNC_ | 149 | 0.021702 | 0.00192 | 0.045102 | 0.3152 | 5359.976 |
| GOMF_STE | 26 | 0.048415 | 0.001796 | 0.10063 | 0.31522 | 5360.316 |
| GOBP_POS | 7 | 0.099938 | 0.001924 | 0.20773 | 0.31523 | 5360.486 |
| PUJANA_BI | 401 | 0.011983 | 0.001728 | 0.02491 | 0.31525 | 5360.826 |
| GOBP_POS | 5 | 0.12138 | 0.001975 | 0.25255 | 0.31539 | 5363.207 |
| PEPPER_CH | 20 | 0.052229 | 0.001699 | 0.10876 | 0.31554 | 5365.758 |
| GOBP_RUF | 41 | 0.037279 | 0.001735 | 0.077663 | 0.31561 | 5366.948 |
| GOBP_LEU | 8 | 0.081556 | 0.001679 | 0.16995 | 0.31566 | 5367.798 |
| BUFFA_HYI | 47 | 0.035276 | 0.001758 | 0.073535 | 0.31572 | 5368.819 |
| PID_SHP2_ | 56 | 0.034111 | 0.001855 | 0.071145 | 0.31581 | 5370.349 |
| GOBP_INO | 18 | 0.051029 | 0.001575 | 0.10644 | 0.31582 | 5370.519 |
| GOMF_MIS | 11 | 0.058626 | 0.001415 | 0.12228 | 0.31582 | 5370.519 |
| GOBP_NEG | 14 | 0.070864 | 0.001929 | 0.14785 | 0.31587 | 5371.369 |
| DARWICHE | 147 | 0.018221 | 0.001602 | 0.03803 | 0.31593 | 5372.39 |
| REACTOME | 6 | 0.11864 | 0.002115 | 0.24776 | 0.31602 | 5373.92 |
| GOBP_HOM | 62 | 0.027157 | 0.001554 | 0.056723 | 0.31606 | 5374.6 |
| GOBP_REG | 5 | 0.12307 | 0.002003 | 0.25715 | 0.31611 | 5375.451 |
| GOBP_REG | 78 | 0.028009 | 0.001797 | 0.058521 | 0.31611 | 5375.451 |
| REACTOME | 86 | 0.024055 | 0.00162 | 0.050261 | 0.31611 | 5375.451 |
| GOBP_EST | 1571 | 0.006187 | 0.001709 | 0.012938 | 0.31626 | 5378.001 |
| GOBP_CHE | 20 | 0.054815 | 0.001783 | 0.11478 | 0.31648 | 5381.742 |
| GOMF_COI | 5 | 0.11691 | 0.001902 | 0.24511 | 0.3167 | 5385.484 |
| JONES_TCC | 5 | 0.1216 | 0.001979 | 0.25496 | 0.31671 | 5385.654 |
| GOBP_NUC | 21 | 0.053962 | 0.001799 | 0.11318 | 0.31676 | 5386.504 |
| GOBP_SEM | 14 | 0.069707 | 0.001898 | 0.14631 | 0.31688 | 5388.544 |
| GOMF_LYS | 21 | 0.056476 | 0.001883 | 0.11854 | 0.31689 | 5388.714 |
| DORN_ADE | 38 | 0.034788 | 0.001559 | 0.073038 | 0.31693 | 5389.395 |
| REACTOME | 54 | 0.032982 | 0.001762 | 0.069281 | 0.31702 | 5390.925 |

| | | | | | | |
|---|---|---|---|---|---|---|
| GOBP_SOD | 54 | 0.035404 | 0.001891 | 0.074384 | 0.31706 | 5391.605 |
| GOBP_REG | 173 | 0.018824 | 0.001794 | 0.039552 | 0.31707 | 5391.775 |
| BAKER_HEI | 14 | 0.068226 | 0.001857 | 0.14338 | 0.3171 | 5392.286 |
| HOLLERN_I | 129 | 0.020767 | 0.001711 | 0.043649 | 0.31712 | 5392.626 |
| GUTIERREZ | 9 | 0.081668 | 0.001783 | 0.17173 | 0.3172 | 5393.986 |
| GOBP_POS | 8 | 0.094084 | 0.001936 | 0.19786 | 0.31722 | 5394.326 |
| GOBP_PUL | 22 | 0.056442 | 0.001926 | 0.11874 | 0.31728 | 5395.346 |
| GOBP_MIT | 11 | 0.075037 | 0.001811 | 0.15788 | 0.3173 | 5395.687 |
| KLEIN_PRIM | 52 | 0.032073 | 0.001681 | 0.067496 | 0.31733 | 5396.197 |
| GOBP_PRIM | 13 | 0.072144 | 0.001893 | 0.152 | 0.31752 | 5399.428 |
| PID_BCR_5 | 60 | 0.02911 | 0.001639 | 0.061329 | 0.31752 | 5399.428 |
| GOBP_HEM | 32 | 0.041127 | 0.001692 | 0.086652 | 0.31753 | 5399.598 |
| GOBP_POS | 5 | 0.14446 | 0.002351 | 0.30446 | 0.31758 | 5400.448 |
| WP_LNCRN | 6 | 0.10725 | 0.001912 | 0.22605 | 0.31759 | 5400.618 |
| SONG_TAR | 54 | 0.031367 | 0.001675 | 0.066123 | 0.31762 | 5401.128 |
| MCBRYAN_ | 117 | 0.022065 | 0.001732 | 0.046521 | 0.31765 | 5401.638 |
| BIOCARTA_ | 11 | 0.060243 | 0.001454 | 0.12704 | 0.31768 | 5402.148 |
| MEISSNER_ | 516 | 0.010704 | 0.001745 | 0.022579 | 0.31772 | 5402.829 |
| REACTOME | 58 | 0.032833 | 0.001817 | 0.06926 | 0.31773 | 5402.999 |
| GOBP_NER | 87 | 0.028397 | 0.001923 | 0.059915 | 0.31777 | 5403.679 |
| GOBP_NEU | 53 | 0.034721 | 0.001837 | 0.073271 | 0.3178 | 5404.189 |
| GOMF_CAL | 24 | 0.041506 | 0.001479 | 0.087727 | 0.31806 | 5408.61 |
| WP_SEROT | 10 | 0.073277 | 0.001686 | 0.15489 | 0.31808 | 5408.95 |
| GOBP_POS | 7 | 0.10214 | 0.001967 | 0.21593 | 0.31809 | 5409.12 |
| GOBP_HOM | 281 | 0.014006 | 0.001696 | 0.029616 | 0.31814 | 5409.971 |
| GOBP_OPT | 7 | 0.087219 | 0.001679 | 0.18449 | 0.31819 | 5410.821 |
| GOBP_REG | 15 | 0.063305 | 0.001784 | 0.1339 | 0.31819 | 5410.821 |
| GOCC_REP | 62 | 0.028634 | 0.001638 | 0.060571 | 0.31821 | 5411.161 |
| MARIADAS | 60 | 0.031551 | 0.001776 | 0.066794 | 0.31834 | 5413.372 |
| GOBP_REG | 64 | 0.027484 | 0.001598 | 0.058192 | 0.31836 | 5413.712 |
| GOBP_THY | 48 | 0.035485 | 0.001787 | 0.075135 | 0.31836 | 5413.712 |
| SEMBA_FH | 11 | 0.061494 | 0.001484 | 0.13024 | 0.31841 | 5414.562 |
| GOBP_DNA | 19 | 0.052212 | 0.001656 | 0.1106 | 0.31843 | 5414.902 |
| WP_DNA_I | 53 | 0.031077 | 0.001644 | 0.065844 | 0.31847 | 5415.582 |
| XU_RESPO | 11 | 0.073909 | 0.001784 | 0.1566 | 0.31848 | 5415.752 |
| BIOCARTA_ | 23 | 0.04661 | 0.001626 | 0.09878 | 0.31852 | 5416.433 |
| BIOCARTA_ | 6 | 0.078976 | 0.001408 | 0.16743 | 0.31858 | 5417.453 |
| GOBP_NEU | 35 | 0.040169 | 0.001728 | 0.085181 | 0.31862 | 5418.133 |
| GOBP_POS | 64 | 0.029167 | 0.001695 | 0.061864 | 0.31866 | 5418.813 |
| GOBP_ASP | 17 | 0.055238 | 0.001657 | 0.11717 | 0.31867 | 5418.983 |
| SANA_TNF | 75 | 0.030293 | 0.001906 | 0.064292 | 0.31876 | 5420.514 |
| SMIRNOV_ | 49 | 0.032885 | 0.001673 | 0.069821 | 0.31883 | 5421.704 |
| REACTOME | 11 | 0.06695 | 0.001616 | 0.14218 | 0.31886 | 5422.214 |
| PASTURAL_ | 5 | 0.11656 | 0.001897 | 0.24755 | 0.31887 | 5422.384 |
| GOMF_NU | 7 | 0.093902 | 0.001808 | 0.19946 | 0.3189 | 5422.895 |

| | | | | | | |
|---|---|---|---|---|---|---|
| WP_METAI | 173 | 0.020007 | 0.001906 | 0.04252 | 0.31899 | 5424.425 |
| WP_PHOSI | 54 | 0.032782 | 0.001751 | 0.069673 | 0.319 | 5424.595 |
| GOBP_POS | 6 | 0.090625 | 0.001615 | 0.19315 | 0.31947 | 5432.587 |
| SCIBETTA_I | 73 | 0.026472 | 0.001643 | 0.056423 | 0.31948 | 5432.757 |
| WP_EGFEG | 156 | 0.01892 | 0.001713 | 0.040329 | 0.31948 | 5432.757 |
| GOBP_REG | 20 | 0.050058 | 0.001629 | 0.10675 | 0.31957 | 5434.288 |
| GOMF_TRA | 5 | 0.089535 | 0.001457 | 0.19102 | 0.31964 | 5435.478 |
| BIOCARTA_ | 36 | 0.043708 | 0.001907 | 0.093362 | 0.31984 | 5438.879 |
| GOMF_TRA | 41 | 0.038338 | 0.001785 | 0.08191 | 0.31988 | 5439.559 |
| GOBP_CELI | 34 | 0.041806 | 0.001773 | 0.089366 | 0.31996 | 5440.92 |
| GOBP_RIB( | 6 | 0.08378 | 0.001493 | 0.17917 | 0.32004 | 5442.28 |
| GOBP_REG | 410 | 0.011512 | 0.001678 | 0.024634 | 0.32014 | 5443.981 |
| GOBP_MEI | 5 | 0.12579 | 0.002047 | 0.26931 | 0.32023 | 5445.511 |
| GOMF_CLA | 10 | 0.067 | 0.001542 | 0.14349 | 0.32028 | 5446.361 |
| GOBP_PEP | 113 | 0.022036 | 0.0017 | 0.04721 | 0.32034 | 5447.382 |
| ZHANG_BR | 409 | 0.011357 | 0.001654 | 0.024332 | 0.32034 | 5447.382 |
| REACTOME | 11 | 0.060631 | 0.001463 | 0.12995 | 0.3204 | 5448.402 |
| GOBP_NK_ | 10 | 0.068135 | 0.001568 | 0.14609 | 0.32047 | 5449.592 |
| IWANAGA_ | 6 | 0.10154 | 0.00181 | 0.21774 | 0.32049 | 5449.932 |
| GOCC_LAT | 62 | 0.028773 | 0.001646 | 0.061842 | 0.32088 | 5456.564 |
| GOMF_CAI | 13 | 0.069678 | 0.001828 | 0.14979 | 0.32091 | 5457.075 |
| BLANCO_M | 167 | 0.01716 | 0.001607 | 0.036892 | 0.32092 | 5457.245 |
| GOMF_STE | 23 | 0.054244 | 0.001892 | 0.11665 | 0.32096 | 5457.925 |
| GOBP_POS | 8 | 0.068752 | 0.001415 | 0.14803 | 0.32117 | 5461.496 |
| GOBP_MA | 50 | 0.030463 | 0.001566 | 0.065629 | 0.32127 | 5463.196 |
| REACTOME | 19 | 0.053436 | 0.001694 | 0.11517 | 0.32133 | 5464.217 |
| GOBP_NEU | 4 | 0.14413 | 0.002098 | 0.31072 | 0.32137 | 5464.897 |
| GOMF_HIS | 106 | 0.021314 | 0.001593 | 0.045953 | 0.32139 | 5465.237 |
| GOBP_NEG | 5 | 0.10714 | 0.001744 | 0.23103 | 0.32141 | 5465.577 |
| GOBP_ORG | 306 | 0.013062 | 0.00165 | 0.028191 | 0.32156 | 5468.128 |
| GOBP_ETH | 25 | 0.04142 | 0.001506 | 0.089432 | 0.32163 | 5469.318 |
| XIE_ST_HS( | 23 | 0.046928 | 0.001637 | 0.10133 | 0.32164 | 5469.488 |
| GOBP_CYC | 13 | 0.063913 | 0.001677 | 0.13802 | 0.32166 | 5469.828 |
| BIOCARTA_ | 18 | 0.050148 | 0.001548 | 0.1083 | 0.32167 | 5469.998 |
| REACTOME | 165 | 0.017804 | 0.001657 | 0.038452 | 0.32167 | 5469.998 |
| MARTINEZ_ | 534 | 0.010231 | 0.001696 | 0.022101 | 0.32171 | 5470.679 |
| GHO_ATF5 | 16 | 0.056062 | 0.001631 | 0.12113 | 0.32175 | 5471.359 |
| GOBP_NEU | 23 | 0.04923 | 0.001717 | 0.10637 | 0.32175 | 5471.359 |
| PEDRIOLI_I | 379 | 0.012088 | 0.001695 | 0.026124 | 0.32179 | 5472.039 |
| GOBP_BUN | 6 | 0.11842 | 0.002111 | 0.25619 | 0.32196 | 5474.93 |
| GOBP_POS | 17 | 0.055546 | 0.001666 | 0.12026 | 0.32208 | 5476.97 |
| BERTUCCI_ | 42 | 0.03815 | 0.001798 | 0.082598 | 0.32209 | 5477.14 |
| GOBP_LIP( | 7 | 0.10127 | 0.00195 | 0.21928 | 0.3221 | 5477.311 |
| GOBP_MY( | 6 | 0.10232 | 0.001824 | 0.22166 | 0.32219 | 5478.841 |
| GOBP_CELI | 213 | 0.015046 | 0.001589 | 0.032599 | 0.3222 | 5479.011 |

| | | | | | | |
|---|---|---|---|---|---|---|
| GOBP_NEG | 12 | 0.058326 | 0.00147 | 0.12637 | 0.3222 | 5479.011 |
| WP_OXYST | 45 | 0.034905 | 0.001702 | 0.075708 | 0.32239 | 5482.242 |
| GOBP_RES | 9 | 0.071031 | 0.001551 | 0.15408 | 0.32241 | 5482.582 |
| SIG_BCR_S | 44 | 0.036619 | 0.001766 | 0.079452 | 0.32244 | 5483.092 |
| SUNG_MET | 52 | 0.028699 | 0.001504 | 0.062306 | 0.32254 | 5484.793 |
| ACOSTA_PI | 81 | 0.024932 | 0.00163 | 0.054152 | 0.32261 | 5485.983 |
| REACTOME | 32 | 0.036234 | 0.001491 | 0.078703 | 0.32262 | 5486.153 |
| GOBP_REG | 310 | 0.013606 | 0.001729 | 0.029558 | 0.32265 | 5486.663 |
| RICKMAN_ | 244 | 0.014214 | 0.001606 | 0.030881 | 0.32265 | 5486.663 |
| MORI_PRE | 74 | 0.025737 | 0.001608 | 0.055914 | 0.32266 | 5486.833 |
| QUINTENS_ | 79 | 0.024269 | 0.001567 | 0.052742 | 0.32271 | 5487.684 |
| REACTOME | 19 | 0.052902 | 0.001677 | 0.115 | 0.32275 | 5488.364 |
| SOTIRIOU_ | 51 | 0.032145 | 0.001669 | 0.069882 | 0.32277 | 5488.704 |
| FIGUEROA_ | 42 | 0.032559 | 0.001534 | 0.070798 | 0.3228 | 5489.214 |
| WARTERS_ | 46 | 0.033502 | 0.001652 | 0.072853 | 0.32282 | 5489.554 |
| GOBP_PRC | 96 | 0.022849 | 0.001625 | 0.049697 | 0.32284 | 5489.894 |
| GOBP_TRA | 6 | 0.09413 | 0.001678 | 0.20488 | 0.32296 | 5491.935 |
| KEGG_FC_I | 77 | 0.026888 | 0.001714 | 0.05858 | 0.32312 | 5494.656 |
| SESTO_RES | 46 | 0.033437 | 0.001649 | 0.072853 | 0.32313 | 5494.826 |
| REACTOME | 18 | 0.060686 | 0.001873 | 0.13224 | 0.32315 | 5495.166 |
| REACTOME | 21 | 0.052185 | 0.00174 | 0.11375 | 0.3232 | 5496.016 |
| GOBP_RES | 9 | 0.070847 | 0.001547 | 0.15443 | 0.32321 | 5496.186 |
| REACTOME | 6 | 0.10168 | 0.001812 | 0.22164 | 0.32321 | 5496.186 |
| GOBP_QUI | 38 | 0.034052 | 0.001526 | 0.074301 | 0.32337 | 5498.907 |
| GOMF_STE | 36 | 0.042019 | 0.001833 | 0.091806 | 0.32359 | 5502.648 |
| REACTOME | 37 | 0.035319 | 0.001562 | 0.077173 | 0.3236 | 5502.818 |
| GOBP_POS | 325 | 0.012573 | 0.001635 | 0.027508 | 0.32381 | 5506.389 |
| REACTOME | 15 | 0.061087 | 0.001721 | 0.13365 | 0.32381 | 5506.389 |
| GOMF_AM | 4 | 0.10541 | 0.001534 | 0.23083 | 0.32396 | 5508.94 |
| FISCHER_D | 911 | 0.007426 | 0.001591 | 0.01628 | 0.32415 | 5512.171 |
| GOMF_ARY | 9 | 0.079955 | 0.001745 | 0.17529 | 0.32415 | 5512.171 |
| GOMF_ENG | 4 | 0.11876 | 0.001729 | 0.26044 | 0.3242 | 5513.021 |
| REACTOME | 7 | 0.081141 | 0.001562 | 0.17794 | 0.3242 | 5513.021 |
| BROWNE_I | 90 | 0.024444 | 0.001684 | 0.053608 | 0.32421 | 5513.191 |
| GOMF_BET | 82 | 0.025041 | 0.001647 | 0.054945 | 0.32429 | 5514.551 |
| EPPERT_CE | 38 | 0.043169 | 0.001935 | 0.094736 | 0.32431 | 5514.892 |
| GOBP_POS | 9 | 0.079167 | 0.001728 | 0.17382 | 0.32439 | 5516.252 |
| AMIT_EGF_ | 65 | 0.025565 | 0.001498 | 0.056136 | 0.32441 | 5516.592 |
| UDAYAKUN | 218 | 0.015565 | 0.001663 | 0.034189 | 0.32446 | 5517.442 |
| GOBP_MIC | 82 | 0.022446 | 0.001476 | 0.049313 | 0.3245 | 5518.123 |
| GOBP_NEG | 23 | 0.044212 | 0.001542 | 0.097146 | 0.32452 | 5518.463 |
| REACTOME | 156 | 0.017387 | 0.001574 | 0.038208 | 0.32454 | 5518.803 |
| GOBP_C_T | 7 | 0.10157 | 0.001956 | 0.2235 | 0.32475 | 5522.374 |
| GOBP_MIC | 5 | 0.10561 | 0.001719 | 0.23245 | 0.3248 | 5523.224 |
| GOBP_AMY | 8 | 0.072507 | 0.001492 | 0.15964 | 0.32485 | 5524.074 |

| | | | | | | |
|---|---|---|---|---|---|---|
| GOBP_DEA | 9 | 0.067841 | 0.001481 | 0.14939 | 0.32488 | 5524.584 |
| GOMF_COI | 40 | 0.035655 | 0.00164 | 0.078635 | 0.32512 | 5528.666 |
| REACTOME | 15 | 0.060845 | 0.001714 | 0.13424 | 0.32519 | 5529.856 |
| GOBP_REG | 99 | 0.021573 | 0.001558 | 0.047625 | 0.32528 | 5531.386 |
| GOBP_MIT | 89 | 0.022309 | 0.001528 | 0.049257 | 0.32531 | 5531.897 |
| GOBP_REG | 543 | 0.010321 | 0.001725 | 0.022791 | 0.32533 | 5532.237 |
| WP_PENTC | 7 | 0.083479 | 0.001607 | 0.18435 | 0.32534 | 5532.407 |
| GOBP_LON | 67 | 0.027089 | 0.001611 | 0.059855 | 0.32543 | 5533.937 |
| PID_EPHB_ | 39 | 0.036542 | 0.001659 | 0.080768 | 0.32548 | 5534.787 |
| GOBP_ASY | 16 | 0.056813 | 0.001653 | 0.1256 | 0.32551 | 5535.298 |
| ROSTY_CEI | 135 | 0.018354 | 0.001547 | 0.040608 | 0.32565 | 5537.678 |
| PID_CERAN | 42 | 0.033793 | 0.001592 | 0.074797 | 0.32571 | 5538.699 |
| WP_7Q112 | 98 | 0.023742 | 0.001706 | 0.052554 | 0.32572 | 5538.869 |
| GOCC_ELO | 6 | 0.10232 | 0.001824 | 0.22671 | 0.32588 | 5541.589 |
| GOBP_ACT | 207 | 0.015952 | 0.001661 | 0.035373 | 0.32601 | 5543.8 |
| GOBP_CELI | 5 | 0.12434 | 0.002023 | 0.2758 | 0.32606 | 5544.65 |
| GOBP_MEI | 4 | 0.11218 | 0.001633 | 0.24898 | 0.32615 | 5546.181 |
| GOMF_VEI | 9 | 0.069858 | 0.001525 | 0.15507 | 0.32618 | 5546.691 |
| GOBP_MEI | 119 | 0.0211 | 0.00167 | 0.046854 | 0.32624 | 5547.711 |
| GOBP_POS | 487 | 0.010182 | 0.001614 | 0.022645 | 0.32649 | 5551.962 |
| GOBP_NEG | 14 | 0.062239 | 0.001694 | 0.13849 | 0.32658 | 5553.493 |
| REACTOME | 6 | 0.097676 | 0.001741 | 0.21744 | 0.32664 | 5554.513 |
| LOPEZ_EPI | 17 | 0.05407 | 0.001622 | 0.12039 | 0.32667 | 5555.023 |
| GOBP_MAI | 25 | 0.045189 | 0.001643 | 0.10064 | 0.32672 | 5555.874 |
| INAMURA_ | 13 | 0.056305 | 0.001477 | 0.12556 | 0.32693 | 5559.445 |
| DUTERTRE_ | 215 | 0.015232 | 0.001616 | 0.033979 | 0.32698 | 5560.295 |
| MACLACHL | 19 | 0.043372 | 0.001375 | 0.096759 | 0.32699 | 5560.465 |
| PID_AR_PA | 58 | 0.031361 | 0.001736 | 0.069966 | 0.327 | 5560.635 |
| WP_MITOC | 18 | 0.052347 | 0.001616 | 0.1168 | 0.32702 | 5560.975 |
| GOBP_5_P | 4 | 0.095113 | 0.001384 | 0.2124 | 0.32715 | 5563.186 |
| VANTVEER_ | 143 | 0.019113 | 0.001657 | 0.042685 | 0.32716 | 5563.356 |
| MIKHAYLO | 6 | 0.094269 | 0.00168 | 0.21068 | 0.32728 | 5565.396 |
| VART_KSHY | 152 | 0.019233 | 0.001719 | 0.042988 | 0.3273 | 5565.737 |
| SHIN_B_CE | 27 | 0.041625 | 0.001573 | 0.093074 | 0.32736 | 5566.757 |
| RHODES_U | 66 | 0.02534 | 0.001496 | 0.056678 | 0.32741 | 5567.607 |
| GOBP_RHO | 119 | 0.019589 | 0.00155 | 0.043816 | 0.32742 | 5567.777 |
| GOBP_REG | 93 | 0.023482 | 0.001644 | 0.052529 | 0.32743 | 5567.947 |
| GENTILE_U | 34 | 0.041791 | 0.001772 | 0.093532 | 0.32751 | 5569.308 |
| REACTOME | 27 | 0.037225 | 0.001407 | 0.083317 | 0.32752 | 5569.478 |
| GOBP_CELI | 22 | 0.05225 | 0.001783 | 0.11696 | 0.32753 | 5569.648 |
| GOMF_EFF | 15 | 0.058762 | 0.001656 | 0.13155 | 0.32755 | 5569.988 |
| GOBP_RES | 5 | 0.093153 | 0.001516 | 0.20861 | 0.32761 | 5571.008 |
| GOCC_MIT | 30 | 0.037623 | 0.001499 | 0.084274 | 0.32764 | 5571.518 |
| GOBP_REG | 48 | 0.034601 | 0.001743 | 0.077518 | 0.32767 | 5572.028 |
| GOMF_MIC | 7 | 0.096103 | 0.00185 | 0.2153 | 0.32767 | 5572.028 |

| | | | | | | |
|---|---|---|---|---|---|---|
| GOCC_MSL | 3 | 0.14615 | 0.001842 | 0.32757 | 0.32774 | 5573.219 |
| GOMF_ACT | 24 | 0.043005 | 0.001532 | 0.096398 | 0.32776 | 5573.559 |
| GOBP_NEG | 141 | 0.019811 | 0.001706 | 0.044415 | 0.32779 | 5574.069 |
| CHESLER_E | 36 | 0.035512 | 0.001549 | 0.079661 | 0.32788 | 5575.599 |
| BRUECKNE | 105 | 0.023996 | 0.001785 | 0.053877 | 0.32803 | 5578.15 |
| BOHN_PRI | 45 | 0.030766 | 0.0015 | 0.069092 | 0.32806 | 5578.66 |
| GOBP_REG | 7 | 0.098186 | 0.00189 | 0.22065 | 0.32817 | 5580.531 |
| GOCC_INSU | 6 | 0.071361 | 0.001272 | 0.16041 | 0.32822 | 5581.381 |
| GARGALOV | 25 | 0.041736 | 0.001518 | 0.093893 | 0.32834 | 5583.422 |
| GOMF_GLU | 19 | 0.048804 | 0.001548 | 0.1098 | 0.32835 | 5583.592 |
| REACTOME | 5 | 0.11595 | 0.001887 | 0.26118 | 0.32854 | 5586.823 |
| REACTOME | 9 | 0.079993 | 0.001746 | 0.18028 | 0.32863 | 5588.353 |
| WP_MFAP5 | 18 | 0.052275 | 0.001613 | 0.11787 | 0.32871 | 5589.714 |
| TESAR_ALK | 6 | 0.1195 | 0.00213 | 0.26953 | 0.32875 | 5590.394 |
| GOMF_BET | 5 | 0.094302 | 0.001535 | 0.21271 | 0.32877 | 5590.734 |
| GOBP_NUC | 35 | 0.037317 | 0.001605 | 0.084207 | 0.32883 | 5591.754 |
| REACTOME | 23 | 0.050467 | 0.001761 | 0.11393 | 0.3289 | 5592.945 |
| REACTOME | 5 | 0.082502 | 0.001343 | 0.18648 | 0.3291 | 5596.346 |
| GOBP_REG | 10 | 0.056604 | 0.001302 | 0.12801 | 0.32919 | 5597.876 |
| MOREIRA_ | 24 | 0.03913 | 0.001394 | 0.0885 | 0.32919 | 5597.876 |
| OUELLET_C | 110 | 0.02059 | 0.001567 | 0.046571 | 0.3292 | 5598.046 |
| GOBP_SPH | 14 | 0.059093 | 0.001609 | 0.13369 | 0.32924 | 5598.726 |
| GOBP_REG | 66 | 0.024861 | 0.001467 | 0.056305 | 0.32941 | 5601.617 |
| REACTOME | 6 | 0.096006 | 0.001711 | 0.21767 | 0.32959 | 5604.678 |
| GOUYER_T | 17 | 0.051234 | 0.001537 | 0.11622 | 0.32966 | 5605.868 |
| GOBP_POS | 6 | 0.094827 | 0.00169 | 0.21515 | 0.3297 | 5606.549 |
| WIEDERSCI | 55 | 0.030252 | 0.001631 | 0.068745 | 0.32995 | 5610.8 |
| GOBP_REG | 37 | 0.036108 | 0.001597 | 0.082097 | 0.33004 | 5612.33 |
| GOBP_REG | 15 | 0.058514 | 0.001649 | 0.13305 | 0.33005 | 5612.5 |
| WEBER_MI | 15 | 0.06387 | 0.0018 | 0.1453 | 0.33013 | 5613.861 |
| NIKOLSKY_ | 8 | 0.10561 | 0.002174 | 0.24045 | 0.33025 | 5615.901 |
| GOBP_POS | 49 | 0.032297 | 0.001643 | 0.073541 | 0.33027 | 5616.241 |
| GOCC_BOX | 3 | 0.13449 | 0.001695 | 0.30628 | 0.33029 | 5616.581 |
| GOBP_PRC | 6 | 0.071981 | 0.001283 | 0.16393 | 0.3303 | 5616.752 |
| GOMF_PRC | 67 | 0.027247 | 0.00162 | 0.062064 | 0.33033 | 5617.262 |
| GOBP_CAR | 24 | 0.040883 | 0.001457 | 0.093142 | 0.33036 | 5617.772 |
| GOCC_OUT | 5 | 0.09797 | 0.001594 | 0.22325 | 0.33039 | 5618.282 |
| GOMF_POI | 26 | 0.040018 | 0.001484 | 0.091204 | 0.33041 | 5618.622 |
| GOBP_DET | 61 | 0.028372 | 0.00161 | 0.064663 | 0.33042 | 5618.792 |
| REACTOME | 349 | 0.011919 | 0.001606 | 0.027167 | 0.33043 | 5618.962 |
| YAN_ESCAI | 19 | 0.053663 | 0.001702 | 0.12242 | 0.33057 | 5621.343 |
| GOBP_MIT | 81 | 0.022392 | 0.001464 | 0.051087 | 0.33058 | 5621.513 |
| HOFFMANI | 57 | 0.027818 | 0.001526 | 0.063512 | 0.3307 | 5623.554 |
| GOBP_EME | 69 | 0.026896 | 0.001623 | 0.061422 | 0.33074 | 5624.234 |
| WP_ENDO | 31 | 0.041533 | 0.001682 | 0.094853 | 0.33075 | 5624.404 |

| | | | | | | |
|---|---|---|---|---|---|---|
| KYNG_DNA | 14 | 0.058122 | 0.001582 | 0.13286 | 0.33089 | 5626.784 |
| ZHAN_LATI | 28 | 0.041916 | 0.001613 | 0.095819 | 0.33089 | 5626.784 |
| REACTOME | 8 | 0.07295 | 0.001501 | 0.16691 | 0.33104 | 5629.335 |
| GOBP_CON | 5 | 0.085791 | 0.001396 | 0.19633 | 0.33107 | 5629.845 |
| GOMF_ALC | 6 | 0.14848 | 0.002647 | 0.34004 | 0.33118 | 5631.716 |
| GOBP_CAR | 14 | 0.056787 | 0.001546 | 0.13005 | 0.33119 | 5631.886 |
| REACTOME | 34 | 0.038751 | 0.001643 | 0.088765 | 0.33122 | 5632.396 |
| REACTOME | 22 | 0.046767 | 0.001596 | 0.10713 | 0.33122 | 5632.396 |
| GOBP_POS | 11 | 0.060845 | 0.001468 | 0.1394 | 0.33125 | 5632.906 |
| GOBP_LAB | 22 | 0.051361 | 0.001752 | 0.11791 | 0.33157 | 5638.348 |
| REACTOME | 70 | 0.025702 | 0.001562 | 0.059042 | 0.33167 | 5640.048 |
| GOBP_NER | 10 | 0.067497 | 0.001553 | 0.15526 | 0.33188 | 5643.619 |
| GOCC_COL | 80 | 0.025683 | 0.001668 | 0.059199 | 0.3322 | 5649.061 |
| GOBP_FAT | 114 | 0.020936 | 0.001622 | 0.048263 | 0.33222 | 5649.401 |
| GOBP_REG | 4 | 0.12596 | 0.001833 | 0.29044 | 0.33226 | 5650.081 |
| REACTOME | 9 | 0.076458 | 0.001669 | 0.1763 | 0.33226 | 5650.081 |
| GOBP_FAT | 5 | 0.10554 | 0.001717 | 0.24338 | 0.33227 | 5650.251 |
| GOMF_NIT | 5 | 0.099246 | 0.001615 | 0.22904 | 0.3324 | 5652.462 |
| GOBP_NEG | 20 | 0.046022 | 0.001497 | 0.10622 | 0.33241 | 5652.632 |
| GOBP_T_C | 54 | 0.028259 | 0.001509 | 0.065235 | 0.33244 | 5653.142 |
| GOBP_RHO | 7 | 0.091465 | 0.001761 | 0.21121 | 0.33249 | 5653.992 |
| KEGG_PRO | 23 | 0.043101 | 0.001504 | 0.099561 | 0.33254 | 5654.843 |
| GOBP_RNA | 99 | 0.020787 | 0.001501 | 0.048023 | 0.33257 | 5655.353 |
| GOBP_POS | 45 | 0.028447 | 0.001387 | 0.065864 | 0.3329 | 5660.965 |
| GOBP_REG | 349 | 0.012144 | 0.001636 | 0.028144 | 0.33306 | 5663.685 |
| GOBP_ETH | 12 | 0.052895 | 0.001333 | 0.12264 | 0.33312 | 5664.706 |
| GOBP_LOW | 16 | 0.051179 | 0.001489 | 0.11875 | 0.33325 | 5666.916 |
| GRAHAM_( | 54 | 0.029924 | 0.001598 | 0.069464 | 0.33331 | 5667.937 |
| GOBP_PER | 9 | 0.070435 | 0.001538 | 0.16355 | 0.33336 | 5668.787 |
| GOBP_DIA( | 9 | 0.083937 | 0.001832 | 0.19491 | 0.33337 | 5668.957 |
| GOMF_NU | 19 | 0.049046 | 0.001555 | 0.1139 | 0.33338 | 5669.127 |
| ROESSLER_ | 49 | 0.028953 | 0.001473 | 0.067284 | 0.33349 | 5670.997 |
| GOCC_CER | 7 | 0.084099 | 0.001619 | 0.19548 | 0.33352 | 5671.508 |
| BIOCARTA_ | 33 | 0.039173 | 0.001636 | 0.09106 | 0.33353 | 5671.678 |
| GOBP_HIST | 39 | 0.033756 | 0.001533 | 0.078468 | 0.33353 | 5671.678 |
| PID_AR_TF | 48 | 0.030791 | 0.001551 | 0.07158 | 0.33354 | 5671.848 |
| GOBP_MEI | 43 | 0.031774 | 0.001515 | 0.073945 | 0.33371 | 5674.739 |
| GOBP_IMM | 12 | 0.075265 | 0.001897 | 0.17524 | 0.33378 | 5675.929 |
| WAKASUGI | 51 | 0.027385 | 0.001421 | 0.063767 | 0.3338 | 5676.269 |
| FEVR_CTNI | 650 | 0.008339 | 0.00152 | 0.019429 | 0.3339 | 5677.97 |
| GOMF_INT | 9 | 0.0741 | 0.001618 | 0.17265 | 0.3339 | 5677.97 |
| GOBP_NEP | 13 | 0.060591 | 0.00159 | 0.14127 | 0.33399 | 5679.5 |
| REACTOME | 7 | 0.084025 | 0.001618 | 0.19593 | 0.33402 | 5680.01 |
| GOBP_NEG | 31 | 0.040328 | 0.001633 | 0.094044 | 0.33403 | 5680.18 |
| GOBP_POS | 85 | 0.023081 | 0.001545 | 0.053823 | 0.33403 | 5680.18 |

| | | | | | | |
|---|---|---|---|---|---|---|
| GOBP_REG | 78 | 0.024936 | 0.0016 | 0.058159 | 0.33406 | 5680.69 |
| GOBP_POS | 1610 | 0.005656 | 0.00158 | 0.013196 | 0.33411 | 5681.541 |
| WP_BLADD | 39 | 0.034893 | 0.001584 | 0.081465 | 0.33421 | 5683.241 |
| WP_FAMIL | 30 | 0.039122 | 0.001558 | 0.091349 | 0.33423 | 5683.581 |
| WANG_BA | 35 | 0.035702 | 0.001536 | 0.083461 | 0.33441 | 5686.642 |
| BIOCARTA_ | 9 | 0.084501 | 0.001845 | 0.19771 | 0.33454 | 5688.853 |
| YAMASHIT/ | 73 | 0.025281 | 0.001569 | 0.059169 | 0.3346 | 5689.873 |
| KRIGE_RES | 680 | 0.008186 | 0.001526 | 0.019162 | 0.33461 | 5690.043 |
| GOBP_REG | 7 | 0.076239 | 0.001468 | 0.17853 | 0.33468 | 5691.233 |
| REACTOME | 30 | 0.034177 | 0.001361 | 0.08003 | 0.33468 | 5691.233 |
| REACTOME | 40 | 0.037405 | 0.00172 | 0.087597 | 0.33469 | 5691.403 |
| WANG_TAI | 42 | 0.033705 | 0.001588 | 0.078952 | 0.33473 | 5692.084 |
| GOBP_RES | 8 | 0.085803 | 0.001766 | 0.20104 | 0.33477 | 5692.764 |
| KIM_ALL_D | 138 | 0.017362 | 0.001479 | 0.040697 | 0.33484 | 5693.954 |
| WATANABI | 14 | 0.049179 | 0.001339 | 0.11534 | 0.33492 | 5695.315 |
| GOBP_NCR | 63 | 0.024067 | 0.001388 | 0.05646 | 0.33496 | 5695.995 |
| GOBP_CYT | 82 | 0.023778 | 0.001564 | 0.055794 | 0.33499 | 5696.505 |
| GOBP_LON | 18 | 0.049445 | 0.001526 | 0.11608 | 0.33507 | 5697.865 |
| GOMF_MA | 209 | 0.01458 | 0.001526 | 0.034229 | 0.33507 | 5697.865 |
| GOMF_HO | 82 | 0.025408 | 0.001671 | 0.059659 | 0.33509 | 5698.205 |
| GOBP_POS | 36 | 0.034777 | 0.001517 | 0.081734 | 0.33524 | 5700.756 |
| GOBP_POS | 22 | 0.044988 | 0.001535 | 0.10573 | 0.33524 | 5700.756 |
| GOBP_CELI | 71 | 0.025177 | 0.001541 | 0.059231 | 0.3354 | 5703.477 |
| GOBP_COT | 22 | 0.042943 | 0.001465 | 0.10103 | 0.33541 | 5703.647 |
| WP_ACQUI | 9 | 0.07318 | 0.001598 | 0.17218 | 0.33541 | 5703.647 |
| MENSE_HY | 95 | 0.021865 | 0.001547 | 0.051521 | 0.33564 | 5707.558 |
| GOBP_RES | 150 | 0.017191 | 0.001526 | 0.040532 | 0.33574 | 5709.259 |
| PID_AP1_P | 68 | 0.027744 | 0.001662 | 0.065418 | 0.33575 | 5709.429 |
| CROONQU | 30 | 0.037611 | 0.001498 | 0.088708 | 0.33579 | 5710.109 |
| SEIDEN_MI | 19 | 0.050631 | 0.001605 | 0.11947 | 0.33586 | 5711.299 |
| KEGG_MET | 65 | 0.032984 | 0.001932 | 0.077874 | 0.33595 | 5712.83 |
| TAYLOR_M | 74 | 0.026719 | 0.00167 | 0.063105 | 0.336 | 5713.68 |
| GOBP_POS | 7 | 0.095563 | 0.00184 | 0.22577 | 0.33605 | 5714.53 |
| GOBP_RES | 5 | 0.10644 | 0.001732 | 0.25152 | 0.33607 | 5714.87 |
| DACOSTA_ | 83 | 0.026231 | 0.001736 | 0.06205 | 0.33625 | 5717.931 |
| GOMF_OXI | 674 | 0.008214 | 0.001524 | 0.019434 | 0.33628 | 5718.441 |
| WP_HEME: | 8 | 0.059975 | 0.001234 | 0.14196 | 0.33634 | 5719.462 |
| WP_HEME_ | 8 | 0.059975 | 0.001234 | 0.14196 | 0.33634 | 5719.462 |
| REACTOME | 6 | 0.074752 | 0.001333 | 0.17708 | 0.33646 | 5721.502 |
| SPIELMAN_ | 561 | 0.008934 | 0.001517 | 0.021167 | 0.3365 | 5722.183 |
| REACTOME | 13 | 0.056226 | 0.001475 | 0.13328 | 0.33656 | 5723.203 |
| GOBP_REG | 11 | 0.064228 | 0.00155 | 0.15225 | 0.33657 | 5723.373 |
| TERAO_AO | 22 | 0.045352 | 0.001547 | 0.1076 | 0.3367 | 5725.584 |
| GOBP_COL | 8 | 0.089696 | 0.001846 | 0.2129 | 0.33677 | 5726.774 |
| GOBP_HIST | 6 | 0.073777 | 0.001315 | 0.17515 | 0.3368 | 5727.284 |

| | | | | | | |
|---|---|---|---|---|---|---|
| GOBP_INTI | 278 | 0.012507 | 0.001507 | 0.029694 | 0.33681 | 5727.454 |
| BLANCO_M | 123 | 0.018851 | 0.001517 | 0.044764 | 0.33684 | 5727.964 |
| GOCC_EKC | 4 | 0.10796 | 0.001571 | 0.25637 | 0.33685 | 5728.134 |
| GOBP_GLC | 22 | 0.050052 | 0.001708 | 0.11892 | 0.33692 | 5729.325 |
| GOBP_PRC | 230 | 0.01394 | 0.001529 | 0.033146 | 0.33704 | 5731.365 |
| GOMF_CGI | 15 | 0.053034 | 0.001494 | 0.12614 | 0.33709 | 5732.215 |
| GOBP_POS | 97 | 0.022185 | 0.001586 | 0.052802 | 0.33719 | 5733.916 |
| WENG_POI | 22 | 0.046899 | 0.0016 | 0.11173 | 0.33733 | 5736.297 |
| GOBP_TYR | 17 | 0.049277 | 0.001478 | 0.11751 | 0.33748 | 5738.847 |
| KOBAYASH | 17 | 0.047867 | 0.001436 | 0.11414 | 0.33748 | 5738.847 |
| GOBP_NEG | 402 | 0.010446 | 0.001508 | 0.024913 | 0.3375 | 5739.188 |
| GOBP_NEG | 6 | 0.097512 | 0.001738 | 0.23257 | 0.33751 | 5739.358 |
| GOCC_AXC | 7 | 0.070622 | 0.00136 | 0.16855 | 0.33761 | 5741.058 |
| GOMF_ISO | 152 | 0.015803 | 0.001412 | 0.037744 | 0.33772 | 5742.929 |
| REACTOME | 33 | 0.035588 | 0.001487 | 0.085041 | 0.3378 | 5744.289 |
| GOBP_CON | 15 | 0.048654 | 0.001371 | 0.11627 | 0.33782 | 5744.629 |
| GOBP_COL | 41 | 0.038374 | 0.001786 | 0.091738 | 0.33787 | 5745.479 |
| GOMF_MII | 30 | 0.037112 | 0.001478 | 0.088803 | 0.33801 | 5747.86 |
| REACTOME | 6 | 0.087294 | 0.001556 | 0.20903 | 0.33812 | 5749.731 |
| GOCC_LYS( | 90 | 0.020965 | 0.001444 | 0.050214 | 0.33815 | 5750.241 |
| MA_RAT_A | 92 | 0.02056 | 0.001432 | 0.049276 | 0.33825 | 5751.941 |
| REACTOME | 27 | 0.040687 | 0.001538 | 0.09752 | 0.33826 | 5752.111 |
| GOBP_REG | 202 | 0.014043 | 0.001445 | 0.033663 | 0.33828 | 5752.451 |
| MANNE_C( | 138 | 0.016647 | 0.001418 | 0.039912 | 0.33831 | 5752.962 |
| GOBP_CAT | 18 | 0.053856 | 0.001662 | 0.12914 | 0.33833 | 5753.302 |
| GOBP_REG | 14 | 0.057533 | 0.001566 | 0.13797 | 0.33834 | 5753.472 |
| DE_YY1_TA | 89 | 0.02255 | 0.001545 | 0.054106 | 0.33842 | 5754.832 |
| GOBP_MO | 147 | 0.017778 | 0.001563 | 0.042685 | 0.33852 | 5756.533 |
| REACTOME | 134 | 0.017098 | 0.001435 | 0.041054 | 0.33853 | 5756.703 |
| GOBP_NEG | 40 | 0.031492 | 0.001448 | 0.075626 | 0.33855 | 5757.043 |
| GOBP_REG | 13 | 0.0567 | 0.001487 | 0.13623 | 0.33863 | 5758.403 |
| REACTOME | 62 | 0.027138 | 0.001553 | 0.065219 | 0.33867 | 5759.083 |
| GOMF_PRE | 4 | 0.096036 | 0.001398 | 0.23089 | 0.33873 | 5760.104 |
| WP_SRF_A | 10 | 0.066505 | 0.00153 | 0.15992 | 0.33876 | 5760.614 |
| BIOCARTA_ | 10 | 0.059207 | 0.001362 | 0.14269 | 0.33909 | 5766.225 |
| REACTOME | 656 | 0.008373 | 0.001533 | 0.020184 | 0.33914 | 5767.076 |
| GOBP_ARC | 31 | 0.038433 | 0.001556 | 0.092691 | 0.33921 | 5768.266 |
| GOBP_POS | 9 | 0.079363 | 0.001732 | 0.19143 | 0.33923 | 5768.606 |
| GOBP_POS | 12 | 0.057849 | 0.001458 | 0.13961 | 0.3393 | 5769.797 |
| PID_NECTII | 30 | 0.037974 | 0.001513 | 0.091666 | 0.33934 | 5770.477 |
| LEE_BMP2_ | 857 | 0.007102 | 0.001479 | 0.017162 | 0.33949 | 5773.027 |
| KYNG_RESI | 17 | 0.048402 | 0.001452 | 0.11699 | 0.33954 | 5773.878 |
| REACTOME | 27 | 0.040731 | 0.001539 | 0.098451 | 0.33954 | 5773.878 |
| CARRILLOR | 37 | 0.032236 | 0.001426 | 0.078018 | 0.33974 | 5777.279 |
| GOMF_LOI | 16 | 0.046572 | 0.001355 | 0.11295 | 0.34006 | 5782.72 |

| | | | | | | |
|---|---|---|---|---|---|---|
| GOBP_REG | 19 | 0.049307 | 0.001564 | 0.11963 | 0.34011 | 5783.571 |
| GOCC_END | 23 | 0.039779 | 0.001388 | 0.096546 | 0.34016 | 5784.421 |
| GOBP_SPE | 133 | 0.01813 | 0.001516 | 0.044018 | 0.34022 | 5785.441 |
| GOMF_THI | 5 | 0.080337 | 0.001307 | 0.19507 | 0.34023 | 5785.611 |
| GOMF_POI | 9 | 0.07469 | 0.00163 | 0.18161 | 0.34044 | 5789.182 |
| GOBP_REG | 50 | 0.029317 | 0.001507 | 0.071321 | 0.34052 | 5790.543 |
| GOMF_OXI | 4 | 0.099441 | 0.001447 | 0.242 | 0.34057 | 5791.393 |
| GOBP_NEG | 117 | 0.018658 | 0.001464 | 0.045439 | 0.34068 | 5793.263 |
| GOMF_TRN | 73 | 0.022149 | 0.001375 | 0.054014 | 0.34088 | 5796.664 |
| GOBP_POS | 5 | 0.10788 | 0.001756 | 0.26312 | 0.3409 | 5797.005 |
| GOBP_NEG | 220 | 0.013738 | 0.001474 | 0.033536 | 0.34104 | 5799.385 |
| REACTOME | 8 | 0.074071 | 0.001525 | 0.18082 | 0.34104 | 5799.385 |
| GOBP_NEG | 5 | 0.075146 | 0.001223 | 0.18348 | 0.34107 | 5799.895 |
| IIZUKA_LIV | 8 | 0.076587 | 0.001576 | 0.18702 | 0.34108 | 5800.065 |
| GOBP_POS | 60 | 0.028877 | 0.001625 | 0.070522 | 0.3411 | 5800.406 |
| GOBP_DOF | 12 | 0.066493 | 0.001676 | 0.16248 | 0.34118 | 5801.766 |
| GOBP_DEV | 215 | 0.014425 | 0.001531 | 0.035253 | 0.3412 | 5802.106 |
| GOBP_FAT | 24 | 0.041111 | 0.001465 | 0.10047 | 0.3412 | 5802.106 |
| GOCC_END | 162 | 0.01577 | 0.001455 | 0.038542 | 0.34121 | 5802.276 |
| SESTO_RES | 69 | 0.025305 | 0.001527 | 0.061847 | 0.34121 | 5802.276 |
| MOOTHA_ | 419 | 0.009873 | 0.001455 | 0.024132 | 0.34123 | 5802.616 |
| GOCC_ACT | 464 | 0.009855 | 0.001526 | 0.024096 | 0.34128 | 5803.466 |
| GOCC_AXC | 58 | 0.025927 | 0.001435 | 0.063439 | 0.34138 | 5805.167 |
| PID_ER_NC | 39 | 0.031594 | 0.001435 | 0.077455 | 0.34167 | 5810.098 |
| GOMF_OXI | 5 | 0.091129 | 0.001483 | 0.22345 | 0.3417 | 5810.609 |
| GOBP_PRC | 20 | 0.040268 | 0.00131 | 0.098776 | 0.34176 | 5811.629 |
| GOBP_SER | 10 | 0.074178 | 0.001707 | 0.18202 | 0.34181 | 5812.479 |
| GOBP_POS | 18 | 0.045995 | 0.00142 | 0.11287 | 0.34182 | 5812.649 |
| CHANGOLK | 46 | 0.029732 | 0.001466 | 0.072973 | 0.34185 | 5813.159 |
| GOBP_PRC | 9 | 0.080973 | 0.001768 | 0.19876 | 0.34186 | 5813.329 |
| GOBP_MEI | 15 | 0.053737 | 0.001514 | 0.13193 | 0.34189 | 5813.839 |
| GOBP_ADF | 10 | 0.055699 | 0.001282 | 0.1369 | 0.34206 | 5816.73 |
| GOMF_POT | 118 | 0.021013 | 0.001656 | 0.051667 | 0.34212 | 5817.751 |
| GOBP_LEN | 14 | 0.059278 | 0.001614 | 0.14582 | 0.34218 | 5818.771 |
| GOMF_PRC | 44 | 0.030655 | 0.001478 | 0.075415 | 0.3422 | 5819.111 |
| GOBP_POS | 25 | 0.040905 | 0.001488 | 0.10067 | 0.34225 | 5819.961 |
| GOBP_DNA | 6 | 0.10113 | 0.001803 | 0.24899 | 0.34232 | 5821.152 |
| GOMF_HEX | 15 | 0.047028 | 0.001325 | 0.11581 | 0.34234 | 5821.492 |
| GOBP_N_T | 7 | 0.06165 | 0.001187 | 0.15183 | 0.34236 | 5821.832 |
| GOBP_CELI | 5 | 0.11463 | 0.001865 | 0.28265 | 0.34254 | 5824.893 |
| GOMF_C_X | 5 | 0.11071 | 0.001801 | 0.27301 | 0.34256 | 5825.233 |
| GOBP_HEN | 131 | 0.018251 | 0.001515 | 0.045034 | 0.34265 | 5826.763 |
| MISIAK_AN | 44 | 0.032118 | 0.001549 | 0.079311 | 0.34276 | 5828.634 |
| GOCC_MYC | 24 | 0.042684 | 0.001521 | 0.10545 | 0.34283 | 5829.824 |
| REACTOME | 89 | 0.021368 | 0.001464 | 0.052802 | 0.34286 | 5830.334 |

| | | | | | | |
|---|---|---|---|---|---|---|
| WAMUNYC | 260 | 0.01205 | 0.001404 | 0.029788 | 0.34292 | 5831.355 |
| GOMF_AN | 77 | 0.022665 | 0.001445 | 0.056102 | 0.34311 | 5834.586 |
| GOBP_NEG | 5 | 0.1039 | 0.001691 | 0.25735 | 0.34321 | 5836.286 |
| GLASS_IGF | 114 | 0.017665 | 0.001369 | 0.043766 | 0.34325 | 5836.966 |
| GOBP_COE | 18 | 0.044922 | 0.001387 | 0.11131 | 0.34327 | 5837.306 |
| REACTOME | 65 | 0.023505 | 0.001377 | 0.058253 | 0.34329 | 5837.646 |
| GOMF_ME | 15 | 0.053664 | 0.001512 | 0.13302 | 0.34332 | 5838.157 |
| GOBP_TRI | 6 | 0.092631 | 0.001651 | 0.22967 | 0.34336 | 5838.837 |
| GOMF_PH | 10 | 0.071837 | 0.001653 | 0.17812 | 0.34336 | 5838.837 |
| GOBP_REV | 16 | 0.061628 | 0.001793 | 0.1529 | 0.34345 | 5840.367 |
| GOBP_MO | 45 | 0.028489 | 0.001389 | 0.070702 | 0.3435 | 5841.218 |
| GOBP_OD | 6 | 0.097013 | 0.001729 | 0.24091 | 0.34359 | 5842.748 |
| MCCOLLUM | 10 | 0.061488 | 0.001415 | 0.1527 | 0.3436 | 5842.918 |
| WP_LUNG_ | 57 | 0.030413 | 0.001669 | 0.075545 | 0.34363 | 5843.428 |
| GOBP_SON | 66 | 0.025719 | 0.001518 | 0.063927 | 0.34373 | 5845.129 |
| ABBUD_LIF | 7 | 0.085862 | 0.001653 | 0.21344 | 0.34374 | 5845.299 |
| COLIN_PIL | 31 | 0.039376 | 0.001594 | 0.097939 | 0.34383 | 5846.829 |
| GOBP_LEU | 571 | 0.008876 | 0.00152 | 0.022083 | 0.34386 | 5847.339 |
| YAO_TEMP | 69 | 0.024534 | 0.001481 | 0.061054 | 0.3439 | 5848.02 |
| GOCC_MIC | 51 | 0.025248 | 0.001311 | 0.062843 | 0.34393 | 5848.53 |
| GOBP_NEG | 8 | 0.088273 | 0.001817 | 0.21974 | 0.34395 | 5848.87 |
| STEIN_ESR | 367 | 0.010135 | 0.001399 | 0.025241 | 0.34402 | 5850.06 |
| GOBP_PRC | 30 | 0.034578 | 0.001377 | 0.086139 | 0.34406 | 5850.74 |
| WP_INTER | 9 | 0.06067 | 0.001324 | 0.15124 | 0.34416 | 5852.441 |
| GOMF_GLU | 40 | 0.030248 | 0.001391 | 0.075439 | 0.34423 | 5853.631 |
| GOBP_NEG | 11 | 0.062005 | 0.001496 | 0.15467 | 0.34425 | 5853.971 |
| GOCC_TEL | 19 | 0.042914 | 0.001361 | 0.10709 | 0.34431 | 5854.992 |
| HOWLIN_C | 32 | 0.034494 | 0.001419 | 0.086083 | 0.34432 | 5855.162 |
| GOBP_MET | 15 | 0.054427 | 0.001534 | 0.13583 | 0.34433 | 5855.332 |
| MULLIGHA | 224 | 0.013408 | 0.001452 | 0.033487 | 0.34444 | 5857.202 |
| WP_DEGRA | 15 | 0.05143 | 0.001449 | 0.1286 | 0.34461 | 5860.093 |
| ALCALA_AI | 83 | 0.020588 | 0.001362 | 0.051484 | 0.34462 | 5860.263 |
| REACTOME | 14 | 0.06327 | 0.001722 | 0.15833 | 0.34472 | 5861.964 |
| GOBP_CELI | 88 | 0.021207 | 0.001445 | 0.053102 | 0.34482 | 5863.664 |
| PEDERSEN_ | 27 | 0.041518 | 0.001569 | 0.10398 | 0.34484 | 5864.004 |
| JECHLINGE | 60 | 0.02456 | 0.001382 | 0.061534 | 0.3449 | 5865.025 |
| CHIN_BREA | 27 | 0.038048 | 0.001438 | 0.095334 | 0.34491 | 5865.195 |
| GOBP_REG | 13 | 0.050027 | 0.001312 | 0.12554 | 0.34514 | 5869.106 |
| GOBP_REG | 43 | 0.027001 | 0.001287 | 0.067765 | 0.34515 | 5869.276 |
| SASSON_FS | 7 | 0.091172 | 0.001755 | 0.22882 | 0.34515 | 5869.276 |
| BANG_VER | 18 | 0.046399 | 0.001432 | 0.11647 | 0.34518 | 5869.786 |
| GOBP_NEG | 32 | 0.032988 | 0.001357 | 0.082838 | 0.34524 | 5870.806 |
| GOBP_REG | 11 | 0.057325 | 0.001383 | 0.14397 | 0.34525 | 5870.976 |
| GOMF_CAI | 11 | 0.058623 | 0.001415 | 0.14739 | 0.34542 | 5873.867 |
| PID_AURO | 31 | 0.031547 | 0.001277 | 0.079323 | 0.34543 | 5874.037 |

| | | | | | | |
|---|---|---|---|---|---|---|
| WP_THYRC | 66 | 0.024898 | 0.00147 | 0.062624 | 0.34548 | 5874.887 |
| GOMF_HEA | 116 | 0.018485 | 0.001445 | 0.046518 | 0.34555 | 5876.078 |
| GOBP_POS | 59 | 0.023874 | 0.001333 | 0.060102 | 0.3456 | 5876.928 |
| GOBP_MES | 6 | 0.070211 | 0.001252 | 0.17677 | 0.34561 | 5877.098 |
| REACTOME | 13 | 0.054231 | 0.001423 | 0.13656 | 0.34564 | 5877.608 |
| GOCC_PAR | 11 | 0.056247 | 0.001357 | 0.14165 | 0.34566 | 5877.948 |
| BENPORAT | 272 | 0.011781 | 0.001404 | 0.029691 | 0.34576 | 5879.649 |
| ZHANG_AD | 14 | 0.054378 | 0.00148 | 0.13711 | 0.34583 | 5880.839 |
| BIOCARTA_ | 37 | 0.032493 | 0.001437 | 0.082009 | 0.34598 | 5883.39 |
| GOBP_POS | 6 | 0.083345 | 0.001486 | 0.21086 | 0.34633 | 5889.342 |
| WP_B_CEL | 93 | 0.02041 | 0.001429 | 0.051647 | 0.34636 | 5889.852 |
| WP_NONC | 6 | 0.081309 | 0.001449 | 0.20586 | 0.34643 | 5891.042 |
| CHIN_BREA | 13 | 0.055188 | 0.001448 | 0.13973 | 0.34644 | 5891.212 |
| REACTOME | 90 | 0.020417 | 0.001406 | 0.051712 | 0.34649 | 5892.062 |
| GOBP_REA | 69 | 0.023572 | 0.001423 | 0.059713 | 0.34652 | 5892.573 |
| VALK_AML | 29 | 0.038583 | 0.001511 | 0.097746 | 0.34652 | 5892.573 |
| NAKAMUR. | 34 | 0.037703 | 0.001599 | 0.095545 | 0.34657 | 5893.423 |
| GOBP_REG | 21 | 0.042927 | 0.001431 | 0.1088 | 0.34659 | 5893.763 |
| GOBP_PRC | 26 | 0.039685 | 0.001472 | 0.10063 | 0.34665 | 5894.783 |
| GOBP_THIA | 7 | 0.083935 | 0.001616 | 0.2129 | 0.3467 | 5895.634 |
| GOBP_CELI | 6 | 0.097898 | 0.001745 | 0.24837 | 0.34673 | 5896.144 |
| REACTOME | 53 | 0.028157 | 0.00149 | 0.071437 | 0.34674 | 5896.314 |
| ALONSO_M | 182 | 0.014282 | 0.001396 | 0.036262 | 0.34684 | 5898.014 |
| RAMJAUN_ | 10 | 0.063889 | 0.00147 | 0.16227 | 0.34689 | 5898.864 |
| GOBP_NEG | 6 | 0.074262 | 0.001324 | 0.18867 | 0.34694 | 5899.715 |
| MOSERLE_ | 31 | 0.03873 | 0.001568 | 0.098429 | 0.34698 | 5900.395 |
| REACTOME | 51 | 0.0271 | 0.001407 | 0.068896 | 0.34703 | 5901.245 |
| GOBP_POS | 18 | 0.049771 | 0.001536 | 0.12655 | 0.34706 | 5901.755 |
| GOBP_LEU | 8 | 0.069757 | 0.001436 | 0.17757 | 0.34722 | 5904.476 |
| SWEET_LU | 400 | 0.010436 | 0.001503 | 0.026588 | 0.34734 | 5906.517 |
| SANSOM_A | 340 | 0.01042 | 0.001386 | 0.02655 | 0.34736 | 5906.857 |
| SCHLOSSEF | 660 | 0.007595 | 0.001395 | 0.019352 | 0.34737 | 5907.027 |
| GOBP_POS | 29 | 0.041382 | 0.001621 | 0.10548 | 0.34741 | 5907.707 |
| REACTOME | 28 | 0.033827 | 0.001302 | 0.086248 | 0.34745 | 5908.387 |
| WP_HIPPO | 22 | 0.03936 | 0.001343 | 0.10038 | 0.3475 | 5909.238 |
| GOBP_REG | 18 | 0.046273 | 0.001428 | 0.11808 | 0.34758 | 5910.598 |
| PID_FAS_P | 36 | 0.034367 | 0.001499 | 0.087746 | 0.34766 | 5911.958 |
| GOBP_NUC | 67 | 0.022779 | 0.001355 | 0.058166 | 0.34767 | 5912.128 |
| GOBP_MO | 581 | 0.00824 | 0.001423 | 0.021057 | 0.34778 | 5913.999 |
| GOBP_SEC | 59 | 0.026872 | 0.0015 | 0.06867 | 0.34778 | 5913.999 |
| WP_INTERA | 16 | 0.044312 | 0.00129 | 0.11331 | 0.34788 | 5915.699 |
| GOMF_SPH | 8 | 0.059188 | 0.001218 | 0.15143 | 0.34796 | 5917.06 |
| GOBP_POS | 27 | 0.042034 | 0.001589 | 0.10758 | 0.348 | 5917.74 |
| GOBP_PUR | 6 | 0.095227 | 0.001697 | 0.24392 | 0.34812 | 5919.781 |
| GOBP_POS | 6 | 0.086483 | 0.001542 | 0.22197 | 0.34841 | 5924.712 |

| | | | | | | |
|---|---|---|---|---|---|---|
| GOMF_PH( | 8 | 0.060156 | 0.001238 | 0.15461 | 0.34861 | 5928.113 |
| GOBP_DEP | 8 | 0.066753 | 0.001374 | 0.17161 | 0.34865 | 5928.793 |
| GOBP_LIPII | 1307 | 0.005678 | 0.001441 | 0.014598 | 0.34865 | 5928.793 |
| REACTOME | 24 | 0.041862 | 0.001492 | 0.10769 | 0.34874 | 5930.324 |
| GOBP_LEU | 6 | 0.077202 | 0.001376 | 0.1988 | 0.34888 | 5932.704 |
| BIOCARTA_ | 11 | 0.069249 | 0.001671 | 0.17851 | 0.34903 | 5935.255 |
| ZIRN_TRET | 23 | 0.045677 | 0.001593 | 0.11775 | 0.34904 | 5935.425 |
| REACTOME | 62 | 0.02634 | 0.001507 | 0.067938 | 0.34912 | 5936.786 |
| REACTOME | 6 | 0.079304 | 0.001414 | 0.20466 | 0.3492 | 5938.146 |
| GOMF_CAI | 5 | 0.07709 | 0.001254 | 0.199 | 0.34924 | 5938.826 |
| GOCC_NEU | 169 | 0.015251 | 0.001437 | 0.039374 | 0.34926 | 5939.166 |
| GOMF_MA | 12 | 0.052998 | 0.001336 | 0.13685 | 0.34928 | 5939.506 |
| WP_NONSI | 69 | 0.023651 | 0.001427 | 0.061117 | 0.34939 | 5941.377 |
| GOCC_PRC | 16 | 0.051693 | 0.001504 | 0.13363 | 0.34944 | 5942.227 |
| GOBP_POS | 64 | 0.025256 | 0.001468 | 0.065336 | 0.34954 | 5943.928 |
| PID_BETA_ | 73 | 0.023532 | 0.001461 | 0.060879 | 0.34955 | 5944.098 |
| GOBP_POS | 8 | 0.058693 | 0.001208 | 0.15189 | 0.34959 | 5944.778 |
| GOBP_NEG | 49 | 0.030419 | 0.001548 | 0.078722 | 0.3496 | 5944.948 |
| GOBP_NEG | 282 | 0.011626 | 0.00141 | 0.030094 | 0.34963 | 5945.458 |
| GOMF_OXI | 9 | 0.077339 | 0.001688 | 0.2002 | 0.34964 | 5945.628 |
| MARIADAS | 7 | 0.053724 | 0.001034 | 0.1391 | 0.34967 | 5946.138 |
| GOMF_ICO | 8 | 0.066161 | 0.001362 | 0.17144 | 0.34978 | 5948.009 |
| GOBP_REG | 10 | 0.059854 | 0.001377 | 0.15529 | 0.34997 | 5951.24 |
| GOMF_TRI | 22 | 0.045462 | 0.001551 | 0.118 | 0.35002 | 5952.09 |
| GOBP_DNA | 54 | 0.025541 | 0.001364 | 0.066305 | 0.35004 | 5952.43 |
| YIH_RESPO | 26 | 0.038727 | 0.001436 | 0.10053 | 0.35004 | 5952.43 |
| GOBP_POS | 30 | 0.032405 | 0.001291 | 0.084151 | 0.35009 | 5953.28 |
| WELCH_GA | 24 | 0.032132 | 0.001145 | 0.083481 | 0.35016 | 5954.471 |
| GOBP_RES | 6 | 0.091513 | 0.001631 | 0.23783 | 0.3502 | 5955.151 |
| WP_EBSTE | 18 | 0.041132 | 0.00127 | 0.10693 | 0.35024 | 5955.831 |
| GOBP_POS | 43 | 0.029873 | 0.001424 | 0.077727 | 0.35037 | 5958.042 |
| MYLLYKAN | 18 | 0.044128 | 0.001362 | 0.11489 | 0.35045 | 5959.402 |
| GOBP_REC | 6 | 0.083365 | 0.001486 | 0.21708 | 0.35048 | 5959.912 |
| GOCC_DOF | 14 | 0.047949 | 0.001305 | 0.1249 | 0.35053 | 5960.763 |
| GOBP_NEU | 53 | 0.027214 | 0.00144 | 0.070915 | 0.35058 | 5961.613 |
| ROSS_ACU | 78 | 0.022044 | 0.001414 | 0.057465 | 0.35064 | 5962.633 |
| GOBP_REG | 5 | 0.080272 | 0.001306 | 0.20957 | 0.35085 | 5966.204 |
| GOBP_NEG | 17 | 0.04561 | 0.001368 | 0.11911 | 0.35089 | 5966.884 |
| WP_IL5_SI( | 38 | 0.034739 | 0.001557 | 0.090734 | 0.35091 | 5967.225 |
| NEMETH_I | 27 | 0.036864 | 0.001393 | 0.096293 | 0.35092 | 5967.395 |
| GOBP_ACE | 5 | 0.07865 | 0.00128 | 0.20547 | 0.35094 | 5967.735 |
| GOBP_RES | 16 | 0.048702 | 0.001417 | 0.12724 | 0.35095 | 5967.905 |
| WP_INTRA | 26 | 0.039208 | 0.001454 | 0.10243 | 0.35095 | 5967.905 |
| GOMF_NA | 19 | 0.050083 | 0.001588 | 0.13086 | 0.35096 | 5968.075 |
| GOBP_MA | 83 | 0.020375 | 0.001348 | 0.053246 | 0.35099 | 5968.585 |

| | | | | | | |
|---|---|---|---|---|---|---|
| WP_TNFAL | 86 | 0.019708 | 0.001327 | 0.051519 | 0.35103 | 5969.265 |
| GOBP_SECI | 7 | 0.064253 | 0.001237 | 0.16802 | 0.35108 | 5970.115 |
| HOSHIDA_I | 211 | 0.013197 | 0.001387 | 0.034519 | 0.35112 | 5970.796 |
| REACTOME | 38 | 0.031179 | 0.001397 | 0.08168 | 0.35134 | 5974.537 |
| WP_NOTCI | 15 | 0.053793 | 0.001516 | 0.14094 | 0.35136 | 5974.877 |
| GOBP_CAP | 7 | 0.058522 | 0.001127 | 0.1534 | 0.35142 | 5975.897 |
| REACTOME | 687 | 0.007486 | 0.001402 | 0.019634 | 0.3515 | 5977.258 |
| GOBP_REG | 20 | 0.043738 | 0.001423 | 0.1149 | 0.35172 | 5980.999 |
| GOCC_BRIS | 5 | 0.087343 | 0.001421 | 0.22958 | 0.35181 | 5982.529 |
| GOCC_PRC | 7 | 0.055261 | 0.001064 | 0.14537 | 0.35193 | 5984.57 |
| WP_TP53_ | 19 | 0.043414 | 0.001377 | 0.11435 | 0.3521 | 5987.461 |
| NAKAMURA | 44 | 0.027502 | 0.001326 | 0.072458 | 0.35214 | 5988.141 |
| GOCC_AZU | 87 | 0.019234 | 0.001303 | 0.050686 | 0.35217 | 5988.651 |
| HINATA_NI | 22 | 0.041914 | 0.00143 | 0.11048 | 0.3522 | 5989.161 |
| GOBP_NEG | 8 | 0.060413 | 0.001243 | 0.15931 | 0.35226 | 5990.181 |
| GOBP_RES | 16 | 0.051434 | 0.001497 | 0.13563 | 0.35226 | 5990.181 |
| MEISSNER_ | 140 | 0.016894 | 0.001449 | 0.04456 | 0.3523 | 5990.862 |
| HE_PTEN_ | 18 | 0.047984 | 0.001481 | 0.12658 | 0.35231 | 5991.032 |
| GOMF_CAI | 11 | 0.048751 | 0.001177 | 0.12869 | 0.35242 | 5992.902 |
| WP_SARSC | 24 | 0.036391 | 0.001297 | 0.096099 | 0.35246 | 5993.582 |
| GOMF_PHO | 104 | 0.018126 | 0.001342 | 0.047889 | 0.35253 | 5994.773 |
| ELVIDGE_H | 87 | 0.019958 | 0.001352 | 0.052731 | 0.35254 | 5994.943 |
| GOCC_DEN | 6 | 0.067083 | 0.001196 | 0.17725 | 0.35255 | 5995.113 |
| GOCC_MES | 12 | 0.056635 | 0.001428 | 0.14971 | 0.35261 | 5996.133 |
| WP_CHOLE | 5 | 0.10241 | 0.001667 | 0.27081 | 0.35265 | 5996.813 |
| GOBP_TRA | 58 | 0.021108 | 0.001168 | 0.055823 | 0.35267 | 5997.153 |
| NICK_RESP | 24 | 0.035948 | 0.001281 | 0.095216 | 0.35289 | 6000.894 |
| REACTOME | 31 | 0.039368 | 0.001594 | 0.10443 | 0.35309 | 6004.295 |
| GOBP_LOC | 12 | 0.057052 | 0.001438 | 0.15149 | 0.35323 | 6006.676 |
| REACTOME | 8 | 0.057262 | 0.001179 | 0.15205 | 0.35324 | 6006.846 |
| GOBP_NEG | 59 | 0.025655 | 0.001432 | 0.068191 | 0.35338 | 6009.227 |
| GOCC_FICO | 176 | 0.013646 | 0.001312 | 0.036288 | 0.35344 | 6010.247 |
| GOBP_POS | 26 | 0.035253 | 0.001307 | 0.093754 | 0.35346 | 6010.587 |
| BIOCARTA_ | 5 | 0.072489 | 0.00118 | 0.19315 | 0.35372 | 6015.009 |
| GOBP_POS | 4 | 0.088863 | 0.001293 | 0.23688 | 0.35378 | 6016.029 |
| GOBP_PRC | 70 | 0.022792 | 0.001385 | 0.06076 | 0.35379 | 6016.199 |
| GOBP_POS | 6 | 0.066586 | 0.001187 | 0.17753 | 0.35381 | 6016.539 |
| MIKHAYLO | 6 | 0.067047 | 0.001195 | 0.17883 | 0.35386 | 6017.389 |
| GOMF_KIN | 44 | 0.026123 | 0.00126 | 0.069687 | 0.35389 | 6017.899 |
| GOBP_PRC | 55 | 0.024959 | 0.001345 | 0.066617 | 0.35396 | 6019.09 |
| GOBP_CHC | 6 | 0.093439 | 0.001666 | 0.24945 | 0.35399 | 6019.6 |
| SHETH_LIV | 156 | 0.014254 | 0.00129 | 0.038111 | 0.3542 | 6023.171 |
| REACTOME | 12 | 0.065071 | 0.00164 | 0.17411 | 0.3543 | 6024.872 |
| TSENG_AD | 27 | 0.037696 | 0.001425 | 0.1009 | 0.35435 | 6025.722 |
| GOBP_L_PI | 6 | 0.071587 | 0.001276 | 0.19165 | 0.35438 | 6026.232 |

| | | | | | | |
|---|---|---|---|---|---|---|
| REACTOME | 11 | 0.061075 | 0.001474 | 0.16352 | 0.35439 | 6026.402 |
| GOBP_POS | 30 | 0.032946 | 0.001312 | 0.088227 | 0.35442 | 6026.912 |
| GOBP_PEP | 5 | 0.095966 | 0.001562 | 0.25719 | 0.35453 | 6028.783 |
| GRAHAM_ | 88 | 0.019736 | 0.001344 | 0.052918 | 0.35459 | 6029.803 |
| GOCC_TET | 10 | 0.053596 | 0.001233 | 0.14373 | 0.35462 | 6030.313 |
| GOMF_BH | 30 | 0.034867 | 0.001389 | 0.093651 | 0.35483 | 6033.884 |
| GOBP_ASP | 46 | 0.026947 | 0.001329 | 0.072418 | 0.35491 | 6035.245 |
| REACTOME | 14 | 0.04975 | 0.001354 | 0.13373 | 0.35494 | 6035.755 |
| GOBP_REG | 41 | 0.024433 | 0.001137 | 0.0657 | 0.35499 | 6036.605 |
| BIOCARTA_ | 19 | 0.03993 | 0.001266 | 0.10739 | 0.35501 | 6036.945 |
| GOMF_TRA | 17 | 0.049383 | 0.001481 | 0.13283 | 0.35503 | 6037.285 |
| GOBP_REL | 32 | 0.036136 | 0.001487 | 0.097284 | 0.35515 | 6039.326 |
| GOBP_RES | 16 | 0.05247 | 0.001527 | 0.14126 | 0.35515 | 6039.326 |
| REACTOME | 16 | 0.044364 | 0.001291 | 0.11944 | 0.35516 | 6039.496 |
| CAIRO_PM | 12 | 0.055491 | 0.001399 | 0.14948 | 0.35524 | 6040.856 |
| MARTENS_ | 440 | 0.008798 | 0.001327 | 0.023701 | 0.35524 | 6040.856 |
| GOBP_TRA | 11 | 0.066199 | 0.001598 | 0.17835 | 0.35525 | 6041.026 |
| GOMF_ME | 65 | 0.023419 | 0.001372 | 0.063126 | 0.35532 | 6042.217 |
| REACTOME | 16 | 0.045688 | 0.00133 | 0.12316 | 0.35533 | 6042.387 |
| LI_WILMS_ | 176 | 0.015078 | 0.001449 | 0.040697 | 0.35551 | 6045.448 |
| GOMF_ATF | 25 | 0.03735 | 0.001358 | 0.10086 | 0.35557 | 6046.468 |
| REACTOME | 42 | 0.02893 | 0.001363 | 0.078212 | 0.35573 | 6049.189 |
| GOBP_IMM | 192 | 0.013868 | 0.001391 | 0.037537 | 0.3559 | 6052.08 |
| GOBP_CAR | 20 | 0.043455 | 0.001414 | 0.11772 | 0.35602 | 6054.12 |
| GOMF_LEU | 6 | 0.062999 | 0.001123 | 0.17073 | 0.35606 | 6054.8 |
| GOBP_CELI | 100 | 0.018928 | 0.001374 | 0.051303 | 0.35609 | 6055.31 |
| GOBP_MYE | 157 | 0.01427 | 0.001296 | 0.038692 | 0.35614 | 6056.161 |
| GOBP_VAS | 28 | 0.038843 | 0.001495 | 0.10536 | 0.35619 | 6057.011 |
| CAFFAREL_ | 9 | 0.062552 | 0.001366 | 0.16974 | 0.35625 | 6058.031 |
| WP_VITAM | 47 | 0.026541 | 0.001323 | 0.072025 | 0.35625 | 6058.031 |
| GOBP_POS | 48 | 0.025352 | 0.001277 | 0.068814 | 0.35628 | 6058.541 |
| REACTOME | 20 | 0.042997 | 0.001399 | 0.11671 | 0.35629 | 6058.711 |
| GOBP_NEG | 46 | 0.026318 | 0.001298 | 0.07151 | 0.35643 | 6061.092 |
| GOBP_REG | 43 | 0.03044 | 0.001451 | 0.082819 | 0.35661 | 6064.153 |
| GOMF_OPS | 4 | 0.09051 | 0.001317 | 0.24633 | 0.35665 | 6064.833 |
| POMEROY_ | 44 | 0.026724 | 0.001289 | 0.072738 | 0.35666 | 6065.003 |
| GOMF_GAI | 7 | 0.079254 | 0.001526 | 0.21576 | 0.35669 | 6065.513 |
| GOBP_RIBO | 64 | 0.021517 | 0.001251 | 0.058597 | 0.35674 | 6066.364 |
| REACTOME | 27 | 0.033883 | 0.001281 | 0.092292 | 0.35676 | 6066.704 |
| GOMF_ATF | 9 | 0.05561 | 0.001214 | 0.1515 | 0.35679 | 6067.214 |
| GOBP_POS | 7 | 0.071174 | 0.00137 | 0.19415 | 0.35696 | 6070.105 |
| GOBP_SPH | 17 | 0.045914 | 0.001377 | 0.1253 | 0.35703 | 6071.295 |
| GOCC_NUC | 43 | 0.026006 | 0.00124 | 0.071018 | 0.35712 | 6072.826 |
| GOMF_MIC | 35 | 0.033969 | 0.001461 | 0.092782 | 0.35714 | 6073.166 |
| GOBP_DEV | 302 | 0.01072 | 0.001345 | 0.029286 | 0.35716 | 6073.506 |

| | | | | | | |
|---|---|---|---|---|---|---|
| GOBP_REG | 12 | 0.061351 | 0.001546 | 0.1676 | 0.35717 | 6073.676 |
| VECCHI_GA | 322 | 0.011146 | 0.001443 | 0.030454 | 0.35718 | 6073.846 |
| TOOKER_G | 77 | 0.01996 | 0.001272 | 0.054544 | 0.3572 | 6074.186 |
| GOCC_HOS | 9 | 0.056207 | 0.001227 | 0.15362 | 0.35723 | 6074.696 |
| GOBP_NUC | 18 | 0.050088 | 0.001546 | 0.13695 | 0.35728 | 6075.546 |
| WP_LEPTIN | 16 | 0.045317 | 0.001319 | 0.12393 | 0.35731 | 6076.057 |
| PID_TNF_P | 43 | 0.025094 | 0.001196 | 0.068719 | 0.3575 | 6079.288 |
| GOBP_REG | 4 | 0.088621 | 0.00129 | 0.2428 | 0.35756 | 6080.308 |
| MEBARKI_I | 74 | 0.02227 | 0.001392 | 0.061032 | 0.3576 | 6080.988 |
| GOBP_NEG | 6 | 0.067866 | 0.00121 | 0.18604 | 0.35764 | 6081.668 |
| GOBP_RNA | 97 | 0.017326 | 0.001239 | 0.047514 | 0.35769 | 6082.518 |
| GOBP_INO | 5 | 0.090718 | 0.001476 | 0.2488 | 0.3577 | 6082.689 |
| GOBP_PUR | 9 | 0.054182 | 0.001183 | 0.14868 | 0.35777 | 6083.879 |
| GOBP_NUC | 10 | 0.081242 | 0.001869 | 0.22305 | 0.35785 | 6085.239 |
| REACTOME | 19 | 0.040285 | 0.001277 | 0.11062 | 0.35786 | 6085.409 |
| GOBP_AMY | 5 | 0.088523 | 0.001441 | 0.2433 | 0.35799 | 6087.62 |
| MANNE_CO | 141 | 0.016386 | 0.001411 | 0.045076 | 0.35811 | 6089.661 |
| GOBP_IRON | 12 | 0.047658 | 0.001201 | 0.13114 | 0.35816 | 6090.511 |
| REACTOME | 14 | 0.059545 | 0.001621 | 0.16388 | 0.35817 | 6090.681 |
| GOBP_FAS_ | 5 | 0.09861 | 0.001605 | 0.27149 | 0.35822 | 6091.531 |
| GOBP_PRC | 10 | 0.053445 | 0.00123 | 0.14717 | 0.35824 | 6091.871 |
| GOMF_U2_ | 6 | 0.081977 | 0.001461 | 0.22587 | 0.35833 | 6093.402 |
| GOCC_END | 11 | 0.055559 | 0.001341 | 0.15322 | 0.35845 | 6095.442 |
| GOBP_SRP_ | 4 | 0.086626 | 0.001261 | 0.23899 | 0.3585 | 6096.293 |
| KIM_ALL_D | 30 | 0.032764 | 0.001305 | 0.090408 | 0.35853 | 6096.803 |
| GOBP_ALP | 86 | 0.018718 | 0.001261 | 0.051655 | 0.35854 | 6096.973 |
| GOBP_REG | 84 | 0.018425 | 0.001226 | 0.050844 | 0.35854 | 6096.973 |
| WATANABE | 89 | 0.018706 | 0.001281 | 0.051687 | 0.35871 | 6099.864 |
| GOMF_AM | 348 | 0.009777 | 0.001315 | 0.027018 | 0.35873 | 6100.204 |
| GOMF_MO | 470 | 0.008324 | 0.001297 | 0.023003 | 0.35874 | 6100.374 |
| GOBP_PHO | 1680 | 0.004596 | 0.001309 | 0.012703 | 0.35875 | 6100.544 |
| GOBP_HOR | 12 | 0.052433 | 0.001322 | 0.14493 | 0.35876 | 6100.714 |
| GOBP_REG | 7 | 0.057852 | 0.001114 | 0.15991 | 0.35876 | 6100.714 |
| HARRIS_HY | 76 | 0.022338 | 0.001415 | 0.061745 | 0.35876 | 6100.714 |
| GOMF_PRI | 7 | 0.080796 | 0.001556 | 0.22335 | 0.35877 | 6100.884 |
| HEIDENBLA | 7 | 0.0672 | 0.001294 | 0.18576 | 0.35877 | 6100.884 |
| GUENTHER | 24 | 0.039341 | 0.001402 | 0.10879 | 0.35882 | 6101.734 |
| GOBP_NEU | 23 | 0.038433 | 0.001341 | 0.10629 | 0.35883 | 6101.904 |
| WP_ANGIO | 24 | 0.04049 | 0.001443 | 0.112 | 0.35885 | 6102.244 |
| GOBP_L_SE | 6 | 0.074593 | 0.00133 | 0.20653 | 0.35899 | 6104.625 |
| BIOCARTA_ | 5 | 0.078412 | 0.001276 | 0.21719 | 0.35904 | 6105.475 |
| GOBP_REG | 52 | 0.027517 | 0.001442 | 0.076236 | 0.35907 | 6105.985 |
| KEGG_REN | 15 | 0.053399 | 0.001505 | 0.14797 | 0.3591 | 6106.496 |
| GOCC_TRA | 6 | 0.055414 | 0.000988 | 0.15357 | 0.35911 | 6106.666 |
| GOBP_DNA | 156 | 0.014085 | 0.001275 | 0.039037 | 0.35912 | 6106.836 |

| | | | | | | |
|---|---|---|---|---|---|---|
| GOBP_NEG | 21 | 0.035926 | 0.001198 | 0.099605 | 0.35917 | 6107.686 |
| REACTOME | 11 | 0.05054 | 0.00122 | 0.14018 | 0.35922 | 6108.536 |
| GOBP_REG | 15 | 0.044474 | 0.001253 | 0.12346 | 0.35934 | 6110.577 |
| GOBP_RES | 19 | 0.036243 | 0.001149 | 0.10063 | 0.35936 | 6110.917 |
| GOBP_CELI | 931 | 0.006012 | 0.001302 | 0.016709 | 0.3595 | 6113.298 |
| GOBP_PHC | 9 | 0.06437 | 0.001405 | 0.17891 | 0.3595 | 6113.298 |
| GOBP_PRC | 6 | 0.069723 | 0.001243 | 0.1939 | 0.35958 | 6114.658 |
| GOBP_COL | 93 | 0.020089 | 0.001407 | 0.055891 | 0.35964 | 6115.678 |
| REACTOME | 6 | 0.086359 | 0.001539 | 0.24054 | 0.35979 | 6118.229 |
| GOBP_TEL( | 9 | 0.055724 | 0.001216 | 0.15544 | 0.35999 | 6121.63 |
| GOBP_RES | 20 | 0.040775 | 0.001327 | 0.11386 | 0.36013 | 6124.011 |
| CHANDRAN | 299 | 0.010692 | 0.001335 | 0.029908 | 0.36036 | 6127.922 |
| GOMF_ATF | 528 | 0.007809 | 0.001288 | 0.021846 | 0.36038 | 6128.262 |
| GOBP_REG | 6 | 0.059634 | 0.001063 | 0.16685 | 0.3604 | 6128.602 |
| FULCHER_I | 419 | 0.00896 | 0.00132 | 0.025088 | 0.3605 | 6130.303 |
| GOMF_EST | 15 | 0.047618 | 0.001342 | 0.13349 | 0.36066 | 6133.023 |
| GOBP_NEG | 30 | 0.034413 | 0.001371 | 0.096485 | 0.36067 | 6133.193 |
| IVANOVA_ | 420 | 0.008535 | 0.001259 | 0.023931 | 0.36069 | 6133.533 |
| GOBP_PEP | 5 | 0.071458 | 0.001163 | 0.20039 | 0.3607 | 6133.704 |
| TOYOTA_T | 421 | 0.008555 | 0.001263 | 0.024021 | 0.36087 | 6136.594 |
| COWLING_ | 39 | 0.028002 | 0.001271 | 0.07873 | 0.36105 | 6139.655 |
| GROSS_HY | 202 | 0.012578 | 0.001294 | 0.035369 | 0.36107 | 6139.995 |
| AKL_HTLV1 | 64 | 0.023071 | 0.001341 | 0.064918 | 0.36115 | 6141.356 |
| GOCC_CHR | 12 | 0.050057 | 0.001262 | 0.14086 | 0.36116 | 6141.526 |
| GOBP_GAE | 16 | 0.052538 | 0.001529 | 0.14791 | 0.36122 | 6142.546 |
| GOMF_DN | 23 | 0.037727 | 0.001316 | 0.10621 | 0.36122 | 6142.546 |
| GOBP_REG | 7 | 0.073809 | 0.001421 | 0.20788 | 0.36127 | 6143.396 |
| GOLDRATH | 329 | 0.009809 | 0.001284 | 0.027632 | 0.3613 | 6143.907 |
| GOCC_TER | 53 | 0.023315 | 0.001234 | 0.0657 | 0.36134 | 6144.587 |
| WEBER_MI | 13 | 0.050651 | 0.001329 | 0.14273 | 0.36134 | 6144.587 |
| GOBP_PEP | 6 | 0.069501 | 0.001239 | 0.1959 | 0.36138 | 6145.267 |
| BIOCARTA_ | 12 | 0.057134 | 0.00144 | 0.16109 | 0.36142 | 6145.947 |
| GOBP_PRII | 6 | 0.070351 | 0.001254 | 0.19837 | 0.36143 | 6146.117 |
| GOBP_VES | 25 | 0.036211 | 0.001317 | 0.10213 | 0.36147 | 6146.797 |
| GOBP_LAC | 37 | 0.029421 | 0.001301 | 0.083147 | 0.36173 | 6151.219 |
| GOBP_POS | 93 | 0.018894 | 0.001323 | 0.053407 | 0.36175 | 6151.559 |
| GOBP_PER | 11 | 0.063275 | 0.001527 | 0.17909 | 0.36193 | 6154.62 |
| GOBP_WO | 7 | 0.08114 | 0.001562 | 0.22969 | 0.36195 | 6154.96 |
| GOBP_REG | 4 | 0.087029 | 0.001267 | 0.24639 | 0.36197 | 6155.3 |
| GOBP_SELI | 12 | 0.057014 | 0.001437 | 0.16152 | 0.36205 | 6156.66 |
| GOBP_REG | 8 | 0.068474 | 0.001409 | 0.1942 | 0.36219 | 6159.041 |
| GOBP_TRA | 165 | 0.012893 | 0.0012 | 0.036563 | 0.36219 | 6159.041 |
| GOBP_RES | 8 | 0.068421 | 0.001408 | 0.19405 | 0.3622 | 6159.211 |
| GOBP_REG | 5 | 0.092298 | 0.001502 | 0.26186 | 0.36225 | 6160.061 |
| GARGALOV | 12 | 0.044932 | 0.001133 | 0.12763 | 0.3624 | 6162.612 |

| | | | | | | |
|---|---|---|---|---|---|---|
| GOBP_MAI | 28 | 0.029495 | 0.001135 | 0.083829 | 0.36248 | 6163.972 |
| DAUER_ST/ | 47 | 0.027881 | 0.001389 | 0.0793 | 0.36258 | 6165.673 |
| GOBP_INTI | 7 | 0.062227 | 0.001198 | 0.17702 | 0.3626 | 6166.013 |
| GOBP_REG | 45 | 0.028627 | 0.001396 | 0.081439 | 0.36261 | 6166.183 |
| GOBP_REG | 26 | 0.030615 | 0.001135 | 0.087142 | 0.36267 | 6167.203 |
| GOBP_GAL | 6 | 0.086405 | 0.00154 | 0.24601 | 0.36271 | 6167.884 |
| GOMF_GLU | 13 | 0.04309 | 0.00113 | 0.12269 | 0.36272 | 6168.054 |
| GOBP_RNA | 268 | 0.010253 | 0.001213 | 0.029198 | 0.36274 | 6168.394 |
| BROWNE_I | 188 | 0.012528 | 0.001244 | 0.035683 | 0.36276 | 6168.734 |
| GOBP_ATP | 14 | 0.04663 | 0.001269 | 0.13283 | 0.36278 | 6169.074 |
| GOBP_CAR | 67 | 0.023998 | 0.001427 | 0.068396 | 0.36285 | 6170.264 |
| GOBP_NEG | 15 | 0.04517 | 0.001273 | 0.1288 | 0.36291 | 6171.285 |
| LEE_LIVER_ | 60 | 0.022436 | 0.001263 | 0.063978 | 0.36292 | 6171.455 |
| REACTOME | 14 | 0.05387 | 0.001467 | 0.15375 | 0.36303 | 6173.325 |
| REACTOME | 141 | 0.013396 | 0.001153 | 0.038279 | 0.36318 | 6175.876 |
| GOBP_INN | 100 | 0.019267 | 0.001399 | 0.055059 | 0.3632 | 6176.216 |
| GOBP_NEG | 16 | 0.045023 | 0.00131 | 0.12866 | 0.3632 | 6176.216 |
| GOBP_POS | 12 | 0.045367 | 0.001143 | 0.12974 | 0.3633 | 6177.917 |
| GOMF_MC | 16 | 0.047464 | 0.001381 | 0.13608 | 0.36363 | 6183.528 |
| GOBP_POS | 23 | 0.03675 | 0.001282 | 0.10539 | 0.36365 | 6183.868 |
| JOHNSTON | 797 | 0.00612 | 0.001231 | 0.017554 | 0.36367 | 6184.208 |
| SPIELMAN_ | 19 | 0.041337 | 0.001311 | 0.11865 | 0.36377 | 6185.909 |
| ONGUSAH/ | 12 | 0.051203 | 0.001291 | 0.14717 | 0.36396 | 6189.14 |
| STAMBOLS | 23 | 0.036663 | 0.001279 | 0.10538 | 0.36396 | 6189.14 |
| GOMF_PYF | 4 | 0.093645 | 0.001363 | 0.27002 | 0.36437 | 6196.112 |
| CHEOK_RE: | 19 | 0.037125 | 0.001177 | 0.10706 | 0.36439 | 6196.452 |
| GOBP_POS | 43 | 0.026556 | 0.001266 | 0.076587 | 0.3644 | 6196.622 |
| REACTOME | 6 | 0.066155 | 0.001179 | 0.19084 | 0.36443 | 6197.132 |
| WP_SMC1! | 9 | 0.064642 | 0.001411 | 0.18666 | 0.36456 | 6199.343 |
| GOCC_SIDE | 506 | 0.008184 | 0.001322 | 0.023635 | 0.36457 | 6199.513 |
| GOBP_DN/ | 111 | 0.016332 | 0.001249 | 0.047171 | 0.36459 | 6199.853 |
| GOBP_HIST | 13 | 0.047838 | 0.001255 | 0.13817 | 0.36459 | 6199.853 |
| REACTOME | 277 | 0.01001 | 0.001204 | 0.028924 | 0.36464 | 6200.703 |
| GOCC_NUC | 81 | 0.019012 | 0.001243 | 0.054984 | 0.36476 | 6202.744 |
| GOBP_RET | 50 | 0.025494 | 0.00131 | 0.073749 | 0.36479 | 6203.254 |
| BIOCARTA_ | 20 | 0.041756 | 0.001358 | 0.1208 | 0.3648 | 6203.424 |
| CLAUS_PGI | 10 | 0.049336 | 0.001135 | 0.14276 | 0.36483 | 6203.934 |
| GOBP_MAI | 6 | 0.10652 | 0.001899 | 0.30842 | 0.3649 | 6205.125 |
| GOBP_MES | 5 | 0.099411 | 0.001618 | 0.28784 | 0.36491 | 6205.295 |
| GOBP_LIPII | 19 | 0.0367 | 0.001164 | 0.10627 | 0.36492 | 6205.465 |
| GOBP_CELI | 84 | 0.019497 | 0.001298 | 0.056509 | 0.36504 | 6207.505 |
| GOMF_DN/ | 12 | 0.038981 | 0.000983 | 0.11307 | 0.36514 | 6209.206 |
| GOBP_NER | 12 | 0.056988 | 0.001436 | 0.16531 | 0.36515 | 6209.376 |
| REACTOME | 26 | 0.034122 | 0.001266 | 0.098986 | 0.36516 | 6209.546 |
| GOCC_CLA | 193 | 0.012134 | 0.001221 | 0.035207 | 0.36519 | 6210.056 |

| | | | | | | |
|---|---|---|---|---|---|---|
| FONTAINE_ | 71 | 0.020381 | 0.001248 | 0.059147 | 0.36521 | 6210.396 |
| GOBP_REG | 666 | 0.006776 | 0.00125 | 0.019674 | 0.36528 | 6211.586 |
| GOMF_EXC | 44 | 0.021393 | 0.001032 | 0.062123 | 0.36529 | 6211.756 |
| GOBP_INSU | 57 | 0.021791 | 0.001196 | 0.063287 | 0.3653 | 6211.927 |
| GOCC_CON | 18 | 0.044182 | 0.001364 | 0.12836 | 0.36535 | 6212.777 |
| STEIN_ESR | 77 | 0.020766 | 0.001324 | 0.06037 | 0.36543 | 6214.137 |
| GOBP_POS | 10 | 0.059932 | 0.001379 | 0.17433 | 0.36551 | 6215.498 |
| GOBP_DIO | 21 | 0.037023 | 0.001234 | 0.1078 | 0.36564 | 6217.708 |
| GOBP_POS | 12 | 0.0494 | 0.001245 | 0.1439 | 0.36569 | 6218.558 |
| GOBP_LIPC | 10 | 0.052004 | 0.001197 | 0.15151 | 0.36571 | 6218.899 |
| GOBP_PUR | 5 | 0.080886 | 0.001316 | 0.23574 | 0.36576 | 6219.749 |
| GOBP_VITA | 5 | 0.060058 | 0.000977 | 0.17508 | 0.36579 | 6220.259 |
| GOBP_ATR | 8 | 0.071872 | 0.001479 | 0.20977 | 0.36594 | 6222.81 |
| REACTOME | 81 | 0.016765 | 0.001096 | 0.048933 | 0.36595 | 6222.98 |
| CHIARADO | 134 | 0.015123 | 0.00127 | 0.0442 | 0.36612 | 6225.871 |
| GOMF_DIC | 29 | 0.031733 | 0.001243 | 0.093168 | 0.3667 | 6235.734 |
| CREIGHTOI | 677 | 0.006822 | 0.001269 | 0.02003 | 0.36671 | 6235.904 |
| GOBP_INTI | 11 | 0.052394 | 0.001264 | 0.15394 | 0.3668 | 6237.434 |
| HOFFMAN_ | 9 | 0.062807 | 0.001371 | 0.18463 | 0.36686 | 6238.454 |
| REACTOME | 5 | 0.074225 | 0.001208 | 0.21837 | 0.36697 | 6240.325 |
| GOBP_INTI | 41 | 0.025638 | 0.001194 | 0.07545 | 0.367 | 6240.835 |
| WIERENGA | 119 | 0.014913 | 0.00118 | 0.043906 | 0.36706 | 6241.855 |
| GOBP_PAR | 7 | 0.068372 | 0.001316 | 0.20145 | 0.36716 | 6243.556 |
| GOBP_POS | 35 | 0.031346 | 0.001349 | 0.092362 | 0.36716 | 6243.556 |
| MYLLYKAN | 9 | 0.08132 | 0.001775 | 0.23989 | 0.36731 | 6246.107 |
| GOCC_GOL | 84 | 0.016185 | 0.001077 | 0.047748 | 0.36732 | 6246.277 |
| GOBP_GLY | 298 | 0.009883 | 0.001232 | 0.029161 | 0.36734 | 6246.617 |
| DELASERNA | 52 | 0.024083 | 0.001262 | 0.071073 | 0.36736 | 6246.957 |
| GOMF_FAT | 10 | 0.051098 | 0.001176 | 0.1508 | 0.36737 | 6247.127 |
| GOBP_ANT | 18 | 0.042098 | 0.001299 | 0.12427 | 0.3674 | 6247.637 |
| GOBP_CAN | 4 | 0.079814 | 0.001162 | 0.23561 | 0.3674 | 6247.637 |
| GOBP_LYM | 17 | 0.042544 | 0.001276 | 0.12566 | 0.36747 | 6248.827 |
| PID_S1P_S | 21 | 0.0347 | 0.001157 | 0.1025 | 0.36748 | 6248.997 |
| GOBP_CD4 | 6 | 0.078955 | 0.001407 | 0.23332 | 0.36753 | 6249.848 |
| GOMF_EXT | 38 | 0.029922 | 0.001341 | 0.088475 | 0.36761 | 6251.208 |
| GOBP_POS | 9 | 0.066827 | 0.001459 | 0.1977 | 0.36768 | 6252.398 |
| REACTOME | 19 | 0.036912 | 0.00117 | 0.10923 | 0.36771 | 6252.909 |
| GOBP_OST | 97 | 0.018201 | 0.001301 | 0.053907 | 0.36782 | 6254.779 |
| WP_T_CELI | 43 | 0.027319 | 0.001302 | 0.080987 | 0.36794 | 6256.82 |
| GOBP_POS | 58 | 0.021393 | 0.001184 | 0.063463 | 0.36802 | 6258.18 |
| GOBP_VAS | 40 | 0.02877 | 0.001323 | 0.085344 | 0.36802 | 6258.18 |
| GOCC_RUF | 175 | 0.012859 | 0.001232 | 0.038189 | 0.36817 | 6260.731 |
| GOMF_LIP | 18 | 0.035666 | 0.001101 | 0.10604 | 0.36831 | 6263.112 |
| GOMF_CEL | 523 | 0.00762 | 0.001251 | 0.022662 | 0.36834 | 6263.622 |
| GOBP_REG | 57 | 0.02131 | 0.001169 | 0.063419 | 0.36843 | 6265.152 |

| | | | | | | |
|---|---|---|---|---|---|---|
| GOMF_HYI | 15 | 0.040327 | 0.001136 | 0.12004 | 0.36846 | 6265.662 |
| GOBP_MEI | 204 | 0.010953 | 0.001133 | 0.032607 | 0.36847 | 6265.832 |
| GOBP_PUL | 18 | 0.044046 | 0.001359 | 0.13115 | 0.36849 | 6266.172 |
| GOBP_PUR | 7 | 0.06068 | 0.001168 | 0.18066 | 0.36849 | 6266.172 |
| GOBP_PHC | 16 | 0.043311 | 0.00126 | 0.12898 | 0.36851 | 6266.513 |
| WANG_SM | 269 | 0.011168 | 0.001324 | 0.033257 | 0.36851 | 6266.513 |
| DESERT_EX | 20 | 0.041472 | 0.001349 | 0.12354 | 0.36855 | 6267.193 |
| GOBP_RNA | 119 | 0.014537 | 0.001151 | 0.043308 | 0.36856 | 6267.363 |
| REACTOME | 9 | 0.050638 | 0.001105 | 0.15099 | 0.36867 | 6269.233 |
| BURTON_A | 38 | 0.026591 | 0.001192 | 0.07935 | 0.36877 | 6270.934 |
| GOBP_RES | 17 | 0.037582 | 0.001127 | 0.11227 | 0.36891 | 6273.315 |
| CHEOK_RE | 18 | 0.040444 | 0.001248 | 0.12084 | 0.36893 | 6273.655 |
| REACTOME | 450 | 0.007849 | 0.001197 | 0.023461 | 0.36898 | 6274.505 |
| GOMF_KET | 5 | 0.087967 | 0.001431 | 0.26315 | 0.36909 | 6276.375 |
| GOBP_POS | 3 | 0.11865 | 0.001496 | 0.35525 | 0.36919 | 6278.076 |
| GOBP_ENS | 138 | 0.014416 | 0.001228 | 0.043166 | 0.36921 | 6278.416 |
| REACTOME | 48 | 0.023317 | 0.001174 | 0.069835 | 0.36924 | 6278.926 |
| REACTOME | 18 | 0.044593 | 0.001376 | 0.13358 | 0.36926 | 6279.266 |
| GOCC_CYT | 13 | 0.041558 | 0.00109 | 0.12458 | 0.36935 | 6280.797 |
| GOBP_VISU | 51 | 0.024744 | 0.001284 | 0.07419 | 0.36937 | 6281.137 |
| PID_ERBB1 | 40 | 0.027297 | 0.001255 | 0.081917 | 0.36948 | 6283.007 |
| MAYBURD_ | 25 | 0.030519 | 0.00111 | 0.091588 | 0.36949 | 6283.177 |
| GOBP_EPIT | 312 | 0.010087 | 0.001286 | 0.030291 | 0.36957 | 6284.538 |
| GOBP_LON | 34 | 0.027626 | 0.001171 | 0.082962 | 0.36957 | 6284.538 |
| GOCC_LAM | 188 | 0.012186 | 0.00121 | 0.036606 | 0.36961 | 6285.218 |
| GOMF_TRA | 11 | 0.058784 | 0.001419 | 0.1766 | 0.36962 | 6285.388 |
| BACOLOD_ | 45 | 0.025537 | 0.001245 | 0.076728 | 0.36964 | 6285.728 |
| GOBP_POS | 9 | 0.05474 | 0.001195 | 0.16449 | 0.36965 | 6285.898 |
| GOBP_CELI | 6 | 0.064163 | 0.001144 | 0.19299 | 0.36977 | 6287.939 |
| REACTOME | 89 | 0.01503 | 0.00103 | 0.045239 | 0.36985 | 6289.299 |
| LOPEZ_ME | 7 | 0.058307 | 0.001123 | 0.17556 | 0.3699 | 6290.15 |
| LIU_LIVER_ | 32 | 0.026597 | 0.001094 | 0.080187 | 0.37006 | 6292.87 |
| GOCC_TRA | 36 | 0.024887 | 0.001086 | 0.075035 | 0.37007 | 6293.04 |
| GOBP_BUN | 14 | 0.052454 | 0.001428 | 0.15841 | 0.37028 | 6296.611 |
| REACTOME | 154 | 0.013828 | 0.001244 | 0.041772 | 0.37031 | 6297.122 |
| GOBP_NEG | 17 | 0.044815 | 0.001344 | 0.13541 | 0.37034 | 6297.632 |
| PALOMERO | 6 | 0.066449 | 0.001184 | 0.20082 | 0.37037 | 6298.142 |
| GOMF_PHO | 4 | 0.098709 | 0.001437 | 0.29835 | 0.37038 | 6298.312 |
| REACTOME | 34 | 0.025391 | 0.001077 | 0.076748 | 0.37038 | 6298.312 |
| GOBP_POS | 5 | 0.09096 | 0.00148 | 0.27507 | 0.37044 | 6299.332 |
| GOBP_CELI | 85 | 0.017901 | 0.001199 | 0.054176 | 0.37054 | 6301.033 |
| WP_MELAT | 32 | 0.027836 | 0.001145 | 0.084248 | 0.37055 | 6301.203 |
| GOBP_POS | 133 | 0.014796 | 0.001238 | 0.044794 | 0.37058 | 6301.713 |
| GOBP_REG | 15 | 0.041914 | 0.001181 | 0.12691 | 0.3706 | 6302.053 |
| JIANG_TIP3 | 43 | 0.026628 | 0.001269 | 0.080692 | 0.3707 | 6303.754 |

| | | | | | | |
|---|---|---|---|---|---|---|
| GOBP_MIT | 20 | 0.035124 | 0.001143 | 0.10654 | 0.37083 | 6305.964 |
| GOBP_EST/ | 10 | 0.05982 | 0.001376 | 0.18151 | 0.37086 | 6306.474 |
| GOBP_RES | 7 | 0.060586 | 0.001166 | 0.18413 | 0.37106 | 6309.875 |
| MA_MYEL( | 37 | 0.025796 | 0.001141 | 0.078441 | 0.37114 | 6311.236 |
| GOBP_RES | 20 | 0.035295 | 0.001148 | 0.10735 | 0.37117 | 6311.746 |
| GOMF_MC | 33 | 0.03248 | 0.001357 | 0.098813 | 0.37119 | 6312.086 |
| KEGG_ALL( | 15 | 0.046585 | 0.001313 | 0.14174 | 0.37121 | 6312.426 |
| GOMF_HYI | 9 | 0.05631 | 0.001229 | 0.17135 | 0.37122 | 6312.596 |
| GOBP_NAT | 6 | 0.068056 | 0.001213 | 0.20717 | 0.37127 | 6313.446 |
| GOBP_ORG | 475 | 0.007364 | 0.001153 | 0.022418 | 0.37128 | 6313.616 |
| REACTOME | 5 | 0.07586 | 0.001234 | 0.23096 | 0.37128 | 6313.616 |
| GOBP_CELI | 542 | 0.007397 | 0.001235 | 0.022528 | 0.37133 | 6314.467 |
| KIM_ALL_D | 708 | 0.006092 | 0.001157 | 0.018565 | 0.37141 | 6315.827 |
| GUENTHER | 20 | 0.042447 | 0.001381 | 0.12938 | 0.37142 | 6315.997 |
| GOBP_CYT | 200 | 0.011007 | 0.001127 | 0.033589 | 0.37157 | 6318.548 |
| REACTOME | 6 | 0.073316 | 0.001307 | 0.22384 | 0.37163 | 6319.568 |
| SOUCEK_M | 8 | 0.06642 | 0.001367 | 0.20279 | 0.37164 | 6319.738 |
| GOBP_MEI | 9 | 0.056159 | 0.001226 | 0.17165 | 0.37177 | 6321.949 |
| GOBP_NAD | 6 | 0.055904 | 0.000996 | 0.17125 | 0.37204 | 6326.54 |
| REACTOME | 80 | 0.02152 | 0.001398 | 0.065944 | 0.37208 | 6327.22 |
| KEGG_AMI | 43 | 0.023012 | 0.001097 | 0.070523 | 0.3721 | 6327.561 |
| GOBP_REG | 65 | 0.021227 | 0.001243 | 0.065069 | 0.37213 | 6328.071 |
| REACTOME | 5 | 0.076398 | 0.001243 | 0.23454 | 0.37231 | 6331.132 |
| GOMF_LIP( | 7 | 0.070469 | 0.001357 | 0.21638 | 0.37234 | 6331.642 |
| GOMF_GLU | 5 | 0.068268 | 0.001111 | 0.20991 | 0.37251 | 6334.533 |
| BEGUM_T/ | 54 | 0.026631 | 0.001422 | 0.08195 | 0.3726 | 6336.063 |
| GOMF_CAT | 120 | 0.013641 | 0.001084 | 0.041985 | 0.37263 | 6336.573 |
| REACTOME | 9 | 0.059005 | 0.001288 | 0.18168 | 0.37268 | 6337.423 |
| GOBP_FAT | 43 | 0.024379 | 0.001162 | 0.07516 | 0.37284 | 6340.144 |
| GOBP_NEG | 15 | 0.033261 | 0.000937 | 0.10258 | 0.37287 | 6340.654 |
| GOBP_ADE | 4 | 0.074628 | 0.001086 | 0.23017 | 0.37288 | 6340.824 |
| GOBP_EST/ | 428 | 0.007632 | 0.001136 | 0.023539 | 0.37288 | 6340.824 |
| SCHUHMA( | 75 | 0.018345 | 0.001154 | 0.05662 | 0.37297 | 6342.355 |
| MATZUK_F | 9 | 0.060133 | 0.001313 | 0.18564 | 0.373 | 6342.865 |
| GOMF_SEC | 12 | 0.060497 | 0.001525 | 0.18691 | 0.37309 | 6344.395 |
| GOBP_REG | 5 | 0.075777 | 0.001233 | 0.23434 | 0.37321 | 6346.436 |
| GOBP_CELI | 612 | 0.006848 | 0.001213 | 0.021193 | 0.37331 | 6348.137 |
| CHYLA_CBI | 230 | 0.010342 | 0.001135 | 0.032022 | 0.37336 | 6348.987 |
| GOBP_REG | 26 | 0.034143 | 0.001266 | 0.10573 | 0.37338 | 6349.327 |
| GOBP_MIT | 49 | 0.020514 | 0.001044 | 0.063531 | 0.37339 | 6349.497 |
| ROYLANCE_ | 61 | 0.029673 | 0.001684 | 0.091905 | 0.3734 | 6349.667 |
| GOBP_VEN | 14 | 0.051815 | 0.001411 | 0.1605 | 0.37342 | 6350.007 |
| GOBP_REG | 11 | 0.039696 | 0.000958 | 0.12316 | 0.37361 | 6353.238 |
| HOLLERN_/ | 15 | 0.036286 | 0.001022 | 0.1126 | 0.37363 | 6353.578 |
| BERENJEN( | 20 | 0.041444 | 0.001348 | 0.12867 | 0.37369 | 6354.598 |

| | | | | | | |
|---|---|---|---|---|---|---|
| GOBP_CYT | 36 | 0.024271 | 0.001059 | 0.075383 | 0.37374 | 6355.449 |
| ZWANG_E( | 30 | 0.028378 | 0.00113 | 0.088197 | 0.37382 | 6356.809 |
| GOMF_TR/ | 97 | 0.014747 | 0.001054 | 0.045909 | 0.37402 | 6360.21 |
| REACTOME | 86 | 0.0172 | 0.001158 | 0.053627 | 0.37421 | 6363.441 |
| GOBP_SPIN | 20 | 0.038364 | 0.001248 | 0.11969 | 0.37428 | 6364.631 |
| GUTIERREZ | 55 | 0.02275 | 0.001226 | 0.07114 | 0.37457 | 6369.563 |
| REACTOME | 11 | 0.054814 | 0.001323 | 0.17148 | 0.37462 | 6370.413 |
| GOBP_RES | 1053 | 0.00514 | 0.00118 | 0.016083 | 0.37464 | 6370.753 |
| GOBP_FAT_ | 41 | 0.024227 | 0.001128 | 0.07587 | 0.37474 | 6372.454 |
| GOCC_U7_ | 7 | 0.084974 | 0.001636 | 0.26611 | 0.37474 | 6372.454 |
| GOBP_AST | 19 | 0.038249 | 0.001213 | 0.1198 | 0.37476 | 6372.794 |
| GOBP_REG | 103 | 0.015362 | 0.001132 | 0.048132 | 0.3748 | 6373.474 |
| GOBP_CON | 7 | 0.07035 | 0.001354 | 0.22042 | 0.37481 | 6373.644 |
| REACTOME | 42 | 0.028576 | 0.001346 | 0.089543 | 0.37481 | 6373.644 |
| WP_PHYSI( | 24 | 0.034992 | 0.001247 | 0.10964 | 0.37481 | 6373.644 |
| GOBP_T_C | 9 | 0.054182 | 0.001183 | 0.1698 | 0.37483 | 6373.984 |
| GOBP_ORG | 129 | 0.013637 | 0.001124 | 0.042747 | 0.37486 | 6374.494 |
| GINESTIER_ | 100 | 0.017755 | 0.001289 | 0.055665 | 0.37488 | 6374.834 |
| GOBP_DET | 18 | 0.036359 | 0.001122 | 0.11404 | 0.37493 | 6375.685 |
| GOBP_REP | 29 | 0.031482 | 0.001233 | 0.098749 | 0.37494 | 6375.855 |
| GOBP_ESTI | 9 | 0.061752 | 0.001348 | 0.19376 | 0.37498 | 6376.535 |
| DAZARD_U | 13 | 0.045691 | 0.001199 | 0.14344 | 0.37505 | 6377.725 |
| GOCC_VAR | 7 | 0.070015 | 0.001348 | 0.22015 | 0.37523 | 6380.786 |
| GOBP_VIR/ | 31 | 0.03011 | 0.001219 | 0.094713 | 0.37528 | 6381.636 |
| TAVOR_CE | 31 | 0.026959 | 0.001092 | 0.084808 | 0.37529 | 6381.806 |
| GOBP_INTI | 10 | 0.058179 | 0.001339 | 0.18314 | 0.37537 | 6383.167 |
| GOBP_G_C | 7 | 0.069342 | 0.001335 | 0.21835 | 0.37541 | 6383.847 |
| GOMF_LIP/ | 127 | 0.013944 | 0.00114 | 0.043914 | 0.37542 | 6384.017 |
| REACTOME | 7 | 0.062017 | 0.001194 | 0.19532 | 0.37543 | 6384.187 |
| REACTOME | 51 | 0.022807 | 0.001184 | 0.071847 | 0.37546 | 6384.697 |
| GOBP_POS | 8 | 0.055014 | 0.001132 | 0.17332 | 0.37547 | 6384.867 |
| GOBP_POS | 5 | 0.082628 | 0.001345 | 0.26044 | 0.37552 | 6385.718 |
| PIONTEK_F | 13 | 0.042062 | 0.001103 | 0.13261 | 0.37555 | 6386.228 |
| HO_LIVER_ | 12 | 0.045274 | 0.001141 | 0.14277 | 0.37558 | 6386.738 |
| FUNG_IL2_ | 8 | 0.058845 | 0.001211 | 0.18567 | 0.37565 | 6387.928 |
| GOBP_NEG | 5 | 0.079365 | 0.001291 | 0.25053 | 0.3757 | 6388.779 |
| HEDENFALI | 16 | 0.036211 | 0.001054 | 0.11431 | 0.37571 | 6388.949 |
| GOBP_HIST | 16 | 0.047215 | 0.001374 | 0.14912 | 0.37577 | 6389.969 |
| REACTOME | 7 | 0.057594 | 0.001109 | 0.18192 | 0.37578 | 6390.139 |
| GOBP_MAI | 6 | 0.083008 | 0.00148 | 0.26227 | 0.37581 | 6390.649 |
| GRADE_CO | 51 | 0.020101 | 0.001043 | 0.063516 | 0.37582 | 6390.819 |
| WP_INHIBI | 17 | 0.035115 | 0.001053 | 0.11109 | 0.37596 | 6393.2 |
| WP_WHITE | 32 | 0.030475 | 0.001254 | 0.09652 | 0.3761 | 6395.581 |
| BIOCARTA_ | 30 | 0.029854 | 0.001189 | 0.094558 | 0.37611 | 6395.751 |
| GOBP_POS | 19 | 0.036577 | 0.00116 | 0.11592 | 0.37618 | 6396.941 |

| | | | | | | |
|---|---|---|---|---|---|---|
| GOMF_PRO | 1202 | 0.004666 | 0.001139 | 0.014793 | 0.37622 | 6397.621 |
| GOBP_SUC | 6 | 0.064723 | 0.001154 | 0.20532 | 0.37629 | 6398.811 |
| GOBP_CAR | 33 | 0.02844 | 0.001188 | 0.090259 | 0.37635 | 6399.832 |
| WU_APOPT | 9 | 0.054022 | 0.001179 | 0.17148 | 0.37637 | 6400.172 |
| REACTOME | 5 | 0.062384 | 0.001015 | 0.19813 | 0.37643 | 6401.192 |
| GOMF_PRO | 21 | 0.032787 | 0.001093 | 0.10419 | 0.3765 | 6402.383 |
| MARTINEZ_ | 583 | 0.006502 | 0.001125 | 0.020676 | 0.37659 | 6403.913 |
| GOBP_REG | 169 | 0.011725 | 0.001104 | 0.037307 | 0.37666 | 6405.103 |
| NABA_PRO | 33 | 0.03181 | 0.001329 | 0.10122 | 0.37666 | 6405.103 |
| TESAR_ALK | 4 | 0.11392 | 0.001658 | 0.36249 | 0.37666 | 6405.103 |
| BREDEMEY | 36 | 0.029598 | 0.001291 | 0.094267 | 0.37677 | 6406.974 |
| MOOTHA_ | 6 | 0.077449 | 0.001381 | 0.2468 | 0.37683 | 6407.994 |
| GOMF_ME | 28 | 0.031689 | 0.00122 | 0.10106 | 0.37693 | 6409.695 |
| GOBP_MIT | 5 | 0.082806 | 0.001347 | 0.26414 | 0.37696 | 6410.205 |
| ZHU_CMV_ | 37 | 0.026555 | 0.001175 | 0.084757 | 0.37703 | 6411.395 |
| CHARAFE_ | 51 | 0.024175 | 0.001255 | 0.077209 | 0.3771 | 6412.586 |
| GOMF_NU | 203 | 0.01035 | 0.001068 | 0.033096 | 0.37724 | 6414.966 |
| GARGALOV | 32 | 0.028317 | 0.001165 | 0.090552 | 0.37725 | 6415.136 |
| GOBP_REG | 327 | 0.008535 | 0.001114 | 0.027295 | 0.37726 | 6415.306 |
| HE_PTEN_ | 7 | 0.069254 | 0.001333 | 0.2215 | 0.37727 | 6415.476 |
| REACTOME | 17 | 0.039204 | 0.001176 | 0.12538 | 0.37727 | 6415.476 |
| GOCC_AZU | 149 | 0.01213 | 0.001073 | 0.038821 | 0.37735 | 6416.837 |
| DAVICIONI | 244 | 0.011035 | 0.001247 | 0.035334 | 0.3774 | 6417.687 |
| KEGG_CYS | 33 | 0.028286 | 0.001182 | 0.090612 | 0.37746 | 6418.707 |
| GOBP_RNA | 52 | 0.021187 | 0.00111 | 0.067913 | 0.37753 | 6419.898 |
| GOBP_RES | 11 | 0.054455 | 0.001314 | 0.17465 | 0.3776 | 6421.088 |
| GOBP_VEN | 13 | 0.052917 | 0.001388 | 0.16989 | 0.37772 | 6423.129 |
| GOBP_VAS | 82 | 0.018589 | 0.001223 | 0.059693 | 0.37774 | 6423.469 |
| KEGG_GLY | 16 | 0.035536 | 0.001034 | 0.11427 | 0.37791 | 6426.36 |
| WP_CLEAR | 82 | 0.017852 | 0.001174 | 0.057419 | 0.37794 | 6426.87 |
| GOBP_DIET | 13 | 0.047184 | 0.001238 | 0.15195 | 0.37809 | 6429.42 |
| BERENJENO | 8 | 0.062433 | 0.001285 | 0.20115 | 0.37814 | 6430.271 |
| GOMF_NEU | 43 | 0.024409 | 0.001164 | 0.078646 | 0.37814 | 6430.271 |
| GOMF_PAL | 9 | 0.047988 | 0.001048 | 0.15474 | 0.37823 | 6431.801 |
| GOBP_AMI | 4 | 0.082583 | 0.001202 | 0.2663 | 0.37824 | 6431.971 |
| BIOCARTA_ | 50 | 0.021959 | 0.001129 | 0.070823 | 0.37826 | 6432.311 |
| SWEET_KR | 81 | 0.018716 | 0.001223 | 0.060388 | 0.37831 | 6433.162 |
| LINDGREN_ | 48 | 0.023862 | 0.001202 | 0.077006 | 0.37833 | 6433.502 |
| REACTOME | 18 | 0.042797 | 0.001321 | 0.13813 | 0.37835 | 6433.842 |
| GOMF_AM | 3 | 0.11046 | 0.001392 | 0.35671 | 0.37841 | 6434.862 |
| NIKOLSKY_ | 15 | 0.055242 | 0.001557 | 0.17855 | 0.37851 | 6436.563 |
| REACTOME | 49 | 0.019366 | 0.000985 | 0.062592 | 0.37851 | 6436.563 |
| GOBP_TNF | 5 | 0.077597 | 0.001263 | 0.25149 | 0.37884 | 6442.174 |
| GARGALOV | 127 | 0.013037 | 0.001066 | 0.042286 | 0.37892 | 6443.535 |
| GOBP_PEP | 15 | 0.046808 | 0.001319 | 0.15181 | 0.37892 | 6443.535 |

| | | | | | | |
|---|---|---|---|---|---|---|
| LEE_AGING | 38 | 0.024707 | 0.001107 | 0.08014 | 0.37893 | 6443.705 |
| REACTOME | 17 | 0.037456 | 0.001124 | 0.12153 | 0.37896 | 6444.215 |
| GOBP_PHC | 14 | 0.048161 | 0.001311 | 0.15633 | 0.37902 | 6445.235 |
| GOBP_RES | 27 | 0.028796 | 0.001088 | 0.093502 | 0.37905 | 6445.745 |
| REACTOME | 85 | 0.015941 | 0.001067 | 0.051765 | 0.37906 | 6445.915 |
| GOCC_CD4 | 11 | 0.042448 | 0.001024 | 0.13788 | 0.37909 | 6446.425 |
| GOBP_ARA | 53 | 0.021157 | 0.001119 | 0.068728 | 0.37911 | 6446.766 |
| GOBP_TRA | 9 | 0.049504 | 0.001081 | 0.16083 | 0.37912 | 6446.936 |
| GOBP_REG | 4 | 0.093931 | 0.001367 | 0.30519 | 0.37913 | 6447.106 |
| GOBP_NEU | 1027 | 0.005052 | 0.001146 | 0.016416 | 0.37914 | 6447.276 |
| GOMF_HIS | 204 | 0.010633 | 0.001099 | 0.034571 | 0.3792 | 6448.296 |
| REACTOME | 36 | 0.02522 | 0.0011 | 0.081991 | 0.3792 | 6448.296 |
| MATZUK_F | 7 | 0.058603 | 0.001128 | 0.19067 | 0.37929 | 6449.826 |
| GOMF_ADI | 12 | 0.051699 | 0.001303 | 0.16845 | 0.37946 | 6452.717 |
| PID_HEDGI | 46 | 0.023295 | 0.001149 | 0.076199 | 0.37991 | 6460.37 |
| SASAI_TAR | 8 | 0.05414 | 0.001114 | 0.17717 | 0.37997 | 6461.39 |
| GOBP_POS | 60 | 0.019355 | 0.001089 | 0.063382 | 0.38004 | 6462.58 |
| ZHAN_MUI | 85 | 0.016739 | 0.001121 | 0.054849 | 0.38011 | 6463.771 |
| GOBP_TYP | 40 | 0.024983 | 0.001149 | 0.081935 | 0.38022 | 6465.641 |
| GOBP_MEI | 126 | 0.012448 | 0.001014 | 0.040862 | 0.38032 | 6467.342 |
| SEKI_INFLA | 72 | 0.018749 | 0.001156 | 0.061605 | 0.38044 | 6469.382 |
| GOMF_D_( | 12 | 0.044555 | 0.001123 | 0.14654 | 0.38055 | 6471.253 |
| SU_LIVER | 55 | 0.020417 | 0.0011 | 0.067198 | 0.38063 | 6472.613 |
| GOBP_AGG | 4 | 0.072496 | 0.001055 | 0.23876 | 0.3807 | 6473.804 |
| REACTOME | 23 | 0.033184 | 0.001158 | 0.10929 | 0.38071 | 6473.974 |
| KARAKAS_ | 18 | 0.040707 | 0.001256 | 0.13411 | 0.38074 | 6474.484 |
| GOBP_RES | 36 | 0.027704 | 0.001209 | 0.091313 | 0.38079 | 6475.334 |
| REACTOME | 40 | 0.021753 | 0.001 | 0.071704 | 0.3808 | 6475.504 |
| XU_HGF_T/ | 22 | 0.028337 | 0.000967 | 0.093423 | 0.38083 | 6476.014 |
| GOCC_PRC | 23 | 0.026247 | 0.000916 | 0.086646 | 0.38098 | 6478.565 |
| GOBP_RET | 13 | 0.045298 | 0.001188 | 0.14957 | 0.381 | 6478.905 |
| GOBP_PRC | 10 | 0.040022 | 0.000921 | 0.1322 | 0.38105 | 6479.755 |
| MURAKAM | 14 | 0.0413 | 0.001124 | 0.13644 | 0.38106 | 6479.925 |
| GOBP_REG | 11 | 0.043124 | 0.001041 | 0.14247 | 0.38107 | 6480.095 |
| GOBP_HIPI | 41 | 0.025989 | 0.00121 | 0.085896 | 0.38111 | 6480.776 |
| GOBP_MAI | 216 | 0.010411 | 0.001107 | 0.034416 | 0.38114 | 6481.286 |
| REACTOME | 61 | 0.018891 | 0.001072 | 0.062664 | 0.38153 | 6487.918 |
| GOBP_BON | 110 | 0.015297 | 0.001164 | 0.050744 | 0.38154 | 6488.088 |
| GOBP_NOI | 8 | 0.064831 | 0.001334 | 0.21515 | 0.38158 | 6488.768 |
| BIOCARTA_ | 20 | 0.031442 | 0.001023 | 0.10459 | 0.38186 | 6493.529 |
| GOBP_DNA | 547 | 0.006123 | 0.001027 | 0.020392 | 0.38198 | 6495.57 |
| MINGUEZ_ | 21 | 0.036212 | 0.001207 | 0.12059 | 0.38198 | 6495.57 |
| GOBP_POS | 21 | 0.034764 | 0.001159 | 0.11579 | 0.382 | 6495.91 |
| GOBP_REG | 482 | 0.00685 | 0.001081 | 0.022835 | 0.3821 | 6497.611 |
| GOBP_REG | 5 | 0.056169 | 0.000914 | 0.18763 | 0.38233 | 6501.522 |

| | | | | | | |
|---|---|---|---|---|---|---|
| GOBP_POS | 5 | 0.081099 | 0.00132 | 0.27102 | 0.38238 | 6502.372 |
| GOCC_PICL | 6 | 0.083852 | 0.001495 | 0.28054 | 0.38251 | 6504.583 |
| GOBP_CELI | 179 | 0.011069 | 0.001073 | 0.037084 | 0.38267 | 6507.303 |
| SIG_CHEM | 40 | 0.023542 | 0.001083 | 0.079049 | 0.38293 | 6511.725 |
| GENTILE_R | 52 | 0.022057 | 0.001156 | 0.074091 | 0.38297 | 6512.405 |
| GOBP_OLI( | 96 | 0.016064 | 0.001143 | 0.053996 | 0.38304 | 6513.595 |
| GOMF_GLY | 7 | 0.065497 | 0.001261 | 0.22021 | 0.38307 | 6514.105 |
| LINDGREN_ | 112 | 0.013752 | 0.001056 | 0.046236 | 0.38307 | 6514.105 |
| CHEBOTAE | 76 | 0.017705 | 0.001121 | 0.059537 | 0.38309 | 6514.445 |
| PID_PI3KCI | 34 | 0.025019 | 0.001061 | 0.084143 | 0.38311 | 6514.786 |
| GOBP_INTI | 50 | 0.020579 | 0.001058 | 0.069237 | 0.38315 | 6515.466 |
| GOMF_DYI | 12 | 0.043701 | 0.001101 | 0.14714 | 0.38324 | 6516.996 |
| ZAIDI_OSTI | 13 | 0.043796 | 0.001149 | 0.14748 | 0.38325 | 6517.166 |
| GOMF_PR( | 23 | 0.02881 | 0.001005 | 0.097084 | 0.38333 | 6518.527 |
| GOBP_DEC | 42 | 0.022462 | 0.001058 | 0.075705 | 0.38335 | 6518.867 |
| GOMF_PH( | 22 | 0.032235 | 0.0011 | 0.10877 | 0.38348 | 6521.077 |
| REACTOMI | 74 | 0.01658 | 0.001036 | 0.055987 | 0.38356 | 6522.438 |
| REACTOMI | 19 | 0.032049 | 0.001016 | 0.10823 | 0.38357 | 6522.608 |
| WP_METH | 11 | 0.044611 | 0.001077 | 0.15065 | 0.38357 | 6522.608 |
| GOBP_CAR | 13 | 0.047181 | 0.001238 | 0.15934 | 0.38358 | 6522.778 |
| GOBP_REG | 157 | 0.011611 | 0.001054 | 0.039222 | 0.38361 | 6523.288 |
| WP_BIOTIN | 8 | 0.054465 | 0.001121 | 0.18404 | 0.38364 | 6523.798 |
| GOBP_MIC | 5 | 0.049165 | 0.0008 | 0.16624 | 0.38371 | 6524.989 |
| GOBP_PYR | 6 | 0.058594 | 0.001044 | 0.19818 | 0.38375 | 6525.669 |
| GOBP_HOI | 300 | 0.008535 | 0.001067 | 0.028881 | 0.3838 | 6526.519 |
| GOBP_PRC | 9 | 0.052628 | 0.001149 | 0.1781 | 0.38381 | 6526.689 |
| GOMF_AZ( | 11 | 0.050343 | 0.001215 | 0.17037 | 0.38381 | 6526.689 |
| GOBP_ENL | 6 | 0.064667 | 0.001153 | 0.21894 | 0.38386 | 6527.539 |
| GOBP_PRE | 7 | 0.048449 | 0.000933 | 0.16417 | 0.38395 | 6529.07 |
| GOBP_REC | 165 | 0.010945 | 0.001019 | 0.0371 | 0.38399 | 6529.75 |
| TAKEDA_T/ | 41 | 0.022712 | 0.001057 | 0.077002 | 0.38402 | 6530.26 |
| GOBP_RAB | 16 | 0.038034 | 0.001107 | 0.12899 | 0.38405 | 6530.77 |
| GOBP_CELI | 4 | 0.074541 | 0.001085 | 0.25355 | 0.38439 | 6536.552 |
| REACTOMI | 29 | 0.027481 | 0.001076 | 0.093628 | 0.38457 | 6539.613 |
| GOCC_PRC | 23 | 0.031374 | 0.001094 | 0.1069 | 0.38458 | 6539.783 |
| GOBP_REG | 18 | 0.037557 | 0.001159 | 0.128 | 0.38461 | 6540.293 |
| LU_TUMOI | 25 | 0.032556 | 0.001184 | 0.11102 | 0.38467 | 6541.313 |
| GEORGES_ | 61 | 0.019417 | 0.001102 | 0.066243 | 0.38472 | 6542.164 |
| GOMF_PR( | 966 | 0.004884 | 0.001076 | 0.016666 | 0.38475 | 6542.674 |
| REACTOMI | 14 | 0.039844 | 0.001085 | 0.13597 | 0.38475 | 6542.674 |
| GOBP_REG | 5 | 0.076582 | 0.001246 | 0.26184 | 0.38496 | 6546.245 |
| GOMF_INC | 7 | 0.051723 | 0.000996 | 0.17698 | 0.38504 | 6547.605 |
| GOBP_CEN | 15 | 0.0357 | 0.001006 | 0.12225 | 0.38514 | 6549.306 |
| GOCC_SEC | 16 | 0.040223 | 0.001171 | 0.13774 | 0.38514 | 6549.306 |
| GOBP_REG | 43 | 0.022333 | 0.001065 | 0.076482 | 0.38515 | 6549.476 |

| | | | | | | |
|---|---|---|---|---|---|---|
| GOBP_REG | 177 | 0.01103 | 0.001063 | 0.037807 | 0.38524 | 6551.006 |
| JAATINEN_ | 286 | 0.008887 | 0.001086 | 0.030517 | 0.38544 | 6554.407 |
| REACTOME | 36 | 0.02569 | 0.001121 | 0.088242 | 0.38548 | 6555.087 |
| GOMF_STF | 5 | 0.078971 | 0.001285 | 0.27157 | 0.38561 | 6557.298 |
| GOBP_MEI | 6 | 0.058416 | 0.001041 | 0.20114 | 0.38575 | 6559.679 |
| GOBP_ATT | 43 | 0.020188 | 0.000962 | 0.069518 | 0.38576 | 6559.849 |
| GOMF_ME | 5 | 0.057491 | 0.000936 | 0.19806 | 0.38581 | 6560.699 |
| GOBP_MEI | 101 | 0.013694 | 0.000999 | 0.047198 | 0.38586 | 6561.549 |
| GOBP_HEN | 13 | 0.03367 | 0.000883 | 0.11612 | 0.38592 | 6562.57 |
| GOBP_NEG | 16 | 0.041613 | 0.001211 | 0.14351 | 0.38592 | 6562.57 |
| GOMF_AM | 10 | 0.046476 | 0.001069 | 0.16031 | 0.38594 | 6562.91 |
| GOBP_APIC | 14 | 0.042839 | 0.001166 | 0.14784 | 0.386 | 6563.93 |
| GOMF_CEL | 60 | 0.020014 | 0.001127 | 0.069092 | 0.38604 | 6564.61 |
| GOBP_REG | 17 | 0.032818 | 0.000984 | 0.11332 | 0.38606 | 6564.95 |
| GOBP_NEG | 22 | 0.030912 | 0.001055 | 0.10695 | 0.38628 | 6568.691 |
| PEDERSEN_ | 101 | 0.015558 | 0.001135 | 0.053857 | 0.38634 | 6569.712 |
| GOBP_EXT | 38 | 0.023326 | 0.001046 | 0.080806 | 0.38642 | 6571.072 |
| GOBP_RES | 10 | 0.046868 | 0.001078 | 0.16259 | 0.38658 | 6573.793 |
| REACTOME | 88 | 0.015176 | 0.001034 | 0.052663 | 0.38661 | 6574.303 |
| GOBP_CAL | 417 | 0.007298 | 0.001073 | 0.025328 | 0.38662 | 6574.473 |
| GOBP_HYD | 5 | 0.070488 | 0.001147 | 0.24465 | 0.38663 | 6574.643 |
| REACTOME | 11 | 0.03819 | 0.000922 | 0.13257 | 0.38664 | 6574.813 |
| CHESLER_E | 6 | 0.051085 | 0.000911 | 0.17734 | 0.38665 | 6574.983 |
| FURUKAWA | 71 | 0.017124 | 0.001048 | 0.059475 | 0.38671 | 6576.004 |
| GOBP_ALP | 194 | 0.010261 | 0.001035 | 0.035644 | 0.38672 | 6576.174 |
| PID_P38_A | 29 | 0.024575 | 0.000962 | 0.085491 | 0.38688 | 6578.894 |
| DORN_ADE | 37 | 0.020697 | 0.000915 | 0.072008 | 0.3869 | 6579.235 |
| GOCC_ISW | 12 | 0.055574 | 0.001401 | 0.1935 | 0.38698 | 6580.595 |
| GOCC_INO | 26 | 0.026275 | 0.000974 | 0.091544 | 0.38705 | 6581.785 |
| GOBP_CELI | 17 | 0.03674 | 0.001102 | 0.12812 | 0.38715 | 6583.486 |
| HAHTOLA_ | 91 | 0.014382 | 0.000996 | 0.050168 | 0.38718 | 6583.996 |
| GOBP_CAR | 160 | 0.01128 | 0.001034 | 0.039396 | 0.38731 | 6586.207 |
| GROSS_ELK | 33 | 0.025375 | 0.00106 | 0.088636 | 0.38733 | 6586.547 |
| NEWMAN_ | 36 | 0.026823 | 0.00117 | 0.09374 | 0.38739 | 6587.567 |
| GOCC_LAN | 9 | 0.054102 | 0.001181 | 0.18921 | 0.38746 | 6588.757 |
| GOBP_NEG | 16 | 0.039582 | 0.001152 | 0.13866 | 0.38765 | 6591.988 |
| ONDER_CD | 128 | 0.012484 | 0.001024 | 0.043795 | 0.38781 | 6594.709 |
| GOBP_POS | 27 | 0.027239 | 0.001029 | 0.095623 | 0.38788 | 6595.899 |
| GOZGIT_ES | 678 | 0.005961 | 0.001109 | 0.020925 | 0.38788 | 6595.899 |
| GOMF_SUI | 4 | 0.092158 | 0.001341 | 0.32376 | 0.38796 | 6597.26 |
| GOBP_FOC | 83 | 0.015736 | 0.001041 | 0.05529 | 0.38798 | 6597.6 |
| GOBP_POS | 401 | 0.007041 | 0.001015 | 0.024769 | 0.3881 | 6599.641 |
| WALLACE_ | 22 | 0.02613 | 0.000891 | 0.091952 | 0.38814 | 6600.321 |
| GOBP_NEG | 5 | 0.077531 | 0.001262 | 0.27291 | 0.38817 | 6600.831 |
| GOBP_REG | 285 | 0.00824 | 0.001005 | 0.02902 | 0.38823 | 6601.851 |

| | | | | | | |
|---|---|---|---|---|---|---|
| WP_IL7_SI( | 24 | 0.034056 | 0.001214 | 0.11997 | 0.38826 | 6602.361 |
| GOBP_INN | 802 | 0.00519 | 0.001047 | 0.018296 | 0.38833 | 6603.552 |
| GOBP_REG | 196 | 0.0099 | 0.001004 | 0.034897 | 0.38833 | 6603.552 |
| NIKOLSKY_ | 10 | 0.070424 | 0.00162 | 0.24836 | 0.38838 | 6604.402 |
| GOBP_NEG | 6 | 0.06921 | 0.001234 | 0.24414 | 0.3884 | 6604.742 |
| DOANE_BR | 44 | 0.023032 | 0.001111 | 0.081332 | 0.38852 | 6606.783 |
| REACTOME | 32 | 0.024868 | 0.001023 | 0.087944 | 0.38868 | 6609.503 |
| REACTOME | 95 | 0.013507 | 0.000956 | 0.047811 | 0.38878 | 6611.204 |
| GOBP_POS | 3 | 0.093842 | 0.001183 | 0.33223 | 0.3888 | 6611.544 |
| GOBP_REG | 14 | 0.040579 | 0.001105 | 0.14368 | 0.38881 | 6611.714 |
| REACTOME | 42 | 0.021948 | 0.001034 | 0.077772 | 0.38889 | 6613.074 |
| REACTOME | 14 | 0.045898 | 0.00125 | 0.16281 | 0.38901 | 6615.115 |
| DER_IFN_A | 67 | 0.016028 | 0.000953 | 0.056938 | 0.38916 | 6617.666 |
| GOBP_REG | 54 | 0.021287 | 0.001137 | 0.075662 | 0.38922 | 6618.686 |
| HUMMERI( | 19 | 0.032701 | 0.001037 | 0.11625 | 0.38924 | 6619.026 |
| GOBP_SUL | 16 | 0.036265 | 0.001055 | 0.12894 | 0.38926 | 6619.366 |
| BLALOCK_A | 1151 | 0.004315 | 0.001032 | 0.015344 | 0.38928 | 6619.706 |
| REACTOME | 29 | 0.026456 | 0.001036 | 0.094088 | 0.38929 | 6619.876 |
| KEGG_AXO | 121 | 0.013055 | 0.001042 | 0.046486 | 0.38942 | 6622.087 |
| REACTOME | 15 | 0.037448 | 0.001055 | 0.13335 | 0.38942 | 6622.087 |
| NGO_MALI | 14 | 0.039461 | 0.001074 | 0.14054 | 0.38944 | 6622.427 |
| BIOCARTA_ | 17 | 0.036403 | 0.001092 | 0.1297 | 0.38948 | 6623.107 |
| GOMF_CAI | 17 | 0.032902 | 0.000987 | 0.11743 | 0.38967 | 6626.338 |
| GAUSSMAI | 16 | 0.039697 | 0.001155 | 0.14178 | 0.38974 | 6627.529 |
| WANG_CLI | 218 | 0.009187 | 0.000982 | 0.032819 | 0.38976 | 6627.869 |
| GOMF_CAI | 51 | 0.021069 | 0.001094 | 0.075462 | 0.39005 | 6632.8 |
| GOBP_DNA | 124 | 0.012498 | 0.00101 | 0.044806 | 0.39015 | 6634.501 |
| GOBP_POS | 544 | 0.006237 | 0.001043 | 0.022359 | 0.39015 | 6634.501 |
| GOBP_RES | 36 | 0.022279 | 0.000972 | 0.079915 | 0.39021 | 6635.521 |
| REACTOME | 11 | 0.041925 | 0.001012 | 0.15055 | 0.39033 | 6637.562 |
| GOBP_NEG | 6 | 0.061691 | 0.0011 | 0.22162 | 0.39037 | 6638.242 |
| GOMF_ACI | 18 | 0.034511 | 0.001065 | 0.12402 | 0.3904 | 6638.752 |
| ACOSTA_PI | 108 | 0.01275 | 0.000962 | 0.045822 | 0.39041 | 6638.922 |
| PAPASPYRI | 49 | 0.018495 | 0.000941 | 0.066585 | 0.3906 | 6642.153 |
| GOBP_MRI | 12 | 0.03343 | 0.000843 | 0.12041 | 0.39065 | 6643.003 |
| PHONG_TN | 8 | 0.047579 | 0.000979 | 0.1717 | 0.39085 | 6646.404 |
| LIAN_LIPA_ | 58 | 0.0181 | 0.001002 | 0.065321 | 0.39086 | 6646.574 |
| GOMF_N_A | 6 | 0.062175 | 0.001108 | 0.22482 | 0.39106 | 6649.975 |
| WEIGEL_O | 33 | 0.022399 | 0.000936 | 0.080997 | 0.39107 | 6650.145 |
| HELLER_HD | 389 | 0.007135 | 0.001014 | 0.025814 | 0.39112 | 6650.996 |
| GOMF_ALK | 4 | 0.062689 | 0.000912 | 0.22683 | 0.39113 | 6651.166 |
| SU_KIDNEY | 11 | 0.044387 | 0.001071 | 0.16072 | 0.39121 | 6652.526 |
| BIOCARTA_ | 8 | 0.041111 | 0.000846 | 0.14937 | 0.39157 | 6658.648 |
| MORI_EMU | 16 | 0.036307 | 0.001057 | 0.13232 | 0.3919 | 6664.26 |
| GOBP_ORG | 555 | 0.005705 | 0.000964 | 0.020794 | 0.39192 | 6664.6 |

| | | | | | | |
|---|---|---|---|---|---|---|
| GOMF_NU | 52 | 0.019205 | 0.001007 | 0.070114 | 0.39208 | 6667.32 |
| GOBP_REG | 6 | 0.053187 | 0.000948 | 0.19428 | 0.39213 | 6668.171 |
| GOCC_INCI | 69 | 0.015138 | 0.000914 | 0.055312 | 0.39217 | 6668.851 |
| GOBP_REG | 7 | 0.048392 | 0.000932 | 0.17694 | 0.39224 | 6670.041 |
| GOBP_REG | 59 | 0.018804 | 0.00105 | 0.068857 | 0.39239 | 6672.592 |
| GOLDRATH | 141 | 0.011637 | 0.001002 | 0.04265 | 0.39249 | 6674.292 |
| GOBP_REG | 7 | 0.044616 | 0.000859 | 0.16371 | 0.39261 | 6676.333 |
| GOBP_NEG | 28 | 0.025353 | 0.000976 | 0.093128 | 0.39272 | 6678.204 |
| GOBP_NEG | 19 | 0.034515 | 0.001094 | 0.12712 | 0.393 | 6682.965 |
| GOBP_PUT | 5 | 0.050832 | 0.000827 | 0.18729 | 0.39304 | 6683.645 |
| GOBP_CELI | 306 | 0.007639 | 0.000965 | 0.028158 | 0.39309 | 6684.495 |
| REACTOME | 53 | 0.019529 | 0.001033 | 0.072022 | 0.39314 | 6685.346 |
| MIKKELSEN | 97 | 0.015325 | 0.001096 | 0.056613 | 0.39332 | 6688.407 |
| GOBP_POS | 14 | 0.030809 | 0.000839 | 0.1139 | 0.3934 | 6689.767 |
| MUNSHI_M | 9 | 0.044505 | 0.000972 | 0.16469 | 0.3935 | 6691.468 |
| GOBP_POS | 78 | 0.015054 | 0.000966 | 0.05572 | 0.39351 | 6691.638 |
| GOMF_HIS | 12 | 0.042315 | 0.001067 | 0.15674 | 0.39359 | 6692.998 |
| MOTAMED | 4 | 0.068726 | 0.001 | 0.25461 | 0.39361 | 6693.338 |
| GOBP_PRC | 9 | 0.043951 | 0.000959 | 0.16284 | 0.39362 | 6693.508 |
| GOCC_CEN | 594 | 0.005507 | 0.000961 | 0.020406 | 0.39363 | 6693.678 |
| GOCC_ZON | 7 | 0.054088 | 0.001041 | 0.20056 | 0.3937 | 6694.869 |
| GOBP_PRC | 84 | 0.014575 | 0.00097 | 0.054063 | 0.39374 | 6695.549 |
| GOMF_PLU | 26 | 0.026156 | 0.00097 | 0.09707 | 0.39379 | 6696.399 |
| CLAUS_PGI | 12 | 0.044614 | 0.001125 | 0.16559 | 0.39381 | 6696.739 |
| RASHI_NFK | 18 | 0.031898 | 0.000984 | 0.11844 | 0.39384 | 6697.249 |
| REACTOME | 7 | 0.072459 | 0.001395 | 0.26937 | 0.39397 | 6699.46 |
| REACTOME | 12 | 0.036548 | 0.000921 | 0.13588 | 0.39398 | 6699.63 |
| GOBP_REG | 13 | 0.034234 | 0.000898 | 0.12766 | 0.39429 | 6704.901 |
| GOBP_MES | 6 | 0.065366 | 0.001165 | 0.24442 | 0.39457 | 6709.663 |
| BOYAULT_I | 27 | 0.027657 | 0.001045 | 0.10347 | 0.39462 | 6710.513 |
| GOMF_RIB | 151 | 0.010743 | 0.000957 | 0.040194 | 0.39463 | 6710.683 |
| GOBP_S_SI | 7 | 0.059751 | 0.00115 | 0.22357 | 0.39464 | 6710.853 |
| GOCC_MIC | 6 | 0.048936 | 0.000872 | 0.18311 | 0.39464 | 6710.853 |
| GOBP_CER | 6 | 0.065269 | 0.001163 | 0.24427 | 0.39466 | 6711.193 |
| GOBP_SPEI | 27 | 0.025369 | 0.000959 | 0.094942 | 0.39466 | 6711.193 |
| GOBP_NEG | 10 | 0.042422 | 0.000976 | 0.15884 | 0.39471 | 6712.044 |
| GOCC_SEC | 4 | 0.063074 | 0.000918 | 0.23627 | 0.39475 | 6712.724 |
| REACTOME | 6 | 0.052885 | 0.000943 | 0.19844 | 0.39492 | 6715.615 |
| STEGMEIEF | 10 | 0.042922 | 0.000988 | 0.16107 | 0.39494 | 6715.955 |
| GOMF_LAC | 5 | 0.061382 | 0.000999 | 0.23043 | 0.39497 | 6716.465 |
| WOOD_EB | 45 | 0.019321 | 0.000942 | 0.072599 | 0.39507 | 6718.165 |
| GOBP_NEG | 13 | 0.034064 | 0.000894 | 0.12817 | 0.39521 | 6720.546 |
| GOMF_STF | 7 | 0.057963 | 0.001116 | 0.2181 | 0.39521 | 6720.546 |
| WP_MAMI | 10 | 0.040241 | 0.000926 | 0.1515 | 0.39527 | 6721.566 |
| SARTIPY_N | 29 | 0.023678 | 0.000927 | 0.089157 | 0.39528 | 6721.736 |

| | | | | | | |
|---|---|---|---|---|---|---|
| GOBP_MIR | 5 | 0.051472 | 0.000838 | 0.19436 | 0.39557 | 6726.668 |
| KUROZUMI | 35 | 0.023758 | 0.001022 | 0.089727 | 0.39559 | 6727.008 |
| BIOCARTA_ | 12 | 0.046322 | 0.001168 | 0.17504 | 0.39564 | 6727.858 |
| GOBP_NEG | 22 | 0.029621 | 0.001011 | 0.11198 | 0.39569 | 6728.708 |
| WP_TYPE_ | 32 | 0.02319 | 0.000954 | 0.087671 | 0.3957 | 6728.879 |
| GOMF_PH( | 62 | 0.016915 | 0.000968 | 0.064007 | 0.39579 | 6730.409 |
| GOBP_RES | 82 | 0.014928 | 0.000982 | 0.056515 | 0.39584 | 6731.259 |
| REACTOME | 14 | 0.0316 | 0.00086 | 0.11965 | 0.39585 | 6731.429 |
| GOBP_REG | 19 | 0.029238 | 0.000927 | 0.11072 | 0.39587 | 6731.769 |
| GOBP_POS | 8 | 0.057258 | 0.001178 | 0.21691 | 0.3959 | 6732.28 |
| REACTOME | 9 | 0.048078 | 0.00105 | 0.18219 | 0.39594 | 6732.96 |
| TURASHVIL | 180 | 0.010471 | 0.001018 | 0.039696 | 0.39598 | 6733.64 |
| STAMBOLS | 49 | 0.017971 | 0.000914 | 0.06815 | 0.39601 | 6734.15 |
| GOBP_HIST | 6 | 0.051437 | 0.000917 | 0.19509 | 0.39603 | 6734.49 |
| PUJANA_BI | 54 | 0.017028 | 0.000909 | 0.064586 | 0.39603 | 6734.49 |
| WP_INFLUI | 34 | 0.021593 | 0.000916 | 0.081979 | 0.39612 | 6736.021 |
| PID_UPA_U | 41 | 0.02338 | 0.001088 | 0.08877 | 0.39613 | 6736.191 |
| GOBP_GLU | 24 | 0.032548 | 0.00116 | 0.1236 | 0.39615 | 6736.531 |
| GOBP_PRC | 7 | 0.058822 | 0.001133 | 0.22337 | 0.39615 | 6736.531 |
| REACTOME | 14 | 0.031662 | 0.000862 | 0.12031 | 0.39621 | 6737.551 |
| GOBP_NEG | 7 | 0.051752 | 0.000996 | 0.19675 | 0.39626 | 6738.401 |
| GOBP_REG | 12 | 0.038425 | 0.000968 | 0.14624 | 0.39637 | 6740.272 |
| GOBP_POS | 7 | 0.055275 | 0.001064 | 0.2104 | 0.39638 | 6740.442 |
| GOBP_LUN | 28 | 0.027297 | 0.001051 | 0.10392 | 0.3964 | 6740.782 |
| PID_INTEG | 33 | 0.021804 | 0.000911 | 0.08301 | 0.39641 | 6740.952 |
| CALVET_IR | 5 | 0.0505 | 0.000822 | 0.19228 | 0.39642 | 6741.122 |
| REACTOME | 5 | 0.0601 | 0.000978 | 0.2289 | 0.39645 | 6741.632 |
| GOMF_GAI | 5 | 0.073049 | 0.001189 | 0.2784 | 0.39651 | 6742.653 |
| GOBP_COL | 25 | 0.030627 | 0.001114 | 0.11675 | 0.39653 | 6742.993 |
| GOBP_REG | 6 | 0.058622 | 0.001045 | 0.22349 | 0.39655 | 6743.333 |
| REACTOME | 14 | 0.035827 | 0.000975 | 0.13659 | 0.39655 | 6743.333 |
| RASHI_RES | 129 | 0.011218 | 0.000924 | 0.04278 | 0.39657 | 6743.673 |
| BIOCARTA_ | 27 | 0.024161 | 0.000913 | 0.092158 | 0.3966 | 6744.183 |
| REACTOME | 33 | 0.019332 | 0.000808 | 0.073762 | 0.39663 | 6744.693 |
| GOBP_RES | 35 | 0.022214 | 0.000956 | 0.084864 | 0.39676 | 6746.904 |
| GOBP_PHC | 49 | 0.020613 | 0.001049 | 0.078829 | 0.39686 | 6748.604 |
| WIERENGA | 178 | 0.009598 | 0.000928 | 0.03671 | 0.39688 | 6748.944 |
| GOBP_POS | 17 | 0.033502 | 0.001005 | 0.12822 | 0.39694 | 6749.965 |
| REACTOME | 22 | 0.03006 | 0.001026 | 0.11518 | 0.39706 | 6752.005 |
| BROWNE_I | 238 | 0.008286 | 0.000924 | 0.031776 | 0.39714 | 6753.366 |
| GOBP_RES | 19 | 0.028429 | 0.000901 | 0.10925 | 0.39735 | 6756.937 |
| GOCC_NUC | 15 | 0.030698 | 0.000865 | 0.11799 | 0.39737 | 6757.277 |
| KEGG_TOLI | 93 | 0.014655 | 0.001026 | 0.056356 | 0.39742 | 6758.127 |
| GOBP_NEG | 15 | 0.034235 | 0.000965 | 0.1318 | 0.39753 | 6759.998 |
| GOBP_CER | 10 | 0.047742 | 0.001099 | 0.18392 | 0.3976 | 6761.188 |

| | | | | | | |
|---|---|---|---|---|---|---|
| GOBP_MO | 13 | 0.03513 | 0.000922 | 0.13535 | 0.3976 | 6761.188 |
| GOBP_REG | 5 | 0.073724 | 0.0012 | 0.28436 | 0.39772 | 6763.229 |
| GOBP_EST/ | 34 | 0.02056 | 0.000872 | 0.079335 | 0.39776 | 6763.909 |
| GOMF_TFII | 5 | 0.062836 | 0.001023 | 0.24281 | 0.3979 | 6766.29 |
| REACTOME | 8 | 0.037488 | 0.000772 | 0.1449 | 0.39793 | 6766.8 |
| GOBP_REG | 87 | 0.014132 | 0.000957 | 0.054631 | 0.39794 | 6766.97 |
| GOBP_ANT | 7 | 0.05814 | 0.001119 | 0.22521 | 0.39814 | 6770.371 |
| GOBP_END | 26 | 0.026178 | 0.000971 | 0.1014 | 0.39814 | 6770.371 |
| GOBP_INTI | 26 | 0.025401 | 0.000942 | 0.098429 | 0.39818 | 6771.051 |
| GOBP_REG | 13 | 0.031241 | 0.00082 | 0.12116 | 0.39826 | 6772.411 |
| REACTOME | 5 | 0.072167 | 0.001174 | 0.27996 | 0.39829 | 6772.921 |
| GOBP_NEG | 48 | 0.017328 | 0.000873 | 0.067253 | 0.39834 | 6773.772 |
| GOBP_CAL | 4 | 0.056324 | 0.00082 | 0.21876 | 0.39841 | 6774.962 |
| GOBP_REG | 4 | 0.076064 | 0.001107 | 0.29542 | 0.39841 | 6774.962 |
| GOBP_POS | 63 | 0.017165 | 0.00099 | 0.066684 | 0.39843 | 6775.302 |
| GOBP_POS | 22 | 0.025883 | 0.000883 | 0.10055 | 0.39843 | 6775.302 |
| AIYAR_COE | 28 | 0.023907 | 0.00092 | 0.092877 | 0.39844 | 6775.472 |
| DAVICIONI_ | 59 | 0.018347 | 0.001024 | 0.071302 | 0.39847 | 6775.982 |
| GOBP_MU | 841 | 0.004503 | 0.000929 | 0.017503 | 0.3985 | 6776.493 |
| MCDOWEL | 44 | 0.019158 | 0.000924 | 0.074473 | 0.3985 | 6776.493 |
| GOBP_POS | 23 | 0.028292 | 0.000987 | 0.11007 | 0.39858 | 6777.853 |
| DER_IFN_G | 10 | 0.035026 | 0.000806 | 0.13634 | 0.39863 | 6778.703 |
| GOBP_DN/ | 21 | 0.028894 | 0.000963 | 0.11253 | 0.39868 | 6779.553 |
| GOMF_SIN | 9 | 0.039053 | 0.000853 | 0.15233 | 0.39883 | 6782.104 |
| MATTIOLI_ | 15 | 0.037996 | 0.001071 | 0.14821 | 0.39883 | 6782.104 |
| GROSS_HIF | 7 | 0.057592 | 0.001109 | 0.22476 | 0.39889 | 6783.124 |
| GOTZMANI | 65 | 0.017148 | 0.001005 | 0.067104 | 0.39915 | 6787.546 |
| GOBP_INN | 70 | 0.015698 | 0.000954 | 0.061461 | 0.39921 | 6788.566 |
| GOBP_STEI | 146 | 0.010584 | 0.000927 | 0.041453 | 0.39924 | 6789.076 |
| GOBP_PRC | 235 | 0.008211 | 0.00091 | 0.032172 | 0.39928 | 6789.756 |
| GOBP_REG | 6 | 0.051755 | 0.000923 | 0.20288 | 0.39933 | 6790.607 |
| GOCC_TRA | 45 | 0.018587 | 0.000906 | 0.073018 | 0.39953 | 6794.008 |
| BURTON_A | 70 | 0.015475 | 0.000941 | 0.060811 | 0.39956 | 6794.518 |
| GOBP_NEG | 11 | 0.044201 | 0.001067 | 0.17391 | 0.39969 | 6796.728 |
| KUROKAW/ | 10 | 0.035528 | 0.000817 | 0.13992 | 0.39978 | 6798.259 |
| DONATO_C | 6 | 0.056267 | 0.001003 | 0.22163 | 0.3998 | 6798.599 |
| GOBP_TRA | 7 | 0.05825 | 0.001122 | 0.22946 | 0.3998 | 6798.599 |
| LOPEZ_ME | 12 | 0.03759 | 0.000947 | 0.14806 | 0.3998 | 6798.599 |
| GOBP_POS | 5 | 0.054662 | 0.000889 | 0.21539 | 0.39983 | 6799.109 |
| ENK_UV_R | 460 | 0.005894 | 0.000909 | 0.023246 | 0.39993 | 6800.81 |
| GOBP_NEG | 24 | 0.025604 | 0.000912 | 0.10099 | 0.39994 | 6800.98 |
| GOBP_POS | 12 | 0.036793 | 0.000927 | 0.14521 | 0.39999 | 6801.83 |
| MURAKAM | 18 | 0.028306 | 0.000874 | 0.11194 | 0.40019 | 6805.231 |
| GOMF_POT | 12 | 0.042311 | 0.001066 | 0.16743 | 0.40025 | 6806.251 |
| GOCC_CILI/ | 47 | 0.019169 | 0.000955 | 0.075889 | 0.40029 | 6806.931 |

| | | | | | | |
|---|---|---|---|---|---|---|
| GOMF_L_G | 14 | 0.037414 | 0.001019 | 0.14821 | 0.40035 | 6807.952 |
| GOBP_XEN | 43 | 0.02067 | 0.000985 | 0.081897 | 0.40037 | 6808.292 |
| REACTOME | 113 | 0.012112 | 0.000934 | 0.047994 | 0.40038 | 6808.462 |
| MARKS_HD | 21 | 0.029373 | 0.000979 | 0.1165 | 0.40047 | 6809.992 |
| SCHUETZ_E | 324 | 0.00755 | 0.000981 | 0.029996 | 0.40064 | 6812.883 |
| LIU_IL13_P | 15 | 0.032439 | 0.000914 | 0.12893 | 0.40068 | 6813.563 |
| GOBP_POS | 7 | 0.051928 | 0.001 | 0.20664 | 0.40079 | 6815.434 |
| GOBP_NEG | 29 | 0.021324 | 0.000835 | 0.084919 | 0.40087 | 6816.794 |
| GOBP_RES | 7 | 0.051993 | 0.001001 | 0.20709 | 0.40088 | 6816.964 |
| GOMF_INS | 13 | 0.031573 | 0.000828 | 0.1258 | 0.40092 | 6817.645 |
| GOBP_RES | 85 | 0.013108 | 0.000878 | 0.052311 | 0.40107 | 6820.195 |
| FRASOR_RI | 69 | 0.014961 | 0.000903 | 0.059709 | 0.40108 | 6820.365 |
| GOBP_CYT | 12 | 0.031168 | 0.000786 | 0.12444 | 0.40111 | 6820.876 |
| GOBP_REG | 10 | 0.033284 | 0.000766 | 0.13301 | 0.40121 | 6822.576 |
| GOMF_PH | 20 | 0.032035 | 0.001042 | 0.12805 | 0.40123 | 6822.916 |
| GOBP_MYE | 214 | 0.009217 | 0.000976 | 0.036849 | 0.40124 | 6823.086 |
| FOURNIER_ | 268 | 0.007682 | 0.000909 | 0.030754 | 0.40138 | 6825.467 |
| GOBP_POS | 5 | 0.074146 | 0.001207 | 0.29694 | 0.40141 | 6825.977 |
| REACTOME | 20 | 0.027058 | 0.00088 | 0.10837 | 0.40142 | 6826.147 |
| GOBP_STEI | 10 | 0.041457 | 0.000954 | 0.16611 | 0.40146 | 6826.827 |
| GOBP_POS | 2 | 0.076922 | 0.000792 | 0.3086 | 0.40158 | 6828.868 |
| FLECHNER_ | 526 | 0.005515 | 0.000908 | 0.022143 | 0.40166 | 6830.228 |
| GOBP_UBI | 19 | 0.024176 | 0.000767 | 0.097122 | 0.40171 | 6831.079 |
| GOBP_NEG | 35 | 0.019296 | 0.00083 | 0.077521 | 0.40172 | 6831.249 |
| GOBP_NEU | 10 | 0.036947 | 0.00085 | 0.14879 | 0.40195 | 6835.16 |
| GOBP_STEI | 55 | 0.01566 | 0.000844 | 0.063096 | 0.40199 | 6835.84 |
| GOBP_INTI | 19 | 0.028634 | 0.000908 | 0.11541 | 0.40203 | 6836.52 |
| GOBP_REG | 10 | 0.040714 | 0.000937 | 0.16413 | 0.40204 | 6836.69 |
| GOMF_DE | 5 | 0.04112 | 0.000669 | 0.16582 | 0.40208 | 6837.37 |
| GOCC_ION | 279 | 0.008026 | 0.000968 | 0.032371 | 0.40209 | 6837.54 |
| GOMF_SUI | 8 | 0.0391 | 0.000805 | 0.15844 | 0.40254 | 6845.193 |
| REACTOME | 8 | 0.038592 | 0.000794 | 0.15652 | 0.40262 | 6846.553 |
| GOBP_NEG | 7 | 0.038078 | 0.000733 | 0.15471 | 0.4028 | 6849.614 |
| GOBP_NEG | 5 | 0.056971 | 0.000927 | 0.2315 | 0.40281 | 6849.784 |
| GOBP_POS | 25 | 0.025888 | 0.000941 | 0.10524 | 0.40284 | 6850.294 |
| GOBP_PRC | 13 | 0.036352 | 0.000954 | 0.14776 | 0.40284 | 6850.294 |
| TURJANSKI | 7 | 0.04464 | 0.000859 | 0.18147 | 0.40284 | 6850.294 |
| GOBP_CON | 7 | 0.049713 | 0.000957 | 0.20215 | 0.40287 | 6850.804 |
| GOBP_POS | 5 | 0.066571 | 0.001083 | 0.27112 | 0.40302 | 6853.355 |
| GOBP_NEU | 24 | 0.024858 | 0.000886 | 0.10135 | 0.40313 | 6855.226 |
| REACTOME | 15 | 0.034381 | 0.000969 | 0.14018 | 0.40313 | 6855.226 |
| REACTOME | 38 | 0.018337 | 0.000822 | 0.074984 | 0.4034 | 6859.817 |
| VERNELL_F | 21 | 0.0266 | 0.000887 | 0.10877 | 0.4034 | 6859.817 |
| ASTON_MA | 45 | 0.018368 | 0.000896 | 0.075131 | 0.40343 | 6860.327 |
| GOBP_DEN | 66 | 0.014147 | 0.000835 | 0.057896 | 0.40348 | 6861.177 |

| | | | | | | |
|---|---|---|---|---|---|---|
| REACTOME | 106 | 0.011463 | 0.000857 | 0.046923 | 0.40351 | 6861.688 |
| REACTOME | 14 | 0.031224 | 0.00085 | 0.12795 | 0.4036 | 6863.218 |
| GOBP_KILL | 43 | 0.019836 | 0.000946 | 0.081285 | 0.40361 | 6863.388 |
| MCCLUNG_ | 47 | 0.018506 | 0.000922 | 0.07596 | 0.40376 | 6865.939 |
| GOBP_SUL | 293 | 0.007223 | 0.000893 | 0.02965 | 0.40377 | 6866.109 |
| GOBP_HIST | 121 | 0.010952 | 0.000874 | 0.044982 | 0.40382 | 6866.959 |
| REACTOME | 21 | 0.026819 | 0.000894 | 0.11019 | 0.40385 | 6867.469 |
| GOBP_POS | 6 | 0.056032 | 0.000999 | 0.23028 | 0.40388 | 6867.979 |
| BERENJENG | 48 | 0.018938 | 0.000954 | 0.077844 | 0.40389 | 6868.149 |
| GOBP_VIRA | 5 | 0.043713 | 0.000711 | 0.18015 | 0.40414 | 6872.401 |
| GOBP_REG | 22 | 0.025388 | 0.000866 | 0.10465 | 0.40416 | 6872.741 |
| CAFFAREL_ | 12 | 0.03433 | 0.000865 | 0.14153 | 0.40417 | 6872.911 |
| GOBP_COE | 9 | 0.037585 | 0.00082 | 0.15503 | 0.40422 | 6873.761 |
| ONDER_CD | 15 | 0.037237 | 0.001049 | 0.1536 | 0.40423 | 6873.931 |
| GOBP_BLO | 109 | 0.01167 | 0.000884 | 0.048145 | 0.40424 | 6874.101 |
| SEMENZA_ | 35 | 0.020379 | 0.000877 | 0.084092 | 0.40426 | 6874.441 |
| REACTOME | 8 | 0.040518 | 0.000834 | 0.16725 | 0.40429 | 6874.951 |
| WP_NAD_I | 15 | 0.030743 | 0.000866 | 0.12713 | 0.40446 | 6877.842 |
| GOMF_COI | 5 | 0.0553 | 0.0009 | 0.22876 | 0.40449 | 6878.352 |
| BIOCARTA_ | 14 | 0.034161 | 0.00093 | 0.14145 | 0.40458 | 6879.883 |
| GOCC_FAS | 9 | 0.042623 | 0.00093 | 0.17655 | 0.40462 | 6880.563 |
| GOBP_PRO | 42 | 0.017897 | 0.000843 | 0.074237 | 0.40475 | 6882.774 |
| MIKKELSEN | 190 | 0.008708 | 0.000869 | 0.036129 | 0.40477 | 6883.114 |
| WORSCHE | 9 | 0.040133 | 0.000876 | 0.16662 | 0.40483 | 6884.134 |
| GOBP_NEG | 5 | 0.054881 | 0.000893 | 0.22789 | 0.40485 | 6884.474 |
| PUJANA_BI | 94 | 0.011767 | 0.000828 | 0.048865 | 0.40485 | 6884.474 |
| LU_AGING_ | 142 | 0.010355 | 0.000895 | 0.04307 | 0.405 | 6887.025 |
| GOBP_ACT | 5 | 0.058955 | 0.000959 | 0.24523 | 0.40501 | 6887.195 |
| GOBP_POS | 102 | 0.011483 | 0.000842 | 0.047807 | 0.40509 | 6888.555 |
| GOBP_CELI | 68 | 0.015631 | 0.000936 | 0.065169 | 0.40522 | 6890.766 |
| ZHOU_CELI | 7 | 0.040913 | 0.000788 | 0.17057 | 0.40522 | 6890.766 |
| GOBP_LIPI | 6 | 0.047295 | 0.000843 | 0.19726 | 0.40526 | 6891.446 |
| KOHN_EMT | 22 | 0.024133 | 0.000823 | 0.10081 | 0.40541 | 6893.997 |
| MATTIOLI_ | 34 | 0.021691 | 0.00092 | 0.090687 | 0.40548 | 6895.187 |
| GOBP_NEG | 6 | 0.052109 | 0.000929 | 0.21791 | 0.4055 | 6895.528 |
| VECCHI_GA | 394 | 0.006024 | 0.000861 | 0.0252 | 0.40554 | 6896.208 |
| IIZUKA_LIV | 24 | 0.022179 | 0.00079 | 0.092862 | 0.40562 | 6897.568 |
| GOCC_CILIA | 13 | 0.030138 | 0.000791 | 0.12632 | 0.40571 | 6899.099 |
| GOBP_GLU | 70 | 0.015509 | 0.000943 | 0.065077 | 0.40582 | 6900.969 |
| GOCC_DEN | 42 | 0.018535 | 0.000873 | 0.07783 | 0.40589 | 6902.159 |
| HEIDENBLA | 37 | 0.018198 | 0.000805 | 0.076442 | 0.40592 | 6902.67 |
| GOMF_PRO | 12 | 0.032743 | 0.000825 | 0.13776 | 0.40606 | 6905.05 |
| WP_REGUO | 30 | 0.02268 | 0.000903 | 0.095692 | 0.40633 | 6909.642 |
| GOMF_SUI | 15 | 0.029708 | 0.000837 | 0.12545 | 0.4064 | 6910.832 |
| TAKEDA_TA | 166 | 0.00964 | 0.0009 | 0.040711 | 0.40642 | 6911.172 |

| | | | | | | |
|---|---|---|---|---|---|---|
| GOBP_PRC | 4 | 0.053759 | 0.000782 | 0.22735 | 0.40654 | 6913.213 |
| ZHAN_MUI | 46 | 0.018585 | 0.000916 | 0.078754 | 0.40672 | 6916.274 |
| GOBP_REG | 11 | 0.034689 | 0.000837 | 0.14707 | 0.40677 | 6917.124 |
| GOCC_INTI | 6 | 0.052383 | 0.000934 | 0.22219 | 0.40681 | 6917.804 |
| GOBP_REG | 13 | 0.027928 | 0.000733 | 0.11856 | 0.40689 | 6919.164 |
| PODAR_RE | 133 | 0.010613 | 0.000888 | 0.045062 | 0.40691 | 6919.505 |
| GOBP_EST/ | 14 | 0.029781 | 0.000811 | 0.1265 | 0.40694 | 6920.015 |
| GOCC_INN | 131 | 0.009344 | 0.000776 | 0.039702 | 0.40697 | 6920.525 |
| SCHRAETS_ | 30 | 0.022764 | 0.000907 | 0.096749 | 0.40699 | 6920.865 |
| GOBP_RAC | 40 | 0.019604 | 0.000901 | 0.083371 | 0.40705 | 6921.885 |
| GOCC_COF | 18 | 0.025046 | 0.000773 | 0.10654 | 0.40707 | 6922.225 |
| BOYLAN_M | 63 | 0.013972 | 0.000806 | 0.059465 | 0.40712 | 6923.076 |
| GOMF_N_/ | 6 | 0.053624 | 0.000956 | 0.22822 | 0.40712 | 6923.076 |
| GOMF_G_I | 7 | 0.033835 | 0.000651 | 0.1441 | 0.40719 | 6924.266 |
| GOMF_EPI | 31 | 0.021707 | 0.000879 | 0.092524 | 0.40726 | 6925.456 |
| GOBP_NEG | 87 | 0.012464 | 0.000844 | 0.053141 | 0.40728 | 6925.796 |
| REACTOME | 9 | 0.029057 | 0.000634 | 0.12402 | 0.40738 | 6927.497 |
| PHESSE_TA | 10 | 0.034001 | 0.000782 | 0.14522 | 0.40744 | 6928.517 |
| PID_S1P_S | 23 | 0.024891 | 0.000868 | 0.10645 | 0.40756 | 6930.558 |
| BIOCARTA_ | 8 | 0.037057 | 0.000763 | 0.1587 | 0.40768 | 6932.598 |
| GOCC_CYT | 14 | 0.031785 | 0.000865 | 0.1361 | 0.40768 | 6932.598 |
| GOCC_NUC | 293 | 0.006548 | 0.000809 | 0.028091 | 0.40784 | 6935.319 |
| GOBP_SPH | 5 | 0.049565 | 0.000807 | 0.21282 | 0.40792 | 6936.68 |
| GOBP_MEI | 5 | 0.060814 | 0.00099 | 0.26115 | 0.40793 | 6936.85 |
| GOBP_PYR | 8 | 0.046781 | 0.000963 | 0.20112 | 0.40804 | 6938.72 |
| GOBP_CELI | 7 | 0.042644 | 0.000821 | 0.18337 | 0.40805 | 6938.89 |
| GOBP_POS | 8 | 0.042806 | 0.000881 | 0.18452 | 0.40828 | 6942.801 |
| GOBP_PYR | 28 | 0.021248 | 0.000818 | 0.091722 | 0.4084 | 6944.842 |
| WP_THERM | 105 | 0.01164 | 0.000866 | 0.050257 | 0.40842 | 6945.182 |
| GOBP_ALD | 11 | 0.03593 | 0.000867 | 0.15518 | 0.40845 | 6945.692 |
| SARTIPY_B | 18 | 0.028724 | 0.000887 | 0.12411 | 0.40849 | 6946.372 |
| GOMF_IMI | 115 | 0.011865 | 0.000923 | 0.051343 | 0.40862 | 6948.583 |
| FOROUTAN | 82 | 0.013142 | 0.000864 | 0.056921 | 0.4087 | 6949.944 |
| GOBP_SYN | 6 | 0.048986 | 0.000873 | 0.21238 | 0.40879 | 6951.474 |
| GOBP_NEG | 76 | 0.013224 | 0.000837 | 0.057381 | 0.40887 | 6952.834 |
| GOBP_RRN | 5 | 0.051277 | 0.000834 | 0.22251 | 0.40887 | 6952.834 |
| LIAO_HAVE | 40 | 0.019119 | 0.000879 | 0.082971 | 0.40888 | 6953.004 |
| YANG_BRE | 45 | 0.0169 | 0.000824 | 0.073439 | 0.409 | 6955.045 |
| GOBP_OO( | 54 | 0.015182 | 0.000811 | 0.065979 | 0.40901 | 6955.215 |
| GOBP_DEN | 51 | 0.01627 | 0.000845 | 0.070734 | 0.40904 | 6955.725 |
| GOBP_UBI( | 5 | 0.062488 | 0.001017 | 0.27203 | 0.40916 | 6957.766 |
| REACTOME | 13 | 0.030936 | 0.000812 | 0.13485 | 0.40928 | 6959.806 |
| GOBP_ACT | 7 | 0.04046 | 0.000779 | 0.17662 | 0.40941 | 6962.017 |
| BIOCARTA_ | 19 | 0.024347 | 0.000772 | 0.10635 | 0.40946 | 6962.867 |
| GOBP_NEG | 66 | 0.012871 | 0.00076 | 0.056229 | 0.40947 | 6963.037 |

| | | | | | | |
|---|---|---|---|---|---|---|
| GOBP_MO | 47 | 0.018205 | 0.000907 | 0.079583 | 0.40953 | 6964.058 |
| HAN_SATB | 364 | 0.006191 | 0.000851 | 0.027107 | 0.40968 | 6966.608 |
| GOMF_MIT | 7 | 0.052514 | 0.001011 | 0.23036 | 0.40984 | 6969.329 |
| SHIRAISHI_ | 9 | 0.03367 | 0.000735 | 0.14773 | 0.40986 | 6969.669 |
| GOBP_AMI | 33 | 0.017918 | 0.000749 | 0.078659 | 0.4099 | 6970.35 |
| GOBP_NEG | 22 | 0.024786 | 0.000846 | 0.10883 | 0.40992 | 6970.69 |
| BIOCARTA_ | 8 | 0.042352 | 0.000872 | 0.186 | 0.40994 | 6971.03 |
| GOBP_RES | 18 | 0.025915 | 0.0008 | 0.11389 | 0.41 | 6972.05 |
| REACTOME | 37 | 0.018112 | 0.000801 | 0.079621 | 0.41003 | 6972.56 |
| LU_TUMOI | 4 | 0.057433 | 0.000836 | 0.25253 | 0.41005 | 6972.9 |
| GOBP_RNA | 9 | 0.038905 | 0.000849 | 0.17158 | 0.41031 | 6977.322 |
| GOBP_RES | 14 | 0.030418 | 0.000828 | 0.1342 | 0.41035 | 6978.002 |
| GOMF_G_I | 4 | 0.062044 | 0.000903 | 0.27389 | 0.4104 | 6978.852 |
| SMID_BRE/ | 6 | 0.058584 | 0.001044 | 0.25881 | 0.41046 | 6979.872 |
| GOBP_PRC | 10 | 0.038378 | 0.000883 | 0.16976 | 0.41057 | 6981.743 |
| MULLIGHA | 304 | 0.00642 | 0.000808 | 0.028405 | 0.41059 | 6982.083 |
| REACTOME | 11 | 0.03394 | 0.000819 | 0.15021 | 0.41062 | 6982.593 |
| WP_CANOI | 26 | 0.024568 | 0.000911 | 0.10874 | 0.41063 | 6982.763 |
| GOBP_PYR | 6 | 0.037553 | 0.000669 | 0.16631 | 0.41068 | 6983.613 |
| GOBP_REG | 11 | 0.038632 | 0.000932 | 0.17117 | 0.41072 | 6984.294 |
| GOBP_REG | 18 | 0.027562 | 0.000851 | 0.12234 | 0.41088 | 6987.014 |
| SMIRNOV_ | 159 | 0.00882 | 0.000806 | 0.039147 | 0.41088 | 6987.014 |
| GOBP_NEG | 21 | 0.021678 | 0.000723 | 0.096333 | 0.41098 | 6988.715 |
| GOMF_DO | 7 | 0.034501 | 0.000664 | 0.15334 | 0.41099 | 6988.885 |
| BIOCARTA_ | 7 | 0.043851 | 0.000844 | 0.19498 | 0.41103 | 6989.565 |
| GOMF_OXI | 165 | 0.00911 | 0.000848 | 0.040512 | 0.41104 | 6989.735 |
| GOBP_CELI | 318 | 0.006316 | 0.000813 | 0.028119 | 0.41114 | 6991.436 |
| WP_FGF23 | 21 | 0.02396 | 0.000799 | 0.10668 | 0.41115 | 6991.606 |
| GOBP_RIB( | 44 | 0.016891 | 0.000814 | 0.075226 | 0.41117 | 6991.946 |
| SU_PLACEI | 23 | 0.023484 | 0.000819 | 0.10462 | 0.41119 | 6992.286 |
| REACTOME | 51 | 0.016461 | 0.000854 | 0.073331 | 0.4112 | 6992.456 |
| GOBP_REG | 5 | 0.046178 | 0.000751 | 0.20581 | 0.41123 | 6992.966 |
| GOCC_NBA | 16 | 0.027395 | 0.000797 | 0.12227 | 0.41136 | 6995.177 |
| GOBP_CHC | 15 | 0.030374 | 0.000856 | 0.13565 | 0.41141 | 6996.027 |
| GOBP_NEG | 25 | 0.022201 | 0.000807 | 0.099191 | 0.41145 | 6996.707 |
| BIOCARTA_ | 7 | 0.032157 | 0.000619 | 0.1437 | 0.41147 | 6997.047 |
| GOBP_UV_ | 12 | 0.030196 | 0.000761 | 0.13498 | 0.41149 | 6997.387 |
| GOBP_REG | 25 | 0.020408 | 0.000742 | 0.091268 | 0.41153 | 6998.068 |
| GRABARCZ | 58 | 0.01497 | 0.000828 | 0.066948 | 0.41154 | 6998.238 |
| KEGG_INO! | 52 | 0.015022 | 0.000787 | 0.067236 | 0.41161 | 6999.428 |
| GOBP_CYT( | 20 | 0.027032 | 0.000879 | 0.12123 | 0.41177 | 7002.149 |
| GOBP_PRC | 74 | 0.013234 | 0.000827 | 0.059367 | 0.4118 | 7002.659 |
| WP_NAD_I | 22 | 0.023134 | 0.000789 | 0.10388 | 0.41189 | 7004.189 |
| REACTOME | 696 | 0.004303 | 0.000811 | 0.019349 | 0.41201 | 7006.23 |
| GOBP_IMN | 15 | 0.032128 | 0.000905 | 0.14451 | 0.41203 | 7006.57 |

| | | | | | | |
|---|---|---|---|---|---|---|
| GOBP_REG | 6 | 0.041493 | 0.00074 | 0.18674 | 0.41208 | 7007.42 |
| GOBP_POS | 36 | 0.017897 | 0.000781 | 0.080568 | 0.41211 | 7007.931 |
| PROVENZA | 186 | 0.008472 | 0.000837 | 0.038146 | 0.41212 | 7008.101 |
| GOCC_DEN | 572 | 0.004799 | 0.000823 | 0.021613 | 0.41214 | 7008.441 |
| GOBP_SUB | 6 | 0.037803 | 0.000674 | 0.17041 | 0.41222 | 7009.801 |
| PARK_OSTI | 12 | 0.030587 | 0.000771 | 0.13788 | 0.41222 | 7009.801 |
| GOBP_REG | 309 | 0.006733 | 0.000854 | 0.030388 | 0.41233 | 7011.672 |
| BIOCARTA_ | 7 | 0.045 | 0.000866 | 0.20336 | 0.41244 | 7013.542 |
| WP_DNA_I | 40 | 0.016323 | 0.000751 | 0.073768 | 0.41244 | 7013.542 |
| GOMF_RIB | 14 | 0.026404 | 0.000719 | 0.11955 | 0.4126 | 7016.263 |
| REACTOME | 15 | 0.024331 | 0.000686 | 0.11019 | 0.41262 | 7016.603 |
| GOBP_REG | 5 | 0.049766 | 0.00081 | 0.22566 | 0.41273 | 7018.474 |
| GOBP_GLY | 18 | 0.025447 | 0.000785 | 0.11544 | 0.41277 | 7019.154 |
| GOBP_NEG | 9 | 0.032043 | 0.000699 | 0.14538 | 0.41278 | 7019.324 |
| GOBP_PHC | 14 | 0.029203 | 0.000795 | 0.13264 | 0.41287 | 7020.854 |
| GOBP_PRC | 5 | 0.059974 | 0.000976 | 0.27242 | 0.41288 | 7021.024 |
| REACTOME | 21 | 0.02431 | 0.00081 | 0.11056 | 0.41298 | 7022.725 |
| GOMF_SM | 5 | 0.041014 | 0.000667 | 0.1867 | 0.41306 | 7024.085 |
| KEGG_TAU | 10 | 0.033382 | 0.000768 | 0.152 | 0.41309 | 7024.595 |
| GOBP_REG | 70 | 0.01378 | 0.000838 | 0.06289 | 0.41328 | 7027.826 |
| GOCC_SMI | 7 | 0.04523 | 0.000871 | 0.20661 | 0.41336 | 7029.187 |
| GOBP_REG | 8 | 0.044531 | 0.000917 | 0.20353 | 0.41341 | 7030.037 |
| BROWNE_I | 49 | 0.016297 | 0.000829 | 0.074499 | 0.41342 | 7030.207 |
| GOBP_RNA | 20 | 0.02578 | 0.000839 | 0.11802 | 0.41354 | 7032.248 |
| GOBP_REG | 15 | 0.029804 | 0.00084 | 0.13646 | 0.41356 | 7032.588 |
| GOBP_MO | 782 | 0.004093 | 0.000816 | 0.018749 | 0.4136 | 7033.268 |
| BYSTRYKH_ | 4 | 0.041533 | 0.000605 | 0.19055 | 0.41373 | 7035.479 |
| GOBP_NEG | 6 | 0.049416 | 0.000881 | 0.2268 | 0.41376 | 7035.989 |
| GOBP_NUC | 7 | 0.044401 | 0.000855 | 0.20396 | 0.41384 | 7037.349 |
| GOCC_ROU | 22 | 0.022077 | 0.000753 | 0.10157 | 0.41397 | 7039.56 |
| LEE_CALOF | 37 | 0.017066 | 0.000755 | 0.078639 | 0.4141 | 7041.771 |
| WANG_TH | 5 | 0.049417 | 0.000804 | 0.2279 | 0.41417 | 7042.961 |
| REACTOME | 8 | 0.03773 | 0.000777 | 0.17408 | 0.41421 | 7043.641 |
| REACTOME | 15 | 0.023558 | 0.000664 | 0.1088 | 0.41429 | 7045.001 |
| GOBP_POS | 5 | 0.044171 | 0.000719 | 0.20418 | 0.41436 | 7046.192 |
| GOBP_LON | 7 | 0.040686 | 0.000783 | 0.1881 | 0.41438 | 7046.532 |
| GOBP_NEG | 2 | 0.077557 | 0.000798 | 0.36013 | 0.41475 | 7052.824 |
| BOYLAN_M | 8 | 0.04072 | 0.000838 | 0.18915 | 0.41478 | 7053.334 |
| WP_TRYPT | 16 | 0.025908 | 0.000754 | 0.12054 | 0.41491 | 7055.545 |
| GOBP_SUP | 36 | 0.017218 | 0.000751 | 0.080116 | 0.41492 | 7055.715 |
| GOBP_DEC | 8 | 0.034332 | 0.000707 | 0.15977 | 0.41493 | 7055.885 |
| REACTOME | 7 | 0.039549 | 0.000761 | 0.18427 | 0.41503 | 7057.585 |
| RAMALHO_ | 186 | 0.007595 | 0.00075 | 0.035399 | 0.41506 | 7058.095 |
| GOBP_REG | 17 | 0.028707 | 0.000861 | 0.13398 | 0.41517 | 7059.966 |
| GOBP_SUB | 10 | 0.038643 | 0.000889 | 0.18091 | 0.41543 | 7064.387 |

| | | | | | | |
|---|---|---|---|---|---|---|
| GOBP_NEG | 4 | 0.051596 | 0.000751 | 0.2418 | 0.41551 | 7065.748 |
| GOBP_REG | 13 | 0.033361 | 0.000875 | 0.15642 | 0.41555 | 7066.428 |
| GOBP_SKEI | 39 | 0.016969 | 0.00077 | 0.079626 | 0.41562 | 7067.618 |
| GOBP_TON | 20 | 0.02526 | 0.000822 | 0.11857 | 0.41565 | 7068.128 |
| GUILLAUM | 27 | 0.019761 | 0.000747 | 0.092831 | 0.41572 | 7069.319 |
| NABA_COL | 40 | 0.018397 | 0.000846 | 0.086462 | 0.41575 | 7069.829 |
| REACTOME | 40 | 0.018397 | 0.000846 | 0.086462 | 0.41575 | 7069.829 |
| BIOCARTA_ | 22 | 0.020601 | 0.000703 | 0.096837 | 0.41576 | 7069.999 |
| GOBP_BAS | 12 | 0.027783 | 0.0007 | 0.13059 | 0.41576 | 7069.999 |
| GOBP_CEN | 82 | 0.012311 | 0.00081 | 0.057886 | 0.41579 | 7070.509 |
| GOBP_CRA | 10 | 0.041859 | 0.000963 | 0.19688 | 0.41582 | 7071.019 |
| REACTOME | 62 | 0.014185 | 0.000812 | 0.066757 | 0.41587 | 7071.869 |
| GOBP_ALK | 8 | 0.039477 | 0.000812 | 0.18591 | 0.41592 | 7072.72 |
| ZHOU_TNF | 46 | 0.013791 | 0.00068 | 0.065047 | 0.41605 | 7074.93 |
| GOMF_7S_ | 7 | 0.036592 | 0.000704 | 0.17286 | 0.41618 | 7077.141 |
| WP_FARNE | 19 | 0.023889 | 0.000757 | 0.11291 | 0.41622 | 7077.821 |
| BIOCARTA_ | 9 | 0.037689 | 0.000823 | 0.17821 | 0.41625 | 7078.331 |
| GOMF_TRA | 4 | 0.052177 | 0.000759 | 0.24673 | 0.41626 | 7078.501 |
| GOCC_DEN | 5 | 0.052909 | 0.000861 | 0.25025 | 0.41628 | 7078.841 |
| ABE_VEGFA | 26 | 0.020721 | 0.000768 | 0.098154 | 0.4164 | 7080.882 |
| WP_NONA | 145 | 0.008274 | 0.000722 | 0.039199 | 0.41642 | 7081.222 |
| GOBP_RES | 8 | 0.037987 | 0.000782 | 0.1801 | 0.41648 | 7082.242 |
| MARTIN_V | 81 | 0.011722 | 0.000766 | 0.055577 | 0.41648 | 7082.242 |
| GOBP_LYSO | 18 | 0.022128 | 0.000683 | 0.10501 | 0.41655 | 7083.433 |
| GOBP_DEN | 10 | 0.041364 | 0.000952 | 0.19639 | 0.41659 | 7084.113 |
| GOCC_NUC | 457 | 0.004821 | 0.000741 | 0.022908 | 0.41667 | 7085.473 |
| GOMF_PHO | 100 | 0.010265 | 0.000745 | 0.048806 | 0.41671 | 7086.154 |
| WP_VITAM | 13 | 0.032183 | 0.000844 | 0.15317 | 0.41679 | 7087.514 |
| REACTOME | 55 | 0.014584 | 0.000786 | 0.069489 | 0.41689 | 7089.214 |
| CHANDRAN | 34 | 0.017465 | 0.000741 | 0.083251 | 0.41692 | 7089.725 |
| ZHANG_RE | 215 | 0.007458 | 0.000791 | 0.035577 | 0.41698 | 7090.745 |
| HOLLERN_S | 168 | 0.008609 | 0.000809 | 0.041226 | 0.41729 | 7096.016 |
| GOBP_REG | 7 | 0.042219 | 0.000813 | 0.20218 | 0.4173 | 7096.187 |
| REACTOME | 5 | 0.059159 | 0.000963 | 0.28352 | 0.41736 | 7097.207 |
| NADERI_BF | 48 | 0.015085 | 0.00076 | 0.072371 | 0.41744 | 7098.567 |
| GOBP_INCI | 21 | 0.024527 | 0.000818 | 0.11783 | 0.41755 | 7100.438 |
| GOCC_SIGI | 6 | 0.038647 | 0.000689 | 0.18578 | 0.41761 | 7101.458 |
| MEBARKI_I | 112 | 0.010731 | 0.000824 | 0.051649 | 0.4177 | 7102.989 |
| GOBP_REG | 73 | 0.011736 | 0.000728 | 0.056532 | 0.41777 | 7104.179 |
| GOBP_BLO | 41 | 0.018514 | 0.000862 | 0.08927 | 0.41785 | 7105.539 |
| GOBP_REG | 1308 | 0.002984 | 0.000758 | 0.014389 | 0.41785 | 7105.539 |
| ZWANG_EG | 85 | 0.011339 | 0.000759 | 0.054696 | 0.41789 | 7106.219 |
| GOBP_HIST | 19 | 0.020194 | 0.00064 | 0.09762 | 0.41806 | 7109.11 |
| GOBP_CELI | 16 | 0.024491 | 0.000713 | 0.11843 | 0.41808 | 7109.45 |
| GOCC_ENZ | 6 | 0.027358 | 0.000488 | 0.13236 | 0.41813 | 7110.301 |

| | | | | | | |
|---|---|---|---|---|---|---|
| GOBP_MRI | 6 | 0.035517 | 0.000633 | 0.17203 | 0.41822 | 7111.831 |
| MYLLYKAN | 8 | 0.043827 | 0.000902 | 0.21229 | 0.41822 | 7111.831 |
| GOBP_CELI | 4 | 0.049722 | 0.000724 | 0.24091 | 0.41824 | 7112.171 |
| GOMF_QU | 15 | 0.024505 | 0.00069 | 0.11873 | 0.41824 | 7112.171 |
| PID_EPHRII | 29 | 0.020915 | 0.000819 | 0.10135 | 0.41826 | 7112.511 |
| GOBP_NEU | 34 | 0.019473 | 0.000826 | 0.094491 | 0.41836 | 7114.212 |
| BILBAN_B_ | 36 | 0.017345 | 0.000757 | 0.084244 | 0.41844 | 7115.572 |
| ONO_FOXF | 21 | 0.024004 | 0.0008 | 0.11669 | 0.41851 | 7116.763 |
| GOBP_LEU | 117 | 0.009677 | 0.000759 | 0.047197 | 0.41877 | 7121.184 |
| GOBP_RES | 5 | 0.052839 | 0.00086 | 0.2578 | 0.4188 | 7121.694 |
| GOMF_GD | 67 | 0.011983 | 0.000713 | 0.058466 | 0.4188 | 7121.694 |
| GOBP_POS | 27 | 0.021457 | 0.000811 | 0.10471 | 0.41882 | 7122.034 |
| GOMF_COI | 8 | 0.036862 | 0.000759 | 0.17993 | 0.41884 | 7122.374 |
| GOBP_TRA | 7 | 0.049965 | 0.000962 | 0.2442 | 0.41894 | 7124.075 |
| WENG_POI | 22 | 0.021823 | 0.000745 | 0.10668 | 0.41896 | 7124.415 |
| GOBP_NEG | 45 | 0.015703 | 0.000766 | 0.076974 | 0.41917 | 7127.986 |
| GOBP_CRA | 5 | 0.048267 | 0.000785 | 0.23688 | 0.41927 | 7129.686 |
| GOBP_RES | 191 | 0.007383 | 0.000739 | 0.036267 | 0.41935 | 7131.047 |
| GOBP_MO | 16 | 0.026929 | 0.000784 | 0.13263 | 0.41955 | 7134.448 |
| GOBP_REG | 5 | 0.045123 | 0.000734 | 0.22224 | 0.41955 | 7134.448 |
| REACTOME | 6 | 0.040802 | 0.000727 | 0.20108 | 0.4196 | 7135.298 |
| BIOCARTA_ | 12 | 0.027927 | 0.000704 | 0.13835 | 0.42002 | 7142.44 |
| GOCC_CCA | 6 | 0.035614 | 0.000635 | 0.17647 | 0.42003 | 7142.61 |
| GOMF_PH( | 5 | 0.050039 | 0.000814 | 0.24814 | 0.42009 | 7143.63 |
| GOBP_POS | 11 | 0.028638 | 0.000691 | 0.14214 | 0.42016 | 7144.821 |
| GOBP_POS | 12 | 0.027444 | 0.000692 | 0.13628 | 0.4202 | 7145.501 |
| GOBP_NEG | 42 | 0.015013 | 0.000707 | 0.074554 | 0.42021 | 7145.671 |
| URS_ADIP( | 64 | 0.012936 | 0.000752 | 0.064463 | 0.42048 | 7150.262 |
| MCBRYAN_ | 122 | 0.009104 | 0.00073 | 0.04538 | 0.4205 | 7150.603 |
| GOBP_POS | 8 | 0.032352 | 0.000666 | 0.16151 | 0.42062 | 7152.643 |
| DACOSTA_ | 83 | 0.012149 | 0.000804 | 0.060679 | 0.42065 | 7153.153 |
| GOMF_XM | 6 | 0.04965 | 0.000885 | 0.24809 | 0.42069 | 7153.833 |
| GOBP_INTI | 55 | 0.014083 | 0.000759 | 0.070425 | 0.42075 | 7154.854 |
| GOBP_POS | 3 | 0.04778 | 0.000602 | 0.23895 | 0.42076 | 7155.024 |
| RICKMAN_ | 52 | 0.013637 | 0.000715 | 0.068227 | 0.42079 | 7155.534 |
| GOBP_PUR | 53 | 0.013452 | 0.000712 | 0.067312 | 0.4208 | 7155.704 |
| MAGRANG | 17 | 0.023122 | 0.000694 | 0.11588 | 0.42092 | 7157.745 |
| IWANAGA_ | 334 | 0.005423 | 0.000715 | 0.02718 | 0.42093 | 7157.915 |
| GOMF_CAI | 14 | 0.031093 | 0.000846 | 0.156 | 0.42101 | 7159.275 |
| GOBP_POS | 388 | 0.00517 | 0.000734 | 0.026068 | 0.42139 | 7165.737 |
| GOBP_NEG | 38 | 0.015867 | 0.000711 | 0.080192 | 0.42158 | 7168.968 |
| GOBP_POS | 31 | 0.020951 | 0.000848 | 0.10593 | 0.42161 | 7169.478 |
| BIOCARTA_ | 16 | 0.022073 | 0.000642 | 0.11169 | 0.42167 | 7170.498 |
| GOBP_HIST | 8 | 0.033332 | 0.000686 | 0.16874 | 0.42171 | 7171.179 |
| GOBP_GRC | 20 | 0.022439 | 0.00073 | 0.11362 | 0.42172 | 7171.349 |

| | | | | | | |
|---|---|---|---|---|---|---|
| MOOTHA_( | 29 | 0.018043 | 0.000707 | 0.091394 | 0.42175 | 7171.859 |
| SOTIRIOU_ | 143 | 0.007714 | 0.000669 | 0.039073 | 0.42175 | 7171.859 |
| GOMF_OR( | 32 | 0.017889 | 0.000736 | 0.090684 | 0.42181 | 7172.879 |
| GOBP_PRII | 16 | 0.020632 | 0.0006 | 0.1046 | 0.42182 | 7173.049 |
| GOBP_REG | 20 | 0.020869 | 0.000679 | 0.10588 | 0.42188 | 7174.069 |
| GOMF_1_F | 6 | 0.039668 | 0.000707 | 0.20128 | 0.42188 | 7174.069 |
| GOBP_HIG | 11 | 0.030755 | 0.000742 | 0.15608 | 0.42189 | 7174.239 |
| GOBP_NEG | 9 | 0.03546 | 0.000774 | 0.18012 | 0.42197 | 7175.6 |
| GOBP_REG | 17 | 0.022228 | 0.000667 | 0.11309 | 0.42209 | 7177.64 |
| GOBP_U2_ | 22 | 0.019777 | 0.000675 | 0.10063 | 0.4221 | 7177.811 |
| GOCC_TRA | 382 | 0.005254 | 0.00074 | 0.026761 | 0.42218 | 7179.171 |
| GOBP_LIPI | 676 | 0.003832 | 0.000712 | 0.019527 | 0.42221 | 7179.681 |
| GOBP_NEG | 161 | 0.00767 | 0.000705 | 0.039101 | 0.42224 | 7180.191 |
| GOBP_POL | 61 | 0.012093 | 0.000686 | 0.061648 | 0.42224 | 7180.191 |
| GOBP_POS | 30 | 0.016205 | 0.000646 | 0.08261 | 0.42224 | 7180.191 |
| GOBP_CAN | 5 | 0.043198 | 0.000703 | 0.22035 | 0.42229 | 7181.041 |
| WU_HBX_T | 18 | 0.024753 | 0.000764 | 0.12638 | 0.42236 | 7182.232 |
| GOBP_FUC | 16 | 0.022251 | 0.000648 | 0.11365 | 0.42239 | 7182.742 |
| GOBP_REG | 146 | 0.007626 | 0.000668 | 0.038954 | 0.4224 | 7182.912 |
| WHITFIELD | 191 | 0.007156 | 0.000716 | 0.036611 | 0.42252 | 7184.953 |
| GOMF_ADI | 11 | 0.033784 | 0.000815 | 0.1729 | 0.42254 | 7185.293 |
| GOCC_U2_ | 23 | 0.01941 | 0.000677 | 0.099344 | 0.42255 | 7185.463 |
| GOBP_OVU | 75 | 0.011957 | 0.000752 | 0.061261 | 0.42262 | 7186.653 |
| GOMF_SPH | 8 | 0.038125 | 0.000785 | 0.19567 | 0.42276 | 7189.034 |
| REACTOME | 35 | 0.015972 | 0.000687 | 0.082001 | 0.42279 | 7189.544 |
| LEIN_PONS | 83 | 0.010137 | 0.000671 | 0.05213 | 0.42291 | 7191.585 |
| REACTOME | 15 | 0.023341 | 0.000658 | 0.12011 | 0.42296 | 7192.435 |
| GOBP_CYC | 20 | 0.025276 | 0.000822 | 0.13017 | 0.42302 | 7193.455 |
| KEGG_ADIF | 62 | 0.011709 | 0.00067 | 0.060337 | 0.42306 | 7194.135 |
| GOBP_POS | 7 | 0.040446 | 0.000779 | 0.20855 | 0.42311 | 7194.986 |
| GOBP_REG | 42 | 0.017873 | 0.000842 | 0.092177 | 0.42313 | 7195.326 |
| GOBP_NEG | 16 | 0.022196 | 0.000646 | 0.11449 | 0.42314 | 7195.496 |
| GOMF_TRA | 41 | 0.014037 | 0.000653 | 0.072405 | 0.42314 | 7195.496 |
| REACTOME | 10 | 0.028312 | 0.000651 | 0.14613 | 0.42319 | 7196.346 |
| GOBP_REG | 957 | 0.003229 | 0.000708 | 0.016672 | 0.42322 | 7196.856 |
| GOCC_CILI | 8 | 0.033882 | 0.000697 | 0.17503 | 0.42325 | 7197.366 |
| GOMF_SPH | 17 | 0.022281 | 0.000668 | 0.11511 | 0.42326 | 7197.536 |
| GOBP_TELI | 25 | 0.01892 | 0.000688 | 0.097855 | 0.42334 | 7198.897 |
| GOBP_GOL | 10 | 0.02866 | 0.000659 | 0.1484 | 0.42343 | 7200.427 |
| REACTOME | 5 | 0.038306 | 0.000623 | 0.19844 | 0.42346 | 7200.937 |
| BIOCARTA_ | 5 | 0.036233 | 0.00059 | 0.18781 | 0.42351 | 7201.788 |
| GOBP_CRA | 8 | 0.036678 | 0.000755 | 0.19058 | 0.42369 | 7204.848 |
| GOCC_GOL | 9 | 0.037096 | 0.00081 | 0.19278 | 0.4237 | 7205.019 |
| REACTOME | 9 | 0.038826 | 0.000848 | 0.20189 | 0.42375 | 7205.869 |
| GOBP_TUB | 6 | 0.04725 | 0.000842 | 0.24595 | 0.42383 | 7207.229 |

| | | | | | | |
|---|---|---|---|---|---|---|
| KOINUMA_ | 798 | 0.003516 | 0.000707 | 0.018313 | 0.42387 | 7207.909 |
| BIOCARTA_ | 15 | 0.023595 | 0.000665 | 0.12291 | 0.42388 | 7208.079 |
| HILLION_H | 83 | 0.009861 | 0.000652 | 0.051392 | 0.42392 | 7208.76 |
| GOBP_LUN | 7 | 0.04463 | 0.000859 | 0.23291 | 0.42402 | 7210.46 |
| CASTELLAN | 66 | 0.01181 | 0.000697 | 0.061652 | 0.42404 | 7210.8 |
| REACTOME | 33 | 0.015943 | 0.000666 | 0.083244 | 0.42406 | 7211.14 |
| BIOCARTA_ | 9 | 0.029342 | 0.000641 | 0.15331 | 0.42411 | 7211.991 |
| REACTOME | 275 | 0.00566 | 0.000678 | 0.029605 | 0.42419 | 7213.351 |
| BIOCARTA_ | 22 | 0.018222 | 0.000622 | 0.095466 | 0.42432 | 7215.562 |
| REACTOME | 16 | 0.023958 | 0.000697 | 0.12552 | 0.42432 | 7215.562 |
| REACTOME | 8 | 0.034081 | 0.000701 | 0.17867 | 0.42436 | 7216.242 |
| GOBP_HEN | 17 | 0.020025 | 0.000601 | 0.10504 | 0.4244 | 7216.922 |
| PID_RAS_P | 28 | 0.018339 | 0.000706 | 0.096262 | 0.42446 | 7217.942 |
| PHONG_TN | 61 | 0.013257 | 0.000752 | 0.069634 | 0.42451 | 7218.793 |
| GOBP_REG | 95 | 0.009727 | 0.000688 | 0.051115 | 0.42454 | 7219.303 |
| GOCC_NAD | 14 | 0.023514 | 0.00064 | 0.12361 | 0.42457 | 7219.813 |
| KYNG_ENV | 47 | 0.014506 | 0.000723 | 0.076276 | 0.42458 | 7219.983 |
| REACTOME | 8 | 0.040872 | 0.000841 | 0.21497 | 0.4246 | 7220.323 |
| GOBP_HIST | 8 | 0.028871 | 0.000594 | 0.15208 | 0.42472 | 7222.364 |
| GOBP_INTI | 6 | 0.038385 | 0.000684 | 0.20233 | 0.42477 | 7223.214 |
| REACTOME | 17 | 0.020578 | 0.000617 | 0.10854 | 0.42482 | 7224.064 |
| IKEDA_MIF | 41 | 0.015658 | 0.000729 | 0.082632 | 0.42486 | 7224.744 |
| GOBP_DEN | 12 | 0.030763 | 0.000775 | 0.16239 | 0.42488 | 7225.084 |
| GOBP_MO | 7 | 0.038692 | 0.000745 | 0.20425 | 0.42488 | 7225.084 |
| REACTOME | 9 | 0.032612 | 0.000712 | 0.17275 | 0.42513 | 7229.336 |
| BAKKER_FC | 51 | 0.013125 | 0.000681 | 0.069619 | 0.42523 | 7231.036 |
| EHLERS_AN | 39 | 0.015307 | 0.000695 | 0.081237 | 0.42528 | 7231.886 |
| GOBP_POS | 5 | 0.041471 | 0.000675 | 0.22032 | 0.42535 | 7233.077 |
| ANDERSEN | 170 | 0.007076 | 0.000668 | 0.037607 | 0.42538 | 7233.587 |
| REACTOME | 171 | 0.007461 | 0.000707 | 0.03967 | 0.42541 | 7234.097 |
| GOBP_NEG | 5 | 0.040909 | 0.000666 | 0.2176 | 0.42544 | 7234.607 |
| KEGG_ACU | 54 | 0.013458 | 0.000719 | 0.071612 | 0.42547 | 7235.117 |
| GOBP_CELI | 1393 | 0.002749 | 0.000719 | 0.014632 | 0.42549 | 7235.457 |
| ACEVEDO_ | 161 | 0.008273 | 0.000761 | 0.04407 | 0.42555 | 7236.478 |
| GOBP_NEG | 12 | 0.028047 | 0.000707 | 0.1494 | 0.42555 | 7236.478 |
| WP_MITOC | 16 | 0.022437 | 0.000653 | 0.11952 | 0.42555 | 7236.478 |
| GOBP_REG | 30 | 0.016709 | 0.000666 | 0.089028 | 0.42556 | 7236.648 |
| GOBP_INTI | 32 | 0.016173 | 0.000665 | 0.086276 | 0.42565 | 7238.178 |
| GOBP_POS | 21 | 0.023381 | 0.000779 | 0.12475 | 0.42567 | 7238.518 |
| GOCC_COF | 69 | 0.011205 | 0.000676 | 0.0598 | 0.42569 | 7238.858 |
| PID_MAPK_ | 33 | 0.016044 | 0.00067 | 0.085648 | 0.4257 | 7239.029 |
| GOBP_POS | 13 | 0.029107 | 0.000764 | 0.15541 | 0.42571 | 7239.199 |
| GOBP_REG | 9 | 0.032942 | 0.000719 | 0.17591 | 0.42572 | 7239.369 |
| REACTOME | 142 | 0.008102 | 0.0007 | 0.043262 | 0.42572 | 7239.369 |
| GOBP_EST/ | 220 | 0.006508 | 0.000698 | 0.034761 | 0.42575 | 7239.879 |

| | | | | | | |
|---|---|---|---|---|---|---|
| GOCC_CIA_ | 5 | 0.040456 | 0.000658 | 0.2161 | 0.42575 | 7239.879 |
| GOMF_CO/ | 5 | 0.043904 | 0.000714 | 0.23522 | 0.42597 | 7243.62 |
| GOBP_VAS | 12 | 0.029572 | 0.000745 | 0.1585 | 0.426 | 7244.13 |
| KIM_GERM | 12 | 0.026677 | 0.000672 | 0.14307 | 0.42604 | 7244.81 |
| GOBP_TRI( | 10 | 0.031458 | 0.000724 | 0.16882 | 0.42609 | 7245.66 |
| GOBP_KINI | 17 | 0.020215 | 0.000606 | 0.10855 | 0.42614 | 7246.511 |
| REACTOME | 122 | 0.008418 | 0.000675 | 0.045275 | 0.42625 | 7248.381 |
| BIOCARTA_ | 16 | 0.021378 | 0.000622 | 0.11507 | 0.42631 | 7249.402 |
| GOMF_IMI | 22 | 0.019358 | 0.00066 | 0.10441 | 0.42646 | 7251.952 |
| GOBP_VIT/ | 50 | 0.013536 | 0.000696 | 0.073077 | 0.42653 | 7253.143 |
| GOBP_REG | 246 | 0.005964 | 0.000676 | 0.032239 | 0.42662 | 7254.673 |
| REACTOME | 8 | 0.032322 | 0.000665 | 0.17489 | 0.42669 | 7255.863 |
| REACTOME | 13 | 0.027947 | 0.000733 | 0.15122 | 0.42669 | 7255.863 |
| REACTOME | 21 | 0.018691 | 0.000623 | 0.10114 | 0.42669 | 7255.863 |
| GOBP_EPIN | 8 | 0.036551 | 0.000752 | 0.19779 | 0.4267 | 7256.034 |
| REACTOME | 16 | 0.02337 | 0.00068 | 0.12647 | 0.4267 | 7256.034 |
| GOBP_MU( | 7 | 0.036921 | 0.000711 | 0.20022 | 0.42685 | 7258.584 |
| REACTOME | 7 | 0.034008 | 0.000655 | 0.18456 | 0.4269 | 7259.435 |
| GOBP_REG | 10 | 0.03282 | 0.000755 | 0.17835 | 0.427 | 7261.135 |
| GOBP_COG | 295 | 0.005549 | 0.000688 | 0.030192 | 0.42709 | 7262.665 |
| PUIFFE_IN\ | 136 | 0.007852 | 0.000664 | 0.042808 | 0.42723 | 7265.046 |
| REACTOME | 24 | 0.018378 | 0.000655 | 0.10022 | 0.42725 | 7265.386 |
| REACTOME | 58 | 0.013324 | 0.000737 | 0.072665 | 0.42726 | 7265.556 |
| PID_MET_I | 78 | 0.010223 | 0.000656 | 0.055774 | 0.42728 | 7265.896 |
| REACTOME | 11 | 0.02974 | 0.000718 | 0.16238 | 0.42734 | 7266.917 |
| GOBP_NUC | 266 | 0.005464 | 0.000644 | 0.029875 | 0.42744 | 7268.617 |
| REACTOME | 34 | 0.016117 | 0.000683 | 0.088161 | 0.42747 | 7269.127 |
| GOMF_HY/ | 8 | 0.030782 | 0.000634 | 0.16863 | 0.42758 | 7270.998 |
| GOBP_AMI | 40 | 0.014665 | 0.000674 | 0.080376 | 0.42761 | 7271.508 |
| HAHTOLA_ | 55 | 0.012381 | 0.000667 | 0.068049 | 0.42781 | 7274.909 |
| CHNG_MU | 30 | 0.015923 | 0.000634 | 0.087543 | 0.42784 | 7275.419 |
| BIOCARTA_ | 29 | 0.0166 | 0.00065 | 0.091285 | 0.42785 | 7275.589 |
| GOBP_REG | 48 | 0.012685 | 0.000639 | 0.069892 | 0.42799 | 7277.97 |
| GOCC_CELI | 791 | 0.003443 | 0.00069 | 0.019039 | 0.42824 | 7282.221 |
| GOBP_POS | 78 | 0.010378 | 0.000666 | 0.057388 | 0.42825 | 7282.391 |
| CHEN_HOX | 38 | 0.014265 | 0.000639 | 0.078899 | 0.42826 | 7282.561 |
| GOBP_CELI | 355 | 0.004798 | 0.000652 | 0.02661 | 0.42845 | 7285.792 |
| GOMF_ENI | 17 | 0.019448 | 0.000583 | 0.10792 | 0.4285 | 7286.643 |
| GOBP_IND | 22 | 0.021069 | 0.000719 | 0.11694 | 0.42851 | 7286.813 |
| KEGG_GAL | 25 | 0.016432 | 0.000598 | 0.091207 | 0.42851 | 7286.813 |
| GOBP_MET | 8 | 0.029339 | 0.000604 | 0.16287 | 0.42853 | 7287.153 |
| GOBP_REG | 19 | 0.018907 | 0.0006 | 0.10511 | 0.42863 | 7288.853 |
| GOBP_PRC | 184 | 0.006236 | 0.000613 | 0.034744 | 0.42878 | 7291.404 |
| BENPORAT | 350 | 0.004633 | 0.000625 | 0.025823 | 0.42881 | 7291.914 |
| REACTOME | 62 | 0.011379 | 0.000651 | 0.063459 | 0.42885 | 7292.594 |

| | | | | | | |
|---|---|---|---|---|---|---|
| GOBP_MU: | 5 | 0.048417 | 0.000788 | 0.27032 | 0.42893 | 7293.955 |
| GOBP_ANI | 42 | 0.013363 | 0.00063 | 0.074643 | 0.42896 | 7294.465 |
| GOCC_U4_ | 9 | 0.035328 | 0.000771 | 0.19746 | 0.42901 | 7295.315 |
| VICENT_MI | 14 | 0.024085 | 0.000656 | 0.13463 | 0.42901 | 7295.315 |
| GOBP_PRC | 31 | 0.016419 | 0.000665 | 0.092169 | 0.42931 | 7300.417 |
| GOCC_PER | 146 | 0.007521 | 0.000659 | 0.042244 | 0.42935 | 7301.097 |
| GOBP_ACE | 4 | 0.058457 | 0.000851 | 0.32935 | 0.42956 | 7304.668 |
| GOBP_DIC/ | 90 | 0.00989 | 0.000681 | 0.055873 | 0.42975 | 7307.899 |
| HEIDENBLA | 34 | 0.015584 | 0.000661 | 0.08807 | 0.42977 | 7308.239 |
| GOBP_PEP | 353 | 0.004924 | 0.000667 | 0.027851 | 0.42984 | 7309.429 |
| MULLIGHA | 393 | 0.004321 | 0.000617 | 0.024477 | 0.42993 | 7310.96 |
| REACTOME | 29 | 0.018926 | 0.000741 | 0.10721 | 0.42994 | 7311.13 |
| GOCC_SON | 787 | 0.003255 | 0.000651 | 0.018446 | 0.42996 | 7311.47 |
| GOBP_UNS | 47 | 0.012086 | 0.000602 | 0.068563 | 0.43004 | 7312.83 |
| GOCC_OLI( | 13 | 0.021152 | 0.000555 | 0.12004 | 0.43007 | 7313.34 |
| GOMF_MC | 97 | 0.010181 | 0.000728 | 0.057819 | 0.43011 | 7314.021 |
| GOBP_NOT | 11 | 0.029531 | 0.000713 | 0.16779 | 0.43015 | 7314.701 |
| GOMF_NU | 13 | 0.027194 | 0.000713 | 0.15463 | 0.4302 | 7315.551 |
| GOBP_CELI | 51 | 0.012756 | 0.000662 | 0.072548 | 0.43021 | 7315.721 |
| GOBP_OPT | 18 | 0.022999 | 0.00071 | 0.1308 | 0.43021 | 7315.721 |
| GOMF_HIS | 8 | 0.031011 | 0.000638 | 0.17638 | 0.43022 | 7315.891 |
| REACTOME | 37 | 0.018891 | 0.000836 | 0.10745 | 0.43022 | 7315.891 |
| BIOCARTA_ | 14 | 0.020853 | 0.000568 | 0.11876 | 0.43031 | 7317.422 |
| GOBP_POS | 8 | 0.027845 | 0.000573 | 0.15866 | 0.43034 | 7317.932 |
| GOBP_CD8 | 7 | 0.02845 | 0.000548 | 0.16214 | 0.43036 | 7318.272 |
| GOBP_DICI | 8 | 0.036763 | 0.000757 | 0.20969 | 0.43041 | 7319.122 |
| GOBP_FEN | 5 | 0.036891 | 0.0006 | 0.21094 | 0.43058 | 7322.013 |
| GOBP_POS | 7 | 0.031757 | 0.000611 | 0.18159 | 0.43059 | 7322.183 |
| KIM_LIVER | 41 | 0.014959 | 0.000696 | 0.085616 | 0.43065 | 7323.203 |
| GOBP_EST/ | 16 | 0.022925 | 0.000667 | 0.13123 | 0.43066 | 7323.373 |
| GOMF_INH | 8 | 0.040174 | 0.000827 | 0.23048 | 0.43081 | 7325.924 |
| GOMF_S_N | 9 | 0.028714 | 0.000627 | 0.16521 | 0.43101 | 7329.325 |
| REACTOME | 49 | 0.011917 | 0.000606 | 0.068599 | 0.43104 | 7329.835 |
| GOBP_REG | 1301 | 0.00258 | 0.000654 | 0.014875 | 0.43115 | 7331.706 |
| KIM_ALL_D | 34 | 0.013838 | 0.000587 | 0.079843 | 0.4312 | 7332.556 |
| GOBP_PUR | 10 | 0.027954 | 0.000643 | 0.16134 | 0.43123 | 7333.066 |
| GOBP_END | 32 | 0.016161 | 0.000665 | 0.09356 | 0.43143 | 7336.467 |
| GARGALOV | 23 | 0.018895 | 0.000659 | 0.10951 | 0.43151 | 7337.828 |
| GOBP_NEG | 11 | 0.027274 | 0.000658 | 0.15808 | 0.43151 | 7337.828 |
| GOBP_SKIN | 117 | 0.008067 | 0.000633 | 0.046809 | 0.43158 | 7339.018 |
| GOMF_OXI | 13 | 0.019978 | 0.000524 | 0.11633 | 0.43182 | 7343.099 |
| GOMF_2_I | 21 | 0.017234 | 0.000574 | 0.10036 | 0.43183 | 7343.269 |
| GOBP_RES | 4 | 0.038116 | 0.000555 | 0.22207 | 0.43186 | 7343.779 |
| GOBP_REG | 4 | 0.039466 | 0.000574 | 0.23007 | 0.4319 | 7344.46 |
| GOBP_EMI | 5 | 0.038592 | 0.000628 | 0.22548 | 0.43205 | 7347.01 |

| | | | | | | |
|---|---|---|---|---|---|---|
| REACTOME | 22 | 0.018277 | 0.000624 | 0.10687 | 0.4321 | 7347.861 |
| GOBP_HIST | 10 | 0.023364 | 0.000538 | 0.13679 | 0.43219 | 7349.391 |
| GOBP_MAI | 41 | 0.012034 | 0.00056 | 0.070497 | 0.43223 | 7350.071 |
| GOBP_REG | 8 | 0.032758 | 0.000674 | 0.19204 | 0.43228 | 7350.921 |
| GOBP_CYS | 12 | 0.025378 | 0.00064 | 0.14899 | 0.43237 | 7352.452 |
| REACTOME | 16 | 0.024579 | 0.000715 | 0.14436 | 0.4324 | 7352.962 |
| REACTOME | 7 | 0.030591 | 0.000589 | 0.17972 | 0.43242 | 7353.302 |
| GOBP_NEG | 95 | 0.009212 | 0.000652 | 0.054155 | 0.43247 | 7354.152 |
| GOBP_NEG | 5 | 0.036367 | 0.000592 | 0.2138 | 0.43247 | 7354.152 |
| GOBP_SIN | 6 | 0.041802 | 0.000745 | 0.24583 | 0.43249 | 7354.492 |
| GOCC_G_P | 8 | 0.032198 | 0.000663 | 0.18942 | 0.43251 | 7354.833 |
| REACTOME | 13 | 0.023664 | 0.000621 | 0.13939 | 0.4326 | 7356.363 |
| GOBP_REG | 37 | 0.015481 | 0.000685 | 0.091206 | 0.43261 | 7356.533 |
| NIKOLSKY_ | 21 | 0.029974 | 0.000999 | 0.17722 | 0.43285 | 7360.614 |
| GOBP_CELI | 97 | 0.008905 | 0.000637 | 0.052677 | 0.43288 | 7361.124 |
| BIOCARTA_ | 12 | 0.024392 | 0.000615 | 0.14464 | 0.43304 | 7363.845 |
| GOBP_EMI | 11 | 0.024636 | 0.000595 | 0.14612 | 0.43306 | 7364.185 |
| GOBP_LIPII | 464 | 0.004057 | 0.000628 | 0.024089 | 0.43313 | 7365.376 |
| BALDWIN_ | 38 | 0.014665 | 0.000657 | 0.087114 | 0.43316 | 7365.886 |
| RUAN_RES | 13 | 0.025628 | 0.000672 | 0.15223 | 0.43316 | 7365.886 |
| GOBP_PRC | 6 | 0.035696 | 0.000636 | 0.2121 | 0.43318 | 7366.226 |
| GOBP_MEI | 43 | 0.014649 | 0.000698 | 0.087053 | 0.43319 | 7366.396 |
| GOCC_PHC | 86 | 0.008782 | 0.000591 | 0.052305 | 0.43333 | 7368.777 |
| ZHOU_CELI | 119 | 0.007294 | 0.000577 | 0.043564 | 0.43351 | 7371.838 |
| BLANCO_M | 91 | 0.008968 | 0.000621 | 0.05359 | 0.43355 | 7372.518 |
| GOBP_REG | 18 | 0.018586 | 0.000574 | 0.11111 | 0.43358 | 7373.028 |
| BIOCARTA_ | 12 | 0.022457 | 0.000566 | 0.13435 | 0.43363 | 7373.878 |
| PEART_HD/ | 52 | 0.011581 | 0.000607 | 0.069314 | 0.43365 | 7374.218 |
| GOBP_APC | 559 | 0.003569 | 0.000605 | 0.021386 | 0.43373 | 7375.579 |
| GOBP_REG | 5 | 0.044583 | 0.000725 | 0.26744 | 0.4338 | 7376.769 |
| GOBP_REG | 22 | 0.016344 | 0.000558 | 0.098037 | 0.4338 | 7376.769 |
| GOBP_CELI | 60 | 0.010086 | 0.000568 | 0.06054 | 0.43384 | 7377.449 |
| GOBP_NEG | 26 | 0.017539 | 0.00065 | 0.10527 | 0.43384 | 7377.449 |
| GOBP_PUR | 19 | 0.017915 | 0.000568 | 0.10769 | 0.43394 | 7379.15 |
| GOMF_MA | 9 | 0.029789 | 0.00065 | 0.17912 | 0.43396 | 7379.49 |
| GOBP_REG | 11 | 0.024203 | 0.000584 | 0.14571 | 0.43404 | 7380.85 |
| GOCC_VAC | 9 | 0.022736 | 0.000496 | 0.13718 | 0.43418 | 7383.231 |
| REACTOME | 13 | 0.021066 | 0.000553 | 0.12717 | 0.43422 | 7383.911 |
| REACTOME | 14 | 0.022663 | 0.000617 | 0.13686 | 0.43424 | 7384.251 |
| PID_IL4_2F | 56 | 0.010996 | 0.000598 | 0.066411 | 0.43425 | 7384.421 |
| RUAN_RES | 77 | 0.009378 | 0.000598 | 0.056705 | 0.43432 | 7385.612 |
| GOBP_DER | 8 | 0.025783 | 0.000531 | 0.15606 | 0.43439 | 7386.802 |
| HEIDENBL/ | 21 | 0.016807 | 0.00056 | 0.10175 | 0.4344 | 7386.972 |
| GOBP_REG | 17 | 0.021418 | 0.000642 | 0.12993 | 0.43454 | 7389.353 |
| GOBP_EMI | 11 | 0.02478 | 0.000598 | 0.15044 | 0.43459 | 7390.203 |

| | | | | | | |
|---|---|---|---|---|---|---|
| REACTOME | 20 | 0.018458 | 0.0006 | 0.11217 | 0.43465 | 7391.223 |
| GOCC_CLE/ | 54 | 0.010984 | 0.000587 | 0.066757 | 0.43466 | 7391.393 |
| REACTOME | 8 | 0.03115 | 0.000641 | 0.18933 | 0.43466 | 7391.393 |
| GOBP_POS | 15 | 0.021784 | 0.000614 | 0.13242 | 0.43467 | 7391.563 |
| GOBP_OXI | 121 | 0.006831 | 0.000545 | 0.04155 | 0.43471 | 7392.244 |
| GOBP_SEQ | 17 | 0.01946 | 0.000584 | 0.11837 | 0.43471 | 7392.244 |
| GOBP_RES | 477 | 0.003699 | 0.00058 | 0.022507 | 0.43474 | 7392.754 |
| WP_HEPAT | 142 | 0.007419 | 0.000641 | 0.045158 | 0.43475 | 7392.924 |
| GOBP_REG | 52 | 0.011642 | 0.00061 | 0.070869 | 0.43476 | 7393.094 |
| GOMF_SPH | 7 | 0.044347 | 0.000854 | 0.27012 | 0.4348 | 7393.774 |
| GOBP_REG | 11 | 0.023057 | 0.000556 | 0.14058 | 0.43486 | 7394.794 |
| GOBP_PRC | 48 | 0.012574 | 0.000633 | 0.076676 | 0.43487 | 7394.964 |
| GOBP_POS | 34 | 0.013611 | 0.000577 | 0.083069 | 0.43493 | 7395.985 |
| GOBP_REG | 40 | 0.013624 | 0.000626 | 0.083217 | 0.43498 | 7396.835 |
| GOBP_ANE | 9 | 0.025131 | 0.000549 | 0.15352 | 0.43499 | 7397.005 |
| GOBP_CELI | 11 | 0.030161 | 0.000728 | 0.18447 | 0.43506 | 7398.195 |
| GOCC_INTI | 30 | 0.014926 | 0.000595 | 0.091539 | 0.43524 | 7401.256 |
| KEGG_PRO | 20 | 0.017593 | 0.000572 | 0.10793 | 0.43526 | 7401.596 |
| REACTOME | 6 | 0.034254 | 0.000611 | 0.21017 | 0.43527 | 7401.766 |
| GOBP_MIT | 128 | 0.006439 | 0.000528 | 0.039566 | 0.43536 | 7403.297 |
| GOBP_INTI | 5 | 0.03155 | 0.000513 | 0.19412 | 0.43545 | 7404.827 |
| GOBP_POS | 287 | 0.004806 | 0.000588 | 0.029571 | 0.43545 | 7404.827 |
| GOBP_POS | 10 | 0.029796 | 0.000686 | 0.18385 | 0.43563 | 7407.888 |
| GOMF_NEU | 8 | 0.034482 | 0.00071 | 0.21291 | 0.43567 | 7408.568 |
| GOBP_HYA | 16 | 0.019007 | 0.000553 | 0.1174 | 0.43569 | 7408.908 |
| REACTOME | 80 | 0.009209 | 0.000598 | 0.05688 | 0.4357 | 7409.079 |
| GOBP_TRO | 19 | 0.017574 | 0.000557 | 0.10862 | 0.43574 | 7409.759 |
| LEE_TARGE | 82 | 0.009314 | 0.000613 | 0.057641 | 0.43581 | 7410.949 |
| GOMF_CCI | 4 | 0.046945 | 0.000683 | 0.29076 | 0.43587 | 7411.969 |
| BIOCARTA_ | 15 | 0.020616 | 0.000581 | 0.12782 | 0.43593 | 7412.99 |
| REACTOME | 11 | 0.024943 | 0.000602 | 0.15488 | 0.43603 | 7414.69 |
| GOMF_CAI | 20 | 0.016276 | 0.00053 | 0.10116 | 0.43609 | 7415.71 |
| GOBP_REG | 5 | 0.042952 | 0.000699 | 0.26702 | 0.4361 | 7415.881 |
| GOBP_PRC | 8 | 0.023217 | 0.000478 | 0.14455 | 0.4362 | 7417.581 |
| GOBP_SNR | 20 | 0.017409 | 0.000566 | 0.10862 | 0.43633 | 7419.792 |
| RUAN_RES | 9 | 0.030181 | 0.000659 | 0.1886 | 0.43643 | 7421.492 |
| BIOCARTA_ | 28 | 0.014792 | 0.000569 | 0.092502 | 0.43648 | 7422.342 |
| GOMF_PEF | 9 | 0.025622 | 0.000559 | 0.16058 | 0.43662 | 7424.723 |
| SANSOM_/ | 121 | 0.007527 | 0.000601 | 0.047238 | 0.4367 | 7426.084 |
| GOMF_SUL | 15 | 0.019742 | 0.000556 | 0.12393 | 0.43672 | 7426.424 |
| REACTOME | 215 | 0.005082 | 0.000539 | 0.031914 | 0.43674 | 7426.764 |
| WP_TRANS | 8 | 0.031499 | 0.000648 | 0.19801 | 0.4368 | 7427.784 |
| LIN_TUMO | 18 | 0.019019 | 0.000587 | 0.11957 | 0.43681 | 7427.954 |
| PETRETTO_ | 10 | 0.031846 | 0.000733 | 0.20026 | 0.43683 | 7428.294 |
| GOBP_POS | 13 | 0.023253 | 0.00061 | 0.14626 | 0.43684 | 7428.464 |

| | | | | | | |
|---|---|---|---|---|---|---|
| GOMF_ME | 21 | 0.016198 | 0.00054 | 0.10222 | 0.43705 | 7432.035 |
| GOBP_RES | 28 | 0.013365 | 0.000514 | 0.084534 | 0.43719 | 7434.416 |
| REACTOME | 11 | 0.023246 | 0.000561 | 0.14707 | 0.43721 | 7434.756 |
| CROSBY_E2 | 6 | 0.026157 | 0.000466 | 0.16575 | 0.4373 | 7436.287 |
| GOBP_POS | 131 | 0.006743 | 0.00056 | 0.042767 | 0.43736 | 7437.307 |
| GOBP_REG | 406 | 0.003887 | 0.000564 | 0.024674 | 0.43741 | 7438.157 |
| GOMF_CO | 10 | 0.027123 | 0.000624 | 0.1723 | 0.43746 | 7439.007 |
| GOBP_REG | 45 | 0.011254 | 0.000549 | 0.071604 | 0.43755 | 7440.538 |
| GOCC_RIBI | 13 | 0.022102 | 0.00058 | 0.14076 | 0.43762 | 7441.728 |
| WP_DNA_I | 114 | 0.00692 | 0.000536 | 0.04408 | 0.43763 | 7441.898 |
| REACTOME | 6 | 0.027563 | 0.000491 | 0.17578 | 0.4377 | 7443.089 |
| GOBP_SER | 12 | 0.024823 | 0.000626 | 0.15839 | 0.43773 | 7443.599 |
| REACTOME | 29 | 0.013329 | 0.000522 | 0.085187 | 0.43783 | 7445.299 |
| BENPORAT | 145 | 0.006121 | 0.000534 | 0.03924 | 0.43802 | 7448.53 |
| GOBP_NEU | 6 | 0.033876 | 0.000604 | 0.21754 | 0.43813 | 7450.401 |
| GOMF_DO | 5 | 0.032765 | 0.000533 | 0.21095 | 0.43828 | 7452.951 |
| GOCC_BRA | 14 | 0.021109 | 0.000575 | 0.13625 | 0.43844 | 7455.672 |
| GOBP_REG | 40 | 0.012574 | 0.000578 | 0.081255 | 0.43851 | 7456.863 |
| GROSS_HY | 101 | 0.007835 | 0.000572 | 0.050686 | 0.43858 | 7458.053 |
| GOBP_MAI | 5 | 0.028519 | 0.000464 | 0.18475 | 0.43866 | 7459.413 |
| GOMF_3_E | 5 | 0.036608 | 0.000596 | 0.23721 | 0.43868 | 7459.753 |
| GOBP_NEG | 56 | 0.010801 | 0.000587 | 0.070002 | 0.43869 | 7459.923 |
| MCBRYAN_ | 180 | 0.00569 | 0.000553 | 0.036892 | 0.43871 | 7460.264 |
| GOBP_REG | 6 | 0.03851 | 0.000686 | 0.24981 | 0.43874 | 7460.774 |
| GOCC_U12 | 25 | 0.014649 | 0.000533 | 0.095019 | 0.43874 | 7460.774 |
| GOBP_POS | 16 | 0.018775 | 0.000546 | 0.12191 | 0.4388 | 7461.794 |
| BIOCARTA_ | 34 | 0.01345 | 0.00057 | 0.08743 | 0.43887 | 7462.984 |
| GOBP_REG | 7 | 0.031527 | 0.000607 | 0.20537 | 0.439 | 7465.195 |
| GOBP_GEP | 5 | 0.030878 | 0.000502 | 0.20125 | 0.43903 | 7465.705 |
| GOMF_PEF | 7 | 0.027252 | 0.000525 | 0.1777 | 0.43906 | 7466.215 |
| GOBP_REG | 12 | 0.023081 | 0.000582 | 0.15055 | 0.43908 | 7466.555 |
| GOBP_REG | 17 | 0.017417 | 0.000522 | 0.11366 | 0.43911 | 7467.066 |
| GOBP_COC | 23 | 0.01659 | 0.000579 | 0.10857 | 0.43928 | 7469.956 |
| GOBP_DET | 33 | 0.014121 | 0.00059 | 0.09244 | 0.43929 | 7470.126 |
| GOBP_ASP | 21 | 0.01635 | 0.000545 | 0.10703 | 0.4393 | 7470.297 |
| GOBP_ISOT | 14 | 0.024352 | 0.000663 | 0.1596 | 0.43936 | 7471.317 |
| GOMF_SUI | 6 | 0.030565 | 0.000545 | 0.20051 | 0.43942 | 7472.337 |
| CHIARADO | 144 | 0.006325 | 0.00055 | 0.041637 | 0.43963 | 7475.908 |
| GOMF_UBI | 13 | 0.022453 | 0.000589 | 0.14797 | 0.4397 | 7477.099 |
| GOBP_CGN | 10 | 0.02842 | 0.000654 | 0.1876 | 0.43979 | 7478.629 |
| GOBP_NAT | 10 | 0.02295 | 0.000528 | 0.15152 | 0.4398 | 7478.799 |
| GOBP_PRC | 12 | 0.020267 | 0.000511 | 0.13382 | 0.43981 | 7478.969 |
| GOMF_HYI | 88 | 0.007412 | 0.000505 | 0.048991 | 0.43987 | 7479.989 |
| GOBP_POS | 6 | 0.029375 | 0.000524 | 0.1945 | 0.43998 | 7481.86 |
| GOBP_REG | 67 | 0.00862 | 0.000513 | 0.057092 | 0.44 | 7482.2 |

| | | | | | | |
|---|---|---|---|---|---|---|
| GOBP_POS | 21 | 0.014245 | 0.000475 | 0.094703 | 0.44022 | 7485.941 |
| GOBP_COR | 7 | 0.02892 | 0.000557 | 0.19237 | 0.44025 | 7486.451 |
| GOCC_TRA | 5 | 0.036394 | 0.000592 | 0.24213 | 0.44026 | 7486.621 |
| BROWNE_I | 145 | 0.005758 | 0.000503 | 0.038327 | 0.44029 | 7487.131 |
| REACTOME | 33 | 0.014113 | 0.00059 | 0.094114 | 0.4404 | 7489.002 |
| GOBP_REG | 65 | 0.009945 | 0.000583 | 0.066446 | 0.44051 | 7490.873 |
| GOBP_REG | 6 | 0.034049 | 0.000607 | 0.22774 | 0.44058 | 7492.063 |
| MULLIGHA | 359 | 0.00387 | 0.000529 | 0.025918 | 0.44065 | 7493.253 |
| TONKS_TA | 149 | 0.006123 | 0.000542 | 0.041068 | 0.44074 | 7494.784 |
| WONG_EN | 24 | 0.014391 | 0.000513 | 0.09658 | 0.44078 | 7495.464 |
| GOBP_PRC | 11 | 0.022622 | 0.000546 | 0.15192 | 0.44082 | 7496.144 |
| WP_NAD_I | 22 | 0.014597 | 0.000498 | 0.098257 | 0.44095 | 7498.355 |
| GOMF_MY | 66 | 0.009034 | 0.000533 | 0.060831 | 0.44097 | 7498.695 |
| GOBP_POS | 8 | 0.029072 | 0.000598 | 0.19578 | 0.44098 | 7498.865 |
| GOMF_ME | 187 | 0.005921 | 0.000586 | 0.03987 | 0.44098 | 7498.865 |
| MOOTHA_ | 393 | 0.003686 | 0.000526 | 0.024823 | 0.44098 | 7498.865 |
| GOMF_CAI | 191 | 0.005419 | 0.000542 | 0.036535 | 0.44105 | 7500.055 |
| HUMMERI( | 20 | 0.016606 | 0.00054 | 0.11198 | 0.44106 | 7500.225 |
| GOBP_SON | 78 | 0.008488 | 0.000544 | 0.057256 | 0.44108 | 7500.565 |
| TORCHIA_T | 253 | 0.004527 | 0.000521 | 0.030607 | 0.44121 | 7502.776 |
| GOBP_UTE | 10 | 0.026892 | 0.000619 | 0.18184 | 0.44122 | 7502.946 |
| PID_ARF6_ | 35 | 0.012244 | 0.000527 | 0.082877 | 0.44127 | 7503.796 |
| GOBP_REG | 130 | 0.006429 | 0.000532 | 0.043543 | 0.44131 | 7504.477 |
| GOCC_SPEI | 3 | 0.041062 | 0.000518 | 0.27835 | 0.44136 | 7505.327 |
| GOBP_AUT | 7 | 0.022999 | 0.000443 | 0.15599 | 0.44139 | 7505.837 |
| WP_SELEN | 82 | 0.008047 | 0.000529 | 0.054746 | 0.44157 | 7508.898 |
| GOCC_SITE | 159 | 0.005723 | 0.000523 | 0.038973 | 0.44162 | 7509.748 |
| HINATA_NI | 75 | 0.00843 | 0.00053 | 0.057544 | 0.44176 | 7512.129 |
| GOBP_REG | 32 | 0.01304 | 0.000536 | 0.089203 | 0.44189 | 7514.339 |
| GOMF_NAI | 5 | 0.031771 | 0.000517 | 0.21733 | 0.44189 | 7514.339 |
| BURTON_A | 27 | 0.014712 | 0.000556 | 0.10071 | 0.44193 | 7515.02 |
| GOMF_PEF | 7 | 0.026011 | 0.000501 | 0.17823 | 0.44198 | 7515.87 |
| GOBP_POS | 31 | 0.012761 | 0.000517 | 0.087807 | 0.44223 | 7520.121 |
| GOBP_NEG | 5 | 0.029814 | 0.000485 | 0.2053 | 0.44227 | 7520.801 |
| REACTOME | 19 | 0.016707 | 0.00053 | 0.11507 | 0.44228 | 7520.971 |
| GOBP_BRA | 20 | 0.01549 | 0.000504 | 0.10686 | 0.44238 | 7522.672 |
| GOBP_RES | 5 | 0.03534 | 0.000575 | 0.24387 | 0.44239 | 7522.842 |
| REACTOME | 19 | 0.014367 | 0.000456 | 0.099314 | 0.44249 | 7524.542 |
| GOMF_TO› | 9 | 0.023624 | 0.000516 | 0.16346 | 0.44254 | 7525.393 |
| GOBP_GLY | 6 | 0.029366 | 0.000523 | 0.20329 | 0.44257 | 7525.903 |
| GOBP_REG | 5 | 0.036799 | 0.000599 | 0.25478 | 0.44258 | 7526.073 |
| GOBP_TRO | 13 | 0.021079 | 0.000553 | 0.1462 | 0.44268 | 7527.773 |
| WP_GLYCC | 36 | 0.011324 | 0.000494 | 0.078602 | 0.44273 | 7528.624 |
| GOBP_ANA | 16 | 0.019471 | 0.000567 | 0.13532 | 0.44279 | 7529.644 |
| GOBP_REG | 129 | 0.006096 | 0.000502 | 0.042421 | 0.44287 | 7531.004 |

| | | | | | | |
|---|---|---|---|---|---|---|
| REACTOME | 264 | 0.004267 | 0.000501 | 0.029695 | 0.44287 | 7531.004 |
| GOBP_CELI | 72 | 0.008497 | 0.000524 | 0.059207 | 0.44294 | 7532.195 |
| GOCC_ZYM | 13 | 0.019598 | 0.000514 | 0.13678 | 0.44303 | 7533.725 |
| GOBP_ASC | 5 | 0.031835 | 0.000518 | 0.2223 | 0.44306 | 7534.235 |
| GOBP_TER | 93 | 0.007514 | 0.000526 | 0.052475 | 0.44307 | 7534.405 |
| MAGRANG | 25 | 0.015273 | 0.000555 | 0.10667 | 0.44307 | 7534.405 |
| FRASOR_TA | 45 | 0.011328 | 0.000552 | 0.07915 | 0.4431 | 7534.916 |
| GOBP_REG | 18 | 0.015172 | 0.000468 | 0.10609 | 0.44314 | 7535.596 |
| GOMF_O_A | 8 | 0.026916 | 0.000554 | 0.1885 | 0.44323 | 7537.126 |
| FOURNIER_ | 19 | 0.015082 | 0.000478 | 0.10573 | 0.44329 | 7538.146 |
| GOBP_SNR | 6 | 0.022796 | 0.000406 | 0.16016 | 0.44341 | 7540.187 |
| GALINDO_I | 70 | 0.0089 | 0.000541 | 0.062583 | 0.44346 | 7541.037 |
| GOBP_MO | 11 | 0.024411 | 0.000589 | 0.17172 | 0.44348 | 7541.377 |
| GOBP_SEG | 16 | 0.018996 | 0.000553 | 0.13375 | 0.44353 | 7542.228 |
| GOBP_REG | 24 | 0.013346 | 0.000476 | 0.094281 | 0.44372 | 7545.459 |
| GOBP_TRA | 65 | 0.008659 | 0.000507 | 0.061227 | 0.44377 | 7546.309 |
| GOMF_UBI | 423 | 0.003458 | 0.000512 | 0.024458 | 0.44378 | 7546.479 |
| GOBP_NEU | 9 | 0.022697 | 0.000495 | 0.16056 | 0.44379 | 7546.649 |
| GOBP_PLA | 8 | 0.027789 | 0.000572 | 0.19656 | 0.44379 | 7546.649 |
| WEIGEL_O | 34 | 0.012559 | 0.000533 | 0.088905 | 0.44383 | 7547.329 |
| NAKAYAMA | 72 | 0.008898 | 0.000548 | 0.063032 | 0.44387 | 7548.009 |
| WP_LIPID_ | 27 | 0.012965 | 0.00049 | 0.091935 | 0.44393 | 7549.03 |
| IKEDA_MIF | 6 | 0.034239 | 0.00061 | 0.24317 | 0.44401 | 7550.39 |
| MULLIGAN | 8 | 0.025854 | 0.000532 | 0.18374 | 0.44405 | 7551.07 |
| REACTOME | 14 | 0.023029 | 0.000627 | 0.16369 | 0.44406 | 7551.24 |
| DUTERTRE_ | 484 | 0.003388 | 0.000536 | 0.0241 | 0.4441 | 7551.921 |
| GOMF_OXI | 9 | 0.018746 | 0.000409 | 0.13357 | 0.44419 | 7553.451 |
| GOBP_CELI | 63 | 0.009182 | 0.00053 | 0.065444 | 0.44421 | 7553.791 |
| GOBP_PRC | 935 | 0.002279 | 0.000495 | 0.016249 | 0.44423 | 7554.131 |
| GOBP_MEI | 7 | 0.03129 | 0.000602 | 0.22316 | 0.44425 | 7554.471 |
| GOCC_LAT | 9 | 0.024962 | 0.000545 | 0.17817 | 0.44429 | 7555.151 |
| GRAHAM_( | 7 | 0.025585 | 0.000493 | 0.18264 | 0.4443 | 7555.322 |
| GOBP_LEU | 349 | 0.003911 | 0.000527 | 0.027926 | 0.44432 | 7555.662 |
| GOBP_CLA | 15 | 0.017588 | 0.000496 | 0.1258 | 0.4444 | 7557.022 |
| GOBP_CELI | 11 | 0.021211 | 0.000512 | 0.15183 | 0.44445 | 7557.872 |
| GOMF_LIG | 104 | 0.00727 | 0.000538 | 0.052058 | 0.44447 | 7558.212 |
| GOBP_TYP | 4 | 0.032139 | 0.000468 | 0.23076 | 0.44462 | 7560.763 |
| GOBP_CAR | 12 | 0.020997 | 0.000529 | 0.15086 | 0.44466 | 7561.443 |
| GOBP_POS | 37 | 0.012446 | 0.00055 | 0.089494 | 0.4447 | 7562.124 |
| GOBP_REG | 13 | 0.017651 | 0.000463 | 0.12693 | 0.4447 | 7562.124 |
| GOBP_FAT | 10 | 0.024583 | 0.000566 | 0.17709 | 0.4448 | 7563.824 |
| GOBP_PHC | 16 | 0.016449 | 0.000479 | 0.11849 | 0.4448 | 7563.824 |
| DUTTA_AP | 29 | 0.014206 | 0.000556 | 0.10237 | 0.44481 | 7563.994 |
| GOCC_VAC | 165 | 0.005223 | 0.000486 | 0.037709 | 0.44492 | 7565.865 |
| REACTOME | 10 | 0.023543 | 0.000542 | 0.17028 | 0.44502 | 7567.565 |

| | | | | | | |
|---|---|---|---|---|---|---|
| LIU_IL13_N | 4 | 0.03036 | 0.000442 | 0.21966 | 0.44504 | 7567.905 |
| GOBP_CER | 67 | 0.008884 | 0.000528 | 0.064454 | 0.44519 | 7570.456 |
| REACTOME | 48 | 0.010005 | 0.000504 | 0.072598 | 0.4452 | 7570.626 |
| GOBP_POS | 58 | 0.009276 | 0.000513 | 0.067401 | 0.44527 | 7571.816 |
| LIU_CDX2_ | 32 | 0.012955 | 0.000533 | 0.094367 | 0.4454 | 7574.027 |
| GRAHAM_( | 178 | 0.005007 | 0.000484 | 0.036561 | 0.44554 | 7576.408 |
| REACTOME | 27 | 0.013545 | 0.000512 | 0.099016 | 0.4456 | 7577.428 |
| GOBP_PYR | 7 | 0.023408 | 0.000451 | 0.17145 | 0.4457 | 7579.129 |
| GOBP_POS | 28 | 0.014133 | 0.000544 | 0.10361 | 0.44575 | 7579.979 |
| GOBP_BLE | 11 | 0.022535 | 0.000544 | 0.16524 | 0.44576 | 7580.149 |
| GOCC_CYT | 5 | 0.039666 | 0.000645 | 0.29083 | 0.44576 | 7580.149 |
| WP_INTER | 20 | 0.014783 | 0.000481 | 0.10839 | 0.44576 | 7580.149 |
| CROONQU | 90 | 0.007208 | 0.000496 | 0.052947 | 0.44586 | 7581.849 |
| WP_KININ | 7 | 0.031103 | 0.000599 | 0.2286 | 0.44589 | 7582.359 |
| GOBP_REG | 26 | 0.013927 | 0.000517 | 0.1024 | 0.44591 | 7582.7 |
| GOBP_POL | 17 | 0.01544 | 0.000463 | 0.11354 | 0.44592 | 7582.87 |
| REACTOME | 38 | 0.010215 | 0.000458 | 0.075341 | 0.44608 | 7585.59 |
| KANG_FLU | 16 | 0.014869 | 0.000433 | 0.1097 | 0.44609 | 7585.76 |
| VANTVEER | 48 | 0.010558 | 0.000532 | 0.077956 | 0.44613 | 7586.441 |
| GOMF_TAU | 22 | 0.014071 | 0.00048 | 0.10404 | 0.44621 | 7587.801 |
| MASSARW | 221 | 0.004833 | 0.00052 | 0.035807 | 0.44632 | 7589.672 |
| GOCC_MIT | 6 | 0.024605 | 0.000439 | 0.18238 | 0.44634 | 7590.012 |
| GOBP_SPLI | 13 | 0.019295 | 0.000506 | 0.14304 | 0.44635 | 7590.182 |
| KEGG_VIBF | 51 | 0.009108 | 0.000473 | 0.067742 | 0.44652 | 7593.073 |
| HOLLEMAN | 27 | 0.013449 | 0.000508 | 0.10044 | 0.44674 | 7596.814 |
| KIM_GLIS2 | 71 | 0.008267 | 0.000506 | 0.061874 | 0.44686 | 7598.854 |
| GOBP_REG | 43 | 0.010472 | 0.000499 | 0.078433 | 0.44689 | 7599.364 |
| REACTOME | 16 | 0.016296 | 0.000474 | 0.12215 | 0.44693 | 7600.045 |
| GOMF_LIG | 6 | 0.028294 | 0.000504 | 0.21213 | 0.44695 | 7600.385 |
| GOCC_AST | 10 | 0.024717 | 0.000569 | 0.18536 | 0.44696 | 7600.555 |
| GOBP_POS | 852 | 0.002352 | 0.000488 | 0.017649 | 0.447 | 7601.235 |
| GOBP_DNA | 16 | 0.014875 | 0.000433 | 0.11166 | 0.44701 | 7601.405 |
| GOBP_POS | 25 | 0.01372 | 0.000499 | 0.10302 | 0.44703 | 7601.745 |
| GOBP_REG | 319 | 0.003753 | 0.000484 | 0.028181 | 0.44703 | 7601.745 |
| DIAZ_CHRO | 1322 | 0.001862 | 0.000475 | 0.014009 | 0.44712 | 7603.276 |
| GOBP_REG | 165 | 0.005243 | 0.000488 | 0.039448 | 0.44713 | 7603.446 |
| GOCC_PLA | 61 | 0.008881 | 0.000504 | 0.06694 | 0.44723 | 7605.146 |
| GOBP_MIR | 90 | 0.007035 | 0.000485 | 0.053125 | 0.44733 | 7606.847 |
| WP_CYSTE | 15 | 0.017358 | 0.000489 | 0.13112 | 0.44734 | 7607.017 |
| GOMF_VAS | 4 | 0.040906 | 0.000595 | 0.30941 | 0.44741 | 7608.207 |
| PID_AJDISS | 47 | 0.009634 | 0.00048 | 0.073004 | 0.44751 | 7609.908 |
| KINSEY_TA | 1211 | 0.001932 | 0.000473 | 0.014667 | 0.44761 | 7611.608 |
| GOBP_DNA | 7 | 0.02404 | 0.000463 | 0.18259 | 0.44763 | 7611.948 |
| GOBP_CELI | 681 | 0.002668 | 0.000498 | 0.020327 | 0.44779 | 7614.669 |
| GOBP_CELI | 6 | 0.030578 | 0.000545 | 0.23301 | 0.4478 | 7614.839 |

| | | | | | | |
|---|---|---|---|---|---|---|
| GOBP_MAI | 11 | 0.021427 | 0.000517 | 0.16341 | 0.44784 | 7615.519 |
| GOBP_REG | 107 | 0.006079 | 0.000456 | 0.046476 | 0.44797 | 7617.73 |
| WP_HYPOT | 7 | 0.0248 | 0.000477 | 0.18991 | 0.44805 | 7619.09 |
| BANDRES_ | 8 | 0.020481 | 0.000422 | 0.15692 | 0.44808 | 7619.6 |
| GOCC_MIT | 13 | 0.015424 | 0.000405 | 0.11849 | 0.44822 | 7621.981 |
| GOBP_DNA | 9 | 0.021727 | 0.000474 | 0.16706 | 0.44826 | 7622.661 |
| GOBP_POS | 112 | 0.006129 | 0.000471 | 0.047152 | 0.44829 | 7623.171 |
| GOBP_ZIN | 8 | 0.021581 | 0.000444 | 0.16659 | 0.44846 | 7626.062 |
| GOBP_AXC | 61 | 0.007779 | 0.000441 | 0.060224 | 0.44862 | 7628.783 |
| GOBP_TRA | 592 | 0.002732 | 0.000476 | 0.021222 | 0.44878 | 7631.504 |
| GOBP_FAT | 73 | 0.006788 | 0.000421 | 0.052798 | 0.44885 | 7632.694 |
| GOMF_KIN | 690 | 0.002459 | 0.000461 | 0.01916 | 0.44895 | 7634.395 |
| REACTOME | 183 | 0.004737 | 0.000464 | 0.036953 | 0.449 | 7635.245 |
| MAINA_VH | 10 | 0.017699 | 0.000407 | 0.13839 | 0.44912 | 7637.286 |
| GOBP_REG | 9 | 0.022703 | 0.000496 | 0.17777 | 0.44919 | 7638.476 |
| GOBP_POS | 42 | 0.009906 | 0.000467 | 0.077736 | 0.4493 | 7640.347 |
| GOBP_POS | 6 | 0.028807 | 0.000513 | 0.22604 | 0.4493 | 7640.347 |
| GOBP_ENE | 296 | 0.003547 | 0.000441 | 0.02784 | 0.44931 | 7640.517 |
| GOBP_JNK_ | 160 | 0.004945 | 0.000453 | 0.03893 | 0.44946 | 7643.067 |
| CROMER_T | 52 | 0.009846 | 0.000516 | 0.07782 | 0.44966 | 7646.468 |
| GOMF_SUL | 53 | 0.009009 | 0.000477 | 0.071201 | 0.44966 | 7646.468 |
| REACTOME | 40 | 0.008776 | 0.000404 | 0.069413 | 0.4497 | 7647.149 |
| SA_FAS_SI | 9 | 0.019487 | 0.000425 | 0.15426 | 0.44974 | 7647.829 |
| GOBP_RES | 9 | 0.019021 | 0.000415 | 0.15062 | 0.44975 | 7647.999 |
| RIZ_ERYTH | 39 | 0.009034 | 0.00041 | 0.071546 | 0.44976 | 7648.169 |
| GOCC_LEW | 8 | 0.017945 | 0.000369 | 0.14217 | 0.44978 | 7648.509 |
| GOBP_MAI | 549 | 0.00266 | 0.000447 | 0.021088 | 0.44982 | 7649.189 |
| WP_DISOR | 20 | 0.014934 | 0.000486 | 0.11878 | 0.44997 | 7651.74 |
| EHRLICH_I | 11 | 0.021123 | 0.00051 | 0.16817 | 0.45002 | 7652.59 |
| ASGHARZA | 40 | 0.009545 | 0.000439 | 0.076 | 0.45003 | 7652.76 |
| LI_CISPLAT | 25 | 0.015996 | 0.000582 | 0.12751 | 0.45008 | 7653.61 |
| MADAN_D | 48 | 0.009156 | 0.000461 | 0.073048 | 0.45012 | 7654.291 |
| GOCC_CELI | 479 | 0.002976 | 0.000468 | 0.023747 | 0.45014 | 7654.631 |
| LOPEZ_ME | 13 | 0.017579 | 0.000461 | 0.14044 | 0.45019 | 7655.481 |
| PID_LKB1_ | 46 | 0.009434 | 0.000465 | 0.07537 | 0.45019 | 7655.481 |
| REACTOME | 5 | 0.034162 | 0.000556 | 0.27379 | 0.45035 | 7658.202 |
| GOBP_NEG | 12 | 0.019273 | 0.000486 | 0.1548 | 0.45046 | 7660.072 |
| GOMF_ENI | 13 | 0.016639 | 0.000436 | 0.13368 | 0.45047 | 7660.242 |
| GOMF_NU | 8 | 0.020998 | 0.000432 | 0.16894 | 0.45054 | 7661.433 |
| WP_FAMIL | 17 | 0.014637 | 0.000439 | 0.11781 | 0.45056 | 7661.773 |
| WP_FAMIL | 15 | 0.016877 | 0.000476 | 0.13616 | 0.45068 | 7663.813 |
| GOBP_CAN | 10 | 0.022345 | 0.000514 | 0.18071 | 0.4508 | 7665.854 |
| WP_NITRIC | 12 | 0.019456 | 0.00049 | 0.15784 | 0.45095 | 7668.405 |
| GOBP_REG | 102 | 0.006444 | 0.000472 | 0.052356 | 0.45102 | 7669.595 |
| GOBP_RES | 23 | 0.011642 | 0.000406 | 0.094811 | 0.45114 | 7671.636 |

| | | | | | | |
|---|---|---|---|---|---|---|
| GOBP_TEL( | 5 | 0.026066 | 0.000424 | 0.21228 | 0.45114 | 7671.636 |
| GOMF_THI | 35 | 0.010191 | 0.000438 | 0.083041 | 0.45116 | 7671.976 |
| GOBP_DET | 5 | 0.039846 | 0.000648 | 0.32469 | 0.45117 | 7672.146 |
| GOBP_NEG | 9 | 0.022054 | 0.000481 | 0.17986 | 0.45121 | 7672.826 |
| GEORGES_ | 842 | 0.002174 | 0.000449 | 0.017745 | 0.45126 | 7673.676 |
| GOBP_ALC | 340 | 0.003341 | 0.000444 | 0.027274 | 0.45126 | 7673.676 |
| GOMF_COI | 5 | 0.031012 | 0.000505 | 0.25334 | 0.45129 | 7674.186 |
| RUTELLA_F | 300 | 0.003568 | 0.000446 | 0.029156 | 0.4513 | 7674.357 |
| GOBP_NEG | 5 | 0.023088 | 0.000376 | 0.18892 | 0.45137 | 7675.547 |
| GOBP_EST/ | 11 | 0.021044 | 0.000508 | 0.17258 | 0.45148 | 7677.417 |
| GOCC_RES | 22 | 0.011127 | 0.00038 | 0.091319 | 0.45151 | 7677.928 |
| GOBP_LIVE | 28 | 0.010837 | 0.000417 | 0.089091 | 0.45159 | 7679.288 |
| GOMF_OXI | 24 | 0.012745 | 0.000454 | 0.10489 | 0.45165 | 7680.308 |
| GOBP_POS | 8 | 0.021976 | 0.000452 | 0.18094 | 0.45166 | 7680.478 |
| GOCC_MIT | 8 | 0.022551 | 0.000464 | 0.18596 | 0.45174 | 7681.839 |
| GOBP_POS | 26 | 0.011069 | 0.000411 | 0.091642 | 0.45193 | 7685.07 |
| GOBP_TOX | 39 | 0.009077 | 0.000412 | 0.075336 | 0.45205 | 7687.11 |
| GOBP_REG | 27 | 0.012132 | 0.000458 | 0.10099 | 0.45219 | 7689.491 |
| GOBP_REG | 921 | 0.00197 | 0.000424 | 0.016416 | 0.45225 | 7690.511 |
| GOBP_CELI | 410 | 0.003132 | 0.000457 | 0.026203 | 0.45243 | 7693.572 |
| ANDERSEN | 60 | 0.008049 | 0.000453 | 0.067417 | 0.45248 | 7694.422 |
| GOMF_PH( | 5 | 0.023254 | 0.000378 | 0.1948 | 0.45249 | 7694.592 |
| GOBP_FEM | 145 | 0.00488 | 0.000426 | 0.040906 | 0.45252 | 7695.103 |
| REACTOME | 30 | 0.011814 | 0.000471 | 0.099171 | 0.45259 | 7696.293 |
| REACTOME | 6 | 0.022747 | 0.000405 | 0.1913 | 0.45268 | 7697.823 |
| GOBP_CELI | 5 | 0.025363 | 0.000413 | 0.21337 | 0.45269 | 7697.993 |
| GOBP_NEG | 90 | 0.006762 | 0.000466 | 0.056982 | 0.45277 | 7699.354 |
| VALK_AML | 23 | 0.012921 | 0.000451 | 0.10909 | 0.45286 | 7700.884 |
| KEGG_SNA | 36 | 0.009913 | 0.000432 | 0.083757 | 0.45289 | 7701.394 |
| GOCC_PHC | 25 | 0.011342 | 0.000412 | 0.09585 | 0.4529 | 7701.565 |
| GOBP_POS | 6 | 0.025613 | 0.000457 | 0.21648 | 0.45291 | 7701.735 |
| GOBP_NU( | 34 | 0.009798 | 0.000415 | 0.082853 | 0.45293 | 7702.075 |
| SHEDDEN_ | 188 | 0.004561 | 0.000453 | 0.038667 | 0.45305 | 7704.115 |
| GOBP_NEG | 78 | 0.006451 | 0.000414 | 0.054744 | 0.4531 | 7704.966 |
| GOBP_SPIN | 10 | 0.021124 | 0.000486 | 0.17935 | 0.45312 | 7705.306 |
| GOBP_CYT | 12 | 0.019471 | 0.000491 | 0.16568 | 0.45322 | 7707.006 |
| GOMF_CYT | 12 | 0.019471 | 0.000491 | 0.16568 | 0.45322 | 7707.006 |
| GOBP_DN/ | 9 | 0.016198 | 0.000354 | 0.13819 | 0.45335 | 7709.217 |
| REACTOME | 11 | 0.021302 | 0.000514 | 0.18209 | 0.45344 | 7710.747 |
| GOBP_NEG | 47 | 0.008884 | 0.000443 | 0.075972 | 0.45346 | 7711.087 |
| GOCC_NU( | 28 | 0.010147 | 0.000391 | 0.08682 | 0.45348 | 7711.427 |
| GOBP_CAL | 20 | 0.012983 | 0.000422 | 0.11112 | 0.4535 | 7711.768 |
| GOBP_PRC | 75 | 0.006789 | 0.000427 | 0.058221 | 0.45359 | 7713.298 |
| REACTOME | 19 | 0.01186 | 0.000376 | 0.10184 | 0.45365 | 7714.318 |
| GOBP_REG | 29 | 0.011011 | 0.000431 | 0.094586 | 0.45366 | 7714.488 |

| | | | | | | |
|---|---|---|---|---|---|---|
| GOCC_AP_ | 9 | 0.018725 | 0.000409 | 0.16093 | 0.45369 | 7714.998 |
| SMID_BREA | 93 | 0.006391 | 0.000447 | 0.055016 | 0.45376 | 7716.189 |
| REACTOME | 11 | 0.018281 | 0.000441 | 0.15775 | 0.45387 | 7718.059 |
| GOCC_OUT | 11 | 0.020425 | 0.000493 | 0.17684 | 0.45402 | 7720.61 |
| GOBP_INTI | 6 | 0.021421 | 0.000382 | 0.18555 | 0.45405 | 7721.12 |
| GOBP_TUB | 148 | 0.005002 | 0.000441 | 0.043368 | 0.45409 | 7721.8 |
| REACTOME | 12 | 0.015519 | 0.000391 | 0.13466 | 0.45413 | 7722.481 |
| GAVIN_FO | 84 | 0.006714 | 0.000447 | 0.058274 | 0.45414 | 7722.651 |
| BIOCARTA_ | 8 | 0.019952 | 0.000411 | 0.17326 | 0.45416 | 7722.991 |
| GOBP_COF | 41 | 0.009111 | 0.000424 | 0.079283 | 0.45426 | 7724.691 |
| WP_INTERI | 45 | 0.008083 | 0.000394 | 0.070672 | 0.45447 | 7728.262 |
| GOBP_ACII | 58 | 0.008393 | 0.000465 | 0.073438 | 0.4545 | 7728.773 |
| MARTORIA | 159 | 0.004618 | 0.000422 | 0.040451 | 0.45456 | 7729.793 |
| GOBP_REG | 14 | 0.018322 | 0.000499 | 0.1607 | 0.45461 | 7730.643 |
| GOBP_ANT | 47 | 0.007762 | 0.000387 | 0.068094 | 0.45463 | 7730.983 |
| GOBP_DRII | 10 | 0.020251 | 0.000466 | 0.17814 | 0.45475 | 7733.024 |
| GOCC_SPE | 9 | 0.019683 | 0.00043 | 0.17315 | 0.45475 | 7733.024 |
| GOMF_COI | 4 | 0.032995 | 0.00048 | 0.29075 | 0.45482 | 7734.214 |
| REACTOME | 28 | 0.010966 | 0.000422 | 0.096686 | 0.45485 | 7734.724 |
| GOBP_HYP | 10 | 0.018367 | 0.000423 | 0.16265 | 0.45505 | 7738.125 |
| GOMF_CAI | 5 | 0.025531 | 0.000415 | 0.22644 | 0.45511 | 7739.146 |
| REACTOME | 17 | 0.0142 | 0.000426 | 0.12634 | 0.45525 | 7741.526 |
| GOBP_INTI | 817 | 0.001916 | 0.00039 | 0.01705 | 0.45528 | 7742.036 |
| GOMF_OXI | 53 | 0.007508 | 0.000397 | 0.066903 | 0.45532 | 7742.717 |
| GOBP_GRC | 10 | 0.018738 | 0.000431 | 0.16734 | 0.45542 | 7744.417 |
| GOBP_NEG | 63 | 0.006792 | 0.000392 | 0.060674 | 0.45544 | 7744.757 |
| GOMF_CILI | 6 | 0.026533 | 0.000473 | 0.23708 | 0.45545 | 7744.927 |
| GOBP_PRC | 90 | 0.005905 | 0.000407 | 0.053009 | 0.45565 | 7748.328 |
| GINESTIER_ | 66 | 0.006868 | 0.000405 | 0.061662 | 0.45566 | 7748.498 |
| GOBP_LOC | 196 | 0.004022 | 0.000408 | 0.036233 | 0.45581 | 7751.049 |
| GOBP_POS | 102 | 0.005293 | 0.000388 | 0.047746 | 0.45586 | 7751.899 |
| GOBP_POS | 25 | 0.009963 | 0.000362 | 0.090079 | 0.45597 | 7753.77 |
| WP_TOLLLI | 93 | 0.006196 | 0.000434 | 0.056061 | 0.456 | 7754.28 |
| GOBP_PRC | 6 | 0.02163 | 0.000386 | 0.19647 | 0.45617 | 7757.171 |
| GOBP_COF | 25 | 0.011272 | 0.00041 | 0.10281 | 0.45635 | 7760.232 |
| REACTOME | 86 | 0.005843 | 0.000394 | 0.053603 | 0.4566 | 7764.483 |
| GOBP_REG | 18 | 0.012862 | 0.000397 | 0.11807 | 0.45663 | 7764.993 |
| BIOCARTA_ | 9 | 0.022504 | 0.000491 | 0.20665 | 0.45664 | 7765.163 |
| GOCC_SHE | 5 | 0.025738 | 0.000419 | 0.23695 | 0.45675 | 7767.034 |
| WP_GLYCC | 23 | 0.010967 | 0.000383 | 0.1012 | 0.45685 | 7768.734 |
| MCBRYAN_ | 204 | 0.004009 | 0.000414 | 0.037028 | 0.4569 | 7769.585 |
| GOBP_REG | 23 | 0.011501 | 0.000401 | 0.1064 | 0.45696 | 7770.605 |
| REACTOME | 14 | 0.015181 | 0.000413 | 0.14045 | 0.45697 | 7770.775 |
| BIOCARTA_ | 12 | 0.018105 | 0.000456 | 0.16807 | 0.45711 | 7773.156 |
| GOCC_ER_ | 3 | 0.030497 | 0.000384 | 0.28315 | 0.45711 | 7773.156 |

| | | | | | | |
|---|---|---|---|---|---|---|
| GOBP_CELI | 6 | 0.017795 | 0.000317 | 0.1659 | 0.45729 | 7776.216 |
| REACTOME | 10 | 0.017734 | 0.000408 | 0.1662 | 0.45751 | 7779.958 |
| BIOCARTA_ | 17 | 0.013324 | 0.0004 | 0.12505 | 0.45757 | 7780.978 |
| REACTOME | 299 | 0.002906 | 0.000363 | 0.027309 | 0.45762 | 7781.828 |
| REACTOME | 83 | 0.005923 | 0.000392 | 0.055746 | 0.45769 | 7783.018 |
| GOBP_NITI | 21 | 0.012181 | 0.000406 | 0.11503 | 0.45783 | 7785.399 |
| BIOCARTA_ | 21 | 0.011628 | 0.000388 | 0.10984 | 0.45785 | 7785.739 |
| GOBP_PEP | 38 | 0.008245 | 0.00037 | 0.077962 | 0.45789 | 7786.419 |
| GOMF_CYC | 28 | 0.008811 | 0.000339 | 0.083555 | 0.45801 | 7788.46 |
| GOBP_NEU | 10 | 0.01322 | 0.000304 | 0.12548 | 0.45805 | 7789.14 |
| WANG_AD | 26 | 0.010383 | 0.000385 | 0.098664 | 0.45809 | 7789.82 |
| GOBP_MIT | 19 | 0.010468 | 0.000332 | 0.099598 | 0.45815 | 7790.841 |
| ZHANG_RE | 62 | 0.006432 | 0.000368 | 0.061211 | 0.45816 | 7791.011 |
| REACTOME | 43 | 0.007509 | 0.000358 | 0.071528 | 0.4582 | 7791.691 |
| GOBP_DNA | 14 | 0.012038 | 0.000328 | 0.11522 | 0.45839 | 7794.922 |
| GOBP_LYM | 22 | 0.011384 | 0.000388 | 0.10893 | 0.45839 | 7794.922 |
| GOBP_REG | 16 | 0.011825 | 0.000344 | 0.11325 | 0.45842 | 7795.432 |
| GOBP_ACT | 17 | 0.012691 | 0.000381 | 0.12169 | 0.45847 | 7796.282 |
| GOBP_PHC | 134 | 0.004831 | 0.000406 | 0.046318 | 0.45847 | 7796.282 |
| CHIBA_RES | 41 | 0.008054 | 0.000375 | 0.077317 | 0.45852 | 7797.133 |
| GOBP_DNA | 37 | 0.007993 | 0.000354 | 0.076896 | 0.45861 | 7798.663 |
| LU_TUMOI | 27 | 0.011108 | 0.00042 | 0.1069 | 0.45862 | 7798.833 |
| REACTOME | 25 | 0.009392 | 0.000342 | 0.090422 | 0.45864 | 7799.173 |
| GOBP_MIC | 9 | 0.018603 | 0.000406 | 0.17942 | 0.45871 | 7800.364 |
| GOBP_POS | 6 | 0.01784 | 0.000318 | 0.17234 | 0.45878 | 7801.554 |
| GOBP_POS | 8 | 0.018335 | 0.000377 | 0.17715 | 0.45878 | 7801.554 |
| GOBP_NEG | 32 | 0.009759 | 0.000401 | 0.094498 | 0.45888 | 7803.254 |
| GOCC_POS | 14 | 0.014219 | 0.000387 | 0.13812 | 0.459 | 7805.295 |
| GOBP_FOR | 7 | 0.023561 | 0.000454 | 0.22899 | 0.45902 | 7805.635 |
| REACTOME | 7 | 0.014367 | 0.000277 | 0.13962 | 0.45902 | 7805.635 |
| GOBP_NEU | 8 | 0.019956 | 0.000411 | 0.194 | 0.45904 | 7805.975 |
| GOMF_ANI | 8 | 0.017897 | 0.000368 | 0.17419 | 0.45908 | 7806.655 |
| GOBP_RAD | 6 | 0.019338 | 0.000345 | 0.18824 | 0.45909 | 7806.825 |
| SHEPARD_I | 188 | 0.003606 | 0.000358 | 0.035148 | 0.45915 | 7807.846 |
| GOBP_MYE | 76 | 0.005639 | 0.000357 | 0.055039 | 0.4592 | 7808.696 |
| GOBP_L_KY | 7 | 0.021506 | 0.000414 | 0.20999 | 0.45921 | 7808.866 |
| BILANGES_ | 29 | 0.010057 | 0.000394 | 0.098263 | 0.45924 | 7809.376 |
| GOBP_POS | 12 | 0.012465 | 0.000314 | 0.12193 | 0.45929 | 7810.226 |
| GOBP_OTC | 9 | 0.018021 | 0.000393 | 0.17649 | 0.45933 | 7810.907 |
| GOCC_VAC | 422 | 0.002412 | 0.000357 | 0.023617 | 0.45933 | 7810.907 |
| REACTOME | 10 | 0.015121 | 0.000348 | 0.14844 | 0.45943 | 7812.607 |
| GOBP_NEG | 5 | 0.02579 | 0.00042 | 0.25382 | 0.45953 | 7814.308 |
| WP_ASPIRI | 10 | 0.016931 | 0.00039 | 0.16689 | 0.4596 | 7815.498 |
| GOBP_OLIC | 9 | 0.014912 | 0.000326 | 0.14707 | 0.45962 | 7815.838 |
| ELVIDGE_H | 39 | 0.008086 | 0.000367 | 0.079851 | 0.45967 | 7816.688 |

| | | | | | | |
|---|---|---|---|---|---|---|
| REACTOME | 41 | 0.008831 | 0.000411 | 0.087255 | 0.45969 | 7817.028 |
| KUNINGER_ | 41 | 0.008453 | 0.000394 | 0.083688 | 0.45977 | 7818.389 |
| GOBP_MO | 6 | 0.022393 | 0.000399 | 0.22217 | 0.45986 | 7819.919 |
| MILI_PSEU | 633 | 0.001961 | 0.000353 | 0.019537 | 0.46002 | 7822.64 |
| GOBP_END | 88 | 0.005106 | 0.000348 | 0.050938 | 0.46008 | 7823.66 |
| GOBP_POS | 16 | 0.013343 | 0.000388 | 0.13363 | 0.46023 | 7826.211 |
| GOMF_PEF | 47 | 0.008301 | 0.000414 | 0.083395 | 0.46036 | 7828.422 |
| REACTOME | 15 | 0.012772 | 0.00036 | 0.12836 | 0.46037 | 7828.592 |
| GOBP_DNA | 123 | 0.004227 | 0.00034 | 0.042548 | 0.46043 | 7829.612 |
| GOBP_GAL | 7 | 0.022615 | 0.000435 | 0.22876 | 0.46063 | 7833.013 |
| REACTOME | 31 | 0.009762 | 0.000395 | 0.09882 | 0.46065 | 7833.353 |
| GOBP_CELI | 67 | 0.005799 | 0.000345 | 0.05875 | 0.46069 | 7834.033 |
| VERHAAK_ | 155 | 0.00411 | 0.000371 | 0.041755 | 0.46079 | 7835.734 |
| GOMF_OLI | 6 | 0.019753 | 0.000352 | 0.20069 | 0.4608 | 7835.904 |
| GOBP_IMP | 159 | 0.003755 | 0.000343 | 0.038224 | 0.46087 | 7837.094 |
| GOCC_ROD | 5 | 0.015677 | 0.000255 | 0.15962 | 0.46088 | 7837.264 |
| GOBP_FAC | 40 | 0.008033 | 0.000369 | 0.081858 | 0.46091 | 7837.775 |
| KERLEY_RE | 42 | 0.007437 | 0.00035 | 0.075915 | 0.46098 | 7838.965 |
| LANDIS_ER | 36 | 0.008293 | 0.000362 | 0.084672 | 0.46099 | 7839.135 |
| GOBP_REG | 11 | 0.014908 | 0.00036 | 0.1523 | 0.46101 | 7839.475 |
| GOBP_FEV | 10 | 0.018388 | 0.000423 | 0.18798 | 0.46104 | 7839.985 |
| GOMF_IMI | 9 | 0.016924 | 0.000369 | 0.17323 | 0.46109 | 7840.835 |
| BYSTRYKH_ | 6 | 0.019941 | 0.000355 | 0.20422 | 0.46111 | 7841.176 |
| GALE_APL_ | 57 | 0.006621 | 0.000363 | 0.068264 | 0.46137 | 7845.597 |
| GOBP_C4_ | 29 | 0.009305 | 0.000364 | 0.096038 | 0.46141 | 7846.277 |
| GOBP_REG | 18 | 0.009312 | 0.000287 | 0.096134 | 0.46142 | 7846.447 |
| GOBP_REG | 7 | 0.017242 | 0.000332 | 0.17827 | 0.46148 | 7847.467 |
| GOBP_CEN | 15 | 0.01021 | 0.000288 | 0.10597 | 0.46162 | 7849.848 |
| GOBP_POS | 15 | 0.011992 | 0.000338 | 0.12464 | 0.46168 | 7850.868 |
| GOCC_DEN | 4 | 0.021076 | 0.000307 | 0.21935 | 0.46173 | 7851.719 |
| TUOMISTO | 16 | 0.012097 | 0.000352 | 0.12591 | 0.46173 | 7851.719 |
| GOBP_CYT | 697 | 0.001848 | 0.000348 | 0.019261 | 0.46179 | 7852.739 |
| WANG_BA | 24 | 0.010313 | 0.000367 | 0.10762 | 0.46183 | 7853.419 |
| GOBP_MAI | 7 | 0.020505 | 0.000395 | 0.21531 | 0.46207 | 7857.5 |
| GOBP_NEG | 27 | 0.009725 | 0.000368 | 0.10218 | 0.46209 | 7857.84 |
| MATZUK_L | 8 | 0.016976 | 0.000349 | 0.17843 | 0.4621 | 7858.011 |
| GOBP_MO | 239 | 0.002928 | 0.000327 | 0.030784 | 0.46212 | 7858.351 |
| GOBP_NUC | 150 | 0.003708 | 0.000329 | 0.03921 | 0.46233 | 7861.922 |
| GOBP_CYT | 12 | 0.012211 | 0.000308 | 0.12928 | 0.46238 | 7862.772 |
| GOBP_PHE | 103 | 0.005078 | 0.000374 | 0.054061 | 0.46258 | 7866.173 |
| GOBP_SINC | 5 | 0.028254 | 0.00046 | 0.30111 | 0.46262 | 7866.853 |
| GOBP_REG | 636 | 0.001891 | 0.000341 | 0.02018 | 0.46267 | 7867.703 |
| GOBP_RES | 10 | 0.012518 | 0.000288 | 0.13382 | 0.46274 | 7868.894 |
| SOGA_COL | 77 | 0.004743 | 0.000302 | 0.050823 | 0.46282 | 7870.254 |
| GOMF_PUI | 1848 | 0.001129 | 0.000335 | 0.012101 | 0.46285 | 7870.764 |

| | | | | | | |
|---|---|---|---|---|---|---|
| GOBP_NEG | 296 | 0.002659 | 0.00033 | 0.02855 | 0.4629 | 7871.615 |
| BILBAN_B_ | 57 | 0.006566 | 0.00036 | 0.07061 | 0.46296 | 7872.635 |
| GOMF_INT | 5 | 0.028441 | 0.000463 | 0.30616 | 0.46299 | 7873.145 |
| GOBP_TRN | 39 | 0.006016 | 0.000273 | 0.064775 | 0.463 | 7873.315 |
| REACTOME | 115 | 0.004139 | 0.000322 | 0.044559 | 0.463 | 7873.315 |
| GOMF_HIS | 5 | 0.019061 | 0.00031 | 0.20643 | 0.46321 | 7876.886 |
| GOBP_REG | 13 | 0.01205 | 0.000316 | 0.13057 | 0.46324 | 7877.396 |
| KEGG_GLY | 14 | 0.011467 | 0.000312 | 0.12453 | 0.46332 | 7878.757 |
| GOCC_CD9 | 6 | 0.024141 | 0.00043 | 0.26236 | 0.46334 | 7879.097 |
| TURASHVIL | 83 | 0.005319 | 0.000352 | 0.057836 | 0.46336 | 7879.437 |
| GOBP_PEP | 45 | 0.006733 | 0.000328 | 0.073331 | 0.46342 | 7880.457 |
| GOBP_CELI | 134 | 0.003929 | 0.00033 | 0.042825 | 0.46345 | 7880.967 |
| GOBP_MIT | 25 | 0.009226 | 0.000336 | 0.10057 | 0.46345 | 7880.967 |
| GOBP_CELI | 37 | 0.00716 | 0.000317 | 0.0781 | 0.46348 | 7881.477 |
| BIOCARTA_ | 43 | 0.007011 | 0.000334 | 0.076564 | 0.46352 | 7882.158 |
| GOMF_EPI | 5 | 0.023138 | 0.000377 | 0.25289 | 0.46355 | 7882.668 |
| GOBP_NEG | 13 | 0.012728 | 0.000334 | 0.13965 | 0.46369 | 7885.048 |
| NIKOLSKY_ | 75 | 0.008857 | 0.000557 | 0.097403 | 0.46377 | 7886.409 |
| GOBP_REG | 4 | 0.027023 | 0.000393 | 0.29787 | 0.46386 | 7887.939 |
| GOCC_MEI | 14 | 0.011339 | 0.000309 | 0.12511 | 0.46389 | 7888.449 |
| KEGG_GLY | 25 | 0.009122 | 0.000332 | 0.10092 | 0.46399 | 7890.15 |
| GOBP_CAN | 28 | 0.009171 | 0.000353 | 0.10161 | 0.46404 | 7891 |
| GOBP_CAR | 14 | 0.014286 | 0.000389 | 0.15829 | 0.46404 | 7891 |
| WP_HEAD_ | 73 | 0.005107 | 0.000317 | 0.056691 | 0.46411 | 7892.191 |
| GOBP_POS | 32 | 0.008718 | 0.000359 | 0.096863 | 0.46414 | 7892.701 |
| LIU_OVARI | 77 | 0.00534 | 0.00034 | 0.059375 | 0.46417 | 7893.211 |
| DACOSTA_ | 52 | 0.006253 | 0.000328 | 0.070067 | 0.46445 | 7897.972 |
| GOMF_CAI | 6 | 0.020496 | 0.000365 | 0.22978 | 0.46446 | 7898.142 |
| KEGG_CELI | 106 | 0.004597 | 0.000343 | 0.051558 | 0.46448 | 7898.482 |
| GOMF_17_ | 11 | 0.013552 | 0.000327 | 0.15222 | 0.46453 | 7899.333 |
| GOMF_CAI | 9 | 0.013893 | 0.000303 | 0.15657 | 0.46465 | 7901.373 |
| GOCC_DEA | 9 | 0.0171 | 0.000373 | 0.19288 | 0.46468 | 7901.883 |
| GOBP_POS | 172 | 0.003512 | 0.000334 | 0.039655 | 0.46472 | 7902.564 |
| VERRECCHI | 19 | 0.010083 | 0.00032 | 0.11386 | 0.46472 | 7902.564 |
| REACTOME | 20 | 0.009915 | 0.000323 | 0.11206 | 0.46475 | 7903.074 |
| GOBP_POS | 27 | 0.008532 | 0.000322 | 0.096553 | 0.46479 | 7903.754 |
| GOBP_RET | 17 | 0.011571 | 0.000347 | 0.13092 | 0.46479 | 7903.754 |
| GOBP_T_C | 83 | 0.004947 | 0.000327 | 0.056033 | 0.46483 | 7904.434 |
| MACLACHL | 14 | 0.010936 | 0.000298 | 0.12439 | 0.46497 | 7906.815 |
| GOBP_REG | 35 | 0.006944 | 0.000299 | 0.079423 | 0.46516 | 7910.046 |
| DAZARD_R | 291 | 0.002567 | 0.000316 | 0.029375 | 0.46519 | 7910.556 |
| GOMF_PRO | 10 | 0.014408 | 0.000332 | 0.16527 | 0.46527 | 7911.916 |
| RODRIGUE | 484 | 0.002061 | 0.000326 | 0.023659 | 0.46529 | 7912.256 |
| GOCC_CILI | 70 | 0.005065 | 0.000308 | 0.058199 | 0.46533 | 7912.937 |
| HESSON_TI | 8 | 0.012351 | 0.000254 | 0.14214 | 0.46538 | 7913.787 |

| | | | | | | |
|---|---|---|---|---|---|---|
| KEGG_ASTI | 14 | 0.011024 | 0.0003 | 0.12804 | 0.4657 | 7919.229 |
| GOBP_ASP | 5 | 0.014806 | 0.000241 | 0.1722 | 0.46574 | 7919.909 |
| GOBP_RES | 5 | 0.023176 | 0.000377 | 0.26977 | 0.46577 | 7920.419 |
| GOBP_SEC | 140 | 0.003583 | 0.000307 | 0.041765 | 0.46582 | 7921.269 |
| GOBP_THY | 19 | 0.00909 | 0.000288 | 0.10626 | 0.46592 | 7922.97 |
| KEGG_LON | 68 | 0.005543 | 0.000332 | 0.064885 | 0.46596 | 7923.65 |
| GOBP_REG | 8 | 0.013461 | 0.000277 | 0.15817 | 0.46609 | 7925.86 |
| PETRETTO_ | 6 | 0.023229 | 0.000414 | 0.27319 | 0.46612 | 7926.371 |
| BLANCO_M | 9 | 0.014687 | 0.000321 | 0.17366 | 0.4663 | 7929.432 |
| FARDIN_HY | 7 | 0.014954 | 0.000288 | 0.17897 | 0.46671 | 7936.404 |
| GOBP_REG | 7 | 0.014254 | 0.000274 | 0.17074 | 0.46673 | 7936.744 |
| REACTOME | 16 | 0.009492 | 0.000276 | 0.1138 | 0.46676 | 7937.254 |
| REACTOME | 6 | 0.019268 | 0.000343 | 0.23203 | 0.46691 | 7939.805 |
| GOBP_CELI | 11 | 0.012673 | 0.000306 | 0.1528 | 0.46695 | 7940.485 |
| GOBP_AXC | 5 | 0.021789 | 0.000355 | 0.26433 | 0.46715 | 7943.886 |
| GOBP_MIT | 24 | 0.007221 | 0.000257 | 0.087708 | 0.46719 | 7944.566 |
| GOBP_REG | 11 | 0.014129 | 0.000341 | 0.17181 | 0.46723 | 7945.246 |
| GOBP_CIRC | 18 | 0.009798 | 0.000302 | 0.11925 | 0.46726 | 7945.756 |
| REACTOME | 12 | 0.012676 | 0.00032 | 0.15459 | 0.46733 | 7946.947 |
| GOBP_NEG | 13 | 0.012878 | 0.000338 | 0.15725 | 0.46737 | 7947.627 |
| GOBP_POS | 5 | 0.018723 | 0.000305 | 0.22892 | 0.46741 | 7948.307 |
| PID_PRL_SI | 23 | 0.008231 | 0.000287 | 0.1009 | 0.46749 | 7949.667 |
| GOBP_POS | 27 | 0.007695 | 0.000291 | 0.09489 | 0.46768 | 7952.898 |
| GOBP_REG | 14 | 0.012412 | 0.000338 | 0.15301 | 0.46768 | 7952.898 |
| ELVIDGE_H | 100 | 0.004123 | 0.000299 | 0.051216 | 0.46792 | 7956.98 |
| GOBP_AXC | 107 | 0.003842 | 0.000288 | 0.048001 | 0.46811 | 7960.211 |
| BIOCARTA_ | 28 | 0.00784 | 0.000302 | 0.098152 | 0.46817 | 7961.231 |
| GOBP_NEG | 15 | 0.011764 | 0.000331 | 0.14854 | 0.46844 | 7965.822 |
| GOBP_REG | 71 | 0.004962 | 0.000304 | 0.062722 | 0.46847 | 7966.332 |
| GOBP_REG | 66 | 0.004753 | 0.000281 | 0.060072 | 0.46847 | 7966.332 |
| GOBP_NEG | 5 | 0.014751 | 0.00024 | 0.18652 | 0.46848 | 7966.502 |
| GOBP_POS | 5 | 0.019339 | 0.000315 | 0.24514 | 0.46856 | 7967.863 |
| GOBP_REG | 33 | 0.007044 | 0.000294 | 0.090204 | 0.46888 | 7973.304 |
| GOBP_HIST | 8 | 0.01456 | 0.0003 | 0.18661 | 0.46891 | 7973.815 |
| GOBP_MO | 5 | 0.019025 | 0.00031 | 0.24384 | 0.46891 | 7973.815 |
| REACTOME | 9 | 0.015607 | 0.000341 | 0.20044 | 0.46897 | 7974.835 |
| REACTOME | 32 | 0.006784 | 0.000279 | 0.087181 | 0.46899 | 7975.175 |
| SHIN_B_CE | 37 | 0.006056 | 0.000268 | 0.077826 | 0.46899 | 7975.175 |
| TSAI_DNAJ | 12 | 0.011371 | 0.000287 | 0.14612 | 0.46899 | 7975.175 |
| NIKOLSKY_ | 146 | 0.005174 | 0.000453 | 0.066537 | 0.46901 | 7975.515 |
| WP_INTEG | 44 | 0.005627 | 0.000271 | 0.072384 | 0.46902 | 7975.685 |
| GOCC_NEU | 69 | 0.004529 | 0.000273 | 0.058273 | 0.46903 | 7975.855 |
| SCHLOSSEF | 155 | 0.003218 | 0.00029 | 0.04149 | 0.46909 | 7976.875 |
| PID_PS1_P | 45 | 0.005592 | 0.000273 | 0.072328 | 0.46919 | 7978.576 |
| GOBP_N_A | 5 | 0.016842 | 0.000274 | 0.21859 | 0.46929 | 7980.276 |

| | | | | | | |
|---|---|---|---|---|---|---|
| REACTOME | 15 | 0.008602 | 0.000242 | 0.11171 | 0.46931 | 7980.617 |
| TSUNODA_ | 14 | 0.0106 | 0.000289 | 0.13796 | 0.46938 | 7981.807 |
| GOBP_REG | 16 | 0.009254 | 0.000269 | 0.12053 | 0.4694 | 7982.147 |
| THEILGAAR | 70 | 0.004743 | 0.000288 | 0.061783 | 0.4694 | 7982.147 |
| GOBP_CRIS | 15 | 0.009046 | 0.000255 | 0.11873 | 0.46963 | 7986.058 |
| REACTOME | 8 | 0.013887 | 0.000286 | 0.18221 | 0.46963 | 7986.058 |
| GOMF_TEL | 7 | 0.014159 | 0.000273 | 0.18611 | 0.46968 | 7986.908 |
| GOBP_POS | 17 | 0.008231 | 0.000247 | 0.10838 | 0.46973 | 7987.759 |
| CHARAFE_I | 116 | 0.003491 | 0.000273 | 0.04605 | 0.46979 | 7988.779 |
| REACTOME | 10 | 0.012757 | 0.000294 | 0.16834 | 0.4698 | 7988.949 |
| GOBP_VAS | 18 | 0.010774 | 0.000333 | 0.14231 | 0.46982 | 7989.289 |
| GOCC_PHA | 26 | 0.00722 | 0.000268 | 0.09542 | 0.46984 | 7989.629 |
| GOBP_POS | 10 | 0.012468 | 0.000287 | 0.16503 | 0.46989 | 7990.479 |
| GOBP_GLU | 5 | 0.018594 | 0.000303 | 0.24767 | 0.47008 | 7993.71 |
| GOCC_SWI | 13 | 0.00938 | 0.000246 | 0.12538 | 0.47018 | 7995.411 |
| RUAN_RES | 15 | 0.009867 | 0.000278 | 0.13186 | 0.47018 | 7995.411 |
| GOBP_POS | 9 | 0.012449 | 0.000272 | 0.16698 | 0.47029 | 7997.281 |
| DARWICHE | 145 | 0.002902 | 0.000253 | 0.038951 | 0.4703 | 7997.452 |
| GOMF_VOI | 11 | 0.012318 | 0.000297 | 0.16534 | 0.47031 | 7997.622 |
| GOBP_NEG | 12 | 0.011004 | 0.000277 | 0.14777 | 0.47032 | 7997.792 |
| KLEIN_TAR | 41 | 0.005525 | 0.000257 | 0.07427 | 0.47035 | 7998.302 |
| GOBP_ADA | 157 | 0.003 | 0.000272 | 0.040408 | 0.47041 | 7999.322 |
| GOBP_DNA | 18 | 0.008004 | 0.000247 | 0.10782 | 0.47041 | 7999.322 |
| AMUNDSO | 201 | 0.002626 | 0.00027 | 0.035439 | 0.47047 | 8000.342 |
| MCBRYAN_ | 60 | 0.004858 | 0.000273 | 0.065886 | 0.47061 | 8002.723 |
| BIOCARTA_ | 7 | 0.014366 | 0.000277 | 0.19512 | 0.47065 | 8003.403 |
| GOMF_IGG | 9 | 0.014772 | 0.000322 | 0.20197 | 0.47085 | 8006.804 |
| YAO_TEMP | 134 | 0.003121 | 0.000262 | 0.042787 | 0.47093 | 8008.165 |
| SALVADOR | 15 | 0.00966 | 0.000272 | 0.13303 | 0.47106 | 8010.375 |
| SMID_BREA | 131 | 0.003184 | 0.000264 | 0.043929 | 0.47111 | 8011.226 |
| KOMMAGA | 9 | 0.013468 | 0.000294 | 0.1859 | 0.47112 | 8011.396 |
| GOBP_REG | 55 | 0.004684 | 0.000252 | 0.065006 | 0.47128 | 8014.116 |
| GOBP_MIT | 171 | 0.002544 | 0.000241 | 0.03531 | 0.47129 | 8014.286 |
| GOBP_HIST | 6 | 0.015713 | 0.00028 | 0.21847 | 0.47133 | 8014.967 |
| GOBP_NEG | 30 | 0.006671 | 0.000266 | 0.093024 | 0.47142 | 8016.497 |
| BIOCARTA_ | 13 | 0.009065 | 0.000238 | 0.12679 | 0.4715 | 8017.858 |
| GOMF_LYS | 6 | 0.017731 | 0.000316 | 0.24826 | 0.47153 | 8018.368 |
| GOBP_COE | 11 | 0.00964 | 0.000233 | 0.13578 | 0.4717 | 8021.259 |
| GOBP_CELI | 112 | 0.0034 | 0.000261 | 0.047968 | 0.47175 | 8022.109 |
| GROSS_ELK | 26 | 0.008138 | 0.000302 | 0.11515 | 0.47183 | 8023.469 |
| GOBP_NEG | 14 | 0.009817 | 0.000267 | 0.13952 | 0.47195 | 8025.51 |
| REACTOME | 32 | 0.005663 | 0.000233 | 0.080626 | 0.472 | 8026.36 |
| MEISSNER_ | 340 | 0.002045 | 0.000272 | 0.02914 | 0.47203 | 8026.87 |
| DAIRKEE_C | 111 | 0.003108 | 0.000238 | 0.044416 | 0.47211 | 8028.231 |
| REACTOME | 139 | 0.002998 | 0.000256 | 0.04299 | 0.47221 | 8029.931 |

| | | | | | | |
|---|---|---|---|---|---|---|
| MORI_PLA! | 34 | 0.006367 | 0.00027 | 0.09179 | 0.47235 | 8032.312 |
| GOBP_RES | 78 | 0.003998 | 0.000256 | 0.057651 | 0.47236 | 8032.482 |
| KYNG_ENV | 11 | 0.011258 | 0.000272 | 0.16253 | 0.47239 | 8032.992 |
| GOBP_OST | 6 | 0.016018 | 0.000286 | 0.23198 | 0.47248 | 8034.522 |
| REACTOMI | 24 | 0.006967 | 0.000248 | 0.10093 | 0.47249 | 8034.692 |
| GOBP_SM/ | 444 | 0.001674 | 0.000254 | 0.024293 | 0.47254 | 8035.543 |
| XU_RESPOI | 14 | 0.009991 | 0.000272 | 0.14528 | 0.47259 | 8036.393 |
| GENTILE_U | 277 | 0.002133 | 0.000257 | 0.031135 | 0.47269 | 8038.093 |
| RATTENBA( | 375 | 0.001782 | 0.000249 | 0.026087 | 0.47277 | 8039.454 |
| REACTOMI | 142 | 0.002996 | 0.000259 | 0.043856 | 0.47277 | 8039.454 |
| GOMF_STF | 9 | 0.011856 | 0.000259 | 0.17382 | 0.47281 | 8040.134 |
| BIOCARTA_ | 12 | 0.011357 | 0.000286 | 0.1667 | 0.47284 | 8040.644 |
| REACTOMI | 24 | 0.006131 | 0.000218 | 0.090162 | 0.47289 | 8041.494 |
| GOBP_POS | 8 | 0.013407 | 0.000276 | 0.19749 | 0.47294 | 8042.345 |
| GOBP_AMI | 5 | 0.015936 | 0.000259 | 0.23537 | 0.47301 | 8043.535 |
| GOBP_IROI | 4 | 0.018501 | 0.000269 | 0.27332 | 0.47302 | 8043.705 |
| GOBP_NAD | 20 | 0.006744 | 0.000219 | 0.099672 | 0.47303 | 8043.875 |
| GOBP_REG | 6 | 0.015634 | 0.000279 | 0.2315 | 0.47308 | 8044.725 |
| GOBP_GLY | 238 | 0.002173 | 0.000242 | 0.03223 | 0.47313 | 8045.576 |
| GOBP_INTI | 6 | 0.012327 | 0.00022 | 0.18374 | 0.47326 | 8047.786 |
| GOBP_RET | 9 | 0.013816 | 0.000302 | 0.2072 | 0.47342 | 8050.507 |
| REACTOMI | 61 | 0.004144 | 0.000235 | 0.062321 | 0.4735 | 8051.868 |
| GOMF_SOI | 75 | 0.004017 | 0.000253 | 0.06046 | 0.47352 | 8052.208 |
| GOBP_CITF | 5 | 0.015256 | 0.000248 | 0.22986 | 0.47354 | 8052.548 |
| MANALO_I | 188 | 0.002647 | 0.000263 | 0.039918 | 0.47357 | 8053.058 |
| WP_RESIST | 31 | 0.005633 | 0.000228 | 0.085616 | 0.47377 | 8056.459 |
| GOMF_GAI | 28 | 0.005738 | 0.000221 | 0.087266 | 0.47379 | 8056.799 |
| REACTOMI | 54 | 0.004556 | 0.000243 | 0.069868 | 0.47401 | 8060.54 |
| PATIL_LIVE | 603 | 0.0013 | 0.000229 | 0.019971 | 0.47406 | 8061.39 |
| REACTOMI | 9 | 0.011525 | 0.000252 | 0.17755 | 0.47412 | 8062.411 |
| GOBP_VIT/ | 9 | 0.009677 | 0.000211 | 0.14911 | 0.47413 | 8062.581 |
| GOMF_SOI | 108 | 0.003175 | 0.000239 | 0.049039 | 0.47419 | 8063.601 |
| GOBP_ANG | 27 | 0.006924 | 0.000262 | 0.10707 | 0.47422 | 8064.111 |
| HEDVAT_E | 12 | 0.009675 | 0.000244 | 0.1497 | 0.47423 | 8064.281 |
| GOMF_SOI | 40 | 0.005405 | 0.000249 | 0.083777 | 0.47428 | 8065.131 |
| HEIDENBL/ | 13 | 0.00957 | 0.000251 | 0.14838 | 0.47429 | 8065.301 |
| GOMF_CL/ | 7 | 0.012379 | 0.000238 | 0.1935 | 0.47449 | 8068.702 |
| WP_CIRCA | 196 | 0.002297 | 0.000233 | 0.036013 | 0.47458 | 8070.233 |
| WAKABAY/ | 29 | 0.006239 | 0.000244 | 0.098152 | 0.47466 | 8071.593 |
| GOBP_RES | 184 | 0.002322 | 0.000228 | 0.036615 | 0.47472 | 8072.614 |
| GOMF_HIG | 10 | 0.011407 | 0.000262 | 0.18052 | 0.47481 | 8074.144 |
| GOMF_LIG | 26 | 0.006102 | 0.000226 | 0.096825 | 0.47488 | 8075.334 |
| REACTOMI | 35 | 0.005075 | 0.000218 | 0.080587 | 0.47489 | 8075.504 |
| GOBP_POS | 339 | 0.001797 | 0.000239 | 0.028549 | 0.47491 | 8075.845 |
| GOBP_CER | 8 | 0.013147 | 0.000271 | 0.20918 | 0.47494 | 8076.355 |

| | | | | | | |
|---|---|---|---|---|---|---|
| GOBP_HET | 434 | 0.001444 | 0.000216 | 0.023251 | 0.47525 | 8081.626 |
| GOMF_TRN | 11 | 0.008565 | 0.000207 | 0.13804 | 0.47526 | 8081.796 |
| REACTOME | 25 | 0.007289 | 0.000265 | 0.1177 | 0.47531 | 8082.647 |
| REACTOME | 51 | 0.004344 | 0.000226 | 0.070191 | 0.47532 | 8082.817 |
| KEGG_HIST | 27 | 0.006618 | 0.00025 | 0.10759 | 0.47548 | 8085.537 |
| REACTOME | 55 | 0.003863 | 0.000208 | 0.062878 | 0.4755 | 8085.878 |
| KEGG_HEM | 77 | 0.003713 | 0.000237 | 0.060481 | 0.47552 | 8086.218 |
| GOBP_HIST | 7 | 0.010437 | 0.000201 | 0.17069 | 0.47562 | 8087.918 |
| WP_LIPID_ | 20 | 0.006611 | 0.000215 | 0.10818 | 0.47563 | 8088.088 |
| GOBP_REG | 272 | 0.001896 | 0.000226 | 0.031265 | 0.47582 | 8091.319 |
| GOBP_REG | 125 | 0.002692 | 0.000218 | 0.044449 | 0.47585 | 8091.829 |
| GOBP_NUC | 9 | 0.010476 | 0.000229 | 0.17303 | 0.47586 | 8091.999 |
| REACTOME | 19 | 0.006023 | 0.000191 | 0.10041 | 0.47608 | 8095.74 |
| REACTOME | 13 | 0.006965 | 0.000183 | 0.11615 | 0.47609 | 8095.91 |
| SHIN_B_CE | 17 | 0.006908 | 0.000207 | 0.11609 | 0.47628 | 8099.141 |
| GOBP_RES | 29 | 0.005682 | 0.000223 | 0.095731 | 0.47633 | 8099.992 |
| GOBP_INTE | 22 | 0.007129 | 0.000243 | 0.12026 | 0.47636 | 8100.502 |
| GOBP_POS | 51 | 0.004581 | 0.000238 | 0.077519 | 0.47644 | 8101.862 |
| GOBP_NUC | 10 | 0.009413 | 0.000217 | 0.15973 | 0.47651 | 8103.053 |
| GOBP_NEG | 10 | 0.007885 | 0.000181 | 0.1342 | 0.47657 | 8104.073 |
| GOCC_PLA | 20 | 0.006433 | 0.000209 | 0.1095 | 0.47658 | 8104.243 |
| REN_BOUN | 57 | 0.003434 | 0.000188 | 0.05847 | 0.47658 | 8104.243 |
| YORDY_RE | 22 | 0.005979 | 0.000204 | 0.10338 | 0.47694 | 8110.365 |
| GOBP_RES | 42 | 0.004422 | 0.000208 | 0.076756 | 0.47703 | 8111.895 |
| PHONG_TN | 325 | 0.001679 | 0.000218 | 0.029651 | 0.47742 | 8118.527 |
| SU_TESTIS | 72 | 0.003059 | 0.000189 | 0.054194 | 0.47749 | 8119.717 |
| ONO_FOXP | 40 | 0.004673 | 0.000215 | 0.083065 | 0.47757 | 8121.078 |
| GOBP_ARC | 16 | 0.007327 | 0.000213 | 0.1307 | 0.47765 | 8122.438 |
| GOMF_SIG | 52 | 0.003592 | 0.000188 | 0.064079 | 0.47765 | 8122.438 |
| GOCC_CYT | 184 | 0.002076 | 0.000204 | 0.037282 | 0.47779 | 8124.819 |
| BIOCARTA_ | 6 | 0.010724 | 0.000191 | 0.19312 | 0.47786 | 8126.009 |
| KOINUMA_ | 15 | 0.007593 | 0.000214 | 0.13716 | 0.47793 | 8127.2 |
| PID_EPHA_ | 33 | 0.004869 | 0.000203 | 0.088183 | 0.47799 | 8128.22 |
| MAHADEVA | 19 | 0.007241 | 0.00023 | 0.13147 | 0.47804 | 8129.07 |
| TAKAO_RES | 76 | 0.003032 | 0.000192 | 0.05511 | 0.47806 | 8129.41 |
| DER_IFN_G | 64 | 0.003225 | 0.000187 | 0.05882 | 0.47814 | 8130.771 |
| GOBP_TELI | 13 | 0.009371 | 0.000246 | 0.17095 | 0.47814 | 8130.771 |
| GOBP_SOM | 4 | 0.016586 | 0.000241 | 0.30388 | 0.47824 | 8132.471 |
| GOBP_POS | 9 | 0.00994 | 0.000217 | 0.18322 | 0.47837 | 8134.682 |
| WP_BLOOD | 20 | 0.006436 | 0.000209 | 0.11905 | 0.47844 | 8135.872 |
| HOELZEL_N | 105 | 0.002985 | 0.000222 | 0.055337 | 0.47849 | 8136.722 |
| GOBP_REG | 29 | 0.00478 | 0.000187 | 0.088783 | 0.47853 | 8137.403 |
| WP_ALTER | 7 | 0.008269 | 0.000159 | 0.15386 | 0.47857 | 8138.083 |
| WANG_TN | 24 | 0.005349 | 0.000191 | 0.099846 | 0.47864 | 8139.273 |
| GOBP_REG | 750 | 0.001008 | 0.000197 | 0.018826 | 0.47865 | 8139.443 |

| | | | | | | |
|---|---|---|---|---|---|---|
| GOBP_REG | 4 | 0.012351 | 0.00018 | 0.2312 | 0.4787 | 8140.294 |
| GOCC_MO | 235 | 0.001711 | 0.00019 | 0.032021 | 0.4787 | 8140.294 |
| REACTOME | 9 | 0.009664 | 0.000211 | 0.18159 | 0.47878 | 8141.654 |
| GOBP_NEG | 145 | 0.002215 | 0.000193 | 0.041727 | 0.47883 | 8142.504 |
| GOBP_POS | 8 | 0.009047 | 0.000186 | 0.17067 | 0.47886 | 8143.014 |
| GOBP_REG | 8 | 0.009047 | 0.000186 | 0.17067 | 0.47886 | 8143.014 |
| GOMF_RET | 4 | 0.013759 | 0.0002 | 0.26036 | 0.47893 | 8144.205 |
| NADERI_BI | 17 | 0.006849 | 0.000205 | 0.12964 | 0.47893 | 8144.205 |
| GOBP_POS | 8 | 0.009031 | 0.000186 | 0.17168 | 0.47902 | 8145.735 |
| REACTOME | 26 | 0.004927 | 0.000183 | 0.093662 | 0.47903 | 8145.905 |
| GOBP_NEG | 9 | 0.008637 | 0.000189 | 0.16454 | 0.47907 | 8146.585 |
| BARRIER_C | 10 | 0.008486 | 0.000195 | 0.1625 | 0.47918 | 8148.456 |
| GOBP_REG | 1667 | 0.000686 | 0.000195 | 0.013146 | 0.47918 | 8148.456 |
| GOBP_POS | 12 | 0.00775 | 0.000195 | 0.14864 | 0.47921 | 8148.966 |
| GOMF_SPH | 7 | 0.012049 | 0.000232 | 0.23121 | 0.47922 | 8149.136 |
| GOBP_EST/ | 49 | 0.003631 | 0.000185 | 0.069748 | 0.47924 | 8149.476 |
| GOMF_STE | 23 | 0.005866 | 0.000205 | 0.11268 | 0.47924 | 8149.476 |
| IVANOVA_ | 128 | 0.002145 | 0.000176 | 0.041372 | 0.47932 | 8150.837 |
| REACTOME | 51 | 0.003531 | 0.000183 | 0.06821 | 0.47936 | 8151.517 |
| GOCC_BET. | 11 | 0.007486 | 0.000181 | 0.14514 | 0.47943 | 8152.707 |
| GOBP_REG | 8 | 0.009763 | 0.000201 | 0.19058 | 0.47957 | 8155.088 |
| REACTOME | 9 | 0.008427 | 0.000184 | 0.16461 | 0.47959 | 8155.428 |
| MCBRYAN_ | 182 | 0.001906 | 0.000186 | 0.037283 | 0.47962 | 8155.938 |
| GOBP_RES | 15 | 0.006688 | 0.000188 | 0.13178 | 0.47976 | 8158.319 |
| LEE_LIVER_ | 169 | 0.001973 | 0.000186 | 0.038918 | 0.47979 | 8158.829 |
| GOBP_AMI | 10 | 0.008778 | 0.000202 | 0.17349 | 0.47982 | 8159.339 |
| REACTOME | 8 | 0.010959 | 0.000226 | 0.21743 | 0.4799 | 8160.7 |
| GOBP_BOD | 86 | 0.002797 | 0.000188 | 0.055583 | 0.47993 | 8161.21 |
| GOBP_HEP | 16 | 0.006604 | 0.000192 | 0.13157 | 0.47998 | 8162.06 |
| GOBP_ACU | 46 | 0.003916 | 0.000193 | 0.078082 | 0.48 | 8162.4 |
| GOBP_EXP | 62 | 0.003539 | 0.000202 | 0.070706 | 0.48004 | 8163.08 |
| GOCC_B_W | 7 | 0.012241 | 0.000236 | 0.24481 | 0.48006 | 8163.42 |
| GOBP_REG | 21 | 0.006032 | 0.000201 | 0.12069 | 0.48007 | 8163.59 |
| REACTOME | 91 | 0.002323 | 0.000161 | 0.046486 | 0.48007 | 8163.59 |
| WP_APOE_ | 7 | 0.008108 | 0.000156 | 0.16303 | 0.48017 | 8165.291 |
| GOMF_DO | 6 | 0.010379 | 0.000185 | 0.2092 | 0.48022 | 8166.141 |
| GOBP_SPH | 20 | 0.005542 | 0.00018 | 0.11227 | 0.48032 | 8167.842 |
| WP_ACETY | 6 | 0.009185 | 0.000164 | 0.18638 | 0.48035 | 8168.352 |
| GOBP_MAI | 7 | 0.01138 | 0.000219 | 0.23125 | 0.48038 | 8168.862 |
| GOBP_FOR | 13 | 0.005835 | 0.000153 | 0.11984 | 0.48059 | 8172.433 |
| LEE_CALOF | 66 | 0.002929 | 0.000173 | 0.060371 | 0.48065 | 8173.453 |
| GOBP_CELI | 193 | 0.001754 | 0.000176 | 0.036473 | 0.48082 | 8176.344 |
| PID_HIF1_T | 65 | 0.00322 | 0.000189 | 0.067043 | 0.48085 | 8176.854 |
| GOBP_REG | 8 | 0.006724 | 0.000138 | 0.14037 | 0.4809 | 8177.705 |
| GOCC_BOF | 8 | 0.006724 | 0.000138 | 0.14037 | 0.4809 | 8177.705 |

| | | | | | | |
|---|---|---|---|---|---|---|
| GOBP_RES | 136 | 0.002014 | 0.00017 | 0.04256 | 0.48113 | 8181.616 |
| GOBP_PRC | 100 | 0.002444 | 0.000177 | 0.051939 | 0.48124 | 8183.486 |
| GOBP_POS | 6 | 0.011264 | 0.000201 | 0.23978 | 0.48127 | 8183.996 |
| GOBP_REG | 159 | 0.001902 | 0.000174 | 0.040495 | 0.48127 | 8183.996 |
| GOBP_REG | 8 | 0.006702 | 0.000138 | 0.14292 | 0.4813 | 8184.507 |
| WP_KISSPE | 38 | 0.003794 | 0.00017 | 0.081588 | 0.48145 | 8187.057 |
| BIOCARTA_ | 9 | 0.008687 | 0.00019 | 0.18767 | 0.48154 | 8188.588 |
| GOBP_G_P | 14 | 0.006047 | 0.000165 | 0.13104 | 0.4816 | 8189.608 |
| GOMF_4_C | 6 | 0.009508 | 0.000169 | 0.20647 | 0.48164 | 8190.288 |
| GOBP_LUT | 7 | 0.009087 | 0.000175 | 0.19831 | 0.48173 | 8191.819 |
| REACTOME | 132 | 0.002021 | 0.000168 | 0.044155 | 0.48175 | 8192.159 |
| GOBP_REG | 32 | 0.003853 | 0.000158 | 0.084547 | 0.48183 | 8193.519 |
| GOBP_NEG | 7 | 0.008412 | 0.000162 | 0.18495 | 0.48186 | 8194.029 |
| GOMF_URI | 7 | 0.008023 | 0.000154 | 0.17655 | 0.48188 | 8194.369 |
| JINESH_BLI | 356 | 0.001236 | 0.000168 | 0.02726 | 0.48192 | 8195.05 |
| GOBP_RES | 6 | 0.010145 | 0.000181 | 0.22447 | 0.48198 | 8196.07 |
| GOCC_MIB | 11 | 0.006768 | 0.000163 | 0.14997 | 0.482 | 8196.41 |
| GOCC_MIC | 148 | 0.001778 | 0.000157 | 0.039776 | 0.48218 | 8199.471 |
| WP_REGUI | 73 | 0.002775 | 0.000172 | 0.062126 | 0.48219 | 8199.641 |
| GOCC_RIB( | 3 | 0.01431 | 0.00018 | 0.32155 | 0.48225 | 8200.661 |
| GOMF_RIB | 3 | 0.01431 | 0.00018 | 0.32155 | 0.48225 | 8200.661 |
| REACTOME | 29 | 0.004043 | 0.000158 | 0.090833 | 0.48225 | 8200.661 |
| WANG_RES | 29 | 0.004381 | 0.000172 | 0.098543 | 0.48227 | 8201.001 |
| CHEOK_RE: | 11 | 0.005171 | 0.000125 | 0.11657 | 0.48231 | 8201.682 |
| GOBP_CELI | 160 | 0.001689 | 0.000155 | 0.038193 | 0.48236 | 8202.532 |
| GOBP_PUR | 6 | 0.008717 | 0.000155 | 0.19757 | 0.4824 | 8203.212 |
| GROSS_HY | 33 | 0.004299 | 0.00018 | 0.097746 | 0.48246 | 8204.232 |
| OHASHI_AU | 5 | 0.008171 | 0.000133 | 0.18641 | 0.48252 | 8205.253 |
| HOWLIN_C | 37 | 0.004078 | 0.00018 | 0.093144 | 0.48254 | 8205.593 |
| GOBP_GRC | 5 | 0.011839 | 0.000193 | 0.27238 | 0.48267 | 8207.803 |
| WP_FOCAL | 288 | 0.001282 | 0.000157 | 0.029512 | 0.48268 | 8207.973 |
| GOMF_DIC | 85 | 0.002176 | 0.000146 | 0.050462 | 0.4828 | 8210.014 |
| GOCC_UNI | 6 | 0.009039 | 0.000161 | 0.21057 | 0.48288 | 8211.374 |
| GOBP_POS | 106 | 0.002273 | 0.00017 | 0.053098 | 0.48293 | 8212.225 |
| GOBP_GLY | 13 | 0.005486 | 0.000144 | 0.12826 | 0.48294 | 8212.395 |
| GOBP_KINI | 22 | 0.004185 | 0.000143 | 0.097982 | 0.48297 | 8212.905 |
| CHEN_PDG | 18 | 0.005332 | 0.000165 | 0.12516 | 0.48301 | 8213.585 |
| GOBP_EMI | 16 | 0.005574 | 0.000162 | 0.13272 | 0.48325 | 8217.666 |
| REACTOME | 12 | 0.006287 | 0.000158 | 0.15001 | 0.48328 | 8218.176 |
| GOMF_ADI | 6 | 0.00889 | 0.000158 | 0.21515 | 0.48352 | 8222.258 |
| GOMF_INS | 8 | 0.008631 | 0.000178 | 0.20951 | 0.48357 | 8223.108 |
| GOBP_OST | 17 | 0.004759 | 0.000143 | 0.11584 | 0.48361 | 8223.788 |
| GOBP_SER( | 19 | 0.0052 | 0.000165 | 0.12693 | 0.48366 | 8224.638 |
| REACTOME | 5 | 0.008982 | 0.000146 | 0.22239 | 0.48389 | 8228.549 |
| WEIGEL_O: | 58 | 0.002846 | 0.000158 | 0.070903 | 0.48399 | 8230.25 |

| | | | | | | |
|---|---|---|---|---|---|---|
| GOBP_POS | 5 | 0.007913 | 0.000129 | 0.19865 | 0.48411 | 8232.291 |
| GOBP_D_A | 5 | 0.011802 | 0.000192 | 0.29657 | 0.48413 | 8232.631 |
| GOMF_ACE | 10 | 0.006425 | 0.000148 | 0.16331 | 0.48431 | 8235.692 |
| GOMF_NAI | 7 | 0.007358 | 0.000142 | 0.18747 | 0.48435 | 8236.372 |
| GOBP_CAL | 173 | 0.001512 | 0.000144 | 0.038604 | 0.48438 | 8236.882 |
| GOBP_CELI | 239 | 0.001231 | 0.000138 | 0.031667 | 0.48449 | 8238.752 |
| GOBP_POL | 98 | 0.001922 | 0.000138 | 0.049764 | 0.48459 | 8240.453 |
| REACTOME | 52 | 0.002603 | 0.000136 | 0.067643 | 0.48465 | 8241.473 |
| BIOCARTA_ | 20 | 0.004038 | 0.000131 | 0.10502 | 0.48466 | 8241.643 |
| REACTOME | 147 | 0.001626 | 0.000143 | 0.042591 | 0.48478 | 8243.684 |
| BIOCARTA_ | 7 | 0.007074 | 0.000136 | 0.18741 | 0.48494 | 8246.405 |
| GOCC_CAL | 5 | 0.008736 | 0.000142 | 0.23149 | 0.48495 | 8246.575 |
| REACTOME | 17 | 0.004529 | 0.000136 | 0.12075 | 0.48504 | 8248.105 |
| GOBP_REG | 8 | 0.005689 | 0.000117 | 0.1526 | 0.48513 | 8249.636 |
| GOBP_DNA | 62 | 0.002464 | 0.000141 | 0.06661 | 0.48525 | 8251.676 |
| GOCC_GEN | 9 | 0.005968 | 0.00013 | 0.16183 | 0.48529 | 8252.356 |
| GOBP_NEG | 22 | 0.003926 | 0.000134 | 0.10651 | 0.4853 | 8252.527 |
| DESERT_PE | 149 | 0.001486 | 0.000131 | 0.040361 | 0.48532 | 8252.867 |
| BHATI_G2N | 110 | 0.001766 | 0.000134 | 0.048224 | 0.48539 | 8254.057 |
| WP_2Q112 | 36 | 0.004005 | 0.000175 | 0.11042 | 0.48553 | 8256.438 |
| GOBP_POS | 5 | 0.007897 | 0.000128 | 0.21782 | 0.48554 | 8256.608 |
| GOBP_MAT | 19 | 0.004178 | 0.000132 | 0.11529 | 0.48555 | 8256.778 |
| GOBP_ESTA | 59 | 0.002394 | 0.000134 | 0.066267 | 0.48559 | 8257.458 |
| GOBP_MEN | 11 | 0.005698 | 0.000138 | 0.16067 | 0.48585 | 8261.879 |
| CEBALLOS_ | 29 | 0.003257 | 0.000128 | 0.09213 | 0.4859 | 8262.73 |
| GOBP_CELI | 19 | 0.004354 | 0.000138 | 0.12319 | 0.4859 | 8262.73 |
| GOBP_NEG | 3 | 0.014277 | 0.00018 | 0.40411 | 0.48591 | 8262.9 |
| GOBP_INTI | 51 | 0.002322 | 0.000121 | 0.066294 | 0.48603 | 8264.94 |
| PARK_OSTI | 6 | 0.007598 | 0.000135 | 0.2169 | 0.48603 | 8264.94 |
| GOBP_REG | 23 | 0.003311 | 0.000116 | 0.095348 | 0.48615 | 8266.981 |
| REACTOME | 36 | 0.002903 | 0.000127 | 0.083878 | 0.48619 | 8267.661 |
| GOBP_NUC | 6 | 0.006126 | 0.000109 | 0.17781 | 0.48626 | 8268.851 |
| GOBP_REG | 27 | 0.003301 | 0.000125 | 0.096306 | 0.48633 | 8270.042 |
| GOCC_NM | 9 | 0.005525 | 0.000121 | 0.16129 | 0.48634 | 8270.212 |
| GOMF_PRE | 33 | 0.002774 | 0.000116 | 0.081774 | 0.48647 | 8272.422 |
| GOBP_TRN | 16 | 0.004262 | 0.000124 | 0.12578 | 0.48648 | 8272.592 |
| GOBP_REG | 16 | 0.00476 | 0.000139 | 0.14185 | 0.48662 | 8274.973 |
| GOBP_TOL | 16 | 0.004313 | 0.000126 | 0.1296 | 0.48673 | 8276.844 |
| GOCC_BLO | 104 | 0.001713 | 0.000127 | 0.051531 | 0.48674 | 8277.014 |
| RADMACHI | 74 | 0.001823 | 0.000114 | 0.054877 | 0.48675 | 8277.184 |
| GOBP_TOL | 4 | 0.009012 | 0.000131 | 0.27172 | 0.48677 | 8277.524 |
| GOCC_CON | 10 | 0.004426 | 0.000102 | 0.13347 | 0.48677 | 8277.524 |
| GOBP_POS | 14 | 0.004524 | 0.000123 | 0.13696 | 0.48682 | 8278.374 |
| GOBP_SNR | 5 | 0.007744 | 0.000126 | 0.23432 | 0.48682 | 8278.374 |
| HOLLEMAN | 19 | 0.003803 | 0.000121 | 0.11512 | 0.48682 | 8278.374 |

| | | | | | | |
|---|---|---|---|---|---|---|
| GOBP_MAI | 722 | 0.000635 | 0.000122 | 0.019305 | 0.48688 | 8279.394 |
| REACTOME | 34 | 0.00271 | 0.000115 | 0.082529 | 0.4869 | 8279.735 |
| GOBP_BON | 32 | 0.003013 | 0.000124 | 0.092844 | 0.48705 | 8282.285 |
| GOBP_MYE | 9 | 0.005324 | 0.000116 | 0.16493 | 0.48712 | 8283.476 |
| HOFFMANI | 36 | 0.002567 | 0.000112 | 0.079917 | 0.48719 | 8284.666 |
| GOCC_DIH | 9 | 0.004617 | 0.000101 | 0.1439 | 0.4872 | 8284.836 |
| GOBP_RES | 34 | 0.002759 | 0.000117 | 0.086291 | 0.48725 | 8285.686 |
| MCGOWAN | 19 | 0.003643 | 0.000116 | 0.11432 | 0.48729 | 8286.366 |
| GOBP_SYN | 25 | 0.003551 | 0.000129 | 0.11177 | 0.48733 | 8287.047 |
| WP_ELECTI | 85 | 0.001489 | 9.97E-05 | 0.046841 | 0.48733 | 8287.047 |
| LIM_MAMI | 104 | 0.001564 | 0.000116 | 0.049638 | 0.48743 | 8288.747 |
| GOBP_POD | 18 | 0.003966 | 0.000122 | 0.12644 | 0.48749 | 8289.767 |
| REACTOME | 14 | 0.004399 | 0.00012 | 0.14175 | 0.48762 | 8291.978 |
| BIOCARTA_ | 9 | 0.005763 | 0.000126 | 0.18749 | 0.48774 | 8294.019 |
| GRUETZMA | 335 | 0.000856 | 0.000113 | 0.027902 | 0.48777 | 8294.529 |
| GOBP_MEI | 8 | 0.006239 | 0.000128 | 0.20497 | 0.48786 | 8296.059 |
| GOBP_CELI | 25 | 0.003212 | 0.000117 | 0.10567 | 0.48788 | 8296.399 |
| GOBP_RISC | 9 | 0.00468 | 0.000102 | 0.15526 | 0.48798 | 8298.1 |
| GOMF_TOI | 5 | 0.008227 | 0.000134 | 0.27306 | 0.48798 | 8298.1 |
| FONTAINE_ | 34 | 0.002735 | 0.000116 | 0.09087 | 0.48799 | 8298.27 |
| GOBP_AMI | 7 | 0.005541 | 0.000107 | 0.18397 | 0.48799 | 8298.27 |
| DASU_IL6_ | 15 | 0.004385 | 0.000124 | 0.1458 | 0.488 | 8298.44 |
| GOCC_HOI | 9 | 0.004364 | 9.53E-05 | 0.14565 | 0.48805 | 8299.29 |
| GOBP_POS | 34 | 0.00248 | 0.000105 | 0.083021 | 0.48809 | 8299.97 |
| GOBP_INFI | 743 | 0.000574 | 0.000112 | 0.019259 | 0.48811 | 8300.311 |
| HECKER_IF | 90 | 0.001677 | 0.000116 | 0.056433 | 0.48815 | 8300.991 |
| GOBP_POS | 7 | 0.005539 | 0.000107 | 0.18768 | 0.48823 | 8302.351 |
| GOBP_PRC | 16 | 0.003215 | 9.35E-05 | 0.10968 | 0.48831 | 8303.712 |
| WP_LDLRA | 7 | 0.006764 | 0.00013 | 0.23074 | 0.48831 | 8303.712 |
| GOBP_REG | 10 | 0.004582 | 0.000105 | 0.15662 | 0.48833 | 8304.052 |
| GOCC_CUL | 35 | 0.002673 | 0.000115 | 0.09264 | 0.48849 | 8306.772 |
| GOBP_KET | 6 | 0.005438 | 9.69E-05 | 0.18879 | 0.48851 | 8307.113 |
| GOBP_NEG | 35 | 0.002343 | 0.000101 | 0.08153 | 0.48854 | 8307.623 |
| REACTOME | 32 | 0.002422 | 9.96E-05 | 0.085359 | 0.48868 | 8310.003 |
| GOBP_OVU | 52 | 0.002125 | 0.000111 | 0.075165 | 0.48873 | 8310.854 |
| POOLA_IN | 254 | 0.000905 | 0.000104 | 0.032144 | 0.48877 | 8311.534 |
| REACTOME | 19 | 0.0031 | 9.83E-05 | 0.11035 | 0.48879 | 8311.874 |
| KANG_IMN | 83 | 0.001536 | 0.000102 | 0.054715 | 0.4888 | 8312.044 |
| GOBP_NEG | 23 | 0.002952 | 0.000103 | 0.10544 | 0.48883 | 8312.554 |
| BIOCARTA_ | 16 | 0.003694 | 0.000108 | 0.13332 | 0.48895 | 8314.595 |
| WP_CELLS_ | 16 | 0.003694 | 0.000108 | 0.13332 | 0.48895 | 8314.595 |
| JOHANSSO | 54 | 0.001726 | 9.22E-05 | 0.062466 | 0.48898 | 8315.105 |
| WP_GPCRS | 14 | 0.004333 | 0.000118 | 0.15844 | 0.48909 | 8316.975 |
| GOBP_CON | 5 | 0.007071 | 0.000115 | 0.2588 | 0.4891 | 8317.146 |
| HUMMERI | 83 | 0.001572 | 0.000104 | 0.057619 | 0.48912 | 8317.486 |

| | | | | | | |
|---|---|---|---|---|---|---|
| REACTOME | 11 | 0.003948 | 9.53E-05 | 0.14587 | 0.4892 | 8318.846 |
| REACTOME | 118 | 0.001211 | 9.54E-05 | 0.044919 | 0.48925 | 8319.696 |
| VERRECCHI | 20 | 0.002922 | 9.50E-05 | 0.10879 | 0.48929 | 8320.376 |
| WANG_NF | 24 | 0.002813 | 0.0001 | 0.1072 | 0.48954 | 8324.628 |
| LEE_METAS | 43 | 0.002031 | 9.68E-05 | 0.077608 | 0.48956 | 8324.968 |
| GOCC_COF | 15 | 0.003288 | 9.27E-05 | 0.12736 | 0.4897 | 8327.349 |
| DESERT_ST | 234 | 0.000868 | 9.61E-05 | 0.033658 | 0.48971 | 8327.519 |
| GOBP_HEP | 18 | 0.002731 | 8.43E-05 | 0.10684 | 0.4898 | 8329.049 |
| GOMF_T_C | 7 | 0.004948 | 9.53E-05 | 0.19491 | 0.48987 | 8330.239 |
| CLASPER_L | 33 | 0.002389 | 9.98E-05 | 0.094712 | 0.48994 | 8331.43 |
| GOBP_INTI | 9 | 0.003727 | 8.14E-05 | 0.14858 | 0.48999 | 8332.28 |
| GOCC_NEU | 357 | 0.000688 | 9.38E-05 | 0.027445 | 0.49 | 8332.45 |
| REACTOME | 5 | 0.005717 | 9.30E-05 | 0.22839 | 0.49002 | 8332.79 |
| GOMF_HIS | 8 | 0.004378 | 9.01E-05 | 0.17515 | 0.49003 | 8332.96 |
| REACTOME | 53 | 0.00171 | 9.05E-05 | 0.068553 | 0.49005 | 8333.3 |
| GOMF_ACI | 11 | 0.003026 | 7.30E-05 | 0.12169 | 0.49008 | 8333.81 |
| CHESLER_E | 69 | 0.001416 | 8.55E-05 | 0.057006 | 0.49009 | 8333.98 |
| GOBP_HIST | 7 | 0.004541 | 8.74E-05 | 0.18313 | 0.49011 | 8334.321 |
| GOBP_L_AS | 6 | 0.005244 | 9.35E-05 | 0.21445 | 0.49025 | 8336.701 |
| REACTOME | 5 | 0.004612 | 7.50E-05 | 0.18862 | 0.49025 | 8336.701 |
| REACTOME | 33 | 0.002189 | 9.14E-05 | 0.089669 | 0.49026 | 8336.871 |
| GOCC_UNC | 8 | 0.004357 | 8.97E-05 | 0.1792 | 0.4903 | 8337.552 |
| LIANG_HEN | 16 | 0.003069 | 8.93E-05 | 0.1262 | 0.4903 | 8337.552 |
| SATO_SILEI | 367 | 0.00064 | 8.83E-05 | 0.026387 | 0.49033 | 8338.062 |
| GOBP_MAC | 6 | 0.004577 | 8.16E-05 | 0.19191 | 0.49049 | 8340.782 |
| ZHU_SKIL_ | 9 | 0.00414 | 9.04E-05 | 0.17531 | 0.49058 | 8342.313 |
| REACTOME | 29 | 0.002504 | 9.81E-05 | 0.10632 | 0.49061 | 8342.823 |
| GOBP_MIT | 159 | 0.000823 | 7.52E-05 | 0.035607 | 0.49078 | 8345.714 |
| REACTOME | 22 | 0.002434 | 8.30E-05 | 0.10528 | 0.49078 | 8345.714 |
| WP_MAJOI | 15 | 0.00345 | 9.72E-05 | 0.14921 | 0.49078 | 8345.714 |
| GOBP_INO | 12 | 0.003524 | 8.88E-05 | 0.15676 | 0.49103 | 8349.965 |
| GOBP_PYR | 14 | 0.003137 | 8.54E-05 | 0.13977 | 0.49105 | 8350.305 |
| GOBP_VITA | 101 | 0.001096 | 8.00E-05 | 0.049121 | 0.4911 | 8351.156 |
| GOBP_APC | 5 | 0.005432 | 8.84E-05 | 0.24558 | 0.49118 | 8352.516 |
| REACTOME | 10 | 0.003339 | 7.68E-05 | 0.1538 | 0.49134 | 8355.237 |
| GOBP_REG | 13 | 0.003337 | 8.75E-05 | 0.15432 | 0.49138 | 8355.917 |
| WP_METAI | 39 | 0.001599 | 7.26E-05 | 0.0741 | 0.4914 | 8356.257 |
| GOBP_ATT | 6 | 0.004165 | 7.42E-05 | 0.19442 | 0.49146 | 8357.277 |
| HOLLMANI | 242 | 0.000679 | 7.64E-05 | 0.032163 | 0.49157 | 8359.148 |
| GOMF_INT | 25 | 0.002115 | 7.69E-05 | 0.10054 | 0.49161 | 8359.828 |
| LU_EZH2_T | 258 | 0.000605 | 7.03E-05 | 0.029081 | 0.49169 | 8361.188 |
| GOBP_POS | 31 | 0.001811 | 7.33E-05 | 0.087286 | 0.49172 | 8361.699 |
| REACTOME | 56 | 0.001371 | 7.46E-05 | 0.066206 | 0.49174 | 8362.039 |
| GOMF_CCI | 31 | 0.002294 | 9.29E-05 | 0.11108 | 0.49176 | 8362.379 |
| GOBP_RES | 7 | 0.004553 | 8.76E-05 | 0.2207 | 0.49177 | 8362.549 |

| | | | | | | |
|---|---|---|---|---|---|---|
| GOBP_MYE | 19 | 0.002485 | 7.88E-05 | 0.12062 | 0.49178 | 8362.719 |
| GOBP_PRC | 8 | 0.004329 | 8.91E-05 | 0.21192 | 0.49185 | 8363.909 |
| GOMF_MR | 5 | 0.005688 | 9.26E-05 | 0.28495 | 0.49204 | 8367.14 |
| GOBP_TRA | 8 | 0.003593 | 7.39E-05 | 0.18076 | 0.49207 | 8367.65 |
| GOBP_POS | 11 | 0.003439 | 8.30E-05 | 0.17363 | 0.4921 | 8368.161 |
| REACTOME | 51 | 0.001473 | 7.64E-05 | 0.076335 | 0.4923 | 8371.562 |
| GOBP_REG | 7 | 0.003261 | 6.28E-05 | 0.17011 | 0.49235 | 8372.412 |
| GOBP_POS | 97 | 0.000983 | 7.03E-05 | 0.051457 | 0.49238 | 8372.922 |
| GOBP_FOR | 23 | 0.002335 | 8.14E-05 | 0.12253 | 0.4924 | 8373.262 |
| GOBP_NEG | 39 | 0.001378 | 6.26E-05 | 0.074106 | 0.49258 | 8376.323 |
| REACTOME | 24 | 0.001855 | 6.61E-05 | 0.10016 | 0.49261 | 8376.833 |
| GOBP_CAR | 8 | 0.003342 | 6.88E-05 | 0.18398 | 0.49275 | 8379.214 |
| ZAMORA_I | 81 | 0.001025 | 6.70E-05 | 0.056425 | 0.49276 | 8379.384 |
| GOMF_3_5 | 16 | 0.002235 | 6.50E-05 | 0.12382 | 0.4928 | 8380.064 |
| GOBP_SPEI | 49 | 0.001389 | 7.07E-05 | 0.077208 | 0.49283 | 8380.574 |
| RODRIGUE | 31 | 0.001707 | 6.91E-05 | 0.094916 | 0.49283 | 8380.574 |
| GOBP_MUS | 19 | 0.00223 | 7.07E-05 | 0.12638 | 0.49296 | 8382.785 |
| REACTOME | 6 | 0.003644 | 6.50E-05 | 0.21284 | 0.49317 | 8386.356 |
| GOBP_LYM | 249 | 0.000567 | 6.47E-05 | 0.033264 | 0.4932 | 8386.866 |
| WP_IL18_S | 255 | 0.000536 | 6.19E-05 | 0.031608 | 0.49324 | 8387.546 |
| GOBP_UDF | 14 | 0.002644 | 7.20E-05 | 0.15769 | 0.49331 | 8388.737 |
| GOBP_NEG | 19 | 0.002063 | 6.54E-05 | 0.12327 | 0.49332 | 8388.907 |
| GOBP_TRA | 10 | 0.002699 | 6.21E-05 | 0.16287 | 0.49339 | 8390.097 |
| THEODORC | 28 | 0.001735 | 6.68E-05 | 0.10495 | 0.49341 | 8390.437 |
| GOMF_3_C | 5 | 0.004186 | 6.81E-05 | 0.25925 | 0.49356 | 8392.988 |
| BOSCO_TH | 111 | 0.000824 | 6.30E-05 | 0.051249 | 0.49359 | 8393.498 |
| GOCC_BBA | 10 | 0.002632 | 6.06E-05 | 0.16388 | 0.49359 | 8393.498 |
| GOBP_NEG | 14 | 0.002064 | 5.62E-05 | 0.13045 | 0.49369 | 8395.198 |
| GOBP_RES | 122 | 0.0007 | 5.61E-05 | 0.04485 | 0.49377 | 8396.559 |
| GOMF_ADI | 6 | 0.002817 | 5.02E-05 | 0.18165 | 0.49381 | 8397.239 |
| GOBP_REG | 5 | 0.002498 | 4.06E-05 | 0.16713 | 0.49404 | 8401.15 |
| GOCC_IROI | 10 | 0.002477 | 5.70E-05 | 0.16906 | 0.49415 | 8403.021 |
| GOCC_MAI | 51 | 0.00098 | 5.09E-05 | 0.067482 | 0.4942 | 8403.871 |
| HESS_TARC | 68 | 0.000867 | 5.20E-05 | 0.059704 | 0.4942 | 8403.871 |
| GOBP_NEG | 43 | 0.001172 | 5.59E-05 | 0.081884 | 0.49429 | 8405.401 |
| GOBP_REG | 144 | 0.00055 | 4.79E-05 | 0.040432 | 0.49457 | 8410.163 |
| REACTOME | 19 | 0.001335 | 4.23E-05 | 0.10004 | 0.49468 | 8412.033 |
| GOBP_OTI | 8 | 0.002983 | 6.14E-05 | 0.22761 | 0.49477 | 8413.564 |
| GOBP_AZC | 12 | 0.002121 | 5.34E-05 | 0.16302 | 0.49481 | 8414.244 |
| GOBP_POS | 7 | 0.003044 | 5.86E-05 | 0.23609 | 0.49486 | 8415.094 |
| GOBP_POS | 6 | 0.002893 | 5.16E-05 | 0.22731 | 0.49492 | 8416.115 |
| NADLER_O | 48 | 0.000859 | 4.33E-05 | 0.067627 | 0.49493 | 8416.285 |
| GOBP_POS | 103 | 0.000643 | 4.74E-05 | 0.05231 | 0.4951 | 8419.176 |
| GOBP_NEG | 349 | 0.000331 | 4.46E-05 | 0.027124 | 0.49513 | 8419.686 |
| LEIN_ASTR | 43 | 0.000895 | 4.27E-05 | 0.075794 | 0.49529 | 8422.406 |

| | | | | | | |
|---|---|---|---|---|---|---|
| REACTOME | 19 | 0.001261 | 4.00E-05 | 0.10748 | 0.49532 | 8422.917 |
| REACTOME | 53 | 0.000765 | 4.05E-05 | 0.066238 | 0.49539 | 8424.107 |
| GOBP_REG | 18 | 0.001293 | 3.99E-05 | 0.11418 | 0.49548 | 8425.637 |
| BROWNE_I | 58 | 0.000712 | 3.94E-05 | 0.063381 | 0.49552 | 8426.318 |
| GOBP_REN | 26 | 0.001267 | 4.70E-05 | 0.11292 | 0.49552 | 8426.318 |
| CHEN_LUN | 24 | 0.001097 | 3.91E-05 | 0.098836 | 0.49557 | 8427.168 |
| WP_DDX1_ | 7 | 0.00188 | 3.62E-05 | 0.17415 | 0.49569 | 8429.208 |
| GOBP_CLE/ | 7 | 0.001929 | 3.71E-05 | 0.17937 | 0.49571 | 8429.549 |
| DORN_ADE | 41 | 0.000729 | 3.39E-05 | 0.06813 | 0.49573 | 8429.889 |
| GOBP_POS | 6 | 0.002269 | 4.04E-05 | 0.21751 | 0.49584 | 8431.759 |
| WP_CHOLE | 19 | 0.001258 | 3.99E-05 | 0.12232 | 0.4959 | 8432.78 |
| GOCC_ANC | 848 | 0.000179 | 3.71E-05 | 0.017671 | 0.49596 | 8433.8 |
| GOBP_GOL | 14 | 0.00117 | 3.19E-05 | 0.11711 | 0.49601 | 8434.65 |
| GOBP_OLIC | 20 | 0.001017 | 3.31E-05 | 0.10449 | 0.49612 | 8436.521 |
| GOBP_RES | 38 | 0.000771 | 3.45E-05 | 0.079429 | 0.49613 | 8436.691 |
| BIOCARTA_ | 16 | 0.001339 | 3.90E-05 | 0.14011 | 0.49619 | 8437.711 |
| GOBP_REG | 8 | 0.00146 | 3.01E-05 | 0.1546 | 0.49623 | 8438.391 |
| GOBP_POS | 11 | 0.001615 | 3.90E-05 | 0.17393 | 0.4963 | 8439.582 |
| CUI_GLUC( | 57 | 0.000637 | 3.50E-05 | 0.069976 | 0.49637 | 8440.772 |
| GOBP_NEG | 17 | 0.00114 | 3.42E-05 | 0.13086 | 0.49652 | 8443.323 |
| GOBP_RES | 13 | 0.000941 | 2.47E-05 | 0.11005 | 0.49659 | 8444.513 |
| BROWNE_I | 54 | 0.000649 | 3.47E-05 | 0.077359 | 0.49665 | 8445.533 |
| PID_CD8_T | 48 | 0.0006 | 3.02E-05 | 0.072024 | 0.49667 | 8445.873 |
| GOMF_FAT | 17 | 0.000931 | 2.79E-05 | 0.1134 | 0.49673 | 8446.894 |
| REACTOME | 11 | 0.001083 | 2.61E-05 | 0.13221 | 0.49673 | 8446.894 |
| MCCABE_H | 26 | 0.000803 | 2.98E-05 | 0.098356 | 0.49674 | 8447.064 |
| GOBP_NEG | 31 | 0.000626 | 2.53E-05 | 0.078091 | 0.4968 | 8448.084 |
| GOBP_DIO | 30 | 0.000752 | 2.99E-05 | 0.095207 | 0.49685 | 8448.934 |
| GOBP_REG | 105 | 0.000376 | 2.80E-05 | 0.048922 | 0.49693 | 8450.295 |
| GOMF_PH( | 9 | 0.00143 | 3.12E-05 | 0.1861 | 0.49693 | 8450.295 |
| REACTOME | 9 | 0.001111 | 2.43E-05 | 0.14561 | 0.49696 | 8450.805 |
| REACTOME | 5 | 0.001595 | 2.60E-05 | 0.20973 | 0.49697 | 8450.975 |
| RUTELLA_F | 393 | 0.000198 | 2.82E-05 | 0.02604 | 0.49697 | 8450.975 |
| JOHNSTON | 6 | 0.001578 | 2.81E-05 | 0.20934 | 0.49699 | 8451.315 |
| CHIANG_LI | 25 | 0.000791 | 2.88E-05 | 0.10797 | 0.49708 | 8452.845 |
| GOCC_POS | 339 | 0.000206 | 2.74E-05 | 0.028268 | 0.49709 | 8453.015 |
| SCHAVOLT_ | 15 | 0.000989 | 2.79E-05 | 0.13557 | 0.49709 | 8453.015 |
| PARK_TRET | 28 | 0.00075 | 2.88E-05 | 0.10405 | 0.49713 | 8453.696 |
| REACTOME | 21 | 0.000734 | 2.45E-05 | 0.10465 | 0.4972 | 8454.886 |
| REACTOME | 26 | 0.000644 | 2.39E-05 | 0.093303 | 0.49725 | 8455.736 |
| WP_INFLAI | 39 | 0.000628 | 2.85E-05 | 0.09231 | 0.49729 | 8456.416 |
| REACTOME | 9 | 0.001046 | 2.28E-05 | 0.15477 | 0.4973 | 8456.587 |
| GOBP_PRC | 5 | 0.001238 | 2.01E-05 | 0.18601 | 0.49734 | 8457.267 |
| GOBP_EST/ | 41 | 0.0005 | 2.33E-05 | 0.077531 | 0.49743 | 8458.797 |
| ONKEN_UV | 735 | 0.000122 | 2.35E-05 | 0.0193 | 0.49749 | 8459.817 |

| | | | | | | |
|---|---|---|---|---|---|---|
| GOBP_CYT | 112 | 0.000292 | 2.25E-05 | 0.046892 | 0.49751 | 8460.158 |
| REACTOME | 7 | 0.00114 | 2.19E-05 | 0.18708 | 0.49757 | 8461.178 |
| BOYLAN_M | 254 | 0.0002 | 2.30E-05 | 0.033313 | 0.49761 | 8461.858 |
| CREIGHTON | 21 | 0.000649 | 2.16E-05 | 0.11293 | 0.49771 | 8463.559 |
| REACTOME | 7 | 0.00123 | 2.37E-05 | 0.21825 | 0.49775 | 8464.239 |
| GOBP_AGG | 10 | 0.000871 | 2.00E-05 | 0.15792 | 0.4978 | 8465.089 |
| GOBP_NEG | 5 | 0.001108 | 1.80E-05 | 0.20614 | 0.49786 | 8466.109 |
| GOBP_MAI | 9 | 0.000762 | 1.66E-05 | 0.15038 | 0.49798 | 8468.15 |
| REACTOME | 638 | 9.47E-05 | 1.71E-05 | 0.019687 | 0.49808 | 8469.85 |
| FERRANDO | 17 | 0.000516 | 1.55E-05 | 0.11811 | 0.49826 | 8472.911 |
| WP_NAD_I | 8 | 0.00067 | 1.38E-05 | 0.15661 | 0.49829 | 8473.421 |
| GOCC_BOX | 7 | 0.000751 | 1.45E-05 | 0.1757 | 0.4983 | 8473.592 |
| GOBP_EST/ | 152 | 0.00015 | 1.34E-05 | 0.040814 | 0.49854 | 8477.673 |
| GOBP_REG | 11 | 0.000676 | 1.63E-05 | 0.18928 | 0.49857 | 8478.183 |
| GOBP_PRE | 44 | 0.000276 | 1.33E-05 | 0.079892 | 0.49862 | 8479.033 |
| GOBP_VES | 141 | 0.000139 | 1.20E-05 | 0.041067 | 0.49865 | 8479.543 |
| GOCC_PER | 17 | 0.000474 | 1.42E-05 | 0.14499 | 0.4987 | 8480.394 |
| BYSTRYKH_ | 7 | 0.000434 | 8.35E-06 | 0.16369 | 0.49894 | 8484.475 |
| REACTOME | 10 | 0.000483 | 1.11E-05 | 0.18356 | 0.49895 | 8484.645 |
| GOBP_POS | 92 | 0.000126 | 8.76E-06 | 0.054792 | 0.49908 | 8486.855 |
| REACTOME | 66 | 0.000134 | 7.90E-06 | 0.05993 | 0.49911 | 8487.366 |
| WP_NEUR | 11 | 0.000287 | 6.93E-06 | 0.14868 | 0.49923 | 8489.406 |
| GOCC_POS | 12 | 0.000227 | 5.72E-06 | 0.14448 | 0.49937 | 8491.787 |
| GOCC_X_C | 9 | 0.000287 | 6.27E-06 | 0.18469 | 0.49938 | 8491.957 |
| GOBP_CITI | 6 | 0.000327 | 5.84E-06 | 0.21644 | 0.4994 | 8492.297 |
| NAKAMUR/ | 13 | 0.000171 | 4.48E-06 | 0.11427 | 0.4994 | 8492.297 |
| BONOME_ | 219 | 3.49E-05 | 3.74E-06 | 0.034352 | 0.49959 | 8495.528 |
| REACTOME | 9 | 0.000182 | 3.97E-06 | 0.18665 | 0.49961 | 8495.868 |
| GOBP_CER | 7 | 0.000147 | 2.82E-06 | 0.15215 | 0.49962 | 8496.038 |
| GOBP_MET | 12 | 0.000134 | 3.39E-06 | 0.15043 | 0.49964 | 8496.378 |
| GOBP_GLA | 72 | 4.50E-05 | 2.78E-06 | 0.063641 | 0.49972 | 8497.739 |
| GOBP_REG | 36 | 5.05E-05 | 2.20E-06 | 0.082012 | 0.49975 | 8498.249 |
| BIOCARTA_ | 11 | 6.40E-05 | 1.54E-06 | 0.14277 | 0.49982 | 8499.439 |
| WP_EXTRA | 67 | 3.74E-06 | 2.22E-07 | 0.063853 | 0.49998 | 8502.16 |
| REACTOME | 24 | -4.16E-06 | -1.48E-07 | 0.098526 | 0.50002 | 8502.84 |
| GOMF_TR/ | 29 | -7.45E-06 | -2.92E-07 | 0.090487 | 0.50003 | 8503.01 |
| REACTOME | 51 | -1.47E-05 | -7.63E-07 | 0.072727 | 0.50008 | 8503.86 |
| SCHAEFFEF | 11 | -5.16E-05 | -1.25E-06 | 0.18559 | 0.50011 | 8504.371 |
| MONNIER_ | 355 | -9.90E-06 | -1.35E-06 | 0.026368 | 0.50015 | 8505.051 |
| GOBP_POS | 20 | -4.67E-05 | -1.52E-06 | 0.11337 | 0.50016 | 8505.221 |
| GOBP_PHA | 24 | -5.17E-05 | -1.84E-06 | 0.11337 | 0.50018 | 8505.561 |
| GOBP_MAI | 9 | -0.00022 | -4.87E-06 | 0.18934 | 0.50047 | 8510.492 |
| GOBP_ZYM | 61 | -8.38E-05 | -4.75E-06 | 0.067509 | 0.5005 | 8511.003 |
| GOBP_NCR | 45 | -0.00013 | -6.46E-06 | 0.076851 | 0.50069 | 8514.233 |
| GOBP_T_C | 75 | -0.00011 | -6.74E-06 | 0.060229 | 0.50071 | 8514.574 |

| | | | | | | |
|---|---|---|---|---|---|---|
| GOBP_REN | 6 | -0.00049 | -8.81E-06 | 0.25688 | 0.50077 | 8515.594 |
| ROY_WOU | 20 | -0.00025 | -8.00E-06 | 0.11492 | 0.50085 | 8516.954 |
| GOMF_ACT | 9 | -0.00046 | -1.01E-05 | 0.21237 | 0.50087 | 8517.294 |
| GOMF_EXT | 5 | -0.00056 | -9.06E-06 | 0.2424 | 0.50092 | 8518.145 |
| GOBP_NEG | 6 | -0.00051 | -9.11E-06 | 0.18805 | 0.50108 | 8520.865 |
| GOBP_REG | 36 | -0.00024 | -1.04E-05 | 0.083623 | 0.50114 | 8521.886 |
| GOCC_CILI | 72 | -0.00017 | -1.02E-05 | 0.057334 | 0.50115 | 8522.056 |
| MCCLUNG_ | 95 | -0.00018 | -1.24E-05 | 0.053048 | 0.50132 | 8524.947 |
| LU_AGING_ | 244 | -0.00012 | -1.32E-05 | 0.033806 | 0.50138 | 8525.967 |
| GOBP_COR | 18 | -0.00055 | -1.70E-05 | 0.15542 | 0.50142 | 8526.647 |
| GOCC_CDC | 7 | -0.00061 | -1.17E-05 | 0.17129 | 0.50142 | 8526.647 |
| GOBP_NEG | 38 | -0.00034 | -1.51E-05 | 0.08903 | 0.50151 | 8528.178 |
| GOBP_OLIG | 28 | -0.00036 | -1.39E-05 | 0.093878 | 0.50154 | 8528.688 |
| GOBP_ANT | 6 | -0.00086 | -1.53E-05 | 0.19367 | 0.50177 | 8532.599 |
| FUJIWARA_ | 8 | -0.00099 | -2.04E-05 | 0.20647 | 0.50191 | 8534.98 |
| GOBP_ANT | 16 | -0.00065 | -1.89E-05 | 0.11682 | 0.50222 | 8540.251 |
| GOBP_NEG | 61 | -0.00039 | -2.21E-05 | 0.065234 | 0.50238 | 8542.972 |
| GOBP_REG | 49 | -0.00047 | -2.39E-05 | 0.072087 | 0.50259 | 8546.543 |
| GOBP_G_P | 21 | -0.00069 | -2.30E-05 | 0.10571 | 0.50261 | 8546.883 |
| GOMF_GAI | 13 | -0.00094 | -2.46E-05 | 0.14172 | 0.50264 | 8547.393 |
| GOBP_PHO | 142 | -0.00027 | -2.33E-05 | 0.040422 | 0.50266 | 8547.733 |
| REACTOME | 91 | -0.00033 | -2.32E-05 | 0.049767 | 0.50268 | 8548.073 |
| GOMF_RNA | 12 | -0.00084 | -2.13E-05 | 0.12431 | 0.50271 | 8548.584 |
| GOMF_ABC | 44 | -0.00053 | -2.55E-05 | 0.077667 | 0.50272 | 8548.754 |
| KIM_RESPO | 8 | -0.00112 | -2.30E-05 | 0.16237 | 0.50274 | 8549.094 |
| GOBP_REG | 12 | -0.00104 | -2.61E-05 | 0.1494 | 0.50277 | 8549.604 |
| GOBP_EXC | 37 | -0.00061 | -2.69E-05 | 0.087488 | 0.50278 | 8549.774 |
| REACTOME | 8 | -0.00144 | -2.97E-05 | 0.18874 | 0.50305 | 8554.365 |
| SUNG_MET | 104 | -0.00036 | -2.69E-05 | 0.047196 | 0.50308 | 8554.875 |
| GOBP_TOL | 17 | -0.00103 | -3.10E-05 | 0.13323 | 0.50309 | 8555.045 |
| GOBP_NEG | 22 | -0.00086 | -2.94E-05 | 0.10777 | 0.50319 | 8556.746 |
| GOBP_POS | 6 | -0.00199 | -3.55E-05 | 0.24897 | 0.50319 | 8556.746 |
| LIAN_LIPA_ | 71 | -0.00049 | -3.02E-05 | 0.061357 | 0.5032 | 8556.916 |
| GOBP_NEU | 5 | -0.00181 | -2.94E-05 | 0.22452 | 0.50321 | 8557.086 |
| BIOCARTA_ | 12 | -0.00136 | -3.42E-05 | 0.16764 | 0.50323 | 8557.426 |
| WP_PPARA | 26 | -0.00079 | -2.91E-05 | 0.095049 | 0.5033 | 8558.617 |
| GOBP_POL | 15 | -0.00113 | -3.19E-05 | 0.13367 | 0.50337 | 8559.807 |
| GOMF_OPI | 7 | -0.00178 | -3.42E-05 | 0.20916 | 0.50339 | 8560.147 |
| REACTOME | 6 | -0.00208 | -3.70E-05 | 0.24102 | 0.50344 | 8560.997 |
| REACTOME | 31 | -0.00085 | -3.43E-05 | 0.092849 | 0.50364 | 8564.398 |
| GOCC_SYM | 8 | -0.00199 | -4.10E-05 | 0.21353 | 0.50372 | 8565.759 |
| SMIRNOV_ | 5 | -0.00168 | -2.74E-05 | 0.17746 | 0.50379 | 8566.949 |
| GOBP_NEG | 6 | -0.00242 | -4.31E-05 | 0.24983 | 0.50386 | 8568.139 |
| GOBP_NEG | 7 | -0.00169 | -3.26E-05 | 0.17459 | 0.50387 | 8568.309 |
| GOCC_PHO | 39 | -0.00077 | -3.51E-05 | 0.079118 | 0.5039 | 8568.82 |

| | | | | | | |
|---|---|---|---|---|---|---|
| GOBP_POS | 10 | -0.00129 | -2.97E-05 | 0.12974 | 0.50397 | 8570.01 |
| WP_PROSU | 14 | -0.00127 | -3.45E-05 | 0.12346 | 0.5041 | 8572.221 |
| DEURIG_T_ | 322 | -0.00028 | -3.69E-05 | 0.027337 | 0.50416 | 8573.241 |
| GOBP_DIA( | 5 | -0.00221 | -3.59E-05 | 0.20797 | 0.50423 | 8574.431 |
| GOBP_ALL/ | 9 | -0.00187 | -4.07E-05 | 0.17492 | 0.50425 | 8574.771 |
| REACTOME | 10 | -0.00156 | -3.59E-05 | 0.14617 | 0.50425 | 8574.771 |
| GOCC_EXT | 6 | -0.00292 | -5.20E-05 | 0.2704 | 0.50431 | 8575.792 |
| GOCC_OXC | 5 | -0.00202 | -3.28E-05 | 0.18437 | 0.50436 | 8576.642 |
| GAVIN_IL2_ | 17 | -0.00139 | -4.18E-05 | 0.12572 | 0.50442 | 8577.662 |
| GOBP_CER | 17 | -0.00122 | -3.67E-05 | 0.11008 | 0.50443 | 8577.832 |
| REACTOME | 39 | -0.00084 | -3.80E-05 | 0.074404 | 0.50448 | 8578.682 |
| DARWICHE | 161 | -0.00041 | -3.78E-05 | 0.03653 | 0.50449 | 8578.852 |
| REACTOME | 14 | -0.00144 | -3.93E-05 | 0.12642 | 0.50455 | 8579.873 |
| GOBP_FRU | 4 | -0.00224 | -3.27E-05 | 0.19566 | 0.50457 | 8580.213 |
| GOMF_FRU | 4 | -0.00224 | -3.27E-05 | 0.19566 | 0.50457 | 8580.213 |
| GOBP_LUN | 6 | -0.00293 | -5.22E-05 | 0.24337 | 0.5048 | 8584.124 |
| GOBP_NEG | 17 | -0.00168 | -5.04E-05 | 0.13425 | 0.50499 | 8587.355 |
| GOBP_NEG | 16 | -0.00154 | -4.47E-05 | 0.12256 | 0.505 | 8587.525 |
| WP_P53_T | 88 | -0.00065 | -4.44E-05 | 0.052074 | 0.505 | 8587.525 |
| WP_GLIAL_ | 5 | -0.00289 | -4.71E-05 | 0.22845 | 0.50505 | 8588.375 |
| GOBP_EST/ | 7 | -0.00275 | -5.30E-05 | 0.21655 | 0.50507 | 8588.715 |
| DEMAGALH | 50 | -0.00086 | -4.40E-05 | 0.066703 | 0.50512 | 8589.566 |
| GOBP_OUT | 7 | -0.00274 | -5.27E-05 | 0.21119 | 0.50517 | 8590.416 |
| GOBP_POS | 238 | -0.00045 | -5.03E-05 | 0.033799 | 0.50532 | 8592.967 |
| GOBP_REG | 11 | -0.002 | -4.83E-05 | 0.14422 | 0.50554 | 8596.708 |
| MATHEW_ | 10 | -0.00237 | -5.46E-05 | 0.17075 | 0.50554 | 8596.708 |
| GOMF_VOI | 15 | -0.00176 | -4.95E-05 | 0.12597 | 0.50557 | 8597.218 |
| GOMF_C_N | 6 | -0.0021 | -3.74E-05 | 0.1496 | 0.5056 | 8597.728 |
| GOCC_CAL | 5 | -0.00308 | -5.02E-05 | 0.2194 | 0.50561 | 8597.898 |
| KEGG_LINC | 29 | -0.00146 | -5.70E-05 | 0.1021 | 0.50569 | 8599.258 |
| GOBP_ON( | 5 | -0.00383 | -6.24E-05 | 0.2678 | 0.50571 | 8599.599 |
| GOBP_BUD | 7 | -0.00282 | -5.43E-05 | 0.19429 | 0.50579 | 8600.959 |
| SASAKI_TA | 12 | -0.00194 | -4.90E-05 | 0.13222 | 0.50586 | 8602.149 |
| WP_PI3KAI | 29 | -0.00141 | -5.53E-05 | 0.095421 | 0.5059 | 8602.83 |
| GOMF_RN/ | 60 | -0.00087 | -4.92E-05 | 0.058624 | 0.50594 | 8603.51 |
| GARCIA_T/ | 46 | -0.001 | -4.91E-05 | 0.066246 | 0.506 | 8604.53 |
| GOMF_SYN | 64 | -0.00086 | -5.00E-05 | 0.056963 | 0.50602 | 8604.87 |
| GOBP_REG | 29 | -0.00143 | -5.60E-05 | 0.09156 | 0.50622 | 8608.271 |
| MIZUKAMI_ | 12 | -0.00246 | -6.19E-05 | 0.15743 | 0.50622 | 8608.271 |
| WP_BMP2' | 13 | -0.00225 | -5.91E-05 | 0.14458 | 0.50622 | 8608.271 |
| SCHAEFFER | 7 | -0.00363 | -7.00E-05 | 0.23047 | 0.50629 | 8609.461 |
| GOBP_TRN | 5 | -0.00364 | -5.92E-05 | 0.22957 | 0.50632 | 8609.972 |
| KEGG_PRIN | 16 | -0.0021 | -6.10E-05 | 0.12877 | 0.5065 | 8613.033 |
| HOLLERN_ | 9 | -0.00327 | -7.14E-05 | 0.19966 | 0.50653 | 8613.543 |
| GOBP_MEI | 172 | -0.00062 | -5.90E-05 | 0.037618 | 0.50658 | 8614.393 |

| | | | | | | |
|---|---|---|---|---|---|---|
| GOBP_POS | 117 | -0.00079 | -6.24E-05 | 0.04768 | 0.50665 | 8615.583 |
| GARGALOV | 15 | -0.00226 | -6.37E-05 | 0.13123 | 0.50687 | 8619.324 |
| GOBP_REG | 11 | -0.00299 | -7.21E-05 | 0.17138 | 0.50695 | 8620.685 |
| BAUS_TFF2 | 10 | -0.00308 | -7.08E-05 | 0.17091 | 0.50718 | 8624.596 |
| GOBP_REG | 7 | -0.00406 | -7.81E-05 | 0.22426 | 0.50721 | 8625.106 |
| GOMF_3_5 | 53 | -0.00108 | -5.73E-05 | 0.058887 | 0.50733 | 8627.147 |
| GOBP_CAR | 10 | -0.00313 | -7.21E-05 | 0.16923 | 0.50739 | 8628.167 |
| KORKOLA_ | 40 | -0.00148 | -6.79E-05 | 0.079457 | 0.50741 | 8628.507 |
| GOBP_REG | 82 | -0.00108 | -7.12E-05 | 0.057144 | 0.50755 | 8630.888 |
| GOBP_REG | 72 | -0.00119 | -7.33E-05 | 0.062804 | 0.50756 | 8631.058 |
| HUMMERI( | 16 | -0.00213 | -6.18E-05 | 0.11186 | 0.50758 | 8631.398 |
| GOBP_REG | 34 | -0.00154 | -6.55E-05 | 0.081106 | 0.50759 | 8631.568 |
| GOBP_REG | 37 | -0.00151 | -6.69E-05 | 0.079197 | 0.50762 | 8632.078 |
| BIOCARTA_ | 17 | -0.00232 | -6.96E-05 | 0.12027 | 0.5077 | 8633.439 |
| GOMF_VOI | 5 | -0.00492 | -8.01E-05 | 0.24714 | 0.50795 | 8637.69 |
| GOCC_SWI | 91 | -0.00109 | -7.57E-05 | 0.054415 | 0.50801 | 8638.71 |
| WP_CLASS | 14 | -0.00286 | -7.78E-05 | 0.14166 | 0.50805 | 8639.39 |
| GOBP_POS | 9 | -0.00374 | -8.16E-05 | 0.18313 | 0.50814 | 8640.921 |
| GOMF_CAI | 59 | -0.00146 | -8.17E-05 | 0.068798 | 0.50849 | 8646.872 |
| REACTOME | 50 | -0.0015 | -7.70E-05 | 0.070049 | 0.50853 | 8647.553 |
| WP_SCFA_ | 6 | -0.0041 | -7.31E-05 | 0.18909 | 0.50865 | 8649.593 |
| GOBP_REG | 3 | -0.00681 | -8.58E-05 | 0.31226 | 0.5087 | 8650.444 |
| GOMF_PR( | 24 | -0.00204 | -7.28E-05 | 0.093356 | 0.50873 | 8650.954 |
| GOBP_PUR | 8 | -0.00428 | -8.81E-05 | 0.19378 | 0.50882 | 8652.484 |
| GOBP_REG | 6 | -0.00533 | -9.49E-05 | 0.23924 | 0.50888 | 8653.504 |
| GOBP_PUT | 7 | -0.00359 | -6.90E-05 | 0.15934 | 0.50898 | 8655.205 |
| GOBP_AN( | 5 | -0.00743 | -0.00012 | 0.32519 | 0.50911 | 8657.416 |
| GOMF_LAI | 7 | -0.00556 | -0.00011 | 0.23847 | 0.5093 | 8660.647 |
| REACTOME | 59 | -0.00146 | -8.13E-05 | 0.061422 | 0.50946 | 8663.367 |
| GOBP_RES | 10 | -0.00345 | -7.95E-05 | 0.14507 | 0.5095 | 8664.048 |
| GOBP_REG | 148 | -0.00098 | -8.66E-05 | 0.040684 | 0.50963 | 8666.258 |
| GOBP_AMI | 75 | -0.00137 | -8.60E-05 | 0.055735 | 0.50979 | 8668.979 |
| GOBP_CAT | 68 | -0.00156 | -9.34E-05 | 0.062819 | 0.5099 | 8670.85 |
| GOBP_NE( | 32 | -0.00231 | -9.50E-05 | 0.093072 | 0.5099 | 8670.85 |
| REACTOME | 19 | -0.00236 | -7.48E-05 | 0.094981 | 0.50991 | 8671.02 |
| GOMF_RN/ | 18 | -0.00294 | -9.07E-05 | 0.1174 | 0.50999 | 8672.38 |
| GOBP_ERR | 5 | -0.00467 | -7.60E-05 | 0.18508 | 0.51006 | 8673.57 |
| GOCC_END | 26 | -0.00238 | -8.83E-05 | 0.094314 | 0.51007 | 8673.74 |
| REACTOME | 80 | -0.00138 | -8.93E-05 | 0.054284 | 0.5101 | 8674.251 |
| REACTOME | 12 | -0.00423 | -0.00011 | 0.1645 | 0.51027 | 8677.141 |
| GOMF_SN/ | 35 | -0.00215 | -9.23E-05 | 0.082656 | 0.51036 | 8678.672 |
| TANG_SEN | 33 | -0.00216 | -9.03E-05 | 0.083167 | 0.51037 | 8678.842 |
| SCHAEFFEF | 8 | -0.00514 | -0.00011 | 0.1968 | 0.51042 | 8679.692 |
| CHEOK_RE: | 17 | -0.00332 | -9.94E-05 | 0.12609 | 0.51049 | 8680.882 |
| GOBP_MO | 56 | -0.00183 | -9.97E-05 | 0.069741 | 0.51049 | 8680.882 |

| | | | | | | |
|---|---|---|---|---|---|---|
| GOBP_POS | 11 | -0.00438 | -0.00011 | 0.16564 | 0.51055 | 8681.903 |
| WP_MIR12 | 6 | -0.00481 | -8.58E-05 | 0.18189 | 0.51056 | 8682.073 |
| KEGG_LEU | 108 | -0.00129 | -9.70E-05 | 0.048036 | 0.51067 | 8683.943 |
| GOBP_SUL | 17 | -0.00303 | -9.09E-05 | 0.11287 | 0.51071 | 8684.624 |
| PID_HIF2P | 31 | -0.0024 | -9.73E-05 | 0.088657 | 0.51081 | 8686.324 |
| BLALOCK_ | 156 | -0.00101 | -9.17E-05 | 0.037104 | 0.51088 | 8687.514 |
| GOBP_CEL | 119 | -0.00126 | -9.99E-05 | 0.045914 | 0.51097 | 8689.045 |
| KEGG_TYR | 39 | -0.00236 | -0.00011 | 0.085031 | 0.51105 | 8690.405 |
| GOBP_REG | 7 | -0.0057 | -0.00011 | 0.20452 | 0.51111 | 8691.426 |
| GOBP_REG | 11 | -0.00453 | -0.00011 | 0.16246 | 0.51112 | 8691.596 |
| GOBP_V_D | 19 | -0.0029 | -9.21E-05 | 0.10356 | 0.51118 | 8692.616 |
| FOSTER_KD | 196 | -0.00095 | -9.61E-05 | 0.033426 | 0.51131 | 8694.827 |
| GOBP_POS | 14 | -0.00372 | -0.0001 | 0.13025 | 0.51138 | 8696.017 |
| GOCC_PHA | 35 | -0.00234 | -0.0001 | 0.081655 | 0.51142 | 8696.697 |
| GOBP_EXT | 211 | -0.00105 | -0.00011 | 0.036422 | 0.51145 | 8697.207 |
| BIOCARTA_ | 9 | -0.00489 | -0.00011 | 0.16841 | 0.51158 | 8699.418 |
| GOBP_MA | 23 | -0.00288 | -0.0001 | 0.099037 | 0.5116 | 8699.758 |
| GOMF_TR | 15 | -0.00436 | -0.00012 | 0.14936 | 0.51163 | 8700.268 |
| WP_MFAP | 13 | -0.00419 | -0.00011 | 0.13984 | 0.51196 | 8705.88 |
| GOBP_DU | 5 | -0.00675 | -0.00011 | 0.22379 | 0.51203 | 8707.07 |
| GOBP_POS | 61 | -0.00195 | -0.00011 | 0.064526 | 0.51207 | 8707.75 |
| GOBP_TEL | 11 | -0.00452 | -0.00011 | 0.1492 | 0.51208 | 8707.92 |
| GOMF_NA | 5 | -0.0069 | -0.00011 | 0.22745 | 0.5121 | 8708.261 |
| GOMF_PH | 4 | -0.0095 | -0.00014 | 0.31086 | 0.5122 | 8709.961 |
| GOBP_REG | 6 | -0.00726 | -0.00013 | 0.23703 | 0.51222 | 8710.301 |
| AMIT_EGF_ | 21 | -0.00403 | -0.00013 | 0.13123 | 0.51225 | 8710.811 |
| GOBP_REG | 13 | -0.00437 | -0.00011 | 0.14215 | 0.51228 | 8711.321 |
| GOCC_TRA | 11 | -0.00458 | -0.00011 | 0.14844 | 0.5123 | 8711.662 |
| GOMF_OX | 10 | -0.00387 | -8.90E-05 | 0.12473 | 0.51237 | 8712.852 |
| GOCC_DEU | 5 | -0.01121 | -0.00018 | 0.36005 | 0.51242 | 8713.702 |
| GOBP_AXI | 7 | -0.00556 | -0.00011 | 0.17777 | 0.51247 | 8714.552 |
| GOCC_CLA | 16 | -0.00382 | -0.00011 | 0.12118 | 0.51257 | 8716.253 |
| GOBP_POS | 35 | -0.00268 | -0.00012 | 0.084645 | 0.51265 | 8717.613 |
| GOMF_AM | 6 | -0.00674 | -0.00012 | 0.21091 | 0.51275 | 8719.314 |
| GOBP_HO | 1558 | -0.00042 | -0.00012 | 0.013283 | 0.51276 | 8719.484 |
| VALK_AML | 24 | -0.00328 | -0.00012 | 0.10172 | 0.51285 | 8721.014 |
| GOBP_CER | 44 | -0.00235 | -0.00011 | 0.072618 | 0.51292 | 8722.205 |
| GOBP_SUC | 10 | -0.00464 | -0.00011 | 0.14268 | 0.51296 | 8722.885 |
| WP_NEUR | 40 | -0.00264 | -0.00012 | 0.08093 | 0.51299 | 8723.395 |
| GOBP_NEG | 9 | -0.00507 | -0.00011 | 0.15483 | 0.51307 | 8724.755 |
| GOMF_HIS | 65 | -0.00202 | -0.00012 | 0.061267 | 0.51318 | 8726.626 |
| PID_MYC_ | 77 | -0.00178 | -0.00011 | 0.053737 | 0.51324 | 8727.646 |
| GOBP_INT | 9 | -0.00548 | -0.00012 | 0.16339 | 0.51337 | 8729.857 |
| GOCC_SEM | 9 | -0.00678 | -0.00015 | 0.20185 | 0.51339 | 8730.197 |
| WP_NEUR | 37 | -0.00281 | -0.00012 | 0.083666 | 0.51339 | 8730.197 |

| | | | | | | |
|---|---|---|---|---|---|---|
| GOCC_EGG | 5 | -0.00664 | -0.00011 | 0.19702 | 0.51344 | 8731.047 |
| REACTOME | 67 | -0.00201 | -0.00012 | 0.059102 | 0.51355 | 8732.918 |
| GOMF_L_P | 7 | -0.00773 | -0.00015 | 0.22708 | 0.51358 | 8733.428 |
| GOBP_HEA | 31 | -0.00354 | -0.00014 | 0.10206 | 0.51382 | 8737.509 |
| GOBP_LIPC | 17 | -0.00388 | -0.00012 | 0.11166 | 0.51385 | 8738.019 |
| VALK_AML | 33 | -0.00308 | -0.00013 | 0.08832 | 0.51391 | 8739.04 |
| GOCC_PRC | 17 | -0.00418 | -0.00013 | 0.11963 | 0.51393 | 8739.38 |
| REACTOME | 5 | -0.00915 | -0.00015 | 0.26088 | 0.51399 | 8740.4 |
| BLANCO_M | 51 | -0.00264 | -0.00014 | 0.075109 | 0.514 | 8740.57 |
| GOBP_REG | 32 | -0.00348 | -0.00014 | 0.098638 | 0.51406 | 8741.59 |
| WP_AMPA | 66 | -0.00217 | -0.00013 | 0.060974 | 0.51421 | 8744.141 |
| GOBP_POS | 12 | -0.0054 | -0.00014 | 0.1511 | 0.51426 | 8744.991 |
| GOBP_END | 5 | -0.00871 | -0.00014 | 0.24012 | 0.51447 | 8748.562 |
| GOBP_POS | 6 | -0.00951 | -0.00017 | 0.26198 | 0.51447 | 8748.562 |
| MEBARKI_I | 176 | -0.00148 | -0.00014 | 0.040724 | 0.51448 | 8748.732 |
| GOBP_NEG | 3 | -0.01111 | -0.00014 | 0.3029 | 0.51464 | 8751.453 |
| GOBP_RES | 83 | -0.00201 | -0.00013 | 0.054262 | 0.51481 | 8754.344 |
| GOMF_TRI | 5 | -0.00712 | -0.00012 | 0.18913 | 0.51501 | 8757.745 |
| HOFFMANI | 27 | -0.00364 | -0.00014 | 0.096676 | 0.51501 | 8757.745 |
| REACTOME | 9 | -0.00735 | -0.00016 | 0.19417 | 0.5151 | 8759.276 |
| GOBP_REG | 8 | -0.0082 | -0.00017 | 0.21354 | 0.51532 | 8763.017 |
| GOBP_NEG | 12 | -0.0065 | -0.00016 | 0.16912 | 0.51534 | 8763.357 |
| NIKOLSKY_ | 21 | -0.00597 | -0.0002 | 0.15452 | 0.5154 | 8764.377 |
| REACTOME | 99 | -0.00217 | -0.00016 | 0.055417 | 0.51561 | 8767.948 |
| GOBP_PYR | 9 | -0.00616 | -0.00013 | 0.15703 | 0.51565 | 8768.628 |
| GOBP_NLS | 18 | -0.0041 | -0.00013 | 0.10428 | 0.51569 | 8769.308 |
| GOBP_PRC | 35 | -0.00374 | -0.00016 | 0.094707 | 0.51575 | 8770.329 |
| GOMF_PRC | 9 | -0.00664 | -0.00014 | 0.16655 | 0.5159 | 8772.88 |
| REACTOME | 192 | -0.00146 | -0.00015 | 0.03647 | 0.51591 | 8773.05 |
| REACTOME | 15 | -0.00514 | -0.00014 | 0.12882 | 0.51592 | 8773.22 |
| GOBP_HEP | 13 | -0.00522 | -0.00014 | 0.12875 | 0.51616 | 8777.301 |
| GOBP_ORG | 117 | -0.0018 | -0.00014 | 0.044411 | 0.51619 | 8777.811 |
| GAZIN_EPI | 25 | -0.00374 | -0.00014 | 0.091763 | 0.51626 | 8779.001 |
| KRASNOSE | 42 | -0.00311 | -0.00015 | 0.076143 | 0.51628 | 8779.341 |
| RODWELL_ | 426 | -0.00099 | -0.00015 | 0.024221 | 0.51628 | 8779.341 |
| GOBP_REG | 17 | -0.00458 | -0.00014 | 0.11186 | 0.51631 | 8779.852 |
| PID_TRAIL_ | 27 | -0.00401 | -0.00015 | 0.097534 | 0.51638 | 8781.042 |
| GOBP_NEG | 88 | -0.00215 | -0.00015 | 0.051829 | 0.51654 | 8783.763 |
| GOBP_NEG | 11 | -0.00679 | -0.00016 | 0.1618 | 0.51674 | 8787.164 |
| AMIT_EGF_ | 11 | -0.00581 | -0.00014 | 0.13706 | 0.51689 | 8789.714 |
| REACTOME | 12 | -0.00675 | -0.00017 | 0.15905 | 0.51693 | 8790.395 |
| WP_MALE_ | 128 | -0.00192 | -0.00016 | 0.045311 | 0.51694 | 8790.565 |
| GOBP_LUN | 11 | -0.00649 | -0.00016 | 0.1524 | 0.51697 | 8791.075 |
| GOBP_SING | 12 | -0.00597 | -0.00015 | 0.13979 | 0.51703 | 8792.095 |
| TAKEDA_TA | 150 | -0.00181 | -0.00016 | 0.042442 | 0.51704 | 8792.265 |

| | | | | | | |
|---|---|---|---|---|---|---|
| GOBP_MEI | 147 | -0.00181 | -0.00016 | 0.042286 | 0.51705 | 8792.435 |
| GOBP_REG | 13 | -0.00781 | -0.0002 | 0.18189 | 0.51712 | 8793.626 |
| GOCC_POS | 255 | -0.00149 | -0.00017 | 0.034533 | 0.51721 | 8795.156 |
| GOBP_NEG | 113 | -0.00205 | -0.00016 | 0.047444 | 0.51724 | 8795.666 |
| BYSTRYKH_ | 724 | -0.0008 | -0.00015 | 0.018553 | 0.51726 | 8796.006 |
| BIOCARTA_ | 15 | -0.00588 | -0.00017 | 0.13544 | 0.51733 | 8797.197 |
| GOMF_THI | 14 | -0.0055 | -0.00015 | 0.12664 | 0.51733 | 8797.197 |
| GOMF_CHI | 8 | -0.00845 | -0.00017 | 0.194 | 0.51737 | 8797.877 |
| GOBP_POS | 29 | -0.00436 | -0.00017 | 0.099751 | 0.51743 | 8798.897 |
| GOBP_LAC | 7 | -0.01002 | -0.00019 | 0.22833 | 0.5175 | 8800.088 |
| GOBP_RES | 33 | -0.00401 | -0.00017 | 0.091196 | 0.51755 | 8800.938 |
| GOBP_POS | 5 | -0.00902 | -0.00015 | 0.20439 | 0.51759 | 8801.618 |
| GOMF_RN/ | 24 | -0.00469 | -0.00017 | 0.10635 | 0.51761 | 8801.958 |
| GOBP_REG | 5 | -0.00927 | -0.00015 | 0.20869 | 0.51772 | 8803.829 |
| GOCC_CUL | 31 | -0.00374 | -0.00015 | 0.083096 | 0.51794 | 8807.57 |
| GOBP_REG | 5 | -0.01228 | -0.0002 | 0.27192 | 0.51801 | 8808.76 |
| GOCC_NON | 9 | -0.00736 | -0.00016 | 0.16274 | 0.51803 | 8809.1 |
| GOBP_CELI | 24 | -0.00458 | -0.00016 | 0.10129 | 0.51804 | 8809.27 |
| GOMF_NIT | 7 | -0.00805 | -0.00015 | 0.17683 | 0.51815 | 8811.141 |
| CROONQU | 93 | -0.00223 | -0.00016 | 0.04886 | 0.51816 | 8811.311 |
| GOBP_POS | 5 | -0.01047 | -0.00017 | 0.22883 | 0.51824 | 8812.671 |
| GOBP_CELI | 19 | -0.00478 | -0.00015 | 0.1035 | 0.51841 | 8815.562 |
| GOBP_PUR | 10 | -0.00759 | -0.00017 | 0.16419 | 0.51844 | 8816.072 |
| GOBP_RRN | 7 | -0.00974 | -0.00019 | 0.21074 | 0.51844 | 8816.072 |
| REACTOME | 9 | -0.00766 | -0.00017 | 0.16551 | 0.51846 | 8816.412 |
| GOBP_MIC | 119 | -0.00194 | -0.00015 | 0.041673 | 0.51855 | 8817.943 |
| GOBP_RIB( | 8 | -0.00719 | -0.00015 | 0.15432 | 0.51859 | 8818.623 |
| CHEOK_RE: | 8 | -0.00836 | -0.00017 | 0.1784 | 0.51869 | 8820.323 |
| GOBP_POS | 148 | -0.00191 | -0.00017 | 0.040349 | 0.51885 | 8823.044 |
| GOMF_ATI | 8 | -0.00999 | -0.00021 | 0.21141 | 0.51885 | 8823.044 |
| GOBP_PRC | 46 | -0.00327 | -0.00016 | 0.068608 | 0.51898 | 8825.255 |
| PETRETTO_ | 30 | -0.00433 | -0.00017 | 0.090563 | 0.51905 | 8826.445 |
| GOBP_NEG | 8 | -0.00776 | -0.00016 | 0.16087 | 0.51923 | 8829.506 |
| GOBP_REG | 32 | -0.00469 | -0.00019 | 0.096684 | 0.51934 | 8831.377 |
| REACTOME | 17 | -0.00564 | -0.00017 | 0.11552 | 0.51947 | 8833.587 |
| FIGUEROA_ | 33 | -0.0044 | -0.00018 | 0.090129 | 0.51948 | 8833.757 |
| BIOCARTA_ | 16 | -0.00599 | -0.00017 | 0.1225 | 0.5195 | 8834.098 |
| ONO_AML: | 23 | -0.00498 | -0.00017 | 0.10184 | 0.5195 | 8834.098 |
| GOBP_CELI | 44 | -0.00374 | -0.00018 | 0.076236 | 0.51959 | 8835.628 |
| GOBP_T_C | 288 | -0.00148 | -0.00018 | 0.030138 | 0.5196 | 8835.798 |
| GOMF_C_X | 5 | -0.0122 | -0.0002 | 0.24822 | 0.5196 | 8835.798 |
| GOBP_REG | 9 | -0.00737 | -0.00016 | 0.14963 | 0.51964 | 8836.478 |
| GOCC_R2T | 5 | -0.01015 | -0.00017 | 0.20394 | 0.51984 | 8839.879 |
| GOBP_CELI | 59 | -0.00335 | -0.00019 | 0.06715 | 0.51986 | 8840.219 |
| DORN_ADE | 10 | -0.0068 | -0.00016 | 0.1365 | 0.51987 | 8840.389 |

| | | | | | | |
|---|---|---|---|---|---|---|
| GOBP_NUC | 20 | -0.00557 | -0.00018 | 0.11149 | 0.51993 | 8841.41 |
| GOBP_ADE | 10 | -0.00929 | -0.00021 | 0.18498 | 0.52002 | 8842.94 |
| GOMF_TRN | 8 | -0.00745 | -0.00015 | 0.14792 | 0.52007 | 8843.79 |
| REACTOME | 30 | -0.00473 | -0.00019 | 0.093601 | 0.52017 | 8845.491 |
| WP_BIOMA | 11 | -0.00683 | -0.00016 | 0.13436 | 0.52028 | 8847.361 |
| SETLUR_PR | 17 | -0.0064 | -0.00019 | 0.12564 | 0.5203 | 8847.702 |
| BLUM_RES | 232 | -0.00158 | -0.00017 | 0.031075 | 0.52033 | 8848.212 |
| GOCC_G_P | 6 | -0.01071 | -0.00019 | 0.2099 | 0.52035 | 8848.552 |
| GOBP_MO | 6 | -0.01169 | -0.00021 | 0.22855 | 0.5204 | 8849.402 |
| GOMF_NU | 10 | -0.00792 | -0.00018 | 0.15375 | 0.52055 | 8851.953 |
| GOBP_REG | 6 | -0.01413 | -0.00025 | 0.27295 | 0.52065 | 8853.653 |
| GOBP_REG | 15 | -0.00732 | -0.00021 | 0.14141 | 0.52065 | 8853.653 |
| CAFFAREL_ | 31 | -0.00443 | -0.00018 | 0.085353 | 0.52071 | 8854.674 |
| TONKS_TA | 7 | -0.01108 | -0.00021 | 0.21291 | 0.52074 | 8855.184 |
| BIOCARTA_ | 51 | -0.00384 | -0.0002 | 0.073716 | 0.52078 | 8855.864 |
| JI_METAST | 25 | -0.0061 | -0.00022 | 0.11704 | 0.52078 | 8855.864 |
| GOBP_RES | 55 | -0.00353 | -0.00019 | 0.067547 | 0.52086 | 8857.224 |
| GOMF_L_A | 61 | -0.00346 | -0.0002 | 0.066123 | 0.52087 | 8857.394 |
| GOBP_POS | 26 | -0.00531 | -0.0002 | 0.10143 | 0.52088 | 8857.564 |
| GOCC_PAR | 17 | -0.00634 | -0.00019 | 0.1204 | 0.521 | 8859.605 |
| GOBP_NEG | 51 | -0.0033 | -0.00017 | 0.062524 | 0.52103 | 8860.115 |
| GOBP_GOL | 283 | -0.00155 | -0.00019 | 0.02937 | 0.52104 | 8860.285 |
| CHEN_LVA | 41 | -0.00433 | -0.0002 | 0.081811 | 0.52109 | 8861.135 |
| GOBP_RES | 7 | -0.00888 | -0.00017 | 0.16691 | 0.52122 | 8863.346 |
| GOBP_STO | 5 | -0.01339 | -0.00022 | 0.2514 | 0.52124 | 8863.686 |
| WP_MAP3 | 16 | -0.00685 | -0.0002 | 0.12852 | 0.52125 | 8863.856 |
| GOMF_APC | 18 | -0.00673 | -0.00021 | 0.12626 | 0.52126 | 8864.026 |
| GOBP_REG | 216 | -0.00188 | -0.0002 | 0.035061 | 0.52133 | 8865.217 |
| GOBP_NEG | 46 | -0.00378 | -0.00019 | 0.070535 | 0.52135 | 8865.557 |
| GOBP_GRC | 4 | -0.01431 | -0.00021 | 0.26513 | 0.52153 | 8868.618 |
| GOBP_TRIC | 36 | -0.00432 | -0.00019 | 0.079908 | 0.52156 | 8869.128 |
| SABATES_C | 10 | -0.00916 | -0.00021 | 0.16865 | 0.52166 | 8870.828 |
| HADDAD_T | 57 | -0.00368 | -0.0002 | 0.067623 | 0.52168 | 8871.168 |
| KAYO_AGII | 207 | -0.00187 | -0.0002 | 0.034113 | 0.52191 | 8875.08 |
| GOBP_GLU | 23 | -0.00557 | -0.00019 | 0.10094 | 0.52202 | 8876.95 |
| GOBP_REG | 1592 | -0.00073 | -0.0002 | 0.013191 | 0.5221 | 8878.311 |
| GOBP_GAS | 22 | -0.00716 | -0.00024 | 0.129 | 0.52213 | 8878.821 |
| GOBP_INTI | 9 | -0.00869 | -0.00019 | 0.15648 | 0.52214 | 8878.991 |
| RANKIN_AI | 5 | -0.0144 | -0.00023 | 0.25889 | 0.52218 | 8879.671 |
| REACTOME | 14 | -0.0066 | -0.00018 | 0.11862 | 0.5222 | 8880.011 |
| GOBP_POL | 10 | -0.00985 | -0.00023 | 0.17684 | 0.52221 | 8880.181 |
| GOBP_CER | 6 | -0.0123 | -0.00022 | 0.22055 | 0.52223 | 8880.521 |
| ZHOU_PAN | 10 | -0.00984 | -0.00023 | 0.17523 | 0.52238 | 8883.072 |
| GOBP_POS | 16 | -0.00652 | -0.00019 | 0.11607 | 0.52239 | 8883.242 |
| GOBP_ADE | 9 | -0.01109 | -0.00024 | 0.19681 | 0.52247 | 8884.602 |

| | | | | | | |
|---|---|---|---|---|---|---|
| GOMF_G_I | 9 | -0.01109 | -0.00024 | 0.19681 | 0.52247 | 8884.602 |
| COLDREN_ | 80 | -0.00298 | -0.00019 | 0.052701 | 0.52255 | 8885.963 |
| BIOCARTA_ | 13 | -0.00796 | -0.00021 | 0.14029 | 0.52263 | 8887.323 |
| GOMF_GU | 8 | -0.00905 | -0.00019 | 0.15911 | 0.52268 | 8888.173 |
| GOBP_REG | 5 | -0.01015 | -0.00017 | 0.17638 | 0.52295 | 8892.765 |
| BIOCARTA_ | 19 | -0.00722 | -0.00023 | 0.12533 | 0.52296 | 8892.935 |
| GOMF_ACI | 4 | -0.01424 | -0.00021 | 0.24697 | 0.52299 | 8893.445 |
| GOBP_POS | 64 | -0.00361 | -0.00021 | 0.062331 | 0.52306 | 8894.635 |
| GOMF_LOI | 4 | -0.01386 | -0.0002 | 0.23952 | 0.52308 | 8894.975 |
| GOBP_REG | 5 | -0.01598 | -0.00026 | 0.27445 | 0.52321 | 8897.186 |
| GOCC_PI_E | 9 | -0.01055 | -0.00023 | 0.17939 | 0.52344 | 8901.097 |
| GOBP_NEG | 171 | -0.00229 | -0.00022 | 0.038854 | 0.52345 | 8901.267 |
| GOBP_REG | 200 | -0.00206 | -0.00021 | 0.034945 | 0.5235 | 8902.118 |
| VISALA_AG | 18 | -0.00735 | -0.00023 | 0.12414 | 0.52361 | 8903.988 |
| GOBP_REG | 10 | -0.00962 | -0.00022 | 0.16162 | 0.52372 | 8905.859 |
| GOBP_LYM | 16 | -0.00714 | -0.00021 | 0.11913 | 0.5239 | 8908.92 |
| FOROUTAN | 73 | -0.00379 | -0.00024 | 0.063204 | 0.52393 | 8909.43 |
| GOBP_INTI | 6 | -0.01366 | -0.00024 | 0.22749 | 0.52394 | 8909.6 |
| GOBP_ACE | 6 | -0.01519 | -0.00027 | 0.25297 | 0.52395 | 8909.77 |
| GOBP_POS | 7 | -0.01006 | -0.00019 | 0.16614 | 0.52414 | 8913.001 |
| GOBP_PUR | 27 | -0.00607 | -0.00023 | 0.10003 | 0.52418 | 8913.681 |
| GOBP_ORN | 9 | -0.00884 | -0.00019 | 0.14569 | 0.52419 | 8913.851 |
| WP_IL1_SI | 51 | -0.0039 | -0.0002 | 0.064137 | 0.52422 | 8914.361 |
| REACTOME | 6 | -0.01549 | -0.00028 | 0.25494 | 0.52423 | 8914.531 |
| BOYAULT_I | 64 | -0.00383 | -0.00022 | 0.062801 | 0.52434 | 8916.402 |
| TIEN_INTES | 28 | -0.00506 | -0.00019 | 0.082886 | 0.52434 | 8916.402 |
| REACTOME | 6 | -0.00989 | -0.00018 | 0.16178 | 0.52436 | 8916.742 |
| WP_ACUTE | 79 | -0.00348 | -0.00022 | 0.056798 | 0.52442 | 8917.762 |
| GOBP_REG | 318 | -0.00172 | -0.00022 | 0.027786 | 0.52474 | 8923.204 |
| GOBP_POS | 8 | -0.01051 | -0.00022 | 0.16929 | 0.52475 | 8923.374 |
| GOBP_REG | 22 | -0.00774 | -0.00026 | 0.12442 | 0.5248 | 8924.224 |
| GOMF_PEF | 269 | -0.00194 | -0.00023 | 0.031181 | 0.5248 | 8924.224 |
| REACTOME | 26 | -0.00558 | -0.00021 | 0.089641 | 0.5248 | 8924.224 |
| NOJIMA_SI | 25 | -0.00613 | -0.00022 | 0.097831 | 0.52497 | 8927.115 |
| BIOCARTA_ | 8 | -0.00954 | -0.0002 | 0.15195 | 0.52503 | 8928.135 |
| GOBP_NEG | 31 | -0.0063 | -0.00026 | 0.10011 | 0.52509 | 8929.155 |
| REACTOME | 9 | -0.01044 | -0.00023 | 0.16532 | 0.52518 | 8930.686 |
| GOBP_PRC | 5 | -0.01282 | -0.00021 | 0.20263 | 0.52522 | 8931.366 |
| GOBP_ORG | 924 | -0.00103 | -0.00022 | 0.016329 | 0.52523 | 8931.536 |
| WP_MEASI | 124 | -0.00302 | -0.00024 | 0.04761 | 0.52527 | 8932.216 |
| GOBP_PRC | 21 | -0.00631 | -0.00021 | 0.098439 | 0.52554 | 8936.808 |
| GOBP_SCA | 4 | -0.0187 | -0.00027 | 0.29169 | 0.52555 | 8936.978 |
| WP_METH | 9 | -0.01058 | -0.00023 | 0.16393 | 0.52573 | 8940.039 |
| GOBP_GLI | 16 | -0.00796 | -0.00023 | 0.12314 | 0.52576 | 8940.549 |
| GOBP_TYP | 8 | -0.01438 | -0.0003 | 0.22201 | 0.52581 | 8941.399 |

| | | | | | | |
|---|---|---|---|---|---|---|
| STAMBOLS | 16 | -0.00855 | -0.00025 | 0.13147 | 0.52594 | 8943.61 |
| GOBP_END | 10 | -0.0107 | -0.00025 | 0.1641 | 0.526 | 8944.63 |
| GOBP_REG | 10 | -0.01099 | -0.00025 | 0.16761 | 0.52614 | 8947.011 |
| GOBP_POS | 27 | -0.00698 | -0.00026 | 0.10623 | 0.52618 | 8947.691 |
| GOBP_NEG | 37 | -0.00563 | -0.00025 | 0.085608 | 0.52622 | 8948.371 |
| KEGG_STEI | 48 | -0.00548 | -0.00028 | 0.083311 | 0.52624 | 8948.711 |
| REACTOME | 72 | -0.00368 | -0.00023 | 0.055917 | 0.52625 | 8948.881 |
| GOBP_PRC | 256 | -0.002 | -0.00023 | 0.030202 | 0.52639 | 8951.262 |
| MATZUK_S | 7 | -0.01426 | -0.00027 | 0.21527 | 0.52641 | 8951.602 |
| GOBP_REG | 19 | -0.00802 | -0.00025 | 0.12041 | 0.52654 | 8953.813 |
| DAZARD_U | 11 | -0.00979 | -0.00024 | 0.14666 | 0.52661 | 8955.003 |
| GOBP_ALC | 50 | -0.00457 | -0.00023 | 0.068408 | 0.52663 | 8955.343 |
| GOBP_GRA | 43 | -0.00478 | -0.00023 | 0.071381 | 0.52672 | 8956.874 |
| GOMF_LEU | 5 | -0.01637 | -0.00027 | 0.24397 | 0.52675 | 8957.384 |
| GOBP_FUN | 6 | -0.01234 | -0.00022 | 0.18291 | 0.52689 | 8959.764 |
| GOMF_SUI | 12 | -0.01072 | -0.00027 | 0.1589 | 0.5269 | 8959.935 |
| GOBP_CELI | 15 | -0.00799 | -0.00023 | 0.11765 | 0.52709 | 8963.165 |
| GOBP_POS | 6 | -0.0144 | -0.00026 | 0.21029 | 0.52729 | 8966.566 |
| GOBP_PRC | 5 | -0.01746 | -0.00028 | 0.25385 | 0.52741 | 8968.607 |
| GOBP_NEG | 21 | -0.00713 | -0.00024 | 0.10346 | 0.52746 | 8969.457 |
| BIOCARTA_ | 12 | -0.00988 | -0.00025 | 0.14301 | 0.52753 | 8970.648 |
| GOBP_REG | 4 | -0.01829 | -0.00027 | 0.26481 | 0.52753 | 8970.648 |
| GOBP_REG | 240 | -0.00232 | -0.00026 | 0.03354 | 0.5276 | 8971.838 |
| GOBP_INFI | 67 | -0.0043 | -0.00026 | 0.0621 | 0.52762 | 8972.178 |
| GOBP_NEU | 236 | -0.00222 | -0.00025 | 0.032023 | 0.52767 | 8973.028 |
| GOCC_WN | 13 | -0.00977 | -0.00026 | 0.14015 | 0.5278 | 8975.239 |
| GOCC_AXC | 153 | -0.00274 | -0.00025 | 0.039229 | 0.52786 | 8976.259 |
| WP_DOPAI | 30 | -0.0073 | -0.00029 | 0.10435 | 0.52788 | 8976.599 |
| WP_TH17_ | 67 | -0.00447 | -0.00027 | 0.063984 | 0.52788 | 8976.599 |
| MONTERO_ | 12 | -0.01079 | -0.00027 | 0.15371 | 0.52798 | 8978.3 |
| GOBP_POS | 18 | -0.00854 | -0.00026 | 0.1215 | 0.52801 | 8978.81 |
| GOBP_MU | 8 | -0.01405 | -0.00029 | 0.19966 | 0.52804 | 8979.32 |
| GOBP_PHY | 22 | -0.00754 | -0.00026 | 0.10704 | 0.52808 | 8980 |
| HANN_RES | 32 | -0.00672 | -0.00028 | 0.095319 | 0.52808 | 8980 |
| GOBP_POS | 52 | -0.00494 | -0.00026 | 0.070062 | 0.52811 | 8980.511 |
| GOBP_CAR | 6 | -0.01481 | -0.00026 | 0.20976 | 0.52815 | 8981.191 |
| GOCC_H3_ | 8 | -0.0113 | -0.00023 | 0.15975 | 0.52819 | 8981.871 |
| GOBP_POS | 45 | -0.00571 | -0.00028 | 0.080628 | 0.52824 | 8982.721 |
| LEIN_OLIG | 74 | -0.00433 | -0.00027 | 0.060873 | 0.52832 | 8984.082 |
| GOBP_SER | 9 | -0.01194 | -0.00026 | 0.16775 | 0.52838 | 8985.102 |
| BARRIER_C | 39 | -0.00557 | -0.00025 | 0.078116 | 0.5284 | 8985.442 |
| BIOCARTA_ | 23 | -0.00772 | -0.00027 | 0.10803 | 0.52847 | 8986.632 |
| GOBP_MES | 7 | -0.0167 | -0.00032 | 0.23365 | 0.52848 | 8986.802 |
| REACTOME | 7 | -0.01731 | -0.00033 | 0.24201 | 0.52851 | 8987.313 |
| EPPERT_HS | 111 | -0.00359 | -0.00027 | 0.050086 | 0.52856 | 8988.163 |

| | | | | | | |
|---|---|---|---|---|---|---|
| PID_P75_N | 61 | -0.00453 | -0.00026 | 0.062658 | 0.5288 | 8992.244 |
| KEGG_B_C | 73 | -0.00418 | -0.00026 | 0.057851 | 0.52881 | 8992.414 |
| GOBP_L_M | 5 | -0.01461 | -0.00024 | 0.20183 | 0.52884 | 8992.924 |
| GOBP_IMN | 12 | -0.0116 | -0.00029 | 0.16027 | 0.52886 | 8993.264 |
| GOCC_APIC | 422 | -0.00182 | -0.00027 | 0.025042 | 0.52889 | 8993.774 |
| GOBP_ARF | 21 | -0.00789 | -0.00026 | 0.10848 | 0.52898 | 8995.305 |
| GOBP_NEG | 10 | -0.01233 | -0.00028 | 0.16947 | 0.52899 | 8995.475 |
| GOBP_WAI | 34 | -0.00609 | -0.00026 | 0.082976 | 0.52925 | 8999.896 |
| REACTOME | 17 | -0.00861 | -0.00026 | 0.11731 | 0.52927 | 9000.236 |
| GOBP_MIT | 3 | -0.01877 | -0.00024 | 0.25442 | 0.52941 | 9002.617 |
| GOBP_MAT | 35 | -0.00681 | -0.00029 | 0.092209 | 0.52945 | 9003.297 |
| GOBP_TRA | 7 | -0.01497 | -0.00029 | 0.20185 | 0.52957 | 9005.338 |
| GOMF_NEU | 16 | -0.00986 | -0.00029 | 0.13272 | 0.52961 | 9006.018 |
| ZWANG_CI | 49 | -0.00557 | -0.00028 | 0.074964 | 0.52962 | 9006.188 |
| GOMF_GAI | 4 | -0.01786 | -0.00026 | 0.23993 | 0.52967 | 9007.038 |
| GOBP_REG | 6 | -0.01475 | -0.00026 | 0.19752 | 0.52976 | 9008.569 |
| RODWELL_ | 200 | -0.00274 | -0.00028 | 0.036577 | 0.52989 | 9010.779 |
| GOBP_REG | 13 | -0.00879 | -0.00023 | 0.11676 | 0.53001 | 9012.82 |
| GOBP_INN | 21 | -0.00889 | -0.0003 | 0.11781 | 0.53007 | 9013.84 |
| GOMF_MR | 4 | -0.02312 | -0.00034 | 0.30635 | 0.53008 | 9014.01 |
| PID_NETRII | 32 | -0.00689 | -0.00028 | 0.09113 | 0.53012 | 9014.691 |
| GOBP_HIST | 19 | -0.00759 | -0.00024 | 0.10028 | 0.53017 | 9015.541 |
| DASU_IL6_ | 7 | -0.01549 | -0.0003 | 0.20332 | 0.53036 | 9018.772 |
| GOBP_CELI | 10 | -0.0111 | -0.00026 | 0.14514 | 0.53049 | 9020.982 |
| GOBP_SPIN | 5 | -0.01762 | -0.00029 | 0.22982 | 0.53055 | 9022.003 |
| STEARMAN | 119 | -0.00346 | -0.00027 | 0.045073 | 0.53056 | 9022.173 |
| GOMF_SEC | 10 | -0.01106 | -0.00025 | 0.14418 | 0.53057 | 9022.343 |
| GERHOLD_ | 60 | -0.00513 | -0.00029 | 0.06686 | 0.53058 | 9022.513 |
| GOBP_AMI | 20 | -0.00789 | -0.00026 | 0.10238 | 0.53073 | 9025.064 |
| MEISSNER_ | 28 | -0.00702 | -0.00027 | 0.090887 | 0.53076 | 9025.574 |
| GOBP_TUN | 159 | -0.00304 | -0.00028 | 0.039291 | 0.5308 | 9026.254 |
| GOBP_ESTA | 27 | -0.00781 | -0.0003 | 0.10104 | 0.53081 | 9026.424 |
| GOBP_REG | 6 | -0.01426 | -0.00025 | 0.18338 | 0.53099 | 9029.485 |
| GOCC_BAS | 5 | -0.01501 | -0.00024 | 0.19294 | 0.53101 | 9029.825 |
| SERVITJA_I | 102 | -0.0039 | -0.00029 | 0.049984 | 0.5311 | 9031.356 |
| FAELT_B_C | 38 | -0.00546 | -0.00024 | 0.070008 | 0.53111 | 9031.526 |
| GOMF_PHO | 14 | -0.01025 | -0.00028 | 0.13113 | 0.53114 | 9032.036 |
| REACTOME | 7 | -0.01195 | -0.00023 | 0.15273 | 0.53119 | 9032.886 |
| GOMF_PHO | 29 | -0.00709 | -0.00028 | 0.090433 | 0.53125 | 9033.906 |
| REACTOME | 36 | -0.00724 | -0.00032 | 0.092247 | 0.53128 | 9034.416 |
| GOMF_CHO | 5 | -0.02245 | -0.00037 | 0.28549 | 0.53134 | 9035.437 |
| REACTOME | 8 | -0.01515 | -0.00031 | 0.19234 | 0.53138 | 9036.117 |
| WIERENGA | 40 | -0.00609 | -0.00028 | 0.077325 | 0.53141 | 9036.627 |
| REACTOME | 5 | -0.01563 | -0.00025 | 0.19736 | 0.53156 | 9039.178 |
| GOBP_PRC | 11 | -0.01283 | -0.00031 | 0.16186 | 0.53159 | 9039.688 |

| | | | | | | |
|---|---|---|---|---|---|---|
| GOMF_SM | 34 | -0.00663 | -0.00028 | 0.083437 | 0.53167 | 9041.048 |
| CAVARD_LI | 27 | -0.0077 | -0.00029 | 0.096839 | 0.53168 | 9041.218 |
| GOBP_GLIA | 47 | -0.00653 | -0.00033 | 0.081879 | 0.53176 | 9042.579 |
| REACTOME | 429 | -0.00203 | -0.0003 | 0.025233 | 0.53206 | 9047.68 |
| REACTOME | 100 | -0.00404 | -0.00029 | 0.050192 | 0.53208 | 9048.02 |
| REACTOME | 41 | -0.00551 | -0.00026 | 0.0683 | 0.53218 | 9049.721 |
| GOBP_HYA | 7 | -0.01739 | -0.00033 | 0.21528 | 0.5322 | 9050.061 |
| WP_SUDDI | 148 | -0.00353 | -0.00031 | 0.043547 | 0.53226 | 9051.081 |
| GOBP_MA( | 56 | -0.00554 | -0.0003 | 0.068205 | 0.53239 | 9053.292 |
| BIOCARTA_ | 6 | -0.01398 | -0.00025 | 0.17197 | 0.5324 | 9053.462 |
| GOBP_REG | 41 | -0.00655 | -0.00031 | 0.080358 | 0.53249 | 9054.992 |
| GOBP_REG | 4 | -0.02414 | -0.00035 | 0.29585 | 0.53251 | 9055.333 |
| GOBP_RES | 31 | -0.00744 | -0.0003 | 0.091161 | 0.53252 | 9055.503 |
| ZHANG_AN | 25 | -0.00848 | -0.00031 | 0.10388 | 0.53252 | 9055.503 |
| TONG_INTI | 53 | -0.00571 | -0.0003 | 0.069882 | 0.53254 | 9055.843 |
| REACTOME | 30 | -0.00741 | -0.00029 | 0.090663 | 0.53255 | 9056.013 |
| GOBP_POS | 22 | -0.00823 | -0.00028 | 0.10058 | 0.53262 | 9057.203 |
| RASHI_RES | 45 | -0.00642 | -0.00031 | 0.078494 | 0.53262 | 9057.203 |
| GOMF_LON | 19 | -0.00911 | -0.00029 | 0.11127 | 0.53264 | 9057.543 |
| GOCC_COS | 16 | -0.01005 | -0.00029 | 0.12261 | 0.53265 | 9057.713 |
| GOBP_MU | 196 | -0.00301 | -0.0003 | 0.036475 | 0.53286 | 9061.284 |
| GOBP_G_P | 11 | -0.0132 | -0.00032 | 0.16004 | 0.53287 | 9061.454 |
| PID_CDC42 | 69 | -0.00492 | -0.0003 | 0.059668 | 0.53289 | 9061.794 |
| BLANCO_M | 71 | -0.00496 | -0.0003 | 0.059981 | 0.53295 | 9062.815 |
| GOBP_CON | 9 | -0.01147 | -0.00025 | 0.1387 | 0.53295 | 9062.815 |
| GOCC_DEN | 38 | -0.00761 | -0.00034 | 0.091979 | 0.53295 | 9062.815 |
| GOBP_PRC | 24 | -0.00707 | -0.00025 | 0.085386 | 0.533 | 9063.665 |
| GOBP_NK_ | 8 | -0.01361 | -0.00028 | 0.16409 | 0.53306 | 9064.685 |
| GOMF_VOI | 11 | -0.01475 | -0.00036 | 0.17761 | 0.5331 | 9065.366 |
| GOBP_POS | 8 | -0.01541 | -0.00032 | 0.18434 | 0.53331 | 9068.937 |
| REACTOME | 32 | -0.00764 | -0.00031 | 0.091229 | 0.53337 | 9069.957 |
| GOCC_CHR | 12 | -0.01107 | -0.00028 | 0.13182 | 0.53345 | 9071.317 |
| MURAKAM | 33 | -0.00657 | -0.00027 | 0.078139 | 0.53351 | 9072.338 |
| GOBP_REG | 298 | -0.00258 | -0.00032 | 0.03057 | 0.53358 | 9073.528 |
| BIOCARTA_ | 7 | -0.01639 | -0.00032 | 0.19439 | 0.5336 | 9073.868 |
| KEGG_PRO | 41 | -0.0066 | -0.00031 | 0.078044 | 0.53367 | 9075.058 |
| RAMJAUN_ | 5 | -0.02187 | -0.00036 | 0.25852 | 0.5337 | 9075.569 |
| GOMF_LYS | 25 | -0.00786 | -0.00029 | 0.09274 | 0.53375 | 9076.419 |
| GOBP_CEN | 9 | -0.01737 | -0.00038 | 0.20498 | 0.53377 | 9076.759 |
| PID_CD40_ | 29 | -0.00792 | -0.00031 | 0.093309 | 0.53382 | 9077.609 |
| GOBP_SPEI | 7 | -0.01289 | -0.00025 | 0.1514 | 0.53391 | 9079.14 |
| GOBP_PHC | 13 | -0.00895 | -0.00023 | 0.10502 | 0.53397 | 9080.16 |
| REACTOME | 5 | -0.02245 | -0.00037 | 0.26321 | 0.53398 | 9080.33 |
| REACTOME | 24 | -0.00893 | -0.00032 | 0.10456 | 0.53403 | 9081.18 |
| VANTVEER_ | 43 | -0.00681 | -0.00032 | 0.079608 | 0.53407 | 9081.86 |

| | | | | | | |
|---|---|---|---|---|---|---|
| GOBP_NEG | 453 | -0.00208 | -0.00032 | 0.024262 | 0.53408 | 9082.03 |
| GOBP_POS | 5 | -0.02264 | -0.00037 | 0.26465 | 0.53409 | 9082.2 |
| GOMF_INS | 13 | -0.01386 | -0.00036 | 0.16201 | 0.53409 | 9082.2 |
| REACTOME | 43 | -0.00668 | -0.00032 | 0.078052 | 0.53409 | 9082.2 |
| SENESE_HI | 417 | -0.00214 | -0.00031 | 0.02497 | 0.53413 | 9082.881 |
| GOBP_NUC | 114 | -0.00379 | -0.00029 | 0.044168 | 0.53416 | 9083.391 |
| GOBP_REG | 494 | -0.0019 | -0.0003 | 0.022127 | 0.53419 | 9083.901 |
| REACTOME | 5 | -0.02505 | -0.00041 | 0.29174 | 0.53422 | 9084.411 |
| GOBP_POS | 129 | -0.00394 | -0.00032 | 0.045822 | 0.53424 | 9084.751 |
| TOMIDA_L | 5 | -0.02207 | -0.00036 | 0.25641 | 0.53429 | 9085.601 |
| HAHTOLA_ | 39 | -0.00734 | -0.00033 | 0.085223 | 0.5343 | 9085.772 |
| BIOCARTA_ | 20 | -0.00952 | -0.00031 | 0.11057 | 0.53431 | 9085.942 |
| GOBP_CON | 5 | -0.02304 | -0.00037 | 0.26703 | 0.53437 | 9086.962 |
| GOBP_POS | 287 | -0.00257 | -0.00031 | 0.02969 | 0.53448 | 9088.832 |
| FARMER_B | 16 | -0.01078 | -0.00031 | 0.12443 | 0.53451 | 9089.343 |
| GOBP_IMN | 573 | -0.00191 | -0.00033 | 0.022076 | 0.53451 | 9089.343 |
| DUTERTRE_ | 305 | -0.0025 | -0.00032 | 0.028774 | 0.53464 | 9091.553 |
| GOBP_NOF | 20 | -0.01117 | -0.00036 | 0.12837 | 0.53466 | 9091.893 |
| REACTOME | 7 | -0.01639 | -0.00032 | 0.18814 | 0.53471 | 9092.744 |
| GOBP_REG | 8 | -0.0158 | -0.00033 | 0.18032 | 0.5349 | 9095.975 |
| REACTOME | 20 | -0.00897 | -0.00029 | 0.10204 | 0.53501 | 9097.845 |
| GOCC_COF | 99 | -0.00444 | -0.00032 | 0.050391 | 0.5351 | 9099.376 |
| NEBEN_AN | 12 | -0.0107 | -0.00027 | 0.12117 | 0.53518 | 9100.736 |
| NABA_SECI | 314 | -0.00286 | -0.00037 | 0.032239 | 0.53537 | 9103.967 |
| MOOTHA_ | 6 | -0.02384 | -0.00042 | 0.26781 | 0.53546 | 9105.497 |
| FIGUEROA_ | 108 | -0.00443 | -0.00033 | 0.049629 | 0.53553 | 9106.688 |
| GOBP_DET | 25 | -0.00859 | -0.00031 | 0.096058 | 0.53562 | 9108.218 |
| GOBP_L_A: | 12 | -0.01322 | -0.00033 | 0.1476 | 0.53568 | 9109.238 |
| CARDOSO_ | 12 | -0.01314 | -0.00033 | 0.1466 | 0.5357 | 9109.579 |
| GOBP_REG | 5 | -0.02264 | -0.00037 | 0.25214 | 0.53577 | 9110.769 |
| GOBP_INTI | 29 | -0.00919 | -0.00036 | 0.10222 | 0.53581 | 9111.449 |
| MULLIGHA | 118 | -0.00437 | -0.00034 | 0.048561 | 0.53582 | 9111.619 |
| GOMF_POT | 152 | -0.00402 | -0.00036 | 0.04469 | 0.53584 | 9111.959 |
| BOYLAN_M | 100 | -0.00465 | -0.00034 | 0.051691 | 0.53587 | 9112.469 |
| WP_AMPLI | 17 | -0.0133 | -0.0004 | 0.14679 | 0.53609 | 9116.21 |
| GOBP_REG | 5 | -0.02167 | -0.00035 | 0.23875 | 0.53616 | 9117.401 |
| GOBP_DE_ | 8 | -0.01363 | -0.00028 | 0.14985 | 0.53622 | 9118.421 |
| GOBP_RES | 21 | -0.01018 | -0.00034 | 0.11184 | 0.53626 | 9119.101 |
| GARGALOV | 8 | -0.01966 | -0.0004 | 0.21509 | 0.53641 | 9121.652 |
| GOCC_RES | 83 | -0.00461 | -0.0003 | 0.05042 | 0.53642 | 9121.822 |
| GOCC_SITE | 106 | -0.00429 | -0.00032 | 0.046884 | 0.53643 | 9121.992 |
| PELLICCIOT | 41 | -0.00753 | -0.00035 | 0.081968 | 0.53659 | 9124.713 |
| GOCC_HAL | 7 | -0.01366 | -0.00026 | 0.14854 | 0.53664 | 9125.563 |
| GOBP_REG | 16 | -0.01099 | -0.00032 | 0.11941 | 0.53665 | 9125.733 |
| RAY_TUMC | 188 | -0.00345 | -0.00034 | 0.037461 | 0.53666 | 9125.903 |

| | | | | | | |
|---|---|---|---|---|---|---|
| YEGNASUB | 105 | -0.00475 | -0.00035 | 0.051645 | 0.53668 | 9126.243 |
| WP_MTHFI | 22 | -0.00943 | -0.00032 | 0.10226 | 0.53675 | 9127.434 |
| GOMF_CAI | 6 | -0.01815 | -0.00032 | 0.19612 | 0.53687 | 9129.474 |
| GOBP_GLU | 16 | -0.01059 | -0.00031 | 0.11421 | 0.53692 | 9130.325 |
| REACTOME | 25 | -0.00985 | -0.00036 | 0.1062 | 0.53694 | 9130.665 |
| DAZARD_R | 112 | -0.00441 | -0.00034 | 0.047557 | 0.53696 | 9131.005 |
| GOBP_REG | 131 | -0.0044 | -0.00037 | 0.047423 | 0.537 | 9131.685 |
| PID_CDC42 | 25 | -0.00909 | -0.00033 | 0.097508 | 0.53713 | 9133.896 |
| HADDAD_E | 258 | -0.00308 | -0.00036 | 0.032965 | 0.53717 | 9134.576 |
| GOBP_REG | 57 | -0.00662 | -0.00036 | 0.070931 | 0.53718 | 9134.746 |
| GOBP_POS | 7 | -0.02208 | -0.00043 | 0.23649 | 0.5372 | 9135.086 |
| GOBP_REG | 7 | -0.01548 | -0.0003 | 0.1656 | 0.53723 | 9135.596 |
| GOMF_U4_ | 5 | -0.02566 | -0.00042 | 0.27454 | 0.53723 | 9135.596 |
| GOBP_POS | 5 | -0.02291 | -0.00037 | 0.24477 | 0.53728 | 9136.446 |
| GOBP_PEP | 20 | -0.01058 | -0.00034 | 0.11282 | 0.53735 | 9137.637 |
| REACTOME | 12 | -0.01092 | -0.00028 | 0.11636 | 0.53738 | 9138.147 |
| GOBP_POS | 28 | -0.00977 | -0.00038 | 0.10385 | 0.53747 | 9139.677 |
| REACTOME | 25 | -0.00887 | -0.00032 | 0.094157 | 0.53753 | 9140.698 |
| GOMF_HIS | 14 | -0.01106 | -0.0003 | 0.11723 | 0.53758 | 9141.548 |
| GOBP_MAI | 6 | -0.02151 | -0.00038 | 0.22758 | 0.53765 | 9142.738 |
| GOBP_REG | 4 | -0.02508 | -0.00036 | 0.26488 | 0.53771 | 9143.759 |
| GOBP_REG | 40 | -0.00878 | -0.0004 | 0.092731 | 0.53773 | 9144.099 |
| GOBP_POS | 13 | -0.01269 | -0.00033 | 0.13347 | 0.53788 | 9146.649 |
| GOBP_RES | 826 | -0.00171 | -0.00035 | 0.017967 | 0.5379 | 9146.99 |
| GOBP_NEG | 7 | -0.02166 | -0.00042 | 0.22681 | 0.53805 | 9149.54 |
| SHARMA_A | 5 | -0.0249 | -0.00041 | 0.2603 | 0.5381 | 9150.391 |
| GOBP_RES | 20 | -0.01081 | -0.00035 | 0.11282 | 0.53816 | 9151.411 |
| JAZAG_TGI | 103 | -0.00429 | -0.00032 | 0.044706 | 0.53819 | 9151.921 |
| WP_PERTU | 16 | -0.01058 | -0.00031 | 0.11031 | 0.53821 | 9152.261 |
| CROONQU | 70 | -0.00556 | -0.00034 | 0.057854 | 0.53826 | 9153.111 |
| GOBP_POS | 11 | -0.01295 | -0.00031 | 0.13448 | 0.53835 | 9154.642 |
| GOBP_RES | 527 | -0.00213 | -0.00035 | 0.022167 | 0.53836 | 9154.812 |
| WP_MYD8 | 15 | -0.01242 | -0.00035 | 0.12841 | 0.53852 | 9157.533 |
| GOBP_ORG | 942 | -0.00158 | -0.00034 | 0.016315 | 0.53854 | 9157.873 |
| GOBP_REG | 25 | -0.00953 | -0.00035 | 0.09805 | 0.53872 | 9160.934 |
| CADWELL_ | 62 | -0.00607 | -0.00035 | 0.06216 | 0.53887 | 9163.484 |
| GOBP_L_C | 6 | -0.01912 | -0.00034 | 0.1957 | 0.53892 | 9164.335 |
| GOBP_POS | 12 | -0.01454 | -0.00037 | 0.14884 | 0.53892 | 9164.335 |
| GOBP_RES | 29 | -0.00965 | -0.00038 | 0.098648 | 0.53898 | 9165.355 |
| GOBP_REG | 103 | -0.00508 | -0.00037 | 0.051814 | 0.53907 | 9166.885 |
| SMIRNOV_ | 150 | -0.00406 | -0.00036 | 0.041199 | 0.53928 | 9170.456 |
| GOBP_CAR | 33 | -0.00976 | -0.00041 | 0.098789 | 0.53934 | 9171.477 |
| GOBP_NEG | 4 | -0.03015 | -0.00044 | 0.30463 | 0.53942 | 9172.837 |
| GOBP_REG | 7 | -0.01917 | -0.00037 | 0.19367 | 0.53942 | 9172.837 |
| GOBP_REG | 80 | -0.00504 | -0.00033 | 0.050922 | 0.53944 | 9173.177 |

| | | | | | | |
|---|---|---|---|---|---|---|
| GOBP_MA( | 38 | -0.00819 | -0.00037 | 0.082658 | 0.53946 | 9173.517 |
| GOBP_REG | 6 | -0.02435 | -0.00043 | 0.24572 | 0.53947 | 9173.687 |
| GOBP_POS | 36 | -0.00888 | -0.00039 | 0.089632 | 0.53948 | 9173.857 |
| GOBP_ORG | 67 | -0.00598 | -0.00036 | 0.06031 | 0.53949 | 9174.027 |
| IVANOVA_ | 20 | -0.01089 | -0.00035 | 0.10945 | 0.53963 | 9176.408 |
| REACTOME | 9 | -0.01423 | -0.00031 | 0.14299 | 0.53964 | 9176.578 |
| SUZUKI_RE | 20 | -0.01275 | -0.00041 | 0.12788 | 0.5397 | 9177.599 |
| REACTOME | 10 | -0.01491 | -0.00034 | 0.14944 | 0.53973 | 9178.109 |
| REACTOME | 66 | -0.0061 | -0.00036 | 0.060991 | 0.53983 | 9179.809 |
| GOBP_RES | 5 | -0.02838 | -0.00046 | 0.2834 | 0.53989 | 9180.829 |
| PID_ATF2_ | 59 | -0.00669 | -0.00037 | 0.066625 | 0.53997 | 9182.19 |
| REACTOME | 12 | -0.01418 | -0.00036 | 0.14118 | 0.53999 | 9182.53 |
| GOBP_CAR | 10 | -0.01834 | -0.00042 | 0.18246 | 0.54003 | 9183.21 |
| JOHNSTON | 398 | -0.00258 | -0.00037 | 0.025649 | 0.54007 | 9183.89 |
| GOBP_ARG | 9 | -0.01675 | -0.00037 | 0.16633 | 0.5401 | 9184.401 |
| GOCC_API( | 9 | -0.0155 | -0.00034 | 0.15359 | 0.54018 | 9185.761 |
| WP_INTEG | 97 | -0.00519 | -0.00037 | 0.051257 | 0.5403 | 9187.802 |
| GOBP_MA( | 12 | -0.01307 | -0.00033 | 0.12857 | 0.54048 | 9190.862 |
| GOBP_LIPI | 189 | -0.00356 | -0.00035 | 0.03491 | 0.54063 | 9193.413 |
| GOBP_SUB | 26 | -0.01201 | -0.00045 | 0.11766 | 0.54064 | 9193.583 |
| GOBP_CHE | 10 | -0.01444 | -0.00033 | 0.14125 | 0.54071 | 9194.774 |
| BIOCARTA_ | 7 | -0.02012 | -0.00039 | 0.19674 | 0.54073 | 9195.114 |
| GOBP_DN/ | 28 | -0.00989 | -0.00038 | 0.096684 | 0.54074 | 9195.284 |
| WP_SULINI | 5 | -0.0259 | -0.00042 | 0.25309 | 0.54075 | 9195.454 |
| GOBP_POS | 10 | -0.01913 | -0.00044 | 0.18651 | 0.54086 | 9197.324 |
| GOCC_BOX | 4 | -0.02328 | -0.00034 | 0.22612 | 0.54101 | 9199.875 |
| GOCC_BOX | 4 | -0.02328 | -0.00034 | 0.22612 | 0.54101 | 9199.875 |
| GOMF_BO) | 4 | -0.02328 | -0.00034 | 0.22612 | 0.54101 | 9199.875 |
| GOCC_SECI | 979 | -0.00166 | -0.00037 | 0.016081 | 0.54103 | 9200.215 |
| GOBP_BINI | 36 | -0.00926 | -0.0004 | 0.089564 | 0.54118 | 9202.766 |
| BIOCARTA_ | 18 | -0.01063 | -0.00033 | 0.10274 | 0.54119 | 9202.936 |
| GOBP_RES | 66 | -0.00667 | -0.00039 | 0.064491 | 0.5412 | 9203.106 |
| GOBP_REG | 6 | -0.02355 | -0.00042 | 0.22671 | 0.54136 | 9205.827 |
| GOBP_POS | 18 | -0.0129 | -0.0004 | 0.12397 | 0.54143 | 9207.017 |
| REACTOME | 56 | -0.00695 | -0.00038 | 0.066704 | 0.54148 | 9207.867 |
| GOBP_MIT | 13 | -0.0156 | -0.00041 | 0.14961 | 0.54153 | 9208.718 |
| GOCC_INN | 6 | -0.01766 | -0.00031 | 0.16929 | 0.54154 | 9208.888 |
| GOBP_REG | 41 | -0.00799 | -0.00037 | 0.076575 | 0.54157 | 9209.398 |
| GOCC_NU( | 6 | -0.0208 | -0.00037 | 0.1989 | 0.54165 | 9210.758 |
| REACTOME | 58 | -0.00674 | -0.00037 | 0.064215 | 0.5418 | 9213.309 |
| GOBP_OVU | 21 | -0.01279 | -0.00043 | 0.12166 | 0.54186 | 9214.329 |
| GOCC_EXT | 8 | -0.02099 | -0.00043 | 0.19956 | 0.54188 | 9214.669 |
| GOBP_CELI | 9 | -0.01669 | -0.00036 | 0.15771 | 0.54214 | 9219.091 |
| GOBP_EPIT | 134 | -0.00478 | -0.0004 | 0.045172 | 0.54215 | 9219.261 |
| GOBP_PRE | 11 | -0.01603 | -0.00039 | 0.15101 | 0.54227 | 9221.301 |

| | | | | | | |
|---|---|---|---|---|---|---|
| GOBP_POS | 9 | -0.01824 | -0.0004 | 0.17154 | 0.54233 | 9222.322 |
| GOBP_ME( | 20 | -0.01173 | -0.00038 | 0.11023 | 0.54236 | 9222.832 |
| WP_METH | 21 | -0.01123 | -0.00037 | 0.10515 | 0.54251 | 9225.383 |
| MANNE_C( | 200 | -0.0041 | -0.00042 | 0.038359 | 0.54259 | 9226.743 |
| GOMF_CAL | 647 | -0.00231 | -0.00042 | 0.02153 | 0.54271 | 9228.784 |
| GOBP_REG | 126 | -0.00472 | -0.00038 | 0.043995 | 0.54274 | 9229.294 |
| GOCC_NPE | 14 | -0.01367 | -0.00037 | 0.12733 | 0.54274 | 9229.294 |
| GOBP_REG | 161 | -0.00455 | -0.00042 | 0.042254 | 0.54288 | 9231.674 |
| GOMF_UD | 10 | -0.01768 | -0.00041 | 0.16409 | 0.54289 | 9231.844 |
| REACTOME | 27 | -0.0115 | -0.00043 | 0.10648 | 0.54299 | 9233.545 |
| GOBP_WA | 56 | -0.00732 | -0.0004 | 0.067652 | 0.54307 | 9234.905 |
| GOBP_FAT | 7 | -0.02622 | -0.0005 | 0.2417 | 0.54319 | 9236.946 |
| GOBP_PRC | 5 | -0.02452 | -0.0004 | 0.22561 | 0.54327 | 9238.306 |
| GOMF_ADI | 7 | -0.02275 | -0.00044 | 0.20881 | 0.54337 | 9240.007 |
| GOTZMANI | 198 | -0.00368 | -0.00037 | 0.033725 | 0.54344 | 9241.197 |
| BIOCARTA_ | 9 | -0.01451 | -0.00032 | 0.13259 | 0.54357 | 9243.408 |
| GOBP_MA! | 56 | -0.00728 | -0.0004 | 0.066563 | 0.54357 | 9243.408 |
| GOBP_MIT | 6 | -0.01665 | -0.0003 | 0.15187 | 0.54365 | 9244.768 |
| WP_IMMU | 36 | -0.00957 | -0.00042 | 0.087079 | 0.54374 | 9246.299 |
| GOBP_RNA | 147 | -0.00416 | -0.00037 | 0.037879 | 0.54375 | 9246.469 |
| GOBP_REG | 10 | -0.01715 | -0.00039 | 0.15581 | 0.54381 | 9247.489 |
| GOMF_INC | 5 | -0.02489 | -0.0004 | 0.22607 | 0.54383 | 9247.829 |
| GOBP_B_C | 260 | -0.00365 | -0.00043 | 0.033141 | 0.5439 | 9249.02 |
| GOBP_NEG | 6 | -0.01843 | -0.00033 | 0.16689 | 0.54398 | 9250.38 |
| GOBP_REG | 5 | -0.02642 | -0.00043 | 0.23905 | 0.544 | 9250.72 |
| HSIAO_LIVI | 234 | -0.00364 | -0.0004 | 0.032911 | 0.544 | 9250.72 |
| WP_ANGIC | 126 | -0.00484 | -0.00039 | 0.043736 | 0.54401 | 9250.89 |
| CHEN_ETV! | 21 | -0.01087 | -0.00036 | 0.098028 | 0.54413 | 9252.931 |
| GOBP_NEG | 31 | -0.00947 | -0.00038 | 0.085458 | 0.54413 | 9252.931 |
| GOBP_STR | 7 | -0.02567 | -0.00049 | 0.23098 | 0.54424 | 9254.801 |
| REACTOME | 9 | -0.01883 | -0.00041 | 0.16927 | 0.5443 | 9255.822 |
| GOBP_PRC | 18 | -0.01347 | -0.00042 | 0.12099 | 0.54433 | 9256.332 |
| GOMF_NU | 27 | -0.01032 | -0.00039 | 0.092652 | 0.54434 | 9256.502 |
| GOCC_CUL | 12 | -0.01601 | -0.0004 | 0.14351 | 0.54442 | 9257.862 |
| REACTOME | 29 | -0.00975 | -0.00038 | 0.087368 | 0.54442 | 9257.862 |
| GOBP_POS | 10 | -0.01764 | -0.00041 | 0.15798 | 0.54445 | 9258.372 |
| GOBP_CAR | 608 | -0.00226 | -0.0004 | 0.020214 | 0.54448 | 9258.882 |
| GOBP_REG | 12 | -0.01519 | -0.00038 | 0.13565 | 0.54459 | 9260.753 |
| GOCC_SPE( | 58 | -0.00718 | -0.0004 | 0.064045 | 0.54461 | 9261.093 |
| PEPPER_CH | 29 | -0.01138 | -0.00045 | 0.1014 | 0.54468 | 9262.283 |
| MOOTHA_ | 22 | -0.01078 | -0.00037 | 0.09557 | 0.54491 | 9266.195 |
| PEREZ_TP5 | 188 | -0.00432 | -0.00043 | 0.038295 | 0.54492 | 9266.365 |
| HOFFMANI | 42 | -0.00874 | -0.00041 | 0.077479 | 0.54493 | 9266.535 |
| GOMF_NAI | 5 | -0.02459 | -0.0004 | 0.21779 | 0.54495 | 9266.875 |
| GOBP_NEG | 10 | -0.01833 | -0.00042 | 0.16225 | 0.54497 | 9267.215 |

| | | | | | | |
|---|---|---|---|---|---|---|
| GOBP_CELI | 172 | -0.00441 | -0.00042 | 0.039059 | 0.54498 | 9267.385 |
| GOBP_EOS | 6 | -0.02319 | -0.00041 | 0.20508 | 0.545 | 9267.725 |
| REACTOME | 45 | -0.00944 | -0.00046 | 0.083459 | 0.54503 | 9268.235 |
| GOBP_GRC | 23 | -0.01248 | -0.00044 | 0.11009 | 0.54511 | 9269.596 |
| CHESLER_E | 34 | -0.0092 | -0.00039 | 0.081112 | 0.54515 | 9270.276 |
| FAELT_B_C | 44 | -0.00908 | -0.00044 | 0.079962 | 0.54519 | 9270.956 |
| GOBP_EXT | 15 | -0.01553 | -0.00044 | 0.13678 | 0.5452 | 9271.126 |
| GOMF_MIT | 16 | -0.01371 | -0.0004 | 0.12055 | 0.54526 | 9272.146 |
| GOMF_LNC | 9 | -0.01815 | -0.0004 | 0.15956 | 0.54529 | 9272.656 |
| MATSUDA_ | 452 | -0.00275 | -0.00042 | 0.024066 | 0.54557 | 9277.418 |
| WP_TCELL_ | 58 | -0.00733 | -0.00041 | 0.063833 | 0.54574 | 9280.309 |
| GOMF_G_I | 11 | -0.0156 | -0.00038 | 0.13575 | 0.54575 | 9280.479 |
| GOBP_SPEI | 36 | -0.00928 | -0.00041 | 0.080707 | 0.54579 | 9281.159 |
| GOBP_PRC | 9 | -0.01836 | -0.0004 | 0.15949 | 0.54581 | 9281.499 |
| WP_TNFRE | 39 | -0.00884 | -0.0004 | 0.076752 | 0.54586 | 9282.349 |
| SCIAN_CEL | 22 | -0.01187 | -0.0004 | 0.103 | 0.54587 | 9282.519 |
| REACTOME | 25 | -0.01206 | -0.00044 | 0.10461 | 0.54588 | 9282.689 |
| REACTOME | 5 | -0.02212 | -0.00036 | 0.19181 | 0.54591 | 9283.2 |
| GOBP_MAI | 10 | -0.01782 | -0.00041 | 0.15439 | 0.54594 | 9283.71 |
| REACTOME | 10 | -0.01917 | -0.00044 | 0.1661 | 0.54594 | 9283.71 |
| FINETTI_BF | 20 | -0.01325 | -0.00043 | 0.11466 | 0.54599 | 9284.56 |
| GOBP_NEG | 6 | -0.02133 | -0.00038 | 0.18459 | 0.54599 | 9284.56 |
| GOBP_CELI | 24 | -0.01251 | -0.00045 | 0.10819 | 0.54602 | 9285.07 |
| GOBP_LEN | 6 | -0.02375 | -0.00042 | 0.20541 | 0.54603 | 9285.24 |
| REACTOME | 20 | -0.01276 | -0.00042 | 0.11035 | 0.54603 | 9285.24 |
| GOBP_END | 61 | -0.00719 | -0.00041 | 0.062179 | 0.54604 | 9285.41 |
| PID_RB_1P | 60 | -0.0076 | -0.00043 | 0.065706 | 0.54605 | 9285.58 |
| GOBP_CELI | 867 | -0.0021 | -0.00044 | 0.018151 | 0.54606 | 9285.75 |
| GOMF_HIS | 9 | -0.01724 | -0.00038 | 0.14847 | 0.54621 | 9288.301 |
| REACTOME | 158 | -0.00415 | -0.00038 | 0.035754 | 0.54622 | 9288.471 |
| GOBP_REG | 341 | -0.00314 | -0.00042 | 0.026941 | 0.54639 | 9291.362 |
| GOBP_CELI | 7 | -0.02765 | -0.00053 | 0.23694 | 0.54644 | 9292.212 |
| GOBP_APC | 1740 | -0.00147 | -0.00043 | 0.012591 | 0.54653 | 9293.743 |
| GOCC_PLA | 14 | -0.01413 | -0.00038 | 0.12087 | 0.54654 | 9293.913 |
| GOBP_POS | 6 | -0.02681 | -0.00048 | 0.22912 | 0.54657 | 9294.423 |
| REACTOME | 16 | -0.01369 | -0.0004 | 0.11683 | 0.54663 | 9295.443 |
| REACTOME | 9 | -0.02016 | -0.00044 | 0.17193 | 0.54666 | 9295.953 |
| GOMF_NAI | 18 | -0.01348 | -0.00042 | 0.11487 | 0.54672 | 9296.974 |
| GOCC_ORC | 507 | -0.00242 | -0.00039 | 0.02058 | 0.54682 | 9298.674 |
| WP_QUERC | 15 | -0.01459 | -0.00041 | 0.12404 | 0.54682 | 9298.674 |
| GOCC_LAT | 5 | -0.02978 | -0.00048 | 0.25288 | 0.54687 | 9299.524 |
| BIOCARTA_ | 13 | -0.01523 | -0.0004 | 0.12931 | 0.54689 | 9299.864 |
| REACTOME | 6 | -0.02136 | -0.00038 | 0.181 | 0.54697 | 9301.225 |
| GOMF_CO_ | 8 | -0.02133 | -0.00044 | 0.18016 | 0.54711 | 9303.606 |
| REN_MIF_ | 5 | -0.02737 | -0.00045 | 0.23114 | 0.54712 | 9303.776 |

| | | | | | | |
|---|---|---|---|---|---|---|
| GOBP_CON | 10 | -0.02118 | -0.00049 | 0.1787 | 0.54716 | 9304.456 |
| GOBP_ALP | 111 | -0.00588 | -0.00045 | 0.049533 | 0.54724 | 9305.816 |
| REACTOME | 15 | -0.0176 | -0.0005 | 0.14782 | 0.54739 | 9308.367 |
| REACTOME | 6 | -0.02815 | -0.0005 | 0.23601 | 0.54748 | 9309.897 |
| GOMF_OLE | 6 | -0.02407 | -0.00043 | 0.20169 | 0.5475 | 9310.238 |
| GOBP_POS | 41 | -0.00936 | -0.00044 | 0.078362 | 0.54753 | 9310.748 |
| GOCC_END | 305 | -0.00329 | -0.00041 | 0.02743 | 0.54769 | 9313.468 |
| GOMF_LON | 14 | -0.01499 | -0.00041 | 0.12508 | 0.54769 | 9313.468 |
| REACTOME | 20 | -0.01385 | -0.00045 | 0.11555 | 0.54769 | 9313.468 |
| GOBP_REG | 66 | -0.00809 | -0.00048 | 0.067071 | 0.548 | 9318.74 |
| REACTOME | 7 | -0.02337 | -0.00045 | 0.19316 | 0.54814 | 9321.121 |
| GOBP_NEG | 11 | -0.01535 | -0.00037 | 0.12664 | 0.54825 | 9322.991 |
| GOBP_NEG | 31 | -0.00967 | -0.00039 | 0.079519 | 0.54838 | 9325.202 |
| GOBP_REG | 29 | -0.01284 | -0.0005 | 0.1054 | 0.54849 | 9327.072 |
| GOBP_REG | 51 | -0.00797 | -0.00041 | 0.065217 | 0.54863 | 9329.453 |
| GOCC_PRO | 17 | -0.0144 | -0.00043 | 0.11767 | 0.54869 | 9330.473 |
| GOBP_NUC | 8 | -0.01982 | -0.00041 | 0.1619 | 0.54872 | 9330.984 |
| GOBP_RES | 6 | -0.02128 | -0.00038 | 0.17377 | 0.54872 | 9330.984 |
| REACTOME | 9 | -0.02409 | -0.00053 | 0.19664 | 0.54874 | 9331.324 |
| MEINHOLD | 19 | -0.01327 | -0.00042 | 0.10821 | 0.54879 | 9332.174 |
| GOBP_REG | 79 | -0.00701 | -0.00045 | 0.057155 | 0.54883 | 9332.854 |
| WIKMAN_A | 29 | -0.0108 | -0.00042 | 0.087914 | 0.54886 | 9333.364 |
| PID_NFKAP | 20 | -0.01374 | -0.00045 | 0.11125 | 0.54914 | 9338.126 |
| REACTOME | 5 | -0.03464 | -0.00056 | 0.27991 | 0.54924 | 9339.826 |
| REACTOME | 73 | -0.00701 | -0.00043 | 0.056467 | 0.54938 | 9342.207 |
| REACTOME | 44 | -0.00951 | -0.00046 | 0.076533 | 0.54945 | 9343.397 |
| WP_TCA_C | 9 | -0.02254 | -0.00049 | 0.18135 | 0.54945 | 9343.397 |
| GOCC_NUF | 5 | -0.04141 | -0.00067 | 0.33316 | 0.54946 | 9343.567 |
| GOBP_POS | 16 | -0.01488 | -0.00043 | 0.11957 | 0.5495 | 9344.248 |
| GOBP_NEG | 7 | -0.02361 | -0.00045 | 0.18976 | 0.54951 | 9344.418 |
| GOBP_L_AS | 8 | -0.02558 | -0.00053 | 0.20554 | 0.54952 | 9344.588 |
| GOBP_POS | 51 | -0.00891 | -0.00046 | 0.071421 | 0.54962 | 9346.288 |
| GOCC_SUM | 13 | -0.01742 | -0.00046 | 0.13957 | 0.54967 | 9347.138 |
| GOBP_EME | 30 | -0.0117 | -0.00047 | 0.093577 | 0.54975 | 9348.499 |
| GOMF_EUI | 9 | -0.02062 | -0.00045 | 0.16486 | 0.54978 | 9349.009 |
| ABBUD_LIF | 40 | -0.01022 | -0.00047 | 0.08162 | 0.54983 | 9349.859 |
| REACTOME | 7 | -0.02157 | -0.00042 | 0.17199 | 0.5499 | 9351.05 |
| GOMF_OXA | 9 | -0.0201 | -0.00044 | 0.16025 | 0.54991 | 9351.22 |
| GOBP_CILIU | 4 | -0.03386 | -0.00049 | 0.26956 | 0.54998 | 9352.41 |
| GOBP_GLY | 15 | -0.01713 | -0.00048 | 0.13633 | 0.54998 | 9352.41 |
| GOMF_LAC | 6 | -0.02446 | -0.00044 | 0.19474 | 0.54998 | 9352.41 |
| LI_DCP2_B | 82 | -0.00668 | -0.00044 | 0.053216 | 0.54998 | 9352.41 |
| GOMF_S_A | 18 | -0.01416 | -0.00044 | 0.11255 | 0.55006 | 9353.77 |
| GOCC_VES | 1130 | -0.00191 | -0.00045 | 0.015203 | 0.55011 | 9354.621 |
| MCCABE_H | 19 | -0.01368 | -0.00043 | 0.1086 | 0.55011 | 9354.621 |

| | | | | | | |
|---|---|---|---|---|---|---|
| GOBP_REG | 28 | -0.01078 | -0.00041 | 0.085417 | 0.55019 | 9355.981 |
| REACTOME | 13 | -0.017 | -0.00045 | 0.13474 | 0.5502 | 9356.151 |
| GOBP_SHC | 10 | -0.0184 | -0.00042 | 0.14568 | 0.55026 | 9357.171 |
| GOMF_BILI | 10 | -0.02095 | -0.00048 | 0.16572 | 0.5503 | 9357.852 |
| REACTOME | 18 | -0.01419 | -0.00044 | 0.1122 | 0.55031 | 9358.022 |
| POS_RESPC | 11 | -0.01766 | -0.00043 | 0.13931 | 0.55043 | 9360.062 |
| GOBP_MIT | 14 | -0.01757 | -0.00048 | 0.13859 | 0.55045 | 9360.402 |
| GOBP_HIST | 5 | -0.03029 | -0.00049 | 0.23806 | 0.55062 | 9363.293 |
| GOMF_LIPI | 164 | -0.00506 | -0.00047 | 0.039766 | 0.55064 | 9363.633 |
| GOMF_RET | 8 | -0.0252 | -0.00052 | 0.19782 | 0.55068 | 9364.313 |
| GOCC_INO | 15 | -0.01558 | -0.00044 | 0.12194 | 0.55082 | 9366.694 |
| GOBP_RES | 11 | -0.02071 | -0.0005 | 0.16174 | 0.55094 | 9368.735 |
| CAFFAREL_ | 52 | -0.00811 | -0.00043 | 0.06332 | 0.55095 | 9368.905 |
| LEE_AGING | 79 | -0.00674 | -0.00043 | 0.05258 | 0.55097 | 9369.245 |
| GOBP_RES | 372 | -0.00331 | -0.00046 | 0.025842 | 0.55102 | 9370.095 |
| GOCC_CYC | 47 | -0.0086 | -0.00043 | 0.066975 | 0.55106 | 9370.775 |
| GOCC_PHA | 6 | -0.02441 | -0.00044 | 0.19008 | 0.55109 | 9371.285 |
| RUIZ_TNC_ | 139 | -0.00566 | -0.00048 | 0.04405 | 0.5511 | 9371.456 |
| CHIBA_RES | 20 | -0.01332 | -0.00043 | 0.10371 | 0.55111 | 9371.626 |
| GOMF_MH | 12 | -0.02342 | -0.00059 | 0.18213 | 0.55116 | 9372.476 |
| WP_NEURC | 56 | -0.00898 | -0.00049 | 0.069711 | 0.55126 | 9374.176 |
| GOBP_REG | 11 | -0.02081 | -0.0005 | 0.16144 | 0.55129 | 9374.686 |
| BIOCARTA_ | 5 | -0.03532 | -0.00057 | 0.27322 | 0.55143 | 9377.067 |
| GOMF_STR | 4 | -0.03148 | -0.00046 | 0.24338 | 0.55146 | 9377.577 |
| GOBP_LYM | 4 | -0.03288 | -0.00048 | 0.25296 | 0.55171 | 9381.829 |
| GOBP_ORG | 547 | -0.00273 | -0.00046 | 0.020931 | 0.5519 | 9385.06 |
| CONCANN( | 222 | -0.00429 | -0.00046 | 0.032877 | 0.55194 | 9385.74 |
| HOUSTIS_F | 32 | -0.0125 | -0.00051 | 0.0953 | 0.55218 | 9389.821 |
| HAN_JNK_S | 36 | -0.01211 | -0.00053 | 0.092156 | 0.55227 | 9391.351 |
| GOCC_TOR | 5 | -0.02815 | -0.00046 | 0.21396 | 0.55233 | 9392.372 |
| GOBP_NUC | 23 | -0.01374 | -0.00048 | 0.10443 | 0.55234 | 9392.542 |
| GOBP_NEG | 5 | -0.03288 | -0.00053 | 0.24973 | 0.55237 | 9393.052 |
| GOBP_NEG | 22 | -0.01426 | -0.00049 | 0.10807 | 0.55248 | 9394.922 |
| REACTOME | 8 | -0.02338 | -0.00048 | 0.17687 | 0.55258 | 9396.623 |
| GOMF_CLA | 67 | -0.00755 | -0.00045 | 0.057054 | 0.55262 | 9397.303 |
| GOMF_FAT | 49 | -0.0092 | -0.00047 | 0.06941 | 0.5527 | 9398.664 |
| GOMF_PYR | 8 | -0.02401 | -0.00049 | 0.18111 | 0.55272 | 9399.004 |
| GOBP_MAT | 5 | -0.02483 | -0.0004 | 0.18731 | 0.55274 | 9399.344 |
| INGA_TP53 | 15 | -0.01586 | -0.00045 | 0.11959 | 0.55274 | 9399.344 |
| GOBP_REG | 17 | -0.01815 | -0.00054 | 0.13676 | 0.55278 | 9400.024 |
| GOBP_CMI | 7 | -0.02493 | -0.00048 | 0.18781 | 0.5528 | 9400.364 |
| GOBP_INSU | 15 | -0.01737 | -0.00049 | 0.13082 | 0.55282 | 9400.704 |
| GOMF_TRA | 105 | -0.00636 | -0.00047 | 0.047824 | 0.55291 | 9402.235 |
| FUJIWARA_ | 6 | -0.02798 | -0.0005 | 0.21027 | 0.55293 | 9402.575 |
| REACTOME | 6 | -0.03039 | -0.00054 | 0.22835 | 0.55293 | 9402.575 |

| | | | | | | |
|---|---|---|---|---|---|---|
| GOBP_POS | 33 | -0.01259 | -0.00053 | 0.09462 | 0.55294 | 9402.745 |
| GOBP_REG | 8 | -0.02688 | -0.00055 | 0.20174 | 0.553 | 9403.765 |
| GOMF_ABC | 14 | -0.01922 | -0.00052 | 0.1442 | 0.55302 | 9404.105 |
| MARKS_AC | 11 | -0.0178 | -0.00043 | 0.13326 | 0.55312 | 9405.806 |
| BIOCARTA_ | 13 | -0.01689 | -0.00044 | 0.12639 | 0.55316 | 9406.486 |
| BEGUM_TA | 7 | -0.0305 | -0.00059 | 0.22814 | 0.55318 | 9406.826 |
| MIKKELSEN | 40 | -0.01022 | -0.00047 | 0.076308 | 0.55325 | 9408.016 |
| GOBP_POS | 9 | -0.02301 | -0.0005 | 0.17168 | 0.55332 | 9409.207 |
| WANG_RES | 332 | -0.00366 | -0.00048 | 0.027244 | 0.5534 | 9410.567 |
| GOBP_POD | 7 | -0.02661 | -0.00051 | 0.19817 | 0.55341 | 9410.737 |
| GOBP_NEG | 42 | -0.01187 | -0.00056 | 0.088131 | 0.55356 | 9413.288 |
| OZEN_MIR | 22 | -0.01412 | -0.00048 | 0.10458 | 0.5537 | 9415.669 |
| HOSHIDA_I | 63 | -0.0082 | -0.00047 | 0.060595 | 0.5538 | 9417.369 |
| ZHOU_INFI | 348 | -0.00352 | -0.00047 | 0.026011 | 0.55385 | 9418.219 |
| GOMF_TUI | 41 | -0.01109 | -0.00052 | 0.081641 | 0.554 | 9420.77 |
| GOBP_PRC | 29 | -0.01295 | -0.00051 | 0.09495 | 0.55424 | 9424.851 |
| PID_SMAD | 75 | -0.00798 | -0.0005 | 0.058445 | 0.55428 | 9425.531 |
| TURASHVIL | 21 | -0.01504 | -0.0005 | 0.11012 | 0.5543 | 9425.872 |
| GOCC_MYC | 52 | -0.00987 | -0.00052 | 0.072237 | 0.55433 | 9426.382 |
| HOLLEMAN | 37 | -0.01158 | -0.00051 | 0.084621 | 0.55443 | 9428.082 |
| GOBP_REG | 17 | -0.01527 | -0.00046 | 0.11151 | 0.55446 | 9428.592 |
| GOBP_NEG | 8 | -0.02506 | -0.00052 | 0.1829 | 0.55448 | 9428.932 |
| GOBP_POS | 54 | -0.01004 | -0.00054 | 0.073282 | 0.55448 | 9428.932 |
| GOBP_REG | 14 | -0.01901 | -0.00052 | 0.13879 | 0.55448 | 9428.932 |
| GOBP_POS | 4 | -0.02902 | -0.00042 | 0.2118 | 0.5545 | 9429.273 |
| GOBP_GLY | 29 | -0.01292 | -0.00051 | 0.093967 | 0.55466 | 9431.993 |
| GOBP_MO | 22 | -0.01531 | -0.00052 | 0.11128 | 0.55469 | 9432.503 |
| GOBP_PRC | 7 | -0.02983 | -0.00057 | 0.21683 | 0.55471 | 9432.844 |
| GOMF_3_H | 5 | -0.03984 | -0.00065 | 0.28928 | 0.55477 | 9433.864 |
| PID_RAC1_ | 36 | -0.01171 | -0.00051 | 0.084978 | 0.55479 | 9434.204 |
| GOBP_PRC | 11 | -0.02018 | -0.00049 | 0.14647 | 0.5548 | 9434.374 |
| REACTOME | 51 | -0.00969 | -0.0005 | 0.070182 | 0.55491 | 9436.245 |
| GOBP_REG | 15 | -0.01812 | -0.00051 | 0.13118 | 0.55493 | 9436.585 |
| GOBP_OPT | 6 | -0.0331 | -0.00059 | 0.23957 | 0.55494 | 9436.755 |
| TESAR_JAK | 6 | -0.03207 | -0.00057 | 0.23215 | 0.55494 | 9436.755 |
| KANNAN_T | 22 | -0.01451 | -0.0005 | 0.10494 | 0.555 | 9437.775 |
| ZHAN_MUI | 42 | -0.01082 | -0.00051 | 0.07825 | 0.555 | 9437.775 |
| GOBP_REG | 60 | -0.00877 | -0.00049 | 0.06333 | 0.55507 | 9438.965 |
| BILANGES_ | 58 | -0.00939 | -0.00052 | 0.067793 | 0.55508 | 9439.135 |
| GOBP_TRIC | 5 | -0.03459 | -0.00056 | 0.24973 | 0.55509 | 9439.305 |
| WANG_ME | 27 | -0.01425 | -0.00054 | 0.10271 | 0.55517 | 9440.666 |
| GOBP_POS | 254 | -0.00463 | -0.00053 | 0.033289 | 0.55536 | 9443.897 |
| GOBP_PAN | 13 | -0.01922 | -0.0005 | 0.13792 | 0.55542 | 9444.917 |
| VERRECCHI | 53 | -0.00955 | -0.00051 | 0.068472 | 0.55546 | 9445.597 |
| GOBP_OLIC | 16 | -0.01569 | -0.00046 | 0.11246 | 0.55547 | 9445.767 |

| | | | | | | |
|---|---|---|---|---|---|---|
| KIM_MYCN | 101 | -0.0076 | -0.00055 | 0.054474 | 0.55547 | 9445.767 |
| GOBP_POS | 210 | -0.00504 | -0.00053 | 0.036059 | 0.55556 | 9447.298 |
| KEGG_REN | 67 | -0.0087 | -0.00052 | 0.062134 | 0.55565 | 9448.828 |
| OSWALD_H | 257 | -0.00432 | -0.0005 | 0.030849 | 0.55575 | 9450.529 |
| GOMF_NM | 8 | -0.02426 | -0.0005 | 0.17277 | 0.55584 | 9452.059 |
| GOBP_POS | 8 | -0.02386 | -0.00049 | 0.16947 | 0.55597 | 9454.27 |
| MIKKELSEN | 7 | -0.02832 | -0.00055 | 0.20088 | 0.55606 | 9455.8 |
| WP_AMYO | 34 | -0.01203 | -0.00051 | 0.085068 | 0.55623 | 9458.691 |
| GOBP_INTI | 18 | -0.01853 | -0.00057 | 0.13095 | 0.55626 | 9459.201 |
| BLANCO_M | 34 | -0.01184 | -0.0005 | 0.083609 | 0.5563 | 9459.882 |
| GOBP_NEG | 7 | -0.02086 | -0.0004 | 0.14725 | 0.55632 | 9460.222 |
| GOBP_TRA | 51 | -0.00908 | -0.00047 | 0.064001 | 0.55639 | 9461.412 |
| GOBP_COL | 47 | -0.01124 | -0.00056 | 0.079128 | 0.5565 | 9463.283 |
| GOMF_INC | 5 | -0.03116 | -0.00051 | 0.21884 | 0.55662 | 9465.323 |
| STONER_ES | 7 | -0.02475 | -0.00048 | 0.17365 | 0.55667 | 9466.173 |
| GOMF_INT | 145 | -0.00619 | -0.00054 | 0.043402 | 0.55668 | 9466.343 |
| GOBP_REG | 16 | -0.01947 | -0.00057 | 0.13586 | 0.55699 | 9471.615 |
| GOBP_CAT | 12 | -0.0228 | -0.00057 | 0.15901 | 0.55701 | 9471.955 |
| MCCABE_B | 383 | -0.00372 | -0.00053 | 0.025908 | 0.55715 | 9474.336 |
| GOBP_POS | 5 | -0.03173 | -0.00052 | 0.22066 | 0.55717 | 9474.676 |
| GOBP_ATP | 193 | -0.0049 | -0.00049 | 0.033965 | 0.55734 | 9477.567 |
| GAZDA_DIA | 30 | -0.01427 | -0.00057 | 0.098669 | 0.55748 | 9479.947 |
| WP_CELL_I | 21 | -0.0189 | -0.00063 | 0.13072 | 0.55749 | 9480.117 |
| REACTOME | 57 | -0.00988 | -0.00054 | 0.068229 | 0.55757 | 9481.478 |
| BLANCO_M | 19 | -0.01693 | -0.00054 | 0.11651 | 0.55776 | 9484.709 |
| GOBP_CAR | 5 | -0.02902 | -0.00047 | 0.19966 | 0.55778 | 9485.049 |
| GOBP_FOR | 5 | -0.0357 | -0.00058 | 0.2456 | 0.55779 | 9485.219 |
| GOCC_GOL | 1521 | -0.00193 | -0.00052 | 0.013246 | 0.55782 | 9485.729 |
| BIOCARTA_ | 24 | -0.01366 | -0.00049 | 0.093777 | 0.55791 | 9487.26 |
| CROMER_T | 45 | -0.01196 | -0.00058 | 0.082085 | 0.55792 | 9487.43 |
| DELYS_THY | 420 | -0.00371 | -0.00055 | 0.025431 | 0.55796 | 9488.11 |
| GOBP_LAC | 7 | -0.02711 | -0.00052 | 0.18551 | 0.55809 | 9490.32 |
| SIG_PIP3_S | 63 | -0.00955 | -0.00055 | 0.065314 | 0.55814 | 9491.171 |
| GOBP_REG | 27 | -0.01327 | -0.0005 | 0.090725 | 0.55815 | 9491.341 |
| REACTOME | 32 | -0.01237 | -0.00051 | 0.084552 | 0.55815 | 9491.341 |
| SCHAEFFER | 20 | -0.01852 | -0.0006 | 0.12643 | 0.55822 | 9492.531 |
| REACTOME | 22 | -0.0143 | -0.00049 | 0.097538 | 0.55828 | 9493.551 |
| GOCC_CUL | 168 | -0.00555 | -0.00052 | 0.037817 | 0.55832 | 9494.232 |
| WHITFIELD | 142 | -0.00596 | -0.00051 | 0.040588 | 0.55834 | 9494.572 |
| GOMF_ORG | 155 | -0.00601 | -0.00054 | 0.0409 | 0.55845 | 9496.442 |
| GOBP_MET | 6 | -0.02828 | -0.0005 | 0.19221 | 0.55849 | 9497.122 |
| GOCC_AXO | 597 | -0.00312 | -0.00055 | 0.021184 | 0.55859 | 9498.823 |
| GOBP_PRO | 5 | -0.0422 | -0.00069 | 0.28601 | 0.55865 | 9499.843 |
| GOCC_EXT | 12 | -0.01749 | -0.00044 | 0.11844 | 0.55869 | 9500.523 |
| REACTOME | 17 | -0.0195 | -0.00059 | 0.13185 | 0.5588 | 9502.394 |

| | | | | | | |
|---|---|---|---|---|---|---|
| BIOCARTA_ | 7 | -0.02947 | -0.00057 | 0.19896 | 0.55887 | 9503.584 |
| WP_7OXO( | 24 | -0.01564 | -0.00056 | 0.10555 | 0.55889 | 9503.924 |
| GOCC_ATP | 139 | -0.00619 | -0.00053 | 0.04168 | 0.55903 | 9506.305 |
| GOBP_REG | 425 | -0.00381 | -0.00056 | 0.025613 | 0.55906 | 9506.815 |
| GOBP_MYE | 105 | -0.0071 | -0.00053 | 0.047711 | 0.55912 | 9507.836 |
| GOBP_CELI | 10 | -0.0248 | -0.00057 | 0.16643 | 0.55922 | 9509.536 |
| GOBP_ACT | 7 | -0.02655 | -0.00051 | 0.17814 | 0.55923 | 9509.706 |
| GOBP_MAT | 35 | -0.01131 | -0.00049 | 0.075874 | 0.55923 | 9509.706 |
| GOBP_REG | 49 | -0.01008 | -0.00051 | 0.067388 | 0.55943 | 9513.107 |
| REACTOME | 22 | -0.01641 | -0.00056 | 0.10934 | 0.55963 | 9516.508 |
| REACTOME | 15 | -0.02235 | -0.00063 | 0.14842 | 0.55984 | 9520.079 |
| GOBP_POS | 172 | -0.00607 | -0.00058 | 0.040272 | 0.55987 | 9520.589 |
| GOBP_PIGI | 74 | -0.00864 | -0.00054 | 0.057269 | 0.55995 | 9521.95 |
| REACTOME | 18 | -0.018 | -0.00056 | 0.11889 | 0.56016 | 9525.521 |
| REACTOME | 6 | -0.02135 | -0.00038 | 0.14099 | 0.56019 | 9526.031 |
| GOBP_INTI | 7 | -0.03137 | -0.0006 | 0.20706 | 0.56022 | 9526.541 |
| GOBP_TYP | 23 | -0.01743 | -0.00061 | 0.11493 | 0.56027 | 9527.391 |
| GOBP_B_C | 58 | -0.01016 | -0.00056 | 0.066815 | 0.56044 | 9530.282 |
| GOBP_RES | 23 | -0.01672 | -0.00058 | 0.1099 | 0.56046 | 9530.622 |
| GOCC_SPEI | 13 | -0.01918 | -0.0005 | 0.12606 | 0.56046 | 9530.622 |
| GOMF_TRA | 13 | -0.02273 | -0.0006 | 0.14938 | 0.56047 | 9530.792 |
| GOBP_NEG | 9 | -0.02797 | -0.00061 | 0.18338 | 0.56062 | 9533.343 |
| GOBP_NEG | 6 | -0.03433 | -0.00061 | 0.22508 | 0.56062 | 9533.343 |
| REACTOME | 56 | -0.01059 | -0.00058 | 0.069331 | 0.56067 | 9534.193 |
| REACTOME | 12 | -0.02564 | -0.00065 | 0.16785 | 0.5607 | 9534.704 |
| REACTOME | 85 | -0.00842 | -0.00056 | 0.055132 | 0.56071 | 9534.874 |
| GOBP_NEG | 6 | -0.02474 | -0.00044 | 0.16185 | 0.56074 | 9535.384 |
| GOBP_REG | 11 | -0.02394 | -0.00058 | 0.15652 | 0.56079 | 9536.234 |
| MANNE_C( | 183 | -0.00617 | -0.0006 | 0.04021 | 0.56097 | 9539.295 |
| KEGG_SYST | 72 | -0.01014 | -0.00063 | 0.066049 | 0.56101 | 9539.975 |
| WP_FOLLIC | 27 | -0.01488 | -0.00056 | 0.096691 | 0.56115 | 9542.356 |
| GOBP_ACT | 58 | -0.0112 | -0.00062 | 0.072591 | 0.56128 | 9544.566 |
| GOMF_PR( | 24 | -0.01815 | -0.00065 | 0.11761 | 0.56133 | 9545.417 |
| MARSHALL | 9 | -0.03003 | -0.00066 | 0.19436 | 0.5614 | 9546.607 |
| GOBP_REG | 75 | -0.00927 | -0.00058 | 0.059934 | 0.56145 | 9547.457 |
| GOBP_GLY | 71 | -0.00912 | -0.00056 | 0.058962 | 0.56147 | 9547.797 |
| REACTOME | 14 | -0.02163 | -0.00059 | 0.13916 | 0.56175 | 9552.559 |
| GOBP_POS | 69 | -0.0096 | -0.00058 | 0.061721 | 0.56182 | 9553.749 |
| GOBP_CEN | 38 | -0.01252 | -0.00056 | 0.080295 | 0.56194 | 9555.79 |
| GOBP_FC_ | 22 | -0.01627 | -0.00055 | 0.10369 | 0.56233 | 9562.422 |
| GOMF_ADI | 1530 | -0.00207 | -0.00056 | 0.013176 | 0.56234 | 9562.592 |
| GEISS_RESI | 35 | -0.01395 | -0.0006 | 0.088866 | 0.56238 | 9563.272 |
| GOBP_MEI | 9 | -0.02946 | -0.00064 | 0.18754 | 0.56242 | 9563.952 |
| GOBP_REG | 93 | -0.00822 | -0.00058 | 0.052321 | 0.56243 | 9564.122 |
| GAZDA_DI/ | 417 | -0.00372 | -0.00055 | 0.02358 | 0.5627 | 9568.714 |

| | | | | | | |
|---|---|---|---|---|---|---|
| GOBP_OLI( | 40 | -0.01221 | -0.00056 | 0.077254 | 0.56276 | 9569.734 |
| REACTOME | 83 | -0.00888 | -0.00059 | 0.056186 | 0.56276 | 9569.734 |
| ISHIDA_TA | 6 | -0.03338 | -0.0006 | 0.21083 | 0.5629 | 9572.115 |
| GOBP_REG | 143 | -0.00643 | -0.00056 | 0.040485 | 0.56306 | 9574.835 |
| GOBP_NEU | 43 | -0.01344 | -0.00064 | 0.084617 | 0.56312 | 9575.856 |
| DEBIASI_AI | 254 | -0.00483 | -0.00056 | 0.030274 | 0.56337 | 9580.107 |
| GOBP_NEG | 25 | -0.01642 | -0.0006 | 0.10289 | 0.56338 | 9580.277 |
| GOBP_PLA | 20 | -0.01613 | -0.00052 | 0.10067 | 0.56363 | 9584.528 |
| GOBP_END | 13 | -0.02302 | -0.0006 | 0.14351 | 0.56371 | 9585.889 |
| GOBP_HIST | 29 | -0.01448 | -0.00057 | 0.090267 | 0.56371 | 9585.889 |
| GOBP_SECI | 43 | -0.01305 | -0.00062 | 0.081358 | 0.56373 | 9586.229 |
| MEINHOLD | 12 | -0.02262 | -0.00057 | 0.14067 | 0.56387 | 9588.609 |
| REACTOME | 9 | -0.02848 | -0.00062 | 0.17712 | 0.56387 | 9588.609 |
| KEGG_MAT | 24 | -0.01858 | -0.00066 | 0.11547 | 0.5639 | 9589.12 |
| KONDO_CC | 3 | -0.05275 | -0.00066 | 0.3274 | 0.564 | 9590.82 |
| REACTOME | 29 | -0.01477 | -0.00058 | 0.091514 | 0.5641 | 9592.521 |
| GOBP_REG | 12 | -0.02044 | -0.00052 | 0.12607 | 0.5644 | 9597.622 |
| LANG_MYE | 28 | -0.01542 | -0.00059 | 0.095039 | 0.56445 | 9598.472 |
| NAKAMUR/ | 6 | -0.03029 | -0.00054 | 0.18658 | 0.56448 | 9598.982 |
| KEGG_TRYI | 37 | -0.01344 | -0.00059 | 0.082651 | 0.56459 | 9600.853 |
| GOBP_PTE | 32 | -0.01344 | -0.00055 | 0.082602 | 0.56463 | 9601.533 |
| GOCC_FLO | 8 | -0.02778 | -0.00057 | 0.17065 | 0.56467 | 9602.213 |
| REACTOME | 14 | -0.02354 | -0.00064 | 0.14438 | 0.56474 | 9603.404 |
| GOMF_PHI | 3 | -0.07125 | -0.0009 | 0.43622 | 0.56487 | 9605.614 |
| HUTTMANI | 56 | -0.01203 | -0.00065 | 0.073635 | 0.56487 | 9605.614 |
| MALONEY_ | 36 | -0.01371 | -0.0006 | 0.083935 | 0.56487 | 9605.614 |
| GOCC_HIPI | 5 | -0.03211 | -0.00052 | 0.1965 | 0.56491 | 9606.295 |
| REACTOME | 29 | -0.01588 | -0.00062 | 0.096969 | 0.56502 | 9608.165 |
| GOBP_POS | 90 | -0.00829 | -0.00057 | 0.050605 | 0.56503 | 9608.335 |
| GOBP_POS | 36 | -0.01512 | -0.00066 | 0.092314 | 0.56504 | 9608.505 |
| GOMF_DO | 68 | -0.00971 | -0.00058 | 0.059178 | 0.56515 | 9610.376 |
| REACTOME | 67 | -0.01023 | -0.00061 | 0.062384 | 0.56515 | 9610.376 |
| GOBP_NEG | 7 | -0.02999 | -0.00058 | 0.18262 | 0.56522 | 9611.566 |
| GOBP_REG | 4 | -0.04276 | -0.00062 | 0.26027 | 0.56524 | 9611.906 |
| GOCC_NEU | 3 | -0.0447 | -0.00056 | 0.27188 | 0.5653 | 9612.927 |
| GOBP_POS | 144 | -0.00693 | -0.0006 | 0.042105 | 0.5654 | 9614.627 |
| GOCC_DN/ | 22 | -0.01798 | -0.00061 | 0.10886 | 0.56558 | 9617.688 |
| GOBP_MEI | 5 | -0.04408 | -0.00072 | 0.26674 | 0.56563 | 9618.538 |
| GOBP_REA | 48 | -0.01152 | -0.00058 | 0.069728 | 0.56563 | 9618.538 |
| STARK_BR/ | 11 | -0.02263 | -0.00055 | 0.1368 | 0.56569 | 9619.558 |
| PID_TRKR_ | 60 | -0.01062 | -0.0006 | 0.064016 | 0.56588 | 9622.789 |
| REACTOME | 11 | -0.02489 | -0.0006 | 0.15001 | 0.56589 | 9622.959 |
| REACTOME | 11 | -0.02471 | -0.0006 | 0.14829 | 0.56617 | 9627.721 |
| GOBP_POS | 4 | -0.03426 | -0.0005 | 0.20555 | 0.56618 | 9627.891 |
| REACTOME | 55 | -0.01124 | -0.00061 | 0.0674 | 0.56619 | 9628.061 |

| | | | | | | |
|---|---|---|---|---|---|---|
| GOBP_NEG | 75 | -0.00999 | -0.00063 | 0.059833 | 0.5663 | 9629.932 |
| WP_TCA_C | 5 | -0.03978 | -0.00065 | 0.23817 | 0.56632 | 9630.272 |
| GOBP_AMI | 22 | -0.02019 | -0.00069 | 0.12084 | 0.56634 | 9630.612 |
| BERENJENG | 25 | -0.01554 | -0.00057 | 0.09293 | 0.56639 | 9631.462 |
| GOBP_IMM | 57 | -0.01188 | -0.00065 | 0.070961 | 0.56647 | 9632.822 |
| GOMF_INO | 11 | -0.02728 | -0.00066 | 0.16242 | 0.56668 | 9636.393 |
| GOBP_REG | 26 | -0.0162 | -0.0006 | 0.09636 | 0.56674 | 9637.414 |
| WANG_RES | 30 | -0.01417 | -0.00056 | 0.084303 | 0.56675 | 9637.584 |
| WHITFIELD | 117 | -0.00739 | -0.00058 | 0.043812 | 0.56697 | 9641.325 |
| GOBP_DOF | 49 | -0.01252 | -0.00064 | 0.07419 | 0.56698 | 9641.495 |
| GOBP_RES | 7 | -0.02781 | -0.00054 | 0.16469 | 0.56704 | 9642.515 |
| GOMF_L_A | 10 | -0.02646 | -0.00061 | 0.15669 | 0.56704 | 9642.515 |
| GOBP_FOR | 41 | -0.01491 | -0.00069 | 0.088123 | 0.56718 | 9644.896 |
| GOMF_NAI | 14 | -0.02959 | -0.00081 | 0.17485 | 0.56719 | 9645.066 |
| LIU_TARGE | 39 | -0.01321 | -0.0006 | 0.077864 | 0.56734 | 9647.617 |
| NIKOLSKY_ | 130 | -0.0112 | -0.00093 | 0.066013 | 0.56738 | 9648.297 |
| GOBP_T_H | 14 | -0.02613 | -0.00071 | 0.15393 | 0.56739 | 9648.467 |
| GOMF_ACE | 42 | -0.01277 | -0.0006 | 0.075219 | 0.56743 | 9649.147 |
| REACTOME | 8 | -0.02644 | -0.00054 | 0.1555 | 0.5675 | 9650.338 |
| GOBP_CELI | 30 | -0.0156 | -0.00062 | 0.091571 | 0.56764 | 9652.718 |
| GRAHAM_I | 87 | -0.00907 | -0.00061 | 0.053214 | 0.56764 | 9652.718 |
| KYNG_DNA | 52 | -0.01119 | -0.00059 | 0.065444 | 0.56785 | 9656.289 |
| GOBP_NEU | 158 | -0.00718 | -0.00065 | 0.041965 | 0.56795 | 9657.99 |
| GOBP_REG | 8 | -0.03247 | -0.00067 | 0.18952 | 0.56801 | 9659.01 |
| GOBP_REG | 71 | -0.01004 | -0.00061 | 0.058595 | 0.56801 | 9659.01 |
| GOBP_SYN | 13 | -0.02784 | -0.00073 | 0.16239 | 0.56806 | 9659.86 |
| FOROUTAN | 107 | -0.00872 | -0.00065 | 0.050812 | 0.56812 | 9660.881 |
| DEN_INTER | 21 | -0.02067 | -0.00069 | 0.1201 | 0.56831 | 9664.112 |
| GOBP_POS | 13 | -0.02573 | -0.00067 | 0.1495 | 0.56832 | 9664.282 |
| WIEMANN_ | 11 | -0.02971 | -0.00072 | 0.17231 | 0.56845 | 9666.492 |
| DIAZ_CHRC | 110 | -0.00844 | -0.00064 | 0.048954 | 0.56847 | 9666.832 |
| REACTOME | 8 | -0.03076 | -0.00063 | 0.17812 | 0.56854 | 9668.023 |
| GOBP_REG | 349 | -0.00472 | -0.00064 | 0.027331 | 0.56862 | 9669.383 |
| GOBP_FOR | 7 | -0.0389 | -0.00075 | 0.22472 | 0.56872 | 9671.084 |
| NIKOLSKY_ | 156 | -0.01214 | -0.0011 | 0.070068 | 0.56879 | 9672.274 |
| GOCC_ASY | 13 | -0.02675 | -0.0007 | 0.15417 | 0.56886 | 9673.464 |
| GOBP_REG | 5 | -0.03584 | -0.00058 | 0.20652 | 0.56889 | 9673.974 |
| REACTOME | 9 | -0.02862 | -0.00062 | 0.16442 | 0.56908 | 9677.205 |
| YAO_TEMP | 96 | -0.00805 | -0.00057 | 0.046252 | 0.56912 | 9677.886 |
| BAKER_HEI | 10 | -0.03274 | -0.00075 | 0.18763 | 0.56926 | 9680.266 |
| GOBP_NEG | 36 | -0.01402 | -0.00061 | 0.080338 | 0.56928 | 9680.606 |
| REACTOME | 170 | -0.00689 | -0.00065 | 0.039449 | 0.56933 | 9681.457 |
| GOMF_AM | 79 | -0.01018 | -0.00066 | 0.058275 | 0.56935 | 9681.797 |
| LEE_METAS | 14 | -0.03009 | -0.00082 | 0.17213 | 0.56938 | 9682.307 |
| GOBP_POS | 79 | -0.00964 | -0.00062 | 0.055065 | 0.56951 | 9684.518 |

| | | | | | | |
|---|---|---|---|---|---|---|
| GOBP_TRA | 32 | -0.01445 | -0.00059 | 0.082501 | 0.56951 | 9684.518 |
| WP_FOLAT | 66 | -0.01029 | -0.00061 | 0.058733 | 0.56956 | 9685.368 |
| REACTOME | 9 | -0.02467 | -0.00054 | 0.14064 | 0.56963 | 9686.558 |
| GOBP_MIC | 23 | -0.01755 | -0.00061 | 0.09966 | 0.56989 | 9690.979 |
| GOMF_MC | 1203 | -0.00269 | -0.00066 | 0.015273 | 0.56989 | 9690.979 |
| GOCC_POL | 5 | -0.04091 | -0.00067 | 0.23218 | 0.56993 | 9691.66 |
| REACTOME | 49 | -0.01287 | -0.00065 | 0.072946 | 0.57003 | 9693.36 |
| GOBP_PRC | 62 | -0.0115 | -0.00066 | 0.06514 | 0.57007 | 9694.04 |
| WP_NUCLE | 33 | -0.01573 | -0.00066 | 0.08888 | 0.57022 | 9696.591 |
| JAIN_NFKB | 69 | -0.01043 | -0.00063 | 0.058866 | 0.57028 | 9697.611 |
| GOMF_CEL | 7 | -0.0336 | -0.00065 | 0.18956 | 0.57035 | 9698.802 |
| GOCC_CYT | 15 | -0.02215 | -0.00062 | 0.12485 | 0.57042 | 9699.992 |
| REACTOME | 18 | -0.02049 | -0.00063 | 0.1154 | 0.57046 | 9700.672 |
| WP_STING_ | 19 | -0.01877 | -0.0006 | 0.10525 | 0.57076 | 9705.774 |
| KAAB_HEA | 248 | -0.00565 | -0.00064 | 0.03166 | 0.57084 | 9707.134 |
| GOBP_LON | 28 | -0.01899 | -0.00073 | 0.10613 | 0.57101 | 9710.025 |
| GOMF_CYC | 34 | -0.01395 | -0.00059 | 0.077954 | 0.57102 | 9710.195 |
| GOBP_SPEI | 30 | -0.01616 | -0.00064 | 0.090277 | 0.57105 | 9710.705 |
| GOBP_AUT | 8 | -0.0308 | -0.00063 | 0.17201 | 0.57106 | 9710.875 |
| REACTOME | 11 | -0.03234 | -0.00078 | 0.18061 | 0.57106 | 9710.875 |
| GOBP_PIGI | 103 | -0.00873 | -0.00064 | 0.048713 | 0.57107 | 9711.045 |
| GOCC_ORG | 88 | -0.00854 | -0.00058 | 0.047676 | 0.57111 | 9711.726 |
| PURBEY_TA | 360 | -0.00461 | -0.00063 | 0.02567 | 0.57124 | 9713.936 |
| GOMF_CHA | 41 | -0.01251 | -0.00058 | 0.069659 | 0.57127 | 9714.446 |
| GOBP_CON | 11 | -0.03121 | -0.00075 | 0.17377 | 0.57128 | 9714.616 |
| KAPOSI_LIV | 5 | -0.05023 | -0.00082 | 0.27907 | 0.57141 | 9716.827 |
| GOBP_NEG | 5 | -0.03804 | -0.00062 | 0.21045 | 0.57172 | 9722.099 |
| SMID_BREA | 16 | -0.01867 | -0.00054 | 0.10331 | 0.57172 | 9722.099 |
| GOBP_REG | 5 | -0.0442 | -0.00072 | 0.24415 | 0.57183 | 9723.969 |
| GOBP_REG | 20 | -0.02122 | -0.00069 | 0.11704 | 0.57193 | 9725.67 |
| GOBP_POS | 13 | -0.02767 | -0.00073 | 0.15209 | 0.57218 | 9729.921 |
| GOBP_VES | 67 | -0.00997 | -0.00059 | 0.054795 | 0.57218 | 9729.921 |
| VISALA_AG | 5 | -0.04418 | -0.00072 | 0.24234 | 0.57233 | 9732.472 |
| GOBP_VIRI | 9 | -0.03118 | -0.00068 | 0.17052 | 0.57255 | 9736.213 |
| GOMF_PHO | 6 | -0.0415 | -0.00074 | 0.22673 | 0.57262 | 9737.403 |
| GOBP_PHO | 53 | -0.01208 | -0.00064 | 0.065991 | 0.57263 | 9737.573 |
| GOUYER_T | 9 | -0.0356 | -0.00078 | 0.1943 | 0.57269 | 9738.593 |
| GOMF_PRC | 541 | -0.00399 | -0.00067 | 0.021753 | 0.57277 | 9739.954 |
| JOHNSTON | 59 | -0.01243 | -0.00069 | 0.067739 | 0.57277 | 9739.954 |
| SEIDEN_ON | 84 | -0.0099 | -0.00066 | 0.053806 | 0.57295 | 9743.015 |
| REACTOME | 14 | -0.02492 | -0.00068 | 0.13525 | 0.57308 | 9745.225 |
| GOBP_CELI | 48 | -0.01302 | -0.00066 | 0.070622 | 0.57316 | 9746.586 |
| GOBP_DET | 25 | -0.01811 | -0.00066 | 0.09821 | 0.57316 | 9746.586 |
| GOBP_REN | 9 | -0.03499 | -0.00076 | 0.18972 | 0.57316 | 9746.586 |
| GOBP_REG | 104 | -0.00874 | -0.00065 | 0.047181 | 0.57345 | 9751.517 |

| | | | | | | |
|---|---|---|---|---|---|---|
| GOBP_GLU | 50 | -0.01322 | -0.00068 | 0.071239 | 0.5736 | 9754.068 |
| REACTOME | 252 | -0.00594 | -0.00068 | 0.032008 | 0.5736 | 9754.068 |
| GOCC_TRA | 765 | -0.00325 | -0.00064 | 0.017526 | 0.57365 | 9754.918 |
| REACTOME | 48 | -0.01236 | -0.00062 | 0.066519 | 0.57372 | 9756.109 |
| GOBP_REG | 32 | -0.01713 | -0.0007 | 0.092092 | 0.57377 | 9756.959 |
| GOBP_INTI | 5 | -0.03595 | -0.00059 | 0.19317 | 0.57382 | 9757.809 |
| GOBP_NUC | 14 | -0.02196 | -0.0006 | 0.11783 | 0.57392 | 9759.51 |
| GOBP_HIST | 92 | -0.00951 | -0.00066 | 0.050992 | 0.57394 | 9759.85 |
| GOCC_FAN | 12 | -0.02648 | -0.00067 | 0.14179 | 0.57407 | 9762.06 |
| GOBP_CELI | 1473 | -0.00255 | -0.00068 | 0.013616 | 0.57423 | 9764.781 |
| WP_RAS_A | 26 | -0.02145 | -0.0008 | 0.11454 | 0.57427 | 9765.461 |
| BILD_MYC_ | 191 | -0.00648 | -0.00065 | 0.034487 | 0.57452 | 9769.713 |
| REACTOME | 8 | -0.03397 | -0.0007 | 0.1807 | 0.57455 | 9770.223 |
| GARGALOV | 17 | -0.02565 | -0.00077 | 0.13601 | 0.5748 | 9774.474 |
| WP_CELLU | 4 | -0.04727 | -0.00069 | 0.25042 | 0.57485 | 9775.324 |
| GOCC_ATG | 6 | -0.04557 | -0.00081 | 0.24139 | 0.57486 | 9775.494 |
| GOBP_NEG | 5 | -0.0548 | -0.00089 | 0.29022 | 0.57489 | 9776.004 |
| GOBP_CYT | 53 | -0.01207 | -0.00064 | 0.063842 | 0.57495 | 9777.025 |
| GOBP_POS | 456 | -0.00459 | -0.00071 | 0.024242 | 0.57516 | 9780.596 |
| GOBP_IND | 5 | -0.0414 | -0.00067 | 0.21807 | 0.57528 | 9782.636 |
| GOBP_MY( | 7 | -0.03295 | -0.00063 | 0.17356 | 0.57529 | 9782.806 |
| YAMASHIT | 16 | -0.02172 | -0.00063 | 0.11438 | 0.5753 | 9782.977 |
| YAO_TEMF | 26 | -0.02119 | -0.00079 | 0.11157 | 0.57533 | 9783.487 |
| GOBP_VER | 6 | -0.02948 | -0.00053 | 0.15475 | 0.57553 | 9786.888 |
| GOMF_COI | 10 | -0.0383 | -0.00088 | 0.20072 | 0.57565 | 9788.928 |
| VANDESLU | 15 | -0.02578 | -0.00073 | 0.13512 | 0.57565 | 9788.928 |
| GOBP_SER | 38 | -0.01477 | -0.00066 | 0.077386 | 0.57568 | 9789.438 |
| GOBP_REG | 28 | -0.01649 | -0.00063 | 0.086286 | 0.57577 | 9790.969 |
| REACTOME | 16 | -0.02558 | -0.00074 | 0.13385 | 0.57578 | 9791.139 |
| GOBP_MUS | 9 | -0.0397 | -0.00087 | 0.20769 | 0.57579 | 9791.309 |
| REACTOME | 16 | -0.02676 | -0.00078 | 0.13996 | 0.57581 | 9791.649 |
| CHESLER_E | 6 | -0.03298 | -0.00059 | 0.17242 | 0.57585 | 9792.329 |
| GOBP_PHC | 18 | -0.01933 | -0.0006 | 0.10104 | 0.57586 | 9792.499 |
| GOMF_CEF | 6 | -0.03247 | -0.00058 | 0.16969 | 0.57587 | 9792.669 |
| GOMF_RN/ | 40 | -0.01449 | -0.00067 | 0.075642 | 0.57594 | 9793.86 |
| BLANCO_M | 352 | -0.00541 | -0.00073 | 0.028241 | 0.57595 | 9794.03 |
| GOBP_MEI | 20 | -0.02471 | -0.0008 | 0.12889 | 0.57602 | 9795.22 |
| LOPEZ_ME | 7 | -0.03777 | -0.00073 | 0.19699 | 0.57602 | 9795.22 |
| REACTOME | 68 | -0.01184 | -0.00071 | 0.06174 | 0.57606 | 9795.9 |
| GOBP_NITI | 26 | -0.02111 | -0.00078 | 0.11 | 0.57607 | 9796.07 |
| GOBP_BEH | 612 | -0.00405 | -0.00072 | 0.021036 | 0.57626 | 9799.301 |
| GOBP_NEU | 92 | -0.0119 | -0.00083 | 0.061839 | 0.5763 | 9799.982 |
| GOBP_MO | 168 | -0.00757 | -0.00071 | 0.039311 | 0.57633 | 9800.492 |
| GOBP_POS | 42 | -0.01527 | -0.00072 | 0.079163 | 0.57648 | 9803.042 |
| GOBP_END | 27 | -0.01779 | -0.00067 | 0.092195 | 0.57649 | 9803.212 |

| | | | | | | |
|---|---|---|---|---|---|---|
| GOBP_OXA | 5 | -0.03691 | -0.0006 | 0.1912 | 0.57653 | 9803.893 |
| GOBP_LEA | 9 | -0.03289 | -0.00072 | 0.17021 | 0.57661 | 9805.253 |
| GOCC_CELI | 5 | -0.04979 | -0.00081 | 0.25762 | 0.57662 | 9805.423 |
| GOBP_PRC | 51 | -0.01341 | -0.0007 | 0.069316 | 0.57672 | 9807.124 |
| GOCC_DYN | 8 | -0.03259 | -0.00067 | 0.16839 | 0.57673 | 9807.294 |
| GOBP_SYN | 5 | -0.03537 | -0.00058 | 0.18256 | 0.57681 | 9808.654 |
| GOBP_RET | 9 | -0.03653 | -0.0008 | 0.18851 | 0.57683 | 9808.994 |
| NABA_MAT | 955 | -0.00357 | -0.00078 | 0.018391 | 0.57687 | 9809.674 |
| GOBP_HON | 13 | -0.0305 | -8.00E-04 | 0.15673 | 0.57714 | 9814.266 |
| GOBP_CELI | 70 | -0.01264 | -0.00077 | 0.064962 | 0.57716 | 9814.606 |
| GRADE_CO | 96 | -0.01006 | -0.00072 | 0.051657 | 0.57719 | 9815.116 |
| GOBP_VAS | 5 | -0.05931 | -0.00097 | 0.30434 | 0.57726 | 9816.306 |
| WP_ESTRO | 22 | -0.02265 | -0.00077 | 0.11619 | 0.57728 | 9816.646 |
| GOCC_MAI | 8 | -0.0313 | -0.00064 | 0.16032 | 0.5774 | 9818.687 |
| GOMF_PH_ | 8 | -0.03433 | -0.00071 | 0.17581 | 0.57741 | 9818.857 |
| GOBP_SMC | 5 | -0.05211 | -0.00085 | 0.26652 | 0.5775 | 9820.388 |
| GOBP_REG | 41 | -0.01443 | -0.00067 | 0.073788 | 0.57754 | 9821.068 |
| GOBP_REG | 10 | -0.02636 | -0.00061 | 0.13454 | 0.57767 | 9823.278 |
| GOBP_CELI | 16 | -0.02643 | -0.00077 | 0.13486 | 0.57768 | 9823.448 |
| JAZAG_TGI | 95 | -0.00944 | -0.00067 | 0.048176 | 0.5777 | 9823.789 |
| RICKMAN_ | 105 | -0.00966 | -0.00072 | 0.04926 | 0.57774 | 9824.469 |
| PID_FOXM | 40 | -0.0162 | -0.00075 | 0.082585 | 0.57777 | 9824.979 |
| CARD_MIR | 75 | -0.0121 | -0.00076 | 0.061651 | 0.5778 | 9825.489 |
| GOBP_GLY | 7 | -0.04607 | -0.00089 | 0.2347 | 0.5778 | 9825.489 |
| GOMF_GLY | 7 | -0.04607 | -0.00089 | 0.2347 | 0.5778 | 9825.489 |
| ROVERSI_G | 54 | -0.01684 | -0.0009 | 0.085673 | 0.57792 | 9827.53 |
| GOBP_DE_ | 8 | -0.03862 | -0.00079 | 0.19632 | 0.57797 | 9828.38 |
| GOCC_MEI | 7 | -0.0364 | -0.0007 | 0.1846 | 0.57815 | 9831.441 |
| WU_ALZHE | 10 | -0.03291 | -0.00076 | 0.16652 | 0.57832 | 9834.332 |
| GOBP_REG | 5 | -0.06409 | -0.00104 | 0.32424 | 0.57834 | 9834.672 |
| REACTOME | 6 | -0.03847 | -0.00069 | 0.1945 | 0.5784 | 9835.692 |
| REACTOME | 25 | -0.01984 | -0.00072 | 0.10009 | 0.57854 | 9838.073 |
| GOBP_NEG | 144 | -0.00828 | -0.00072 | 0.041766 | 0.57855 | 9838.243 |
| GOBP_REG | 91 | -0.01076 | -0.00075 | 0.054276 | 0.57857 | 9838.583 |
| GOBP_REG | 47 | -0.0143 | -0.00071 | 0.071981 | 0.57871 | 9840.964 |
| FINETTI_BI | 15 | -0.02353 | -0.00066 | 0.11843 | 0.57873 | 9841.304 |
| GOBP_SER | 17 | -0.02316 | -0.00069 | 0.11657 | 0.57875 | 9841.644 |
| REACTOME | 19 | -0.02289 | -0.00073 | 0.11511 | 0.57879 | 9842.324 |
| GOMF_INT | 50 | -0.0126 | -0.00065 | 0.063294 | 0.57888 | 9843.854 |
| GOBP_PEP | 8 | -0.03553 | -0.00073 | 0.1785 | 0.57889 | 9844.024 |
| GOBP_RIB | 208 | -0.00671 | -0.0007 | 0.033687 | 0.57892 | 9844.535 |
| GOCC_INTI | 5 | -0.05108 | -0.00083 | 0.25647 | 0.57893 | 9844.705 |
| GOBP_SIN | 150 | -0.0084 | -0.00075 | 0.042027 | 0.57917 | 9848.786 |
| GOCC_MIT | 461 | -0.00451 | -0.0007 | 0.02255 | 0.57925 | 9850.146 |
| GOBP_MY | 4 | -0.05741 | -0.00084 | 0.28665 | 0.57937 | 9852.187 |

| | | | | | | |
|---|---|---|---|---|---|---|
| GOBP_B_C | 8 | -0.04095 | -0.00084 | 0.20443 | 0.57939 | 9852.527 |
| REACTOME | 5 | -0.03806 | -0.00062 | 0.18996 | 0.57939 | 9852.527 |
| GOBP_NEG | 13 | -0.02627 | -0.00069 | 0.13112 | 0.5794 | 9852.697 |
| GOMF_PRO | 10 | -0.03469 | -0.0008 | 0.17291 | 0.5795 | 9854.398 |
| GOBP_BRO | 4 | -0.05631 | -0.00082 | 0.28025 | 0.57962 | 9856.438 |
| GOBP_HAII | 9 | -0.03037 | -0.00066 | 0.15103 | 0.57969 | 9857.628 |
| WP_MIRNA | 24 | -0.02019 | -0.00072 | 0.10031 | 0.57976 | 9858.819 |
| GOBP_LIPO | 101 | -0.00992 | -0.00072 | 0.049239 | 0.5798 | 9859.499 |
| GOBP_POS | 2 | -0.08393 | -0.00086 | 0.41631 | 0.57988 | 9860.859 |
| GOCC_PRO | 4 | -0.06306 | -0.00092 | 0.31264 | 0.57992 | 9861.54 |
| WP_ECTOD | 124 | -0.00967 | -0.00078 | 0.04793 | 0.57992 | 9861.54 |
| GOMF_PRO | 21 | -0.0212 | -0.00071 | 0.10494 | 0.58005 | 9863.75 |
| FARMER_B | 38 | -0.01727 | -0.00077 | 0.085335 | 0.58017 | 9865.791 |
| GOBP_REG | 270 | -0.00635 | -0.00075 | 0.031317 | 0.58037 | 9869.192 |
| GOBP_REG | 28 | -0.0232 | -0.00089 | 0.11436 | 0.58039 | 9869.532 |
| GOBP_NEG | 5 | -0.05335 | -0.00087 | 0.2625 | 0.58053 | 9871.913 |
| GOCC_BRO | 7 | -0.04085 | -0.00079 | 0.20085 | 0.58057 | 9872.593 |
| REACTOME | 76 | -0.01122 | -0.00071 | 0.05501 | 0.58077 | 9875.994 |
| VANHARAN | 63 | -0.01377 | -0.00079 | 0.067543 | 0.58077 | 9875.994 |
| GOBP_REG | 9 | -0.0405 | -0.00088 | 0.19853 | 0.58082 | 9876.844 |
| GOBP_REG | 6 | -0.03766 | -0.00067 | 0.18459 | 0.58082 | 9876.844 |
| GOBP_REG | 8 | -0.03757 | -0.00077 | 0.18406 | 0.58087 | 9877.694 |
| GOBP_VES | 205 | -0.00678 | -0.0007 | 0.033185 | 0.58089 | 9878.034 |
| GOBP_GLU | 7 | -0.03706 | -0.00071 | 0.1815 | 0.5809 | 9878.205 |
| GOMF_SEN | 10 | -0.03887 | -0.00089 | 0.19018 | 0.58096 | 9879.225 |
| BANDRES_ | 30 | -0.01895 | -0.00075 | 0.092636 | 0.58102 | 9880.245 |
| GOCC_LAM | 22 | -0.02313 | -0.00079 | 0.11304 | 0.58107 | 9881.095 |
| GOBP_BLO | 23 | -0.0234 | -0.00082 | 0.114 | 0.58131 | 9885.177 |
| GOBP_PHO | 239 | -0.00654 | -0.00073 | 0.031841 | 0.58131 | 9885.177 |
| GOCC_NEU | 28 | -0.01985 | -0.00076 | 0.09651 | 0.58148 | 9888.067 |
| REACTOME | 32 | -0.01711 | -0.0007 | 0.083198 | 0.58148 | 9888.067 |
| GOCC_CILI | 695 | -0.00386 | -0.00073 | 0.018764 | 0.5815 | 9888.408 |
| REACTOME | 643 | -0.00409 | -0.00074 | 0.01982 | 0.58169 | 9891.638 |
| GOCC_TIM | 7 | -0.03121 | -0.0006 | 0.15129 | 0.58171 | 9891.979 |
| DITTMER_I | 110 | -0.00989 | -0.00075 | 0.047904 | 0.58177 | 9892.999 |
| GOBP_ADH | 7 | -0.03987 | -0.00077 | 0.19314 | 0.58177 | 9892.999 |
| PID_CONE_ | 21 | -0.02132 | -0.00071 | 0.10318 | 0.58186 | 9894.529 |
| BIOCARTA_ | 24 | -0.02207 | -0.00079 | 0.10679 | 0.58188 | 9894.869 |
| CASTELLAN | 5 | -0.04167 | -0.00068 | 0.20153 | 0.58191 | 9895.38 |
| REACTOME | 6 | -0.04557 | -0.00081 | 0.22028 | 0.58194 | 9895.89 |
| LY_AGING_ | 55 | -0.01306 | -0.0007 | 0.063068 | 0.582 | 9896.91 |
| MUNSHI_M | 71 | -0.01165 | -0.00071 | 0.056269 | 0.58202 | 9897.25 |
| GOBP_REG | 238 | -0.00645 | -0.00072 | 0.031118 | 0.58206 | 9897.93 |
| GOMF_ORG | 217 | -0.00715 | -0.00076 | 0.034471 | 0.58217 | 9899.801 |
| GOBP_NEG | 8 | -0.033 | -0.00068 | 0.15902 | 0.58218 | 9899.971 |

| | | | | | | |
|---|---|---|---|---|---|---|
| GOBP_RIB( | 8 | -0.03609 | -0.00074 | 0.17385 | 0.58222 | 9900.651 |
| GOBP_SPEI | 6 | -0.04539 | -0.00081 | 0.21866 | 0.58222 | 9900.651 |
| GOBP_AXC | 7 | -0.0456 | -0.00088 | 0.21958 | 0.58225 | 9901.161 |
| GOBP_TRO | 14 | -0.02986 | -0.00081 | 0.14364 | 0.58233 | 9902.522 |
| MATTIOLI_ | 93 | -0.01041 | -0.00073 | 0.050024 | 0.5824 | 9903.712 |
| PETRETTO_ | 33 | -0.01939 | -0.00081 | 0.09309 | 0.58248 | 9905.072 |
| GOBP_AMI | 8 | -0.04545 | -0.00094 | 0.21818 | 0.58251 | 9905.583 |
| GOMF_PH( | 261 | -0.00651 | -0.00076 | 0.031188 | 0.58267 | 9908.303 |
| GOBP_SOD | 16 | -0.02965 | -0.00086 | 0.14195 | 0.58271 | 9908.984 |
| GOCC_INTI | 16 | -0.02263 | -0.00066 | 0.10833 | 0.58273 | 9909.324 |
| GOBP_NAT | 13 | -0.02603 | -0.00068 | 0.12454 | 0.58278 | 9910.174 |
| GOCC_EXC | 56 | -0.0146 | -0.00079 | 0.069798 | 0.58283 | 9911.024 |
| GOMF_NU | 22 | -0.02211 | -0.00075 | 0.10525 | 0.5832 | 9917.316 |
| TIAN_BHLH | 16 | -0.027 | -0.00079 | 0.12852 | 0.5832 | 9917.316 |
| GOBP_COE | 9 | -0.03765 | -0.00082 | 0.17919 | 0.58321 | 9917.486 |
| WP_SEROT | 32 | -0.01868 | -0.00077 | 0.088835 | 0.58327 | 9918.506 |
| GOCC_PHC | 8 | -0.03677 | -0.00076 | 0.17454 | 0.58342 | 9921.057 |
| REACTOME | 156 | -0.00865 | -0.00078 | 0.041061 | 0.58342 | 9921.057 |
| GOBP_NEG | 21 | -0.02379 | -0.00079 | 0.1129 | 0.58343 | 9921.227 |
| REACTOME | 8 | -0.04687 | -0.00096 | 0.22243 | 0.58345 | 9921.567 |
| KORKOLA_ | 29 | -0.02028 | -0.00079 | 0.096197 | 0.58348 | 9922.077 |
| STEIN_ESRI | 495 | -0.00462 | -0.00074 | 0.021906 | 0.58357 | 9923.608 |
| REACTOME | 108 | -0.01 | -0.00075 | 0.047384 | 0.58359 | 9923.948 |
| GOCC_CUT | 9 | -0.03327 | -0.00073 | 0.15749 | 0.58364 | 9924.798 |
| HAHTOLA_ | 23 | -0.02041 | -0.00071 | 0.096623 | 0.58364 | 9924.798 |
| WP_ALTER | 10 | -0.03079 | -0.00071 | 0.14576 | 0.58364 | 9924.798 |
| GOBP_NAD | 51 | -0.01392 | -0.00072 | 0.065902 | 0.58365 | 9924.968 |
| IKEDA_MIF | 24 | -0.02311 | -0.00082 | 0.10932 | 0.58369 | 9925.648 |
| GOMF_ACT | 402 | -0.00536 | -0.00077 | 0.025296 | 0.58384 | 9928.199 |
| REACTOME | 5 | -0.05402 | -0.00088 | 0.25473 | 0.58396 | 9930.24 |
| GOMF_BET | 5 | -0.03694 | -0.0006 | 0.17403 | 0.58405 | 9931.77 |
| KEGG_P53_ | 67 | -0.01322 | -0.00079 | 0.062261 | 0.58407 | 9932.11 |
| WP_TRIACY | 22 | -0.02545 | -0.00087 | 0.11974 | 0.58414 | 9933.301 |
| GOBP_MA( | 11 | -0.03391 | -0.00082 | 0.15948 | 0.58419 | 9934.151 |
| GOBP_REG | 7 | -0.0515 | -0.00099 | 0.24205 | 0.58424 | 9935.001 |
| ZIRN_TRET | 18 | -0.0261 | -0.00081 | 0.12268 | 0.58425 | 9935.171 |
| GOBP_INTI | 5 | -0.05576 | -0.00091 | 0.26199 | 0.58427 | 9935.511 |
| GOBP_PRC | 12 | -0.02947 | -0.00074 | 0.1382 | 0.58441 | 9937.892 |
| GOBP_MEI | 62 | -0.01284 | -0.00073 | 0.060199 | 0.58442 | 9938.062 |
| GOBP_EXO | 5 | -0.05686 | -0.00093 | 0.2664 | 0.58451 | 9939.593 |
| GOBP_NEG | 8 | -0.03899 | -0.0008 | 0.18264 | 0.58452 | 9939.763 |
| GOBP_REG | 456 | -0.00515 | -0.00079 | 0.02411 | 0.58454 | 9940.103 |
| GOBP_LYM | 9 | -0.03558 | -0.00078 | 0.16658 | 0.58457 | 9940.613 |
| IRITANI_M. | 42 | -0.01693 | -0.0008 | 0.079254 | 0.58459 | 9940.953 |
| WP_G1_TC | 63 | -0.01271 | -0.00073 | 0.059436 | 0.58463 | 9941.633 |

| | | | | | | |
|---|---|---|---|---|---|---|
| GOMF_GR( | 151 | -0.00906 | -0.00081 | 0.042347 | 0.58466 | 9942.143 |
| GOBP_GEN | 457 | -0.00479 | -0.00074 | 0.022401 | 0.58467 | 9942.313 |
| ELLWOOD_ | 42 | -0.01532 | -0.00072 | 0.071587 | 0.58473 | 9943.334 |
| GOMF_PR( | 396 | -0.00526 | -0.00075 | 0.024557 | 0.58488 | 9945.884 |
| BIOCARTA_ | 5 | -0.06706 | -0.00109 | 0.31231 | 0.58501 | 9948.095 |
| GOBP_COM | 13 | -0.03121 | -0.00082 | 0.14524 | 0.58508 | 9949.285 |
| GOBP_TRA | 26 | -0.02197 | -0.00081 | 0.1022 | 0.58511 | 9949.796 |
| GAVIN_FO) | 152 | -0.00864 | -0.00077 | 0.040187 | 0.58513 | 9950.136 |
| GOBP_POS | 21 | -0.0205 | -0.00068 | 0.095285 | 0.58515 | 9950.476 |
| GOBP_CELI | 180 | -0.00822 | -0.0008 | 0.038177 | 0.58519 | 9951.156 |
| ZHAN_MUI | 42 | -0.0168 | -0.00079 | 0.078081 | 0.58519 | 9951.156 |
| WANG_ES( | 9 | -0.03929 | -0.00086 | 0.18216 | 0.58538 | 9954.387 |
| GOBP_NEG | 9 | -0.03885 | -0.00085 | 0.17987 | 0.5855 | 9956.428 |
| MARTINELI | 12 | -0.02933 | -0.00074 | 0.13573 | 0.58554 | 9957.108 |
| REACTOME | 54 | -0.0149 | -0.0008 | 0.068948 | 0.58557 | 9957.618 |
| GOCC_SITE | 75 | -0.01213 | -0.00076 | 0.056124 | 0.58558 | 9957.788 |
| GOBP_HOF | 177 | -0.00815 | -0.00079 | 0.03769 | 0.58561 | 9958.298 |
| WANG_RES | 331 | -0.00598 | -0.00078 | 0.027572 | 0.58586 | 9962.549 |
| GOMF_MC | 11 | -0.03499 | -0.00084 | 0.1613 | 0.58587 | 9962.719 |
| GOBP_GLY | 32 | -0.01883 | -0.00077 | 0.086666 | 0.58602 | 9965.27 |
| GOMF_DIP | 4 | -0.06213 | -0.0009 | 0.28553 | 0.58612 | 9966.971 |
| GOMF_DN/ | 7 | -0.03228 | -0.00062 | 0.14828 | 0.58616 | 9967.651 |
| BIOCARTA_ | 19 | -0.02874 | -0.00091 | 0.13176 | 0.58634 | 9970.712 |
| GOBP_MA( | 11 | -0.03441 | -0.00083 | 0.15775 | 0.58634 | 9970.712 |
| GOCC_APC | 5 | -0.05454 | -0.00089 | 0.24997 | 0.58636 | 9971.052 |
| VANHARAN | 59 | -0.01518 | -0.00085 | 0.069533 | 0.58639 | 9971.562 |
| GOBP_RES | 9 | -0.03979 | -0.00087 | 0.18227 | 0.5864 | 9971.732 |
| GOMF_RN/ | 56 | -0.01487 | -0.00081 | 0.068073 | 0.58643 | 9972.242 |
| GOBP_POS | 443 | -0.00515 | -0.00078 | 0.023558 | 0.58654 | 9974.113 |
| GOCC_AGG | 31 | -0.01809 | -0.00073 | 0.082658 | 0.58661 | 9975.303 |
| GOMF_SPE | 26 | -0.02183 | -0.00081 | 0.099661 | 0.58671 | 9977.004 |
| GOBP_NEG | 43 | -0.01704 | -0.00081 | 0.077645 | 0.58685 | 9979.384 |
| GOBP_POS | 122 | -0.0096 | -0.00077 | 0.043713 | 0.58693 | 9980.745 |
| GOBP_REG | 8 | -0.04822 | -0.00099 | 0.21947 | 0.58695 | 9981.085 |
| GOMF_CCF | 5 | -0.05172 | -0.00084 | 0.23513 | 0.58705 | 9982.785 |
| MOROSETT | 12 | -0.0241 | -0.00061 | 0.10957 | 0.58705 | 9982.785 |
| GOCC_BRU | 54 | -0.01529 | -0.00082 | 0.069487 | 0.5871 | 9983.636 |
| GOBP_REG | 7 | -0.05236 | -0.00101 | 0.2375 | 0.58724 | 9986.016 |
| GOBP_POS | 3 | -0.06444 | -0.00081 | 0.29207 | 0.5873 | 9987.037 |
| GOBP_RET | 31 | -0.02155 | -0.00087 | 0.097606 | 0.58736 | 9988.057 |
| MARSON_I | 953 | -0.00373 | -0.00082 | 0.016893 | 0.58739 | 9988.567 |
| FLECHNER_ | 48 | -0.01555 | -0.00078 | 0.070407 | 0.58742 | 9989.077 |
| GOBP_CELI | 12 | -0.02799 | -0.00071 | 0.12661 | 0.58747 | 9989.927 |
| GOBP_XEN | 25 | -0.0224 | -0.00081 | 0.10131 | 0.58748 | 9990.097 |
| GOBP_NEG | 15 | -0.02615 | -0.00074 | 0.11809 | 0.58763 | 9992.648 |

| | | | | | | |
|---|---|---|---|---|---|---|
| GOBP_EMI | 6 | -0.03381 | -0.0006 | 0.15249 | 0.58773 | 9994.349 |
| GOBP_NEU | 4 | -0.05343 | -0.00078 | 0.24074 | 0.58782 | 9995.879 |
| GOBP_PHC | 356 | -0.00581 | -0.00079 | 0.026155 | 0.58793 | 9997.75 |
| GOBP_TYP | 90 | -0.01132 | -0.00078 | 0.050867 | 0.58802 | 9999.28 |
| GOBP_REL | 9 | -0.03633 | -0.00079 | 0.16322 | 0.58807 | 10000.13 |
| GOBP_MH | 8 | -0.04593 | -0.00095 | 0.20582 | 0.5883 | 10004.04 |
| GOBP_POS | 5 | -0.04974 | -0.00081 | 0.22281 | 0.58832 | 10004.38 |
| GOBP_MU | 719 | -0.00426 | -0.00082 | 0.019074 | 0.58839 | 10005.57 |
| COLLER_M | 24 | -0.02391 | -0.00085 | 0.1068 | 0.58856 | 10008.46 |
| WP_PYRIM | 19 | -0.02784 | -0.00088 | 0.12426 | 0.58864 | 10009.82 |
| SPIELMAN_ | 12 | -0.03105 | -0.00078 | 0.1385 | 0.5887 | 10010.84 |
| GOBP_COF | 15 | -0.0253 | -0.00071 | 0.11283 | 0.58871 | 10011.01 |
| SCHMAHL_ | 9 | -0.04082 | -0.00089 | 0.18194 | 0.58875 | 10011.69 |
| GOBP_BILE | 34 | -0.02118 | -0.0009 | 0.094404 | 0.58877 | 10012.03 |
| GOBP_MEI | 5 | -0.04925 | -0.0008 | 0.21948 | 0.58877 | 10012.03 |
| GOMF_RIB | 126 | -0.00938 | -0.00076 | 0.041752 | 0.58888 | 10013.9 |
| LANDIS_ER | 143 | -0.00972 | -0.00084 | 0.043245 | 0.58888 | 10013.9 |
| WANG_ES( | 97 | -0.01118 | -0.0008 | 0.049645 | 0.58905 | 10016.8 |
| REACTOME | 12 | -0.03447 | -0.00087 | 0.1528 | 0.58923 | 10019.86 |
| LANDIS_ER | 22 | -0.02899 | -0.00099 | 0.12838 | 0.58932 | 10021.39 |
| REACTOME | 34 | -0.01962 | -0.00083 | 0.086886 | 0.58934 | 10021.73 |
| REACTOME | 49 | -0.01628 | -0.00083 | 0.072051 | 0.58935 | 10021.9 |
| PASQUALU | 275 | -0.00722 | -0.00087 | 0.031962 | 0.58937 | 10022.24 |
| GOBP_DEF | 97 | -0.01159 | -0.00083 | 0.051268 | 0.58942 | 10023.09 |
| GOMF_VIT | 34 | -0.02094 | -0.00089 | 0.092613 | 0.58942 | 10023.09 |
| GOMF_PR( | 341 | -0.00603 | -0.0008 | 0.026659 | 0.58947 | 10023.94 |
| REACTOME | 53 | -0.01834 | -0.00097 | 0.081091 | 0.58948 | 10024.11 |
| HALMOS_( | 47 | -0.01724 | -0.00086 | 0.076182 | 0.58952 | 10024.79 |
| GOBP_CHC | 4 | -0.05596 | -0.00081 | 0.24724 | 0.58953 | 10024.96 |
| GOBP_VIT/ | 13 | -0.03671 | -0.00096 | 0.16218 | 0.58954 | 10025.13 |
| LEE_LIVER_ | 61 | -0.01502 | -0.00085 | 0.066363 | 0.58955 | 10025.3 |
| GOBP_REG | 102 | -0.01204 | -0.00088 | 0.053128 | 0.58963 | 10026.66 |
| REACTOME | 16 | -0.0289 | -0.00084 | 0.1275 | 0.58967 | 10027.34 |
| GOBP_REN | 29 | -0.02229 | -0.00087 | 0.098284 | 0.58972 | 10028.19 |
| GOMF_VOI | 9 | -0.04332 | -0.00095 | 0.19067 | 0.58985 | 10030.4 |
| GEISS_RESI | 16 | -0.02817 | -0.00082 | 0.12375 | 0.59005 | 10033.8 |
| GOBP_MIT | 17 | -0.02903 | -0.00087 | 0.12744 | 0.59008 | 10034.31 |
| GOBP_NEG | 7 | -0.0468 | -0.0009 | 0.20541 | 0.59011 | 10034.82 |
| KIM_GERM | 58 | -0.01506 | -0.00083 | 0.065767 | 0.59054 | 10042.13 |
| GOBP_SCH | 4 | -0.07617 | -0.00111 | 0.3326 | 0.59057 | 10042.64 |
| PID_INTEG | 28 | -0.0221 | -0.00085 | 0.096397 | 0.59065 | 10044 |
| GOBP_END | 10 | -0.03229 | -0.00074 | 0.1408 | 0.59069 | 10044.68 |
| REACTOME | 13 | -0.03399 | -0.00089 | 0.14811 | 0.59075 | 10045.7 |
| GOBP_POS | 23 | -0.02585 | -0.0009 | 0.11254 | 0.59081 | 10046.72 |
| GOBP_NEU | 82 | -0.01213 | -0.0008 | 0.052735 | 0.59094 | 10048.93 |

| | | | | | | |
|---|---|---|---|---|---|---|
| MIKKELSEN | 663 | -0.00432 | -0.00079 | 0.018769 | 0.59098 | 10049.61 |
| PID_IL23_F | 36 | -0.01961 | -0.00086 | 0.08525 | 0.59098 | 10049.61 |
| REICHERT_ | 8 | -0.04033 | -0.00083 | 0.17524 | 0.59101 | 10050.13 |
| GOBP_CELI | 5 | -0.05751 | -0.00094 | 0.24978 | 0.59105 | 10050.81 |
| GOBP_GOL | 10 | -0.03486 | -0.0008 | 0.15136 | 0.59109 | 10051.49 |
| GOBP_REG | 33 | -0.02235 | -0.00093 | 0.09703 | 0.59109 | 10051.49 |
| GOBP_RES | 51 | -0.01499 | -0.00078 | 0.065069 | 0.59112 | 10052 |
| GOBP_GLU | 5 | -0.05222 | -0.00085 | 0.22628 | 0.59126 | 10054.38 |
| GOBP_MA( | 302 | -0.00641 | -0.0008 | 0.027774 | 0.59132 | 10055.4 |
| GOBP_FOR | 50 | -0.01841 | -0.00095 | 0.079704 | 0.59135 | 10055.91 |
| GOBP_POS | 54 | -0.01682 | -0.0009 | 0.072772 | 0.59137 | 10056.25 |
| GOMF_AM | 7 | -0.05539 | -0.00107 | 0.23927 | 0.59154 | 10059.14 |
| REACTOME | 29 | -0.02208 | -0.00086 | 0.095114 | 0.59178 | 10063.22 |
| GOBP_REG | 19 | -0.02473 | -0.00078 | 0.10584 | 0.59237 | 10073.25 |
| GOCC_AST | 8 | -0.04899 | -0.00101 | 0.20968 | 0.59237 | 10073.25 |
| GOCC_ALV | 7 | -0.04395 | -0.00085 | 0.18784 | 0.5925 | 10075.46 |
| GOBP_REG | 14 | -0.03159 | -0.00086 | 0.13499 | 0.59251 | 10075.63 |
| REACTOME | 13 | -0.02955 | -0.00078 | 0.12627 | 0.59251 | 10075.63 |
| PID_PDGFF | 126 | -0.01019 | -0.00083 | 0.043532 | 0.59252 | 10075.8 |
| KEGG_REG | 198 | -0.00823 | -0.00084 | 0.035184 | 0.59253 | 10075.97 |
| GOMF_SM | 10 | -0.03623 | -0.00083 | 0.15467 | 0.59259 | 10076.99 |
| GOBP_REG | 9 | -0.03924 | -0.00086 | 0.1675 | 0.59261 | 10077.33 |
| LASTOWSK | 38 | -0.02194 | -0.00098 | 0.093577 | 0.59266 | 10078.18 |
| GOBP_POS | 41 | -0.01952 | -0.00091 | 0.08325 | 0.59269 | 10078.69 |
| WAMUNY( | 62 | -0.015 | -0.00086 | 0.063883 | 0.59282 | 10080.9 |
| GOBP_ADII | 6 | -0.04975 | -0.00089 | 0.2118 | 0.59285 | 10081.41 |
| XIE_LT_HS( | 28 | -0.02324 | -0.00089 | 0.098909 | 0.59287 | 10081.75 |
| WP_GENES | 17 | -0.0301 | -0.0009 | 0.12801 | 0.59295 | 10083.11 |
| GOBP_NU( | 124 | -0.01005 | -0.00081 | 0.042652 | 0.59309 | 10085.5 |
| GOBP_REG | 38 | -0.01914 | -0.00086 | 0.081203 | 0.59318 | 10087.03 |
| GOMF_INT | 6 | -0.04539 | -0.00081 | 0.19235 | 0.59326 | 10088.39 |
| ROYLANCE_ | 24 | -0.03698 | -0.00132 | 0.15668 | 0.59329 | 10088.9 |
| GOBP_ER_ | 4 | -0.05302 | -0.00077 | 0.22457 | 0.59332 | 10089.41 |
| GOBP_GAN | 16 | -0.02953 | -0.00086 | 0.12507 | 0.59332 | 10089.41 |
| HELLER_SIL | 102 | -0.01166 | -0.00085 | 0.049285 | 0.59346 | 10091.79 |
| REACTOME | 72 | -0.01382 | -0.00085 | 0.05844 | 0.59348 | 10092.13 |
| ELVIDGE_H | 57 | -0.01526 | -0.00084 | 0.064501 | 0.5935 | 10092.47 |
| GOBP_LAN | 64 | -0.01511 | -0.00088 | 0.063813 | 0.59357 | 10093.66 |
| GOCC_RIP( | 6 | -0.06111 | -0.00109 | 0.25792 | 0.59365 | 10095.02 |
| GOBP_NEG | 64 | -0.0148 | -0.00086 | 0.062438 | 0.59367 | 10095.36 |
| GOBP_RNA | 78 | -0.01319 | -0.00085 | 0.055408 | 0.5941 | 10102.67 |
| GAVIN_PDI | 21 | -0.02814 | -0.00094 | 0.11806 | 0.59419 | 10104.2 |
| GOBP_CAR | 12 | -0.03883 | -0.00098 | 0.16289 | 0.59419 | 10104.2 |
| GOBP_MIT | 23 | -0.02508 | -0.00087 | 0.10516 | 0.59426 | 10105.39 |
| GOBP_SON | 4 | -0.07347 | -0.00107 | 0.30795 | 0.59428 | 10105.73 |

| | | | | | | |
|---|---|---|---|---|---|---|
| GOMF_SOI | 4 | -0.07347 | -0.00107 | 0.30795 | 0.59428 | 10105.73 |
| GOCC_MIC | 28 | -0.0215 | -0.00083 | 0.090109 | 0.59429 | 10105.9 |
| REACTOME | 34 | -0.02042 | -0.00087 | 0.085508 | 0.59436 | 10107.09 |
| GOBP_PER | 22 | -0.0315 | -0.00107 | 0.13183 | 0.59441 | 10107.94 |
| HOFMANN | 22 | -0.02423 | -0.00083 | 0.10139 | 0.59444 | 10108.45 |
| BLANCO_M | 71 | -0.0152 | -0.00093 | 0.063475 | 0.59461 | 10111.34 |
| GOCC_IMN | 41 | -0.01971 | -0.00092 | 0.082332 | 0.59461 | 10111.34 |
| NELSON_R | 80 | -0.01424 | -0.00093 | 0.059476 | 0.59462 | 10111.51 |
| GOBP_REG | 5 | -0.05099 | -0.00083 | 0.21239 | 0.59485 | 10115.42 |
| GOMF_ORI | 5 | -0.05099 | -0.00083 | 0.21239 | 0.59485 | 10115.42 |
| REACTOME | 40 | -0.01801 | -0.00083 | 0.074915 | 0.59498 | 10117.63 |
| REACTOME | 90 | -0.01217 | -0.00084 | 0.050584 | 0.59508 | 10119.34 |
| GOBP_HIST | 6 | -0.05301 | -0.00094 | 0.22021 | 0.59511 | 10119.85 |
| GOBP_AV_ | 9 | -0.04798 | -0.00105 | 0.19926 | 0.59514 | 10120.36 |
| LUI_THYRC | 50 | -0.01824 | -0.00094 | 0.075568 | 0.59536 | 10124.1 |
| RODWELL_ | 133 | -0.01073 | -0.0009 | 0.044441 | 0.59537 | 10124.27 |
| GOBP_PRC | 14 | -0.03845 | -0.00105 | 0.15908 | 0.59551 | 10126.65 |
| GOCC_ARP | 9 | -0.03975 | -0.00087 | 0.16436 | 0.59556 | 10127.5 |
| GOBP_SEM | 9 | -0.04969 | -0.00108 | 0.20517 | 0.59568 | 10129.54 |
| GOBP_CELI | 5 | -0.0647 | -0.00105 | 0.26712 | 0.59569 | 10129.71 |
| GOBP_DOF | 7 | -0.05838 | -0.00112 | 0.24098 | 0.59571 | 10130.05 |
| GOBP_POS | 5 | -0.05808 | -0.00095 | 0.23969 | 0.59574 | 10130.56 |
| GOBP_POS | 56 | -0.01782 | -0.00097 | 0.073524 | 0.59575 | 10130.73 |
| GOBP_REG | 104 | -0.01144 | -0.00085 | 0.047085 | 0.59597 | 10134.47 |
| GOBP_SUP | 60 | -0.01528 | -0.00086 | 0.062912 | 0.59597 | 10134.47 |
| KUMAMOT | 9 | -0.03591 | -0.00078 | 0.1478 | 0.59597 | 10134.47 |
| GOBP_NEG | 7 | -0.03677 | -0.00071 | 0.15128 | 0.59603 | 10135.49 |
| GOBP_RES | 140 | -0.0106 | -0.00091 | 0.043579 | 0.59606 | 10136 |
| GOBP_NEU | 15 | -0.02985 | -0.00084 | 0.12272 | 0.59607 | 10136.17 |
| TOMLINS_I | 20 | -0.02639 | -0.00086 | 0.10834 | 0.59622 | 10138.72 |
| GOBP_END | 5 | -0.0472 | -0.00077 | 0.19355 | 0.59632 | 10140.42 |
| CROONQU | 11 | -0.03539 | -0.00085 | 0.14511 | 0.59634 | 10140.76 |
| REACTOME | 69 | -0.01509 | -0.00091 | 0.061886 | 0.59634 | 10140.76 |
| WP_TAMO | 21 | -0.02741 | -0.00091 | 0.11231 | 0.5964 | 10141.78 |
| PARK_APL_ | 45 | -0.01952 | -0.00095 | 0.079944 | 0.59644 | 10142.46 |
| REACTOME | 5 | -0.04674 | -0.00076 | 0.19124 | 0.59654 | 10144.16 |
| GOMF_MY | 10 | -0.03162 | -0.00073 | 0.12931 | 0.59659 | 10145.01 |
| MYLLYKAN | 10 | -0.04128 | -0.00095 | 0.16877 | 0.5966 | 10145.18 |
| GOBP_PEP | 1115 | -0.00375 | -0.00088 | 0.015296 | 0.5967 | 10146.88 |
| GOMF_VAS | 9 | -0.05128 | -0.00112 | 0.20905 | 0.59688 | 10149.94 |
| ONDER_CD | 162 | -0.0097 | -0.0009 | 0.039566 | 0.59688 | 10149.94 |
| REACTOME | 5 | -0.04656 | -0.00076 | 0.18977 | 0.5969 | 10150.28 |
| GOMF_GLU | 5 | -0.0641 | -0.00104 | 0.26086 | 0.59705 | 10152.84 |
| GOBP_CELI | 29 | -0.0247 | -0.00097 | 0.10043 | 0.59712 | 10154.03 |
| BOYAULT_I | 112 | -0.01169 | -0.0009 | 0.047485 | 0.59726 | 10156.41 |

| | | | | | | |
|---|---|---|---|---|---|---|
| GOBP_PYR | 11 | -0.03569 | -0.00086 | 0.14482 | 0.59732 | 10157.43 |
| PID_ALPHA | 31 | -0.02385 | -0.00097 | 0.09679 | 0.59733 | 10157.6 |
| REACTOME | 30 | -0.02246 | -0.00089 | 0.091116 | 0.59734 | 10157.77 |
| GOBP_BAS | 9 | -0.03789 | -0.00083 | 0.15362 | 0.5974 | 10158.79 |
| GOMF_EPH | 18 | -0.0327 | -0.00101 | 0.13244 | 0.59751 | 10160.66 |
| BARRIER_C | 15 | -0.03834 | -0.00108 | 0.15521 | 0.59755 | 10161.34 |
| GOBP_DEN | 7 | -0.04709 | -0.00091 | 0.19061 | 0.59756 | 10161.51 |
| RADAEVA_ | 43 | -0.01884 | -0.0009 | 0.076247 | 0.59756 | 10161.51 |
| GOCC_REC | 373 | -0.00678 | -0.00094 | 0.027422 | 0.5977 | 10163.89 |
| WP_BIOSYI | 8 | -0.05002 | -0.00103 | 0.20186 | 0.59785 | 10166.44 |
| PID_AR_NC | 30 | -0.01931 | -0.00077 | 0.077768 | 0.59804 | 10169.67 |
| GOBP_MAS | 68 | -0.0146 | -0.00087 | 0.058785 | 0.59809 | 10170.52 |
| GOCC_PRO | 127 | -0.0109 | -0.00089 | 0.04387 | 0.59809 | 10170.52 |
| MCCABE_H | 21 | -0.03004 | -0.001 | 0.12089 | 0.59812 | 10171.03 |
| GOBP_NEG | 23 | -0.0265 | -0.00092 | 0.1066 | 0.59814 | 10171.37 |
| GOBP_ENT | 35 | -0.0224 | -0.00096 | 0.090081 | 0.59817 | 10171.88 |
| GOMF_PLA | 13 | -0.04121 | -0.00108 | 0.16571 | 0.59819 | 10172.22 |
| RICKMAN_ | 79 | -0.01468 | -0.00095 | 0.058987 | 0.59829 | 10173.92 |
| WP_INTERA | 24 | -0.02753 | -0.00098 | 0.11049 | 0.59838 | 10175.45 |
| GOBP_EMB | 49 | -0.01786 | -0.00091 | 0.071615 | 0.59848 | 10177.15 |
| GOBP_CAL | 42 | -0.02209 | -0.00104 | 0.088541 | 0.59851 | 10177.66 |
| GOMF_ARI | 10 | -0.03316 | -0.00076 | 0.13281 | 0.59857 | 10178.68 |
| GOBP_CELI | 44 | -0.01878 | -0.00091 | 0.075172 | 0.59866 | 10180.21 |
| GOMF_NEI | 7 | -0.04651 | -0.0009 | 0.18592 | 0.59877 | 10182.08 |
| GOBP_MIC | 202 | -0.00835 | -0.00086 | 0.033348 | 0.59881 | 10182.76 |
| GOCC_GID_ | 5 | -0.05921 | -0.00096 | 0.2365 | 0.59884 | 10183.27 |
| ABBUD_LIF | 23 | -0.0264 | -0.00092 | 0.10525 | 0.59903 | 10186.51 |
| REACTOME | 65 | -0.01531 | -0.0009 | 0.060988 | 0.59909 | 10187.53 |
| GOCC_VES | 55 | -0.01743 | -0.00094 | 0.069414 | 0.59911 | 10187.87 |
| SCHAEFFER | 5 | -0.06005 | -0.00098 | 0.23898 | 0.5992 | 10189.4 |
| RUTELLA_F | 229 | -0.00839 | -0.00092 | 0.033353 | 0.59927 | 10190.59 |
| GOBP_CAR | 8 | -0.05414 | -0.00111 | 0.21523 | 0.5993 | 10191.1 |
| GOBP_3_P | 15 | -0.03613 | -0.00102 | 0.1435 | 0.5994 | 10192.8 |
| WORSCHE( | 55 | -0.01759 | -0.00095 | 0.06985 | 0.59943 | 10193.31 |
| WP_UREA_ | 8 | -0.0415 | -0.00085 | 0.16462 | 0.59952 | 10194.84 |
| GOMF_NU | 9 | -0.04636 | -0.00101 | 0.18356 | 0.5997 | 10197.9 |
| GOBP_REG | 8 | -0.04525 | -0.00093 | 0.17891 | 0.59982 | 10199.94 |
| GOBP_REG | 71 | -0.01548 | -0.00095 | 0.061197 | 0.59984 | 10200.28 |
| GOBP_NEG | 15 | -0.03013 | -0.00085 | 0.11909 | 0.59987 | 10200.79 |
| GOMF_NU | 9 | -0.0401 | -0.00088 | 0.15842 | 0.5999 | 10201.3 |
| GOMF_SER | 10 | -0.043 | -0.00099 | 0.16986 | 0.59992 | 10201.64 |
| BIOCARTA_ | 11 | -0.03077 | -0.00074 | 0.12151 | 0.59995 | 10202.15 |
| GOBP_REG | 183 | -0.00955 | -0.00094 | 0.037708 | 0.59997 | 10202.49 |
| GOMF_NAI | 8 | -0.03514 | -0.00072 | 0.13864 | 0.60004 | 10203.68 |
| GOBP_NEG | 5 | -0.06028 | -0.00098 | 0.23775 | 0.60007 | 10204.19 |

| | | | | | | |
|---|---|---|---|---|---|---|
| GOBP_CELI | 5 | -0.05368 | -0.00087 | 0.21144 | 0.60019 | 10206.23 |
| ZWANG_TI | 1414 | -0.00353 | -0.00093 | 0.013906 | 0.60024 | 10207.08 |
| GOMF_SEF | 5 | -0.05736 | -0.00093 | 0.22584 | 0.60025 | 10207.25 |
| YOSHIMUR | 1193 | -0.00389 | -0.00095 | 0.015281 | 0.60036 | 10209.12 |
| GOBP_REG | 5 | -0.05687 | -0.00093 | 0.22331 | 0.60051 | 10211.67 |
| GOBP_REG | 14 | -0.0344 | -0.00094 | 0.13502 | 0.60055 | 10212.35 |
| GOBP_CELI | 18 | -0.02899 | -0.00089 | 0.1136 | 0.60069 | 10214.73 |
| GOBP_INO | 8 | -0.04505 | -0.00093 | 0.17646 | 0.60075 | 10215.75 |
| GOCC_PER | 4 | -0.07591 | -0.0011 | 0.29681 | 0.60093 | 10218.81 |
| GOBP_G_P | 9 | -0.05157 | -0.00113 | 0.2016 | 0.60095 | 10219.15 |
| GOMF_FAT | 5 | -0.05733 | -0.00093 | 0.22389 | 0.60105 | 10220.86 |
| GOBP_POS | 8 | -0.04981 | -0.00103 | 0.19422 | 0.60121 | 10223.58 |
| BIOCARTA_ | 26 | -0.02466 | -0.00091 | 0.096127 | 0.60122 | 10223.75 |
| GOBP_REG | 9 | -0.04799 | -0.00105 | 0.18694 | 0.60129 | 10224.94 |
| GOBP_POS | 34 | -0.0214 | -0.00091 | 0.083348 | 0.6013 | 10225.11 |
| GOBP_REG | 17 | -0.03176 | -0.00095 | 0.12367 | 0.60134 | 10225.79 |
| PID_PI3K_I | 36 | -0.02191 | -0.00096 | 0.085217 | 0.60143 | 10227.32 |
| GOBP_REG | 9 | -0.03418 | -0.00075 | 0.1327 | 0.60162 | 10230.55 |
| GOMF_PRE | 16 | -0.02957 | -0.00086 | 0.11472 | 0.6017 | 10231.91 |
| GARGALOV | 11 | -0.04134 | -0.001 | 0.16027 | 0.60176 | 10232.93 |
| BENITEZ_G | 47 | -0.01967 | -0.00098 | 0.076134 | 0.60192 | 10235.65 |
| SOGA_COL | 69 | -0.01593 | -0.00096 | 0.061658 | 0.60192 | 10235.65 |
| REACTOME | 76 | -0.01455 | -0.00092 | 0.056257 | 0.60202 | 10237.35 |
| GOBP_NCR | 9 | -0.03936 | -0.00086 | 0.15216 | 0.60205 | 10237.86 |
| REACTOME | 240 | -0.00826 | -0.00092 | 0.031909 | 0.60207 | 10238.2 |
| GOBP_INTI | 58 | -0.01725 | -0.00095 | 0.066645 | 0.60213 | 10239.22 |
| GOBP_FER | 184 | -0.00974 | -0.00096 | 0.037637 | 0.60214 | 10239.39 |
| GOBP_ACY | 7 | -0.05568 | -0.00107 | 0.21482 | 0.60225 | 10241.26 |
| GOCC_AXC | 20 | -0.03167 | -0.00103 | 0.1222 | 0.60226 | 10241.43 |
| GOMF_SYN | 140 | -0.01138 | -0.00098 | 0.043899 | 0.6023 | 10242.11 |
| GOCC_BAS | 78 | -0.01578 | -0.00101 | 0.060816 | 0.60237 | 10243.3 |
| REACTOME | 37 | -0.02035 | -0.0009 | 0.078426 | 0.60237 | 10243.3 |
| REACTOME | 124 | -0.01139 | -0.00092 | 0.04388 | 0.60238 | 10243.47 |
| GOBP_REG | 5 | -0.05111 | -0.00083 | 0.19691 | 0.6024 | 10243.81 |
| BIOCARTA_ | 14 | -0.03646 | -0.00099 | 0.14045 | 0.60242 | 10244.15 |
| STREICHER | 19 | -0.0304 | -0.00096 | 0.11709 | 0.60242 | 10244.15 |
| GOBP_TRN | 7 | -0.05487 | -0.00106 | 0.21108 | 0.60255 | 10246.36 |
| GOBP_CELI | 16 | -0.0345 | -0.001 | 0.1326 | 0.60262 | 10247.55 |
| WP_GPR14 | 29 | -0.02521 | -0.00099 | 0.096885 | 0.60265 | 10248.06 |
| GOBP_CIR( | 23 | -0.02826 | -0.00099 | 0.10856 | 0.60271 | 10249.08 |
| TOMLINS_I | 37 | -0.02209 | -0.00098 | 0.084813 | 0.60273 | 10249.42 |
| GOBP_MEI | 24 | -0.03081 | -0.0011 | 0.11827 | 0.60276 | 10249.93 |
| GOBP_MA( | 6 | -0.0486 | -0.00087 | 0.18645 | 0.60282 | 10250.95 |
| GOBP_POL | 25 | -0.02338 | -0.00085 | 0.089683 | 0.60284 | 10251.29 |
| IWANAGA_ | 174 | -0.00945 | -0.0009 | 0.036263 | 0.60284 | 10251.29 |

| | | | | | | |
|---|---|---|---|---|---|---|
| GOMF_TRA | 994 | -0.00425 | -0.00095 | 0.016275 | 0.6029 | 10252.31 |
| BILD_CTNN | 74 | -0.01571 | -0.00098 | 0.060195 | 0.60292 | 10252.65 |
| GOMF_OPS | 5 | -0.06067 | -0.00099 | 0.2325 | 0.60293 | 10252.82 |
| GOBP_PRC | 27 | -0.02792 | -0.00106 | 0.10688 | 0.60304 | 10254.7 |
| ROVERSI_G | 97 | -0.01674 | -0.0012 | 0.064065 | 0.60306 | 10255.04 |
| REACTOME | 15 | -0.03552 | -0.001 | 0.13584 | 0.60313 | 10256.23 |
| GOMF_PR( | 30 | -0.02265 | -0.0009 | 0.086592 | 0.60317 | 10256.91 |
| HUNSBERG | 28 | -0.02585 | -0.00099 | 0.098807 | 0.6032 | 10257.42 |
| GOMF_DEA | 17 | -0.03347 | -0.001 | 0.12778 | 0.60332 | 10259.46 |
| KAYO_CAL( | 76 | -0.01384 | -0.00088 | 0.052813 | 0.60334 | 10259.8 |
| GOBP_RES | 4 | -0.07295 | -0.00106 | 0.27788 | 0.60354 | 10263.2 |
| GOMF_PEF | 7 | -0.04731 | -0.00091 | 0.1802 | 0.60355 | 10263.37 |
| REACTOME | 10 | -0.04679 | -0.00108 | 0.17811 | 0.60361 | 10264.39 |
| POMEROY_ | 52 | -0.01828 | -0.00096 | 0.069579 | 0.60362 | 10264.56 |
| WP_SARSC | 6 | -0.0654 | -0.00117 | 0.24859 | 0.60376 | 10266.94 |
| KUMAMOT | 9 | -0.0397 | -0.00087 | 0.15078 | 0.60383 | 10268.13 |
| GOBP_POS | 5 | -0.06011 | -0.00098 | 0.22783 | 0.60405 | 10271.87 |
| GOCC_NU( | 136 | -0.01117 | -0.00094 | 0.042305 | 0.60408 | 10272.38 |
| REACTOME | 15 | -0.03332 | -0.00094 | 0.12622 | 0.6041 | 10272.72 |
| GOBP_ATF | 7 | -0.05178 | -0.001 | 0.19606 | 0.60414 | 10273.4 |
| GOMF_K48 | 9 | -0.03931 | -0.00086 | 0.14881 | 0.60418 | 10274.08 |
| KEGG_GLY( | 44 | -0.02287 | -0.0011 | 0.086402 | 0.60436 | 10277.14 |
| GRANDVAL | 14 | -0.03292 | -0.0009 | 0.12438 | 0.60438 | 10277.48 |
| SWEET_KR. | 59 | -0.01702 | -0.00095 | 0.064203 | 0.60452 | 10279.86 |
| GOBP_VEN | 24 | -0.03052 | -0.00109 | 0.11511 | 0.60453 | 10280.03 |
| SCHAEFFEF | 147 | -0.01092 | -0.00096 | 0.041167 | 0.6046 | 10281.22 |
| GOBP_END | 9 | -0.04315 | -0.00094 | 0.1626 | 0.60464 | 10281.9 |
| REACTOME | 599 | -0.0054 | -0.00095 | 0.020277 | 0.60492 | 10286.66 |
| GOBP_POS | 26 | -0.02466 | -0.00091 | 0.092625 | 0.60498 | 10287.68 |
| GOBP_MIT | 16 | -0.03298 | -0.00096 | 0.12375 | 0.60508 | 10289.39 |
| GOMF_PYF | 6 | -0.044 | -0.00078 | 0.16508 | 0.60509 | 10289.56 |
| WP_1Q211 | 24 | -0.03186 | -0.00114 | 0.11948 | 0.60512 | 10290.07 |
| GOBP_TYP | 30 | -0.02576 | -0.00103 | 0.096522 | 0.60521 | 10291.6 |
| GOBP_RES | 6 | -0.06552 | -0.00117 | 0.24543 | 0.60525 | 10292.28 |
| GOMF_ASF | 7 | -0.05705 | -0.0011 | 0.21359 | 0.6053 | 10293.13 |
| GOBP_RES | 32 | -0.02458 | -0.00101 | 0.091998 | 0.60532 | 10293.47 |
| REACTOME | 24 | -0.02608 | -0.00093 | 0.097584 | 0.60534 | 10293.81 |
| GOBP_HAII | 4 | -0.06234 | -0.00091 | 0.23325 | 0.60537 | 10294.32 |
| BIOCARTA_ | 25 | -0.02351 | -0.00086 | 0.087934 | 0.60542 | 10295.17 |
| GOBP_OO( | 92 | -0.01395 | -0.00097 | 0.052171 | 0.60543 | 10295.34 |
| BIOCARTA_ | 17 | -0.03083 | -0.00092 | 0.11522 | 0.60547 | 10296.02 |
| GOBP_UBI( | 13 | -0.03229 | -0.00085 | 0.12066 | 0.6055 | 10296.53 |
| GOBP_CYT | 12 | -0.04424 | -0.00112 | 0.16526 | 0.60553 | 10297.04 |
| GOBP_MAI | 8 | -0.05257 | -0.00108 | 0.19614 | 0.60565 | 10299.08 |
| GOBP_DNA | 6 | -0.05626 | -0.001 | 0.20988 | 0.60568 | 10299.59 |

| | | | | | | |
|---|---|---|---|---|---|---|
| GOBP_REG | 343 | -0.0074 | -0.00099 | 0.027603 | 0.6057 | 10299.93 |
| GOBP_RET | 151 | -0.01135 | -0.00101 | 0.04231 | 0.60578 | 10301.29 |
| PID_VEGFR | 69 | -0.01626 | -0.00098 | 0.060511 | 0.60592 | 10303.67 |
| GOBP_VES | 64 | -0.0177 | -0.00103 | 0.065848 | 0.60598 | 10304.69 |
| GOBP_PRC | 10 | -0.03319 | -0.00076 | 0.12331 | 0.60608 | 10306.39 |
| JI_RESPON | 50 | -0.01938 | -0.001 | 0.072018 | 0.60608 | 10306.39 |
| KEGG_PRIC | 31 | -0.0275 | -0.00111 | 0.10206 | 0.60619 | 10308.26 |
| GOCC_CYT | 272 | -0.00796 | -0.00095 | 0.029509 | 0.60629 | 10309.96 |
| GOBP_LIPII | 316 | -0.00743 | -0.00095 | 0.027512 | 0.60648 | 10313.19 |
| SU_SALIVA | 13 | -0.04109 | -0.00108 | 0.15192 | 0.6066 | 10315.23 |
| RODRIGUE | 150 | -0.01111 | -0.00099 | 0.041069 | 0.60661 | 10315.4 |
| GOBP_GAN | 6 | -0.04993 | -0.00089 | 0.18455 | 0.60662 | 10315.57 |
| GOMF_PRC | 20 | -0.02866 | -0.00093 | 0.10572 | 0.60683 | 10319.14 |
| GOMF_UN | 105 | -0.01263 | -0.00094 | 0.046563 | 0.6069 | 10320.33 |
| GOBP_SUC | 5 | -0.07122 | -0.00116 | 0.26256 | 0.60691 | 10320.5 |
| GOCC_PRC | 11 | -0.03315 | -0.0008 | 0.12208 | 0.607 | 10322.04 |
| REACTOME | 5 | -0.05551 | -0.0009 | 0.20379 | 0.60734 | 10327.82 |
| OHGUCHI_ | 45 | -0.02106 | -0.00103 | 0.077275 | 0.60739 | 10328.67 |
| WP_IL3_SI | 48 | -0.02066 | -0.00104 | 0.075757 | 0.60745 | 10329.69 |
| TUOMISTO | 19 | -0.03349 | -0.00106 | 0.12276 | 0.60749 | 10330.37 |
| REACTOME | 5 | -0.06405 | -0.00104 | 0.2346 | 0.60757 | 10331.73 |
| REACTOME | 29 | -0.03112 | -0.00122 | 0.11381 | 0.60774 | 10334.62 |
| REACTOME | 100 | -0.01278 | -0.00093 | 0.046717 | 0.6078 | 10335.64 |
| GOBP_NEG | 6 | -0.05963 | -0.00106 | 0.21793 | 0.60781 | 10335.81 |
| GOBP_REG | 37 | -0.02155 | -0.00095 | 0.078616 | 0.60798 | 10338.7 |
| REACTOME | 11 | -0.0687 | -0.00166 | 0.25068 | 0.60798 | 10338.7 |
| GOBP_DEV | 207 | -0.0098 | -0.00102 | 0.03574 | 0.60805 | 10339.89 |
| GOBP_PYR | 42 | -0.0228 | -0.00107 | 0.083118 | 0.60805 | 10339.89 |
| WP_GLYCC | 20 | -0.03094 | -0.00101 | 0.11282 | 0.60806 | 10340.06 |
| REACTOME | 17 | -0.04025 | -0.00121 | 0.14669 | 0.6081 | 10340.74 |
| GOMF_HIS | 12 | -0.03304 | -0.00083 | 0.12042 | 0.60811 | 10340.91 |
| WHITESIDE | 12 | -0.03459 | -0.00087 | 0.12598 | 0.60816 | 10341.76 |
| GOBP_CON | 9 | -0.04135 | -0.0009 | 0.1506 | 0.60818 | 10342.1 |
| GOBP_REG | 19 | -0.03468 | -0.0011 | 0.12628 | 0.60818 | 10342.1 |
| KEGG_BET | 21 | -0.03049 | -0.00102 | 0.11086 | 0.60836 | 10345.16 |
| GOCC_RPA | 9 | -0.03958 | -0.00086 | 0.14383 | 0.6084 | 10345.84 |
| GOBP_MO | 79 | -0.01605 | -0.00104 | 0.058261 | 0.60851 | 10347.71 |
| REACTOME | 6 | -0.04698 | -0.00084 | 0.17054 | 0.60853 | 10348.05 |
| GOBP_NEG | 205 | -0.00986 | -0.00102 | 0.035757 | 0.60859 | 10349.07 |
| GOBP_SIAL | 21 | -0.03128 | -0.00104 | 0.11333 | 0.60871 | 10351.11 |
| GOMF_SIA | 21 | -0.03128 | -0.00104 | 0.11333 | 0.60871 | 10351.11 |
| GOBP_PHE | 5 | -0.06706 | -0.00109 | 0.24282 | 0.60878 | 10352.3 |
| WILSON_P | 19 | -0.03041 | -0.00096 | 0.1101 | 0.60878 | 10352.3 |
| GOMF_OXI | 8 | -0.05272 | -0.00109 | 0.1907 | 0.6089 | 10354.34 |
| GOCC_BLO | 8 | -0.04651 | -0.00096 | 0.16818 | 0.60895 | 10355.19 |

| | | | | | | |
|---|---|---|---|---|---|---|
| YAMASHIT/ | 44 | -0.0227 | -0.00109 | 0.082065 | 0.60896 | 10355.36 |
| GOBP_CHC | 9 | -0.05171 | -0.00113 | 0.18688 | 0.60899 | 10355.87 |
| GOBP_POS | 32 | -0.02313 | -0.00095 | 0.08358 | 0.609 | 10356.05 |
| PID_TCR_JI | 14 | -0.04094 | -0.00111 | 0.14788 | 0.60906 | 10357.07 |
| VALK_AML | 33 | -0.02612 | -0.00109 | 0.094345 | 0.60906 | 10357.07 |
| GOCC_9PLI | 4 | -0.05319 | -0.00077 | 0.19193 | 0.60915 | 10358.6 |
| REACTOME | 249 | -0.00927 | -0.00106 | 0.033425 | 0.60921 | 10359.62 |
| LEE_LIVER_ | 15 | -0.04039 | -0.00114 | 0.14561 | 0.60925 | 10360.3 |
| GOCC_FLEI | 35 | -0.02587 | -0.00111 | 0.093258 | 0.60927 | 10360.64 |
| REACTOME | 25 | -0.02655 | -0.00097 | 0.095575 | 0.60941 | 10363.02 |
| GOBP_OXII | 18 | -0.0286 | -0.00088 | 0.10292 | 0.60945 | 10363.7 |
| TORCHIA_T | 297 | -0.00825 | -0.00103 | 0.029689 | 0.60945 | 10363.7 |
| GOBP_MEC | 66 | -0.01749 | -0.00103 | 0.062903 | 0.60951 | 10364.72 |
| GOBP_GLI/ | 5 | -0.07442 | -0.00121 | 0.26761 | 0.60953 | 10365.06 |
| GOCC_HIPI | 41 | -0.02171 | -0.00101 | 0.078055 | 0.60955 | 10365.4 |
| REACTOME | 130 | -0.0127 | -0.00105 | 0.045669 | 0.60956 | 10365.57 |
| BIOCARTA_ | 15 | -0.0342 | -0.00096 | 0.12277 | 0.60971 | 10368.12 |
| KEGG_GLY( | 20 | -0.02735 | -0.00089 | 0.098067 | 0.60983 | 10370.16 |
| WP_2Q211 | 17 | -0.03786 | -0.00114 | 0.13568 | 0.60989 | 10371.18 |
| GOMF_PEF | 131 | -0.01352 | -0.00112 | 0.048418 | 0.60995 | 10372.2 |
| LEE_LIVER_ | 61 | -0.01967 | -0.00112 | 0.07032 | 0.61013 | 10375.26 |
| GOMF_TR/ | 58 | -0.01785 | -0.00099 | 0.063756 | 0.61024 | 10377.13 |
| GOBP_REG | 80 | -0.01649 | -0.00107 | 0.058884 | 0.61026 | 10377.47 |
| GOMF_INC | 5 | -0.05154 | -0.00084 | 0.18396 | 0.61032 | 10378.49 |
| GOBP_POS | 1003 | -0.00468 | -0.00105 | 0.016701 | 0.61037 | 10379.34 |
| GOBP_HIST | 4 | -0.05402 | -0.00079 | 0.19271 | 0.61038 | 10379.51 |
| GOMF_EX( | 99 | -0.0145 | -0.00105 | 0.051687 | 0.6105 | 10381.55 |
| GOBP_REG | 146 | -0.0126 | -0.0011 | 0.044894 | 0.61052 | 10381.89 |
| GOBP_REG | 18 | -0.03645 | -0.00113 | 0.12983 | 0.61054 | 10382.23 |
| GOBP_TRI( | 24 | -0.03162 | -0.00113 | 0.11264 | 0.61055 | 10382.4 |
| VANLOO_S | 8 | -0.03939 | -0.00081 | 0.14026 | 0.61058 | 10382.91 |
| GOBP_ACT | 28 | -0.02745 | -0.00106 | 0.097668 | 0.61065 | 10384.1 |
| GOBP_NEG | 11 | -0.03776 | -0.00091 | 0.13437 | 0.61066 | 10384.27 |
| GOBP_TEL( | 7 | -0.05348 | -0.00103 | 0.19028 | 0.61067 | 10384.44 |
| HAHTOLA_ | 13 | -0.04042 | -0.00106 | 0.14381 | 0.61067 | 10384.44 |
| GOCC_END | 12 | -0.03901 | -0.00098 | 0.13864 | 0.61078 | 10386.31 |
| WP_REGUI | 22 | -0.02929 | -0.001 | 0.10396 | 0.61094 | 10389.03 |
| GOBP_ERK | 5 | -0.08183 | -0.00133 | 0.29036 | 0.61095 | 10389.2 |
| GOMF_PH( | 8 | -0.05452 | -0.00112 | 0.19344 | 0.61096 | 10389.37 |
| BIOCARTA_ | 22 | -0.0342 | -0.00117 | 0.12097 | 0.61131 | 10395.33 |
| GOBP_REG | 16 | -0.034 | -0.00099 | 0.1202 | 0.61134 | 10395.84 |
| GOMF_CAT | 11 | -0.04875 | -0.00118 | 0.17233 | 0.61137 | 10396.35 |
| GOBP_NEG | 5 | -0.06056 | -0.00099 | 0.21401 | 0.61141 | 10397.03 |
| KOHOUTEK | 55 | -0.0184 | -0.00099 | 0.065019 | 0.61142 | 10397.2 |
| GOBP_ORG | 152 | -0.01103 | -0.00099 | 0.03895 | 0.61145 | 10397.71 |

| | | | | | | |
|---|---|---|---|---|---|---|
| REACTOME | 9 | -0.04882 | -0.00107 | 0.17245 | 0.61145 | 10397.71 |
| CHEMNITZ_ | 360 | -0.00787 | -0.00108 | 0.027794 | 0.61146 | 10397.88 |
| REACTOME | 8 | -0.0513 | -0.00106 | 0.18115 | 0.61149 | 10398.39 |
| GOBP_POS | 187 | -0.01006 | -0.001 | 0.035531 | 0.6115 | 10398.56 |
| GOBP_REG | 19 | -0.03113 | -0.00099 | 0.10975 | 0.61165 | 10401.11 |
| VALK_AML | 4 | -0.08826 | -0.00128 | 0.31109 | 0.61169 | 10401.79 |
| REACTOME | 342 | -0.00741 | -0.00099 | 0.026089 | 0.61173 | 10402.47 |
| YAGI_AML_ | 110 | -0.0129 | -0.00098 | 0.045403 | 0.61188 | 10405.02 |
| GOBP_VES | 45 | -0.01792 | -0.00087 | 0.063004 | 0.61196 | 10406.38 |
| GOBP_SKE | 21 | -0.03062 | -0.00102 | 0.1076 | 0.612 | 10407.06 |
| GARGALOV | 11 | -0.04879 | -0.00118 | 0.17146 | 0.61201 | 10407.23 |
| GOBP_ORG | 85 | -0.01387 | -0.00093 | 0.048716 | 0.61202 | 10407.4 |
| GOBP_RES | 8 | -0.05134 | -0.00106 | 0.18018 | 0.61215 | 10409.61 |
| GOBP_REG | 7 | -0.06495 | -0.00125 | 0.22791 | 0.61216 | 10409.78 |
| GOBP_VAS | 247 | -0.00983 | -0.00112 | 0.034467 | 0.61222 | 10410.8 |
| GOBP_RES | 26 | -0.02796 | -0.00104 | 0.097995 | 0.6123 | 10412.16 |
| GOMF_ACI | 5 | -0.0553 | -0.0009 | 0.19377 | 0.61233 | 10412.67 |
| ANDERSEN | 30 | -0.02492 | -0.00099 | 0.087237 | 0.61243 | 10414.37 |
| GOBP_IMN | 15 | -0.04098 | -0.00115 | 0.14346 | 0.61243 | 10414.37 |
| GOCC_ACR | 133 | -0.01185 | -0.00099 | 0.041464 | 0.61244 | 10414.54 |
| HUTTMANI | 254 | -0.00873 | -0.00101 | 0.030567 | 0.61244 | 10414.54 |
| GOBP_CER | 17 | -0.03434 | -0.00103 | 0.12014 | 0.61248 | 10415.22 |
| GOBP_REG | 5 | -0.06365 | -0.00104 | 0.2226 | 0.61254 | 10416.24 |
| GOBP_WN | 5 | -0.06365 | -0.00104 | 0.2226 | 0.61254 | 10416.24 |
| GOMF_ME | 16 | -0.03911 | -0.00114 | 0.13675 | 0.61255 | 10416.41 |
| GOBP_POS | 5 | -0.06623 | -0.00108 | 0.2315 | 0.61259 | 10417.09 |
| GOMF_ION | 7 | -0.06535 | -0.00126 | 0.22834 | 0.61264 | 10417.94 |
| GOBP_FOR | 22 | -0.03376 | -0.00115 | 0.11785 | 0.61275 | 10419.81 |
| SUBTIL_PR | 33 | -0.02487 | -0.00104 | 0.086639 | 0.61296 | 10423.38 |
| GOBP_POS | 83 | -0.0156 | -0.00103 | 0.054304 | 0.61307 | 10425.26 |
| GOBP_PHA | 46 | -0.02254 | -0.00111 | 0.07841 | 0.61313 | 10426.28 |
| GOMF_PH | 26 | -0.02919 | -0.00108 | 0.10154 | 0.61313 | 10426.28 |
| HOEBEKE_I | 91 | -0.01608 | -0.00111 | 0.055922 | 0.61313 | 10426.28 |
| REACTOME | 7 | -0.06818 | -0.00131 | 0.23711 | 0.61314 | 10426.45 |
| GOBP_RES | 45 | -0.02248 | -0.0011 | 0.078161 | 0.61315 | 10426.62 |
| GOBP_POS | 28 | -0.03024 | -0.00116 | 0.10511 | 0.61322 | 10427.81 |
| GOMF_MC | 638 | -0.00579 | -0.00105 | 0.020094 | 0.61331 | 10429.34 |
| BURTON_A | 72 | -0.01731 | -0.00107 | 0.060104 | 0.61332 | 10429.51 |
| GOBP_STR | 20 | -0.03803 | -0.00124 | 0.13194 | 0.61342 | 10431.21 |
| REACTOME | 37 | -0.02207 | -0.00098 | 0.076549 | 0.61344 | 10431.55 |
| GOMF_PH | 11 | -0.03975 | -0.00096 | 0.13785 | 0.61345 | 10431.72 |
| PID_IL27_F | 25 | -0.03032 | -0.0011 | 0.10518 | 0.61345 | 10431.72 |
| WP_GASTF | 113 | -0.01351 | -0.00104 | 0.046837 | 0.61353 | 10433.08 |
| GOMF_LAN | 15 | -0.03342 | -0.00094 | 0.1158 | 0.61354 | 10433.25 |
| GOBP_HIN | 5 | -0.08272 | -0.00135 | 0.28654 | 0.61359 | 10434.1 |

| | | | | | | |
|---|---|---|---|---|---|---|
| KEGG_ALD | 40 | -0.02422 | -0.00111 | 0.083854 | 0.61365 | 10435.12 |
| GOBP_ANT | 3 | -0.07347 | -0.00093 | 0.25421 | 0.61372 | 10436.31 |
| GOBP_TRA | 10 | -0.04384 | -0.00101 | 0.15161 | 0.61376 | 10436.99 |
| REACTOME | 11 | -0.05003 | -0.00121 | 0.17299 | 0.61378 | 10437.33 |
| GOBP_PEP | 22 | -0.029 | -0.00099 | 0.10017 | 0.6139 | 10439.37 |
| GOBP_NEG | 31 | -0.025 | -0.00101 | 0.086319 | 0.61392 | 10439.71 |
| GOBP_EPIT | 10 | -0.05508 | -0.00127 | 0.19006 | 0.61401 | 10441.24 |
| FLORIO_NE | 362 | -0.00799 | -0.0011 | 0.027547 | 0.61404 | 10441.75 |
| BIOCARTA_ | 8 | -0.05358 | -0.0011 | 0.18467 | 0.61413 | 10443.28 |
| WANG_LM | 334 | -0.00787 | -0.00104 | 0.0271 | 0.61418 | 10444.13 |
| NABA_ECM | 158 | -0.01281 | -0.00117 | 0.044103 | 0.61423 | 10444.98 |
| GOBP_ACT | 8 | -0.05002 | -0.00103 | 0.17202 | 0.61438 | 10447.53 |
| GOBP_ACR | 9 | -0.05447 | -0.00119 | 0.1873 | 0.61441 | 10448.04 |
| GOBP_PRC | 13 | -0.04441 | -0.00117 | 0.15248 | 0.61457 | 10450.76 |
| GOBP_T_C | 34 | -0.02397 | -0.00102 | 0.082286 | 0.61461 | 10451.44 |
| GOBP_BEH | 19 | -0.03312 | -0.00105 | 0.11359 | 0.61468 | 10452.63 |
| GOBP_DOL | 5 | -0.05834 | -0.00095 | 0.20011 | 0.61468 | 10452.63 |
| BIOCARTA_ | 9 | -0.04454 | -0.00097 | 0.15266 | 0.61476 | 10453.99 |
| GOMF_5S_ | 10 | -0.03726 | -0.00086 | 0.12763 | 0.61483 | 10455.18 |
| OUYANG_F | 19 | -0.03673 | -0.00116 | 0.12561 | 0.615 | 10458.08 |
| BIOCARTA_ | 17 | -0.03358 | -0.00101 | 0.11481 | 0.61503 | 10458.59 |
| GOBP_POS | 13 | -0.04296 | -0.00113 | 0.14684 | 0.61506 | 10459.1 |
| GOBP_REG | 22 | -0.03354 | -0.00114 | 0.11462 | 0.61509 | 10459.61 |
| REACTOME | 6 | -0.05457 | -0.00097 | 0.18631 | 0.61519 | 10461.31 |
| GOBP_S_A | 16 | -0.03797 | -0.00111 | 0.12962 | 0.61522 | 10461.82 |
| GOBP_POS | 52 | -0.02148 | -0.00113 | 0.073279 | 0.61527 | 10462.67 |
| GOMF_PH( | 13 | -0.04159 | -0.00109 | 0.1419 | 0.61527 | 10462.67 |
| GOBP_POS | 37 | -0.02392 | -0.00106 | 0.081538 | 0.61536 | 10464.2 |
| GOBP_DNA | 308 | -0.00808 | -0.00102 | 0.027541 | 0.61544 | 10465.56 |
| GOMF_FLA | 83 | -0.01608 | -0.00106 | 0.054742 | 0.6155 | 10466.58 |
| GOBP_VES | 1447 | -0.00397 | -0.00106 | 0.013519 | 0.61551 | 10466.75 |
| GOBP_ICOS | 48 | -0.02256 | -0.00114 | 0.076761 | 0.61557 | 10467.77 |
| GOBP_DEN | 129 | -0.01322 | -0.00109 | 0.044962 | 0.61564 | 10468.96 |
| ROSS_LEUK | 74 | -0.01816 | -0.00113 | 0.061705 | 0.61573 | 10470.49 |
| GOBP_LEU | 7 | -0.06045 | -0.00116 | 0.20525 | 0.61581 | 10471.85 |
| GOBP_POL | 13 | -0.03625 | -0.00095 | 0.12294 | 0.61594 | 10474.06 |
| GOBP_ACT | 104 | -0.01482 | -0.0011 | 0.050164 | 0.61613 | 10477.29 |
| REACTOME | 9 | -0.05023 | -0.0011 | 0.16994 | 0.61622 | 10478.82 |
| GOBP_NEG | 11 | -0.04114 | -0.00099 | 0.13918 | 0.61623 | 10478.99 |
| BIOCARTA_ | 10 | -0.0495 | -0.00114 | 0.16738 | 0.61628 | 10479.84 |
| REACTOME | 5 | -0.0609 | -0.00099 | 0.20584 | 0.61632 | 10480.52 |
| IRITANI_M | 12 | -0.04533 | -0.00114 | 0.1529 | 0.61657 | 10484.77 |
| KEGG_ALA | 31 | -0.02706 | -0.0011 | 0.091239 | 0.6166 | 10485.28 |
| GOCC_PRE | 5 | -0.06687 | -0.00109 | 0.22512 | 0.61678 | 10488.34 |
| BOYLAN_M | 53 | -0.02256 | -0.00119 | 0.075905 | 0.61685 | 10489.53 |

| | | | | | | |
|---|---|---|---|---|---|---|
| GOBP_REG | 5 | -0.08029 | -0.00131 | 0.26996 | 0.61692 | 10490.72 |
| GOBP_T_H | 42 | -0.02201 | -0.00104 | 0.073968 | 0.61699 | 10491.91 |
| GOBP_ACU | 23 | -0.03448 | -0.0012 | 0.11584 | 0.61701 | 10492.26 |
| REACTOME | 22 | -0.03036 | -0.00104 | 0.10199 | 0.61702 | 10492.43 |
| REACTOME | 8 | -0.05376 | -0.00111 | 0.18053 | 0.61707 | 10493.28 |
| GOBP_SOD | 168 | -0.01246 | -0.00117 | 0.04176 | 0.61726 | 10496.51 |
| MARZEC_IL | 36 | -0.02459 | -0.00107 | 0.082408 | 0.61728 | 10496.85 |
| GOBP_ORN | 6 | -0.06382 | -0.00114 | 0.21379 | 0.61734 | 10497.87 |
| GOMF_OD | 95 | -0.01838 | -0.0013 | 0.061544 | 0.61739 | 10498.72 |
| GOCC_T_C | 13 | -0.04934 | -0.00129 | 0.16513 | 0.61744 | 10499.57 |
| GOMF_TES | 8 | -0.0527 | -0.00108 | 0.17623 | 0.61753 | 10501.1 |
| GHO_ATF5 | 12 | -0.04251 | -0.00107 | 0.14193 | 0.61772 | 10504.33 |
| HUANG_FC | 36 | -0.02299 | -0.001 | 0.076696 | 0.61779 | 10505.52 |
| GOBP_REG | 16 | -0.03684 | -0.00107 | 0.12289 | 0.61782 | 10506.03 |
| GOBP_POS | 10 | -0.05259 | -0.00121 | 0.17537 | 0.61786 | 10506.71 |
| GOBP_NEG | 5 | -0.06209 | -0.00101 | 0.20703 | 0.61788 | 10507.05 |
| REACTOME | 9 | -0.04907 | -0.00107 | 0.16353 | 0.61793 | 10507.9 |
| GOBP_SYN | 15 | -0.04247 | -0.0012 | 0.14147 | 0.61798 | 10508.75 |
| FUNG_IL2_ | 11 | -0.04662 | -0.00113 | 0.15522 | 0.61805 | 10509.94 |
| GOBP_MY( | 120 | -0.01414 | -0.00112 | 0.047086 | 0.61805 | 10509.94 |
| KEGG_ABC | 40 | -0.02473 | -0.00114 | 0.082252 | 0.61816 | 10511.81 |
| GOBP_T_C | 104 | -0.01539 | -0.00114 | 0.051143 | 0.61823 | 10513 |
| GOBP_ADH | 11 | -0.0473 | -0.00114 | 0.15719 | 0.61825 | 10513.34 |
| HENDRICKS | 44 | -0.02146 | -0.00103 | 0.071312 | 0.61826 | 10513.51 |
| GOMF_PH( | 19 | -0.03868 | -0.00123 | 0.12849 | 0.61831 | 10514.36 |
| GOCC_SNA | 45 | -0.02248 | -0.0011 | 0.074647 | 0.61836 | 10515.21 |
| WP_FOLAT | 9 | -0.06656 | -0.00145 | 0.22098 | 0.61836 | 10515.21 |
| GOBP_CELI | 8 | -0.062 | -0.00128 | 0.20579 | 0.6184 | 10515.89 |
| GOBP_POS | 44 | -0.02359 | -0.00114 | 0.078201 | 0.61854 | 10518.27 |
| JOHNSTON | 323 | -0.00876 | -0.00114 | 0.029018 | 0.61859 | 10519.12 |
| GOBP_MO | 4 | -0.07628 | -0.00111 | 0.25247 | 0.61872 | 10521.33 |
| KEGG_RNA | 54 | -0.02007 | -0.00107 | 0.066429 | 0.61873 | 10521.5 |
| GOMF_PH( | 6 | -0.05058 | -0.0009 | 0.16734 | 0.61878 | 10522.35 |
| WP_AFLAT | 7 | -0.05302 | -0.00102 | 0.17527 | 0.61887 | 10523.88 |
| GOMF_HIS | 7 | -0.06187 | -0.00119 | 0.20429 | 0.61899 | 10525.92 |
| GOBP_ACR | 40 | -0.02506 | -0.00115 | 0.082643 | 0.61914 | 10528.48 |
| GOBP_CELI | 6 | -0.06795 | -0.00121 | 0.22362 | 0.61937 | 10532.39 |
| GOBP_SKEI | 8 | -0.05398 | -0.00111 | 0.17764 | 0.61938 | 10532.56 |
| GOBP_TISS | 163 | -0.01251 | -0.00116 | 0.04116 | 0.61943 | 10533.41 |
| GOBP_CELI | 211 | -0.00977 | -0.00103 | 0.032132 | 0.61948 | 10534.26 |
| GOBP_RET | 22 | -0.0351 | -0.0012 | 0.11541 | 0.61949 | 10534.43 |
| KIM_TIAL1 | 31 | -0.0275 | -0.00111 | 0.09031 | 0.61961 | 10536.47 |
| GOBP_POS | 4 | -0.09253 | -0.00135 | 0.3039 | 0.61962 | 10536.64 |
| GOMF_CAI | 15 | -0.03364 | -0.00095 | 0.11048 | 0.61963 | 10536.81 |
| GOBP_NEU | 59 | -0.02001 | -0.00112 | 0.0657 | 0.61965 | 10537.15 |

| | | | | | | |
|---|---|---|---|---|---|---|
| SUZUKI_RE | 15 | -0.03857 | -0.00109 | 0.12651 | 0.61975 | 10538.85 |
| GOBP_NEG | 28 | -0.03001 | -0.00115 | 0.09825 | 0.61998 | 10542.76 |
| GOBP_LEU | 12 | -0.05133 | -0.00129 | 0.16793 | 0.62007 | 10544.29 |
| GOCC_CYT | 18 | -0.0374 | -0.00115 | 0.12232 | 0.62009 | 10544.63 |
| GOBP_SPEI | 27 | -0.02911 | -0.0011 | 0.095163 | 0.62014 | 10545.48 |
| WEBER_MI | 16 | -0.03908 | -0.00114 | 0.12771 | 0.62018 | 10546.16 |
| GOBP_AXC | 21 | -0.0382 | -0.00127 | 0.12471 | 0.62032 | 10548.54 |
| BOYAULT_I | 25 | -0.03349 | -0.00122 | 0.1093 | 0.62036 | 10549.22 |
| GOBP_REG | 6 | -0.07305 | -0.0013 | 0.23804 | 0.62053 | 10552.11 |
| GOBP_REG | 28 | -0.03323 | -0.00128 | 0.10819 | 0.62062 | 10553.64 |
| REACTOME | 70 | -0.01856 | -0.00113 | 0.060408 | 0.62067 | 10554.49 |
| PID_CASPA | 49 | -0.02221 | -0.00113 | 0.072233 | 0.62077 | 10556.19 |
| EHLERS_AN | 12 | -0.04748 | -0.0012 | 0.15424 | 0.62089 | 10558.23 |
| GOBP_DET | 4 | -0.07037 | -0.00102 | 0.22836 | 0.62101 | 10560.28 |
| GOMF_S10 | 13 | -0.04137 | -0.00109 | 0.13417 | 0.62107 | 10561.3 |
| GOBP_PRC | 78 | -0.01699 | -0.00109 | 0.055098 | 0.62109 | 10561.64 |
| DASU_IL6_ | 30 | -0.02788 | -0.00111 | 0.090317 | 0.6212 | 10563.51 |
| GOCC_CYT | 13 | -0.03998 | -0.00105 | 0.12954 | 0.62121 | 10563.68 |
| BIOCARTA_ | 8 | -0.06045 | -0.00124 | 0.19548 | 0.62142 | 10567.25 |
| GOBP_NEG | 43 | -0.02312 | -0.0011 | 0.074751 | 0.62143 | 10567.42 |
| KANG_DOX | 52 | -0.0207 | -0.00109 | 0.066931 | 0.62146 | 10567.93 |
| REACTOME | 57 | -0.01996 | -0.0011 | 0.064467 | 0.62155 | 10569.46 |
| GOBP_GLU | 9 | -0.0497 | -0.00109 | 0.16048 | 0.62161 | 10570.48 |
| GOBP_REG | 330 | -0.00855 | -0.00112 | 0.027592 | 0.62164 | 10570.99 |
| GOCC_CYT | 152 | -0.01278 | -0.00114 | 0.041253 | 0.62165 | 10571.16 |
| GOBP_PRC | 28 | -0.03324 | -0.00128 | 0.10727 | 0.62167 | 10571.5 |
| REACTOME | 38 | -0.02538 | -0.00114 | 0.081916 | 0.62167 | 10571.5 |
| GOBP_NEG | 13 | -0.04261 | -0.00112 | 0.13739 | 0.62177 | 10573.2 |
| GOMF_SPL | 5 | -0.06636 | -0.00108 | 0.21392 | 0.6218 | 10573.71 |
| GOBP_RIB | 20 | -0.04035 | -0.00131 | 0.12998 | 0.62189 | 10575.24 |
| GOBP_RES | 30 | -0.03186 | -0.00127 | 0.10255 | 0.62198 | 10576.77 |
| GOBP_IMN | 382 | -0.00829 | -0.00117 | 0.026691 | 0.62199 | 10576.94 |
| GOBP_POS | 73 | -0.01752 | -0.00109 | 0.056325 | 0.62208 | 10578.47 |
| GOBP_T_H | 18 | -0.0392 | -0.00121 | 0.12598 | 0.62215 | 10579.66 |
| GOBP_T_TI | 6 | -0.05568 | -0.00099 | 0.1789 | 0.62219 | 10580.34 |
| GOBP_ADA | 20 | -0.03616 | -0.00118 | 0.11606 | 0.62231 | 10582.38 |
| GOBP_MIT | 11 | -0.04207 | -0.00102 | 0.13491 | 0.62241 | 10584.08 |
| REACTOME | 45 | -0.02389 | -0.00116 | 0.076555 | 0.62248 | 10585.27 |
| MIKKELSEN | 408 | -0.00755 | -0.0011 | 0.024185 | 0.62257 | 10586.8 |
| GOBP_REG | 60 | -0.02284 | -0.00129 | 0.073112 | 0.62263 | 10587.82 |
| GOMF_RRI | 8 | -0.05095 | -0.00105 | 0.16305 | 0.62266 | 10588.33 |
| GOBP_ALD | 21 | -0.03213 | -0.00107 | 0.10278 | 0.6227 | 10589.01 |
| LINDSTEDT | 44 | -0.02447 | -0.00118 | 0.078232 | 0.62275 | 10589.86 |
| GOBP_CAR | 3 | -0.08239 | -0.00104 | 0.2634 | 0.62278 | 10590.37 |
| DIERICK_SE | 6 | -0.06059 | -0.00108 | 0.19316 | 0.62312 | 10596.16 |

| | | | | | | |
|---|---|---|---|---|---|---|
| GOBP_VER | 11 | -0.04588 | -0.00111 | 0.14624 | 0.62314 | 10596.5 |
| GOBP_NEG | 8 | -0.04645 | -0.00096 | 0.14798 | 0.62319 | 10597.35 |
| LEE_INTRA | 18 | -0.04009 | -0.00124 | 0.12769 | 0.62323 | 10598.03 |
| GOBP_TRA | 27 | -0.0291 | -0.0011 | 0.092623 | 0.62329 | 10599.05 |
| GOMF_NA | 13 | -0.04166 | -0.00109 | 0.13258 | 0.62332 | 10599.56 |
| GOBP_DE_ | 5 | -0.11263 | -0.00183 | 0.35832 | 0.62336 | 10600.24 |
| GOBP_DET | 120 | -0.01461 | -0.00116 | 0.04646 | 0.62344 | 10601.6 |
| BIOCARTA_ | 8 | -0.05306 | -0.00109 | 0.16857 | 0.62353 | 10603.13 |
| GOBP_DNA | 86 | -0.01612 | -0.00109 | 0.051223 | 0.62353 | 10603.13 |
| GOBP_RES | 13 | -0.04662 | -0.00122 | 0.1479 | 0.62369 | 10605.85 |
| GOBP_REG | 74 | -0.01826 | -0.00114 | 0.057872 | 0.62379 | 10607.55 |
| GUILLAUM | 50 | -0.02062 | -0.00106 | 0.065361 | 0.62379 | 10607.55 |
| GOMF_VO | 5 | -0.0862 | -0.0014 | 0.27322 | 0.62381 | 10607.89 |
| GOMF_CAL | 53 | -0.02194 | -0.00116 | 0.069498 | 0.6239 | 10609.42 |
| MATZUK_M | 37 | -0.02312 | -0.00102 | 0.073215 | 0.62391 | 10609.59 |
| GOBP_NEG | 171 | -0.01166 | -0.0011 | 0.036904 | 0.62392 | 10609.76 |
| GAL_LEUKE | 220 | -0.01043 | -0.00112 | 0.033011 | 0.62394 | 10610.1 |
| GOBP_TRA | 18 | -0.0427 | -0.00132 | 0.13517 | 0.62395 | 10610.27 |
| GOBP_KET | 10 | -0.0488 | -0.00112 | 0.15449 | 0.62396 | 10610.44 |
| GOBP_COL | 42 | -0.02483 | -0.00117 | 0.078565 | 0.62399 | 10610.95 |
| PID_AMB2_ | 39 | -0.02514 | -0.00114 | 0.079501 | 0.62407 | 10612.31 |
| GOBP_TEM | 175 | -0.0123 | -0.00118 | 0.03887 | 0.62415 | 10613.67 |
| GOCC_CYT | 22 | -0.03653 | -0.00125 | 0.11544 | 0.62415 | 10613.67 |
| GOBP_POL | 66 | -0.01944 | -0.00115 | 0.061411 | 0.62419 | 10614.35 |
| GOBP_MAT | 5 | -0.08756 | -0.00142 | 0.27648 | 0.62426 | 10615.54 |
| GOBP_RES | 209 | -0.01127 | -0.00118 | 0.035573 | 0.62434 | 10616.9 |
| WP_4HYDF | 18 | -0.03589 | -0.00111 | 0.11307 | 0.62452 | 10619.96 |
| GOBP_TUB | 4 | -0.09525 | -0.00139 | 0.30004 | 0.62455 | 10620.47 |
| GOMF_CAF | 127 | -0.01439 | -0.00118 | 0.045324 | 0.62458 | 10620.98 |
| GOBP_REG | 4 | -0.09717 | -0.00141 | 0.30585 | 0.62464 | 10622 |
| GOBP_GLY | 5 | -0.07068 | -0.00115 | 0.22242 | 0.62468 | 10622.68 |
| LEE_BMP2_ | 729 | -0.00597 | -0.00115 | 0.018786 | 0.6247 | 10623.02 |
| GOMF_HEX | 185 | -0.01161 | -0.00114 | 0.03652 | 0.62476 | 10624.04 |
| WP_DOPAI | 11 | -0.04969 | -0.0012 | 0.15614 | 0.62484 | 10625.4 |
| GOBP_LAC | 15 | -0.04064 | -0.00115 | 0.12746 | 0.62507 | 10629.32 |
| GOBP_NEG | 20 | -0.0342 | -0.00111 | 0.10699 | 0.62537 | 10634.42 |
| WP_CREAT | 4 | -0.07466 | -0.00109 | 0.23354 | 0.6254 | 10634.93 |
| REACTOME | 19 | -0.04092 | -0.0013 | 0.12785 | 0.62553 | 10637.14 |
| CALVET_IR | 4 | -0.06658 | -0.00097 | 0.20785 | 0.62563 | 10638.84 |
| GOBP_PHE | 8 | -0.05828 | -0.0012 | 0.18187 | 0.62568 | 10639.69 |
| GOMF_11_ | 8 | -0.05479 | -0.00113 | 0.17083 | 0.62579 | 10641.56 |
| GOBP_MEL | 18 | -0.0357 | -0.0011 | 0.11122 | 0.62589 | 10643.26 |
| GOBP_REG | 84 | -0.01905 | -0.00127 | 0.05932 | 0.62592 | 10643.77 |
| GOBP_GLA | 35 | -0.0293 | -0.00126 | 0.091232 | 0.62593 | 10643.94 |
| KOYAMA_S | 230 | -0.01075 | -0.00118 | 0.03347 | 0.62593 | 10643.94 |

| | | | | | | |
|---|---|---|---|---|---|---|
| GOBP_LYM | 51 | -0.0238 | -0.00124 | 0.074013 | 0.62612 | 10647.17 |
| FARDIN_HY | 30 | -0.02829 | -0.00113 | 0.087935 | 0.62614 | 10647.51 |
| BIOCARTA_ | 18 | -0.03775 | -0.00117 | 0.11732 | 0.62618 | 10648.19 |
| GOMF_ME | 9 | -0.07197 | -0.00157 | 0.22365 | 0.6262 | 10648.53 |
| RANKIN_AI | 8 | -0.06031 | -0.00124 | 0.1874 | 0.6262 | 10648.53 |
| GOCC_MIT | 10 | -0.05066 | -0.00117 | 0.15741 | 0.62621 | 10648.7 |
| WP_DISOR | 14 | -0.04276 | -0.00116 | 0.13282 | 0.62624 | 10649.21 |
| GOMF_PRO | 17 | -0.04164 | -0.00125 | 0.12933 | 0.62627 | 10649.72 |
| GOBP_REG | 11 | -0.04263 | -0.00103 | 0.13231 | 0.62635 | 10651.08 |
| GAUSSMAI | 226 | -0.0109 | -0.00119 | 0.033828 | 0.62638 | 10651.59 |
| GOBP_STR | 229 | -0.01063 | -0.00116 | 0.032965 | 0.62647 | 10653.12 |
| GOMF_LEU | 10 | -0.04946 | -0.00114 | 0.15324 | 0.62656 | 10654.65 |
| GOMF_NEU | 29 | -0.02925 | -0.00115 | 0.090579 | 0.62661 | 10655.5 |
| GOBP_CER | 24 | -0.03421 | -0.00122 | 0.10591 | 0.62666 | 10656.35 |
| GOCC_CAL | 19 | -0.03832 | -0.00122 | 0.11842 | 0.62687 | 10659.92 |
| PURBEY_TA | 150 | -0.01285 | -0.00114 | 0.039704 | 0.62687 | 10659.92 |
| GOBP_NEU | 12 | -0.0481 | -0.00121 | 0.14861 | 0.62691 | 10660.6 |
| MIDORIKAV | 36 | -0.03722 | -0.00162 | 0.11494 | 0.62696 | 10661.45 |
| BIOCARTA_ | 14 | -0.04097 | -0.00112 | 0.12638 | 0.62708 | 10663.5 |
| GOBP_PYR | 9 | -0.05718 | -0.00125 | 0.17639 | 0.6271 | 10663.84 |
| GOBP_SNO | 5 | -0.06979 | -0.00114 | 0.21522 | 0.62712 | 10664.18 |
| GOBP_NEG | 5 | -0.09625 | -0.00157 | 0.2965 | 0.62726 | 10666.56 |
| KEGG_NUC | 40 | -0.02215 | -0.00102 | 0.068183 | 0.62733 | 10667.75 |
| WP_NUCLE | 40 | -0.02215 | -0.00102 | 0.068183 | 0.62733 | 10667.75 |
| GOCC_ALP | 5 | -0.09099 | -0.00148 | 0.27998 | 0.6274 | 10668.94 |
| GOBP_AMI | 17 | -0.04247 | -0.00127 | 0.13059 | 0.62749 | 10670.47 |
| GOBP_REG | 8 | -0.05267 | -0.00108 | 0.16183 | 0.62759 | 10672.17 |
| GOBP_NEG | 13 | -0.04396 | -0.00115 | 0.13485 | 0.62779 | 10675.57 |
| GOBP_L_A | 14 | -0.04837 | -0.00132 | 0.14836 | 0.6278 | 10675.74 |
| MYLLYKAN | 9 | -0.05104 | -0.00111 | 0.15648 | 0.62786 | 10676.76 |
| GOBP_ACT | 10 | -0.0484 | -0.00111 | 0.14813 | 0.62806 | 10680.16 |
| GOCC_POS | 7 | -0.06464 | -0.00124 | 0.19774 | 0.62813 | 10681.35 |
| GOBP_POS | 5 | -0.07973 | -0.0013 | 0.24384 | 0.62816 | 10681.86 |
| GOBP_XYLU | 5 | -0.05896 | -0.00096 | 0.18027 | 0.62819 | 10682.37 |
| REACTOME | 5 | -0.05896 | -0.00096 | 0.18027 | 0.62819 | 10682.37 |
| GOMF_FOU | 6 | -0.07261 | -0.00129 | 0.22197 | 0.62821 | 10682.71 |
| GOBP_MU | 92 | -0.01712 | -0.00119 | 0.052303 | 0.62828 | 10683.9 |
| LEE_LIVER_ | 59 | -0.02168 | -0.00121 | 0.066207 | 0.62833 | 10684.75 |
| GOBP_REG | 43 | -0.02626 | -0.00125 | 0.080049 | 0.62857 | 10688.83 |
| GOBP_NEG | 14 | -0.05102 | -0.00139 | 0.15531 | 0.62874 | 10691.72 |
| AMIT_SERU | 55 | -0.02387 | -0.00129 | 0.072651 | 0.62876 | 10692.06 |
| GOBP_POS | 46 | -0.02429 | -0.0012 | 0.073931 | 0.62876 | 10692.06 |
| GOMF_GRO | 135 | -0.01504 | -0.00127 | 0.045749 | 0.62879 | 10692.57 |
| GOBP_EPIT | 123 | -0.01477 | -0.00119 | 0.044939 | 0.6288 | 10692.74 |
| WP_RENIN | 41 | -0.02659 | -0.00124 | 0.080771 | 0.62899 | 10695.97 |

| | | | | | | |
|---|---|---|---|---|---|---|
| SANSOM_A | 215 | -0.01088 | -0.00115 | 0.033026 | 0.62908 | 10697.51 |
| QI_PLASMA | 93 | -0.01762 | -0.00123 | 0.053497 | 0.62909 | 10697.68 |
| GOBP_RES | 314 | -0.00945 | -0.00121 | 0.028675 | 0.62913 | 10698.36 |
| GOBP_VES | 43 | -0.0253 | -0.00121 | 0.076735 | 0.62918 | 10699.21 |
| GOCC_SPEI | 30 | -0.02833 | -0.00113 | 0.085911 | 0.6292 | 10699.55 |
| GOMF_TRN | 31 | -0.02413 | -0.00098 | 0.073142 | 0.62927 | 10700.74 |
| GOMF_MC | 307 | -0.01008 | -0.00128 | 0.030527 | 0.62939 | 10702.78 |
| GOBP_ADE | 76 | -0.02079 | -0.00132 | 0.062921 | 0.62947 | 10704.14 |
| DAZARD_U | 19 | -0.03552 | -0.00113 | 0.10732 | 0.62968 | 10707.71 |
| GOBP_NEG | 24 | -0.03449 | -0.00123 | 0.10418 | 0.6297 | 10708.05 |
| DOANE_RE | 165 | -0.01283 | -0.00119 | 0.03874 | 0.62977 | 10709.24 |
| KYNG_NOF | 14 | -0.04547 | -0.00124 | 0.13724 | 0.6298 | 10709.75 |
| MOREAUX_ | 330 | -0.0088 | -0.00115 | 0.026575 | 0.6298 | 10709.75 |
| GOCC_TRA | 5 | -0.07322 | -0.00119 | 0.22088 | 0.62986 | 10710.77 |
| GOBP_REG | 17 | -0.04391 | -0.00132 | 0.13238 | 0.62993 | 10711.96 |
| GOBP_G0_ | 36 | -0.02464 | -0.00107 | 0.074273 | 0.62994 | 10712.13 |
| BYSTROEM | 56 | -0.02159 | -0.00117 | 0.065041 | 0.63005 | 10714 |
| WP_DISOR | 12 | -0.04304 | -0.00108 | 0.12959 | 0.63009 | 10714.68 |
| GENTILE_U | 34 | -0.02991 | -0.00127 | 0.090047 | 0.63011 | 10715.02 |
| PID_P38_G | 11 | -0.05557 | -0.00134 | 0.16731 | 0.63011 | 10715.02 |
| GOBP_NOF | 12 | -0.05314 | -0.00134 | 0.15991 | 0.63018 | 10716.21 |
| MORI_MAT | 82 | -0.01848 | -0.00122 | 0.055602 | 0.63018 | 10716.21 |
| GOBP_T_H | 28 | -0.03127 | -0.0012 | 0.094053 | 0.63022 | 10716.89 |
| REACTOME | 75 | -0.01979 | -0.00124 | 0.059487 | 0.63028 | 10717.91 |
| GOBP_LATI | 8 | -0.05586 | -0.00115 | 0.16771 | 0.63047 | 10721.14 |
| FARMER_B | 19 | -0.0431 | -0.00137 | 0.12924 | 0.63062 | 10723.69 |
| GOBP_CAT | 54 | -0.02338 | -0.00125 | 0.070006 | 0.6308 | 10726.75 |
| GOMF_OXI | 11 | -0.05144 | -0.00124 | 0.15403 | 0.6308 | 10726.75 |
| GOBP_ALP | 158 | -0.01397 | -0.00127 | 0.041791 | 0.63088 | 10728.11 |
| GOMF_TES | 6 | -0.06438 | -0.00115 | 0.19263 | 0.63088 | 10728.11 |
| GOBP_LYTI | 96 | -0.01714 | -0.00122 | 0.051184 | 0.63112 | 10732.2 |
| HOSHIDA_I | 103 | -0.01542 | -0.00114 | 0.046034 | 0.63118 | 10733.22 |
| GOBP_NEG | 6 | -0.07415 | -0.00132 | 0.22115 | 0.63129 | 10735.09 |
| GOBP_NEG | 21 | -0.03974 | -0.00132 | 0.11848 | 0.63132 | 10735.6 |
| REACTOME | 5 | -0.0671 | -0.00109 | 0.20004 | 0.63135 | 10736.11 |
| WP_IL1_AN | 22 | -0.03547 | -0.00121 | 0.10572 | 0.63139 | 10736.79 |
| GOBP_PRO | 112 | -0.01668 | -0.00128 | 0.049667 | 0.63149 | 10738.49 |
| WP_CANCE | 13 | -0.04935 | -0.00129 | 0.14694 | 0.63151 | 10738.83 |
| GOBP_ICOS | 65 | -0.02255 | -0.00132 | 0.067138 | 0.63152 | 10739 |
| GREGORY_ | 131 | -0.01555 | -0.00129 | 0.046293 | 0.63154 | 10739.34 |
| REACTOME | 12 | -0.04504 | -0.00114 | 0.13406 | 0.63154 | 10739.34 |
| GOCC_ORI | 9 | -0.04884 | -0.00107 | 0.14531 | 0.63161 | 10740.53 |
| GOBP_CYT | 17 | -0.03939 | -0.00118 | 0.11713 | 0.63168 | 10741.72 |
| GOMF_HIS | 50 | -0.02341 | -0.0012 | 0.06957 | 0.63174 | 10742.74 |
| GOBP_PRO | 6 | -0.08311 | -0.00148 | 0.24693 | 0.63178 | 10743.42 |

| | | | | | | |
|---|---|---|---|---|---|---|
| GOMF_ENZ | 331 | -0.00955 | -0.00125 | 0.028355 | 0.63184 | 10744.44 |
| GOCC_RNA | 6 | -0.07873 | -0.0014 | 0.23329 | 0.63211 | 10749.03 |
| GOBP_NEG | 7 | -0.06264 | -0.00121 | 0.18541 | 0.63225 | 10751.41 |
| GOMF_FAT | 21 | -0.03748 | -0.00125 | 0.11092 | 0.63228 | 10751.92 |
| LIEN_BREA | 108 | -0.01566 | -0.00118 | 0.046293 | 0.6324 | 10753.96 |
| GOBP_FILO | 64 | -0.02267 | -0.00132 | 0.06702 | 0.63242 | 10754.3 |
| GOBP_GAS | 18 | -0.04221 | -0.0013 | 0.12477 | 0.63243 | 10754.47 |
| GOMF_POI | 22 | -0.04137 | -0.00141 | 0.12226 | 0.63244 | 10754.64 |
| WEBER_MI | 10 | -0.05303 | -0.00122 | 0.15668 | 0.6325 | 10755.66 |
| GOBP_PRC | 5 | -0.07694 | -0.00125 | 0.22721 | 0.63255 | 10756.51 |
| GOBP_NEG | 3 | -0.09992 | -0.00126 | 0.29504 | 0.63257 | 10756.85 |
| GOMF_INC | 5 | -0.06906 | -0.00112 | 0.2034 | 0.63289 | 10762.29 |
| GOBP_POS | 10 | -0.05294 | -0.00122 | 0.15562 | 0.63315 | 10766.72 |
| BIOCARTA_ | 21 | -0.03454 | -0.00115 | 0.10151 | 0.63317 | 10767.06 |
| PID_INTEG | 11 | -0.05736 | -0.00138 | 0.16859 | 0.63317 | 10767.06 |
| GOMF_HIS | 8 | -0.05316 | -0.00109 | 0.15618 | 0.63322 | 10767.91 |
| GOBP_B_C | 85 | -0.02047 | -0.00137 | 0.060099 | 0.63327 | 10768.76 |
| GOMF_CAT | 21 | -0.03717 | -0.00124 | 0.10905 | 0.63337 | 10770.46 |
| GOCC_SIN3 | 20 | -0.03828 | -0.00125 | 0.11226 | 0.63344 | 10771.65 |
| GOBP_POS | 41 | -0.02566 | -0.00119 | 0.075212 | 0.63349 | 10772.5 |
| REACTOME | 27 | -0.03323 | -0.00126 | 0.097367 | 0.63356 | 10773.69 |
| GOBP_FEEI | 102 | -0.01797 | -0.00132 | 0.052583 | 0.63374 | 10776.75 |
| GOBP_REG | 5 | -0.06782 | -0.0011 | 0.19832 | 0.63381 | 10777.94 |
| GOMF_NEU | 18 | -0.04043 | -0.00125 | 0.11816 | 0.63388 | 10779.13 |
| PID_CXCR3 | 42 | -0.02619 | -0.00123 | 0.076352 | 0.63421 | 10784.74 |
| PYEON_CA | 18 | -0.04253 | -0.00131 | 0.12395 | 0.63426 | 10785.59 |
| GOBP_PYR | 5 | -0.06405 | -0.00104 | 0.18649 | 0.63436 | 10787.29 |
| GOBP_FEM | 17 | -0.04431 | -0.00133 | 0.12897 | 0.63442 | 10788.31 |
| GOMF_CAI | 10 | -0.04875 | -0.00112 | 0.14183 | 0.63448 | 10789.33 |
| GOBP_RIBO | 15 | -0.04066 | -0.00115 | 0.11822 | 0.63456 | 10790.69 |
| BANDRES_ | 18 | -0.03702 | -0.00114 | 0.10762 | 0.63458 | 10791.03 |
| GOBP_REG | 11 | -0.04905 | -0.00118 | 0.14255 | 0.6346 | 10791.37 |
| GOBP_CAD | 5 | -0.07805 | -0.00127 | 0.22672 | 0.63466 | 10792.39 |
| GOBP_XEN | 118 | -0.01721 | -0.00136 | 0.049974 | 0.63471 | 10793.24 |
| GOBP_TYP | 4 | -0.08087 | -0.00118 | 0.23436 | 0.63498 | 10797.83 |
| WINZEN_D | 97 | -0.01973 | -0.00141 | 0.057173 | 0.63499 | 10798 |
| GOBP_INO | 124 | -0.01675 | -0.00135 | 0.04852 | 0.635 | 10798.18 |
| GOBP_SNR | 25 | -0.03211 | -0.00117 | 0.09303 | 0.635 | 10798.18 |
| GOBP_POS | 5 | -0.09197 | -0.0015 | 0.26644 | 0.63502 | 10798.52 |
| MOTAMED | 6 | -0.06159 | -0.0011 | 0.1779 | 0.63541 | 10805.15 |
| WP_HOSTF | 18 | -0.04219 | -0.0013 | 0.12182 | 0.63544 | 10805.66 |
| GOBP_CELI | 579 | -0.00754 | -0.0013 | 0.021753 | 0.63556 | 10807.7 |
| GOMF_PHO | 9 | -0.05215 | -0.00114 | 0.1503 | 0.63569 | 10809.91 |
| GOCC_ZYM | 16 | -0.03884 | -0.00113 | 0.1119 | 0.63575 | 10810.93 |
| FISCHER_D | 302 | -0.00976 | -0.00122 | 0.028111 | 0.63581 | 10811.95 |

| | | | | | | |
|---|---|---|---|---|---|---|
| REACTOME | 115 | -0.0174 | -0.00135 | 0.050075 | 0.63587 | 10812.97 |
| WP_FATTY_ | 32 | -0.02909 | -0.0012 | 0.083649 | 0.636 | 10815.18 |
| GOBP_MYE | 16 | -0.04163 | -0.00121 | 0.11968 | 0.63602 | 10815.52 |
| WP_MIRNA | 30 | -0.0307 | -0.00122 | 0.088228 | 0.63605 | 10816.03 |
| GOMF_CAI | 319 | -0.00986 | -0.00127 | 0.028337 | 0.63607 | 10816.37 |
| LANDIS_ER | 142 | -0.01525 | -0.00132 | 0.043807 | 0.6361 | 10816.88 |
| BRUINS_UV | 834 | -0.00628 | -0.00129 | 0.01804 | 0.63622 | 10818.92 |
| GOBP_REG | 7 | -0.07086 | -0.00136 | 0.20338 | 0.63623 | 10819.09 |
| GOBP_NEG | 10 | -0.05793 | -0.00133 | 0.16627 | 0.63624 | 10819.26 |
| SUZUKI_CT | 12 | -0.05088 | -0.00128 | 0.14589 | 0.63636 | 10821.3 |
| GOMF_ACY | 19 | -0.03743 | -0.00119 | 0.10728 | 0.6364 | 10821.98 |
| GOBP_REG | 22 | -0.03586 | -0.00122 | 0.10273 | 0.63647 | 10823.17 |
| GOCC_SER | 17 | -0.04757 | -0.00143 | 0.13624 | 0.6365 | 10823.68 |
| GOBP_NEG | 16 | -0.04055 | -0.00118 | 0.11593 | 0.63675 | 10827.93 |
| LEE_LIVER_ | 53 | -0.02445 | -0.00129 | 0.069797 | 0.63692 | 10830.82 |
| GOMF_CHO | 10 | -0.06525 | -0.0015 | 0.18613 | 0.63703 | 10832.7 |
| WILLIAMS_ | 11 | -0.06239 | -0.00151 | 0.17792 | 0.63708 | 10833.55 |
| GOBP_PHO | 42 | -0.0263 | -0.00124 | 0.074937 | 0.63719 | 10835.42 |
| GOMF_TRA | 6 | -0.05872 | -0.00105 | 0.16728 | 0.63721 | 10835.76 |
| WP_COVID | 5 | -0.08123 | -0.00132 | 0.23142 | 0.63722 | 10835.93 |
| GOCC_PHO | 8 | -0.06818 | -0.0014 | 0.19416 | 0.63725 | 10836.44 |
| MIKKELSEN | 56 | -0.02377 | -0.00129 | 0.067685 | 0.63727 | 10836.78 |
| GOBP_SRP_ | 8 | -0.05655 | -0.00116 | 0.16085 | 0.63742 | 10839.33 |
| REACTOME | 9 | -0.06129 | -0.00134 | 0.1742 | 0.63751 | 10840.86 |
| REACTOME | 57 | -0.02101 | -0.00115 | 0.059684 | 0.63755 | 10841.54 |
| WP_TRYPT | 31 | -0.03239 | -0.00131 | 0.091967 | 0.63766 | 10843.41 |
| PID_IFNG_ | 40 | -0.02744 | -0.00126 | 0.077737 | 0.63794 | 10848.17 |
| GOBP_SEN | 28 | -0.03245 | -0.00125 | 0.091904 | 0.63797 | 10848.68 |
| NIKOLSKY_ | 21 | -0.04154 | -0.00138 | 0.11759 | 0.63806 | 10850.21 |
| GOBP_POS | 3 | -0.12093 | -0.00152 | 0.34216 | 0.63812 | 10851.23 |
| GOMF_AN | 3 | -0.1243 | -0.00157 | 0.3516 | 0.63816 | 10851.91 |
| GOBP_CELI | 11 | -0.06565 | -0.00158 | 0.18568 | 0.63817 | 10852.08 |
| GOBP_RRN | 8 | -0.06051 | -0.00125 | 0.17111 | 0.63819 | 10852.42 |
| REACTOME | 28 | -0.03238 | -0.00125 | 0.09153 | 0.63822 | 10852.93 |
| HOLLERN_I | 17 | -0.04865 | -0.00146 | 0.13754 | 0.63823 | 10853.1 |
| GOCC_SPE | 9 | -0.05832 | -0.00127 | 0.16476 | 0.63833 | 10854.8 |
| GOBP_INTI | 5 | -0.08966 | -0.00146 | 0.25323 | 0.63835 | 10855.14 |
| XU_RESPO | 14 | -0.04714 | -0.00128 | 0.13313 | 0.63837 | 10855.48 |
| REACTOME | 80 | -0.02085 | -0.00135 | 0.058857 | 0.63843 | 10856.5 |
| GOBP_MO | 970 | -0.00593 | -0.00131 | 0.016732 | 0.63845 | 10856.84 |
| GOBP_IMN | 22 | -0.0398 | -0.00136 | 0.11221 | 0.63859 | 10859.22 |
| GOBP_END | 6 | -0.08846 | -0.00158 | 0.24932 | 0.63863 | 10859.9 |
| GOBP_MAS | 7 | -0.0696 | -0.00134 | 0.19605 | 0.63871 | 10861.26 |
| GOMF_AM | 39 | -0.03119 | -0.00142 | 0.087834 | 0.63876 | 10862.11 |
| GOBP_PUR | 5 | -0.09381 | -0.00153 | 0.26405 | 0.63881 | 10862.96 |

| | | | | | | |
|---|---|---|---|---|---|---|
| REACTOME | 14 | -0.04884 | -0.00133 | 0.13737 | 0.63889 | 10864.32 |
| GOCC_ESC | 11 | -0.0438 | -0.00106 | 0.12307 | 0.63904 | 10866.88 |
| GOBP_HYP | 6 | -0.0902 | -0.00161 | 0.25326 | 0.63913 | 10868.41 |
| GOBP_AMI | 6 | -0.0954 | -0.0017 | 0.26767 | 0.63923 | 10870.11 |
| GOMF_AM | 6 | -0.0954 | -0.0017 | 0.26767 | 0.63923 | 10870.11 |
| GOBP_PRC | 15 | -0.04933 | -0.00139 | 0.13835 | 0.63929 | 10871.13 |
| GOMF_UR/ | 6 | -0.05922 | -0.00106 | 0.16606 | 0.6393 | 10871.3 |
| GOBP_ADE | 5 | -0.08524 | -0.00139 | 0.23881 | 0.63942 | 10873.34 |
| RAMASWA | 60 | -0.02377 | -0.00134 | 0.066579 | 0.63947 | 10874.19 |
| GOBP_POS | 6 | -0.07983 | -0.00142 | 0.22349 | 0.63953 | 10875.21 |
| GOBP_NEG | 26 | -0.03508 | -0.0013 | 0.098171 | 0.63959 | 10876.23 |
| KEGG_PUR | 151 | -0.01396 | -0.00124 | 0.039055 | 0.63962 | 10876.74 |
| GOBP_URE | 5 | -0.09064 | -0.00147 | 0.25339 | 0.63971 | 10878.27 |
| GOBP_RES | 203 | -0.01303 | -0.00134 | 0.036408 | 0.63981 | 10879.97 |
| GOBP_LEU | 8 | -0.06046 | -0.00124 | 0.16868 | 0.63997 | 10882.69 |
| GOBP_POS | 5 | -0.05649 | -0.00092 | 0.1576 | 0.63999 | 10883.03 |
| GOMF_OXI | 11 | -0.06053 | -0.00146 | 0.16888 | 0.63999 | 10883.03 |
| PARK_HSC_ | 17 | -0.04275 | -0.00128 | 0.11921 | 0.64006 | 10884.22 |
| GOBP_EPIL | 361 | -0.01038 | -0.00142 | 0.028929 | 0.64016 | 10885.92 |
| TOMLINS_I | 14 | -0.04352 | -0.00118 | 0.12127 | 0.64016 | 10885.92 |
| ROETH_TEI | 9 | -0.0498 | -0.00109 | 0.13872 | 0.64021 | 10886.77 |
| REACTOME | 40 | -0.03011 | -0.00138 | 0.083822 | 0.64029 | 10888.13 |
| WP_EFFECT | 25 | -0.0335 | -0.00122 | 0.093119 | 0.64047 | 10891.19 |
| WP_FATTY_ | 17 | -0.04117 | -0.00123 | 0.11437 | 0.64055 | 10892.55 |
| GOCC_PHC | 68 | -0.02154 | -0.00129 | 0.059817 | 0.64063 | 10893.91 |
| DANG_REG | 64 | -0.02251 | -0.00131 | 0.062486 | 0.64065 | 10894.25 |
| GOMF_NEU | 29 | -0.03163 | -0.00124 | 0.087801 | 0.64066 | 10894.42 |
| MATZUK_C | 8 | -0.07508 | -0.00155 | 0.20837 | 0.6407 | 10895.1 |
| GOBP_POS | 125 | -0.01627 | -0.00132 | 0.045113 | 0.64079 | 10896.63 |
| GOBP_COF | 8 | -0.08297 | -0.00171 | 0.22986 | 0.64093 | 10899.01 |
| KEGG_OXII | 111 | -0.01541 | -0.00118 | 0.042705 | 0.64093 | 10899.01 |
| MOHANKU | 381 | -0.00902 | -0.00127 | 0.024988 | 0.64095 | 10899.35 |
| GOBP_CD4 | 82 | -0.02079 | -0.00137 | 0.057559 | 0.64102 | 10900.55 |
| GOBP_CELI | 7 | -0.06899 | -0.00133 | 0.19088 | 0.64112 | 10902.25 |
| GOBP_REG | 8 | -0.07793 | -0.0016 | 0.2155 | 0.64118 | 10903.27 |
| WP_SARS_ | 29 | -0.04591 | -0.0018 | 0.12693 | 0.64119 | 10903.44 |
| YAMANAK/ | 7 | -0.07245 | -0.00139 | 0.20031 | 0.64121 | 10903.78 |
| GOMF_HIS | 6 | -0.07363 | -0.00131 | 0.20343 | 0.6413 | 10905.31 |
| GOBP_T_C | 5 | -0.08295 | -0.00135 | 0.22908 | 0.64135 | 10906.16 |
| GOMF_PH( | 14 | -0.05172 | -0.00141 | 0.14283 | 0.64136 | 10906.33 |
| GOMF_POI | 5 | -0.07009 | -0.00114 | 0.19324 | 0.64158 | 10910.07 |
| GOBP_CHE | 6 | -0.07858 | -0.0014 | 0.21665 | 0.64159 | 10910.24 |
| BIOCARTA_ | 7 | -0.06495 | -0.00125 | 0.17898 | 0.64165 | 10911.26 |
| REACTOME | 35 | -0.03306 | -0.00142 | 0.091098 | 0.64167 | 10911.6 |
| GOBP_MIC | 609 | -0.00715 | -0.00126 | 0.019697 | 0.64168 | 10911.77 |

| | | | | | | |
|---|---|---|---|---|---|---|
| GOCC_BRU | 98 | -0.01883 | -0.00135 | 0.051849 | 0.64179 | 10913.64 |
| SERVITJA_I | 130 | -0.01576 | -0.0013 | 0.043383 | 0.64179 | 10913.64 |
| GOBP_POS | 4 | -0.11003 | -0.0016 | 0.30287 | 0.6418 | 10913.81 |
| GOBP_TETI | 14 | -0.04211 | -0.00115 | 0.11585 | 0.64187 | 10915 |
| GOMF_FAT | 7 | -0.08911 | -0.00172 | 0.24516 | 0.64188 | 10915.17 |
| GOBP_PLA | 21 | -0.03593 | -0.0012 | 0.098833 | 0.6419 | 10915.51 |
| GOMF_N_I | 5 | -0.07563 | -0.00123 | 0.20799 | 0.64192 | 10915.85 |
| WP_CATAL | 5 | -0.07563 | -0.00123 | 0.20799 | 0.64192 | 10915.85 |
| GOBP_REG | 6 | -0.08058 | -0.00144 | 0.22142 | 0.64203 | 10917.72 |
| GOBP_MO | 33 | -0.03478 | -0.00145 | 0.095545 | 0.64207 | 10918.4 |
| REACTOME | 47 | -0.02625 | -0.00131 | 0.072109 | 0.64209 | 10918.74 |
| GOBP_MAI | 21 | -0.04397 | -0.00147 | 0.12074 | 0.64214 | 10919.59 |
| GOCC_LIPC | 4 | -0.0876 | -0.00128 | 0.24047 | 0.64218 | 10920.27 |
| GOBP_IROI | 9 | -0.05781 | -0.00126 | 0.15865 | 0.64221 | 10920.78 |
| GOBP_RES | 94 | -0.01962 | -0.00138 | 0.053836 | 0.64225 | 10921.46 |
| GOBP_G_P | 11 | -0.05617 | -0.00136 | 0.15405 | 0.6423 | 10922.31 |
| GOMF_CAI | 5 | -0.09942 | -0.00162 | 0.27254 | 0.64236 | 10923.33 |
| REACTOME | 14 | -0.0494 | -0.00134 | 0.13533 | 0.64245 | 10924.86 |
| GOBP_LYM | 30 | -0.03598 | -0.00143 | 0.098507 | 0.64252 | 10926.05 |
| REACTOME | 53 | -0.02534 | -0.00134 | 0.069368 | 0.64255 | 10926.56 |
| GOMF_CYS | 6 | -0.07228 | -0.00129 | 0.19786 | 0.64256 | 10926.73 |
| GOBP_RES | 211 | -0.01244 | -0.00131 | 0.034019 | 0.64274 | 10929.79 |
| REACTOME | 22 | -0.0386 | -0.00132 | 0.1055 | 0.64278 | 10930.47 |
| BIOCARTA_ | 17 | -0.05032 | -0.00151 | 0.13737 | 0.64294 | 10933.19 |
| GOBP_REN | 22 | -0.04226 | -0.00144 | 0.11517 | 0.64316 | 10936.94 |
| GOBP_LOV | 27 | -0.03711 | -0.0014 | 0.10112 | 0.64317 | 10937.11 |
| YAGI_AML_ | 33 | -0.02788 | -0.00116 | 0.075912 | 0.64329 | 10939.15 |
| GOBP_NEG | 281 | -0.01176 | -0.00142 | 0.03201 | 0.64336 | 10940.34 |
| WP_DISOR | 5 | -0.07639 | -0.00124 | 0.2079 | 0.64336 | 10940.34 |
| GOBP_NITI | 5 | -0.08613 | -0.0014 | 0.23438 | 0.64337 | 10940.51 |
| GOCC_NUC | 9 | -0.05958 | -0.0013 | 0.16211 | 0.64338 | 10940.68 |
| GOMF_L_A | 19 | -0.04118 | -0.00131 | 0.11201 | 0.64342 | 10941.36 |
| KINNEY_DI | 14 | -0.05665 | -0.00154 | 0.15407 | 0.64345 | 10941.87 |
| ZHAN_MUI | 42 | -0.0327 | -0.00154 | 0.088876 | 0.64354 | 10943.4 |
| GOBP_SUL | 142 | -0.01565 | -0.00135 | 0.042503 | 0.64365 | 10945.27 |
| WP_OSX_A | 14 | -0.05759 | -0.00157 | 0.15639 | 0.64365 | 10945.27 |
| REACTOME | 8 | -0.07703 | -0.00159 | 0.20916 | 0.64366 | 10945.44 |
| GOBP_LAN | 82 | -0.02075 | -0.00136 | 0.056329 | 0.64367 | 10945.61 |
| LIU_COMM | 60 | -0.02246 | -0.00126 | 0.060975 | 0.64369 | 10945.95 |
| GOBP_REG | 6 | -0.07293 | -0.0013 | 0.19795 | 0.64372 | 10946.46 |
| GOBP_DITI | 11 | -0.05417 | -0.00131 | 0.14695 | 0.6438 | 10947.82 |
| GOBP_LYM | 715 | -0.00727 | -0.00139 | 0.019712 | 0.64387 | 10949.01 |
| GAURNIER_ | 45 | -0.03115 | -0.00152 | 0.084341 | 0.64404 | 10951.9 |
| GOBP_REG | 27 | -0.03019 | -0.00114 | 0.081693 | 0.64415 | 10953.77 |
| GOBP_RES | 61 | -0.02204 | -0.00125 | 0.059636 | 0.64415 | 10953.77 |

| | | | | | | |
|---|---|---|---|---|---|---|
| WP_GLUTA | 18 | -0.04408 | -0.00136 | 0.11925 | 0.64417 | 10954.11 |
| GOBP_MA1 | 30 | -0.03807 | -0.00152 | 0.10299 | 0.64418 | 10954.28 |
| GOMF_AM | 81 | -0.02102 | -0.00137 | 0.056852 | 0.64422 | 10954.96 |
| GOBP_SIGI | 8 | -0.07204 | -0.00148 | 0.19478 | 0.64425 | 10955.47 |
| GOMF_R_S | 24 | -0.03776 | -0.00135 | 0.10206 | 0.64431 | 10956.49 |
| GOBP_REG | 8 | -0.05892 | -0.00121 | 0.15908 | 0.64444 | 10958.7 |
| GOCC_EXT | 11 | -0.05732 | -0.00138 | 0.15476 | 0.64444 | 10958.7 |
| GOMF_BAS | 17 | -0.0513 | -0.00154 | 0.13851 | 0.64445 | 10958.87 |
| GOMF_DN. | 9 | -0.05608 | -0.00122 | 0.1514 | 0.64446 | 10959.04 |
| GOMF_BAS | 59 | -0.02378 | -0.00133 | 0.064181 | 0.64452 | 10960.06 |
| GOBP_UV_ | 12 | -0.04709 | -0.00119 | 0.12703 | 0.64456 | 10960.74 |
| GOBP_EPIT | 16 | -0.05044 | -0.00147 | 0.13606 | 0.64458 | 10961.08 |
| GOBP_RES | 5 | -0.07806 | -0.00127 | 0.21027 | 0.64477 | 10964.31 |
| GOCC_PRC | 44 | -0.0246 | -0.00119 | 0.06623 | 0.64484 | 10965.5 |
| GOMF_MU | 7 | -0.05562 | -0.00107 | 0.14966 | 0.64492 | 10966.86 |
| GOBP_NEU | 65 | -0.02327 | -0.00136 | 0.062614 | 0.64494 | 10967.2 |
| ZHAN_MUI | 42 | -0.02879 | -0.00136 | 0.077425 | 0.64498 | 10967.88 |
| GOBP_ETH | 8 | -0.08961 | -0.00184 | 0.24099 | 0.64499 | 10968.05 |
| KORKOLA_ | 7 | -0.08058 | -0.00155 | 0.21671 | 0.64499 | 10968.05 |
| GOBP_POS | 51 | -0.02628 | -0.00136 | 0.070657 | 0.64503 | 10968.74 |
| WP_BIOM/ | 15 | -0.04737 | -0.00133 | 0.12731 | 0.64508 | 10969.59 |
| GOBP_NEG | 5 | -0.07872 | -0.00128 | 0.21152 | 0.6451 | 10969.93 |
| KEGG_SULI | 11 | -0.05621 | -0.00136 | 0.15104 | 0.6451 | 10969.93 |
| GOBP_NEG | 15 | -0.04696 | -0.00132 | 0.12614 | 0.64516 | 10970.95 |
| GOMF_LIP( | 35 | -0.0295 | -0.00127 | 0.079033 | 0.64553 | 10977.24 |
| GOBP_POS | 398 | -0.00983 | -0.00141 | 0.026309 | 0.64559 | 10978.26 |
| GOBP_NEG | 27 | -0.0324 | -0.00122 | 0.086711 | 0.64565 | 10979.28 |
| YAMASHIT/ | 7 | -0.07272 | -0.0014 | 0.19431 | 0.64589 | 10983.36 |
| GOBP_EPIC | 234 | -0.014 | -0.00155 | 0.037389 | 0.64597 | 10984.72 |
| GOBP_REG | 9 | -0.05302 | -0.00116 | 0.14147 | 0.64607 | 10986.42 |
| REACTOME | 5 | -0.10408 | -0.00169 | 0.27759 | 0.64615 | 10987.78 |
| REACTOME | 26 | -0.04417 | -0.00164 | 0.11757 | 0.64643 | 10992.54 |
| GOBP_PHC | 15 | -0.04423 | -0.00125 | 0.1177 | 0.64646 | 10993.05 |
| GOMF_ALF | 6 | -0.06291 | -0.00112 | 0.16737 | 0.6465 | 10993.73 |
| GOBP_SIRI | 8 | -0.06716 | -0.00138 | 0.17865 | 0.64651 | 10993.9 |
| BIOCARTA_ | 15 | -0.04927 | -0.00139 | 0.13101 | 0.64658 | 10995.09 |
| GOBP_REG | 2 | -0.13007 | -0.00134 | 0.34572 | 0.64663 | 10995.94 |
| WP_COMP | 90 | -0.02047 | -0.00141 | 0.054376 | 0.64667 | 10996.62 |
| HOFFMANI | 72 | -0.02196 | -0.00135 | 0.058271 | 0.64685 | 10999.68 |
| LIU_CMYB_ | 153 | -0.01564 | -0.0014 | 0.04149 | 0.64693 | 11001.04 |
| REACTOME | 7 | -0.06275 | -0.00121 | 0.16632 | 0.64701 | 11002.41 |
| GOBP_REG | 11 | -0.05209 | -0.00126 | 0.13804 | 0.64703 | 11002.75 |
| GOBP_MO | 137 | -0.01686 | -0.00143 | 0.044676 | 0.64705 | 11003.09 |
| GOMF_GLY | 12 | -0.06237 | -0.00157 | 0.16527 | 0.64705 | 11003.09 |
| GOBP_NEU | 158 | -0.01505 | -0.00137 | 0.039877 | 0.64709 | 11003.77 |

| | | | | | | |
|---|---|---|---|---|---|---|
| GOBP_NEG | 7 | -0.0841 | -0.00162 | 0.22262 | 0.64721 | 11005.81 |
| KEGG_HUN | 164 | -0.01331 | -0.00124 | 0.035224 | 0.64726 | 11006.66 |
| REACTOME | 49 | -0.02787 | -0.00142 | 0.073714 | 0.64733 | 11007.85 |
| GOMF_PH( | 47 | -0.02879 | -0.00143 | 0.076075 | 0.64744 | 11009.72 |
| GOBP_POS | 71 | -0.02266 | -0.00139 | 0.05986 | 0.64746 | 11010.06 |
| WP_STERO | 10 | -0.06661 | -0.00153 | 0.17586 | 0.64756 | 11011.76 |
| MCBRYAN_ | 159 | -0.01513 | -0.00138 | 0.039925 | 0.64764 | 11013.12 |
| VALK_AML | 29 | -0.03741 | -0.00147 | 0.098705 | 0.64767 | 11013.63 |
| WENG_PO | 42 | -0.02986 | -0.00141 | 0.078761 | 0.64769 | 11013.97 |
| GOBP_CER | 34 | -0.03102 | -0.00132 | 0.081801 | 0.64774 | 11014.82 |
| GOBP_TRI( | 13 | -0.05239 | -0.00137 | 0.13813 | 0.64775 | 11014.99 |
| GOBP_REG | 80 | -0.02154 | -0.0014 | 0.056795 | 0.64776 | 11015.16 |
| GOBP_REG | 13 | -0.04519 | -0.00119 | 0.11913 | 0.64778 | 11015.5 |
| REACTOME | 35 | -0.03145 | -0.00135 | 0.082898 | 0.64778 | 11015.5 |
| TERAMOT( | 13 | -0.04833 | -0.00127 | 0.12735 | 0.64783 | 11016.35 |
| GOBP_AG( | 8 | -0.08012 | -0.00165 | 0.21099 | 0.64791 | 11017.71 |
| GOCC_CO/ | 278 | -0.01099 | -0.00132 | 0.028949 | 0.64792 | 11017.88 |
| GOBP_GLY | 20 | -0.03739 | -0.00122 | 0.098444 | 0.64795 | 11018.39 |
| GOBP_DEC | 23 | -0.04061 | -0.00142 | 0.10681 | 0.6481 | 11020.94 |
| GOMF_ATI | 26 | -0.04178 | -0.00155 | 0.10988 | 0.6481 | 11020.94 |
| GOBP_POS | 24 | -0.04589 | -0.00164 | 0.12052 | 0.64832 | 11024.68 |
| REACTOME | 67 | -0.02289 | -0.00136 | 0.060088 | 0.64835 | 11025.19 |
| GOBP_NEG | 646 | -0.00751 | -0.00136 | 0.019702 | 0.64838 | 11025.7 |
| PID_IL1_P/ | 32 | -0.03346 | -0.00138 | 0.087805 | 0.6484 | 11026.04 |
| REACTOME | 8 | -0.06181 | -0.00127 | 0.16222 | 0.6484 | 11026.04 |
| GOBP_NEG | 48 | -0.02743 | -0.00138 | 0.071988 | 0.64842 | 11026.38 |
| GOMF_TR/ | 77 | -0.01935 | -0.00123 | 0.050716 | 0.6486 | 11029.44 |
| GOMF_NEI | 11 | -0.06233 | -0.0015 | 0.16327 | 0.64867 | 11030.63 |
| KEGG_NAT | 119 | -0.01903 | -0.00151 | 0.049846 | 0.6487 | 11031.14 |
| LIANG_SILI | 45 | -0.02855 | -0.00139 | 0.074761 | 0.6487 | 11031.14 |
| HU_GENOT | 45 | -0.0272 | -0.00133 | 0.071203 | 0.64875 | 11031.99 |
| GOCC_CLA | 117 | -0.01742 | -0.00137 | 0.045581 | 0.64884 | 11033.52 |
| GOBP_OR( | 57 | -0.02776 | -0.00152 | 0.072622 | 0.64885 | 11033.69 |
| GOBP_KET | 14 | -0.05098 | -0.00139 | 0.13328 | 0.64894 | 11035.22 |
| BIOCARTA_ | 7 | -0.09271 | -0.00178 | 0.24213 | 0.6491 | 11037.95 |
| IIZUKA_LIV | 10 | -0.0562 | -0.00129 | 0.14673 | 0.64913 | 11038.46 |
| LU_TUMOI | 8 | -0.07389 | -0.00152 | 0.19272 | 0.64929 | 11041.18 |
| GOBP_SEQ | 8 | -0.05388 | -0.00111 | 0.14045 | 0.64936 | 11042.37 |
| GOBP_REG | 20 | -0.04505 | -0.00147 | 0.11729 | 0.64952 | 11045.09 |
| XU_HGF_T. | 18 | -0.0473 | -0.00146 | 0.12313 | 0.64957 | 11045.94 |
| GOBP_PRC | 33 | -0.03106 | -0.0013 | 0.080848 | 0.64958 | 11046.11 |
| DORN_ADE | 13 | -0.04671 | -0.00123 | 0.1215 | 0.64965 | 11047.3 |
| GOBP_REN | 12 | -0.0588 | -0.00148 | 0.15294 | 0.64968 | 11047.81 |
| GOBP_NEG | 22 | -0.04167 | -0.00142 | 0.10838 | 0.64969 | 11047.98 |
| LIN_MELAI | 70 | -0.0262 | -0.00159 | 0.068137 | 0.64972 | 11048.49 |

| | | | | | | |
|---|---|---|---|---|---|---|
| GOBP_PHC | 5 | -0.07876 | -0.00128 | 0.2048 | 0.64973 | 11048.66 |
| WP_NICOT | 6 | -0.08634 | -0.00154 | 0.22444 | 0.64976 | 11049.17 |
| GOCC_TRN | 8 | -0.0624 | -0.00128 | 0.16209 | 0.64987 | 11051.04 |
| GOBP_NUC | 11 | -0.05295 | -0.00128 | 0.13749 | 0.64991 | 11051.72 |
| GOBP_MAI | 60 | -0.02381 | -0.00134 | 0.06178 | 0.64999 | 11053.08 |
| BIOCARTA_ | 7 | -0.077 | -0.00148 | 0.19983 | 0.65001 | 11053.42 |
| REACTOME | 31 | -0.0347 | -0.00141 | 0.090013 | 0.65006 | 11054.27 |
| GOBP_SYN | 22 | -0.04593 | -0.00157 | 0.1191 | 0.65011 | 11055.12 |
| GOMF_TYF | 6 | -0.10213 | -0.00182 | 0.26454 | 0.65027 | 11057.84 |
| GOBP_POL | 109 | -0.01795 | -0.00136 | 0.046457 | 0.65035 | 11059.2 |
| REACTOME | 34 | -0.03683 | -0.00156 | 0.095309 | 0.65039 | 11059.88 |
| GOMF_PH( | 9 | -0.06183 | -0.00135 | 0.15991 | 0.65049 | 11061.58 |
| BIOCARTA_ | 23 | -0.04441 | -0.00155 | 0.11485 | 0.6505 | 11061.75 |
| BIOCARTA_ | 31 | -0.03413 | -0.00138 | 0.088228 | 0.65055 | 11062.6 |
| GOMF_NEI | 4 | -0.1237 | -0.0018 | 0.31977 | 0.65056 | 11062.77 |
| GOBP_CELI | 456 | -0.00913 | -0.0014 | 0.023603 | 0.65061 | 11063.62 |
| BIOCARTA_ | 18 | -0.04452 | -0.00137 | 0.11504 | 0.65063 | 11063.96 |
| GOBP_DER | 11 | -0.04938 | -0.00119 | 0.12753 | 0.65069 | 11064.98 |
| GOBP_7_M | 8 | -0.06334 | -0.0013 | 0.16354 | 0.65073 | 11065.66 |
| REACTOME | 20 | -0.04061 | -0.00132 | 0.10482 | 0.65077 | 11066.34 |
| GOBP_MO | 18 | -0.04237 | -0.00131 | 0.10927 | 0.65089 | 11068.38 |
| GOBP_POS | 89 | -0.02126 | -0.00146 | 0.054754 | 0.65112 | 11072.3 |
| GOBP_REV | 4 | -0.10451 | -0.00152 | 0.26912 | 0.65112 | 11072.3 |
| GAVIN_FO | 96 | -0.02153 | -0.00153 | 0.05535 | 0.65133 | 11075.87 |
| HOLLERN_ | 116 | -0.01732 | -0.00135 | 0.044504 | 0.65138 | 11076.72 |
| GOBP_POS | 4 | -0.09875 | -0.00144 | 0.2537 | 0.65144 | 11077.74 |
| CHIARETTI_ | 27 | -0.03665 | -0.00138 | 0.094136 | 0.65147 | 11078.25 |
| WP_PPAR_ | 65 | -0.02345 | -0.00137 | 0.060179 | 0.65162 | 11080.8 |
| GOCC_RNA | 12 | -0.05328 | -0.00134 | 0.13671 | 0.65163 | 11080.97 |
| BIOCARTA_ | 20 | -0.04453 | -0.00145 | 0.11421 | 0.65167 | 11081.65 |
| GOBP_PEP | 241 | -0.01255 | -0.00141 | 0.032183 | 0.65167 | 11081.65 |
| CERVERA_S | 113 | -0.01976 | -0.00152 | 0.050684 | 0.65168 | 11081.82 |
| GOBP_NUC | 9 | -0.07012 | -0.00153 | 0.17986 | 0.65168 | 11081.82 |
| GOCC_RNA | 5 | -0.10929 | -0.00178 | 0.28014 | 0.65177 | 11083.35 |
| REACTOME | 10 | -0.0683 | -0.00157 | 0.17501 | 0.65183 | 11084.37 |
| GOCC_SM( | 7 | -0.08218 | -0.00158 | 0.21046 | 0.6519 | 11085.56 |
| GOBP_LATI | 12 | -0.0631 | -0.00159 | 0.16153 | 0.65197 | 11086.75 |
| REACTOME | 71 | -0.02317 | -0.00142 | 0.059303 | 0.65202 | 11087.6 |
| GOBP_NEG | 7 | -0.07137 | -0.00137 | 0.18256 | 0.65209 | 11088.79 |
| GOBP_DNA | 7 | -0.05298 | -0.00102 | 0.13529 | 0.65233 | 11092.87 |
| GOBP_REG | 11 | -0.05912 | -0.00143 | 0.15097 | 0.65233 | 11092.87 |
| LINDGREN_ | 8 | -0.06125 | -0.00126 | 0.15614 | 0.65257 | 11096.95 |
| DAVICIONI_ | 53 | -0.0293 | -0.00155 | 0.074678 | 0.65262 | 11097.8 |
| GOMF_ANI | 19 | -0.04411 | -0.0014 | 0.1124 | 0.65264 | 11098.14 |
| GOMF_FLC | 15 | -0.05318 | -0.0015 | 0.1355 | 0.65265 | 11098.31 |

| | | | | | | |
|---|---|---|---|---|---|---|
| BIOCARTA_ | 17 | -0.04886 | -0.00147 | 0.12448 | 0.65267 | 11098.65 |
| WP_VITAN | 11 | -0.05184 | -0.00125 | 0.13203 | 0.65271 | 11099.33 |
| GOBP_ACT | 44 | -0.02971 | -0.00143 | 0.075565 | 0.65289 | 11102.39 |
| REACTOME | 43 | -0.02921 | -0.00139 | 0.074286 | 0.65289 | 11102.39 |
| MATTIOLI_ | 11 | -0.06508 | -0.00157 | 0.16552 | 0.6529 | 11102.56 |
| WP_ACE_II | 14 | -0.06401 | -0.00174 | 0.16274 | 0.65296 | 11103.58 |
| GOBP_TETI | 8 | -0.06877 | -0.00142 | 0.1748 | 0.653 | 11104.27 |
| GOBP_CAR | 69 | -0.02699 | -0.00163 | 0.068561 | 0.65306 | 11105.29 |
| GOBP_REG | 4 | -0.11803 | -0.00172 | 0.29983 | 0.65308 | 11105.63 |
| GOBP_POS | 12 | -0.06252 | -0.00158 | 0.15882 | 0.65309 | 11105.8 |
| REACTOME | 114 | -0.01808 | -0.0014 | 0.045925 | 0.65311 | 11106.14 |
| REACTOME | 16 | -0.05519 | -0.00161 | 0.14011 | 0.65317 | 11107.16 |
| GOCC_MIT | 5 | -0.07133 | -0.00116 | 0.18105 | 0.65321 | 11107.84 |
| GOBP_REG | 6 | -0.07512 | -0.00134 | 0.19019 | 0.65357 | 11113.96 |
| GOMF_COI | 4 | -0.10168 | -0.00148 | 0.25744 | 0.65357 | 11113.96 |
| GOBP_POS | 7 | -0.07782 | -0.0015 | 0.19698 | 0.6536 | 11114.47 |
| BASSO_B_I | 137 | -0.01707 | -0.00145 | 0.043191 | 0.65363 | 11114.98 |
| GOMF_PEI | 5 | -0.09093 | -0.00148 | 0.23009 | 0.65365 | 11115.32 |
| GOBP_FOL | 25 | -0.03674 | -0.00134 | 0.092945 | 0.65367 | 11115.66 |
| GOBP_PYR | 5 | -0.09168 | -0.00149 | 0.2318 | 0.65376 | 11117.19 |
| GOMF_URI | 5 | -0.09168 | -0.00149 | 0.2318 | 0.65376 | 11117.19 |
| GOBP_REN | 20 | -0.04862 | -0.00158 | 0.12287 | 0.65384 | 11118.55 |
| GOBP_PYR | 23 | -0.0387 | -0.00135 | 0.097645 | 0.65405 | 11122.12 |
| GOCC_MYI | 6 | -0.09089 | -0.00162 | 0.22925 | 0.65412 | 11123.31 |
| REACTOME | 8 | -0.0768 | -0.00158 | 0.19368 | 0.65413 | 11123.48 |
| GOBP_GLU | 6 | -0.08303 | -0.00148 | 0.20925 | 0.65423 | 11125.18 |
| GOBP_MIT | 23 | -0.04065 | -0.00142 | 0.10245 | 0.65424 | 11125.35 |
| GOBP_PRC | 18 | -0.04294 | -0.00133 | 0.1082 | 0.65427 | 11125.86 |
| BIOCARTA_ | 19 | -0.04319 | -0.00137 | 0.10877 | 0.65434 | 11127.05 |
| GOBP_TRN | 14 | -0.06557 | -0.00178 | 0.16512 | 0.65435 | 11127.22 |
| GOBP_NEG | 11 | -0.06857 | -0.00165 | 0.17248 | 0.65451 | 11129.94 |
| REACTOME | 24 | -0.04385 | -0.00156 | 0.11024 | 0.65459 | 11131.3 |
| GOBP_HAII | 15 | -0.05118 | -0.00144 | 0.12858 | 0.65469 | 11133 |
| GOBP_REG | 11 | -0.07229 | -0.00174 | 0.18163 | 0.65469 | 11133 |
| GOCC_END | 71 | -0.0229 | -0.0014 | 0.057529 | 0.65469 | 11133 |
| GOMF_L_C | 4 | -0.11637 | -0.00169 | 0.29225 | 0.65474 | 11133.85 |
| GOBP_CAR | 6 | -0.10382 | -0.00185 | 0.26072 | 0.65475 | 11134.02 |
| GOBP_NEG | 40 | -0.03148 | -0.00145 | 0.079053 | 0.65475 | 11134.02 |
| FOURNIER_ | 6 | -0.08566 | -0.00153 | 0.21497 | 0.65485 | 11135.72 |
| GOCC_TRA | 5 | -0.12922 | -0.0021 | 0.324 | 0.65498 | 11137.93 |
| WP_MAPK_ | 10 | -0.06299 | -0.00145 | 0.15794 | 0.65498 | 11137.93 |
| MCBRYAN_ | 12 | -0.05861 | -0.00148 | 0.14692 | 0.65501 | 11138.45 |
| GOBP_AMY | 6 | -0.09139 | -0.00163 | 0.22908 | 0.65502 | 11138.62 |
| KIM_WT1_ | 115 | -0.01763 | -0.00137 | 0.044201 | 0.65502 | 11138.62 |
| GOCC_END | 85 | -0.02199 | -0.00147 | 0.05512 | 0.65504 | 11138.96 |

| | | | | | | |
|---|---|---|---|---|---|---|
| GOBP_LYM | 198 | -0.01565 | -0.00159 | 0.039202 | 0.65509 | 11139.81 |
| GOBP_POS | 929 | -0.00676 | -0.00146 | 0.016925 | 0.65513 | 11140.49 |
| GOBP_PHE | 46 | -0.03128 | -0.00154 | 0.078299 | 0.65524 | 11142.36 |
| GOBP_REG | 220 | -0.01407 | -0.00151 | 0.035171 | 0.65546 | 11146.1 |
| WP_AUTO: | 11 | -0.06159 | -0.00149 | 0.15391 | 0.65547 | 11146.27 |
| GOBP_CEN | 28 | -0.03402 | -0.00131 | 0.084969 | 0.65556 | 11147.8 |
| WP_HAIR_ | 87 | -0.02342 | -0.00159 | 0.058473 | 0.65563 | 11148.99 |
| GOBP_NEG | 5 | -0.09678 | -0.00157 | 0.24148 | 0.6557 | 11150.18 |
| GOBP_MAI | 7 | -0.09124 | -0.00176 | 0.22753 | 0.65578 | 11151.54 |
| HENDRICKS | 52 | -0.0305 | -0.0016 | 0.076053 | 0.65581 | 11152.05 |
| GOMF_CEF | 6 | -0.07196 | -0.00128 | 0.17926 | 0.65595 | 11154.43 |
| MOOTHA_ | 14 | -0.05452 | -0.00148 | 0.13567 | 0.65611 | 11157.15 |
| ZHENG_IL2 | 33 | -0.03355 | -0.0014 | 0.083414 | 0.65625 | 11159.53 |
| GOBP_HYP | 7 | -0.09398 | -0.00181 | 0.23359 | 0.65628 | 11160.04 |
| GOBP_POS | 9 | -0.07272 | -0.00159 | 0.18075 | 0.65629 | 11160.21 |
| GOBP_INTI | 46 | -0.02911 | -0.00144 | 0.0723 | 0.65637 | 11161.57 |
| NAKAMUR. | 60 | -0.0291 | -0.00164 | 0.072278 | 0.65637 | 11161.57 |
| GOBP_NUC | 11 | -0.0574 | -0.00139 | 0.14243 | 0.65653 | 11164.29 |
| GOMF_CYS | 19 | -0.04439 | -0.00141 | 0.11014 | 0.65653 | 11164.29 |
| GOBP_NEG | 5 | -0.10911 | -0.00178 | 0.27068 | 0.65656 | 11164.8 |
| GOBP_POT | 34 | -0.03582 | -0.00152 | 0.088834 | 0.65662 | 11165.82 |
| ZHAN_VAR | 16 | -0.04644 | -0.00135 | 0.11517 | 0.65662 | 11165.82 |
| GOBP_INH | 16 | -0.05785 | -0.00168 | 0.14335 | 0.65674 | 11167.86 |
| REACTOME | 6 | -0.09237 | -0.00165 | 0.22872 | 0.65684 | 11169.56 |
| GOMF_RN/ | 11 | -0.05842 | -0.00141 | 0.14452 | 0.65698 | 11171.94 |
| GOBP_CHE | 5 | -0.11208 | -0.00182 | 0.27722 | 0.657 | 11172.29 |
| RUTELLA_F | 387 | -0.01054 | -0.00149 | 0.026078 | 0.657 | 11172.29 |
| EBAUER_T/ | 49 | -0.03005 | -0.00153 | 0.074317 | 0.65703 | 11172.8 |
| GOBP_CELI | 11 | -0.06404 | -0.00155 | 0.15833 | 0.65707 | 11173.48 |
| WP_NAD_I | 11 | -0.06601 | -0.00159 | 0.16318 | 0.65708 | 11173.65 |
| XU_RESPOI | 12 | -0.05838 | -0.00147 | 0.14429 | 0.65711 | 11174.16 |
| REACTOME | 52 | -0.02628 | -0.00138 | 0.06485 | 0.65734 | 11178.07 |
| GOBP_REG | 15 | -0.05283 | -0.00149 | 0.13035 | 0.65738 | 11178.75 |
| GOBP_PRC | 12 | -0.04775 | -0.0012 | 0.11778 | 0.6574 | 11179.09 |
| GENTLES_L | 17 | -0.05056 | -0.00152 | 0.12446 | 0.6577 | 11184.19 |
| MOREIRA_ | 18 | -0.04408 | -0.00136 | 0.10851 | 0.6577 | 11184.19 |
| GOMF_BU | 9 | -0.09037 | -0.00197 | 0.22236 | 0.65778 | 11185.55 |
| REACTOME | 12 | -0.05828 | -0.00147 | 0.14337 | 0.6578 | 11185.89 |
| GOBP_END | 6 | -0.08125 | -0.00145 | 0.19971 | 0.65793 | 11188.1 |
| GOBP_TRY | 10 | -0.07199 | -0.00166 | 0.17688 | 0.65799 | 11189.12 |
| GOBP_POS | 8 | -0.07348 | -0.00151 | 0.18048 | 0.65804 | 11189.97 |
| GOBP_MAI | 320 | -0.01145 | -0.00148 | 0.02812 | 0.65809 | 11190.82 |
| GOBP_HIST | 57 | -0.02554 | -0.0014 | 0.062685 | 0.65816 | 11192.01 |
| ROSS_AML | 48 | -0.0311 | -0.00157 | 0.0763 | 0.65819 | 11192.52 |
| PID_S1P_N | 21 | -0.04772 | -0.00159 | 0.11706 | 0.65824 | 11193.37 |

| | | | | | | |
|---|---|---|---|---|---|---|
| WP_MYON | 154 | -0.01629 | -0.00147 | 0.039928 | 0.65838 | 11195.75 |
| GOBP_POS | 15 | -0.0569 | -0.0016 | 0.1392 | 0.65865 | 11200.34 |
| GOBP_ORG | 333 | -0.01162 | -0.00153 | 0.028431 | 0.65866 | 11200.51 |
| WEINMAN | 23 | -0.04659 | -0.00163 | 0.11392 | 0.65871 | 11201.36 |
| VERRECCHI | 14 | -0.05464 | -0.00149 | 0.13359 | 0.65872 | 11201.53 |
| GOCC_PRC | 16 | -0.0457 | -0.00133 | 0.11169 | 0.65881 | 11203.06 |
| GOBP_ACT | 7 | -0.09986 | -0.00192 | 0.24396 | 0.65885 | 11203.74 |
| WP_BILE_A | 12 | -0.06102 | -0.00154 | 0.14902 | 0.65889 | 11204.42 |
| JAZAERI_BI | 47 | -0.02712 | -0.00135 | 0.066189 | 0.659 | 11206.3 |
| GOBP_RES | 40 | -0.03311 | -0.00152 | 0.080809 | 0.65901 | 11206.47 |
| GOBP_REG | 35 | -0.03325 | -0.00143 | 0.081074 | 0.65914 | 11208.68 |
| GOMF_GAI | 5 | -0.08689 | -0.00141 | 0.21181 | 0.65917 | 11209.19 |
| GOBP_REG | 15 | -0.05258 | -0.00148 | 0.12792 | 0.65948 | 11214.46 |
| GOBP_GLY | 11 | -0.05839 | -0.00141 | 0.14203 | 0.65951 | 11214.97 |
| WP_SIGNA | 30 | -0.03708 | -0.00148 | 0.090017 | 0.65978 | 11219.56 |
| KEGG_CIRC | 13 | -0.05354 | -0.0014 | 0.12977 | 0.66004 | 11223.98 |
| RUAN_RES | 37 | -0.0347 | -0.00153 | 0.084085 | 0.66007 | 11224.49 |
| TANAKA_M | 98 | -0.02474 | -0.00178 | 0.059948 | 0.6601 | 11225 |
| GOMF_N_A | 5 | -0.10891 | -0.00177 | 0.26382 | 0.66013 | 11225.51 |
| RAMALHO_ | 65 | -0.02493 | -0.00146 | 0.060377 | 0.66018 | 11226.36 |
| GOMF_CYC | 21 | -0.04404 | -0.00147 | 0.10665 | 0.66019 | 11226.53 |
| WP_PEPTII | 67 | -0.02899 | -0.00172 | 0.070154 | 0.66026 | 11227.72 |
| GHANDHI_ | 74 | -0.02658 | -0.00166 | 0.064222 | 0.6605 | 11231.8 |
| REACTOME | 51 | -0.03269 | -0.0017 | 0.078855 | 0.66078 | 11236.56 |
| GOCC_CHR | 13 | -0.07092 | -0.00186 | 0.17099 | 0.66083 | 11237.41 |
| GOCC_INTI | 48 | -0.02991 | -0.00151 | 0.072121 | 0.66084 | 11237.58 |
| GOBP_DEF | 51 | -0.02684 | -0.00139 | 0.064703 | 0.66086 | 11237.92 |
| GOMF_CYT | 7 | -0.05731 | -0.0011 | 0.13809 | 0.66094 | 11239.28 |
| RODRIGUE | 111 | -0.02066 | -0.00158 | 0.049786 | 0.66095 | 11239.45 |
| GOBP_RET | 18 | -0.05043 | -0.00156 | 0.12146 | 0.66099 | 11240.13 |
| YAO_TEMP | 49 | -0.02981 | -0.00152 | 0.071782 | 0.66102 | 11240.65 |
| UEDA_CEN | 82 | -0.02341 | -0.00154 | 0.056383 | 0.66103 | 11240.82 |
| GOMF_ENI | 33 | -0.03619 | -0.00151 | 0.08711 | 0.6611 | 11242.01 |
| GOBP_PHA | 9 | -0.07939 | -0.00173 | 0.19081 | 0.66131 | 11245.58 |
| GOBP_PRC | 530 | -0.00895 | -0.00148 | 0.021509 | 0.66141 | 11247.28 |
| WENDT_CC | 32 | -0.03857 | -0.00159 | 0.09265 | 0.66141 | 11247.28 |
| GOMF_ALD | 12 | -0.07053 | -0.00178 | 0.16938 | 0.66142 | 11247.45 |
| GRADE_CO | 261 | -0.01247 | -0.00146 | 0.029937 | 0.66153 | 11249.32 |
| GOMF_LIPI | 14 | -0.05731 | -0.00156 | 0.13747 | 0.6616 | 11250.51 |
| ZHAN_MUI | 41 | -0.0303 | -0.00141 | 0.072686 | 0.66161 | 11250.68 |
| GOBP_DEN | 41 | -0.03489 | -0.00162 | 0.083689 | 0.66162 | 11250.85 |
| GOCC_IGA_ | 2 | -0.09557 | -0.00098 | 0.22919 | 0.66165 | 11251.36 |
| GOCC_IGA_ | 2 | -0.09557 | -0.00098 | 0.22919 | 0.66165 | 11251.36 |
| GOCC_IMN | 2 | -0.09557 | -0.00098 | 0.22919 | 0.66165 | 11251.36 |
| REACTOME | 13 | -0.05964 | -0.00156 | 0.14294 | 0.66176 | 11253.23 |

| | | | | | | |
|---|---|---|---|---|---|---|
| GOBP_POS | 8 | -0.09544 | -0.00196 | 0.22852 | 0.6619 | 11255.61 |
| GOBP_RIB( | 6 | -0.08808 | -0.00157 | 0.21078 | 0.66197 | 11256.8 |
| GOMF_RIB | 6 | -0.08808 | -0.00157 | 0.21078 | 0.66197 | 11256.8 |
| GOBP_KYN | 11 | -0.06584 | -0.00159 | 0.15755 | 0.66199 | 11257.14 |
| HUANG_D/ | 75 | -0.02692 | -0.00169 | 0.064351 | 0.66212 | 11259.35 |
| GOBP_GLY | 355 | -0.0112 | -0.00152 | 0.026744 | 0.6623 | 11262.41 |
| WP_ATR_S | 9 | -0.05895 | -0.00129 | 0.14064 | 0.66243 | 11264.62 |
| GOBP_CYT | 5 | -0.07833 | -0.00127 | 0.18687 | 0.66245 | 11264.96 |
| GOCC_AP_ | 9 | -0.05683 | -0.00124 | 0.13557 | 0.66245 | 11264.96 |
| ITO_PTTG1 | 9 | -0.06179 | -0.00135 | 0.14737 | 0.66249 | 11265.64 |
| GOBP_AXC | 90 | -0.02079 | -0.00143 | 0.049562 | 0.66256 | 11266.83 |
| GOMF_PR( | 6 | -0.08317 | -0.00148 | 0.19813 | 0.66268 | 11268.87 |
| GOBP_PAN | 10 | -0.07945 | -0.00183 | 0.18921 | 0.66272 | 11269.55 |
| GOCC_AMI | 11 | -0.07672 | -0.00185 | 0.18255 | 0.66286 | 11271.93 |
| GOBP_CELI | 7 | -0.08771 | -0.00169 | 0.20859 | 0.66292 | 11272.95 |
| GOBP_REG | 27 | -0.044 | -0.00166 | 0.10464 | 0.66294 | 11273.29 |
| GOBP_REG | 58 | -0.0307 | -0.0017 | 0.072958 | 0.66304 | 11275 |
| ALONSO_M | 5 | -0.11304 | -0.00184 | 0.26857 | 0.66309 | 11275.85 |
| WEBER_MI | 8 | -0.06566 | -0.00135 | 0.15597 | 0.66312 | 11276.36 |
| KEGG_T_CI | 103 | -0.02096 | -0.00154 | 0.04978 | 0.66316 | 11277.04 |
| GOBP_NK_ | 12 | -0.05854 | -0.00148 | 0.13895 | 0.66323 | 11278.23 |
| GOBP_REG | 114 | -0.01965 | -0.00152 | 0.04663 | 0.66323 | 11278.23 |
| GOBP_RES | 8 | -0.08807 | -0.00181 | 0.20904 | 0.66323 | 11278.23 |
| GOBP_RES | 6 | -0.09693 | -0.00173 | 0.23006 | 0.66324 | 11278.4 |
| REACTOME | 16 | -0.05352 | -0.00156 | 0.12699 | 0.66327 | 11278.91 |
| GOMF_TR/ | 22 | -0.04534 | -0.00155 | 0.10757 | 0.66332 | 11279.76 |
| GOBP_NEG | 20 | -0.04656 | -0.00151 | 0.11039 | 0.66339 | 11280.95 |
| GOBP_NEG | 22 | -0.04543 | -0.00155 | 0.10767 | 0.66346 | 11282.14 |
| GOBP_NEG | 17 | -0.0531 | -0.00159 | 0.12579 | 0.66352 | 11283.16 |
| WP_TRANS | 15 | -0.05713 | -0.00161 | 0.13531 | 0.66356 | 11283.84 |
| PID_SYNDE | 32 | -0.04104 | -0.00169 | 0.097027 | 0.66384 | 11288.6 |
| GOBP_MID | 7 | -0.09153 | -0.00176 | 0.21635 | 0.66387 | 11289.11 |
| GOBP_PRC | 5 | -0.08431 | -0.00137 | 0.1993 | 0.66387 | 11289.11 |
| GOCC_FHF | 5 | -0.08051 | -0.00131 | 0.19031 | 0.66388 | 11289.28 |
| GOBP_SYN | 74 | -0.02391 | -0.00149 | 0.056472 | 0.66401 | 11291.49 |
| MIKKELSEN | 202 | -0.01561 | -0.00161 | 0.036854 | 0.66408 | 11292.68 |
| GOBP_LEN | 20 | -0.05233 | -0.0017 | 0.12351 | 0.6641 | 11293.02 |
| GOBP_END | 46 | -0.03295 | -0.00162 | 0.07767 | 0.66427 | 11295.91 |
| GOBP_SNA | 22 | -0.03927 | -0.00134 | 0.092511 | 0.6644 | 11298.12 |
| GOBP_NEG | 18 | -0.05262 | -0.00162 | 0.12391 | 0.66444 | 11298.8 |
| GOBP_REG | 12 | -0.05999 | -0.00151 | 0.14115 | 0.66457 | 11301.01 |
| GOBP_LEU | 218 | -0.01531 | -0.00164 | 0.035959 | 0.66482 | 11305.26 |
| WP_FLUOF | 33 | -0.03895 | -0.00163 | 0.091439 | 0.66492 | 11306.96 |
| GOMF_AP_ | 5 | -0.10746 | -0.00175 | 0.25223 | 0.66495 | 11307.47 |
| CHEN_HOX | 5 | -0.0896 | -0.00146 | 0.21025 | 0.66501 | 11308.5 |

| | | | | | | |
|---|---|---|---|---|---|---|
| GERY_CEBI | 119 | -0.02034 | -0.00161 | 0.047709 | 0.66506 | 11309.35 |
| GOBP_PRC | 11 | -0.06311 | -0.00152 | 0.148 | 0.6651 | 11310.03 |
| REACTOME | 434 | -0.01117 | -0.00167 | 0.02618 | 0.66517 | 11311.22 |
| JOHNSTON | 129 | -0.01789 | -0.00147 | 0.041932 | 0.66519 | 11311.56 |
| REACTOME | 7 | -0.08567 | -0.00165 | 0.20032 | 0.66555 | 11317.68 |
| GOBP_RES | 30 | -0.04234 | -0.00169 | 0.098905 | 0.66569 | 11320.06 |
| WP_15Q13 | 14 | -0.07195 | -0.00196 | 0.16807 | 0.6657 | 11320.23 |
| KAMMING. | 38 | -0.03513 | -0.00157 | 0.082017 | 0.66579 | 11321.76 |
| GOMF_THY | 6 | -0.09833 | -0.00175 | 0.22944 | 0.66588 | 11323.29 |
| REACTOME | 28 | -0.03837 | -0.00148 | 0.089525 | 0.66591 | 11323.8 |
| GOBP_RELI | 56 | -0.02769 | -0.00151 | 0.064577 | 0.66593 | 11324.14 |
| REACTOME | 15 | -0.05714 | -0.00161 | 0.13318 | 0.66604 | 11326.01 |
| GOBP_REG | 23 | -0.05071 | -0.00177 | 0.11814 | 0.66613 | 11327.54 |
| REACTOME | 44 | -0.03246 | -0.00157 | 0.075573 | 0.66623 | 11329.24 |
| SEMBA_FH | 9 | -0.07771 | -0.0017 | 0.18091 | 0.66624 | 11329.41 |
| WP_KYNUI | 22 | -0.04458 | -0.00152 | 0.10376 | 0.66625 | 11329.58 |
| PEREZ_TP5 | 1093 | -0.00672 | -0.00157 | 0.015628 | 0.66635 | 11331.28 |
| LINSLEY_M | 197 | -0.01411 | -0.00143 | 0.032815 | 0.66636 | 11331.45 |
| HAHTOLA_ | 24 | -0.04275 | -0.00152 | 0.099409 | 0.6664 | 11332.13 |
| VERHAAK_ | 120 | -0.01958 | -0.00156 | 0.04552 | 0.66641 | 11332.3 |
| TAKADA_G | 7 | -0.09669 | -0.00186 | 0.22455 | 0.66661 | 11335.7 |
| GOBP_RAD | 14 | -0.06634 | -0.00181 | 0.15407 | 0.66662 | 11335.87 |
| GOBP_PYR | 12 | -0.06346 | -0.0016 | 0.14737 | 0.66664 | 11336.21 |
| GOBP_HEX | 7 | -0.07212 | -0.00139 | 0.16745 | 0.66665 | 11336.38 |
| RASHI_RES | 46 | -0.03263 | -0.00161 | 0.07575 | 0.66668 | 11336.89 |
| GOCC_INTI | 5 | -0.08498 | -0.00138 | 0.19718 | 0.66675 | 11338.08 |
| GOBP_REG | 143 | -0.01939 | -0.00168 | 0.044976 | 0.66684 | 11339.61 |
| GOBP_REN | 4 | -0.13044 | -0.0019 | 0.30234 | 0.66693 | 11341.14 |
| GOCC_TIGI | 121 | -0.01996 | -0.00159 | 0.046224 | 0.66703 | 11342.85 |
| GOBP_NEG | 11 | -0.05745 | -0.00139 | 0.13304 | 0.66707 | 11343.53 |
| BUCKANO\ | 20 | -0.05214 | -0.0017 | 0.12067 | 0.66715 | 11344.89 |
| GOCC_KER | 94 | -0.03173 | -0.00223 | 0.073391 | 0.66723 | 11346.25 |
| GOBP_REC | 7 | -0.09162 | -0.00176 | 0.21193 | 0.66725 | 11346.59 |
| GOBP_FIBI | 13 | -0.06861 | -0.0018 | 0.15855 | 0.66739 | 11348.97 |
| GOCC_SM( | 9 | -0.07689 | -0.00168 | 0.17759 | 0.66748 | 11350.5 |
| GOBP_REG | 19 | -0.05252 | -0.00167 | 0.12128 | 0.66752 | 11351.18 |
| GOMF_HO | 117 | -0.0197 | -0.00155 | 0.045491 | 0.66752 | 11351.18 |
| GOBP_NEG | 5 | -0.08943 | -0.00146 | 0.20639 | 0.6676 | 11352.54 |
| KIM_WT1_ | 439 | -0.01051 | -0.00158 | 0.024259 | 0.66761 | 11352.71 |
| PID_INSULI | 44 | -0.03535 | -0.0017 | 0.081477 | 0.66779 | 11355.77 |
| GOBP_DGN | 5 | -0.09353 | -0.00152 | 0.2155 | 0.66785 | 11356.79 |
| FINETTI_BI | 14 | -0.06253 | -0.0017 | 0.14389 | 0.66807 | 11360.53 |
| GOMF_3_5 | 15 | -0.06099 | -0.00172 | 0.14026 | 0.66815 | 11361.89 |
| WP_GALAN | 21 | -0.05629 | -0.00188 | 0.12941 | 0.6682 | 11362.74 |
| GOBP_COE | 6 | -0.07894 | -0.00141 | 0.18136 | 0.66832 | 11364.78 |

| | | | | | | |
|---|---|---|---|---|---|---|
| GOBP_REG | 47 | -0.03427 | -0.00171 | 0.078691 | 0.66838 | 11365.8 |
| WP_STERO | 7 | -0.08439 | -0.00162 | 0.19379 | 0.66838 | 11365.8 |
| DELPUECH_ | 61 | -0.02947 | -0.00167 | 0.06763 | 0.66851 | 11368.01 |
| GOBP_REG | 33 | -0.03386 | -0.00141 | 0.077693 | 0.66853 | 11368.35 |
| WP_PRION | 31 | -0.04183 | -0.00169 | 0.095944 | 0.66858 | 11369.2 |
| REACTOME | 11 | -0.06918 | -0.00167 | 0.15855 | 0.66869 | 11371.07 |
| GOBP_PLA | 8 | -0.07354 | -0.00151 | 0.16849 | 0.66875 | 11372.09 |
| GOMF_ALC | 23 | -0.04604 | -0.00161 | 0.10545 | 0.66879 | 11372.77 |
| GOBP_SEQ | 23 | -0.03965 | -0.00138 | 0.090804 | 0.66881 | 11373.11 |
| REACTOME | 55 | -0.03137 | -0.00169 | 0.071757 | 0.66898 | 11376 |
| HIRSCH_CE | 235 | -0.0141 | -0.00156 | 0.032255 | 0.66902 | 11376.69 |
| GOBP_POS | 16 | -0.05517 | -0.00161 | 0.12618 | 0.66903 | 11376.86 |
| GOBP_VITA | 8 | -0.07347 | -0.00151 | 0.16795 | 0.6691 | 11378.05 |
| TERAMOTO | 8 | -0.06733 | -0.00139 | 0.1539 | 0.66912 | 11378.39 |
| GOBP_IND | 12 | -0.06527 | -0.00165 | 0.14911 | 0.6692 | 11379.75 |
| MAHADEVA | 17 | -0.06465 | -0.00194 | 0.14744 | 0.66948 | 11384.51 |
| CASORELLI_ | 148 | -0.01933 | -0.0017 | 0.044044 | 0.66962 | 11386.89 |
| GOBP_PEN | 11 | -0.06402 | -0.00155 | 0.14585 | 0.66965 | 11387.4 |
| JAEGER_M | 39 | -0.03823 | -0.00174 | 0.087074 | 0.66969 | 11388.08 |
| GOCC_EPSI | 5 | -0.13028 | -0.00212 | 0.29662 | 0.66974 | 11388.93 |
| GOBP_GAL | 5 | -0.08572 | -0.00139 | 0.19506 | 0.66983 | 11390.46 |
| KEGG_ECM | 80 | -0.02565 | -0.00167 | 0.058341 | 0.66988 | 11391.31 |
| GOBP_RES | 5 | -0.09765 | -0.00159 | 0.22205 | 0.66995 | 11392.5 |
| PARK_HSC_ | 41 | -0.03453 | -0.00161 | 0.078449 | 0.67007 | 11394.54 |
| GOBP_PYR | 83 | -0.02256 | -0.00149 | 0.051245 | 0.67014 | 11395.73 |
| GOCC_B_C | 5 | -0.09582 | -0.00156 | 0.21751 | 0.67022 | 11397.09 |
| GOCC_BBS | 8 | -0.10571 | -0.00218 | 0.23988 | 0.67027 | 11397.94 |
| GOMF_ADI | 4 | -0.1053 | -0.00153 | 0.23877 | 0.67039 | 11399.98 |
| GOMF_ATF | 115 | -0.01985 | -0.00154 | 0.044995 | 0.67041 | 11400.32 |
| LUI_THYRO | 13 | -0.06292 | -0.00165 | 0.14262 | 0.67045 | 11401 |
| GOMF_L_A | 12 | -0.06981 | -0.00176 | 0.15822 | 0.67046 | 11401.17 |
| REACTOME | 232 | -0.01461 | -0.00161 | 0.033111 | 0.67046 | 11401.17 |
| GOBP_NEG | 7 | -0.10209 | -0.00197 | 0.23127 | 0.67055 | 11402.7 |
| GOBP_POS | 5 | -0.09333 | -0.00152 | 0.21142 | 0.67056 | 11402.87 |
| GOBP_DOF | 36 | -0.03798 | -0.00166 | 0.086005 | 0.67061 | 11403.72 |
| POMEROY_ | 57 | -0.02846 | -0.00156 | 0.064444 | 0.67061 | 11403.72 |
| GOMF_CAF | 16 | -0.06135 | -0.00179 | 0.13879 | 0.67077 | 11406.44 |
| NAKAMUR | 42 | -0.03533 | -0.00166 | 0.079885 | 0.67084 | 11407.63 |
| KEGG_GLU | 46 | -0.03494 | -0.00172 | 0.078962 | 0.67093 | 11409.16 |
| GOCC_TRA | 11 | -0.07044 | -0.0017 | 0.15914 | 0.67099 | 11410.18 |
| GOMF_HEL | 143 | -0.01783 | -0.00155 | 0.040284 | 0.67099 | 11410.18 |
| SEKI_INFLA | 21 | -0.05183 | -0.00173 | 0.11706 | 0.67103 | 11410.87 |
| GOBP_SON | 52 | -0.03368 | -0.00177 | 0.076053 | 0.67105 | 11411.21 |
| KEGG_JAK_ | 146 | -0.02056 | -0.0018 | 0.04632 | 0.67141 | 11417.33 |
| WP_ETHAN | 27 | -0.04477 | -0.00169 | 0.10086 | 0.67141 | 11417.33 |

| | | | | | | |
|---|---|---|---|---|---|---|
| GOBP_REG | 13 | -0.06465 | -0.0017 | 0.14561 | 0.67146 | 11418.18 |
| GOCC_EXT | 5 | -0.0838 | -0.00136 | 0.18871 | 0.6715 | 11418.86 |
| GOMF_COI | 45 | -0.03574 | -0.00174 | 0.080432 | 0.67161 | 11420.73 |
| GOBP_RET | 48 | -0.0306 | -0.00154 | 0.068853 | 0.67163 | 11421.07 |
| GOBP_SUL | 22 | -0.05515 | -0.00188 | 0.12399 | 0.67176 | 11423.28 |
| GOMF_K48 | 11 | -0.06441 | -0.00155 | 0.1448 | 0.67176 | 11423.28 |
| GOBP_NAT | 33 | -0.04772 | -0.00199 | 0.10714 | 0.67198 | 11427.02 |
| REACTOME | 31 | -0.03577 | -0.00145 | 0.080295 | 0.67202 | 11427.7 |
| TAVAZOIE_ | 90 | -0.02337 | -0.00161 | 0.052452 | 0.67202 | 11427.7 |
| GOCC_INFI | 15 | -0.05695 | -0.0016 | 0.1278 | 0.67206 | 11428.38 |
| REACTOME | 37 | -0.03626 | -0.0016 | 0.081334 | 0.67213 | 11429.57 |
| REACTOME | 87 | -0.02341 | -0.00159 | 0.052475 | 0.67226 | 11431.78 |
| GOBP_L_A | 77 | -0.02622 | -0.00167 | 0.058753 | 0.6723 | 11432.46 |
| GOBP_CELI | 8 | -0.06534 | -0.00134 | 0.14639 | 0.67232 | 11432.8 |
| REACTOME | 17 | -0.04593 | -0.00138 | 0.10285 | 0.67238 | 11433.82 |
| WP_NIPBL_ | 7 | -0.08388 | -0.00162 | 0.18782 | 0.67243 | 11434.67 |
| GOBP_PHC | 56 | -0.02749 | -0.0015 | 0.061539 | 0.67246 | 11435.18 |
| WP_GPCRS | 222 | -0.01644 | -0.00177 | 0.0368 | 0.67248 | 11435.52 |
| GOBP_LAYI | 14 | -0.05504 | -0.0015 | 0.12307 | 0.67262 | 11437.9 |
| GOBP_REG | 11 | -0.07252 | -0.00175 | 0.16211 | 0.67268 | 11438.92 |
| GOBP_SKIN | 298 | -0.01425 | -0.00178 | 0.031801 | 0.67293 | 11443.17 |
| GOBP_DET | 5 | -0.10966 | -0.00178 | 0.24473 | 0.67294 | 11443.34 |
| GOBP_LIPII | 13 | -0.05336 | -0.0014 | 0.11894 | 0.67312 | 11446.41 |
| REACTOME | 9 | -0.07314 | -0.0016 | 0.16305 | 0.67313 | 11446.58 |
| GOBP_END | 16 | -0.05488 | -0.0016 | 0.12228 | 0.67321 | 11447.94 |
| JIANG_HYP | 18 | -0.04388 | -0.00135 | 0.097769 | 0.67322 | 11448.11 |
| GOBP_POR | 49 | -0.02929 | -0.00149 | 0.065068 | 0.67369 | 11456.1 |
| WEINMAN | 33 | -0.04647 | -0.00194 | 0.10323 | 0.67369 | 11456.1 |
| REACTOME | 12 | -0.07111 | -0.00179 | 0.15794 | 0.67371 | 11456.44 |
| RAMJAUN_ | 8 | -0.07245 | -0.00149 | 0.16074 | 0.6739 | 11459.67 |
| GOBP_PEP | 22 | -0.05082 | -0.00173 | 0.1127 | 0.67398 | 11461.03 |
| GOBP_REG | 75 | -0.02657 | -0.00167 | 0.058914 | 0.67398 | 11461.03 |
| GOBP_MIC | 42 | -0.03294 | -0.00155 | 0.073032 | 0.67402 | 11461.71 |
| GOBP_NEU | 109 | -0.02493 | -0.00189 | 0.055208 | 0.67418 | 11464.43 |
| GOBP_LYSO | 19 | -0.05509 | -0.00175 | 0.12193 | 0.67428 | 11466.13 |
| KYNG_WEF | 168 | -0.01663 | -0.00156 | 0.036811 | 0.67432 | 11466.81 |
| REACTOME | 75 | -0.02625 | -0.00165 | 0.058096 | 0.67432 | 11466.81 |
| RHODES_C | 61 | -0.02786 | -0.00158 | 0.061647 | 0.67432 | 11466.81 |
| GOBP_REG | 20 | -0.04127 | -0.00134 | 0.09132 | 0.67434 | 11467.15 |
| GOBP_PRC | 1612 | -0.00601 | -0.00168 | 0.013292 | 0.67435 | 11467.32 |
| BLANCO_M | 135 | -0.01919 | -0.00162 | 0.042396 | 0.67461 | 11471.74 |
| CHIANG_LI | 70 | -0.029 | -0.00176 | 0.064049 | 0.67462 | 11471.91 |
| GOBP_NEG | 29 | -0.04686 | -0.00184 | 0.10346 | 0.67469 | 11473.1 |
| GOBP_POS | 473 | -0.01036 | -0.00162 | 0.022856 | 0.67488 | 11476.33 |
| GOCC_UBI | 6 | -0.0713 | -0.00127 | 0.15724 | 0.67488 | 11476.33 |

| | | | | | | |
|---|---|---|---|---|---|---|
| GOBP_TRA | 11 | -0.07353 | -0.00177 | 0.16207 | 0.67497 | 11477.86 |
| GOBP_POS | 41 | -0.03723 | -0.00173 | 0.082029 | 0.67504 | 11479.06 |
| VIETOR_IFI | 22 | -0.04894 | -0.00167 | 0.10781 | 0.67505 | 11479.23 |
| WP_SPHIN | 26 | -0.05246 | -0.00195 | 0.11548 | 0.67519 | 11481.61 |
| GOMF_INT | 7 | -0.08997 | -0.00173 | 0.19802 | 0.67521 | 11481.95 |
| PID_ANGIC | 46 | -0.03347 | -0.00165 | 0.073656 | 0.67522 | 11482.12 |
| CONRAD_G | 9 | -0.08243 | -0.0018 | 0.18137 | 0.67526 | 11482.8 |
| SHI_SPARC | 23 | -0.04944 | -0.00172 | 0.10861 | 0.67552 | 11487.22 |
| WP_PROST | 31 | -0.04752 | -0.00192 | 0.10439 | 0.67552 | 11487.22 |
| GOMF_EST | 5 | -0.1072 | -0.00174 | 0.23533 | 0.67563 | 11489.09 |
| GOBP_NEG | 8 | -0.10163 | -0.00209 | 0.22309 | 0.67564 | 11489.26 |
| GOBP_RIBO | 107 | -0.02125 | -0.0016 | 0.046592 | 0.6758 | 11491.98 |
| GOBP_GRO | 18 | -0.05591 | -0.00173 | 0.12254 | 0.6759 | 11493.68 |
| BIOCARTA_ | 14 | -0.06067 | -0.00165 | 0.13289 | 0.67599 | 11495.21 |
| GOBP_NEG | 21 | -0.0516 | -0.00172 | 0.11291 | 0.67617 | 11498.27 |
| GOBP_DET | 8 | -0.069 | -0.00142 | 0.15081 | 0.67636 | 11501.5 |
| GOBP_NEG | 44 | -0.03781 | -0.00182 | 0.082638 | 0.67637 | 11501.67 |
| GOBP_GLY | 60 | -0.02943 | -0.00166 | 0.064286 | 0.67643 | 11502.69 |
| REACTOME | 33 | -0.03272 | -0.00137 | 0.071468 | 0.67646 | 11503.2 |
| GOBP_CDP | 6 | -0.07501 | -0.00134 | 0.1638 | 0.67649 | 11503.71 |
| WP_BRAIN | 136 | -0.02068 | -0.00175 | 0.045117 | 0.67667 | 11506.77 |
| REACTOME | 9 | -0.07284 | -0.00159 | 0.15884 | 0.67672 | 11507.62 |
| XU_GH1_E | 48 | -0.03356 | -0.00169 | 0.073184 | 0.67674 | 11507.96 |
| GOBP_MAT | 13 | -0.0588 | -0.00154 | 0.12817 | 0.6768 | 11508.98 |
| GOBP_RHO | 12 | -0.0529 | -0.00133 | 0.11531 | 0.67681 | 11509.15 |
| LUDWICZEI | 7 | -0.08108 | -0.00156 | 0.17668 | 0.67685 | 11509.83 |
| GAJATE_RE | 62 | -0.02895 | -0.00166 | 0.063078 | 0.67687 | 11510.17 |
| LEE_LIVER_ | 63 | -0.03028 | -0.00175 | 0.065962 | 0.67691 | 11510.85 |
| GOBP_RET | 12 | -0.06655 | -0.00168 | 0.14494 | 0.67692 | 11511.02 |
| GOCC_PHO | 6 | -0.09094 | -0.00162 | 0.19787 | 0.67709 | 11513.92 |
| MODY_HIP | 54 | -0.03288 | -0.00176 | 0.071518 | 0.67714 | 11514.77 |
| GOBP_POS | 13 | -0.06432 | -0.00169 | 0.13982 | 0.67725 | 11516.64 |
| GOBP_NEG | 30 | -0.04371 | -0.00174 | 0.095006 | 0.67728 | 11517.15 |
| KANG_AR_ | 16 | -0.06462 | -0.00188 | 0.14038 | 0.67734 | 11518.17 |
| GOBP_LOC | 1240 | -0.00699 | -0.00173 | 0.015186 | 0.67739 | 11519.02 |
| GOBP_NUC | 12 | -0.06014 | -0.00152 | 0.13058 | 0.67745 | 11520.04 |
| WP_MIRNA | 69 | -0.02734 | -0.00165 | 0.059364 | 0.67745 | 11520.04 |
| REACTOME | 10 | -0.06948 | -0.0016 | 0.15084 | 0.67746 | 11520.21 |
| REACTOME | 24 | -0.04715 | -0.00168 | 0.10231 | 0.67754 | 11521.57 |
| GRAHAM_( | 22 | -0.05353 | -0.00183 | 0.11613 | 0.67757 | 11522.08 |
| GOBP_NEG | 4 | -0.13604 | -0.00198 | 0.29477 | 0.67778 | 11525.65 |
| BIOCARTA_ | 11 | -0.07458 | -0.0018 | 0.16152 | 0.67787 | 11527.18 |
| GOBP_SKEI | 8 | -0.09674 | -0.00199 | 0.20946 | 0.67791 | 11527.86 |
| GOCC_CHO | 10 | -0.07868 | -0.00181 | 0.1703 | 0.67795 | 11528.54 |
| SHIPP_DLB | 37 | -0.03765 | -0.00167 | 0.081484 | 0.67798 | 11529.05 |

| | | | | | | |
|---|---|---|---|---|---|---|
| GOMF_PH( | 22 | -0.05519 | -0.00188 | 0.11936 | 0.6781 | 11531.09 |
| BIOCARTA_ | 18 | -0.05244 | -0.00162 | 0.11339 | 0.67813 | 11531.6 |
| GOCC_SPH | 7 | -0.10535 | -0.00203 | 0.22775 | 0.67816 | 11532.11 |
| ROETH_TEI | 14 | -0.07197 | -0.00196 | 0.15536 | 0.67841 | 11536.36 |
| LI_CYTIDIN | 14 | -0.05451 | -0.00148 | 0.11759 | 0.6785 | 11537.89 |
| MEBARKI_I | 511 | -0.01035 | -0.00168 | 0.022312 | 0.67861 | 11539.76 |
| GOBP_POL | 16 | -0.05308 | -0.00154 | 0.11429 | 0.67883 | 11543.5 |
| KEGG_O_G | 29 | -0.04443 | -0.00174 | 0.095668 | 0.67883 | 11543.5 |
| GOBP_REG | 7 | -0.10659 | -0.00205 | 0.22949 | 0.67884 | 11543.67 |
| GOBP_REG | 39 | -0.03516 | -0.0016 | 0.075685 | 0.67885 | 11543.84 |
| REACTOME | 93 | -0.02369 | -0.00166 | 0.050973 | 0.67896 | 11545.71 |
| GOBP_LIPII | 32 | -0.03775 | -0.00155 | 0.081196 | 0.67902 | 11546.74 |
| WP_MITOC | 8 | -0.07196 | -0.00148 | 0.15473 | 0.67906 | 11547.42 |
| GOBP_NEG | 6 | -0.09645 | -0.00172 | 0.20717 | 0.67923 | 11550.31 |
| GOBP_CELI | 7 | -0.09505 | -0.00183 | 0.2041 | 0.67929 | 11551.33 |
| GOBP_CELI | 7 | -0.09325 | -0.0018 | 0.20023 | 0.6793 | 11551.5 |
| GOBP_PHC | 8 | -0.09273 | -0.00191 | 0.19894 | 0.67944 | 11553.88 |
| GOBP_REG | 32 | -0.04214 | -0.00173 | 0.090365 | 0.67949 | 11554.73 |
| GOBP_SM( | 7 | -0.10251 | -0.00197 | 0.2198 | 0.67954 | 11555.58 |
| AMIT_SERU | 30 | -0.04448 | -0.00177 | 0.095255 | 0.67971 | 11558.47 |
| GOMF_GAI | 29 | -0.03959 | -0.00155 | 0.08476 | 0.67979 | 11559.83 |
| GOMF_PO1 | 12 | -0.06674 | -0.00168 | 0.14258 | 0.68014 | 11565.78 |
| GOBP_REG | 8 | -0.08286 | -0.00171 | 0.177 | 0.68015 | 11565.95 |
| GOBP_NEU | 8 | -0.08638 | -0.00178 | 0.1845 | 0.68016 | 11566.12 |
| GOBP_PRE | 32 | -0.0441 | -0.00181 | 0.094195 | 0.68016 | 11566.12 |
| REACTOME | 23 | -0.04608 | -0.00161 | 0.098373 | 0.68024 | 11567.48 |
| GOMF_ADI | 9 | -0.07444 | -0.00163 | 0.15886 | 0.68032 | 11568.84 |
| RAO_BOUN | 214 | -0.01654 | -0.00175 | 0.035286 | 0.68041 | 11570.37 |
| GOBP_CEN | 73 | -0.02669 | -0.00166 | 0.056912 | 0.68045 | 11571.05 |
| GOMF_XEN | 30 | -0.04691 | -0.00187 | 0.10001 | 0.68047 | 11571.39 |
| GOMF_PRE | 8 | -0.09937 | -0.00205 | 0.2118 | 0.68053 | 11572.41 |
| MIKI_COEX | 6 | -0.10137 | -0.00181 | 0.21598 | 0.68059 | 11573.43 |
| SA_G1_AN | 15 | -0.0584 | -0.00165 | 0.12429 | 0.68077 | 11576.49 |
| PID_E2F_P. | 70 | -0.02825 | -0.00172 | 0.060061 | 0.68096 | 11579.72 |
| GOCC_MH( | 2 | -0.12107 | -0.00125 | 0.25729 | 0.68101 | 11580.58 |
| GOCC_NEU | 5 | -0.13304 | -0.00216 | 0.28252 | 0.68114 | 11582.79 |
| BROWNE_I | 162 | -0.01859 | -0.00171 | 0.039468 | 0.68119 | 11583.64 |
| GOBP_POS | 20 | -0.05062 | -0.00165 | 0.10735 | 0.68138 | 11586.87 |
| GOBP_POS | 6 | -0.11454 | -0.00204 | 0.24222 | 0.68184 | 11594.69 |
| AMIT_EGF_ | 160 | -0.01879 | -0.00172 | 0.039736 | 0.68186 | 11595.03 |
| GOBP_REG | 4 | -0.13247 | -0.00193 | 0.27989 | 0.68199 | 11597.24 |
| JIANG_TIP3 | 22 | -0.04884 | -0.00167 | 0.10315 | 0.68205 | 11598.26 |
| GOBP_SIGI | 13 | -0.0629 | -0.00165 | 0.13285 | 0.68206 | 11598.43 |
| GOBP_REG | 387 | -0.01203 | -0.0017 | 0.025364 | 0.68228 | 11602.17 |
| GOBP_TYP | 25 | -0.04845 | -0.00176 | 0.10219 | 0.68228 | 11602.17 |

| | | | | | | |
|---|---|---|---|---|---|---|
| GOBP_REG | 4 | -0.1148 | -0.00167 | 0.24196 | 0.68241 | 11604.38 |
| GOMF_DIA | 14 | -0.0515 | -0.0014 | 0.10848 | 0.68253 | 11606.42 |
| GOBP_NEG | 229 | -0.01489 | -0.00163 | 0.031343 | 0.68257 | 11607.1 |
| GOMF_RET | 5 | -0.13127 | -0.00214 | 0.27643 | 0.68257 | 11607.1 |
| BIOCARTA_ | 7 | -0.08869 | -0.00171 | 0.18674 | 0.68258 | 11607.27 |
| GOBP_MO | 61 | -0.02735 | -0.00155 | 0.057581 | 0.68263 | 11608.12 |
| REACTOME | 28 | -0.04418 | -0.0017 | 0.092922 | 0.68275 | 11610.16 |
| GOBP_REG | 37 | -0.04141 | -0.00183 | 0.087094 | 0.68278 | 11610.67 |
| ZHONG_RE | 151 | -0.01794 | -0.0016 | 0.03771 | 0.68287 | 11612.2 |
| GOBP_CHR | 6 | -0.09374 | -0.00167 | 0.19692 | 0.68298 | 11614.07 |
| GOBP_CELI | 28 | -0.05034 | -0.00194 | 0.10568 | 0.68309 | 11615.95 |
| GOBP_NEU | 93 | -0.02595 | -0.00182 | 0.054444 | 0.68317 | 11617.31 |
| WP_ESTRO | 21 | -0.05423 | -0.00181 | 0.11376 | 0.68321 | 11617.99 |
| GOBP_INTI | 32 | -0.0448 | -0.00184 | 0.093966 | 0.68324 | 11618.5 |
| SHETH_LIV | 52 | -0.03228 | -0.00169 | 0.067674 | 0.68332 | 11619.86 |
| GOBP_REF | 21 | -0.05254 | -0.00175 | 0.11014 | 0.68333 | 11620.03 |
| REACTOME | 17 | -0.05521 | -0.00166 | 0.11561 | 0.6835 | 11622.92 |
| SA_CASPAS | 18 | -0.05665 | -0.00175 | 0.11861 | 0.68354 | 11623.6 |
| GOMF_STR | 12 | -0.07086 | -0.00179 | 0.14833 | 0.68358 | 11624.28 |
| GOCC_PRC | 6 | -0.09243 | -0.00165 | 0.19344 | 0.6836 | 11624.62 |
| GOBP_REG | 22 | -0.05241 | -0.00179 | 0.10965 | 0.68366 | 11625.64 |
| KEGG_VALI | 8 | -0.10652 | -0.00219 | 0.22264 | 0.68382 | 11628.36 |
| GOBP_NOF | 7 | -0.10877 | -0.00209 | 0.2273 | 0.68386 | 11629.04 |
| GOBP_AGII | 161 | -0.01934 | -0.00178 | 0.040403 | 0.6839 | 11629.72 |
| GOBP_CELI | 1551 | -0.00646 | -0.00177 | 0.013501 | 0.6839 | 11629.72 |
| GOBP_GAN | 6 | -0.08194 | -0.00146 | 0.17113 | 0.68397 | 11630.91 |
| GOBP_NEG | 8 | -0.0785 | -0.00162 | 0.16391 | 0.68399 | 11631.25 |
| GOBP_END | 6 | -0.10876 | -0.00194 | 0.22704 | 0.68403 | 11631.93 |
| GOMF_BET | 7 | -0.0751 | -0.00145 | 0.15676 | 0.68406 | 11632.44 |
| OSADA_AS | 43 | -0.03859 | -0.00184 | 0.080537 | 0.6841 | 11633.12 |
| GOBP_REG | 238 | -0.01559 | -0.00174 | 0.032478 | 0.68433 | 11637.03 |
| GOBP_NAT | 92 | -0.02715 | -0.00189 | 0.056548 | 0.68441 | 11638.39 |
| GOBP_TEL( | 14 | -0.06329 | -0.00172 | 0.13181 | 0.68444 | 11638.9 |
| GOBP_TRA | 4 | -0.10901 | -0.00159 | 0.22693 | 0.68452 | 11640.26 |
| GOBP_CELI | 33 | -0.04226 | -0.00177 | 0.087947 | 0.68455 | 11640.77 |
| GHANDHI_ | 96 | -0.02632 | -0.00187 | 0.054772 | 0.68457 | 11641.11 |
| GOBP_ELE( | 152 | -0.01805 | -0.00161 | 0.03752 | 0.68472 | 11643.66 |
| GOBP_PLA | 144 | -0.02 | -0.00174 | 0.041579 | 0.68478 | 11644.68 |
| GOBP_NEG | 31 | -0.04356 | -0.00176 | 0.090488 | 0.68486 | 11646.04 |
| GOBP_PRC | 4 | -0.09089 | -0.00132 | 0.18872 | 0.68496 | 11647.74 |
| WP_PDGFF | 28 | -0.04214 | -0.00162 | 0.087478 | 0.68498 | 11648.08 |
| GOCC_HIST | 52 | -0.03144 | -0.00165 | 0.065258 | 0.68501 | 11648.6 |
| GOCC_NU( | 12 | -0.06543 | -0.00165 | 0.13577 | 0.68508 | 11649.79 |
| GOBP_NEG | 9 | -0.06869 | -0.0015 | 0.14251 | 0.68509 | 11649.96 |
| SIG_REGUL | 31 | -0.04117 | -0.00167 | 0.085366 | 0.68518 | 11651.49 |

| | | | | | | |
|---|---|---|---|---|---|---|
| GOBP_ERY | 15 | -0.056 | -0.00158 | 0.11612 | 0.6852 | 11651.83 |
| GOMF_PH( | 12 | -0.072 | -0.00181 | 0.14918 | 0.68532 | 11653.87 |
| WEST_ADR | 17 | -0.06401 | -0.00192 | 0.13253 | 0.68543 | 11655.74 |
| GOMF_GA | 290 | -0.01555 | -0.00191 | 0.032168 | 0.68558 | 11658.29 |
| WP_COVID | 7 | -0.12412 | -0.00239 | 0.25679 | 0.68558 | 11658.29 |
| KEGG_LIM( | 10 | -0.07417 | -0.00171 | 0.15336 | 0.68566 | 11659.65 |
| GOBP_NEG | 83 | -0.0271 | -0.00179 | 0.056018 | 0.68572 | 11660.67 |
| GOCC_COF | 22 | -0.04593 | -0.00157 | 0.094939 | 0.68573 | 11660.84 |
| GOBP_BEN | 26 | -0.05189 | -0.00192 | 0.10725 | 0.68575 | 11661.18 |
| KYNG_ENV | 27 | -0.0474 | -0.00179 | 0.097909 | 0.68586 | 11663.05 |
| SCIAN_CEL | 9 | -0.07473 | -0.00163 | 0.15435 | 0.68586 | 11663.05 |
| GOBP_POS | 8 | -0.09509 | -0.00196 | 0.19631 | 0.68593 | 11664.24 |
| REACTOME | 7 | -0.09718 | -0.00187 | 0.20061 | 0.68595 | 11664.58 |
| GOBP_SPH | 109 | -0.02424 | -0.00184 | 0.050035 | 0.68599 | 11665.26 |
| REACTOME | 24 | -0.04702 | -0.00168 | 0.097043 | 0.686 | 11665.43 |
| GOCC_END | 41 | -0.03527 | -0.00164 | 0.072764 | 0.68604 | 11666.11 |
| WP_GLUC( | 68 | -0.03096 | -0.00185 | 0.063879 | 0.68605 | 11666.28 |
| GOBP_MU( | 8 | -0.09102 | -0.00187 | 0.18765 | 0.68617 | 11668.32 |
| REACTOME | 106 | -0.02375 | -0.00177 | 0.048964 | 0.68618 | 11668.49 |
| GOBP_P_B | 20 | -0.04696 | -0.00153 | 0.096798 | 0.68619 | 11668.66 |
| GOBP_ANT | 6 | -0.12047 | -0.00215 | 0.2483 | 0.68621 | 11669 |
| GOBP_ART | 5 | -0.10027 | -0.00163 | 0.20661 | 0.68626 | 11669.85 |
| GOCC_PRC | 10 | -0.09273 | -0.00213 | 0.19102 | 0.68632 | 11670.87 |
| GOBP_BON | 8 | -0.08884 | -0.00183 | 0.18288 | 0.68644 | 11672.91 |
| HOLLEMAN | 10 | -0.08263 | -0.0019 | 0.16996 | 0.68659 | 11675.46 |
| GOBP_PLA | 23 | -0.05422 | -0.00189 | 0.11147 | 0.68666 | 11676.65 |
| GOBP_NEG | 18 | -0.05521 | -0.0017 | 0.11344 | 0.68675 | 11678.18 |
| GOCC_PSE | 15 | -0.05736 | -0.00162 | 0.11783 | 0.68679 | 11678.86 |
| GOBP_TAC | 6 | -0.10282 | -0.00183 | 0.2112 | 0.6868 | 11679.03 |
| GOBP_POS | 14 | -0.06176 | -0.00168 | 0.12681 | 0.68687 | 11680.22 |
| WP_INITIA | 30 | -0.04572 | -0.00182 | 0.093861 | 0.6869 | 11680.73 |
| IBRAHIM_N | 507 | -0.01055 | -0.00171 | 0.02166 | 0.68696 | 11681.75 |
| GOBP_POS | 9 | -0.07771 | -0.0017 | 0.15944 | 0.687 | 11682.44 |
| BIOCARTA_ | 24 | -0.04586 | -0.00163 | 0.094065 | 0.68705 | 11683.29 |
| REACTOME | 8 | -0.0848 | -0.00175 | 0.17391 | 0.68707 | 11683.63 |
| MIKKELSEN | 57 | -0.0327 | -0.00179 | 0.067065 | 0.68708 | 11683.8 |
| REACTOME | 43 | -0.03654 | -0.00174 | 0.074917 | 0.68715 | 11684.99 |
| GOBP_NEG | 7 | -0.08751 | -0.00168 | 0.17927 | 0.68728 | 11687.2 |
| GOBP_GLY | 7 | -0.07237 | -0.00139 | 0.14821 | 0.68733 | 11688.05 |
| CHUANG_( | 26 | -0.04963 | -0.00184 | 0.10163 | 0.68736 | 11688.56 |
| GOBP_REG | 19 | -0.05245 | -0.00166 | 0.10739 | 0.68736 | 11688.56 |
| GOBP_NU( | 81 | -0.0243 | -0.00159 | 0.049719 | 0.68749 | 11690.77 |
| REACTOME | 51 | -0.0334 | -0.00173 | 0.068333 | 0.68751 | 11691.11 |
| ENK_UV_R | 493 | -0.01141 | -0.00182 | 0.023328 | 0.68761 | 11692.81 |
| GOBP_VAG | 9 | -0.08573 | -0.00187 | 0.17523 | 0.68766 | 11693.66 |

| | | | | | | |
|---|---|---|---|---|---|---|
| KEGG_PAR | 107 | -0.02109 | -0.00158 | 0.043089 | 0.68769 | 11694.17 |
| GOBP_MO | 934 | -0.00853 | -0.00185 | 0.017432 | 0.68772 | 11694.68 |
| GOCC_COF | 70 | -0.0284 | -0.00173 | 0.057995 | 0.68782 | 11696.38 |
| GOMF_GPI | 8 | -0.09199 | -0.00189 | 0.18771 | 0.68795 | 11698.59 |
| GOBP_DNA | 39 | -0.03788 | -0.00172 | 0.077203 | 0.68818 | 11702.5 |
| GOBP_POS | 18 | -0.06233 | -0.00192 | 0.1269 | 0.68835 | 11705.39 |
| GOBP_NEG | 5 | -0.11488 | -0.00187 | 0.23385 | 0.68837 | 11705.73 |
| GOCC_GOL | 605 | -0.00995 | -0.00175 | 0.02024 | 0.68845 | 11707.09 |
| GOMF_TRA | 10 | -0.06671 | -0.00154 | 0.13573 | 0.68846 | 11707.26 |
| GOMF_ARA | 10 | -0.07031 | -0.00162 | 0.143 | 0.68853 | 11708.45 |
| ZIRN_TRET | 5 | -0.10693 | -0.00174 | 0.21747 | 0.68853 | 11708.45 |
| GOBP_SAT | 5 | -0.10094 | -0.00164 | 0.20528 | 0.68854 | 11708.62 |
| GOBP_REN | 28 | -0.04929 | -0.0019 | 0.10018 | 0.68863 | 11710.15 |
| SHAFFER_I | 33 | -0.04842 | -0.00202 | 0.098402 | 0.68866 | 11710.66 |
| GOCC_PRE | 12 | -0.07701 | -0.00194 | 0.15643 | 0.68873 | 11711.85 |
| JOSEPH_RE | 59 | -0.03088 | -0.00172 | 0.062727 | 0.68873 | 11711.85 |
| GAUSSMAN | 10 | -0.07199 | -0.00166 | 0.14615 | 0.68885 | 11713.89 |
| GOBP_POS | 60 | -0.03075 | -0.00173 | 0.062412 | 0.68887 | 11714.23 |
| REACTOME | 125 | -0.02393 | -0.00194 | 0.048545 | 0.689 | 11716.45 |
| GOBP_NEG | 16 | -0.06122 | -0.00178 | 0.12412 | 0.68906 | 11717.47 |
| GOBP_NEG | 2 | -0.25457 | -0.00262 | 0.51596 | 0.68912 | 11718.49 |
| GOMF_2_C | 53 | -0.03048 | -0.00161 | 0.061732 | 0.68925 | 11720.7 |
| GOMF_OLI | 6 | -0.09298 | -0.00166 | 0.18826 | 0.6893 | 11721.55 |
| GOBP_REG | 29 | -0.04214 | -0.00165 | 0.085264 | 0.6894 | 11723.25 |
| GOMF_SUI | 24 | -0.04861 | -0.00173 | 0.098349 | 0.68944 | 11723.93 |
| GOBP_DET | 3 | -0.12444 | -0.00157 | 0.25174 | 0.68946 | 11724.27 |
| REACTOME | 20 | -0.05474 | -0.00178 | 0.11069 | 0.68953 | 11725.46 |
| REACTOME | 107 | -0.02085 | -0.00157 | 0.042161 | 0.68954 | 11725.63 |
| GOMF_UD | 11 | -0.07051 | -0.0017 | 0.14251 | 0.68961 | 11726.82 |
| PID_AVB3_ | 30 | -0.04651 | -0.00185 | 0.093996 | 0.68962 | 11726.99 |
| BIOCARTA_ | 18 | -0.05572 | -0.00172 | 0.11256 | 0.68971 | 11728.52 |
| GOBP_INSU | 186 | -0.01797 | -0.00177 | 0.036293 | 0.68971 | 11728.52 |
| GOBP_POS | 37 | -0.04333 | -0.00192 | 0.087505 | 0.68974 | 11729.03 |
| GOMF_LIP | 10 | -0.10024 | -0.00231 | 0.20241 | 0.68977 | 11729.54 |
| WP_HYPER | 19 | -0.06305 | -0.002 | 0.12731 | 0.68977 | 11729.54 |
| GOBP_TETI | 58 | -0.03032 | -0.00168 | 0.061221 | 0.6898 | 11730.05 |
| GOBP_GLU | 8 | -0.08487 | -0.00175 | 0.17131 | 0.68983 | 11730.56 |
| GOBP_AXC | 23 | -0.04913 | -0.00171 | 0.099162 | 0.68986 | 11731.07 |
| WP_SELECT | 25 | -0.05768 | -0.0021 | 0.11624 | 0.69013 | 11735.66 |
| GOBP_CELI | 8 | -0.09276 | -0.00191 | 0.18687 | 0.69018 | 11736.51 |
| GOMF_MC | 6 | -0.10568 | -0.00188 | 0.21284 | 0.69023 | 11737.36 |
| SMID_BREA | 22 | -0.05259 | -0.00179 | 0.10589 | 0.69026 | 11737.87 |
| GOBP_EPIT | 199 | -0.01832 | -0.00187 | 0.036881 | 0.69028 | 11738.21 |
| GOBP_POS | 11 | -0.08298 | -0.002 | 0.16699 | 0.69036 | 11739.57 |
| MULLIGAN | 21 | -0.05314 | -0.00177 | 0.10695 | 0.69036 | 11739.57 |

| | | | | | | |
|---|---|---|---|---|---|---|
| BIOCARTA_ | 4 | -0.10745 | -0.00156 | 0.21607 | 0.69051 | 11742.12 |
| GOCC_CELI | 292 | -0.01485 | -0.00183 | 0.02985 | 0.69056 | 11742.97 |
| GOBP_TRY | 9 | -0.09064 | -0.00198 | 0.1821 | 0.69066 | 11744.67 |
| GOCC_TIM | 12 | -0.07264 | -0.00183 | 0.14594 | 0.69066 | 11744.67 |
| GOBP_HIST | 58 | -0.03259 | -0.0018 | 0.065442 | 0.69074 | 11746.03 |
| TOMLINS_I | 39 | -0.04175 | -0.0019 | 0.083822 | 0.6908 | 11747.05 |
| GOBP_MIT | 7 | -0.07628 | -0.00147 | 0.15307 | 0.69088 | 11748.41 |
| GOBP_TOR | 125 | -0.0226 | -0.00183 | 0.045343 | 0.69088 | 11748.41 |
| PUIFFE_INV | 69 | -0.02928 | -0.00177 | 0.058662 | 0.69114 | 11752.84 |
| GOBP_REG | 5 | -0.14456 | -0.00235 | 0.2893 | 0.69135 | 11756.41 |
| RODRIGUE | 16 | -0.0644 | -0.00187 | 0.12886 | 0.69139 | 11757.09 |
| REACTOME | 11 | -0.06946 | -0.00168 | 0.13892 | 0.69145 | 11758.11 |
| REACTOME | 32 | -0.04522 | -0.00186 | 0.090344 | 0.69164 | 11761.34 |
| REACTOME | 5 | -0.0961 | -0.00156 | 0.19197 | 0.69167 | 11761.85 |
| CONCANN( | 152 | -0.02068 | -0.00185 | 0.041296 | 0.69176 | 11763.38 |
| GOBP_REG | 134 | -0.02118 | -0.00178 | 0.042288 | 0.69179 | 11763.89 |
| GOBP_PRE | 8 | -0.11251 | -0.00232 | 0.22454 | 0.69183 | 11764.57 |
| KHETCHOU | 7 | -0.09604 | -0.00185 | 0.19167 | 0.69184 | 11764.74 |
| HOSHIDA_I | 70 | -0.03124 | -0.0019 | 0.062315 | 0.69193 | 11766.27 |
| GOBP_REG | 6 | -0.10274 | -0.00183 | 0.2048 | 0.69204 | 11768.14 |
| GOBP_INTI | 108 | -0.02432 | -0.00183 | 0.04844 | 0.69216 | 11770.18 |
| GOBP_REG | 56 | -0.03843 | -0.00209 | 0.076546 | 0.6922 | 11770.86 |
| GOMF_ME | 6 | -0.11272 | -0.00201 | 0.22445 | 0.69224 | 11771.54 |
| LIM_MAMI | 58 | -0.03246 | -0.0018 | 0.064588 | 0.69236 | 11773.58 |
| PETROVA_ | 128 | -0.02266 | -0.00186 | 0.045079 | 0.69238 | 11773.92 |
| GOBP_CLA | 7 | -0.08266 | -0.00159 | 0.16439 | 0.69245 | 11775.11 |
| GOBP_MIC | 8 | -0.09256 | -0.0019 | 0.18398 | 0.69254 | 11776.64 |
| GOMF_TYF | 17 | -0.11232 | -0.00337 | 0.22326 | 0.69254 | 11776.64 |
| GOBP_NEG | 146 | -0.01997 | -0.00175 | 0.039668 | 0.69262 | 11778 |
| LE_NEURO | 16 | -0.06073 | -0.00177 | 0.12066 | 0.69262 | 11778 |
| LANDIS_ER | 56 | -0.03194 | -0.00174 | 0.063433 | 0.69267 | 11778.85 |
| GOBP_REG | 94 | -0.02804 | -0.00197 | 0.05568 | 0.6927 | 11779.36 |
| TORCHIA_T | 18 | -0.06037 | -0.00186 | 0.11988 | 0.69272 | 11779.7 |
| GOBP_REG | 7 | -0.11144 | -0.00215 | 0.22121 | 0.69278 | 11780.72 |
| GOBP_SKEI | 37 | -0.04083 | -0.00181 | 0.081027 | 0.69283 | 11781.57 |
| GOBP_NEU | 42 | -0.0382 | -0.0018 | 0.075748 | 0.69295 | 11783.61 |
| GOBP_EYEI | 14 | -0.07479 | -0.00204 | 0.1483 | 0.69297 | 11783.95 |
| KEGG_EPIT | 63 | -0.0302 | -0.00174 | 0.059855 | 0.69308 | 11785.83 |
| GOMF_ADI | 13 | -0.07257 | -0.0019 | 0.14369 | 0.69324 | 11788.55 |
| GOBP_REG | 378 | -0.01361 | -0.00191 | 0.02695 | 0.69327 | 11789.06 |
| GOBP_NEG | 7 | -0.09262 | -0.00178 | 0.18328 | 0.69334 | 11790.25 |
| GOMF_VIN | 11 | -0.07775 | -0.00188 | 0.1538 | 0.6934 | 11791.27 |
| GOBP_DEN | 20 | -0.05563 | -0.00181 | 0.11001 | 0.69345 | 11792.12 |
| SHETH_LIV | 98 | -0.02685 | -0.00193 | 0.053095 | 0.69348 | 11792.63 |
| GOBP_POS | 103 | -0.02476 | -0.00182 | 0.048949 | 0.6935 | 11792.97 |

| | | | | | | |
|---|---|---|---|---|---|---|
| GOMF_OXI | 5 | -0.12919 | -0.0021 | 0.25518 | 0.69366 | 11795.69 |
| GOBP_POL | 7 | -0.08416 | -0.00162 | 0.1662 | 0.6937 | 11796.37 |
| GOBP_INTI | 139 | -0.02141 | -0.00183 | 0.042276 | 0.69374 | 11797.05 |
| MATZUK_C | 29 | -0.04952 | -0.00194 | 0.097773 | 0.69374 | 11797.05 |
| GOBP_MIT | 7 | -0.08575 | -0.00165 | 0.16928 | 0.69377 | 11797.56 |
| GOBP_B_C | 25 | -0.04909 | -0.00179 | 0.096873 | 0.69382 | 11798.41 |
| GOBP_LON | 10 | -0.07787 | -0.00179 | 0.15368 | 0.69382 | 11798.41 |
| GOCC_CIS_ | 7 | -0.08672 | -0.00167 | 0.171 | 0.69397 | 11800.96 |
| GOBP_RIBO | 8 | -0.09416 | -0.00194 | 0.18551 | 0.69411 | 11803.34 |
| GOBP_REG | 10 | -0.09071 | -0.00209 | 0.17854 | 0.69429 | 11806.4 |
| GOBP_RES | 103 | -0.02293 | -0.00169 | 0.045125 | 0.69429 | 11806.4 |
| GOBP_AMI | 327 | -0.01433 | -0.00187 | 0.02821 | 0.6943 | 11806.57 |
| GOBP_CAL | 45 | -0.04146 | -0.00202 | 0.081586 | 0.69432 | 11806.91 |
| BLANCO_M | 14 | -0.07554 | -0.00206 | 0.14849 | 0.69452 | 11810.31 |
| REACTOME | 33 | -0.04363 | -0.00182 | 0.085764 | 0.69453 | 11810.48 |
| MARKEY_R | 112 | -0.024 | -0.00184 | 0.047153 | 0.69459 | 11811.5 |
| GOBP_CDC | 16 | -0.06465 | -0.00188 | 0.12699 | 0.69465 | 11812.52 |
| GOBP_BON | 88 | -0.02785 | -0.0019 | 0.054667 | 0.69479 | 11814.9 |
| GOBP_APC | 5 | -0.12746 | -0.00207 | 0.25013 | 0.69482 | 11815.41 |
| GOBP_CAL | 61 | -0.03307 | -0.00188 | 0.064855 | 0.69493 | 11817.28 |
| GOBP_ION_ | 10 | -0.09711 | -0.00223 | 0.19041 | 0.69496 | 11817.79 |
| REACTOME | 8 | -0.09037 | -0.00186 | 0.17709 | 0.69509 | 11820.01 |
| GOBP_MEI | 202 | -0.0187 | -0.00192 | 0.036625 | 0.69517 | 11821.37 |
| REACTOME | 4 | -0.14612 | -0.00213 | 0.28617 | 0.69518 | 11821.54 |
| FERRARI_R | 21 | -0.06432 | -0.00214 | 0.12597 | 0.69519 | 11821.71 |
| GOBP_REG | 56 | -0.03079 | -0.00167 | 0.060292 | 0.6952 | 11821.88 |
| REACTOME | 13 | -0.06942 | -0.00182 | 0.13592 | 0.69522 | 11822.22 |
| GOMF_FUC | 13 | -0.07006 | -0.00184 | 0.13714 | 0.69528 | 11823.24 |
| GOBP_NEG | 144 | -0.02135 | -0.00186 | 0.041789 | 0.69532 | 11823.92 |
| GOBP_N_A | 9 | -0.10004 | -0.00218 | 0.19577 | 0.69532 | 11823.92 |
| ELVIDGE_H | 65 | -0.0309 | -0.00181 | 0.0604 | 0.69552 | 11827.32 |
| WP_SEROT | 15 | -0.07818 | -0.0022 | 0.15282 | 0.69552 | 11827.32 |
| GOMF_ACY | 13 | -0.06106 | -0.0016 | 0.11933 | 0.69557 | 11828.17 |
| SA_REG_CA | 13 | -0.06714 | -0.00176 | 0.1312 | 0.69559 | 11828.51 |
| GOMF_SPH | 7 | -0.08692 | -0.00167 | 0.16976 | 0.69567 | 11829.87 |
| CORONEL_ | 40 | -0.04209 | -0.00194 | 0.082197 | 0.6957 | 11830.38 |
| GOMF_RRM | 4 | -0.11748 | -0.00171 | 0.22921 | 0.69587 | 11833.27 |
| GOMF_3_5 | 11 | -0.07178 | -0.00173 | 0.13998 | 0.69594 | 11834.46 |
| GOBP_POS | 62 | -0.03502 | -0.002 | 0.068287 | 0.69595 | 11834.63 |
| GOBP_CAR | 19 | -0.05578 | -0.00177 | 0.10873 | 0.69602 | 11835.82 |
| GOBP_POS | 144 | -0.02194 | -0.00191 | 0.042776 | 0.69602 | 11835.82 |
| TIAN_TNF_ | 26 | -0.05524 | -0.00205 | 0.10766 | 0.69605 | 11836.33 |
| GOBP_TRA | 10 | -0.06598 | -0.00152 | 0.12857 | 0.69608 | 11836.84 |
| KEGG_AMY | 50 | -0.03619 | -0.00186 | 0.070507 | 0.69612 | 11837.52 |
| GOBP_REN | 14 | -0.06955 | -0.00189 | 0.13536 | 0.6963 | 11840.58 |

| | | | | | | |
|---|---|---|---|---|---|---|
| GOBP_APC | 16 | -0.06633 | -0.00193 | 0.12908 | 0.69632 | 11840.92 |
| GOMF_CAI | 133 | -0.02294 | -0.00192 | 0.04461 | 0.69642 | 11842.62 |
| GOBP_CELI | 183 | -0.0204 | -0.002 | 0.039665 | 0.69645 | 11843.13 |
| GOBP_PUR | 4 | -0.12803 | -0.00186 | 0.24889 | 0.69652 | 11844.32 |
| GOBP_NEG | 6 | -0.11626 | -0.00207 | 0.22591 | 0.69659 | 11845.51 |
| GOBP_NEG | 9 | -0.08371 | -0.00183 | 0.16263 | 0.69663 | 11846.19 |
| GOBP_REG | 48 | -0.0372 | -0.00187 | 0.072225 | 0.69674 | 11848.06 |
| GOMF_HIS | 6 | -0.10105 | -0.0018 | 0.19618 | 0.69675 | 11848.23 |
| GOBP_I_KA | 16 | -0.06453 | -0.00188 | 0.12527 | 0.69677 | 11848.57 |
| GOBP_REG | 32 | -0.04471 | -0.00184 | 0.086678 | 0.69699 | 11852.31 |
| NUNODA_I | 11 | -0.07526 | -0.00182 | 0.14591 | 0.697 | 11852.49 |
| MEBARKI_I | 18 | -0.06856 | -0.00212 | 0.13288 | 0.69705 | 11853.34 |
| GOMF_NAI | 15 | -0.07492 | -0.00211 | 0.14514 | 0.69715 | 11855.04 |
| PELLICCIOT | 60 | -0.03155 | -0.00178 | 0.061087 | 0.69721 | 11856.06 |
| KIM_MYCN | 86 | -0.03064 | -0.00206 | 0.059271 | 0.69738 | 11858.95 |
| GOBP_NEG | 5 | -0.09999 | -0.00163 | 0.19343 | 0.69739 | 11859.12 |
| GOBP_SYN | 54 | -0.03656 | -0.00195 | 0.070679 | 0.69751 | 11861.16 |
| REACTOME | 14 | -0.06524 | -0.00178 | 0.12608 | 0.69757 | 11862.18 |
| GOMF_TFII | 10 | -0.07968 | -0.00183 | 0.15393 | 0.69765 | 11863.54 |
| GOBP_POL | 9 | -0.09479 | -0.00207 | 0.18307 | 0.69768 | 11864.05 |
| KEGG_BAS | 29 | -0.04757 | -0.00186 | 0.09185 | 0.69772 | 11864.73 |
| GOCC_TER | 44 | -0.041 | -0.00198 | 0.079177 | 0.69773 | 11864.9 |
| GOCC_ORG | 1372 | -0.00712 | -0.00185 | 0.013757 | 0.69774 | 11865.07 |
| GOMF_LYS | 4 | -0.11643 | -0.00169 | 0.22461 | 0.6979 | 11867.79 |
| GOBP_RES | 27 | -0.0566 | -0.00214 | 0.10906 | 0.69812 | 11871.53 |
| GOBP_ANA | 5 | -0.11243 | -0.00183 | 0.21648 | 0.69823 | 11873.4 |
| WP_SYNAF | 48 | -0.03708 | -0.00187 | 0.07138 | 0.69828 | 11874.25 |
| PID_CIRCA | 14 | -0.06437 | -0.00175 | 0.12392 | 0.69829 | 11874.42 |
| GOBP_POS | 12 | -0.09192 | -0.00232 | 0.17682 | 0.69841 | 11876.46 |
| GOBP_NEG | 46 | -0.0399 | -0.00197 | 0.076707 | 0.69852 | 11878.33 |
| GOMF_DN | 28 | -0.0485 | -0.00187 | 0.093182 | 0.69863 | 11880.2 |
| GOBP_ACT | 5 | -0.13326 | -0.00217 | 0.256 | 0.69865 | 11880.54 |
| GOBP_FAT | 19 | -0.05499 | -0.00174 | 0.10551 | 0.69886 | 11884.11 |
| LY_AGING_ | 16 | -0.06006 | -0.00175 | 0.11523 | 0.69889 | 11884.62 |
| REACTOME | 21 | -0.05788 | -0.00193 | 0.11101 | 0.69893 | 11885.3 |
| REACTOME | 6 | -0.097 | -0.00173 | 0.18604 | 0.69896 | 11885.81 |
| GOBP_T_C | 113 | -0.02637 | -0.00203 | 0.050574 | 0.69898 | 11886.15 |
| GOBP_POS | 9 | -0.11837 | -0.00258 | 0.2267 | 0.69922 | 11890.24 |
| REACTOME | 80 | -0.0266 | -0.00173 | 0.050927 | 0.69925 | 11890.75 |
| GOCC_CAT | 1561 | -0.00669 | -0.00184 | 0.012797 | 0.69934 | 11892.28 |
| GOBP_NEG | 16 | -0.06598 | -0.00192 | 0.12624 | 0.69939 | 11893.13 |
| GOBP_POS | 19 | -0.06623 | -0.0021 | 0.12657 | 0.69961 | 11896.87 |
| GOBP_MO | 11 | -0.07231 | -0.00174 | 0.13816 | 0.69962 | 11897.04 |
| VISALA_RE | 12 | -0.07725 | -0.00195 | 0.14755 | 0.69968 | 11898.06 |
| KEGG_GLY | 58 | -0.0375 | -0.00208 | 0.071629 | 0.69971 | 11898.57 |

| | | | | | | |
|---|---|---|---|---|---|---|
| PID_P38_M | 21 | -0.05795 | -0.00193 | 0.11067 | 0.69971 | 11898.57 |
| WONG_EN | 8 | -0.09028 | -0.00186 | 0.17242 | 0.69972 | 11898.74 |
| GOBP_REN | 23 | -0.05629 | -0.00196 | 0.10746 | 0.6998 | 11900.1 |
| GOBP_REG | 38 | -0.04059 | -0.00182 | 0.077481 | 0.69982 | 11900.44 |
| DORN_ADE | 27 | -0.04561 | -0.00172 | 0.086971 | 0.70002 | 11903.84 |
| BILD_SRC_ | 57 | -0.03415 | -0.00187 | 0.065058 | 0.70016 | 11906.22 |
| REACTOME | 6 | -0.09861 | -0.00176 | 0.18786 | 0.70017 | 11906.39 |
| GOMF_ORG | 31 | -0.05278 | -0.00214 | 0.10052 | 0.70022 | 11907.24 |
| PID_RHOA_ | 44 | -0.03849 | -0.00186 | 0.073292 | 0.70026 | 11907.92 |
| BOYAULT_I | 39 | -0.04046 | -0.00184 | 0.076993 | 0.70038 | 11909.96 |
| GOBP_TRA | 16 | -0.05347 | -0.00156 | 0.10175 | 0.7004 | 11910.3 |
| GOMF_SCA | 62 | -0.03525 | -0.00202 | 0.066951 | 0.70071 | 11915.57 |
| REACTOME | 10 | -0.0852 | -0.00196 | 0.16175 | 0.70081 | 11917.27 |
| GOBP_AMI | 122 | -0.02384 | -0.00191 | 0.045258 | 0.70083 | 11917.61 |
| GOBP_TAX | 561 | -0.01189 | -0.00202 | 0.022559 | 0.70084 | 11917.78 |
| RIZKI_TUM | 196 | -0.01969 | -0.002 | 0.037359 | 0.70093 | 11919.31 |
| LINDGREN_ | 301 | -0.01529 | -0.00192 | 0.028976 | 0.70111 | 11922.38 |
| GOBP_GOL | 5 | -0.10667 | -0.00174 | 0.20211 | 0.70117 | 11923.4 |
| GOBP_FOL | 15 | -0.06187 | -0.00174 | 0.11723 | 0.70118 | 11923.57 |
| ZHENG_FO | 26 | -0.05714 | -0.00212 | 0.10821 | 0.70128 | 11925.27 |
| GOBP_CELI | 410 | -0.01308 | -0.00191 | 0.024749 | 0.70142 | 11927.65 |
| AUNG_GAS | 47 | -0.03754 | -0.00187 | 0.07098 | 0.70155 | 11929.86 |
| SUZUKI_AM | 17 | -0.07093 | -0.00213 | 0.13406 | 0.70163 | 11931.22 |
| GOMF_NF_ | 30 | -0.04264 | -0.0017 | 0.080562 | 0.7017 | 11932.41 |
| GOMF_SOI | 12 | -0.07199 | -0.00181 | 0.13574 | 0.70205 | 11938.36 |
| GOBP_NOF | 11 | -0.09628 | -0.00232 | 0.18147 | 0.70214 | 11939.89 |
| WP_PHOTO | 23 | -0.05994 | -0.00209 | 0.11297 | 0.70214 | 11939.89 |
| GOBP_REG | 6 | -0.10044 | -0.00179 | 0.18928 | 0.70217 | 11940.4 |
| REACTOME | 71 | -0.02959 | -0.00181 | 0.055762 | 0.70217 | 11940.4 |
| BAUS_TFF2 | 26 | -0.04825 | -0.00179 | 0.09081 | 0.70238 | 11943.97 |
| ALONSO_M | 16 | -0.07171 | -0.00209 | 0.13497 | 0.7024 | 11944.31 |
| WP_COVID | 12 | -0.08675 | -0.00219 | 0.16325 | 0.70242 | 11944.65 |
| BROWNE_I | 270 | -0.01643 | -0.00195 | 0.030903 | 0.7025 | 11946.01 |
| GOBP_REG | 8 | -0.09349 | -0.00192 | 0.17583 | 0.70254 | 11946.69 |
| PILON_KLF | 457 | -0.01244 | -0.00191 | 0.02339 | 0.70258 | 11947.37 |
| IVANOVA_ | 278 | -0.01531 | -0.00184 | 0.028778 | 0.70264 | 11948.39 |
| WP_NICOT | 21 | -0.05966 | -0.00199 | 0.11214 | 0.70264 | 11948.39 |
| REACTOME | 5 | -0.13849 | -0.00225 | 0.26008 | 0.7028 | 11951.11 |
| GOBP_POS | 5 | -0.1326 | -0.00216 | 0.24895 | 0.70285 | 11951.96 |
| GOBP_POS | 5 | -0.15215 | -0.00248 | 0.28532 | 0.70307 | 11955.71 |
| KEGG_ETH | 33 | -0.04696 | -0.00196 | 0.088051 | 0.70309 | 11956.05 |
| REACTOME | 99 | -0.02629 | -0.0019 | 0.049279 | 0.70314 | 11956.9 |
| ONDER_CD | 58 | -0.03744 | -0.00207 | 0.070169 | 0.70315 | 11957.07 |
| GOBP_PAR | 6 | -0.09675 | -0.00172 | 0.18133 | 0.70317 | 11957.41 |
| GOBP_MYC | 39 | -0.04087 | -0.00186 | 0.076591 | 0.70319 | 11957.75 |

| | | | | | | |
|---|---|---|---|---|---|---|
| GOBP_ACT | 410 | -0.01346 | -0.00196 | 0.02522 | 0.70325 | 11958.77 |
| FRASOR_RI | 22 | -0.06314 | -0.00215 | 0.11823 | 0.70334 | 11960.3 |
| BIOCARTA_ | 4 | -0.15679 | -0.00228 | 0.29356 | 0.70336 | 11960.64 |
| BEIER_GLIO | 55 | -0.03505 | -0.00189 | 0.065623 | 0.70337 | 11960.81 |
| GOBP_MIC | 118 | -0.02291 | -0.00181 | 0.042836 | 0.70361 | 11964.89 |
| LA_MEN1_ | 21 | -0.05653 | -0.00188 | 0.10566 | 0.70368 | 11966.08 |
| GOBP_RES | 404 | -0.01356 | -0.00196 | 0.02532 | 0.70378 | 11967.78 |
| ONDER_CD | 233 | -0.01916 | -0.00212 | 0.035769 | 0.70386 | 11969.14 |
| WP_MICRO | 38 | -0.0453 | -0.00203 | 0.084554 | 0.70393 | 11970.33 |
| GOBP_NEG | 7 | -0.12292 | -0.00237 | 0.2294 | 0.70395 | 11970.67 |
| GOMF_OXI | 10 | -0.08942 | -0.00206 | 0.16687 | 0.70398 | 11971.18 |
| REACTOME | 9 | -0.10135 | -0.00221 | 0.18903 | 0.70407 | 11972.71 |
| REACTOME | 139 | -0.02209 | -0.00189 | 0.04117 | 0.70419 | 11974.75 |
| REACTOME | 4 | -0.11774 | -0.00171 | 0.21947 | 0.70419 | 11974.75 |
| GOBP_MO | 34 | -0.04064 | -0.00172 | 0.075725 | 0.70426 | 11975.94 |
| OUYANG_P | 20 | -0.0615 | -0.002 | 0.11453 | 0.70436 | 11977.64 |
| GOBP_CHO | 5 | -0.09838 | -0.0016 | 0.18289 | 0.70469 | 11983.25 |
| GOBP_PEP | 26 | -0.05746 | -0.00213 | 0.1068 | 0.70472 | 11983.76 |
| GOMF_CHA | 145 | -0.02266 | -0.00198 | 0.042084 | 0.70488 | 11986.48 |
| GOBP_T_H | 45 | -0.04264 | -0.00208 | 0.07917 | 0.7049 | 11986.82 |
| GOBP_SEM | 7 | -0.12525 | -0.00241 | 0.23255 | 0.70491 | 11986.99 |
| GOBP_SYN | 26 | -0.04957 | -0.00184 | 0.092015 | 0.70494 | 11987.5 |
| GOCC_SOD | 19 | -0.06612 | -0.0021 | 0.12272 | 0.70497 | 11988.01 |
| GOCC_GOL | 54 | -0.03159 | -0.00169 | 0.058605 | 0.70506 | 11989.55 |
| MA_MYELO | 41 | -0.0432 | -0.00201 | 0.080105 | 0.70515 | 11991.08 |
| GOBP_MAT | 56 | -0.0359 | -0.00195 | 0.066508 | 0.70531 | 11993.8 |
| GOBP_NEU | 102 | -0.02743 | -0.00201 | 0.050763 | 0.70551 | 11997.2 |
| WP_CHOLE | 44 | -0.03963 | -0.00191 | 0.073346 | 0.70552 | 11997.37 |
| FOURNIER_ | 22 | -0.05906 | -0.00202 | 0.10927 | 0.70558 | 11998.39 |
| GOBP_NEU | 10 | -0.08736 | -0.00201 | 0.16153 | 0.70568 | 12000.09 |
| GOBP_DNA | 9 | -0.06838 | -0.00149 | 0.12642 | 0.7057 | 12000.43 |
| GOBP_NEG | 49 | -0.03816 | -0.00194 | 0.070539 | 0.70575 | 12001.28 |
| REACTOME | 22 | -0.05435 | -0.00185 | 0.10044 | 0.70578 | 12001.79 |
| GOMF_GTP | 12 | -0.07541 | -0.0019 | 0.13935 | 0.70579 | 12001.96 |
| GOBP_REG | 10 | -0.09579 | -0.0022 | 0.17699 | 0.70581 | 12002.3 |
| BIOCARTA_ | 9 | -0.08165 | -0.00178 | 0.15052 | 0.70624 | 12009.61 |
| WP_TCELL_ | 87 | -0.03027 | -0.00205 | 0.055768 | 0.70635 | 12011.48 |
| REACTOME | 15 | -0.07668 | -0.00216 | 0.14124 | 0.70639 | 12012.16 |
| GOBP_MIT | 61 | -0.03408 | -0.00193 | 0.062776 | 0.7064 | 12012.33 |
| CHASSOT_S | 9 | -0.10264 | -0.00224 | 0.18899 | 0.70646 | 12013.35 |
| GOBP_DET | 130 | -0.024 | -0.00199 | 0.044191 | 0.70649 | 12013.86 |
| GOBP_REG | 6 | -0.11375 | -0.00203 | 0.20938 | 0.70653 | 12014.54 |
| GOBP_DUN | 6 | -0.10881 | -0.00194 | 0.20018 | 0.70662 | 12016.07 |
| GARGALOV | 9 | -0.09805 | -0.00214 | 0.18034 | 0.70667 | 12016.92 |
| GOBP_NEG | 107 | -0.02468 | -0.00185 | 0.045356 | 0.70678 | 12018.79 |

| | | | | | | |
|---|---|---|---|---|---|---|
| REACTOME | 15 | -0.0733 | -0.00207 | 0.13471 | 0.70682 | 12019.47 |
| REACTOME | 10 | -0.0976 | -0.00225 | 0.17935 | 0.70684 | 12019.81 |
| GOMF_PYF | 26 | -0.04874 | -0.00181 | 0.089567 | 0.70685 | 12019.98 |
| GOMF_G_I | 5 | -0.10279 | -0.00167 | 0.18887 | 0.70686 | 12020.15 |
| GOBP_ACT | 130 | -0.02529 | -0.00209 | 0.046448 | 0.70695 | 12021.68 |
| GOBP_CYC | 9 | -0.07727 | -0.00169 | 0.14177 | 0.70713 | 12024.75 |
| GOBP_POS | 15 | -0.06686 | -0.00188 | 0.12266 | 0.70715 | 12025.09 |
| REACTOME | 208 | -0.02092 | -0.00218 | 0.03837 | 0.70717 | 12025.43 |
| GOBP_REG | 6 | -0.0867 | -0.00155 | 0.15901 | 0.7072 | 12025.94 |
| GOBP_WAT | 36 | -0.04975 | -0.00217 | 0.091223 | 0.70724 | 12026.62 |
| GOBP_IMN | 12 | -0.08426 | -0.00212 | 0.15448 | 0.70726 | 12026.96 |
| GOMF_NU | 24 | -0.05093 | -0.00181 | 0.093298 | 0.70743 | 12029.85 |
| WP_PKCGA | 21 | -0.06307 | -0.0021 | 0.11552 | 0.70743 | 12029.85 |
| LIANG_SILE | 29 | -0.05618 | -0.0022 | 0.1029 | 0.70744 | 12030.02 |
| GOBP_INN | 36 | -0.04023 | -0.00176 | 0.07366 | 0.70752 | 12031.38 |
| IZADPANAI | 119 | -0.0247 | -0.00195 | 0.045208 | 0.70757 | 12032.23 |
| GOMF_ORG | 210 | -0.01913 | -0.00201 | 0.035021 | 0.70758 | 12032.4 |
| WP_WNT_ | 98 | -0.02767 | -0.00199 | 0.050617 | 0.70767 | 12033.93 |
| GOBP_PHE | 7 | -0.12398 | -0.00239 | 0.22673 | 0.70775 | 12035.29 |
| GOMF_ATF | 7 | -0.09934 | -0.00191 | 0.18164 | 0.70776 | 12035.46 |
| GOBP_FAT | 13 | -0.08044 | -0.00211 | 0.14705 | 0.70782 | 12036.48 |
| KUROKAWA | 4 | -0.10979 | -0.0016 | 0.20049 | 0.70801 | 12039.71 |
| GOBP_ORG | 35 | -0.04882 | -0.0021 | 0.089016 | 0.7083 | 12044.64 |
| GOBP_COC | 49 | -0.04093 | -0.00208 | 0.074621 | 0.70834 | 12045.32 |
| TURASHVIL | 4 | -0.13806 | -0.00201 | 0.25166 | 0.70835 | 12045.49 |
| REACTOME | 4 | -0.11878 | -0.00173 | 0.2165 | 0.70837 | 12045.83 |
| GOBP_POS | 9 | -0.08606 | -0.00188 | 0.15685 | 0.70838 | 12046 |
| GOBP_AUT | 18 | -0.06989 | -0.00216 | 0.12734 | 0.70845 | 12047.19 |
| GOBP_POS | 24 | -0.0564 | -0.00201 | 0.1027 | 0.70855 | 12048.89 |
| KUROZUMI | 19 | -0.06195 | -0.00196 | 0.11281 | 0.70856 | 12049.06 |
| REACTOME | 26 | -0.04848 | -0.0018 | 0.088179 | 0.70877 | 12052.63 |
| GOBP_VES | 8 | -0.11019 | -0.00227 | 0.20041 | 0.70878 | 12052.8 |
| GOBP_TOL | 4 | -0.15626 | -0.00227 | 0.28412 | 0.70883 | 12053.65 |
| GOBP_NUC | 10 | -0.09946 | -0.00229 | 0.18083 | 0.70884 | 12053.82 |
| WP_NODLI | 8 | -0.09083 | -0.00187 | 0.16506 | 0.70892 | 12055.18 |
| GOBP_REG | 17 | -0.06384 | -0.00191 | 0.11584 | 0.7092 | 12059.95 |
| REACTOME | 285 | -0.01751 | -0.00213 | 0.03176 | 0.70923 | 12060.46 |
| GOBP_REG | 19 | -0.06093 | -0.00193 | 0.11052 | 0.70927 | 12061.14 |
| LE_NEURO | 18 | -0.05506 | -0.0017 | 0.099869 | 0.7093 | 12061.65 |
| ZHAN_MUI | 44 | -0.04288 | -0.00207 | 0.07772 | 0.70942 | 12063.69 |
| WEBER_MI | 38 | -0.04554 | -0.00204 | 0.082484 | 0.70954 | 12065.73 |
| GOBP_REG | 8 | -0.10409 | -0.00214 | 0.18846 | 0.70962 | 12067.09 |
| RIZ_ERYTH | 26 | -0.05718 | -0.00212 | 0.10353 | 0.70962 | 12067.09 |
| GOMF_SEC | 15 | -0.06747 | -0.0019 | 0.12213 | 0.70967 | 12067.94 |
| GOBP_POS | 38 | -0.04906 | -0.0022 | 0.088748 | 0.70981 | 12070.32 |

| | | | | | | |
|---|---|---|---|---|---|---|
| GOMF_DEI | 34 | -0.04067 | -0.00172 | 0.073562 | 0.70983 | 12070.66 |
| GOBP_REG | 656 | -0.01144 | -0.0021 | 0.020684 | 0.70991 | 12072.02 |
| REACTOME | 12 | -0.07013 | -0.00177 | 0.12674 | 0.70996 | 12072.87 |
| GOBP_POS | 5 | -0.11842 | -0.00193 | 0.21403 | 0.70997 | 12073.04 |
| REACTOME | 67 | -0.03375 | -0.00201 | 0.060975 | 0.71001 | 12073.72 |
| MCCABE_H | 9 | -0.10684 | -0.00233 | 0.19298 | 0.71008 | 12074.91 |
| GOBP_REG | 6 | -0.13599 | -0.00242 | 0.24559 | 0.71011 | 12075.42 |
| GOBP_CELI | 67 | -0.03337 | -0.00198 | 0.060245 | 0.71019 | 12076.78 |
| RICKMAN_ | 367 | -0.01456 | -0.00201 | 0.026252 | 0.71037 | 12079.84 |
| GOBP_EAR | 13 | -0.06782 | -0.00178 | 0.12226 | 0.71045 | 12081.2 |
| REACTOME | 4 | -0.13107 | -0.00191 | 0.23619 | 0.71053 | 12082.56 |
| GOCC_AMI | 23 | -0.05793 | -0.00202 | 0.10436 | 0.71057 | 12083.24 |
| GOBP_RES | 11 | -0.08949 | -0.00216 | 0.1612 | 0.7106 | 12083.75 |
| GOCC_POS | 121 | -0.02737 | -0.00218 | 0.049284 | 0.71066 | 12084.77 |
| GOBP_RES | 34 | -0.04736 | -0.00201 | 0.085229 | 0.71077 | 12086.64 |
| GOBP_GAN | 25 | -0.05407 | -0.00197 | 0.097061 | 0.71125 | 12094.81 |
| GOMF_OLI | 15 | -0.08418 | -0.00237 | 0.15092 | 0.71149 | 12098.89 |
| ZHANG_PR | 49 | -0.04049 | -0.00206 | 0.072569 | 0.71153 | 12099.57 |
| KYNG_ENV | 33 | -0.05296 | -0.00221 | 0.094857 | 0.71167 | 12101.95 |
| BANDRES_ | 31 | -0.04713 | -0.00191 | 0.084414 | 0.7117 | 12102.46 |
| MCBRYAN_ | 6 | -0.12911 | -0.0023 | 0.23117 | 0.71174 | 12103.14 |
| GOBP_REG | 102 | -0.0288 | -0.00211 | 0.051563 | 0.71175 | 12103.31 |
| MARIADAS | 23 | -0.05742 | -0.002 | 0.1028 | 0.71175 | 12103.31 |
| PID_RETIN | 29 | -0.04974 | -0.00195 | 0.088997 | 0.71188 | 12105.52 |
| ABDULRAH | 13 | -0.09506 | -0.00249 | 0.17006 | 0.7119 | 12105.86 |
| GOBP_SER | 19 | -0.05869 | -0.00186 | 0.10497 | 0.71196 | 12106.88 |
| REACTOME | 7 | -0.14653 | -0.00282 | 0.26202 | 0.71199 | 12107.39 |
| ZHANG_RE | 14 | -0.07899 | -0.00215 | 0.14124 | 0.71202 | 12107.9 |
| MCLACHLA | 208 | -0.01993 | -0.00208 | 0.035604 | 0.71215 | 12110.11 |
| GOBP_CELI | 53 | -0.04057 | -0.00215 | 0.072465 | 0.71221 | 12111.13 |
| MAINA_HY | 6 | -0.1203 | -0.00214 | 0.21483 | 0.71224 | 12111.64 |
| REACTOME | 83 | -0.03204 | -0.00212 | 0.05721 | 0.71224 | 12111.64 |
| GOBP_NEG | 22 | -0.05893 | -0.00201 | 0.10523 | 0.71227 | 12112.15 |
| GOBP_TRN | 13 | -0.06981 | -0.00183 | 0.12459 | 0.71236 | 12113.68 |
| GOBP_DEN | 215 | -0.01942 | -0.00206 | 0.034661 | 0.71238 | 12114.02 |
| BIOCARTA_ | 16 | -0.07389 | -0.00215 | 0.1318 | 0.71248 | 12115.72 |
| GOCC_TOR | 10 | -0.09464 | -0.00218 | 0.1687 | 0.71258 | 12117.42 |
| APPIERTO_ | 37 | -0.04655 | -0.00206 | 0.082955 | 0.71263 | 12118.27 |
| WP_GLYCE | 22 | -0.05595 | -0.00191 | 0.099692 | 0.71266 | 12118.78 |
| GOBP_REG | 8 | -0.12308 | -0.00253 | 0.21911 | 0.71284 | 12121.84 |
| BIOCARTA_ | 8 | -0.11859 | -0.00244 | 0.21111 | 0.71286 | 12122.18 |
| FIGUEROA_ | 48 | -0.04015 | -0.00202 | 0.071424 | 0.713 | 12124.57 |
| ZHOU_PAN | 12 | -0.08446 | -0.00213 | 0.15018 | 0.71307 | 12125.76 |
| REACTOME | 277 | -0.01686 | -0.00203 | 0.02997 | 0.71308 | 12125.93 |
| REACTOME | 51 | -0.03932 | -0.00204 | 0.069912 | 0.7131 | 12126.27 |

| | | | | | | |
|---|---|---|---|---|---|---|
| GOMF_DE( | 5 | -0.12373 | -0.00201 | 0.21975 | 0.71329 | 12129.5 |
| SCHMIDT_I | 8 | -0.1161 | -0.00239 | 0.2062 | 0.71329 | 12129.5 |
| REACTOME | 9 | -0.08439 | -0.00184 | 0.14986 | 0.71331 | 12129.84 |
| ZHAN_MUI | 43 | -0.04239 | -0.00202 | 0.075272 | 0.71333 | 12130.18 |
| GOBP_NEG | 10 | -0.09131 | -0.0021 | 0.16209 | 0.71339 | 12131.2 |
| GOBP_HOI | 5 | -0.13407 | -0.00218 | 0.2379 | 0.71347 | 12132.56 |
| BOYLAN_M | 34 | -0.05259 | -0.00223 | 0.093277 | 0.71353 | 12133.58 |
| GOCC_ACR | 31 | -0.04977 | -0.00202 | 0.088265 | 0.71356 | 12134.09 |
| GOCC_PIGI | 105 | -0.02681 | -0.00199 | 0.04755 | 0.71359 | 12134.6 |
| GOBP_SM( | 107 | -0.02867 | -0.00215 | 0.050828 | 0.71361 | 12134.94 |
| GOBP_NEU | 12 | -0.07873 | -0.00198 | 0.13958 | 0.71364 | 12135.45 |
| GOBP_PRC | 4 | -0.20215 | -0.00294 | 0.35834 | 0.71366 | 12135.79 |
| REACTOME | 8 | -0.08001 | -0.00165 | 0.14181 | 0.7137 | 12136.47 |
| BROWN_M | 122 | -0.02519 | -0.00202 | 0.044599 | 0.71386 | 12139.19 |
| GOMF_ENZ | 540 | -0.01244 | -0.00207 | 0.022015 | 0.71399 | 12141.4 |
| GOBP_REG | 11 | -0.08111 | -0.00196 | 0.14348 | 0.71406 | 12142.59 |
| GOBP_ME( | 14 | -0.07817 | -0.00213 | 0.13827 | 0.71408 | 12142.93 |
| GOMF_RIB | 69 | -0.03381 | -0.00204 | 0.059779 | 0.71413 | 12143.78 |
| GOBP_POS | 33 | -0.05396 | -0.00225 | 0.09539 | 0.71418 | 12144.63 |
| LEE_AGING | 38 | -0.04434 | -0.00199 | 0.078391 | 0.71419 | 12144.8 |
| GOMF_CAI | 23 | -0.0535 | -0.00187 | 0.094559 | 0.71422 | 12145.31 |
| GOBP_THY | 6 | -0.12334 | -0.0022 | 0.21797 | 0.71424 | 12145.65 |
| GOCC_TRA | 378 | -0.01419 | -0.00199 | 0.025061 | 0.7143 | 12146.67 |
| ZEILSTRA_( | 8 | -0.11066 | -0.00228 | 0.19549 | 0.71433 | 12147.18 |
| REACTOME | 19 | -0.05785 | -0.00183 | 0.10209 | 0.71452 | 12150.41 |
| GOBP_MIT | 31 | -0.05155 | -0.00209 | 0.090962 | 0.71456 | 12151.09 |
| GOBP_REG | 80 | -0.02866 | -0.00186 | 0.050558 | 0.71461 | 12151.94 |
| GOBP_REG | 13 | -0.08491 | -0.00223 | 0.14967 | 0.71474 | 12154.15 |
| BLANCO_M | 191 | -0.02129 | -0.00213 | 0.037527 | 0.71478 | 12154.83 |
| MARCHINI_ | 17 | -0.0669 | -0.00201 | 0.11787 | 0.71483 | 12155.68 |
| GOMF_STF | 22 | -0.06404 | -0.00218 | 0.11277 | 0.71493 | 12157.38 |
| GOBP_POS | 62 | -0.03506 | -0.00201 | 0.061693 | 0.71507 | 12159.77 |
| WP_SMALI | 18 | -0.06909 | -0.00213 | 0.12154 | 0.71512 | 12160.62 |
| GOBP_POS | 7 | -0.11228 | -0.00216 | 0.19748 | 0.71517 | 12161.47 |
| GOBP_POS | 41 | -0.05297 | -0.00247 | 0.093152 | 0.7152 | 12161.98 |
| REACTOME | 7 | -0.08124 | -0.00156 | 0.14285 | 0.71521 | 12162.15 |
| GOMF_CO/ | 21 | -0.06061 | -0.00202 | 0.10646 | 0.7154 | 12165.38 |
| GOBP_NU( | 79 | -0.03121 | -0.00201 | 0.054769 | 0.71558 | 12168.44 |
| GOBP_PIGI | 11 | -0.08525 | -0.00206 | 0.14958 | 0.71564 | 12169.46 |
| GOBP_RNA | 5 | -0.11034 | -0.0018 | 0.19357 | 0.71567 | 12169.97 |
| GOCC_PHC | 13 | -0.0966 | -0.00253 | 0.16942 | 0.7157 | 12170.48 |
| GOBP_DEL | 6 | -0.11867 | -0.00212 | 0.20809 | 0.71576 | 12171.5 |
| GOBP_CELI | 14 | -0.07332 | -0.002 | 0.12854 | 0.71578 | 12171.84 |
| GOMF_SUI | 8 | -0.10046 | -0.00207 | 0.17605 | 0.71587 | 12173.37 |
| PARK_TRET | 22 | -0.06899 | -0.00235 | 0.12085 | 0.71596 | 12174.9 |

| | | | | | | |
|---|---|---|---|---|---|---|
| KORKOLA_ | 20 | -0.06076 | -0.00198 | 0.10639 | 0.71605 | 12176.43 |
| GOCC_PRE | 10 | -0.10209 | -0.00235 | 0.17872 | 0.71606 | 12176.6 |
| GOBP_PRC | 18 | -0.07224 | -0.00223 | 0.12646 | 0.7161 | 12177.28 |
| HUANG_G/ | 142 | -0.02425 | -0.0021 | 0.042437 | 0.71612 | 12177.62 |
| BIOCARTA_ | 11 | -0.1029 | -0.00248 | 0.17988 | 0.71635 | 12181.53 |
| GOCC_TER | 155 | -0.02292 | -0.00207 | 0.040069 | 0.71637 | 12181.87 |
| MAGRANG | 36 | -0.05043 | -0.0022 | 0.088155 | 0.71637 | 12181.87 |
| GOMF_PH( | 15 | -0.08378 | -0.00236 | 0.14639 | 0.71644 | 12183.06 |
| GOBP_CAN | 6 | -0.09628 | -0.00172 | 0.16814 | 0.71653 | 12184.59 |
| REACTOME | 29 | -0.05844 | -0.00229 | 0.10203 | 0.7166 | 12185.78 |
| PID_FANC( | 45 | -0.04066 | -0.00198 | 0.070926 | 0.71674 | 12188.16 |
| GOBP_MES | 11 | -0.08742 | -0.00211 | 0.15248 | 0.71678 | 12188.84 |
| GOCC_PHC | 3 | -0.17081 | -0.00215 | 0.29788 | 0.71681 | 12189.35 |
| GOMF_HIS | 21 | -0.05565 | -0.00186 | 0.096959 | 0.71701 | 12192.76 |
| GOBP_CHC | 7 | -0.12755 | -0.00246 | 0.22219 | 0.71702 | 12192.93 |
| PID_DELTA | 47 | -0.04151 | -0.00207 | 0.072282 | 0.71708 | 12193.95 |
| GOBP_LYM | 8 | -0.10287 | -0.00212 | 0.17911 | 0.71712 | 12194.63 |
| TARTE_PLA | 74 | -0.03322 | -0.00208 | 0.057833 | 0.71714 | 12194.97 |
| REACTOME | 64 | -0.0353 | -0.00205 | 0.061436 | 0.71718 | 12195.65 |
| WP_GANG | 12 | -0.07684 | -0.00194 | 0.13369 | 0.71725 | 12196.84 |
| BOYAULT_I | 51 | -0.03952 | -0.00205 | 0.068724 | 0.71736 | 12198.71 |
| GOMF_INT | 19 | -0.07864 | -0.00249 | 0.13675 | 0.71737 | 12198.88 |
| KYNG_ENV | 21 | -0.06875 | -0.00229 | 0.11949 | 0.71747 | 12200.58 |
| GOBP_NAI | 11 | -0.0911 | -0.0022 | 0.15833 | 0.71748 | 12200.75 |
| REACTOME | 40 | -0.04308 | -0.00198 | 0.074816 | 0.71762 | 12203.13 |
| GOBP_REG | 129 | -0.0254 | -0.00209 | 0.044096 | 0.71768 | 12204.15 |
| GOMF_ELE | 104 | -0.02555 | -0.00189 | 0.044321 | 0.71784 | 12206.87 |
| GOMF_POI | 7 | -0.09206 | -0.00177 | 0.15966 | 0.71791 | 12208.06 |
| GOBP_DIC/ | 94 | -0.02852 | -0.00201 | 0.049449 | 0.71796 | 12208.91 |
| GOBP_PSEI | 16 | -0.07143 | -0.00208 | 0.12371 | 0.71817 | 12212.48 |
| GOMF_CYT | 9 | -0.09198 | -0.00201 | 0.15928 | 0.71818 | 12212.65 |
| IVANOVA_ | 202 | -0.02084 | -0.00214 | 0.036074 | 0.71824 | 12213.67 |
| HOLLEMAN | 8 | -0.09422 | -0.00194 | 0.1631 | 0.71825 | 12213.84 |
| BIOCARTA_ | 8 | -0.09417 | -0.00194 | 0.16296 | 0.71833 | 12215.2 |
| GOBP_SYN | 51 | -0.03743 | -0.00194 | 0.064752 | 0.71837 | 12215.88 |
| AMBROSIN | 104 | -0.02973 | -0.0022 | 0.051422 | 0.71844 | 12217.07 |
| GOBP_POS | 19 | -0.06113 | -0.00194 | 0.10565 | 0.71858 | 12219.45 |
| WP_NOTCI | 57 | -0.03899 | -0.00214 | 0.067334 | 0.71871 | 12221.66 |
| VETTER_TA | 15 | -0.0711 | -0.002 | 0.12262 | 0.71897 | 12226.08 |
| KEGG_DOF | 23 | -0.06654 | -0.00232 | 0.11475 | 0.71899 | 12226.42 |
| GOMF_RN/ | 69 | -0.03466 | -0.00209 | 0.059749 | 0.71905 | 12227.45 |
| GOBP_REG | 5 | -0.14265 | -0.00232 | 0.24592 | 0.71906 | 12227.62 |
| GOBP_RES | 18 | -0.06569 | -0.00203 | 0.1131 | 0.71929 | 12231.53 |
| SCHAEFFEF | 7 | -0.11008 | -0.00212 | 0.18954 | 0.71929 | 12231.53 |
| BRIDEAU_I | 61 | -0.04378 | -0.00248 | 0.075362 | 0.71935 | 12232.55 |

| | | | | | | |
|---|---|---|---|---|---|---|
| GOMF_SUI | 12 | -0.08452 | -0.00213 | 0.14542 | 0.71944 | 12234.08 |
| WP_MRNA | 7 | -0.0905 | -0.00174 | 0.15571 | 0.71944 | 12234.08 |
| GOBP_REG | 28 | -0.05583 | -0.00215 | 0.096031 | 0.7195 | 12235.1 |
| GOBP_POS | 161 | -0.02326 | -0.00214 | 0.03998 | 0.71961 | 12236.97 |
| GOBP_SPIN | 61 | -0.03667 | -0.00208 | 0.063031 | 0.71961 | 12236.97 |
| GOBP_ANG | 12 | -0.09029 | -0.00228 | 0.15518 | 0.71965 | 12237.65 |
| HOLLEMAN | 12 | -0.07568 | -0.00191 | 0.13007 | 0.71965 | 12237.65 |
| LANDIS_BR | 43 | -0.0472 | -0.00225 | 0.081113 | 0.71968 | 12238.16 |
| GOMF_ALF | 5 | -0.15392 | -0.0025 | 0.26442 | 0.71975 | 12239.35 |
| GOBP_POS | 16 | -0.07363 | -0.00214 | 0.12647 | 0.71977 | 12239.69 |
| GOBP_REG | 24 | -0.06255 | -0.00223 | 0.10739 | 0.71985 | 12241.05 |
| GOMF_TOX | 10 | -0.11403 | -0.00262 | 0.19578 | 0.71986 | 12241.22 |
| SLEBOS_HE | 70 | -0.03433 | -0.00209 | 0.058933 | 0.71987 | 12241.39 |
| GOBP_MIC | 149 | -0.02321 | -0.00205 | 0.039832 | 0.71997 | 12243.09 |
| WP_AMINO | 7 | -0.1088 | -0.00209 | 0.18669 | 0.71997 | 12243.09 |
| GOBP_REC | 10 | -0.0847 | -0.00195 | 0.14529 | 0.72004 | 12244.28 |
| GOBP_SYN | 6 | -0.09848 | -0.00176 | 0.16892 | 0.72005 | 12244.45 |
| GOMF_ACE | 4 | -0.17856 | -0.0026 | 0.30622 | 0.72009 | 12245.13 |
| WP_NEPHF | 44 | -0.04504 | -0.00217 | 0.077225 | 0.72012 | 12245.64 |
| GOBP_REG | 331 | -0.01649 | -0.00216 | 0.028262 | 0.72026 | 12248.02 |
| WP_UREA_ | 23 | -0.05871 | -0.00205 | 0.10059 | 0.72026 | 12248.02 |
| GOBP_SECI | 12 | -0.07979 | -0.00201 | 0.13669 | 0.72029 | 12248.53 |
| REACTOME | 139 | -0.02495 | -0.00213 | 0.042742 | 0.72029 | 12248.53 |
| GOBP_REG | 18 | -0.07017 | -0.00217 | 0.1202 | 0.72031 | 12248.87 |
| REACTOME | 14 | -0.08078 | -0.0022 | 0.13832 | 0.72038 | 12250.06 |
| REACTOME | 20 | -0.06232 | -0.00203 | 0.10665 | 0.72049 | 12251.93 |
| GOBP_INTE | 20 | -0.06742 | -0.00219 | 0.11538 | 0.7205 | 12252.1 |
| SHAFFER_I | 61 | -0.039 | -0.00221 | 0.066737 | 0.72051 | 12252.27 |
| ABRAHAM_ | 20 | -0.0609 | -0.00198 | 0.1041 | 0.72072 | 12255.84 |
| GOMF_CAF | 44 | -0.04592 | -0.00221 | 0.078462 | 0.72079 | 12257.03 |
| GOBP_NAD | 43 | -0.04264 | -0.00203 | 0.072847 | 0.72085 | 12258.05 |
| GOBP_CELI | 27 | -0.05422 | -0.00205 | 0.092607 | 0.72088 | 12258.56 |
| GOBP_POS | 18 | -0.07782 | -0.0024 | 0.1329 | 0.7209 | 12258.9 |
| GOBP_END | 20 | -0.06059 | -0.00197 | 0.1034 | 0.72105 | 12261.46 |
| GOBP_RES | 12 | -0.09863 | -0.00249 | 0.16826 | 0.72112 | 12262.65 |
| GOMF_MC | 11 | -0.08069 | -0.00195 | 0.13765 | 0.72112 | 12262.65 |
| WANG_PRO | 59 | -0.03718 | -0.00208 | 0.063428 | 0.72112 | 12262.65 |
| GOCC_PRE | 9 | -0.07823 | -0.00171 | 0.13342 | 0.72118 | 12263.67 |
| CHARAFE_I | 428 | -0.01533 | -0.00228 | 0.026126 | 0.72125 | 12264.86 |
| GOBP_HEN | 9 | -0.10095 | -0.0022 | 0.17207 | 0.72128 | 12265.37 |
| REACTOME | 75 | -0.03285 | -0.00207 | 0.055969 | 0.7214 | 12267.41 |
| GOBP_SNR | 8 | -0.10229 | -0.00211 | 0.17408 | 0.7216 | 12270.81 |
| XU_GH1_E | 95 | -0.03014 | -0.00213 | 0.051266 | 0.72169 | 12272.34 |
| GOCC_SIGI | 5 | -0.11806 | -0.00192 | 0.20081 | 0.7217 | 12272.51 |
| INAMURA_ | 15 | -0.08482 | -0.00239 | 0.14427 | 0.72172 | 12272.85 |

| | | | | | | |
|---|---|---|---|---|---|---|
| YAMASHIT/ | 60 | -0.03722 | -0.00209 | 0.063299 | 0.72172 | 12272.85 |
| GOCC_CHI | 19 | -0.07188 | -0.00228 | 0.12223 | 0.72175 | 12273.36 |
| ALCALAY_/ | 130 | -0.02771 | -0.00229 | 0.047119 | 0.72177 | 12273.7 |
| GOBP_NEU | 19 | -0.05698 | -0.00181 | 0.096843 | 0.72186 | 12275.23 |
| GOBP_PRC | 24 | -0.05864 | -0.00209 | 0.099644 | 0.72189 | 12275.74 |
| GOBP_RES | 231 | -0.01974 | -0.00217 | 0.033551 | 0.7219 | 12275.91 |
| GOBP_PRC | 62 | -0.03635 | -0.00208 | 0.06175 | 0.72196 | 12276.93 |
| LEE_LIVER_ | 61 | -0.03882 | -0.0022 | 0.065931 | 0.72198 | 12277.27 |
| GOBP_REG | 19 | -0.07548 | -0.00239 | 0.12801 | 0.72227 | 12282.2 |
| GOBP_HIST | 6 | -0.121 | -0.00216 | 0.20519 | 0.7223 | 12282.71 |
| GOCC_COF | 13 | -0.0719 | -0.00189 | 0.12182 | 0.72245 | 12285.26 |
| HOLLERN_I | 43 | -0.04465 | -0.00213 | 0.075643 | 0.72247 | 12285.6 |
| GOCC_SNC | 23 | -0.05811 | -0.00203 | 0.098452 | 0.72248 | 12285.77 |
| GOMF_ACY | 5 | -0.15542 | -0.00253 | 0.26326 | 0.72253 | 12286.62 |
| KORKOLA_ | 12 | -0.08316 | -0.0021 | 0.14075 | 0.72267 | 12289 |
| GOMF_ALF | 35 | -0.05065 | -0.00218 | 0.085707 | 0.72272 | 12289.85 |
| GOBP_CELI | 7 | -0.09274 | -0.00179 | 0.15678 | 0.72291 | 12293.08 |
| GOBP_REG | 6 | -0.14733 | -0.00263 | 0.24905 | 0.72293 | 12293.42 |
| WP_INTERI | 44 | -0.04483 | -0.00216 | 0.075776 | 0.72294 | 12293.59 |
| GOBP_NEU | 8 | -0.08205 | -0.00169 | 0.13864 | 0.723 | 12294.62 |
| BIOCARTA_ | 14 | -0.07752 | -0.00211 | 0.13073 | 0.7234 | 12301.42 |
| GOBP_HEN | 36 | -0.04899 | -0.00214 | 0.082611 | 0.72341 | 12301.59 |
| GOBP_PRC | 14 | -0.0837 | -0.00228 | 0.14101 | 0.7236 | 12304.82 |
| WALLACE_ | 252 | -0.01963 | -0.00225 | 0.033056 | 0.72371 | 12306.69 |
| TAKEDA_T/ | 30 | -0.05632 | -0.00224 | 0.094807 | 0.72375 | 12307.37 |
| GOBP_MIT | 31 | -0.05362 | -0.00217 | 0.090257 | 0.72377 | 12307.71 |
| HOLLEMAN | 16 | -0.07046 | -0.00205 | 0.11849 | 0.72396 | 12310.94 |
| KEGG_PRO | 31 | -0.05764 | -0.00233 | 0.096912 | 0.72401 | 12311.79 |
| REACTOME | 5 | -0.12392 | -0.00202 | 0.20831 | 0.72403 | 12312.13 |
| GOBP_MU | 26 | -0.07004 | -0.0026 | 0.11763 | 0.72421 | 12315.19 |
| GOBP_CHR | 79 | -0.03204 | -0.00207 | 0.053789 | 0.72426 | 12316.04 |
| WP_MITOC | 48 | -0.03897 | -0.00196 | 0.065381 | 0.72441 | 12318.59 |
| GOBP_ANT | 5 | -0.22422 | -0.00365 | 0.37591 | 0.72456 | 12321.14 |
| GOMF_LIPI | 5 | -0.22422 | -0.00365 | 0.37591 | 0.72456 | 12321.14 |
| KUMAR_AU | 64 | -0.036 | -0.00209 | 0.060315 | 0.72468 | 12323.18 |
| GOBP_ANI | 61 | -0.03958 | -0.00225 | 0.066319 | 0.7247 | 12323.52 |
| GOMF_AN( | 7 | -0.09611 | -0.00185 | 0.16095 | 0.72479 | 12325.05 |
| WP_ALLOG | 64 | -0.04178 | -0.00243 | 0.06996 | 0.72479 | 12325.05 |
| BASSO_CD | 87 | -0.03192 | -0.00216 | 0.053403 | 0.725 | 12328.63 |
| GOBP_UDF | 6 | -0.13259 | -0.00236 | 0.22168 | 0.72511 | 12330.5 |
| GOMF_ACI | 30 | -0.05558 | -0.00221 | 0.092897 | 0.72519 | 12331.86 |
| GOBP_REG | 7 | -0.10481 | -0.00202 | 0.17509 | 0.72528 | 12333.39 |
| KEGG_DRU | 47 | -0.04898 | -0.00244 | 0.081798 | 0.72535 | 12334.58 |
| REACTOME | 10 | -0.0712 | -0.00164 | 0.11883 | 0.72547 | 12336.62 |
| GOMF_NEU | 39 | -0.05586 | -0.00254 | 0.093197 | 0.72554 | 12337.81 |

| | | | | | | |
|---|---|---|---|---|---|---|
| GOBP_NEG | 5 | -0.13338 | -0.00217 | 0.22239 | 0.72566 | 12339.85 |
| GOBP_REG | 43 | -0.04449 | -0.00212 | 0.07412 | 0.72583 | 12342.74 |
| RICKMAN_ | 95 | -0.03448 | -0.00244 | 0.057411 | 0.72594 | 12344.61 |
| GOBP_REG | 4 | -0.12122 | -0.00176 | 0.20175 | 0.72603 | 12346.14 |
| FARMER_B | 19 | -0.0665 | -0.00211 | 0.11055 | 0.72625 | 12349.88 |
| GOMF_INC | 6 | -0.10265 | -0.00183 | 0.17053 | 0.7264 | 12352.43 |
| WU_SILEN | 49 | -0.04429 | -0.00225 | 0.073548 | 0.72647 | 12353.62 |
| GOBP_NEG | 6 | -0.14578 | -0.0026 | 0.24177 | 0.72673 | 12358.04 |
| WP_NEURO | 133 | -0.02759 | -0.00231 | 0.045743 | 0.7268 | 12359.23 |
| BIOCARTA_ | 7 | -0.10021 | -0.00193 | 0.16612 | 0.72682 | 12359.57 |
| BAELDE_DI | 80 | -0.03432 | -0.00223 | 0.056879 | 0.72688 | 12360.59 |
| GOBP_NEG | 3 | -0.18411 | -0.00232 | 0.305 | 0.72695 | 12361.78 |
| GOBP_POS | 8 | -0.08984 | -0.00185 | 0.14883 | 0.72696 | 12361.95 |
| MEISSNER_ | 28 | -0.05284 | -0.00203 | 0.087536 | 0.72696 | 12361.95 |
| GOBP_NEG | 25 | -0.06689 | -0.00243 | 0.11078 | 0.72701 | 12362.81 |
| GOBP_NEG | 14 | -0.08528 | -0.00232 | 0.1412 | 0.72707 | 12363.83 |
| GOBP_QUA | 27 | -0.06563 | -0.00248 | 0.10865 | 0.7271 | 12364.34 |
| REACTOME | 101 | -0.03016 | -0.0022 | 0.049911 | 0.72713 | 12364.85 |
| SANDERSO | 15 | -0.07903 | -0.00223 | 0.13063 | 0.7274 | 12369.44 |
| WP_OSTEC | 12 | -0.08614 | -0.00217 | 0.14237 | 0.72741 | 12369.61 |
| PID_PDGFF | 21 | -0.06195 | -0.00207 | 0.10236 | 0.72748 | 12370.8 |
| GOBP_NEG | 5 | -0.15772 | -0.00257 | 0.26057 | 0.7275 | 12371.14 |
| GOMF_PRO | 58 | -0.03993 | -0.00221 | 0.065957 | 0.72753 | 12371.65 |
| KORKOLA_ | 11 | -0.0904 | -0.00218 | 0.14931 | 0.72754 | 12371.82 |
| GOBP_REG | 97 | -0.0313 | -0.00224 | 0.051686 | 0.72758 | 12372.5 |
| GOBP_REG | 10 | -0.0784 | -0.0018 | 0.12947 | 0.72758 | 12372.5 |
| WP_THYRO | 91 | -0.0323 | -0.00224 | 0.053343 | 0.72758 | 12372.5 |
| GOMF_STR | 22 | -0.06562 | -0.00224 | 0.10837 | 0.72759 | 12372.67 |
| GOCC_CAL | 6 | -0.13658 | -0.00243 | 0.2254 | 0.72773 | 12375.05 |
| NAGY_PCA | 8 | -0.09975 | -0.00205 | 0.1646 | 0.72774 | 12375.22 |
| GOBP_POS | 21 | -0.06215 | -0.00207 | 0.10252 | 0.72779 | 12376.07 |
| GOCC_IMN | 7 | -0.0957 | -0.00184 | 0.15777 | 0.72793 | 12378.45 |
| GOBP_NEG | 25 | -0.06054 | -0.0022 | 0.099806 | 0.72794 | 12378.62 |
| GOBP_POS | 7 | -0.097 | -0.00187 | 0.1599 | 0.72795 | 12378.79 |
| GOMF_PRO | 7 | -0.11268 | -0.00217 | 0.18573 | 0.72796 | 12378.96 |
| GOBP_REG | 7 | -0.08639 | -0.00166 | 0.14238 | 0.72798 | 12379.3 |
| BIOCARTA_ | 19 | -0.06198 | -0.00197 | 0.10206 | 0.72818 | 12382.7 |
| GOBP_REP | 43 | -0.05193 | -0.00248 | 0.085407 | 0.7284 | 12386.44 |
| REACTOME | 15 | -0.082 | -0.00231 | 0.13482 | 0.72847 | 12387.63 |
| GOBP_ENT | 151 | -0.02522 | -0.00225 | 0.041439 | 0.72858 | 12389.5 |
| GOBP_PUL | 7 | -0.12178 | -0.00234 | 0.20005 | 0.72865 | 12390.69 |
| GOBP_INTI | 50 | -0.04127 | -0.00212 | 0.06779 | 0.72868 | 12391.2 |
| GOBP_CELI | 86 | -0.03577 | -0.00241 | 0.058756 | 0.72869 | 12391.37 |
| REACTOME | 6 | -0.12817 | -0.00228 | 0.21049 | 0.72869 | 12391.37 |
| GOMF_1_F | 7 | -0.12185 | -0.00235 | 0.20006 | 0.72876 | 12392.56 |

| | | | | | | |
|---|---|---|---|---|---|---|
| GOBP_L_LY | 8 | -0.1241 | -0.00255 | 0.20358 | 0.72893 | 12395.45 |
| REACTOME | 15 | -0.08142 | -0.00229 | 0.13356 | 0.72893 | 12395.45 |
| WP_SUPRE | 7 | -0.10382 | -0.002 | 0.17025 | 0.72899 | 12396.47 |
| KEGG_RIG_ | 65 | -0.04209 | -0.00247 | 0.068987 | 0.72909 | 12398.18 |
| GOBP_DET | 20 | -0.06333 | -0.00206 | 0.10377 | 0.72915 | 12399.2 |
| GOBP_CELI | 149 | -0.02763 | -0.00244 | 0.045261 | 0.72921 | 12400.22 |
| GOBP_EPH | 49 | -0.04603 | -0.00234 | 0.075397 | 0.72921 | 12400.22 |
| GOBP_RES | 22 | -0.06833 | -0.00233 | 0.11194 | 0.72921 | 12400.22 |
| REACTOME | 10 | -0.12272 | -0.00282 | 0.20101 | 0.72923 | 12400.56 |
| GOMF_LIG | 158 | -0.02387 | -0.00217 | 0.039094 | 0.72929 | 12401.58 |
| GOBP_MEI | 89 | -0.03053 | -0.00209 | 0.049988 | 0.72931 | 12401.92 |
| GOCC_TRA | 11 | -0.10718 | -0.00259 | 0.17531 | 0.72952 | 12405.49 |
| GOBP_CRA | 9 | -0.10637 | -0.00232 | 0.17397 | 0.72955 | 12406 |
| BIOCARTA_ | 8 | -0.12081 | -0.00249 | 0.19755 | 0.72957 | 12406.34 |
| WP_METAI | 21 | -0.06958 | -0.00232 | 0.11373 | 0.72967 | 12408.04 |
| GOBP_DTT | 5 | -0.11752 | -0.00191 | 0.19194 | 0.72981 | 12410.42 |
| WP_PHOT( | 33 | -0.05071 | -0.00212 | 0.082786 | 0.72992 | 12412.29 |
| GOBP_INTI | 10 | -0.10679 | -0.00246 | 0.17424 | 0.73003 | 12414.16 |
| GOBP_SNR | 52 | -0.03837 | -0.00201 | 0.062593 | 0.73006 | 12414.67 |
| WP_MOLYI | 7 | -0.11551 | -0.00222 | 0.18838 | 0.73011 | 12415.52 |
| GOCC_PER | 4 | -0.14289 | -0.00208 | 0.23295 | 0.73019 | 12416.88 |
| KEGG_OTH | 15 | -0.06418 | -0.00181 | 0.10461 | 0.73021 | 12417.22 |
| GOBP_MO | 448 | -0.01523 | -0.00232 | 0.024823 | 0.73028 | 12418.41 |
| GOBP_MIT | 79 | -0.0332 | -0.00214 | 0.054063 | 0.73043 | 12420.96 |
| WP_LDL_IN | 24 | -0.06002 | -0.00214 | 0.097725 | 0.73044 | 12421.13 |
| GOBP_REG | 19 | -0.06374 | -0.00202 | 0.10378 | 0.73045 | 12421.3 |
| XU_AKT1_ | 9 | -0.11823 | -0.00258 | 0.19245 | 0.7305 | 12422.15 |
| WP_AGER/ | 63 | -0.03754 | -0.00217 | 0.061075 | 0.73061 | 12424.02 |
| KYNG_DNA | 70 | -0.03761 | -0.00229 | 0.061174 | 0.73067 | 12425.04 |
| GOBP_POS | 83 | -0.03302 | -0.00218 | 0.053704 | 0.73069 | 12425.38 |
| GOCC_CUL | 22 | -0.07116 | -0.00243 | 0.11568 | 0.73077 | 12426.74 |
| REACTOME | 33 | -0.05304 | -0.00222 | 0.086223 | 0.73077 | 12426.74 |
| PID_TAP63 | 53 | -0.04324 | -0.00229 | 0.07027 | 0.73081 | 12427.42 |
| IWANAGA_ | 24 | -0.07258 | -0.00259 | 0.11793 | 0.73086 | 12428.27 |
| WP_SMALI | 92 | -0.03143 | -0.00219 | 0.051038 | 0.73096 | 12429.97 |
| GOMF_MIT | 5 | -0.127 | -0.00207 | 0.20622 | 0.73099 | 12430.48 |
| WP_HERED | 18 | -0.07581 | -0.00234 | 0.1231 | 0.73099 | 12430.48 |
| GOCC_GRA | 7 | -0.12091 | -0.00233 | 0.19627 | 0.73107 | 12431.85 |
| PID_IL12_S | 31 | -0.06364 | -0.00258 | 0.10329 | 0.73109 | 12432.19 |
| GOBP_SOD | 237 | -0.02115 | -0.00235 | 0.034316 | 0.73112 | 12432.7 |
| GOBP_POS | 147 | -0.02415 | -0.00212 | 0.039179 | 0.7312 | 12434.06 |
| GOBP_NEU | 15 | -0.07873 | -0.00222 | 0.12769 | 0.73124 | 12434.74 |
| GOBP_REG | 22 | -0.07394 | -0.00252 | 0.11988 | 0.73129 | 12435.59 |
| GOBP_REG | 31 | -0.05365 | -0.00217 | 0.08698 | 0.73131 | 12435.93 |
| GOBP_REG | 15 | -0.08752 | -0.00247 | 0.14187 | 0.73136 | 12436.78 |

| | | | | | | |
|---|---|---|---|---|---|---|
| GOMF_DN/ | 15 | -0.08032 | -0.00226 | 0.13014 | 0.73144 | 12438.14 |
| GOBP_FLO | 11 | -0.09573 | -0.00231 | 0.15506 | 0.73151 | 12439.33 |
| GOMF_OXI | 55 | -0.0365 | -0.00197 | 0.059089 | 0.73163 | 12441.37 |
| GOBP_NEG | 203 | -0.02411 | -0.00249 | 0.03899 | 0.73186 | 12445.28 |
| GOBP_CHE | 15 | -0.08404 | -0.00237 | 0.13586 | 0.73189 | 12445.79 |
| GOMF_ACI | 33 | -0.0563 | -0.00235 | 0.091005 | 0.73191 | 12446.13 |
| LIANG_HEI | 42 | -0.04953 | -0.00233 | 0.080044 | 0.73198 | 12447.32 |
| GOMF_PRC | 47 | -0.04692 | -0.00234 | 0.075815 | 0.732 | 12447.66 |
| GOBP_SLEI | 29 | -0.06125 | -0.0024 | 0.098933 | 0.73207 | 12448.85 |
| GOBP_INO | 50 | -0.04135 | -0.00213 | 0.066748 | 0.73217 | 12450.55 |
| GOBP_PHC | 33 | -0.05543 | -0.00232 | 0.089412 | 0.73233 | 12453.27 |
| REACTOMI | 34 | -0.05159 | -0.00219 | 0.083187 | 0.73243 | 12454.97 |
| GOBP_POS | 12 | -0.08786 | -0.00221 | 0.14159 | 0.73253 | 12456.67 |
| GOBP_REG | 68 | -0.03498 | -0.0021 | 0.056352 | 0.7326 | 12457.86 |
| RICKMAN_ | 289 | -0.01852 | -0.00227 | 0.029839 | 0.73261 | 12458.03 |
| GOBP_CELI | 869 | -0.01151 | -0.00241 | 0.018532 | 0.73269 | 12459.39 |
| GOMF_EXT | 10 | -0.10159 | -0.00234 | 0.16358 | 0.7327 | 12459.56 |
| GOBP_MO | 17 | -0.07475 | -0.00224 | 0.1203 | 0.7328 | 12461.26 |
| GOBP_CER | 5 | -0.16755 | -0.00273 | 0.26948 | 0.73294 | 12463.64 |
| GOBP_GEN | 11 | -0.11119 | -0.00268 | 0.17884 | 0.73294 | 12463.64 |
| GOBP_POS | 6 | -0.15238 | -0.00272 | 0.24508 | 0.73295 | 12463.81 |
| KEGG_INTE | 32 | -0.05449 | -0.00224 | 0.087633 | 0.73296 | 12463.98 |
| GOBP_NEU | 7 | -0.16434 | -0.00316 | 0.26416 | 0.73307 | 12465.86 |
| GOBP_CELI | 17 | -0.07475 | -0.00224 | 0.12007 | 0.73319 | 12467.9 |
| VALK_AML | 38 | -0.05076 | -0.00228 | 0.081522 | 0.73323 | 12468.58 |
| KEGG_DRU | 65 | -0.04785 | -0.0028 | 0.076797 | 0.73337 | 12470.96 |
| GOBP_PHC | 101 | -0.0346 | -0.00252 | 0.055486 | 0.73356 | 12474.19 |
| GOBP_TYP | 103 | -0.03295 | -0.00243 | 0.052833 | 0.73357 | 12474.36 |
| REACTOMI | 47 | -0.04472 | -0.00223 | 0.071713 | 0.73357 | 12474.36 |
| GOBP_POS | 124 | -0.02988 | -0.00241 | 0.047899 | 0.7336 | 12474.87 |
| GOBP_NEG | 7 | -0.12708 | -0.00245 | 0.20367 | 0.73367 | 12476.06 |
| MYLLYKAN | 9 | -0.10508 | -0.00229 | 0.16835 | 0.73374 | 12477.25 |
| GOBP_RES | 39 | -0.05146 | -0.00234 | 0.08244 | 0.73375 | 12477.42 |
| GOBP_END | 7 | -0.12397 | -0.00239 | 0.19854 | 0.73381 | 12478.44 |
| GOMF_PSE | 11 | -0.09008 | -0.00217 | 0.14426 | 0.73382 | 12478.61 |
| GOBP_POS | 2 | -0.2446 | -0.00252 | 0.3917 | 0.73383 | 12478.78 |
| GOBP_MAT | 33 | -0.05104 | -0.00213 | 0.0817 | 0.7339 | 12479.97 |
| WP_DEVEL | 13 | -0.09953 | -0.00261 | 0.15927 | 0.73399 | 12481.5 |
| NATSUME_ | 66 | -0.04123 | -0.00243 | 0.065959 | 0.73401 | 12481.84 |
| KOBAYASH | 91 | -0.03169 | -0.00219 | 0.050695 | 0.73404 | 12482.35 |
| REACTOMI | 74 | -0.03612 | -0.00226 | 0.057763 | 0.73412 | 12483.71 |
| ELLWOOD_ | 12 | -0.09487 | -0.00239 | 0.15168 | 0.73416 | 12484.39 |
| GOBP_BRU | 6 | -0.13404 | -0.00239 | 0.21419 | 0.73427 | 12486.26 |
| WP_PRADI | 59 | -0.04274 | -0.00239 | 0.068291 | 0.73427 | 12486.26 |
| GOCC_NUC | 59 | -0.03795 | -0.00212 | 0.060638 | 0.7343 | 12486.77 |

| | | | | | | |
|---|---|---|---|---|---|---|
| STEIN_ESRI | 39 | -0.05322 | -0.00242 | 0.085012 | 0.73434 | 12487.45 |
| AZARE_NE( | 18 | -0.07636 | -0.00236 | 0.12193 | 0.73443 | 12488.98 |
| SANSOM_/ | 200 | -0.02286 | -0.00234 | 0.036506 | 0.73444 | 12489.15 |
| SHAFFER_I | 78 | -0.03589 | -0.0023 | 0.057244 | 0.73463 | 12492.38 |
| GOBP_REG | 176 | -0.02325 | -0.00223 | 0.03707 | 0.73471 | 12493.74 |
| GOMF_MC | 58 | -0.04001 | -0.00221 | 0.063792 | 0.73473 | 12494.08 |
| REACTOME | 11 | -0.09406 | -0.00227 | 0.14986 | 0.73487 | 12496.46 |
| GOBP_PRC | 20 | -0.08097 | -0.00263 | 0.12899 | 0.7349 | 12496.97 |
| GOBP_POS | 11 | -0.10289 | -0.00248 | 0.16381 | 0.73502 | 12499.02 |
| DAVIES_MI | 23 | -0.06521 | -0.00227 | 0.1036 | 0.73544 | 12506.16 |
| GOBP_STEI | 82 | -0.03708 | -0.00244 | 0.058907 | 0.73549 | 12507.01 |
| BIOCARTA_ | 17 | -0.07516 | -0.00225 | 0.11938 | 0.73552 | 12507.52 |
| GOBP_OLI( | 63 | -0.03858 | -0.00223 | 0.061273 | 0.73553 | 12507.69 |
| GOBP_REG | 7 | -0.10905 | -0.0021 | 0.1732 | 0.73553 | 12507.69 |
| GOBP_MRI | 17 | -0.06358 | -0.00191 | 0.10097 | 0.73555 | 12508.03 |
| GOBP_PRC | 67 | -0.03884 | -0.00231 | 0.061683 | 0.73555 | 12508.03 |
| GOMF_GLU | 32 | -0.06642 | -0.00273 | 0.10545 | 0.73559 | 12508.71 |
| GOBP_PEN | 11 | -0.10863 | -0.00262 | 0.17241 | 0.73566 | 12509.9 |
| GOMF_INT | 6 | -0.10285 | -0.00183 | 0.16323 | 0.73568 | 12510.24 |
| GOMF_SIR | 8 | -0.13067 | -0.00269 | 0.20736 | 0.7357 | 12510.58 |
| GOBP_TOL | 9 | -0.10501 | -0.00229 | 0.16658 | 0.73578 | 12511.94 |
| BIOCARTA_ | 9 | -0.10965 | -0.00239 | 0.1738 | 0.73594 | 12514.66 |
| GOMF_ANI | 6 | -0.09249 | -0.00165 | 0.14656 | 0.73599 | 12515.51 |
| GOBP_REG | 129 | -0.02749 | -0.00226 | 0.043556 | 0.73601 | 12515.85 |
| DAVICIONI_ | 310 | -0.01933 | -0.00246 | 0.03061 | 0.73608 | 12517.04 |
| GOBP_MIT | 7 | -0.14478 | -0.00279 | 0.2293 | 0.73611 | 12517.55 |
| REACTOME | 15 | -0.08264 | -0.00233 | 0.13086 | 0.73615 | 12518.23 |
| GOBP_REG | 399 | -0.01667 | -0.0024 | 0.026382 | 0.73624 | 12519.76 |
| GOBP_NEU | 14 | -0.07653 | -0.00208 | 0.12108 | 0.73631 | 12520.95 |
| GOBP_PYR | 9 | -0.10439 | -0.00228 | 0.16505 | 0.73646 | 12523.5 |
| SHIN_B_CE | 31 | -0.05579 | -0.00226 | 0.088134 | 0.73663 | 12526.39 |
| GOBP_OPS | 15 | -0.08815 | -0.00248 | 0.13921 | 0.73671 | 12527.75 |
| GOBP_CITI | 4 | -0.15767 | -0.00229 | 0.24884 | 0.73683 | 12529.79 |
| GOCC_SECI | 299 | -0.01867 | -0.00233 | 0.029457 | 0.73683 | 12529.79 |
| GOBP_REG | 10 | -0.10374 | -0.00239 | 0.16371 | 0.73685 | 12530.13 |
| BIOCARTA_ | 15 | -0.09208 | -0.00259 | 0.14524 | 0.73695 | 12531.83 |
| GOBP_REG | 10 | -0.08432 | -0.00194 | 0.13295 | 0.73703 | 12533.2 |
| GOBP_REG | 36 | -0.05571 | -0.00243 | 0.087837 | 0.73705 | 12533.54 |
| GOMF_HIS | 12 | -0.09891 | -0.00249 | 0.15593 | 0.73706 | 12533.71 |
| GOCC_TRA | 10 | -0.07719 | -0.00178 | 0.12168 | 0.73709 | 12534.22 |
| GOBP_L_A | 4 | -0.12887 | -0.00188 | 0.20275 | 0.73748 | 12540.85 |
| BRUINS_U\ | 510 | -0.01422 | -0.00231 | 0.022353 | 0.73762 | 12543.23 |
| GOMF_VAS | 6 | -0.1408 | -0.00251 | 0.22135 | 0.73763 | 12543.4 |
| RAO_BOUI | 492 | -0.01503 | -0.00239 | 0.023626 | 0.73767 | 12544.08 |
| GOBP_TEL( | 6 | -0.09637 | -0.00172 | 0.1513 | 0.73791 | 12548.16 |

| | | | | | | |
|---|---|---|---|---|---|---|
| GOMF_LIP( | 6 | -0.10794 | -0.00192 | 0.16946 | 0.73792 | 12548.33 |
| GOMF_5_3 | 8 | -0.10076 | -0.00207 | 0.15804 | 0.73812 | 12551.73 |
| GOBP_REG | 14 | -0.07085 | -0.00193 | 0.11111 | 0.73816 | 12552.41 |
| GOMF_BILI | 5 | -0.10448 | -0.0017 | 0.16376 | 0.73827 | 12554.28 |
| GOBP_POS | 46 | -0.05022 | -0.00248 | 0.078674 | 0.73837 | 12555.98 |
| KEGG_SMA | 80 | -0.03583 | -0.00233 | 0.056099 | 0.7385 | 12558.19 |
| GOBP_EPIT | 23 | -0.07049 | -0.00246 | 0.11035 | 0.73852 | 12558.53 |
| GOBP_PRC | 25 | -0.06192 | -0.00225 | 0.096915 | 0.73854 | 12558.87 |
| GOMF_RA( | 10 | -0.09215 | -0.00212 | 0.1442 | 0.73861 | 12560.06 |
| GOBP_GLIA | 47 | -0.04588 | -0.00229 | 0.071775 | 0.73865 | 12560.74 |
| JISON_SICK | 166 | -0.02468 | -0.0023 | 0.038585 | 0.73876 | 12562.61 |
| GOCC_MLL | 28 | -0.05541 | -0.00213 | 0.08663 | 0.73877 | 12562.78 |
| REACTOME | 11 | -0.08716 | -0.0021 | 0.13627 | 0.73879 | 12563.12 |
| GOBP_REG | 6 | -0.12966 | -0.00231 | 0.20265 | 0.73884 | 12563.97 |
| WP_SPHIN | 28 | -0.05845 | -0.00225 | 0.091334 | 0.73888 | 12564.65 |
| REACTOME | 10 | -0.07686 | -0.00177 | 0.1201 | 0.73889 | 12564.82 |
| ACEVEDO_ | 117 | -0.03334 | -0.00262 | 0.052093 | 0.73892 | 12565.33 |
| BOGNI_TRI | 26 | -0.05234 | -0.00194 | 0.081736 | 0.73902 | 12567.04 |
| GOBP_REG | 725 | -0.01239 | -0.00238 | 0.019349 | 0.73909 | 12568.23 |
| GOMF_CY( | 12 | -0.09236 | -0.00233 | 0.14418 | 0.7391 | 12568.4 |
| GOBP_SOM | 5 | -0.1583 | -0.00258 | 0.24706 | 0.73915 | 12569.25 |
| GOBP_MIT | 13 | -0.08896 | -0.00233 | 0.13876 | 0.73927 | 12571.29 |
| GOBP_OUT | 22 | -0.06618 | -0.00226 | 0.10317 | 0.73937 | 12572.99 |
| GOCC_VAC | 777 | -0.01126 | -0.00224 | 0.017553 | 0.7394 | 12573.5 |
| GOBP_REG | 7 | -0.1224 | -0.00236 | 0.1908 | 0.73941 | 12573.67 |
| MOHANKU | 155 | -0.0269 | -0.00243 | 0.041933 | 0.73943 | 12574.01 |
| GOMF_PRE | 12 | -0.08386 | -0.00211 | 0.13065 | 0.73951 | 12575.37 |
| GOBP_POS | 5 | -0.15057 | -0.00245 | 0.23451 | 0.73958 | 12576.56 |
| GOBP_NEG | 12 | -0.08861 | -0.00223 | 0.13786 | 0.7398 | 12580.3 |
| GOBP_CEN | 29 | -0.06102 | -0.00239 | 0.094882 | 0.73993 | 12582.51 |
| GOBP_UDF | 5 | -0.13996 | -0.00228 | 0.21759 | 0.73996 | 12583.02 |
| REACTOME | 71 | -0.0377 | -0.00231 | 0.058595 | 0.74002 | 12584.04 |
| GENTILE_U | 17 | -0.08278 | -0.00248 | 0.12865 | 0.74003 | 12584.21 |
| SCHWAB_T | 14 | -0.08597 | -0.00234 | 0.13355 | 0.74011 | 12585.57 |
| REACTOME | 31 | -0.05561 | -0.00225 | 0.086383 | 0.74013 | 12585.91 |
| REACTOME | 12 | -0.08583 | -0.00216 | 0.1332 | 0.74032 | 12589.14 |
| GOBP_NEG | 4 | -0.1397 | -0.00203 | 0.2168 | 0.74033 | 12589.31 |
| GOMF_PAL | 35 | -0.05421 | -0.00233 | 0.084107 | 0.74037 | 12589.99 |
| GOBP_POS | 99 | -0.03251 | -0.00235 | 0.050399 | 0.74055 | 12593.05 |
| WP_SECRE | 5 | -0.13353 | -0.00217 | 0.20699 | 0.74056 | 12593.22 |
| GOBP_END | 128 | -0.02789 | -0.00229 | 0.043219 | 0.74066 | 12594.92 |
| GOMF_TUI | 5 | -0.14241 | -0.00232 | 0.22054 | 0.74077 | 12596.79 |
| GOMF_AM | 9 | -0.10327 | -0.00225 | 0.15984 | 0.74089 | 12598.83 |
| REACTOME | 21 | -0.06741 | -0.00225 | 0.10432 | 0.7409 | 12599 |
| GOMF_PR( | 58 | -0.04391 | -0.00243 | 0.067943 | 0.74096 | 12600.02 |

| | | | | | | |
|---|---|---|---|---|---|---|
| GOBP_NEG | 4 | -0.1875 | -0.00273 | 0.29 | 0.74103 | 12601.22 |
| GOBP_REG | 33 | -0.05308 | -0.00222 | 0.08203 | 0.7412 | 12604.11 |
| GOBP_VES | 64 | -0.03929 | -0.00228 | 0.06072 | 0.7412 | 12604.11 |
| MIKKELSEN | 14 | -0.0834 | -0.00227 | 0.12881 | 0.74132 | 12606.15 |
| GOMF_SNI | 44 | -0.05033 | -0.00243 | 0.077705 | 0.74141 | 12607.68 |
| ZHANG_GA | 13 | -0.09446 | -0.00248 | 0.14583 | 0.74142 | 12607.85 |
| GOMF_INT | 41 | -0.05695 | -0.00265 | 0.087906 | 0.74146 | 12608.53 |
| HOFFMANI | 36 | -0.0517 | -0.00226 | 0.079776 | 0.74152 | 12609.55 |
| GOBP_REG | 28 | -0.0591 | -0.00227 | 0.091167 | 0.7416 | 12610.91 |
| KEGG_MTC | 49 | -0.04637 | -0.00236 | 0.071512 | 0.74165 | 12611.76 |
| GOBP_REG | 18 | -0.08205 | -0.00253 | 0.12653 | 0.74166 | 12611.93 |
| GOMF_SOI | 15 | -0.09728 | -0.00274 | 0.14991 | 0.74179 | 12614.14 |
| WP_T_CELI | 28 | -0.05932 | -0.00228 | 0.091425 | 0.74179 | 12614.14 |
| GOBP_REG | 10 | -0.08719 | -0.00201 | 0.13436 | 0.7418 | 12614.31 |
| BROWNE_I | 63 | -0.04005 | -0.00231 | 0.061708 | 0.74184 | 12614.99 |
| GOBP_INO | 170 | -0.02598 | -0.00245 | 0.040023 | 0.74189 | 12615.84 |
| GOBP_POS | 12 | -0.08485 | -0.00214 | 0.13067 | 0.74192 | 12616.35 |
| GOBP_NEG | 8 | -0.12844 | -0.00264 | 0.19778 | 0.74197 | 12617.2 |
| GOBP_INN | 44 | -0.05281 | -0.00255 | 0.081306 | 0.742 | 12617.71 |
| MEBARKI_I | 58 | -0.04234 | -0.00234 | 0.065163 | 0.74204 | 12618.39 |
| GOBP_TON | 11 | -0.12083 | -0.00292 | 0.18595 | 0.74208 | 12619.07 |
| KEGG_FAT | 40 | -0.04925 | -0.00226 | 0.075718 | 0.74227 | 12622.3 |
| GOBP_NEG | 6 | -0.18046 | -0.00322 | 0.2774 | 0.74232 | 12623.15 |
| REACTOME | 102 | -0.03566 | -0.00261 | 0.054793 | 0.74243 | 12625.02 |
| PID_REG_G | 82 | -0.03581 | -0.00236 | 0.055017 | 0.74246 | 12625.53 |
| GOBP_PAR | 17 | -0.08182 | -0.00245 | 0.12557 | 0.74265 | 12628.76 |
| GOBP_POS | 27 | -0.06859 | -0.00259 | 0.10527 | 0.74266 | 12628.93 |
| GOBP_MEL | 24 | -0.07736 | -0.00276 | 0.11862 | 0.74284 | 12631.99 |
| GOBP_REG | 88 | -0.0342 | -0.00233 | 0.052425 | 0.74289 | 12632.84 |
| GOBP_DNA | 13 | -0.13686 | -0.00359 | 0.2098 | 0.74291 | 12633.18 |
| GOBP_TRA | 92 | -0.03334 | -0.00232 | 0.051101 | 0.74293 | 12633.52 |
| GOBP_REG | 7 | -0.12155 | -0.00234 | 0.18628 | 0.74296 | 12634.03 |
| DER_IFN_B | 95 | -0.03116 | -0.0022 | 0.047719 | 0.7431 | 12636.42 |
| HOOI_ST7_ | 87 | -0.04051 | -0.00274 | 0.062016 | 0.74316 | 12637.44 |
| GOBP_SPR | 123 | -0.03065 | -0.00247 | 0.046925 | 0.74318 | 12637.78 |
| GOBP_RET | 8 | -0.11983 | -0.00247 | 0.18343 | 0.74321 | 12638.29 |
| GOBP_DEF | 80 | -0.0382 | -0.00248 | 0.058455 | 0.74327 | 12639.31 |
| GOBP_POS | 14 | -0.09079 | -0.00247 | 0.13891 | 0.74332 | 12640.16 |
| GOCC_COF | 5 | -0.13952 | -0.00227 | 0.21334 | 0.74344 | 12642.2 |
| TAGHAVI_I | 11 | -0.08142 | -0.00196 | 0.12447 | 0.74347 | 12642.71 |
| REACTOME | 5 | -0.13458 | -0.00219 | 0.20573 | 0.74348 | 12642.88 |
| GOBP_REG | 45 | -0.04388 | -0.00214 | 0.067073 | 0.74351 | 12643.39 |
| WP_GLYCE | 29 | -0.06029 | -0.00236 | 0.092086 | 0.74367 | 12646.11 |
| GOBP_END | 24 | -0.07105 | -0.00253 | 0.10848 | 0.74376 | 12647.64 |
| GOBP_NUC | 97 | -0.03132 | -0.00224 | 0.047786 | 0.74388 | 12649.68 |

| | | | | | | |
|---|---|---|---|---|---|---|
| GOBP_RES | 22 | -0.07399 | -0.00252 | 0.1129 | 0.74388 | 12649.68 |
| GOBP_REG | 6 | -0.12207 | -0.00218 | 0.18624 | 0.74391 | 12650.19 |
| GOBP_POS | 14 | -0.10692 | -0.00291 | 0.16311 | 0.74392 | 12650.36 |
| GOBP_RIB( | 14 | -0.09349 | -0.00255 | 0.14258 | 0.744 | 12651.72 |
| GOCC_GAE | 75 | -0.04259 | -0.00268 | 0.064939 | 0.74401 | 12651.89 |
| GOCC_CYT( | 79 | -0.0366 | -0.00236 | 0.055773 | 0.74415 | 12654.27 |
| GOMF_DE( | 9 | -0.15325 | -0.00335 | 0.23342 | 0.74425 | 12655.97 |
| REACTOME | 5 | -0.18111 | -0.00295 | 0.27586 | 0.74425 | 12655.97 |
| GOCC_BAS | 220 | -0.0225 | -0.00241 | 0.034215 | 0.74461 | 12662.09 |
| FONTAINE_ | 25 | -0.06426 | -0.00234 | 0.097683 | 0.74466 | 12662.94 |
| REACTOME | 18 | -0.08677 | -0.00268 | 0.13192 | 0.74466 | 12662.94 |
| GOBP_PYR | 6 | -0.14146 | -0.00252 | 0.21502 | 0.7447 | 12663.62 |
| GOBP_NU( | 18 | -0.08614 | -0.00266 | 0.13086 | 0.74481 | 12665.49 |
| GOBP_CELI | 17 | -0.07412 | -0.00222 | 0.11259 | 0.74482 | 12665.66 |
| GOBP_REP | 10 | -0.10703 | -0.00246 | 0.16257 | 0.74484 | 12666 |
| GOBP_AMI | 14 | -0.09369 | -0.00255 | 0.14227 | 0.7449 | 12667.02 |
| GOBP_SYN | 32 | -0.05567 | -0.00229 | 0.084528 | 0.7449 | 12667.02 |
| GOBP_REG | 19 | -0.08765 | -0.00278 | 0.13304 | 0.745 | 12668.73 |
| KAMIKUBO | 18 | -0.071 | -0.00219 | 0.10775 | 0.74502 | 12669.07 |
| GOMF_TRA | 120 | -0.03125 | -0.00248 | 0.047418 | 0.74503 | 12669.24 |
| GOBP_NEU | 33 | -0.05712 | -0.00239 | 0.086662 | 0.74508 | 12670.09 |
| REACTOME | 10 | -0.09322 | -0.00214 | 0.14141 | 0.74512 | 12670.77 |
| GOBP_NEG | 19 | -0.06768 | -0.00215 | 0.10263 | 0.74519 | 12671.96 |
| GOBP_NEG | 6 | -0.16343 | -0.00291 | 0.24775 | 0.74525 | 12672.98 |
| REACTOME | 39 | -0.05517 | -0.00251 | 0.083625 | 0.74528 | 12673.49 |
| GOMF_CEN | 6 | -0.15302 | -0.00273 | 0.23192 | 0.74531 | 12674 |
| GOBP_LYSI | 8 | -0.10684 | -0.0022 | 0.16183 | 0.74542 | 12675.87 |
| GOBP_POS | 13 | -0.10429 | -0.00274 | 0.15792 | 0.74549 | 12677.06 |
| GOMF_SER | 180 | -0.02759 | -0.00268 | 0.041784 | 0.7455 | 12677.23 |
| GOBP_POS | 3 | -0.24797 | -0.00313 | 0.37537 | 0.74556 | 12678.25 |
| GOBP_DEC | 14 | -0.09644 | -0.00263 | 0.14599 | 0.74557 | 12678.42 |
| GOBP_CHO | 35 | -0.05562 | -0.00239 | 0.084193 | 0.74558 | 12678.59 |
| BIOCARTA_ | 16 | -0.08573 | -0.00249 | 0.1297 | 0.74569 | 12680.46 |
| GOBP_SPEI | 17 | -0.0773 | -0.00232 | 0.1168 | 0.74593 | 12684.54 |
| GOBP_NEG | 9 | -0.1351 | -0.00295 | 0.20413 | 0.74596 | 12685.05 |
| GOCC_BAS | 270 | -0.02028 | -0.00241 | 0.030635 | 0.74596 | 12685.05 |
| GOBP_NEG | 17 | -0.07303 | -0.00219 | 0.11017 | 0.74628 | 12690.49 |
| GOBP_POS | 7 | -0.13302 | -0.00256 | 0.20067 | 0.7463 | 12690.83 |
| GOBP_POS | 18 | -0.08146 | -0.00251 | 0.12286 | 0.74634 | 12691.51 |
| GOBP_URA | 8 | -0.1142 | -0.00235 | 0.17219 | 0.7464 | 12692.53 |
| GOCC_DNA | 18 | -0.0815 | -0.00252 | 0.12287 | 0.74643 | 12693.04 |
| REACTOME | 76 | -0.03845 | -0.00243 | 0.057956 | 0.74645 | 12693.38 |
| GOBP_NEG | 13 | -0.10193 | -0.00267 | 0.15363 | 0.74647 | 12693.72 |
| ZHOU_TNF | 42 | -0.05156 | -0.00243 | 0.077679 | 0.74657 | 12695.42 |
| GOBP_POS | 6 | -0.17004 | -0.00303 | 0.25614 | 0.74661 | 12696.1 |

| | | | | | | |
|---|---|---|---|---|---|---|
| EINAV_INT | 27 | -0.0643 | -0.00243 | 0.096802 | 0.74673 | 12698.14 |
| GOBP_CYT | 9 | -0.1085 | -0.00237 | 0.16327 | 0.74681 | 12699.5 |
| GOBP_INTI | 106 | -0.03296 | -0.00246 | 0.049599 | 0.74682 | 12699.67 |
| GOBP_POS | 13 | -0.11569 | -0.00304 | 0.17409 | 0.74684 | 12700.01 |
| GOBP_ADH | 15 | -0.09531 | -0.00269 | 0.14331 | 0.74698 | 12702.39 |
| PETROVA_ | 155 | -0.02645 | -0.00239 | 0.039771 | 0.74702 | 12703.08 |
| GOBP_HET | 58 | -0.04787 | -0.00265 | 0.071957 | 0.74706 | 12703.76 |
| REACTOME | 39 | -0.05046 | -0.00229 | 0.075845 | 0.74707 | 12703.93 |
| GOBP_RES | 23 | -0.06451 | -0.00225 | 0.096949 | 0.7471 | 12704.44 |
| LINDGREN_ | 130 | -0.03115 | -0.00258 | 0.046816 | 0.74711 | 12704.61 |
| GOMF_HEI | 9 | -0.11942 | -0.00261 | 0.17935 | 0.74724 | 12706.82 |
| LEE_LIVER_ | 64 | -0.04237 | -0.00246 | 0.063625 | 0.74724 | 12706.82 |
| TURASHVIL | 61 | -0.04194 | -0.00238 | 0.062983 | 0.74726 | 12707.16 |
| STANHILL_ | 4 | -0.22619 | -0.00329 | 0.33958 | 0.74732 | 12708.18 |
| GALI_TP53_ | 8 | -0.11048 | -0.00227 | 0.16585 | 0.74734 | 12708.52 |
| PID_CXCR4 | 97 | -0.03224 | -0.00231 | 0.048394 | 0.74734 | 12708.52 |
| GOBP_NEG | 137 | -0.03102 | -0.00263 | 0.046562 | 0.74735 | 12708.69 |
| WP_NRF2A | 23 | -0.07456 | -0.0026 | 0.1119 | 0.74738 | 12709.2 |
| GOBP_REG | 7 | -0.09446 | -0.00182 | 0.14169 | 0.74749 | 12711.07 |
| GOCC_DIST | 260 | -0.02097 | -0.00244 | 0.031417 | 0.74771 | 12714.81 |
| GOMF_HEI | 5 | -0.20533 | -0.00334 | 0.30753 | 0.74782 | 12716.68 |
| GOBP_PLA | 28 | -0.06422 | -0.00247 | 0.096038 | 0.74815 | 12722.29 |
| LUI_THYRO | 46 | -0.04664 | -0.0023 | 0.069743 | 0.74815 | 12722.29 |
| GOMF_CYS | 50 | -0.04992 | -0.00257 | 0.074642 | 0.7482 | 12723.14 |
| GOBP_POS | 68 | -0.04211 | -0.00252 | 0.062951 | 0.74822 | 12723.48 |
| GOMF_DN | 7 | -0.11186 | -0.00215 | 0.16723 | 0.74822 | 12723.48 |
| BANDRES_ | 12 | -0.0884 | -0.00223 | 0.13203 | 0.74841 | 12726.71 |
| GOBP_EPIT | 27 | -0.06772 | -0.00256 | 0.10114 | 0.74841 | 12726.71 |
| GOBP_POS | 7 | -0.15593 | -0.003 | 0.23291 | 0.74841 | 12726.71 |
| GOBP_REG | 9 | -0.10771 | -0.00235 | 0.16087 | 0.74842 | 12726.88 |
| GOMF_RN | 7 | -0.09927 | -0.00191 | 0.14824 | 0.74844 | 12727.22 |
| GOMF_PH | 12 | -0.10366 | -0.00261 | 0.15476 | 0.74851 | 12728.41 |
| REACTOME | 43 | -0.04793 | -0.00228 | 0.071531 | 0.74858 | 12729.6 |
| GOBP_NEG | 299 | -0.02039 | -0.00255 | 0.030421 | 0.74865 | 12730.79 |
| SILIGAN_T | 15 | -0.07476 | -0.00211 | 0.11147 | 0.74878 | 12733 |
| ITO_PTTG1 | 12 | -0.10713 | -0.0027 | 0.15972 | 0.7488 | 12733.34 |
| GOBP_GLA | 9 | -0.10939 | -0.00239 | 0.16304 | 0.74887 | 12734.53 |
| GOBP_PAR | 6 | -0.14443 | -0.00257 | 0.21525 | 0.74888 | 12734.7 |
| GOBP_SNR | 18 | -0.06487 | -0.002 | 0.096666 | 0.74892 | 12735.38 |
| HOWLIN_P | 60 | -0.04575 | -0.00258 | 0.068161 | 0.74896 | 12736.06 |
| GOBP_POS | 38 | -0.05277 | -0.00236 | 0.078574 | 0.74905 | 12737.6 |
| GOMF_INC | 6 | -0.12983 | -0.00231 | 0.19321 | 0.74918 | 12739.81 |
| SIMBULAN_ | 9 | -0.11471 | -0.0025 | 0.17067 | 0.74924 | 12740.83 |
| GOBP_EPIT | 10 | -0.09642 | -0.00222 | 0.14344 | 0.74926 | 12741.17 |
| GOBP_POS | 5 | -0.14494 | -0.00236 | 0.2155 | 0.74939 | 12743.38 |

| | | | | | | |
|---|---|---|---|---|---|---|
| HU_GENOT | 32 | -0.06032 | -0.00248 | 0.089666 | 0.74942 | 12743.89 |
| GOMF_UBI | 9 | -0.09906 | -0.00216 | 0.14721 | 0.74948 | 12744.91 |
| GOBP_ROS | 10 | -0.12843 | -0.00296 | 0.19059 | 0.7498 | 12750.35 |
| REACTOME | 168 | -0.02548 | -0.00239 | 0.0378 | 0.74982 | 12750.69 |
| GOMF_LIP/ | 5 | -0.13658 | -0.00222 | 0.20265 | 0.74983 | 12750.86 |
| GOBP_REG | 4 | -0.20089 | -0.00292 | 0.29805 | 0.74985 | 12751.2 |
| GOBP_REG | 6 | -0.14151 | -0.00252 | 0.20993 | 0.74987 | 12751.54 |
| REACTOME | 28 | -0.06504 | -0.0025 | 0.09647 | 0.7499 | 12752.05 |
| GOBP_ADF | 88 | -0.03387 | -0.00231 | 0.050217 | 0.74996 | 12753.07 |
| GOBP_RES | 118 | -0.03335 | -0.00263 | 0.049456 | 0.74997 | 12753.24 |
| GOBP_POS | 5 | -0.11655 | -0.0019 | 0.17281 | 0.74998 | 12753.41 |
| GOBP_DIS/ | 12 | -0.09201 | -0.00232 | 0.13642 | 0.75 | 12753.75 |
| GOBP_SPH | 8 | -0.11013 | -0.00227 | 0.16321 | 0.75009 | 12755.28 |
| GOMF_G_I | 20 | -0.07982 | -0.0026 | 0.11829 | 0.75009 | 12755.28 |
| GOBP_CELI | 99 | -0.03594 | -0.0026 | 0.05323 | 0.75022 | 12757.49 |
| PIEPOLI_LC | 13 | -0.09169 | -0.00241 | 0.13577 | 0.75027 | 12758.34 |
| GOBP_INN | 33 | -0.05594 | -0.00234 | 0.082822 | 0.75028 | 12758.51 |
| GOBP_REG | 19 | -0.0734 | -0.00233 | 0.10868 | 0.75029 | 12758.68 |
| WP_TYPE_ | 28 | -0.06681 | -0.00257 | 0.098892 | 0.75033 | 12759.36 |
| GOMF_CHI | 8 | -0.11463 | -0.00236 | 0.16966 | 0.75036 | 12759.87 |
| GOMF_RN/ | 7 | -0.11543 | -0.00222 | 0.17082 | 0.75039 | 12760.38 |
| GOMF_SYN | 20 | -0.07074 | -0.0023 | 0.10468 | 0.7504 | 12760.55 |
| RYAN_MAI | 146 | -0.02975 | -0.00261 | 0.044026 | 0.75041 | 12760.72 |
| GOBP_APC | 5 | -0.1558 | -0.00254 | 0.23049 | 0.75045 | 12761.4 |
| PID_ALK2_ | 11 | -0.11021 | -0.00266 | 0.16303 | 0.75048 | 12761.91 |
| GOBP_REG | 8 | -0.13369 | -0.00275 | 0.1976 | 0.75066 | 12764.97 |
| GOBP_POS | 19 | -0.08315 | -0.00264 | 0.12288 | 0.75067 | 12765.14 |
| GOBP_T_C | 12 | -0.09218 | -0.00232 | 0.13623 | 0.75068 | 12765.31 |
| GOBP_LUN | 5 | -0.15748 | -0.00256 | 0.23267 | 0.75075 | 12766.5 |
| HOWLIN_C | 16 | -0.0944 | -0.00275 | 0.13943 | 0.7508 | 12767.35 |
| GOMF_MA | 5 | -0.13574 | -0.00221 | 0.20036 | 0.75094 | 12769.73 |
| GOBP_REG | 952 | -0.01157 | -0.00253 | 0.017064 | 0.75107 | 12771.95 |
| GOBP_NEU | 19 | -0.07133 | -0.00226 | 0.10519 | 0.75112 | 12772.8 |
| GOCC_ANN | 5 | -0.11036 | -0.0018 | 0.16273 | 0.75117 | 12773.65 |
| GOMF_APC | 6 | -0.13545 | -0.00241 | 0.19961 | 0.75128 | 12775.52 |
| GOMF_GU. | 7 | -0.14963 | -0.00288 | 0.22045 | 0.75135 | 12776.71 |
| REACTOME | 20 | -0.07467 | -0.00243 | 0.11 | 0.75135 | 12776.71 |
| GOCC_CIS_ | 59 | -0.04246 | -0.00237 | 0.06255 | 0.75137 | 12777.05 |
| GOBP_REG | 32 | -0.06925 | -0.00285 | 0.102 | 0.7514 | 12777.56 |
| RAGHAVAC | 63 | -0.04321 | -0.00249 | 0.063651 | 0.7514 | 12777.56 |
| WINTER_H | 89 | -0.03358 | -0.0023 | 0.049437 | 0.75152 | 12779.6 |
| GOBP_RET | 5 | -0.17667 | -0.00287 | 0.26001 | 0.75158 | 12780.62 |
| SERVITJA_I | 156 | -0.02823 | -0.00256 | 0.04153 | 0.75166 | 12781.98 |
| GOBP_DER | 5 | -0.13408 | -0.00218 | 0.19722 | 0.75169 | 12782.49 |
| GOBP_RES | 6 | -0.1268 | -0.00226 | 0.18649 | 0.75172 | 12783 |

| | | | | | | |
|---|---|---|---|---|---|---|
| GOBP_NUC | 14 | -0.08558 | -0.00233 | 0.12586 | 0.75173 | 12783.17 |
| ISSAEVA_M | 53 | -0.05237 | -0.00277 | 0.07697 | 0.75187 | 12785.55 |
| GOBP_REG | 117 | -0.0316 | -0.00248 | 0.046431 | 0.75193 | 12786.57 |
| REACTOME | 7 | -0.13554 | -0.00261 | 0.19906 | 0.75203 | 12788.27 |
| GOBP_POS | 49 | -0.04981 | -0.00253 | 0.073149 | 0.75205 | 12788.61 |
| GOBP_POS | 28 | -0.06702 | -0.00258 | 0.098408 | 0.75206 | 12788.78 |
| GOBP_NEG | 24 | -0.07447 | -0.00265 | 0.10925 | 0.75226 | 12792.18 |
| GOBP_MET | 14 | -0.08358 | -0.00228 | 0.12261 | 0.75227 | 12792.35 |
| GOBP_REG | 18 | -0.08052 | -0.00249 | 0.11811 | 0.75229 | 12792.69 |
| GOCC_CBM | 5 | -0.11997 | -0.00195 | 0.17597 | 0.7523 | 12792.86 |
| GOBP_REG | 12 | -0.11728 | -0.00296 | 0.172 | 0.75234 | 12793.54 |
| REACTOME | 90 | -0.03412 | -0.00235 | 0.050001 | 0.7525 | 12796.26 |
| GOBP_IND | 11 | -0.10917 | -0.00263 | 0.15997 | 0.75251 | 12796.43 |
| GOBP_POS | 136 | -0.03104 | -0.00262 | 0.045467 | 0.75257 | 12797.45 |
| WP_COMP | 17 | -0.09009 | -0.0027 | 0.13194 | 0.75262 | 12798.3 |
| ROSS_AML | 69 | -0.03957 | -0.00239 | 0.057951 | 0.75263 | 12798.47 |
| GOBP_NEU | 35 | -0.05722 | -0.00246 | 0.083789 | 0.75267 | 12799.15 |
| REACTOME | 11 | -0.09507 | -0.00229 | 0.1392 | 0.75269 | 12799.49 |
| GOBP_PYR | 16 | -0.09898 | -0.00288 | 0.14491 | 0.7527 | 12799.66 |
| FRASOR_RI | 35 | -0.06371 | -0.00274 | 0.093277 | 0.75271 | 12799.83 |
| GOBP_NEG | 6 | -0.13673 | -0.00244 | 0.20015 | 0.75273 | 12800.17 |
| REACTOME | 29 | -0.08397 | -0.00329 | 0.12291 | 0.75274 | 12800.34 |
| GOBP_STEI | 87 | -0.03884 | -0.00263 | 0.056848 | 0.75276 | 12800.68 |
| GOBP_RES | 9 | -0.0944 | -0.00206 | 0.13812 | 0.75282 | 12801.7 |
| RODWELL_ | 136 | -0.03065 | -0.00259 | 0.044833 | 0.75292 | 12803.4 |
| WP_MINEF | 9 | -0.12703 | -0.00277 | 0.18572 | 0.753 | 12804.77 |
| GOBP_LON | 83 | -0.03981 | -0.00263 | 0.058184 | 0.75306 | 12805.79 |
| GOBP_INTI | 11 | -0.08545 | -0.00206 | 0.12483 | 0.75316 | 12807.49 |
| BIOCARTA_ | 9 | -0.13447 | -0.00294 | 0.19645 | 0.75317 | 12807.66 |
| SIG_INSULI | 48 | -0.04926 | -0.00248 | 0.071941 | 0.75323 | 12808.68 |
| GOBP_ASP | 20 | -0.08192 | -0.00267 | 0.11963 | 0.75325 | 12809.02 |
| GOBP_PRC | 48 | -0.04642 | -0.00234 | 0.067747 | 0.7534 | 12811.57 |
| GOBP_BAS | 20 | -0.07976 | -0.00259 | 0.11638 | 0.75343 | 12812.08 |
| GOBP_POS | 28 | -0.06902 | -0.00266 | 0.10068 | 0.75349 | 12813.1 |
| GOBP_REG | 207 | -0.02385 | -0.00248 | 0.034782 | 0.75357 | 12814.46 |
| GOBP_PRC | 61 | -0.04276 | -0.00243 | 0.062344 | 0.75362 | 12815.31 |
| GOMF_PH( | 5 | -0.17952 | -0.00292 | 0.26169 | 0.75364 | 12815.65 |
| GOBP_AXC | 9 | -0.13123 | -0.00286 | 0.19101 | 0.75397 | 12821.26 |
| REACTOME | 9 | -0.10876 | -0.00237 | 0.1583 | 0.75397 | 12821.26 |
| GOBP_INTI | 7 | -0.156 | -0.003 | 0.22698 | 0.75404 | 12822.45 |
| MIKKELSEN | 8 | -0.11577 | -0.00238 | 0.16842 | 0.75407 | 12822.96 |
| GOBP_DEC | 24 | -0.06427 | -0.00229 | 0.093471 | 0.75415 | 12824.32 |
| DACOSTA_ | 26 | -0.06806 | -0.00252 | 0.098929 | 0.75425 | 12826.02 |
| BIOCARTA_ | 11 | -0.11797 | -0.00285 | 0.17135 | 0.75442 | 12828.91 |
| GOBP_MEI | 13 | -0.11002 | -0.00289 | 0.15981 | 0.75442 | 12828.91 |

| | | | | | | |
|---|---|---|---|---|---|---|
| HUPER_BR | 54 | -0.05273 | -0.00282 | 0.076568 | 0.75446 | 12829.59 |
| GOBP_TRN | 6 | -0.11846 | -0.00211 | 0.17202 | 0.75447 | 12829.76 |
| LIN_NPAS4 | 161 | -0.02715 | -0.0025 | 0.039429 | 0.75448 | 12829.93 |
| GOBP_MO | 6 | -0.11548 | -0.00206 | 0.16767 | 0.75451 | 12830.44 |
| GOCC_POS | 15 | -0.08774 | -0.00247 | 0.1272 | 0.75483 | 12835.88 |
| GOBP_REG | 67 | -0.04076 | -0.00242 | 0.059061 | 0.75494 | 12837.75 |
| GOBP_REG | 26 | -0.06636 | -0.00246 | 0.096136 | 0.75499 | 12838.6 |
| GOBP_REG | 4 | -0.23734 | -0.00345 | 0.3438 | 0.75501 | 12838.95 |
| GOBP_POS | 7 | -0.12439 | -0.00239 | 0.18018 | 0.75502 | 12839.12 |
| GOBP_POS | 26 | -0.07153 | -0.00265 | 0.1036 | 0.75504 | 12839.46 |
| REACTOME | 103 | -0.03738 | -0.00275 | 0.054114 | 0.75512 | 12840.82 |
| GOBP_POS | 12 | -0.09904 | -0.0025 | 0.14337 | 0.75514 | 12841.16 |
| GOMF_PH( | 40 | -0.05497 | -0.00253 | 0.079572 | 0.75514 | 12841.16 |
| GOCC_MAI | 16 | -0.08473 | -0.00247 | 0.12264 | 0.75518 | 12841.84 |
| HOELZEL_N | 122 | -0.0337 | -0.0027 | 0.048769 | 0.75521 | 12842.35 |
| GOMF_E_E | 47 | -0.05627 | -0.0028 | 0.081423 | 0.75523 | 12842.69 |
| GOMF_UBI | 16 | -0.07615 | -0.00222 | 0.11018 | 0.75527 | 12843.37 |
| REACTOME | 17 | -0.08623 | -0.00259 | 0.12471 | 0.75536 | 12844.9 |
| DARWICHE | 138 | -0.02824 | -0.00241 | 0.040821 | 0.75548 | 12846.94 |
| REACTOME | 101 | -0.03445 | -0.00251 | 0.049786 | 0.75551 | 12847.45 |
| GOBP_NEG | 10 | -0.12657 | -0.00291 | 0.18288 | 0.75555 | 12848.13 |
| GOBP_POS | 15 | -0.10032 | -0.00283 | 0.14494 | 0.75557 | 12848.47 |
| GOCC_RISC | 8 | -0.1262 | -0.0026 | 0.18228 | 0.75563 | 12849.49 |
| NAKAYAMA | 67 | -0.04843 | -0.00288 | 0.069916 | 0.75572 | 12851.02 |
| REACTOME | 102 | -0.03713 | -0.00272 | 0.053593 | 0.75577 | 12851.87 |
| NIKOLSKY_ | 93 | -0.03701 | -0.00259 | 0.053399 | 0.75587 | 12853.57 |
| HALMOS_C | 43 | -0.05706 | -0.00272 | 0.082311 | 0.75589 | 12853.91 |
| REACTOME | 43 | -0.04827 | -0.0023 | 0.069636 | 0.75591 | 12854.25 |
| GOBP_CELI | 40 | -0.05787 | -0.00266 | 0.083471 | 0.75594 | 12854.76 |
| YAGI_AML_ | 330 | -0.01884 | -0.00247 | 0.027162 | 0.75604 | 12856.46 |
| WILENSKY_ | 24 | -0.06214 | -0.00221 | 0.089581 | 0.75606 | 12856.8 |
| PYEON_HP | 8 | -0.12375 | -0.00255 | 0.17838 | 0.75608 | 12857.14 |
| GOMF_GLU | 10 | -0.12881 | -0.00296 | 0.18559 | 0.75616 | 12858.5 |
| KEGG_GRA | 17 | -0.09105 | -0.00273 | 0.13119 | 0.75618 | 12858.84 |
| GOBP_RES | 32 | -0.05786 | -0.00238 | 0.083317 | 0.7563 | 12860.88 |
| REACTOME | 187 | -0.02458 | -0.00243 | 0.035384 | 0.75636 | 12861.9 |
| GOBP_POS | 7 | -0.13504 | -0.0026 | 0.19429 | 0.75648 | 12863.94 |
| LEE_LIVER_ | 62 | -0.04478 | -0.00256 | 0.064419 | 0.7565 | 12864.28 |
| GOBP_NEG | 7 | -0.11184 | -0.00215 | 0.16083 | 0.7566 | 12865.98 |
| GOBP_DUN | 7 | -0.12903 | -0.00248 | 0.18526 | 0.75693 | 12871.59 |
| WP_MIRNA | 6 | -0.14486 | -0.00258 | 0.20799 | 0.75693 | 12871.59 |
| GOBP_VAS | 73 | -0.0462 | -0.00287 | 0.066318 | 0.75698 | 12872.44 |
| GOCC_MIC | 7 | -0.15684 | -0.00302 | 0.22509 | 0.75703 | 12873.3 |
| GOMF_RET | 9 | -0.11539 | -0.00252 | 0.16558 | 0.75704 | 12873.47 |
| GOBP_NEG | 22 | -0.07563 | -0.00258 | 0.10849 | 0.75713 | 12875 |

| | | | | | | |
|---|---|---|---|---|---|---|
| REACTOME | 31 | -0.0585 | -0.00237 | 0.083881 | 0.75723 | 12876.7 |
| PETRETTO_ | 12 | -0.08765 | -0.00221 | 0.12554 | 0.75747 | 12880.78 |
| GOBP_GAL | 8 | -0.11747 | -0.00242 | 0.16824 | 0.75749 | 12881.12 |
| GOBP_RES | 225 | -0.02383 | -0.00259 | 0.034114 | 0.75758 | 12882.65 |
| KONDO_PF | 181 | -0.02743 | -0.00267 | 0.039256 | 0.75765 | 12883.84 |
| RAY_TUMC | 256 | -0.0224 | -0.00259 | 0.032048 | 0.7577 | 12884.69 |
| GOBP_IMN | 275 | -0.02193 | -0.00263 | 0.031368 | 0.75773 | 12885.2 |
| GOMF_PRC | 138 | -0.03199 | -0.00272 | 0.045747 | 0.75778 | 12886.05 |
| WP_ESTRO | 14 | -0.08646 | -0.00235 | 0.12361 | 0.75787 | 12887.58 |
| GOBP_RES | 10 | -0.12567 | -0.00289 | 0.17964 | 0.7579 | 12888.09 |
| WP_GLYCC | 16 | -0.07127 | -0.00207 | 0.10187 | 0.75793 | 12888.6 |
| KIM_RESPC | 115 | -0.03017 | -0.00235 | 0.043115 | 0.75799 | 12889.62 |
| REACTOME | 44 | -0.05591 | -0.0027 | 0.079885 | 0.75799 | 12889.62 |
| GOBP_POS | 78 | -0.03864 | -0.00248 | 0.055196 | 0.75804 | 12890.47 |
| SETLUR_PF | 66 | -0.04342 | -0.00256 | 0.062015 | 0.75808 | 12891.15 |
| GOBP_REG | 53 | -0.04846 | -0.00256 | 0.069198 | 0.75812 | 12891.83 |
| GOBP_INN | 44 | -0.0521 | -0.00251 | 0.074399 | 0.75813 | 12892 |
| SMID_BREA | 160 | -0.02893 | -0.00265 | 0.041298 | 0.75822 | 12893.53 |
| GOBP_REG | 38 | -0.05452 | -0.00244 | 0.077797 | 0.75827 | 12894.38 |
| GOBP_OXA | 4 | -0.14082 | -0.00205 | 0.20094 | 0.75829 | 12894.72 |
| REACTOME | 12 | -0.09948 | -0.00251 | 0.1419 | 0.75835 | 12895.74 |
| GOBP_SPEI | 4 | -0.13222 | -0.00192 | 0.18859 | 0.75837 | 12896.08 |
| ROSS_AML | 72 | -0.04155 | -0.00256 | 0.059263 | 0.7584 | 12896.59 |
| GOBP_REG | 19 | -0.08439 | -0.00268 | 0.12027 | 0.75856 | 12899.31 |
| GILDEA_M | 29 | -0.06596 | -0.00258 | 0.093985 | 0.7586 | 12899.99 |
| GOBP_CLA | 46 | -0.04988 | -0.00246 | 0.071052 | 0.75866 | 12901.01 |
| REACTOME | 12 | -0.11319 | -0.00285 | 0.16124 | 0.75867 | 12901.18 |
| REACTOME | 6 | -0.14539 | -0.00259 | 0.20705 | 0.75872 | 12902.03 |
| FIRESTEIN_ | 154 | -0.02752 | -0.00248 | 0.039183 | 0.75875 | 12902.54 |
| GOMF_ENZ | 38 | -0.06241 | -0.0028 | 0.088855 | 0.75878 | 12903.05 |
| PID_FOXO_ | 47 | -0.0541 | -0.0027 | 0.077009 | 0.7588 | 12903.39 |
| WP_ZINC_I | 35 | -0.05878 | -0.00253 | 0.083679 | 0.7588 | 12903.39 |
| PID_HIF1A_ | 18 | -0.08221 | -0.00254 | 0.11701 | 0.75883 | 12903.9 |
| GOBP_TRA | 46 | -0.05671 | -0.0028 | 0.080716 | 0.75885 | 12904.24 |
| GOMF_RNA | 14 | -0.08521 | -0.00232 | 0.12127 | 0.75886 | 12904.41 |
| VANTVEER_ | 39 | -0.05896 | -0.00268 | 0.08389 | 0.75892 | 12905.43 |
| GOBP_DEC | 27 | -0.06683 | -0.00253 | 0.095084 | 0.75893 | 12905.6 |
| TSUNODA_ | 43 | -0.05323 | -0.00254 | 0.075723 | 0.75894 | 12905.77 |
| GOBP_RES | 146 | -0.0304 | -0.00266 | 0.04325 | 0.75896 | 12906.11 |
| WP_DNA_I | 67 | -0.04248 | -0.00253 | 0.060399 | 0.75905 | 12907.65 |
| GOBP_POS | 5 | -0.14292 | -0.00233 | 0.2032 | 0.75907 | 12907.99 |
| GOBP_PRC | 10 | -0.10615 | -0.00244 | 0.15086 | 0.75916 | 12909.52 |
| GOBP_CELI | 11 | -0.0956 | -0.00231 | 0.13581 | 0.75925 | 12911.05 |
| GOMF_MA | 15 | -0.08609 | -0.00243 | 0.12229 | 0.75926 | 12911.22 |
| REACTOME | 48 | -0.04941 | -0.00249 | 0.070194 | 0.75926 | 12911.22 |

| | | | | | | |
|---|---|---|---|---|---|---|
| GOBP_RES | 14 | -0.11513 | -0.00313 | 0.16349 | 0.75934 | 12912.58 |
| GOBP_REG | 58 | -0.04559 | -0.00252 | 0.064721 | 0.75938 | 12913.26 |
| SPIRA_SM( | 14 | -0.08439 | -0.0023 | 0.1198 | 0.75941 | 12913.77 |
| GOBP_ANT | 72 | -0.04698 | -0.0029 | 0.066684 | 0.75944 | 12914.28 |
| GOMF_CAI | 51 | -0.04812 | -0.0025 | 0.06824 | 0.75964 | 12917.68 |
| WP_IL17_S | 30 | -0.05829 | -0.00232 | 0.082641 | 0.75968 | 12918.36 |
| GOBP_REG | 40 | -0.05797 | -0.00267 | 0.082178 | 0.75972 | 12919.04 |
| GOBP_NEU | 6 | -0.14532 | -0.00259 | 0.20588 | 0.75985 | 12921.25 |
| GOMF_STE | 7 | -0.14871 | -0.00286 | 0.21069 | 0.75985 | 12921.25 |
| KOBAYASH | 18 | -0.08633 | -0.00266 | 0.12229 | 0.75988 | 12921.76 |
| GOBP_PRC | 7 | -0.12039 | -0.00232 | 0.17045 | 0.76 | 12923.8 |
| GOMF_ENZ | 1114 | -0.01097 | -0.00259 | 0.015535 | 0.76 | 12923.8 |
| REACTOME | 5 | -0.17587 | -0.00286 | 0.24887 | 0.76011 | 12925.67 |
| GOBP_MO | 9 | -0.11918 | -0.0026 | 0.16848 | 0.76033 | 12929.41 |
| GOCC_ALP | 4 | -0.15653 | -0.00228 | 0.22124 | 0.76037 | 12930.09 |
| GOBP_CON | 8 | -0.12342 | -0.00254 | 0.17443 | 0.76039 | 12930.43 |
| GOBP_MAI | 32 | -0.06671 | -0.00274 | 0.094284 | 0.76039 | 12930.43 |
| GOMF_WA | 16 | -0.10033 | -0.00292 | 0.14179 | 0.76039 | 12930.43 |
| GOBP_PAR | 11 | -0.126 | -0.00304 | 0.17805 | 0.76043 | 12931.11 |
| GOBP_NEG | 17 | -0.08568 | -0.00257 | 0.12104 | 0.76048 | 12931.96 |
| GOBP_NEG | 6 | -0.15374 | -0.00274 | 0.21707 | 0.7606 | 12934 |
| GOBP_PRC | 64 | -0.04508 | -0.00262 | 0.063648 | 0.76063 | 12934.51 |
| GOCC_CELI | 7 | -0.12154 | -0.00234 | 0.17158 | 0.76063 | 12934.51 |
| HORIUCHI_ | 282 | -0.02205 | -0.00267 | 0.03111 | 0.76073 | 12936.21 |
| GOBP_NEG | 16 | -0.08801 | -0.00256 | 0.12415 | 0.76079 | 12937.23 |
| GOBP_PIRI | 21 | -0.07148 | -0.00238 | 0.10082 | 0.76083 | 12937.91 |
| GOBP_REG | 15 | -0.08451 | -0.00238 | 0.11918 | 0.76086 | 12938.42 |
| GOBP_ARA | 32 | -0.06407 | -0.00264 | 0.09032 | 0.76094 | 12939.78 |
| GOMF_POS | 49 | -0.0554 | -0.00282 | 0.078042 | 0.76111 | 12942.68 |
| GOMF_ADI | 6 | -0.14622 | -0.00261 | 0.20595 | 0.76114 | 12943.19 |
| GOBP_RES | 5 | -0.13027 | -0.00212 | 0.18347 | 0.76115 | 12943.36 |
| REACTOME | 8 | -0.11788 | -0.00243 | 0.16599 | 0.7612 | 12944.21 |
| STEIN_ESRI | 25 | -0.07237 | -0.00263 | 0.10191 | 0.7612 | 12944.21 |
| GOBP_POS | 7 | -0.14692 | -0.00283 | 0.20684 | 0.76124 | 12944.89 |
| GOBP_MEI | 34 | -0.0576 | -0.00244 | 0.081055 | 0.76135 | 12946.76 |
| GOBP_REG | 3 | -0.28527 | -0.0036 | 0.40138 | 0.76137 | 12947.1 |
| GOBP_VAS | 23 | -0.09274 | -0.00324 | 0.1304 | 0.76151 | 12949.48 |
| GOBP_L_G | 28 | -0.07085 | -0.00273 | 0.09959 | 0.76157 | 12950.5 |
| GOBP_NU( | 6 | -0.14442 | -0.00257 | 0.2029 | 0.76169 | 12952.54 |
| GOBP_PYR | 7 | -0.15295 | -0.00294 | 0.21487 | 0.7617 | 12952.71 |
| PID_ILK_PA | 44 | -0.05288 | -0.00255 | 0.074282 | 0.76174 | 12953.39 |
| REACTOME | 15 | -0.08567 | -0.00241 | 0.12031 | 0.76179 | 12954.24 |
| VERNOCHE | 19 | -0.07925 | -0.00251 | 0.11124 | 0.7619 | 12956.11 |
| SATO_SILEI | 39 | -0.06008 | -0.00273 | 0.08431 | 0.76195 | 12956.96 |
| GOBP_REG | 14 | -0.10117 | -0.00275 | 0.14196 | 0.76197 | 12957.3 |

| | | | | | | |
|---|---|---|---|---|---|---|
| GOBP_T_H | 8 | -0.14676 | -0.00302 | 0.20584 | 0.76206 | 12958.83 |
| GOMF_FLII | 15 | -0.10897 | -0.00307 | 0.15281 | 0.76211 | 12959.68 |
| GOMF_RNA | 7 | -0.10937 | -0.00211 | 0.15324 | 0.76229 | 12962.74 |
| GOBP_NEG | 112 | -0.03328 | -0.00256 | 0.046602 | 0.76245 | 12965.46 |
| GOBP_POS | 13 | -0.09205 | -0.00241 | 0.12883 | 0.76253 | 12966.82 |
| WP_SEROT | 12 | -0.12664 | -0.00319 | 0.17723 | 0.76255 | 12967.16 |
| GOBP_REG | 10 | -0.10488 | -0.00241 | 0.14673 | 0.76262 | 12968.35 |
| GOBP_REG | 17 | -0.09589 | -0.00288 | 0.13415 | 0.76263 | 12968.52 |
| FLECHNER_ | 38 | -0.05423 | -0.00243 | 0.075824 | 0.76276 | 12970.73 |
| GOBP_MIT | 10 | -0.10853 | -0.0025 | 0.15174 | 0.76277 | 12970.9 |
| GOBP_THIO | 85 | -0.0401 | -0.00268 | 0.056019 | 0.76294 | 12973.79 |
| GOMF_EXO | 75 | -0.04373 | -0.00275 | 0.061055 | 0.76307 | 12976.01 |
| REACTOME | 19 | -0.07461 | -0.00237 | 0.10417 | 0.76307 | 12976.01 |
| REACTOME | 8 | -0.12253 | -0.00252 | 0.17106 | 0.76309 | 12976.35 |
| GOBP_ENA | 18 | -0.08379 | -0.00259 | 0.11696 | 0.76312 | 12976.86 |
| GOMF_PRO | 81 | -0.03751 | -0.00245 | 0.052331 | 0.76321 | 12978.39 |
| NABA_MAT | 699 | -0.01543 | -0.00291 | 0.02152 | 0.76325 | 12979.07 |
| GOBP_REG | 4 | -0.12203 | -0.00178 | 0.1702 | 0.76331 | 12980.09 |
| BHAT_ESR1 | 271 | -0.02236 | -0.00266 | 0.031178 | 0.76332 | 12980.26 |
| GOBP_NEG | 32 | -0.06802 | -0.0028 | 0.094833 | 0.7634 | 12981.62 |
| GOBP_NEG | 4 | -0.18283 | -0.00266 | 0.25466 | 0.76361 | 12985.19 |
| GOMF_MH | 7 | -0.14552 | -0.0028 | 0.20264 | 0.76365 | 12985.87 |
| GOMF_CAI | 6 | -0.15694 | -0.0028 | 0.21837 | 0.76383 | 12988.93 |
| GOBP_RES | 86 | -0.041 | -0.00276 | 0.057011 | 0.76396 | 12991.14 |
| GOMF_MA | 14 | -0.10611 | -0.00289 | 0.14752 | 0.76401 | 12991.99 |
| LOPES_ME | 28 | -0.07168 | -0.00276 | 0.099595 | 0.76415 | 12994.37 |
| GOMF_L_L | 7 | -0.15797 | -0.00304 | 0.21947 | 0.76416 | 12994.54 |
| GOBP_SEQ | 6 | -0.15175 | -0.0027 | 0.21078 | 0.76421 | 12995.39 |
| GOBP_REG | 93 | -0.03667 | -0.00257 | 0.050925 | 0.76423 | 12995.73 |
| GOMF_PEF | 17 | -0.09844 | -0.00295 | 0.13664 | 0.76435 | 12997.77 |
| REACTOME | 15 | -0.08752 | -0.00247 | 0.12146 | 0.76441 | 12998.79 |
| ACEVEDO_ | 710 | -0.01464 | -0.00278 | 0.020293 | 0.7646 | 13002.02 |
| LEONARD_ | 40 | -0.05423 | -0.00249 | 0.075124 | 0.76479 | 13005.25 |
| SAMOLS_T | 49 | -0.05269 | -0.00268 | 0.072992 | 0.76479 | 13005.25 |
| MARSON_I | 29 | -0.0703 | -0.00275 | 0.097384 | 0.76481 | 13005.59 |
| GOBP_POS | 4 | -0.14331 | -0.00209 | 0.19851 | 0.76482 | 13005.76 |
| GOBP_PER | 9 | -0.11776 | -0.00257 | 0.16309 | 0.76486 | 13006.44 |
| GAVIN_FOX | 92 | -0.03551 | -0.00247 | 0.049148 | 0.76499 | 13008.65 |
| GOBP_OXII | 5 | -0.11412 | -0.00186 | 0.15796 | 0.76499 | 13008.65 |
| GOMF_OXI | 5 | -0.11412 | -0.00186 | 0.15796 | 0.76499 | 13008.65 |
| GOBP_POS | 22 | -0.06548 | -0.00223 | 0.090619 | 0.76502 | 13009.17 |
| GOBP_REG | 66 | -0.04623 | -0.00273 | 0.063985 | 0.76502 | 13009.17 |
| GOBP_BRO | 11 | -0.11797 | -0.00285 | 0.16324 | 0.76506 | 13009.85 |
| GOMF_MO | 85 | -0.03989 | -0.00267 | 0.055192 | 0.76509 | 13010.36 |
| WP_EV_RE | 6 | -0.16039 | -0.00286 | 0.22191 | 0.76509 | 13010.36 |

| | | | | | | |
|---|---|---|---|---|---|---|
| BIOCARTA_ | 13 | -0.12261 | -0.00322 | 0.16956 | 0.7652 | 13012.23 |
| GOMF_THI | 6 | -0.16261 | -0.0029 | 0.22472 | 0.76534 | 13014.61 |
| GOBP_AUT | 8 | -0.12957 | -0.00267 | 0.179 | 0.76541 | 13015.8 |
| GOBP_NEG | 11 | -0.11073 | -0.00267 | 0.15281 | 0.76565 | 13019.88 |
| GOMF_CAI | 46 | -0.0531 | -0.00262 | 0.073247 | 0.76576 | 13021.75 |
| SHIRAISHI_ | 9 | -0.10979 | -0.0024 | 0.15144 | 0.76577 | 13021.92 |
| REACTOME | 28 | -0.06794 | -0.00261 | 0.093669 | 0.76585 | 13023.28 |
| ACEVEDO_ | 240 | -0.0266 | -0.00298 | 0.036663 | 0.76593 | 13024.64 |
| GOBP_ADU | 79 | -0.04138 | -0.00267 | 0.057026 | 0.76596 | 13025.15 |
| REACTOME | 12 | -0.11029 | -0.00278 | 0.15199 | 0.76596 | 13025.15 |
| GOBP_SYN | 9 | -0.10291 | -0.00225 | 0.14178 | 0.76603 | 13026.34 |
| GOBP_NEG | 24 | -0.06764 | -0.00241 | 0.093121 | 0.76618 | 13028.89 |
| BERTUCCI_ | 163 | -0.02908 | -0.00269 | 0.03998 | 0.76649 | 13034.16 |
| GOBP_CHE | 934 | -0.01235 | -0.00268 | 0.016982 | 0.76653 | 13034.84 |
| GOCC_SAR | 9 | -0.1211 | -0.00264 | 0.1664 | 0.76662 | 13036.37 |
| GOBP_VEN | 32 | -0.07477 | -0.00308 | 0.1027 | 0.76669 | 13037.56 |
| GOBP_POS | 89 | -0.03894 | -0.00267 | 0.053466 | 0.76678 | 13039.09 |
| LUI_TARGE | 33 | -0.05755 | -0.0024 | 0.078993 | 0.76684 | 13040.11 |
| GOBP_PIGI | 11 | -0.10934 | -0.00264 | 0.15003 | 0.76693 | 13041.64 |
| GOBP_MYE | 5 | -0.19262 | -0.00313 | 0.26426 | 0.76696 | 13042.15 |
| GOBP_CAN | 5 | -0.17853 | -0.00291 | 0.24491 | 0.76699 | 13042.66 |
| GOBP_REG | 4 | -0.24305 | -0.00354 | 0.33341 | 0.76699 | 13042.66 |
| REACTOME | 29 | -0.07062 | -0.00277 | 0.096836 | 0.76707 | 13044.03 |
| WP_OXIDA | 50 | -0.04563 | -0.00235 | 0.062551 | 0.76712 | 13044.88 |
| XU_HGF_SI | 32 | -0.06306 | -0.00259 | 0.086452 | 0.76712 | 13044.88 |
| LEE_NAIVE | 16 | -0.09339 | -0.00272 | 0.12801 | 0.76717 | 13045.73 |
| GOBP_DOU | 12 | -0.10205 | -0.00257 | 0.13981 | 0.76727 | 13047.43 |
| GOBP_INTI | 46 | -0.05383 | -0.00265 | 0.073705 | 0.76742 | 13049.98 |
| REACTOME | 16 | -0.08761 | -0.00255 | 0.11992 | 0.76746 | 13050.66 |
| GOCC_HET | 25 | -0.07038 | -0.00256 | 0.096251 | 0.76766 | 13054.06 |
| GOBP_FRU | 13 | -0.09245 | -0.00243 | 0.12639 | 0.76775 | 13055.59 |
| GOBP_NEG | 4 | -0.21282 | -0.0031 | 0.29091 | 0.76778 | 13056.1 |
| GOMF_INC | 17 | -0.07903 | -0.00237 | 0.10802 | 0.76778 | 13056.1 |
| GOCC_AST | 19 | -0.08844 | -0.0028 | 0.12081 | 0.76793 | 13058.65 |
| GOBP_PRC | 15 | -0.08512 | -0.0024 | 0.11627 | 0.76794 | 13058.82 |
| GOBP_ESTA | 22 | -0.06945 | -0.00237 | 0.094753 | 0.76819 | 13063.07 |
| GOBP_BRA | 3 | -0.24975 | -0.00315 | 0.34074 | 0.7682 | 13063.24 |
| GOBP_MA( | 18 | -0.08965 | -0.00277 | 0.12229 | 0.76824 | 13063.92 |
| GOBP_MIC | 13 | -0.0953 | -0.0025 | 0.13 | 0.76824 | 13063.92 |
| GOBP_POS | 10 | -0.11543 | -0.00266 | 0.15744 | 0.76826 | 13064.26 |
| GOBP_PRIN | 13 | -0.10991 | -0.00288 | 0.14986 | 0.76834 | 13065.62 |
| LIU_VAV3_ | 16 | -0.08648 | -0.00252 | 0.11789 | 0.76839 | 13066.47 |
| GOBP_CELI | 69 | -0.04329 | -0.00261 | 0.058998 | 0.76844 | 13067.32 |
| GOBP_VES | 8 | -0.1662 | -0.00342 | 0.22649 | 0.76846 | 13067.66 |
| GOBP_POS | 41 | -0.05635 | -0.00262 | 0.076769 | 0.76853 | 13068.85 |

| | | | | | | |
|---|---|---|---|---|---|---|
| GOCC_TRA | 16 | -0.07662 | -0.00223 | 0.10436 | 0.76855 | 13069.19 |
| GOBP_POS | 12 | -0.10241 | -0.00258 | 0.1395 | 0.76856 | 13069.36 |
| GOBP_N_T | 29 | -0.06158 | -0.00241 | 0.083867 | 0.76859 | 13069.87 |
| JAATINEN_ | 218 | -0.02512 | -0.00268 | 0.034166 | 0.76885 | 13074.29 |
| GOMF_HIS | 11 | -0.10715 | -0.00259 | 0.14572 | 0.76892 | 13075.48 |
| REACTOME | 28 | -0.06487 | -0.0025 | 0.088222 | 0.76892 | 13075.48 |
| GOBP_PRC | 64 | -0.04214 | -0.00245 | 0.057291 | 0.769 | 13076.85 |
| GOBP_NEU | 10 | -0.1317 | -0.00303 | 0.17902 | 0.76903 | 13077.36 |
| GOBP_REG | 20 | -0.0894 | -0.00291 | 0.12145 | 0.76916 | 13079.57 |
| GOBP_NUC | 26 | -0.0804 | -0.00298 | 0.10917 | 0.76927 | 13081.44 |
| GOBP_REG | 7 | -0.11775 | -0.00227 | 0.1598 | 0.76939 | 13083.48 |
| REACTOME | 49 | -0.05004 | -0.00255 | 0.06789 | 0.76947 | 13084.84 |
| GOBP_SKE | 14 | -0.09052 | -0.00246 | 0.12276 | 0.76954 | 13086.03 |
| REACTOME | 5 | -0.12771 | -0.00208 | 0.17315 | 0.76961 | 13087.22 |
| WP_CONST | 30 | -0.07017 | -0.0028 | 0.095134 | 0.76961 | 13087.22 |
| GOBP_REG | 19 | -0.07703 | -0.00244 | 0.10443 | 0.76962 | 13087.39 |
| REACTOME | 8 | -0.12148 | -0.0025 | 0.16465 | 0.76968 | 13088.41 |
| GOMF_OXI | 27 | -0.06609 | -0.0025 | 0.089533 | 0.76978 | 13090.11 |
| GOBP_MEI | 8 | -0.13715 | -0.00282 | 0.18574 | 0.76986 | 13091.47 |
| GOBP_RES | 326 | -0.02105 | -0.00274 | 0.028504 | 0.76993 | 13092.66 |
| SCHAEFFEF | 14 | -0.09734 | -0.00265 | 0.13175 | 0.76999 | 13093.68 |
| GOBP_POS | 7 | -0.15306 | -0.00295 | 0.20714 | 0.77002 | 13094.19 |
| NIKOLSKY_ | 10 | -0.1924 | -0.00443 | 0.26031 | 0.77007 | 13095.04 |
| GOCC_COA | 173 | -0.02713 | -0.00259 | 0.036699 | 0.77013 | 13096.06 |
| GOMF_2_A | 9 | -0.11494 | -0.00251 | 0.15545 | 0.77015 | 13096.4 |
| GOBP_NEG | 14 | -0.12517 | -0.00341 | 0.16925 | 0.7702 | 13097.25 |
| GOBP_INSU | 9 | -0.1276 | -0.00279 | 0.1724 | 0.77039 | 13100.48 |
| GOBP_POS | 9 | -0.10735 | -0.00234 | 0.14504 | 0.77039 | 13100.48 |
| PEREZ_TP6 | 323 | -0.02131 | -0.00276 | 0.028767 | 0.7706 | 13104.05 |
| GOBP_REG | 38 | -0.06136 | -0.00275 | 0.082797 | 0.77067 | 13105.24 |
| GOBP_REG | 207 | -0.02656 | -0.00277 | 0.035826 | 0.77075 | 13106.6 |
| GOBP_POS | 33 | -0.06692 | -0.0028 | 0.090223 | 0.77085 | 13108.3 |
| GOMF_PRC | 9 | -0.13651 | -0.00298 | 0.18405 | 0.77086 | 13108.47 |
| GOBP_CAR | 5 | -0.17342 | -0.00282 | 0.2338 | 0.77087 | 13108.64 |
| GOBP_REP | 16 | -0.08291 | -0.00241 | 0.11178 | 0.77088 | 13108.81 |
| KEGG_PPA | 66 | -0.0444 | -0.00262 | 0.05984 | 0.77094 | 13109.83 |
| REACTOME | 16 | -0.09034 | -0.00263 | 0.12173 | 0.77099 | 13110.68 |
| GOMF_HO | 30 | -0.06819 | -0.00272 | 0.091877 | 0.77102 | 13111.2 |
| GOBP_CELI | 5 | -0.17343 | -0.00282 | 0.23342 | 0.77125 | 13115.11 |
| GOBP_COF | 13 | -0.08921 | -0.00234 | 0.12006 | 0.77128 | 13115.62 |
| GOBP_POS | 37 | -0.05517 | -0.00244 | 0.074232 | 0.77131 | 13116.13 |
| MARIADAS | 9 | -0.13906 | -0.00304 | 0.18708 | 0.77136 | 13116.98 |
| BERGER_M | 4 | -0.19874 | -0.00289 | 0.2672 | 0.77149 | 13119.19 |
| REACTOME | 191 | -0.0266 | -0.00266 | 0.035759 | 0.77149 | 13119.19 |
| BIOCARTA_ | 11 | -0.11451 | -0.00276 | 0.15392 | 0.77154 | 13120.04 |

| | | | | | | |
|---|---|---|---|---|---|---|
| GOBP_REG | 7 | -0.12767 | -0.00246 | 0.17157 | 0.77159 | 13120.89 |
| GOCC_CHE | 5 | -0.13818 | -0.00225 | 0.18554 | 0.77177 | 13123.95 |
| GOCC_EXT | 7 | -0.11938 | -0.0023 | 0.16028 | 0.7718 | 13124.46 |
| GOBP_ORG | 407 | -0.01877 | -0.00273 | 0.025193 | 0.77185 | 13125.31 |
| GOBP_UM | 5 | -0.15783 | -0.00257 | 0.21178 | 0.77193 | 13126.67 |
| GOMF_RET | 19 | -0.07971 | -0.00253 | 0.10696 | 0.77193 | 13126.67 |
| GOBP_CITF | 5 | -0.18687 | -0.00304 | 0.25072 | 0.77196 | 13127.18 |
| GOBP_REG | 9 | -0.14326 | -0.00313 | 0.19221 | 0.77196 | 13127.18 |
| GOBP_REN | 4 | -0.17877 | -0.0026 | 0.23977 | 0.77204 | 13128.54 |
| GOMF_CAI | 9 | -0.11821 | -0.00258 | 0.15853 | 0.77206 | 13128.88 |
| GOMF_HIS | 19 | -0.08831 | -0.0028 | 0.11842 | 0.77208 | 13129.22 |
| REACTOME | 55 | -0.04983 | -0.00269 | 0.066785 | 0.77219 | 13131.09 |
| GOBP_REG | 181 | -0.02718 | -0.00265 | 0.036412 | 0.77228 | 13132.62 |
| GOBP_INTI | 5 | -0.23645 | -0.00385 | 0.31676 | 0.77231 | 13133.13 |
| BIOCARTA_ | 12 | -0.10618 | -0.00268 | 0.1422 | 0.77237 | 13134.15 |
| GOBP_PHA | 7 | -0.14147 | -0.00272 | 0.18932 | 0.77254 | 13137.04 |
| WP_SPHIN | 23 | -0.08883 | -0.0031 | 0.1187 | 0.77286 | 13142.48 |
| HASLINGEF | 20 | -0.08117 | -0.00264 | 0.10842 | 0.77296 | 13144.18 |
| GOBP_REG | 8 | -0.14669 | -0.00302 | 0.19592 | 0.77298 | 13144.52 |
| GOCC_9PLU | 155 | -0.02902 | -0.00262 | 0.038756 | 0.77299 | 13144.69 |
| IYENGAR_F | 10 | -0.13778 | -0.00317 | 0.18378 | 0.77327 | 13149.46 |
| GOBP_RES | 6 | -0.15512 | -0.00277 | 0.20688 | 0.77331 | 13150.14 |
| GOBP_REG | 60 | -0.04726 | -0.00266 | 0.062992 | 0.77345 | 13152.52 |
| GOBP_POS | 48 | -0.05214 | -0.00263 | 0.069467 | 0.77352 | 13153.71 |
| GOBP_NEG | 7 | -0.13539 | -0.00261 | 0.18033 | 0.77361 | 13155.24 |
| WP_AMINO | 85 | -0.04016 | -0.00269 | 0.053483 | 0.77362 | 13155.41 |
| GOBP_NEU | 8 | -0.12343 | -0.00254 | 0.16436 | 0.77366 | 13156.09 |
| GOCC_END | 122 | -0.03282 | -0.00263 | 0.043691 | 0.77373 | 13157.28 |
| GOMF_TRA | 14 | -0.11113 | -0.00303 | 0.14793 | 0.77373 | 13157.28 |
| GOBP_POS | 10 | -0.11268 | -0.00259 | 0.14998 | 0.77376 | 13157.79 |
| GOBP_VES | 5 | -0.17955 | -0.00292 | 0.23897 | 0.77377 | 13157.96 |
| REACTOME | 206 | -0.02703 | -0.00281 | 0.035978 | 0.77379 | 13158.3 |
| VANASSE_I | 28 | -0.07787 | -0.003 | 0.10362 | 0.77381 | 13158.64 |
| GAVIN_FO | 86 | -0.04011 | -0.0027 | 0.053378 | 0.77382 | 13158.81 |
| GOCC_CTF | 7 | -0.11738 | -0.00226 | 0.15614 | 0.7739 | 13160.17 |
| GOCC_CYT | 8 | -0.15975 | -0.00329 | 0.21251 | 0.7739 | 13160.17 |
| GOBP_NEG | 18 | -0.08895 | -0.00275 | 0.1183 | 0.77394 | 13160.85 |
| REACTOME | 30 | -0.07174 | -0.00286 | 0.095409 | 0.77395 | 13161.02 |
| GOBP_TRA | 7 | -0.15727 | -0.00303 | 0.20905 | 0.77406 | 13162.89 |
| GOBP_REG | 35 | -0.05832 | -0.00251 | 0.0775 | 0.77413 | 13164.08 |
| REACTOME | 11 | -0.13538 | -0.00327 | 0.17969 | 0.77439 | 13168.5 |
| CAIRO_LIVI | 211 | -0.02559 | -0.00269 | 0.033965 | 0.77444 | 13169.35 |
| GOBP_REG | 18 | -0.10552 | -0.00326 | 0.14 | 0.7745 | 13170.37 |
| GOMF_DO | 5 | -0.13062 | -0.00213 | 0.17316 | 0.77467 | 13173.26 |
| GOBP_HIST | 66 | -0.04256 | -0.00251 | 0.05641 | 0.77471 | 13173.94 |

| | | | | | | |
|---|---|---|---|---|---|---|
| GOBP_INTI | 5 | -0.13136 | -0.00214 | 0.17407 | 0.77475 | 13174.62 |
| OHGUCHI_ | 157 | -0.03058 | -0.00278 | 0.040516 | 0.77481 | 13175.64 |
| GOBP_POS | 118 | -0.03625 | -0.00286 | 0.048023 | 0.77484 | 13176.15 |
| WP_MIRNA | 15 | -0.09721 | -0.00274 | 0.12874 | 0.77489 | 13177 |
| WP_NOTCI | 44 | -0.05173 | -0.00249 | 0.068494 | 0.77495 | 13178.02 |
| VALK_AML | 17 | -0.09852 | -0.00296 | 0.13045 | 0.77496 | 13178.19 |
| GOCC_PHA | 5 | -0.14183 | -0.00231 | 0.18772 | 0.77503 | 13179.39 |
| ZHONG_RE | 63 | -0.04669 | -0.00269 | 0.061752 | 0.77519 | 13182.11 |
| GOBP_POS | 23 | -0.07601 | -0.00265 | 0.10052 | 0.77522 | 13182.62 |
| GOBP_PYR | 6 | -0.13517 | -0.00241 | 0.17867 | 0.77532 | 13184.32 |
| SESTO_RES | 67 | -0.04781 | -0.00284 | 0.063188 | 0.77536 | 13185 |
| GOBP_AMI | 44 | -0.05817 | -0.00281 | 0.076876 | 0.77537 | 13185.17 |
| REACTOME | 63 | -0.04687 | -0.0027 | 0.061941 | 0.77538 | 13185.34 |
| YAGUE_PR | 7 | -0.15667 | -0.00302 | 0.20704 | 0.77539 | 13185.51 |
| GOBP_POS | 24 | -0.07329 | -0.00261 | 0.096839 | 0.77543 | 13186.19 |
| GOBP_POS | 6 | -0.17004 | -0.00303 | 0.22465 | 0.77543 | 13186.19 |
| GOBP_RES | 30 | -0.07477 | -0.00298 | 0.098791 | 0.77543 | 13186.19 |
| GOCC_NLS | 10 | -0.10625 | -0.00244 | 0.14034 | 0.77549 | 13187.21 |
| GOBP_FOR | 62 | -0.04797 | -0.00274 | 0.063315 | 0.77564 | 13189.76 |
| GOBP_NEG | 5 | -0.1504 | -0.00245 | 0.19832 | 0.77588 | 13193.84 |
| KEGG_ANT | 54 | -0.05908 | -0.00316 | 0.077886 | 0.77592 | 13194.52 |
| GOBP_POS | 11 | -0.12106 | -0.00292 | 0.15954 | 0.776 | 13195.88 |
| REACTOME | 29 | -0.07315 | -0.00287 | 0.096406 | 0.77601 | 13196.05 |
| PID_INTEG | 17 | -0.10249 | -0.00307 | 0.13506 | 0.77602 | 13196.22 |
| GOBP_NEG | 12 | -0.11589 | -0.00292 | 0.15264 | 0.77615 | 13198.43 |
| GOBP_HIST | 14 | -0.09613 | -0.00262 | 0.12659 | 0.77618 | 13198.94 |
| GOBP_NITI | 6 | -0.15484 | -0.00276 | 0.2039 | 0.77618 | 13198.94 |
| GOMF_HIS | 38 | -0.05958 | -0.00267 | 0.078449 | 0.77622 | 13199.62 |
| GOBP_NEG | 157 | -0.03056 | -0.00278 | 0.040231 | 0.77624 | 13199.96 |
| GOBP_ALP | 42 | -0.06031 | -0.00284 | 0.079391 | 0.77626 | 13200.3 |
| KEGG_SPH | 37 | -0.06435 | -0.00285 | 0.084697 | 0.77629 | 13200.81 |
| WP_VITAN | 171 | -0.03053 | -0.00289 | 0.04018 | 0.77631 | 13201.15 |
| GOBP_RES | 851 | -0.01374 | -0.00285 | 0.018078 | 0.77635 | 13201.83 |
| FARMER_B | 307 | -0.02184 | -0.00276 | 0.028699 | 0.77662 | 13206.42 |
| MARTORIA | 147 | -0.03232 | -0.00284 | 0.042442 | 0.77682 | 13209.82 |
| GOCC_MRI | 18 | -0.07957 | -0.00246 | 0.10443 | 0.77696 | 13212.2 |
| GOBP_REG | 441 | -0.01839 | -0.00278 | 0.024114 | 0.77709 | 13214.42 |
| VANLOO_S | 84 | -0.04034 | -0.00268 | 0.052874 | 0.77725 | 13217.14 |
| GOBP_GLU | 5 | -0.15368 | -0.0025 | 0.20133 | 0.77736 | 13219.01 |
| ZHENG_FO | 12 | -0.11819 | -0.00298 | 0.15483 | 0.77737 | 13219.18 |
| GILMORE_ | 11 | -0.09963 | -0.0024 | 0.13049 | 0.77741 | 13219.86 |
| WP_IMATI | 19 | -0.09525 | -0.00302 | 0.12471 | 0.77749 | 13221.22 |
| GOMF_HIS | 6 | -0.1293 | -0.0023 | 0.16921 | 0.7776 | 13223.09 |
| GOMF_REC | 41 | -0.05884 | -0.00274 | 0.076995 | 0.77763 | 13223.6 |
| GOMF_BEN | 2 | -0.24127 | -0.00248 | 0.31564 | 0.77767 | 13224.28 |

| | | | | | | |
|---|---|---|---|---|---|---|
| CHIARETTI_ | 67 | -0.04635 | -0.00276 | 0.060629 | 0.77772 | 13225.13 |
| VANDESLU | 20 | -0.08495 | -0.00276 | 0.11099 | 0.77798 | 13229.55 |
| GOBP_POS | 9 | -0.15625 | -0.00341 | 0.20399 | 0.77816 | 13232.61 |
| REACTOME | 12 | -0.1033 | -0.0026 | 0.13483 | 0.77819 | 13233.12 |
| GOBP_VAS | 52 | -0.05544 | -0.00291 | 0.072346 | 0.77825 | 13234.14 |
| REACTOME | 30 | -0.0706 | -0.00281 | 0.092129 | 0.77825 | 13234.14 |
| GOBP_RNA | 16 | -0.0911 | -0.00265 | 0.11884 | 0.77833 | 13235.5 |
| KEGG_STAI | 46 | -0.05745 | -0.00283 | 0.074935 | 0.77835 | 13235.84 |
| REACTOME | 6 | -0.1397 | -0.00249 | 0.18217 | 0.77841 | 13236.86 |
| GOBP_NEG | 4 | -0.23765 | -0.00346 | 0.30989 | 0.77842 | 13237.03 |
| ZHAN_MUI | 46 | -0.05621 | -0.00277 | 0.073288 | 0.77842 | 13237.03 |
| GOBP_REC | 4 | -0.20634 | -0.003 | 0.26902 | 0.77846 | 13237.71 |
| REACTOME | 24 | -0.07448 | -0.00265 | 0.097065 | 0.77855 | 13239.24 |
| GOBP_END | 8 | -0.1317 | -0.00271 | 0.17161 | 0.7786 | 13240.09 |
| GOBP_INTI | 55 | -0.05259 | -0.00283 | 0.068512 | 0.77861 | 13240.26 |
| GOBP_ETH | 7 | -0.14975 | -0.00288 | 0.19509 | 0.77863 | 13240.6 |
| GOBP_REG | 9 | -0.13161 | -0.00287 | 0.17141 | 0.77869 | 13241.62 |
| GOCC_PEP | 11 | -0.11614 | -0.0028 | 0.15115 | 0.77888 | 13244.85 |
| GENTILE_U | 18 | -0.08752 | -0.0027 | 0.11389 | 0.77889 | 13245.02 |
| GOCC_NLR | 7 | -0.15316 | -0.00295 | 0.19931 | 0.77889 | 13245.02 |
| GOMF_RET | 15 | -0.08986 | -0.00253 | 0.11693 | 0.77891 | 13245.36 |
| GOMF_HYI | 7 | -0.15887 | -0.00306 | 0.20665 | 0.77898 | 13246.55 |
| GOBP_PRC | 6 | -0.15872 | -0.00283 | 0.20646 | 0.77899 | 13246.72 |
| GOMF_GTI | 299 | -0.02241 | -0.0028 | 0.029142 | 0.77904 | 13247.58 |
| GOBP_REG | 122 | -0.03761 | -0.00301 | 0.048898 | 0.77906 | 13247.92 |
| GOBP_POS | 5 | -0.1417 | -0.00231 | 0.18424 | 0.77908 | 13248.26 |
| GOMF_MC | 23 | -0.07904 | -0.00276 | 0.10269 | 0.77924 | 13250.98 |
| GOBP_INH | 16 | -0.10466 | -0.00305 | 0.13595 | 0.77931 | 13252.17 |
| GOBP_REG | 6 | -0.15022 | -0.00268 | 0.19512 | 0.77931 | 13252.17 |
| BIOCARTA_ | 14 | -0.11368 | -0.00309 | 0.1476 | 0.77941 | 13253.87 |
| GOMF_NU | 13 | -0.10352 | -0.00272 | 0.13438 | 0.77944 | 13254.38 |
| HASINA_N | 13 | -0.12701 | -0.00333 | 0.16486 | 0.77947 | 13254.89 |
| GOMF_HIS | 12 | -0.09599 | -0.00242 | 0.12447 | 0.77969 | 13258.63 |
| MANNE_CO | 122 | -0.03243 | -0.0026 | 0.042033 | 0.77981 | 13260.67 |
| BIOCARTA_ | 28 | -0.07529 | -0.0029 | 0.097569 | 0.77983 | 13261.01 |
| RUTELLA_F | 223 | -0.02552 | -0.00276 | 0.033072 | 0.77986 | 13261.52 |
| GOBP_REG | 7 | -0.17586 | -0.00339 | 0.22778 | 0.77995 | 13263.05 |
| BIOCARTA_ | 10 | -0.10502 | -0.00242 | 0.13595 | 0.78008 | 13265.26 |
| REACTOME | 46 | -0.05551 | -0.00274 | 0.071844 | 0.78011 | 13265.77 |
| AMUNDSO | 16 | -0.1064 | -0.0031 | 0.13769 | 0.78016 | 13266.62 |
| GOMF_G_I | 21 | -0.09057 | -0.00302 | 0.11719 | 0.78018 | 13266.96 |
| GOBP_EOS | 22 | -0.0953 | -0.00325 | 0.12328 | 0.78024 | 13267.98 |
| ZERBINI_RI | 9 | -0.15475 | -0.00338 | 0.20017 | 0.78026 | 13268.32 |
| GOBP_MIT | 39 | -0.06006 | -0.00273 | 0.077643 | 0.78038 | 13270.36 |
| REACTOME | 7 | -0.1847 | -0.00356 | 0.23878 | 0.78038 | 13270.36 |

| | | | | | | |
|---|---|---|---|---|---|---|
| YU_BAP1_T | 29 | -0.07079 | -0.00277 | 0.09148 | 0.78048 | 13272.06 |
| GOBP_CELI | 202 | -0.02648 | -0.00272 | 0.034214 | 0.7805 | 13272.4 |
| GOBP_REG | 15 | -0.09554 | -0.00269 | 0.12344 | 0.78053 | 13272.91 |
| GOMF_TRA | 853 | -0.01334 | -0.00277 | 0.017217 | 0.78071 | 13275.97 |
| GOBP_ANT | 7 | -0.16521 | -0.00318 | 0.21326 | 0.78074 | 13276.48 |
| GOBP_REG | 31 | -0.0763 | -0.00309 | 0.098443 | 0.78083 | 13278.01 |
| WP_NUCLE | 39 | -0.05982 | -0.00272 | 0.077169 | 0.78088 | 13278.86 |
| GOBP_REG | 5 | -0.13619 | -0.00222 | 0.17565 | 0.78092 | 13279.54 |
| BIOCARTA_ | 10 | -0.12498 | -0.00288 | 0.16109 | 0.78107 | 13282.1 |
| GOBP_AER | 166 | -0.02803 | -0.00262 | 0.036114 | 0.78113 | 13283.12 |
| GOBP_GLU | 40 | -0.06023 | -0.00277 | 0.077546 | 0.78131 | 13286.18 |
| WEI_MIR34 | 141 | -0.03321 | -0.00286 | 0.042756 | 0.78134 | 13286.69 |
| GOBP_HUN | 41 | -0.06976 | -0.00325 | 0.089797 | 0.78138 | 13287.37 |
| GOBP_HYP | 3 | -0.24956 | -0.00315 | 0.32107 | 0.7815 | 13289.41 |
| REACTOME | 5 | -0.17096 | -0.00278 | 0.21992 | 0.78153 | 13289.92 |
| GOBP_AMY | 67 | -0.0471 | -0.0028 | 0.060559 | 0.78163 | 13291.62 |
| REACTOME | 3 | -0.28612 | -0.00361 | 0.3679 | 0.78163 | 13291.62 |
| MIKKELSEN | 37 | -0.06543 | -0.00289 | 0.084114 | 0.78166 | 13292.13 |
| GOBP_MU | 9 | -0.1097 | -0.00239 | 0.14096 | 0.78178 | 13294.17 |
| GOBP_MYE | 23 | -0.07606 | -0.00265 | 0.097729 | 0.7818 | 13294.51 |
| GOBP_TOC | 27 | -0.07791 | -0.00294 | 0.1001 | 0.7818 | 13294.51 |
| REACTOME | 9 | -0.13004 | -0.00284 | 0.167 | 0.78192 | 13296.55 |
| GOCC_INTE | 234 | -0.03362 | -0.00372 | 0.043172 | 0.78195 | 13297.06 |
| GOBP_IMP | 243 | -0.02651 | -0.00299 | 0.034026 | 0.78208 | 13299.27 |
| GOBP_SEN | 7 | -0.13733 | -0.00264 | 0.17623 | 0.78208 | 13299.27 |
| KANG_GLIS | 34 | -0.07397 | -0.00314 | 0.094871 | 0.7822 | 13301.31 |
| GOBP_RES | 6 | -0.13267 | -0.00236 | 0.17011 | 0.78228 | 13302.67 |
| GOCC_FILO | 98 | -0.04074 | -0.00293 | 0.052228 | 0.78228 | 13302.67 |
| GOCC_PAR | 6 | -0.16647 | -0.00297 | 0.21344 | 0.78229 | 13302.84 |
| TERAMOTO | 14 | -0.08869 | -0.00241 | 0.11365 | 0.78242 | 13305.05 |
| GOBP_ANT | 52 | -0.06003 | -0.00315 | 0.076908 | 0.78245 | 13305.56 |
| GOBP_POS | 63 | -0.05094 | -0.00294 | 0.065254 | 0.78249 | 13306.24 |
| GOMF_ME | 115 | -0.03973 | -0.00309 | 0.05089 | 0.78252 | 13306.75 |
| REACTOME | 18 | -0.09604 | -0.00296 | 0.12299 | 0.78256 | 13307.43 |
| GOBP_INTE | 271 | -0.02413 | -0.00287 | 0.030883 | 0.78267 | 13309.3 |
| BEIER_GLIO | 37 | -0.06346 | -0.00281 | 0.081189 | 0.78278 | 13311.17 |
| CERVERA_S | 33 | -0.07275 | -0.00304 | 0.093072 | 0.78278 | 13311.17 |
| WANG_ME | 13 | -0.12268 | -0.00322 | 0.15696 | 0.78278 | 13311.17 |
| HOLLEMAN | 11 | -0.10741 | -0.00259 | 0.13737 | 0.78285 | 13312.36 |
| GOBP_REG | 75 | -0.04879 | -0.00307 | 0.062388 | 0.78289 | 13313.04 |
| GOBP_LEU | 359 | -0.02176 | -0.00297 | 0.027823 | 0.7829 | 13313.21 |
| GOBP_REG | 6 | -0.13357 | -0.00238 | 0.17077 | 0.78295 | 13314.06 |
| RUTELLA_F | 316 | -0.02249 | -0.00288 | 0.02873 | 0.78307 | 13316.11 |
| GOBP_REG | 7 | -0.14187 | -0.00273 | 0.18121 | 0.78315 | 13317.47 |
| GOBP_MO | 21 | -0.08244 | -0.00275 | 0.1053 | 0.78317 | 13317.81 |

| | | | | | | |
|---|---|---|---|---|---|---|
| REACTOME | 52 | -0.06091 | -0.00319 | 0.077727 | 0.78338 | 13321.38 |
| YAGI_AML_ | 178 | -0.03027 | -0.00293 | 0.038608 | 0.78351 | 13323.59 |
| GOBP_INTI | 29 | -0.06401 | -0.00251 | 0.081595 | 0.78362 | 13325.46 |
| GOBP_CELI | 70 | -0.05143 | -0.00313 | 0.065549 | 0.78367 | 13326.31 |
| KEGG_STEI | 15 | -0.09421 | -0.00265 | 0.12005 | 0.78371 | 13326.99 |
| WP_SPINA | 112 | -0.03927 | -0.00302 | 0.050004 | 0.78384 | 13329.2 |
| GOBP_REG | 39 | -0.06446 | -0.00293 | 0.082087 | 0.78386 | 13329.54 |
| GOBP_REG | 57 | -0.04938 | -0.00271 | 0.062868 | 0.78392 | 13330.56 |
| GOMF_REC | 19 | -0.1009 | -0.0032 | 0.12844 | 0.78393 | 13330.73 |
| GOBP_GLU | 47 | -0.05883 | -0.00293 | 0.074883 | 0.78395 | 13331.07 |
| WEBER_MI | 20 | -0.08447 | -0.00275 | 0.10748 | 0.78402 | 13332.26 |
| GOBP_NEG | 5 | -0.16638 | -0.00271 | 0.21165 | 0.78409 | 13333.45 |
| NIKOLSKY_ | 117 | -0.07357 | -0.00577 | 0.09359 | 0.78409 | 13333.45 |
| GOBP_POS | 14 | -0.09937 | -0.00271 | 0.12637 | 0.78417 | 13334.81 |
| GOMF_CXC | 18 | -0.11901 | -0.00367 | 0.15134 | 0.78418 | 13334.98 |
| GOBP_MO | 29 | -0.06264 | -0.00245 | 0.079599 | 0.78432 | 13337.36 |
| GOBP_COF | 9 | -0.16854 | -0.00368 | 0.21416 | 0.78435 | 13337.87 |
| LEE_LIVER_ | 56 | -0.05466 | -0.00297 | 0.06942 | 0.78448 | 13340.08 |
| GOMF_NEU | 5 | -0.18537 | -0.00302 | 0.23536 | 0.78453 | 13340.93 |
| WP_NOVEI | 54 | -0.0532 | -0.00284 | 0.067548 | 0.78453 | 13340.93 |
| GOBP_POS | 7 | -0.17033 | -0.00328 | 0.21621 | 0.78459 | 13341.95 |
| TURASHVIL | 5 | -0.23707 | -0.00386 | 0.30085 | 0.78464 | 13342.8 |
| GOMF_CAI | 7 | -0.14071 | -0.00271 | 0.17856 | 0.78465 | 13342.97 |
| GOMF_OXI | 11 | -0.12948 | -0.00312 | 0.1643 | 0.78467 | 13343.31 |
| WP_S1P_R | 25 | -0.0737 | -0.00268 | 0.093493 | 0.78473 | 13344.33 |
| GOBP_DEN | 10 | -0.13821 | -0.00318 | 0.17528 | 0.78479 | 13345.35 |
| KORKOLA_ | 11 | -0.11394 | -0.00275 | 0.14449 | 0.78483 | 13346.03 |
| GOMF_DO | 6 | -0.17978 | -0.0032 | 0.22784 | 0.78495 | 13348.07 |
| GOBP_REG | 4 | -0.20981 | -0.00305 | 0.26579 | 0.78506 | 13349.95 |
| GOCC_CLA | 8 | -0.17779 | -0.00366 | 0.22518 | 0.7851 | 13350.63 |
| BIOCARTA_ | 20 | -0.08111 | -0.00264 | 0.10269 | 0.78519 | 13352.16 |
| PID_TOLL_ | 21 | -0.08573 | -0.00286 | 0.10853 | 0.7852 | 13352.33 |
| GOBP_NUC | 111 | -0.03861 | -0.00295 | 0.04887 | 0.78526 | 13353.35 |
| GOMF_P_T | 22 | -0.08368 | -0.00286 | 0.10579 | 0.78553 | 13357.94 |
| GOBP_SEQ | 5 | -0.19531 | -0.00318 | 0.24686 | 0.78557 | 13358.62 |
| GOBP_RES | 25 | -0.08368 | -0.00304 | 0.10574 | 0.78565 | 13359.98 |
| GOMF_GD | 9 | -0.13175 | -0.00288 | 0.16645 | 0.78567 | 13360.32 |
| GOBP_REG | 39 | -0.063 | -0.00286 | 0.079545 | 0.7858 | 13362.53 |
| GOBP_POS | 7 | -0.17488 | -0.00337 | 0.22075 | 0.78587 | 13363.72 |
| GOBP_REG | 23 | -0.0916 | -0.0032 | 0.11563 | 0.78587 | 13363.72 |
| GOBP_TOR | 11 | -0.12282 | -0.00296 | 0.15502 | 0.7859 | 13364.23 |
| GOBP_REG | 14 | -0.09982 | -0.00272 | 0.12595 | 0.78598 | 13365.59 |
| KEGG_PAN | 16 | -0.09587 | -0.00279 | 0.12091 | 0.78607 | 13367.12 |
| GOBP_IMP | 193 | -0.03039 | -0.00306 | 0.038309 | 0.7862 | 13369.33 |
| GOBP_POS | 18 | -0.08195 | -0.00253 | 0.10328 | 0.78624 | 13370.01 |

| | | | | | | |
|---|---|---|---|---|---|---|
| REACTOME | 4 | -0.16209 | -0.00236 | 0.20423 | 0.7863 | 13371.03 |
| KEGG_TIGH | 128 | -0.03671 | -0.00301 | 0.046237 | 0.7864 | 13372.73 |
| GOBP_POS | 163 | -0.0319 | -0.00295 | 0.040157 | 0.78651 | 13374.6 |
| BRACHAT_ | 20 | -0.08779 | -0.00286 | 0.11046 | 0.78662 | 13376.47 |
| GOBP_RES | 259 | -0.02444 | -0.00284 | 0.030744 | 0.78665 | 13376.98 |
| GOBP_HIST | 6 | -0.15271 | -0.00272 | 0.19207 | 0.78671 | 13378 |
| REACTOME | 31 | -0.06958 | -0.00282 | 0.087495 | 0.78674 | 13378.51 |
| GOBP_REG | 47 | -0.05775 | -0.00288 | 0.072578 | 0.78687 | 13380.72 |
| SMID_BREA | 5 | -0.21479 | -0.0035 | 0.26994 | 0.7869 | 13381.23 |
| REACTOME | 23 | -0.0766 | -0.00267 | 0.096213 | 0.78703 | 13383.45 |
| REACTOME | 8 | -0.14392 | -0.00296 | 0.18072 | 0.78709 | 13384.47 |
| REACTOME | 26 | -0.08386 | -0.00311 | 0.10522 | 0.78728 | 13387.7 |
| GOBP_BON | 10 | -0.11833 | -0.00272 | 0.14846 | 0.78729 | 13387.87 |
| GOBP_NEG | 5 | -0.18574 | -0.00302 | 0.23292 | 0.7874 | 13389.74 |
| GOMF_HIS | 14 | -0.09074 | -0.00247 | 0.11379 | 0.7874 | 13389.74 |
| GOMF_PHO | 27 | -0.08159 | -0.00308 | 0.10231 | 0.7874 | 13389.74 |
| GOBP_TUM | 96 | -0.04137 | -0.00294 | 0.051878 | 0.78741 | 13389.91 |
| GOBP_EPIT | 11 | -0.12216 | -0.00295 | 0.15299 | 0.7877 | 13394.84 |
| GAVIN_FOX | 76 | -0.04783 | -0.00303 | 0.059902 | 0.78771 | 13395.01 |
| GOBP_GLY | 19 | -0.08322 | -0.00264 | 0.10422 | 0.78771 | 13395.01 |
| REACTOME | 72 | -0.0453 | -0.00279 | 0.056713 | 0.78775 | 13395.69 |
| GOBP_REG | 19 | -0.09581 | -0.00304 | 0.11995 | 0.78778 | 13396.2 |
| GOBP_REG | 12 | -0.13107 | -0.0033 | 0.164 | 0.7879 | 13398.24 |
| REACTOME | 69 | -0.04758 | -0.00287 | 0.059526 | 0.78796 | 13399.26 |
| GOMF_CAF | 41 | -0.06788 | -0.00316 | 0.084891 | 0.78801 | 13400.11 |
| GOCC_FICO | 58 | -0.05415 | -0.003 | 0.067703 | 0.78808 | 13401.3 |
| REACTOME | 37 | -0.06928 | -0.00306 | 0.08662 | 0.78808 | 13401.3 |
| REACTOME | 12 | -0.11767 | -0.00297 | 0.14711 | 0.78809 | 13401.47 |
| WP_ROLES | 22 | -0.08823 | -0.00301 | 0.11031 | 0.7881 | 13401.64 |
| DEBIASI_AI | 281 | -0.02406 | -0.00291 | 0.030079 | 0.78813 | 13402.15 |
| GOMF_SHO | 6 | -0.17312 | -0.00309 | 0.21641 | 0.78813 | 13402.15 |
| GOCC_POS | 9 | -0.12397 | -0.00271 | 0.15492 | 0.78821 | 13403.51 |
| CASTELLAN | 5 | -0.18647 | -0.00303 | 0.23296 | 0.78826 | 13404.36 |
| GOBP_POS | 7 | -0.13749 | -0.00265 | 0.17177 | 0.78826 | 13404.36 |
| GOBP_MYO | 107 | -0.0382 | -0.00287 | 0.047698 | 0.78837 | 13406.23 |
| GOBP_NEG | 152 | -0.03397 | -0.00304 | 0.042397 | 0.78846 | 13407.76 |
| GOCC_EXT | 6 | -0.19776 | -0.00353 | 0.24676 | 0.78855 | 13409.29 |
| GOMF_CYC | 5 | -0.14882 | -0.00242 | 0.18567 | 0.78858 | 13409.8 |
| REACTOME | 52 | -0.05683 | -0.00298 | 0.070902 | 0.78859 | 13409.97 |
| GOMF_ENI | 169 | -0.0335 | -0.00316 | 0.041786 | 0.78861 | 13410.31 |
| GOBP_DEF | 273 | -0.02571 | -0.00307 | 0.032058 | 0.78867 | 13411.33 |
| GOBP_POS | 24 | -0.08296 | -0.00296 | 0.10346 | 0.78868 | 13411.5 |
| REACTOME | 25 | -0.08235 | -0.00299 | 0.10268 | 0.78872 | 13412.18 |
| REACTOME | 6 | -0.18466 | -0.00329 | 0.23024 | 0.78873 | 13412.35 |
| GOMF_MR | 12 | -0.09757 | -0.00246 | 0.12164 | 0.78875 | 13412.69 |

| | | | | | | |
|---|---|---|---|---|---|---|
| GOMF_PRO | 33 | -0.06776 | -0.00283 | 0.084437 | 0.78884 | 13414.22 |
| WIELAND_ | 82 | -0.0431 | -0.00283 | 0.053639 | 0.78915 | 13419.5 |
| GOBP_POS | 4 | -0.23423 | -0.00341 | 0.2915 | 0.78916 | 13419.67 |
| BURTON_A | 187 | -0.02811 | -0.00278 | 0.034978 | 0.78921 | 13420.52 |
| TONKS_TA | 54 | -0.05487 | -0.00293 | 0.068266 | 0.78922 | 13420.69 |
| GOBP_REG | 186 | -0.02941 | -0.00291 | 0.036576 | 0.78933 | 13422.56 |
| GOBP_ACR | 27 | -0.08299 | -0.00314 | 0.10314 | 0.78947 | 13424.94 |
| GOCC_INTI | 18 | -0.0875 | -0.0027 | 0.10874 | 0.78948 | 13425.11 |
| GOBP_POS | 138 | -0.035 | -0.00298 | 0.043484 | 0.78953 | 13425.96 |
| BIOCARTA_ | 10 | -0.13342 | -0.00307 | 0.16569 | 0.78965 | 13428 |
| BLUM_RES | 321 | -0.02256 | -0.00292 | 0.027982 | 0.78992 | 13432.59 |
| TESAR_ALK | 5 | -0.23394 | -0.00381 | 0.29019 | 0.78992 | 13432.59 |
| WP_AIRWA | 15 | -0.10178 | -0.00287 | 0.12624 | 0.78994 | 13432.93 |
| GOBP_HEP | 9 | -0.165 | -0.0036 | 0.20463 | 0.78997 | 13433.44 |
| WP_PTF1A | 11 | -0.14507 | -0.0035 | 0.1799 | 0.78998 | 13433.61 |
| GOBP_SUR | 17 | -0.10723 | -0.00322 | 0.13298 | 0.78999 | 13433.78 |
| REACTOME | 38 | -0.06633 | -0.00297 | 0.082247 | 0.79001 | 13434.12 |
| ZHAN_MUI | 51 | -0.05831 | -0.00303 | 0.072289 | 0.79005 | 13434.8 |
| GOBP_POS | 9 | -0.12904 | -0.00282 | 0.15997 | 0.79007 | 13435.14 |
| REACTOME | 17 | -0.08796 | -0.00264 | 0.10899 | 0.79018 | 13437.01 |
| GOBP_MIT | 6 | -0.12463 | -0.00222 | 0.15438 | 0.79025 | 13438.2 |
| GOBP_MAI | 12 | -0.17224 | -0.00434 | 0.21328 | 0.79033 | 13439.56 |
| GOBP_POS | 6 | -0.19785 | -0.00353 | 0.2446 | 0.79069 | 13445.68 |
| GOBP_ALA | 16 | -0.1107 | -0.00322 | 0.13685 | 0.7907 | 13445.85 |
| REACTOME | 14 | -0.0997 | -0.00271 | 0.12325 | 0.79072 | 13446.19 |
| GOBP_HIST | 4 | -0.17904 | -0.00261 | 0.22114 | 0.79091 | 13449.42 |
| GOBP_GLY | 8 | -0.15323 | -0.00315 | 0.18922 | 0.79096 | 13450.27 |
| GOCC_F_A | 8 | -0.15023 | -0.00309 | 0.18542 | 0.79108 | 13452.32 |
| MILICIC_FA | 9 | -0.14971 | -0.00327 | 0.18476 | 0.79112 | 13453 |
| GOMF_3_F | 9 | -0.14742 | -0.00322 | 0.18192 | 0.79113 | 13453.17 |
| GOBP_ANT | 164 | -0.03204 | -0.00297 | 0.039521 | 0.79123 | 13454.87 |
| GOMF_ACI | 23 | -0.08989 | -0.00314 | 0.11083 | 0.79132 | 13456.4 |
| TENEDINI_ | 59 | -0.05282 | -0.00295 | 0.065116 | 0.79137 | 13457.25 |
| GOMF_ALK | 6 | -0.24038 | -0.00428 | 0.29618 | 0.79148 | 13459.12 |
| GOBP_BRA | 7 | -0.16787 | -0.00323 | 0.20674 | 0.7916 | 13461.16 |
| GOBP_NUC | 6 | -0.19817 | -0.00353 | 0.24404 | 0.7916 | 13461.16 |
| GOBP_MEI | 8 | -0.1542 | -0.00317 | 0.18986 | 0.79164 | 13461.84 |
| PID_S1P_S | 29 | -0.0777 | -0.00304 | 0.095663 | 0.79167 | 13462.35 |
| GOCC_MRI | 14 | -0.09334 | -0.00254 | 0.11488 | 0.79175 | 13463.71 |
| GOCC_PRC | 55 | -0.0566 | -0.00305 | 0.069649 | 0.79176 | 13463.88 |
| GOBP_POS | 243 | -0.02536 | -0.00286 | 0.031208 | 0.79178 | 13464.22 |
| GOBP_FIBF | 5 | -0.19243 | -0.00313 | 0.23677 | 0.79181 | 13464.73 |
| REACTOME | 24 | -0.07406 | -0.00264 | 0.091079 | 0.79194 | 13466.94 |
| GOCC_TUB | 5 | -0.15924 | -0.00259 | 0.19574 | 0.79204 | 13468.64 |
| GOCC_PRC | 56 | -0.05456 | -0.00297 | 0.067044 | 0.79209 | 13469.49 |

| | | | | | | |
|---|---|---|---|---|---|---|
| GOBP_REG | 7 | -0.15008 | -0.00289 | 0.18441 | 0.79214 | 13470.34 |
| GOBP_MO | 23 | -0.08528 | -0.00298 | 0.10477 | 0.79216 | 13470.68 |
| GOBP_CELI | 571 | -0.01737 | -0.00298 | 0.021332 | 0.79227 | 13472.55 |
| GOMF_ACT | 191 | -0.03016 | -0.00302 | 0.037034 | 0.79227 | 13472.55 |
| WP_BIOSYI | 8 | -0.13889 | -0.00286 | 0.17049 | 0.79235 | 13473.91 |
| GOBP_POS | 5 | -0.1391 | -0.00226 | 0.17074 | 0.79237 | 13474.25 |
| GOCC_POS | 12 | -0.1046 | -0.00264 | 0.12837 | 0.79241 | 13474.93 |
| GOBP_REG | 46 | -0.05817 | -0.00287 | 0.071389 | 0.79242 | 13475.1 |
| GOBP_NEG | 3 | -0.16516 | -0.00208 | 0.20264 | 0.79247 | 13475.95 |
| REACTOME | 9 | -0.1615 | -0.00353 | 0.19813 | 0.79249 | 13476.29 |
| PID_NFKAF | 15 | -0.09429 | -0.00266 | 0.11565 | 0.79253 | 13476.97 |
| GOBP_POS | 14 | -0.10378 | -0.00283 | 0.12728 | 0.79255 | 13477.31 |
| MEISSNER_ | 17 | -0.09464 | -0.00284 | 0.11605 | 0.7926 | 13478.16 |
| GOBP_INTI | 38 | -0.06199 | -0.00278 | 0.075983 | 0.7927 | 13479.86 |
| GOBP_B_C | 119 | -0.03898 | -0.00309 | 0.047754 | 0.79282 | 13481.9 |
| GOBP_GRA | 18 | -0.08918 | -0.00275 | 0.10924 | 0.79286 | 13482.58 |
| GOBP_REG | 25 | -0.08934 | -0.00325 | 0.10943 | 0.79286 | 13482.58 |
| GOBP_PHC | 47 | -0.06269 | -0.00312 | 0.07677 | 0.79291 | 13483.43 |
| KYNG_ENV | 16 | -0.1086 | -0.00316 | 0.13295 | 0.79298 | 13484.62 |
| GOMF_CYC | 21 | -0.1006 | -0.00335 | 0.12313 | 0.79303 | 13485.48 |
| GOBP_REG | 34 | -0.07236 | -0.00307 | 0.088548 | 0.79307 | 13486.16 |
| GOMF_SH3 | 114 | -0.03785 | -0.00293 | 0.046314 | 0.79313 | 13487.18 |
| GOBP_LAM | 18 | -0.10116 | -0.00312 | 0.12375 | 0.79316 | 13487.69 |
| WINTER_H | 45 | -0.06341 | -0.00309 | 0.077507 | 0.79335 | 13490.92 |
| STEIN_ESTI | 17 | -0.11941 | -0.00358 | 0.14595 | 0.79336 | 13491.09 |
| WP_GLYCC | 9 | -0.12308 | -0.00269 | 0.15041 | 0.79339 | 13491.6 |
| GOBP_END | 5 | -0.21307 | -0.00347 | 0.26036 | 0.79342 | 13492.11 |
| GOCC_ER_ | 42 | -0.06066 | -0.00286 | 0.074114 | 0.79344 | 13492.45 |
| GOBP_VES | 5 | -0.25955 | -0.00422 | 0.31709 | 0.79347 | 13492.96 |
| GOBP_NEG | 58 | -0.05538 | -0.00306 | 0.067649 | 0.7935 | 13493.47 |
| GOCC_POS | 96 | -0.04545 | -0.00323 | 0.055513 | 0.7935 | 13493.47 |
| GOBP_PLA | 54 | -0.05446 | -0.00291 | 0.066506 | 0.79357 | 13494.66 |
| GOBP_CELI | 16 | -0.10929 | -0.00318 | 0.1334 | 0.79368 | 13496.53 |
| GOBP_REG | 42 | -0.06447 | -0.00304 | 0.07864 | 0.79383 | 13499.08 |
| GOMF_CAI | 17 | -0.113 | -0.00339 | 0.13775 | 0.79398 | 13501.63 |
| KEGG_NOT | 46 | -0.05464 | -0.00269 | 0.066611 | 0.79398 | 13501.63 |
| REACTOME | 52 | -0.05509 | -0.00289 | 0.067151 | 0.79398 | 13501.63 |
| OKUMURA | 164 | -0.03078 | -0.00286 | 0.037491 | 0.79415 | 13504.52 |
| PID_ENDO | 62 | -0.05481 | -0.00314 | 0.06676 | 0.79415 | 13504.52 |
| GOCC_PER | 29 | -0.07106 | -0.00278 | 0.086545 | 0.7942 | 13505.37 |
| GOMF_CHI | 55 | -0.0666 | -0.00359 | 0.081104 | 0.79422 | 13505.71 |
| GOBP_PRE | 67 | -0.04893 | -0.00291 | 0.059574 | 0.79426 | 13506.39 |
| WANG_CLA | 27 | -0.08072 | -0.00305 | 0.098257 | 0.79431 | 13507.24 |
| IVANOVSK | 87 | -0.04187 | -0.00284 | 0.050967 | 0.79434 | 13507.75 |
| GOBP_POS | 24 | -0.09574 | -0.00341 | 0.1165 | 0.7944 | 13508.77 |

| | | | | | | |
|---|---|---|---|---|---|---|
| GOBP_INO | 114 | -0.03926 | -0.00304 | 0.047763 | 0.79444 | 13509.45 |
| GOBP_REG | 353 | -0.02213 | -0.003 | 0.026911 | 0.7946 | 13512.17 |
| GOBP_THR | 5 | -0.12849 | -0.00209 | 0.15621 | 0.79461 | 13512.34 |
| GOMF_OM | 13 | -0.11309 | -0.00297 | 0.13734 | 0.79486 | 13516.59 |
| REACTOME | 38 | -0.08907 | -0.00399 | 0.10817 | 0.79486 | 13516.59 |
| GOMF_PRI | 9 | -0.12141 | -0.00265 | 0.14744 | 0.79487 | 13516.76 |
| MARIADAS | 15 | -0.098 | -0.00276 | 0.11899 | 0.7949 | 13517.27 |
| WP_CONTI | 13 | -0.12976 | -0.0034 | 0.15753 | 0.79495 | 13518.12 |
| GOMF_PEF | 18 | -0.09217 | -0.00284 | 0.11189 | 0.79496 | 13518.29 |
| WP_16P11 | 31 | -0.08363 | -0.00339 | 0.1015 | 0.79501 | 13519.15 |
| GOMF_PAT | 25 | -0.0891 | -0.00324 | 0.10801 | 0.79527 | 13523.57 |
| GOBP_POS | 8 | -0.14515 | -0.00299 | 0.17591 | 0.79536 | 13525.1 |
| GOMF_DN/ | 57 | -0.05419 | -0.00297 | 0.065669 | 0.79537 | 13525.27 |
| BROWNE_I | 168 | -0.03194 | -0.003 | 0.038697 | 0.79545 | 13526.63 |
| FUJIWARA_ | 10 | -0.12725 | -0.00293 | 0.15416 | 0.79545 | 13526.63 |
| PETROVA_ | 55 | -0.05648 | -0.00304 | 0.068349 | 0.79568 | 13530.54 |
| BROWNE_I | 43 | -0.06821 | -0.00325 | 0.082546 | 0.7957 | 13530.88 |
| GOBP_TRIC | 30 | -0.08037 | -0.0032 | 0.097241 | 0.79574 | 13531.56 |
| GOBP_REG | 5 | -0.20983 | -0.00341 | 0.25381 | 0.7958 | 13532.58 |
| GOBP_ALD | 6 | -0.14744 | -0.00263 | 0.17833 | 0.79581 | 13532.75 |
| GOBP_REE | 7 | -0.1549 | -0.00298 | 0.18733 | 0.79583 | 13533.09 |
| SENGUPTA | 150 | -0.03396 | -0.00302 | 0.041065 | 0.79589 | 13534.11 |
| GOBP_INTI | 9 | -0.1594 | -0.00348 | 0.19264 | 0.79601 | 13536.15 |
| KEGG_GLY( | 21 | -0.08447 | -0.00282 | 0.10205 | 0.7961 | 13537.68 |
| GALE_APL_ | 12 | -0.10171 | -0.00256 | 0.12276 | 0.7963 | 13541.08 |
| GOBP_SNR | 16 | -0.08759 | -0.00255 | 0.10571 | 0.79632 | 13541.42 |
| SHI_SPARC | 11 | -0.13199 | -0.00319 | 0.1593 | 0.79632 | 13541.42 |
| REACTOME | 860 | -0.0141 | -0.00294 | 0.01701 | 0.79646 | 13543.8 |
| GOMF_VIT | 6 | -0.17918 | -0.00319 | 0.2161 | 0.79649 | 13544.31 |
| GOBP_HIST | 12 | -0.11388 | -0.00287 | 0.13723 | 0.79668 | 13547.54 |
| GOBP_REG | 83 | -0.04717 | -0.00312 | 0.056836 | 0.79668 | 13547.54 |
| GOBP_SEC( | 25 | -0.08603 | -0.00313 | 0.10366 | 0.79672 | 13548.22 |
| GOMF_PH( | 9 | -0.12423 | -0.00271 | 0.14963 | 0.79678 | 13549.24 |
| GOBP_POS | 8 | -0.12947 | -0.00266 | 0.15586 | 0.79692 | 13551.62 |
| ODONNELL | 70 | -0.0524 | -0.00318 | 0.063056 | 0.797 | 13552.99 |
| GOMF_NU | 43 | -0.06537 | -0.00312 | 0.078655 | 0.79703 | 13553.5 |
| REACTOME | 132 | -0.0385 | -0.00321 | 0.046319 | 0.79705 | 13553.84 |
| VALK_AML | 30 | -0.07527 | -0.003 | 0.090549 | 0.79708 | 13554.35 |
| GOMF_NU | 11 | -0.14008 | -0.00338 | 0.16841 | 0.79722 | 13556.73 |
| GOBP_HEN | 5 | -0.17588 | -0.00286 | 0.21145 | 0.79723 | 13556.9 |
| GOBP_SPH | 29 | -0.07224 | -0.00283 | 0.086842 | 0.79724 | 13557.07 |
| MILICIC_FA | 9 | -0.14374 | -0.00314 | 0.1728 | 0.79724 | 13557.07 |
| GOCC_INH | 17 | -0.10717 | -0.00321 | 0.12883 | 0.79725 | 13557.24 |
| GOBP_REG | 7 | -0.16545 | -0.00319 | 0.19884 | 0.79731 | 13558.26 |
| GOCC_DER | 8 | -0.11977 | -0.00247 | 0.14391 | 0.79737 | 13559.28 |

| | | | | | | |
|---|---|---|---|---|---|---|
| GOBP_HIST | 20 | -0.08348 | -0.00272 | 0.10014 | 0.79774 | 13565.57 |
| GOBP_CHA | 20 | -0.09918 | -0.00323 | 0.11897 | 0.79776 | 13565.91 |
| GOBP_NEG | 3 | -0.21439 | -0.0027 | 0.25701 | 0.7979 | 13568.29 |
| GOBP_AXC | 37 | -0.06323 | -0.0028 | 0.075772 | 0.798 | 13569.99 |
| VERHAAK_ | 209 | -0.02839 | -0.00297 | 0.03402 | 0.79803 | 13570.5 |
| GOMF_UN | 7 | -0.14658 | -0.00282 | 0.17559 | 0.79808 | 13571.35 |
| REACTOME | 218 | -0.02723 | -0.00291 | 0.03262 | 0.79809 | 13571.52 |
| GOMF_PRC | 10 | -0.13029 | -0.003 | 0.15604 | 0.79813 | 13572.2 |
| GOMF_TRA | 10 | -0.13049 | -0.003 | 0.15627 | 0.79814 | 13572.37 |
| GOBP_REG | 12 | -0.13308 | -0.00335 | 0.15932 | 0.79821 | 13573.56 |
| REACTOME | 90 | -0.04336 | -0.00299 | 0.051887 | 0.79833 | 13575.6 |
| REACTOME | 41 | -0.08359 | -0.00389 | 0.09996 | 0.79847 | 13577.98 |
| BASSO_HA | 5 | -0.17273 | -0.00281 | 0.20651 | 0.79854 | 13579.17 |
| GOMF_PYF | 5 | -0.19391 | -0.00316 | 0.23183 | 0.79854 | 13579.17 |
| GOBP_GLU | 72 | -0.04974 | -0.00307 | 0.059445 | 0.79863 | 13580.7 |
| GOMF_POI | 5 | -0.20546 | -0.00334 | 0.24549 | 0.79868 | 13581.55 |
| GOBP_PRC | 5 | -0.22972 | -0.00374 | 0.27444 | 0.79871 | 13582.06 |
| BIOCARTA_ | 8 | -0.13443 | -0.00277 | 0.16059 | 0.79873 | 13582.4 |
| GOBP_CAR | 78 | -0.0458 | -0.00294 | 0.054686 | 0.79886 | 13584.61 |
| GOBP_RES | 616 | -0.01779 | -0.00316 | 0.021244 | 0.79887 | 13584.78 |
| GOMF_RIB | 13 | -0.1233 | -0.00323 | 0.1472 | 0.79889 | 13585.12 |
| YIH_RESPO | 10 | -0.17819 | -0.0041 | 0.21272 | 0.79889 | 13585.12 |
| IWANAGA_ | 165 | -0.03313 | -0.00308 | 0.03953 | 0.79904 | 13587.68 |
| GOMF_PHC | 17 | -0.10237 | -0.00307 | 0.1221 | 0.79911 | 13588.87 |
| GOBP_UBIC | 5 | -0.26977 | -0.00439 | 0.32166 | 0.79917 | 13589.89 |
| GOMF_SNA | 102 | -0.0388 | -0.00284 | 0.046257 | 0.7992 | 13590.4 |
| SMID_BREA | 425 | -0.02149 | -0.00319 | 0.025601 | 0.79932 | 13592.44 |
| GOBP_VEST | 11 | -0.16172 | -0.0039 | 0.19269 | 0.79934 | 13592.78 |
| DACOSTA_ | 24 | -0.08106 | -0.00289 | 0.096557 | 0.79939 | 13593.63 |
| GOMF_OXI | 76 | -0.04461 | -0.00282 | 0.053103 | 0.79954 | 13596.18 |
| GOBP_NEG | 17 | -0.09467 | -0.00284 | 0.11268 | 0.79958 | 13596.86 |
| GOCC_LIPI | 94 | -0.04332 | -0.00305 | 0.051541 | 0.79965 | 13598.05 |
| REACTOME | 21 | -0.09712 | -0.00324 | 0.11554 | 0.79969 | 13598.73 |
| GOCC_NF_ | 6 | -0.13871 | -0.00247 | 0.165 | 0.79972 | 13599.24 |
| WP_TCELL_ | 83 | -0.04685 | -0.0031 | 0.055728 | 0.79974 | 13599.58 |
| GOBP_EXT | 41 | -0.06429 | -0.00299 | 0.076442 | 0.79981 | 13600.77 |
| GOBP_NEG | 6 | -0.19813 | -0.00353 | 0.23553 | 0.79988 | 13601.96 |
| LIU_VMYB_ | 116 | -0.04023 | -0.00314 | 0.047779 | 0.80007 | 13605.19 |
| GOCC_NSL | 9 | -0.14988 | -0.00327 | 0.17801 | 0.80009 | 13605.53 |
| CHEOK_RE: | 5 | -0.13906 | -0.00226 | 0.16504 | 0.80026 | 13608.42 |
| GOMF_PYF | 7 | -0.17109 | -0.00329 | 0.20305 | 0.80027 | 13608.59 |
| GOMF_C3H | 4 | -0.18534 | -0.0027 | 0.21982 | 0.80041 | 13610.97 |
| REACTOME | 21 | -0.09578 | -0.00319 | 0.1136 | 0.80041 | 13610.97 |
| PID_MYC_I | 24 | -0.07818 | -0.00279 | 0.092724 | 0.80043 | 13611.31 |
| REACTOME | 76 | -0.04844 | -0.00307 | 0.057439 | 0.80047 | 13611.99 |

| | | | | | | |
|---|---|---|---|---|---|---|
| WEBER_MI | 28 | -0.08223 | -0.00316 | 0.097485 | 0.80053 | 13613.01 |
| GOMF_PH( | 11 | -0.12776 | -0.00308 | 0.15139 | 0.80063 | 13614.71 |
| GOCC_GAN | 3 | -0.24587 | -0.0031 | 0.29125 | 0.80071 | 13616.07 |
| GOBP_PRC | 11 | -0.13914 | -0.00336 | 0.1647 | 0.80089 | 13619.13 |
| GOBP_ZIN( | 8 | -0.11893 | -0.00245 | 0.14076 | 0.80092 | 13619.64 |
| REACTOME | 662 | -0.01756 | -0.00323 | 0.020781 | 0.80099 | 13620.83 |
| REACTOME | 5 | -0.17457 | -0.00284 | 0.20612 | 0.80148 | 13629.17 |
| GOBP_POS | 517 | -0.01906 | -0.00311 | 0.022502 | 0.80153 | 13630.02 |
| GOBP_NEG | 3 | -0.24736 | -0.00312 | 0.292 | 0.80154 | 13630.19 |
| REACTOME | 28 | -0.07845 | -0.00302 | 0.092594 | 0.80157 | 13630.7 |
| AMIT_EGF_ | 59 | -0.05842 | -0.00326 | 0.068862 | 0.80186 | 13635.63 |
| GOBP_POS | 27 | -0.09317 | -0.00352 | 0.10982 | 0.80187 | 13635.8 |
| GOBP_ACT | 29 | -0.08407 | -0.00329 | 0.09909 | 0.8019 | 13636.31 |
| LANDIS_ER | 17 | -0.10673 | -0.0032 | 0.12578 | 0.80192 | 13636.65 |
| GOBP_HIST | 82 | -0.04337 | -0.00285 | 0.0511 | 0.80196 | 13637.33 |
| GOBP_NEU | 75 | -0.05269 | -0.00331 | 0.06206 | 0.80207 | 13639.2 |
| GOBP_DEF | 12 | -0.12934 | -0.00326 | 0.15218 | 0.80232 | 13643.45 |
| KOHOUTEK | 49 | -0.05612 | -0.00286 | 0.066005 | 0.80241 | 13644.98 |
| GOMF_POI | 11 | -0.14136 | -0.00341 | 0.16604 | 0.80271 | 13650.08 |
| GOBP_REG | 17 | -0.09316 | -0.00279 | 0.1094 | 0.80276 | 13650.93 |
| GOBP_RES | 408 | -0.02241 | -0.00326 | 0.026302 | 0.80286 | 13652.63 |
| GOBP_PRC | 12 | -0.13908 | -0.00351 | 0.1632 | 0.80294 | 13653.99 |
| GOMF_ATF | 33 | -0.0726 | -0.00303 | 0.085188 | 0.80294 | 13653.99 |
| GOCC_NEU | 43 | -0.0662 | -0.00316 | 0.077652 | 0.80302 | 13655.36 |
| REACTOME | 181 | -0.03015 | -0.00294 | 0.035363 | 0.80304 | 13655.7 |
| REACTOME | 112 | -0.04011 | -0.00308 | 0.047047 | 0.80304 | 13655.7 |
| GOBP_REG | 9 | -0.13826 | -0.00302 | 0.16213 | 0.8031 | 13656.72 |
| AKL_HTLV1 | 25 | -0.08266 | -0.00301 | 0.096924 | 0.80311 | 13656.89 |
| WP_INCLU | 11 | -0.12215 | -0.00295 | 0.14321 | 0.80316 | 13657.74 |
| GOBP_CON | 11 | -0.17025 | -0.00411 | 0.19941 | 0.80338 | 13661.48 |
| GRANDVAU | 18 | -0.10499 | -0.00324 | 0.12287 | 0.80359 | 13665.05 |
| NIKOLSKY_ | 25 | -0.13675 | -0.00497 | 0.16002 | 0.8036 | 13665.22 |
| GOBP_END | 6 | -0.19843 | -0.00354 | 0.2321 | 0.80371 | 13667.09 |
| REACTOME | 25 | -0.07829 | -0.00285 | 0.091564 | 0.80373 | 13667.43 |
| REACTOME | 14 | -0.10229 | -0.00278 | 0.11963 | 0.80375 | 13667.77 |
| GOBP_RES | 296 | -0.02512 | -0.00312 | 0.029358 | 0.80387 | 13669.81 |
| GOCC_POL | 60 | -0.0495 | -0.00279 | 0.057849 | 0.80391 | 13670.49 |
| GOBP_POS | 75 | -0.04968 | -0.00312 | 0.058028 | 0.80401 | 13672.19 |
| GOMF_ATF | 4 | -0.18929 | -0.00276 | 0.22111 | 0.80402 | 13672.36 |
| REACTOME | 7 | -0.12995 | -0.0025 | 0.15178 | 0.80404 | 13672.7 |
| REACTOME | 83 | -0.04586 | -0.00303 | 0.053564 | 0.80405 | 13672.87 |
| CREIGHTOI | 31 | -0.07279 | -0.00295 | 0.085015 | 0.80406 | 13673.04 |
| GOBP_CELI | 94 | -0.04699 | -0.00331 | 0.054876 | 0.80407 | 13673.21 |
| REACTOME | 85 | -0.04984 | -0.00334 | 0.058172 | 0.80419 | 13675.25 |
| GOBP_CAL | 12 | -0.1218 | -0.00307 | 0.14216 | 0.80421 | 13675.59 |

| | | | | | | |
|---|---|---|---|---|---|---|
| GOBP_NEU | 6 | -0.19439 | -0.00347 | 0.22684 | 0.80426 | 13676.44 |
| PID_IL8_CX | 27 | -0.07864 | -0.00297 | 0.091671 | 0.80451 | 13680.69 |
| REACTOME | 5 | -0.20376 | -0.00332 | 0.23745 | 0.80457 | 13681.71 |
| GOBP_DNA | 17 | -0.10221 | -0.00307 | 0.1191 | 0.8046 | 13682.22 |
| GOBP_REG | 7 | -0.19245 | -0.00371 | 0.22389 | 0.80498 | 13688.68 |
| GOMF_ALL | 6 | -0.19781 | -0.00353 | 0.23002 | 0.8051 | 13690.73 |
| GOBP_POS | 16 | -0.12301 | -0.00358 | 0.14301 | 0.80513 | 13691.24 |
| GOBP_POS | 435 | -0.02106 | -0.00316 | 0.024476 | 0.80524 | 13693.11 |
| GOMF_GLY | 5 | -0.22821 | -0.00371 | 0.26506 | 0.80537 | 13695.32 |
| GOMF_GLY | 29 | -0.07946 | -0.00311 | 0.09225 | 0.80546 | 13696.85 |
| GOMF_POI | 11 | -0.14961 | -0.00361 | 0.1737 | 0.80546 | 13696.85 |
| GOBP_REG | 8 | -0.12904 | -0.00266 | 0.14979 | 0.8055 | 13697.53 |
| GOCC_P_B | 95 | -0.04378 | -0.0031 | 0.050811 | 0.80556 | 13698.55 |
| REACTOME | 5 | -0.19089 | -0.00311 | 0.22129 | 0.80582 | 13702.97 |
| GOBP_PHO | 14 | -0.1118 | -0.00304 | 0.12955 | 0.80593 | 13704.84 |
| GOBP_AMY | 52 | -0.05945 | -0.00312 | 0.068859 | 0.80602 | 13706.37 |
| GOBP_IRON | 16 | -0.10428 | -0.00303 | 0.12071 | 0.80616 | 13708.75 |
| REACTOME | 12 | -0.12751 | -0.00321 | 0.14754 | 0.80626 | 13710.45 |
| WP_PROXI | 56 | -0.05685 | -0.00309 | 0.065743 | 0.80639 | 13712.66 |
| GOBP_KER | 55 | -0.05903 | -0.00318 | 0.06826 | 0.80643 | 13713.34 |
| REACTOME | 97 | -0.04118 | -0.00294 | 0.047611 | 0.80644 | 13713.51 |
| GOMF_LIG | 39 | -0.06797 | -0.00309 | 0.078568 | 0.80652 | 13714.87 |
| REACTOME | 5 | -0.20112 | -0.00327 | 0.23247 | 0.80652 | 13714.87 |
| GOBP_RES | 7 | -0.13592 | -0.00262 | 0.15707 | 0.80657 | 13715.72 |
| DITTMER_I | 70 | -0.05046 | -0.00307 | 0.058289 | 0.80665 | 13717.08 |
| GOBP_POS | 13 | -0.13774 | -0.00361 | 0.15907 | 0.80672 | 13718.27 |
| GOBP_ATP | 83 | -0.04355 | -0.00288 | 0.050279 | 0.80678 | 13719.29 |
| GOBP_END | 557 | -0.01823 | -0.00309 | 0.02105 | 0.80678 | 13719.29 |
| REACTOME | 71 | -0.04863 | -0.00298 | 0.056126 | 0.80687 | 13720.82 |
| SHAFFER_I | 97 | -0.04521 | -0.00323 | 0.052171 | 0.80692 | 13721.67 |
| GOBP_POS | 8 | -0.16662 | -0.00343 | 0.19211 | 0.80711 | 13724.91 |
| GOBP_REG | 66 | -0.05257 | -0.0031 | 0.060587 | 0.8072 | 13726.44 |
| GOBP_T_H | 21 | -0.09927 | -0.00331 | 0.11428 | 0.80747 | 13731.03 |
| GOCC_CYT | 13 | -0.11623 | -0.00305 | 0.1338 | 0.80749 | 13731.37 |
| KEGG_NITF | 21 | -0.09653 | -0.00322 | 0.1111 | 0.80752 | 13731.88 |
| GOBP_POS | 7 | -0.13856 | -0.00267 | 0.15947 | 0.80753 | 13732.05 |
| JIANG_VHL | 84 | -0.04425 | -0.00294 | 0.050918 | 0.80756 | 13732.56 |
| GOBP_LEU | 6 | -0.17446 | -0.00311 | 0.20074 | 0.80759 | 13733.07 |
| GOMF_GLU | 6 | -0.17446 | -0.00311 | 0.20074 | 0.80759 | 13733.07 |
| GOBP_MIT | 134 | -0.03755 | -0.00315 | 0.043198 | 0.80763 | 13733.75 |
| REACTOME | 18 | -0.10795 | -0.00333 | 0.12419 | 0.80764 | 13733.92 |
| REACTOME | 21 | -0.09554 | -0.00318 | 0.10991 | 0.80764 | 13733.92 |
| GOBP_POS | 20 | -0.10839 | -0.00353 | 0.12469 | 0.80765 | 13734.09 |
| GOBP_DNA | 9 | -0.22146 | -0.00483 | 0.25474 | 0.80766 | 13734.26 |
| GOBP_REG | 5 | -0.21146 | -0.00344 | 0.24319 | 0.80772 | 13735.28 |

| | | | | | | |
|---|---|---|---|---|---|---|
| GOCC_MM | 4 | -0.20317 | -0.00296 | 0.23361 | 0.80776 | 13735.96 |
| WP_KETOG | 8 | -0.15456 | -0.00318 | 0.17771 | 0.80777 | 13736.13 |
| WP_MATR | 25 | -0.1051 | -0.00382 | 0.12082 | 0.8078 | 13736.64 |
| GOBP_POS | 14 | -0.10813 | -0.00294 | 0.12423 | 0.80796 | 13739.36 |
| VANTVEER_ | 104 | -0.04342 | -0.00321 | 0.049834 | 0.80819 | 13743.27 |
| REACTOME | 8 | -0.17676 | -0.00364 | 0.20286 | 0.8082 | 13743.44 |
| ONO_AML | 37 | -0.07812 | -0.00346 | 0.089639 | 0.80826 | 13744.46 |
| WP_MAPK_ | 15 | -0.11501 | -0.00324 | 0.13187 | 0.80843 | 13747.35 |
| GOBP_RES | 16 | -0.10843 | -0.00316 | 0.1242 | 0.80867 | 13751.43 |
| GOCC_NUC | 2 | -0.26991 | -0.00278 | 0.30895 | 0.80883 | 13754.15 |
| KEGG_GLY | 13 | -0.11082 | -0.00291 | 0.1268 | 0.80893 | 13755.85 |
| CHIARETTI_ | 19 | -0.10545 | -0.00334 | 0.12065 | 0.80894 | 13756.02 |
| REACTOME | 29 | -0.07259 | -0.00284 | 0.083009 | 0.80905 | 13757.9 |
| GOBP_POS | 15 | -0.11564 | -0.00326 | 0.13217 | 0.8092 | 13760.45 |
| GOBP_MES | 59 | -0.05869 | -0.00328 | 0.067068 | 0.80921 | 13760.62 |
| BIOCARTA_ | 23 | -0.08336 | -0.00291 | 0.095185 | 0.80941 | 13764.02 |
| GOBP_MIN | 5 | -0.17977 | -0.00293 | 0.20519 | 0.80951 | 13765.72 |
| WP_PROTE | 50 | -0.06356 | -0.00327 | 0.07253 | 0.80955 | 13766.4 |
| GOMF_MA | 11 | -0.13102 | -0.00316 | 0.14951 | 0.80957 | 13766.74 |
| WP_ARYL_ | 45 | -0.06267 | -0.00306 | 0.071494 | 0.80964 | 13767.93 |
| GOBP_POS | 6 | -0.22756 | -0.00406 | 0.25954 | 0.80968 | 13768.61 |
| GOBP_NEG | 184 | -0.03358 | -0.0033 | 0.038292 | 0.80974 | 13769.63 |
| GOCC_POD | 30 | -0.09037 | -0.0036 | 0.10305 | 0.80974 | 13769.63 |
| GOBP_SER | 26 | -0.11482 | -0.00426 | 0.13092 | 0.80977 | 13770.14 |
| GOMF_1_A | 7 | -0.16645 | -0.0032 | 0.18976 | 0.8098 | 13770.65 |
| REACTOME | 22 | -0.09543 | -0.00326 | 0.10878 | 0.80983 | 13771.16 |
| REACTOME | 81 | -0.04802 | -0.00314 | 0.054727 | 0.80986 | 13771.67 |
| GOMF_ATF | 6 | -0.19497 | -0.00348 | 0.2222 | 0.80987 | 13771.84 |
| COLIN_PIL | 27 | -0.08876 | -0.00335 | 0.10112 | 0.80996 | 13773.37 |
| GOBP_PHA | 13 | -0.12503 | -0.00328 | 0.14241 | 0.81001 | 13774.22 |
| GOBP_NAT | 6 | -0.20896 | -0.00372 | 0.23798 | 0.81005 | 13774.9 |
| WAGNER_A | 26 | -0.08806 | -0.00327 | 0.10026 | 0.8101 | 13775.75 |
| GOMF_RNA | 18 | -0.08578 | -0.00265 | 0.097584 | 0.8103 | 13779.15 |
| GOMF_DO | 5 | -0.22246 | -0.00362 | 0.25302 | 0.81035 | 13780 |
| REACTOME | 5 | -0.22246 | -0.00362 | 0.25302 | 0.81035 | 13780 |
| GOMF_POI | 19 | -0.10887 | -0.00345 | 0.12383 | 0.81036 | 13780.17 |
| YAO_TEMP | 72 | -0.05022 | -0.0031 | 0.057114 | 0.81037 | 13780.34 |
| GOBP_HEP | 8 | -0.13047 | -0.00269 | 0.14835 | 0.81042 | 13781.19 |
| GOCC_SAG | 21 | -0.07999 | -0.00267 | 0.090883 | 0.81061 | 13784.42 |
| GOBP_CAR | 126 | -0.04142 | -0.00337 | 0.047046 | 0.81065 | 13785.1 |
| BIOCARTA_ | 8 | -0.17076 | -0.00351 | 0.1939 | 0.81075 | 13786.8 |
| GOCC_GRC | 8 | -0.15599 | -0.00321 | 0.17711 | 0.81077 | 13787.14 |
| GOBP_DNA | 35 | -0.07337 | -0.00316 | 0.08327 | 0.81085 | 13788.5 |
| GOBP_HIST | 43 | -0.05932 | -0.00283 | 0.06733 | 0.81086 | 13788.67 |
| GOMF_MC | 79 | -0.05419 | -0.0035 | 0.0615 | 0.81086 | 13788.67 |

| | | | | | | |
|---|---|---|---|---|---|---|
| REACTOME | 27 | -0.08974 | -0.00339 | 0.1018 | 0.81097 | 13790.54 |
| GOBP_REG | 75 | -0.0548 | -0.00345 | 0.062146 | 0.81105 | 13791.91 |
| NAKAJIMA_ | 22 | -0.09834 | -0.00336 | 0.11144 | 0.81122 | 13794.8 |
| GOBP_NEG | 133 | -0.03981 | -0.00333 | 0.045107 | 0.81125 | 13795.31 |
| GOBP_GLY | 33 | -0.07758 | -0.00324 | 0.0879 | 0.81126 | 13795.48 |
| REACTOME | 37 | -0.06673 | -0.00295 | 0.07559 | 0.81132 | 13796.5 |
| GOBP_GLY | 278 | -0.02649 | -0.00319 | 0.030003 | 0.81133 | 13796.67 |
| PURBEY_TA | 272 | -0.02677 | -0.00319 | 0.030316 | 0.81136 | 13797.18 |
| GOBP_POS | 10 | -0.15944 | -0.00367 | 0.18057 | 0.81137 | 13797.35 |
| GOBP_DEN | 33 | -0.07908 | -0.0033 | 0.089532 | 0.81145 | 13798.71 |
| YAMANAKA | 11 | -0.13271 | -0.0032 | 0.15022 | 0.8115 | 13799.56 |
| GOBP_CHC | 17 | -0.1032 | -0.0031 | 0.11681 | 0.81151 | 13799.73 |
| GOBP_INTI | 7 | -0.1884 | -0.00363 | 0.2132 | 0.81157 | 13800.75 |
| GOBP_PEP | 9 | -0.12347 | -0.0027 | 0.13966 | 0.81167 | 13802.45 |
| GOBP_PIGI | 24 | -0.08123 | -0.00289 | 0.091868 | 0.81169 | 13802.79 |
| GOBP_ER_ | 10 | -0.13112 | -0.00302 | 0.14826 | 0.81176 | 13803.98 |
| GOCC_DEN | 4 | -0.20859 | -0.00304 | 0.23574 | 0.81187 | 13805.85 |
| GOBP_ACT | 111 | -0.04161 | -0.00318 | 0.047001 | 0.81197 | 13807.55 |
| GOBP_FIBI | 95 | -0.04663 | -0.0033 | 0.05267 | 0.81202 | 13808.4 |
| GOBP_NEG | 9 | -0.15532 | -0.00339 | 0.1753 | 0.8122 | 13811.46 |
| GOMF_NAI | 19 | -0.10254 | -0.00325 | 0.11569 | 0.81227 | 13812.65 |
| GOBP_REG | 22 | -0.09787 | -0.00334 | 0.11041 | 0.81229 | 13812.99 |
| CREIGHTOI | 423 | -0.02079 | -0.00308 | 0.023437 | 0.81245 | 13815.71 |
| GOBP_REG | 11 | -0.14654 | -0.00354 | 0.1652 | 0.81247 | 13816.05 |
| REACTOME | 49 | -0.0564 | -0.00287 | 0.063548 | 0.81261 | 13818.43 |
| MCGOWAI | 5 | -0.17135 | -0.00279 | 0.19303 | 0.81263 | 13818.77 |
| RIEGE_DEL | 134 | -0.03764 | -0.00316 | 0.042399 | 0.81267 | 13819.45 |
| GOCC_SYN | 1361 | -0.0127 | -0.00329 | 0.014305 | 0.81274 | 13820.64 |
| KASLER_HI | 6 | -0.1789 | -0.00319 | 0.20125 | 0.81297 | 13824.55 |
| WP_LET7_I | 5 | -0.24167 | -0.00393 | 0.27188 | 0.81297 | 13824.55 |
| GOBP_MRI | 56 | -0.05517 | -0.003 | 0.062052 | 0.81301 | 13825.24 |
| GOBP_RES | 46 | -0.06546 | -0.00323 | 0.073618 | 0.81304 | 13825.75 |
| REACTOME | 4 | -0.2214 | -0.00322 | 0.24887 | 0.81316 | 13827.79 |
| GOBP_PHC | 9 | -0.15481 | -0.00338 | 0.174 | 0.81318 | 13828.13 |
| GOBP_REG | 7 | -0.15286 | -0.00294 | 0.17178 | 0.81322 | 13828.81 |
| GOBP_REG | 15 | -0.13944 | -0.00393 | 0.15668 | 0.81324 | 13829.15 |
| REACTOME | 51 | -0.05118 | -0.00266 | 0.057499 | 0.81329 | 13830 |
| LUI_THYRC | 28 | -0.07279 | -0.0028 | 0.081766 | 0.81333 | 13830.68 |
| WP_DRAVI | 25 | -0.09313 | -0.00339 | 0.1046 | 0.81333 | 13830.68 |
| WP_FOXA2 | 20 | -0.09131 | -0.00297 | 0.10256 | 0.81333 | 13830.68 |
| GOBP_POS | 13 | -0.14814 | -0.00389 | 0.16639 | 0.81335 | 13831.02 |
| YOKOE_CA | 32 | -0.08634 | -0.00355 | 0.096959 | 0.8134 | 13831.87 |
| GOBP_ACT | 421 | -0.02267 | -0.00335 | 0.025451 | 0.81345 | 13832.72 |
| GOBP_POS | 4 | -0.24567 | -0.00358 | 0.27573 | 0.81352 | 13833.91 |
| YIH_RESPO | 21 | -0.08724 | -0.00291 | 0.097918 | 0.81352 | 13833.91 |

| | | | | | | |
|---|---|---|---|---|---|---|
| LEE_EARLY_ | 51 | -0.06258 | -0.00325 | 0.070166 | 0.81375 | 13837.82 |
| GOCC_PHA | 18 | -0.10162 | -0.00314 | 0.11394 | 0.81377 | 13838.16 |
| GOBP_NEG | 5 | -0.21495 | -0.0035 | 0.24059 | 0.81418 | 13845.13 |
| GOBP_POS | 23 | -0.09246 | -0.00323 | 0.10338 | 0.81444 | 13849.55 |
| ZHAN_VAR | 29 | -0.07382 | -0.00289 | 0.0825 | 0.81456 | 13851.59 |
| GOBP_PYR | 5 | -0.21314 | -0.00347 | 0.23818 | 0.81457 | 13851.76 |
| GOBP_NEG | 16 | -0.11963 | -0.00348 | 0.13365 | 0.81464 | 13852.95 |
| NABA_ECM | 227 | -0.03346 | -0.00365 | 0.037358 | 0.81475 | 13854.82 |
| GOBP_POS | 10 | -0.15203 | -0.0035 | 0.16967 | 0.81488 | 13857.03 |
| GOBP_SUL | 51 | -0.06299 | -0.00327 | 0.070278 | 0.81493 | 13857.88 |
| GOBP_PRC | 7 | -0.12889 | -0.00248 | 0.1438 | 0.81495 | 13858.22 |
| GOCC_CLA | 12 | -0.15377 | -0.00388 | 0.17153 | 0.81498 | 13858.73 |
| GOBP_UTE | 6 | -0.15707 | -0.0028 | 0.17519 | 0.81503 | 13859.59 |
| REACTOME | 51 | -0.06396 | -0.00332 | 0.071317 | 0.81507 | 13860.27 |
| GOMF_SUC | 30 | -0.08656 | -0.00345 | 0.09648 | 0.81519 | 13862.31 |
| GOBP_POS | 25 | -0.09104 | -0.00331 | 0.10143 | 0.81528 | 13863.84 |
| ABE_INNEF | 41 | -0.06405 | -0.00298 | 0.071351 | 0.81531 | 13864.35 |
| STARK_HYF | 49 | -0.06214 | -0.00316 | 0.069204 | 0.81538 | 13865.54 |
| GOBP_NEG | 30 | -0.08182 | -0.00326 | 0.091056 | 0.81555 | 13868.43 |
| GOBP_REG | 53 | -0.05639 | -0.00298 | 0.062711 | 0.81573 | 13871.49 |
| GOBP_ISOI | 5 | -0.18204 | -0.00296 | 0.2024 | 0.81578 | 13872.34 |
| GOBP_LEU | 18 | -0.10059 | -0.0031 | 0.11181 | 0.81584 | 13873.36 |
| CAIRO_HEF | 14 | -0.12753 | -0.00347 | 0.14173 | 0.81587 | 13873.87 |
| GAUSSMAI | 34 | -0.08056 | -0.00342 | 0.089501 | 0.81595 | 13875.23 |
| GOTTWEIN | 62 | -0.05793 | -0.00331 | 0.064326 | 0.81608 | 13877.44 |
| GOBP_CER | 13 | -0.12414 | -0.00326 | 0.13785 | 0.81609 | 13877.61 |
| REACTOME | 14 | -0.1181 | -0.00322 | 0.13112 | 0.81612 | 13878.12 |
| MUELLER_( | 13 | -0.13358 | -0.0035 | 0.14827 | 0.81618 | 13879.14 |
| REACTOME | 21 | -0.09923 | -0.00331 | 0.1101 | 0.81627 | 13880.67 |
| GOBP_REG | 28 | -0.07825 | -0.00301 | 0.086733 | 0.8165 | 13884.58 |
| BIOCARTA_ | 15 | -0.10747 | -0.00303 | 0.1191 | 0.81656 | 13885.6 |
| GOBP_XEN | 14 | -0.11981 | -0.00326 | 0.13277 | 0.81657 | 13885.77 |
| GOBP_NEG | 5 | -0.21843 | -0.00355 | 0.24199 | 0.81664 | 13886.96 |
| GOBP_GLY | 78 | -0.05144 | -0.0033 | 0.056978 | 0.81666 | 13887.3 |
| GOBP_LUT | 12 | -0.13496 | -0.0034 | 0.14946 | 0.81672 | 13888.32 |
| WP_SLEEP_ | 38 | -0.07922 | -0.00355 | 0.08772 | 0.81675 | 13888.83 |
| GOCC_GTP | 30 | -0.08015 | -0.00319 | 0.088746 | 0.81677 | 13889.17 |
| GOMF_RNA | 9 | -0.12396 | -0.00271 | 0.13714 | 0.81696 | 13892.4 |
| DAZARD_R | 199 | -0.03138 | -0.0032 | 0.034704 | 0.81706 | 13894.11 |
| GOBP_ENT | 9 | -0.15112 | -0.0033 | 0.16704 | 0.81718 | 13896.15 |
| GOBP_RES | 6 | -0.16746 | -0.00298 | 0.1851 | 0.81718 | 13896.15 |
| GOBP_NEG | 6 | -0.18803 | -0.00335 | 0.20772 | 0.81732 | 13898.53 |
| GOBP_ADH | 57 | -0.06612 | -0.00363 | 0.07302 | 0.81741 | 13900.06 |
| GOBP_REG | 12 | -0.12322 | -0.00311 | 0.13601 | 0.81753 | 13902.1 |
| GOBP_INO | 28 | -0.08337 | -0.00321 | 0.092006 | 0.81756 | 13902.61 |

| | | | | | | |
|---|---|---|---|---|---|---|
| GOBP_MEI | 8 | -0.1866 | -0.00384 | 0.20593 | 0.81756 | 13902.61 |
| REACTOME | 50 | -0.06412 | -0.0033 | 0.070745 | 0.8176 | 13903.29 |
| WP_GABA_ | 27 | -0.09841 | -0.00372 | 0.10858 | 0.81761 | 13903.46 |
| GOMF_RN/ | 3 | -0.23742 | -0.00299 | 0.26189 | 0.81767 | 13904.48 |
| REACTOME | 12 | -0.123 | -0.0031 | 0.13564 | 0.81773 | 13905.5 |
| WP_IL10_/ | 12 | -0.13011 | -0.00328 | 0.14337 | 0.81791 | 13908.56 |
| GOBP_REG | 56 | -0.05844 | -0.00318 | 0.064376 | 0.81799 | 13909.92 |
| GOBP_CHC | 22 | -0.09525 | -0.00325 | 0.10492 | 0.81801 | 13910.26 |
| GOBP_CER | 7 | -0.2098 | -0.00404 | 0.23104 | 0.81808 | 13911.45 |
| MIZUSHIM | 12 | -0.1235 | -0.00311 | 0.13599 | 0.8181 | 13911.79 |
| GOBP_POS | 15 | -0.10705 | -0.00302 | 0.11787 | 0.81813 | 13912.3 |
| GOBP_GLU | 9 | -0.17406 | -0.0038 | 0.19162 | 0.81815 | 13912.64 |
| KEGG_CON | 63 | -0.06121 | -0.00353 | 0.067363 | 0.81822 | 13913.83 |
| GOBP_POS | 67 | -0.05783 | -0.00344 | 0.063618 | 0.81832 | 13915.53 |
| BIOCARTA_ | 9 | -0.18662 | -0.00407 | 0.20526 | 0.81836 | 13916.21 |
| REACTOME | 11 | -0.15178 | -0.00366 | 0.16687 | 0.81847 | 13918.08 |
| GOBP_REG | 11 | -0.16193 | -0.00391 | 0.17799 | 0.81852 | 13918.93 |
| GOMF_GTI | 14 | -0.10756 | -0.00293 | 0.1182 | 0.81859 | 13920.12 |
| GOBP_POS | 63 | -0.06047 | -0.00349 | 0.066375 | 0.81885 | 13924.54 |
| IBRAHIM_I | 405 | -0.02203 | -0.00319 | 0.024183 | 0.81888 | 13925.05 |
| GOBP_GAN | 21 | -0.09516 | -0.00317 | 0.10438 | 0.81903 | 13927.61 |
| WP_COMP | 53 | -0.06557 | -0.00347 | 0.071911 | 0.81907 | 13928.29 |
| BIOCARTA_ | 33 | -0.07646 | -0.00319 | 0.083849 | 0.81909 | 13928.63 |
| GOMF_PR( | 4 | -0.23036 | -0.00335 | 0.25249 | 0.8192 | 13930.5 |
| KANG_IMN | 97 | -0.04742 | -0.00339 | 0.051951 | 0.81931 | 13932.37 |
| REACTOME | 5 | -0.22159 | -0.00361 | 0.24272 | 0.81937 | 13933.39 |
| NEWMAN_ | 25 | -0.10853 | -0.00395 | 0.11881 | 0.8195 | 13935.6 |
| GOBP_ONE | 23 | -0.08682 | -0.00303 | 0.094988 | 0.81963 | 13937.81 |
| LEE_AGING | 62 | -0.05414 | -0.0031 | 0.059225 | 0.81966 | 13938.32 |
| GOMF_RN/ | 35 | -0.06837 | -0.00294 | 0.074796 | 0.81967 | 13938.49 |
| GOBP_INO | 10 | -0.13805 | -0.00318 | 0.151 | 0.8197 | 13939 |
| GOBP_RES | 20 | -0.10393 | -0.00338 | 0.11362 | 0.81981 | 13940.87 |
| GOBP_REG | 8 | -0.19456 | -0.004 | 0.21269 | 0.81982 | 13941.04 |
| REACTOME | 10 | -0.16873 | -0.00388 | 0.18443 | 0.81986 | 13941.72 |
| BIOCARTA_ | 19 | -0.09732 | -0.00309 | 0.10634 | 0.81993 | 13942.91 |
| GOCC_PLA | 1212 | -0.0139 | -0.00341 | 0.015187 | 0.81997 | 13943.59 |
| GOBP_LEU | 45 | -0.06368 | -0.00311 | 0.069524 | 0.82013 | 13946.31 |
| KESHELAV/ | 71 | -0.04939 | -0.00302 | 0.053918 | 0.82016 | 13946.82 |
| WP_OMEG | 13 | -0.13088 | -0.00343 | 0.14285 | 0.82021 | 13947.67 |
| CASTELLAN | 14 | -0.12471 | -0.00339 | 0.13609 | 0.82026 | 13948.52 |
| GOBP_CELI | 7 | -0.19916 | -0.00383 | 0.21723 | 0.82038 | 13950.56 |
| JAZAG_TGI | 59 | -0.06127 | -0.00342 | 0.066811 | 0.82045 | 13951.75 |
| WATANABI | 28 | -0.08633 | -0.00332 | 0.094102 | 0.82053 | 13953.11 |
| GOBP_REG | 49 | -0.06922 | -0.00352 | 0.075415 | 0.82066 | 13955.32 |
| GOBP_POS | 16 | -0.13105 | -0.00381 | 0.1427 | 0.82077 | 13957.19 |

| | | | | | | |
|---|---|---|---|---|---|---|
| REACTOME | 6 | -0.15969 | -0.00285 | 0.17385 | 0.82083 | 13958.21 |
| GOMF_PH( | 18 | -0.12394 | -0.00383 | 0.13483 | 0.82101 | 13961.28 |
| GOBP_CAL | 45 | -0.06675 | -0.00326 | 0.072592 | 0.82109 | 13962.64 |
| GOBP_REG | 400 | -0.02272 | -0.00327 | 0.024701 | 0.82111 | 13962.98 |
| GOBP_REG | 10 | -0.1755 | -0.00404 | 0.19064 | 0.82136 | 13967.23 |
| GOBP_NEG | 15 | -0.10642 | -0.003 | 0.11555 | 0.82146 | 13968.93 |
| GOBP_GLU | 33 | -0.07884 | -0.00329 | 0.085598 | 0.82147 | 13969.1 |
| GOBP_ADU | 9 | -0.13581 | -0.00296 | 0.14744 | 0.8215 | 13969.61 |
| GOBP_MEI | 6 | -0.19818 | -0.00353 | 0.215 | 0.82166 | 13972.33 |
| GOBP_REG | 33 | -0.08146 | -0.0034 | 0.088338 | 0.82175 | 13973.86 |
| GOBP_INTI | 17 | -0.10681 | -0.0032 | 0.1158 | 0.82183 | 13975.22 |
| GOMF_CAI | 11 | -0.14157 | -0.00342 | 0.15346 | 0.82186 | 13975.73 |
| GOBP_EXT | 4 | -0.26687 | -0.00388 | 0.28928 | 0.82187 | 13975.9 |
| GOBP_DIG | 133 | -0.04361 | -0.00365 | 0.047254 | 0.82194 | 13977.09 |
| REACTOME | 22 | -0.1049 | -0.00358 | 0.11366 | 0.82197 | 13977.6 |
| GOBP_CELI | 6 | -0.15513 | -0.00277 | 0.16793 | 0.82219 | 13981.34 |
| GOBP_REG | 22 | -0.09139 | -0.00312 | 0.098889 | 0.82229 | 13983.04 |
| GINESTIER_ | 152 | -0.03393 | -0.00303 | 0.036712 | 0.82232 | 13983.55 |
| PID_ATM_I | 32 | -0.07825 | -0.00322 | 0.084653 | 0.82235 | 13984.06 |
| GOBP_AMY | 50 | -0.06425 | -0.0033 | 0.069497 | 0.82236 | 13984.23 |
| SESTO_RES | 68 | -0.05417 | -0.00325 | 0.058553 | 0.82254 | 13987.29 |
| WP_INTRA | 26 | -0.10626 | -0.00394 | 0.11484 | 0.82258 | 13987.97 |
| WP_OVERV | 115 | -0.05266 | -0.0041 | 0.056851 | 0.82284 | 13992.39 |
| REACTOME | 11 | -0.12623 | -0.00305 | 0.13622 | 0.82294 | 13994.09 |
| NAKAMUR. | 45 | -0.06959 | -0.00339 | 0.075095 | 0.82295 | 13994.26 |
| REACTOME | 7 | -0.18534 | -0.00357 | 0.19998 | 0.82297 | 13994.6 |
| REACTOME | 11 | -0.12061 | -0.00291 | 0.13012 | 0.82302 | 13995.46 |
| GOBP_RES | 7 | -0.22639 | -0.00436 | 0.24419 | 0.82306 | 13996.14 |
| GOBP_MYE | 223 | -0.03117 | -0.00337 | 0.033603 | 0.82314 | 13997.5 |
| GOCC_SPE( | 151 | -0.0377 | -0.00336 | 0.040651 | 0.82314 | 13997.5 |
| GOBP_NEG | 60 | -0.06096 | -0.00343 | 0.065727 | 0.82315 | 13997.67 |
| GOBP_POS | 41 | -0.07496 | -0.00349 | 0.080822 | 0.82315 | 13997.67 |
| KLEIN_PRIN | 46 | -0.07157 | -0.00353 | 0.077157 | 0.82319 | 13998.35 |
| GOBP_INTI | 26 | -0.08982 | -0.00333 | 0.096803 | 0.82326 | 13999.54 |
| GOBP_DET | 7 | -0.19801 | -0.00381 | 0.21332 | 0.82336 | 14001.24 |
| GOBP_NEG | 10 | -0.15301 | -0.00352 | 0.16473 | 0.82351 | 14003.79 |
| GOBP_NEG | 13 | -0.12704 | -0.00333 | 0.13674 | 0.82357 | 14004.81 |
| HUMMERI( | 18 | -0.11204 | -0.00346 | 0.12059 | 0.82358 | 14004.98 |
| GOCC_NUC | 5 | -0.19336 | -0.00315 | 0.20801 | 0.82369 | 14006.85 |
| GOCC_SPEI | 4 | -0.24716 | -0.0036 | 0.26589 | 0.82369 | 14006.85 |
| GOBP_ZYG | 6 | -0.26268 | -0.00468 | 0.28246 | 0.8238 | 14008.72 |
| GOBP_POS | 53 | -0.06777 | -0.00359 | 0.072826 | 0.82394 | 14011.1 |
| GOBP_NEG | 10 | -0.1447 | -0.00333 | 0.15543 | 0.82405 | 14012.97 |
| GOMF_SOI | 147 | -0.04031 | -0.00354 | 0.043292 | 0.82408 | 14013.48 |
| GOBP_REG | 912 | -0.01562 | -0.00335 | 0.016779 | 0.82409 | 14013.65 |

| | | | | | | |
|---|---|---|---|---|---|---|
| GOBP_PRC | 77 | -0.05279 | -0.00336 | 0.056694 | 0.82411 | 14013.99 |
| GOBP_L_A | 6 | -0.19396 | -0.00346 | 0.20825 | 0.82417 | 14015.01 |
| PID_TCR_C | 27 | -0.09521 | -0.0036 | 0.10221 | 0.82419 | 14015.35 |
| GOMF_CAI | 18 | -0.09919 | -0.00306 | 0.10649 | 0.8242 | 14015.52 |
| GOBP_AMI | 92 | -0.05033 | -0.00351 | 0.054007 | 0.82431 | 14017.39 |
| GOMF_TBF | 24 | -0.08783 | -0.00313 | 0.094241 | 0.82432 | 14017.56 |
| GOMF_THY | 6 | -0.21447 | -0.00382 | 0.23008 | 0.82436 | 14018.24 |
| GOBP_FAC | 9 | -0.1482 | -0.00324 | 0.15894 | 0.82444 | 14019.6 |
| VALK_AML | 24 | -0.09578 | -0.00341 | 0.10268 | 0.82453 | 14021.13 |
| GOCC_END | 30 | -0.0879 | -0.0035 | 0.094215 | 0.82457 | 14021.81 |
| WALLACE_ | 75 | -0.05498 | -0.00346 | 0.058887 | 0.82477 | 14025.21 |
| GOBP_SUB | 8 | -0.15919 | -0.00328 | 0.17045 | 0.82483 | 14026.23 |
| GOBP_POS | 7 | -0.17667 | -0.0034 | 0.18914 | 0.82486 | 14026.74 |
| PUJANA_BI | 8 | -0.20993 | -0.00432 | 0.22473 | 0.82488 | 14027.08 |
| WP_NANO | 22 | -0.1034 | -0.00353 | 0.1106 | 0.82508 | 14030.49 |
| GOBP_STR | 44 | -0.07398 | -0.00357 | 0.079124 | 0.82509 | 14030.66 |
| GOCC_NET | 6 | -0.22122 | -0.00394 | 0.2366 | 0.82509 | 14030.66 |
| GOBP_NEG | 86 | -0.05147 | -0.00347 | 0.055008 | 0.82527 | 14033.72 |
| GOMF_ARY | 6 | -0.20295 | -0.00362 | 0.21685 | 0.82533 | 14034.74 |
| GOBP_RES | 4 | -0.20871 | -0.00304 | 0.22299 | 0.82534 | 14034.91 |
| DOUGLAS_ | 301 | -0.0279 | -0.0035 | 0.02981 | 0.82536 | 14035.25 |
| GOCC_CYC | 6 | -0.21457 | -0.00382 | 0.22924 | 0.82536 | 14035.25 |
| HOFMANN | 45 | -0.06984 | -0.00341 | 0.074619 | 0.82536 | 14035.25 |
| LINDGREN_ | 114 | -0.04799 | -0.00372 | 0.051246 | 0.82547 | 14037.12 |
| GOMF_ICO | 13 | -0.15236 | -0.004 | 0.16264 | 0.82556 | 14038.65 |
| GOCC_API( | 22 | -0.10375 | -0.00354 | 0.1107 | 0.82568 | 14040.69 |
| GOBP_REG | 18 | -0.1137 | -0.00351 | 0.12122 | 0.82586 | 14043.75 |
| GOBP_POS | 7 | -0.1529 | -0.00294 | 0.16297 | 0.82593 | 14044.94 |
| FRASOR_RI | 49 | -0.06431 | -0.00327 | 0.068537 | 0.82594 | 14045.11 |
| HOEGERKC | 25 | -0.10187 | -0.0037 | 0.10858 | 0.82594 | 14045.11 |
| LEE_LIVER_ | 72 | -0.05635 | -0.00347 | 0.059992 | 0.82619 | 14049.36 |
| REACTOME | 34 | -0.07129 | -0.00302 | 0.075879 | 0.82626 | 14050.55 |
| GOBP_3_U | 20 | -0.11213 | -0.00365 | 0.11932 | 0.82631 | 14051.4 |
| GOBP_DET | 13 | -0.12941 | -0.00339 | 0.13761 | 0.8265 | 14054.63 |
| GOMF_1_F | 10 | -0.1435 | -0.0033 | 0.15258 | 0.82651 | 14054.8 |
| GOMF_HIS | 8 | -0.16568 | -0.00341 | 0.17613 | 0.82655 | 14055.48 |
| GOBP_PRC | 5 | -0.19693 | -0.0032 | 0.20922 | 0.82671 | 14058.2 |
| GOBP_REG | 56 | -0.06446 | -0.00351 | 0.068437 | 0.82688 | 14061.09 |
| GOMF_CAI | 9 | -0.13993 | -0.00305 | 0.14852 | 0.82694 | 14062.11 |
| GOMF_CYT | 22 | -0.0968 | -0.0033 | 0.10269 | 0.82706 | 14064.16 |
| GOBP_GLY | 5 | -0.20153 | -0.00328 | 0.21354 | 0.82734 | 14068.92 |
| GOBP_VES | 9 | -0.18381 | -0.00401 | 0.19473 | 0.82738 | 14069.6 |
| GOMF_PH( | 8 | -0.19728 | -0.00406 | 0.20895 | 0.82744 | 14070.62 |
| GOBP_POS | 74 | -0.05385 | -0.00336 | 0.057031 | 0.82745 | 14070.79 |
| GOBP_PHA | 17 | -0.12208 | -0.00366 | 0.12915 | 0.82772 | 14075.38 |

| | | | | | | |
|---|---|---|---|---|---|---|
| GOBP_BILE | 29 | -0.08161 | -0.0032 | 0.086326 | 0.82775 | 14075.89 |
| GOBP_NEU | 27 | -0.0887 | -0.00335 | 0.093801 | 0.82782 | 14077.08 |
| CHENG_RE | 40 | -0.07322 | -0.00337 | 0.077363 | 0.82804 | 14080.82 |
| GOBP_AMI | 73 | -0.05475 | -0.0034 | 0.057838 | 0.82806 | 14081.16 |
| GOBP_REG | 215 | -0.03175 | -0.00337 | 0.033535 | 0.82814 | 14082.52 |
| GOCC_TRIC | 15 | -0.11351 | -0.0032 | 0.11986 | 0.82818 | 14083.2 |
| REACTOME | 19 | -0.09991 | -0.00317 | 0.10549 | 0.8282 | 14083.54 |
| JAERVINEN | 40 | -0.07272 | -0.00334 | 0.076774 | 0.82822 | 14083.88 |
| GARGALOV | 32 | -0.09153 | -0.00377 | 0.096627 | 0.82824 | 14084.22 |
| GOBP_REG | 5 | -0.17858 | -0.00291 | 0.1885 | 0.82827 | 14084.73 |
| GOBP_POS | 30 | -0.09408 | -0.00375 | 0.099302 | 0.82828 | 14084.9 |
| GOBP_CHC | 6 | -0.22722 | -0.00405 | 0.23981 | 0.8283 | 14085.24 |
| REACTOME | 7 | -0.19243 | -0.0037 | 0.2031 | 0.8283 | 14085.24 |
| VANHARAN | 34 | -0.08504 | -0.00361 | 0.089655 | 0.82856 | 14089.66 |
| GOMF_BIC | 14 | -0.11979 | -0.00326 | 0.12628 | 0.82857 | 14089.83 |
| REACTOME | 70 | -0.05673 | -0.00345 | 0.059751 | 0.82878 | 14093.4 |
| GOBP_NUC | 11 | -0.13378 | -0.00323 | 0.14089 | 0.82883 | 14094.25 |
| NGUYEN_N | 73 | -0.05596 | -0.00347 | 0.05892 | 0.82887 | 14094.93 |
| KEGG_CITR | 28 | -0.08704 | -0.00335 | 0.091602 | 0.82899 | 14096.97 |
| GOBP_NEG | 3 | -0.24683 | -0.00311 | 0.25965 | 0.8291 | 14098.85 |
| KYNG_DNA | 76 | -0.05538 | -0.00351 | 0.058252 | 0.8291 | 14098.85 |
| GOBP_CD4 | 105 | -0.04925 | -0.00366 | 0.051771 | 0.82927 | 14101.74 |
| GOBP_POS | 17 | -0.10772 | -0.00323 | 0.11323 | 0.82928 | 14101.91 |
| GOCC_MIC | 91 | -0.05037 | -0.00349 | 0.052945 | 0.82929 | 14102.08 |
| GOBP_NEG | 12 | -0.13045 | -0.00329 | 0.13705 | 0.8294 | 14103.95 |
| GOBP_CAR | 7 | -0.18075 | -0.00348 | 0.18988 | 0.82943 | 14104.46 |
| MAEKAWA | 22 | -0.09144 | -0.00312 | 0.096006 | 0.82954 | 14106.33 |
| GOBP_AUT | 7 | -0.19276 | -0.00371 | 0.20238 | 0.82956 | 14106.67 |
| GOBP_THI( | 41 | -0.07516 | -0.0035 | 0.078898 | 0.8296 | 14107.35 |
| GOBP_GOL | 143 | -0.04144 | -0.00359 | 0.043491 | 0.82968 | 14108.71 |
| GOBP_POS | 5 | -0.25297 | -0.00412 | 0.26547 | 0.82968 | 14108.71 |
| REACTOME | 12 | -0.12655 | -0.00319 | 0.13278 | 0.8297 | 14109.05 |
| WP_PATH( | 17 | -0.12654 | -0.0038 | 0.13259 | 0.83006 | 14115.17 |
| FREDERICK | 9 | -0.16418 | -0.00358 | 0.17197 | 0.83012 | 14116.19 |
| GOBP_REG | 15 | -0.14621 | -0.00412 | 0.15314 | 0.83014 | 14116.53 |
| REACTOME | 72 | -0.05014 | -0.00309 | 0.052507 | 0.83016 | 14116.87 |
| REACTOME | 4 | -0.32937 | -0.00479 | 0.34486 | 0.83022 | 14117.89 |
| GOBP_REG | 5 | -0.24456 | -0.00398 | 0.25581 | 0.83047 | 14122.14 |
| GOMF_DEI | 6 | -0.1548 | -0.00276 | 0.16192 | 0.83047 | 14122.14 |
| GOBP_POS | 6 | -0.19442 | -0.00347 | 0.20334 | 0.83048 | 14122.31 |
| GOBP_COL | 5 | -0.2244 | -0.00365 | 0.23465 | 0.83054 | 14123.33 |
| LIAO_MET/ | 495 | -0.02114 | -0.00338 | 0.0221 | 0.83062 | 14124.69 |
| REACTOME | 89 | -0.05046 | -0.00346 | 0.05273 | 0.83071 | 14126.22 |
| GOMF_PH( | 35 | -0.08066 | -0.00347 | 0.084282 | 0.83072 | 14126.39 |
| GOBP_POS | 5 | -0.21856 | -0.00356 | 0.2283 | 0.8308 | 14127.75 |

| | | | | | | |
|---|---|---|---|---|---|---|
| GOBP_PYR | 104 | -0.04496 | -0.00333 | 0.046937 | 0.8309 | 14129.45 |
| REACTOME | 24 | -0.10658 | -0.0038 | 0.11126 | 0.83093 | 14129.96 |
| GOBP_NEG | 14 | -0.14036 | -0.00382 | 0.14649 | 0.831 | 14131.16 |
| HAHTOLA_ | 108 | -0.04377 | -0.0033 | 0.045674 | 0.83102 | 14131.5 |
| GOBP_MU | 55 | -0.06696 | -0.00361 | 0.06986 | 0.83108 | 14132.52 |
| GOBP_NEG | 8 | -0.1658 | -0.00341 | 0.17298 | 0.8311 | 14132.86 |
| GOBP_SUL | 13 | -0.12253 | -0.00321 | 0.12779 | 0.83117 | 14134.05 |
| GOMF_PO | 49 | -0.0686 | -0.00349 | 0.071526 | 0.83124 | 14135.24 |
| IIZUKA_LIV | 20 | -0.10045 | -0.00327 | 0.1047 | 0.83133 | 14136.77 |
| GOBP_CEL | 148 | -0.04184 | -0.00369 | 0.043598 | 0.83139 | 14137.79 |
| GOMF_ALC | 9 | -0.20687 | -0.00452 | 0.21548 | 0.83148 | 14139.32 |
| GOBP_PEP | 340 | -0.02486 | -0.00331 | 0.025895 | 0.8315 | 14139.66 |
| GOMF_ATI | 9 | -0.16804 | -0.00367 | 0.17488 | 0.83169 | 14142.89 |
| GOBP_REG | 18 | -0.11448 | -0.00353 | 0.1191 | 0.83177 | 14144.25 |
| BIOCARTA_ | 16 | -0.13043 | -0.0038 | 0.13567 | 0.83181 | 14144.93 |
| GOBP_SEC | 890 | -0.01643 | -0.00348 | 0.017081 | 0.83186 | 14145.78 |
| GOBP_PRC | 29 | -0.08666 | -0.00339 | 0.090108 | 0.83191 | 14146.63 |
| WILLIAMS_ | 25 | -0.09895 | -0.0036 | 0.10283 | 0.83203 | 14148.67 |
| GOBP_CHE | 88 | -0.05202 | -0.00354 | 0.05405 | 0.83206 | 14149.18 |
| GOBP_EMI | 9 | -0.15993 | -0.00349 | 0.16619 | 0.83206 | 14149.18 |
| GOMF_ACI | 9 | -0.15518 | -0.00339 | 0.16118 | 0.83216 | 14150.88 |
| GOBP_REG | 42 | -0.07433 | -0.0035 | 0.077206 | 0.83217 | 14151.05 |
| GOMF_PH( | 11 | -0.17115 | -0.00413 | 0.17772 | 0.83223 | 14152.07 |
| BIOCARTA_ | 5 | -0.23957 | -0.0039 | 0.24868 | 0.83231 | 14153.43 |
| REACTOME | 32 | -0.07363 | -0.00303 | 0.076409 | 0.83238 | 14154.62 |
| GOBP_DISI | 5 | -0.29904 | -0.00487 | 0.31008 | 0.83257 | 14157.85 |
| REACTOME | 13 | -0.12024 | -0.00315 | 0.12466 | 0.8326 | 14158.36 |
| GOBP_POS | 303 | -0.02795 | -0.00351 | 0.028974 | 0.83267 | 14159.55 |
| REACTOME | 19 | -0.12636 | -0.00401 | 0.13096 | 0.83268 | 14159.72 |
| GOBP_REG | 593 | -0.02062 | -0.0036 | 0.021366 | 0.83274 | 14160.74 |
| UZONYI_RI | 36 | -0.08964 | -0.00391 | 0.09285 | 0.83283 | 14162.27 |
| GOBP_REG | 48 | -0.06844 | -0.00345 | 0.070881 | 0.83285 | 14162.61 |
| GOBP_LEU | 32 | -0.08019 | -0.0033 | 0.083022 | 0.83294 | 14164.14 |
| GOBP_NEG | 9 | -0.19379 | -0.00423 | 0.2006 | 0.83298 | 14164.82 |
| GOBP_REG | 13 | -0.12435 | -0.00326 | 0.12857 | 0.83327 | 14169.76 |
| WANG_TH( | 17 | -0.12908 | -0.00387 | 0.13344 | 0.83329 | 14170.1 |
| GOBP_HAI | 51 | -0.07391 | -0.00384 | 0.076407 | 0.83331 | 14170.44 |
| GOCC_RET | 11 | -0.12819 | -0.00309 | 0.13248 | 0.83337 | 14171.46 |
| GOBP_MAI | 5 | -0.22492 | -0.00366 | 0.23228 | 0.83355 | 14174.52 |
| GOCC_CLA | 5 | -0.27707 | -0.00451 | 0.28611 | 0.83357 | 14174.86 |
| LIAN_NEUT | 24 | -0.09533 | -0.0034 | 0.098349 | 0.83379 | 14178.6 |
| GOBP_REG | 69 | -0.05945 | -0.00359 | 0.061334 | 0.8338 | 14178.77 |
| WP_ULCER | 14 | -0.15171 | -0.00413 | 0.15648 | 0.83384 | 14179.45 |
| GOBP_S_A | 7 | -0.17023 | -0.00328 | 0.17549 | 0.83397 | 14181.66 |
| GOBP_REC | 167 | -0.03993 | -0.00374 | 0.041159 | 0.83401 | 14182.34 |

| | | | | | | |
|---|---|---|---|---|---|---|
| GOBP_REG | 8 | -0.13768 | -0.00283 | 0.1419 | 0.83404 | 14182.85 |
| GOBP_POL | 5 | -0.24561 | -0.004 | 0.25293 | 0.83423 | 14186.08 |
| GOMF_TOI | 14 | -0.13809 | -0.00376 | 0.14213 | 0.83436 | 14188.29 |
| GOCC_DNA | 5 | -0.18794 | -0.00306 | 0.19341 | 0.8344 | 14188.97 |
| GOBP_CAR | 7 | -0.19808 | -0.00381 | 0.2038 | 0.83444 | 14189.65 |
| FLOTHO_PI | 27 | -0.09102 | -0.00344 | 0.093648 | 0.83445 | 14189.82 |
| GOBP_HIST | 20 | -0.11371 | -0.0037 | 0.11699 | 0.83446 | 14189.99 |
| DODD_NAS | 1592 | -0.01259 | -0.0035 | 0.012947 | 0.83453 | 14191.18 |
| GOBP_MO | 1139 | -0.01527 | -0.00364 | 0.0157 | 0.83468 | 14193.73 |
| GOBP_ACT | 70 | -0.06161 | -0.00374 | 0.063328 | 0.83469 | 14193.9 |
| GOBP_REG | 15 | -0.10857 | -0.00306 | 0.11157 | 0.83473 | 14194.58 |
| GOMF_LIPO | 5 | -0.22176 | -0.00361 | 0.22789 | 0.83473 | 14194.58 |
| GOBP_REG | 62 | -0.06169 | -0.00353 | 0.063376 | 0.8348 | 14195.77 |
| GOMF_PRO | 11 | -0.11578 | -0.00279 | 0.11895 | 0.83481 | 14195.94 |
| GOBP_ORG | 146 | -0.041 | -0.00359 | 0.042114 | 0.83484 | 14196.45 |
| GOCC_CAT | 31 | -0.09654 | -0.00391 | 0.099162 | 0.83487 | 14196.96 |
| REACTOME | 18 | -0.10074 | -0.00311 | 0.10347 | 0.83488 | 14197.13 |
| KEGG_LYSI | 41 | -0.06975 | -0.00325 | 0.071631 | 0.83491 | 14197.64 |
| GOCC_CELI | 68 | -0.05971 | -0.00358 | 0.061311 | 0.83492 | 14197.81 |
| SCHWAB_T | 12 | -0.14283 | -0.0036 | 0.14667 | 0.83493 | 14197.98 |
| GOBP_REG | 147 | -0.03863 | -0.0034 | 0.039666 | 0.83495 | 14198.32 |
| LOPEZ_ME | 10 | -0.19246 | -0.00443 | 0.19761 | 0.83495 | 14198.32 |
| GOMF_GU | 217 | -0.0335 | -0.00357 | 0.034362 | 0.83522 | 14202.92 |
| GOBP_VAC | 200 | -0.03375 | -0.00346 | 0.034596 | 0.83535 | 14205.13 |
| REACTOME | 25 | -0.09414 | -0.00342 | 0.096403 | 0.8356 | 14209.38 |
| SMID_BREA | 656 | -0.02014 | -0.00369 | 0.020617 | 0.83561 | 14209.55 |
| GOBP_NEG | 3 | -0.35033 | -0.00442 | 0.35871 | 0.83562 | 14209.72 |
| GOBP_REG | 8 | -0.17664 | -0.00364 | 0.18072 | 0.83581 | 14212.95 |
| GOCC_LSM | 6 | -0.25377 | -0.00452 | 0.25957 | 0.83587 | 14213.97 |
| GOCC_U6_ | 6 | -0.25377 | -0.00452 | 0.25957 | 0.83587 | 14213.97 |
| REACTOME | 7 | -0.2943 | -0.00567 | 0.30102 | 0.83587 | 14213.97 |
| GOMF_TET | 138 | -0.04407 | -0.00375 | 0.045064 | 0.83592 | 14214.82 |
| GOBP_COL | 13 | -0.13847 | -0.00363 | 0.14159 | 0.83594 | 14215.16 |
| GOBP_COF | 13 | -0.11287 | -0.00296 | 0.11538 | 0.83602 | 14216.52 |
| GOMF_PEN | 53 | -0.06809 | -0.0036 | 0.069602 | 0.83603 | 14216.69 |
| GOBP_APIC | 70 | -0.06205 | -0.00377 | 0.063415 | 0.83606 | 14217.2 |
| GOBP_REG | 6 | -0.21814 | -0.00389 | 0.22292 | 0.83609 | 14217.71 |
| GOMF_LIG | 49 | -0.07101 | -0.00361 | 0.072556 | 0.83614 | 14218.56 |
| GOCC_BLO | 13 | -0.12714 | -0.00334 | 0.12986 | 0.83621 | 14219.75 |
| GOCC_SET1 | 12 | -0.13694 | -0.00345 | 0.13986 | 0.83623 | 14220.09 |
| REACTOME | 24 | -0.10915 | -0.00389 | 0.11142 | 0.83635 | 14222.13 |
| GOBP_RES | 194 | -0.03372 | -0.0034 | 0.034424 | 0.83636 | 14222.3 |
| WONG_PR | 45 | -0.06719 | -0.00328 | 0.068577 | 0.83638 | 14222.64 |
| GOBP_RES | 118 | -0.04457 | -0.00351 | 0.045477 | 0.83644 | 14223.66 |
| GOMF_NU | 24 | -0.09094 | -0.00324 | 0.092783 | 0.83647 | 14224.17 |

| | | | | | | |
|---|---|---|---|---|---|---|
| WP_NEURO | 40 | -0.07497 | -0.00345 | 0.076447 | 0.83662 | 14226.72 |
| GOBP_PLA | 23 | -0.09434 | -0.00329 | 0.096131 | 0.83679 | 14229.61 |
| ODONNELL | 21 | -0.09873 | -0.00329 | 0.10056 | 0.83691 | 14231.65 |
| GOBP_END | 14 | -0.13475 | -0.00367 | 0.13723 | 0.83694 | 14232.16 |
| BIOCARTA_ | 18 | -0.12803 | -0.00395 | 0.13037 | 0.83697 | 14232.67 |
| MIKKELSEN | 195 | -0.03526 | -0.00357 | 0.035901 | 0.837 | 14233.19 |
| GOMF_INC | 133 | -0.04417 | -0.00369 | 0.044959 | 0.83705 | 14234.04 |
| REACTOME | 9 | -0.15756 | -0.00344 | 0.16035 | 0.83709 | 14234.72 |
| GOBP_REG | 5 | -0.18928 | -0.00308 | 0.19259 | 0.83715 | 14235.74 |
| MEBARKI_I | 14 | -0.12794 | -0.00348 | 0.13018 | 0.83715 | 14235.74 |
| GOBP_RES | 36 | -0.08671 | -0.00378 | 0.088219 | 0.83717 | 14236.08 |
| GOBP_CER | 101 | -0.05212 | -0.0038 | 0.052975 | 0.83739 | 14239.82 |
| GOBP_POS | 18 | -0.12746 | -0.00393 | 0.12954 | 0.83741 | 14240.16 |
| REACTOME | 7 | -0.16633 | -0.0032 | 0.16906 | 0.83741 | 14240.16 |
| GOMF_SER | 83 | -0.06009 | -0.00398 | 0.061052 | 0.83748 | 14241.35 |
| DESERT_PE | 146 | -0.04118 | -0.00361 | 0.041838 | 0.83751 | 14241.86 |
| WEBER_MI | 6 | -0.22248 | -0.00397 | 0.226 | 0.83754 | 14242.37 |
| REACTOME | 17 | -0.11883 | -0.00356 | 0.12068 | 0.8376 | 14243.39 |
| GOBP_FAC | 7 | -0.18675 | -0.0036 | 0.18952 | 0.83777 | 14246.28 |
| GOBP_REG | 44 | -0.07573 | -0.00365 | 0.076851 | 0.83777 | 14246.28 |
| WP_DEVEL | 31 | -0.10256 | -0.00415 | 0.10402 | 0.83791 | 14248.66 |
| GOBP_CON | 5 | -0.21531 | -0.0035 | 0.21829 | 0.83802 | 14250.53 |
| REACTOME | 68 | -0.05751 | -0.00345 | 0.058267 | 0.83817 | 14253.08 |
| REACTOME | 9 | -0.21017 | -0.00459 | 0.21284 | 0.83827 | 14254.78 |
| WP_EBOLA | 107 | -0.04749 | -0.00357 | 0.048095 | 0.83829 | 14255.12 |
| GOBP_CILI | 5 | -0.26707 | -0.00435 | 0.27043 | 0.83831 | 14255.46 |
| GOMF_GAI | 17 | -0.13981 | -0.00419 | 0.14152 | 0.83839 | 14256.82 |
| GOBP_POS | 14 | -0.11387 | -0.0031 | 0.11524 | 0.83844 | 14257.67 |
| GOBP_REG | 72 | -0.06758 | -0.00417 | 0.068333 | 0.83865 | 14261.24 |
| GOBP_CELI | 8 | -0.20073 | -0.00413 | 0.20296 | 0.83866 | 14261.41 |
| GOCC_HEN | 7 | -0.17264 | -0.00332 | 0.17455 | 0.83868 | 14261.75 |
| GOBP_NEG | 6 | -0.19421 | -0.00346 | 0.19621 | 0.83886 | 14264.81 |
| REACTOME | 54 | -0.06295 | -0.00336 | 0.063587 | 0.83891 | 14265.66 |
| GRAHAM_( | 80 | -0.06046 | -0.00393 | 0.060964 | 0.83931 | 14272.47 |
| SCHEIDERE | 51 | -0.06529 | -0.00339 | 0.065822 | 0.83938 | 14273.66 |
| GOCC_EXT | 49 | -0.06763 | -0.00344 | 0.068173 | 0.83942 | 14274.34 |
| GOBP_REG | 26 | -0.10183 | -0.00378 | 0.1026 | 0.83951 | 14275.87 |
| WP_IRINOT | 12 | -0.14163 | -0.00357 | 0.14263 | 0.83963 | 14277.91 |
| GOBP_POS | 438 | -0.02464 | -0.00371 | 0.024803 | 0.8397 | 14279.1 |
| GOBP_PYR | 8 | -0.14911 | -0.00307 | 0.1501 | 0.83974 | 14279.78 |
| BIOCARTA_ | 8 | -0.18797 | -0.00387 | 0.18921 | 0.83976 | 14280.12 |
| GOBP_REG | 12 | -0.17647 | -0.00445 | 0.17763 | 0.83976 | 14280.12 |
| GOBP_GER | 301 | -0.0286 | -0.00358 | 0.028789 | 0.83977 | 14280.29 |
| REACTOME | 28 | -0.08076 | -0.00311 | 0.081267 | 0.83983 | 14281.31 |
| REACTOME | 16 | -0.13473 | -0.00392 | 0.13555 | 0.83987 | 14281.99 |

| | | | | | | |
|---|---|---|---|---|---|---|
| GOBP_NUC | 54 | -0.06957 | -0.00372 | 0.069963 | 0.83996 | 14283.52 |
| BIOCARTA_ | 21 | -0.10899 | -0.00363 | 0.10959 | 0.84001 | 14284.37 |
| GOBP_NEG | 5 | -0.21939 | -0.00357 | 0.22057 | 0.84005 | 14285.05 |
| GOCC_RAG | 5 | -0.26745 | -0.00435 | 0.26881 | 0.84011 | 14286.07 |
| GOBP_GOL | 17 | -0.13449 | -0.00403 | 0.13517 | 0.84012 | 14286.24 |
| GOBP_OM | 11 | -0.16192 | -0.00391 | 0.16267 | 0.84023 | 14288.11 |
| GOBP_TETI | 13 | -0.13231 | -0.00347 | 0.1329 | 0.84026 | 14288.62 |
| RAY_TARG | 14 | -0.13367 | -0.00364 | 0.13419 | 0.84039 | 14290.83 |
| GOMF_BIC | 5 | -0.2261 | -0.00368 | 0.22694 | 0.84044 | 14291.68 |
| GOBP_EYE_ | 4 | -0.38645 | -0.00562 | 0.38784 | 0.84047 | 14292.19 |
| GOBP_RES | 33 | -0.0888 | -0.00371 | 0.089101 | 0.84052 | 14293.04 |
| WAKABAYA | 829 | -0.01746 | -0.00358 | 0.017522 | 0.84053 | 14293.21 |
| PEDERSEN_ | 18 | -0.13141 | -0.00406 | 0.13185 | 0.84054 | 14293.38 |
| GOBP_ESTA | 9 | -0.16999 | -0.00371 | 0.1705 | 0.8406 | 14294.4 |
| REACTOME | 16 | -0.13404 | -0.0039 | 0.13441 | 0.84068 | 14295.76 |
| GOBP_REG | 31 | -0.09528 | -0.00386 | 0.095488 | 0.84081 | 14297.97 |
| REACTOME | 21 | -0.12122 | -0.00404 | 0.12149 | 0.84081 | 14297.97 |
| GOBP_POS | 13 | -0.16351 | -0.00429 | 0.16385 | 0.84083 | 14298.31 |
| REACTOME | 8 | -0.16102 | -0.00331 | 0.16135 | 0.84085 | 14298.65 |
| KEGG_ALPI | 19 | -0.11571 | -0.00367 | 0.11589 | 0.84097 | 14300.69 |
| GOCC_CAR | 16 | -0.10739 | -0.00313 | 0.10748 | 0.84115 | 14303.76 |
| GOBP_REG | 186 | -0.03633 | -0.00359 | 0.036344 | 0.84121 | 14304.78 |
| GOBP_POS | 12 | -0.14679 | -0.0037 | 0.14675 | 0.8414 | 14308.01 |
| GOBP_POS | 219 | -0.03509 | -0.00376 | 0.035059 | 0.84152 | 14310.05 |
| GOBP_REG | 40 | -0.08304 | -0.00382 | 0.082972 | 0.84154 | 14310.39 |
| GOBP_GLY | 33 | -0.08893 | -0.00371 | 0.088726 | 0.84189 | 14316.34 |
| REACTOME | 8 | -0.18718 | -0.00385 | 0.18675 | 0.8419 | 14316.51 |
| WP_TYPE_ | 10 | -0.18028 | -0.00415 | 0.17982 | 0.84196 | 14317.53 |
| IBRAHIM_N | 5 | -0.23699 | -0.00386 | 0.23638 | 0.84197 | 14317.7 |
| GOBP_POS | 594 | -0.02141 | -0.00374 | 0.021346 | 0.84202 | 14318.55 |
| PID_FRA_P | 36 | -0.08505 | -0.00371 | 0.084737 | 0.84224 | 14322.29 |
| GOMF_LIPA | 20 | -0.12468 | -0.00406 | 0.12419 | 0.84228 | 14322.97 |
| WP_PREGN | 31 | -0.10336 | -0.00418 | 0.10293 | 0.84233 | 14323.82 |
| GOBP_GRA | 7 | -0.17746 | -0.00342 | 0.17672 | 0.84235 | 14324.16 |
| GOMF_CUI | 5 | -0.2512 | -0.00409 | 0.25003 | 0.84247 | 14326.2 |
| GOBP_REG | 10 | -0.17184 | -0.00395 | 0.17095 | 0.84261 | 14328.58 |
| GOBP_REG | 55 | -0.06315 | -0.0034 | 0.062812 | 0.84262 | 14328.75 |
| GOBP_RES | 14 | -0.15311 | -0.00417 | 0.15223 | 0.84273 | 14330.62 |
| REACTOME | 34 | -0.07752 | -0.00329 | 0.07706 | 0.84276 | 14331.13 |
| REACTOME | 100 | -0.05034 | -0.00365 | 0.050042 | 0.84278 | 14331.47 |
| GOBP_REA | 207 | -0.03512 | -0.00366 | 0.034906 | 0.84283 | 14332.32 |
| REACTOME | 45 | -0.08119 | -0.00396 | 0.080635 | 0.84298 | 14334.87 |
| WP_CODEI | 14 | -0.14186 | -0.00386 | 0.14088 | 0.84302 | 14335.56 |
| GOBP_BON | 13 | -0.13873 | -0.00364 | 0.13777 | 0.84303 | 14335.73 |
| WP_TARGE | 35 | -0.0806 | -0.00347 | 0.080002 | 0.84312 | 14337.26 |

| | | | | | | |
|---|---|---|---|---|---|---|
| GOBP_T_C | 506 | -0.02345 | -0.00379 | 0.023276 | 0.84317 | 14338.11 |
| GOCC_MIC | 3 | -0.29844 | -0.00376 | 0.29611 | 0.84323 | 14339.13 |
| VALK_AML | 24 | -0.1125 | -0.00401 | 0.1116 | 0.84329 | 14340.15 |
| ZHAN_MUI | 20 | -0.10726 | -0.00349 | 0.10636 | 0.84337 | 14341.51 |
| GOMF_PH( | 6 | -0.22273 | -0.00397 | 0.22079 | 0.84345 | 14342.87 |
| GOBERT_O | 1017 | -0.01632 | -0.00369 | 0.016175 | 0.84356 | 14344.74 |
| GOCC_CLA | 6 | -0.17745 | -0.00316 | 0.17583 | 0.84356 | 14344.74 |
| WP_CANOI | 7 | -0.16837 | -0.00324 | 0.16675 | 0.84368 | 14346.78 |
| NUYTTEN_ | 720 | -0.019 | -0.00364 | 0.018814 | 0.84371 | 14347.29 |
| GOCC_PEP | 115 | -0.04614 | -0.00359 | 0.045667 | 0.84381 | 14348.99 |
| GOCC_CHL | 47 | -0.07934 | -0.00395 | 0.078528 | 0.84383 | 14349.33 |
| GOBP_MU | 9 | -0.19881 | -0.00434 | 0.19676 | 0.84385 | 14349.67 |
| GOBP_P38 | 52 | -0.07003 | -0.00367 | 0.06929 | 0.84391 | 14350.69 |
| GOBP_STEI | 124 | -0.04445 | -0.00359 | 0.043969 | 0.84395 | 14351.37 |
| GOBP_PHC | 11 | -0.17592 | -0.00425 | 0.174 | 0.84399 | 14352.05 |
| GOBP_CAR | 150 | -0.03865 | -0.00343 | 0.038216 | 0.84407 | 14353.41 |
| GOMF_CHI | 20 | -0.13482 | -0.00439 | 0.1332 | 0.84426 | 14356.64 |
| GOBP_NEG | 327 | -0.02732 | -0.00356 | 0.026983 | 0.84431 | 14357.49 |
| GOBP_ADH | 13 | -0.14645 | -0.00384 | 0.14455 | 0.84448 | 14360.38 |
| GOBP_TISS | 249 | -0.03252 | -0.00371 | 0.032103 | 0.84449 | 14360.55 |
| GOBP_POS | 7 | -0.21712 | -0.00418 | 0.21426 | 0.84455 | 14361.57 |
| GOMF_ANI | 8 | -0.16041 | -0.0033 | 0.15828 | 0.84457 | 14361.91 |
| GOBP_REG | 124 | -0.04254 | -0.00344 | 0.041967 | 0.8446 | 14362.42 |
| GOMF_CCI | 5 | -0.24779 | -0.00403 | 0.24438 | 0.84469 | 14363.95 |
| GOBP_POS | 28 | -0.10015 | -0.00385 | 0.098745 | 0.84477 | 14365.31 |
| REACTOME | 9 | -0.17282 | -0.00377 | 0.17028 | 0.84492 | 14367.86 |
| GOBP_PHC | 22 | -0.10079 | -0.00344 | 0.099252 | 0.84506 | 14370.25 |
| REACTOME | 7 | -0.19946 | -0.00384 | 0.19637 | 0.84512 | 14371.27 |
| GOBP_REG | 38 | -0.09147 | -0.0041 | 0.090021 | 0.84521 | 14372.8 |
| NGUYEN_N | 25 | -0.10478 | -0.00381 | 0.10308 | 0.8453 | 14374.33 |
| GOBP_VES | 43 | -0.07592 | -0.00362 | 0.074677 | 0.84532 | 14374.67 |
| GOBP_CAN | 7 | -0.191 | -0.00368 | 0.18787 | 0.84533 | 14374.84 |
| GOBP_NEG | 62 | -0.06466 | -0.0037 | 0.063584 | 0.84539 | 14375.86 |
| GABRIELY_ | 272 | -0.0315 | -0.00375 | 0.030972 | 0.84543 | 14376.54 |
| GOBP_AMY | 12 | -0.1336 | -0.00337 | 0.13135 | 0.84545 | 14376.88 |
| BIOCARTA_ | 19 | -0.10291 | -0.00326 | 0.10109 | 0.84566 | 14380.45 |
| GOBP_CELI | 20 | -0.1274 | -0.00414 | 0.12506 | 0.84582 | 14383.17 |
| GOBP_REG | 23 | -0.10342 | -0.00361 | 0.1015 | 0.84587 | 14384.02 |
| GOBP_CAT | 4 | -0.29834 | -0.00434 | 0.29278 | 0.8459 | 14384.53 |
| SU_THYMU | 18 | -0.13499 | -0.00417 | 0.13242 | 0.84598 | 14385.89 |
| HOLLEMAN | 12 | -0.13607 | -0.00343 | 0.13336 | 0.84621 | 14389.8 |
| STEIN_ESRI | 94 | -0.05034 | -0.00354 | 0.049333 | 0.84621 | 14389.8 |
| GOBP_TYP | 10 | -0.19297 | -0.00444 | 0.18911 | 0.84623 | 14390.14 |
| GOBP_2_O | 17 | -0.11074 | -0.00332 | 0.10849 | 0.84632 | 14391.67 |
| GOBP_NEG | 5 | -0.24669 | -0.00401 | 0.24158 | 0.8464 | 14393.03 |

| | | | | | | |
|---|---|---|---|---|---|---|
| GOBP_DET | 49 | -0.07245 | -0.00369 | 0.07092 | 0.84651 | 14394.9 |
| WP_CELL_I | 13 | -0.13308 | -0.00349 | 0.13026 | 0.84652 | 14395.07 |
| WP_3Q29_ | 62 | -0.07218 | -0.00413 | 0.070629 | 0.84659 | 14396.26 |
| SU_PANCR | 39 | -0.07998 | -0.00363 | 0.078241 | 0.84667 | 14397.62 |
| GOBP_XEN | 14 | -0.15079 | -0.0041 | 0.14747 | 0.84671 | 14398.3 |
| GOCC_PRC | 8 | -0.17929 | -0.00369 | 0.17531 | 0.84677 | 14399.32 |
| KEGG_POR | 36 | -0.08777 | -0.00383 | 0.085805 | 0.8468 | 14399.83 |
| GOMF_CAI | 15 | -0.13509 | -0.00381 | 0.13205 | 0.84685 | 14400.68 |
| GOBP_SPEI | 191 | -0.03568 | -0.00357 | 0.034855 | 0.84701 | 14403.41 |
| REACTOME | 6 | -0.21823 | -0.00389 | 0.21309 | 0.8471 | 14404.94 |
| GOBP_MY( | 25 | -0.09626 | -0.0035 | 0.093949 | 0.8472 | 14406.64 |
| REACTOME | 76 | -0.04981 | -0.00315 | 0.048615 | 0.8472 | 14406.64 |
| GOBP_SYN | 711 | -0.01983 | -0.00378 | 0.019351 | 0.84728 | 14408 |
| GOCC_TRA | 16 | -0.13481 | -0.00392 | 0.13147 | 0.84742 | 14410.38 |
| GOMF_OXI | 15 | -0.14613 | -0.00412 | 0.14242 | 0.84756 | 14412.76 |
| ALONSO_M | 24 | -0.09779 | -0.00348 | 0.095293 | 0.8476 | 14413.44 |
| GOBP_REG | 6 | -0.2462 | -0.00439 | 0.23986 | 0.84765 | 14414.29 |
| SMITH_TEI | 81 | -0.05455 | -0.00357 | 0.053142 | 0.84765 | 14414.29 |
| DAZARD_U | 19 | -0.12929 | -0.0041 | 0.12595 | 0.84767 | 14414.63 |
| GOBP_NEG | 93 | -0.05435 | -0.00381 | 0.052929 | 0.84773 | 14415.65 |
| BOSCO_IN | 73 | -0.06029 | -0.00374 | 0.058681 | 0.84789 | 14418.37 |
| GOBP_MO | 5 | -0.27736 | -0.00451 | 0.26981 | 0.84801 | 14420.41 |
| CERVERA_S | 108 | -0.05255 | -0.00396 | 0.051115 | 0.84802 | 14420.58 |
| GOBP_AMI | 136 | -0.04672 | -0.00395 | 0.045443 | 0.84805 | 14421.09 |
| GOBP_PRC | 26 | -0.10119 | -0.00375 | 0.098413 | 0.84807 | 14421.43 |
| GOMF_ISO | 4 | -0.23462 | -0.00341 | 0.22816 | 0.8481 | 14421.94 |
| PID_RAC1_ | 52 | -0.06516 | -0.00342 | 0.063352 | 0.84814 | 14422.62 |
| GOBP_ACT | 179 | -0.03947 | -0.00382 | 0.03835 | 0.84828 | 14425 |
| GOBP_VITA | 19 | -0.13278 | -0.00421 | 0.12902 | 0.84829 | 14425.17 |
| PID_NFAT_ | 53 | -0.07838 | -0.00415 | 0.076151 | 0.84832 | 14425.68 |
| GOBP_POS | 4 | -0.28214 | -0.00411 | 0.27397 | 0.84845 | 14427.89 |
| BIOCARTA_ | 9 | -0.16933 | -0.0037 | 0.16441 | 0.84848 | 14428.4 |
| SASSON_RI | 85 | -0.0621 | -0.00416 | 0.060235 | 0.84871 | 14432.31 |
| GOBP_PRC | 6 | -0.19581 | -0.00349 | 0.18991 | 0.84874 | 14432.82 |
| REACTOME | 35 | -0.09232 | -0.00397 | 0.089515 | 0.84881 | 14434.01 |
| GOBP_POS | 124 | -0.04563 | -0.00369 | 0.044215 | 0.84893 | 14436.05 |
| GOMF_DN/ | 3 | -0.26599 | -0.00335 | 0.2577 | 0.84899 | 14437.07 |
| REACTOME | 5 | -0.18284 | -0.00298 | 0.17703 | 0.84914 | 14439.63 |
| SCHLOSSEF | 32 | -0.08285 | -0.00341 | 0.080173 | 0.84929 | 14442.18 |
| WP_FATTY_ | 14 | -0.16597 | -0.00452 | 0.16054 | 0.84939 | 14443.88 |
| GOBP_POS | 57 | -0.07469 | -0.0041 | 0.072245 | 0.8494 | 14444.05 |
| GOBP_EST/ | 5 | -0.18417 | -0.003 | 0.17809 | 0.84945 | 14444.9 |
| SCIAN_INV | 28 | -0.0918 | -0.00353 | 0.08875 | 0.84952 | 14446.09 |
| BILANGES_ | 71 | -0.06168 | -0.00378 | 0.059609 | 0.8496 | 14447.45 |
| GOMF_AP_ | 10 | -0.18534 | -0.00426 | 0.17909 | 0.84963 | 14447.96 |

| | | | | | | |
|---|---|---|---|---|---|---|
| GOBP_NEG | 8 | -0.19129 | -0.00394 | 0.18467 | 0.84985 | 14451.7 |
| WP_TCA_C | 17 | -0.12286 | -0.00369 | 0.11859 | 0.84989 | 14452.38 |
| FOSTER_TC | 156 | -0.04082 | -0.00369 | 0.039382 | 0.84998 | 14453.91 |
| GOBP_PRC | 62 | -0.06182 | -0.00354 | 0.05962 | 0.85009 | 14455.78 |
| GOBP_MRI | 8 | -0.18994 | -0.00391 | 0.18317 | 0.85012 | 14456.29 |
| GOBP_PYR | 8 | -0.19187 | -0.00395 | 0.18495 | 0.85022 | 14457.99 |
| GOBP_CEN | 7 | -0.18884 | -0.00364 | 0.18194 | 0.85035 | 14460.2 |
| GOBP_ANT | 25 | -0.09525 | -0.00346 | 0.09175 | 0.8504 | 14461.05 |
| WP_TOLLLI | 26 | -0.09845 | -0.00365 | 0.094775 | 0.85053 | 14463.26 |
| GOMF_ASF | 26 | -0.10291 | -0.00382 | 0.099065 | 0.85055 | 14463.6 |
| GOMF_MH | 12 | -0.13466 | -0.00339 | 0.12954 | 0.85071 | 14466.32 |
| GOBP_PRC | 29 | -0.1073 | -0.0042 | 0.1032 | 0.85076 | 14467.17 |
| REACTOME | 11 | -0.14564 | -0.00351 | 0.14007 | 0.85078 | 14467.51 |
| CHOW_RAS | 30 | -0.09961 | -0.00397 | 0.095759 | 0.85088 | 14469.21 |
| CAMPS_CC | 47 | -0.09286 | -0.00463 | 0.08925 | 0.85093 | 14470.06 |
| REACTOME | 5 | -0.28134 | -0.00458 | 0.27035 | 0.85097 | 14470.74 |
| GOBP_APC | 101 | -0.04767 | -0.00348 | 0.0458 | 0.85103 | 14471.77 |
| GOBP_REG | 10 | -0.17761 | -0.00409 | 0.17053 | 0.85118 | 14474.32 |
| GOMF_PH( | 7 | -0.17283 | -0.00333 | 0.16591 | 0.85122 | 14475 |
| GOBP_DNA | 130 | -0.04611 | -0.00381 | 0.044257 | 0.85124 | 14475.34 |
| GOCC_GLIA | 36 | -0.09351 | -0.00408 | 0.089755 | 0.85124 | 14475.34 |
| GOCC_POL | 21 | -0.11454 | -0.00382 | 0.10983 | 0.85149 | 14479.59 |
| GOBP_NEG | 14 | -0.14319 | -0.0039 | 0.1373 | 0.8515 | 14479.76 |
| GOBP_POS | 21 | -0.13491 | -0.0045 | 0.12926 | 0.85167 | 14482.65 |
| GOBP_GRC | 8 | -0.18984 | -0.00391 | 0.18188 | 0.85169 | 14482.99 |
| GOBP_PRC | 9 | -0.15066 | -0.00329 | 0.1443 | 0.85178 | 14484.52 |
| GOBP_ESTA | 8 | -0.16378 | -0.00337 | 0.15684 | 0.85181 | 14485.03 |
| GUO_HEX_ | 63 | -0.06444 | -0.00372 | 0.061657 | 0.85202 | 14488.6 |
| GOMF_ACI | 83 | -0.0545 | -0.00361 | 0.052128 | 0.85209 | 14489.79 |
| GOBP_NEG | 137 | -0.04432 | -0.00376 | 0.042388 | 0.85214 | 14490.64 |
| GOMF_PH( | 27 | -0.10458 | -0.00395 | 0.099995 | 0.85219 | 14491.49 |
| GOBP_COL | 12 | -0.14614 | -0.00368 | 0.13972 | 0.8522 | 14491.66 |
| GOMF_POT | 9 | -0.15159 | -0.00331 | 0.14489 | 0.85226 | 14492.68 |
| GOBP_EYE_ | 46 | -0.08461 | -0.00417 | 0.080798 | 0.85247 | 14496.25 |
| FINAK_BRE | 22 | -0.11849 | -0.00404 | 0.11315 | 0.85249 | 14496.59 |
| GOCC_ROU | 80 | -0.0583 | -0.00379 | 0.055673 | 0.85249 | 14496.59 |
| GOBP_ERY | 35 | -0.07815 | -0.00336 | 0.074619 | 0.85252 | 14497.1 |
| GOBP_NEG | 16 | -0.1335 | -0.00388 | 0.12743 | 0.85259 | 14498.29 |
| YAO_TEMF | 67 | -0.06358 | -0.00378 | 0.060673 | 0.85267 | 14499.65 |
| GOMF_VIR | 11 | -0.18099 | -0.00437 | 0.17266 | 0.85273 | 14500.67 |
| GOMF_PR( | 15 | -0.12911 | -0.00364 | 0.12316 | 0.85276 | 14501.18 |
| PID_P73PA | 78 | -0.06139 | -0.00394 | 0.058529 | 0.85289 | 14503.39 |
| GOMF_THI | 10 | -0.13848 | -0.00319 | 0.13201 | 0.8529 | 14503.56 |
| ZHENG_FO | 12 | -0.15406 | -0.00388 | 0.14676 | 0.85306 | 14506.29 |
| GOCC_RIB( | 254 | -0.03242 | -0.00373 | 0.030876 | 0.8531 | 14506.97 |

| | | | | | | |
|---|---|---|---|---|---|---|
| GOBP_POS | 79 | -0.06616 | -0.00427 | 0.062997 | 0.85316 | 14507.99 |
| SMID_BREA | 9 | -0.19616 | -0.00428 | 0.18672 | 0.85326 | 14509.69 |
| GOBP_INTI | 8 | -0.21331 | -0.00439 | 0.20301 | 0.8533 | 14510.37 |
| GOBP_T_C | 121 | -0.04861 | -0.00388 | 0.046254 | 0.85333 | 14510.88 |
| GOCC_NEU | 5 | -0.24371 | -0.00397 | 0.23187 | 0.85338 | 14511.73 |
| GOBP_POS | 21 | -0.12214 | -0.00407 | 0.11619 | 0.8534 | 14512.07 |
| GOMF_SOI | 17 | -0.13589 | -0.00408 | 0.12926 | 0.85343 | 14512.58 |
| LEE_SP4_T | 14 | -0.17521 | -0.00477 | 0.16666 | 0.85343 | 14512.58 |
| GOBP_CAR | 47 | -0.08805 | -0.00439 | 0.083738 | 0.85349 | 14513.6 |
| MEBARKI_I | 26 | -0.11458 | -0.00425 | 0.10896 | 0.85349 | 14513.6 |
| REACTOME | 46 | -0.0833 | -0.00411 | 0.079204 | 0.85353 | 14514.28 |
| REACTOME | 5 | -0.29632 | -0.00482 | 0.28164 | 0.85362 | 14515.81 |
| GOBP_CLU | 6 | -0.25191 | -0.00449 | 0.23937 | 0.85368 | 14516.83 |
| JAEGER_M | 244 | -0.03433 | -0.00388 | 0.032625 | 0.85368 | 14516.83 |
| CHOI_ATL_ | 44 | -0.07526 | -0.00363 | 0.071506 | 0.85369 | 14517 |
| GOBP_ZIN | 27 | -0.09486 | -0.00359 | 0.090139 | 0.85369 | 14517 |
| GOMF_COI | 10 | -0.22208 | -0.00511 | 0.21088 | 0.85385 | 14519.72 |
| GOBP_EXO | 328 | -0.02907 | -0.0038 | 0.027597 | 0.85387 | 14520.06 |
| GOMF_HYA | 24 | -0.10551 | -0.00376 | 0.10017 | 0.85389 | 14520.4 |
| GOBP_CILI | 402 | -0.02596 | -0.00375 | 0.024643 | 0.85394 | 14521.25 |
| GOBP_LEU | 17 | -0.13403 | -0.00402 | 0.12722 | 0.85395 | 14521.42 |
| GOBP_PRC | 5 | -0.21622 | -0.00352 | 0.20519 | 0.854 | 14522.27 |
| GOCC_EXO | 209 | -0.03499 | -0.00366 | 0.033196 | 0.85409 | 14523.8 |
| MUELLER_ | 35 | -0.08693 | -0.00374 | 0.082451 | 0.85414 | 14524.65 |
| GOBP_RIB | 7 | -0.18185 | -0.0035 | 0.1724 | 0.85424 | 14526.35 |
| GOBP_CHL | 103 | -0.05454 | -0.00402 | 0.051687 | 0.85433 | 14527.88 |
| GOCC_OXI | 108 | -0.04937 | -0.00372 | 0.046785 | 0.85435 | 14528.22 |
| GOBP_TOL | 26 | -0.109 | -0.00404 | 0.10322 | 0.85449 | 14530.6 |
| REACTOME | 24 | -0.09526 | -0.00339 | 0.090174 | 0.85459 | 14532.3 |
| REACTOME | 4 | -0.29631 | -0.00431 | 0.28041 | 0.85468 | 14533.83 |
| CHAUHAN_ | 46 | -0.06949 | -0.00343 | 0.065747 | 0.85471 | 14534.34 |
| GOMF_GLY | 10 | -0.17544 | -0.00404 | 0.16598 | 0.85474 | 14534.85 |
| GOBP_NEG | 20 | -0.12148 | -0.00395 | 0.11492 | 0.85476 | 14535.19 |
| WP_LTF_D | 16 | -0.14245 | -0.00415 | 0.13475 | 0.85476 | 14535.19 |
| GOBP_PRC | 16 | -0.12883 | -0.00375 | 0.12184 | 0.85481 | 14536.04 |
| MISSIAGLI | 108 | -0.04962 | -0.00374 | 0.046931 | 0.85482 | 14536.21 |
| GOBP_RES | 6 | -0.26796 | -0.00478 | 0.2532 | 0.85504 | 14539.96 |
| IIZUKA_LIV | 24 | -0.10275 | -0.00366 | 0.097084 | 0.85505 | 14540.13 |
| GOBP_REG | 29 | -0.11306 | -0.00443 | 0.10682 | 0.85506 | 14540.3 |
| GOBP_DNA | 27 | -0.08827 | -0.00334 | 0.083386 | 0.8551 | 14540.98 |
| PID_ERBB1 | 103 | -0.05274 | -0.00388 | 0.049789 | 0.85524 | 14543.36 |
| GOCC_EAR | 12 | -0.16677 | -0.0042 | 0.15741 | 0.85529 | 14544.21 |
| GOCC_EXT | 10 | -0.21244 | -0.00489 | 0.20043 | 0.85541 | 14546.25 |
| GOBP_NEG | 20 | -0.11617 | -0.00378 | 0.10957 | 0.85548 | 14547.44 |
| NIKOLSKY_ | 21 | -0.14623 | -0.00487 | 0.1379 | 0.8555 | 14547.78 |

| | | | | | | |
|---|---|---|---|---|---|---|
| SAKAI_CHR | 35 | -0.08386 | -0.00361 | 0.079082 | 0.85551 | 14547.95 |
| BOQUEST_ | 200 | -0.03968 | -0.00406 | 0.037419 | 0.85554 | 14548.46 |
| GOBP_EYE_ | 33 | -0.10072 | -0.00421 | 0.094955 | 0.85558 | 14549.14 |
| GOBP_POS | 13 | -0.13617 | -0.00357 | 0.12836 | 0.8556 | 14549.48 |
| GOBP_NEG | 12 | -0.12328 | -0.00311 | 0.11621 | 0.85561 | 14549.65 |
| REACTOME | 417 | -0.02618 | -0.00385 | 0.024664 | 0.85579 | 14552.71 |
| GOBP_ESTA | 16 | -0.15175 | -0.00442 | 0.14294 | 0.8558 | 14552.88 |
| SHEDDEN_ | 65 | -0.06452 | -0.00378 | 0.060738 | 0.85594 | 14555.26 |
| GOBP_REG | 6 | -0.17298 | -0.00308 | 0.1628 | 0.856 | 14556.28 |
| GOBP_NAD | 10 | -0.17111 | -0.00394 | 0.16103 | 0.85601 | 14556.45 |
| WATANABE | 55 | -0.07155 | -0.00386 | 0.067319 | 0.85606 | 14557.3 |
| REACTOME | 13 | -0.13824 | -0.00363 | 0.13003 | 0.85612 | 14558.32 |
| GOBP_NEU | 31 | -0.09957 | -0.00403 | 0.093654 | 0.85613 | 14558.49 |
| GROSS_HIF | 25 | -0.10736 | -0.0039 | 0.10094 | 0.85624 | 14560.36 |
| GOBP_NEG | 4 | -0.29781 | -0.00433 | 0.27976 | 0.85645 | 14563.93 |
| REACTOME | 93 | -0.0535 | -0.00375 | 0.050249 | 0.85647 | 14564.27 |
| GOBP_REG | 30 | -0.1018 | -0.00406 | 0.095442 | 0.85692 | 14571.92 |
| GOMF_LIG | 132 | -0.05038 | -0.0042 | 0.047222 | 0.85698 | 14572.94 |
| ICHIBA_GR | 53 | -0.0782 | -0.00414 | 0.073296 | 0.85698 | 14572.94 |
| GOBP_AMI | 23 | -0.11118 | -0.00388 | 0.10419 | 0.85702 | 14573.63 |
| NIKOLSKY_ | 8 | -0.22713 | -0.00467 | 0.21283 | 0.85705 | 14574.14 |
| GOCC_PHO | 16 | -0.11511 | -0.00335 | 0.10786 | 0.85706 | 14574.31 |
| REACTOME | 233 | -0.0357 | -0.00394 | 0.033431 | 0.85722 | 14577.03 |
| REACTOME | 11 | -0.1771 | -0.00427 | 0.16582 | 0.85723 | 14577.2 |
| GOBP_NEG | 14 | -0.13128 | -0.00357 | 0.12286 | 0.85736 | 14579.41 |
| GOBP_MAC | 99 | -0.05335 | -0.00385 | 0.04992 | 0.85739 | 14579.92 |
| GOBP_NEG | 17 | -0.10978 | -0.00329 | 0.1027 | 0.85745 | 14580.94 |
| WORSCHEC | 28 | -0.10691 | -0.00411 | 0.099962 | 0.85757 | 14582.98 |
| GOBP_REG | 30 | -0.10061 | -0.00401 | 0.094029 | 0.85769 | 14585.02 |
| GOBP_POS | 84 | -0.05297 | -0.00353 | 0.049471 | 0.85786 | 14587.91 |
| GOBP_POS | 120 | -0.05054 | -0.00402 | 0.047195 | 0.85788 | 14588.25 |
| GOBP_NEG | 2 | -0.44198 | -0.00455 | 0.41262 | 0.85794 | 14589.27 |
| GAVIN_FOX | 91 | -0.05898 | -0.00408 | 0.05505 | 0.85798 | 14589.95 |
| GOBP_POS | 5 | -0.28304 | -0.00461 | 0.26412 | 0.85804 | 14590.97 |
| GOMF_G_F | 6 | -0.25562 | -0.00456 | 0.23847 | 0.85812 | 14592.33 |
| REACTOME | 24 | -0.09738 | -0.00347 | 0.090839 | 0.85813 | 14592.5 |
| GOBP_DIG | 106 | -0.0558 | -0.00417 | 0.051986 | 0.85842 | 14597.43 |
| REACTOME | 13 | -0.14749 | -0.00387 | 0.13733 | 0.85856 | 14599.81 |
| COULOUAR | 101 | -0.0541 | -0.00395 | 0.050352 | 0.85866 | 14601.51 |
| GOBP_NEU | 26 | -0.10933 | -0.00405 | 0.10175 | 0.8587 | 14602.19 |
| GOMF_GLY | 260 | -0.03351 | -0.00391 | 0.03117 | 0.8588 | 14603.89 |
| REACTOME | 29 | -0.0885 | -0.00347 | 0.08232 | 0.85881 | 14604.06 |
| KYNG_WER | 13 | -0.14744 | -0.00387 | 0.13711 | 0.85888 | 14605.25 |
| REACTOME | 121 | -0.0491 | -0.00392 | 0.045654 | 0.8589 | 14605.59 |
| REACTOME | 506 | -0.02421 | -0.00391 | 0.022516 | 0.8589 | 14605.59 |

| | | | | | | |
|---|---|---|---|---|---|---|
| GOBP_REG | 8 | -0.16503 | -0.0034 | 0.15345 | 0.85891 | 14605.76 |
| GOMF_ZIN | 22 | -0.10746 | -0.00367 | 0.099887 | 0.85899 | 14607.12 |
| REACTOME | 17 | -0.12156 | -0.00365 | 0.11295 | 0.85908 | 14608.66 |
| GOBP_REG | 37 | -0.08934 | -0.00395 | 0.083006 | 0.8591 | 14609 |
| REACTOME | 14 | -0.14053 | -0.00383 | 0.1305 | 0.85922 | 14611.04 |
| GOMF_RN/ | 12 | -0.13441 | -0.00339 | 0.12477 | 0.85931 | 14612.57 |
| GOBP_POS | 12 | -0.13883 | -0.0035 | 0.12883 | 0.85939 | 14613.93 |
| BANDRES_ | 17 | -0.10504 | -0.00315 | 0.097433 | 0.85949 | 14615.63 |
| BROWNE_I | 36 | -0.09239 | -0.00403 | 0.085704 | 0.85949 | 14615.63 |
| GOMF_PR( | 6 | -0.22642 | -0.00404 | 0.20999 | 0.85953 | 14616.31 |
| TOMIDA_N | 21 | -0.10621 | -0.00354 | 0.098494 | 0.85956 | 14616.82 |
| GOBP_OXY | 16 | -0.1636 | -0.00476 | 0.15169 | 0.8596 | 14617.5 |
| GOMF_GEI | 45 | -0.07664 | -0.00374 | 0.070979 | 0.85987 | 14622.09 |
| GOCC_DEN | 36 | -0.08933 | -0.0039 | 0.082657 | 0.86007 | 14625.49 |
| GOBP_ACT | 149 | -0.04499 | -0.00398 | 0.041623 | 0.86011 | 14626.17 |
| BIOCARTA_ | 5 | -0.24163 | -0.00393 | 0.22338 | 0.8603 | 14629.4 |
| WP_PARKII | 37 | -0.08497 | -0.00376 | 0.078552 | 0.86031 | 14629.57 |
| GRAHAM_( | 35 | -0.09668 | -0.00416 | 0.089367 | 0.86032 | 14629.74 |
| REACTOME | 102 | -0.05411 | -0.00397 | 0.050018 | 0.86032 | 14629.74 |
| GOMF_HE> | 25 | -0.11073 | -0.00403 | 0.10235 | 0.86034 | 14630.08 |
| GOCC_GOL | 131 | -0.04611 | -0.00383 | 0.042604 | 0.86043 | 14631.61 |
| GOBP_HYD | 14 | -0.13248 | -0.00361 | 0.12235 | 0.86054 | 14633.48 |
| TURJANSKI | 7 | -0.19994 | -0.00385 | 0.18463 | 0.86057 | 14633.99 |
| MATZUK_S | 109 | -0.05487 | -0.00416 | 0.050658 | 0.86061 | 14634.67 |
| REACTOME | 12 | -0.16551 | -0.00417 | 0.15275 | 0.8607 | 14636.2 |
| GOBP_T_C | 63 | -0.06861 | -0.00396 | 0.06331 | 0.86074 | 14636.88 |
| GOBP_NEG | 51 | -0.0786 | -0.00408 | 0.072529 | 0.86075 | 14637.05 |
| WP_DYRK1 | 61 | -0.06991 | -0.00397 | 0.064496 | 0.8608 | 14637.9 |
| GOBP_REG | 7 | -0.18955 | -0.00365 | 0.17481 | 0.86088 | 14639.26 |
| GOBP_REG | 502 | -0.02416 | -0.00389 | 0.022269 | 0.86104 | 14641.99 |
| GOCC_MO | 5 | -0.25561 | -0.00416 | 0.23554 | 0.86107 | 14642.5 |
| BLANCO_N | 389 | -0.0282 | -0.00401 | 0.025979 | 0.86116 | 14644.03 |
| WP_PI3KAI | 20 | -0.11507 | -0.00374 | 0.10599 | 0.86118 | 14644.37 |
| GOBP_REG | 8 | -0.20521 | -0.00422 | 0.18899 | 0.86121 | 14644.88 |
| WP_AUTOI | 30 | -0.09427 | -0.00375 | 0.086809 | 0.86123 | 14645.22 |
| GOBP_INO | 501 | -0.02512 | -0.00404 | 0.023132 | 0.86126 | 14645.73 |
| GOBP_NEG | 13 | -0.16927 | -0.00444 | 0.15586 | 0.86126 | 14645.73 |
| WP_FATTY_ | 20 | -0.11258 | -0.00366 | 0.10361 | 0.86139 | 14647.94 |
| GOBP_POS | 30 | -0.09728 | -0.00388 | 0.089491 | 0.86149 | 14649.64 |
| REACTOME | 11 | -0.15735 | -0.0038 | 0.14471 | 0.86155 | 14650.66 |
| GOBP_HYA | 14 | -0.14499 | -0.00395 | 0.13331 | 0.8616 | 14651.51 |
| REACTOME | 41 | -0.0856 | -0.00399 | 0.07866 | 0.86175 | 14654.06 |
| TOOKER_G | 120 | -0.04834 | -0.00384 | 0.044402 | 0.86185 | 14655.76 |
| GRATIAS_R | 18 | -0.16224 | -0.00501 | 0.14893 | 0.86199 | 14658.14 |
| GOBP_DIC/ | 18 | -0.11413 | -0.00352 | 0.10476 | 0.86201 | 14658.48 |

| | | | | | | |
|---|---|---|---|---|---|---|
| GOMF_PEF | 43 | -0.07985 | -0.00381 | 0.073264 | 0.86212 | 14660.35 |
| ZHENG_BC | 473 | -0.02713 | -0.00424 | 0.024878 | 0.8622 | 14661.71 |
| GOBP_POS | 21 | -0.12584 | -0.00419 | 0.11541 | 0.86222 | 14662.05 |
| GOBP_REG | 9 | -0.2085 | -0.00455 | 0.19113 | 0.86233 | 14663.92 |
| REACTOME | 63 | -0.05869 | -0.00338 | 0.053779 | 0.86241 | 14665.28 |
| GOBP_GRA | 137 | -0.05126 | -0.00435 | 0.046962 | 0.86245 | 14665.96 |
| YAUCH_HE | 129 | -0.04995 | -0.00412 | 0.045736 | 0.86263 | 14669.02 |
| GOBP_POS | 18 | -0.13785 | -0.00425 | 0.1262 | 0.86264 | 14669.19 |
| GOMF_BILI | 19 | -0.11401 | -0.00362 | 0.10437 | 0.86266 | 14669.53 |
| GOMF_ENI | 114 | -0.05074 | -0.00393 | 0.046449 | 0.86266 | 14669.53 |
| FUJII_YBX1 | 180 | -0.03994 | -0.00388 | 0.036541 | 0.8628 | 14671.91 |
| GOBP_NEG | 6 | -0.24166 | -0.00431 | 0.22107 | 0.86283 | 14672.42 |
| DELYS_THY | 214 | -0.04119 | -0.00436 | 0.03767 | 0.86287 | 14673.1 |
| GOCC_AP_ | 35 | -0.07922 | -0.00341 | 0.072423 | 0.86299 | 14675.14 |
| GOMF_NAI | 6 | -0.24213 | -0.00432 | 0.22123 | 0.86312 | 14677.36 |
| GOMF_RNA | 8 | -0.19517 | -0.00402 | 0.17818 | 0.86331 | 14680.59 |
| GOCC_EXO | 13 | -0.17871 | -0.00469 | 0.16315 | 0.86332 | 14680.76 |
| BIOCARTA_ | 18 | -0.13529 | -0.00418 | 0.12349 | 0.86336 | 14681.44 |
| GOBP_POS | 4 | -0.32489 | -0.00473 | 0.29637 | 0.8635 | 14683.82 |
| GOBP_NEG | 4 | -0.24123 | -0.00351 | 0.22005 | 0.86351 | 14683.99 |
| GOBP_POS | 5 | -0.27477 | -0.00447 | 0.25064 | 0.86351 | 14683.99 |
| GOBP_POS | 5 | -0.27477 | -0.00447 | 0.25064 | 0.86351 | 14683.99 |
| GOBP_XEN | 6 | -0.25553 | -0.00455 | 0.23308 | 0.86352 | 14684.16 |
| REACTOME | 5 | -0.22421 | -0.00365 | 0.20435 | 0.86372 | 14687.56 |
| GOCC_DNA | 12 | -0.17022 | -0.00429 | 0.15503 | 0.86388 | 14690.28 |
| GOBP_NEG | 14 | -0.12481 | -0.0034 | 0.11365 | 0.86393 | 14691.13 |
| GOBP_NEG | 30 | -0.09754 | -0.00389 | 0.088811 | 0.86395 | 14691.47 |
| GOMF_FAT | 8 | -0.20486 | -0.00422 | 0.18646 | 0.86403 | 14692.83 |
| LI_WILMS_ | 17 | -0.113 | -0.00339 | 0.10282 | 0.86411 | 14694.19 |
| GOBP_INTI | 7 | -0.23229 | -0.00447 | 0.21135 | 0.86413 | 14694.53 |
| LIU_OVARI | 33 | -0.10561 | -0.00441 | 0.096052 | 0.86423 | 14696.23 |
| GOBP_NEG | 2 | -0.47906 | -0.00493 | 0.43561 | 0.86427 | 14696.91 |
| GOMF_INT | 17 | -0.17431 | -0.00523 | 0.15834 | 0.8645 | 14700.82 |
| WP_HOSTF | 30 | -0.10585 | -0.00422 | 0.096115 | 0.8646 | 14702.52 |
| WAGSCHAI | 13 | -0.16836 | -0.00442 | 0.15278 | 0.86476 | 14705.24 |
| GOBP_PIGI | 10 | -0.17039 | -0.00392 | 0.15459 | 0.86481 | 14706.09 |
| GOMF_POI | 6 | -0.23016 | -0.0041 | 0.20874 | 0.8649 | 14707.62 |
| GOBP_SEB/ | 10 | -0.18773 | -0.00432 | 0.17022 | 0.86496 | 14708.64 |
| GOBP_MUS | 23 | -0.11977 | -0.00418 | 0.10856 | 0.86503 | 14709.84 |
| GOBP_THY | 9 | -0.16861 | -0.00368 | 0.15278 | 0.8651 | 14711.03 |
| FERREIRA_ | 90 | -0.0559 | -0.00385 | 0.050642 | 0.86516 | 14712.05 |
| GOBP_REG | 16 | -0.145 | -0.00422 | 0.13131 | 0.86525 | 14713.58 |
| GOBP_POS | 42 | -0.09528 | -0.00449 | 0.086274 | 0.86527 | 14713.92 |
| GOMF_CYC | 11 | -0.14935 | -0.0036 | 0.1352 | 0.86534 | 14715.11 |
| GOBP_POS | 13 | -0.17596 | -0.00462 | 0.15918 | 0.8655 | 14717.83 |

| | | | | | | |
|---|---|---|---|---|---|---|
| GOBP_REG | 40 | -0.08505 | -0.00391 | 0.076911 | 0.86559 | 14719.36 |
| GOBP_POS | 67 | -0.07136 | -0.00424 | 0.064519 | 0.86563 | 14720.04 |
| GOMF_LYS | 4 | -0.20298 | -0.00295 | 0.1835 | 0.86566 | 14720.55 |
| GOMF_PH( | 12 | -0.14117 | -0.00356 | 0.12761 | 0.86568 | 14720.89 |
| REACTOME | 26 | -0.11117 | -0.00412 | 0.10049 | 0.86569 | 14721.06 |
| GOBP_PRC | 12 | -0.14407 | -0.00363 | 0.13017 | 0.8658 | 14722.93 |
| GOBP_MU: | 84 | -0.06506 | -0.00433 | 0.058782 | 0.86581 | 14723.1 |
| GOBP_STR | 27 | -0.10582 | -0.004 | 0.095606 | 0.86581 | 14723.1 |
| GOBP_GU/ | 7 | -0.24679 | -0.00475 | 0.22284 | 0.86594 | 14725.31 |
| GOBP_POS | 5 | -0.28679 | -0.00467 | 0.25897 | 0.86594 | 14725.31 |
| BIOCARTA_ | 14 | -0.12811 | -0.00349 | 0.11567 | 0.86595 | 14725.48 |
| GOBP_REG | 37 | -0.08717 | -0.00386 | 0.078688 | 0.86602 | 14726.67 |
| GOMF_OXI | 27 | -0.09764 | -0.00369 | 0.088126 | 0.86605 | 14727.18 |
| BHAT_ESR1 | 203 | -0.04088 | -0.00422 | 0.036893 | 0.86609 | 14727.86 |
| GOMF_S_/ | 28 | -0.09738 | -0.00375 | 0.087872 | 0.8661 | 14728.03 |
| GOBP_POS | 79 | -0.06125 | -0.00395 | 0.055234 | 0.86625 | 14730.58 |
| REACTOME | 7 | -0.20393 | -0.00393 | 0.18389 | 0.86627 | 14730.92 |
| GOBP_CAL | 49 | -0.09056 | -0.00461 | 0.081635 | 0.86636 | 14732.45 |
| GOBP_SALI | 13 | -0.17826 | -0.00468 | 0.16066 | 0.86639 | 14732.96 |
| CHENG_TA | 6 | -0.2279 | -0.00406 | 0.2054 | 0.8664 | 14733.13 |
| GOBP_DER | 8 | -0.16346 | -0.00336 | 0.14722 | 0.86657 | 14736.02 |
| GOBP_REG | 6 | -0.22065 | -0.00393 | 0.1986 | 0.86671 | 14738.4 |
| REACTOME | 142 | -0.04472 | -0.00386 | 0.040249 | 0.86671 | 14738.4 |
| GOBP_RES | 11 | -0.19497 | -0.0047 | 0.17547 | 0.86673 | 14738.74 |
| GOBP_REG | 198 | -0.04126 | -0.0042 | 0.037127 | 0.86676 | 14739.25 |
| GOBP_REG | 6 | -0.27871 | -0.00497 | 0.25079 | 0.86678 | 14739.59 |
| GOCC_CLA | 23 | -0.1018 | -0.00355 | 0.091572 | 0.86686 | 14740.95 |
| PID_HDAC_ | 24 | -0.11632 | -0.00414 | 0.10461 | 0.86691 | 14741.8 |
| GOBP_ACE | 8 | -0.2161 | -0.00445 | 0.19401 | 0.86732 | 14748.78 |
| GOBP_REG | 11 | -0.18689 | -0.00451 | 0.16778 | 0.86733 | 14748.95 |
| GOBP_ANT | 109 | -0.05827 | -0.00442 | 0.052309 | 0.86735 | 14749.29 |
| WP_VITAM | 9 | -0.17462 | -0.00381 | 0.15673 | 0.86737 | 14749.63 |
| BIOCARTA_ | 13 | -0.15541 | -0.00408 | 0.13929 | 0.86774 | 14755.92 |
| GOBP_TYP | 28 | -0.10224 | -0.00393 | 0.091625 | 0.86776 | 14756.26 |
| WP_2Q37_ | 131 | -0.05766 | -0.00479 | 0.051658 | 0.86781 | 14757.11 |
| GOBP_REG | 11 | -0.16642 | -0.00402 | 0.14908 | 0.86785 | 14757.79 |
| GOBP_LIG/ | 30 | -0.10967 | -0.00437 | 0.098123 | 0.86813 | 14762.55 |
| GOMF_PH( | 8 | -0.21195 | -0.00436 | 0.18949 | 0.86833 | 14765.95 |
| GOBP_REG | 6 | -0.23687 | -0.00422 | 0.21174 | 0.86835 | 14766.29 |
| GOBP_VES | 7 | -0.28771 | -0.00554 | 0.25719 | 0.86835 | 14766.29 |
| GOBP_CYT | 7 | -0.21919 | -0.00422 | 0.19592 | 0.86837 | 14766.63 |
| GOBP_NEG | 12 | -0.18502 | -0.00466 | 0.16535 | 0.86842 | 14767.48 |
| REACTOME | 62 | -0.07432 | -0.00425 | 0.066412 | 0.86844 | 14767.82 |
| GOBP_POS | 42 | -0.09052 | -0.00427 | 0.080857 | 0.86854 | 14769.52 |
| GOMF_GU | 7 | -0.22136 | -0.00426 | 0.19751 | 0.8688 | 14773.94 |

| | | | | | | |
|---|---|---|---|---|---|---|
| GOBP_CELI | 275 | -0.03561 | -0.00427 | 0.031775 | 0.86881 | 14774.11 |
| GOMF_APC | 5 | -0.31823 | -0.00518 | 0.28387 | 0.86886 | 14774.96 |
| MCCLUNG_ | 71 | -0.06798 | -0.00416 | 0.060612 | 0.86895 | 14776.49 |
| GOBP_HEP | 32 | -0.10114 | -0.00416 | 0.09013 | 0.86909 | 14778.88 |
| GOBP_GAN | 13 | -0.14909 | -0.00391 | 0.1328 | 0.8692 | 14780.75 |
| GUTIERREZ | 9 | -0.21519 | -0.0047 | 0.1916 | 0.8693 | 14782.45 |
| RICKMAN_ | 228 | -0.0349 | -0.00381 | 0.031065 | 0.86936 | 14783.47 |
| GOBP_POS | 6 | -0.24634 | -0.00439 | 0.21923 | 0.86941 | 14784.32 |
| GOMF_N6_ | 10 | -0.18177 | -0.00418 | 0.16168 | 0.86955 | 14786.7 |
| GOMF_CUI | 8 | -0.188 | -0.00387 | 0.16713 | 0.86966 | 14788.57 |
| GOBP_L_G | 16 | -0.15108 | -0.0044 | 0.13428 | 0.86972 | 14789.59 |
| GOBP_NEG | 57 | -0.07471 | -0.0041 | 0.06639 | 0.86976 | 14790.27 |
| WP_SARSC | 8 | -0.1939 | -0.00399 | 0.17228 | 0.86981 | 14791.12 |
| REACTOME | 20 | -0.12543 | -0.00408 | 0.11142 | 0.86986 | 14791.97 |
| SAFFORD_ | 83 | -0.06261 | -0.00414 | 0.055611 | 0.86988 | 14792.31 |
| REACTOME | 302 | -0.03548 | -0.00445 | 0.031489 | 0.87004 | 14795.03 |
| GOBP_DEN | 9 | -0.19473 | -0.00425 | 0.17284 | 0.87005 | 14795.2 |
| GOBP_NEG | 16 | -0.14186 | -0.00413 | 0.12588 | 0.87011 | 14796.22 |
| GOBP_NEG | 41 | -0.09831 | -0.00458 | 0.087213 | 0.87016 | 14797.07 |
| PID_HES_H | 46 | -0.08576 | -0.00423 | 0.076075 | 0.87018 | 14797.41 |
| GOMF_MH | 5 | -0.28784 | -0.00468 | 0.25522 | 0.87029 | 14799.28 |
| GOBP_CELI | 294 | -0.03308 | -0.0041 | 0.029326 | 0.8703 | 14799.45 |
| GOBP_PHO | 61 | -0.07748 | -0.0044 | 0.068696 | 0.8703 | 14799.45 |
| GOBP_REG | 95 | -0.05688 | -0.00402 | 0.0504 | 0.87045 | 14802 |
| GOBP_REG | 74 | -0.06912 | -0.00432 | 0.061242 | 0.87046 | 14802.17 |
| ZHENG_FO | 6 | -0.23249 | -0.00414 | 0.20584 | 0.87064 | 14805.23 |
| GOBP_SMC | 4 | -0.32371 | -0.00471 | 0.28648 | 0.87074 | 14806.93 |
| BRACHAT_ | 24 | -0.10449 | -0.00372 | 0.092469 | 0.87076 | 14807.27 |
| GOBP_FOR | 5 | -0.27939 | -0.00455 | 0.24722 | 0.87078 | 14807.61 |
| GOBP_REG | 17 | -0.1356 | -0.00407 | 0.11998 | 0.87079 | 14807.78 |
| REACTOME | 66 | -0.06484 | -0.00383 | 0.057357 | 0.87084 | 14808.63 |
| GOMF_INT | 6 | -0.31511 | -0.00562 | 0.27869 | 0.87089 | 14809.48 |
| GOMF_MC | 552 | -0.02475 | -0.00417 | 0.021871 | 0.87106 | 14812.38 |
| GOBP_NEG | 17 | -0.15213 | -0.00456 | 0.13438 | 0.8712 | 14814.76 |
| GOBP_ACE | 29 | -0.10198 | -0.00399 | 0.090015 | 0.87136 | 14817.48 |
| GOCC_TRA | 19 | -0.1102 | -0.00349 | 0.097259 | 0.8714 | 14818.16 |
| GOBP_SOD | 14 | -0.17316 | -0.00471 | 0.15282 | 0.87141 | 14818.33 |
| GOBP_INTI | 2 | -0.49869 | -0.00513 | 0.4395 | 0.87173 | 14823.77 |
| GOMF_ALA | 13 | -0.16984 | -0.00446 | 0.14965 | 0.87178 | 14824.62 |
| GOBP_END | 52 | -0.08352 | -0.00438 | 0.073561 | 0.87187 | 14826.15 |
| GOBP_REG | 89 | -0.06384 | -0.00437 | 0.056223 | 0.87191 | 14826.83 |
| GOCC_NEU | 1253 | -0.01676 | -0.00417 | 0.014751 | 0.87199 | 14828.19 |
| GOBP_ACT | 8 | -0.18858 | -0.00388 | 0.16599 | 0.87203 | 14828.87 |
| KEGG_LYSC | 114 | -0.04793 | -0.00371 | 0.042171 | 0.87213 | 14830.57 |
| GOBP_PTE | 12 | -0.15525 | -0.00391 | 0.13656 | 0.8722 | 14831.76 |

| | | | | | | |
|---|---|---|---|---|---|---|
| GOMF_POT | 6 | -0.22592 | -0.00403 | 0.19861 | 0.87233 | 14833.97 |
| GOBP_GLY | 228 | -0.03698 | -0.00404 | 0.032502 | 0.87239 | 14834.99 |
| GOBP_BAR | 25 | -0.12576 | -0.00457 | 0.11048 | 0.8725 | 14836.86 |
| KEGG_RETI | 58 | -0.09046 | -0.00501 | 0.079428 | 0.87262 | 14838.9 |
| WP_NRF2_ | 134 | -0.05327 | -0.00447 | 0.046747 | 0.87275 | 14841.11 |
| GOBP_POS | 7 | -0.24423 | -0.0047 | 0.21431 | 0.87277 | 14841.45 |
| GOBP_REG | 38 | -0.09315 | -0.00418 | 0.081684 | 0.87292 | 14844 |
| GOMF_L_H | 3 | -0.4051 | -0.00511 | 0.35515 | 0.87298 | 14845.02 |
| GOBP_RES | 14 | -0.13425 | -0.00365 | 0.11768 | 0.87301 | 14845.54 |
| REACTOME | 13 | -0.14244 | -0.00374 | 0.12486 | 0.87301 | 14845.54 |
| REACTOME | 5 | -0.30233 | -0.00492 | 0.26478 | 0.87322 | 14849.11 |
| GOMF_ATF | 91 | -0.05988 | -0.00415 | 0.052442 | 0.87324 | 14849.45 |
| GOCC_CCR | 6 | -0.20674 | -0.00369 | 0.18104 | 0.87325 | 14849.62 |
| GOBP_REG | 179 | -0.04105 | -0.00398 | 0.035932 | 0.87335 | 14851.32 |
| GOMF_IMI | 5 | -0.22682 | -0.00369 | 0.19853 | 0.87336 | 14851.49 |
| GOCC_STEI | 5 | -0.22224 | -0.00362 | 0.19448 | 0.87342 | 14852.51 |
| GOBP_INSI | 13 | -0.18615 | -0.00488 | 0.1629 | 0.87343 | 14852.68 |
| KRISHNAN_ | 8 | -0.21245 | -0.00437 | 0.18576 | 0.87361 | 14855.74 |
| GOBP_POS | 308 | -0.03183 | -0.00403 | 0.027828 | 0.87362 | 14855.91 |
| RAMPON_I | 10 | -0.17887 | -0.00412 | 0.15639 | 0.87364 | 14856.25 |
| LUCAS_HN | 6 | -0.29003 | -0.00517 | 0.25353 | 0.87368 | 14856.93 |
| GOCC_GLU | 385 | -0.02982 | -0.00422 | 0.026066 | 0.8737 | 14857.27 |
| GOBP_POS | 19 | -0.1351 | -0.00428 | 0.11808 | 0.87371 | 14857.44 |
| GOCC_LAM | 5 | -0.23431 | -0.00381 | 0.20477 | 0.87373 | 14857.78 |
| LU_TUMOI | 20 | -0.1492 | -0.00485 | 0.13037 | 0.87376 | 14858.29 |
| GOBP_NAD | 40 | -0.07851 | -0.00361 | 0.06853 | 0.87402 | 14862.71 |
| REACTOME | 13 | -0.15887 | -0.00417 | 0.13868 | 0.87402 | 14862.71 |
| GOMF_GPI | 7 | -0.24801 | -0.00477 | 0.21645 | 0.87405 | 14863.22 |
| GOBP_CELI | 12 | -0.15919 | -0.00401 | 0.13891 | 0.87408 | 14863.73 |
| GOCC_MKS | 13 | -0.14326 | -0.00376 | 0.12501 | 0.87409 | 14863.9 |
| GOBP_COR | 47 | -0.07949 | -0.00396 | 0.069348 | 0.87414 | 14864.75 |
| GOBP_REG | 5 | -0.20685 | -0.00337 | 0.18044 | 0.87417 | 14865.26 |
| NOURUZI_ | 125 | -0.05583 | -0.00453 | 0.048688 | 0.87424 | 14866.45 |
| GOBP_GAM | 6 | -0.27651 | -0.00493 | 0.24082 | 0.87455 | 14871.72 |
| KEGG_BUT | 33 | -0.10334 | -0.00432 | 0.090003 | 0.87455 | 14871.72 |
| BIOCARTA_ | 11 | -0.18395 | -0.00444 | 0.16018 | 0.87459 | 14872.4 |
| RASHI_RES | 66 | -0.06897 | -0.00407 | 0.060027 | 0.87469 | 14874.1 |
| GOBP_POS | 59 | -0.07944 | -0.00443 | 0.069135 | 0.87473 | 14874.78 |
| GOBP_ISOI | 5 | -0.20737 | -0.00337 | 0.18039 | 0.87483 | 14876.48 |
| GOBP_CELI | 9 | -0.19422 | -0.00424 | 0.16882 | 0.87502 | 14879.72 |
| REACTOME | 126 | -0.06797 | -0.00553 | 0.059078 | 0.87504 | 14880.06 |
| GOBP_G2_ | 5 | -0.21278 | -0.00346 | 0.18475 | 0.87527 | 14883.97 |
| REACTOME | 10 | -0.17243 | -0.00397 | 0.1497 | 0.87528 | 14884.14 |
| BIOCARTA_ | 17 | -0.13916 | -0.00417 | 0.1208 | 0.87534 | 14885.16 |
| REACTOME | 9 | -0.20304 | -0.00443 | 0.17616 | 0.87545 | 14887.03 |

| | | | | | | |
|---|---|---|---|---|---|---|
| GOMF_ME | 411 | -0.02969 | -0.00433 | 0.025746 | 0.87561 | 14889.75 |
| GOCC_EXT | 271 | -0.03597 | -0.00428 | 0.031176 | 0.87569 | 14891.11 |
| GOMF_PH( | 43 | -0.09323 | -0.00444 | 0.080754 | 0.87585 | 14893.83 |
| SA_G2_AN | 8 | -0.15446 | -0.00318 | 0.13373 | 0.87594 | 14895.36 |
| BLANCO_M | 447 | -0.02812 | -0.00428 | 0.024348 | 0.87596 | 14895.7 |
| OUILLETTE_ | 50 | -0.08176 | -0.0042 | 0.070761 | 0.87603 | 14896.89 |
| GOBP_REG | 8 | -0.24139 | -0.00497 | 0.20891 | 0.87605 | 14897.23 |
| GOBP_SYN | 6 | -0.28764 | -0.00513 | 0.24889 | 0.87609 | 14897.91 |
| GOMF_PAS | 453 | -0.02908 | -0.00445 | 0.025164 | 0.87609 | 14897.91 |
| WP_GANG | 12 | -0.15099 | -0.00381 | 0.13064 | 0.87611 | 14898.25 |
| GOBP_G_P | 10 | -0.22736 | -0.00523 | 0.1967 | 0.87613 | 14898.59 |
| GOMF_G_I | 10 | -0.22736 | -0.00523 | 0.1967 | 0.87613 | 14898.59 |
| REACTOME | 27 | -0.09473 | -0.00358 | 0.08195 | 0.87613 | 14898.59 |
| ROVERSI_G | 45 | -0.10667 | -0.0052 | 0.092175 | 0.8764 | 14903.18 |
| GOBP_NAT | 11 | -0.19083 | -0.00461 | 0.16481 | 0.87653 | 14905.39 |
| GOBP_COF | 5 | -0.20754 | -0.00338 | 0.17922 | 0.87655 | 14905.73 |
| GOBP_REG | 3 | -0.35536 | -0.00448 | 0.30679 | 0.87663 | 14907.09 |
| GOBP_REG | 3 | -0.35536 | -0.00448 | 0.30679 | 0.87663 | 14907.09 |
| GOMF_CAT | 20 | -0.1401 | -0.00456 | 0.12092 | 0.87668 | 14907.94 |
| GOBP_RIB( | 5 | -0.28541 | -0.00464 | 0.24623 | 0.87679 | 14909.81 |
| WP_TYPE_ | 33 | -0.10371 | -0.00433 | 0.089411 | 0.87694 | 14912.36 |
| GOBP_GLU | 23 | -0.11468 | -0.004 | 0.098871 | 0.87696 | 14912.7 |
| GOBP_POS | 58 | -0.07304 | -0.00404 | 0.062968 | 0.87696 | 14912.7 |
| GOBP_GER | 14 | -0.15229 | -0.00415 | 0.13128 | 0.87697 | 14912.87 |
| GOMF_SAL | 529 | -0.02637 | -0.00435 | 0.022724 | 0.87702 | 14913.73 |
| GOCC_POS | 5 | -0.25069 | -0.00408 | 0.21595 | 0.87714 | 14915.77 |
| DE_YY1_TA | 16 | -0.16589 | -0.00483 | 0.14278 | 0.87735 | 14919.34 |
| GOBP_GLY | 195 | -0.04086 | -0.00413 | 0.035164 | 0.87736 | 14919.51 |
| NIKOLSKY_ | 14 | -0.25465 | -0.00693 | 0.21915 | 0.87737 | 14919.68 |
| VALK_AML | 31 | -0.10196 | -0.00413 | 0.087739 | 0.8774 | 14920.19 |
| GOBP_IND | 7 | -0.26389 | -0.00508 | 0.22703 | 0.87745 | 14921.04 |
| GOMF_N_A | 56 | -0.07438 | -0.00404 | 0.063986 | 0.87745 | 14921.04 |
| GOBP_POS | 61 | -0.08673 | -0.00492 | 0.074582 | 0.87754 | 14922.57 |
| REACTOME | 13 | -0.15835 | -0.00415 | 0.13614 | 0.8776 | 14923.59 |
| CHOI_ATL_ | 4 | -0.25804 | -0.00376 | 0.22185 | 0.87761 | 14923.76 |
| GOBP_DNA | 20 | -0.12765 | -0.00415 | 0.10974 | 0.87761 | 14923.76 |
| GOBP_REG | 15 | -0.14236 | -0.00401 | 0.12232 | 0.87774 | 14925.97 |
| GOBP_REG | 40 | -0.0859 | -0.00395 | 0.073771 | 0.87787 | 14928.18 |
| GOCC_RSC_ | 15 | -0.14535 | -0.0041 | 0.12476 | 0.878 | 14930.39 |
| GOBP_CHC | 18 | -0.15198 | -0.00469 | 0.13037 | 0.87814 | 14932.77 |
| GOBP_POS | 110 | -0.05809 | -0.00442 | 0.049814 | 0.87821 | 14933.96 |
| GOBP_CELI | 11 | -0.20996 | -0.00507 | 0.17981 | 0.87852 | 14939.23 |
| REACTOME | 6 | -0.1909 | -0.0034 | 0.16345 | 0.87858 | 14940.25 |
| REACTOME | 17 | -0.14977 | -0.00449 | 0.12818 | 0.87869 | 14942.12 |
| STEINER_E | 14 | -0.15758 | -0.00429 | 0.13484 | 0.87872 | 14942.63 |

| | | | | | | |
|---|---|---|---|---|---|---|
| REACTOME | 11 | -0.19177 | -0.00463 | 0.16407 | 0.87876 | 14943.31 |
| GOCC_MEI | 90 | -0.05516 | -0.0038 | 0.04715 | 0.87895 | 14946.54 |
| REACTOME | 5 | -0.25953 | -0.00422 | 0.22185 | 0.87896 | 14946.71 |
| GOBP_POS | 10 | -0.22739 | -0.00523 | 0.19435 | 0.87898 | 14947.05 |
| LI_WILMS_ | 5 | -0.28553 | -0.00465 | 0.24402 | 0.87901 | 14947.57 |
| GOBP_REG | 3 | -0.35876 | -0.00452 | 0.30649 | 0.8791 | 14949.1 |
| GOCC_CELI | 39 | -0.08854 | -0.00402 | 0.075614 | 0.87919 | 14950.63 |
| GOBP_POS | 71 | -0.07083 | -0.00434 | 0.060436 | 0.87939 | 14954.03 |
| GOBP_AUT | 104 | -0.0543 | -0.00402 | 0.046303 | 0.87953 | 14956.41 |
| WP_MODU | 58 | -0.07727 | -0.00428 | 0.065882 | 0.87956 | 14956.92 |
| GOBP_THC | 5 | -0.37188 | -0.00605 | 0.31703 | 0.8796 | 14957.6 |
| GOBP_PRC | 350 | -0.0317 | -0.00428 | 0.027017 | 0.87969 | 14959.13 |
| BOGNI_TRI | 24 | -0.11683 | -0.00416 | 0.099515 | 0.87979 | 14960.83 |
| GOBP_BEH | 13 | -0.15707 | -0.00412 | 0.13375 | 0.87987 | 14962.19 |
| GOBP_ADE | 9 | -0.19453 | -0.00425 | 0.16563 | 0.87989 | 14962.53 |
| WP_PROTE | 18 | -0.13026 | -0.00402 | 0.11089 | 0.87993 | 14963.21 |
| GOBP_GRA | 116 | -0.06129 | -0.00479 | 0.052116 | 0.88022 | 14968.14 |
| GOBP_CELI | 203 | -0.0438 | -0.00452 | 0.037241 | 0.88023 | 14968.31 |
| XU_CREBBI | 36 | -0.0934 | -0.00407 | 0.079412 | 0.88023 | 14968.31 |
| GOMF_PH( | 4 | -0.30734 | -0.00447 | 0.26113 | 0.88039 | 14971.03 |
| REACTOME | 108 | -0.05525 | -0.00417 | 0.046942 | 0.88041 | 14971.37 |
| GOBP_NEG | 6 | -0.24031 | -0.00428 | 0.20413 | 0.88044 | 14971.88 |
| SCHAEFFEF | 10 | -0.16157 | -0.00372 | 0.13716 | 0.88058 | 14974.26 |
| GOBP_PHC | 43 | -0.08936 | -0.00426 | 0.075859 | 0.88059 | 14974.43 |
| GOBP_TIGI | 75 | -0.07135 | -0.00449 | 0.06056 | 0.88063 | 14975.11 |
| WP_TYPE_ | 32 | -0.10404 | -0.00428 | 0.088306 | 0.88064 | 14975.28 |
| REACTOME | 25 | -0.11955 | -0.00435 | 0.10132 | 0.88098 | 14981.06 |
| GOBP_POS | 18 | -0.12747 | -0.00393 | 0.10797 | 0.88111 | 14983.28 |
| KEGG_GLY( | 25 | -0.11989 | -0.00436 | 0.10155 | 0.88111 | 14983.28 |
| GOMF_GEI | 51 | -0.0797 | -0.00414 | 0.067468 | 0.88124 | 14985.49 |
| GOMF_PAI | 14 | -0.14842 | -0.00404 | 0.12563 | 0.88126 | 14985.83 |
| GOBP_RES | 44 | -0.09046 | -0.00436 | 0.076538 | 0.88136 | 14987.53 |
| GOBP_INTI | 5 | -0.23342 | -0.0038 | 0.19743 | 0.88144 | 14988.89 |
| GOBP_CELI | 723 | -0.02242 | -0.0043 | 0.018965 | 0.88146 | 14989.23 |
| DAVIES_MI | 12 | -0.15196 | -0.00383 | 0.12851 | 0.88148 | 14989.57 |
| GOBP_NEG | 7 | -0.27254 | -0.00525 | 0.23047 | 0.8815 | 14989.91 |
| GOMF_NEI | 89 | -0.07052 | -0.00483 | 0.05961 | 0.8816 | 14991.61 |
| ZHANG_IN | 23 | -0.13842 | -0.00483 | 0.11693 | 0.88175 | 14994.16 |
| GOMF_CY( | 46 | -0.08245 | -0.00406 | 0.069624 | 0.88182 | 14995.35 |
| GALI_TP53 | 6 | -0.20326 | -0.00362 | 0.17161 | 0.88187 | 14996.2 |
| GOBP_POS | 899 | -0.02055 | -0.00438 | 0.017341 | 0.88201 | 14998.58 |
| REACTOME | 81 | -0.06307 | -0.00412 | 0.053198 | 0.88208 | 14999.77 |
| GOBP_LUN | 9 | -0.17752 | -0.00388 | 0.14973 | 0.88211 | 15000.28 |
| GOMF_PH( | 36 | -0.10365 | -0.00452 | 0.087395 | 0.88219 | 15001.64 |
| LINDSTEDT | 56 | -0.08164 | -0.00444 | 0.068818 | 0.88224 | 15002.49 |

| | | | | | | |
|---|---|---|---|---|---|---|
| GOBP_POS | 64 | -0.0754 | -0.00438 | 0.063558 | 0.88225 | 15002.66 |
| GOBP_MUS | 99 | -0.06414 | -0.00463 | 0.054053 | 0.88229 | 15003.34 |
| GOMF_ENI | 381 | -0.03271 | -0.0046 | 0.02756 | 0.88237 | 15004.7 |
| GOBP_INSU | 121 | -0.05257 | -0.0042 | 0.044284 | 0.88239 | 15005.04 |
| GOCC_ARY | 8 | -0.19183 | -0.00395 | 0.16157 | 0.88243 | 15005.72 |
| GOMF_ARY | 9 | -0.22186 | -0.00484 | 0.18685 | 0.88244 | 15005.89 |
| GOBP_MEI | 14 | -0.15703 | -0.00427 | 0.13222 | 0.8825 | 15006.91 |
| GOCC_H4_ | 25 | -0.10828 | -0.00394 | 0.091029 | 0.88286 | 15013.03 |
| GOBP_PRC | 7 | -0.19905 | -0.00383 | 0.16726 | 0.88298 | 15015.07 |
| KIM_LIVER_ | 22 | -0.12825 | -0.00438 | 0.10776 | 0.88299 | 15015.24 |
| REACTOME | 5 | -0.26952 | -0.00439 | 0.22641 | 0.88304 | 15016.1 |
| GOMF_INC | 621 | -0.02482 | -0.00443 | 0.020843 | 0.88315 | 15017.97 |
| GOBP_HYA | 34 | -0.10287 | -0.00436 | 0.086367 | 0.88317 | 15018.31 |
| GOBP_ANT | 84 | -0.0621 | -0.00413 | 0.052124 | 0.88323 | 15019.33 |
| GOBP_NEG | 13 | -0.14748 | -0.00387 | 0.12373 | 0.88335 | 15021.37 |
| REACTOME | 7 | -0.21736 | -0.00418 | 0.18224 | 0.8835 | 15023.92 |
| FULCHER_I | 555 | -0.02645 | -0.00447 | 0.022173 | 0.88351 | 15024.09 |
| CHANG_PC | 14 | -0.16862 | -0.00459 | 0.14133 | 0.88357 | 15025.11 |
| GOBP_SECI | 62 | -0.07719 | -0.00442 | 0.064696 | 0.88357 | 15025.11 |
| GOMF_MR | 3 | -0.26798 | -0.00338 | 0.22458 | 0.8836 | 15025.62 |
| GOCC_LSM | 4 | -0.29387 | -0.00428 | 0.24628 | 0.88361 | 15025.79 |
| GOBP_PEP | 23 | -0.12914 | -0.0045 | 0.10822 | 0.88363 | 15026.13 |
| GOCC_PRC | 24 | -0.11537 | -0.00411 | 0.096656 | 0.88367 | 15026.81 |
| PID_IL8_CX | 33 | -0.10221 | -0.00427 | 0.085638 | 0.88367 | 15026.81 |
| GOBP_FC_ | 52 | -0.08407 | -0.00441 | 0.070417 | 0.88374 | 15028 |
| GOMF_OXY | 37 | -0.10547 | -0.00466 | 0.088332 | 0.88376 | 15028.34 |
| REACTOME | 40 | -0.08528 | -0.00392 | 0.07142 | 0.88377 | 15028.51 |
| GOBP_POS | 176 | -0.04398 | -0.00423 | 0.036823 | 0.8838 | 15029.02 |
| GOBP_CELI | 23 | -0.13513 | -0.00471 | 0.11308 | 0.88393 | 15031.23 |
| GOBP_VAC | 31 | -0.10676 | -0.00432 | 0.089279 | 0.88411 | 15034.29 |
| REACTOME | 19 | -0.14622 | -0.00464 | 0.12225 | 0.88416 | 15035.14 |
| GOBP_REG | 6 | -0.25202 | -0.00449 | 0.21069 | 0.88417 | 15035.31 |
| REACTOME | 11 | -0.22534 | -0.00544 | 0.18839 | 0.88417 | 15035.31 |
| REACTOME | 12 | -0.17711 | -0.00446 | 0.14801 | 0.88426 | 15036.84 |
| GOBP_L_A | 10 | -0.19188 | -0.00442 | 0.16033 | 0.8843 | 15037.52 |
| GOMF_OXI | 5 | -0.3397 | -0.00553 | 0.28353 | 0.88456 | 15041.94 |
| GOBP_TYR | 77 | -0.0693 | -0.00442 | 0.057729 | 0.88501 | 15049.6 |
| GOBP_REG | 91 | -0.06591 | -0.00457 | 0.054877 | 0.88513 | 15051.64 |
| GOMF_L_L | 7 | -0.20169 | -0.00388 | 0.16784 | 0.88525 | 15053.68 |
| REACTOME | 142 | -0.04893 | -0.00423 | 0.040706 | 0.88534 | 15055.21 |
| GOCC_RNA | 16 | -0.16957 | -0.00493 | 0.141 | 0.88543 | 15056.74 |
| GOBP_SEN | 7 | -0.20568 | -0.00396 | 0.17101 | 0.88545 | 15057.08 |
| GOMF_HIS | 21 | -0.11556 | -0.00385 | 0.096028 | 0.88558 | 15059.29 |
| GOBP_SER | 12 | -0.16685 | -0.00421 | 0.13862 | 0.88562 | 15059.97 |
| GOBP_MIT | 47 | -0.08835 | -0.0044 | 0.073403 | 0.88563 | 15060.14 |

| | | | | | | |
|---|---|---|---|---|---|---|
| GOCC_SPE( | 86 | -0.06651 | -0.00448 | 0.055206 | 0.88584 | 15063.71 |
| GOBP_REG | 18 | -0.1631 | -0.00503 | 0.13531 | 0.88597 | 15065.92 |
| BIOCARTA_ | 9 | -0.16394 | -0.00358 | 0.13595 | 0.88605 | 15067.28 |
| PID_SYNDE | 16 | -0.14931 | -0.00434 | 0.12377 | 0.88614 | 15068.81 |
| GOBP_MIC | 398 | -0.02925 | -0.0042 | 0.024244 | 0.8862 | 15069.83 |
| GOMF_MY | 11 | -0.15516 | -0.00374 | 0.12854 | 0.88629 | 15071.36 |
| GOBP_REG | 9 | -0.27386 | -0.00598 | 0.22685 | 0.88631 | 15071.7 |
| GOBP_REG | 30 | -0.12146 | -0.00484 | 0.10059 | 0.88637 | 15072.72 |
| REACTOME | 19 | -0.11776 | -0.00373 | 0.097503 | 0.88641 | 15073.4 |
| REACTOME | 16 | -0.13667 | -0.00398 | 0.1131 | 0.88655 | 15075.78 |
| GOBP_REG | 142 | -0.05213 | -0.0045 | 0.043131 | 0.88658 | 15076.29 |
| GOBP_OLI( | 7 | -0.25292 | -0.00487 | 0.20917 | 0.88669 | 15078.16 |
| GOBP_CELI | 285 | -0.03838 | -0.00468 | 0.031733 | 0.88676 | 15079.35 |
| GOBP_REG | 27 | -0.12248 | -0.00463 | 0.10124 | 0.8868 | 15080.03 |
| GOBP_INTI | 5 | -0.25624 | -0.00417 | 0.2117 | 0.88693 | 15082.24 |
| WP_THYM | 45 | -0.09108 | -0.00444 | 0.075237 | 0.88697 | 15082.92 |
| GOBP_POS | 63 | -0.07722 | -0.00445 | 0.063778 | 0.88699 | 15083.26 |
| THILLAINAI | 41 | -0.09303 | -0.00433 | 0.076829 | 0.88702 | 15083.78 |
| GOBP_NU( | 52 | -0.08257 | -0.00433 | 0.068172 | 0.88709 | 15084.97 |
| GOBP_REG | 22 | -0.13672 | -0.00466 | 0.11285 | 0.88716 | 15086.16 |
| YANG_BRE | 22 | -0.13614 | -0.00464 | 0.11224 | 0.88741 | 15090.41 |
| GOBP_MIT | 19 | -0.12886 | -0.00409 | 0.10618 | 0.88753 | 15092.45 |
| GOCC_VES | 58 | -0.07243 | -0.00401 | 0.059666 | 0.8876 | 15093.64 |
| GOBP_REG | 3 | -0.35449 | -0.00447 | 0.29201 | 0.88762 | 15093.98 |
| REACTOME | 8 | -0.19495 | -0.00401 | 0.16054 | 0.88768 | 15095 |
| GOBP_GAN | 8 | -0.25262 | -0.0052 | 0.208 | 0.88771 | 15095.51 |
| GOBP_CELI | 556 | -0.0273 | -0.00462 | 0.022476 | 0.88777 | 15096.53 |
| DACOSTA_ | 831 | -0.02269 | -0.00466 | 0.018673 | 0.88786 | 15098.06 |
| GOBP_POS | 28 | -0.10164 | -0.00391 | 0.083622 | 0.8879 | 15098.74 |
| GOBP_PIGI | 34 | -0.11466 | -0.00486 | 0.094293 | 0.88799 | 15100.27 |
| GOBP_RES | 29 | -0.10748 | -0.00421 | 0.088389 | 0.888 | 15100.44 |
| GOBP_CHI | 6 | -0.25517 | -0.00455 | 0.20977 | 0.88808 | 15101.8 |
| GOMF_CHI | 6 | -0.25517 | -0.00455 | 0.20977 | 0.88808 | 15101.8 |
| GOBP_NEU | 14 | -0.18319 | -0.00499 | 0.15053 | 0.88819 | 15103.67 |
| REACTOME | 3 | -0.41124 | -0.00518 | 0.33788 | 0.88821 | 15104.01 |
| GOBP_MEI | 10 | -0.16177 | -0.00372 | 0.13285 | 0.88831 | 15105.71 |
| REACTOME | 9 | -0.19523 | -0.00426 | 0.16016 | 0.88857 | 15110.13 |
| PID_MTOR | 67 | -0.07234 | -0.0043 | 0.059306 | 0.88871 | 15112.51 |
| GOCC_MIT | 7 | -0.25026 | -0.00482 | 0.20508 | 0.88882 | 15114.38 |
| GOBP_CAL | 22 | -0.13853 | -0.00473 | 0.11346 | 0.88894 | 15116.42 |
| GOMF_PH( | 11 | -0.19019 | -0.00459 | 0.15571 | 0.88903 | 15117.96 |
| COATES_M | 82 | -0.06469 | -0.00425 | 0.052931 | 0.88915 | 15120 |
| GOCC_NEU | 8 | -0.22911 | -0.00472 | 0.18733 | 0.88933 | 15123.06 |
| GOBP_PEP | 14 | -0.15698 | -0.00427 | 0.12833 | 0.88937 | 15123.74 |
| GOBP_POS | 7 | -0.22684 | -0.00437 | 0.18541 | 0.88941 | 15124.42 |

| | | | | | | |
|---|---|---|---|---|---|---|
| GOBP_POS | 5 | -0.26531 | -0.00432 | 0.21679 | 0.88948 | 15125.61 |
| GOBP_POS | 162 | -0.05052 | -0.00466 | 0.041233 | 0.88974 | 15130.03 |
| GOMF_PH( | 10 | -0.19895 | -0.00458 | 0.16233 | 0.88982 | 15131.39 |
| CHEMELLO | 4 | -0.27835 | -0.00405 | 0.22693 | 0.89 | 15134.45 |
| MEISSNER_ | 35 | -0.09918 | -0.00427 | 0.080858 | 0.89001 | 15134.62 |
| GOBP_POS | 15 | -0.13906 | -0.00392 | 0.1133 | 0.89016 | 15137.17 |
| WU_HBX_T | 14 | -0.17056 | -0.00464 | 0.1389 | 0.89025 | 15138.7 |
| GOBP_REG | 7 | -0.25885 | -0.00498 | 0.2108 | 0.89026 | 15138.87 |
| LY_AGING_ | 30 | -0.10182 | -0.00406 | 0.082894 | 0.89033 | 15140.06 |
| GOMF_DYI | 15 | -0.17003 | -0.00479 | 0.13841 | 0.89035 | 15140.4 |
| GOBP_POS | 139 | -0.05144 | -0.0044 | 0.041866 | 0.89037 | 15140.74 |
| GOBP_ACT | 10 | -0.18105 | -0.00417 | 0.14734 | 0.89041 | 15141.42 |
| GOMF_DYI | 23 | -0.12334 | -0.0043 | 0.10036 | 0.89045 | 15142.1 |
| GOBP_REG | 16 | -0.14501 | -0.00422 | 0.11799 | 0.89047 | 15142.44 |
| DELLA_RES | 18 | -0.14624 | -0.00451 | 0.11896 | 0.89053 | 15143.46 |
| GOBP_DES | 10 | -0.21262 | -0.00489 | 0.1729 | 0.89059 | 15144.48 |
| SALVADOR | 22 | -0.13325 | -0.00455 | 0.10831 | 0.8907 | 15146.35 |
| GOMF_FAI | 38 | -0.10431 | -0.00468 | 0.084745 | 0.8908 | 15148.05 |
| GOMF_PH( | 88 | -0.06627 | -0.00451 | 0.053823 | 0.89088 | 15149.41 |
| GOBP_COF | 5 | -0.19185 | -0.00312 | 0.1558 | 0.89089 | 15149.58 |
| GOBP_CRA | 18 | -0.16989 | -0.00524 | 0.13791 | 0.89099 | 15151.28 |
| STARK_HYI | 21 | -0.18088 | -0.00603 | 0.14672 | 0.89118 | 15154.52 |
| HILLION_H | 80 | -0.06613 | -0.0043 | 0.053607 | 0.89132 | 15156.9 |
| GOBP_MEI | 8 | -0.17762 | -0.00366 | 0.14397 | 0.89134 | 15157.24 |
| REACTOME | 20 | -0.12946 | -0.00421 | 0.1049 | 0.89141 | 15158.43 |
| GOBP_REG | 11 | -0.19169 | -0.00463 | 0.15531 | 0.89143 | 15158.77 |
| SASSON_RI | 84 | -0.0742 | -0.00494 | 0.060121 | 0.89143 | 15158.77 |
| GOBP_POS | 15 | -0.14693 | -0.00414 | 0.11903 | 0.89145 | 15159.11 |
| GOBP_POS | 13 | -0.1825 | -0.00479 | 0.14784 | 0.89146 | 15159.28 |
| KEGG_CHE | 173 | -0.04786 | -0.00456 | 0.038766 | 0.89151 | 15160.13 |
| REACTOME | 7 | -0.26094 | -0.00502 | 0.21134 | 0.89152 | 15160.3 |
| YANG_BRE | 26 | -0.1131 | -0.00419 | 0.091574 | 0.89159 | 15161.49 |
| GOBP_NEG | 42 | -0.09607 | -0.00453 | 0.077775 | 0.89161 | 15161.83 |
| GOBP_FLU | 32 | -0.11337 | -0.00466 | 0.091775 | 0.89163 | 15162.17 |
| GOMF_DYI | 10 | -0.18456 | -0.00425 | 0.14939 | 0.89164 | 15162.34 |
| GOBP_CELI | 135 | -0.05215 | -0.00439 | 0.042203 | 0.89169 | 15163.19 |
| GOMF_CYS | 5 | -0.28737 | -0.00468 | 0.23252 | 0.89175 | 15164.21 |
| GOBP_REG | 9 | -0.18975 | -0.00414 | 0.1534 | 0.89195 | 15167.61 |
| GOBP_EPIT | 31 | -0.11294 | -0.00457 | 0.091285 | 0.892 | 15168.46 |
| GOBP_NOT | 9 | -0.18587 | -0.00406 | 0.15008 | 0.89223 | 15172.37 |
| WP_EFFEC | 7 | -0.24115 | -0.00464 | 0.1947 | 0.89224 | 15172.54 |
| GOBP_NEG | 10 | -0.18542 | -0.00427 | 0.14968 | 0.89227 | 15173.05 |
| WP_METAI | 41 | -0.09939 | -0.00463 | 0.08021 | 0.89233 | 15174.07 |
| GOCC_SWI | 30 | -0.11093 | -0.00442 | 0.089502 | 0.8924 | 15175.26 |
| GOBP_INTI | 51 | -0.08973 | -0.00466 | 0.072394 | 0.89241 | 15175.43 |

| | | | | | | |
|---|---|---|---|---|---|---|
| GOBP_DEV | 44 | -0.10196 | -0.00492 | 0.082244 | 0.89244 | 15175.94 |
| POOLA_IN\ | 126 | -0.0549 | -0.00447 | 0.044233 | 0.8927 | 15180.36 |
| GOBP_POS | 27 | -0.11701 | -0.00442 | 0.094272 | 0.89273 | 15180.87 |
| WOOD_EB | 104 | -0.05919 | -0.00438 | 0.047656 | 0.89287 | 15183.25 |
| REACTOME | 13 | -0.16547 | -0.00434 | 0.13313 | 0.89304 | 15186.15 |
| REACTOME | 40 | -0.09618 | -0.00442 | 0.07737 | 0.89307 | 15186.66 |
| GOBP_MEI | 12 | -0.15902 | -0.00401 | 0.12787 | 0.89318 | 15188.53 |
| GOMF_N_I | 4 | -0.44462 | -0.00647 | 0.3574 | 0.89325 | 15189.72 |
| GOCC_HEN | 12 | -0.20257 | -0.00511 | 0.16283 | 0.89326 | 15189.89 |
| TURJANSKI | 4 | -0.29976 | -0.00436 | 0.2408 | 0.8934 | 15192.27 |
| GOBP_NEG | 8 | -0.2461 | -0.00507 | 0.19769 | 0.89341 | 15192.44 |
| GOBP_RES | 5 | -0.25216 | -0.0041 | 0.20255 | 0.89341 | 15192.44 |
| GOBP_POS | 5 | -0.31697 | -0.00516 | 0.25454 | 0.89347 | 15193.46 |
| FLORIO_HU | 38 | -0.09485 | -0.00425 | 0.076132 | 0.89358 | 15195.33 |
| GOBP_COF | 22 | -0.15251 | -0.0052 | 0.12241 | 0.89359 | 15195.5 |
| MARTIN_V | 47 | -0.08983 | -0.00448 | 0.072099 | 0.89359 | 15195.5 |
| GOBP_REG | 73 | -0.0761 | -0.00472 | 0.061057 | 0.89367 | 15196.86 |
| BARIS_THY | 23 | -0.13934 | -0.00486 | 0.1118 | 0.89368 | 15197.03 |
| LIU_CDX2_ | 8 | -0.24037 | -0.00495 | 0.1927 | 0.89386 | 15200.09 |
| MANN_RES | 19 | -0.13497 | -0.00428 | 0.10817 | 0.89392 | 15201.11 |
| GOBP_CHI/ | 5 | -0.2316 | -0.00377 | 0.1856 | 0.89395 | 15201.62 |
| MCCLUNG_ | 73 | -0.07447 | -0.00462 | 0.059658 | 0.89403 | 15202.98 |
| REACTOME | 8 | -0.25593 | -0.00527 | 0.205 | 0.89405 | 15203.32 |
| KEGG_GLY( | 74 | -0.07203 | -0.0045 | 0.057674 | 0.89416 | 15205.19 |
| REACTOME | 24 | -0.13876 | -0.00494 | 0.11109 | 0.89418 | 15205.53 |
| GOBP_BILE | 12 | -0.19318 | -0.00487 | 0.15459 | 0.89427 | 15207.06 |
| GOBP_CELI | 25 | -0.12979 | -0.00472 | 0.10383 | 0.89434 | 15208.25 |
| GOBP_RES | 61 | -0.08051 | -0.00457 | 0.064403 | 0.89436 | 15208.59 |
| GOMF_OXY | 14 | -0.2021 | -0.0055 | 0.16159 | 0.89447 | 15210.46 |
| ALCALAY_A | 171 | -0.04887 | -0.00463 | 0.039061 | 0.89453 | 15211.48 |
| GOBP_NEG | 5 | -0.31464 | -0.00512 | 0.25151 | 0.89453 | 15211.48 |
| WP_ID_SIG | 15 | -0.14989 | -0.00422 | 0.11979 | 0.89457 | 15212.16 |
| REACTOME | 9 | -0.19778 | -0.00432 | 0.15798 | 0.8947 | 15214.37 |
| GOBP_POS | 37 | -0.11313 | -0.005 | 0.090327 | 0.89479 | 15215.9 |
| GOMF_HIS | 10 | -0.17434 | -0.00401 | 0.13916 | 0.89486 | 15217.09 |
| GOBP_MIC | 881 | -0.0207 | -0.00437 | 0.016512 | 0.89497 | 15218.96 |
| WP_FLUOF | 6 | -0.301 | -0.00537 | 0.24012 | 0.89499 | 15219.3 |
| GOBP_AUT | 44 | -0.09981 | -0.00481 | 0.079589 | 0.89508 | 15220.84 |
| GOBP_GEN | 12 | -0.15775 | -0.00398 | 0.12571 | 0.89523 | 15223.39 |
| WP_GASTF | 11 | -0.18231 | -0.0044 | 0.14521 | 0.89534 | 15225.26 |
| REACTOME | 52 | -0.08939 | -0.00469 | 0.071138 | 0.89554 | 15228.66 |
| GOBP_REG | 183 | -0.04854 | -0.00476 | 0.038623 | 0.89557 | 15229.17 |
| GOCC_CLA | 31 | -0.10175 | -0.00412 | 0.080955 | 0.89559 | 15229.51 |
| ZHENG_IL2 | 54 | -0.08761 | -0.00468 | 0.069692 | 0.89564 | 15230.36 |
| GOBP_ISO( | 5 | -0.25674 | -0.00418 | 0.20417 | 0.8957 | 15231.38 |

| | | | | | | |
|---|---|---|---|---|---|---|
| GOBP_WYI | 5 | -0.26009 | -0.00423 | 0.20682 | 0.89572 | 15231.72 |
| GOMF_INT | 5 | -0.24095 | -0.00392 | 0.1915 | 0.89584 | 15233.76 |
| GOBP_POS | 16 | -0.1703 | -0.00496 | 0.13532 | 0.89589 | 15234.61 |
| WP_DUAL_ | 8 | -0.25043 | -0.00515 | 0.1989 | 0.89599 | 15236.31 |
| WP_CHROI | 40 | -0.11609 | -0.00534 | 0.092137 | 0.89615 | 15239.03 |
| GOMF_RN/ | 34 | -0.10885 | -0.00462 | 0.086386 | 0.89616 | 15239.2 |
| GOBP_ADA | 403 | -0.03346 | -0.00484 | 0.026553 | 0.8962 | 15239.88 |
| GOMF_P_T | 18 | -0.14715 | -0.00454 | 0.11668 | 0.89636 | 15242.6 |
| GOMF_PEF | 11 | -0.19315 | -0.00466 | 0.15314 | 0.89638 | 15242.94 |
| GOBP_POS | 24 | -0.13041 | -0.00465 | 0.10339 | 0.89639 | 15243.11 |
| GOBP_NEG | 4 | -0.35102 | -0.00511 | 0.27819 | 0.89648 | 15244.64 |
| GOMF_MH | 13 | -0.15741 | -0.00413 | 0.12469 | 0.89658 | 15246.34 |
| REACTOME | 47 | -0.09388 | -0.00468 | 0.074352 | 0.89662 | 15247.02 |
| GOBP_NEU | 12 | -0.15369 | -0.00387 | 0.12169 | 0.89668 | 15248.04 |
| GOBP_CAR | 148 | -0.05399 | -0.00476 | 0.042739 | 0.89673 | 15248.89 |
| GOBP_NET | 9 | -0.20055 | -0.00438 | 0.15875 | 0.89675 | 15249.23 |
| GOCC_PLA | 11 | -0.1838 | -0.00444 | 0.1455 | 0.89675 | 15249.23 |
| GOBP_POS | 9 | -0.20055 | -0.00438 | 0.15874 | 0.89678 | 15249.74 |
| GOCC_END | 5 | -0.29504 | -0.0048 | 0.23344 | 0.89686 | 15251.1 |
| EPPERT_PF | 132 | -0.05499 | -0.00458 | 0.043494 | 0.89691 | 15251.95 |
| GOBP_CHO | 8 | -0.25492 | -0.00525 | 0.20162 | 0.89695 | 15252.63 |
| HATADA_N | 359 | -0.03461 | -0.00473 | 0.027375 | 0.89695 | 15252.63 |
| GOMF_BIC | 6 | -0.20323 | -0.00362 | 0.16071 | 0.89698 | 15253.14 |
| GOBP_NEG | 19 | -0.16411 | -0.0052 | 0.12975 | 0.89702 | 15253.83 |
| WP_OVERV | 18 | -0.1473 | -0.00455 | 0.11645 | 0.89704 | 15254.17 |
| WP_RIBOF | 14 | -0.16214 | -0.00441 | 0.12816 | 0.89708 | 15254.85 |
| WP_REGUI | 140 | -0.05387 | -0.00462 | 0.04258 | 0.8971 | 15255.19 |
| ROY_WOU | 46 | -0.09345 | -0.00461 | 0.073793 | 0.89731 | 15258.76 |
| PID_LYSOP | 64 | -0.07884 | -0.00458 | 0.062251 | 0.89732 | 15258.93 |
| DELACROIX | 26 | -0.12903 | -0.00479 | 0.10186 | 0.89738 | 15259.95 |
| REACTOME | 52 | -0.0815 | -0.00427 | 0.064331 | 0.89738 | 15259.95 |
| GOCC_PER | 5 | -0.2983 | -0.00485 | 0.23533 | 0.89752 | 15262.33 |
| BIOCARTA_ | 18 | -0.15191 | -0.00469 | 0.1198 | 0.8976 | 15263.69 |
| SATO_SILEI | 39 | -0.10364 | -0.00471 | 0.081688 | 0.89771 | 15265.56 |
| WP_SREBF | 16 | -0.14331 | -0.00417 | 0.11296 | 0.89772 | 15265.73 |
| GOBP_RES | 6 | -0.24834 | -0.00443 | 0.19565 | 0.89783 | 15267.6 |
| GOMF_PRI | 146 | -0.04999 | -0.00438 | 0.039383 | 0.89783 | 15267.6 |
| GOBP_CELI | 571 | -0.02665 | -0.00456 | 0.020991 | 0.89789 | 15268.62 |
| GOBP_POS | 140 | -0.05467 | -0.00469 | 0.043047 | 0.89794 | 15269.47 |
| GOBP_REG | 5 | -0.24009 | -0.00391 | 0.18904 | 0.89796 | 15269.81 |
| LEE_LIVER_ | 59 | -0.08439 | -0.00471 | 0.066423 | 0.89804 | 15271.17 |
| SUZUKI_RE | 16 | -0.18205 | -0.0053 | 0.14324 | 0.89812 | 15272.53 |
| BOHN_PRII | 35 | -0.11641 | -0.00501 | 0.091582 | 0.89815 | 15273.04 |
| GOMF_HIS | 5 | -0.3412 | -0.00555 | 0.26838 | 0.89818 | 15273.55 |
| OUELLET_( | 63 | -0.07372 | -0.00425 | 0.057962 | 0.89828 | 15275.25 |

| | | | | | | |
|---|---|---|---|---|---|---|
| GOBP_DICA | 13 | -0.18491 | -0.00485 | 0.1452 | 0.89857 | 15280.18 |
| GOBP_MDA | 10 | -0.18989 | -0.00437 | 0.1491 | 0.89858 | 15280.35 |
| GOMF_PEF | 39 | -0.09779 | -0.00444 | 0.076774 | 0.8986 | 15280.69 |
| WP_ATM_S | 37 | -0.10724 | -0.00474 | 0.084187 | 0.89863 | 15281.2 |
| WEST_ADR | 19 | -0.13029 | -0.00413 | 0.10222 | 0.89877 | 15283.58 |
| ZWANG_EC | 198 | -0.0445 | -0.00453 | 0.034913 | 0.89878 | 15283.75 |
| GOMF_NAI | 28 | -0.12802 | -0.00493 | 0.10041 | 0.89883 | 15284.6 |
| GOCC_INTI | 197 | -0.06862 | -0.00697 | 0.053818 | 0.89886 | 15285.11 |
| GOBP_PYR | 24 | -0.13417 | -0.00478 | 0.10518 | 0.89896 | 15286.81 |
| GOBP_OLF | 6 | -0.2931 | -0.00522 | 0.22974 | 0.89898 | 15287.15 |
| GOBP_POS | 39 | -0.11237 | -0.0051 | 0.088074 | 0.89899 | 15287.32 |
| GOUYER_T | 9 | -0.23722 | -0.00518 | 0.1859 | 0.89902 | 15287.84 |
| GOBP_DOF | 37 | -0.1026 | -0.00454 | 0.080396 | 0.89905 | 15288.35 |
| GOBP_ETH | 18 | -0.17695 | -0.00546 | 0.13865 | 0.89906 | 15288.52 |
| GOBP_SPH | 157 | -0.05394 | -0.0049 | 0.04226 | 0.89909 | 15289.03 |
| HOFMANN | 20 | -0.14812 | -0.00482 | 0.116 | 0.89917 | 15290.39 |
| GAUSSMAI | 90 | -0.07095 | -0.00489 | 0.055528 | 0.8993 | 15292.6 |
| GOBP_POS | 7 | -0.18436 | -0.00355 | 0.14428 | 0.89934 | 15293.28 |
| GOBP_BLO | 8 | -0.24592 | -0.00506 | 0.19243 | 0.89936 | 15293.62 |
| KEGG_TAS | 50 | -0.10619 | -0.00546 | 0.083091 | 0.89936 | 15293.62 |
| GOBP_TER | 8 | -0.24019 | -0.00494 | 0.18791 | 0.8994 | 15294.3 |
| GOMF_DEU | 88 | -0.06239 | -0.00425 | 0.048792 | 0.89948 | 15295.66 |
| BILANGES_ | 35 | -0.09631 | -0.00414 | 0.075296 | 0.89956 | 15297.02 |
| SCHAEFFEF | 6 | -0.25233 | -0.0045 | 0.19724 | 0.89959 | 15297.53 |
| GOBP_REG | 47 | -0.09617 | -0.00479 | 0.075114 | 0.89978 | 15300.76 |
| WANG_ME | 34 | -0.1069 | -0.00453 | 0.083464 | 0.89985 | 15301.95 |
| REACTOME | 12 | -0.18122 | -0.00457 | 0.14148 | 0.89988 | 15302.46 |
| WEI_MYCN | 757 | -0.02409 | -0.00473 | 0.018802 | 0.89988 | 15302.46 |
| GOBP_MAI | 9 | -0.1766 | -0.00386 | 0.13783 | 0.89994 | 15303.48 |
| GOBP_N_G | 20 | -0.14269 | -0.00464 | 0.11135 | 0.89998 | 15304.16 |
| BIOCARTA_ | 8 | -0.26105 | -0.00537 | 0.2037 | 0.89999 | 15304.33 |
| HOEGERKC | 12 | -0.1848 | -0.00466 | 0.14416 | 0.90004 | 15305.18 |
| GOBP_POS | 14 | -0.1712 | -0.00466 | 0.1335 | 0.90013 | 15306.71 |
| GOBP_REG | 4 | -0.28409 | -0.00413 | 0.22149 | 0.90018 | 15307.56 |
| GOBP_PHC | 5 | -0.23608 | -0.00384 | 0.184 | 0.90025 | 15308.75 |
| FUNG_IL2_ | 8 | -0.29826 | -0.00614 | 0.23246 | 0.90026 | 15308.92 |
| GOMF_PRC | 36 | -0.10274 | -0.00448 | 0.080036 | 0.90036 | 15310.62 |
| GOMF_NAI | 50 | -0.08902 | -0.00458 | 0.069323 | 0.90044 | 15311.98 |
| GOBP_NEU | 5 | -0.25168 | -0.0041 | 0.19598 | 0.90046 | 15312.32 |
| GOBP_GLU | 4 | -0.27442 | -0.00399 | 0.21366 | 0.90048 | 15312.66 |
| GOBP_PYR | 21 | -0.13998 | -0.00467 | 0.10899 | 0.90049 | 15312.83 |
| GOMF_DEI | 10 | -0.23284 | -0.00536 | 0.18127 | 0.9005 | 15313 |
| GOBP_NEU | 22 | -0.12334 | -0.00421 | 0.096011 | 0.90052 | 15313.34 |
| GOBP_REG | 47 | -0.0955 | -0.00476 | 0.074332 | 0.90056 | 15314.02 |
| GOBP_NOI | 16 | -0.16695 | -0.00486 | 0.12991 | 0.90062 | 15315.04 |

| | | | | | | |
|---|---|---|---|---|---|---|
| WP_PYRIM | 78 | -0.06766 | -0.00434 | 0.052646 | 0.90064 | 15315.38 |
| PID_TXA2P | 54 | -0.08697 | -0.00464 | 0.067632 | 0.90075 | 15317.25 |
| GOMF_OXI | 49 | -0.08708 | -0.00443 | 0.067703 | 0.90081 | 15318.27 |
| GOBP_REG | 9 | -0.23267 | -0.00508 | 0.18089 | 0.90082 | 15318.44 |
| MOREAUX_ | 74 | -0.07376 | -0.00461 | 0.057318 | 0.90091 | 15319.97 |
| GOBP_REG | 5 | -0.26312 | -0.00428 | 0.20438 | 0.90101 | 15321.68 |
| GOMF_EXT | 7 | -0.25115 | -0.00484 | 0.19506 | 0.90104 | 15322.19 |
| GOBP_LEU | 76 | -0.07346 | -0.00465 | 0.057042 | 0.90108 | 15322.87 |
| GOBP_POS | 14 | -0.17396 | -0.00474 | 0.13487 | 0.90143 | 15328.82 |
| GOBP_PHO | 9 | -0.22576 | -0.00493 | 0.17495 | 0.90154 | 15330.69 |
| GOMF_NU | 15 | -0.16218 | -0.00457 | 0.12564 | 0.90161 | 15331.88 |
| BIOCARTA_ | 7 | -0.23902 | -0.0046 | 0.18512 | 0.90167 | 15332.9 |
| GOBP_ADA | 266 | -0.04086 | -0.00482 | 0.031631 | 0.90179 | 15334.94 |
| CHUNG_BL | 42 | -0.10466 | -0.00493 | 0.080979 | 0.90188 | 15336.47 |
| GOBP_DET | 5 | -0.25589 | -0.00416 | 0.19791 | 0.90198 | 15338.17 |
| UDAYAKUN | 129 | -0.05376 | -0.00443 | 0.041578 | 0.90198 | 15338.17 |
| GOBP_CAT | 21 | -0.14958 | -0.00499 | 0.11561 | 0.90213 | 15340.72 |
| HU_ANGIO | 20 | -0.13935 | -0.00453 | 0.10768 | 0.90218 | 15341.57 |
| WP_VITAM | 41 | -0.10408 | -0.00485 | 0.080428 | 0.90218 | 15341.57 |
| TONKS_TA | 49 | -0.09695 | -0.00493 | 0.074906 | 0.9022 | 15341.91 |
| GOBP_TOL | 7 | -0.27509 | -0.0053 | 0.21247 | 0.90229 | 15343.44 |
| COATES_M | 75 | -0.07937 | -0.00499 | 0.061288 | 0.90233 | 15344.12 |
| GOCC_SER | 6 | -0.22426 | -0.004 | 0.17294 | 0.90263 | 15349.22 |
| GOBP_POS | 13 | -0.17557 | -0.00461 | 0.13524 | 0.90287 | 15353.3 |
| GOBP_ETH | 10 | -0.22013 | -0.00507 | 0.1695 | 0.90297 | 15355 |
| GOBP_PHO | 27 | -0.11903 | -0.0045 | 0.091639 | 0.903 | 15355.52 |
| GOMF_SIN | 5 | -0.27665 | -0.0045 | 0.21285 | 0.90314 | 15357.9 |
| GOBP_FOL | 5 | -0.32266 | -0.00525 | 0.24813 | 0.90326 | 15359.94 |
| KONDO_HY | 7 | -0.25787 | -0.00496 | 0.19803 | 0.90356 | 15365.04 |
| GOMF_PRO | 7 | -0.22898 | -0.00441 | 0.17584 | 0.90357 | 15365.21 |
| GOBP_RES | 34 | -0.12546 | -0.00532 | 0.096329 | 0.9036 | 15365.72 |
| GOBP_TOL | 2 | -0.41961 | -0.00432 | 0.32202 | 0.90371 | 15367.59 |
| BLANCO_M | 70 | -0.08147 | -0.00495 | 0.062496 | 0.90381 | 15369.29 |
| REACTOME | 10 | -0.2175 | -0.005 | 0.16682 | 0.90384 | 15369.8 |
| BIOCARTA_ | 17 | -0.14234 | -0.00427 | 0.10908 | 0.90404 | 15373.2 |
| GOBP_INTI | 14 | -0.18951 | -0.00516 | 0.1452 | 0.90406 | 15373.54 |
| GOCC_NUC | 10 | -0.17246 | -0.00397 | 0.13207 | 0.90419 | 15375.75 |
| SA_B_CELL | 22 | -0.13947 | -0.00476 | 0.10678 | 0.90423 | 15376.43 |
| GOBP_KER | 169 | -0.0591 | -0.00557 | 0.04525 | 0.90424 | 15376.6 |
| GOBP_REG | 4 | -0.36978 | -0.00538 | 0.28309 | 0.90425 | 15376.77 |
| GOBP_REG | 30 | -0.1122 | -0.00447 | 0.085857 | 0.90435 | 15378.47 |
| GOBP_ION | 22 | -0.14442 | -0.00493 | 0.11046 | 0.90445 | 15380.17 |
| GOBP_POS | 73 | -0.07552 | -0.00469 | 0.057748 | 0.90452 | 15381.36 |
| GOBP_END | 78 | -0.07553 | -0.00484 | 0.057739 | 0.90456 | 15382.04 |
| XU_CREBBI | 21 | -0.13736 | -0.00458 | 0.105 | 0.90459 | 15382.55 |

| | | | | | | |
|---|---|---|---|---|---|---|
| DACOSTA_ | 32 | -0.12289 | -0.00506 | 0.093934 | 0.9046 | 15382.72 |
| GOBP_RES | 186 | -0.04683 | -0.00463 | 0.035796 | 0.90462 | 15383.06 |
| GOBP_LIPI | 80 | -0.07336 | -0.00477 | 0.056049 | 0.9047 | 15384.42 |
| GOBP_ANT | 4 | -0.2271 | -0.00331 | 0.17349 | 0.90472 | 15384.76 |
| SCHAEFFER | 11 | -0.25448 | -0.00614 | 0.19442 | 0.90472 | 15384.76 |
| GOBP_DNA | 5 | -0.29039 | -0.00473 | 0.22168 | 0.90489 | 15387.65 |
| TONKS_TA | 15 | -0.16497 | -0.00465 | 0.12573 | 0.90525 | 15393.78 |
| REACTOME | 76 | -0.07582 | -0.0048 | 0.057778 | 0.90527 | 15394.12 |
| WP_SARSC | 6 | -0.21394 | -0.00381 | 0.16301 | 0.90529 | 15394.46 |
| BRACHAT_ | 21 | -0.12788 | -0.00426 | 0.097398 | 0.9054 | 15396.33 |
| GOBP_POS | 5 | -0.34597 | -0.00563 | 0.26347 | 0.90542 | 15396.67 |
| GOBP_LEU | 4 | -0.36148 | -0.00526 | 0.27488 | 0.90575 | 15402.28 |
| GOMF_GLY | 4 | -0.38316 | -0.00558 | 0.29134 | 0.90577 | 15402.62 |
| GOBP_T_C | 45 | -0.10003 | -0.00488 | 0.076041 | 0.90582 | 15403.47 |
| GOMF_ISO | 43 | -0.10292 | -0.00491 | 0.078211 | 0.90589 | 15404.66 |
| GOBP_NEG | 6 | -0.233 | -0.00415 | 0.17705 | 0.9059 | 15404.83 |
| KYNG_DNA | 191 | -0.04835 | -0.00484 | 0.036741 | 0.90591 | 15405 |
| KEGG_SELE | 26 | -0.1182 | -0.00438 | 0.089757 | 0.90605 | 15407.38 |
| GOMF_RET | 23 | -0.151 | -0.00527 | 0.11466 | 0.90606 | 15407.55 |
| GOCC_BAS | 6 | -0.30963 | -0.00552 | 0.23495 | 0.90622 | 15410.27 |
| GOMF_TRN | 15 | -0.16218 | -0.00457 | 0.12306 | 0.90622 | 15410.27 |
| MARTENS_ | 795 | -0.02444 | -0.00491 | 0.018521 | 0.90651 | 15415.2 |
| GOBP_PRO | 8 | -0.1928 | -0.00397 | 0.14605 | 0.90659 | 15416.56 |
| GOBP_REG | 5 | -0.41706 | -0.00679 | 0.31586 | 0.90664 | 15417.41 |
| GOMF_ME | 63 | -0.07855 | -0.00453 | 0.059491 | 0.90664 | 15417.41 |
| GRAHAM_ | 12 | -0.16554 | -0.00417 | 0.12537 | 0.90664 | 15417.41 |
| RAY_TARG | 14 | -0.17889 | -0.00487 | 0.13546 | 0.90668 | 15418.09 |
| CREIGHTON | 453 | -0.0318 | -0.00487 | 0.024075 | 0.90674 | 15419.11 |
| GOMF_SOL | 8 | -0.25284 | -0.0052 | 0.19138 | 0.90677 | 15419.62 |
| GOBP_NEG | 70 | -0.08527 | -0.00518 | 0.064529 | 0.90681 | 15420.3 |
| GOMF_FER | 24 | -0.13046 | -0.00465 | 0.098707 | 0.90686 | 15421.15 |
| GOBP_LOC | 12 | -0.19228 | -0.00485 | 0.14529 | 0.90714 | 15425.92 |
| GOBP_REG | 35 | -0.10049 | -0.00432 | 0.075915 | 0.90719 | 15426.77 |
| GOBP_POS | 89 | -0.06954 | -0.00476 | 0.052519 | 0.90725 | 15427.79 |
| GOBP_REG | 12 | -0.18386 | -0.00463 | 0.1388 | 0.90734 | 15429.32 |
| KYNG_ENV | 27 | -0.13433 | -0.00508 | 0.10139 | 0.90737 | 15429.83 |
| GOMF_ACY | 26 | -0.13101 | -0.00486 | 0.098877 | 0.90741 | 15430.51 |
| GOBP_REG | 8 | -0.24819 | -0.00511 | 0.18727 | 0.90745 | 15431.19 |
| TARTE_PLA | 34 | -0.1168 | -0.00495 | 0.088056 | 0.90764 | 15434.42 |
| GOBP_REG | 206 | -0.04646 | -0.00483 | 0.035019 | 0.9077 | 15435.44 |
| GOCC_GOL | 77 | -0.07404 | -0.00472 | 0.055787 | 0.90777 | 15436.63 |
| WP_CALCIU | 147 | -0.05669 | -0.00498 | 0.042711 | 0.9078 | 15437.14 |
| GOBP_NEG | 9 | -0.22574 | -0.00493 | 0.17005 | 0.90781 | 15437.31 |
| GOBP_REG | 42 | -0.10077 | -0.00475 | 0.075894 | 0.90787 | 15438.33 |
| GOBP_DCN | 6 | -0.26634 | -0.00475 | 0.20045 | 0.90802 | 15440.88 |

| | | | | | | |
|---|---|---|---|---|---|---|
| GOBP_BEH | 8 | -0.24539 | -0.00505 | 0.18453 | 0.9082 | 15443.94 |
| GOCC_TRA | 262 | -0.03984 | -0.00466 | 0.029955 | 0.90823 | 15444.45 |
| REACTOME | 27 | -0.12348 | -0.00467 | 0.092821 | 0.90828 | 15445.3 |
| GOBP_BILE | 9 | -0.1903 | -0.00415 | 0.14301 | 0.90834 | 15446.32 |
| GOBP_RES | 38 | -0.11887 | -0.00533 | 0.089328 | 0.90834 | 15446.32 |
| GOCC_SYN | 6 | -0.28583 | -0.00509 | 0.21473 | 0.90842 | 15447.68 |
| GOMF_UBI | 33 | -0.11396 | -0.00476 | 0.085562 | 0.90855 | 15449.89 |
| GOMF_ATF | 50 | -0.09049 | -0.00465 | 0.067937 | 0.90856 | 15450.06 |
| GOMF_CAI | 5 | -0.28837 | -0.00469 | 0.21644 | 0.90862 | 15451.08 |
| GOBP_LEU | 5 | -0.24275 | -0.00395 | 0.18213 | 0.9087 | 15452.44 |
| GOBP_REG | 44 | -0.10719 | -0.00517 | 0.080399 | 0.90877 | 15453.63 |
| GOBP_ATP | 88 | -0.06722 | -0.00458 | 0.050393 | 0.90888 | 15455.5 |
| GOBP_NEU | 9 | -0.17985 | -0.00393 | 0.13477 | 0.90897 | 15457.03 |
| GOBP_POS | 14 | -0.18419 | -0.00501 | 0.13801 | 0.90898 | 15457.2 |
| GOBP_CYC | 5 | -0.34926 | -0.00568 | 0.26148 | 0.90917 | 15460.44 |
| MEBARKI_I | 77 | -0.07585 | -0.00483 | 0.056784 | 0.90918 | 15460.61 |
| RAMASWA | 50 | -0.09212 | -0.00473 | 0.068965 | 0.90918 | 15460.61 |
| MAGRANG | 21 | -0.13739 | -0.00458 | 0.10284 | 0.90921 | 15461.12 |
| GOBP_PLA | 8 | -0.2591 | -0.00533 | 0.19381 | 0.90936 | 15463.67 |
| GOBP_RNA | 41 | -0.09361 | -0.00436 | 0.069941 | 0.90962 | 15468.09 |
| GOBP_REG | 7 | -0.2554 | -0.00492 | 0.1908 | 0.90964 | 15468.43 |
| GOMF_PEF | 555 | -0.02977 | -0.00503 | 0.022232 | 0.90973 | 15469.96 |
| GOBP_GLY | 58 | -0.08798 | -0.00487 | 0.065671 | 0.90981 | 15471.32 |
| GOBP_LYN | 6 | -0.22212 | -0.00396 | 0.16579 | 0.90983 | 15471.66 |
| GOMF_OXI | 5 | -0.38557 | -0.00627 | 0.28769 | 0.9099 | 15472.85 |
| GOBP_REG | 33 | -0.11191 | -0.00467 | 0.083452 | 0.91002 | 15474.89 |
| GOBP_TRY | 5 | -0.29838 | -0.00486 | 0.22251 | 0.91003 | 15475.06 |
| KIM_MYC_ | 179 | -0.05128 | -0.00497 | 0.038229 | 0.91007 | 15475.74 |
| GOMF_SOI | 6 | -0.28783 | -0.00513 | 0.21459 | 0.91009 | 15476.08 |
| GOBP_CON | 51 | -0.10772 | -0.00559 | 0.080302 | 0.9101 | 15476.25 |
| REACTOME | 84 | -0.07614 | -0.00507 | 0.05675 | 0.91013 | 15476.76 |
| CHOW_RAS | 22 | -0.1419 | -0.00484 | 0.10576 | 0.91015 | 15477.1 |
| WP_EXRNA | 6 | -0.30051 | -0.00536 | 0.22369 | 0.91043 | 15481.86 |
| KEGG_NOD | 57 | -0.08794 | -0.00482 | 0.06542 | 0.91055 | 15483.9 |
| GOCC_ATA | 13 | -0.18732 | -0.00491 | 0.13933 | 0.91058 | 15484.41 |
| GOBP_PRC | 29 | -0.11456 | -0.00449 | 0.085181 | 0.91066 | 15485.77 |
| GOBP_NEG | 27 | -0.12828 | -0.00485 | 0.095301 | 0.91085 | 15489 |
| GOMF_PH( | 11 | -0.18365 | -0.00443 | 0.13643 | 0.91085 | 15489 |
| GOMF_MA | 28 | -0.1236 | -0.00476 | 0.091776 | 0.91097 | 15491.04 |
| WP_CYTOK | 23 | -0.1579 | -0.00551 | 0.11717 | 0.9111 | 15493.26 |
| GOBP_POS | 3 | -0.38553 | -0.00486 | 0.28601 | 0.91116 | 15494.28 |
| GOCC_GBA | 14 | -0.17169 | -0.00467 | 0.12737 | 0.91116 | 15494.28 |
| GOBP_REG | 9 | -0.2617 | -0.00571 | 0.1941 | 0.9112 | 15494.96 |
| BIOCARTA_ | 13 | -0.15191 | -0.00398 | 0.11261 | 0.91132 | 15497 |
| GOBP_NEG | 9 | -0.21287 | -0.00465 | 0.15779 | 0.91134 | 15497.34 |

| | | | | | | |
|---|---|---|---|---|---|---|
| GOBP_REG | 18 | -0.16619 | -0.00513 | 0.12317 | 0.91137 | 15497.85 |
| GOMF_ALD | 29 | -0.12892 | -0.00505 | 0.095522 | 0.91142 | 15498.7 |
| GOBP_POS | 6 | -0.25771 | -0.00459 | 0.19083 | 0.91156 | 15501.08 |
| GUTIERREZ | 11 | -0.21043 | -0.00508 | 0.15578 | 0.9116 | 15501.76 |
| GOBP_NEG | 10 | -0.23396 | -0.00538 | 0.17319 | 0.91162 | 15502.1 |
| GOMF_HSF | 46 | -0.10311 | -0.00508 | 0.076288 | 0.91173 | 15503.97 |
| GOBP_REG | 17 | -0.16308 | -0.00489 | 0.12062 | 0.9118 | 15505.16 |
| GOCC_KIN | 10 | -0.19703 | -0.00453 | 0.14572 | 0.91183 | 15505.67 |
| BASAKI_YB | 363 | -0.03674 | -0.00505 | 0.027158 | 0.91192 | 15507.2 |
| GOCC_H4_ | 37 | -0.10388 | -0.00459 | 0.076795 | 0.91192 | 15507.2 |
| GOBP_POS | 9 | -0.23752 | -0.00518 | 0.17546 | 0.91207 | 15509.75 |
| REACTOME | 83 | -0.06994 | -0.00463 | 0.051648 | 0.91214 | 15510.94 |
| GOBP_ANT | 21 | -0.12848 | -0.00428 | 0.094872 | 0.91216 | 15511.28 |
| REACTOME | 12 | -0.20133 | -0.00507 | 0.14866 | 0.91217 | 15511.45 |
| GOBP_POS | 10 | -0.22133 | -0.00509 | 0.16327 | 0.91238 | 15515.02 |
| GRAHAM_I | 59 | -0.09449 | -0.00527 | 0.069638 | 0.91259 | 15518.59 |
| GOMF_CAS | 12 | -0.19486 | -0.00491 | 0.14352 | 0.91272 | 15520.8 |
| GOMF_NEU | 26 | -0.1577 | -0.00585 | 0.11614 | 0.91272 | 15520.8 |
| GOBP_NEG | 36 | -0.10963 | -0.00478 | 0.080715 | 0.9128 | 15522.16 |
| BIOCARTA_ | 7 | -0.24333 | -0.00468 | 0.17907 | 0.9129 | 15523.86 |
| GOBP_INO | 55 | -0.09112 | -0.00491 | 0.067021 | 0.913 | 15525.57 |
| TING_SILEN | 29 | -0.12881 | -0.00504 | 0.094718 | 0.91307 | 15526.76 |
| GOBP_ARP | 41 | -0.09963 | -0.00464 | 0.073223 | 0.91316 | 15528.29 |
| REACTOME | 31 | -0.11772 | -0.00477 | 0.08648 | 0.91327 | 15530.16 |
| GOBP_POS | 15 | -0.19068 | -0.00537 | 0.13995 | 0.91347 | 15533.56 |
| RAMPON_I | 20 | -0.15055 | -0.0049 | 0.11048 | 0.91351 | 15534.24 |
| GOBP_REG | 63 | -0.08801 | -0.00508 | 0.064555 | 0.9136 | 15535.77 |
| GOCC_CELI | 529 | -0.0303 | -0.005 | 0.022213 | 0.91371 | 15537.64 |
| GOBP_ACE | 15 | -0.17813 | -0.00502 | 0.13059 | 0.91372 | 15537.81 |
| GOBP_REG | 15 | -0.16909 | -0.00476 | 0.12396 | 0.91372 | 15537.81 |
| GOBP_POS | 10 | -0.21359 | -0.00491 | 0.15658 | 0.91373 | 15537.98 |
| GOMF_ACT | 216 | -0.04483 | -0.00477 | 0.03286 | 0.91375 | 15538.32 |
| GOBP_THY | 7 | -0.27989 | -0.00539 | 0.205 | 0.91392 | 15541.21 |
| GOBP_N_T | 8 | -0.22887 | -0.00471 | 0.16759 | 0.91397 | 15542.06 |
| GOBP_GAN | 17 | -0.17716 | -0.00531 | 0.12971 | 0.91399 | 15542.4 |
| WP_ENERG | 47 | -0.09375 | -0.00467 | 0.068618 | 0.91406 | 15543.59 |
| GOMF_FER | 9 | -0.2374 | -0.00518 | 0.17358 | 0.91429 | 15547.5 |
| GOMF_N_A | 80 | -0.07551 | -0.00491 | 0.055213 | 0.91429 | 15547.5 |
| GOBP_NUC | 31 | -0.13936 | -0.00564 | 0.10187 | 0.91433 | 15548.18 |
| GOCC_PHC | 19 | -0.12692 | -0.00402 | 0.092772 | 0.91434 | 15548.35 |
| SAGIV_CD2 | 22 | -0.13626 | -0.00465 | 0.099523 | 0.91451 | 15551.24 |
| GOCC_CEN | 7 | -0.22009 | -0.00424 | 0.16068 | 0.91461 | 15552.94 |
| GOBP_NEG | 66 | -0.08841 | -0.00522 | 0.064501 | 0.91474 | 15555.15 |
| GOCC_CLA | 70 | -0.07745 | -0.00471 | 0.056498 | 0.91477 | 15555.66 |
| GOMF_RAC | 10 | -0.17538 | -0.00404 | 0.12793 | 0.91479 | 15556 |

| | | | | | | |
|---|---|---|---|---|---|---|
| GRAHAM_( | 7 | -0.26444 | -0.00509 | 0.19289 | 0.91479 | 15556 |
| XU_HGF_SI | 23 | -0.15072 | -0.00526 | 0.10991 | 0.91485 | 15557.02 |
| GOBP_PEP | 160 | -0.05181 | -0.00475 | 0.03777 | 0.91489 | 15557.7 |
| BAKKER_F( | 160 | -0.05529 | -0.00507 | 0.040304 | 0.91494 | 15558.55 |
| GOBP_LEU | 10 | -0.19446 | -0.00447 | 0.14169 | 0.91503 | 15560.09 |
| GOBP_URE | 11 | -0.2498 | -0.00603 | 0.18181 | 0.91526 | 15564 |
| GOBP_MO | 8 | -0.25572 | -0.00526 | 0.18595 | 0.91545 | 15567.23 |
| GOCC_SAG | 34 | -0.10456 | -0.00443 | 0.076016 | 0.91549 | 15567.91 |
| GOBP_POS | 45 | -0.11114 | -0.00542 | 0.080802 | 0.9155 | 15568.08 |
| GOBP_CON | 30 | -0.15044 | -0.00599 | 0.10937 | 0.91551 | 15568.25 |
| GOBP_NEG | 20 | -0.15309 | -0.00498 | 0.11129 | 0.91551 | 15568.25 |
| BARRIER_C | 15 | -0.15253 | -0.0043 | 0.11084 | 0.9156 | 15569.78 |
| LUND_SILE | 16 | -0.18216 | -0.0053 | 0.13223 | 0.91582 | 15573.52 |
| GOBP_POS | 90 | -0.07732 | -0.00533 | 0.056115 | 0.91586 | 15574.2 |
| NEBEN_AN | 8 | -0.19355 | -0.00398 | 0.14046 | 0.91588 | 15574.54 |
| GOBP_PHO | 72 | -0.07946 | -0.0049 | 0.057658 | 0.9159 | 15574.88 |
| REACTOME | 5 | -0.31125 | -0.00506 | 0.22577 | 0.91599 | 15576.41 |
| GOBP_POS | 24 | -0.14084 | -0.00502 | 0.10214 | 0.91603 | 15577.09 |
| GOBP_INTI | 41 | -0.10988 | -0.00511 | 0.079672 | 0.91606 | 15577.6 |
| GOBP_MA( | 115 | -0.05996 | -0.00467 | 0.043474 | 0.91609 | 15578.11 |
| PASINI_SU | 96 | -0.0662 | -0.00471 | 0.047991 | 0.91612 | 15578.62 |
| GOBP_MAS | 7 | -0.27478 | -0.00529 | 0.19903 | 0.91629 | 15581.51 |
| GOMF_GLU | 10 | -0.22325 | -0.00514 | 0.16167 | 0.91633 | 15582.19 |
| MARIADAS | 13 | -0.17052 | -0.00447 | 0.12348 | 0.91634 | 15582.36 |
| WP_CHEM | 157 | -0.05432 | -0.00493 | 0.039285 | 0.91661 | 15586.95 |
| GOBP_GAN | 11 | -0.21718 | -0.00524 | 0.15703 | 0.91667 | 15587.97 |
| GOBP_NEG | 11 | -0.24353 | -0.00588 | 0.17603 | 0.91672 | 15588.82 |
| WP_TLR4_ | 24 | -0.15331 | -0.00546 | 0.11077 | 0.91682 | 15590.52 |
| GOBP_GER | 9 | -0.26675 | -0.00582 | 0.19259 | 0.91697 | 15593.07 |
| GOBP_TRY | 5 | -0.25823 | -0.0042 | 0.18637 | 0.91705 | 15594.44 |
| GOBP_REG | 15 | -0.17981 | -0.00507 | 0.12969 | 0.91718 | 15596.65 |
| WP_BARDE | 84 | -0.07804 | -0.00519 | 0.056214 | 0.91746 | 15601.41 |
| HAHTOLA_ | 14 | -0.20382 | -0.00555 | 0.14671 | 0.91762 | 15604.13 |
| GENTILE_U | 35 | -0.12355 | -0.00531 | 0.088888 | 0.91771 | 15605.66 |
| GOMF_LIP/ | 16 | -0.18326 | -0.00533 | 0.13178 | 0.91782 | 15607.53 |
| GOBP_REG | 28 | -0.11752 | -0.00452 | 0.084493 | 0.91786 | 15608.21 |
| GOBP_REG | 142 | -0.0596 | -0.00515 | 0.042831 | 0.91796 | 15609.91 |
| WANG_NE | 16 | -0.19973 | -0.00581 | 0.1435 | 0.91801 | 15610.76 |
| BIOCARTA_ | 9 | -0.24399 | -0.00533 | 0.17528 | 0.91803 | 15611.1 |
| BOYAULT_ | 50 | -0.09411 | -0.00484 | 0.067582 | 0.9181 | 15612.29 |
| STAEGE_EV | 25 | -0.16288 | -0.00592 | 0.11693 | 0.91818 | 15613.65 |
| GOBP_REG | 5 | -0.24001 | -0.00391 | 0.17229 | 0.91819 | 15613.82 |
| GOBP_MYE | 28 | -0.14022 | -0.0054 | 0.10059 | 0.91832 | 15616.03 |
| GOMF_AC | 229 | -0.04696 | -0.00514 | 0.033681 | 0.91839 | 15617.22 |
| GOBP_REG | 169 | -0.05557 | -0.00523 | 0.039831 | 0.9185 | 15619.09 |

| | | | | | | |
|---|---|---|---|---|---|---|
| GOCC_AXC | 22 | -0.14882 | -0.00508 | 0.10667 | 0.9185 | 15619.09 |
| GOBP_DIAI | 5 | -0.30708 | -0.005 | 0.21961 | 0.91898 | 15627.25 |
| GOBP_INTI | 14 | -0.22322 | -0.00608 | 0.15952 | 0.91913 | 15629.81 |
| GOBP_UBI( | 9 | -0.25441 | -0.00555 | 0.18181 | 0.91913 | 15629.81 |
| GOBP_ANT | 22 | -0.14619 | -0.00499 | 0.10445 | 0.91918 | 15630.66 |
| RHEIN_ALL | 67 | -0.08607 | -0.00512 | 0.061453 | 0.91933 | 15633.21 |
| PID_ECADH | 21 | -0.13208 | -0.0044 | 0.094236 | 0.91948 | 15635.76 |
| GOBP_NEG | 5 | -0.38955 | -0.00634 | 0.27789 | 0.91951 | 15636.27 |
| GOCC_PRC | 8 | -0.23824 | -0.0049 | 0.16994 | 0.91952 | 15636.44 |
| GOBP_LEU | 43 | -0.10641 | -0.00507 | 0.075898 | 0.91954 | 15636.78 |
| REACTOME | 29 | -0.13446 | -0.00527 | 0.095895 | 0.91956 | 15637.12 |
| KEGG_CYT( | 50 | -0.10422 | -0.00536 | 0.074319 | 0.91957 | 15637.29 |
| WP_GPCRS | 85 | -0.08524 | -0.00571 | 0.060767 | 0.91965 | 15638.65 |
| WUNDER_I | 48 | -0.09631 | -0.00485 | 0.068571 | 0.91991 | 15643.07 |
| REACTOME | 5 | -0.37773 | -0.00615 | 0.26883 | 0.92 | 15644.6 |
| GOBP_REG | 24 | -0.1638 | -0.00584 | 0.11653 | 0.92007 | 15645.79 |
| GOBP_NEG | 7 | -0.2084 | -0.00401 | 0.14821 | 0.92014 | 15646.98 |
| HOEBEKE_I | 81 | -0.0766 | -0.00501 | 0.054479 | 0.92014 | 15646.98 |
| PID_INSULI | 26 | -0.13072 | -0.00485 | 0.092945 | 0.92019 | 15647.83 |
| GOBP_REG | 83 | -0.07436 | -0.00492 | 0.052843 | 0.92029 | 15649.53 |
| GOBP_NEG | 25 | -0.16141 | -0.00587 | 0.1147 | 0.92031 | 15649.87 |
| REACTOME | 33 | -0.13653 | -0.0057 | 0.097004 | 0.92034 | 15650.38 |
| GOBP_NEG | 6 | -0.21888 | -0.0039 | 0.15542 | 0.92047 | 15652.59 |
| GOCC_SYN | 361 | -0.0398 | -0.00545 | 0.028259 | 0.92047 | 15652.59 |
| GOBP_PLA | 8 | -0.19603 | -0.00403 | 0.13903 | 0.92071 | 15656.67 |
| GOBP_DED | 8 | -0.28913 | -0.00595 | 0.20505 | 0.92072 | 15656.84 |
| GOBP_NEU | 35 | -0.10682 | -0.0046 | 0.075702 | 0.92087 | 15659.39 |
| WP_HIJACI | 8 | -0.24473 | -0.00504 | 0.17338 | 0.92095 | 15660.75 |
| GOCC_AP_ | 10 | -0.19693 | -0.00453 | 0.13941 | 0.9211 | 15663.31 |
| GOBP_GAN | 24 | -0.16502 | -0.00588 | 0.1168 | 0.92115 | 15664.16 |
| MORI_PLA: | 49 | -0.09786 | -0.00498 | 0.069204 | 0.92133 | 15667.22 |
| ZHAN_EAR | 39 | -0.11328 | -0.00514 | 0.080101 | 0.92134 | 15667.39 |
| GOBP_REG | 7 | -0.28841 | -0.00555 | 0.20369 | 0.92159 | 15671.64 |
| RICKMAN_ | 102 | -0.06974 | -0.00511 | 0.049238 | 0.92165 | 15672.66 |
| SCHEIDERE | 17 | -0.15658 | -0.0047 | 0.11051 | 0.92174 | 15674.19 |
| GOMF_CAI | 33 | -0.12547 | -0.00524 | 0.088532 | 0.92179 | 15675.04 |
| GOBP_MYE | 93 | -0.07266 | -0.00509 | 0.05125 | 0.92185 | 15676.06 |
| WP_JOUBE | 71 | -0.08342 | -0.00511 | 0.05882 | 0.92194 | 15677.59 |
| REACTOME | 29 | -0.14335 | -0.00561 | 0.10105 | 0.92199 | 15678.44 |
| GOBP_PRC | 6 | -0.26886 | -0.00479 | 0.1895 | 0.92201 | 15678.78 |
| WP_DRUG_ | 17 | -0.16356 | -0.00491 | 0.11528 | 0.92201 | 15678.78 |
| GOBP_POS | 6 | -0.34875 | -0.00622 | 0.24576 | 0.92205 | 15679.46 |
| GOBP_POS | 533 | -0.03187 | -0.00528 | 0.022453 | 0.92212 | 15680.65 |
| GOBP_ACU | 108 | -0.06936 | -0.00523 | 0.048849 | 0.92217 | 15681.5 |
| GOBP_COF | 12 | -0.19352 | -0.00488 | 0.13628 | 0.9222 | 15682.01 |

| | | | | | | |
|---|---|---|---|---|---|---|
| GOBP_INN | 27 | -0.15831 | -0.00598 | 0.11146 | 0.92224 | 15682.69 |
| GOBP_REG | 7 | -0.31853 | -0.00613 | 0.22426 | 0.92225 | 15682.86 |
| GOBP_CELI | 5 | -0.29447 | -0.00479 | 0.20722 | 0.92234 | 15684.39 |
| GOCC_OUT | 13 | -0.20955 | -0.0055 | 0.14743 | 0.92238 | 15685.07 |
| GOBP_CELI | 234 | -0.04887 | -0.00541 | 0.034378 | 0.9224 | 15685.41 |
| GOMF_OXI | 47 | -0.1058 | -0.00527 | 0.074428 | 0.92241 | 15685.58 |
| SHIN_B_CE | 12 | -0.18067 | -0.00455 | 0.12707 | 0.92244 | 15686.09 |
| GOBP_VES | 31 | -0.12118 | -0.00491 | 0.085212 | 0.92249 | 15686.94 |
| GOCC_CHR | 9 | -0.29199 | -0.00637 | 0.20525 | 0.92256 | 15688.13 |
| GOBP_VAC | 83 | -0.07469 | -0.00494 | 0.052496 | 0.92257 | 15688.3 |
| PID_IL12_2 | 57 | -0.09696 | -0.00532 | 0.06814 | 0.92262 | 15689.15 |
| SANSOM_A | 229 | -0.04559 | -0.00499 | 0.032029 | 0.92268 | 15690.17 |
| WP_OVERV | 36 | -0.15707 | -0.00685 | 0.11027 | 0.92283 | 15692.72 |
| KEGG_PYRI | 92 | -0.06968 | -0.00485 | 0.048893 | 0.92294 | 15694.59 |
| GOCC_INTI | 25 | -0.13788 | -0.00501 | 0.09668 | 0.92307 | 15696.81 |
| WP_WNTB | 26 | -0.1417 | -0.00526 | 0.099332 | 0.92313 | 15697.83 |
| GOCC_PRC | 5 | -0.33549 | -0.00546 | 0.23516 | 0.92315 | 15698.17 |
| CHANDRAN | 41 | -0.10454 | -0.00487 | 0.073234 | 0.92327 | 15700.21 |
| REACTOME | 27 | -0.14246 | -0.00538 | 0.0998 | 0.92327 | 15700.21 |
| WP_PURIN | 28 | -0.1398 | -0.00538 | 0.097896 | 0.92335 | 15701.57 |
| GOBP_MAI | 5 | -0.2673 | -0.00435 | 0.1871 | 0.92344 | 15703.1 |
| GOBP_INTI | 473 | -0.03396 | -0.00531 | 0.023759 | 0.92353 | 15704.63 |
| GOBP_REG | 40 | -0.12259 | -0.00564 | 0.08576 | 0.92355 | 15704.97 |
| GOBP_RES | 7 | -0.26599 | -0.00512 | 0.18605 | 0.92359 | 15705.65 |
| GOBP_RES | 120 | -0.06878 | -0.00547 | 0.04809 | 0.92367 | 15707.01 |
| GOBP_TRA | 33 | -0.13396 | -0.0056 | 0.093623 | 0.92376 | 15708.54 |
| PEDERSEN_ | 72 | -0.08483 | -0.00523 | 0.059264 | 0.92384 | 15709.9 |
| GOCC_ORG | 44 | -0.10769 | -0.00519 | 0.075205 | 0.92391 | 15711.09 |
| GOBP_REG | 586 | -0.02981 | -0.00517 | 0.020811 | 0.92398 | 15712.28 |
| GOMF_DN/ | 5 | -0.21118 | -0.00344 | 0.1474 | 0.92402 | 15712.96 |
| GOBP_POS | 210 | -0.05201 | -0.00545 | 0.036297 | 0.92403 | 15713.13 |
| REACTOME | 196 | -0.05164 | -0.00523 | 0.036029 | 0.92411 | 15714.49 |
| GOMF_PRC | 22 | -0.15185 | -0.00518 | 0.10583 | 0.92433 | 15718.23 |
| WP_FIBRIN | 39 | -0.12175 | -0.00553 | 0.084784 | 0.9245 | 15721.12 |
| GOBP_TUB | 19 | -0.1565 | -0.00496 | 0.10896 | 0.92453 | 15721.63 |
| DAZARD_R | 90 | -0.0721 | -0.00497 | 0.050185 | 0.92459 | 15722.65 |
| GOCC_CEN | 102 | -0.07291 | -0.00534 | 0.050745 | 0.9246 | 15722.82 |
| CAIRO_HEF | 203 | -0.05003 | -0.00516 | 0.034806 | 0.92467 | 15724.01 |
| GOMF_ADI | 5 | -0.32555 | -0.0053 | 0.22651 | 0.92467 | 15724.01 |
| GOCC_NEU | 122 | -0.06907 | -0.00553 | 0.048055 | 0.92468 | 15724.18 |
| GOBP_REG | 43 | -0.11803 | -0.00563 | 0.082068 | 0.9248 | 15726.22 |
| GOBP_RET | 75 | -0.07855 | -0.00494 | 0.05461 | 0.92483 | 15726.73 |
| REACTOME | 22 | -0.13266 | -0.00453 | 0.092218 | 0.92485 | 15727.07 |
| GOMF_ACI | 5 | -0.34566 | -0.00562 | 0.24027 | 0.92486 | 15727.24 |
| WP_HORM | 5 | -0.28297 | -0.0046 | 0.19662 | 0.92494 | 15728.6 |

| | | | | | | |
|---|---|---|---|---|---|---|
| GOBP_IROI | 50 | -0.09745 | -0.00501 | 0.067642 | 0.92514 | 15732.01 |
| DEURIG_T_ | 290 | -0.0415 | -0.0051 | 0.028802 | 0.92519 | 15732.86 |
| MARSON_I | 51 | -0.09862 | -0.00512 | 0.068439 | 0.92521 | 15733.2 |
| GOBP_POS | 25 | -0.1409 | -0.00512 | 0.097768 | 0.92523 | 15733.54 |
| GOCC_CELI | 12 | -0.18887 | -0.00476 | 0.131 | 0.92531 | 15734.9 |
| GOCC_GAN | 7 | -0.24993 | -0.00481 | 0.1733 | 0.92537 | 15735.92 |
| GOMF_OXI | 17 | -0.1604 | -0.00481 | 0.1112 | 0.9254 | 15736.43 |
| GOBP_NEU | 164 | -0.05891 | -0.00547 | 0.040837 | 0.92542 | 15736.77 |
| WP_NUCLE | 301 | -0.04362 | -0.00546 | 0.030231 | 0.92545 | 15737.28 |
| GOBP_LIPII | 11 | -0.18291 | -0.00441 | 0.12664 | 0.92567 | 15741.02 |
| WP_NANO | 6 | -0.3466 | -0.00618 | 0.23996 | 0.92568 | 15741.19 |
| REACTOME | 23 | -0.14936 | -0.00521 | 0.10338 | 0.92573 | 15742.04 |
| REACTOME | 20 | -0.17572 | -0.00572 | 0.12153 | 0.92588 | 15744.59 |
| NAKAMUR | 5 | -0.25075 | -0.00408 | 0.17326 | 0.92607 | 15747.82 |
| GOBP_POS | 5 | -0.33742 | -0.00549 | 0.23312 | 0.9261 | 15748.33 |
| GOMF_RA( | 17 | -0.16453 | -0.00494 | 0.11352 | 0.92637 | 15752.92 |
| GOMF_FEF | 5 | -0.33409 | -0.00544 | 0.23048 | 0.9264 | 15753.43 |
| BIOCARTA_ | 12 | -0.19341 | -0.00487 | 0.13341 | 0.92643 | 15753.94 |
| GOBP_ISG1 | 6 | -0.29893 | -0.00533 | 0.20601 | 0.92661 | 15757 |
| GOBP_REG | 43 | -0.10199 | -0.00486 | 0.070263 | 0.92668 | 15758.19 |
| GOCC_MCI | 8 | -0.21625 | -0.00445 | 0.14897 | 0.92669 | 15758.36 |
| REACTOME | 170 | -0.05887 | -0.00556 | 0.040524 | 0.92682 | 15760.57 |
| DAZARD_U | 53 | -0.09756 | -0.00516 | 0.06714 | 0.9269 | 15761.93 |
| GOBP_PEP | 13 | -0.20474 | -0.00537 | 0.14088 | 0.92692 | 15762.27 |
| GOBP_NEG | 9 | -0.19678 | -0.0043 | 0.13538 | 0.92696 | 15762.95 |
| WP_ALSTR | 31 | -0.12833 | -0.0052 | 0.088266 | 0.92701 | 15763.81 |
| GOBP_REG | 7 | -0.26795 | -0.00516 | 0.18422 | 0.92709 | 15765.17 |
| REACTOME | 37 | -0.10448 | -0.00462 | 0.071833 | 0.92709 | 15765.17 |
| GOBP_POS | 184 | -0.0513 | -0.00504 | 0.035258 | 0.92715 | 15766.19 |
| GOMF_URI | 7 | -0.31413 | -0.00605 | 0.2159 | 0.92715 | 15766.19 |
| GOCC_VCP | 8 | -0.26943 | -0.00555 | 0.18508 | 0.92727 | 15768.23 |
| GOBP_NEU | 17 | -0.15101 | -0.00453 | 0.10371 | 0.9273 | 15768.74 |
| GOBP_PRC | 17 | -0.16257 | -0.00488 | 0.11161 | 0.92737 | 15769.93 |
| REACTOME | 33 | -0.13428 | -0.00561 | 0.092155 | 0.92745 | 15771.29 |
| GOMF_INC | 5 | -0.2783 | -0.00453 | 0.1909 | 0.92754 | 15772.82 |
| GOMF_SOI | 8 | -0.29231 | -0.00602 | 0.20045 | 0.92761 | 15774.01 |
| GOMF_AN | 37 | -0.12221 | -0.0054 | 0.083795 | 0.92762 | 15774.18 |
| REACTOME | 27 | -0.13094 | -0.00495 | 0.089754 | 0.9277 | 15775.54 |
| WP_MBDN | 35 | -0.12632 | -0.00543 | 0.086563 | 0.92774 | 15776.22 |
| BIOCARTA_ | 18 | -0.15192 | -0.00469 | 0.10408 | 0.92779 | 15777.07 |
| GOBP_DEN | 32 | -0.13269 | -0.00546 | 0.090896 | 0.92782 | 15777.58 |
| GOBP_L_H | 4 | -0.43815 | -0.00638 | 0.30006 | 0.92788 | 15778.6 |
| REACTOME | 46 | -0.10033 | -0.00495 | 0.068688 | 0.92793 | 15779.45 |
| GOBP_GU/ | 8 | -0.2898 | -0.00596 | 0.19832 | 0.92802 | 15780.98 |
| GOMF_D_I | 5 | -0.38233 | -0.00622 | 0.26155 | 0.92809 | 15782.17 |

| | | | | | | |
|---|---|---|---|---|---|---|
| GOBP_ANT | 7 | -0.21397 | -0.00412 | 0.1463 | 0.9282 | 15784.04 |
| HASLINGEF | 16 | -0.19723 | -0.00574 | 0.13481 | 0.92826 | 15785.06 |
| BLANCO_M | 79 | -0.07526 | -0.00486 | 0.051435 | 0.92828 | 15785.4 |
| FAELT_B_C | 44 | -0.11654 | -0.00562 | 0.079653 | 0.92828 | 15785.4 |
| GOBP_POS | 5 | -0.37615 | -0.00612 | 0.25704 | 0.92831 | 15785.91 |
| GOMF_MC | 657 | -0.02975 | -0.00545 | 0.020319 | 0.92842 | 15787.78 |
| GOBP_POS | 21 | -0.15377 | -0.00513 | 0.10495 | 0.92855 | 15789.99 |
| IVANOV_M | 11 | -0.18854 | -0.00455 | 0.1286 | 0.92868 | 15792.2 |
| GOBP_MEI | 43 | -0.11303 | -0.00539 | 0.077089 | 0.9287 | 15792.54 |
| REACTOME | 55 | -0.09445 | -0.00509 | 0.064414 | 0.9287 | 15792.54 |
| GOBP_REG | 12 | -0.21329 | -0.00538 | 0.1454 | 0.92878 | 15793.9 |
| GOMF_INC | 11 | -0.18611 | -0.00449 | 0.12682 | 0.92886 | 15795.26 |
| FIGUEROA_ | 155 | -0.06084 | -0.00549 | 0.041432 | 0.929 | 15797.65 |
| REACTOME | 5 | -0.409 | -0.00666 | 0.27848 | 0.92903 | 15798.16 |
| GOBP_FUC | 17 | -0.1711 | -0.00513 | 0.11647 | 0.92908 | 15799.01 |
| GOBP_RES | 5 | -0.39221 | -0.00638 | 0.26695 | 0.92911 | 15799.52 |
| GOBP_HUM | 220 | -0.0558 | -0.00599 | 0.037972 | 0.92914 | 15800.03 |
| GOBP_REG | 5 | -0.26364 | -0.00429 | 0.17939 | 0.92916 | 15800.37 |
| MIYAGAWA | 246 | -0.04968 | -0.00563 | 0.033794 | 0.92923 | 15801.56 |
| GOCC_PLA | 9 | -0.24319 | -0.00531 | 0.16531 | 0.92937 | 15803.94 |
| CHANG_IM | 66 | -0.09175 | -0.00542 | 0.062343 | 0.92944 | 15805.13 |
| GOBP_HIST | 5 | -0.33106 | -0.00539 | 0.22495 | 0.92944 | 15805.13 |
| GOBP_REG | 9 | -0.23574 | -0.00515 | 0.16013 | 0.9295 | 15806.15 |
| GOBP_POS | 35 | -0.12586 | -0.00541 | 0.085454 | 0.92959 | 15807.68 |
| GOBP_POS | 7 | -0.25379 | -0.00489 | 0.17226 | 0.92966 | 15808.87 |
| GOBP_OSM | 5 | -0.29796 | -0.00485 | 0.2022 | 0.9297 | 15809.55 |
| SINGH_NFE | 13 | -0.18957 | -0.00497 | 0.12854 | 0.92985 | 15812.1 |
| GOMF_TRA | 5 | -0.44089 | -0.00717 | 0.29877 | 0.92998 | 15814.31 |
| BIOCARTA_ | 5 | -0.29888 | -0.00486 | 0.20239 | 0.93012 | 15816.69 |
| REACTOME | 12 | -0.20815 | -0.00525 | 0.14094 | 0.93014 | 15817.03 |
| GOBP_REG | 96 | -0.08573 | -0.0061 | 0.057987 | 0.93034 | 15820.43 |
| PID_RXR_V | 24 | -0.1605 | -0.00572 | 0.10854 | 0.93038 | 15821.11 |
| GOBP_GLU | 6 | -0.2673 | -0.00476 | 0.18075 | 0.93039 | 15821.28 |
| GOBP_POS | 6 | -0.29299 | -0.00522 | 0.19801 | 0.93051 | 15823.32 |
| GOMF_PEF | 207 | -0.05529 | -0.00576 | 0.037361 | 0.93054 | 15823.83 |
| GOBP_REG | 9 | -0.2917 | -0.00637 | 0.19702 | 0.93062 | 15825.19 |
| BIOCARTA_ | 8 | -0.23954 | -0.00493 | 0.16179 | 0.93063 | 15825.36 |
| WP_CYTOS | 68 | -0.09209 | -0.00552 | 0.062188 | 0.93066 | 15825.87 |
| GOBP_REG | 56 | -0.10661 | -0.0058 | 0.071992 | 0.93067 | 15826.04 |
| GOBP_NEU | 7 | -0.26437 | -0.00509 | 0.17852 | 0.93068 | 15826.21 |
| GOBP_EST/ | 29 | -0.1363 | -0.00534 | 0.091941 | 0.93089 | 15829.78 |
| GOMF_ARI | 14 | -0.16253 | -0.00442 | 0.10962 | 0.93091 | 15830.12 |
| KANG_CISF | 19 | -0.15407 | -0.00489 | 0.10384 | 0.93105 | 15832.51 |
| GOBP_CD4 | 19 | -0.18142 | -0.00575 | 0.12226 | 0.93106 | 15832.68 |
| GOMF_CAI | 5 | -0.45461 | -0.0074 | 0.30616 | 0.9312 | 15835.06 |

| | | | | | | |
|---|---|---|---|---|---|---|
| GOBP_NEG | 8 | -0.25239 | -0.00519 | 0.1699 | 0.93128 | 15836.42 |
| GOBP_DNA | 18 | -0.15765 | -0.00487 | 0.10611 | 0.93132 | 15837.1 |
| REACTOME | 42 | -0.10238 | -0.00482 | 0.068907 | 0.93132 | 15837.1 |
| GOBP_AMI | 37 | -0.11873 | -0.00525 | 0.079858 | 0.93145 | 15839.31 |
| RASHI_RES | 37 | -0.12095 | -0.00535 | 0.081307 | 0.93156 | 15841.18 |
| SATO_SILEI | 41 | -0.11606 | -0.0054 | 0.077981 | 0.93166 | 15842.88 |
| REACTOME | 103 | -0.06973 | -0.00514 | 0.046778 | 0.93197 | 15848.15 |
| GOBP_TRA | 8 | -0.28418 | -0.00585 | 0.19053 | 0.93208 | 15850.02 |
| WP_ARYLA | 6 | -0.36051 | -0.00643 | 0.2417 | 0.93208 | 15850.02 |
| GOBP_KER | 79 | -0.10811 | -0.00698 | 0.072438 | 0.9322 | 15852.06 |
| GOBP_HIST | 9 | -0.25501 | -0.00557 | 0.17077 | 0.93232 | 15854.1 |
| GOBP_SIGI | 444 | -0.03585 | -0.00543 | 0.024008 | 0.93232 | 15854.1 |
| WANG_TAI | 43 | -0.11559 | -0.00551 | 0.077365 | 0.93242 | 15855.8 |
| GOCC_TRA | 234 | -0.04706 | -0.00521 | 0.031486 | 0.93247 | 15856.65 |
| REACTOME | 8 | -0.32794 | -0.00675 | 0.21938 | 0.93251 | 15857.33 |
| GOMF_RNA | 16 | -0.16294 | -0.00474 | 0.10896 | 0.93259 | 15858.69 |
| WP_GAMM | 6 | -0.28953 | -0.00516 | 0.19359 | 0.9326 | 15858.86 |
| GOMF_CYS | 92 | -0.07798 | -0.00543 | 0.052074 | 0.93285 | 15863.11 |
| GOBP_REG | 7 | -0.26569 | -0.00512 | 0.17741 | 0.93288 | 15863.62 |
| GOBP_NEG | 25 | -0.14743 | -0.00536 | 0.098389 | 0.93298 | 15865.32 |
| BIOCARTA_ | 14 | -0.21243 | -0.00578 | 0.14173 | 0.93303 | 15866.18 |
| WP_EFFECT | 8 | -0.26015 | -0.00535 | 0.17357 | 0.93303 | 15866.18 |
| KEGG_REG | 33 | -0.14865 | -0.00621 | 0.099119 | 0.93315 | 15868.22 |
| WP_CILIAR | 208 | -0.05245 | -0.00548 | 0.03497 | 0.93318 | 15868.73 |
| ABE_VEGFA | 18 | -0.18673 | -0.00576 | 0.12447 | 0.93321 | 15869.24 |
| BOYLAN_M | 28 | -0.15718 | -0.00605 | 0.10475 | 0.93325 | 15869.92 |
| STAMBOLS | 77 | -0.08879 | -0.00566 | 0.059159 | 0.93329 | 15870.6 |
| GOBP_CELI | 45 | -0.12258 | -0.00598 | 0.081665 | 0.93331 | 15870.94 |
| REACTOME | 15 | -0.20419 | -0.00575 | 0.13592 | 0.93347 | 15873.66 |
| GOBP_POS | 347 | -0.04233 | -0.00569 | 0.02818 | 0.93348 | 15873.83 |
| WP_GENES | 102 | -0.07293 | -0.00535 | 0.04852 | 0.93358 | 15875.53 |
| GOCC_SYN | 116 | -0.06924 | -0.00541 | 0.046059 | 0.93361 | 15876.04 |
| RIGGI_EWI | 408 | -0.04164 | -0.00606 | 0.027691 | 0.93367 | 15877.06 |
| GOBP_ACT | 14 | -0.19756 | -0.00538 | 0.13136 | 0.93368 | 15877.23 |
| GOMF_CHI | 38 | -0.15983 | -0.00716 | 0.10627 | 0.9337 | 15877.57 |
| GOBP_REG | 68 | -0.09883 | -0.00592 | 0.065705 | 0.93371 | 15877.74 |
| LIU_NASOI | 61 | -0.08388 | -0.00476 | 0.05577 | 0.93371 | 15877.74 |
| PID_ATR_P | 38 | -0.11134 | -0.00499 | 0.074019 | 0.93373 | 15878.08 |
| GOMF_MC | 114 | -0.07367 | -0.00571 | 0.048973 | 0.93374 | 15878.25 |
| REACTOME | 45 | -0.10921 | -0.00533 | 0.072587 | 0.93377 | 15878.76 |
| GOBP_REG | 5 | -0.28245 | -0.0046 | 0.18738 | 0.93414 | 15885.05 |
| GOBP_REG | 39 | -0.12907 | -0.00586 | 0.085589 | 0.93422 | 15886.41 |
| GOBP_COR | 39 | -0.1124 | -0.0051 | 0.074515 | 0.93427 | 15887.26 |
| GOBP_FC_ | 5 | -0.34816 | -0.00567 | 0.2308 | 0.93427 | 15887.26 |
| GOCC_PHA | 59 | -0.08893 | -0.00496 | 0.058955 | 0.93427 | 15887.26 |

| | | | | | | |
|---|---|---|---|---|---|---|
| GOMF_NU | 19 | -0.14662 | -0.00465 | 0.097161 | 0.93434 | 15888.45 |
| GOBP_CD4 | 20 | -0.19149 | -0.00623 | 0.12688 | 0.93437 | 15888.96 |
| GOBP_INTI | 9 | -0.19083 | -0.00417 | 0.12634 | 0.93453 | 15891.68 |
| GOCC_TRA | 29 | -0.14461 | -0.00566 | 0.095717 | 0.93457 | 15892.36 |
| GOBP_REG | 49 | -0.11217 | -0.00571 | 0.074221 | 0.93463 | 15893.38 |
| KATSANOU | 138 | -0.06523 | -0.00556 | 0.043124 | 0.9348 | 15896.27 |
| JINESH_BLI | 254 | -0.04586 | -0.00528 | 0.030316 | 0.93481 | 15896.44 |
| GOBP_PHC | 6 | -0.32447 | -0.00578 | 0.21448 | 0.93483 | 15896.78 |
| TURJANSKI | 9 | -0.27113 | -0.00592 | 0.17921 | 0.93484 | 15896.95 |
| GOBP_T_H | 13 | -0.21624 | -0.00567 | 0.1429 | 0.93487 | 15897.46 |
| GOBP_T_F( | 6 | -0.25998 | -0.00463 | 0.1716 | 0.9351 | 15901.38 |
| GOBP_PRC | 4 | -0.45179 | -0.00658 | 0.29816 | 0.93513 | 15901.89 |
| IGARASHI_ | 92 | -0.08261 | -0.00575 | 0.054514 | 0.93515 | 15902.23 |
| GOBP_POS | 4 | -0.29019 | -0.00422 | 0.1914 | 0.93525 | 15903.93 |
| GOBP_REG | 12 | -0.19476 | -0.00491 | 0.12833 | 0.93545 | 15907.33 |
| WU_HBX_T | 14 | -0.20599 | -0.00561 | 0.13571 | 0.93546 | 15907.5 |
| GOMF_RIB | 11 | -0.22505 | -0.00543 | 0.14818 | 0.93557 | 15909.37 |
| GOBP_REG | 21 | -0.16521 | -0.00551 | 0.10871 | 0.9357 | 15911.58 |
| GOMF_RNA | 9 | -0.22437 | -0.0049 | 0.1475 | 0.93588 | 15914.64 |
| GOMF_CY( | 8 | -0.28606 | -0.00589 | 0.18795 | 0.93599 | 15916.51 |
| SMID_BREA | 22 | -0.16604 | -0.00566 | 0.10907 | 0.93603 | 15917.19 |
| GOBP_WAT | 23 | -0.1669 | -0.00582 | 0.10962 | 0.93604 | 15917.36 |
| GOBP_SEN | 45 | -0.13074 | -0.00638 | 0.085833 | 0.93613 | 15918.89 |
| GOMF_BIC | 23 | -0.1498 | -0.00523 | 0.098317 | 0.93619 | 15919.91 |
| GOBP_POS | 23 | -0.17357 | -0.00605 | 0.11382 | 0.93636 | 15922.8 |
| LINDSTEDT | 61 | -0.09982 | -0.00567 | 0.0654 | 0.93653 | 15925.69 |
| BIOCARTA_ | 7 | -0.34436 | -0.00663 | 0.22555 | 0.93658 | 15926.54 |
| GOBP_PYR | 8 | -0.26363 | -0.00543 | 0.17266 | 0.9366 | 15926.88 |
| GOMF_ADI | 30 | -0.15237 | -0.00607 | 0.099785 | 0.93661 | 15927.05 |
| GOBP_PRC | 57 | -0.09974 | -0.00547 | 0.065288 | 0.9367 | 15928.58 |
| GOBP_MY( | 6 | -0.36402 | -0.00649 | 0.23826 | 0.93671 | 15928.75 |
| GOBP_INTI | 76 | -0.09114 | -0.00577 | 0.059594 | 0.9369 | 15931.98 |
| GOBP_REG | 326 | -0.04255 | -0.00554 | 0.027815 | 0.93695 | 15932.83 |
| GOBP_NEG | 12 | -0.23357 | -0.00589 | 0.15254 | 0.93713 | 15935.9 |
| LEIN_NEUF | 60 | -0.10144 | -0.00571 | 0.066241 | 0.93715 | 15936.24 |
| PID_THROI | 15 | -0.1961 | -0.00553 | 0.12797 | 0.93727 | 15938.28 |
| GOMF_UBI | 5 | -0.26456 | -0.00431 | 0.17256 | 0.93737 | 15939.98 |
| GOBP_POS | 144 | -0.06319 | -0.0055 | 0.041204 | 0.93741 | 15940.66 |
| GOBP_POS | 12 | -0.20636 | -0.0052 | 0.1345 | 0.93752 | 15942.53 |
| SHARMA_F | 7 | -0.3478 | -0.0067 | 0.2264 | 0.93775 | 15946.44 |
| BIOCARTA_ | 5 | -0.36015 | -0.00586 | 0.23426 | 0.9379 | 15948.99 |
| GOCC_SPEI | 6 | -0.34918 | -0.00622 | 0.227 | 0.93799 | 15950.52 |
| WP_TRANS | 63 | -0.09449 | -0.00545 | 0.061429 | 0.93799 | 15950.52 |
| GOBP_AUD | 22 | -0.17155 | -0.00585 | 0.11152 | 0.93801 | 15950.86 |
| GOBP_REG | 4 | -0.3227 | -0.0047 | 0.20977 | 0.93801 | 15950.86 |

| | | | | | | |
|---|---|---|---|---|---|---|
| GOBP_GLIA | 11 | -0.26881 | -0.00649 | 0.17459 | 0.93817 | 15953.58 |
| GOCC_CLU | 153 | -0.06252 | -0.00561 | 0.040602 | 0.9382 | 15954.09 |
| REACTOME | 8 | -0.25401 | -0.00523 | 0.16491 | 0.93824 | 15954.77 |
| MINGUEZ_ | 13 | -0.20575 | -0.0054 | 0.13354 | 0.9383 | 15955.79 |
| GOBP_NEG | 44 | -0.11439 | -0.00552 | 0.074196 | 0.93843 | 15958 |
| GOMF_ALP | 8 | -0.2487 | -0.00512 | 0.16118 | 0.93858 | 15960.55 |
| GOMF_IRO | 139 | -0.06989 | -0.00597 | 0.04528 | 0.93863 | 15961.4 |
| GOBP_GLU | 7 | -0.25812 | -0.00497 | 0.16714 | 0.93873 | 15963.1 |
| GOBP_RIBO | 8 | -0.20956 | -0.00431 | 0.13565 | 0.9388 | 15964.29 |
| GOBP_REG | 196 | -0.0576 | -0.00584 | 0.037242 | 0.93901 | 15967.87 |
| GOBP_MH | 20 | -0.17256 | -0.00561 | 0.11155 | 0.93905 | 15968.55 |
| GOMF_SUP | 4 | -0.27267 | -0.00397 | 0.17622 | 0.9391 | 15969.4 |
| GOBP_POS | 9 | -0.28484 | -0.00622 | 0.18398 | 0.93921 | 15971.27 |
| GOBP_NEG | 5 | -0.27146 | -0.00442 | 0.17511 | 0.93945 | 15975.35 |
| GOMF_LIG | 5 | -0.26819 | -0.00436 | 0.17296 | 0.9395 | 15976.2 |
| REACTOME | 15 | -0.20671 | -0.00582 | 0.13325 | 0.93958 | 15977.56 |
| GOBP_MYO | 8 | -0.30163 | -0.00621 | 0.19417 | 0.93983 | 15981.81 |
| GOBP_DOU | 6 | -0.28552 | -0.00509 | 0.18363 | 0.94001 | 15984.87 |
| GOBP_VES | 30 | -0.15907 | -0.00634 | 0.10229 | 0.94004 | 15985.38 |
| REACTOME | 62 | -0.09332 | -0.00534 | 0.059987 | 0.94009 | 15986.23 |
| GOBP_NEU | 7 | -0.34122 | -0.00657 | 0.21932 | 0.94011 | 15986.57 |
| GOBP_RES | 30 | -0.15969 | -0.00636 | 0.10256 | 0.94026 | 15989.12 |
| GOMF_IRO | 8 | -0.27278 | -0.00561 | 0.17518 | 0.94027 | 15989.29 |
| GOBP_SPIN | 11 | -0.26599 | -0.00642 | 0.17076 | 0.94033 | 15990.31 |
| GOBP_ANT | 19 | -0.15198 | -0.00482 | 0.097558 | 0.94035 | 15990.65 |
| GOMF_COI | 4 | -0.29884 | -0.00435 | 0.19181 | 0.94037 | 15990.99 |
| GOCC_COL | 4 | -0.44914 | -0.00654 | 0.2881 | 0.94049 | 15993.03 |
| GOMF_L_A | 9 | -0.25882 | -0.00565 | 0.16599 | 0.94052 | 15993.54 |
| GOMF_14_ | 33 | -0.13429 | -0.00561 | 0.0859 | 0.94101 | 16001.88 |
| GOMF_HIS | 8 | -0.27326 | -0.00562 | 0.17474 | 0.94106 | 16002.73 |
| GOCC_DYN | 53 | -0.11167 | -0.00591 | 0.071202 | 0.94158 | 16011.57 |
| REACTOME | 16 | -0.2152 | -0.00626 | 0.13716 | 0.94166 | 16012.93 |
| GOBP_CELI | 4 | -0.33156 | -0.00483 | 0.21131 | 0.94168 | 16013.27 |
| REACTOME | 131 | -0.06758 | -0.00561 | 0.043072 | 0.94168 | 16013.27 |
| GOBP_ANT | 30 | -0.13152 | -0.00524 | 0.083797 | 0.94173 | 16014.12 |
| GOBP_NEG | 17 | -0.20284 | -0.00608 | 0.12905 | 0.94199 | 16018.54 |
| GOBP_PRO | 144 | -0.05928 | -0.00516 | 0.037674 | 0.94219 | 16021.94 |
| GOBP_CELI | 10 | -0.22363 | -0.00515 | 0.1421 | 0.94222 | 16022.45 |
| GOBP_EXP | 821 | -0.02823 | -0.00576 | 0.017934 | 0.94223 | 16022.62 |
| GOBP_CAL | 9 | -0.25459 | -0.00556 | 0.16171 | 0.94229 | 16023.64 |
| GOMF_SEC | 242 | -0.05051 | -0.00568 | 0.032074 | 0.94233 | 16024.32 |
| GOBP_REG | 118 | -0.07139 | -0.00563 | 0.045328 | 0.94236 | 16024.83 |
| KEGG_FC_ | 92 | -0.0849 | -0.00591 | 0.053898 | 0.94238 | 16025.17 |
| GOCC_APIC | 139 | -0.06649 | -0.00568 | 0.042202 | 0.94242 | 16025.85 |
| GOBP_REG | 11 | -0.21464 | -0.00518 | 0.13613 | 0.94255 | 16028.06 |

| | | | | | | |
|---|---|---|---|---|---|---|
| BROWNE_I | 48 | -0.12341 | -0.00622 | 0.078215 | 0.94269 | 16030.44 |
| GOBP_RES | 10 | -0.24447 | -0.00563 | 0.15476 | 0.94289 | 16033.84 |
| GOBP_CAT | 48 | -0.12473 | -0.00628 | 0.078947 | 0.94292 | 16034.35 |
| GOMF_CYS | 5 | -0.26215 | -0.00427 | 0.16592 | 0.94293 | 16034.52 |
| GOBP_REG | 45 | -0.10546 | -0.00514 | 0.066715 | 0.94302 | 16036.06 |
| GOBP_URC | 23 | -0.19832 | -0.00692 | 0.12537 | 0.94315 | 16038.27 |
| GOBP_LOB | 6 | -0.33977 | -0.00606 | 0.2146 | 0.9433 | 16040.82 |
| GOCC_DEN | 7 | -0.31057 | -0.00598 | 0.19615 | 0.94332 | 16041.16 |
| GOBP_RES | 113 | -0.07145 | -0.00551 | 0.045118 | 0.94335 | 16041.67 |
| CERIBELLI_ | 35 | -0.1398 | -0.00601 | 0.088267 | 0.94337 | 16042.01 |
| WP_CELL_I | 19 | -0.17936 | -0.00569 | 0.11318 | 0.94348 | 16043.88 |
| GOBP_DEC | 6 | -0.38697 | -0.0069 | 0.24415 | 0.9435 | 16044.22 |
| GOBP_TRN | 13 | -0.218 | -0.00572 | 0.13749 | 0.94358 | 16045.58 |
| GOBP_SYN | 106 | -0.082 | -0.00613 | 0.051711 | 0.9436 | 16045.92 |
| FIGUEROA_ | 44 | -0.14285 | -0.00689 | 0.090074 | 0.94361 | 16046.09 |
| SHAFFER_I | 64 | -0.10569 | -0.00614 | 0.066628 | 0.94365 | 16046.77 |
| GOBP_THE | 8 | -0.26934 | -0.00554 | 0.16975 | 0.9437 | 16047.62 |
| GOMF_GR( | 9 | -0.23471 | -0.00512 | 0.14792 | 0.94371 | 16047.79 |
| REACTOME | 83 | -0.08747 | -0.00579 | 0.055117 | 0.94374 | 16048.3 |
| GOMF_PR( | 6 | -0.2734 | -0.00487 | 0.17226 | 0.94375 | 16048.47 |
| GOBP_REG | 378 | -0.04469 | -0.00626 | 0.028157 | 0.94376 | 16048.64 |
| REACTOME | 13 | -0.20176 | -0.00529 | 0.12711 | 0.94377 | 16048.81 |
| GOCC_PAR | 6 | -0.3427 | -0.00611 | 0.21587 | 0.94379 | 16049.15 |
| BAE_BRCA: | 65 | -0.09216 | -0.0054 | 0.058054 | 0.9438 | 16049.32 |
| GOBP_REG | 155 | -0.06326 | -0.00571 | 0.039825 | 0.9439 | 16051.02 |
| GOBP_HIST | 431 | -0.03668 | -0.00548 | 0.023076 | 0.944 | 16052.72 |
| GOCC_PIP_ | 5 | -0.34534 | -0.00562 | 0.21724 | 0.94403 | 16053.23 |
| GOBP_NEG | 23 | -0.16425 | -0.00573 | 0.1033 | 0.94407 | 16053.91 |
| KEGG_LEIS | 54 | -0.11087 | -0.00592 | 0.069703 | 0.94414 | 16055.1 |
| WEBER_MI | 13 | -0.21478 | -0.00563 | 0.13503 | 0.94414 | 16055.1 |
| GOMF_GLU | 26 | -0.16916 | -0.00627 | 0.10633 | 0.94418 | 16055.78 |
| GOBP_NAD | 13 | -0.19549 | -0.00513 | 0.12284 | 0.94424 | 16056.8 |
| GOBP_RNA | 5 | -0.30193 | -0.00491 | 0.18968 | 0.94428 | 16057.48 |
| REACTOME | 40 | -0.13068 | -0.00601 | 0.082057 | 0.94436 | 16058.84 |
| BIOCARTA_ | 6 | -0.31683 | -0.00565 | 0.19875 | 0.94453 | 16061.73 |
| SUMI_HNF | 29 | -0.14521 | -0.00569 | 0.091088 | 0.94454 | 16061.9 |
| GOBP_REG | 217 | -0.05769 | -0.00615 | 0.036155 | 0.94469 | 16064.45 |
| GOBP_G_P | 1080 | -0.02826 | -0.00656 | 0.017701 | 0.94479 | 16066.15 |
| REACTOME | 50 | -0.10963 | -0.00563 | 0.068623 | 0.94492 | 16068.36 |
| GOCC_NAT | 4 | -0.65282 | -0.0095 | 0.4085 | 0.94498 | 16069.38 |
| GOBP_VAC | 5 | -0.33542 | -0.00546 | 0.20979 | 0.94506 | 16070.75 |
| BROWNE_I | 207 | -0.05574 | -0.0058 | 0.034856 | 0.9451 | 16071.43 |
| GOBP_FC_ | 32 | -0.15047 | -0.00619 | 0.094019 | 0.94523 | 16073.64 |
| GOBP_GUA | 4 | -0.50105 | -0.00729 | 0.31304 | 0.94526 | 16074.15 |
| WP_NICOT | 4 | -0.45211 | -0.00658 | 0.2824 | 0.9453 | 16074.83 |

| | | | | | | |
|---|---|---|---|---|---|---|
| GOBP_DNA | 4 | -0.30231 | -0.0044 | 0.18878 | 0.94534 | 16075.51 |
| TERAO_AO | 25 | -0.16598 | -0.00604 | 0.10363 | 0.94537 | 16076.02 |
| GOMF_DYI | 35 | -0.14452 | -0.00622 | 0.090094 | 0.94565 | 16080.78 |
| REACTOME | 19 | -0.19879 | -0.0063 | 0.12383 | 0.94578 | 16082.99 |
| GOCC_EUK | 4 | -0.57592 | -0.00838 | 0.35861 | 0.94585 | 16084.18 |
| KEGG_PYRI | 38 | -0.13636 | -0.00611 | 0.084893 | 0.94589 | 16084.86 |
| GOBP_REG | 44 | -0.12153 | -0.00586 | 0.075642 | 0.94593 | 16085.54 |
| GOCC_CLA | 45 | -0.11019 | -0.00537 | 0.068567 | 0.94597 | 16086.22 |
| REACTOME | 9 | -0.30268 | -0.00661 | 0.18833 | 0.94598 | 16086.39 |
| GOBP_PRC | 226 | -0.05154 | -0.00561 | 0.032063 | 0.94602 | 16087.07 |
| REACTOME | 22 | -0.15374 | -0.00525 | 0.095635 | 0.94602 | 16087.07 |
| DACOSTA_ | 450 | -0.03984 | -0.00608 | 0.024766 | 0.94616 | 16089.45 |
| WP_GLUCU | 23 | -0.2243 | -0.00782 | 0.1392 | 0.94644 | 16094.21 |
| GOMF_ACE | 16 | -0.23309 | -0.00678 | 0.14445 | 0.94668 | 16098.29 |
| GOBP_POS | 17 | -0.17394 | -0.00522 | 0.10772 | 0.94681 | 16100.5 |
| GOCC_CER | 7 | -0.33922 | -0.00653 | 0.21005 | 0.94683 | 16100.84 |
| GOBP_INN | 14 | -0.19989 | -0.00544 | 0.12371 | 0.94692 | 16102.37 |
| GOBP_RES | 6 | -0.41077 | -0.00732 | 0.25413 | 0.94698 | 16103.39 |
| WALLACE_ | 19 | -0.18448 | -0.00585 | 0.11399 | 0.94721 | 16107.31 |
| REACTOME | 5 | -0.41691 | -0.00678 | 0.25755 | 0.94724 | 16107.82 |
| REACTOME | 17 | -0.20066 | -0.00602 | 0.12389 | 0.94734 | 16109.52 |
| MARSON_I | 17 | -0.1826 | -0.00548 | 0.1127 | 0.94739 | 16110.37 |
| GOMF_LYS | 7 | -0.31814 | -0.00613 | 0.19626 | 0.94748 | 16111.9 |
| GOBP_SKIN | 12 | -0.23679 | -0.00597 | 0.146 | 0.94757 | 16113.43 |
| REACTOME | 17 | -0.20019 | -0.006 | 0.12339 | 0.94763 | 16114.45 |
| GOBP_CDP | 13 | -0.24989 | -0.00656 | 0.154 | 0.94766 | 16114.96 |
| GOBP_MA( | 16 | -0.21582 | -0.00628 | 0.13299 | 0.94767 | 16115.13 |
| GOMF_CLA | 7 | -0.23708 | -0.00456 | 0.14608 | 0.94769 | 16115.47 |
| ASTIER_INT | 53 | -0.11728 | -0.00621 | 0.072206 | 0.94782 | 16117.68 |
| REACTOME | 72 | -0.10092 | -0.00622 | 0.062081 | 0.94798 | 16120.4 |
| VERHAAK_ | 168 | -0.06611 | -0.00621 | 0.040664 | 0.94798 | 16120.4 |
| WP_TCA_C | 15 | -0.20318 | -0.00573 | 0.12493 | 0.94806 | 16121.76 |
| HAEGERSTI | 9 | -0.31004 | -0.00677 | 0.19054 | 0.94814 | 16123.12 |
| GOBP_PHC | 22 | -0.16512 | -0.00563 | 0.10139 | 0.94828 | 16125.5 |
| LIN_NPAS4 | 68 | -0.1037 | -0.00621 | 0.063658 | 0.94834 | 16126.52 |
| GOBP_REG | 6 | -0.37052 | -0.0066 | 0.22703 | 0.94865 | 16131.79 |
| REACTOME | 15 | -0.20019 | -0.00564 | 0.12262 | 0.94872 | 16132.98 |
| GOBP_POS | 7 | -0.3178 | -0.00612 | 0.19456 | 0.9488 | 16134.34 |
| DAZARD_U | 27 | -0.17215 | -0.00651 | 0.10535 | 0.94887 | 16135.53 |
| GOBP_REG | 45 | -0.12739 | -0.00621 | 0.07795 | 0.94888 | 16135.7 |
| GOBP_REG | 26 | -0.17015 | -0.00631 | 0.1041 | 0.94891 | 16136.21 |
| REACTOME | 10 | -0.25257 | -0.00581 | 0.15451 | 0.94893 | 16136.55 |
| GOCC_END | 34 | -0.12923 | -0.00548 | 0.079022 | 0.94901 | 16137.92 |
| DARWICHE | 72 | -0.0913 | -0.00563 | 0.055801 | 0.94909 | 16139.28 |
| GOBP_REG | 298 | -0.05157 | -0.00643 | 0.031518 | 0.9491 | 16139.45 |

| | | | | | | |
|---|---|---|---|---|---|---|
| GOBP_POS | 27 | -0.16621 | -0.00628 | 0.10156 | 0.94913 | 16139.96 |
| GOBP_PHC | 36 | -0.13083 | -0.00571 | 0.07992 | 0.94918 | 16140.81 |
| GOCC_GOL | 100 | -0.08056 | -0.00585 | 0.049211 | 0.94918 | 16140.81 |
| REACTOME | 22 | -0.22355 | -0.00763 | 0.13643 | 0.94934 | 16143.53 |
| GOBP_SEN | 205 | -0.05911 | -0.00613 | 0.036067 | 0.94938 | 16144.21 |
| REACTOME | 18 | -0.19523 | -0.00603 | 0.11896 | 0.94961 | 16148.12 |
| GOBP_POS | 58 | -0.10595 | -0.00586 | 0.064455 | 0.94988 | 16152.71 |
| GOBP_POS | 18 | -0.19113 | -0.0059 | 0.11626 | 0.9499 | 16153.05 |
| GOMF_BIT | 24 | -0.2545 | -0.00907 | 0.15478 | 0.94994 | 16153.73 |
| GOBP_RES | 103 | -0.08387 | -0.00618 | 0.050977 | 0.95003 | 16155.26 |
| RAO_BOUN | 178 | -0.06343 | -0.00613 | 0.038545 | 0.95006 | 16155.77 |
| PID_ECADH | 38 | -0.12422 | -0.00557 | 0.075399 | 0.95027 | 16159.34 |
| GOBP_ADU | 137 | -0.07238 | -0.00614 | 0.043901 | 0.95039 | 16161.38 |
| ZHAN_LATI | 17 | -0.18345 | -0.0055 | 0.11121 | 0.95047 | 16162.74 |
| WP_GASTF | 27 | -0.14439 | -0.00546 | 0.087513 | 0.95051 | 16163.42 |
| GOMF_SUL | 6 | -0.37296 | -0.00665 | 0.22602 | 0.95053 | 16163.76 |
| GOCC_CUL | 8 | -0.28292 | -0.00582 | 0.1712 | 0.95079 | 16168.18 |
| GOBP_DOF | 19 | -0.21826 | -0.00692 | 0.13204 | 0.95082 | 16168.69 |
| BORCZUK_ | 87 | -0.08666 | -0.00587 | 0.052386 | 0.95095 | 16170.9 |
| GOBP_RES | 17 | -0.20633 | -0.00619 | 0.12471 | 0.95098 | 16171.41 |
| GOBP_REG | 17 | -0.18683 | -0.0056 | 0.11289 | 0.95103 | 16172.27 |
| GOBP_MES | 6 | -0.41846 | -0.00746 | 0.25281 | 0.95105 | 16172.61 |
| GOBP_URE | 9 | -0.3201 | -0.00699 | 0.19327 | 0.95116 | 16174.48 |
| GOBP_REG | 5 | -0.4267 | -0.00694 | 0.25761 | 0.95117 | 16174.65 |
| GOBP_FLA\ | 6 | -0.3508 | -0.00625 | 0.21167 | 0.95126 | 16176.18 |
| GOCC_TER | 67 | -0.10561 | -0.00628 | 0.063698 | 0.95134 | 16177.54 |
| GOBP_DEN | 25 | -0.17291 | -0.00629 | 0.10424 | 0.95142 | 16178.9 |
| GOBP_POS | 6 | -0.35979 | -0.00641 | 0.21684 | 0.95146 | 16179.58 |
| GOCC_INTI | 8 | -0.2886 | -0.00594 | 0.17392 | 0.95147 | 16179.75 |
| GOBP_BIO | 86 | -0.09303 | -0.00626 | 0.056049 | 0.95151 | 16180.43 |
| WP_FTO_C | 8 | -0.37081 | -0.00763 | 0.22333 | 0.95157 | 16181.45 |
| GOBP_REG | 65 | -0.09669 | -0.00566 | 0.058203 | 0.95165 | 16182.81 |
| GOBP_SYN | 26 | -0.15116 | -0.00561 | 0.090924 | 0.95178 | 16185.02 |
| GREENBAU | 18 | -0.18078 | -0.00558 | 0.10869 | 0.95185 | 16186.21 |
| WP_NCRN/ | 13 | -0.21411 | -0.00562 | 0.12874 | 0.95185 | 16186.21 |
| GOBP_SYN | 62 | -0.1118 | -0.0064 | 0.067214 | 0.95187 | 16186.55 |
| GOBP_BRA | 14 | -0.21174 | -0.00576 | 0.12722 | 0.95197 | 16188.25 |
| GOMF_SIA | 21 | -0.22577 | -0.00753 | 0.13565 | 0.95197 | 16188.25 |
| CAIRO_PM | 22 | -0.17154 | -0.00585 | 0.10304 | 0.95201 | 16188.93 |
| GOMF_GLY | 7 | -0.36012 | -0.00693 | 0.21631 | 0.95201 | 16188.93 |
| REACTOME | 11 | -0.28818 | -0.00695 | 0.17304 | 0.95207 | 16189.95 |
| GOMF_CYS | 38 | -0.13941 | -0.00625 | 0.083679 | 0.95213 | 16190.97 |
| GOMF_BR/ | 10 | -0.24927 | -0.00574 | 0.14955 | 0.95221 | 16192.33 |
| GOBP_ACII | 44 | -0.12365 | -0.00596 | 0.074177 | 0.95223 | 16192.67 |
| REACTOME | 33 | -0.13064 | -0.00546 | 0.07835 | 0.95227 | 16193.35 |

| | | | | | | |
|---|---|---|---|---|---|---|
| WHITEHUR | 34 | -0.14277 | -0.00605 | 0.085573 | 0.95238 | 16195.22 |
| ZHENG_RE | 15 | -0.22244 | -0.00627 | 0.13331 | 0.95239 | 16195.39 |
| ZHAN_EAR | 6 | -0.42638 | -0.0076 | 0.25547 | 0.95243 | 16196.07 |
| REACTOME | 29 | -0.14937 | -0.00585 | 0.089485 | 0.95246 | 16196.58 |
| BIOCARTA_ | 24 | -0.17008 | -0.00606 | 0.10188 | 0.95247 | 16196.75 |
| WP_SULFA | 14 | -0.25592 | -0.00697 | 0.15326 | 0.95251 | 16197.43 |
| GOCC_NUC | 10 | -0.24282 | -0.00559 | 0.14541 | 0.95253 | 16197.77 |
| GOBP_10_ | 6 | -0.29067 | -0.00518 | 0.17403 | 0.95255 | 16198.11 |
| ZHAN_V1_ | 30 | -0.15222 | -0.00606 | 0.091082 | 0.95266 | 16199.98 |
| GOCC_NAD | 40 | -0.12923 | -0.00594 | 0.077298 | 0.95271 | 16200.83 |
| GOBP_MYC | 6 | -0.35859 | -0.00639 | 0.21441 | 0.95277 | 16201.85 |
| REACTOME | 102 | -0.08126 | -0.00596 | 0.04857 | 0.95283 | 16202.87 |
| REACTOME | 31 | -0.12336 | -0.00499 | 0.073716 | 0.95286 | 16203.38 |
| REACTOME | 39 | -0.14542 | -0.0066 | 0.086837 | 0.95299 | 16205.59 |
| GOMF_MA | 9 | -0.27137 | -0.00592 | 0.16201 | 0.95302 | 16206.11 |
| HESS_TARC | 81 | -0.09297 | -0.00608 | 0.055485 | 0.95309 | 16207.3 |
| KEGG_PEN | 25 | -0.19484 | -0.00709 | 0.11624 | 0.95315 | 16208.32 |
| WP_CEREB | 8 | -0.28655 | -0.0059 | 0.17086 | 0.95323 | 16209.68 |
| GOBP_COR | 14 | -0.18877 | -0.00514 | 0.11248 | 0.95333 | 16211.38 |
| GOMF_UD | 136 | -0.07349 | -0.00622 | 0.043734 | 0.95355 | 16215.12 |
| GOBP_SER | 5 | -0.3907 | -0.00636 | 0.23223 | 0.95374 | 16218.35 |
| GOCC_CCR | 14 | -0.20951 | -0.0057 | 0.12452 | 0.95376 | 16218.69 |
| GOBP_END | 12 | -0.24375 | -0.00614 | 0.1448 | 0.95384 | 16220.05 |
| GOCC_RIBO | 9 | -0.24199 | -0.00528 | 0.14368 | 0.95392 | 16221.41 |
| GOBP_REG | 458 | -0.04162 | -0.0064 | 0.024703 | 0.95396 | 16222.09 |
| GOBP_POS | 43 | -0.14397 | -0.00686 | 0.085457 | 0.95397 | 16222.26 |
| GOMF_PRO | 16 | -0.20642 | -0.00601 | 0.12252 | 0.95398 | 16222.43 |
| GOBP_PHA | 220 | -0.05819 | -0.00624 | 0.03452 | 0.95406 | 16223.79 |
| LIU_PROST | 89 | -0.08939 | -0.00612 | 0.05301 | 0.95412 | 16224.81 |
| REACTOME | 19 | -0.18753 | -0.00595 | 0.11117 | 0.95418 | 16225.83 |
| GOBP_AUD | 18 | -0.20182 | -0.00623 | 0.11962 | 0.9542 | 16226.17 |
| GOBP_REG | 58 | -0.10315 | -0.00571 | 0.061134 | 0.95422 | 16226.51 |
| BIOCARTA_ | 6 | -0.42487 | -0.00757 | 0.25175 | 0.95425 | 16227.02 |
| GOBP_REG | 11 | -0.24994 | -0.00603 | 0.14807 | 0.95428 | 16227.53 |
| GOBP_CAL | 5 | -0.41613 | -0.00677 | 0.24651 | 0.95429 | 16227.7 |
| GOBP_PHA | 27 | -0.17572 | -0.00664 | 0.10399 | 0.95445 | 16230.42 |
| GOMF_COI | 4 | -0.28352 | -0.00413 | 0.16764 | 0.95459 | 16232.8 |
| GOMF_O_I | 5 | -0.38329 | -0.00624 | 0.22662 | 0.9546 | 16232.97 |
| GOBP_POS | 11 | -0.27768 | -0.0067 | 0.16415 | 0.95463 | 16233.48 |
| CHEOK_RES | 23 | -0.16989 | -0.00593 | 0.10042 | 0.95466 | 16233.99 |
| GOMF_MC | 85 | -0.08527 | -0.00571 | 0.050339 | 0.95484 | 16237.05 |
| GOBP_BEH | 9 | -0.28357 | -0.00619 | 0.1673 | 0.95495 | 16238.92 |
| GOBP_RIGI | 8 | -0.29681 | -0.00611 | 0.17492 | 0.95512 | 16241.82 |
| GOMF_NAI | 37 | -0.12962 | -0.00573 | 0.076361 | 0.95518 | 16242.84 |
| GOBP_REG | 5 | -0.41666 | -0.00678 | 0.24533 | 0.95527 | 16244.37 |

| | | | | | | |
|---|---|---|---|---|---|---|
| STREICHER | 40 | -0.14418 | -0.00663 | 0.084893 | 0.95528 | 16244.54 |
| GOBP_NEG | 30 | -0.16253 | -0.00647 | 0.095666 | 0.95533 | 16245.39 |
| GOBP_REG | 10 | -0.29688 | -0.00683 | 0.17464 | 0.95543 | 16247.09 |
| REACTOME | 22 | -0.16378 | -0.00559 | 0.096265 | 0.95555 | 16249.13 |
| REACTOME | 28 | -0.12811 | -0.00493 | 0.07529 | 0.95557 | 16249.47 |
| SENGUPTA | 332 | -0.04582 | -0.00602 | 0.02689 | 0.9558 | 16253.38 |
| GOBP_BICA | 29 | -0.14349 | -0.00562 | 0.084169 | 0.95588 | 16254.74 |
| GOMF_ALP | 6 | -0.42957 | -0.00766 | 0.25196 | 0.95588 | 16254.74 |
| GOCC_PHA | 119 | -0.07047 | -0.00558 | 0.04133 | 0.9559 | 16255.08 |
| WP_COPPE | 49 | -0.11029 | -0.00561 | 0.064682 | 0.95591 | 16255.25 |
| GOBP_SYN | 18 | -0.17874 | -0.00552 | 0.10456 | 0.9563 | 16261.88 |
| GOBP_TRN | 15 | -0.22196 | -0.00625 | 0.12976 | 0.95641 | 16263.75 |
| GOBP_POS | 8 | -0.35487 | -0.0073 | 0.20744 | 0.95642 | 16263.92 |
| REACTOME | 47 | -0.13821 | -0.00689 | 0.080784 | 0.95643 | 16264.09 |
| GOBP_REG | 17 | -0.21461 | -0.00644 | 0.12539 | 0.9565 | 16265.28 |
| WP_IRON_ | 12 | -0.22335 | -0.00563 | 0.13049 | 0.95651 | 16265.45 |
| GOMF_ADI | 31 | -0.15175 | -0.00614 | 0.088469 | 0.95685 | 16271.23 |
| REACTOME | 22 | -0.19869 | -0.00678 | 0.11581 | 0.95687 | 16271.57 |
| GOMF_PEF | 21 | -0.19757 | -0.00659 | 0.11516 | 0.95688 | 16271.74 |
| GOBP_HYP | 12 | -0.22546 | -0.00568 | 0.13131 | 0.957 | 16273.79 |
| GOMF_GLU | 7 | -0.34265 | -0.0066 | 0.19957 | 0.957 | 16273.79 |
| GOBP_TAU | 7 | -0.40703 | -0.00784 | 0.23704 | 0.95701 | 16273.96 |
| GOMF_TAS | 28 | -0.22995 | -0.00885 | 0.1338 | 0.95715 | 16276.34 |
| REACTOME | 16 | -0.21391 | -0.00622 | 0.12445 | 0.95717 | 16276.68 |
| GOBP_CELI | 26 | -0.16132 | -0.00598 | 0.093849 | 0.95718 | 16276.85 |
| GOBP_N_T | 16 | -0.2077 | -0.00604 | 0.12081 | 0.95721 | 16277.36 |
| REACTOME | 988 | -0.02851 | -0.00635 | 0.016576 | 0.95728 | 16278.55 |
| GOBP_SEN | 170 | -0.06945 | -0.00656 | 0.040369 | 0.95731 | 16279.06 |
| GOBP_URII | 10 | -0.29072 | -0.00669 | 0.1689 | 0.95739 | 16280.42 |
| GOMF_SOI | 21 | -0.17462 | -0.00582 | 0.10138 | 0.95749 | 16282.12 |
| GOBP_POS | 132 | -0.08054 | -0.00671 | 0.046758 | 0.95751 | 16282.46 |
| GOCC_RIBO | 12 | -0.24021 | -0.00605 | 0.13942 | 0.95754 | 16282.97 |
| GOMF_NU | 30 | -0.145 | -0.00578 | 0.084162 | 0.95754 | 16282.97 |
| GOCC_MU | 9 | -0.26312 | -0.00574 | 0.15261 | 0.95765 | 16284.84 |
| GOMF_RIB | 9 | -0.26312 | -0.00574 | 0.15261 | 0.95765 | 16284.84 |
| REACTOME | 33 | -0.146 | -0.0061 | 0.084581 | 0.95783 | 16287.9 |
| GOBP_REG | 83 | -0.09495 | -0.00628 | 0.055004 | 0.95784 | 16288.07 |
| REACTOME | 11 | -0.25585 | -0.00617 | 0.14811 | 0.95795 | 16289.94 |
| GOBP_MEI | 54 | -0.11954 | -0.00638 | 0.069166 | 0.95802 | 16291.13 |
| GOMF_NU | 17 | -0.23938 | -0.00718 | 0.1385 | 0.95802 | 16291.13 |
| MARIADAS | 6 | -0.31475 | -0.00561 | 0.18208 | 0.95805 | 16291.64 |
| GOBP_MRI | 47 | -0.12164 | -0.00606 | 0.070332 | 0.95813 | 16293 |
| GOBP_REG | 242 | -0.05786 | -0.00651 | 0.033449 | 0.95817 | 16293.68 |
| GOBP_NEG | 7 | -0.35557 | -0.00685 | 0.20544 | 0.95824 | 16294.87 |
| GOBP_NEG | 3 | -0.39115 | -0.00493 | 0.22599 | 0.95825 | 16295.04 |

| | | | | | | |
|---|---|---|---|---|---|---|
| GOMF_4_I | 40 | -0.12779 | -0.00588 | 0.073826 | 0.95827 | 16295.38 |
| GOBP_POS | 10 | -0.3396 | -0.00781 | 0.19608 | 0.95835 | 16296.74 |
| GOBP_LOB | 5 | -0.39642 | -0.00645 | 0.22867 | 0.95849 | 16299.12 |
| GOMF_RNA | 5 | -0.3365 | -0.00548 | 0.1941 | 0.9585 | 16299.29 |
| GOMF_TEL | 5 | -0.3365 | -0.00548 | 0.1941 | 0.9585 | 16299.29 |
| GOBP_NEG | 12 | -0.21806 | -0.0055 | 0.12577 | 0.95852 | 16299.63 |
| REACTOME | 295 | -0.05538 | -0.00687 | 0.031916 | 0.95864 | 16301.67 |
| GOMF_OPS | 19 | -0.22944 | -0.00728 | 0.13215 | 0.95873 | 16303.2 |
| GOBP_POS | 18 | -0.21245 | -0.00656 | 0.12233 | 0.95877 | 16303.88 |
| GOCC_SCF_ | 62 | -0.11052 | -0.00632 | 0.063602 | 0.95885 | 16305.24 |
| GOMF_OXY | 5 | -0.31767 | -0.00517 | 0.18268 | 0.95897 | 16307.28 |
| GOMF_OXI | 4 | -0.43644 | -0.00635 | 0.25079 | 0.95908 | 16309.16 |
| GOMF_ORG | 49 | -0.12208 | -0.00621 | 0.070046 | 0.95931 | 16313.07 |
| GOBP_SYN | 39 | -0.12356 | -0.00561 | 0.070889 | 0.95932 | 16313.24 |
| GOMF_CHA | 14 | -0.25544 | -0.00695 | 0.14647 | 0.95941 | 16314.77 |
| GOMF_ME | 10 | -0.24346 | -0.0056 | 0.13957 | 0.95944 | 16315.28 |
| GOMF_STC | 8 | -0.31162 | -0.00641 | 0.1786 | 0.95948 | 16315.96 |
| GOBP_SCF_ | 45 | -0.13283 | -0.00648 | 0.076069 | 0.9596 | 16318 |
| GOMF_ARG | 8 | -0.28808 | -0.00593 | 0.16495 | 0.95962 | 16318.34 |
| REACTOME | 38 | -0.14042 | -0.00629 | 0.080386 | 0.95965 | 16318.85 |
| REACTOME | 33 | -0.14246 | -0.00595 | 0.081507 | 0.95974 | 16320.38 |
| GOBP_PRC | 236 | -0.05833 | -0.00648 | 0.033354 | 0.95982 | 16321.74 |
| GOBP_ACT | 52 | -0.11481 | -0.00602 | 0.06564 | 0.95986 | 16322.42 |
| GOBP_NEG | 23 | -0.1635 | -0.0057 | 0.093475 | 0.95986 | 16322.42 |
| GOBP_POS | 5 | -0.4341 | -0.00706 | 0.24815 | 0.95987 | 16322.59 |
| GOBP_IMM | 9 | -0.26922 | -0.00588 | 0.15377 | 0.96 | 16324.8 |
| KYNG_ENV | 11 | -0.27481 | -0.00663 | 0.15681 | 0.96015 | 16327.35 |
| GOBP_NEG | 32 | -0.16868 | -0.00694 | 0.09624 | 0.96016 | 16327.52 |
| GOBP_CELI | 13 | -0.26457 | -0.00694 | 0.15086 | 0.96025 | 16329.05 |
| REACTOME | 6 | -0.43757 | -0.0078 | 0.24949 | 0.96027 | 16329.39 |
| GOMF_BEN | 9 | -0.36495 | -0.00797 | 0.20807 | 0.96028 | 16329.56 |
| GOMF_RNA | 10 | -0.23745 | -0.00546 | 0.13535 | 0.9603 | 16329.9 |
| NOUSHME | 50 | -0.12547 | -0.00645 | 0.071524 | 0.9603 | 16329.9 |
| GNATENKC | 33 | -0.13596 | -0.00568 | 0.077482 | 0.96033 | 16330.41 |
| GOBP_FC_( | 33 | -0.16385 | -0.00684 | 0.093382 | 0.96033 | 16330.41 |
| GOCC_N_T | 9 | -0.29327 | -0.0064 | 0.16698 | 0.96047 | 16332.79 |
| GOBP_POS | 11 | -0.25107 | -0.00606 | 0.14292 | 0.96051 | 16333.47 |
| VILLANUEV | 164 | -0.06814 | -0.00632 | 0.038789 | 0.96051 | 16333.47 |
| GOBP_MIT | 4 | -0.37337 | -0.00543 | 0.21239 | 0.96061 | 16335.17 |
| EHRLICH_I( | 12 | -0.24509 | -0.00618 | 0.1394 | 0.96063 | 16335.51 |
| GOCC_EXT | 77 | -0.10287 | -0.00656 | 0.058502 | 0.96065 | 16335.85 |
| FLECHNER_ | 56 | -0.11011 | -0.00599 | 0.06261 | 0.96068 | 16336.36 |
| WP_PEROX | 8 | -0.31296 | -0.00644 | 0.17789 | 0.96072 | 16337.04 |
| GOBP_FLA\ | 11 | -0.25331 | -0.00611 | 0.14398 | 0.96073 | 16337.21 |
| GOMF_SOI | 34 | -0.17897 | -0.00759 | 0.10172 | 0.96074 | 16337.38 |

| | | | | | | |
|---|---|---|---|---|---|---|
| GOBP_REG | 16 | -0.22486 | -0.00654 | 0.12776 | 0.96078 | 16338.06 |
| GOBP_NEG | 7 | -0.29284 | -0.00564 | 0.16625 | 0.96091 | 16340.27 |
| GOBP_REG | 8 | -0.43383 | -0.00893 | 0.24614 | 0.961 | 16341.81 |
| REACTOME | 212 | -0.09306 | -0.00981 | 0.052782 | 0.96104 | 16342.49 |
| GOBP_POS | 285 | -0.05428 | -0.00662 | 0.030778 | 0.9611 | 16343.51 |
| REACTOME | 155 | -0.07062 | -0.00637 | 0.040024 | 0.96115 | 16344.36 |
| GOMF_ME | 35 | -0.14288 | -0.00615 | 0.080933 | 0.96124 | 16345.89 |
| GOBP_POS | 238 | -0.05885 | -0.00657 | 0.033319 | 0.96131 | 16347.08 |
| GOBP_CELI | 317 | -0.0491 | -0.00631 | 0.027771 | 0.96146 | 16349.63 |
| GOBP_POS | 15 | -0.22125 | -0.00623 | 0.12509 | 0.96152 | 16350.65 |
| GOMF_PH( | 25 | -0.16437 | -0.00598 | 0.092921 | 0.96154 | 16350.99 |
| GOBP_POS | 39 | -0.14899 | -0.00676 | 0.084215 | 0.96155 | 16351.16 |
| GOCC_RNA | 18 | -0.18429 | -0.00569 | 0.10408 | 0.96168 | 16353.37 |
| REACTOME | 18 | -0.18429 | -0.00569 | 0.10408 | 0.96168 | 16353.37 |
| GOMF_NU | 450 | -0.04286 | -0.00654 | 0.024193 | 0.96176 | 16354.73 |
| TSENG_IRS | 107 | -0.08291 | -0.00622 | 0.046785 | 0.9618 | 16355.41 |
| GOBP_MAI | 10 | -0.27783 | -0.00639 | 0.15677 | 0.96181 | 16355.58 |
| MALTA_CU | 20 | -0.2325 | -0.00756 | 0.13117 | 0.96184 | 16356.09 |
| GOMF_PEF | 47 | -0.12724 | -0.00634 | 0.071695 | 0.96202 | 16359.15 |
| GOCC_DYN | 20 | -0.16369 | -0.00533 | 0.09213 | 0.96219 | 16362.04 |
| REACTOME | 20 | -0.18223 | -0.00593 | 0.10254 | 0.96222 | 16362.55 |
| REACTOME | 40 | -0.13768 | -0.00633 | 0.077427 | 0.96231 | 16364.08 |
| PAL_PRMT | 27 | -0.18304 | -0.00692 | 0.1029 | 0.96235 | 16364.76 |
| NAGY_STA | 12 | -0.20523 | -0.00517 | 0.11534 | 0.9624 | 16365.61 |
| GOBP_POS | 8 | -0.34211 | -0.00704 | 0.19219 | 0.96246 | 16366.63 |
| REACTOME | 26 | -0.18551 | -0.00688 | 0.10417 | 0.96252 | 16367.65 |
| GOBP_DET | 46 | -0.15231 | -0.00751 | 0.085505 | 0.96256 | 16368.33 |
| GOBP_ONE | 53 | -0.12359 | -0.00654 | 0.06935 | 0.96262 | 16369.35 |
| BLANCO_M | 148 | -0.07576 | -0.00668 | 0.04244 | 0.96287 | 16373.6 |
| GOBP_LYM | 104 | -0.09108 | -0.00674 | 0.05102 | 0.96288 | 16373.77 |
| GOBP_REG | 17 | -0.22257 | -0.00668 | 0.12455 | 0.96302 | 16376.16 |
| PACHER_TA | 35 | -0.15541 | -0.00669 | 0.086882 | 0.96316 | 16378.54 |
| REACTOME | 56 | -0.10461 | -0.00569 | 0.058485 | 0.96316 | 16378.54 |
| REACTOME | 180 | -0.07377 | -0.00717 | 0.041229 | 0.9632 | 16379.22 |
| GOMF_PYF | 5 | -0.46283 | -0.00753 | 0.25857 | 0.96326 | 16380.24 |
| GOBP_PUR | 28 | -0.18702 | -0.0072 | 0.10447 | 0.96327 | 16380.41 |
| GOBP_NEG | 11 | -0.23155 | -0.00559 | 0.12918 | 0.96346 | 16383.64 |
| GOBP_CELI | 18 | -0.23616 | -0.00729 | 0.13169 | 0.96353 | 16384.83 |
| GOBP_CELI | 81 | -0.09724 | -0.00636 | 0.05421 | 0.96357 | 16385.51 |
| GOBP_POS | 27 | -0.1796 | -0.00679 | 0.10012 | 0.96357 | 16385.51 |
| PID_ARF6_ | 48 | -0.13409 | -0.00675 | 0.074712 | 0.96364 | 16386.7 |
| GOBP_REG | 8 | -0.29479 | -0.00607 | 0.16404 | 0.96383 | 16389.93 |
| GOMF_NAI | 6 | -0.4375 | -0.0078 | 0.24336 | 0.96388 | 16390.78 |
| GOBP_REG | 341 | -0.05076 | -0.00676 | 0.028221 | 0.96396 | 16392.14 |
| WP_ARYL_ | 44 | -0.13091 | -0.00631 | 0.072749 | 0.96401 | 16392.99 |

| | | | | | | |
|---|---|---|---|---|---|---|
| REACTOME | 89 | -0.10169 | -0.00697 | 0.056462 | 0.96414 | 16395.2 |
| REACTOME | 15 | -0.21554 | -0.00607 | 0.11967 | 0.96415 | 16395.37 |
| GOBP_REG | 27 | -0.16389 | -0.00619 | 0.090969 | 0.96418 | 16395.88 |
| NAKAYAMA | 63 | -0.11356 | -0.00655 | 0.063022 | 0.96421 | 16396.39 |
| GOBP_NEG | 17 | -0.21696 | -0.00651 | 0.1204 | 0.96422 | 16396.56 |
| BIOCARTA_ | 21 | -0.1817 | -0.00606 | 0.10068 | 0.96444 | 16400.3 |
| GOMF_MH | 26 | -0.17687 | -0.00656 | 0.097915 | 0.96456 | 16402.34 |
| GOBP_NEG | 7 | -0.38299 | -0.00737 | 0.212 | 0.96457 | 16402.51 |
| BILD_HRAS | 237 | -0.06026 | -0.00671 | 0.033346 | 0.96461 | 16403.19 |
| GAUSSMAI | 37 | -0.15859 | -0.00701 | 0.087764 | 0.96461 | 16403.19 |
| REACTOME | 10 | -0.25258 | -0.00581 | 0.1397 | 0.96469 | 16404.55 |
| REACTOME | 26 | -0.16731 | -0.0062 | 0.092517 | 0.96472 | 16405.06 |
| GOBP_POS | 21 | -0.18101 | -0.00603 | 0.10007 | 0.96475 | 16405.57 |
| GOBP_POS | 23 | -0.18772 | -0.00655 | 0.10375 | 0.9648 | 16406.42 |
| GOMF_JUN | 9 | -0.29967 | -0.00654 | 0.1655 | 0.96489 | 16407.95 |
| GOCC_RNA | 72 | -0.09637 | -0.00594 | 0.053125 | 0.96515 | 16412.38 |
| PID_RHOD | 22 | -0.16377 | -0.00559 | 0.090215 | 0.96526 | 16414.25 |
| GOMF_BM | 10 | -0.31534 | -0.00726 | 0.17367 | 0.96529 | 16414.76 |
| GOBP_URI | 8 | -0.33922 | -0.00698 | 0.18679 | 0.96531 | 16415.1 |
| JINESH_BL | 325 | -0.04961 | -0.00645 | 0.027306 | 0.96536 | 16415.95 |
| NAGY_TFT | 17 | -0.19408 | -0.00582 | 0.10681 | 0.96539 | 16416.46 |
| GOBP_LUN | 45 | -0.139 | -0.00678 | 0.076492 | 0.9654 | 16416.63 |
| IIZUKA_LIV | 12 | -0.2288 | -0.00577 | 0.1258 | 0.96552 | 16418.67 |
| JI_CARCIN | 12 | -0.27952 | -0.00705 | 0.1534 | 0.96578 | 16423.09 |
| GOBP_NEG | 6 | -0.37568 | -0.0067 | 0.20612 | 0.96581 | 16423.6 |
| WP_KENNE | 13 | -0.20989 | -0.00551 | 0.11513 | 0.96584 | 16424.11 |
| GOBP_POS | 6 | -0.37725 | -0.00672 | 0.20673 | 0.96598 | 16426.49 |
| BOYAULT_ | 11 | -0.29854 | -0.0072 | 0.16334 | 0.9662 | 16430.23 |
| GOBP_NAD | 5 | -0.32804 | -0.00534 | 0.17941 | 0.96625 | 16431.08 |
| GOBP_NEG | 5 | -0.45376 | -0.00738 | 0.24767 | 0.96653 | 16435.84 |
| KEGG_NEU | 248 | -0.0655 | -0.00746 | 0.035716 | 0.96666 | 16438.05 |
| GOBP_BLA | 21 | -0.1841 | -0.00614 | 0.10033 | 0.96673 | 16439.24 |
| GOBP_REG | 11 | -0.26169 | -0.00632 | 0.1426 | 0.96675 | 16439.58 |
| GOMF_QU | 17 | -0.2646 | -0.00794 | 0.14404 | 0.96689 | 16441.96 |
| WP_GABA_ | 10 | -0.27053 | -0.00622 | 0.14726 | 0.9669 | 16442.13 |
| GOBP_NEU | 189 | -0.06992 | -0.00696 | 0.038003 | 0.96709 | 16445.37 |
| REACTOME | 7 | -0.36825 | -0.00709 | 0.1998 | 0.96733 | 16449.45 |
| GOCC_PRE | 531 | -0.04128 | -0.00682 | 0.022369 | 0.96749 | 16452.17 |
| GOBP_TRA | 64 | -0.11629 | -0.00676 | 0.062996 | 0.96754 | 16453.02 |
| HERNANDE | 5 | -0.48939 | -0.00796 | 0.2651 | 0.96755 | 16453.19 |
| GOBP_POS | 10 | -0.30377 | -0.00699 | 0.16454 | 0.96756 | 16453.36 |
| ZHOU_PAN | 11 | -0.30726 | -0.00741 | 0.16637 | 0.96761 | 16454.21 |
| GOBP_POS | 45 | -0.13222 | -0.00645 | 0.071584 | 0.96762 | 16454.38 |
| GOMF_O_ | 47 | -0.13869 | -0.00691 | 0.075089 | 0.96762 | 16454.38 |
| GOBP_IMN | 7 | -0.31617 | -0.00609 | 0.17111 | 0.96767 | 16455.23 |

| | | | | | | |
|---|---|---|---|---|---|---|
| GOBP_POS | 169 | -0.07557 | -0.00712 | 0.040889 | 0.9677 | 16455.74 |
| GOBP_REG | 28 | -0.17727 | -0.00682 | 0.095888 | 0.96774 | 16456.42 |
| GOCC_HAF | 11 | -0.376 | -0.00907 | 0.20334 | 0.96777 | 16456.93 |
| GOMF_DIN | 8 | -0.34336 | -0.00707 | 0.18546 | 0.96793 | 16459.65 |
| GOLUB_AL | 24 | -0.17032 | -0.00607 | 0.091991 | 0.96794 | 16459.82 |
| LEE_DIFFEI | 173 | -0.0694 | -0.00661 | 0.037471 | 0.96798 | 16460.5 |
| GOBP_CHC | 8 | -0.34118 | -0.00702 | 0.18419 | 0.968 | 16460.84 |
| REACTOME | 7 | -0.3949 | -0.0076 | 0.21307 | 0.96807 | 16462.03 |
| GOBP_T_C | 41 | -0.14238 | -0.00663 | 0.076745 | 0.96821 | 16464.41 |
| GOBP_INTI | 42 | -0.13135 | -0.00619 | 0.070755 | 0.96829 | 16465.77 |
| GRABARCZ | 71 | -0.10201 | -0.00624 | 0.054932 | 0.96833 | 16466.45 |
| BIOCARTA_ | 18 | -0.22333 | -0.00689 | 0.12023 | 0.96837 | 16467.13 |
| GOBP_REG | 22 | -0.32066 | -0.01094 | 0.17258 | 0.96841 | 16467.81 |
| REACTOME | 5 | -0.48134 | -0.00783 | 0.259 | 0.96844 | 16468.32 |
| GOMF_AN | 96 | -0.08938 | -0.00636 | 0.048053 | 0.96855 | 16470.19 |
| GOBP_FLA | 6 | -0.38345 | -0.00684 | 0.20594 | 0.96869 | 16472.57 |
| GOBP_REG | 8 | -0.31776 | -0.00654 | 0.1705 | 0.96881 | 16474.61 |
| GOMF_MY | 12 | -0.26809 | -0.00676 | 0.14383 | 0.96883 | 16474.95 |
| GOBP_POS | 151 | -0.075 | -0.00668 | 0.04021 | 0.96891 | 16476.31 |
| TONKS_TAI | 18 | -0.22401 | -0.00691 | 0.1201 | 0.96891 | 16476.31 |
| GOBP_MA | 10 | -0.32928 | -0.00758 | 0.17631 | 0.96909 | 16479.38 |
| GOBP_REG | 28 | -0.18789 | -0.00723 | 0.10058 | 0.96912 | 16479.89 |
| REACTOME | 10 | -0.28312 | -0.00651 | 0.15156 | 0.96912 | 16479.89 |
| GOCC_CHR | 12 | -0.25264 | -0.00637 | 0.13521 | 0.96914 | 16480.23 |
| GOBP_POS | 25 | -0.18626 | -0.00677 | 0.099599 | 0.96925 | 16482.1 |
| GOBP_PRC | 187 | -0.06634 | -0.00657 | 0.035443 | 0.96937 | 16484.14 |
| GOBP_TRA | 1434 | -0.02646 | -0.00701 | 0.014135 | 0.96937 | 16484.14 |
| GOBP_POS | 13 | -0.23711 | -0.00622 | 0.12663 | 0.96942 | 16484.99 |
| KEGG_RIBC | 15 | -0.22234 | -0.00626 | 0.1187 | 0.96947 | 16485.84 |
| GOBP_REG | 382 | -0.04643 | -0.00654 | 0.02475 | 0.96966 | 16489.07 |
| GOBP_NEG | 12 | -0.24449 | -0.00616 | 0.13023 | 0.96975 | 16490.6 |
| REACTOME | 16 | -0.24398 | -0.0071 | 0.12996 | 0.96975 | 16490.6 |
| GOBP_LEU | 320 | -0.05494 | -0.00709 | 0.029258 | 0.96977 | 16490.94 |
| REACTOME | 9 | -0.36075 | -0.00788 | 0.19214 | 0.96977 | 16490.94 |
| GOMF_OXI | 38 | -0.15752 | -0.00706 | 0.083871 | 0.96981 | 16491.62 |
| GOBP_PHC | 35 | -0.16997 | -0.00731 | 0.090355 | 0.97002 | 16495.19 |
| GOBP_I_KA | 262 | -0.05807 | -0.00679 | 0.030857 | 0.97007 | 16496.04 |
| GOBP_RET | 32 | -0.17581 | -0.00723 | 0.093377 | 0.97013 | 16497.06 |
| GOBP_CELI | 18 | -0.32876 | -0.01015 | 0.17444 | 0.97025 | 16499.1 |
| DACOSTA_ | 12 | -0.26535 | -0.00669 | 0.14056 | 0.97046 | 16502.67 |
| BIOCARTA_ | 15 | -0.22556 | -0.00636 | 0.11945 | 0.9705 | 16503.35 |
| SMIRNOV_ | 54 | -0.13251 | -0.00708 | 0.070169 | 0.97051 | 16503.52 |
| GOBP_REG | 13 | -0.29055 | -0.00762 | 0.15375 | 0.9706 | 16505.05 |
| SENGUPTA | 137 | -0.07613 | -0.00646 | 0.040285 | 0.9706 | 16505.05 |
| GOBP_PRC | 12 | -0.27036 | -0.00681 | 0.14303 | 0.97063 | 16505.56 |

| | | | | | | |
|---|---|---|---|---|---|---|
| GOBP_AMI | 99 | -0.09931 | -0.00717 | 0.052532 | 0.97064 | 16505.73 |
| GOMF_UBI | 103 | -0.0835 | -0.00615 | 0.044121 | 0.97079 | 16508.28 |
| GOBP_POS | 57 | -0.1327 | -0.00728 | 0.070067 | 0.97087 | 16509.64 |
| GOBP_PER | 12 | -0.32353 | -0.00815 | 0.17066 | 0.971 | 16511.86 |
| GOBP_REG | 11 | -0.3131 | -0.00756 | 0.16516 | 0.971 | 16511.86 |
| GOMF_P_T | 8 | -0.33013 | -0.00679 | 0.17386 | 0.9712 | 16515.26 |
| GOBP_POS | 36 | -0.16191 | -0.00706 | 0.085259 | 0.97121 | 16515.43 |
| PID_NFAT_ | 41 | -0.15088 | -0.00702 | 0.079391 | 0.97131 | 16517.13 |
| GOCC_INTI | 825 | -0.03231 | -0.00661 | 0.016991 | 0.97139 | 16518.49 |
| GOBP_MIT | 8 | -0.3643 | -0.0075 | 0.19106 | 0.97171 | 16523.93 |
| GOBP_REG | 441 | -0.04747 | -0.00717 | 0.024888 | 0.97176 | 16524.78 |
| GOBP_NEG | 7 | -0.32669 | -0.00629 | 0.17119 | 0.97182 | 16525.8 |
| GOBP_T_C | 192 | -0.071 | -0.00712 | 0.037154 | 0.97199 | 16528.69 |
| GOMF_ALF | 10 | -0.29055 | -0.00669 | 0.15186 | 0.97214 | 16531.24 |
| GOBP_REG | 97 | -0.09486 | -0.00678 | 0.049502 | 0.97232 | 16534.3 |
| FLECHNER_ | 22 | -0.19305 | -0.00659 | 0.10071 | 0.97236 | 16534.98 |
| REACTOME | 14 | -0.30385 | -0.00827 | 0.15851 | 0.97236 | 16534.98 |
| REACTOME | 12 | -0.27667 | -0.00697 | 0.14433 | 0.97237 | 16535.15 |
| TAVOR_CE | 46 | -0.15088 | -0.00744 | 0.078679 | 0.97242 | 16536 |
| GOBP_ANT | 5 | -0.42319 | -0.00689 | 0.22055 | 0.97249 | 16537.19 |
| WP_VITAM | 19 | -0.23939 | -0.00759 | 0.12463 | 0.97261 | 16539.23 |
| ELVIDGE_H | 6 | -0.34773 | -0.0062 | 0.18085 | 0.97273 | 16541.27 |
| CHEN_ETV! | 22 | -0.19924 | -0.0068 | 0.10336 | 0.97304 | 16546.55 |
| GOBP_REG | 11 | -0.33293 | -0.00803 | 0.17259 | 0.97313 | 16548.08 |
| SCIBETTA_ | 18 | -0.2123 | -0.00655 | 0.11004 | 0.97315 | 16548.42 |
| WP_RETIN | 15 | -0.23921 | -0.00674 | 0.12397 | 0.97316 | 16548.59 |
| GOBP_CELI | 148 | -0.07772 | -0.00685 | 0.040251 | 0.97324 | 16549.95 |
| REACTOME | 93 | -0.09835 | -0.00689 | 0.050938 | 0.97324 | 16549.95 |
| GOBP_ACT | 15 | -0.23061 | -0.0065 | 0.1193 | 0.97337 | 16552.16 |
| GOBP_REG | 2 | -0.69062 | -0.00711 | 0.35726 | 0.97338 | 16552.33 |
| GOMF_TRA | 19 | -0.18597 | -0.0059 | 0.096055 | 0.97356 | 16555.39 |
| GOBP_POS | 8 | -0.3019 | -0.00621 | 0.15588 | 0.97361 | 16556.24 |
| GOCC_TRA | 5 | -0.62067 | -0.0101 | 0.32044 | 0.97362 | 16556.41 |
| GOCC_GUA | 17 | -0.23669 | -0.0071 | 0.12215 | 0.97366 | 16557.09 |
| KIM_GLIS2 | 6 | -0.31666 | -0.00564 | 0.1631 | 0.97389 | 16561 |
| NABA_BAS | 37 | -0.17931 | -0.00793 | 0.092361 | 0.97389 | 16561 |
| GOBP_MIT | 83 | -0.10349 | -0.00685 | 0.053243 | 0.97403 | 16563.38 |
| GOMF_SIN | 23 | -0.18599 | -0.00649 | 0.095598 | 0.97414 | 16565.25 |
| GOMF_OXI | 4 | -0.59498 | -0.00866 | 0.30563 | 0.97421 | 16566.44 |
| GOBP_ARC | 9 | -0.30834 | -0.00673 | 0.15832 | 0.97426 | 16567.29 |
| GOBP_REG | 15 | -0.26134 | -0.00736 | 0.13395 | 0.97446 | 16570.69 |
| GOBP_NEU | 132 | -0.08695 | -0.00725 | 0.044552 | 0.9745 | 16571.37 |
| BYSTRYKH_ | 26 | -0.17774 | -0.00659 | 0.091069 | 0.97451 | 16571.54 |
| GOBP_POS | 7 | -0.45238 | -0.00871 | 0.23152 | 0.97464 | 16573.75 |
| GOMF_ATI | 8 | -0.32019 | -0.00659 | 0.16384 | 0.97466 | 16574.09 |

| | | | | | | |
|---|---|---|---|---|---|---|
| GOCC_TRA | 87 | -0.09781 | -0.00662 | 0.050037 | 0.97468 | 16574.43 |
| GOBP_PRC | 9 | -0.25294 | -0.00552 | 0.12928 | 0.97479 | 16576.3 |
| REACTOME | 21 | -0.22038 | -0.00735 | 0.11262 | 0.97481 | 16576.64 |
| CERIBELLI_ | 31 | -0.17868 | -0.00723 | 0.091254 | 0.97488 | 16577.83 |
| GOBP_COM | 13 | -0.25402 | -0.00666 | 0.12966 | 0.97494 | 16578.85 |
| REACTOME | 6 | -0.3498 | -0.00624 | 0.17845 | 0.975 | 16579.88 |
| GOMF_ACT | 391 | -0.0488 | -0.00695 | 0.024868 | 0.97513 | 16582.09 |
| GOBP_REG | 61 | -0.12809 | -0.00727 | 0.065256 | 0.97516 | 16582.6 |
| BURTON_A | 42 | -0.15043 | -0.00709 | 0.076627 | 0.97517 | 16582.77 |
| GOCC_RNA | 13 | -0.24376 | -0.00639 | 0.12408 | 0.97525 | 16584.13 |
| REACTOME | 11 | -0.38414 | -0.00927 | 0.19542 | 0.97532 | 16585.32 |
| GOMF_CAF | 258 | -0.06456 | -0.0075 | 0.03283 | 0.97537 | 16586.17 |
| GOCC_STEI | 61 | -0.12125 | -0.00688 | 0.061453 | 0.97575 | 16592.63 |
| KEGG_ARG | 48 | -0.13084 | -0.00659 | 0.066286 | 0.9758 | 16593.48 |
| GOBP_NEG | 9 | -0.38858 | -0.00848 | 0.19683 | 0.97581 | 16593.65 |
| GOBP_REG | 9 | -0.36464 | -0.00796 | 0.18466 | 0.97584 | 16594.16 |
| GOCC_EXT | 141 | -0.087 | -0.00749 | 0.044047 | 0.97587 | 16594.67 |
| GOMF_COI | 22 | -0.22438 | -0.00766 | 0.11359 | 0.97588 | 16594.84 |
| GOBP_CELI | 210 | -0.07165 | -0.00751 | 0.03626 | 0.97591 | 16595.35 |
| GOBP_REG | 45 | -0.15134 | -0.00738 | 0.076543 | 0.97598 | 16596.54 |
| GOBP_ENT | 11 | -0.29754 | -0.00718 | 0.15038 | 0.97606 | 16597.9 |
| GOBP_NEG | 4 | -0.41668 | -0.00606 | 0.21026 | 0.97624 | 16600.96 |
| GOBP_NEG | 9 | -0.31298 | -0.00683 | 0.1578 | 0.97633 | 16602.49 |
| GOMF_1_A | 8 | -0.31508 | -0.00648 | 0.1588 | 0.97637 | 16603.17 |
| REACTOME | 24 | -0.20962 | -0.00747 | 0.10559 | 0.97643 | 16604.19 |
| GOBP_RES | 44 | -0.1586 | -0.00765 | 0.079825 | 0.97652 | 16605.72 |
| GOBP_FLAV | 4 | -0.58437 | -0.00851 | 0.29401 | 0.97656 | 16606.4 |
| KYNG_RESI | 34 | -0.16906 | -0.00717 | 0.085032 | 0.9766 | 16607.08 |
| GOBP_REG | 28 | -0.19176 | -0.00738 | 0.096393 | 0.97666 | 16608.1 |
| GOCC_ACE | 15 | -0.2931 | -0.00826 | 0.14726 | 0.97672 | 16609.12 |
| GOMF_COI | 54 | -0.13215 | -0.00706 | 0.066323 | 0.97684 | 16611.16 |
| REACTOME | 6 | -0.45101 | -0.00804 | 0.22628 | 0.97687 | 16611.67 |
| GOBP_REG | 51 | -0.12986 | -0.00674 | 0.065105 | 0.97695 | 16613.03 |
| PARK_TRET | 12 | -0.35467 | -0.00894 | 0.17776 | 0.97698 | 16613.54 |
| GOCC_GOL | 339 | -0.05248 | -0.00697 | 0.026278 | 0.97708 | 16615.25 |
| SESTO_RES | 18 | -0.23373 | -0.00721 | 0.11704 | 0.97708 | 16615.25 |
| REACTOME | 6 | -0.43736 | -0.0078 | 0.21885 | 0.97716 | 16616.61 |
| GOBP_REG | 41 | -0.15562 | -0.00724 | 0.07781 | 0.97724 | 16617.97 |
| GOCC_OM | 5 | -0.40041 | -0.00652 | 0.20008 | 0.97731 | 16619.16 |
| REACTOME | 7 | -0.43908 | -0.00845 | 0.21933 | 0.97734 | 16619.67 |
| GOMF_C_C | 8 | -0.4131 | -0.0085 | 0.20635 | 0.97735 | 16619.84 |
| GOBP_REG | 12 | -0.29676 | -0.00748 | 0.14804 | 0.97748 | 16622.05 |
| REACTOME | 10 | -0.36665 | -0.00844 | 0.18289 | 0.9775 | 16622.39 |
| GOBP_RES | 19 | -0.25418 | -0.00806 | 0.12678 | 0.97751 | 16622.56 |
| REACTOME | 23 | -0.19468 | -0.00679 | 0.097087 | 0.97752 | 16622.73 |

| | | | | | | |
|---|---|---|---|---|---|---|
| GOBP_RNA | 22 | -0.20862 | -0.00712 | 0.10398 | 0.97759 | 16623.92 |
| GOMF_MC | 1288 | -0.03312 | -0.00835 | 0.016506 | 0.9776 | 16624.09 |
| GOBP_PHA | 28 | -0.18605 | -0.00716 | 0.092685 | 0.97764 | 16624.77 |
| GOBP_REG | 94 | -0.10439 | -0.00735 | 0.052 | 0.97764 | 16624.77 |
| GOMF_ION | 18 | -0.2488 | -0.00768 | 0.12379 | 0.97777 | 16626.98 |
| GOBP_ANT | 45 | -0.1441 | -0.00703 | 0.071666 | 0.97782 | 16627.83 |
| SHIPP_DLB | 34 | -0.181 | -0.00767 | 0.089988 | 0.97785 | 16628.34 |
| GOBP_BON | 64 | -0.13018 | -0.00757 | 0.064703 | 0.97788 | 16628.85 |
| GOBP_MO | 565 | -0.04382 | -0.00747 | 0.021778 | 0.97789 | 16629.02 |
| REACTOME | 39 | -0.14833 | -0.00673 | 0.073717 | 0.97789 | 16629.02 |
| GOBP_MO | 178 | -0.07712 | -0.00745 | 0.038305 | 0.97795 | 16630.04 |
| LU_EZH2_T | 352 | -0.05412 | -0.00732 | 0.026848 | 0.97808 | 16632.25 |
| GOBP_TET | 5 | -0.41578 | -0.00677 | 0.20625 | 0.97809 | 16632.42 |
| GOMF_HAI | 10 | -0.42333 | -0.00974 | 0.20961 | 0.97828 | 16635.65 |
| BIOCARTA_ | 25 | -0.21438 | -0.0078 | 0.1061 | 0.97833 | 16636.5 |
| GOBP_REG | 250 | -0.06376 | -0.00729 | 0.03154 | 0.97837 | 16637.18 |
| GOBP_PHC | 20 | -0.21469 | -0.00698 | 0.1061 | 0.97847 | 16638.88 |
| REACTOME | 29 | -0.19979 | -0.00782 | 0.098643 | 0.97858 | 16640.75 |
| YAO_TEMP | 72 | -0.11548 | -0.00712 | 0.056974 | 0.97865 | 16641.94 |
| GOBP_REG | 111 | -0.09559 | -0.00731 | 0.047139 | 0.9787 | 16642.79 |
| GOBP_ACT | 19 | -0.22875 | -0.00725 | 0.1126 | 0.97889 | 16646.02 |
| GOBP_REG | 24 | -0.20755 | -0.0074 | 0.10201 | 0.97904 | 16648.58 |
| WP_KREBS | 13 | -0.26643 | -0.00699 | 0.1309 | 0.97908 | 16649.26 |
| GOCC_PRE | 18 | -0.25179 | -0.00777 | 0.12344 | 0.9793 | 16653 |
| GOCC_TRA | 199 | -0.07057 | -0.00721 | 0.034554 | 0.97943 | 16655.21 |
| GOBP_MIT | 10 | -0.28947 | -0.00666 | 0.1414 | 0.97967 | 16659.29 |
| GOBP_LYM | 43 | -0.16437 | -0.00784 | 0.08027 | 0.9797 | 16659.8 |
| GOBP_POS | 10 | -0.31101 | -0.00716 | 0.15185 | 0.97972 | 16660.14 |
| BIOCARTA_ | 8 | -0.34963 | -0.0072 | 0.17069 | 0.97973 | 16660.31 |
| MATZUK_S | 37 | -0.15653 | -0.00692 | 0.076378 | 0.97978 | 16661.16 |
| REACTOME | 29 | -0.16811 | -0.00658 | 0.081989 | 0.97983 | 16662.01 |
| GOMF_CAI | 16 | -0.27288 | -0.00794 | 0.13302 | 0.97987 | 16662.69 |
| TERAMOTO | 6 | -0.40784 | -0.00727 | 0.19821 | 0.98018 | 16667.96 |
| PID_THRON | 43 | -0.15248 | -0.00727 | 0.074055 | 0.98025 | 16669.15 |
| PID_ECADH | 41 | -0.15574 | -0.00725 | 0.075612 | 0.98028 | 16669.66 |
| ONDER_CD | 451 | -0.05112 | -0.00781 | 0.024792 | 0.98039 | 16671.53 |
| GOBP_POS | 10 | -0.34829 | -0.00801 | 0.1686 | 0.98057 | 16674.59 |
| GOBP_VES | 23 | -0.21002 | -0.00733 | 0.10159 | 0.98065 | 16675.95 |
| KENNY_CTI | 50 | -0.15505 | -0.00797 | 0.074974 | 0.98067 | 16676.29 |
| AIYAR_COE | 38 | -0.19346 | -0.00867 | 0.093486 | 0.98074 | 16677.48 |
| GOBP_MY( | 46 | -0.16727 | -0.00825 | 0.080636 | 0.98097 | 16681.39 |
| ZHANG_TA | 82 | -0.12019 | -0.0079 | 0.057861 | 0.9811 | 16683.61 |
| GOBP_SPEI | 124 | -0.08684 | -0.00702 | 0.041791 | 0.98114 | 16684.29 |
| REACTOME | 7 | -0.4605 | -0.00887 | 0.22159 | 0.98115 | 16684.46 |
| GOBP_TRA | 89 | -0.10153 | -0.00695 | 0.048849 | 0.98116 | 16684.63 |

| | | | | | | |
|---|---|---|---|---|---|---|
| GOBP_POS | 129 | -0.08961 | -0.00738 | 0.043094 | 0.9812 | 16685.31 |
| REACTOME | 23 | -0.21013 | -0.00733 | 0.10095 | 0.9813 | 16687.01 |
| REACTOME | 6 | -0.34154 | -0.00609 | 0.164 | 0.98135 | 16687.86 |
| ZHAN_MUI | 44 | -0.15661 | -0.00755 | 0.075165 | 0.98139 | 16688.54 |
| GOBP_REG | 11 | -0.303 | -0.00731 | 0.14513 | 0.98158 | 16691.77 |
| GOBP_REG | 35 | -0.16783 | -0.00722 | 0.080273 | 0.98171 | 16693.98 |
| REACTOME | 17 | -0.26509 | -0.00795 | 0.12673 | 0.98176 | 16694.83 |
| GOCC_SMC | 30 | -0.19 | -0.00757 | 0.090798 | 0.9818 | 16695.51 |
| SABATES_C | 130 | -0.09784 | -0.00809 | 0.046698 | 0.98192 | 16697.55 |
| GOBP_POS | 18 | -0.23852 | -0.00736 | 0.11378 | 0.98197 | 16698.4 |
| BOYLAN_M | 69 | -0.13785 | -0.00832 | 0.065581 | 0.98222 | 16702.65 |
| REACTOME | 64 | -0.14485 | -0.00842 | 0.068911 | 0.98222 | 16702.65 |
| GOMF_G_I | 691 | -0.04954 | -0.0093 | 0.023562 | 0.98223 | 16702.82 |
| GOBP_DOF | 12 | -0.31165 | -0.00785 | 0.1482 | 0.98226 | 16703.33 |
| GOBP_PRC | 310 | -0.06041 | -0.00768 | 0.028714 | 0.9823 | 16704.01 |
| WP_LIVER_ | 10 | -0.31612 | -0.00727 | 0.15019 | 0.98234 | 16704.69 |
| GOBP_REG | 9 | -0.31207 | -0.00681 | 0.14812 | 0.98243 | 16706.22 |
| GOBP_NEG | 9 | -0.29854 | -0.00652 | 0.14167 | 0.98245 | 16706.56 |
| GOBP_POS | 16 | -0.23868 | -0.00695 | 0.11326 | 0.98245 | 16706.56 |
| GOMF_INH | 12 | -0.35459 | -0.00894 | 0.16825 | 0.98245 | 16706.56 |
| GOBP_CHE | 7 | -0.36009 | -0.00693 | 0.17083 | 0.98247 | 16706.9 |
| REACTOME | 41 | -0.13951 | -0.00649 | 0.066143 | 0.98253 | 16707.92 |
| GOCC_RNA | 100 | -0.09565 | -0.00694 | 0.045318 | 0.98259 | 16708.94 |
| GOMF_GTI | 247 | -0.06984 | -0.00794 | 0.033062 | 0.98266 | 16710.13 |
| GOBP_REG | 26 | -0.20858 | -0.00774 | 0.098701 | 0.9827 | 16710.81 |
| GOBP_ICOS | 54 | -0.13885 | -0.00742 | 0.065673 | 0.98274 | 16711.49 |
| WP_OXIDA | 29 | -0.20462 | -0.00801 | 0.096788 | 0.98274 | 16711.49 |
| LIANG_SILE | 10 | -0.28692 | -0.0066 | 0.13554 | 0.98285 | 16713.36 |
| GOBP_REG | 74 | -0.12636 | -0.0079 | 0.059689 | 0.98286 | 16713.53 |
| GOBP_POS | 15 | -0.25758 | -0.00726 | 0.12122 | 0.9832 | 16719.32 |
| GOBP_PRC | 7 | -0.39742 | -0.00765 | 0.18693 | 0.98324 | 16720 |
| GOBP_LEU | 96 | -0.10801 | -0.00768 | 0.050738 | 0.98335 | 16721.87 |
| GOBP_POS | 148 | -0.09182 | -0.0081 | 0.043125 | 0.98337 | 16722.21 |
| GOBP_HYD | 29 | -0.20458 | -0.00801 | 0.095982 | 0.98347 | 16723.91 |
| HOSHIDA_I | 252 | -0.06604 | -0.00758 | 0.030977 | 0.98349 | 16724.25 |
| GOBP_PHC | 10 | -0.31799 | -0.00732 | 0.14908 | 0.98353 | 16724.93 |
| GOBP_CEN | 45 | -0.144 | -0.00702 | 0.067494 | 0.98355 | 16725.27 |
| VALK_AML | 27 | -0.21148 | -0.00799 | 0.099071 | 0.9836 | 16726.12 |
| GOBP_CAL | 21 | -0.24065 | -0.00802 | 0.11241 | 0.98385 | 16730.37 |
| GOBP_DOF | 34 | -0.20116 | -0.00853 | 0.093961 | 0.98385 | 16730.37 |
| GOMF_PRC | 7 | -0.42084 | -0.0081 | 0.1962 | 0.98402 | 16733.26 |
| GOBP_POS | 133 | -0.09543 | -0.00798 | 0.044447 | 0.9841 | 16734.62 |
| GOBP_ESTA | 6 | -0.3957 | -0.00705 | 0.1842 | 0.98414 | 16735.3 |
| GOBP_VEN | 32 | -0.18452 | -0.00759 | 0.085889 | 0.98415 | 16735.47 |
| GOBP_CELI | 84 | -0.11648 | -0.00775 | 0.054185 | 0.9842 | 16736.32 |

| | | | | | | |
|---|---|---|---|---|---|---|
| GOBP_REG | 27 | -0.20829 | -0.00787 | 0.096835 | 0.98426 | 16737.34 |
| GOBP_SEN | 67 | -0.14155 | -0.00842 | 0.065684 | 0.98441 | 16739.89 |
| GOCC_INN | 5 | -0.38878 | -0.00633 | 0.18003 | 0.98459 | 16742.95 |
| REACTOME | 9 | -0.39121 | -0.00854 | 0.18115 | 0.98459 | 16742.95 |
| GOMF_U6_ | 12 | -0.31896 | -0.00804 | 0.14741 | 0.98475 | 16745.67 |
| REACTOME | 24 | -0.20674 | -0.00737 | 0.095526 | 0.98477 | 16746.01 |
| GOMF_MI | 17 | -0.28128 | -0.00844 | 0.1299 | 0.98482 | 16746.86 |
| DORMOY_ | 17 | -0.25853 | -0.00775 | 0.11921 | 0.98494 | 16748.9 |
| GOMF_TR | 1126 | -0.03369 | -0.00798 | 0.015529 | 0.98498 | 16749.58 |
| RICKMAN_ | 30 | -0.20362 | -0.00811 | 0.093806 | 0.98502 | 16750.27 |
| REACTOME | 74 | -0.11554 | -0.00722 | 0.05313 | 0.98517 | 16752.82 |
| GOMF_LYS | 20 | -0.24578 | -0.008 | 0.11281 | 0.98532 | 16755.37 |
| GOBP_NEU | 95 | -0.11926 | -0.00844 | 0.054724 | 0.98534 | 16755.71 |
| WP_SARSC | 60 | -0.15587 | -0.00877 | 0.071488 | 0.98538 | 16756.39 |
| GOBP_NEG | 15 | -0.28084 | -0.00791 | 0.12875 | 0.98542 | 16757.07 |
| GOBP_REG | 43 | -0.15835 | -0.00755 | 0.072536 | 0.98548 | 16758.09 |
| GOBP_REG | 7 | -0.35703 | -0.00687 | 0.16322 | 0.98564 | 16760.81 |
| GOBP_POS | 292 | -0.06774 | -0.00836 | 0.030958 | 0.98566 | 16761.15 |
| GOBP_RES | 10 | -0.29361 | -0.00676 | 0.13416 | 0.98567 | 16761.32 |
| GOBP_ACT | 100 | -0.10972 | -0.00796 | 0.050046 | 0.98581 | 16763.7 |
| GOBP_RES | 14 | -0.28057 | -0.00764 | 0.12789 | 0.98587 | 16764.72 |
| GOBP_MIT | 42 | -0.15614 | -0.00736 | 0.071068 | 0.98599 | 16766.76 |
| GOBP_REG | 62 | -0.14964 | -0.00856 | 0.067983 | 0.98613 | 16769.14 |
| MYLLYKAN | 9 | -0.40264 | -0.00879 | 0.18277 | 0.9862 | 16770.33 |
| GOCC_MIT | 18 | -0.2509 | -0.00774 | 0.11382 | 0.98624 | 16771.01 |
| GOBP_CILI | 148 | -0.08569 | -0.00756 | 0.03887 | 0.98625 | 16771.18 |
| GOMF_CYS | 154 | -0.08401 | -0.00756 | 0.038072 | 0.98632 | 16772.37 |
| GOMF_RN | 5 | -0.51979 | -0.00846 | 0.23547 | 0.98635 | 16772.88 |
| GOBP_LEU | 365 | -0.05998 | -0.00826 | 0.02715 | 0.98641 | 16773.9 |
| GOMF_MH | 6 | -0.47188 | -0.00841 | 0.21326 | 0.98653 | 16775.94 |
| GOMF_PIR | 5 | -0.42375 | -0.0069 | 0.19147 | 0.98655 | 16776.28 |
| GOBP_NEG | 4 | -0.67509 | -0.00983 | 0.30487 | 0.98659 | 16776.96 |
| GOBP_REG | 15 | -0.26841 | -0.00756 | 0.12118 | 0.98661 | 16777.3 |
| GOBP_FOL | 7 | -0.41872 | -0.00806 | 0.18889 | 0.98667 | 16778.32 |
| GOBP_RRN | 5 | -0.40343 | -0.00656 | 0.18199 | 0.98668 | 16778.49 |
| GOBP_REG | 27 | -0.22421 | -0.00847 | 0.10106 | 0.98674 | 16779.51 |
| XU_GH1_A | 171 | -0.0874 | -0.00828 | 0.039364 | 0.98679 | 16780.36 |
| GOBP_ZYM | 7 | -0.4105 | -0.0079 | 0.18451 | 0.98694 | 16782.91 |
| PARENT_M | 42 | -0.17166 | -0.00809 | 0.077116 | 0.98699 | 16783.76 |
| GOBP_REG | 16 | -0.27858 | -0.00811 | 0.12508 | 0.98702 | 16784.28 |
| GOMF_GT | 286 | -0.06321 | -0.00772 | 0.028381 | 0.98703 | 16784.45 |
| GOBP_IMM | 26 | -0.23192 | -0.0086 | 0.10398 | 0.98713 | 16786.15 |
| REACTOME | 24 | -0.22757 | -0.00811 | 0.10203 | 0.98714 | 16786.32 |
| GOBP_REG | 6 | -0.4829 | -0.00861 | 0.21639 | 0.98717 | 16786.83 |
| WP_FERRC | 59 | -0.14234 | -0.00795 | 0.063733 | 0.98723 | 16787.85 |

| | | | | | | |
|---|---|---|---|---|---|---|
| GOMF_POT | 8 | -0.40657 | -0.00837 | 0.18181 | 0.98732 | 16789.38 |
| GOBP_REG | 26 | -0.1935 | -0.00718 | 0.086439 | 0.9874 | 16790.74 |
| GOBP_REG | 6 | -0.40531 | -0.00722 | 0.18083 | 0.98749 | 16792.27 |
| DEBOSSCH | 19 | -0.27812 | -0.00882 | 0.12402 | 0.98753 | 16792.95 |
| GOBP_REG | 49 | -0.16377 | -0.00833 | 0.072945 | 0.98761 | 16794.31 |
| FONTAINE_ | 72 | -0.13676 | -0.00843 | 0.060901 | 0.98763 | 16794.65 |
| GOBP_B_C | 30 | -0.21105 | -0.00841 | 0.093968 | 0.98764 | 16794.82 |
| GOBP_REG | 6 | -0.45737 | -0.00815 | 0.20345 | 0.98771 | 16796.01 |
| GOCC_ACT | 211 | -0.07705 | -0.0081 | 0.034271 | 0.98771 | 16796.01 |
| GOBP_AMI | 13 | -0.31956 | -0.00838 | 0.14194 | 0.98781 | 16797.71 |
| GOBP_GLU | 23 | -0.22174 | -0.00774 | 0.098295 | 0.98795 | 16800.09 |
| GOBP_PEP | 4 | -0.4731 | -0.00689 | 0.20934 | 0.98808 | 16802.3 |
| GOCC_MH | 4 | -0.4731 | -0.00689 | 0.20934 | 0.98808 | 16802.3 |
| GOMF_DES | 7 | -0.31004 | -0.00597 | 0.13703 | 0.98816 | 16803.66 |
| GOMF_CH | 6 | -0.51599 | -0.0092 | 0.22765 | 0.98828 | 16805.7 |
| GOBP_IRO | 77 | -0.12902 | -0.00822 | 0.05689 | 0.98833 | 16806.55 |
| GOBP_INFL | 31 | -0.20394 | -0.00826 | 0.089772 | 0.98844 | 16808.42 |
| GOMF_LIG | 16 | -0.32596 | -0.00949 | 0.14343 | 0.98847 | 16808.93 |
| GOMF_DYI | 27 | -0.21722 | -0.00821 | 0.095562 | 0.98848 | 16809.1 |
| GOBP_NEG | 13 | -0.34775 | -0.00912 | 0.1526 | 0.98865 | 16811.99 |
| REACTOME | 32 | -0.19425 | -0.00799 | 0.085155 | 0.98872 | 16813.18 |
| GOBP_CELI | 260 | -0.06943 | -0.00809 | 0.030363 | 0.98888 | 16815.9 |
| GOBP_VES | 204 | -0.08177 | -0.00845 | 0.035757 | 0.98889 | 16816.07 |
| FURUKAW/ | 67 | -0.13966 | -0.00831 | 0.060866 | 0.98912 | 16819.99 |
| GOBP_C_X | 5 | -0.54674 | -0.0089 | 0.23825 | 0.98912 | 16819.99 |
| GOBP_REG | 6 | -0.49885 | -0.00889 | 0.21738 | 0.98912 | 16819.99 |
| GOMF_MH | 21 | -0.23818 | -0.00794 | 0.10372 | 0.98917 | 16820.84 |
| REACTOME | 12 | -0.35619 | -0.00898 | 0.15511 | 0.98917 | 16820.84 |
| WP_NETW | 202 | -0.08426 | -0.00867 | 0.036639 | 0.98926 | 16822.37 |
| REACTOME | 38 | -0.20339 | -0.00912 | 0.088407 | 0.98929 | 16822.88 |
| GOBP_KER | 12 | -0.32479 | -0.00819 | 0.14095 | 0.98939 | 16824.58 |
| REACTOME | 41 | -0.1621 | -0.00755 | 0.070274 | 0.98946 | 16825.77 |
| GOCC_NU | 5 | -0.41087 | -0.00669 | 0.17797 | 0.98951 | 16826.62 |
| GOBP_STO | 20 | -0.23986 | -0.0078 | 0.10387 | 0.98953 | 16826.96 |
| GOBP_REG | 63 | -0.14679 | -0.00847 | 0.063471 | 0.98962 | 16828.49 |
| REACTOME | 17 | -0.29787 | -0.00893 | 0.12829 | 0.98987 | 16832.74 |
| GOBP_GLC | 5 | -0.64316 | -0.01047 | 0.27665 | 0.98995 | 16834.1 |
| REACTOME | 78 | -0.13714 | -0.0088 | 0.058956 | 0.98999 | 16834.78 |
| GOBP_POS | 21 | -0.25299 | -0.00843 | 0.10817 | 0.99032 | 16840.39 |
| GOMF_PR | 12 | -0.37534 | -0.00946 | 0.16049 | 0.99032 | 16840.39 |
| GOBP_PYR | 5 | -0.41079 | -0.00668 | 0.17516 | 0.99049 | 16843.28 |
| GOBP_POS | 6 | -0.42853 | -0.00764 | 0.18259 | 0.99053 | 16843.96 |
| GOBP_XEN | 6 | -0.57689 | -0.01028 | 0.24536 | 0.99064 | 16845.83 |
| GOMF_COI | 7 | -0.46012 | -0.00886 | 0.19543 | 0.99072 | 16847.19 |
| GOBP_AMI | 105 | -0.12272 | -0.00913 | 0.052113 | 0.99073 | 16847.36 |

| | | | | | | |
|---|---|---|---|---|---|---|
| FIGUEROA_ | 5 | -0.61247 | -0.00997 | 0.25967 | 0.99082 | 16848.89 |
| GOBP_POS | 36 | -0.20059 | -0.00875 | 0.084777 | 0.991 | 16851.96 |
| WP_CILIOP | 175 | -0.08824 | -0.00846 | 0.03707 | 0.99134 | 16857.74 |
| GOBP_REG | 319 | -0.06887 | -0.00888 | 0.028933 | 0.99135 | 16857.91 |
| GOBP_POS | 7 | -0.47849 | -0.00921 | 0.20092 | 0.99137 | 16858.25 |
| LIN_SILENC | 103 | -0.12028 | -0.00886 | 0.050478 | 0.9914 | 16858.76 |
| GOBP_REG | 8 | -0.4077 | -0.00839 | 0.17103 | 0.99143 | 16859.27 |
| REACTOME | 11 | -0.2884 | -0.00696 | 0.12078 | 0.99152 | 16860.8 |
| GOCC_ACE | 90 | -0.11545 | -0.00795 | 0.04823 | 0.99166 | 16863.18 |
| GOBP_POS | 6 | -0.3899 | -0.00695 | 0.1628 | 0.99168 | 16863.52 |
| REACTOME | 13 | -0.35076 | -0.0092 | 0.14644 | 0.99169 | 16863.69 |
| GOBP_NEG | 40 | -0.17638 | -0.00811 | 0.073616 | 0.99171 | 16864.03 |
| REACTOME | 8 | -0.36114 | -0.00743 | 0.15069 | 0.99172 | 16864.2 |
| GOBP_EXO | 83 | -0.12902 | -0.00854 | 0.053771 | 0.99178 | 16865.22 |
| GOMF_CEL | 5 | -0.55542 | -0.00904 | 0.23063 | 0.99198 | 16868.62 |
| GOMF_MH | 7 | -0.48116 | -0.00926 | 0.19953 | 0.99205 | 16869.81 |
| GOBP_POS | 31 | -0.22071 | -0.00894 | 0.091392 | 0.99213 | 16871.17 |
| GOCC_FILO | 17 | -0.29978 | -0.00899 | 0.12405 | 0.99216 | 16871.68 |
| GOBP_REG | 136 | -0.11062 | -0.00936 | 0.045622 | 0.99234 | 16874.74 |
| GOBP_POS | 135 | -0.104 | -0.00876 | 0.042638 | 0.99263 | 16879.67 |
| GOMF_OLF | 254 | -0.10368 | -0.01195 | 0.042378 | 0.99278 | 16882.22 |
| GOBP_STR | 5 | -0.45024 | -0.00733 | 0.18356 | 0.99291 | 16884.43 |
| GOMF_GTF | 22 | -0.28376 | -0.00968 | 0.11565 | 0.99293 | 16884.77 |
| GOBP_DEC | 11 | -0.34939 | -0.00843 | 0.14202 | 0.99305 | 16886.82 |
| GOBP_AM/ | 12 | -0.38216 | -0.00963 | 0.15524 | 0.99308 | 16887.33 |
| GOBP_POS | 37 | -0.19133 | -0.00846 | 0.077539 | 0.99319 | 16889.2 |
| GOBP_POS | 4 | -0.79426 | -0.01156 | 0.32117 | 0.9933 | 16891.07 |
| GOBP_RES | 11 | -0.39771 | -0.0096 | 0.16029 | 0.99345 | 16893.62 |
| GOBP_NEG | 5 | -0.46802 | -0.00762 | 0.18855 | 0.99347 | 16893.96 |
| VALK_AML | 25 | -0.2749 | -0.01 | 0.11067 | 0.9935 | 16894.47 |
| SCHAEFFER | 33 | -0.21789 | -0.0091 | 0.087668 | 0.99353 | 16894.98 |
| GOBP_CHE | 122 | -0.11841 | -0.00949 | 0.047554 | 0.99361 | 16896.34 |
| REACTOME | 47 | -0.20509 | -0.01022 | 0.082207 | 0.99369 | 16897.7 |
| GOBP_POS | 8 | -0.41123 | -0.00846 | 0.16443 | 0.9938 | 16899.57 |
| GOCC_KAI | 5 | -0.57825 | -0.00941 | 0.23075 | 0.99389 | 16901.1 |
| GOMF_KAI | 5 | -0.57825 | -0.00941 | 0.23075 | 0.99389 | 16901.1 |
| GOBP_NEG | 13 | -0.35716 | -0.00937 | 0.14241 | 0.99392 | 16901.61 |
| GOBP_REG | 9 | -0.37844 | -0.00826 | 0.15066 | 0.99399 | 16902.8 |
| REACTOME | 16 | -0.28067 | -0.00817 | 0.11151 | 0.99408 | 16904.33 |
| GOBP_RES | 5 | -0.55614 | -0.00905 | 0.22082 | 0.9941 | 16904.67 |
| GOBP_LYM | 66 | -0.15664 | -0.00925 | 0.062182 | 0.99411 | 16904.84 |
| GOBP_NEG | 9 | -0.41746 | -0.00911 | 0.16535 | 0.99421 | 16906.54 |
| GOCC_STE | 20 | -0.2768 | -0.009 | 0.1096 | 0.99422 | 16906.71 |
| GOBP_SYN | 11 | -0.37371 | -0.00902 | 0.14791 | 0.99424 | 16907.05 |
| GOBP_CEL | 49 | -0.17236 | -0.00877 | 0.06803 | 0.99435 | 16908.92 |

| | | | | | | |
|---|---|---|---|---|---|---|
| GOMF_EX( | 29 | -0.26103 | -0.01022 | 0.10287 | 0.99441 | 16909.94 |
| GOMF_ALD | 17 | -0.3304 | -0.00991 | 0.13016 | 0.99443 | 16910.28 |
| REACTOME | 63 | -0.14704 | -0.00848 | 0.057573 | 0.99467 | 16914.36 |
| GOBP_POS | 238 | -0.08626 | -0.00962 | 0.03377 | 0.99468 | 16914.53 |
| KEGG_RNA | 26 | -0.21271 | -0.00789 | 0.083071 | 0.99477 | 16916.06 |
| GOBP_PHC | 28 | -0.23732 | -0.00913 | 0.09254 | 0.99483 | 16917.08 |
| GOBP_CILI | 183 | -0.0905 | -0.00887 | 0.03526 | 0.99486 | 16917.59 |
| GOMF_PR( | 8 | -0.46753 | -0.00962 | 0.18175 | 0.99494 | 16918.95 |
| GOCC_LUN | 15 | -0.30494 | -0.00859 | 0.11842 | 0.99498 | 16919.63 |
| GOMF_NU | 141 | -0.10507 | -0.00905 | 0.040777 | 0.99501 | 16920.15 |
| BOYAULT_ | 39 | -0.21441 | -0.00974 | 0.082957 | 0.99512 | 16922.02 |
| IM_SREBF1 | 5 | -0.46917 | -0.00763 | 0.18104 | 0.99522 | 16923.72 |
| GOMF_ASI | 7 | -0.41663 | -0.00802 | 0.16072 | 0.99523 | 16923.89 |
| GOBP_POS | 64 | -0.15851 | -0.00921 | 0.061101 | 0.99526 | 16924.4 |
| REACTOME | 83 | -0.13657 | -0.00904 | 0.052639 | 0.99526 | 16924.4 |
| REACTOME | 11 | -0.45391 | -0.01095 | 0.1749 | 0.99527 | 16924.57 |
| REACTOME | 59 | -0.15445 | -0.00862 | 0.059472 | 0.99529 | 16924.91 |
| GOBP_REG | 213 | -0.08876 | -0.00937 | 0.034105 | 0.99537 | 16926.27 |
| GOMF_GAI | 11 | -0.49677 | -0.01199 | 0.19074 | 0.99539 | 16926.61 |
| GOBP_RES | 397 | -0.06437 | -0.00924 | 0.024527 | 0.99566 | 16931.2 |
| GOMF_TRA | 37 | -0.19687 | -0.00871 | 0.074878 | 0.99572 | 16932.22 |
| WU_CELL_ | 172 | -0.10505 | -0.00998 | 0.039919 | 0.99574 | 16932.56 |
| GOMF_NEU | 34 | -0.25086 | -0.01064 | 0.095106 | 0.99582 | 16933.92 |
| GOBP_NEG | 78 | -0.15738 | -0.0101 | 0.05954 | 0.99589 | 16935.11 |
| GOMF_BET | 36 | -0.21225 | -0.00926 | 0.08024 | 0.99591 | 16935.45 |
| REACTOME | 56 | -0.17845 | -0.0097 | 0.067346 | 0.99597 | 16936.47 |
| GOBP_HYD | 48 | -0.18931 | -0.00953 | 0.071381 | 0.996 | 16936.98 |
| KEGG_FOL | 11 | -0.53085 | -0.01281 | 0.20002 | 0.99602 | 16937.32 |
| GOBP_REG | 874 | -0.04791 | -0.01007 | 0.018045 | 0.99603 | 16937.49 |
| THILLAINAI | 8 | -0.51345 | -0.01057 | 0.19335 | 0.99604 | 16937.66 |
| GOBP_B_C | 99 | -0.15065 | -0.01088 | 0.056259 | 0.99629 | 16941.91 |
| GOCC_GAE | 16 | -0.42296 | -0.01231 | 0.15704 | 0.99646 | 16944.8 |
| GOMF_GAI | 17 | -0.40819 | -0.01224 | 0.15104 | 0.99656 | 16946.5 |
| GOBP_LIPC | 60 | -0.18371 | -0.01034 | 0.067842 | 0.99661 | 16947.35 |
| GOMF_PEF | 8 | -0.47399 | -0.00976 | 0.17483 | 0.99664 | 16947.86 |
| GOBP_AMI | 12 | -0.39888 | -0.01005 | 0.14601 | 0.99685 | 16951.43 |
| GOMF_AM | 12 | -0.39888 | -0.01005 | 0.14601 | 0.99685 | 16951.43 |
| WP_ACTIV | 7 | -0.51411 | -0.0099 | 0.18778 | 0.9969 | 16952.28 |
| GOMF_NAI | 53 | -0.17684 | -0.00936 | 0.064515 | 0.99693 | 16952.79 |
| GOCC_PRE | 77 | -0.15301 | -0.00975 | 0.055765 | 0.99696 | 16953.3 |
| WP_EUKAF | 39 | -0.18759 | -0.00852 | 0.068262 | 0.997 | 16953.99 |
| SPIRA_SM( | 36 | -0.23245 | -0.01014 | 0.084008 | 0.99717 | 16956.88 |
| GOBP_INN | 15 | -0.33633 | -0.00948 | 0.12135 | 0.99721 | 16957.56 |
| GOCC_GOL | 18 | -0.32154 | -0.00992 | 0.11602 | 0.99721 | 16957.56 |
| GOBP_DET | 543 | -0.07307 | -0.01221 | 0.026313 | 0.99725 | 16958.24 |

| | | | | | | |
|---|---|---|---|---|---|---|
| GOBP_POS | 15 | -0.37639 | -0.01061 | 0.1354 | 0.99728 | 16958.75 |
| GOBP_SEN | 343 | -0.10564 | -0.01411 | 0.037765 | 0.99742 | 16961.13 |
| GOMF_LIG | 15 | -0.38612 | -0.01088 | 0.13759 | 0.99749 | 16962.32 |
| MISHRA_C. | 21 | -0.27683 | -0.00923 | 0.098207 | 0.99759 | 16964.02 |
| GOMF_BUI | 8 | -0.62659 | -0.0129 | 0.22075 | 0.99773 | 16966.4 |
| GOMF_1_F | 15 | -0.33414 | -0.00942 | 0.11739 | 0.99779 | 16967.42 |
| GOBP_REG | 20 | -0.34257 | -0.01114 | 0.12015 | 0.99782 | 16967.93 |
| CROMER_I | 77 | -0.16098 | -0.01026 | 0.056406 | 0.99784 | 16968.27 |
| GOBP_PRC | 15 | -0.38132 | -0.01074 | 0.13362 | 0.99784 | 16968.27 |
| GOBP_PEP | 9 | -0.47397 | -0.01035 | 0.16593 | 0.99786 | 16968.61 |
| GOMF_PH( | 9 | -0.47306 | -0.01033 | 0.16483 | 0.99794 | 16969.97 |
| GOBP_POS | 7 | -0.53884 | -0.01037 | 0.18718 | 0.998 | 16970.99 |
| GOBP_SYN | 19 | -0.34213 | -0.01085 | 0.11876 | 0.99802 | 16971.33 |
| GOCC_ZON | 8 | -0.47155 | -0.00971 | 0.1627 | 0.99812 | 16973.03 |
| GOCC_PRE | 32 | -0.25939 | -0.01067 | 0.086529 | 0.99864 | 16981.87 |
| GOBP_REG | 3 | -1.0283 | -0.01296 | 0.3417 | 0.99869 | 16982.72 |
| GOMF_PR( | 5 | -0.628 | -0.01022 | 0.20846 | 0.9987 | 16982.89 |
| GOCC_LUM | 8 | -0.46249 | -0.00952 | 0.15314 | 0.99873 | 16983.4 |
| GOMF_TR/ | 51 | -0.24409 | -0.01267 | 0.080544 | 0.99878 | 16984.25 |
| GOCC_SCA | 11 | -0.39423 | -0.00951 | 0.12903 | 0.99887 | 16985.78 |
| GOBP_SYN | 92 | -0.1615 | -0.01125 | 0.052684 | 0.99891 | 16986.46 |
| GOBP_POS | 512 | -0.07142 | -0.0116 | 0.023254 | 0.99893 | 16986.8 |
| GOBP_DET | 430 | -0.0975 | -0.01455 | 0.031709 | 0.99895 | 16987.14 |
| WP_ANTIV | 27 | -0.31723 | -0.01199 | 0.10029 | 0.99922 | 16991.74 |
| KEGG_ASC | 22 | -0.45689 | -0.01559 | 0.14365 | 0.99926 | 16992.42 |
| FLECHNER_ | 79 | -0.17622 | -0.01138 | 0.054193 | 0.99943 | 16995.31 |
| GOBP_DET | 399 | -0.109 | -0.01568 | 0.033465 | 0.99944 | 16995.48 |
| REACTOME | 10 | -0.50367 | -0.01159 | 0.15396 | 0.99946 | 16995.82 |
| GOBP_CYT | 4 | -0.82154 | -0.01196 | 0.25098 | 0.99947 | 16995.99 |
| GOMF_EXT | 62 | -0.23577 | -0.01349 | 0.071633 | 0.9995 | 16996.5 |
| GOCC_PRE | 154 | -0.14244 | -0.01281 | 0.042094 | 0.99964 | 16998.88 |
| GOBP_NER | 1348 | -0.05229 | -0.01346 | 0.01536 | 0.99967 | 16999.39 |
| GOBP_AMI | 5 | -0.92679 | -0.01508 | 0.26761 | 0.99973 | 17000.41 |
| KEGG_OLF. | 326 | -0.12834 | -0.01672 | 0.03705 | 0.99973 | 17000.41 |
| REACTOME | 326 | -0.13726 | -0.01788 | 0.039421 | 0.99975 | 17000.75 |
| GOBP_SEN | 418 | -0.11679 | -0.01719 | 0.03251 | 0.99984 | 17002.28 |
| REACTOME | 5 | -0.73306 | -0.01193 | 0.20335 | 0.99984 | 17002.28 |
| GOMF_NO | 41 | -0.29114 | -0.01355 | 0.080152 | 0.99986 | 17002.62 |
| GOBP_SEN | 854 | -0.07347 | -0.01527 | 0.020122 | 0.99987 | 17002.79 |
| GOCC_IKAI | 5 | -0.80566 | -0.01311 | 0.20443 | 0.99996 | 17004.32 |
| REACTOME | 530 | -0.10456 | -0.01727 | 0.026601 | 0.99996 | 17004.32 |
