## Supplementary Table 8 for "Hundreds of cardiac MRI traits derived using 3D diffusion autoencoders share a common genetic architecture"

| Gene_set | Term | Overlap | P-value | Adjusted P | Old P-value | Old Adjuste | Odds Ratio | Combined |
|---|---|---|---|---|---|---|---|---|
| UK_Bioban | Pulse rate 4 | 13/71 | 3.71E-18 | 1.06E-15 | 0 | 0 | 60.13863 | 2413.658 |
| UK_Bioban | ECG heart r | 8-May | 7.68E-11 | 1.09E-08 | 0 | 0 | 404.6748 | 9424.694 |
| GeDiPNet_ | Gout | 12/183 | 2.34E-11 | 1.36E-08 | 0 | 0 | 18.47205 | 452.1736 |
| PhenGenI_ | Heart Rate | 14/314 | 8.00E-11 | 1.85E-08 | 0 | 0 | 12.53799 | 291.5024 |
| GeDiPNet_ | Gouty Arth | 11/159 | 9.42E-11 | 2.75E-08 | 0 | 0 | 19.32921 | 446.2307 |
| dbGaP | Metabolic S | 23-Jun | 5.41E-10 | 3.52E-08 | 0 | 0 | 86.69281 | 1849.841 |
| GWAS_Cat | Resting He | Aug-63 | 2.94E-10 | 3.59E-07 | 0 | 0 | 36.56249 | 802.4455 |
| UK_Bioban | Automatec | 12/304 | 7.86E-09 | 7.46E-07 | 0 | 0 | 10.75123 | 200.6377 |
| UK_Bioban | Forced vita | 14/480 | 1.90E-08 | 1.35E-06 | 0 | 0 | 8.003351 | 142.2894 |
| dbGaP | Alzheimer I | Jul-80 | 5.66E-08 | 1.84E-06 | 0 | 0 | 23.78082 | 396.8205 |
| DisGeNET | Uric acid m | 9/112 | 1.39E-09 | 3.65E-06 | 0 | 0 | 22.19193 | 452.5446 |
| UK_Bioban | Forced exp | 13/447 | 6.65E-08 | 3.79E-06 | 0 | 0 | 7.884761 | 130.3062 |
| DisGeNET | RESTING H | Jul-53 | 3.02E-09 | 3.96E-06 | 0 | 0 | 37.79049 | 741.3632 |
| ClinVar_20 | primary dil | May-34 | 3.49E-07 | 4.37E-06 | 0 | 0 | 41.80824 | 621.5792 |
| ClinVar_20 | familial dila | 30-May | 1.81E-07 | 4.37E-06 | 0 | 0 | 48.50732 | 752.9731 |
| GWAS_Cat | Waist Circu | 7-Apr | 1.16E-08 | 7.06E-06 | 0 | 0 | 319.8394 | 5844.973 |
| UK_Bioban | Comparativ | 16/760 | 1.54E-07 | 7.31E-06 | 0 | 0 | 5.806149 | 91.0798 |
| ClinVar_20 | cardiomyop | May-48 | 2.05E-06 | 1.44E-05 | 0 | 0 | 28.1764 | 369.0584 |
| ClinVar_20 | primary far | 22-Apr | 2.30E-06 | 1.44E-05 | 0 | 0 | 53.2664 | 691.5122 |
| OMIM_Dis | cardiomyop | May-42 | 1.04E-06 | 1.56E-05 | 0 | 0 | 32.75544 | 451.2791 |
| ClinVar_20 | limb-girdle | 25-Apr | 3.94E-06 | 1.97E-05 | 0 | 0 | 45.65003 | 568.0859 |
| PhenGenI_ | Metabolic S | Jun-60 | 2.36E-07 | 2.72E-05 | 0 | 0 | 27.24143 | 415.7011 |
| ClinVar_20 | limb-girdle | 9-Mar | 6.55E-06 | 2.73E-05 | 0 | 0 | 118.494 | 1414.295 |
| UK_Bioban | Forced exp | 12/463 | 7.57E-07 | 3.08E-05 | 0 | 0 | 6.904479 | 97.3099 |
| Jensen_DIS | Dilated car | May-32 | 2.55E-07 | 4.96E-05 | 0 | 0 | 44.90967 | 681.9048 |
| UK_Bioban | Forced vita | 12/496 | 1.56E-06 | 4.99E-05 | 0 | 0 | 6.42281 | 85.89631 |
| UK_Bioban | Maximum I | 6-Mar | 1.58E-06 | 4.99E-05 | 0 | 0 | 237.0238 | 3166.928 |
| GWAS_Cat | Triglyceride | 16/774 | 1.97E-07 | 5.26E-05 | 0 | 0 | 5.694749 | 87.92652 |
| GWAS_Cat | Descending | 31-May | 2.15E-07 | 5.26E-05 | 0 | 0 | 46.63931 | 715.9321 |
| GWAS_Cat | Fasting Blo | 12-Apr | 1.61E-07 | 5.26E-05 | 0 | 0 | 119.9096 | 1875.637 |
| UK_Bioban | Right arm p | 15/815 | 2.13E-06 | 6.06E-05 | 0 | 0 | 4.977344 | 65.01177 |
| PhenGenI_ | Amyloid be | 20-Apr | 1.53E-06 | 8.86E-05 | 0 | 0 | 59.93072 | 802.322 |
| PhenGenI_ | Cerebral Co | 19-Apr | 1.23E-06 | 8.86E-05 | 0 | 0 | 63.92932 | 869.9064 |
| OMIM_Dis | cardiomyop | Apr-33 | 1.24E-05 | 9.31E-05 | 0 | 0 | 33.04362 | 373.2919 |
| Jensen_DIS | Cardiovasc | 9/246 | 1.24E-06 | 0.000121 | 0 | 0 | 9.579357 | 130.2875 |
| Human_Ph | Dilated car | Jun-65 | 3.82E-07 | 0.000146 | 0 | 0 | 24.92655 | 368.3283 |
| DisGeNET | heart rate | 30-May | 1.81E-07 | 0.000158 | 0 | 0 | 48.50732 | 752.9731 |
| GWAS_Cat | Predicted H | 5-Mar | 7.90E-07 | 0.000161 | 0 | 0 | 355.5536 | 4995.963 |
| UK_Bioban | Alzheimer's | 9-Mar | 6.55E-06 | 0.00017 | 0 | 0 | 118.494 | 1414.295 |
| GeDiPNet_ | Atrial Fibril | 9/248 | 1.33E-06 | 0.000193 | 0 | 0 | 9.49823 | 128.5474 |
| GeDiPNet_ | Dilated Car | May-44 | 1.32E-06 | 0.000193 | 0 | 0 | 31.07255 | 420.7164 |
| Human_Ph | Palmoplant | 7/122 | 1.03E-06 | 0.000197 | 0 | 0 | 15.0637 | 207.6539 |
| GWAS_Cat | Fat-free Ma | 7/124 | 1.15E-06 | 0.000201 | 0 | 0 | 14.8047 | 202.4596 |
| GWAS_Cat | Fasting Blo | 6-Mar | 1.58E-06 | 0.000214 | 0 | 0 | 237.0238 | 3166.928 |

| | | | | | | | | |
|---|---|---|---|---|---|---|---|---|
| GWAS_Cat | Vertex-wis | 11/405 | 1.44E-06 | 0.000214 | 0 | 0 | 7.170351 | 96.42315 |
| UK_Bioban | Forced exp | 10/400 | 9.30E-06 | 0.000217 | 0 | 0 | 6.501166 | 75.32108 |
| UK_Bioban | Right arm f | 14/810 | 9.90E-06 | 0.000217 | 0 | 0 | 4.605872 | 53.07164 |
| GeDiPNet_ | Arrhythmo | 21-Apr | 1.89E-06 | 0.00022 | 0 | 0 | 56.40255 | 743.3574 |
| GWAS_Cat | Body Comp | May-48 | 2.05E-06 | 0.00025 | 0 | 0 | 28.1764 | 369.0584 |
| OMIM_Dis | cardiomyo | 17-Mar | 5.17E-05 | 0.000259 | 0 | 0 | 50.76276 | 501.0024 |
| UK_Bioban | Basal meta | 14/834 | 1.38E-05 | 0.00028 | 0 | 0 | 4.465453 | 49.98808 |
| Rare_Disea | Arrhythmo | May-37 | 5.42E-07 | 0.000305 | 0 | 0 | 37.883 | 546.605 |
| Rare_Disea | Myotubula | Jun-79 | 1.23E-06 | 0.000305 | 0 | 0 | 20.13191 | 274.0283 |
| Rare_Disea | Primary pr | 19-Apr | 1.23E-06 | 0.000305 | 0 | 0 | 63.92932 | 869.9064 |
| GeDiPNet_ | Palmoplan | 7/145 | 3.28E-06 | 0.000318 | 0 | 0 | 12.5385 | 158.3405 |
| PhenGenI_ | Breast | May-62 | 7.38E-06 | 0.000341 | 0 | 0 | 21.24091 | 250.9915 |
| UK_Bioban | Whole bod | 14/880 | 2.50E-05 | 0.00042 | 0 | 0 | 4.218071 | 44.70252 |
| UK_Bioban | Left arm pr | 13/763 | 2.47E-05 | 0.00042 | 0 | 0 | 4.488631 | 47.61088 |
| UK_Bioban | Right leg pr | 13/764 | 2.51E-05 | 0.00042 | 0 | 0 | 4.48242 | 47.48381 |
| DisGeNET | Cardiomyo | Jun-73 | 7.67E-07 | 0.000502 | 0 | 0 | 21.9414 | 308.9603 |
| UK_Bioban | Trunk pred | 14/904 | 3.36E-05 | 0.000514 | 0 | 0 | 4.099153 | 42.23103 |
| UK_Bioban | Sitting heig | 15/1029 | 3.43E-05 | 0.000514 | 0 | 0 | 3.882931 | 39.92304 |
| Rare_Disea | Familial dil | May-51 | 2.78E-06 | 0.000518 | 0 | 0 | 26.33484 | 336.8806 |
| Human_Ph | Lipoatroph | May-55 | 4.06E-06 | 0.000518 | 0 | 0 | 24.22317 | 300.6869 |
| UK_Bioban | Trunk fat-fr | 14/917 | 3.92E-05 | 0.000559 | 0 | 0 | 4.037379 | 40.96551 |
| GeDiPNet_ | Paroxysma | 7/163 | 7.10E-06 | 0.000591 | 0 | 0 | 11.08165 | 131.3814 |
| UK_Bioban | Standing he | 18/1456 | 4.55E-05 | 0.000618 | 0 | 0 | 3.351575 | 33.50593 |
| Rare_Disea | Familial hy | May-61 | 6.81E-06 | 0.000633 | 0 | 0 | 21.6213 | 257.2306 |
| Rare_Disea | Single vent | 6/104 | 6.14E-06 | 0.000633 | 0 | 0 | 14.97732 | 179.7443 |
| Rare_Disea | Idiopathic ( | 7/159 | 6.03E-06 | 0.000633 | 0 | 0 | 11.37558 | 136.7233 |
| Rare_Disea | Dilated car | 7/159 | 6.03E-06 | 0.000633 | 0 | 0 | 11.37558 | 136.7233 |
| Rare_Disea | Progressive | 10-Mar | 9.33E-06 | 0.000771 | 0 | 0 | 101.5612 | 1176.288 |
| GWAS_Cat | Triglyceride | 8/236 | 8.75E-06 | 0.000773 | 0 | 0 | 8.74306 | 101.8287 |
| GWAS_Cat | Breast Size | 30-Apr | 8.39E-06 | 0.000773 | 0 | 0 | 36.86191 | 430.8378 |
| GWAS_Cat | Cortical Thi | 10/401 | 9.50E-06 | 0.000773 | 0 | 0 | 6.484206 | 74.9837 |
| GWAS_Cat | Diastolic Bl | 14/803 | 8.98E-06 | 0.000773 | 0 | 0 | 4.648437 | 54.0179 |
| GWAS_Cat | Alzheimer's | 10-Mar | 9.33E-06 | 0.000773 | 0 | 0 | 101.5612 | 1176.288 |
| UK_Bioban | Left 6mm s | 6/157 | 6.36E-05 | 0.000824 | 0 | 0 | 9.694383 | 93.68059 |
| UK_Bioban | Diastolic bl | 8/314 | 6.73E-05 | 0.000834 | 0 | 0 | 6.488624 | 62.33267 |
| UK_Bioban | Left arm fa | 14/977 | 7.76E-05 | 0.000885 | 0 | 0 | 3.77388 | 35.71358 |
| UK_Bioban | Left leg fat- | 12/733 | 7.66E-05 | 0.000885 | 0 | 0 | 4.258974 | 40.35977 |
| UK_Bioban | Sodium in u | Apr-54 | 8.95E-05 | 0.000981 | 0 | 0 | 19.14506 | 178.4623 |
| UK_Bioban | Whole bod | 13/873 | 9.81E-05 | 0.001035 | 0 | 0 | 3.892033 | 35.92206 |
| GWAS_Cat | Systolic Blo | 16/1073 | 1.36E-05 | 0.00104 | 0 | 0 | 4.020094 | 45.03364 |
| DisGeNET | Intracrania | 22-Apr | 2.30E-06 | 0.001056 | 0 | 0 | 53.2664 | 691.5122 |
| DisGeNET | Waist Circu | Jun-91 | 2.82E-06 | 0.001056 | 0 | 0 | 17.2793 | 220.8056 |
| DisGeNET | response to | 23-Apr | 2.78E-06 | 0.001056 | 0 | 0 | 50.46037 | 645.6106 |
| Jensen_DIS | Acquired m | 9/338 | 1.63E-05 | 0.001059 | 0 | 0 | 6.868366 | 75.72351 |
| UK_Bioban | Right leg fa | 12/764 | 0.000113 | 0.001114 | 0 | 0 | 4.076809 | 37.03732 |

| | | | | | | | | |
|---|---|---|---|---|---|---|---|---|
| UK_Bioban | Hip circum | 10/537 | 0.000112 | 0.001114 | 0 | 0 | 4.777348 | 43.44769 |
| GWAS_Cat | Atrial Fibril | 7/186 | 1.68E-05 | 0.001204 | 0 | 0 | 9.646508 | 106.0721 |
| Jensen_DIS | Cardiomyo | May-80 | 2.58E-05 | 0.001257 | 0 | 0 | 16.12846 | 170.4037 |
| CellMarker | Cardiomyo | 26-Apr | 4.64E-06 | 0.001272 | 0 | 0 | 43.57284 | 535.1021 |
| CellMarker | Cardiomyo | 10-Mar | 9.33E-06 | 0.001278 | 0 | 0 | 101.5612 | 1176.288 |
| Rare_Disea | Congenital | May-74 | 1.76E-05 | 0.001312 | 0 | 0 | 17.53623 | 191.9395 |
| DisGeNET | CARDIOMY | 8-Mar | 4.38E-06 | 0.001314 | 0 | 0 | 142.2 | 1754.449 |
| DisGeNET | Atrial Fibril | 9/288 | 4.51E-06 | 0.001314 | 0 | 0 | 8.119934 | 99.94322 |
| GeDiPNet_ | Neuromusc | 13-Mar | 2.20E-05 | 0.001415 | 0 | 0 | 71.08214 | 762.2138 |
| GeDiPNet_ | Metabolic | May-79 | 2.43E-05 | 0.001415 | 0 | 0 | 16.34723 | 173.7145 |
| GeDiPNet_ | Lipoatroph | May-77 | 2.14E-05 | 0.001415 | 0 | 0 | 16.80301 | 180.6552 |
| GWAS_Cat | Alzheimer's | May-79 | 2.43E-05 | 0.001645 | 0 | 0 | 16.34723 | 173.7145 |
| DisGeNET | Centronucl | 28-Apr | 6.31E-06 | 0.001654 | 0 | 0 | 39.93775 | 478.1657 |
| GWAS_Cat | Family Hist | Apr-40 | 2.71E-05 | 0.001739 | 0 | 0 | 26.6091 | 279.838 |
| UK_Bioban | Medicatior | 26-Mar | 0.000192 | 0.001827 | 0 | 0 | 30.88509 | 264.2718 |
| HuBMAP_A | Cardiac Mu | 6/105 | 6.49E-06 | 0.001868 | 0 | 0 | 14.82529 | 177.0988 |
| GWAS_Cat | Urinary Alb | May-83 | 3.08E-05 | 0.001881 | 0 | 0 | 15.50578 | 161.0565 |
| UK_Bioban | Eats other | 27-Mar | 0.000216 | 0.001982 | 0 | 0 | 29.59673 | 249.8547 |
| GWAS_Cat | Descending | Apr-43 | 3.62E-05 | 0.002008 | 0 | 0 | 24.55854 | 251.1403 |
| GWAS_Cat | RR Interval | 15-Mar | 3.48E-05 | 0.002008 | 0 | 0 | 59.22917 | 607.9868 |
| UK_Bioban | None of th | 7/285 | 0.000245 | 0.002181 | 0 | 0 | 6.180081 | 51.38491 |
| GWAS_Cat | Cerebral Ar | 16-Mar | 4.27E-05 | 0.002267 | 0 | 0 | 54.67033 | 550.0107 |
| UK_Bioban | Any ICDMA | 6-Feb | 0.000277 | 0.002396 | 0 | 0 | 117.1118 | 959.145 |
| Jensen_DIS | Hypertroph | 18-Mar | 6.19E-05 | 0.002413 | 0 | 0 | 47.37619 | 459.0901 |
| SysMyo_M | MGS1341 I | 7/151 | 4.29E-06 | 0.002427 | 0 | 0 | 12.01241 | 148.4686 |
| GeDiPNet_ | Congestive | 8/300 | 4.89E-05 | 0.002513 | 0 | 0 | 6.804578 | 67.54476 |
| GeDiPNet_ | Centronucl | 17-Mar | 5.17E-05 | 0.002513 | 0 | 0 | 50.76276 | 501.0024 |
| GWAS_Cat | C-reactive | 9/393 | 5.29E-05 | 0.002582 | 0 | 0 | 5.868089 | 57.78177 |
| GWAS_Cat | Pulse Rate | 17-Mar | 5.17E-05 | 0.002582 | 0 | 0 | 50.76276 | 501.0024 |
| PheWeb_2 | Hyperlipide | Apr-46 | 4.74E-05 | 0.002892 | 0 | 0 | 22.80092 | 227.0231 |
| PheWeb_2 | Disorders c | Apr-45 | 4.34E-05 | 0.002892 | 0 | 0 | 23.35821 | 234.6195 |
| UK_Bioban | Left leg pre | 11/738 | 0.00035 | 0.002931 | 0 | 0 | 3.819699 | 30.39897 |
| GeDiPNet_ | Myofibrilla | 19-Mar | 7.33E-05 | 0.003285 | 0 | 0 | 44.41295 | 422.8826 |
| OMIM_Dis | myopathy | Mar-44 | 0.000926 | 0.003471 | 0 | 0 | 17.3101 | 120.912 |
| CellMarker | Contractile | 16-Mar | 4.27E-05 | 0.003903 | 0 | 0 | 54.67033 | 550.0107 |
| GWAS_Cat | Estimated | 10/518 | 8.35E-05 | 0.003917 | 0 | 0 | 4.960886 | 46.58681 |
| GWAS_Cat | Creatinine | 8/326 | 8.74E-05 | 0.003932 | 0 | 0 | 6.239949 | 58.31387 |
| GWAS_Cat | Drinks Per | 9/426 | 9.80E-05 | 0.003932 | 0 | 0 | 5.394577 | 49.79307 |
| GWAS_Cat | Nonalcoho | 21-Mar | 9.99E-05 | 0.003932 | 0 | 0 | 39.47421 | 363.6059 |
| GWAS_Cat | L-selectin L | 21-Mar | 9.99E-05 | 0.003932 | 0 | 0 | 39.47421 | 363.6059 |
| GWAS_Cat | Uterine Lei | 21-Mar | 9.99E-05 | 0.003932 | 0 | 0 | 39.47421 | 363.6059 |
| dbGaP | Blood Sedi | 5-Feb | 0.000185 | 0.004018 | 0 | 0 | 156.1569 | 1341.798 |
| UK_Bioban | Soft cheese | Apr-84 | 0.000494 | 0.004026 | 0 | 0 | 11.94759 | 90.94771 |
| Rare_Disea | Wilson dise | 7/229 | 6.32E-05 | 0.004277 | 0 | 0 | 7.761092 | 75.04001 |
| UK_Bioban | Right 6mm | 5/153 | 0.000547 | 0.004334 | 0 | 0 | 8.143128 | 61.15618 |

| | | | | | | | | |
|---|---|---|---|---|---|---|---|---|
| GWAS_Cat | Heart Rate | 22-Mar | 0.000115 | 0.004397 | 0 | 0 | 37.39474 | 339.0868 |
| GeDiPNet_ | Lipodystro| | 5/108 | 0.000109 | 0.004527 | 0 | 0 | 11.72744 | 107.0331 |
| UK_Bioban | Dried fruit | Mar-38 | 0.000601 | 0.004628 | 0 | 0 | 20.28367 | 150.4498 |
| DisGeNET | Familial dil | May-76 | 2.01E-05 | 0.004786 | 0 | 0 | 17.04054 | 184.2948 |
| GWAS_Cat | Whole Brai | 6/180 | 0.000135 | 0.004844 | 0 | 0 | 8.40315 | 74.87375 |
| GWAS_Cat | Lipid Traits | 23-Mar | 0.000132 | 0.004844 | 0 | 0 | 35.52321 | 317.2632 |
| GWAS_Cat | Apolipopro | 5-Feb | 0.000185 | 0.004941 | 0 | 0 | 156.1569 | 1341.798 |
| GWAS_Cat | Postprandi | 5-Feb | 0.000185 | 0.004941 | 0 | 0 | 156.1569 | 1341.798 |
| GWAS_Cat | Sick Sinus S | 5-Feb | 0.000185 | 0.004941 | 0 | 0 | 156.1569 | 1341.798 |
| GWAS_Cat | C-reactive | 9/464 | 0.000186 | 0.004941 | 0 | 0 | 4.934404 | 42.37714 |
| GWAS_Cat | Brain Morp | 9/463 | 0.000183 | 0.004941 | 0 | 0 | 4.945527 | 42.55211 |
| GWAS_Cat | Triglyceride | 8/361 | 0.000176 | 0.004941 | 0 | 0 | 5.611217 | 48.50279 |
| GWAS_Cat | Electrocarc | 7/269 | 0.000172 | 0.004941 | 0 | 0 | 6.562834 | 56.88435 |
| GWAS_Cat | Gallstone D | Apr-62 | 0.000154 | 0.004941 | 0 | 0 | 16.49771 | 144.8582 |
| GWAS_Cat | Erythrocyte | 5-Feb | 0.000185 | 0.004941 | 0 | 0 | 156.1569 | 1341.798 |
| GWAS_Cat | Fasting Fre | 5-Feb | 0.000185 | 0.004941 | 0 | 0 | 156.1569 | 1341.798 |
| GWAS_Cat | Fasting Mo | 5-Feb | 0.000185 | 0.004941 | 0 | 0 | 156.1569 | 1341.798 |
| GWAS_Cat | Body Mass | 5-Feb | 0.000185 | 0.004941 | 0 | 0 | 156.1569 | 1341.798 |
| CellMarker | Cardiomyo | 19-Mar | 7.33E-05 | 0.005018 | 0 | 0 | 44.41295 | 422.8826 |
| UK_Bioban | Fresh fruit | Mar-40 | 0.000699 | 0.00511 | 0 | 0 | 19.18533 | 139.392 |
| UK_Bioban | Left 6mm v | 5/161 | 0.00069 | 0.00511 | 0 | 0 | 7.722405 | 56.21249 |
| UK_Bioban | High light s | 8/446 | 0.00072 | 0.005133 | 0 | 0 | 4.50263 | 32.57994 |
| UK_Bioban | Weight 23( | 10/683 | 0.000757 | 0.005134 | 0 | 0 | 3.712781 | 26.68271 |
| UK_Bioban | Right 6mm | 5/164 | 0.00075 | 0.005134 | 0 | 0 | 7.575548 | 54.51247 |
| DisGeNET | Conductior | May-79 | 2.43E-05 | 0.005298 | 0 | 0 | 16.34723 | 173.7145 |
| UK_Bioban | None of the | 10-Feb | 0.000823 | 0.005454 | 0 | 0 | 58.54412 | 415.8217 |
| GWAS_Cat | Histidine Le | 27-Mar | 0.000216 | 0.005544 | 0 | 0 | 29.59673 | 249.8547 |
| GWAS_Cat | Alzheimer's | 6-Feb | 0.000277 | 0.005544 | 0 | 0 | 117.1118 | 959.145 |
| GWAS_Cat | Transmeml | 6-Feb | 0.000277 | 0.005544 | 0 | 0 | 117.1118 | 959.145 |
| GWAS_Cat | Inactive Ga | 6-Feb | 0.000277 | 0.005544 | 0 | 0 | 117.1118 | 959.145 |
| GWAS_Cat | Compleme | 6-Feb | 0.000277 | 0.005544 | 0 | 0 | 117.1118 | 959.145 |
| GWAS_Cat | Gamma Gl | 7/284 | 0.00024 | 0.005544 | 0 | 0 | 6.202708 | 51.70564 |
| GWAS_Cat | Appendicu | 12/836 | 0.000261 | 0.005544 | 0 | 0 | 3.706602 | 30.5814 |
| GWAS_Cat | Subcortical | 5/131 | 0.000268 | 0.005544 | 0 | 0 | 9.57559 | 78.7391 |
| GWAS_Cat | 1-linoleoyl- | 6-Feb | 0.000277 | 0.005544 | 0 | 0 | 117.1118 | 959.145 |
| GWAS_Cat | Fasting Om | 6-Feb | 0.000277 | 0.005544 | 0 | 0 | 117.1118 | 959.145 |
| GWAS_Cat | Postprandi | 6-Feb | 0.000277 | 0.005544 | 0 | 0 | 117.1118 | 959.145 |
| GWAS_Cat | Tumor Prot | 6-Feb | 0.000277 | 0.005544 | 0 | 0 | 117.1118 | 959.145 |
| GWAS_Cat | Endothelia | 6-Feb | 0.000277 | 0.005544 | 0 | 0 | 117.1118 | 959.145 |
| GWAS_Cat | Total Chole | 12/843 | 0.000282 | 0.005544 | 0 | 0 | 3.674031 | 30.03336 |
| GWAS_Cat | Trihexosylc | 6-Feb | 0.000277 | 0.005544 | 0 | 0 | 117.1118 | 959.145 |
| GWAS_Cat | Excessive H | 6-Feb | 0.000277 | 0.005544 | 0 | 0 | 117.1118 | 959.145 |
| DisGeNET | Congenital | 8/277 | 2.78E-05 | 0.005597 | 0 | 0 | 7.39504 | 77.58536 |
| DisGeNET | Hypoplasti | Apr-41 | 2.99E-05 | 0.005598 | 0 | 0 | 25.88864 | 269.6893 |
| DisGeNET | Arrhythmo | Apr-42 | 3.30E-05 | 0.005755 | 0 | 0 | 25.20609 | 260.1398 |

| | | | | | | | | |
|---|---|---|---|---|---|---|---|---|
| GWAS_Cat | Aspartate / | 5/134 | 0.000298 | 0.005772 | 0 | 0 | 9.351484 | 75.91653 |
| GWAS_Cat | Height (Sta | 8/392 | 0.000307 | 0.005859 | 0 | 0 | 5.150053 | 41.65087 |
| dbGaP | Cholestero | 7/304 | 0.000362 | 0.005879 | 0 | 0 | 5.779125 | 45.79638 |
| KEGG_202: | Dilated car | May-96 | 6.21E-05 | 0.0059 | 0 | 0 | 13.28196 | 128.6587 |
| Human_Ph | Abnormalit | 18-Mar | 6.19E-05 | 0.00591 | 0 | 0 | 47.37619 | 459.0901 |
| Azimuth_2 | Lung V2 (H | 9-Feb | 0.00066 | 0.006015 | 0 | 0 | 66.91092 | 489.9901 |
| Azimuth_2 | Fetal Devel | 9-Feb | 0.00066 | 0.006015 | 0 | 0 | 66.91092 | 489.9901 |
| Azimuth_2 | Heart-L2-A | 9-Feb | 0.00066 | 0.006015 | 0 | 0 | 66.91092 | 489.9901 |
| Azimuth_2 | Fetal Devel | 9-Feb | 0.00066 | 0.006015 | 0 | 0 | 66.91092 | 489.9901 |
| Azimuth_2 | Lung V2 (H | 9-Feb | 0.00066 | 0.006015 | 0 | 0 | 66.91092 | 489.9901 |
| Azimuth_2 | Motor Cort | 9-Feb | 0.00066 | 0.006015 | 0 | 0 | 66.91092 | 489.9901 |
| Azimuth_2 | Lung v1-L1- | 9-Feb | 0.00066 | 0.006015 | 0 | 0 | 66.91092 | 489.9901 |
| Azimuth_2 | Heart-L2-V | 9-Feb | 0.00066 | 0.006015 | 0 | 0 | 66.91092 | 489.9901 |
| Azimuth_2 | Motor Cort | 9-Feb | 0.00066 | 0.006015 | 0 | 0 | 66.91092 | 489.9901 |
| GWAS_Cat | Circulating | 31-Mar | 0.000327 | 0.006142 | 0 | 0 | 25.36352 | 203.5388 |
| UK_Bioban | Left heel qu | 5/178 | 0.001082 | 0.006168 | 0 | 0 | 6.957564 | 47.51185 |
| UK_Bioban | Left heel bo | 5/178 | 0.001082 | 0.006168 | 0 | 0 | 6.957564 | 47.51185 |
| UK_Bioban | Left 3mm s | 5/177 | 0.001055 | 0.006168 | 0 | 0 | 6.998369 | 47.9662 |
| UK_Bioban | Heel bone | 7/363 | 0.001031 | 0.006168 | 0 | 0 | 4.806847 | 33.0571 |
| UK_Bioban | Medicatior | 11-Feb | 0.001003 | 0.006168 | 0 | 0 | 52.0366 | 359.3058 |
| UK_Bioban | Number of | 11-Feb | 0.001003 | 0.006168 | 0 | 0 | 52.0366 | 359.3058 |
| UK_Bioban | Right 3mm | 5/173 | 0.000953 | 0.006168 | 0 | 0 | 7.166449 | 49.8497 |
| dbGaP | Triglyceride | 6/244 | 0.000687 | 0.006376 | 0 | 0 | 6.123561 | 44.60198 |
| dbGaP | Lipoprotein | Mar-36 | 0.000512 | 0.006376 | 0 | 0 | 21.51515 | 163.0377 |
| dbGaP | C-Reactive | Apr-88 | 0.00059 | 0.006376 | 0 | 0 | 11.37636 | 84.59459 |
| GWAS_Cat | Descending | Mar-33 | 0.000395 | 0.00642 | 0 | 0 | 23.67024 | 185.5129 |
| GWAS_Cat | tRNA (guar | 7-Feb | 0.000387 | 0.00642 | 0 | 0 | 93.68471 | 736.0204 |
| GWAS_Cat | Cerebrospi | 7-Feb | 0.000387 | 0.00642 | 0 | 0 | 93.68471 | 736.0204 |
| GWAS_Cat | Cholesteryl | Mar-32 | 0.00036 | 0.00642 | 0 | 0 | 24.48768 | 194.1766 |
| GWAS_Cat | Liver Proto | 7-Feb | 0.000387 | 0.00642 | 0 | 0 | 93.68471 | 736.0204 |
| GWAS_Cat | Cardioresp | 7-Feb | 0.000387 | 0.00642 | 0 | 0 | 93.68471 | 736.0204 |
| GWAS_Cat | Left Ventric | 7-Feb | 0.000387 | 0.00642 | 0 | 0 | 93.68471 | 736.0204 |
| GWAS_Cat | Metabolica | 7-Feb | 0.000387 | 0.00642 | 0 | 0 | 93.68471 | 736.0204 |
| GWAS_Cat | Triglyceride | Mar-32 | 0.00036 | 0.00642 | 0 | 0 | 24.48768 | 194.1766 |
| GWAS_Cat | Fruit Consu | Mar-33 | 0.000395 | 0.00642 | 0 | 0 | 23.67024 | 185.5129 |
| DisGeNET | Disorder of | 16-Mar | 4.27E-05 | 0.006586 | 0 | 0 | 54.67033 | 550.0107 |
| DisGeNET | Facial musc | May-88 | 4.09E-05 | 0.006586 | 0 | 0 | 14.56803 | 147.2001 |
| DisGeNET | Cardiomyo | 9/388 | 4.79E-05 | 0.006635 | 0 | 0 | 5.947027 | 59.14671 |
| DisGeNET | Facial Pare | May-92 | 5.06E-05 | 0.006635 | 0 | 0 | 13.89543 | 137.4338 |
| DisGeNET | Bell Palsy | May-92 | 5.06E-05 | 0.006635 | 0 | 0 | 13.89543 | 137.4338 |
| GeDiPNet_ | Distal Musc | 5-Feb | 0.000185 | 0.006758 | 0 | 0 | 156.1569 | 1341.798 |
| GeDiPNet_ | Welander I | 5-Feb | 0.000185 | 0.006758 | 0 | 0 | 156.1569 | 1341.798 |
| Jensen_DIS | Alzheimer's | 6/195 | 0.000209 | 0.006778 | 0 | 0 | 7.730355 | 65.5167 |
| UK_Bioban | Relative ag | 4/107 | 0.001226 | 0.00685 | 0 | 0 | 9.26892 | 62.14076 |
| UK_Bioban | Left heel bo | 5/184 | 0.001254 | 0.006872 | 0 | 0 | 6.722305 | 44.91504 |

| | | | | | | | | |
|---|---|---|---|---|---|---|---|---|
| GWAS_Cat | Triglyceride | Mar-34 | 0.000432 | 0.006893 | 0 | 0 | 22.90553 | 177.4756 |
| GWAS_Cat | Apolipopro | 8/413 | 0.000435 | 0.006893 | 0 | 0 | 4.877762 | 37.75395 |
| GeDiPNet_ | Myopathy | 7/278 | 0.00021 | 0.007218 | 0 | 0 | 6.341974 | 53.69199 |
| GWAS_Cat | C-reactive | 5/148 | 0.000471 | 0.00736 | 0 | 0 | 8.429985 | 64.58697 |
| DisGeNET | Autosomal | 18-Mar | 6.19E-05 | 0.007369 | 0 | 0 | 47.37619 | 459.0901 |
| DisGeNET | Primary Pro | 18-Mar | 6.19E-05 | 0.007369 | 0 | 0 | 47.37619 | 459.0901 |
| GWAS_Cat | Threonine | 8-Feb | 0.000515 | 0.007569 | 0 | 0 | 78.06667 | 591.0822 |
| GWAS_Cat | Heart Rate | 8-Feb | 0.000515 | 0.007569 | 0 | 0 | 78.06667 | 591.0822 |
| GWAS_Cat | PH And SEC | 8-Feb | 0.000515 | 0.007569 | 0 | 0 | 78.06667 | 591.0822 |
| GWAS_Cat | Intrahepati | 8-Feb | 0.000515 | 0.007569 | 0 | 0 | 78.06667 | 591.0822 |
| GWAS_Cat | Bipolar Dis | 8-Feb | 0.000515 | 0.007569 | 0 | 0 | 78.06667 | 591.0822 |
| GWAS_Cat | Serum Tota | 7/324 | 0.000529 | 0.007688 | 0 | 0 | 5.408991 | 40.80467 |
| GWAS_Cat | LDL Choles | 6/233 | 0.000539 | 0.007739 | 0 | 0 | 6.423886 | 48.34267 |
| CellMarker | Exhausted | 7/261 | 0.000143 | 0.00783 | 0 | 0 | 6.772293 | 59.95841 |
| GWAS_Cat | Orbital Telo | Mar-37 | 0.000555 | 0.007874 | 0 | 0 | 20.8813 | 156.5343 |
| DisGeNET | C-reactive | Apr-51 | 7.14E-05 | 0.007997 | 0 | 0 | 20.37016 | 194.4766 |
| DisGeNET | Congenital | 19-Mar | 7.33E-05 | 0.007997 | 0 | 0 | 44.41295 | 422.8826 |
| dbGaP | Glomerulon | 11-Feb | 0.001003 | 0.008149 | 0 | 0 | 52.0366 | 359.3058 |
| GWAS_Cat | Brain Shape | Apr-88 | 0.00059 | 0.008269 | 0 | 0 | 11.37636 | 84.59459 |
| GWAS_Cat | Atrial Fibril | Apr-89 | 0.000615 | 0.008531 | 0 | 0 | 11.24196 | 83.11525 |
| GWAS_Cat | Serum Alka | 9/548 | 0.000627 | 0.008595 | 0 | 0 | 4.147424 | 30.58513 |
| PhenGenI_ | Triglyceride | 14/1082 | 0.000226 | 0.008712 | 0 | 0 | 3.383998 | 28.40441 |
| GWAS_Cat | Phosphatic | 9-Feb | 0.00066 | 0.008754 | 0 | 0 | 66.91092 | 489.9901 |
| GWAS_Cat | Frontal QRS | 9-Feb | 0.00066 | 0.008754 | 0 | 0 | 66.91092 | 489.9901 |
| GWAS_Cat | High Light S | 8/439 | 0.00065 | 0.008754 | 0 | 0 | 4.577403 | 33.59251 |
| UK_Bioban | Butter/spre | 14-Feb | 0.001645 | 0.008847 | 0 | 0 | 39.02157 | 250.1213 |
| GWAS_Cat | Glutamine | Mar-40 | 0.000699 | 0.009172 | 0 | 0 | 19.18533 | 139.392 |
| GO_Molec | Phosphatic | 5-Feb | 0.000185 | 0.009273 | 0 | 0 | 156.1569 | 1341.798 |
| GO_Molec | Protein Ser | Apr-55 | 9.62E-05 | 0.009273 | 0 | 0 | 18.76872 | 173.5991 |
| DisGeNET | Protein C a | Apr-55 | 9.62E-05 | 0.009691 | 0 | 0 | 18.76872 | 173.5991 |
| DisGeNET | Protein C n | Apr-55 | 9.62E-05 | 0.009691 | 0 | 0 | 18.76872 | 173.5991 |
| GWAS_Cat | Total Lipid | Mar-41 | 0.000752 | 0.009761 | 0 | 0 | 18.67951 | 134.3561 |
| GWAS_Cat | Height (We | Apr-95 | 0.000787 | 0.010102 | 0 | 0 | 10.49755 | 75.0339 |
| UK_Bioban | Left ankle s | 5/204 | 0.001974 | 0.010417 | 0 | 0 | 6.040569 | 37.61947 |
| UK_Bioban | Peak expira | 5/205 | 0.002016 | 0.010448 | 0 | 0 | 6.010061 | 37.30138 |
| GWAS_Cat | Alzheimer's | 10-Feb | 0.000823 | 0.010457 | 0 | 0 | 58.54412 | 415.8217 |
| Rare_Disea | Myofibrilla | 25-Mar | 0.000171 | 0.010578 | 0 | 0 | 32.29058 | 280.1555 |
| UK_Bioban | Right leg fa | 8/528 | 0.002106 | 0.010716 | 0 | 0 | 3.776631 | 23.27589 |
| UK_Bioban | Self-report | 16-Feb | 0.002157 | 0.010787 | 0 | 0 | 33.4437 | 205.3055 |
| DisGeNET | Restrictive | 22-Mar | 0.000115 | 0.010791 | 0 | 0 | 37.39474 | 339.0868 |
| DisGeNET | Sudden dea | 22-Mar | 0.000115 | 0.010791 | 0 | 0 | 37.39474 | 339.0868 |
| Human_Ph | Neck musc | 24-Mar | 0.000151 | 0.010863 | 0 | 0 | 33.82993 | 297.7292 |
| Human_Ph | EMG: myop | 25-Mar | 0.000171 | 0.010863 | 0 | 0 | 32.29058 | 280.1555 |
| GWAS_Cat | Cholestero | Mar-43 | 0.000865 | 0.010882 | 0 | 0 | 17.74375 | 125.1385 |
| PheWeb_2 | Open wour | 29-Mar | 0.000268 | 0.010886 | 0 | 0 | 27.31731 | 224.7047 |

| | | | | | | | | |
|---|---|---|---|---|---|---|---|---|
| GWAS_Cat | Alcohol Co | Apr-98 | 0.000884 | 0.011002 | 0 | 0 | 10.16098 | 71.44486 |
| ARCHS4_Ti | ATRIUM | 23/2316 | 0.000104 | 0.011196 | 0 | 0 | 2.76153 | 25.33511 |
| GeDiPNet_ | Obstructive | 7-Feb | 0.000387 | 0.011289 | 0 | 0 | 93.68471 | 736.0204 |
| GeDiPNet_ | Hypertropl | 7-Feb | 0.000387 | 0.011289 | 0 | 0 | 93.68471 | 736.0204 |
| GeDiPNet_ | Ventricular | 7-Feb | 0.000387 | 0.011289 | 0 | 0 | 93.68471 | 736.0204 |
| GWAS_Cat | Lung Funct | 5/172 | 0.000929 | 0.011328 | 0 | 0 | 7.209727 | 50.33777 |
| GWAS_Cat | Cholesteryl | Mar-44 | 0.000926 | 0.011328 | 0 | 0 | 17.3101 | 120.912 |
| Jensen_DIS | Heart cond | Apr-82 | 0.000451 | 0.011469 | 0 | 0 | 12.25517 | 94.41149 |
| Jensen_DIS | Lipid metal | Mar-35 | 0.000471 | 0.011469 | 0 | 0 | 22.18862 | 170.0021 |
| GWAS_Cat | Cholestero | 11-Feb | 0.001003 | 0.011765 | 0 | 0 | 52.0366 | 359.3058 |
| GWAS_Cat | Left Ventri | 11-Feb | 0.001003 | 0.011765 | 0 | 0 | 52.0366 | 359.3058 |
| GWAS_Cat | Alzheimer's | 11-Feb | 0.001003 | 0.011765 | 0 | 0 | 52.0366 | 359.3058 |
| GWAS_Cat | Breast Can | 8/469 | 0.000997 | 0.011765 | 0 | 0 | 4.272934 | 29.53043 |
| UK_Bioban | No major d | 17-Feb | 0.002438 | 0.011778 | 0 | 0 | 31.21255 | 187.7904 |
| UK_Bioban | Doctor diag | 17-Feb | 0.002438 | 0.011778 | 0 | 0 | 31.21255 | 187.7904 |
| DisGeNET | Lipids mea: | 23-Mar | 0.000132 | 0.011944 | 0 | 0 | 35.52321 | 317.2632 |
| ClinVar_20 | acute myel | 20-Feb | 0.003378 | 0.012063 | 0 | 0 | 26.00654 | 147.9928 |
| GWAS_Cat | Waist Circu | Mar-46 | 0.001054 | 0.012134 | 0 | 0 | 16.50332 | 113.1284 |
| GWAS_Cat | Insulin-like | 6/265 | 0.001053 | 0.012134 | 0 | 0 | 5.62105 | 38.53642 |
| UK_Bioban | Left hand g | 5/218 | 0.002632 | 0.012502 | 0 | 0 | 5.639528 | 33.49894 |
| UK_Bioban | Ankle spaci | 7/430 | 0.002698 | 0.012588 | 0 | 0 | 4.031619 | 23.84858 |
| UK_Bioban | Left leg fat | 8/551 | 0.002739 | 0.012588 | 0 | 0 | 3.612374 | 21.31424 |
| GWAS_Cat | Testostero | 5/179 | 0.001109 | 0.01265 | 0 | 0 | 6.917227 | 47.06385 |
| CellMarker | Smooth Mu | 6-Feb | 0.000277 | 0.012669 | 0 | 0 | 117.1118 | 959.145 |
| Gene_Pert | Rbm15 KO | 9/304 | 6.99E-06 | 0.012675 | 0 | 0 | 7.673272 | 91.09283 |
| GWAS_Cat | Familial Co | Mar-47 | 0.001123 | 0.012682 | 0 | 0 | 16.12744 | 109.5385 |
| DisGeNET | Behavioral | 24-Mar | 0.000151 | 0.01273 | 0 | 0 | 33.82993 | 297.7292 |
| DisGeNET | X-linked ce | 24-Mar | 0.000151 | 0.01273 | 0 | 0 | 33.82993 | 297.7292 |
| Human_Ph | Congestive | 5/128 | 0.000241 | 0.012782 | 0 | 0 | 9.810629 | 81.72619 |
| Human_Ph | Abnormalit | 29-Mar | 0.000268 | 0.012782 | 0 | 0 | 27.31731 | 224.7047 |
| GeDiPNet_ | Peroneal M | 8-Feb | 0.000515 | 0.013025 | 0 | 0 | 78.06667 | 591.0822 |
| GeDiPNet_ | Distal Myo | 8-Feb | 0.000515 | 0.013025 | 0 | 0 | 78.06667 | 591.0822 |
| GeDiPNet_ | Sphenoid W | 12-Feb | 0.0012 | 0.013025 | 0 | 0 | 46.83059 | 314.9532 |
| GeDiPNet_ | Septal Hyp | 12-Feb | 0.0012 | 0.013025 | 0 | 0 | 46.83059 | 314.9532 |
| GeDiPNet_ | Hemangiob | 12-Feb | 0.0012 | 0.013025 | 0 | 0 | 46.83059 | 314.9532 |
| GeDiPNet_ | Psammoma | 12-Feb | 0.0012 | 0.013025 | 0 | 0 | 46.83059 | 314.9532 |
| GeDiPNet_ | Fossa Men | 12-Feb | 0.0012 | 0.013025 | 0 | 0 | 46.83059 | 314.9532 |
| GeDiPNet_ | Transitiona | 12-Feb | 0.0012 | 0.013025 | 0 | 0 | 46.83059 | 314.9532 |
| GeDiPNet_ | Intraventri | 12-Feb | 0.0012 | 0.013025 | 0 | 0 | 46.83059 | 314.9532 |
| GeDiPNet_ | Fibrous Me | 12-Feb | 0.0012 | 0.013025 | 0 | 0 | 46.83059 | 314.9532 |
| GeDiPNet_ | Parasagitta | 12-Feb | 0.0012 | 0.013025 | 0 | 0 | 46.83059 | 314.9532 |
| GeDiPNet_ | Papillary M | 12-Feb | 0.0012 | 0.013025 | 0 | 0 | 46.83059 | 314.9532 |
| GeDiPNet_ | Ovarian Ad | Mar-41 | 0.000752 | 0.013025 | 0 | 0 | 18.67951 | 134.3561 |
| GeDiPNet_ | Wolff-Park | Mar-43 | 0.000865 | 0.013025 | 0 | 0 | 17.74375 | 125.1385 |
| GeDiPNet_ | Left Ventri | Apr-95 | 0.000787 | 0.013025 | 0 | 0 | 10.49755 | 75.0339 |

| | | | | | | | | |
|---|---|---|---|---|---|---|---|---|
| GeDiPNet_ | Facial Para | 5/166 | 0.000792 | 0.013025 | 0 | 0 | 7.480685 | 53.42157 |
| GeDiPNet_ | Lung Cance | 7/324 | 0.000529 | 0.013025 | 0 | 0 | 5.408991 | 40.80467 |
| GeDiPNet_ | Angioblasti | 12-Feb | 0.0012 | 0.013025 | 0 | 0 | 46.83059 | 314.9532 |
| GeDiPNet_ | Combined | Mar-44 | 0.000926 | 0.013025 | 0 | 0 | 17.3101 | 120.912 |
| GeDiPNet_ | Malignant | 11-Feb | 0.001003 | 0.013025 | 0 | 0 | 52.0366 | 359.3058 |
| GeDiPNet_ | Malignant | 12-Feb | 0.0012 | 0.013025 | 0 | 0 | 46.83059 | 314.9532 |
| GeDiPNet_ | Cerebral Co | 12-Feb | 0.0012 | 0.013025 | 0 | 0 | 46.83059 | 314.9532 |
| GeDiPNet_ | Benign Me | 12-Feb | 0.0012 | 0.013025 | 0 | 0 | 46.83059 | 314.9532 |
| GeDiPNet_ | Senile Dem | Apr-99 | 0.000918 | 0.013025 | 0 | 0 | 10.05352 | 70.30771 |
| GeDiPNet_ | Microstom | 5/171 | 0.000905 | 0.013025 | 0 | 0 | 7.253526 | 50.83298 |
| GeDiPNet_ | Cardiomyo | 7/373 | 0.001206 | 0.013025 | 0 | 0 | 4.673122 | 31.40388 |
| GeDiPNet_ | Secretory M | 12-Feb | 0.0012 | 0.013025 | 0 | 0 | 46.83059 | 314.9532 |
| GeDiPNet_ | Hemangiop | 12-Feb | 0.0012 | 0.013025 | 0 | 0 | 46.83059 | 314.9532 |
| GeDiPNet_ | Xanthomat | 12-Feb | 0.0012 | 0.013025 | 0 | 0 | 46.83059 | 314.9532 |
| GeDiPNet_ | Intraorbita | 12-Feb | 0.0012 | 0.013025 | 0 | 0 | 46.83059 | 314.9532 |
| GeDiPNet_ | Olfactory G | 12-Feb | 0.0012 | 0.013025 | 0 | 0 | 46.83059 | 314.9532 |
| GeDiPNet_ | Angiomato | 12-Feb | 0.0012 | 0.013025 | 0 | 0 | 46.83059 | 314.9532 |
| GeDiPNet_ | Meningoth | 12-Feb | 0.0012 | 0.013025 | 0 | 0 | 46.83059 | 314.9532 |
| GeDiPNet_ | Microcystic | 12-Feb | 0.0012 | 0.013025 | 0 | 0 | 46.83059 | 314.9532 |
| GWAS_Cat | Cognitive D | 12-Feb | 0.0012 | 0.01319 | 0 | 0 | 46.83059 | 314.9532 |
| GWAS_Cat | Left Ventri | 12-Feb | 0.0012 | 0.01319 | 0 | 0 | 46.83059 | 314.9532 |
| GWAS_Cat | Cognitive In | 12-Feb | 0.0012 | 0.01319 | 0 | 0 | 46.83059 | 314.9532 |
| GeDiPNet_ | Alopecia | 6/277 | 0.001322 | 0.01401 | 0 | 0 | 5.368867 | 35.58941 |
| GWAS_Cat | Sex Hormo | 10/735 | 0.001318 | 0.014354 | 0 | 0 | 3.43717 | 22.79464 |
| GWAS_Cat | Electrocarc | Mar-50 | 0.001344 | 0.014387 | 0 | 0 | 15.09574 | 99.8109 |
| GWAS_Cat | Phospholip | Mar-50 | 0.001344 | 0.014387 | 0 | 0 | 15.09574 | 99.8109 |
| Rare_Disea | Subvalvula | 6/202 | 0.000252 | 0.014436 | 0 | 0 | 7.451625 | 61.73755 |
| GWAS_Cat | Heart Rate | 13-Feb | 0.001414 | 0.014622 | 0 | 0 | 42.57112 | 279.3154 |
| GWAS_Cat | Uterine Lei | 13-Feb | 0.001414 | 0.014622 | 0 | 0 | 42.57112 | 279.3154 |
| GWAS_Cat | Urinary Alb | 13-Feb | 0.001414 | 0.014622 | 0 | 0 | 42.57112 | 279.3154 |
| GWAS_Cat | Body Mass | 13-Feb | 0.001414 | 0.014622 | 0 | 0 | 42.57112 | 279.3154 |
| GeDiPNet_ | Heart Failu | 5/189 | 0.001412 | 0.014698 | 0 | 0 | 6.537977 | 42.90806 |
| Jensen_DIS | Macular de | Mar-40 | 0.000699 | 0.01515 | 0 | 0 | 19.18533 | 139.392 |
| UK_Bioban | Trunk fat m | 8/573 | 0.003475 | 0.01572 | 0 | 0 | 3.467772 | 19.63512 |
| DisGeNET | Amino acid | 26-Mar | 0.000192 | 0.015742 | 0 | 0 | 30.88509 | 264.2718 |
| Jensen_DIS | Cavernous | 10-Feb | 0.000823 | 0.016046 | 0 | 0 | 58.54412 | 415.8217 |
| Rare_Disea | Coats disea | Apr-74 | 0.000305 | 0.016193 | 0 | 0 | 13.66127 | 110.604 |
| GWAS_Cat | Low Densit | 10/754 | 0.001593 | 0.016196 | 0 | 0 | 3.346076 | 21.55574 |
| GWAS_Cat | Waist Circu | Mar-53 | 0.001592 | 0.016196 | 0 | 0 | 14.18786 | 91.4113 |
| GeDiPNet_ | Androgene | Mar-53 | 0.001592 | 0.01628 | 0 | 0 | 14.18786 | 91.4113 |
| GeneSigDB | 19061838-T | 7/203 | 2.94E-05 | 0.016393 | 0 | 0 | 8.802232 | 91.83778 |
| GWAS_Cat | Waist-hip F | 9/628 | 0.001626 | 0.016399 | 0 | 0 | 3.596496 | 23.09438 |
| UK_Bioban | Left heel bı | 4/145 | 0.003704 | 0.016494 | 0 | 0 | 6.757925 | 37.83344 |
| GeDiPNet_ | Congenital | 14-Feb | 0.001645 | 0.016538 | 0 | 0 | 39.02157 | 250.1213 |
| UK_Bioban | Hot drink t | Mar-72 | 0.003819 | 0.016745 | 0 | 0 | 10.27122 | 57.1878 |

| | | | | | | | | |
|---|---|---|---|---|---|---|---|---|
| Rare_Disea | Frontotem | 6/215 | 0.000352 | 0.017004 | 0 | 0 | 6.983519 | 55.52746 |
| Rare_Disea | Semantic d | 6/219 | 0.000389 | 0.017004 | 0 | 0 | 6.850982 | 53.80163 |
| Rare_Disea | Inclusion b | 5/141 | 0.000377 | 0.017004 | 0 | 0 | 8.867019 | 69.90321 |
| GWAS_Cat | Reticulocyt | 7/397 | 0.001723 | 0.01723 | 0 | 0 | 4.38016 | 27.87396 |
| GeDiPNet_ | Atrioventri | Mar-55 | 0.001771 | 0.017505 | 0 | 0 | 13.6408 | 86.42716 |
| Jensen_DIS | Croup | 11-Feb | 0.001003 | 0.017779 | 0 | 0 | 52.0366 | 359.3058 |
| UK_Bioban | Impedance | 8/590 | 0.004143 | 0.01789 | 0 | 0 | 3.363522 | 18.45342 |
| GeDiPNet_ | Spinal Men | 15-Feb | 0.001893 | 0.018093 | 0 | 0 | 36.0181 | 225.8172 |
| GeDiPNet_ | Urolithiasis | 15-Feb | 0.001893 | 0.018093 | 0 | 0 | 36.0181 | 225.8172 |
| DisGeNET | Calcificatio | 8/375 | 0.000228 | 0.018109 | 0 | 0 | 5.393302 | 45.22684 |
| UK_Bioban | Salt added | Mar-75 | 0.004284 | 0.018222 | 0 | 0 | 9.841766 | 53.66662 |
| GWAS_Cat | Waist Circu | Mar-56 | 0.001866 | 0.018358 | 0 | 0 | 13.38275 | 84.09717 |
| GWAS_Cat | Spatial QRS | Mar-56 | 0.001866 | 0.018358 | 0 | 0 | 13.38275 | 84.09717 |
| GWAS_Cat | Waist-to-hi | 15-Feb | 0.001893 | 0.018477 | 0 | 0 | 36.0181 | 225.8172 |
| DisGeNET | Benign Me | 28-Mar | 0.000241 | 0.018551 | 0 | 0 | 28.41143 | 236.7179 |
| UK_Bioban | Attenpted | Mar-77 | 0.004612 | 0.019328 | 0 | 0 | 9.574807 | 51.50443 |
| GWAS_Cat | Non-album | 5/206 | 0.002059 | 0.019673 | 0 | 0 | 5.979857 | 36.9872 |
| GWAS_Cat | Triglyceride | Mar-58 | 0.002064 | 0.019673 | 0 | 0 | 12.89481 | 79.72951 |
| GWAS_Cat | Serum Met | Mar-58 | 0.002064 | 0.019673 | 0 | 0 | 12.89481 | 79.72951 |
| DisGeNET | Myocardial | 13/966 | 0.000266 | 0.01969 | 0 | 0 | 3.49508 | 28.77274 |
| DisGeNET | Blood Sedi | 6-Feb | 0.000277 | 0.01969 | 0 | 0 | 117.1118 | 959.145 |
| DisGeNET | Cardiac Arr | 5/132 | 0.000278 | 0.01969 | 0 | 0 | 9.499712 | 77.78059 |
| UK_Bioban | Cardiac arr | 24-Feb | 0.004851 | 0.019752 | 0 | 0 | 21.2738 | 113.3575 |
| UK_Bioban | Disorders c | 24-Feb | 0.004851 | 0.019752 | 0 | 0 | 21.2738 | 113.3575 |
| Human_Ge | Heart | 8/415 | 0.000449 | 0.019765 | 0 | 0 | 4.853295 | 37.40947 |
| GWAS_Cat | Whole Brai | 4/124 | 0.002107 | 0.019928 | 0 | 0 | 7.948996 | 48.98517 |
| GWAS_Cat | Waist-to-hi | 16-Feb | 0.002157 | 0.01994 | 0 | 0 | 33.4437 | 205.3055 |
| GWAS_Cat | 3-Hydroxyb | 16-Feb | 0.002157 | 0.01994 | 0 | 0 | 33.4437 | 205.3055 |
| GWAS_Cat | Lung Cance | 16-Feb | 0.002157 | 0.01994 | 0 | 0 | 33.4437 | 205.3055 |
| CellMarker | Myocyte U | 8-Feb | 0.000515 | 0.020155 | 0 | 0 | 78.06667 | 591.0822 |
| HumanCyc_ | 3-phospho | 20-Feb | 0.003378 | 0.020265 | 0 | 0 | 26.00654 | 147.9928 |
| GeDiPNet_ | Alopecia, N | Mar-59 | 0.002168 | 0.020385 | 0 | 0 | 12.6639 | 77.68052 |
| GWAS_Cat | Lung Funct | 5/210 | 0.002239 | 0.020385 | 0 | 0 | 5.861987 | 35.76819 |
| GWAS_Cat | Tonsillecto | 4/126 | 0.002233 | 0.020385 | 0 | 0 | 7.817895 | 47.72214 |
| GWAS_Cat | Glycated H | 6/308 | 0.002258 | 0.020401 | 0 | 0 | 4.810155 | 29.31062 |
| UK_Bioban | Impedance | 9/747 | 0.005149 | 0.020667 | 0 | 0 | 2.997967 | 15.79642 |
| UK_Bioban | Heel quant | 6/367 | 0.005319 | 0.020765 | 0 | 0 | 4.011901 | 21.00841 |
| UK_Bioban | Heel bone | 6/367 | 0.005319 | 0.020765 | 0 | 0 | 4.011901 | 21.00841 |
| GWAS_Cat | Cholesteryl | Mar-61 | 0.002385 | 0.021395 | 0 | 0 | 12.22599 | 73.82703 |
| GWAS_Cat | Vaginal Mi | 17-Feb | 0.002438 | 0.021712 | 0 | 0 | 31.21255 | 187.7904 |
| UK_Bioban | Heart disea | 26-Feb | 0.005681 | 0.021878 | 0 | 0 | 19.49902 | 100.8238 |
| GWAS_Cat | LDL Choles | 6/314 | 0.002485 | 0.02197 | 0 | 0 | 4.715007 | 28.27783 |
| DisGeNET | Charcot-M | 7/298 | 0.000321 | 0.022123 | 0 | 0 | 5.900086 | 47.46306 |
| GeDiPNet_ | Intracrania | 17-Feb | 0.002438 | 0.02221 | 0 | 0 | 31.21255 | 187.7904 |
| GeDiPNet_ | Subaortic S | 17-Feb | 0.002438 | 0.02221 | 0 | 0 | 31.21255 | 187.7904 |

| | | | | | | | | |
|---|---|---|---|---|---|---|---|---|
| GeneSigDB | 19643001-: | 7/238 | 8.05E-05 | 0.022429 | 0 | 0 | 7.455303 | 70.27976 |
| GWAS_Cat | Chronic Ele | Mar-63 | 0.002615 | 0.022936 | 0 | 0 | 11.81726 | 70.27008 |
| GWAS_Cat | Low-densit | 5/218 | 0.002632 | 0.022936 | 0 | 0 | 5.639528 | 33.49894 |
| UK_Bioban | Time to co | Mar-85 | 0.006074 | 0.02308 | 0 | 0 | 8.637195 | 44.08228 |
| Jensen_DIS | Ovarian ca | Mar-51 | 0.001424 | 0.023138 | 0 | 0 | 14.78051 | 96.87677 |
| GWAS_Cat | Cerebrospi | 18-Feb | 0.002735 | 0.023336 | 0 | 0 | 29.26029 | 172.6809 |
| GWAS_Cat | Cholestero | Mar-64 | 0.002735 | 0.023336 | 0 | 0 | 11.62295 | 68.5929 |
| GWAS_Cat | Edge-level | 4/133 | 0.002717 | 0.023336 | 0 | 0 | 7.391053 | 43.6691 |
| UK_Bioban | Medicatior | Mar-86 | 0.006274 | 0.023527 | 0 | 0 | 8.532702 | 43.27254 |
| DisGeNET | Welander I | 7-Feb | 0.000387 | 0.023597 | 0 | 0 | 93.68471 | 736.0204 |
| DisGeNET | Myopathy, | 7-Feb | 0.000387 | 0.023597 | 0 | 0 | 93.68471 | 736.0204 |
| DisGeNET | Familial res | 7-Feb | 0.000387 | 0.023597 | 0 | 0 | 93.68471 | 736.0204 |
| DisGeNET | Reduced sy | 7-Feb | 0.000387 | 0.023597 | 0 | 0 | 93.68471 | 736.0204 |
| DisGeNET | Memory pe | 7-Feb | 0.000387 | 0.023597 | 0 | 0 | 93.68471 | 736.0204 |
| UK_Bioban | ICD10 atria | 28-Feb | 0.00657 | 0.024005 | 0 | 0 | 17.99729 | 90.44125 |
| UK_Bioban | Narcolepsy | 28-Feb | 0.00657 | 0.024005 | 0 | 0 | 17.99729 | 90.44125 |
| GWAS_Cat | Alzheimer's | Mar-65 | 0.002859 | 0.024053 | 0 | 0 | 11.43491 | 66.97852 |
| GWAS_Cat | Triglyceride | Mar-65 | 0.002859 | 0.024053 | 0 | 0 | 11.43491 | 66.97852 |
| PhenGenI_ | Insulin-Like | 5/168 | 0.000836 | 0.024126 | 0 | 0 | 7.388149 | 52.36302 |
| PhenGenI_ | Memory | Apr-95 | 0.000787 | 0.024126 | 0 | 0 | 10.49755 | 75.0339 |
| Rare_Disea | Laing distal | Mar-39 | 0.000649 | 0.02425 | 0 | 0 | 19.71925 | 144.7467 |
| Rare_Disea | Alzheimer ( | 5/159 | 0.000652 | 0.02425 | 0 | 0 | 7.823487 | 57.39044 |
| Rare_Disea | Distal myo | Mar-39 | 0.000649 | 0.02425 | 0 | 0 | 19.71925 | 144.7467 |
| UK_Bioban | Left 3mm v | 4/173 | 0.006895 | 0.024875 | 0 | 0 | 5.630284 | 28.02158 |
| GWAS_Cat | Triglyceride | Mar-66 | 0.002986 | 0.024948 | 0 | 0 | 11.25283 | 65.42369 |
| GWAS_Cat | Cardiovasc | 19-Feb | 0.003048 | 0.025129 | 0 | 0 | 27.53772 | 159.5298 |
| GWAS_Cat | Kidney Stor | 19-Feb | 0.003048 | 0.025129 | 0 | 0 | 27.53772 | 159.5298 |
| GWAS_Cat | Subcortical | Mar-67 | 0.003116 | 0.025512 | 0 | 0 | 11.07645 | 63.92535 |
| UK_Bioban | Right 3mm | 4/175 | 0.007175 | 0.025563 | 0 | 0 | 5.56387 | 27.46932 |
| DisGeNET | Left ventric | Mar-34 | 0.000432 | 0.025696 | 0 | 0 | 22.90553 | 177.4756 |
| PhenGenI_ | Ossificatior | 11-Feb | 0.001003 | 0.025741 | 0 | 0 | 52.0366 | 359.3058 |
| GWAS_Cat | Dyslipidem | 20-Feb | 0.003378 | 0.027289 | 0 | 0 | 26.00654 | 147.9928 |
| GWAS_Cat | Triacylglyce | 20-Feb | 0.003378 | 0.027289 | 0 | 0 | 26.00654 | 147.9928 |
| UK_Bioban | Forced exp | Mar-93 | 0.007784 | 0.027388 | 0 | 0 | 7.86627 | 38.19617 |
| UK_Bioban | Coffee inta | Feb-31 | 0.008014 | 0.027518 | 0 | 0 | 16.13306 | 77.8673 |
| UK_Bioban | Enxymatic | Feb-31 | 0.008014 | 0.027518 | 0 | 0 | 16.13306 | 77.8673 |
| DisGeNET | Obstructive | 8-Feb | 0.000515 | 0.027532 | 0 | 0 | 78.06667 | 591.0822 |
| DisGeNET | 5q-syndror | Mar-36 | 0.000512 | 0.027532 | 0 | 0 | 21.51515 | 163.0377 |
| DisGeNET | Ovarian Mu | Mar-36 | 0.000512 | 0.027532 | 0 | 0 | 21.51515 | 163.0377 |
| DisGeNET | Cardiomyo | 8/419 | 0.000479 | 0.027532 | 0 | 0 | 4.805076 | 36.73337 |
| DisGeNET | Breast size | 8-Feb | 0.000515 | 0.027532 | 0 | 0 | 78.06667 | 591.0822 |
| UK_Bioban | Trunk fat p | 7/528 | 0.008156 | 0.027671 | 0 | 0 | 3.256814 | 15.66213 |
| Rare_Disea | Fallot tetra | Apr-95 | 0.000787 | 0.027869 | 0 | 0 | 10.49755 | 75.0339 |
| DisGeNET | Heart Disea | 7/325 | 0.000539 | 0.02825 | 0 | 0 | 5.391706 | 40.57567 |
| GWAS_Cat | Triglyceride | Mar-70 | 0.003527 | 0.02831 | 0 | 0 | 10.57889 | 59.74182 |

| | | | | | | | | |
|---|---|---|---|---|---|---|---|---|
| DisGeNET | Hemorrhag | Mar-37 | 0.000555 | 0.028516 | 0 | 0 | 20.8813 | 156.5343 |
| UK_Bioban | Self-report | 5/289 | 0.008583 | 0.028778 | 0 | 0 | 4.214402 | 20.05212 |
| GWAS_Cat | Total Trigly | Mar-71 | 0.003671 | 0.028927 | 0 | 0 | 10.42279 | 58.44282 |
| GWAS_Cat | Blood Urea | 5/236 | 0.003696 | 0.028927 | 0 | 0 | 5.195333 | 29.09659 |
| GWAS_Cat | Cerebrospi | 21-Feb | 0.003723 | 0.028927 | 0 | 0 | 24.63653 | 137.8005 |
| GWAS_Cat | Left Ventri | 21-Feb | 0.003723 | 0.028927 | 0 | 0 | 24.63653 | 137.8005 |
| GWAS_Cat | Lipid Metal | 21-Feb | 0.003723 | 0.028927 | 0 | 0 | 24.63653 | 137.8005 |
| GeDiPNet_ | Charcot-Ma | Mar-68 | 0.003249 | 0.029145 | 0 | 0 | 10.90549 | 62.48067 |
| GWAS_Cat | Serum Crea | 6/342 | 0.003783 | 0.029214 | 0 | 0 | 4.315917 | 24.07034 |
| GO_Biologi | Regulation | Mar-34 | 0.000432 | 0.02982 | 0 | 0 | 22.90553 | 177.4756 |
| GO_Biologi | Adult Hear | 8-Feb | 0.000515 | 0.02982 | 0 | 0 | 78.06667 | 591.0822 |
| GO_Biologi | Regulation | 8-Feb | 0.000515 | 0.02982 | 0 | 0 | 78.06667 | 591.0822 |
| GO_Biologi | Cardiac Mu | 27-Mar | 0.000216 | 0.02982 | 0 | 0 | 29.59673 | 249.8547 |
| GO_Biologi | Cardiac Mu | 31-Mar | 0.000327 | 0.02982 | 0 | 0 | 25.36352 | 203.5388 |
| GO_Biologi | Cholestero | 6-Feb | 0.000277 | 0.02982 | 0 | 0 | 117.1118 | 959.145 |
| GO_Biologi | Sarcomere | 29-Mar | 0.000268 | 0.02982 | 0 | 0 | 27.31731 | 224.7047 |
| GO_Biologi | Negative R | 7-Feb | 0.000387 | 0.02982 | 0 | 0 | 93.68471 | 736.0204 |
| GO_Biologi | Purine Ribo | Mar-32 | 0.00036 | 0.02982 | 0 | 0 | 24.48768 | 194.1766 |
| GO_Biologi | Negative R | 9-Feb | 0.00066 | 0.02982 | 0 | 0 | 66.91092 | 489.9901 |
| GO_Biologi | Regulation | 9-Feb | 0.00066 | 0.02982 | 0 | 0 | 66.91092 | 489.9901 |
| GO_Biologi | Heart Cont | Mar-41 | 0.000752 | 0.02982 | 0 | 0 | 18.67951 | 134.3561 |
| GO_Biologi | Alcohol Cat | 8-Feb | 0.000515 | 0.02982 | 0 | 0 | 78.06667 | 591.0822 |
| GO_Biologi | Acylglycero | Mar-41 | 0.000752 | 0.02982 | 0 | 0 | 18.67951 | 134.3561 |
| GO_Biologi | Sterol Cata | 6-Feb | 0.000277 | 0.02982 | 0 | 0 | 117.1118 | 959.145 |
| GO_Biologi | Regulation | 9-Feb | 0.00066 | 0.02982 | 0 | 0 | 66.91092 | 489.9901 |
| GO_Biologi | Cardiac Mu | Mar-33 | 0.000395 | 0.02982 | 0 | 0 | 23.67024 | 185.5129 |
| GO_Biologi | Muscle Cor | Apr-94 | 0.000756 | 0.02982 | 0 | 0 | 10.61473 | 76.29308 |
| GO_Molec | Phosphatic | 12-Feb | 0.0012 | 0.030002 | 0 | 0 | 46.83059 | 314.9532 |
| GO_Molec | Phosphatic | 11-Feb | 0.001003 | 0.030002 | 0 | 0 | 52.0366 | 359.3058 |
| KEGG_202 | Hypertroph | Apr-90 | 0.000642 | 0.030488 | 0 | 0 | 11.11068 | 81.67616 |
| GO_Biologi | Muscle Fila | 10-Feb | 0.000823 | 0.030749 | 0 | 0 | 58.54412 | 415.8217 |
| DisGeNET | Triglyceride | 7/336 | 0.000656 | 0.030886 | 0 | 0 | 5.208511 | 38.17198 |
| DisGeNET | Spindle Cel | 9-Feb | 0.00066 | 0.030886 | 0 | 0 | 66.91092 | 489.9901 |
| DisGeNET | TIBIAL MUS | 9-Feb | 0.00066 | 0.030886 | 0 | 0 | 66.91092 | 489.9901 |
| DisGeNET | Cardiomyo | 9-Feb | 0.00066 | 0.030886 | 0 | 0 | 66.91092 | 489.9901 |
| DisGeNET | Myofibrilla | Mar-39 | 0.000649 | 0.030886 | 0 | 0 | 19.71925 | 144.7467 |
| GWAS_Cat | Low Testos | 22-Feb | 0.004083 | 0.031135 | 0 | 0 | 23.40353 | 128.7392 |
| GWAS_Cat | IgA Nephro | 22-Feb | 0.004083 | 0.031135 | 0 | 0 | 23.40353 | 128.7392 |
| UK_Bioban | White bloo | 8/679 | 0.0094 | 0.03115 | 0 | 0 | 2.90396 | 13.553 |
| UK_Bioban | Takes chole | Feb-34 | 0.009587 | 0.031212 | 0 | 0 | 14.61838 | 67.9362 |
| UK_Bioban | Weight 210 | 8/682 | 0.009637 | 0.031212 | 0 | 0 | 2.890583 | 13.41836 |
| UK_Bioban | Neutrophil | 6/418 | 0.009801 | 0.031384 | 0 | 0 | 3.506113 | 16.21685 |
| Rare_Disea | Myopathy | 4/102 | 0.001026 | 0.031666 | 0 | 0 | 9.744283 | 67.06001 |
| Rare_Disea | Hypoplasti | Mar-46 | 0.001054 | 0.031666 | 0 | 0 | 16.50332 | 113.1284 |
| Rare_Disea | Buerger dis | Mar-46 | 0.001054 | 0.031666 | 0 | 0 | 16.50332 | 113.1284 |

| | | | | | | | | |
|---|---|---|---|---|---|---|---|---|
| Rare_Disea | Atrial septa | 4/103 | 0.001064 | 0.031666 | 0 | 0 | 9.645369 | 66.029 |
| GWAS_Cat | Lung Funct | 7/466 | 0.004195 | 0.03179 | 0 | 0 | 3.708551 | 20.29987 |
| Jensen_DIS | Gastrointes | 16-Feb | 0.002157 | 0.032361 | 0 | 0 | 33.4437 | 205.3055 |
| GeDiPNet_ | Arterioscle | 21-Feb | 0.003723 | 0.032392 | 0 | 0 | 24.63653 | 137.8005 |
| GeDiPNet_ | Ventricular | 21-Feb | 0.003723 | 0.032392 | 0 | 0 | 24.63653 | 137.8005 |
| GeDiPNet_ | Malocclusi | 4/147 | 0.003889 | 0.033067 | 0 | 0 | 6.662735 | 36.97528 |
| GeDiPNet_ | Amyotroph | Mar-73 | 0.00397 | 0.033067 | 0 | 0 | 10.12398 | 55.97469 |
| GeDiPNet_ | Bladder Ca | Mar-73 | 0.00397 | 0.033067 | 0 | 0 | 10.12398 | 55.97469 |
| Rare_Disea | Wildervanc | 12-Feb | 0.0012 | 0.033069 | 0 | 0 | 46.83059 | 314.9532 |
| Rare_Disea | Holt-Oram | 12-Feb | 0.0012 | 0.033069 | 0 | 0 | 46.83059 | 314.9532 |
| GWAS_Cat | Pyruvate Le | 23-Feb | 0.00446 | 0.033175 | 0 | 0 | 22.28796 | 120.6379 |
| GWAS_Cat | Direct Bilir | Mar-76 | 0.004446 | 0.033175 | 0 | 0 | 9.706458 | 52.56811 |
| GWAS_Cat | Cerebrospi | 23-Feb | 0.00446 | 0.033175 | 0 | 0 | 22.28796 | 120.6379 |
| UK_Bioban | Doctor diag | 5/304 | 0.010529 | 0.033343 | 0 | 0 | 3.999918 | 18.214 |
| UK_Bioban | Water intal | Feb-36 | 0.010706 | 0.03353 | 0 | 0 | 13.75709 | 62.41496 |
| Jensen_DIS | Coffin-Siris | 17-Feb | 0.002438 | 0.03396 | 0 | 0 | 31.21255 | 187.7904 |
| GWAS_Cat | Estimated ( | 9/734 | 0.004598 | 0.033995 | 0 | 0 | 3.053793 | 16.43614 |
| GWAS_Cat | Chronic Ob | 5/249 | 0.004636 | 0.034069 | 0 | 0 | 4.915284 | 26.41466 |
| GWAS_Cat | Very Low D | 24-Feb | 0.004851 | 0.034612 | 0 | 0 | 21.2738 | 113.3575 |
| GWAS_Cat | Descending | 24-Feb | 0.004851 | 0.034612 | 0 | 0 | 21.2738 | 113.3575 |
| GWAS_Cat | Chin Dimpl | 24-Feb | 0.004851 | 0.034612 | 0 | 0 | 21.2738 | 113.3575 |
| GWAS_Cat | Alzheimer's | 4/156 | 0.004801 | 0.034612 | 0 | 0 | 6.265377 | 33.4505 |
| GWAS_Cat | Optic Cup / | 24-Feb | 0.004851 | 0.034612 | 0 | 0 | 21.2738 | 113.3575 |
| Human_Ph | Centrally n | 10-Feb | 0.000823 | 0.034926 | 0 | 0 | 58.54412 | 415.8217 |
| UK_Bioban | Hair/baldir | 4/200 | 0.011332 | 0.035105 | 0 | 0 | 4.848045 | 21.71979 |
| RNA-Seq_D | Ezh2 Heart | 10/498 | 6.02E-05 | 0.035147 | 0 | 0 | 5.169523 | 50.23512 |
| RNA-Seq_D | Casz1 Hear | 10/499 | 6.12E-05 | 0.035147 | 0 | 0 | 5.158686 | 50.04358 |
| DisGeNET | Erythroker | 10-Feb | 0.000823 | 0.035343 | 0 | 0 | 58.54412 | 415.8217 |
| DisGeNET | EMG: myop | Mar-42 | 0.000807 | 0.035343 | 0 | 0 | 18.19963 | 129.6128 |
| DisGeNET | Idiopathic I | 10-Feb | 0.000823 | 0.035343 | 0 | 0 | 58.54412 | 415.8217 |
| DisGeNET | Heart failu | 11/815 | 0.000799 | 0.035343 | 0 | 0 | 3.44002 | 24.53302 |
| DisGeNET | Hallermanr | 10-Feb | 0.000823 | 0.035343 | 0 | 0 | 58.54412 | 415.8217 |
| GO_Biologi | Actin-Myos | 12-Feb | 0.0012 | 0.035502 | 0 | 0 | 46.83059 | 314.9532 |
| GO_Biologi | Phospholip | 11-Feb | 0.001003 | 0.035502 | 0 | 0 | 52.0366 | 359.3058 |
| GO_Biologi | Negative R | 12-Feb | 0.0012 | 0.035502 | 0 | 0 | 46.83059 | 314.9532 |
| GO_Biologi | Myofibril A | Mar-46 | 0.001054 | 0.035502 | 0 | 0 | 16.50332 | 113.1284 |
| GO_Biologi | Atrial Cardi | 12-Feb | 0.0012 | 0.035502 | 0 | 0 | 46.83059 | 314.9532 |
| GWAS_Cat | Brain Morp | Mar-80 | 0.005132 | 0.03598 | 0 | 0 | 9.200371 | 48.50752 |
| GWAS_Cat | Hip Index | 5/255 | 0.005122 | 0.03598 | 0 | 0 | 4.795854 | 25.29419 |
| GWAS_Cat | Heel Bone | 11/1034 | 0.005127 | 0.03598 | 0 | 0 | 2.672609 | 14.09343 |
| UK_Bioban | Birth weigh | 3/109 | 0.011983 | 0.036333 | 0 | 0 | 6.673518 | 29.52517 |
| UK_Bioban | Right ankle | 4/203 | 0.011916 | 0.036333 | 0 | 0 | 4.774233 | 21.14924 |
| CellMarker | Alveolar M | 12-Feb | 0.0012 | 0.036536 | 0 | 0 | 46.83059 | 314.9532 |
| CellMarker | Astrocyte C | 12-Feb | 0.0012 | 0.036536 | 0 | 0 | 46.83059 | 314.9532 |
| GWAS_Cat | Fresh Fruit | 25-Feb | 0.005258 | 0.036658 | 0 | 0 | 20.34783 | 106.7841 |

| | | | | | | | | |
|---|---|---|---|---|---|---|---|---|
| Rare_Disea | Peripartum | 13-Feb | 0.001414 | 0.037579 | 0 | 0 | 42.57112 | 279.3154 |
| Azimuth_2 | Fetal Devel | 9-Jan | 0.038483 | 0.038483 | 0 | 0 | 28.93169 | 94.24608 |
| Azimuth_2 | Motor Cort | 8-Jan | 0.03428 | 0.038483 | 0 | 0 | 33.06645 | 111.5391 |
| Azimuth_2 | Lung V2 (H | 9-Jan | 0.038483 | 0.038483 | 0 | 0 | 28.93169 | 94.24608 |
| Azimuth_2 | Fetal Devel | 9-Jan | 0.038483 | 0.038483 | 0 | 0 | 28.93169 | 94.24608 |
| Azimuth_2 | Fetal Devel | 9-Jan | 0.038483 | 0.038483 | 0 | 0 | 28.93169 | 94.24608 |
| Azimuth_2 | Mouse Mo | 9-Jan | 0.038483 | 0.038483 | 0 | 0 | 28.93169 | 94.24608 |
| Azimuth_2 | Mouse Mo | 9-Jan | 0.038483 | 0.038483 | 0 | 0 | 28.93169 | 94.24608 |
| Azimuth_2 | Mouse Mo | 9-Jan | 0.038483 | 0.038483 | 0 | 0 | 28.93169 | 94.24608 |
| Azimuth_2 | Motor Cort | 9-Jan | 0.038483 | 0.038483 | 0 | 0 | 28.93169 | 94.24608 |
| Azimuth_2 | Fetal Devel | 9-Jan | 0.038483 | 0.038483 | 0 | 0 | 28.93169 | 94.24608 |
| Azimuth_2 | Lung v1-L2- | 9-Jan | 0.038483 | 0.038483 | 0 | 0 | 28.93169 | 94.24608 |
| Azimuth_2 | Mouse Mo | 9-Jan | 0.038483 | 0.038483 | 0 | 0 | 28.93169 | 94.24608 |
| Azimuth_2 | Lung v1-L2- | 9-Jan | 0.038483 | 0.038483 | 0 | 0 | 28.93169 | 94.24608 |
| Azimuth_2 | Lung V2 (H | 9-Jan | 0.038483 | 0.038483 | 0 | 0 | 28.93169 | 94.24608 |
| Azimuth_2 | Fetal Devel | 9-Jan | 0.038483 | 0.038483 | 0 | 0 | 28.93169 | 94.24608 |
| Azimuth_2 | PBMC-L2-H | 9-Jan | 0.038483 | 0.038483 | 0 | 0 | 28.93169 | 94.24608 |
| Azimuth_2 | Tonsil-L2-C | 9-Jan | 0.038483 | 0.038483 | 0 | 0 | 28.93169 | 94.24608 |
| Azimuth_2 | Lung v1-L2- | 9-Jan | 0.038483 | 0.038483 | 0 | 0 | 28.93169 | 94.24608 |
| Azimuth_2 | Fetal Devel | 9-Jan | 0.038483 | 0.038483 | 0 | 0 | 28.93169 | 94.24608 |
| Azimuth_2 | PBMC-L1-N | 9-Jan | 0.038483 | 0.038483 | 0 | 0 | 28.93169 | 94.24608 |
| Azimuth_2 | Bone Marr | 9-Jan | 0.038483 | 0.038483 | 0 | 0 | 28.93169 | 94.24608 |
| Azimuth_2 | Fetal Devel | 9-Jan | 0.038483 | 0.038483 | 0 | 0 | 28.93169 | 94.24608 |
| Azimuth_2 | Motor Cort | 7-Jan | 0.03006 | 0.038483 | 0 | 0 | 38.57946 | 135.2043 |
| Azimuth_2 | Liver-L2-He | 9-Jan | 0.038483 | 0.038483 | 0 | 0 | 28.93169 | 94.24608 |
| Azimuth_2 | Lung v1-L1- | 9-Jan | 0.038483 | 0.038483 | 0 | 0 | 28.93169 | 94.24608 |
| Azimuth_2 | Lung V2 (H | 9-Jan | 0.038483 | 0.038483 | 0 | 0 | 28.93169 | 94.24608 |
| Azimuth_2 | Bone Marr | 9-Jan | 0.038483 | 0.038483 | 0 | 0 | 28.93169 | 94.24608 |
| Azimuth_2 | Bone Marr | 9-Jan | 0.038483 | 0.038483 | 0 | 0 | 28.93169 | 94.24608 |
| Azimuth_2 | Lung V2 (H | 9-Jan | 0.038483 | 0.038483 | 0 | 0 | 28.93169 | 94.24608 |
| Azimuth_2 | PBMC-L2-C | 9-Jan | 0.038483 | 0.038483 | 0 | 0 | 28.93169 | 94.24608 |
| Azimuth_2 | Mouse Mo | 9-Jan | 0.038483 | 0.038483 | 0 | 0 | 28.93169 | 94.24608 |
| Azimuth_2 | Mouse Mo | 9-Jan | 0.038483 | 0.038483 | 0 | 0 | 28.93169 | 94.24608 |
| Azimuth_2 | Mouse Mo | 9-Jan | 0.038483 | 0.038483 | 0 | 0 | 28.93169 | 94.24608 |
| Azimuth_2 | Mouse Mo | 9-Jan | 0.038483 | 0.038483 | 0 | 0 | 28.93169 | 94.24608 |
| Azimuth_2 | Mouse Mo | 9-Jan | 0.038483 | 0.038483 | 0 | 0 | 28.93169 | 94.24608 |
| Azimuth_2 | Lung V2 (H | 9-Jan | 0.038483 | 0.038483 | 0 | 0 | 28.93169 | 94.24608 |
| Azimuth_2 | Fetal Devel | 9-Jan | 0.038483 | 0.038483 | 0 | 0 | 28.93169 | 94.24608 |
| Azimuth_2 | Bone Marr | 9-Jan | 0.038483 | 0.038483 | 0 | 0 | 28.93169 | 94.24608 |
| Azimuth_2 | Bone Marr | 9-Jan | 0.038483 | 0.038483 | 0 | 0 | 28.93169 | 94.24608 |
| Azimuth_2 | Kidney-L2-/ | 9-Jan | 0.038483 | 0.038483 | 0 | 0 | 28.93169 | 94.24608 |
| Azimuth_2 | Fetal Devel | 9-Jan | 0.038483 | 0.038483 | 0 | 0 | 28.93169 | 94.24608 |
| Azimuth_2 | Lung v1-L1- | 9-Jan | 0.038483 | 0.038483 | 0 | 0 | 28.93169 | 94.24608 |
| Azimuth_2 | Pancreas-L | 9-Jan | 0.038483 | 0.038483 | 0 | 0 | 28.93169 | 94.24608 |
| Azimuth_2 | Lung V2 (H | 9-Jan | 0.038483 | 0.038483 | 0 | 0 | 28.93169 | 94.24608 |

| | | | | | | | | |
|---|---|---|---|---|---|---|---|---|
| Azimuth_2 | Mouse Mo | 9-Jan | 0.038483 | 0.038483 | 0 | 0 | 28.93169 | 94.24608 |
| Azimuth_2 | Bone Marr | 9-Jan | 0.038483 | 0.038483 | 0 | 0 | 28.93169 | 94.24608 |
| Azimuth_2 | Lung V2 (H | 9-Jan | 0.038483 | 0.038483 | 0 | 0 | 28.93169 | 94.24608 |
| Azimuth_2 | Bone Marr | 9-Jan | 0.038483 | 0.038483 | 0 | 0 | 28.93169 | 94.24608 |
| Azimuth_2 | Fetal Devel | 9-Jan | 0.038483 | 0.038483 | 0 | 0 | 28.93169 | 94.24608 |
| Azimuth_2 | Fetal Devel | 9-Jan | 0.038483 | 0.038483 | 0 | 0 | 28.93169 | 94.24608 |
| Azimuth_2 | Pancreas-L | 9-Jan | 0.038483 | 0.038483 | 0 | 0 | 28.93169 | 94.24608 |
| Azimuth_2 | Fetal Devel | 9-Jan | 0.038483 | 0.038483 | 0 | 0 | 28.93169 | 94.24608 |
| Azimuth_2 | Liver-L1-He | 9-Jan | 0.038483 | 0.038483 | 0 | 0 | 28.93169 | 94.24608 |
| Azimuth_2 | Motor Cort | 9-Jan | 0.038483 | 0.038483 | 0 | 0 | 28.93169 | 94.24608 |
| Azimuth_2 | Motor Cort | 9-Jan | 0.038483 | 0.038483 | 0 | 0 | 28.93169 | 94.24608 |
| Azimuth_2 | Fetal Devel | 9-Jan | 0.038483 | 0.038483 | 0 | 0 | 28.93169 | 94.24608 |
| Azimuth_2 | Motor Cort | 9-Jan | 0.038483 | 0.038483 | 0 | 0 | 28.93169 | 94.24608 |
| Azimuth_2 | Pancreas-L | 9-Jan | 0.038483 | 0.038483 | 0 | 0 | 28.93169 | 94.24608 |
| Azimuth_2 | PBMC-L2-C | 9-Jan | 0.038483 | 0.038483 | 0 | 0 | 28.93169 | 94.24608 |
| Azimuth_2 | Lung V2 (H | 9-Jan | 0.038483 | 0.038483 | 0 | 0 | 28.93169 | 94.24608 |
| Azimuth_2 | Fetal Devel | 9-Jan | 0.038483 | 0.038483 | 0 | 0 | 28.93169 | 94.24608 |
| Azimuth_2 | Lung v1-L2 | 9-Jan | 0.038483 | 0.038483 | 0 | 0 | 28.93169 | 94.24608 |
| Azimuth_2 | Tonsil-L2-D | 9-Jan | 0.038483 | 0.038483 | 0 | 0 | 28.93169 | 94.24608 |
| Azimuth_2 | Mouse Mo | 9-Jan | 0.038483 | 0.038483 | 0 | 0 | 28.93169 | 94.24608 |
| Azimuth_2 | Fetal Devel | 9-Jan | 0.038483 | 0.038483 | 0 | 0 | 28.93169 | 94.24608 |
| Azimuth_2 | Pancreas-L | 9-Jan | 0.038483 | 0.038483 | 0 | 0 | 28.93169 | 94.24608 |
| Azimuth_2 | Fetal Devel | 9-Jan | 0.038483 | 0.038483 | 0 | 0 | 28.93169 | 94.24608 |
| Azimuth_2 | Pancreas-L | 9-Jan | 0.038483 | 0.038483 | 0 | 0 | 28.93169 | 94.24608 |
| Azimuth_2 | Tonsil-L2-M | 9-Jan | 0.038483 | 0.038483 | 0 | 0 | 28.93169 | 94.24608 |
| Azimuth_2 | Kidney-L3- | 9-Jan | 0.038483 | 0.038483 | 0 | 0 | 28.93169 | 94.24608 |
| Azimuth_2 | Lung V2 (H | 9-Jan | 0.038483 | 0.038483 | 0 | 0 | 28.93169 | 94.24608 |
| Azimuth_2 | Fetal Devel | 9-Jan | 0.038483 | 0.038483 | 0 | 0 | 28.93169 | 94.24608 |
| Azimuth_2 | Heart-L1-C | 9-Jan | 0.038483 | 0.038483 | 0 | 0 | 28.93169 | 94.24608 |
| GO_Biologi | Outer Mito | 13-Feb | 0.001414 | 0.03862 | 0 | 0 | 42.57112 | 279.3154 |
| GO_Biologi | Protein Ins | 13-Feb | 0.001414 | 0.03862 | 0 | 0 | 42.57112 | 279.3154 |
| DisGeNET | Young onse | Mar-45 | 0.000989 | 0.038679 | 0 | 0 | 16.89711 | 116.9144 |
| DisGeNET | LATERAL M | Mar-45 | 0.000989 | 0.038679 | 0 | 0 | 16.89711 | 116.9144 |
| DisGeNET | Distal Musc | Mar-45 | 0.000989 | 0.038679 | 0 | 0 | 16.89711 | 116.9144 |
| DisGeNET | ECG abnor | 4/100 | 0.000953 | 0.038679 | 0 | 0 | 9.948293 | 69.19848 |
| DisGeNET | Neuromusc | 4/101 | 0.000989 | 0.038679 | 0 | 0 | 9.845237 | 68.11622 |
| DisGeNET | Abnormal | 4/100 | 0.000953 | 0.038679 | 0 | 0 | 9.948293 | 69.19848 |
| GWAS_Cat | Node-level | Mar-83 | 0.005685 | 0.039188 | 0 | 0 | 8.854018 | 45.77397 |
| GWAS_Cat | Mental Hea | 26-Feb | 0.005681 | 0.039188 | 0 | 0 | 19.49902 | 100.8238 |
| SysMyo_M | MGS1343 | 5/114 | 0.00014 | 0.03935 | 0 | 0 | 11.07854 | 98.2864 |
| SysMyo_M | MGS1331 | 6/195 | 0.000209 | 0.03935 | 0 | 0 | 7.730355 | 65.5167 |
| DisGeNET | Cardiac co | 4/102 | 0.001026 | 0.039535 | 0 | 0 | 9.744283 | 67.06001 |
| DisGeNET | Obstructive | Mar-46 | 0.001054 | 0.039826 | 0 | 0 | 16.50332 | 113.1284 |
| DisGeNET | Electrocard | 4/103 | 0.001064 | 0.039826 | 0 | 0 | 9.645369 | 66.029 |
| GeDiPNet_ | Dyslipidem | 24-Feb | 0.004851 | 0.039836 | 0 | 0 | 21.2738 | 113.3575 |

| | | | | | | | | |
|---|---|---|---|---|---|---|---|---|
| GWAS_Cat | Waist-hip I | 8/625 | 0.005828 | 0.039948 | 0 | 0 | 3.166978 | 16.29414 |
| GWAS_Cat | Lobe Attac | Mar-84 | 0.005878 | 0.04006 | 0 | 0 | 8.744268 | 44.91577 |
| DisGeNET | Electrocarc | 4/104 | 0.001103 | 0.040293 | 0 | 0 | 9.548434 | 65.02235 |
| DisGeNET | Protein me | Mar-47 | 0.001123 | 0.040293 | 0 | 0 | 16.12744 | 109.5385 |
| DisGeNET | LIMB-MAN | Mar-47 | 0.001123 | 0.040293 | 0 | 0 | 16.12744 | 109.5385 |
| Rare_Disea | Thalassemi | 6/288 | 0.001611 | 0.040804 | 0 | 0 | 5.156554 | 33.16158 |
| Rare_Disea | Hypoplasti | 14-Feb | 0.001645 | 0.040804 | 0 | 0 | 39.02157 | 250.1213 |
| DisGeNET | Cardiomyo | 8/480 | 0.001156 | 0.04093 | 0 | 0 | 4.170993 | 28.20744 |
| GWAS_Cat | Metabolic ! | Mar-85 | 0.006074 | 0.041167 | 0 | 0 | 8.637195 | 44.08228 |
| UK_Bioban | Medicatior | Feb-41 | 0.013741 | 0.041223 | 0 | 0 | 11.99035 | 51.40702 |
| GeDiPNet_ | Motor Dela | 6/365 | 0.005181 | 0.041427 | 0 | 0 | 4.034664 | 21.23315 |
| GeDiPNet_ | Atrial Septa | 5/256 | 0.005207 | 0.041427 | 0 | 0 | 4.776504 | 25.114 |
| GeDiPNet_ | Left Ventri | 25-Feb | 0.005258 | 0.041427 | 0 | 0 | 20.34783 | 106.7841 |
| DisGeNET | Intraventri | 13-Feb | 0.001414 | 0.041633 | 0 | 0 | 42.57112 | 279.3154 |
| DisGeNET | obsolete C | 13-Feb | 0.001414 | 0.041633 | 0 | 0 | 42.57112 | 279.3154 |
| DisGeNET | Posterior F | 13-Feb | 0.001414 | 0.041633 | 0 | 0 | 42.57112 | 279.3154 |
| DisGeNET | Olfactory G | 13-Feb | 0.001414 | 0.041633 | 0 | 0 | 42.57112 | 279.3154 |
| DisGeNET | Parasagitta | 13-Feb | 0.001414 | 0.041633 | 0 | 0 | 42.57112 | 279.3154 |
| DisGeNET | Microcystic | 13-Feb | 0.001414 | 0.041633 | 0 | 0 | 42.57112 | 279.3154 |
| DisGeNET | Hemangiot | 13-Feb | 0.001414 | 0.041633 | 0 | 0 | 42.57112 | 279.3154 |
| DisGeNET | Subaortic s | 12-Feb | 0.0012 | 0.041633 | 0 | 0 | 46.83059 | 314.9532 |
| DisGeNET | Congenital | 4/110 | 0.001358 | 0.041633 | 0 | 0 | 9.005228 | 59.45169 |
| DisGeNET | Angioblasti | 13-Feb | 0.001414 | 0.041633 | 0 | 0 | 42.57112 | 279.3154 |
| DisGeNET | Xanthomat | 13-Feb | 0.001414 | 0.041633 | 0 | 0 | 42.57112 | 279.3154 |
| DisGeNET | Hemangiop | 13-Feb | 0.001414 | 0.041633 | 0 | 0 | 42.57112 | 279.3154 |
| DisGeNET | Intraorbita | 13-Feb | 0.001414 | 0.041633 | 0 | 0 | 42.57112 | 279.3154 |
| DisGeNET | Cerebral C | 13-Feb | 0.001414 | 0.041633 | 0 | 0 | 42.57112 | 279.3154 |
| DisGeNET | Angiomato | 13-Feb | 0.001414 | 0.041633 | 0 | 0 | 42.57112 | 279.3154 |
| UK_Bioban | Monocyte | 5/329 | 0.014413 | 0.042127 | 0 | 0 | 3.686578 | 15.62979 |
| UK_Bioban | Time spent | 3/117 | 0.014486 | 0.042127 | 0 | 0 | 6.202694 | 26.26582 |
| UK_Bioban | Both eyes p | 7/589 | 0.014257 | 0.042127 | 0 | 0 | 2.906293 | 12.35329 |
| DisGeNET | Low densit | 6/283 | 0.001474 | 0.04291 | 0 | 0 | 5.250969 | 34.23508 |
| GO_Biologi | Cardiac Cel | 14-Feb | 0.001645 | 0.043266 | 0 | 0 | 39.02157 | 250.1213 |
| DisGeNET | Myeloid Le | 8/501 | 0.001517 | 0.043662 | 0 | 0 | 3.989011 | 25.89404 |
| GeDiPNet_ | Chronic Ob | 5/261 | 0.005644 | 0.043872 | 0 | 0 | 4.682022 | 24.23965 |
| GWAS_Cat | Cortical Su | 7/506 | 0.006523 | 0.043964 | 0 | 0 | 3.404259 | 17.13187 |
| GWAS_Cat | Average Di | 28-Feb | 0.00657 | 0.044039 | 0 | 0 | 17.99729 | 90.44125 |
| UK_Bioban | Takes chole | Feb-44 | 0.01572 | 0.044258 | 0 | 0 | 11.13221 | 46.2302 |
| UK_Bioban | Right intra- | 3/121 | 0.01584 | 0.044258 | 0 | 0 | 5.991223 | 24.83499 |
| UK_Bioban | Hair/baldir | 4/220 | 0.01559 | 0.044258 | 0 | 0 | 4.39469 | 18.28685 |
| UK_Bioban | Systolic blc | 5/336 | 0.015652 | 0.044258 | 0 | 0 | 3.607324 | 14.9962 |
| GWAS_Cat | Late-onset | Mar-88 | 0.006685 | 0.04457 | 0 | 0 | 8.331092 | 41.72057 |
| Rare_Disea | Locked-in s | Mar-56 | 0.001866 | 0.044782 | 0 | 0 | 13.38275 | 84.09717 |
| GWAS_Cat | Alanine Am | 6/386 | 0.00676 | 0.044822 | 0 | 0 | 3.807602 | 19.02552 |
| dbGaP | Cholestero | 5/268 | 0.006299 | 0.045492 | 0 | 0 | 4.555782 | 23.08587 |

| | | | | | | | | |
|---|---|---|---|---|---|---|---|---|
| UK_Bioban | Left intra-o | 3/123 | 0.016543 | 0.045773 | 0 | 0 | 5.890774 | 24.16285 |
| DisGeNET | Transitiona | 14-Feb | 0.001645 | 0.046181 | 0 | 0 | 39.02157 | 250.1213 |
| DisGeNET | Meningoth | 14-Feb | 0.001645 | 0.046181 | 0 | 0 | 39.02157 | 250.1213 |
| DisGeNET | Spinal Mus | 5/196 | 0.001657 | 0.046181 | 0 | 0 | 6.296131 | 40.31296 |
| DisGeNET | Diabetic Ca | Mar-54 | 0.00168 | 0.046336 | 0 | 0 | 13.90896 | 88.8629 |
| GTEx_Tissu | GTEX-XQ3S | 13/732 | 1.60E-05 | 0.046339 | 0 | 0 | 4.689734 | 51.78321 |
| GO_Biologi | Mitotic Chr | 15-Feb | 0.001893 | 0.046348 | 0 | 0 | 36.0181 | 225.8172 |
| GO_Biologi | High-Densi | 15-Feb | 0.001893 | 0.046348 | 0 | 0 | 36.0181 | 225.8172 |
| GWAS_Cat | Male Pube | Feb-29 | 0.007037 | 0.046404 | 0 | 0 | 17.32985 | 85.89758 |
| GO_Biologi | Muscle Org | Mar-58 | 0.002064 | 0.046417 | 0 | 0 | 12.89481 | 79.72951 |
| GO_Biologi | Striated Mu | Mar-57 | 0.001963 | 0.046417 | 0 | 0 | 13.13426 | 81.86658 |
| GO_Biologi | Phosphatic | 16-Feb | 0.002157 | 0.046417 | 0 | 0 | 33.4437 | 205.3055 |
| GO_Biologi | Lymphocyt | 16-Feb | 0.002157 | 0.046417 | 0 | 0 | 33.4437 | 205.3055 |
| SysMyo_M | MGS1175 I | 5/137 | 0.00033 | 0.046717 | 0 | 0 | 9.137565 | 73.24614 |
| UK_Bioban | Other joint | 4-Jan | 0.017288 | 0.047376 | 0 | 0 | 77.17054 | 313.1385 |
| GeDiPNet_ | Alzheimer I | 6/380 | 0.006277 | 0.048155 | 0 | 0 | 3.869875 | 19.62333 |
| GWAS_Cat | Serum Albu | 5/279 | 0.007434 | 0.048758 | 0 | 0 | 4.370438 | 21.42276 |
| Disease_Pe | Myocardial | 7/264 | 0.000153 | 0.049213 | 0 | 0 | 6.692218 | 58.77796 |
| Disease_Pe | MELAS - M | 6/169 | 9.55E-05 | 0.049213 | 0 | 0 | 8.975233 | 83.07519 |
| GWAS_Cat | Immature I | 5/281 | 0.007654 | 0.049937 | 0 | 0 | 4.338326 | 21.13844 |
| DisGeNET | Psammoma | 15-Feb | 0.001893 | 0.050611 | 0 | 0 | 36.0181 | 225.8172 |
| DisGeNET | Sphenoid V | 15-Feb | 0.001893 | 0.050611 | 0 | 0 | 36.0181 | 225.8172 |
| DisGeNET | Clear Cell N | 15-Feb | 0.001893 | 0.050611 | 0 | 0 | 36.0181 | 225.8172 |
| GTEx_Tissu | GTEX-S3XE | 19/1567 | 3.50E-05 | 0.050643 | 0 | 0 | 3.314856 | 34.00997 |
| GeDiPNet_ | Hypertroph | 5/272 | 0.006696 | 0.050701 | 0 | 0 | 4.486617 | 22.46089 |
| GWAS_Cat | Creatine Ki | Feb-31 | 0.008014 | 0.052006 | 0 | 0 | 16.13306 | 77.8673 |
| DisGeNET | Acute Meg | 4/122 | 0.001986 | 0.052554 | 0 | 0 | 8.084542 | 50.29973 |
| GeDiPNet_ | Congenital | Feb-29 | 0.007037 | 0.052594 | 0 | 0 | 17.32985 | 85.89758 |
| DisGeNET | Adenocarci | 4/125 | 0.00217 | 0.052634 | 0 | 0 | 7.882904 | 48.34739 |
| DisGeNET | Papillary M | 16-Feb | 0.002157 | 0.052634 | 0 | 0 | 33.4437 | 205.3055 |
| DisGeNET | Secretory r | 16-Feb | 0.002157 | 0.052634 | 0 | 0 | 33.4437 | 205.3055 |
| DisGeNET | Fibrous Me | 16-Feb | 0.002157 | 0.052634 | 0 | 0 | 33.4437 | 205.3055 |
| DisGeNET | Vital Capac | 16-Feb | 0.002157 | 0.052634 | 0 | 0 | 33.4437 | 205.3055 |
| DisGeNET | Spinal Men | 16-Feb | 0.002157 | 0.052634 | 0 | 0 | 33.4437 | 205.3055 |
| DisGeNET | Autosomal | 16-Feb | 0.002157 | 0.052634 | 0 | 0 | 33.4437 | 205.3055 |
| DisGeNET | Tetralogy c | 4/125 | 0.00217 | 0.052634 | 0 | 0 | 7.882904 | 48.34739 |
| DisGeNET | Neurofibril | 16-Feb | 0.002157 | 0.052634 | 0 | 0 | 33.4437 | 205.3055 |
| PheWeb_2 | Hx of malig | 19-Feb | 0.003048 | 0.053129 | 0 | 0 | 27.53772 | 159.5298 |
| PheWeb_2 | Open wour | 19-Feb | 0.003048 | 0.053129 | 0 | 0 | 27.53772 | 159.5298 |
| PheWeb_2 | Dementias | 17-Feb | 0.002438 | 0.053129 | 0 | 0 | 31.21255 | 187.7904 |
| PheWeb_2 | Optic atrop | 16-Feb | 0.002157 | 0.053129 | 0 | 0 | 33.4437 | 205.3055 |
| ClinVar_20 | congenital | 6-Jan | 0.025821 | 0.053793 | 0 | 0 | 46.29767 | 169.2909 |
| ClinVar_20 | meningiom | 6-Jan | 0.025821 | 0.053793 | 0 | 0 | 46.29767 | 169.2909 |
| ClinVar_20 | myofibrilla | 6-Jan | 0.025821 | 0.053793 | 0 | 0 | 46.29767 | 169.2909 |
| ClinVar_20 | leukoencep | 5-Jan | 0.021564 | 0.053793 | 0 | 0 | 57.875 | 222.0519 |

| | | | | | | | | |
|---|---|---|---|---|---|---|---|---|
| ClinVar_20 | atrial septa | 5-Jan | 0.021564 | 0.053793 | 0 | 0 | 57.875 | 222.0519 |
| UK_Bioban | Right arm f | 6/489 | 0.019824 | 0.053808 | 0 | 0 | 2.979833 | 11.68352 |
| GeDiPNet_ | Non-Alcoh | Mar-91 | 0.007333 | 0.054113 | 0 | 0 | 8.04586 | 39.5488 |
| DisGeNET | Androgene | Mar-60 | 0.002275 | 0.05468 | 0 | 0 | 12.4411 | 75.71458 |
| UK_Bioban | ICD10 anky | 5-Jan | 0.021564 | 0.054872 | 0 | 0 | 57.875 | 222.0519 |
| UK_Bioban | Father still | 5-Jan | 0.021564 | 0.054872 | 0 | 0 | 57.875 | 222.0519 |
| UK_Bioban | Hard chees | 5-Jan | 0.021564 | 0.054872 | 0 | 0 | 57.875 | 222.0519 |
| UK_Bioban | Right hand | 4/240 | 0.020741 | 0.054872 | 0 | 0 | 4.018174 | 15.57293 |
| UK_Bioban | Takes iron | 5-Jan | 0.021564 | 0.054872 | 0 | 0 | 57.875 | 222.0519 |
| UK_Bioban | Immature r | 5/364 | 0.02132 | 0.054872 | 0 | 0 | 3.321217 | 12.78047 |
| UK_Bioban | None of th | 5-Jan | 0.021564 | 0.054872 | 0 | 0 | 57.875 | 222.0519 |
| GWAS_Cat | Urolithiasis | Feb-32 | 0.008524 | 0.055024 | 0 | 0 | 15.59451 | 74.30533 |
| dbGaP | Phospholip | Feb-32 | 0.008524 | 0.055408 | 0 | 0 | 15.59451 | 74.30533 |
| UK_Bioban | Platelet cri | 7/647 | 0.022597 | 0.056493 | 0 | 0 | 2.63498 | 9.986389 |
| UK_Bioban | Mean retic | 6/503 | 0.02241 | 0.056493 | 0 | 0 | 2.893807 | 10.99139 |
| GO_Biologi | Cardiac Atr | 18-Feb | 0.002735 | 0.057118 | 0 | 0 | 29.26029 | 172.6809 |
| DisGeNET | Myopathy, | 18-Feb | 0.002735 | 0.057793 | 0 | 0 | 29.26029 | 172.6809 |
| DisGeNET | precancerc | 4/132 | 0.002644 | 0.057793 | 0 | 0 | 7.449172 | 44.21552 |
| DisGeNET | Metabolic S | 8/548 | 0.002648 | 0.057793 | 0 | 0 | 3.633005 | 21.5575 |
| DisGeNET | Centrally n | 18-Feb | 0.002735 | 0.057793 | 0 | 0 | 29.26029 | 172.6809 |
| DisGeNET | Primary cu | 18-Feb | 0.002735 | 0.057793 | 0 | 0 | 29.26029 | 172.6809 |
| DisGeNET | Diastrophic | 18-Feb | 0.002735 | 0.057793 | 0 | 0 | 29.26029 | 172.6809 |
| DisGeNET | Lewy Body | 4/133 | 0.002717 | 0.057793 | 0 | 0 | 7.391053 | 43.6691 |
| DisGeNET | PACHYONY | 4/133 | 0.002717 | 0.057793 | 0 | 0 | 7.391053 | 43.6691 |
| DisGeNET | Refractory | 18-Feb | 0.002735 | 0.057793 | 0 | 0 | 29.26029 | 172.6809 |
| DisGeNET | Niemann-P | 18-Feb | 0.002735 | 0.057793 | 0 | 0 | 29.26029 | 172.6809 |
| DisGeNET | Opposition | 18-Feb | 0.002735 | 0.057793 | 0 | 0 | 29.26029 | 172.6809 |
| DisGeNET | Alagille Syr | 17-Feb | 0.002438 | 0.057793 | 0 | 0 | 31.21255 | 187.7904 |
| DisGeNET | Mitochond | 18-Feb | 0.002735 | 0.057793 | 0 | 0 | 29.26029 | 172.6809 |
| DisGeNET | Body Heigh | 6/314 | 0.002485 | 0.057793 | 0 | 0 | 4.715007 | 28.27783 |
| DisGeNET | Frontal bos | 5/216 | 0.002529 | 0.057793 | 0 | 0 | 5.693561 | 34.04628 |
| GWAS_Cat | Parental Lc | Feb-33 | 0.009049 | 0.057798 | 0 | 0 | 15.0907 | 71.00369 |
| GWAS_Cat | Docosahex | Feb-33 | 0.009049 | 0.057798 | 0 | 0 | 15.0907 | 71.00369 |
| GeDiPNet_ | Palmoplant | Feb-31 | 0.008014 | 0.058402 | 0 | 0 | 16.13306 | 77.8673 |
| GeDiPNet_ | Lung Neop | 5/286 | 0.008226 | 0.059206 | 0 | 0 | 4.260047 | 20.45021 |
| DisGeNET | Neurilemm | 4/135 | 0.002867 | 0.060089 | 0 | 0 | 7.277476 | 42.60636 |
| GWAS_Cat | Phospholip | Feb-34 | 0.009587 | 0.060291 | 0 | 0 | 14.61838 | 67.9362 |
| GWAS_Cat | Phospholip | Feb-34 | 0.009587 | 0.060291 | 0 | 0 | 14.61838 | 67.9362 |
| GWAS_Cat | Leisure Sec | 3/100 | 0.009491 | 0.060291 | 0 | 0 | 7.296024 | 33.98049 |
| UK_Bioban | Alzheimer's | 6-Jan | 0.025821 | 0.060818 | 0 | 0 | 46.29767 | 169.2909 |
| UK_Bioban | Chronic bro | 6-Jan | 0.025821 | 0.060818 | 0 | 0 | 46.29767 | 169.2909 |
| UK_Bioban | Headache l | Feb-56 | 0.024747 | 0.060818 | 0 | 0 | 8.653159 | 32.00857 |
| UK_Bioban | Self-report | 3/145 | 0.025421 | 0.060818 | 0 | 0 | 4.972586 | 18.26021 |
| UK_Bioban | Fractured v | 6-Jan | 0.025821 | 0.060818 | 0 | 0 | 46.29767 | 169.2909 |
| UK_Bioban | Shortness c | 6-Jan | 0.025821 | 0.060818 | 0 | 0 | 46.29767 | 169.2909 |

| | | | | | | | | |
|---|---|---|---|---|---|---|---|---|
| UK_Bioban | Alzheimer's | 6-Jan | 0.025821 | 0.060818 | 0 | 0 | 46.29767 | 169.2909 |
| DisGeNET | Acute Erytl | 5/224 | 0.002958 | 0.061506 | 0 | 0 | 5.48335 | 31.93106 |
| Rare_Disea | Central cor | 5/219 | 0.002684 | 0.061669 | 0 | 0 | 5.61289 | 33.23002 |
| Rare_Disea | Moyamoya | Mar-64 | 0.002735 | 0.061669 | 0 | 0 | 11.62295 | 68.5929 |
| DisGeNET | Mixed phei | 19-Feb | 0.003048 | 0.061913 | 0 | 0 | 27.53772 | 159.5298 |
| DisGeNET | Generalize | 19-Feb | 0.003048 | 0.061913 | 0 | 0 | 27.53772 | 159.5298 |
| DisGeNET | Asymmetri | 19-Feb | 0.003048 | 0.061913 | 0 | 0 | 27.53772 | 159.5298 |
| DisGeNET | Hypertropl | 6/328 | 0.003083 | 0.062142 | 0 | 0 | 4.506786 | 26.05698 |
| Jensen_DIS | Vascular di | Mar-78 | 0.004781 | 0.062156 | 0 | 0 | 9.446667 | 50.47405 |
| GWAS_Cat | Cholestero | 3/102 | 0.010016 | 0.062663 | 0 | 0 | 7.147908 | 32.90605 |
| PhenGenI_ | Amino Acid | Mar-70 | 0.003527 | 0.062675 | 0 | 0 | 10.57889 | 59.74182 |
| PhenGenI_ | Amyloid Be | Mar-67 | 0.003116 | 0.062675 | 0 | 0 | 11.07645 | 63.92535 |
| PhenGenI_ | Neuroimag | 20-Feb | 0.003378 | 0.062675 | 0 | 0 | 26.00654 | 147.9928 |
| PhenGenI_ | Metabolisr | 6/329 | 0.00313 | 0.062675 | 0 | 0 | 4.492604 | 25.90773 |
| Human_Ph | Sudden dea | 14-Feb | 0.001645 | 0.062851 | 0 | 0 | 39.02157 | 250.1213 |
| UK_Bioban | Birth weigh | Feb-59 | 0.027266 | 0.062881 | 0 | 0 | 8.196491 | 29.5248 |
| UK_Bioban | Mean sphe | 6/527 | 0.027359 | 0.062881 | 0 | 0 | 2.757091 | 9.922011 |
| UK_Bioban | Hair/baldir | Feb-59 | 0.027266 | 0.062881 | 0 | 0 | 8.196491 | 29.5248 |
| GWAS_Cat | Urinary Alb | Feb-35 | 0.01014 | 0.063115 | 0 | 0 | 14.17469 | 65.07997 |
| GWAS_Cat | Sex Hormo | 7/552 | 0.010263 | 0.063559 | 0 | 0 | 3.109541 | 14.23919 |
| GeDiPNet_ | Hip Contra | Feb-33 | 0.009049 | 0.063674 | 0 | 0 | 15.0907 | 71.00369 |
| GeDiPNet_ | Gastric Car | 6/411 | 0.009065 | 0.063674 | 0 | 0 | 3.567993 | 16.78144 |
| GWAS_Cat | Cortical Su | Feb-36 | 0.010706 | 0.064636 | 0 | 0 | 13.75709 | 62.41496 |
| GWAS_Cat | Sex Hormo | Feb-36 | 0.010706 | 0.064636 | 0 | 0 | 13.75709 | 62.41496 |
| GWAS_Cat | Cholesteryl | Feb-36 | 0.010706 | 0.064636 | 0 | 0 | 13.75709 | 62.41496 |
| GWAS_Cat | Cholestero | Feb-36 | 0.010706 | 0.064636 | 0 | 0 | 13.75709 | 62.41496 |
| GWAS_Cat | Medicatior | 3/104 | 0.010557 | 0.064636 | 0 | 0 | 7.005658 | 31.88247 |
| GWAS_Cat | Vertex-wis | 6/427 | 0.010808 | 0.064636 | 0 | 0 | 3.429577 | 15.52732 |
| GWAS_Cat | Cystatin C I | 4/197 | 0.010767 | 0.064636 | 0 | 0 | 4.924153 | 22.31264 |
| dbGaP | Carcinoma, | 3-Jan | 0.012994 | 0.064969 | 0 | 0 | 115.7616 | 502.7851 |
| dbGaP | Hypertrigly | 3-Jan | 0.012994 | 0.064969 | 0 | 0 | 115.7616 | 502.7851 |
| dbGaP | Amyloid be | 3-Jan | 0.012994 | 0.064969 | 0 | 0 | 115.7616 | 502.7851 |
| DisGeNET | Short palpe | Mar-68 | 0.003249 | 0.064988 | 0 | 0 | 10.90549 | 62.48067 |
| GWAS_Cat | Endometric | Feb-37 | 0.011286 | 0.065568 | 0 | 0 | 13.36336 | 59.92353 |
| GWAS_Cat | High Light ! | 6/429 | 0.011041 | 0.065568 | 0 | 0 | 3.413011 | 15.37938 |
| GWAS_Cat | Total Lipid | Feb-37 | 0.011286 | 0.065568 | 0 | 0 | 13.36336 | 59.92353 |
| GWAS_Cat | Phospholip | Feb-37 | 0.011286 | 0.065568 | 0 | 0 | 13.36336 | 59.92353 |
| GWAS_Cat | Cardioemb | Feb-37 | 0.011286 | 0.065568 | 0 | 0 | 13.36336 | 59.92353 |
| GWAS_Cat | Concentrat | Feb-37 | 0.011286 | 0.065568 | 0 | 0 | 13.36336 | 59.92353 |
| GWAS_Cat | Total Body | 3/107 | 0.0114 | 0.065916 | 0 | 0 | 6.802541 | 30.43536 |
| ClinVar_20 | hyperlipop | 8-Jan | 0.03428 | 0.065924 | 0 | 0 | 33.06645 | 111.5391 |
| GWAS_Cat | Cholesteryl | Feb-38 | 0.01188 | 0.06618 | 0 | 0 | 12.9915 | 57.59009 |
| GWAS_Cat | Cholesteryl | Feb-38 | 0.01188 | 0.06618 | 0 | 0 | 12.9915 | 57.59009 |
| GWAS_Cat | Urea Levels | 3/108 | 0.01169 | 0.06618 | 0 | 0 | 6.737415 | 29.97504 |
| GWAS_Cat | Ratio Of Po | Feb-38 | 0.01188 | 0.06618 | 0 | 0 | 12.9915 | 57.59009 |

| | | | | | | | | |
|---|---|---|---|---|---|---|---|---|
| GWAS_Cat | Phospholip | Feb-38 | 0.01188 | 0.06618 | 0 | 0 | 12.9915 | 57.59009 |
| GWAS_Cat | Phospholip | Feb-38 | 0.01188 | 0.06618 | 0 | 0 | 12.9915 | 57.59009 |
| GWAS_Cat | Waist-to-hi | Feb-38 | 0.01188 | 0.06618 | 0 | 0 | 12.9915 | 57.59009 |
| GWAS_Cat | Whole Brai | 4/202 | 0.011719 | 0.06618 | 0 | 0 | 4.798588 | 21.33702 |
| DisGeNET | Underdeve | Mar-69 | 0.003387 | 0.066712 | 0 | 0 | 10.73972 | 61.08699 |
| DisGeNET | Solitary fib | 20-Feb | 0.003378 | 0.066712 | 0 | 0 | 26.00654 | 147.9928 |
| GO_Biologi | T Cell Medi | 20-Feb | 0.003378 | 0.06679 | 0 | 0 | 26.00654 | 147.9928 |
| GO_Biologi | Regulation | Mar-69 | 0.003387 | 0.06679 | 0 | 0 | 10.73972 | 61.08699 |
| UK_Bioban | ICD10 fract | 7-Jan | 0.03006 | 0.06693 | 0 | 0 | 38.57946 | 135.2043 |
| UK_Bioban | General ho | 7-Jan | 0.03006 | 0.06693 | 0 | 0 | 38.57946 | 135.2043 |
| UK_Bioban | Right 3mm | 7-Jan | 0.03006 | 0.06693 | 0 | 0 | 38.57946 | 135.2043 |
| UK_Bioban | Medicatior | 7-Jan | 0.03006 | 0.06693 | 0 | 0 | 38.57946 | 135.2043 |
| SysMyo_M | MGS914 M | 5/156 | 0.000598 | 0.067691 | 0 | 0 | 7.980132 | 59.22823 |
| GWAS_Cat | Bone Mine | Feb-39 | 0.012487 | 0.06801 | 0 | 0 | 12.63975 | 55.40083 |
| GWAS_Cat | Body Surfa | 5/317 | 0.012445 | 0.06801 | 0 | 0 | 3.830715 | 16.80313 |
| GWAS_Cat | Phospholip | Feb-39 | 0.012487 | 0.06801 | 0 | 0 | 12.63975 | 55.40083 |
| GWAS_Cat | HDL Choles | Feb-39 | 0.012487 | 0.06801 | 0 | 0 | 12.63975 | 55.40083 |
| GWAS_Cat | Free Chole: | Feb-39 | 0.012487 | 0.06801 | 0 | 0 | 12.63975 | 55.40083 |
| GWAS_Cat | Estimated ( | 3/111 | 0.012584 | 0.068231 | 0 | 0 | 6.549272 | 28.65547 |
| UK_Bioban | Neutrophil | 5/402 | 0.030946 | 0.068368 | 0 | 0 | 2.997481 | 10.41781 |
| ClinVar_20 | malignant t | 9-Jan | 0.038483 | 0.06872 | 0 | 0 | 28.93169 | 94.24608 |
| GWAS_Cat | Vertex-wis | 7/576 | 0.012744 | 0.068797 | 0 | 0 | 2.974692 | 12.97757 |
| WikiPathw | Striated Mu | Mar-38 | 0.000601 | 0.069089 | 0 | 0 | 20.28367 | 150.4498 |
| GWAS_Cat | Bone Mine | Feb-40 | 0.013107 | 0.070136 | 0 | 0 | 12.3065 | 53.34346 |
| GWAS_Cat | Cholestero | Feb-40 | 0.013107 | 0.070136 | 0 | 0 | 12.3065 | 53.34346 |
| GeDiPNet_ | Hypoglycer | 4/194 | 0.010221 | 0.070935 | 0 | 0 | 5.002663 | 22.92898 |
| ClinVar_20 | alzheimer's | 10-Jan | 0.042667 | 0.071112 | 0 | 0 | 25.71576 | 81.1157 |
| DisGeNET | Pathologica | 21-Feb | 0.003723 | 0.071714 | 0 | 0 | 24.63653 | 137.8005 |
| DisGeNET | Siderosis | 21-Feb | 0.003723 | 0.071714 | 0 | 0 | 24.63653 | 137.8005 |
| DisGeNET | Meningiom | 21-Feb | 0.003723 | 0.071714 | 0 | 0 | 24.63653 | 137.8005 |
| Rare_Disea | Endomyoca | 20-Feb | 0.003378 | 0.071797 | 0 | 0 | 26.00654 | 147.9928 |
| Rare_Disea | Endocardia | 20-Feb | 0.003378 | 0.071797 | 0 | 0 | 26.00654 | 147.9928 |
| GWAS_Cat | Noncogniti | 3/114 | 0.013515 | 0.072004 | 0 | 0 | 6.3713 | 27.42156 |
| GWAS_Cat | Total Lipid | Feb-41 | 0.013741 | 0.072259 | 0 | 0 | 11.99035 | 51.40702 |
| GWAS_Cat | Cholesteryl | Feb-41 | 0.013741 | 0.072259 | 0 | 0 | 11.99035 | 51.40702 |
| GWAS_Cat | Triglyceride | Feb-41 | 0.013741 | 0.072259 | 0 | 0 | 11.99035 | 51.40702 |
| GWAS_Cat | Weight | 7/587 | 0.014016 | 0.073388 | 0 | 0 | 2.916616 | 12.44683 |
| GWAS_Cat | Integral Me | 5-Jan | 0.021564 | 0.073601 | 0 | 0 | 57.875 | 222.0519 |
| GWAS_Cat | Hippocamp | 5-Jan | 0.021564 | 0.073601 | 0 | 0 | 57.875 | 222.0519 |
| GWAS_Cat | X-24870 Le | 5-Jan | 0.021564 | 0.073601 | 0 | 0 | 57.875 | 222.0519 |
| GWAS_Cat | Serum Leve | 6-Jan | 0.025821 | 0.073601 | 0 | 0 | 46.29767 | 169.2909 |
| GWAS_Cat | Cerebral Ar | 6-Jan | 0.025821 | 0.073601 | 0 | 0 | 46.29767 | 169.2909 |
| GWAS_Cat | Gout (Com | 6-Jan | 0.025821 | 0.073601 | 0 | 0 | 46.29767 | 169.2909 |
| GWAS_Cat | Postprandi | 6-Jan | 0.025821 | 0.073601 | 0 | 0 | 46.29767 | 169.2909 |
| GWAS_Cat | Postprandi | 6-Jan | 0.025821 | 0.073601 | 0 | 0 | 46.29767 | 169.2909 |

| | | | | | | | |
|---|---|---|---|---|---|---|---|
| GWAS_Cat Triglyceride | Feb-56 | 0.024747 | 0.073601 | 0 | 0 | 8.653159 | 32.00857 |
| GWAS_Cat Cholesteryl | Feb-56 | 0.024747 | 0.073601 | 0 | 0 | 8.653159 | 32.00857 |
| GWAS_Cat Free Choles | Feb-57 | 0.025575 | 0.073601 | 0 | 0 | 8.495401 | 31.14526 |
| GWAS_Cat HLA Class I | 5-Jan | 0.021564 | 0.073601 | 0 | 0 | 57.875 | 222.0519 |
| GWAS_Cat Trihexosylc | 5-Jan | 0.021564 | 0.073601 | 0 | 0 | 57.875 | 222.0519 |
| GWAS_Cat Circulating | 5-Jan | 0.021564 | 0.073601 | 0 | 0 | 57.875 | 222.0519 |
| GWAS_Cat Myocardial | 5-Jan | 0.021564 | 0.073601 | 0 | 0 | 57.875 | 222.0519 |
| GWAS_Cat Monohexo | 5-Jan | 0.021564 | 0.073601 | 0 | 0 | 57.875 | 222.0519 |
| GWAS_Cat Drusen Are | 6-Jan | 0.025821 | 0.073601 | 0 | 0 | 46.29767 | 169.2909 |
| GWAS_Cat Phosphatic | 6-Jan | 0.025821 | 0.073601 | 0 | 0 | 46.29767 | 169.2909 |
| GWAS_Cat Total Lipid | Feb-55 | 0.02393 | 0.073601 | 0 | 0 | 8.81687 | 32.91021 |
| GWAS_Cat Body Mass | 8/807 | 0.024119 | 0.073601 | 0 | 0 | 2.422522 | 9.023298 |
| GWAS_Cat Glomerular | Feb-56 | 0.024747 | 0.073601 | 0 | 0 | 8.653159 | 32.00857 |
| GWAS_Cat Sex Hormo | 3/120 | 0.015495 | 0.073601 | 0 | 0 | 6.042735 | 25.18157 |
| GWAS_Cat Triglyceride | Feb-44 | 0.01572 | 0.073601 | 0 | 0 | 11.13221 | 46.2302 |
| GWAS_Cat Cerebrospi | Feb-44 | 0.01572 | 0.073601 | 0 | 0 | 11.13221 | 46.2302 |
| GWAS_Cat Phospholip | Feb-43 | 0.015047 | 0.073601 | 0 | 0 | 11.4043 | 47.85875 |
| GWAS_Cat Breast Can | 6-Jan | 0.025821 | 0.073601 | 0 | 0 | 46.29767 | 169.2909 |
| GWAS_Cat Retinol (Vit | 5-Jan | 0.021564 | 0.073601 | 0 | 0 | 57.875 | 222.0519 |
| GWAS_Cat Killer Cell Ir | 5-Jan | 0.021564 | 0.073601 | 0 | 0 | 57.875 | 222.0519 |
| GWAS_Cat Phosphatic | 5-Jan | 0.021564 | 0.073601 | 0 | 0 | 57.875 | 222.0519 |
| GWAS_Cat 1-palmitoy | 5-Jan | 0.021564 | 0.073601 | 0 | 0 | 57.875 | 222.0519 |
| GWAS_Cat Aspartate T | 5-Jan | 0.021564 | 0.073601 | 0 | 0 | 57.875 | 222.0519 |
| GWAS_Cat Serum Leve | 5-Jan | 0.021564 | 0.073601 | 0 | 0 | 57.875 | 222.0519 |
| GWAS_Cat Glucose-de | 5-Jan | 0.021564 | 0.073601 | 0 | 0 | 57.875 | 222.0519 |
| GWAS_Cat Lithium Cle | 6-Jan | 0.025821 | 0.073601 | 0 | 0 | 46.29767 | 169.2909 |
| GWAS_Cat Right Ventr | 6-Jan | 0.025821 | 0.073601 | 0 | 0 | 46.29767 | 169.2909 |
| GWAS_Cat Fear Of Sev | 6-Jan | 0.025821 | 0.073601 | 0 | 0 | 46.29767 | 169.2909 |
| GWAS_Cat Dehydroch | 6-Jan | 0.025821 | 0.073601 | 0 | 0 | 46.29767 | 169.2909 |
| GWAS_Cat Albumin Le | 3/120 | 0.015495 | 0.073601 | 0 | 0 | 6.042735 | 25.18157 |
| GWAS_Cat Chronic Lyr | 5-Jan | 0.021564 | 0.073601 | 0 | 0 | 57.875 | 222.0519 |
| GWAS_Cat Alkenylpho | 5-Jan | 0.021564 | 0.073601 | 0 | 0 | 57.875 | 222.0519 |
| GWAS_Cat Entorhinal | 5-Jan | 0.021564 | 0.073601 | 0 | 0 | 57.875 | 222.0519 |
| GWAS_Cat Fasting Trig | 6-Jan | 0.025821 | 0.073601 | 0 | 0 | 46.29767 | 169.2909 |
| GWAS_Cat Postprandi | 6-Jan | 0.025821 | 0.073601 | 0 | 0 | 46.29767 | 169.2909 |
| GWAS_Cat Diacylglyce | 6-Jan | 0.025821 | 0.073601 | 0 | 0 | 46.29767 | 169.2909 |
| GWAS_Cat Glycemic T | 6-Jan | 0.025821 | 0.073601 | 0 | 0 | 46.29767 | 169.2909 |
| GWAS_Cat Postprandi | 6-Jan | 0.025821 | 0.073601 | 0 | 0 | 46.29767 | 169.2909 |
| GWAS_Cat Left Ventric | 6-Jan | 0.025821 | 0.073601 | 0 | 0 | 46.29767 | 169.2909 |
| GWAS_Cat Age Of Ons | 6-Jan | 0.025821 | 0.073601 | 0 | 0 | 46.29767 | 169.2909 |
| GWAS_Cat Heart Rate | 6-Jan | 0.025821 | 0.073601 | 0 | 0 | 46.29767 | 169.2909 |
| GWAS_Cat Neurogenic | 5-Jan | 0.021564 | 0.073601 | 0 | 0 | 57.875 | 222.0519 |
| GWAS_Cat F-fatty Foo | 6-Jan | 0.025821 | 0.073601 | 0 | 0 | 46.29767 | 169.2909 |
| GWAS_Cat Vaginal Mic | 6-Jan | 0.025821 | 0.073601 | 0 | 0 | 46.29767 | 169.2909 |
| GWAS_Cat Psychosis A | 6-Jan | 0.025821 | 0.073601 | 0 | 0 | 46.29767 | 169.2909 |

| | | | | | | | |
|---|---|---|---|---|---|---|---|
| GWAS_Cat Spread Typ | 6-Jan | 0.025821 | 0.073601 | 0 | 0 | 46.29767 | 169.2909 |
| GWAS_Cat P Wave Ter | 6-Jan | 0.025821 | 0.073601 | 0 | 0 | 46.29767 | 169.2909 |
| GWAS_Cat 1-(1-Enyl-P | 6-Jan | 0.025821 | 0.073601 | 0 | 0 | 46.29767 | 169.2909 |
| GWAS_Cat C-C Motif C | 6-Jan | 0.025821 | 0.073601 | 0 | 0 | 46.29767 | 169.2909 |
| GWAS_Cat 1-stearoyl- | 6-Jan | 0.025821 | 0.073601 | 0 | 0 | 46.29767 | 169.2909 |
| GWAS_Cat Palmitic Ac | 5-Jan | 0.021564 | 0.073601 | 0 | 0 | 57.875 | 222.0519 |
| GWAS_Cat Cadherin-3 | 5-Jan | 0.021564 | 0.073601 | 0 | 0 | 57.875 | 222.0519 |
| GWAS_Cat Globe Artic | 5-Jan | 0.021564 | 0.073601 | 0 | 0 | 57.875 | 222.0519 |
| GWAS_Cat Alkenylpho | 6-Jan | 0.025821 | 0.073601 | 0 | 0 | 46.29767 | 169.2909 |
| GWAS_Cat Cardiovasc | 4/227 | 0.017289 | 0.073601 | 0 | 0 | 4.255227 | 17.26642 |
| GWAS_Cat Fasting Trig | 5-Jan | 0.021564 | 0.073601 | 0 | 0 | 57.875 | 222.0519 |
| GWAS_Cat Fasting Trig | 5-Jan | 0.021564 | 0.073601 | 0 | 0 | 57.875 | 222.0519 |
| GWAS_Cat Postprandi | 5-Jan | 0.021564 | 0.073601 | 0 | 0 | 57.875 | 222.0519 |
| GWAS_Cat Repulsive C | 5-Jan | 0.021564 | 0.073601 | 0 | 0 | 57.875 | 222.0519 |
| GWAS_Cat Coagulatio | 5-Jan | 0.021564 | 0.073601 | 0 | 0 | 57.875 | 222.0519 |
| GWAS_Cat Emotional | 5-Jan | 0.021564 | 0.073601 | 0 | 0 | 57.875 | 222.0519 |
| GWAS_Cat Osteoarthr | 5-Jan | 0.021564 | 0.073601 | 0 | 0 | 57.875 | 222.0519 |
| GWAS_Cat Triglyceride | 5-Jan | 0.021564 | 0.073601 | 0 | 0 | 57.875 | 222.0519 |
| GWAS_Cat Biomedical | 5-Jan | 0.021564 | 0.073601 | 0 | 0 | 57.875 | 222.0519 |
| GWAS_Cat Postprandi | 5-Jan | 0.021564 | 0.073601 | 0 | 0 | 57.875 | 222.0519 |
| GWAS_Cat Longevity ( | 6-Jan | 0.025821 | 0.073601 | 0 | 0 | 46.29767 | 169.2909 |
| GWAS_Cat Body Mass | 15/1912 | 0.017397 | 0.073601 | 0 | 0 | 1.978563 | 8.016064 |
| GWAS_Cat Free Chole | Feb-51 | 0.020776 | 0.073601 | 0 | 0 | 9.538535 | 36.95177 |
| GWAS_Cat Free Chole | Feb-51 | 0.020776 | 0.073601 | 0 | 0 | 9.538535 | 36.95177 |
| GWAS_Cat Cholestero | Feb-52 | 0.021547 | 0.073601 | 0 | 0 | 9.347294 | 35.8704 |
| GWAS_Cat Triglyceride | Feb-52 | 0.021547 | 0.073601 | 0 | 0 | 9.347294 | 35.8704 |
| GWAS_Cat N-acetylglu | 5-Jan | 0.021564 | 0.073601 | 0 | 0 | 57.875 | 222.0519 |
| GWAS_Cat Cholesteryl | Feb-50 | 0.020017 | 0.073601 | 0 | 0 | 9.737745 | 38.08585 |
| GWAS_Cat Concentrat | Feb-50 | 0.020017 | 0.073601 | 0 | 0 | 9.737745 | 38.08585 |
| GWAS_Cat Free Chole | Feb-50 | 0.020017 | 0.073601 | 0 | 0 | 9.737745 | 38.08585 |
| GWAS_Cat Free Chole | Feb-50 | 0.020017 | 0.073601 | 0 | 0 | 9.737745 | 38.08585 |
| GWAS_Cat Male Pube | Feb-51 | 0.020776 | 0.073601 | 0 | 0 | 9.538535 | 36.95177 |
| GWAS_Cat Total Free C | Feb-51 | 0.020776 | 0.073601 | 0 | 0 | 9.538535 | 36.95177 |
| GWAS_Cat Two-hour C | 5-Jan | 0.021564 | 0.073601 | 0 | 0 | 57.875 | 222.0519 |
| GWAS_Cat Cholelithia | Feb-47 | 0.017813 | 0.073601 | 0 | 0 | 10.3885 | 41.84306 |
| GWAS_Cat Familial Co | Feb-47 | 0.017813 | 0.073601 | 0 | 0 | 10.3885 | 41.84306 |
| GWAS_Cat Central Co | 3/128 | 0.018376 | 0.073601 | 0 | 0 | 5.653714 | 22.5964 |
| GWAS_Cat Fish- And P | Feb-48 | 0.018536 | 0.073601 | 0 | 0 | 10.16215 | 40.52728 |
| GWAS_Cat Body Fat D | Feb-48 | 0.018536 | 0.073601 | 0 | 0 | 10.16215 | 40.52728 |
| GWAS_Cat Free Chole | Feb-49 | 0.01927 | 0.073601 | 0 | 0 | 9.945432 | 39.27635 |
| GWAS_Cat Endometri | Feb-50 | 0.020017 | 0.073601 | 0 | 0 | 9.737745 | 38.08585 |
| GWAS_Cat Fasting Insu | Feb-45 | 0.016405 | 0.073601 | 0 | 0 | 10.87278 | 44.68894 |
| GWAS_Cat Total Lipid | Feb-45 | 0.016405 | 0.073601 | 0 | 0 | 10.87278 | 44.68894 |
| GWAS_Cat Triglyceride | Feb-46 | 0.017103 | 0.073601 | 0 | 0 | 10.62513 | 43.22851 |
| GWAS_Cat Cholestero | Feb-46 | 0.017103 | 0.073601 | 0 | 0 | 10.62513 | 43.22851 |

| | | | | | | | |
|---|---|---|---|---|---|---|---|
| GWAS_Cat Cerebral M | 6-Jan | 0.025821 | 0.073601 | 0 | 0 | 46.29767 | 169.2909 |
| GWAS_Cat Total Alkylo | 6-Jan | 0.025821 | 0.073601 | 0 | 0 | 46.29767 | 169.2909 |
| GWAS_Cat Vaginal Mi | 6-Jan | 0.025821 | 0.073601 | 0 | 0 | 46.29767 | 169.2909 |
| GWAS_Cat Serum Tota | 6-Jan | 0.025821 | 0.073601 | 0 | 0 | 46.29767 | 169.2909 |
| GWAS_Cat ADP-ribosy | 6-Jan | 0.025821 | 0.073601 | 0 | 0 | 46.29767 | 169.2909 |
| GWAS_Cat Free Chole | Feb-42 | 0.014388 | 0.073601 | 0 | 0 | 11.69 | 49.58172 |
| GWAS_Cat Free Chole | Feb-43 | 0.015047 | 0.073601 | 0 | 0 | 11.4043 | 47.85875 |
| GWAS_Cat Bone Mine | Feb-43 | 0.015047 | 0.073601 | 0 | 0 | 11.4043 | 47.85875 |
| GWAS_Cat Ratio Of Or | Feb-43 | 0.015047 | 0.073601 | 0 | 0 | 11.4043 | 47.85875 |
| GWAS_Cat Cholesteryl | Feb-43 | 0.015047 | 0.073601 | 0 | 0 | 11.4043 | 47.85875 |
| GWAS_Cat Total Chole | Feb-43 | 0.015047 | 0.073601 | 0 | 0 | 11.4043 | 47.85875 |
| GWAS_Cat Cholestero | Feb-42 | 0.014388 | 0.073601 | 0 | 0 | 11.69 | 49.58172 |
| GWAS_Cat BMI X Envi | 6-Jan | 0.025821 | 0.073601 | 0 | 0 | 46.29767 | 169.2909 |
| GWAS_Cat Tripeptidyl | 6-Jan | 0.025821 | 0.073601 | 0 | 0 | 46.29767 | 169.2909 |
| GWAS_Cat Gout Vs As | 6-Jan | 0.025821 | 0.073601 | 0 | 0 | 46.29767 | 169.2909 |
| GWAS_Cat Fasting Fre | 5-Jan | 0.021564 | 0.073601 | 0 | 0 | 57.875 | 222.0519 |
| GWAS_Cat Postprandi | 5-Jan | 0.021564 | 0.073601 | 0 | 0 | 57.875 | 222.0519 |
| GWAS_Cat Cholesteryl | 5-Jan | 0.021564 | 0.073601 | 0 | 0 | 57.875 | 222.0519 |
| GWAS_Cat Alkenylpho | 5-Jan | 0.021564 | 0.073601 | 0 | 0 | 57.875 | 222.0519 |
| GWAS_Cat Alkyldiacyl | 5-Jan | 0.021564 | 0.073601 | 0 | 0 | 57.875 | 222.0519 |
| GWAS_Cat Diabetic M | 5-Jan | 0.021564 | 0.073601 | 0 | 0 | 57.875 | 222.0519 |
| GWAS_Cat Longevity ( | 5-Jan | 0.021564 | 0.073601 | 0 | 0 | 57.875 | 222.0519 |
| GWAS_Cat Fasting Pho | 5-Jan | 0.021564 | 0.073601 | 0 | 0 | 57.875 | 222.0519 |
| GWAS_Cat Fasting Fre | 5-Jan | 0.021564 | 0.073601 | 0 | 0 | 57.875 | 222.0519 |
| GWAS_Cat Fasting Tot | 5-Jan | 0.021564 | 0.073601 | 0 | 0 | 57.875 | 222.0519 |
| GWAS_Cat Phosphatic | 6-Jan | 0.025821 | 0.073601 | 0 | 0 | 46.29767 | 169.2909 |
| GWAS_Cat Fasting Me | 5-Jan | 0.021564 | 0.073601 | 0 | 0 | 57.875 | 222.0519 |
| GWAS_Cat Plasma Am | 5-Jan | 0.021564 | 0.073601 | 0 | 0 | 57.875 | 222.0519 |
| GWAS_Cat Postprandi | 5-Jan | 0.021564 | 0.073601 | 0 | 0 | 57.875 | 222.0519 |
| GWAS_Cat Fasting Trig | 5-Jan | 0.021564 | 0.073601 | 0 | 0 | 57.875 | 222.0519 |
| GWAS_Cat Trihexosylc | 6-Jan | 0.025821 | 0.073601 | 0 | 0 | 46.29767 | 169.2909 |
| GWAS_Cat Trihexosylc | 6-Jan | 0.025821 | 0.073601 | 0 | 0 | 46.29767 | 169.2909 |
| GWAS_Cat Protein FAI | 6-Jan | 0.025821 | 0.073601 | 0 | 0 | 46.29767 | 169.2909 |
| GWAS_Cat lysoPhosph | 5-Jan | 0.021564 | 0.073601 | 0 | 0 | 57.875 | 222.0519 |
| GWAS_Cat Alkenylpho | 6-Jan | 0.025821 | 0.073601 | 0 | 0 | 46.29767 | 169.2909 |
| GWAS_Cat Phosphatic | 6-Jan | 0.025821 | 0.073601 | 0 | 0 | 46.29767 | 169.2909 |
| GWAS_Cat Osteosarcc | 6-Jan | 0.025821 | 0.073601 | 0 | 0 | 46.29767 | 169.2909 |
| GWAS_Cat Multiple Sc | 5-Jan | 0.021564 | 0.073601 | 0 | 0 | 57.875 | 222.0519 |
| GWAS_Cat HSP 70 Lev | 6-Jan | 0.025821 | 0.073601 | 0 | 0 | 46.29767 | 169.2909 |
| GWAS_Cat Circulating | 6-Jan | 0.025821 | 0.073601 | 0 | 0 | 46.29767 | 169.2909 |
| GWAS_Cat Neutrophil | 5-Jan | 0.021564 | 0.073601 | 0 | 0 | 57.875 | 222.0519 |
| GWAS_Cat S-methylm | 6-Jan | 0.025821 | 0.073601 | 0 | 0 | 46.29767 | 169.2909 |
| GWAS_Cat Triacylglyce | 6-Jan | 0.025821 | 0.073601 | 0 | 0 | 46.29767 | 169.2909 |
| GWAS_Cat Total Lipid | Feb-57 | 0.025575 | 0.073601 | 0 | 0 | 8.495401 | 31.14526 |
| GWAS_Cat Concentrat | Feb-57 | 0.025575 | 0.073601 | 0 | 0 | 8.495401 | 31.14526 |

| | | | | | | | |
|---|---|---|---|---|---|---|---|
| GWAS_Cat Asparagine | 6-Jan | 0.025821 | 0.073601 | 0 | 0 | 46.29767 | 169.2909 |
| GWAS_Cat Psychosis P | 6-Jan | 0.025821 | 0.073601 | 0 | 0 | 46.29767 | 169.2909 |
| GWAS_Cat 1-(1-enyl-p | 6-Jan | 0.025821 | 0.073601 | 0 | 0 | 46.29767 | 169.2909 |
| GWAS_Cat N-acetylate | 6-Jan | 0.025821 | 0.073601 | 0 | 0 | 46.29767 | 169.2909 |
| GWAS_Cat Triglyceride | Feb-53 | 0.02233 | 0.073601 | 0 | 0 | 9.163552 | 34.83838 |
| GWAS_Cat Total Esteri | Feb-55 | 0.02393 | 0.073601 | 0 | 0 | 8.81687 | 32.91021 |
| GWAS_Cat Cholesteryl | Feb-55 | 0.02393 | 0.073601 | 0 | 0 | 8.81687 | 32.91021 |
| GWAS_Cat Sphingomy | Feb-55 | 0.02393 | 0.073601 | 0 | 0 | 8.81687 | 32.91021 |
| GWAS_Cat Cholesteryl | Feb-55 | 0.02393 | 0.073601 | 0 | 0 | 8.81687 | 32.91021 |
| GWAS_Cat Cholesteryl | Feb-55 | 0.02393 | 0.073601 | 0 | 0 | 8.81687 | 32.91021 |
| GWAS_Cat Central Cer | 5-Jan | 0.021564 | 0.073601 | 0 | 0 | 57.875 | 222.0519 |
| GWAS_Cat Metabolic S | 5-Jan | 0.021564 | 0.073601 | 0 | 0 | 57.875 | 222.0519 |
| GWAS_Cat Pallidum Vo | 5-Jan | 0.021564 | 0.073601 | 0 | 0 | 57.875 | 222.0519 |
| GWAS_Cat Mitral Valv | 5-Jan | 0.021564 | 0.073601 | 0 | 0 | 57.875 | 222.0519 |
| GWAS_Cat Adult Body | 5/368 | 0.022225 | 0.073601 | 0 | 0 | 3.283948 | 12.50041 |
| GWAS_Cat Free Chole: | Feb-53 | 0.02233 | 0.073601 | 0 | 0 | 9.163552 | 34.83838 |
| GWAS_Cat Triglyceride | Feb-53 | 0.02233 | 0.073601 | 0 | 0 | 9.163552 | 34.83838 |
| GWAS_Cat Alcohol Co | 3/121 | 0.01584 | 0.073601 | 0 | 0 | 5.991223 | 24.83499 |
| GWAS_Cat Chronic Ob | 3/138 | 0.022367 | 0.073601 | 0 | 0 | 5.232275 | 19.88343 |
| GWAS_Cat Cholestero | Feb-54 | 0.023124 | 0.073601 | 0 | 0 | 8.986878 | 33.8526 |
| GWAS_Cat FEV1 | 3/141 | 0.02365 | 0.073601 | 0 | 0 | 5.117754 | 19.16288 |
| GWAS_Cat Non-oily Fi: | 5-Jan | 0.021564 | 0.073601 | 0 | 0 | 57.875 | 222.0519 |
| GWAS_Cat Postprandi | 5-Jan | 0.021564 | 0.073601 | 0 | 0 | 57.875 | 222.0519 |
| GWAS_Cat Weight Los | 5-Jan | 0.021564 | 0.073601 | 0 | 0 | 57.875 | 222.0519 |
| GWAS_Cat Compleme | 5-Jan | 0.021564 | 0.073601 | 0 | 0 | 57.875 | 222.0519 |
| GWAS_Cat Sphingomy | 5-Jan | 0.021564 | 0.073601 | 0 | 0 | 57.875 | 222.0519 |
| GWAS_Cat Sphingomy | 5-Jan | 0.021564 | 0.073601 | 0 | 0 | 57.875 | 222.0519 |
| GWAS_Cat 1-palmitoy | 5-Jan | 0.021564 | 0.073601 | 0 | 0 | 57.875 | 222.0519 |
| GWAS_Cat Glucocortic | 5-Jan | 0.021564 | 0.073601 | 0 | 0 | 57.875 | 222.0519 |
| GWAS_Cat Lateral Ver | 5-Jan | 0.021564 | 0.073601 | 0 | 0 | 57.875 | 222.0519 |
| GWAS_Cat Liver Carbo | 5-Jan | 0.021564 | 0.073601 | 0 | 0 | 57.875 | 222.0519 |
| GWAS_Cat Surfeit Locu | 5-Jan | 0.021564 | 0.073601 | 0 | 0 | 57.875 | 222.0519 |
| GWAS_Cat Parental Lo | 5-Jan | 0.021564 | 0.073601 | 0 | 0 | 57.875 | 222.0519 |
| GWAS_Cat Total Lysoa | 5-Jan | 0.021564 | 0.073601 | 0 | 0 | 57.875 | 222.0519 |
| GWAS_Cat Total Lysop | 5-Jan | 0.021564 | 0.073601 | 0 | 0 | 57.875 | 222.0519 |
| GWAS_Cat Cerebrospi | 5-Jan | 0.021564 | 0.073601 | 0 | 0 | 57.875 | 222.0519 |
| GWAS_Cat Total Trihe: | 5-Jan | 0.021564 | 0.073601 | 0 | 0 | 57.875 | 222.0519 |
| GWAS_Cat Hip Circum | 9/900 | 0.016229 | 0.073601 | 0 | 0 | 2.463351 | 10.15136 |
| GWAS_Cat Waist-to-hi | 9/900 | 0.016229 | 0.073601 | 0 | 0 | 2.463351 | 10.15136 |
| GWAS_Cat Phospholip | Feb-45 | 0.016405 | 0.073601 | 0 | 0 | 10.87278 | 44.68894 |
| GWAS_Cat Lamb Liking | 5-Jan | 0.021564 | 0.073601 | 0 | 0 | 57.875 | 222.0519 |
| GWAS_Cat Physical Ac | 5-Jan | 0.021564 | 0.073601 | 0 | 0 | 57.875 | 222.0519 |
| GWAS_Cat Compleme | 5-Jan | 0.021564 | 0.073601 | 0 | 0 | 57.875 | 222.0519 |
| GWAS_Cat Pyridoxine- | 5-Jan | 0.021564 | 0.073601 | 0 | 0 | 57.875 | 222.0519 |
| GWAS_Cat Beta-defen | 5-Jan | 0.021564 | 0.073601 | 0 | 0 | 57.875 | 222.0519 |

| | | | | | | | |
|---|---|---|---|---|---|---|---|
| GWAS_Cat Lysoalkeny | 5-Jan | 0.021564 | 0.073601 | 0 | 0 | 57.875 | 222.0519 |
| GWAS_Cat 40S Riboso | 5-Jan | 0.021564 | 0.073601 | 0 | 0 | 57.875 | 222.0519 |
| GWAS_Cat Ceramide ( | 5-Jan | 0.021564 | 0.073601 | 0 | 0 | 57.875 | 222.0519 |
| GWAS_Cat Alanine Lev | Feb-44 | 0.01572 | 0.073601 | 0 | 0 | 11.13221 | 46.2302 |
| GWAS_Cat Heart Failu | Feb-44 | 0.01572 | 0.073601 | 0 | 0 | 11.13221 | 46.2302 |
| GWAS_Cat COVID-19 ( | Feb-44 | 0.01572 | 0.073601 | 0 | 0 | 11.13221 | 46.2302 |
| GWAS_Cat QRS Durati | 3/121 | 0.01584 | 0.073601 | 0 | 0 | 5.991223 | 24.83499 |
| GWAS_Cat Post Bronc | 4/258 | 0.026179 | 0.074449 | 0 | 0 | 3.730007 | 13.58763 |
| GWAS_Cat Alcohol Co | 7-Jan | 0.03006 | 0.074538 | 0 | 0 | 38.57946 | 135.2043 |
| GWAS_Cat Cathepsin I | 7-Jan | 0.03006 | 0.074538 | 0 | 0 | 38.57946 | 135.2043 |
| GWAS_Cat Electrocarc | 7-Jan | 0.03006 | 0.074538 | 0 | 0 | 38.57946 | 135.2043 |
| GWAS_Cat Verbal Lear | 7-Jan | 0.03006 | 0.074538 | 0 | 0 | 38.57946 | 135.2043 |
| GWAS_Cat Sphingomy | 7-Jan | 0.03006 | 0.074538 | 0 | 0 | 38.57946 | 135.2043 |
| GWAS_Cat Mean Arte | 4/261 | 0.027161 | 0.074538 | 0 | 0 | 3.685903 | 13.29124 |
| GWAS_Cat Liver Liking | 7-Jan | 0.03006 | 0.074538 | 0 | 0 | 38.57946 | 135.2043 |
| GWAS_Cat F-savour/ca | 7-Jan | 0.03006 | 0.074538 | 0 | 0 | 38.57946 | 135.2043 |
| GWAS_Cat Linoleoyl-a | 7-Jan | 0.03006 | 0.074538 | 0 | 0 | 38.57946 | 135.2043 |
| GWAS_Cat Right Ventr | 7-Jan | 0.03006 | 0.074538 | 0 | 0 | 38.57946 | 135.2043 |
| GWAS_Cat Fasting Blo | 7-Jan | 0.03006 | 0.074538 | 0 | 0 | 38.57946 | 135.2043 |
| GWAS_Cat Lumbar Spi | 7-Jan | 0.03006 | 0.074538 | 0 | 0 | 38.57946 | 135.2043 |
| GWAS_Cat Heart Rate | 7-Jan | 0.03006 | 0.074538 | 0 | 0 | 38.57946 | 135.2043 |
| GWAS_Cat Beta-endor | 7-Jan | 0.03006 | 0.074538 | 0 | 0 | 38.57946 | 135.2043 |
| GWAS_Cat Antineutro | 7-Jan | 0.03006 | 0.074538 | 0 | 0 | 38.57946 | 135.2043 |
| GWAS_Cat Fasting Lin | 7-Jan | 0.03006 | 0.074538 | 0 | 0 | 38.57946 | 135.2043 |
| GWAS_Cat Circulating | 7-Jan | 0.03006 | 0.074538 | 0 | 0 | 38.57946 | 135.2043 |
| GWAS_Cat Relative Pr | 7-Jan | 0.03006 | 0.074538 | 0 | 0 | 38.57946 | 135.2043 |
| GWAS_Cat 1-stearoyl- | 7-Jan | 0.03006 | 0.074538 | 0 | 0 | 38.57946 | 135.2043 |
| GWAS_Cat 1-stearoyl- | 7-Jan | 0.03006 | 0.074538 | 0 | 0 | 38.57946 | 135.2043 |
| GWAS_Cat Bone Mine | 7-Jan | 0.03006 | 0.074538 | 0 | 0 | 38.57946 | 135.2043 |
| GWAS_Cat Phosphatic | 7-Jan | 0.03006 | 0.074538 | 0 | 0 | 38.57946 | 135.2043 |
| GWAS_Cat Triglyceride | Feb-62 | 0.029883 | 0.074538 | 0 | 0 | 7.78549 | 27.33058 |
| GWAS_Cat Non-HDL C | 5/396 | 0.029271 | 0.074538 | 0 | 0 | 3.044414 | 10.75027 |
| GWAS_Cat Triglyceride | Feb-61 | 0.029 | 0.074538 | 0 | 0 | 7.917846 | 28.0328 |
| GWAS_Cat RAC-beta S | 7-Jan | 0.03006 | 0.074538 | 0 | 0 | 38.57946 | 135.2043 |
| GWAS_Cat Alkyldiacylg | 7-Jan | 0.03006 | 0.074538 | 0 | 0 | 38.57946 | 135.2043 |
| GWAS_Cat Atrial Fibril | Feb-60 | 0.028127 | 0.074538 | 0 | 0 | 8.054767 | 28.76365 |
| GWAS_Cat Ratio Of M | Feb-60 | 0.028127 | 0.074538 | 0 | 0 | 8.054767 | 28.76365 |
| GWAS_Cat Glycoprote | Feb-59 | 0.027266 | 0.074538 | 0 | 0 | 8.196491 | 29.5248 |
| GWAS_Cat Vigorous Pl | 7-Jan | 0.03006 | 0.074538 | 0 | 0 | 38.57946 | 135.2043 |
| GWAS_Cat Myocardial | 7-Jan | 0.03006 | 0.074538 | 0 | 0 | 38.57946 | 135.2043 |
| GWAS_Cat Myocardial | 7-Jan | 0.03006 | 0.074538 | 0 | 0 | 38.57946 | 135.2043 |
| GWAS_Cat Sphingomy | 7-Jan | 0.03006 | 0.074538 | 0 | 0 | 38.57946 | 135.2043 |
| GWAS_Cat Sphingomy | 7-Jan | 0.03006 | 0.074538 | 0 | 0 | 38.57946 | 135.2043 |
| GWAS_Cat F-bitter Foo | 7-Jan | 0.03006 | 0.074538 | 0 | 0 | 38.57946 | 135.2043 |
| GWAS_Cat GDH/6PGL | 7-Jan | 0.03006 | 0.074538 | 0 | 0 | 38.57946 | 135.2043 |

| | | | | | | | | |
|---|---|---|---|---|---|---|---|---|
| GWAS_Cat | Apolipoprc | 7-Jan | 0.03006 | 0.074538 | 0 | 0 | 38.57946 | 135.2043 |
| GWAS_Cat | Cortisol Le | 7-Jan | 0.03006 | 0.074538 | 0 | 0 | 38.57946 | 135.2043 |
| GWAS_Cat | Triglyceride | Feb-61 | 0.029 | 0.074538 | 0 | 0 | 7.917846 | 28.0328 |
| GWAS_Cat | Cholesteryl | Feb-61 | 0.029 | 0.074538 | 0 | 0 | 7.917846 | 28.0328 |
| GWAS_Cat | Waist Circu | Feb-60 | 0.028127 | 0.074538 | 0 | 0 | 8.054767 | 28.76365 |
| GWAS_Cat | Free Chole: | Feb-59 | 0.027266 | 0.074538 | 0 | 0 | 8.196491 | 29.5248 |
| GWAS_Cat | Average Di | Feb-59 | 0.027266 | 0.074538 | 0 | 0 | 8.196491 | 29.5248 |
| GWAS_Cat | Cholestero | Feb-59 | 0.027266 | 0.074538 | 0 | 0 | 8.196491 | 29.5248 |
| GWAS_Cat | TB-LM Or T | 7-Jan | 0.03006 | 0.074538 | 0 | 0 | 38.57946 | 135.2043 |
| GWAS_Cat | Gamma Gl | 4/260 | 0.026831 | 0.074538 | 0 | 0 | 3.700489 | 13.38904 |
| GWAS_Cat | Waist Circu | Feb-58 | 0.026415 | 0.074538 | 0 | 0 | 8.343277 | 30.31803 |
| GWAS_Cat | Left Ventri | 7-Jan | 0.03006 | 0.074538 | 0 | 0 | 38.57946 | 135.2043 |
| GWAS_Cat | 1-myristoy | 7-Jan | 0.03006 | 0.074538 | 0 | 0 | 38.57946 | 135.2043 |
| GWAS_Cat | Phospholip | Feb-58 | 0.026415 | 0.074538 | 0 | 0 | 8.343277 | 30.31803 |
| GWAS_Cat | Lipoprotei | 7-Jan | 0.03006 | 0.074538 | 0 | 0 | 38.57946 | 135.2043 |
| GWAS_Cat | Ratio Of Ap | Feb-58 | 0.026415 | 0.074538 | 0 | 0 | 8.343277 | 30.31803 |
| GWAS_Cat | Total Chole | 7-Jan | 0.03006 | 0.074538 | 0 | 0 | 38.57946 | 135.2043 |
| GWAS_Cat | Fasting Trig | 7-Jan | 0.03006 | 0.074538 | 0 | 0 | 38.57946 | 135.2043 |
| GWAS_Cat | Free Chole: | Feb-59 | 0.027266 | 0.074538 | 0 | 0 | 8.196491 | 29.5248 |
| GWAS_Cat | Linoleic Aci | Feb-59 | 0.027266 | 0.074538 | 0 | 0 | 8.196491 | 29.5248 |
| GWAS_Cat | Sphingomy | 7-Jan | 0.03006 | 0.074538 | 0 | 0 | 38.57946 | 135.2043 |
| GWAS_Cat | Number Of | 7-Jan | 0.03006 | 0.074538 | 0 | 0 | 38.57946 | 135.2043 |
| GWAS_Cat | Liver Volun | 7-Jan | 0.03006 | 0.074538 | 0 | 0 | 38.57946 | 135.2043 |
| GWAS_Cat | BMI At 2 Y | 7-Jan | 0.03006 | 0.074538 | 0 | 0 | 38.57946 | 135.2043 |
| GWAS_Cat | Alkenylpho | 7-Jan | 0.03006 | 0.074538 | 0 | 0 | 38.57946 | 135.2043 |
| GWAS_Cat | Alkenylpho | 7-Jan | 0.03006 | 0.074538 | 0 | 0 | 38.57946 | 135.2043 |
| UK_Bioban | Leisure/soc | 8-Jan | 0.03428 | 0.07458 | 0 | 0 | 33.06645 | 111.5391 |
| UK_Bioban | None of th | 8-Jan | 0.03428 | 0.07458 | 0 | 0 | 33.06645 | 111.5391 |
| DisGeNET | Long QT Sy | Mar-73 | 0.00397 | 0.075378 | 0 | 0 | 10.12398 | 55.97469 |
| DisGeNET | Attention D | Mar-73 | 0.00397 | 0.075378 | 0 | 0 | 10.12398 | 55.97469 |
| GWAS_Cat | Lipoprotei | Feb-63 | 0.030777 | 0.07555 | 0 | 0 | 7.657473 | 26.65545 |
| GWAS_Cat | Cholestero | Feb-63 | 0.030777 | 0.07555 | 0 | 0 | 7.657473 | 26.65545 |
| GWAS_Cat | Triglyceride | Feb-63 | 0.030777 | 0.07555 | 0 | 0 | 7.657473 | 26.65545 |
| GWAS_Cat | Triglyceride | Feb-63 | 0.030777 | 0.07555 | 0 | 0 | 7.657473 | 26.65545 |
| GWAS_Cat | Leisure Scr | Feb-63 | 0.030777 | 0.07555 | 0 | 0 | 7.657473 | 26.65545 |
| PheWeb_2 | Osteoarthr | Feb-30 | 0.007518 | 0.076434 | 0 | 0 | 16.71008 | 81.71977 |
| PheWeb_2 | Open wour | 25-Feb | 0.005258 | 0.076434 | 0 | 0 | 20.34783 | 106.7841 |
| PheWeb_2 | Osteoarthr | 27-Feb | 0.006118 | 0.076434 | 0 | 0 | 18.71812 | 95.39811 |
| PheWeb_2 | Dysmenorr | Feb-30 | 0.007518 | 0.076434 | 0 | 0 | 16.71008 | 81.71977 |
| PheWeb_2 | Syncope ar | Feb-30 | 0.007518 | 0.076434 | 0 | 0 | 16.71008 | 81.71977 |
| DisGeNET | Protein Mi: | 22-Feb | 0.004083 | 0.076966 | 0 | 0 | 23.40353 | 128.7392 |
| GeDiPNet_ | Meningiom | Feb-37 | 0.011286 | 0.077411 | 0 | 0 | 13.36336 | 59.92353 |
| GWAS_Cat | Free Chole: | Feb-64 | 0.031682 | 0.077459 | 0 | 0 | 7.533586 | 26.00597 |
| GWAS_Cat | Cholestero | Feb-64 | 0.031682 | 0.077459 | 0 | 0 | 7.533586 | 26.00597 |
| GWAS_Cat | Plateletcrit | 6/546 | 0.031757 | 0.077487 | 0 | 0 | 2.657476 | 9.167359 |

| | | | | | | | | |
|---|---|---|---|---|---|---|---|---|
| GWAS_Cat | IGF-1 And I | 8-Jan | 0.03428 | 0.077994 | 0 | 0 | 33.06645 | 111.5391 |
| GWAS_Cat | Phosphatic | 8-Jan | 0.03428 | 0.077994 | 0 | 0 | 33.06645 | 111.5391 |
| GWAS_Cat | Phosphatic | 8-Jan | 0.03428 | 0.077994 | 0 | 0 | 33.06645 | 111.5391 |
| GWAS_Cat | Triglyceride | Feb-67 | 0.034458 | 0.077994 | 0 | 0 | 7.184796 | 24.19848 |
| GWAS_Cat | Lamb Cons | 8-Jan | 0.03428 | 0.077994 | 0 | 0 | 33.06645 | 111.5391 |
| GWAS_Cat | Vitamin D L | Feb-67 | 0.034458 | 0.077994 | 0 | 0 | 7.184796 | 24.19848 |
| GWAS_Cat | Alcohol Co | 8-Jan | 0.03428 | 0.077994 | 0 | 0 | 33.06645 | 111.5391 |
| GWAS_Cat | Rapid Auto | 8-Jan | 0.03428 | 0.077994 | 0 | 0 | 33.06645 | 111.5391 |
| GWAS_Cat | Oral Cavity | 8-Jan | 0.03428 | 0.077994 | 0 | 0 | 33.06645 | 111.5391 |
| GWAS_Cat | Autism Spe | 8-Jan | 0.03428 | 0.077994 | 0 | 0 | 33.06645 | 111.5391 |
| GWAS_Cat | Cerebellum | 8-Jan | 0.03428 | 0.077994 | 0 | 0 | 33.06645 | 111.5391 |
| GWAS_Cat | Alzheimer's | 8-Jan | 0.03428 | 0.077994 | 0 | 0 | 33.06645 | 111.5391 |
| GWAS_Cat | Alkenylpho | 8-Jan | 0.03428 | 0.077994 | 0 | 0 | 33.06645 | 111.5391 |
| GWAS_Cat | Lactate Lev | 8-Jan | 0.03428 | 0.077994 | 0 | 0 | 33.06645 | 111.5391 |
| GWAS_Cat | Type 2 Dial | Feb-67 | 0.034458 | 0.077994 | 0 | 0 | 7.184796 | 24.19848 |
| GWAS_Cat | Alkenylpho | 8-Jan | 0.03428 | 0.077994 | 0 | 0 | 33.06645 | 111.5391 |
| GWAS_Cat | Urinary Alb | 8-Jan | 0.03428 | 0.077994 | 0 | 0 | 33.06645 | 111.5391 |
| GWAS_Cat | Phenyleph | 8-Jan | 0.03428 | 0.077994 | 0 | 0 | 33.06645 | 111.5391 |
| GWAS_Cat | BMI At 5 Ye | 8-Jan | 0.03428 | 0.077994 | 0 | 0 | 33.06645 | 111.5391 |
| GWAS_Cat | Free Chole | Feb-65 | 0.032597 | 0.077994 | 0 | 0 | 7.413632 | 25.38081 |
| GWAS_Cat | Phosphatic | 8-Jan | 0.03428 | 0.077994 | 0 | 0 | 33.06645 | 111.5391 |
| GWAS_Cat | Protein C L | 8-Jan | 0.03428 | 0.077994 | 0 | 0 | 33.06645 | 111.5391 |
| GWAS_Cat | Serine Leve | 8-Jan | 0.03428 | 0.077994 | 0 | 0 | 33.06645 | 111.5391 |
| GWAS_Cat | High-densi | 8-Jan | 0.03428 | 0.077994 | 0 | 0 | 33.06645 | 111.5391 |
| GWAS_Cat | Beer Or Cic | 8-Jan | 0.03428 | 0.077994 | 0 | 0 | 33.06645 | 111.5391 |
| GWAS_Cat | Abdominal | 8-Jan | 0.03428 | 0.077994 | 0 | 0 | 33.06645 | 111.5391 |
| GWAS_Cat | Diacylglyce | 8-Jan | 0.03428 | 0.077994 | 0 | 0 | 33.06645 | 111.5391 |
| GWAS_Cat | Triglyceride | Feb-65 | 0.032597 | 0.077994 | 0 | 0 | 7.413632 | 25.38081 |
| GWAS_Cat | Relative Ca | 8-Jan | 0.03428 | 0.077994 | 0 | 0 | 33.06645 | 111.5391 |
| GWAS_Cat | TNF-relate | 8-Jan | 0.03428 | 0.077994 | 0 | 0 | 33.06645 | 111.5391 |
| GWAS_Cat | Monounsa | Feb-66 | 0.033522 | 0.077994 | 0 | 0 | 7.297426 | 24.7787 |
| GWAS_Cat | Coronary A | Feb-66 | 0.033522 | 0.077994 | 0 | 0 | 7.297426 | 24.7787 |
| GWAS_Cat | Free Chole | Feb-65 | 0.032597 | 0.077994 | 0 | 0 | 7.413632 | 25.38081 |
| GWAS_Cat | Waist-to-hi | Feb-65 | 0.032597 | 0.077994 | 0 | 0 | 7.413632 | 25.38081 |
| GWAS_Cat | Nonalcoho | 8-Jan | 0.03428 | 0.077994 | 0 | 0 | 33.06645 | 111.5391 |
| GWAS_Cat | Monohexo | 8-Jan | 0.03428 | 0.077994 | 0 | 0 | 33.06645 | 111.5391 |
| GWAS_Cat | Osteoarthr | 8-Jan | 0.03428 | 0.077994 | 0 | 0 | 33.06645 | 111.5391 |
| GWAS_Cat | Post Bronc | 8-Jan | 0.03428 | 0.077994 | 0 | 0 | 33.06645 | 111.5391 |
| GWAS_Cat | Chitinase-3 | 8-Jan | 0.03428 | 0.077994 | 0 | 0 | 33.06645 | 111.5391 |
| DisGeNET | Uterine Fib | 6/349 | 0.004175 | 0.07813 | 0 | 0 | 4.226325 | 23.15465 |
| DisGeNET | Pulmonary | 4/151 | 0.004278 | 0.079037 | 0 | 0 | 6.480125 | 35.34375 |
| DisGeNET | Endometri | Mar-75 | 0.004284 | 0.079037 | 0 | 0 | 9.841766 | 53.66662 |
| GWAS_Cat | Hematocrit | 6/559 | 0.035019 | 0.079118 | 0 | 0 | 2.593262 | 8.692246 |
| GWAS_Cat | Bipolar Dis | Feb-68 | 0.035404 | 0.079544 | 0 | 0 | 7.075579 | 23.63906 |
| GWAS_Cat | Bitter Alcol | Feb-68 | 0.035404 | 0.079544 | 0 | 0 | 7.075579 | 23.63906 |

| | | | | | | | | |
|---|---|---|---|---|---|---|---|---|
| GWAS_Cat | Body Fat D | Feb-68 | 0.035404 | 0.079544 | 0 | 0 | 7.075579 | 23.63906 |
| PhenGenI_ | 1-Alkyl-2-a | 24-Feb | 0.004851 | 0.080047 | 0 | 0 | 21.2738 | 113.3575 |
| DisGeNET | Aneurysm | 4/152 | 0.00438 | 0.080243 | 0 | 0 | 6.436014 | 34.9526 |
| GWAS_Cat | Blood Pres: | 4/285 | 0.035811 | 0.080312 | 0 | 0 | 3.366977 | 11.21034 |
| GeDiPNet_ | Ventricular | Feb-38 | 0.01188 | 0.080535 | 0 | 0 | 12.9915 | 57.59009 |
| GWAS_Cat | Reticulocyt | 5/419 | 0.036014 | 0.080618 | 0 | 0 | 2.871892 | 9.545744 |
| GWAS_Cat | Cholesteryl | Feb-69 | 0.03636 | 0.081094 | 0 | 0 | 6.969622 | 23.09942 |
| GWAS_Cat | Problemati | Feb-69 | 0.03636 | 0.081094 | 0 | 0 | 6.969622 | 23.09942 |
| GO_Molec | Low-Densit | 22-Feb | 0.004083 | 0.081666 | 0 | 0 | 23.40353 | 128.7392 |
| GWAS_Cat | Systolic Blc | 9-Jan | 0.038483 | 0.082079 | 0 | 0 | 28.93169 | 94.24608 |
| GWAS_Cat | Hearing Fu | 9-Jan | 0.038483 | 0.082079 | 0 | 0 | 28.93169 | 94.24608 |
| GWAS_Cat | Biological A | 9-Jan | 0.038483 | 0.082079 | 0 | 0 | 28.93169 | 94.24608 |
| GWAS_Cat | Diacylglyce | 9-Jan | 0.038483 | 0.082079 | 0 | 0 | 28.93169 | 94.24608 |
| GWAS_Cat | Plasma Ant | 9-Jan | 0.038483 | 0.082079 | 0 | 0 | 28.93169 | 94.24608 |
| GWAS_Cat | Gut Microb | 9-Jan | 0.038483 | 0.082079 | 0 | 0 | 28.93169 | 94.24608 |
| GWAS_Cat | Insulin Leve | 9-Jan | 0.038483 | 0.082079 | 0 | 0 | 28.93169 | 94.24608 |
| GWAS_Cat | Hypertrigly | 9-Jan | 0.038483 | 0.082079 | 0 | 0 | 28.93169 | 94.24608 |
| GWAS_Cat | Central Cer | 9-Jan | 0.038483 | 0.082079 | 0 | 0 | 28.93169 | 94.24608 |
| GWAS_Cat | Tumor Nec | 9-Jan | 0.038483 | 0.082079 | 0 | 0 | 28.93169 | 94.24608 |
| GWAS_Cat | Caudal Ant | 9-Jan | 0.038483 | 0.082079 | 0 | 0 | 28.93169 | 94.24608 |
| GWAS_Cat | Heart Rate | 9-Jan | 0.038483 | 0.082079 | 0 | 0 | 28.93169 | 94.24608 |
| GWAS_Cat | Sphingomy | 9-Jan | 0.038483 | 0.082079 | 0 | 0 | 28.93169 | 94.24608 |
| GWAS_Cat | Sporadic N | 9-Jan | 0.038483 | 0.082079 | 0 | 0 | 28.93169 | 94.24608 |
| GWAS_Cat | Exploratory | 9-Jan | 0.038483 | 0.082079 | 0 | 0 | 28.93169 | 94.24608 |
| GWAS_Cat | Herring Lik | 9-Jan | 0.038483 | 0.082079 | 0 | 0 | 28.93169 | 94.24608 |
| GWAS_Cat | Parkinson's | 9-Jan | 0.038483 | 0.082079 | 0 | 0 | 28.93169 | 94.24608 |
| GWAS_Cat | Homeostas | 9-Jan | 0.038483 | 0.082079 | 0 | 0 | 28.93169 | 94.24608 |
| GWAS_Cat | Renal Over | 9-Jan | 0.038483 | 0.082079 | 0 | 0 | 28.93169 | 94.24608 |
| GWAS_Cat | Lewy Body | 9-Jan | 0.038483 | 0.082079 | 0 | 0 | 28.93169 | 94.24608 |
| GWAS_Cat | Fasting Insu | 9-Jan | 0.038483 | 0.082079 | 0 | 0 | 28.93169 | 94.24608 |
| GWAS_Cat | Caffeine M | 9-Jan | 0.038483 | 0.082079 | 0 | 0 | 28.93169 | 94.24608 |
| GWAS_Cat | Diacylglyce | 9-Jan | 0.038483 | 0.082079 | 0 | 0 | 28.93169 | 94.24608 |
| GWAS_Cat | Mucinous A | 9-Jan | 0.038483 | 0.082079 | 0 | 0 | 28.93169 | 94.24608 |
| GWAS_Cat | Bone Ultra: | 9-Jan | 0.038483 | 0.082079 | 0 | 0 | 28.93169 | 94.24608 |
| Gene_Pert | Pitx2 KO m | 7/243 | 9.17E-05 | 0.082216 | 0 | 0 | 7.295498 | 67.8255 |
| Gene_Pert | Lama2 Defi | 8/301 | 5.00E-05 | 0.082216 | 0 | 0 | 6.781008 | 67.15197 |
| ClinVar_20 | hypogonad | 14-Jan | 0.059226 | 0.082258 | 0 | 0 | 17.79964 | 50.30878 |
| ClinVar_20 | age-related | 13-Jan | 0.055113 | 0.082258 | 0 | 0 | 19.28391 | 55.89185 |
| ClinVar_20 | limb-girdle | 14-Jan | 0.059226 | 0.082258 | 0 | 0 | 17.79964 | 50.30878 |
| Jensen_DIS | Heart disea | Mar-94 | 0.008016 | 0.082266 | 0 | 0 | 7.779435 | 37.54635 |
| Jensen_DIS | DOID:9917 | Feb-30 | 0.007518 | 0.082266 | 0 | 0 | 16.71008 | 81.71977 |
| Jensen_DIS | Metabolic : | Feb-31 | 0.008014 | 0.082266 | 0 | 0 | 16.13306 | 77.8673 |
| Jensen_DIS | limb-girdle | Feb-30 | 0.007518 | 0.082266 | 0 | 0 | 16.71008 | 81.71977 |
| GTEx_Tissu | GTEX-R53T | 14/986 | 8.56E-05 | 0.082558 | 0 | 0 | 3.737161 | 35.00108 |
| UK_Bioban | Pork intake | 9-Jan | 0.038483 | 0.082643 | 0 | 0 | 28.93169 | 94.24608 |

| | | | | | | | | |
|---|---|---|---|---|---|---|---|---|
| UK_Bioban | Haematocr | 5/427 | 0.038567 | 0.082643 | 0 | 0 | 2.816293 | 9.168059 |
| GO_Biologi | Cholestero | 23-Feb | 0.00446 | 0.083324 | 0 | 0 | 22.28796 | 120.6379 |
| GO_Biologi | Adaptive Ir | 23-Feb | 0.00446 | 0.083324 | 0 | 0 | 22.28796 | 120.6379 |
| Rare_Disea | Gall bladde | 4/150 | 0.004179 | 0.08333 | 0 | 0 | 6.524839 | 35.74144 |
| Rare_Disea | Marek dise | Mar-74 | 0.004125 | 0.08333 | 0 | 0 | 9.980885 | 54.80158 |
| Rare_Disea | Pediatric T- | 5/244 | 0.004256 | 0.08333 | 0 | 0 | 5.01939 | 27.40285 |
| CellMarker | Stromal Ce | 19-Feb | 0.003048 | 0.083526 | 0 | 0 | 27.53772 | 159.5298 |
| GWAS_Cat | Blood Gluc | Feb-72 | 0.039286 | 0.083645 | 0 | 0 | 6.669916 | 21.58979 |
| GO_Biologi | Actomyosii | Mar-77 | 0.004612 | 0.083956 | 0 | 0 | 9.574807 | 51.50443 |
| GO_Biologi | Response T | 24-Feb | 0.004851 | 0.084011 | 0 | 0 | 21.2738 | 113.3575 |
| GO_Biologi | Acylglycerc | 24-Feb | 0.004851 | 0.084011 | 0 | 0 | 21.2738 | 113.3575 |
| GTEx_Tissu | GTEX-X62C | 12/766 | 0.000116 | 0.084031 | 0 | 0 | 4.06557 | 36.83541 |
| GeDiPNet_ | Micrognatl | 7/575 | 0.012633 | 0.084657 | 0 | 0 | 2.980084 | 13.02723 |
| GeDiPNet_ | Coronary H | 4/208 | 0.012932 | 0.084709 | 0 | 0 | 4.656036 | 20.24485 |
| GeDiPNet_ | Frontal Bos | 5/320 | 0.012919 | 0.084709 | 0 | 0 | 3.793651 | 16.49886 |
| GWAS_Cat | Apolipoprc | 5/432 | 0.040218 | 0.08548 | 0 | 0 | 2.782601 | 8.941757 |
| DisGeNET | Pulmonary | Mar-78 | 0.004781 | 0.0858 | 0 | 0 | 9.446667 | 50.47405 |
| DisGeNET | Sudden infa | Mar-78 | 0.004781 | 0.0858 | 0 | 0 | 9.446667 | 50.47405 |
| DisGeNET | Pleural Effu | Mar-78 | 0.004781 | 0.0858 | 0 | 0 | 9.446667 | 50.47405 |
| DisGeNET | Carotid art | 24-Feb | 0.004851 | 0.085882 | 0 | 0 | 21.2738 | 113.3575 |
| DisGeNET | Cognitive c | 24-Feb | 0.004851 | 0.085882 | 0 | 0 | 21.2738 | 113.3575 |
| GWAS_Cat | Gestationa | 10-Jan | 0.042667 | 0.086326 | 0 | 0 | 25.71576 | 81.1157 |
| GWAS_Cat | Granulocyt | 10-Jan | 0.042667 | 0.086326 | 0 | 0 | 25.71576 | 81.1157 |
| GWAS_Cat | Bone Mine | 10-Jan | 0.042667 | 0.086326 | 0 | 0 | 25.71576 | 81.1157 |
| GWAS_Cat | Executive F | 10-Jan | 0.042667 | 0.086326 | 0 | 0 | 25.71576 | 81.1157 |
| GWAS_Cat | BMI At 1.5 | 10-Jan | 0.042667 | 0.086326 | 0 | 0 | 25.71576 | 81.1157 |
| GWAS_Cat | Alkenylpho | 10-Jan | 0.042667 | 0.086326 | 0 | 0 | 25.71576 | 81.1157 |
| GWAS_Cat | Alkenylpho | 10-Jan | 0.042667 | 0.086326 | 0 | 0 | 25.71576 | 81.1157 |
| GWAS_Cat | Dental Dev | 10-Jan | 0.042667 | 0.086326 | 0 | 0 | 25.71576 | 81.1157 |
| GWAS_Cat | Alcohol Us | 10-Jan | 0.042667 | 0.086326 | 0 | 0 | 25.71576 | 81.1157 |
| GWAS_Cat | Body Fat P | 10-Jan | 0.042667 | 0.086326 | 0 | 0 | 25.71576 | 81.1157 |
| GWAS_Cat | Alanine Tra | 10-Jan | 0.042667 | 0.086326 | 0 | 0 | 25.71576 | 81.1157 |
| GWAS_Cat | Cortical Ar | 10-Jan | 0.042667 | 0.086326 | 0 | 0 | 25.71576 | 81.1157 |
| GWAS_Cat | Parental Li | 10-Jan | 0.042667 | 0.086326 | 0 | 0 | 25.71576 | 81.1157 |
| GWAS_Cat | Osteoprote | 10-Jan | 0.042667 | 0.086326 | 0 | 0 | 25.71576 | 81.1157 |
| GWAS_Cat | Lipoprotei | 10-Jan | 0.042667 | 0.086326 | 0 | 0 | 25.71576 | 81.1157 |
| GWAS_Cat | Factor VII L | 10-Jan | 0.042667 | 0.086326 | 0 | 0 | 25.71576 | 81.1157 |
| GWAS_Cat | Gout | Feb-74 | 0.041285 | 0.086326 | 0 | 0 | 6.483987 | 20.66613 |
| GWAS_Cat | Waist Circu | 10-Jan | 0.042667 | 0.086326 | 0 | 0 | 25.71576 | 81.1157 |
| GWAS_Cat | Pulse Press | 7/735 | 0.040934 | 0.086326 | 0 | 0 | 2.305889 | 7.369133 |
| GWAS_Cat | Acetate Le | 10-Jan | 0.042667 | 0.086326 | 0 | 0 | 25.71576 | 81.1157 |
| GWAS_Cat | Familial Co | 10-Jan | 0.042667 | 0.086326 | 0 | 0 | 25.71576 | 81.1157 |
| GWAS_Cat | P Wave Du | 10-Jan | 0.042667 | 0.086326 | 0 | 0 | 25.71576 | 81.1157 |
| GWAS_Cat | Adiponecti | 10-Jan | 0.042667 | 0.086326 | 0 | 0 | 25.71576 | 81.1157 |
| GWAS_Cat | Posterior C | 10-Jan | 0.042667 | 0.086326 | 0 | 0 | 25.71576 | 81.1157 |

| | | | | | | | | |
|---|---|---|---|---|---|---|---|---|
| GWAS_Cat | Neuropath | 10-Jan | 0.042667 | 0.086326 | 0 | 0 | 25.71576 | 81.1157 |
| GWAS_Cat | Body Fat P | 10-Jan | 0.042667 | 0.086326 | 0 | 0 | 25.71576 | 81.1157 |
| GWAS_Cat | Hospital Co | 10-Jan | 0.042667 | 0.086326 | 0 | 0 | 25.71576 | 81.1157 |
| GWAS_Cat | Physical Ac | 10-Jan | 0.042667 | 0.086326 | 0 | 0 | 25.71576 | 81.1157 |
| GWAS_Cat | Age-relatec | 10-Jan | 0.042667 | 0.086326 | 0 | 0 | 25.71576 | 81.1157 |
| DisGeNET | Glioblastor | Mar-79 | 0.004955 | 0.086493 | 0 | 0 | 9.321898 | 49.47554 |
| DisGeNET | Chronic gra | Mar-79 | 0.004955 | 0.086493 | 0 | 0 | 9.321898 | 49.47554 |
| DisGeNET | Congestive | 10/884 | 0.004985 | 0.086493 | 0 | 0 | 2.829059 | 14.99782 |
| GO_Biologi | Cardiac Cor | 25-Feb | 0.005258 | 0.086824 | 0 | 0 | 20.34783 | 106.7841 |
| GO_Biologi | Ventricular | 25-Feb | 0.005258 | 0.086824 | 0 | 0 | 20.34783 | 106.7841 |
| GWAS_Cat | Lung Funct | 6/589 | 0.043357 | 0.087576 | 0 | 0 | 2.456007 | 7.70764 |
| UK_Bioban | Oily fish int | Feb-74 | 0.041285 | 0.087808 | 0 | 0 | 6.483987 | 20.66613 |
| UK_Bioban | Self-report | 10-Jan | 0.042667 | 0.088118 | 0 | 0 | 25.71576 | 81.1157 |
| UK_Bioban | Frequency | 10-Jan | 0.042667 | 0.088118 | 0 | 0 | 25.71576 | 81.1157 |
| UK_Bioban | Number of | 10-Jan | 0.042667 | 0.088118 | 0 | 0 | 25.71576 | 81.1157 |
| UK_Bioban | Self-report | 10-Jan | 0.042667 | 0.088118 | 0 | 0 | 25.71576 | 81.1157 |
| GWAS_Cat | PR Interval | 3/180 | 0.043866 | 0.088457 | 0 | 0 | 3.982244 | 12.45094 |
| ClinVar_20 | susceptibili | 16-Jan | 0.067399 | 0.088683 | 0 | 0 | 15.42481 | 41.60265 |
| GWAS_Cat | Blood Pres | 4/305 | 0.044109 | 0.088801 | 0 | 0 | 3.140055 | 9.800381 |
| SysMyo_M | MGS1329 I | 6/277 | 0.001322 | 0.088941 | 0 | 0 | 5.368867 | 35.58941 |
| SysMyo_M | MGS864 M | 5/183 | 0.001224 | 0.088941 | 0 | 0 | 6.760414 | 45.33303 |
| SysMyo_M | MGS61 Hu | 13-Feb | 0.001414 | 0.088941 | 0 | 0 | 42.57112 | 279.3154 |
| SysMyo_M | MGS1402 I | Mar-47 | 0.001123 | 0.088941 | 0 | 0 | 16.12744 | 109.5385 |
| GWAS_Cat | Corneal Re | 3/181 | 0.04447 | 0.089379 | 0 | 0 | 3.959671 | 12.32626 |
| DisGeNET | Hydroceph | 25-Feb | 0.005258 | 0.090045 | 0 | 0 | 20.34783 | 106.7841 |
| DisGeNET | Tooth Attri | 25-Feb | 0.005258 | 0.090045 | 0 | 0 | 20.34783 | 106.7841 |
| GWAS_Cat | Type 2 Dial | 8/913 | 0.044998 | 0.090293 | 0 | 0 | 2.126918 | 6.595852 |
| DisGeNET | Acute myel | Mar-81 | 0.005312 | 0.090379 | 0 | 0 | 9.08196 | 47.56873 |
| GWAS_Cat | LDL Choles | 11-Jan | 0.046834 | 0.090551 | 0 | 0 | 23.14302 | 70.84419 |
| GWAS_Cat | Cirrhosis (A | 11-Jan | 0.046834 | 0.090551 | 0 | 0 | 23.14302 | 70.84419 |
| GWAS_Cat | Lung Funct | 11-Jan | 0.046834 | 0.090551 | 0 | 0 | 23.14302 | 70.84419 |
| GWAS_Cat | Serum Glut | 11-Jan | 0.046834 | 0.090551 | 0 | 0 | 23.14302 | 70.84419 |
| GWAS_Cat | Alcohol Cor | 11-Jan | 0.046834 | 0.090551 | 0 | 0 | 23.14302 | 70.84419 |
| GWAS_Cat | Body Fat D | Feb-78 | 0.045395 | 0.090551 | 0 | 0 | 6.141486 | 18.99159 |
| GWAS_Cat | Aspartate A | 5/447 | 0.045426 | 0.090551 | 0 | 0 | 2.6861 | 8.304515 |
| GWAS_Cat | Calcium Le | 4/308 | 0.04544 | 0.090551 | 0 | 0 | 3.108592 | 9.609793 |
| GWAS_Cat | Liver Enzyn | 3/184 | 0.046305 | 0.090551 | 0 | 0 | 3.893449 | 11.96266 |
| GWAS_Cat | Body Fat P | 11-Jan | 0.046834 | 0.090551 | 0 | 0 | 23.14302 | 70.84419 |
| GWAS_Cat | Left Atrial A | 11-Jan | 0.046834 | 0.090551 | 0 | 0 | 23.14302 | 70.84419 |
| GWAS_Cat | Right Ventr | 11-Jan | 0.046834 | 0.090551 | 0 | 0 | 23.14302 | 70.84419 |
| GWAS_Cat | Palmitoleic | 11-Jan | 0.046834 | 0.090551 | 0 | 0 | 23.14302 | 70.84419 |
| GWAS_Cat | Primary To | 11-Jan | 0.046834 | 0.090551 | 0 | 0 | 23.14302 | 70.84419 |
| GWAS_Cat | Lipoprotein | 11-Jan | 0.046834 | 0.090551 | 0 | 0 | 23.14302 | 70.84419 |
| GWAS_Cat | Pediatric B | 11-Jan | 0.046834 | 0.090551 | 0 | 0 | 23.14302 | 70.84419 |
| GWAS_Cat | Supranorm | 11-Jan | 0.046834 | 0.090551 | 0 | 0 | 23.14302 | 70.84419 |

| | | | | | | | | |
|---|---|---|---|---|---|---|---|---|
| GWAS_Cat | Medicatior | 11-Jan | 0.046834 | 0.090551 | 0 | 0 | 23.14302 | 70.84419 |
| GWAS_Cat | Moderate | 11-Jan | 0.046834 | 0.090551 | 0 | 0 | 23.14302 | 70.84419 |
| GWAS_Cat | Sphingomy | 11-Jan | 0.046834 | 0.090551 | 0 | 0 | 23.14302 | 70.84419 |
| GWAS_Cat | Facial Mor | 11-Jan | 0.046834 | 0.090551 | 0 | 0 | 23.14302 | 70.84419 |
| GWAS_Cat | Hernia | 11-Jan | 0.046834 | 0.090551 | 0 | 0 | 23.14302 | 70.84419 |
| GWAS_Cat | Dilated Car | 11-Jan | 0.046834 | 0.090551 | 0 | 0 | 23.14302 | 70.84419 |
| Rare_Disea | Charcot-M | 4/158 | 0.005021 | 0.090902 | 0 | 0 | 6.183383 | 32.73506 |
| Rare_Disea | Roussy Lev | 4/158 | 0.005021 | 0.090902 | 0 | 0 | 6.183383 | 32.73506 |
| Rare_Disea | Anthrax | Mar-80 | 0.005132 | 0.090902 | 0 | 0 | 9.200371 | 48.50752 |
| Rare_Disea | Ventricular | 4/156 | 0.004801 | 0.090902 | 0 | 0 | 6.265377 | 33.4505 |
| UK_Bioban | High light s | 27/4539 | 0.045089 | 0.091137 | 0 | 0 | 1.536004 | 4.760268 |
| UK_Bioban | Heel Broad | 4/306 | 0.04455 | 0.091137 | 0 | 0 | 3.129498 | 9.736294 |
| UK_Bioban | Dark browr | 3/182 | 0.045077 | 0.091137 | 0 | 0 | 3.93735 | 12.20334 |
| GO_Biologi | Cardiac Mu | 26-Feb | 0.005681 | 0.091663 | 0 | 0 | 19.49902 | 100.8238 |
| GWAS_Cat | Endometri | Feb-80 | 0.047505 | 0.091703 | 0 | 0 | 5.983409 | 18.23095 |
| DisGeNET | Body mass | 6/370 | 0.00553 | 0.092279 | 0 | 0 | 3.978225 | 20.67731 |
| DisGeNET | Finding of I | 6/370 | 0.00553 | 0.092279 | 0 | 0 | 3.978225 | 20.67731 |
| DisGeNET | Nephrobla | 6/370 | 0.00553 | 0.092279 | 0 | 0 | 3.978225 | 20.67731 |
| GO_Biologi | Purine Ribc | Mar-84 | 0.005878 | 0.092737 | 0 | 0 | 8.744268 | 44.91577 |
| UK_Bioban | Diabetes di | Feb-79 | 0.046446 | 0.093219 | 0 | 0 | 6.061421 | 18.60534 |
| UK_Bioban | Glaucoma ( | 11-Jan | 0.046834 | 0.09334 | 0 | 0 | 23.14302 | 70.84419 |
| GWAS_Cat | High Densi | 7/764 | 0.048647 | 0.093759 | 0 | 0 | 2.214201 | 6.693901 |
| DisGeNET | Tangier Dis | 26-Feb | 0.005681 | 0.094196 | 0 | 0 | 19.49902 | 100.8238 |
| PhenGenI_ | Breast Neo | 7/500 | 0.006123 | 0.094296 | 0 | 0 | 3.446755 | 17.56359 |
| Rare_Disea | Sudden inf | 4/162 | 0.005483 | 0.094863 | 0 | 0 | 6.025621 | 31.37037 |
| Human_Ph | Abnormalit | 18-Feb | 0.002735 | 0.094987 | 0 | 0 | 29.26029 | 172.6809 |
| GeDiPNet_ | Hypothyroi | 5/332 | 0.014935 | 0.096032 | 0 | 0 | 3.652197 | 15.35391 |
| GeDiPNet_ | Skeletal Dy | 3/119 | 0.015154 | 0.096032 | 0 | 0 | 6.095135 | 25.53542 |
| GeDiPNet_ | Ventricular | Feb-43 | 0.015047 | 0.096032 | 0 | 0 | 11.4043 | 47.85875 |
| Rare_Disea | Rod myopa | 26-Feb | 0.005681 | 0.096053 | 0 | 0 | 19.49902 | 100.8238 |
| DisGeNET | Atrial Septa | 4/165 | 0.005847 | 0.096247 | 0 | 0 | 5.912445 | 30.40115 |
| DisGeNET | Distal musc | Mar-84 | 0.005878 | 0.096247 | 0 | 0 | 8.744268 | 44.91577 |
| GWAS_Cat | Protein Z-d | 12-Jan | 0.050982 | 0.096582 | 0 | 0 | 21.03805 | 62.61499 |
| GWAS_Cat | Fasting Blo | 12-Jan | 0.050982 | 0.096582 | 0 | 0 | 21.03805 | 62.61499 |
| GWAS_Cat | Surgical Kn | 12-Jan | 0.050982 | 0.096582 | 0 | 0 | 21.03805 | 62.61499 |
| GWAS_Cat | Glaucoma ( | 12-Jan | 0.050982 | 0.096582 | 0 | 0 | 21.03805 | 62.61499 |
| GWAS_Cat | Brain Shap | 12-Jan | 0.050982 | 0.096582 | 0 | 0 | 21.03805 | 62.61499 |
| GWAS_Cat | Alzheimer' | 12-Jan | 0.050982 | 0.096582 | 0 | 0 | 21.03805 | 62.61499 |
| GWAS_Cat | BMI At 6 M | 12-Jan | 0.050982 | 0.096582 | 0 | 0 | 21.03805 | 62.61499 |
| GWAS_Cat | Abdominal | 12-Jan | 0.050982 | 0.096582 | 0 | 0 | 21.03805 | 62.61499 |
| GWAS_Cat | Response T | 12-Jan | 0.050982 | 0.096582 | 0 | 0 | 21.03805 | 62.61499 |
| GWAS_Cat | Body Mass | 12-Jan | 0.050982 | 0.096582 | 0 | 0 | 21.03805 | 62.61499 |
| GWAS_Cat | Cholecystit | 12-Jan | 0.050982 | 0.096582 | 0 | 0 | 21.03805 | 62.61499 |
| Human_Ph | Exostoses ( | 19-Feb | 0.003048 | 0.097041 | 0 | 0 | 27.53772 | 159.5298 |
| GeDiPNet_ | Bulbar Pals | Feb-44 | 0.01572 | 0.097206 | 0 | 0 | 11.13221 | 46.2302 |

| | | | | | | | | |
|---|---|---|---|---|---|---|---|---|
| GeDiPNet_ | Osteochon | 3/121 | 0.01584 | 0.097206 | 0 | 0 | 5.991223 | 24.83499 |
| GeDiPNet_ | Microphth | 4/221 | 0.015826 | 0.097206 | 0 | 0 | 4.374216 | 18.13595 |
| GeDiPNet_ | Macroceph | 5/338 | 0.016019 | 0.097281 | 0 | 0 | 3.585293 | 14.82156 |
| GO_Biologi | Negative R | 28-Feb | 0.00657 | 0.097953 | 0 | 0 | 17.99729 | 90.44125 |
| GO_Biologi | Protein Ins | 28-Feb | 0.00657 | 0.097953 | 0 | 0 | 17.99729 | 90.44125 |
| GO_Biologi | Heart Deve | 4/171 | 0.006622 | 0.097953 | 0 | 0 | 5.69829 | 28.5902 |
| UK_Bioban | Average to | Feb-82 | 0.04965 | 0.098266 | 0 | 0 | 5.833235 | 17.51576 |
| DisGeNET | Neck musc | 27-Feb | 0.006118 | 0.098334 | 0 | 0 | 18.71812 | 95.39811 |
| DisGeNET | Corneal Dis | 27-Feb | 0.006118 | 0.098334 | 0 | 0 | 18.71812 | 95.39811 |
| DisGeNET | Bulbar pals | 27-Feb | 0.006118 | 0.098334 | 0 | 0 | 18.71812 | 95.39811 |
| DisGeNET | Distal limb | Mar-86 | 0.006274 | 0.098427 | 0 | 0 | 8.532702 | 43.27254 |
| DisGeNET | Myocardial | Mar-86 | 0.006274 | 0.098427 | 0 | 0 | 8.532702 | 43.27254 |
| DisGeNET | Coronary h | 10/913 | 0.006225 | 0.098427 | 0 | 0 | 2.734032 | 13.88675 |
| DisGeNET | Herpes NO | Mar-86 | 0.006274 | 0.098427 | 0 | 0 | 8.532702 | 43.27254 |
| UK_Bioban | ICD10 acut | 12-Jan | 0.050982 | 0.098844 | 0 | 0 | 21.03805 | 62.61499 |
| UK_Bioban | Medicatior | 12-Jan | 0.050982 | 0.098844 | 0 | 0 | 21.03805 | 62.61499 |
| UK_Bioban | Takes aspir | 12-Jan | 0.050982 | 0.098844 | 0 | 0 | 21.03805 | 62.61499 |
| dbGaP | Dupuytren | 5-Jan | 0.021564 | 0.100117 | 0 | 0 | 57.875 | 222.0519 |
| Human_Ph | Arterial thr | 22-Feb | 0.004083 | 0.10021 | 0 | 0 | 23.40353 | 128.7392 |
| Human_Ph | Steppage g | 23-Feb | 0.00446 | 0.10021 | 0 | 0 | 22.28796 | 120.6379 |
| Human_Ph | Scapular w | 23-Feb | 0.00446 | 0.10021 | 0 | 0 | 22.28796 | 120.6379 |
| Human_Ph | Short palpe | 22-Feb | 0.004083 | 0.10021 | 0 | 0 | 23.40353 | 128.7392 |
| Human_Ph | Acute myel | 23-Feb | 0.00446 | 0.10021 | 0 | 0 | 22.28796 | 120.6379 |
| HumanCyc | superpathv | Feb-68 | 0.035404 | 0.100242 | 0 | 0 | 7.075579 | 23.63906 |
| HumanCyc | superpathv | 20-Jan | 0.083535 | 0.100242 | 0 | 0 | 12.17503 | 30.22444 |
| HumanCyc | D-myo-inos | 15-Jan | 0.063321 | 0.100242 | 0 | 0 | 16.52741 | 45.60793 |
| HumanCyc | D-myo-inos | 19-Jan | 0.079527 | 0.100242 | 0 | 0 | 12.85207 | 32.53709 |
| Rare_Disea | Atrial fibrill | Mar-85 | 0.006074 | 0.10042 | 0 | 0 | 8.637195 | 44.08228 |
| DisGeNET | Cytomegal | 5/270 | 0.006495 | 0.10066 | 0 | 0 | 4.520939 | 22.77038 |
| DisGeNET | Granulocyt | 28-Feb | 0.00657 | 0.10066 | 0 | 0 | 17.99729 | 90.44125 |
| DisGeNET | Multiple tu | Mar-87 | 0.006478 | 0.10066 | 0 | 0 | 8.430697 | 42.48559 |
| DisGeNET | Tumor Exp | 28-Feb | 0.00657 | 0.10066 | 0 | 0 | 17.99729 | 90.44125 |
| Rare_Disea | Myocarditi | 4/168 | 0.006226 | 0.100705 | 0 | 0 | 5.803409 | 29.4753 |
| GWAS_Cat | Total Body | 13-Jan | 0.055113 | 0.10111 | 0 | 0 | 19.28391 | 55.89185 |
| GWAS_Cat | Sphingomy | 13-Jan | 0.055113 | 0.10111 | 0 | 0 | 19.28391 | 55.89185 |
| GWAS_Cat | Ischemic St | 13-Jan | 0.055113 | 0.10111 | 0 | 0 | 19.28391 | 55.89185 |
| GWAS_Cat | Dementia \ | 13-Jan | 0.055113 | 0.10111 | 0 | 0 | 19.28391 | 55.89185 |
| GWAS_Cat | Fasting Glu | 13-Jan | 0.055113 | 0.10111 | 0 | 0 | 19.28391 | 55.89185 |
| GWAS_Cat | Alzheimer's | 13-Jan | 0.055113 | 0.10111 | 0 | 0 | 19.28391 | 55.89185 |
| GWAS_Cat | Percent Liv | 13-Jan | 0.055113 | 0.10111 | 0 | 0 | 19.28391 | 55.89185 |
| GWAS_Cat | Weight Gai | 13-Jan | 0.055113 | 0.10111 | 0 | 0 | 19.28391 | 55.89185 |
| GWAS_Cat | Coffee Inta | 13-Jan | 0.055113 | 0.10111 | 0 | 0 | 19.28391 | 55.89185 |
| GWAS_Cat | Male-patte | 4/328 | 0.054882 | 0.10111 | 0 | 0 | 2.913729 | 8.457304 |
| GWAS_Cat | C-reactive | 13-Jan | 0.055113 | 0.10111 | 0 | 0 | 19.28391 | 55.89185 |
| GWAS_Cat | Total Body | 13-Jan | 0.055113 | 0.10111 | 0 | 0 | 19.28391 | 55.89185 |

| | | | | | | | | |
|---|---|---|---|---|---|---|---|---|
| GWAS_Cat | Sense Of Sı | 13-Jan | 0.055113 | 0.10111 | 0 | 0 | 19.28391 | 55.89185 |
| GWAS_Cat | Blood Meta | 13-Jan | 0.055113 | 0.10111 | 0 | 0 | 19.28391 | 55.89185 |
| GWAS_Cat | Phosphatic | 13-Jan | 0.055113 | 0.10111 | 0 | 0 | 19.28391 | 55.89185 |
| GWAS_Cat | Platelet Co | 8/949 | 0.054198 | 0.10111 | 0 | 0 | 2.041674 | 5.951689 |
| GWAS_Cat | Isoleucine I | 13-Jan | 0.055113 | 0.10111 | 0 | 0 | 19.28391 | 55.89185 |
| GWAS_Cat | Plasma Om | 13-Jan | 0.055113 | 0.10111 | 0 | 0 | 19.28391 | 55.89185 |
| GWAS_Cat | Cerebrospi | 13-Jan | 0.055113 | 0.10111 | 0 | 0 | 19.28391 | 55.89185 |
| GWAS_Cat | Neuroblast | 13-Jan | 0.055113 | 0.10111 | 0 | 0 | 19.28391 | 55.89185 |
| GWAS_Cat | Benign Pro | 13-Jan | 0.055113 | 0.10111 | 0 | 0 | 19.28391 | 55.89185 |
| Rare_Disea | Emery-Dre | 28-Feb | 0.00657 | 0.101832 | 0 | 0 | 17.99729 | 90.44125 |
| Rare_Disea | Acute myel | 6/382 | 0.006435 | 0.101832 | 0 | 0 | 3.848897 | 19.42128 |
| GO_Molecı | Lipoproteir | 27-Feb | 0.006118 | 0.101962 | 0 | 0 | 18.71812 | 95.39811 |
| HuBMAP_/ | PI16+ Fibrc | 4/103 | 0.001064 | 0.102149 | 0 | 0 | 9.645369 | 66.029 |
| HuBMAP_/ | Detrusor Sı | 4/102 | 0.001026 | 0.102149 | 0 | 0 | 9.744283 | 67.06001 |
| DisGeNET | High Grade | Feb-29 | 0.007037 | 0.103573 | 0 | 0 | 17.32985 | 85.89758 |
| DisGeNET | Congenital | Feb-29 | 0.007037 | 0.103573 | 0 | 0 | 17.32985 | 85.89758 |
| DisGeNET | Aspartate a | Feb-29 | 0.007037 | 0.103573 | 0 | 0 | 17.32985 | 85.89758 |
| DisGeNET | Exostoses | Feb-29 | 0.007037 | 0.103573 | 0 | 0 | 17.32985 | 85.89758 |
| DisGeNET | Bulbar wea | Feb-29 | 0.007037 | 0.103573 | 0 | 0 | 17.32985 | 85.89758 |
| DisGeNET | Keratosis F | Feb-29 | 0.007037 | 0.103573 | 0 | 0 | 17.32985 | 85.89758 |
| DisGeNET | Acute mon | Feb-29 | 0.007037 | 0.103573 | 0 | 0 | 17.32985 | 85.89758 |
| UK_Bioban | Impedance | 6/623 | 0.054211 | 0.104393 | 0 | 0 | 2.316586 | 6.752548 |
| GTEx_Tissu | Heart - Left | Mar-66 | 0.002986 | 0.10508 | 0 | 0 | 11.25283 | 65.42369 |
| GTEx_Tissu | Heart - Left | 3/100 | 0.009491 | 0.10508 | 0 | 0 | 7.296024 | 33.98049 |
| GTEx_Tissu | Heart - Left | 3/100 | 0.009491 | 0.10508 | 0 | 0 | 7.296024 | 33.98049 |
| GTEx_Tissu | Heart - Left | 3/100 | 0.009491 | 0.10508 | 0 | 0 | 7.296024 | 33.98049 |
| GTEx_Tissu | Heart - Left | 3/100 | 0.009491 | 0.10508 | 0 | 0 | 7.296024 | 33.98049 |
| GTEx_Tissu | Heart - Left | 3/100 | 0.009491 | 0.10508 | 0 | 0 | 7.296024 | 33.98049 |
| GTEx_Tissu | Liver Fema | 3/100 | 0.009491 | 0.10508 | 0 | 0 | 7.296024 | 33.98049 |
| GTEx_Tissu | Colon - Sigı | 3/100 | 0.009491 | 0.10508 | 0 | 0 | 7.296024 | 33.98049 |
| GTEx_Tissu | Heart - Left | 3/100 | 0.009491 | 0.10508 | 0 | 0 | 7.296024 | 33.98049 |
| GTEx_Tissu | Liver Male | 3/100 | 0.009491 | 0.10508 | 0 | 0 | 7.296024 | 33.98049 |
| GTEx_Tissu | Brain - Hyp | 3/100 | 0.009491 | 0.10508 | 0 | 0 | 7.296024 | 33.98049 |
| GTEx_Tissu | Heart - Left | 3/100 | 0.009491 | 0.10508 | 0 | 0 | 7.296024 | 33.98049 |
| GTEx_Tissu | Heart - Left | 3/100 | 0.009491 | 0.10508 | 0 | 0 | 7.296024 | 33.98049 |
| GTEx_Tissu | Heart - Left | 3/100 | 0.009491 | 0.10508 | 0 | 0 | 7.296024 | 33.98049 |
| UK_Bioban | Medicatior | 13-Jan | 0.055113 | 0.105418 | 0 | 0 | 19.28391 | 55.89185 |
| GWAS_Cat | Mammogra | 14-Jan | 0.059226 | 0.105483 | 0 | 0 | 17.79964 | 50.30878 |
| GWAS_Cat | F-small Fisl | 14-Jan | 0.059226 | 0.105483 | 0 | 0 | 17.79964 | 50.30878 |
| GWAS_Cat | F-strong Ve | 14-Jan | 0.059226 | 0.105483 | 0 | 0 | 17.79964 | 50.30878 |
| GWAS_Cat | Lactate Del | 14-Jan | 0.059226 | 0.105483 | 0 | 0 | 17.79964 | 50.30878 |
| GWAS_Cat | Benign Pro | 14-Jan | 0.059226 | 0.105483 | 0 | 0 | 17.79964 | 50.30878 |
| GWAS_Cat | Cerebrospi | 14-Jan | 0.059226 | 0.105483 | 0 | 0 | 17.79964 | 50.30878 |
| GWAS_Cat | Renal Unde | 14-Jan | 0.059226 | 0.105483 | 0 | 0 | 17.79964 | 50.30878 |
| GWAS_Cat | Plasma Ant | 14-Jan | 0.059226 | 0.105483 | 0 | 0 | 17.79964 | 50.30878 |

| | | | | | | | | |
|---|---|---|---|---|---|---|---|---|
| GWAS_Cat | Blood Suga | 14-Jan | 0.059226 | 0.105483 | 0 | 0 | 17.79964 | 50.30878 |
| GWAS_Cat | BMI At 1 Ye | 14-Jan | 0.059226 | 0.105483 | 0 | 0 | 17.79964 | 50.30878 |
| GWAS_Cat | Waist Circu | 8/966 | 0.058938 | 0.105483 | 0 | 0 | 2.003647 | 5.672872 |
| GWAS_Cat | Waist-to-hi | 14-Jan | 0.059226 | 0.105483 | 0 | 0 | 17.79964 | 50.30878 |
| GWAS_Cat | C-reactive I | 14-Jan | 0.059226 | 0.105483 | 0 | 0 | 17.79964 | 50.30878 |
| GWAS_Cat | C-reactive I | 14-Jan | 0.059226 | 0.105483 | 0 | 0 | 17.79964 | 50.30878 |
| GWAS_Cat | Prostate-sp | 14-Jan | 0.059226 | 0.105483 | 0 | 0 | 17.79964 | 50.30878 |
| GWAS_Cat | Posterior T | 14-Jan | 0.059226 | 0.105483 | 0 | 0 | 17.79964 | 50.30878 |
| GWAS_Cat | Response T | 14-Jan | 0.059226 | 0.105483 | 0 | 0 | 17.79964 | 50.30878 |
| GWAS_Cat | Ultradistal | 14-Jan | 0.059226 | 0.105483 | 0 | 0 | 17.79964 | 50.30878 |
| GWAS_Cat | Macular Te | 14-Jan | 0.059226 | 0.105483 | 0 | 0 | 17.79964 | 50.30878 |
| GWAS_Cat | BMI At 8 M | 14-Jan | 0.059226 | 0.105483 | 0 | 0 | 17.79964 | 50.30878 |
| PheWeb_2 | Chronic air | Feb-37 | 0.011286 | 0.105918 | 0 | 0 | 13.36336 | 59.92353 |
| GeDiPNet_ | Microcorne | 3/126 | 0.017629 | 0.105958 | 0 | 0 | 5.746225 | 23.20435 |
| GeDiPNet_ | Cognitive D | Feb-47 | 0.017813 | 0.105969 | 0 | 0 | 10.3885 | 41.84306 |
| GeDiPNet_ | Glomerulo | 3/127 | 0.018 | 0.106002 | 0 | 0 | 5.699597 | 22.89738 |
| UK_Bioban | Left cornea | 4/331 | 0.056384 | 0.106419 | 0 | 0 | 2.886555 | 8.300512 |
| UK_Bioban | Self-report | Feb-88 | 0.05629 | 0.106419 | 0 | 0 | 5.424624 | 15.60794 |
| DisGeNET | Optic Atrop | Feb-30 | 0.007518 | 0.106697 | 0 | 0 | 16.71008 | 81.71977 |
| DisGeNET | Decreased | 6/395 | 0.007534 | 0.106697 | 0 | 0 | 3.717795 | 18.17384 |
| DisGeNET | Decreased | 6/395 | 0.007534 | 0.106697 | 0 | 0 | 3.717795 | 18.17384 |
| DisGeNET | Ventricular | Feb-30 | 0.007518 | 0.106697 | 0 | 0 | 16.71008 | 81.71977 |
| DisGeNET | Retrusion c | 6/395 | 0.007534 | 0.106697 | 0 | 0 | 3.717795 | 18.17384 |
| DisGeNET | Hypoplasti | 6/395 | 0.007534 | 0.106697 | 0 | 0 | 3.717795 | 18.17384 |
| DisGeNET | X-Linked Er | Feb-30 | 0.007518 | 0.106697 | 0 | 0 | 16.71008 | 81.71977 |
| Rare_Disea | Dihydropte | Feb-29 | 0.007037 | 0.106842 | 0 | 0 | 17.32985 | 85.89758 |
| Rare_Disea | Edwards sy | Mar-91 | 0.007333 | 0.10697 | 0 | 0 | 8.04586 | 39.5488 |
| Rare_Disea | Stress card | 4/176 | 0.007318 | 0.10697 | 0 | 0 | 5.531241 | 27.19912 |
| Disease_Pe | melanoma | 7/337 | 0.000668 | 0.107209 | 0 | 0 | 5.192462 | 37.96347 |
| Disease_Pe | Senescence | 6/236 | 0.000577 | 0.107209 | 0 | 0 | 6.33913 | 47.27858 |
| GO_Biologi | Phospholip | Feb-30 | 0.007518 | 0.108061 | 0 | 0 | 16.71008 | 81.71977 |
| GO_Biologi | Negative R | 4/178 | 0.00761 | 0.108061 | 0 | 0 | 5.46711 | 26.67023 |
| DisGeNET | Aplasia/Hy | 6/397 | 0.007714 | 0.108663 | 0 | 0 | 3.698399 | 17.99155 |
| Jensen_TIS | Longissimu | 9-Feb | 0.00066 | 0.10874 | 0 | 0 | 66.91092 | 489.9901 |
| Jensen_TIS | Fiber | 6/263 | 0.001013 | 0.10874 | 0 | 0 | 5.66537 | 39.06122 |
| Jensen_TIS | Whole plar | Mar-44 | 0.000926 | 0.10874 | 0 | 0 | 17.3101 | 120.912 |
| GWAS_Cat | Height | 34/6159 | 0.061348 | 0.109103 | 0 | 0 | 1.444103 | 4.03077 |
| GeDiPNet_ | CACH Synd | 5-Jan | 0.021564 | 0.109318 | 0 | 0 | 57.875 | 222.0519 |
| GeDiPNet_ | Megakaryo | 5-Jan | 0.021564 | 0.109318 | 0 | 0 | 57.875 | 222.0519 |
| GeDiPNet_ | Sea-Blue H | 5-Jan | 0.021564 | 0.109318 | 0 | 0 | 57.875 | 222.0519 |
| GeDiPNet_ | Hearing Lo: | 8/775 | 0.019476 | 0.109318 | 0 | 0 | 2.527817 | 9.956017 |
| GeDiPNet_ | Oral Cavity | 5-Jan | 0.021564 | 0.109318 | 0 | 0 | 57.875 | 222.0519 |
| GeDiPNet_ | Hemangiob | 5-Jan | 0.021564 | 0.109318 | 0 | 0 | 57.875 | 222.0519 |
| GeDiPNet_ | Common C | 5-Jan | 0.021564 | 0.109318 | 0 | 0 | 57.875 | 222.0519 |
| GeDiPNet_ | Cree Leuko | 5-Jan | 0.021564 | 0.109318 | 0 | 0 | 57.875 | 222.0519 |

| | | | | | | | | |
|---|---|---|---|---|---|---|---|---|
| GeDiPNet_ | Internal Ca | 5-Jan | 0.021564 | 0.109318 | 0 | 0 | 57.875 | 222.0519 |
| GeDiPNet_ | Complete / | 5-Jan | 0.021564 | 0.109318 | 0 | 0 | 57.875 | 222.0519 |
| GeDiPNet_ | Angioma | 5-Jan | 0.021564 | 0.109318 | 0 | 0 | 57.875 | 222.0519 |
| GeDiPNet_ | Patchy Palr | 5-Jan | 0.021564 | 0.109318 | 0 | 0 | 57.875 | 222.0519 |
| GeDiPNet_ | Erythroker | 5-Jan | 0.021564 | 0.109318 | 0 | 0 | 57.875 | 222.0519 |
| GeDiPNet_ | Endometri | 5-Jan | 0.021564 | 0.109318 | 0 | 0 | 57.875 | 222.0519 |
| GeDiPNet_ | External Ca | 5-Jan | 0.021564 | 0.109318 | 0 | 0 | 57.875 | 222.0519 |
| GeDiPNet_ | Fibroather | 5-Jan | 0.021564 | 0.109318 | 0 | 0 | 57.875 | 222.0519 |
| UK_Bioban | Myocardial | 14-Jan | 0.059226 | 0.109607 | 0 | 0 | 17.79964 | 50.30878 |
| UK_Bioban | Fractured/ | 14-Jan | 0.059226 | 0.109607 | 0 | 0 | 17.79964 | 50.30878 |
| UK_Bioban | Myocardial | 14-Jan | 0.059226 | 0.109607 | 0 | 0 | 17.79964 | 50.30878 |
| GeDiPNet_ | Congenital | 4/244 | 0.021883 | 0.109982 | 0 | 0 | 3.950402 | 15.09859 |
| DisGeNET | Progressive | Feb-31 | 0.008014 | 0.110509 | 0 | 0 | 16.13306 | 77.8673 |
| DisGeNET | Micrognatl | 6/400 | 0.007991 | 0.110509 | 0 | 0 | 3.669675 | 17.72267 |
| DisGeNET | Longevity | Feb-31 | 0.008014 | 0.110509 | 0 | 0 | 16.13306 | 77.8673 |
| DisGeNET | Lipomatosi | Feb-31 | 0.008014 | 0.110509 | 0 | 0 | 16.13306 | 77.8673 |
| GWAS_Cat | Male Pube | 15-Jan | 0.063321 | 0.110518 | 0 | 0 | 16.52741 | 45.60793 |
| GWAS_Cat | Decaffeina | 15-Jan | 0.063321 | 0.110518 | 0 | 0 | 16.52741 | 45.60793 |
| GWAS_Cat | Low High D | 15-Jan | 0.063321 | 0.110518 | 0 | 0 | 16.52741 | 45.60793 |
| GWAS_Cat | Cardiovasc | 15-Jan | 0.063321 | 0.110518 | 0 | 0 | 16.52741 | 45.60793 |
| GWAS_Cat | F-sharp Fla | 15-Jan | 0.063321 | 0.110518 | 0 | 0 | 16.52741 | 45.60793 |
| GWAS_Cat | Parental Lc | 15-Jan | 0.063321 | 0.110518 | 0 | 0 | 16.52741 | 45.60793 |
| GWAS_Cat | Hernia (An | 15-Jan | 0.063321 | 0.110518 | 0 | 0 | 16.52741 | 45.60793 |
| GWAS_Cat | Cigarettes | Feb-94 | 0.063221 | 0.110518 | 0 | 0 | 5.069309 | 13.99697 |
| GWAS_Cat | Response T | 15-Jan | 0.063321 | 0.110518 | 0 | 0 | 16.52741 | 45.60793 |
| GWAS_Cat | Parkinson's | 15-Jan | 0.063321 | 0.110518 | 0 | 0 | 16.52741 | 45.60793 |
| GWAS_Cat | Psoriasis O | 15-Jan | 0.063321 | 0.110518 | 0 | 0 | 16.52741 | 45.60793 |
| GWAS_Cat | Glaucoma ( | 15-Jan | 0.063321 | 0.110518 | 0 | 0 | 16.52741 | 45.60793 |
| GWAS_Cat | Glucagon L | 15-Jan | 0.063321 | 0.110518 | 0 | 0 | 16.52741 | 45.60793 |
| Jensen_TIS | Peripheral | 22-Feb | 0.004083 | 0.110692 | 0 | 0 | 23.40353 | 128.7392 |
| Jensen_TIS | Heart | 73/14216 | 0.00408 | 0.110692 | 0 | 0 | 2.127302 | 11.70372 |
| Jensen_TIS | Spindle cel | Mar-66 | 0.002986 | 0.110692 | 0 | 0 | 11.25283 | 65.42369 |
| Jensen_TIS | U-937 cell | Mar-74 | 0.004125 | 0.110692 | 0 | 0 | 9.980885 | 54.80158 |
| Jensen_TIS | Fascicle | 21-Feb | 0.003723 | 0.110692 | 0 | 0 | 24.63653 | 137.8005 |
| Jensen_TIS | Hepatoblas | 20-Feb | 0.003378 | 0.110692 | 0 | 0 | 26.00654 | 147.9928 |
| Jensen_TIS | Stomach | 81/16255 | 0.001546 | 0.110692 | 0 | 0 | 3.120842 | 20.19919 |
| Jensen_TIS | Interstitial | 17-Feb | 0.002438 | 0.110692 | 0 | 0 | 31.21255 | 187.7904 |
| Jensen_TIS | Bile | 18-Feb | 0.002735 | 0.110692 | 0 | 0 | 29.26029 | 172.6809 |
| Jensen_DIS | Oropharyn | 6-Jan | 0.025821 | 0.111412 | 0 | 0 | 46.29767 | 169.2909 |
| Jensen_DIS | biotin-resp | 6-Jan | 0.025821 | 0.111412 | 0 | 0 | 46.29767 | 169.2909 |
| Jensen_DIS | Intestinal a | 6-Jan | 0.025821 | 0.111412 | 0 | 0 | 46.29767 | 169.2909 |
| Jensen_DIS | Lymphangi | 6-Jan | 0.025821 | 0.111412 | 0 | 0 | 46.29767 | 169.2909 |
| Jensen_DIS | Motor peri | 7-Jan | 0.03006 | 0.111412 | 0 | 0 | 38.57946 | 135.2043 |
| Jensen_DIS | Erythroker | 7-Jan | 0.03006 | 0.111412 | 0 | 0 | 38.57946 | 135.2043 |
| Jensen_DIS | Distal arthr | Feb-45 | 0.016405 | 0.111412 | 0 | 0 | 10.87278 | 44.68894 |

| | | | | | | | | |
|---|---|---|---|---|---|---|---|---|
| Jensen_DIS | Atheroscle | Feb-52 | 0.021547 | 0.111412 | 0 | 0 | 9.347294 | 35.8704 |
| Jensen_DIS | Head and r | 6-Jan | 0.025821 | 0.111412 | 0 | 0 | 46.29767 | 169.2909 |
| Jensen_DIS | extra-adrer | 7-Jan | 0.03006 | 0.111412 | 0 | 0 | 38.57946 | 135.2043 |
| Jensen_DIS | Large cell c | 5-Jan | 0.021564 | 0.111412 | 0 | 0 | 57.875 | 222.0519 |
| Jensen_DIS | Avian influ | 7-Jan | 0.03006 | 0.111412 | 0 | 0 | 38.57946 | 135.2043 |
| Jensen_DIS | Myelodysp | 8-Jan | 0.03428 | 0.111412 | 0 | 0 | 33.06645 | 111.5391 |
| Jensen_DIS | Abdominal | 8-Jan | 0.03428 | 0.111412 | 0 | 0 | 33.06645 | 111.5391 |
| Jensen_DIS | Pneumonia | Feb-64 | 0.031682 | 0.111412 | 0 | 0 | 7.533586 | 26.00597 |
| Jensen_DIS | Leishmania | Feb-40 | 0.013107 | 0.111412 | 0 | 0 | 12.3065 | 53.34346 |
| Jensen_DIS | Necrotizing | 8-Jan | 0.03428 | 0.111412 | 0 | 0 | 33.06645 | 111.5391 |
| Jensen_DIS | Skin diseas | Feb-41 | 0.013741 | 0.111412 | 0 | 0 | 11.99035 | 51.40702 |
| Jensen_DIS | Large intes | 3/122 | 0.016189 | 0.111412 | 0 | 0 | 5.940576 | 24.49549 |
| Jensen_DIS | Bannayan-I | 7-Jan | 0.03006 | 0.111412 | 0 | 0 | 38.57946 | 135.2043 |
| Jensen_DIS | Ovarian ser | 8-Jan | 0.03428 | 0.111412 | 0 | 0 | 33.06645 | 111.5391 |
| Jensen_DIS | Skin cancer | 6/454 | 0.01427 | 0.111412 | 0 | 0 | 3.218419 | 13.67709 |
| Jensen_DIS | Breast canc | 6/434 | 0.011641 | 0.111412 | 0 | 0 | 3.372274 | 15.01756 |
| Jensen_DIS | Juvenile pc | 5-Jan | 0.021564 | 0.111412 | 0 | 0 | 57.875 | 222.0519 |
| Jensen_DIS | Proteus syr | 5-Jan | 0.021564 | 0.111412 | 0 | 0 | 57.875 | 222.0519 |
| Jensen_DIS | Osteosarcc | 5-Jan | 0.021564 | 0.111412 | 0 | 0 | 57.875 | 222.0519 |
| Jensen_DIS | Atrial heart | 7-Jan | 0.03006 | 0.111412 | 0 | 0 | 38.57946 | 135.2043 |
| Jensen_DIS | Swine influ | 7-Jan | 0.03006 | 0.111412 | 0 | 0 | 38.57946 | 135.2043 |
| Jensen_DIS | Osteopoiki | 5-Jan | 0.021564 | 0.111412 | 0 | 0 | 57.875 | 222.0519 |
| Jensen_DIS | Food allerg | 5-Jan | 0.021564 | 0.111412 | 0 | 0 | 57.875 | 222.0519 |
| Jensen_DIS | Cognitive d | Feb-52 | 0.021547 | 0.111412 | 0 | 0 | 9.347294 | 35.8704 |
| Jensen_DIS | Biotin defic | 8-Jan | 0.03428 | 0.111412 | 0 | 0 | 33.06645 | 111.5391 |
| Jensen_DIS | Pulmonary | 7-Jan | 0.03006 | 0.111412 | 0 | 0 | 38.57946 | 135.2043 |
| Jensen_DIS | Lung cance | 5/400 | 0.030381 | 0.111412 | 0 | 0 | 3.012967 | 10.52712 |
| Jensen_DIS | Type 1 diat | 3/158 | 0.031658 | 0.111412 | 0 | 0 | 4.552535 | 15.71884 |
| Jensen_DIS | Lung large | 5-Jan | 0.021564 | 0.111412 | 0 | 0 | 57.875 | 222.0519 |
| Jensen_DIS | Syndactyly | Feb-53 | 0.02233 | 0.111412 | 0 | 0 | 9.163552 | 34.83838 |
| Jensen_DIS | Melanoma | 6/505 | 0.022797 | 0.111412 | 0 | 0 | 2.881912 | 10.89684 |
| Jensen_DIS | Crohn's dis | 4/248 | 0.023063 | 0.111412 | 0 | 0 | 3.884851 | 14.64409 |
| Jensen_DIS | Myopathy | 6-Jan | 0.025821 | 0.111412 | 0 | 0 | 46.29767 | 169.2909 |
| Jensen_DIS | Optic atrop | Feb-40 | 0.013107 | 0.111412 | 0 | 0 | 12.3065 | 53.34346 |
| GO_Biologi | Triglyceride | Feb-31 | 0.008014 | 0.111567 | 0 | 0 | 16.13306 | 77.8673 |
| GO_Biologi | Amide Trar | Feb-32 | 0.008524 | 0.112078 | 0 | 0 | 15.59451 | 74.30533 |
| GO_Biologi | Muscle Cel | Feb-32 | 0.008524 | 0.112078 | 0 | 0 | 15.59451 | 74.30533 |
| GO_Biologi | Chromosor | Feb-32 | 0.008524 | 0.112078 | 0 | 0 | 15.59451 | 74.30533 |
| Jensen_DIS | Scarlet feve | 9-Jan | 0.038483 | 0.1137 | 0 | 0 | 28.93169 | 94.24608 |
| Jensen_DIS | Mood disor | 9-Jan | 0.038483 | 0.1137 | 0 | 0 | 28.93169 | 94.24608 |
| Jensen_DIS | Alpha 1-an | 9-Jan | 0.038483 | 0.1137 | 0 | 0 | 28.93169 | 94.24608 |
| Jensen_DIS | IgA glomer | 9-Jan | 0.038483 | 0.1137 | 0 | 0 | 28.93169 | 94.24608 |
| Jensen_DIS | Biotinidase | 9-Jan | 0.038483 | 0.1137 | 0 | 0 | 28.93169 | 94.24608 |
| Jensen_DIS | Clouston sy | 9-Jan | 0.038483 | 0.1137 | 0 | 0 | 28.93169 | 94.24608 |
| Jensen_DIS | protein-los | 10-Jan | 0.042667 | 0.114158 | 0 | 0 | 25.71576 | 81.1157 |

| | | | | | | | | |
|---|---|---|---|---|---|---|---|---|
| Jensen_DIS | Hemophilia | 11-Jan | 0.046834 | 0.114158 | 0 | 0 | 23.14302 | 70.84419 |
| Jensen_DIS | Histiocytos | 11-Jan | 0.046834 | 0.114158 | 0 | 0 | 23.14302 | 70.84419 |
| Jensen_DIS | Lipomatosi | 11-Jan | 0.046834 | 0.114158 | 0 | 0 | 23.14302 | 70.84419 |
| Jensen_DIS | Thymoma | 10-Jan | 0.042667 | 0.114158 | 0 | 0 | 25.71576 | 81.1157 |
| Jensen_DIS | Cancer | 4/300 | 0.041941 | 0.114158 | 0 | 0 | 3.193911 | 10.12943 |
| Jensen_DIS | Hydranenc | 10-Jan | 0.042667 | 0.114158 | 0 | 0 | 25.71576 | 81.1157 |
| Jensen_DIS | Liver cance | 6/597 | 0.045775 | 0.114158 | 0 | 0 | 2.421758 | 7.468727 |
| Jensen_DIS | Brain disea | Feb-77 | 0.044354 | 0.114158 | 0 | 0 | 6.223686 | 19.39022 |
| Jensen_DIS | Acute myel | 11-Jan | 0.046834 | 0.114158 | 0 | 0 | 23.14302 | 70.84419 |
| Jensen_DIS | Myoepithe | 11-Jan | 0.046834 | 0.114158 | 0 | 0 | 23.14302 | 70.84419 |
| Jensen_DIS | Dental ena | 10-Jan | 0.042667 | 0.114158 | 0 | 0 | 25.71576 | 81.1157 |
| Jensen_DIS | Lysosomal | 10-Jan | 0.042667 | 0.114158 | 0 | 0 | 25.71576 | 81.1157 |
| Jensen_DIS | Capillary he | 11-Jan | 0.046834 | 0.114158 | 0 | 0 | 23.14302 | 70.84419 |
| GWAS_Cat | Longevity | Feb-96 | 0.065593 | 0.114319 | 0 | 0 | 4.960951 | 13.51507 |
| Jensen_TIS | Adipose tis | 65/12174 | 0.004625 | 0.11457 | 0 | 0 | 1.904143 | 10.237 |
| GWAS_Cat | Brain Regic | 3/213 | 0.065919 | 0.114724 | 0 | 0 | 3.35085 | 9.112059 |
| GO_Biologi | Ventricular | Feb-33 | 0.009049 | 0.114725 | 0 | 0 | 15.0907 | 71.00369 |
| GO_Biologi | Negative R | Feb-33 | 0.009049 | 0.114725 | 0 | 0 | 15.0907 | 71.00369 |
| GWAS_Cat | Invasive Ep | 16-Jan | 0.067399 | 0.115002 | 0 | 0 | 15.42481 | 41.60265 |
| GWAS_Cat | Glycemic T | 16-Jan | 0.067399 | 0.115002 | 0 | 0 | 15.42481 | 41.60265 |
| GWAS_Cat | Factor VII A | 16-Jan | 0.067399 | 0.115002 | 0 | 0 | 15.42481 | 41.60265 |
| GWAS_Cat | Attention D | 16-Jan | 0.067399 | 0.115002 | 0 | 0 | 15.42481 | 41.60265 |
| GWAS_Cat | Alanine Am | 16-Jan | 0.067399 | 0.115002 | 0 | 0 | 15.42481 | 41.60265 |
| GWAS_Cat | White Mat | 16-Jan | 0.067399 | 0.115002 | 0 | 0 | 15.42481 | 41.60265 |
| GWAS_Cat | Cognitive F | 16-Jan | 0.067399 | 0.115002 | 0 | 0 | 15.42481 | 41.60265 |
| GWAS_Cat | Facial Morp | 16-Jan | 0.067399 | 0.115002 | 0 | 0 | 15.42481 | 41.60265 |
| GWAS_Cat | HDL Choles | 4/351 | 0.066957 | 0.115002 | 0 | 0 | 2.717406 | 7.347065 |
| GWAS_Cat | Lung Funct | 16-Jan | 0.067399 | 0.115002 | 0 | 0 | 15.42481 | 41.60265 |
| GWAS_Cat | Medicatior | 16-Jan | 0.067399 | 0.115002 | 0 | 0 | 15.42481 | 41.60265 |
| GWAS_Cat | Electrocarc | 16-Jan | 0.067399 | 0.115002 | 0 | 0 | 15.42481 | 41.60265 |
| GWAS_Cat | Low Hand ( | 16-Jan | 0.067399 | 0.115002 | 0 | 0 | 15.42481 | 41.60265 |
| GWAS_Cat | Bitter Beve | 16-Jan | 0.067399 | 0.115002 | 0 | 0 | 15.42481 | 41.60265 |
| DisGeNET | Giant Cell F | Feb-32 | 0.008524 | 0.116321 | 0 | 0 | 15.59451 | 74.30533 |
| DisGeNET | Dermatofik | Feb-32 | 0.008524 | 0.116321 | 0 | 0 | 15.59451 | 74.30533 |
| GO_Biologi | Regulation | 11/1127 | 0.009559 | 0.117362 | 0 | 0 | 2.43783 | 11.33652 |
| GO_Biologi | ATP Metab | Feb-34 | 0.009587 | 0.117362 | 0 | 0 | 14.61838 | 67.9362 |
| Jensen_TIS | Mesenchyr | 5/264 | 0.005918 | 0.117417 | 0 | 0 | 4.627084 | 23.73548 |
| Jensen_TIS | Adult | 10/918 | 0.006461 | 0.117417 | 0 | 0 | 2.718262 | 13.70539 |
| Jensen_TIS | Promastigc | Feb-29 | 0.007037 | 0.117417 | 0 | 0 | 17.32985 | 85.89758 |
| Jensen_TIS | Lymphocyt | Mar-84 | 0.005878 | 0.117417 | 0 | 0 | 8.744268 | 44.91577 |
| Jensen_TIS | Spleen | 75/14937 | 0.00667 | 0.117417 | 0 | 0 | 2.124125 | 10.64224 |
| Jensen_TIS | Connective | 55/9893 | 0.006678 | 0.117417 | 0 | 0 | 1.760155 | 8.816421 |
| Jensen_TIS | Smooth mu | 53/9462 | 0.007293 | 0.117417 | 0 | 0 | 1.740236 | 8.563422 |
| GWAS_Cat | Red Blood | 8/1001 | 0.069509 | 0.118437 | 0 | 0 | 1.929456 | 5.14451 |
| GWAS_Cat | Body Fat Pe | 17-Jan | 0.071459 | 0.118936 | 0 | 0 | 14.46003 | 38.15469 |

| | | | | | | | | |
|---|---|---|---|---|---|---|---|---|
| GWAS_Cat | Raw Veget | 17-Jan | 0.071459 | 0.118936 | 0 | 0 | 14.46003 | 38.15469 |
| GWAS_Cat | Triglyceride | 17-Jan | 0.071459 | 0.118936 | 0 | 0 | 14.46003 | 38.15469 |
| GWAS_Cat | Alzheimer's | 17-Jan | 0.071459 | 0.118936 | 0 | 0 | 14.46003 | 38.15469 |
| GWAS_Cat | Facial Morp | 17-Jan | 0.071459 | 0.118936 | 0 | 0 | 14.46003 | 38.15469 |
| GWAS_Cat | Chronic Ob | 17-Jan | 0.071459 | 0.118936 | 0 | 0 | 14.46003 | 38.15469 |
| GWAS_Cat | Pubertal Ar | 17-Jan | 0.071459 | 0.118936 | 0 | 0 | 14.46003 | 38.15469 |
| GWAS_Cat | Chloride Le | 17-Jan | 0.071459 | 0.118936 | 0 | 0 | 14.46003 | 38.15469 |
| GWAS_Cat | Decaffeina | 17-Jan | 0.071459 | 0.118936 | 0 | 0 | 14.46003 | 38.15469 |
| GWAS_Cat | Recurrent I | 17-Jan | 0.071459 | 0.118936 | 0 | 0 | 14.46003 | 38.15469 |
| GWAS_Cat | Serous Inva | 17-Jan | 0.071459 | 0.118936 | 0 | 0 | 14.46003 | 38.15469 |
| GWAS_Cat | Anthropom | 17-Jan | 0.071459 | 0.118936 | 0 | 0 | 14.46003 | 38.15469 |
| GWAS_Cat | Two-hour ( | 17-Jan | 0.071459 | 0.118936 | 0 | 0 | 14.46003 | 38.15469 |
| GWAS_Cat | Problemati | 17-Jan | 0.071459 | 0.118936 | 0 | 0 | 14.46003 | 38.15469 |
| GWAS_Cat | Sleep Apne | 17-Jan | 0.071459 | 0.118936 | 0 | 0 | 14.46003 | 38.15469 |
| GWAS_Cat | F-seafood I | 17-Jan | 0.071459 | 0.118936 | 0 | 0 | 14.46003 | 38.15469 |
| GWAS_Cat | Percent Ma | 17-Jan | 0.071459 | 0.118936 | 0 | 0 | 14.46003 | 38.15469 |
| ClinVar_20 | neoplasm c | 23-Jan | 0.095455 | 0.119319 | 0 | 0 | 10.51321 | 24.6966 |
| GeDiPNet_ | Multiple Cc | 4/251 | 0.023972 | 0.119452 | 0 | 0 | 3.837081 | 14.31558 |
| Jensen_DIS | Ebstein anc | 12-Jan | 0.050982 | 0.119778 | 0 | 0 | 21.03805 | 62.61499 |
| Jensen_DIS | Hard palate | 12-Jan | 0.050982 | 0.119778 | 0 | 0 | 21.03805 | 62.61499 |
| Jensen_DIS | Lysinuric pı | 12-Jan | 0.050982 | 0.119778 | 0 | 0 | 21.03805 | 62.61499 |
| GO_Biologi | Regulation | 3/102 | 0.010016 | 0.120529 | 0 | 0 | 7.147908 | 32.90605 |
| DisGeNET | Delusions | Feb-33 | 0.009049 | 0.121578 | 0 | 0 | 15.0907 | 71.00369 |
| DisGeNET | Angiomyoli | Feb-33 | 0.009049 | 0.121578 | 0 | 0 | 15.0907 | 71.00369 |
| DisGeNET | Scapular w | Feb-33 | 0.009049 | 0.121578 | 0 | 0 | 15.0907 | 71.00369 |
| Jensen_DIS | Oculodento | 13-Jan | 0.055113 | 0.122126 | 0 | 0 | 19.28391 | 55.89185 |
| Jensen_DIS | Cryptospor | 13-Jan | 0.055113 | 0.122126 | 0 | 0 | 19.28391 | 55.89185 |
| Jensen_DIS | Upper resp | 13-Jan | 0.055113 | 0.122126 | 0 | 0 | 19.28391 | 55.89185 |
| Jensen_DIS | Fibroma | 13-Jan | 0.055113 | 0.122126 | 0 | 0 | 19.28391 | 55.89185 |
| Jensen_DIS | Limb ischer | 13-Jan | 0.055113 | 0.122126 | 0 | 0 | 19.28391 | 55.89185 |
| GeDiPNet_ | Ovarian Se | Feb-56 | 0.024747 | 0.122266 | 0 | 0 | 8.653159 | 32.00857 |
| GeDiPNet_ | Complete A | 6-Jan | 0.025821 | 0.122387 | 0 | 0 | 46.29767 | 169.2909 |
| GeDiPNet_ | Basal Gang | 6-Jan | 0.025821 | 0.122387 | 0 | 0 | 46.29767 | 169.2909 |
| GeDiPNet_ | Partial Atri | 6-Jan | 0.025821 | 0.122387 | 0 | 0 | 46.29767 | 169.2909 |
| GeDiPNet_ | Intussuscep | 6-Jan | 0.025821 | 0.122387 | 0 | 0 | 46.29767 | 169.2909 |
| GeDiPNet_ | Ovarioleuk | 6-Jan | 0.025821 | 0.122387 | 0 | 0 | 46.29767 | 169.2909 |
| dbGaP | Heart Rate | 3/155 | 0.030154 | 0.122499 | 0 | 0 | 4.643092 | 16.25756 |
| dbGaP | 1-Alkyl-2-a | 7-Jan | 0.03006 | 0.122499 | 0 | 0 | 38.57946 | 135.2043 |
| DisGeNET | Attention c | 6/412 | 0.009168 | 0.122547 | 0 | 0 | 3.559022 | 16.6992 |
| Disease_Pe | atheroscle | 6/271 | 0.001182 | 0.122746 | 0 | 0 | 5.492103 | 37.02115 |
| Disease_Pe | cerebral pa | 6/309 | 0.002294 | 0.122746 | 0 | 0 | 4.794035 | 29.13487 |
| Disease_Pe | Transplant | 6/292 | 0.001727 | 0.122746 | 0 | 0 | 5.083398 | 32.33671 |
| Disease_Pe | Neurologic | 7/370 | 0.001152 | 0.122746 | 0 | 0 | 4.712466 | 31.88762 |
| Disease_Pe | Polymyosit | 6/279 | 0.001371 | 0.122746 | 0 | 0 | 5.328992 | 35.12965 |
| Disease_Pe | Duchenne | 6/287 | 0.001583 | 0.122746 | 0 | 0 | 5.175168 | 33.37244 |

| | | | | | | | | |
|---|---|---|---|---|---|---|---|---|
| Disease_Pe | cerebral pa | 6/309 | 0.002294 | 0.122746 | 0 | 0 | 4.794035 | 29.13487 |
| Disease_Pe | idiopathic | 6/300 | 0.001979 | 0.122746 | 0 | 0 | 4.943059 | 30.77152 |
| DisGeNET | Palmoplan | Mar-99 | 0.009235 | 0.122821 | 0 | 0 | 7.372396 | 34.53788 |
| PhenGenI_ | Intra-Abdo | Feb-33 | 0.009049 | 0.122957 | 0 | 0 | 15.0907 | 71.00369 |
| PhenGenI_ | Tau Protei | Mar-97 | 0.008735 | 0.122957 | 0 | 0 | 7.530015 | 35.6954 |
| Rare_Disea | Visceral ste | 5/291 | 0.008827 | 0.123905 | 0 | 0 | 4.184505 | 19.79266 |
| Rare_Disea | Leishmania | Mar-97 | 0.008735 | 0.123905 | 0 | 0 | 7.530015 | 35.6954 |
| UK_Bioban | Alcohol int | 16-Jan | 0.067399 | 0.123927 | 0 | 0 | 15.42481 | 41.60265 |
| CellMarker | Stem Cell A | 8-Jan | 0.03428 | 0.124051 | 0 | 0 | 33.06645 | 111.5391 |
| CellMarker | Dental Pulp | 5-Jan | 0.021564 | 0.124051 | 0 | 0 | 57.875 | 222.0519 |
| CellMarker | Cardiomyo | 5-Jan | 0.021564 | 0.124051 | 0 | 0 | 57.875 | 222.0519 |
| CellMarker | Fibroblast | 8-Jan | 0.03428 | 0.124051 | 0 | 0 | 33.06645 | 111.5391 |
| CellMarker | Multipoter | 8-Jan | 0.03428 | 0.124051 | 0 | 0 | 33.06645 | 111.5391 |
| CellMarker | Multipoter | 8-Jan | 0.03428 | 0.124051 | 0 | 0 | 33.06645 | 111.5391 |
| CellMarker | Mesenchyr | 7-Jan | 0.03006 | 0.124051 | 0 | 0 | 38.57946 | 135.2043 |
| CellMarker | Smooth Mu | 7-Jan | 0.03006 | 0.124051 | 0 | 0 | 38.57946 | 135.2043 |
| CellMarker | Myofibrobl | 9-Jan | 0.038483 | 0.124051 | 0 | 0 | 28.93169 | 94.24608 |
| CellMarker | Meiotic Pro | 9/1049 | 0.038472 | 0.124051 | 0 | 0 | 2.093898 | 6.821558 |
| CellMarker | Ciliated Cel | 4/283 | 0.035036 | 0.124051 | 0 | 0 | 3.391458 | 11.36607 |
| CellMarker | Mesenchyr | 5-Jan | 0.021564 | 0.124051 | 0 | 0 | 57.875 | 222.0519 |
| CellMarker | Mesenchyr | 8-Jan | 0.03428 | 0.124051 | 0 | 0 | 33.06645 | 111.5391 |
| CellMarker | Endothelia | 8-Jan | 0.03428 | 0.124051 | 0 | 0 | 33.06645 | 111.5391 |
| CellMarker | Microglial | 9-Jan | 0.038483 | 0.124051 | 0 | 0 | 28.93169 | 94.24608 |
| CellMarker | Stem Cell B | 9-Jan | 0.038483 | 0.124051 | 0 | 0 | 28.93169 | 94.24608 |
| CellMarker | Dermal Cel | 7-Jan | 0.03006 | 0.124051 | 0 | 0 | 38.57946 | 135.2043 |
| CellMarker | Macrophag | 6-Jan | 0.025821 | 0.124051 | 0 | 0 | 46.29767 | 169.2909 |
| CellMarker | Progenitor | 6-Jan | 0.025821 | 0.124051 | 0 | 0 | 46.29767 | 169.2909 |
| CellMarker | Cardiomyo | 6-Jan | 0.025821 | 0.124051 | 0 | 0 | 46.29767 | 169.2909 |
| CellMarker | Hematopoi | 6-Jan | 0.025821 | 0.124051 | 0 | 0 | 46.29767 | 169.2909 |
| CellMarker | Mesangial | 9-Jan | 0.038483 | 0.124051 | 0 | 0 | 28.93169 | 94.24608 |
| CellMarker | Stem Cell N | 27-Feb | 0.006118 | 0.124051 | 0 | 0 | 18.71812 | 95.39811 |
| CellMarker | Macrophag | 3/141 | 0.02365 | 0.124051 | 0 | 0 | 5.117754 | 19.16288 |
| CellMarker | Mitotic Arr | 9/956 | 0.023017 | 0.124051 | 0 | 0 | 2.31086 | 8.715432 |
| CellMarker | Smooth Mu | 5-Jan | 0.021564 | 0.124051 | 0 | 0 | 57.875 | 222.0519 |
| CellMarker | Mesenchyr | 5-Jan | 0.021564 | 0.124051 | 0 | 0 | 57.875 | 222.0519 |
| CellMarker | Mesenchyr | 6-Jan | 0.025821 | 0.124051 | 0 | 0 | 46.29767 | 169.2909 |
| CellMarker | Mesenchyr | 6-Jan | 0.025821 | 0.124051 | 0 | 0 | 46.29767 | 169.2909 |
| CellMarker | Myofibrobl | 5-Jan | 0.021564 | 0.124051 | 0 | 0 | 57.875 | 222.0519 |
| CellMarker | Stem Cell L | 6-Jan | 0.025821 | 0.124051 | 0 | 0 | 46.29767 | 169.2909 |
| CellMarker | Endothelia | 9-Jan | 0.038483 | 0.124051 | 0 | 0 | 28.93169 | 94.24608 |
| CellMarker | Hematopoi | 7/565 | 0.011559 | 0.124051 | 0 | 0 | 3.035058 | 13.53735 |
| CellMarker | Quiescent | 9-Jan | 0.038483 | 0.124051 | 0 | 0 | 28.93169 | 94.24608 |
| CellMarker | Hepatocyte | Feb-31 | 0.008014 | 0.124051 | 0 | 0 | 16.13306 | 77.8673 |
| CellMarker | Sensory Ne | 5-Jan | 0.021564 | 0.124051 | 0 | 0 | 57.875 | 222.0519 |
| CellMarker | Mesenchyr | 5-Jan | 0.021564 | 0.124051 | 0 | 0 | 57.875 | 222.0519 |

| | | | | | | | |
|---|---|---|---|---|---|---|---|
| CellMarker Cancer Ste | 3/110 | 0.012281 | 0.124051 | 0 | 0 | 6.610814 | 29.08541 |
| CellMarker Mesenchyr | 7-Jan | 0.03006 | 0.124051 | 0 | 0 | 38.57946 | 135.2043 |
| CellMarker Hematopoi | 9-Jan | 0.038483 | 0.124051 | 0 | 0 | 28.93169 | 94.24608 |
| CellMarker P53-like Ce | 8-Jan | 0.03428 | 0.124051 | 0 | 0 | 33.06645 | 111.5391 |
| CellMarker Stem Cell D | 9-Jan | 0.038483 | 0.124051 | 0 | 0 | 28.93169 | 94.24608 |
| CellMarker Cancer Ste | 8-Jan | 0.03428 | 0.124051 | 0 | 0 | 33.06645 | 111.5391 |
| CellMarker Multipoter | 6-Jan | 0.025821 | 0.124051 | 0 | 0 | 46.29767 | 169.2909 |
| CellMarker T Helper 2( | 7-Jan | 0.03006 | 0.124051 | 0 | 0 | 38.57946 | 135.2043 |
| CellMarker Epithelial C | 5-Jan | 0.021564 | 0.124051 | 0 | 0 | 57.875 | 222.0519 |
| CellMarker Regulatory | Feb-49 | 0.01927 | 0.124051 | 0 | 0 | 9.945432 | 39.27635 |
| CellMarker Neutrophil | Feb-49 | 0.01927 | 0.124051 | 0 | 0 | 9.945432 | 39.27635 |
| CellMarker Macrophag | Feb-45 | 0.016405 | 0.124051 | 0 | 0 | 10.87278 | 44.68894 |
| CellMarker Hematopoi | Feb-40 | 0.013107 | 0.124051 | 0 | 0 | 12.3065 | 53.34346 |
| CellMarker Hepatic Ce | 8-Jan | 0.03428 | 0.124051 | 0 | 0 | 33.06645 | 111.5391 |
| CellMarker Bone Marr | 7-Jan | 0.03006 | 0.124051 | 0 | 0 | 38.57946 | 135.2043 |
| CellMarker Mesenchyr | 5-Jan | 0.021564 | 0.124051 | 0 | 0 | 57.875 | 222.0519 |
| CellMarker Hematopoi | 5-Jan | 0.021564 | 0.124051 | 0 | 0 | 57.875 | 222.0519 |
| CellMarker Glutamater | 5-Jan | 0.021564 | 0.124051 | 0 | 0 | 57.875 | 222.0519 |
| CellMarker Mural Cell | 5-Jan | 0.021564 | 0.124051 | 0 | 0 | 57.875 | 222.0519 |
| CellMarker Regulatory | 6-Jan | 0.025821 | 0.124051 | 0 | 0 | 46.29767 | 169.2909 |
| CellMarker Hepatocyte | Feb-35 | 0.01014 | 0.124051 | 0 | 0 | 14.17469 | 65.07997 |
| CellMarker Hematopoi | Feb-37 | 0.011286 | 0.124051 | 0 | 0 | 13.36336 | 59.92353 |
| CellMarker Adipocyte I | 6-Jan | 0.025821 | 0.124051 | 0 | 0 | 46.29767 | 169.2909 |
| CellMarker Interstitial | 7-Jan | 0.03006 | 0.124051 | 0 | 0 | 38.57946 | 135.2043 |
| CellMarker Epithelial C | 7-Jan | 0.03006 | 0.124051 | 0 | 0 | 38.57946 | 135.2043 |
| CellMarker Muscle Cel | 6-Jan | 0.025821 | 0.124051 | 0 | 0 | 46.29767 | 169.2909 |
| CellMarker Hematopoi | Feb-62 | 0.029883 | 0.124051 | 0 | 0 | 7.78549 | 27.33058 |
| CellMarker Epidermal S | 6-Jan | 0.025821 | 0.124051 | 0 | 0 | 46.29767 | 169.2909 |
| CellMarker Mesenchyr | 7-Jan | 0.03006 | 0.124051 | 0 | 0 | 38.57946 | 135.2043 |
| CellMarker Mesenchyr | 7-Jan | 0.03006 | 0.124051 | 0 | 0 | 38.57946 | 135.2043 |
| CellMarker Adipose-de | 7-Jan | 0.03006 | 0.124051 | 0 | 0 | 38.57946 | 135.2043 |
| CellMarker Dendritic C | 7-Jan | 0.03006 | 0.124051 | 0 | 0 | 38.57946 | 135.2043 |
| CellMarker Myoblast M | 7-Jan | 0.03006 | 0.124051 | 0 | 0 | 38.57946 | 135.2043 |
| CellMarker Cancer Ste | 7-Jan | 0.03006 | 0.124051 | 0 | 0 | 38.57946 | 135.2043 |
| CellMarker Hematopoi | 7-Jan | 0.03006 | 0.124051 | 0 | 0 | 38.57946 | 135.2043 |
| CellMarker Hemangiok | 7-Jan | 0.03006 | 0.124051 | 0 | 0 | 38.57946 | 135.2043 |
| CellMarker Fibroblast V | 7-Jan | 0.03006 | 0.124051 | 0 | 0 | 38.57946 | 135.2043 |
| CellMarker B Cell Splee | 6-Jan | 0.025821 | 0.124051 | 0 | 0 | 46.29767 | 169.2909 |
| GWAS_Cat Inflammato | 18-Jan | 0.075502 | 0.124308 | 0 | 0 | 13.60876 | 35.1596 |
| GWAS_Cat Hypertroph | 18-Jan | 0.075502 | 0.124308 | 0 | 0 | 13.60876 | 35.1596 |
| GWAS_Cat Alcohol Fre | 18-Jan | 0.075502 | 0.124308 | 0 | 0 | 13.60876 | 35.1596 |
| GWAS_Cat Essential H | 18-Jan | 0.075502 | 0.124308 | 0 | 0 | 13.60876 | 35.1596 |
| GWAS_Cat Aortic Root | 18-Jan | 0.075502 | 0.124308 | 0 | 0 | 13.60876 | 35.1596 |
| GWAS_Cat Periodontit | 18-Jan | 0.075502 | 0.124308 | 0 | 0 | 13.60876 | 35.1596 |
| GWAS_Cat Alcohol Us | 18-Jan | 0.075502 | 0.124308 | 0 | 0 | 13.60876 | 35.1596 |

| | | | | | | | | |
|---|---|---|---|---|---|---|---|---|
| GWAS_Cat | Osteoarthr | 18-Jan | 0.075502 | 0.124308 | 0 | 0 | 13.60876 | 35.1596 |
| GWAS_Cat | HDL Choles | 5/519 | 0.075844 | 0.124703 | 0 | 0 | 2.301295 | 5.935216 |
| DisGeNET | Myopathy | 6/415 | 0.00948 | 0.124969 | 0 | 0 | 3.532373 | 16.45568 |
| DisGeNET | Winged sca | Feb-34 | 0.009587 | 0.124969 | 0 | 0 | 14.61838 | 67.9362 |
| DisGeNET | Wolff-Park | Feb-34 | 0.009587 | 0.124969 | 0 | 0 | 14.61838 | 67.9362 |
| DisGeNET | Myelodysp | Feb-34 | 0.009587 | 0.124969 | 0 | 0 | 14.61838 | 67.9362 |
| GWAS_Cat | Body Fat Pe | 3/227 | 0.076549 | 0.125693 | 0 | 0 | 3.13919 | 8.067162 |
| DisGeNET | Transient Is | 4/191 | 0.009692 | 0.125713 | 0 | 0 | 5.083693 | 23.5701 |
| Jensen_DIS | Takayasu's | 17-Jan | 0.071459 | 0.125836 | 0 | 0 | 14.46003 | 38.15469 |
| Jensen_DIS | Cutaneous | 15-Jan | 0.063321 | 0.125836 | 0 | 0 | 16.52741 | 45.60793 |
| Jensen_DIS | Stomach ca | 14-Jan | 0.059226 | 0.125836 | 0 | 0 | 17.79964 | 50.30878 |
| Jensen_DIS | Fibromyalg | 16-Jan | 0.067399 | 0.125836 | 0 | 0 | 15.42481 | 41.60265 |
| Jensen_DIS | Kidney dise | Feb-91 | 0.05972 | 0.125836 | 0 | 0 | 5.240978 | 14.76951 |
| Jensen_DIS | Exophthalm | 14-Jan | 0.059226 | 0.125836 | 0 | 0 | 17.79964 | 50.30878 |
| Jensen_DIS | Colorectal | 17-Jan | 0.071459 | 0.125836 | 0 | 0 | 14.46003 | 38.15469 |
| Jensen_DIS | Asthma | Feb-92 | 0.060879 | 0.125836 | 0 | 0 | 5.182484 | 14.50504 |
| Jensen_DIS | Cleft palate | Feb-98 | 0.067994 | 0.125836 | 0 | 0 | 4.857108 | 13.05753 |
| Jensen_DIS | Left ventric | 16-Jan | 0.067399 | 0.125836 | 0 | 0 | 15.42481 | 41.60265 |
| Jensen_DIS | Pneumotho | 16-Jan | 0.067399 | 0.125836 | 0 | 0 | 15.42481 | 41.60265 |
| Jensen_DIS | Compleme | 16-Jan | 0.067399 | 0.125836 | 0 | 0 | 15.42481 | 41.60265 |
| Jensen_DIS | Opposition | 16-Jan | 0.067399 | 0.125836 | 0 | 0 | 15.42481 | 41.60265 |
| Jensen_DIS | Lymphatic | 18-Jan | 0.075502 | 0.125836 | 0 | 0 | 13.60876 | 35.1596 |
| Jensen_DIS | Lipoma | 18-Jan | 0.075502 | 0.125836 | 0 | 0 | 13.60876 | 35.1596 |
| Jensen_DIS | Pearson sy | 18-Jan | 0.075502 | 0.125836 | 0 | 0 | 13.60876 | 35.1596 |
| Jensen_DIS | Adams-Oliv | 15-Jan | 0.063321 | 0.125836 | 0 | 0 | 16.52741 | 45.60793 |
| Jensen_DIS | Coronary a | 3/205 | 0.060177 | 0.125836 | 0 | 0 | 3.484972 | 9.794417 |
| Jensen_DIS | Thyroid car | 18-Jan | 0.075502 | 0.125836 | 0 | 0 | 13.60876 | 35.1596 |
| Jensen_DIS | Neurilemm | 16-Jan | 0.067399 | 0.125836 | 0 | 0 | 15.42481 | 41.60265 |
| Jensen_DIS | Ophthalmo | 16-Jan | 0.067399 | 0.125836 | 0 | 0 | 15.42481 | 41.60265 |
| Jensen_DIS | Papilloma | 17-Jan | 0.071459 | 0.125836 | 0 | 0 | 14.46003 | 38.15469 |
| Jensen_DIS | Bronchiect | 18-Jan | 0.075502 | 0.125836 | 0 | 0 | 13.60876 | 35.1596 |
| Jensen_DIS | Phenylketo | 17-Jan | 0.071459 | 0.125836 | 0 | 0 | 14.46003 | 38.15469 |
| Jensen_DIS | Obesity | 2/103 | 0.074122 | 0.125836 | 0 | 0 | 4.615492 | 12.00969 |
| Jensen_DIS | Angiomyoli | 18-Jan | 0.075502 | 0.125836 | 0 | 0 | 13.60876 | 35.1596 |
| Jensen_DIS | Cowden dis | 17-Jan | 0.071459 | 0.125836 | 0 | 0 | 14.46003 | 38.15469 |
| Jensen_DIS | Myasthenia | 15-Jan | 0.063321 | 0.125836 | 0 | 0 | 16.52741 | 45.60793 |
| Jensen_DIS | Variant Cre | 16-Jan | 0.067399 | 0.125836 | 0 | 0 | 15.42481 | 41.60265 |
| ClinVar_20 | neoplasm o | 27-Jan | 0.111111 | 0.126262 | 0 | 0 | 8.894007 | 19.54216 |
| ClinVar_20 | familial car | 27-Jan | 0.111111 | 0.126262 | 0 | 0 | 8.894007 | 19.54216 |
| Rare_Disea | Acute myel | 4/188 | 0.009182 | 0.126513 | 0 | 0 | 5.167365 | 24.23737 |
| Rare_Disea | Craniofacia | 3/102 | 0.010016 | 0.127865 | 0 | 0 | 7.147908 | 32.90605 |
| Rare_Disea | Klinefelter | Feb-35 | 0.01014 | 0.127865 | 0 | 0 | 14.17469 | 65.07997 |
| Rare_Disea | Neuroblast | 3/102 | 0.010016 | 0.127865 | 0 | 0 | 7.147908 | 32.90605 |
| Rare_Disea | Tetrahydro | Feb-35 | 0.01014 | 0.127865 | 0 | 0 | 14.17469 | 65.07997 |
| Rare_Disea | Leukodystr | 3/101 | 0.009751 | 0.127865 | 0 | 0 | 7.22121 | 33.4367 |

| | | | | | | | | |
|---|---|---|---|---|---|---|---|---|
| Jensen_TIS | Seminifero | Feb-34 | 0.009587 | 0.12863 | 0 | 0 | 14.61838 | 67.9362 |
| Jensen_TIS | Hypothalar | 67/12937 | 0.009132 | 0.12863 | 0 | 0 | 1.83326 | 8.608992 |
| Jensen_TIS | Myoblast | Mar-99 | 0.009235 | 0.12863 | 0 | 0 | 7.372396 | 34.53788 |
| Jensen_TIS | Corpus call | 51/9077 | 0.008837 | 0.12863 | 0 | 0 | 1.708758 | 8.080446 |
| GWAS_Cat | High-grade | 19-Jan | 0.079527 | 0.129191 | 0 | 0 | 12.85207 | 32.53709 |
| GWAS_Cat | Free Chole: | 19-Jan | 0.079527 | 0.129191 | 0 | 0 | 12.85207 | 32.53709 |
| GWAS_Cat | Visceral Ad | 19-Jan | 0.079527 | 0.129191 | 0 | 0 | 12.85207 | 32.53709 |
| GWAS_Cat | Liver Enzyn | 3/230 | 0.07892 | 0.129191 | 0 | 0 | 3.097231 | 7.864854 |
| GWAS_Cat | Diverticula | 2/107 | 0.079148 | 0.129191 | 0 | 0 | 4.438768 | 11.25866 |
| GWAS_Cat | Alcohol Us | 19-Jan | 0.079527 | 0.129191 | 0 | 0 | 12.85207 | 32.53709 |
| GWAS_Cat | Response T | 19-Jan | 0.079527 | 0.129191 | 0 | 0 | 12.85207 | 32.53709 |
| GWAS_Cat | Estradiol Le | 19-Jan | 0.079527 | 0.129191 | 0 | 0 | 12.85207 | 32.53709 |
| DisGeNET | PARAGANG | Feb-35 | 0.01014 | 0.129592 | 0 | 0 | 14.17469 | 65.07997 |
| DisGeNET | Gait, Drop | Feb-35 | 0.01014 | 0.129592 | 0 | 0 | 14.17469 | 65.07997 |
| DisGeNET | Acinar Cell | Feb-35 | 0.01014 | 0.129592 | 0 | 0 | 14.17469 | 65.07997 |
| CellMarker | Stellate Cel | 10-Jan | 0.042667 | 0.129621 | 0 | 0 | 25.71576 | 81.1157 |
| CellMarker | Rheaume E | 10-Jan | 0.042667 | 0.129621 | 0 | 0 | 25.71576 | 81.1157 |
| CellMarker | Cardiac Prc | 11-Jan | 0.046834 | 0.129621 | 0 | 0 | 23.14302 | 70.84419 |
| CellMarker | Endothelia | Feb-79 | 0.046446 | 0.129621 | 0 | 0 | 6.061421 | 18.60534 |
| CellMarker | Endothelia | 11-Jan | 0.046834 | 0.129621 | 0 | 0 | 23.14302 | 70.84419 |
| CellMarker | Stem Cell E | 11-Jan | 0.046834 | 0.129621 | 0 | 0 | 23.14302 | 70.84419 |
| CellMarker | Urine-deriv | 11-Jan | 0.046834 | 0.129621 | 0 | 0 | 23.14302 | 70.84419 |
| CellMarker | Glutamate | 11-Jan | 0.046834 | 0.129621 | 0 | 0 | 23.14302 | 70.84419 |
| CellMarker | Adipose-de | 10-Jan | 0.042667 | 0.129621 | 0 | 0 | 25.71576 | 81.1157 |
| CellMarker | Mesenchyr | 11-Jan | 0.046834 | 0.129621 | 0 | 0 | 23.14302 | 70.84419 |
| CellMarker | Hematopoi | 10-Jan | 0.042667 | 0.129621 | 0 | 0 | 25.71576 | 81.1157 |
| CellMarker | Ciliated Cel | 11-Jan | 0.046834 | 0.129621 | 0 | 0 | 23.14302 | 70.84419 |
| CellMarker | Cytotoxic T | 11-Jan | 0.046834 | 0.129621 | 0 | 0 | 23.14302 | 70.84419 |
| CellMarker | Rheaume E | 10-Jan | 0.042667 | 0.129621 | 0 | 0 | 25.71576 | 81.1157 |
| Human_Ph | Foot dorsif | 27-Feb | 0.006118 | 0.129832 | 0 | 0 | 18.71812 | 95.39811 |
| UK_Bioban | Year endec | 17-Jan | 0.071459 | 0.13055 | 0 | 0 | 14.46003 | 38.15469 |
| dbGaP | Glucose Tra | 8-Jan | 0.03428 | 0.131072 | 0 | 0 | 33.06645 | 111.5391 |
| Rare_Disea | Aortic valv | 4/196 | 0.010583 | 0.131227 | 0 | 0 | 4.95005 | 22.51542 |
| GO_Biologi | Negative R | Feb-37 | 0.011286 | 0.131365 | 0 | 0 | 13.36336 | 59.92353 |
| GO_Biologi | Regulation | Feb-37 | 0.011286 | 0.131365 | 0 | 0 | 13.36336 | 59.92353 |
| Jensen_DIS | Centronucl | 19-Jan | 0.079527 | 0.131421 | 0 | 0 | 12.85207 | 32.53709 |
| GeDiPNet_ | Cardiovasc | 4/266 | 0.028847 | 0.131766 | 0 | 0 | 3.614642 | 12.81666 |
| GeDiPNet_ | Diabetic M | 7-Jan | 0.03006 | 0.131766 | 0 | 0 | 38.57946 | 135.2043 |
| GeDiPNet_ | Madelung | 7-Jan | 0.03006 | 0.131766 | 0 | 0 | 38.57946 | 135.2043 |
| GeDiPNet_ | Thymus Ne | 7-Jan | 0.03006 | 0.131766 | 0 | 0 | 38.57946 | 135.2043 |
| GeDiPNet_ | Retinal Nor | 7-Jan | 0.03006 | 0.131766 | 0 | 0 | 38.57946 | 135.2043 |
| GeDiPNet_ | Biventricul | 7-Jan | 0.03006 | 0.131766 | 0 | 0 | 38.57946 | 135.2043 |
| GeDiPNet_ | Frontonasa | 7-Jan | 0.03006 | 0.131766 | 0 | 0 | 38.57946 | 135.2043 |
| GeDiPNet_ | Congenital | 7-Jan | 0.03006 | 0.131766 | 0 | 0 | 38.57946 | 135.2043 |
| GeDiPNet_ | Salaam Sei: | Feb-62 | 0.029883 | 0.131766 | 0 | 0 | 7.78549 | 27.33058 |

| | | | | | | | | |
|---|---|---|---|---|---|---|---|---|
| GeDiPNet_ | Emery-Dre | 7-Jan | 0.03006 | 0.131766 | 0 | 0 | 38.57946 | 135.2043 |
| OMIM_Dis | thyroid car | 17-Jan | 0.071459 | 0.132545 | 0 | 0 | 14.46003 | 38.15469 |
| OMIM_Dis | autism | 19-Jan | 0.079527 | 0.132545 | 0 | 0 | 12.85207 | 32.53709 |
| OMIM_Dis | malaria | 14-Jan | 0.059226 | 0.132545 | 0 | 0 | 17.79964 | 50.30878 |
| OMIM_Dis | myocardial | 16-Jan | 0.067399 | 0.132545 | 0 | 0 | 15.42481 | 41.60265 |
| OMIM_Dis | macular de | 18-Jan | 0.075502 | 0.132545 | 0 | 0 | 13.60876 | 35.1596 |
| GWAS_Cat | Alanine Tra | 20-Jan | 0.083535 | 0.133045 | 0 | 0 | 12.17503 | 30.22444 |
| GWAS_Cat | Total Conc | 20-Jan | 0.083535 | 0.133045 | 0 | 0 | 12.17503 | 30.22444 |
| GWAS_Cat | Drink Temp | 20-Jan | 0.083535 | 0.133045 | 0 | 0 | 12.17503 | 30.22444 |
| GWAS_Cat | White Mat | 20-Jan | 0.083535 | 0.133045 | 0 | 0 | 12.17503 | 30.22444 |
| GWAS_Cat | Femur Tota | 20-Jan | 0.083535 | 0.133045 | 0 | 0 | 12.17503 | 30.22444 |
| GWAS_Cat | Bladder Ca | 20-Jan | 0.083535 | 0.133045 | 0 | 0 | 12.17503 | 30.22444 |
| GWAS_Cat | F-strong Fl | 20-Jan | 0.083535 | 0.133045 | 0 | 0 | 12.17503 | 30.22444 |
| GWAS_Cat | Crohn's Dis | 20-Jan | 0.083535 | 0.133045 | 0 | 0 | 12.17503 | 30.22444 |
| GWAS_Cat | Aging Trait | 20-Jan | 0.083535 | 0.133045 | 0 | 0 | 12.17503 | 30.22444 |
| GWAS_Cat | Familial Co | 20-Jan | 0.083535 | 0.133045 | 0 | 0 | 12.17503 | 30.22444 |
| GWAS_Cat | Psoriasis O | 20-Jan | 0.083535 | 0.133045 | 0 | 0 | 12.17503 | 30.22444 |
| GWAS_Cat | Urate Leve | 20-Jan | 0.083535 | 0.133045 | 0 | 0 | 12.17503 | 30.22444 |
| GWAS_Cat | Urate Leve | 20-Jan | 0.083535 | 0.133045 | 0 | 0 | 12.17503 | 30.22444 |
| GWAS_Cat | Interleukin | 20-Jan | 0.083535 | 0.133045 | 0 | 0 | 12.17503 | 30.22444 |
| GWAS_Cat | Age-relate | 20-Jan | 0.083535 | 0.133045 | 0 | 0 | 12.17503 | 30.22444 |
| CellMarker | Schwalie Et | 3/188 | 0.04881 | 0.133738 | 0 | 0 | 3.808494 | 11.501 |
| GO_Biologi | Glycerophc | 3/108 | 0.01169 | 0.133867 | 0 | 0 | 6.737415 | 29.97504 |
| GWAS_Cat | Hemoglobi | 5/536 | 0.084327 | 0.133957 | 0 | 0 | 2.225667 | 5.504199 |
| GWAS_Cat | Alzheimer's | 2/111 | 0.084277 | 0.133957 | 0 | 0 | 4.275013 | 10.57487 |
| GWAS_Cat | Alzheimer? | 3/237 | 0.084577 | 0.134179 | 0 | 0 | 3.00351 | 7.418958 |
| DisGeNET | Ischemia | 3/104 | 0.010557 | 0.134269 | 0 | 0 | 7.005658 | 31.88247 |
| UK_Bioban | Major coro | 18-Jan | 0.075502 | 0.134487 | 0 | 0 | 13.60876 | 35.1596 |
| UK_Bioban | Lamb/mutt | 18-Jan | 0.075502 | 0.134487 | 0 | 0 | 13.60876 | 35.1596 |
| UK_Bioban | Self-report | 18-Jan | 0.075502 | 0.134487 | 0 | 0 | 13.60876 | 35.1596 |
| UK_Bioban | Major coro | 18-Jan | 0.075502 | 0.134487 | 0 | 0 | 13.60876 | 35.1596 |
| Jensen_DIS | Craniosyno | 20-Jan | 0.083535 | 0.134622 | 0 | 0 | 12.17503 | 30.22444 |
| Jensen_DIS | Aphasia | 20-Jan | 0.083535 | 0.134622 | 0 | 0 | 12.17503 | 30.22444 |
| Jensen_DIS | Steatorrhe | 20-Jan | 0.083535 | 0.134622 | 0 | 0 | 12.17503 | 30.22444 |
| Rare_Disea | Phenylketc | Feb-37 | 0.011286 | 0.135436 | 0 | 0 | 13.36336 | 59.92353 |
| Rare_Disea | Beta-thalas | 4/199 | 0.011142 | 0.135436 | 0 | 0 | 4.873154 | 21.91487 |
| DisGeNET | Peripheral | Feb-36 | 0.010706 | 0.135509 | 0 | 0 | 13.75709 | 62.41496 |
| CellMarker | Dendritic C | 12-Jan | 0.050982 | 0.136953 | 0 | 0 | 21.03805 | 62.61499 |
| CellMarker | Epithelial C | 12-Jan | 0.050982 | 0.136953 | 0 | 0 | 21.03805 | 62.61499 |
| Rare_Disea | Familial ve | Feb-38 | 0.01188 | 0.137164 | 0 | 0 | 12.9915 | 57.59009 |
| Rare_Disea | Pigment-di | 3/108 | 0.01169 | 0.137164 | 0 | 0 | 6.737415 | 29.97504 |
| Rare_Disea | Sudden Arr | 3/109 | 0.011983 | 0.137164 | 0 | 0 | 6.673518 | 29.52517 |
| Rare_Disea | Plasma thr | Feb-42 | 0.014388 | 0.137237 | 0 | 0 | 11.69 | 49.58172 |
| Rare_Disea | Acute myel | 5/328 | 0.014241 | 0.137237 | 0 | 0 | 3.69818 | 15.72324 |
| Rare_Disea | Congenital | 3/115 | 0.013835 | 0.137237 | 0 | 0 | 6.314094 | 27.02797 |

| | | | | | | | | |
|---|---|---|---|---|---|---|---|---|
| Rare_Disea | Bednar's tu | Feb-40 | 0.013107 | 0.137237 | 0 | 0 | 12.3065 | 53.34346 |
| Rare_Disea | Oculootofa | 3/113 | 0.013201 | 0.137237 | 0 | 0 | 6.429545 | 27.82383 |
| Rare_Disea | Dermatofik | Feb-40 | 0.013107 | 0.137237 | 0 | 0 | 12.3065 | 53.34346 |
| Rare_Disea | Opthalmor | 3/113 | 0.013201 | 0.137237 | 0 | 0 | 6.429545 | 27.82383 |
| Rare_Disea | Hemochroı | Feb-42 | 0.014388 | 0.137237 | 0 | 0 | 11.69 | 49.58172 |
| Rare_Disea | Myxoma-sı | 3/115 | 0.013835 | 0.137237 | 0 | 0 | 6.314094 | 27.02797 |
| Rare_Disea | Glaucoma i | 4/211 | 0.013567 | 0.137237 | 0 | 0 | 4.587859 | 19.72845 |
| Rare_Disea | Hemochroı | Feb-41 | 0.013741 | 0.137237 | 0 | 0 | 11.99035 | 51.40702 |
| Rare_Disea | Transpositi | Feb-41 | 0.013741 | 0.137237 | 0 | 0 | 11.99035 | 51.40702 |
| Rare_Disea | Infective m | 3/110 | 0.012281 | 0.137237 | 0 | 0 | 6.610814 | 29.08541 |
| PheWeb_2 | Seborrheic | 30-Jan | 0.122676 | 0.137308 | 0 | 0 | 7.972735 | 16.72843 |
| PheWeb_2 | Ulcerative | 27-Jan | 0.111111 | 0.137308 | 0 | 0 | 8.894007 | 19.54216 |
| PheWeb_2 | Encounter | 21-Jan | 0.087525 | 0.137308 | 0 | 0 | 11.5657 | 28.17205 |
| PheWeb_2 | Other disea | 26-Jan | 0.107222 | 0.137308 | 0 | 0 | 9.250233 | 20.65441 |
| PheWeb_2 | Premenstru | 21-Jan | 0.087525 | 0.137308 | 0 | 0 | 11.5657 | 28.17205 |
| PheWeb_2 | Noninflamı | 21-Jan | 0.087525 | 0.137308 | 0 | 0 | 11.5657 | 28.17205 |
| PheWeb_2 | Neutropen | 21-Jan | 0.087525 | 0.137308 | 0 | 0 | 11.5657 | 28.17205 |
| PheWeb_2 | Erectile dys | 21-Jan | 0.087525 | 0.137308 | 0 | 0 | 11.5657 | 28.17205 |
| PheWeb_2 | Osteoporo: | 25-Jan | 0.103317 | 0.137308 | 0 | 0 | 9.636143 | 21.87363 |
| PheWeb_2 | Degenerati | 26-Jan | 0.107222 | 0.137308 | 0 | 0 | 9.250233 | 20.65441 |
| PheWeb_2 | Delirium de | 26-Jan | 0.107222 | 0.137308 | 0 | 0 | 9.250233 | 20.65441 |
| PheWeb_2 | Hyperhidro | 30-Jan | 0.122676 | 0.137308 | 0 | 0 | 7.972735 | 16.72843 |
| PheWeb_2 | Otalgia | 25-Jan | 0.103317 | 0.137308 | 0 | 0 | 9.636143 | 21.87363 |
| PheWeb_2 | Papanicola | 22-Jan | 0.091499 | 0.137308 | 0 | 0 | 11.0144 | 26.34017 |
| PheWeb_2 | Diseases of | 26-Jan | 0.107222 | 0.137308 | 0 | 0 | 9.250233 | 20.65441 |
| PheWeb_2 | Known or s | 26-Jan | 0.107222 | 0.137308 | 0 | 0 | 9.250233 | 20.65441 |
| PheWeb_2 | Disorders c | 26-Jan | 0.107222 | 0.137308 | 0 | 0 | 9.250233 | 20.65441 |
| PheWeb_2 | Restless leg | 26-Jan | 0.107222 | 0.137308 | 0 | 0 | 9.250233 | 20.65441 |
| PheWeb_2 | Osteoporo: | 26-Jan | 0.107222 | 0.137308 | 0 | 0 | 9.250233 | 20.65441 |
| PheWeb_2 | Primary an | 19-Jan | 0.079527 | 0.137308 | 0 | 0 | 12.85207 | 32.53709 |
| PheWeb_2 | Cardiac shu | 19-Jan | 0.079527 | 0.137308 | 0 | 0 | 12.85207 | 32.53709 |
| PheWeb_2 | Other disoı | 20-Jan | 0.083535 | 0.137308 | 0 | 0 | 12.17503 | 30.22444 |
| PheWeb_2 | Aphasia | 20-Jan | 0.083535 | 0.137308 | 0 | 0 | 12.17503 | 30.22444 |
| PheWeb_2 | Retinal det | 13-Jan | 0.055113 | 0.137308 | 0 | 0 | 19.28391 | 55.89185 |
| PheWeb_2 | Abnormal f | 14-Jan | 0.059226 | 0.137308 | 0 | 0 | 17.79964 | 50.30878 |
| PheWeb_2 | Epistaxis oı | 20-Jan | 0.083535 | 0.137308 | 0 | 0 | 12.17503 | 30.22444 |
| PheWeb_2 | Rhesus isoi | 20-Jan | 0.083535 | 0.137308 | 0 | 0 | 12.17503 | 30.22444 |
| PheWeb_2 | Hyperpotas | 15-Jan | 0.063321 | 0.137308 | 0 | 0 | 16.52741 | 45.60793 |
| PheWeb_2 | Neuralgia, | 15-Jan | 0.063321 | 0.137308 | 0 | 0 | 16.52741 | 45.60793 |
| PheWeb_2 | Hereditary | 9-Jan | 0.038483 | 0.137308 | 0 | 0 | 28.93169 | 94.24608 |
| PheWeb_2 | Other endc | 9-Jan | 0.038483 | 0.137308 | 0 | 0 | 28.93169 | 94.24608 |
| PheWeb_2 | Epilepsy, re | 10-Jan | 0.042667 | 0.137308 | 0 | 0 | 25.71576 | 81.1157 |
| PheWeb_2 | Otitis medi | 13-Jan | 0.055113 | 0.137308 | 0 | 0 | 19.28391 | 55.89185 |
| PheWeb_2 | Polyneurop | 19-Jan | 0.079527 | 0.137308 | 0 | 0 | 12.85207 | 32.53709 |
| PheWeb_2 | Musculosk | 19-Jan | 0.079527 | 0.137308 | 0 | 0 | 12.85207 | 32.53709 |

| | | | | | | | |
|---|---|---|---|---|---|---|---|
| PheWeb_2 Arthropath | 20-Jan | 0.083535 | 0.137308 | 0 | 0 | 12.17503 | 30.22444 |
| PheWeb_2 Viral warts | 20-Jan | 0.083535 | 0.137308 | 0 | 0 | 12.17503 | 30.22444 |
| PheWeb_2 Liver replac | 14-Jan | 0.059226 | 0.137308 | 0 | 0 | 17.79964 | 50.30878 |
| PheWeb_2 Congenital | 18-Jan | 0.075502 | 0.137308 | 0 | 0 | 13.60876 | 35.1596 |
| PheWeb_2 Other local | 18-Jan | 0.075502 | 0.137308 | 0 | 0 | 13.60876 | 35.1596 |
| PheWeb_2 Other disor | 25-Jan | 0.103317 | 0.137308 | 0 | 0 | 9.636143 | 21.87363 |
| PheWeb_2 Cystitis anc | 25-Jan | 0.103317 | 0.137308 | 0 | 0 | 9.636143 | 21.87363 |
| PheWeb_2 Paralytic ile | 21-Jan | 0.087525 | 0.137308 | 0 | 0 | 11.5657 | 28.17205 |
| PheWeb_2 Bacterial in | 21-Jan | 0.087525 | 0.137308 | 0 | 0 | 11.5657 | 28.17205 |
| PheWeb_2 Peptic ulce | 21-Jan | 0.087525 | 0.137308 | 0 | 0 | 11.5657 | 28.17205 |
| PheWeb_2 Benign nec | 21-Jan | 0.087525 | 0.137308 | 0 | 0 | 11.5657 | 28.17205 |
| PheWeb_2 Cancer of k | 21-Jan | 0.087525 | 0.137308 | 0 | 0 | 11.5657 | 28.17205 |
| PheWeb_2 Portal hype | 17-Jan | 0.071459 | 0.137308 | 0 | 0 | 14.46003 | 38.15469 |
| PheWeb_2 Carcinoma | 27-Jan | 0.111111 | 0.137308 | 0 | 0 | 8.894007 | 19.54216 |
| PheWeb_2 Hidradeniti | 28-Jan | 0.114983 | 0.137308 | 0 | 0 | 8.564169 | 18.52408 |
| PheWeb_2 Other pulm | 19-Jan | 0.079527 | 0.137308 | 0 | 0 | 12.85207 | 32.53709 |
| PheWeb_2 Iron deficie | 25-Jan | 0.103317 | 0.137308 | 0 | 0 | 9.636143 | 21.87363 |
| PheWeb_2 Nontoxic n | 25-Jan | 0.103317 | 0.137308 | 0 | 0 | 9.636143 | 21.87363 |
| PheWeb_2 Retinal det | 25-Jan | 0.103317 | 0.137308 | 0 | 0 | 9.636143 | 21.87363 |
| PheWeb_2 Hyperplasia | 26-Jan | 0.107222 | 0.137308 | 0 | 0 | 9.250233 | 20.65441 |
| PheWeb_2 Lupus (loca | 17-Jan | 0.071459 | 0.137308 | 0 | 0 | 14.46003 | 38.15469 |
| PheWeb_2 Disorders c | 29-Jan | 0.118838 | 0.137308 | 0 | 0 | 8.25789 | 17.58927 |
| PheWeb_2 Other chro | 30-Jan | 0.122676 | 0.137308 | 0 | 0 | 7.972735 | 16.72843 |
| PheWeb_2 Bone cance | 23-Jan | 0.095455 | 0.137308 | 0 | 0 | 10.51321 | 24.6966 |
| PheWeb_2 Pneumonit | 23-Jan | 0.095455 | 0.137308 | 0 | 0 | 10.51321 | 24.6966 |
| PheWeb_2 Chronic Kic | 23-Jan | 0.095455 | 0.137308 | 0 | 0 | 10.51321 | 24.6966 |
| PheWeb_2 Chronic de | 23-Jan | 0.095455 | 0.137308 | 0 | 0 | 10.51321 | 24.6966 |
| PheWeb_2 Anorexia | 24-Jan | 0.099394 | 0.137308 | 0 | 0 | 10.05561 | 23.215 |
| PheWeb_2 Nonspecifi | 24-Jan | 0.099394 | 0.137308 | 0 | 0 | 10.05561 | 23.215 |
| PheWeb_2 Malaise an | 23-Jan | 0.095455 | 0.137308 | 0 | 0 | 10.51321 | 24.6966 |
| PheWeb_2 Disorders c | 23-Jan | 0.095455 | 0.137308 | 0 | 0 | 10.51321 | 24.6966 |
| PheWeb_2 Lipoma of s | 16-Jan | 0.067399 | 0.137308 | 0 | 0 | 15.42481 | 41.60265 |
| PheWeb_2 Symptoms | 17-Jan | 0.071459 | 0.137308 | 0 | 0 | 14.46003 | 38.15469 |
| PheWeb_2 Otitis medi | 17-Jan | 0.071459 | 0.137308 | 0 | 0 | 14.46003 | 38.15469 |
| PheWeb_2 Contractur | 16-Jan | 0.067399 | 0.137308 | 0 | 0 | 15.42481 | 41.60265 |
| PheWeb_2 Antepartur | 21-Jan | 0.087525 | 0.137308 | 0 | 0 | 11.5657 | 28.17205 |
| PheWeb_2 Iron deficie | 21-Jan | 0.087525 | 0.137308 | 0 | 0 | 11.5657 | 28.17205 |
| PheWeb_2 Temporom | 23-Jan | 0.095455 | 0.137308 | 0 | 0 | 10.51321 | 24.6966 |
| PheWeb_2 Uterine/Ut | 23-Jan | 0.095455 | 0.137308 | 0 | 0 | 10.51321 | 24.6966 |
| PheWeb_2 Oliguria an | 20-Jan | 0.083535 | 0.137308 | 0 | 0 | 12.17503 | 30.22444 |
| PheWeb_2 Chronic cys | 20-Jan | 0.083535 | 0.137308 | 0 | 0 | 12.17503 | 30.22444 |
| PheWeb_2 Sleep relat | 21-Jan | 0.087525 | 0.137308 | 0 | 0 | 11.5657 | 28.17205 |
| PheWeb_2 Iatrogenic I | 21-Jan | 0.087525 | 0.137308 | 0 | 0 | 11.5657 | 28.17205 |
| PheWeb_2 Disorders c | 19-Jan | 0.079527 | 0.137308 | 0 | 0 | 12.85207 | 32.53709 |
| PheWeb_2 Dysthymic | 19-Jan | 0.079527 | 0.137308 | 0 | 0 | 12.85207 | 32.53709 |

| | | | | | | | | |
|---|---|---|---|---|---|---|---|---|
| PheWeb_2 | Lipoma | 28-Jan | 0.114983 | 0.137308 | 0 | 0 | 8.564169 | 18.52408 |
| PheWeb_2 | Disorders c | 28-Jan | 0.114983 | 0.137308 | 0 | 0 | 8.564169 | 18.52408 |
| PheWeb_2 | Cirrhosis of | 29-Jan | 0.118838 | 0.137308 | 0 | 0 | 8.25789 | 17.58927 |
| PheWeb_2 | Coagulatio | 29-Jan | 0.118838 | 0.137308 | 0 | 0 | 8.25789 | 17.58927 |
| PheWeb_2 | Cystitis | 29-Jan | 0.118838 | 0.137308 | 0 | 0 | 8.25789 | 17.58927 |
| PheWeb_2 | Hypothyroi | 25-Jan | 0.103317 | 0.137308 | 0 | 0 | 9.636143 | 21.87363 |
| PheWeb_2 | Diseases of | 30-Jan | 0.122676 | 0.137308 | 0 | 0 | 7.972735 | 16.72843 |
| PheWeb_2 | Bursitis | 30-Jan | 0.122676 | 0.137308 | 0 | 0 | 7.972735 | 16.72843 |
| PheWeb_2 | Disorders c | 22-Jan | 0.091499 | 0.137308 | 0 | 0 | 11.0144 | 26.34017 |
| PheWeb_2 | Malignant | 22-Jan | 0.091499 | 0.137308 | 0 | 0 | 11.0144 | 26.34017 |
| PheWeb_2 | Chronic ulc | 23-Jan | 0.095455 | 0.137308 | 0 | 0 | 10.51321 | 24.6966 |
| PheWeb_2 | Staphyloco | 23-Jan | 0.095455 | 0.137308 | 0 | 0 | 10.51321 | 24.6966 |
| PheWeb_2 | Kidney rep | 22-Jan | 0.091499 | 0.137308 | 0 | 0 | 11.0144 | 26.34017 |
| PheWeb_2 | Abnormal r | 22-Jan | 0.091499 | 0.137308 | 0 | 0 | 11.0144 | 26.34017 |
| PheWeb_2 | Retinal vas | 21-Jan | 0.087525 | 0.137308 | 0 | 0 | 11.5657 | 28.17205 |
| PheWeb_2 | Liver absce | 26-Jan | 0.107222 | 0.137308 | 0 | 0 | 9.250233 | 20.65441 |
| GWAS_Cat | Maximum | 21-Jan | 0.087525 | 0.137604 | 0 | 0 | 11.5657 | 28.17205 |
| GWAS_Cat | Medicatior | 21-Jan | 0.087525 | 0.137604 | 0 | 0 | 11.5657 | 28.17205 |
| GWAS_Cat | HDL Choles | 21-Jan | 0.087525 | 0.137604 | 0 | 0 | 11.5657 | 28.17205 |
| GWAS_Cat | Ratio Of Sa | 21-Jan | 0.087525 | 0.137604 | 0 | 0 | 11.5657 | 28.17205 |
| GWAS_Cat | Optic Disc / | 21-Jan | 0.087525 | 0.137604 | 0 | 0 | 11.5657 | 28.17205 |
| GWAS_Cat | F-acquired | 21-Jan | 0.087525 | 0.137604 | 0 | 0 | 11.5657 | 28.17205 |
| GWAS_Cat | Valine Leve | 21-Jan | 0.087525 | 0.137604 | 0 | 0 | 11.5657 | 28.17205 |
| DisGeNET | Serum tota | 5/307 | 0.010952 | 0.137954 | 0 | 0 | 3.959578 | 17.87442 |
| dbGaP | Lipid Metal | 9-Jan | 0.038483 | 0.138966 | 0 | 0 | 28.93169 | 94.24608 |
| Disease_Pe | Congestive | 6/328 | 0.003083 | 0.139145 | 0 | 0 | 4.506786 | 26.05698 |
| Disease_Pe | facioscapul | 7/438 | 0.002988 | 0.139145 | 0 | 0 | 3.955162 | 22.99233 |
| Disease_Pe | Obesity C0 | 5/229 | 0.003251 | 0.139145 | 0 | 0 | 5.359593 | 30.70392 |
| GWAS_Cat | Mean Sphe | 5/545 | 0.089014 | 0.139765 | 0 | 0 | 2.187556 | 5.291603 |
| Jensen_DIS | Peutz-Jegh | 21-Jan | 0.087525 | 0.139897 | 0 | 0 | 11.5657 | 28.17205 |
| DisGeNET | Microstom | 3/107 | 0.0114 | 0.140229 | 0 | 0 | 6.802541 | 30.43536 |
| DisGeNET | Fibroid Tur | 5/310 | 0.011386 | 0.140229 | 0 | 0 | 3.920032 | 17.54347 |
| DisGeNET | Tubular Ag | Feb-37 | 0.011286 | 0.140229 | 0 | 0 | 13.36336 | 59.92353 |
| DisGeNET | Leukoaraio | Feb-37 | 0.011286 | 0.140229 | 0 | 0 | 13.36336 | 59.92353 |
| DisGeNET | Kidney Fail | 5/310 | 0.011386 | 0.140229 | 0 | 0 | 3.920032 | 17.54347 |
| PheWeb_2 | Degenerati | 31-Jan | 0.126498 | 0.140298 | 0 | 0 | 7.706589 | 15.93356 |
| GWAS_Cat | Eosinophil | 2/115 | 0.089505 | 0.140355 | 0 | 0 | 4.122853 | 9.950336 |
| GeDiPNet_ | Developme | 8-Jan | 0.03428 | 0.140743 | 0 | 0 | 33.06645 | 111.5391 |
| GeDiPNet_ | Hydranenc | 8-Jan | 0.03428 | 0.140743 | 0 | 0 | 33.06645 | 111.5391 |
| GeDiPNet_ | Coronary S | 8-Jan | 0.03428 | 0.140743 | 0 | 0 | 33.06645 | 111.5391 |
| GeDiPNet_ | Erythroker | 8-Jan | 0.03428 | 0.140743 | 0 | 0 | 33.06645 | 111.5391 |
| GeDiPNet_ | Shagreen P | 8-Jan | 0.03428 | 0.140743 | 0 | 0 | 33.06645 | 111.5391 |
| GeDiPNet_ | Motor Skill | 8-Jan | 0.03428 | 0.140743 | 0 | 0 | 33.06645 | 111.5391 |
| GeDiPNet_ | Sirenomeli | 8-Jan | 0.03428 | 0.140743 | 0 | 0 | 33.06645 | 111.5391 |
| GeDiPNet_ | Breast Carc | 8/859 | 0.033251 | 0.140743 | 0 | 0 | 2.268307 | 7.720554 |

| | | | | | | | | |
|---|---|---|---|---|---|---|---|---|
| GeDiPNet_ | Cerebral Ar | 8-Jan | 0.03428 | 0.140743 | 0 | 0 | 33.06645 | 111.5391 |
| CellMarker | FGFR1High | 3/195 | 0.05335 | 0.141131 | 0 | 0 | 3.668341 | 10.75147 |
| CellMarker | Fibroblast I | 13-Jan | 0.055113 | 0.141131 | 0 | 0 | 19.28391 | 55.89185 |
| CellMarker | Stem Cell E | 13-Jan | 0.055113 | 0.141131 | 0 | 0 | 19.28391 | 55.89185 |
| CellMarker | Sinoatrial N | 13-Jan | 0.055113 | 0.141131 | 0 | 0 | 19.28391 | 55.89185 |
| CellMarker | Stem Cell B | 13-Jan | 0.055113 | 0.141131 | 0 | 0 | 19.28391 | 55.89185 |
| Human_Ph | Craniofacia | Feb-29 | 0.007037 | 0.141473 | 0 | 0 | 17.32985 | 85.89758 |
| DisGeNET | Abnormalit | Feb-38 | 0.01188 | 0.141479 | 0 | 0 | 12.9915 | 57.59009 |
| DisGeNET | Coffin-Siris | Feb-38 | 0.01188 | 0.141479 | 0 | 0 | 12.9915 | 57.59009 |
| DisGeNET | Disorder of | Feb-38 | 0.01188 | 0.141479 | 0 | 0 | 12.9915 | 57.59009 |
| DisGeNET | Gross motc | Feb-38 | 0.01188 | 0.141479 | 0 | 0 | 12.9915 | 57.59009 |
| DisGeNET | Milroy Dise | Feb-38 | 0.01188 | 0.141479 | 0 | 0 | 12.9915 | 57.59009 |
| DisGeNET | Reticulocyt | 4/202 | 0.011719 | 0.141479 | 0 | 0 | 4.798588 | 21.33702 |
| DisGeNET | Arterioven | Feb-38 | 0.01188 | 0.141479 | 0 | 0 | 12.9915 | 57.59009 |
| GWAS_Cat | High-densi | 3/244 | 0.090403 | 0.141581 | 0 | 0 | 2.915234 | 7.00671 |
| PheWeb_2 | Symptoma | Jan-32 | 0.130304 | 0.141938 | 0 | 0 | 7.457614 | 15.19775 |
| PheWeb_2 | Hypercoag | Jan-32 | 0.130304 | 0.141938 | 0 | 0 | 7.457614 | 15.19775 |
| GWAS_Cat | Dupuytren | 22-Jan | 0.091499 | 0.142021 | 0 | 0 | 11.0144 | 26.34017 |
| GWAS_Cat | Prostate-sp | 22-Jan | 0.091499 | 0.142021 | 0 | 0 | 11.0144 | 26.34017 |
| GWAS_Cat | Osteoporo: | 22-Jan | 0.091499 | 0.142021 | 0 | 0 | 11.0144 | 26.34017 |
| GWAS_Cat | Birth Lengt | 22-Jan | 0.091499 | 0.142021 | 0 | 0 | 11.0144 | 26.34017 |
| GWAS_Cat | Glycated H | 22-Jan | 0.091499 | 0.142021 | 0 | 0 | 11.0144 | 26.34017 |
| GWAS_Cat | Phospholip | 22-Jan | 0.091499 | 0.142021 | 0 | 0 | 11.0144 | 26.34017 |
| GWAS_Cat | Facial Mor | 22-Jan | 0.091499 | 0.142021 | 0 | 0 | 11.0144 | 26.34017 |
| PhenGenI_ | Atrial Fibril | 3/108 | 0.01169 | 0.142123 | 0 | 0 | 6.737415 | 29.97504 |
| PhenGenI_ | Glomerulo | Feb-37 | 0.011286 | 0.142123 | 0 | 0 | 13.36336 | 59.92353 |
| Rare_Disea | Human T-c | 3/119 | 0.015154 | 0.142718 | 0 | 0 | 6.095135 | 25.53542 |
| PhenGenI_ | Proteins | 3/116 | 0.014158 | 0.143031 | 0 | 0 | 6.257901 | 26.64281 |
| PhenGenI_ | Macular De | 5/328 | 0.014241 | 0.143031 | 0 | 0 | 3.69818 | 15.72324 |
| PhenGenI_ | Paliperidor | 4/209 | 0.013141 | 0.143031 | 0 | 0 | 4.633088 | 20.0706 |
| PhenGenI_ | Neoplasms | 3/113 | 0.013201 | 0.143031 | 0 | 0 | 6.429545 | 27.82383 |
| CellMarker | Conventior | 15-Jan | 0.063321 | 0.143389 | 0 | 0 | 16.52741 | 45.60793 |
| CellMarker | Leukemic S | 14-Jan | 0.059226 | 0.143389 | 0 | 0 | 17.79964 | 50.30878 |
| CellMarker | Oviduct-de | 15-Jan | 0.063321 | 0.143389 | 0 | 0 | 16.52741 | 45.60793 |
| CellMarker | Endothelia | 14-Jan | 0.059226 | 0.143389 | 0 | 0 | 17.79964 | 50.30878 |
| CellMarker | Macrophag | 14-Jan | 0.059226 | 0.143389 | 0 | 0 | 17.79964 | 50.30878 |
| CellMarker | Epithelial C | 15-Jan | 0.063321 | 0.143389 | 0 | 0 | 16.52741 | 45.60793 |
| CellMarker | Pericyte Br | 15-Jan | 0.063321 | 0.143389 | 0 | 0 | 16.52741 | 45.60793 |
| CellMarker | Astrocyte E | 4/343 | 0.062611 | 0.143389 | 0 | 0 | 2.782671 | 7.710271 |
| CellMarker | Axin2+ Cell | 15-Jan | 0.063321 | 0.143389 | 0 | 0 | 16.52741 | 45.60793 |
| CellMarker | Endothelia | 3/207 | 0.061589 | 0.143389 | 0 | 0 | 3.450455 | 9.617355 |
| CellMarker | Monocyte | 15-Jan | 0.063321 | 0.143389 | 0 | 0 | 16.52741 | 45.60793 |
| CellMarker | Astrocyte E | 3/204 | 0.059476 | 0.143389 | 0 | 0 | 3.502488 | 9.88465 |
| CellMarker | Endothelia | 14-Jan | 0.059226 | 0.143389 | 0 | 0 | 17.79964 | 50.30878 |
| CellMarker | Stem Cell C | 15-Jan | 0.063321 | 0.143389 | 0 | 0 | 16.52741 | 45.60793 |

| | | | | | | | | |
|---|---|---|---|---|---|---|---|---|
| PheWeb_2 | Hearing los | Jan-33 | 0.134093 | 0.143503 | 0 | 0 | 7.224201 | 14.515 |
| PheWeb_2 | Abnormal s | Jan-33 | 0.134093 | 0.143503 | 0 | 0 | 7.224201 | 14.515 |
| Human_Ph | Generalize | Feb-30 | 0.007518 | 0.143595 | 0 | 0 | 16.71008 | 81.71977 |
| GWAS_Cat | QT Interval | 3/247 | 0.09295 | 0.14409 | 0 | 0 | 2.878952 | 6.839504 |
| GeDiPNet_ | Malignant | 9-Jan | 0.038483 | 0.144343 | 0 | 0 | 28.93169 | 94.24608 |
| GeDiPNet_ | Epidermal | 9-Jan | 0.038483 | 0.144343 | 0 | 0 | 28.93169 | 94.24608 |
| GeDiPNet_ | Malignant | 9-Jan | 0.038483 | 0.144343 | 0 | 0 | 28.93169 | 94.24608 |
| GeDiPNet_ | Malignant | 9-Jan | 0.038483 | 0.144343 | 0 | 0 | 28.93169 | 94.24608 |
| GeDiPNet_ | Cowden Sy | 9-Jan | 0.038483 | 0.144343 | 0 | 0 | 28.93169 | 94.24608 |
| GeDiPNet_ | Malignant | 9-Jan | 0.038483 | 0.144343 | 0 | 0 | 28.93169 | 94.24608 |
| GeDiPNet_ | Lymphangi | 9-Jan | 0.038483 | 0.144343 | 0 | 0 | 28.93169 | 94.24608 |
| GeDiPNet_ | Liver Failur | 3/171 | 0.038623 | 0.144343 | 0 | 0 | 4.197491 | 13.65819 |
| GeDiPNet_ | Carotid Art | 9-Jan | 0.038483 | 0.144343 | 0 | 0 | 28.93169 | 94.24608 |
| GeDiPNet_ | Limb Amyc | 9-Jan | 0.038483 | 0.144343 | 0 | 0 | 28.93169 | 94.24608 |
| GeDiPNet_ | Xanthoma | 9-Jan | 0.038483 | 0.144343 | 0 | 0 | 28.93169 | 94.24608 |
| GeDiPNet_ | Polymicrog | 3/168 | 0.036952 | 0.144343 | 0 | 0 | 4.274459 | 14.09775 |
| GeDiPNet_ | Tumors Of | 9-Jan | 0.038483 | 0.144343 | 0 | 0 | 28.93169 | 94.24608 |
| GeDiPNet_ | Nephrobla | Feb-69 | 0.03636 | 0.144343 | 0 | 0 | 6.969622 | 23.09942 |
| DisGeNET | Bone Dens | 3/110 | 0.012281 | 0.145598 | 0 | 0 | 6.610814 | 29.08541 |
| DisGeNET | Anosmia | Feb-39 | 0.012487 | 0.146054 | 0 | 0 | 12.63975 | 55.40083 |
| DisGeNET | Sense of sn | Feb-39 | 0.012487 | 0.146054 | 0 | 0 | 12.63975 | 55.40083 |
| DisGeNET | Alcohol cor | Feb-39 | 0.012487 | 0.146054 | 0 | 0 | 12.63975 | 55.40083 |
| GWAS_Cat | Caffeine Cc | 23-Jan | 0.095455 | 0.146117 | 0 | 0 | 10.51321 | 24.6966 |
| GWAS_Cat | Parental Lc | 23-Jan | 0.095455 | 0.146117 | 0 | 0 | 10.51321 | 24.6966 |
| GWAS_Cat | Neuritic Pla | 23-Jan | 0.095455 | 0.146117 | 0 | 0 | 10.51321 | 24.6966 |
| GWAS_Cat | Leucine Lev | 23-Jan | 0.095455 | 0.146117 | 0 | 0 | 10.51321 | 24.6966 |
| GWAS_Cat | Alzheimer's | 23-Jan | 0.095455 | 0.146117 | 0 | 0 | 10.51321 | 24.6966 |
| GWAS_Cat | Abdominal | 23-Jan | 0.095455 | 0.146117 | 0 | 0 | 10.51321 | 24.6966 |
| GWAS_Cat | Ratio Of Or | 23-Jan | 0.095455 | 0.146117 | 0 | 0 | 10.51321 | 24.6966 |
| GWAS_Cat | Verbal Lear | 23-Jan | 0.095455 | 0.146117 | 0 | 0 | 10.51321 | 24.6966 |
| GWAS_Cat | Polycystic ( | 23-Jan | 0.095455 | 0.146117 | 0 | 0 | 10.51321 | 24.6966 |
| GWAS_Cat | Male Pube | 23-Jan | 0.095455 | 0.146117 | 0 | 0 | 10.51321 | 24.6966 |
| KEGG_202 | Compleme | Mar-85 | 0.006074 | 0.14622 | 0 | 0 | 8.637195 | 44.08228 |
| KEGG_202 | Phosphatic | Mar-97 | 0.008735 | 0.14622 | 0 | 0 | 7.530015 | 35.6954 |
| KEGG_202 | Cardiac mu | Mar-87 | 0.006478 | 0.14622 | 0 | 0 | 8.430697 | 42.48559 |
| KEGG_202 | Hematopoi | Mar-99 | 0.009235 | 0.14622 | 0 | 0 | 7.372396 | 34.53788 |
| GTEx_Tissu | GTEX-U4B1 | 18/1711 | 0.000349 | 0.146409 | 0 | 0 | 2.807468 | 22.34652 |
| GTEx_Tissu | GTEX-VUS( | 18/1713 | 0.000354 | 0.146409 | 0 | 0 | 2.803848 | 22.27795 |
| GTEx_Tissu | GTEX-WFG | 17/1530 | 0.000278 | 0.146409 | 0 | 0 | 2.953451 | 24.17891 |
| DisGeNET | Physical Ac | 3/111 | 0.012584 | 0.146528 | 0 | 0 | 6.549272 | 28.65547 |
| Jensen_DIS | Ankylosis | 23-Jan | 0.095455 | 0.146565 | 0 | 0 | 10.51321 | 24.6966 |
| Jensen_DIS | Meniere's ( | 23-Jan | 0.095455 | 0.146565 | 0 | 0 | 10.51321 | 24.6966 |
| Jensen_DIS | Atypical au | 23-Jan | 0.095455 | 0.146565 | 0 | 0 | 10.51321 | 24.6966 |
| Jensen_DIS | Omphaloce | 23-Jan | 0.095455 | 0.146565 | 0 | 0 | 10.51321 | 24.6966 |
| Jensen_DIS | Polycythen | 23-Jan | 0.095455 | 0.146565 | 0 | 0 | 10.51321 | 24.6966 |

| | | | | | | | | |
|---|---|---|---|---|---|---|---|---|
| Jensen_DIS | Hypotricho | 24-Jan | 0.099394 | 0.146832 | 0 | 0 | 10.05561 | 23.215 |
| Jensen_DIS | Mastocytos | 24-Jan | 0.099394 | 0.146832 | 0 | 0 | 10.05561 | 23.215 |
| Jensen_DIS | Colitis | 24-Jan | 0.099394 | 0.146832 | 0 | 0 | 10.05561 | 23.215 |
| Jensen_DIS | Leiomyoma | 24-Jan | 0.099394 | 0.146832 | 0 | 0 | 10.05561 | 23.215 |
| Jensen_DIS | Connective | 24-Jan | 0.099394 | 0.146832 | 0 | 0 | 10.05561 | 23.215 |
| UK_Bioban | Medicatior | 20-Jan | 0.083535 | 0.146959 | 0 | 0 | 12.17503 | 30.22444 |
| UK_Bioban | Diabetes in | 20-Jan | 0.083535 | 0.146959 | 0 | 0 | 12.17503 | 30.22444 |
| CellMarker | B Cell Unde | Feb-96 | 0.065593 | 0.147315 | 0 | 0 | 4.960951 | 13.51507 |
| PheWeb_2 | Primary hy | Jan-35 | 0.141623 | 0.147675 | 0 | 0 | 6.798564 | 13.28839 |
| PheWeb_2 | Diseases of | Jan-35 | 0.141623 | 0.147675 | 0 | 0 | 6.798564 | 13.28839 |
| PheWeb_2 | Symptoms | Jan-35 | 0.141623 | 0.147675 | 0 | 0 | 6.798564 | 13.28839 |
| Jensen_TIS | ROS cell | 5-Jan | 0.021564 | 0.147733 | 0 | 0 | 57.875 | 222.0519 |
| Jensen_TIS | HTC cell | 5-Jan | 0.021564 | 0.147733 | 0 | 0 | 57.875 | 222.0519 |
| Jensen_TIS | Nerve | 3/118 | 0.014818 | 0.147733 | 0 | 0 | 6.148447 | 25.89677 |
| Jensen_TIS | H9 cell | 5-Jan | 0.021564 | 0.147733 | 0 | 0 | 57.875 | 222.0519 |
| Jensen_TIS | Cerebral co | 77/15695 | 0.011661 | 0.147733 | 0 | 0 | 2.117525 | 9.4261 |
| Jensen_TIS | Vascular ce | Feb-43 | 0.015047 | 0.147733 | 0 | 0 | 11.4043 | 47.85875 |
| Jensen_TIS | MC-3T3 cel | 5-Jan | 0.021564 | 0.147733 | 0 | 0 | 57.875 | 222.0519 |
| Jensen_TIS | Breast | 63/12084 | 0.013154 | 0.147733 | 0 | 0 | 1.723359 | 7.463916 |
| Jensen_TIS | pa317 | 5-Jan | 0.021564 | 0.147733 | 0 | 0 | 57.875 | 222.0519 |
| Jensen_TIS | Esophagus | 76/15560 | 0.017231 | 0.147733 | 0 | 0 | 1.976257 | 8.025639 |
| Jensen_TIS | Skeletal mu | 75/15243 | 0.015398 | 0.147733 | 0 | 0 | 1.955185 | 8.160011 |
| Jensen_TIS | Blood vess | 3/124 | 0.016901 | 0.147733 | 0 | 0 | 5.841795 | 23.83691 |
| Jensen_TIS | Osteocyte | Feb-43 | 0.015047 | 0.147733 | 0 | 0 | 11.4043 | 47.85875 |
| Jensen_TIS | HIT-T15 cel | 5-Jan | 0.021564 | 0.147733 | 0 | 0 | 57.875 | 222.0519 |
| Jensen_TIS | Tube foot | 5-Jan | 0.021564 | 0.147733 | 0 | 0 | 57.875 | 222.0519 |
| Jensen_TIS | Medial lon | 5-Jan | 0.021564 | 0.147733 | 0 | 0 | 57.875 | 222.0519 |
| Jensen_TIS | Yolk sac | Feb-43 | 0.015047 | 0.147733 | 0 | 0 | 11.4043 | 47.85875 |
| Jensen_TIS | Mouth | 10/1073 | 0.017937 | 0.147733 | 0 | 0 | 2.302965 | 9.260013 |
| Jensen_TIS | Cerebrospi | Feb-44 | 0.01572 | 0.147733 | 0 | 0 | 11.13221 | 46.2302 |
| Jensen_TIS | Rosette | Feb-44 | 0.01572 | 0.147733 | 0 | 0 | 11.13221 | 46.2302 |
| Jensen_TIS | Acute lymp | Feb-51 | 0.020776 | 0.147733 | 0 | 0 | 9.538535 | 36.95177 |
| Jensen_TIS | Lymph nod | 73/14834 | 0.021332 | 0.147733 | 0 | 0 | 1.819931 | 7.002233 |
| Jensen_TIS | Uterine gla | 5-Jan | 0.021564 | 0.147733 | 0 | 0 | 57.875 | 222.0519 |
| CellMarker | Pit Cell Sto | 16-Jan | 0.067399 | 0.147738 | 0 | 0 | 15.42481 | 41.60265 |
| CellMarker | Endothelia | 16-Jan | 0.067399 | 0.147738 | 0 | 0 | 15.42481 | 41.60265 |
| CellMarker | Cardiomyo | 16-Jan | 0.067399 | 0.147738 | 0 | 0 | 15.42481 | 41.60265 |
| PheWeb_2 | Actinic ker | Jan-36 | 0.145363 | 0.147786 | 0 | 0 | 6.603987 | 12.73591 |
| PheWeb_2 | Cancer of p | Jan-36 | 0.145363 | 0.147786 | 0 | 0 | 6.603987 | 12.73591 |
| PheWeb_2 | Hypothyroi | Jan-36 | 0.145363 | 0.147786 | 0 | 0 | 6.603987 | 12.73591 |
| CellMarker | Conventior | 17-Jan | 0.071459 | 0.148332 | 0 | 0 | 14.46003 | 38.15469 |
| CellMarker | Delta Cell F | 17-Jan | 0.071459 | 0.148332 | 0 | 0 | 14.46003 | 38.15469 |
| CellMarker | Adipose-de | 17-Jan | 0.071459 | 0.148332 | 0 | 0 | 14.46003 | 38.15469 |
| CellMarker | Stromal Ce | 17-Jan | 0.071459 | 0.148332 | 0 | 0 | 14.46003 | 38.15469 |
| CellMarker | Mesenchyr | 17-Jan | 0.071459 | 0.148332 | 0 | 0 | 14.46003 | 38.15469 |

| | | | | | | | | |
|---|---|---|---|---|---|---|---|---|
| CellMarker | Osteocyte | 17-Jan | 0.071459 | 0.148332 | 0 | 0 | 14.46003 | 38.15469 |
| CellMarker | Endothelia | 17-Jan | 0.071459 | 0.148332 | 0 | 0 | 14.46003 | 38.15469 |
| GTEx_Tissu | GTEX-PWO | 17/1582 | 0.000411 | 0.148621 | 0 | 0 | 2.847248 | 22.19988 |
| DisGeNET | Muscular D | 3/112 | 0.01289 | 0.148773 | 0 | 0 | 6.48886 | 28.23504 |
| DisGeNET | Glomerulo: | 3/112 | 0.01289 | 0.148773 | 0 | 0 | 6.48886 | 28.23504 |
| GWAS_Cat | Medicatior | 2/121 | 0.097522 | 0.149094 | 0 | 0 | 3.913791 | 9.110048 |
| Jensen_DIS | Leukodystr | 25-Jan | 0.103317 | 0.149235 | 0 | 0 | 9.636143 | 21.87363 |
| Jensen_DIS | Familial ad | 25-Jan | 0.103317 | 0.149235 | 0 | 0 | 9.636143 | 21.87363 |
| Jensen_DIS | Wiskott-Al | 25-Jan | 0.103317 | 0.149235 | 0 | 0 | 9.636143 | 21.87363 |
| Jensen_DIS | Toxic ence | 2/126 | 0.104352 | 0.149623 | 0 | 0 | 3.755028 | 8.486292 |
| DisGeNET | Motor sym | Feb-40 | 0.013107 | 0.149963 | 0 | 0 | 12.3065 | 53.34346 |
| DisGeNET | Psychiatric | Feb-40 | 0.013107 | 0.149963 | 0 | 0 | 12.3065 | 53.34346 |
| Rare_Disea | Kimura dis | 3/123 | 0.016543 | 0.150135 | 0 | 0 | 5.890774 | 24.16285 |
| Rare_Disea | Encephaloc | Feb-45 | 0.016405 | 0.150135 | 0 | 0 | 10.87278 | 44.68894 |
| Rare_Disea | Non-alcoho | 4/224 | 0.016547 | 0.150135 | 0 | 0 | 4.31391 | 17.69367 |
| CellMarker | CD141+CLE | 2/102 | 0.072883 | 0.150149 | 0 | 0 | 4.661882 | 12.20902 |
| GWAS_Cat | Depression | 24-Jan | 0.099394 | 0.150261 | 0 | 0 | 10.05561 | 23.215 |
| GWAS_Cat | Subcutane | 24-Jan | 0.099394 | 0.150261 | 0 | 0 | 10.05561 | 23.215 |
| GWAS_Cat | Premature | 24-Jan | 0.099394 | 0.150261 | 0 | 0 | 10.05561 | 23.215 |
| GWAS_Cat | HIV-1 Susc | 24-Jan | 0.099394 | 0.150261 | 0 | 0 | 10.05561 | 23.215 |
| GWAS_Cat | Hematolog | 24-Jan | 0.099394 | 0.150261 | 0 | 0 | 10.05561 | 23.215 |
| GWAS_Cat | Medicatior | 24-Jan | 0.099394 | 0.150261 | 0 | 0 | 10.05561 | 23.215 |
| GWAS_Cat | Renal Cell ( | 24-Jan | 0.099394 | 0.150261 | 0 | 0 | 10.05561 | 23.215 |
| GWAS_Cat | COVID-19 [ | 24-Jan | 0.099394 | 0.150261 | 0 | 0 | 10.05561 | 23.215 |
| GWAS_Cat | COVID-19 [ | 24-Jan | 0.099394 | 0.150261 | 0 | 0 | 10.05561 | 23.215 |
| PheWeb_2 | Early or thr | Jan-37 | 0.149088 | 0.15032 | 0 | 0 | 6.42022 | 12.21909 |
| UK_Bioban | Salad/raw | 21-Jan | 0.087525 | 0.151018 | 0 | 0 | 11.5657 | 28.17205 |
| UK_Bioban | Pulse wave | 23/3995 | 0.087421 | 0.151018 | 0 | 0 | 1.442295 | 3.514898 |
| UK_Bioban | Diabetes in | 21-Jan | 0.087525 | 0.151018 | 0 | 0 | 11.5657 | 28.17205 |
| UK_Bioban | Whole bod | 5/543 | 0.087961 | 0.151018 | 0 | 0 | 2.195915 | 5.337964 |
| DisGeNET | Glomerulo | 4/210 | 0.013353 | 0.152105 | 0 | 0 | 4.610364 | 19.89848 |
| dbGaP | Apolipopro | 11-Jan | 0.046834 | 0.15221 | 0 | 0 | 23.14302 | 70.84419 |
| dbGaP | Memory | 11-Jan | 0.046834 | 0.15221 | 0 | 0 | 23.14302 | 70.84419 |
| DisGeNET | Myocardial | 6/448 | 0.013441 | 0.152449 | 0 | 0 | 3.263114 | 14.06218 |
| GeDiPNet_ | Oropharyn | 10-Jan | 0.042667 | 0.152608 | 0 | 0 | 25.71576 | 81.1157 |
| GeDiPNet_ | Conjunctiv | 10-Jan | 0.042667 | 0.152608 | 0 | 0 | 25.71576 | 81.1157 |
| GeDiPNet_ | Polydactyl | 3/176 | 0.041494 | 0.152608 | 0 | 0 | 4.075145 | 12.96795 |
| GeDiPNet_ | Ramer Lad | 10-Jan | 0.042667 | 0.152608 | 0 | 0 | 25.71576 | 81.1157 |
| GeDiPNet_ | Subcutane | 10-Jan | 0.042667 | 0.152608 | 0 | 0 | 25.71576 | 81.1157 |
| GeDiPNet_ | Angioid Str | 10-Jan | 0.042667 | 0.152608 | 0 | 0 | 25.71576 | 81.1157 |
| GeDiPNet_ | Brachydact | 4/300 | 0.041941 | 0.152608 | 0 | 0 | 3.193911 | 10.12943 |
| HumanCyc | tRNA charg | Jan-38 | 0.152796 | 0.152796 | 0 | 0 | 6.246386 | 11.73478 |
| UK_Bioban | Left spheri | 2/117 | 0.092155 | 0.152797 | 0 | 0 | 4.050742 | 9.658129 |
| UK_Bioban | Body fat pe | 5/551 | 0.092214 | 0.152797 | 0 | 0 | 2.162847 | 5.155447 |
| UK_Bioban | Bread intal | 22-Jan | 0.091499 | 0.152797 | 0 | 0 | 11.0144 | 26.34017 |

| | | | | | | | | |
|---|---|---|---|---|---|---|---|---|
| UK_Bioban | High blood | 22-Jan | 0.091499 | 0.152797 | 0 | 0 | 11.0144 | 26.34017 |
| UK_Bioban | Self-report | 22-Jan | 0.091499 | 0.152797 | 0 | 0 | 11.0144 | 26.34017 |
| UK_Bioban | Self-report | 22-Jan | 0.091499 | 0.152797 | 0 | 0 | 11.0144 | 26.34017 |
| DisGeNET | High densit | 5/324 | 0.013569 | 0.153237 | 0 | 0 | 3.745317 | 16.10472 |
| CellMarker | Endometri | 18-Jan | 0.075502 | 0.15324 | 0 | 0 | 13.60876 | 35.1596 |
| CellMarker | Plasmacyto | 18-Jan | 0.075502 | 0.15324 | 0 | 0 | 13.60876 | 35.1596 |
| OMIM_Dis | systemic lu | 28-Jan | 0.114983 | 0.153346 | 0 | 0 | 8.564169 | 18.52408 |
| OMIM_Dis | prostate ca | 30-Jan | 0.122676 | 0.153346 | 0 | 0 | 7.972735 | 16.72843 |
| OMIM_Dis | alzheimer c | 26-Jan | 0.107222 | 0.153346 | 0 | 0 | 9.250233 | 20.65441 |
| DisGeNET | Nephrotic S | Feb-41 | 0.013741 | 0.153588 | 0 | 0 | 11.99035 | 51.40702 |
| DisGeNET | Malignant | Feb-41 | 0.013741 | 0.153588 | 0 | 0 | 11.99035 | 51.40702 |
| DisGeNET | Carcinoma, | 3/115 | 0.013835 | 0.153588 | 0 | 0 | 6.314094 | 27.02797 |
| DisGeNET | Carcinoma, | 3/115 | 0.013835 | 0.153588 | 0 | 0 | 6.314094 | 27.02797 |
| CellMarker | Oligodendr | 2/110 | 0.082985 | 0.153614 | 0 | 0 | 4.314815 | 10.73997 |
| CellMarker | Axin2-Palpl | 19-Jan | 0.079527 | 0.153614 | 0 | 0 | 12.85207 | 32.53709 |
| CellMarker | Red Blood | 20-Jan | 0.083535 | 0.153614 | 0 | 0 | 12.17503 | 30.22444 |
| CellMarker | MKI67+ Pro | 2/110 | 0.082985 | 0.153614 | 0 | 0 | 4.314815 | 10.73997 |
| CellMarker | Macrophag | 19-Jan | 0.079527 | 0.153614 | 0 | 0 | 12.85207 | 32.53709 |
| CellMarker | Mesenchyr | 19-Jan | 0.079527 | 0.153614 | 0 | 0 | 12.85207 | 32.53709 |
| CellMarker | Macrophag | 20-Jan | 0.083535 | 0.153614 | 0 | 0 | 12.17503 | 30.22444 |
| CellMarker | Mesenchyr | 20-Jan | 0.083535 | 0.153614 | 0 | 0 | 12.17503 | 30.22444 |
| CellMarker | Stem Cell U | 19-Jan | 0.079527 | 0.153614 | 0 | 0 | 12.85207 | 32.53709 |
| CellMarker | Endothelia | 20-Jan | 0.083535 | 0.153614 | 0 | 0 | 12.17503 | 30.22444 |
| CellMarker | Astrocyte H | 19-Jan | 0.079527 | 0.153614 | 0 | 0 | 12.85207 | 32.53709 |
| CellMarker | Mesenchyr | 20-Jan | 0.083535 | 0.153614 | 0 | 0 | 12.17503 | 30.22444 |
| CellMarker | Mesenchyr | 20-Jan | 0.083535 | 0.153614 | 0 | 0 | 12.17503 | 30.22444 |
| CellMarker | Mast Cell U | 20-Jan | 0.083535 | 0.153614 | 0 | 0 | 12.17503 | 30.22444 |
| DisGeNET | Big calvaria | 4/213 | 0.014001 | 0.154127 | 0 | 0 | 4.543495 | 19.39453 |
| DisGeNET | Increased s | 4/213 | 0.014001 | 0.154127 | 0 | 0 | 4.543495 | 19.39453 |
| Jensen_DIS | Osteoporos | 2/130 | 0.109909 | 0.154632 | 0 | 0 | 3.636949 | 8.030761 |
| Jensen_DIS | ENSP0000C | 28-Jan | 0.114983 | 0.154632 | 0 | 0 | 8.564169 | 18.52408 |
| Jensen_DIS | Age relatec | 28-Jan | 0.114983 | 0.154632 | 0 | 0 | 8.564169 | 18.52408 |
| Jensen_DIS | Amyloidosi | 27-Jan | 0.111111 | 0.154632 | 0 | 0 | 8.894007 | 19.54216 |
| Jensen_DIS | Hyperhom | 27-Jan | 0.111111 | 0.154632 | 0 | 0 | 8.894007 | 19.54216 |
| Jensen_DIS | Primary cu | 28-Jan | 0.114983 | 0.154632 | 0 | 0 | 8.564169 | 18.52408 |
| Jensen_DIS | maturity-o | 28-Jan | 0.114983 | 0.154632 | 0 | 0 | 8.564169 | 18.52408 |
| Jensen_DIS | Turner syn | 28-Jan | 0.114983 | 0.154632 | 0 | 0 | 8.564169 | 18.52408 |
| Jensen_DIS | Glycine enc | 27-Jan | 0.111111 | 0.154632 | 0 | 0 | 8.894007 | 19.54216 |
| OMIM_Dis | muscular d | Jan-33 | 0.134093 | 0.154723 | 0 | 0 | 7.224201 | 14.515 |
| DisGeNET | Adenocarci | 3/117 | 0.014486 | 0.15491 | 0 | 0 | 6.202694 | 26.26582 |
| DisGeNET | Diastolic bl | 3/117 | 0.014486 | 0.15491 | 0 | 0 | 6.202694 | 26.26582 |
| DisGeNET | Hyperhom | 3/117 | 0.014486 | 0.15491 | 0 | 0 | 6.202694 | 26.26582 |
| DisGeNET | Reduced bo | Feb-42 | 0.014388 | 0.15491 | 0 | 0 | 11.69 | 49.58172 |
| DisGeNET | Ischemic ca | 3/116 | 0.014158 | 0.15491 | 0 | 0 | 6.257901 | 26.64281 |
| DisGeNET | Increased h | 4/215 | 0.014444 | 0.15491 | 0 | 0 | 4.499971 | 19.06857 |

| | | | | | | | | |
|---|---|---|---|---|---|---|---|---|
| DisGeNET | Parkinsonia | 4/214 | 0.014221 | 0.15491 | 0 | 0 | 4.521629 | 19.23057 |
| GeDiPNet_ | Fibroadenc | 11-Jan | 0.046834 | 0.155137 | 0 | 0 | 23.14302 | 70.84419 |
| GeDiPNet_ | Noncompa | 11-Jan | 0.046834 | 0.155137 | 0 | 0 | 23.14302 | 70.84419 |
| GeDiPNet_ | Language D | 11-Jan | 0.046834 | 0.155137 | 0 | 0 | 23.14302 | 70.84419 |
| GeDiPNet_ | Fingernail D | 11-Jan | 0.046834 | 0.155137 | 0 | 0 | 23.14302 | 70.84419 |
| GeDiPNet_ | Parietal For | 11-Jan | 0.046834 | 0.155137 | 0 | 0 | 23.14302 | 70.84419 |
| GeDiPNet_ | Hyperhom | 11-Jan | 0.046834 | 0.155137 | 0 | 0 | 23.14302 | 70.84419 |
| GeDiPNet_ | Renal Steat | 11-Jan | 0.046834 | 0.155137 | 0 | 0 | 23.14302 | 70.84419 |
| GeDiPNet_ | Manic Diso | Feb-79 | 0.046446 | 0.155137 | 0 | 0 | 6.061421 | 18.60534 |
| GeDiPNet_ | Mitral Valv | Feb-79 | 0.046446 | 0.155137 | 0 | 0 | 6.061421 | 18.60534 |
| GeDiPNet_ | Ductal Carc | 11-Jan | 0.046834 | 0.155137 | 0 | 0 | 23.14302 | 70.84419 |
| GeDiPNet_ | Skin Diseas | 11-Jan | 0.046834 | 0.155137 | 0 | 0 | 23.14302 | 70.84419 |
| GeDiPNet_ | Arcus Senil | 11-Jan | 0.046834 | 0.155137 | 0 | 0 | 23.14302 | 70.84419 |
| GeDiPNet_ | Cortical Dy | Feb-78 | 0.045395 | 0.155137 | 0 | 0 | 6.141486 | 18.99159 |
| GWAS_Cat | Cryptic Phe | 25-Jan | 0.103317 | 0.155613 | 0 | 0 | 9.636143 | 21.87363 |
| GWAS_Cat | Parental Lo | 25-Jan | 0.103317 | 0.155613 | 0 | 0 | 9.636143 | 21.87363 |
| GWAS_Cat | Meat-relat | 25-Jan | 0.103317 | 0.155613 | 0 | 0 | 9.636143 | 21.87363 |
| OMIM_Dis | blood | Jan-36 | 0.145363 | 0.155747 | 0 | 0 | 6.603987 | 12.73591 |
| Jensen_TIS | Filament | 3/140 | 0.023218 | 0.155755 | 0 | 0 | 5.15537 | 19.39875 |
| dbGaP | Lung Neop | 12-Jan | 0.050982 | 0.155755 | 0 | 0 | 21.03805 | 62.61499 |
| dbGaP | Glucose To | 13-Jan | 0.055113 | 0.155755 | 0 | 0 | 19.28391 | 55.89185 |
| dbGaP | Blood Prot | 13-Jan | 0.055113 | 0.155755 | 0 | 0 | 19.28391 | 55.89185 |
| PhenGenI_ | Mortality | 3/122 | 0.016189 | 0.15582 | 0 | 0 | 5.940576 | 24.49549 |
| Rare_Disea | Idiopathic | 3/126 | 0.017629 | 0.156146 | 0 | 0 | 5.746225 | 23.20435 |
| Rare_Disea | Muscular d | 3/126 | 0.017629 | 0.156146 | 0 | 0 | 5.746225 | 23.20435 |
| Jensen_DIS | Chronic ob | 2/135 | 0.116962 | 0.156216 | 0 | 0 | 3.499337 | 7.509252 |
| Human_Ph | Abnormalit | Feb-38 | 0.01188 | 0.156487 | 0 | 0 | 12.9915 | 57.59009 |
| Human_Ph | Blepharopl | 3/104 | 0.010557 | 0.156487 | 0 | 0 | 7.005658 | 31.88247 |
| Human_Ph | Cranial hyp | Feb-36 | 0.010706 | 0.156487 | 0 | 0 | 13.75709 | 62.41496 |
| Human_Ph | Ventricular | Feb-36 | 0.010706 | 0.156487 | 0 | 0 | 13.75709 | 62.41496 |
| Human_Ph | Increased i | Feb-38 | 0.01188 | 0.156487 | 0 | 0 | 12.9915 | 57.59009 |
| Human_Ph | Slow progr | Mar-98 | 0.008983 | 0.156487 | 0 | 0 | 7.450376 | 35.10935 |
| Human_Ph | Abnormalit | Feb-36 | 0.010706 | 0.156487 | 0 | 0 | 13.75709 | 62.41496 |
| Human_Ph | Abnormalit | Feb-38 | 0.01188 | 0.156487 | 0 | 0 | 12.9915 | 57.59009 |
| Human_Ph | Autosomal | 11/1134 | 0.009986 | 0.156487 | 0 | 0 | 2.421732 | 11.1559 |
| GWAS_Cat | Migraine | 2/126 | 0.104352 | 0.156979 | 0 | 0 | 3.755028 | 8.486292 |
| GO_Biologi | Substantia | Feb-42 | 0.014388 | 0.157158 | 0 | 0 | 11.69 | 49.58172 |
| GO_Biologi | Cardiac Ve | Feb-42 | 0.014388 | 0.157158 | 0 | 0 | 11.69 | 49.58172 |
| GO_Biologi | Translation | Feb-42 | 0.014388 | 0.157158 | 0 | 0 | 11.69 | 49.58172 |
| Jensen_TIS | Embryoid b | Feb-55 | 0.02393 | 0.157251 | 0 | 0 | 8.81687 | 32.91021 |
| Jensen_DIS | Toxic shock | 29-Jan | 0.118838 | 0.157642 | 0 | 0 | 8.25789 | 17.58927 |
| DisGeNET | Erythroker | Feb-43 | 0.015047 | 0.157697 | 0 | 0 | 11.4043 | 47.85875 |
| DisGeNET | Becker Mu | Feb-43 | 0.015047 | 0.157697 | 0 | 0 | 11.4043 | 47.85875 |
| DisGeNET | Transient C | Feb-43 | 0.015047 | 0.157697 | 0 | 0 | 11.4043 | 47.85875 |
| DisGeNET | Dissection | Feb-43 | 0.015047 | 0.157697 | 0 | 0 | 11.4043 | 47.85875 |

| | | | | | | | | |
|---|---|---|---|---|---|---|---|---|
| DisGeNET | Sarcoma, E | Feb-43 | 0.015047 | 0.157697 | 0 | 0 | 11.4043 | 47.85875 |
| Jensen_DIS | Immune sy | 2/137 | 0.119815 | 0.157864 | 0 | 0 | 3.447146 | 7.31418 |
| Jensen_DIS | Nephrolith | 30-Jan | 0.122676 | 0.158423 | 0 | 0 | 7.972735 | 16.72843 |
| Jensen_DIS | Hemangior | 30-Jan | 0.122676 | 0.158423 | 0 | 0 | 7.972735 | 16.72843 |
| Jensen_DIS | Respiratory | 30-Jan | 0.122676 | 0.158423 | 0 | 0 | 7.972735 | 16.72843 |
| GWAS_Cat | Total Testo | 4/413 | 0.105701 | 0.158666 | 0 | 0 | 2.298171 | 5.164304 |
| GWAS_Cat | Inguinal He | 2/127 | 0.105734 | 0.158666 | 0 | 0 | 3.7248 | 8.368983 |
| GWAS_Cat | Neonatal A | 26-Jan | 0.107222 | 0.15972 | 0 | 0 | 9.250233 | 20.65441 |
| GWAS_Cat | Fasting Blo | 26-Jan | 0.107222 | 0.15972 | 0 | 0 | 9.250233 | 20.65441 |
| GWAS_Cat | Time To Co | 26-Jan | 0.107222 | 0.15972 | 0 | 0 | 9.250233 | 20.65441 |
| GWAS_Cat | Forns Index | 26-Jan | 0.107222 | 0.15972 | 0 | 0 | 9.250233 | 20.65441 |
| GWAS_Cat | Parental Lc | 26-Jan | 0.107222 | 0.15972 | 0 | 0 | 9.250233 | 20.65441 |
| GWAS_Cat | Systolic Blc | 26-Jan | 0.107222 | 0.15972 | 0 | 0 | 9.250233 | 20.65441 |
| CellMarker | Mesenchyr | 21-Jan | 0.087525 | 0.159879 | 0 | 0 | 11.5657 | 28.17205 |
| Jensen_TIS | Metacyclic | 8-Jan | 0.03428 | 0.159975 | 0 | 0 | 33.06645 | 111.5391 |
| Jensen_TIS | Palisade pa | 7-Jan | 0.03006 | 0.159975 | 0 | 0 | 38.57946 | 135.2043 |
| Jensen_TIS | LNCaP-C4-2 | 6-Jan | 0.025821 | 0.159975 | 0 | 0 | 46.29767 | 169.2909 |
| Jensen_TIS | CH-1 cell | 6-Jan | 0.025821 | 0.159975 | 0 | 0 | 46.29767 | 169.2909 |
| Jensen_TIS | Neointima | Feb-58 | 0.026415 | 0.159975 | 0 | 0 | 8.343277 | 30.31803 |
| Jensen_TIS | Neuroepith | 7-Jan | 0.03006 | 0.159975 | 0 | 0 | 38.57946 | 135.2043 |
| Jensen_TIS | Thyroid gla | 68/13652 | 0.027655 | 0.159975 | 0 | 0 | 1.667488 | 5.982836 |
| Jensen_TIS | phi1 | 7-Jan | 0.03006 | 0.159975 | 0 | 0 | 38.57946 | 135.2043 |
| Jensen_TIS | Ppes | 8-Jan | 0.03428 | 0.159975 | 0 | 0 | 33.06645 | 111.5391 |
| Jensen_TIS | Blood clot | Feb-63 | 0.030777 | 0.159975 | 0 | 0 | 7.657473 | 26.65545 |
| Jensen_TIS | Epididymis | 63/12472 | 0.03173 | 0.159975 | 0 | 0 | 1.587396 | 5.477315 |
| Jensen_TIS | Intercalate | 7-Jan | 0.03006 | 0.159975 | 0 | 0 | 38.57946 | 135.2043 |
| Jensen_TIS | Brain | 66/13237 | 0.033457 | 0.159975 | 0 | 0 | 1.608773 | 5.465786 |
| Jensen_TIS | Mesocarp | 8-Jan | 0.03428 | 0.159975 | 0 | 0 | 33.06645 | 111.5391 |
| Jensen_TIS | Clonei | 8-Jan | 0.03428 | 0.159975 | 0 | 0 | 33.06645 | 111.5391 |
| Jensen_TIS | Sternocleic | 7-Jan | 0.03006 | 0.159975 | 0 | 0 | 38.57946 | 135.2043 |
| Jensen_TIS | Sinus node | 7-Jan | 0.03006 | 0.159975 | 0 | 0 | 38.57946 | 135.2043 |
| Jensen_TIS | Epithelial li | 8-Jan | 0.03428 | 0.159975 | 0 | 0 | 33.06645 | 111.5391 |
| Jensen_TIS | Digestive c | 8-Jan | 0.03428 | 0.159975 | 0 | 0 | 33.06645 | 111.5391 |
| Jensen_TIS | pg13 | 7-Jan | 0.03006 | 0.159975 | 0 | 0 | 38.57946 | 135.2043 |
| CellMarker | Natural Kill | 2/114 | 0.088189 | 0.160025 | 0 | 0 | 4.159874 | 10.1013 |
| GTEx_Tissu | GTEX-S32V | 21/2242 | 0.0005 | 0.16057 | 0 | 0 | 2.534567 | 19.2674 |
| GeDiPNet_ | Protein-los | 12-Jan | 0.050982 | 0.160664 | 0 | 0 | 21.03805 | 62.61499 |
| GeDiPNet_ | Carotid Art | 12-Jan | 0.050982 | 0.160664 | 0 | 0 | 21.03805 | 62.61499 |
| GeDiPNet_ | Metaphyse | 12-Jan | 0.050982 | 0.160664 | 0 | 0 | 21.03805 | 62.61499 |
| GeDiPNet_ | Emphysem | 12-Jan | 0.050982 | 0.160664 | 0 | 0 | 21.03805 | 62.61499 |
| GeDiPNet_ | Semantic D | 12-Jan | 0.050982 | 0.160664 | 0 | 0 | 21.03805 | 62.61499 |
| GeDiPNet_ | Eyelid Xant | 12-Jan | 0.050982 | 0.160664 | 0 | 0 | 21.03805 | 62.61499 |
| GeDiPNet_ | Keratosis P | 12-Jan | 0.050982 | 0.160664 | 0 | 0 | 21.03805 | 62.61499 |
| GeDiPNet_ | Hyperostos | 12-Jan | 0.050982 | 0.160664 | 0 | 0 | 21.03805 | 62.61499 |
| GeDiPNet_ | Panacinar I | 12-Jan | 0.050982 | 0.160664 | 0 | 0 | 21.03805 | 62.61499 |

| | | | | | | | | |
|---|---|---|---|---|---|---|---|---|
| GWAS_Cat | Educationa | 13/2058 | 0.108352 | 0.161206 | 0 | 0 | 1.53495 | 3.411229 |
| Jensen_DIS | Palmoplant | 31-Jan | 0.126498 | 0.161224 | 0 | 0 | 7.706589 | 15.93356 |
| Jensen_DIS | Bronchitis | 31-Jan | 0.126498 | 0.161224 | 0 | 0 | 7.706589 | 15.93356 |
| Jensen_TIS | Respiratory | 9-Jan | 0.038483 | 0.161634 | 0 | 0 | 28.93169 | 94.24608 |
| Jensen_TIS | Vermiform | 9-Jan | 0.038483 | 0.161634 | 0 | 0 | 28.93169 | 94.24608 |
| Jensen_TIS | Sessile cell | 9-Jan | 0.038483 | 0.161634 | 0 | 0 | 28.93169 | 94.24608 |
| Jensen_TIS | Spinal cord | 58/11398 | 0.041635 | 0.161634 | 0 | 0 | 1.511993 | 4.806328 |
| Jensen_TIS | K-562 cell | Feb-75 | 0.042299 | 0.161634 | 0 | 0 | 6.394843 | 20.22688 |
| Jensen_TIS | Mature cel | 10-Jan | 0.042667 | 0.161634 | 0 | 0 | 25.71576 | 81.1157 |
| Jensen_TIS | Oral mucos | 55/10627 | 0.036734 | 0.161634 | 0 | 0 | 1.518619 | 5.017609 |
| Jensen_TIS | Myofibrobl | Feb-74 | 0.041285 | 0.161634 | 0 | 0 | 6.483987 | 20.66613 |
| Jensen_TIS | Sertoli cell | Feb-72 | 0.039286 | 0.161634 | 0 | 0 | 6.669916 | 21.58979 |
| Jensen_TIS | U-251MG c | 9-Jan | 0.038483 | 0.161634 | 0 | 0 | 28.93169 | 94.24608 |
| Jensen_TIS | Nerve shea | 10-Jan | 0.042667 | 0.161634 | 0 | 0 | 25.71576 | 81.1157 |
| Jensen_TIS | Endometria | 10-Jan | 0.042667 | 0.161634 | 0 | 0 | 25.71576 | 81.1157 |
| Jensen_TIS | Te | 10-Jan | 0.042667 | 0.161634 | 0 | 0 | 25.71576 | 81.1157 |
| Jensen_TIS | Leaf disc | 9-Jan | 0.038483 | 0.161634 | 0 | 0 | 28.93169 | 94.24608 |
| Jensen_TIS | Plant cuticl | 10-Jan | 0.042667 | 0.161634 | 0 | 0 | 25.71576 | 81.1157 |
| Jensen_TIS | Laryngeal r | 10-Jan | 0.042667 | 0.161634 | 0 | 0 | 25.71576 | 81.1157 |
| ClinVar_20 | combined c | Jan-37 | 0.149088 | 0.162052 | 0 | 0 | 6.42022 | 12.21909 |
| Rare_Disea | Cohen Hay | 5-Jan | 0.021564 | 0.162054 | 0 | 0 | 57.875 | 222.0519 |
| Rare_Disea | Insulin-resi | 4/238 | 0.020185 | 0.162054 | 0 | 0 | 4.05293 | 15.8179 |
| Rare_Disea | Mitochond | Feb-50 | 0.020017 | 0.162054 | 0 | 0 | 9.737745 | 38.08585 |
| Rare_Disea | Oliver synd | Feb-49 | 0.01927 | 0.162054 | 0 | 0 | 9.945432 | 39.27635 |
| Rare_Disea | Cardiomyo | 5-Jan | 0.021564 | 0.162054 | 0 | 0 | 57.875 | 222.0519 |
| Rare_Disea | Oculodento | 5-Jan | 0.021564 | 0.162054 | 0 | 0 | 57.875 | 222.0519 |
| Rare_Disea | Anaplastic | 5-Jan | 0.021564 | 0.162054 | 0 | 0 | 57.875 | 222.0519 |
| Rare_Disea | Arthrogryp | 5-Jan | 0.021564 | 0.162054 | 0 | 0 | 57.875 | 222.0519 |
| Rare_Disea | Benign aut | 5-Jan | 0.021564 | 0.162054 | 0 | 0 | 57.875 | 222.0519 |
| Rare_Disea | Ruvalcaba | 5-Jan | 0.021564 | 0.162054 | 0 | 0 | 57.875 | 222.0519 |
| Rare_Disea | Holocarbox | 5-Jan | 0.021564 | 0.162054 | 0 | 0 | 57.875 | 222.0519 |
| Rare_Disea | Proteus syr | 5-Jan | 0.021564 | 0.162054 | 0 | 0 | 57.875 | 222.0519 |
| Rare_Disea | Dermatofib | Feb-51 | 0.020776 | 0.162054 | 0 | 0 | 9.538535 | 36.95177 |
| Rare_Disea | Osteochon | Feb-48 | 0.018536 | 0.162054 | 0 | 0 | 10.16215 | 40.52728 |
| Rare_Disea | Alpha-Thal | 3/135 | 0.021124 | 0.162054 | 0 | 0 | 5.352002 | 20.6445 |
| GeDiPNet_ | Absence Of | Feb-84 | 0.05183 | 0.162456 | 0 | 0 | 5.690387 | 16.84234 |
| DisGeNET | Nonorganic | 4/220 | 0.01559 | 0.162733 | 0 | 0 | 4.39469 | 18.28685 |
| CellMarker | Fibroblast I | 2/116 | 0.090827 | 0.162796 | 0 | 0 | 4.086481 | 9.802643 |
| CellMarker | Stem Cell S | 22-Jan | 0.091499 | 0.162796 | 0 | 0 | 11.0144 | 26.34017 |
| CellMarker | Cancer Ster | 22-Jan | 0.091499 | 0.162796 | 0 | 0 | 11.0144 | 26.34017 |
| UK_Bioban | Right spher | 2/122 | 0.098877 | 0.162801 | 0 | 0 | 3.88098 | 8.980101 |
| UK_Bioban | Doctor diag | 24-Jan | 0.099394 | 0.162801 | 0 | 0 | 10.05561 | 23.215 |
| Jensen_TIS | Cerebellum | 72/14879 | 0.043485 | 0.162817 | 0 | 0 | 1.655217 | 5.189657 |
| GWAS_Cat | Hip Circum | 2/130 | 0.109909 | 0.163324 | 0 | 0 | 3.636949 | 8.030761 |
| DisGeNET | Sickle Cell | Feb-44 | 0.01572 | 0.163436 | 0 | 0 | 11.13221 | 46.2302 |

| | | | | | | | | |
|---|---|---|---|---|---|---|---|---|
| PheWeb_2 | Excessive c | Jan-41 | 0.163825 | 0.163825 | 0 | 0 | 5.777035 | 10.45039 |
| Jensen_TIS | Temporom | 11-Jan | 0.046834 | 0.163919 | 0 | 0 | 23.14302 | 70.84419 |
| Jensen_TIS | Motor cort | 11-Jan | 0.046834 | 0.163919 | 0 | 0 | 23.14302 | 70.84419 |
| Jensen_TIS | COS-1 cell | 11-Jan | 0.046834 | 0.163919 | 0 | 0 | 23.14302 | 70.84419 |
| Jensen_TIS | Cancer ster | Feb-78 | 0.045395 | 0.163919 | 0 | 0 | 6.141486 | 18.99159 |
| Jensen_TIS | Craniofacia | 11-Jan | 0.046834 | 0.163919 | 0 | 0 | 23.14302 | 70.84419 |
| Jensen_TIS | hutu80 | 11-Jan | 0.046834 | 0.163919 | 0 | 0 | 23.14302 | 70.84419 |
| GWAS_Cat | QTc Interva | 27-Jan | 0.111111 | 0.164008 | 0 | 0 | 8.894007 | 19.54216 |
| GWAS_Cat | Waist-to-hi | 27-Jan | 0.111111 | 0.164008 | 0 | 0 | 8.894007 | 19.54216 |
| GWAS_Cat | Photic Snee | 27-Jan | 0.111111 | 0.164008 | 0 | 0 | 8.894007 | 19.54216 |
| GWAS_Cat | Diastolic Bl | 27-Jan | 0.111111 | 0.164008 | 0 | 0 | 8.894007 | 19.54216 |
| GWAS_Cat | Pulmonary | 27-Jan | 0.111111 | 0.164008 | 0 | 0 | 8.894007 | 19.54216 |
| GWAS_Cat | Fasting Glu | 2/131 | 0.11131 | 0.164008 | 0 | 0 | 3.608573 | 7.922388 |
| GWAS_Cat | Liver Enzyn | 2/131 | 0.11131 | 0.164008 | 0 | 0 | 3.608573 | 7.922388 |
| DisGeNET | Adult Acute | 3/121 | 0.01584 | 0.164033 | 0 | 0 | 5.991223 | 24.83499 |
| Jensen_DIS | Chagas dise | Jan-32 | 0.130304 | 0.164995 | 0 | 0 | 7.457614 | 15.19775 |
| UK_Bioban | Body mass | 5/568 | 0.101601 | 0.165465 | 0 | 0 | 2.095698 | 4.79223 |
| Jensen_DIS | Hypertensi | 3/289 | 0.131539 | 0.165484 | 0 | 0 | 2.450924 | 4.971583 |
| DisGeNET | Microphtha | 4/222 | 0.016064 | 0.165701 | 0 | 0 | 4.353929 | 17.98681 |
| GeDiPNet_ | Congenital | 14-Jan | 0.059226 | 0.166004 | 0 | 0 | 17.79964 | 50.30878 |
| GeDiPNet_ | Lung Aden | 4/336 | 0.058935 | 0.166004 | 0 | 0 | 2.842357 | 8.047601 |
| GeDiPNet_ | Peritoneal | 14-Jan | 0.059226 | 0.166004 | 0 | 0 | 17.79964 | 50.30878 |
| GeDiPNet_ | Facioscapu | 13-Jan | 0.055113 | 0.166004 | 0 | 0 | 19.28391 | 55.89185 |
| GeDiPNet_ | Hamartom | 13-Jan | 0.055113 | 0.166004 | 0 | 0 | 19.28391 | 55.89185 |
| GeDiPNet_ | Senile Plaq | 14-Jan | 0.059226 | 0.166004 | 0 | 0 | 17.79964 | 50.30878 |
| GeDiPNet_ | Leukoence | 14-Jan | 0.059226 | 0.166004 | 0 | 0 | 17.79964 | 50.30878 |
| GeDiPNet_ | Venous Ins | 14-Jan | 0.059226 | 0.166004 | 0 | 0 | 17.79964 | 50.30878 |
| GeDiPNet_ | Carotid Ath | 14-Jan | 0.059226 | 0.166004 | 0 | 0 | 17.79964 | 50.30878 |
| GeDiPNet_ | Hemimega | 14-Jan | 0.059226 | 0.166004 | 0 | 0 | 17.79964 | 50.30878 |
| GeDiPNet_ | Reactive H | 14-Jan | 0.059226 | 0.166004 | 0 | 0 | 17.79964 | 50.30878 |
| GeDiPNet_ | Tubular Ag | 14-Jan | 0.059226 | 0.166004 | 0 | 0 | 17.79964 | 50.30878 |
| GeDiPNet_ | Thyroid Dis | 14-Jan | 0.059226 | 0.166004 | 0 | 0 | 17.79964 | 50.30878 |
| GeDiPNet_ | Subcutane | 13-Jan | 0.055113 | 0.166004 | 0 | 0 | 19.28391 | 55.89185 |
| GeDiPNet_ | Auditory P | 13-Jan | 0.055113 | 0.166004 | 0 | 0 | 19.28391 | 55.89185 |
| GeDiPNet_ | Hypoplasia | 14-Jan | 0.059226 | 0.166004 | 0 | 0 | 17.79964 | 50.30878 |
| GeDiPNet_ | Dysarthria | 5/470 | 0.054167 | 0.166004 | 0 | 0 | 2.550223 | 7.435639 |
| GeDiPNet_ | Immunoglo | 14-Jan | 0.059226 | 0.166004 | 0 | 0 | 17.79964 | 50.30878 |
| GeDiPNet_ | Centronucl | 13-Jan | 0.055113 | 0.166004 | 0 | 0 | 19.28391 | 55.89185 |
| GeDiPNet_ | Congenital | 13-Jan | 0.055113 | 0.166004 | 0 | 0 | 19.28391 | 55.89185 |
| GeDiPNet_ | Atresia Of | 14-Jan | 0.059226 | 0.166004 | 0 | 0 | 17.79964 | 50.30878 |
| GeDiPNet_ | Asbestosis | 14-Jan | 0.059226 | 0.166004 | 0 | 0 | 17.79964 | 50.30878 |
| CellMarker | Neutrophil | 24-Jan | 0.099394 | 0.166061 | 0 | 0 | 10.05561 | 23.215 |
| CellMarker | Myeloid Ce | 24-Jan | 0.099394 | 0.166061 | 0 | 0 | 10.05561 | 23.215 |
| CellMarker | Mesenchyr | 24-Jan | 0.099394 | 0.166061 | 0 | 0 | 10.05561 | 23.215 |
| CellMarker | Eosinophil | 24-Jan | 0.099394 | 0.166061 | 0 | 0 | 10.05561 | 23.215 |

| | | | | | | | | |
|---|---|---|---|---|---|---|---|---|
| CellMarker | CD4+ T Cel | 24-Jan | 0.099394 | 0.166061 | 0 | 0 | 10.05561 | 23.215 |
| CellMarker | Macrophag | 23-Jan | 0.095455 | 0.166061 | 0 | 0 | 10.51321 | 24.6966 |
| CellMarker | Endothelia | 23-Jan | 0.095455 | 0.166061 | 0 | 0 | 10.51321 | 24.6966 |
| CellMarker | GABAergic | 24-Jan | 0.099394 | 0.166061 | 0 | 0 | 10.05561 | 23.215 |
| CellMarker | Mesenchyr | 23-Jan | 0.095455 | 0.166061 | 0 | 0 | 10.51321 | 24.6966 |
| CellMarker | Myeloid Ce | 2/120 | 0.096172 | 0.166061 | 0 | 0 | 3.947159 | 9.242743 |
| SysMyo_M | MGS64 Mc | Mar-68 | 0.003249 | 0.166246 | 0 | 0 | 10.90549 | 62.48067 |
| SysMyo_M | MGS12 Rat | 4/143 | 0.003525 | 0.166246 | 0 | 0 | 6.855855 | 38.72168 |
| SysMyo_M | MGS1396 I | 4/142 | 0.003437 | 0.166246 | 0 | 0 | 6.905884 | 39.17754 |
| DisGeNET | Adenocarci | 3/122 | 0.016189 | 0.166335 | 0 | 0 | 5.940576 | 24.49549 |
| UK_Bioban | Hernia K11 | 25-Jan | 0.103317 | 0.166357 | 0 | 0 | 9.636143 | 21.87363 |
| UK_Bioban | Number of | 25-Jan | 0.103317 | 0.166357 | 0 | 0 | 9.636143 | 21.87363 |
| GeDiPNet_ | Gynecoma: | Feb-91 | 0.05972 | 0.166588 | 0 | 0 | 5.240978 | 14.76951 |
| DisGeNET | Abetalipop | Feb-45 | 0.016405 | 0.166594 | 0 | 0 | 10.87278 | 44.68894 |
| DisGeNET | Granuloma | Feb-45 | 0.016405 | 0.166594 | 0 | 0 | 10.87278 | 44.68894 |
| DisGeNET | Liposarcon | Feb-45 | 0.016405 | 0.166594 | 0 | 0 | 10.87278 | 44.68894 |
| GO_Biologi | Positive Re | 6-Jan | 0.025821 | 0.166738 | 0 | 0 | 46.29767 | 169.2909 |
| GO_Biologi | Positive Re | 5-Jan | 0.021564 | 0.166738 | 0 | 0 | 57.875 | 222.0519 |
| GO_Biologi | Lipoprotein | 5-Jan | 0.021564 | 0.166738 | 0 | 0 | 57.875 | 222.0519 |
| GO_Biologi | Locomotor | 6-Jan | 0.025821 | 0.166738 | 0 | 0 | 46.29767 | 169.2909 |
| GO_Biologi | Negative R | Feb-54 | 0.023124 | 0.166738 | 0 | 0 | 8.986878 | 33.8526 |
| GO_Biologi | Negative R | 6-Jan | 0.025821 | 0.166738 | 0 | 0 | 46.29767 | 169.2909 |
| GO_Biologi | Positive Re | 7-Jan | 0.03006 | 0.166738 | 0 | 0 | 38.57946 | 135.2043 |
| GO_Biologi | Detection ( | 6-Jan | 0.025821 | 0.166738 | 0 | 0 | 46.29767 | 169.2909 |
| GO_Biologi | Regulation | 7-Jan | 0.03006 | 0.166738 | 0 | 0 | 38.57946 | 135.2043 |
| GO_Biologi | Skeletal Mu | 6-Jan | 0.025821 | 0.166738 | 0 | 0 | 46.29767 | 169.2909 |
| GO_Biologi | Dephosphc | 3/147 | 0.026333 | 0.166738 | 0 | 0 | 4.903026 | 17.83201 |
| GO_Biologi | Regulation | 6-Jan | 0.025821 | 0.166738 | 0 | 0 | 46.29767 | 169.2909 |
| GO_Biologi | Regulation | Feb-47 | 0.017813 | 0.166738 | 0 | 0 | 10.3885 | 41.84306 |
| GO_Biologi | Protein De | 3/124 | 0.016901 | 0.166738 | 0 | 0 | 5.841795 | 23.83691 |
| GO_Biologi | Negative R | Feb-50 | 0.020017 | 0.166738 | 0 | 0 | 9.737745 | 38.08585 |
| GO_Biologi | Negative R | 5-Jan | 0.021564 | 0.166738 | 0 | 0 | 57.875 | 222.0519 |
| GO_Biologi | Cardiac Mu | 7-Jan | 0.03006 | 0.166738 | 0 | 0 | 38.57946 | 135.2043 |
| GO_Biologi | Glomerulus | 5-Jan | 0.021564 | 0.166738 | 0 | 0 | 57.875 | 222.0519 |
| GO_Biologi | Regulation | 5-Jan | 0.021564 | 0.166738 | 0 | 0 | 57.875 | 222.0519 |
| GO_Biologi | Cardiac Pac | 6-Jan | 0.025821 | 0.166738 | 0 | 0 | 46.29767 | 169.2909 |
| GO_Biologi | Keratinocy | 6-Jan | 0.025821 | 0.166738 | 0 | 0 | 46.29767 | 169.2909 |
| GO_Biologi | Regulation | 7-Jan | 0.03006 | 0.166738 | 0 | 0 | 38.57946 | 135.2043 |
| GO_Biologi | Regulation | 7-Jan | 0.03006 | 0.166738 | 0 | 0 | 38.57946 | 135.2043 |
| GO_Biologi | Regulation | 7-Jan | 0.03006 | 0.166738 | 0 | 0 | 38.57946 | 135.2043 |
| GO_Biologi | Negative R | 5-Jan | 0.021564 | 0.166738 | 0 | 0 | 57.875 | 222.0519 |
| GO_Biologi | Positive Re | 7-Jan | 0.03006 | 0.166738 | 0 | 0 | 38.57946 | 135.2043 |
| GO_Biologi | Prepulse In | 5-Jan | 0.021564 | 0.166738 | 0 | 0 | 57.875 | 222.0519 |
| GO_Biologi | Regulation | 5-Jan | 0.021564 | 0.166738 | 0 | 0 | 57.875 | 222.0519 |
| GO_Biologi | Triglyceride | Feb-45 | 0.016405 | 0.166738 | 0 | 0 | 10.87278 | 44.68894 |

| | | | | | | | | |
|---|---|---|---|---|---|---|---|---|
| GO_Biologi | Postsynapt | 5-Jan | 0.021564 | 0.166738 | 0 | 0 | 57.875 | 222.0519 |
| GO_Biologi | Regulation | 5-Jan | 0.021564 | 0.166738 | 0 | 0 | 57.875 | 222.0519 |
| GO_Biologi | Mitochond | 5-Jan | 0.021564 | 0.166738 | 0 | 0 | 57.875 | 222.0519 |
| GO_Biologi | Skeletal Sy | 3/149 | 0.027262 | 0.166738 | 0 | 0 | 4.835372 | 17.41828 |
| GO_Biologi | Positive Re | 5-Jan | 0.021564 | 0.166738 | 0 | 0 | 57.875 | 222.0519 |
| GO_Biologi | Regulation | 7-Jan | 0.03006 | 0.166738 | 0 | 0 | 38.57946 | 135.2043 |
| GO_Biologi | Very-Low-D | 7-Jan | 0.03006 | 0.166738 | 0 | 0 | 38.57946 | 135.2043 |
| GO_Biologi | Atrial Cardi | 7-Jan | 0.03006 | 0.166738 | 0 | 0 | 38.57946 | 135.2043 |
| GO_Biologi | Detection ( | 5-Jan | 0.021564 | 0.166738 | 0 | 0 | 57.875 | 222.0519 |
| GO_Biologi | Negative R | 5-Jan | 0.021564 | 0.166738 | 0 | 0 | 57.875 | 222.0519 |
| GO_Biologi | Cardiac Mu | Feb-52 | 0.021547 | 0.166738 | 0 | 0 | 9.347294 | 35.8704 |
| GO_Biologi | Negative R | 5-Jan | 0.021564 | 0.166738 | 0 | 0 | 57.875 | 222.0519 |
| GO_Biologi | Striated Mu | 5-Jan | 0.021564 | 0.166738 | 0 | 0 | 57.875 | 222.0519 |
| GO_Biologi | Positive Re | 6-Jan | 0.025821 | 0.166738 | 0 | 0 | 46.29767 | 169.2909 |
| GO_Biologi | CENP-A Co | 6-Jan | 0.025821 | 0.166738 | 0 | 0 | 46.29767 | 169.2909 |
| GO_Biologi | Negative R | 6-Jan | 0.025821 | 0.166738 | 0 | 0 | 46.29767 | 169.2909 |
| GO_Biologi | Positive Re | 6-Jan | 0.025821 | 0.166738 | 0 | 0 | 46.29767 | 169.2909 |
| GO_Biologi | Vascular W | 6-Jan | 0.025821 | 0.166738 | 0 | 0 | 46.29767 | 169.2909 |
| GO_Biologi | Postsynapt | 7-Jan | 0.03006 | 0.166738 | 0 | 0 | 38.57946 | 135.2043 |
| GO_Biologi | Positive Re | 6-Jan | 0.025821 | 0.166738 | 0 | 0 | 46.29767 | 169.2909 |
| GO_Biologi | Sinoatrial N | 7-Jan | 0.03006 | 0.166738 | 0 | 0 | 38.57946 | 135.2043 |
| GO_Biologi | tRNA Amin | 7-Jan | 0.03006 | 0.166738 | 0 | 0 | 38.57946 | 135.2043 |
| GO_Biologi | Protein Tar | Feb-59 | 0.027266 | 0.166738 | 0 | 0 | 8.196491 | 29.5248 |
| GO_Biologi | Phospholip | Feb-59 | 0.027266 | 0.166738 | 0 | 0 | 8.196491 | 29.5248 |
| GO_Biologi | Regulation | 7-Jan | 0.03006 | 0.166738 | 0 | 0 | 38.57946 | 135.2043 |
| GO_Biologi | Chromatin | 6-Jan | 0.025821 | 0.166738 | 0 | 0 | 46.29767 | 169.2909 |
| GO_Biologi | Cholestero | Feb-51 | 0.020776 | 0.166738 | 0 | 0 | 9.538535 | 36.95177 |
| GO_Biologi | DNA Dealky | 7-Jan | 0.03006 | 0.166738 | 0 | 0 | 38.57946 | 135.2043 |
| GO_Biologi | Regulation | 6-Jan | 0.025821 | 0.166738 | 0 | 0 | 46.29767 | 169.2909 |
| GO_Biologi | Positive Re | 6-Jan | 0.025821 | 0.166738 | 0 | 0 | 46.29767 | 169.2909 |
| GO_Biologi | DNA Dealky | 7-Jan | 0.03006 | 0.166738 | 0 | 0 | 38.57946 | 135.2043 |
| GO_Biologi | Negative R | 5-Jan | 0.021564 | 0.166738 | 0 | 0 | 57.875 | 222.0519 |
| GO_Biologi | Negative R | 7-Jan | 0.03006 | 0.166738 | 0 | 0 | 38.57946 | 135.2043 |
| GO_Biologi | Stress-Indu | 7-Jan | 0.03006 | 0.166738 | 0 | 0 | 38.57946 | 135.2043 |
| GeDiPNet_ | Penis Agen | 3/205 | 0.060177 | 0.167062 | 0 | 0 | 3.484972 | 9.794417 |
| DisGeNET | Hyperplasia | 3/123 | 0.016543 | 0.167343 | 0 | 0 | 5.890774 | 24.16285 |
| GWAS_Cat | Fibrinogen | 28-Jan | 0.114983 | 0.167397 | 0 | 0 | 8.564169 | 18.52408 |
| GWAS_Cat | Venous Thr | 28-Jan | 0.114983 | 0.167397 | 0 | 0 | 8.564169 | 18.52408 |
| GWAS_Cat | Cholestero | 28-Jan | 0.114983 | 0.167397 | 0 | 0 | 8.564169 | 18.52408 |
| GWAS_Cat | Parental Lo | 28-Jan | 0.114983 | 0.167397 | 0 | 0 | 8.564169 | 18.52408 |
| GWAS_Cat | Migraine O | 28-Jan | 0.114983 | 0.167397 | 0 | 0 | 8.564169 | 18.52408 |
| GWAS_Cat | Epstein-Bar | 28-Jan | 0.114983 | 0.167397 | 0 | 0 | 8.564169 | 18.52408 |
| GWAS_Cat | Spleen Volu | 28-Jan | 0.114983 | 0.167397 | 0 | 0 | 8.564169 | 18.52408 |
| GWAS_Cat | Phospholip | 28-Jan | 0.114983 | 0.167397 | 0 | 0 | 8.564169 | 18.52408 |
| GWAS_Cat | Gut Microb | 28-Jan | 0.114983 | 0.167397 | 0 | 0 | 8.564169 | 18.52408 |

| | | | | | | | | |
|---|---|---|---|---|---|---|---|---|
| GWAS_Cat | Coffee Con | 28-Jan | 0.114983 | 0.167397 | 0 | 0 | 8.564169 | 18.52408 |
| Jensen_DIS | Endometri | 3/292 | 0.134486 | 0.168108 | 0 | 0 | 2.425111 | 4.86548 |
| DisGeNET | CNS disord | 3/124 | 0.016901 | 0.168456 | 0 | 0 | 5.841795 | 23.83691 |
| DisGeNET | Hyperthyrc | 3/124 | 0.016901 | 0.168456 | 0 | 0 | 5.841795 | 23.83691 |
| DisGeNET | Exanthema | 4/225 | 0.016792 | 0.168456 | 0 | 0 | 4.294172 | 17.54962 |
| DisGeNET | Ventricular | 4/226 | 0.017039 | 0.168456 | 0 | 0 | 4.274612 | 17.40721 |
| DisGeNET | Single tum | Feb-46 | 0.017103 | 0.168456 | 0 | 0 | 10.62513 | 43.22851 |
| DisGeNET | Dysautono | Feb-46 | 0.017103 | 0.168456 | 0 | 0 | 10.62513 | 43.22851 |
| DisGeNET | Foot-drop | Feb-46 | 0.017103 | 0.168456 | 0 | 0 | 10.62513 | 43.22851 |
| CellMarker | Macrophag | 25-Jan | 0.103317 | 0.168504 | 0 | 0 | 9.636143 | 21.87363 |
| CellMarker | Macrophag | 25-Jan | 0.103317 | 0.168504 | 0 | 0 | 9.636143 | 21.87363 |
| CellMarker | Hematopoi | 25-Jan | 0.103317 | 0.168504 | 0 | 0 | 9.636143 | 21.87363 |
| CellMarker | Endothelia | 25-Jan | 0.103317 | 0.168504 | 0 | 0 | 9.636143 | 21.87363 |
| Jensen_TIS | Oviduct | 61/12240 | 0.053065 | 0.169014 | 0 | 0 | 1.489872 | 4.374618 |
| Jensen_TIS | Stratum co | 12-Jan | 0.050982 | 0.169014 | 0 | 0 | 21.03805 | 62.61499 |
| Jensen_TIS | Stellate cel | 12-Jan | 0.050982 | 0.169014 | 0 | 0 | 21.03805 | 62.61499 |
| Jensen_TIS | Endometri | 13-Jan | 0.055113 | 0.169014 | 0 | 0 | 19.28391 | 55.89185 |
| Jensen_TIS | BTO:00054 | 39/7164 | 0.051597 | 0.169014 | 0 | 0 | 1.458281 | 4.322769 |
| Jensen_TIS | Interstitial | 3/194 | 0.052689 | 0.169014 | 0 | 0 | 3.687734 | 10.85427 |
| Jensen_TIS | Subcutane | 13-Jan | 0.055113 | 0.169014 | 0 | 0 | 19.28391 | 55.89185 |
| Jensen_TIS | Aedesalbo | 12-Jan | 0.050982 | 0.169014 | 0 | 0 | 21.03805 | 62.61499 |
| Jensen_TIS | T-cell acute | 13-Jan | 0.055113 | 0.169014 | 0 | 0 | 19.28391 | 55.89185 |
| Jensen_TIS | Inflorescen | 13-Jan | 0.055113 | 0.169014 | 0 | 0 | 19.28391 | 55.89185 |
| Jensen_TIS | CACO-2 cel | 13-Jan | 0.055113 | 0.169014 | 0 | 0 | 19.28391 | 55.89185 |
| Jensen_TIS | Sertoli cell | 13-Jan | 0.055113 | 0.169014 | 0 | 0 | 19.28391 | 55.89185 |
| Jensen_TIS | Bone marr | 13-Jan | 0.055113 | 0.169014 | 0 | 0 | 19.28391 | 55.89185 |
| Jensen_TIS | Colon | 79/16859 | 0.056859 | 0.169135 | 0 | 0 | 1.843765 | 5.286421 |
| Jensen_TIS | Rectum | 71/14744 | 0.056348 | 0.169135 | 0 | 0 | 1.584713 | 4.557965 |
| Jensen_TIS | Hippocamp | 57/11325 | 0.057254 | 0.169135 | 0 | 0 | 1.457712 | 4.169441 |
| Jensen_TIS | Uterine en | 57/11318 | 0.056505 | 0.169135 | 0 | 0 | 1.459799 | 4.194627 |
| SysMyo_M | MGS1039 I | 5/244 | 0.004256 | 0.169141 | 0 | 0 | 5.01939 | 27.40285 |
| SysMyo_M | MGS976 M | 4/153 | 0.004483 | 0.169141 | 0 | 0 | 6.392496 | 34.56784 |
| SysMyo_M | MGS1006 I | 4/147 | 0.003889 | 0.169141 | 0 | 0 | 6.662735 | 36.97528 |
| DisGeNET | Myotonic D | 3/125 | 0.017263 | 0.169395 | 0 | 0 | 5.793618 | 23.51747 |
| GO_Biologi | Phosphatic | Feb-63 | 0.030777 | 0.169395 | 0 | 0 | 7.657473 | 26.65545 |
| Rare_Disea | Hypergona | 3/139 | 0.022791 | 0.169562 | 0 | 0 | 5.19354 | 19.6389 |
| GO_Biologi | Negative R | 8-Jan | 0.03428 | 0.169897 | 0 | 0 | 33.06645 | 111.5391 |
| GO_Biologi | Presynapti | 8-Jan | 0.03428 | 0.169897 | 0 | 0 | 33.06645 | 111.5391 |
| GO_Biologi | Negative R | 8-Jan | 0.03428 | 0.169897 | 0 | 0 | 33.06645 | 111.5391 |
| GO_Biologi | Triglyceride | 8-Jan | 0.03428 | 0.169897 | 0 | 0 | 33.06645 | 111.5391 |
| GO_Biologi | Skeletal My | 8-Jan | 0.03428 | 0.169897 | 0 | 0 | 33.06645 | 111.5391 |
| GO_Biologi | Regulation | 8-Jan | 0.03428 | 0.169897 | 0 | 0 | 33.06645 | 111.5391 |
| GO_Biologi | Regulation | 8-Jan | 0.03428 | 0.169897 | 0 | 0 | 33.06645 | 111.5391 |
| GO_Biologi | Negative R | 8-Jan | 0.03428 | 0.169897 | 0 | 0 | 33.06645 | 111.5391 |
| GO_Biologi | Cholestero | Feb-67 | 0.034458 | 0.169897 | 0 | 0 | 7.184796 | 24.19848 |

| | | | | | | | | |
|---|---|---|---|---|---|---|---|---|
| GO_Biologi | Regulation | 3/162 | 0.033724 | 0.169897 | 0 | 0 | 4.437107 | 15.03977 |
| GO_Biologi | Canonical \ | Feb-65 | 0.032597 | 0.169897 | 0 | 0 | 7.413632 | 25.38081 |
| GO_Biologi | Negative R | Feb-66 | 0.033522 | 0.169897 | 0 | 0 | 7.297426 | 24.7787 |
| GO_Biologi | Regulation | 8-Jan | 0.03428 | 0.169897 | 0 | 0 | 33.06645 | 111.5391 |
| GO_Biologi | Positive Re | 8-Jan | 0.03428 | 0.169897 | 0 | 0 | 33.06645 | 111.5391 |
| GO_Biologi | Regulation | Feb-66 | 0.033522 | 0.169897 | 0 | 0 | 7.297426 | 24.7787 |
| GeDiPNet_ | Flexion Cor | 15-Jan | 0.063321 | 0.170121 | 0 | 0 | 16.52741 | 45.60793 |
| GeDiPNet_ | Endocrine ( | 15-Jan | 0.063321 | 0.170121 | 0 | 0 | 16.52741 | 45.60793 |
| GeDiPNet_ | Digestive S | 15-Jan | 0.063321 | 0.170121 | 0 | 0 | 16.52741 | 45.60793 |
| GeDiPNet_ | Secundum | 15-Jan | 0.063321 | 0.170121 | 0 | 0 | 16.52741 | 45.60793 |
| GeDiPNet_ | Nephrosis | 15-Jan | 0.063321 | 0.170121 | 0 | 0 | 16.52741 | 45.60793 |
| GeDiPNet_ | Congenital | 15-Jan | 0.063321 | 0.170121 | 0 | 0 | 16.52741 | 45.60793 |
| GeDiPNet_ | Ankyloglos | 15-Jan | 0.063321 | 0.170121 | 0 | 0 | 16.52741 | 45.60793 |
| GWAS_Cat | Ratio Of Lir | 29-Jan | 0.118838 | 0.170768 | 0 | 0 | 8.25789 | 17.58927 |
| GWAS_Cat | Large Arter | 29-Jan | 0.118838 | 0.170768 | 0 | 0 | 8.25789 | 17.58927 |
| GWAS_Cat | Cognitive F | 29-Jan | 0.118838 | 0.170768 | 0 | 0 | 8.25789 | 17.58927 |
| GWAS_Cat | Response T | 29-Jan | 0.118838 | 0.170768 | 0 | 0 | 8.25789 | 17.58927 |
| GWAS_Cat | Iron Status | 29-Jan | 0.118838 | 0.170768 | 0 | 0 | 8.25789 | 17.58927 |
| GWAS_Cat | Carotid Inti | 29-Jan | 0.118838 | 0.170768 | 0 | 0 | 8.25789 | 17.58927 |
| GWAS_Cat | Microalbur | 29-Jan | 0.118838 | 0.170768 | 0 | 0 | 8.25789 | 17.58927 |
| GWAS_Cat | Age-relate( | 29-Jan | 0.118838 | 0.170768 | 0 | 0 | 8.25789 | 17.58927 |
| GWAS_Cat | Tyrosine Le | 29-Jan | 0.118838 | 0.170768 | 0 | 0 | 8.25789 | 17.58927 |
| GWAS_Cat | Chronic Inf | 2/136 | 0.118386 | 0.170768 | 0 | 0 | 3.473047 | 7.410799 |
| GWAS_Cat | Waist-to-hi | 29-Jan | 0.118838 | 0.170768 | 0 | 0 | 8.25789 | 17.58927 |
| Jensen_TIS | Groin | 16-Jan | 0.067399 | 0.170939 | 0 | 0 | 15.42481 | 41.60265 |
| Jensen_TIS | Immature ( | 16-Jan | 0.067399 | 0.170939 | 0 | 0 | 15.42481 | 41.60265 |
| Jensen_TIS | Human aor | 15-Jan | 0.063321 | 0.170939 | 0 | 0 | 16.52741 | 45.60793 |
| Jensen_TIS | Regio parie | 16-Jan | 0.067399 | 0.170939 | 0 | 0 | 15.42481 | 41.60265 |
| Jensen_TIS | Claw | 16-Jan | 0.067399 | 0.170939 | 0 | 0 | 15.42481 | 41.60265 |
| Jensen_TIS | Salivary gla | 73/15335 | 0.066349 | 0.170939 | 0 | 0 | 1.589021 | 4.310746 |
| Jensen_TIS | Renal proxi | 16-Jan | 0.067399 | 0.170939 | 0 | 0 | 15.42481 | 41.60265 |
| Jensen_TIS | Gastric ulc( | 15-Jan | 0.063321 | 0.170939 | 0 | 0 | 16.52741 | 45.60793 |
| Jensen_TIS | Gbm cell | 15-Jan | 0.063321 | 0.170939 | 0 | 0 | 16.52741 | 45.60793 |
| Jensen_TIS | Cervical car | Feb-96 | 0.065593 | 0.170939 | 0 | 0 | 4.960951 | 13.51507 |
| Jensen_TIS | Pt-K2 cell | 15-Jan | 0.063321 | 0.170939 | 0 | 0 | 16.52741 | 45.60793 |
| Jensen_TIS | Myelocyte | 15-Jan | 0.063321 | 0.170939 | 0 | 0 | 16.52741 | 45.60793 |
| Jensen_TIS | Frontal lob | 58/11591 | 0.060435 | 0.170939 | 0 | 0 | 1.453221 | 4.078001 |
| Jensen_TIS | Erythroleul | 25/4265 | 0.06295 | 0.170939 | 0 | 0 | 1.490509 | 4.121881 |
| Jensen_TIS | Microspori | 14-Jan | 0.059226 | 0.170939 | 0 | 0 | 17.79964 | 50.30878 |
| Jensen_TIS | Skin | 74/15601 | 0.06742 | 0.170939 | 0 | 0 | 1.607939 | 4.336308 |
| Jensen_TIS | SW-1116 c( | 15-Jan | 0.063321 | 0.170939 | 0 | 0 | 16.52741 | 45.60793 |
| Jensen_TIS | Gall bladde | 77/16366 | 0.06392 | 0.170939 | 0 | 0 | 1.713107 | 4.711252 |
| Jensen_TIS | Marrow | Feb-98 | 0.067994 | 0.171048 | 0 | 0 | 4.857108 | 13.05753 |
| PhenGenI_ | Lipids | 5/358 | 0.020007 | 0.171259 | 0 | 0 | 3.378705 | 13.21642 |
| PhenGenI_ | Tonsillecto | Feb-50 | 0.020017 | 0.171259 | 0 | 0 | 9.737745 | 38.08585 |

| | | | | | | | | |
|---|---|---|---|---|---|---|---|---|
| PhenGenI_ | Hypertrigly | Feb-50 | 0.020017 | 0.171259 | 0 | 0 | 9.737745 | 38.08585 |
| Jensen_TIS | Temporal l | 44/8425 | 0.068666 | 0.171398 | 0 | 0 | 1.407969 | 3.771256 |
| Rare_Disea | Malignant | Feb-57 | 0.025575 | 0.171524 | 0 | 0 | 8.495401 | 31.14526 |
| Rare_Disea | Langer mes | 6-Jan | 0.025821 | 0.171524 | 0 | 0 | 46.29767 | 169.2909 |
| Rare_Disea | Meningiom | 3/145 | 0.025421 | 0.171524 | 0 | 0 | 4.972586 | 18.26021 |
| Rare_Disea | Adenomyo | 3/145 | 0.025421 | 0.171524 | 0 | 0 | 4.972586 | 18.26021 |
| Rare_Disea | Autoimmu | Feb-56 | 0.024747 | 0.171524 | 0 | 0 | 8.653159 | 32.00857 |
| Rare_Disea | Precocious | 6-Jan | 0.025821 | 0.171524 | 0 | 0 | 46.29767 | 169.2909 |
| Rare_Disea | Rectal canc | 6-Jan | 0.025821 | 0.171524 | 0 | 0 | 46.29767 | 169.2909 |
| Rare_Disea | Logopenic | 6-Jan | 0.025821 | 0.171524 | 0 | 0 | 46.29767 | 169.2909 |
| Rare_Disea | Megalence | 6-Jan | 0.025821 | 0.171524 | 0 | 0 | 46.29767 | 169.2909 |
| Rare_Disea | Pleomorph | Feb-57 | 0.025575 | 0.171524 | 0 | 0 | 8.495401 | 31.14526 |
| Rare_Disea | Limb-girdle | 3/143 | 0.024527 | 0.171524 | 0 | 0 | 5.044133 | 18.70358 |
| Rare_Disea | Oculodento | 6-Jan | 0.025821 | 0.171524 | 0 | 0 | 46.29767 | 169.2909 |
| PhenGenI_ | Diarrhea | 5-Jan | 0.021564 | 0.171765 | 0 | 0 | 57.875 | 222.0519 |
| PhenGenI_ | 25-Hydroxy | Feb-52 | 0.021547 | 0.171765 | 0 | 0 | 9.347294 | 35.8704 |
| DisGeNET | Memory Lo | 3/126 | 0.017629 | 0.172347 | 0 | 0 | 5.746225 | 23.20435 |
| Rare_Disea | Cold agglut | Feb-58 | 0.026415 | 0.172392 | 0 | 0 | 8.343277 | 30.31803 |
| Rare_Disea | Intracrania | Feb-58 | 0.026415 | 0.172392 | 0 | 0 | 8.343277 | 30.31803 |
| DisGeNET | Foot dorsif | Feb-47 | 0.017813 | 0.172852 | 0 | 0 | 10.3885 | 41.84306 |
| DisGeNET | Phospholip | Feb-47 | 0.017813 | 0.172852 | 0 | 0 | 10.3885 | 41.84306 |
| Jensen_TIS | Adrenal gla | 76/16140 | 0.06979 | 0.172864 | 0 | 0 | 1.655446 | 4.407242 |
| Rare_Disea | Cytokine d | 3/148 | 0.026795 | 0.173353 | 0 | 0 | 4.868966 | 17.62338 |
| Rare_Disea | Ramon Syn | Feb-59 | 0.027266 | 0.173382 | 0 | 0 | 8.196491 | 29.5248 |
| Rare_Disea | Brugada sy | Feb-59 | 0.027266 | 0.173382 | 0 | 0 | 8.196491 | 29.5248 |
| CellMarker | Endothelia | 26-Jan | 0.107222 | 0.173839 | 0 | 0 | 9.250233 | 20.65441 |
| GeDiPNet_ | Mitral Valv | 16-Jan | 0.067399 | 0.173865 | 0 | 0 | 15.42481 | 41.60265 |
| GeDiPNet_ | Melanoma | 4/350 | 0.066405 | 0.173865 | 0 | 0 | 2.725399 | 7.391226 |
| GeDiPNet_ | Blepharopl | Feb-96 | 0.065593 | 0.173865 | 0 | 0 | 4.960951 | 13.51507 |
| GeDiPNet_ | Pyle Metap | 16-Jan | 0.067399 | 0.173865 | 0 | 0 | 15.42481 | 41.60265 |
| GeDiPNet_ | Hypoplasia | 4/351 | 0.066957 | 0.173865 | 0 | 0 | 2.717406 | 7.347065 |
| GeDiPNet_ | Synostotic | 16-Jan | 0.067399 | 0.173865 | 0 | 0 | 15.42481 | 41.60265 |
| GeDiPNet_ | Adenocarci | 3/214 | 0.066654 | 0.173865 | 0 | 0 | 3.3348 | 9.031434 |
| GeDiPNet_ | Testicular I | 16-Jan | 0.067399 | 0.173865 | 0 | 0 | 15.42481 | 41.60265 |
| GeDiPNet_ | Sinus Node | 16-Jan | 0.067399 | 0.173865 | 0 | 0 | 15.42481 | 41.60265 |
| GWAS_Cat | Hypothyroi | 2/138 | 0.121248 | 0.174026 | 0 | 0 | 3.421626 | 7.219351 |
| Jensen_TIS | Ovum | 17-Jan | 0.071459 | 0.174317 | 0 | 0 | 14.46003 | 38.15469 |
| Jensen_TIS | RAJI cell | 17-Jan | 0.071459 | 0.174317 | 0 | 0 | 14.46003 | 38.15469 |
| PhenGenI_ | Cholestero | 9/961 | 0.02371 | 0.174338 | 0 | 0 | 2.298117 | 8.599252 |
| PhenGenI_ | Triple Nega | 6-Jan | 0.025821 | 0.174338 | 0 | 0 | 46.29767 | 169.2909 |
| PhenGenI_ | Cold-Shock | 6-Jan | 0.025821 | 0.174338 | 0 | 0 | 46.29767 | 169.2909 |
| PhenGenI_ | Vascular Ca | 3/142 | 0.024086 | 0.174338 | 0 | 0 | 5.080678 | 18.9312 |
| PhenGenI_ | Zeaxanthin | Feb-58 | 0.026415 | 0.174338 | 0 | 0 | 8.343277 | 30.31803 |
| PhenGenI_ | Blood Sedi | Feb-56 | 0.024747 | 0.174338 | 0 | 0 | 8.653159 | 32.00857 |
| Jensen_TIS | Bronchus | 62/12644 | 0.072013 | 0.174348 | 0 | 0 | 1.444991 | 3.801631 |

| | | | | | | | | |
|---|---|---|---|---|---|---|---|---|
| GeDiPNet_ | Congenital | Feb-98 | 0.067994 | 0.174628 | 0 | 0 | 4.857108 | 13.05753 |
| GWAS_Cat | Endometri | 30-Jan | 0.122676 | 0.174843 | 0 | 0 | 7.972735 | 16.72843 |
| GWAS_Cat | Familial Co | 30-Jan | 0.122676 | 0.174843 | 0 | 0 | 7.972735 | 16.72843 |
| GWAS_Cat | Age-relate | 30-Jan | 0.122676 | 0.174843 | 0 | 0 | 7.972735 | 16.72843 |
| GWAS_Cat | Maximum | 30-Jan | 0.122676 | 0.174843 | 0 | 0 | 7.972735 | 16.72843 |
| GWAS_Cat | Age-relate | 30-Jan | 0.122676 | 0.174843 | 0 | 0 | 7.972735 | 16.72843 |
| GWAS_Cat | Age-relate | 30-Jan | 0.122676 | 0.174843 | 0 | 0 | 7.972735 | 16.72843 |
| GTEx_Tissu | GTEX-XLM4 | 9/551 | 0.000652 | 0.174922 | 0 | 0 | 4.123829 | 30.25059 |
| GTEx_Tissu | GTEX-Q2AI | 17/1650 | 0.000665 | 0.174922 | 0 | 0 | 2.718572 | 19.8879 |
| Human_Ph | Childhood | Feb-41 | 0.013741 | 0.174969 | 0 | 0 | 11.99035 | 51.40702 |
| GO_Biologi | Myelin Ma | 9-Jan | 0.038483 | 0.175147 | 0 | 0 | 28.93169 | 94.24608 |
| GO_Biologi | Telomere M | 9-Jan | 0.038483 | 0.175147 | 0 | 0 | 28.93169 | 94.24608 |
| GO_Biologi | Regulation | 9-Jan | 0.038483 | 0.175147 | 0 | 0 | 28.93169 | 94.24608 |
| GO_Biologi | Negative R | 9-Jan | 0.038483 | 0.175147 | 0 | 0 | 28.93169 | 94.24608 |
| GO_Biologi | Excitatory | 9-Jan | 0.038483 | 0.175147 | 0 | 0 | 28.93169 | 94.24608 |
| GO_Biologi | Regulation | 9-Jan | 0.038483 | 0.175147 | 0 | 0 | 28.93169 | 94.24608 |
| GO_Biologi | High-Densi | 9-Jan | 0.038483 | 0.175147 | 0 | 0 | 28.93169 | 94.24608 |
| GO_Biologi | Kinetochor | 9-Jan | 0.038483 | 0.175147 | 0 | 0 | 28.93169 | 94.24608 |
| GO_Biologi | Negative R | 9-Jan | 0.038483 | 0.175147 | 0 | 0 | 28.93169 | 94.24608 |
| GO_Biologi | Presynapti | 9-Jan | 0.038483 | 0.175147 | 0 | 0 | 28.93169 | 94.24608 |
| GO_Biologi | Lipid Phosp | 9-Jan | 0.038483 | 0.175147 | 0 | 0 | 28.93169 | 94.24608 |
| GO_Biologi | Positive Re | 9-Jan | 0.038483 | 0.175147 | 0 | 0 | 28.93169 | 94.24608 |
| Jensen_DIS | Lymphoid | 2/152 | 0.141732 | 0.176037 | 0 | 0 | 3.100078 | 6.056983 |
| Jensen_TIS | Cardiac mu | 72/15135 | 0.074355 | 0.17617 | 0 | 0 | 1.545509 | 4.016628 |
| Jensen_TIS | Myotube | 2/103 | 0.074122 | 0.17617 | 0 | 0 | 4.615492 | 12.00969 |
| Jensen_TIS | Vagina | 51/10073 | 0.075251 | 0.17617 | 0 | 0 | 1.398149 | 3.616903 |
| Jensen_TIS | Jaw | 18-Jan | 0.075502 | 0.17617 | 0 | 0 | 13.60876 | 35.1596 |
| Jensen_TIS | Limb bud | 18-Jan | 0.075502 | 0.17617 | 0 | 0 | 13.60876 | 35.1596 |
| Human_Ph | Carious tee | 2/119 | 0.094827 | 0.176598 | 0 | 0 | 3.981096 | 9.378266 |
| Human_Ph | Testicular | 16-Jan | 0.067399 | 0.176598 | 0 | 0 | 15.42481 | 41.60265 |
| Human_Ph | Asymmetry | 8-Jan | 0.03428 | 0.176598 | 0 | 0 | 33.06645 | 111.5391 |
| Human_Ph | Enlarged p | 8-Jan | 0.03428 | 0.176598 | 0 | 0 | 33.06645 | 111.5391 |
| Human_Ph | Aplasia inv | 8-Jan | 0.03428 | 0.176598 | 0 | 0 | 33.06645 | 111.5391 |
| Human_Ph | Aplasia inv | 8-Jan | 0.03428 | 0.176598 | 0 | 0 | 33.06645 | 111.5391 |
| Human_Ph | Aplasia/Hy | 18-Jan | 0.075502 | 0.176598 | 0 | 0 | 13.60876 | 35.1596 |
| Human_Ph | Hypoplasti | 18-Jan | 0.075502 | 0.176598 | 0 | 0 | 13.60876 | 35.1596 |
| Human_Ph | Bone cyst ( | 18-Jan | 0.075502 | 0.176598 | 0 | 0 | 13.60876 | 35.1596 |
| Human_Ph | Absent thu | 18-Jan | 0.075502 | 0.176598 | 0 | 0 | 13.60876 | 35.1596 |
| Human_Ph | Breast carc | 22-Jan | 0.091499 | 0.176598 | 0 | 0 | 11.0144 | 26.34017 |
| Human_Ph | Limited hip | 22-Jan | 0.091499 | 0.176598 | 0 | 0 | 11.0144 | 26.34017 |
| Human_Ph | Neoplasm | 16-Jan | 0.067399 | 0.176598 | 0 | 0 | 15.42481 | 41.60265 |
| Human_Ph | Squamous | 17-Jan | 0.071459 | 0.176598 | 0 | 0 | 14.46003 | 38.15469 |
| Human_Ph | Thyroid car | 18-Jan | 0.075502 | 0.176598 | 0 | 0 | 13.60876 | 35.1596 |
| Human_Ph | CNS hypom | 22-Jan | 0.091499 | 0.176598 | 0 | 0 | 11.0144 | 26.34017 |
| Human_Ph | Abnormalit | 20-Jan | 0.083535 | 0.176598 | 0 | 0 | 12.17503 | 30.22444 |

| | | | | | | | | |
|---|---|---|---|---|---|---|---|---|
| Human_Ph | Abnormalit | 20-Jan | 0.083535 | 0.176598 | 0 | 0 | 12.17503 | 30.22444 |
| Human_Ph | Basal gangl | 20-Jan | 0.083535 | 0.176598 | 0 | 0 | 12.17503 | 30.22444 |
| Human_Ph | Poor eye co | 18-Jan | 0.075502 | 0.176598 | 0 | 0 | 13.60876 | 35.1596 |
| Human_Ph | Skull defec | 18-Jan | 0.075502 | 0.176598 | 0 | 0 | 13.60876 | 35.1596 |
| Human_Ph | Delusions ( | 18-Jan | 0.075502 | 0.176598 | 0 | 0 | 13.60876 | 35.1596 |
| Human_Ph | Intention tı | 22-Jan | 0.091499 | 0.176598 | 0 | 0 | 11.0144 | 26.34017 |
| Human_Ph | Hyperchole | 17-Jan | 0.071459 | 0.176598 | 0 | 0 | 14.46003 | 38.15469 |
| Human_Ph | Hyperthyro | 24-Jan | 0.099394 | 0.176598 | 0 | 0 | 10.05561 | 23.215 |
| Human_Ph | Optic atrop | 4/374 | 0.080303 | 0.176598 | 0 | 0 | 2.54549 | 6.419589 |
| Human_Ph | Abnormal g | 14-Jan | 0.059226 | 0.176598 | 0 | 0 | 17.79964 | 50.30878 |
| Human_Ph | Abnormalit | 14-Jan | 0.059226 | 0.176598 | 0 | 0 | 17.79964 | 50.30878 |
| Human_Ph | Amblyopia | 14-Jan | 0.059226 | 0.176598 | 0 | 0 | 17.79964 | 50.30878 |
| Human_Ph | Narrow mo | Feb-93 | 0.062046 | 0.176598 | 0 | 0 | 5.125275 | 14.24761 |
| Human_Ph | Abnormalit | 19-Jan | 0.079527 | 0.176598 | 0 | 0 | 12.85207 | 32.53709 |
| Human_Ph | Cutaneous | 24-Jan | 0.099394 | 0.176598 | 0 | 0 | 10.05561 | 23.215 |
| Human_Ph | Stillbirth (H | 24-Jan | 0.099394 | 0.176598 | 0 | 0 | 10.05561 | 23.215 |
| Human_Ph | Aplasia/Hy | 24-Jan | 0.099394 | 0.176598 | 0 | 0 | 10.05561 | 23.215 |
| Human_Ph | Bladder car | 24-Jan | 0.099394 | 0.176598 | 0 | 0 | 10.05561 | 23.215 |
| Human_Ph | Short midd | 24-Jan | 0.099394 | 0.176598 | 0 | 0 | 10.05561 | 23.215 |
| Human_Ph | Adenoma s | 17-Jan | 0.071459 | 0.176598 | 0 | 0 | 14.46003 | 38.15469 |
| Human_Ph | Aplasia/Hy | 16-Jan | 0.067399 | 0.176598 | 0 | 0 | 15.42481 | 41.60265 |
| Human_Ph | Heterochro | 16-Jan | 0.067399 | 0.176598 | 0 | 0 | 15.42481 | 41.60265 |
| Human_Ph | Mixed hear | 16-Jan | 0.067399 | 0.176598 | 0 | 0 | 15.42481 | 41.60265 |
| Human_Ph | Neurofibril | 8-Jan | 0.03428 | 0.176598 | 0 | 0 | 33.06645 | 111.5391 |
| Human_Ph | Hypoplastic | 16-Jan | 0.067399 | 0.176598 | 0 | 0 | 15.42481 | 41.60265 |
| Human_Ph | Wide nasal | 3/217 | 0.068882 | 0.176598 | 0 | 0 | 3.28755 | 8.795386 |
| Human_Ph | Premature | 17-Jan | 0.071459 | 0.176598 | 0 | 0 | 14.46003 | 38.15469 |
| Human_Ph | Severe visu | 8-Jan | 0.03428 | 0.176598 | 0 | 0 | 33.06645 | 111.5391 |
| Human_Ph | Mesangial | 8-Jan | 0.03428 | 0.176598 | 0 | 0 | 33.06645 | 111.5391 |
| Human_Ph | Mandibula | Feb-97 | 0.06679 | 0.176598 | 0 | 0 | 4.908483 | 13.28336 |
| Human_Ph | Hip contrac | 16-Jan | 0.067399 | 0.176598 | 0 | 0 | 15.42481 | 41.60265 |
| Human_Ph | Diaphragm | 9-Jan | 0.038483 | 0.176598 | 0 | 0 | 28.93169 | 94.24608 |
| Human_Ph | Abnormal s | 23-Jan | 0.095455 | 0.176598 | 0 | 0 | 10.51321 | 24.6966 |
| Human_Ph | Slow-growi | 24-Jan | 0.099394 | 0.176598 | 0 | 0 | 10.05561 | 23.215 |
| Human_Ph | Mediastina | 20-Jan | 0.083535 | 0.176598 | 0 | 0 | 12.17503 | 30.22444 |
| Human_Ph | Hematoche | 8-Jan | 0.03428 | 0.176598 | 0 | 0 | 33.06645 | 111.5391 |
| Human_Ph | Bell-shaped | 10-Jan | 0.042667 | 0.176598 | 0 | 0 | 25.71576 | 81.1157 |
| Human_Ph | Asymmetri | 10-Jan | 0.042667 | 0.176598 | 0 | 0 | 25.71576 | 81.1157 |
| Human_Ph | Abnormalit | 10-Jan | 0.042667 | 0.176598 | 0 | 0 | 25.71576 | 81.1157 |
| Human_Ph | Broad alve | 10-Jan | 0.042667 | 0.176598 | 0 | 0 | 25.71576 | 81.1157 |
| Human_Ph | Aplasia/Hy | 9-Jan | 0.038483 | 0.176598 | 0 | 0 | 28.93169 | 94.24608 |
| Human_Ph | Muscle hyp | 9-Jan | 0.038483 | 0.176598 | 0 | 0 | 28.93169 | 94.24608 |
| Human_Ph | Conjunctiva | 9-Jan | 0.038483 | 0.176598 | 0 | 0 | 28.93169 | 94.24608 |
| Human_Ph | Blepharitis | 23-Jan | 0.095455 | 0.176598 | 0 | 0 | 10.51321 | 24.6966 |
| Human_Ph | Abnormalit | 9-Jan | 0.038483 | 0.176598 | 0 | 0 | 28.93169 | 94.24608 |

| | | | | | | | | |
|---|---|---|---|---|---|---|---|---|
| Human_Ph | Right ventr | 9-Jan | 0.038483 | 0.176598 | 0 | 0 | 28.93169 | 94.24608 |
| Human_Ph | Cutaneous | 8-Jan | 0.03428 | 0.176598 | 0 | 0 | 33.06645 | 111.5391 |
| Human_Ph | Subaortic s | 8-Jan | 0.03428 | 0.176598 | 0 | 0 | 33.06645 | 111.5391 |
| Human_Ph | Aplasia cut | 10-Jan | 0.042667 | 0.176598 | 0 | 0 | 25.71576 | 81.1157 |
| Human_Ph | Neoplasm ( | 10-Jan | 0.042667 | 0.176598 | 0 | 0 | 25.71576 | 81.1157 |
| Human_Ph | Aplasia/Hy | 13-Jan | 0.055113 | 0.176598 | 0 | 0 | 19.28391 | 55.89185 |
| Human_Ph | Absent pha | 12-Jan | 0.050982 | 0.176598 | 0 | 0 | 21.03805 | 62.61499 |
| Human_Ph | Atrioventri | 12-Jan | 0.050982 | 0.176598 | 0 | 0 | 21.03805 | 62.61499 |
| Human_Ph | Calf muscle | 8-Jan | 0.03428 | 0.176598 | 0 | 0 | 33.06645 | 111.5391 |
| Human_Ph | Abnormalit | 12-Jan | 0.050982 | 0.176598 | 0 | 0 | 21.03805 | 62.61499 |
| Human_Ph | Abnormalit | 20-Jan | 0.083535 | 0.176598 | 0 | 0 | 12.17503 | 30.22444 |
| Human_Ph | Abnormalit | 17-Jan | 0.071459 | 0.176598 | 0 | 0 | 14.46003 | 38.15469 |
| Human_Ph | Microcorne | Feb-97 | 0.06679 | 0.176598 | 0 | 0 | 4.908483 | 13.28336 |
| Human_Ph | Progressive | 21-Jan | 0.087525 | 0.176598 | 0 | 0 | 11.5657 | 28.17205 |
| Human_Ph | Upslanted | 2/108 | 0.080421 | 0.176598 | 0 | 0 | 4.39667 | 11.08174 |
| Human_Ph | Dolichocep | 2/109 | 0.0817 | 0.176598 | 0 | 0 | 4.35536 | 10.90889 |
| Human_Ph | Hepatocell | 20-Jan | 0.083535 | 0.176598 | 0 | 0 | 12.17503 | 30.22444 |
| Human_Ph | Lower limb | 17-Jan | 0.071459 | 0.176598 | 0 | 0 | 14.46003 | 38.15469 |
| Human_Ph | Median cle | 17-Jan | 0.071459 | 0.176598 | 0 | 0 | 14.46003 | 38.15469 |
| Human_Ph | Hypoplasia | 2/103 | 0.074122 | 0.176598 | 0 | 0 | 4.615492 | 12.00969 |
| Human_Ph | 2-3 toe syn | 22-Jan | 0.091499 | 0.176598 | 0 | 0 | 11.0144 | 26.34017 |
| Human_Ph | Absent hair | 20-Jan | 0.083535 | 0.176598 | 0 | 0 | 12.17503 | 30.22444 |
| Human_Ph | Diminished | 19-Jan | 0.079527 | 0.176598 | 0 | 0 | 12.85207 | 32.53709 |
| Human_Ph | EMG: neur | 20-Jan | 0.083535 | 0.176598 | 0 | 0 | 12.17503 | 30.22444 |
| Human_Ph | Dislocated | 20-Jan | 0.083535 | 0.176598 | 0 | 0 | 12.17503 | 30.22444 |
| Human_Ph | Bladder ne | 24-Jan | 0.099394 | 0.176598 | 0 | 0 | 10.05561 | 23.215 |
| Human_Ph | Severe mus | 24-Jan | 0.099394 | 0.176598 | 0 | 0 | 10.05561 | 23.215 |
| Human_Ph | Coarctation | 24-Jan | 0.099394 | 0.176598 | 0 | 0 | 10.05561 | 23.215 |
| Human_Ph | Abnormalit | 24-Jan | 0.099394 | 0.176598 | 0 | 0 | 10.05561 | 23.215 |
| Human_Ph | Palmar hyp | 24-Jan | 0.099394 | 0.176598 | 0 | 0 | 10.05561 | 23.215 |
| Human_Ph | Chest pain | 24-Jan | 0.099394 | 0.176598 | 0 | 0 | 10.05561 | 23.215 |
| Human_Ph | Unsteady g | 24-Jan | 0.099394 | 0.176598 | 0 | 0 | 10.05561 | 23.215 |
| Human_Ph | Atrioventri | 24-Jan | 0.099394 | 0.176598 | 0 | 0 | 10.05561 | 23.215 |
| Human_Ph | Impaired so | 23-Jan | 0.095455 | 0.176598 | 0 | 0 | 10.51321 | 24.6966 |
| Human_Ph | Radial devi | 3/229 | 0.078126 | 0.176598 | 0 | 0 | 3.111094 | 7.931512 |
| Human_Ph | Aplasia/Hy | 19-Jan | 0.079527 | 0.176598 | 0 | 0 | 12.85207 | 32.53709 |
| Human_Ph | Bowel dive | 19-Jan | 0.079527 | 0.176598 | 0 | 0 | 12.85207 | 32.53709 |
| Human_Ph | Taurodonti | 19-Jan | 0.079527 | 0.176598 | 0 | 0 | 12.85207 | 32.53709 |
| Human_Ph | Bony spicu | 19-Jan | 0.079527 | 0.176598 | 0 | 0 | 12.85207 | 32.53709 |
| Human_Ph | Dry hair (H | 9-Jan | 0.038483 | 0.176598 | 0 | 0 | 28.93169 | 94.24608 |
| Human_Ph | Midline de | 14-Jan | 0.059226 | 0.176598 | 0 | 0 | 17.79964 | 50.30878 |
| Human_Ph | Abnormalit | 8-Jan | 0.03428 | 0.176598 | 0 | 0 | 33.06645 | 111.5391 |
| Human_Ph | Hypoplastic | 8-Jan | 0.03428 | 0.176598 | 0 | 0 | 33.06645 | 111.5391 |
| Human_Ph | Macroorch | 13-Jan | 0.055113 | 0.176598 | 0 | 0 | 19.28391 | 55.89185 |
| Human_Ph | Hypoplastic | 18-Jan | 0.075502 | 0.176598 | 0 | 0 | 13.60876 | 35.1596 |

| | | | | | | | | |
|---|---|---|---|---|---|---|---|---|
| Human_Ph | Depressed | 3/228 | 0.077336 | 0.176598 | 0 | 0 | 3.125079 | 7.998944 |
| Human_Ph | Hamartom | 13-Jan | 0.055113 | 0.176598 | 0 | 0 | 19.28391 | 55.89185 |
| Human_Ph | Abnormal a | 19-Jan | 0.079527 | 0.176598 | 0 | 0 | 12.85207 | 32.53709 |
| Human_Ph | Limb-girdle | 11-Jan | 0.046834 | 0.176598 | 0 | 0 | 23.14302 | 70.84419 |
| Human_Ph | Lymphangi | 11-Jan | 0.046834 | 0.176598 | 0 | 0 | 23.14302 | 70.84419 |
| Human_Ph | Rimmed va | 11-Jan | 0.046834 | 0.176598 | 0 | 0 | 23.14302 | 70.84419 |
| Human_Ph | High palate | 3/191 | 0.050731 | 0.176598 | 0 | 0 | 3.74715 | 11.17106 |
| Human_Ph | Secondary | 19-Jan | 0.079527 | 0.176598 | 0 | 0 | 12.85207 | 32.53709 |
| Human_Ph | Supernume | 19-Jan | 0.079527 | 0.176598 | 0 | 0 | 12.85207 | 32.53709 |
| Human_Ph | Genu recur | 18-Jan | 0.075502 | 0.176598 | 0 | 0 | 13.60876 | 35.1596 |
| Human_Ph | Heart block | 18-Jan | 0.075502 | 0.176598 | 0 | 0 | 13.60876 | 35.1596 |
| Human_Ph | Hemangior | 17-Jan | 0.071459 | 0.176598 | 0 | 0 | 14.46003 | 38.15469 |
| Human_Ph | Xanthomat | 14-Jan | 0.059226 | 0.176598 | 0 | 0 | 17.79964 | 50.30878 |
| Human_Ph | Hyporeflex | 12-Jan | 0.050982 | 0.176598 | 0 | 0 | 21.03805 | 62.61499 |
| Human_Ph | Buphthalm | 12-Jan | 0.050982 | 0.176598 | 0 | 0 | 21.03805 | 62.61499 |
| Human_Ph | Clinodactyl | 3/192 | 0.05138 | 0.176598 | 0 | 0 | 3.727135 | 11.06404 |
| Human_Ph | Ragged-rec | 12-Jan | 0.050982 | 0.176598 | 0 | 0 | 21.03805 | 62.61499 |
| Human_Ph | Fine hair (H | Feb-54 | 0.023124 | 0.176598 | 0 | 0 | 8.986878 | 33.8526 |
| Human_Ph | Wrist flexic | 8-Jan | 0.03428 | 0.176598 | 0 | 0 | 33.06645 | 111.5391 |
| Human_Ph | Short midd | 8-Jan | 0.03428 | 0.176598 | 0 | 0 | 33.06645 | 111.5391 |
| Human_Ph | Hypoplastic | 13-Jan | 0.055113 | 0.176598 | 0 | 0 | 19.28391 | 55.89185 |
| Human_Ph | Sudden car | Feb-88 | 0.05629 | 0.176598 | 0 | 0 | 5.424624 | 15.60794 |
| Human_Ph | Juvenile on | Feb-88 | 0.05629 | 0.176598 | 0 | 0 | 5.424624 | 15.60794 |
| Human_Ph | Abnormalit | 19-Jan | 0.079527 | 0.176598 | 0 | 0 | 12.85207 | 32.53709 |
| Human_Ph | Long nose | 12-Jan | 0.050982 | 0.176598 | 0 | 0 | 21.03805 | 62.61499 |
| Human_Ph | Tetrapares | 9-Jan | 0.038483 | 0.176598 | 0 | 0 | 28.93169 | 94.24608 |
| Human_Ph | Microgloss | 9-Jan | 0.038483 | 0.176598 | 0 | 0 | 28.93169 | 94.24608 |
| Human_Ph | Aqueducta | 10-Jan | 0.042667 | 0.176598 | 0 | 0 | 25.71576 | 81.1157 |
| Human_Ph | Mesomelia | 10-Jan | 0.042667 | 0.176598 | 0 | 0 | 25.71576 | 81.1157 |
| Human_Ph | Global brai | 10-Jan | 0.042667 | 0.176598 | 0 | 0 | 25.71576 | 81.1157 |
| Human_Ph | Abnormal r | 13-Jan | 0.055113 | 0.176598 | 0 | 0 | 19.28391 | 55.89185 |
| Human_Ph | Abnormalit | 11-Jan | 0.046834 | 0.176598 | 0 | 0 | 23.14302 | 70.84419 |
| Human_Ph | Long clavic | 9-Jan | 0.038483 | 0.176598 | 0 | 0 | 28.93169 | 94.24608 |
| Human_Ph | Type 1 mus | 9-Jan | 0.038483 | 0.176598 | 0 | 0 | 28.93169 | 94.24608 |
| Human_Ph | Premature | 15-Jan | 0.063321 | 0.176598 | 0 | 0 | 16.52741 | 45.60793 |
| Human_Ph | Ovoid verte | 21-Jan | 0.087525 | 0.176598 | 0 | 0 | 11.5657 | 28.17205 |
| Human_Ph | Delayed gr | 21-Jan | 0.087525 | 0.176598 | 0 | 0 | 11.5657 | 28.17205 |
| Human_Ph | Abnormalit | 21-Jan | 0.087525 | 0.176598 | 0 | 0 | 11.5657 | 28.17205 |
| Human_Ph | Abnormalit | 21-Jan | 0.087525 | 0.176598 | 0 | 0 | 11.5657 | 28.17205 |
| Human_Ph | Generalize | 22-Jan | 0.091499 | 0.176598 | 0 | 0 | 11.0144 | 26.34017 |
| Human_Ph | Palmoplant | 22-Jan | 0.091499 | 0.176598 | 0 | 0 | 11.0144 | 26.34017 |
| Human_Ph | Mildly elev | 15-Jan | 0.063321 | 0.176598 | 0 | 0 | 16.52741 | 45.60793 |
| Human_Ph | Short hallu | 15-Jan | 0.063321 | 0.176598 | 0 | 0 | 16.52741 | 45.60793 |
| Human_Ph | Maternal d | 15-Jan | 0.063321 | 0.176598 | 0 | 0 | 16.52741 | 45.60793 |
| Human_Ph | Abnormalit | 15-Jan | 0.063321 | 0.176598 | 0 | 0 | 16.52741 | 45.60793 |

| | | | | | | | | |
|---|---|---|---|---|---|---|---|---|
| Human_Ph | Selective to | 11-Jan | 0.046834 | 0.176598 | 0 | 0 | 23.14302 | 70.84419 |
| Human_Ph | Short 5th fi | 15-Jan | 0.063321 | 0.176598 | 0 | 0 | 16.52741 | 45.60793 |
| Human_Ph | Abnormal f | 14-Jan | 0.059226 | 0.176598 | 0 | 0 | 17.79964 | 50.30878 |
| Human_Ph | Acute hepa | 14-Jan | 0.059226 | 0.176598 | 0 | 0 | 17.79964 | 50.30878 |
| Human_Ph | Fragile nail | 14-Jan | 0.059226 | 0.176598 | 0 | 0 | 17.79964 | 50.30878 |
| Human_Ph | Fibular apla | 8-Jan | 0.03428 | 0.176598 | 0 | 0 | 33.06645 | 111.5391 |
| Human_Ph | Aplasia of t | 8-Jan | 0.03428 | 0.176598 | 0 | 0 | 33.06645 | 111.5391 |
| Human_Ph | Narrow nas | 15-Jan | 0.063321 | 0.176598 | 0 | 0 | 16.52741 | 45.60793 |
| Human_Ph | Abnormalit | 15-Jan | 0.063321 | 0.176598 | 0 | 0 | 16.52741 | 45.60793 |
| Human_Ph | Abnormalit | 19-Jan | 0.079527 | 0.176598 | 0 | 0 | 12.85207 | 32.53709 |
| Human_Ph | Abnormalit | 19-Jan | 0.079527 | 0.176598 | 0 | 0 | 12.85207 | 32.53709 |
| Human_Ph | Paraparesis | 22-Jan | 0.091499 | 0.176598 | 0 | 0 | 11.0144 | 26.34017 |
| Human_Ph | Angiofibror | 17-Jan | 0.071459 | 0.176598 | 0 | 0 | 14.46003 | 38.15469 |
| Human_Ph | Hypokinesi | 11-Jan | 0.046834 | 0.176598 | 0 | 0 | 23.14302 | 70.84419 |
| Human_Ph | Bulbar pals | 19-Jan | 0.079527 | 0.176598 | 0 | 0 | 12.85207 | 32.53709 |
| Human_Ph | Congenital | 19-Jan | 0.079527 | 0.176598 | 0 | 0 | 12.85207 | 32.53709 |
| Human_Ph | Late onset | 11-Jan | 0.046834 | 0.176598 | 0 | 0 | 23.14302 | 70.84419 |
| Human_Ph | Cerebral hy | 11-Jan | 0.046834 | 0.176598 | 0 | 0 | 23.14302 | 70.84419 |
| Human_Ph | Obstructive | 22-Jan | 0.091499 | 0.176598 | 0 | 0 | 11.0144 | 26.34017 |
| Human_Ph | Transitiona | 22-Jan | 0.091499 | 0.176598 | 0 | 0 | 11.0144 | 26.34017 |
| Human_Ph | Cubitus val | 22-Jan | 0.091499 | 0.176598 | 0 | 0 | 11.0144 | 26.34017 |
| Human_Ph | Bicuspid ac | 21-Jan | 0.087525 | 0.176598 | 0 | 0 | 11.5657 | 28.17205 |
| Human_Ph | Telecanthu | Feb-83 | 0.050736 | 0.176598 | 0 | 0 | 5.76093 | 17.17404 |
| Human_Ph | Aplasia/Hy | 12-Jan | 0.050982 | 0.176598 | 0 | 0 | 21.03805 | 62.61499 |
| Human_Ph | Personality | 17-Jan | 0.071459 | 0.176598 | 0 | 0 | 14.46003 | 38.15469 |
| Human_Ph | Meningiom | 17-Jan | 0.071459 | 0.176598 | 0 | 0 | 14.46003 | 38.15469 |
| Human_Ph | Cerebral in | 11-Jan | 0.046834 | 0.176598 | 0 | 0 | 23.14302 | 70.84419 |
| Human_Ph | Aplasia/Hy | 16-Jan | 0.067399 | 0.176598 | 0 | 0 | 15.42481 | 41.60265 |
| Human_Ph | Attenuatio | 21-Jan | 0.087525 | 0.176598 | 0 | 0 | 11.5657 | 28.17205 |
| Human_Ph | Cavernous | 22-Jan | 0.091499 | 0.176598 | 0 | 0 | 11.0144 | 26.34017 |
| Human_Ph | Chronic ob | 22-Jan | 0.091499 | 0.176598 | 0 | 0 | 11.0144 | 26.34017 |
| Human_Ph | Visceral an | 16-Jan | 0.067399 | 0.176598 | 0 | 0 | 15.42481 | 41.60265 |
| Human_Ph | Abnormalit | 11-Jan | 0.046834 | 0.176598 | 0 | 0 | 23.14302 | 70.84419 |
| HuBMAP_/ | Bronchial S | 3/102 | 0.010016 | 0.176776 | 0 | 0 | 7.147908 | 32.90605 |
| HuBMAP_/ | Lymphatic | 3/111 | 0.012584 | 0.176776 | 0 | 0 | 6.549272 | 28.65547 |
| HuBMAP_/ | Inh L1 LAM | 3/103 | 0.010284 | 0.176776 | 0 | 0 | 7.076071 | 32.38812 |
| HuBMAP_/ | B Cell Zone | 3/105 | 0.010834 | 0.176776 | 0 | 0 | 6.936625 | 31.38872 |
| HuBMAP_/ | T Cell Zone | 3/105 | 0.010834 | 0.176776 | 0 | 0 | 6.936625 | 31.38872 |
| HuBMAP_/ | Medullary | 3/105 | 0.010834 | 0.176776 | 0 | 0 | 6.936625 | 31.38872 |
| HuBMAP_/ | Marginal R | 3/106 | 0.011115 | 0.176776 | 0 | 0 | 6.868932 | 30.90647 |
| HuBMAP_/ | Splenic Wh | 3/101 | 0.009751 | 0.176776 | 0 | 0 | 7.22121 | 33.4367 |
| HuBMAP_/ | Erythroid L | 3/108 | 0.01169 | 0.176776 | 0 | 0 | 6.737415 | 29.97504 |
| HuBMAP_/ | Pericyte ce | 3/102 | 0.010016 | 0.176776 | 0 | 0 | 7.147908 | 32.90605 |
| HuBMAP_/ | Prostate pe | 3/102 | 0.010016 | 0.176776 | 0 | 0 | 7.147908 | 32.90605 |
| HuBMAP_/ | Splenic Ma | 3/101 | 0.009751 | 0.176776 | 0 | 0 | 7.22121 | 33.4367 |

| | | | | | | | | |
|---|---|---|---|---|---|---|---|---|
| HuBMAP_/ | Oligo L2-6 ( | 3/102 | 0.010016 | 0.176776 | 0 | 0 | 7.147908 | 32.90605 |
| HuBMAP_/ | Lymphatic | 3/111 | 0.012584 | 0.176776 | 0 | 0 | 6.549272 | 28.65547 |
| HuBMAP_/ | Smooth Mu | 3/102 | 0.010016 | 0.176776 | 0 | 0 | 7.147908 | 32.90605 |
| HuBMAP_/ | DPT+ Fibro | 3/101 | 0.009751 | 0.176776 | 0 | 0 | 7.22121 | 33.4367 |
| HuBMAP_/ | Neuro-Mec | 3/110 | 0.012281 | 0.176776 | 0 | 0 | 6.610814 | 29.08541 |
| HuBMAP_/ | Lymphatic | 3/112 | 0.01289 | 0.176776 | 0 | 0 | 6.48886 | 28.23504 |
| CellMarker | Astrocyte E | 5/582 | 0.109681 | 0.17678 | 0 | 0 | 2.04337 | 4.516217 |
| GeDiPNet_ | Myocardial | 4/355 | 0.069188 | 0.176914 | 0 | 0 | 2.685889 | 7.173822 |
| CellMarker | Macrophag | 27-Jan | 0.111111 | 0.177002 | 0 | 0 | 8.894007 | 19.54216 |
| CellMarker | CD4+ T Cel | 27-Jan | 0.111111 | 0.177002 | 0 | 0 | 8.894007 | 19.54216 |
| DisGeNET | Intracrania | 6-Jan | 0.025821 | 0.17715 | 0 | 0 | 46.29767 | 169.2909 |
| DisGeNET | Townes syr | 8-Jan | 0.03428 | 0.17715 | 0 | 0 | 33.06645 | 111.5391 |
| DisGeNET | Tubular ad | 8-Jan | 0.03428 | 0.17715 | 0 | 0 | 33.06645 | 111.5391 |
| DisGeNET | Congenital | 7-Jan | 0.03006 | 0.17715 | 0 | 0 | 38.57946 | 135.2043 |
| DisGeNET | Papillary cy | 7-Jan | 0.03006 | 0.17715 | 0 | 0 | 38.57946 | 135.2043 |
| DisGeNET | Aplasia/Hy | 7-Jan | 0.03006 | 0.17715 | 0 | 0 | 38.57946 | 135.2043 |
| DisGeNET | Arcus Senil | 7-Jan | 0.03006 | 0.17715 | 0 | 0 | 38.57946 | 135.2043 |
| DisGeNET | Asymmetry | 7-Jan | 0.03006 | 0.17715 | 0 | 0 | 38.57946 | 135.2043 |
| DisGeNET | Malignant | 7-Jan | 0.03006 | 0.17715 | 0 | 0 | 38.57946 | 135.2043 |
| DisGeNET | MINICORE | 7-Jan | 0.03006 | 0.17715 | 0 | 0 | 38.57946 | 135.2043 |
| DisGeNET | Broad alve | 7-Jan | 0.03006 | 0.17715 | 0 | 0 | 38.57946 | 135.2043 |
| DisGeNET | Colitis, Mic | 6-Jan | 0.025821 | 0.17715 | 0 | 0 | 46.29767 | 169.2909 |
| DisGeNET | Scaly skin | 6-Jan | 0.025821 | 0.17715 | 0 | 0 | 46.29767 | 169.2909 |
| DisGeNET | Sea-Blue H | 6-Jan | 0.025821 | 0.17715 | 0 | 0 | 46.29767 | 169.2909 |
| DisGeNET | Idiopathic r | 6-Jan | 0.025821 | 0.17715 | 0 | 0 | 46.29767 | 169.2909 |
| DisGeNET | HYPERALPH | 8-Jan | 0.03428 | 0.17715 | 0 | 0 | 33.06645 | 111.5391 |
| DisGeNET | Speech Sou | 6-Jan | 0.025821 | 0.17715 | 0 | 0 | 46.29767 | 169.2909 |
| DisGeNET | Neoplasms | 6-Jan | 0.025821 | 0.17715 | 0 | 0 | 46.29767 | 169.2909 |
| DisGeNET | Stunned M | 6-Jan | 0.025821 | 0.17715 | 0 | 0 | 46.29767 | 169.2909 |
| DisGeNET | Aristolochi | 6-Jan | 0.025821 | 0.17715 | 0 | 0 | 46.29767 | 169.2909 |
| DisGeNET | Axonal reg | 6-Jan | 0.025821 | 0.17715 | 0 | 0 | 46.29767 | 169.2909 |
| DisGeNET | Tension He | 6-Jan | 0.025821 | 0.17715 | 0 | 0 | 46.29767 | 169.2909 |
| DisGeNET | Hyperplasi | 6-Jan | 0.025821 | 0.17715 | 0 | 0 | 46.29767 | 169.2909 |
| DisGeNET | Trichilemm | 6-Jan | 0.025821 | 0.17715 | 0 | 0 | 46.29767 | 169.2909 |
| DisGeNET | Trichinellos | 6-Jan | 0.025821 | 0.17715 | 0 | 0 | 46.29767 | 169.2909 |
| DisGeNET | Abnormal l | 6-Jan | 0.025821 | 0.17715 | 0 | 0 | 46.29767 | 169.2909 |
| DisGeNET | Unconsciou | 6-Jan | 0.025821 | 0.17715 | 0 | 0 | 46.29767 | 169.2909 |
| DisGeNET | NONAKA M | Feb-51 | 0.020776 | 0.17715 | 0 | 0 | 9.538535 | 36.95177 |
| DisGeNET | Presenile d | 5/362 | 0.020876 | 0.17715 | 0 | 0 | 3.340165 | 12.92363 |
| DisGeNET | Iodine defi | 6-Jan | 0.025821 | 0.17715 | 0 | 0 | 46.29767 | 169.2909 |
| DisGeNET | Familial rer | 8-Jan | 0.03428 | 0.17715 | 0 | 0 | 33.06645 | 111.5391 |
| DisGeNET | Epilepsy | 10/1177 | 0.031495 | 0.17715 | 0 | 0 | 2.086157 | 7.213798 |
| DisGeNET | Temporom | 7-Jan | 0.03006 | 0.17715 | 0 | 0 | 38.57946 | 135.2043 |
| DisGeNET | Toxic multi | 7-Jan | 0.03006 | 0.17715 | 0 | 0 | 38.57946 | 135.2043 |
| DisGeNET | Anxiety Dis | 5/396 | 0.029271 | 0.17715 | 0 | 0 | 3.044414 | 10.75027 |

| | | | | | | | | |
|---|---|---|---|---|---|---|---|---|
| DisGeNET | Hypothyroi | 4/268 | 0.029538 | 0.17715 | 0 | 0 | 3.586893 | 12.63333 |
| DisGeNET | Multiple pc | Feb-62 | 0.029883 | 0.17715 | 0 | 0 | 7.78549 | 27.33058 |
| DisGeNET | Precursor c | Feb-62 | 0.029883 | 0.17715 | 0 | 0 | 7.78549 | 27.33058 |
| DisGeNET | Myopia | 4/269 | 0.029887 | 0.17715 | 0 | 0 | 3.573176 | 12.54301 |
| DisGeNET | Enlargemer | 7-Jan | 0.03006 | 0.17715 | 0 | 0 | 38.57946 | 135.2043 |
| DisGeNET | Macular De | 8-Jan | 0.03428 | 0.17715 | 0 | 0 | 33.06645 | 111.5391 |
| DisGeNET | Broad colu | 6-Jan | 0.025821 | 0.17715 | 0 | 0 | 46.29767 | 169.2909 |
| DisGeNET | Amyloidosi | 8-Jan | 0.03428 | 0.17715 | 0 | 0 | 33.06645 | 111.5391 |
| DisGeNET | Aphasia, Pr | 8-Jan | 0.03428 | 0.17715 | 0 | 0 | 33.06645 | 111.5391 |
| DisGeNET | Atrophy, D | 8-Jan | 0.03428 | 0.17715 | 0 | 0 | 33.06645 | 111.5391 |
| DisGeNET | Keratitis-Ic | 6-Jan | 0.025821 | 0.17715 | 0 | 0 | 46.29767 | 169.2909 |
| DisGeNET | Scalp defec | 8-Jan | 0.03428 | 0.17715 | 0 | 0 | 33.06645 | 111.5391 |
| DisGeNET | Secondary | 8-Jan | 0.03428 | 0.17715 | 0 | 0 | 33.06645 | 111.5391 |
| DisGeNET | Urethral Ol | 8-Jan | 0.03428 | 0.17715 | 0 | 0 | 33.06645 | 111.5391 |
| DisGeNET | Uterine car | 8-Jan | 0.03428 | 0.17715 | 0 | 0 | 33.06645 | 111.5391 |
| DisGeNET | Sensory de | 8-Jan | 0.03428 | 0.17715 | 0 | 0 | 33.06645 | 111.5391 |
| DisGeNET | Flexion cor | 8-Jan | 0.03428 | 0.17715 | 0 | 0 | 33.06645 | 111.5391 |
| DisGeNET | Focal abser | 8-Jan | 0.03428 | 0.17715 | 0 | 0 | 33.06645 | 111.5391 |
| DisGeNET | Frontometa | 8-Jan | 0.03428 | 0.17715 | 0 | 0 | 33.06645 | 111.5391 |
| DisGeNET | Simple End | 8-Jan | 0.03428 | 0.17715 | 0 | 0 | 33.06645 | 111.5391 |
| DisGeNET | Glaucoma c | 8-Jan | 0.03428 | 0.17715 | 0 | 0 | 33.06645 | 111.5391 |
| DisGeNET | Broad alve | 7-Jan | 0.03006 | 0.17715 | 0 | 0 | 38.57946 | 135.2043 |
| DisGeNET | Broad long | 7-Jan | 0.03006 | 0.17715 | 0 | 0 | 38.57946 | 135.2043 |
| DisGeNET | Spindle Cel | 8-Jan | 0.03428 | 0.17715 | 0 | 0 | 33.06645 | 111.5391 |
| DisGeNET | Sprains anc | 8-Jan | 0.03428 | 0.17715 | 0 | 0 | 33.06645 | 111.5391 |
| DisGeNET | Childhood | Feb-50 | 0.020017 | 0.17715 | 0 | 0 | 9.737745 | 38.08585 |
| DisGeNET | Carcinoma, | 6-Jan | 0.025821 | 0.17715 | 0 | 0 | 46.29767 | 169.2909 |
| DisGeNET | Severe mus | Feb-54 | 0.023124 | 0.17715 | 0 | 0 | 8.986878 | 33.8526 |
| DisGeNET | Syndactyly | Feb-65 | 0.032597 | 0.17715 | 0 | 0 | 7.413632 | 25.38081 |
| DisGeNET | Paroxysma | Feb-65 | 0.032597 | 0.17715 | 0 | 0 | 7.413632 | 25.38081 |
| DisGeNET | Adenocarci | 10/1187 | 0.033102 | 0.17715 | 0 | 0 | 2.067329 | 7.045775 |
| DisGeNET | Congenital | Feb-66 | 0.033522 | 0.17715 | 0 | 0 | 7.297426 | 24.7787 |
| DisGeNET | Disorder ch | 8-Jan | 0.03428 | 0.17715 | 0 | 0 | 33.06645 | 111.5391 |
| DisGeNET | Prodrome | 8-Jan | 0.03428 | 0.17715 | 0 | 0 | 33.06645 | 111.5391 |
| DisGeNET | contracture | Feb-48 | 0.018536 | 0.17715 | 0 | 0 | 10.16215 | 40.52728 |
| DisGeNET | Epidemic k | 6-Jan | 0.025821 | 0.17715 | 0 | 0 | 46.29767 | 169.2909 |
| DisGeNET | Epithelial h | Feb-61 | 0.029 | 0.17715 | 0 | 0 | 7.917846 | 28.0328 |
| DisGeNET | Mydriasis | 6-Jan | 0.025821 | 0.17715 | 0 | 0 | 46.29767 | 169.2909 |
| DisGeNET | Myeloid M | 6-Jan | 0.025821 | 0.17715 | 0 | 0 | 46.29767 | 169.2909 |
| DisGeNET | Cessation c | 6-Jan | 0.025821 | 0.17715 | 0 | 0 | 46.29767 | 169.2909 |
| DisGeNET | Skin Diseas | 6-Jan | 0.025821 | 0.17715 | 0 | 0 | 46.29767 | 169.2909 |
| DisGeNET | Vesicular S | 3/133 | 0.020317 | 0.17715 | 0 | 0 | 5.43489 | 21.17595 |
| DisGeNET | beta Thalas | 3/133 | 0.020317 | 0.17715 | 0 | 0 | 5.43489 | 21.17595 |
| DisGeNET | beta^+^ Th | 3/128 | 0.018376 | 0.17715 | 0 | 0 | 5.653714 | 22.5964 |
| DisGeNET | Hyperlipop | 3/153 | 0.029172 | 0.17715 | 0 | 0 | 4.705476 | 16.63166 |

| | | | | | | | | |
|---|---|---|---|---|---|---|---|---|
| DisGeNET | Cervical Sq | 3/153 | 0.029172 | 0.17715 | 0 | 0 | 4.705476 | 16.63166 |
| DisGeNET | Memory in | 4/267 | 0.029191 | 0.17715 | 0 | 0 | 3.600715 | 12.72455 |
| DisGeNET | gliosarcom | Feb-63 | 0.030777 | 0.17715 | 0 | 0 | 7.657473 | 26.65545 |
| DisGeNET | Skeletal ma | 6-Jan | 0.025821 | 0.17715 | 0 | 0 | 46.29767 | 169.2909 |
| DisGeNET | Sleep-wake | 6-Jan | 0.025821 | 0.17715 | 0 | 0 | 46.29767 | 169.2909 |
| DisGeNET | Small cell c | 6-Jan | 0.025821 | 0.17715 | 0 | 0 | 46.29767 | 169.2909 |
| DisGeNET | Ventricular | Feb-60 | 0.028127 | 0.17715 | 0 | 0 | 8.054767 | 28.76365 |
| DisGeNET | Common v | 8-Jan | 0.03428 | 0.17715 | 0 | 0 | 33.06645 | 111.5391 |
| DisGeNET | Common w | 8-Jan | 0.03428 | 0.17715 | 0 | 0 | 33.06645 | 111.5391 |
| DisGeNET | Xanthoma | 8-Jan | 0.03428 | 0.17715 | 0 | 0 | 33.06645 | 111.5391 |
| DisGeNET | tumor vasc | 3/129 | 0.018755 | 0.17715 | 0 | 0 | 5.60856 | 22.30125 |
| DisGeNET | LIPOPROTE | 6-Jan | 0.025821 | 0.17715 | 0 | 0 | 46.29767 | 169.2909 |
| DisGeNET | Fingernail ( | 8-Jan | 0.03428 | 0.17715 | 0 | 0 | 33.06645 | 111.5391 |
| DisGeNET | Dermatolo | 5/406 | 0.032095 | 0.17715 | 0 | 0 | 2.966973 | 10.20359 |
| DisGeNET | Multiple, su | Feb-65 | 0.032597 | 0.17715 | 0 | 0 | 7.413632 | 25.38081 |
| DisGeNET | Hepatic me | Feb-65 | 0.032597 | 0.17715 | 0 | 0 | 7.413632 | 25.38081 |
| DisGeNET | Bifid nasal | 6-Jan | 0.025821 | 0.17715 | 0 | 0 | 46.29767 | 169.2909 |
| DisGeNET | Melanoma | 6-Jan | 0.025821 | 0.17715 | 0 | 0 | 46.29767 | 169.2909 |
| DisGeNET | Birth lengtl | 6-Jan | 0.025821 | 0.17715 | 0 | 0 | 46.29767 | 169.2909 |
| DisGeNET | Myocardial | 8-Jan | 0.03428 | 0.17715 | 0 | 0 | 33.06645 | 111.5391 |
| DisGeNET | Myofibrom | 8-Jan | 0.03428 | 0.17715 | 0 | 0 | 33.06645 | 111.5391 |
| DisGeNET | Proximal w | 8-Jan | 0.03428 | 0.17715 | 0 | 0 | 33.06645 | 111.5391 |
| DisGeNET | Unipolar D | 6/517 | 0.025216 | 0.17715 | 0 | 0 | 2.812495 | 10.35077 |
| DisGeNET | Abnormally | 3/145 | 0.025421 | 0.17715 | 0 | 0 | 4.972586 | 18.26021 |
| DisGeNET | Ewings sarc | 5/352 | 0.018748 | 0.17715 | 0 | 0 | 3.438181 | 13.67242 |
| DisGeNET | Solitary sca | 8-Jan | 0.03428 | 0.17715 | 0 | 0 | 33.06645 | 111.5391 |
| DisGeNET | Chronic my | 4/233 | 0.018834 | 0.17715 | 0 | 0 | 4.142474 | 16.45435 |
| DisGeNET | Low grade | 6-Jan | 0.025821 | 0.17715 | 0 | 0 | 46.29767 | 169.2909 |
| DisGeNET | Diaphragm | 8-Jan | 0.03428 | 0.17715 | 0 | 0 | 33.06645 | 111.5391 |
| DisGeNET | Diffuse Lar | 8-Jan | 0.03428 | 0.17715 | 0 | 0 | 33.06645 | 111.5391 |
| DisGeNET | Diminished | 8-Jan | 0.03428 | 0.17715 | 0 | 0 | 33.06645 | 111.5391 |
| DisGeNET | Direct Cont | 8-Jan | 0.03428 | 0.17715 | 0 | 0 | 33.06645 | 111.5391 |
| DisGeNET | cardiac syn | 8-Jan | 0.03428 | 0.17715 | 0 | 0 | 33.06645 | 111.5391 |
| DisGeNET | Meningiom | 4/241 | 0.021023 | 0.17715 | 0 | 0 | 4.001017 | 15.45242 |
| DisGeNET | Middle Cer | 3/135 | 0.021124 | 0.17715 | 0 | 0 | 5.352002 | 20.6445 |
| DisGeNET | Cholelithia | 3/136 | 0.021534 | 0.17715 | 0 | 0 | 5.311493 | 20.38612 |
| DisGeNET | Localized a | 6-Jan | 0.025821 | 0.17715 | 0 | 0 | 46.29767 | 169.2909 |
| DisGeNET | Familial me | 8-Jan | 0.03428 | 0.17715 | 0 | 0 | 33.06645 | 111.5391 |
| DisGeNET | Subcortical | 6-Jan | 0.025821 | 0.17715 | 0 | 0 | 46.29767 | 169.2909 |
| DisGeNET | Myositis | 3/143 | 0.024527 | 0.17715 | 0 | 0 | 5.044133 | 18.70358 |
| DisGeNET | Axial muscl | 8-Jan | 0.03428 | 0.17715 | 0 | 0 | 33.06645 | 111.5391 |
| DisGeNET | Respiratory | Feb-56 | 0.024747 | 0.17715 | 0 | 0 | 8.653159 | 32.00857 |
| DisGeNET | Psychotic s | Feb-48 | 0.018536 | 0.17715 | 0 | 0 | 10.16215 | 40.52728 |
| DisGeNET | leukemia | 15/1941 | 0.019688 | 0.17715 | 0 | 0 | 1.945634 | 7.641996 |
| DisGeNET | IGA Glome | 4/237 | 0.01991 | 0.17715 | 0 | 0 | 4.070531 | 15.94241 |

| | | | | | | | | |
|---|---|---|---|---|---|---|---|---|
| DisGeNET | Small Lymp | Feb-50 | 0.020017 | 0.17715 | 0 | 0 | 9.737745 | 38.08585 |
| DisGeNET | Other spec | Feb-53 | 0.02233 | 0.17715 | 0 | 0 | 9.163552 | 34.83838 |
| DisGeNET | Childhood | Feb-53 | 0.02233 | 0.17715 | 0 | 0 | 9.163552 | 34.83838 |
| DisGeNET | NEUROTICI | Feb-53 | 0.02233 | 0.17715 | 0 | 0 | 9.163552 | 34.83838 |
| DisGeNET | Endometri | 6-Jan | 0.025821 | 0.17715 | 0 | 0 | 46.29767 | 169.2909 |
| DisGeNET | Nodular gl | Feb-53 | 0.02233 | 0.17715 | 0 | 0 | 9.163552 | 34.83838 |
| DisGeNET | Brain Neop | 7/646 | 0.02243 | 0.17715 | 0 | 0 | 2.639241 | 10.02216 |
| DisGeNET | Tachycardi | Feb-51 | 0.020776 | 0.17715 | 0 | 0 | 9.538535 | 36.95177 |
| DisGeNET | Adult Lymp | Feb-51 | 0.020776 | 0.17715 | 0 | 0 | 9.538535 | 36.95177 |
| DisGeNET | Lafora Dise | Feb-51 | 0.020776 | 0.17715 | 0 | 0 | 9.538535 | 36.95177 |
| DisGeNET | Lymphede | 8-Jan | 0.03428 | 0.17715 | 0 | 0 | 33.06645 | 111.5391 |
| DisGeNET | Alzheimer | 4/253 | 0.024591 | 0.17715 | 0 | 0 | 3.805874 | 14.1022 |
| DisGeNET | Aplasia cut | 8-Jan | 0.03428 | 0.17715 | 0 | 0 | 33.06645 | 111.5391 |
| DisGeNET | Treatment | Feb-56 | 0.024747 | 0.17715 | 0 | 0 | 8.653159 | 32.00857 |
| DisGeNET | Amyotroph | 7/660 | 0.024853 | 0.17715 | 0 | 0 | 2.580781 | 9.535457 |
| DisGeNET | Flexion cor | Feb-52 | 0.021547 | 0.17715 | 0 | 0 | 9.347294 | 35.8704 |
| DisGeNET | Malignant | Feb-52 | 0.021547 | 0.17715 | 0 | 0 | 9.347294 | 35.8704 |
| DisGeNET | Hypertrigly | 3/157 | 0.031152 | 0.17715 | 0 | 0 | 4.582328 | 15.89551 |
| DisGeNET | Decreased | 3/145 | 0.025421 | 0.17715 | 0 | 0 | 4.972586 | 18.26021 |
| DisGeNET | Cataract | 6/519 | 0.025635 | 0.17715 | 0 | 0 | 2.801242 | 10.26317 |
| DisGeNET | OSSIFICATI | 8-Jan | 0.03428 | 0.17715 | 0 | 0 | 33.06645 | 111.5391 |
| DisGeNET | Xanthelasn | 8-Jan | 0.03428 | 0.17715 | 0 | 0 | 33.06645 | 111.5391 |
| DisGeNET | Wernicke-I | 8-Jan | 0.03428 | 0.17715 | 0 | 0 | 33.06645 | 111.5391 |
| DisGeNET | Old myoca | 8-Jan | 0.03428 | 0.17715 | 0 | 0 | 33.06645 | 111.5391 |
| DisGeNET | Brain Disea | 4/250 | 0.023667 | 0.17715 | 0 | 0 | 3.852875 | 14.42394 |
| DisGeNET | Adenoma c | 4/258 | 0.026179 | 0.17715 | 0 | 0 | 3.730007 | 13.58763 |
| DisGeNET | Mixed dem | 6-Jan | 0.025821 | 0.17715 | 0 | 0 | 46.29767 | 169.2909 |
| DisGeNET | Calf muscle | 6-Jan | 0.025821 | 0.17715 | 0 | 0 | 46.29767 | 169.2909 |
| DisGeNET | Waddling g | Feb-59 | 0.027266 | 0.17715 | 0 | 0 | 8.196491 | 29.5248 |
| DisGeNET | Drug Resist | Feb-59 | 0.027266 | 0.17715 | 0 | 0 | 8.196491 | 29.5248 |
| DisGeNET | ACTIVATED | 7-Jan | 0.03006 | 0.17715 | 0 | 0 | 38.57946 | 135.2043 |
| DisGeNET | Decreased | 6-Jan | 0.025821 | 0.17715 | 0 | 0 | 46.29767 | 169.2909 |
| DisGeNET | Dementia i | 6-Jan | 0.025821 | 0.17715 | 0 | 0 | 46.29767 | 169.2909 |
| DisGeNET | Numerous | 6-Jan | 0.025821 | 0.17715 | 0 | 0 | 46.29767 | 169.2909 |
| DisGeNET | Congenital | 6-Jan | 0.025821 | 0.17715 | 0 | 0 | 46.29767 | 169.2909 |
| DisGeNET | Patchy palı | 6-Jan | 0.025821 | 0.17715 | 0 | 0 | 46.29767 | 169.2909 |
| DisGeNET | Bipolar Dis | 8/837 | 0.029138 | 0.17715 | 0 | 0 | 2.331191 | 8.242451 |
| DisGeNET | Familial lic | 3/153 | 0.029172 | 0.17715 | 0 | 0 | 4.705476 | 16.63166 |
| DisGeNET | Primary Cu | 6-Jan | 0.025821 | 0.17715 | 0 | 0 | 46.29767 | 169.2909 |
| DisGeNET | Proteus Syı | 6-Jan | 0.025821 | 0.17715 | 0 | 0 | 46.29767 | 169.2909 |
| DisGeNET | Coronary S | Feb-52 | 0.021547 | 0.17715 | 0 | 0 | 9.347294 | 35.8704 |
| DisGeNET | SCHIZOPHI | Feb-58 | 0.026415 | 0.17715 | 0 | 0 | 8.343277 | 30.31803 |
| DisGeNET | Amnesia | Feb-58 | 0.026415 | 0.17715 | 0 | 0 | 8.343277 | 30.31803 |
| DisGeNET | Forgetful | Feb-59 | 0.027266 | 0.17715 | 0 | 0 | 8.196491 | 29.5248 |
| DisGeNET | Secondary | 6-Jan | 0.025821 | 0.17715 | 0 | 0 | 46.29767 | 169.2909 |

| | | | | | | | | |
|---|---|---|---|---|---|---|---|---|
| DisGeNET | First degre | 6-Jan | 0.025821 | 0.17715 | 0 | 0 | 46.29767 | 169.2909 |
| DisGeNET | Mild cognit | 3/150 | 0.027733 | 0.17715 | 0 | 0 | 4.802235 | 17.21664 |
| DisGeNET | Malignant | 4/263 | 0.027828 | 0.17715 | 0 | 0 | 3.657068 | 13.09856 |
| DisGeNET | AURAL ATF | Feb-60 | 0.028127 | 0.17715 | 0 | 0 | 8.054767 | 28.76365 |
| DisGeNET | Encephalo | 4/234 | 0.019099 | 0.17715 | 0 | 0 | 4.124254 | 16.32424 |
| DisGeNET | Sparse eye | Feb-49 | 0.01927 | 0.17715 | 0 | 0 | 9.945432 | 39.27635 |
| DisGeNET | COWDEN-L | 8-Jan | 0.03428 | 0.17715 | 0 | 0 | 33.06645 | 111.5391 |
| DisGeNET | Disease Ex | 3/144 | 0.024972 | 0.17715 | 0 | 0 | 5.008105 | 18.47995 |
| DisGeNET | Complex E | 8-Jan | 0.03428 | 0.17715 | 0 | 0 | 33.06645 | 111.5391 |
| DisGeNET | Bile duct c | Feb-61 | 0.029 | 0.17715 | 0 | 0 | 7.917846 | 28.0328 |
| DisGeNET | Pericarditis | 8-Jan | 0.03428 | 0.17715 | 0 | 0 | 33.06645 | 111.5391 |
| DisGeNET | NADH cyto | 7-Jan | 0.03006 | 0.17715 | 0 | 0 | 38.57946 | 135.2043 |
| DisGeNET | Myelolipor | 7-Jan | 0.03006 | 0.17715 | 0 | 0 | 38.57946 | 135.2043 |
| DisGeNET | Calvarial h | 7-Jan | 0.03006 | 0.17715 | 0 | 0 | 38.57946 | 135.2043 |
| DisGeNET | Vascular Di | 5/401 | 0.030663 | 0.17715 | 0 | 0 | 3.005204 | 10.47227 |
| DisGeNET | Other spec | 3/155 | 0.030154 | 0.17715 | 0 | 0 | 4.643092 | 16.25756 |
| DisGeNET | Lymphede | Feb-50 | 0.020017 | 0.17715 | 0 | 0 | 9.737745 | 38.08585 |
| DisGeNET | Sudden Ca | Feb-50 | 0.020017 | 0.17715 | 0 | 0 | 9.737745 | 38.08585 |
| DisGeNET | Subcutane | Feb-65 | 0.032597 | 0.17715 | 0 | 0 | 7.413632 | 25.38081 |
| DisGeNET | Cerebral A | 7-Jan | 0.03006 | 0.17715 | 0 | 0 | 38.57946 | 135.2043 |
| DisGeNET | Panniculitis | 8-Jan | 0.03428 | 0.17715 | 0 | 0 | 33.06645 | 111.5391 |
| DisGeNET | OVARIOLEU | 8-Jan | 0.03428 | 0.17715 | 0 | 0 | 33.06645 | 111.5391 |
| DisGeNET | Endocardia | 8-Jan | 0.03428 | 0.17715 | 0 | 0 | 33.06645 | 111.5391 |
| DisGeNET | Proximal m | 8-Jan | 0.03428 | 0.17715 | 0 | 0 | 33.06645 | 111.5391 |
| DisGeNET | Amnestic D | 7-Jan | 0.03006 | 0.17715 | 0 | 0 | 38.57946 | 135.2043 |
| DisGeNET | Adenoid cy | 7-Jan | 0.03006 | 0.17715 | 0 | 0 | 38.57946 | 135.2043 |
| DisGeNET | Infectious | 7-Jan | 0.03006 | 0.17715 | 0 | 0 | 38.57946 | 135.2043 |
| DisGeNET | Proximal lo | 7-Jan | 0.03006 | 0.17715 | 0 | 0 | 38.57946 | 135.2043 |
| DisGeNET | Lymphangi | Feb-63 | 0.030777 | 0.17715 | 0 | 0 | 7.657473 | 26.65545 |
| DisGeNET | Arcus juver | 6-Jan | 0.025821 | 0.17715 | 0 | 0 | 46.29767 | 169.2909 |
| DisGeNET | Inclusion B | Feb-63 | 0.030777 | 0.17715 | 0 | 0 | 7.657473 | 26.65545 |
| DisGeNET | Fibroadeno | Feb-63 | 0.030777 | 0.17715 | 0 | 0 | 7.657473 | 26.65545 |
| DisGeNET | Selective to | 7-Jan | 0.03006 | 0.17715 | 0 | 0 | 38.57946 | 135.2043 |
| DisGeNET | Fat redistri | 7-Jan | 0.03006 | 0.17715 | 0 | 0 | 38.57946 | 135.2043 |
| DisGeNET | Abnormalit | 3/155 | 0.030154 | 0.17715 | 0 | 0 | 4.643092 | 16.25756 |
| DisGeNET | Pupillary al | 7-Jan | 0.03006 | 0.17715 | 0 | 0 | 38.57946 | 135.2043 |
| DisGeNET | Alcohol Wi | 8-Jan | 0.03428 | 0.17715 | 0 | 0 | 33.06645 | 111.5391 |
| DisGeNET | Postoperat | 7-Jan | 0.03006 | 0.17715 | 0 | 0 | 38.57946 | 135.2043 |
| DisGeNET | Peroneal m | 7-Jan | 0.03006 | 0.17715 | 0 | 0 | 38.57946 | 135.2043 |
| DisGeNET | Congenital | 7-Jan | 0.03006 | 0.17715 | 0 | 0 | 38.57946 | 135.2043 |
| DisGeNET | Muscle bio | Feb-64 | 0.031682 | 0.17715 | 0 | 0 | 7.533586 | 26.00597 |
| DisGeNET | Fibrosis, Li | 6/545 | 0.031514 | 0.17715 | 0 | 0 | 2.662544 | 9.205238 |
| DisGeNET | Congenital | 7-Jan | 0.03006 | 0.17715 | 0 | 0 | 38.57946 | 135.2043 |
| DisGeNET | Nasal obstr | 7-Jan | 0.03006 | 0.17715 | 0 | 0 | 38.57946 | 135.2043 |
| DisGeNET | MYOPATHY | 7-Jan | 0.03006 | 0.17715 | 0 | 0 | 38.57946 | 135.2043 |

| | | | | | | | | |
|---|---|---|---|---|---|---|---|---|
| DisGeNET | Absence of | 7-Jan | 0.03006 | 0.17715 | 0 | 0 | 38.57946 | 135.2043 |
| DisGeNET | Type 1 fibe | 7-Jan | 0.03006 | 0.17715 | 0 | 0 | 38.57946 | 135.2043 |
| DisGeNET | Abnormal s | 7-Jan | 0.03006 | 0.17715 | 0 | 0 | 38.57946 | 135.2043 |
| DisGeNET | Transitiona | 7-Jan | 0.03006 | 0.17715 | 0 | 0 | 38.57946 | 135.2043 |
| DisGeNET | Angiosarco | 7-Jan | 0.03006 | 0.17715 | 0 | 0 | 38.57946 | 135.2043 |
| DisGeNET | Angiokerat | 7-Jan | 0.03006 | 0.17715 | 0 | 0 | 38.57946 | 135.2043 |
| DisGeNET | PROSTATE | 7-Jan | 0.03006 | 0.17715 | 0 | 0 | 38.57946 | 135.2043 |
| DisGeNET | Nemaline M | 7-Jan | 0.03006 | 0.17715 | 0 | 0 | 38.57946 | 135.2043 |
| DisGeNET | Infantile m | 7-Jan | 0.03006 | 0.17715 | 0 | 0 | 38.57946 | 135.2043 |
| DisGeNET | GIANT AXC | 7-Jan | 0.03006 | 0.17715 | 0 | 0 | 38.57946 | 135.2043 |
| DisGeNET | Gastrointes | 7-Jan | 0.03006 | 0.17715 | 0 | 0 | 38.57946 | 135.2043 |
| DisGeNET | Flat Ductal | 7-Jan | 0.03006 | 0.17715 | 0 | 0 | 38.57946 | 135.2043 |
| DisGeNET | Ventricular | 7-Jan | 0.03006 | 0.17715 | 0 | 0 | 38.57946 | 135.2043 |
| DisGeNET | Increased c | 7-Jan | 0.03006 | 0.17715 | 0 | 0 | 38.57946 | 135.2043 |
| DisGeNET | Leg Length | 8-Jan | 0.03428 | 0.17715 | 0 | 0 | 33.06645 | 111.5391 |
| DisGeNET | Clear-cell n | 3/142 | 0.024086 | 0.17715 | 0 | 0 | 5.080678 | 18.9312 |
| DisGeNET | Squamous | 8-Jan | 0.03428 | 0.17715 | 0 | 0 | 33.06645 | 111.5391 |
| DisGeNET | Absent axil | 7-Jan | 0.03006 | 0.17715 | 0 | 0 | 38.57946 | 135.2043 |
| DisGeNET | Venous Ins | 7-Jan | 0.03006 | 0.17715 | 0 | 0 | 38.57946 | 135.2043 |
| DisGeNET | Nephrogen | 6-Jan | 0.025821 | 0.17715 | 0 | 0 | 46.29767 | 169.2909 |
| DisGeNET | Notched na | 6-Jan | 0.025821 | 0.17715 | 0 | 0 | 46.29767 | 169.2909 |
| DisGeNET | Acral Lentig | 7-Jan | 0.03006 | 0.17715 | 0 | 0 | 38.57946 | 135.2043 |
| GO_Biologi | Meiotic Sis | 10-Jan | 0.042667 | 0.177157 | 0 | 0 | 25.71576 | 81.1157 |
| GO_Biologi | Fatty Acid I | 10-Jan | 0.042667 | 0.177157 | 0 | 0 | 25.71576 | 81.1157 |
| GO_Biologi | Protein Kin | 10-Jan | 0.042667 | 0.177157 | 0 | 0 | 25.71576 | 81.1157 |
| GO_Biologi | Lipoprotein | 10-Jan | 0.042667 | 0.177157 | 0 | 0 | 25.71576 | 81.1157 |
| GO_Biologi | Hematopoi | 10-Jan | 0.042667 | 0.177157 | 0 | 0 | 25.71576 | 81.1157 |
| GO_Biologi | Very-Low-D | 10-Jan | 0.042667 | 0.177157 | 0 | 0 | 25.71576 | 81.1157 |
| GO_Biologi | Acidic Amir | 10-Jan | 0.042667 | 0.177157 | 0 | 0 | 25.71576 | 81.1157 |
| GO_Biologi | Positive Re | 10-Jan | 0.042667 | 0.177157 | 0 | 0 | 25.71576 | 81.1157 |
| GO_Biologi | Angiogenes | 10-Jan | 0.042667 | 0.177157 | 0 | 0 | 25.71576 | 81.1157 |
| GO_Biologi | Regulation | Feb-74 | 0.041285 | 0.177157 | 0 | 0 | 6.483987 | 20.66613 |
| GO_Biologi | Kinetochor | 10-Jan | 0.042667 | 0.177157 | 0 | 0 | 25.71576 | 81.1157 |
| GO_Biologi | Negative R | 10-Jan | 0.042667 | 0.177157 | 0 | 0 | 25.71576 | 81.1157 |
| GO_Biologi | Negative R | Feb-74 | 0.041285 | 0.177157 | 0 | 0 | 6.483987 | 20.66613 |
| GO_Biologi | Negative R | 10-Jan | 0.042667 | 0.177157 | 0 | 0 | 25.71576 | 81.1157 |
| GO_Biologi | Positive Re | 10-Jan | 0.042667 | 0.177157 | 0 | 0 | 25.71576 | 81.1157 |
| GeDiPNet_ | Attention D | 4/359 | 0.071457 | 0.177279 | 0 | 0 | 2.655082 | 7.005859 |
| GeDiPNet_ | Follicular T | 17-Jan | 0.071459 | 0.177279 | 0 | 0 | 14.46003 | 38.15469 |
| GeDiPNet_ | Peg-Shaped | 17-Jan | 0.071459 | 0.177279 | 0 | 0 | 14.46003 | 38.15469 |
| GeDiPNet_ | Delirium | 17-Jan | 0.071459 | 0.177279 | 0 | 0 | 14.46003 | 38.15469 |
| GeDiPNet_ | Microgloss | 17-Jan | 0.071459 | 0.177279 | 0 | 0 | 14.46003 | 38.15469 |
| GeDiPNet_ | Xanthoma | 17-Jan | 0.071459 | 0.177279 | 0 | 0 | 14.46003 | 38.15469 |
| GeDiPNet_ | Visceral An | 17-Jan | 0.071459 | 0.177279 | 0 | 0 | 14.46003 | 38.15469 |
| Rare_Disea | Hemochroi | Feb-60 | 0.028127 | 0.177346 | 0 | 0 | 8.054767 | 28.76365 |

| | | | | | | | | |
|---|---|---|---|---|---|---|---|---|
| Rare_Disea | Salla diseas | 7-Jan | 0.03006 | 0.177496 | 0 | 0 | 38.57946 | 135.2043 |
| Rare_Disea | Frontonasa | 7-Jan | 0.03006 | 0.177496 | 0 | 0 | 38.57946 | 135.2043 |
| Rare_Disea | Atkin syndr | 7-Jan | 0.03006 | 0.177496 | 0 | 0 | 38.57946 | 135.2043 |
| Rare_Disea | Angiosarco | 7-Jan | 0.03006 | 0.177496 | 0 | 0 | 38.57946 | 135.2043 |
| Rare_Disea | Idiopathic l | 7-Jan | 0.03006 | 0.177496 | 0 | 0 | 38.57946 | 135.2043 |
| Rare_Disea | Optic atrop | Feb-62 | 0.029883 | 0.177496 | 0 | 0 | 7.78549 | 27.33058 |
| Rare_Disea | Localized s | Feb-62 | 0.029883 | 0.177496 | 0 | 0 | 7.78549 | 27.33058 |
| Rare_Disea | Wallenberg | 7-Jan | 0.03006 | 0.177496 | 0 | 0 | 38.57946 | 135.2043 |
| SysMyo_M | MGS1345 I | Mar-84 | 0.005878 | 0.177548 | 0 | 0 | 8.744268 | 44.91577 |
| SysMyo_M | MGS1089 I | 4/168 | 0.006226 | 0.177548 | 0 | 0 | 5.803409 | 29.4753 |
| SysMyo_M | MGS1277 I | 4/165 | 0.005847 | 0.177548 | 0 | 0 | 5.912445 | 30.40115 |
| SysMyo_M | MGS1047 I | 4/159 | 0.005134 | 0.177548 | 0 | 0 | 6.143179 | 32.3858 |
| SysMyo_M | MGS772 M | Mar-86 | 0.006274 | 0.177548 | 0 | 0 | 8.532702 | 43.27254 |
| GeDiPNet_ | Movement | 3/221 | 0.071905 | 0.177629 | 0 | 0 | 3.226573 | 8.493677 |
| DisGeNET | Fine hair | Feb-67 | 0.034458 | 0.177717 | 0 | 0 | 7.184796 | 24.19848 |
| UK_Bioban | ICD10 chol | 27-Jan | 0.111111 | 0.177902 | 0 | 0 | 8.894007 | 19.54216 |
| CellMarker | Myeloid Ce | 28-Jan | 0.114983 | 0.177996 | 0 | 0 | 8.564169 | 18.52408 |
| CellMarker | Stem Cell L | 28-Jan | 0.114983 | 0.177996 | 0 | 0 | 8.564169 | 18.52408 |
| CellMarker | Fibroblast U | 28-Jan | 0.114983 | 0.177996 | 0 | 0 | 8.564169 | 18.52408 |
| CellMarker | T Helper Ce | 28-Jan | 0.114983 | 0.177996 | 0 | 0 | 8.564169 | 18.52408 |
| CellMarker | Endothelia | 28-Jan | 0.114983 | 0.177996 | 0 | 0 | 8.564169 | 18.52408 |
| GWAS_Cat | Diastolic Bl | 31-Jan | 0.126498 | 0.178414 | 0 | 0 | 7.706589 | 15.93356 |
| GWAS_Cat | Bone Mine | 31-Jan | 0.126498 | 0.178414 | 0 | 0 | 7.706589 | 15.93356 |
| GWAS_Cat | Subjective | 31-Jan | 0.126498 | 0.178414 | 0 | 0 | 7.706589 | 15.93356 |
| GWAS_Cat | Response T | 31-Jan | 0.126498 | 0.178414 | 0 | 0 | 7.706589 | 15.93356 |
| GWAS_Cat | Systolic Blc | 31-Jan | 0.126498 | 0.178414 | 0 | 0 | 7.706589 | 15.93356 |
| GWAS_Cat | Ratio Of Or | 31-Jan | 0.126498 | 0.178414 | 0 | 0 | 7.706589 | 15.93356 |
| GWAS_Cat | Glomerular | 31-Jan | 0.126498 | 0.178414 | 0 | 0 | 7.706589 | 15.93356 |
| GWAS_Cat | Birth Weigl | 31-Jan | 0.126498 | 0.178414 | 0 | 0 | 7.706589 | 15.93356 |
| GWAS_Cat | Left-hande | 31-Jan | 0.126498 | 0.178414 | 0 | 0 | 7.706589 | 15.93356 |
| GO_Biologi | Wnt Signal | Feb-76 | 0.043322 | 0.178828 | 0 | 0 | 6.308108 | 19.80178 |
| Rare_Disea | N syndrom | 5/401 | 0.030663 | 0.178894 | 0 | 0 | 3.005204 | 10.47227 |
| Rare_Disea | Morphea | Feb-63 | 0.030777 | 0.178894 | 0 | 0 | 7.657473 | 26.65545 |
| GWAS_Cat | Spherical E | 2/142 | 0.127022 | 0.178946 | 0 | 0 | 3.323193 | 6.857052 |
| Jensen_TIS | Melanocyt | 19-Jan | 0.079527 | 0.179074 | 0 | 0 | 12.85207 | 32.53709 |
| Jensen_TIS | A7r5 cell | 19-Jan | 0.079527 | 0.179074 | 0 | 0 | 12.85207 | 32.53709 |
| Jensen_TIS | Trophozoit | 19-Jan | 0.079527 | 0.179074 | 0 | 0 | 12.85207 | 32.53709 |
| Jensen_TIS | Oocyst | 19-Jan | 0.079527 | 0.179074 | 0 | 0 | 12.85207 | 32.53709 |
| Jensen_TIS | Mesoneph | 19-Jan | 0.079527 | 0.179074 | 0 | 0 | 12.85207 | 32.53709 |
| Human_Ph | Flared met | 25-Jan | 0.103317 | 0.179395 | 0 | 0 | 9.636143 | 21.87363 |
| Human_Ph | Onion bulb | 25-Jan | 0.103317 | 0.179395 | 0 | 0 | 9.636143 | 21.87363 |
| Human_Ph | Decreased | 25-Jan | 0.103317 | 0.179395 | 0 | 0 | 9.636143 | 21.87363 |
| Human_Ph | Acanthosis | 25-Jan | 0.103317 | 0.179395 | 0 | 0 | 9.636143 | 21.87363 |
| Human_Ph | Patent duc | 2/125 | 0.102976 | 0.179395 | 0 | 0 | 3.785748 | 8.60599 |
| DisGeNET | Long clavic | 9-Jan | 0.038483 | 0.179405 | 0 | 0 | 28.93169 | 94.24608 |

| | | | | | | | | |
|---|---|---|---|---|---|---|---|---|
| DisGeNET | Pediatric O | Feb-71 | 0.038301 | 0.179405 | 0 | 0 | 6.766922 | 22.07565 |
| DisGeNET | LOW DENS | 9-Jan | 0.038483 | 0.179405 | 0 | 0 | 28.93169 | 94.24608 |
| DisGeNET | alpha-Thala | Feb-70 | 0.037325 | 0.179405 | 0 | 0 | 6.866782 | 22.57858 |
| DisGeNET | Proximal m | Feb-71 | 0.038301 | 0.179405 | 0 | 0 | 6.766922 | 22.07565 |
| DisGeNET | Chromosor | 9-Jan | 0.038483 | 0.179405 | 0 | 0 | 28.93169 | 94.24608 |
| DisGeNET | Deposits in | 9-Jan | 0.038483 | 0.179405 | 0 | 0 | 28.93169 | 94.24608 |
| DisGeNET | Diseases of | 9-Jan | 0.038483 | 0.179405 | 0 | 0 | 28.93169 | 94.24608 |
| DisGeNET | proliferativ | 9-Jan | 0.038483 | 0.179405 | 0 | 0 | 28.93169 | 94.24608 |
| DisGeNET | Aortic Anei | 4/288 | 0.036992 | 0.179405 | 0 | 0 | 3.330901 | 10.98213 |
| DisGeNET | Glaucoma, | Feb-70 | 0.037325 | 0.179405 | 0 | 0 | 6.866782 | 22.57858 |
| DisGeNET | Nephrogen | 9-Jan | 0.038483 | 0.179405 | 0 | 0 | 28.93169 | 94.24608 |
| DisGeNET | Cerebral Th | 9-Jan | 0.038483 | 0.179405 | 0 | 0 | 28.93169 | 94.24608 |
| DisGeNET | Nasal cong | 9-Jan | 0.038483 | 0.179405 | 0 | 0 | 28.93169 | 94.24608 |
| DisGeNET | Carcinoma | 17/2476 | 0.036723 | 0.179405 | 0 | 0 | 1.723802 | 5.696065 |
| DisGeNET | Hereditary | 3/167 | 0.036403 | 0.179405 | 0 | 0 | 4.30074 | 14.24877 |
| DisGeNET | Abnormal s | 9-Jan | 0.038483 | 0.179405 | 0 | 0 | 28.93169 | 94.24608 |
| DisGeNET | Abnormal s | 9-Jan | 0.038483 | 0.179405 | 0 | 0 | 28.93169 | 94.24608 |
| DisGeNET | Hypertroph | 9-Jan | 0.038483 | 0.179405 | 0 | 0 | 28.93169 | 94.24608 |
| DisGeNET | Renal medu | 9-Jan | 0.038483 | 0.179405 | 0 | 0 | 28.93169 | 94.24608 |
| DisGeNET | Hypoplastic | 9-Jan | 0.038483 | 0.179405 | 0 | 0 | 28.93169 | 94.24608 |
| DisGeNET | Sclerosing | 9-Jan | 0.038483 | 0.179405 | 0 | 0 | 28.93169 | 94.24608 |
| DisGeNET | Diabetic Ne | 6/561 | 0.03554 | 0.179405 | 0 | 0 | 2.58365 | 8.621909 |
| DisGeNET | Fibular apla | 9-Jan | 0.038483 | 0.179405 | 0 | 0 | 28.93169 | 94.24608 |
| DisGeNET | Serum albu | 9-Jan | 0.038483 | 0.179405 | 0 | 0 | 28.93169 | 94.24608 |
| DisGeNET | Frontonasa | 9-Jan | 0.038483 | 0.179405 | 0 | 0 | 28.93169 | 94.24608 |
| DisGeNET | Keratitis, H | 9-Jan | 0.038483 | 0.179405 | 0 | 0 | 28.93169 | 94.24608 |
| DisGeNET | Steatohepa | 6/569 | 0.037672 | 0.179405 | 0 | 0 | 2.545885 | 8.347555 |
| DisGeNET | Metastatic | 5/417 | 0.035393 | 0.179405 | 0 | 0 | 2.88613 | 9.643295 |
| DisGeNET | Broad philt | 9-Jan | 0.038483 | 0.179405 | 0 | 0 | 28.93169 | 94.24608 |
| DisGeNET | Motor Skill | 9-Jan | 0.038483 | 0.179405 | 0 | 0 | 28.93169 | 94.24608 |
| DisGeNET | Carbamoyl- | 9-Jan | 0.038483 | 0.179405 | 0 | 0 | 28.93169 | 94.24608 |
| DisGeNET | Gout | Feb-71 | 0.038301 | 0.179405 | 0 | 0 | 6.766922 | 22.07565 |
| DisGeNET | Abnormalit | Feb-71 | 0.038301 | 0.179405 | 0 | 0 | 6.766922 | 22.07565 |
| DisGeNET | Thyroid car | 9-Jan | 0.038483 | 0.179405 | 0 | 0 | 28.93169 | 94.24608 |
| DisGeNET | eyelids (syr | 9-Jan | 0.038483 | 0.179405 | 0 | 0 | 28.93169 | 94.24608 |
| DisGeNET | Argyrophili | 9-Jan | 0.038483 | 0.179405 | 0 | 0 | 28.93169 | 94.24608 |
| DisGeNET | Proximal ne | Feb-71 | 0.038301 | 0.179405 | 0 | 0 | 6.766922 | 22.07565 |
| DisGeNET | Enlargemer | 9-Jan | 0.038483 | 0.179405 | 0 | 0 | 28.93169 | 94.24608 |
| DisGeNET | Hepatolent | Feb-69 | 0.03636 | 0.179405 | 0 | 0 | 6.969622 | 23.09942 |
| DisGeNET | Glycogen S | 9-Jan | 0.038483 | 0.179405 | 0 | 0 | 28.93169 | 94.24608 |
| DisGeNET | Thick skull | 9-Jan | 0.038483 | 0.179405 | 0 | 0 | 28.93169 | 94.24608 |
| DisGeNET | Autosomal | 9-Jan | 0.038483 | 0.179405 | 0 | 0 | 28.93169 | 94.24608 |
| DisGeNET | Glycogen st | 9-Jan | 0.038483 | 0.179405 | 0 | 0 | 28.93169 | 94.24608 |
| DisGeNET | Dysembryo | 9-Jan | 0.038483 | 0.179405 | 0 | 0 | 28.93169 | 94.24608 |
| DisGeNET | Partial atric | 9-Jan | 0.038483 | 0.179405 | 0 | 0 | 28.93169 | 94.24608 |

| | | | | | | | | |
|---|---|---|---|---|---|---|---|---|
| DisGeNET | Conjunctiva | 9-Jan | 0.038483 | 0.179405 | 0 | 0 | 28.93169 | 94.24608 |
| DisGeNET | Cranial hyp | 9-Jan | 0.038483 | 0.179405 | 0 | 0 | 28.93169 | 94.24608 |
| DisGeNET | Delayed er | 9-Jan | 0.038483 | 0.179405 | 0 | 0 | 28.93169 | 94.24608 |
| DisGeNET | Abnormalit | 9-Jan | 0.038483 | 0.179405 | 0 | 0 | 28.93169 | 94.24608 |
| DisGeNET | Excessive g | 9-Jan | 0.038483 | 0.179405 | 0 | 0 | 28.93169 | 94.24608 |
| DisGeNET | Cooley's ar | Feb-70 | 0.037325 | 0.179405 | 0 | 0 | 6.866782 | 22.57858 |
| DisGeNET | Philadelphi | Feb-70 | 0.037325 | 0.179405 | 0 | 0 | 6.866782 | 22.57858 |
| DisGeNET | Duodenal U | Feb-71 | 0.038301 | 0.179405 | 0 | 0 | 6.766922 | 22.07565 |
| DisGeNET | Anterior m | 10-Jan | 0.042667 | 0.180305 | 0 | 0 | 25.71576 | 81.1157 |
| DisGeNET | cortex bon | 10-Jan | 0.042667 | 0.180305 | 0 | 0 | 25.71576 | 81.1157 |
| DisGeNET | Radiohume | 10-Jan | 0.042667 | 0.180305 | 0 | 0 | 25.71576 | 81.1157 |
| DisGeNET | Ramer Lad | 10-Jan | 0.042667 | 0.180305 | 0 | 0 | 25.71576 | 81.1157 |
| DisGeNET | Subcutane | 10-Jan | 0.042667 | 0.180305 | 0 | 0 | 25.71576 | 81.1157 |
| DisGeNET | B Acute Lyr | 10-Jan | 0.042667 | 0.180305 | 0 | 0 | 25.71576 | 81.1157 |
| DisGeNET | Ambiguous | 10-Jan | 0.042667 | 0.180305 | 0 | 0 | 25.71576 | 81.1157 |
| DisGeNET | Arthus Rea | 10-Jan | 0.042667 | 0.180305 | 0 | 0 | 25.71576 | 81.1157 |
| DisGeNET | Lymphangi | 10-Jan | 0.042667 | 0.180305 | 0 | 0 | 25.71576 | 81.1157 |
| DisGeNET | Xanthoma | 10-Jan | 0.042667 | 0.180305 | 0 | 0 | 25.71576 | 81.1157 |
| DisGeNET | Increased s | 10-Jan | 0.042667 | 0.180305 | 0 | 0 | 25.71576 | 81.1157 |
| DisGeNET | Neck flexor | 10-Jan | 0.042667 | 0.180305 | 0 | 0 | 25.71576 | 81.1157 |
| DisGeNET | Low set ear | 4/293 | 0.039011 | 0.180305 | 0 | 0 | 3.272439 | 10.61551 |
| DisGeNET | Hallucinati | Feb-72 | 0.039286 | 0.180305 | 0 | 0 | 6.669916 | 21.58979 |
| DisGeNET | Hyperurice | Feb-72 | 0.039286 | 0.180305 | 0 | 0 | 6.669916 | 21.58979 |
| DisGeNET | Leukemoge | 7/730 | 0.039691 | 0.180305 | 0 | 0 | 2.322441 | 7.493635 |
| DisGeNET | Dysphoric r | 10-Jan | 0.042667 | 0.180305 | 0 | 0 | 25.71576 | 81.1157 |
| DisGeNET | Low-grade | 10-Jan | 0.042667 | 0.180305 | 0 | 0 | 25.71576 | 81.1157 |
| DisGeNET | Precursor T | 10-Jan | 0.042667 | 0.180305 | 0 | 0 | 25.71576 | 81.1157 |
| DisGeNET | Hibernoma | 10-Jan | 0.042667 | 0.180305 | 0 | 0 | 25.71576 | 81.1157 |
| DisGeNET | Bone marr | 10-Jan | 0.042667 | 0.180305 | 0 | 0 | 25.71576 | 81.1157 |
| DisGeNET | Mild deme | 10-Jan | 0.042667 | 0.180305 | 0 | 0 | 25.71576 | 81.1157 |
| DisGeNET | Canavan Di | 10-Jan | 0.042667 | 0.180305 | 0 | 0 | 25.71576 | 81.1157 |
| DisGeNET | Decreased | Feb-74 | 0.041285 | 0.180305 | 0 | 0 | 6.483987 | 20.66613 |
| DisGeNET | Meningiom | 5/438 | 0.042255 | 0.180305 | 0 | 0 | 2.743198 | 8.679586 |
| DisGeNET | Malignant | Feb-75 | 0.042299 | 0.180305 | 0 | 0 | 6.394843 | 20.22688 |
| DisGeNET | Porphyria, | 10-Jan | 0.042667 | 0.180305 | 0 | 0 | 25.71576 | 81.1157 |
| DisGeNET | Ambiguous | 10-Jan | 0.042667 | 0.180305 | 0 | 0 | 25.71576 | 81.1157 |
| DisGeNET | Ambiguous | 10-Jan | 0.042667 | 0.180305 | 0 | 0 | 25.71576 | 81.1157 |
| DisGeNET | Bannayan-I | 10-Jan | 0.042667 | 0.180305 | 0 | 0 | 25.71576 | 81.1157 |
| DisGeNET | Bile Duct N | 10-Jan | 0.042667 | 0.180305 | 0 | 0 | 25.71576 | 81.1157 |
| DisGeNET | Angioid Str | 10-Jan | 0.042667 | 0.180305 | 0 | 0 | 25.71576 | 81.1157 |
| DisGeNET | Variegate F | 10-Jan | 0.042667 | 0.180305 | 0 | 0 | 25.71576 | 81.1157 |
| DisGeNET | PARIETAL F | 10-Jan | 0.042667 | 0.180305 | 0 | 0 | 25.71576 | 81.1157 |
| DisGeNET | FRAXE Sync | 10-Jan | 0.042667 | 0.180305 | 0 | 0 | 25.71576 | 81.1157 |
| DisGeNET | Histiocytoi | 10-Jan | 0.042667 | 0.180305 | 0 | 0 | 25.71576 | 81.1157 |
| DisGeNET | Stage 0 Ski | 10-Jan | 0.042667 | 0.180305 | 0 | 0 | 25.71576 | 81.1157 |

| | | | | | | | | |
|---|---|---|---|---|---|---|---|---|
| DisGeNET | Gliomatosi | 10-Jan | 0.042667 | 0.180305 | 0 | 0 | 25.71576 | 81.1157 |
| DisGeNET | Quebec pla | 10-Jan | 0.042667 | 0.180305 | 0 | 0 | 25.71576 | 81.1157 |
| DisGeNET | Developme | 10-Jan | 0.042667 | 0.180305 | 0 | 0 | 25.71576 | 81.1157 |
| DisGeNET | Dementia c | 10-Jan | 0.042667 | 0.180305 | 0 | 0 | 25.71576 | 81.1157 |
| DisGeNET | Dementia c | 10-Jan | 0.042667 | 0.180305 | 0 | 0 | 25.71576 | 81.1157 |
| DisGeNET | Polygenic h | 10-Jan | 0.042667 | 0.180305 | 0 | 0 | 25.71576 | 81.1157 |
| DisGeNET | FAMILIAL A | 10-Jan | 0.042667 | 0.180305 | 0 | 0 | 25.71576 | 81.1157 |
| DisGeNET | clinical dep | 10-Jan | 0.042667 | 0.180305 | 0 | 0 | 25.71576 | 81.1157 |
| DisGeNET | Vanishing v | 10-Jan | 0.042667 | 0.180305 | 0 | 0 | 25.71576 | 81.1157 |
| DisGeNET | Tunnel visu | 10-Jan | 0.042667 | 0.180305 | 0 | 0 | 25.71576 | 81.1157 |
| DisGeNET | Thiamine D | 10-Jan | 0.042667 | 0.180305 | 0 | 0 | 25.71576 | 81.1157 |
| DisGeNET | Giardiasis | 10-Jan | 0.042667 | 0.180305 | 0 | 0 | 25.71576 | 81.1157 |
| DisGeNET | Shoulder gi | 10-Jan | 0.042667 | 0.180305 | 0 | 0 | 25.71576 | 81.1157 |
| DisGeNET | Sex hormo | 10-Jan | 0.042667 | 0.180305 | 0 | 0 | 25.71576 | 81.1157 |
| DisGeNET | Syndactyly | Feb-73 | 0.040281 | 0.180305 | 0 | 0 | 6.575642 | 21.1202 |
| DisGeNET | Hamartom | Feb-74 | 0.041285 | 0.180305 | 0 | 0 | 6.483987 | 20.66613 |
| DisGeNET | Epitheliom 4/298 | | 0.041092 | 0.180305 | 0 | 0 | 3.215966 | 10.2652 |
| DisGeNET | Diabetes m | 10-Jan | 0.042667 | 0.180305 | 0 | 0 | 25.71576 | 81.1157 |
| DisGeNET | Cancer Cell | 10-Jan | 0.042667 | 0.180305 | 0 | 0 | 25.71576 | 81.1157 |
| DisGeNET | Severe den | 10-Jan | 0.042667 | 0.180305 | 0 | 0 | 25.71576 | 81.1157 |
| DisGeNET | COPD exac | 10-Jan | 0.042667 | 0.180305 | 0 | 0 | 25.71576 | 81.1157 |
| GWAS_Cat | Vascular Er | Jan-32 | 0.130304 | 0.181889 | 0 | 0 | 7.457614 | 15.19775 |
| GWAS_Cat | Acceptance | Jan-32 | 0.130304 | 0.181889 | 0 | 0 | 7.457614 | 15.19775 |
| GWAS_Cat | Total Omeg | Jan-32 | 0.130304 | 0.181889 | 0 | 0 | 7.457614 | 15.19775 |
| GWAS_Cat | TRAIL Leve | Jan-32 | 0.130304 | 0.181889 | 0 | 0 | 7.457614 | 15.19775 |
| GWAS_Cat | COVID-19 ( 2/144 | | 0.129934 | 0.181889 | 0 | 0 | 3.276056 | 6.685543 |
| GWAS_Cat | Number Of | Jan-32 | 0.130304 | 0.181889 | 0 | 0 | 7.457614 | 15.19775 |
| GWAS_Cat | Albumin-gl | Jan-32 | 0.130304 | 0.181889 | 0 | 0 | 7.457614 | 15.19775 |
| GWAS_Cat | Dentures | Jan-32 | 0.130304 | 0.181889 | 0 | 0 | 7.457614 | 15.19775 |
| ClinVar_20 | charcot-ma | Jan-44 | 0.174713 | 0.181993 | 0 | 0 | 5.373175 | 9.3741 |
| DisGeNET | Microcorne | Feb-76 | 0.043322 | 0.182481 | 0 | 0 | 6.308108 | 19.80178 |
| DisGeNET | Machado-J | Feb-76 | 0.043322 | 0.182481 | 0 | 0 | 6.308108 | 19.80178 |
| GeDiPNet_ | Coronary A | 18-Jan | 0.075502 | 0.182645 | 0 | 0 | 13.60876 | 35.1596 |
| GeDiPNet_ | Mediastina | 18-Jan | 0.075502 | 0.182645 | 0 | 0 | 13.60876 | 35.1596 |
| GeDiPNet_ | Optic Atro 5/518 | | 0.07536 | 0.182645 | 0 | 0 | 2.3059 | 5.961845 |
| GeDiPNet_ | Congenital | 18-Jan | 0.075502 | 0.182645 | 0 | 0 | 13.60876 | 35.1596 |
| GeDiPNet_ | Enlarged Pe | 18-Jan | 0.075502 | 0.182645 | 0 | 0 | 13.60876 | 35.1596 |
| GO_Biologi | Negative R | 11-Jan | 0.046834 | 0.182704 | 0 | 0 | 23.14302 | 70.84419 |
| GO_Biologi | Central Ne | 11-Jan | 0.046834 | 0.182704 | 0 | 0 | 23.14302 | 70.84419 |
| GO_Biologi | Central Ne | 11-Jan | 0.046834 | 0.182704 | 0 | 0 | 23.14302 | 70.84419 |
| GO_Biologi | Brain Morp | 11-Jan | 0.046834 | 0.182704 | 0 | 0 | 23.14302 | 70.84419 |
| GO_Biologi | Regulation | 11-Jan | 0.046834 | 0.182704 | 0 | 0 | 23.14302 | 70.84419 |
| GO_Biologi | Golgi To Ly | 11-Jan | 0.046834 | 0.182704 | 0 | 0 | 23.14302 | 70.84419 |
| GO_Biologi | Positive Re | 11-Jan | 0.046834 | 0.182704 | 0 | 0 | 23.14302 | 70.84419 |
| GO_Biologi | Negative R | 11-Jan | 0.046834 | 0.182704 | 0 | 0 | 23.14302 | 70.84419 |

| | | | | | | | | |
|---|---|---|---|---|---|---|---|---|
| GO_Biologi | Regulation | 11-Jan | 0.046834 | 0.182704 | 0 | 0 | 23.14302 | 70.84419 |
| GO_Biologi | Compleme | 11-Jan | 0.046834 | 0.182704 | 0 | 0 | 23.14302 | 70.84419 |
| Rare_Disea | Follicular d | Feb-64 | 0.031682 | 0.182724 | 0 | 0 | 7.533586 | 26.00597 |
| Jensen_DIS | Kidney can | 15/2584 | 0.148401 | 0.182844 | 0 | 0 | 1.406514 | 2.6834 |
| Jensen_DIS | Idiopathic s | Jan-37 | 0.149088 | 0.182844 | 0 | 0 | 6.42022 | 12.21909 |
| Jensen_TIS | af1 | 20-Jan | 0.083535 | 0.182981 | 0 | 0 | 12.17503 | 30.22444 |
| Jensen_TIS | Adult liver | 20-Jan | 0.083535 | 0.182981 | 0 | 0 | 12.17503 | 30.22444 |
| Jensen_TIS | Hepatopan | 20-Jan | 0.083535 | 0.182981 | 0 | 0 | 12.17503 | 30.22444 |
| Jensen_TIS | Egg yolk | 20-Jan | 0.083535 | 0.182981 | 0 | 0 | 12.17503 | 30.22444 |
| UK_Bioban | Self-report | 28-Jan | 0.114983 | 0.183073 | 0 | 0 | 8.564169 | 18.52408 |
| DisGeNET | Fetal Grow | 4/304 | 0.043671 | 0.183655 | 0 | 0 | 3.150683 | 9.865033 |
| GO_Biologi | Negative R | Feb-80 | 0.047505 | 0.18431 | 0 | 0 | 5.983409 | 18.23095 |
| Jensen_TIS | Immune sy | 8/1046 | 0.084731 | 0.184347 | 0 | 0 | 1.841419 | 4.54513 |
| Human_Ph | Thyroiditis | 26-Jan | 0.107222 | 0.184499 | 0 | 0 | 9.250233 | 20.65441 |
| Human_Ph | Mucosal te | 26-Jan | 0.107222 | 0.184499 | 0 | 0 | 9.250233 | 20.65441 |
| DisGeNET | Fatty Liver | 5/443 | 0.044 | 0.184742 | 0 | 0 | 2.711187 | 8.468599 |
| DisGeNET | Hydranenc | 11-Jan | 0.046834 | 0.184797 | 0 | 0 | 23.14302 | 70.84419 |
| DisGeNET | Hypersensi | 11-Jan | 0.046834 | 0.184797 | 0 | 0 | 23.14302 | 70.84419 |
| DisGeNET | Epilepsy, T | 3/181 | 0.04447 | 0.184797 | 0 | 0 | 3.959671 | 12.32626 |
| DisGeNET | Transient Is | 11-Jan | 0.046834 | 0.184797 | 0 | 0 | 23.14302 | 70.84419 |
| DisGeNET | Transient Is | 11-Jan | 0.046834 | 0.184797 | 0 | 0 | 23.14302 | 70.84419 |
| DisGeNET | Malignant | 11-Jan | 0.046834 | 0.184797 | 0 | 0 | 23.14302 | 70.84419 |
| DisGeNET | Malignant | 11-Jan | 0.046834 | 0.184797 | 0 | 0 | 23.14302 | 70.84419 |
| DisGeNET | Lean body | 11-Jan | 0.046834 | 0.184797 | 0 | 0 | 23.14302 | 70.84419 |
| DisGeNET | Angioma | 11-Jan | 0.046834 | 0.184797 | 0 | 0 | 23.14302 | 70.84419 |
| DisGeNET | Malignant | 11-Jan | 0.046834 | 0.184797 | 0 | 0 | 23.14302 | 70.84419 |
| DisGeNET | malignant r | 11-Jan | 0.046834 | 0.184797 | 0 | 0 | 23.14302 | 70.84419 |
| DisGeNET | Leukemic In | 11-Jan | 0.046834 | 0.184797 | 0 | 0 | 23.14302 | 70.84419 |
| DisGeNET | Arterial an | 11-Jan | 0.046834 | 0.184797 | 0 | 0 | 23.14302 | 70.84419 |
| DisGeNET | Apraxia of | 11-Jan | 0.046834 | 0.184797 | 0 | 0 | 23.14302 | 70.84419 |
| DisGeNET | Redundant | 11-Jan | 0.046834 | 0.184797 | 0 | 0 | 23.14302 | 70.84419 |
| DisGeNET | Localized C | 11-Jan | 0.046834 | 0.184797 | 0 | 0 | 23.14302 | 70.84419 |
| DisGeNET | Localized N | 11-Jan | 0.046834 | 0.184797 | 0 | 0 | 23.14302 | 70.84419 |
| DisGeNET | insulinoma | 3/181 | 0.04447 | 0.184797 | 0 | 0 | 3.959671 | 12.32626 |
| DisGeNET | Crohn Dise | 8/912 | 0.044759 | 0.184797 | 0 | 0 | 2.129383 | 6.614865 |
| DisGeNET | Subcutane | 11-Jan | 0.046834 | 0.184797 | 0 | 0 | 23.14302 | 70.84419 |
| DisGeNET | Granular ce | 11-Jan | 0.046834 | 0.184797 | 0 | 0 | 23.14302 | 70.84419 |
| DisGeNET | Cerebral ar | Feb-78 | 0.045395 | 0.184797 | 0 | 0 | 6.141486 | 18.99159 |
| DisGeNET | Acute flacc | 11-Jan | 0.046834 | 0.184797 | 0 | 0 | 23.14302 | 70.84419 |
| DisGeNET | CARDIOMY | 11-Jan | 0.046834 | 0.184797 | 0 | 0 | 23.14302 | 70.84419 |
| DisGeNET | Brain Stem | 11-Jan | 0.046834 | 0.184797 | 0 | 0 | 23.14302 | 70.84419 |
| DisGeNET | Carotid Cir | 11-Jan | 0.046834 | 0.184797 | 0 | 0 | 23.14302 | 70.84419 |
| DisGeNET | Eyelid Xant | 11-Jan | 0.046834 | 0.184797 | 0 | 0 | 23.14302 | 70.84419 |
| DisGeNET | Median cle | 11-Jan | 0.046834 | 0.184797 | 0 | 0 | 23.14302 | 70.84419 |
| DisGeNET | Right bund | 11-Jan | 0.046834 | 0.184797 | 0 | 0 | 23.14302 | 70.84419 |

| | | | | | | | | |
|---|---|---|---|---|---|---|---|---|
| DisGeNET | Postherpet | 11-Jan | 0.046834 | 0.184797 | 0 | 0 | 23.14302 | 70.84419 |
| DisGeNET | Primary go | 11-Jan | 0.046834 | 0.184797 | 0 | 0 | 23.14302 | 70.84419 |
| DisGeNET | Dry hair | 11-Jan | 0.046834 | 0.184797 | 0 | 0 | 23.14302 | 70.84419 |
| DisGeNET | Primary Pr | 11-Jan | 0.046834 | 0.184797 | 0 | 0 | 23.14302 | 70.84419 |
| DisGeNET | Electrocarc | 11-Jan | 0.046834 | 0.184797 | 0 | 0 | 23.14302 | 70.84419 |
| DisGeNET | Crescendo | 11-Jan | 0.046834 | 0.184797 | 0 | 0 | 23.14302 | 70.84419 |
| DisGeNET | Posterior C | 11-Jan | 0.046834 | 0.184797 | 0 | 0 | 23.14302 | 70.84419 |
| DisGeNET | Microgloss | 11-Jan | 0.046834 | 0.184797 | 0 | 0 | 23.14302 | 70.84419 |
| DisGeNET | Lysosomal | Feb-79 | 0.046446 | 0.184797 | 0 | 0 | 6.061421 | 18.60534 |
| DisGeNET | Abnormal s | Feb-78 | 0.045395 | 0.184797 | 0 | 0 | 6.141486 | 18.99159 |
| DisGeNET | Type 1 mus | 11-Jan | 0.046834 | 0.184797 | 0 | 0 | 23.14302 | 70.84419 |
| DisGeNET | Respiratory | 3/185 | 0.046925 | 0.184877 | 0 | 0 | 3.87186 | 11.84483 |
| Human_Ph | Talipes equ | 2/129 | 0.108512 | 0.185052 | 0 | 0 | 3.665771 | 8.141279 |
| Human_Ph | Pes cavus ( | 2/129 | 0.108512 | 0.185052 | 0 | 0 | 3.665771 | 8.141279 |
| GWAS_Cat | Fibrinogen | Jan-33 | 0.134093 | 0.185691 | 0 | 0 | 7.224201 | 14.515 |
| GWAS_Cat | Corneal Cu | Jan-33 | 0.134093 | 0.185691 | 0 | 0 | 7.224201 | 14.515 |
| GWAS_Cat | Iron Status | Jan-33 | 0.134093 | 0.185691 | 0 | 0 | 7.224201 | 14.515 |
| GWAS_Cat | Mean Corp | 3/291 | 0.133501 | 0.185691 | 0 | 0 | 2.433656 | 4.900515 |
| GWAS_Cat | Response T | Jan-33 | 0.134093 | 0.185691 | 0 | 0 | 7.224201 | 14.515 |
| GWAS_Cat | Monobrow | Jan-33 | 0.134093 | 0.185691 | 0 | 0 | 7.224201 | 14.515 |
| GWAS_Cat | Cholesteryl | Jan-33 | 0.134093 | 0.185691 | 0 | 0 | 7.224201 | 14.515 |
| GWAS_Cat | Mean Retic | 4/452 | 0.134305 | 0.185774 | 0 | 0 | 2.093911 | 4.203817 |
| UK_Bioban | Body mass | 5/595 | 0.117458 | 0.185976 | 0 | 0 | 1.997003 | 4.276924 |
| GO_Biologi | Positive Re | Feb-83 | 0.050736 | 0.186585 | 0 | 0 | 5.76093 | 17.17404 |
| GO_Biologi | Endotheliu | 12-Jan | 0.050982 | 0.186585 | 0 | 0 | 21.03805 | 62.61499 |
| GO_Biologi | Mitochond | 12-Jan | 0.050982 | 0.186585 | 0 | 0 | 21.03805 | 62.61499 |
| GO_Biologi | Relaxation | 12-Jan | 0.050982 | 0.186585 | 0 | 0 | 21.03805 | 62.61499 |
| GO_Biologi | Detection ( | 12-Jan | 0.050982 | 0.186585 | 0 | 0 | 21.03805 | 62.61499 |
| GO_Biologi | Negative R | 12-Jan | 0.050982 | 0.186585 | 0 | 0 | 21.03805 | 62.61499 |
| GO_Biologi | Ribosome I | 12-Jan | 0.050982 | 0.186585 | 0 | 0 | 21.03805 | 62.61499 |
| GO_Biologi | Atrial Cardi | 12-Jan | 0.050982 | 0.186585 | 0 | 0 | 21.03805 | 62.61499 |
| GO_Biologi | Regulation | 12-Jan | 0.050982 | 0.186585 | 0 | 0 | 21.03805 | 62.61499 |
| GO_Biologi | Axonogene | 3/188 | 0.04881 | 0.186585 | 0 | 0 | 3.808494 | 11.501 |
| GO_Biologi | Negative R | 12-Jan | 0.050982 | 0.186585 | 0 | 0 | 21.03805 | 62.61499 |
| DisGeNET | Dementia, | Feb-80 | 0.047505 | 0.186602 | 0 | 0 | 5.983409 | 18.23095 |
| DisGeNET | Prion Disea | Feb-80 | 0.047505 | 0.186602 | 0 | 0 | 5.983409 | 18.23095 |
| UK_Bioban | Left arm fa | 4/431 | 0.118527 | 0.186631 | 0 | 0 | 2.199261 | 4.690176 |
| Jensen_TIS | Retina | 55/11092 | 0.087632 | 0.18687 | 0 | 0 | 1.382226 | 3.365186 |
| Jensen_TIS | HUVEC cell | 2/113 | 0.086879 | 0.18687 | 0 | 0 | 4.197562 | 10.25564 |
| Jensen_TIS | Calcaneal b | 21-Jan | 0.087525 | 0.18687 | 0 | 0 | 11.5657 | 28.17205 |
| Human_Ph | Anencepha | 27-Jan | 0.111111 | 0.186979 | 0 | 0 | 8.894007 | 19.54216 |
| Human_Ph | Venous ins | 27-Jan | 0.111111 | 0.186979 | 0 | 0 | 8.894007 | 19.54216 |
| Human_Ph | Hammerto | 27-Jan | 0.111111 | 0.186979 | 0 | 0 | 8.894007 | 19.54216 |
| DisGeNET | External Ca | 12-Jan | 0.050982 | 0.187079 | 0 | 0 | 21.03805 | 62.61499 |
| DisGeNET | Cholecysto | Feb-82 | 0.04965 | 0.187079 | 0 | 0 | 5.833235 | 17.51576 |

| | | | | | | | | |
|---|---|---|---|---|---|---|---|---|
| DisGeNET | Platelet Co | 3/191 | 0.050731 | 0.187079 | 0 | 0 | 3.74715 | 11.17106 |
| DisGeNET | bone destr | Feb-83 | 0.050736 | 0.187079 | 0 | 0 | 5.76093 | 17.17404 |
| DisGeNET | Migraine V | 12-Jan | 0.050982 | 0.187079 | 0 | 0 | 21.03805 | 62.61499 |
| DisGeNET | Multiple Sc | 9/1106 | 0.050858 | 0.187079 | 0 | 0 | 1.979104 | 5.895174 |
| DisGeNET | Mucopolys | 12-Jan | 0.050982 | 0.187079 | 0 | 0 | 21.03805 | 62.61499 |
| DisGeNET | Panacinar E | 12-Jan | 0.050982 | 0.187079 | 0 | 0 | 21.03805 | 62.61499 |
| DisGeNET | Arterial Dis | 12-Jan | 0.050982 | 0.187079 | 0 | 0 | 21.03805 | 62.61499 |
| DisGeNET | Decreased | 12-Jan | 0.050982 | 0.187079 | 0 | 0 | 21.03805 | 62.61499 |
| DisGeNET | Progressive | 12-Jan | 0.050982 | 0.187079 | 0 | 0 | 21.03805 | 62.61499 |
| DisGeNET | PTEN Hama | 12-Jan | 0.050982 | 0.187079 | 0 | 0 | 21.03805 | 62.61499 |
| DisGeNET | Craniofacia | 3/187 | 0.048177 | 0.187079 | 0 | 0 | 3.829387 | 11.61403 |
| DisGeNET | Auditory Pr | 12-Jan | 0.050982 | 0.187079 | 0 | 0 | 21.03805 | 62.61499 |
| DisGeNET | Sensory ne | Feb-81 | 0.048573 | 0.187079 | 0 | 0 | 5.907372 | 17.86791 |
| DisGeNET | Vascular le | Feb-81 | 0.048573 | 0.187079 | 0 | 0 | 5.907372 | 17.86791 |
| DisGeNET | Short 5th fi | 12-Jan | 0.050982 | 0.187079 | 0 | 0 | 21.03805 | 62.61499 |
| DisGeNET | Pelizaeus M | 12-Jan | 0.050982 | 0.187079 | 0 | 0 | 21.03805 | 62.61499 |
| DisGeNET | Peripheral | 12-Jan | 0.050982 | 0.187079 | 0 | 0 | 21.03805 | 62.61499 |
| DisGeNET | Persistent I | 12-Jan | 0.050982 | 0.187079 | 0 | 0 | 21.03805 | 62.61499 |
| DisGeNET | Dysplastic I | 12-Jan | 0.050982 | 0.187079 | 0 | 0 | 21.03805 | 62.61499 |
| DisGeNET | Early Hepa | 12-Jan | 0.050982 | 0.187079 | 0 | 0 | 21.03805 | 62.61499 |
| DisGeNET | Psychotic e | 12-Jan | 0.050982 | 0.187079 | 0 | 0 | 21.03805 | 62.61499 |
| DisGeNET | Serum Alar | 12-Jan | 0.050982 | 0.187079 | 0 | 0 | 21.03805 | 62.61499 |
| DisGeNET | Heterochro | 12-Jan | 0.050982 | 0.187079 | 0 | 0 | 21.03805 | 62.61499 |
| DisGeNET | Glucose tol | 12-Jan | 0.050982 | 0.187079 | 0 | 0 | 21.03805 | 62.61499 |
| DisGeNET | Hamartom | 12-Jan | 0.050982 | 0.187079 | 0 | 0 | 21.03805 | 62.61499 |
| DisGeNET | NEVUS, EPI | 12-Jan | 0.050982 | 0.187079 | 0 | 0 | 21.03805 | 62.61499 |
| DisGeNET | Alport Sync | 12-Jan | 0.050982 | 0.187079 | 0 | 0 | 21.03805 | 62.61499 |
| DisGeNET | Hypergranu | 12-Jan | 0.050982 | 0.187079 | 0 | 0 | 21.03805 | 62.61499 |
| DisGeNET | Hyporeflex | 12-Jan | 0.050982 | 0.187079 | 0 | 0 | 21.03805 | 62.61499 |
| DisGeNET | Abnormalit | 12-Jan | 0.050982 | 0.187079 | 0 | 0 | 21.03805 | 62.61499 |
| DisGeNET | Cirrhosis, C | 12-Jan | 0.050982 | 0.187079 | 0 | 0 | 21.03805 | 62.61499 |
| DisGeNET | Acute mou | 12-Jan | 0.050982 | 0.187079 | 0 | 0 | 21.03805 | 62.61499 |
| DisGeNET | Internal Ca | 12-Jan | 0.050982 | 0.187079 | 0 | 0 | 21.03805 | 62.61499 |
| DisGeNET | Adenocarci | 12-Jan | 0.050982 | 0.187079 | 0 | 0 | 21.03805 | 62.61499 |
| DisGeNET | Large head | 12-Jan | 0.050982 | 0.187079 | 0 | 0 | 21.03805 | 62.61499 |
| DisGeNET | Leri-Weill c | 12-Jan | 0.050982 | 0.187079 | 0 | 0 | 21.03805 | 62.61499 |
| DisGeNET | Focal Emph | 12-Jan | 0.050982 | 0.187079 | 0 | 0 | 21.03805 | 62.61499 |
| DisGeNET | Abnormal l | 12-Jan | 0.050982 | 0.187079 | 0 | 0 | 21.03805 | 62.61499 |
| DisGeNET | Liver disea | 6/606 | 0.048595 | 0.187079 | 0 | 0 | 2.384321 | 7.210741 |
| DisGeNET | Dental Fluc | 12-Jan | 0.050982 | 0.187079 | 0 | 0 | 21.03805 | 62.61499 |
| DisGeNET | Semantic-P | 12-Jan | 0.050982 | 0.187079 | 0 | 0 | 21.03805 | 62.61499 |
| DisGeNET | Senile dem | 12-Jan | 0.050982 | 0.187079 | 0 | 0 | 21.03805 | 62.61499 |
| DisGeNET | Psychotic D | 4/317 | 0.049566 | 0.187079 | 0 | 0 | 3.017822 | 9.066892 |
| DisGeNET | Familial Alz | 3/188 | 0.04881 | 0.187079 | 0 | 0 | 3.808494 | 11.501 |
| DisGeNET | Thalassemi | Feb-82 | 0.04965 | 0.187079 | 0 | 0 | 5.833235 | 17.51576 |

| | | | | | | | | |
|---|---|---|---|---|---|---|---|---|
| GTEx_Tissu | GTEX-Q2AI | 17/1673 | 0.000777 | 0.187416 | 0 | 0 | 2.677441 | 19.16932 |
| GWAS_Cat | Educationa | 6/801 | 0.135767 | 0.187583 | 0 | 0 | 1.781318 | 3.556959 |
| PhenGenI_ | Hernia, Ing | 7-Jan | 0.03006 | 0.18767 | 0 | 0 | 38.57946 | 135.2043 |
| PhenGenI_ | Gallstones | 7-Jan | 0.03006 | 0.18767 | 0 | 0 | 38.57946 | 135.2043 |
| DisGeNET | Hallucinati | 13-Jan | 0.055113 | 0.18794 | 0 | 0 | 19.28391 | 55.89185 |
| DisGeNET | Language D | 13-Jan | 0.055113 | 0.18794 | 0 | 0 | 19.28391 | 55.89185 |
| DisGeNET | Hemangios | Feb-87 | 0.055163 | 0.18794 | 0 | 0 | 5.48872 | 15.9034 |
| DisGeNET | Lipoma | Feb-87 | 0.055163 | 0.18794 | 0 | 0 | 5.48872 | 15.9034 |
| DisGeNET | Gynecoma: | Feb-87 | 0.055163 | 0.18794 | 0 | 0 | 5.48872 | 15.9034 |
| DisGeNET | Paraparesi: | 13-Jan | 0.055113 | 0.18794 | 0 | 0 | 19.28391 | 55.89185 |
| DisGeNET | Diabetes in | 13-Jan | 0.055113 | 0.18794 | 0 | 0 | 19.28391 | 55.89185 |
| DisGeNET | Proximal sp | 13-Jan | 0.055113 | 0.18794 | 0 | 0 | 19.28391 | 55.89185 |
| DisGeNET | Generalize | Feb-87 | 0.055163 | 0.18794 | 0 | 0 | 5.48872 | 15.9034 |
| DisGeNET | Lysinuric Pr | 13-Jan | 0.055113 | 0.18794 | 0 | 0 | 19.28391 | 55.89185 |
| DisGeNET | Macular re | 13-Jan | 0.055113 | 0.18794 | 0 | 0 | 19.28391 | 55.89185 |
| DisGeNET | Axenfeld-R | 13-Jan | 0.055113 | 0.18794 | 0 | 0 | 19.28391 | 55.89185 |
| DisGeNET | Mediastina | 13-Jan | 0.055113 | 0.18794 | 0 | 0 | 19.28391 | 55.89185 |
| DisGeNET | Calcium me | 13-Jan | 0.055113 | 0.18794 | 0 | 0 | 19.28391 | 55.89185 |
| DisGeNET | Multi-core | 13-Jan | 0.055113 | 0.18794 | 0 | 0 | 19.28391 | 55.89185 |
| DisGeNET | Neonatal h | 13-Jan | 0.055113 | 0.18794 | 0 | 0 | 19.28391 | 55.89185 |
| DisGeNET | Hypoplasia | 13-Jan | 0.055113 | 0.18794 | 0 | 0 | 19.28391 | 55.89185 |
| DisGeNET | Infantile m | 13-Jan | 0.055113 | 0.18794 | 0 | 0 | 19.28391 | 55.89185 |
| DisGeNET | Age-relate | 13-Jan | 0.055113 | 0.18794 | 0 | 0 | 19.28391 | 55.89185 |
| DisGeNET | Oxidative F | 13-Jan | 0.055113 | 0.18794 | 0 | 0 | 19.28391 | 55.89185 |
| DisGeNET | Gastrointe: | 4/323 | 0.052429 | 0.18794 | 0 | 0 | 2.960154 | 8.727434 |
| DisGeNET | Alzheimer': | Feb-85 | 0.052932 | 0.18794 | 0 | 0 | 5.621545 | 16.52025 |
| DisGeNET | Acute Conf | Feb-85 | 0.052932 | 0.18794 | 0 | 0 | 5.621545 | 16.52025 |
| DisGeNET | Lesion of b | Feb-85 | 0.052932 | 0.18794 | 0 | 0 | 5.621545 | 16.52025 |
| DisGeNET | Benign sym | 13-Jan | 0.055113 | 0.18794 | 0 | 0 | 19.28391 | 55.89185 |
| DisGeNET | Angina Pec | 13-Jan | 0.055113 | 0.18794 | 0 | 0 | 19.28391 | 55.89185 |
| DisGeNET | Epithelioid | 13-Jan | 0.055113 | 0.18794 | 0 | 0 | 19.28391 | 55.89185 |
| DisGeNET | Restrictive | 13-Jan | 0.055113 | 0.18794 | 0 | 0 | 19.28391 | 55.89185 |
| DisGeNET | Cerebral Ar | 13-Jan | 0.055113 | 0.18794 | 0 | 0 | 19.28391 | 55.89185 |
| DisGeNET | Hypotroph | Feb-84 | 0.05183 | 0.18794 | 0 | 0 | 5.690387 | 16.84234 |
| DisGeNET | Maxillary r | Feb-84 | 0.05183 | 0.18794 | 0 | 0 | 5.690387 | 16.84234 |
| DisGeNET | Decreased | Feb-84 | 0.05183 | 0.18794 | 0 | 0 | 5.690387 | 16.84234 |
| DisGeNET | Deficiency | Feb-84 | 0.05183 | 0.18794 | 0 | 0 | 5.690387 | 16.84234 |
| DisGeNET | Childhood . | 13-Jan | 0.055113 | 0.18794 | 0 | 0 | 19.28391 | 55.89185 |
| DisGeNET | Neurogeni | 13-Jan | 0.055113 | 0.18794 | 0 | 0 | 19.28391 | 55.89185 |
| DisGeNET | Non-toxic r | 13-Jan | 0.055113 | 0.18794 | 0 | 0 | 19.28391 | 55.89185 |
| DisGeNET | Retrusion c | Feb-84 | 0.05183 | 0.18794 | 0 | 0 | 5.690387 | 16.84234 |
| DisGeNET | Hemochroi | Feb-84 | 0.05183 | 0.18794 | 0 | 0 | 5.690387 | 16.84234 |
| DisGeNET | Thrombopl | Feb-84 | 0.05183 | 0.18794 | 0 | 0 | 5.690387 | 16.84234 |
| DisGeNET | Hypoplasia | Feb-84 | 0.05183 | 0.18794 | 0 | 0 | 5.690387 | 16.84234 |
| DisGeNET | Gastrointe: | 13-Jan | 0.055113 | 0.18794 | 0 | 0 | 19.28391 | 55.89185 |

| | | | | | | | | |
|---|---|---|---|---|---|---|---|---|
| DisGeNET | Hamartom | 13-Jan | 0.055113 | 0.18794 | 0 | 0 | 19.28391 | 55.89185 |
| DisGeNET | MYOPATHY | 13-Jan | 0.055113 | 0.18794 | 0 | 0 | 19.28391 | 55.89185 |
| DisGeNET | Human Pap | 13-Jan | 0.055113 | 0.18794 | 0 | 0 | 19.28391 | 55.89185 |
| DisGeNET | Hyperactiv | 13-Jan | 0.055113 | 0.18794 | 0 | 0 | 19.28391 | 55.89185 |
| DisGeNET | Lecithin Ac | 13-Jan | 0.055113 | 0.18794 | 0 | 0 | 19.28391 | 55.89185 |
| DisGeNET | Liver and Ir | 13-Jan | 0.055113 | 0.18794 | 0 | 0 | 19.28391 | 55.89185 |
| DisGeNET | Liver and Ir | 13-Jan | 0.055113 | 0.18794 | 0 | 0 | 19.28391 | 55.89185 |
| DisGeNET | Low densit | 13-Jan | 0.055113 | 0.18794 | 0 | 0 | 19.28391 | 55.89185 |
| DisGeNET | Ceroid lipo | 13-Jan | 0.055113 | 0.18794 | 0 | 0 | 19.28391 | 55.89185 |
| DisGeNET | Eosinophili | 13-Jan | 0.055113 | 0.18794 | 0 | 0 | 19.28391 | 55.89185 |
| DisGeNET | Disseminat | 3/195 | 0.05335 | 0.18794 | 0 | 0 | 3.668341 | 10.75147 |
| DisGeNET | Headache | Feb-86 | 0.054043 | 0.18794 | 0 | 0 | 5.554342 | 16.20739 |
| DisGeNET | Leukoence | Feb-86 | 0.054043 | 0.18794 | 0 | 0 | 5.554342 | 16.20739 |
| DisGeNET | Chronic Lyr | 9/1120 | 0.054265 | 0.18794 | 0 | 0 | 1.952711 | 5.689966 |
| GeDiPNet_ | Morgagni D | 19-Jan | 0.079527 | 0.188472 | 0 | 0 | 12.85207 | 32.53709 |
| GeDiPNet_ | Septo-Opti | 19-Jan | 0.079527 | 0.188472 | 0 | 0 | 12.85207 | 32.53709 |
| GeDiPNet_ | Posterolate | 19-Jan | 0.079527 | 0.188472 | 0 | 0 | 12.85207 | 32.53709 |
| GeDiPNet_ | Osteopetrc | 19-Jan | 0.079527 | 0.188472 | 0 | 0 | 12.85207 | 32.53709 |
| GeDiPNet_ | Coronal Cra | 19-Jan | 0.079527 | 0.188472 | 0 | 0 | 12.85207 | 32.53709 |
| Human_Ph | Postnatal n | 28-Jan | 0.114983 | 0.188512 | 0 | 0 | 8.564169 | 18.52408 |
| Human_Ph | Furrowed t | 28-Jan | 0.114983 | 0.188512 | 0 | 0 | 8.564169 | 18.52408 |
| Human_Ph | Abnormalit | 28-Jan | 0.114983 | 0.188512 | 0 | 0 | 8.564169 | 18.52408 |
| Human_Ph | Abnormalit | 28-Jan | 0.114983 | 0.188512 | 0 | 0 | 8.564169 | 18.52408 |
| Human_Ph | Generalize | 28-Jan | 0.114983 | 0.188512 | 0 | 0 | 8.564169 | 18.52408 |
| Human_Ph | Plantar hyp | 28-Jan | 0.114983 | 0.188512 | 0 | 0 | 8.564169 | 18.52408 |
| GeDiPNet_ | Communic | 20-Jan | 0.083535 | 0.188663 | 0 | 0 | 12.17503 | 30.22444 |
| GeDiPNet_ | Mental Ret | 12/1766 | 0.080175 | 0.188663 | 0 | 0 | 1.656465 | 4.180168 |
| GeDiPNet_ | Adenoma S | 20-Jan | 0.083535 | 0.188663 | 0 | 0 | 12.17503 | 30.22444 |
| GeDiPNet_ | Atrioventri | 20-Jan | 0.083535 | 0.188663 | 0 | 0 | 12.17503 | 30.22444 |
| GeDiPNet_ | Bipolar Dis | 6/697 | 0.083122 | 0.188663 | 0 | 0 | 2.060567 | 5.125552 |
| GeDiPNet_ | Cystic Fibrc | 20-Jan | 0.083535 | 0.188663 | 0 | 0 | 12.17503 | 30.22444 |
| GeDiPNet_ | Dupuytren | 20-Jan | 0.083535 | 0.188663 | 0 | 0 | 12.17503 | 30.22444 |
| GeDiPNet_ | Restrictive | 20-Jan | 0.083535 | 0.188663 | 0 | 0 | 12.17503 | 30.22444 |
| GeDiPNet_ | Hereditary | 20-Jan | 0.083535 | 0.188663 | 0 | 0 | 12.17503 | 30.22444 |
| GeDiPNet_ | Carcinoma | 5/535 | 0.083814 | 0.188663 | 0 | 0 | 2.229982 | 5.528463 |
| GeDiPNet_ | Thoracolur | 20-Jan | 0.083535 | 0.188663 | 0 | 0 | 12.17503 | 30.22444 |
| GeDiPNet_ | Limb Musc | 20-Jan | 0.083535 | 0.188663 | 0 | 0 | 12.17503 | 30.22444 |
| GeDiPNet_ | Benign Pro | 20-Jan | 0.083535 | 0.188663 | 0 | 0 | 12.17503 | 30.22444 |
| Rare_Disea | Auditory p | 8-Jan | 0.03428 | 0.188923 | 0 | 0 | 33.06645 | 111.5391 |
| Rare_Disea | Antley Bixl | 8-Jan | 0.03428 | 0.188923 | 0 | 0 | 33.06645 | 111.5391 |
| Rare_Disea | Ebstein's a | 8-Jan | 0.03428 | 0.188923 | 0 | 0 | 33.06645 | 111.5391 |
| Rare_Disea | Hypoalpha | 8-Jan | 0.03428 | 0.188923 | 0 | 0 | 33.06645 | 111.5391 |
| Rare_Disea | Double ute | 4/280 | 0.033892 | 0.188923 | 0 | 0 | 3.428846 | 11.6052 |
| Rare_Disea | Pantothen | 8-Jan | 0.03428 | 0.188923 | 0 | 0 | 33.06645 | 111.5391 |
| Jensen_DIS | Night blind | Jan-39 | 0.156488 | 0.189536 | 0 | 0 | 6.081701 | 11.28018 |

| | | | | | | | | |
|---|---|---|---|---|---|---|---|---|
| Jensen_DIS | Alopecia | Jan-39 | 0.156488 | 0.189536 | 0 | 0 | 6.081701 | 11.28018 |
| GeDiPNet_ | Infiltrating | 21-Jan | 0.087525 | 0.189692 | 0 | 0 | 11.5657 | 28.17205 |
| GeDiPNet_ | Synostotic | 21-Jan | 0.087525 | 0.189692 | 0 | 0 | 11.5657 | 28.17205 |
| GeDiPNet_ | Patent Duc | 3/239 | 0.086224 | 0.189692 | 0 | 0 | 2.977754 | 7.297891 |
| GeDiPNet_ | Hypoplasia | 2/112 | 0.085575 | 0.189692 | 0 | 0 | 4.235936 | 10.41346 |
| GeDiPNet_ | Thumb Apl | 21-Jan | 0.087525 | 0.189692 | 0 | 0 | 11.5657 | 28.17205 |
| GeDiPNet_ | Pulmonary | 21-Jan | 0.087525 | 0.189692 | 0 | 0 | 11.5657 | 28.17205 |
| GeDiPNet_ | Paraganglio | 21-Jan | 0.087525 | 0.189692 | 0 | 0 | 11.5657 | 28.17205 |
| GeDiPNet_ | Short Femu | 21-Jan | 0.087525 | 0.189692 | 0 | 0 | 11.5657 | 28.17205 |
| GeDiPNet_ | Speech Del | 21-Jan | 0.087525 | 0.189692 | 0 | 0 | 11.5657 | 28.17205 |
| GeDiPNet_ | Infantile Sp | 21-Jan | 0.087525 | 0.189692 | 0 | 0 | 11.5657 | 28.17205 |
| GWAS_Cat | Citrate Lev | Jan-34 | 0.137866 | 0.189838 | 0 | 0 | 7.004933 | 13.88007 |
| GWAS_Cat | Cholestero | Jan-34 | 0.137866 | 0.189838 | 0 | 0 | 7.004933 | 13.88007 |
| GWAS_Cat | Free Chole: | Jan-34 | 0.137866 | 0.189838 | 0 | 0 | 7.004933 | 13.88007 |
| DisGeNET | Secondary | 4/330 | 0.055881 | 0.190139 | 0 | 0 | 2.895558 | 8.352346 |
| GeDiPNet_ | Liver Cirrho | 2/114 | 0.088189 | 0.190423 | 0 | 0 | 4.159874 | 10.1013 |
| Human_Ph | Hamartom | 29-Jan | 0.118838 | 0.19074 | 0 | 0 | 8.25789 | 17.58927 |
| Human_Ph | Papilloma ( | 29-Jan | 0.118838 | 0.19074 | 0 | 0 | 8.25789 | 17.58927 |
| Human_Ph | Rhizomelia | 29-Jan | 0.118838 | 0.19074 | 0 | 0 | 8.25789 | 17.58927 |
| Human_Ph | Pulmonary | 29-Jan | 0.118838 | 0.19074 | 0 | 0 | 8.25789 | 17.58927 |
| Human_Ph | Verrucae (I | 29-Jan | 0.118838 | 0.19074 | 0 | 0 | 8.25789 | 17.58927 |
| DisGeNET | Abnormalit | Feb-88 | 0.05629 | 0.190789 | 0 | 0 | 5.424624 | 15.60794 |
| DisGeNET | Retrovirida | Feb-88 | 0.05629 | 0.190789 | 0 | 0 | 5.424624 | 15.60794 |
| DisGeNET | Blepharoph | Feb-88 | 0.05629 | 0.190789 | 0 | 0 | 5.424624 | 15.60794 |
| DisGeNET | Adult B Acu | 14-Jan | 0.059226 | 0.191808 | 0 | 0 | 17.79964 | 50.30878 |
| DisGeNET | Renal fibro | 3/202 | 0.058087 | 0.191808 | 0 | 0 | 3.538047 | 10.06862 |
| DisGeNET | Hypoplastic | 14-Jan | 0.059226 | 0.191808 | 0 | 0 | 17.79964 | 50.30878 |
| DisGeNET | Vacuolar m | 14-Jan | 0.059226 | 0.191808 | 0 | 0 | 17.79964 | 50.30878 |
| DisGeNET | Visual sym | 14-Jan | 0.059226 | 0.191808 | 0 | 0 | 17.79964 | 50.30878 |
| DisGeNET | Adams Oliv | 14-Jan | 0.059226 | 0.191808 | 0 | 0 | 17.79964 | 50.30878 |
| DisGeNET | Hip circum | Feb-90 | 0.058569 | 0.191808 | 0 | 0 | 5.300802 | 15.0413 |
| DisGeNET | Adenovirus | Feb-90 | 0.058569 | 0.191808 | 0 | 0 | 5.300802 | 15.0413 |
| DisGeNET | Cardiac Arr | 3/201 | 0.057398 | 0.191808 | 0 | 0 | 3.556097 | 10.16239 |
| DisGeNET | Genital ulc | 14-Jan | 0.059226 | 0.191808 | 0 | 0 | 17.79964 | 50.30878 |
| DisGeNET | Germinoma | 14-Jan | 0.059226 | 0.191808 | 0 | 0 | 17.79964 | 50.30878 |
| DisGeNET | Skin tag | 14-Jan | 0.059226 | 0.191808 | 0 | 0 | 17.79964 | 50.30878 |
| DisGeNET | hemangiop | 14-Jan | 0.059226 | 0.191808 | 0 | 0 | 17.79964 | 50.30878 |
| DisGeNET | Subcortical | 14-Jan | 0.059226 | 0.191808 | 0 | 0 | 17.79964 | 50.30878 |
| DisGeNET | Myocardial | Feb-89 | 0.057425 | 0.191808 | 0 | 0 | 5.362001 | 15.32067 |
| DisGeNET | Left Ventri | 3/203 | 0.05878 | 0.191808 | 0 | 0 | 3.520179 | 9.976043 |
| DisGeNET | Mood Diso | 4/336 | 0.058935 | 0.191808 | 0 | 0 | 2.842357 | 8.047601 |
| DisGeNET | Familial hy | 14-Jan | 0.059226 | 0.191808 | 0 | 0 | 17.79964 | 50.30878 |
| DisGeNET | Histiocytor | 14-Jan | 0.059226 | 0.191808 | 0 | 0 | 17.79964 | 50.30878 |
| DisGeNET | Gastric Gas | 14-Jan | 0.059226 | 0.191808 | 0 | 0 | 17.79964 | 50.30878 |
| DisGeNET | Mastitis-m | 14-Jan | 0.059226 | 0.191808 | 0 | 0 | 17.79964 | 50.30878 |

| | | | | | | | | |
|---|---|---|---|---|---|---|---|---|
| DisGeNET | Mucopolys | 14-Jan | 0.059226 | 0.191808 | 0 | 0 | 17.79964 | 50.30878 |
| DisGeNET | Centriacina | 14-Jan | 0.059226 | 0.191808 | 0 | 0 | 17.79964 | 50.30878 |
| DisGeNET | Poikiloderr | 14-Jan | 0.059226 | 0.191808 | 0 | 0 | 17.79964 | 50.30878 |
| DisGeNET | Pleomorph | 14-Jan | 0.059226 | 0.191808 | 0 | 0 | 17.79964 | 50.30878 |
| DisGeNET | Congenital | 14-Jan | 0.059226 | 0.191808 | 0 | 0 | 17.79964 | 50.30878 |
| DisGeNET | ARTERIAL T | 14-Jan | 0.059226 | 0.191808 | 0 | 0 | 17.79964 | 50.30878 |
| DisGeNET | Corticobas | 14-Jan | 0.059226 | 0.191808 | 0 | 0 | 17.79964 | 50.30878 |
| DisGeNET | Lone atrial | 14-Jan | 0.059226 | 0.191808 | 0 | 0 | 17.79964 | 50.30878 |
| DisGeNET | Aqueducta | 14-Jan | 0.059226 | 0.191808 | 0 | 0 | 17.79964 | 50.30878 |
| DisGeNET | ALZHEIMEI | 14-Jan | 0.059226 | 0.191808 | 0 | 0 | 17.79964 | 50.30878 |
| DisGeNET | Semantic D | 14-Jan | 0.059226 | 0.191808 | 0 | 0 | 17.79964 | 50.30878 |
| DisGeNET | Hereditary | 14-Jan | 0.059226 | 0.191808 | 0 | 0 | 17.79964 | 50.30878 |
| DisGeNET | Creutzfeldt | Feb-89 | 0.057425 | 0.191808 | 0 | 0 | 5.362001 | 15.32067 |
| DisGeNET | Testicular I | 14-Jan | 0.059226 | 0.191808 | 0 | 0 | 17.79964 | 50.30878 |
| DisGeNET | Testicular D | 14-Jan | 0.059226 | 0.191808 | 0 | 0 | 17.79964 | 50.30878 |
| GO_Biologi | Regulation | 13-Jan | 0.055113 | 0.191816 | 0 | 0 | 19.28391 | 55.89185 |
| GO_Biologi | ATP Transp | 13-Jan | 0.055113 | 0.191816 | 0 | 0 | 19.28391 | 55.89185 |
| GO_Biologi | Regulation | 13-Jan | 0.055113 | 0.191816 | 0 | 0 | 19.28391 | 55.89185 |
| GO_Biologi | Regulation | 13-Jan | 0.055113 | 0.191816 | 0 | 0 | 19.28391 | 55.89185 |
| GO_Biologi | Negative R | 13-Jan | 0.055113 | 0.191816 | 0 | 0 | 19.28391 | 55.89185 |
| GO_Biologi | GTP Metab | 13-Jan | 0.055113 | 0.191816 | 0 | 0 | 19.28391 | 55.89185 |
| GO_Biologi | Positive Re | 13-Jan | 0.055113 | 0.191816 | 0 | 0 | 19.28391 | 55.89185 |
| GO_Biologi | Relaxation | 13-Jan | 0.055113 | 0.191816 | 0 | 0 | 19.28391 | 55.89185 |
| GO_Biologi | Regulation | 13-Jan | 0.055113 | 0.191816 | 0 | 0 | 19.28391 | 55.89185 |
| GO_Biologi | Regulation | 13-Jan | 0.055113 | 0.191816 | 0 | 0 | 19.28391 | 55.89185 |
| GTEx_Tissu | GTEX-WH7 | 9/607 | 0.001287 | 0.191902 | 0 | 0 | 3.726846 | 24.80414 |
| GTEx_Tissu | GTEX-X4XX | 9/614 | 0.001393 | 0.191902 | 0 | 0 | 3.68239 | 24.21649 |
| GTEx_Tissu | GTEX-OHPI | 15/1446 | 0.001345 | 0.191902 | 0 | 0 | 2.690717 | 17.78869 |
| GTEx_Tissu | GTEX-NFK9 | 8/489 | 0.001301 | 0.191902 | 0 | 0 | 4.091055 | 27.1835 |
| GTEx_Tissu | GTEX-XMK | 16/1579 | 0.001169 | 0.191902 | 0 | 0 | 2.645689 | 17.86232 |
| GTEx_Tissu | GTEX-WY7 | 10/725 | 0.001189 | 0.191902 | 0 | 0 | 3.487058 | 23.48307 |
| GTEx_Tissu | GTEX-RUSC | 23/2728 | 0.001117 | 0.191902 | 0 | 0 | 2.286183 | 15.53877 |
| GTEx_Tissu | GTEX-QLQ\ | 20/2193 | 0.000982 | 0.191902 | 0 | 0 | 2.436964 | 16.87805 |
| GTEx_Tissu | GTEX-PLZ5 | 8/489 | 0.001301 | 0.191902 | 0 | 0 | 4.091055 | 27.1835 |
| DisGeNET | Turridolich | Feb-91 | 0.05972 | 0.191984 | 0 | 0 | 5.240978 | 14.76951 |
| DisGeNET | Narrow hea | Feb-91 | 0.05972 | 0.191984 | 0 | 0 | 5.240978 | 14.76951 |
| DisGeNET | Papilloma | 3/204 | 0.059476 | 0.191984 | 0 | 0 | 3.502488 | 9.88465 |
| DisGeNET | Narrow sku | Feb-91 | 0.05972 | 0.191984 | 0 | 0 | 5.240978 | 14.76951 |
| DisGeNET | Leukodystr | 3/204 | 0.059476 | 0.191984 | 0 | 0 | 3.502488 | 9.88465 |
| DisGeNET | Narrow cra | Feb-91 | 0.05972 | 0.191984 | 0 | 0 | 5.240978 | 14.76951 |
| UK_Bioban | Ease of skir | 2/139 | 0.122685 | 0.192117 | 0 | 0 | 3.396479 | 7.126265 |
| GeDiPNet_ | Craniosyno | 2/115 | 0.089505 | 0.192552 | 0 | 0 | 4.122853 | 9.950336 |
| Jensen_TIS | Reticulum | 22-Jan | 0.091499 | 0.192566 | 0 | 0 | 11.0144 | 26.34017 |
| Jensen_TIS | Myeloid ce | 22-Jan | 0.091499 | 0.192566 | 0 | 0 | 11.0144 | 26.34017 |
| DisGeNET | Congenital | 15-Jan | 0.063321 | 0.193134 | 0 | 0 | 16.52741 | 45.60793 |

| | | | | | | | | |
|---|---|---|---|---|---|---|---|---|
| DisGeNET | Heart Failu | 15-Jan | 0.063321 | 0.193134 | 0 | 0 | 16.52741 | 45.60793 |
| DisGeNET | Muscular D | Feb-93 | 0.062046 | 0.193134 | 0 | 0 | 5.125275 | 14.24761 |
| DisGeNET | Systolic hy | 15-Jan | 0.063321 | 0.193134 | 0 | 0 | 16.52741 | 45.60793 |
| DisGeNET | Hypoplasti | 15-Jan | 0.063321 | 0.193134 | 0 | 0 | 16.52741 | 45.60793 |
| DisGeNET | INCLUSION | 15-Jan | 0.063321 | 0.193134 | 0 | 0 | 16.52741 | 45.60793 |
| DisGeNET | Large earlo | 15-Jan | 0.063321 | 0.193134 | 0 | 0 | 16.52741 | 45.60793 |
| DisGeNET | Left ventric | 15-Jan | 0.063321 | 0.193134 | 0 | 0 | 16.52741 | 45.60793 |
| DisGeNET | Recurrent l | 15-Jan | 0.063321 | 0.193134 | 0 | 0 | 16.52741 | 45.60793 |
| DisGeNET | Benign sca | 15-Jan | 0.063321 | 0.193134 | 0 | 0 | 16.52741 | 45.60793 |
| DisGeNET | Mesomelia | 15-Jan | 0.063321 | 0.193134 | 0 | 0 | 16.52741 | 45.60793 |
| DisGeNET | Cardiomyo | Feb-92 | 0.060879 | 0.193134 | 0 | 0 | 5.182484 | 14.50504 |
| DisGeNET | Thyroid ass | Feb-92 | 0.060879 | 0.193134 | 0 | 0 | 5.182484 | 14.50504 |
| DisGeNET | Malignant | 7/805 | 0.061053 | 0.193134 | 0 | 0 | 2.095943 | 5.860289 |
| DisGeNET | Hyperglyce | 5/488 | 0.061648 | 0.193134 | 0 | 0 | 2.452911 | 6.834599 |
| DisGeNET | Short midd | 15-Jan | 0.063321 | 0.193134 | 0 | 0 | 16.52741 | 45.60793 |
| DisGeNET | Small wing | 15-Jan | 0.063321 | 0.193134 | 0 | 0 | 16.52741 | 45.60793 |
| DisGeNET | Hematoche | 15-Jan | 0.063321 | 0.193134 | 0 | 0 | 16.52741 | 45.60793 |
| DisGeNET | Pulmonary | 15-Jan | 0.063321 | 0.193134 | 0 | 0 | 16.52741 | 45.60793 |
| DisGeNET | Limb-girdle | 15-Jan | 0.063321 | 0.193134 | 0 | 0 | 16.52741 | 45.60793 |
| DisGeNET | Metaphyse | 15-Jan | 0.063321 | 0.193134 | 0 | 0 | 16.52741 | 45.60793 |
| DisGeNET | Thymoma | 3/205 | 0.060177 | 0.193134 | 0 | 0 | 3.484972 | 9.794417 |
| DisGeNET | Peg-shape | 15-Jan | 0.063321 | 0.193134 | 0 | 0 | 16.52741 | 45.60793 |
| DisGeNET | Pointed to | 15-Jan | 0.063321 | 0.193134 | 0 | 0 | 16.52741 | 45.60793 |
| DisGeNET | Manic | Feb-93 | 0.062046 | 0.193134 | 0 | 0 | 5.125275 | 14.24761 |
| DisGeNET | Smoking B | 3/209 | 0.063017 | 0.193134 | 0 | 0 | 3.416609 | 9.444704 |
| DisGeNET | Myxoid cys | 3/209 | 0.063017 | 0.193134 | 0 | 0 | 3.416609 | 9.444704 |
| DisGeNET | Long narro | Feb-94 | 0.063221 | 0.193134 | 0 | 0 | 5.069309 | 13.99697 |
| DisGeNET | Myelodysp | Feb-94 | 0.063221 | 0.193134 | 0 | 0 | 5.069309 | 13.99697 |
| DisGeNET | Diabetes M | 11/1507 | 0.061872 | 0.193134 | 0 | 0 | 1.78183 | 4.958287 |
| DisGeNET | Chromosor | 15-Jan | 0.063321 | 0.193134 | 0 | 0 | 16.52741 | 45.60793 |
| DisGeNET | Nevi and M | 15-Jan | 0.063321 | 0.193134 | 0 | 0 | 16.52741 | 45.60793 |
| DisGeNET | Oestrogen | 15-Jan | 0.063321 | 0.193134 | 0 | 0 | 16.52741 | 45.60793 |
| DisGeNET | Congenital | 15-Jan | 0.063321 | 0.193134 | 0 | 0 | 16.52741 | 45.60793 |
| DisGeNET | PARKINSOI | Feb-93 | 0.062046 | 0.193134 | 0 | 0 | 5.125275 | 14.24761 |
| DisGeNET | Pulmonary | 15-Jan | 0.063321 | 0.193134 | 0 | 0 | 16.52741 | 45.60793 |
| DisGeNET | Genital He | 15-Jan | 0.063321 | 0.193134 | 0 | 0 | 16.52741 | 45.60793 |
| DisGeNET | Brachydact | 15-Jan | 0.063321 | 0.193134 | 0 | 0 | 16.52741 | 45.60793 |
| DisGeNET | Lens Opaci | 4/339 | 0.060496 | 0.193134 | 0 | 0 | 2.816472 | 7.900705 |
| DisGeNET | Muscle We | 4/339 | 0.060496 | 0.193134 | 0 | 0 | 2.816472 | 7.900705 |
| DisGeNET | Gaucher Di | 15-Jan | 0.063321 | 0.193134 | 0 | 0 | 16.52741 | 45.60793 |
| DisGeNET | Executive c | 15-Jan | 0.063321 | 0.193134 | 0 | 0 | 16.52741 | 45.60793 |
| DisGeNET | Autosomal | 15-Jan | 0.063321 | 0.193134 | 0 | 0 | 16.52741 | 45.60793 |
| DisGeNET | Reiter Sync | 15-Jan | 0.063321 | 0.193134 | 0 | 0 | 16.52741 | 45.60793 |
| GWAS_Cat | Post Bronc | 2/152 | 0.141732 | 0.193199 | 0 | 0 | 3.100078 | 6.056983 |
| GWAS_Cat | Intake Of T | Jan-35 | 0.141623 | 0.193199 | 0 | 0 | 6.798564 | 13.28839 |

| | | | | | | | | |
|---|---|---|---|---|---|---|---|---|
| GWAS_Cat | Age-relate | Jan-35 | 0.141623 | 0.193199 | 0 | 0 | 6.798564 | 13.28839 |
| GWAS_Cat | Scarlet Fev | Jan-35 | 0.141623 | 0.193199 | 0 | 0 | 6.798564 | 13.28839 |
| GWAS_Cat | LDL Choles | Jan-35 | 0.141623 | 0.193199 | 0 | 0 | 6.798564 | 13.28839 |
| GWAS_Cat | Alcohol Co | Jan-35 | 0.141623 | 0.193199 | 0 | 0 | 6.798564 | 13.28839 |
| GWAS_Cat | Cholesteryl | Jan-35 | 0.141623 | 0.193199 | 0 | 0 | 6.798564 | 13.28839 |
| GWAS_Cat | Neurofibril | Jan-35 | 0.141623 | 0.193199 | 0 | 0 | 6.798564 | 13.28839 |
| GWAS_Cat | Pork Consu | Jan-35 | 0.141623 | 0.193199 | 0 | 0 | 6.798564 | 13.28839 |
| CellMarker | Mast Cell L | 31-Jan | 0.126498 | 0.193635 | 0 | 0 | 7.706589 | 15.93356 |
| CellMarker | Microglial | 31-Jan | 0.126498 | 0.193635 | 0 | 0 | 7.706589 | 15.93356 |
| Rare_Disea | Cytoplasmi | Feb-68 | 0.035404 | 0.193679 | 0 | 0 | 7.075579 | 23.63906 |
| HuBMAP_ | Endothelia | 3/118 | 0.014818 | 0.19398 | 0 | 0 | 6.148447 | 25.89677 |
| DisGeNET | Germ cell t | 3/210 | 0.063737 | 0.194175 | 0 | 0 | 3.399931 | 9.359987 |
| GeDiPNet_ | Central Vis | 2/116 | 0.090827 | 0.194355 | 0 | 0 | 4.086481 | 9.802643 |
| GeDiPNet_ | LEOPARD S | 22-Jan | 0.091499 | 0.194355 | 0 | 0 | 11.0144 | 26.34017 |
| GeDiPNet_ | Hamartom | 22-Jan | 0.091499 | 0.194355 | 0 | 0 | 11.0144 | 26.34017 |
| GeDiPNet_ | Hypertensi | 5/550 | 0.091677 | 0.194355 | 0 | 0 | 2.166928 | 5.17784 |
| GO_Biologi | Negative R | Feb-88 | 0.05629 | 0.194956 | 0 | 0 | 5.424624 | 15.60794 |
| Human_Ph | Hypertrigly | 30-Jan | 0.122676 | 0.19526 | 0 | 0 | 7.972735 | 16.72843 |
| Human_Ph | Emphysem | 30-Jan | 0.122676 | 0.19526 | 0 | 0 | 7.972735 | 16.72843 |
| GO_Biologi | Negative R | 14-Jan | 0.059226 | 0.195584 | 0 | 0 | 17.79964 | 50.30878 |
| GO_Biologi | Striated M | 14-Jan | 0.059226 | 0.195584 | 0 | 0 | 17.79964 | 50.30878 |
| GO_Biologi | Formation | 14-Jan | 0.059226 | 0.195584 | 0 | 0 | 17.79964 | 50.30878 |
| GO_Biologi | Positive Re | 14-Jan | 0.059226 | 0.195584 | 0 | 0 | 17.79964 | 50.30878 |
| GO_Biologi | Lipoprotei | 14-Jan | 0.059226 | 0.195584 | 0 | 0 | 17.79964 | 50.30878 |
| GO_Biologi | Response T | 14-Jan | 0.059226 | 0.195584 | 0 | 0 | 17.79964 | 50.30878 |
| GO_Biologi | Centromer | 14-Jan | 0.059226 | 0.195584 | 0 | 0 | 17.79964 | 50.30878 |
| GO_Biologi | Mitotic Re | 14-Jan | 0.059226 | 0.195584 | 0 | 0 | 17.79964 | 50.30878 |
| GO_Biologi | Regulation | Feb-90 | 0.058569 | 0.195584 | 0 | 0 | 5.300802 | 15.0413 |
| GO_Biologi | Positive Re | 5/480 | 0.058253 | 0.195584 | 0 | 0 | 2.49525 | 7.093875 |
| DisGeNET | Rheumatis | Feb-95 | 0.064403 | 0.195977 | 0 | 0 | 5.014548 | 13.75286 |
| CellMarker | Epithelial C | Jan-32 | 0.130304 | 0.196172 | 0 | 0 | 7.457614 | 15.19775 |
| CellMarker | Endothelia | 2/144 | 0.129934 | 0.196172 | 0 | 0 | 3.276056 | 6.685543 |
| CellMarker | Neutrophil | Jan-32 | 0.130304 | 0.196172 | 0 | 0 | 7.457614 | 15.19775 |
| GO_Molec | Translation | Feb-41 | 0.013741 | 0.196301 | 0 | 0 | 11.99035 | 51.40702 |
| Human_Ph | Abnormalit | 31-Jan | 0.126498 | 0.196433 | 0 | 0 | 7.706589 | 15.93356 |
| Human_Ph | Fibroma (H | 31-Jan | 0.126498 | 0.196433 | 0 | 0 | 7.706589 | 15.93356 |
| Human_Ph | Aplasia/Hy | 31-Jan | 0.126498 | 0.196433 | 0 | 0 | 7.706589 | 15.93356 |
| Human_Ph | Prenatal m | 31-Jan | 0.126498 | 0.196433 | 0 | 0 | 7.706589 | 15.93356 |
| Human_Ph | Hypoplasti | 31-Jan | 0.126498 | 0.196433 | 0 | 0 | 7.706589 | 15.93356 |
| Human_Ph | Melanoma | 31-Jan | 0.126498 | 0.196433 | 0 | 0 | 7.706589 | 15.93356 |
| Human_Ph | Congenital | 2/142 | 0.127022 | 0.196447 | 0 | 0 | 3.323193 | 6.857052 |
| GeDiPNet_ | Defect Of S | 23-Jan | 0.095455 | 0.196644 | 0 | 0 | 10.51321 | 24.6966 |
| GeDiPNet_ | Congenital | 23-Jan | 0.095455 | 0.196644 | 0 | 0 | 10.51321 | 24.6966 |
| GeDiPNet_ | Hashimoto | 23-Jan | 0.095455 | 0.196644 | 0 | 0 | 10.51321 | 24.6966 |
| GeDiPNet_ | Social Com | 23-Jan | 0.095455 | 0.196644 | 0 | 0 | 10.51321 | 24.6966 |

| | | | | | | | | |
|---|---|---|---|---|---|---|---|---|
| GeDiPNet_ | Polyp Of La | 23-Jan | 0.095455 | 0.196644 | 0 | 0 | 10.51321 | 24.6966 |
| GeDiPNet_ | Rectal Prol | 23-Jan | 0.095455 | 0.196644 | 0 | 0 | 10.51321 | 24.6966 |
| GeDiPNet_ | Parkinson I | 2/119 | 0.094827 | 0.196644 | 0 | 0 | 3.981096 | 9.378266 |
| GeDiPNet_ | Physiologic | 2/118 | 0.093488 | 0.196644 | 0 | 0 | 4.015619 | 9.516699 |
| GWAS_Cat | Serum 25-I | 2/154 | 0.144717 | 0.197048 | 0 | 0 | 3.058978 | 5.912923 |
| GWAS_Cat | Free Chole | Jan-36 | 0.145363 | 0.197048 | 0 | 0 | 6.603987 | 12.73591 |
| GWAS_Cat | Clinical LDL | Jan-36 | 0.145363 | 0.197048 | 0 | 0 | 6.603987 | 12.73591 |
| GWAS_Cat | Phospholip | Jan-36 | 0.145363 | 0.197048 | 0 | 0 | 6.603987 | 12.73591 |
| GWAS_Cat | Thiazide-in | Jan-36 | 0.145363 | 0.197048 | 0 | 0 | 6.603987 | 12.73591 |
| PhenGenI_ | Cerebral Ar | 9-Jan | 0.038483 | 0.197283 | 0 | 0 | 28.93169 | 94.24608 |
| PhenGenI_ | Alzheimer I | 9/1037 | 0.036158 | 0.197283 | 0 | 0 | 2.119687 | 7.037071 |
| PhenGenI_ | Saccades | 8-Jan | 0.03428 | 0.197283 | 0 | 0 | 33.06645 | 111.5391 |
| PhenGenI_ | Vital Capac | 3/170 | 0.038062 | 0.197283 | 0 | 0 | 4.22284 | 13.80251 |
| PhenGenI_ | Lipoprotein | Feb-70 | 0.037325 | 0.197283 | 0 | 0 | 6.866782 | 22.57858 |
| PhenGenI_ | Glucose To | 9-Jan | 0.038483 | 0.197283 | 0 | 0 | 28.93169 | 94.24608 |
| PhenGenI_ | Eyebrows | Feb-72 | 0.039286 | 0.197283 | 0 | 0 | 6.669916 | 21.58979 |
| PhenGenI_ | C-Peptide | 9-Jan | 0.038483 | 0.197283 | 0 | 0 | 28.93169 | 94.24608 |
| PhenGenI_ | Hematocrit | 4/292 | 0.038602 | 0.197283 | 0 | 0 | 3.283969 | 10.6875 |
| DisGeNET | Abnormalit | 16-Jan | 0.067399 | 0.197302 | 0 | 0 | 15.42481 | 41.60265 |
| DisGeNET | Uterine Co | 16-Jan | 0.067399 | 0.197302 | 0 | 0 | 15.42481 | 41.60265 |
| DisGeNET | Visceral an | 16-Jan | 0.067399 | 0.197302 | 0 | 0 | 15.42481 | 41.60265 |
| DisGeNET | Adenosine | 16-Jan | 0.067399 | 0.197302 | 0 | 0 | 15.42481 | 41.60265 |
| DisGeNET | Telecanthu | Feb-96 | 0.065593 | 0.197302 | 0 | 0 | 4.960951 | 13.51507 |
| DisGeNET | IMMUNE S | Feb-96 | 0.065593 | 0.197302 | 0 | 0 | 4.960951 | 13.51507 |
| DisGeNET | Quadripare | 16-Jan | 0.067399 | 0.197302 | 0 | 0 | 15.42481 | 41.60265 |
| DisGeNET | Juvenile pc | 16-Jan | 0.067399 | 0.197302 | 0 | 0 | 15.42481 | 41.60265 |
| DisGeNET | Lhermitte-I | 16-Jan | 0.067399 | 0.197302 | 0 | 0 | 15.42481 | 41.60265 |
| DisGeNET | Macroence | 16-Jan | 0.067399 | 0.197302 | 0 | 0 | 15.42481 | 41.60265 |
| DisGeNET | Severe [Du | 16-Jan | 0.067399 | 0.197302 | 0 | 0 | 15.42481 | 41.60265 |
| DisGeNET | Globoid ce | 16-Jan | 0.067399 | 0.197302 | 0 | 0 | 15.42481 | 41.60265 |
| DisGeNET | Dermatofik | 16-Jan | 0.067399 | 0.197302 | 0 | 0 | 15.42481 | 41.60265 |
| DisGeNET | Desmoplas | 16-Jan | 0.067399 | 0.197302 | 0 | 0 | 15.42481 | 41.60265 |
| DisGeNET | Disproport | 16-Jan | 0.067399 | 0.197302 | 0 | 0 | 15.42481 | 41.60265 |
| DisGeNET | Pyle metap | 16-Jan | 0.067399 | 0.197302 | 0 | 0 | 15.42481 | 41.60265 |
| DisGeNET | Large for g | 16-Jan | 0.067399 | 0.197302 | 0 | 0 | 15.42481 | 41.60265 |
| DisGeNET | Lepidic Pre | 16-Jan | 0.067399 | 0.197302 | 0 | 0 | 15.42481 | 41.60265 |
| DisGeNET | Ovarian Gr | 16-Jan | 0.067399 | 0.197302 | 0 | 0 | 15.42481 | 41.60265 |
| DisGeNET | Pleomorph | 16-Jan | 0.067399 | 0.197302 | 0 | 0 | 15.42481 | 41.60265 |
| DisGeNET | Craniosync | 3/214 | 0.066654 | 0.197302 | 0 | 0 | 3.3348 | 9.031434 |
| DisGeNET | Ovarian Se | Feb-97 | 0.06679 | 0.197302 | 0 | 0 | 4.908483 | 13.28336 |
| DisGeNET | Adult T-Cel | 5/500 | 0.066947 | 0.197302 | 0 | 0 | 2.391968 | 6.467549 |
| DisGeNET | Secondary | 16-Jan | 0.067399 | 0.197302 | 0 | 0 | 15.42481 | 41.60265 |
| DisGeNET | Gaucher Di | 16-Jan | 0.067399 | 0.197302 | 0 | 0 | 15.42481 | 41.60265 |
| DisGeNET | Auricular s | 16-Jan | 0.067399 | 0.197302 | 0 | 0 | 15.42481 | 41.60265 |
| DisGeNET | Ocular mus | 16-Jan | 0.067399 | 0.197302 | 0 | 0 | 15.42481 | 41.60265 |

| | | | | | | | | |
|---|---|---|---|---|---|---|---|---|
| DisGeNET | Cerebellar | 16-Jan | 0.067399 | 0.197302 | 0 | 0 | 15.42481 | 41.60265 |
| DisGeNET | Cerebral hy | 16-Jan | 0.067399 | 0.197302 | 0 | 0 | 15.42481 | 41.60265 |
| DisGeNET | Glioblastor | 16-Jan | 0.067399 | 0.197302 | 0 | 0 | 15.42481 | 41.60265 |
| DisGeNET | Reactive hy | 16-Jan | 0.067399 | 0.197302 | 0 | 0 | 15.42481 | 41.60265 |
| DisGeNET | Carcinoma | 9/1162 | 0.065375 | 0.197302 | 0 | 0 | 1.877377 | 5.120762 |
| DisGeNET | Lymphoid l | 3/213 | 0.065919 | 0.197302 | 0 | 0 | 3.35085 | 9.112059 |
| DisGeNET | Hand polyc | 16-Jan | 0.067399 | 0.197302 | 0 | 0 | 15.42481 | 41.60265 |
| Rare_Disea | Alpha 1-an | Feb-69 | 0.03636 | 0.197456 | 0 | 0 | 6.969622 | 23.09942 |
| Human_Ph | Hallux valg | Jan-32 | 0.130304 | 0.197525 | 0 | 0 | 7.457614 | 15.19775 |
| Human_Ph | Aplasia/Hy | Jan-32 | 0.130304 | 0.197525 | 0 | 0 | 7.457614 | 15.19775 |
| Human_Ph | Atrial fibrill | Jan-32 | 0.130304 | 0.197525 | 0 | 0 | 7.457614 | 15.19775 |
| Human_Ph | Fibrous tiss | Jan-32 | 0.130304 | 0.197525 | 0 | 0 | 7.457614 | 15.19775 |
| Human_Ph | Renal hypo | Jan-32 | 0.130304 | 0.197525 | 0 | 0 | 7.457614 | 15.19775 |
| GO_Biologi | Negative R | 15-Jan | 0.063321 | 0.198054 | 0 | 0 | 16.52741 | 45.60793 |
| GO_Biologi | Negative R | 15-Jan | 0.063321 | 0.198054 | 0 | 0 | 16.52741 | 45.60793 |
| GO_Biologi | Keratan Su | 15-Jan | 0.063321 | 0.198054 | 0 | 0 | 16.52741 | 45.60793 |
| GO_Biologi | Presynapse | 15-Jan | 0.063321 | 0.198054 | 0 | 0 | 16.52741 | 45.60793 |
| GO_Biologi | Regulation | 15-Jan | 0.063321 | 0.198054 | 0 | 0 | 16.52741 | 45.60793 |
| GO_Biologi | Regulation | 15-Jan | 0.063321 | 0.198054 | 0 | 0 | 16.52741 | 45.60793 |
| GO_Biologi | Negative R | 15-Jan | 0.063321 | 0.198054 | 0 | 0 | 16.52741 | 45.60793 |
| GO_Biologi | Regulation | 15-Jan | 0.063321 | 0.198054 | 0 | 0 | 16.52741 | 45.60793 |
| GO_Biologi | Regulation | 15-Jan | 0.063321 | 0.198054 | 0 | 0 | 16.52741 | 45.60793 |
| GO_Biologi | Negative R | 15-Jan | 0.063321 | 0.198054 | 0 | 0 | 16.52741 | 45.60793 |
| GO_Biologi | Dendritic S | 15-Jan | 0.063321 | 0.198054 | 0 | 0 | 16.52741 | 45.60793 |
| GO_Biologi | Cardiac My | 15-Jan | 0.063321 | 0.198054 | 0 | 0 | 16.52741 | 45.60793 |
| Jensen_TIS | Fascia | 23-Jan | 0.095455 | 0.1983 | 0 | 0 | 10.51321 | 24.6966 |
| Jensen_TIS | ChaGo-K-1 | 23-Jan | 0.095455 | 0.1983 | 0 | 0 | 10.51321 | 24.6966 |
| GeDiPNet_ | Glossoptos | 24-Jan | 0.099394 | 0.198448 | 0 | 0 | 10.05561 | 23.215 |
| GeDiPNet_ | Hepatocell | 24-Jan | 0.099394 | 0.198448 | 0 | 0 | 10.05561 | 23.215 |
| GeDiPNet_ | Hepatolent | 24-Jan | 0.099394 | 0.198448 | 0 | 0 | 10.05561 | 23.215 |
| GeDiPNet_ | Anhedonia | 24-Jan | 0.099394 | 0.198448 | 0 | 0 | 10.05561 | 23.215 |
| GeDiPNet_ | Tracheal St | 24-Jan | 0.099394 | 0.198448 | 0 | 0 | 10.05561 | 23.215 |
| GeDiPNet_ | Intrahepati | 24-Jan | 0.099394 | 0.198448 | 0 | 0 | 10.05561 | 23.215 |
| GeDiPNet_ | Metopic Sy | 24-Jan | 0.099394 | 0.198448 | 0 | 0 | 10.05561 | 23.215 |
| GeDiPNet_ | Lung Carcir | 4/403 | 0.098869 | 0.198448 | 0 | 0 | 2.356977 | 5.453938 |
| GeDiPNet_ | Pulmonary | 2/122 | 0.098877 | 0.198448 | 0 | 0 | 3.88098 | 8.980101 |
| GO_Molecu | Mitogen-Ac | 6-Jan | 0.025821 | 0.198622 | 0 | 0 | 46.29767 | 169.2909 |
| GO_Molecu | Telomerase | 5-Jan | 0.021564 | 0.198622 | 0 | 0 | 57.875 | 222.0519 |
| GO_Molecu | Protease B | 3/125 | 0.017263 | 0.198622 | 0 | 0 | 5.793618 | 23.51747 |
| GO_Molecu | RNA-direct | 5-Jan | 0.021564 | 0.198622 | 0 | 0 | 57.875 | 222.0519 |
| GO_Molecu | Phosphatic | 6-Jan | 0.025821 | 0.198622 | 0 | 0 | 46.29767 | 169.2909 |
| GO_Molecu | Telethonin | 6-Jan | 0.025821 | 0.198622 | 0 | 0 | 46.29767 | 169.2909 |
| DisGeNET | Dyslipidem | 3/216 | 0.068136 | 0.198792 | 0 | 0 | 3.303152 | 8.873113 |
| DisGeNET | Alopecia | 3/216 | 0.068136 | 0.198792 | 0 | 0 | 3.303152 | 8.873113 |
| DisGeNET | Muscle Spa | 4/353 | 0.068068 | 0.198792 | 0 | 0 | 2.701557 | 7.25977 |

| | | | | | | | | |
|---|---|---|---|---|---|---|---|---|
| UK_Bioban | ICD10 fibro | Jan-32 | 0.130304 | 0.198848 | 0 | 0 | 7.457614 | 15.19775 |
| UK_Bioban | Cereal inta | Jan-32 | 0.130304 | 0.198848 | 0 | 0 | 7.457614 | 15.19775 |
| UK_Bioban | Had menop | Jan-32 | 0.130304 | 0.198848 | 0 | 0 | 7.457614 | 15.19775 |
| UK_Bioban | Recent cha | Jan-32 | 0.130304 | 0.198848 | 0 | 0 | 7.457614 | 15.19775 |
| UK_Bioban | Eosinophill | 4/447 | 0.130472 | 0.198848 | 0 | 0 | 2.118089 | 4.313691 |
| GO_Biologi | Regulation | 16-Jan | 0.067399 | 0.198964 | 0 | 0 | 15.42481 | 41.60265 |
| GO_Biologi | Golgi To Va | 17-Jan | 0.071459 | 0.198964 | 0 | 0 | 14.46003 | 38.15469 |
| GO_Biologi | Negative R | 17-Jan | 0.071459 | 0.198964 | 0 | 0 | 14.46003 | 38.15469 |
| GO_Biologi | Peptidyl-Th | 17-Jan | 0.071459 | 0.198964 | 0 | 0 | 14.46003 | 38.15469 |
| GO_Biologi | Regulation | 17-Jan | 0.071459 | 0.198964 | 0 | 0 | 14.46003 | 38.15469 |
| GO_Biologi | Regulation | 17-Jan | 0.071459 | 0.198964 | 0 | 0 | 14.46003 | 38.15469 |
| GO_Biologi | Central Ne | 17-Jan | 0.071459 | 0.198964 | 0 | 0 | 14.46003 | 38.15469 |
| GO_Biologi | Muscle Tiss | 17-Jan | 0.071459 | 0.198964 | 0 | 0 | 14.46003 | 38.15469 |
| GO_Biologi | Negative R | 17-Jan | 0.071459 | 0.198964 | 0 | 0 | 14.46003 | 38.15469 |
| GO_Biologi | Regulation | Feb-96 | 0.065593 | 0.198964 | 0 | 0 | 4.960951 | 13.51507 |
| GO_Biologi | Synaptic Tr | 16-Jan | 0.067399 | 0.198964 | 0 | 0 | 15.42481 | 41.60265 |
| GO_Biologi | Negative R | 16-Jan | 0.067399 | 0.198964 | 0 | 0 | 15.42481 | 41.60265 |
| GO_Biologi | Alternative | 16-Jan | 0.067399 | 0.198964 | 0 | 0 | 15.42481 | 41.60265 |
| GO_Biologi | Positive Re | 16-Jan | 0.067399 | 0.198964 | 0 | 0 | 15.42481 | 41.60265 |
| GO_Biologi | Keratan Su | 17-Jan | 0.071459 | 0.198964 | 0 | 0 | 14.46003 | 38.15469 |
| GO_Biologi | Protein Kin | 16-Jan | 0.067399 | 0.198964 | 0 | 0 | 15.42481 | 41.60265 |
| GO_Biologi | Positive Re | 17-Jan | 0.071459 | 0.198964 | 0 | 0 | 14.46003 | 38.15469 |
| GO_Biologi | Positive Re | 17-Jan | 0.071459 | 0.198964 | 0 | 0 | 14.46003 | 38.15469 |
| GO_Biologi | Amyloid Pr | 17-Jan | 0.071459 | 0.198964 | 0 | 0 | 14.46003 | 38.15469 |
| GO_Biologi | Negative R | 17-Jan | 0.071459 | 0.198964 | 0 | 0 | 14.46003 | 38.15469 |
| GO_Biologi | Oligodendr | 16-Jan | 0.067399 | 0.198964 | 0 | 0 | 15.42481 | 41.60265 |
| GO_Biologi | Cell Comm | 16-Jan | 0.067399 | 0.198964 | 0 | 0 | 15.42481 | 41.60265 |
| GO_Biologi | Negative R | 16-Jan | 0.067399 | 0.198964 | 0 | 0 | 15.42481 | 41.60265 |
| GO_Biologi | Negative R | 16-Jan | 0.067399 | 0.198964 | 0 | 0 | 15.42481 | 41.60265 |
| GO_Biologi | Negative R | 17-Jan | 0.071459 | 0.198964 | 0 | 0 | 14.46003 | 38.15469 |
| GO_Biologi | Reverse Ch | 16-Jan | 0.067399 | 0.198964 | 0 | 0 | 15.42481 | 41.60265 |
| GO_Biologi | RNA Splicir | Feb-98 | 0.067994 | 0.198964 | 0 | 0 | 4.857108 | 13.05753 |
| GO_Biologi | Phosphatic | Feb-98 | 0.067994 | 0.198964 | 0 | 0 | 4.857108 | 13.05753 |
| DisGeNET | Herpes Sim | 5/503 | 0.06831 | 0.19908 | 0 | 0 | 2.377192 | 6.379661 |
| UK_Bioban | Waist circu | 4/451 | 0.133535 | 0.19936 | 0 | 0 | 2.098704 | 4.225511 |
| UK_Bioban | ICD10 ingu | Jan-33 | 0.134093 | 0.19936 | 0 | 0 | 7.224201 | 14.515 |
| UK_Bioban | Longest pe | 8/1166 | 0.134246 | 0.19936 | 0 | 0 | 1.640104 | 3.293463 |
| UK_Bioban | Right arm f | 4/452 | 0.134305 | 0.19936 | 0 | 0 | 2.093911 | 4.203817 |
| UK_Bioban | Fibroblastic | Jan-33 | 0.134093 | 0.19936 | 0 | 0 | 7.224201 | 14.515 |
| Human_Ph | Abnormalit | Jan-33 | 0.134093 | 0.199488 | 0 | 0 | 7.224201 | 14.515 |
| Human_Ph | Asymmetri | Jan-34 | 0.137866 | 0.199488 | 0 | 0 | 7.004933 | 13.88007 |
| Human_Ph | Scrotal hyp | Jan-34 | 0.137866 | 0.199488 | 0 | 0 | 7.004933 | 13.88007 |
| Human_Ph | Cutis marm | Jan-34 | 0.137866 | 0.199488 | 0 | 0 | 7.004933 | 13.88007 |
| Human_Ph | Abnormalit | Jan-33 | 0.134093 | 0.199488 | 0 | 0 | 7.224201 | 14.515 |
| Human_Ph | Muscular d | Jan-34 | 0.137866 | 0.199488 | 0 | 0 | 7.004933 | 13.88007 |

| | | | | | | | | |
|---|---|---|---|---|---|---|---|---|
| Human_Ph | Venous abı | Jan-34 | 0.137866 | 0.199488 | 0 | 0 | 7.004933 | 13.88007 |
| Human_Ph | Deviation c | Jan-34 | 0.137866 | 0.199488 | 0 | 0 | 7.004933 | 13.88007 |
| Human_Ph | Short nose | 2/146 | 0.132861 | 0.199488 | 0 | 0 | 3.230229 | 6.520055 |
| Human_Ph | Elbow flexi | Jan-34 | 0.137866 | 0.199488 | 0 | 0 | 7.004933 | 13.88007 |
| Human_Ph | Neoplasm ( | Jan-34 | 0.137866 | 0.199488 | 0 | 0 | 7.004933 | 13.88007 |
| Human_Ph | Absent toe | Jan-34 | 0.137866 | 0.199488 | 0 | 0 | 7.004933 | 13.88007 |
| GWAS_Cat | Snoring | Jan-37 | 0.149088 | 0.199876 | 0 | 0 | 6.42022 | 12.21909 |
| GWAS_Cat | Advanced / | Jan-37 | 0.149088 | 0.199876 | 0 | 0 | 6.42022 | 12.21909 |
| GWAS_Cat | Triglyceride | Jan-37 | 0.149088 | 0.199876 | 0 | 0 | 6.42022 | 12.21909 |
| GWAS_Cat | Acute Lymp | Jan-37 | 0.149088 | 0.199876 | 0 | 0 | 6.42022 | 12.21909 |
| GWAS_Cat | White Mat | Jan-37 | 0.149088 | 0.199876 | 0 | 0 | 6.42022 | 12.21909 |
| GWAS_Cat | Otosclerosi | Jan-37 | 0.149088 | 0.199876 | 0 | 0 | 6.42022 | 12.21909 |
| GWAS_Cat | Lumbar Spi | Jan-37 | 0.149088 | 0.199876 | 0 | 0 | 6.42022 | 12.21909 |
| GWAS_Cat | Urinary Soc | Jan-37 | 0.149088 | 0.199876 | 0 | 0 | 6.42022 | 12.21909 |
| GWAS_Cat | Nonsyndro | Jan-37 | 0.149088 | 0.199876 | 0 | 0 | 6.42022 | 12.21909 |
| GWAS_Cat | BMI In Nor | Jan-37 | 0.149088 | 0.199876 | 0 | 0 | 6.42022 | 12.21909 |
| GTEx_Tissu | GTEX-OIZG | 7/416 | 0.002243 | 0.200017 | 0 | 0 | 4.172616 | 25.45329 |
| GTEx_Tissu | GTEX-SUCS | 17/1838 | 0.002172 | 0.200017 | 0 | 0 | 2.412834 | 14.79526 |
| GTEx_Tissu | GTEX-OXRK | 19/2159 | 0.002047 | 0.200017 | 0 | 0 | 2.320554 | 14.3675 |
| GTEx_Tissu | GTEX-X5EB | 9/644 | 0.001931 | 0.200017 | 0 | 0 | 3.502968 | 21.89288 |
| GTEx_Tissu | GTEX-WYJK | 7/403 | 0.001876 | 0.200017 | 0 | 0 | 4.312468 | 27.07695 |
| GTEx_Tissu | GTEX-U3ZN | 12/1072 | 0.002282 | 0.200017 | 0 | 0 | 2.845736 | 17.31031 |
| GTEx_Tissu | GTEX-WH7 | 9/648 | 0.002014 | 0.200017 | 0 | 0 | 3.480318 | 21.60516 |
| GTEx_Tissu | GTEX-S33H | 15/1486 | 0.001763 | 0.200017 | 0 | 0 | 2.611885 | 16.56155 |
| GTEx_Tissu | GTEX-XUZC | 16/1653 | 0.001882 | 0.200017 | 0 | 0 | 2.515904 | 15.78861 |
| GTEx_Tissu | GTEX-QEG5 | 11/926 | 0.002218 | 0.200017 | 0 | 0 | 3.005148 | 18.36511 |
| GTEx_Tissu | GTEX-XV7C | 17/1841 | 0.00221 | 0.200017 | 0 | 0 | 2.408466 | 14.7268 |
| GTEx_Tissu | GTEX-R45C | 8/504 | 0.001575 | 0.200017 | 0 | 0 | 3.964271 | 25.58445 |
| DisGeNET | Seizures | 9/1174 | 0.068799 | 0.200281 | 0 | 0 | 1.85685 | 4.96999 |
| DisGeNET | synovial sa | 3/217 | 0.068882 | 0.2003 | 0 | 0 | 3.28755 | 8.795386 |
| Human_Ph | Abnormal ( | Jan-35 | 0.141623 | 0.20037 | 0 | 0 | 6.798564 | 13.28839 |
| Human_Ph | Benign nec | Jan-35 | 0.141623 | 0.20037 | 0 | 0 | 6.798564 | 13.28839 |
| Human_Ph | Primary atı | Jan-35 | 0.141623 | 0.20037 | 0 | 0 | 6.798564 | 13.28839 |
| Human_Ph | Intestinal p | Jan-35 | 0.141623 | 0.20037 | 0 | 0 | 6.798564 | 13.28839 |
| Human_Ph | Hypoplasia | Jan-35 | 0.141623 | 0.20037 | 0 | 0 | 6.798564 | 13.28839 |
| Human_Ph | Abnormalit | Jan-35 | 0.141623 | 0.20037 | 0 | 0 | 6.798564 | 13.28839 |
| GeDiPNet_ | Kidney Fail | 3/256 | 0.100769 | 0.200506 | 0 | 0 | 2.775268 | 6.369029 |
| GO_Biologi | Negative R | 18-Jan | 0.075502 | 0.200772 | 0 | 0 | 13.60876 | 35.1596 |
| GO_Biologi | Maintenan | 18-Jan | 0.075502 | 0.200772 | 0 | 0 | 13.60876 | 35.1596 |
| GO_Biologi | Negative R | 18-Jan | 0.075502 | 0.200772 | 0 | 0 | 13.60876 | 35.1596 |
| GO_Biologi | Mitochond | 18-Jan | 0.075502 | 0.200772 | 0 | 0 | 13.60876 | 35.1596 |
| GO_Biologi | Protein Imp | 18-Jan | 0.075502 | 0.200772 | 0 | 0 | 13.60876 | 35.1596 |
| GO_Biologi | Nitric Oxide | 18-Jan | 0.075502 | 0.200772 | 0 | 0 | 13.60876 | 35.1596 |
| GO_Biologi | Maintenan | 18-Jan | 0.075502 | 0.200772 | 0 | 0 | 13.60876 | 35.1596 |
| GO_Biologi | Positive Re | 4/364 | 0.074347 | 0.200772 | 0 | 0 | 2.617537 | 6.803013 |

| | | | | | | | | |
|---|---|---|---|---|---|---|---|---|
| GO_Biologi | Positive Re | 18-Jan | 0.075502 | 0.200772 | 0 | 0 | 13.60876 | 35.1596 |
| GO_Biologi | Negative R | 18-Jan | 0.075502 | 0.200772 | 0 | 0 | 13.60876 | 35.1596 |
| GO_Biologi | Detection ( | 18-Jan | 0.075502 | 0.200772 | 0 | 0 | 13.60876 | 35.1596 |
| GO_Biologi | Endothelia | 18-Jan | 0.075502 | 0.200772 | 0 | 0 | 13.60876 | 35.1596 |
| CellMarker | Ciliated Cel | Jan-33 | 0.134093 | 0.200774 | 0 | 0 | 7.224201 | 14.515 |
| DisGeNET | Adenoma | 8/1000 | 0.069191 | 0.200977 | 0 | 0 | 1.931503 | 5.158807 |
| Jensen_TIS | Vasculatur | 2/121 | 0.097522 | 0.201076 | 0 | 0 | 3.913791 | 9.110048 |
| Jensen_TIS | Ovary | 75/16042 | 0.09804 | 0.201076 | 0 | 0 | 1.544592 | 3.587126 |
| DisGeNET | Hearing Lo: | 17-Jan | 0.071459 | 0.20142 | 0 | 0 | 14.46003 | 38.15469 |
| DisGeNET | Psychologi | 17-Jan | 0.071459 | 0.20142 | 0 | 0 | 14.46003 | 38.15469 |
| DisGeNET | Forced exp | 2/101 | 0.07165 | 0.20142 | 0 | 0 | 4.70921 | 12.4133 |
| DisGeNET | Hemangior | 2/101 | 0.07165 | 0.20142 | 0 | 0 | 4.70921 | 12.4133 |
| DisGeNET | Genu recur | 17-Jan | 0.071459 | 0.20142 | 0 | 0 | 14.46003 | 38.15469 |
| DisGeNET | Macroangi | 17-Jan | 0.071459 | 0.20142 | 0 | 0 | 14.46003 | 38.15469 |
| DisGeNET | Lipoprotei | 17-Jan | 0.071459 | 0.20142 | 0 | 0 | 14.46003 | 38.15469 |
| DisGeNET | Childhood | 2/100 | 0.070424 | 0.20142 | 0 | 0 | 4.757503 | 12.62269 |
| DisGeNET | Mucosal te | 17-Jan | 0.071459 | 0.20142 | 0 | 0 | 14.46003 | 38.15469 |
| DisGeNET | Multiple cc | 17-Jan | 0.071459 | 0.20142 | 0 | 0 | 14.46003 | 38.15469 |
| DisGeNET | Colon diver | 17-Jan | 0.071459 | 0.20142 | 0 | 0 | 14.46003 | 38.15469 |
| DisGeNET | Cranial ner | 17-Jan | 0.071459 | 0.20142 | 0 | 0 | 14.46003 | 38.15469 |
| DisGeNET | T-lymphoc | 17-Jan | 0.071459 | 0.20142 | 0 | 0 | 14.46003 | 38.15469 |
| DisGeNET | Hematocrit | 2/101 | 0.07165 | 0.20142 | 0 | 0 | 4.70921 | 12.4133 |
| DisGeNET | Atypical En | 17-Jan | 0.071459 | 0.20142 | 0 | 0 | 14.46003 | 38.15469 |
| DisGeNET | Synostotic | 17-Jan | 0.071459 | 0.20142 | 0 | 0 | 14.46003 | 38.15469 |
| DisGeNET | Stomach N | 7/835 | 0.071269 | 0.20142 | 0 | 0 | 2.016833 | 5.327045 |
| DisGeNET | Oral Cavity | 2/100 | 0.070424 | 0.20142 | 0 | 0 | 4.757503 | 12.62269 |
| DisGeNET | Neoplasm, | 3/220 | 0.071143 | 0.20142 | 0 | 0 | 3.241606 | 8.567754 |
| DisGeNET | Craniofacia | 17-Jan | 0.071459 | 0.20142 | 0 | 0 | 14.46003 | 38.15469 |
| DisGeNET | Thick crani | 17-Jan | 0.071459 | 0.20142 | 0 | 0 | 14.46003 | 38.15469 |
| DisGeNET | Hyperkerat | 17-Jan | 0.071459 | 0.20142 | 0 | 0 | 14.46003 | 38.15469 |
| DisGeNET | Hypertroph | 17-Jan | 0.071459 | 0.20142 | 0 | 0 | 14.46003 | 38.15469 |
| DisGeNET | Acute mon | 17-Jan | 0.071459 | 0.20142 | 0 | 0 | 14.46003 | 38.15469 |
| DisGeNET | Enlargeme | 17-Jan | 0.071459 | 0.20142 | 0 | 0 | 14.46003 | 38.15469 |
| DisGeNET | Curly hair ( | 17-Jan | 0.071459 | 0.20142 | 0 | 0 | 14.46003 | 38.15469 |
| DisGeNET | Diverticula | 17-Jan | 0.071459 | 0.20142 | 0 | 0 | 14.46003 | 38.15469 |
| DisGeNET | Memory dy | 17-Jan | 0.071459 | 0.20142 | 0 | 0 | 14.46003 | 38.15469 |
| DisGeNET | Schistosor | 17-Jan | 0.071459 | 0.20142 | 0 | 0 | 14.46003 | 38.15469 |
| DisGeNET | Familial De | 17-Jan | 0.071459 | 0.20142 | 0 | 0 | 14.46003 | 38.15469 |
| GWAS_Cat | Waist Circu | 2/158 | 0.150727 | 0.201852 | 0 | 0 | 2.97994 | 5.638896 |
| GO_Biologi | Regulation | 2/105 | 0.076622 | 0.202236 | 0 | 0 | 4.525414 | 11.62522 |
| GO_Biologi | Negative R | 2/105 | 0.076622 | 0.202236 | 0 | 0 | 4.525414 | 11.62522 |
| UK_Bioban | Number of | Jan-34 | 0.137866 | 0.202535 | 0 | 0 | 7.004933 | 13.88007 |
| UK_Bioban | Takes none | Jan-34 | 0.137866 | 0.202535 | 0 | 0 | 7.004933 | 13.88007 |
| Jensen_TIS | Kidney can | 24-Jan | 0.099394 | 0.202563 | 0 | 0 | 10.05561 | 23.215 |
| DisGeNET | Impaired c | 6/671 | 0.072135 | 0.202565 | 0 | 0 | 2.144027 | 5.637115 |

| | | | | | | | | |
|---|---|---|---|---|---|---|---|---|
| GeDiPNet_ | Mesomelia | 25-Jan | 0.103317 | 0.202807 | 0 | 0 | 9.636143 | 21.87363 |
| GeDiPNet_ | Lipoma | 25-Jan | 0.103317 | 0.202807 | 0 | 0 | 9.636143 | 21.87363 |
| GeDiPNet_ | Stenosis Of | 25-Jan | 0.103317 | 0.202807 | 0 | 0 | 9.636143 | 21.87363 |
| GeDiPNet_ | Hyperlipop | 25-Jan | 0.103317 | 0.202807 | 0 | 0 | 9.636143 | 21.87363 |
| GWAS_Cat | Bioavailabl | 3/309 | 0.151612 | 0.202815 | 0 | 0 | 2.288399 | 4.316902 |
| Jensen_TIS | Eggshell | 25-Jan | 0.103317 | 0.202853 | 0 | 0 | 9.636143 | 21.87363 |
| Jensen_TIS | Flower | 25-Jan | 0.103317 | 0.202853 | 0 | 0 | 9.636143 | 21.87363 |
| Jensen_TIS | Epithelium | 25-Jan | 0.103317 | 0.202853 | 0 | 0 | 9.636143 | 21.87363 |
| Jensen_TIS | Junctional | 25-Jan | 0.103317 | 0.202853 | 0 | 0 | 9.636143 | 21.87363 |
| Jensen_TIS | Female puc | 25-Jan | 0.103317 | 0.202853 | 0 | 0 | 9.636143 | 21.87363 |
| Jensen_TIS | Muscle | 12/1841 | 0.101527 | 0.202853 | 0 | 0 | 1.581979 | 3.618672 |
| Rare_Disea | Congenital | 9-Jan | 0.038483 | 0.203059 | 0 | 0 | 28.93169 | 94.24608 |
| Rare_Disea | Halo nevi | 3/170 | 0.038062 | 0.203059 | 0 | 0 | 4.22284 | 13.80251 |
| Rare_Disea | Von Hippel | 3/170 | 0.038062 | 0.203059 | 0 | 0 | 4.22284 | 13.80251 |
| Rare_Disea | Neurogenic | 9-Jan | 0.038483 | 0.203059 | 0 | 0 | 28.93169 | 94.24608 |
| GeDiPNet_ | Diabetes M | 8/1096 | 0.103803 | 0.203077 | 0 | 0 | 1.752141 | 3.969062 |
| CellMarker | Natural Kill | Jan-34 | 0.137866 | 0.203093 | 0 | 0 | 7.004933 | 13.88007 |
| CellMarker | Gonadal M | 3/295 | 0.137457 | 0.203093 | 0 | 0 | 2.399829 | 4.762324 |
| CellMarker | Monocyte | Jan-34 | 0.137866 | 0.203093 | 0 | 0 | 7.004933 | 13.88007 |
| DisGeNET | mandibular | 2/102 | 0.072883 | 0.203141 | 0 | 0 | 4.661882 | 12.20902 |
| DisGeNET | Mandibula | 2/102 | 0.072883 | 0.203141 | 0 | 0 | 4.661882 | 12.20902 |
| DisGeNET | Precursor E | 2/102 | 0.072883 | 0.203141 | 0 | 0 | 4.661882 | 12.20902 |
| DisGeNET | Hepatitis B | 7/839 | 0.072705 | 0.203141 | 0 | 0 | 2.006716 | 5.260305 |
| DisGeNET | Hypertroph | 2/102 | 0.072883 | 0.203141 | 0 | 0 | 4.661882 | 12.20902 |
| DisGeNET | Downward | 3/222 | 0.07267 | 0.203141 | 0 | 0 | 3.211676 | 8.420478 |
| DisGeNET | Intervertek | 2/102 | 0.072883 | 0.203141 | 0 | 0 | 4.661882 | 12.20902 |
| DisGeNET | Major Depr | 5/513 | 0.072967 | 0.203161 | 0 | 0 | 2.329196 | 6.097235 |
| DisGeNET | Solid Neop | 7/840 | 0.073066 | 0.20322 | 0 | 0 | 2.004202 | 5.243772 |
| GWAS_Cat | Cholesteryl | Jan-38 | 0.152796 | 0.203284 | 0 | 0 | 6.246386 | 11.73478 |
| GWAS_Cat | White Mat | Jan-38 | 0.152796 | 0.203284 | 0 | 0 | 6.246386 | 11.73478 |
| GWAS_Cat | Familial Sq | Jan-38 | 0.152796 | 0.203284 | 0 | 0 | 6.246386 | 11.73478 |
| GWAS_Cat | Lymphocyt | 2/159 | 0.152237 | 0.203284 | 0 | 0 | 2.960809 | 5.573176 |
| GWAS_Cat | Cataracts | Jan-38 | 0.152796 | 0.203284 | 0 | 0 | 6.246386 | 11.73478 |
| Human_Ph | Lymphedem | Jan-37 | 0.149088 | 0.203398 | 0 | 0 | 6.42022 | 12.21909 |
| Human_Ph | Supraventr | Jan-36 | 0.145363 | 0.203398 | 0 | 0 | 6.603987 | 12.73591 |
| Human_Ph | Abnormalit | Jan-37 | 0.149088 | 0.203398 | 0 | 0 | 6.42022 | 12.21909 |
| Human_Ph | Hypoteloris | Jan-37 | 0.149088 | 0.203398 | 0 | 0 | 6.42022 | 12.21909 |
| Human_Ph | Supraventr | Jan-37 | 0.149088 | 0.203398 | 0 | 0 | 6.42022 | 12.21909 |
| Human_Ph | Non-midlin | Jan-37 | 0.149088 | 0.203398 | 0 | 0 | 6.42022 | 12.21909 |
| Human_Ph | Neoplasm | Jan-37 | 0.149088 | 0.203398 | 0 | 0 | 6.42022 | 12.21909 |
| Human_Ph | Intestinal p | Jan-37 | 0.149088 | 0.203398 | 0 | 0 | 6.42022 | 12.21909 |
| Human_Ph | Abnormalit | Jan-36 | 0.145363 | 0.203398 | 0 | 0 | 6.603987 | 12.73591 |
| Human_Ph | Abnormalit | Jan-36 | 0.145363 | 0.203398 | 0 | 0 | 6.603987 | 12.73591 |
| SysMyo_M | MGS1337 I | Mar-92 | 0.007556 | 0.20366 | 0 | 0 | 7.955056 | 38.86343 |
| Gene_Pert | Map2k3 Se | 7/301 | 0.000341 | 0.203707 | 0 | 0 | 5.838988 | 46.61909 |

| | | | | | | | | |
|---|---|---|---|---|---|---|---|---|
| DisGeNET | Compressi | 18-Jan | 0.075502 | 0.204143 | 0 | 0 | 13.60876 | 35.1596 |
| DisGeNET | Increased s | 2/104 | 0.075369 | 0.204143 | 0 | 0 | 4.570012 | 11.81514 |
| DisGeNET | Narrowing | 18-Jan | 0.075502 | 0.204143 | 0 | 0 | 13.60876 | 35.1596 |
| DisGeNET | Alexander | 18-Jan | 0.075502 | 0.204143 | 0 | 0 | 13.60876 | 35.1596 |
| DisGeNET | Prostate sp | 18-Jan | 0.075502 | 0.204143 | 0 | 0 | 13.60876 | 35.1596 |
| DisGeNET | Papillomat | 18-Jan | 0.075502 | 0.204143 | 0 | 0 | 13.60876 | 35.1596 |
| DisGeNET | Congenital | 18-Jan | 0.075502 | 0.204143 | 0 | 0 | 13.60876 | 35.1596 |
| DisGeNET | Congenital | 18-Jan | 0.075502 | 0.204143 | 0 | 0 | 13.60876 | 35.1596 |
| DisGeNET | Heart murr | 18-Jan | 0.075502 | 0.204143 | 0 | 0 | 13.60876 | 35.1596 |
| DisGeNET | Carcinoid t | 2/104 | 0.075369 | 0.204143 | 0 | 0 | 4.570012 | 11.81514 |
| DisGeNET | Acute panc | 2/104 | 0.075369 | 0.204143 | 0 | 0 | 4.570012 | 11.81514 |
| DisGeNET | Precocious | 18-Jan | 0.075502 | 0.204143 | 0 | 0 | 13.60876 | 35.1596 |
| DisGeNET | Dental Plac | 18-Jan | 0.075502 | 0.204143 | 0 | 0 | 13.60876 | 35.1596 |
| DisGeNET | Malignant | 18-Jan | 0.075502 | 0.204143 | 0 | 0 | 13.60876 | 35.1596 |
| DisGeNET | Bilateral m | 18-Jan | 0.075502 | 0.204143 | 0 | 0 | 13.60876 | 35.1596 |
| DisGeNET | Heat Strok | 18-Jan | 0.075502 | 0.204143 | 0 | 0 | 13.60876 | 35.1596 |
| DisGeNET | Rimmed va | 18-Jan | 0.075502 | 0.204143 | 0 | 0 | 13.60876 | 35.1596 |
| DisGeNET | Infarction, | 18-Jan | 0.075502 | 0.204143 | 0 | 0 | 13.60876 | 35.1596 |
| DisGeNET | Smith-Lem | 18-Jan | 0.075502 | 0.204143 | 0 | 0 | 13.60876 | 35.1596 |
| DisGeNET | Stenosis | 18-Jan | 0.075502 | 0.204143 | 0 | 0 | 13.60876 | 35.1596 |
| DisGeNET | Teratoma, | 18-Jan | 0.075502 | 0.204143 | 0 | 0 | 13.60876 | 35.1596 |
| DisGeNET | Hyperlipop | 18-Jan | 0.075502 | 0.204143 | 0 | 0 | 13.60876 | 35.1596 |
| DisGeNET | Hypoplasti | 18-Jan | 0.075502 | 0.204143 | 0 | 0 | 13.60876 | 35.1596 |
| DisGeNET | Cafe au lait | 18-Jan | 0.075502 | 0.204143 | 0 | 0 | 13.60876 | 35.1596 |
| DisGeNET | Xanthomat | 18-Jan | 0.075502 | 0.204143 | 0 | 0 | 13.60876 | 35.1596 |
| DisGeNET | Adult Burki | 18-Jan | 0.075502 | 0.204143 | 0 | 0 | 13.60876 | 35.1596 |
| DisGeNET | Severe Sep | 2/103 | 0.074122 | 0.204143 | 0 | 0 | 4.615492 | 12.00969 |
| Jensen_TIS | Fetal meml | 26-Jan | 0.107222 | 0.204293 | 0 | 0 | 9.250233 | 20.65441 |
| Jensen_TIS | Erythroid p | 26-Jan | 0.107222 | 0.204293 | 0 | 0 | 9.250233 | 20.65441 |
| Jensen_TIS | Seminal ve | 57/11672 | 0.105216 | 0.204293 | 0 | 0 | 1.3574 | 3.05651 |
| Jensen_TIS | Rhabdomy | 26-Jan | 0.107222 | 0.204293 | 0 | 0 | 9.250233 | 20.65441 |
| Jensen_TIS | Urine | 26-Jan | 0.107222 | 0.204293 | 0 | 0 | 9.250233 | 20.65441 |
| dbGaP | Kidney Dise | 18-Jan | 0.075502 | 0.204483 | 0 | 0 | 13.60876 | 35.1596 |
| Jensen_TIS | Lateral ven | 57/11697 | 0.10957 | 0.204569 | 0 | 0 | 1.350404 | 2.986 |
| Jensen_TIS | Blastocyst | 2/129 | 0.108512 | 0.204569 | 0 | 0 | 3.665771 | 8.141279 |
| Jensen_TIS | Vegetative | 27-Jan | 0.111111 | 0.204569 | 0 | 0 | 8.894007 | 19.54216 |
| Jensen_TIS | Neurofibril | 27-Jan | 0.111111 | 0.204569 | 0 | 0 | 8.894007 | 19.54216 |
| Jensen_TIS | Bladder wa | 27-Jan | 0.111111 | 0.204569 | 0 | 0 | 8.894007 | 19.54216 |
| Jensen_TIS | Root nodul | 2/131 | 0.11131 | 0.204569 | 0 | 0 | 3.608573 | 7.922388 |
| Jensen_TIS | Gut | 40/7807 | 0.111814 | 0.204569 | 0 | 0 | 1.33089 | 2.915867 |
| GO_Biologi | Oligodendr | 19-Jan | 0.079527 | 0.20458 | 0 | 0 | 12.85207 | 32.53709 |
| GO_Biologi | Regulation | 19-Jan | 0.079527 | 0.20458 | 0 | 0 | 12.85207 | 32.53709 |
| GO_Biologi | Negative R | 19-Jan | 0.079527 | 0.20458 | 0 | 0 | 12.85207 | 32.53709 |
| GO_Biologi | Negative R | 19-Jan | 0.079527 | 0.20458 | 0 | 0 | 12.85207 | 32.53709 |
| GO_Biologi | Negative R | 19-Jan | 0.079527 | 0.20458 | 0 | 0 | 12.85207 | 32.53709 |

| | | | | | | | | |
|---|---|---|---|---|---|---|---|---|
| GO_Biologi | Skeletal Mu | 19-Jan | 0.079527 | 0.20458 | 0 | 0 | 12.85207 | 32.53709 |
| GO_Biologi | Negative R | 19-Jan | 0.079527 | 0.20458 | 0 | 0 | 12.85207 | 32.53709 |
| UK_Bioban | Haemoglob | 4/460 | 0.140535 | 0.204886 | 0 | 0 | 2.056331 | 4.035136 |
| UK_Bioban | Palmar fasc | Jan-35 | 0.141623 | 0.204886 | 0 | 0 | 6.798564 | 13.28839 |
| UK_Bioban | Impedance | 5/632 | 0.140982 | 0.204886 | 0 | 0 | 1.875559 | 3.674455 |
| GeDiPNet_ | Ulcerative | 3/261 | 0.105224 | 0.20517 | 0 | 0 | 2.720792 | 6.1263 |
| GeDiPNet_ | Achoo Sync | 26-Jan | 0.107222 | 0.206305 | 0 | 0 | 9.250233 | 20.65441 |
| GeDiPNet_ | Congenital | 26-Jan | 0.107222 | 0.206305 | 0 | 0 | 9.250233 | 20.65441 |
| GeDiPNet_ | Malignant | 26-Jan | 0.107222 | 0.206305 | 0 | 0 | 9.250233 | 20.65441 |
| GeDiPNet_ | Short Clavi | 26-Jan | 0.107222 | 0.206305 | 0 | 0 | 9.250233 | 20.65441 |
| GWAS_Cat | Age At Firs | 2/161 | 0.155267 | 0.206346 | 0 | 0 | 2.92327 | 5.444911 |
| DisGeNET | M5b Acute | 2/105 | 0.076622 | 0.206745 | 0 | 0 | 4.525414 | 11.62522 |
| DisGeNET | Pick Diseas | 2/105 | 0.076622 | 0.206745 | 0 | 0 | 4.525414 | 11.62522 |
| DisGeNET | Orbital sep | 4/368 | 0.076701 | 0.206746 | 0 | 0 | 2.588243 | 6.646184 |
| Human_Ph | Delayed my | Jan-38 | 0.152796 | 0.206846 | 0 | 0 | 6.246386 | 11.73478 |
| Human_Ph | Uterine ne | Jan-38 | 0.152796 | 0.206846 | 0 | 0 | 6.246386 | 11.73478 |
| Human_Ph | Broad long | Jan-39 | 0.156488 | 0.206846 | 0 | 0 | 6.081701 | 11.28018 |
| Human_Ph | Skin tags (H | Jan-39 | 0.156488 | 0.206846 | 0 | 0 | 6.081701 | 11.28018 |
| Human_Ph | Neoplasm | Jan-39 | 0.156488 | 0.206846 | 0 | 0 | 6.081701 | 11.28018 |
| Human_Ph | Neoplasm | Jan-39 | 0.156488 | 0.206846 | 0 | 0 | 6.081701 | 11.28018 |
| Human_Ph | CNS demye | Jan-39 | 0.156488 | 0.206846 | 0 | 0 | 6.081701 | 11.28018 |
| Human_Ph | Anal atresi | Jan-39 | 0.156488 | 0.206846 | 0 | 0 | 6.081701 | 11.28018 |
| Human_Ph | Melanocyti | Jan-39 | 0.156488 | 0.206846 | 0 | 0 | 6.081701 | 11.28018 |
| GeDiPNet_ | Mental De | 8/1106 | 0.107887 | 0.206903 | 0 | 0 | 1.735261 | 3.863848 |
| GeDiPNet_ | Central Ne | 27-Jan | 0.111111 | 0.206957 | 0 | 0 | 8.894007 | 19.54216 |
| GeDiPNet_ | Dislocated | 27-Jan | 0.111111 | 0.206957 | 0 | 0 | 8.894007 | 19.54216 |
| GeDiPNet_ | Strawberry | 27-Jan | 0.111111 | 0.206957 | 0 | 0 | 8.894007 | 19.54216 |
| GeDiPNet_ | Neurogenic | 27-Jan | 0.111111 | 0.206957 | 0 | 0 | 8.894007 | 19.54216 |
| GeDiPNet_ | Chromoph | 2/129 | 0.108512 | 0.206957 | 0 | 0 | 3.665771 | 8.141279 |
| GeDiPNet_ | Liver Carcir | 5/581 | 0.109093 | 0.206957 | 0 | 0 | 2.047023 | 4.535284 |
| GeDiPNet_ | Sick Sinus S | 27-Jan | 0.111111 | 0.206957 | 0 | 0 | 8.894007 | 19.54216 |
| GeDiPNet_ | Lewy Body | 27-Jan | 0.111111 | 0.206957 | 0 | 0 | 8.894007 | 19.54216 |
| GeDiPNet_ | Peripheral | 27-Jan | 0.111111 | 0.206957 | 0 | 0 | 8.894007 | 19.54216 |
| MAGMA_D | ATC Code E | 6-Jan | 0.025821 | 0.207377 | 0 | 0 | 46.29767 | 169.2909 |
| MAGMA_D | 11 Beta Hy | 25-Jan | 0.103317 | 0.207377 | 0 | 0 | 9.636143 | 21.87363 |
| MAGMA_D | ATC Code A | 25-Jan | 0.103317 | 0.207377 | 0 | 0 | 9.636143 | 21.87363 |
| MAGMA_D | Carbenoxo | 25-Jan | 0.103317 | 0.207377 | 0 | 0 | 9.636143 | 21.87363 |
| MAGMA_D | Ritonavir | 25-Jan | 0.103317 | 0.207377 | 0 | 0 | 9.636143 | 21.87363 |
| MAGMA_D | Ifosfamide | 17-Jan | 0.071459 | 0.207377 | 0 | 0 | 14.46003 | 38.15469 |
| MAGMA_D | Graves Dise | 17-Jan | 0.071459 | 0.207377 | 0 | 0 | 14.46003 | 38.15469 |
| MAGMA_D | Pioglitazon | 16-Jan | 0.067399 | 0.207377 | 0 | 0 | 15.42481 | 41.60265 |
| MAGMA_D | Propylthiou | 17-Jan | 0.071459 | 0.207377 | 0 | 0 | 14.46003 | 38.15469 |
| MAGMA_D | Hemorrhag | 17-Jan | 0.071459 | 0.207377 | 0 | 0 | 14.46003 | 38.15469 |
| MAGMA_D | Goiter | 17-Jan | 0.071459 | 0.207377 | 0 | 0 | 14.46003 | 38.15469 |
| MAGMA_D | Labetalol | 13-Jan | 0.055113 | 0.207377 | 0 | 0 | 19.28391 | 55.89185 |

| | | | | | | | | |
|---|---|---|---|---|---|---|---|---|
| MAGMA_D | Thyroid Pe | 17-Jan | 0.071459 | 0.207377 | 0 | 0 | 14.46003 | 38.15469 |
| MAGMA_D | Deep Vein | 6-Jan | 0.025821 | 0.207377 | 0 | 0 | 46.29767 | 169.2909 |
| MAGMA_D | Irbesartan | 12-Jan | 0.050982 | 0.207377 | 0 | 0 | 21.03805 | 62.61499 |
| MAGMA_D | PARP Inhib | 9-Jan | 0.038483 | 0.207377 | 0 | 0 | 28.93169 | 94.24608 |
| MAGMA_D | Niraparib | 9-Jan | 0.038483 | 0.207377 | 0 | 0 | 28.93169 | 94.24608 |
| MAGMA_D | Tryptophar | 9-Jan | 0.038483 | 0.207377 | 0 | 0 | 28.93169 | 94.24608 |
| MAGMA_D | Serotonin F | 9-Jan | 0.038483 | 0.207377 | 0 | 0 | 28.93169 | 94.24608 |
| MAGMA_D | Acenocoun | 6-Jan | 0.025821 | 0.207377 | 0 | 0 | 46.29767 | 169.2909 |
| MAGMA_D | Abirateron | 6-Jan | 0.025821 | 0.207377 | 0 | 0 | 46.29767 | 169.2909 |
| MAGMA_D | BCL Inhibit | 15-Jan | 0.063321 | 0.207377 | 0 | 0 | 16.52741 | 45.60793 |
| MAGMA_D | ATC Code D | 12-Jan | 0.050982 | 0.207377 | 0 | 0 | 21.03805 | 62.61499 |
| MAGMA_D | Cyclitis | 26-Jan | 0.107222 | 0.207377 | 0 | 0 | 9.250233 | 20.65441 |
| MAGMA_D | Urea | 12-Jan | 0.050982 | 0.207377 | 0 | 0 | 21.03805 | 62.61499 |
| MAGMA_D | Nimesulide | 15-Jan | 0.063321 | 0.207377 | 0 | 0 | 16.52741 | 45.60793 |
| MAGMA_D | Temsirolim | 15-Jan | 0.063321 | 0.207377 | 0 | 0 | 16.52741 | 45.60793 |
| MAGMA_D | Cervical Ca | 15-Jan | 0.063321 | 0.207377 | 0 | 0 | 16.52741 | 45.60793 |
| MAGMA_D | Fulvestrant | 14-Jan | 0.059226 | 0.207377 | 0 | 0 | 17.79964 | 50.30878 |
| MAGMA_D | Phenylephi | 14-Jan | 0.059226 | 0.207377 | 0 | 0 | 17.79964 | 50.30878 |
| MAGMA_D | Binimetinik | 18-Jan | 0.075502 | 0.207377 | 0 | 0 | 13.60876 | 35.1596 |
| MAGMA_D | Ichthyosis V | 12-Jan | 0.050982 | 0.207377 | 0 | 0 | 21.03805 | 62.61499 |
| MAGMA_D | Bladder Ca | 2/146 | 0.132861 | 0.207377 | 0 | 0 | 3.230229 | 6.520055 |
| MAGMA_D | Vitamin K A | 6-Jan | 0.025821 | 0.207377 | 0 | 0 | 46.29767 | 169.2909 |
| MAGMA_D | Idelalisib | 6-Jan | 0.025821 | 0.207377 | 0 | 0 | 46.29767 | 169.2909 |
| MAGMA_D | Prostate Ca | 6-Jan | 0.025821 | 0.207377 | 0 | 0 | 46.29767 | 169.2909 |
| MAGMA_D | Carvedilol | 19-Jan | 0.079527 | 0.207377 | 0 | 0 | 12.85207 | 32.53709 |
| MAGMA_D | MEK Inhibi | 18-Jan | 0.075502 | 0.207377 | 0 | 0 | 13.60876 | 35.1596 |
| MAGMA_D | Follicular Ly | 18-Jan | 0.075502 | 0.207377 | 0 | 0 | 13.60876 | 35.1596 |
| MAGMA_D | Copanlisib | 18-Jan | 0.075502 | 0.207377 | 0 | 0 | 13.60876 | 35.1596 |
| MAGMA_D | Bloating | 9-Jan | 0.038483 | 0.207377 | 0 | 0 | 28.93169 | 94.24608 |
| MAGMA_D | Primary Pe | 9-Jan | 0.038483 | 0.207377 | 0 | 0 | 28.93169 | 94.24608 |
| MAGMA_D | Encorafeni | 8-Jan | 0.03428 | 0.207377 | 0 | 0 | 33.06645 | 111.5391 |
| MAGMA_D | Androgen E | 6-Jan | 0.025821 | 0.207377 | 0 | 0 | 46.29767 | 169.2909 |
| MAGMA_D | Melanoma | 18-Jan | 0.075502 | 0.207377 | 0 | 0 | 13.60876 | 35.1596 |
| MAGMA_D | Venetoclax | 15-Jan | 0.063321 | 0.207377 | 0 | 0 | 16.52741 | 45.60793 |
| MAGMA_D | Triamcinolo | 23-Jan | 0.095455 | 0.207377 | 0 | 0 | 10.51321 | 24.6966 |
| MAGMA_D | Allopurinol | 27-Jan | 0.111111 | 0.207377 | 0 | 0 | 8.894007 | 19.54216 |
| MAGMA_D | Kidney Stor | 27-Jan | 0.111111 | 0.207377 | 0 | 0 | 8.894007 | 19.54216 |
| MAGMA_D | ATC Code N | 27-Jan | 0.111111 | 0.207377 | 0 | 0 | 8.894007 | 19.54216 |
| MAGMA_D | PI3K Inhibit | Jan-33 | 0.134093 | 0.207377 | 0 | 0 | 7.224201 | 14.515 |
| MAGMA_D | Ovarian Ca | 2/146 | 0.132861 | 0.207377 | 0 | 0 | 3.230229 | 6.520055 |
| MAGMA_D | Cisplatin | 2/146 | 0.132861 | 0.207377 | 0 | 0 | 3.230229 | 6.520055 |
| MAGMA_D | Hydroxy Ra | 12-Jan | 0.050982 | 0.207377 | 0 | 0 | 21.03805 | 62.61499 |
| MAGMA_D | Talazoparik | 11-Jan | 0.046834 | 0.207377 | 0 | 0 | 23.14302 | 70.84419 |
| MAGMA_D | Vemurafen | 30-Jan | 0.122676 | 0.207377 | 0 | 0 | 7.972735 | 16.72843 |
| MAGMA_D | Gout | 27-Jan | 0.111111 | 0.207377 | 0 | 0 | 8.894007 | 19.54216 |

| | | | | | | | | |
|---|---|---|---|---|---|---|---|---|
| MAGMA_D | Testicular ( | 2/146 | 0.132861 | 0.207377 | 0 | 0 | 3.230229 | 6.520055 |
| MAGMA_D | Capecitabi | Jan-32 | 0.130304 | 0.207377 | 0 | 0 | 7.457614 | 15.19775 |
| MAGMA_D | Prednisone | 31-Jan | 0.126498 | 0.207377 | 0 | 0 | 7.706589 | 15.93356 |
| MAGMA_D | Congenital | 31-Jan | 0.126498 | 0.207377 | 0 | 0 | 7.706589 | 15.93356 |
| MAGMA_D | Temozolon | 20-Jan | 0.083535 | 0.207377 | 0 | 0 | 12.17503 | 30.22444 |
| MAGMA_D | Ketanserin | 20-Jan | 0.083535 | 0.207377 | 0 | 0 | 12.17503 | 30.22444 |
| MAGMA_D | Alpelisib | Jan-33 | 0.134093 | 0.207377 | 0 | 0 | 7.224201 | 14.515 |
| MAGMA_D | Epicondylit | 31-Jan | 0.126498 | 0.207377 | 0 | 0 | 7.706589 | 15.93356 |
| MAGMA_D | Dermatom | 31-Jan | 0.126498 | 0.207377 | 0 | 0 | 7.706589 | 15.93356 |
| MAGMA_D | Enalapril | 20-Jan | 0.083535 | 0.207377 | 0 | 0 | 12.17503 | 30.22444 |
| MAGMA_D | Topotecan | 15-Jan | 0.063321 | 0.207377 | 0 | 0 | 16.52741 | 45.60793 |
| MAGMA_D | Mesothelic | 14-Jan | 0.059226 | 0.207377 | 0 | 0 | 17.79964 | 50.30878 |
| MAGMA_D | Pemetrexe | 14-Jan | 0.059226 | 0.207377 | 0 | 0 | 17.79964 | 50.30878 |
| MAGMA_D | Hemorrhoi | 14-Jan | 0.059226 | 0.207377 | 0 | 0 | 17.79964 | 50.30878 |
| MAGMA_D | Ranitidine | 14-Jan | 0.059226 | 0.207377 | 0 | 0 | 17.79964 | 50.30878 |
| MAGMA_D | ATC Code E | 12-Jan | 0.050982 | 0.207377 | 0 | 0 | 21.03805 | 62.61499 |
| MAGMA_D | Xerosis Cut | 12-Jan | 0.050982 | 0.207377 | 0 | 0 | 21.03805 | 62.61499 |
| MAGMA_D | Fluvastatin | 21-Jan | 0.087525 | 0.207377 | 0 | 0 | 11.5657 | 28.17205 |
| MAGMA_D | Left Ventri | 20-Jan | 0.083535 | 0.207377 | 0 | 0 | 12.17503 | 30.22444 |
| MAGMA_D | Hypertrigly | 30-Jan | 0.122676 | 0.207377 | 0 | 0 | 7.972735 | 16.72843 |
| MAGMA_D | Xanthine O | 27-Jan | 0.111111 | 0.207377 | 0 | 0 | 8.894007 | 19.54216 |
| MAGMA_D | Prednisolo | 26-Jan | 0.107222 | 0.207377 | 0 | 0 | 9.250233 | 20.65441 |
| MAGMA_D | Iritis | 26-Jan | 0.107222 | 0.207377 | 0 | 0 | 9.250233 | 20.65441 |
| MAGMA_D | Midostauri | 26-Jan | 0.107222 | 0.207377 | 0 | 0 | 9.250233 | 20.65441 |
| MAGMA_D | Punctate K | 26-Jan | 0.107222 | 0.207377 | 0 | 0 | 9.250233 | 20.65441 |
| MAGMA_D | Peptic Ulce | 25-Jan | 0.103317 | 0.207377 | 0 | 0 | 9.636143 | 21.87363 |
| MAGMA_D | Mouth Infl | 25-Jan | 0.103317 | 0.207377 | 0 | 0 | 9.636143 | 21.87363 |
| MAGMA_D | Fenofibrate | 30-Jan | 0.122676 | 0.207377 | 0 | 0 | 7.972735 | 16.72843 |
| MAGMA_D | Olaparib | 30-Jan | 0.122676 | 0.207377 | 0 | 0 | 7.972735 | 16.72843 |
| CellMarker | Cancer Ste | Jan-35 | 0.141623 | 0.207512 | 0 | 0 | 6.798564 | 13.28839 |
| GWAS_Cat | Stable Angi | Jan-39 | 0.156488 | 0.207517 | 0 | 0 | 6.081701 | 11.28018 |
| GWAS_Cat | Cholestero | Jan-39 | 0.156488 | 0.207517 | 0 | 0 | 6.081701 | 11.28018 |
| DisGeNET | MYELODYS | 6/683 | 0.077094 | 0.207593 | 0 | 0 | 2.10471 | 5.39379 |
| DisGeNET | Lobular Ne | 19-Jan | 0.079527 | 0.207737 | 0 | 0 | 12.85207 | 32.53709 |
| DisGeNET | Aortic Rupt | 19-Jan | 0.079527 | 0.207737 | 0 | 0 | 12.85207 | 32.53709 |
| DisGeNET | No develop | 3/229 | 0.078126 | 0.207737 | 0 | 0 | 3.111094 | 7.931512 |
| DisGeNET | Refractory | 19-Jan | 0.079527 | 0.207737 | 0 | 0 | 12.85207 | 32.53709 |
| DisGeNET | THROMBO | 19-Jan | 0.079527 | 0.207737 | 0 | 0 | 12.85207 | 32.53709 |
| DisGeNET | Thrombosi | 19-Jan | 0.079527 | 0.207737 | 0 | 0 | 12.85207 | 32.53709 |
| DisGeNET | Lipidemias | 19-Jan | 0.079527 | 0.207737 | 0 | 0 | 12.85207 | 32.53709 |
| DisGeNET | Depressed | 19-Jan | 0.079527 | 0.207737 | 0 | 0 | 12.85207 | 32.53709 |
| DisGeNET | Defect of s | 19-Jan | 0.079527 | 0.207737 | 0 | 0 | 12.85207 | 32.53709 |
| DisGeNET | Cerebral Sr | 19-Jan | 0.079527 | 0.207737 | 0 | 0 | 12.85207 | 32.53709 |
| DisGeNET | Amyloid ar | 19-Jan | 0.079527 | 0.207737 | 0 | 0 | 12.85207 | 32.53709 |
| DisGeNET | Aortopulm | 19-Jan | 0.079527 | 0.207737 | 0 | 0 | 12.85207 | 32.53709 |

| | | | | | | | | |
|---|---|---|---|---|---|---|---|---|
| DisGeNET | Communic | 19-Jan | 0.079527 | 0.207737 | 0 | 0 | 12.85207 | 32.53709 |
| DisGeNET | Decreased | 19-Jan | 0.079527 | 0.207737 | 0 | 0 | 12.85207 | 32.53709 |
| DisGeNET | Carcinoma | 19-Jan | 0.079527 | 0.207737 | 0 | 0 | 12.85207 | 32.53709 |
| DisGeNET | Calvarial de | 19-Jan | 0.079527 | 0.207737 | 0 | 0 | 12.85207 | 32.53709 |
| DisGeNET | Pulmonary | 19-Jan | 0.079527 | 0.207737 | 0 | 0 | 12.85207 | 32.53709 |
| DisGeNET | CADASIL Sy | 19-Jan | 0.079527 | 0.207737 | 0 | 0 | 12.85207 | 32.53709 |
| DisGeNET | Massive Os | 19-Jan | 0.079527 | 0.207737 | 0 | 0 | 12.85207 | 32.53709 |
| DisGeNET | Bell-shapec | 19-Jan | 0.079527 | 0.207737 | 0 | 0 | 12.85207 | 32.53709 |
| DisGeNET | ABLEPHAR( | 19-Jan | 0.079527 | 0.207737 | 0 | 0 | 12.85207 | 32.53709 |
| DisGeNET | RETINITIS F | 19-Jan | 0.079527 | 0.207737 | 0 | 0 | 12.85207 | 32.53709 |
| DisGeNET | Aneurysm, | 19-Jan | 0.079527 | 0.207737 | 0 | 0 | 12.85207 | 32.53709 |
| DisGeNET | Chronic gra | 2/106 | 0.077881 | 0.207737 | 0 | 0 | 4.481674 | 11.43977 |
| DisGeNET | Thumb apl | 19-Jan | 0.079527 | 0.207737 | 0 | 0 | 12.85207 | 32.53709 |
| DisGeNET | Somnolenc | 19-Jan | 0.079527 | 0.207737 | 0 | 0 | 12.85207 | 32.53709 |
| DisGeNET | Global brai | 19-Jan | 0.079527 | 0.207737 | 0 | 0 | 12.85207 | 32.53709 |
| DisGeNET | Histiocytor | 19-Jan | 0.079527 | 0.207737 | 0 | 0 | 12.85207 | 32.53709 |
| DisGeNET | Fragile nail | 19-Jan | 0.079527 | 0.207737 | 0 | 0 | 12.85207 | 32.53709 |
| DisGeNET | Hypertroph | 19-Jan | 0.079527 | 0.207737 | 0 | 0 | 12.85207 | 32.53709 |
| SysMyo_M | MGS251 C | 5/300 | 0.009983 | 0.207891 | 0 | 0 | 4.054981 | 18.68068 |
| SysMyo_M | MGS249 C | 5/300 | 0.009983 | 0.207891 | 0 | 0 | 4.054981 | 18.68068 |
| SysMyo_M | MGS1061 I | Mar-99 | 0.009235 | 0.207891 | 0 | 0 | 7.372396 | 34.53788 |
| SysMyo_M | MGS551 H | 5/293 | 0.009075 | 0.207891 | 0 | 0 | 4.155022 | 19.53781 |
| SysMyo_M | MGS261 H | 5/300 | 0.009983 | 0.207891 | 0 | 0 | 4.054981 | 18.68068 |
| SysMyo_M | MGS333 C | 5/300 | 0.009983 | 0.207891 | 0 | 0 | 4.054981 | 18.68068 |
| SysMyo_M | MGS1347 I | 3/103 | 0.010284 | 0.207891 | 0 | 0 | 7.076071 | 32.38812 |
| UK_Bioban | Relative ag | Jan-36 | 0.145363 | 0.208184 | 0 | 0 | 6.603987 | 12.73591 |
| UK_Bioban | ICD10 chro | Jan-36 | 0.145363 | 0.208184 | 0 | 0 | 6.603987 | 12.73591 |
| GeDiPNet_ | Leukemia | 7/935 | 0.112312 | 0.208529 | 0 | 0 | 1.790073 | 3.913941 |
| DisGeNET | Hypoglycer | 3/232 | 0.080519 | 0.208871 | 0 | 0 | 3.069869 | 7.733813 |
| DisGeNET | Hyperlipide | 3/232 | 0.080519 | 0.208871 | 0 | 0 | 3.069869 | 7.733813 |
| DisGeNET | Motor dela | 3/232 | 0.080519 | 0.208871 | 0 | 0 | 3.069869 | 7.733813 |
| DisGeNET | RDW - Red | 2/108 | 0.080421 | 0.208871 | 0 | 0 | 4.39667 | 11.08174 |
| DisGeNET | Red cell dis | 2/108 | 0.080421 | 0.208871 | 0 | 0 | 4.39667 | 11.08174 |
| DisGeNET | Leukemia, | 2/108 | 0.080421 | 0.208871 | 0 | 0 | 4.39667 | 11.08174 |
| DisGeNET | Class III ma | 2/108 | 0.080421 | 0.208871 | 0 | 0 | 4.39667 | 11.08174 |
| GTEx_Tissu | GTEX-WFG | 10/800 | 0.002457 | 0.209066 | 0 | 0 | 3.143679 | 18.88972 |
| DisGeNET | Reperfusio | 2/110 | 0.082985 | 0.209236 | 0 | 0 | 4.314815 | 10.73997 |
| DisGeNET | Hyperplast | 2/110 | 0.082985 | 0.209236 | 0 | 0 | 4.314815 | 10.73997 |
| DisGeNET | Autosomal | 20-Jan | 0.083535 | 0.209236 | 0 | 0 | 12.17503 | 30.22444 |
| DisGeNET | Bone spicu | 20-Jan | 0.083535 | 0.209236 | 0 | 0 | 12.17503 | 30.22444 |
| DisGeNET | Tonsillar Ca | 20-Jan | 0.083535 | 0.209236 | 0 | 0 | 12.17503 | 30.22444 |
| DisGeNET | Calf muscle | 20-Jan | 0.083535 | 0.209236 | 0 | 0 | 12.17503 | 30.22444 |
| DisGeNET | Calvarial sk | 20-Jan | 0.083535 | 0.209236 | 0 | 0 | 12.17503 | 30.22444 |
| DisGeNET | Carcinoma | 20-Jan | 0.083535 | 0.209236 | 0 | 0 | 12.17503 | 30.22444 |
| DisGeNET | Multiple ve | 20-Jan | 0.083535 | 0.209236 | 0 | 0 | 12.17503 | 30.22444 |

| | | | | | | | | |
|---|---|---|---|---|---|---|---|---|
| DisGeNET | Squamous | 20-Jan | 0.083535 | 0.209236 | 0 | 0 | 12.17503 | 30.22444 |
| DisGeNET | Stenosis of | 20-Jan | 0.083535 | 0.209236 | 0 | 0 | 12.17503 | 30.22444 |
| DisGeNET | stage, gastı | 20-Jan | 0.083535 | 0.209236 | 0 | 0 | 12.17503 | 30.22444 |
| DisGeNET | Cholesteryl | 20-Jan | 0.083535 | 0.209236 | 0 | 0 | 12.17503 | 30.22444 |
| DisGeNET | Abnormal v | 20-Jan | 0.083535 | 0.209236 | 0 | 0 | 12.17503 | 30.22444 |
| DisGeNET | Abnormal v | 20-Jan | 0.083535 | 0.209236 | 0 | 0 | 12.17503 | 30.22444 |
| DisGeNET | Idiopathic I | 20-Jan | 0.083535 | 0.209236 | 0 | 0 | 12.17503 | 30.22444 |
| DisGeNET | Arterioscle | 20-Jan | 0.083535 | 0.209236 | 0 | 0 | 12.17503 | 30.22444 |
| DisGeNET | Acral pseud | 20-Jan | 0.083535 | 0.209236 | 0 | 0 | 12.17503 | 30.22444 |
| DisGeNET | Dyslipopro | 20-Jan | 0.083535 | 0.209236 | 0 | 0 | 12.17503 | 30.22444 |
| DisGeNET | Coronal cra | 20-Jan | 0.083535 | 0.209236 | 0 | 0 | 12.17503 | 30.22444 |
| DisGeNET | Accessory r | 20-Jan | 0.083535 | 0.209236 | 0 | 0 | 12.17503 | 30.22444 |
| DisGeNET | Cerebral Va | 20-Jan | 0.083535 | 0.209236 | 0 | 0 | 12.17503 | 30.22444 |
| DisGeNET | Observatio | 20-Jan | 0.083535 | 0.209236 | 0 | 0 | 12.17503 | 30.22444 |
| DisGeNET | Polymicrog | 2/110 | 0.082985 | 0.209236 | 0 | 0 | 4.314815 | 10.73997 |
| DisGeNET | Retinoic ac | 20-Jan | 0.083535 | 0.209236 | 0 | 0 | 12.17503 | 30.22444 |
| DisGeNET | Plaque, Am | 3/235 | 0.082943 | 0.209236 | 0 | 0 | 3.029711 | 7.542769 |
| DisGeNET | Hyperinsul | 4/377 | 0.082135 | 0.209236 | 0 | 0 | 2.524629 | 6.310023 |
| DisGeNET | Fissured to | 20-Jan | 0.083535 | 0.209236 | 0 | 0 | 12.17503 | 30.22444 |
| DisGeNET | Endophtha | 20-Jan | 0.083535 | 0.209236 | 0 | 0 | 12.17503 | 30.22444 |
| DisGeNET | Idiopathic I | 4/379 | 0.083368 | 0.209236 | 0 | 0 | 2.510908 | 6.238313 |
| DisGeNET | Cutaneous | 20-Jan | 0.083535 | 0.209236 | 0 | 0 | 12.17503 | 30.22444 |
| DisGeNET | Cutaneous | 20-Jan | 0.083535 | 0.209236 | 0 | 0 | 12.17503 | 30.22444 |
| DisGeNET | Peyronie D | 20-Jan | 0.083535 | 0.209236 | 0 | 0 | 12.17503 | 30.22444 |
| DisGeNET | Sick Sinus S | 20-Jan | 0.083535 | 0.209236 | 0 | 0 | 12.17503 | 30.22444 |
| DisGeNET | Prostate ca | 19/3145 | 0.081524 | 0.209236 | 0 | 0 | 1.500475 | 3.761486 |
| DisGeNET | Hamman-R | 3/233 | 0.081323 | 0.209236 | 0 | 0 | 3.056366 | 7.669407 |
| GTEx_Tissu | GTEX-XV7C | 8/544 | 0.002532 | 0.209282 | 0 | 0 | 3.660873 | 21.88754 |
| DisGeNET | Chronic Kic | 4/380 | 0.083988 | 0.210172 | 0 | 0 | 2.504102 | 6.202851 |
| Human_Ph | Arterioven | Jan-40 | 0.160165 | 0.210251 | 0 | 0 | 5.925462 | 10.85279 |
| Human_Ph | Atresia of t | Jan-40 | 0.160165 | 0.210251 | 0 | 0 | 5.925462 | 10.85279 |
| Jensen_DIS | Candidiasis | Jan-44 | 0.174713 | 0.210303 | 0 | 0 | 5.373175 | 9.3741 |
| GO_Biologi | Xenobiotic | 20-Jan | 0.083535 | 0.210318 | 0 | 0 | 12.17503 | 30.22444 |
| GO_Biologi | Regulation | 20-Jan | 0.083535 | 0.210318 | 0 | 0 | 12.17503 | 30.22444 |
| GO_Biologi | Positive Re | 20-Jan | 0.083535 | 0.210318 | 0 | 0 | 12.17503 | 30.22444 |
| GO_Biologi | Negative R | 20-Jan | 0.083535 | 0.210318 | 0 | 0 | 12.17503 | 30.22444 |
| GO_Biologi | Sensory Pe | 20-Jan | 0.083535 | 0.210318 | 0 | 0 | 12.17503 | 30.22444 |
| GO_Biologi | Negative R | 20-Jan | 0.083535 | 0.210318 | 0 | 0 | 12.17503 | 30.22444 |
| GTEx_Tissu | GTEX-S3XE | 8/547 | 0.002619 | 0.210458 | 0 | 0 | 3.639933 | 21.6394 |
| GO_Biologi | Cytoskelet | 2/111 | 0.084277 | 0.210693 | 0 | 0 | 4.275013 | 10.57487 |
| GO_Biologi | Mitotic Sist | 2/111 | 0.084277 | 0.210693 | 0 | 0 | 4.275013 | 10.57487 |
| CellMarker | Fibroblast S | Jan-36 | 0.145363 | 0.210738 | 0 | 0 | 6.603987 | 12.73591 |
| CellMarker | Morula Cel | Jan-36 | 0.145363 | 0.210738 | 0 | 0 | 6.603987 | 12.73591 |
| GWAS_Cat | Degree Of | Jan-40 | 0.160165 | 0.210789 | 0 | 0 | 5.925462 | 10.85279 |
| GWAS_Cat | Concentrat | Jan-40 | 0.160165 | 0.210789 | 0 | 0 | 5.925462 | 10.85279 |

| | | | | | | | | |
|---|---|---|---|---|---|---|---|---|
| GWAS_Cat | Parental Lc | Jan-40 | 0.160165 | 0.210789 | 0 | 0 | 5.925462 | 10.85279 |
| GWAS_Cat | Fasting Blo | Jan-40 | 0.160165 | 0.210789 | 0 | 0 | 5.925462 | 10.85279 |
| GWAS_Cat | Phospholip | Jan-40 | 0.160165 | 0.210789 | 0 | 0 | 5.925462 | 10.85279 |
| GWAS_Cat | LDL Choles | Jan-40 | 0.160165 | 0.210789 | 0 | 0 | 5.925462 | 10.85279 |
| GWAS_Cat | Hippocamp | Jan-40 | 0.160165 | 0.210789 | 0 | 0 | 5.925462 | 10.85279 |
| Rare_Disea | Long QT sy | Feb-73 | 0.040281 | 0.211048 | 0 | 0 | 6.575642 | 21.1202 |
| UK_Bioban | Impedance | 5/645 | 0.14971 | 0.211283 | 0 | 0 | 1.836223 | 3.487092 |
| UK_Bioban | Coronary a | Jan-37 | 0.149088 | 0.211283 | 0 | 0 | 6.42022 | 12.21909 |
| UK_Bioban | Lymphocyt | 15/2588 | 0.149751 | 0.211283 | 0 | 0 | 1.404003 | 2.665891 |
| KEGG_202 | Malaria | Feb-50 | 0.020017 | 0.211294 | 0 | 0 | 9.737745 | 38.08585 |
| KEGG_202 | Cholestero | Feb-50 | 0.020017 | 0.211294 | 0 | 0 | 9.737745 | 38.08585 |
| KEGG_202 | Thyroid ho | 3/121 | 0.01584 | 0.211294 | 0 | 0 | 5.991223 | 24.83499 |
| UK_Bioban | Right corne | 3/308 | 0.150586 | 0.211414 | 0 | 0 | 2.296019 | 4.346874 |
| GeDiPNet_ | Long QT Sy | 28-Jan | 0.114983 | 0.211466 | 0 | 0 | 8.564169 | 18.52408 |
| GeDiPNet_ | Testicular I | 28-Jan | 0.114983 | 0.211466 | 0 | 0 | 8.564169 | 18.52408 |
| GeDiPNet_ | Sezary Syn | 28-Jan | 0.114983 | 0.211466 | 0 | 0 | 8.564169 | 18.52408 |
| GO_Molec | Translation | 14-Jan | 0.059226 | 0.211522 | 0 | 0 | 17.79964 | 50.30878 |
| GO_Molec | Guanyl-Nu | 3/203 | 0.05878 | 0.211522 | 0 | 0 | 3.520179 | 9.976043 |
| GO_Molec | Compleme | 10-Jan | 0.042667 | 0.211522 | 0 | 0 | 25.71576 | 81.1157 |
| GO_Molec | Lipase Inhil | 9-Jan | 0.038483 | 0.211522 | 0 | 0 | 28.93169 | 94.24608 |
| GO_Molec | Diacylglyce | 10-Jan | 0.042667 | 0.211522 | 0 | 0 | 25.71576 | 81.1157 |
| GO_Molec | Protein Tyr | Feb-76 | 0.043322 | 0.211522 | 0 | 0 | 6.308108 | 19.80178 |
| GO_Molec | Metalloenc | 13-Jan | 0.055113 | 0.211522 | 0 | 0 | 19.28391 | 55.89185 |
| GO_Molec | Beta-1,3-G | 14-Jan | 0.059226 | 0.211522 | 0 | 0 | 17.79964 | 50.30878 |
| GO_Molec | Histone Me | 11-Jan | 0.046834 | 0.211522 | 0 | 0 | 23.14302 | 70.84419 |
| GO_Molec | Phosphatic | 9-Jan | 0.038483 | 0.211522 | 0 | 0 | 28.93169 | 94.24608 |
| GO_Molec | Phosphatic | 13-Jan | 0.055113 | 0.211522 | 0 | 0 | 19.28391 | 55.89185 |
| GO_Molec | UDP-galact | 11-Jan | 0.046834 | 0.211522 | 0 | 0 | 23.14302 | 70.84419 |
| GO_Molec | Muscle Alp | 14-Jan | 0.059226 | 0.211522 | 0 | 0 | 17.79964 | 50.30878 |
| GO_Molec | Compleme | 11-Jan | 0.046834 | 0.211522 | 0 | 0 | 23.14302 | 70.84419 |
| GO_Molec | Phospholip | 9-Jan | 0.038483 | 0.211522 | 0 | 0 | 28.93169 | 94.24608 |
| GTEx_Tissu | GTEX-UPK5 | 9/685 | 0.002923 | 0.211873 | 0 | 0 | 3.283512 | 19.16003 |
| GTEx_Tissu | GTEX-QCQ | 15/1566 | 0.002929 | 0.211873 | 0 | 0 | 2.46642 | 14.38647 |
| GTEx_Tissu | GTEX-OHPI | 8/556 | 0.002894 | 0.211873 | 0 | 0 | 3.57849 | 20.91669 |
| GTEx_Tissu | GTEX-QDV. | 14/1406 | 0.002882 | 0.211873 | 0 | 0 | 2.551704 | 14.92572 |
| PhenGenI_ | Electrocarc | 8/909 | 0.044045 | 0.211966 | 0 | 0 | 2.13681 | 6.672286 |
| PhenGenI_ | Gout | Feb-76 | 0.043322 | 0.211966 | 0 | 0 | 6.308108 | 19.80178 |
| Human_Ph | Underdeve | Jan-41 | 0.163825 | 0.212862 | 0 | 0 | 5.777035 | 10.45039 |
| Human_Ph | Abnormalit | Jan-41 | 0.163825 | 0.212862 | 0 | 0 | 5.777035 | 10.45039 |
| Human_Ph | Waddling g | Jan-41 | 0.163825 | 0.212862 | 0 | 0 | 5.777035 | 10.45039 |
| Jensen_DIS | Thalassemi | Jan-45 | 0.178311 | 0.213317 | 0 | 0 | 5.250793 | 9.053559 |
| UK_Bioban | Ischaemic I | Jan-38 | 0.152796 | 0.213465 | 0 | 0 | 6.246386 | 11.73478 |
| DisGeNET | Anaplastic | 21-Jan | 0.087525 | 0.213715 | 0 | 0 | 11.5657 | 28.17205 |
| DisGeNET | Basal gangl | 21-Jan | 0.087525 | 0.213715 | 0 | 0 | 11.5657 | 28.17205 |
| DisGeNET | Mild global | 21-Jan | 0.087525 | 0.213715 | 0 | 0 | 11.5657 | 28.17205 |

| | | | | | | | | |
|---|---|---|---|---|---|---|---|---|
| DisGeNET | Nodular Go | 21-Jan | 0.087525 | 0.213715 | 0 | 0 | 11.5657 | 28.17205 |
| DisGeNET | Oculo-dent | 21-Jan | 0.087525 | 0.213715 | 0 | 0 | 11.5657 | 28.17205 |
| DisGeNET | Pancreatic | 21-Jan | 0.087525 | 0.213715 | 0 | 0 | 11.5657 | 28.17205 |
| DisGeNET | Fibrous ski | 21-Jan | 0.087525 | 0.213715 | 0 | 0 | 11.5657 | 28.17205 |
| DisGeNET | Shared Par | 21-Jan | 0.087525 | 0.213715 | 0 | 0 | 11.5657 | 28.17205 |
| DisGeNET | Sinus Node | 21-Jan | 0.087525 | 0.213715 | 0 | 0 | 11.5657 | 28.17205 |
| DisGeNET | Merkel cell | 2/112 | 0.085575 | 0.213715 | 0 | 0 | 4.235936 | 10.41346 |
| DisGeNET | Myeloid Le | 7/876 | 0.086802 | 0.213715 | 0 | 0 | 1.917549 | 4.686721 |
| DisGeNET | Alzheimer's | 13/1982 | 0.086803 | 0.213715 | 0 | 0 | 1.600977 | 3.91297 |
| DisGeNET | Paresis | 2/113 | 0.086879 | 0.213715 | 0 | 0 | 4.197562 | 10.25564 |
| DisGeNET | Regurgitati | 21-Jan | 0.087525 | 0.213715 | 0 | 0 | 11.5657 | 28.17205 |
| DisGeNET | Feeding dif | 3/239 | 0.086224 | 0.213715 | 0 | 0 | 2.977754 | 7.297891 |
| DisGeNET | EMG: neur | 21-Jan | 0.087525 | 0.213715 | 0 | 0 | 11.5657 | 28.17205 |
| DisGeNET | Xanthoma | 21-Jan | 0.087525 | 0.213715 | 0 | 0 | 11.5657 | 28.17205 |
| DisGeNET | Amyloid ne | 21-Jan | 0.087525 | 0.213715 | 0 | 0 | 11.5657 | 28.17205 |
| DisGeNET | Premature | 21-Jan | 0.087525 | 0.213715 | 0 | 0 | 11.5657 | 28.17205 |
| DisGeNET | Early tooth | 21-Jan | 0.087525 | 0.213715 | 0 | 0 | 11.5657 | 28.17205 |
| DisGeNET | Gastric Ade | 4/383 | 0.085862 | 0.213715 | 0 | 0 | 2.483899 | 6.098006 |
| DisGeNET | Glucose me | 21-Jan | 0.087525 | 0.213715 | 0 | 0 | 11.5657 | 28.17205 |
| DisGeNET | Stillbirth | 21-Jan | 0.087525 | 0.213715 | 0 | 0 | 11.5657 | 28.17205 |
| DisGeNET | Hereditary | 21-Jan | 0.087525 | 0.213715 | 0 | 0 | 11.5657 | 28.17205 |
| DisGeNET | Hypoalpha | 21-Jan | 0.087525 | 0.213715 | 0 | 0 | 11.5657 | 28.17205 |
| DisGeNET | Intestinal P | 21-Jan | 0.087525 | 0.213715 | 0 | 0 | 11.5657 | 28.17205 |
| GeDiPNet_ | Stereotype | 2/135 | 0.116962 | 0.213758 | 0 | 0 | 3.499337 | 7.509252 |
| GeDiPNet_ | Papillary Re | 2/135 | 0.116962 | 0.213758 | 0 | 0 | 3.499337 | 7.509252 |
| Jensen_TIS | Myeloid pr | 29-Jan | 0.118838 | 0.213775 | 0 | 0 | 8.25789 | 17.58927 |
| Jensen_TIS | Larynx | 3/275 | 0.118102 | 0.213775 | 0 | 0 | 2.578913 | 5.509085 |
| Jensen_TIS | Keloid | 29-Jan | 0.118838 | 0.213775 | 0 | 0 | 8.25789 | 17.58927 |
| SysMyo_M | MGS1064 I | 4/198 | 0.010953 | 0.213779 | 0 | 0 | 4.898522 | 22.11248 |
| PhenGenI_ | Chin | Feb-78 | 0.045395 | 0.214007 | 0 | 0 | 6.141486 | 18.99159 |
| Human_Ph | Microtia (H | Jan-42 | 0.16747 | 0.214677 | 0 | 0 | 5.635848 | 10.07097 |
| Human_Ph | Prominent | Jan-42 | 0.16747 | 0.214677 | 0 | 0 | 5.635848 | 10.07097 |
| Human_Ph | Goiter (HP: | Jan-42 | 0.16747 | 0.214677 | 0 | 0 | 5.635848 | 10.07097 |
| Human_Ph | Left ventric | Jan-42 | 0.16747 | 0.214677 | 0 | 0 | 5.635848 | 10.07097 |
| GWAS_Cat | Smoking Be | Jan-41 | 0.163825 | 0.21468 | 0 | 0 | 5.777035 | 10.45039 |
| GWAS_Cat | Type 2 Dial | Jan-41 | 0.163825 | 0.21468 | 0 | 0 | 5.777035 | 10.45039 |
| GWAS_Cat | Concentrat | Jan-41 | 0.163825 | 0.21468 | 0 | 0 | 5.777035 | 10.45039 |
| GWAS_Cat | Uric Acid Le | Jan-41 | 0.163825 | 0.21468 | 0 | 0 | 5.777035 | 10.45039 |
| DisGeNET | leiomyosar | 2/114 | 0.088189 | 0.214935 | 0 | 0 | 4.159874 | 10.1013 |
| DisGeNET | Intracrania | 2/114 | 0.088189 | 0.214935 | 0 | 0 | 4.159874 | 10.1013 |
| CellMarker | Mesenchyr | Jan-37 | 0.149088 | 0.215 | 0 | 0 | 6.42022 | 12.21909 |
| GO_Biologi | Negative R | 21-Jan | 0.087525 | 0.215027 | 0 | 0 | 11.5657 | 28.17205 |
| GO_Biologi | Regulation | 21-Jan | 0.087525 | 0.215027 | 0 | 0 | 11.5657 | 28.17205 |
| GO_Biologi | Chaperone | 21-Jan | 0.087525 | 0.215027 | 0 | 0 | 11.5657 | 28.17205 |
| GO_Biologi | Regulation | 21-Jan | 0.087525 | 0.215027 | 0 | 0 | 11.5657 | 28.17205 |

| | | | | | | | | |
|---|---|---|---|---|---|---|---|---|
| GO_Biologi | Sulfur Com | 21-Jan | 0.087525 | 0.215027 | 0 | 0 | 11.5657 | 28.17205 |
| GeDiPNet_ | Kallmann S | 29-Jan | 0.118838 | 0.215163 | 0 | 0 | 8.25789 | 17.58927 |
| GeDiPNet_ | Amyloidosi | 29-Jan | 0.118838 | 0.215163 | 0 | 0 | 8.25789 | 17.58927 |
| GeDiPNet_ | Delusions | 29-Jan | 0.118838 | 0.215163 | 0 | 0 | 8.25789 | 17.58927 |
| Disease_Pe | ulcerative ( | 7/386 | 0.001468 | 0.215544 | 0 | 0 | 4.509828 | 29.42044 |
| Disease_Pe | Insulin Resi | 6/290 | 0.001668 | 0.215544 | 0 | 0 | 5.119718 | 32.74548 |
| Disease_Pe | Type 2 diak | 7/315 | 0.000448 | 0.215544 | 0 | 0 | 5.569602 | 42.95028 |
| Disease_Pe | Muscular D | 7/362 | 0.001015 | 0.215544 | 0 | 0 | 4.820634 | 33.22899 |
| Disease_Pe | systemic lu | 6/287 | 0.001583 | 0.215544 | 0 | 0 | 5.175168 | 33.37244 |
| UK_Bioban | Age catara | 2/161 | 0.155267 | 0.215859 | 0 | 0 | 2.92327 | 5.444911 |
| Rare_Disea | Renal cell c | 3/176 | 0.041494 | 0.215885 | 0 | 0 | 4.075145 | 12.96795 |
| GO_Biologi | Regulation | 2/114 | 0.088189 | 0.215911 | 0 | 0 | 4.159874 | 10.1013 |
| UK_Bioban | North co-o | Jan-39 | 0.156488 | 0.216501 | 0 | 0 | 6.081701 | 11.28018 |
| MAGMA_D | Pravastatin | Jan-35 | 0.141623 | 0.216504 | 0 | 0 | 6.798564 | 13.28839 |
| UK_Bioban | Right leg fa | 4/481 | 0.157425 | 0.216744 | 0 | 0 | 1.963679 | 3.630464 |
| Rare_Disea | Defective a | 10-Jan | 0.042667 | 0.217429 | 0 | 0 | 25.71576 | 81.1157 |
| Rare_Disea | Pontocerek | 10-Jan | 0.042667 | 0.217429 | 0 | 0 | 25.71576 | 81.1157 |
| Rare_Disea | Crouzon sy | Feb-75 | 0.042299 | 0.217429 | 0 | 0 | 6.394843 | 20.22688 |
| Human_Ph | Emotional | Jan-43 | 0.171099 | 0.217688 | 0 | 0 | 5.501384 | 9.712751 |
| Human_Ph | Hypertelor | 3/328 | 0.171529 | 0.217688 | 0 | 0 | 2.152527 | 3.794916 |
| Human_Ph | Multiple lip | Jan-43 | 0.171099 | 0.217688 | 0 | 0 | 5.501384 | 9.712751 |
| CellMarker | Microglial ( | 4/476 | 0.153335 | 0.217688 | 0 | 0 | 1.984991 | 3.722114 |
| CellMarker | Specialist A | Jan-38 | 0.152796 | 0.217688 | 0 | 0 | 6.246386 | 11.73478 |
| CellMarker | B Cell Bone | Jan-38 | 0.152796 | 0.217688 | 0 | 0 | 6.246386 | 11.73478 |
| SysMyo_M | MGS115 M | 4/205 | 0.012316 | 0.217836 | 0 | 0 | 4.726248 | 20.7807 |
| SysMyo_M | MGS1333 I | 3/108 | 0.01169 | 0.217836 | 0 | 0 | 6.737415 | 29.97504 |
| SysMyo_M | MGS800 M | 4/204 | 0.012115 | 0.217836 | 0 | 0 | 4.75012 | 20.96382 |
| GeDiPNet_ | Psoriasis | 3/278 | 0.120936 | 0.217969 | 0 | 0 | 2.55039 | 5.387681 |
| GeDiPNet_ | Cataract | 5/601 | 0.121135 | 0.217969 | 0 | 0 | 1.976285 | 4.171633 |
| Jensen_TIS | Blood vess | 30-Jan | 0.122676 | 0.218242 | 0 | 0 | 7.972735 | 16.72843 |
| Jensen_TIS | Vascular tis | 30-Jan | 0.122676 | 0.218242 | 0 | 0 | 7.972735 | 16.72843 |
| GWAS_Cat | Oily Fish Cc | Jan-42 | 0.16747 | 0.218284 | 0 | 0 | 5.635848 | 10.07097 |
| GWAS_Cat | Hemoglobi | 4/493 | 0.167405 | 0.218284 | 0 | 0 | 1.914308 | 3.421514 |
| GWAS_Cat | Omega-3 F | Jan-42 | 0.16747 | 0.218284 | 0 | 0 | 5.635848 | 10.07097 |
| GWAS_Cat | Free Chole: | Jan-42 | 0.16747 | 0.218284 | 0 | 0 | 5.635848 | 10.07097 |
| GWAS_Cat | LDL Choles | Jan-42 | 0.16747 | 0.218284 | 0 | 0 | 5.635848 | 10.07097 |
| Human_Ph | Hepatome | 3/329 | 0.172598 | 0.218319 | 0 | 0 | 2.145815 | 3.769752 |
| GTEx_Tissu | GTEX-WHP | 10/832 | 0.003256 | 0.218356 | 0 | 0 | 3.016242 | 17.27501 |
| GTEx_Tissu | GTEX-TKQ2 | 16/1746 | 0.003268 | 0.218356 | 0 | 0 | 2.368542 | 13.55647 |
| GTEx_Tissu | GTEX-U3ZH | 10/827 | 0.003119 | 0.218356 | 0 | 0 | 3.035496 | 17.5157 |
| GTEx_Tissu | GTEX-P4QS | 17/1914 | 0.003321 | 0.218356 | 0 | 0 | 2.306439 | 13.16397 |
| CellMarker | Cancer Cell | Jan-39 | 0.156488 | 0.218764 | 0 | 0 | 6.081701 | 11.28018 |
| CellMarker | Monocyte | Jan-39 | 0.156488 | 0.218764 | 0 | 0 | 6.081701 | 11.28018 |
| CellMarker | Lymphoid ( | Jan-39 | 0.156488 | 0.218764 | 0 | 0 | 6.081701 | 11.28018 |
| UK_Bioban | Right heel I | 2/164 | 0.159833 | 0.219003 | 0 | 0 | 2.8687 | 5.260115 |

| | | | | | | | | |
|---|---|---|---|---|---|---|---|---|
| PhenGenI_ | Cerebrospi | 3/186 | 0.047549 | 0.219676 | 0 | 0 | 3.850507 | 11.72863 |
| MAGMA_D | Warfarin | Jan-36 | 0.145363 | 0.219697 | 0 | 0 | 6.603987 | 12.73591 |
| DisGeNET | Hypothalar | 22-Jan | 0.091499 | 0.219933 | 0 | 0 | 11.0144 | 26.34017 |
| DisGeNET | Personality | 22-Jan | 0.091499 | 0.219933 | 0 | 0 | 11.0144 | 26.34017 |
| DisGeNET | Psychomot | 22-Jan | 0.091499 | 0.219933 | 0 | 0 | 11.0144 | 26.34017 |
| DisGeNET | Dystrophic | 22-Jan | 0.091499 | 0.219933 | 0 | 0 | 11.0144 | 26.34017 |
| DisGeNET | Lower resp | 22-Jan | 0.091499 | 0.219933 | 0 | 0 | 11.0144 | 26.34017 |
| DisGeNET | Hypoplasti | 22-Jan | 0.091499 | 0.219933 | 0 | 0 | 11.0144 | 26.34017 |
| DisGeNET | Infiltrating | 22-Jan | 0.091499 | 0.219933 | 0 | 0 | 11.0144 | 26.34017 |
| DisGeNET | Flexion cor | 22-Jan | 0.091499 | 0.219933 | 0 | 0 | 11.0144 | 26.34017 |
| DisGeNET | Alzheimer | 2/116 | 0.090827 | 0.219933 | 0 | 0 | 4.086481 | 9.802643 |
| DisGeNET | Cluster Hea | 22-Jan | 0.091499 | 0.219933 | 0 | 0 | 11.0144 | 26.34017 |
| DisGeNET | Conduct Di | 22-Jan | 0.091499 | 0.219933 | 0 | 0 | 11.0144 | 26.34017 |
| DisGeNET | Severe dep | 22-Jan | 0.091499 | 0.219933 | 0 | 0 | 11.0144 | 26.34017 |
| DisGeNET | Weak cry | 22-Jan | 0.091499 | 0.219933 | 0 | 0 | 11.0144 | 26.34017 |
| DisGeNET | Herpes zos | 22-Jan | 0.091499 | 0.219933 | 0 | 0 | 11.0144 | 26.34017 |
| DisGeNET | Leukemia, | 22-Jan | 0.091499 | 0.219933 | 0 | 0 | 11.0144 | 26.34017 |
| PhenGenI_ | Hydroxyme | Feb-81 | 0.048573 | 0.220009 | 0 | 0 | 5.907372 | 17.86791 |
| GeDiPNet_ | Papilloma | 30-Jan | 0.122676 | 0.220063 | 0 | 0 | 7.972735 | 16.72843 |
| GO_Biologi | Positive Re | 22-Jan | 0.091499 | 0.220217 | 0 | 0 | 11.0144 | 26.34017 |
| GO_Biologi | Positive Re | 22-Jan | 0.091499 | 0.220217 | 0 | 0 | 11.0144 | 26.34017 |
| GO_Biologi | Negative R | 22-Jan | 0.091499 | 0.220217 | 0 | 0 | 11.0144 | 26.34017 |
| GO_Biologi | Regulation | 22-Jan | 0.091499 | 0.220217 | 0 | 0 | 11.0144 | 26.34017 |
| GO_Biologi | tRNA Amin | 22-Jan | 0.091499 | 0.220217 | 0 | 0 | 11.0144 | 26.34017 |
| Human_Ph | Pulmonary | Jan-44 | 0.174713 | 0.220265 | 0 | 0 | 5.373175 | 9.3741 |
| dbGaP | Body Heigh | 4/385 | 0.087122 | 0.220275 | 0 | 0 | 2.470607 | 6.029369 |
| dbGaP | gamma-Glu | 22-Jan | 0.091499 | 0.220275 | 0 | 0 | 11.0144 | 26.34017 |
| dbGaP | Alcohol Dri | 22-Jan | 0.091499 | 0.220275 | 0 | 0 | 11.0144 | 26.34017 |
| Jensen_DIS | Anorexia n | Jan-47 | 0.18546 | 0.220517 | 0 | 0 | 5.021992 | 8.46163 |
| GWAS_Cat | Intelligence | 3/326 | 0.169397 | 0.220559 | 0 | 0 | 2.166077 | 3.845893 |
| UK_Bioban | Chest pain | Jan-41 | 0.163825 | 0.221281 | 0 | 0 | 5.777035 | 10.45039 |
| UK_Bioban | Medication | Jan-41 | 0.163825 | 0.221281 | 0 | 0 | 5.777035 | 10.45039 |
| UK_Bioban | Snoring 12 | Jan-41 | 0.163825 | 0.221281 | 0 | 0 | 5.777035 | 10.45039 |
| DisGeNET | Teratoma | 2/117 | 0.092155 | 0.221306 | 0 | 0 | 4.050742 | 9.658129 |
| GWAS_Cat | Femoral Ne | Jan-43 | 0.171099 | 0.221359 | 0 | 0 | 5.501384 | 9.712751 |
| GWAS_Cat | Cholestero | Jan-43 | 0.171099 | 0.221359 | 0 | 0 | 5.501384 | 9.712751 |
| GWAS_Cat | Total Lipid | Jan-43 | 0.171099 | 0.221359 | 0 | 0 | 5.501384 | 9.712751 |
| GWAS_Cat | Depressive | 2/171 | 0.170583 | 0.221359 | 0 | 0 | 2.748904 | 4.86152 |
| GWAS_Cat | Ovarian Ca | Jan-43 | 0.171099 | 0.221359 | 0 | 0 | 5.501384 | 9.712751 |
| GWAS_Cat | Free Chole | Jan-43 | 0.171099 | 0.221359 | 0 | 0 | 5.501384 | 9.712751 |
| Jensen_TIS | Cranium | 2/141 | 0.125573 | 0.222167 | 0 | 0 | 3.34727 | 6.945157 |
| GO_Biologi | Regulation | 23-Jan | 0.095455 | 0.222207 | 0 | 0 | 10.51321 | 24.6966 |
| GO_Biologi | Diacylglyce | 23-Jan | 0.095455 | 0.222207 | 0 | 0 | 10.51321 | 24.6966 |
| GO_Biologi | Immune Re | 23-Jan | 0.095455 | 0.222207 | 0 | 0 | 10.51321 | 24.6966 |
| GO_Biologi | Rescue Of | 23-Jan | 0.095455 | 0.222207 | 0 | 0 | 10.51321 | 24.6966 |

| | | | | | | | | |
|---|---|---|---|---|---|---|---|---|
| GO_Biologi | Long-Chain | 23-Jan | 0.095455 | 0.222207 | 0 | 0 | 10.51321 | 24.6966 |
| GO_Biologi | Regulation | 23-Jan | 0.095455 | 0.222207 | 0 | 0 | 10.51321 | 24.6966 |
| GO_Biologi | Positive Re | 23-Jan | 0.095455 | 0.222207 | 0 | 0 | 10.51321 | 24.6966 |
| GO_Biologi | Glial Cell D | 23-Jan | 0.095455 | 0.222207 | 0 | 0 | 10.51321 | 24.6966 |
| GO_Biologi | Regulation | 2/119 | 0.094827 | 0.222207 | 0 | 0 | 3.981096 | 9.378266 |
| GO_Biologi | Regulation | 23-Jan | 0.095455 | 0.222207 | 0 | 0 | 10.51321 | 24.6966 |
| KEGG_202 | Viral myoca | Feb-60 | 0.028127 | 0.222675 | 0 | 0 | 8.054767 | 28.76365 |
| KEGG_202 | Legionellos | Feb-57 | 0.025575 | 0.222675 | 0 | 0 | 8.495401 | 31.14526 |
| KEGG_202 | Adrenergic | 3/150 | 0.027733 | 0.222675 | 0 | 0 | 4.802235 | 17.21664 |
| MAGMA_D | Metformin | Jan-37 | 0.149088 | 0.222794 | 0 | 0 | 6.42022 | 12.21909 |
| DisGeNET | Acute myo | 3/247 | 0.09295 | 0.223012 | 0 | 0 | 2.878952 | 6.839504 |
| DisGeNET | Hypobetali | 23-Jan | 0.095455 | 0.223296 | 0 | 0 | 10.51321 | 24.6966 |
| DisGeNET | Hypoxic-Isc | 23-Jan | 0.095455 | 0.223296 | 0 | 0 | 10.51321 | 24.6966 |
| DisGeNET | Leukemia s | 23-Jan | 0.095455 | 0.223296 | 0 | 0 | 10.51321 | 24.6966 |
| DisGeNET | Andersen S | 23-Jan | 0.095455 | 0.223296 | 0 | 0 | 10.51321 | 24.6966 |
| DisGeNET | Camurati-E | 23-Jan | 0.095455 | 0.223296 | 0 | 0 | 10.51321 | 24.6966 |
| DisGeNET | Petechiae | 23-Jan | 0.095455 | 0.223296 | 0 | 0 | 10.51321 | 24.6966 |
| DisGeNET | Porphyria C | 23-Jan | 0.095455 | 0.223296 | 0 | 0 | 10.51321 | 24.6966 |
| DisGeNET | Convulsion | 2/118 | 0.093488 | 0.223296 | 0 | 0 | 4.015619 | 9.516699 |
| DisGeNET | Acute-On-C | 23-Jan | 0.095455 | 0.223296 | 0 | 0 | 10.51321 | 24.6966 |
| DisGeNET | Macule | 23-Jan | 0.095455 | 0.223296 | 0 | 0 | 10.51321 | 24.6966 |
| DisGeNET | Systolic Pre | 2/118 | 0.093488 | 0.223296 | 0 | 0 | 4.015619 | 9.516699 |
| DisGeNET | Varicocele | 23-Jan | 0.095455 | 0.223296 | 0 | 0 | 10.51321 | 24.6966 |
| DisGeNET | Multi-infar | 23-Jan | 0.095455 | 0.223296 | 0 | 0 | 10.51321 | 24.6966 |
| DisGeNET | Cellulitis of | 23-Jan | 0.095455 | 0.223296 | 0 | 0 | 10.51321 | 24.6966 |
| DisGeNET | Chromosor | 23-Jan | 0.095455 | 0.223296 | 0 | 0 | 10.51321 | 24.6966 |
| DisGeNET | Pre B-cell a | 23-Jan | 0.095455 | 0.223296 | 0 | 0 | 10.51321 | 24.6966 |
| DisGeNET | Chronic urt | 23-Jan | 0.095455 | 0.223296 | 0 | 0 | 10.51321 | 24.6966 |
| DisGeNET | cocaine use | 23-Jan | 0.095455 | 0.223296 | 0 | 0 | 10.51321 | 24.6966 |
| DisGeNET | Retinal Vei | 23-Jan | 0.095455 | 0.223296 | 0 | 0 | 10.51321 | 24.6966 |
| DisGeNET | Exfoliation | 23-Jan | 0.095455 | 0.223296 | 0 | 0 | 10.51321 | 24.6966 |
| DisGeNET | Rhabdomy | 4/397 | 0.094874 | 0.223296 | 0 | 0 | 2.393697 | 5.637647 |
| DisGeNET | Muscle Rig | 2/119 | 0.094827 | 0.223296 | 0 | 0 | 3.981096 | 9.378266 |
| DisGeNET | Hemimega | 23-Jan | 0.095455 | 0.223296 | 0 | 0 | 10.51321 | 24.6966 |
| DisGeNET | Acquired cu | 23-Jan | 0.095455 | 0.223296 | 0 | 0 | 10.51321 | 24.6966 |
| DisGeNET | Pseudoaph | 23-Jan | 0.095455 | 0.223296 | 0 | 0 | 10.51321 | 24.6966 |
| DisGeNET | Congenital | 23-Jan | 0.095455 | 0.223296 | 0 | 0 | 10.51321 | 24.6966 |
| DisGeNET | Abnormalit | 23-Jan | 0.095455 | 0.223296 | 0 | 0 | 10.51321 | 24.6966 |
| DisGeNET | Infarction, | 2/119 | 0.094827 | 0.223296 | 0 | 0 | 3.981096 | 9.378266 |
| CellMarker | Epithelial C | 3/318 | 0.160951 | 0.223861 | 0 | 0 | 2.221995 | 4.058822 |
| UK_Bioban | Age startec | Jan-42 | 0.16747 | 0.224068 | 0 | 0 | 5.635848 | 10.07097 |
| UK_Bioban | Duration tc | 9/1429 | 0.167972 | 0.224068 | 0 | 0 | 1.502681 | 2.680722 |
| UK_Bioban | Mean corp | 4/494 | 0.168247 | 0.224068 | 0 | 0 | 1.910302 | 3.404772 |
| GTEx_Tissu | GTEX-V1D1 | 11/985 | 0.003564 | 0.22417 | 0 | 0 | 2.814344 | 15.86377 |
| GTEx_Tissu | GTEX-QDVI | 14/1438 | 0.003532 | 0.22417 | 0 | 0 | 2.490053 | 14.05872 |

| | | | | | | | | |
|---|---|---|---|---|---|---|---|---|
| DisGeNET | Neurologic | 2/120 | 0.096172 | 0.224773 | 0 | 0 | 3.947159 | 9.242743 |
| GWAS_Cat | Cholesteryl | Jan-44 | 0.174713 | 0.224842 | 0 | 0 | 5.373175 | 9.3741 |
| GWAS_Cat | Total Phos | Jan-44 | 0.174713 | 0.224842 | 0 | 0 | 5.373175 | 9.3741 |
| GWAS_Cat | Cholesteryl | Jan-44 | 0.174713 | 0.224842 | 0 | 0 | 5.373175 | 9.3741 |
| GWAS_Cat | Phospholip | Jan-44 | 0.174713 | 0.224842 | 0 | 0 | 5.373175 | 9.3741 |
| GWAS_Cat | Remnant C | Jan-44 | 0.174713 | 0.224842 | 0 | 0 | 5.373175 | 9.3741 |
| DisGeNET | Heart Block | 24-Jan | 0.099394 | 0.225271 | 0 | 0 | 10.05561 | 23.215 |
| DisGeNET | Synostotic | 24-Jan | 0.099394 | 0.225271 | 0 | 0 | 10.05561 | 23.215 |
| DisGeNET | Lyme Arthr | 24-Jan | 0.099394 | 0.225271 | 0 | 0 | 10.05561 | 23.215 |
| DisGeNET | Sebaceous | 24-Jan | 0.099394 | 0.225271 | 0 | 0 | 10.05561 | 23.215 |
| DisGeNET | Fetal Alcoh | 24-Jan | 0.099394 | 0.225271 | 0 | 0 | 10.05561 | 23.215 |
| DisGeNET | Broad cran | 2/121 | 0.097522 | 0.225271 | 0 | 0 | 3.913791 | 9.110048 |
| DisGeNET | Juvenile M | 2/121 | 0.097522 | 0.225271 | 0 | 0 | 3.913791 | 9.110048 |
| DisGeNET | Severe con | 24-Jan | 0.099394 | 0.225271 | 0 | 0 | 10.05561 | 23.215 |
| DisGeNET | Congenital | 2/122 | 0.098877 | 0.225271 | 0 | 0 | 3.88098 | 8.980101 |
| DisGeNET | Basophilic | 24-Jan | 0.099394 | 0.225271 | 0 | 0 | 10.05561 | 23.215 |
| DisGeNET | bilateral br | 24-Jan | 0.099394 | 0.225271 | 0 | 0 | 10.05561 | 23.215 |
| DisGeNET | Waist-Hip I | 2/122 | 0.098877 | 0.225271 | 0 | 0 | 3.88098 | 8.980101 |
| DisGeNET | Acute myel | 24-Jan | 0.099394 | 0.225271 | 0 | 0 | 10.05561 | 23.215 |
| DisGeNET | Pilocytic As | 2/122 | 0.098877 | 0.225271 | 0 | 0 | 3.88098 | 8.980101 |
| DisGeNET | Chronic atr | 24-Jan | 0.099394 | 0.225271 | 0 | 0 | 10.05561 | 23.215 |
| DisGeNET | Child Devel | 24-Jan | 0.099394 | 0.225271 | 0 | 0 | 10.05561 | 23.215 |
| DisGeNET | Erectile abı | 24-Jan | 0.099394 | 0.225271 | 0 | 0 | 10.05561 | 23.215 |
| DisGeNET | Asymptom | 24-Jan | 0.099394 | 0.225271 | 0 | 0 | 10.05561 | 23.215 |
| DisGeNET | Signs and S | 24-Jan | 0.099394 | 0.225271 | 0 | 0 | 10.05561 | 23.215 |
| DisGeNET | Wide skull | 2/121 | 0.097522 | 0.225271 | 0 | 0 | 3.913791 | 9.110048 |
| DisGeNET | Chronic thr | 24-Jan | 0.099394 | 0.225271 | 0 | 0 | 10.05561 | 23.215 |
| DisGeNET | Inattention | 24-Jan | 0.099394 | 0.225271 | 0 | 0 | 10.05561 | 23.215 |
| DisGeNET | Hyperlipop | 24-Jan | 0.099394 | 0.225271 | 0 | 0 | 10.05561 | 23.215 |
| DisGeNET | Norrie dise | 24-Jan | 0.099394 | 0.225271 | 0 | 0 | 10.05561 | 23.215 |
| DisGeNET | Poor eye co | 24-Jan | 0.099394 | 0.225271 | 0 | 0 | 10.05561 | 23.215 |
| DisGeNET | Precursor T | 24-Jan | 0.099394 | 0.225271 | 0 | 0 | 10.05561 | 23.215 |
| DisGeNET | Endemic Cr | 24-Jan | 0.099394 | 0.225271 | 0 | 0 | 10.05561 | 23.215 |
| DisGeNET | Leukemia, | 24-Jan | 0.099394 | 0.225271 | 0 | 0 | 10.05561 | 23.215 |
| DisGeNET | Ostium sec | 24-Jan | 0.099394 | 0.225271 | 0 | 0 | 10.05561 | 23.215 |
| DisGeNET | Pinched bri | 24-Jan | 0.099394 | 0.225271 | 0 | 0 | 10.05561 | 23.215 |
| DisGeNET | Pinched na | 24-Jan | 0.099394 | 0.225271 | 0 | 0 | 10.05561 | 23.215 |
| DisGeNET | Child Devel | 24-Jan | 0.099394 | 0.225271 | 0 | 0 | 10.05561 | 23.215 |
| DisGeNET | Long nose | 24-Jan | 0.099394 | 0.225271 | 0 | 0 | 10.05561 | 23.215 |
| DisGeNET | Intestinal P | 24-Jan | 0.099394 | 0.225271 | 0 | 0 | 10.05561 | 23.215 |
| DisGeNET | Infant Leuk | 24-Jan | 0.099394 | 0.225271 | 0 | 0 | 10.05561 | 23.215 |
| DisGeNET | Diabetes | 9/1268 | 0.099525 | 0.225372 | 0 | 0 | 1.709599 | 3.944635 |
| GeDiPNet_ | Bright Dise | 31-Jan | 0.126498 | 0.225531 | 0 | 0 | 7.706589 | 15.93356 |
| GeDiPNet_ | Hemangior | 31-Jan | 0.126498 | 0.225531 | 0 | 0 | 7.706589 | 15.93356 |
| Jensen_TIS | High endot | Jan-32 | 0.130304 | 0.22558 | 0 | 0 | 7.457614 | 15.19775 |

| | | | | | | | | |
|---|---|---|---|---|---|---|---|---|
| Jensen_TIS | Cob | Jan-32 | 0.130304 | 0.22558 | 0 | 0 | 7.457614 | 15.19775 |
| Jensen_TIS | BMMC cell | Jan-32 | 0.130304 | 0.22558 | 0 | 0 | 7.457614 | 15.19775 |
| Jensen_TIS | Plasma cell | 3/286 | 0.128615 | 0.22558 | 0 | 0 | 2.477284 | 5.080743 |
| DisGeNET | Acute lymp | 7/907 | 0.099747 | 0.225679 | 0 | 0 | 1.848486 | 4.260985 |
| GO_Biologi | Protein Loc | 2/122 | 0.098877 | 0.226186 | 0 | 0 | 3.88098 | 8.980101 |
| GO_Biologi | Dicarboxyli | 24-Jan | 0.099394 | 0.226186 | 0 | 0 | 10.05561 | 23.215 |
| GO_Biologi | Negative R | 24-Jan | 0.099394 | 0.226186 | 0 | 0 | 10.05561 | 23.215 |
| GO_Biologi | Negative R | 24-Jan | 0.099394 | 0.226186 | 0 | 0 | 10.05561 | 23.215 |
| GO_Biologi | Protein Kin | 24-Jan | 0.099394 | 0.226186 | 0 | 0 | 10.05561 | 23.215 |
| GO_Biologi | Positive Re | 24-Jan | 0.099394 | 0.226186 | 0 | 0 | 10.05561 | 23.215 |
| GO_Biologi | Regulation | 24-Jan | 0.099394 | 0.226186 | 0 | 0 | 10.05561 | 23.215 |
| DisGeNET | Arterioscle | 8/1087 | 0.100203 | 0.2264 | 0 | 0 | 1.7676 | 4.066467 |
| DisGeNET | Substance- | 2/123 | 0.100238 | 0.2264 | 0 | 0 | 3.848712 | 8.852824 |
| Rare_Disea | Leri Weill d | 11-Jan | 0.046834 | 0.226702 | 0 | 0 | 23.14302 | 70.84419 |
| Rare_Disea | Hyperinsul | 3/184 | 0.046305 | 0.226702 | 0 | 0 | 3.893449 | 11.96266 |
| Rare_Disea | Peyronie d | 11-Jan | 0.046834 | 0.226702 | 0 | 0 | 23.14302 | 70.84419 |
| Rare_Disea | Spongiforn | 3/185 | 0.046925 | 0.226702 | 0 | 0 | 3.87186 | 11.84483 |
| Rare_Disea | Enlarged ve | 11-Jan | 0.046834 | 0.226702 | 0 | 0 | 23.14302 | 70.84419 |
| Rare_Disea | Japanese e | Feb-79 | 0.046446 | 0.226702 | 0 | 0 | 6.061421 | 18.60534 |
| Rare_Disea | Watson syr | 3/185 | 0.046925 | 0.226702 | 0 | 0 | 3.87186 | 11.84483 |
| Rare_Disea | Acetyl-carr | 11-Jan | 0.046834 | 0.226702 | 0 | 0 | 23.14302 | 70.84419 |
| CellMarker | Macrophag | Jan-41 | 0.163825 | 0.226708 | 0 | 0 | 5.777035 | 10.45039 |
| UK_Bioban | Right 6mm | Jan-43 | 0.171099 | 0.226806 | 0 | 0 | 5.501384 | 9.712751 |
| MAGMA_D | Everolimus | Jan-41 | 0.163825 | 0.226967 | 0 | 0 | 5.777035 | 10.45039 |
| MAGMA_D | Renal Angi | Jan-41 | 0.163825 | 0.226967 | 0 | 0 | 5.777035 | 10.45039 |
| MAGMA_D | Subependy | Jan-41 | 0.163825 | 0.226967 | 0 | 0 | 5.777035 | 10.45039 |
| MAGMA_D | Tuberous S | Jan-41 | 0.163825 | 0.226967 | 0 | 0 | 5.777035 | 10.45039 |
| MAGMA_D | Neuroendo | Jan-41 | 0.163825 | 0.226967 | 0 | 0 | 5.777035 | 10.45039 |
| MAGMA_D | mTOR Inhil | Jan-41 | 0.163825 | 0.226967 | 0 | 0 | 5.777035 | 10.45039 |
| MAGMA_D | Cytarabine | Jan-41 | 0.163825 | 0.226967 | 0 | 0 | 5.777035 | 10.45039 |
| CellMarker | B Cell Kidn | 5/667 | 0.164993 | 0.227176 | 0 | 0 | 1.773174 | 3.195 |
| Jensen_TIS | Amastigote | Jan-33 | 0.134093 | 0.227253 | 0 | 0 | 7.224201 | 14.515 |
| Jensen_TIS | adipose-de | Jan-33 | 0.134093 | 0.227253 | 0 | 0 | 7.224201 | 14.515 |
| Jensen_TIS | Uterus | 61/12771 | 0.133725 | 0.227253 | 0 | 0 | 1.32961 | 2.675131 |
| Jensen_TIS | Merozoite | Jan-33 | 0.134093 | 0.227253 | 0 | 0 | 7.224201 | 14.515 |
| GWAS_Cat | Ratio Of Pc | Jan-45 | 0.178311 | 0.227551 | 0 | 0 | 5.250793 | 9.053559 |
| GWAS_Cat | Phospholip | Jan-45 | 0.178311 | 0.227551 | 0 | 0 | 5.250793 | 9.053559 |
| GWAS_Cat | Major Depi | Jan-45 | 0.178311 | 0.227551 | 0 | 0 | 5.250793 | 9.053559 |
| GWAS_Cat | Saturated F | Jan-45 | 0.178311 | 0.227551 | 0 | 0 | 5.250793 | 9.053559 |
| GWAS_Cat | Concentrat | Jan-45 | 0.178311 | 0.227551 | 0 | 0 | 5.250793 | 9.053559 |
| GWAS_Cat | Testostero | Jan-45 | 0.178311 | 0.227551 | 0 | 0 | 5.250793 | 9.053559 |
| GWAS_Cat | Cholestero | Jan-45 | 0.178311 | 0.227551 | 0 | 0 | 5.250793 | 9.053559 |
| GWAS_Cat | Triglyceride | Jan-45 | 0.178311 | 0.227551 | 0 | 0 | 5.250793 | 9.053559 |
| Jensen_DIS | Dental cari | Jan-49 | 0.192548 | 0.227557 | 0 | 0 | 4.812258 | 7.927759 |
| DisGeNET | Malignant | 19/3239 | 0.102144 | 0.227853 | 0 | 0 | 1.448516 | 3.304603 |

| | | | | | | | | |
|---|---|---|---|---|---|---|---|---|
| DisGeNET | Dystonia, P | 25-Jan | 0.103317 | 0.227853 | 0 | 0 | 9.636143 | 21.87363 |
| DisGeNET | Peripheral | 2/125 | 0.102976 | 0.227853 | 0 | 0 | 3.785748 | 8.60599 |
| DisGeNET | Malignant | 12/1846 | 0.103063 | 0.227853 | 0 | 0 | 1.57723 | 3.584114 |
| DisGeNET | Hypokinesi | 25-Jan | 0.103317 | 0.227853 | 0 | 0 | 9.636143 | 21.87363 |
| DisGeNET | Lymphoma | 25-Jan | 0.103317 | 0.227853 | 0 | 0 | 9.636143 | 21.87363 |
| DisGeNET | Goiter | 25-Jan | 0.103317 | 0.227853 | 0 | 0 | 9.636143 | 21.87363 |
| DisGeNET | Perianal Sq | 25-Jan | 0.103317 | 0.227853 | 0 | 0 | 9.636143 | 21.87363 |
| DisGeNET | Temporom | 25-Jan | 0.103317 | 0.227853 | 0 | 0 | 9.636143 | 21.87363 |
| DisGeNET | Hemangior | 25-Jan | 0.103317 | 0.227853 | 0 | 0 | 9.636143 | 21.87363 |
| DisGeNET | Hallervord | 25-Jan | 0.103317 | 0.227853 | 0 | 0 | 9.636143 | 21.87363 |
| DisGeNET | Hepatic Fo | 25-Jan | 0.103317 | 0.227853 | 0 | 0 | 9.636143 | 21.87363 |
| DisGeNET | Hyperostos | 25-Jan | 0.103317 | 0.227853 | 0 | 0 | 9.636143 | 21.87363 |
| DisGeNET | Excessive v | 25-Jan | 0.103317 | 0.227853 | 0 | 0 | 9.636143 | 21.87363 |
| DisGeNET | Onion bulb | 25-Jan | 0.103317 | 0.227853 | 0 | 0 | 9.636143 | 21.87363 |
| DisGeNET | Atrioventri | 25-Jan | 0.103317 | 0.227853 | 0 | 0 | 9.636143 | 21.87363 |
| DisGeNET | Pulmonary | 25-Jan | 0.103317 | 0.227853 | 0 | 0 | 9.636143 | 21.87363 |
| DisGeNET | Encephaliti | 2/124 | 0.101605 | 0.227853 | 0 | 0 | 3.816972 | 8.728145 |
| DisGeNET | CNS metas | 25-Jan | 0.103317 | 0.227853 | 0 | 0 | 9.636143 | 21.87363 |
| DisGeNET | Parathyroi | 25-Jan | 0.103317 | 0.227853 | 0 | 0 | 9.636143 | 21.87363 |
| DisGeNET | Sclerosis | 25-Jan | 0.103317 | 0.227853 | 0 | 0 | 9.636143 | 21.87363 |
| DisGeNET | Uterine Ca | 25-Jan | 0.103317 | 0.227853 | 0 | 0 | 9.636143 | 21.87363 |
| DisGeNET | Acrocephal | 25-Jan | 0.103317 | 0.227853 | 0 | 0 | 9.636143 | 21.87363 |
| DisGeNET | Inflammat | 7/911 | 0.101491 | 0.227853 | 0 | 0 | 1.83992 | 4.209334 |
| DisGeNET | Cyanosis | 25-Jan | 0.103317 | 0.227853 | 0 | 0 | 9.636143 | 21.87363 |
| DisGeNET | Quadripleg | 25-Jan | 0.103317 | 0.227853 | 0 | 0 | 9.636143 | 21.87363 |
| DisGeNET | Diabetic fo | 25-Jan | 0.103317 | 0.227853 | 0 | 0 | 9.636143 | 21.87363 |
| DisGeNET | Progressive | 25-Jan | 0.103317 | 0.227853 | 0 | 0 | 9.636143 | 21.87363 |
| GeDiPNet_ | Celiac Dise | 2/143 | 0.128476 | 0.22791 | 0 | 0 | 3.299458 | 6.770526 |
| GeDiPNet_ | Camptodac | 3/286 | 0.128615 | 0.22791 | 0 | 0 | 2.477284 | 5.080743 |
| Human_Ph | Ovarian ne | Jan-46 | 0.181893 | 0.228563 | 0 | 0 | 5.13385 | 8.749803 |
| DisGeNET | Astrocyton | 6/741 | 0.103734 | 0.228581 | 0 | 0 | 1.932779 | 4.379539 |
| CellMarker | Fibroblast | Jan-42 | 0.16747 | 0.229434 | 0 | 0 | 5.635848 | 10.07097 |
| GeDiPNet_ | Cholangioc | Jan-32 | 0.130304 | 0.229508 | 0 | 0 | 7.457614 | 15.19775 |
| GeDiPNet_ | Mirror Mov | Jan-32 | 0.130304 | 0.229508 | 0 | 0 | 7.457614 | 15.19775 |
| DisGeNET | Left-Sided | 2/126 | 0.104352 | 0.229558 | 0 | 0 | 3.755028 | 8.486292 |
| DisGeNET | Laryngeal S | 2/126 | 0.104352 | 0.229558 | 0 | 0 | 3.755028 | 8.486292 |
| MAGMA_D | Tetanus | Jan-43 | 0.171099 | 0.229861 | 0 | 0 | 5.501384 | 9.712751 |
| MAGMA_D | Acute Inter | Jan-43 | 0.171099 | 0.229861 | 0 | 0 | 5.501384 | 9.712751 |
| MAGMA_D | Chlorprom | Jan-43 | 0.171099 | 0.229861 | 0 | 0 | 5.501384 | 9.712751 |
| GWAS_Cat | Concentrat | Jan-46 | 0.181893 | 0.230436 | 0 | 0 | 5.13385 | 8.749803 |
| GWAS_Cat | Uterine Lei | Jan-46 | 0.181893 | 0.230436 | 0 | 0 | 5.13385 | 8.749803 |
| GWAS_Cat | Triglycerid | Jan-46 | 0.181893 | 0.230436 | 0 | 0 | 5.13385 | 8.749803 |
| GWAS_Cat | Total Omeg | Jan-46 | 0.181893 | 0.230436 | 0 | 0 | 5.13385 | 8.749803 |
| GWAS_Cat | Refractive | 3/337 | 0.181217 | 0.230436 | 0 | 0 | 2.093563 | 3.575931 |
| GWAS_Cat | Total Lipid | Jan-46 | 0.181893 | 0.230436 | 0 | 0 | 5.13385 | 8.749803 |

| | | | | | | | | |
|---|---|---|---|---|---|---|---|---|
| GWAS_Cat | Phospholip | Jan-46 | 0.181893 | 0.230436 | 0 | 0 | 5.13385 | 8.749803 |
| GO_Biologi | Neuron Prc | 2/124 | 0.101605 | 0.230477 | 0 | 0 | 3.816972 | 8.728145 |
| UK_Bioban | North co-o | Jan-44 | 0.174713 | 0.230524 | 0 | 0 | 5.373175 | 9.3741 |
| Human_Ph | Increased ( | Jan-47 | 0.18546 | 0.230768 | 0 | 0 | 5.021992 | 8.46163 |
| Human_Ph | Hypoplasia | Jan-47 | 0.18546 | 0.230768 | 0 | 0 | 5.021992 | 8.46163 |
| Human_Ph | Preaxial ha | Jan-47 | 0.18546 | 0.230768 | 0 | 0 | 5.021992 | 8.46163 |
| GTEx_Tissu | GTEX-U3ZN | 11/992 | 0.00376 | 0.230798 | 0 | 0 | 2.793229 | 15.59518 |
| GTEx_Tissu | GTEX-XUZC | 15/1611 | 0.003829 | 0.230798 | 0 | 0 | 2.391004 | 13.30608 |
| DisGeNET | Gastritis, A | 2/127 | 0.105734 | 0.231402 | 0 | 0 | 3.7248 | 8.368983 |
| DisGeNET | Pituitary ca | 26-Jan | 0.107222 | 0.231402 | 0 | 0 | 9.250233 | 20.65441 |
| DisGeNET | Epidermoly | 26-Jan | 0.107222 | 0.231402 | 0 | 0 | 9.250233 | 20.65441 |
| DisGeNET | Sarcomato | 2/128 | 0.107121 | 0.231402 | 0 | 0 | 3.695051 | 8.253999 |
| DisGeNET | Blepharitis | 26-Jan | 0.107222 | 0.231402 | 0 | 0 | 9.250233 | 20.65441 |
| DisGeNET | Interstitial | 2/127 | 0.105734 | 0.231402 | 0 | 0 | 3.7248 | 8.368983 |
| DisGeNET | ATRIOVEN | 26-Jan | 0.107222 | 0.231402 | 0 | 0 | 9.250233 | 20.65441 |
| DisGeNET | Malignant | 26-Jan | 0.107222 | 0.231402 | 0 | 0 | 9.250233 | 20.65441 |
| DisGeNET | Osteoarthr | 26-Jan | 0.107222 | 0.231402 | 0 | 0 | 9.250233 | 20.65441 |
| DisGeNET | Adult T Acu | 26-Jan | 0.107222 | 0.231402 | 0 | 0 | 9.250233 | 20.65441 |
| DisGeNET | Malignant | 15/2449 | 0.106967 | 0.231402 | 0 | 0 | 1.49608 | 3.344091 |
| DisGeNET | Multiple lip | 26-Jan | 0.107222 | 0.231402 | 0 | 0 | 9.250233 | 20.65441 |
| DisGeNET | Cyclic neut | 26-Jan | 0.107222 | 0.231402 | 0 | 0 | 9.250233 | 20.65441 |
| DisGeNET | Familial Aty | 26-Jan | 0.107222 | 0.231402 | 0 | 0 | 9.250233 | 20.65441 |
| DisGeNET | Personality | 26-Jan | 0.107222 | 0.231402 | 0 | 0 | 9.250233 | 20.65441 |
| DisGeNET | Hereditary | 2/128 | 0.107121 | 0.231402 | 0 | 0 | 3.695051 | 8.253999 |
| DisGeNET | Fatty acid r | 26-Jan | 0.107222 | 0.231402 | 0 | 0 | 9.250233 | 20.65441 |
| DisGeNET | Schizophre | 26-Jan | 0.107222 | 0.231402 | 0 | 0 | 9.250233 | 20.65441 |
| DisGeNET | Severe Apl | 26-Jan | 0.107222 | 0.231402 | 0 | 0 | 9.250233 | 20.65441 |
| DisGeNET | Generalize | 26-Jan | 0.107222 | 0.231402 | 0 | 0 | 9.250233 | 20.65441 |
| DisGeNET | MUTYH-As | 26-Jan | 0.107222 | 0.231402 | 0 | 0 | 9.250233 | 20.65441 |
| DisGeNET | Carotid Art | 26-Jan | 0.107222 | 0.231402 | 0 | 0 | 9.250233 | 20.65441 |
| DisGeNET | Borna Dise | 26-Jan | 0.107222 | 0.231402 | 0 | 0 | 9.250233 | 20.65441 |
| GO_Biologi | Regulation | 25-Jan | 0.103317 | 0.231403 | 0 | 0 | 9.636143 | 21.87363 |
| GO_Biologi | Regulation | 25-Jan | 0.103317 | 0.231403 | 0 | 0 | 9.636143 | 21.87363 |
| GO_Biologi | Negative R | 25-Jan | 0.103317 | 0.231403 | 0 | 0 | 9.636143 | 21.87363 |
| GO_Biologi | Negative R | 25-Jan | 0.103317 | 0.231403 | 0 | 0 | 9.636143 | 21.87363 |
| GWAS_Cat | Birth Weigl | 2/179 | 0.183014 | 0.231615 | 0 | 0 | 2.623596 | 4.455375 |
| DisGeNET | Cervix carc | 8/1105 | 0.107475 | 0.231757 | 0 | 0 | 1.736935 | 3.874229 |
| DisGeNET | Myeloproli | 3/264 | 0.107935 | 0.232365 | 0 | 0 | 2.689108 | 5.986573 |
| DisGeNET | Brain Ische | 3/264 | 0.107935 | 0.232365 | 0 | 0 | 2.689108 | 5.986573 |
| Jensen_TIS | Cerebral w | Jan-34 | 0.137866 | 0.232424 | 0 | 0 | 7.004933 | 13.88007 |
| CellMarker | Schwalie Et | 2/171 | 0.170583 | 0.232537 | 0 | 0 | 2.748904 | 4.86152 |
| DisGeNET | Tauopathie | 2/130 | 0.109909 | 0.232581 | 0 | 0 | 3.636949 | 8.030761 |
| DisGeNET | Flared met | 27-Jan | 0.111111 | 0.232581 | 0 | 0 | 8.894007 | 19.54216 |
| DisGeNET | Mucopolys | 27-Jan | 0.111111 | 0.232581 | 0 | 0 | 8.894007 | 19.54216 |
| DisGeNET | Aphasia | 27-Jan | 0.111111 | 0.232581 | 0 | 0 | 8.894007 | 19.54216 |

| | | | | | | | | |
|---|---|---|---|---|---|---|---|---|
| DisGeNET | Pseudohyp | 2/131 | 0.11131 | 0.232581 | 0 | 0 | 3.608573 | 7.922388 |
| DisGeNET | Metabolic I | 4/421 | 0.111319 | 0.232581 | 0 | 0 | 2.253157 | 4.946473 |
| DisGeNET | early pregr | 2/129 | 0.108512 | 0.232581 | 0 | 0 | 3.665771 | 8.141279 |
| DisGeNET | Acquired s | 4/418 | 0.109197 | 0.232581 | 0 | 0 | 2.269833 | 5.026777 |
| DisGeNET | Pervasive D | 2/129 | 0.108512 | 0.232581 | 0 | 0 | 3.665771 | 8.141279 |
| DisGeNET | Cervical dy | 27-Jan | 0.111111 | 0.232581 | 0 | 0 | 8.894007 | 19.54216 |
| DisGeNET | Chronic live | 2/129 | 0.108512 | 0.232581 | 0 | 0 | 3.665771 | 8.141279 |
| DisGeNET | Congenital | 27-Jan | 0.111111 | 0.232581 | 0 | 0 | 8.894007 | 19.54216 |
| DisGeNET | Aortic Valv | 2/130 | 0.109909 | 0.232581 | 0 | 0 | 3.636949 | 8.030761 |
| DisGeNET | Hereditary | 27-Jan | 0.111111 | 0.232581 | 0 | 0 | 8.894007 | 19.54216 |
| DisGeNET | Idiopathic I | 27-Jan | 0.111111 | 0.232581 | 0 | 0 | 8.894007 | 19.54216 |
| DisGeNET | Uveitis | 2/130 | 0.109909 | 0.232581 | 0 | 0 | 3.636949 | 8.030761 |
| DisGeNET | Hearing Lo: | 27-Jan | 0.111111 | 0.232581 | 0 | 0 | 8.894007 | 19.54216 |
| DisGeNET | Gastroente | 27-Jan | 0.111111 | 0.232581 | 0 | 0 | 8.894007 | 19.54216 |
| DisGeNET | Disorder of | 2/131 | 0.11131 | 0.232581 | 0 | 0 | 3.608573 | 7.922388 |
| DisGeNET | Neoplastic | 2/131 | 0.11131 | 0.232581 | 0 | 0 | 3.608573 | 7.922388 |
| DisGeNET | Primary Ma | 27-Jan | 0.111111 | 0.232581 | 0 | 0 | 8.894007 | 19.54216 |
| DisGeNET | Coronary M | 27-Jan | 0.111111 | 0.232581 | 0 | 0 | 8.894007 | 19.54216 |
| DisGeNET | Adenocarci | 3/267 | 0.110672 | 0.232581 | 0 | 0 | 2.658144 | 5.851062 |
| DisGeNET | Trichohepa | 3/267 | 0.110672 | 0.232581 | 0 | 0 | 2.658144 | 5.851062 |
| DisGeNET | Benign Pro | 4/420 | 0.11061 | 0.232581 | 0 | 0 | 2.258689 | 4.97306 |
| DisGeNET | Pancreatic | 12/1869 | 0.110315 | 0.232581 | 0 | 0 | 1.555714 | 3.429439 |
| DisGeNET | Congenital | 2/130 | 0.109909 | 0.232581 | 0 | 0 | 3.636949 | 8.030761 |
| DisGeNET | Brachycepł | 2/130 | 0.109909 | 0.232581 | 0 | 0 | 3.636949 | 8.030761 |
| DisGeNET | Mildly elev | 27-Jan | 0.111111 | 0.232581 | 0 | 0 | 8.894007 | 19.54216 |
| DisGeNET | Apathy | 27-Jan | 0.111111 | 0.232581 | 0 | 0 | 8.894007 | 19.54216 |
| DisGeNET | Metopic sy | 27-Jan | 0.111111 | 0.232581 | 0 | 0 | 8.894007 | 19.54216 |
| DisGeNET | Anterior uv | 27-Jan | 0.111111 | 0.232581 | 0 | 0 | 8.894007 | 19.54216 |
| DisGeNET | Leg Ulcer | 27-Jan | 0.111111 | 0.232581 | 0 | 0 | 8.894007 | 19.54216 |
| DisGeNET | Extramedu | 27-Jan | 0.111111 | 0.232581 | 0 | 0 | 8.894007 | 19.54216 |
| DisGeNET | Attenuatio | 27-Jan | 0.111111 | 0.232581 | 0 | 0 | 8.894007 | 19.54216 |
| DisGeNET | Extraskelet | 27-Jan | 0.111111 | 0.232581 | 0 | 0 | 8.894007 | 19.54216 |
| DisGeNET | Mucinous r | 27-Jan | 0.111111 | 0.232581 | 0 | 0 | 8.894007 | 19.54216 |
| SysMyo_M | MGS436 Hı | 4/211 | 0.013567 | 0.232688 | 0 | 0 | 4.587859 | 19.72845 |
| Jensen_TIS | Testis | 80/17517 | 0.138836 | 0.232839 | 0 | 0 | 1.622822 | 3.204202 |
| GWAS_Cat | Age At Firs | Jan-47 | 0.18546 | 0.233019 | 0 | 0 | 5.021992 | 8.46163 |
| GWAS_Cat | Metabolic I | 2/180 | 0.184577 | 0.233019 | 0 | 0 | 2.608724 | 4.407929 |
| GWAS_Cat | White Mat | Jan-47 | 0.18546 | 0.233019 | 0 | 0 | 5.021992 | 8.46163 |
| GWAS_Cat | Cholesteryl | Jan-47 | 0.18546 | 0.233019 | 0 | 0 | 5.021992 | 8.46163 |
| GWAS_Cat | COVID-19 ( | Jan-47 | 0.18546 | 0.233019 | 0 | 0 | 5.021992 | 8.46163 |
| GWAS_Cat | Angina Pec | Jan-47 | 0.18546 | 0.233019 | 0 | 0 | 5.021992 | 8.46163 |
| GWAS_Cat | Phosphogly | Jan-47 | 0.18546 | 0.233019 | 0 | 0 | 5.021992 | 8.46163 |
| UK_Bioban | Red blood ( | 3/334 | 0.177971 | 0.23374 | 0 | 0 | 2.112861 | 3.647087 |
| DisGeNET | Primary ma | 14/2268 | 0.112138 | 0.234104 | 0 | 0 | 1.50251 | 3.287533 |
| GO_Biologi | Regulation | 26-Jan | 0.107222 | 0.234239 | 0 | 0 | 9.250233 | 20.65441 |

| | | | | | | | | |
|---|---|---|---|---|---|---|---|---|
| GO_Biologi | Dendritic S | 26-Jan | 0.107222 | 0.234239 | 0 | 0 | 9.250233 | 20.65441 |
| GO_Biologi | Negative R | 26-Jan | 0.107222 | 0.234239 | 0 | 0 | 9.250233 | 20.65441 |
| GO_Biologi | Regulation | 26-Jan | 0.107222 | 0.234239 | 0 | 0 | 9.250233 | 20.65441 |
| GO_Biologi | Organelle F | 26-Jan | 0.107222 | 0.234239 | 0 | 0 | 9.250233 | 20.65441 |
| GO_Biologi | Negative R | 26-Jan | 0.107222 | 0.234239 | 0 | 0 | 9.250233 | 20.65441 |
| GO_Biologi | Negative R | 26-Jan | 0.107222 | 0.234239 | 0 | 0 | 9.250233 | 20.65441 |
| GO_Biologi | Positive Re | 26-Jan | 0.107222 | 0.234239 | 0 | 0 | 9.250233 | 20.65441 |
| GWAS_Cat | Neutrophil | 3/342 | 0.186664 | 0.23429 | 0 | 0 | 2.062158 | 3.461222 |
| Human_Ph | Respiratory | Jan-48 | 0.189012 | 0.234424 | 0 | 0 | 4.914894 | 8.187947 |
| Jensen_DIS | Charcot-Ma | Jan-51 | 0.199575 | 0.23444 | 0 | 0 | 4.619302 | 7.444308 |
| GeDiPNet_ | Scaphocep | Jan-33 | 0.134093 | 0.234764 | 0 | 0 | 7.224201 | 14.515 |
| GeDiPNet_ | Prostate Ad | Jan-33 | 0.134093 | 0.234764 | 0 | 0 | 7.224201 | 14.515 |
| DisGeNET | Mouth Nec | 2/132 | 0.112716 | 0.234937 | 0 | 0 | 3.580633 | 7.816104 |
| DisGeNET | Sleep Apne | 2/132 | 0.112716 | 0.234937 | 0 | 0 | 3.580633 | 7.816104 |
| UK_Bioban | Left arm fa | 4/508 | 0.180193 | 0.235573 | 0 | 0 | 1.8559 | 3.180508 |
| GeDiPNet_ | Multiple Sc | 3/293 | 0.135474 | 0.235765 | 0 | 0 | 2.416626 | 4.830773 |
| GeDiPNet_ | Crohn Dise | 3/293 | 0.135474 | 0.235765 | 0 | 0 | 2.416626 | 4.830773 |
| DisGeNET | Coronary A | 6/760 | 0.1134 | 0.236175 | 0 | 0 | 1.882208 | 4.097254 |
| GWAS_Cat | Cholestero | Jan-48 | 0.189012 | 0.236265 | 0 | 0 | 4.914894 | 8.187947 |
| GWAS_Cat | Phospholip | Jan-48 | 0.189012 | 0.236265 | 0 | 0 | 4.914894 | 8.187947 |
| GWAS_Cat | Triglyceride | Jan-48 | 0.189012 | 0.236265 | 0 | 0 | 4.914894 | 8.187947 |
| GWAS_Cat | Phospholip | Jan-48 | 0.189012 | 0.236265 | 0 | 0 | 4.914894 | 8.187947 |
| Jensen_TIS | Amyloid pla | Jan-35 | 0.141623 | 0.236283 | 0 | 0 | 6.798564 | 13.28839 |
| Gene_Pert | Adrb2 KO r | 7/288 | 0.000261 | 0.236726 | 0 | 0 | 6.113167 | 50.4397 |
| PhenGenI_ | Longevity | 4/326 | 0.053893 | 0.236881 | 0 | 0 | 2.932126 | 8.56401 |
| PhenGenI_ | Waist-Hip I | 5/473 | 0.055375 | 0.236881 | 0 | 0 | 2.533484 | 7.330969 |
| PhenGenI_ | Carcinoma, | 13-Jan | 0.055113 | 0.236881 | 0 | 0 | 19.28391 | 55.89185 |
| Rare_Disea | Long QT sy | 12-Jan | 0.050982 | 0.237069 | 0 | 0 | 21.03805 | 62.61499 |
| Rare_Disea | Juvenile pc | 12-Jan | 0.050982 | 0.237069 | 0 | 0 | 21.03805 | 62.61499 |
| Rare_Disea | Whooping | 12-Jan | 0.050982 | 0.237069 | 0 | 0 | 21.03805 | 62.61499 |
| Rare_Disea | Transcobal | 12-Jan | 0.050982 | 0.237069 | 0 | 0 | 21.03805 | 62.61499 |
| Rare_Disea | Nonseminc | 12-Jan | 0.050982 | 0.237069 | 0 | 0 | 21.03805 | 62.61499 |
| Rare_Disea | Pineocyton | 12-Jan | 0.050982 | 0.237069 | 0 | 0 | 21.03805 | 62.61499 |
| UK_Bioban | Right heel I | 2/179 | 0.183014 | 0.237086 | 0 | 0 | 2.623596 | 4.455375 |
| UK_Bioban | Right heel ( | 2/179 | 0.183014 | 0.237086 | 0 | 0 | 2.623596 | 4.455375 |
| DisGeNET | Childhood | 28-Jan | 0.114983 | 0.237208 | 0 | 0 | 8.564169 | 18.52408 |
| DisGeNET | CRANIOME | 28-Jan | 0.114983 | 0.237208 | 0 | 0 | 8.564169 | 18.52408 |
| DisGeNET | Chronic col | 28-Jan | 0.114983 | 0.237208 | 0 | 0 | 8.564169 | 18.52408 |
| DisGeNET | Extrinsic all | 28-Jan | 0.114983 | 0.237208 | 0 | 0 | 8.564169 | 18.52408 |
| DisGeNET | Agranulocy | 28-Jan | 0.114983 | 0.237208 | 0 | 0 | 8.564169 | 18.52408 |
| DisGeNET | Lymphoma | 28-Jan | 0.114983 | 0.237208 | 0 | 0 | 8.564169 | 18.52408 |
| DisGeNET | Serum gam | 28-Jan | 0.114983 | 0.237208 | 0 | 0 | 8.564169 | 18.52408 |
| DisGeNET | Liposarcon | 28-Jan | 0.114983 | 0.237208 | 0 | 0 | 8.564169 | 18.52408 |
| DisGeNET | Lung Disea | 28-Jan | 0.114983 | 0.237208 | 0 | 0 | 8.564169 | 18.52408 |
| DisGeNET | Seborrheic | 28-Jan | 0.114983 | 0.237208 | 0 | 0 | 8.564169 | 18.52408 |

| | | | | | | | | |
|---|---|---|---|---|---|---|---|---|
| DisGeNET | Hyperemia | 28-Jan | 0.114983 | 0.237208 | 0 | 0 | 8.564169 | 18.52408 |
| DisGeNET | Fetal Death | 28-Jan | 0.114983 | 0.237208 | 0 | 0 | 8.564169 | 18.52408 |
| DisGeNET | Macrotia | 2/134 | 0.115542 | 0.237614 | 0 | 0 | 3.526025 | 7.609588 |
| DisGeNET | Large auric | 2/134 | 0.115542 | 0.237614 | 0 | 0 | 3.526025 | 7.609588 |
| DisGeNET | Large pinna | 2/134 | 0.115542 | 0.237614 | 0 | 0 | 3.526025 | 7.609588 |
| DisGeNET | Thyroid Gla | 2/134 | 0.115542 | 0.237614 | 0 | 0 | 3.526025 | 7.609588 |
| GO_Biologi | Positive Re | 27-Jan | 0.111111 | 0.237616 | 0 | 0 | 8.894007 | 19.54216 |
| GO_Biologi | Regulation | 27-Jan | 0.111111 | 0.237616 | 0 | 0 | 8.894007 | 19.54216 |
| GO_Biologi | Monoubiqu | 27-Jan | 0.111111 | 0.237616 | 0 | 0 | 8.894007 | 19.54216 |
| GO_Biologi | In Utero Er | 27-Jan | 0.111111 | 0.237616 | 0 | 0 | 8.894007 | 19.54216 |
| GO_Biologi | Negative R | 27-Jan | 0.111111 | 0.237616 | 0 | 0 | 8.894007 | 19.54216 |
| GO_Biologi | Monoubiqu | 27-Jan | 0.111111 | 0.237616 | 0 | 0 | 8.894007 | 19.54216 |
| GO_Biologi | Organelle ( | 4/418 | 0.109197 | 0.237616 | 0 | 0 | 2.269833 | 5.026777 |
| UK_Bioban | Skin colour | 2/180 | 0.184577 | 0.238029 | 0 | 0 | 2.608724 | 4.407929 |
| GeDiPNet_ | Nevus | Jan-34 | 0.137866 | 0.238505 | 0 | 0 | 7.004933 | 13.88007 |
| GeDiPNet_ | Taurodonti | Jan-34 | 0.137866 | 0.238505 | 0 | 0 | 7.004933 | 13.88007 |
| GO_Biologi | Nuclear-Tra | 28-Jan | 0.114983 | 0.238707 | 0 | 0 | 8.564169 | 18.52408 |
| GO_Biologi | Regulation | 28-Jan | 0.114983 | 0.238707 | 0 | 0 | 8.564169 | 18.52408 |
| GO_Biologi | Hematopoi | 28-Jan | 0.114983 | 0.238707 | 0 | 0 | 8.564169 | 18.52408 |
| GO_Biologi | Regulation | 28-Jan | 0.114983 | 0.238707 | 0 | 0 | 8.564169 | 18.52408 |
| GO_Biologi | Regulation | 28-Jan | 0.114983 | 0.238707 | 0 | 0 | 8.564169 | 18.52408 |
| GO_Biologi | Regulation | 28-Jan | 0.114983 | 0.238707 | 0 | 0 | 8.564169 | 18.52408 |
| GO_Biologi | Limb Devel | 28-Jan | 0.114983 | 0.238707 | 0 | 0 | 8.564169 | 18.52408 |
| GO_Biologi | Regulation | 28-Jan | 0.114983 | 0.238707 | 0 | 0 | 8.564169 | 18.52408 |
| GO_Biologi | Negative R | 28-Jan | 0.114983 | 0.238707 | 0 | 0 | 8.564169 | 18.52408 |
| GO_Biologi | Circulatory | 2/132 | 0.112716 | 0.238707 | 0 | 0 | 3.580633 | 7.816104 |
| DisGeNET | Dysgermin | 29-Jan | 0.118838 | 0.238952 | 0 | 0 | 8.25789 | 17.58927 |
| DisGeNET | LEOPARD S | 29-Jan | 0.118838 | 0.238952 | 0 | 0 | 8.25789 | 17.58927 |
| DisGeNET | Cholestero | 29-Jan | 0.118838 | 0.238952 | 0 | 0 | 8.25789 | 17.58927 |
| DisGeNET | Collagen Di | 29-Jan | 0.118838 | 0.238952 | 0 | 0 | 8.25789 | 17.58927 |
| DisGeNET | Charcot-Ma | 29-Jan | 0.118838 | 0.238952 | 0 | 0 | 8.25789 | 17.58927 |
| DisGeNET | alpha-1-An | 29-Jan | 0.118838 | 0.238952 | 0 | 0 | 8.25789 | 17.58927 |
| DisGeNET | Salivary du | 29-Jan | 0.118838 | 0.238952 | 0 | 0 | 8.25789 | 17.58927 |
| DisGeNET | Open Bite | 29-Jan | 0.118838 | 0.238952 | 0 | 0 | 8.25789 | 17.58927 |
| DisGeNET | Generalize | 29-Jan | 0.118838 | 0.238952 | 0 | 0 | 8.25789 | 17.58927 |
| DisGeNET | Gall Bladde | 29-Jan | 0.118838 | 0.238952 | 0 | 0 | 8.25789 | 17.58927 |
| DisGeNET | Social disin | 29-Jan | 0.118838 | 0.238952 | 0 | 0 | 8.25789 | 17.58927 |
| DisGeNET | Cortical wh | 29-Jan | 0.118838 | 0.238952 | 0 | 0 | 8.25789 | 17.58927 |
| DisGeNET | Coronary A | 29-Jan | 0.118838 | 0.238952 | 0 | 0 | 8.25789 | 17.58927 |
| DisGeNET | Contiguous | 29-Jan | 0.118838 | 0.238952 | 0 | 0 | 8.25789 | 17.58927 |
| DisGeNET | Acquired C | 29-Jan | 0.118838 | 0.238952 | 0 | 0 | 8.25789 | 17.58927 |
| DisGeNET | HMN (Here | 29-Jan | 0.118838 | 0.238952 | 0 | 0 | 8.25789 | 17.58927 |
| DisGeNET | Left ventric | 29-Jan | 0.118838 | 0.238952 | 0 | 0 | 8.25789 | 17.58927 |
| DisGeNET | Follicular a | 2/136 | 0.118386 | 0.238952 | 0 | 0 | 3.473047 | 7.410799 |
| DisGeNET | Asbestosis | 29-Jan | 0.118838 | 0.238952 | 0 | 0 | 8.25789 | 17.58927 |

| | | | | | | | | |
|---|---|---|---|---|---|---|---|---|
| DisGeNET | Anterior hy | 29-Jan | 0.118838 | 0.238952 | 0 | 0 | 8.25789 | 17.58927 |
| DisGeNET | Multiple er | 29-Jan | 0.118838 | 0.238952 | 0 | 0 | 8.25789 | 17.58927 |
| DisGeNET | Disease of | 29-Jan | 0.118838 | 0.238952 | 0 | 0 | 8.25789 | 17.58927 |
| DisGeNET | Primary pe | 29-Jan | 0.118838 | 0.238952 | 0 | 0 | 8.25789 | 17.58927 |
| DisGeNET | Intermitter | 29-Jan | 0.118838 | 0.238952 | 0 | 0 | 8.25789 | 17.58927 |
| DisGeNET | Cardiac Lyr | 29-Jan | 0.118838 | 0.238952 | 0 | 0 | 8.25789 | 17.58927 |
| DisGeNET | Concave br | 3/275 | 0.118102 | 0.238952 | 0 | 0 | 2.578913 | 5.509085 |
| DisGeNET | Hormone r | 2/135 | 0.116962 | 0.238952 | 0 | 0 | 3.499337 | 7.509252 |
| DisGeNET | Central Cor | 29-Jan | 0.118838 | 0.238952 | 0 | 0 | 8.25789 | 17.58927 |
| DisGeNET | Depressed | 3/275 | 0.118102 | 0.238952 | 0 | 0 | 2.578913 | 5.509085 |
| DisGeNET | Bacterial Ir | 3/276 | 0.119044 | 0.239184 | 0 | 0 | 2.569335 | 5.468219 |
| GWAS_Cat | Concentrat | Jan-49 | 0.192548 | 0.239214 | 0 | 0 | 4.812258 | 7.927759 |
| GWAS_Cat | Medicatior | Jan-49 | 0.192548 | 0.239214 | 0 | 0 | 4.812258 | 7.927759 |
| GWAS_Cat | Triglyceride | Jan-49 | 0.192548 | 0.239214 | 0 | 0 | 4.812258 | 7.927759 |
| GWAS_Cat | Phospholip | Jan-49 | 0.192548 | 0.239214 | 0 | 0 | 4.812258 | 7.927759 |
| GWAS_Cat | Waist-to-hi | Jan-49 | 0.192548 | 0.239214 | 0 | 0 | 4.812258 | 7.927759 |
| GWAS_Cat | Cholesteryl | Jan-49 | 0.192548 | 0.239214 | 0 | 0 | 4.812258 | 7.927759 |
| Rare_Disea | Nijmegen b | Feb-84 | 0.05183 | 0.239479 | 0 | 0 | 5.690387 | 16.84234 |
| Rare_Disea | Cerebral pa | Feb-86 | 0.054043 | 0.239479 | 0 | 0 | 5.554342 | 16.20739 |
| Rare_Disea | Hyperinsul | 13-Jan | 0.055113 | 0.239479 | 0 | 0 | 19.28391 | 55.89185 |
| Rare_Disea | Internal ca | 14-Jan | 0.059226 | 0.239479 | 0 | 0 | 17.79964 | 50.30878 |
| Rare_Disea | Hypobetali | 14-Jan | 0.059226 | 0.239479 | 0 | 0 | 17.79964 | 50.30878 |
| Rare_Disea | Acute myel | 4/335 | 0.05842 | 0.239479 | 0 | 0 | 2.85109 | 8.097363 |
| Rare_Disea | Protein R d | 3/198 | 0.055356 | 0.239479 | 0 | 0 | 3.611355 | 10.45114 |
| Rare_Disea | Guttate ps | 13-Jan | 0.055113 | 0.239479 | 0 | 0 | 19.28391 | 55.89185 |
| Rare_Disea | Caroli disea | 14-Jan | 0.059226 | 0.239479 | 0 | 0 | 17.79964 | 50.30878 |
| Rare_Disea | Kerion cels | 14-Jan | 0.059226 | 0.239479 | 0 | 0 | 17.79964 | 50.30878 |
| Rare_Disea | Multiple er | 14-Jan | 0.059226 | 0.239479 | 0 | 0 | 17.79964 | 50.30878 |
| Rare_Disea | Pineoblast | 13-Jan | 0.055113 | 0.239479 | 0 | 0 | 19.28391 | 55.89185 |
| Rare_Disea | Pendred sy | 14-Jan | 0.059226 | 0.239479 | 0 | 0 | 17.79964 | 50.30878 |
| Rare_Disea | Norum dise | 14-Jan | 0.059226 | 0.239479 | 0 | 0 | 17.79964 | 50.30878 |
| Rare_Disea | Coffin synd | 14-Jan | 0.059226 | 0.239479 | 0 | 0 | 17.79964 | 50.30878 |
| Rare_Disea | Mycetoma | 14-Jan | 0.059226 | 0.239479 | 0 | 0 | 17.79964 | 50.30878 |
| Rare_Disea | Canavan di | 13-Jan | 0.055113 | 0.239479 | 0 | 0 | 19.28391 | 55.89185 |
| Rare_Disea | Athetosis | 13-Jan | 0.055113 | 0.239479 | 0 | 0 | 19.28391 | 55.89185 |
| Rare_Disea | North Caro | 13-Jan | 0.055113 | 0.239479 | 0 | 0 | 19.28391 | 55.89185 |
| Rare_Disea | Multiple re | 13-Jan | 0.055113 | 0.239479 | 0 | 0 | 19.28391 | 55.89185 |
| Rare_Disea | EAF | 14-Jan | 0.059226 | 0.239479 | 0 | 0 | 17.79964 | 50.30878 |
| Rare_Disea | Pleuropulm | 14-Jan | 0.059226 | 0.239479 | 0 | 0 | 17.79964 | 50.30878 |
| Rare_Disea | Afibrinoger | 14-Jan | 0.059226 | 0.239479 | 0 | 0 | 17.79964 | 50.30878 |
| Rare_Disea | Schwannor | 14-Jan | 0.059226 | 0.239479 | 0 | 0 | 17.79964 | 50.30878 |
| GO_Biologi | RNA Biosyr | 29-Jan | 0.118838 | 0.240384 | 0 | 0 | 8.25789 | 17.58927 |
| GO_Biologi | Regulation | 29-Jan | 0.118838 | 0.240384 | 0 | 0 | 8.25789 | 17.58927 |
| GO_Biologi | Phosphatic | 29-Jan | 0.118838 | 0.240384 | 0 | 0 | 8.25789 | 17.58927 |
| GO_Biologi | Modified A | 29-Jan | 0.118838 | 0.240384 | 0 | 0 | 8.25789 | 17.58927 |

| | | | | | | | | |
|---|---|---|---|---|---|---|---|---|
| GO_Biologi | Establishm | 29-Jan | 0.118838 | 0.240384 | 0 | 0 | 8.25789 | 17.58927 |
| GO_Biologi | Positive Re | 29-Jan | 0.118838 | 0.240384 | 0 | 0 | 8.25789 | 17.58927 |
| GO_Biologi | Regulation | 29-Jan | 0.118838 | 0.240384 | 0 | 0 | 8.25789 | 17.58927 |
| GO_Biologi | Maintenan | 29-Jan | 0.118838 | 0.240384 | 0 | 0 | 8.25789 | 17.58927 |
| GO_Biologi | Non-Memb | 2/135 | 0.116962 | 0.240384 | 0 | 0 | 3.499337 | 7.509252 |
| CellMarker | Multipoter | Jan-45 | 0.178311 | 0.240676 | 0 | 0 | 5.250793 | 9.053559 |
| CellMarker | Macrophag | Jan-45 | 0.178311 | 0.240676 | 0 | 0 | 5.250793 | 9.053559 |
| DisGeNET | Depressed | 3/277 | 0.119989 | 0.240897 | 0 | 0 | 2.559828 | 5.427752 |
| MAGMA_D | Palbociclib | Jan-47 | 0.18546 | 0.241826 | 0 | 0 | 5.021992 | 8.46163 |
| MAGMA_D | Vorinostat | Jan-47 | 0.18546 | 0.241826 | 0 | 0 | 5.021992 | 8.46163 |
| MAGMA_D | Verapamil | Jan-47 | 0.18546 | 0.241826 | 0 | 0 | 5.021992 | 8.46163 |
| GeDiPNet_ | Cubitus Val | Jan-35 | 0.141623 | 0.24213 | 0 | 0 | 6.798564 | 13.28839 |
| GeDiPNet_ | Hypopigme | Jan-35 | 0.141623 | 0.24213 | 0 | 0 | 6.798564 | 13.28839 |
| GeDiPNet_ | Aplasia Cut | Jan-35 | 0.141623 | 0.24213 | 0 | 0 | 6.798564 | 13.28839 |
| GeDiPNet_ | Acrocephal | Jan-35 | 0.141623 | 0.24213 | 0 | 0 | 6.798564 | 13.28839 |
| GWAS_Cat | White Mat | Jan-50 | 0.196069 | 0.242355 | 0 | 0 | 4.713811 | 7.680157 |
| GWAS_Cat | Total Conc | Jan-50 | 0.196069 | 0.242355 | 0 | 0 | 4.713811 | 7.680157 |
| GWAS_Cat | Triglyceride | Jan-50 | 0.196069 | 0.242355 | 0 | 0 | 4.713811 | 7.680157 |
| GWAS_Cat | Glucose Ho | Jan-50 | 0.196069 | 0.242355 | 0 | 0 | 4.713811 | 7.680157 |
| GWAS_Cat | Total Lipid | Jan-50 | 0.196069 | 0.242355 | 0 | 0 | 4.713811 | 7.680157 |
| Human_Ph | Chorioretir | Jan-50 | 0.196069 | 0.24239 | 0 | 0 | 4.713811 | 7.680157 |
| DisGeNET | familial atr | 30-Jan | 0.122676 | 0.242942 | 0 | 0 | 7.972735 | 16.72843 |
| DisGeNET | Delirium, D | 30-Jan | 0.122676 | 0.242942 | 0 | 0 | 7.972735 | 16.72843 |
| DisGeNET | Carney Cor | 30-Jan | 0.122676 | 0.242942 | 0 | 0 | 7.972735 | 16.72843 |
| DisGeNET | Fasting Hyp | 30-Jan | 0.122676 | 0.242942 | 0 | 0 | 7.972735 | 16.72843 |
| DisGeNET | Serum chol | 30-Jan | 0.122676 | 0.242942 | 0 | 0 | 7.972735 | 16.72843 |
| DisGeNET | Stomach D | 30-Jan | 0.122676 | 0.242942 | 0 | 0 | 7.972735 | 16.72843 |
| DisGeNET | Horseshoe | 30-Jan | 0.122676 | 0.242942 | 0 | 0 | 7.972735 | 16.72843 |
| DisGeNET | TUBEROUS | 30-Jan | 0.122676 | 0.242942 | 0 | 0 | 7.972735 | 16.72843 |
| DisGeNET | Lymphader | 30-Jan | 0.122676 | 0.242942 | 0 | 0 | 7.972735 | 16.72843 |
| DisGeNET | Overlap syı | 30-Jan | 0.122676 | 0.242942 | 0 | 0 | 7.972735 | 16.72843 |
| DisGeNET | Collecting I | 2/138 | 0.121248 | 0.242942 | 0 | 0 | 3.421626 | 7.219351 |
| DisGeNET | nervous sy: | 4/436 | 0.122206 | 0.242942 | 0 | 0 | 2.173249 | 4.568268 |
| DisGeNET | Dementia | 4/436 | 0.122206 | 0.242942 | 0 | 0 | 2.173249 | 4.568268 |
| DisGeNET | Carcinoma, | 30-Jan | 0.122676 | 0.242942 | 0 | 0 | 7.972735 | 16.72843 |
| DisGeNET | Noncancer | 30-Jan | 0.122676 | 0.242942 | 0 | 0 | 7.972735 | 16.72843 |
| DisGeNET | Cornelia De | 30-Jan | 0.122676 | 0.242942 | 0 | 0 | 7.972735 | 16.72843 |
| DisGeNET | Persistent a | 30-Jan | 0.122676 | 0.242942 | 0 | 0 | 7.972735 | 16.72843 |
| DisGeNET | Hamartom | 2/138 | 0.121248 | 0.242942 | 0 | 0 | 3.421626 | 7.219351 |
| DisGeNET | Kidney Fail | 4/437 | 0.122948 | 0.242974 | 0 | 0 | 2.168118 | 4.544364 |
| DisGeNET | Byzanthine | 3/280 | 0.122839 | 0.242974 | 0 | 0 | 2.531717 | 5.308703 |
| DisGeNET | Colonic Ne | 6/778 | 0.122971 | 0.242974 | 0 | 0 | 1.836596 | 3.849154 |
| GO_Biologi | Regulation | 30-Jan | 0.122676 | 0.243297 | 0 | 0 | 7.972735 | 16.72843 |
| GO_Biologi | Negative R | 30-Jan | 0.122676 | 0.243297 | 0 | 0 | 7.972735 | 16.72843 |
| GO_Biologi | Cytoplasmi | 30-Jan | 0.122676 | 0.243297 | 0 | 0 | 7.972735 | 16.72843 |

| | | | | | | | | |
|---|---|---|---|---|---|---|---|---|
| GO_Biologi | Cardiac Mu | 30-Jan | 0.122676 | 0.243297 | 0 | 0 | 7.972735 | 16.72843 |
| GO_Biologi | DNA Geom | 30-Jan | 0.122676 | 0.243297 | 0 | 0 | 7.972735 | 16.72843 |
| GO_Biologi | Negative R | 30-Jan | 0.122676 | 0.243297 | 0 | 0 | 7.972735 | 16.72843 |
| GO_Biologi | Negative R | 30-Jan | 0.122676 | 0.243297 | 0 | 0 | 7.972735 | 16.72843 |
| CellMarker | Type IB Spi | Jan-46 | 0.181893 | 0.244308 | 0 | 0 | 5.13385 | 8.749803 |
| DisGeNET | Sebaceous | 31-Jan | 0.126498 | 0.244415 | 0 | 0 | 7.706589 | 15.93356 |
| DisGeNET | Deformity | 31-Jan | 0.126498 | 0.244415 | 0 | 0 | 7.706589 | 15.93356 |
| DisGeNET | Adenocarci | 31-Jan | 0.126498 | 0.244415 | 0 | 0 | 7.706589 | 15.93356 |
| DisGeNET | Protein C D | 31-Jan | 0.126498 | 0.244415 | 0 | 0 | 7.706589 | 15.93356 |
| DisGeNET | Slow-growi | 31-Jan | 0.126498 | 0.244415 | 0 | 0 | 7.706589 | 15.93356 |
| DisGeNET | Major depr | 31-Jan | 0.126498 | 0.244415 | 0 | 0 | 7.706589 | 15.93356 |
| DisGeNET | Cholangitis | 2/140 | 0.124127 | 0.244415 | 0 | 0 | 3.371697 | 7.034881 |
| DisGeNET | HIV Encepł | 31-Jan | 0.126498 | 0.244415 | 0 | 0 | 7.706589 | 15.93356 |
| DisGeNET | Onset of ly | 31-Jan | 0.126498 | 0.244415 | 0 | 0 | 7.706589 | 15.93356 |
| DisGeNET | Terminal tr | 31-Jan | 0.126498 | 0.244415 | 0 | 0 | 7.706589 | 15.93356 |
| DisGeNET | Plasma Cel | 31-Jan | 0.126498 | 0.244415 | 0 | 0 | 7.706589 | 15.93356 |
| DisGeNET | Lymphatic | 31-Jan | 0.126498 | 0.244415 | 0 | 0 | 7.706589 | 15.93356 |
| DisGeNET | Bright Dise | 31-Jan | 0.126498 | 0.244415 | 0 | 0 | 7.706589 | 15.93356 |
| DisGeNET | Experimen | 3/282 | 0.124754 | 0.244415 | 0 | 0 | 2.513313 | 5.231246 |
| DisGeNET | Opportunis | 31-Jan | 0.126498 | 0.244415 | 0 | 0 | 7.706589 | 15.93356 |
| DisGeNET | Liver and Ir | 5/607 | 0.124867 | 0.244415 | 0 | 0 | 1.95598 | 4.069431 |
| DisGeNET | Hyperdiplo | 31-Jan | 0.126498 | 0.244415 | 0 | 0 | 7.706589 | 15.93356 |
| DisGeNET | Antevertec | 3/283 | 0.125715 | 0.244415 | 0 | 0 | 2.504209 | 5.193076 |
| DisGeNET | Substance | 2/140 | 0.124127 | 0.244415 | 0 | 0 | 3.371697 | 7.034881 |
| DisGeNET | Invasive as | 31-Jan | 0.126498 | 0.244415 | 0 | 0 | 7.706589 | 15.93356 |
| DisGeNET | AIDS-Assoc | 31-Jan | 0.126498 | 0.244415 | 0 | 0 | 7.706589 | 15.93356 |
| DisGeNET | Atheroscle | 31-Jan | 0.126498 | 0.244415 | 0 | 0 | 7.706589 | 15.93356 |
| DisGeNET | Alopecia, N | 31-Jan | 0.126498 | 0.244415 | 0 | 0 | 7.706589 | 15.93356 |
| DisGeNET | Reduced nu | 31-Jan | 0.126498 | 0.244415 | 0 | 0 | 7.706589 | 15.93356 |
| DisGeNET | Hepatitis C | 6/783 | 0.125699 | 0.244415 | 0 | 0 | 1.8243 | 3.783352 |
| DisGeNET | Thyroid Dis | 2/140 | 0.124127 | 0.244415 | 0 | 0 | 3.371697 | 7.034881 |
| DisGeNET | Histiocytic | 31-Jan | 0.126498 | 0.244415 | 0 | 0 | 7.706589 | 15.93356 |
| DisGeNET | Refractory | 31-Jan | 0.126498 | 0.244415 | 0 | 0 | 7.706589 | 15.93356 |
| DisGeNET | Childhood | 31-Jan | 0.126498 | 0.244415 | 0 | 0 | 7.706589 | 15.93356 |
| DisGeNET | Anaplasia | 3/283 | 0.125715 | 0.244415 | 0 | 0 | 2.504209 | 5.193076 |
| GWAS_Cat | Total Lipid | Jan-51 | 0.199575 | 0.245198 | 0 | 0 | 4.619302 | 7.444308 |
| GWAS_Cat | Multisite C | Jan-51 | 0.199575 | 0.245198 | 0 | 0 | 4.619302 | 7.444308 |
| GWAS_Cat | Concentrat | Jan-51 | 0.199575 | 0.245198 | 0 | 0 | 4.619302 | 7.444308 |
| GWAS_Cat | Total Choli | Jan-51 | 0.199575 | 0.245198 | 0 | 0 | 4.619302 | 7.444308 |
| GWAS_Cat | Nonalcoho | Jan-51 | 0.199575 | 0.245198 | 0 | 0 | 4.619302 | 7.444308 |
| GWAS_Cat | VLDL Chole | Jan-51 | 0.199575 | 0.245198 | 0 | 0 | 4.619302 | 7.444308 |
| Human_Ph | Teleangiec | Jan-51 | 0.199575 | 0.245928 | 0 | 0 | 4.619302 | 7.444308 |
| GeDiPNet_ | Nystagmus | 6/818 | 0.145621 | 0.246079 | 0 | 0 | 1.742474 | 3.357306 |
| GeDiPNet_ | Dolichocep | 2/154 | 0.144717 | 0.246079 | 0 | 0 | 3.058978 | 5.912923 |
| GeDiPNet_ | Papillary A | Jan-36 | 0.145363 | 0.246079 | 0 | 0 | 6.603987 | 12.73591 |

| | | | | | | | | |
|---|---|---|---|---|---|---|---|---|
| GeDiPNet_ | Necrotizing | Jan-36 | 0.145363 | 0.246079 | 0 | 0 | 6.603987 | 12.73591 |
| UK_Bioban | Monocyte | 11/1881 | 0.19262 | 0.246173 | 0 | 0 | 1.396517 | 2.300116 |
| UK_Bioban | Unsalted n | Jan-49 | 0.192548 | 0.246173 | 0 | 0 | 4.812258 | 7.927759 |
| DisGeNET | polyps | 3/285 | 0.127646 | 0.246267 | 0 | 0 | 2.486196 | 5.117829 |
| DisGeNET | Congenital | 3/285 | 0.127646 | 0.246267 | 0 | 0 | 2.486196 | 5.117829 |
| Rare_Disea | Dementia f | 15-Jan | 0.063321 | 0.246655 | 0 | 0 | 16.52741 | 45.60793 |
| Rare_Disea | Deafness c | 15-Jan | 0.063321 | 0.246655 | 0 | 0 | 16.52741 | 45.60793 |
| Rare_Disea | Twin twin t | 15-Jan | 0.063321 | 0.246655 | 0 | 0 | 16.52741 | 45.60793 |
| Rare_Disea | Multiple cc | Feb-94 | 0.063221 | 0.246655 | 0 | 0 | 5.069309 | 13.99697 |
| Rare_Disea | Hereditary | 15-Jan | 0.063321 | 0.246655 | 0 | 0 | 16.52741 | 45.60793 |
| Rare_Disea | Congenital | 15-Jan | 0.063321 | 0.246655 | 0 | 0 | 16.52741 | 45.60793 |
| Rare_Disea | Fibromuscu | 15-Jan | 0.063321 | 0.246655 | 0 | 0 | 16.52741 | 45.60793 |
| GO_Biologi | Modulation | 31-Jan | 0.126498 | 0.246742 | 0 | 0 | 7.706589 | 15.93356 |
| GO_Biologi | Negative R | 31-Jan | 0.126498 | 0.246742 | 0 | 0 | 7.706589 | 15.93356 |
| GO_Biologi | Negative R | 31-Jan | 0.126498 | 0.246742 | 0 | 0 | 7.706589 | 15.93356 |
| GO_Biologi | Positive Re | 31-Jan | 0.126498 | 0.246742 | 0 | 0 | 7.706589 | 15.93356 |
| GO_Biologi | Positive Re | 31-Jan | 0.126498 | 0.246742 | 0 | 0 | 7.706589 | 15.93356 |
| GO_Biologi | Regulation | 31-Jan | 0.126498 | 0.246742 | 0 | 0 | 7.706589 | 15.93356 |
| UK_Bioban | Right heel | 2/186 | 0.193998 | 0.246828 | 0 | 0 | 2.52289 | 4.137304 |
| GTEx_Tissu | GTEX-OIZH | 15/1628 | 0.004224 | 0.247093 | 0 | 0 | 2.363608 | 12.92185 |
| GTEx_Tissu | GTEX-P4PP | 8/593 | 0.004271 | 0.247093 | 0 | 0 | 3.345754 | 18.25448 |
| GTEx_Tissu | GTEX-R55G | 10/868 | 0.00439 | 0.247313 | 0 | 0 | 2.884237 | 15.65708 |
| GTEx_Tissu | GTEX-WFO | 8/597 | 0.004445 | 0.247313 | 0 | 0 | 3.322344 | 17.9935 |
| GTEx_Tissu | GTEX-SIU7- | 8/599 | 0.004535 | 0.247528 | 0 | 0 | 3.310758 | 17.86481 |
| Human_Ph | Gynecomas | Jan-52 | 0.203066 | 0.247547 | 0 | 0 | 4.5285 | 7.219451 |
| Human_Ph | Brachydact | 2/192 | 0.203481 | 0.247547 | 0 | 0 | 2.442477 | 3.888868 |
| Human_Ph | Hydroceph | 2/191 | 0.201897 | 0.247547 | 0 | 0 | 2.455524 | 3.928837 |
| Human_Ph | Glomerulo | Jan-52 | 0.203066 | 0.247547 | 0 | 0 | 4.5285 | 7.219451 |
| DisGeNET | Increased i | Jan-32 | 0.130304 | 0.247928 | 0 | 0 | 7.457614 | 15.19775 |
| DisGeNET | Verrucous | Jan-32 | 0.130304 | 0.247928 | 0 | 0 | 7.457614 | 15.19775 |
| DisGeNET | Lipomatosi | Jan-32 | 0.130304 | 0.247928 | 0 | 0 | 7.457614 | 15.19775 |
| DisGeNET | Aortic valve | Jan-32 | 0.130304 | 0.247928 | 0 | 0 | 7.457614 | 15.19775 |
| DisGeNET | Cardiovasc | Jan-32 | 0.130304 | 0.247928 | 0 | 0 | 7.457614 | 15.19775 |
| DisGeNET | Juvenile pc | Jan-32 | 0.130304 | 0.247928 | 0 | 0 | 7.457614 | 15.19775 |
| DisGeNET | African Try | Jan-32 | 0.130304 | 0.247928 | 0 | 0 | 7.457614 | 15.19775 |
| DisGeNET | Cataplexy | Jan-32 | 0.130304 | 0.247928 | 0 | 0 | 7.457614 | 15.19775 |
| DisGeNET | Lipid Metal | Jan-32 | 0.130304 | 0.247928 | 0 | 0 | 7.457614 | 15.19775 |
| DisGeNET | Anhedonia | Jan-32 | 0.130304 | 0.247928 | 0 | 0 | 7.457614 | 15.19775 |
| DisGeNET | Fibrillation | Jan-32 | 0.130304 | 0.247928 | 0 | 0 | 7.457614 | 15.19775 |
| DisGeNET | Columnar C | Jan-32 | 0.130304 | 0.247928 | 0 | 0 | 7.457614 | 15.19775 |
| DisGeNET | Bowen's Di | Jan-32 | 0.130304 | 0.247928 | 0 | 0 | 7.457614 | 15.19775 |
| DisGeNET | Ophthalmc | 2/144 | 0.129934 | 0.247928 | 0 | 0 | 3.276056 | 6.685543 |
| DisGeNET | Myopathie | Jan-32 | 0.130304 | 0.247928 | 0 | 0 | 7.457614 | 15.19775 |
| DisGeNET | Amblyopia | Jan-32 | 0.130304 | 0.247928 | 0 | 0 | 7.457614 | 15.19775 |
| DisGeNET | HIV encepl | Jan-32 | 0.130304 | 0.247928 | 0 | 0 | 7.457614 | 15.19775 |

| | | | | | | | | |
|---|---|---|---|---|---|---|---|---|
| DisGeNET | Squamous | Jan-32 | 0.130304 | 0.247928 | 0 | 0 | 7.457614 | 15.19775 |
| DisGeNET | Increased I | Jan-32 | 0.130304 | 0.247928 | 0 | 0 | 7.457614 | 15.19775 |
| GWAS_Cat | Free Chole: | Jan-52 | 0.203066 | 0.248237 | 0 | 0 | 4.5285 | 7.219451 |
| GWAS_Cat | Metabolic ' | Jan-52 | 0.203066 | 0.248237 | 0 | 0 | 4.5285 | 7.219451 |
| GWAS_Cat | Total Lipid | Jan-52 | 0.203066 | 0.248237 | 0 | 0 | 4.5285 | 7.219451 |
| GWAS_Cat | Total Lipid | Jan-52 | 0.203066 | 0.248237 | 0 | 0 | 4.5285 | 7.219451 |
| GWAS_Cat | Polyunsatu | Jan-52 | 0.203066 | 0.248237 | 0 | 0 | 4.5285 | 7.219451 |
| dbGaP | Body Weig | Jan-34 | 0.137866 | 0.24848 | 0 | 0 | 7.004933 | 13.88007 |
| dbGaP | Thyrotropi | Jan-33 | 0.134093 | 0.24848 | 0 | 0 | 7.224201 | 14.515 |
| dbGaP | HIV-1 | Jan-37 | 0.149088 | 0.24848 | 0 | 0 | 6.42022 | 12.21909 |
| dbGaP | Inflammat | Jan-35 | 0.141623 | 0.24848 | 0 | 0 | 6.798564 | 13.28839 |
| dbGaP | Monocytes | Jan-37 | 0.149088 | 0.24848 | 0 | 0 | 6.42022 | 12.21909 |
| dbGaP | Sleep | Jan-35 | 0.141623 | 0.24848 | 0 | 0 | 6.798564 | 13.28839 |
| dbGaP | Uric Acid | Jan-37 | 0.149088 | 0.24848 | 0 | 0 | 6.42022 | 12.21909 |
| dbGaP | Inflammati | 27-Jan | 0.111111 | 0.24848 | 0 | 0 | 8.894007 | 19.54216 |
| dbGaP | Body Comp | Jan-36 | 0.145363 | 0.24848 | 0 | 0 | 6.603987 | 12.73591 |
| dbGaP | Longevity | Jan-32 | 0.130304 | 0.24848 | 0 | 0 | 7.457614 | 15.19775 |
| dbGaP | Respiratory | 2/151 | 0.140245 | 0.24848 | 0 | 0 | 3.121042 | 6.130869 |
| dbGaP | Aorta | Jan-34 | 0.137866 | 0.24848 | 0 | 0 | 7.004933 | 13.88007 |
| GWAS_Cat | Crohn's Dis | 2/192 | 0.203481 | 0.248495 | 0 | 0 | 2.442477 | 3.888868 |
| GO_Biologi | Negative R | Jan-32 | 0.130304 | 0.248699 | 0 | 0 | 7.457614 | 15.19775 |
| GO_Biologi | Regulation | Dec-22 | 0.128161 | 0.248699 | 0 | 0 | 1.508105 | 3.09835 |
| GO_Biologi | Regulation | Jan-32 | 0.130304 | 0.248699 | 0 | 0 | 7.457614 | 15.19775 |
| GO_Biologi | Protein Tra | Jan-32 | 0.130304 | 0.248699 | 0 | 0 | 7.457614 | 15.19775 |
| GO_Biologi | Regulation | Jan-32 | 0.130304 | 0.248699 | 0 | 0 | 7.457614 | 15.19775 |
| GO_Biologi | Histone Lys | Jan-32 | 0.130304 | 0.248699 | 0 | 0 | 7.457614 | 15.19775 |
| GO_Biologi | Phosphatic | Jan-32 | 0.130304 | 0.248699 | 0 | 0 | 7.457614 | 15.19775 |
| GO_Biologi | Negative R | Jan-32 | 0.130304 | 0.248699 | 0 | 0 | 7.457614 | 15.19775 |
| PhenGenI_ | Fractures, I | 14-Jan | 0.059226 | 0.24875 | 0 | 0 | 17.79964 | 50.30878 |
| GWAS_Cat | Cholestero | Jan-53 | 0.206541 | 0.248993 | 0 | 0 | 4.44119 | 7.004886 |
| GWAS_Cat | Omega-6 F | Jan-53 | 0.206541 | 0.248993 | 0 | 0 | 4.44119 | 7.004886 |
| GWAS_Cat | Free Chole: | Jan-53 | 0.206541 | 0.248993 | 0 | 0 | 4.44119 | 7.004886 |
| GWAS_Cat | Body Mass | Jan-53 | 0.206541 | 0.248993 | 0 | 0 | 4.44119 | 7.004886 |
| GWAS_Cat | Adiponecti | Jan-53 | 0.206541 | 0.248993 | 0 | 0 | 4.44119 | 7.004886 |
| GWAS_Cat | Concentrat | Jan-53 | 0.206541 | 0.248993 | 0 | 0 | 4.44119 | 7.004886 |
| GWAS_Cat | LDL Choles | Jan-53 | 0.206541 | 0.248993 | 0 | 0 | 4.44119 | 7.004886 |
| GWAS_Cat | Cortical Th | Jan-53 | 0.206541 | 0.248993 | 0 | 0 | 4.44119 | 7.004886 |
| GWAS_Cat | Cholesteryl | Jan-53 | 0.206541 | 0.248993 | 0 | 0 | 4.44119 | 7.004886 |
| GWAS_Cat | Vertical Cu | 2/193 | 0.205067 | 0.248993 | 0 | 0 | 2.429566 | 3.84945 |
| GWAS_Cat | Total Biliru | 2/193 | 0.205067 | 0.248993 | 0 | 0 | 2.429566 | 3.84945 |
| GWAS_Cat | Phospholip | Jan-53 | 0.206541 | 0.248993 | 0 | 0 | 4.44119 | 7.004886 |
| GWAS_Cat | Triglyceride | Jan-53 | 0.206541 | 0.248993 | 0 | 0 | 4.44119 | 7.004886 |
| GeDiPNet_ | Endometri | Jan-37 | 0.149088 | 0.249049 | 0 | 0 | 6.42022 | 12.21909 |
| GeDiPNet_ | Intestinal P | Jan-37 | 0.149088 | 0.249049 | 0 | 0 | 6.42022 | 12.21909 |
| GeDiPNet_ | Breast Hyp | Jan-37 | 0.149088 | 0.249049 | 0 | 0 | 6.42022 | 12.21909 |

| | | | | | | | | |
|---|---|---|---|---|---|---|---|---|
| GeDiPNet_ | Adenoma | Jan-37 | 0.149088 | 0.249049 | 0 | 0 | 6.42022 | 12.21909 |
| GeDiPNet_ | Hydroceph | 3/307 | 0.149562 | 0.249127 | 0 | 0 | 2.303689 | 4.377116 |
| DisGeNET | Thrombocy | 2/145 | 0.131396 | 0.249642 | 0 | 0 | 3.252982 | 6.602064 |
| DisGeNET | Frontotem | 2/145 | 0.131396 | 0.249642 | 0 | 0 | 3.252982 | 6.602064 |
| GO_Biologi | Regulation | Jan-33 | 0.134093 | 0.249672 | 0 | 0 | 7.224201 | 14.515 |
| GO_Biologi | Negative R | Jan-33 | 0.134093 | 0.249672 | 0 | 0 | 7.224201 | 14.515 |
| GO_Biologi | Negative R | 2/147 | 0.134331 | 0.249672 | 0 | 0 | 3.207789 | 6.43948 |
| GO_Biologi | Negative R | 2/146 | 0.132861 | 0.249672 | 0 | 0 | 3.230229 | 6.520055 |
| GO_Biologi | Phosphatic | Jan-33 | 0.134093 | 0.249672 | 0 | 0 | 7.224201 | 14.515 |
| GO_Biologi | Positive Re | Jan-33 | 0.134093 | 0.249672 | 0 | 0 | 7.224201 | 14.515 |
| GO_Biologi | Positive Re | Jan-33 | 0.134093 | 0.249672 | 0 | 0 | 7.224201 | 14.515 |
| GO_Biologi | Regulation | 4/452 | 0.134305 | 0.249672 | 0 | 0 | 2.093911 | 4.203817 |
| GO_Biologi | Monoubiqu | Jan-33 | 0.134093 | 0.249672 | 0 | 0 | 7.224201 | 14.515 |
| GO_Biologi | Negative R | Jan-33 | 0.134093 | 0.249672 | 0 | 0 | 7.224201 | 14.515 |
| Jensen_TIS | Nasophary | 59/12374 | 0.150446 | 0.249709 | 0 | 0 | 1.300046 | 2.462486 |
| Jensen_TIS | Notochord | Jan-38 | 0.152796 | 0.249748 | 0 | 0 | 6.246386 | 11.73478 |
| Jensen_TIS | Middle ear | Jan-38 | 0.152796 | 0.249748 | 0 | 0 | 6.246386 | 11.73478 |
| Jensen_TIS | Mesenchyr | 2/159 | 0.152237 | 0.249748 | 0 | 0 | 2.960809 | 5.573176 |
| DisGeNET | Precursor ( | 5/618 | 0.131846 | 0.250317 | 0 | 0 | 1.919787 | 3.889717 |
| GeDiPNet_ | Nyctalopia | 2/158 | 0.150727 | 0.250353 | 0 | 0 | 2.97994 | 5.638896 |
| Human_Ph | Muscular h | Jan-53 | 0.206541 | 0.250472 | 0 | 0 | 4.44119 | 7.004886 |
| GWAS_Cat | Concentrat | Jan-54 | 0.210002 | 0.250933 | 0 | 0 | 4.357174 | 6.79997 |
| GWAS_Cat | BMI (Adjus | Jan-54 | 0.210002 | 0.250933 | 0 | 0 | 4.357174 | 6.79997 |
| GWAS_Cat | Cholestero | Jan-54 | 0.210002 | 0.250933 | 0 | 0 | 4.357174 | 6.79997 |
| GWAS_Cat | Platelet-to- | 2/196 | 0.209833 | 0.250933 | 0 | 0 | 2.391631 | 3.734399 |
| GWAS_Cat | Blood Prote | Jan-54 | 0.210002 | 0.250933 | 0 | 0 | 4.357174 | 6.79997 |
| GWAS_Cat | Inflammato | 2/196 | 0.209833 | 0.250933 | 0 | 0 | 2.391631 | 3.734399 |
| GWAS_Cat | Triglyceride | Jan-54 | 0.210002 | 0.250933 | 0 | 0 | 4.357174 | 6.79997 |
| GWAS_Cat | Walking Pa | Jan-54 | 0.210002 | 0.250933 | 0 | 0 | 4.357174 | 6.79997 |
| GWAS_Cat | Concentrat | Jan-54 | 0.210002 | 0.250933 | 0 | 0 | 4.357174 | 6.79997 |
| DisGeNET | Acute biph | Jan-33 | 0.134093 | 0.251211 | 0 | 0 | 7.224201 | 14.515 |
| DisGeNET | X-Linked Co | Jan-33 | 0.134093 | 0.251211 | 0 | 0 | 7.224201 | 14.515 |
| DisGeNET | High altituc | Jan-33 | 0.134093 | 0.251211 | 0 | 0 | 7.224201 | 14.515 |
| DisGeNET | Mild Menta | 2/147 | 0.134331 | 0.251211 | 0 | 0 | 3.207789 | 6.43948 |
| DisGeNET | Chromosor | Jan-33 | 0.134093 | 0.251211 | 0 | 0 | 7.224201 | 14.515 |
| DisGeNET | Severe visu | Jan-33 | 0.134093 | 0.251211 | 0 | 0 | 7.224201 | 14.515 |
| DisGeNET | Depressive | 2/147 | 0.134331 | 0.251211 | 0 | 0 | 3.207789 | 6.43948 |
| DisGeNET | Albinism, C | Jan-33 | 0.134093 | 0.251211 | 0 | 0 | 7.224201 | 14.515 |
| DisGeNET | Gerstmann | Jan-33 | 0.134093 | 0.251211 | 0 | 0 | 7.224201 | 14.515 |
| DisGeNET | Soluble Inte | Jan-33 | 0.134093 | 0.251211 | 0 | 0 | 7.224201 | 14.515 |
| DisGeNET | Monosomy | 2/147 | 0.134331 | 0.251211 | 0 | 0 | 3.207789 | 6.43948 |
| DisGeNET | Agitation | Jan-33 | 0.134093 | 0.251211 | 0 | 0 | 7.224201 | 14.515 |
| DisGeNET | Nephritis, I | Jan-33 | 0.134093 | 0.251211 | 0 | 0 | 7.224201 | 14.515 |
| DisGeNET | Hypoplasia | Jan-33 | 0.134093 | 0.251211 | 0 | 0 | 7.224201 | 14.515 |
| DisGeNET | Refractory | Jan-33 | 0.134093 | 0.251211 | 0 | 0 | 7.224201 | 14.515 |

| | | | | | | | | |
|---|---|---|---|---|---|---|---|---|
| DisGeNET | Spastic | Jan-33 | 0.134093 | 0.251211 | 0 | 0 | 7.224201 | 14.515 |
| DisGeNET | Leukocytos | Jan-33 | 0.134093 | 0.251211 | 0 | 0 | 7.224201 | 14.515 |
| DisGeNET | Atrioventri | Jan-33 | 0.134093 | 0.251211 | 0 | 0 | 7.224201 | 14.515 |
| DisGeNET | Chronic Air | Jan-33 | 0.134093 | 0.251211 | 0 | 0 | 7.224201 | 14.515 |
| DisGeNET | Polydactyly | 2/147 | 0.134331 | 0.251211 | 0 | 0 | 3.207789 | 6.43948 |
| DisGeNET | Glaucoma, | 2/147 | 0.134331 | 0.251211 | 0 | 0 | 3.207789 | 6.43948 |
| GO_Biologi | Axon Guida | 2/149 | 0.13728 | 0.251633 | 0 | 0 | 3.163826 | 6.282501 |
| GO_Biologi | Cellular Re: | Jan-34 | 0.137866 | 0.251633 | 0 | 0 | 7.004933 | 13.88007 |
| GO_Biologi | Regulation | Jan-34 | 0.137866 | 0.251633 | 0 | 0 | 7.004933 | 13.88007 |
| GO_Biologi | Establishm | Jan-34 | 0.137866 | 0.251633 | 0 | 0 | 7.004933 | 13.88007 |
| GO_Biologi | Negative R | 2/149 | 0.13728 | 0.251633 | 0 | 0 | 3.163826 | 6.282501 |
| GO_Biologi | Regulation | Jan-34 | 0.137866 | 0.251633 | 0 | 0 | 7.004933 | 13.88007 |
| GO_Biologi | Histone De | Jan-34 | 0.137866 | 0.251633 | 0 | 0 | 7.004933 | 13.88007 |
| Jensen_TIS | Blood plate | 29/5583 | 0.156364 | 0.251946 | 0 | 0 | 1.292672 | 2.398639 |
| Jensen_TIS | Colorectum | Jan-39 | 0.156488 | 0.251946 | 0 | 0 | 6.081701 | 11.28018 |
| Jensen_TIS | Satellite ce | Jan-39 | 0.156488 | 0.251946 | 0 | 0 | 6.081701 | 11.28018 |
| GWAS_Cat | Lymphocyt | 3/364 | 0.211116 | 0.252017 | 0 | 0 | 1.934309 | 3.008525 |
| GO_Biologi | Regulation | Jan-35 | 0.141623 | 0.252644 | 0 | 0 | 6.798564 | 13.28839 |
| GO_Biologi | Regulation | 3/297 | 0.13945 | 0.252644 | 0 | 0 | 2.38326 | 4.695137 |
| GO_Biologi | Establishm | Jan-35 | 0.141623 | 0.252644 | 0 | 0 | 6.798564 | 13.28839 |
| GO_Biologi | Regulation | Jan-35 | 0.141623 | 0.252644 | 0 | 0 | 6.798564 | 13.28839 |
| GO_Biologi | Regulation | Jan-35 | 0.141623 | 0.252644 | 0 | 0 | 6.798564 | 13.28839 |
| GO_Biologi | tRNA Amin | Jan-35 | 0.141623 | 0.252644 | 0 | 0 | 6.798564 | 13.28839 |
| GO_Biologi | Dendrite M | Jan-35 | 0.141623 | 0.252644 | 0 | 0 | 6.798564 | 13.28839 |
| GO_Biologi | Negative R | Jan-35 | 0.141623 | 0.252644 | 0 | 0 | 6.798564 | 13.28839 |
| GO_Biologi | Skeletal Mu | Jan-35 | 0.141623 | 0.252644 | 0 | 0 | 6.798564 | 13.28839 |
| GWAS_Cat | Free Chole: | Jan-55 | 0.213448 | 0.253067 | 0 | 0 | 4.27627 | 6.604112 |
| GWAS_Cat | Phospholip | Jan-55 | 0.213448 | 0.253067 | 0 | 0 | 4.27627 | 6.604112 |
| GWAS_Cat | Phosphatic | Jan-55 | 0.213448 | 0.253067 | 0 | 0 | 4.27627 | 6.604112 |
| GWAS_Cat | Phospholip | Jan-55 | 0.213448 | 0.253067 | 0 | 0 | 4.27627 | 6.604112 |
| GWAS_Cat | Phospholip | Jan-55 | 0.213448 | 0.253067 | 0 | 0 | 4.27627 | 6.604112 |
| GWAS_Cat | Phospholip | Jan-55 | 0.213448 | 0.253067 | 0 | 0 | 4.27627 | 6.604112 |
| GWAS_Cat | Phospholip | Jan-55 | 0.213448 | 0.253067 | 0 | 0 | 4.27627 | 6.604112 |
| GeDiPNet_ | Paraplegia | Jan-38 | 0.152796 | 0.253069 | 0 | 0 | 6.246386 | 11.73478 |
| GTEx_Tissu | Breast - Ma | Feb-86 | 0.054043 | 0.253855 | 0 | 0 | 5.554342 | 16.20739 |
| GTEx_Tissu | Liver Fema | 2/100 | 0.070424 | 0.253855 | 0 | 0 | 4.757503 | 12.62269 |
| GTEx_Tissu | Heart - Atri | 2/100 | 0.070424 | 0.253855 | 0 | 0 | 4.757503 | 12.62269 |
| GTEx_Tissu | Heart - Atri | 2/100 | 0.070424 | 0.253855 | 0 | 0 | 4.757503 | 12.62269 |
| GTEx_Tissu | Colon - Sigi | 2/100 | 0.070424 | 0.253855 | 0 | 0 | 4.757503 | 12.62269 |
| GTEx_Tissu | Liver Male | 2/100 | 0.070424 | 0.253855 | 0 | 0 | 4.757503 | 12.62269 |
| GTEx_Tissu | Heart - Atri | 2/100 | 0.070424 | 0.253855 | 0 | 0 | 4.757503 | 12.62269 |
| GTEx_Tissu | Heart - Atri | 2/100 | 0.070424 | 0.253855 | 0 | 0 | 4.757503 | 12.62269 |
| GTEx_Tissu | Liver Male | 2/100 | 0.070424 | 0.253855 | 0 | 0 | 4.757503 | 12.62269 |
| GTEx_Tissu | Esophagus | Feb-89 | 0.057425 | 0.253855 | 0 | 0 | 5.362001 | 15.32067 |
| GTEx_Tissu | Muscle - Sk | 2/100 | 0.070424 | 0.253855 | 0 | 0 | 4.757503 | 12.62269 |

| | | | | | | | | |
|---|---|---|---|---|---|---|---|---|
| GTEx_Tissu | Breast - Ma | Feb-90 | 0.058569 | 0.253855 | 0 | 0 | 5.300802 | 15.0413 |
| GTEx_Tissu | Colon - Sigi | 2/100 | 0.070424 | 0.253855 | 0 | 0 | 4.757503 | 12.62269 |
| GTEx_Tissu | Heart - Atri | 2/100 | 0.070424 | 0.253855 | 0 | 0 | 4.757503 | 12.62269 |
| GTEx_Tissu | Heart - Atri | 2/100 | 0.070424 | 0.253855 | 0 | 0 | 4.757503 | 12.62269 |
| GTEx_Tissu | Muscle - Sk | 2/100 | 0.070424 | 0.253855 | 0 | 0 | 4.757503 | 12.62269 |
| GTEx_Tissu | Artery - Tib | 2/100 | 0.070424 | 0.253855 | 0 | 0 | 4.757503 | 12.62269 |
| GTEx_Tissu | Colon - Sigi | 2/100 | 0.070424 | 0.253855 | 0 | 0 | 4.757503 | 12.62269 |
| GTEx_Tissu | Muscle - Sk | 2/100 | 0.070424 | 0.253855 | 0 | 0 | 4.757503 | 12.62269 |
| GTEx_Tissu | Heart - Atri | 2/100 | 0.070424 | 0.253855 | 0 | 0 | 4.757503 | 12.62269 |
| GTEx_Tissu | Heart - Atri | 2/100 | 0.070424 | 0.253855 | 0 | 0 | 4.757503 | 12.62269 |
| GTEx_Tissu | Brain - Hip | 2/100 | 0.070424 | 0.253855 | 0 | 0 | 4.757503 | 12.62269 |
| GTEx_Tissu | Muscle - Sk | 2/100 | 0.070424 | 0.253855 | 0 | 0 | 4.757503 | 12.62269 |
| GTEx_Tissu | Heart - Atri | 2/100 | 0.070424 | 0.253855 | 0 | 0 | 4.757503 | 12.62269 |
| GTEx_Tissu | Heart - Atri | 2/100 | 0.070424 | 0.253855 | 0 | 0 | 4.757503 | 12.62269 |
| GTEx_Tissu | Liver Male | 2/100 | 0.070424 | 0.253855 | 0 | 0 | 4.757503 | 12.62269 |
| GTEx_Tissu | Heart - Atri | 2/100 | 0.070424 | 0.253855 | 0 | 0 | 4.757503 | 12.62269 |
| GTEx_Tissu | Brain - Hip | 2/100 | 0.070424 | 0.253855 | 0 | 0 | 4.757503 | 12.62269 |
| GTEx_Tissu | Brain - Sub | 2/100 | 0.070424 | 0.253855 | 0 | 0 | 4.757503 | 12.62269 |
| GWAS_Cat | Cholesteryl | Jan-56 | 0.216879 | 0.254171 | 0 | 0 | 4.198309 | 6.416764 |
| GWAS_Cat | Adverse Re | Jan-56 | 0.216879 | 0.254171 | 0 | 0 | 4.198309 | 6.416764 |
| GWAS_Cat | Free Chole | Jan-56 | 0.216879 | 0.254171 | 0 | 0 | 4.198309 | 6.416764 |
| GWAS_Cat | Phospholip | Jan-56 | 0.216879 | 0.254171 | 0 | 0 | 4.198309 | 6.416764 |
| GWAS_Cat | Total Lipid | Jan-56 | 0.216879 | 0.254171 | 0 | 0 | 4.198309 | 6.416764 |
| GWAS_Cat | Cholestero | Jan-56 | 0.216879 | 0.254171 | 0 | 0 | 4.198309 | 6.416764 |
| GWAS_Cat | Free Chole | Jan-56 | 0.216879 | 0.254171 | 0 | 0 | 4.198309 | 6.416764 |
| GWAS_Cat | Cholestero | Jan-56 | 0.216879 | 0.254171 | 0 | 0 | 4.198309 | 6.416764 |
| GWAS_Cat | LDL Choles | Jan-56 | 0.216879 | 0.254171 | 0 | 0 | 4.198309 | 6.416764 |
| GWAS_Cat | Free Chole | Jan-56 | 0.216879 | 0.254171 | 0 | 0 | 4.198309 | 6.416764 |
| GWAS_Cat | Phospholip | Jan-56 | 0.216879 | 0.254171 | 0 | 0 | 4.198309 | 6.416764 |
| GWAS_Cat | Lymphocyt | 5/736 | 0.216628 | 0.254171 | 0 | 0 | 1.600047 | 2.447387 |
| GTEx_Tissu | GTEX-V1D1 | 8/606 | 0.004859 | 0.254322 | 0 | 0 | 3.270818 | 17.42357 |
| GTEx_Tissu | GTEX-PWO | 8/605 | 0.004811 | 0.254322 | 0 | 0 | 3.276467 | 17.48574 |
| GTEx_Tissu | GTEX-X261 | 7/480 | 0.004923 | 0.254322 | 0 | 0 | 3.596195 | 19.10964 |
| Diabetes_F | GSE14946E | 5/162 | 0.000709 | 0.254651 | 0 | 0 | 7.672829 | 55.63708 |
| DisGeNET | hearing im | 3/294 | 0.136464 | 0.254837 | 0 | 0 | 2.408198 | 4.796389 |
| DisGeNET | Pituitary A | 3/294 | 0.136464 | 0.254837 | 0 | 0 | 2.408198 | 4.796389 |
| GO_Biologi | Regulation | 2/153 | 0.143223 | 0.254858 | 0 | 0 | 3.079392 | 5.984343 |
| DisGeNET | Cholangioc | 4/455 | 0.136628 | 0.254961 | 0 | 0 | 2.079662 | 4.139557 |
| DisGeNET | Malignant | Jan-34 | 0.137866 | 0.255272 | 0 | 0 | 7.004933 | 13.88007 |
| DisGeNET | Autosomal | Jan-34 | 0.137866 | 0.255272 | 0 | 0 | 7.004933 | 13.88007 |
| DisGeNET | Trigonocep | Jan-34 | 0.137866 | 0.255272 | 0 | 0 | 7.004933 | 13.88007 |
| DisGeNET | Generalize | Jan-34 | 0.137866 | 0.255272 | 0 | 0 | 7.004933 | 13.88007 |
| DisGeNET | Chromoph | 2/149 | 0.13728 | 0.255272 | 0 | 0 | 3.163826 | 6.282501 |
| DisGeNET | Secondary | Jan-34 | 0.137866 | 0.255272 | 0 | 0 | 7.004933 | 13.88007 |
| DisGeNET | Broad-base | Jan-34 | 0.137866 | 0.255272 | 0 | 0 | 7.004933 | 13.88007 |

| | | | | | | | | |
|---|---|---|---|---|---|---|---|---|
| DisGeNET | Amyotroph | Jan-34 | 0.137866 | 0.255272 | 0 | 0 | 7.004933 | 13.88007 |
| DisGeNET | Narrow no: | Jan-34 | 0.137866 | 0.255272 | 0 | 0 | 7.004933 | 13.88007 |
| DisGeNET | Carcinoma, | Jan-34 | 0.137866 | 0.255272 | 0 | 0 | 7.004933 | 13.88007 |
| DisGeNET | Anti-Neutr | Jan-34 | 0.137866 | 0.255272 | 0 | 0 | 7.004933 | 13.88007 |
| Jensen_TIS | Pancreas | 74/16051 | 0.160273 | 0.255485 | 0 | 0 | 1.402324 | 2.567481 |
| Jensen_TIS | Chondrosa | Jan-40 | 0.160165 | 0.255485 | 0 | 0 | 5.925462 | 10.85279 |
| GTEx_Tissu | GTEX-RUSC | 8/611 | 0.005101 | 0.255718 | 0 | 0 | 3.242857 | 17.11691 |
| GTEx_Tissu | GTEX-O5YT | 11/1034 | 0.005127 | 0.255718 | 0 | 0 | 2.672609 | 14.09343 |
| GWAS_Cat | Cholesteryl | Jan-57 | 0.220295 | 0.256695 | 0 | 0 | 4.123131 | 6.237423 |
| GWAS_Cat | Cholestero | Jan-57 | 0.220295 | 0.256695 | 0 | 0 | 4.123131 | 6.237423 |
| GWAS_Cat | Hyperopia | Jan-57 | 0.220295 | 0.256695 | 0 | 0 | 4.123131 | 6.237423 |
| GWAS_Cat | Phospholip | Jan-57 | 0.220295 | 0.256695 | 0 | 0 | 4.123131 | 6.237423 |
| GWAS_Cat | COVID-19 ( | Jan-57 | 0.220295 | 0.256695 | 0 | 0 | 4.123131 | 6.237423 |
| GWAS_Cat | Phospholip | Jan-57 | 0.220295 | 0.256695 | 0 | 0 | 4.123131 | 6.237423 |
| GO_Biologi | Negative R | Jan-36 | 0.145363 | 0.256736 | 0 | 0 | 6.603987 | 12.73591 |
| GO_Biologi | Negative R | Jan-36 | 0.145363 | 0.256736 | 0 | 0 | 6.603987 | 12.73591 |
| GO_Biologi | Histone H3 | Jan-36 | 0.145363 | 0.256736 | 0 | 0 | 6.603987 | 12.73591 |
| Rare_Disea | Nocardiosis | 16-Jan | 0.067399 | 0.257153 | 0 | 0 | 15.42481 | 41.60265 |
| Rare_Disea | Mycobacte | 16-Jan | 0.067399 | 0.257153 | 0 | 0 | 15.42481 | 41.60265 |
| Rare_Disea | Symphalan | 16-Jan | 0.067399 | 0.257153 | 0 | 0 | 15.42481 | 41.60265 |
| Rare_Disea | Brain tumo | 3/215 | 0.067393 | 0.257153 | 0 | 0 | 3.318902 | 8.951791 |
| GO_Molecu | Ubiquitin-S | 20-Jan | 0.083535 | 0.257427 | 0 | 0 | 12.17503 | 30.22444 |
| GO_Molecu | Endopeptic | 2/112 | 0.085575 | 0.257427 | 0 | 0 | 4.235936 | 10.41346 |
| GO_Molecu | Telomerase | 21-Jan | 0.087525 | 0.257427 | 0 | 0 | 11.5657 | 28.17205 |
| GO_Molecu | Lipid Kinase | 21-Jan | 0.087525 | 0.257427 | 0 | 0 | 11.5657 | 28.17205 |
| GO_Molecu | UDP-galact | 19-Jan | 0.079527 | 0.257427 | 0 | 0 | 12.85207 | 32.53709 |
| GO_Molecu | Actinin Bin | 20-Jan | 0.083535 | 0.257427 | 0 | 0 | 12.17503 | 30.22444 |
| GTEx_Tissu | GTEX-U3ZN | 14/1504 | 0.005256 | 0.257709 | 0 | 0 | 2.37126 | 12.44541 |
| GeDiPNet_ | Degenerati | Jan-39 | 0.156488 | 0.25772 | 0 | 0 | 6.081701 | 11.28018 |
| GeDiPNet_ | Neurodege | Jan-39 | 0.156488 | 0.25772 | 0 | 0 | 6.081701 | 11.28018 |
| DisGeNET | Lung Neop | 8/1177 | 0.139414 | 0.257956 | 0 | 0 | 1.623718 | 3.199221 |
| GO_Molecu | N-acetyltra | 23-Jan | 0.095455 | 0.257986 | 0 | 0 | 10.51321 | 24.6966 |
| GO_Molecu | Protein Tyr | 23-Jan | 0.095455 | 0.257986 | 0 | 0 | 10.51321 | 24.6966 |
| GO_Molecu | Alpha-Actir | 23-Jan | 0.095455 | 0.257986 | 0 | 0 | 10.51321 | 24.6966 |
| PhenGenI_ | Mouth Nec | 15-Jan | 0.063321 | 0.258083 | 0 | 0 | 16.52741 | 45.60793 |
| PhenGenI_ | Coronary D | 4/345 | 0.063683 | 0.258083 | 0 | 0 | 2.766067 | 7.617307 |
| Rare_Disea | Hereditary | Feb-98 | 0.067994 | 0.2581 | 0 | 0 | 4.857108 | 13.05753 |
| Jensen_TIS | Gill | Jan-41 | 0.163825 | 0.258587 | 0 | 0 | 5.777035 | 10.45039 |
| Jensen_TIS | Orbit | Jan-41 | 0.163825 | 0.258587 | 0 | 0 | 5.777035 | 10.45039 |
| Human_Ph | Autosomal | 10/1722 | 0.214135 | 0.258859 | 0 | 0 | 1.380705 | 2.127872 |
| Rare_Disea | Hereditary | 18-Jan | 0.075502 | 0.259017 | 0 | 0 | 13.60876 | 35.1596 |
| Rare_Disea | Cushing's s | 2/105 | 0.076622 | 0.259017 | 0 | 0 | 4.525414 | 11.62522 |
| Rare_Disea | Chronic my | 2/105 | 0.076622 | 0.259017 | 0 | 0 | 4.525414 | 11.62522 |
| Rare_Disea | Mycobacte | 2/108 | 0.080421 | 0.259017 | 0 | 0 | 4.39667 | 11.08174 |
| Rare_Disea | Extragonac | 18-Jan | 0.075502 | 0.259017 | 0 | 0 | 13.60876 | 35.1596 |

| | | | | | | | | |
|---|---|---|---|---|---|---|---|---|
| Rare_Disea | Polysyndac | 19-Jan | 0.079527 | 0.259017 | 0 | 0 | 12.85207 | 32.53709 |
| Rare_Disea | Van Benthe | 2/107 | 0.079148 | 0.259017 | 0 | 0 | 4.438768 | 11.25866 |
| Rare_Disea | Birt-Hogg-D | 19-Jan | 0.079527 | 0.259017 | 0 | 0 | 12.85207 | 32.53709 |
| Rare_Disea | Dysferlinop | 18-Jan | 0.075502 | 0.259017 | 0 | 0 | 13.60876 | 35.1596 |
| Rare_Disea | Ovarian ca | 19-Jan | 0.079527 | 0.259017 | 0 | 0 | 12.85207 | 32.53709 |
| Rare_Disea | Cerebral fo | 18-Jan | 0.075502 | 0.259017 | 0 | 0 | 13.60876 | 35.1596 |
| Rare_Disea | Mercury po | 18-Jan | 0.075502 | 0.259017 | 0 | 0 | 13.60876 | 35.1596 |
| Rare_Disea | Axenfeld-R | 18-Jan | 0.075502 | 0.259017 | 0 | 0 | 13.60876 | 35.1596 |
| Rare_Disea | Typhus | 18-Jan | 0.075502 | 0.259017 | 0 | 0 | 13.60876 | 35.1596 |
| Rare_Disea | Peripheral | 2/103 | 0.074122 | 0.259017 | 0 | 0 | 4.615492 | 12.00969 |
| Rare_Disea | Intracrania | 2/108 | 0.080421 | 0.259017 | 0 | 0 | 4.39667 | 11.08174 |
| Rare_Disea | Leiomyoma | 19-Jan | 0.079527 | 0.259017 | 0 | 0 | 12.85207 | 32.53709 |
| Rare_Disea | Dysostosis | 19-Jan | 0.079527 | 0.259017 | 0 | 0 | 12.85207 | 32.53709 |
| Rare_Disea | Siderosis | 19-Jan | 0.079527 | 0.259017 | 0 | 0 | 12.85207 | 32.53709 |
| Rare_Disea | Binswange | 3/219 | 0.070386 | 0.259017 | 0 | 0 | 3.256779 | 8.642721 |
| Rare_Disea | Trisomy 6 | 2/103 | 0.074122 | 0.259017 | 0 | 0 | 4.615492 | 12.00969 |
| Rare_Disea | Choriocarc | 2/103 | 0.074122 | 0.259017 | 0 | 0 | 4.615492 | 12.00969 |
| Rare_Disea | Amyotroph | 2/108 | 0.080421 | 0.259017 | 0 | 0 | 4.39667 | 11.08174 |
| Rare_Disea | Optic neuri | 2/106 | 0.077881 | 0.259017 | 0 | 0 | 4.481674 | 11.43977 |
| Rare_Disea | Familial col | 2/101 | 0.07165 | 0.259017 | 0 | 0 | 4.70921 | 12.4133 |
| Rare_Disea | Plasma cell | 2/101 | 0.07165 | 0.259017 | 0 | 0 | 4.70921 | 12.4133 |
| Rare_Disea | Sarcoidosis | 3/222 | 0.07267 | 0.259017 | 0 | 0 | 3.211676 | 8.420478 |
| Rare_Disea | Van Bogaer | 2/107 | 0.079148 | 0.259017 | 0 | 0 | 4.438768 | 11.25866 |
| Rare_Disea | Acromegal | 2/108 | 0.080421 | 0.259017 | 0 | 0 | 4.39667 | 11.08174 |
| Rare_Disea | Mucolipido | 18-Jan | 0.075502 | 0.259017 | 0 | 0 | 13.60876 | 35.1596 |
| Rare_Disea | Acute resp | 3/221 | 0.071905 | 0.259017 | 0 | 0 | 3.226573 | 8.493677 |
| Rare_Disea | Oculophary | 17-Jan | 0.071459 | 0.259017 | 0 | 0 | 14.46003 | 38.15469 |
| Rare_Disea | Amyotonia | 4/358 | 0.070886 | 0.259017 | 0 | 0 | 2.662719 | 7.047366 |
| Rare_Disea | Leukoence | 2/100 | 0.070424 | 0.259017 | 0 | 0 | 4.757503 | 12.62269 |
| Rare_Disea | Chronic my | 2/101 | 0.07165 | 0.259017 | 0 | 0 | 4.70921 | 12.4133 |
| DisGeNET | Prostate Ca | Jan-35 | 0.141623 | 0.259115 | 0 | 0 | 6.798564 | 13.28839 |
| DisGeNET | Limb defec | Jan-35 | 0.141623 | 0.259115 | 0 | 0 | 6.798564 | 13.28839 |
| DisGeNET | Neoplasm | Jan-35 | 0.141623 | 0.259115 | 0 | 0 | 6.798564 | 13.28839 |
| DisGeNET | Ventricular | Jan-35 | 0.141623 | 0.259115 | 0 | 0 | 6.798564 | 13.28839 |
| DisGeNET | Malignant | 2/151 | 0.140245 | 0.259115 | 0 | 0 | 3.121042 | 6.130869 |
| DisGeNET | Intrahepati | Jan-35 | 0.141623 | 0.259115 | 0 | 0 | 6.798564 | 13.28839 |
| DisGeNET | Therapy-re | Jan-35 | 0.141623 | 0.259115 | 0 | 0 | 6.798564 | 13.28839 |
| DisGeNET | Intracrania | Jan-35 | 0.141623 | 0.259115 | 0 | 0 | 6.798564 | 13.28839 |
| DisGeNET | Follicular n | Jan-35 | 0.141623 | 0.259115 | 0 | 0 | 6.798564 | 13.28839 |
| DisGeNET | Fibrosarcor | 3/298 | 0.14045 | 0.259115 | 0 | 0 | 2.375061 | 4.662008 |
| DisGeNET | COLORBLIN | Jan-35 | 0.141623 | 0.259115 | 0 | 0 | 6.798564 | 13.28839 |
| DisGeNET | Acanthosis | Jan-35 | 0.141623 | 0.259115 | 0 | 0 | 6.798564 | 13.28839 |
| DisGeNET | Esophagea | Jan-35 | 0.141623 | 0.259115 | 0 | 0 | 6.798564 | 13.28839 |
| DisGeNET | Sparse axill | Jan-35 | 0.141623 | 0.259115 | 0 | 0 | 6.798564 | 13.28839 |
| DisGeNET | Tooth ager | Jan-35 | 0.141623 | 0.259115 | 0 | 0 | 6.798564 | 13.28839 |

| | | | | | | | | |
|---|---|---|---|---|---|---|---|---|
| DisGeNET | Acanthosis | Jan-35 | 0.141623 | 0.259115 | 0 | 0 | 6.798564 | 13.28839 |
| DisGeNET | Osteochon | 2/152 | 0.141732 | 0.259134 | 0 | 0 | 3.100078 | 6.056983 |
| GWAS_Cat | Total Lipid | Jan-58 | 0.223696 | 0.259667 | 0 | 0 | 4.050592 | 6.065622 |
| GWAS_Cat | Vertical Cu | Jan-58 | 0.223696 | 0.259667 | 0 | 0 | 4.050592 | 6.065622 |
| GWAS_Cat | Total Fatty | Jan-58 | 0.223696 | 0.259667 | 0 | 0 | 4.050592 | 6.065622 |
| GWAS_Cat | Triglyceride | Jan-58 | 0.223696 | 0.259667 | 0 | 0 | 4.050592 | 6.065622 |
| DisGeNET | Hereditary | 3/300 | 0.142458 | 0.259917 | 0 | 0 | 2.358826 | 4.596658 |
| DisGeNET | Choriocarc | 3/300 | 0.142458 | 0.259917 | 0 | 0 | 2.358826 | 4.596658 |
| DisGeNET | Obesity | Dec-61 | 0.142292 | 0.259917 | 0 | 0 | 1.474726 | 2.875524 |
| GeDiPNet_ | Anaplastic | 2/163 | 0.158308 | 0.259983 | 0 | 0 | 2.886664 | 5.320729 |
| GO_Biologi | Regulation | Jan-37 | 0.149088 | 0.260079 | 0 | 0 | 6.42022 | 12.21909 |
| GO_Biologi | Melanosom | Jan-37 | 0.149088 | 0.260079 | 0 | 0 | 6.42022 | 12.21909 |
| GO_Biologi | Negative R | Jan-37 | 0.149088 | 0.260079 | 0 | 0 | 6.42022 | 12.21909 |
| GO_Biologi | Negative R | Jan-37 | 0.149088 | 0.260079 | 0 | 0 | 6.42022 | 12.21909 |
| GO_Biologi | Muscle Cel | Jan-37 | 0.149088 | 0.260079 | 0 | 0 | 6.42022 | 12.21909 |
| dbGaP | Leukocyte | Jan-40 | 0.160165 | 0.260268 | 0 | 0 | 5.925462 | 10.85279 |
| Human_Ph | Hydroneph | Jan-56 | 0.216879 | 0.260527 | 0 | 0 | 4.198309 | 6.416764 |
| Human_Ph | Optic disc p | Jan-56 | 0.216879 | 0.260527 | 0 | 0 | 4.198309 | 6.416764 |
| GO_Molecı | Active Mor | 24-Jan | 0.099394 | 0.261564 | 0 | 0 | 10.05561 | 23.215 |
| UK_Bioban | Age comple | Jan-53 | 0.206541 | 0.261619 | 0 | 0 | 4.44119 | 7.004886 |
| Jensen_TIS | Ileal mucos | Jan-42 | 0.16747 | 0.261774 | 0 | 0 | 5.635848 | 10.07097 |
| Jensen_TIS | Biofilm | Jan-42 | 0.16747 | 0.261774 | 0 | 0 | 5.635848 | 10.07097 |
| GWAS_Cat | Glaucoma | Jan-59 | 0.227083 | 0.261854 | 0 | 0 | 3.980553 | 5.900928 |
| GWAS_Cat | Concentrat | Jan-59 | 0.227083 | 0.261854 | 0 | 0 | 3.980553 | 5.900928 |
| GWAS_Cat | Triglyceride | Jan-59 | 0.227083 | 0.261854 | 0 | 0 | 3.980553 | 5.900928 |
| GWAS_Cat | Concentrat | Jan-59 | 0.227083 | 0.261854 | 0 | 0 | 3.980553 | 5.900928 |
| GWAS_Cat | Total Lipid | Jan-59 | 0.227083 | 0.261854 | 0 | 0 | 3.980553 | 5.900928 |
| GWAS_Cat | Coronary A | Jan-59 | 0.227083 | 0.261854 | 0 | 0 | 3.980553 | 5.900928 |
| GWAS_Cat | Triglyceride | Jan-59 | 0.227083 | 0.261854 | 0 | 0 | 3.980553 | 5.900928 |
| DisGeNET | Anxiety | 4/464 | 0.143693 | 0.261987 | 0 | 0 | 2.03803 | 3.953934 |
| CellMarker | Fibroblast | Jan-50 | 0.196069 | 0.262063 | 0 | 0 | 4.713811 | 7.680157 |
| DisGeNET | Inflammato | Jan-36 | 0.145363 | 0.262838 | 0 | 0 | 6.603987 | 12.73591 |
| DisGeNET | Mesenchyr | Jan-36 | 0.145363 | 0.262838 | 0 | 0 | 6.603987 | 12.73591 |
| DisGeNET | High-grade | Jan-36 | 0.145363 | 0.262838 | 0 | 0 | 6.603987 | 12.73591 |
| DisGeNET | Hepatitis E | Jan-36 | 0.145363 | 0.262838 | 0 | 0 | 6.603987 | 12.73591 |
| DisGeNET | Oestrogen | Jan-36 | 0.145363 | 0.262838 | 0 | 0 | 6.603987 | 12.73591 |
| DisGeNET | Myoclonus | 2/154 | 0.144717 | 0.262838 | 0 | 0 | 3.058978 | 5.912923 |
| DisGeNET | Plexiform N | Jan-36 | 0.145363 | 0.262838 | 0 | 0 | 6.603987 | 12.73591 |
| DisGeNET | DEAFNESS, | Jan-36 | 0.145363 | 0.262838 | 0 | 0 | 6.603987 | 12.73591 |
| DisGeNET | Low Tensic | Jan-36 | 0.145363 | 0.262838 | 0 | 0 | 6.603987 | 12.73591 |
| DisGeNET | Hammer To | Jan-36 | 0.145363 | 0.262838 | 0 | 0 | 6.603987 | 12.73591 |
| DisGeNET | Scaphycepl | Jan-36 | 0.145363 | 0.262838 | 0 | 0 | 6.603987 | 12.73591 |
| DisGeNET | Invasive Ca | 2/154 | 0.144717 | 0.262838 | 0 | 0 | 3.058978 | 5.912923 |
| DisGeNET | Kidney Dise | 5/639 | 0.145653 | 0.263179 | 0 | 0 | 1.854178 | 3.57213 |
| KEGG_202 | Arrhythmo | Feb-77 | 0.044354 | 0.263352 | 0 | 0 | 6.223686 | 19.39022 |

| | | | | | | | | |
|---|---|---|---|---|---|---|---|---|
| KEGG_202: | Leishmania | Feb-77 | 0.044354 | 0.263352 | 0 | 0 | 6.223686 | 19.39022 |
| KEGG_202: | Inositol pho | Feb-73 | 0.040281 | 0.263352 | 0 | 0 | 6.575642 | 21.1202 |
| KEGG_202: | cGMP-PKG | 3/167 | 0.036403 | 0.263352 | 0 | 0 | 4.30074 | 14.24877 |
| GWAS_Cat | Total Lipid | Jan-60 | 0.230455 | 0.263748 | 0 | 0 | 3.912889 | 5.742942 |
| GWAS_Cat | Telomere L | Jan-60 | 0.230455 | 0.263748 | 0 | 0 | 3.912889 | 5.742942 |
| GWAS_Cat | Cholesteryl | Jan-60 | 0.230455 | 0.263748 | 0 | 0 | 3.912889 | 5.742942 |
| GWAS_Cat | Concentrat | Jan-60 | 0.230455 | 0.263748 | 0 | 0 | 3.912889 | 5.742942 |
| GWAS_Cat | Triglyceride | Jan-60 | 0.230455 | 0.263748 | 0 | 0 | 3.912889 | 5.742942 |
| GWAS_Cat | Educationa | 5/753 | 0.23009 | 0.263748 | 0 | 0 | 1.562296 | 2.295461 |
| GWAS_Cat | Total Lipid | Jan-60 | 0.230455 | 0.263748 | 0 | 0 | 3.912889 | 5.742942 |
| GWAS_Cat | Medicatior | Jan-60 | 0.230455 | 0.263748 | 0 | 0 | 3.912889 | 5.742942 |
| GO_Biologi | Membrane | Jan-38 | 0.152796 | 0.263954 | 0 | 0 | 6.246386 | 11.73478 |
| GO_Biologi | Fatty Acid T | Jan-38 | 0.152796 | 0.263954 | 0 | 0 | 6.246386 | 11.73478 |
| GO_Biologi | Import Into | Jan-38 | 0.152796 | 0.263954 | 0 | 0 | 6.246386 | 11.73478 |
| GO_Biologi | Response T | Jan-38 | 0.152796 | 0.263954 | 0 | 0 | 6.246386 | 11.73478 |
| SysMyo_M | MGS1398 I | 4/222 | 0.016064 | 0.264074 | 0 | 0 | 4.353929 | 17.98681 |
| SysMyo_M | MGS450 H | 4/225 | 0.016792 | 0.264074 | 0 | 0 | 4.294172 | 17.54962 |
| SysMyo_M | MGS1046 I | 3/125 | 0.017263 | 0.264074 | 0 | 0 | 5.793618 | 23.51747 |
| SysMyo_M | MGS7 Rat ( | Feb-46 | 0.017103 | 0.264074 | 0 | 0 | 10.62513 | 43.22851 |
| CellMarker | Immune Ce | Jan-51 | 0.199575 | 0.264172 | 0 | 0 | 4.619302 | 7.444308 |
| CellMarker | Endothelia | Jan-51 | 0.199575 | 0.264172 | 0 | 0 | 4.619302 | 7.444308 |
| GO_Molec | Wide Pore | 25-Jan | 0.103317 | 0.264914 | 0 | 0 | 9.636143 | 21.87363 |
| GO_Biologi | Nuclear Tra | Jan-39 | 0.156488 | 0.265043 | 0 | 0 | 6.081701 | 11.28018 |
| GO_Biologi | Mitochond | 2/162 | 0.156786 | 0.265043 | 0 | 0 | 2.904853 | 5.382321 |
| GO_Biologi | Negative R | Jan-39 | 0.156488 | 0.265043 | 0 | 0 | 6.081701 | 11.28018 |
| GO_Biologi | Vitamin Tra | Jan-39 | 0.156488 | 0.265043 | 0 | 0 | 6.081701 | 11.28018 |
| GO_Biologi | Carboxylic | Jan-39 | 0.156488 | 0.265043 | 0 | 0 | 6.081701 | 11.28018 |
| GO_Biologi | Regulation | Jan-39 | 0.156488 | 0.265043 | 0 | 0 | 6.081701 | 11.28018 |
| GO_Biologi | Negative R | Jan-39 | 0.156488 | 0.265043 | 0 | 0 | 6.081701 | 11.28018 |
| GO_Biologi | Negative R | Jan-39 | 0.156488 | 0.265043 | 0 | 0 | 6.081701 | 11.28018 |
| GO_Biologi | Positive Re | Jan-39 | 0.156488 | 0.265043 | 0 | 0 | 6.081701 | 11.28018 |
| DisGeNET | Immunolog | 5/641 | 0.147 | 0.26543 | 0 | 0 | 1.848155 | 3.543515 |
| GO_Biologi | Negative R | 2/163 | 0.158308 | 0.265694 | 0 | 0 | 2.886664 | 5.320729 |
| GO_Biologi | Nucleic Aci | Jan-40 | 0.160165 | 0.265694 | 0 | 0 | 5.925462 | 10.85279 |
| GO_Biologi | Cyclic-Nucl | Jan-40 | 0.160165 | 0.265694 | 0 | 0 | 5.925462 | 10.85279 |
| GO_Biologi | Regulation | Jan-40 | 0.160165 | 0.265694 | 0 | 0 | 5.925462 | 10.85279 |
| GO_Biologi | Regulation | Jan-40 | 0.160165 | 0.265694 | 0 | 0 | 5.925462 | 10.85279 |
| GO_Biologi | Regulation | Jan-40 | 0.160165 | 0.265694 | 0 | 0 | 5.925462 | 10.85279 |
| GO_Biologi | Negative R | Jan-40 | 0.160165 | 0.265694 | 0 | 0 | 5.925462 | 10.85279 |
| GO_Biologi | Negative R | Jan-40 | 0.160165 | 0.265694 | 0 | 0 | 5.925462 | 10.85279 |
| CellMarker | Schwalie Et | 2/191 | 0.201897 | 0.26596 | 0 | 0 | 2.455524 | 3.928837 |
| Jensen_TIS | Pelvis | Jan-43 | 0.171099 | 0.266155 | 0 | 0 | 5.501384 | 9.712751 |
| PhenGenI_ | Phospholip | Feb-98 | 0.067994 | 0.266214 | 0 | 0 | 4.857108 | 13.05753 |
| PhenGenI_ | Serum Albu | 16-Jan | 0.067399 | 0.266214 | 0 | 0 | 15.42481 | 41.60265 |
| CellMarker | Mural Cell | Jan-52 | 0.203066 | 0.26622 | 0 | 0 | 4.5285 | 7.219451 |

| | | | | | | | | |
|---|---|---|---|---|---|---|---|---|
| Rare_Disea | Pseudopela | 3/236 | 0.083758 | 0.266308 | 0 | 0 | 3.016554 | 7.480516 |
| Rare_Disea | Peters ano | 20-Jan | 0.083535 | 0.266308 | 0 | 0 | 12.17503 | 30.22444 |
| Rare_Disea | Hereditary | 20-Jan | 0.083535 | 0.266308 | 0 | 0 | 12.17503 | 30.22444 |
| GTEx_Tissu | GTEX-O5YT | 16/1846 | 0.005618 | 0.266447 | 0 | 0 | 2.226799 | 11.53873 |
| GTEx_Tissu | GTEX-Q734 | 14/1514 | 0.005568 | 0.266447 | 0 | 0 | 2.354174 | 12.21971 |
| DisGeNET | Elevated cr | 2/156 | 0.147716 | 0.266539 | 0 | 0 | 3.018946 | 5.773631 |
| DisGeNET | Vitelliform | Jan-37 | 0.149088 | 0.266628 | 0 | 0 | 6.42022 | 12.21909 |
| DisGeNET | Oculocereb | Jan-37 | 0.149088 | 0.266628 | 0 | 0 | 6.42022 | 12.21909 |
| DisGeNET | Pelvic Orga | Jan-37 | 0.149088 | 0.266628 | 0 | 0 | 6.42022 | 12.21909 |
| DisGeNET | Adenoma, | Jan-37 | 0.149088 | 0.266628 | 0 | 0 | 6.42022 | 12.21909 |
| DisGeNET | Juvenile Pil | Jan-37 | 0.149088 | 0.266628 | 0 | 0 | 6.42022 | 12.21909 |
| DisGeNET | Escherichia | Jan-37 | 0.149088 | 0.266628 | 0 | 0 | 6.42022 | 12.21909 |
| DisGeNET | Glomerulor | Jan-37 | 0.149088 | 0.266628 | 0 | 0 | 6.42022 | 12.21909 |
| DisGeNET | Paget Disea | Jan-37 | 0.149088 | 0.266628 | 0 | 0 | 6.42022 | 12.21909 |
| DisGeNET | Congenital | Jan-37 | 0.149088 | 0.266628 | 0 | 0 | 6.42022 | 12.21909 |
| DisGeNET | Adenoma, | Jan-37 | 0.149088 | 0.266628 | 0 | 0 | 6.42022 | 12.21909 |
| DisGeNET | Blood Plate | Jan-37 | 0.149088 | 0.266628 | 0 | 0 | 6.42022 | 12.21909 |
| DisGeNET | Abscess | Jan-37 | 0.149088 | 0.266628 | 0 | 0 | 6.42022 | 12.21909 |
| DisGeNET | Simple syn | Jan-37 | 0.149088 | 0.266628 | 0 | 0 | 6.42022 | 12.21909 |
| DisGeNET | Curvature c | 2/157 | 0.14922 | 0.266682 | 0 | 0 | 2.999317 | 5.705706 |
| MAGMA_D | Erlotinib | Jan-53 | 0.206541 | 0.266699 | 0 | 0 | 4.44119 | 7.004886 |
| GeDiPNet_ | Hypoplastic | Jan-41 | 0.163825 | 0.266788 | 0 | 0 | 5.777035 | 10.45039 |
| GeDiPNet_ | Glaucoma, | Jan-41 | 0.163825 | 0.266788 | 0 | 0 | 5.777035 | 10.45039 |
| GeDiPNet_ | Breast Ade | Jan-41 | 0.163825 | 0.266788 | 0 | 0 | 5.777035 | 10.45039 |
| GWAS_Cat | Triglyceride | Jan-61 | 0.233813 | 0.26684 | 0 | 0 | 3.847481 | 5.591291 |
| GWAS_Cat | AFP Levels | Jan-61 | 0.233813 | 0.26684 | 0 | 0 | 3.847481 | 5.591291 |
| GWAS_Cat | Cholestero | Jan-61 | 0.233813 | 0.26684 | 0 | 0 | 3.847481 | 5.591291 |
| DisGeNET | Arthrogryp | 3/307 | 0.149562 | 0.266929 | 0 | 0 | 2.303689 | 4.377116 |
| DisGeNET | Malignant | 3/307 | 0.149562 | 0.266929 | 0 | 0 | 2.303689 | 4.377116 |
| Rare_Disea | Acral lentig | 22-Jan | 0.091499 | 0.266987 | 0 | 0 | 11.0144 | 26.34017 |
| Rare_Disea | Mental reta | 22-Jan | 0.091499 | 0.266987 | 0 | 0 | 11.0144 | 26.34017 |
| Rare_Disea | Collagenou | 22-Jan | 0.091499 | 0.266987 | 0 | 0 | 11.0144 | 26.34017 |
| Rare_Disea | Myoepithe | 2/115 | 0.089505 | 0.266987 | 0 | 0 | 4.122853 | 9.950336 |
| Rare_Disea | Poland syn | 2/117 | 0.092155 | 0.266987 | 0 | 0 | 4.050742 | 9.658129 |
| Rare_Disea | Gordon syr | 2/117 | 0.092155 | 0.266987 | 0 | 0 | 4.050742 | 9.658129 |
| Rare_Disea | AIDS Deme | 2/118 | 0.093488 | 0.266987 | 0 | 0 | 4.015619 | 9.516699 |
| Rare_Disea | Malignant g | 2/118 | 0.093488 | 0.266987 | 0 | 0 | 4.015619 | 9.516699 |
| Rare_Disea | Factor 2 de | 2/118 | 0.093488 | 0.266987 | 0 | 0 | 4.015619 | 9.516699 |
| Rare_Disea | Blastoma | 23-Jan | 0.095455 | 0.266987 | 0 | 0 | 10.51321 | 24.6966 |
| Rare_Disea | Encephalop | 23-Jan | 0.095455 | 0.266987 | 0 | 0 | 10.51321 | 24.6966 |
| Rare_Disea | West syndr | 2/117 | 0.092155 | 0.266987 | 0 | 0 | 4.050742 | 9.658129 |
| Rare_Disea | Kikuchi dise | 3/249 | 0.094665 | 0.266987 | 0 | 0 | 2.855256 | 6.731013 |
| Rare_Disea | Cytokine re | 2/117 | 0.092155 | 0.266987 | 0 | 0 | 4.050742 | 9.658129 |
| Rare_Disea | Metastatic | 21-Jan | 0.087525 | 0.266987 | 0 | 0 | 11.5657 | 28.17205 |
| Rare_Disea | Anaplastic | 2/114 | 0.088189 | 0.266987 | 0 | 0 | 4.159874 | 10.1013 |

| | | | | | | | | |
|---|---|---|---|---|---|---|---|---|
| Rare_Disea | Berger dise | 2/114 | 0.088189 | 0.266987 | 0 | 0 | 4.159874 | 10.1013 |
| Rare_Disea | Myxopapill | 2/114 | 0.088189 | 0.266987 | 0 | 0 | 4.159874 | 10.1013 |
| Rare_Disea | Ependymo | 2/114 | 0.088189 | 0.266987 | 0 | 0 | 4.159874 | 10.1013 |
| Rare_Disea | Heart tumc | 2/112 | 0.085575 | 0.266987 | 0 | 0 | 4.235936 | 10.41346 |
| Rare_Disea | Syncampto | 3/239 | 0.086224 | 0.266987 | 0 | 0 | 2.977754 | 7.297891 |
| Rare_Disea | Ollier disea | 2/113 | 0.086879 | 0.266987 | 0 | 0 | 4.197562 | 10.25564 |
| Rare_Disea | Hypertropł | 3/249 | 0.094665 | 0.266987 | 0 | 0 | 2.855256 | 6.731013 |
| Rare_Disea | Nasophary | 2/119 | 0.094827 | 0.266987 | 0 | 0 | 3.981096 | 9.378266 |
| Rare_Disea | Wolff-Park | 23-Jan | 0.095455 | 0.266987 | 0 | 0 | 10.51321 | 24.6966 |
| Rare_Disea | Polysyndac | 22-Jan | 0.091499 | 0.266987 | 0 | 0 | 11.0144 | 26.34017 |
| Rare_Disea | Post-traum | 22-Jan | 0.091499 | 0.266987 | 0 | 0 | 11.0144 | 26.34017 |
| Rare_Disea | Leigh syndı | 3/246 | 0.092098 | 0.266987 | 0 | 0 | 2.890947 | 6.894635 |
| Rare_Disea | HELLP sync | 2/113 | 0.086879 | 0.266987 | 0 | 0 | 4.197562 | 10.25564 |
| Rare_Disea | Lymphocyt | 21-Jan | 0.087525 | 0.266987 | 0 | 0 | 11.5657 | 28.17205 |
| Rare_Disea | Leukemia I | 2/119 | 0.094827 | 0.266987 | 0 | 0 | 3.981096 | 9.378266 |
| Rare_Disea | Fibrosarcoı | 2/116 | 0.090827 | 0.266987 | 0 | 0 | 4.086481 | 9.802643 |
| GeDiPNet_ | Dyssomnia | 2/167 | 0.164425 | 0.267019 | 0 | 0 | 2.816114 | 5.083934 |
| Human_Ph | Respiratory | Jan-58 | 0.223696 | 0.267875 | 0 | 0 | 4.050592 | 6.065622 |
| Rare_Disea | Myelodysp | 2/120 | 0.096172 | 0.267984 | 0 | 0 | 3.947159 | 9.242743 |
| SysMyo_M | MGS1033 I | 3/127 | 0.018 | 0.268109 | 0 | 0 | 5.699597 | 22.89738 |
| GTEx_Tissu | GTEX-X3Y1 | 9/760 | 0.00575 | 0.268284 | 0 | 0 | 2.944075 | 15.18738 |
| DisGeNET | Infection c | 3/308 | 0.150586 | 0.268574 | 0 | 0 | 2.296019 | 4.346874 |
| GeDiPNet_ | Cholangitis | 2/168 | 0.165961 | 0.268764 | 0 | 0 | 2.799008 | 5.027029 |
| DisGeNET | Lip and Ora | 4/473 | 0.150901 | 0.268954 | 0 | 0 | 1.997996 | 3.778467 |
| GeDiPNet_ | Cerebral At | 3/324 | 0.167273 | 0.268967 | 0 | 0 | 2.179795 | 3.897752 |
| GeDiPNet_ | Congenital | Jan-42 | 0.16747 | 0.268967 | 0 | 0 | 5.635848 | 10.07097 |
| GeDiPNet_ | Ovarian Ca | Jan-42 | 0.16747 | 0.268967 | 0 | 0 | 5.635848 | 10.07097 |
| Disease_Pe | Dermatom | 5/280 | 0.007543 | 0.269048 | 0 | 0 | 4.354324 | 21.27994 |
| Disease_Pe | Duchenne | 5/276 | 0.007111 | 0.269048 | 0 | 0 | 4.419494 | 21.8593 |
| Disease_Pe | Tendonopa | 5/278 | 0.007325 | 0.269048 | 0 | 0 | 4.38667 | 21.56691 |
| GO_Biologi | Negative R | Jan-41 | 0.163825 | 0.26925 | 0 | 0 | 5.777035 | 10.45039 |
| GO_Biologi | Regulation | Jan-41 | 0.163825 | 0.26925 | 0 | 0 | 5.777035 | 10.45039 |
| GO_Biologi | O-glycan Pı | Jan-41 | 0.163825 | 0.26925 | 0 | 0 | 5.777035 | 10.45039 |
| GO_Biologi | DNA Duple | Jan-41 | 0.163825 | 0.26925 | 0 | 0 | 5.777035 | 10.45039 |
| GWAS_Cat | Waist-to-hi | Jan-62 | 0.237156 | 0.269395 | 0 | 0 | 3.784217 | 5.445629 |
| GWAS_Cat | Verbal-nun | Jan-62 | 0.237156 | 0.269395 | 0 | 0 | 3.784217 | 5.445629 |
| GWAS_Cat | White Mat | Jan-62 | 0.237156 | 0.269395 | 0 | 0 | 3.784217 | 5.445629 |
| GWAS_Cat | Triglyceride | Jan-62 | 0.237156 | 0.269395 | 0 | 0 | 3.784217 | 5.445629 |
| GWAS_Cat | Glycine Lev | Jan-62 | 0.237156 | 0.269395 | 0 | 0 | 3.784217 | 5.445629 |
| DisGeNET | Common a | Jan-38 | 0.152796 | 0.269398 | 0 | 0 | 6.246386 | 11.73478 |
| DisGeNET | Papillary ac | Jan-38 | 0.152796 | 0.269398 | 0 | 0 | 6.246386 | 11.73478 |
| DisGeNET | Prostatic D | Jan-38 | 0.152796 | 0.269398 | 0 | 0 | 6.246386 | 11.73478 |
| DisGeNET | Pelvic Infla | Jan-38 | 0.152796 | 0.269398 | 0 | 0 | 6.246386 | 11.73478 |
| DisGeNET | End Stage I | Jan-38 | 0.152796 | 0.269398 | 0 | 0 | 6.246386 | 11.73478 |
| DisGeNET | Nephritis, T | Jan-38 | 0.152796 | 0.269398 | 0 | 0 | 6.246386 | 11.73478 |

| | | | | | | | | |
|---|---|---|---|---|---|---|---|---|
| DisGeNET | Vitamin D I | Jan-38 | 0.152796 | 0.269398 | 0 | 0 | 6.246386 | 11.73478 |
| DisGeNET | Autosomal | Jan-38 | 0.152796 | 0.269398 | 0 | 0 | 6.246386 | 11.73478 |
| DisGeNET | Chronic eo: | Jan-38 | 0.152796 | 0.269398 | 0 | 0 | 6.246386 | 11.73478 |
| DisGeNET | Clinodactyl | 2/159 | 0.152237 | 0.269398 | 0 | 0 | 2.960809 | 5.573176 |
| DisGeNET | Down Sync | 4/475 | 0.152522 | 0.269398 | 0 | 0 | 1.989308 | 3.740783 |
| DisGeNET | Adenoma, | Jan-38 | 0.152796 | 0.269398 | 0 | 0 | 6.246386 | 11.73478 |
| DisGeNET | Salmonella | Jan-38 | 0.152796 | 0.269398 | 0 | 0 | 6.246386 | 11.73478 |
| DisGeNET | Bladder ne | Jan-38 | 0.152796 | 0.269398 | 0 | 0 | 6.246386 | 11.73478 |
| DisGeNET | Creatine pl | 2/159 | 0.152237 | 0.269398 | 0 | 0 | 2.960809 | 5.573176 |
| DisGeNET | Neurobeha | Jan-38 | 0.152796 | 0.269398 | 0 | 0 | 6.246386 | 11.73478 |
| Human_Ph | Dysarthria | 2/206 | 0.225797 | 0.269545 | 0 | 0 | 2.273241 | 3.382851 |
| GeDiPNet_ | High Palate | 4/496 | 0.169936 | 0.269597 | 0 | 0 | 1.902341 | 3.371586 |
| GeDiPNet_ | Geographic | Jan-43 | 0.171099 | 0.269597 | 0 | 0 | 5.501384 | 9.712751 |
| GeDiPNet_ | Dysgraphia | Jan-43 | 0.171099 | 0.269597 | 0 | 0 | 5.501384 | 9.712751 |
| GeDiPNet_ | Ovarian Ca | 2/170 | 0.16904 | 0.269597 | 0 | 0 | 2.765406 | 4.91584 |
| GeDiPNet_ | Developme | 3/326 | 0.169397 | 0.269597 | 0 | 0 | 2.166077 | 3.845893 |
| GeDiPNet_ | Diabetes Ir | Jan-43 | 0.171099 | 0.269597 | 0 | 0 | 5.501384 | 9.712751 |
| GeDiPNet_ | Bundle Bra | Jan-43 | 0.171099 | 0.269597 | 0 | 0 | 5.501384 | 9.712751 |
| Rare_Disea | Respiratory | 2/121 | 0.097522 | 0.269726 | 0 | 0 | 3.913791 | 9.110048 |
| Rare_Disea | Growth ho | 3/252 | 0.097262 | 0.269726 | 0 | 0 | 2.820425 | 6.572577 |
| Rare_Disea | Granulosa | 24-Jan | 0.099394 | 0.269888 | 0 | 0 | 10.05561 | 23.215 |
| Rare_Disea | Brain stem | 2/122 | 0.098877 | 0.269888 | 0 | 0 | 3.88098 | 8.980101 |
| Rare_Disea | Hereditary | 24-Jan | 0.099394 | 0.269888 | 0 | 0 | 10.05561 | 23.215 |
| Rare_Disea | Tangier dis | 24-Jan | 0.099394 | 0.269888 | 0 | 0 | 10.05561 | 23.215 |
| Rare_Disea | Pelizaeus-M | 24-Jan | 0.099394 | 0.269888 | 0 | 0 | 10.05561 | 23.215 |
| GO_Molecu | Acetyltrans | 27-Jan | 0.111111 | 0.270087 | 0 | 0 | 8.894007 | 19.54216 |
| GO_Molecu | GTPase Act | 3/265 | 0.108844 | 0.270087 | 0 | 0 | 2.678708 | 5.940942 |
| GO_Molecu | Ribonuclec | 3/270 | 0.113436 | 0.270087 | 0 | 0 | 2.627876 | 5.719604 |
| Rare_Disea | Usual inter | 2/123 | 0.100238 | 0.270208 | 0 | 0 | 3.848712 | 8.852824 |
| Rare_Disea | Common v | 2/123 | 0.100238 | 0.270208 | 0 | 0 | 3.848712 | 8.852824 |
| Human_Ph | Incomplete | Jan-59 | 0.227083 | 0.270236 | 0 | 0 | 3.980553 | 5.900928 |
| MAGMA_D | Vandetanib | Jan-55 | 0.213448 | 0.270367 | 0 | 0 | 4.27627 | 6.604112 |
| MAGMA_D | Simvastatin | Jan-55 | 0.213448 | 0.270367 | 0 | 0 | 4.27627 | 6.604112 |
| Jensen_TIS | BTO:00029 | Jan-44 | 0.174713 | 0.270469 | 0 | 0 | 5.373175 | 9.3741 |
| GeDiPNet_ | Ankylosing | 2/172 | 0.172129 | 0.270489 | 0 | 0 | 2.732595 | 4.808027 |
| Rare_Disea | Subependy | 2/126 | 0.104352 | 0.270517 | 0 | 0 | 3.755028 | 8.486292 |
| Rare_Disea | Fibrosing a | 2/126 | 0.104352 | 0.270517 | 0 | 0 | 3.755028 | 8.486292 |
| Rare_Disea | Pilocytic as | 2/126 | 0.104352 | 0.270517 | 0 | 0 | 3.755028 | 8.486292 |
| Rare_Disea | Anaplastic | 2/126 | 0.104352 | 0.270517 | 0 | 0 | 3.755028 | 8.486292 |
| Rare_Disea | Diffuse ast | 2/126 | 0.104352 | 0.270517 | 0 | 0 | 3.755028 | 8.486292 |
| Rare_Disea | Restless leg | 25-Jan | 0.103317 | 0.270517 | 0 | 0 | 9.636143 | 21.87363 |
| Rare_Disea | Thyroid car | 25-Jan | 0.103317 | 0.270517 | 0 | 0 | 9.636143 | 21.87363 |
| Rare_Disea | SARS | 2/125 | 0.102976 | 0.270517 | 0 | 0 | 3.785748 | 8.60599 |
| Rare_Disea | Pulmonary | 25-Jan | 0.103317 | 0.270517 | 0 | 0 | 9.636143 | 21.87363 |
| Rare_Disea | Compleme | 2/124 | 0.101605 | 0.270517 | 0 | 0 | 3.816972 | 8.728145 |

| | | | | | | | | |
|---|---|---|---|---|---|---|---|---|
| Rare_Disea | Fazio Londe | 25-Jan | 0.103317 | 0.270517 | 0 | 0 | 9.636143 | 21.87363 |
| UK_Bioban | Mean plate | 5/734 | 0.215062 | 0.270994 | 0 | 0 | 1.604604 | 2.466002 |
| UK_Bioban | Platelet co | 5/735 | 0.215845 | 0.270994 | 0 | 0 | 1.602322 | 2.456674 |
| UK_Bioban | Takes none | Jan-56 | 0.216879 | 0.271099 | 0 | 0 | 4.198309 | 6.416764 |
| MAGMA_D | Lymphangi | Jan-57 | 0.220295 | 0.271289 | 0 | 0 | 4.123131 | 6.237423 |
| MAGMA_D | Nifedipine | Jan-57 | 0.220295 | 0.271289 | 0 | 0 | 4.123131 | 6.237423 |
| MAGMA_D | Sirolimus | Jan-57 | 0.220295 | 0.271289 | 0 | 0 | 4.123131 | 6.237423 |
| GeDiPNet_ | Liver Fibros | 2/173 | 0.173677 | 0.271458 | 0 | 0 | 2.716477 | 4.755345 |
| GeDiPNet_ | Rod-cone D | 2/173 | 0.173677 | 0.271458 | 0 | 0 | 2.716477 | 4.755345 |
| Rare_Disea | Cartilagino | 3/261 | 0.105224 | 0.271829 | 0 | 0 | 2.720792 | 6.1263 |
| GWAS_Cat | Triglyceride | Jan-63 | 0.240485 | 0.27191 | 0 | 0 | 3.722993 | 5.305634 |
| GWAS_Cat | Cholesteryl | Jan-63 | 0.240485 | 0.27191 | 0 | 0 | 3.722993 | 5.305634 |
| GWAS_Cat | DNA Methy | 2/215 | 0.240243 | 0.27191 | 0 | 0 | 2.176194 | 3.103476 |
| GWAS_Cat | Cognitive F | Jan-63 | 0.240485 | 0.27191 | 0 | 0 | 3.722993 | 5.305634 |
| GWAS_Cat | Concentrat | Jan-63 | 0.240485 | 0.27191 | 0 | 0 | 3.722993 | 5.305634 |
| GO_Biologi | Cellular Re: | Jan-42 | 0.16747 | 0.272091 | 0 | 0 | 5.635848 | 10.07097 |
| GO_Biologi | Lipid Modif | Jan-42 | 0.16747 | 0.272091 | 0 | 0 | 5.635848 | 10.07097 |
| GO_Biologi | Regulation | Jan-42 | 0.16747 | 0.272091 | 0 | 0 | 5.635848 | 10.07097 |
| GO_Biologi | Regulation | Jan-42 | 0.16747 | 0.272091 | 0 | 0 | 5.635848 | 10.07097 |
| GO_Biologi | Keratinocy | Jan-42 | 0.16747 | 0.272091 | 0 | 0 | 5.635848 | 10.07097 |
| GeDiPNet_ | Ovarian Ep | Jan-44 | 0.174713 | 0.272347 | 0 | 0 | 5.373175 | 9.3741 |
| DisGeNET | Amyloidosi | 6/833 | 0.154585 | 0.272368 | 0 | 0 | 1.709526 | 3.191707 |
| DisGeNET | Celiac Dise | 3/312 | 0.154705 | 0.272398 | 0 | 0 | 2.265834 | 4.228577 |
| GeDiPNet_ | Fatty Liver | 2/174 | 0.175228 | 0.272421 | 0 | 0 | 2.700547 | 4.703455 |
| PhenGenI_ | Diet | 18-Jan | 0.075502 | 0.272513 | 0 | 0 | 13.60876 | 35.1596 |
| PhenGenI_ | Seizures, Fe | 18-Jan | 0.075502 | 0.272513 | 0 | 0 | 13.60876 | 35.1596 |
| PhenGenI_ | Cholestero | 9/1192 | 0.074145 | 0.272513 | 0 | 0 | 1.826842 | 4.752958 |
| PhenGenI_ | Thyrotropi | 3/225 | 0.074986 | 0.272513 | 0 | 0 | 3.167793 | 8.206005 |
| PhenGenI_ | Frontotem | 17-Jan | 0.071459 | 0.272513 | 0 | 0 | 14.46003 | 38.15469 |
| DisGeNET | Primary ma | Jan-39 | 0.156488 | 0.273333 | 0 | 0 | 6.081701 | 11.28018 |
| DisGeNET | Orbital sep | Jan-39 | 0.156488 | 0.273333 | 0 | 0 | 6.081701 | 11.28018 |
| DisGeNET | Optic Neur | Jan-39 | 0.156488 | 0.273333 | 0 | 0 | 6.081701 | 11.28018 |
| DisGeNET | Abnormal c | Jan-39 | 0.156488 | 0.273333 | 0 | 0 | 6.081701 | 11.28018 |
| DisGeNET | Klinefelter' | Jan-39 | 0.156488 | 0.273333 | 0 | 0 | 6.081701 | 11.28018 |
| DisGeNET | heavy drinl | Jan-39 | 0.156488 | 0.273333 | 0 | 0 | 6.081701 | 11.28018 |
| DisGeNET | Ovarian fai | Jan-39 | 0.156488 | 0.273333 | 0 | 0 | 6.081701 | 11.28018 |
| DisGeNET | Growth Dis | Jan-39 | 0.156488 | 0.273333 | 0 | 0 | 6.081701 | 11.28018 |
| DisGeNET | Gowers sig | Jan-39 | 0.156488 | 0.273333 | 0 | 0 | 6.081701 | 11.28018 |
| DisGeNET | Atypical He | Jan-39 | 0.156488 | 0.273333 | 0 | 0 | 6.081701 | 11.28018 |
| DisGeNET | Fasting blo | Jan-39 | 0.156488 | 0.273333 | 0 | 0 | 6.081701 | 11.28018 |
| DisGeNET | Lymphatic | Jan-39 | 0.156488 | 0.273333 | 0 | 0 | 6.081701 | 11.28018 |
| DisGeNET | Nephritis | 2/162 | 0.156786 | 0.273671 | 0 | 0 | 2.904853 | 5.382321 |
| CellMarker | Myeloid Ce | Jan-54 | 0.210002 | 0.274003 | 0 | 0 | 4.357174 | 6.79997 |
| UK_Bioban | Age at mer | Jan-57 | 0.220295 | 0.274166 | 0 | 0 | 4.123131 | 6.237423 |
| GeDiPNet_ | Stomach N | 3/333 | 0.176892 | 0.274277 | 0 | 0 | 2.119372 | 3.671207 |

| | | | | | | | | |
|---|---|---|---|---|---|---|---|---|
| CellMarker | Type II Spir | 4/543 | 0.211234 | 0.274304 | 0 | 0 | 1.732257 | 2.69329 |
| Jensen_TIS | Mammary | Jan-45 | 0.178311 | 0.274718 | 0 | 0 | 5.250793 | 9.053559 |
| Rare_Disea | Astroblastc | 2/128 | 0.107121 | 0.27482 | 0 | 0 | 3.695051 | 8.253999 |
| Rare_Disea | Gliomatosi | 2/128 | 0.107121 | 0.27482 | 0 | 0 | 3.695051 | 8.253999 |
| GeDiPNet_ | Thyroiditis | Jan-45 | 0.178311 | 0.275014 | 0 | 0 | 5.250793 | 9.053559 |
| GeDiPNet_ | Kidney Can | Jan-45 | 0.178311 | 0.275014 | 0 | 0 | 5.250793 | 9.053559 |
| GWAS_Cat | Phospholip | Jan-64 | 0.243799 | 0.275148 | 0 | 0 | 3.663714 | 5.171007 |
| GWAS_Cat | Autoimmu | Jan-64 | 0.243799 | 0.275148 | 0 | 0 | 3.663714 | 5.171007 |
| CellMarker | Mesangial | 2/198 | 0.213017 | 0.275314 | 0 | 0 | 2.366987 | 3.660273 |
| GO_Biologi | Response T | Jan-43 | 0.171099 | 0.275466 | 0 | 0 | 5.501384 | 9.712751 |
| GO_Biologi | Regulation | Jan-43 | 0.171099 | 0.275466 | 0 | 0 | 5.501384 | 9.712751 |
| GO_Biologi | Regulation | Jan-43 | 0.171099 | 0.275466 | 0 | 0 | 5.501384 | 9.712751 |
| GO_Biologi | Energy Der | Jan-43 | 0.171099 | 0.275466 | 0 | 0 | 5.501384 | 9.712751 |
| GeDiPNet_ | Cerebellar | Jan-46 | 0.181893 | 0.276156 | 0 | 0 | 5.13385 | 8.749803 |
| GeDiPNet_ | Exudative I | Jan-46 | 0.181893 | 0.276156 | 0 | 0 | 5.13385 | 8.749803 |
| GeDiPNet_ | Ovarian Cy | Jan-46 | 0.181893 | 0.276156 | 0 | 0 | 5.13385 | 8.749803 |
| GeDiPNet_ | Myeloid Le | 2/177 | 0.179893 | 0.276156 | 0 | 0 | 2.653849 | 4.552394 |
| GeDiPNet_ | Language D | Jan-46 | 0.181893 | 0.276156 | 0 | 0 | 5.13385 | 8.749803 |
| GeDiPNet_ | Dysmorphi | 3/337 | 0.181217 | 0.276156 | 0 | 0 | 2.093563 | 3.575931 |
| Jensen_TIS | Artery | 3/336 | 0.180133 | 0.276204 | 0 | 0 | 2.099957 | 3.599451 |
| GO_Biologi | Generatior | 2/172 | 0.172129 | 0.276497 | 0 | 0 | 2.732595 | 4.808027 |
| Human_Ph | Retinal det | Jan-61 | 0.233813 | 0.276522 | 0 | 0 | 3.847481 | 5.591291 |
| Human_Ph | Metabolic a | Jan-61 | 0.233813 | 0.276522 | 0 | 0 | 3.847481 | 5.591291 |
| UK_Bioban | Right mear | Jan-58 | 0.223696 | 0.277189 | 0 | 0 | 4.050592 | 6.065622 |
| Rare_Disea | Narcolepsy | 3/265 | 0.108844 | 0.277329 | 0 | 0 | 2.678708 | 5.940942 |
| Rare_Disea | Primary lat | 2/129 | 0.108512 | 0.277329 | 0 | 0 | 3.665771 | 8.141279 |
| DisGeNET | Malignant | Jan-40 | 0.160165 | 0.277351 | 0 | 0 | 5.925462 | 10.85279 |
| DisGeNET | Muscular D | Jan-40 | 0.160165 | 0.277351 | 0 | 0 | 5.925462 | 10.85279 |
| DisGeNET | Anxiety Sta | Jan-40 | 0.160165 | 0.277351 | 0 | 0 | 5.925462 | 10.85279 |
| DisGeNET | AIDS Deme | Jan-40 | 0.160165 | 0.277351 | 0 | 0 | 5.925462 | 10.85279 |
| DisGeNET | Anemia, se | Jan-40 | 0.160165 | 0.277351 | 0 | 0 | 5.925462 | 10.85279 |
| DisGeNET | Sparse pub | Jan-40 | 0.160165 | 0.277351 | 0 | 0 | 5.925462 | 10.85279 |
| DisGeNET | Hepatitis | 3/317 | 0.159904 | 0.277351 | 0 | 0 | 2.229186 | 4.086495 |
| DisGeNET | Islet Cell Tu | Jan-40 | 0.160165 | 0.277351 | 0 | 0 | 5.925462 | 10.85279 |
| DisGeNET | Pneumonia | Jan-40 | 0.160165 | 0.277351 | 0 | 0 | 5.925462 | 10.85279 |
| DisGeNET | Rhizomelia | Jan-40 | 0.160165 | 0.277351 | 0 | 0 | 5.925462 | 10.85279 |
| DisGeNET | Degenerati | Jan-40 | 0.160165 | 0.277351 | 0 | 0 | 5.925462 | 10.85279 |
| DisGeNET | Contractur | Jan-40 | 0.160165 | 0.277351 | 0 | 0 | 5.925462 | 10.85279 |
| GWAS_Cat | Cholestero | Jan-65 | 0.247099 | 0.277844 | 0 | 0 | 3.606286 | 5.041467 |
| GWAS_Cat | Total Lipid | Jan-65 | 0.247099 | 0.277844 | 0 | 0 | 3.606286 | 5.041467 |
| GWAS_Cat | Cholestero | Jan-65 | 0.247099 | 0.277844 | 0 | 0 | 3.606286 | 5.041467 |
| GWAS_Cat | Ascending | Jan-65 | 0.247099 | 0.277844 | 0 | 0 | 3.606286 | 5.041467 |
| GO_Biologi | Establishm | Jan-44 | 0.174713 | 0.27813 | 0 | 0 | 5.373175 | 9.3741 |
| GO_Biologi | Response T | Jan-44 | 0.174713 | 0.27813 | 0 | 0 | 5.373175 | 9.3741 |
| GO_Biologi | Positive Re | Jan-44 | 0.174713 | 0.27813 | 0 | 0 | 5.373175 | 9.3741 |

| | | | | | | | | |
|---|---|---|---|---|---|---|---|---|
| GO_Biologi | Bone Deve | Jan-44 | 0.174713 | 0.27813 | 0 | 0 | 5.373175 | 9.3741 |
| GO_Biologi | Regulation | 2/174 | 0.175228 | 0.278326 | 0 | 0 | 2.700547 | 4.703455 |
| PhenGenI_ | Subcutane | 2/107 | 0.079148 | 0.278344 | 0 | 0 | 4.438768 | 11.25866 |
| PhenGenI_ | Diagnostic | 19-Jan | 0.079527 | 0.278344 | 0 | 0 | 12.85207 | 32.53709 |
| UK_Bioban | Reticulocyt | 6/941 | 0.225721 | 0.278487 | 0 | 0 | 1.503506 | 2.2379 |
| GTEx_Tissu | GTEX-PWC | 14/1529 | 0.006065 | 0.278528 | 0 | 0 | 2.328966 | 11.88972 |
| GeDiPNet_ | Trigonocep | Jan-47 | 0.18546 | 0.278668 | 0 | 0 | 5.021992 | 8.46163 |
| GeDiPNet_ | Syndactyly | 2/180 | 0.184577 | 0.278668 | 0 | 0 | 2.608724 | 4.407929 |
| GeDiPNet_ | Hyperlipide | Jan-47 | 0.18546 | 0.278668 | 0 | 0 | 5.021992 | 8.46163 |
| GeDiPNet_ | Uterine Ce | Jan-47 | 0.18546 | 0.278668 | 0 | 0 | 5.021992 | 8.46163 |
| Human_Ph | Frontal bos | 2/213 | 0.237028 | 0.278749 | 0 | 0 | 2.197045 | 3.162813 |
| Human_Ph | Distal sens | Jan-62 | 0.237156 | 0.278749 | 0 | 0 | 3.784217 | 5.445629 |
| Rare_Disea | Factor XIII | 27-Jan | 0.111111 | 0.279278 | 0 | 0 | 8.894007 | 19.54216 |
| Rare_Disea | Conjunctiva | 27-Jan | 0.111111 | 0.279278 | 0 | 0 | 8.894007 | 19.54216 |
| Rare_Disea | Syndactyly | 27-Jan | 0.111111 | 0.279278 | 0 | 0 | 8.894007 | 19.54216 |
| Rare_Disea | Microphtha | 27-Jan | 0.111111 | 0.279278 | 0 | 0 | 8.894007 | 19.54216 |
| ClinVar_20 | retinitis pig | Jan-75 | 0.279326 | 0.279326 | 0 | 0 | 3.117379 | 3.975827 |
| Disease_Pe | juvenile de | Mar-97 | 0.008735 | 0.280394 | 0 | 0 | 7.530015 | 35.6954 |
| Disease_Pe | cerebral pa | 5/288 | 0.008463 | 0.280394 | 0 | 0 | 4.22951 | 20.18361 |
| Jensen_DIS | Carcinoma | 53/11318 | 0.240267 | 0.280552 | 0 | 0 | 1.196689 | 1.706482 |
| MAGMA_D | Primary Dy | Jan-63 | 0.240485 | 0.280565 | 0 | 0 | 3.722993 | 5.305634 |
| MAGMA_D | Heart Attac | Jan-62 | 0.237156 | 0.280565 | 0 | 0 | 3.784217 | 5.445629 |
| MAGMA_D | ATC Code C | Jan-62 | 0.237156 | 0.280565 | 0 | 0 | 3.784217 | 5.445629 |
| MAGMA_D | Celecoxib | Jan-63 | 0.240485 | 0.280565 | 0 | 0 | 3.722993 | 5.305634 |
| MAGMA_D | HMGCR Inh | Jan-62 | 0.237156 | 0.280565 | 0 | 0 | 3.784217 | 5.445629 |
| MAGMA_D | Atorvastati | Jan-62 | 0.237156 | 0.280565 | 0 | 0 | 3.784217 | 5.445629 |
| GWAS_Cat | Intracrania | Jan-66 | 0.250385 | 0.280762 | 0 | 0 | 3.550626 | 4.916752 |
| GWAS_Cat | Coronary A | Jan-66 | 0.250385 | 0.280762 | 0 | 0 | 3.550626 | 4.916752 |
| GWAS_Cat | Central Cor | Jan-66 | 0.250385 | 0.280762 | 0 | 0 | 3.550626 | 4.916752 |
| Jensen_TIS | Uterine cer | 56/11814 | 0.185001 | 0.280992 | 0 | 0 | 1.252901 | 2.11414 |
| Jensen_TIS | Placenta | 73/15885 | 0.184241 | 0.280992 | 0 | 0 | 1.352377 | 2.287561 |
| DisGeNET | Alcohol wit | Jan-41 | 0.163825 | 0.281642 | 0 | 0 | 5.777035 | 10.45039 |
| DisGeNET | Malignant | 2/166 | 0.162892 | 0.281642 | 0 | 0 | 2.833429 | 5.141737 |
| DisGeNET | Endometri | Jan-41 | 0.163825 | 0.281642 | 0 | 0 | 5.777035 | 10.45039 |
| DisGeNET | Anaplastic | 2/166 | 0.162892 | 0.281642 | 0 | 0 | 2.833429 | 5.141737 |
| DisGeNET | AIDS relate | Jan-41 | 0.163825 | 0.281642 | 0 | 0 | 5.777035 | 10.45039 |
| DisGeNET | Human imr | Jan-41 | 0.163825 | 0.281642 | 0 | 0 | 5.777035 | 10.45039 |
| DisGeNET | Pancreatic | 5/665 | 0.163578 | 0.281642 | 0 | 0 | 1.778732 | 3.220336 |
| DisGeNET | Irregular hy | Jan-41 | 0.163825 | 0.281642 | 0 | 0 | 5.777035 | 10.45039 |
| DisGeNET | Alcoholic Ir | Jan-41 | 0.163825 | 0.281642 | 0 | 0 | 5.777035 | 10.45039 |
| DisGeNET | Chikungun | Jan-41 | 0.163825 | 0.281642 | 0 | 0 | 5.777035 | 10.45039 |
| DisGeNET | alpha 1-An | Jan-41 | 0.163825 | 0.281642 | 0 | 0 | 5.777035 | 10.45039 |
| GeDiPNet_ | Microceph | 6/886 | 0.188142 | 0.281972 | 0 | 0 | 1.602104 | 2.676405 |
| CellMarker | Cycling Bas | 2/202 | 0.219398 | 0.282231 | 0 | 0 | 2.319176 | 3.517879 |
| GWAS_Cat | Eosinophil | 5/780 | 0.251961 | 0.28227 | 0 | 0 | 1.505744 | 2.075639 |

| | | | | | | | | |
|---|---|---|---|---|---|---|---|---|
| GeDiPNet_ | Cardiovasc | Jan-48 | 0.189012 | 0.282548 | 0 | 0 | 4.914894 | 8.187947 |
| GO_Biologi | Chromosor | Jan-45 | 0.178311 | 0.282591 | 0 | 0 | 5.250793 | 9.053559 |
| GeneSigDB | 16373702-1 | 19-Feb | 0.003048 | 0.282993 | 0 | 0 | 27.53772 | 159.5298 |
| GeneSigDB | 20035825-1 | 19-Feb | 0.003048 | 0.282993 | 0 | 0 | 27.53772 | 159.5298 |
| GeneSigDB | 16404635-1 | 19-Feb | 0.003048 | 0.282993 | 0 | 0 | 27.53772 | 159.5298 |
| GeneSigDB | 20035825-1 | 17-Feb | 0.002438 | 0.282993 | 0 | 0 | 31.21255 | 187.7904 |
| UK_Bioban | Self-report | Jan-60 | 0.230455 | 0.283102 | 0 | 0 | 3.912889 | 5.742942 |
| GWAS_Cat | Ratio Of Tr | Jan-67 | 0.253656 | 0.283389 | 0 | 0 | 3.496653 | 4.79662 |
| GWAS_Cat | White Bloo | Jan-67 | 0.253656 | 0.283389 | 0 | 0 | 3.496653 | 4.79662 |
| GWAS_Cat | Free Chole: | Jan-67 | 0.253656 | 0.283389 | 0 | 0 | 3.496653 | 4.79662 |
| Jensen_TIS | Stromal cel | 2/182 | 0.18771 | 0.283768 | 0 | 0 | 2.579477 | 4.315096 |
| PhenGenI_ | Dupuytren | 20-Jan | 0.083535 | 0.283772 | 0 | 0 | 12.17503 | 30.22444 |
| PhenGenI_ | Caffeine | 20-Jan | 0.083535 | 0.283772 | 0 | 0 | 12.17503 | 30.22444 |
| GTEx_Tissu | GTEX-Q2AI | 15/1709 | 0.006591 | 0.284171 | 0 | 0 | 2.240629 | 11.25267 |
| GTEx_Tissu | GTEX-PW2 | 14/1543 | 0.006561 | 0.284171 | 0 | 0 | 2.305885 | 11.59072 |
| GTEx_Tissu | GTEX-S4Z8 | 6/383 | 0.006516 | 0.284171 | 0 | 0 | 3.838491 | 19.32131 |
| GTEx_Tissu | GTEX-X3Y1 | 8/635 | 0.006395 | 0.284171 | 0 | 0 | 3.114853 | 15.73705 |
| GTEx_Tissu | GTEX-SNM | 11/1072 | 0.006679 | 0.284171 | 0 | 0 | 2.571705 | 12.88095 |
| Rare_Disea | Light chain | 28-Jan | 0.114983 | 0.284209 | 0 | 0 | 8.564169 | 18.52408 |
| Rare_Disea | CADASIL | 28-Jan | 0.114983 | 0.284209 | 0 | 0 | 8.564169 | 18.52408 |
| Rare_Disea | Urethral ca | 28-Jan | 0.114983 | 0.284209 | 0 | 0 | 8.564169 | 18.52408 |
| Rare_Disea | Autoimmu | 28-Jan | 0.114983 | 0.284209 | 0 | 0 | 8.564169 | 18.52408 |
| Rare_Disea | Ossificatior | 28-Jan | 0.114983 | 0.284209 | 0 | 0 | 8.564169 | 18.52408 |
| DisGeNET | Dyskeratos | Jan-42 | 0.16747 | 0.284412 | 0 | 0 | 5.635848 | 10.07097 |
| DisGeNET | Diffuse Scle | Jan-42 | 0.16747 | 0.284412 | 0 | 0 | 5.635848 | 10.07097 |
| DisGeNET | Degenerati | Jan-42 | 0.16747 | 0.284412 | 0 | 0 | 5.635848 | 10.07097 |
| DisGeNET | Hernia | Jan-42 | 0.16747 | 0.284412 | 0 | 0 | 5.635848 | 10.07097 |
| DisGeNET | Aortic aneu | Jan-42 | 0.16747 | 0.284412 | 0 | 0 | 5.635848 | 10.07097 |
| DisGeNET | Infection b | Jan-42 | 0.16747 | 0.284412 | 0 | 0 | 5.635848 | 10.07097 |
| DisGeNET | Acute leuk | 4/492 | 0.166565 | 0.284412 | 0 | 0 | 1.918329 | 3.438356 |
| DisGeNET | Breast ade | 2/168 | 0.165961 | 0.284412 | 0 | 0 | 2.799008 | 5.027029 |
| DisGeNET | Malignant | 4/492 | 0.166565 | 0.284412 | 0 | 0 | 1.918329 | 3.438356 |
| DisGeNET | Clear Cell S | Jan-42 | 0.16747 | 0.284412 | 0 | 0 | 5.635848 | 10.07097 |
| DisGeNET | T-Cell Lymp | 3/324 | 0.167273 | 0.284412 | 0 | 0 | 2.179795 | 3.897752 |
| DisGeNET | Optic Neur | Jan-42 | 0.16747 | 0.284412 | 0 | 0 | 5.635848 | 10.07097 |
| DisGeNET | Dystrophic | Jan-42 | 0.16747 | 0.284412 | 0 | 0 | 5.635848 | 10.07097 |
| DisGeNET | Ragged-rec | Jan-42 | 0.16747 | 0.284412 | 0 | 0 | 5.635848 | 10.07097 |
| DisGeNET | Defective t | Jan-42 | 0.16747 | 0.284412 | 0 | 0 | 5.635848 | 10.07097 |
| DisGeNET | Cyst | 2/169 | 0.167499 | 0.284412 | 0 | 0 | 2.782106 | 4.971004 |
| DisGeNET | Papillary Re | 2/169 | 0.167499 | 0.284412 | 0 | 0 | 2.782106 | 4.971004 |
| DisGeNET | exudative r | Jan-42 | 0.16747 | 0.284412 | 0 | 0 | 5.635848 | 10.07097 |
| DisGeNET | Cleft upper | 2/169 | 0.167499 | 0.284412 | 0 | 0 | 2.782106 | 4.971004 |
| GeDiPNet_ | Brachycepl | 2/184 | 0.19085 | 0.284567 | 0 | 0 | 2.550873 | 4.224921 |
| Rare_Disea | Glioma | 2/134 | 0.115542 | 0.284647 | 0 | 0 | 3.526025 | 7.609588 |
| Human_Ph | Hypermela | Jan-64 | 0.243799 | 0.285679 | 0 | 0 | 3.663714 | 5.171007 |

| | | | | | | | | |
|---|---|---|---|---|---|---|---|---|
| GO_Biologi | Regulation | Jan-46 | 0.181893 | 0.285717 | 0 | 0 | 5.13385 | 8.749803 |
| GO_Biologi | Negative R | Jan-46 | 0.181893 | 0.285717 | 0 | 0 | 5.13385 | 8.749803 |
| GO_Biologi | Response T | Jan-46 | 0.181893 | 0.285717 | 0 | 0 | 5.13385 | 8.749803 |
| GO_Biologi | Intracellula | Jan-46 | 0.181893 | 0.285717 | 0 | 0 | 5.13385 | 8.749803 |
| UK_Bioban | Nervous fe | Jan-61 | 0.233813 | 0.285994 | 0 | 0 | 3.847481 | 5.591291 |
| GeDiPNet_ | Cholestasis | 2/185 | 0.192423 | 0.286181 | 0 | 0 | 2.536805 | 4.180799 |
| Jensen_DIS | Multiple sc | 2/219 | 0.246681 | 0.286326 | 0 | 0 | 2.135647 | 2.989182 |
| GWAS_Cat | Concentrat | Jan-68 | 0.256914 | 0.286504 | 0 | 0 | 3.44429 | 4.68084 |
| GWAS_Cat | Cholesteryl | Jan-68 | 0.256914 | 0.286504 | 0 | 0 | 3.44429 | 4.68084 |
| DisGeNET | Respiratory | 2/170 | 0.16904 | 0.286657 | 0 | 0 | 2.765406 | 4.91584 |
| DisGeNET | Developme | 2/170 | 0.16904 | 0.286657 | 0 | 0 | 2.765406 | 4.91584 |
| Gene_Pert | Cav3 Defici | 6/275 | 0.001274 | 0.286902 | 0 | 0 | 5.409335 | 36.05776 |
| Gene_Pert | BMI1 Defic | 6/303 | 0.00208 | 0.286902 | 0 | 0 | 4.892381 | 30.21198 |
| Gene_Pert | Dmd Defici | 5/191 | 0.001479 | 0.286902 | 0 | 0 | 6.467021 | 42.14213 |
| Gene_Pert | Akt2 knock | 7/368 | 0.001116 | 0.286902 | 0 | 0 | 4.739058 | 32.21584 |
| Gene_Pert | Lama2 Defi | 7/362 | 0.001015 | 0.286902 | 0 | 0 | 4.820634 | 33.22899 |
| Gene_Pert | NET1 knocl | 5/159 | 0.000652 | 0.286902 | 0 | 0 | 7.823487 | 57.39044 |
| Gene_Pert | Hras mous | 7/403 | 0.001876 | 0.286902 | 0 | 0 | 4.312468 | 27.07695 |
| Gene_Pert | NET1 knocl | 6/300 | 0.001979 | 0.286902 | 0 | 0 | 4.943059 | 30.77152 |
| Gene_Pert | INSR mutat | 6/290 | 0.001668 | 0.286902 | 0 | 0 | 5.119718 | 32.74548 |
| Gene_Pert | Cstb KO mc | 7/387 | 0.00149 | 0.286902 | 0 | 0 | 4.49773 | 29.27507 |
| DisGeNET | Depressive | Jan-43 | 0.171099 | 0.287544 | 0 | 0 | 5.501384 | 9.712751 |
| DisGeNET | Dysplasia c | Jan-43 | 0.171099 | 0.287544 | 0 | 0 | 5.501384 | 9.712751 |
| DisGeNET | Coinfectior | 2/171 | 0.170583 | 0.287544 | 0 | 0 | 2.748904 | 4.86152 |
| DisGeNET | Blood Coag | 2/171 | 0.170583 | 0.287544 | 0 | 0 | 2.748904 | 4.86152 |
| DisGeNET | Secondary | Jan-43 | 0.171099 | 0.287544 | 0 | 0 | 5.501384 | 9.712751 |
| DisGeNET | Thin denta | Jan-43 | 0.171099 | 0.287544 | 0 | 0 | 5.501384 | 9.712751 |
| DisGeNET | Primary Ca | Jan-43 | 0.171099 | 0.287544 | 0 | 0 | 5.501384 | 9.712751 |
| DisGeNET | Hyperlipop | Jan-43 | 0.171099 | 0.287544 | 0 | 0 | 5.501384 | 9.712751 |
| DisGeNET | Tooth Abnc | Jan-43 | 0.171099 | 0.287544 | 0 | 0 | 5.501384 | 9.712751 |
| DisGeNET | Autoimmu | Jan-43 | 0.171099 | 0.287544 | 0 | 0 | 5.501384 | 9.712751 |
| DisGeNET | Macroceph | Jan-43 | 0.171099 | 0.287544 | 0 | 0 | 5.501384 | 9.712751 |
| DisGeNET | Gemistocyt | Jan-43 | 0.171099 | 0.287544 | 0 | 0 | 5.501384 | 9.712751 |
| DisGeNET | Defective e | Jan-43 | 0.171099 | 0.287544 | 0 | 0 | 5.501384 | 9.712751 |
| DisGeNET | Acute resp | Jan-43 | 0.171099 | 0.287544 | 0 | 0 | 5.501384 | 9.712751 |
| GeDiPNet_ | Glaucoma | 3/349 | 0.194361 | 0.288327 | 0 | 0 | 2.019715 | 3.308368 |
| GeDiPNet_ | Secondary | Jan-50 | 0.196069 | 0.288657 | 0 | 0 | 4.713811 | 7.680157 |
| GeDiPNet_ | Thyroid Gla | Jan-50 | 0.196069 | 0.288657 | 0 | 0 | 4.713811 | 7.680157 |
| GeDiPNet_ | Seizure | 3/350 | 0.195467 | 0.288657 | 0 | 0 | 2.013792 | 3.287236 |
| UK_Bioban | Blonde hai | 2/213 | 0.237028 | 0.288688 | 0 | 0 | 2.197045 | 3.162813 |
| OMIM_Dis | leukemia | Jan-78 | 0.288726 | 0.288726 | 0 | 0 | 2.99547 | 3.721207 |
| GO_Biologi | Secondary | Jan-47 | 0.18546 | 0.288765 | 0 | 0 | 5.021992 | 8.46163 |
| GO_Biologi | Regulation | Jan-47 | 0.18546 | 0.288765 | 0 | 0 | 5.021992 | 8.46163 |
| GO_Biologi | RNA Splicir | 2/180 | 0.184577 | 0.288765 | 0 | 0 | 2.608724 | 4.407929 |
| GO_Biologi | Positive Re | Jan-47 | 0.18546 | 0.288765 | 0 | 0 | 5.021992 | 8.46163 |

| | | | | | | | | |
|---|---|---|---|---|---|---|---|---|
| GWAS_Cat | Autism Spe | Jan-69 | 0.260157 | 0.289327 | 0 | 0 | 3.393468 | 4.569198 |
| GWAS_Cat | Haemorrho | Jan-69 | 0.260157 | 0.289327 | 0 | 0 | 3.393468 | 4.569198 |
| GWAS_Cat | Celiac Dise | Jan-69 | 0.260157 | 0.289327 | 0 | 0 | 3.393468 | 4.569198 |
| GeDiPNet_ | Macrotia | 2/188 | 0.197153 | 0.289521 | 0 | 0 | 2.495509 | 4.05215 |
| DisGeNET | Neurocogn | Jan-44 | 0.174713 | 0.289714 | 0 | 0 | 5.373175 | 9.3741 |
| DisGeNET | Confusion | Jan-44 | 0.174713 | 0.289714 | 0 | 0 | 5.373175 | 9.3741 |
| DisGeNET | Birth Weig | Jan-44 | 0.174713 | 0.289714 | 0 | 0 | 5.373175 | 9.3741 |
| DisGeNET | Abnormal f | Jan-44 | 0.174713 | 0.289714 | 0 | 0 | 5.373175 | 9.3741 |
| DisGeNET | Acquired a | Jan-44 | 0.174713 | 0.289714 | 0 | 0 | 5.373175 | 9.3741 |
| DisGeNET | HELLP Sync | Jan-44 | 0.174713 | 0.289714 | 0 | 0 | 5.373175 | 9.3741 |
| DisGeNET | Thyroiditis | Jan-44 | 0.174713 | 0.289714 | 0 | 0 | 5.373175 | 9.3741 |
| DisGeNET | Strawberry | Jan-44 | 0.174713 | 0.289714 | 0 | 0 | 5.373175 | 9.3741 |
| DisGeNET | Apraxias | Jan-44 | 0.174713 | 0.289714 | 0 | 0 | 5.373175 | 9.3741 |
| DisGeNET | Memory Di | Jan-44 | 0.174713 | 0.289714 | 0 | 0 | 5.373175 | 9.3741 |
| DisGeNET | Osteopenia | 3/329 | 0.172598 | 0.289714 | 0 | 0 | 2.145815 | 3.769752 |
| DisGeNET | CARCINOM | Jan-44 | 0.174713 | 0.289714 | 0 | 0 | 5.373175 | 9.3741 |
| DisGeNET | Broad flat r | 3/330 | 0.173668 | 0.289714 | 0 | 0 | 2.139144 | 3.744801 |
| DisGeNET | Neoplasm c | Jan-44 | 0.174713 | 0.289714 | 0 | 0 | 5.373175 | 9.3741 |
| DisGeNET | Muscular s | Jan-44 | 0.174713 | 0.289714 | 0 | 0 | 5.373175 | 9.3741 |
| DisGeNET | Birthmark | Jan-44 | 0.174713 | 0.289714 | 0 | 0 | 5.373175 | 9.3741 |
| DisGeNET | Japanese E | Jan-44 | 0.174713 | 0.289714 | 0 | 0 | 5.373175 | 9.3741 |
| DisGeNET | Acute ante | Jan-44 | 0.174713 | 0.289714 | 0 | 0 | 5.373175 | 9.3741 |
| DisGeNET | Multiple Sc | Jan-44 | 0.174713 | 0.289714 | 0 | 0 | 5.373175 | 9.3741 |
| DisGeNET | Neoplasm I | 21/3920 | 0.174247 | 0.289714 | 0 | 0 | 1.306839 | 2.283416 |
| DisGeNET | Muscle dar | Jan-44 | 0.174713 | 0.289714 | 0 | 0 | 5.373175 | 9.3741 |
| Rare_Disea | Alexander ( | 2/136 | 0.118386 | 0.289735 | 0 | 0 | 3.473047 | 7.410799 |
| Rare_Disea | Glioblastor | 2/136 | 0.118386 | 0.289735 | 0 | 0 | 3.473047 | 7.410799 |
| Rare_Disea | Refsum dis | 29-Jan | 0.118838 | 0.289886 | 0 | 0 | 8.25789 | 17.58927 |
| Jensen_TIS | Vascular sy | 6/894 | 0.193446 | 0.290546 | 0 | 0 | 1.587004 | 2.607063 |
| Jensen_TIS | Ear | 2/186 | 0.193998 | 0.290546 | 0 | 0 | 2.52289 | 4.137304 |
| Diabetes_F | GSE150679 | 5/197 | 0.001694 | 0.290755 | 0 | 0 | 6.263021 | 39.96113 |
| Diabetes_F | GSE149468 | 5/214 | 0.00243 | 0.290755 | 0 | 0 | 5.748629 | 34.60665 |
| DisGeNET | melanoma | 14/2454 | 0.17547 | 0.290785 | 0 | 0 | 1.373355 | 2.390034 |
| DisGeNET | Cerebral Ar | Jan-45 | 0.178311 | 0.290934 | 0 | 0 | 5.250793 | 9.053559 |
| DisGeNET | Malignant ( | 4/504 | 0.17675 | 0.290934 | 0 | 0 | 1.871133 | 3.242707 |
| DisGeNET | Nasal bridg | 3/333 | 0.176892 | 0.290934 | 0 | 0 | 2.119372 | 3.671207 |
| DisGeNET | Obesity, Vi | Jan-45 | 0.178311 | 0.290934 | 0 | 0 | 5.250793 | 9.053559 |
| DisGeNET | Skin Diseas | Jan-45 | 0.178311 | 0.290934 | 0 | 0 | 5.250793 | 9.053559 |
| DisGeNET | Carcinoma | 2/176 | 0.178336 | 0.290934 | 0 | 0 | 2.669236 | 4.601995 |
| DisGeNET | Delirium | Jan-45 | 0.178311 | 0.290934 | 0 | 0 | 5.250793 | 9.053559 |
| DisGeNET | Congenital | Jan-45 | 0.178311 | 0.290934 | 0 | 0 | 5.250793 | 9.053559 |
| DisGeNET | Leukemia, | Jan-45 | 0.178311 | 0.290934 | 0 | 0 | 5.250793 | 9.053559 |
| DisGeNET | Cafe-au-Lai | Jan-45 | 0.178311 | 0.290934 | 0 | 0 | 5.250793 | 9.053559 |
| DisGeNET | Lipoatroph | Jan-45 | 0.178311 | 0.290934 | 0 | 0 | 5.250793 | 9.053559 |
| DisGeNET | Age-relatec | Jan-45 | 0.178311 | 0.290934 | 0 | 0 | 5.250793 | 9.053559 |

| | | | | | | | | |
|---|---|---|---|---|---|---|---|---|
| DisGeNET | Impulsive c | Jan-45 | 0.178311 | 0.290934 | 0 | 0 | 5.250793 | 9.053559 |
| DisGeNET | Well Differ | 2/176 | 0.178336 | 0.290934 | 0 | 0 | 2.669236 | 4.601995 |
| DisGeNET | Small nose | 2/176 | 0.178336 | 0.290934 | 0 | 0 | 2.669236 | 4.601995 |
| DisGeNET | Endometri | Jan-45 | 0.178311 | 0.290934 | 0 | 0 | 5.250793 | 9.053559 |
| DisGeNET | MYELOPRC | Jan-45 | 0.178311 | 0.290934 | 0 | 0 | 5.250793 | 9.053559 |
| DisGeNET | Liver Failur | 2/176 | 0.178336 | 0.290934 | 0 | 0 | 2.669236 | 4.601995 |
| DisGeNET | Mammary | 2/176 | 0.178336 | 0.290934 | 0 | 0 | 2.669236 | 4.601995 |
| DisGeNET | Hypopituit | Jan-45 | 0.178311 | 0.290934 | 0 | 0 | 5.250793 | 9.053559 |
| DisGeNET | Non-alcoho | 3/334 | 0.177971 | 0.290934 | 0 | 0 | 2.112861 | 3.647087 |
| DisGeNET | Autoimmu | 7/1060 | 0.177899 | 0.290934 | 0 | 0 | 1.567189 | 2.705815 |
| DisGeNET | Malignant | 4/505 | 0.177609 | 0.290934 | 0 | 0 | 1.867302 | 3.22702 |
| DisGeNET | Liver Cirrho | 5/683 | 0.176491 | 0.290934 | 0 | 0 | 1.729891 | 3.000474 |
| DisGeNET | Adenoma, | Jan-45 | 0.178311 | 0.290934 | 0 | 0 | 5.250793 | 9.053559 |
| GeDiPNet_ | Sleep Disor | 2/189 | 0.198732 | 0.291108 | 0 | 0 | 2.482038 | 4.010467 |
| Rare_Disea | Testicular c | 2/137 | 0.119815 | 0.291315 | 0 | 0 | 3.447146 | 7.31418 |
| GeDiPNet_ | Patent Fora | Jan-51 | 0.199575 | 0.29161 | 0 | 0 | 4.619302 | 7.444308 |
| Jensen_TIS | Pronephros | 5/709 | 0.19581 | 0.291902 | 0 | 0 | 1.663751 | 2.71293 |
| DisGeNET | Graves Dise | 3/335 | 0.179051 | 0.291919 | 0 | 0 | 2.10639 | 3.623169 |
| GWAS_Cat | Basophil Co | 2/229 | 0.262799 | 0.291999 | 0 | 0 | 2.040529 | 2.726893 |
| GeneSigDB | 18410693- | Mar-71 | 0.003671 | 0.29213 | 0 | 0 | 10.42279 | 58.44282 |
| DisGeNET | Genetic Dis | 4/507 | 0.17933 | 0.292192 | 0 | 0 | 1.859685 | 3.195922 |
| GWAS_Cat | Cholestero | Jan-70 | 0.263387 | 0.292386 | 0 | 0 | 3.344119 | 4.461494 |
| Human_Ph | Intellectual | Jan-66 | 0.250385 | 0.292498 | 0 | 0 | 3.550626 | 4.916752 |
| GO_Biologi | RNA Proces | 2/183 | 0.189279 | 0.292785 | 0 | 0 | 2.565096 | 4.269683 |
| GO_Biologi | Myelinatio | Jan-48 | 0.189012 | 0.292785 | 0 | 0 | 4.914894 | 8.187947 |
| GO_Biologi | Negative R | Jan-48 | 0.189012 | 0.292785 | 0 | 0 | 4.914894 | 8.187947 |
| Rare_Disea | Corpus call | 2/138 | 0.121248 | 0.292885 | 0 | 0 | 3.421626 | 7.219351 |
| Rare_Disea | Basal cell c | 2/138 | 0.121248 | 0.292885 | 0 | 0 | 3.421626 | 7.219351 |
| DisGeNET | Refractory | 2/177 | 0.179893 | 0.292927 | 0 | 0 | 2.653849 | 4.552394 |
| GWAS_Cat | A Body Sha | 3/410 | 0.264211 | 0.293034 | 0 | 0 | 1.711653 | 2.278224 |
| DisGeNET | Secondary | 4/508 | 0.180193 | 0.293233 | 0 | 0 | 1.8559 | 3.180508 |
| DisGeNET | Disseminat | Jan-46 | 0.181893 | 0.293448 | 0 | 0 | 5.13385 | 8.749803 |
| DisGeNET | Moyamoya | Jan-46 | 0.181893 | 0.293448 | 0 | 0 | 5.13385 | 8.749803 |
| DisGeNET | Malignant | Jan-46 | 0.181893 | 0.293448 | 0 | 0 | 5.13385 | 8.749803 |
| DisGeNET | Adult Diffu | Jan-46 | 0.181893 | 0.293448 | 0 | 0 | 5.13385 | 8.749803 |
| DisGeNET | Childhood | Jan-46 | 0.181893 | 0.293448 | 0 | 0 | 5.13385 | 8.749803 |
| DisGeNET | Sleep Apne | Jan-46 | 0.181893 | 0.293448 | 0 | 0 | 5.13385 | 8.749803 |
| DisGeNET | Early gastri | Jan-46 | 0.181893 | 0.293448 | 0 | 0 | 5.13385 | 8.749803 |
| DisGeNET | Decreased | Jan-46 | 0.181893 | 0.293448 | 0 | 0 | 5.13385 | 8.749803 |
| DisGeNET | Abnormalit | Jan-46 | 0.181893 | 0.293448 | 0 | 0 | 5.13385 | 8.749803 |
| DisGeNET | Thrombast | Jan-46 | 0.181893 | 0.293448 | 0 | 0 | 5.13385 | 8.749803 |
| DisGeNET | Telangiecta | Jan-46 | 0.181893 | 0.293448 | 0 | 0 | 5.13385 | 8.749803 |
| DisGeNET | Drusen | Jan-46 | 0.181893 | 0.293448 | 0 | 0 | 5.13385 | 8.749803 |
| DisGeNET | Hereditary | Jan-46 | 0.181893 | 0.293448 | 0 | 0 | 5.13385 | 8.749803 |
| DisGeNET | Cerebrovas | 2/178 | 0.181452 | 0.293448 | 0 | 0 | 2.638636 | 4.503525 |

| | | | | | | | | |
|---|---|---|---|---|---|---|---|---|
| Rare_Disea | Syndactyly | 30-Jan | 0.122676 | 0.293498 | 0 | 0 | 7.972735 | 16.72843 |
| Rare_Disea | Spina bifida | 2/139 | 0.122685 | 0.293498 | 0 | 0 | 3.396479 | 7.126265 |
| Rare_Disea | Gynandrob | 2/139 | 0.122685 | 0.293498 | 0 | 0 | 3.396479 | 7.126265 |
| Human_Ph | Hirsutism ( | Jan-67 | 0.253656 | 0.293626 | 0 | 0 | 3.496653 | 4.79662 |
| Human_Ph | Oligohydra | Jan-67 | 0.253656 | 0.293626 | 0 | 0 | 3.496653 | 4.79662 |
| Human_Ph | Cafe-au-lai | Jan-67 | 0.253656 | 0.293626 | 0 | 0 | 3.496653 | 4.79662 |
| GO_Molec | SH2 Domai | 31-Jan | 0.126498 | 0.294183 | 0 | 0 | 7.706589 | 15.93356 |
| UK_Bioban | Dentures 6 | Jan-64 | 0.243799 | 0.294418 | 0 | 0 | 3.663714 | 5.171007 |
| UK_Bioban | Microalbur | 9/1571 | 0.242914 | 0.294418 | 0 | 0 | 1.355585 | 1.918219 |
| Jensen_TIS | Sputum | Jan-51 | 0.199575 | 0.294785 | 0 | 0 | 4.619302 | 7.444308 |
| Jensen_TIS | Submucosa | Jan-51 | 0.199575 | 0.294785 | 0 | 0 | 4.619302 | 7.444308 |
| GWAS_Cat | Glomerular | Jan-71 | 0.266602 | 0.294882 | 0 | 0 | 3.296179 | 4.35754 |
| GWAS_Cat | Coronary A | Jan-71 | 0.266602 | 0.294882 | 0 | 0 | 3.296179 | 4.35754 |
| GWAS_Cat | Dyslexia | Jan-71 | 0.266602 | 0.294882 | 0 | 0 | 3.296179 | 4.35754 |
| GTEx_Tissu | GTEX-T2IS- | 12/1239 | 0.00724 | 0.294999 | 0 | 0 | 2.436642 | 12.00814 |
| GTEx_Tissu | GTEX-R53T | 16/1894 | 0.007161 | 0.294999 | 0 | 0 | 2.164124 | 10.68879 |
| GTEx_Tissu | GTEX-NFK9 | 13/1398 | 0.007237 | 0.294999 | 0 | 0 | 2.350122 | 11.5828 |
| CellMarker | Mesenchyr | Jan-60 | 0.230455 | 0.295069 | 0 | 0 | 3.912889 | 5.742942 |
| DisGeNET | Short nose | 2/179 | 0.183014 | 0.295074 | 0 | 0 | 2.623596 | 4.455375 |
| PhenGenI_ | Optic Disk | 2/115 | 0.089505 | 0.295367 | 0 | 0 | 4.122853 | 9.950336 |
| PhenGenI_ | Hydrochlor | 2/115 | 0.089505 | 0.295367 | 0 | 0 | 4.122853 | 9.950336 |
| Jensen_DIS | Cerebrovas | 2/225 | 0.256348 | 0.295787 | 0 | 0 | 2.077552 | 2.828 |
| GeDiPNet_ | Dyskinetic | 2/192 | 0.203481 | 0.295834 | 0 | 0 | 2.442477 | 3.888868 |
| GeDiPNet_ | Arnold-Chia | Jan-52 | 0.203066 | 0.295834 | 0 | 0 | 4.5285 | 7.219451 |
| Rare_Disea | Angiomato | 31-Jan | 0.126498 | 0.295959 | 0 | 0 | 7.706589 | 15.93356 |
| Rare_Disea | Esophagea | 31-Jan | 0.126498 | 0.295959 | 0 | 0 | 7.706589 | 15.93356 |
| Rare_Disea | Hepatocell | 31-Jan | 0.126498 | 0.295959 | 0 | 0 | 7.706589 | 15.93356 |
| Rare_Disea | Angiofollicu | 31-Jan | 0.126498 | 0.295959 | 0 | 0 | 7.706589 | 15.93356 |
| Rare_Disea | Tuberculou | 31-Jan | 0.126498 | 0.295959 | 0 | 0 | 7.706589 | 15.93356 |
| Rare_Disea | Malignant | 31-Jan | 0.126498 | 0.295959 | 0 | 0 | 7.706589 | 15.93356 |
| Rare_Disea | Cluster hea | 31-Jan | 0.126498 | 0.295959 | 0 | 0 | 7.706589 | 15.93356 |
| GO_Molec | Monocarbo | Jan-32 | 0.130304 | 0.296146 | 0 | 0 | 7.457614 | 15.19775 |
| Rare_Disea | Gamma he | 2/142 | 0.127022 | 0.296253 | 0 | 0 | 3.323193 | 6.857052 |
| Human_Ph | Postnatal g | Jan-68 | 0.256914 | 0.296499 | 0 | 0 | 3.44429 | 4.68084 |
| Jensen_DIS | Gout | Jan-69 | 0.260157 | 0.296671 | 0 | 0 | 3.393468 | 4.569198 |
| Jensen_DIS | Human imr | Jan-69 | 0.260157 | 0.296671 | 0 | 0 | 3.393468 | 4.569198 |
| SysMyo_M | MGS440 Hu | 4/239 | 0.020462 | 0.29696 | 0 | 0 | 4.035478 | 15.69475 |
| MAGMA_D | Gefitinib | Jan-68 | 0.256914 | 0.297127 | 0 | 0 | 3.44429 | 4.68084 |
| Rare_Disea | Juvenile Hu | 2/143 | 0.128476 | 0.297138 | 0 | 0 | 3.299458 | 6.770526 |
| Rare_Disea | Supraglotti | Jan-32 | 0.130304 | 0.297138 | 0 | 0 | 7.457614 | 15.19775 |
| Rare_Disea | Complex re | Jan-32 | 0.130304 | 0.297138 | 0 | 0 | 7.457614 | 15.19775 |
| Rare_Disea | Diabetes m | Jan-32 | 0.130304 | 0.297138 | 0 | 0 | 7.457614 | 15.19775 |
| Rare_Disea | Pediatric C | 2/145 | 0.131396 | 0.297138 | 0 | 0 | 3.252982 | 6.602064 |
| Rare_Disea | Eclampsia | 2/145 | 0.131396 | 0.297138 | 0 | 0 | 3.252982 | 6.602064 |
| Rare_Disea | Acute pron | 2/145 | 0.131396 | 0.297138 | 0 | 0 | 3.252982 | 6.602064 |

| | | | | | | | | |
|---|---|---|---|---|---|---|---|---|
| Rare_Disea | Opisthorch | Jan-32 | 0.130304 | 0.297138 | 0 | 0 | 7.457614 | 15.19775 |
| Rare_Disea | Huntingtor | 2/143 | 0.128476 | 0.297138 | 0 | 0 | 3.299458 | 6.770526 |
| Rare_Disea | Acute myel | 2/144 | 0.129934 | 0.297138 | 0 | 0 | 3.276056 | 6.685543 |
| GO_Biologi | Negative R | Jan-49 | 0.192548 | 0.297194 | 0 | 0 | 4.812258 | 7.927759 |
| DisGeNET | Hypertensi | Jan-47 | 0.18546 | 0.297372 | 0 | 0 | 5.021992 | 8.46163 |
| DisGeNET | Disseminat | Jan-47 | 0.18546 | 0.297372 | 0 | 0 | 5.021992 | 8.46163 |
| DisGeNET | Abnormalit | Jan-47 | 0.18546 | 0.297372 | 0 | 0 | 5.021992 | 8.46163 |
| DisGeNET | FRAGILE X | Jan-47 | 0.18546 | 0.297372 | 0 | 0 | 5.021992 | 8.46163 |
| DisGeNET | Diabetic Fo | Jan-47 | 0.18546 | 0.297372 | 0 | 0 | 5.021992 | 8.46163 |
| DisGeNET | Abnormalit | Jan-47 | 0.18546 | 0.297372 | 0 | 0 | 5.021992 | 8.46163 |
| DisGeNET | Lumbar hyp | Jan-47 | 0.18546 | 0.297372 | 0 | 0 | 5.021992 | 8.46163 |
| DisGeNET | Severe con | Jan-47 | 0.18546 | 0.297372 | 0 | 0 | 5.021992 | 8.46163 |
| DisGeNET | Serum trigl | Jan-47 | 0.18546 | 0.297372 | 0 | 0 | 5.021992 | 8.46163 |
| GWAS_Cat | Primary Bil | Jan-72 | 0.269804 | 0.297398 | 0 | 0 | 3.249591 | 4.257158 |
| GWAS_Cat | Severe CO\ | 3/415 | 0.270096 | 0.297398 | 0 | 0 | 1.690447 | 2.212759 |
| GWAS_Cat | Varicose Ve | Jan-72 | 0.269804 | 0.297398 | 0 | 0 | 3.249591 | 4.257158 |
| GWAS_Cat | Cholesteryl | Jan-72 | 0.269804 | 0.297398 | 0 | 0 | 3.249591 | 4.257158 |
| GWAS_Cat | Smoking In | 7/1205 | 0.269971 | 0.297398 | 0 | 0 | 1.366914 | 1.789893 |
| GTEx_Tissu | GTEX-XUJ4 | 15/1734 | 0.007507 | 0.297514 | 0 | 0 | 2.205013 | 10.78666 |
| GTEx_Tissu | GTEX-XOT4 | 11/1089 | 0.007484 | 0.297514 | 0 | 0 | 2.528867 | 12.37864 |
| CellMarker | Ependymal | Jan-61 | 0.233813 | 0.297975 | 0 | 0 | 3.847481 | 5.591291 |
| GO_Molecu | Frizzled Bir | Jan-33 | 0.134093 | 0.297985 | 0 | 0 | 7.224201 | 14.515 |
| DisGeNET | Conductive | 2/181 | 0.186143 | 0.298284 | 0 | 0 | 2.594019 | 4.361174 |
| Jensen_TIS | Amniotic fl | Jan-52 | 0.203066 | 0.298571 | 0 | 0 | 4.5285 | 7.219451 |
| Reactome_ | Translation | 4/281 | 0.034271 | 0.298672 | 0 | 0 | 3.416293 | 11.52474 |
| Reactome_ | Chylomicro | 9-Jan | 0.038483 | 0.298672 | 0 | 0 | 28.93169 | 94.24608 |
| Reactome_ | RHOT2 GTI | 7-Jan | 0.03006 | 0.298672 | 0 | 0 | 38.57946 | 135.2043 |
| Reactome_ | Mitophagy | 28-Feb | 0.00657 | 0.298672 | 0 | 0 | 17.99729 | 90.44125 |
| Reactome_ | Selective A | Feb-61 | 0.029 | 0.298672 | 0 | 0 | 7.917846 | 28.0328 |
| Reactome_ | PINK1-PRKI | 22-Feb | 0.004083 | 0.298672 | 0 | 0 | 23.40353 | 128.7392 |
| Reactome_ | VLDL Cleara | 5-Jan | 0.021564 | 0.298672 | 0 | 0 | 57.875 | 222.0519 |
| Reactome_ | Synthesis C | Feb-51 | 0.020776 | 0.298672 | 0 | 0 | 9.538535 | 36.95177 |
| Reactome_ | NR1H3 And | Feb-36 | 0.010706 | 0.298672 | 0 | 0 | 13.75709 | 62.41496 |
| Reactome_ | Plasma Lip | Feb-35 | 0.01014 | 0.298672 | 0 | 0 | 14.17469 | 65.07997 |
| Reactome_ | Macroauto | 3/114 | 0.013515 | 0.298672 | 0 | 0 | 6.3713 | 27.42156 |
| Reactome_ | NR1H2 And | Feb-46 | 0.017103 | 0.298672 | 0 | 0 | 10.62513 | 43.22851 |
| Reactome_ | Striated Mu | Feb-33 | 0.009049 | 0.298672 | 0 | 0 | 15.0907 | 71.00369 |
| Reactome_ | DNA Dama | 8-Jan | 0.03428 | 0.298672 | 0 | 0 | 33.06645 | 111.5391 |
| Reactome_ | Recycling C | 8-Jan | 0.03428 | 0.298672 | 0 | 0 | 33.06645 | 111.5391 |
| Reactome_ | Condensati | 10-Jan | 0.042667 | 0.298672 | 0 | 0 | 25.71576 | 81.1157 |
| Reactome_ | Miro GTPa: | 8-Jan | 0.03428 | 0.298672 | 0 | 0 | 33.06645 | 111.5391 |
| Reactome_ | Plasma Lip | Feb-71 | 0.038301 | 0.298672 | 0 | 0 | 6.766922 | 22.07565 |
| Reactome_ | RUNX1 And | 10-Jan | 0.042667 | 0.298672 | 0 | 0 | 25.71576 | 81.1157 |
| Reactome_ | Platelet Ac | 4/254 | 0.024904 | 0.298672 | 0 | 0 | 3.790458 | 13.99716 |
| Reactome_ | SARS-CoV-: | 5-Jan | 0.021564 | 0.298672 | 0 | 0 | 57.875 | 222.0519 |

| | | | | | | | | |
|---|---|---|---|---|---|---|---|---|
| Reactome_ | Chylomicrc | 5-Jan | 0.021564 | 0.298672 | 0 | 0 | 57.875 | 222.0519 |
| Reactome_ | Formation | 9-Jan | 0.038483 | 0.298672 | 0 | 0 | 28.93169 | 94.24608 |
| Reactome_ | HDL Remoc | 10-Jan | 0.042667 | 0.298672 | 0 | 0 | 25.71576 | 81.1157 |
| Reactome_ | Reversal O | 7-Jan | 0.03006 | 0.298672 | 0 | 0 | 38.57946 | 135.2043 |
| Reactome_ | Chylomicrc | 9-Jan | 0.038483 | 0.298672 | 0 | 0 | 28.93169 | 94.24608 |
| Reactome_ | Regulation | 9-Jan | 0.038483 | 0.298672 | 0 | 0 | 28.93169 | 94.24608 |
| Reactome_ | Gap Junctic | 10-Jan | 0.042667 | 0.298672 | 0 | 0 | 25.71576 | 81.1157 |
| Reactome_ | Plasma Lip | 16-Feb | 0.002157 | 0.298672 | 0 | 0 | 33.4437 | 205.3055 |
| Reactome_ | Autophagy 3/128 | | 0.018376 | 0.298672 | 0 | 0 | 5.653714 | 22.5964 |
| Reactome_ | Scavenging | 6-Jan | 0.025821 | 0.298672 | 0 | 0 | 46.29767 | 169.2909 |
| Reactome_ | Binding An | Feb-41 | 0.013741 | 0.298672 | 0 | 0 | 11.99035 | 51.40702 |
| GO_Biologi | Negative R 2/186 | | 0.193998 | 0.298782 | 0 | 0 | 2.52289 | 4.137304 |
| GeDiPNet_ | Fundus Col | Jan-53 | 0.206541 | 0.299536 | 0 | 0 | 4.44119 | 7.004886 |
| Rare_Disea | Omenn syr 3/292 | | 0.134486 | 0.299575 | 0 | 0 | 2.425111 | 4.86548 |
| Rare_Disea | Anencepha | Jan-33 | 0.134093 | 0.299575 | 0 | 0 | 7.224201 | 14.515 |
| Rare_Disea | Dysembryc | Jan-33 | 0.134093 | 0.299575 | 0 | 0 | 7.224201 | 14.515 |
| Rare_Disea | Aortic supr | Jan-33 | 0.134093 | 0.299575 | 0 | 0 | 7.224201 | 14.515 |
| Rare_Disea | Severe con 3/292 | | 0.134486 | 0.299575 | 0 | 0 | 2.425111 | 4.86548 |
| GO_Molecı | Solute:Sod | Jan-34 | 0.137866 | 0.299709 | 0 | 0 | 7.004933 | 13.88007 |
| Reactome_ | Biotin Tran | 11-Jan | 0.046834 | 0.299737 | 0 | 0 | 23.14302 | 70.84419 |
| Reactome_ | G Alpha (1 | Feb-78 | 0.045395 | 0.299737 | 0 | 0 | 6.141486 | 18.99159 |
| Reactome_ | Regulation | 11-Jan | 0.046834 | 0.299737 | 0 | 0 | 23.14302 | 70.84419 |
| GWAS_Cat | Worry | Jan-73 | 0.272992 | 0.299775 | 0 | 0 | 3.204296 | 4.160181 |
| GWAS_Cat | Depressive | Jan-73 | 0.272992 | 0.299775 | 0 | 0 | 3.204296 | 4.160181 |
| GWAS_Cat | Average Di | Jan-73 | 0.272992 | 0.299775 | 0 | 0 | 3.204296 | 4.160181 |
| GO_Biologi | Positive Re | Jan-50 | 0.196069 | 0.30002 | 0 | 0 | 4.713811 | 7.680157 |
| GO_Biologi | Blood Circu | Jan-50 | 0.196069 | 0.30002 | 0 | 0 | 4.713811 | 7.680157 |
| GO_Biologi | Nucleocytc | Jan-50 | 0.196069 | 0.30002 | 0 | 0 | 4.713811 | 7.680157 |
| GeDiPNet_ | Rhizomelia | Jan-54 | 0.210002 | 0.300077 | 0 | 0 | 4.357174 | 6.79997 |
| GeDiPNet_ | Heart Sept | Jan-54 | 0.210002 | 0.300077 | 0 | 0 | 4.357174 | 6.79997 |
| GeDiPNet_ | Retinal Col | Jan-54 | 0.210002 | 0.300077 | 0 | 0 | 4.357174 | 6.79997 |
| GeDiPNet_ | Transient I | Jan-54 | 0.210002 | 0.300077 | 0 | 0 | 4.357174 | 6.79997 |
| GeDiPNet_ | Syndactyly 2/195 | | 0.208243 | 0.300077 | 0 | 0 | 2.404145 | 3.772225 |
| GeDiPNet_ | Metastatic | Jan-54 | 0.210002 | 0.300077 | 0 | 0 | 4.357174 | 6.79997 |
| DisGeNET | Fabry Disea | Jan-48 | 0.189012 | 0.300553 | 0 | 0 | 4.914894 | 8.187947 |
| DisGeNET | oligodendr 2/183 | | 0.189279 | 0.300553 | 0 | 0 | 2.565096 | 4.269683 |
| DisGeNET | Dupuytren | Jan-48 | 0.189012 | 0.300553 | 0 | 0 | 4.914894 | 8.187947 |
| DisGeNET | Hepatitis, ( 2/183 | | 0.189279 | 0.300553 | 0 | 0 | 2.565096 | 4.269683 |
| DisGeNET | Decreased | Jan-48 | 0.189012 | 0.300553 | 0 | 0 | 4.914894 | 8.187947 |
| DisGeNET | Corneal Ne | Jan-48 | 0.189012 | 0.300553 | 0 | 0 | 4.914894 | 8.187947 |
| DisGeNET | Renal tubu | Jan-48 | 0.189012 | 0.300553 | 0 | 0 | 4.914894 | 8.187947 |
| DisGeNET | Manic Diso | Jan-48 | 0.189012 | 0.300553 | 0 | 0 | 4.914894 | 8.187947 |
| DisGeNET | Neoplasm I 2/183 | | 0.189279 | 0.300553 | 0 | 0 | 2.565096 | 4.269683 |
| DisGeNET | Right Ventı | Jan-48 | 0.189012 | 0.300553 | 0 | 0 | 4.914894 | 8.187947 |
| DisGeNET | Sleep Disor | Jan-48 | 0.189012 | 0.300553 | 0 | 0 | 4.914894 | 8.187947 |

| | | | | | | | | |
|---|---|---|---|---|---|---|---|---|
| DisGeNET | Synovial Cy | Jan-48 | 0.189012 | 0.300553 | 0 | 0 | 4.914894 | 8.187947 |
| DisGeNET | Alcoholic S | Jan-48 | 0.189012 | 0.300553 | 0 | 0 | 4.914894 | 8.187947 |
| DisGeNET | Delayed my | Jan-48 | 0.189012 | 0.300553 | 0 | 0 | 4.914894 | 8.187947 |
| DisGeNET | Respiratory | 2/182 | 0.18771 | 0.300553 | 0 | 0 | 2.579477 | 4.315096 |
| Rare_Disea | Optic atrop | 2/148 | 0.135804 | 0.300708 | 0 | 0 | 3.185657 | 6.360306 |
| Rare_Disea | Bare lymph | 3/293 | 0.135474 | 0.300708 | 0 | 0 | 2.416626 | 4.830773 |
| Rare_Disea | Mitral atre | Jan-34 | 0.137866 | 0.300799 | 0 | 0 | 7.004933 | 13.88007 |
| Rare_Disea | Cardiospas | Jan-34 | 0.137866 | 0.300799 | 0 | 0 | 7.004933 | 13.88007 |
| Rare_Disea | Bubonic pla | Jan-34 | 0.137866 | 0.300799 | 0 | 0 | 7.004933 | 13.88007 |
| Rare_Disea | Hydroceph | 2/149 | 0.13728 | 0.300799 | 0 | 0 | 3.163826 | 6.282501 |
| Rare_Disea | Arginase de | Jan-34 | 0.137866 | 0.300799 | 0 | 0 | 7.004933 | 13.88007 |
| CellMarker | Macrophag | Jan-62 | 0.237156 | 0.300837 | 0 | 0 | 3.784217 | 5.445629 |
| CellMarker | Secretory C | Jan-63 | 0.240485 | 0.30088 | 0 | 0 | 3.722993 | 5.305634 |
| CellMarker | Epithelial C | Jan-63 | 0.240485 | 0.30088 | 0 | 0 | 3.722993 | 5.305634 |
| CellMarker | Megakaryo | Jan-63 | 0.240485 | 0.30088 | 0 | 0 | 3.722993 | 5.305634 |
| GTEx_Tissu | GTEX-QDVI | 8/656 | 0.00772 | 0.301818 | 0 | 0 | 3.010627 | 14.64343 |
| Rare_Disea | Reiter synd | 2/150 | 0.138761 | 0.301866 | 0 | 0 | 3.142289 | 6.206032 |
| Human_Ph | Gingival ov | Jan-70 | 0.263387 | 0.302143 | 0 | 0 | 3.344119 | 4.461494 |
| Human_Ph | Memory in | Jan-70 | 0.263387 | 0.302143 | 0 | 0 | 3.344119 | 4.461494 |
| GWAS_Cat | Estimated ( | Jan-74 | 0.276166 | 0.302715 | 0 | 0 | 3.160242 | 4.066453 |
| GWAS_Cat | Parkinson's | Jan-74 | 0.276166 | 0.302715 | 0 | 0 | 3.160242 | 4.066453 |
| Jensen_TIS | Occipital lo | 48/10045 | 0.206911 | 0.302843 | 0 | 0 | 1.220797 | 1.923324 |
| DisGeNET | Proteinuria | 2/184 | 0.19085 | 0.302864 | 0 | 0 | 2.550873 | 4.224921 |
| MAGMA_D | Tourettes D | Jan-71 | 0.266602 | 0.303061 | 0 | 0 | 3.296179 | 4.35754 |
| MAGMA_D | Haloperido | Jan-71 | 0.266602 | 0.303061 | 0 | 0 | 3.296179 | 4.35754 |
| Gene_Pert | Ncoa2 KO r | 7/367 | 0.001099 | 0.303096 | 0 | 0 | 4.752465 | 32.3817 |
| Gene_Pert | Lmna KO m | 7/322 | 0.00051 | 0.303096 | 0 | 0 | 5.443889 | 41.26816 |
| Gene_Pert | Hras Select | 6/265 | 0.001053 | 0.303096 | 0 | 0 | 5.62105 | 38.53642 |
| Gene_Pert | Ppara OE m | 7/371 | 0.00117 | 0.303096 | 0 | 0 | 4.699279 | 31.72523 |
| Gene_Pert | Atf6 tamox | 6/243 | 0.000672 | 0.303096 | 0 | 0 | 6.149711 | 44.92436 |
| DisGeNET | Intracrania | Jan-49 | 0.192548 | 0.303353 | 0 | 0 | 4.812258 | 7.927759 |
| DisGeNET | Dental Ena | Jan-49 | 0.192548 | 0.303353 | 0 | 0 | 4.812258 | 7.927759 |
| DisGeNET | MIXED LINI | 2/185 | 0.192423 | 0.303353 | 0 | 0 | 2.536805 | 4.180799 |
| DisGeNET | Malignant | 2/185 | 0.192423 | 0.303353 | 0 | 0 | 2.536805 | 4.180799 |
| DisGeNET | Recurrent t | 3/347 | 0.192154 | 0.303353 | 0 | 0 | 2.031665 | 3.351151 |
| DisGeNET | Cardiovasc | Jan-49 | 0.192548 | 0.303353 | 0 | 0 | 4.812258 | 7.927759 |
| DisGeNET | cardiac eve | Jan-49 | 0.192548 | 0.303353 | 0 | 0 | 4.812258 | 7.927759 |
| DisGeNET | Female Bre | Jan-49 | 0.192548 | 0.303353 | 0 | 0 | 4.812258 | 7.927759 |
| DisGeNET | Central ner | Jan-49 | 0.192548 | 0.303353 | 0 | 0 | 4.812258 | 7.927759 |
| DisGeNET | Airway Obs | Jan-49 | 0.192548 | 0.303353 | 0 | 0 | 4.812258 | 7.927759 |
| DisGeNET | Hypermela | Jan-49 | 0.192548 | 0.303353 | 0 | 0 | 4.812258 | 7.927759 |
| DisGeNET | Thromboar | Jan-49 | 0.192548 | 0.303353 | 0 | 0 | 4.812258 | 7.927759 |
| GWAS_Cat | Urate Leve | 3/421 | 0.277179 | 0.303553 | 0 | 0 | 1.66567 | 2.13721 |
| Human_Ph | Retinopath | Jan-71 | 0.266602 | 0.304006 | 0 | 0 | 3.296179 | 4.35754 |
| Human_Ph | Polycystic ( | Jan-71 | 0.266602 | 0.304006 | 0 | 0 | 3.296179 | 4.35754 |

| | | | | | | | | |
|---|---|---|---|---|---|---|---|---|
| GO_Biologi | Negative R | Jan-51 | 0.199575 | 0.304073 | 0 | 0 | 4.619302 | 7.444308 |
| GO_Biologi | Sensory Pe | Jan-51 | 0.199575 | 0.304073 | 0 | 0 | 4.619302 | 7.444308 |
| DisGeNET | Encephalor | 3/348 | 0.193257 | 0.304103 | 0 | 0 | 2.025673 | 3.329673 |
| DisGeNET | Ankylosing | 3/348 | 0.193257 | 0.304103 | 0 | 0 | 2.025673 | 3.329673 |
| Human_Ph | Intrauterin | 2/232 | 0.267638 | 0.304279 | 0 | 0 | 2.013606 | 2.654174 |
| Jensen_TIS | Bone marr | 67/14508 | 0.208981 | 0.304489 | 0 | 0 | 1.269386 | 1.987237 |
| Rare_Disea | Ectopia pu | Jan-35 | 0.141623 | 0.30453 | 0 | 0 | 6.798564 | 13.28839 |
| Rare_Disea | Wallerian c | Jan-35 | 0.141623 | 0.30453 | 0 | 0 | 6.798564 | 13.28839 |
| Rare_Disea | Herpes sim | Jan-35 | 0.141623 | 0.30453 | 0 | 0 | 6.798564 | 13.28839 |
| Rare_Disea | Hereditary | Jan-35 | 0.141623 | 0.30453 | 0 | 0 | 6.798564 | 13.28839 |
| Jensen_TIS | ae3 | Jan-54 | 0.210002 | 0.304598 | 0 | 0 | 4.357174 | 6.79997 |
| Human_Ph | External ea | Jan-72 | 0.269804 | 0.304926 | 0 | 0 | 3.249591 | 4.257158 |
| Human_Ph | Microdonti | Jan-72 | 0.269804 | 0.304926 | 0 | 0 | 3.249591 | 4.257158 |
| UK_Bioban | Usual walk | Jan-67 | 0.253656 | 0.30503 | 0 | 0 | 3.496653 | 4.79662 |
| DisGeNET | Respiratory | 2/186 | 0.193998 | 0.305087 | 0 | 0 | 2.52289 | 4.137304 |
| DisGeNET | Leishmania | Jan-50 | 0.196069 | 0.305229 | 0 | 0 | 4.713811 | 7.680157 |
| DisGeNET | Erythrobla | Jan-50 | 0.196069 | 0.305229 | 0 | 0 | 4.713811 | 7.680157 |
| DisGeNET | Atypical Du | Jan-50 | 0.196069 | 0.305229 | 0 | 0 | 4.713811 | 7.680157 |
| DisGeNET | Congenital | Jan-50 | 0.196069 | 0.305229 | 0 | 0 | 4.713811 | 7.680157 |
| DisGeNET | Decreased | Jan-50 | 0.196069 | 0.305229 | 0 | 0 | 4.713811 | 7.680157 |
| DisGeNET | Biparietal r | Jan-50 | 0.196069 | 0.305229 | 0 | 0 | 4.713811 | 7.680157 |
| DisGeNET | Childhood | 2/187 | 0.195575 | 0.305229 | 0 | 0 | 2.509126 | 4.094425 |
| DisGeNET | Turcot sync | Jan-50 | 0.196069 | 0.305229 | 0 | 0 | 4.713811 | 7.680157 |
| DisGeNET | Giant Cell T | Jan-50 | 0.196069 | 0.305229 | 0 | 0 | 4.713811 | 7.680157 |
| DisGeNET | Li-Fraumer | Jan-50 | 0.196069 | 0.305229 | 0 | 0 | 4.713811 | 7.680157 |
| DisGeNET | Varicella zc | Jan-50 | 0.196069 | 0.305229 | 0 | 0 | 4.713811 | 7.680157 |
| DisGeNET | Nephrosis | Jan-50 | 0.196069 | 0.305229 | 0 | 0 | 4.713811 | 7.680157 |
| DisGeNET | Moyamoya | Jan-50 | 0.196069 | 0.305229 | 0 | 0 | 4.713811 | 7.680157 |
| DisGeNET | Malignant | 3/349 | 0.194361 | 0.305229 | 0 | 0 | 2.019715 | 3.308368 |
| DisGeNET | Candidiasis | Jan-50 | 0.196069 | 0.305229 | 0 | 0 | 4.713811 | 7.680157 |
| DisGeNET | Hypoplasia | 2/187 | 0.195575 | 0.305229 | 0 | 0 | 2.509126 | 4.094425 |
| DisGeNET | Carcinosar | Jan-50 | 0.196069 | 0.305229 | 0 | 0 | 4.713811 | 7.680157 |
| Disease_Pe | ulcerative | 6/329 | 0.00313 | 0.305517 | 0 | 0 | 4.492604 | 25.90773 |
| Disease_Pe | Malignant | 6/342 | 0.003783 | 0.305517 | 0 | 0 | 4.315917 | 24.07034 |
| Disease_Pe | Emery-Dre | 6/335 | 0.00342 | 0.305517 | 0 | 0 | 4.409321 | 25.03652 |
| Jensen_DIS | Neutropen | Jan-72 | 0.269804 | 0.305882 | 0 | 0 | 3.249591 | 4.257158 |
| DisGeNET | Malignant | 2/188 | 0.197153 | 0.30637 | 0 | 0 | 2.495509 | 4.05215 |
| DisGeNET | Acquired Ir | 2/188 | 0.197153 | 0.30637 | 0 | 0 | 2.495509 | 4.05215 |
| DisGeNET | Carcinoma | 2/188 | 0.197153 | 0.30637 | 0 | 0 | 2.495509 | 4.05215 |
| Human_Ph | Increased s | Jan-73 | 0.272992 | 0.306714 | 0 | 0 | 3.204296 | 4.160181 |
| Human_Ph | Hypermetr | Jan-73 | 0.272992 | 0.306714 | 0 | 0 | 3.204296 | 4.160181 |
| GO_Biologi | Regulation | Jan-52 | 0.203066 | 0.308042 | 0 | 0 | 4.5285 | 7.219451 |
| GO_Biologi | Neuron Pro | 2/192 | 0.203481 | 0.308042 | 0 | 0 | 2.442477 | 3.888868 |
| GO_Biologi | Negative R | Jan-52 | 0.203066 | 0.308042 | 0 | 0 | 4.5285 | 7.219451 |
| DisGeNET | Malignant | Jan-51 | 0.199575 | 0.308124 | 0 | 0 | 4.619302 | 7.444308 |

| | | | | | | | | |
|---|---|---|---|---|---|---|---|---|
| DisGeNET | Tumors of . | Jan-51 | 0.199575 | 0.308124 | 0 | 0 | 4.619302 | 7.444308 |
| DisGeNET | Tardive Dy: | Jan-51 | 0.199575 | 0.308124 | 0 | 0 | 4.619302 | 7.444308 |
| DisGeNET | Premature | Jan-51 | 0.199575 | 0.308124 | 0 | 0 | 4.619302 | 7.444308 |
| DisGeNET | Adult Gliob | 2/189 | 0.198732 | 0.308124 | 0 | 0 | 2.482038 | 4.010467 |
| DisGeNET | Cockayne S | Jan-51 | 0.199575 | 0.308124 | 0 | 0 | 4.619302 | 7.444308 |
| DisGeNET | Pauci-immu | Jan-51 | 0.199575 | 0.308124 | 0 | 0 | 4.619302 | 7.444308 |
| DisGeNET | Autosomal | Jan-51 | 0.199575 | 0.308124 | 0 | 0 | 4.619302 | 7.444308 |
| DisGeNET | Breast Dise | Jan-51 | 0.199575 | 0.308124 | 0 | 0 | 4.619302 | 7.444308 |
| DisGeNET | Drug-induc | Jan-51 | 0.199575 | 0.308124 | 0 | 0 | 4.619302 | 7.444308 |
| DisGeNET | Cystadenoı | Jan-51 | 0.199575 | 0.308124 | 0 | 0 | 4.619302 | 7.444308 |
| PhenGenI_ | Smell | 23-Jan | 0.095455 | 0.308206 | 0 | 0 | 10.51321 | 24.6966 |
| PhenGenI_ | Body Heigh | 11/1632 | 0.096064 | 0.308206 | 0 | 0 | 1.633267 | 3.826319 |
| GWAS_Cat | Coronary A | Jan-76 | 0.282473 | 0.308244 | 0 | 0 | 3.075659 | 3.888164 |
| GWAS_Cat | Fasting Pla: | Jan-76 | 0.282473 | 0.308244 | 0 | 0 | 3.075659 | 3.888164 |
| GWAS_Cat | Myopia | Jan-76 | 0.282473 | 0.308244 | 0 | 0 | 3.075659 | 3.888164 |
| GWAS_Cat | Insomnia | 5/816 | 0.281911 | 0.308244 | 0 | 0 | 1.436197 | 1.818461 |
| GeDiPNet_ | Melanocyti | Jan-56 | 0.216879 | 0.308391 | 0 | 0 | 4.198309 | 6.416764 |
| GeDiPNet_ | Coronary S | Jan-56 | 0.216879 | 0.308391 | 0 | 0 | 4.198309 | 6.416764 |
| GO_Biologi | Organelle I | Jan-54 | 0.210002 | 0.308699 | 0 | 0 | 4.357174 | 6.79997 |
| GO_Biologi | Regulation | Jan-54 | 0.210002 | 0.308699 | 0 | 0 | 4.357174 | 6.79997 |
| GO_Biologi | Regulation | Jan-54 | 0.210002 | 0.308699 | 0 | 0 | 4.357174 | 6.79997 |
| GO_Biologi | Endomemb | 2/196 | 0.209833 | 0.308699 | 0 | 0 | 2.391631 | 3.734399 |
| GO_Biologi | Epidermal ( | Jan-54 | 0.210002 | 0.308699 | 0 | 0 | 4.357174 | 6.79997 |
| GO_Biologi | Regulation | Jan-53 | 0.206541 | 0.308699 | 0 | 0 | 4.44119 | 7.004886 |
| GO_Biologi | Mitochond | Jan-53 | 0.206541 | 0.308699 | 0 | 0 | 4.44119 | 7.004886 |
| GO_Biologi | Negative R | 4/537 | 0.205803 | 0.308699 | 0 | 0 | 1.7523 | 2.770095 |
| GO_Biologi | Regulation | Jan-54 | 0.210002 | 0.308699 | 0 | 0 | 4.357174 | 6.79997 |
| GO_Biologi | Regulation | Jan-53 | 0.206541 | 0.308699 | 0 | 0 | 4.44119 | 7.004886 |
| GO_Biologi | Negative R | Jan-53 | 0.206541 | 0.308699 | 0 | 0 | 4.44119 | 7.004886 |
| GO_Biologi | Positive Re | 2/196 | 0.209833 | 0.308699 | 0 | 0 | 2.391631 | 3.734399 |
| GO_Biologi | Regulation | Jan-54 | 0.210002 | 0.308699 | 0 | 0 | 4.357174 | 6.79997 |
| GO_Biologi | Monoatom | Jan-54 | 0.210002 | 0.308699 | 0 | 0 | 4.357174 | 6.79997 |
| GeDiPNet_ | Hyperthyro | Jan-57 | 0.220295 | 0.308731 | 0 | 0 | 4.123131 | 6.237423 |
| GeDiPNet_ | Brain Neop | Jan-57 | 0.220295 | 0.308731 | 0 | 0 | 4.123131 | 6.237423 |
| GeDiPNet_ | Palmoplant | Jan-57 | 0.220295 | 0.308731 | 0 | 0 | 4.123131 | 6.237423 |
| GeDiPNet_ | Mouth Neo | Jan-57 | 0.220295 | 0.308731 | 0 | 0 | 4.123131 | 6.237423 |
| GeDiPNet_ | Obesity | 4/551 | 0.21854 | 0.308731 | 0 | 0 | 1.706218 | 2.594793 |
| GeDiPNet_ | Aphasia | Jan-57 | 0.220295 | 0.308731 | 0 | 0 | 4.123131 | 6.237423 |
| DisGeNET | Muscular D | 2/190 | 0.200314 | 0.308901 | 0 | 0 | 2.468711 | 3.969367 |
| DisGeNET | Long philtr | 2/190 | 0.200314 | 0.308901 | 0 | 0 | 2.468711 | 3.969367 |
| Reactome_ | PI Metabol | Feb-82 | 0.04965 | 0.308935 | 0 | 0 | 5.833235 | 17.51576 |
| GWAS_Cat | General Co | 4/620 | 0.284002 | 0.309636 | 0 | 0 | 1.509701 | 1.900371 |
| Rare_Disea | Hypomelar | 2/154 | 0.144717 | 0.309886 | 0 | 0 | 3.058978 | 5.912923 |
| Rare_Disea | Achalasia | Jan-36 | 0.145363 | 0.309886 | 0 | 0 | 6.603987 | 12.73591 |
| Rare_Disea | Cervical dy | Jan-36 | 0.145363 | 0.309886 | 0 | 0 | 6.603987 | 12.73591 |

| | | | | | | | | |
|---|---|---|---|---|---|---|---|---|
| Rare_Disea | Rabies | Jan-37 | 0.149088 | 0.31011 | 0 | 0 | 6.42022 | 12.21909 |
| Rare_Disea | Amyloidosi | 2/157 | 0.14922 | 0.31011 | 0 | 0 | 2.999317 | 5.705706 |
| Rare_Disea | Hemiplegia | Jan-37 | 0.149088 | 0.31011 | 0 | 0 | 6.42022 | 12.21909 |
| Rare_Disea | Idiopathic | 2/155 | 0.146215 | 0.31011 | 0 | 0 | 3.038831 | 5.842694 |
| Rare_Disea | Hm syndro | Jan-37 | 0.149088 | 0.31011 | 0 | 0 | 6.42022 | 12.21909 |
| Rare_Disea | Imperforat | Jan-37 | 0.149088 | 0.31011 | 0 | 0 | 6.42022 | 12.21909 |
| Rare_Disea | Pheochron | 2/157 | 0.14922 | 0.31011 | 0 | 0 | 2.999317 | 5.705706 |
| Rare_Disea | Colorectal | Jan-37 | 0.149088 | 0.31011 | 0 | 0 | 6.42022 | 12.21909 |
| Rare_Disea | Lhermitte-I | Jan-37 | 0.149088 | 0.31011 | 0 | 0 | 6.42022 | 12.21909 |
| GO_Molecu | N-acyltrans | Jan-37 | 0.149088 | 0.3106 | 0 | 0 | 6.42022 | 12.21909 |
| GO_Molecu | aminoacyl- | Jan-37 | 0.149088 | 0.3106 | 0 | 0 | 6.42022 | 12.21909 |
| MAGMA_D | Progestero | Jan-74 | 0.276166 | 0.311272 | 0 | 0 | 3.160242 | 4.066453 |
| Jensen_DIS | Cataract | Jan-74 | 0.276166 | 0.311285 | 0 | 0 | 3.160242 | 4.066453 |
| CellMarker | Endothelia | Jan-66 | 0.250385 | 0.311843 | 0 | 0 | 3.550626 | 4.916752 |
| DisGeNET | Drug usage | Jan-52 | 0.203066 | 0.311859 | 0 | 0 | 4.5285 | 7.219451 |
| DisGeNET | Range of jo | Jan-52 | 0.203066 | 0.311859 | 0 | 0 | 4.5285 | 7.219451 |
| DisGeNET | Gait, Unste | Jan-52 | 0.203066 | 0.311859 | 0 | 0 | 4.5285 | 7.219451 |
| DisGeNET | Hereditary | Jan-52 | 0.203066 | 0.311859 | 0 | 0 | 4.5285 | 7.219451 |
| DisGeNET | Hippocamp | Jan-52 | 0.203066 | 0.311859 | 0 | 0 | 4.5285 | 7.219451 |
| DisGeNET | Invasive Lo | Jan-52 | 0.203066 | 0.311859 | 0 | 0 | 4.5285 | 7.219451 |
| DisGeNET | X-linked ag | Jan-52 | 0.203066 | 0.311859 | 0 | 0 | 4.5285 | 7.219451 |
| GeDiPNet_ | Premature | Jan-59 | 0.227083 | 0.312309 | 0 | 0 | 3.980553 | 5.900928 |
| GeDiPNet_ | Thyroid Ne | Jan-59 | 0.227083 | 0.312309 | 0 | 0 | 3.980553 | 5.900928 |
| GeDiPNet_ | Inflammato | 2/205 | 0.224196 | 0.312309 | 0 | 0 | 2.284555 | 3.415945 |
| GeDiPNet_ | Hypopituita | Jan-59 | 0.227083 | 0.312309 | 0 | 0 | 3.980553 | 5.900928 |
| GeDiPNet_ | Skin Cance | Jan-59 | 0.227083 | 0.312309 | 0 | 0 | 3.980553 | 5.900928 |
| GeDiPNet_ | Malignant | 2/205 | 0.224196 | 0.312309 | 0 | 0 | 2.284555 | 3.415945 |
| GeDiPNet_ | Colorectal | 6/943 | 0.227134 | 0.312309 | 0 | 0 | 1.500138 | 2.223528 |
| GeDiPNet_ | Cafe-au-lai | Jan-59 | 0.227083 | 0.312309 | 0 | 0 | 3.980553 | 5.900928 |
| DisGeNET | Hematopoi | 2/192 | 0.203481 | 0.312314 | 0 | 0 | 2.442477 | 3.888868 |
| Rare_Disea | Cervical int | 2/158 | 0.150727 | 0.31237 | 0 | 0 | 2.97994 | 5.638896 |
| Human_Ph | Splenomeg | 2/239 | 0.278929 | 0.312465 | 0 | 0 | 1.953438 | 2.494149 |
| GO_Biologi | Neuron Mi | Jan-55 | 0.213448 | 0.31247 | 0 | 0 | 4.27627 | 6.604112 |
| GO_Biologi | Response T | Jan-55 | 0.213448 | 0.31247 | 0 | 0 | 4.27627 | 6.604112 |
| Rare_Disea | Ganglioglic | Jan-38 | 0.152796 | 0.313169 | 0 | 0 | 6.246386 | 11.73478 |
| Rare_Disea | Bannayan-I | Jan-38 | 0.152796 | 0.313169 | 0 | 0 | 6.246386 | 11.73478 |
| Rare_Disea | Hemangiop | Jan-38 | 0.152796 | 0.313169 | 0 | 0 | 6.246386 | 11.73478 |
| Rare_Disea | Syndactyly | Jan-38 | 0.152796 | 0.313169 | 0 | 0 | 6.246386 | 11.73478 |
| UK_Bioban | Red blood | 4/597 | 0.261768 | 0.313462 | 0 | 0 | 1.570125 | 2.104431 |
| GO_Biologi | DNA-templ | 2/199 | 0.21461 | 0.313525 | 0 | 0 | 2.354852 | 3.623955 |
| GWAS_Cat | Common E | Jan-78 | 0.288726 | 0.313665 | 0 | 0 | 2.99547 | 3.721207 |
| GWAS_Cat | Alcohol Co | Jan-78 | 0.288726 | 0.313665 | 0 | 0 | 2.99547 | 3.721207 |
| GWAS_Cat | Lifetime Sn | Jan-78 | 0.288726 | 0.313665 | 0 | 0 | 2.99547 | 3.721207 |
| GWAS_Cat | Subjective | Jan-78 | 0.288726 | 0.313665 | 0 | 0 | 2.99547 | 3.721207 |
| DisGeNET | Leukemia, | 10/1703 | 0.204576 | 0.313811 | 0 | 0 | 1.397657 | 2.217823 |

| | | | | | | | | |
|---|---|---|---|---|---|---|---|---|
| GeDiPNet_ | Diverticula | Jan-60 | 0.230455 | 0.313915 | 0 | 0 | 3.912889 | 5.742942 |
| GeDiPNet_ | Hypoalbum | Jan-60 | 0.230455 | 0.313915 | 0 | 0 | 3.912889 | 5.742942 |
| GeDiPNet_ | Leigh Syndı | Jan-60 | 0.230455 | 0.313915 | 0 | 0 | 3.912889 | 5.742942 |
| GeDiPNet_ | Dental Ena | Jan-60 | 0.230455 | 0.313915 | 0 | 0 | 3.912889 | 5.742942 |
| UK_Bioban | Fluid intelli | Jan-70 | 0.263387 | 0.31408 | 0 | 0 | 3.344119 | 4.461494 |
| Rare_Disea | Membranc | 2/160 | 0.153751 | 0.314259 | 0 | 0 | 2.941921 | 5.508522 |
| DisGeNET | Developme | 2/193 | 0.205067 | 0.31438 | 0 | 0 | 2.429566 | 3.84945 |
| Human_Ph | Cerebral is | Jan-76 | 0.282473 | 0.314591 | 0 | 0 | 3.075659 | 3.888164 |
| Human_Ph | Premature | Jan-76 | 0.282473 | 0.314591 | 0 | 0 | 3.075659 | 3.888164 |
| DisGeNET | Situs ambig | Jan-53 | 0.206541 | 0.314787 | 0 | 0 | 4.44119 | 7.004886 |
| DisGeNET | Prostate ca | Jan-53 | 0.206541 | 0.314787 | 0 | 0 | 4.44119 | 7.004886 |
| DisGeNET | Cerebellar | Jan-53 | 0.206541 | 0.314787 | 0 | 0 | 4.44119 | 7.004886 |
| DisGeNET | Eclampsia | 2/194 | 0.206654 | 0.314787 | 0 | 0 | 2.416789 | 3.810572 |
| DisGeNET | Leukemia, | Jan-53 | 0.206541 | 0.314787 | 0 | 0 | 4.44119 | 7.004886 |
| DisGeNET | granulosa c | Jan-53 | 0.206541 | 0.314787 | 0 | 0 | 4.44119 | 7.004886 |
| DisGeNET | Turner Syn | Jan-53 | 0.206541 | 0.314787 | 0 | 0 | 4.44119 | 7.004886 |
| DisGeNET | Tumor-Ass | Jan-53 | 0.206541 | 0.314787 | 0 | 0 | 4.44119 | 7.004886 |
| DisGeNET | Language D | Jan-53 | 0.206541 | 0.314787 | 0 | 0 | 4.44119 | 7.004886 |
| DisGeNET | Anxiety neı | Jan-53 | 0.206541 | 0.314787 | 0 | 0 | 4.44119 | 7.004886 |
| DisGeNET | Mental det | Jan-53 | 0.206541 | 0.314787 | 0 | 0 | 4.44119 | 7.004886 |
| SysMyo_M | MGS719 M | 7-Jan | 0.03006 | 0.314984 | 0 | 0 | 38.57946 | 135.2043 |
| SysMyo_M | MGS1060 I | 3/158 | 0.031658 | 0.314984 | 0 | 0 | 4.552535 | 15.71884 |
| SysMyo_M | MGS121 H | Feb-64 | 0.031682 | 0.314984 | 0 | 0 | 7.533586 | 26.00597 |
| SysMyo_M | MGS237 M | 4/300 | 0.041941 | 0.314984 | 0 | 0 | 3.193911 | 10.12943 |
| SysMyo_M | MGS245 C | 4/300 | 0.041941 | 0.314984 | 0 | 0 | 3.193911 | 10.12943 |
| SysMyo_M | MGS275 M | 4/300 | 0.041941 | 0.314984 | 0 | 0 | 3.193911 | 10.12943 |
| SysMyo_M | MGS281 M | 4/300 | 0.041941 | 0.314984 | 0 | 0 | 3.193911 | 10.12943 |
| SysMyo_M | MGS289 C | 4/300 | 0.041941 | 0.314984 | 0 | 0 | 3.193911 | 10.12943 |
| SysMyo_M | MGS1279 I | 3/165 | 0.035319 | 0.314984 | 0 | 0 | 4.354277 | 14.55783 |
| SysMyo_M | MGS1223 I | 3/176 | 0.041494 | 0.314984 | 0 | 0 | 4.075145 | 12.96795 |
| SysMyo_M | MGS213 C | 4/300 | 0.041941 | 0.314984 | 0 | 0 | 3.193911 | 10.12943 |
| SysMyo_M | MGS852 M | 3/169 | 0.037505 | 0.314984 | 0 | 0 | 4.248494 | 13.94901 |
| SysMyo_M | MGS1317 I | 3/178 | 0.042672 | 0.314984 | 0 | 0 | 4.028163 | 12.70571 |
| SysMyo_M | MGS329 M | 4/300 | 0.041941 | 0.314984 | 0 | 0 | 3.193911 | 10.12943 |
| SysMyo_M | MGS1243 I | 3/181 | 0.04447 | 0.314984 | 0 | 0 | 3.959671 | 12.32626 |
| SysMyo_M | MGS102 M | Feb-77 | 0.044354 | 0.314984 | 0 | 0 | 6.223686 | 19.39022 |
| SysMyo_M | MGS857 M | Feb-62 | 0.029883 | 0.314984 | 0 | 0 | 7.78549 | 27.33058 |
| SysMyo_M | MGS862 M | Feb-54 | 0.023124 | 0.314984 | 0 | 0 | 8.986878 | 33.8526 |
| SysMyo_M | MGS247 C | 3/154 | 0.029661 | 0.314984 | 0 | 0 | 4.674078 | 16.44307 |
| SysMyo_M | MGS1480 I | 3/152 | 0.028688 | 0.314984 | 0 | 0 | 4.737296 | 16.82341 |
| SysMyo_M | MGS1365 I | 4/251 | 0.023972 | 0.314984 | 0 | 0 | 3.837081 | 14.31558 |
| SysMyo_M | MGS1326 I | 3/180 | 0.043866 | 0.314984 | 0 | 0 | 3.982244 | 12.45094 |
| SysMyo_M | MGS239 M | 4/300 | 0.041941 | 0.314984 | 0 | 0 | 3.193911 | 10.12943 |
| SysMyo_M | MGS553 H | 4/251 | 0.023972 | 0.314984 | 0 | 0 | 3.837081 | 14.31558 |
| SysMyo_M | MGS1043 I | 3/139 | 0.022791 | 0.314984 | 0 | 0 | 5.19354 | 19.6389 |

| | | | | | | | | |
|---|---|---|---|---|---|---|---|---|
| SysMyo_M | MGS642 Hu | 8-Jan | 0.03428 | 0.314984 | 0 | 0 | 33.06645 | 111.5391 |
| SysMyo_M | MGS1450 M | 8-Jan | 0.03428 | 0.314984 | 0 | 0 | 33.06645 | 111.5391 |
| SysMyo_M | MGS171 M | Feb-66 | 0.033522 | 0.314984 | 0 | 0 | 7.297426 | 24.7787 |
| SysMyo_M | MGS438 Hu | 3/161 | 0.033201 | 0.314984 | 0 | 0 | 4.465416 | 15.20552 |
| SysMyo_M | MGS731 M | 3/159 | 0.032168 | 0.314984 | 0 | 0 | 4.523123 | 15.545 |
| SysMyo_M | MGS1351 M | 4/264 | 0.028165 | 0.314984 | 0 | 0 | 3.642817 | 13.00366 |
| SysMyo_M | MGS877 M | 4/261 | 0.027161 | 0.314984 | 0 | 0 | 3.685903 | 13.29124 |
| SysMyo_M | MGS29 Mu | Feb-70 | 0.037325 | 0.314984 | 0 | 0 | 6.866782 | 22.57858 |
| SysMyo_M | MGS1069 M | 3/168 | 0.036952 | 0.314984 | 0 | 0 | 4.274459 | 14.09775 |
| SysMyo_M | MGS1034 M | 3/173 | 0.039759 | 0.314984 | 0 | 0 | 4.147689 | 13.37596 |
| SysMyo_M | MGS1045 M | 3/175 | 0.040912 | 0.314984 | 0 | 0 | 4.099045 | 13.10196 |
| SysMyo_M | MGS1377 M | 3/182 | 0.045077 | 0.314984 | 0 | 0 | 3.93735 | 12.20334 |
| SysMyo_M | MGS542 Hu | 3/182 | 0.045077 | 0.314984 | 0 | 0 | 3.93735 | 12.20334 |
| SysMyo_M | MGS695 M | 8-Jan | 0.03428 | 0.314984 | 0 | 0 | 33.06645 | 111.5391 |
| SysMyo_M | MGS1192 M | 3/165 | 0.035319 | 0.314984 | 0 | 0 | 4.354277 | 14.55783 |
| SysMyo_M | MGS548 Hu | 3/170 | 0.038062 | 0.314984 | 0 | 0 | 4.22284 | 13.80251 |
| SysMyo_M | MGS1311 M | 3/170 | 0.038062 | 0.314984 | 0 | 0 | 4.22284 | 13.80251 |
| Human_Ph | Dementia ( | Jan-77 | 0.285606 | 0.315322 | 0 | 0 | 3.035037 | 3.803331 |
| Human_Ph | Broad forel | Jan-77 | 0.285606 | 0.315322 | 0 | 0 | 3.035037 | 3.803331 |
| Human_Ph | Blindness ( | Jan-77 | 0.285606 | 0.315322 | 0 | 0 | 3.035037 | 3.803331 |
| SysMyo_M | MGS587 Hu | 3/183 | 0.045689 | 0.315365 | 0 | 0 | 3.915278 | 12.08215 |
| SysMyo_M | MGS574 Hu | 11-Jan | 0.046834 | 0.315572 | 0 | 0 | 23.14302 | 70.84419 |
| SysMyo_M | MGS1095 M | Feb-79 | 0.046446 | 0.315572 | 0 | 0 | 6.061421 | 18.60534 |
| DisGeNET | Ulcerative | 6/915 | 0.207639 | 0.315812 | 0 | 0 | 1.548629 | 2.434374 |
| DisGeNET | Lymphoma | 4/539 | 0.207609 | 0.315812 | 0 | 0 | 1.745569 | 2.744206 |
| DisGeNET | Lymphoma | 8/1307 | 0.207688 | 0.315812 | 0 | 0 | 1.451087 | 2.280698 |
| GO_Biologi | Cellular Re | Jan-56 | 0.216879 | 0.316189 | 0 | 0 | 4.198309 | 6.416764 |
| SysMyo_M | MGS657 Hu | Feb-80 | 0.047505 | 0.316329 | 0 | 0 | 5.983409 | 18.23095 |
| DisGeNET | Gait abnor | 2/195 | 0.208243 | 0.316471 | 0 | 0 | 2.404145 | 3.772225 |
| HuBMAP_A | Articular cl | 2/103 | 0.074122 | 0.316591 | 0 | 0 | 4.615492 | 12.00969 |
| HuBMAP_A | Luminal Ce | 2/100 | 0.070424 | 0.316591 | 0 | 0 | 4.757503 | 12.62269 |
| HuBMAP_A | Periepithel | 2/100 | 0.070424 | 0.316591 | 0 | 0 | 4.757503 | 12.62269 |
| HuBMAP_A | Interstitial | 2/100 | 0.070424 | 0.316591 | 0 | 0 | 4.757503 | 12.62269 |
| HuBMAP_A | pericyte - H | 2/100 | 0.070424 | 0.316591 | 0 | 0 | 4.757503 | 12.62269 |
| HuBMAP_A | osteoblast | 2/102 | 0.072883 | 0.316591 | 0 | 0 | 4.661882 | 12.20902 |
| HuBMAP_A | osteocyte - | 2/102 | 0.072883 | 0.316591 | 0 | 0 | 4.661882 | 12.20902 |
| HuBMAP_A | Endo L2-5 I | 2/101 | 0.07165 | 0.316591 | 0 | 0 | 4.70921 | 12.4133 |
| HuBMAP_A | Lens Epithe | 2/101 | 0.07165 | 0.316591 | 0 | 0 | 4.70921 | 12.4133 |
| HuBMAP_A | Smooth Mu | 2/101 | 0.07165 | 0.316591 | 0 | 0 | 4.70921 | 12.4133 |
| HuBMAP_A | Endothelia | 2/101 | 0.07165 | 0.316591 | 0 | 0 | 4.70921 | 12.4133 |
| HuBMAP_A | B cell - Lym | 2/101 | 0.07165 | 0.316591 | 0 | 0 | 4.70921 | 12.4133 |
| HuBMAP_A | myofibrobl | 2/102 | 0.072883 | 0.316591 | 0 | 0 | 4.661882 | 12.20902 |
| HuBMAP_A | Kidney Effe | 2/103 | 0.074122 | 0.316591 | 0 | 0 | 4.615492 | 12.00969 |
| HuBMAP_A | Vasa Recta | 2/103 | 0.074122 | 0.316591 | 0 | 0 | 4.615492 | 12.00969 |
| HuBMAP_A | Hepatic Ste | 2/103 | 0.074122 | 0.316591 | 0 | 0 | 4.615492 | 12.00969 |

| | | | | | | | | |
|---|---|---|---|---|---|---|---|---|
| HuBMAP_A | Exc L3 ROR | 2/102 | 0.072883 | 0.316591 | 0 | 0 | 4.661882 | 12.20902 |
| HuBMAP_A | Endothelia | 2/103 | 0.074122 | 0.316591 | 0 | 0 | 4.615492 | 12.00969 |
| HuBMAP_A | Megakaryc | 2/103 | 0.074122 | 0.316591 | 0 | 0 | 4.615492 | 12.00969 |
| HuBMAP_A | Vasa Recta | 2/102 | 0.072883 | 0.316591 | 0 | 0 | 4.661882 | 12.20902 |
| HuBMAP_A | Renal Inter | 2/102 | 0.072883 | 0.316591 | 0 | 0 | 4.661882 | 12.20902 |
| HuBMAP_A | Pericyte ce | 2/102 | 0.072883 | 0.316591 | 0 | 0 | 4.661882 | 12.20902 |
| HuBMAP_A | CD141-pos | 2/104 | 0.075369 | 0.316591 | 0 | 0 | 4.570012 | 11.81514 |
| HuBMAP_A | Myofibrobl | 2/103 | 0.074122 | 0.316591 | 0 | 0 | 4.615492 | 12.00969 |
| HuBMAP_A | Mature Na | 2/102 | 0.072883 | 0.316591 | 0 | 0 | 4.661882 | 12.20902 |
| HuBMAP_A | Splenic Tin | 2/102 | 0.072883 | 0.316591 | 0 | 0 | 4.661882 | 12.20902 |
| HuBMAP_A | CD14-low, | 2/106 | 0.077881 | 0.316591 | 0 | 0 | 4.481674 | 11.43977 |
| HuBMAP_A | Splenic Enc | 2/106 | 0.077881 | 0.316591 | 0 | 0 | 4.481674 | 11.43977 |
| HuBMAP_A | Descending | 2/107 | 0.079148 | 0.316591 | 0 | 0 | 4.438768 | 11.25866 |
| HuBMAP_A | macrophag | 2/107 | 0.079148 | 0.316591 | 0 | 0 | 4.438768 | 11.25866 |
| HuBMAP_A | Matrix Fibr | 2/100 | 0.070424 | 0.316591 | 0 | 0 | 4.757503 | 12.62269 |
| HuBMAP_A | Kidney Affe | 2/105 | 0.076622 | 0.316591 | 0 | 0 | 4.525414 | 11.62522 |
| HuBMAP_A | Capsule M | 2/102 | 0.072883 | 0.316591 | 0 | 0 | 4.661882 | 12.20902 |
| HuBMAP_A | Luminal Ep | 2/103 | 0.074122 | 0.316591 | 0 | 0 | 4.615492 | 12.00969 |
| HuBMAP_A | Prostate Gl | 2/103 | 0.074122 | 0.316591 | 0 | 0 | 4.615492 | 12.00969 |
| HuBMAP_A | Splenic Per | 2/103 | 0.074122 | 0.316591 | 0 | 0 | 4.615492 | 12.00969 |
| HuBMAP_A | Dendritic C | 2/105 | 0.076622 | 0.316591 | 0 | 0 | 4.525414 | 11.62522 |
| HuBMAP_A | Endothelia | 2/103 | 0.074122 | 0.316591 | 0 | 0 | 4.615492 | 12.00969 |
| HuBMAP_A | Uterine ma | 2/104 | 0.075369 | 0.316591 | 0 | 0 | 4.570012 | 11.81514 |
| HuBMAP_A | Oligo L3-6 | 2/104 | 0.075369 | 0.316591 | 0 | 0 | 4.570012 | 11.81514 |
| HuBMAP_A | basophil - | 2/102 | 0.072883 | 0.316591 | 0 | 0 | 4.661882 | 12.20902 |
| HuBMAP_A | Stromal cel | 2/104 | 0.075369 | 0.316591 | 0 | 0 | 4.570012 | 11.81514 |
| HuBMAP_A | CAP1 - Res | 2/104 | 0.075369 | 0.316591 | 0 | 0 | 4.570012 | 11.81514 |
| HuBMAP_A | Pulmonary | 2/105 | 0.076622 | 0.316591 | 0 | 0 | 4.525414 | 11.62522 |
| HuBMAP_A | Effector M | 2/105 | 0.076622 | 0.316591 | 0 | 0 | 4.525414 | 11.62522 |
| HuBMAP_A | Lympho-M | 2/106 | 0.077881 | 0.316591 | 0 | 0 | 4.481674 | 11.43977 |
| HuBMAP_A | myofibrobl | 2/102 | 0.072883 | 0.316591 | 0 | 0 | 4.661882 | 12.20902 |
| HuBMAP_A | Inh L1-6 PV | 2/102 | 0.072883 | 0.316591 | 0 | 0 | 4.661882 | 12.20902 |
| HuBMAP_A | Portal fibro | 2/102 | 0.072883 | 0.316591 | 0 | 0 | 4.661882 | 12.20902 |
| HuBMAP_A | Uterine Sm | 2/105 | 0.076622 | 0.316591 | 0 | 0 | 4.525414 | 11.62522 |
| Jensen_TIS | Acute myel | Jan-57 | 0.220295 | 0.316674 | 0 | 0 | 4.123131 | 6.237423 |
| Jensen_TIS | Tonsil | 64/13827 | 0.219546 | 0.316674 | 0 | 0 | 1.243409 | 1.885249 |
| GWAS_Cat | White Mat | Jan-79 | 0.291832 | 0.316757 | 0 | 0 | 2.956917 | 3.641673 |
| GeDiPNet_ | Polycystic | 2/211 | 0.233816 | 0.31701 | 0 | 0 | 2.218294 | 3.223675 |
| GeDiPNet_ | Hypotricho | Jan-61 | 0.233813 | 0.31701 | 0 | 0 | 3.847481 | 5.591291 |
| Jensen_TIS | Kidney | 77/16989 | 0.221605 | 0.317142 | 0 | 0 | 1.366349 | 2.058893 |
| DisGeNET | Action Trer | Jan-54 | 0.210002 | 0.317671 | 0 | 0 | 4.357174 | 6.79997 |
| DisGeNET | Autistic bel | Jan-54 | 0.210002 | 0.317671 | 0 | 0 | 4.357174 | 6.79997 |
| DisGeNET | Malignant | Jan-54 | 0.210002 | 0.317671 | 0 | 0 | 4.357174 | 6.79997 |
| DisGeNET | Cerebral Ar | Jan-54 | 0.210002 | 0.317671 | 0 | 0 | 4.357174 | 6.79997 |
| DisGeNET | Optic Atrop | 3/363 | 0.209989 | 0.317671 | 0 | 0 | 1.939782 | 3.027421 |

| | | | | | | | | |
|---|---|---|---|---|---|---|---|---|
| DisGeNET | Hepatocell | Jan-54 | 0.210002 | 0.317671 | 0 | 0 | 4.357174 | 6.79997 |
| DisGeNET | Hypertensi | 8/1310 | 0.209407 | 0.317671 | 0 | 0 | 1.44751 | 2.263148 |
| DisGeNET | Carcinoma, | 2/196 | 0.209833 | 0.317671 | 0 | 0 | 2.391631 | 3.734399 |
| Rare_Disea | Essential th | 2/162 | 0.156786 | 0.318713 | 0 | 0 | 2.904853 | 5.382321 |
| Rare_Disea | Cowden's d | Jan-39 | 0.156488 | 0.318713 | 0 | 0 | 6.081701 | 11.28018 |
| Jensen_TIS | Somite | Jan-58 | 0.223696 | 0.318718 | 0 | 0 | 4.050592 | 6.065622 |
| GO_Biologi | Post-Trans | Jan-57 | 0.220295 | 0.319203 | 0 | 0 | 4.123131 | 6.237423 |
| GO_Biologi | Regulation | 2/202 | 0.219398 | 0.319203 | 0 | 0 | 2.319176 | 3.517879 |
| GO_Biologi | Regulation | Jan-57 | 0.220295 | 0.319203 | 0 | 0 | 4.123131 | 6.237423 |
| GWAS_Cat | Coffee Con | Jan-80 | 0.294925 | 0.319262 | 0 | 0 | 2.919341 | 3.564619 |
| GWAS_Cat | Respiratory | Jan-80 | 0.294925 | 0.319262 | 0 | 0 | 2.919341 | 3.564619 |
| GWAS_Cat | Chronic Kic | Jan-80 | 0.294925 | 0.319262 | 0 | 0 | 2.919341 | 3.564619 |
| GeDiPNet_ | Vasculitis | Jan-62 | 0.237156 | 0.319312 | 0 | 0 | 3.784217 | 5.445629 |
| GeDiPNet_ | Neuronal H | Jan-62 | 0.237156 | 0.319312 | 0 | 0 | 3.784217 | 5.445629 |
| GeDiPNet_ | Oral Cleft | Jan-62 | 0.237156 | 0.319312 | 0 | 0 | 3.784217 | 5.445629 |
| SysMyo_M | MGS1121 I | 3/189 | 0.049446 | 0.319341 | 0 | 0 | 3.787826 | 11.38952 |
| SysMyo_M | MGS571 Hu | Feb-82 | 0.04965 | 0.319341 | 0 | 0 | 5.833235 | 17.51576 |
| SysMyo_M | MGS536 Hu | Feb-82 | 0.04965 | 0.319341 | 0 | 0 | 5.833235 | 17.51576 |
| GO_Molecu | Tau Protein | Jan-39 | 0.156488 | 0.319364 | 0 | 0 | 6.081701 | 11.28018 |
| DisGeNET | Tuberous S | 2/197 | 0.211424 | 0.319453 | 0 | 0 | 2.379246 | 3.697085 |
| DisGeNET | Allergic ast | 2/197 | 0.211424 | 0.319453 | 0 | 0 | 2.379246 | 3.697085 |
| GTEx_Tissu | GTEX-V955 | 16/1924 | 0.008289 | 0.319748 | 0 | 0 | 2.126554 | 10.19211 |
| Jensen_DIS | Essential hy | Jan-78 | 0.288726 | 0.319895 | 0 | 0 | 2.99547 | 3.721207 |
| Jensen_DIS | Cleft lip | Jan-77 | 0.285606 | 0.319895 | 0 | 0 | 3.035037 | 3.803331 |
| Jensen_DIS | Acquired ir | Jan-78 | 0.288726 | 0.319895 | 0 | 0 | 2.99547 | 3.721207 |
| Rare_Disea | Shigellosis | Jan-40 | 0.160165 | 0.32033 | 0 | 0 | 5.925462 | 10.85279 |
| GO_Molecu | Acetylgluco | Jan-40 | 0.160165 | 0.32033 | 0 | 0 | 5.925462 | 10.85279 |
| Rare_Disea | Fish-eye di | Jan-40 | 0.160165 | 0.32033 | 0 | 0 | 5.925462 | 10.85279 |
| Rare_Disea | Carcinoid s | Jan-40 | 0.160165 | 0.32033 | 0 | 0 | 5.925462 | 10.85279 |
| Rare_Disea | Li Fraumen | Jan-40 | 0.160165 | 0.32033 | 0 | 0 | 5.925462 | 10.85279 |
| Rare_Disea | Carney con | Jan-40 | 0.160165 | 0.32033 | 0 | 0 | 5.925462 | 10.85279 |
| Rare_Disea | Neurofibro | 2/163 | 0.158308 | 0.32033 | 0 | 0 | 2.886664 | 5.320729 |
| GeDiPNet_ | Anxiety Dis | 3/388 | 0.238547 | 0.320445 | 0 | 0 | 1.811503 | 2.596225 |
| DisGeNET | Brugada Sy | Jan-55 | 0.213448 | 0.320845 | 0 | 0 | 4.27627 | 6.604112 |
| DisGeNET | Ganglioneu | Jan-55 | 0.213448 | 0.320845 | 0 | 0 | 4.27627 | 6.604112 |
| DisGeNET | Atherogeni | Jan-55 | 0.213448 | 0.320845 | 0 | 0 | 4.27627 | 6.604112 |
| DisGeNET | Hallucinatio | Jan-55 | 0.213448 | 0.320845 | 0 | 0 | 4.27627 | 6.604112 |
| DisGeNET | Osteoporos | Jan-55 | 0.213448 | 0.320845 | 0 | 0 | 4.27627 | 6.604112 |
| DisGeNET | Adenosqua | Jan-55 | 0.213448 | 0.320845 | 0 | 0 | 4.27627 | 6.604112 |
| DisGeNET | Congenital | 2/198 | 0.213017 | 0.320845 | 0 | 0 | 2.366987 | 3.660273 |
| DisGeNET | Insulin Sen | Jan-55 | 0.213448 | 0.320845 | 0 | 0 | 4.27627 | 6.604112 |
| DisGeNET | Scoliosis, u | Jan-55 | 0.213448 | 0.320845 | 0 | 0 | 4.27627 | 6.604112 |
| GeDiPNet_ | Myopia | 3/389 | 0.239704 | 0.321259 | 0 | 0 | 1.806717 | 2.580624 |
| Human_Ph | Depressed | Jan-79 | 0.291832 | 0.321267 | 0 | 0 | 2.956917 | 3.641673 |
| GeDiPNet_ | Ptosis | 4/575 | 0.240857 | 0.322063 | 0 | 0 | 1.632477 | 2.323919 |

| | | | | | | | | |
|---|---|---|---|---|---|---|---|---|
| dbGaP | Cholestero | 3/357 | 0.203255 | 0.322234 | 0 | 0 | 1.973265 | 3.143989 |
| DisGeNET | Precursor E | 3/367 | 0.214506 | 0.32225 | 0 | 0 | 1.918073 | 2.952718 |
| GWAS_Cat | Photorecep | Jan-81 | 0.298004 | 0.322309 | 0 | 0 | 2.882703 | 3.48994 |
| GO_Biologi | Memory (G | Jan-58 | 0.223696 | 0.322814 | 0 | 0 | 4.050592 | 6.065622 |
| GO_Biologi | Glycosamir | Jan-58 | 0.223696 | 0.322814 | 0 | 0 | 4.050592 | 6.065622 |
| UK_Bioban | Mean corp | 4/608 | 0.27236 | 0.323428 | 0 | 0 | 1.540653 | 2.003818 |
| GO_Biologi | Heart Morp | Jan-59 | 0.227083 | 0.323554 | 0 | 0 | 3.980553 | 5.900928 |
| GO_Biologi | Positive Re | Jan-59 | 0.227083 | 0.323554 | 0 | 0 | 3.980553 | 5.900928 |
| GO_Biologi | Chordate E | Jan-59 | 0.227083 | 0.323554 | 0 | 0 | 3.980553 | 5.900928 |
| GO_Biologi | Regulation | Jan-59 | 0.227083 | 0.323554 | 0 | 0 | 3.980553 | 5.900928 |
| GO_Biologi | Negative R | 2/207 | 0.227399 | 0.323554 | 0 | 0 | 2.262037 | 3.350186 |
| GO_Biologi | Positive Re | Jan-59 | 0.227083 | 0.323554 | 0 | 0 | 3.980553 | 5.900928 |
| GO_Biologi | DNA-templ | Jan-59 | 0.227083 | 0.323554 | 0 | 0 | 3.980553 | 5.900928 |
| Human_Ph | Pulmonary | Jan-80 | 0.294925 | 0.323739 | 0 | 0 | 2.919341 | 3.564619 |
| GO_Biologi | Negative R | 3/379 | 0.228181 | 0.324017 | 0 | 0 | 1.855718 | 2.74204 |
| GeDiPNet_ | Agenesis O | 2/217 | 0.243461 | 0.324059 | 0 | 0 | 2.155732 | 3.045615 |
| GeDiPNet_ | Renal Carci | 2/217 | 0.243461 | 0.324059 | 0 | 0 | 2.155732 | 3.045615 |
| Human_Ph | Deeply set | Jan-81 | 0.298004 | 0.324323 | 0 | 0 | 2.882703 | 3.48994 |
| Human_Ph | Dental mal | Jan-81 | 0.298004 | 0.324323 | 0 | 0 | 2.882703 | 3.48994 |
| Human_Ph | Lethargy (H | Jan-81 | 0.298004 | 0.324323 | 0 | 0 | 2.882703 | 3.48994 |
| DisGeNET | Speech imp | 2/200 | 0.216205 | 0.324618 | 0 | 0 | 2.34284 | 3.588123 |
| DisGeNET | Carotid Ste | Jan-56 | 0.216879 | 0.324699 | 0 | 0 | 4.198309 | 6.416764 |
| DisGeNET | HER2 gene | Jan-56 | 0.216879 | 0.324699 | 0 | 0 | 4.198309 | 6.416764 |
| DisGeNET | Autoimmu | Jan-56 | 0.216879 | 0.324699 | 0 | 0 | 4.198309 | 6.416764 |
| DisGeNET | Congenital | Jan-56 | 0.216879 | 0.324699 | 0 | 0 | 4.198309 | 6.416764 |
| DisGeNET | Adenoviral | Jan-56 | 0.216879 | 0.324699 | 0 | 0 | 4.198309 | 6.416764 |
| GO_Biologi | Sterol Meta | Jan-60 | 0.230455 | 0.325295 | 0 | 0 | 3.912889 | 5.742942 |
| GO_Biologi | Negative R | Jan-60 | 0.230455 | 0.325295 | 0 | 0 | 3.912889 | 5.742942 |
| GO_Biologi | Histone Ac | Jan-60 | 0.230455 | 0.325295 | 0 | 0 | 3.912889 | 5.742942 |
| DisGeNET | Eosinophil | 2/201 | 0.217801 | 0.325707 | 0 | 0 | 2.330949 | 3.552767 |
| DisGeNET | Eosinophil | 2/201 | 0.217801 | 0.325707 | 0 | 0 | 2.330949 | 3.552767 |
| Jensen_TIS | Colonic car | 4/563 | 0.229627 | 0.325726 | 0 | 0 | 1.668556 | 2.454945 |
| PhenGenI_ | Docosapen | 26-Jan | 0.107222 | 0.325899 | 0 | 0 | 9.250233 | 20.65441 |
| PhenGenI_ | Smoking Ce | 26-Jan | 0.107222 | 0.325899 | 0 | 0 | 9.250233 | 20.65441 |
| PhenGenI_ | Lewy Body | 25-Jan | 0.103317 | 0.325899 | 0 | 0 | 9.636143 | 21.87363 |
| PhenGenI_ | gamma-Glu | 2/127 | 0.105734 | 0.325899 | 0 | 0 | 3.7248 | 8.368983 |
| Rare_Disea | Amyloid ne | Jan-41 | 0.163825 | 0.325899 | 0 | 0 | 5.777035 | 10.45039 |
| Rare_Disea | Kallmann s | Jan-41 | 0.163825 | 0.325899 | 0 | 0 | 5.777035 | 10.45039 |
| SysMyo_M | MGS820 M | Feb-84 | 0.05183 | 0.325953 | 0 | 0 | 5.690387 | 16.84234 |
| SysMyo_M | MGS1213 I | 3/192 | 0.05138 | 0.325953 | 0 | 0 | 3.727135 | 11.06404 |
| MAGMA_D | Gemcitabir | Jan-79 | 0.291832 | 0.326165 | 0 | 0 | 2.956917 | 3.641673 |
| Rare_Disea | Lupus neph | 2/167 | 0.164425 | 0.326219 | 0 | 0 | 2.816114 | 5.083934 |
| UK_Bioban | Left intra-o | Jan-74 | 0.276166 | 0.326586 | 0 | 0 | 3.160242 | 4.066453 |
| Reactome_ | Muscle Cor | 3/196 | 0.054015 | 0.327008 | 0 | 0 | 3.649149 | 10.65003 |
| Jensen_TIS | Urinary bla | 73/16036 | 0.233657 | 0.327342 | 0 | 0 | 1.290261 | 1.875913 |

| | | | | | | | | |
|---|---|---|---|---|---|---|---|---|
| Jensen_TIS | Brain corte | 2/211 | 0.233816 | 0.327342 | 0 | 0 | 2.218294 | 3.223675 |
| Jensen_TIS | Duodenum | 72/15789 | 0.232407 | 0.327342 | 0 | 0 | 1.281466 | 1.870001 |
| DisGeNET | Abnormal I | 3/372 | 0.220182 | 0.327381 | 0 | 0 | 1.891599 | 2.862559 |
| DisGeNET | Geographic | Jan-57 | 0.220295 | 0.327381 | 0 | 0 | 4.123131 | 6.237423 |
| DisGeNET | Peutz-Jegh | Jan-57 | 0.220295 | 0.327381 | 0 | 0 | 4.123131 | 6.237423 |
| DisGeNET | Spondylart | Jan-57 | 0.220295 | 0.327381 | 0 | 0 | 4.123131 | 6.237423 |
| DisGeNET | Pneumocys | Jan-57 | 0.220295 | 0.327381 | 0 | 0 | 4.123131 | 6.237423 |
| DisGeNET | Carotid Art | Jan-57 | 0.220295 | 0.327381 | 0 | 0 | 4.123131 | 6.237423 |
| DisGeNET | Hyperaldos | Jan-57 | 0.220295 | 0.327381 | 0 | 0 | 4.123131 | 6.237423 |
| DisGeNET | Broad nasa | Jan-57 | 0.220295 | 0.327381 | 0 | 0 | 4.123131 | 6.237423 |
| DisGeNET | Abnormalit | Jan-57 | 0.220295 | 0.327381 | 0 | 0 | 4.123131 | 6.237423 |
| DisGeNET | Respiratory | Jan-57 | 0.220295 | 0.327381 | 0 | 0 | 4.123131 | 6.237423 |
| DisGeNET | Atrophic | Jan-57 | 0.220295 | 0.327381 | 0 | 0 | 4.123131 | 6.237423 |
| HuBMAP_A | Follicular D | 2/110 | 0.082985 | 0.327394 | 0 | 0 | 4.314815 | 10.73997 |
| MAGMA_D | Sorafenib | Jan-85 | 0.310189 | 0.327422 | 0 | 0 | 2.744878 | 3.21308 |
| MAGMA_D | Polar Auxir | Jan-84 | 0.307163 | 0.327422 | 0 | 0 | 2.778089 | 3.279195 |
| MAGMA_D | Hepatocell | Jan-85 | 0.310189 | 0.327422 | 0 | 0 | 2.744878 | 3.21308 |
| MAGMA_D | Proteasom | Jan-84 | 0.307163 | 0.327422 | 0 | 0 | 2.778089 | 3.279195 |
| MAGMA_D | Bortezomib | Jan-84 | 0.307163 | 0.327422 | 0 | 0 | 2.778089 | 3.279195 |
| MAGMA_D | Quercetin | Jan-84 | 0.307163 | 0.327422 | 0 | 0 | 2.778089 | 3.279195 |
| MAGMA_D | Multiple M | Jan-84 | 0.307163 | 0.327422 | 0 | 0 | 2.778089 | 3.279195 |
| CellMarker | Regulatory | 3/411 | 0.265386 | 0.327574 | 0 | 0 | 1.70737 | 2.264943 |
| CellMarker | Natural Kill | Jan-71 | 0.266602 | 0.327574 | 0 | 0 | 3.296179 | 4.35754 |
| CellMarker | Macrophag | Jan-71 | 0.266602 | 0.327574 | 0 | 0 | 3.296179 | 4.35754 |
| HuBMAP_A | Medullary | 2/111 | 0.084277 | 0.327997 | 0 | 0 | 4.275013 | 10.57487 |
| DisGeNET | Tooth Crov | Jan-58 | 0.223696 | 0.32801 | 0 | 0 | 4.050592 | 6.065622 |
| DisGeNET | Intestinal C | Jan-58 | 0.223696 | 0.32801 | 0 | 0 | 4.050592 | 6.065622 |
| DisGeNET | Carcinoma | Jan-58 | 0.223696 | 0.32801 | 0 | 0 | 4.050592 | 6.065622 |
| DisGeNET | Anaplastic | 2/204 | 0.222596 | 0.32801 | 0 | 0 | 2.295981 | 3.449476 |
| DisGeNET | Melanocyti | 2/203 | 0.220997 | 0.32801 | 0 | 0 | 2.307521 | 3.483452 |
| DisGeNET | Hepatomeg | 3/373 | 0.221321 | 0.32801 | 0 | 0 | 1.88639 | 2.844943 |
| DisGeNET | Muscular D | 2/203 | 0.220997 | 0.32801 | 0 | 0 | 2.307521 | 3.483452 |
| DisGeNET | Brachydact | 2/204 | 0.222596 | 0.32801 | 0 | 0 | 2.295981 | 3.449476 |
| DisGeNET | Respiratory | Jan-58 | 0.223696 | 0.32801 | 0 | 0 | 4.050592 | 6.065622 |
| DisGeNET | Neurodege | 5/745 | 0.223723 | 0.32801 | 0 | 0 | 1.579845 | 2.365576 |
| DisGeNET | Cutis Laxa | Jan-58 | 0.223696 | 0.32801 | 0 | 0 | 4.050592 | 6.065622 |
| DisGeNET | Oncocytic I | Jan-58 | 0.223696 | 0.32801 | 0 | 0 | 4.050592 | 6.065622 |
| DisGeNET | Nerve Shea | Jan-58 | 0.223696 | 0.32801 | 0 | 0 | 4.050592 | 6.065622 |
| DisGeNET | Mycoplasm | Jan-58 | 0.223696 | 0.32801 | 0 | 0 | 4.050592 | 6.065622 |
| DisGeNET | Tooth size | Jan-58 | 0.223696 | 0.32801 | 0 | 0 | 4.050592 | 6.065622 |
| DisGeNET | Tooth mass | Jan-58 | 0.223696 | 0.32801 | 0 | 0 | 4.050592 | 6.065622 |
| DisGeNET | Heart valve | Jan-58 | 0.223696 | 0.32801 | 0 | 0 | 4.050592 | 6.065622 |
| DisGeNET | Atheroscle | 7/1134 | 0.223116 | 0.32801 | 0 | 0 | 1.45854 | 2.187903 |
| DisGeNET | Sparse and | Jan-58 | 0.223696 | 0.32801 | 0 | 0 | 4.050592 | 6.065622 |
| DisGeNET | Carcinoma, | Jan-58 | 0.223696 | 0.32801 | 0 | 0 | 4.050592 | 6.065622 |

| | | | | | | | | |
|---|---|---|---|---|---|---|---|---|
| DisGeNET | Inclusion B | Jan-58 | 0.223696 | 0.32801 | 0 | 0 | 4.050592 | 6.065622 |
| DisGeNET | Large cell c | Jan-58 | 0.223696 | 0.32801 | 0 | 0 | 4.050592 | 6.065622 |
| DisGeNET | Maternal o | Jan-58 | 0.223696 | 0.32801 | 0 | 0 | 4.050592 | 6.065622 |
| DisGeNET | Inadequate | Jan-58 | 0.223696 | 0.32801 | 0 | 0 | 4.050592 | 6.065622 |
| Rare_Disea | Testotoxic | 3/323 | 0.166214 | 0.32802 | 0 | 0 | 2.186719 | 3.924018 |
| Rare_Disea | Precocious | 3/323 | 0.166214 | 0.32802 | 0 | 0 | 2.186719 | 3.924018 |
| GeDiPNet_ | Acne | Jan-65 | 0.247099 | 0.328152 | 0 | 0 | 3.606286 | 5.041467 |
| DisGeNET | Embryonal | 2/205 | 0.224196 | 0.328153 | 0 | 0 | 2.284555 | 3.415945 |
| DisGeNET | Hematolog | 4/557 | 0.224065 | 0.328153 | 0 | 0 | 1.687183 | 2.523721 |
| DisGeNET | Hypogonac | 2/205 | 0.224196 | 0.328153 | 0 | 0 | 2.284555 | 3.415945 |
| SysMyo_M | MGS1264 I | 3/195 | 0.05335 | 0.328218 | 0 | 0 | 3.668341 | 10.75147 |
| SysMyo_M | MGS1504 I | Feb-85 | 0.052932 | 0.328218 | 0 | 0 | 5.621545 | 16.52025 |
| KEGG_202 | Diabetic ca | 3/203 | 0.05878 | 0.328475 | 0 | 0 | 3.520179 | 9.976043 |
| Human_Ph | Infantile or | 2/254 | 0.303079 | 0.32891 | 0 | 0 | 1.835761 | 2.191461 |
| Jensen_TIS | Cyst | Jan-62 | 0.237156 | 0.329156 | 0 | 0 | 3.784217 | 5.445629 |
| Jensen_TIS | Ovary canc | Jan-62 | 0.237156 | 0.329156 | 0 | 0 | 3.784217 | 5.445629 |
| GO_Biologi | mRNA Splic | 2/211 | 0.233816 | 0.329383 | 0 | 0 | 2.218294 | 3.223675 |
| SysMyo_M | MGS733 M | 3/198 | 0.055356 | 0.329807 | 0 | 0 | 3.611355 | 10.45114 |
| SysMyo_M | MGS1421 I | 3/197 | 0.054684 | 0.329807 | 0 | 0 | 3.630155 | 10.54993 |
| SysMyo_M | MGS1166 I | 3/198 | 0.055356 | 0.329807 | 0 | 0 | 3.611355 | 10.45114 |
| DisGeNET | Radial club | Jan-59 | 0.227083 | 0.329982 | 0 | 0 | 3.980553 | 5.900928 |
| DisGeNET | Chronic lur | Jan-59 | 0.227083 | 0.329982 | 0 | 0 | 3.980553 | 5.900928 |
| DisGeNET | Body fluid r | Jan-59 | 0.227083 | 0.329982 | 0 | 0 | 3.980553 | 5.900928 |
| DisGeNET | Progeria | Jan-59 | 0.227083 | 0.329982 | 0 | 0 | 3.980553 | 5.900928 |
| DisGeNET | Anaplastic | Jan-59 | 0.227083 | 0.329982 | 0 | 0 | 3.980553 | 5.900928 |
| DisGeNET | Language D | 2/206 | 0.225797 | 0.329982 | 0 | 0 | 2.273241 | 3.382851 |
| DisGeNET | Skin Abnor | Jan-59 | 0.227083 | 0.329982 | 0 | 0 | 3.980553 | 5.900928 |
| DisGeNET | Moderate r | Jan-59 | 0.227083 | 0.329982 | 0 | 0 | 3.980553 | 5.900928 |
| DisGeNET | Sleep Apne | Jan-59 | 0.227083 | 0.329982 | 0 | 0 | 3.980553 | 5.900928 |
| DisGeNET | Squamous | Jan-59 | 0.227083 | 0.329982 | 0 | 0 | 3.980553 | 5.900928 |
| DisGeNET | Paraparesis | Jan-59 | 0.227083 | 0.329982 | 0 | 0 | 3.980553 | 5.900928 |
| DisGeNET | AICARDI-G | Jan-59 | 0.227083 | 0.329982 | 0 | 0 | 3.980553 | 5.900928 |
| DisGeNET | Angina Pec | Jan-59 | 0.227083 | 0.329982 | 0 | 0 | 3.980553 | 5.900928 |
| CellMarker | Macrophag | Jan-72 | 0.269804 | 0.330028 | 0 | 0 | 3.249591 | 4.257158 |
| DisGeNET | Visual seizu | 2/207 | 0.227399 | 0.330258 | 0 | 0 | 2.262037 | 3.350186 |
| Jensen_TIS | Femur | Jan-63 | 0.240485 | 0.330923 | 0 | 0 | 3.722993 | 5.305634 |
| Jensen_TIS | Synovia | Jan-63 | 0.240485 | 0.330923 | 0 | 0 | 3.722993 | 5.305634 |
| GWAS_Cat | Mitochond | Jan-84 | 0.307163 | 0.331627 | 0 | 0 | 2.778089 | 3.279195 |
| GWAS_Cat | Coronary A | Jan-84 | 0.307163 | 0.331627 | 0 | 0 | 2.778089 | 3.279195 |
| GTEx_Tissu | GTEX-XYKS | 15/1766 | 0.008828 | 0.331683 | 0 | 0 | 2.160908 | 10.2207 |
| GTEx_Tissu | GTEX-Q2AI | 9/812 | 0.008716 | 0.331683 | 0 | 0 | 2.745953 | 13.02298 |
| GeDiPNet_ | Diabetes | Jan-66 | 0.250385 | 0.33176 | 0 | 0 | 3.550626 | 4.916752 |
| DisGeNET | Systemic Sc | 4/562 | 0.228697 | 0.33196 | 0 | 0 | 1.671633 | 2.466252 |
| DisGeNET | Ataxia | 2/208 | 0.229002 | 0.332034 | 0 | 0 | 2.250942 | 3.317943 |
| DisGeNET | Addictive B | 2/208 | 0.229002 | 0.332034 | 0 | 0 | 2.250942 | 3.317943 |

| | | | | | | | | |
|---|---|---|---|---|---|---|---|---|
| GO_Biologi | Glycerophc | Jan-62 | 0.237156 | 0.332112 | 0 | 0 | 3.784217 | 5.445629 |
| GO_Biologi | Phosphatic | Jan-62 | 0.237156 | 0.332112 | 0 | 0 | 3.784217 | 5.445629 |
| GO_Biologi | Negative R | Jan-62 | 0.237156 | 0.332112 | 0 | 0 | 3.784217 | 5.445629 |
| GWAS_Cat | Prostate Ca | 2/257 | 0.307896 | 0.332125 | 0 | 0 | 1.813887 | 2.136747 |
| DisGeNET | Hypotricho | Jan-60 | 0.230455 | 0.332333 | 0 | 0 | 3.912889 | 5.742942 |
| DisGeNET | MAJOR AFI | Jan-60 | 0.230455 | 0.332333 | 0 | 0 | 3.912889 | 5.742942 |
| DisGeNET | Malignant | 7/1144 | 0.22953 | 0.332333 | 0 | 0 | 1.444943 | 2.126551 |
| DisGeNET | MAJOR AFI | Jan-60 | 0.230455 | 0.332333 | 0 | 0 | 3.912889 | 5.742942 |
| DisGeNET | Fatty Liver, | Jan-60 | 0.230455 | 0.332333 | 0 | 0 | 3.912889 | 5.742942 |
| DisGeNET | Autoimmu | Jan-60 | 0.230455 | 0.332333 | 0 | 0 | 3.912889 | 5.742942 |
| DisGeNET | Cerebral ca | Jan-60 | 0.230455 | 0.332333 | 0 | 0 | 3.912889 | 5.742942 |
| DisGeNET | Glaucoma | 3/381 | 0.230477 | 0.332333 | 0 | 0 | 1.845711 | 2.708774 |
| DisGeNET | Cavernous | Jan-60 | 0.230455 | 0.332333 | 0 | 0 | 3.912889 | 5.742942 |
| DisGeNET | Factor V Le | Jan-60 | 0.230455 | 0.332333 | 0 | 0 | 3.912889 | 5.742942 |
| DisGeNET | Nonalcoho | 2/209 | 0.230606 | 0.332336 | 0 | 0 | 2.239955 | 3.286115 |
| Human_Ph | Nephrotic s | Jan-84 | 0.307163 | 0.332397 | 0 | 0 | 2.778089 | 3.279195 |
| GWAS_Cat | Monocyte | 4/645 | 0.308493 | 0.332475 | 0 | 0 | 1.448941 | 1.704035 |
| Rare_Disea | Pulmonary | Jan-43 | 0.171099 | 0.333241 | 0 | 0 | 5.501384 | 9.712751 |
| Rare_Disea | C syndrom | 2/171 | 0.170583 | 0.333241 | 0 | 0 | 2.748904 | 4.86152 |
| Rare_Disea | Nephrotic s | Jan-43 | 0.171099 | 0.333241 | 0 | 0 | 5.501384 | 9.712751 |
| Rare_Disea | Oral cancer | 3/327 | 0.170462 | 0.333241 | 0 | 0 | 2.159281 | 3.820295 |
| Rare_Disea | Infective er | Jan-43 | 0.171099 | 0.333241 | 0 | 0 | 5.501384 | 9.712751 |
| DisGeNET | Age relatec | 3/382 | 0.231626 | 0.333306 | 0 | 0 | 1.840746 | 2.692329 |
| DisGeNET | Renal Cell ( | 8/1348 | 0.231661 | 0.333306 | 0 | 0 | 1.40359 | 2.052724 |
| DisGeNET | Cerebellar | 4/565 | 0.231489 | 0.333306 | 0 | 0 | 1.662436 | 2.432513 |
| PhenGenI_ | Neurofibril | 27-Jan | 0.111111 | 0.333332 | 0 | 0 | 8.894007 | 19.54216 |
| GO_Biologi | mRNA Proc | 2/214 | 0.238636 | 0.333526 | 0 | 0 | 2.18657 | 3.132957 |
| SysMyo_M | MGS160 H | 5/476 | 0.056598 | 0.333693 | 0 | 0 | 2.516959 | 7.228153 |
| DisGeNET | Lupus Nepl | 2/210 | 0.23221 | 0.333731 | 0 | 0 | 2.229072 | 3.254694 |
| DisGeNET | Neurofibro | 2/210 | 0.23221 | 0.333731 | 0 | 0 | 2.229072 | 3.254694 |
| GWAS_Cat | White Mat | Jan-85 | 0.310189 | 0.334008 | 0 | 0 | 2.744878 | 3.21308 |
| DisGeNET | Mesothelic | 3/383 | 0.232777 | 0.334362 | 0 | 0 | 1.835808 | 2.676009 |
| DisGeNET | Ganglioglic | Jan-61 | 0.233813 | 0.334367 | 0 | 0 | 3.847481 | 5.591291 |
| DisGeNET | Bradykines | Jan-61 | 0.233813 | 0.334367 | 0 | 0 | 3.847481 | 5.591291 |
| DisGeNET | Depression | Jan-61 | 0.233813 | 0.334367 | 0 | 0 | 3.847481 | 5.591291 |
| DisGeNET | MYELOPRC | Jan-61 | 0.233813 | 0.334367 | 0 | 0 | 3.847481 | 5.591291 |
| DisGeNET | Mitochond | 3/384 | 0.233929 | 0.334367 | 0 | 0 | 1.830896 | 2.659811 |
| DisGeNET | Noonan Sy | Jan-61 | 0.233813 | 0.334367 | 0 | 0 | 3.847481 | 5.591291 |
| DisGeNET | Leukoplaki | Jan-61 | 0.233813 | 0.334367 | 0 | 0 | 3.847481 | 5.591291 |
| DisGeNET | Increased ( | Jan-61 | 0.233813 | 0.334367 | 0 | 0 | 3.847481 | 5.591291 |
| DisGeNET | MAJOR AFI | Jan-61 | 0.233813 | 0.334367 | 0 | 0 | 3.847481 | 5.591291 |
| Gene_Pert | MTDH DEP | 6/327 | 0.003037 | 0.334385 | 0 | 0 | 4.521057 | 26.20743 |
| Gene_Pert | Lmna LMN. | 6/335 | 0.00342 | 0.334385 | 0 | 0 | 4.409321 | 25.03652 |
| Gene_Pert | Ovol2 OE n | 6/340 | 0.003677 | 0.334385 | 0 | 0 | 4.342204 | 24.34108 |
| Gene_Pert | Map2k3 OI | 6/329 | 0.00313 | 0.334385 | 0 | 0 | 4.492604 | 25.90773 |

| | | | | | | | | |
|---|---|---|---|---|---|---|---|---|
| Gene_Pert | Tgfbr2 kno | 6/332 | 0.003273 | 0.334385 | 0 | 0 | 4.450579 | 25.46698 |
| Gene_Pert | Map2k3 m | 6/341 | 0.00373 | 0.334385 | 0 | 0 | 4.329022 | 24.20519 |
| Gene_Pert | Ncoa2 KO r | 5/233 | 0.0035 | 0.334385 | 0 | 0 | 5.264495 | 29.77034 |
| GeDiPNet_ | Hereditary | Jan-67 | 0.253656 | 0.334574 | 0 | 0 | 3.496653 | 4.79662 |
| GeDiPNet_ | Cerebral Is | Jan-67 | 0.253656 | 0.334574 | 0 | 0 | 3.496653 | 4.79662 |
| GO_Biologi | Inorganic A | Jan-63 | 0.240485 | 0.334792 | 0 | 0 | 3.722993 | 5.305634 |
| GO_Biologi | Regulation | Jan-63 | 0.240485 | 0.334792 | 0 | 0 | 3.722993 | 5.305634 |
| SysMyo_M | MGS1062 I | 3/201 | 0.057398 | 0.334923 | 0 | 0 | 3.556097 | 10.16239 |
| Rare_Disea | Subacute s | Jan-44 | 0.174713 | 0.335017 | 0 | 0 | 5.373175 | 9.3741 |
| Rare_Disea | Oral leukop | 2/173 | 0.173677 | 0.335017 | 0 | 0 | 2.716477 | 4.755345 |
| Rare_Disea | Rectal neo | 3/329 | 0.172598 | 0.335017 | 0 | 0 | 2.145815 | 3.769752 |
| Rare_Disea | Polycythem | 2/173 | 0.173677 | 0.335017 | 0 | 0 | 2.716477 | 4.755345 |
| Rare_Disea | Neonatal n | Jan-44 | 0.174713 | 0.335017 | 0 | 0 | 5.373175 | 9.3741 |
| Rare_Disea | Keratosis s | Jan-44 | 0.174713 | 0.335017 | 0 | 0 | 5.373175 | 9.3741 |
| Jensen_DIS | Retinitis pi | Jan-83 | 0.304123 | 0.335051 | 0 | 0 | 2.81211 | 3.34732 |
| GTEx_Tissu | GTEX-OXRI | 9/832 | 0.010125 | 0.33531 | 0 | 0 | 2.676418 | 12.29216 |
| GTEx_Tissu | GTEX-OHPI | 8/687 | 0.010044 | 0.33531 | 0 | 0 | 2.868552 | 13.19765 |
| GTEx_Tissu | GTEX-XXEK | 7/549 | 0.00998 | 0.33531 | 0 | 0 | 3.127237 | 14.40772 |
| GTEx_Tissu | GTEX-XBEC | 9/827 | 0.009757 | 0.33531 | 0 | 0 | 2.693483 | 12.47009 |
| GTEx_Tissu | GTEX-S4Q7 | 14/1622 | 0.010006 | 0.33531 | 0 | 0 | 2.183177 | 10.05257 |
| GTEx_Tissu | GTEX-RWS | 7/542 | 0.009342 | 0.33531 | 0 | 0 | 3.169299 | 14.81101 |
| GTEx_Tissu | GTEX-P4PC | 8/686 | 0.009961 | 0.33531 | 0 | 0 | 2.872932 | 13.24143 |
| GTEx_Tissu | GTEX-RU1J | 9/833 | 0.0102 | 0.33531 | 0 | 0 | 2.67303 | 12.25695 |
| GTEx_Tissu | GTEX-XPVG | 7/539 | 0.009077 | 0.33531 | 0 | 0 | 3.187664 | 14.98828 |
| GTEx_Tissu | GTEX-T6M | 13/1458 | 0.010127 | 0.33531 | 0 | 0 | 2.245245 | 10.31144 |
| GTEx_Tissu | GTEX-PWC | 8/677 | 0.009244 | 0.33531 | 0 | 0 | 2.912944 | 13.6437 |
| GO_Biologi | Negative R | Jan-64 | 0.243799 | 0.336111 | 0 | 0 | 3.663714 | 5.171007 |
| GO_Biologi | Endocytic F | Jan-64 | 0.243799 | 0.336111 | 0 | 0 | 3.663714 | 5.171007 |
| GO_Biologi | Regulation | Jan-64 | 0.243799 | 0.336111 | 0 | 0 | 3.663714 | 5.171007 |
| GO_Biologi | Positive Re | Jan-64 | 0.243799 | 0.336111 | 0 | 0 | 3.663714 | 5.171007 |
| GO_Biologi | Response T | Jan-64 | 0.243799 | 0.336111 | 0 | 0 | 3.663714 | 5.171007 |
| PhenGenI_ | Neurodege | 28-Jan | 0.114983 | 0.336215 | 0 | 0 | 8.564169 | 18.52408 |
| PhenGenI_ | Forced Exp | 6/761 | 0.113921 | 0.336215 | 0 | 0 | 1.879617 | 4.082993 |
| Jensen_DIS | Pancreatiti | Jan-84 | 0.307163 | 0.336498 | 0 | 0 | 2.778089 | 3.279195 |
| Disease_Pe | schizophre | 5/311 | 0.011534 | 0.336575 | 0 | 0 | 3.907022 | 17.43501 |
| Disease_Pe | COPD - Chr | 5/311 | 0.011534 | 0.336575 | 0 | 0 | 3.907022 | 17.43501 |
| DisGeNET | Macular de | Jan-62 | 0.237156 | 0.337323 | 0 | 0 | 3.784217 | 5.445629 |
| DisGeNET | Xerostomia | Jan-62 | 0.237156 | 0.337323 | 0 | 0 | 3.784217 | 5.445629 |
| DisGeNET | Speech Del | 2/213 | 0.237028 | 0.337323 | 0 | 0 | 2.197045 | 3.162813 |
| DisGeNET | Carcinoid T | 2/213 | 0.237028 | 0.337323 | 0 | 0 | 2.197045 | 3.162813 |
| DisGeNET | Hereditary | Jan-62 | 0.237156 | 0.337323 | 0 | 0 | 3.784217 | 5.445629 |
| DisGeNET | Abnormalit | Jan-62 | 0.237156 | 0.337323 | 0 | 0 | 3.784217 | 5.445629 |
| DisGeNET | Cancer of N | Jan-62 | 0.237156 | 0.337323 | 0 | 0 | 3.784217 | 5.445629 |
| DisGeNET | Diabetes M | 6/956 | 0.236389 | 0.337323 | 0 | 0 | 1.478596 | 2.132546 |
| DisGeNET | Nijmegen E | Jan-62 | 0.237156 | 0.337323 | 0 | 0 | 3.784217 | 5.445629 |

| | | | | | | | | |
|---|---|---|---|---|---|---|---|---|
| GTEx_Tissu | GTEX-WL4( | 8/691 | 0.010378 | 0.337338 | 0 | 0 | 2.851159 | 13.02433 |
| GeDiPNet_ | Ovarian Ne | 2/225 | 0.256348 | 0.337362 | 0 | 0 | 2.077552 | 2.828 |
| GTEx_Tissu | GTEX-WFO | 9/837 | 0.010503 | 0.337601 | 0 | 0 | 2.65956 | 12.1173 |
| dbGaP | Insulin Resi | Jan-58 | 0.223696 | 0.33773 | 0 | 0 | 4.050592 | 6.065622 |
| dbGaP | Hematocrit | Jan-61 | 0.233813 | 0.33773 | 0 | 0 | 3.847481 | 5.591291 |
| dbGaP | Calcium | Jan-61 | 0.233813 | 0.33773 | 0 | 0 | 3.847481 | 5.591291 |
| dbGaP | monocyte ( | Jan-58 | 0.223696 | 0.33773 | 0 | 0 | 4.050592 | 6.065622 |
| Human_Ph | Delayed er | Jan-86 | 0.313203 | 0.337976 | 0 | 0 | 2.712449 | 3.148895 |
| DisGeNET | Thin face | Jan-63 | 0.240485 | 0.338565 | 0 | 0 | 3.722993 | 5.305634 |
| DisGeNET | Aortic Valv | 2/214 | 0.238636 | 0.338565 | 0 | 0 | 2.18657 | 3.132957 |
| DisGeNET | Familial ap | Jan-63 | 0.240485 | 0.338565 | 0 | 0 | 3.722993 | 5.305634 |
| DisGeNET | Idiopathic l | Jan-63 | 0.240485 | 0.338565 | 0 | 0 | 3.722993 | 5.305634 |
| DisGeNET | Kidney Calc | Jan-63 | 0.240485 | 0.338565 | 0 | 0 | 3.722993 | 5.305634 |
| DisGeNET | Monocytic | Jan-63 | 0.240485 | 0.338565 | 0 | 0 | 3.722993 | 5.305634 |
| DisGeNET | Malignant | 13/2398 | 0.239921 | 0.338565 | 0 | 0 | 1.291087 | 1.842958 |
| DisGeNET | Pallor of op | Jan-63 | 0.240485 | 0.338565 | 0 | 0 | 3.722993 | 5.305634 |
| DisGeNET | Bulbo-Spin | Jan-63 | 0.240485 | 0.338565 | 0 | 0 | 3.722993 | 5.305634 |
| DisGeNET | Neuro-dege | Jan-63 | 0.240485 | 0.338565 | 0 | 0 | 3.722993 | 5.305634 |
| DisGeNET | Blast Phase | 2/214 | 0.238636 | 0.338565 | 0 | 0 | 2.18657 | 3.132957 |
| DisGeNET | Rheumatic | Jan-63 | 0.240485 | 0.338565 | 0 | 0 | 3.722993 | 5.305634 |
| DisGeNET | Epilepsy, C | Jan-63 | 0.240485 | 0.338565 | 0 | 0 | 3.722993 | 5.305634 |
| DisGeNET | Narrow fac | Jan-63 | 0.240485 | 0.338565 | 0 | 0 | 3.722993 | 5.305634 |
| DisGeNET | Simple ear | Jan-63 | 0.240485 | 0.338565 | 0 | 0 | 3.722993 | 5.305634 |
| DisGeNET | Restenosis | 2/215 | 0.240243 | 0.338565 | 0 | 0 | 2.176194 | 3.103476 |
| DisGeNET | Viral hepat | Jan-63 | 0.240485 | 0.338565 | 0 | 0 | 3.722993 | 5.305634 |
| DisGeNET | Chronic kid | 3/388 | 0.238547 | 0.338565 | 0 | 0 | 1.811503 | 2.596225 |
| DisGeNET | Papule | Jan-63 | 0.240485 | 0.338565 | 0 | 0 | 3.722993 | 5.305634 |
| CellMarker | Fibroblast l | Jan-75 | 0.279326 | 0.338652 | 0 | 0 | 3.117379 | 3.975827 |
| CellMarker | Lymphoid-p | Jan-75 | 0.279326 | 0.338652 | 0 | 0 | 3.117379 | 3.975827 |
| DisGeNET | Mental Dep | 4/575 | 0.240857 | 0.338907 | 0 | 0 | 1.632477 | 2.323919 |
| RNA-Seq_D | Baf60c Car | 8/498 | 0.00146 | 0.33952 | 0 | 0 | 4.014053 | 26.20863 |
| RNA-Seq_D | Epac1-PLN | 8/499 | 0.001479 | 0.33952 | 0 | 0 | 4.005672 | 26.1032 |
| RNA-Seq_D | Obesity On | 8/498 | 0.00146 | 0.33952 | 0 | 0 | 4.014053 | 26.20863 |
| DisGeNET | Mastocytos | Jan-64 | 0.243799 | 0.339582 | 0 | 0 | 3.663714 | 5.171007 |
| DisGeNET | Congenital | Jan-64 | 0.243799 | 0.339582 | 0 | 0 | 3.663714 | 5.171007 |
| DisGeNET | Osteopetro | Jan-64 | 0.243799 | 0.339582 | 0 | 0 | 3.663714 | 5.171007 |
| DisGeNET | Cytogeneti | Jan-64 | 0.243799 | 0.339582 | 0 | 0 | 3.663714 | 5.171007 |
| DisGeNET | Short meta | Jan-64 | 0.243799 | 0.339582 | 0 | 0 | 3.663714 | 5.171007 |
| DisGeNET | Cerebrovas | 5/768 | 0.242171 | 0.339582 | 0 | 0 | 1.530384 | 2.170258 |
| DisGeNET | Anus, Impe | Jan-64 | 0.243799 | 0.339582 | 0 | 0 | 3.663714 | 5.171007 |
| DisGeNET | Uterine Ca | Jan-64 | 0.243799 | 0.339582 | 0 | 0 | 3.663714 | 5.171007 |
| DisGeNET | Multiple Er | Jan-64 | 0.243799 | 0.339582 | 0 | 0 | 3.663714 | 5.171007 |
| DisGeNET | Oligohydra | Jan-64 | 0.243799 | 0.339582 | 0 | 0 | 3.663714 | 5.171007 |
| DisGeNET | Adenomato | 3/392 | 0.243181 | 0.339582 | 0 | 0 | 1.792508 | 2.534513 |
| DisGeNET | Adenocarci | Jan-64 | 0.243799 | 0.339582 | 0 | 0 | 3.663714 | 5.171007 |

| | | | | | | | | |
|---|---|---|---|---|---|---|---|---|
| DisGeNET | Polyp of lar | Jan-64 | 0.243799 | 0.339582 | 0 | 0 | 3.663714 | 5.171007 |
| DisGeNET | Diaphragm | Jan-64 | 0.243799 | 0.339582 | 0 | 0 | 3.663714 | 5.171007 |
| DisGeNET | Cortical Dy | Jan-64 | 0.243799 | 0.339582 | 0 | 0 | 3.663714 | 5.171007 |
| DisGeNET | Carcinoma | 3/392 | 0.243181 | 0.339582 | 0 | 0 | 1.792508 | 2.534513 |
| DisGeNET | Febrile Cor | Jan-64 | 0.243799 | 0.339582 | 0 | 0 | 3.663714 | 5.171007 |
| DisGeNET | Extrapyram | Jan-64 | 0.243799 | 0.339582 | 0 | 0 | 3.663714 | 5.171007 |
| DisGeNET | Carcinoma | 2/216 | 0.241852 | 0.339582 | 0 | 0 | 2.165915 | 3.074364 |
| GO_Biologi | Regulation | Jan-65 | 0.247099 | 0.340001 | 0 | 0 | 3.606286 | 5.041467 |
| Rare_Disea | Hepatic en | Jan-47 | 0.18546 | 0.340061 | 0 | 0 | 5.021992 | 8.46163 |
| Rare_Disea | Corticobas | Jan-47 | 0.18546 | 0.340061 | 0 | 0 | 5.021992 | 8.46163 |
| Rare_Disea | Microscopi | Jan-47 | 0.18546 | 0.340061 | 0 | 0 | 5.021992 | 8.46163 |
| Rare_Disea | Renal tubu | Jan-47 | 0.18546 | 0.340061 | 0 | 0 | 5.021992 | 8.46163 |
| Rare_Disea | Endomyoca | Jan-47 | 0.18546 | 0.340061 | 0 | 0 | 5.021992 | 8.46163 |
| Rare_Disea | Oligodendr | 2/180 | 0.184577 | 0.340061 | 0 | 0 | 2.608724 | 4.407929 |
| Rare_Disea | Malignant | Jan-47 | 0.18546 | 0.340061 | 0 | 0 | 5.021992 | 8.46163 |
| Rare_Disea | Anaplastic | 2/180 | 0.184577 | 0.340061 | 0 | 0 | 2.608724 | 4.407929 |
| Rare_Disea | Cytomegali | 3/341 | 0.185571 | 0.340061 | 0 | 0 | 2.068364 | 3.483785 |
| Rare_Disea | Chagas dis | 2/179 | 0.183014 | 0.340061 | 0 | 0 | 2.623596 | 4.455375 |
| Rare_Disea | Coal worke | Jan-46 | 0.181893 | 0.340061 | 0 | 0 | 5.13385 | 8.749803 |
| Rare_Disea | Neonatal h | Jan-46 | 0.181893 | 0.340061 | 0 | 0 | 5.13385 | 8.749803 |
| Rare_Disea | Oropharyn | 2/180 | 0.184577 | 0.340061 | 0 | 0 | 2.608724 | 4.407929 |
| Rare_Disea | Holoprosen | Jan-47 | 0.18546 | 0.340061 | 0 | 0 | 5.021992 | 8.46163 |
| Rare_Disea | Hyperlipop | Jan-47 | 0.18546 | 0.340061 | 0 | 0 | 5.021992 | 8.46163 |
| Rare_Disea | Thymic hyp | Jan-45 | 0.178311 | 0.340061 | 0 | 0 | 5.250793 | 9.053559 |
| Rare_Disea | Myasthenia | 2/177 | 0.179893 | 0.340061 | 0 | 0 | 2.653849 | 4.552394 |
| Rare_Disea | Mental reta | 2/177 | 0.179893 | 0.340061 | 0 | 0 | 2.653849 | 4.552394 |
| DisGeNET | Lung diseas | 3/393 | 0.244342 | 0.340158 | 0 | 0 | 1.787821 | 2.51937 |
| CellMarker | Brush Cell ( | Jan-76 | 0.282473 | 0.340959 | 0 | 0 | 3.075659 | 3.888164 |
| DisGeNET | Invasive Du | 2/218 | 0.245071 | 0.34099 | 0 | 0 | 2.145643 | 3.017223 |
| Disease_Pe | Adenocarci | 5/319 | 0.01276 | 0.341319 | 0 | 0 | 3.805927 | 16.59944 |
| Disease_Pe | juvenile de | 3/111 | 0.012584 | 0.341319 | 0 | 0 | 6.549272 | 28.65547 |
| GeDiPNet_ | Nonorganic | Jan-69 | 0.260157 | 0.341603 | 0 | 0 | 3.393468 | 4.569198 |
| GO_Molecu | Signaling R | Jan-44 | 0.174713 | 0.342363 | 0 | 0 | 5.373175 | 9.3741 |
| GO_Molecu | Proton Tran | Jan-45 | 0.178311 | 0.342363 | 0 | 0 | 5.250793 | 9.053559 |
| GO_Molecu | Cytokine A | 2/178 | 0.181452 | 0.342363 | 0 | 0 | 2.638636 | 4.503525 |
| Jensen_TIS | Breast canc | 2/221 | 0.249902 | 0.342419 | 0 | 0 | 2.115928 | 2.934131 |
| DisGeNET | Lymphocyt | Jan-65 | 0.247099 | 0.342539 | 0 | 0 | 3.606286 | 5.041467 |
| DisGeNET | Systemic In | Jan-65 | 0.247099 | 0.342539 | 0 | 0 | 3.606286 | 5.041467 |
| DisGeNET | Adult Liver | Jan-65 | 0.247099 | 0.342539 | 0 | 0 | 3.606286 | 5.041467 |
| DisGeNET | Stress, Psyc | Jan-65 | 0.247099 | 0.342539 | 0 | 0 | 3.606286 | 5.041467 |
| DisGeNET | Glomerular | Jan-65 | 0.247099 | 0.342539 | 0 | 0 | 3.606286 | 5.041467 |
| DisGeNET | Small hand | Jan-65 | 0.247099 | 0.342539 | 0 | 0 | 3.606286 | 5.041467 |
| DisGeNET | HEMOCHRO | Jan-65 | 0.247099 | 0.342539 | 0 | 0 | 3.606286 | 5.041467 |
| GTEx_Tissu | GTEX-T2IS- | 3/105 | 0.010834 | 0.342584 | 0 | 0 | 6.936625 | 31.38872 |
| GTEx_Tissu | GTEX-XGQ4 | 8/697 | 0.010894 | 0.342584 | 0 | 0 | 2.825449 | 12.76961 |

| | | | | | | | | |
|---|---|---|---|---|---|---|---|---|
| Rare_Disea | Asbestosis | Jan-48 | 0.189012 | 0.342987 | 0 | 0 | 4.914894 | 8.187947 |
| Rare_Disea | Hypolipopr | Jan-48 | 0.189012 | 0.342987 | 0 | 0 | 4.914894 | 8.187947 |
| Rare_Disea | Leukoplaki | 2/182 | 0.18771 | 0.342987 | 0 | 0 | 2.579477 | 4.315096 |
| Rare_Disea | Diamond-B | Jan-48 | 0.189012 | 0.342987 | 0 | 0 | 4.914894 | 8.187947 |
| GO_Biologi | Negative R | Jan-66 | 0.250385 | 0.343191 | 0 | 0 | 3.550626 | 4.916752 |
| GO_Biologi | Response T | Jan-66 | 0.250385 | 0.343191 | 0 | 0 | 3.550626 | 4.916752 |
| CellMarker | Stem Cell E | Jan-77 | 0.285606 | 0.343228 | 0 | 0 | 3.035037 | 3.803331 |
| UK_Bioban | Facial ageir | Jan-79 | 0.291832 | 0.343686 | 0 | 0 | 2.956917 | 3.641673 |
| GTEx_Tissu | GTEX-TKQ2 | 9/844 | 0.011049 | 0.343718 | 0 | 0 | 2.636297 | 11.87753 |
| DisGeNET | Delayed sp | 2/220 | 0.248291 | 0.34401 | 0 | 0 | 2.125742 | 2.961486 |
| DisGeNET | Glycogen s | Jan-66 | 0.250385 | 0.344904 | 0 | 0 | 3.550626 | 4.916752 |
| DisGeNET | Paroxysma | Jan-66 | 0.250385 | 0.344904 | 0 | 0 | 3.550626 | 4.916752 |
| DisGeNET | GRN-relate | Jan-66 | 0.250385 | 0.344904 | 0 | 0 | 3.550626 | 4.916752 |
| DisGeNET | Bronchitis, | Jan-66 | 0.250385 | 0.344904 | 0 | 0 | 3.550626 | 4.916752 |
| DisGeNET | Giant Cell T | Jan-66 | 0.250385 | 0.344904 | 0 | 0 | 3.550626 | 4.916752 |
| DisGeNET | Hip Disloca | Jan-66 | 0.250385 | 0.344904 | 0 | 0 | 3.550626 | 4.916752 |
| DisGeNET | Round, full | Jan-66 | 0.250385 | 0.344904 | 0 | 0 | 3.550626 | 4.916752 |
| DisGeNET | Arachnoda | Jan-66 | 0.250385 | 0.344904 | 0 | 0 | 3.550626 | 4.916752 |
| DisGeNET | stage, non- | Jan-66 | 0.250385 | 0.344904 | 0 | 0 | 3.550626 | 4.916752 |
| DisGeNET | Hypertroph | Jan-66 | 0.250385 | 0.344904 | 0 | 0 | 3.550626 | 4.916752 |
| DisGeNET | Round face | Jan-66 | 0.250385 | 0.344904 | 0 | 0 | 3.550626 | 4.916752 |
| CellMarker | Neuroendo | Jan-78 | 0.288726 | 0.345462 | 0 | 0 | 2.99547 | 3.721207 |
| Rare_Disea | Minimal ch | 2/184 | 0.19085 | 0.345481 | 0 | 0 | 2.550873 | 4.224921 |
| PhenGenI_ | Glaucoma, | 30-Jan | 0.122676 | 0.345588 | 0 | 0 | 7.972735 | 16.72843 |
| PhenGenI_ | Coffee | 30-Jan | 0.122676 | 0.345588 | 0 | 0 | 7.972735 | 16.72843 |
| PhenGenI_ | Birth Weigl | 30-Jan | 0.122676 | 0.345588 | 0 | 0 | 7.972735 | 16.72843 |
| DisGeNET | Tongue Car | Jan-67 | 0.253656 | 0.345955 | 0 | 0 | 3.496653 | 4.79662 |
| DisGeNET | Abnormal l | Jan-67 | 0.253656 | 0.345955 | 0 | 0 | 3.496653 | 4.79662 |
| DisGeNET | Aplasia/Hy | Jan-67 | 0.253656 | 0.345955 | 0 | 0 | 3.496653 | 4.79662 |
| DisGeNET | Gestationa | Jan-67 | 0.253656 | 0.345955 | 0 | 0 | 3.496653 | 4.79662 |
| DisGeNET | Mastocyto: | Jan-67 | 0.253656 | 0.345955 | 0 | 0 | 3.496653 | 4.79662 |
| DisGeNET | Skin blister | Jan-67 | 0.253656 | 0.345955 | 0 | 0 | 3.496653 | 4.79662 |
| DisGeNET | Parasitemi | Jan-67 | 0.253656 | 0.345955 | 0 | 0 | 3.496653 | 4.79662 |
| DisGeNET | Cushing Sy | Jan-67 | 0.253656 | 0.345955 | 0 | 0 | 3.496653 | 4.79662 |
| DisGeNET | neurofibro | Jan-67 | 0.253656 | 0.345955 | 0 | 0 | 3.496653 | 4.79662 |
| DisGeNET | Oxyphilic A | Jan-67 | 0.253656 | 0.345955 | 0 | 0 | 3.496653 | 4.79662 |
| DisGeNET | Congenital | Jan-67 | 0.253656 | 0.345955 | 0 | 0 | 3.496653 | 4.79662 |
| DisGeNET | Schistosom | Jan-67 | 0.253656 | 0.345955 | 0 | 0 | 3.496653 | 4.79662 |
| DisGeNET | Arteriopatl | Jan-67 | 0.253656 | 0.345955 | 0 | 0 | 3.496653 | 4.79662 |
| DisGeNET | Metastatic | Jan-67 | 0.253656 | 0.345955 | 0 | 0 | 3.496653 | 4.79662 |
| DisGeNET | Nail dyspla | Jan-67 | 0.253656 | 0.345955 | 0 | 0 | 3.496653 | 4.79662 |
| DisGeNET | Pustulosis ( | Jan-67 | 0.253656 | 0.345955 | 0 | 0 | 3.496653 | 4.79662 |
| DisGeNET | Cervical Ad | Jan-67 | 0.253656 | 0.345955 | 0 | 0 | 3.496653 | 4.79662 |
| DisGeNET | Short dista | Jan-67 | 0.253656 | 0.345955 | 0 | 0 | 3.496653 | 4.79662 |
| DisGeNET | Cerebral H | Jan-67 | 0.253656 | 0.345955 | 0 | 0 | 3.496653 | 4.79662 |

| | | | | | | | | |
|---|---|---|---|---|---|---|---|---|
| CellMarker | FOXN4+ Ce | Jan-79 | 0.291832 | 0.346156 | 0 | 0 | 2.956917 | 3.641673 |
| CellMarker | T Cell Unde | Jan-79 | 0.291832 | 0.346156 | 0 | 0 | 2.956917 | 3.641673 |
| Jensen_TIS | Breast canc | 2/227 | 0.259573 | 0.34616 | 0 | 0 | 2.058876 | 2.776839 |
| Jensen_TIS | Atheroscle | Jan-69 | 0.260157 | 0.34616 | 0 | 0 | 3.393468 | 4.569198 |
| Jensen_TIS | Colorectal | Jan-68 | 0.256914 | 0.34616 | 0 | 0 | 3.44429 | 4.68084 |
| Jensen_TIS | Palate | Jan-69 | 0.260157 | 0.34616 | 0 | 0 | 3.393468 | 4.569198 |
| Jensen_TIS | Lung | 75/16585 | 0.256273 | 0.34616 | 0 | 0 | 1.288234 | 1.753946 |
| Jensen_TIS | Teratocarc | 4/592 | 0.256982 | 0.34616 | 0 | 0 | 1.583887 | 2.152104 |
| Jensen_TIS | Uroepithel | Jan-68 | 0.256914 | 0.34616 | 0 | 0 | 3.44429 | 4.68084 |
| GTEx_Tissu | GTEX-X4XY | 12/1314 | 0.011273 | 0.346937 | 0 | 0 | 2.287066 | 10.25833 |
| GO_Biologi | Negative R | Jan-67 | 0.253656 | 0.347006 | 0 | 0 | 3.496653 | 4.79662 |
| Jensen_DIS | Celiac disea | Jan-88 | 0.319191 | 0.347084 | 0 | 0 | 2.649826 | 3.026014 |
| Jensen_DIS | Down synd | Jan-89 | 0.322165 | 0.347084 | 0 | 0 | 2.619582 | 2.967177 |
| Jensen_DIS | Neuropath | Jan-89 | 0.322165 | 0.347084 | 0 | 0 | 2.619582 | 2.967177 |
| DisGeNET | Blood baso | 2/224 | 0.254736 | 0.347247 | 0 | 0 | 2.087016 | 2.85405 |
| GWAS_Cat | Mean Corp | 5/864 | 0.322877 | 0.347363 | 0 | 0 | 1.352537 | 1.529022 |
| GTEx_Tissu | GTEX-T5JC- | 18/1603 | 0.000156 | 0.347535 | 0 | 0 | 3.016541 | 26.43818 |
| Rare_Disea | Apo A-I def | Jan-49 | 0.192548 | 0.347708 | 0 | 0 | 4.812258 | 7.927759 |
| DisGeNET | Skin bullae | Jan-68 | 0.256914 | 0.347775 | 0 | 0 | 3.44429 | 4.68084 |
| DisGeNET | Angle class | Jan-68 | 0.256914 | 0.347775 | 0 | 0 | 3.44429 | 4.68084 |
| DisGeNET | Pulmonary | 2/225 | 0.256348 | 0.347775 | 0 | 0 | 2.077552 | 2.828 |
| DisGeNET | Alveolar rh | Jan-68 | 0.256914 | 0.347775 | 0 | 0 | 3.44429 | 4.68084 |
| DisGeNET | Angle class | Jan-68 | 0.256914 | 0.347775 | 0 | 0 | 3.44429 | 4.68084 |
| DisGeNET | Leishmania | Jan-68 | 0.256914 | 0.347775 | 0 | 0 | 3.44429 | 4.68084 |
| DisGeNET | Hodgkin Di | 4/592 | 0.256982 | 0.347775 | 0 | 0 | 1.583887 | 2.152104 |
| DisGeNET | Migraine w | Jan-68 | 0.256914 | 0.347775 | 0 | 0 | 3.44429 | 4.68084 |
| DisGeNET | Low poster | Jan-68 | 0.256914 | 0.347775 | 0 | 0 | 3.44429 | 4.68084 |
| DisGeNET | Gastrointes | 2/225 | 0.256348 | 0.347775 | 0 | 0 | 2.077552 | 2.828 |
| DisGeNET | Malnutritic | 2/225 | 0.256348 | 0.347775 | 0 | 0 | 2.077552 | 2.828 |
| DisGeNET | Adenomato | Jan-68 | 0.256914 | 0.347775 | 0 | 0 | 3.44429 | 4.68084 |
| DisGeNET | Undifferen | Jan-68 | 0.256914 | 0.347775 | 0 | 0 | 3.44429 | 4.68084 |
| DisGeNET | Wiskott-Al | Jan-68 | 0.256914 | 0.347775 | 0 | 0 | 3.44429 | 4.68084 |
| DisGeNET | Strabismus | 3/404 | 0.257172 | 0.347852 | 0 | 0 | 1.737798 | 2.359949 |
| UK_Bioban | Right 3mm | 2/250 | 0.296648 | 0.347921 | 0 | 0 | 1.86575 | 2.267274 |
| DisGeNET | Tremor | 2/226 | 0.257961 | 0.34838 | 0 | 0 | 2.068172 | 2.802265 |
| DisGeNET | Graft-vs-Hc | 2/226 | 0.257961 | 0.34838 | 0 | 0 | 2.068172 | 2.802265 |
| DisGeNET | Cardiovasc | 5/787 | 0.25772 | 0.34838 | 0 | 0 | 1.491719 | 2.022596 |
| GeDiPNet_ | Severe Con | Jan-71 | 0.266602 | 0.348496 | 0 | 0 | 3.296179 | 4.35754 |
| GeDiPNet_ | Elbow Flexi | Jan-71 | 0.266602 | 0.348496 | 0 | 0 | 3.296179 | 4.35754 |
| GO_Biologi | Telomere M | Jan-68 | 0.256914 | 0.348774 | 0 | 0 | 3.44429 | 4.68084 |
| GO_Biologi | Aerobic Ele | Jan-68 | 0.256914 | 0.348774 | 0 | 0 | 3.44429 | 4.68084 |
| GO_Biologi | Intermedia | Jan-68 | 0.256914 | 0.348774 | 0 | 0 | 3.44429 | 4.68084 |
| GO_Biologi | Skin Develo | Jan-68 | 0.256914 | 0.348774 | 0 | 0 | 3.44429 | 4.68084 |
| GWAS_Cat | Systolic Blo | Jan-90 | 0.325127 | 0.349476 | 0 | 0 | 2.590018 | 2.909988 |
| Rare_Disea | Hansen's d | 2/186 | 0.193998 | 0.349478 | 0 | 0 | 2.52289 | 4.137304 |

| | | | | | | | | |
|---|---|---|---|---|---|---|---|---|
| GeDiPNet_ | Nail Dyspla | Jan-72 | 0.269804 | 0.349546 | 0 | 0 | 3.249591 | 4.257158 |
| GeDiPNet_ | Nail Diseas | Jan-72 | 0.269804 | 0.349546 | 0 | 0 | 3.249591 | 4.257158 |
| GeDiPNet_ | Carcinoma | Jan-72 | 0.269804 | 0.349546 | 0 | 0 | 3.249591 | 4.257158 |
| GeDiPNet_ | Talipes | Jan-72 | 0.269804 | 0.349546 | 0 | 0 | 3.249591 | 4.257158 |
| DisGeNET | Diabetic Ar | Jan-69 | 0.260157 | 0.349724 | 0 | 0 | 3.393468 | 4.569198 |
| DisGeNET | Thin lips | Jan-69 | 0.260157 | 0.349724 | 0 | 0 | 3.393468 | 4.569198 |
| DisGeNET | Histiocytos | Jan-69 | 0.260157 | 0.349724 | 0 | 0 | 3.393468 | 4.569198 |
| DisGeNET | Colorectal | 17/3298 | 0.260098 | 0.349724 | 0 | 0 | 1.231088 | 1.657905 |
| DisGeNET | hiv-infectic | Jan-69 | 0.260157 | 0.349724 | 0 | 0 | 3.393468 | 4.569198 |
| DisGeNET | Creatinine | Jan-69 | 0.260157 | 0.349724 | 0 | 0 | 3.393468 | 4.569198 |
| DisGeNET | Williams Sy | Jan-69 | 0.260157 | 0.349724 | 0 | 0 | 3.393468 | 4.569198 |
| DisGeNET | Progressive | Jan-69 | 0.260157 | 0.349724 | 0 | 0 | 3.393468 | 4.569198 |
| DisGeNET | Kallmann S | Jan-69 | 0.260157 | 0.349724 | 0 | 0 | 3.393468 | 4.569198 |
| GTEx_Tissu | GTEX-UJM( | 9/852 | 0.0117 | 0.35039 | 0 | 0 | 2.610183 | 11.61048 |
| GTEx_Tissu | GTEX-POM | 10/1002 | 0.011581 | 0.35039 | 0 | 0 | 2.477089 | 11.04388 |
| GTEx_Tissu | GTEX-XAJ8 | 10/1006 | 0.011885 | 0.35039 | 0 | 0 | 2.46662 | 10.93326 |
| GTEx_Tissu | GTEX-W5W | 14/1658 | 0.011991 | 0.35039 | 0 | 0 | 2.13117 | 9.427524 |
| GTEx_Tissu | GTEX-NPJ8 | 7/568 | 0.011874 | 0.35039 | 0 | 0 | 3.01836 | 13.38169 |
| Jensen_DIS | Type 2 diak | 2/269 | 0.327097 | 0.350461 | 0 | 0 | 1.731306 | 1.934732 |
| WikiPathw | Phosphoin | Feb-49 | 0.01927 | 0.350483 | 0 | 0 | 9.945432 | 39.27635 |
| WikiPathw | Fatty Acids | Feb-35 | 0.01014 | 0.350483 | 0 | 0 | 14.17469 | 65.07997 |
| WikiPathw | 11P11 2 Cc | Feb-56 | 0.024747 | 0.350483 | 0 | 0 | 8.653159 | 32.00857 |
| WikiPathw | Translation | Feb-50 | 0.020017 | 0.350483 | 0 | 0 | 9.737745 | 38.08585 |
| WikiPathw | Somatroph | 6-Jan | 0.025821 | 0.350483 | 0 | 0 | 46.29767 | 169.2909 |
| WikiPathw | Urotensin I | Feb-69 | 0.03636 | 0.350483 | 0 | 0 | 6.969622 | 23.09942 |
| WikiPathw | ApoE And r | 8-Jan | 0.03428 | 0.350483 | 0 | 0 | 33.06645 | 111.5391 |
| WikiPathw | Lipid Partic | 10-Jan | 0.042667 | 0.350483 | 0 | 0 | 25.71576 | 81.1157 |
| WikiPathw | Arrhythmo | Feb-74 | 0.041285 | 0.350483 | 0 | 0 | 6.483987 | 20.66613 |
| WikiPathw | Cholestero | Feb-72 | 0.039286 | 0.350483 | 0 | 0 | 6.669916 | 21.58979 |
| WikiPathw | Statin Inhik | Feb-29 | 0.007037 | 0.350483 | 0 | 0 | 17.32985 | 85.89758 |
| WikiPathw | Calcium Re | 3/151 | 0.028208 | 0.350483 | 0 | 0 | 4.769546 | 17.01838 |
| WikiPathw | Compleme | Feb-58 | 0.026415 | 0.350483 | 0 | 0 | 8.343277 | 30.31803 |
| Rare_Disea | Penile canc | Jan-50 | 0.196069 | 0.350667 | 0 | 0 | 4.713811 | 7.680157 |
| Rare_Disea | Signet ring | 2/187 | 0.195575 | 0.350667 | 0 | 0 | 2.509126 | 4.094425 |
| Rare_Disea | Oligoastroc | 2/190 | 0.200314 | 0.350667 | 0 | 0 | 2.468711 | 3.969367 |
| Rare_Disea | Lewy body | 2/190 | 0.200314 | 0.350667 | 0 | 0 | 2.468711 | 3.969367 |
| Rare_Disea | Ovarian ins | Jan-51 | 0.199575 | 0.350667 | 0 | 0 | 4.619302 | 7.444308 |
| Rare_Disea | Children's i | Jan-51 | 0.199575 | 0.350667 | 0 | 0 | 4.619302 | 7.444308 |
| Rare_Disea | Cerebral ca | Jan-51 | 0.199575 | 0.350667 | 0 | 0 | 4.619302 | 7.444308 |
| Rare_Disea | Membranc | Jan-51 | 0.199575 | 0.350667 | 0 | 0 | 4.619302 | 7.444308 |
| Rare_Disea | Hirschsprui | 2/190 | 0.200314 | 0.350667 | 0 | 0 | 2.468711 | 3.969367 |
| Rare_Disea | Long QT sy | Jan-51 | 0.199575 | 0.350667 | 0 | 0 | 4.619302 | 7.444308 |
| Rare_Disea | Patent duc | 2/190 | 0.200314 | 0.350667 | 0 | 0 | 2.468711 | 3.969367 |
| Rare_Disea | Spinal shoc | Jan-51 | 0.199575 | 0.350667 | 0 | 0 | 4.619302 | 7.444308 |
| Jensen_DIS | Polydactyly | Jan-92 | 0.331012 | 0.350801 | 0 | 0 | 2.532839 | 2.800307 |

| | | | | | | | | |
|---|---|---|---|---|---|---|---|---|
| Jensen_DIS | DOID:2627 | Jan-92 | 0.331012 | 0.350801 | 0 | 0 | 2.532839 | 2.800307 |
| DisGeNET | Ki-1+ Anap | 2/228 | 0.261186 | 0.350927 | 0 | 0 | 2.049662 | 2.751716 |
| DisGeNET | Precursor T | 3/408 | 0.261862 | 0.351474 | 0 | 0 | 1.720282 | 2.305074 |
| DisGeNET | Curvature ( | 3/408 | 0.261862 | 0.351474 | 0 | 0 | 1.720282 | 2.305074 |
| DisGeNET | Stromal Ne | Jan-70 | 0.263387 | 0.35154 | 0 | 0 | 3.344119 | 4.461494 |
| DisGeNET | Metabolic a | Jan-70 | 0.263387 | 0.35154 | 0 | 0 | 3.344119 | 4.461494 |
| DisGeNET | Deformity ( | Jan-70 | 0.263387 | 0.35154 | 0 | 0 | 3.344119 | 4.461494 |
| DisGeNET | Squamous | Jan-70 | 0.263387 | 0.35154 | 0 | 0 | 3.344119 | 4.461494 |
| DisGeNET | Distortion ( | Jan-70 | 0.263387 | 0.35154 | 0 | 0 | 3.344119 | 4.461494 |
| DisGeNET | Funny look | Jan-70 | 0.263387 | 0.35154 | 0 | 0 | 3.344119 | 4.461494 |
| DisGeNET | Anemia, Di | Jan-70 | 0.263387 | 0.35154 | 0 | 0 | 3.344119 | 4.461494 |
| DisGeNET | Seminoma | 2/229 | 0.262799 | 0.35154 | 0 | 0 | 2.040529 | 2.726893 |
| DisGeNET | Pulmonary | Jan-70 | 0.263387 | 0.35154 | 0 | 0 | 3.344119 | 4.461494 |
| DisGeNET | Angina, Un | Jan-70 | 0.263387 | 0.35154 | 0 | 0 | 3.344119 | 4.461494 |
| DisGeNET | Chromosor | Jan-70 | 0.263387 | 0.35154 | 0 | 0 | 3.344119 | 4.461494 |
| GO_Biologi | Positive Re | Jan-69 | 0.260157 | 0.351832 | 0 | 0 | 3.393468 | 4.569198 |
| GO_Biologi | Regulation | Jan-69 | 0.260157 | 0.351832 | 0 | 0 | 3.393468 | 4.569198 |
| DisGeNET | Retinitis Pi | 3/410 | 0.264211 | 0.35246 | 0 | 0 | 1.711653 | 2.278224 |
| CellMarker | Oligodendr | 3/439 | 0.298533 | 0.352578 | 0 | 0 | 1.595429 | 1.928672 |
| GO_Biologi | Sulfur Com | Jan-70 | 0.263387 | 0.352839 | 0 | 0 | 3.344119 | 4.461494 |
| GO_Biologi | Mitochond | Jan-70 | 0.263387 | 0.352839 | 0 | 0 | 3.344119 | 4.461494 |
| GO_Biologi | Negative R | Jan-70 | 0.263387 | 0.352839 | 0 | 0 | 3.344119 | 4.461494 |
| GO_Biologi | Positive Re | Jan-70 | 0.263387 | 0.352839 | 0 | 0 | 3.344119 | 4.461494 |
| GO_Biologi | Regulation | Jan-70 | 0.263387 | 0.352839 | 0 | 0 | 3.344119 | 4.461494 |
| GeDiPNet_ | Exotropia | Jan-73 | 0.272992 | 0.352892 | 0 | 0 | 3.204296 | 4.160181 |
| Rare_Disea | Pitt syndro | Jan-52 | 0.203066 | 0.352993 | 0 | 0 | 4.5285 | 7.219451 |
| Rare_Disea | Thyroid car | Jan-52 | 0.203066 | 0.352993 | 0 | 0 | 4.5285 | 7.219451 |
| Rare_Disea | Gardner sy | Jan-52 | 0.203066 | 0.352993 | 0 | 0 | 4.5285 | 7.219451 |
| GTEx_Tissu | Breast - Ma | 1/100 | 0.35405 | 0.35405 | 0 | 0 | 2.327226 | 2.416399 |
| GTEx_Tissu | Breast - Ma | Jan-59 | 0.227083 | 0.35405 | 0 | 0 | 3.980553 | 5.900928 |
| GTEx_Tissu | Brain - Cort | 1/100 | 0.35405 | 0.35405 | 0 | 0 | 2.327226 | 2.416399 |
| GTEx_Tissu | Skin - Not S | 1/100 | 0.35405 | 0.35405 | 0 | 0 | 2.327226 | 2.416399 |
| GTEx_Tissu | Brain - Hipp | 1/100 | 0.35405 | 0.35405 | 0 | 0 | 2.327226 | 2.416399 |
| GTEx_Tissu | Brain - Cort | 1/100 | 0.35405 | 0.35405 | 0 | 0 | 2.327226 | 2.416399 |
| GTEx_Tissu | Brain - Fror | 1/100 | 0.35405 | 0.35405 | 0 | 0 | 2.327226 | 2.416399 |
| GTEx_Tissu | Esophagus | 1/100 | 0.35405 | 0.35405 | 0 | 0 | 2.327226 | 2.416399 |
| GTEx_Tissu | Small Intes | 1/100 | 0.35405 | 0.35405 | 0 | 0 | 2.327226 | 2.416399 |
| GTEx_Tissu | Bladder Ma | 1/100 | 0.35405 | 0.35405 | 0 | 0 | 2.327226 | 2.416399 |
| GTEx_Tissu | Liver Male | 1/100 | 0.35405 | 0.35405 | 0 | 0 | 2.327226 | 2.416399 |
| GTEx_Tissu | Muscle - Sk | 1/100 | 0.35405 | 0.35405 | 0 | 0 | 2.327226 | 2.416399 |
| GTEx_Tissu | Small Intes | 1/100 | 0.35405 | 0.35405 | 0 | 0 | 2.327226 | 2.416399 |
| GTEx_Tissu | Esophagus | 1/100 | 0.35405 | 0.35405 | 0 | 0 | 2.327226 | 2.416399 |
| GTEx_Tissu | Brain - Spir | 1/100 | 0.35405 | 0.35405 | 0 | 0 | 2.327226 | 2.416399 |
| GTEx_Tissu | Skin - Not S | 1/100 | 0.35405 | 0.35405 | 0 | 0 | 2.327226 | 2.416399 |
| GTEx_Tissu | Esophagus | Jan-60 | 0.230455 | 0.35405 | 0 | 0 | 3.912889 | 5.742942 |

| | | | | | | | | |
|---|---|---|---|---|---|---|---|---|
| GTEx_Tissu | Brain - Ant | 1/100 | 0.35405 | 0.35405 | 0 | 0 | 2.327226 | 2.416399 |
| GTEx_Tissu | Brain - Hip | 1/100 | 0.35405 | 0.35405 | 0 | 0 | 2.327226 | 2.416399 |
| GTEx_Tissu | Brain - Ant | 1/100 | 0.35405 | 0.35405 | 0 | 0 | 2.327226 | 2.416399 |
| GTEx_Tissu | Adipose - S | 1/100 | 0.35405 | 0.35405 | 0 | 0 | 2.327226 | 2.416399 |
| GTEx_Tissu | Adrenal Gl | 1/100 | 0.35405 | 0.35405 | 0 | 0 | 2.327226 | 2.416399 |
| GTEx_Tissu | Colon - Sig | 1/100 | 0.35405 | 0.35405 | 0 | 0 | 2.327226 | 2.416399 |
| GTEx_Tissu | Small Intes | 1/100 | 0.35405 | 0.35405 | 0 | 0 | 2.327226 | 2.416399 |
| GTEx_Tissu | Adrenal Gl | 1/100 | 0.35405 | 0.35405 | 0 | 0 | 2.327226 | 2.416399 |
| GTEx_Tissu | Heart - Atri | 1/100 | 0.35405 | 0.35405 | 0 | 0 | 2.327226 | 2.416399 |
| GTEx_Tissu | Brain - Cau | 1/100 | 0.35405 | 0.35405 | 0 | 0 | 2.327226 | 2.416399 |
| GTEx_Tissu | Adipose - S | 1/100 | 0.35405 | 0.35405 | 0 | 0 | 2.327226 | 2.416399 |
| GTEx_Tissu | Artery - Tib | 1/100 | 0.35405 | 0.35405 | 0 | 0 | 2.327226 | 2.416399 |
| GTEx_Tissu | Colon - Sig | 1/100 | 0.35405 | 0.35405 | 0 | 0 | 2.327226 | 2.416399 |
| GTEx_Tissu | Colon - Sig | 1/100 | 0.35405 | 0.35405 | 0 | 0 | 2.327226 | 2.416399 |
| GTEx_Tissu | Skin - Not S | 1/100 | 0.35405 | 0.35405 | 0 | 0 | 2.327226 | 2.416399 |
| GTEx_Tissu | Esophagus | 1/100 | 0.35405 | 0.35405 | 0 | 0 | 2.327226 | 2.416399 |
| GTEx_Tissu | Small Intes | 1/100 | 0.35405 | 0.35405 | 0 | 0 | 2.327226 | 2.416399 |
| GTEx_Tissu | Adipose - V | 1/100 | 0.35405 | 0.35405 | 0 | 0 | 2.327226 | 2.416399 |
| GTEx_Tissu | Brain - Sub | 1/100 | 0.35405 | 0.35405 | 0 | 0 | 2.327226 | 2.416399 |
| GTEx_Tissu | Ovary Fem | 1/100 | 0.35405 | 0.35405 | 0 | 0 | 2.327226 | 2.416399 |
| GTEx_Tissu | Skin - Not S | 1/100 | 0.35405 | 0.35405 | 0 | 0 | 2.327226 | 2.416399 |
| GTEx_Tissu | Brain - Hip | 1/100 | 0.35405 | 0.35405 | 0 | 0 | 2.327226 | 2.416399 |
| GTEx_Tissu | Colon - Sig | 1/100 | 0.35405 | 0.35405 | 0 | 0 | 2.327226 | 2.416399 |
| GTEx_Tissu | Small Intes | 1/100 | 0.35405 | 0.35405 | 0 | 0 | 2.327226 | 2.416399 |
| GTEx_Tissu | Adrenal Gl | 1/100 | 0.35405 | 0.35405 | 0 | 0 | 2.327226 | 2.416399 |
| GTEx_Tissu | Brain - Sub | 1/100 | 0.35405 | 0.35405 | 0 | 0 | 2.327226 | 2.416399 |
| GTEx_Tissu | Adipose - V | 1/100 | 0.35405 | 0.35405 | 0 | 0 | 2.327226 | 2.416399 |
| GTEx_Tissu | Adrenal Gl | 1/100 | 0.35405 | 0.35405 | 0 | 0 | 2.327226 | 2.416399 |
| GTEx_Tissu | Brain - Hip | 1/100 | 0.35405 | 0.35405 | 0 | 0 | 2.327226 | 2.416399 |
| GTEx_Tissu | Adipose - V | Jan-74 | 0.276166 | 0.35405 | 0 | 0 | 3.160242 | 4.066453 |
| GTEx_Tissu | Esophagus | Jan-57 | 0.220295 | 0.35405 | 0 | 0 | 4.123131 | 6.237423 |
| GTEx_Tissu | Breast - Ma | Jan-44 | 0.174713 | 0.35405 | 0 | 0 | 5.373175 | 9.3741 |
| GTEx_Tissu | Adipose - V | Jan-41 | 0.163825 | 0.35405 | 0 | 0 | 5.777035 | 10.45039 |
| GTEx_Tissu | Skin - Not S | 1/100 | 0.35405 | 0.35405 | 0 | 0 | 2.327226 | 2.416399 |
| GTEx_Tissu | Heart - Left | 1/100 | 0.35405 | 0.35405 | 0 | 0 | 2.327226 | 2.416399 |
| GTEx_Tissu | Artery - Tib | 1/100 | 0.35405 | 0.35405 | 0 | 0 | 2.327226 | 2.416399 |
| GTEx_Tissu | Muscle - Sk | 1/100 | 0.35405 | 0.35405 | 0 | 0 | 2.327226 | 2.416399 |
| GTEx_Tissu | Esophagus | 1/100 | 0.35405 | 0.35405 | 0 | 0 | 2.327226 | 2.416399 |
| GTEx_Tissu | Muscle - Sk | 1/100 | 0.35405 | 0.35405 | 0 | 0 | 2.327226 | 2.416399 |
| GTEx_Tissu | Testis Male | 1/100 | 0.35405 | 0.35405 | 0 | 0 | 2.327226 | 2.416399 |
| GTEx_Tissu | Brain - Ant | 1/100 | 0.35405 | 0.35405 | 0 | 0 | 2.327226 | 2.416399 |
| GTEx_Tissu | Brain - Cort | 1/100 | 0.35405 | 0.35405 | 0 | 0 | 2.327226 | 2.416399 |
| GTEx_Tissu | Adipose - S | Jan-69 | 0.260157 | 0.35405 | 0 | 0 | 3.393468 | 4.569198 |
| GTEx_Tissu | Breast - Ma | Jan-65 | 0.247099 | 0.35405 | 0 | 0 | 3.606286 | 5.041467 |
| GTEx_Tissu | Ovary Fem | 1/100 | 0.35405 | 0.35405 | 0 | 0 | 2.327226 | 2.416399 |

| | | | | | | | | |
|---|---|---|---|---|---|---|---|---|
| GTEx_Tissu | Uterus Fem | 1/100 | 0.35405 | 0.35405 | 0 | 0 | 2.327226 | 2.416399 |
| GTEx_Tissu | Muscle - Sk | 1/100 | 0.35405 | 0.35405 | 0 | 0 | 2.327226 | 2.416399 |
| GTEx_Tissu | Brain - Spir | 1/100 | 0.35405 | 0.35405 | 0 | 0 | 2.327226 | 2.416399 |
| GTEx_Tissu | Brain - Sub | 1/100 | 0.35405 | 0.35405 | 0 | 0 | 2.327226 | 2.416399 |
| GTEx_Tissu | Brain - Spir | 1/100 | 0.35405 | 0.35405 | 0 | 0 | 2.327226 | 2.416399 |
| GTEx_Tissu | Small Intes | 1/100 | 0.35405 | 0.35405 | 0 | 0 | 2.327226 | 2.416399 |
| GTEx_Tissu | Skin - Not S | 1/100 | 0.35405 | 0.35405 | 0 | 0 | 2.327226 | 2.416399 |
| GTEx_Tissu | Testis Male | 1/100 | 0.35405 | 0.35405 | 0 | 0 | 2.327226 | 2.416399 |
| GTEx_Tissu | Small Intes | 1/100 | 0.35405 | 0.35405 | 0 | 0 | 2.327226 | 2.416399 |
| GTEx_Tissu | Small Intes | 1/100 | 0.35405 | 0.35405 | 0 | 0 | 2.327226 | 2.416399 |
| GTEx_Tissu | Brain - Ant | 1/100 | 0.35405 | 0.35405 | 0 | 0 | 2.327226 | 2.416399 |
| GTEx_Tissu | Brain - Spir | 1/100 | 0.35405 | 0.35405 | 0 | 0 | 2.327226 | 2.416399 |
| GTEx_Tissu | Uterus Fem | 1/100 | 0.35405 | 0.35405 | 0 | 0 | 2.327226 | 2.416399 |
| GTEx_Tissu | Muscle - Sk | 1/100 | 0.35405 | 0.35405 | 0 | 0 | 2.327226 | 2.416399 |
| GTEx_Tissu | Brain - Spir | 1/100 | 0.35405 | 0.35405 | 0 | 0 | 2.327226 | 2.416399 |
| GTEx_Tissu | Brain - Fror | 1/100 | 0.35405 | 0.35405 | 0 | 0 | 2.327226 | 2.416399 |
| GTEx_Tissu | Artery - Tib | 1/100 | 0.35405 | 0.35405 | 0 | 0 | 2.327226 | 2.416399 |
| GTEx_Tissu | Muscle - Sk | 1/100 | 0.35405 | 0.35405 | 0 | 0 | 2.327226 | 2.416399 |
| GTEx_Tissu | Artery - Tib | 1/100 | 0.35405 | 0.35405 | 0 | 0 | 2.327226 | 2.416399 |
| GTEx_Tissu | Adipose - V | Jan-79 | 0.291832 | 0.35405 | 0 | 0 | 2.956917 | 3.641673 |
| GTEx_Tissu | Uterus Fem | 1/100 | 0.35405 | 0.35405 | 0 | 0 | 2.327226 | 2.416399 |
| GTEx_Tissu | Adipose - V | 1/100 | 0.35405 | 0.35405 | 0 | 0 | 2.327226 | 2.416399 |
| GTEx_Tissu | Esophagus | 1/100 | 0.35405 | 0.35405 | 0 | 0 | 2.327226 | 2.416399 |
| GTEx_Tissu | Brain - Spir | 1/100 | 0.35405 | 0.35405 | 0 | 0 | 2.327226 | 2.416399 |
| GTEx_Tissu | Brain - Cort | 1/100 | 0.35405 | 0.35405 | 0 | 0 | 2.327226 | 2.416399 |
| GTEx_Tissu | Whole Bloc | 1/100 | 0.35405 | 0.35405 | 0 | 0 | 2.327226 | 2.416399 |
| GTEx_Tissu | Brain - Hip | 1/100 | 0.35405 | 0.35405 | 0 | 0 | 2.327226 | 2.416399 |
| GTEx_Tissu | Artery - Tib | 1/100 | 0.35405 | 0.35405 | 0 | 0 | 2.327226 | 2.416399 |
| GTEx_Tissu | Esophagus | 1/100 | 0.35405 | 0.35405 | 0 | 0 | 2.327226 | 2.416399 |
| GTEx_Tissu | Ovary Fem | 1/100 | 0.35405 | 0.35405 | 0 | 0 | 2.327226 | 2.416399 |
| GTEx_Tissu | Small Intes | 1/100 | 0.35405 | 0.35405 | 0 | 0 | 2.327226 | 2.416399 |
| GTEx_Tissu | Skin - Not S | 1/100 | 0.35405 | 0.35405 | 0 | 0 | 2.327226 | 2.416399 |
| GTEx_Tissu | Skin - Not S | 1/100 | 0.35405 | 0.35405 | 0 | 0 | 2.327226 | 2.416399 |
| GTEx_Tissu | Brain - Ant | 1/100 | 0.35405 | 0.35405 | 0 | 0 | 2.327226 | 2.416399 |
| GTEx_Tissu | Brain - Spir | 1/100 | 0.35405 | 0.35405 | 0 | 0 | 2.327226 | 2.416399 |
| GTEx_Tissu | Liver Fema | 1/100 | 0.35405 | 0.35405 | 0 | 0 | 2.327226 | 2.416399 |
| GTEx_Tissu | Breast - Ma | Jan-71 | 0.266602 | 0.35405 | 0 | 0 | 3.296179 | 4.35754 |
| GTEx_Tissu | Brain - Spir | 1/100 | 0.35405 | 0.35405 | 0 | 0 | 2.327226 | 2.416399 |
| GTEx_Tissu | Brain - Cort | 1/100 | 0.35405 | 0.35405 | 0 | 0 | 2.327226 | 2.416399 |
| GTEx_Tissu | Brain - Hip | 1/100 | 0.35405 | 0.35405 | 0 | 0 | 2.327226 | 2.416399 |
| GTEx_Tissu | Adipose - S | Jan-86 | 0.313203 | 0.35405 | 0 | 0 | 2.712449 | 3.148895 |
| GTEx_Tissu | Adrenal Gl | 1/100 | 0.35405 | 0.35405 | 0 | 0 | 2.327226 | 2.416399 |
| GTEx_Tissu | Artery - Tib | 1/100 | 0.35405 | 0.35405 | 0 | 0 | 2.327226 | 2.416399 |
| GTEx_Tissu | Cells - Cult | 1/100 | 0.35405 | 0.35405 | 0 | 0 | 2.327226 | 2.416399 |
| GTEx_Tissu | Artery - Tib | 1/100 | 0.35405 | 0.35405 | 0 | 0 | 2.327226 | 2.416399 |

| | | | | | | | | |
|---|---|---|---|---|---|---|---|---|
| GTEx_Tissu | Colon - Sigı | 1/100 | 0.35405 | 0.35405 | 0 | 0 | 2.327226 | 2.416399 |
| GTEx_Tissu | Esophagus | Jan-82 | 0.30107 | 0.35405 | 0 | 0 | 2.846971 | 3.417539 |
| GTEx_Tissu | Esophagus | Jan-89 | 0.322165 | 0.35405 | 0 | 0 | 2.619582 | 2.967177 |
| GTEx_Tissu | Small Intes | 1/100 | 0.35405 | 0.35405 | 0 | 0 | 2.327226 | 2.416399 |
| GTEx_Tissu | Liver Male | 1/100 | 0.35405 | 0.35405 | 0 | 0 | 2.327226 | 2.416399 |
| GO_Biologi | Negative R | 2/231 | 0.266025 | 0.354471 | 0 | 0 | 2.022502 | 2.678127 |
| GO_Biologi | Synapse As | Jan-71 | 0.266602 | 0.354471 | 0 | 0 | 3.296179 | 4.35754 |
| GO_Biologi | Negative R | Jan-71 | 0.266602 | 0.354471 | 0 | 0 | 3.296179 | 4.35754 |
| GO_Biologi | Cell-Cell Ju | Jan-71 | 0.266602 | 0.354471 | 0 | 0 | 3.296179 | 4.35754 |
| GeDiPNet_ | Thyroid Ca | Jan-74 | 0.276166 | 0.354636 | 0 | 0 | 3.160242 | 4.066453 |
| GeDiPNet_ | Congenital | Jan-74 | 0.276166 | 0.354636 | 0 | 0 | 3.160242 | 4.066453 |
| GeDiPNet_ | Non-Syndr | Jan-74 | 0.276166 | 0.354636 | 0 | 0 | 3.160242 | 4.066453 |
| GWAS_Cat | Multiple Sc | 2/271 | 0.330285 | 0.354707 | 0 | 0 | 1.718259 | 1.903487 |
| GWAS_Cat | Glaucoma ( | Jan-92 | 0.331012 | 0.354864 | 0 | 0 | 2.532839 | 2.800307 |
| GWAS_Cat | Offspring B | Jan-92 | 0.331012 | 0.354864 | 0 | 0 | 2.532839 | 2.800307 |
| DisGeNET | Rhabdoid T | Jan-71 | 0.266602 | 0.355108 | 0 | 0 | 3.296179 | 4.35754 |
| DisGeNET | Leukemia, | Jan-71 | 0.266602 | 0.355108 | 0 | 0 | 3.296179 | 4.35754 |
| DisGeNET | Malocclusi | Jan-71 | 0.266602 | 0.355108 | 0 | 0 | 3.296179 | 4.35754 |
| GTEx_Tissu | GTEX-N7M | 7/574 | 0.012523 | 0.355174 | 0 | 0 | 2.985494 | 13.07713 |
| GTEx_Tissu | GTEX-XPT6 | 7/574 | 0.012523 | 0.355174 | 0 | 0 | 2.985494 | 13.07713 |
| GTEx_Tissu | GTEX-WI4N | 7/572 | 0.012303 | 0.355174 | 0 | 0 | 2.996372 | 13.17766 |
| PhenGenI_ | Alcohol Dri | 2/151 | 0.140245 | 0.355358 | 0 | 0 | 3.121042 | 6.130869 |
| PhenGenI_ | Plaque, Am | Jan-35 | 0.141623 | 0.355358 | 0 | 0 | 6.798564 | 13.28839 |
| PhenGenI_ | Kidney Dise | Jan-33 | 0.134093 | 0.355358 | 0 | 0 | 7.224201 | 14.515 |
| PhenGenI_ | Bladder Ex | Jan-36 | 0.145363 | 0.355358 | 0 | 0 | 6.603987 | 12.73591 |
| PhenGenI_ | Urinary Bla | Jan-35 | 0.141623 | 0.355358 | 0 | 0 | 6.798564 | 13.28839 |
| PhenGenI_ | Dementia | Jan-34 | 0.137866 | 0.355358 | 0 | 0 | 7.004933 | 13.88007 |
| PhenGenI_ | Blood Circu | Jan-36 | 0.145363 | 0.355358 | 0 | 0 | 6.603987 | 12.73591 |
| PhenGenI_ | Glucocortic | Jan-36 | 0.145363 | 0.355358 | 0 | 0 | 6.603987 | 12.73591 |
| PhenGenI_ | Apolipopro | Jan-33 | 0.134093 | 0.355358 | 0 | 0 | 7.224201 | 14.515 |
| PhenGenI_ | Osteonecro | Jan-33 | 0.134093 | 0.355358 | 0 | 0 | 7.224201 | 14.515 |
| PhenGenI_ | Body Mass | Dec-71 | 0.14605 | 0.355358 | 0 | 0 | 1.466381 | 2.821033 |
| PhenGenI_ | Adiponecti | 2/156 | 0.147716 | 0.355358 | 0 | 0 | 3.018946 | 5.773631 |
| PhenGenI_ | Telomere | Jan-37 | 0.149088 | 0.355358 | 0 | 0 | 6.42022 | 12.21909 |
| PhenGenI_ | Cognition [ | 2/157 | 0.14922 | 0.355358 | 0 | 0 | 2.999317 | 5.705706 |
| PhenGenI_ | Neuropsycl | 3/287 | 0.129587 | 0.355358 | 0 | 0 | 2.468436 | 5.04401 |
| GO_Biologi | Negative R | Jan-72 | 0.269804 | 0.3554 | 0 | 0 | 3.249591 | 4.257158 |
| GO_Biologi | Positive Re | 2/233 | 0.269251 | 0.3554 | 0 | 0 | 2.004787 | 2.630503 |
| GO_Biologi | Cholestero | Jan-72 | 0.269804 | 0.3554 | 0 | 0 | 3.249591 | 4.257158 |
| GO_Biologi | Plasma Me | Jan-72 | 0.269804 | 0.3554 | 0 | 0 | 3.249591 | 4.257158 |
| GO_Biologi | Second-Me | Jan-72 | 0.269804 | 0.3554 | 0 | 0 | 3.249591 | 4.257158 |
| Disease_Si | Carcinoma, | 5/300 | 0.009983 | 0.356069 | 0 | 0 | 4.054981 | 18.68068 |
| Disease_Si | Huntingtor | 5/300 | 0.009983 | 0.356069 | 0 | 0 | 4.054981 | 18.68068 |
| Disease_Si | Muscular D | 5/300 | 0.009983 | 0.356069 | 0 | 0 | 4.054981 | 18.68068 |
| GTEx_Tissu | GTEX-T6MI | 13/1501 | 0.012707 | 0.356917 | 0 | 0 | 2.175285 | 9.496363 |

| | | | | | | | | |
|---|---|---|---|---|---|---|---|---|
| GO_Biologi | Monocarbo | Jan-73 | 0.272992 | 0.356951 | 0 | 0 | 3.204296 | 4.160181 |
| GO_Biologi | Regulation | Jan-73 | 0.272992 | 0.356951 | 0 | 0 | 3.204296 | 4.160181 |
| GO_Biologi | Positive Re | Jan-73 | 0.272992 | 0.356951 | 0 | 0 | 3.204296 | 4.160181 |
| GO_Biologi | Sterol Hom | Jan-73 | 0.272992 | 0.356951 | 0 | 0 | 3.204296 | 4.160181 |
| GWAS_Cat | Spherical E | Jan-93 | 0.333936 | 0.357057 | 0 | 0 | 2.505182 | 2.747701 |
| GWAS_Cat | Sleep Dura | Jan-93 | 0.333936 | 0.357057 | 0 | 0 | 2.505182 | 2.747701 |
| GWAS_Cat | Cancer (Ple | Jan-93 | 0.333936 | 0.357057 | 0 | 0 | 2.505182 | 2.747701 |
| UK_Bioban | North co-o | 2/256 | 0.306291 | 0.357312 | 0 | 0 | 1.821121 | 2.154787 |
| UK_Bioban | Sweets inta | Jan-84 | 0.307163 | 0.357312 | 0 | 0 | 2.778089 | 3.279195 |
| Jensen_TIS | SH-SY5Y ce | Jan-72 | 0.269804 | 0.357518 | 0 | 0 | 3.249591 | 4.257158 |
| Rare_Disea | Temporal e | Jan-53 | 0.206541 | 0.35756 | 0 | 0 | 4.44119 | 7.004886 |
| Rare_Disea | Neurotoxic | 2/194 | 0.206654 | 0.35756 | 0 | 0 | 2.416789 | 3.810572 |
| CellMarker | Oligodendr | Jan-83 | 0.304123 | 0.357638 | 0 | 0 | 2.81211 | 3.34732 |
| DisGeNET | Cryptorchi | 3/414 | 0.268918 | 0.35801 | 0 | 0 | 1.694647 | 2.225665 |
| DisGeNET | MAJOR AFI | Jan-72 | 0.269804 | 0.358098 | 0 | 0 | 3.249591 | 4.257158 |
| DisGeNET | alpha^+^ T | Jan-72 | 0.269804 | 0.358098 | 0 | 0 | 3.249591 | 4.257158 |
| DisGeNET | Spinocereb | Jan-72 | 0.269804 | 0.358098 | 0 | 0 | 3.249591 | 4.257158 |
| DisGeNET | Gallbladde | 2/233 | 0.269251 | 0.358098 | 0 | 0 | 2.004787 | 2.630503 |
| DisGeNET | secondary | Jan-72 | 0.269804 | 0.358098 | 0 | 0 | 3.249591 | 4.257158 |
| DisGeNET | MAJOR AFI | Jan-72 | 0.269804 | 0.358098 | 0 | 0 | 3.249591 | 4.257158 |
| Jensen_DIS | Thrombocy | Jan-95 | 0.339745 | 0.35811 | 0 | 0 | 2.451633 | 2.646685 |
| SysMyo_M | MGS684 Hu | 3/212 | 0.065188 | 0.358217 | 0 | 0 | 3.367054 | 9.193681 |
| SysMyo_M | MGS674 Hu | Feb-95 | 0.064403 | 0.358217 | 0 | 0 | 5.014548 | 13.75286 |
| SysMyo_M | MGS1016 I | 3/210 | 0.063737 | 0.358217 | 0 | 0 | 3.399931 | 9.359987 |
| SysMyo_M | MGS1186 I | 3/210 | 0.063737 | 0.358217 | 0 | 0 | 3.399931 | 9.359987 |
| SysMyo_M | MGS1357 I | 3/212 | 0.065188 | 0.358217 | 0 | 0 | 3.367054 | 9.193681 |
| SysMyo_M | MGS78 Rat | Feb-94 | 0.063221 | 0.358217 | 0 | 0 | 5.069309 | 13.99697 |
| Disease_Pe | Emery-Dre | 5/266 | 0.006107 | 0.358623 | 0 | 0 | 4.59116 | 23.40751 |
| Disease_Pe | Treacher C | 5/258 | 0.005379 | 0.358623 | 0 | 0 | 4.738263 | 24.75901 |
| Disease_Pe | nemaline n | 6/374 | 0.00582 | 0.358623 | 0 | 0 | 3.934179 | 20.24674 |
| Reactome_ | Class B/2 (S | Feb-92 | 0.060879 | 0.358869 | 0 | 0 | 5.182484 | 14.50504 |
| HuBMAP_A | Lymphatic | 2/118 | 0.093488 | 0.358994 | 0 | 0 | 4.015619 | 9.516699 |
| ARCHS4_Ti | CARDIAC M | 18/2316 | 0.009974 | 0.359062 | 0 | 0 | 1.999659 | 9.213991 |
| ARCHS4_Ti | HEART (BU | 18/2316 | 0.009974 | 0.359062 | 0 | 0 | 1.999659 | 9.213991 |
| Jensen_DIS | Azoosperm | Jan-96 | 0.342631 | 0.35921 | 0 | 0 | 2.425704 | 2.598176 |
| DisGeNET | Epstein-Ba | 2/234 | 0.270864 | 0.359324 | 0 | 0 | 1.996045 | 2.607109 |
| Human_Ph | Hypopigme | Jan-93 | 0.333936 | 0.359334 | 0 | 0 | 2.505182 | 2.747701 |
| Rare_Disea | Multiple sy | 2/195 | 0.208243 | 0.359473 | 0 | 0 | 2.404145 | 3.772225 |
| DisGeNET | Congenital | 4/607 | 0.271394 | 0.359844 | 0 | 0 | 1.543288 | 2.01273 |
| Rare_Disea | Metaplasti | Jan-54 | 0.210002 | 0.360004 | 0 | 0 | 4.357174 | 6.79997 |
| Rare_Disea | Intravenou | Jan-54 | 0.210002 | 0.360004 | 0 | 0 | 4.357174 | 6.79997 |
| Rare_Disea | Heavy met | Jan-54 | 0.210002 | 0.360004 | 0 | 0 | 4.357174 | 6.79997 |
| Jensen_TIS | Prostate gl | 72/15901 | 0.273002 | 0.360274 | 0 | 0 | 1.238436 | 1.60783 |
| GO_Biologi | Import Into | Jan-74 | 0.276166 | 0.360437 | 0 | 0 | 3.160242 | 4.066453 |
| DisGeNET | Hypopigme | Jan-73 | 0.272992 | 0.360685 | 0 | 0 | 3.204296 | 4.160181 |

| | | | | | | | | |
|---|---|---|---|---|---|---|---|---|
| DisGeNET | Inborn Errc | Jan-73 | 0.272992 | 0.360685 | 0 | 0 | 3.204296 | 4.160181 |
| DisGeNET | mixed glior | Jan-73 | 0.272992 | 0.360685 | 0 | 0 | 3.204296 | 4.160181 |
| DisGeNET | Photosensi | Jan-73 | 0.272992 | 0.360685 | 0 | 0 | 3.204296 | 4.160181 |
| DisGeNET | Guillain-Ba | Jan-73 | 0.272992 | 0.360685 | 0 | 0 | 3.204296 | 4.160181 |
| DisGeNET | Marfan Syr | Jan-73 | 0.272992 | 0.360685 | 0 | 0 | 3.204296 | 4.160181 |
| DisGeNET | Dysplastic I | Jan-73 | 0.272992 | 0.360685 | 0 | 0 | 3.204296 | 4.160181 |
| DisGeNET | Rheumatoi | 10/1833 | 0.273442 | 0.361009 | 0 | 0 | 1.288728 | 1.671048 |
| DisGeNET | HIV Infectio | 5/806 | 0.273513 | 0.361009 | 0 | 0 | 1.454889 | 1.886128 |
| GeDiPNet_ | Hemangior | Jan-76 | 0.282473 | 0.361144 | 0 | 0 | 3.075659 | 3.888164 |
| GeDiPNet_ | Diabetic Ne | Jan-76 | 0.282473 | 0.361144 | 0 | 0 | 3.075659 | 3.888164 |
| GTEx_Tissu | GTEX-WHS | 9/867 | 0.012998 | 0.361567 | 0 | 0 | 2.562534 | 11.129 |
| DisGeNET | Sarcoidosis | 2/236 | 0.27409 | 0.361589 | 0 | 0 | 1.978783 | 2.561135 |
| Rare_Disea | Pure red ce | Jan-55 | 0.213448 | 0.361743 | 0 | 0 | 4.27627 | 6.604112 |
| Rare_Disea | Peutz Jegh | Jan-55 | 0.213448 | 0.361743 | 0 | 0 | 4.27627 | 6.604112 |
| Rare_Disea | Dextrocard | Jan-55 | 0.213448 | 0.361743 | 0 | 0 | 4.27627 | 6.604112 |
| Rare_Disea | Juvenile my | Jan-55 | 0.213448 | 0.361743 | 0 | 0 | 4.27627 | 6.604112 |
| Rare_Disea | Fetal macro | Jan-55 | 0.213448 | 0.361743 | 0 | 0 | 4.27627 | 6.604112 |
| Rare_Disea | Batten Dise | Jan-56 | 0.216879 | 0.361789 | 0 | 0 | 4.198309 | 6.416764 |
| Rare_Disea | Teratoma | 2/199 | 0.21461 | 0.361789 | 0 | 0 | 2.354852 | 3.623955 |
| Rare_Disea | Germinoma | 2/199 | 0.21461 | 0.361789 | 0 | 0 | 2.354852 | 3.623955 |
| Rare_Disea | Compartme | 2/200 | 0.216205 | 0.361789 | 0 | 0 | 2.34284 | 3.588123 |
| Rare_Disea | Sydenham' | 3/369 | 0.216772 | 0.361789 | 0 | 0 | 1.907397 | 2.916234 |
| Rare_Disea | Chorea mir | 3/369 | 0.216772 | 0.361789 | 0 | 0 | 1.907397 | 2.916234 |
| Rare_Disea | Ocular mel | Jan-56 | 0.216879 | 0.361789 | 0 | 0 | 4.198309 | 6.416764 |
| Rare_Disea | Portal hype | 2/201 | 0.217801 | 0.362515 | 0 | 0 | 2.330949 | 3.552767 |
| Rare_Disea | Congenital | Jan-57 | 0.220295 | 0.362609 | 0 | 0 | 4.123131 | 6.237423 |
| Rare_Disea | Gangliocyt | Jan-57 | 0.220295 | 0.362609 | 0 | 0 | 4.123131 | 6.237423 |
| Rare_Disea | Mental reta | Jan-57 | 0.220295 | 0.362609 | 0 | 0 | 4.123131 | 6.237423 |
| Rare_Disea | Progressive | Jan-57 | 0.220295 | 0.362609 | 0 | 0 | 4.123131 | 6.237423 |
| Rare_Disea | Glaucoma ( | Jan-57 | 0.220295 | 0.362609 | 0 | 0 | 4.123131 | 6.237423 |
| GWAS_Cat | Cortical Sur | Jan-95 | 0.339745 | 0.362717 | 0 | 0 | 2.451633 | 2.646685 |
| GWAS_Cat | Hair Color | 2/277 | 0.339825 | 0.362717 | 0 | 0 | 1.680257 | 1.813544 |
| GeDiPNet_ | Dermatolo | Jan-77 | 0.285606 | 0.362763 | 0 | 0 | 3.035037 | 3.803331 |
| GeDiPNet_ | Febrile Seiz | Jan-77 | 0.285606 | 0.362763 | 0 | 0 | 3.035037 | 3.803331 |
| GeDiPNet_ | Cutis Marm | Jan-77 | 0.285606 | 0.362763 | 0 | 0 | 3.035037 | 3.803331 |
| Human_Ph | Low-set, po | 2/276 | 0.338237 | 0.36294 | 0 | 0 | 1.686475 | 1.828152 |
| GO_Biologi | Regulation | Jan-75 | 0.279326 | 0.363226 | 0 | 0 | 3.117379 | 3.975827 |
| GO_Biologi | Negative R | Jan-75 | 0.279326 | 0.363226 | 0 | 0 | 3.117379 | 3.975827 |
| SysMyo_M | MGS688 M | 16-Jan | 0.067399 | 0.363312 | 0 | 0 | 15.42481 | 41.60265 |
| SysMyo_M | MGS1482 I | Feb-97 | 0.06679 | 0.363312 | 0 | 0 | 4.908483 | 13.28336 |
| CellMarker | Neuroendo | 3/449 | 0.310446 | 0.363513 | 0 | 0 | 1.558857 | 1.823466 |
| DisGeNET | Undifferen | 2/237 | 0.275703 | 0.363534 | 0 | 0 | 1.970263 | 2.538546 |
| Human_Ph | Long face ( | Jan-95 | 0.339745 | 0.363537 | 0 | 0 | 2.451633 | 2.646685 |
| DisGeNET | MAJOR AFI | Jan-74 | 0.276166 | 0.363595 | 0 | 0 | 3.160242 | 4.066453 |
| DisGeNET | Acoustic Ne | Jan-74 | 0.276166 | 0.363595 | 0 | 0 | 3.160242 | 4.066453 |

| | | | | | | | | |
|---|---|---|---|---|---|---|---|---|
| DisGeNET | Necrotizing | Jan-74 | 0.276166 | 0.363595 | 0 | 0 | 3.160242 | 4.066453 |
| SysMyo_M | MGS1339 I | 3/216 | 0.068136 | 0.363818 | 0 | 0 | 3.303152 | 8.873113 |
| GeDiPNet_ | Obsessive-( | Jan-78 | 0.288726 | 0.364344 | 0 | 0 | 2.99547 | 3.721207 |
| GeDiPNet_ | Neck Webb | Jan-78 | 0.288726 | 0.364344 | 0 | 0 | 2.99547 | 3.721207 |
| GeDiPNet_ | Lymphopei | Jan-78 | 0.288726 | 0.364344 | 0 | 0 | 2.99547 | 3.721207 |
| DisGeNET | Overweigh | 2/238 | 0.277316 | 0.364926 | 0 | 0 | 1.961815 | 2.516219 |
| GWAS_Cat | Neutrophil | Jan-96 | 0.342631 | 0.365074 | 0 | 0 | 2.425704 | 2.598176 |
| GWAS_Cat | Anorexia N | Jan-96 | 0.342631 | 0.365074 | 0 | 0 | 2.425704 | 2.598176 |
| GTEx_Tissu | GTEX-QESD | 8/722 | 0.013255 | 0.365215 | 0 | 0 | 2.722973 | 11.77239 |
| MAGMA_D | Thymidylat | 1/104 | 0.365273 | 0.365273 | 0 | 0 | 2.236396 | 2.252299 |
| MAGMA_D | Pancreatic | 1/104 | 0.365273 | 0.365273 | 0 | 0 | 2.236396 | 2.252299 |
| MAGMA_D | Colorectal | 1/104 | 0.365273 | 0.365273 | 0 | 0 | 2.236396 | 2.252299 |
| MAGMA_D | 5 Fluoroura | 1/104 | 0.365273 | 0.365273 | 0 | 0 | 2.236396 | 2.252299 |
| MAGMA_D | Breast Can | 1/104 | 0.365273 | 0.365273 | 0 | 0 | 2.236396 | 2.252299 |
| MAGMA_D | Paclitaxel | 1/101 | 0.356874 | 0.365273 | 0 | 0 | 2.303837 | 2.373811 |
| MAGMA_D | Gastric Ade | 1/104 | 0.365273 | 0.365273 | 0 | 0 | 2.236396 | 2.252299 |
| Rare_Disea | Acute myel | 2/204 | 0.222596 | 0.365588 | 0 | 0 | 2.295981 | 3.449476 |
| Jensen_TIS | Branchial a | Jan-75 | 0.279326 | 0.365622 | 0 | 0 | 3.117379 | 3.975827 |
| Jensen_TIS | Schwann ce | Jan-75 | 0.279326 | 0.365622 | 0 | 0 | 3.117379 | 3.975827 |
| DisGeNET | Bladder Ne | 7/1217 | 0.278161 | 0.365855 | 0 | 0 | 1.35249 | 1.730584 |
| DisGeNET | Comatose | Jan-75 | 0.279326 | 0.3661 | 0 | 0 | 3.117379 | 3.975827 |
| DisGeNET | Shy-Drager | Jan-75 | 0.279326 | 0.3661 | 0 | 0 | 3.117379 | 3.975827 |
| DisGeNET | Polycythen | Jan-75 | 0.279326 | 0.3661 | 0 | 0 | 3.117379 | 3.975827 |
| DisGeNET | Ovarian Cy | Jan-75 | 0.279326 | 0.3661 | 0 | 0 | 3.117379 | 3.975827 |
| DisGeNET | Medullary | 2/239 | 0.278929 | 0.3661 | 0 | 0 | 1.953438 | 2.494149 |
| DisGeNET | Mitral Valv | Jan-75 | 0.279326 | 0.3661 | 0 | 0 | 3.117379 | 3.975827 |
| DisGeNET | Ewing Sarc | Jan-75 | 0.279326 | 0.3661 | 0 | 0 | 3.117379 | 3.975827 |
| Jensen_DIS | Parkinson's | Jan-99 | 0.351214 | 0.366239 | 0 | 0 | 2.351092 | 2.46009 |
| UK_Bioban | A/AS levels | Jan-87 | 0.316203 | 0.366333 | 0 | 0 | 2.680773 | 3.086563 |
| Rare_Disea | Myxoid lipo | Jan-58 | 0.223696 | 0.366586 | 0 | 0 | 4.050592 | 6.065622 |
| Rare_Disea | Dengue fev | 2/205 | 0.224196 | 0.366598 | 0 | 0 | 2.284555 | 3.415945 |
| GO_Biologi | Positive Re | Jan-76 | 0.282473 | 0.366647 | 0 | 0 | 3.075659 | 3.888164 |
| GTEx_Tissu | GTEX-W5X | 8/725 | 0.013562 | 0.366681 | 0 | 0 | 2.711156 | 11.65928 |
| GTEx_Tissu | GTEX-QVJC | 13/1512 | 0.013443 | 0.366681 | 0 | 0 | 2.158033 | 9.299528 |
| Gene_Pert | Il13 OE mo | 5/195 | 0.00162 | 0.367355 | 0 | 0 | 6.329589 | 40.66935 |
| GeDiPNet_ | Muscular D | Jan-79 | 0.291832 | 0.367469 | 0 | 0 | 2.956917 | 3.641673 |
| GWAS_Cat | Hay Fever / | Jan-97 | 0.345504 | 0.367814 | 0 | 0 | 2.400315 | 2.550937 |
| DisGeNET | Granuloma | Jan-76 | 0.282473 | 0.368199 | 0 | 0 | 3.075659 | 3.888164 |
| DisGeNET | Varicosity | Jan-76 | 0.282473 | 0.368199 | 0 | 0 | 3.075659 | 3.888164 |
| DisGeNET | Acromegal | Jan-76 | 0.282473 | 0.368199 | 0 | 0 | 3.075659 | 3.888164 |
| DisGeNET | Blood pres | Jan-76 | 0.282473 | 0.368199 | 0 | 0 | 3.075659 | 3.888164 |
| DisGeNET | Early-Stage | Jan-76 | 0.282473 | 0.368199 | 0 | 0 | 3.075659 | 3.888164 |
| DisGeNET | Blood Pres | Jan-76 | 0.282473 | 0.368199 | 0 | 0 | 3.075659 | 3.888164 |
| DisGeNET | Chest Pain | Jan-76 | 0.282473 | 0.368199 | 0 | 0 | 3.075659 | 3.888164 |
| DisGeNET | Respiratior | Jan-76 | 0.282473 | 0.368199 | 0 | 0 | 3.075659 | 3.888164 |

| | | | | | | | | |
|---|---|---|---|---|---|---|---|---|
| DisGeNET | Systemic ar | Jan-76 | 0.282473 | 0.368199 | 0 | 0 | 3.075659 | 3.888164 |
| DisGeNET | Acute Undi | Jan-76 | 0.282473 | 0.368199 | 0 | 0 | 3.075659 | 3.888164 |
| DisGeNET | Stable angi | Jan-76 | 0.282473 | 0.368199 | 0 | 0 | 3.075659 | 3.888164 |
| GO_Biologi | Positive Re | Jan-77 | 0.285606 | 0.369363 | 0 | 0 | 3.035037 | 3.803331 |
| GO_Biologi | Regulation | Jan-77 | 0.285606 | 0.369363 | 0 | 0 | 3.035037 | 3.803331 |
| DisGeNET | Psoriasis | 5/818 | 0.283597 | 0.36948 | 0 | 0 | 1.432514 | 1.805254 |
| CellMarker | Podocyte K | 2/263 | 0.317511 | 0.370204 | 0 | 0 | 1.771648 | 2.03251 |
| GWAS_Cat | Adolescent | 4/685 | 0.3481 | 0.370255 | 0 | 0 | 1.361003 | 1.436219 |
| DisGeNET | Congenital | Jan-77 | 0.285606 | 0.370256 | 0 | 0 | 3.035037 | 3.803331 |
| DisGeNET | PANIC DISC | Jan-77 | 0.285606 | 0.370256 | 0 | 0 | 3.035037 | 3.803331 |
| DisGeNET | Neoplastic | Jan-77 | 0.285606 | 0.370256 | 0 | 0 | 3.035037 | 3.803331 |
| DisGeNET | Vitiligo | 2/243 | 0.285377 | 0.370256 | 0 | 0 | 1.920625 | 2.408358 |
| DisGeNET | High-Grade | Jan-77 | 0.285606 | 0.370256 | 0 | 0 | 3.035037 | 3.803331 |
| DisGeNET | Non-small | Jan-77 | 0.285606 | 0.370256 | 0 | 0 | 3.035037 | 3.803331 |
| DisGeNET | Niemann-P | Jan-77 | 0.285606 | 0.370256 | 0 | 0 | 3.035037 | 3.803331 |
| DisGeNET | aggressive | Jan-77 | 0.285606 | 0.370256 | 0 | 0 | 3.035037 | 3.803331 |
| DisGeNET | Progressive | Jan-77 | 0.285606 | 0.370256 | 0 | 0 | 3.035037 | 3.803331 |
| DisGeNET | Decreased | Jan-77 | 0.285606 | 0.370256 | 0 | 0 | 3.035037 | 3.803331 |
| GO_Biologi | Regulation | 2/244 | 0.286988 | 0.370475 | 0 | 0 | 1.912591 | 2.387516 |
| GeDiPNet_ | Developme | Jan-80 | 0.294925 | 0.370563 | 0 | 0 | 2.919341 | 3.564619 |
| GeDiPNet_ | Renal Cyst | Jan-81 | 0.298004 | 0.371231 | 0 | 0 | 2.882703 | 3.48994 |
| GeDiPNet_ | Retinal Det | Jan-81 | 0.298004 | 0.371231 | 0 | 0 | 2.882703 | 3.48994 |
| GeDiPNet_ | Cranial Ner | Jan-81 | 0.298004 | 0.371231 | 0 | 0 | 2.882703 | 3.48994 |
| GeDiPNet_ | Spastic Qua | Jan-81 | 0.298004 | 0.371231 | 0 | 0 | 2.882703 | 3.48994 |
| DisGeNET | Inflammati | 3/429 | 0.286652 | 0.371429 | 0 | 0 | 1.633719 | 2.041308 |
| UK_Bioban | Right intra- | Jan-89 | 0.322165 | 0.371729 | 0 | 0 | 2.619582 | 2.967177 |
| GO_Biologi | Negative R | Jan-78 | 0.288726 | 0.372042 | 0 | 0 | 2.99547 | 3.721207 |
| DisGeNET | Peripheral | Jan-78 | 0.288726 | 0.372641 | 0 | 0 | 2.99547 | 3.721207 |
| DisGeNET | Dyskinetic | Jan-78 | 0.288726 | 0.372641 | 0 | 0 | 2.99547 | 3.721207 |
| DisGeNET | Giant Cell C | Jan-78 | 0.288726 | 0.372641 | 0 | 0 | 2.99547 | 3.721207 |
| DisGeNET | Bronchial H | Jan-78 | 0.288726 | 0.372641 | 0 | 0 | 2.99547 | 3.721207 |
| DisGeNET | Marinesco- | Jan-78 | 0.288726 | 0.372641 | 0 | 0 | 2.99547 | 3.721207 |
| DisGeNET | Klinefelter | Jan-78 | 0.288726 | 0.372641 | 0 | 0 | 2.99547 | 3.721207 |
| DisGeNET | Acute mon | Jan-78 | 0.288726 | 0.372641 | 0 | 0 | 2.99547 | 3.721207 |
| DisGeNET | Multiple Sc | Jan-78 | 0.288726 | 0.372641 | 0 | 0 | 2.99547 | 3.721207 |
| WikiPathw | Familial Hy | 13-Jan | 0.055113 | 0.372825 | 0 | 0 | 19.28391 | 55.89185 |
| WikiPathw | Genes Targ | 13-Jan | 0.055113 | 0.372825 | 0 | 0 | 19.28391 | 55.89185 |
| WikiPathw | PtdIns 4 5 I | 12-Jan | 0.050982 | 0.372825 | 0 | 0 | 21.03805 | 62.61499 |
| GO_Biologi | Positive Re | Jan-79 | 0.291832 | 0.373334 | 0 | 0 | 2.956917 | 3.641673 |
| GO_Biologi | Protein Imp | Jan-79 | 0.291832 | 0.373334 | 0 | 0 | 2.956917 | 3.641673 |
| GO_Biologi | Regulation | Jan-79 | 0.291832 | 0.373334 | 0 | 0 | 2.956917 | 3.641673 |
| GO_Biologi | Positive Re | Jan-79 | 0.291832 | 0.373334 | 0 | 0 | 2.956917 | 3.641673 |
| GeDiPNet_ | Oculovestil | Jan-82 | 0.30107 | 0.374058 | 0 | 0 | 2.846971 | 3.417539 |
| GeDiPNet_ | Alveolitis | Jan-83 | 0.304123 | 0.374058 | 0 | 0 | 2.81211 | 3.34732 |
| GeDiPNet_ | Apraxia | Jan-83 | 0.304123 | 0.374058 | 0 | 0 | 2.81211 | 3.34732 |

| | | | | | | | | |
|---|---|---|---|---|---|---|---|---|
| GeDiPNet_ | Dwarfism | 6/1046 | 0.303309 | 0.374058 | 0 | 0 | 1.344231 | 1.603673 |
| GeDiPNet_ | Nephrolith | Jan-83 | 0.304123 | 0.374058 | 0 | 0 | 2.81211 | 3.34732 |
| GeDiPNet_ | Respiratory | Jan-83 | 0.304123 | 0.374058 | 0 | 0 | 2.81211 | 3.34732 |
| DisGeNET | Intellectual | 13/2503 | 0.290331 | 0.374345 | 0 | 0 | 1.229236 | 1.520235 |
| DisGeNET | Pulmonary | 3/432 | 0.290213 | 0.374345 | 0 | 0 | 1.622045 | 2.006699 |
| GO_Molec | Serine-Typ | Jan-53 | 0.206541 | 0.375004 | 0 | 0 | 4.44119 | 7.004886 |
| GO_Molec | Polyubiqui | Jan-53 | 0.206541 | 0.375004 | 0 | 0 | 4.44119 | 7.004886 |
| GO_Molec | Oxidoredu | Jan-54 | 0.210002 | 0.375004 | 0 | 0 | 4.357174 | 6.79997 |
| Rare_Disea | Familial hy | Jan-60 | 0.230455 | 0.375428 | 0 | 0 | 3.912889 | 5.742942 |
| Rare_Disea | Acute myel | 2/209 | 0.230606 | 0.375428 | 0 | 0 | 2.239955 | 3.286115 |
| DisGeNET | Osteitis De | Jan-79 | 0.291832 | 0.37554 | 0 | 0 | 2.956917 | 3.641673 |
| DisGeNET | Polyendocr | Jan-79 | 0.291832 | 0.37554 | 0 | 0 | 2.956917 | 3.641673 |
| DisGeNET | Distal sens | Jan-79 | 0.291832 | 0.37554 | 0 | 0 | 2.956917 | 3.641673 |
| DisGeNET | Diabetic Re | 2/247 | 0.29182 | 0.37554 | 0 | 0 | 1.888884 | 2.326383 |
| Gene_Pert | Idh2 R140C | 6/306 | 0.002185 | 0.375676 | 0 | 0 | 4.842716 | 29.66659 |
| Gene_Pert | DNMT3B K | 5/212 | 0.002333 | 0.375676 | 0 | 0 | 5.80476 | 35.18047 |
| Gene_Pert | Klf15 KO m | 6/314 | 0.002485 | 0.375676 | 0 | 0 | 4.715007 | 28.27783 |
| Gene_Pert | BMI1 KO m | 6/303 | 0.00208 | 0.375676 | 0 | 0 | 4.892381 | 30.21198 |
| DisGeNET | Central neu | 9/1655 | 0.292151 | 0.375766 | 0 | 0 | 1.280517 | 1.575657 |
| GeDiPNet_ | Nonsyndro | Jan-84 | 0.307163 | 0.37621 | 0 | 0 | 2.778089 | 3.279195 |
| GeDiPNet_ | Renal Glom | Jan-84 | 0.307163 | 0.37621 | 0 | 0 | 2.778089 | 3.279195 |
| GWAS_Cat | Macular Th | 1/100 | 0.35405 | 0.376255 | 0 | 0 | 2.327226 | 2.416399 |
| GO_Biologi | Regulation | Jan-80 | 0.294925 | 0.376612 | 0 | 0 | 2.919341 | 3.564619 |
| Jensen_TIS | B-lymphoc | 39/8270 | 0.289441 | 0.377328 | 0 | 0 | 1.153156 | 1.429686 |
| DisGeNET | Arthritis | 4/630 | 0.293764 | 0.377656 | 0 | 0 | 1.484815 | 1.818864 |
| DisGeNET | Bloom Syn | Jan-80 | 0.294925 | 0.37785 | 0 | 0 | 2.919341 | 3.564619 |
| DisGeNET | Abnormalit | Jan-80 | 0.294925 | 0.37785 | 0 | 0 | 2.919341 | 3.564619 |
| DisGeNET | Depressed | Jan-80 | 0.294925 | 0.37785 | 0 | 0 | 2.919341 | 3.564619 |
| DisGeNET | Abnormalit | Jan-80 | 0.294925 | 0.37785 | 0 | 0 | 2.919341 | 3.564619 |
| DisGeNET | Extrahepat | Jan-80 | 0.294925 | 0.37785 | 0 | 0 | 2.919341 | 3.564619 |
| DisGeNET | Paroxysma | Jan-80 | 0.294925 | 0.37785 | 0 | 0 | 2.919341 | 3.564619 |
| DisGeNET | Microdonti | Jan-80 | 0.294925 | 0.37785 | 0 | 0 | 2.919341 | 3.564619 |
| SysMyo_M | MGS166 Hu | 17-Jan | 0.071459 | 0.377998 | 0 | 0 | 14.46003 | 38.15469 |
| GeDiPNet_ | Diffuse Lyn | Jan-85 | 0.310189 | 0.378327 | 0 | 0 | 2.744878 | 3.21308 |
| GeDiPNet_ | Atheroscle | Jan-85 | 0.310189 | 0.378327 | 0 | 0 | 2.744878 | 3.21308 |
| Jensen_DIS | Neurodege | 2/293 | 0.365056 | 0.378648 | 0 | 0 | 1.586578 | 1.598802 |
| Jensen_TIS | Needle | Jan-79 | 0.291832 | 0.378911 | 0 | 0 | 2.956917 | 3.641673 |
| GWAS_Cat | Medicatior | 1/101 | 0.356874 | 0.378926 | 0 | 0 | 2.303837 | 2.373811 |
| Rare_Disea | Encephalor | Jan-61 | 0.233813 | 0.379818 | 0 | 0 | 3.847481 | 5.591291 |
| HuBMAP_A | Blood Vess | 2/123 | 0.100238 | 0.379851 | 0 | 0 | 3.848712 | 8.852824 |
| GO_Biologi | Regulation | Jan-81 | 0.298004 | 0.379861 | 0 | 0 | 2.882703 | 3.48994 |
| GTEx_Tissu | GTEX-XUW | 10/1034 | 0.014188 | 0.380042 | 0 | 0 | 2.395622 | 10.19431 |
| DisGeNET | Insulin Resi | Jan-81 | 0.298004 | 0.380864 | 0 | 0 | 2.882703 | 3.48994 |
| DisGeNET | Reduced fe | Jan-81 | 0.298004 | 0.380864 | 0 | 0 | 2.882703 | 3.48994 |
| DisGeNET | Neoplasms | Jan-81 | 0.298004 | 0.380864 | 0 | 0 | 2.882703 | 3.48994 |

| | | | | | | | | |
|---|---|---|---|---|---|---|---|---|
| DisGeNET | Congenital | Jan-81 | 0.298004 | 0.380864 | 0 | 0 | 2.882703 | 3.48994 |
| DisGeNET | Granulocyt | Jan-81 | 0.298004 | 0.380864 | 0 | 0 | 2.882703 | 3.48994 |
| DisGeNET | Patent duc | 2/251 | 0.298257 | 0.381001 | 0 | 0 | 1.858162 | 2.248005 |
| GeDiPNet_ | Autism | 4/650 | 0.313422 | 0.381472 | 0 | 0 | 1.437353 | 1.667624 |
| SysMyo_M | MGS1063 I | 2/102 | 0.072883 | 0.381959 | 0 | 0 | 4.661882 | 12.20902 |
| GTEx_Tissu | GTEX-XUZC | 9/882 | 0.014401 | 0.382209 | 0 | 0 | 2.516521 | 10.67128 |
| GWAS_Cat | Serum Uric | 2/290 | 0.36035 | 0.382285 | 0 | 0 | 1.60335 | 1.636504 |
| GTEx_Tissu | GTEX-OIZG | 10/1038 | 0.014542 | 0.382459 | 0 | 0 | 2.385795 | 10.09359 |
| Reactome_ | ADP Signali | 25-Jan | 0.103317 | 0.382905 | 0 | 0 | 9.636143 | 21.87363 |
| Reactome_ | Regulation | 20-Jan | 0.083535 | 0.382905 | 0 | 0 | 12.17503 | 30.22444 |
| Reactome_ | ADP Signali | 22-Jan | 0.091499 | 0.382905 | 0 | 0 | 11.0144 | 26.34017 |
| Reactome_ | Cap-depen | 2/116 | 0.090827 | 0.382905 | 0 | 0 | 4.086481 | 9.802643 |
| Reactome_ | Presynapti | 21-Jan | 0.087525 | 0.382905 | 0 | 0 | 11.5657 | 28.17205 |
| Reactome_ | Mitochond | 21-Jan | 0.087525 | 0.382905 | 0 | 0 | 11.5657 | 28.17205 |
| Reactome_ | Scavenging | 19-Jan | 0.079527 | 0.382905 | 0 | 0 | 12.85207 | 32.53709 |
| Reactome_ | Response T | 2/130 | 0.109909 | 0.382905 | 0 | 0 | 3.636949 | 8.030761 |
| Reactome_ | Gap Junctio | 27-Jan | 0.111111 | 0.382905 | 0 | 0 | 8.894007 | 19.54216 |
| Reactome_ | G Beta:Gar | 25-Jan | 0.103317 | 0.382905 | 0 | 0 | 9.636143 | 21.87363 |
| Reactome_ | Prostacycli | 19-Jan | 0.079527 | 0.382905 | 0 | 0 | 12.85207 | 32.53709 |
| Reactome_ | Platelet De | 2/125 | 0.102976 | 0.382905 | 0 | 0 | 3.785748 | 8.60599 |
| Reactome_ | Thromboxa | 24-Jan | 0.099394 | 0.382905 | 0 | 0 | 10.05561 | 23.215 |
| Reactome_ | Regulation | 2/123 | 0.100238 | 0.382905 | 0 | 0 | 3.848712 | 8.852824 |
| Reactome_ | Post-transl | 2/106 | 0.077881 | 0.382905 | 0 | 0 | 4.481674 | 11.43977 |
| Reactome_ | Gap Junctio | 16-Jan | 0.067399 | 0.382905 | 0 | 0 | 15.42481 | 41.60265 |
| Reactome_ | Synthesis C | 26-Jan | 0.107222 | 0.382905 | 0 | 0 | 9.250233 | 20.65441 |
| Reactome_ | WNT Ligan | 26-Jan | 0.107222 | 0.382905 | 0 | 0 | 9.250233 | 20.65441 |
| Reactome_ | G-protein A | 24-Jan | 0.099394 | 0.382905 | 0 | 0 | 10.05561 | 23.215 |
| Reactome_ | G Beta:Gar | 18-Jan | 0.075502 | 0.382905 | 0 | 0 | 13.60876 | 35.1596 |
| Reactome_ | Vitamin B5 | 17-Jan | 0.071459 | 0.382905 | 0 | 0 | 14.46003 | 38.15469 |
| Reactome_ | Effects Of F | 27-Jan | 0.111111 | 0.382905 | 0 | 0 | 8.894007 | 19.54216 |
| Reactome_ | Signaling B | 3/260 | 0.104327 | 0.382905 | 0 | 0 | 2.731518 | 6.173843 |
| Reactome_ | Hemostasis | 5/576 | 0.10618 | 0.382905 | 0 | 0 | 2.065482 | 4.632088 |
| Reactome_ | G Beta:Gar | 20-Jan | 0.083535 | 0.382905 | 0 | 0 | 12.17503 | 30.22444 |
| Reactome_ | G Beta:Gar | 20-Jan | 0.083535 | 0.382905 | 0 | 0 | 12.17503 | 30.22444 |
| Reactome_ | Gap Junctio | 25-Jan | 0.103317 | 0.382905 | 0 | 0 | 9.636143 | 21.87363 |
| GTEx_Tissu | GTEX-SIU8- | 13/1530 | 0.014719 | 0.383624 | 0 | 0 | 2.130343 | 8.987086 |
| DisGeNET | Salivary Gla | Jan-82 | 0.30107 | 0.383659 | 0 | 0 | 2.846971 | 3.417539 |
| DisGeNET | Decreased | Jan-82 | 0.30107 | 0.383659 | 0 | 0 | 2.846971 | 3.417539 |
| DisGeNET | Malignant | Jan-82 | 0.30107 | 0.383659 | 0 | 0 | 2.846971 | 3.417539 |
| DisGeNET | Hyperlipide | Jan-82 | 0.30107 | 0.383659 | 0 | 0 | 2.846971 | 3.417539 |
| DisGeNET | Non-medu | Jan-82 | 0.30107 | 0.383659 | 0 | 0 | 2.846971 | 3.417539 |
| GWAS_Cat | Granulocyt | 1/103 | 0.362485 | 0.383882 | 0 | 0 | 2.258436 | 2.291796 |
| GWAS_Cat | Sum Neutr | 1/103 | 0.362485 | 0.383882 | 0 | 0 | 2.258436 | 2.291796 |
| DisGeNET | Hydroceph | 2/253 | 0.301472 | 0.383985 | 0 | 0 | 1.843169 | 2.210102 |
| GeDiPNet_ | Oculomotc | Jan-87 | 0.316203 | 0.384055 | 0 | 0 | 2.680773 | 3.086563 |

| | | | | | | | | |
|---|---|---|---|---|---|---|---|---|
| Rare_Disea | Mycobacte | Jan-62 | 0.237156 | 0.38441 | 0 | 0 | 3.784217 | 5.445629 |
| Reactome_ | Keratan Su | 28-Jan | 0.114983 | 0.384419 | 0 | 0 | 8.564169 | 18.52408 |
| Reactome_ | Adrenaline | 28-Jan | 0.114983 | 0.384419 | 0 | 0 | 8.564169 | 18.52408 |
| GO_Biologi | Vesicle-Me | Jan-83 | 0.304123 | 0.384897 | 0 | 0 | 2.81211 | 3.34732 |
| GO_Biologi | Regulation | Jan-83 | 0.304123 | 0.384897 | 0 | 0 | 2.81211 | 3.34732 |
| GO_Biologi | Vascular Tr | Jan-83 | 0.304123 | 0.384897 | 0 | 0 | 2.81211 | 3.34732 |
| GO_Biologi | Positive Re | Jan-83 | 0.304123 | 0.384897 | 0 | 0 | 2.81211 | 3.34732 |
| Reactome_ | Signal Amp | Jan-33 | 0.134093 | 0.385089 | 0 | 0 | 7.224201 | 14.515 |
| Reactome_ | Cargo Conc | Jan-32 | 0.130304 | 0.385089 | 0 | 0 | 7.457614 | 15.19775 |
| Reactome_ | Glucagon S | Jan-32 | 0.130304 | 0.385089 | 0 | 0 | 7.457614 | 15.19775 |
| Reactome_ | Plasma Lip | Jan-33 | 0.134093 | 0.385089 | 0 | 0 | 7.224201 | 14.515 |
| Reactome_ | Keratan Su | Jan-33 | 0.134093 | 0.385089 | 0 | 0 | 7.224201 | 14.515 |
| Reactome_ | Glucagon-t | Jan-33 | 0.134093 | 0.385089 | 0 | 0 | 7.224201 | 14.515 |
| Reactome_ | Nuclear Sig | Jan-32 | 0.130304 | 0.385089 | 0 | 0 | 7.457614 | 15.19775 |
| Reactome_ | Activation | 29-Jan | 0.118838 | 0.385089 | 0 | 0 | 8.25789 | 17.58927 |
| Reactome_ | Activation | 29-Jan | 0.118838 | 0.385089 | 0 | 0 | 8.25789 | 17.58927 |
| Reactome_ | G-protein E | Jan-32 | 0.130304 | 0.385089 | 0 | 0 | 7.457614 | 15.19775 |
| Reactome_ | Thrombin S | Jan-32 | 0.130304 | 0.385089 | 0 | 0 | 7.457614 | 15.19775 |
| dbGaP | Bone Dens | Jan-73 | 0.272992 | 0.385749 | 0 | 0 | 3.204296 | 4.160181 |
| PhenGenI_ | Anti-Neutr | Jan-41 | 0.163825 | 0.38616 | 0 | 0 | 5.777035 | 10.45039 |
| GWAS_Cat | Sum Basop | 1/104 | 0.365273 | 0.386164 | 0 | 0 | 2.236396 | 2.252299 |
| GWAS_Cat | Serum Pho | 1/104 | 0.365273 | 0.386164 | 0 | 0 | 2.236396 | 2.252299 |
| PhenGenI_ | Diabetes M | 3/327 | 0.170462 | 0.386387 | 0 | 0 | 2.159281 | 3.820295 |
| PhenGenI_ | Overweigh | Jan-48 | 0.189012 | 0.386387 | 0 | 0 | 4.914894 | 8.187947 |
| PhenGenI_ | Bipolar Dis | 5/675 | 0.170703 | 0.386387 | 0 | 0 | 1.751274 | 3.095957 |
| PhenGenI_ | Abacavir | Jan-43 | 0.171099 | 0.386387 | 0 | 0 | 5.501384 | 9.712751 |
| PhenGenI_ | Binge-Eatir | Jan-48 | 0.189012 | 0.386387 | 0 | 0 | 4.914894 | 8.187947 |
| PhenGenI_ | Leukocyte | 2/176 | 0.178336 | 0.386387 | 0 | 0 | 2.669236 | 4.601995 |
| PhenGenI_ | C-Reactive | 4/492 | 0.166565 | 0.386387 | 0 | 0 | 1.918329 | 3.438356 |
| PhenGenI_ | Behavior | Jan-48 | 0.189012 | 0.386387 | 0 | 0 | 4.914894 | 8.187947 |
| PhenGenI_ | Vitamin K | Jan-47 | 0.18546 | 0.386387 | 0 | 0 | 5.021992 | 8.46163 |
| PhenGenI_ | Ghrelin | Jan-47 | 0.18546 | 0.386387 | 0 | 0 | 5.021992 | 8.46163 |
| PhenGenI_ | Rhinitis, All | Jan-47 | 0.18546 | 0.386387 | 0 | 0 | 5.021992 | 8.46163 |
| PhenGenI_ | Blood Prot | Jan-45 | 0.178311 | 0.386387 | 0 | 0 | 5.250793 | 9.053559 |
| PhenGenI_ | Leucine | Jan-44 | 0.174713 | 0.386387 | 0 | 0 | 5.373175 | 9.3741 |
| PhenGenI_ | Antibodies | Jan-44 | 0.174713 | 0.386387 | 0 | 0 | 5.373175 | 9.3741 |
| PhenGenI_ | Receptors, | Jan-46 | 0.181893 | 0.386387 | 0 | 0 | 5.13385 | 8.749803 |
| DisGeNET | Drug toxici | Jan-83 | 0.304123 | 0.386422 | 0 | 0 | 2.81211 | 3.34732 |
| DisGeNET | Protein De | Jan-83 | 0.304123 | 0.386422 | 0 | 0 | 2.81211 | 3.34732 |
| DisGeNET | Maturity o | Jan-83 | 0.304123 | 0.386422 | 0 | 0 | 2.81211 | 3.34732 |
| DisGeNET | Adverse re | Jan-83 | 0.304123 | 0.386422 | 0 | 0 | 2.81211 | 3.34732 |
| DisGeNET | Testicular I | Jan-83 | 0.304123 | 0.386422 | 0 | 0 | 2.81211 | 3.34732 |
| GeneSigDB | 19567819-: | 5/260 | 0.005554 | 0.386729 | 0 | 0 | 4.700622 | 24.41105 |
| GeDiPNet_ | Endometri | Jan-88 | 0.319191 | 0.386878 | 0 | 0 | 2.649826 | 3.026014 |
| UK_Bioban | Eosinophill | 3/471 | 0.336703 | 0.386937 | 0 | 0 | 1.483898 | 1.615303 |

| | | | | | | | | |
|---|---|---|---|---|---|---|---|---|
| Reactome_ | Signaling B | 3/294 | 0.136464 | 0.386937 | 0 | 0 | 2.408198 | 4.796389 |
| DisGeNET | Congenital | 2/255 | 0.304685 | 0.386949 | 0 | 0 | 1.828412 | 2.173024 |
| GWAS_Cat | Serum Met | 2/294 | 0.366622 | 0.387254 | 0 | 0 | 1.581064 | 1.586479 |
| CellMarker | Endothelia | Jan-93 | 0.333936 | 0.387705 | 0 | 0 | 2.505182 | 2.747701 |
| GO_Biologi | Establishm | Jan-85 | 0.310189 | 0.387736 | 0 | 0 | 2.744878 | 3.21308 |
| GO_Biologi | Negative R | Jan-85 | 0.310189 | 0.387736 | 0 | 0 | 2.744878 | 3.21308 |
| GO_Biologi | Regulation | Jan-85 | 0.310189 | 0.387736 | 0 | 0 | 2.744878 | 3.21308 |
| GO_Biologi | Transport / | Jan-85 | 0.310189 | 0.387736 | 0 | 0 | 2.744878 | 3.21308 |
| GO_Biologi | Nuclear-Tr | Jan-85 | 0.310189 | 0.387736 | 0 | 0 | 2.744878 | 3.21308 |
| GO_Biologi | Cellular Re | Jan-85 | 0.310189 | 0.387736 | 0 | 0 | 2.744878 | 3.21308 |
| GO_Biologi | Positive Re | Jan-85 | 0.310189 | 0.387736 | 0 | 0 | 2.744878 | 3.21308 |
| GWAS_Cat | Glucose Le | 1/105 | 0.368048 | 0.387754 | 0 | 0 | 2.214781 | 2.213764 |
| GWAS_Cat | Myeloid W | 1/105 | 0.368048 | 0.387754 | 0 | 0 | 2.214781 | 2.213764 |
| GWAS_Cat | Epithelial C | 1/105 | 0.368048 | 0.387754 | 0 | 0 | 2.214781 | 2.213764 |
| Jensen_DIS | Hyperglyce | 1/108 | 0.376303 | 0.388249 | 0 | 0 | 2.152358 | 2.10363 |
| HuBMAP_/ | CD8 T Cell | 1/102 | 0.359686 | 0.388491 | 0 | 0 | 2.280912 | 2.332289 |
| HuBMAP_/ | Endothelia | 1/104 | 0.365273 | 0.388491 | 0 | 0 | 2.236396 | 2.252299 |
| HuBMAP_/ | Vein Endot | 1/104 | 0.365273 | 0.388491 | 0 | 0 | 2.236396 | 2.252299 |
| HuBMAP_/ | Inh L1-6 LA | 1/104 | 0.365273 | 0.388491 | 0 | 0 | 2.236396 | 2.252299 |
| HuBMAP_/ | Plasma cell | 1/103 | 0.362485 | 0.388491 | 0 | 0 | 2.258436 | 2.291796 |
| HuBMAP_/ | Granulocyt | 1/103 | 0.362485 | 0.388491 | 0 | 0 | 2.258436 | 2.291796 |
| HuBMAP_/ | Splenic Rec | 1/103 | 0.362485 | 0.388491 | 0 | 0 | 2.258436 | 2.291796 |
| HuBMAP_/ | Intercalary | 1/103 | 0.362485 | 0.388491 | 0 | 0 | 2.258436 | 2.291796 |
| HuBMAP_/ | Intercalary | 1/103 | 0.362485 | 0.388491 | 0 | 0 | 2.258436 | 2.291796 |
| HuBMAP_/ | Capillary Er | 1/103 | 0.362485 | 0.388491 | 0 | 0 | 2.258436 | 2.291796 |
| HuBMAP_/ | Inh L1 LAM | 1/102 | 0.359686 | 0.388491 | 0 | 0 | 2.280912 | 2.332289 |
| HuBMAP_/ | Vascular As | 1/102 | 0.359686 | 0.388491 | 0 | 0 | 2.280912 | 2.332289 |
| HuBMAP_/ | Endothelia | 1/102 | 0.359686 | 0.388491 | 0 | 0 | 2.280912 | 2.332289 |
| HuBMAP_/ | Perineural | 1/102 | 0.359686 | 0.388491 | 0 | 0 | 2.280912 | 2.332289 |
| HuBMAP_/ | Perivascula | 1/102 | 0.359686 | 0.388491 | 0 | 0 | 2.280912 | 2.332289 |
| HuBMAP_/ | lymphocyte | 1/102 | 0.359686 | 0.388491 | 0 | 0 | 2.280912 | 2.332289 |
| HuBMAP_/ | Naive Thyn | 1/104 | 0.365273 | 0.388491 | 0 | 0 | 2.236396 | 2.252299 |
| HuBMAP_/ | Vascular Sr | 1/102 | 0.359686 | 0.388491 | 0 | 0 | 2.280912 | 2.332289 |
| HuBMAP_/ | Monocyte- | 1/102 | 0.359686 | 0.388491 | 0 | 0 | 2.280912 | 2.332289 |
| HuBMAP_/ | Cardiac Pac | 1/102 | 0.359686 | 0.388491 | 0 | 0 | 2.280912 | 2.332289 |
| HuBMAP_/ | Telocyte (Ir | 1/102 | 0.359686 | 0.388491 | 0 | 0 | 2.280912 | 2.332289 |
| HuBMAP_/ | Heart Valve | 1/102 | 0.359686 | 0.388491 | 0 | 0 | 2.280912 | 2.332289 |
| HuBMAP_/ | Mast cell - | 1/102 | 0.359686 | 0.388491 | 0 | 0 | 2.280912 | 2.332289 |
| HuBMAP_/ | Sympathet | 1/102 | 0.359686 | 0.388491 | 0 | 0 | 2.280912 | 2.332289 |
| HuBMAP_/ | Inh L1 LAM | 1/102 | 0.359686 | 0.388491 | 0 | 0 | 2.280912 | 2.332289 |
| HuBMAP_/ | Exc L5 FEZF | 1/103 | 0.362485 | 0.388491 | 0 | 0 | 2.258436 | 2.291796 |
| HuBMAP_/ | Mononucle | 1/103 | 0.362485 | 0.388491 | 0 | 0 | 2.258436 | 2.291796 |
| HuBMAP_/ | Inh L2-5 PV | 1/103 | 0.362485 | 0.388491 | 0 | 0 | 2.258436 | 2.291796 |
| HuBMAP_/ | Exc L5 ROR | 1/103 | 0.362485 | 0.388491 | 0 | 0 | 2.258436 | 2.291796 |
| HuBMAP_/ | Inh L1-6 VI | 1/102 | 0.359686 | 0.388491 | 0 | 0 | 2.280912 | 2.332289 |

| | | | | | | | | |
|---|---|---|---|---|---|---|---|---|
| HuBMAP_/ | Thymic Inte | 1/108 | 0.376303 | 0.388491 | 0 | 0 | 2.152358 | 2.10363 |
| HuBMAP_/ | enterocyte | 1/104 | 0.365273 | 0.388491 | 0 | 0 | 2.236396 | 2.252299 |
| HuBMAP_/ | Erythroid L | 1/104 | 0.365273 | 0.388491 | 0 | 0 | 2.236396 | 2.252299 |
| HuBMAP_/ | CD1c-posit | 1/104 | 0.365273 | 0.388491 | 0 | 0 | 2.236396 | 2.252299 |
| HuBMAP_/ | Inh L5-6 SS | 1/104 | 0.365273 | 0.388491 | 0 | 0 | 2.236396 | 2.252299 |
| HuBMAP_/ | Double Neg | 1/107 | 0.373563 | 0.388491 | 0 | 0 | 2.172773 | 2.139459 |
| HuBMAP_/ | Epithelial S | 1/105 | 0.368048 | 0.388491 | 0 | 0 | 2.214781 | 2.213764 |
| HuBMAP_/ | Hepatic Ov | 1/105 | 0.368048 | 0.388491 | 0 | 0 | 2.214781 | 2.213764 |
| HuBMAP_/ | Venous End | 1/104 | 0.365273 | 0.388491 | 0 | 0 | 2.236396 | 2.252299 |
| HuBMAP_/ | CAP2 - Resp | 1/103 | 0.362485 | 0.388491 | 0 | 0 | 2.258436 | 2.291796 |
| HuBMAP_/ | Peritubular | 1/103 | 0.362485 | 0.388491 | 0 | 0 | 2.258436 | 2.291796 |
| HuBMAP_/ | Exc L5 THEI | 1/103 | 0.362485 | 0.388491 | 0 | 0 | 2.258436 | 2.291796 |
| HuBMAP_/ | Exc L5 THEI | 1/103 | 0.362485 | 0.388491 | 0 | 0 | 2.258436 | 2.291796 |
| HuBMAP_/ | Lens Fiber | 1/102 | 0.359686 | 0.388491 | 0 | 0 | 2.280912 | 2.332289 |
| HuBMAP_/ | macrophag | 1/103 | 0.362485 | 0.388491 | 0 | 0 | 2.258436 | 2.291796 |
| HuBMAP_/ | Fat cell - Pa | 1/103 | 0.362485 | 0.388491 | 0 | 0 | 2.258436 | 2.291796 |
| HuBMAP_/ | Naive Thym | 1/104 | 0.365273 | 0.388491 | 0 | 0 | 2.236396 | 2.252299 |
| HuBMAP_/ | Medullary | 1/108 | 0.376303 | 0.388491 | 0 | 0 | 2.152358 | 2.10363 |
| HuBMAP_/ | Exc L2-3 RC | 1/104 | 0.365273 | 0.388491 | 0 | 0 | 2.236396 | 2.252299 |
| HuBMAP_/ | Exc L3 THEI | 1/104 | 0.365273 | 0.388491 | 0 | 0 | 2.236396 | 2.252299 |
| HuBMAP_/ | Exc L5-6 FE | 1/104 | 0.365273 | 0.388491 | 0 | 0 | 2.236396 | 2.252299 |
| HuBMAP_/ | Retinal Gar | 1/104 | 0.365273 | 0.388491 | 0 | 0 | 2.236396 | 2.252299 |
| HuBMAP_/ | Stem cell - | 1/105 | 0.368048 | 0.388491 | 0 | 0 | 2.214781 | 2.213764 |
| HuBMAP_/ | Tracheobro | 1/104 | 0.365273 | 0.388491 | 0 | 0 | 2.236396 | 2.252299 |
| HuBMAP_/ | Interstitial | 1/104 | 0.365273 | 0.388491 | 0 | 0 | 2.236396 | 2.252299 |
| HuBMAP_/ | Kidney Inte | 1/104 | 0.365273 | 0.388491 | 0 | 0 | 2.236396 | 2.252299 |
| HuBMAP_/ | keratinocyt | 1/104 | 0.365273 | 0.388491 | 0 | 0 | 2.236396 | 2.252299 |
| HuBMAP_/ | Mast cell - | 1/104 | 0.365273 | 0.388491 | 0 | 0 | 2.236396 | 2.252299 |
| HuBMAP_/ | CD14-posit | 1/102 | 0.359686 | 0.388491 | 0 | 0 | 2.280912 | 2.332289 |
| HuBMAP_/ | Hematopoi | 1/103 | 0.362485 | 0.388491 | 0 | 0 | 2.258436 | 2.291796 |
| HuBMAP_/ | CD14-posit | 1/103 | 0.362485 | 0.388491 | 0 | 0 | 2.258436 | 2.291796 |
| HuBMAP_/ | CD16-nega | 1/103 | 0.362485 | 0.388491 | 0 | 0 | 2.258436 | 2.291796 |
| HuBMAP_/ | Endothelia | 1/102 | 0.359686 | 0.388491 | 0 | 0 | 2.280912 | 2.332289 |
| HuBMAP_/ | Astro L1-6 | 1/102 | 0.359686 | 0.388491 | 0 | 0 | 2.280912 | 2.332289 |
| HuBMAP_/ | VLMC L1-5 | 1/102 | 0.359686 | 0.388491 | 0 | 0 | 2.280912 | 2.332289 |
| HuBMAP_/ | endoethlia | 1/102 | 0.359686 | 0.388491 | 0 | 0 | 2.280912 | 2.332289 |
| HuBMAP_/ | hepatocyte | 1/102 | 0.359686 | 0.388491 | 0 | 0 | 2.280912 | 2.332289 |
| HuBMAP_/ | Periportal I | 1/102 | 0.359686 | 0.388491 | 0 | 0 | 2.280912 | 2.332289 |
| HuBMAP_/ | Vein Endot | 1/102 | 0.359686 | 0.388491 | 0 | 0 | 2.280912 | 2.332289 |
| HuBMAP_/ | neutrophil | 1/100 | 0.35405 | 0.388491 | 0 | 0 | 2.327226 | 2.416399 |
| HuBMAP_/ | Endothelia | 1/103 | 0.362485 | 0.388491 | 0 | 0 | 2.258436 | 2.291796 |
| HuBMAP_/ | Inh L5-6 PV | 1/103 | 0.362485 | 0.388491 | 0 | 0 | 2.258436 | 2.291796 |
| HuBMAP_/ | Inh L5-6 PV | 1/103 | 0.362485 | 0.388491 | 0 | 0 | 2.258436 | 2.291796 |
| HuBMAP_/ | Cytotoxic T | 1/102 | 0.359686 | 0.388491 | 0 | 0 | 2.280912 | 2.332289 |
| HuBMAP_/ | Exc L5 FEZF | 1/102 | 0.359686 | 0.388491 | 0 | 0 | 2.280912 | 2.332289 |

| | | | | | | | | |
|---|---|---|---|---|---|---|---|---|
| HuBMAP_A | B Cell (Inclu | 1/102 | 0.359686 | 0.388491 | 0 | 0 | 2.280912 | 2.332289 |
| HuBMAP_A | Naive Thym | 1/102 | 0.359686 | 0.388491 | 0 | 0 | 2.280912 | 2.332289 |
| HuBMAP_A | Inh L5-6 SS | 1/102 | 0.359686 | 0.388491 | 0 | 0 | 2.280912 | 2.332289 |
| HuBMAP_A | Oligo L2-6 ( | 1/102 | 0.359686 | 0.388491 | 0 | 0 | 2.280912 | 2.332289 |
| HuBMAP_A | Inh L6 SST ' | 1/102 | 0.359686 | 0.388491 | 0 | 0 | 2.280912 | 2.332289 |
| HuBMAP_A | Connective | 1/102 | 0.359686 | 0.388491 | 0 | 0 | 2.280912 | 2.332289 |
| HuBMAP_A | macrophag | 1/102 | 0.359686 | 0.388491 | 0 | 0 | 2.280912 | 2.332289 |
| HuBMAP_A | Gastrin Sec | 1/101 | 0.356874 | 0.388491 | 0 | 0 | 2.303837 | 2.373811 |
| HuBMAP_A | Invaginatin | 1/101 | 0.356874 | 0.388491 | 0 | 0 | 2.303837 | 2.373811 |
| HuBMAP_A | Dendritic c | 1/101 | 0.356874 | 0.388491 | 0 | 0 | 2.303837 | 2.373811 |
| HuBMAP_A | megakaryo | 1/101 | 0.356874 | 0.388491 | 0 | 0 | 2.303837 | 2.373811 |
| HuBMAP_A | Endothelia | 1/101 | 0.356874 | 0.388491 | 0 | 0 | 2.303837 | 2.373811 |
| HuBMAP_A | Mucosal Ty | 1/101 | 0.356874 | 0.388491 | 0 | 0 | 2.303837 | 2.373811 |
| HuBMAP_A | Kidney Coll | 1/101 | 0.356874 | 0.388491 | 0 | 0 | 2.303837 | 2.373811 |
| HuBMAP_A | Adult Endo | 1/101 | 0.356874 | 0.388491 | 0 | 0 | 2.303837 | 2.373811 |
| HuBMAP_A | Kidney Loo | 1/101 | 0.356874 | 0.388491 | 0 | 0 | 2.303837 | 2.373811 |
| HuBMAP_A | Lung Neuro | 1/101 | 0.356874 | 0.388491 | 0 | 0 | 2.303837 | 2.373811 |
| HuBMAP_A | B cell - Sple | 1/100 | 0.35405 | 0.388491 | 0 | 0 | 2.327226 | 2.416399 |
| HuBMAP_A | Skin fibrob | 1/103 | 0.362485 | 0.388491 | 0 | 0 | 2.258436 | 2.291796 |
| HuBMAP_A | Naive B cel | 1/108 | 0.376303 | 0.388491 | 0 | 0 | 2.152358 | 2.10363 |
| HuBMAP_A | CD141-pos | 1/108 | 0.376303 | 0.388491 | 0 | 0 | 2.152358 | 2.10363 |
| HuBMAP_A | Hematopoi | 1/109 | 0.379031 | 0.388491 | 0 | 0 | 2.132321 | 2.068645 |
| HuBMAP_A | Endothelia | 1/102 | 0.359686 | 0.388491 | 0 | 0 | 2.280912 | 2.332289 |
| HuBMAP_A | Endometria | 1/102 | 0.359686 | 0.388491 | 0 | 0 | 2.280912 | 2.332289 |
| HuBMAP_A | Exc L6 FEZI | 1/102 | 0.359686 | 0.388491 | 0 | 0 | 2.280912 | 2.332289 |
| HuBMAP_A | CD141-pos | 1/103 | 0.362485 | 0.388491 | 0 | 0 | 2.258436 | 2.291796 |
| HuBMAP_A | CD14-posit | 1/103 | 0.362485 | 0.388491 | 0 | 0 | 2.258436 | 2.291796 |
| HuBMAP_A | Serous Cell | 1/103 | 0.362485 | 0.388491 | 0 | 0 | 2.258436 | 2.291796 |
| HuBMAP_A | Myofibrobl | 1/103 | 0.362485 | 0.388491 | 0 | 0 | 2.258436 | 2.291796 |
| HuBMAP_A | Pericyte ce | 1/103 | 0.362485 | 0.388491 | 0 | 0 | 2.258436 | 2.291796 |
| HuBMAP_A | Parietal Ep | 1/102 | 0.359686 | 0.388491 | 0 | 0 | 2.280912 | 2.332289 |
| HuBMAP_A | Blood Vess | 1/108 | 0.376303 | 0.388491 | 0 | 0 | 2.152358 | 2.10363 |
| HuBMAP_A | Oligo L5-6 ( | 1/103 | 0.362485 | 0.388491 | 0 | 0 | 2.258436 | 2.291796 |
| HuBMAP_A | Superficial | 1/103 | 0.362485 | 0.388491 | 0 | 0 | 2.258436 | 2.291796 |
| HuBMAP_A | Extravillous | 1/103 | 0.362485 | 0.388491 | 0 | 0 | 2.258436 | 2.291796 |
| HuBMAP_A | Mast cell - | 1/103 | 0.362485 | 0.388491 | 0 | 0 | 2.258436 | 2.291796 |
| HuBMAP_A | Immature ( | 1/103 | 0.362485 | 0.388491 | 0 | 0 | 2.258436 | 2.291796 |
| HuBMAP_A | Lipofibrobl | 1/102 | 0.359686 | 0.388491 | 0 | 0 | 2.280912 | 2.332289 |
| HuBMAP_A | Pericyte Ce | 1/102 | 0.359686 | 0.388491 | 0 | 0 | 2.280912 | 2.332289 |
| HuBMAP_A | Blue Cone I | 1/101 | 0.356874 | 0.388491 | 0 | 0 | 2.303837 | 2.373811 |
| HuBMAP_A | Diffuse Bip | 1/101 | 0.356874 | 0.388491 | 0 | 0 | 2.303837 | 2.373811 |
| HuBMAP_A | Smooth Mu | 1/102 | 0.359686 | 0.388491 | 0 | 0 | 2.280912 | 2.332289 |
| HuBMAP_A | Pulmonary | 1/102 | 0.359686 | 0.388491 | 0 | 0 | 2.280912 | 2.332289 |
| HuBMAP_A | Endothelia | 1/102 | 0.359686 | 0.388491 | 0 | 0 | 2.280912 | 2.332289 |
| HuBMAP_A | Inh L5-6 PV | 1/103 | 0.362485 | 0.388491 | 0 | 0 | 2.258436 | 2.291796 |

| | | | | | | | | |
|---|---|---|---|---|---|---|---|---|
| HuBMAP_A | Exc L2-3 RC | 1/103 | 0.362485 | 0.388491 | 0 | 0 | 2.258436 | 2.291796 |
| HuBMAP_A | Exc L3-5 RC | 1/103 | 0.362485 | 0.388491 | 0 | 0 | 2.258436 | 2.291796 |
| HuBMAP_A | Exc L3-5 RC | 1/103 | 0.362485 | 0.388491 | 0 | 0 | 2.258436 | 2.291796 |
| HuBMAP_A | Hofbauer c | 1/102 | 0.359686 | 0.388491 | 0 | 0 | 2.280912 | 2.332289 |
| HuBMAP_A | Inh L5-6 SS | 1/103 | 0.362485 | 0.388491 | 0 | 0 | 2.258436 | 2.291796 |
| HuBMAP_A | Capillary La | 1/100 | 0.35405 | 0.388491 | 0 | 0 | 2.327226 | 2.416399 |
| HuBMAP_A | neutrophil | 1/100 | 0.35405 | 0.388491 | 0 | 0 | 2.327226 | 2.416399 |
| HuBMAP_A | Perineurial | 1/102 | 0.359686 | 0.388491 | 0 | 0 | 2.280912 | 2.332289 |
| HuBMAP_A | Pericyte ce | 1/103 | 0.362485 | 0.388491 | 0 | 0 | 2.258436 | 2.291796 |
| HuBMAP_A | Monocytoi | 1/105 | 0.368048 | 0.388491 | 0 | 0 | 2.214781 | 2.213764 |
| HuBMAP_A | Endothelia | 1/102 | 0.359686 | 0.388491 | 0 | 0 | 2.280912 | 2.332289 |
| HuBMAP_A | Mesothelia | 1/102 | 0.359686 | 0.388491 | 0 | 0 | 2.280912 | 2.332289 |
| HuBMAP_A | Inh L3-6 VII | 1/103 | 0.362485 | 0.388491 | 0 | 0 | 2.258436 | 2.291796 |
| HuBMAP_A | Inh L1 PAX( | 1/103 | 0.362485 | 0.388491 | 0 | 0 | 2.258436 | 2.291796 |
| HuBMAP_A | Inh L3-5 SS | 1/103 | 0.362485 | 0.388491 | 0 | 0 | 2.258436 | 2.291796 |
| HuBMAP_A | B cell - Sma | 1/103 | 0.362485 | 0.388491 | 0 | 0 | 2.258436 | 2.291796 |
| HuBMAP_A | fibroblasts | 1/100 | 0.35405 | 0.388491 | 0 | 0 | 2.327226 | 2.416399 |
| HuBMAP_A | Tuft cells - | 1/103 | 0.362485 | 0.388491 | 0 | 0 | 2.258436 | 2.291796 |
| HuBMAP_A | Stratified C | 1/102 | 0.359686 | 0.388491 | 0 | 0 | 2.280912 | 2.332289 |
| HuBMAP_A | CD1c-posit | 1/110 | 0.381747 | 0.388491 | 0 | 0 | 2.112652 | 2.034479 |
| HuBMAP_A | CD14-low, | 1/106 | 0.370812 | 0.388491 | 0 | 0 | 2.193577 | 2.176161 |
| HuBMAP_A | neutrophil | 1/106 | 0.370812 | 0.388491 | 0 | 0 | 2.193577 | 2.176161 |
| HuBMAP_A | Myo-Medu | 1/110 | 0.381747 | 0.388491 | 0 | 0 | 2.112652 | 2.034479 |
| HuBMAP_A | Plasma cell | 1/110 | 0.381747 | 0.388491 | 0 | 0 | 2.112652 | 2.034479 |
| HuBMAP_A | Endothelia | 1/103 | 0.362485 | 0.388491 | 0 | 0 | 2.258436 | 2.291796 |
| HuBMAP_A | plasmablas | 1/109 | 0.379031 | 0.388491 | 0 | 0 | 2.132321 | 2.068645 |
| HuBMAP_A | Hematopoi | 1/106 | 0.370812 | 0.388491 | 0 | 0 | 2.193577 | 2.176161 |
| HuBMAP_A | Semilunar | 1/101 | 0.356874 | 0.388491 | 0 | 0 | 2.303837 | 2.373811 |
| HuBMAP_A | Endothelia | 1/103 | 0.362485 | 0.388491 | 0 | 0 | 2.258436 | 2.291796 |
| HuBMAP_A | Schwann c | 1/101 | 0.356874 | 0.388491 | 0 | 0 | 2.303837 | 2.373811 |
| HuBMAP_A | Inh L1-5 VII | 1/102 | 0.359686 | 0.388491 | 0 | 0 | 2.280912 | 2.332289 |
| HuBMAP_A | basophil - E | 1/101 | 0.356874 | 0.388491 | 0 | 0 | 2.303837 | 2.373811 |
| HuBMAP_A | Secretory c | 1/101 | 0.356874 | 0.388491 | 0 | 0 | 2.303837 | 2.373811 |
| HuBMAP_A | myelocyte | 1/101 | 0.356874 | 0.388491 | 0 | 0 | 2.303837 | 2.373811 |
| HuBMAP_A | Detrusor Sı | 1/101 | 0.356874 | 0.388491 | 0 | 0 | 2.303837 | 2.373811 |
| HuBMAP_A | B cell - Fall | 1/101 | 0.356874 | 0.388491 | 0 | 0 | 2.303837 | 2.373811 |
| HuBMAP_A | Perineural | 1/101 | 0.356874 | 0.388491 | 0 | 0 | 2.303837 | 2.373811 |
| HuBMAP_A | Skeletal Mu | 1/101 | 0.356874 | 0.388491 | 0 | 0 | 2.303837 | 2.373811 |
| HuBMAP_A | macrophag | 1/101 | 0.356874 | 0.388491 | 0 | 0 | 2.303837 | 2.373811 |
| HuBMAP_A | Tuft cell - S | 1/103 | 0.362485 | 0.388491 | 0 | 0 | 2.258436 | 2.291796 |
| HuBMAP_A | Inh L1-6 SS | 1/101 | 0.356874 | 0.388491 | 0 | 0 | 2.303837 | 2.373811 |
| HuBMAP_A | macrophag | 1/101 | 0.356874 | 0.388491 | 0 | 0 | 2.303837 | 2.373811 |
| HuBMAP_A | Quiescent | 1/101 | 0.356874 | 0.388491 | 0 | 0 | 2.303837 | 2.373811 |
| HuBMAP_A | Secondary | 1/101 | 0.356874 | 0.388491 | 0 | 0 | 2.303837 | 2.373811 |
| HuBMAP_A | Corneal En | 1/101 | 0.356874 | 0.388491 | 0 | 0 | 2.303837 | 2.373811 |

| | | | | | | | | |
|---|---|---|---|---|---|---|---|---|
| HuBMAP_A | Dense-Core | 1/101 | 0.356874 | 0.388491 | 0 | 0 | 2.303837 | 2.373811 |
| HuBMAP_A | eosinophil | 1/101 | 0.356874 | 0.388491 | 0 | 0 | 2.303837 | 2.373811 |
| HuBMAP_A | promyeloc | 1/100 | 0.35405 | 0.388491 | 0 | 0 | 2.327226 | 2.416399 |
| HuBMAP_A | Uterine NK | 1/102 | 0.359686 | 0.388491 | 0 | 0 | 2.280912 | 2.332289 |
| HuBMAP_A | fibroblast - | 1/100 | 0.35405 | 0.388491 | 0 | 0 | 2.327226 | 2.416399 |
| HuBMAP_A | Common L | 1/106 | 0.370812 | 0.388491 | 0 | 0 | 2.193577 | 2.176161 |
| HuBMAP_A | Descending | 1/106 | 0.370812 | 0.388491 | 0 | 0 | 2.193577 | 2.176161 |
| HuBMAP_A | Kidney Loo | 1/106 | 0.370812 | 0.388491 | 0 | 0 | 2.193577 | 2.176161 |
| HuBMAP_A | Myoepithe | 1/106 | 0.370812 | 0.388491 | 0 | 0 | 2.193577 | 2.176161 |
| HuBMAP_A | Inh L1 PVA | 1/103 | 0.362485 | 0.388491 | 0 | 0 | 2.258436 | 2.291796 |
| HuBMAP_A | Thymic ma | 1/106 | 0.370812 | 0.388491 | 0 | 0 | 2.193577 | 2.176161 |
| HuBMAP_A | Thymic Cor | 1/106 | 0.370812 | 0.388491 | 0 | 0 | 2.193577 | 2.176161 |
| HuBMAP_A | Endothelia | 1/103 | 0.362485 | 0.388491 | 0 | 0 | 2.258436 | 2.291796 |
| HuBMAP_A | Endothelia | 1/104 | 0.365273 | 0.388491 | 0 | 0 | 2.236396 | 2.252299 |
| HuBMAP_A | Corneal Ep | 1/104 | 0.365273 | 0.388491 | 0 | 0 | 2.236396 | 2.252299 |
| HuBMAP_A | monocyte - | 1/107 | 0.373563 | 0.388491 | 0 | 0 | 2.172773 | 2.139459 |
| HuBMAP_A | Centroblas | 1/107 | 0.373563 | 0.388491 | 0 | 0 | 2.172773 | 2.139459 |
| HuBMAP_A | Lymph Noc | 1/107 | 0.373563 | 0.388491 | 0 | 0 | 2.172773 | 2.139459 |
| HuBMAP_A | CD8 T Cell | 1/106 | 0.370812 | 0.388491 | 0 | 0 | 2.193577 | 2.176161 |
| HuBMAP_A | Megakaryc | 1/106 | 0.370812 | 0.388491 | 0 | 0 | 2.193577 | 2.176161 |
| HuBMAP_A | Thymic Cap | 1/107 | 0.373563 | 0.388491 | 0 | 0 | 2.172773 | 2.139459 |
| HuBMAP_A | CD1c-posit | 1/106 | 0.370812 | 0.388491 | 0 | 0 | 2.193577 | 2.176161 |
| HuBMAP_A | Centrocyte | 1/106 | 0.370812 | 0.388491 | 0 | 0 | 2.193577 | 2.176161 |
| HuBMAP_A | Lymph Noc | 1/106 | 0.370812 | 0.388491 | 0 | 0 | 2.193577 | 2.176161 |
| HuBMAP_A | Keratinocy | 1/103 | 0.362485 | 0.388491 | 0 | 0 | 2.258436 | 2.291796 |
| HuBMAP_A | Innate Lym | 1/106 | 0.370812 | 0.388491 | 0 | 0 | 2.193577 | 2.176161 |
| HuBMAP_A | Kidney Res | 1/103 | 0.362485 | 0.388491 | 0 | 0 | 2.258436 | 2.291796 |
| HuBMAP_A | Double Neg | 1/106 | 0.370812 | 0.388491 | 0 | 0 | 2.193577 | 2.176161 |
| HuBMAP_A | Early T Line | 1/106 | 0.370812 | 0.388491 | 0 | 0 | 2.193577 | 2.176161 |
| HuBMAP_A | macrophag | 1/106 | 0.370812 | 0.388491 | 0 | 0 | 2.193577 | 2.176161 |
| HuBMAP_A | Medullary | 1/106 | 0.370812 | 0.388491 | 0 | 0 | 2.193577 | 2.176161 |
| HuBMAP_A | Glomerular | 1/105 | 0.368048 | 0.388491 | 0 | 0 | 2.214781 | 2.213764 |
| HuBMAP_A | neuronal - | 1/103 | 0.362485 | 0.388491 | 0 | 0 | 2.258436 | 2.291796 |
| HuBMAP_A | macrophag | 1/103 | 0.362485 | 0.388491 | 0 | 0 | 2.258436 | 2.291796 |
| HuBMAP_A | Vein Endot | 1/103 | 0.362485 | 0.388491 | 0 | 0 | 2.258436 | 2.291796 |
| HuBMAP_A | Descending | 1/105 | 0.368048 | 0.388491 | 0 | 0 | 2.214781 | 2.213764 |
| HuBMAP_A | CD8-positiv | 1/105 | 0.368048 | 0.388491 | 0 | 0 | 2.214781 | 2.213764 |
| HuBMAP_A | CD4-positiv | 1/105 | 0.368048 | 0.388491 | 0 | 0 | 2.214781 | 2.213764 |
| HuBMAP_A | enterocyte | 1/104 | 0.365273 | 0.388491 | 0 | 0 | 2.236396 | 2.252299 |
| HuBMAP_A | Dendritic C | 1/106 | 0.370812 | 0.388491 | 0 | 0 | 2.193577 | 2.176161 |
| HuBMAP_A | Glomerular | 1/106 | 0.370812 | 0.388491 | 0 | 0 | 2.193577 | 2.176161 |
| HuBMAP_A | Thymic Me | 1/106 | 0.370812 | 0.388491 | 0 | 0 | 2.193577 | 2.176161 |
| GO_Molecu | PDZ Domai | Jan-62 | 0.237156 | 0.38878 | 0 | 0 | 3.784217 | 5.445629 |
| GO_Molecu | UDP-glycos | Jan-61 | 0.233813 | 0.38878 | 0 | 0 | 3.847481 | 5.591291 |
| GO_Molecu | SH3 Domai | Jan-59 | 0.227083 | 0.38878 | 0 | 0 | 3.980553 | 5.900928 |

| | | | | | | | | |
|---|---|---|---|---|---|---|---|---|
| GO_Molecı | Carboxylic | Jan-59 | 0.227083 | 0.38878 | 0 | 0 | 3.980553 | 5.900928 |
| GO_Molecı | Nucleosom | Jan-62 | 0.237156 | 0.38878 | 0 | 0 | 3.784217 | 5.445629 |
| DisGeNET | Skin Neopl | 2/256 | 0.306291 | 0.3888 | 0 | 0 | 1.821121 | 2.154787 |
| DisGeNET | Short philtı | Jan-84 | 0.307163 | 0.389152 | 0 | 0 | 2.778089 | 3.279195 |
| DisGeNET | Soft Tissue | Jan-84 | 0.307163 | 0.389152 | 0 | 0 | 2.778089 | 3.279195 |
| DisGeNET | Late tooth | Jan-84 | 0.307163 | 0.389152 | 0 | 0 | 2.778089 | 3.279195 |
| DisGeNET | Mitochond | Jan-84 | 0.307163 | 0.389152 | 0 | 0 | 2.778089 | 3.279195 |
| CellMarker | Paneth Cel | 2/276 | 0.338237 | 0.389399 | 0 | 0 | 1.686475 | 1.828152 |
| CellMarker | Macrophag | Jan-94 | 0.336847 | 0.389399 | 0 | 0 | 2.47812 | 2.69651 |
| CellMarker | Primitive E | Jan-95 | 0.339745 | 0.389498 | 0 | 0 | 2.451633 | 2.646685 |
| DisGeNET | Neuroendc | 2/257 | 0.307896 | 0.389704 | 0 | 0 | 1.813887 | 2.136747 |
| DisGeNET | Urothelial ( | 2/257 | 0.307896 | 0.389704 | 0 | 0 | 1.813887 | 2.136747 |
| GTEx_Tissu | GTEX-UPK5 | 9/889 | 0.015092 | 0.389842 | 0 | 0 | 2.495586 | 10.4654 |
| HuBMAP_A | Splenic Litt | 1/112 | 0.387144 | 0.389851 | 0 | 0 | 2.074377 | 1.968499 |
| HuBMAP_A | Splenic Strı | 1/112 | 0.387144 | 0.389851 | 0 | 0 | 2.074377 | 1.968499 |
| HuBMAP_A | Transitiona | 1/112 | 0.387144 | 0.389851 | 0 | 0 | 2.074377 | 1.968499 |
| PhenGenI_ | Insulin-Like | Jan-49 | 0.192548 | 0.390163 | 0 | 0 | 4.812258 | 7.927759 |
| DisGeNET | Syndactyly | Jan-85 | 0.310189 | 0.390719 | 0 | 0 | 2.744878 | 3.21308 |
| DisGeNET | Prominent | Jan-85 | 0.310189 | 0.390719 | 0 | 0 | 2.744878 | 3.21308 |
| DisGeNET | Downturne | Jan-85 | 0.310189 | 0.390719 | 0 | 0 | 2.744878 | 3.21308 |
| DisGeNET | Primary ph | Jan-85 | 0.310189 | 0.390719 | 0 | 0 | 2.744878 | 3.21308 |
| DisGeNET | Convex nas | Jan-85 | 0.310189 | 0.390719 | 0 | 0 | 2.744878 | 3.21308 |
| DisGeNET | Hurthle Cel | Jan-85 | 0.310189 | 0.390719 | 0 | 0 | 2.744878 | 3.21308 |
| DisGeNET | Abnormalit | Jan-85 | 0.310189 | 0.390719 | 0 | 0 | 2.744878 | 3.21308 |
| DisGeNET | Hip Disloca | Jan-85 | 0.310189 | 0.390719 | 0 | 0 | 2.744878 | 3.21308 |
| DisGeNET | Severe Acu | Jan-85 | 0.310189 | 0.390719 | 0 | 0 | 2.744878 | 3.21308 |
| DisGeNET | Cicatrix, Hy | Jan-85 | 0.310189 | 0.390719 | 0 | 0 | 2.744878 | 3.21308 |
| GO_Biologi | Negative R | Jan-86 | 0.313203 | 0.390815 | 0 | 0 | 2.712449 | 3.148895 |
| Human_Ph | Macrotia (I | 1/105 | 0.368048 | 0.391628 | 0 | 0 | 2.214781 | 2.213764 |
| Human_Ph | Migraine (I | 1/105 | 0.368048 | 0.391628 | 0 | 0 | 2.214781 | 2.213764 |
| SysMyo_M | MGS103 M | 18-Jan | 0.075502 | 0.392054 | 0 | 0 | 13.60876 | 35.1596 |
| Jensen_TIS | Bladder | 3/443 | 0.303295 | 0.392213 | 0 | 0 | 1.580601 | 1.885733 |
| PhenGenI_ | Bronchodil | 7/1095 | 0.198766 | 0.392435 | 0 | 0 | 1.513959 | 2.445995 |
| PhenGenI_ | Anthropon | 2/188 | 0.197153 | 0.392435 | 0 | 0 | 2.495509 | 4.05215 |
| PhenGenI_ | Follicle Stir | Jan-50 | 0.196069 | 0.392435 | 0 | 0 | 4.713811 | 7.680157 |
| GeDiPNet_ | Ambiguous | Jan-90 | 0.325127 | 0.392441 | 0 | 0 | 2.590018 | 2.909988 |
| GeDiPNet_ | Thyroid Ca | Jan-90 | 0.325127 | 0.392441 | 0 | 0 | 2.590018 | 2.909988 |
| DisGeNET | Malaria, Ce | Jan-86 | 0.313203 | 0.392627 | 0 | 0 | 2.712449 | 3.148895 |
| DisGeNET | Postnatal g | Jan-86 | 0.313203 | 0.392627 | 0 | 0 | 2.712449 | 3.148895 |
| DisGeNET | Diabetic Ne | Jan-86 | 0.313203 | 0.392627 | 0 | 0 | 2.712449 | 3.148895 |
| DisGeNET | Curvature ( | Jan-86 | 0.313203 | 0.392627 | 0 | 0 | 2.712449 | 3.148895 |
| DisGeNET | Leishmania | Jan-86 | 0.313203 | 0.392627 | 0 | 0 | 2.712449 | 3.148895 |
| DisGeNET | Cystadenoı | Jan-86 | 0.313203 | 0.392627 | 0 | 0 | 2.712449 | 3.148895 |
| DisGeNET | Lymphoma | 2/260 | 0.312707 | 0.392627 | 0 | 0 | 1.792522 | 2.083787 |
| DisGeNET | Hearing Lo: | 2/260 | 0.312707 | 0.392627 | 0 | 0 | 1.792522 | 2.083787 |

| | | | | | | | | |
|---|---|---|---|---|---|---|---|---|
| DisGeNET | Muscular h | Jan-86 | 0.313203 | 0.392627 | 0 | 0 | 2.712449 | 3.148895 |
| DisGeNET | Acquired H | Jan-86 | 0.313203 | 0.392627 | 0 | 0 | 2.712449 | 3.148895 |
| CellMarker | Naive CD8+ | Jan-97 | 0.345504 | 0.392814 | 0 | 0 | 2.400315 | 2.550937 |
| CellMarker | Acinar Cell | Jan-97 | 0.345504 | 0.392814 | 0 | 0 | 2.400315 | 2.550937 |
| GTEx_Tissu | GTEX-XPT6 | 7/599 | 0.015505 | 0.392998 | 0 | 0 | 2.855722 | 11.8987 |
| GTEx_Tissu | GTEX-XXEK | 10/1052 | 0.015836 | 0.392998 | 0 | 0 | 2.351995 | 9.750097 |
| GTEx_Tissu | GTEX-WFG | 8/746 | 0.015857 | 0.392998 | 0 | 0 | 2.631128 | 10.90373 |
| GTEx_Tissu | GTEX-XPVG | 7/602 | 0.015894 | 0.392998 | 0 | 0 | 2.840882 | 11.76644 |
| GTEx_Tissu | GTEX-OXRI | 8/745 | 0.015742 | 0.392998 | 0 | 0 | 2.634835 | 10.93833 |
| GTEx_Tissu | GTEX-S4Z8 | 14/1723 | 0.016347 | 0.393138 | 0 | 0 | 2.042819 | 8.403613 |
| GTEx_Tissu | GTEX-RM2 | 14/1720 | 0.016122 | 0.393138 | 0 | 0 | 2.046749 | 8.448119 |
| GTEx_Tissu | GTEX-REY6 | 12/1381 | 0.01619 | 0.393138 | 0 | 0 | 2.167305 | 8.936564 |
| GTEx_Tissu | GTEX-XBED | 8/751 | 0.016443 | 0.393138 | 0 | 0 | 2.61274 | 10.73275 |
| GO_Biologi | Regulation | Jan-87 | 0.316203 | 0.393177 | 0 | 0 | 2.680773 | 3.086563 |
| GO_Biologi | Glucose Hc | Jan-87 | 0.316203 | 0.393177 | 0 | 0 | 2.680773 | 3.086563 |
| GeDiPNet_ | Mood Diso | 2/269 | 0.327097 | 0.393191 | 0 | 0 | 1.731306 | 1.934732 |
| GeDiPNet_ | Hypogonac | 2/269 | 0.327097 | 0.393191 | 0 | 0 | 1.731306 | 1.934732 |
| GWAS_Cat | Medicatior | 1/107 | 0.373563 | 0.393225 | 0 | 0 | 2.172773 | 2.139459 |
| HuBMAP_A | Follicular D | 1/114 | 0.392494 | 0.393862 | 0 | 0 | 2.037456 | 1.905499 |
| Rare_Disea | Variant Cre | Jan-64 | 0.243799 | 0.394318 | 0 | 0 | 3.663714 | 5.171007 |
| CellMarker | Basal Cell T | Jan-98 | 0.348365 | 0.39443 | 0 | 0 | 2.37545 | 2.504922 |
| DisGeNET | Adult type | Jan-87 | 0.316203 | 0.395254 | 0 | 0 | 2.680773 | 3.086563 |
| DisGeNET | Actinic kera | Jan-87 | 0.316203 | 0.395254 | 0 | 0 | 2.680773 | 3.086563 |
| DisGeNET | Nevus | Jan-87 | 0.316203 | 0.395254 | 0 | 0 | 2.680773 | 3.086563 |
| DisGeNET | Dysmorphi | Jan-87 | 0.316203 | 0.395254 | 0 | 0 | 2.680773 | 3.086563 |
| DisGeNET | Chronic act | Jan-87 | 0.316203 | 0.395254 | 0 | 0 | 2.680773 | 3.086563 |
| DisGeNET | Endothelia | 2/262 | 0.31591 | 0.395254 | 0 | 0 | 1.778552 | 2.04942 |
| GO_Biologi | Regulation | Jan-88 | 0.319191 | 0.395507 | 0 | 0 | 2.649826 | 3.026014 |
| GO_Biologi | Negative R | Jan-88 | 0.319191 | 0.395507 | 0 | 0 | 2.649826 | 3.026014 |
| Rare_Disea | Thyroid car | 2/218 | 0.245071 | 0.395515 | 0 | 0 | 2.145643 | 3.017223 |
| GWAS_Cat | Blond Vs. B | 1/108 | 0.376303 | 0.395767 | 0 | 0 | 2.152358 | 2.10363 |
| Jensen_TIS | Parietal lob | 46/9921 | 0.307308 | 0.395813 | 0 | 0 | 1.140471 | 1.345645 |
| CellMarker | Basal Cell L | Jan-99 | 0.351214 | 0.396019 | 0 | 0 | 2.351092 | 2.46009 |
| DisGeNET | Parkinson I | 6/1064 | 0.317153 | 0.396252 | 0 | 0 | 1.320101 | 1.515964 |
| GeDiPNet_ | Skin Neopla | Jan-92 | 0.331012 | 0.396263 | 0 | 0 | 2.532839 | 2.800307 |
| GeDiPNet_ | Leukoence | Jan-92 | 0.331012 | 0.396263 | 0 | 0 | 2.532839 | 2.800307 |
| DisGeNET | Renal Insuf | 2/263 | 0.317511 | 0.39651 | 0 | 0 | 1.771648 | 2.03251 |
| Reactome_ | Inwardly Re | Jan-35 | 0.141623 | 0.396544 | 0 | 0 | 6.798564 | 13.28839 |
| GO_Biologi | Regulation | 2/265 | 0.32071 | 0.396601 | 0 | 0 | 1.757996 | 1.999226 |
| GO_Biologi | Positive Re | 2/266 | 0.322308 | 0.396601 | 0 | 0 | 1.751248 | 1.982846 |
| GO_Biologi | Regulation | Jan-89 | 0.322165 | 0.396601 | 0 | 0 | 2.619582 | 2.967177 |
| GO_Biologi | Sodium Ior | Jan-89 | 0.322165 | 0.396601 | 0 | 0 | 2.619582 | 2.967177 |
| DisGeNET | Neuroblast | 9/1698 | 0.31838 | 0.397406 | 0 | 0 | 1.244979 | 1.42489 |
| DisGeNET | Brain Tumc | Jan-88 | 0.319191 | 0.397471 | 0 | 0 | 2.649826 | 3.026014 |
| DisGeNET | Alcohol Us | Jan-88 | 0.319191 | 0.397471 | 0 | 0 | 2.649826 | 3.026014 |

| | | | | | | | | |
|---|---|---|---|---|---|---|---|---|
| DisGeNET | Atypical sca | Jan-88 | 0.319191 | 0.397471 | 0 | 0 | 2.649826 | 3.026014 |
| DisGeNET | Iron Overlo | Jan-88 | 0.319191 | 0.397471 | 0 | 0 | 2.649826 | 3.026014 |
| DisGeNET | Chordoma | Jan-88 | 0.319191 | 0.397471 | 0 | 0 | 2.649826 | 3.026014 |
| PhenGenI_ | Cytokine Re | Jan-53 | 0.206541 | 0.397592 | 0 | 0 | 4.44119 | 7.004886 |
| PhenGenI_ | Atrial Press | Jan-53 | 0.206541 | 0.397592 | 0 | 0 | 4.44119 | 7.004886 |
| PhenGenI_ | Macrophag | Jan-53 | 0.206541 | 0.397592 | 0 | 0 | 4.44119 | 7.004886 |
| PhenGenI_ | Amyloidosi | Jan-54 | 0.210002 | 0.397633 | 0 | 0 | 4.357174 | 6.79997 |
| PhenGenI_ | Efavirenz | Jan-55 | 0.213448 | 0.397633 | 0 | 0 | 4.27627 | 6.604112 |
| PhenGenI_ | Blood Press | 15/2756 | 0.21249 | 0.397633 | 0 | 0 | 1.305181 | 2.021545 |
| PhenGenI_ | Chlorine | Jan-55 | 0.213448 | 0.397633 | 0 | 0 | 4.27627 | 6.604112 |
| Rare_Disea | Ehlers-Dan | Jan-65 | 0.247099 | 0.397926 | 0 | 0 | 3.606286 | 5.041467 |
| GWAS_Cat | Neutrophil | 1/109 | 0.379031 | 0.398293 | 0 | 0 | 2.132321 | 2.068645 |
| Rare_Disea | Insulinoma | 2/220 | 0.248291 | 0.398982 | 0 | 0 | 2.125742 | 2.961486 |
| Human_Ph | High foreh | 1/108 | 0.376303 | 0.399299 | 0 | 0 | 2.152358 | 2.10363 |
| GO_Biologi | Positive Re | Jan-90 | 0.325127 | 0.399377 | 0 | 0 | 2.590018 | 2.909988 |
| DisGeNET | Bronchiolo | Jan-89 | 0.322165 | 0.399455 | 0 | 0 | 2.619582 | 2.967177 |
| DisGeNET | Acute Kidn | Jan-89 | 0.322165 | 0.399455 | 0 | 0 | 2.619582 | 2.967177 |
| DisGeNET | Tactile Allo | Jan-89 | 0.322165 | 0.399455 | 0 | 0 | 2.619582 | 2.967177 |
| DisGeNET | Refractory | 2/266 | 0.322308 | 0.399455 | 0 | 0 | 1.751248 | 1.982846 |
| DisGeNET | Congenital | Jan-89 | 0.322165 | 0.399455 | 0 | 0 | 2.619582 | 2.967177 |
| DisGeNET | Sarcoma | 4/658 | 0.321324 | 0.399455 | 0 | 0 | 1.419181 | 1.611202 |
| DisGeNET | Chagas Dis | Jan-89 | 0.322165 | 0.399455 | 0 | 0 | 2.619582 | 2.967177 |
| DisGeNET | Nasophary | Jan-89 | 0.322165 | 0.399455 | 0 | 0 | 2.619582 | 2.967177 |
| DisGeNET | Malaria, Fa | Jan-89 | 0.322165 | 0.399455 | 0 | 0 | 2.619582 | 2.967177 |
| DisGeNET | Cerebral Pa | Jan-89 | 0.322165 | 0.399455 | 0 | 0 | 2.619582 | 2.967177 |
| UK_Bioban | Inverse dis | 7/1318 | 0.349283 | 0.399782 | 0 | 0 | 1.241552 | 1.305954 |
| CellMarker | Macrophag | 1/101 | 0.356874 | 0.400752 | 0 | 0 | 2.303837 | 2.373811 |
| Human_Ph | Autism (HP | 1/109 | 0.379031 | 0.40108 | 0 | 0 | 2.132321 | 2.068645 |
| Rare_Disea | Breast canc | Jan-66 | 0.250385 | 0.401479 | 0 | 0 | 3.550626 | 4.916752 |
| GO_Biologi | Regulation | Jan-91 | 0.328076 | 0.40161 | 0 | 0 | 2.561111 | 2.854384 |
| GO_Biologi | Positive Re | Jan-91 | 0.328076 | 0.40161 | 0 | 0 | 2.561111 | 2.854384 |
| GTEx_Tissu | GTEX-XQ8I | 7/610 | 0.016966 | 0.401684 | 0 | 0 | 2.802032 | 11.42266 |
| GTEx_Tissu | GTEX-S32W | 11/1226 | 0.017078 | 0.401684 | 0 | 0 | 2.227399 | 9.065409 |
| Reactome_ | Transcripti | Jan-36 | 0.145363 | 0.401993 | 0 | 0 | 6.603987 | 12.73591 |
| DisGeNET | Gaucher Di | Jan-90 | 0.325127 | 0.402377 | 0 | 0 | 2.590018 | 2.909988 |
| DisGeNET | Cystic kidn | Jan-90 | 0.325127 | 0.402377 | 0 | 0 | 2.590018 | 2.909988 |
| DisGeNET | Periodontit | Jan-90 | 0.325127 | 0.402377 | 0 | 0 | 2.590018 | 2.909988 |
| GeDiPNet_ | Syndactyly | Jan-94 | 0.336847 | 0.402421 | 0 | 0 | 2.47812 | 2.69651 |
| dbGaP | Metabolisr | Jan-83 | 0.304123 | 0.403004 | 0 | 0 | 2.81211 | 3.34732 |
| dbGaP | Coronary D | Jan-82 | 0.30107 | 0.403004 | 0 | 0 | 2.846971 | 3.417539 |
| dbGaP | Body Weig | Jan-87 | 0.316203 | 0.403004 | 0 | 0 | 2.680773 | 3.086563 |
| dbGaP | Electrocard | Jan-87 | 0.316203 | 0.403004 | 0 | 0 | 2.680773 | 3.086563 |
| dbGaP | Lipids | Jan-87 | 0.316203 | 0.403004 | 0 | 0 | 2.680773 | 3.086563 |
| SysMyo_M | MGS1501 I | 3/232 | 0.080519 | 0.403306 | 0 | 0 | 3.069869 | 7.733813 |
| SysMyo_M | MGS1363 I | 3/232 | 0.080519 | 0.403306 | 0 | 0 | 3.069869 | 7.733813 |

| | | | | | | | | |
|---|---|---|---|---|---|---|---|---|
| SysMyo_M | MGS555 H | 3/232 | 0.080519 | 0.403306 | 0 | 0 | 3.069869 | 7.733813 |
| SysMyo_M | MGS1262 I | 3/231 | 0.079718 | 0.403306 | 0 | 0 | 3.08349 | 7.798958 |
| GWAS_Cat | Age Of Sm | 1/111 | 0.384451 | 0.403641 | 0 | 0 | 2.09334 | 2.001105 |
| GO_Biologi | Positive Re | Jan-92 | 0.331012 | 0.403812 | 0 | 0 | 2.532839 | 2.800307 |
| GO_Biologi | Positive Re | Jan-92 | 0.331012 | 0.403812 | 0 | 0 | 2.532839 | 2.800307 |
| GO_Biologi | Protein O-l | Jan-93 | 0.333936 | 0.403909 | 0 | 0 | 2.505182 | 2.747701 |
| GO_Biologi | Negative R | Jan-93 | 0.333936 | 0.403909 | 0 | 0 | 2.505182 | 2.747701 |
| GO_Biologi | Chemical S | 2/273 | 0.333469 | 0.403909 | 0 | 0 | 1.705405 | 1.872885 |
| GO_Biologi | Regulation | Jan-93 | 0.333936 | 0.403909 | 0 | 0 | 2.505182 | 2.747701 |
| GO_Biologi | Regulation | Jan-93 | 0.333936 | 0.403909 | 0 | 0 | 2.505182 | 2.747701 |
| GeDiPNet_ | Hyperchole | Jan-95 | 0.339745 | 0.404227 | 0 | 0 | 2.451633 | 2.646685 |
| GeDiPNet_ | Neuroblast | Jan-95 | 0.339745 | 0.404227 | 0 | 0 | 2.451633 | 2.646685 |
| WikiPathw | Nsp1 From | 16-Jan | 0.067399 | 0.404706 | 0 | 0 | 15.42481 | 41.60265 |
| WikiPathw | Mitochond | 16-Jan | 0.067399 | 0.404706 | 0 | 0 | 15.42481 | 41.60265 |
| WikiPathw | DNA Dama | 2/110 | 0.082985 | 0.404706 | 0 | 0 | 4.314815 | 10.73997 |
| WikiPathw | Metabolic I | 17-Jan | 0.071459 | 0.404706 | 0 | 0 | 14.46003 | 38.15469 |
| WikiPathw | Extracellul | 19-Jan | 0.079527 | 0.404706 | 0 | 0 | 12.85207 | 32.53709 |
| WikiPathw | Embryonic | 2/116 | 0.090827 | 0.404706 | 0 | 0 | 4.086481 | 9.802643 |
| WikiPathw | Synthesis C | 22-Jan | 0.091499 | 0.404706 | 0 | 0 | 11.0144 | 26.34017 |
| WikiPathw | Familial Hy | 22-Jan | 0.091499 | 0.404706 | 0 | 0 | 11.0144 | 26.34017 |
| WikiPathw | Hematopoi | 20-Jan | 0.083535 | 0.404706 | 0 | 0 | 12.17503 | 30.22444 |
| Jensen_TIS | T-cell chror | Dec-44 | 0.319383 | 0.404887 | 0 | 0 | 1.206244 | 1.376763 |
| Jensen_TIS | Embryonic | Jan-88 | 0.319191 | 0.404887 | 0 | 0 | 2.649826 | 3.026014 |
| Jensen_TIS | Neonate | Jan-87 | 0.316203 | 0.404887 | 0 | 0 | 2.680773 | 3.086563 |
| Jensen_TIS | Germ cell | 2/264 | 0.319111 | 0.404887 | 0 | 0 | 1.764796 | 2.015779 |
| DisGeNET | Acute Cere | Jan-91 | 0.328076 | 0.405452 | 0 | 0 | 2.561111 | 2.854384 |
| DisGeNET | Embryonal | Jan-91 | 0.328076 | 0.405452 | 0 | 0 | 2.561111 | 2.854384 |
| DisGeNET | Nonsyndro | Jan-91 | 0.328076 | 0.405452 | 0 | 0 | 2.561111 | 2.854384 |
| Jensen_DIS | Ulcerative | 1/115 | 0.395152 | 0.40555 | 0 | 0 | 2.019482 | 1.875059 |
| PhenGenI_ | Waist Circu | 5/742 | 0.22135 | 0.405808 | 0 | 0 | 1.586524 | 2.392495 |
| PhenGenI_ | Interleukin | Jan-57 | 0.220295 | 0.405808 | 0 | 0 | 4.123131 | 6.237423 |
| Rare_Disea | Eosinophili | Jan-67 | 0.253656 | 0.40585 | 0 | 0 | 3.496653 | 4.79662 |
| GTEx_Tissu | GTEX-WHV | 10/1068 | 0.017419 | 0.40639 | 0 | 0 | 2.314462 | 9.374057 |
| GeneSigDB | 17786710- | 28-Feb | 0.00657 | 0.406598 | 0 | 0 | 17.99729 | 90.44125 |
| Rare_Disea | Ocular mot | 2/224 | 0.254736 | 0.406704 | 0 | 0 | 2.087016 | 2.85405 |
| GO_Biologi | Vesicle Org | Jan-94 | 0.336847 | 0.406737 | 0 | 0 | 2.47812 | 2.69651 |
| Jensen_TIS | Prostate ca | Jan-89 | 0.322165 | 0.406813 | 0 | 0 | 2.619582 | 2.967177 |
| GeDiPNet_ | Aortic Aneu | Jan-96 | 0.342631 | 0.40683 | 0 | 0 | 2.425704 | 2.598176 |
| Reactome_ | Cooperatio | Jan-37 | 0.149088 | 0.407264 | 0 | 0 | 6.42022 | 12.21909 |
| Human_Ph | Type II diat | 1/112 | 0.387144 | 0.407407 | 0 | 0 | 2.074377 | 1.968499 |
| Human_Ph | Elevated he | 1/112 | 0.387144 | 0.407407 | 0 | 0 | 2.074377 | 1.968499 |
| Rare_Disea | PROSTAGL | Jan-68 | 0.256914 | 0.407556 | 0 | 0 | 3.44429 | 4.68084 |
| Rare_Disea | Schwannor | 2/225 | 0.256348 | 0.407556 | 0 | 0 | 2.077552 | 2.828 |
| Rare_Disea | Apraxia | Jan-68 | 0.256914 | 0.407556 | 0 | 0 | 3.44429 | 4.68084 |
| SysMyo_M | MGS1274 I | 2/111 | 0.084277 | 0.407699 | 0 | 0 | 4.275013 | 10.57487 |

| | | | | | | | | |
|---|---|---|---|---|---|---|---|---|
| SysMyo_M | MGS423 H | 20-Jan | 0.083535 | 0.407699 | 0 | 0 | 12.17503 | 30.22444 |
| SysMyo_M | MGS1122 I | 2/111 | 0.084277 | 0.407699 | 0 | 0 | 4.275013 | 10.57487 |
| SysMyo_M | MGS1141 I | 2/111 | 0.084277 | 0.407699 | 0 | 0 | 4.275013 | 10.57487 |
| Disease_Si | Squamous | 4/300 | 0.041941 | 0.407975 | 0 | 0 | 3.193911 | 10.12943 |
| Disease_Si | COPD - Chr | 5/300 | 0.009983 | 0.407975 | 0 | 0 | 4.054981 | 18.68068 |
| Disease_Si | Astrocyton | 4/300 | 0.041941 | 0.407975 | 0 | 0 | 3.193911 | 10.12943 |
| Disease_Si | Bipolar Dis | 4/300 | 0.041941 | 0.407975 | 0 | 0 | 3.193911 | 10.12943 |
| Disease_Si | Uterine lei | 4/300 | 0.041941 | 0.407975 | 0 | 0 | 3.193911 | 10.12943 |
| Disease_Si | Diabetic Ne | 4/300 | 0.041941 | 0.407975 | 0 | 0 | 3.193911 | 10.12943 |
| Disease_Si | Inclusion B | 4/300 | 0.041941 | 0.407975 | 0 | 0 | 3.193911 | 10.12943 |
| Disease_Si | Leukemia, . | 4/300 | 0.041941 | 0.407975 | 0 | 0 | 3.193911 | 10.12943 |
| Disease_Si | Adenocarci | 4/300 | 0.041941 | 0.407975 | 0 | 0 | 3.193911 | 10.12943 |
| Disease_Si | Obstructive | 4/300 | 0.041941 | 0.407975 | 0 | 0 | 3.193911 | 10.12943 |
| Disease_Si | Huntingtor | 5/300 | 0.009983 | 0.407975 | 0 | 0 | 4.054981 | 18.68068 |
| GWAS_Cat | Metabolite | 5/940 | 0.388948 | 0.408011 | 0 | 0 | 1.237642 | 1.168716 |
| DisGeNET | Acute kidn | Jan-92 | 0.331012 | 0.408503 | 0 | 0 | 2.532839 | 2.800307 |
| DisGeNET | Cholestasis | Jan-92 | 0.331012 | 0.408503 | 0 | 0 | 2.532839 | 2.800307 |
| DisGeNET | Premature | Jan-92 | 0.331012 | 0.408503 | 0 | 0 | 2.532839 | 2.800307 |
| GWAS_Cat | Positive Af | 1/113 | 0.389825 | 0.408579 | 0 | 0 | 2.055752 | 1.936638 |
| GO_Biologi | T Cell Rece | Jan-95 | 0.339745 | 0.408846 | 0 | 0 | 2.451633 | 2.646685 |
| GO_Biologi | Cell Junctic | Jan-95 | 0.339745 | 0.408846 | 0 | 0 | 2.451633 | 2.646685 |
| GeDiPNet_ | Strabismus | 4/682 | 0.34512 | 0.408953 | 0 | 0 | 1.367239 | 1.454555 |
| dbGaP | Creatinine | Jan-92 | 0.331012 | 0.409544 | 0 | 0 | 2.532839 | 2.800307 |
| dbGaP | Waist-Hip I | Jan-93 | 0.333936 | 0.409544 | 0 | 0 | 2.505182 | 2.747701 |
| Jensen_TIS | BTO:00044 | 43/9290 | 0.325772 | 0.40976 | 0 | 0 | 1.12724 | 1.264266 |
| PhenGenI_ | Beta-Globu | Jan-59 | 0.227083 | 0.409814 | 0 | 0 | 3.980553 | 5.900928 |
| PhenGenI_ | Wet Macul | Jan-59 | 0.227083 | 0.409814 | 0 | 0 | 3.980553 | 5.900928 |
| Jensen_TIS | Tongue | 3/463 | 0.327152 | 0.409895 | 0 | 0 | 1.510326 | 1.687532 |
| DisGeNET | Virus Disea | 5/875 | 0.332381 | 0.409999 | 0 | 0 | 1.334665 | 1.470099 |
| Rare_Disea | Tuberous s | Jan-69 | 0.260157 | 0.410079 | 0 | 0 | 3.393468 | 4.569198 |
| Rare_Disea | Cystosarco | Jan-69 | 0.260157 | 0.410079 | 0 | 0 | 3.393468 | 4.569198 |
| Rare_Disea | Silicosis | Jan-69 | 0.260157 | 0.410079 | 0 | 0 | 3.393468 | 4.569198 |
| SysMyo_M | MGS1066 I | 2/112 | 0.085575 | 0.410109 | 0 | 0 | 4.235936 | 10.41346 |
| SysMyo_M | MGS651 H | 3/239 | 0.086224 | 0.410109 | 0 | 0 | 2.977754 | 7.297891 |
| DisGeNET | Pregnancy | Jan-93 | 0.333936 | 0.410179 | 0 | 0 | 2.505182 | 2.747701 |
| DisGeNET | Panic Disor | Jan-93 | 0.333936 | 0.410179 | 0 | 0 | 2.505182 | 2.747701 |
| DisGeNET | Subarachn | Jan-93 | 0.333936 | 0.410179 | 0 | 0 | 2.505182 | 2.747701 |
| DisGeNET | Glioblastor | Oct-37 | 0.333237 | 0.410179 | 0 | 0 | 1.212166 | 1.332051 |
| DisGeNET | Edema | Jan-93 | 0.333936 | 0.410179 | 0 | 0 | 2.505182 | 2.747701 |
| DisGeNET | Liver Cirrhc | Jan-93 | 0.333936 | 0.410179 | 0 | 0 | 2.505182 | 2.747701 |
| DisGeNET | Deafness | 2/273 | 0.333469 | 0.410179 | 0 | 0 | 1.705405 | 1.872885 |
| DisGeNET | Epileptic er | 3/468 | 0.333122 | 0.410179 | 0 | 0 | 1.493702 | 1.641948 |
| DisGeNET | Mucinous / | Jan-93 | 0.333936 | 0.410179 | 0 | 0 | 2.505182 | 2.747701 |
| DisGeNET | Malaria | 3/469 | 0.334315 | 0.410453 | 0 | 0 | 1.49042 | 1.633009 |
| UK_Bioban | Reticulocyt | 7/1333 | 0.360081 | 0.410493 | 0 | 0 | 1.226518 | 1.252797 |

| | | | | | | | | |
|---|---|---|---|---|---|---|---|---|
| GWAS_Cat | Medicatior | 1/114 | 0.392494 | 0.411024 | 0 | 0 | 2.037456 | 1.905499 |
| DisGeNET | Uranostapl | 2/274 | 0.335059 | 0.411174 | 0 | 0 | 1.699048 | 1.85782 |
| DisGeNET | Malignant | 3/470 | 0.335509 | 0.411533 | 0 | 0 | 1.487152 | 1.624127 |
| UK_Bioban | East co-orc | 1/103 | 0.362485 | 0.411587 | 0 | 0 | 2.258436 | 2.291796 |
| GO_Biologi | Positive Re | Jan-96 | 0.342631 | 0.411621 | 0 | 0 | 2.425704 | 2.598176 |
| GeDiPNet_ | Pulmonary | Jan-98 | 0.348365 | 0.411961 | 0 | 0 | 2.37545 | 2.504922 |
| DisGeNET | Malignant | 4/673 | 0.336185 | 0.412168 | 0 | 0 | 1.386281 | 1.511177 |
| DisGeNET | Cerebral Is | 2/275 | 0.336649 | 0.412352 | 0 | 0 | 1.692739 | 1.842909 |
| DisGeNET | Infant, Sma | 2/275 | 0.336649 | 0.412352 | 0 | 0 | 1.692739 | 1.842909 |
| DisGeNET | Macroglos | Jan-94 | 0.336847 | 0.412401 | 0 | 0 | 2.47812 | 2.69651 |
| SysMyo_M | MGS953 M | 21-Jan | 0.087525 | 0.412827 | 0 | 0 | 11.5657 | 28.17205 |
| UK_Bioban | Overall hea | 1/104 | 0.365273 | 0.413106 | 0 | 0 | 2.236396 | 2.252299 |
| DisGeNET | Stomach Ca | Dec-78 | 0.337594 | 0.413123 | 0 | 0 | 1.18661 | 1.288552 |
| Rare_Disea | Marinesco- | 2/229 | 0.262799 | 0.413367 | 0 | 0 | 2.040529 | 2.726893 |
| Rare_Disea | Tuberous s | Jan-70 | 0.263387 | 0.413417 | 0 | 0 | 3.344119 | 4.461494 |
| GWAS_Cat | Smoking St | 1/115 | 0.395152 | 0.413452 | 0 | 0 | 2.019482 | 1.875059 |
| Human_Ph | Proteinuria | 1/115 | 0.395152 | 0.413556 | 0 | 0 | 2.019482 | 1.875059 |
| Human_Ph | Recurrent f | 1/115 | 0.395152 | 0.413556 | 0 | 0 | 2.019482 | 1.875059 |
| GeDiPNet_ | Papillary Th | Jan-99 | 0.351214 | 0.413652 | 0 | 0 | 2.351092 | 2.46009 |
| GeDiPNet_ | Renal Ager | Jan-99 | 0.351214 | 0.413652 | 0 | 0 | 2.351092 | 2.46009 |
| DisGeNET | Rectal Carc | 2/276 | 0.338237 | 0.413717 | 0 | 0 | 1.686475 | 1.828152 |
| Jensen_DIS | Dementia | 1/119 | 0.405668 | 0.414164 | 0 | 0 | 1.950631 | 1.759896 |
| GO_Biologi | Regulation | Jan-97 | 0.345504 | 0.414371 | 0 | 0 | 2.400315 | 2.550937 |
| DisGeNET | Sciatic Neu | Jan-95 | 0.339745 | 0.4144 | 0 | 0 | 2.451633 | 2.646685 |
| DisGeNET | Congenital | Jan-95 | 0.339745 | 0.4144 | 0 | 0 | 2.451633 | 2.646685 |
| DisGeNET | Membranc | Jan-95 | 0.339745 | 0.4144 | 0 | 0 | 2.451633 | 2.646685 |
| DisGeNET | Rotting tee | Jan-95 | 0.339745 | 0.4144 | 0 | 0 | 2.451633 | 2.646685 |
| DisGeNET | Renal cyst | Jan-95 | 0.339745 | 0.4144 | 0 | 0 | 2.451633 | 2.646685 |
| DisGeNET | Fragile X Sy | Jan-95 | 0.339745 | 0.4144 | 0 | 0 | 2.451633 | 2.646685 |
| UK_Bioban | Alcohol int | 1/105 | 0.368048 | 0.4146 | 0 | 0 | 2.214781 | 2.213764 |
| DisGeNET | Autistic Dis | 4/677 | 0.340155 | 0.414707 | 0 | 0 | 1.377755 | 1.485709 |
| UK_Bioban | Longest pe | 2/296 | 0.369749 | 0.414875 | 0 | 0 | 1.570148 | 1.56219 |
| DisGeNET | Cardiac fibr | Jan-96 | 0.342631 | 0.414878 | 0 | 0 | 2.425704 | 2.598176 |
| DisGeNET | Mammary | 3/476 | 0.34267 | 0.414878 | 0 | 0 | 1.467834 | 1.572031 |
| DisGeNET | MAJOR AFI | Jan-96 | 0.342631 | 0.414878 | 0 | 0 | 2.425704 | 2.598176 |
| DisGeNET | Steroid Sul | Jan-96 | 0.342631 | 0.414878 | 0 | 0 | 2.425704 | 2.598176 |
| DisGeNET | Sparse hair | Jan-96 | 0.342631 | 0.414878 | 0 | 0 | 2.425704 | 2.598176 |
| DisGeNET | Thin, spars | Jan-96 | 0.342631 | 0.414878 | 0 | 0 | 2.425704 | 2.598176 |
| DisGeNET | PARKINSOI | Jan-96 | 0.342631 | 0.414878 | 0 | 0 | 2.425704 | 2.598176 |
| DisGeNET | Aortic Anei | Jan-96 | 0.342631 | 0.414878 | 0 | 0 | 2.425704 | 2.598176 |
| DisGeNET | Idiopathic p | Jan-96 | 0.342631 | 0.414878 | 0 | 0 | 2.425704 | 2.598176 |
| DisGeNET | Broad forel | Jan-96 | 0.342631 | 0.414878 | 0 | 0 | 2.425704 | 2.598176 |
| DisGeNET | Essential H | 2/278 | 0.341411 | 0.414878 | 0 | 0 | 1.674084 | 1.799086 |
| DisGeNET | Intrauterin | 2/278 | 0.341411 | 0.414878 | 0 | 0 | 1.674084 | 1.799086 |
| DisGeNET | Mammary | Dec-87 | 0.342456 | 0.414878 | 0 | 0 | 1.181507 | 1.266119 |

| | | | | | | | | |
|---|---|---|---|---|---|---|---|---|
| DisGeNET | Renal failu | Jan-96 | 0.342631 | 0.414878 | 0 | 0 | 2.425704 | 2.598176 |
| DisGeNET | Retinal Det | Jan-96 | 0.342631 | 0.414878 | 0 | 0 | 2.425704 | 2.598176 |
| Rare_Disea | Palmoplan | Jan-71 | 0.266602 | 0.414963 | 0 | 0 | 3.296179 | 4.35754 |
| Rare_Disea | Anaplastic | 2/231 | 0.266025 | 0.414963 | 0 | 0 | 2.022502 | 2.678127 |
| Rare_Disea | Anaplastic | Jan-71 | 0.266602 | 0.414963 | 0 | 0 | 3.296179 | 4.35754 |
| Rare_Disea | Non-Hodgk | Jan-71 | 0.266602 | 0.414963 | 0 | 0 | 3.296179 | 4.35754 |
| GO_Biologi | Mitochond | Jan-98 | 0.348365 | 0.416396 | 0 | 0 | 2.37545 | 2.504922 |
| GO_Biologi | Positive Re | Jan-98 | 0.348365 | 0.416396 | 0 | 0 | 2.37545 | 2.504922 |
| Jensen_TIS | Liver | 73/16290 | 0.333871 | 0.416691 | 0 | 0 | 1.188383 | 1.303658 |
| GO_Biologi | Positive Re | 2/283 | 0.349324 | 0.41684 | 0 | 0 | 1.643877 | 1.728956 |
| SysMyo_M | MGS915 M | 3/245 | 0.091249 | 0.417277 | 0 | 0 | 2.90304 | 6.950367 |
| SysMyo_M | MGS718 M | 2/117 | 0.092155 | 0.417277 | 0 | 0 | 4.050742 | 9.658129 |
| SysMyo_M | MGS811 M | 2/115 | 0.089505 | 0.417277 | 0 | 0 | 4.122853 | 9.950336 |
| SysMyo_M | MGS1024 I | 2/117 | 0.092155 | 0.417277 | 0 | 0 | 4.050742 | 9.658129 |
| SysMyo_M | MGS393 H | 3/244 | 0.090403 | 0.417277 | 0 | 0 | 2.915234 | 7.00671 |
| GeDiPNet_ | Neurodeve | 1/101 | 0.356874 | 0.417786 | 0 | 0 | 2.303837 | 2.373811 |
| GeDiPNet_ | Acute Kidn | 1/101 | 0.356874 | 0.417786 | 0 | 0 | 2.303837 | 2.373811 |
| GeDiPNet_ | Jacksonian | 1/101 | 0.356874 | 0.417786 | 0 | 0 | 2.303837 | 2.373811 |
| DisGeNET | Neoplasms | Jan-97 | 0.345504 | 0.417923 | 0 | 0 | 2.400315 | 2.550937 |
| DisGeNET | Adult Hepa | Jan-97 | 0.345504 | 0.417923 | 0 | 0 | 2.400315 | 2.550937 |
| GTEx_Tissu | GTEX-U3ZH | 13/1572 | 0.018059 | 0.417965 | 0 | 0 | 2.068218 | 8.302017 |
| GO_Biologi | Positive Re | 3/483 | 0.351019 | 0.418161 | 0 | 0 | 1.445908 | 1.513741 |
| WikiPathw | miRNA Reg | 24-Jan | 0.099394 | 0.41837 | 0 | 0 | 10.05561 | 23.215 |
| WikiPathw | mBDNF An | Jan-38 | 0.152796 | 0.41837 | 0 | 0 | 6.246386 | 11.73478 |
| WikiPathw | EGF EGFR S | 2/159 | 0.152237 | 0.41837 | 0 | 0 | 2.960809 | 5.573176 |
| WikiPathw | 1Q21 1 Co | 27-Jan | 0.111111 | 0.41837 | 0 | 0 | 8.894007 | 19.54216 |
| WikiPathw | Amyotroph | Jan-38 | 0.152796 | 0.41837 | 0 | 0 | 6.246386 | 11.73478 |
| WikiPathw | Myometria | 2/156 | 0.147716 | 0.41837 | 0 | 0 | 3.018946 | 5.773631 |
| WikiPathw | Cardiomyo | Jan-33 | 0.134093 | 0.41837 | 0 | 0 | 7.224201 | 14.515 |
| WikiPathw | Hepatocyte | Jan-34 | 0.137866 | 0.41837 | 0 | 0 | 7.004933 | 13.88007 |
| WikiPathw | 16P11 2 Di | 31-Jan | 0.126498 | 0.41837 | 0 | 0 | 7.706589 | 15.93356 |
| WikiPathw | PI3K AKT m | Jan-32 | 0.130304 | 0.41837 | 0 | 0 | 7.457614 | 15.19775 |
| WikiPathw | Breast Can | 2/154 | 0.144717 | 0.41837 | 0 | 0 | 3.058978 | 5.912923 |
| WikiPathw | Melatonin | Jan-37 | 0.149088 | 0.41837 | 0 | 0 | 6.42022 | 12.21909 |
| WikiPathw | Wnt Signal | Jan-36 | 0.145363 | 0.41837 | 0 | 0 | 6.603987 | 12.73591 |
| WikiPathw | Factors And | Jan-36 | 0.145363 | 0.41837 | 0 | 0 | 6.603987 | 12.73591 |
| WikiPathw | NRF2 Pathw | 2/141 | 0.125573 | 0.41837 | 0 | 0 | 3.34727 | 6.945157 |
| WikiPathw | T Cell Rece | 28-Jan | 0.114983 | 0.41837 | 0 | 0 | 8.564169 | 18.52408 |
| DisGeNET | Secondary | 4/683 | 0.346113 | 0.418466 | 0 | 0 | 1.365154 | 1.448414 |
| GeneSigDB | 16964388- | Feb-30 | 0.007518 | 0.418756 | 0 | 0 | 16.71008 | 81.71977 |
| Jensen_TIS | Germ | Jan-94 | 0.336847 | 0.418782 | 0 | 0 | 2.47812 | 2.69651 |
| UK_Bioban | Left leg fat | 3/503 | 0.374809 | 0.418904 | 0 | 0 | 1.386643 | 1.360766 |
| Rare_Disea | Meleda dis | Jan-72 | 0.269804 | 0.419069 | 0 | 0 | 3.249591 | 4.257158 |
| CellMarker | Morula Cel | 1/108 | 0.376303 | 0.419134 | 0 | 0 | 2.152358 | 2.10363 |
| CellMarker | Mast Cell K | 1/108 | 0.376303 | 0.419134 | 0 | 0 | 2.152358 | 2.10363 |

| | | | | | | | | |
|---|---|---|---|---|---|---|---|---|
| DisGeNET | Frontotem | Jan-98 | 0.348365 | 0.420026 | 0 | 0 | 2.37545 | 2.504922 |
| DisGeNET | Dysarthria | 2/282 | 0.347744 | 0.420026 | 0 | 0 | 1.649832 | 1.7427 |
| DisGeNET | Myelofibro | Jan-98 | 0.348365 | 0.420026 | 0 | 0 | 2.37545 | 2.504922 |
| DisGeNET | Mycobacte | Jan-98 | 0.348365 | 0.420026 | 0 | 0 | 2.37545 | 2.504922 |
| DisGeNET | Henoch-Scl | Jan-98 | 0.348365 | 0.420026 | 0 | 0 | 2.37545 | 2.504922 |
| DisGeNET | Central Ser | Jan-98 | 0.348365 | 0.420026 | 0 | 0 | 2.37545 | 2.504922 |
| DisGeNET | Cerebral In | 2/283 | 0.349324 | 0.420795 | 0 | 0 | 1.643877 | 1.728956 |
| DisGeNET | Squamous | 2/283 | 0.349324 | 0.420795 | 0 | 0 | 1.643877 | 1.728956 |
| GO_Molecu | RNA Polym | 2/228 | 0.261186 | 0.421268 | 0 | 0 | 2.049662 | 2.751716 |
| GWAS_Cat | Cerebrospi | 1/118 | 0.403056 | 0.421361 | 0 | 0 | 1.967402 | 1.787737 |
| Rare_Disea | Chromosor | Jan-73 | 0.272992 | 0.422201 | 0 | 0 | 3.204296 | 4.160181 |
| Rare_Disea | Poncet-Spi | 2/236 | 0.27409 | 0.422201 | 0 | 0 | 1.978783 | 2.561135 |
| Rare_Disea | Seminoma | 2/236 | 0.27409 | 0.422201 | 0 | 0 | 1.978783 | 2.561135 |
| Rare_Disea | Diffuse scle | 2/236 | 0.27409 | 0.422201 | 0 | 0 | 1.978783 | 2.561135 |
| Reactome_ | Ovarian Tu | Jan-39 | 0.156488 | 0.42233 | 0 | 0 | 6.081701 | 11.28018 |
| GTEx_Tissu | GTEX-X585 | 13/1576 | 0.018405 | 0.422594 | 0 | 0 | 2.062475 | 8.239818 |
| DisGeNET | Leigh Disea | Jan-99 | 0.351214 | 0.422682 | 0 | 0 | 2.351092 | 2.46009 |
| DisGeNET | Nephrolith | Jan-99 | 0.351214 | 0.422682 | 0 | 0 | 2.351092 | 2.46009 |
| DisGeNET | Liver neopl | 7/1321 | 0.35144 | 0.42276 | 0 | 0 | 1.238518 | 1.29514 |
| PhenGenI_ | Hip | 6/956 | 0.236389 | 0.423301 | 0 | 0 | 1.478596 | 2.132546 |
| Gene_Pert | Sod2 KO m | 6/328 | 0.003083 | 0.423319 | 0 | 0 | 4.506786 | 26.05698 |
| Gene_Pert | Hes1 OE m | 4/140 | 0.003267 | 0.423319 | 0 | 0 | 7.00815 | 40.11369 |
| GeDiPNet_ | Microdonti | 1/103 | 0.362485 | 0.423505 | 0 | 0 | 2.258436 | 2.291796 |
| Rare_Disea | Gata1 Gen | 2/237 | 0.275703 | 0.423644 | 0 | 0 | 1.970263 | 2.538546 |
| Rare_Disea | Extracrania | Jan-74 | 0.276166 | 0.423644 | 0 | 0 | 3.160242 | 4.066453 |
| GWAS_Cat | Type 1 Dial | 1/119 | 0.405668 | 0.423729 | 0 | 0 | 1.950631 | 1.759896 |
| Jensen_TIS | Peripheral | Jan-96 | 0.342631 | 0.424335 | 0 | 0 | 2.425704 | 2.598176 |
| GO_Biologi | Hemopoies | 1/101 | 0.356874 | 0.424423 | 0 | 0 | 2.303837 | 2.373811 |
| DisGeNET | Rhinitis | 1/100 | 0.35405 | 0.425004 | 0 | 0 | 2.327226 | 2.416399 |
| DisGeNET | Seizures, A | 1/101 | 0.356874 | 0.425004 | 0 | 0 | 2.303837 | 2.373811 |
| DisGeNET | Mood swin | 1/100 | 0.35405 | 0.425004 | 0 | 0 | 2.327226 | 2.416399 |
| DisGeNET | Myalgia | 1/100 | 0.35405 | 0.425004 | 0 | 0 | 2.327226 | 2.416399 |
| DisGeNET | Gustatory s | 1/101 | 0.356874 | 0.425004 | 0 | 0 | 2.303837 | 2.373811 |
| DisGeNET | Sleep distu | 1/101 | 0.356874 | 0.425004 | 0 | 0 | 2.303837 | 2.373811 |
| DisGeNET | Atonic Abs | 1/101 | 0.356874 | 0.425004 | 0 | 0 | 2.303837 | 2.373811 |
| DisGeNET | Generalize | 1/101 | 0.356874 | 0.425004 | 0 | 0 | 2.303837 | 2.373811 |
| DisGeNET | Nonepilept | 1/101 | 0.356874 | 0.425004 | 0 | 0 | 2.303837 | 2.373811 |
| DisGeNET | Olfactory s | 1/101 | 0.356874 | 0.425004 | 0 | 0 | 2.303837 | 2.373811 |
| DisGeNET | Non-epilep | 1/101 | 0.356874 | 0.425004 | 0 | 0 | 2.303837 | 2.373811 |
| DisGeNET | Lupus Erytl | 6/1113 | 0.35532 | 0.425004 | 0 | 0 | 1.258389 | 1.302102 |
| DisGeNET | Noninfiltra | 2/287 | 0.355633 | 0.425004 | 0 | 0 | 1.620475 | 1.675339 |
| DisGeNET | Seizures, C | 1/101 | 0.356874 | 0.425004 | 0 | 0 | 2.303837 | 2.373811 |
| DisGeNET | Seizures, S | 1/101 | 0.356874 | 0.425004 | 0 | 0 | 2.303837 | 2.373811 |
| DisGeNET | Seizures, S | 1/101 | 0.356874 | 0.425004 | 0 | 0 | 2.303837 | 2.373811 |
| DisGeNET | Xeroderma | 1/101 | 0.356874 | 0.425004 | 0 | 0 | 2.303837 | 2.373811 |

| | | | | | | | | |
|---|---|---|---|---|---|---|---|---|
| DisGeNET | Hypertensi | 1/101 | 0.356874 | 0.425004 | 0 | 0 | 2.303837 | 2.373811 |
| DisGeNET | Jacksonian | 1/101 | 0.356874 | 0.425004 | 0 | 0 | 2.303837 | 2.373811 |
| DisGeNET | Vertiginous | 1/101 | 0.356874 | 0.425004 | 0 | 0 | 2.303837 | 2.373811 |
| DisGeNET | Single Seizu | 1/101 | 0.356874 | 0.425004 | 0 | 0 | 2.303837 | 2.373811 |
| DisGeNET | Rash and D | 1/100 | 0.35405 | 0.425004 | 0 | 0 | 2.327226 | 2.416399 |
| dbGaP | Bipolar Dis | 1/102 | 0.359686 | 0.425083 | 0 | 0 | 2.280912 | 2.332289 |
| dbGaP | Neuroblast | 1/101 | 0.356874 | 0.425083 | 0 | 0 | 2.303837 | 2.373811 |
| Jensen_DIS | Bipolar disc | 1/124 | 0.418561 | 0.425101 | 0 | 0 | 1.870864 | 1.629399 |
| Diabetes_P | GSE125856 | 4/214 | 0.014221 | 0.425453 | 0 | 0 | 4.521629 | 19.23057 |
| Diabetes_P | GSE149468 | 4/194 | 0.010221 | 0.425453 | 0 | 0 | 5.002663 | 22.92898 |
| Diabetes_P | GSE154198 | 4/200 | 0.011332 | 0.425453 | 0 | 0 | 4.848045 | 21.71979 |
| Diabetes_P | GSE106295 | 4/169 | 0.006357 | 0.425453 | 0 | 0 | 5.767945 | 29.17586 |
| Diabetes_P | GSE127251 | 4/202 | 0.011719 | 0.425453 | 0 | 0 | 4.798588 | 21.33702 |
| Diabetes_P | GSE150679 | 4/189 | 0.00935 | 0.425453 | 0 | 0 | 5.139173 | 24.01195 |
| Diabetes_P | GSE41203 | 5/317 | 0.012445 | 0.425453 | 0 | 0 | 3.830715 | 16.80313 |
| Diabetes_P | GSE124621 | 4/210 | 0.013353 | 0.425453 | 0 | 0 | 4.610364 | 19.89848 |
| Diabetes_P | GSE128361 | 4/158 | 0.005021 | 0.425453 | 0 | 0 | 6.183383 | 32.73506 |
| GO_Biologi | Calcium Ior | 1/102 | 0.359686 | 0.425628 | 0 | 0 | 2.280912 | 2.332289 |
| GO_Biologi | Regulation | 1/102 | 0.359686 | 0.425628 | 0 | 0 | 2.280912 | 2.332289 |
| GO_Biologi | Protein Cat | 1/102 | 0.359686 | 0.425628 | 0 | 0 | 2.280912 | 2.332289 |
| Reactome_ | Transport ( | Jan-43 | 0.171099 | 0.425847 | 0 | 0 | 5.501384 | 9.712751 |
| Reactome_ | Retinoid M | Jan-43 | 0.171099 | 0.425847 | 0 | 0 | 5.501384 | 9.712751 |
| Reactome_ | Vasopressi | Jan-42 | 0.16747 | 0.425847 | 0 | 0 | 5.635848 | 10.07097 |
| Reactome_ | Glucagon-l | Jan-42 | 0.16747 | 0.425847 | 0 | 0 | 5.635848 | 10.07097 |
| Reactome_ | GPER1 Sigr | Jan-43 | 0.171099 | 0.425847 | 0 | 0 | 5.501384 | 9.712751 |
| Reactome_ | Activation ( | Jan-42 | 0.16747 | 0.425847 | 0 | 0 | 5.635848 | 10.07097 |
| Reactome_ | tRNA Amin | Jan-42 | 0.16747 | 0.425847 | 0 | 0 | 5.635848 | 10.07097 |
| GeDiPNet_ | Autism Spe | 1/104 | 0.365273 | 0.425908 | 0 | 0 | 2.236396 | 2.252299 |
| Human_Ph | Hypothyroi | 1/120 | 0.408269 | 0.426117 | 0 | 0 | 1.934141 | 1.732658 |
| PhenGenI_ | Folic Acid | Jan-64 | 0.243799 | 0.426648 | 0 | 0 | 3.663714 | 5.171007 |
| PhenGenI_ | Taste Perce | Jan-64 | 0.243799 | 0.426648 | 0 | 0 | 3.663714 | 5.171007 |
| PhenGenI_ | Fructose-B | Jan-63 | 0.240485 | 0.426648 | 0 | 0 | 3.722993 | 5.305634 |
| Gene_Pert | Klf15 Defic | 7/458 | 0.003818 | 0.426735 | 0 | 0 | 3.775887 | 21.02429 |
| Gene_Pert | Srf NULL M | 5/257 | 0.005292 | 0.426735 | 0 | 0 | 4.757307 | 24.93561 |
| Gene_Pert | Ncoa2 KO r | 6/352 | 0.004351 | 0.426735 | 0 | 0 | 4.189039 | 22.77679 |
| Gene_Pert | DNMT1 KO | 5/238 | 0.003831 | 0.426735 | 0 | 0 | 5.150215 | 28.65946 |
| Gene_Pert | Pitx2 KO m | 6/356 | 0.004596 | 0.426735 | 0 | 0 | 4.140317 | 22.28598 |
| Gene_Pert | Hras Select | 4/161 | 0.005365 | 0.426735 | 0 | 0 | 6.064308 | 31.70357 |
| Gene_Pert | Sirt1 OE mc | 5/263 | 0.005826 | 0.426735 | 0 | 0 | 4.645254 | 23.90189 |
| Gene_Pert | Hras OE mc | 5/262 | 0.005734 | 0.426735 | 0 | 0 | 4.663566 | 24.06993 |
| Gene_Pert | EGFR druga | 5/252 | 0.004875 | 0.426735 | 0 | 0 | 4.854843 | 25.84584 |
| Gene_Pert | TBK1 drugi | 5/274 | 0.006901 | 0.426735 | 0 | 0 | 4.452806 | 22.15724 |
| Gene_Pert | IRS4 KO mc | 5/285 | 0.008109 | 0.426735 | 0 | 0 | 4.275479 | 20.58534 |
| Gene_Pert | Dmd Defici | 5/283 | 0.00788 | 0.426735 | 0 | 0 | 4.306677 | 20.85935 |
| Gene_Pert | Cdkn1a DN | 5/283 | 0.00788 | 0.426735 | 0 | 0 | 4.306677 | 20.85935 |

| | | | | | | | | |
|---|---|---|---|---|---|---|---|---|
| Gene_Pert | Tgfb2 KD n | 5/283 | 0.00788 | 0.426735 | 0 | 0 | 4.306677 | 20.85935 |
| Gene_Pert | Dmd Defici | 5/269 | 0.006397 | 0.426735 | 0 | 0 | 4.538295 | 22.92736 |
| Gene_Pert | Stub1 KO n | 5/272 | 0.006696 | 0.426735 | 0 | 0 | 4.486617 | 22.46089 |
| Gene_Pert | Rybp KO m | 5/288 | 0.008463 | 0.426735 | 0 | 0 | 4.22951 | 20.18361 |
| Gene_Pert | Dmd Defici | 6/402 | 0.008179 | 0.426735 | 0 | 0 | 3.650767 | 17.5464 |
| Gene_Pert | MIR124-1 ( | 4/185 | 0.00869 | 0.426735 | 0 | 0 | 5.253811 | 24.93226 |
| Gene_Pert | Map2k7 Se | 4/177 | 0.007463 | 0.426735 | 0 | 0 | 5.49899 | 26.93279 |
| Gene_Pert | CMAHP KO | 5/276 | 0.007111 | 0.426735 | 0 | 0 | 4.419494 | 21.8593 |
| Gene_Pert | Vldlr KO m | 5/290 | 0.008704 | 0.426735 | 0 | 0 | 4.199401 | 19.9218 |
| Gene_Pert | Mnat1 Defi | 5/289 | 0.008583 | 0.426735 | 0 | 0 | 4.214402 | 20.05212 |
| DisGeNET | Congenital | 2/289 | 0.358779 | 0.427079 | 0 | 0 | 1.609018 | 1.649321 |
| Gene_Pert | Vax2 KO m | 5/292 | 0.00895 | 0.427257 | 0 | 0 | 4.169712 | 19.66466 |
| DisGeNET | Childhood . | 1/102 | 0.359686 | 0.427382 | 0 | 0 | 2.280912 | 2.332289 |
| DisGeNET | Heart Deco | 1/102 | 0.359686 | 0.427382 | 0 | 0 | 2.280912 | 2.332289 |
| DisGeNET | Inflammato | 1/102 | 0.359686 | 0.427382 | 0 | 0 | 2.280912 | 2.332289 |
| DisGeNET | Adult Acute | 1/102 | 0.359686 | 0.427382 | 0 | 0 | 2.280912 | 2.332289 |
| Rare_Disea | Cholestasis | Jan-75 | 0.279326 | 0.42761 | 0 | 0 | 3.117379 | 3.975827 |
| Human_Ph | Camptodac | 1/121 | 0.410859 | 0.427651 | 0 | 0 | 1.917926 | 1.706006 |
| DisGeNET | Migraine D | 2/290 | 0.36035 | 0.427977 | 0 | 0 | 1.60335 | 1.636504 |
| UK_Bioban | Dark choco | 1/111 | 0.384451 | 0.428002 | 0 | 0 | 2.09334 | 2.001105 |
| GeDiPNet_ | Congenital | 1/105 | 0.368048 | 0.428288 | 0 | 0 | 2.214781 | 2.213764 |
| SysMyo_M | MGS1112 I | 2/120 | 0.096172 | 0.428608 | 0 | 0 | 3.947159 | 9.242743 |
| SysMyo_M | MGS54 Mu | 23-Jan | 0.095455 | 0.428608 | 0 | 0 | 10.51321 | 24.6966 |
| Rare_Disea | Maturity-o | Jan-76 | 0.282473 | 0.428898 | 0 | 0 | 3.075659 | 3.888164 |
| Rare_Disea | Paroxysma | Jan-76 | 0.282473 | 0.428898 | 0 | 0 | 3.075659 | 3.888164 |
| Rare_Disea | Myeloid sa | Jan-76 | 0.282473 | 0.428898 | 0 | 0 | 3.075659 | 3.888164 |
| Rare_Disea | Paroxysma | Jan-76 | 0.282473 | 0.428898 | 0 | 0 | 3.075659 | 3.888164 |
| DisGeNET | Infection | 3/492 | 0.361739 | 0.429345 | 0 | 0 | 1.418639 | 1.442516 |
| DisGeNET | Hypoplastic | 1/103 | 0.362485 | 0.429345 | 0 | 0 | 2.258436 | 2.291796 |
| DisGeNET | HIV Coinfe | 1/103 | 0.362485 | 0.429345 | 0 | 0 | 2.258436 | 2.291796 |
| DisGeNET | Epileptic Se | 1/103 | 0.362485 | 0.429345 | 0 | 0 | 2.258436 | 2.291796 |
| DisGeNET | Ichthyosis, | 1/103 | 0.362485 | 0.429345 | 0 | 0 | 2.258436 | 2.291796 |
| DisGeNET | Sunken eye | 1/103 | 0.362485 | 0.429345 | 0 | 0 | 2.258436 | 2.291796 |
| SysMyo_M | MGS946 M | 3/252 | 0.097262 | 0.430079 | 0 | 0 | 2.820425 | 6.572577 |
| Rare_Disea | Situs invers | Jan-77 | 0.285606 | 0.430485 | 0 | 0 | 3.035037 | 3.803331 |
| Rare_Disea | Guillain-Ba | Jan-78 | 0.288726 | 0.430485 | 0 | 0 | 2.99547 | 3.721207 |
| Rare_Disea | Diffuse sys | 2/243 | 0.285377 | 0.430485 | 0 | 0 | 1.920625 | 2.408358 |
| Rare_Disea | Cholesteato | Jan-77 | 0.285606 | 0.430485 | 0 | 0 | 3.035037 | 3.803331 |
| Rare_Disea | Castleman' | Jan-78 | 0.288726 | 0.430485 | 0 | 0 | 2.99547 | 3.721207 |
| Rare_Disea | IgM deficie | Jan-78 | 0.288726 | 0.430485 | 0 | 0 | 2.99547 | 3.721207 |
| Rare_Disea | Acute idiop | Jan-78 | 0.288726 | 0.430485 | 0 | 0 | 2.99547 | 3.721207 |
| Rare_Disea | Hyperlipop | Jan-78 | 0.288726 | 0.430485 | 0 | 0 | 2.99547 | 3.721207 |
| Rare_Disea | Congenital | Jan-78 | 0.288726 | 0.430485 | 0 | 0 | 2.99547 | 3.721207 |
| GO_Biologi | Positive Re | 1/104 | 0.365273 | 0.430803 | 0 | 0 | 2.236396 | 2.252299 |
| GO_Biologi | Macroauto | 1/104 | 0.365273 | 0.430803 | 0 | 0 | 2.236396 | 2.252299 |

| | | | | | | | | |
|---|---|---|---|---|---|---|---|---|
| GWAS_Cat | Alzheimer's | 1/122 | 0.413437 | 0.431474 | 0 | 0 | 1.90198 | 1.679923 |
| DisGeNET | Tonic Seizu | 1/104 | 0.365273 | 0.432061 | 0 | 0 | 2.236396 | 2.252299 |
| DisGeNET | Enophthalr | 1/104 | 0.365273 | 0.432061 | 0 | 0 | 2.236396 | 2.252299 |
| DisGeNET | Xerosis | 1/104 | 0.365273 | 0.432061 | 0 | 0 | 2.236396 | 2.252299 |
| GO_Biologi | Lysosomal | 1/105 | 0.368048 | 0.43264 | 0 | 0 | 2.214781 | 2.213764 |
| GO_Biologi | Positive Re | 1/105 | 0.368048 | 0.43264 | 0 | 0 | 2.214781 | 2.213764 |
| UK_Bioban | College/un | 2/312 | 0.394543 | 0.433147 | 0 | 0 | 1.487894 | 1.383781 |
| UK_Bioban | Never smo | 1/115 | 0.395152 | 0.433147 | 0 | 0 | 2.019482 | 1.875059 |
| UK_Bioban | Right 3mm | 2/310 | 0.391467 | 0.433147 | 0 | 0 | 1.497708 | 1.404633 |
| UK_Bioban | Night shift | 1/114 | 0.392494 | 0.433147 | 0 | 0 | 2.037456 | 1.905499 |
| GO_Molecı | ATPase Bin | Jan-73 | 0.272992 | 0.43332 | 0 | 0 | 3.204296 | 4.160181 |
| HuBMAP_A | Reticular C | 1/130 | 0.433666 | 0.433666 | 0 | 0 | 1.783306 | 1.489916 |
| GeDiPNet_ | Non-Syndrı | 1/107 | 0.373563 | 0.43384 | 0 | 0 | 2.172773 | 2.139459 |
| Rare_Disea | Arthrogryp | Jan-79 | 0.291832 | 0.434246 | 0 | 0 | 2.956917 | 3.641673 |
| DisGeNET | Ehlers-Dan | 1/105 | 0.368048 | 0.434363 | 0 | 0 | 2.214781 | 2.213764 |
| DisGeNET | Carotid Ath | 1/105 | 0.368048 | 0.434363 | 0 | 0 | 2.214781 | 2.213764 |
| DisGeNET | Mixed Saliv | 1/105 | 0.368048 | 0.434363 | 0 | 0 | 2.214781 | 2.213764 |
| DisGeNET | Biliary Atre | 1/105 | 0.368048 | 0.434363 | 0 | 0 | 2.214781 | 2.213764 |
| DisGeNET | Congenital | 3/497 | 0.367685 | 0.434363 | 0 | 0 | 1.403918 | 1.404659 |
| GO_Biologi | Regulation | 1/106 | 0.370812 | 0.434449 | 0 | 0 | 2.193577 | 2.176161 |
| GO_Biologi | Response T | 1/106 | 0.370812 | 0.434449 | 0 | 0 | 2.193577 | 2.176161 |
| SysMyo_M | MGS445 Hı | 3/256 | 0.100769 | 0.435384 | 0 | 0 | 2.775268 | 6.369029 |
| SysMyo_M | MGS873 M | 2/123 | 0.100238 | 0.435384 | 0 | 0 | 3.848712 | 8.852824 |
| SysMyo_M | MGS682 Hı | 2/123 | 0.100238 | 0.435384 | 0 | 0 | 3.848712 | 8.852824 |
| SysMyo_M | MGS14 Rat | 2/124 | 0.101605 | 0.435668 | 0 | 0 | 3.816972 | 8.728145 |
| Rare_Disea | Cholera | 2/248 | 0.29343 | 0.435752 | 0 | 0 | 1.88111 | 2.306459 |
| SysMyo_M | MGS128 C. | 31-Jan | 0.126498 | 0.435829 | 0 | 0 | 7.706589 | 15.93356 |
| SysMyo_M | MGS1312 I | 2/130 | 0.109909 | 0.435829 | 0 | 0 | 3.636949 | 8.030761 |
| SysMyo_M | MGS732 M | 2/141 | 0.125573 | 0.435829 | 0 | 0 | 3.34727 | 6.945157 |
| SysMyo_M | MGS502 Hı | 2/141 | 0.125573 | 0.435829 | 0 | 0 | 3.34727 | 6.945157 |
| SysMyo_M | MGS274 M | 3/300 | 0.142458 | 0.435829 | 0 | 0 | 2.358826 | 4.596658 |
| SysMyo_M | MGS256 M | 3/300 | 0.142458 | 0.435829 | 0 | 0 | 2.358826 | 4.596658 |
| SysMyo_M | MGS283 C. | 3/300 | 0.142458 | 0.435829 | 0 | 0 | 2.358826 | 4.596658 |
| SysMyo_M | MGS279 M | 3/300 | 0.142458 | 0.435829 | 0 | 0 | 2.358826 | 4.596658 |
| SysMyo_M | MGS276 M | 3/300 | 0.142458 | 0.435829 | 0 | 0 | 2.358826 | 4.596658 |
| SysMyo_M | MGS277 M | 3/300 | 0.142458 | 0.435829 | 0 | 0 | 2.358826 | 4.596658 |
| SysMyo_M | MGS1290 I | 2/145 | 0.131396 | 0.435829 | 0 | 0 | 3.252982 | 6.602064 |
| SysMyo_M | MGS923 M | 2/147 | 0.134331 | 0.435829 | 0 | 0 | 3.207789 | 6.43948 |
| SysMyo_M | MGS531 Hı | 2/134 | 0.115542 | 0.435829 | 0 | 0 | 3.526025 | 7.609588 |
| SysMyo_M | MGS989 M | 2/149 | 0.13728 | 0.435829 | 0 | 0 | 3.163826 | 6.282501 |
| SysMyo_M | MGS138 M | 6/800 | 0.135198 | 0.435829 | 0 | 0 | 1.783655 | 3.569121 |
| SysMyo_M | MGS975 M | 2/147 | 0.134331 | 0.435829 | 0 | 0 | 3.207789 | 6.43948 |
| SysMyo_M | MGS1160 I | 2/139 | 0.122685 | 0.435829 | 0 | 0 | 3.396479 | 7.126265 |
| SysMyo_M | MGS879 M | 3/280 | 0.122839 | 0.435829 | 0 | 0 | 2.531717 | 5.308703 |
| SysMyo_M | MGS362 Hı | 2/141 | 0.125573 | 0.435829 | 0 | 0 | 3.34727 | 6.945157 |

| | | | | | | | | |
|---|---|---|---|---|---|---|---|---|
| SysMyo_M | MGS816 M | 2/131 | 0.11131 | 0.435829 | 0 | 0 | 3.608573 | 7.922388 |
| SysMyo_M | MGS742 M | 2/137 | 0.119815 | 0.435829 | 0 | 0 | 3.447146 | 7.31418 |
| SysMyo_M | MGS911 M | 25-Jan | 0.103317 | 0.435829 | 0 | 0 | 9.636143 | 21.87363 |
| SysMyo_M | MGS1506 I | 2/138 | 0.121248 | 0.435829 | 0 | 0 | 3.421626 | 7.219351 |
| SysMyo_M | MGS185 M | 30-Jan | 0.122676 | 0.435829 | 0 | 0 | 7.972735 | 16.72843 |
| SysMyo_M | MGS860 M | 3/270 | 0.113436 | 0.435829 | 0 | 0 | 2.627876 | 5.719604 |
| SysMyo_M | MGS940 M | 2/134 | 0.115542 | 0.435829 | 0 | 0 | 3.526025 | 7.609588 |
| SysMyo_M | MGS1195 I | 2/135 | 0.116962 | 0.435829 | 0 | 0 | 3.499337 | 7.509252 |
| SysMyo_M | MGS1280 I | 2/135 | 0.116962 | 0.435829 | 0 | 0 | 3.499337 | 7.509252 |
| SysMyo_M | MGS1226 I | 2/129 | 0.108512 | 0.435829 | 0 | 0 | 3.665771 | 8.141279 |
| SysMyo_M | MGS118 M | 26-Jan | 0.107222 | 0.435829 | 0 | 0 | 9.250233 | 20.65441 |
| SysMyo_M | MGS173 M | 29-Jan | 0.118838 | 0.435829 | 0 | 0 | 8.25789 | 17.58927 |
| SysMyo_M | MGS42 Hu | 29-Jan | 0.118838 | 0.435829 | 0 | 0 | 8.25789 | 17.58927 |
| SysMyo_M | MGS318 M | 3/300 | 0.142458 | 0.435829 | 0 | 0 | 2.358826 | 4.596658 |
| SysMyo_M | MGS194 M | 3/300 | 0.142458 | 0.435829 | 0 | 0 | 2.358826 | 4.596658 |
| SysMyo_M | MGS206 C | 3/300 | 0.142458 | 0.435829 | 0 | 0 | 2.358826 | 4.596658 |
| SysMyo_M | MGS426 H | 3/283 | 0.125715 | 0.435829 | 0 | 0 | 2.504209 | 5.193076 |
| SysMyo_M | MGS875 M | 31-Jan | 0.126498 | 0.435829 | 0 | 0 | 7.706589 | 15.93356 |
| SysMyo_M | MGS1059 I | 2/142 | 0.127022 | 0.435829 | 0 | 0 | 3.323193 | 6.857052 |
| SysMyo_M | MGS1056 I | 2/151 | 0.140245 | 0.435829 | 0 | 0 | 3.121042 | 6.130869 |
| SysMyo_M | MGS1342 I | 2/149 | 0.13728 | 0.435829 | 0 | 0 | 3.163826 | 6.282501 |
| SysMyo_M | MGS1005 I | 2/153 | 0.143223 | 0.435829 | 0 | 0 | 3.079392 | 5.984343 |
| SysMyo_M | MGS924 M | 2/153 | 0.143223 | 0.435829 | 0 | 0 | 3.079392 | 5.984343 |
| SysMyo_M | MGS1446 I | 2/131 | 0.11131 | 0.435829 | 0 | 0 | 3.608573 | 7.922388 |
| SysMyo_M | MGS722 M | 2/131 | 0.11131 | 0.435829 | 0 | 0 | 3.608573 | 7.922388 |
| SysMyo_M | MGS1055 I | 2/149 | 0.13728 | 0.435829 | 0 | 0 | 3.163826 | 6.282501 |
| SysMyo_M | MGS273 M | 3/288 | 0.130562 | 0.435829 | 0 | 0 | 2.459649 | 5.007625 |
| SysMyo_M | MGS1275 I | 2/144 | 0.129934 | 0.435829 | 0 | 0 | 3.276056 | 6.685543 |
| SysMyo_M | MGS913 M | 2/144 | 0.129934 | 0.435829 | 0 | 0 | 3.276056 | 6.685543 |
| SysMyo_M | MGS238 M | 3/300 | 0.142458 | 0.435829 | 0 | 0 | 2.358826 | 4.596658 |
| SysMyo_M | MGS215 C | 3/300 | 0.142458 | 0.435829 | 0 | 0 | 2.358826 | 4.596658 |
| SysMyo_M | MGS291 C | 3/300 | 0.142458 | 0.435829 | 0 | 0 | 2.358826 | 4.596658 |
| SysMyo_M | MGS293 C | 3/300 | 0.142458 | 0.435829 | 0 | 0 | 2.358826 | 4.596658 |
| SysMyo_M | MGS56 Mu | 31-Jan | 0.126498 | 0.435829 | 0 | 0 | 7.706589 | 15.93356 |
| SysMyo_M | MGS285 C | 3/300 | 0.142458 | 0.435829 | 0 | 0 | 2.358826 | 4.596658 |
| GO_Biologi | Negative R | 1/107 | 0.373563 | 0.43596 | 0 | 0 | 2.172773 | 2.139459 |
| GO_Biologi | Protein Mc | 4/711 | 0.373943 | 0.43596 | 0 | 0 | 1.30918 | 1.287776 |
| GO_Biologi | Regulation | 1/107 | 0.373563 | 0.43596 | 0 | 0 | 2.172773 | 2.139459 |
| PhenGenI_ | Glucose | 3/399 | 0.251327 | 0.436376 | 0 | 0 | 1.760191 | 2.430822 |
| PhenGenI_ | Crohn Dise | 3/403 | 0.256001 | 0.436376 | 0 | 0 | 1.742232 | 2.373918 |
| PhenGenI_ | Heart Failu | 5/785 | 0.256071 | 0.436376 | 0 | 0 | 1.4957 | 2.037594 |
| PhenGenI_ | Gestationa | Jan-68 | 0.256914 | 0.436376 | 0 | 0 | 3.44429 | 4.68084 |
| GO_Biologi | Positive Re | 1/108 | 0.376303 | 0.436561 | 0 | 0 | 2.152358 | 2.10363 |
| GO_Biologi | Positive Re | 1/108 | 0.376303 | 0.436561 | 0 | 0 | 2.152358 | 2.10363 |
| GO_Biologi | Lipid Trans | 1/108 | 0.376303 | 0.436561 | 0 | 0 | 2.152358 | 2.10363 |

| | | | | | | | | |
|---|---|---|---|---|---|---|---|---|
| DisGeNET | Atrial Prem | 1/106 | 0.370812 | 0.436639 | 0 | 0 | 2.193577 | 2.176161 |
| DisGeNET | Nystagmus | 3/500 | 0.371249 | 0.436639 | 0 | 0 | 1.395229 | 1.382508 |
| DisGeNET | Idiopathic | 2/297 | 0.37131 | 0.436639 | 0 | 0 | 1.564746 | 1.550222 |
| DisGeNET | Adenocarci | 2/297 | 0.37131 | 0.436639 | 0 | 0 | 1.564746 | 1.550222 |
| DisGeNET | Kidney Fail | 2/297 | 0.37131 | 0.436639 | 0 | 0 | 1.564746 | 1.550222 |
| DisGeNET | Malignant | 1/106 | 0.370812 | 0.436639 | 0 | 0 | 2.193577 | 2.176161 |
| DisGeNET | Teratocarc | 1/106 | 0.370812 | 0.436639 | 0 | 0 | 2.193577 | 2.176161 |
| DisGeNET | Heart Failu | 1/106 | 0.370812 | 0.436639 | 0 | 0 | 2.193577 | 2.176161 |
| Jensen_TIS | Bronchoalv | 1/100 | 0.35405 | 0.436797 | 0 | 0 | 2.327226 | 2.416399 |
| GO_Biologi | Regulation | 1/109 | 0.379031 | 0.43687 | 0 | 0 | 2.132321 | 2.068645 |
| GO_Biologi | Positive Re | 1/109 | 0.379031 | 0.43687 | 0 | 0 | 2.132321 | 2.068645 |
| GO_Biologi | Cell-Matrix | 1/109 | 0.379031 | 0.43687 | 0 | 0 | 2.132321 | 2.068645 |
| GO_Biologi | Regulation | 1/109 | 0.379031 | 0.43687 | 0 | 0 | 2.132321 | 2.068645 |
| Rare_Disea | Pick's disea | Jan-80 | 0.294925 | 0.437099 | 0 | 0 | 2.919341 | 3.564619 |
| Human_Ph | Abdominal | 1/125 | 0.421106 | 0.437126 | 0 | 0 | 1.855683 | 1.604928 |
| DisGeNET | Hyperactiv | 4/709 | 0.371956 | 0.437203 | 0 | 0 | 1.313031 | 1.298561 |
| SysMyo_M | MGS1042 | 2/154 | 0.144717 | 0.438021 | 0 | 0 | 3.058978 | 5.912923 |
| Gene_Pert | Tcof1 KD m | 5/258 | 0.005379 | 0.438353 | 0 | 0 | 4.738263 | 24.75901 |
| Gene_Pert | Prkag3 KO | 6/365 | 0.005181 | 0.438353 | 0 | 0 | 4.034664 | 21.23315 |
| GeDiPNet_ | Nasophary | 1/109 | 0.379031 | 0.438442 | 0 | 0 | 2.132321 | 2.068645 |
| GeDiPNet_ | Miscarriage | 1/109 | 0.379031 | 0.438442 | 0 | 0 | 2.132321 | 2.068645 |
| DisGeNET | Pleural effu | 1/107 | 0.373563 | 0.438502 | 0 | 0 | 2.172773 | 2.139459 |
| DisGeNET | Alcohol abu | 1/107 | 0.373563 | 0.438502 | 0 | 0 | 2.172773 | 2.139459 |
| DisGeNET | White Bloo | 1/107 | 0.373563 | 0.438502 | 0 | 0 | 2.172773 | 2.139459 |
| GTEx_Tissu | GTEX-POM | 9/928 | 0.019407 | 0.438625 | 0 | 0 | 2.384783 | 9.401123 |
| GTEx_Tissu | GTEX-OIZF- | 9/927 | 0.019286 | 0.438625 | 0 | 0 | 2.387506 | 9.426763 |
| GO_Molecu | GTPase Reg | 3/424 | 0.280728 | 0.438637 | 0 | 0 | 1.653546 | 2.100616 |
| Reactome_ | Regulation | Jan-45 | 0.178311 | 0.438919 | 0 | 0 | 5.250793 | 9.053559 |
| GWAS_Cat | Hand Grip | 1/125 | 0.421106 | 0.439102 | 0 | 0 | 1.855683 | 1.604928 |
| Reactome_ | Ion Homeo | Jan-50 | 0.196069 | 0.439195 | 0 | 0 | 4.713811 | 7.680157 |
| Reactome_ | Metabolism | 2/186 | 0.193998 | 0.439195 | 0 | 0 | 2.52289 | 4.137304 |
| Reactome_ | mRNA Splic | Jan-49 | 0.192548 | 0.439195 | 0 | 0 | 4.812258 | 7.927759 |
| Reactome_ | Aquaporin- | Jan-50 | 0.196069 | 0.439195 | 0 | 0 | 4.713811 | 7.680157 |
| Reactome_ | Transport ( | 5/706 | 0.193543 | 0.439195 | 0 | 0 | 1.671132 | 2.744429 |
| Reactome_ | G Alpha (Z) | Jan-47 | 0.18546 | 0.439195 | 0 | 0 | 5.021992 | 8.46163 |
| Reactome_ | Metabolism | Jan-47 | 0.18546 | 0.439195 | 0 | 0 | 5.021992 | 8.46163 |
| Reactome_ | Formation | Jan-50 | 0.196069 | 0.439195 | 0 | 0 | 4.713811 | 7.680157 |
| Reactome_ | Inositol Ph | Jan-48 | 0.189012 | 0.439195 | 0 | 0 | 4.914894 | 8.187947 |
| GO_Biologi | Regulation | 1/110 | 0.381747 | 0.439287 | 0 | 0 | 2.112652 | 2.034479 |
| GO_Molecu | Phosphotra | Jan-77 | 0.285606 | 0.439394 | 0 | 0 | 3.035037 | 3.803331 |
| GeneSigDB | 12495447- | 2-Jan | 0.008681 | 0.439584 | 0 | 0 | 231.5349 | 1099.002 |
| Gene_Pert | OGT siOGT | 5/296 | 0.009457 | 0.439872 | 0 | 0 | 4.111558 | 19.16397 |
| SysMyo_M | MGS1108 | 2/155 | 0.146215 | 0.4402 | 0 | 0 | 3.038831 | 5.842694 |
| Gene_Pert | Rps6ka3 KC | 5/298 | 0.009718 | 0.440696 | 0 | 0 | 4.083077 | 18.9202 |
| GeDiPNet_ | Bronchiect | 1/110 | 0.381747 | 0.44071 | 0 | 0 | 2.112652 | 2.034479 |

| | | | | | | | | |
|---|---|---|---|---|---|---|---|---|
| Reactome_ | mRNA Splic | 2/189 | 0.198732 | 0.440753 | 0 | 0 | 2.482038 | 4.010467 |
| Rare_Disea | Creutzfeldt | Jan-81 | 0.298004 | 0.440785 | 0 | 0 | 2.882703 | 3.48994 |
| DisGeNET | Vesico-Ure | 1/108 | 0.376303 | 0.441125 | 0 | 0 | 2.152358 | 2.10363 |
| DisGeNET | Long face | 1/108 | 0.376303 | 0.441125 | 0 | 0 | 2.152358 | 2.10363 |
| DisGeNET | Myocardial | 1/108 | 0.376303 | 0.441125 | 0 | 0 | 2.152358 | 2.10363 |
| CellMarker | Exhausted | 1/116 | 0.397798 | 0.441282 | 0 | 0 | 2.00182 | 1.845299 |
| GWAS_Cat | Smoking Ce | 1/126 | 0.42364 | 0.441367 | 0 | 0 | 1.840744 | 1.580964 |
| GO_Biologi | Negative R | 1/111 | 0.384451 | 0.441683 | 0 | 0 | 2.09334 | 2.001105 |
| GeDiPNet_ | Macroglos | 1/111 | 0.384451 | 0.442081 | 0 | 0 | 2.09334 | 2.001105 |
| GeDiPNet_ | Keratoconu | 1/111 | 0.384451 | 0.442081 | 0 | 0 | 2.09334 | 2.001105 |
| GO_Molecu | Amyloid-Be | Jan-79 | 0.291832 | 0.442169 | 0 | 0 | 2.956917 | 3.641673 |
| DisGeNET | Senile Plaq | 2/301 | 0.37754 | 0.44218 | 0 | 0 | 1.543498 | 1.503487 |
| DisGeNET | Hyperrefle: | 2/301 | 0.37754 | 0.44218 | 0 | 0 | 1.543498 | 1.503487 |
| SysMyo_M | MGS264 Hu | 2/156 | 0.147716 | 0.442365 | 0 | 0 | 3.018946 | 5.773631 |
| GeDiPNet_ | Hemiplegia | 1/112 | 0.387144 | 0.442558 | 0 | 0 | 2.074377 | 1.968499 |
| GeDiPNet_ | Vitiligo | 1/112 | 0.387144 | 0.442558 | 0 | 0 | 2.074377 | 1.968499 |
| GeDiPNet_ | Congenital | 1/112 | 0.387144 | 0.442558 | 0 | 0 | 2.074377 | 1.968499 |
| Reactome_ | RHOQ GTP. | Jan-59 | 0.227083 | 0.443147 | 0 | 0 | 3.980553 | 5.900928 |
| Reactome_ | Deposition | Jan-53 | 0.206541 | 0.443147 | 0 | 0 | 4.44119 | 7.004886 |
| Reactome_ | NRAGE Sigr | Jan-57 | 0.220295 | 0.443147 | 0 | 0 | 4.123131 | 6.237423 |
| Reactome_ | Nuclear Re | Jan-53 | 0.206541 | 0.443147 | 0 | 0 | 4.44119 | 7.004886 |
| Reactome_ | RHOJ GTPa | Jan-54 | 0.210002 | 0.443147 | 0 | 0 | 4.357174 | 6.79997 |
| Reactome_ | Ca2+ Pathv | Jan-61 | 0.233813 | 0.443147 | 0 | 0 | 3.847481 | 5.591291 |
| Reactome_ | Ion Transpo | Jan-53 | 0.206541 | 0.443147 | 0 | 0 | 4.44119 | 7.004886 |
| Reactome_ | Translation | Jan-57 | 0.220295 | 0.443147 | 0 | 0 | 4.123131 | 6.237423 |
| Reactome_ | Ribosomal | Jan-57 | 0.220295 | 0.443147 | 0 | 0 | 4.123131 | 6.237423 |
| Reactome_ | O-linked Gl | Jan-59 | 0.227083 | 0.443147 | 0 | 0 | 3.980553 | 5.900928 |
| Reactome_ | mRNA Acti | Jan-58 | 0.223696 | 0.443147 | 0 | 0 | 4.050592 | 6.065622 |
| Reactome_ | Transcripti | Jan-60 | 0.230455 | 0.443147 | 0 | 0 | 3.912889 | 5.742942 |
| Reactome_ | GABA Rece | Jan-59 | 0.227083 | 0.443147 | 0 | 0 | 3.980553 | 5.900928 |
| Reactome_ | G Alpha (Q | 2/212 | 0.235422 | 0.443147 | 0 | 0 | 2.207619 | 3.19305 |
| Reactome_ | Phospholip | 2/208 | 0.229002 | 0.443147 | 0 | 0 | 2.250942 | 3.317943 |
| Reactome_ | Regulation | Jan-61 | 0.233813 | 0.443147 | 0 | 0 | 3.847481 | 5.591291 |
| Reactome_ | Signaling B | Jan-57 | 0.220295 | 0.443147 | 0 | 0 | 4.123131 | 6.237423 |
| Reactome_ | Compleme | Jan-55 | 0.213448 | 0.443147 | 0 | 0 | 4.27627 | 6.604112 |
| GO_Biologi | Regulation | 1/112 | 0.387144 | 0.443342 | 0 | 0 | 2.074377 | 1.968499 |
| GO_Biologi | Negative R | 1/112 | 0.387144 | 0.443342 | 0 | 0 | 2.074377 | 1.968499 |
| DisGeNET | Convulsive | 1/109 | 0.379031 | 0.443405 | 0 | 0 | 2.132321 | 2.068645 |
| DisGeNET | Flexion cor | 1/109 | 0.379031 | 0.443405 | 0 | 0 | 2.132321 | 2.068645 |
| DisGeNET | Pancreatiti | 2/302 | 0.379094 | 0.443405 | 0 | 0 | 1.538275 | 1.492081 |
| Jensen_TIS | Eye | 26/5536 | 0.360909 | 0.44356 | 0 | 0 | 1.114153 | 1.135466 |
| Jensen_TIS | Capillary | 1/103 | 0.362485 | 0.443803 | 0 | 0 | 2.258436 | 2.291796 |
| SysMyo_M | MGS126 Hu | Jan-37 | 0.149088 | 0.444125 | 0 | 0 | 6.42022 | 12.21909 |
| DisGeNET | Dry skin | 1/110 | 0.381747 | 0.444918 | 0 | 0 | 2.112652 | 2.034479 |
| DisGeNET | Post-Traum | 1/110 | 0.381747 | 0.444918 | 0 | 0 | 2.112652 | 2.034479 |

| | | | | | | | | |
|---|---|---|---|---|---|---|---|---|
| DisGeNET | Lethargy | 1/110 | 0.381747 | 0.444918 | 0 | 0 | 2.112652 | 2.034479 |
| DisGeNET | Neutrophil | 1/110 | 0.381747 | 0.444918 | 0 | 0 | 2.112652 | 2.034479 |
| DisGeNET | Borderline | 1/110 | 0.381747 | 0.444918 | 0 | 0 | 2.112652 | 2.034479 |
| DisGeNET | Abortion, T | 1/110 | 0.381747 | 0.444918 | 0 | 0 | 2.112652 | 2.034479 |
| DisGeNET | Small testic | 1/110 | 0.381747 | 0.444918 | 0 | 0 | 2.112652 | 2.034479 |
| DisGeNET | Luminal A I | 1/110 | 0.381747 | 0.444918 | 0 | 0 | 2.112652 | 2.034479 |
| Disease_Pe | Hepatitis C | 6/403 | 0.008274 | 0.445415 | 0 | 0 | 3.641384 | 17.45914 |
| GO_Biologi | Positive Re | 1/113 | 0.389825 | 0.445693 | 0 | 0 | 2.055752 | 1.936638 |
| GWAS_Cat | Smoking St | 1/128 | 0.428675 | 0.446231 | 0 | 0 | 1.811573 | 1.534505 |
| Rare_Disea | Craniorach | 2/255 | 0.304685 | 0.446232 | 0 | 0 | 1.828412 | 2.173024 |
| Rare_Disea | Diastemato | 2/255 | 0.304685 | 0.446232 | 0 | 0 | 1.828412 | 2.173024 |
| Rare_Disea | Iniencepha | 2/255 | 0.304685 | 0.446232 | 0 | 0 | 1.828412 | 2.173024 |
| Rare_Disea | Andersen T | Jan-83 | 0.304123 | 0.446232 | 0 | 0 | 2.81211 | 3.34732 |
| Rare_Disea | Occult spin | 2/255 | 0.304685 | 0.446232 | 0 | 0 | 1.828412 | 2.173024 |
| Human_Ph | Coarse faci | 1/129 | 0.431176 | 0.446367 | 0 | 0 | 1.797329 | 1.511983 |
| GeneSigDB | 17312329- | 5/461 | 0.050638 | 0.446473 | 0 | 0 | 2.60176 | 7.761203 |
| GeneSigDB | 12446012- | 10-Jan | 0.042667 | 0.446473 | 0 | 0 | 25.71576 | 81.1157 |
| GeneSigDB | 16818684- | 3/179 | 0.043267 | 0.446473 | 0 | 0 | 4.005073 | 12.57741 |
| GeneSigDB | 17471573- | Feb-88 | 0.05629 | 0.446473 | 0 | 0 | 5.424624 | 15.60794 |
| GeneSigDB | 18704457- | Feb-89 | 0.057425 | 0.446473 | 0 | 0 | 5.362001 | 15.32067 |
| GeneSigDB | 19808871- | 9/1092 | 0.047598 | 0.446473 | 0 | 0 | 2.006179 | 6.10875 |
| GeneSigDB | 18535662- | 6/605 | 0.048277 | 0.446473 | 0 | 0 | 2.388425 | 7.23886 |
| GeneSigDB | 11823860- | 14/1995 | 0.048798 | 0.446473 | 0 | 0 | 1.735999 | 5.24284 |
| GeneSigDB | 14766275- | Feb-82 | 0.04965 | 0.446473 | 0 | 0 | 5.833235 | 17.51576 |
| GeneSigDB | 18006812- | Feb-75 | 0.042299 | 0.446473 | 0 | 0 | 6.394843 | 20.22688 |
| GeneSigDB | 16467079- | 12-Jan | 0.050982 | 0.446473 | 0 | 0 | 21.03805 | 62.61499 |
| GeneSigDB | 14556934- | 14-Jan | 0.059226 | 0.446473 | 0 | 0 | 17.79964 | 50.30878 |
| GeneSigDB | 17597811- | 3/208 | 0.062301 | 0.446473 | 0 | 0 | 3.433449 | 9.530487 |
| GeneSigDB | 19118092- | 15-Jan | 0.063321 | 0.446473 | 0 | 0 | 16.52741 | 45.60793 |
| GeneSigDB | 18508997- | 15-Jan | 0.063321 | 0.446473 | 0 | 0 | 16.52741 | 45.60793 |
| GeneSigDB | 18714348- | 22-Jan | 0.091499 | 0.446473 | 0 | 0 | 11.0144 | 26.34017 |
| GeneSigDB | 19249242- | 22-Jan | 0.091499 | 0.446473 | 0 | 0 | 11.0144 | 26.34017 |
| GeneSigDB | 17312329- | Feb-66 | 0.033522 | 0.446473 | 0 | 0 | 7.297426 | 24.7787 |
| GeneSigDB | 15941854- | 24-Jan | 0.099394 | 0.446473 | 0 | 0 | 10.05561 | 23.215 |
| GeneSigDB | 14768006- | 24-Jan | 0.099394 | 0.446473 | 0 | 0 | 10.05561 | 23.215 |
| GeneSigDB | 16421595- | 16-Jan | 0.067399 | 0.446473 | 0 | 0 | 15.42481 | 41.60265 |
| GeneSigDB | 16948840- | 19-Jan | 0.079527 | 0.446473 | 0 | 0 | 12.85207 | 32.53709 |
| GeneSigDB | 12471243- | 4/369 | 0.077296 | 0.446473 | 0 | 0 | 2.58102 | 6.60771 |
| GeneSigDB | 20421987- | 15-Jan | 0.063321 | 0.446473 | 0 | 0 | 16.52741 | 45.60793 |
| GeneSigDB | 18397756- | 7-Jan | 0.03006 | 0.446473 | 0 | 0 | 38.57946 | 135.2043 |
| GeneSigDB | 17785550- | Feb-64 | 0.031682 | 0.446473 | 0 | 0 | 7.533586 | 26.00597 |
| GeneSigDB | 14990357- | 18-Jan | 0.075502 | 0.446473 | 0 | 0 | 13.60876 | 35.1596 |
| GeneSigDB | 18704457- | 8-Jan | 0.03428 | 0.446473 | 0 | 0 | 33.06645 | 111.5391 |
| GeneSigDB | 19061838- | Feb-68 | 0.035404 | 0.446473 | 0 | 0 | 7.075579 | 23.63906 |
| GeneSigDB | 12456497- | 12-Jan | 0.050982 | 0.446473 | 0 | 0 | 21.03805 | 62.61499 |

| | | | | | | | |
|---|---|---|---|---|---|---|---|
| GeneSigDB 17009876- | 8/939 | 0.05153 | 0.446473 | 0 | 0 | 2.064692 | 6.123038 |
| GeneSigDB 15064725- | 9-Jan | 0.038483 | 0.446473 | 0 | 0 | 28.93169 | 94.24608 |
| GeneSigDB 12917207- | 4/297 | 0.040671 | 0.446473 | 0 | 0 | 3.227106 | 10.33401 |
| GeneSigDB 18160667- | Feb-74 | 0.041285 | 0.446473 | 0 | 0 | 6.483987 | 20.66613 |
| GeneSigDB 15668952- | 14-Jan | 0.059226 | 0.446473 | 0 | 0 | 17.79964 | 50.30878 |
| GeneSigDB 17495134- | Feb-97 | 0.06679 | 0.446473 | 0 | 0 | 4.908483 | 13.28336 |
| GeneSigDB 15796909- | 16-Jan | 0.067399 | 0.446473 | 0 | 0 | 15.42481 | 41.60265 |
| GeneSigDB 19658182- | 13-Jan | 0.055113 | 0.446473 | 0 | 0 | 19.28391 | 55.89185 |
| GeneSigDB 15073102- | 3-Jan | 0.012994 | 0.446473 | 0 | 0 | 115.7616 | 502.7851 |
| GeneSigDB 15300801- | Feb-41 | 0.013741 | 0.446473 | 0 | 0 | 11.99035 | 51.40702 |
| GeneSigDB 15793299- | 8/967 | 0.059225 | 0.446473 | 0 | 0 | 2.001452 | 5.656943 |
| GeneSigDB 14969821- | 14-Jan | 0.059226 | 0.446473 | 0 | 0 | 17.79964 | 50.30878 |
| GeneSigDB 18927307- | 14-Jan | 0.059226 | 0.446473 | 0 | 0 | 17.79964 | 50.30878 |
| GeneSigDB 15051756- | 20-Jan | 0.083535 | 0.446473 | 0 | 0 | 12.17503 | 30.22444 |
| GeneSigDB 16414199- | 14-Jan | 0.059226 | 0.446473 | 0 | 0 | 17.79964 | 50.30878 |
| GeneSigDB 15256063- | 14-Jan | 0.059226 | 0.446473 | 0 | 0 | 17.79964 | 50.30878 |
| GeneSigDB 19038878- | 4/359 | 0.071457 | 0.446473 | 0 | 0 | 2.655082 | 7.005859 |
| GeneSigDB 12935928- | 17-Jan | 0.071459 | 0.446473 | 0 | 0 | 14.46003 | 38.15469 |
| GeneSigDB 18006812- | 3/209 | 0.063017 | 0.446473 | 0 | 0 | 3.416609 | 9.444704 |
| GeneSigDB 18794102- | 4/344 | 0.063146 | 0.446473 | 0 | 0 | 2.774344 | 7.663604 |
| GeneSigDB 12782714- | 15-Jan | 0.063321 | 0.446473 | 0 | 0 | 16.52741 | 45.60793 |
| GeneSigDB 19050837- | 24-Jan | 0.099394 | 0.446473 | 0 | 0 | 10.05561 | 23.215 |
| GeneSigDB 19303107- | 20-Jan | 0.083535 | 0.446473 | 0 | 0 | 12.17503 | 30.22444 |
| GeneSigDB 16143142- | Feb-99 | 0.069206 | 0.446473 | 0 | 0 | 4.806792 | 12.83737 |
| GeneSigDB 18662380- | 4/355 | 0.069188 | 0.446473 | 0 | 0 | 2.685889 | 7.173822 |
| GeneSigDB 18397756- | 16-Jan | 0.067399 | 0.446473 | 0 | 0 | 15.42481 | 41.60265 |
| GeneSigDB 19199503- | 16-Jan | 0.067399 | 0.446473 | 0 | 0 | 15.42481 | 41.60265 |
| GeneSigDB 19061838- | 16-Jan | 0.067399 | 0.446473 | 0 | 0 | 15.42481 | 41.60265 |
| GeneSigDB 19408105- | 5/559 | 0.096573 | 0.446473 | 0 | 0 | 2.130734 | 4.980494 |
| GeneSigDB 15950496- | 20-Jan | 0.083535 | 0.446473 | 0 | 0 | 12.17503 | 30.22444 |
| GeneSigDB 16207381- | 14/2159 | 0.082672 | 0.446473 | 0 | 0 | 1.588607 | 3.960206 |
| GeneSigDB 18362358- | 20/3353 | 0.082566 | 0.446473 | 0 | 0 | 1.484925 | 3.70364 |
| GeneSigDB 15260889- | 4/375 | 0.080912 | 0.446473 | 0 | 0 | 2.538499 | 6.382797 |
| GeneSigDB 12446012- | 19-Jan | 0.079527 | 0.446473 | 0 | 0 | 12.85207 | 32.53709 |
| GeneSigDB 16373702- | 19-Jan | 0.079527 | 0.446473 | 0 | 0 | 12.85207 | 32.53709 |
| GeneSigDB 16380157- | 11-Jan | 0.046834 | 0.446473 | 0 | 0 | 23.14302 | 70.84419 |
| GeneSigDB 18410693- | Feb-76 | 0.043322 | 0.446473 | 0 | 0 | 6.308108 | 19.80178 |
| GeneSigDB 19880498- | 4/242 | 0.021308 | 0.446473 | 0 | 0 | 3.984003 | 15.33321 |
| GeneSigDB 15917480- | 18-Jan | 0.075502 | 0.446473 | 0 | 0 | 13.60876 | 35.1596 |
| GeneSigDB 11282888- | 18-Jan | 0.075502 | 0.446473 | 0 | 0 | 13.60876 | 35.1596 |
| GeneSigDB 20490655- | 4/363 | 0.073764 | 0.446473 | 0 | 0 | 2.624962 | 6.842967 |
| GeneSigDB 16391795- | 17-Jan | 0.071459 | 0.446473 | 0 | 0 | 14.46003 | 38.15469 |
| GeneSigDB 15832406- | 13-Jan | 0.055113 | 0.446473 | 0 | 0 | 19.28391 | 55.89185 |
| GeneSigDB 19837975- | 6/623 | 0.054211 | 0.446473 | 0 | 0 | 2.316586 | 6.752548 |
| GeneSigDB 16207381- | 17/2585 | 0.051944 | 0.446473 | 0 | 0 | 1.640326 | 4.851395 |

| | | | | | | | | |
|---|---|---|---|---|---|---|---|---|
| GeneSigDB | 16627760-] | Feb-84 | 0.05183 | 0.446473 | 0 | 0 | 5.690387 | 16.84234 |
| GeneSigDB | 18006806-] | Feb-84 | 0.05183 | 0.446473 | 0 | 0 | 5.690387 | 16.84234 |
| GeneSigDB | 11834723-] | Feb-54 | 0.023124 | 0.446473 | 0 | 0 | 8.986878 | 33.8526 |
| GeneSigDB | 16627760-] | Feb-53 | 0.02233 | 0.446473 | 0 | 0 | 9.163552 | 34.83838 |
| GeneSigDB | 20369051-] | 9-Jan | 0.038483 | 0.446473 | 0 | 0 | 28.93169 | 94.24608 |
| GeneSigDB | 18565887-] | 9-Jan | 0.038483 | 0.446473 | 0 | 0 | 28.93169 | 94.24608 |
| GeneSigDB | 18378853-] | 24-Jan | 0.099394 | 0.446473 | 0 | 0 | 10.05561 | 23.215 |
| GeneSigDB | 15831697-] | 3/134 | 0.020718 | 0.446473 | 0 | 0 | 5.39313 | 20.90773 |
| GeneSigDB | 17072343-] | Feb-46 | 0.017103 | 0.446473 | 0 | 0 | 10.62513 | 43.22851 |
| GeneSigDB | 18547747-] | 3/117 | 0.014486 | 0.446473 | 0 | 0 | 6.202694 | 26.26582 |
| GeneSigDB | 16228831-] | Feb-42 | 0.014388 | 0.446473 | 0 | 0 | 11.69 | 49.58172 |
| GeneSigDB | 18338247-: | 5/328 | 0.014241 | 0.446473 | 0 | 0 | 3.69818 | 15.72324 |
| GeneSigDB | 16849555- | Feb-60 | 0.028127 | 0.446473 | 0 | 0 | 8.054767 | 28.76365 |
| GeneSigDB | 17304354-] | Feb-60 | 0.028127 | 0.446473 | 0 | 0 | 8.054767 | 28.76365 |
| GeneSigDB | 18637760-: | Feb-37 | 0.011286 | 0.446473 | 0 | 0 | 13.36336 | 59.92353 |
| GeneSigDB | 19893615-] | 3/103 | 0.010284 | 0.446473 | 0 | 0 | 7.076071 | 32.38812 |
| GeneSigDB | 20103679-] | 5-Jan | 0.021564 | 0.446473 | 0 | 0 | 57.875 | 222.0519 |
| GeneSigDB | 15486989-] | 5-Jan | 0.021564 | 0.446473 | 0 | 0 | 57.875 | 222.0519 |
| GeneSigDB | 17069663-] | Feb-55 | 0.02393 | 0.446473 | 0 | 0 | 8.81687 | 32.91021 |
| GeneSigDB | 17617570-] | Feb-56 | 0.024747 | 0.446473 | 0 | 0 | 8.653159 | 32.00857 |
| GeneSigDB | 18606719-] | 18-Jan | 0.075502 | 0.446473 | 0 | 0 | 13.60876 | 35.1596 |
| GeneSigDB | 16813654-: | 3/250 | 0.095527 | 0.446473 | 0 | 0 | 2.843551 | 6.677635 |
| GeneSigDB | 14555994-] | 24-Jan | 0.099394 | 0.446473 | 0 | 0 | 10.05561 | 23.215 |
| GeneSigDB | 11493696-] | 23-Jan | 0.095455 | 0.446473 | 0 | 0 | 10.51321 | 24.6966 |
| GeneSigDB | 15885264-] | 23-Jan | 0.095455 | 0.446473 | 0 | 0 | 10.51321 | 24.6966 |
| GeneSigDB | 12429807-] | 23-Jan | 0.095455 | 0.446473 | 0 | 0 | 10.51321 | 24.6966 |
| GeneSigDB | 17150101-] | 23-Jan | 0.095455 | 0.446473 | 0 | 0 | 10.51321 | 24.6966 |
| GeneSigDB | 16849555- | 22-Jan | 0.091499 | 0.446473 | 0 | 0 | 11.0144 | 26.34017 |
| GeneSigDB | 17002788-] | 22-Jan | 0.091499 | 0.446473 | 0 | 0 | 11.0144 | 26.34017 |
| GeneSigDB | 16651414-: | 4/386 | 0.087756 | 0.446473 | 0 | 0 | 2.464013 | 5.995422 |
| GeneSigDB | 17555561-] | 8/1054 | 0.08763 | 0.446473 | 0 | 0 | 1.82656 | 4.447005 |
| GeneSigDB | 18676831-] | 21-Jan | 0.087525 | 0.446473 | 0 | 0 | 11.5657 | 28.17205 |
| GeneSigDB | 18714348-] | 21-Jan | 0.087525 | 0.446473 | 0 | 0 | 11.5657 | 28.17205 |
| GeneSigDB | 16705683-] | 21-Jan | 0.087525 | 0.446473 | 0 | 0 | 11.5657 | 28.17205 |
| GeneSigDB | 17483295-] | 2/122 | 0.098877 | 0.446473 | 0 | 0 | 3.88098 | 8.980101 |
| GeneSigDB | 17012836-] | 7-Jan | 0.03006 | 0.446473 | 0 | 0 | 38.57946 | 135.2043 |
| GeneSigDB | 15897907-: | 21/3159 | 0.027906 | 0.446473 | 0 | 0 | 1.700924 | 6.087458 |
| GeneSigDB | 18927307-] | 4/263 | 0.027828 | 0.446473 | 0 | 0 | 3.657068 | 13.09856 |
| SysMyo_M | MGS904 M | 2/158 | 0.150727 | 0.446657 | 0 | 0 | 2.97994 | 5.638896 |
| DisGeNET | Plasmacyto | 1/111 | 0.384451 | 0.447076 | 0 | 0 | 2.09334 | 2.001105 |
| DisGeNET | Thyroid car | 4/721 | 0.383874 | 0.447076 | 0 | 0 | 1.290249 | 1.235337 |
| DisGeNET | Tumor Ang | 3/511 | 0.384285 | 0.447076 | 0 | 0 | 1.364244 | 1.304722 |
| DisGeNET | Diabetes, A | 1/111 | 0.384451 | 0.447076 | 0 | 0 | 2.09334 | 2.001105 |
| DisGeNET | Hydroneph | 1/111 | 0.384451 | 0.447076 | 0 | 0 | 2.09334 | 2.001105 |
| WikiPathw | Synaptic Si | Jan-50 | 0.196069 | 0.447645 | 0 | 0 | 4.713811 | 7.680157 |

| | | | | | | | | |
|---|---|---|---|---|---|---|---|---|
| WikiPathw | Parkin Ubi( | Jan-71 | 0.266602 | 0.447645 | 0 | 0 | 3.296179 | 4.35754 |
| WikiPathw | 15Q11Q13 | Jan-58 | 0.223696 | 0.447645 | 0 | 0 | 4.050592 | 6.065622 |
| WikiPathw | Primary Fo | Jan-72 | 0.269804 | 0.447645 | 0 | 0 | 3.249591 | 4.257158 |
| WikiPathw | T Cell Activ | Jan-88 | 0.319191 | 0.447645 | 0 | 0 | 2.649826 | 3.026014 |
| WikiPathw | Hair Follicle | Jan-87 | 0.316203 | 0.447645 | 0 | 0 | 2.680773 | 3.086563 |
| WikiPathw | Vitamin B1 | Jan-50 | 0.196069 | 0.447645 | 0 | 0 | 4.713811 | 7.680157 |
| WikiPathw | Non Genor | Jan-75 | 0.279326 | 0.447645 | 0 | 0 | 3.117379 | 3.975827 |
| WikiPathw | Pathways A | Jan-65 | 0.247099 | 0.447645 | 0 | 0 | 3.606286 | 5.041467 |
| WikiPathw | Physico Ch | Jan-66 | 0.250385 | 0.447645 | 0 | 0 | 3.550626 | 4.916752 |
| WikiPathw | Melanoma | Jan-68 | 0.256914 | 0.447645 | 0 | 0 | 3.44429 | 4.68084 |
| WikiPathw | Leptin Sign | Jan-71 | 0.266602 | 0.447645 | 0 | 0 | 3.296179 | 4.35754 |
| WikiPathw | Alzheimer : | 2/260 | 0.312707 | 0.447645 | 0 | 0 | 1.792522 | 2.083787 |
| WikiPathw | IL 18 Signal | Jan-56 | 0.216879 | 0.447645 | 0 | 0 | 4.198309 | 6.416764 |
| WikiPathw | Hematopoi | Jan-54 | 0.210002 | 0.447645 | 0 | 0 | 4.357174 | 6.79997 |
| WikiPathw | Apoptosis I | Jan-53 | 0.206541 | 0.447645 | 0 | 0 | 4.44119 | 7.004886 |
| WikiPathw | Cardiac Pro | Jan-53 | 0.206541 | 0.447645 | 0 | 0 | 4.44119 | 7.004886 |
| WikiPathw | Copper Ho | Jan-51 | 0.199575 | 0.447645 | 0 | 0 | 4.619302 | 7.444308 |
| WikiPathw | IL 24 Signal | 2/232 | 0.267638 | 0.447645 | 0 | 0 | 2.013606 | 2.654174 |
| WikiPathw | Alzheimer : | 2/260 | 0.312707 | 0.447645 | 0 | 0 | 1.792522 | 2.083787 |
| WikiPathw | Lung Fibros | Jan-63 | 0.240485 | 0.447645 | 0 | 0 | 3.722993 | 5.305634 |
| WikiPathw | ncRNAs Inv | Jan-85 | 0.310189 | 0.447645 | 0 | 0 | 2.744878 | 3.21308 |
| WikiPathw | MECP2 And | Jan-73 | 0.272992 | 0.447645 | 0 | 0 | 3.204296 | 4.160181 |
| WikiPathw | Integrated | Jan-44 | 0.174713 | 0.447645 | 0 | 0 | 5.373175 | 9.3741 |
| WikiPathw | TROP2 Reg | Jan-45 | 0.178311 | 0.447645 | 0 | 0 | 5.250793 | 9.053559 |
| WikiPathw | Microtubul | Jan-46 | 0.181893 | 0.447645 | 0 | 0 | 5.13385 | 8.749803 |
| WikiPathw | Wnt Signal | Jan-47 | 0.18546 | 0.447645 | 0 | 0 | 5.021992 | 8.46163 |
| WikiPathw | Endometria | Jan-63 | 0.240485 | 0.447645 | 0 | 0 | 3.722993 | 5.305634 |
| WikiPathw | Extrafollicu | Jan-74 | 0.276166 | 0.447645 | 0 | 0 | 3.160242 | 4.066453 |
| WikiPathw | 16P11 2 Pr | Jan-74 | 0.276166 | 0.447645 | 0 | 0 | 3.160242 | 4.066453 |
| WikiPathw | Head And I | Jan-73 | 0.272992 | 0.447645 | 0 | 0 | 3.204296 | 4.160181 |
| WikiPathw | Primary Ov | 2/170 | 0.16904 | 0.447645 | 0 | 0 | 2.765406 | 4.91584 |
| WikiPathw | Androgen I | Jan-87 | 0.316203 | 0.447645 | 0 | 0 | 2.680773 | 3.086563 |
| WikiPathw | Clear Cell R | Jan-85 | 0.310189 | 0.447645 | 0 | 0 | 2.744878 | 3.21308 |
| WikiPathw | EGFR Tyros | Jan-84 | 0.307163 | 0.447645 | 0 | 0 | 2.778089 | 3.279195 |
| WikiPathw | Acute Viral | Jan-84 | 0.307163 | 0.447645 | 0 | 0 | 2.778089 | 3.279195 |
| WikiPathw | A Network | Jan-82 | 0.30107 | 0.447645 | 0 | 0 | 2.846971 | 3.417539 |
| WikiPathw | Glioblastor | Jan-82 | 0.30107 | 0.447645 | 0 | 0 | 2.846971 | 3.417539 |
| WikiPathw | Clock Contı | Jan-80 | 0.294925 | 0.447645 | 0 | 0 | 2.919341 | 3.564619 |
| WikiPathw | Markers Of | Jan-59 | 0.227083 | 0.447645 | 0 | 0 | 3.980553 | 5.900928 |
| Rare_Disea | Exencepha | 2/256 | 0.306291 | 0.447702 | 0 | 0 | 1.821121 | 2.154787 |
| Human_Ph | Ptosis (HP:( | 2/338 | 0.433875 | 0.447947 | 0 | 0 | 1.370938 | 1.144732 |
| GO_Biologi | Negative R | 1/114 | 0.392494 | 0.448024 | 0 | 0 | 2.037456 | 1.905499 |
| Rare_Disea | Still's disea | Jan-84 | 0.307163 | 0.448096 | 0 | 0 | 2.778089 | 3.279195 |
| UK_Bioban | Self-report | 1/121 | 0.410859 | 0.448639 | 0 | 0 | 1.917926 | 1.706006 |
| SysMyo_M | MGS994 M | 2/159 | 0.152237 | 0.448783 | 0 | 0 | 2.960809 | 5.573176 |

| | | | | | | | | |
|---|---|---|---|---|---|---|---|---|
| DisGeNET | Malignant | Oct-26 | 0.386418 | 0.449011 | 0 | 0 | 1.15292 | 1.096237 |
| DisGeNET | Cholera | 1/112 | 0.387144 | 0.449011 | 0 | 0 | 2.074377 | 1.968499 |
| DisGeNET | Neonatal H | 1/112 | 0.387144 | 0.449011 | 0 | 0 | 2.074377 | 1.968499 |
| DisGeNET | Bronchiect | 1/112 | 0.387144 | 0.449011 | 0 | 0 | 2.074377 | 1.968499 |
| DisGeNET | Bilateral ca | 1/112 | 0.387144 | 0.449011 | 0 | 0 | 2.074377 | 1.968499 |
| DisGeNET | Infectious ( | 1/112 | 0.387144 | 0.449011 | 0 | 0 | 2.074377 | 1.968499 |
| WikiPathw | lncRNA In ( | Jan-93 | 0.333936 | 0.449088 | 0 | 0 | 2.505182 | 2.747701 |
| WikiPathw | Compleme | Jan-95 | 0.339745 | 0.449088 | 0 | 0 | 2.451633 | 2.646685 |
| WikiPathw | Small Cell L | Jan-95 | 0.339745 | 0.449088 | 0 | 0 | 2.451633 | 2.646685 |
| WikiPathw | P53 Transc | Jan-91 | 0.328076 | 0.449088 | 0 | 0 | 2.561111 | 2.854384 |
| WikiPathw | Corticotrop | Jan-93 | 0.333936 | 0.449088 | 0 | 0 | 2.505182 | 2.747701 |
| UK_Bioban | Doctor diag | 1/122 | 0.413437 | 0.449731 | 0 | 0 | 1.90198 | 1.679923 |
| GO_Biologi | Endoplasm | 1/115 | 0.395152 | 0.450333 | 0 | 0 | 2.019482 | 1.875059 |
| Disease_Pe | facioscapul | 4/228 | 0.017541 | 0.450441 | 0 | 0 | 4.236015 | 17.12723 |
| GO_Biologi | Protein Tra | 2/313 | 0.396079 | 0.450667 | 0 | 0 | 1.483034 | 1.3735 |
| SysMyo_M | MGS929 M | 2/160 | 0.153751 | 0.450896 | 0 | 0 | 2.941921 | 5.508522 |
| GWAS_Cat | Predicted \ | 1/130 | 0.433666 | 0.451043 | 0 | 0 | 1.783306 | 1.489916 |
| GeneSigDB | 17671232- | 2/182 | 0.18771 | 0.451283 | 0 | 0 | 2.579477 | 4.315096 |
| GeneSigDB | 17885619- | Jan-48 | 0.189012 | 0.451283 | 0 | 0 | 4.914894 | 8.187947 |
| GeneSigDB | 16872506- | 10/1548 | 0.134448 | 0.451283 | 0 | 0 | 1.551602 | 3.113413 |
| GeneSigDB | 17349526- | Jan-34 | 0.137866 | 0.451283 | 0 | 0 | 7.004933 | 13.88007 |
| GeneSigDB | 17213801- | Jan-34 | 0.137866 | 0.451283 | 0 | 0 | 7.004933 | 13.88007 |
| GeneSigDB | 19377508- | Jan-47 | 0.18546 | 0.451283 | 0 | 0 | 5.021992 | 8.46163 |
| GeneSigDB | 16597596- | Jan-47 | 0.18546 | 0.451283 | 0 | 0 | 5.021992 | 8.46163 |
| GeneSigDB | 15618438- | Jan-49 | 0.192548 | 0.451283 | 0 | 0 | 4.812258 | 7.927759 |
| GeneSigDB | 15161681- | Jan-49 | 0.192548 | 0.451283 | 0 | 0 | 4.812258 | 7.927759 |
| GeneSigDB | 19204204- | 11/1746 | 0.135699 | 0.451283 | 0 | 0 | 1.516442 | 3.028811 |
| GeneSigDB | 12917207- | 4/517 | 0.188021 | 0.451283 | 0 | 0 | 1.822495 | 3.045753 |
| GeneSigDB | 15260889- | 4/517 | 0.188021 | 0.451283 | 0 | 0 | 1.822495 | 3.045753 |
| GeneSigDB | 20407444- | Jan-48 | 0.189012 | 0.451283 | 0 | 0 | 4.914894 | 8.187947 |
| GeneSigDB | 15618438- | Jan-45 | 0.178311 | 0.451283 | 0 | 0 | 5.250793 | 9.053559 |
| GeneSigDB | 18778486- | Jan-45 | 0.178311 | 0.451283 | 0 | 0 | 5.250793 | 9.053559 |
| GeneSigDB | 20215539- | Jan-48 | 0.189012 | 0.451283 | 0 | 0 | 4.914894 | 8.187947 |
| GeneSigDB | 19373655- | Jan-48 | 0.189012 | 0.451283 | 0 | 0 | 4.914894 | 8.187947 |
| GeneSigDB | 12900505- | Jan-35 | 0.141623 | 0.451283 | 0 | 0 | 6.798564 | 13.28839 |
| GeneSigDB | 14517214- | Jan-33 | 0.134093 | 0.451283 | 0 | 0 | 7.224201 | 14.515 |
| GeneSigDB | 20436685- | 6/802 | 0.136338 | 0.451283 | 0 | 0 | 1.778988 | 3.544846 |
| GeneSigDB | 15757864- | 30-Jan | 0.122676 | 0.451283 | 0 | 0 | 7.972735 | 16.72843 |
| GeneSigDB | 17430594- | 2/141 | 0.125573 | 0.451283 | 0 | 0 | 3.34727 | 6.945157 |
| GeneSigDB | 12598313- | 31-Jan | 0.126498 | 0.451283 | 0 | 0 | 7.706589 | 15.93356 |
| GeneSigDB | 18403781- | 31-Jan | 0.126498 | 0.451283 | 0 | 0 | 7.706589 | 15.93356 |
| GeneSigDB | 16717027- | 31-Jan | 0.126498 | 0.451283 | 0 | 0 | 7.706589 | 15.93356 |
| GeneSigDB | 18427120- | 2/142 | 0.127022 | 0.451283 | 0 | 0 | 3.323193 | 6.857052 |
| GeneSigDB | 19043454- | 2/176 | 0.178336 | 0.451283 | 0 | 0 | 2.669236 | 4.601995 |
| GeneSigDB | 18662380- | 3/286 | 0.128615 | 0.451283 | 0 | 0 | 2.477284 | 5.080743 |

| | | | | | | | |
|---|---|---|---|---|---|---|---|
| GeneSigDB 17206280-` | 2/133 | 0.114127 | 0.451283 | 0 | 0 | 3.553121 | 7.711855 |
| GeneSigDB 20421987-` | 6/765 | 0.116019 | 0.451283 | 0 | 0 | 1.869321 | 4.026525 |
| GeneSigDB 15467184-` | 29-Jan | 0.118838 | 0.451283 | 0 | 0 | 8.25789 | 17.58927 |
| GeneSigDB 19061838-` | 29-Jan | 0.118838 | 0.451283 | 0 | 0 | 8.25789 | 17.58927 |
| GeneSigDB 17170073-` | 29-Jan | 0.118838 | 0.451283 | 0 | 0 | 8.25789 | 17.58927 |
| GeneSigDB 20166207-` | 30-Jan | 0.122676 | 0.451283 | 0 | 0 | 7.972735 | 16.72843 |
| GeneSigDB 16597596-` | Jan-49 | 0.192548 | 0.451283 | 0 | 0 | 4.812258 | 7.927759 |
| GeneSigDB 15980968-` | Jan-42 | 0.16747 | 0.451283 | 0 | 0 | 5.635848 | 10.07097 |
| GeneSigDB 14633610-` | Jan-42 | 0.16747 | 0.451283 | 0 | 0 | 5.635848 | 10.07097 |
| GeneSigDB 14997935-` | 27-Jan | 0.111111 | 0.451283 | 0 | 0 | 8.894007 | 19.54216 |
| GeneSigDB 18510698-` | 25-Jan | 0.103317 | 0.451283 | 0 | 0 | 9.636143 | 21.87363 |
| GeneSigDB 15120971-` | 27-Jan | 0.111111 | 0.451283 | 0 | 0 | 8.894007 | 19.54216 |
| GeneSigDB 20081105-` | 25-Jan | 0.103317 | 0.451283 | 0 | 0 | 9.636143 | 21.87363 |
| GeneSigDB 17096850-` | Jan-50 | 0.196069 | 0.451283 | 0 | 0 | 4.713811 | 7.680157 |
| GeneSigDB 14767469-` | Jan-48 | 0.189012 | 0.451283 | 0 | 0 | 4.914894 | 8.187947 |
| GeneSigDB 15817677-` | Jan-42 | 0.16747 | 0.451283 | 0 | 0 | 5.635848 | 10.07097 |
| GeneSigDB 16597596-` | Jan-49 | 0.192548 | 0.451283 | 0 | 0 | 4.812258 | 7.927759 |
| GeneSigDB 18398820-` | Jan-49 | 0.192548 | 0.451283 | 0 | 0 | 4.812258 | 7.927759 |
| GeneSigDB 16597596-` | Jan-49 | 0.192548 | 0.451283 | 0 | 0 | 4.812258 | 7.927759 |
| GeneSigDB 19088021-: | 5/708 | 0.195053 | 0.451283 | 0 | 0 | 1.666204 | 2.723383 |
| GeneSigDB 12782714-` | Jan-50 | 0.196069 | 0.451283 | 0 | 0 | 4.713811 | 7.680157 |
| GeneSigDB 14735469-` | Jan-50 | 0.196069 | 0.451283 | 0 | 0 | 4.713811 | 7.680157 |
| GeneSigDB 20035825-` | Jan-50 | 0.196069 | 0.451283 | 0 | 0 | 4.713811 | 7.680157 |
| GeneSigDB 19112514-` | Jan-33 | 0.134093 | 0.451283 | 0 | 0 | 7.224201 | 14.515 |
| GeneSigDB 16597596-` | Jan-42 | 0.16747 | 0.451283 | 0 | 0 | 5.635848 | 10.07097 |
| GeneSigDB 16581771-` | 30-Jan | 0.122676 | 0.451283 | 0 | 0 | 7.972735 | 16.72843 |
| GeneSigDB 12640676-` | 30-Jan | 0.122676 | 0.451283 | 0 | 0 | 7.972735 | 16.72843 |
| GeneSigDB 12734205-` | 2/167 | 0.164425 | 0.451283 | 0 | 0 | 2.816114 | 5.083934 |
| GeneSigDB 16936776-( | 2/175 | 0.176781 | 0.451283 | 0 | 0 | 2.684801 | 4.652344 |
| GeneSigDB 16909099-: | Jan-44 | 0.174713 | 0.451283 | 0 | 0 | 5.373175 | 9.3741 |
| GeneSigDB 14722241-` | Jan-44 | 0.174713 | 0.451283 | 0 | 0 | 5.373175 | 9.3741 |
| GeneSigDB 18086899-` | Jan-44 | 0.174713 | 0.451283 | 0 | 0 | 5.373175 | 9.3741 |
| GeneSigDB 18692832-` | Jan-47 | 0.18546 | 0.451283 | 0 | 0 | 5.021992 | 8.46163 |
| GeneSigDB 15240595-` | Jan-47 | 0.18546 | 0.451283 | 0 | 0 | 5.021992 | 8.46163 |
| GeneSigDB 15908647-: | Jan-43 | 0.171099 | 0.451283 | 0 | 0 | 5.501384 | 9.712751 |
| GeneSigDB 15608061-` | Jan-42 | 0.16747 | 0.451283 | 0 | 0 | 5.635848 | 10.07097 |
| GeneSigDB 16919171-` | Jan-46 | 0.181893 | 0.451283 | 0 | 0 | 5.13385 | 8.749803 |
| GeneSigDB 19840643-` | Jan-46 | 0.181893 | 0.451283 | 0 | 0 | 5.13385 | 8.749803 |
| GeneSigDB 15817677-` | Jan-46 | 0.181893 | 0.451283 | 0 | 0 | 5.13385 | 8.749803 |
| GeneSigDB 18698033-` | 4/469 | 0.14768 | 0.451283 | 0 | 0 | 2.015598 | 3.855244 |
| GeneSigDB 16557592-` | 26-Jan | 0.107222 | 0.451283 | 0 | 0 | 9.250233 | 20.65441 |
| GeneSigDB 16965636- | 26-Jan | 0.107222 | 0.451283 | 0 | 0 | 9.250233 | 20.65441 |
| GeneSigDB 14585372-` | Jan-36 | 0.145363 | 0.451283 | 0 | 0 | 6.603987 | 12.73591 |
| GeneSigDB 18992152-` | Jan-36 | 0.145363 | 0.451283 | 0 | 0 | 6.603987 | 12.73591 |
| GeneSigDB 12972399-` | Jan-40 | 0.160165 | 0.451283 | 0 | 0 | 5.925462 | 10.85279 |

| | | | | | | | |
|---|---|---|---|---|---|---|---|
| GeneSigDB 17210693-‹ | Jan-40 | 0.160165 | 0.451283 | 0 | 0 | 5.925462 | 10.85279 |
| GeneSigDB 15136595-‹ | Jan-35 | 0.141623 | 0.451283 | 0 | 0 | 6.798564 | 13.28839 |
| GeneSigDB 15705876-‹ | Jan-35 | 0.141623 | 0.451283 | 0 | 0 | 6.798564 | 13.28839 |
| GeneSigDB 16760442-‹ | Jan-32 | 0.130304 | 0.451283 | 0 | 0 | 7.457614 | 15.19775 |
| GeneSigDB 12086872-‹ | Jan-34 | 0.137866 | 0.451283 | 0 | 0 | 7.004933 | 13.88007 |
| GeneSigDB 15548366-‹ | Jan-37 | 0.149088 | 0.451283 | 0 | 0 | 6.42022 | 12.21909 |
| GeneSigDB 15843827-‹ | Jan-37 | 0.149088 | 0.451283 | 0 | 0 | 6.42022 | 12.21909 |
| GeneSigDB 20060365-‹ | Jan-37 | 0.149088 | 0.451283 | 0 | 0 | 6.42022 | 12.21909 |
| GeneSigDB 19204204-‹ | 4/459 | 0.13975 | 0.451283 | 0 | 0 | 2.060956 | 4.055757 |
| GeneSigDB 16188652-‹ | Jan-49 | 0.192548 | 0.451283 | 0 | 0 | 4.812258 | 7.927759 |
| GeneSigDB 20194811- | Jan-45 | 0.178311 | 0.451283 | 0 | 0 | 5.250793 | 9.053559 |
| GeneSigDB 19840643-‹ | Jan-45 | 0.178311 | 0.451283 | 0 | 0 | 5.250793 | 9.053559 |
| GeneSigDB 15817677-‹ | Jan-50 | 0.196069 | 0.451283 | 0 | 0 | 4.713811 | 7.680157 |
| GeneSigDB 18955564-‹ | Jan-50 | 0.196069 | 0.451283 | 0 | 0 | 4.713811 | 7.680157 |
| GeneSigDB 20103679-‹ | 3/308 | 0.150586 | 0.451283 | 0 | 0 | 2.296019 | 4.346874 |
| GeneSigDB 18438415-‹ | 2/159 | 0.152237 | 0.451283 | 0 | 0 | 2.960809 | 5.573176 |
| GeneSigDB 16707422-‹ | Jan-48 | 0.189012 | 0.451283 | 0 | 0 | 4.914894 | 8.187947 |
| GeneSigDB 19061838-‹ | 3/345 | 0.189953 | 0.451283 | 0 | 0 | 2.043755 | 3.394639 |
| GeneSigDB 17174972-‹ | 3/346 | 0.191052 | 0.451283 | 0 | 0 | 2.037693 | 3.372806 |
| GeneSigDB 11694447- | Jan-49 | 0.192548 | 0.451283 | 0 | 0 | 4.812258 | 7.927759 |
| GeneSigDB 17122647-‹ | Jan-49 | 0.192548 | 0.451283 | 0 | 0 | 4.812258 | 7.927759 |
| GeneSigDB 19755675-‹ | 4/417 | 0.108494 | 0.451283 | 0 | 0 | 2.275446 | 5.053909 |
| GeneSigDB 15150091-‹ | Jan-38 | 0.152796 | 0.451283 | 0 | 0 | 6.246386 | 11.73478 |
| GeneSigDB 12890387-‹ | Jan-39 | 0.156488 | 0.451283 | 0 | 0 | 6.081701 | 11.28018 |
| GeneSigDB 16518402-‹ | Jan-39 | 0.156488 | 0.451283 | 0 | 0 | 6.081701 | 11.28018 |
| GeneSigDB 15073113-‹ | 26-Jan | 0.107222 | 0.451283 | 0 | 0 | 9.250233 | 20.65441 |
| GeneSigDB 15305381-‹ | Jan-38 | 0.152796 | 0.451283 | 0 | 0 | 6.246386 | 11.73478 |
| GeneSigDB 16289389-‹ | 2/162 | 0.156786 | 0.451283 | 0 | 0 | 2.904853 | 5.382321 |
| GeneSigDB 16488994-‹ | 4/484 | 0.159898 | 0.451283 | 0 | 0 | 1.951104 | 3.576799 |
| GeneSigDB 16919171-‹ | Jan-43 | 0.171099 | 0.451283 | 0 | 0 | 5.501384 | 9.712751 |
| GeneSigDB 15220918-‹ | 8/1230 | 0.165704 | 0.451283 | 0 | 0 | 1.548903 | 2.784232 |
| GeneSigDB 20386565-‹ | 5/635 | 0.142975 | 0.451283 | 0 | 0 | 1.866338 | 3.630182 |
| GeneSigDB 15486989-‹ | 2/144 | 0.129934 | 0.451283 | 0 | 0 | 3.276056 | 6.685543 |
| GeneSigDB 19306436-‹ | 2/168 | 0.165961 | 0.451283 | 0 | 0 | 2.799008 | 5.027029 |
| GeneSigDB 18032706-‹ | 4/408 | 0.102259 | 0.451283 | 0 | 0 | 2.32721 | 5.30662 |
| GeneSigDB 16488994-‹ | 4/489 | 0.164053 | 0.451283 | 0 | 0 | 1.930493 | 3.489493 |
| GeneSigDB 11983058-‹ | Jan-41 | 0.163825 | 0.451283 | 0 | 0 | 5.777035 | 10.45039 |
| GeneSigDB 15757864-‹ | Jan-41 | 0.163825 | 0.451283 | 0 | 0 | 5.777035 | 10.45039 |
| GeneSigDB 15492233-‹ | 3/318 | 0.160951 | 0.451283 | 0 | 0 | 2.221995 | 4.058822 |
| GeneSigDB 17510386-‹ | Jan-40 | 0.160165 | 0.451283 | 0 | 0 | 5.925462 | 10.85279 |
| GeneSigDB 12586635- | Jan-32 | 0.130304 | 0.451283 | 0 | 0 | 7.457614 | 15.19775 |
| GeneSigDB 18535662-‹ | 5/679 | 0.173587 | 0.451283 | 0 | 0 | 1.740519 | 3.04778 |
| GeneSigDB 19204204-‹ | 2/143 | 0.128476 | 0.451283 | 0 | 0 | 3.299458 | 6.770526 |
| GeneSigDB 12644218-‹ | 27-Jan | 0.111111 | 0.451283 | 0 | 0 | 8.894007 | 19.54216 |
| GeneSigDB 17935615-‹ | 27-Jan | 0.111111 | 0.451283 | 0 | 0 | 8.894007 | 19.54216 |

| | | | | | | | | |
|---|---|---|---|---|---|---|---|---|
| DisGeNET | Adenocarci | 1/113 | 0.389825 | 0.451521 | 0 | 0 | 2.055752 | 1.936638 |
| DisGeNET | Erythema | 1/113 | 0.389825 | 0.451521 | 0 | 0 | 2.055752 | 1.936638 |
| DisGeNET | Lipodystro| | 1/113 | 0.389825 | 0.451521 | 0 | 0 | 2.055752 | 1.936638 |
| Rare_Disea | Idiopathic a | Jan-85 | 0.310189 | 0.451626 | 0 | 0 | 2.744878 | 3.21308 |
| GTEx_Tissu | GTEX-R55E | 9/934 | 0.020143 | 0.451736 | 0 | 0 | 2.368565 | 9.248999 |
| DisGeNET | Malignant | 3/516 | 0.390193 | 0.451748 | 0 | 0 | 1.350599 | 1.271068 |
| GeDiPNet_ | Coronary A | 2/313 | 0.396079 | 0.451886 | 0 | 0 | 1.483034 | 1.3735 |
| GeDiPNet_ | Ventricular | 2/314 | 0.397613 | 0.452078 | 0 | 0 | 1.478205 | 1.363314 |
| GeDiPNet_ | Endometria | 1/116 | 0.397798 | 0.452078 | 0 | 0 | 2.00182 | 1.845299 |
| DisGeNET | Mental Ret | 6/1158 | 0.390721 | 0.452159 | 0 | 0 | 1.20634 | 1.133672 |
| GWAS_Cat | Externalizir | 2/339 | 0.435362 | 0.45242 | 0 | 0 | 1.3668 | 1.136601 |
| GeDiPNet_ | Kidney Dise | 2/315 | 0.399145 | 0.452427 | 0 | 0 | 1.473407 | 1.353222 |
| GeDiPNet_ | Clonic Seizu | 1/117 | 0.400433 | 0.452427 | 0 | 0 | 1.984463 | 1.816198 |
| GeDiPNet_ | Pancreatic | 1/117 | 0.400433 | 0.452427 | 0 | 0 | 1.984463 | 1.816198 |
| SysMyo_M | MGS1257 I | 2/163 | 0.158308 | 0.452538 | 0 | 0 | 2.886664 | 5.320729 |
| SysMyo_M | MGS1285 I | 2/163 | 0.158308 | 0.452538 | 0 | 0 | 2.886664 | 5.320729 |
| SysMyo_M | MGS444 Hu | 2/161 | 0.155267 | 0.452538 | 0 | 0 | 2.92327 | 5.444911 |
| SysMyo_M | MGS48 Hur | Jan-39 | 0.156488 | 0.452538 | 0 | 0 | 6.081701 | 11.28018 |
| SysMyo_M | MGS1003 I | 2/162 | 0.156786 | 0.452538 | 0 | 0 | 2.904853 | 5.382321 |
| DisGeNET | Gingival Di | 1/114 | 0.392494 | 0.453011 | 0 | 0 | 2.037456 | 1.905499 |
| DisGeNET | Testicular ( | 1/114 | 0.392494 | 0.453011 | 0 | 0 | 2.037456 | 1.905499 |
| DisGeNET | Urticaria | 1/114 | 0.392494 | 0.453011 | 0 | 0 | 2.037456 | 1.905499 |
| DisGeNET | Carcinoma, | 1/114 | 0.392494 | 0.453011 | 0 | 0 | 2.037456 | 1.905499 |
| DisGeNET | Dermatom | 1/114 | 0.392494 | 0.453011 | 0 | 0 | 2.037456 | 1.905499 |
| DisGeNET | Lymphocyt | 1/114 | 0.392494 | 0.453011 | 0 | 0 | 2.037456 | 1.905499 |
| SysMyo_M | MGS175 M | Jan-40 | 0.160165 | 0.453267 | 0 | 0 | 5.925462 | 10.85279 |
| SysMyo_M | MGS925 M | 2/164 | 0.159833 | 0.453267 | 0 | 0 | 2.8687 | 5.260115 |
| UK_Bioban | QRS durati | 1/124 | 0.418561 | 0.453573 | 0 | 0 | 1.870864 | 1.629399 |
| PhenGenI_ | Ankle Bracl | 2/234 | 0.270864 | 0.453677 | 0 | 0 | 1.996045 | 2.607109 |
| PhenGenI_ | Autosomal | Jan-73 | 0.272992 | 0.453677 | 0 | 0 | 3.204296 | 4.160181 |
| PhenGenI_ | Varicose Ve | Jan-73 | 0.272992 | 0.453677 | 0 | 0 | 3.204296 | 4.160181 |
| GeneSigDB | 16280042- | Jan-51 | 0.199575 | 0.453728 | 0 | 0 | 4.619302 | 7.444308 |
| GeneSigDB | 15318932- | Jan-51 | 0.199575 | 0.453728 | 0 | 0 | 4.619302 | 7.444308 |
| GeneSigDB | 18033314- | Jan-51 | 0.199575 | 0.453728 | 0 | 0 | 4.619302 | 7.444308 |
| Rare_Disea | Hypopituita | Jan-86 | 0.313203 | 0.454235 | 0 | 0 | 2.712449 | 3.148895 |
| Rare_Disea | Primary am | Jan-86 | 0.313203 | 0.454235 | 0 | 0 | 2.712449 | 3.148895 |
| SysMyo_M | MGS1146 I | 2/165 | 0.161361 | 0.45438 | 0 | 0 | 2.850956 | 5.200458 |
| GO_Biologi | Cellular Re | 1/117 | 0.400433 | 0.45444 | 0 | 0 | 1.984463 | 1.816198 |
| GO_Biologi | Supramole | 2/316 | 0.400676 | 0.45444 | 0 | 0 | 1.46864 | 1.343223 |
| GO_Biologi | Regulation | 1/118 | 0.403056 | 0.45496 | 0 | 0 | 1.967402 | 1.787737 |
| GO_Biologi | Positive Re | 1/118 | 0.403056 | 0.45496 | 0 | 0 | 1.967402 | 1.787737 |
| GO_Biologi | Positive Re | 1/118 | 0.403056 | 0.45496 | 0 | 0 | 1.967402 | 1.787737 |
| GWAS_Cat | Well-being | 1/132 | 0.438615 | 0.455413 | 0 | 0 | 1.755903 | 1.447098 |
| Jensen_DIS | Pancreatic | 1/137 | 0.4508 | 0.455472 | 0 | 0 | 1.69092 | 1.347208 |
| DisGeNET | Epileptic di | 1/115 | 0.395152 | 0.455877 | 0 | 0 | 2.019482 | 1.875059 |

| | | | | | | | | |
|---|---|---|---|---|---|---|---|---|
| GO_Biologi | Positive Re | 1/119 | 0.405668 | 0.456457 | 0 | 0 | 1.950631 | 1.759896 |
| GO_Biologi | Regulation | 1/119 | 0.405668 | 0.456457 | 0 | 0 | 1.950631 | 1.759896 |
| Diabetes_F | GSE143319 | 3/123 | 0.016543 | 0.456832 | 0 | 0 | 5.890774 | 24.16285 |
| GeneSigDB | 19096012-1 | 3/357 | 0.203255 | 0.457012 | 0 | 0 | 1.973265 | 3.143989 |
| GeneSigDB | 15525570-1 | 2/192 | 0.203481 | 0.457012 | 0 | 0 | 2.442477 | 3.888868 |
| GeneSigDB | 11823860-1 | 2/191 | 0.201897 | 0.457012 | 0 | 0 | 2.455524 | 3.928837 |
| GTEx_Tissu | GTEX-RNOI | 8/792 | 0.021854 | 0.45702 | 0 | 0 | 2.470809 | 9.446786 |
| GTEx_Tissu | GTEX-WY7( | 7/637 | 0.020962 | 0.45702 | 0 | 0 | 2.678194 | 10.35129 |
| GTEx_Tissu | GTEX-QV44 | 9/941 | 0.021028 | 0.45702 | 0 | 0 | 2.349909 | 9.075169 |
| GTEx_Tissu | GTEX-WL4( | 7/636 | 0.020804 | 0.45702 | 0 | 0 | 2.682591 | 10.38868 |
| GTEx_Tissu | GTEX-PLZ5 | 15/1961 | 0.021398 | 0.45702 | 0 | 0 | 1.923497 | 7.394787 |
| GTEx_Tissu | GTEX-T6MI | 9/940 | 0.020899 | 0.45702 | 0 | 0 | 2.352557 | 9.099765 |
| GTEx_Tissu | GTEX-RNOI | 12/1442 | 0.021958 | 0.45702 | 0 | 0 | 2.068028 | 7.896981 |
| GTEx_Tissu | GTEX-XXEK | 6/497 | 0.021275 | 0.45702 | 0 | 0 | 2.930075 | 11.28141 |
| GTEx_Tissu | GTEX-P4Q1 | 11/1272 | 0.021805 | 0.45702 | 0 | 0 | 2.140866 | 8.190152 |
| GTEx_Tissu | GTEX-QDVI | 9/948 | 0.02194 | 0.45702 | 0 | 0 | 2.331531 | 8.905167 |
| GWAS_Cat | Cigarettes : | 1/133 | 0.441073 | 0.457187 | 0 | 0 | 1.742512 | 1.426323 |
| GWAS_Cat | Lung Cance | 1/133 | 0.441073 | 0.457187 | 0 | 0 | 1.742512 | 1.426323 |
| Rare_Disea | Central ner | Jan-87 | 0.316203 | 0.457695 | 0 | 0 | 2.680773 | 3.086563 |
| Human_Ph | Pectus exca | 1/135 | 0.445958 | 0.457946 | 0 | 0 | 1.716331 | 1.38599 |
| Human_Ph | Abnormalit | 1/135 | 0.445958 | 0.457946 | 0 | 0 | 1.716331 | 1.38599 |
| DisGeNET | Prescriptio | 1/116 | 0.397798 | 0.458123 | 0 | 0 | 2.00182 | 1.845299 |
| DisGeNET | Organic Me | 1/116 | 0.397798 | 0.458123 | 0 | 0 | 2.00182 | 1.845299 |
| DisGeNET | Complex pa | 1/116 | 0.397798 | 0.458123 | 0 | 0 | 2.00182 | 1.845299 |
| DisGeNET | Pancytopei | 1/116 | 0.397798 | 0.458123 | 0 | 0 | 2.00182 | 1.845299 |
| CellMarker | Multilymph | 3/537 | 0.414851 | 0.458343 | 0 | 0 | 1.296081 | 1.140339 |
| Disease_Pe | Dermatom | 4/232 | 0.01857 | 0.458547 | 0 | 0 | 4.160854 | 16.58593 |
| dbGaP | Erythrocyte | 1/115 | 0.395152 | 0.458658 | 0 | 0 | 2.019482 | 1.875059 |
| GTEx_Tissu | GTEX-WZT( | 9/950 | 0.022206 | 0.458865 | 0 | 0 | 2.32633 | 8.857283 |
| SysMyo_M | MGS962 M | Jan-41 | 0.163825 | 0.459036 | 0 | 0 | 5.777035 | 10.45039 |
| GWAS_Cat | JT Interval | 1/134 | 0.443521 | 0.459334 | 0 | 0 | 1.729323 | 1.405958 |
| Rare_Disea | Tuberous s | 2/264 | 0.319111 | 0.459338 | 0 | 0 | 1.764796 | 2.015779 |
| Rare_Disea | Nodular m | Jan-88 | 0.319191 | 0.459338 | 0 | 0 | 2.649826 | 3.026014 |
| Rare_Disea | Bourneville | 2/264 | 0.319111 | 0.459338 | 0 | 0 | 1.764796 | 2.015779 |
| DisGeNET | Cholestasis | 2/315 | 0.399145 | 0.459473 | 0 | 0 | 1.473407 | 1.353222 |
| DisGeNET | Leukemia, | 1/117 | 0.400433 | 0.460147 | 0 | 0 | 1.984463 | 1.816198 |
| DisGeNET | Muscle Hyp | 1/117 | 0.400433 | 0.460147 | 0 | 0 | 1.984463 | 1.816198 |
| DisGeNET | Hypogonac | 1/117 | 0.400433 | 0.460147 | 0 | 0 | 1.984463 | 1.816198 |
| DisGeNET | Carcinoma, | 1/117 | 0.400433 | 0.460147 | 0 | 0 | 1.984463 | 1.816198 |
| GeneSigDB | 16365062-1 | Jan-53 | 0.206541 | 0.460174 | 0 | 0 | 4.44119 | 7.004886 |
| GeneSigDB | 17885619-1 | Jan-53 | 0.206541 | 0.460174 | 0 | 0 | 4.44119 | 7.004886 |
| GeneSigDB | 15150091-1 | Jan-54 | 0.210002 | 0.460516 | 0 | 0 | 4.357174 | 6.79997 |
| GeneSigDB | 15609325-1 | Jan-54 | 0.210002 | 0.460516 | 0 | 0 | 4.357174 | 6.79997 |
| GeneSigDB | 18537973-1 | Jan-54 | 0.210002 | 0.460516 | 0 | 0 | 4.357174 | 6.79997 |
| GeneSigDB | 20521089-1 | Jan-54 | 0.210002 | 0.460516 | 0 | 0 | 4.357174 | 6.79997 |

| | | | | | | | | | |
|---|---|---|---|---|---|---|---|---|---|
| CellMarker | Enteroend( | 1/124 | | 0.418561 | 0.460585 | 0 | 0 | 1.870864 | 1.629399 |
| Reactome_ | Mitochond | | Jan-65 | 0.247099 | 0.461251 | 0 | 0 | 3.606286 | 5.041467 |
| DisGeNET | Red Blood | 1/118 | | 0.403056 | 0.46134 | 0 | 0 | 1.967402 | 1.787737 |
| DisGeNET | Breast Carc | 23/4963 | | 0.402806 | 0.46134 | 0 | 0 | 1.089255 | 0.99046 |
| DisGeNET | Transamin | 1/118 | | 0.403056 | 0.46134 | 0 | 0 | 1.967402 | 1.787737 |
| DisGeNET | Elevated liv | 1/118 | | 0.403056 | 0.46134 | 0 | 0 | 1.967402 | 1.787737 |
| DisGeNET | Erectile dys | 1/118 | | 0.403056 | 0.46134 | 0 | 0 | 1.967402 | 1.787737 |
| DisGeNET | Subclinical | 1/118 | | 0.403056 | 0.46134 | 0 | 0 | 1.967402 | 1.787737 |
| DisGeNET | Liver functi | 1/118 | | 0.403056 | 0.46134 | 0 | 0 | 1.967402 | 1.787737 |
| DisGeNET | Increased l | 1/118 | | 0.403056 | 0.46134 | 0 | 0 | 1.967402 | 1.787737 |
| DisGeNET | Liver enzyn | 1/118 | | 0.403056 | 0.46134 | 0 | 0 | 1.967402 | 1.787737 |
| Rare_Disea | Congenital | | Jan-89 | 0.322165 | 0.461832 | 0 | 0 | 2.619582 | 2.967177 |
| Rare_Disea | Sheehan sy | | Jan-89 | 0.322165 | 0.461832 | 0 | 0 | 2.619582 | 2.967177 |
| DisGeNET | Depressive | 4/741 | | 0.403686 | 0.461859 | 0 | 0 | 1.253928 | 1.137459 |
| GeDiPNet_ | Impaired C | 2/322 | | 0.409821 | 0.462138 | 0 | 0 | 1.440662 | 1.285121 |
| Jensen_TIS | NIH-3T3 ce | 1/109 | | 0.379031 | 0.462303 | 0 | 0 | 2.132321 | 2.068645 |
| GeneSigDB | 14993899- | 3/366 | | 0.213374 | 0.462609 | 0 | 0 | 1.923455 | 2.971175 |
| GeneSigDB | 17683608- | 5/731 | | 0.212719 | 0.462609 | 0 | 0 | 1.611486 | 2.49423 |
| GeneSigDB | 18310505- | | Jan-55 | 0.213448 | 0.462609 | 0 | 0 | 4.27627 | 6.604112 |
| UK_Bioban | Age startec | 1/128 | | 0.428675 | 0.462774 | 0 | 0 | 1.811573 | 1.534505 |
| DisGeNET | Elevated he | 1/119 | | 0.405668 | 0.462914 | 0 | 0 | 1.950631 | 1.759896 |
| DisGeNET | Von Hippel | 1/119 | | 0.405668 | 0.462914 | 0 | 0 | 1.950631 | 1.759896 |
| DisGeNET | Premature | 1/119 | | 0.405668 | 0.462914 | 0 | 0 | 1.950631 | 1.759896 |
| DisGeNET | Increased s | 1/119 | | 0.405668 | 0.462914 | 0 | 0 | 1.950631 | 1.759896 |
| DisGeNET | Retrognath | 1/119 | | 0.405668 | 0.462914 | 0 | 0 | 1.950631 | 1.759896 |
| DisGeNET | Drug Use D | 1/119 | | 0.405668 | 0.462914 | 0 | 0 | 1.950631 | 1.759896 |
| GO_Biologi | Receptor-N | 1/122 | | 0.413437 | 0.462997 | 0 | 0 | 1.90198 | 1.679923 |
| GO_Biologi | Positive Re | 1/122 | | 0.413437 | 0.462997 | 0 | 0 | 1.90198 | 1.679923 |
| GO_Biologi | Regulation | 1/122 | | 0.413437 | 0.462997 | 0 | 0 | 1.90198 | 1.679923 |
| GeneSigDB | 16643655- | 8/1319 | | 0.214597 | 0.463297 | 0 | 0 | 1.436878 | 2.211345 |
| GO_Biologi | Intracellula | 2/325 | | 0.414369 | 0.46331 | 0 | 0 | 1.427062 | 1.257241 |
| Reactome_ | COPII-medi | | Jan-66 | 0.250385 | 0.463522 | 0 | 0 | 3.550626 | 4.916752 |
| GO_Biologi | Organonitr | 1/123 | | 0.416004 | 0.463678 | 0 | 0 | 1.886294 | 1.654392 |
| GO_Biologi | Modulatior | 1/123 | | 0.416004 | 0.463678 | 0 | 0 | 1.886294 | 1.654392 |
| GeneSigDB | 16168083- | | Jan-56 | 0.216879 | 0.464621 | 0 | 0 | 4.198309 | 6.416764 |
| GeneSigDB | 19249085- | | Jan-56 | 0.216879 | 0.464621 | 0 | 0 | 4.198309 | 6.416764 |
| GO_Biologi | Regulation | 1/124 | | 0.418561 | 0.464716 | 0 | 0 | 1.870864 | 1.629399 |
| GO_Biologi | Negative R | 1/124 | | 0.418561 | 0.464716 | 0 | 0 | 1.870864 | 1.629399 |
| GO_Biologi | Protein-Co | 2/328 | | 0.418899 | 0.464716 | 0 | 0 | 1.413713 | 1.230107 |
| Rare_Disea | Gaucher di | | Jan-90 | 0.325127 | 0.465182 | 0 | 0 | 2.590018 | 2.909988 |
| GeDiPNet_ | Glomerulo: | 1/122 | | 0.413437 | 0.465316 | 0 | 0 | 1.90198 | 1.679923 |
| DisGeNET | Liver Failur | 1/120 | | 0.408269 | 0.465679 | 0 | 0 | 1.934141 | 1.732658 |
| Reactome_ | Regulation | | Jan-67 | 0.253656 | 0.46573 | 0 | 0 | 3.496653 | 4.79662 |
| WikiPathw | Wnt Signal | 1/101 | | 0.356874 | 0.466369 | 0 | 0 | 2.303837 | 2.373811 |
| GTEx_Tissu | GTEX-WEY! | 7/648 | | 0.022765 | 0.466373 | 0 | 0 | 2.630733 | 9.950781 |

| | | | | | | | | |
|---|---|---|---|---|---|---|---|---|
| GTEx_Tissu | GTEX-RNOI | 8/799 | 0.022891 | 0.466373 | 0 | 0 | 2.448047 | 9.246255 |
| GO_Biologi | Negative R | 1/125 | 0.421106 | 0.466435 | 0 | 0 | 1.855683 | 1.604928 |
| SysMyo_M | MGS1327 I | 2/169 | 0.167499 | 0.467017 | 0 | 0 | 2.782106 | 4.971004 |
| Jensen_TIS | Tendon | 1/111 | 0.384451 | 0.467144 | 0 | 0 | 2.09334 | 2.001105 |
| GeDiPNet_ | Marfan Syr | 3/538 | 0.416018 | 0.467319 | 0 | 0 | 1.293591 | 1.134515 |
| PhenGenI_ | Eye Moven | Jan-77 | 0.285606 | 0.467908 | 0 | 0 | 3.035037 | 3.803331 |
| PhenGenI_ | Virus Disea | Jan-77 | 0.285606 | 0.467908 | 0 | 0 | 3.035037 | 3.803331 |
| GeneSigDB | 15067324-‍ | 2/202 | 0.219398 | 0.468218 | 0 | 0 | 2.319176 | 3.517879 |
| Rare_Disea | Transverse | Jan-91 | 0.328076 | 0.4685 | 0 | 0 | 2.561111 | 2.854384 |
| GO_Biologi | Cellular Re | 1/126 | 0.42364 | 0.468511 | 0 | 0 | 1.840744 | 1.580964 |
| WikiPathw | CKAP4 Sigr | 1/109 | 0.379031 | 0.468772 | 0 | 0 | 2.132321 | 2.068645 |
| WikiPathw | Androgen I | 1/109 | 0.379031 | 0.468772 | 0 | 0 | 2.132321 | 2.068645 |
| WikiPathw | Electron Tr | 1/105 | 0.368048 | 0.468772 | 0 | 0 | 2.214781 | 2.213764 |
| WikiPathw | Focal Adhe | 2/302 | 0.379094 | 0.468772 | 0 | 0 | 1.538275 | 1.492081 |
| WikiPathw | Compleme | 1/106 | 0.370812 | 0.468772 | 0 | 0 | 2.193577 | 2.176161 |
| DisGeNET | Kaposi Sarc | 2/323 | 0.411339 | 0.468976 | 0 | 0 | 1.4361 | 1.275744 |
| Jensen_TIS | Intestine | 25/5377 | 0.387443 | 0.46901 | 0 | 0 | 1.097042 | 1.040202 |
| DisGeNET | Endometri | 4/749 | 0.411585 | 0.469053 | 0 | 0 | 1.239945 | 1.100749 |
| GeDiPNet_ | Imperforat | 1/124 | 0.418561 | 0.469271 | 0 | 0 | 1.870864 | 1.629399 |
| PhenGenI_ | Sex Hormo | Jan-78 | 0.288726 | 0.469688 | 0 | 0 | 2.99547 | 3.721207 |
| DisGeNET | Short statu | 4/750 | 0.412571 | 0.469972 | 0 | 0 | 1.238218 | 1.096253 |
| Rare_Disea | Tetanus | Jan-92 | 0.331012 | 0.469987 | 0 | 0 | 2.532839 | 2.800307 |
| Rare_Disea | TAU syndro | Jan-92 | 0.331012 | 0.469987 | 0 | 0 | 2.532839 | 2.800307 |
| Rare_Disea | Malignant | Jan-92 | 0.331012 | 0.469987 | 0 | 0 | 2.532839 | 2.800307 |
| DisGeNET | Congenital | 1/122 | 0.413437 | 0.47055 | 0 | 0 | 1.90198 | 1.679923 |
| DisGeNET | Gastrointe: | 1/122 | 0.413437 | 0.47055 | 0 | 0 | 1.90198 | 1.679923 |
| GO_Biologi | Cytosolic T | 1/127 | 0.426163 | 0.470568 | 0 | 0 | 1.826043 | 1.557494 |
| SysMyo_M | MGS241 C: | 2/172 | 0.172129 | 0.470653 | 0 | 0 | 2.732595 | 4.808027 |
| SysMyo_M | MGS1412 I | 2/172 | 0.172129 | 0.470653 | 0 | 0 | 2.732595 | 4.808027 |
| SysMyo_M | MGS543 Hı | 2/171 | 0.170583 | 0.470653 | 0 | 0 | 2.748904 | 4.86152 |
| SysMyo_M | MGS113 M | Jan-43 | 0.171099 | 0.470653 | 0 | 0 | 5.501384 | 9.712751 |
| GTEx_Tissu | GTEX-WEY! | 7/651 | 0.023275 | 0.470878 | 0 | 0 | 2.618071 | 9.844893 |
| GTEx_Tissu | GTEX-O5YV | 7/652 | 0.023447 | 0.471057 | 0 | 0 | 2.613876 | 9.809906 |
| GeDiPNet_ | Glioma | 1/125 | 0.421106 | 0.471218 | 0 | 0 | 1.855683 | 1.604928 |
| GWAS_Cat | Sum Eosinc | 1/139 | 0.455601 | 0.471445 | 0 | 0 | 1.666245 | 1.309899 |
| GeneSigDB | 19204204-: | 7/1132 | 0.221841 | 0.471624 | 0 | 0 | 1.461289 | 2.2004 |
| GeneSigDB | 18081427-‍ | 4/556 | 0.223141 | 0.471965 | 0 | 0 | 1.690327 | 2.535405 |
| GeneSigDB | 11823860- | Jan-58 | 0.223696 | 0.471965 | 0 | 0 | 4.050592 | 6.065622 |
| Rare_Disea | Neuromyel | Jan-93 | 0.333936 | 0.472335 | 0 | 0 | 2.505182 | 2.747701 |
| Rare_Disea | Rett syndro | Jan-93 | 0.333936 | 0.472335 | 0 | 0 | 2.505182 | 2.747701 |
| DisGeNET | Involuntary | 1/123 | 0.416004 | 0.47265 | 0 | 0 | 1.886294 | 1.654392 |
| DisGeNET | Absence Se | 1/123 | 0.416004 | 0.47265 | 0 | 0 | 1.886294 | 1.654392 |
| DisGeNET | Impaired g | 2/326 | 0.415881 | 0.47265 | 0 | 0 | 1.422585 | 1.248115 |
| DisGeNET | Spasmodic | 1/123 | 0.416004 | 0.47265 | 0 | 0 | 1.886294 | 1.654392 |
| GeDiPNet_ | Microtia | 1/126 | 0.42364 | 0.473145 | 0 | 0 | 1.840744 | 1.580964 |

| | | | | | | | | |
|---|---|---|---|---|---|---|---|---|
| GWAS_Cat | Medicatior | 1/140 | 0.457986 | 0.473511 | 0 | 0 | 1.654174 | 1.291774 |
| GO_Biologi | Regulation | 1/129 | 0.431176 | 0.473893 | 0 | 0 | 1.797329 | 1.511983 |
| GO_Biologi | Negative R | 2/336 | 0.430896 | 0.473893 | 0 | 0 | 1.379288 | 1.161207 |
| GO_Biologi | Protein Tar | 1/129 | 0.431176 | 0.473893 | 0 | 0 | 1.797329 | 1.511983 |
| GeneSigDB | 18338247-: | 2/207 | 0.227399 | 0.474387 | 0 | 0 | 2.262037 | 3.350186 |
| GeneSigDB | 20165955-' | Jan-59 | 0.227083 | 0.474387 | 0 | 0 | 3.980553 | 5.900928 |
| GeneSigDB | 15474998-' | 4/559 | 0.225915 | 0.474387 | 0 | 0 | 1.680929 | 2.500545 |
| Rare_Disea | Kohler dise | Jan-94 | 0.336847 | 0.474648 | 0 | 0 | 2.47812 | 2.69651 |
| Rare_Disea | Jensen syn | Jan-94 | 0.336847 | 0.474648 | 0 | 0 | 2.47812 | 2.69651 |
| Gene_Pert | MIR196A2 | 4/169 | 0.006357 | 0.474884 | 0 | 0 | 5.767945 | 29.17586 |
| Gene_Pert | Lmna KO m | 5/266 | 0.006107 | 0.474884 | 0 | 0 | 4.59116 | 23.40751 |
| DisGeNET | Nicotine De | 1/124 | 0.418561 | 0.474937 | 0 | 0 | 1.870864 | 1.629399 |
| DisGeNET | Upward sla | 1/124 | 0.418561 | 0.474937 | 0 | 0 | 1.870864 | 1.629399 |
| DisGeNET | Uterine Cei | 1/124 | 0.418561 | 0.474937 | 0 | 0 | 1.870864 | 1.629399 |
| GeDiPNet_ | Hallucinati | 1/127 | 0.426163 | 0.475053 | 0 | 0 | 1.826043 | 1.557494 |
| UK_Bioban | Frequency | 9/1894 | 0.442388 | 0.475776 | 0 | 0 | 1.10353 | 0.900003 |
| GO_Biologi | Golgi Orgai | 1/130 | 0.433666 | 0.475894 | 0 | 0 | 1.783306 | 1.489916 |
| DisGeNET | Mental diso | 2/329 | 0.420406 | 0.476411 | 0 | 0 | 1.409318 | 1.221224 |
| DisGeNET | Diarrhea | 2/329 | 0.420406 | 0.476411 | 0 | 0 | 1.409318 | 1.221224 |
| DisGeNET | Primary bil | 2/329 | 0.420406 | 0.476411 | 0 | 0 | 1.409318 | 1.221224 |
| UK_Bioban | Time spent | 3/563 | 0.444949 | 0.476731 | 0 | 0 | 1.234247 | 0.999487 |
| Rare_Disea | Indolent B | Jan-95 | 0.339745 | 0.476925 | 0 | 0 | 2.451633 | 2.646685 |
| Rare_Disea | Optic atrop | Jan-95 | 0.339745 | 0.476925 | 0 | 0 | 2.451633 | 2.646685 |
| DisGeNET | Congenital | 1/125 | 0.421106 | 0.476998 | 0 | 0 | 1.855683 | 1.604928 |
| GO_Biologi | Synapse Or | 1/131 | 0.436146 | 0.477876 | 0 | 0 | 1.769499 | 1.468291 |
| PhenGenI_ | Ovarian Ne | Jan-81 | 0.298004 | 0.478454 | 0 | 0 | 2.882703 | 3.48994 |
| PhenGenI_ | Calcium | 2/251 | 0.298257 | 0.478454 | 0 | 0 | 1.858162 | 2.248005 |
| DisGeNET | Generalize | 1/126 | 0.42364 | 0.478627 | 0 | 0 | 1.840744 | 1.580964 |
| DisGeNET | Hypogonac | 1/126 | 0.42364 | 0.478627 | 0 | 0 | 1.840744 | 1.580964 |
| DisGeNET | Myocarditi | 1/126 | 0.42364 | 0.478627 | 0 | 0 | 1.840744 | 1.580964 |
| DisGeNET | Congenital | 1/126 | 0.42364 | 0.478627 | 0 | 0 | 1.840744 | 1.580964 |
| DisGeNET | Spastic Par | 1/126 | 0.42364 | 0.478627 | 0 | 0 | 1.840744 | 1.580964 |
| DisGeNET | Difficulties | 1/126 | 0.42364 | 0.478627 | 0 | 0 | 1.840744 | 1.580964 |
| GeneSigDB | 17069664-: | Jan-60 | 0.230455 | 0.478969 | 0 | 0 | 3.912889 | 5.742942 |
| GO_Biologi | Positive Re | 3/557 | 0.438052 | 0.479103 | 0 | 0 | 1.248002 | 1.030122 |
| GO_Biologi | Plasma Me | 1/132 | 0.438615 | 0.479103 | 0 | 0 | 1.755903 | 1.447098 |
| Rare_Disea | Hyperlipop | Jan-96 | 0.342631 | 0.479168 | 0 | 0 | 2.425704 | 2.598176 |
| Rare_Disea | Devic disea | Jan-96 | 0.342631 | 0.479168 | 0 | 0 | 2.425704 | 2.598176 |
| DisGeNET | Coronary A | 5/981 | 0.424617 | 0.479525 | 0 | 0 | 1.183089 | 1.013395 |
| GeDiPNet_ | Hyperopia | 1/129 | 0.431176 | 0.479724 | 0 | 0 | 1.797329 | 1.511983 |
| Jensen_TIS | Neural ster | 1/116 | 0.397798 | 0.479741 | 0 | 0 | 2.00182 | 1.845299 |
| DisGeNET | Tumor Pro | Oct-90 | 0.425191 | 0.479966 | 0 | 0 | 1.113449 | 0.95224 |
| GeDiPNet_ | Mammary | 3/552 | 0.432281 | 0.480038 | 0 | 0 | 1.259693 | 1.056478 |
| WikiPathw | Wnt Signal | 1/114 | 0.392494 | 0.480179 | 0 | 0 | 2.037456 | 1.905499 |
| Rare_Disea | Cerebellar | 2/280 | 0.34458 | 0.480477 | 0 | 0 | 1.661871 | 1.770607 |

| | | | | | | | | |
|---|---|---|---|---|---|---|---|---|
| Rare_Disea | Autoimmu | Jan-97 | 0.345504 | 0.480477 | 0 | 0 | 2.400315 | 2.550937 |
| Rare_Disea | Yolk sac tu | Jan-97 | 0.345504 | 0.480477 | 0 | 0 | 2.400315 | 2.550937 |
| Rare_Disea | Anal cance | 2/281 | 0.346162 | 0.480494 | 0 | 0 | 1.65583 | 1.756583 |
| DisGeNET | Hyperkerat | 1/127 | 0.426163 | 0.480648 | 0 | 0 | 1.826043 | 1.557494 |
| DisGeNET | Renal glom | 1/127 | 0.426163 | 0.480648 | 0 | 0 | 1.826043 | 1.557494 |
| WikiPathw | Osteoblast | 1/118 | 0.403056 | 0.480947 | 0 | 0 | 1.967402 | 1.787737 |
| WikiPathw | Nuclear Re | 2/314 | 0.397613 | 0.480947 | 0 | 0 | 1.478205 | 1.363314 |
| WikiPathw | Spinal Corc | 1/119 | 0.405668 | 0.480947 | 0 | 0 | 1.950631 | 1.759896 |
| GO_Biologi | Negative R | 1/133 | 0.441073 | 0.481048 | 0 | 0 | 1.742512 | 1.426323 |
| Human_Ph | Progressive | 1/145 | 0.469755 | 0.48109 | 0 | 0 | 1.596334 | 1.2061 |
| GeneSigDB | 18427120- | 3/383 | 0.232777 | 0.481996 | 0 | 0 | 1.835808 | 2.676009 |
| Rare_Disea | Haemophil | Jan-98 | 0.348365 | 0.482651 | 0 | 0 | 2.37545 | 2.504922 |
| DisGeNET | Ependymo | 1/128 | 0.428675 | 0.482858 | 0 | 0 | 1.811573 | 1.534505 |
| DisGeNET | Peritonitis | 1/128 | 0.428675 | 0.482858 | 0 | 0 | 1.811573 | 1.534505 |
| DisGeNET | Biliary Trac | 1/128 | 0.428675 | 0.482858 | 0 | 0 | 1.811573 | 1.534505 |
| GO_Biologi | Antigen Re | 1/134 | 0.443521 | 0.482975 | 0 | 0 | 1.729323 | 1.405958 |
| UK_Bioban | Mean corp | 1/138 | 0.453206 | 0.483219 | 0 | 0 | 1.678493 | 1.328374 |
| UK_Bioban | Workplace | 1/139 | 0.455601 | 0.483219 | 0 | 0 | 1.666245 | 1.309899 |
| UK_Bioban | Distance be | 17/3725 | 0.456091 | 0.483219 | 0 | 0 | 1.061354 | 0.833229 |
| GeDiPNet_ | Mood Swir | 1/132 | 0.438615 | 0.483389 | 0 | 0 | 1.755903 | 1.447098 |
| GeDiPNet_ | Neoplasms | 1/132 | 0.438615 | 0.483389 | 0 | 0 | 1.755903 | 1.447098 |
| GeDiPNet_ | Lymphocyt | 1/132 | 0.438615 | 0.483389 | 0 | 0 | 1.755903 | 1.447098 |
| GeDiPNet_ | Speech Dis | 1/132 | 0.438615 | 0.483389 | 0 | 0 | 1.755903 | 1.447098 |
| DisGeNET | Malignant | 2/335 | 0.429403 | 0.483471 | 0 | 0 | 1.383501 | 1.169555 |
| Disease_Pe | prostate ca | 5/361 | 0.020656 | 0.483852 | 0 | 0 | 3.349719 | 12.99601 |
| Disease_Pe | Parkinson's | 5/365 | 0.021544 | 0.483852 | 0 | 0 | 3.311822 | 12.70968 |
| Disease_Pe | Adenoma ( | 5/371 | 0.022921 | 0.483852 | 0 | 0 | 3.256531 | 12.29569 |
| Disease_Pe | Type 1 diak | 4/248 | 0.023063 | 0.483852 | 0 | 0 | 3.884851 | 14.64409 |
| Disease_Pe | Fracture of | 4/249 | 0.023364 | 0.483852 | 0 | 0 | 3.868798 | 14.53344 |
| PhenGenI_ | Depression | Jan-83 | 0.304123 | 0.484499 | 0 | 0 | 2.81211 | 3.34732 |
| Human_Ph | Photophob | 1/147 | 0.474392 | 0.484539 | 0 | 0 | 1.574307 | 1.173995 |
| Rare_Disea | Familial int | Jan-99 | 0.351214 | 0.484792 | 0 | 0 | 2.351092 | 2.46009 |
| Rare_Disea | Lambdoid s | Jan-99 | 0.351214 | 0.484792 | 0 | 0 | 2.351092 | 2.46009 |
| DisGeNET | Seizures, F | 1/129 | 0.431176 | 0.484842 | 0 | 0 | 1.797329 | 1.511983 |
| DisGeNET | Endometri | 1/129 | 0.431176 | 0.484842 | 0 | 0 | 1.797329 | 1.511983 |
| DisGeNET | liposarcom | 1/129 | 0.431176 | 0.484842 | 0 | 0 | 1.797329 | 1.511983 |
| WikiPathw | Fragile X Sy | 1/122 | 0.413437 | 0.485156 | 0 | 0 | 1.90198 | 1.679923 |
| UK_Bioban | East co-orc | 3/576 | 0.459782 | 0.485325 | 0 | 0 | 1.205435 | 0.936627 |
| DisGeNET | Blepharopt | 2/337 | 0.432387 | 0.485994 | 0 | 0 | 1.375101 | 1.152933 |
| Jensen_DIS | Diarrhea | 1/151 | 0.483545 | 0.486038 | 0 | 0 | 1.532016 | 1.113178 |
| GeneSigDB | 20436685-: | 3/387 | 0.237391 | 0.486128 | 0 | 0 | 1.816313 | 2.611943 |
| GeneSigDB | 18310659-1 | Jan-62 | 0.237156 | 0.486128 | 0 | 0 | 3.784217 | 5.445629 |
| GeneSigDB | 16457687-( | Jan-62 | 0.237156 | 0.486128 | 0 | 0 | 3.784217 | 5.445629 |
| SysMyo_M | MGS1023 I | 2/183 | 0.189279 | 0.486964 | 0 | 0 | 2.565096 | 4.269683 |
| SysMyo_M | MGS1029 I | 2/180 | 0.184577 | 0.486964 | 0 | 0 | 2.608724 | 4.407929 |

| | | | | | | | | |
|---|---|---|---|---|---|---|---|---|
| SysMyo_M | MGS45 C2( | 3/339 | 0.18339 | 0.486964 | 0 | 0 | 2.080889 | 3.529475 |
| SysMyo_M | MGS950 M | Jan-46 | 0.181893 | 0.486964 | 0 | 0 | 5.13385 | 8.749803 |
| SysMyo_M | MGS1315 I | 2/180 | 0.184577 | 0.486964 | 0 | 0 | 2.608724 | 4.407929 |
| SysMyo_M | MGS23 Mu | Jan-47 | 0.18546 | 0.486964 | 0 | 0 | 5.021992 | 8.46163 |
| SysMyo_M | MGS243 C2 | 2/179 | 0.183014 | 0.486964 | 0 | 0 | 2.623596 | 4.455375 |
| SysMyo_M | MGS481 H | 2/179 | 0.183014 | 0.486964 | 0 | 0 | 2.623596 | 4.455375 |
| SysMyo_M | MGS717 M | 2/183 | 0.189279 | 0.486964 | 0 | 0 | 2.565096 | 4.269683 |
| SysMyo_M | MGS945 M | Jan-48 | 0.189012 | 0.486964 | 0 | 0 | 4.914894 | 8.187947 |
| SysMyo_M | MGS979 M | 2/182 | 0.18771 | 0.486964 | 0 | 0 | 2.579477 | 4.315096 |
| SysMyo_M | MGS46 Hu | 3/340 | 0.18448 | 0.486964 | 0 | 0 | 2.074608 | 3.506535 |
| SysMyo_M | MGS715 M | 2/181 | 0.186143 | 0.486964 | 0 | 0 | 2.594019 | 4.361174 |
| GeneSigDB | 20156340- | Jan-63 | 0.240485 | 0.487091 | 0 | 0 | 3.722993 | 5.305634 |
| GeneSigDB | 19014521- | Jan-63 | 0.240485 | 0.487091 | 0 | 0 | 3.722993 | 5.305634 |
| GeneSigDB | 19047173- | Jan-63 | 0.240485 | 0.487091 | 0 | 0 | 3.722993 | 5.305634 |
| GeneSigDB | 17683608- | 5/767 | 0.24136 | 0.487092 | 0 | 0 | 1.532472 | 2.17836 |
| DisGeNET | Delayed Pu | 1/130 | 0.433666 | 0.487224 | 0 | 0 | 1.783306 | 1.489916 |
| DisGeNET | Cerebral at | 2/338 | 0.433875 | 0.487249 | 0 | 0 | 1.370938 | 1.144732 |
| GO_Biologi | Regulation | 1/136 | 0.448384 | 0.487524 | 0 | 0 | 1.703531 | 1.36641 |
| Rare_Disea | Gastrointes | 2/286 | 0.354057 | 0.487812 | 0 | 0 | 1.626263 | 1.688543 |
| GeDiPNet_ | Multicystic | 1/134 | 0.443521 | 0.487873 | 0 | 0 | 1.729323 | 1.405958 |
| SysMyo_M | MGS1038 I | 2/184 | 0.19085 | 0.488784 | 0 | 0 | 2.550873 | 4.224921 |
| Rare_Disea | Retinoblast | 1/101 | 0.356874 | 0.488976 | 0 | 0 | 2.303837 | 2.373811 |
| Rare_Disea | Neisseria m | 1/101 | 0.356874 | 0.488976 | 0 | 0 | 2.303837 | 2.373811 |
| Rare_Disea | Mantle cell | 2/287 | 0.355633 | 0.488976 | 0 | 0 | 1.620475 | 1.675339 |
| WikiPathw | mRNA Proc | 1/125 | 0.421106 | 0.489163 | 0 | 0 | 1.855683 | 1.604928 |
| Rare_Disea | Sickle cell a | 1/105 | 0.368048 | 0.489315 | 0 | 0 | 2.214781 | 2.213764 |
| Rare_Disea | Rhabdoid t | 1/105 | 0.368048 | 0.489315 | 0 | 0 | 2.214781 | 2.213764 |
| Rare_Disea | Hemorrhag | 1/103 | 0.362485 | 0.489315 | 0 | 0 | 2.258436 | 2.291796 |
| Rare_Disea | Mitochond | 1/104 | 0.365273 | 0.489315 | 0 | 0 | 2.236396 | 2.252299 |
| Rare_Disea | Nerve shea | 2/294 | 0.366622 | 0.489315 | 0 | 0 | 1.581064 | 1.586479 |
| Rare_Disea | Spinocereb | 1/105 | 0.368048 | 0.489315 | 0 | 0 | 2.214781 | 2.213764 |
| Rare_Disea | Wilms' tum | 1/104 | 0.365273 | 0.489315 | 0 | 0 | 2.236396 | 2.252299 |
| Rare_Disea | Oncocytom | 1/105 | 0.368048 | 0.489315 | 0 | 0 | 2.214781 | 2.213764 |
| Rare_Disea | Toni-Debre | 1/107 | 0.373563 | 0.489315 | 0 | 0 | 2.172773 | 2.139459 |
| Rare_Disea | Breast canc | 1/107 | 0.373563 | 0.489315 | 0 | 0 | 2.172773 | 2.139459 |
| Rare_Disea | Hereditary | 1/106 | 0.370812 | 0.489315 | 0 | 0 | 2.193577 | 2.176161 |
| Rare_Disea | Cystic fibro | 2/295 | 0.368186 | 0.489315 | 0 | 0 | 1.575587 | 1.574275 |
| Rare_Disea | Cutaneous | 1/105 | 0.368048 | 0.489315 | 0 | 0 | 2.214781 | 2.213764 |
| Rare_Disea | Neurofibro | 2/298 | 0.37287 | 0.489315 | 0 | 0 | 1.55938 | 1.538369 |
| Rare_Disea | Bile duct ca | 1/105 | 0.368048 | 0.489315 | 0 | 0 | 2.214781 | 2.213764 |
| Rare_Disea | Progressive | 1/107 | 0.373563 | 0.489315 | 0 | 0 | 2.172773 | 2.139459 |
| Rare_Disea | Chordoma | 1/107 | 0.373563 | 0.489315 | 0 | 0 | 2.172773 | 2.139459 |
| Rare_Disea | Congenital | 1/107 | 0.373563 | 0.489315 | 0 | 0 | 2.172773 | 2.139459 |
| Rare_Disea | Congenital | 1/107 | 0.373563 | 0.489315 | 0 | 0 | 2.172773 | 2.139459 |
| Rare_Disea | Focal dystc | 1/107 | 0.373563 | 0.489315 | 0 | 0 | 2.172773 | 2.139459 |

| | | | | | | | | |
|---|---|---|---|---|---|---|---|---|
| Rare_Disea | Fragile X sy | 1/107 | 0.373563 | 0.489315 | 0 | 0 | 2.172773 | 2.139459 |
| Rare_Disea | Fanconi Bic | 1/107 | 0.373563 | 0.489315 | 0 | 0 | 2.172773 | 2.139459 |
| Rare_Disea | Fanconi rer | 1/107 | 0.373563 | 0.489315 | 0 | 0 | 2.172773 | 2.139459 |
| Rare_Disea | Nonmedull | 1/107 | 0.373563 | 0.489315 | 0 | 0 | 2.172773 | 2.139459 |
| Rare_Disea | Ewing's fan | 1/107 | 0.373563 | 0.489315 | 0 | 0 | 2.172773 | 2.139459 |
| GO_Biologi | Negative R | 1/137 | 0.4508 | 0.489401 | 0 | 0 | 1.69092 | 1.347208 |
| GeDiPNet_ | Hypogonac | 1/135 | 0.445958 | 0.48963 | 0 | 0 | 1.716331 | 1.38599 |
| SysMyo_M | MGS723 M | 2/185 | 0.192423 | 0.490192 | 0 | 0 | 2.536805 | 4.180799 |
| SysMyo_M | MGS767 M | 2/186 | 0.193998 | 0.490192 | 0 | 0 | 2.52289 | 4.137304 |
| SysMyo_M | MGS972 M | 2/186 | 0.193998 | 0.490192 | 0 | 0 | 2.52289 | 4.137304 |
| GTEx_Tissu | GTEX-WYB: | 5/379 | 0.024845 | 0.490226 | 0 | 0 | 3.185568 | 11.77102 |
| GTEx_Tissu | GTEX-WZT( | 8/812 | 0.02491 | 0.490226 | 0 | 0 | 2.406827 | 8.887219 |
| GTEx_Tissu | GTEX-R55C | 7/659 | 0.024674 | 0.490226 | 0 | 0 | 2.584873 | 9.569242 |
| GWAS_Cat | Monocyte | 2/366 | 0.474693 | 0.490369 | 0 | 0 | 1.263672 | 0.941545 |
| Diabetes_F | GSE149468 | 3/181 | 0.04447 | 0.490631 | 0 | 0 | 3.959671 | 12.32626 |
| Diabetes_F | GSE109285 | 3/197 | 0.054684 | 0.490631 | 0 | 0 | 3.630155 | 10.54993 |
| Diabetes_F | GSE100067 | 3/198 | 0.055356 | 0.490631 | 0 | 0 | 3.611355 | 10.45114 |
| Diabetes_F | GSE41203 : | 3/193 | 0.052032 | 0.490631 | 0 | 0 | 3.707331 | 10.95845 |
| Diabetes_F | GSE137678 | 3/192 | 0.05138 | 0.490631 | 0 | 0 | 3.727135 | 11.06404 |
| Diabetes_F | GSE127251 | 3/198 | 0.055356 | 0.490631 | 0 | 0 | 3.611355 | 10.45114 |
| Diabetes_F | GSE41203 : | 3/195 | 0.05335 | 0.490631 | 0 | 0 | 3.668341 | 10.75147 |
| Diabetes_F | GSE79164 ( | 3/194 | 0.052689 | 0.490631 | 0 | 0 | 3.687734 | 10.85427 |
| Diabetes_F | GSE156067 | 3/181 | 0.04447 | 0.490631 | 0 | 0 | 3.959671 | 12.32626 |
| Diabetes_F | GSE134846 | 3/183 | 0.045689 | 0.490631 | 0 | 0 | 3.915278 | 12.08215 |
| Diabetes_F | GSE41203 : | 3/190 | 0.050087 | 0.490631 | 0 | 0 | 3.76738 | 11.27955 |
| Diabetes_F | GSE41203 : | 3/199 | 0.056033 | 0.490631 | 0 | 0 | 3.592748 | 10.35363 |
| Diabetes_F | GSE153222 | 3/159 | 0.032168 | 0.490631 | 0 | 0 | 4.523123 | 15.545 |
| Diabetes_F | GSE115421 | 3/169 | 0.037505 | 0.490631 | 0 | 0 | 4.248494 | 13.94901 |
| Diabetes_F | GSE87854 I | 3/168 | 0.036952 | 0.490631 | 0 | 0 | 4.274459 | 14.09775 |
| Diabetes_F | GSE87854 I | 3/166 | 0.035859 | 0.490631 | 0 | 0 | 4.327344 | 14.40211 |
| Diabetes_F | GSE155973 | 3/147 | 0.026333 | 0.490631 | 0 | 0 | 4.903026 | 17.83201 |
| Diabetes_F | GSE101657 | 3/138 | 0.022367 | 0.490631 | 0 | 0 | 5.232275 | 19.88343 |
| Diabetes_F | GSE155064 | 3/142 | 0.024086 | 0.490631 | 0 | 0 | 5.080678 | 18.9312 |
| Diabetes_F | GSE122348 | 3/170 | 0.038062 | 0.490631 | 0 | 0 | 4.22284 | 13.80251 |
| Diabetes_F | GSE143319 | 3/173 | 0.039759 | 0.490631 | 0 | 0 | 4.147689 | 13.37596 |
| Diabetes_F | GSE125856 | 3/170 | 0.038062 | 0.490631 | 0 | 0 | 4.22284 | 13.80251 |
| Diabetes_F | GSE41203 ( | 3/176 | 0.041494 | 0.490631 | 0 | 0 | 4.075145 | 12.96795 |
| Diabetes_F | GSE40439 I | 3/179 | 0.043267 | 0.490631 | 0 | 0 | 4.005073 | 12.57741 |
| Diabetes_F | GSE149294 | 3/152 | 0.028688 | 0.490631 | 0 | 0 | 4.737296 | 16.82341 |
| Diabetes_F | GSE156774 | 3/151 | 0.028208 | 0.490631 | 0 | 0 | 4.769546 | 17.01838 |
| Diabetes_F | GSE101657 | 3/157 | 0.031152 | 0.490631 | 0 | 0 | 4.582328 | 15.89551 |
| Diabetes_F | GSE107894 | 3/145 | 0.025421 | 0.490631 | 0 | 0 | 4.972586 | 18.26021 |
| GO_Biologi | Protein Loc | 2/351 | 0.453038 | 0.491079 | 0 | 0 | 1.318995 | 1.044354 |
| Human_Ph | Developme | 1/151 | 0.483545 | 0.491262 | 0 | 0 | 1.532016 | 1.113178 |
| Human_Ph | Umbilical h | 1/151 | 0.483545 | 0.491262 | 0 | 0 | 1.532016 | 1.113178 |

| | | | | | | | | |
|---|---|---|---|---|---|---|---|---|
| GeDiPNet_ | Retinal Dis | 1/136 | 0.448384 | 0.491369 | 0 | 0 | 1.703531 | 1.36641 |
| DisGeNET | Nausea and | 1/132 | 0.438615 | 0.49194 | 0 | 0 | 1.755903 | 1.447098 |
| DisGeNET | Thrombocy | 2/341 | 0.438329 | 0.49194 | 0 | 0 | 1.358598 | 1.120553 |
| DisGeNET | Night Blind | 1/132 | 0.438615 | 0.49194 | 0 | 0 | 1.755903 | 1.447098 |
| Rare_Disea | Transitiona | 1/108 | 0.376303 | 0.492038 | 0 | 0 | 2.152358 | 2.10363 |
| PhenGenI_ | Emotions | 2/259 | 0.311104 | 0.492226 | 0 | 0 | 1.799588 | 2.10125 |
| Human_Ph | Long philtr | 1/152 | 0.485809 | 0.492252 | 0 | 0 | 1.521793 | 1.098642 |
| GWAS_Cat | Hemoglobi | 2/368 | 0.477542 | 0.492895 | 0 | 0 | 1.256638 | 0.928785 |
| GO_Biologi | Intracellula | 1/139 | 0.455601 | 0.493105 | 0 | 0 | 1.666245 | 1.309899 |
| Reactome_ | RHOC GTPa | Jan-73 | 0.272992 | 0.493146 | 0 | 0 | 3.204296 | 4.160181 |
| Reactome_ | Extra-nucle | Jan-73 | 0.272992 | 0.493146 | 0 | 0 | 3.204296 | 4.160181 |
| SysMyo_M | MGS96 Mu | Jan-50 | 0.196069 | 0.493223 | 0 | 0 | 4.713811 | 7.680157 |
| GeneSigDB | 12917485-1 | 3/394 | 0.245504 | 0.493667 | 0 | 0 | 1.783157 | 2.504339 |
| SysMyo_M | MGS1065 I | 2/188 | 0.197153 | 0.493754 | 0 | 0 | 2.495509 | 4.05215 |
| Rare_Disea | CD4 deficie | 1/109 | 0.379031 | 0.493869 | 0 | 0 | 2.132321 | 2.068645 |
| Rare_Disea | Primary bil | 1/109 | 0.379031 | 0.493869 | 0 | 0 | 2.132321 | 2.068645 |
| GWAS_Cat | Ulcerative | 1/149 | 0.478989 | 0.49397 | 0 | 0 | 1.552876 | 1.143039 |
| Jensen_TIS | Ganglion | 2/323 | 0.411339 | 0.49422 | 0 | 0 | 1.4361 | 1.275744 |
| SysMyo_M | MGS1181 I | 2/190 | 0.200314 | 0.494293 | 0 | 0 | 2.468711 | 3.969367 |
| SysMyo_M | MGS1134 I | Jan-52 | 0.203066 | 0.494293 | 0 | 0 | 4.5285 | 7.219451 |
| SysMyo_M | MGS1085 I | 2/191 | 0.201897 | 0.494293 | 0 | 0 | 2.455524 | 3.928837 |
| SysMyo_M | MGS347 Hu | Jan-52 | 0.203066 | 0.494293 | 0 | 0 | 4.5285 | 7.219451 |
| SysMyo_M | MGS981 M | 2/191 | 0.201897 | 0.494293 | 0 | 0 | 2.455524 | 3.928837 |
| SysMyo_M | MGS698 M | 2/192 | 0.203481 | 0.494293 | 0 | 0 | 2.442477 | 3.888868 |
| SysMyo_M | MGS1103 I | 2/190 | 0.200314 | 0.494293 | 0 | 0 | 2.468711 | 3.969367 |
| Rare_Disea | Hydatidifor | 1/110 | 0.381747 | 0.494808 | 0 | 0 | 2.112652 | 2.034479 |
| Rare_Disea | Hypertroph | 1/110 | 0.381747 | 0.494808 | 0 | 0 | 2.112652 | 2.034479 |
| Rare_Disea | Vaginal car | 1/110 | 0.381747 | 0.494808 | 0 | 0 | 2.112652 | 2.034479 |
| Reactome_ | Cell Death | Jan-74 | 0.276166 | 0.494889 | 0 | 0 | 3.160242 | 4.066453 |
| GO_Biologi | Negative R | 1/140 | 0.457986 | 0.494931 | 0 | 0 | 1.654174 | 1.291774 |
| GeneSigDB | 16042785-1 | Jan-65 | 0.247099 | 0.495087 | 0 | 0 | 3.606286 | 5.041467 |
| GeDiPNet_ | Hypotonic | 1/138 | 0.453206 | 0.49572 | 0 | 0 | 1.678493 | 1.328374 |
| Jensen_TIS | HeLa cell | 1/123 | 0.416004 | 0.496124 | 0 | 0 | 1.886294 | 1.654392 |
| Jensen_TIS | Prostate gl | 1/123 | 0.416004 | 0.496124 | 0 | 0 | 1.886294 | 1.654392 |
| Diabetes_F | GSE154198 | 3/205 | 0.060177 | 0.496732 | 0 | 0 | 3.484972 | 9.794417 |
| Diabetes_F | GSE110935 | 3/206 | 0.060881 | 0.496732 | 0 | 0 | 3.467628 | 9.705325 |
| Diabetes_F | GSE154198 | 3/204 | 0.059476 | 0.496732 | 0 | 0 | 3.502488 | 9.88465 |
| DisGeNET | Absent ten | 1/134 | 0.443521 | 0.496793 | 0 | 0 | 1.729323 | 1.405958 |
| DisGeNET | Diabetes M | 8/1672 | 0.443701 | 0.496793 | 0 | 0 | 1.110577 | 0.90246 |
| DisGeNET | Skin Carcin | 1/134 | 0.443521 | 0.496793 | 0 | 0 | 1.729323 | 1.405958 |
| DisGeNET | High foreh | 1/134 | 0.443521 | 0.496793 | 0 | 0 | 1.729323 | 1.405958 |
| DisGeNET | Nasophary | 4/782 | 0.443931 | 0.496839 | 0 | 0 | 1.185307 | 0.96257 |
| GeDiPNet_ | Pancreatiti | 1/139 | 0.455601 | 0.497407 | 0 | 0 | 1.666245 | 1.309899 |
| Rare_Disea | Ovarian lov | 1/111 | 0.384451 | 0.497446 | 0 | 0 | 2.09334 | 2.001105 |
| DisGeNET | Dystonia D | 1/135 | 0.445958 | 0.498256 | 0 | 0 | 1.716331 | 1.38599 |

| | | | | | | | | |
|---|---|---|---|---|---|---|---|---|
| DisGeNET | Flexion cor | 1/135 | 0.445958 | 0.498256 | 0 | 0 | 1.716331 | 1.38599 |
| DisGeNET | Motor Neu | 1/135 | 0.445958 | 0.498256 | 0 | 0 | 1.716331 | 1.38599 |
| DisGeNET | Hirschsprui | 1/135 | 0.445958 | 0.498256 | 0 | 0 | 1.716331 | 1.38599 |
| Disease_Sig | Synovial sa | 4/300 | 0.041941 | 0.498636 | 0 | 0 | 3.193911 | 10.12943 |
| Disease_Sig | Down Sync | 4/300 | 0.041941 | 0.498636 | 0 | 0 | 3.193911 | 10.12943 |
| Disease_Sig | Adenocarci | 4/300 | 0.041941 | 0.498636 | 0 | 0 | 3.193911 | 10.12943 |
| Disease_Sig | Urothelial ( | 4/300 | 0.041941 | 0.498636 | 0 | 0 | 3.193911 | 10.12943 |
| Disease_Sig | LGLL - Larg | 4/300 | 0.041941 | 0.498636 | 0 | 0 | 3.193911 | 10.12943 |
| Disease_Sig | Lymphatic | 4/300 | 0.041941 | 0.498636 | 0 | 0 | 3.193911 | 10.12943 |
| WikiPathw | PI3K Akt Sig | 2/338 | 0.433875 | 0.498956 | 0 | 0 | 1.370938 | 1.144732 |
| Rare_Disea | Polydactyly | 1/112 | 0.387144 | 0.499194 | 0 | 0 | 2.074377 | 1.968499 |
| Rare_Disea | Renal canc | 1/112 | 0.387144 | 0.499194 | 0 | 0 | 2.074377 | 1.968499 |
| GeneSigDB | 19096012-1 | Jan-66 | 0.250385 | 0.499872 | 0 | 0 | 3.550626 | 4.916752 |
| DisGeNET | Amyotroph | 1/136 | 0.448384 | 0.500114 | 0 | 0 | 1.703531 | 1.36641 |
| DisGeNET | Vasculitis | 1/136 | 0.448384 | 0.500114 | 0 | 0 | 1.703531 | 1.36641 |
| DisGeNET | Melanoma | 1/136 | 0.448384 | 0.500114 | 0 | 0 | 1.703531 | 1.36641 |
| DisGeNET | Flexion cor | 1/136 | 0.448384 | 0.500114 | 0 | 0 | 1.703531 | 1.36641 |
| dbGaP | Parkinson I | 1/132 | 0.438615 | 0.500175 | 0 | 0 | 1.755903 | 1.447098 |
| GWAS_Cat | Eczema | 1/152 | 0.485809 | 0.50058 | 0 | 0 | 1.521793 | 1.098642 |
| Rare_Disea | Niemann-P | 1/113 | 0.389825 | 0.500915 | 0 | 0 | 2.055752 | 1.936638 |
| Rare_Disea | Cholecystit | 1/113 | 0.389825 | 0.500915 | 0 | 0 | 2.055752 | 1.936638 |
| Rare_Disea | Hyperlipop | 1/114 | 0.392494 | 0.501745 | 0 | 0 | 2.037456 | 1.905499 |
| Rare_Disea | Wegener's | 1/114 | 0.392494 | 0.501745 | 0 | 0 | 2.037456 | 1.905499 |
| Rare_Disea | Sertoli cell- | 1/114 | 0.392494 | 0.501745 | 0 | 0 | 2.037456 | 1.905499 |
| GO_Biologi | Positive Re | 1/143 | 0.465078 | 0.501832 | 0 | 0 | 1.618981 | 1.239412 |
| DisGeNET | Periodonta | 1/137 | 0.4508 | 0.501954 | 0 | 0 | 1.69092 | 1.347208 |
| DisGeNET | Reflex, Dee | 1/137 | 0.4508 | 0.501954 | 0 | 0 | 1.69092 | 1.347208 |
| DisGeNET | Absent refl | 1/137 | 0.4508 | 0.501954 | 0 | 0 | 1.69092 | 1.347208 |
| DisGeNET | Pruritus | 1/137 | 0.4508 | 0.501954 | 0 | 0 | 1.69092 | 1.347208 |
| GeneSigDB | 19249242-1 | Jan-68 | 0.256914 | 0.502109 | 0 | 0 | 3.44429 | 4.68084 |
| GeneSigDB | 14737219-( | 4/591 | 0.256027 | 0.502109 | 0 | 0 | 1.586667 | 2.161789 |
| GeneSigDB | 16546165-1 | Jan-68 | 0.256914 | 0.502109 | 0 | 0 | 3.44429 | 4.68084 |
| GeneSigDB | 17893858-1 | Jan-68 | 0.256914 | 0.502109 | 0 | 0 | 3.44429 | 4.68084 |
| GeneSigDB | 20436685-1 | 2/225 | 0.256348 | 0.502109 | 0 | 0 | 2.077552 | 2.828 |
| GeneSigDB | 18231641-1 | Jan-67 | 0.253656 | 0.502109 | 0 | 0 | 3.496653 | 4.79662 |
| Diabetes_P | GSE154198 | 3/209 | 0.063017 | 0.502736 | 0 | 0 | 3.416609 | 9.444704 |
| Disease_Pe | liver cirrho | 5/301 | 0.010118 | 0.502781 | 0 | 0 | 4.041076 | 18.56248 |
| DisGeNET | Drug habitu | 1/138 | 0.453206 | 0.503135 | 0 | 0 | 1.678493 | 1.328374 |
| DisGeNET | Ureteral ob | 1/138 | 0.453206 | 0.503135 | 0 | 0 | 1.678493 | 1.328374 |
| DisGeNET | Spontaneo | 1/138 | 0.453206 | 0.503135 | 0 | 0 | 1.678493 | 1.328374 |
| DisGeNET | Mental Ret | 1/138 | 0.453206 | 0.503135 | 0 | 0 | 1.678493 | 1.328374 |
| DisGeNET | Psoriasis vu | 1/138 | 0.453206 | 0.503135 | 0 | 0 | 1.678493 | 1.328374 |
| DisGeNET | Small head | 3/570 | 0.452956 | 0.503135 | 0 | 0 | 1.218569 | 0.965059 |
| DisGeNET | Measles | 1/138 | 0.453206 | 0.503135 | 0 | 0 | 1.678493 | 1.328374 |
| GWAS_Cat | White Bloo | 5/1056 | 0.48881 | 0.503247 | 0 | 0 | 1.094312 | 0.783289 |

| | | | | | | | | |
|---|---|---|---|---|---|---|---|---|
| Disease_Pe | Niemann-P | 5/310 | 0.011386 | 0.503439 | 0 | 0 | 3.920032 | 17.54347 |
| Disease_Pe | polycystic ( | 3/108 | 0.01169 | 0.503439 | 0 | 0 | 6.737415 | 29.97504 |
| GO_Biologi | Actin Filam | 1/144 | 0.467422 | 0.503595 | 0 | 0 | 1.607578 | 1.222601 |
| Reactome_ | Regulation | Jan-77 | 0.285606 | 0.503746 | 0 | 0 | 3.035037 | 3.803331 |
| Reactome_ | Processing | 2/242 | 0.283765 | 0.503746 | 0 | 0 | 1.928725 | 2.429439 |
| GeDiPNet_ | Ichthyosis | 1/142 | 0.462724 | 0.50424 | 0 | 0 | 1.630546 | 1.256538 |
| SysMyo_M | MGS663 H | 2/196 | 0.209833 | 0.505385 | 0 | 0 | 2.391631 | 3.734399 |
| SysMyo_M | MGS1204 I | 2/196 | 0.209833 | 0.505385 | 0 | 0 | 2.391631 | 3.734399 |
| DisGeNET | Differentia | 1/139 | 0.455601 | 0.50558 | 0 | 0 | 1.666245 | 1.309899 |
| Rare_Disea | Craniosync | 1/120 | 0.408269 | 0.506254 | 0 | 0 | 1.934141 | 1.732658 |
| Rare_Disea | Acute myel | 1/120 | 0.408269 | 0.506254 | 0 | 0 | 1.934141 | 1.732658 |
| Rare_Disea | Cardiac rup | 1/119 | 0.405668 | 0.506254 | 0 | 0 | 1.950631 | 1.759896 |
| Rare_Disea | Myelodysp | 1/117 | 0.400433 | 0.506254 | 0 | 0 | 1.984463 | 1.816198 |
| Rare_Disea | Schistosom | 1/120 | 0.408269 | 0.506254 | 0 | 0 | 1.934141 | 1.732658 |
| Rare_Disea | Hodgkin lyı | 1/117 | 0.400433 | 0.506254 | 0 | 0 | 1.984463 | 1.816198 |
| Rare_Disea | Xeroderma | 1/120 | 0.408269 | 0.506254 | 0 | 0 | 1.934141 | 1.732658 |
| Rare_Disea | Placenta di | 1/116 | 0.397798 | 0.506254 | 0 | 0 | 2.00182 | 1.845299 |
| Rare_Disea | Stomach ca | 1/120 | 0.408269 | 0.506254 | 0 | 0 | 1.934141 | 1.732658 |
| Rare_Disea | Oral squam | 1/117 | 0.400433 | 0.506254 | 0 | 0 | 1.984463 | 1.816198 |
| Rare_Disea | Lymphobla | 1/120 | 0.408269 | 0.506254 | 0 | 0 | 1.934141 | 1.732658 |
| Rare_Disea | Mitochond | 1/117 | 0.400433 | 0.506254 | 0 | 0 | 1.984463 | 1.816198 |
| Rare_Disea | Acute myel | 1/120 | 0.408269 | 0.506254 | 0 | 0 | 1.934141 | 1.732658 |
| Rare_Disea | Allergic enc | 2/317 | 0.402204 | 0.506254 | 0 | 0 | 1.463903 | 1.333316 |
| Rare_Disea | Acute myel | 1/118 | 0.403056 | 0.506254 | 0 | 0 | 1.967402 | 1.787737 |
| Rare_Disea | Bloom sync | 1/118 | 0.403056 | 0.506254 | 0 | 0 | 1.967402 | 1.787737 |
| Rare_Disea | Cutaneous | 1/116 | 0.397798 | 0.506254 | 0 | 0 | 2.00182 | 1.845299 |
| Rare_Disea | Esophagea | 1/118 | 0.403056 | 0.506254 | 0 | 0 | 1.967402 | 1.787737 |
| PhenGenI_ | Leptin | Jan-89 | 0.322165 | 0.50626 | 0 | 0 | 2.619582 | 2.967177 |
| GO_Biologi | Neuron Prc | 1/146 | 0.472079 | 0.506308 | 0 | 0 | 1.585245 | 1.1899 |
| GO_Biologi | Positive Re | 1/146 | 0.472079 | 0.506308 | 0 | 0 | 1.585245 | 1.1899 |
| GO_Biologi | Regulation | 1/146 | 0.472079 | 0.506308 | 0 | 0 | 1.585245 | 1.1899 |
| GeneSigDB | 12919955- | Jan-69 | 0.260157 | 0.50667 | 0 | 0 | 3.393468 | 4.569198 |
| GeDiPNet_ | Arthrogryp | 1/144 | 0.467422 | 0.507461 | 0 | 0 | 1.607578 | 1.222601 |
| GeDiPNet_ | Pancreatic | 1/144 | 0.467422 | 0.507461 | 0 | 0 | 1.607578 | 1.222601 |
| DisGeNET | Contractur | 1/140 | 0.457986 | 0.508011 | 0 | 0 | 1.654174 | 1.291774 |
| GO_Biologi | Protein Gly | 1/147 | 0.474392 | 0.508021 | 0 | 0 | 1.574307 | 1.173995 |
| GTEx_Tissu | GTEX-PLZ6 | 13/1652 | 0.02599 | 0.508031 | 0 | 0 | 1.958693 | 7.149327 |
| GTEx_Tissu | GTEX-WK1 | 9/978 | 0.026177 | 0.508247 | 0 | 0 | 2.255775 | 8.217544 |
| DisGeNET | Diffuse Lar | 3/575 | 0.458647 | 0.508529 | 0 | 0 | 1.207605 | 0.941298 |
| GeneSigDB | 18410693- | Jan-71 | 0.266602 | 0.508553 | 0 | 0 | 3.296179 | 4.35754 |
| GeneSigDB | 20096135- | Jan-71 | 0.266602 | 0.508553 | 0 | 0 | 3.296179 | 4.35754 |
| GeneSigDB | 18490921- | Jan-71 | 0.266602 | 0.508553 | 0 | 0 | 3.296179 | 4.35754 |
| GeneSigDB | 18006812- | Jan-71 | 0.266602 | 0.508553 | 0 | 0 | 3.296179 | 4.35754 |
| GeneSigDB | 16699793- | Jan-71 | 0.266602 | 0.508553 | 0 | 0 | 3.296179 | 4.35754 |
| GeneSigDB | 17671232- | Jan-70 | 0.263387 | 0.508553 | 0 | 0 | 3.344119 | 4.461494 |

| | | | | | | | | |
|---|---|---|---|---|---|---|---|---|
| GO_Molecı | DNA-bindir | 2/282 | 0.347744 | 0.509005 | 0 | 0 | 1.649832 | 1.7427 |
| GO_Molecı | Protein Tyr | Jan-99 | 0.351214 | 0.509005 | 0 | 0 | 2.351092 | 2.46009 |
| GO_Molecı | Hexosyltrai | Jan-96 | 0.342631 | 0.509005 | 0 | 0 | 2.425704 | 2.598176 |
| GeDiPNet_ | Colonic Ne | 2/363 | 0.470402 | 0.509748 | 0 | 0 | 1.274369 | 0.961087 |
| SysMyo_M | MGS372 Hı | Jan-55 | 0.213448 | 0.509753 | 0 | 0 | 4.27627 | 6.604112 |
| SysMyo_M | MGS916 M | Jan-55 | 0.213448 | 0.509753 | 0 | 0 | 4.27627 | 6.604112 |
| DisGeNET | Pectus exca | 1/141 | 0.46036 | 0.509997 | 0 | 0 | 1.642276 | 1.27399 |
| DisGeNET | Electroretir | 1/141 | 0.46036 | 0.509997 | 0 | 0 | 1.642276 | 1.27399 |
| SysMyo_M | MGS1259 I | 2/199 | 0.21461 | 0.510376 | 0 | 0 | 2.354852 | 3.623955 |
| Reactome_ | Amyloid Fil | Jan-79 | 0.291832 | 0.510706 | 0 | 0 | 2.956917 | 3.641673 |
| GTEx_Tissu | GTEX-XYKS | 15/2014 | 0.026483 | 0.510774 | 0 | 0 | 1.866975 | 6.779438 |
| GWAS_Cat | Cognitive A | 1/157 | 0.496981 | 0.510798 | 0 | 0 | 1.472645 | 1.029678 |
| GWAS_Cat | Peak Expira | 1/157 | 0.496981 | 0.510798 | 0 | 0 | 1.472645 | 1.029678 |
| CellMarker | Microglial ( | 2/360 | 0.466091 | 0.510836 | 0 | 0 | 1.285245 | 0.981122 |
| Reactome_ | Mitochond | Jan-82 | 0.30107 | 0.510907 | 0 | 0 | 2.846971 | 3.417539 |
| Reactome_ | Regulation | Jan-81 | 0.298004 | 0.510907 | 0 | 0 | 2.882703 | 3.48994 |
| Reactome_ | TP53 Regul | Jan-81 | 0.298004 | 0.510907 | 0 | 0 | 2.882703 | 3.48994 |
| Reactome_ | Vesicle-me | 4/637 | 0.300626 | 0.510907 | 0 | 0 | 1.467862 | 1.764205 |
| Reactome_ | Platelet Ho | Jan-83 | 0.304123 | 0.511208 | 0 | 0 | 2.81211 | 3.34732 |
| Reactome_ | Metabolisr | 10/1890 | 0.305812 | 0.511208 | 0 | 0 | 1.245717 | 1.475906 |
| GTEx_Tissu | GTEX-WYJk | 8/825 | 0.02705 | 0.511477 | 0 | 0 | 2.366918 | 8.544727 |
| GTEx_Tissu | GTEX-PVO\ | 6/525 | 0.026921 | 0.511477 | 0 | 0 | 2.768001 | 10.00594 |
| GTEx_Tissu | GTEX-OHPI | 6/524 | 0.026704 | 0.511477 | 0 | 0 | 2.773488 | 10.04824 |
| Rare_Disea | Conversion | 1/122 | 0.413437 | 0.511809 | 0 | 0 | 1.90198 | 1.679923 |
| PhenGenI_ | Atheroscle | Jan-92 | 0.331012 | 0.511918 | 0 | 0 | 2.532839 | 2.800307 |
| PhenGenI_ | Job Satisfa | Jan-94 | 0.336847 | 0.511918 | 0 | 0 | 2.47812 | 2.69651 |
| PhenGenI_ | Liver Cirrho | Jan-94 | 0.336847 | 0.511918 | 0 | 0 | 2.47812 | 2.69651 |
| PhenGenI_ | Prostate-Sp | Jan-93 | 0.333936 | 0.511918 | 0 | 0 | 2.505182 | 2.747701 |
| PhenGenI_ | Hypertensi | 2/270 | 0.328691 | 0.511918 | 0 | 0 | 1.724759 | 1.919028 |
| DisGeNET | Vascular in | 1/142 | 0.462724 | 0.512183 | 0 | 0 | 1.630546 | 1.256538 |
| DisGeNET | Mycosis Fu | 1/142 | 0.462724 | 0.512183 | 0 | 0 | 1.630546 | 1.256538 |
| Rare_Disea | Mental reta | 2/326 | 0.415881 | 0.512429 | 0 | 0 | 1.422585 | 1.248115 |
| Rare_Disea | Chronic my | 1/123 | 0.416004 | 0.512429 | 0 | 0 | 1.886294 | 1.654392 |
| Rare_Disea | Intrahepati | 1/123 | 0.416004 | 0.512429 | 0 | 0 | 1.886294 | 1.654392 |
| DisGeNET | Uterine Co | 3/579 | 0.463182 | 0.512473 | 0 | 0 | 1.198971 | 0.92277 |
| GeneSigDB | 17150101- | Jan-72 | 0.269804 | 0.512904 | 0 | 0 | 3.249591 | 4.257158 |
| PhenGenI_ | Platelet Co | 2/277 | 0.339825 | 0.513068 | 0 | 0 | 1.680257 | 1.813544 |
| GeDiPNet_ | Pulmonary | 1/147 | 0.474392 | 0.513118 | 0 | 0 | 1.574307 | 1.173995 |
| SysMyo_M | MGS646 Hı | 2/202 | 0.219398 | 0.513139 | 0 | 0 | 2.319176 | 3.517879 |
| SysMyo_M | MGS817 M | 2/201 | 0.217801 | 0.513139 | 0 | 0 | 2.330949 | 3.552767 |
| SysMyo_M | MGS774 M | 2/201 | 0.217801 | 0.513139 | 0 | 0 | 2.330949 | 3.552767 |
| SysMyo_M | MGS1147 I | 2/202 | 0.219398 | 0.513139 | 0 | 0 | 2.319176 | 3.517879 |
| WikiPathw | Ectoderm I | 1/137 | 0.4508 | 0.513288 | 0 | 0 | 1.69092 | 1.347208 |
| GeneSigDB | 18849563- | Jan-75 | 0.279326 | 0.513481 | 0 | 0 | 3.117379 | 3.975827 |
| GeneSigDB | 16705683- | Jan-75 | 0.279326 | 0.513481 | 0 | 0 | 3.117379 | 3.975827 |

| | | | | | | | | |
|---|---|---|---|---|---|---|---|---|
| GeneSigDB | 18691415-' | Jan-74 | 0.276166 | 0.513481 | 0 | 0 | 3.160242 | 4.066453 |
| GeneSigDB | 18849563-: | Jan-74 | 0.276166 | 0.513481 | 0 | 0 | 3.160242 | 4.066453 |
| GeneSigDB | 18849563-: | Jan-75 | 0.279326 | 0.513481 | 0 | 0 | 3.117379 | 3.975827 |
| GeneSigDB | 18516279-( | Jan-74 | 0.276166 | 0.513481 | 0 | 0 | 3.160242 | 4.066453 |
| GeneSigDB | 19799973-' | Jan-74 | 0.276166 | 0.513481 | 0 | 0 | 3.160242 | 4.066453 |
| GeneSigDB | 18849563-: | Jan-75 | 0.279326 | 0.513481 | 0 | 0 | 3.117379 | 3.975827 |
| GeneSigDB | 20166207-' | Jan-73 | 0.272992 | 0.513481 | 0 | 0 | 3.204296 | 4.160181 |
| GeneSigDB | 12917207-' | 2/236 | 0.27409 | 0.513481 | 0 | 0 | 1.978783 | 2.561135 |
| KOMP2_M | Abnormal I | Jan-46 | 0.181893 | 0.513582 | 0 | 0 | 5.13385 | 8.749803 |
| KOMP2_M | Abnormal \ | Jan-46 | 0.181893 | 0.513582 | 0 | 0 | 5.13385 | 8.749803 |
| KOMP2_M | Embryonic | 4/510 | 0.181923 | 0.513582 | 0 | 0 | 1.848374 | 3.149951 |
| KOMP2_M | Abnormal I | Jan-47 | 0.18546 | 0.513582 | 0 | 0 | 5.021992 | 8.46163 |
| KOMP2_M | Enlarged O | 29-Jan | 0.118838 | 0.513582 | 0 | 0 | 8.25789 | 17.58927 |
| KOMP2_M | Abnormal I | 2/161 | 0.155267 | 0.513582 | 0 | 0 | 2.92327 | 5.444911 |
| KOMP2_M | Impaired G | 2/164 | 0.159833 | 0.513582 | 0 | 0 | 2.8687 | 5.260115 |
| KOMP2_M | Enlarged Ly | 3/286 | 0.128615 | 0.513582 | 0 | 0 | 2.477284 | 5.080743 |
| KOMP2_M | Abnormal S | Feb-84 | 0.05183 | 0.513582 | 0 | 0 | 5.690387 | 16.84234 |
| KOMP2_M | Abnormal \ | Jan-46 | 0.181893 | 0.513582 | 0 | 0 | 5.13385 | 8.749803 |
| KOMP2_M | Increased ( | Jan-34 | 0.137866 | 0.513582 | 0 | 0 | 7.004933 | 13.88007 |
| KOMP2_M | Trunk Curl | Jan-37 | 0.149088 | 0.513582 | 0 | 0 | 6.42022 | 12.21909 |
| KOMP2_M | Preweanin | 11/1785 | 0.151074 | 0.513582 | 0 | 0 | 1.479922 | 2.79703 |
| KOMP2_M | Abnormal / | 3/309 | 0.151612 | 0.513582 | 0 | 0 | 2.288399 | 4.316902 |
| KOMP2_M | Decreased | 16-Jan | 0.067399 | 0.513582 | 0 | 0 | 15.42481 | 41.60265 |
| KOMP2_M | Enlarged C | Jan-33 | 0.134093 | 0.513582 | 0 | 0 | 7.224201 | 14.515 |
| KOMP2_M | Enlarged S | 4/360 | 0.07203 | 0.513582 | 0 | 0 | 2.647489 | 6.964668 |
| KOMP2_M | Embryonic | 3/311 | 0.153672 | 0.513582 | 0 | 0 | 2.273307 | 4.257756 |
| KOMP2_M | Hyperactiv | 5/614 | 0.129288 | 0.513582 | 0 | 0 | 1.932797 | 3.953951 |
| KOMP2_M | Facial Cleft | 17-Jan | 0.071459 | 0.513582 | 0 | 0 | 14.46003 | 38.15469 |
| KOMP2_M | Increased ( | 7-Jan | 0.03006 | 0.513582 | 0 | 0 | 38.57946 | 135.2043 |
| KOMP2_M | Corneal De | 11-Jan | 0.046834 | 0.513582 | 0 | 0 | 23.14302 | 70.84419 |
| KOMP2_M | Hypertroph | 11-Jan | 0.046834 | 0.513582 | 0 | 0 | 23.14302 | 70.84419 |
| KOMP2_M | Abnormal I | 30-Jan | 0.122676 | 0.513582 | 0 | 0 | 7.972735 | 16.72843 |
| KOMP2_M | Abnormal I | Jan-41 | 0.163825 | 0.513582 | 0 | 0 | 5.777035 | 10.45039 |
| KOMP2_M | Increased ( | 3/328 | 0.171529 | 0.513582 | 0 | 0 | 2.152527 | 3.794916 |
| KOMP2_M | Abnormal ( | Jan-32 | 0.130304 | 0.513582 | 0 | 0 | 7.457614 | 15.19775 |
| KOMP2_M | Increased ( | 2/145 | 0.131396 | 0.513582 | 0 | 0 | 3.252982 | 6.602064 |
| KOMP2_M | Abnormal S | 11-Jan | 0.046834 | 0.513582 | 0 | 0 | 23.14302 | 70.84419 |
| KOMP2_M | Decreased | 2/176 | 0.178336 | 0.513582 | 0 | 0 | 2.669236 | 4.601995 |
| KOMP2_M | Extramedu | 13-Jan | 0.055113 | 0.513582 | 0 | 0 | 19.28391 | 55.89185 |
| KOMP2_M | Abnormal ( | 13-Jan | 0.055113 | 0.513582 | 0 | 0 | 19.28391 | 55.89185 |
| KOMP2_M | Decreased | Feb-88 | 0.05629 | 0.513582 | 0 | 0 | 5.424624 | 15.60794 |
| KOMP2_M | Increased ( | 14-Jan | 0.059226 | 0.513582 | 0 | 0 | 17.79964 | 50.30878 |
| KOMP2_M | Decreased | 15-Jan | 0.063321 | 0.513582 | 0 | 0 | 16.52741 | 45.60793 |
| KOMP2_M | Increased ( | 2/110 | 0.082985 | 0.513582 | 0 | 0 | 4.314815 | 10.73997 |
| KOMP2_M | Increased / | 21-Jan | 0.087525 | 0.513582 | 0 | 0 | 11.5657 | 28.17205 |

| | | | | | | | | |
|---|---|---|---|---|---|---|---|---|
| KOMP2_M | Abnormal ( | 22-Jan | 0.091499 | 0.513582 | 0 | 0 | 11.0144 | 26.34017 |
| KOMP2_M | Abnormal I | 22-Jan | 0.091499 | 0.513582 | 0 | 0 | 11.0144 | 26.34017 |
| GeDiPNet_ | Congenital | 1/148 | 0.476695 | 0.513703 | 0 | 0 | 1.563518 | 1.158376 |
| GeDiPNet_ | Hyperinsul | 1/148 | 0.476695 | 0.513703 | 0 | 0 | 1.563518 | 1.158376 |
| GO_Biologi | Neuron De | 1/150 | 0.481272 | 0.513839 | 0 | 0 | 1.542376 | 1.127974 |
| GO_Biologi | protein-RN | 1/150 | 0.481272 | 0.513839 | 0 | 0 | 1.542376 | 1.127974 |
| DisGeNET | Lymphoma | 2/359 | 0.46465 | 0.51388 | 0 | 0 | 1.288911 | 0.987914 |
| DisGeNET | Substance | 1/143 | 0.465078 | 0.514137 | 0 | 0 | 1.618981 | 1.239412 |
| SysMyo_M | MGS1503 I | 2/204 | 0.222596 | 0.514242 | 0 | 0 | 2.295981 | 3.449476 |
| SysMyo_M | MGS1071 I | 2/204 | 0.222596 | 0.514242 | 0 | 0 | 2.295981 | 3.449476 |
| SysMyo_M | MGS1310 I | 2/203 | 0.220997 | 0.514242 | 0 | 0 | 2.307521 | 3.483452 |
| GeneSigDB | 17571080-: | 2/242 | 0.283765 | 0.51483 | 0 | 0 | 1.928725 | 2.429439 |
| GeneSigDB | 20038735-' | Jan-76 | 0.282473 | 0.51483 | 0 | 0 | 3.075659 | 3.888164 |
| GeneSigDB | 20157477-' | 2/243 | 0.285377 | 0.51483 | 0 | 0 | 1.920625 | 2.408358 |
| GeneSigDB | 17205517-' | Jan-77 | 0.285606 | 0.51483 | 0 | 0 | 3.035037 | 3.803331 |
| GeneSigDB | 12213815-' | Jan-76 | 0.282473 | 0.51483 | 0 | 0 | 3.075659 | 3.888164 |
| GeneSigDB | 15972966-' | Jan-77 | 0.285606 | 0.51483 | 0 | 0 | 3.035037 | 3.803331 |
| GeneSigDB | 18394172-: | 2/244 | 0.286988 | 0.515449 | 0 | 0 | 1.912591 | 2.387516 |
| GeneSigDB | 18410693-' | Jan-78 | 0.288726 | 0.515449 | 0 | 0 | 2.99547 | 3.721207 |
| GeneSigDB | 20038735-' | Jan-78 | 0.288726 | 0.515449 | 0 | 0 | 2.99547 | 3.721207 |
| Jensen_TIS | Ophthalmi( | 16/3453 | 0.433879 | 0.515532 | 0 | 0 | 1.080274 | 0.902017 |
| SysMyo_M | MGS673 H( | 2/205 | 0.224196 | 0.515833 | 0 | 0 | 2.284555 | 3.415945 |
| GeneSigDB | 17597811-: | 4/627 | 0.290831 | 0.516033 | 0 | 0 | 1.492197 | 1.842884 |
| GeneSigDB | 19798054-: | 2/247 | 0.29182 | 0.516033 | 0 | 0 | 1.888884 | 2.326383 |
| GeneSigDB | 19192944-' | Jan-79 | 0.291832 | 0.516033 | 0 | 0 | 2.956917 | 3.641673 |
| DisGeNET | Encephaliti | 1/144 | 0.467422 | 0.51651 | 0 | 0 | 1.607578 | 1.222601 |
| SysMyo_M | MGS1098 I | 2/206 | 0.225797 | 0.517414 | 0 | 0 | 2.273241 | 3.382851 |
| GeneSigDB | 19061838-' | 5/830 | 0.293757 | 0.517793 | 0 | 0 | 1.410791 | 1.728222 |
| Rare_Disea | Chronic lyn | 1/125 | 0.421106 | 0.517855 | 0 | 0 | 1.855683 | 1.604928 |
| GTEx_Tissu | GTEX-VJYA | 11/1319 | 0.027574 | 0.51799 | 0 | 0 | 2.058738 | 7.392714 |
| DisGeNET | Mammary | 1/145 | 0.469755 | 0.518432 | 0 | 0 | 1.596334 | 1.2061 |
| DisGeNET | Animal Ma | 1/145 | 0.469755 | 0.518432 | 0 | 0 | 1.596334 | 1.2061 |
| DisGeNET | Nerve Dege | 1/145 | 0.469755 | 0.518432 | 0 | 0 | 1.596334 | 1.2061 |
| DisGeNET | Adenocarci | 8/1712 | 0.470646 | 0.519197 | 0 | 0 | 1.08213 | 0.815546 |
| Rare_Disea | Lipodystro| | 1/126 | 0.42364 | 0.519255 | 0 | 0 | 1.840744 | 1.580964 |
| Rare_Disea | Malignant | 1/126 | 0.42364 | 0.519255 | 0 | 0 | 1.840744 | 1.580964 |
| GWAS_Cat | General Ris | 1/161 | 0.505746 | 0.519368 | 0 | 0 | 1.435538 | 0.978637 |
| DisGeNET | Leukemia, ' | 2/364 | 0.471835 | 0.520289 | 0 | 0 | 1.270783 | 0.954519 |
| GeneSigDB | 20068109-: | 2/252 | 0.299865 | 0.520326 | 0 | 0 | 1.850635 | 2.228949 |
| GeneSigDB | 19185848-' | 2/252 | 0.299865 | 0.520326 | 0 | 0 | 1.850635 | 2.228949 |
| GeneSigDB | 20077526-' | Jan-81 | 0.298004 | 0.520326 | 0 | 0 | 2.882703 | 3.48994 |
| GeneSigDB | 17389037-' | 2/251 | 0.298257 | 0.520326 | 0 | 0 | 1.858162 | 2.248005 |
| GeneSigDB | 18425577-' | Jan-81 | 0.298004 | 0.520326 | 0 | 0 | 2.882703 | 3.48994 |
| Rare_Disea | Sezary sync | 1/127 | 0.426163 | 0.521488 | 0 | 0 | 1.826043 | 1.557494 |
| GWAS_Cat | Eosinophil | 2/390 | 0.508263 | 0.521514 | 0 | 0 | 1.184051 | 0.801314 |

| | | | | | | | | |
|---|---|---|---|---|---|---|---|---|
| GeneSigDB | 11108479-: | Jan-83 | 0.304123 | 0.521843 | 0 | 0 | 2.81211 | 3.34732 |
| GeneSigDB | 19171046-: | Jan-83 | 0.304123 | 0.521843 | 0 | 0 | 2.81211 | 3.34732 |
| GeneSigDB | 17638894-: | Jan-83 | 0.304123 | 0.521843 | 0 | 0 | 2.81211 | 3.34732 |
| GeneSigDB | 20460173- | 3/444 | 0.304486 | 0.521843 | 0 | 0 | 1.576936 | 1.875179 |
| PhenGenI_ | Lung Volun | 5/893 | 0.347994 | 0.521992 | 0 | 0 | 1.306375 | 1.378968 |
| SysMyo_M | MGS737 M | 2/208 | 0.229002 | 0.522642 | 0 | 0 | 2.250942 | 3.317943 |
| GO_Biologi | Epithelium | 1/154 | 0.490307 | 0.5227 | 0 | 0 | 1.501748 | 1.070331 |
| Rare_Disea | Acute non | 1/128 | 0.428675 | 0.522843 | 0 | 0 | 1.811573 | 1.534505 |
| Rare_Disea | Hypothalar | 1/128 | 0.428675 | 0.522843 | 0 | 0 | 1.811573 | 1.534505 |
| DisGeNET | Hematolog | 1/147 | 0.474392 | 0.522889 | 0 | 0 | 1.574307 | 1.173995 |
| Rare_Disea | Medullobla | 1/134 | 0.443521 | 0.522947 | 0 | 0 | 1.729323 | 1.405958 |
| Rare_Disea | Hepatocell | 1/134 | 0.443521 | 0.522947 | 0 | 0 | 1.729323 | 1.405958 |
| Rare_Disea | Distomatos | 1/132 | 0.438615 | 0.522947 | 0 | 0 | 1.755903 | 1.447098 |
| Rare_Disea | Homologou | 1/134 | 0.443521 | 0.522947 | 0 | 0 | 1.729323 | 1.405958 |
| Rare_Disea | Familial pr | 1/132 | 0.438615 | 0.522947 | 0 | 0 | 1.755903 | 1.447098 |
| Rare_Disea | German sy | 2/343 | 0.441287 | 0.522947 | 0 | 0 | 1.350492 | 1.104782 |
| Rare_Disea | Lymphatic | 1/134 | 0.443521 | 0.522947 | 0 | 0 | 1.729323 | 1.405958 |
| Rare_Disea | Acoustic ne | 1/129 | 0.431176 | 0.522947 | 0 | 0 | 1.797329 | 1.511983 |
| Rare_Disea | Acute lymp | 1/133 | 0.441073 | 0.522947 | 0 | 0 | 1.742512 | 1.426323 |
| Rare_Disea | Pemphigus | 1/134 | 0.443521 | 0.522947 | 0 | 0 | 1.729323 | 1.405958 |
| Rare_Disea | Aplastic an | 1/134 | 0.443521 | 0.522947 | 0 | 0 | 1.729323 | 1.405958 |
| Rare_Disea | Brown sync | 2/337 | 0.432387 | 0.522947 | 0 | 0 | 1.375101 | 1.152933 |
| Rare_Disea | MYH-assoc | 1/130 | 0.433666 | 0.522947 | 0 | 0 | 1.783306 | 1.489916 |
| Rare_Disea | Chromosor | 2/344 | 0.442764 | 0.522947 | 0 | 0 | 1.346474 | 1.096999 |
| Rare_Disea | Thyroid car | 2/338 | 0.433875 | 0.522947 | 0 | 0 | 1.370938 | 1.144732 |
| Rare_Disea | Ataxia-tela | 1/131 | 0.436146 | 0.522947 | 0 | 0 | 1.769499 | 1.468291 |
| Rare_Disea | Vulvar canc | 1/133 | 0.441073 | 0.522947 | 0 | 0 | 1.742512 | 1.426323 |
| Rare_Disea | Ependymol | 1/132 | 0.438615 | 0.522947 | 0 | 0 | 1.755903 | 1.447098 |
| Rare_Disea | Radiation i | 1/133 | 0.441073 | 0.522947 | 0 | 0 | 1.742512 | 1.426323 |
| Rare_Disea | Merkel cell | 1/129 | 0.431176 | 0.522947 | 0 | 0 | 1.797329 | 1.511983 |
| Rare_Disea | Metagonin | 1/132 | 0.438615 | 0.522947 | 0 | 0 | 1.755903 | 1.447098 |
| SysMyo_M | MGS1179 I | Jan-61 | 0.233813 | 0.523082 | 0 | 0 | 3.847481 | 5.591291 |
| SysMyo_M | MGS1372 I | 2/211 | 0.233816 | 0.523082 | 0 | 0 | 2.218294 | 3.223675 |
| SysMyo_M | MGS146 M | Jan-61 | 0.233813 | 0.523082 | 0 | 0 | 3.847481 | 5.591291 |
| SysMyo_M | MGS565 H | 2/210 | 0.23221 | 0.523082 | 0 | 0 | 2.229072 | 3.254694 |
| SysMyo_M | MGS428 H | 2/209 | 0.230606 | 0.523082 | 0 | 0 | 2.239955 | 3.286115 |
| SysMyo_M | MGS27 Mu | Jan-62 | 0.237156 | 0.523518 | 0 | 0 | 3.784217 | 5.445629 |
| SysMyo_M | MGS1119 I | 2/214 | 0.238636 | 0.523518 | 0 | 0 | 2.18657 | 3.132957 |
| SysMyo_M | MGS65 Mc | Jan-62 | 0.237156 | 0.523518 | 0 | 0 | 3.784217 | 5.445629 |
| SysMyo_M | MGS448 H | 2/214 | 0.238636 | 0.523518 | 0 | 0 | 2.18657 | 3.132957 |
| SysMyo_M | MGS538 H | 2/214 | 0.238636 | 0.523518 | 0 | 0 | 2.18657 | 3.132957 |
| DisGeNET | Medullobla | 3/590 | 0.475572 | 0.523969 | 0 | 0 | 1.175834 | 0.873923 |
| Rare_Disea | Antiphosph | 1/135 | 0.445958 | 0.524159 | 0 | 0 | 1.716331 | 1.38599 |
| Rare_Disea | Fallopian tu | 1/135 | 0.445958 | 0.524159 | 0 | 0 | 1.716331 | 1.38599 |
| Rare_Disea | Familial ad | 1/136 | 0.448384 | 0.524525 | 0 | 0 | 1.703531 | 1.36641 |

| | | | | | | | | |
|---|---|---|---|---|---|---|---|---|
| Rare_Disea | Tongue car | 1/136 | 0.448384 | 0.524525 | 0 | 0 | 1.703531 | 1.36641 |
| Rare_Disea | Laryngeal c | 1/136 | 0.448384 | 0.524525 | 0 | 0 | 1.703531 | 1.36641 |
| KEGG_202 | Vitamin dig | 24-Jan | 0.099394 | 0.524581 | 0 | 0 | 10.05561 | 23.215 |
| GeneSigDB | 17284527-T | Jan-84 | 0.307163 | 0.524815 | 0 | 0 | 2.778089 | 3.279195 |
| GeDiPNet_ | Glioblastor | 1/153 | 0.488063 | 0.524983 | 0 | 0 | 1.511704 | 1.084362 |
| DisGeNET | Chronic Pe | 1/148 | 0.476695 | 0.524986 | 0 | 0 | 1.563518 | 1.158376 |
| SysMyo_M | MGS122 Hu | Jan-63 | 0.240485 | 0.525538 | 0 | 0 | 3.722993 | 5.305634 |
| Rare_Disea | Liver cance | 2/349 | 0.450113 | 0.525698 | 0 | 0 | 1.326733 | 1.059074 |
| Rare_Disea | Uveal disea | 1/137 | 0.4508 | 0.525698 | 0 | 0 | 1.69092 | 1.347208 |
| Reactome_ | Opioid Sign | Jan-88 | 0.319191 | 0.525726 | 0 | 0 | 2.649826 | 3.026014 |
| Reactome_ | Mitochond | Jan-88 | 0.319191 | 0.525726 | 0 | 0 | 2.649826 | 3.026014 |
| GeDiPNet_ | Autoimmu | 1/154 | 0.490307 | 0.526425 | 0 | 0 | 1.501748 | 1.070331 |
| SysMyo_M | MGS1183 I | 2/216 | 0.241852 | 0.526493 | 0 | 0 | 2.165915 | 3.074364 |
| DisGeNET | Adrenoleul | 1/149 | 0.478989 | 0.52729 | 0 | 0 | 1.552876 | 1.143039 |
| PhenGenI_ | Precursor C | 1/100 | 0.35405 | 0.527648 | 0 | 0 | 2.327226 | 2.416399 |
| Reactome_ | Respiratory | Jan-90 | 0.325127 | 0.527742 | 0 | 0 | 2.590018 | 2.909988 |
| Reactome_ | Chaperonir | Jan-90 | 0.325127 | 0.527742 | 0 | 0 | 2.590018 | 2.909988 |
| DisGeNET | Psychomot | 1/150 | 0.481272 | 0.528029 | 0 | 0 | 1.542376 | 1.127974 |
| DisGeNET | Psychomot | 1/150 | 0.481272 | 0.528029 | 0 | 0 | 1.542376 | 1.127974 |
| DisGeNET | Mental det | 1/150 | 0.481272 | 0.528029 | 0 | 0 | 1.542376 | 1.127974 |
| DisGeNET | Loss of dev | 1/150 | 0.481272 | 0.528029 | 0 | 0 | 1.542376 | 1.127974 |
| DisGeNET | Cystic Fibro | 3/594 | 0.480046 | 0.528029 | 0 | 0 | 1.167634 | 0.856895 |
| DisGeNET | Monoclona | 1/150 | 0.481272 | 0.528029 | 0 | 0 | 1.542376 | 1.127974 |
| DisGeNET | Neurodeve | 1/150 | 0.481272 | 0.528029 | 0 | 0 | 1.542376 | 1.127974 |
| DisGeNET | Psychomot | 1/150 | 0.481272 | 0.528029 | 0 | 0 | 1.542376 | 1.127974 |
| GeneSigDB | 16536878-1 | 2/259 | 0.311104 | 0.528307 | 0 | 0 | 1.799588 | 2.10125 |
| GeneSigDB | 19061838-T | Jan-85 | 0.310189 | 0.528307 | 0 | 0 | 2.744878 | 3.21308 |
| PhenGenI_ | Cognition | 1/101 | 0.356874 | 0.528448 | 0 | 0 | 2.303837 | 2.373811 |
| dbGaP | Waist Circu | 1/146 | 0.472079 | 0.529054 | 0 | 0 | 1.585245 | 1.1899 |
| GeDiPNet_ | Dementia | 1/156 | 0.494766 | 0.529264 | 0 | 0 | 1.482221 | 1.042994 |
| GeDiPNet_ | Cirrhosis | 1/156 | 0.494766 | 0.529264 | 0 | 0 | 1.482221 | 1.042994 |
| WikiPathw | Male Infert | 1/145 | 0.469755 | 0.529626 | 0 | 0 | 1.596334 | 1.2061 |
| GO_Molecu | Phosphatic | 1/106 | 0.370812 | 0.529731 | 0 | 0 | 2.193577 | 2.176161 |
| DisGeNET | Primary Sj? | 1/151 | 0.483545 | 0.529857 | 0 | 0 | 1.532016 | 1.113178 |
| DisGeNET | Lung Disea | 1/151 | 0.483545 | 0.529857 | 0 | 0 | 1.532016 | 1.113178 |
| DisGeNET | Thickened | 1/151 | 0.483545 | 0.529857 | 0 | 0 | 1.532016 | 1.113178 |
| GO_Molecu | Protein Kin | 3/511 | 0.384285 | 0.530334 | 0 | 0 | 1.364244 | 1.304722 |
| GO_Molecu | Phosphatas | 1/112 | 0.387144 | 0.530334 | 0 | 0 | 2.074377 | 1.968499 |
| GO_Molecu | Sequence-S | 4/715 | 0.377917 | 0.530334 | 0 | 0 | 1.301544 | 1.266507 |
| Rare_Disea | Weber syn | 1/139 | 0.455601 | 0.530465 | 0 | 0 | 1.666245 | 1.309899 |
| Human_Ph | Constipatic | 1/170 | 0.524918 | 0.530473 | 0 | 0 | 1.35847 | 0.875552 |
| Rare_Disea | Gestationa | 1/140 | 0.457986 | 0.53075 | 0 | 0 | 1.654174 | 1.291774 |
| Rare_Disea | Lymphoma | 1/140 | 0.457986 | 0.53075 | 0 | 0 | 1.654174 | 1.291774 |
| Rare_Disea | Hyperadrei | 1/140 | 0.457986 | 0.53075 | 0 | 0 | 1.654174 | 1.291774 |
| Gene_Pert | Nfe2l2 KO | 6/439 | 0.012263 | 0.53081 | 0 | 0 | 3.332478 | 14.66685 |

| | | | | | | | | |
|---|---|---|---|---|---|---|---|---|
| Gene_Pert | Pten KD mo | 5/316 | 0.01229 | 0.53081 | 0 | 0 | 3.843228 | 16.90625 |
| DisGeNET | Pre-Eclamp | 1/152 | 0.485809 | 0.531449 | 0 | 0 | 1.521793 | 1.098642 |
| DisGeNET | Common V | 1/152 | 0.485809 | 0.531449 | 0 | 0 | 1.521793 | 1.098642 |
| DisGeNET | Decreased | 1/152 | 0.485809 | 0.531449 | 0 | 0 | 1.521793 | 1.098642 |
| DisGeNET | Coarse faci | 1/152 | 0.485809 | 0.531449 | 0 | 0 | 1.521793 | 1.098642 |
| SysMyo_M | MGS413 H | 2/218 | 0.245071 | 0.531456 | 0 | 0 | 2.145643 | 3.017223 |
| GWAS_Cat | Hypertensi | 1/167 | 0.51861 | 0.531684 | 0 | 0 | 1.383231 | 0.908233 |
| Disease_Pe | atheroscle | 4/259 | 0.026504 | 0.531739 | 0 | 0 | 3.71519 | 13.48783 |
| Rare_Disea | Kidney can | 1/141 | 0.46036 | 0.531844 | 0 | 0 | 1.642276 | 1.27399 |
| Rare_Disea | Hepadnavi | 1/141 | 0.46036 | 0.531844 | 0 | 0 | 1.642276 | 1.27399 |
| Rare_Disea | Kienbock's | 1/142 | 0.462724 | 0.532097 | 0 | 0 | 1.630546 | 1.256538 |
| Rare_Disea | Squamous | 1/142 | 0.462724 | 0.532097 | 0 | 0 | 1.630546 | 1.256538 |
| Rare_Disea | Biliary trac | 1/142 | 0.462724 | 0.532097 | 0 | 0 | 1.630546 | 1.256538 |
| GeneSigDB | 20460173- | Jan-87 | 0.316203 | 0.5321 | 0 | 0 | 2.680773 | 3.086563 |
| GeneSigDB | 15892885- | Jan-87 | 0.316203 | 0.5321 | 0 | 0 | 2.680773 | 3.086563 |
| GeneSigDB | 17760805- | Jan-87 | 0.316203 | 0.5321 | 0 | 0 | 2.680773 | 3.086563 |
| PhenGenI_ | Arterial Pre | 2/291 | 0.36192 | 0.532507 | 0 | 0 | 1.59772 | 1.623813 |
| DisGeNET | Squamous | 1/153 | 0.488063 | 0.533024 | 0 | 0 | 1.511704 | 1.084362 |
| DisGeNET | Sezary Syn | 1/153 | 0.488063 | 0.533024 | 0 | 0 | 1.511704 | 1.084362 |
| DisGeNET | Psychomot | 1/153 | 0.488063 | 0.533024 | 0 | 0 | 1.511704 | 1.084362 |
| DisGeNET | Cachexia | 1/153 | 0.488063 | 0.533024 | 0 | 0 | 1.511704 | 1.084362 |
| GTEx_Tissu | GTEX-XUJ4 | 6/534 | 0.028929 | 0.53306 | 0 | 0 | 2.719557 | 9.635186 |
| GTEx_Tissu | GTEX-QLQ7 | 8/835 | 0.028782 | 0.53306 | 0 | 0 | 2.337073 | 8.291937 |
| GTEx_Tissu | GTEX-U8XE | 8/834 | 0.028606 | 0.53306 | 0 | 0 | 2.340025 | 8.316812 |
| Rare_Disea | Osteonecro | 1/143 | 0.465078 | 0.533978 | 0 | 0 | 1.618981 | 1.239412 |
| GO_Molec | Protein Ph | 1/115 | 0.395152 | 0.533989 | 0 | 0 | 2.019482 | 1.875059 |
| DisGeNET | Premature | 1/154 | 0.490307 | 0.534361 | 0 | 0 | 1.501748 | 1.070331 |
| DisGeNET | Breast Can | 1/154 | 0.490307 | 0.534361 | 0 | 0 | 1.501748 | 1.070331 |
| DisGeNET | Hyperchole | 1/154 | 0.490307 | 0.534361 | 0 | 0 | 1.501748 | 1.070331 |
| DisGeNET | Icterus | 1/154 | 0.490307 | 0.534361 | 0 | 0 | 1.501748 | 1.070331 |
| DisGeNET | Developme | 1/154 | 0.490307 | 0.534361 | 0 | 0 | 1.501748 | 1.070331 |
| Gene_Pert | Psap hypor | 5/292 | 0.00895 | 0.534927 | 0 | 0 | 4.169712 | 19.66466 |
| Gene_Pert | Smarca4 K | 4/185 | 0.00869 | 0.534927 | 0 | 0 | 5.253811 | 24.93226 |
| Gene_Pert | MIR124-1 ( | 4/185 | 0.00869 | 0.534927 | 0 | 0 | 5.253811 | 24.93226 |
| Gene_Pert | Dbh KO mo | 5/291 | 0.008827 | 0.534927 | 0 | 0 | 4.184505 | 19.79266 |
| Gene_Pert | Nfe2l2 KO | 4/177 | 0.007463 | 0.534927 | 0 | 0 | 5.49899 | 26.93279 |
| Gene_Pert | Emd KO mo | 5/289 | 0.008583 | 0.534927 | 0 | 0 | 4.214402 | 20.05212 |
| RNA-Seq_D | TGFB3 Pala | 7/498 | 0.005994 | 0.534997 | 0 | 0 | 3.461151 | 17.71064 |
| RNA-Seq_D | Mrpp3 Hea | 7/498 | 0.005994 | 0.534997 | 0 | 0 | 3.461151 | 17.71064 |
| RNA-Seq_D | Hif1a Hear | 7/498 | 0.005994 | 0.534997 | 0 | 0 | 3.461151 | 17.71064 |
| RNA-Seq_D | Dexametha | 7/497 | 0.00593 | 0.534997 | 0 | 0 | 3.468393 | 17.78478 |
| RNA-Seq_D | Parkin Hea | 7/499 | 0.006058 | 0.534997 | 0 | 0 | 3.453938 | 17.63691 |
| RNA-Seq_D | TNFR2 Colo | 7/497 | 0.00593 | 0.534997 | 0 | 0 | 3.468393 | 17.78478 |
| RNA-Seq_D | Barx2 Skele | 7/499 | 0.006058 | 0.534997 | 0 | 0 | 3.453938 | 17.63691 |
| RNA-Seq_D | HnRNP U H | 7/498 | 0.005994 | 0.534997 | 0 | 0 | 3.461151 | 17.71064 |

| | | | | | | | | |
|---|---|---|---|---|---|---|---|---|
| Rare_Disea | Hypophary | 1/144 | 0.467422 | 0.535143 | 0 | 0 | 1.607578 | 1.222601 |
| Rare_Disea | Renal carci | 2/361 | 0.46753 | 0.535143 | 0 | 0 | 1.281599 | 0.974388 |
| GeneSigDB | 18362358-1 | 4/657 | 0.320336 | 0.535817 | 0 | 0 | 1.421428 | 1.618134 |
| GeneSigDB | 19096012-1 | 3/457 | 0.31999 | 0.535817 | 0 | 0 | 1.530758 | 1.744247 |
| DisGeNET | Aplastic An | 1/155 | 0.492541 | 0.535905 | 0 | 0 | 1.491921 | 1.056544 |
| DisGeNET | Substance | 1/155 | 0.492541 | 0.535905 | 0 | 0 | 1.491921 | 1.056544 |
| DisGeNET | Focal glom | 1/155 | 0.492541 | 0.535905 | 0 | 0 | 1.491921 | 1.056544 |
| DisGeNET | Platelet he | 1/155 | 0.492541 | 0.535905 | 0 | 0 | 1.491921 | 1.056544 |
| SysMyo_M | MGS1113 I | 2/220 | 0.248291 | 0.536384 | 0 | 0 | 2.125742 | 2.961486 |
| GO_Biologi | Positive Re | 2/387 | 0.504142 | 0.536643 | 0 | 0 | 1.193461 | 0.817399 |
| DisGeNET | Ovarian Ca | Oct-03 | 0.493474 | 0.536696 | 0 | 0 | 1.049384 | 0.741164 |
| GTEx_Tissu | GTEX-OOB. | 8/838 | 0.029316 | 0.536788 | 0 | 0 | 2.32826 | 8.21784 |
| SysMyo_M | MGS1096 I | 2/221 | 0.249902 | 0.537811 | 0 | 0 | 2.115928 | 2.934131 |
| DisGeNET | Myoclonic | 1/156 | 0.494766 | 0.537878 | 0 | 0 | 1.482221 | 1.042994 |
| PhenGenI_ | Plaque, Atl | 1/105 | 0.368048 | 0.538096 | 0 | 0 | 2.214781 | 2.213764 |
| GTEx_Tissu | GTEX-OHPI | 11/1334 | 0.02963 | 0.539112 | 0 | 0 | 2.033755 | 7.156739 |
| SysMyo_M | MGS1497 I | 2/222 | 0.251513 | 0.539229 | 0 | 0 | 2.106203 | 2.90711 |
| Rare_Disea | Dyskinesia | 1/146 | 0.472079 | 0.539518 | 0 | 0 | 1.585245 | 1.1899 |
| GeneSigDB | 16790086-1 | 2/268 | 0.325502 | 0.539596 | 0 | 0 | 1.737904 | 1.950602 |
| GeneSigDB | 15897907-1 | 2/267 | 0.323905 | 0.539596 | 0 | 0 | 1.74455 | 1.966639 |
| GeneSigDB | 15843827-1 | Jan-90 | 0.325127 | 0.539596 | 0 | 0 | 2.590018 | 2.909988 |
| DisGeNET | Hyperkyph | 1/157 | 0.496981 | 0.540062 | 0 | 0 | 1.472645 | 1.029678 |
| CellMarker | Matrix Fibr | 1/156 | 0.494766 | 0.540103 | 0 | 0 | 1.482221 | 1.042994 |
| GO_Molecu | Receptor Li | 2/319 | 0.405256 | 0.540342 | 0 | 0 | 1.454518 | 1.313773 |
| GeneSigDB | 15459216-1 | Jan-91 | 0.328076 | 0.540646 | 0 | 0 | 2.561111 | 2.854384 |
| GeneSigDB | 12086872-1 | Jan-91 | 0.328076 | 0.540646 | 0 | 0 | 2.561111 | 2.854384 |
| WikiPathw | Nonalcoho | 1/154 | 0.490307 | 0.541219 | 0 | 0 | 1.501748 | 1.070331 |
| WikiPathw | Smith Mag | 1/155 | 0.492541 | 0.541219 | 0 | 0 | 1.491921 | 1.056544 |
| WikiPathw | Sudden Inf | 1/157 | 0.496981 | 0.541219 | 0 | 0 | 1.472645 | 1.029678 |
| WikiPathw | Insulin Sigr | 1/160 | 0.503569 | 0.541219 | 0 | 0 | 1.444639 | 0.991073 |
| WikiPathw | Epithelial T | 1/160 | 0.503569 | 0.541219 | 0 | 0 | 1.444639 | 0.991073 |
| Human_Ph | Ventricular | 1/176 | 0.537289 | 0.541542 | 0 | 0 | 1.311495 | 0.814726 |
| SysMyo_M | MGS58 Hu | Jan-67 | 0.253656 | 0.541772 | 0 | 0 | 3.496653 | 4.79662 |
| CellMarker | Leydig Cell | 2/383 | 0.498613 | 0.542143 | 0 | 0 | 1.206237 | 0.839451 |
| DisGeNET | Blindness, I | 1/158 | 0.499187 | 0.542234 | 0 | 0 | 1.463191 | 1.016589 |
| GeneSigDB | 15892885-1 | Jan-92 | 0.331012 | 0.542276 | 0 | 0 | 2.532839 | 2.800307 |
| GeneSigDB | 15572690-1 | Jan-92 | 0.331012 | 0.542276 | 0 | 0 | 2.532839 | 2.800307 |
| DisGeNET | Endometri | 4/840 | 0.499444 | 0.542289 | 0 | 0 | 1.099729 | 0.763498 |
| GeDiPNet_ | Lymphoma | 1/162 | 0.507913 | 0.542332 | 0 | 0 | 1.426549 | 0.966408 |
| Reactome_ | Downstrea | Jan-94 | 0.336847 | 0.542832 | 0 | 0 | 2.47812 | 2.69651 |
| GeneSigDB | 15744745-1 | Jan-93 | 0.333936 | 0.543252 | 0 | 0 | 2.505182 | 2.747701 |
| GeneSigDB | 16166618-1 | 9/1725 | 0.335121 | 0.543252 | 0 | 0 | 1.223575 | 1.337688 |
| GeneSigDB | 20215539-1 | Jan-93 | 0.333936 | 0.543252 | 0 | 0 | 2.505182 | 2.747701 |
| GeneSigDB | 20077526-1 | 3/470 | 0.335509 | 0.543252 | 0 | 0 | 1.487152 | 1.624127 |
| GeDiPNet_ | Cryptorchi | 3/621 | 0.509778 | 0.543329 | 0 | 0 | 1.11506 | 0.751305 |

| | | | | | | | | |
|---|---|---|---|---|---|---|---|---|
| Reactome_ | P75 NTR Re | Jan-95 | 0.339745 | 0.543592 | 0 | 0 | 2.451633 | 2.646685 |
| Reactome_ | Cellular Re: | Jan-99 | 0.351214 | 0.543896 | 0 | 0 | 2.351092 | 2.46009 |
| Reactome_ | Protein Fol | Jan-96 | 0.342631 | 0.543896 | 0 | 0 | 2.425704 | 2.598176 |
| Reactome_ | Signaling B | 4/689 | 0.352075 | 0.543896 | 0 | 0 | 1.352775 | 1.412175 |
| Reactome_ | Formation | Jan-98 | 0.348365 | 0.543896 | 0 | 0 | 2.37545 | 2.504922 |
| Reactome_ | Visual Phot | Jan-98 | 0.348365 | 0.543896 | 0 | 0 | 2.37545 | 2.504922 |
| DisGeNET | Sporadic Bı | 1/159 | 0.501382 | 0.544168 | 0 | 0 | 1.453856 | 1.003722 |
| Disease_Si | Septic Shoc | 3/300 | 0.142458 | 0.544394 | 0 | 0 | 2.358826 | 4.596658 |
| Disease_Si | Polycystic ( | 3/300 | 0.142458 | 0.544394 | 0 | 0 | 2.358826 | 4.596658 |
| Disease_Si | Schizophre | 3/300 | 0.142458 | 0.544394 | 0 | 0 | 2.358826 | 4.596658 |
| Disease_Si | Appendicit | 3/300 | 0.142458 | 0.544394 | 0 | 0 | 2.358826 | 4.596658 |
| Disease_Si | Alzheimer's | 3/300 | 0.142458 | 0.544394 | 0 | 0 | 2.358826 | 4.596658 |
| Disease_Si | Anaplasmo | 3/300 | 0.142458 | 0.544394 | 0 | 0 | 2.358826 | 4.596658 |
| Disease_Si | Epilepsy GS | 3/300 | 0.142458 | 0.544394 | 0 | 0 | 2.358826 | 4.596658 |
| Disease_Si | Huntingtor | 3/300 | 0.142458 | 0.544394 | 0 | 0 | 2.358826 | 4.596658 |
| Disease_Si | Malignant | 3/300 | 0.142458 | 0.544394 | 0 | 0 | 2.358826 | 4.596658 |
| Disease_Si | Lung trans | 3/300 | 0.142458 | 0.544394 | 0 | 0 | 2.358826 | 4.596658 |
| Disease_Si | Bipolar Dis | 3/300 | 0.142458 | 0.544394 | 0 | 0 | 2.358826 | 4.596658 |
| Disease_Si | Acute Lung | 3/300 | 0.142458 | 0.544394 | 0 | 0 | 2.358826 | 4.596658 |
| Disease_Si | Actinic ker | 3/300 | 0.142458 | 0.544394 | 0 | 0 | 2.358826 | 4.596658 |
| Disease_Si | Infantile ne | 3/300 | 0.142458 | 0.544394 | 0 | 0 | 2.358826 | 4.596658 |
| Disease_Si | Obesity GS | 3/300 | 0.142458 | 0.544394 | 0 | 0 | 2.358826 | 4.596658 |
| Disease_Si | Obesity GS | 3/300 | 0.142458 | 0.544394 | 0 | 0 | 2.358826 | 4.596658 |
| Disease_Si | Overexerti | 3/300 | 0.142458 | 0.544394 | 0 | 0 | 2.358826 | 4.596658 |
| Disease_Si | Leukemia, | 3/300 | 0.142458 | 0.544394 | 0 | 0 | 2.358826 | 4.596658 |
| Disease_Si | NASH GSE2 | 3/300 | 0.142458 | 0.544394 | 0 | 0 | 2.358826 | 4.596658 |
| SysMyo_M | MGS1031 I | Jan-68 | 0.256914 | 0.544619 | 0 | 0 | 3.44429 | 4.68084 |
| SysMyo_M | MGS1502 I | Jan-68 | 0.256914 | 0.544619 | 0 | 0 | 3.44429 | 4.68084 |
| Jensen_TIS | Fibroblast | 1/141 | 0.46036 | 0.544985 | 0 | 0 | 1.642276 | 1.27399 |
| DisGeNET | Hepatitis C | 2/386 | 0.502763 | 0.545441 | 0 | 0 | 1.19663 | 0.822846 |
| SysMyo_M | MGS159 Hı | 3/407 | 0.260688 | 0.545503 | 0 | 0 | 1.724629 | 2.318644 |
| SysMyo_M | MGS1170 I | 2/228 | 0.261186 | 0.545503 | 0 | 0 | 2.049662 | 2.751716 |
| SysMyo_M | MGS182 Hı | Jan-69 | 0.260157 | 0.545503 | 0 | 0 | 3.393468 | 4.569198 |
| SysMyo_M | MGS756 M | 2/228 | 0.261186 | 0.545503 | 0 | 0 | 2.049662 | 2.751716 |
| Gene_Pert | CTNNB1 VF | 4/209 | 0.013141 | 0.5459 | 0 | 0 | 4.633088 | 20.0706 |
| Gene_Pert | Tfam KO m | 5/322 | 0.013241 | 0.5459 | 0 | 0 | 3.769331 | 16.30017 |
| DisGeNET | Mammary | 1/160 | 0.503569 | 0.546089 | 0 | 0 | 1.444639 | 0.991073 |
| GeDiPNet_ | Nephrotic ! | 1/165 | 0.514359 | 0.546214 | 0 | 0 | 1.400241 | 0.930927 |
| GeDiPNet_ | Hypotonia | 1/165 | 0.514359 | 0.546214 | 0 | 0 | 1.400241 | 0.930927 |
| PhenGenI_ | HIV-1 | 2/300 | 0.375985 | 0.546242 | 0 | 0 | 1.548756 | 1.515003 |
| PhenGenI_ | Glomerulaı | 3/510 | 0.383102 | 0.546275 | 0 | 0 | 1.367005 | 1.311577 |
| PhenGenI_ | Attention [ | 3/510 | 0.383102 | 0.546275 | 0 | 0 | 1.367005 | 1.311577 |
| PhenGenI_ | Anatomy | 1/109 | 0.379031 | 0.546275 | 0 | 0 | 2.132321 | 2.068645 |
| GTEx_Tissu | GTEX-WHV | 7/688 | 0.030229 | 0.546571 | 0 | 0 | 2.471072 | 8.646196 |
| Rare_Disea | Bacterial m | 1/149 | 0.478989 | 0.546576 | 0 | 0 | 1.552876 | 1.143039 |

| | | | | | | | | |
|---|---|---|---|---|---|---|---|---|
| GeneSigDB | 12829800- | 3/473 | 0.33909 | 0.54693 | 0 | 0 | 1.477432 | 1.597826 |
| GeneSigDB | 18698033- | Jan-95 | 0.339745 | 0.54693 | 0 | 0 | 2.451633 | 2.646685 |
| GWAS_Cat | Highest Ma | 3/644 | 0.534413 | 0.547425 | 0 | 0 | 1.073769 | 0.67281 |
| dbGaP | Hemoglobi | 1/157 | 0.496981 | 0.547522 | 0 | 0 | 1.472645 | 1.029678 |
| DisGeNET | Weight dec | 1/161 | 0.505746 | 0.547543 | 0 | 0 | 1.435538 | 0.978637 |
| DisGeNET | Polycystic I | 1/161 | 0.505746 | 0.547543 | 0 | 0 | 1.435538 | 0.978637 |
| DisGeNET | Familial mu | 1/161 | 0.505746 | 0.547543 | 0 | 0 | 1.435538 | 0.978637 |
| DisGeNET | Eosinophili | 1/161 | 0.505746 | 0.547543 | 0 | 0 | 1.435538 | 0.978637 |
| WikiPathw | Chemokine | 1/165 | 0.514359 | 0.547697 | 0 | 0 | 1.400241 | 0.930927 |
| Rare_Disea | Inflammato | 1/150 | 0.481272 | 0.54834 | 0 | 0 | 1.542376 | 1.127974 |
| GeneSigDB | 19171046- | Jan-96 | 0.342631 | 0.548406 | 0 | 0 | 2.425704 | 2.598176 |
| GeneSigDB | 18497862- | Jan-96 | 0.342631 | 0.548406 | 0 | 0 | 2.425704 | 2.598176 |
| PhenGenI_ | Cleft Lip | 1/112 | 0.387144 | 0.548651 | 0 | 0 | 2.074377 | 1.968499 |
| Rare_Disea | Small intes | 2/372 | 0.483213 | 0.54971 | 0 | 0 | 1.242798 | 0.903885 |
| SysMyo_M | MGS1266 I | 2/230 | 0.264412 | 0.55021 | 0 | 0 | 2.031476 | 2.702365 |
| Rare_Disea | Primary eff | 1/152 | 0.485809 | 0.550979 | 0 | 0 | 1.521793 | 1.098642 |
| Rare_Disea | Hemeralop | 1/152 | 0.485809 | 0.550979 | 0 | 0 | 1.521793 | 1.098642 |
| GO_Biologi | Membrane | 1/167 | 0.51861 | 0.551217 | 0 | 0 | 1.383231 | 0.908233 |
| Jensen_TIS | Synovium | 1/144 | 0.467422 | 0.551318 | 0 | 0 | 1.607578 | 1.222601 |
| SysMyo_M | MGS1250 I | 2/231 | 0.266025 | 0.551539 | 0 | 0 | 2.022502 | 2.678127 |
| DisGeNET | androgen i | 1/163 | 0.510071 | 0.55177 | 0 | 0 | 1.417672 | 0.954384 |
| DisGeNET | Metastatic | 1/163 | 0.510071 | 0.55177 | 0 | 0 | 1.417672 | 0.954384 |
| Reactome_ | Potassium | 1/102 | 0.359686 | 0.551846 | 0 | 0 | 2.280912 | 2.332289 |
| Rare_Disea | Microceph | 1/153 | 0.488063 | 0.551852 | 0 | 0 | 1.511704 | 1.084362 |
| Rare_Disea | Microence | 1/153 | 0.488063 | 0.551852 | 0 | 0 | 1.511704 | 1.084362 |
| KOMP2_M | Decreased | Jan-57 | 0.220295 | 0.551868 | 0 | 0 | 4.123131 | 6.237423 |
| KOMP2_M | Decreased | 2/219 | 0.246681 | 0.551868 | 0 | 0 | 2.135647 | 2.989182 |
| KOMP2_M | Abnormal I | Jan-65 | 0.247099 | 0.551868 | 0 | 0 | 3.606286 | 5.041467 |
| KOMP2_M | Impaired P | Jan-55 | 0.213448 | 0.551868 | 0 | 0 | 4.27627 | 6.604112 |
| KOMP2_M | Decreased | Jan-58 | 0.223696 | 0.551868 | 0 | 0 | 4.050592 | 6.065622 |
| KOMP2_M | Abnormal I | Jan-62 | 0.237156 | 0.551868 | 0 | 0 | 3.784217 | 5.445629 |
| KOMP2_M | Hemorrhag | Jan-60 | 0.230455 | 0.551868 | 0 | 0 | 3.912889 | 5.742942 |
| KOMP2_M | Abnormal ( | Jan-66 | 0.250385 | 0.551868 | 0 | 0 | 3.550626 | 4.916752 |
| KOMP2_M | Abnormal I | Jan-57 | 0.220295 | 0.551868 | 0 | 0 | 4.123131 | 6.237423 |
| KOMP2_M | Abnormal I | 2/214 | 0.238636 | 0.551868 | 0 | 0 | 2.18657 | 3.132957 |
| DisGeNET | Multiple M | 6/1312 | 0.510489 | 0.551994 | 0 | 0 | 1.055357 | 0.709607 |
| CellMarker | Neural Pro | 1/163 | 0.510071 | 0.552409 | 0 | 0 | 1.417672 | 0.954384 |
| DisGeNET | Asthma | 6/1313 | 0.511246 | 0.552584 | 0 | 0 | 1.054493 | 0.707464 |
| GTEx_Tissu | GTEX-XUJ4 | 8/846 | 0.030775 | 0.552995 | 0 | 0 | 2.305066 | 8.024054 |
| DisGeNET | Blindness | 1/164 | 0.51222 | 0.553409 | 0 | 0 | 1.408903 | 0.942558 |
| GeneSigDB | 17183660- | Jan-99 | 0.351214 | 0.554181 | 0 | 0 | 2.351092 | 2.46009 |
| GeneSigDB | 19061838- | 3/483 | 0.351019 | 0.554181 | 0 | 0 | 1.445908 | 1.513741 |
| GeneSigDB | 20081812- | Jan-99 | 0.351214 | 0.554181 | 0 | 0 | 2.351092 | 2.46009 |
| GeneSigDB | 20015407- | Jan-98 | 0.348365 | 0.554181 | 0 | 0 | 2.37545 | 2.504922 |
| GeneSigDB | 18425577- | 3/481 | 0.348635 | 0.554181 | 0 | 0 | 1.452107 | 1.530129 |

| | | | | | | | | |
|---|---|---|---|---|---|---|---|---|
| GeDiPNet_ | Lymphobla | 1/169 | 0.522824 | 0.554194 | 0 | 0 | 1.366625 | 0.886269 |
| GTEx_Tissu | GTEX-RWS. | 7/692 | 0.031056 | 0.554603 | 0 | 0 | 2.456131 | 8.527581 |
| DisGeNET | Ischemic st | 2/394 | 0.513724 | 0.554805 | 0 | 0 | 1.171729 | 0.780453 |
| GO_Biologi | Brain Deve | 1/169 | 0.522824 | 0.554866 | 0 | 0 | 1.366625 | 0.886269 |
| Diabetes_F | GSE14946‹ | 3/224 | 0.07421 | 0.555032 | 0 | 0 | 3.182288 | 8.276655 |
| Diabetes_F | GSE12029‹ | 3/223 | 0.073438 | 0.555032 | 0 | 0 | 3.196916 | 8.348142 |
| Diabetes_F | GSE37248 | 3/221 | 0.071905 | 0.555032 | 0 | 0 | 3.226573 | 8.493677 |
| DisGeNET | TARSAL-CA | 1/165 | 0.514359 | 0.555033 | 0 | 0 | 1.400241 | 0.930927 |
| DisGeNET | Bone Disea | 1/165 | 0.514359 | 0.555033 | 0 | 0 | 1.400241 | 0.930927 |
| Rare_Disea | Pituitary ca | 1/157 | 0.496981 | 0.555186 | 0 | 0 | 1.472645 | 1.029678 |
| Rare_Disea | Wisconsin : | 1/157 | 0.496981 | 0.555186 | 0 | 0 | 1.472645 | 1.029678 |
| Rare_Disea | Mycosis fu | 1/156 | 0.494766 | 0.555186 | 0 | 0 | 1.482221 | 1.042994 |
| Rare_Disea | Pancreatic | 1/157 | 0.496981 | 0.555186 | 0 | 0 | 1.472645 | 1.029678 |
| Rare_Disea | Lymphocyt | 1/157 | 0.496981 | 0.555186 | 0 | 0 | 1.472645 | 1.029678 |
| Rare_Disea | Dystonia 1 | 1/155 | 0.492541 | 0.555186 | 0 | 0 | 1.491921 | 1.056544 |
| Rare_Disea | Turner syn | 1/157 | 0.496981 | 0.555186 | 0 | 0 | 1.472645 | 1.029678 |
| Rare_Disea | Good synd | 1/156 | 0.494766 | 0.555186 | 0 | 0 | 1.482221 | 1.042994 |
| SysMyo_M | MGS525 H | Jan-72 | 0.269804 | 0.555306 | 0 | 0 | 3.249591 | 4.257158 |
| SysMyo_M | MGS847 M | 2/233 | 0.269251 | 0.555306 | 0 | 0 | 2.004787 | 2.630503 |
| GeDiPNet_ | Clinodactyl | 1/170 | 0.524918 | 0.555403 | 0 | 0 | 1.35847 | 0.875552 |
| SysMyo_M | MGS1416 I | 2/234 | 0.270864 | 0.555468 | 0 | 0 | 1.996045 | 2.607109 |
| SysMyo_M | MGS814 M | Jan-73 | 0.272992 | 0.555803 | 0 | 0 | 3.204296 | 4.160181 |
| SysMyo_M | MGS1017 I | Jan-73 | 0.272992 | 0.555803 | 0 | 0 | 3.204296 | 4.160181 |
| DisGeNET | Keratoconu | 1/166 | 0.516489 | 0.555958 | 0 | 0 | 1.391684 | 0.919487 |
| DisGeNET | VITILIGO-A | 1/166 | 0.516489 | 0.555958 | 0 | 0 | 1.391684 | 0.919487 |
| DisGeNET | invasive ca | 1/166 | 0.516489 | 0.555958 | 0 | 0 | 1.391684 | 0.919487 |
| DisGeNET | Huntingtor | 3/627 | 0.516269 | 0.555958 | 0 | 0 | 1.103995 | 0.729882 |
| DisGeNET | Ataxia Tela | 2/396 | 0.516439 | 0.555958 | 0 | 0 | 1.165661 | 0.770266 |
| DisGeNET | Contractur | 1/166 | 0.516489 | 0.555958 | 0 | 0 | 1.391684 | 0.919487 |
| Rare_Disea | Primary scl | 1/158 | 0.499187 | 0.55598 | 0 | 0 | 1.463191 | 1.016589 |
| Rare_Disea | Osteosarco | 1/158 | 0.499187 | 0.55598 | 0 | 0 | 1.463191 | 1.016589 |
| Jensen_TIS | Monocyte | 34/7640 | 0.473129 | 0.556013 | 0 | 0 | 1.038004 | 0.776828 |
| PhenGenI_ | Cleft Palate | 1/115 | 0.395152 | 0.556586 | 0 | 0 | 2.019482 | 1.875059 |
| DisGeNET | Transitiona | 1/167 | 0.51861 | 0.55664 | 0 | 0 | 1.383231 | 0.908233 |
| DisGeNET | Follicular tl | 1/167 | 0.51861 | 0.55664 | 0 | 0 | 1.383231 | 0.908233 |
| DisGeNET | Retinal pigı | 1/167 | 0.51861 | 0.55664 | 0 | 0 | 1.383231 | 0.908233 |
| DisGeNET | Ataxias, He | 1/167 | 0.51861 | 0.55664 | 0 | 0 | 1.383231 | 0.908233 |
| DisGeNET | Progressive | 1/167 | 0.51861 | 0.55664 | 0 | 0 | 1.383231 | 0.908233 |
| DisGeNET | Irritable Bo | 1/167 | 0.51861 | 0.55664 | 0 | 0 | 1.383231 | 0.908233 |
| DisGeNET | Abnormalit | 1/167 | 0.51861 | 0.55664 | 0 | 0 | 1.383231 | 0.908233 |
| PhenGenI_ | Estradiol | 1/116 | 0.397798 | 0.556917 | 0 | 0 | 2.00182 | 1.845299 |
| Reactome_ | Integration | 1/105 | 0.368048 | 0.557046 | 0 | 0 | 2.214781 | 2.213764 |
| Reactome_ | PI3K/AKT S | 1/105 | 0.368048 | 0.557046 | 0 | 0 | 2.214781 | 2.213764 |
| Jensen_DIS | Arthritis | 1/186 | 0.557204 | 0.557204 | 0 | 0 | 1.239975 | 0.725168 |
| PhenGenI_ | Body Weig | 1/117 | 0.400433 | 0.557229 | 0 | 0 | 1.984463 | 1.816198 |

| | | | | | | | | |
|---|---|---|---|---|---|---|---|---|
| Reactome_ | HATs Acety | 1/106 | 0.370812 | 0.557462 | 0 | 0 | 2.193577 | 2.176161 |
| PhenGenI_ | Alopecia | 1/118 | 0.403056 | 0.557521 | 0 | 0 | 1.967402 | 1.787737 |
| Rare_Disea | Duchenne | 1/159 | 0.501382 | 0.557591 | 0 | 0 | 1.453856 | 1.003722 |
| GWAS_Cat | Balding Typ | 1/180 | 0.545359 | 0.557701 | 0 | 0 | 1.281928 | 0.777247 |
| GWAS_Cat | Myocardial | 1/180 | 0.545359 | 0.557701 | 0 | 0 | 1.281928 | 0.777247 |
| Reactome_ | O-linked Gl | 1/107 | 0.373563 | 0.557855 | 0 | 0 | 2.172773 | 2.139459 |
| Disease_Pe | Duchenne | 4/271 | 0.030593 | 0.557913 | 0 | 0 | 3.546049 | 12.365 |
| Disease_Pe | Duchenne | 4/268 | 0.029538 | 0.557913 | 0 | 0 | 3.586893 | 12.63333 |
| Disease_Pe | esophagus | 4/278 | 0.033142 | 0.557913 | 0 | 0 | 3.454226 | 11.76843 |
| Disease_Pe | Atheroscle | 4/274 | 0.031671 | 0.557913 | 0 | 0 | 3.506113 | 12.1044 |
| Disease_Pe | polycystic c | 4/275 | 0.032035 | 0.557913 | 0 | 0 | 3.492998 | 12.01919 |
| Disease_Pe | Teratosper | 4/276 | 0.032401 | 0.557913 | 0 | 0 | 3.479979 | 11.93481 |
| Disease_Pe | Ventricular | 4/280 | 0.033892 | 0.557913 | 0 | 0 | 3.428846 | 11.6052 |
| DisGeNET | Conventior | 3/631 | 0.520571 | 0.557992 | 0 | 0 | 1.096736 | 0.715981 |
| DisGeNET | Carcinoma, | 2/399 | 0.520494 | 0.557992 | 0 | 0 | 1.156675 | 0.755281 |
| DisGeNET | Childhood | 1/168 | 0.520722 | 0.557992 | 0 | 0 | 1.374878 | 0.897162 |
| DisGeNET | Kyphosis de | 1/168 | 0.520722 | 0.557992 | 0 | 0 | 1.374878 | 0.897162 |
| Reactome_ | L13a-media | 1/108 | 0.376303 | 0.558224 | 0 | 0 | 2.152358 | 2.10363 |
| GO_Biologi | Metal Ion T | 1/171 | 0.527002 | 0.558465 | 0 | 0 | 1.35041 | 0.865006 |
| Reactome_ | GTP Hydrol | 1/109 | 0.379031 | 0.558572 | 0 | 0 | 2.132321 | 2.068645 |
| DisGeNET | Triple Nega | 3/632 | 0.521643 | 0.558751 | 0 | 0 | 1.094935 | 0.712553 |
| GeDiPNet_ | Respiratory | 1/172 | 0.529077 | 0.55879 | 0 | 0 | 1.342445 | 0.854628 |
| DisGeNET | Generalize | 3/633 | 0.522714 | 0.55933 | 0 | 0 | 1.093141 | 0.709142 |
| DisGeNET | Tuberculos | 1/169 | 0.522824 | 0.55933 | 0 | 0 | 1.366625 | 0.886269 |
| DisGeNET | Progressive | 1/169 | 0.522824 | 0.55933 | 0 | 0 | 1.366625 | 0.886269 |
| GTEx_Tissu | GTEX-S32V | 8/850 | 0.031523 | 0.559483 | 0 | 0 | 2.293635 | 7.929192 |
| GeneSigDB | 12782714- | 1/101 | 0.356874 | 0.55994 | 0 | 0 | 2.303837 | 2.373811 |
| GeneSigDB | 16140871- | 1/101 | 0.356874 | 0.55994 | 0 | 0 | 2.303837 | 2.373811 |
| ARCHS4_Ti | SKELETAL M | 17/2316 | 0.020747 | 0.56016 | 0 | 0 | 1.860672 | 7.210793 |
| DisGeNET | Complete a | 1/170 | 0.524918 | 0.561112 | 0 | 0 | 1.35847 | 0.875552 |
| DisGeNET | Dental cari | 1/170 | 0.524918 | 0.561112 | 0 | 0 | 1.35847 | 0.875552 |
| GO_Biologi | Neuron Dif | 1/173 | 0.531144 | 0.562015 | 0 | 0 | 1.334573 | 0.844414 |
| DisGeNET | Keloid | 1/171 | 0.527002 | 0.56288 | 0 | 0 | 1.35041 | 0.865006 |
| DisGeNET | Esophagea | 3/637 | 0.526986 | 0.56288 | 0 | 0 | 1.086018 | 0.695683 |
| Rare_Disea | Becker mus | 1/162 | 0.507913 | 0.56317 | 0 | 0 | 1.426549 | 0.966408 |
| Rare_Disea | Cluttering | 1/162 | 0.507913 | 0.56317 | 0 | 0 | 1.426549 | 0.966408 |
| GO_Biologi | Positive Re | 1/174 | 0.533201 | 0.563352 | 0 | 0 | 1.326791 | 0.834362 |
| GeneSigDB | 19123479- | 1/103 | 0.362485 | 0.563984 | 0 | 0 | 2.258436 | 2.291796 |
| GeneSigDB | 17597811- | 4/701 | 0.364004 | 0.563984 | 0 | 0 | 1.328655 | 1.342724 |
| GeneSigDB | 10445855- | 1/103 | 0.362485 | 0.563984 | 0 | 0 | 2.258436 | 2.291796 |
| GeneSigDB | 20032505- | 1/104 | 0.365273 | 0.563984 | 0 | 0 | 2.236396 | 2.252299 |
| GeneSigDB | 20032505- | 1/104 | 0.365273 | 0.563984 | 0 | 0 | 2.236396 | 2.252299 |
| GeneSigDB | 18786252- | 6/1126 | 0.365527 | 0.563984 | 0 | 0 | 1.242923 | 1.250898 |
| Human_Ph | Downslant | 1/188 | 0.561084 | 0.564037 | 0 | 0 | 1.226589 | 0.708828 |
| Disease_Pe | actinic kera | 5/430 | 0.039552 | 0.564277 | 0 | 0 | 2.795983 | 9.031405 |

| | | | | | | | | |
|---|---|---|---|---|---|---|---|---|
| Disease_Pe | hepatitis C | 4/294 | 0.039422 | 0.564277 | 0 | 0 | 3.260989 | 10.54418 |
| Disease_Pe | Polycystic ( | 4/294 | 0.039422 | 0.564277 | 0 | 0 | 3.260989 | 10.54418 |
| Disease_Pe | Actinic kera | 5/429 | 0.039222 | 0.564277 | 0 | 0 | 2.802721 | 9.076663 |
| Disease_Pe | acute myoc | 4/292 | 0.038602 | 0.564277 | 0 | 0 | 3.283969 | 10.6875 |
| Disease_Pe | West Nile f | Feb-68 | 0.035404 | 0.564277 | 0 | 0 | 7.075579 | 23.63906 |
| GTEx_Tissu | GTEX-WHP | 6/547 | 0.032 | 0.564494 | 0 | 0 | 2.652427 | 9.129675 |
| Disease_Sig | Urothelial ( | 3/300 | 0.142458 | 0.564557 | 0 | 0 | 2.358826 | 4.596658 |
| Disease_Sig | Whipple's I | 3/300 | 0.142458 | 0.564557 | 0 | 0 | 2.358826 | 4.596658 |
| Disease_Sig | Hypoxia GS | 3/300 | 0.142458 | 0.564557 | 0 | 0 | 2.358826 | 4.596658 |
| Disease_Sig | HIV - Huma | 3/300 | 0.142458 | 0.564557 | 0 | 0 | 2.358826 | 4.596658 |
| Disease_Sig | Simian Acq | 3/300 | 0.142458 | 0.564557 | 0 | 0 | 2.358826 | 4.596658 |
| Disease_Sig | Huntingtor | 3/300 | 0.142458 | 0.564557 | 0 | 0 | 2.358826 | 4.596658 |
| Disease_Sig | Dehydratio | 3/300 | 0.142458 | 0.564557 | 0 | 0 | 2.358826 | 4.596658 |
| Disease_Sig | Tachycardi | 3/300 | 0.142458 | 0.564557 | 0 | 0 | 2.358826 | 4.596658 |
| Disease_Sig | Meningocc | 3/300 | 0.142458 | 0.564557 | 0 | 0 | 2.358826 | 4.596658 |
| Disease_Sig | Papillary Ca | 3/300 | 0.142458 | 0.564557 | 0 | 0 | 2.358826 | 4.596658 |
| Disease_Sig | Premature | 3/300 | 0.142458 | 0.564557 | 0 | 0 | 2.358826 | 4.596658 |
| Disease_Sig | Lung transp | 3/300 | 0.142458 | 0.564557 | 0 | 0 | 2.358826 | 4.596658 |
| Disease_Sig | Cancer of ( | 3/300 | 0.142458 | 0.564557 | 0 | 0 | 2.358826 | 4.596658 |
| Disease_Sig | Cardiac Hyp | 3/300 | 0.142458 | 0.564557 | 0 | 0 | 2.358826 | 4.596658 |
| Disease_Sig | Alzheimer's | 3/300 | 0.142458 | 0.564557 | 0 | 0 | 2.358826 | 4.596658 |
| Disease_Sig | B-cell chron | 3/300 | 0.142458 | 0.564557 | 0 | 0 | 2.358826 | 4.596658 |
| GO_Biologi | Positive Re | 1/175 | 0.535249 | 0.564676 | 0 | 0 | 1.319099 | 0.824467 |
| PhenGenI_ | Lung Neop | 1/121 | 0.410859 | 0.564931 | 0 | 0 | 1.917926 | 1.706006 |
| UK_Bioban | Phosphate | 2/412 | 0.537807 | 0.565591 | 0 | 0 | 1.119254 | 0.694223 |
| Jensen_TIS | Spermatid | 1/151 | 0.483545 | 0.566188 | 0 | 0 | 1.532016 | 1.113178 |
| Rare_Disea | Di Guglielm | 1/164 | 0.51222 | 0.566258 | 0 | 0 | 1.408903 | 0.942558 |
| Rare_Disea | Lymph nod | 1/164 | 0.51222 | 0.566258 | 0 | 0 | 1.408903 | 0.942558 |
| GeneSigDB | 17683608-1 | 4/707 | 0.369968 | 0.566275 | 0 | 0 | 1.316903 | 1.309447 |
| GeneSigDB | 17597811-: | 3/499 | 0.370061 | 0.566275 | 0 | 0 | 1.398113 | 1.389846 |
| GeneSigDB | 15870693-: | 1/105 | 0.368048 | 0.566275 | 0 | 0 | 2.214781 | 2.213764 |
| SysMyo_M | MGS1390 I | Jan-75 | 0.279326 | 0.566662 | 0 | 0 | 3.117379 | 3.975827 |
| GTEx_Tissu | GTEX-WL4( | 7/698 | 0.032326 | 0.566785 | 0 | 0 | 2.434045 | 8.353348 |
| Reactome_ | Respiratory | 1/112 | 0.387144 | 0.566799 | 0 | 0 | 2.074377 | 1.968499 |
| DisGeNET | Burkitt Lym | 2/407 | 0.531199 | 0.5669 | 0 | 0 | 1.133362 | 0.716987 |
| DisGeNET | Tonic - clor | 1/173 | 0.531144 | 0.5669 | 0 | 0 | 1.334573 | 0.844414 |
| Gene_Pert | Por Global | 5/301 | 0.010118 | 0.566918 | 0 | 0 | 4.041076 | 18.56248 |
| Gene_Pert | Mstn DEPL | 5/301 | 0.010118 | 0.566918 | 0 | 0 | 4.041076 | 18.56248 |
| Reactome_ | Negative R | 1/113 | 0.389825 | 0.567018 | 0 | 0 | 2.055752 | 1.936638 |
| Disease_Pe | acute myoc | 4/297 | 0.040671 | 0.567081 | 0 | 0 | 3.227106 | 10.33401 |
| Disease_Pe | acute myoc | 4/299 | 0.041515 | 0.567081 | 0 | 0 | 3.204901 | 10.19702 |
| GeDiPNet_ | Congenital | 2/413 | 0.539122 | 0.568369 | 0 | 0 | 1.116473 | 0.689773 |
| DisGeNET | Prostatic Ir | 1/174 | 0.533201 | 0.568574 | 0 | 0 | 1.326791 | 0.834362 |
| DisGeNET | Azoosperm | 1/174 | 0.533201 | 0.568574 | 0 | 0 | 1.326791 | 0.834362 |
| GeDiPNet_ | Lung Disea | 1/178 | 0.541341 | 0.569679 | 0 | 0 | 1.296544 | 0.795696 |

| | | | | | | | | |
|---|---|---|---|---|---|---|---|---|
| DisGeNET | Drug Depe | 1/175 | 0.535249 | 0.570526 | 0 | 0 | 1.319099 | 0.824467 |
| CellMarker | Neuron Bra | 1/172 | 0.529077 | 0.570737 | 0 | 0 | 1.342445 | 0.854628 |
| UK_Bioban | Age when | 1/180 | 0.545359 | 0.571424 | 0 | 0 | 1.281928 | 0.777247 |
| Rare_Disea | Mutagen s | 1/167 | 0.51861 | 0.571624 | 0 | 0 | 1.383231 | 0.908233 |
| Rare_Disea | Liposarcom | 1/167 | 0.51861 | 0.571624 | 0 | 0 | 1.383231 | 0.908233 |
| CellMarker | Ciliated Cel | 3/642 | 0.532297 | 0.571959 | 0 | 0 | 1.077241 | 0.679258 |
| Jensen_TIS | Mast cell | 1/154 | 0.490307 | 0.572025 | 0 | 0 | 1.501748 | 1.070331 |
| Rare_Disea | Sinus cance | 1/169 | 0.522824 | 0.572031 | 0 | 0 | 1.366625 | 0.886269 |
| Rare_Disea | Ectopic pre | 1/168 | 0.520722 | 0.572031 | 0 | 0 | 1.374878 | 0.897162 |
| Rare_Disea | Glomerulo | 1/169 | 0.522824 | 0.572031 | 0 | 0 | 1.366625 | 0.886269 |
| Rare_Disea | Thyroid car | 1/169 | 0.522824 | 0.572031 | 0 | 0 | 1.366625 | 0.886269 |
| Rare_Disea | Measles | 1/168 | 0.520722 | 0.572031 | 0 | 0 | 1.374878 | 0.897162 |
| DisGeNET | Anaplastic | 1/176 | 0.537289 | 0.572235 | 0 | 0 | 1.311495 | 0.814726 |
| DisGeNET | Multiple m | 1/176 | 0.537289 | 0.572235 | 0 | 0 | 1.311495 | 0.814726 |
| GO_Biologi | Positive Re | 1/179 | 0.543355 | 0.572376 | 0 | 0 | 1.289195 | 0.7864 |
| GTEx_Tissu | GTEX-WHS | 8/860 | 0.033447 | 0.572561 | 0 | 0 | 2.265526 | 7.697774 |
| GTEx_Tissu | GTEX-S4UY | 9/1021 | 0.033225 | 0.572561 | 0 | 0 | 2.155024 | 7.33667 |
| GTEx_Tissu | GTEX-U4B1 | 7/701 | 0.032974 | 0.572561 | 0 | 0 | 2.423145 | 8.267853 |
| GTEx_Tissu | GTEX-WXY | 10/1189 | 0.033431 | 0.572561 | 0 | 0 | 2.063602 | 7.012701 |
| GO_Biologi | Endosomal | 1/180 | 0.545359 | 0.573637 | 0 | 0 | 1.281928 | 0.777247 |
| DisGeNET | Carcinoma | 1/177 | 0.53932 | 0.573931 | 0 | 0 | 1.303977 | 0.805137 |
| DisGeNET | Intrahepati | 1/177 | 0.53932 | 0.573931 | 0 | 0 | 1.303977 | 0.805137 |
| Gene_Pert | Nlgn1 KD n | 5/328 | 0.014241 | 0.574079 | 0 | 0 | 3.69818 | 15.72324 |
| DisGeNET | Malignant | 4/884 | 0.539997 | 0.574418 | 0 | 0 | 1.042333 | 0.642278 |
| Reactome_ | TCR Signali | 1/116 | 0.397798 | 0.574882 | 0 | 0 | 2.00182 | 1.845299 |
| Rare_Disea | Lymphosar | 1/171 | 0.527002 | 0.574911 | 0 | 0 | 1.35041 | 0.865006 |
| Rare_Disea | Adrenocor | 1/171 | 0.527002 | 0.574911 | 0 | 0 | 1.35041 | 0.865006 |
| GTEx_Tissu | GTEX-SNO | Apr-85 | 0.000517 | 0.575248 | 0 | 0 | 11.79949 | 89.29053 |
| DisGeNET | Neutropen | 1/178 | 0.541341 | 0.575381 | 0 | 0 | 1.296544 | 0.795696 |
| DisGeNET | Dyschezia | 1/178 | 0.541341 | 0.575381 | 0 | 0 | 1.296544 | 0.795696 |
| Rare_Disea | Small non- | 1/172 | 0.529077 | 0.575488 | 0 | 0 | 1.342445 | 0.854628 |
| Rare_Disea | Lymphocyt | 2/405 | 0.528537 | 0.575488 | 0 | 0 | 1.139104 | 0.72634 |
| PhenGenI_ | Receptors, | 1/125 | 0.421106 | 0.575594 | 0 | 0 | 1.855683 | 1.604928 |
| GTEx_Tissu | GTEX-T5JW | 9/1025 | 0.033942 | 0.575891 | 0 | 0 | 2.146086 | 7.260419 |
| GTEx_Tissu | GTEX-WRH | 8/863 | 0.03404 | 0.575891 | 0 | 0 | 2.257221 | 7.629909 |
| GTEx_Tissu | GTEX-O5Y | 8/866 | 0.03464 | 0.575936 | 0 | 0 | 2.248975 | 7.562747 |
| GTEx_Tissu | GTEX-PX3G | 7/708 | 0.03452 | 0.575936 | 0 | 0 | 2.398074 | 8.072454 |
| GTEx_Tissu | GTEX-S7SF | 8/866 | 0.03464 | 0.575936 | 0 | 0 | 2.248975 | 7.562747 |
| Jensen_TIS | Natural kill | 42/9530 | 0.495513 | 0.576011 | 0 | 0 | 1.025506 | 0.720072 |
| GTEx_Tissu | GTEX-PWO | 9/1031 | 0.035038 | 0.576608 | 0 | 0 | 2.132809 | 7.147744 |
| GTEx_Tissu | GTEX-POM | 8/873 | 0.036068 | 0.576608 | 0 | 0 | 2.229955 | 7.40872 |
| GTEx_Tissu | GTEX-QEG | 12/1553 | 0.036215 | 0.576608 | 0 | 0 | 1.907541 | 6.329737 |
| GTEx_Tissu | GTEX-U4B1 | 7/713 | 0.035653 | 0.576608 | 0 | 0 | 2.380471 | 7.936299 |
| GTEx_Tissu | GTEX-VUS | 8/869 | 0.035247 | 0.576608 | 0 | 0 | 2.240786 | 7.496279 |
| GTEx_Tissu | GTEX-OIZH | 8/874 | 0.036275 | 0.576608 | 0 | 0 | 2.227263 | 7.387019 |

| | | | | | | | | |
|---|---|---|---|---|---|---|---|---|
| GTEx_Tissu | GTEX-T6MI | 8/873 | 0.036068 | 0.576608 | 0 | 0 | 2.229955 | 7.40872 |
| GTEx_Tissu | GTEX-TSE9 | 12/1548 | 0.035458 | 0.576608 | 0 | 0 | 1.914271 | 6.392547 |
| Rare_Disea | Paraganglio | 1/173 | 0.531144 | 0.576892 | 0 | 0 | 1.334573 | 0.844414 |
| CellMarker | Ciliated Epi | 2/413 | 0.539122 | 0.577028 | 0 | 0 | 1.116473 | 0.689773 |
| DisGeNET | Myasthenia | 1/179 | 0.543355 | 0.577053 | 0 | 0 | 1.289195 | 0.7864 |
| DisGeNET | Scleroderm | 1/179 | 0.543355 | 0.577053 | 0 | 0 | 1.289195 | 0.7864 |
| CellMarker | Schwalie Et | 1/178 | 0.541341 | 0.57715 | 0 | 0 | 1.296544 | 0.795696 |
| GeneSigDB | 16478745- | 1/110 | 0.381747 | 0.577195 | 0 | 0 | 2.112652 | 2.034479 |
| GeneSigDB | 12917207- | 3/511 | 0.384285 | 0.577195 | 0 | 0 | 1.364244 | 1.304722 |
| GeneSigDB | 16288205- | 5/930 | 0.380227 | 0.577195 | 0 | 0 | 1.251681 | 1.210358 |
| GeneSigDB | 17150101- | 1/111 | 0.384451 | 0.577195 | 0 | 0 | 2.09334 | 2.001105 |
| GeneSigDB | 18304324- | 1/110 | 0.381747 | 0.577195 | 0 | 0 | 2.112652 | 2.034479 |
| GeneSigDB | 15474998- | Nov-41 | 0.383331 | 0.577195 | 0 | 0 | 1.147705 | 1.100483 |
| GeneSigDB | 17683608- | 3/508 | 0.380735 | 0.577195 | 0 | 0 | 1.37256 | 1.325415 |
| GWAS_Cat | Depression | 1/190 | 0.56493 | 0.577232 | 0 | 0 | 1.213486 | 0.692965 |
| Rare_Disea | Insulin-like | 1/174 | 0.533201 | 0.57744 | 0 | 0 | 1.326791 | 0.834362 |
| Rare_Disea | Paraplegia | 1/174 | 0.533201 | 0.57744 | 0 | 0 | 1.326791 | 0.834362 |
| GeneSigDB | 16617318- | 1/112 | 0.387144 | 0.578121 | 0 | 0 | 2.074377 | 1.968499 |
| GeneSigDB | 16707422- | 1/112 | 0.387144 | 0.578121 | 0 | 0 | 2.074377 | 1.968499 |
| WikiPathw | Ras Signali | 1/184 | 0.55329 | 0.578439 | 0 | 0 | 1.253654 | 0.742005 |
| WikiPathw | Metapathw | 1/184 | 0.55329 | 0.578439 | 0 | 0 | 1.253654 | 0.742005 |
| GO_Molecu | Protein Ser | 2/342 | 0.439809 | 0.578696 | 0 | 0 | 1.354533 | 1.112633 |
| GWAS_Cat | Body Size A | 1/191 | 0.566841 | 0.578699 | 0 | 0 | 1.207038 | 0.685208 |
| Reactome_ | Chromosor | 1/118 | 0.403056 | 0.578748 | 0 | 0 | 1.967402 | 1.787737 |
| CellMarker | Microglial ( | 3/658 | 0.549072 | 0.578921 | 0 | 0 | 1.050055 | 0.629536 |
| CellMarker | Type IC Spi | 1/182 | 0.549341 | 0.578921 | 0 | 0 | 1.267635 | 0.759358 |
| CellMarker | Oligodendr | 1/180 | 0.545359 | 0.578921 | 0 | 0 | 1.281928 | 0.777247 |
| GO_Biologi | Macromole | 1/183 | 0.55132 | 0.579049 | 0 | 0 | 1.260606 | 0.750615 |
| Diabetes_P | GSE135391 | 2/202 | 0.219398 | 0.579147 | 0 | 0 | 2.319176 | 3.517879 |
| Diabetes_P | GSE149468 | 2/167 | 0.164425 | 0.579147 | 0 | 0 | 2.816114 | 5.083934 |
| Diabetes_P | GSE110569 | 2/186 | 0.193998 | 0.579147 | 0 | 0 | 2.52289 | 4.137304 |
| Diabetes_P | GSE149468 | 2/186 | 0.193998 | 0.579147 | 0 | 0 | 2.52289 | 4.137304 |
| Diabetes_P | GSE135445 | 2/184 | 0.19085 | 0.579147 | 0 | 0 | 2.550873 | 4.224921 |
| Diabetes_P | GSE149468 | 2/193 | 0.205067 | 0.579147 | 0 | 0 | 2.429566 | 3.84945 |
| Diabetes_P | GSE154198 | 2/192 | 0.203481 | 0.579147 | 0 | 0 | 2.442477 | 3.888868 |
| Diabetes_P | GSE80780 | 2/188 | 0.197153 | 0.579147 | 0 | 0 | 2.495509 | 4.05215 |
| Diabetes_P | GSE149468 | 2/187 | 0.195575 | 0.579147 | 0 | 0 | 2.509126 | 4.094425 |
| Diabetes_P | GSE155612 | 2/176 | 0.178336 | 0.579147 | 0 | 0 | 2.669236 | 4.601995 |
| Diabetes_P | GSE106295 | 2/145 | 0.131396 | 0.579147 | 0 | 0 | 3.252982 | 6.602064 |
| Diabetes_P | GSE131320 | 2/202 | 0.219398 | 0.579147 | 0 | 0 | 2.319176 | 3.517879 |
| Diabetes_P | GSE143735 | 2/168 | 0.165961 | 0.579147 | 0 | 0 | 2.799008 | 5.027029 |
| Diabetes_P | GSE84759 | 2/202 | 0.219398 | 0.579147 | 0 | 0 | 2.319176 | 3.517879 |
| Diabetes_P | GSE114192 | 2/201 | 0.217801 | 0.579147 | 0 | 0 | 2.330949 | 3.552767 |
| Diabetes_P | GSE40439 | 2/178 | 0.181452 | 0.579147 | 0 | 0 | 2.638636 | 4.503525 |
| Diabetes_P | GSE144061 | 2/178 | 0.181452 | 0.579147 | 0 | 0 | 2.638636 | 4.503525 |

| | | | | | | | | |
|---|---|---|---|---|---|---|---|---|
| Diabetes_F | GSE89035 | 2/177 | 0.179893 | 0.579147 | 0 | 0 | 2.653849 | 4.552394 |
| Diabetes_F | GSE153222 | 2/182 | 0.18771 | 0.579147 | 0 | 0 | 2.579477 | 4.315096 |
| Diabetes_F | GSE131320 | 2/182 | 0.18771 | 0.579147 | 0 | 0 | 2.579477 | 4.315096 |
| Diabetes_F | GSE120429 | 2/181 | 0.186143 | 0.579147 | 0 | 0 | 2.594019 | 4.361174 |
| Diabetes_F | GSE131320 | 2/180 | 0.184577 | 0.579147 | 0 | 0 | 2.608724 | 4.407929 |
| Diabetes_F | GSE41203 | 2/180 | 0.184577 | 0.579147 | 0 | 0 | 2.608724 | 4.407929 |
| Diabetes_F | GSE86949 | 2/164 | 0.159833 | 0.579147 | 0 | 0 | 2.8687 | 5.260115 |
| Diabetes_F | GSE155064 | 2/171 | 0.170583 | 0.579147 | 0 | 0 | 2.748904 | 4.86152 |
| Diabetes_F | GSE110528 | 2/140 | 0.124127 | 0.579147 | 0 | 0 | 3.371697 | 7.034881 |
| Diabetes_F | GSE40439 | 2/171 | 0.170583 | 0.579147 | 0 | 0 | 2.748904 | 4.86152 |
| Diabetes_F | GSE143735 | 2/140 | 0.124127 | 0.579147 | 0 | 0 | 3.371697 | 7.034881 |
| Diabetes_F | GSE129020 | 2/166 | 0.162892 | 0.579147 | 0 | 0 | 2.833429 | 5.141737 |
| Diabetes_F | GSE115421 | 2/164 | 0.159833 | 0.579147 | 0 | 0 | 2.8687 | 5.260115 |
| Diabetes_F | GSE107894 | 2/124 | 0.101605 | 0.579147 | 0 | 0 | 3.816972 | 8.728145 |
| Diabetes_F | GSE149468 | 2/167 | 0.164425 | 0.579147 | 0 | 0 | 2.816114 | 5.083934 |
| Diabetes_F | GSE138252 | 2/166 | 0.162892 | 0.579147 | 0 | 0 | 2.833429 | 5.141737 |
| Diabetes_F | GSE106295 | 2/164 | 0.159833 | 0.579147 | 0 | 0 | 2.8687 | 5.260115 |
| Diabetes_F | GSE144061 | 2/179 | 0.183014 | 0.579147 | 0 | 0 | 2.623596 | 4.455375 |
| Diabetes_F | GSE112213 | 2/161 | 0.155267 | 0.579147 | 0 | 0 | 2.92327 | 5.444911 |
| Diabetes_F | GSE149468 | 2/161 | 0.155267 | 0.579147 | 0 | 0 | 2.92327 | 5.444911 |
| Diabetes_F | GSE127056 | 2/160 | 0.153751 | 0.579147 | 0 | 0 | 2.941921 | 5.508522 |
| Diabetes_F | GSE156903 | 2/159 | 0.152237 | 0.579147 | 0 | 0 | 2.960809 | 5.573176 |
| Diabetes_F | GSE7196 E | 2/175 | 0.176781 | 0.579147 | 0 | 0 | 2.684801 | 4.652344 |
| Diabetes_F | GSE158359 | 2/175 | 0.176781 | 0.579147 | 0 | 0 | 2.684801 | 4.652344 |
| Diabetes_F | GSE127056 | 2/175 | 0.176781 | 0.579147 | 0 | 0 | 2.684801 | 4.652344 |
| Diabetes_F | GSE41203 | 2/172 | 0.172129 | 0.579147 | 0 | 0 | 2.732595 | 4.808027 |
| Diabetes_F | GSE48009 | 2/170 | 0.16904 | 0.579147 | 0 | 0 | 2.765406 | 4.91584 |
| Diabetes_F | GSE41203 | 2/170 | 0.16904 | 0.579147 | 0 | 0 | 2.765406 | 4.91584 |
| Diabetes_F | GSE127251 | 2/128 | 0.107121 | 0.579147 | 0 | 0 | 3.695051 | 8.253999 |
| Diabetes_F | GSE156067 | 2/199 | 0.21461 | 0.579147 | 0 | 0 | 2.354852 | 3.623955 |
| Diabetes_F | GSE107894 | 2/147 | 0.134331 | 0.579147 | 0 | 0 | 3.207789 | 6.43948 |
| Diabetes_F | GSE101537 | 2/197 | 0.211424 | 0.579147 | 0 | 0 | 2.379246 | 3.697085 |
| Diabetes_F | GSE138252 | 2/196 | 0.209833 | 0.579147 | 0 | 0 | 2.391631 | 3.734399 |
| Diabetes_F | GSE143482 | 2/199 | 0.21461 | 0.579147 | 0 | 0 | 2.354852 | 3.623955 |
| Diabetes_F | GSE154198 | 2/198 | 0.213017 | 0.579147 | 0 | 0 | 2.366987 | 3.660273 |
| Diabetes_F | GSE87354 | 2/146 | 0.132861 | 0.579147 | 0 | 0 | 3.230229 | 6.520055 |
| Diabetes_F | GSE156903 | 2/146 | 0.132861 | 0.579147 | 0 | 0 | 3.230229 | 6.520055 |
| Diabetes_F | GSE129369 | 2/147 | 0.134331 | 0.579147 | 0 | 0 | 3.207789 | 6.43948 |
| Diabetes_F | GSE125856 | 2/126 | 0.104352 | 0.579147 | 0 | 0 | 3.755028 | 8.486292 |
| Diabetes_F | GSE109285 | 2/201 | 0.217801 | 0.579147 | 0 | 0 | 2.330949 | 3.552767 |
| Diabetes_F | GSE142025 | 2/132 | 0.112716 | 0.579147 | 0 | 0 | 3.580633 | 7.816104 |
| Diabetes_F | GSE149468 | 2/133 | 0.114127 | 0.579147 | 0 | 0 | 3.553121 | 7.711855 |
| Diabetes_F | GSE127056 | 2/134 | 0.115542 | 0.579147 | 0 | 0 | 3.526025 | 7.609588 |
| Diabetes_F | GSE149468 | 2/184 | 0.19085 | 0.579147 | 0 | 0 | 2.550873 | 4.224921 |
| Diabetes_F | GSE107894 | 2/136 | 0.118386 | 0.579147 | 0 | 0 | 3.473047 | 7.410799 |

| | | | | | | | | |
|---|---|---|---|---|---|---|---|---|
| Diabetes_F | GSE143482 | 3/238 | 0.085399 | 0.579147 | 0 | 0 | 2.990578 | 7.358087 |
| Diabetes_F | GSE12585€ | 2/157 | 0.14922 | 0.579147 | 0 | 0 | 2.999317 | 5.705706 |
| Diabetes_F | GSE128361 | 2/157 | 0.14922 | 0.579147 | 0 | 0 | 2.999317 | 5.705706 |
| Diabetes_F | GSE149468 | 2/158 | 0.150727 | 0.579147 | 0 | 0 | 2.97994 | 5.638896 |
| Diabetes_F | GSE142209 | 2/202 | 0.219398 | 0.579147 | 0 | 0 | 2.319176 | 3.517879 |
| Diabetes_F | GSE136134 | 2/194 | 0.206654 | 0.579147 | 0 | 0 | 2.416789 | 3.810572 |
| Diabetes_F | GSE149468 | 2/179 | 0.183014 | 0.579147 | 0 | 0 | 2.623596 | 4.455375 |
| Diabetes_F | GSE149468 | 2/202 | 0.219398 | 0.579147 | 0 | 0 | 2.319176 | 3.517879 |
| Diabetes_F | GSE148997 | 2/189 | 0.198732 | 0.579147 | 0 | 0 | 2.482038 | 4.010467 |
| Diabetes_F | GSE153222 | 2/189 | 0.198732 | 0.579147 | 0 | 0 | 2.482038 | 4.010467 |
| Diabetes_F | GSE149468 | 2/188 | 0.197153 | 0.579147 | 0 | 0 | 2.495509 | 4.05215 |
| Diabetes_F | GSE14507( | 2/151 | 0.140245 | 0.579147 | 0 | 0 | 3.121042 | 6.130869 |
| Diabetes_F | GSE151588 | 2/148 | 0.135804 | 0.579147 | 0 | 0 | 3.185657 | 6.360306 |
| Diabetes_F | GSE162493 | 2/187 | 0.195575 | 0.579147 | 0 | 0 | 2.509126 | 4.094425 |
| Diabetes_F | GSE144061 | 2/186 | 0.193998 | 0.579147 | 0 | 0 | 2.52289 | 4.137304 |
| Diabetes_F | GSE128361 | 2/186 | 0.193998 | 0.579147 | 0 | 0 | 2.52289 | 4.137304 |
| Diabetes_F | GSE67991 | 2/194 | 0.206654 | 0.579147 | 0 | 0 | 2.416789 | 3.810572 |
| Diabetes_F | GSE74113 | 2/193 | 0.205067 | 0.579147 | 0 | 0 | 2.429566 | 3.84945 |
| Diabetes_F | GSE100067 | 2/159 | 0.152237 | 0.579147 | 0 | 0 | 2.960809 | 5.573176 |
| Diabetes_F | GSE73036 | 2/158 | 0.150727 | 0.579147 | 0 | 0 | 2.97994 | 5.638896 |
| Diabetes_F | GSE86611 | 2/191 | 0.201897 | 0.579147 | 0 | 0 | 2.455524 | 3.928837 |
| Diabetes_F | GSE162493 | 2/190 | 0.200314 | 0.579147 | 0 | 0 | 2.468711 | 3.969367 |
| Diabetes_F | GSE101657 | 2/189 | 0.198732 | 0.579147 | 0 | 0 | 2.482038 | 4.010467 |
| Diabetes_F | GSE12585€ | 2/154 | 0.144717 | 0.579147 | 0 | 0 | 3.058978 | 5.912923 |
| Diabetes_F | GSE149468 | 2/152 | 0.141732 | 0.579147 | 0 | 0 | 3.100078 | 6.056983 |
| Diabetes_F | GSE153222 | 2/151 | 0.140245 | 0.579147 | 0 | 0 | 3.121042 | 6.130869 |
| KOMP2_M | Decreased | Jan-73 | 0.272992 | 0.57929 | 0 | 0 | 3.204296 | 4.160181 |
| KOMP2_M | Abnormal ( | Jan-74 | 0.276166 | 0.57929 | 0 | 0 | 3.160242 | 4.066453 |
| KOMP2_M | Abnormal 1 | Jan-76 | 0.282473 | 0.57929 | 0 | 0 | 3.075659 | 3.888164 |
| KOMP2_M | Enlarged H | 3/427 | 0.284281 | 0.57929 | 0 | 0 | 1.641594 | 2.064784 |
| PhenGenI_ | Trypanosoı | 2/333 | 0.426412 | 0.579419 | 0 | 0 | 1.392003 | 1.186473 |
| GeneSigDB | 20436685-: | 3/516 | 0.390193 | 0.579566 | 0 | 0 | 1.350599 | 1.271068 |
| GeneSigDB | 18062813-( | 1/113 | 0.389825 | 0.579566 | 0 | 0 | 2.055752 | 1.936638 |
| Jensen_TIS | Pineal glan | 26/5881 | 0.501296 | 0.580638 | 0 | 0 | 1.023388 | 0.70671 |
| Rare_Disea | Ovarian cal | 1/176 | 0.537289 | 0.581022 | 0 | 0 | 1.311495 | 0.814726 |
| DisGeNET | Photodyspl | 1/181 | 0.547354 | 0.581065 | 0 | 0 | 1.274742 | 0.768234 |
| GeDiPNet_ | Immunolog | 1/184 | 0.55329 | 0.581203 | 0 | 0 | 1.253654 | 0.742005 |
| WikiPathw | VEGFA VEG | 2/430 | 0.561074 | 0.581293 | 0 | 0 | 1.071193 | 0.619045 |
| SysMyo_M | MGS1498 I | Jan-78 | 0.288726 | 0.581561 | 0 | 0 | 2.99547 | 3.721207 |
| SysMyo_M | MGS1397 I | Jan-78 | 0.288726 | 0.581561 | 0 | 0 | 2.99547 | 3.721207 |
| Jensen_TIS | Dendritic c | 25/5660 | 0.504636 | 0.581608 | 0 | 0 | 1.021696 | 0.698756 |
| Jensen_TIS | Trophoblas | 1/161 | 0.505746 | 0.581608 | 0 | 0 | 1.435538 | 0.978637 |
| Rare_Disea | Cough hea | 1/177 | 0.53932 | 0.582371 | 0 | 0 | 1.303977 | 0.805137 |
| DisGeNET | Mental imp | 1/182 | 0.549341 | 0.582466 | 0 | 0 | 1.267635 | 0.759358 |
| DisGeNET | Nephrotic ! | 1/182 | 0.549341 | 0.582466 | 0 | 0 | 1.267635 | 0.759358 |

| | | | | | | | | |
|---|---|---|---|---|---|---|---|---|
| DisGeNET | Primary My | 1/182 | 0.549341 | 0.582466 | 0 | 0 | 1.267635 | 0.759358 |
| SysMyo_M | MGS861 M | 2/246 | 0.29021 | 0.582478 | 0 | 0 | 1.896721 | 2.346531 |
| Reactome_ | Glycosamir | 1/120 | 0.408269 | 0.582499 | 0 | 0 | 1.934141 | 1.732658 |
| Rare_Disea | Non-small | 1/178 | 0.541341 | 0.582863 | 0 | 0 | 1.296544 | 0.795696 |
| Rare_Disea | Lymphoma | 1/178 | 0.541341 | 0.582863 | 0 | 0 | 1.296544 | 0.795696 |
| GTEx_Tissu | GTEX-WK1 | 8/877 | 0.036901 | 0.583361 | 0 | 0 | 2.219225 | 7.32236 |
| SysMyo_M | MGS591 H | 2/247 | 0.29182 | 0.58364 | 0 | 0 | 1.888884 | 2.326383 |
| PhenGenI_ | Bone Dens | 3/552 | 0.432281 | 0.583959 | 0 | 0 | 1.259693 | 1.056478 |
| GWAS_Cat | Neutrophil | 3/681 | 0.572573 | 0.584063 | 0 | 0 | 1.013222 | 0.564988 |
| DisGeNET | Immune th | 1/183 | 0.55132 | 0.584091 | 0 | 0 | 1.260606 | 0.750615 |
| DisGeNET | Delayed bc | 1/183 | 0.55132 | 0.584091 | 0 | 0 | 1.260606 | 0.750615 |
| Rare_Disea | B-cell lymp | 1/179 | 0.543355 | 0.584185 | 0 | 0 | 1.289195 | 0.7864 |
| WikiPathw | Pleural Me | 2/437 | 0.569898 | 0.585163 | 0 | 0 | 1.053577 | 0.592424 |
| Reactome_ | Metabolisr | 1/122 | 0.413437 | 0.586139 | 0 | 0 | 1.90198 | 1.679923 |
| GTEx_Tissu | GTEX-V955 | 7/720 | 0.037281 | 0.586164 | 0 | 0 | 2.356241 | 7.750311 |
| Rare_Disea | Idiopathic | 1/181 | 0.547354 | 0.586789 | 0 | 0 | 1.274742 | 0.768234 |
| Rare_Disea | Multiple m | 1/181 | 0.547354 | 0.586789 | 0 | 0 | 1.274742 | 0.768234 |
| GO_Biologi | Phosphoryl | 2/429 | 0.559803 | 0.58709 | 0 | 0 | 1.073757 | 0.622962 |
| Diabetes_F | GSE136134 | 2/207 | 0.227399 | 0.587145 | 0 | 0 | 2.262037 | 3.350186 |
| Diabetes_F | GSE148997 | 2/209 | 0.230606 | 0.587145 | 0 | 0 | 2.239955 | 3.286115 |
| Diabetes_F | GSE80780 | 2/208 | 0.229002 | 0.587145 | 0 | 0 | 2.250942 | 3.317943 |
| Diabetes_F | GSE47504 | 2/209 | 0.230606 | 0.587145 | 0 | 0 | 2.239955 | 3.286115 |
| Diabetes_F | GSE155482 | 2/207 | 0.227399 | 0.587145 | 0 | 0 | 2.262037 | 3.350186 |
| GWAS_Cat | Attention D | 1/196 | 0.576271 | 0.587343 | 0 | 0 | 1.17579 | 0.64807 |
| Gene_Pert | Mnat1 Defi | 5/332 | 0.014935 | 0.588975 | 0 | 0 | 3.652197 | 15.35391 |
| GTEx_Tissu | GTEX-QEL4 | 12/1565 | 0.038081 | 0.589141 | 0 | 0 | 1.891565 | 6.18169 |
| GTEx_Tissu | GTEX-XGQ4 | 6/570 | 0.037944 | 0.589141 | 0 | 0 | 2.54124 | 8.31403 |
| GTEx_Tissu | GTEX-S4UY | 11/1387 | 0.037786 | 0.589141 | 0 | 0 | 1.949845 | 6.38731 |
| GO_Biologi | Endocytosi | 1/189 | 0.563011 | 0.589584 | 0 | 0 | 1.220002 | 0.700838 |
| DisGeNET | Hashimoto | 1/186 | 0.557204 | 0.589848 | 0 | 0 | 1.239975 | 0.725168 |
| DisGeNET | Eosinophili | 1/186 | 0.557204 | 0.589848 | 0 | 0 | 1.239975 | 0.725168 |
| GO_Biologi | Nervous Sy | 2/433 | 0.564871 | 0.589853 | 0 | 0 | 1.063573 | 0.607468 |
| GO_Biologi | Intracellula | 1/190 | 0.56493 | 0.589853 | 0 | 0 | 1.213486 | 0.692965 |
| GeneSigDB | 15257931- | 1/117 | 0.400433 | 0.590056 | 0 | 0 | 1.984463 | 1.816198 |
| GeneSigDB | 17804718- | 1/117 | 0.400433 | 0.590056 | 0 | 0 | 1.984463 | 1.816198 |
| GeneSigDB | 16014681- | 1/117 | 0.400433 | 0.590056 | 0 | 0 | 1.984463 | 1.816198 |
| Gene_Pert | PARK2 OE I | 5/334 | 0.015291 | 0.590164 | 0 | 0 | 3.629624 | 15.17363 |
| WikiPathw | Focal Adhe | 1/199 | 0.581831 | 0.590633 | 0 | 0 | 1.157799 | 0.627036 |
| WikiPathw | Circadian R | 1/201 | 0.585497 | 0.590633 | 0 | 0 | 1.146105 | 0.613503 |
| GeneSigDB | 19328798- | 1/118 | 0.403056 | 0.590796 | 0 | 0 | 1.967402 | 1.787737 |
| GeneSigDB | 19306436- | 2/317 | 0.402204 | 0.590796 | 0 | 0 | 1.463903 | 1.333316 |
| CellMarker | Schwalie Et | 1/189 | 0.563011 | 0.591054 | 0 | 0 | 1.220002 | 0.700838 |
| GWAS_Cat | Mean Corp | 4/930 | 0.580614 | 0.591277 | 0 | 0 | 0.98816 | 0.537232 |
| GTEx_Tissu | GTEX-P78B | 8/888 | 0.039261 | 0.591573 | 0 | 0 | 2.190219 | 7.090886 |
| GTEx_Tissu | GTEX-REY6 | 11/1395 | 0.039143 | 0.591573 | 0 | 0 | 1.937738 | 6.279285 |

| | | | | | | | | |
|---|---|---|---|---|---|---|---|---|
| GTEx_Tissu | GTEX-OXRI | 9/1052 | 0.039066 | 0.591573 | 0 | 0 | 2.087543 | 6.768865 |
| GTEx_Tissu | GTEX-UJHI- | 9/1050 | 0.038669 | 0.591573 | 0 | 0 | 2.091776 | 6.803942 |
| GTEx_Tissu | GTEX-WQL | 6/572 | 0.038492 | 0.591573 | 0 | 0 | 2.531998 | 8.247479 |
| Rare_Disea | Thundercla | 1/184 | 0.55329 | 0.592299 | 0 | 0 | 1.253654 | 0.742005 |
| Rare_Disea | Hypnic hea | 1/185 | 0.555251 | 0.592692 | 0 | 0 | 1.246777 | 0.733523 |
| Rare_Disea | Primary ma | 1/185 | 0.555251 | 0.592692 | 0 | 0 | 1.246777 | 0.733523 |
| DisGeNET | Chronic Ob | 3/669 | 0.560403 | 0.592995 | 0 | 0 | 1.032121 | 0.5977 |
| DisGeNET | Photophob | 1/188 | 0.561084 | 0.592997 | 0 | 0 | 1.226589 | 0.708828 |
| DisGeNET | Dyspnea | 1/188 | 0.561084 | 0.592997 | 0 | 0 | 1.226589 | 0.708828 |
| DisGeNET | Bronchopu | 1/188 | 0.561084 | 0.592997 | 0 | 0 | 1.226589 | 0.708828 |
| Rare_Disea | Spinal mus | 1/186 | 0.557204 | 0.593075 | 0 | 0 | 1.239975 | 0.725168 |
| Rare_Disea | Acute myel | 1/186 | 0.557204 | 0.593075 | 0 | 0 | 1.239975 | 0.725168 |
| UK_Bioban | Home large | 1/192 | 0.568743 | 0.593743 | 0 | 0 | 1.200657 | 0.677563 |
| SysMyo_M | MGS675 H | Jan-82 | 0.30107 | 0.593748 | 0 | 0 | 2.846971 | 3.417539 |
| SysMyo_M | MGS1014 I | Jan-82 | 0.30107 | 0.593748 | 0 | 0 | 2.846971 | 3.417539 |
| SysMyo_M | MGS1410 I | Jan-82 | 0.30107 | 0.593748 | 0 | 0 | 2.846971 | 3.417539 |
| SysMyo_M | MGS168 Ra | Jan-81 | 0.298004 | 0.593748 | 0 | 0 | 2.882703 | 3.48994 |
| GWAS_Cat | Cognitive P | 1/200 | 0.583668 | 0.593891 | 0 | 0 | 1.151922 | 0.620221 |
| Diabetes_F | GSE15464S | 2/218 | 0.245071 | 0.594462 | 0 | 0 | 2.145643 | 3.017223 |
| Diabetes_F | GSE11376 | 2/216 | 0.241852 | 0.594462 | 0 | 0 | 2.165915 | 3.074364 |
| Diabetes_F | GSE10841 | 2/214 | 0.238636 | 0.594462 | 0 | 0 | 2.18657 | 3.132957 |
| Diabetes_F | GSE13132 | 2/212 | 0.235422 | 0.594462 | 0 | 0 | 2.207619 | 3.19305 |
| Diabetes_F | GSE47504 | 2/213 | 0.237028 | 0.594462 | 0 | 0 | 2.197045 | 3.162813 |
| Diabetes_F | GSE14946 | 2/217 | 0.243461 | 0.594462 | 0 | 0 | 2.155732 | 3.045615 |
| Diabetes_F | GSE13713 | 2/216 | 0.241852 | 0.594462 | 0 | 0 | 2.165915 | 3.074364 |
| GO_Molec | G Protein-( | 1/140 | 0.457986 | 0.594786 | 0 | 0 | 1.654174 | 1.291774 |
| GTEx_Tissu | GTEX-UJM | 11/1402 | 0.040359 | 0.595819 | 0 | 0 | 1.927258 | 6.186402 |
| GTEx_Tissu | GTEX-QDV. | 8/893 | 0.040367 | 0.595819 | 0 | 0 | 2.177272 | 6.988506 |
| GTEx_Tissu | GTEX-WZT | 11/1401 | 0.040183 | 0.595819 | 0 | 0 | 1.928749 | 6.199579 |
| GTEx_Tissu | GTEX-VUS | 10/1226 | 0.039929 | 0.595819 | 0 | 0 | 1.99686 | 6.431197 |
| GTEx_Tissu | GTEX-V955 | 8/895 | 0.040815 | 0.596347 | 0 | 0 | 2.172135 | 6.948039 |
| GTEx_Tissu | GTEX-V1D1 | 6/580 | 0.040736 | 0.596347 | 0 | 0 | 2.495677 | 7.987797 |
| Rare_Disea | Leiomyosa | 1/188 | 0.561084 | 0.596352 | 0 | 0 | 1.226589 | 0.708828 |
| GeDiPNet_ | Hydroneph | 1/192 | 0.568743 | 0.596362 | 0 | 0 | 1.200657 | 0.677563 |
| GTEx_Tissu | GTEX-PWN | 8/902 | 0.042409 | 0.596371 | 0 | 0 | 2.154334 | 6.808537 |
| GTEx_Tissu | GTEX-PSD | 7/741 | 0.042459 | 0.596371 | 0 | 0 | 2.286325 | 7.222987 |
| GTEx_Tissu | GTEX-Q73 | 9/1069 | 0.042552 | 0.596371 | 0 | 0 | 2.052213 | 6.47889 |
| GTEx_Tissu | GTEX-WFG | 6/589 | 0.043357 | 0.596371 | 0 | 0 | 2.456007 | 7.70764 |
| GTEx_Tissu | GTEX-XBEC | 10/1233 | 0.041251 | 0.596371 | 0 | 0 | 1.984687 | 6.327358 |
| GTEx_Tissu | GTEX-SNM | 11/1409 | 0.0416 | 0.596371 | 0 | 0 | 1.916883 | 6.095018 |
| GTEx_Tissu | GTEX-SIU8- | 7/745 | 0.043496 | 0.596371 | 0 | 0 | 2.273459 | 7.127483 |
| GTEx_Tissu | GTEX-VUS | 8/899 | 0.041721 | 0.596371 | 0 | 0 | 2.161929 | 6.86792 |
| GTEx_Tissu | GTEX-UJHI- | 8/904 | 0.042872 | 0.596371 | 0 | 0 | 2.149299 | 6.769281 |
| GTEx_Tissu | GTEX-R55E | 10/1242 | 0.042994 | 0.596371 | 0 | 0 | 1.96924 | 6.196583 |
| GTEx_Tissu | GTEX-SE5C | 8/906 | 0.043339 | 0.596371 | 0 | 0 | 2.144287 | 6.730287 |

| | | | | | | | | |
|---|---|---|---|---|---|---|---|---|
| GTEx_Tissu | GTEX-T2IS- | 13/1773 | 0.042556 | 0.596371 | 0 | 0 | 1.811955 | 5.720214 |
| GTEx_Tissu | GTEX-WRH | 8/904 | 0.042872 | 0.596371 | 0 | 0 | 2.149299 | 6.769281 |
| DisGeNET | Cirrhosis | 2/433 | 0.564871 | 0.596759 | 0 | 0 | 1.063573 | 0.607468 |
| Reactome_ | Cardiac Co | 1/126 | 0.42364 | 0.596826 | 0 | 0 | 1.840744 | 1.580964 |
| GeneSigDB | 14522937- | 1/120 | 0.408269 | 0.596866 | 0 | 0 | 1.934141 | 1.732658 |
| GTEx_Tissu | GTEX-WRH | 12/1599 | 0.043741 | 0.596893 | 0 | 0 | 1.847612 | 5.782065 |
| SysMyo_M | MGS94 Hu | Jan-84 | 0.307163 | 0.597437 | 0 | 0 | 2.778089 | 3.279195 |
| SysMyo_M | MGS1340 I | Jan-84 | 0.307163 | 0.597437 | 0 | 0 | 2.778089 | 3.279195 |
| SysMyo_M | MGS484 Hu | Jan-84 | 0.307163 | 0.597437 | 0 | 0 | 2.778089 | 3.279195 |
| SysMyo_M | MGS409 Hu | Jan-83 | 0.304123 | 0.597437 | 0 | 0 | 2.81211 | 3.34732 |
| Rare_Disea | Acinic cell | 1/189 | 0.563011 | 0.597547 | 0 | 0 | 1.220002 | 0.700838 |
| GeneSigDB | 20203266- | 2/322 | 0.409821 | 0.597566 | 0 | 0 | 1.440662 | 1.285121 |
| Diabetes_F | GSE155482 | 2/224 | 0.254736 | 0.597715 | 0 | 0 | 2.087016 | 2.85405 |
| Diabetes_F | GSE157585 | 2/224 | 0.254736 | 0.597715 | 0 | 0 | 2.087016 | 2.85405 |
| Diabetes_F | GSE134431 | 2/224 | 0.254736 | 0.597715 | 0 | 0 | 2.087016 | 2.85405 |
| Diabetes_F | GSE141891 | 2/220 | 0.248291 | 0.597715 | 0 | 0 | 2.125742 | 2.961486 |
| Diabetes_F | GSE108654 | 2/221 | 0.249902 | 0.597715 | 0 | 0 | 2.115928 | 2.934131 |
| GeneSigDB | 18667080- | 3/534 | 0.411345 | 0.598222 | 0 | 0 | 1.303605 | 1.158022 |
| GeDiPNet_ | Stroke | 1/194 | 0.572523 | 0.598248 | 0 | 0 | 1.188095 | 0.662603 |
| GeDiPNet_ | Breast Can | 5/1159 | 0.572594 | 0.598248 | 0 | 0 | 0.991197 | 0.55267 |
| DisGeNET | Deglutition | 1/191 | 0.566841 | 0.598598 | 0 | 0 | 1.207038 | 0.685208 |
| Rare_Disea | Cutaneous | 1/190 | 0.56493 | 0.598729 | 0 | 0 | 1.213486 | 0.692965 |
| GeDiPNet_ | Developme | 1/195 | 0.574401 | 0.599062 | 0 | 0 | 1.181911 | 0.655284 |
| KEGG_202 | Autophagy | 2/137 | 0.119815 | 0.599074 | 0 | 0 | 3.447146 | 7.31418 |
| UK_Bioban | Left 6mm r | 1/197 | 0.578132 | 0.60036 | 0 | 0 | 1.169732 | 0.640958 |
| UK_Bioban | Right 6mm | 8/1879 | 0.579294 | 0.60036 | 0 | 0 | 0.976503 | 0.533117 |
| GTEx_Tissu | GTEX-WRH | 6/592 | 0.044254 | 0.601069 | 0 | 0 | 2.443054 | 7.61696 |
| Diabetes_F | GSE144274 | 2/226 | 0.257961 | 0.60135 | 0 | 0 | 2.068172 | 2.802265 |
| Rare_Disea | Sphingolipi | 1/192 | 0.568743 | 0.601913 | 0 | 0 | 1.200657 | 0.677563 |
| GO_Biologi | Golgi Vesic | 1/197 | 0.578132 | 0.602751 | 0 | 0 | 1.169732 | 0.640958 |
| GTEx_Tissu | GTEX-WFG | 5/445 | 0.04471 | 0.602788 | 0 | 0 | 2.698586 | 8.386044 |
| GTEx_Tissu | GTEX-XQ3S | 12/1605 | 0.044798 | 0.602788 | 0 | 0 | 1.84005 | 5.714461 |
| SysMyo_M | MGS447 Hu | Jan-86 | 0.313203 | 0.602968 | 0 | 0 | 2.712449 | 3.148895 |
| SysMyo_M | MGS1209 I | Jan-86 | 0.313203 | 0.602968 | 0 | 0 | 2.712449 | 3.148895 |
| SysMyo_M | MGS537 Hu | Jan-86 | 0.313203 | 0.602968 | 0 | 0 | 2.712449 | 3.148895 |
| GTEx_Tissu | GTEX-S341 | 11/1428 | 0.045105 | 0.604113 | 0 | 0 | 1.88924 | 5.85431 |
| DisGeNET | Mean Corp | 1/194 | 0.572523 | 0.604356 | 0 | 0 | 1.188095 | 0.662603 |
| SysMyo_M | MGS910 M | Jan-87 | 0.316203 | 0.604632 | 0 | 0 | 2.680773 | 3.086563 |
| SysMyo_M | MGS676 Hu | Jan-87 | 0.316203 | 0.604632 | 0 | 0 | 2.680773 | 3.086563 |
| Diabetes_F | GSE110528 | 2/228 | 0.261186 | 0.604941 | 0 | 0 | 2.049662 | 2.751716 |
| GTEx_Tissu | GTEX-XPT6 | 10/1255 | 0.045602 | 0.605087 | 0 | 0 | 1.947322 | 6.012953 |
| GTEx_Tissu | GTEX-X4EP | 7/755 | 0.04616 | 0.605087 | 0 | 0 | 2.241895 | 6.895244 |
| GTEx_Tissu | GTEX-P44H | 7/756 | 0.046433 | 0.605087 | 0 | 0 | 2.238785 | 6.87252 |
| GTEx_Tissu | GTEX-SIU8- | 5/448 | 0.045787 | 0.605087 | 0 | 0 | 2.679899 | 8.26413 |
| GTEx_Tissu | GTEX-OHPI | 7/754 | 0.045889 | 0.605087 | 0 | 0 | 2.245013 | 6.918057 |

| | | | | | | | | |
|---|---|---|---|---|---|---|---|---|
| GTEx_Tissu | GTEX-QEL4 | 9/1086 | 0.046245 | 0.605087 | 0 | 0 | 2.017999 | 6.202946 |
| dbGaP | Heart Failu | 1/187 | 0.559148 | 0.605743 | 0 | 0 | 1.233246 | 0.716937 |
| GeneSigDB | 19185848-' | 2/328 | 0.418899 | 0.606044 | 0 | 0 | 1.413713 | 1.230107 |
| GeneSigDB | 15831697-' | 2/328 | 0.418899 | 0.606044 | 0 | 0 | 1.413713 | 1.230107 |
| DisGeNET | Decreased | 1/195 | 0.574401 | 0.606094 | 0 | 0 | 1.181911 | 0.655284 |
| DisGeNET | Osteoporo: | 2/441 | 0.574883 | 0.606358 | 0 | 0 | 1.043763 | 0.577815 |
| GeneSigDB | 20418243-: | 2/329 | 0.420406 | 0.606648 | 0 | 0 | 1.409318 | 1.221224 |
| SysMyo_M | MGS783 M | 2/288 | 0.357207 | 0.606802 | 0 | 0 | 1.614726 | 1.662266 |
| SysMyo_M | MGS209 C: | 2/300 | 0.375985 | 0.606802 | 0 | 0 | 1.548756 | 1.515003 |
| SysMyo_M | MGS210 C: | 2/300 | 0.375985 | 0.606802 | 0 | 0 | 1.548756 | 1.515003 |
| SysMyo_M | MGS912 M | 2/275 | 0.336649 | 0.606802 | 0 | 0 | 1.692739 | 1.842909 |
| SysMyo_M | MGS1097 I | Jan-94 | 0.336847 | 0.606802 | 0 | 0 | 2.47812 | 2.69651 |
| SysMyo_M | MGS1211 I | Jan-94 | 0.336847 | 0.606802 | 0 | 0 | 2.47812 | 2.69651 |
| SysMyo_M | MGS252 C: | 2/300 | 0.375985 | 0.606802 | 0 | 0 | 1.548756 | 1.515003 |
| SysMyo_M | MGS270 M | 2/300 | 0.375985 | 0.606802 | 0 | 0 | 1.548756 | 1.515003 |
| SysMyo_M | MGS272 M | 2/300 | 0.375985 | 0.606802 | 0 | 0 | 1.548756 | 1.515003 |
| SysMyo_M | MGS278 M | 2/300 | 0.375985 | 0.606802 | 0 | 0 | 1.548756 | 1.515003 |
| SysMyo_M | MGS280 M | 2/300 | 0.375985 | 0.606802 | 0 | 0 | 1.548756 | 1.515003 |
| SysMyo_M | MGS282 M | 2/300 | 0.375985 | 0.606802 | 0 | 0 | 1.548756 | 1.515003 |
| SysMyo_M | MGS818 M | Jan-99 | 0.351214 | 0.606802 | 0 | 0 | 2.351092 | 2.46009 |
| SysMyo_M | MGS575 Hu | 1/100 | 0.35405 | 0.606802 | 0 | 0 | 2.327226 | 2.416399 |
| SysMyo_M | MGS10 Rat | 1/101 | 0.356874 | 0.606802 | 0 | 0 | 2.303837 | 2.373811 |
| SysMyo_M | MGS599 Hu | 1/101 | 0.356874 | 0.606802 | 0 | 0 | 2.303837 | 2.373811 |
| SysMyo_M | MGS246 C: | 2/300 | 0.375985 | 0.606802 | 0 | 0 | 1.548756 | 1.515003 |
| SysMyo_M | MGS734 M | 1/102 | 0.359686 | 0.606802 | 0 | 0 | 2.280912 | 2.332289 |
| SysMyo_M | MGS1422 I | 1/103 | 0.362485 | 0.606802 | 0 | 0 | 2.258436 | 2.291796 |
| SysMyo_M | MGS973 M | 1/103 | 0.362485 | 0.606802 | 0 | 0 | 2.258436 | 2.291796 |
| SysMyo_M | MGS287 C: | 2/300 | 0.375985 | 0.606802 | 0 | 0 | 1.548756 | 1.515003 |
| SysMyo_M | MGS294 C: | 2/300 | 0.375985 | 0.606802 | 0 | 0 | 1.548756 | 1.515003 |
| SysMyo_M | MGS208 C: | 2/300 | 0.375985 | 0.606802 | 0 | 0 | 1.548756 | 1.515003 |
| SysMyo_M | MGS526 Hu | Jan-97 | 0.345504 | 0.606802 | 0 | 0 | 2.400315 | 2.550937 |
| SysMyo_M | MGS1336 I | 1/104 | 0.365273 | 0.606802 | 0 | 0 | 2.236396 | 2.252299 |
| SysMyo_M | MGS1332 I | 1/105 | 0.368048 | 0.606802 | 0 | 0 | 2.214781 | 2.213764 |
| SysMyo_M | MGS1361 I | 2/295 | 0.368186 | 0.606802 | 0 | 0 | 1.575587 | 1.574275 |
| SysMyo_M | MGS869 M | 1/107 | 0.373563 | 0.606802 | 0 | 0 | 2.172773 | 2.139459 |
| SysMyo_M | MGS1241 I | 1/107 | 0.373563 | 0.606802 | 0 | 0 | 2.172773 | 2.139459 |
| SysMyo_M | MGS187 M | 2/300 | 0.375985 | 0.606802 | 0 | 0 | 1.548756 | 1.515003 |
| SysMyo_M | MGS207 C: | 2/300 | 0.375985 | 0.606802 | 0 | 0 | 1.548756 | 1.515003 |
| SysMyo_M | MGS1216 I | 2/293 | 0.365056 | 0.606802 | 0 | 0 | 1.586578 | 1.598802 |
| SysMyo_M | MGS697 M | 1/108 | 0.376303 | 0.606802 | 0 | 0 | 2.152358 | 2.10363 |
| SysMyo_M | MGS334 C: | 2/300 | 0.375985 | 0.606802 | 0 | 0 | 1.548756 | 1.515003 |
| SysMyo_M | MGS244 C: | 2/300 | 0.375985 | 0.606802 | 0 | 0 | 1.548756 | 1.515003 |
| SysMyo_M | MGS93 Hu | Jan-89 | 0.322165 | 0.606802 | 0 | 0 | 2.619582 | 2.967177 |
| SysMyo_M | MGS435 Hu | Jan-89 | 0.322165 | 0.606802 | 0 | 0 | 2.619582 | 2.967177 |
| SysMyo_M | MGS332 C: | 2/300 | 0.375985 | 0.606802 | 0 | 0 | 1.548756 | 1.515003 |

| | | | | | | | | |
|---|---|---|---|---|---|---|---|---|
| SysMyo_M | MGS948 M | Jan-89 | 0.322165 | 0.606802 | 0 | 0 | 2.619582 | 2.967177 |
| SysMyo_M | MGS773 M | Jan-99 | 0.351214 | 0.606802 | 0 | 0 | 2.351092 | 2.46009 |
| SysMyo_M | MGS330 M | 2/300 | 0.375985 | 0.606802 | 0 | 0 | 1.548756 | 1.515003 |
| SysMyo_M | MGS328 M | 2/300 | 0.375985 | 0.606802 | 0 | 0 | 1.548756 | 1.515003 |
| SysMyo_M | MGS799 M | Jan-96 | 0.342631 | 0.606802 | 0 | 0 | 2.425704 | 2.598176 |
| SysMyo_M | MGS1072 I | Jan-96 | 0.342631 | 0.606802 | 0 | 0 | 2.425704 | 2.598176 |
| SysMyo_M | MGS211 H | 2/300 | 0.375985 | 0.606802 | 0 | 0 | 1.548756 | 1.515003 |
| SysMyo_M | MGS219 C | 2/300 | 0.375985 | 0.606802 | 0 | 0 | 1.548756 | 1.515003 |
| SysMyo_M | MGS240 M | 2/300 | 0.375985 | 0.606802 | 0 | 0 | 1.548756 | 1.515003 |
| SysMyo_M | MGS566 H | Jan-90 | 0.325127 | 0.606802 | 0 | 0 | 2.590018 | 2.909988 |
| SysMyo_M | MGS876 M | 2/269 | 0.327097 | 0.606802 | 0 | 0 | 1.731306 | 1.934732 |
| SysMyo_M | MGS1302 I | Jan-92 | 0.331012 | 0.606802 | 0 | 0 | 2.532839 | 2.800307 |
| SysMyo_M | MGS412 H | Jan-93 | 0.333936 | 0.606802 | 0 | 0 | 2.505182 | 2.747701 |
| SysMyo_M | MGS500 H | Jan-93 | 0.333936 | 0.606802 | 0 | 0 | 2.505182 | 2.747701 |
| SysMyo_M | MGS295 C | 2/300 | 0.375985 | 0.606802 | 0 | 0 | 1.548756 | 1.515003 |
| SysMyo_M | MGS297 C | 2/300 | 0.375985 | 0.606802 | 0 | 0 | 1.548756 | 1.515003 |
| SysMyo_M | MGS665 H | Jan-98 | 0.348365 | 0.606802 | 0 | 0 | 2.37545 | 2.504922 |
| GeneSigDB | 18535662- | 5/978 | 0.422016 | 0.607397 | 0 | 0 | 1.186925 | 1.023975 |
| DisGeNET | Profound N | 1/196 | 0.576271 | 0.607577 | 0 | 0 | 1.17579 | 0.64807 |
| GeDiPNet_ | Proptosis | 1/200 | 0.583668 | 0.60764 | 0 | 0 | 1.151922 | 0.620221 |
| DisGeNET | Esophagea | 3/685 | 0.576584 | 0.607663 | 0 | 0 | 1.00707 | 0.554527 |
| DisGeNET | Squamous | 8/1876 | 0.577422 | 0.608057 | 0 | 0 | 0.978234 | 0.537228 |
| DisGeNET | Childhood | 2/443 | 0.577361 | 0.608057 | 0 | 0 | 1.038922 | 0.570668 |
| SysMyo_M | MGS74 Hu | 1/110 | 0.381747 | 0.608644 | 0 | 0 | 2.112652 | 2.034479 |
| SysMyo_M | MGS1086 I | 1/109 | 0.379031 | 0.608644 | 0 | 0 | 2.132321 | 2.068645 |
| SysMyo_M | MGS740 M | 1/110 | 0.381747 | 0.608644 | 0 | 0 | 2.112652 | 2.034479 |
| SysMyo_M | MGS786 M | 1/110 | 0.381747 | 0.608644 | 0 | 0 | 2.112652 | 2.034479 |
| Jensen_TIS | Adrenal co | 20/4574 | 0.531289 | 0.608808 | 0 | 0 | 1.006758 | 0.636724 |
| Disease_Pe | Duchenne | 5/333 | 0.015112 | 0.610166 | 0 | 0 | 3.640876 | 15.26341 |
| Reactome_ | ADORA2B I | 1/131 | 0.436146 | 0.610605 | 0 | 0 | 1.769499 | 1.468291 |
| DisGeNET | Precancerc | 2/446 | 0.581057 | 0.611639 | 0 | 0 | 1.031744 | 0.56014 |
| GeneSigDB | 16735486- | 1/127 | 0.426163 | 0.611785 | 0 | 0 | 1.826043 | 1.557494 |
| Jensen_TIS | Ascites | 4/884 | 0.539997 | 0.61225 | 0 | 0 | 1.042333 | 0.642278 |
| Jensen_TIS | Trachea | 4/882 | 0.538187 | 0.61225 | 0 | 0 | 1.044817 | 0.647315 |
| Jensen_TIS | Blood plasr | 1/176 | 0.537289 | 0.61225 | 0 | 0 | 1.311495 | 0.814726 |
| GWAS_Cat | Highest Ma | 1/211 | 0.603359 | 0.613415 | 0 | 0 | 1.090975 | 0.551207 |
| GeneSigDB | 17676974- | 6/1206 | 0.42851 | 0.613574 | 0 | 0 | 1.155123 | 0.978898 |
| Reactome_ | ER To Golg | 1/133 | 0.441073 | 0.613667 | 0 | 0 | 1.742512 | 1.426323 |
| Rare_Disea | Synovial sa | 1/199 | 0.581831 | 0.614017 | 0 | 0 | 1.157799 | 0.627036 |
| Rare_Disea | Focal or mu | 1/199 | 0.581831 | 0.614017 | 0 | 0 | 1.157799 | 0.627036 |
| GeneSigDB | 18614019- | 8/1652 | 0.430171 | 0.614372 | 0 | 0 | 1.12532 | 0.94929 |
| WikiPathw | Network M | 1/218 | 0.615408 | 0.615408 | 0 | 0 | 1.055407 | 0.512369 |
| SysMyo_M | MGS790 M | 1/112 | 0.387144 | 0.615515 | 0 | 0 | 2.074377 | 1.968499 |
| Diabetes_F | GSE14427 | 2/232 | 0.267638 | 0.615911 | 0 | 0 | 2.013606 | 2.654174 |
| DisGeNET | Arthritis, P | 1/201 | 0.585497 | 0.616065 | 0 | 0 | 1.146105 | 0.613503 |

| | | | | | | | | |
|---|---|---|---|---|---|---|---|---|
| GWAS_Cat | IgG Glycosy | 1/213 | 0.606839 | 0.616439 | 0 | 0 | 1.080573 | 0.539737 |
| GeneSigDB | 19808870-: | 4/772 | 0.434175 | 0.616928 | 0 | 0 | 1.201368 | 1.002309 |
| GeneSigDB | 17096850-' | 1/130 | 0.433666 | 0.616928 | 0 | 0 | 1.783306 | 1.489916 |
| GO_Biologi | Regulation | 1/205 | 0.592736 | 0.617071 | 0 | 0 | 1.123404 | 0.587548 |
| GeneSigDB | 18757322-' | 1/131 | 0.436146 | 0.617572 | 0 | 0 | 1.769499 | 1.468291 |
| GeneSigDB | 17571080-: | 2/340 | 0.436846 | 0.617572 | 0 | 0 | 1.362687 | 1.128542 |
| GeneSigDB | 18378853-' | 1/132 | 0.438615 | 0.617603 | 0 | 0 | 1.755903 | 1.447098 |
| GeneSigDB | 19797726-: | 1/134 | 0.443521 | 0.617603 | 0 | 0 | 1.729323 | 1.405958 |
| GeneSigDB | 17304354-' | 1/134 | 0.443521 | 0.617603 | 0 | 0 | 1.729323 | 1.405958 |
| GeneSigDB | 18974375-' | 1/134 | 0.443521 | 0.617603 | 0 | 0 | 1.729323 | 1.405958 |
| GeneSigDB | 11988840-' | 1/134 | 0.443521 | 0.617603 | 0 | 0 | 1.729323 | 1.405958 |
| GeneSigDB | 19805619-' | 1/133 | 0.441073 | 0.617603 | 0 | 0 | 1.742512 | 1.426323 |
| SysMyo_M | MGS971 M | 1/114 | 0.392494 | 0.617834 | 0 | 0 | 2.037456 | 1.905499 |
| SysMyo_M | MGS1217 I | 1/115 | 0.395152 | 0.617834 | 0 | 0 | 2.019482 | 1.875059 |
| SysMyo_M | MGS724 M | 1/115 | 0.395152 | 0.617834 | 0 | 0 | 2.019482 | 1.875059 |
| SysMyo_M | MGS1492 I | 1/115 | 0.395152 | 0.617834 | 0 | 0 | 2.019482 | 1.875059 |
| SysMyo_M | MGS73 Hu | 1/114 | 0.392494 | 0.617834 | 0 | 0 | 2.037456 | 1.905499 |
| SysMyo_M | MGS768 M | 1/114 | 0.392494 | 0.617834 | 0 | 0 | 2.037456 | 1.905499 |
| DisGeNET | ovarian ne | 4/938 | 0.587474 | 0.617897 | 0 | 0 | 0.979283 | 0.520903 |
| Diabetes_F | GSE73433 | 2/235 | 0.272477 | 0.61911 | 0 | 0 | 1.987377 | 2.583987 |
| Diabetes_F | GSE73433 | 2/234 | 0.270864 | 0.61911 | 0 | 0 | 1.996045 | 2.607109 |
| dbGaP | Blood Pres | 2/454 | 0.5908 | 0.619387 | 0 | 0 | 1.013066 | 0.533155 |
| dbGaP | Hip | 1/202 | 0.587319 | 0.619387 | 0 | 0 | 1.140345 | 0.606877 |
| DisGeNET | Malignant | 1/203 | 0.589132 | 0.619393 | 0 | 0 | 1.134642 | 0.600344 |
| GeneSigDB | 14749371-' | 1/135 | 0.445958 | 0.619448 | 0 | 0 | 1.716331 | 1.38599 |
| GO_Biologi | Regulation | 1/207 | 0.596308 | 0.619881 | 0 | 0 | 1.112384 | 0.575101 |
| CellMarker | Vascular Er | 1/205 | 0.592736 | 0.619884 | 0 | 0 | 1.123404 | 0.587548 |
| Reactome_ | Factors Inv | 1/136 | 0.448384 | 0.619988 | 0 | 0 | 1.703531 | 1.36641 |
| GeneSigDB | 19505326-: | Sep-02 | 0.44749 | 0.62003 | 0 | 0 | 1.098379 | 0.883207 |
| GeneSigDB | 16489042-: | 2/348 | 0.448647 | 0.620091 | 0 | 0 | 1.330636 | 1.066529 |
| GTEx_Tissu | GTEX-TMM | 7/761 | 0.047809 | 0.620227 | 0 | 0 | 2.223359 | 6.760228 |
| KOMP2_M | Abnormal ( | Jan-85 | 0.310189 | 0.620378 | 0 | 0 | 2.744878 | 3.21308 |
| DisGeNET | Osteogene | 1/204 | 0.590938 | 0.620544 | 0 | 0 | 1.128995 | 0.593902 |
| DisGeNET | Eczema | 2/454 | 0.5908 | 0.620544 | 0 | 0 | 1.013066 | 0.533155 |
| DisGeNET | Short neck | 1/204 | 0.590938 | 0.620544 | 0 | 0 | 1.128995 | 0.593902 |
| GO_Biologi | Protein Sta | 1/208 | 0.598082 | 0.620816 | 0 | 0 | 1.106954 | 0.569005 |
| SysMyo_M | MGS1378 I | 1/118 | 0.403056 | 0.621184 | 0 | 0 | 1.967402 | 1.787737 |
| SysMyo_M | MGS716 M | 1/119 | 0.405668 | 0.621184 | 0 | 0 | 1.950631 | 1.759896 |
| SysMyo_M | MGS1325 I | 1/120 | 0.408269 | 0.621184 | 0 | 0 | 1.934141 | 1.732658 |
| SysMyo_M | MGS659 H | 1/119 | 0.405668 | 0.621184 | 0 | 0 | 1.950631 | 1.759896 |
| SysMyo_M | MGS597 H | 1/119 | 0.405668 | 0.621184 | 0 | 0 | 1.950631 | 1.759896 |
| SysMyo_M | MGS1244 I | 1/119 | 0.405668 | 0.621184 | 0 | 0 | 1.950631 | 1.759896 |
| SysMyo_M | MGS1030 I | 1/120 | 0.408269 | 0.621184 | 0 | 0 | 1.934141 | 1.732658 |
| SysMyo_M | MGS810 M | 1/117 | 0.400433 | 0.621184 | 0 | 0 | 1.984463 | 1.816198 |
| SysMyo_M | MGS1316 I | 1/120 | 0.408269 | 0.621184 | 0 | 0 | 1.934141 | 1.732658 |

| | | | | | | | | |
|---|---|---|---|---|---|---|---|---|
| SysMyo_M | MGS120 H | 1/120 | 0.408269 | 0.621184 | 0 | 0 | 1.934141 | 1.732658 |
| DisGeNET | Pancreatic | 3/701 | 0.592398 | 0.621828 | 0 | 0 | 0.983166 | 0.514763 |
| DisGeNET | Cervical Int | 1/205 | 0.592736 | 0.621933 | 0 | 0 | 1.123404 | 0.587548 |
| Reactome_ | PTEN Regu | 1/139 | 0.455601 | 0.622284 | 0 | 0 | 1.666245 | 1.309899 |
| Reactome_ | Death Rece | 1/139 | 0.455601 | 0.622284 | 0 | 0 | 1.666245 | 1.309899 |
| GTEx_Tissu | GTEX-U8T8 | 8/926 | 0.048191 | 0.622396 | 0 | 0 | 2.095364 | 6.354362 |
| Rare_Disea | Myelofibro | 1/204 | 0.590938 | 0.622745 | 0 | 0 | 1.128995 | 0.593902 |
| GWAS_Cat | Bipolar Dis | 1/217 | 0.613709 | 0.622899 | 0 | 0 | 1.060347 | 0.517698 |
| DisGeNET | Cytopenia | 1/206 | 0.594525 | 0.623562 | 0 | 0 | 1.117867 | 0.581282 |
| DisGeNET | Low intellig | 4/947 | 0.595116 | 0.623681 | 0 | 0 | 0.969477 | 0.503158 |
| DisGeNET | Dull intellig | 4/947 | 0.595116 | 0.623681 | 0 | 0 | 0.969477 | 0.503158 |
| GeneSigDB | 18667080- | 4/791 | 0.452671 | 0.624104 | 0 | 0 | 1.171201 | 0.928281 |
| DisGeNET | Liver Dysfu | 1/207 | 0.596308 | 0.624431 | 0 | 0 | 1.112384 | 0.575101 |
| DisGeNET | Polycythen | 1/207 | 0.596308 | 0.624431 | 0 | 0 | 1.112384 | 0.575101 |
| SysMyo_M | MGS1328 I | 1/131 | 0.436146 | 0.624959 | 0 | 0 | 1.769499 | 1.468291 |
| SysMyo_M | MGS834 M | 1/130 | 0.433666 | 0.624959 | 0 | 0 | 1.783306 | 1.489916 |
| SysMyo_M | MGS580 H | 1/130 | 0.433666 | 0.624959 | 0 | 0 | 1.783306 | 1.489916 |
| SysMyo_M | MGS1109 I | 1/131 | 0.436146 | 0.624959 | 0 | 0 | 1.769499 | 1.468291 |
| SysMyo_M | MGS922 M | 1/127 | 0.426163 | 0.624959 | 0 | 0 | 1.826043 | 1.557494 |
| SysMyo_M | MGS1423 I | 1/128 | 0.428675 | 0.624959 | 0 | 0 | 1.811573 | 1.534505 |
| SysMyo_M | MGS1473 I | 1/127 | 0.426163 | 0.624959 | 0 | 0 | 1.826043 | 1.557494 |
| SysMyo_M | MGS898 M | 1/130 | 0.433666 | 0.624959 | 0 | 0 | 1.783306 | 1.489916 |
| SysMyo_M | MGS851 M | 1/131 | 0.436146 | 0.624959 | 0 | 0 | 1.769499 | 1.468291 |
| SysMyo_M | MGS1298 I | 1/125 | 0.421106 | 0.624959 | 0 | 0 | 1.855683 | 1.604928 |
| SysMyo_M | MGS269 M | 1/126 | 0.42364 | 0.624959 | 0 | 0 | 1.840744 | 1.580964 |
| SysMyo_M | MGS558 H | 1/126 | 0.42364 | 0.624959 | 0 | 0 | 1.840744 | 1.580964 |
| SysMyo_M | MGS660 H | 1/126 | 0.42364 | 0.624959 | 0 | 0 | 1.840744 | 1.580964 |
| SysMyo_M | MGS761 M | 1/126 | 0.42364 | 0.624959 | 0 | 0 | 1.840744 | 1.580964 |
| SysMyo_M | MGS544 H | 1/129 | 0.431176 | 0.624959 | 0 | 0 | 1.797329 | 1.511983 |
| SysMyo_M | MGS407 H | 1/128 | 0.428675 | 0.624959 | 0 | 0 | 1.811573 | 1.534505 |
| SysMyo_M | MGS1228 I | 1/122 | 0.413437 | 0.624959 | 0 | 0 | 1.90198 | 1.679923 |
| SysMyo_M | MGS1477 I | 1/125 | 0.421106 | 0.624959 | 0 | 0 | 1.855683 | 1.604928 |
| SysMyo_M | MGS900 M | 1/124 | 0.418561 | 0.624959 | 0 | 0 | 1.870864 | 1.629399 |
| SysMyo_M | MGS1224 I | 1/124 | 0.418561 | 0.624959 | 0 | 0 | 1.870864 | 1.629399 |
| SysMyo_M | MGS547 H | 1/130 | 0.433666 | 0.624959 | 0 | 0 | 1.783306 | 1.489916 |
| SysMyo_M | MGS1282 I | 1/129 | 0.431176 | 0.624959 | 0 | 0 | 1.797329 | 1.511983 |
| SysMyo_M | MGS1237 I | 1/131 | 0.436146 | 0.624959 | 0 | 0 | 1.769499 | 1.468291 |
| DisGeNET | Neurodeve | 1/208 | 0.598082 | 0.62504 | 0 | 0 | 1.106954 | 0.569005 |
| DisGeNET | Colorectal | 1/208 | 0.598082 | 0.62504 | 0 | 0 | 1.106954 | 0.569005 |
| DisGeNET | Adenoid Cy | 1/208 | 0.598082 | 0.62504 | 0 | 0 | 1.106954 | 0.569005 |
| DisGeNET | Constipatic | 1/208 | 0.598082 | 0.62504 | 0 | 0 | 1.106954 | 0.569005 |
| DisGeNET | Congenital | 1/208 | 0.598082 | 0.62504 | 0 | 0 | 1.106954 | 0.569005 |
| GeneSigDB | 15489886- | 2/353 | 0.455954 | 0.625416 | 0 | 0 | 1.311346 | 1.029883 |
| GeneSigDB | 17150101- | 2/353 | 0.455954 | 0.625416 | 0 | 0 | 1.311346 | 1.029883 |
| GeneSigDB | 17009876- | 6/1245 | 0.459011 | 0.625416 | 0 | 0 | 1.116432 | 0.869344 |

| | | | | | | | | |
|---|---|---|---|---|---|---|---|---|
| GeneSigDB | 20077526-1 | 1/140 | 0.457986 | 0.625416 | 0 | 0 | 1.654174 | 1.291774 |
| GeneSigDB | 20460173- | 1/141 | 0.46036 | 0.625416 | 0 | 0 | 1.642276 | 1.27399 |
| GeneSigDB | 17676974-1 | 2/356 | 0.460312 | 0.625416 | 0 | 0 | 1.300033 | 1.008633 |
| SysMyo_M | MGS1070 I | 1/132 | 0.438615 | 0.626333 | 0 | 0 | 1.755903 | 1.447098 |
| SysMyo_M | MGS476 Hu | 1/133 | 0.441073 | 0.626333 | 0 | 0 | 1.742512 | 1.426323 |
| SysMyo_M | MGS888 M | 1/135 | 0.445958 | 0.626333 | 0 | 0 | 1.716331 | 1.38599 |
| SysMyo_M | MGS559 Hu | 1/134 | 0.443521 | 0.626333 | 0 | 0 | 1.729323 | 1.405958 |
| SysMyo_M | MGS1158 I | 1/134 | 0.443521 | 0.626333 | 0 | 0 | 1.729323 | 1.405958 |
| SysMyo_M | MGS849 M | 1/134 | 0.443521 | 0.626333 | 0 | 0 | 1.729323 | 1.405958 |
| SysMyo_M | MGS1151 I | 1/135 | 0.445958 | 0.626333 | 0 | 0 | 1.716331 | 1.38599 |
| SysMyo_M | MGS562 Hu | 1/135 | 0.445958 | 0.626333 | 0 | 0 | 1.716331 | 1.38599 |
| Jensen_TIS | Parathyroid | 45/10379 | 0.555938 | 0.626616 | 0 | 0 | 0.99315 | 0.583077 |
| Jensen_TIS | Bone | 6/1374 | 0.55656 | 0.626616 | 0 | 0 | 1.004169 | 0.588424 |
| SysMyo_M | MGS926 M | 1/136 | 0.448384 | 0.626631 | 0 | 0 | 1.703531 | 1.36641 |
| SysMyo_M | MGS1084 I | 1/136 | 0.448384 | 0.626631 | 0 | 0 | 1.703531 | 1.36641 |
| DisGeNET | Trisomy | 1/209 | 0.599849 | 0.626636 | 0 | 0 | 1.101576 | 0.562991 |
| KOMP2_M | Increased E | Jan-88 | 0.319191 | 0.626774 | 0 | 0 | 2.649826 | 3.026014 |
| SysMyo_M | MGS1258 I | 1/137 | 0.4508 | 0.626912 | 0 | 0 | 1.69092 | 1.347208 |
| SysMyo_M | MGS1286 I | 1/137 | 0.4508 | 0.626912 | 0 | 0 | 1.69092 | 1.347208 |
| Gene_Pert | TAL1 KD hu | 5/342 | 0.01677 | 0.627237 | 0 | 0 | 3.542013 | 14.4804 |
| Gene_Pert | Kras Knock | 4/227 | 0.017289 | 0.627237 | 0 | 0 | 4.255227 | 17.26642 |
| Gene_Pert | Thrb d337T | 5/343 | 0.016961 | 0.627237 | 0 | 0 | 3.531354 | 14.39677 |
| Jensen_TIS | Neuroblast | 1/187 | 0.559148 | 0.627337 | 0 | 0 | 1.233246 | 0.716937 |
| SysMyo_M | MGS1036 I | 1/150 | 0.481272 | 0.62765 | 0 | 0 | 1.542376 | 1.127974 |
| SysMyo_M | MGS1475 I | 1/140 | 0.457986 | 0.62765 | 0 | 0 | 1.654174 | 1.291774 |
| SysMyo_M | MGS692 M | 1/140 | 0.457986 | 0.62765 | 0 | 0 | 1.654174 | 1.291774 |
| SysMyo_M | MGS1387 I | 1/141 | 0.46036 | 0.62765 | 0 | 0 | 1.642276 | 1.27399 |
| SysMyo_M | MGS1048 I | 1/141 | 0.46036 | 0.62765 | 0 | 0 | 1.642276 | 1.27399 |
| SysMyo_M | MGS903 M | 1/142 | 0.462724 | 0.62765 | 0 | 0 | 1.630546 | 1.256538 |
| SysMyo_M | MGS141 M | 1/142 | 0.462724 | 0.62765 | 0 | 0 | 1.630546 | 1.256538 |
| SysMyo_M | MGS586 Hu | 1/142 | 0.462724 | 0.62765 | 0 | 0 | 1.630546 | 1.256538 |
| SysMyo_M | MGS823 M | 1/139 | 0.455601 | 0.62765 | 0 | 0 | 1.666245 | 1.309899 |
| SysMyo_M | MGS1057 I | 1/147 | 0.474392 | 0.62765 | 0 | 0 | 1.574307 | 1.173995 |
| SysMyo_M | MGS1479 I | 1/148 | 0.476695 | 0.62765 | 0 | 0 | 1.563518 | 1.158376 |
| SysMyo_M | MGS1004 I | 1/138 | 0.453206 | 0.62765 | 0 | 0 | 1.678493 | 1.328374 |
| SysMyo_M | MGS941 M | 1/145 | 0.469755 | 0.62765 | 0 | 0 | 1.596334 | 1.2061 |
| SysMyo_M | MGS902 M | 1/145 | 0.469755 | 0.62765 | 0 | 0 | 1.596334 | 1.2061 |
| SysMyo_M | MGS493 Hu | 1/145 | 0.469755 | 0.62765 | 0 | 0 | 1.596334 | 1.2061 |
| SysMyo_M | MGS1078 I | 1/143 | 0.465078 | 0.62765 | 0 | 0 | 1.618981 | 1.239412 |
| SysMyo_M | MGS694 M | 1/147 | 0.474392 | 0.62765 | 0 | 0 | 1.574307 | 1.173995 |
| SysMyo_M | MGS1035 I | 1/150 | 0.481272 | 0.62765 | 0 | 0 | 1.542376 | 1.127974 |
| SysMyo_M | MGS713 M | 1/150 | 0.481272 | 0.62765 | 0 | 0 | 1.542376 | 1.127974 |
| SysMyo_M | MGS142 M | 1/150 | 0.481272 | 0.62765 | 0 | 0 | 1.542376 | 1.127974 |
| SysMyo_M | MGS835 M | 1/149 | 0.478989 | 0.62765 | 0 | 0 | 1.552876 | 1.143039 |
| SysMyo_M | MGS943 M | 1/149 | 0.478989 | 0.62765 | 0 | 0 | 1.552876 | 1.143039 |

| | | | | | | | | |
|---|---|---|---|---|---|---|---|---|
| SysMyo_M | MGS782 M | 1/147 | 0.474392 | 0.62765 | 0 | 0 | 1.574307 | 1.173995 |
| SysMyo_M | MGS1283 I | 1/148 | 0.476695 | 0.62765 | 0 | 0 | 1.563518 | 1.158376 |
| SysMyo_M | MGS1041 I | 1/146 | 0.472079 | 0.62765 | 0 | 0 | 1.585245 | 1.1899 |
| SysMyo_M | MGS1167 I | 1/146 | 0.472079 | 0.62765 | 0 | 0 | 1.585245 | 1.1899 |
| SysMyo_M | MGS1107 I | 1/145 | 0.469755 | 0.62765 | 0 | 0 | 1.596334 | 1.2061 |
| SysMyo_M | MGS944 M | 1/151 | 0.483545 | 0.627722 | 0 | 0 | 1.532016 | 1.113178 |
| SysMyo_M | MGS990 M | 1/151 | 0.483545 | 0.627722 | 0 | 0 | 1.532016 | 1.113178 |
| SysMyo_M | MGS312 C: | 1/152 | 0.485809 | 0.627781 | 0 | 0 | 1.521793 | 1.098642 |
| SysMyo_M | MGS1284 I | 1/152 | 0.485809 | 0.627781 | 0 | 0 | 1.521793 | 1.098642 |
| Rare_Disea | Dermatom | 1/209 | 0.599849 | 0.627831 | 0 | 0 | 1.101576 | 0.562991 |
| Rare_Disea | Collecting ( | 1/211 | 0.603359 | 0.627831 | 0 | 0 | 1.090975 | 0.551207 |
| Rare_Disea | Papillary re | 1/211 | 0.603359 | 0.627831 | 0 | 0 | 1.090975 | 0.551207 |
| Rare_Disea | Juvenile de | 1/209 | 0.599849 | 0.627831 | 0 | 0 | 1.101576 | 0.562991 |
| Rare_Disea | Autoimmu | 1/209 | 0.599849 | 0.627831 | 0 | 0 | 1.101576 | 0.562991 |
| Rare_Disea | Basilar mig | 1/210 | 0.601608 | 0.627831 | 0 | 0 | 1.09625 | 0.557059 |
| Rare_Disea | Chromophi | 1/211 | 0.603359 | 0.627831 | 0 | 0 | 1.090975 | 0.551207 |
| Rare_Disea | Chromoph | 1/211 | 0.603359 | 0.627831 | 0 | 0 | 1.090975 | 0.551207 |
| Rare_Disea | Clear cell r | 1/211 | 0.603359 | 0.627831 | 0 | 0 | 1.090975 | 0.551207 |
| Rare_Disea | Soft tissue | 1/213 | 0.606839 | 0.627939 | 0 | 0 | 1.080573 | 0.539737 |
| Rare_Disea | Splenomeg | 1/212 | 0.605103 | 0.627939 | 0 | 0 | 1.085749 | 0.545433 |
| Rare_Disea | Epithelioid | 1/213 | 0.606839 | 0.627939 | 0 | 0 | 1.080573 | 0.539737 |
| Rare_Disea | Oral phary | 1/213 | 0.606839 | 0.627939 | 0 | 0 | 1.080573 | 0.539737 |
| Reactome_ | Beta-caten | 1/142 | 0.462724 | 0.628183 | 0 | 0 | 1.630546 | 1.256538 |
| DisGeNET | Malignant | 6/1438 | 0.602029 | 0.628663 | 0 | 0 | 0.95598 | 0.485112 |
| Jensen_TIS | Cervical ca | 21/4876 | 0.562338 | 0.628725 | 0 | 0 | 0.986855 | 0.568085 |
| GeDiPNet_ | Disorder O | 1/212 | 0.605103 | 0.628833 | 0 | 0 | 1.085749 | 0.545433 |
| Rare_Disea | Polymyosit | 1/214 | 0.608568 | 0.628853 | 0 | 0 | 1.075445 | 0.534116 |
| Rare_Disea | Hepatoblas | 1/215 | 0.610289 | 0.629757 | 0 | 0 | 1.070365 | 0.528571 |
| GeneSigDB | 19168792- | 1/143 | 0.465078 | 0.630288 | 0 | 0 | 1.618981 | 1.239412 |
| Rare_Disea | Gonadal dy | 1/216 | 0.612003 | 0.630651 | 0 | 0 | 1.065333 | 0.523098 |
| SysMyo_M | MGS1168 I | 1/154 | 0.490307 | 0.630713 | 0 | 0 | 1.501748 | 1.070331 |
| SysMyo_M | MGS346 H | 1/154 | 0.490307 | 0.630713 | 0 | 0 | 1.501748 | 1.070331 |
| SysMyo_M | MGS901 M | 1/155 | 0.492541 | 0.63072 | 0 | 0 | 1.491921 | 1.056544 |
| SysMyo_M | MGS942 M | 1/155 | 0.492541 | 0.63072 | 0 | 0 | 1.491921 | 1.056544 |
| Gene_Pert | BST2 OE hu | 5/347 | 0.017741 | 0.631013 | 0 | 0 | 3.489338 | 14.06864 |
| Disease_Pe | idiopathic | 4/312 | 0.047249 | 0.631954 | 0 | 0 | 3.067595 | 9.3633 |
| GeneSigDB | 19168792- | 4/810 | 0.470982 | 0.632086 | 0 | 0 | 1.142456 | 0.860195 |
| GeneSigDB | 19429869- | 2/364 | 0.471835 | 0.632086 | 0 | 0 | 1.270783 | 0.954519 |
| GeneSigDB | 17389037- | 1/146 | 0.472079 | 0.632086 | 0 | 0 | 1.585245 | 1.1899 |
| GeneSigDB | 11823860- | 2/362 | 0.468968 | 0.632086 | 0 | 0 | 1.277974 | 0.96771 |
| GeneSigDB | 12734205- | 2/362 | 0.468968 | 0.632086 | 0 | 0 | 1.277974 | 0.96771 |
| KOMP2_M | Hyperplasi | Jan-91 | 0.328076 | 0.632718 | 0 | 0 | 2.561111 | 2.854384 |
| GTEx_Tissu | GTEX-XYKS | 6/609 | 0.049559 | 0.634385 | 0 | 0 | 2.37209 | 7.127183 |
| GTEx_Tissu | GTEX-X4EP | 12/1632 | 0.049777 | 0.634385 | 0 | 0 | 1.806716 | 5.420506 |
| GTEx_Tissu | GTEX-TML8 | 14/2000 | 0.049654 | 0.634385 | 0 | 0 | 1.731145 | 5.198057 |

| | | | | | | | | |
|---|---|---|---|---|---|---|---|---|
| SysMyo_M | MGS844 M | 1/162 | 0.507913 | 0.634506 | 0 | 0 | 1.426549 | 0.966408 |
| SysMyo_M | MGS741 M | 1/163 | 0.510071 | 0.634506 | 0 | 0 | 1.417672 | 0.954384 |
| SysMyo_M | MGS1269 I | 1/163 | 0.510071 | 0.634506 | 0 | 0 | 1.417672 | 0.954384 |
| SysMyo_M | MGS585 Hu | 1/158 | 0.499187 | 0.634506 | 0 | 0 | 1.463191 | 1.016589 |
| SysMyo_M | MGS47 Hu | 1/157 | 0.496981 | 0.634506 | 0 | 0 | 1.472645 | 1.029678 |
| SysMyo_M | MGS1388 I | 1/159 | 0.501382 | 0.634506 | 0 | 0 | 1.453856 | 1.003722 |
| SysMyo_M | MGS1176 I | 1/163 | 0.510071 | 0.634506 | 0 | 0 | 1.417672 | 0.954384 |
| SysMyo_M | MGS501 Hu | 1/159 | 0.501382 | 0.634506 | 0 | 0 | 1.453856 | 1.003722 |
| SysMyo_M | MGS189 M | 1/161 | 0.505746 | 0.634506 | 0 | 0 | 1.435538 | 0.978637 |
| SysMyo_M | MGS1044 I | 1/161 | 0.505746 | 0.634506 | 0 | 0 | 1.435538 | 0.978637 |
| SysMyo_M | MGS1505 I | 1/162 | 0.507913 | 0.634506 | 0 | 0 | 1.426549 | 0.966408 |
| SysMyo_M | MGS144 M | 1/161 | 0.505746 | 0.634506 | 0 | 0 | 1.435538 | 0.978637 |
| SysMyo_M | MGS691 M | 1/160 | 0.503569 | 0.634506 | 0 | 0 | 1.444639 | 0.991073 |
| Gene_Pert | Adrb2 KO r | 5/312 | 0.011682 | 0.634741 | 0 | 0 | 3.894097 | 17.32747 |
| GeneSigDB | 18180321- | 1/149 | 0.478989 | 0.63523 | 0 | 0 | 1.552876 | 1.143039 |
| GeneSigDB | 17344918- | 1/149 | 0.478989 | 0.63523 | 0 | 0 | 1.552876 | 1.143039 |
| GeneSigDB | 16818684- | 1/149 | 0.478989 | 0.63523 | 0 | 0 | 1.552876 | 1.143039 |
| GeneSigDB | 18338247- | 1/149 | 0.478989 | 0.63523 | 0 | 0 | 1.552876 | 1.143039 |
| GeneSigDB | 18497862- | 1/150 | 0.481272 | 0.635233 | 0 | 0 | 1.542376 | 1.127974 |
| GeneSigDB | 19061838- | 1/150 | 0.481272 | 0.635233 | 0 | 0 | 1.542376 | 1.127974 |
| DisGeNET | cervical car | 4/965 | 0.610152 | 0.636892 | 0 | 0 | 0.950416 | 0.469549 |
| SysMyo_M | MGS434 Hu | 1/165 | 0.514359 | 0.63704 | 0 | 0 | 1.400241 | 0.930927 |
| SysMyo_M | MGS967 M | 1/165 | 0.514359 | 0.63704 | 0 | 0 | 1.400241 | 0.930927 |
| GeDiPNet_ | Colorectal | 2/474 | 0.614429 | 0.637269 | 0 | 0 | 0.969143 | 0.472033 |
| GeDiPNet_ | Congenital | 1/218 | 0.615408 | 0.637269 | 0 | 0 | 1.055407 | 0.512369 |
| DisGeNET | Sensorineu | 2/471 | 0.610951 | 0.637472 | 0 | 0 | 0.975492 | 0.480663 |
| GWAS_Cat | Major Depr | 1/226 | 0.628735 | 0.63762 | 0 | 0 | 1.017468 | 0.472152 |
| KEGG_202 | mTOR signa | 2/154 | 0.144717 | 0.638016 | 0 | 0 | 3.058978 | 5.912923 |
| KEGG_202 | Pathways c | 4/475 | 0.152522 | 0.638016 | 0 | 0 | 1.989308 | 3.740783 |
| KEGG_202 | Cholinergic | 1/113 | 0.389825 | 0.638016 | 0 | 0 | 2.055752 | 1.936638 |
| KEGG_202 | Wnt signali | 2/166 | 0.162892 | 0.638016 | 0 | 0 | 2.833429 | 5.141737 |
| KEGG_202 | Cellular ser | 1/156 | 0.494766 | 0.638016 | 0 | 0 | 1.482221 | 1.042994 |
| KEGG_202 | Hepatitis C | 1/157 | 0.496981 | 0.638016 | 0 | 0 | 1.472645 | 1.029678 |
| KEGG_202 | Hepatocell | 2/168 | 0.165961 | 0.638016 | 0 | 0 | 2.799008 | 5.027029 |
| KEGG_202 | Antigen pro | Jan-78 | 0.288726 | 0.638016 | 0 | 0 | 2.99547 | 3.721207 |
| KEGG_202 | Neutrophil | 2/189 | 0.198732 | 0.638016 | 0 | 0 | 2.482038 | 4.010467 |
| KEGG_202 | Glycerolipi | Jan-61 | 0.233813 | 0.638016 | 0 | 0 | 3.847481 | 5.591291 |
| KEGG_202 | Tuberculos | 2/180 | 0.184577 | 0.638016 | 0 | 0 | 2.608724 | 4.407929 |
| KEGG_202 | RNA transp | 2/186 | 0.193998 | 0.638016 | 0 | 0 | 2.52289 | 4.137304 |
| KEGG_202 | Pathways i | 3/531 | 0.407833 | 0.638016 | 0 | 0 | 1.311215 | 1.176024 |
| KEGG_202 | Human pap | 2/331 | 0.423413 | 0.638016 | 0 | 0 | 1.400608 | 1.203693 |
| KEGG_202 | Mitophagy | Jan-68 | 0.256914 | 0.638016 | 0 | 0 | 3.44429 | 4.68084 |
| KEGG_202 | Aminoacyl- | Jan-66 | 0.250385 | 0.638016 | 0 | 0 | 3.550626 | 4.916752 |
| KEGG_202 | Retrograde | 1/148 | 0.476695 | 0.638016 | 0 | 0 | 1.563518 | 1.158376 |
| KEGG_202 | Gastric can | 1/149 | 0.478989 | 0.638016 | 0 | 0 | 1.552876 | 1.143039 |

| | | | | | | | | |
|---|---|---|---|---|---|---|---|---|
| KEGG_202: | Central car | Jan-70 | 0.263387 | 0.638016 | 0 | 0 | 3.344119 | 4.461494 |
| KEGG_202: | Phospholip | 1/148 | 0.476695 | 0.638016 | 0 | 0 | 1.563518 | 1.158376 |
| KEGG_202: | Alzheimer ( | 3/369 | 0.216772 | 0.638016 | 0 | 0 | 1.907397 | 2.916234 |
| KEGG_202: | Endometria | Jan-58 | 0.223696 | 0.638016 | 0 | 0 | 4.050592 | 6.065622 |
| KEGG_202: | Prostate ca | Jan-97 | 0.345504 | 0.638016 | 0 | 0 | 2.400315 | 2.550937 |
| KEGG_202: | Basal cell c | Jan-63 | 0.240485 | 0.638016 | 0 | 0 | 3.722993 | 5.305634 |
| KEGG_202: | Breast canc | 2/147 | 0.134331 | 0.638016 | 0 | 0 | 3.207789 | 6.43948 |
| KEGG_202: | Parkinson ( | 2/249 | 0.295039 | 0.638016 | 0 | 0 | 1.873398 | 2.286757 |
| KEGG_202: | Non-alcoho | 1/155 | 0.492541 | 0.638016 | 0 | 0 | 1.491921 | 1.056544 |
| KEGG_202: | Cushing syı | 1/155 | 0.492541 | 0.638016 | 0 | 0 | 1.491921 | 1.056544 |
| KEGG_202: | Gap junctic | Jan-88 | 0.319191 | 0.638016 | 0 | 0 | 2.649826 | 3.026014 |
| KEGG_202: | PD-L1 expr | Jan-89 | 0.322165 | 0.638016 | 0 | 0 | 2.619582 | 2.967177 |
| KEGG_202: | Glycosamir | Jan-53 | 0.206541 | 0.638016 | 0 | 0 | 4.44119 | 7.004886 |
| KEGG_202: | GABAergic | Jan-89 | 0.322165 | 0.638016 | 0 | 0 | 2.619582 | 2.967177 |
| KEGG_202: | Amyotroph | 2/364 | 0.471835 | 0.638016 | 0 | 0 | 1.270783 | 0.954519 |
| KEGG_202: | Cell adhesi | 1/148 | 0.476695 | 0.638016 | 0 | 0 | 1.563518 | 1.158376 |
| KEGG_202: | Relaxin sigı | 1/129 | 0.431176 | 0.638016 | 0 | 0 | 1.797329 | 1.511983 |
| KEGG_202: | FoxO signa | 1/131 | 0.436146 | 0.638016 | 0 | 0 | 1.769499 | 1.468291 |
| KEGG_202: | Morphine a | Jan-91 | 0.328076 | 0.638016 | 0 | 0 | 2.561111 | 2.854384 |
| KEGG_202: | Small cell lı | Jan-92 | 0.331012 | 0.638016 | 0 | 0 | 2.532839 | 2.800307 |
| KEGG_202: | Circadian e | Jan-97 | 0.345504 | 0.638016 | 0 | 0 | 2.400315 | 2.550937 |
| KEGG_202: | Choline me | Jan-98 | 0.348365 | 0.638016 | 0 | 0 | 2.37545 | 2.504922 |
| KEGG_202: | Glyceropho | Jan-98 | 0.348365 | 0.638016 | 0 | 0 | 2.37545 | 2.504922 |
| KEGG_202: | Melanoger | 1/101 | 0.356874 | 0.638016 | 0 | 0 | 2.303837 | 2.373811 |
| KEGG_202: | Insulin resi | 1/108 | 0.376303 | 0.638016 | 0 | 0 | 2.152358 | 2.10363 |
| KEGG_202: | Melanoma | Jan-72 | 0.269804 | 0.638016 | 0 | 0 | 3.249591 | 4.257158 |
| KEGG_202: | p53 signalii | Jan-73 | 0.272992 | 0.638016 | 0 | 0 | 3.204296 | 4.160181 |
| KEGG_202: | Glioma | Jan-75 | 0.279326 | 0.638016 | 0 | 0 | 3.117379 | 3.975827 |
| KEGG_202: | Transcripti | 2/192 | 0.203481 | 0.638016 | 0 | 0 | 2.442477 | 3.888868 |
| KEGG_202: | Glutamater | 1/114 | 0.392494 | 0.638016 | 0 | 0 | 2.037456 | 1.905499 |
| KEGG_202: | Sphingolipi | 1/119 | 0.405668 | 0.638016 | 0 | 0 | 1.950631 | 1.759896 |
| KEGG_202: | PI3K-Akt si | 2/354 | 0.457409 | 0.638016 | 0 | 0 | 1.307553 | 1.02274 |
| KEGG_202: | Dopaminer | 1/132 | 0.438615 | 0.638016 | 0 | 0 | 1.755903 | 1.447098 |
| KEGG_202: | Oxidative p | 1/133 | 0.441073 | 0.638016 | 0 | 0 | 1.742512 | 1.426323 |
| KEGG_202: | Apelin sign | 1/137 | 0.4508 | 0.638016 | 0 | 0 | 1.69092 | 1.347208 |
| KEGG_202: | Serotonerg | 1/113 | 0.389825 | 0.638016 | 0 | 0 | 2.055752 | 1.936638 |
| KEGG_202: | Signaling p | 1/143 | 0.465078 | 0.638016 | 0 | 0 | 1.618981 | 1.239412 |
| DisGeNET | Adenocarci | 1/216 | 0.612003 | 0.638061 | 0 | 0 | 1.065333 | 0.523098 |
| DisGeNET | Respiratory | 1/216 | 0.612003 | 0.638061 | 0 | 0 | 1.065333 | 0.523098 |
| SysMyo_M | MGS560 Hı | 1/166 | 0.516489 | 0.638282 | 0 | 0 | 1.391684 | 0.919487 |
| Disease_Si | Cardiac Hyp | 2/300 | 0.375985 | 0.638578 | 0 | 0 | 1.548756 | 1.515003 |
| Disease_Si | Alzheimer's | 2/300 | 0.375985 | 0.638578 | 0 | 0 | 1.548756 | 1.515003 |
| Disease_Si | Barrett Eso | 2/300 | 0.375985 | 0.638578 | 0 | 0 | 1.548756 | 1.515003 |
| Disease_Si | Bipolar Dis | 2/300 | 0.375985 | 0.638578 | 0 | 0 | 1.548756 | 1.515003 |
| Disease_Si | Huntingtor | 2/300 | 0.375985 | 0.638578 | 0 | 0 | 1.548756 | 1.515003 |

| | | | | | | | | |
|---|---|---|---|---|---|---|---|---|
| Disease_Si | Arrhythmo | 2/300 | 0.375985 | 0.638578 | 0 | 0 | 1.548756 | 1.515003 |
| Disease_Si | Leukemia, . | 2/300 | 0.375985 | 0.638578 | 0 | 0 | 1.548756 | 1.515003 |
| Disease_Si | Leukemia, . | 2/300 | 0.375985 | 0.638578 | 0 | 0 | 1.548756 | 1.515003 |
| Disease_Si | Dermatom | 2/300 | 0.375985 | 0.638578 | 0 | 0 | 1.548756 | 1.515003 |
| Disease_Si | Eczema GS | 2/300 | 0.375985 | 0.638578 | 0 | 0 | 1.548756 | 1.515003 |
| Disease_Si | Endometri | 2/300 | 0.375985 | 0.638578 | 0 | 0 | 1.548756 | 1.515003 |
| Disease_Si | Dermatom | 2/300 | 0.375985 | 0.638578 | 0 | 0 | 1.548756 | 1.515003 |
| Disease_Si | Carcinoma, | 2/300 | 0.375985 | 0.638578 | 0 | 0 | 1.548756 | 1.515003 |
| Disease_Si | Cancer of C | 2/300 | 0.375985 | 0.638578 | 0 | 0 | 1.548756 | 1.515003 |
| Disease_Si | COPD - Chr | 2/300 | 0.375985 | 0.638578 | 0 | 0 | 1.548756 | 1.515003 |
| Disease_Si | CML - Chro | 2/300 | 0.375985 | 0.638578 | 0 | 0 | 1.548756 | 1.515003 |
| Disease_Si | Nephroblas | 2/300 | 0.375985 | 0.638578 | 0 | 0 | 1.548756 | 1.515003 |
| Disease_Si | Familial co | 2/300 | 0.375985 | 0.638578 | 0 | 0 | 1.548756 | 1.515003 |
| Disease_Si | Viral cardic | 2/300 | 0.375985 | 0.638578 | 0 | 0 | 1.548756 | 1.515003 |
| Disease_Si | Malignant | 2/300 | 0.375985 | 0.638578 | 0 | 0 | 1.548756 | 1.515003 |
| Disease_Si | Huntingtor | 2/300 | 0.375985 | 0.638578 | 0 | 0 | 1.548756 | 1.515003 |
| Disease_Si | Tachycardi | 2/300 | 0.375985 | 0.638578 | 0 | 0 | 1.548756 | 1.515003 |
| Disease_Si | Premature | 2/300 | 0.375985 | 0.638578 | 0 | 0 | 1.548756 | 1.515003 |
| Disease_Si | MDS - Mye | 2/300 | 0.375985 | 0.638578 | 0 | 0 | 1.548756 | 1.515003 |
| Disease_Si | Parkinson's | 2/300 | 0.375985 | 0.638578 | 0 | 0 | 1.548756 | 1.515003 |
| Disease_Si | Papillary Ca | 2/300 | 0.375985 | 0.638578 | 0 | 0 | 1.548756 | 1.515003 |
| Disease_Si | Cardiomyo | 2/300 | 0.375985 | 0.638578 | 0 | 0 | 1.548756 | 1.515003 |
| Disease_Si | Cardiomyo | 2/300 | 0.375985 | 0.638578 | 0 | 0 | 1.548756 | 1.515003 |
| Disease_Si | Multiple be | 2/300 | 0.375985 | 0.638578 | 0 | 0 | 1.548756 | 1.515003 |
| Disease_Si | Osteoarthr | 2/300 | 0.375985 | 0.638578 | 0 | 0 | 1.548756 | 1.515003 |
| Disease_Si | Obstructive | 2/300 | 0.375985 | 0.638578 | 0 | 0 | 1.548756 | 1.515003 |
| Disease_Si | Neurologic | 2/300 | 0.375985 | 0.638578 | 0 | 0 | 1.548756 | 1.515003 |
| Disease_Si | Monoclona | 2/300 | 0.375985 | 0.638578 | 0 | 0 | 1.548756 | 1.515003 |
| Disease_Si | MS (Multip | 2/300 | 0.375985 | 0.638578 | 0 | 0 | 1.548756 | 1.515003 |
| Disease_Si | Uterine lei | 2/300 | 0.375985 | 0.638578 | 0 | 0 | 1.548756 | 1.515003 |
| SysMyo_M | MGS191 M | 1/171 | 0.527002 | 0.638722 | 0 | 0 | 1.35041 | 0.865006 |
| SysMyo_M | MGS831 M | 1/171 | 0.527002 | 0.638722 | 0 | 0 | 1.35041 | 0.865006 |
| SysMyo_M | MGS1420 I | 1/167 | 0.51861 | 0.638722 | 0 | 0 | 1.383231 | 0.908233 |
| SysMyo_M | MGS271 M | 1/171 | 0.527002 | 0.638722 | 0 | 0 | 1.35041 | 0.865006 |
| SysMyo_M | MGS1281 I | 1/171 | 0.527002 | 0.638722 | 0 | 0 | 1.35041 | 0.865006 |
| SysMyo_M | MGS707 M | 1/170 | 0.524918 | 0.638722 | 0 | 0 | 1.35847 | 0.875552 |
| SysMyo_M | MGS1445 I | 1/169 | 0.522824 | 0.638722 | 0 | 0 | 1.366625 | 0.886269 |
| SysMyo_M | MGS815 M | 1/169 | 0.522824 | 0.638722 | 0 | 0 | 1.366625 | 0.886269 |
| SysMyo_M | MGS721 M | 1/169 | 0.522824 | 0.638722 | 0 | 0 | 1.366625 | 0.886269 |
| GTEx_Tissu | GTEX-XUJ4 | 9/1104 | 0.050384 | 0.639299 | 0 | 0 | 1.982929 | 5.925164 |
| GTEx_Tissu | GTEX-T6MI | 11/1457 | 0.050842 | 0.639503 | 0 | 0 | 1.848448 | 5.50659 |
| GTEx_Tissu | GTEX-WRH | Feb-83 | 0.050736 | 0.639503 | 0 | 0 | 5.76093 | 17.17404 |
| DisGeNET | Severe Con | 1/217 | 0.613709 | 0.639585 | 0 | 0 | 1.060347 | 0.517698 |
| SysMyo_M | MGS751 M | 1/172 | 0.529077 | 0.639867 | 0 | 0 | 1.342445 | 0.854628 |
| UK_Bioban | Left 6mm a | 1/222 | 0.622129 | 0.640097 | 0 | 0 | 1.036094 | 0.491738 |

| | | | | | | | | |
|---|---|---|---|---|---|---|---|---|
| UK_Bioban | Basophill p | 1/221 | 0.62046 | 0.640097 | 0 | 0 | 1.040856 | 0.496795 |
| Reactome_ | RHOA GTPa | 1/147 | 0.474392 | 0.640143 | 0 | 0 | 1.574307 | 1.173995 |
| DisGeNET | Carcinogen | 17/4065 | 0.61468 | 0.640343 | 0 | 0 | 0.95181 | 0.463201 |
| GWAS_Cat | Triglyceride | 1/228 | 0.631995 | 0.640393 | 0 | 0 | 1.008401 | 0.462729 |
| Diabetes_P | GSE136396 | 1/201 | 0.585497 | 0.640399 | 0 | 0 | 1.146105 | 0.613503 |
| Diabetes_P | GSE149662 | 1/202 | 0.587319 | 0.640399 | 0 | 0 | 1.140345 | 0.606877 |
| Diabetes_P | GSE73036 | 1/203 | 0.589132 | 0.640399 | 0 | 0 | 1.134642 | 0.600344 |
| Diabetes_P | GSE109285 | 1/203 | 0.589132 | 0.640399 | 0 | 0 | 1.134642 | 0.600344 |
| Diabetes_P | GSE138252 | 1/204 | 0.590938 | 0.640399 | 0 | 0 | 1.128995 | 0.593902 |
| Diabetes_P | GSE125856 | 1/198 | 0.579985 | 0.640399 | 0 | 0 | 1.163735 | 0.633948 |
| Diabetes_P | GSE70681 | 1/120 | 0.408269 | 0.640399 | 0 | 0 | 1.934141 | 1.732658 |
| Diabetes_P | GSE125856 | 1/125 | 0.421106 | 0.640399 | 0 | 0 | 1.855683 | 1.604928 |
| Diabetes_P | GSE67991 | 1/125 | 0.421106 | 0.640399 | 0 | 0 | 1.855683 | 1.604928 |
| Diabetes_P | GSE70681 | 1/159 | 0.501382 | 0.640399 | 0 | 0 | 1.453856 | 1.003722 |
| Diabetes_P | GSE107894 | 1/159 | 0.501382 | 0.640399 | 0 | 0 | 1.453856 | 1.003722 |
| Diabetes_P | GSE89035 | 1/151 | 0.483545 | 0.640399 | 0 | 0 | 1.532016 | 1.113178 |
| Diabetes_P | GSE108971 | 1/133 | 0.441073 | 0.640399 | 0 | 0 | 1.742512 | 1.426323 |
| Diabetes_P | GSE140230 | 1/134 | 0.443521 | 0.640399 | 0 | 0 | 1.729323 | 1.405958 |
| Diabetes_P | GSE120290 | 1/136 | 0.448384 | 0.640399 | 0 | 0 | 1.703531 | 1.36641 |
| Diabetes_P | GSE94846 | 1/121 | 0.410859 | 0.640399 | 0 | 0 | 1.917926 | 1.706006 |
| Diabetes_P | GSE149468 | 1/167 | 0.51861 | 0.640399 | 0 | 0 | 1.383231 | 0.908233 |
| Diabetes_P | GSE115421 | 1/168 | 0.520722 | 0.640399 | 0 | 0 | 1.374878 | 0.897162 |
| Diabetes_P | GSE115421 | 1/159 | 0.501382 | 0.640399 | 0 | 0 | 1.453856 | 1.003722 |
| Diabetes_P | GSE114192 | 1/160 | 0.503569 | 0.640399 | 0 | 0 | 1.444639 | 0.991073 |
| Diabetes_P | GSE90531 | 1/160 | 0.503569 | 0.640399 | 0 | 0 | 1.444639 | 0.991073 |
| Diabetes_P | GSE153222 | 1/161 | 0.505746 | 0.640399 | 0 | 0 | 1.435538 | 0.978637 |
| Diabetes_P | GSE137187 | 1/164 | 0.51222 | 0.640399 | 0 | 0 | 1.408903 | 0.942558 |
| Diabetes_P | GSE115421 | 1/212 | 0.605103 | 0.640399 | 0 | 0 | 1.085749 | 0.545433 |
| Diabetes_P | GSE127251 | 1/213 | 0.606839 | 0.640399 | 0 | 0 | 1.080573 | 0.539737 |
| Diabetes_P | GSE138782 | 1/206 | 0.594525 | 0.640399 | 0 | 0 | 1.117867 | 0.581282 |
| Diabetes_P | GSE26978 | 1/206 | 0.594525 | 0.640399 | 0 | 0 | 1.117867 | 0.581282 |
| Diabetes_P | GSE135391 | 1/207 | 0.596308 | 0.640399 | 0 | 0 | 1.112384 | 0.575101 |
| Diabetes_P | GSE59141 | 1/183 | 0.55132 | 0.640399 | 0 | 0 | 1.260606 | 0.750615 |
| Diabetes_P | GSE149468 | 1/183 | 0.55132 | 0.640399 | 0 | 0 | 1.260606 | 0.750615 |
| Diabetes_P | GSE149468 | 1/183 | 0.55132 | 0.640399 | 0 | 0 | 1.260606 | 0.750615 |
| Diabetes_P | GSE59141 | 1/184 | 0.55329 | 0.640399 | 0 | 0 | 1.253654 | 0.742005 |
| Diabetes_P | GSE115421 | 1/210 | 0.601608 | 0.640399 | 0 | 0 | 1.09625 | 0.557059 |
| Diabetes_P | GSE43106 | 1/204 | 0.590938 | 0.640399 | 0 | 0 | 1.128995 | 0.593902 |
| Diabetes_P | GSE131320 | 1/204 | 0.590938 | 0.640399 | 0 | 0 | 1.128995 | 0.593902 |
| Diabetes_P | GSE138252 | 1/204 | 0.590938 | 0.640399 | 0 | 0 | 1.128995 | 0.593902 |
| Diabetes_P | GSE115421 | 1/204 | 0.590938 | 0.640399 | 0 | 0 | 1.128995 | 0.593902 |
| Diabetes_P | GSE127251 | 1/204 | 0.590938 | 0.640399 | 0 | 0 | 1.128995 | 0.593902 |
| Diabetes_P | GSE151030 | 1/125 | 0.421106 | 0.640399 | 0 | 0 | 1.855683 | 1.604928 |
| Diabetes_P | GSE153222 | 1/151 | 0.483545 | 0.640399 | 0 | 0 | 1.532016 | 1.113178 |
| Diabetes_P | GSE149468 | 1/151 | 0.483545 | 0.640399 | 0 | 0 | 1.532016 | 1.113178 |

| | | | | | | | | |
|---|---|---|---|---|---|---|---|---|
| Diabetes_F | GSE143482 | 1/111 | 0.384451 | 0.640399 | 0 | 0 | 2.09334 | 2.001105 |
| Diabetes_F | GSE150679 | 1/164 | 0.51222 | 0.640399 | 0 | 0 | 1.408903 | 0.942558 |
| Diabetes_F | GSE86949 | 1/166 | 0.516489 | 0.640399 | 0 | 0 | 1.391684 | 0.919487 |
| Diabetes_F | GSE149468 | 1/169 | 0.522824 | 0.640399 | 0 | 0 | 1.366625 | 0.886269 |
| Diabetes_F | GSE70681 | 1/132 | 0.438615 | 0.640399 | 0 | 0 | 1.755903 | 1.447098 |
| Diabetes_F | GSE101537 | 1/133 | 0.441073 | 0.640399 | 0 | 0 | 1.742512 | 1.426323 |
| Diabetes_F | GSE123658 | 1/133 | 0.441073 | 0.640399 | 0 | 0 | 1.742512 | 1.426323 |
| Diabetes_F | GSE114192 | 1/169 | 0.522824 | 0.640399 | 0 | 0 | 1.366625 | 0.886269 |
| Diabetes_F | GSE153222 | 1/170 | 0.524918 | 0.640399 | 0 | 0 | 1.35847 | 0.875552 |
| Diabetes_F | GSE101207 | 1/170 | 0.524918 | 0.640399 | 0 | 0 | 1.35847 | 0.875552 |
| Diabetes_F | GSE152576 | 1/163 | 0.510071 | 0.640399 | 0 | 0 | 1.417672 | 0.954384 |
| Diabetes_F | GSE120290 | 1/163 | 0.510071 | 0.640399 | 0 | 0 | 1.417672 | 0.954384 |
| Diabetes_F | GSE26978 | 1/122 | 0.413437 | 0.640399 | 0 | 0 | 1.90198 | 1.679923 |
| Diabetes_F | GSE149468 | 1/122 | 0.413437 | 0.640399 | 0 | 0 | 1.90198 | 1.679923 |
| Diabetes_F | GSE149468 | 1/122 | 0.413437 | 0.640399 | 0 | 0 | 1.90198 | 1.679923 |
| Diabetes_F | GSE134870 | 1/142 | 0.462724 | 0.640399 | 0 | 0 | 1.630546 | 1.256538 |
| Diabetes_F | GSE149294 | 1/143 | 0.465078 | 0.640399 | 0 | 0 | 1.618981 | 1.239412 |
| Diabetes_F | GSE114192 | 1/145 | 0.469755 | 0.640399 | 0 | 0 | 1.596334 | 1.2061 |
| Diabetes_F | GSE153222 | 1/130 | 0.433666 | 0.640399 | 0 | 0 | 1.783306 | 1.489916 |
| Diabetes_F | GSE153222 | 1/130 | 0.433666 | 0.640399 | 0 | 0 | 1.783306 | 1.489916 |
| Diabetes_F | GSE149468 | 1/170 | 0.524918 | 0.640399 | 0 | 0 | 1.35847 | 0.875552 |
| Diabetes_F | GSE114192 | 1/179 | 0.543355 | 0.640399 | 0 | 0 | 1.289195 | 0.7864 |
| Diabetes_F | GSE87354 | 1/180 | 0.545359 | 0.640399 | 0 | 0 | 1.281928 | 0.777247 |
| Diabetes_F | GSE106295 | 1/180 | 0.545359 | 0.640399 | 0 | 0 | 1.281928 | 0.777247 |
| Diabetes_F | GSE149468 | 1/180 | 0.545359 | 0.640399 | 0 | 0 | 1.281928 | 0.777247 |
| Diabetes_F | GSE157585 | 1/181 | 0.547354 | 0.640399 | 0 | 0 | 1.274742 | 0.768234 |
| Diabetes_F | GSE149468 | 1/181 | 0.547354 | 0.640399 | 0 | 0 | 1.274742 | 0.768234 |
| Diabetes_F | GSE80780 | 1/183 | 0.55132 | 0.640399 | 0 | 0 | 1.260606 | 0.750615 |
| Diabetes_F | GSE112213 | 1/175 | 0.535249 | 0.640399 | 0 | 0 | 1.319099 | 0.824467 |
| Diabetes_F | GSE144441 | 1/226 | 0.628735 | 0.640399 | 0 | 0 | 1.017468 | 0.472152 |
| Diabetes_F | GSE131320 | 1/226 | 0.628735 | 0.640399 | 0 | 0 | 1.017468 | 0.472152 |
| Diabetes_F | GSE144441 | 1/227 | 0.630368 | 0.640399 | 0 | 0 | 1.012914 | 0.46741 |
| Diabetes_F | GSE131320 | 1/230 | 0.635226 | 0.640399 | 0 | 0 | 0.999492 | 0.453544 |
| Diabetes_F | GSE73433 | 1/218 | 0.615408 | 0.640399 | 0 | 0 | 1.055407 | 0.512369 |
| Diabetes_F | GSE137631 | 1/218 | 0.615408 | 0.640399 | 0 | 0 | 1.055407 | 0.512369 |
| Diabetes_F | GSE137136 | 1/218 | 0.615408 | 0.640399 | 0 | 0 | 1.055407 | 0.512369 |
| Diabetes_F | GSE44035 | 1/219 | 0.617099 | 0.640399 | 0 | 0 | 1.050512 | 0.507109 |
| Diabetes_F | GSE136134 | 1/231 | 0.636831 | 0.640399 | 0 | 0 | 0.995096 | 0.449038 |
| Diabetes_F | GSE70681 | 1/224 | 0.625446 | 0.640399 | 0 | 0 | 1.026697 | 0.481818 |
| Diabetes_F | GSE133903 | 1/213 | 0.606839 | 0.640399 | 0 | 0 | 1.080573 | 0.539737 |
| Diabetes_F | GSE142209 | 1/214 | 0.608568 | 0.640399 | 0 | 0 | 1.075445 | 0.534116 |
| Diabetes_F | GSE109285 | 1/214 | 0.608568 | 0.640399 | 0 | 0 | 1.075445 | 0.534116 |
| Diabetes_F | GSE135391 | 1/214 | 0.608568 | 0.640399 | 0 | 0 | 1.075445 | 0.534116 |
| Diabetes_F | GSE149468 | 1/227 | 0.630368 | 0.640399 | 0 | 0 | 1.012914 | 0.46741 |
| Diabetes_F | GSE124774 | 1/228 | 0.631995 | 0.640399 | 0 | 0 | 1.008401 | 0.462729 |

| | | | | | | | | |
|---|---|---|---|---|---|---|---|---|
| Diabetes_F | GSE13132( | 1/229 | 0.633614 | 0.640399 | 0 | 0 | 1.003927 | 0.458108 |
| Diabetes_F | GSE156067 | 1/211 | 0.603359 | 0.640399 | 0 | 0 | 1.090975 | 0.551207 |
| Diabetes_F | GSE94846 | 1/176 | 0.537289 | 0.640399 | 0 | 0 | 1.311495 | 0.814726 |
| Diabetes_F | GSE14946[illegible] | 1/176 | 0.537289 | 0.640399 | 0 | 0 | 1.311495 | 0.814726 |
| Diabetes_F | GSE40439 | 1/176 | 0.537289 | 0.640399 | 0 | 0 | 1.311495 | 0.814726 |
| Diabetes_F | GSE14095[illegible] | 1/176 | 0.537289 | 0.640399 | 0 | 0 | 1.311495 | 0.814726 |
| Diabetes_F | GSE15758[illegible] | 1/178 | 0.541341 | 0.640399 | 0 | 0 | 1.296544 | 0.795696 |
| Diabetes_F | GSE26978 | 1/178 | 0.541341 | 0.640399 | 0 | 0 | 1.296544 | 0.795696 |
| Diabetes_F | GSE15322[illegible] | 1/179 | 0.543355 | 0.640399 | 0 | 0 | 1.289195 | 0.7864 |
| Diabetes_F | GSE15758[illegible] | 1/172 | 0.529077 | 0.640399 | 0 | 0 | 1.342445 | 0.854628 |
| Diabetes_F | GSE10629[illegible] | 1/174 | 0.533201 | 0.640399 | 0 | 0 | 1.326791 | 0.834362 |
| Diabetes_F | GSE14946[illegible] | 1/174 | 0.533201 | 0.640399 | 0 | 0 | 1.326791 | 0.834362 |
| Diabetes_F | GSE10789[illegible] | 1/174 | 0.533201 | 0.640399 | 0 | 0 | 1.326791 | 0.834362 |
| Diabetes_F | GSE14946[illegible] | 1/175 | 0.535249 | 0.640399 | 0 | 0 | 1.319099 | 0.824467 |
| Diabetes_F | GSE73036 | 1/161 | 0.505746 | 0.640399 | 0 | 0 | 1.435538 | 0.978637 |
| Diabetes_F | GSE14946[illegible] | 1/162 | 0.507913 | 0.640399 | 0 | 0 | 1.426549 | 0.966408 |
| Diabetes_F | GSE12902( | 1/162 | 0.507913 | 0.640399 | 0 | 0 | 1.426549 | 0.966408 |
| Diabetes_F | GSE162037 | 1/163 | 0.510071 | 0.640399 | 0 | 0 | 1.417672 | 0.954384 |
| Diabetes_F | GSE14929[illegible] | 1/166 | 0.516489 | 0.640399 | 0 | 0 | 1.391684 | 0.919487 |
| Diabetes_F | GSE15506[illegible] | 1/166 | 0.516489 | 0.640399 | 0 | 0 | 1.391684 | 0.919487 |
| Diabetes_F | GSE89035 | 1/167 | 0.51861 | 0.640399 | 0 | 0 | 1.383231 | 0.908233 |
| Diabetes_F | GSE14946[illegible] | 1/191 | 0.566841 | 0.640399 | 0 | 0 | 1.207038 | 0.685208 |
| Diabetes_F | GSE15322[illegible] | 1/191 | 0.566841 | 0.640399 | 0 | 0 | 1.207038 | 0.685208 |
| Diabetes_F | GSE10172[illegible] | 1/191 | 0.566841 | 0.640399 | 0 | 0 | 1.207038 | 0.685208 |
| Diabetes_F | GSE41203 | 1/191 | 0.566841 | 0.640399 | 0 | 0 | 1.207038 | 0.685208 |
| Diabetes_F | GSE26978 | 1/185 | 0.555251 | 0.640399 | 0 | 0 | 1.246777 | 0.733523 |
| Diabetes_F | GSE14946[illegible] | 1/185 | 0.555251 | 0.640399 | 0 | 0 | 1.246777 | 0.733523 |
| Diabetes_F | GSE15322[illegible] | 1/193 | 0.570637 | 0.640399 | 0 | 0 | 1.194344 | 0.670029 |
| Diabetes_F | GSE73036 | 1/193 | 0.570637 | 0.640399 | 0 | 0 | 1.194344 | 0.670029 |
| Diabetes_F | GSE40439 | 1/186 | 0.557204 | 0.640399 | 0 | 0 | 1.239975 | 0.725168 |
| Diabetes_F | GSE14067[illegible] | 1/187 | 0.559148 | 0.640399 | 0 | 0 | 1.233246 | 0.716937 |
| Diabetes_F | GSE80780 | 1/187 | 0.559148 | 0.640399 | 0 | 0 | 1.233246 | 0.716937 |
| Diabetes_F | GSE43106 | 1/187 | 0.559148 | 0.640399 | 0 | 0 | 1.233246 | 0.716937 |
| Diabetes_F | GSE14946[illegible] | 1/187 | 0.559148 | 0.640399 | 0 | 0 | 1.233246 | 0.716937 |
| Diabetes_F | GSE41203 | 1/151 | 0.483545 | 0.640399 | 0 | 0 | 1.532016 | 1.113178 |
| Diabetes_F | GSE14946[illegible] | 1/189 | 0.563011 | 0.640399 | 0 | 0 | 1.220002 | 0.700838 |
| Diabetes_F | GSE14067[illegible] | 1/190 | 0.56493 | 0.640399 | 0 | 0 | 1.213486 | 0.692965 |
| Diabetes_F | GSE15419[illegible] | 1/179 | 0.543355 | 0.640399 | 0 | 0 | 1.289195 | 0.7864 |
| Diabetes_F | GSE48009 | 1/195 | 0.574401 | 0.640399 | 0 | 0 | 1.181911 | 0.655284 |
| Diabetes_F | GSE13132( | 1/195 | 0.574401 | 0.640399 | 0 | 0 | 1.181911 | 0.655284 |
| Diabetes_F | GSE80780 | 1/176 | 0.537289 | 0.640399 | 0 | 0 | 1.311495 | 0.814726 |
| Diabetes_F | GSE15158[illegible] | 1/152 | 0.485809 | 0.640399 | 0 | 0 | 1.521793 | 1.098642 |
| Diabetes_F | GSE10120[illegible] | 1/152 | 0.485809 | 0.640399 | 0 | 0 | 1.521793 | 1.098642 |
| Diabetes_F | GSE15067[illegible] | 1/152 | 0.485809 | 0.640399 | 0 | 0 | 1.521793 | 1.098642 |
| Diabetes_F | GSE11221[illegible] | 1/184 | 0.55329 | 0.640399 | 0 | 0 | 1.253654 | 0.742005 |

| | | | | | | | | |
|---|---|---|---|---|---|---|---|---|
| Diabetes_F | GSE59143 | 1/184 | 0.55329 | 0.640399 | 0 | 0 | 1.253654 | 0.742005 |
| Diabetes_F | GSE154198 | 1/184 | 0.55329 | 0.640399 | 0 | 0 | 1.253654 | 0.742005 |
| Diabetes_F | GSE80780 | 1/185 | 0.555251 | 0.640399 | 0 | 0 | 1.246777 | 0.733523 |
| Diabetes_F | GSE37248 | 1/210 | 0.601608 | 0.640399 | 0 | 0 | 1.09625 | 0.557059 |
| Diabetes_F | GSE67543 | 1/214 | 0.608568 | 0.640399 | 0 | 0 | 1.075445 | 0.534116 |
| Diabetes_F | GSE149468 | 1/215 | 0.610289 | 0.640399 | 0 | 0 | 1.070365 | 0.528571 |
| Diabetes_F | GSE120299 | 1/215 | 0.610289 | 0.640399 | 0 | 0 | 1.070365 | 0.528571 |
| Diabetes_F | GSE37248 | 1/215 | 0.610289 | 0.640399 | 0 | 0 | 1.070365 | 0.528571 |
| Diabetes_F | GSE73433 | 1/219 | 0.617099 | 0.640399 | 0 | 0 | 1.050512 | 0.507109 |
| Diabetes_F | GSE40439 | 1/220 | 0.618783 | 0.640399 | 0 | 0 | 1.045662 | 0.501918 |
| Diabetes_F | GSE137631 | 1/221 | 0.62046 | 0.640399 | 0 | 0 | 1.040856 | 0.496795 |
| Diabetes_F | GSE115421 | 1/197 | 0.578132 | 0.640399 | 0 | 0 | 1.169732 | 0.640958 |
| Diabetes_F | GSE115421 | 1/207 | 0.596308 | 0.640399 | 0 | 0 | 1.112384 | 0.575101 |
| Diabetes_F | GSE142209 | 1/207 | 0.596308 | 0.640399 | 0 | 0 | 1.112384 | 0.575101 |
| Diabetes_F | GSE137631 | 1/209 | 0.599849 | 0.640399 | 0 | 0 | 1.101576 | 0.562991 |
| Diabetes_F | GSE149468 | 1/197 | 0.578132 | 0.640399 | 0 | 0 | 1.169732 | 0.640958 |
| Diabetes_F | GSE149468 | 1/197 | 0.578132 | 0.640399 | 0 | 0 | 1.169732 | 0.640958 |
| Diabetes_F | GSE157585 | 1/198 | 0.579985 | 0.640399 | 0 | 0 | 1.163735 | 0.633948 |
| Diabetes_F | GSE109285 | 1/198 | 0.579985 | 0.640399 | 0 | 0 | 1.163735 | 0.633948 |
| Diabetes_F | GSE149468 | 1/192 | 0.568743 | 0.640399 | 0 | 0 | 1.200657 | 0.677563 |
| Diabetes_F | GSE124774 | 1/193 | 0.570637 | 0.640399 | 0 | 0 | 1.194344 | 0.670029 |
| Diabetes_F | GSE109285 | 1/171 | 0.527002 | 0.640399 | 0 | 0 | 1.35041 | 0.865006 |
| Diabetes_F | GSE133903 | 1/196 | 0.576271 | 0.640399 | 0 | 0 | 1.17579 | 0.64807 |
| Diabetes_F | GSE101537 | 1/194 | 0.572523 | 0.640399 | 0 | 0 | 1.188095 | 0.662603 |
| Diabetes_F | GSE153222 | 1/194 | 0.572523 | 0.640399 | 0 | 0 | 1.188095 | 0.662603 |
| Diabetes_F | GSE136134 | 1/194 | 0.572523 | 0.640399 | 0 | 0 | 1.188095 | 0.662603 |
| Diabetes_F | GSE134068 | 1/195 | 0.574401 | 0.640399 | 0 | 0 | 1.181911 | 0.655284 |
| Diabetes_F | GSE73436 | 1/187 | 0.559148 | 0.640399 | 0 | 0 | 1.233246 | 0.716937 |
| Diabetes_F | GSE110569 | 1/188 | 0.561084 | 0.640399 | 0 | 0 | 1.226589 | 0.708828 |
| Diabetes_F | GSE87854 | 1/168 | 0.520722 | 0.640399 | 0 | 0 | 1.374878 | 0.897162 |
| Diabetes_F | GSE149468 | 1/169 | 0.522824 | 0.640399 | 0 | 0 | 1.366625 | 0.886269 |
| Diabetes_F | GSE144061 | 1/224 | 0.625446 | 0.640399 | 0 | 0 | 1.026697 | 0.481818 |
| Diabetes_F | GSE162037 | 1/156 | 0.494766 | 0.640399 | 0 | 0 | 1.482221 | 1.042994 |
| Diabetes_F | GSE153222 | 1/156 | 0.494766 | 0.640399 | 0 | 0 | 1.482221 | 1.042994 |
| Diabetes_F | GSE122348 | 1/156 | 0.494766 | 0.640399 | 0 | 0 | 1.482221 | 1.042994 |
| Diabetes_F | GSE127251 | 1/156 | 0.494766 | 0.640399 | 0 | 0 | 1.482221 | 1.042994 |
| Diabetes_F | GSE86611 | 1/157 | 0.496981 | 0.640399 | 0 | 0 | 1.472645 | 1.029678 |
| Diabetes_F | GSE107894 | 1/155 | 0.492541 | 0.640399 | 0 | 0 | 1.491921 | 1.056544 |
| Diabetes_F | GSE127251 | 1/146 | 0.472079 | 0.640399 | 0 | 0 | 1.585245 | 1.1899 |
| Diabetes_F | GSE145070 | 1/146 | 0.472079 | 0.640399 | 0 | 0 | 1.585245 | 1.1899 |
| Diabetes_F | GSE143319 | 1/147 | 0.474392 | 0.640399 | 0 | 0 | 1.574307 | 1.173995 |
| Diabetes_F | GSE149468 | 1/149 | 0.478989 | 0.640399 | 0 | 0 | 1.552876 | 1.143039 |
| Diabetes_F | GSE149468 | 1/149 | 0.478989 | 0.640399 | 0 | 0 | 1.552876 | 1.143039 |
| Diabetes_F | GSE114192 | 1/152 | 0.485809 | 0.640399 | 0 | 0 | 1.521793 | 1.098642 |
| Diabetes_F | GSE138252 | 1/137 | 0.4508 | 0.640399 | 0 | 0 | 1.69092 | 1.347208 |

| | | | | | | | | |
|---|---|---|---|---|---|---|---|---|
| Diabetes_F | GSE14946E | 1/138 | 0.453206 | 0.640399 | 0 | 0 | 1.678493 | 1.328374 |
| Diabetes_F | GSE15758E | 1/138 | 0.453206 | 0.640399 | 0 | 0 | 1.678493 | 1.328374 |
| Diabetes_F | GSE14946E | 1/142 | 0.462724 | 0.640399 | 0 | 0 | 1.630546 | 1.256538 |
| Diabetes_F | GSE14946E | 1/142 | 0.462724 | 0.640399 | 0 | 0 | 1.630546 | 1.256538 |
| Diabetes_F | GSE7196 El | 1/153 | 0.488063 | 0.640399 | 0 | 0 | 1.511704 | 1.084362 |
| Diabetes_F | GSE15322 | 1/154 | 0.490307 | 0.640399 | 0 | 0 | 1.501748 | 1.070331 |
| Diabetes_F | GSE14946E | 1/201 | 0.585497 | 0.640399 | 0 | 0 | 1.146105 | 0.613503 |
| Diabetes_F | GSE10006 | 1/201 | 0.585497 | 0.640399 | 0 | 0 | 1.146105 | 0.613503 |
| Diabetes_F | GSE10006 | 1/201 | 0.585497 | 0.640399 | 0 | 0 | 1.146105 | 0.613503 |
| Diabetes_F | GSE13763 | 1/198 | 0.579985 | 0.640399 | 0 | 0 | 1.163735 | 0.633948 |
| Diabetes_F | GSE14946E | 1/198 | 0.579985 | 0.640399 | 0 | 0 | 1.163735 | 0.633948 |
| Diabetes_F | GSE15606 | 1/199 | 0.581831 | 0.640399 | 0 | 0 | 1.157799 | 0.627036 |
| Diabetes_F | GSE12477 | 1/199 | 0.581831 | 0.640399 | 0 | 0 | 1.157799 | 0.627036 |
| Diabetes_F | GSE80780 | 1/200 | 0.583668 | 0.640399 | 0 | 0 | 1.151922 | 0.620221 |
| Diabetes_F | GSE10165 | 1/200 | 0.583668 | 0.640399 | 0 | 0 | 1.151922 | 0.620221 |
| Diabetes_F | GSE15322 | 1/200 | 0.583668 | 0.640399 | 0 | 0 | 1.151922 | 0.620221 |
| Diabetes_F | GSE14495 | 1/128 | 0.428675 | 0.640399 | 0 | 0 | 1.811573 | 1.534505 |
| Diabetes_F | GSE10032 | Jan-89 | 0.322165 | 0.640399 | 0 | 0 | 2.619582 | 2.967177 |
| Diabetes_F | GSE14373 | 1/105 | 0.368048 | 0.640399 | 0 | 0 | 2.214781 | 2.213764 |
| Diabetes_F | GSE59141 | 1/108 | 0.376303 | 0.640399 | 0 | 0 | 2.152358 | 2.10363 |
| Diabetes_F | GSE14023 | 1/110 | 0.381747 | 0.640399 | 0 | 0 | 2.112652 | 2.034479 |
| Diabetes_F | GSE15067 | 1/185 | 0.555251 | 0.640399 | 0 | 0 | 1.246777 | 0.733523 |
| Diabetes_F | GSE12042 | 1/157 | 0.496981 | 0.640399 | 0 | 0 | 1.472645 | 1.029678 |
| SysMyo_M | MGS769 M | 1/173 | 0.531144 | 0.640489 | 0 | 0 | 1.334573 | 0.844414 |
| SysMyo_M | MGS557 H | 1/174 | 0.533201 | 0.640489 | 0 | 0 | 1.326791 | 0.834362 |
| SysMyo_M | MGS1441 I | 1/175 | 0.535249 | 0.640489 | 0 | 0 | 1.319099 | 0.824467 |
| SysMyo_M | MGS357 H | 1/175 | 0.535249 | 0.640489 | 0 | 0 | 1.319099 | 0.824467 |
| SysMyo_M | MGS1400 I | 1/174 | 0.533201 | 0.640489 | 0 | 0 | 1.326791 | 0.834362 |
| DisGeNET | Acute Pron | 2/475 | 0.615583 | 0.641028 | 0 | 0 | 0.967044 | 0.469196 |
| Human_Ph | Epicanthus | 1/233 | 0.640021 | 0.641701 | 0 | 0 | 0.986417 | 0.440193 |
| DisGeNET | Thyroid Ne | 2/476 | 0.616734 | 0.641972 | 0 | 0 | 0.964954 | 0.466379 |
| SysMyo_M | MGS825 M | 1/177 | 0.53932 | 0.642642 | 0 | 0 | 1.303977 | 0.805137 |
| SysMyo_M | MGS1028 I | 1/177 | 0.53932 | 0.642642 | 0 | 0 | 1.303977 | 0.805137 |
| KOMP2_M | Prolonged | Jan-96 | 0.342631 | 0.643352 | 0 | 0 | 2.425704 | 2.598176 |
| KOMP2_M | Decreased | Jan-97 | 0.345504 | 0.643352 | 0 | 0 | 2.400315 | 2.550937 |
| Rare_Disea | Lynch synd | 1/224 | 0.625446 | 0.643613 | 0 | 0 | 1.026697 | 0.481818 |
| GeneSigDB | 15980968- | 1/154 | 0.490307 | 0.644106 | 0 | 0 | 1.501748 | 1.070331 |
| GeneSigDB | 19014521- | 1/154 | 0.490307 | 0.644106 | 0 | 0 | 1.501748 | 1.070331 |
| DisGeNET | Degenerati | 4/976 | 0.619177 | 0.644259 | 0 | 0 | 0.939114 | 0.450178 |
| SysMyo_M | MGS91 Hu | 1/179 | 0.543355 | 0.644735 | 0 | 0 | 1.289195 | 0.7864 |
| SysMyo_M | MGS757 M | 1/179 | 0.543355 | 0.644735 | 0 | 0 | 1.289195 | 0.7864 |
| GeDiPNet_ | Osteopenia | 1/223 | 0.623791 | 0.644806 | 0 | 0 | 1.031374 | 0.486746 |
| Diabetes_F | GSE14946E | 1/235 | 0.643183 | 0.644979 | 0 | 0 | 0.977887 | 0.431568 |
| GTEx_Tissu | GTEX-TML8 | 6/615 | 0.051521 | 0.645238 | 0 | 0 | 2.34799 | 6.963594 |
| DisGeNET | Status Epile | 1/221 | 0.62046 | 0.645337 | 0 | 0 | 1.040856 | 0.496795 |

| | | | | | | | | |
|---|---|---|---|---|---|---|---|---|
| KEGG_202: | Hippo signa | 1/163 | 0.510071 | 0.64609 | 0 | 0 | 1.417672 | 0.954384 |
| Rare_Disea | Malignant | 1/226 | 0.628735 | 0.646103 | 0 | 0 | 1.017468 | 0.472152 |
| Jensen_TIS | CL-48 cell | 1/198 | 0.579985 | 0.646212 | 0 | 0 | 1.163735 | 0.633948 |
| GO_Biologi | Negative R | 2/482 | 0.623589 | 0.646348 | 0 | 0 | 0.952598 | 0.449878 |
| PhenGenI_ | Blood Coag | 1/150 | 0.481272 | 0.646359 | 0 | 0 | 1.542376 | 1.127974 |
| CellMarker | Leydig Prec | 1/221 | 0.62046 | 0.646411 | 0 | 0 | 1.040856 | 0.496795 |
| GO_Molecu | DNA Bindir | 4/846 | 0.505062 | 0.647515 | 0 | 0 | 1.091549 | 0.745609 |
| SysMyo_M | MGS44 C2c | 2/421 | 0.549544 | 0.648004 | 0 | 0 | 1.094707 | 0.655365 |
| SysMyo_M | MGS112 M | 1/181 | 0.547354 | 0.648004 | 0 | 0 | 1.274742 | 0.768234 |
| SysMyo_M | MGS883 M | 1/182 | 0.549341 | 0.648004 | 0 | 0 | 1.267635 | 0.759358 |
| GeneSigDB | 16140871-: | 1/156 | 0.494766 | 0.648435 | 0 | 0 | 1.482221 | 1.042994 |
| Disease_Sig | Endometri | 2/300 | 0.375985 | 0.648877 | 0 | 0 | 1.548756 | 1.515003 |
| Disease_Sig | Leber cong | 2/300 | 0.375985 | 0.648877 | 0 | 0 | 1.548756 | 1.515003 |
| Disease_Sig | Adenoma ( | 2/300 | 0.375985 | 0.648877 | 0 | 0 | 1.548756 | 1.515003 |
| Disease_Sig | Infantile ne | 2/300 | 0.375985 | 0.648877 | 0 | 0 | 1.548756 | 1.515003 |
| Disease_Sig | JRA - Juven | 2/300 | 0.375985 | 0.648877 | 0 | 0 | 1.548756 | 1.515003 |
| Disease_Sig | Familial co | 2/300 | 0.375985 | 0.648877 | 0 | 0 | 1.548756 | 1.515003 |
| Disease_Sig | Gastrointe: | 2/300 | 0.375985 | 0.648877 | 0 | 0 | 1.548756 | 1.515003 |
| Disease_Sig | Hepatitis C | 2/300 | 0.375985 | 0.648877 | 0 | 0 | 1.548756 | 1.515003 |
| Disease_Sig | Huntingtor | 2/300 | 0.375985 | 0.648877 | 0 | 0 | 1.548756 | 1.515003 |
| Disease_Sig | Actinic kera | 2/300 | 0.375985 | 0.648877 | 0 | 0 | 1.548756 | 1.515003 |
| Disease_Sig | Cystic Fibro | 2/300 | 0.375985 | 0.648877 | 0 | 0 | 1.548756 | 1.515003 |
| Disease_Sig | Dermatom | 2/300 | 0.375985 | 0.648877 | 0 | 0 | 1.548756 | 1.515003 |
| Disease_Sig | Multiple be | 2/300 | 0.375985 | 0.648877 | 0 | 0 | 1.548756 | 1.515003 |
| Disease_Sig | Nephrobla: | 2/300 | 0.375985 | 0.648877 | 0 | 0 | 1.548756 | 1.515003 |
| Disease_Sig | Neurologic | 2/300 | 0.375985 | 0.648877 | 0 | 0 | 1.548756 | 1.515003 |
| Disease_Sig | Obesity GS | 2/300 | 0.375985 | 0.648877 | 0 | 0 | 1.548756 | 1.515003 |
| Disease_Sig | Polycystic ( | 2/300 | 0.375985 | 0.648877 | 0 | 0 | 1.548756 | 1.515003 |
| Disease_Sig | SCID - Seve | 2/300 | 0.375985 | 0.648877 | 0 | 0 | 1.548756 | 1.515003 |
| Disease_Sig | Septic Shoc | 2/300 | 0.375985 | 0.648877 | 0 | 0 | 1.548756 | 1.515003 |
| Disease_Sig | Synovial sa | 2/300 | 0.375985 | 0.648877 | 0 | 0 | 1.548756 | 1.515003 |
| Disease_Sig | Type 1 diat | 2/300 | 0.375985 | 0.648877 | 0 | 0 | 1.548756 | 1.515003 |
| Disease_Sig | Ventilator- | 2/300 | 0.375985 | 0.648877 | 0 | 0 | 1.548756 | 1.515003 |
| Disease_Sig | Viral cardic | 2/300 | 0.375985 | 0.648877 | 0 | 0 | 1.548756 | 1.515003 |
| Disease_Sig | Pseudomoi | 2/300 | 0.375985 | 0.648877 | 0 | 0 | 1.548756 | 1.515003 |
| Disease_Sig | Bipolar Dis | 2/300 | 0.375985 | 0.648877 | 0 | 0 | 1.548756 | 1.515003 |
| Disease_Sig | Breast Can | 2/300 | 0.375985 | 0.648877 | 0 | 0 | 1.548756 | 1.515003 |
| Disease_Sig | Leukemia, . | 2/300 | 0.375985 | 0.648877 | 0 | 0 | 1.548756 | 1.515003 |
| Disease_Sig | Lung trans | 2/300 | 0.375985 | 0.648877 | 0 | 0 | 1.548756 | 1.515003 |
| Disease_Sig | MDS - Mye | 2/300 | 0.375985 | 0.648877 | 0 | 0 | 1.548756 | 1.515003 |
| Disease_Sig | MELAS - M | 2/300 | 0.375985 | 0.648877 | 0 | 0 | 1.548756 | 1.515003 |
| Disease_Sig | Cancer of t | 2/300 | 0.375985 | 0.648877 | 0 | 0 | 1.548756 | 1.515003 |
| Disease_Sig | Adenoma ( | 2/300 | 0.375985 | 0.648877 | 0 | 0 | 1.548756 | 1.515003 |
| Disease_Sig | Bipolar Dis | 2/300 | 0.375985 | 0.648877 | 0 | 0 | 1.548756 | 1.515003 |
| Disease_Sig | Arrhythmo | 2/300 | 0.375985 | 0.648877 | 0 | 0 | 1.548756 | 1.515003 |

| | | | | | | | | |
|---|---|---|---|---|---|---|---|---|
| Disease_Si{ | In situ mela | 2/300 | 0.375985 | 0.648877 | 0 | 0 | 1.548756 | 1.515003 |
| Diabetes_F | GSE15464S | 1/239 | 0.649424 | 0.649424 | 0 | 0 | 0.961257 | 0.414945 |
| Disease_Pe | Cardiac Fai | 4/275 | 0.032035 | 0.649525 | 0 | 0 | 3.492998 | 12.01919 |
| Disease_Pe | cerebral pa | 4/292 | 0.038602 | 0.649525 | 0 | 0 | 3.283969 | 10.6875 |
| Disease_Pe | Heart Injur | 4/264 | 0.028165 | 0.649525 | 0 | 0 | 3.642817 | 13.00366 |
| Disease_Pe | Oligodendr | 4/257 | 0.025857 | 0.649525 | 0 | 0 | 3.74494 | 13.68845 |
| Disease_Pe | astrocytom | 4/299 | 0.041515 | 0.649525 | 0 | 0 | 3.204901 | 10.19702 |
| Disease_Pe | astrocytom | 4/301 | 0.04237 | 0.649525 | 0 | 0 | 3.182995 | 10.06245 |
| Disease_Pe | Gamma-hy | 4/292 | 0.038602 | 0.649525 | 0 | 0 | 3.283969 | 10.6875 |
| Disease_Pe | nasophary | 4/303 | 0.043235 | 0.649525 | 0 | 0 | 3.161381 | 9.930257 |
| Disease_Pe | Anxiety Dis | 5/358 | 0.020007 | 0.649525 | 0 | 0 | 3.378705 | 13.21642 |
| Disease_Pe | oligodendr | 4/287 | 0.036596 | 0.649525 | 0 | 0 | 3.342841 | 11.05749 |
| Disease_Pe | nasophary | 4/227 | 0.017289 | 0.649525 | 0 | 0 | 4.255227 | 17.26642 |
| Disease_Pe | Alzheimer's | 4/231 | 0.01831 | 0.649525 | 0 | 0 | 4.179396 | 16.71898 |
| Disease_Pe | obesity DO | 5/426 | 0.038242 | 0.649525 | 0 | 0 | 2.823127 | 9.214202 |
| Disease_Pe | Huntingtor | 4/281 | 0.034271 | 0.649525 | 0 | 0 | 3.416293 | 11.52474 |
| Disease_Pe | non-systen | 5/359 | 0.020222 | 0.649525 | 0 | 0 | 3.368989 | 13.1424 |
| Disease_Pe | Breast Can | 4/276 | 0.032401 | 0.649525 | 0 | 0 | 3.479979 | 11.93481 |
| Disease_Pe | Nemaline M | 5/419 | 0.036014 | 0.649525 | 0 | 0 | 2.871892 | 9.545744 |
| Disease_Pe | Tachycardi | 5/392 | 0.028188 | 0.649525 | 0 | 0 | 3.076511 | 10.97965 |
| Disease_Pe | Duchenne | 4/285 | 0.035811 | 0.649525 | 0 | 0 | 3.366977 | 11.21034 |
| Disease_Pe | severe asth | 4/247 | 0.022764 | 0.649525 | 0 | 0 | 3.901036 | 14.75592 |
| Disease_Pe | cerebral pa | 5/381 | 0.025341 | 0.649525 | 0 | 0 | 3.168299 | 11.64449 |
| Disease_Pe | Spinal Mus | 5/432 | 0.040218 | 0.649525 | 0 | 0 | 2.782601 | 8.941757 |
| Disease_Pe | Cardiac Hy | 4/287 | 0.036596 | 0.649525 | 0 | 0 | 3.342841 | 11.05749 |
| Disease_Pe | Emery-Dre | 4/253 | 0.024591 | 0.649525 | 0 | 0 | 3.805874 | 14.1022 |
| Disease_Pe | Dystonia C | 4/283 | 0.035036 | 0.649525 | 0 | 0 | 3.391458 | 11.36607 |
| Disease_Pe | glioblaston | 4/284 | 0.035422 | 0.649525 | 0 | 0 | 3.379174 | 11.28784 |
| Disease_Pe | Hepatic Cir | 4/297 | 0.040671 | 0.649525 | 0 | 0 | 3.227106 | 10.33401 |
| GeneSigDB | 16760443-: | 1/157 | 0.496981 | 0.649642 | 0 | 0 | 1.472645 | 1.029678 |
| GeneSigDB | 16081686-: | 1/158 | 0.499187 | 0.649642 | 0 | 0 | 1.463191 | 1.016589 |
| GeneSigDB | 18199711-: | 1/158 | 0.499187 | 0.649642 | 0 | 0 | 1.463191 | 1.016589 |
| KEGG_202: | Tight juncti | 1/169 | 0.522824 | 0.650197 | 0 | 0 | 1.366625 | 0.886269 |
| KEGG_202: | Protein prc | 1/171 | 0.527002 | 0.650197 | 0 | 0 | 1.35041 | 0.865006 |
| Rare_Disea | Limb dysto | 1/229 | 0.633614 | 0.650219 | 0 | 0 | 1.003927 | 0.458108 |
| Jensen_TIS | Prostate gl | 2/450 | 0.585949 | 0.650606 | 0 | 0 | 1.022321 | 0.546454 |
| DisGeNET | Dermatitis, | 2/484 | 0.625853 | 0.650689 | 0 | 0 | 0.948548 | 0.444527 |
| SysMyo_M | MGS1002 I | 1/184 | 0.55329 | 0.651064 | 0 | 0 | 1.253654 | 0.742005 |
| DisGeNET | Poor schoc | 4/985 | 0.626465 | 0.651067 | 0 | 0 | 0.930057 | 0.434952 |
| DisGeNET | Pain | 2/485 | 0.626981 | 0.651345 | 0 | 0 | 0.946535 | 0.441879 |
| GeneSigDB | 16135788-: | 2/386 | 0.502763 | 0.651524 | 0 | 0 | 1.19663 | 0.822846 |
| GeneSigDB | 19797726-: | 2/387 | 0.504142 | 0.651524 | 0 | 0 | 1.193461 | 0.817399 |
| GeneSigDB | 19861896-: | 2/387 | 0.504142 | 0.651524 | 0 | 0 | 1.193461 | 0.817399 |
| GeneSigDB | 17177833-: | 4/847 | 0.505996 | 0.652407 | 0 | 0 | 1.090197 | 0.742672 |
| GO_Biologi | Regulation | 1/227 | 0.630368 | 0.652422 | 0 | 0 | 1.012914 | 0.46741 |

| | | | | | | | | |
|---|---|---|---|---|---|---|---|---|
| GeneSigDB | 15474998-‹ | 4/850 | 0.508793 | 0.65299 | 0 | 0 | 1.08616 | 0.733934 |
| GeneSigDB | 16651414-: | 2/390 | 0.508263 | 0.65299 | 0 | 0 | 1.184051 | 0.801314 |
| GO_Biologi | Apoptotic I | 1/228 | 0.631995 | 0.653153 | 0 | 0 | 1.008401 | 0.462729 |
| GTEx_Tissu | GTEX-RN64 | 7/778 | 0.052683 | 0.654128 | 0 | 0 | 2.172406 | 6.394397 |
| GTEx_Tissu | GTEX-QEL4 | 7/778 | 0.052683 | 0.654128 | 0 | 0 | 2.172406 | 6.394397 |
| SysMyo_M | MGS262 H | 1/186 | 0.557204 | 0.65431 | 0 | 0 | 1.239975 | 0.725168 |
| SysMyo_M | MGS1193 I | 1/189 | 0.563011 | 0.654341 | 0 | 0 | 1.220002 | 0.700838 |
| SysMyo_M | MGS747 M | 1/189 | 0.563011 | 0.654341 | 0 | 0 | 1.220002 | 0.700838 |
| SysMyo_M | MGS1208 I | 1/188 | 0.561084 | 0.654341 | 0 | 0 | 1.226589 | 0.708828 |
| SysMyo_M | MGS110 M | 1/187 | 0.559148 | 0.654341 | 0 | 0 | 1.233246 | 0.716937 |
| SysMyo_M | MGS983 M | 1/189 | 0.563011 | 0.654341 | 0 | 0 | 1.220002 | 0.700838 |
| Reactome_ | G Alpha (S) | 1/153 | 0.488063 | 0.654647 | 0 | 0 | 1.511704 | 1.084362 |
| PhenGenI_ | Celiac Dise | 1/154 | 0.490307 | 0.654687 | 0 | 0 | 1.501748 | 1.070331 |
| DisGeNET | Neoplasms | 5/1236 | 0.630498 | 0.654739 | 0 | 0 | 0.925383 | 0.426828 |
| SysMyo_M | MGS1440 I | 1/190 | 0.56493 | 0.655226 | 0 | 0 | 1.213486 | 0.692965 |
| GeneSigDB | 17597811-: | 2/393 | 0.512362 | 0.656059 | 0 | 0 | 1.174786 | 0.785607 |
| SysMyo_M | MGS837 M | 1/192 | 0.568743 | 0.656465 | 0 | 0 | 1.200657 | 0.677563 |
| SysMyo_M | MGS870 M | 1/193 | 0.570637 | 0.656465 | 0 | 0 | 1.194344 | 0.670029 |
| SysMyo_M | MGS167 Ra | 1/193 | 0.570637 | 0.656465 | 0 | 0 | 1.194344 | 0.670029 |
| SysMyo_M | MGS1454 I | 1/193 | 0.570637 | 0.656465 | 0 | 0 | 1.194344 | 0.670029 |
| DisGeNET | Anemia | 2/490 | 0.632583 | 0.656643 | 0 | 0 | 0.936596 | 0.428908 |
| SysMyo_M | MGS1455 I | 1/197 | 0.578132 | 0.657074 | 0 | 0 | 1.169732 | 0.640958 |
| SysMyo_M | MGS158 H | 1/197 | 0.578132 | 0.657074 | 0 | 0 | 1.169732 | 0.640958 |
| SysMyo_M | MGS1348 I | 1/197 | 0.578132 | 0.657074 | 0 | 0 | 1.169732 | 0.640958 |
| SysMyo_M | MGS1417 I | 1/195 | 0.574401 | 0.657074 | 0 | 0 | 1.181911 | 0.655284 |
| SysMyo_M | MGS1485 I | 1/197 | 0.578132 | 0.657074 | 0 | 0 | 1.169732 | 0.640958 |
| SysMyo_M | MGS1335 I | 1/196 | 0.576271 | 0.657074 | 0 | 0 | 1.17579 | 0.64807 |
| GTEx_Tissu | GTEX-WY7( | 10/1291 | 0.053383 | 0.657185 | 0 | 0 | 1.888946 | 5.535092 |
| GTEx_Tissu | GTEX-R55G | 11/1469 | 0.053356 | 0.657185 | 0 | 0 | 1.832043 | 5.369297 |
| DisGeNET | Drug abuse | 1/229 | 0.633614 | 0.657193 | 0 | 0 | 1.003927 | 0.458108 |
| DisGeNET | Endometri | 1/229 | 0.633614 | 0.657193 | 0 | 0 | 1.003927 | 0.458108 |
| GO_Biologi | Negative R | 1/231 | 0.636831 | 0.657195 | 0 | 0 | 0.995096 | 0.449038 |
| KOMP2_M | Decreased | 1/102 | 0.359686 | 0.657491 | 0 | 0 | 2.280912 | 2.332289 |
| KOMP2_M | Increased I | 1/104 | 0.365273 | 0.657491 | 0 | 0 | 2.236396 | 2.252299 |
| DisGeNET | Glioma | Sep-11 | 0.634531 | 0.657884 | 0 | 0 | 0.928055 | 0.422143 |
| GO_Biologi | Regulation | 1/232 | 0.63843 | 0.657888 | 0 | 0 | 0.990738 | 0.444588 |
| DisGeNET | Ductal Carc | 1/230 | 0.635226 | 0.658343 | 0 | 0 | 0.999492 | 0.453544 |
| Disease_Pe | Schizophre | 4/319 | 0.05051 | 0.658741 | 0 | 0 | 2.998355 | 8.951822 |
| Disease_Pe | Huntingtor | 4/307 | 0.044994 | 0.658741 | 0 | 0 | 3.119011 | 9.672767 |
| Disease_Pe | Huntingtor | 4/315 | 0.048632 | 0.658741 | 0 | 0 | 3.037539 | 9.183935 |
| Disease_Pe | glioblaston | 4/315 | 0.048632 | 0.658741 | 0 | 0 | 3.037539 | 9.183935 |
| Disease_Pe | Osteoarthr | 4/317 | 0.049566 | 0.658741 | 0 | 0 | 3.017822 | 9.066892 |
| Disease_Pe | Ventricular | 4/320 | 0.050986 | 0.658741 | 0 | 0 | 2.988714 | 8.895015 |
| Disease_Pe | Cushing Sy | 4/316 | 0.049098 | 0.658741 | 0 | 0 | 3.027649 | 9.125164 |
| DisGeNET | Colorectal | Dec-31 | 0.636184 | 0.659075 | 0 | 0 | 0.931497 | 0.421286 |

| | | | | | | | | |
|---|---|---|---|---|---|---|---|---|
| DisGeNET | Malignant | 21/5054 | 0.636488 | 0.65913 | 0 | 0 | 0.9407 | 0.424998 |
| DisGeNET | Anemia, Si | 1/231 | 0.636831 | 0.659225 | 0 | 0 | 0.995096 | 0.449038 |
| Disease_Pe | basal cell c | 4/320 | 0.050986 | 0.659316 | 0 | 0 | 2.988714 | 8.895015 |
| Disease_Pe | Cardiac Hy | 5/467 | 0.052975 | 0.659316 | 0 | 0 | 2.567179 | 7.542205 |
| Disease_Pe | Cardiac Fai | 4/325 | 0.053403 | 0.659316 | 0 | 0 | 2.941411 | 8.618031 |
| Disease_Pe | NASH C04C | 4/325 | 0.053403 | 0.659316 | 0 | 0 | 2.941411 | 8.618031 |
| GO_Biologi | Translation | 1/234 | 0.641605 | 0.660202 | 0 | 0 | 0.982134 | 0.435854 |
| DisGeNET | Metastasis | 1/232 | 0.63843 | 0.660618 | 0 | 0 | 0.990738 | 0.444588 |
| GWAS_Cat | Intraocular | 1/241 | 0.652504 | 0.660627 | 0 | 0 | 0.953149 | 0.406935 |
| Rare_Disea | Immune th | 1/236 | 0.644753 | 0.660739 | 0 | 0 | 0.973676 | 0.427334 |
| GTEx_Tissu | GTEX-UJM( | 12/1654 | 0.05411 | 0.661587 | 0 | 0 | 1.780365 | 5.192871 |
| GTEx_Tissu | GTEX-S4Z8 | 8/949 | 0.054198 | 0.661587 | 0 | 0 | 2.041674 | 5.951689 |
| GeDiPNet_ | Esophagus | 1/234 | 0.641605 | 0.662046 | 0 | 0 | 0.982134 | 0.435854 |
| DisGeNET | Pneumonia | 2/497 | 0.640317 | 0.66231 | 0 | 0 | 0.923018 | 0.411474 |
| Disease_Pe | glaucoma a | 4/332 | 0.056889 | 0.66238 | 0 | 0 | 2.877608 | 8.249101 |
| Disease_Pe | Down synd | 4/335 | 0.05842 | 0.66238 | 0 | 0 | 2.85109 | 8.097363 |
| Disease_Pe | Epilepsy D( | 3/203 | 0.05878 | 0.66238 | 0 | 0 | 3.520179 | 9.976043 |
| Disease_Pe | nemaline n | 5/472 | 0.05497 | 0.66238 | 0 | 0 | 2.53904 | 7.365651 |
| Disease_Pe | autism spe | 4/336 | 0.058935 | 0.66238 | 0 | 0 | 2.842357 | 8.047601 |
| Disease_Pe | AML - Acut | 4/339 | 0.060496 | 0.66238 | 0 | 0 | 2.816472 | 7.900705 |
| Disease_Pe | polycystic ( | 4/325 | 0.053403 | 0.66238 | 0 | 0 | 2.941411 | 8.618031 |
| Disease_Pe | Infantile ne | 4/338 | 0.059973 | 0.66238 | 0 | 0 | 2.825049 | 7.949276 |
| Disease_Pe | Arthropath | 3/199 | 0.056033 | 0.66238 | 0 | 0 | 3.592748 | 10.35363 |
| PhenGenI_ | Hemoglobi | 1/158 | 0.499187 | 0.662713 | 0 | 0 | 1.463191 | 1.016589 |
| Rare_Disea | Pheochrom | 1/238 | 0.647874 | 0.663024 | 0 | 0 | 0.965362 | 0.419024 |
| DisGeNET | Tumor Initi | 1/234 | 0.641605 | 0.66338 | 0 | 0 | 0.982134 | 0.435854 |
| SysMyo_M | MGS1067 I | 1/201 | 0.585497 | 0.664111 | 0 | 0 | 1.146105 | 0.613503 |
| DisGeNET | Hepatoblas | 1/235 | 0.643183 | 0.664487 | 0 | 0 | 0.977887 | 0.431568 |
| DisGeNET | Microceph | 1/235 | 0.643183 | 0.664487 | 0 | 0 | 0.977887 | 0.431568 |
| SysMyo_M | MGS1313 I | 1/202 | 0.587319 | 0.664845 | 0 | 0 | 1.140345 | 0.606877 |
| GeneSigDB | 20421987- | 2/399 | 0.520494 | 0.664943 | 0 | 0 | 1.156675 | 0.755281 |
| GTEx_Tissu | GTEX-SUCS | 52/9613 | 0.018559 | 0.665396 | 0 | 0 | 1.60863 | 6.413291 |
| GTEx_Tissu | GTEX-R55C | 4/215 | 0.014444 | 0.665396 | 0 | 0 | 4.499971 | 19.06857 |
| GTEx_Tissu | GTEX-PX3G | 3/114 | 0.013515 | 0.665396 | 0 | 0 | 6.3713 | 27.42156 |
| GTEx_Tissu | GTEX-QDT8 | 3/113 | 0.013201 | 0.665396 | 0 | 0 | 6.429545 | 27.82383 |
| GTEx_Tissu | GTEX-XYKS | 20/2979 | 0.029136 | 0.665396 | 0 | 0 | 1.71034 | 6.047385 |
| GTEx_Tissu | GTEX-OXR( | 53/10043 | 0.028786 | 0.665396 | 0 | 0 | 1.548369 | 5.493417 |
| GTEx_Tissu | GTEX-R55E | 4/209 | 0.013141 | 0.665396 | 0 | 0 | 4.633088 | 20.0706 |
| GTEx_Tissu | GTEX-XLM4 | 6/445 | 0.01304 | 0.665396 | 0 | 0 | 3.285919 | 14.26005 |
| GTEx_Tissu | GTEX-Y8E5 | 59/11105 | 0.012931 | 0.665396 | 0 | 0 | 1.691475 | 7.354784 |
| GTEx_Tissu | GTEX-WHP | 19/2765 | 0.027092 | 0.665396 | 0 | 0 | 1.746781 | 6.303298 |
| GTEx_Tissu | GTEX-Q2A( | 9/998 | 0.029307 | 0.665396 | 0 | 0 | 2.207825 | 7.79344 |
| GTEx_Tissu | GTEX-OHPI | 6/446 | 0.013173 | 0.665396 | 0 | 0 | 3.278283 | 14.19369 |
| GTEx_Tissu | GTEX-PX3G | 6/488 | 0.019647 | 0.665396 | 0 | 0 | 2.986169 | 11.73507 |
| GTEx_Tissu | GTEX-U3ZH | Feb-48 | 0.018536 | 0.665396 | 0 | 0 | 10.16215 | 40.52728 |

| | | | | | | | | |
|---|---|---|---|---|---|---|---|---|
| GTEx_Tissu | GTEX-RWS/ | 4/229 | 0.017795 | 0.665396 | 0 | 0 | 4.216975 | 16.98961 |
| GTEx_Tissu | GTEX-S4Q7 | 3/141 | 0.02365 | 0.665396 | 0 | 0 | 5.117754 | 19.16288 |
| GTEx_Tissu | GTEX-U8T8 | 51/9570 | 0.028151 | 0.665396 | 0 | 0 | 1.546889 | 5.52268 |
| GTEx_Tissu | GTEX-X4XY | 39/6345 | 0.006968 | 0.665396 | 0 | 0 | 1.753201 | 8.707255 |
| GTEx_Tissu | GTEX-T2IS- | 5/260 | 0.005554 | 0.665396 | 0 | 0 | 4.700622 | 24.41105 |
| GTEx_Tissu | GTEX-X4EC | Mar-81 | 0.005312 | 0.665396 | 0 | 0 | 9.08196 | 47.56873 |
| GTEx_Tissu | GTEX-QEG4 | 37/5769 | 0.004344 | 0.665396 | 0 | 0 | 1.830764 | 9.957328 |
| GTEx_Tissu | GTEX-XXEK | 30/4841 | 0.019838 | 0.665396 | 0 | 0 | 1.652135 | 6.47666 |
| GTEx_Tissu | GTEX-XQ3S | 7/626 | 0.019262 | 0.665396 | 0 | 0 | 2.727342 | 10.77198 |
| GTEx_Tissu | GTEX-T5JC- | 9/967 | 0.02456 | 0.665396 | 0 | 0 | 2.283001 | 8.462216 |
| GTEx_Tissu | GTEX-XUJ4 | 41/7227 | 0.022629 | 0.665396 | 0 | 0 | 1.578574 | 5.980449 |
| GTEx_Tissu | GTEX-W5W | 43/7492 | 0.014867 | 0.665396 | 0 | 0 | 1.635216 | 6.881999 |
| GTEx_Tissu | GTEX-XGQ4 | 23/3191 | 0.008383 | 0.665396 | 0 | 0 | 1.899537 | 9.082728 |
| GTEx_Tissu | GTEX-QDVI | 19/2179 | 0.002276 | 0.665396 | 0 | 0 | 2.29648 | 13.97503 |
| GTEx_Tissu | GTEX-TMZS | 21/3045 | 0.01925 | 0.665396 | 0 | 0 | 1.777041 | 7.019724 |
| GTEx_Tissu | GTEX-PW2 | 58/10853 | 0.012539 | 0.665396 | 0 | 0 | 1.689301 | 7.39726 |
| GTEx_Tissu | GTEX-OXRI | 7/561 | 0.011148 | 0.665396 | 0 | 0 | 3.057604 | 13.74851 |
| GTEx_Tissu | GTEX-WH7 | 22/3070 | 0.010757 | 0.665396 | 0 | 0 | 1.872754 | 8.487666 |
| GTEx_Tissu | GTEX-U8XE | 34/5422 | 0.00997 | 0.665396 | 0 | 0 | 1.729385 | 7.969261 |
| GTEx_Tissu | GTEX-XLM4 | 22/3355 | 0.027978 | 0.665396 | 0 | 0 | 1.683676 | 6.02142 |
| GTEx_Tissu | GTEX-VJYA | 19/2678 | 0.020049 | 0.665396 | 0 | 0 | 1.813077 | 7.088387 |
| GTEx_Tissu | GTEX-TML8 | 20/2914 | 0.023562 | 0.665396 | 0 | 0 | 1.755459 | 6.579668 |
| GTEx_Tissu | GTEX-TML8 | 43/7588 | 0.018792 | 0.665396 | 0 | 0 | 1.601976 | 6.366813 |
| GTEx_Tissu | GTEX-X88G | 21/3088 | 0.022222 | 0.665396 | 0 | 0 | 1.747666 | 6.652794 |
| GTEx_Tissu | GTEX-RN64 | 25/3869 | 0.022115 | 0.665396 | 0 | 0 | 1.685597 | 6.424686 |
| GTEx_Tissu | GTEX-R3RS | 3/137 | 0.021949 | 0.665396 | 0 | 0 | 5.271588 | 20.13247 |
| GTEx_Tissu | GTEX-WFO | 29/4770 | 0.028474 | 0.665396 | 0 | 0 | 1.600084 | 5.694301 |
| GTEx_Tissu | GTEX-QESD | 27/4359 | 0.028363 | 0.665396 | 0 | 0 | 1.618525 | 5.766256 |
| GTEx_Tissu | GTEX-WFJC | 19/2776 | 0.028103 | 0.665396 | 0 | 0 | 1.738697 | 6.210395 |
| GTEx_Tissu | GTEX-SNOS | 3/150 | 0.027733 | 0.665396 | 0 | 0 | 4.802235 | 17.21664 |
| GTEx_Tissu | GTEX-RWS | 19/2772 | 0.027732 | 0.665396 | 0 | 0 | 1.741629 | 6.24402 |
| GTEx_Tissu | GTEX-U4B1 | 6/527 | 0.027359 | 0.665396 | 0 | 0 | 2.757091 | 9.922011 |
| GTEx_Tissu | GTEX-PX3G | 29/4754 | 0.027304 | 0.665396 | 0 | 0 | 1.607196 | 5.78709 |
| GTEx_Tissu | GTEX-WEY! | 19/2787 | 0.029144 | 0.665396 | 0 | 0 | 1.730677 | 6.118846 |
| GTEx_Tissu | GTEX-P78B | 24/3404 | 0.009139 | 0.665396 | 0 | 0 | 1.863398 | 8.749053 |
| GTEx_Tissu | GTEX-R55G | 3/143 | 0.024527 | 0.665396 | 0 | 0 | 5.044133 | 18.70358 |
| GTEx_Tissu | GTEX-XBEV | 24/3088 | 0.002614 | 0.665396 | 0 | 0 | 2.094865 | 12.45756 |
| GTEx_Tissu | GTEX-U3ZN | 5/215 | 0.002479 | 0.665396 | 0 | 0 | 5.720964 | 34.32481 |
| GTEx_Tissu | GTEX-QVJC | 4/128 | 0.002365 | 0.665396 | 0 | 0 | 7.691022 | 46.50787 |
| GTEx_Tissu | GTEX-UTH( | 4/127 | 0.002298 | 0.665396 | 0 | 0 | 7.753943 | 47.10907 |
| GTEx_Tissu | GTEX-PWO | 23/3404 | 0.017659 | 0.665396 | 0 | 0 | 1.757228 | 7.093051 |
| GTEx_Tissu | GTEX-REY6 | 37/6257 | 0.017453 | 0.665396 | 0 | 0 | 1.629071 | 6.594906 |
| GTEx_Tissu | GTEX-S4UY | Feb-58 | 0.026415 | 0.665396 | 0 | 0 | 8.343277 | 30.31803 |
| GTEx_Tissu | GTEX-NL4V | 15/1876 | 0.014848 | 0.665396 | 0 | 0 | 2.020867 | 8.50765 |
| GTEx_Tissu | GTEX-XBEC | 22/3156 | 0.014635 | 0.665396 | 0 | 0 | 1.812076 | 7.654771 |

| | | | | | | | | |
|---|---|---|---|---|---|---|---|---|
| GTEx_Tissu | GTEX-RTLS- | 27/4128 | 0.014526 | 0.665396 | 0 | 0 | 1.73504 | 7.342327 |
| GTEx_Tissu | GTEX-WHS | 45/8191 | 0.027098 | 0.665396 | 0 | 0 | 1.547692 | 5.584507 |
| GTEx_Tissu | GTEX-QCQ( | 4/187 | 0.009016 | 0.665396 | 0 | 0 | 5.195865 | 24.46586 |
| GTEx_Tissu | GTEX-U3ZN | Mar-96 | 0.008491 | 0.665396 | 0 | 0 | 7.611367 | 36.29655 |
| GTEx_Tissu | GTEX-T5JW | 5/387 | 0.02687 | 0.665396 | 0 | 0 | 3.117578 | 11.2755 |
| GTEx_Tissu | GTEX-TML8 | Mar-96 | 0.008491 | 0.665396 | 0 | 0 | 7.611367 | 36.29655 |
| GTEx_Tissu | GTEX-R55D | 3/124 | 0.016901 | 0.665396 | 0 | 0 | 5.841795 | 23.83691 |
| GTEx_Tissu | GTEX-XBED | 21/3006 | 0.016836 | 0.665396 | 0 | 0 | 1.804416 | 7.369636 |
| GTEx_Tissu | GTEX-TMM | 5/342 | 0.01677 | 0.665396 | 0 | 0 | 3.542013 | 14.4804 |
| GTEx_Tissu | GTEX-Q2AI | 4/223 | 0.016305 | 0.665396 | 0 | 0 | 4.333828 | 17.83939 |
| GTEx_Tissu | GTEX-XLM4 | 22/3184 | 0.016118 | 0.665396 | 0 | 0 | 1.793033 | 7.40133 |
| GTEx_Tissu | GTEX-QDV. | 3/121 | 0.01584 | 0.665396 | 0 | 0 | 5.991223 | 24.83499 |
| GTEx_Tissu | GTEX-T6M( | 5/319 | 0.01276 | 0.665396 | 0 | 0 | 3.805927 | 16.59944 |
| GTEx_Tissu | GTEX-REY6 | 25/3781 | 0.016902 | 0.665396 | 0 | 0 | 1.734537 | 7.077505 |
| GTEx_Tissu | GTEX-Q2AI | 5/352 | 0.018748 | 0.665396 | 0 | 0 | 3.438181 | 13.67242 |
| GTEx_Tissu | GTEX-XMD | 11/1295 | 0.024503 | 0.665396 | 0 | 0 | 2.099924 | 7.788502 |
| GTEx_Tissu | GTEX-R55G | 8/809 | 0.024433 | 0.665396 | 0 | 0 | 2.41622 | 8.968569 |
| GTEx_Tissu | GTEX-T6M( | 52/9730 | 0.02414 | 0.665396 | 0 | 0 | 1.571222 | 5.851028 |
| GTEx_Tissu | GTEX-S7SE- | 20/2919 | 0.023959 | 0.665396 | 0 | 0 | 1.751917 | 6.53714 |
| GTEx_Tissu | GTEX-XAJ8- | Feb-55 | 0.02393 | 0.665396 | 0 | 0 | 8.81687 | 32.91021 |
| GTEx_Tissu | GTEX-U3ZN | 5/284 | 0.007994 | 0.665396 | 0 | 0 | 4.291022 | 20.72172 |
| GTEx_Tissu | GTEX-XOT( | 4/260 | 0.026831 | 0.665396 | 0 | 0 | 3.700489 | 13.38904 |
| GTEx_Tissu | GTEX-WK1 | Mar-88 | 0.006685 | 0.665396 | 0 | 0 | 8.331092 | 41.72057 |
| GTEx_Tissu | GTEX-RTLS- | 45/7408 | 0.003591 | 0.665396 | 0 | 0 | 1.826216 | 10.28047 |
| GTEx_Tissu | GTEX-XOT( | 22/3203 | 0.017191 | 0.665396 | 0 | 0 | 1.780301 | 7.23404 |
| GTEx_Tissu | GTEX-QLQ7 | 43/7435 | 0.012882 | 0.665396 | 0 | 0 | 1.655361 | 7.203989 |
| GTEx_Tissu | GTEX-XMD | 9/617 | 0.001441 | 0.665396 | 0 | 0 | 3.663651 | 23.97019 |
| GTEx_Tissu | GTEX-SJXC- | 31/4539 | 0.004141 | 0.665396 | 0 | 0 | 1.891697 | 10.37933 |
| GTEx_Tissu | GTEX-SNO! | 4/144 | 0.003613 | 0.665396 | 0 | 0 | 6.80654 | 38.27371 |
| GTEx_Tissu | GTEX-V955 | 46/8177 | 0.015625 | 0.665396 | 0 | 0 | 1.625732 | 6.761212 |
| GTEx_Tissu | GTEX-TSE9 | 8/741 | 0.015287 | 0.665396 | 0 | 0 | 2.649766 | 11.07805 |
| GTEx_Tissu | GTEX-NPJ7 | 9/979 | 0.026327 | 0.665396 | 0 | 0 | 2.253331 | 8.195712 |
| GTEx_Tissu | GTEX-T5JC- | 7/665 | 0.02576 | 0.665396 | 0 | 0 | 2.560505 | 9.368718 |
| GTEx_Tissu | GTEX-U3ZN | 19/2744 | 0.025239 | 0.665396 | 0 | 0 | 1.762396 | 6.484496 |
| GTEx_Tissu | GTEX-XUZ( | 49/9061 | 0.025194 | 0.665396 | 0 | 0 | 1.55976 | 5.741729 |
| GTEx_Tissu | GTEX-Q2A( | Feb-56 | 0.024747 | 0.665396 | 0 | 0 | 8.653159 | 32.00857 |
| GTEx_Tissu | GTEX-QCQ( | 5/378 | 0.024599 | 0.665396 | 0 | 0 | 3.194272 | 11.83497 |
| SysMyo_M | MGS1481 I | 1/203 | 0.589132 | 0.665566 | 0 | 0 | 1.134642 | 0.600344 |
| GeDiPNet_ | Congenital | 1/237 | 0.646317 | 0.665729 | 0 | 0 | 0.969501 | 0.423154 |
| DisGeNET | Uveal mela | 1/236 | 0.644753 | 0.665847 | 0 | 0 | 0.973676 | 0.427334 |
| KEGG_202 | NOD-like re | 1/181 | 0.547354 | 0.66665 | 0 | 0 | 1.274742 | 0.768234 |
| GeneSigDB | 12509443-T | 2/401 | 0.523185 | 0.666852 | 0 | 0 | 1.150759 | 0.745484 |
| DisGeNET | Cognition I | 1/237 | 0.646317 | 0.666936 | 0 | 0 | 0.969501 | 0.423154 |
| DisGeNET | Periodontit | 1/237 | 0.646317 | 0.666936 | 0 | 0 | 0.969501 | 0.423154 |
| Jensen_TIS | Superior ce | 20/4740 | 0.603335 | 0.667608 | 0 | 0 | 0.960853 | 0.485501 |

| | | | | | | | | |
|---|---|---|---|---|---|---|---|---|
| SysMyo_M | MGS669 H | 1/207 | 0.596308 | 0.667681 | 0 | 0 | 1.112384 | 0.575101 |
| SysMyo_M | MGS1212 I | 1/206 | 0.594525 | 0.667681 | 0 | 0 | 1.117867 | 0.581282 |
| SysMyo_M | MGS1301 I | 1/208 | 0.598082 | 0.667681 | 0 | 0 | 1.106954 | 0.569005 |
| SysMyo_M | MGS1437 I | 1/206 | 0.594525 | 0.667681 | 0 | 0 | 1.117867 | 0.581282 |
| SysMyo_M | MGS1338 I | 1/208 | 0.598082 | 0.667681 | 0 | 0 | 1.106954 | 0.569005 |
| SysMyo_M | MGS411 H | 1/207 | 0.596308 | 0.667681 | 0 | 0 | 1.112384 | 0.575101 |
| DisGeNET | Hepatitis B | 1/238 | 0.647874 | 0.668016 | 0 | 0 | 0.965362 | 0.419024 |
| DisGeNET | Endometri | 1/238 | 0.647874 | 0.668016 | 0 | 0 | 0.965362 | 0.419024 |
| Jensen_TIS | Cytotoxic T | 22/5212 | 0.605905 | 0.668155 | 0 | 0 | 0.960148 | 0.481066 |
| SysMyo_M | MGS1251 I | 1/209 | 0.599849 | 0.668335 | 0 | 0 | 1.101576 | 0.562991 |
| GO_Biologi | Regulation | 3/766 | 0.652706 | 0.668718 | 0 | 0 | 0.896368 | 0.382416 |
| GO_Biologi | Regulation | 1/240 | 0.650968 | 0.668718 | 0 | 0 | 0.957186 | 0.410916 |
| GO_Biologi | Regulation | 1/241 | 0.652504 | 0.668718 | 0 | 0 | 0.953149 | 0.406935 |
| KEGG_202 | Chemokine | 1/192 | 0.568743 | 0.669266 | 0 | 0 | 1.200657 | 0.677563 |
| KEGG_202 | Kaposi sarc | 1/193 | 0.570637 | 0.669266 | 0 | 0 | 1.194344 | 0.670029 |
| KEGG_202 | Alcoholism | 1/186 | 0.557204 | 0.669266 | 0 | 0 | 1.239975 | 0.725168 |
| GeneSigDB | 16141321- | 4/869 | 0.526346 | 0.669348 | 0 | 0 | 1.061244 | 0.681103 |
| GTEx_Tissu | GTEX-PW2 | 7/786 | 0.055082 | 0.669548 | 0 | 0 | 2.149198 | 6.230375 |
| KOMP2_M | Prenatal Le | 1/109 | 0.379031 | 0.669689 | 0 | 0 | 2.132321 | 2.068645 |
| KOMP2_M | Increased E | 1/111 | 0.384451 | 0.669689 | 0 | 0 | 2.09334 | 2.001105 |
| Reactome_ | GPCR Dowi | 3/619 | 0.507605 | 0.669863 | 0 | 0 | 1.118796 | 0.758602 |
| Reactome_ | Anti-inflam | 1/165 | 0.514359 | 0.669863 | 0 | 0 | 1.400241 | 0.930927 |
| Reactome_ | Protein Loc | 1/164 | 0.51222 | 0.669863 | 0 | 0 | 1.408903 | 0.942558 |
| Reactome_ | Transport T | 1/164 | 0.51222 | 0.669863 | 0 | 0 | 1.408903 | 0.942558 |
| Reactome_ | Citric Acid | 1/163 | 0.510071 | 0.669863 | 0 | 0 | 1.417672 | 0.954384 |
| Rare_Disea | Absent T ly | 1/243 | 0.655558 | 0.669966 | 0 | 0 | 0.945176 | 0.399118 |
| DisGeNET | Pituitary Di | 1/240 | 0.650968 | 0.670678 | 0 | 0 | 0.957186 | 0.410916 |
| DisGeNET | Hyperchole | 1/240 | 0.650968 | 0.670678 | 0 | 0 | 0.957186 | 0.410916 |
| Disease_Pe | Muscular D | 4/329 | 0.05538 | 0.67083 | 0 | 0 | 2.904615 | 8.404609 |
| DisGeNET | Metastatic | 1/241 | 0.652504 | 0.671206 | 0 | 0 | 0.953149 | 0.406935 |
| DisGeNET | Non-Small | Sep-43 | 0.652053 | 0.671206 | 0 | 0 | 0.913109 | 0.390473 |
| DisGeNET | Malignant | Aug-01 | 0.652092 | 0.671206 | 0 | 0 | 0.910529 | 0.389314 |
| DisGeNET | Chondrosa | 1/241 | 0.652504 | 0.671206 | 0 | 0 | 0.953149 | 0.406935 |
| Disease_Pe | Type 2 diak | 4/346 | 0.064222 | 0.671636 | 0 | 0 | 2.757838 | 7.571375 |
| Disease_Pe | asthma DO | 3/211 | 0.06446 | 0.671636 | 0 | 0 | 3.383413 | 9.276318 |
| Disease_Pe | Cancer of t | 4/346 | 0.064222 | 0.671636 | 0 | 0 | 2.757838 | 7.571375 |
| RNA-Seq_I | PSMD13 H | 6/497 | 0.021275 | 0.671706 | 0 | 0 | 2.930075 | 11.28141 |
| RNA-Seq_I | AMPK R29 | 6/496 | 0.02109 | 0.671706 | 0 | 0 | 2.936206 | 11.33069 |
| RNA-Seq_I | WZ4002 N | 6/497 | 0.021275 | 0.671706 | 0 | 0 | 2.930075 | 11.28141 |
| RNA-Seq_I | SMYD1 Tib | 6/498 | 0.021462 | 0.671706 | 0 | 0 | 2.923969 | 11.2324 |
| RNA-Seq_I | Rb1 Lung k | 6/498 | 0.021462 | 0.671706 | 0 | 0 | 2.923969 | 11.2324 |
| RNA-Seq_I | PSMD13 H | 6/498 | 0.021462 | 0.671706 | 0 | 0 | 2.923969 | 11.2324 |
| RNA-Seq_I | CDK12 HCT | 6/498 | 0.021462 | 0.671706 | 0 | 0 | 2.923969 | 11.2324 |
| RNA-Seq_I | YAP Oocyt | 6/497 | 0.021275 | 0.671706 | 0 | 0 | 2.930075 | 11.28141 |
| RNA-Seq_I | Vitamin E k | 6/498 | 0.021462 | 0.671706 | 0 | 0 | 2.923969 | 11.2324 |

| | | | | | | | | |
|---|---|---|---|---|---|---|---|---|
| RNA-Seq_D | Vemurafen | 6/498 | 0.021462 | 0.671706 | 0 | 0 | 2.923969 | 11.2324 |
| RNA-Seq_D | Hif1a Heart | 6/499 | 0.021649 | 0.671706 | 0 | 0 | 2.917887 | 11.18366 |
| RNA-Seq_D | Sarcoma Sa | 6/497 | 0.021275 | 0.671706 | 0 | 0 | 2.930075 | 11.28141 |
| RNA-Seq_D | TGFB3 Pala | 6/496 | 0.02109 | 0.671706 | 0 | 0 | 2.936206 | 11.33069 |
| RNA-Seq_D | Dioxin Live | 6/496 | 0.02109 | 0.671706 | 0 | 0 | 2.936206 | 11.33069 |
| RNA-Seq_D | LPS Astrocy | 6/497 | 0.021275 | 0.671706 | 0 | 0 | 2.930075 | 11.28141 |
| RNA-Seq_D | CFTR Color | 6/497 | 0.021275 | 0.671706 | 0 | 0 | 2.930075 | 11.28141 |
| RNA-Seq_D | Panobinost | 6/497 | 0.021275 | 0.671706 | 0 | 0 | 2.930075 | 11.28141 |
| RNA-Seq_D | Dgcr8 Cone | 6/498 | 0.021462 | 0.671706 | 0 | 0 | 2.923969 | 11.2324 |
| RNA-Seq_D | Gtf2ird1 Li | 6/499 | 0.021649 | 0.671706 | 0 | 0 | 2.917887 | 11.18366 |
| RNA-Seq_D | MEF2 Skele | 6/499 | 0.021649 | 0.671706 | 0 | 0 | 2.917887 | 11.18366 |
| RNA-Seq_D | Bmal1 Live | 6/497 | 0.021275 | 0.671706 | 0 | 0 | 2.930075 | 11.28141 |
| RNA-Seq_D | miR-92a-1 | 6/496 | 0.02109 | 0.671706 | 0 | 0 | 2.936206 | 11.33069 |
| RNA-Seq_D | CDK10 HCT | 6/495 | 0.020906 | 0.671706 | 0 | 0 | 2.942362 | 11.38025 |
| RNA-Seq_D | NIPBL Mou | 6/495 | 0.020906 | 0.671706 | 0 | 0 | 2.942362 | 11.38025 |
| GTEx_Tissu | GTEX-OHPI | 9/1125 | 0.055517 | 0.67201 | 0 | 0 | 1.943445 | 5.618631 |
| UK_Bioban | Left cornea | 1/243 | 0.655558 | 0.672065 | 0 | 0 | 0.945176 | 0.399118 |
| SysMyo_M | MGS819 M | 1/216 | 0.612003 | 0.672434 | 0 | 0 | 1.065333 | 0.523098 |
| SysMyo_M | MGS483 H | 1/216 | 0.612003 | 0.672434 | 0 | 0 | 1.065333 | 0.523098 |
| SysMyo_M | MGS909 M | 1/213 | 0.606839 | 0.672434 | 0 | 0 | 1.080573 | 0.539737 |
| SysMyo_M | MGS140 M | 2/468 | 0.607449 | 0.672434 | 0 | 0 | 0.981924 | 0.489476 |
| SysMyo_M | MGS410 H | 1/217 | 0.613709 | 0.672434 | 0 | 0 | 1.060347 | 0.517698 |
| SysMyo_M | MGS316 M | 1/212 | 0.605103 | 0.672434 | 0 | 0 | 1.085749 | 0.545433 |
| SysMyo_M | MGS1374 I | 1/216 | 0.612003 | 0.672434 | 0 | 0 | 1.065333 | 0.523098 |
| SysMyo_M | MGS1210 I | 1/214 | 0.608568 | 0.672434 | 0 | 0 | 1.075445 | 0.534116 |
| SysMyo_M | MGS1013 I | 1/218 | 0.615408 | 0.672434 | 0 | 0 | 1.055407 | 0.512369 |
| SysMyo_M | MGS1295 I | 1/218 | 0.615408 | 0.672434 | 0 | 0 | 1.055407 | 0.512369 |
| GeDiPNet_ | Psychosis | 1/242 | 0.654034 | 0.67249 | 0 | 0 | 0.949146 | 0.403003 |
| Gene_Pert | Rps6ka3 K( | 5/319 | 0.01276 | 0.67288 | 0 | 0 | 3.805927 | 16.59944 |
| GeneSigDB | 18285497- | 2/410 | 0.535171 | 0.672989 | 0 | 0 | 1.124856 | 0.703224 |
| GeneSigDB | 18849563- | 1/174 | 0.533201 | 0.672989 | 0 | 0 | 1.326791 | 0.834362 |
| GeneSigDB | 16536878- | 1/175 | 0.535249 | 0.672989 | 0 | 0 | 1.319099 | 0.824467 |
| GeneSigDB | 18849563- | 1/174 | 0.533201 | 0.672989 | 0 | 0 | 1.326791 | 0.834362 |
| GeneSigDB | 17389037- | 1/174 | 0.533201 | 0.672989 | 0 | 0 | 1.326791 | 0.834362 |
| GTEx_Tissu | GTEX-SUCS | 8/955 | 0.055842 | 0.673128 | 0 | 0 | 2.028097 | 5.851526 |
| CellMarker | SLC16A7+ ( | 4/1013 | 0.648585 | 0.673153 | 0 | 0 | 0.90291 | 0.390925 |
| PhenGenI_ | Citalopram | 1/163 | 0.510071 | 0.673294 | 0 | 0 | 1.417672 | 0.954384 |
| KEGG_202 | Focal adhe | 1/201 | 0.585497 | 0.673844 | 0 | 0 | 1.146105 | 0.613503 |
| KEGG_202 | Proteoglyc | 1/205 | 0.592736 | 0.673844 | 0 | 0 | 1.123404 | 0.587548 |
| KEGG_202 | cAMP signa | 1/216 | 0.612003 | 0.673844 | 0 | 0 | 1.065333 | 0.523098 |
| KEGG_202 | Human T-c | 1/219 | 0.617099 | 0.673844 | 0 | 0 | 1.050512 | 0.507109 |
| KEGG_202 | Lipid and a | 1/215 | 0.610289 | 0.673844 | 0 | 0 | 1.070365 | 0.528571 |
| KEGG_202 | Human imr | 1/212 | 0.605103 | 0.673844 | 0 | 0 | 1.085749 | 0.545433 |
| KEGG_202 | Thermoger | 1/232 | 0.63843 | 0.673898 | 0 | 0 | 0.990738 | 0.444588 |
| KEGG_202 | Ras signalir | 1/232 | 0.63843 | 0.673898 | 0 | 0 | 0.990738 | 0.444588 |

| | | | | | | | | |
|---|---|---|---|---|---|---|---|---|
| KEGG_202: | Human cyt | 1/225 | 0.627094 | 0.673898 | 0 | 0 | 1.022062 | 0.476954 |
| GeneSigDB | 17676974-' | 6/1348 | 0.537457 | 0.674243 | 0 | 0 | 1.025059 | 0.636466 |
| GeDiPNet_ | Epilepsy | 1/244 | 0.657075 | 0.674427 | 0 | 0 | 0.941238 | 0.39528 |
| GeneSigDB | 18435859-: | 2/414 | 0.540433 | 0.674557 | 0 | 0 | 1.113706 | 0.685357 |
| GeneSigDB | 16288205-' | 1/178 | 0.541341 | 0.674557 | 0 | 0 | 1.296544 | 0.795696 |
| GeneSigDB | 20068086-: | 1/177 | 0.53932 | 0.674557 | 0 | 0 | 1.303977 | 0.805137 |
| GeDiPNet_ | Anemia | 2/514 | 0.658575 | 0.674779 | 0 | 0 | 0.89159 | 0.372397 |
| KOMP2_M | Decreased | 1/156 | 0.494766 | 0.674894 | 0 | 0 | 1.482221 | 1.042994 |
| KOMP2_M | Edema (MI | 1/135 | 0.445958 | 0.674894 | 0 | 0 | 1.716331 | 1.38599 |
| KOMP2_M | Increased E | 1/141 | 0.46036 | 0.674894 | 0 | 0 | 1.642276 | 1.27399 |
| KOMP2_M | Persistence | 1/160 | 0.503569 | 0.674894 | 0 | 0 | 1.444639 | 0.991073 |
| KOMP2_M | Decreased | 1/139 | 0.455601 | 0.674894 | 0 | 0 | 1.666245 | 1.309899 |
| KOMP2_M | Preweaning | 4/801 | 0.462334 | 0.674894 | 0 | 0 | 1.155901 | 0.891741 |
| KOMP2_M | Abnormal ( | 1/162 | 0.507913 | 0.674894 | 0 | 0 | 1.426549 | 0.966408 |
| KOMP2_M | Decreased | 1/132 | 0.438615 | 0.674894 | 0 | 0 | 1.755903 | 1.447098 |
| KOMP2_M | Decreased | 1/127 | 0.426163 | 0.674894 | 0 | 0 | 1.826043 | 1.557494 |
| KOMP2_M | Increased I | 1/152 | 0.485809 | 0.674894 | 0 | 0 | 1.521793 | 1.098642 |
| KOMP2_M | Microphtha | 1/145 | 0.469755 | 0.674894 | 0 | 0 | 1.596334 | 1.2061 |
| KOMP2_M | Enlarged Ki | 1/146 | 0.472079 | 0.674894 | 0 | 0 | 1.585245 | 1.1899 |
| KOMP2_M | Enlarged Tl | 1/146 | 0.472079 | 0.674894 | 0 | 0 | 1.585245 | 1.1899 |
| KOMP2_M | Decreased | 1/121 | 0.410859 | 0.674894 | 0 | 0 | 1.917926 | 1.706006 |
| KOMP2_M | Small Splee | 1/126 | 0.42364 | 0.674894 | 0 | 0 | 1.840744 | 1.580964 |
| KOMP2_M | Decreased | 1/129 | 0.431176 | 0.674894 | 0 | 0 | 1.797329 | 1.511983 |
| KOMP2_M | Increased ( | 2/391 | 0.509632 | 0.674894 | 0 | 0 | 1.180947 | 0.796037 |
| KOMP2_M | Increased I | 1/169 | 0.522824 | 0.674894 | 0 | 0 | 1.366625 | 0.886269 |
| KOMP2_M | Decreased | 1/170 | 0.524918 | 0.674894 | 0 | 0 | 1.35847 | 0.875552 |
| KOMP2_M | Decreased | 1/156 | 0.494766 | 0.674894 | 0 | 0 | 1.482221 | 1.042994 |
| KOMP2_M | Increased ( | 1/133 | 0.441073 | 0.674894 | 0 | 0 | 1.742512 | 1.426323 |
| KOMP2_M | Decreased | 1/170 | 0.524918 | 0.674894 | 0 | 0 | 1.35847 | 0.875552 |
| GeneSigDB | 17229949-' | 1/180 | 0.545359 | 0.674912 | 0 | 0 | 1.281928 | 0.777247 |
| GeneSigDB | 20215513-' | 1/182 | 0.549341 | 0.674912 | 0 | 0 | 1.267635 | 0.759358 |
| GeneSigDB | 20368555-' | 1/182 | 0.549341 | 0.674912 | 0 | 0 | 1.267635 | 0.759358 |
| GeneSigDB | 17284527-' | 1/183 | 0.55132 | 0.674912 | 0 | 0 | 1.260606 | 0.750615 |
| GeneSigDB | 15762987-: | 1/181 | 0.547354 | 0.674912 | 0 | 0 | 1.274742 | 0.768234 |
| GeneSigDB | 18498629-' | 1/181 | 0.547354 | 0.674912 | 0 | 0 | 1.274742 | 0.768234 |
| GeneSigDB | 18691415-' | 1/183 | 0.55132 | 0.674912 | 0 | 0 | 1.260606 | 0.750615 |
| GeneSigDB | 20215513-' | 1/181 | 0.547354 | 0.674912 | 0 | 0 | 1.274742 | 0.768234 |
| GeDiPNet_ | Cerebral Cc | 1/246 | 0.660088 | 0.675143 | 0 | 0 | 0.93346 | 0.387742 |
| GO_Biologi | Positive Re | 1/246 | 0.660088 | 0.675306 | 0 | 0 | 0.93346 | 0.387742 |
| Gene_Pert | Lat2 KD mc | 5/355 | 0.019371 | 0.675746 | 0 | 0 | 3.408188 | 13.44184 |
| GTEx_Tissu | GTEX-XV7C | 8/957 | 0.056397 | 0.676996 | 0 | 0 | 2.023609 | 5.818569 |
| GeDiPNet_ | Scoliosis | 3/778 | 0.663132 | 0.677068 | 0 | 0 | 0.881935 | 0.362283 |
| GTEx_Tissu | GTEX-TMZ: | 38/6697 | 0.030136 | 0.677291 | 0 | 0 | 1.543567 | 5.405644 |
| DisGeNET | Kidney Nec | 1/246 | 0.660088 | 0.678208 | 0 | 0 | 0.93346 | 0.387742 |
| DisGeNET | Gastroesop | 1/246 | 0.660088 | 0.678208 | 0 | 0 | 0.93346 | 0.387742 |

| | | | | | | | | |
|---|---|---|---|---|---|---|---|---|
| DisGeNET | Leukopenia | 1/246 | 0.660088 | 0.678208 | 0 | 0 | 0.93346 | 0.387742 |
| Rare_Disea | Salivary gla | 1/249 | 0.66456 | 0.678234 | 0 | 0 | 0.922027 | 0.376768 |
| Gene_Pert | Stat3 defici | 5/322 | 0.013241 | 0.678329 | 0 | 0 | 3.769331 | 16.30017 |
| DisGeNET | Splenomeg | 1/247 | 0.661585 | 0.67948 | 0 | 0 | 0.929618 | 0.384041 |
| KEGG_202 | Calcium sig | 1/240 | 0.650968 | 0.679582 | 0 | 0 | 0.957186 | 0.410916 |
| GeDiPNet_ | Retinitis Pi | 1/251 | 0.667508 | 0.680345 | 0 | 0 | 0.914558 | 0.369667 |
| DisGeNET | Primary ma | 4/1032 | 0.663107 | 0.68051 | 0 | 0 | 0.885331 | 0.36371 |
| DisGeNET | Mental def | 4/1032 | 0.663107 | 0.68051 | 0 | 0 | 0.885331 | 0.36371 |
| GeneSigDB | 19225562- | 2/427 | 0.557254 | 0.68068 | 0 | 0 | 1.07892 | 0.630882 |
| GTEx_Tissu | GTEX-WOF | 4/271 | 0.030593 | 0.680697 | 0 | 0 | 3.546049 | 12.365 |
| Disease_Pe | Malignant | 4/350 | 0.066405 | 0.680917 | 0 | 0 | 2.725399 | 7.391226 |
| GeneSigDB | 20460173- | 7/1612 | 0.558778 | 0.681049 | 0 | 0 | 0.9981 | 0.580897 |
| GeneSigDB | 18974375- | 1/188 | 0.561084 | 0.681358 | 0 | 0 | 1.226589 | 0.708828 |
| GeneSigDB | 19139136- | 7/1616 | 0.561478 | 0.681358 | 0 | 0 | 0.995401 | 0.574528 |
| Gene_Pert | Dmd Defici | 4/237 | 0.01991 | 0.681442 | 0 | 0 | 4.070531 | 15.94241 |
| DisGeNET | FANCONI A | 1/249 | 0.66456 | 0.681733 | 0 | 0 | 0.922027 | 0.376768 |
| GTEx_Tissu | GTEX-P4PC | 9/1132 | 0.057302 | 0.682197 | 0 | 0 | 1.930612 | 5.520438 |
| GTEx_Tissu | GTEX-XBED | 7/793 | 0.057237 | 0.682197 | 0 | 0 | 2.129278 | 6.090915 |
| KOMP2_M | Increased ( | 1/176 | 0.537289 | 0.682673 | 0 | 0 | 1.311495 | 0.814726 |
| GTEx_Tissu | GTEX-TKQ1 | 18/2615 | 0.030992 | 0.682739 | 0 | 0 | 1.739398 | 6.04273 |
| DisGeNET | Immune Sy | 1/250 | 0.666037 | 0.682982 | 0 | 0 | 0.918278 | 0.373197 |
| Rare_Disea | Brenner tu | 1/253 | 0.670431 | 0.683289 | 0 | 0 | 0.907207 | 0.362732 |
| GTEx_Tissu | GTEX-XQ3S | 7/795 | 0.057862 | 0.68378 | 0 | 0 | 2.123652 | 6.051746 |
| GTEx_Tissu | GTEX-OXR( | 4/334 | 0.057907 | 0.68378 | 0 | 0 | 2.859876 | 8.147531 |
| SysMyo_M | MGS1389 I | 1/225 | 0.627094 | 0.683883 | 0 | 0 | 1.022062 | 0.476954 |
| DisGeNET | Visual Impa | 1/251 | 0.667508 | 0.683955 | 0 | 0 | 0.914558 | 0.369667 |
| DisGeNET | Dystonia | 1/251 | 0.667508 | 0.683955 | 0 | 0 | 0.914558 | 0.369667 |
| GeneSigDB | 16365179- | 1/190 | 0.56493 | 0.684057 | 0 | 0 | 1.213486 | 0.692965 |
| Gene_Pert | ESRRA Defi | 5/325 | 0.013735 | 0.684082 | 0 | 0 | 3.733422 | 16.00818 |
| GTEx_Tissu | GTEX-S33H | 11/1491 | 0.058182 | 0.684234 | 0 | 0 | 1.802658 | 5.127072 |
| SysMyo_M | MGS114 M | 1/226 | 0.628735 | 0.684354 | 0 | 0 | 1.017468 | 0.472152 |
| GO_Biologi | Positive Re | 2/525 | 0.669995 | 0.684455 | 0 | 0 | 0.872343 | 0.34936 |
| GO_Molec | Double-Str | 3/650 | 0.540727 | 0.684464 | 0 | 0 | 1.06348 | 0.653871 |
| GWAS_Cat | Red Cell Di | 3/794 | 0.676685 | 0.68454 | 0 | 0 | 0.863374 | 0.33719 |
| GeDiPNet_ | Eczema | 1/255 | 0.673329 | 0.68508 | 0 | 0 | 0.899973 | 0.355958 |
| GTEx_Tissu | GTEX-QVU | 45/8263 | 0.031722 | 0.685234 | 0 | 0 | 1.524745 | 5.261525 |
| GTEx_Tissu | GTEX-S7SE | 3/160 | 0.032682 | 0.685234 | 0 | 0 | 4.494086 | 15.37391 |
| GTEx_Tissu | GTEX-REY6 | 6/551 | 0.032987 | 0.685234 | 0 | 0 | 2.632416 | 8.980866 |
| GTEx_Tissu | GTEX-WXY | 43/7824 | 0.03222 | 0.685234 | 0 | 0 | 1.523747 | 5.234339 |
| GTEx_Tissu | GTEX-XMK | 19/2817 | 0.032131 | 0.685234 | 0 | 0 | 1.709125 | 5.87588 |
| GTEx_Tissu | GTEX-U3ZC | 58/11290 | 0.033402 | 0.685234 | 0 | 0 | 1.545762 | 5.254257 |
| GTEx_Tissu | GTEX-QESD | 5/411 | 0.033569 | 0.685234 | 0 | 0 | 2.929683 | 9.943809 |
| GTEx_Tissu | GTEX-TML8 | 3/160 | 0.032682 | 0.685234 | 0 | 0 | 4.494086 | 15.37391 |
| UK_Bioban | Right corne | 1/254 | 0.671883 | 0.686213 | 0 | 0 | 0.903576 | 0.359325 |
| UK_Bioban | Longest pe | 3/791 | 0.674174 | 0.686213 | 0 | 0 | 0.866797 | 0.34175 |

| | | | | | | | | |
|---|---|---|---|---|---|---|---|---|
| DisGeNET | Osteosarcc | 4/1042 | 0.67059 | 0.686843 | 0 | 0 | 0.876338 | 0.350183 |
| DisGeNET | Polycystic ( | 2/526 | 0.671018 | 0.687013 | 0 | 0 | 0.870633 | 0.347347 |
| GeneSigDB | 19168792-1 | 2/436 | 0.568645 | 0.687061 | 0 | 0 | 1.056059 | 0.596144 |
| GTEx_Tissu | GTEX-NFK9 | 8/967 | 0.059225 | 0.687228 | 0 | 0 | 2.001452 | 5.656943 |
| GTEx_Tissu | GTEX-P4PP | 9/1140 | 0.059387 | 0.687228 | 0 | 0 | 1.91614 | 5.41056 |
| GTEx_Tissu | GTEX-TKQ2 | 8/966 | 0.058938 | 0.687228 | 0 | 0 | 2.003647 | 5.672872 |
| GTEx_Tissu | GTEX-SIU7- | 8/966 | 0.058938 | 0.687228 | 0 | 0 | 2.003647 | 5.672872 |
| GeneSigDB | 20460173- | 5/1156 | 0.570248 | 0.687507 | 0 | 0 | 0.99394 | 0.55828 |
| DisGeNET | Hepatocarc | 2/527 | 0.672038 | 0.687789 | 0 | 0 | 0.86893 | 0.345347 |
| PhenGenI_ | Renal Insuf | 1/171 | 0.527002 | 0.688097 | 0 | 0 | 1.35041 | 0.865006 |
| PhenGenI_ | Aspartate A | 1/174 | 0.533201 | 0.688097 | 0 | 0 | 1.326791 | 0.834362 |
| PhenGenI_ | Adipose Tis | 1/174 | 0.533201 | 0.688097 | 0 | 0 | 1.326791 | 0.834362 |
| PhenGenI_ | Pancreatic | 1/174 | 0.533201 | 0.688097 | 0 | 0 | 1.326791 | 0.834362 |
| SysMyo_M | MGS1353 I | 1/229 | 0.633614 | 0.688341 | 0 | 0 | 1.003927 | 0.458108 |
| Disease_Pe | Emery-Dre | 4/334 | 0.057907 | 0.688454 | 0 | 0 | 2.859876 | 8.147531 |
| SysMyo_M | MGS546 Hu | 1/230 | 0.635226 | 0.68877 | 0 | 0 | 0.999492 | 0.453544 |
| DisGeNET | Acute Coro | 1/255 | 0.673329 | 0.688841 | 0 | 0 | 0.899973 | 0.355958 |
| DisGeNET | Undergrow | 2/529 | 0.674072 | 0.689063 | 0 | 0 | 0.865543 | 0.341386 |
| DisGeNET | Sepsis | 2/529 | 0.674072 | 0.689063 | 0 | 0 | 0.865543 | 0.341386 |
| Disease_Pe | adrenoleuk | 4/342 | 0.062078 | 0.689609 | 0 | 0 | 2.791046 | 7.75731 |
| Disease_Pe | retinoblast | 3/207 | 0.061589 | 0.689609 | 0 | 0 | 3.450455 | 9.617355 |
| Disease_Pe | glaucoma a | 4/341 | 0.061549 | 0.689609 | 0 | 0 | 2.799471 | 7.804726 |
| Disease_Pe | Tachycardi | 3/208 | 0.062301 | 0.689609 | 0 | 0 | 3.433449 | 9.530487 |
| GeDiPNet_ | Congenital | 1/259 | 0.679049 | 0.689696 | 0 | 0 | 0.885839 | 0.342875 |
| Disease_Pe | Hypoxia CC | 4/349 | 0.065856 | 0.690297 | 0 | 0 | 2.733438 | 7.435733 |
| Disease_Pe | Meningocc | 3/213 | 0.065919 | 0.690297 | 0 | 0 | 3.35085 | 9.112059 |
| Disease_Pe | Gastrointes | 3/224 | 0.07421 | 0.690297 | 0 | 0 | 3.182288 | 8.276655 |
| Disease_Pe | prostate ca | 4/362 | 0.073184 | 0.690297 | 0 | 0 | 2.632429 | 6.883225 |
| Disease_Pe | Asthma, all | 3/217 | 0.068882 | 0.690297 | 0 | 0 | 3.28755 | 8.795386 |
| Disease_Pe | dengue hei | 3/226 | 0.075766 | 0.690297 | 0 | 0 | 3.153427 | 8.136178 |
| Disease_Pe | Huntingtor | 4/371 | 0.078492 | 0.690297 | 0 | 0 | 2.566692 | 6.531619 |
| Disease_Pe | Huntingtor | 4/371 | 0.078492 | 0.690297 | 0 | 0 | 2.566692 | 6.531619 |
| Disease_Pe | Gastrointes | 3/220 | 0.071143 | 0.690297 | 0 | 0 | 3.241606 | 8.567754 |
| Disease_Pe | osteoarthri | 3/226 | 0.075766 | 0.690297 | 0 | 0 | 3.153427 | 8.136178 |
| Disease_Pe | cerebral pa | 3/219 | 0.070386 | 0.690297 | 0 | 0 | 3.256779 | 8.642721 |
| Disease_Pe | Huntingtor | 4/356 | 0.069752 | 0.690297 | 0 | 0 | 2.678122 | 7.131345 |
| Disease_Pe | diabetes m | 4/346 | 0.064222 | 0.690297 | 0 | 0 | 2.757838 | 7.571375 |
| Disease_Pe | Acute Lung | 3/228 | 0.077336 | 0.690297 | 0 | 0 | 3.125079 | 7.998944 |
| Disease_Pe | dengue dis | 3/225 | 0.074986 | 0.690297 | 0 | 0 | 3.167793 | 8.206005 |
| GTEx_Tissu | GTEX-O5YV | 5/484 | 0.059937 | 0.690824 | 0 | 0 | 2.473904 | 6.962721 |
| SysMyo_M | MGS1032 I | 1/232 | 0.63843 | 0.69092 | 0 | 0 | 0.990738 | 0.444588 |
| GeneSigDB | 19838208-1 | 1/195 | 0.574401 | 0.691018 | 0 | 0 | 1.181911 | 0.655284 |
| GTEx_Tissu | GTEX-T5JW | 4/284 | 0.035422 | 0.691356 | 0 | 0 | 3.379174 | 11.28784 |
| GTEx_Tissu | GTEX-X88G | 34/5901 | 0.034898 | 0.691356 | 0 | 0 | 1.535818 | 5.153187 |
| GTEx_Tissu | GTEX-TKQ1 | 6/560 | 0.035279 | 0.691356 | 0 | 0 | 2.588448 | 8.656988 |

| | | | | | | | | |
|---|---|---|---|---|---|---|---|---|
| GTEx_Tissu | GTEX-OOB. | 35/6117 | 0.035089 | 0.691356 | 0 | 0 | 1.530636 | 5.127425 |
| GTEx_Tissu | GTEX-Q2A( | 20/3032 | 0.03439 | 0.691356 | 0 | 0 | 1.674992 | 5.644713 |
| GO_Molecı | Protein Ho | 3/662 | 0.553211 | 0.691514 | 0 | 0 | 1.043464 | 0.617747 |
| DisGeNET | Narcolepsy | 1/258 | 0.677628 | 0.692158 | 0 | 0 | 0.889331 | 0.346089 |
| DisGeNET | Behcet Syn | 1/258 | 0.677628 | 0.692158 | 0 | 0 | 0.889331 | 0.346089 |
| Reactome_ | Ion Channe | 1/175 | 0.535249 | 0.69304 | 0 | 0 | 1.319099 | 0.824467 |
| PhenGenI_ | Diabetes M | 2/416 | 0.543049 | 0.693062 | 0 | 0 | 1.108213 | 0.676626 |
| PhenGenI_ | Mental Cor | 1/178 | 0.541341 | 0.693062 | 0 | 0 | 1.296544 | 0.795696 |
| Gene_Pert | EZH2 KO hı | 5/361 | 0.020656 | 0.693901 | 0 | 0 | 3.349719 | 12.99601 |
| Disease_Pe | Duchenne | 3/237 | 0.084577 | 0.694 | 0 | 0 | 3.00351 | 7.418958 |
| Disease_Pe | amyotroph | 2/109 | 0.0817 | 0.694 | 0 | 0 | 4.35536 | 10.90889 |
| Disease_Pe | polycystic ( | 4/378 | 0.082751 | 0.694 | 0 | 0 | 2.51775 | 6.274037 |
| Disease_Pe | Muscular D | 3/238 | 0.085399 | 0.694 | 0 | 0 | 2.990578 | 7.358087 |
| Disease_Pe | Alzheimer's | 4/380 | 0.083988 | 0.694 | 0 | 0 | 2.504102 | 6.202851 |
| Disease_Pe | Sepsis C02 | 4/374 | 0.080303 | 0.694 | 0 | 0 | 2.54549 | 6.419589 |
| Disease_Pe | Pompe dise | 3/242 | 0.088721 | 0.694622 | 0 | 0 | 2.939928 | 7.121261 |
| Disease_Pe | Bone fractı | 3/241 | 0.087885 | 0.694622 | 0 | 0 | 2.952431 | 7.179488 |
| Disease_Pe | Lewy body | 3/242 | 0.088721 | 0.694622 | 0 | 0 | 2.939928 | 7.121261 |
| SysMyo_M | MGS1255 I | 1/235 | 0.643183 | 0.694735 | 0 | 0 | 0.977887 | 0.431568 |
| DisGeNET | Sjogren's S | 1/260 | 0.680464 | 0.694783 | 0 | 0 | 0.882374 | 0.339697 |
| GTEx_Tissu | GTEX-X638 | 18/2660 | 0.035941 | 0.695388 | 0 | 0 | 1.705329 | 5.671694 |
| Rare_Disea | Williams sy | 1/262 | 0.683274 | 0.695425 | 0 | 0 | 0.875523 | 0.333451 |
| DisGeNET | Low Vision | 1/261 | 0.681872 | 0.695679 | 0 | 0 | 0.878936 | 0.336556 |
| DisGeNET | Hepatitis A | 1/261 | 0.681872 | 0.695679 | 0 | 0 | 0.878936 | 0.336556 |
| GeneSigDB | 17284527-1 | 3/688 | 0.579577 | 0.695743 | 0 | 0 | 1.002503 | 0.546821 |
| GeneSigDB | 18631401-1 | 5/1171 | 0.581913 | 0.697044 | 0 | 0 | 0.980369 | 0.530805 |
| SysMyo_M | MGS139 M | 2/503 | 0.646846 | 0.697142 | 0 | 0 | 0.911683 | 0.397172 |
| SysMyo_M | MGS858 M | 1/238 | 0.647874 | 0.697142 | 0 | 0 | 0.965362 | 0.419024 |
| KOMP2_M | Decreased | 1/185 | 0.555251 | 0.697292 | 0 | 0 | 1.246777 | 0.733523 |
| DisGeNET | Failure to g | 2/539 | 0.684088 | 0.697397 | 0 | 0 | 0.848987 | 0.322334 |
| DisGeNET | Retinoblast | 2/539 | 0.684088 | 0.697397 | 0 | 0 | 0.848987 | 0.322334 |
| Jensen_TIS | B-lymphob | 25/5983 | 0.634593 | 0.697402 | 0 | 0 | 0.944447 | 0.429508 |
| GWAS_Cat | Colorectal | 1/267 | 0.690194 | 0.697627 | 0 | 0 | 0.858848 | 0.318446 |
| Gene_Pert | B4galnt2 O | 5/329 | 0.014413 | 0.69843 | 0 | 0 | 3.686578 | 15.62979 |
| PhenGenI_ | Alanine Tra | 1/183 | 0.55132 | 0.699752 | 0 | 0 | 1.260606 | 0.750615 |
| UK_Bioban | Rarely/nev | 2/545 | 0.689978 | 0.6998 | 0 | 0 | 0.839346 | 0.311477 |
| GeneSigDB | 18768393-1 | 1/201 | 0.585497 | 0.699833 | 0 | 0 | 1.146105 | 0.613503 |
| SysMyo_M | MGS1174 I | 1/241 | 0.652504 | 0.700792 | 0 | 0 | 0.953149 | 0.406935 |
| DisGeNET | Pediatric fa | 2/543 | 0.688025 | 0.701138 | 0 | 0 | 0.842536 | 0.31505 |
| GTEx_Tissu | GTEX-PX3G | 12/1687 | 0.061083 | 0.701245 | 0 | 0 | 1.742137 | 4.870175 |
| KOMP2_M | Increased I | 1/190 | 0.56493 | 0.701292 | 0 | 0 | 1.213486 | 0.692965 |
| CellMarker | Monocyte | 3/796 | 0.678351 | 0.701389 | 0 | 0 | 0.861106 | 0.334187 |
| Reactome_ | Neurotrans | 1/182 | 0.549341 | 0.701456 | 0 | 0 | 1.267635 | 0.759358 |
| Reactome_ | Signaling B | 3/660 | 0.551144 | 0.701456 | 0 | 0 | 1.046749 | 0.62361 |
| Reactome_ | mRNA Splic | 1/181 | 0.547354 | 0.701456 | 0 | 0 | 1.274742 | 0.768234 |

| | | | | | | | | |
|---|---|---|---|---|---|---|---|---|
| UK_Bioban | Fortified w | 1/270 | 0.694273 | 0.701659 | 0 | 0 | 0.84914 | 0.309842 |
| DisGeNET | Metastatic | 2/544 | 0.689003 | 0.701861 | 0 | 0 | 0.840938 | 0.313258 |
| Rare_Disea | Leukemia T | 1/268 | 0.691559 | 0.702897 | 0 | 0 | 0.855587 | 0.315546 |
| GeDiPNet_ | Asthma | 2/549 | 0.693855 | 0.703509 | 0 | 0 | 0.833036 | 0.304468 |
| GTEx_Tissu | GTEX-WY7( | 43/7940 | 0.041225 | 0.703983 | 0 | 0 | 1.487009 | 4.741652 |
| GTEx_Tissu | GTEX-PX3G | 23/3672 | 0.03942 | 0.703983 | 0 | 0 | 1.601774 | 5.179291 |
| GTEx_Tissu | GTEX-R55C | 52/9964 | 0.039545 | 0.703983 | 0 | 0 | 1.499055 | 4.842431 |
| GTEx_Tissu | GTEX-XQ3S | 4/289 | 0.037391 | 0.703983 | 0 | 0 | 3.319045 | 10.90745 |
| GTEx_Tissu | GTEX-T5JW | 5/437 | 0.041911 | 0.703983 | 0 | 0 | 2.749689 | 8.722589 |
| GTEx_Tissu | GTEX-XXEK | 3/170 | 0.038062 | 0.703983 | 0 | 0 | 4.22284 | 13.80251 |
| GTEx_Tissu | GTEX-X638 | 35/6172 | 0.039753 | 0.703983 | 0 | 0 | 1.510886 | 4.872711 |
| GTEx_Tissu | GTEX-WFG | 20/3081 | 0.03986 | 0.703983 | 0 | 0 | 1.6434 | 5.295662 |
| GTEx_Tissu | GTEX-T6MI | 4/296 | 0.040252 | 0.703983 | 0 | 0 | 3.238323 | 10.40343 |
| GTEx_Tissu | GTEX-QDVI | 54/10424 | 0.039132 | 0.703983 | 0 | 0 | 1.505865 | 4.880222 |
| GTEx_Tissu | GTEX-PLZ4 | 3/177 | 0.042081 | 0.703983 | 0 | 0 | 4.051519 | 12.83588 |
| GTEx_Tissu | GTEX-T5JC- | 7/738 | 0.041692 | 0.703983 | 0 | 0 | 2.296067 | 7.295622 |
| GTEx_Tissu | GTEX-R53T | 3/176 | 0.041494 | 0.703983 | 0 | 0 | 4.075145 | 12.96795 |
| GTEx_Tissu | GTEX-WCD | 17/2513 | 0.04146 | 0.703983 | 0 | 0 | 1.694649 | 5.394096 |
| GTEx_Tissu | GTEX-XBEC | 44/8135 | 0.038874 | 0.703983 | 0 | 0 | 1.495109 | 4.855278 |
| GTEx_Tissu | GTEX-N7M | 8/890 | 0.039701 | 0.703983 | 0 | 0 | 2.185023 | 7.049725 |
| GTEx_Tissu | GTEX-Q2AH | Feb-74 | 0.041285 | 0.703983 | 0 | 0 | 6.483987 | 20.66613 |
| GTEx_Tissu | GTEX-TKQ2 | 4/289 | 0.037391 | 0.703983 | 0 | 0 | 3.319045 | 10.90745 |
| Jensen_TIS | Blood | 25/6006 | 0.643356 | 0.704629 | 0 | 0 | 0.939265 | 0.414269 |
| Rare_Disea | Embryonal | 1/270 | 0.694273 | 0.704692 | 0 | 0 | 0.84914 | 0.309842 |
| KOMP2_M | Abnormal S | 2/446 | 0.581057 | 0.705103 | 0 | 0 | 1.031744 | 0.56014 |
| KOMP2_M | Embryonic | 1/198 | 0.579985 | 0.705103 | 0 | 0 | 1.163735 | 0.633948 |
| Reactome_ | Mitotic Pro | 1/186 | 0.557204 | 0.705162 | 0 | 0 | 1.239975 | 0.725168 |
| GeDiPNet_ | Renal Insuf | 1/272 | 0.696964 | 0.705434 | 0 | 0 | 0.842787 | 0.304265 |
| GeneSigDB | 16478745- | 1/205 | 0.592736 | 0.705457 | 0 | 0 | 1.123404 | 0.587548 |
| GeneSigDB | 14768006- | 1/205 | 0.592736 | 0.705457 | 0 | 0 | 1.123404 | 0.587548 |
| DisGeNET | Pancreatiti | 1/269 | 0.692919 | 0.705459 | 0 | 0 | 0.852352 | 0.312678 |
| DisGeNET | Colorectal | 4/1073 | 0.693073 | 0.705459 | 0 | 0 | 0.849527 | 0.311454 |
| GeDiPNet_ | Cerebellar | 1/273 | 0.6983 | 0.705561 | 0 | 0 | 0.839646 | 0.301522 |
| GeneSigDB | 16440291- | 1/206 | 0.594525 | 0.70596 | 0 | 0 | 1.117867 | 0.581282 |
| GeneSigDB | 18818702- | 5/1189 | 0.595694 | 0.70596 | 0 | 0 | 0.964537 | 0.499658 |
| SysMyo_M | MGS371 Hu | 1/245 | 0.658585 | 0.705983 | 0 | 0 | 0.937333 | 0.391489 |
| DisGeNET | Epithelial o | 5/1329 | 0.693938 | 0.706065 | 0 | 0 | 0.856099 | 0.312796 |
| Reactome_ | ESR-mediat | 1/188 | 0.561084 | 0.706083 | 0 | 0 | 1.226589 | 0.708828 |
| SysMyo_M | MGS765 M | 1/246 | 0.660088 | 0.706257 | 0 | 0 | 0.93346 | 0.387742 |
| SysMyo_M | MGS1220 N | 1/247 | 0.661585 | 0.706523 | 0 | 0 | 0.929618 | 0.384041 |
| Disease_Pe | Alzheimer's | 3/220 | 0.071143 | 0.707056 | 0 | 0 | 3.241606 | 8.567754 |
| Disease_Pe | nasophary | 4/358 | 0.070886 | 0.707056 | 0 | 0 | 2.662719 | 7.047366 |
| GeneSigDB | 19204204- | 2/460 | 0.597998 | 0.707186 | 0 | 0 | 0.999486 | 0.513904 |
| Rare_Disea | Spleen neo | 1/273 | 0.6983 | 0.707814 | 0 | 0 | 0.839646 | 0.301522 |
| PhenGenI_ | Periodontit | 1/188 | 0.561084 | 0.708253 | 0 | 0 | 1.226589 | 0.708828 |

| | | | | | | | | |
|---|---|---|---|---|---|---|---|---|
| Disease_Pe | Neurogenic | 4/392 | 0.091605 | 0.70856 | 0 | 0 | 2.425165 | 5.796793 |
| KOMP2_M | Decreased | 1/204 | 0.590938 | 0.709125 | 0 | 0 | 1.128995 | 0.593902 |
| GTEx_Tissu | GTEX-S341 | 9/1150 | 0.062063 | 0.709674 | 0 | 0 | 1.898335 | 5.27663 |
| GTEx_Tissu | GTEX-UPJH | 7/809 | 0.062359 | 0.710255 | 0 | 0 | 2.085053 | 5.785702 |
| GeneSigDB | 16365179- | 1/211 | 0.603359 | 0.71051 | 0 | 0 | 1.090975 | 0.551207 |
| GeneSigDB | 17894856- | 1/211 | 0.603359 | 0.71051 | 0 | 0 | 1.090975 | 0.551207 |
| SysMyo_M | MGS1459 M | 1/251 | 0.667508 | 0.711506 | 0 | 0 | 0.914558 | 0.369667 |
| Jensen_TIS | Hepatoma | 1/241 | 0.652504 | 0.712225 | 0 | 0 | 0.953149 | 0.406935 |
| GeneSigDB | 15902281- | 2/467 | 0.606277 | 0.712439 | 0 | 0 | 0.984086 | 0.492455 |
| GeneSigDB | 18338247- | 1/214 | 0.608568 | 0.713626 | 0 | 0 | 1.075445 | 0.534116 |
| GeneSigDB | 18667080- | 1/215 | 0.610289 | 0.714141 | 0 | 0 | 1.070365 | 0.528571 |
| SysMyo_M | MGS949 M | 1/254 | 0.671883 | 0.714823 | 0 | 0 | 0.903576 | 0.359325 |
| Disease_Pe | colorectal | 3/249 | 0.094665 | 0.714999 | 0 | 0 | 2.855256 | 6.731013 |
| Disease_Pe | acute myo | 3/248 | 0.093806 | 0.714999 | 0 | 0 | 2.867055 | 6.784966 |
| GeneSigDB | 20460173- | 19/4528 | 0.612426 | 0.715139 | 0 | 0 | 0.954548 | 0.468041 |
| DisGeNET | Dwarfism | 1/277 | 0.703588 | 0.715605 | 0 | 0 | 0.827309 | 0.290851 |
| GeneSigDB | 16728581- | 4/970 | 0.61427 | 0.715792 | 0 | 0 | 0.945247 | 0.460638 |
| SysMyo_M | MGS549 H | 1/256 | 0.674769 | 0.716546 | 0 | 0 | 0.896398 | 0.35263 |
| SysMyo_M | MGS856 M | 1/257 | 0.676202 | 0.716723 | 0 | 0 | 0.892851 | 0.34934 |
| Reactome_ | SARS-CoV- | 1/196 | 0.576271 | 0.717137 | 0 | 0 | 1.17579 | 0.64807 |
| Reactome_ | RHO GTPas | 2/441 | 0.574883 | 0.717137 | 0 | 0 | 1.043763 | 0.577815 |
| Reactome_ | TCF Depen | 1/198 | 0.579985 | 0.717772 | 0 | 0 | 1.163735 | 0.633948 |
| GTEx_Tissu | GTEX-PX3G | Feb-76 | 0.043322 | 0.719335 | 0 | 0 | 6.308108 | 19.80178 |
| dbGaP | Echocardio | 1/273 | 0.6983 | 0.720468 | 0 | 0 | 0.839646 | 0.301522 |
| KOMP2_M | Increased M | 1/219 | 0.617099 | 0.720534 | 0 | 0 | 1.050512 | 0.507109 |
| KOMP2_M | Increased ( | 1/221 | 0.62046 | 0.720534 | 0 | 0 | 1.040856 | 0.496795 |
| KOMP2_M | Abnormal I | 1/214 | 0.608568 | 0.720534 | 0 | 0 | 1.075445 | 0.534116 |
| GTEx_Tissu | GTEX-O5YV | 44/8192 | 0.043719 | 0.720548 | 0 | 0 | 1.477492 | 4.624521 |
| Reactome_ | Ub-specific | 1/201 | 0.585497 | 0.720612 | 0 | 0 | 1.146105 | 0.613503 |
| KEGG_202 | Prion disea | 1/273 | 0.6983 | 0.721071 | 0 | 0 | 0.839646 | 0.301522 |
| GeneSigDB | 12631704- | 2/479 | 0.620173 | 0.721162 | 0 | 0 | 0.958737 | 0.458043 |
| GeneSigDB | 20103679- | 1/222 | 0.622129 | 0.721929 | 0 | 0 | 1.036094 | 0.491738 |
| GeneSigDB | 19904269- | 1/223 | 0.623791 | 0.722353 | 0 | 0 | 1.031374 | 0.486746 |
| GeneSigDB | 17975224- | 1/224 | 0.625446 | 0.722767 | 0 | 0 | 1.026697 | 0.481818 |
| SysMyo_M | MGS156 C | 1/262 | 0.683274 | 0.722866 | 0 | 0 | 0.875523 | 0.333451 |
| SysMyo_M | MGS937 M | 1/263 | 0.68467 | 0.722991 | 0 | 0 | 0.872137 | 0.330382 |
| GeneSigDB | 18440302- | 2/485 | 0.626981 | 0.72304 | 0 | 0 | 0.946535 | 0.441879 |
| Reactome_ | Transcripti | 1/204 | 0.590938 | 0.723334 | 0 | 0 | 1.128995 | 0.593902 |
| GTEx_Tissu | GTEX-WYV | 11/1515 | 0.063774 | 0.723518 | 0 | 0 | 1.771583 | 4.876132 |
| GTEx_Tissu | GTEX-OIZH | 51/9791 | 0.044413 | 0.723523 | 0 | 0 | 1.479646 | 4.607938 |
| GTEx_Tissu | GTEX-Q2A( | 7/749 | 0.044549 | 0.723523 | 0 | 0 | 2.260731 | 7.033484 |
| DisGeNET | Neuropath | 1/284 | 0.712621 | 0.724512 | 0 | 0 | 0.806558 | 0.273266 |
| SysMyo_M | MGS854 M | 1/265 | 0.687444 | 0.724568 | 0 | 0 | 0.865442 | 0.324346 |
| DisGeNET | Global dev | 4/1102 | 0.713124 | 0.724742 | 0 | 0 | 0.825817 | 0.279208 |
| Reactome_ | GPCR Ligar | 2/458 | 0.595609 | 0.725089 | 0 | 0 | 1.003973 | 0.52023 |

| | | | | | | | | |
|---|---|---|---|---|---|---|---|---|
| DisGeNET | Osteosarcc | 4/1103 | 0.713799 | 0.725146 | 0 | 0 | 0.825022 | 0.27816 |
| Rare_Disea | Ovarian ge | 1/288 | 0.717661 | 0.725461 | 0 | 0 | 0.795154 | 0.263799 |
| Rare_Disea | Acute mon | 1/288 | 0.717661 | 0.725461 | 0 | 0 | 0.795154 | 0.263799 |
| DisGeNET | Papillary th | 2/571 | 0.714479 | 0.725556 | 0 | 0 | 0.799917 | 0.268934 |
| GO_Biologi | Central Ne | 1/283 | 0.711348 | 0.725656 | 0 | 0 | 0.809459 | 0.275697 |
| KOMP2_M | Abnormal I | 1/228 | 0.631995 | 0.726121 | 0 | 0 | 1.008401 | 0.462729 |
| PhenGenI_ | Basophils | 1/199 | 0.581831 | 0.726502 | 0 | 0 | 1.157799 | 0.627036 |
| PhenGenI_ | Age Of Ons | 2/446 | 0.581057 | 0.726502 | 0 | 0 | 1.031744 | 0.56014 |
| GeneSigDB | 16877703- | 1/230 | 0.635226 | 0.726532 | 0 | 0 | 0.999492 | 0.453544 |
| GeneSigDB | 18160665- | 17/4109 | 0.633933 | 0.726532 | 0 | 0 | 0.938965 | 0.427991 |
| GeneSigDB | 14695196- | 1/230 | 0.635226 | 0.726532 | 0 | 0 | 0.999492 | 0.453544 |
| GeneSigDB | 16318415- | 1/230 | 0.635226 | 0.726532 | 0 | 0 | 0.999492 | 0.453544 |
| Jensen_TIS | Lymph | 3/784 | 0.668261 | 0.726959 | 0 | 0 | 0.874886 | 0.352646 |
| GeneSigDB | 18535662- | 4/999 | 0.637632 | 0.727788 | 0 | 0 | 0.916292 | 0.412327 |
| Disease_Pe | Epilepsy CC | 4/401 | 0.097529 | 0.728065 | 0 | 0 | 2.369094 | 5.514317 |
| DisGeNET | Anoxia | 1/288 | 0.717661 | 0.728505 | 0 | 0 | 0.795154 | 0.263799 |
| Disease_Pe | Diabetic Re | 3/254 | 0.099009 | 0.728724 | 0 | 0 | 2.797666 | 6.469729 |
| Disease_Pe | colitis DOID | 3/255 | 0.099887 | 0.728724 | 0 | 0 | 2.786423 | 6.419114 |
| GTEx_Tissu | GTEX-R55G | 11/1518 | 0.064497 | 0.728864 | 0 | 0 | 1.767768 | 4.8457 |
| ARCHS4_Ti | VENTRICLE | 16/2316 | 0.0405 | 0.729006 | 0 | 0 | 1.725707 | 5.533386 |
| ARCHS4_Ti | OSTEOBLAS | 16/2316 | 0.0405 | 0.729006 | 0 | 0 | 1.725707 | 5.533386 |
| GeneSigDB | 18537972- | 4/1008 | 0.644698 | 0.729974 | 0 | 0 | 0.907647 | 0.398433 |
| GeneSigDB | 17683608- | 2/503 | 0.646846 | 0.729974 | 0 | 0 | 0.911683 | 0.397172 |
| GeneSigDB | 19074828- | 3/760 | 0.64741 | 0.729974 | 0 | 0 | 0.903755 | 0.392931 |
| GeneSigDB | 20479250- | 1/237 | 0.646317 | 0.729974 | 0 | 0 | 0.969501 | 0.423154 |
| GeneSigDB | 17555561- | 5/1252 | 0.641939 | 0.729974 | 0 | 0 | 0.912727 | 0.404577 |
| GeneSigDB | 20479250- | 1/237 | 0.646317 | 0.729974 | 0 | 0 | 0.969501 | 0.423154 |
| dbGaP | Stroke | 1/289 | 0.718907 | 0.73014 | 0 | 0 | 0.792353 | 0.261495 |
| DisGeNET | Fanconi An | 1/290 | 0.720147 | 0.730463 | 0 | 0 | 0.789571 | 0.259216 |
| DisGeNET | Malignant | 2/577 | 0.719901 | 0.730463 | 0 | 0 | 0.791325 | 0.260062 |
| Disease_Si | Epilepsy GS | 1/300 | 0.73226 | 0.73226 | 0 | 0 | 0.762775 | 0.237696 |
| Disease_Si | Preeclamps | 1/300 | 0.73226 | 0.73226 | 0 | 0 | 0.762775 | 0.237696 |
| Disease_Si | Papillary Ca | 1/300 | 0.73226 | 0.73226 | 0 | 0 | 0.762775 | 0.237696 |
| Disease_Si | Overexerti | 1/300 | 0.73226 | 0.73226 | 0 | 0 | 0.762775 | 0.237696 |
| Disease_Si | Barrett Eso | 1/300 | 0.73226 | 0.73226 | 0 | 0 | 0.762775 | 0.237696 |
| Disease_Si | Asthma GS | 1/300 | 0.73226 | 0.73226 | 0 | 0 | 0.762775 | 0.237696 |
| Disease_Si | Asthma, all | 1/300 | 0.73226 | 0.73226 | 0 | 0 | 0.762775 | 0.237696 |
| Disease_Si | Uterine leic | 1/300 | 0.73226 | 0.73226 | 0 | 0 | 0.762775 | 0.237696 |
| Disease_Si | Uterine leic | 1/300 | 0.73226 | 0.73226 | 0 | 0 | 0.762775 | 0.237696 |
| Disease_Si | Hepatic Cir | 1/300 | 0.73226 | 0.73226 | 0 | 0 | 0.762775 | 0.237696 |
| Disease_Si | Huntingtor | 1/300 | 0.73226 | 0.73226 | 0 | 0 | 0.762775 | 0.237696 |
| Disease_Si | Familial co | 1/300 | 0.73226 | 0.73226 | 0 | 0 | 0.762775 | 0.237696 |
| Disease_Si | Essential Tl | 1/300 | 0.73226 | 0.73226 | 0 | 0 | 0.762775 | 0.237696 |
| Disease_Si | Cardiomyo | 1/300 | 0.73226 | 0.73226 | 0 | 0 | 0.762775 | 0.237696 |
| Disease_Si | Dermatom | 1/300 | 0.73226 | 0.73226 | 0 | 0 | 0.762775 | 0.237696 |

| | | | | | | | | |
|---|---|---|---|---|---|---|---|---|
| Disease_Si | Huntingtor | 1/300 | 0.73226 | 0.73226 | 0 | 0 | 0.762775 | 0.237696 |
| Disease_Si | Hepatitis C | 1/300 | 0.73226 | 0.73226 | 0 | 0 | 0.762775 | 0.237696 |
| Disease_Si | Familial co | 1/300 | 0.73226 | 0.73226 | 0 | 0 | 0.762775 | 0.237696 |
| Disease_Si | Dehydratio | 1/300 | 0.73226 | 0.73226 | 0 | 0 | 0.762775 | 0.237696 |
| Disease_Si | MS (Multip | 1/300 | 0.73226 | 0.73226 | 0 | 0 | 0.762775 | 0.237696 |
| Disease_Si | Lymphatic | 1/300 | 0.73226 | 0.73226 | 0 | 0 | 0.762775 | 0.237696 |
| Disease_Si | Leukemia, | 1/300 | 0.73226 | 0.73226 | 0 | 0 | 0.762775 | 0.237696 |
| Disease_Si | Bipolar Dis | 1/300 | 0.73226 | 0.73226 | 0 | 0 | 0.762775 | 0.237696 |
| Disease_Si | Type 1 diak | 1/300 | 0.73226 | 0.73226 | 0 | 0 | 0.762775 | 0.237696 |
| Disease_Si | Pseudomo | 1/300 | 0.73226 | 0.73226 | 0 | 0 | 0.762775 | 0.237696 |
| Disease_Si | Cystic Fibr | 1/300 | 0.73226 | 0.73226 | 0 | 0 | 0.762775 | 0.237696 |
| Disease_Si | Squamous | 1/300 | 0.73226 | 0.73226 | 0 | 0 | 0.762775 | 0.237696 |
| Disease_Si | Nephrolith | 1/300 | 0.73226 | 0.73226 | 0 | 0 | 0.762775 | 0.237696 |
| Disease_Si | Muscular D | 1/300 | 0.73226 | 0.73226 | 0 | 0 | 0.762775 | 0.237696 |
| Disease_Si | Malignant | 1/300 | 0.73226 | 0.73226 | 0 | 0 | 0.762775 | 0.237696 |
| Disease_Si | Bipolar Dis | 1/300 | 0.73226 | 0.73226 | 0 | 0 | 0.762775 | 0.237696 |
| Disease_Si | B-cell chro | 1/300 | 0.73226 | 0.73226 | 0 | 0 | 0.762775 | 0.237696 |
| Disease_Si | Adenoma ( | 1/300 | 0.73226 | 0.73226 | 0 | 0 | 0.762775 | 0.237696 |
| Disease_Si | Acute Lung | 1/300 | 0.73226 | 0.73226 | 0 | 0 | 0.762775 | 0.237696 |
| Disease_Si | Dehydratio | 1/300 | 0.73226 | 0.73226 | 0 | 0 | 0.762775 | 0.237696 |
| Disease_Si | Breast Can | 1/300 | 0.73226 | 0.73226 | 0 | 0 | 0.762775 | 0.237696 |
| Disease_Si | Ulcerative | 1/300 | 0.73226 | 0.73226 | 0 | 0 | 0.762775 | 0.237696 |
| Disease_Si | Anaplasmo | 1/300 | 0.73226 | 0.73226 | 0 | 0 | 0.762775 | 0.237696 |
| Disease_Si | Cardiomyo | 1/300 | 0.73226 | 0.73226 | 0 | 0 | 0.762775 | 0.237696 |
| Disease_Si | Cancer of t | 1/300 | 0.73226 | 0.73226 | 0 | 0 | 0.762775 | 0.237696 |
| Disease_Si | Cancer of p | 1/300 | 0.73226 | 0.73226 | 0 | 0 | 0.762775 | 0.237696 |
| Disease_Si | COPD - Chr | 1/300 | 0.73226 | 0.73226 | 0 | 0 | 0.762775 | 0.237696 |
| Disease_Si | Breast Can | 1/300 | 0.73226 | 0.73226 | 0 | 0 | 0.762775 | 0.237696 |
| Disease_Si | Idiopathic | 1/300 | 0.73226 | 0.73226 | 0 | 0 | 0.762775 | 0.237696 |
| Disease_Si | Bipolar Dis | 1/300 | 0.73226 | 0.73226 | 0 | 0 | 0.762775 | 0.237696 |
| Disease_Si | LGLL - Larg | 1/300 | 0.73226 | 0.73226 | 0 | 0 | 0.762775 | 0.237696 |
| Disease_Si | Huntingtor | 1/300 | 0.73226 | 0.73226 | 0 | 0 | 0.762775 | 0.237696 |
| Disease_Si | Dehydratio | 1/300 | 0.73226 | 0.73226 | 0 | 0 | 0.762775 | 0.237696 |
| Disease_Si | Cardiomyo | 1/300 | 0.73226 | 0.73226 | 0 | 0 | 0.762775 | 0.237696 |
| Disease_Si | Cardiomyo | 1/300 | 0.73226 | 0.73226 | 0 | 0 | 0.762775 | 0.237696 |
| Disease_Si | Lung trans | 1/300 | 0.73226 | 0.73226 | 0 | 0 | 0.762775 | 0.237696 |
| Disease_Si | Leber cong | 1/300 | 0.73226 | 0.73226 | 0 | 0 | 0.762775 | 0.237696 |
| Disease_Si | Ulcerative | 1/300 | 0.73226 | 0.73226 | 0 | 0 | 0.762775 | 0.237696 |
| Disease_Si | Type 1 diak | 1/300 | 0.73226 | 0.73226 | 0 | 0 | 0.762775 | 0.237696 |
| Disease_Si | Sepsis GSE | 1/300 | 0.73226 | 0.73226 | 0 | 0 | 0.762775 | 0.237696 |
| Disease_Si | Schizophre | 1/300 | 0.73226 | 0.73226 | 0 | 0 | 0.762775 | 0.237696 |
| Disease_Si | Psoriasis vu | 1/300 | 0.73226 | 0.73226 | 0 | 0 | 0.762775 | 0.237696 |
| Disease_Si | Sickle Cell | 1/300 | 0.73226 | 0.73226 | 0 | 0 | 0.762775 | 0.237696 |
| Disease_Si | Setleis syn | 1/300 | 0.73226 | 0.73226 | 0 | 0 | 0.762775 | 0.237696 |
| Disease_Si | Serous carc | 1/300 | 0.73226 | 0.73226 | 0 | 0 | 0.762775 | 0.237696 |

| | | | | | | | | |
|---|---|---|---|---|---|---|---|---|
| Disease_Sig | Ventricular | 1/300 | 0.73226 | 0.73226 | 0 | 0 | 0.762775 | 0.237696 |
| Disease_Sig | Osteoarthr | 1/300 | 0.73226 | 0.73226 | 0 | 0 | 0.762775 | 0.237696 |
| Disease_Sig | Nutritional | 1/300 | 0.73226 | 0.73226 | 0 | 0 | 0.762775 | 0.237696 |
| Disease_Sig | Alzheimer's | 1/300 | 0.73226 | 0.73226 | 0 | 0 | 0.762775 | 0.237696 |
| Disease_Sig | Carcinoma, | 1/300 | 0.73226 | 0.73226 | 0 | 0 | 0.762775 | 0.237696 |
| Disease_Sig | Carcinoma, | 1/300 | 0.73226 | 0.73226 | 0 | 0 | 0.762775 | 0.237696 |
| Disease_Sig | COPD - Chr | 1/300 | 0.73226 | 0.73226 | 0 | 0 | 0.762775 | 0.237696 |
| Disease_Sig | Ventricular | 1/300 | 0.73226 | 0.73226 | 0 | 0 | 0.762775 | 0.237696 |
| Disease_Sig | Streptococ | 1/300 | 0.73226 | 0.73226 | 0 | 0 | 0.762775 | 0.237696 |
| Disease_Sig | Appendicit | 1/300 | 0.73226 | 0.73226 | 0 | 0 | 0.762775 | 0.237696 |
| Disease_Sig | SCID - Seve | 1/300 | 0.73226 | 0.73226 | 0 | 0 | 0.762775 | 0.237696 |
| Disease_Sig | Acute Lung | 1/300 | 0.73226 | 0.73226 | 0 | 0 | 0.762775 | 0.237696 |
| Disease_Sig | Infection b | 1/300 | 0.73226 | 0.73226 | 0 | 0 | 0.762775 | 0.237696 |
| Disease_Sig | Type 1 diab | 1/300 | 0.73226 | 0.73226 | 0 | 0 | 0.762775 | 0.237696 |
| Disease_Sig | Ventilator- | 1/300 | 0.73226 | 0.73226 | 0 | 0 | 0.762775 | 0.237696 |
| Disease_Sig | Huntingtor | 1/300 | 0.73226 | 0.73226 | 0 | 0 | 0.762775 | 0.237696 |
| Disease_Sig | JRA - Juven | 1/300 | 0.73226 | 0.73226 | 0 | 0 | 0.762775 | 0.237696 |
| Disease_Sig | Polycystic ( | 1/300 | 0.73226 | 0.73226 | 0 | 0 | 0.762775 | 0.237696 |
| Disease_Sig | Inclusion B | 1/300 | 0.73226 | 0.73226 | 0 | 0 | 0.762775 | 0.237696 |
| Disease_Sig | Sepsis GSE | 1/300 | 0.73226 | 0.73226 | 0 | 0 | 0.762775 | 0.237696 |
| Disease_Sig | Rhinovirus | 1/300 | 0.73226 | 0.73226 | 0 | 0 | 0.762775 | 0.237696 |
| Disease_Sig | MDS - Mye | 1/300 | 0.73226 | 0.73226 | 0 | 0 | 0.762775 | 0.237696 |
| Disease_Sig | Retinoschis | 1/300 | 0.73226 | 0.73226 | 0 | 0 | 0.762775 | 0.237696 |
| Disease_Sig | Psoriasis vu | 1/300 | 0.73226 | 0.73226 | 0 | 0 | 0.762775 | 0.237696 |
| Disease_Sig | Psoriasis vu | 1/300 | 0.73226 | 0.73226 | 0 | 0 | 0.762775 | 0.237696 |
| Disease_Sig | Ewing's sar | 1/300 | 0.73226 | 0.73226 | 0 | 0 | 0.762775 | 0.237696 |
| Disease_Sig | Diabetic Ne | 1/300 | 0.73226 | 0.73226 | 0 | 0 | 0.762775 | 0.237696 |
| Disease_Sig | Nicotine ac | 1/300 | 0.73226 | 0.73226 | 0 | 0 | 0.762775 | 0.237696 |
| Disease_Sig | Nephrolith | 1/300 | 0.73226 | 0.73226 | 0 | 0 | 0.762775 | 0.237696 |
| Disease_Pe | Cardiac Hy | 4/365 | 0.074932 | 0.732608 | 0 | 0 | 2.610153 | 6.763361 |
| Disease_Pe | Viral cardic | 4/373 | 0.079697 | 0.732608 | 0 | 0 | 2.552519 | 6.456655 |
| Disease_Pe | osteoarthri | 3/229 | 0.078126 | 0.732608 | 0 | 0 | 3.111094 | 7.931512 |
| Disease_Pe | asthma DO | 3/231 | 0.079718 | 0.732608 | 0 | 0 | 3.08349 | 7.798958 |
| Disease_Pe | allergic cor | 3/232 | 0.080519 | 0.732608 | 0 | 0 | 3.069869 | 7.733813 |
| Disease_Pe | Dehydratio | 3/230 | 0.07892 | 0.732608 | 0 | 0 | 3.097231 | 7.864854 |
| GeneSigDB | 19204204- | 1/241 | 0.652504 | 0.732752 | 0 | 0 | 0.953149 | 0.406935 |
| GeneSigDB | 15902281- | 1/241 | 0.652504 | 0.732752 | 0 | 0 | 0.953149 | 0.406935 |
| GeneSigDB | 16643655- | 1/242 | 0.654034 | 0.732992 | 0 | 0 | 0.949146 | 0.403003 |
| GO_Biologi | DNA Repai | 1/291 | 0.721383 | 0.734837 | 0 | 0 | 0.786808 | 0.25696 |
| Reactome_ | Neutrophil | 2/468 | 0.607449 | 0.735506 | 0 | 0 | 0.981924 | 0.489476 |
| PhenGenI_ | Tunica Me | 1/207 | 0.596308 | 0.735813 | 0 | 0 | 1.112384 | 0.575101 |
| PhenGenI_ | Cholestero | 6/1438 | 0.602029 | 0.735813 | 0 | 0 | 0.95598 | 0.485112 |
| PhenGenI_ | Fetal Hem | 1/210 | 0.601608 | 0.735813 | 0 | 0 | 1.09625 | 0.557059 |
| PhenGenI_ | Inflammat | 2/463 | 0.601562 | 0.735813 | 0 | 0 | 0.992829 | 0.504582 |
| Jensen_TIS | Parenchym | 21/5163 | 0.679145 | 0.736312 | 0 | 0 | 0.914015 | 0.353651 |

| | | | | | | | | |
|---|---|---|---|---|---|---|---|---|
| GTEx_Tissu | GTEX-TMM | 8/988 | 0.065454 | 0.7368 | 0 | 0 | 1.956394 | 5.333936 |
| GTEx_Tissu | GTEX-XLM4 | 7/757 | 0.046706 | 0.737023 | 0 | 0 | 2.235683 | 6.849885 |
| GTEx_Tissu | GTEX-T5JW | 7/757 | 0.046706 | 0.737023 | 0 | 0 | 2.235683 | 6.849885 |
| GTEx_Tissu | GTEX-X4XX | 39/7110 | 0.046169 | 0.737023 | 0 | 0 | 1.475622 | 4.53821 |
| GTEx_Tissu | GTEX-O5YT | 44/8221 | 0.046361 | 0.737023 | 0 | 0 | 1.468623 | 4.510571 |
| GTEx_Tissu | GTEX-WEY! | 8/989 | 0.06576 | 0.737381 | 0 | 0 | 1.954296 | 5.319087 |
| SysMyo_M | MGS296 C | 1/300 | 0.73226 | 0.737472 | 0 | 0 | 0.762775 | 0.237696 |
| SysMyo_M | MGS331 C | 1/300 | 0.73226 | 0.737472 | 0 | 0 | 0.762775 | 0.237696 |
| SysMyo_M | MGS327 M | 1/300 | 0.73226 | 0.737472 | 0 | 0 | 0.762775 | 0.237696 |
| SysMyo_M | MGS326 M | 1/300 | 0.73226 | 0.737472 | 0 | 0 | 0.762775 | 0.237696 |
| SysMyo_M | MGS563 H | 1/286 | 0.715152 | 0.737472 | 0 | 0 | 0.800816 | 0.268482 |
| SysMyo_M | MGS1368 I | 1/298 | 0.72988 | 0.737472 | 0 | 0 | 0.76799 | 0.241821 |
| SysMyo_M | MGS1359 I | 1/296 | 0.727479 | 0.737472 | 0 | 0 | 0.773276 | 0.246034 |
| SysMyo_M | MGS188 M | 1/300 | 0.73226 | 0.737472 | 0 | 0 | 0.762775 | 0.237696 |
| SysMyo_M | MGS933 M | 1/278 | 0.704895 | 0.737472 | 0 | 0 | 0.82428 | 0.288256 |
| SysMyo_M | MGS193 M | 1/300 | 0.73226 | 0.737472 | 0 | 0 | 0.762775 | 0.237696 |
| SysMyo_M | MGS196 M | 1/300 | 0.73226 | 0.737472 | 0 | 0 | 0.762775 | 0.237696 |
| SysMyo_M | MGS68 Mu | 4/1085 | 0.701485 | 0.737472 | 0 | 0 | 0.839562 | 0.297671 |
| SysMyo_M | MGS424 H | 1/280 | 0.707493 | 0.737472 | 0 | 0 | 0.818288 | 0.28315 |
| SysMyo_M | MGS1405 I | 1/280 | 0.707493 | 0.737472 | 0 | 0 | 0.818288 | 0.28315 |
| SysMyo_M | MGS539 H | 1/283 | 0.711348 | 0.737472 | 0 | 0 | 0.809459 | 0.275697 |
| SysMyo_M | MGS322 M | 1/300 | 0.73226 | 0.737472 | 0 | 0 | 0.762775 | 0.237696 |
| SysMyo_M | MGS320 M | 1/300 | 0.73226 | 0.737472 | 0 | 0 | 0.762775 | 0.237696 |
| SysMyo_M | MGS324 M | 1/300 | 0.73226 | 0.737472 | 0 | 0 | 0.762775 | 0.237696 |
| SysMyo_M | MGS205 C | 1/300 | 0.73226 | 0.737472 | 0 | 0 | 0.762775 | 0.237696 |
| SysMyo_M | MGS200 H | 1/300 | 0.73226 | 0.737472 | 0 | 0 | 0.762775 | 0.237696 |
| SysMyo_M | MGS221 C | 1/300 | 0.73226 | 0.737472 | 0 | 0 | 0.762775 | 0.237696 |
| SysMyo_M | MGS259 H | 1/300 | 0.73226 | 0.737472 | 0 | 0 | 0.762775 | 0.237696 |
| SysMyo_M | MGS257 H | 1/300 | 0.73226 | 0.737472 | 0 | 0 | 0.762775 | 0.237696 |
| SysMyo_M | MGS242 C | 1/300 | 0.73226 | 0.737472 | 0 | 0 | 0.762775 | 0.237696 |
| SysMyo_M | MGS290 C | 1/300 | 0.73226 | 0.737472 | 0 | 0 | 0.762775 | 0.237696 |
| DisGeNET | Pituitary N | 1/296 | 0.727479 | 0.737614 | 0 | 0 | 0.773276 | 0.246034 |
| GTEx_Tissu | GTEX-WFO | 8/990 | 0.066068 | 0.737968 | 0 | 0 | 1.952203 | 5.304285 |
| GO_Molec | Kinase Binc | 2/460 | 0.597998 | 0.738269 | 0 | 0 | 0.999486 | 0.513904 |
| GTEx_Tissu | GTEX-WY7 | 7/821 | 0.066381 | 0.738615 | 0 | 0 | 2.053025 | 5.568516 |
| GTEx_Tissu | GTEX-T2IS- | 7/824 | 0.067411 | 0.738707 | 0 | 0 | 2.045165 | 5.515717 |
| GTEx_Tissu | GTEX-XOT( | 7/824 | 0.067411 | 0.738707 | 0 | 0 | 2.045165 | 5.515717 |
| GTEx_Tissu | GTEX-Q2AI | 7/823 | 0.067066 | 0.738707 | 0 | 0 | 2.047779 | 5.533251 |
| GTEx_Tissu | GTEX-VJYA | 9/1168 | 0.067073 | 0.738707 | 0 | 0 | 1.86706 | 5.044751 |
| GO_Molec | Transcripti | 2/474 | 0.614429 | 0.740276 | 0 | 0 | 0.969143 | 0.472033 |
| GO_Molec | Sequence-! | 3/717 | 0.607836 | 0.740276 | 0 | 0 | 0.960334 | 0.478103 |
| GTEx_Tissu | GTEX-Q2AI | 5/452 | 0.047249 | 0.74034 | 0 | 0 | 2.655372 | 8.105074 |
| Rare_Disea | Peripheral | 1/301 | 0.733443 | 0.740409 | 0 | 0 | 0.760194 | 0.235665 |
| GWAS_Cat | Hemoglobi | 1/301 | 0.733443 | 0.740728 | 0 | 0 | 0.760194 | 0.235665 |
| Jensen_TIS | Trigeminal | 17/4239 | 0.68818 | 0.741117 | 0 | 0 | 0.902575 | 0.337297 |

| | | | | | | | | |
|---|---|---|---|---|---|---|---|---|
| Jensen_TIS | Pharynx | 2/541 | 0.686061 | 0.741117 | 0 | 0 | 0.845749 | 0.318668 |
| Jensen_TIS | Olfactory b | 17/4257 | 0.695349 | 0.742949 | 0 | 0 | 0.897712 | 0.326175 |
| Jensen_TIS | Pancreatic | 20/4972 | 0.696228 | 0.742949 | 0 | 0 | 0.901852 | 0.326541 |
| Jensen_TIS | Small intes | 25/6152 | 0.696803 | 0.742949 | 0 | 0 | 0.907275 | 0.327755 |
| DisGeNET | Cutaneous | 1/301 | 0.733443 | 0.743373 | 0 | 0 | 0.760194 | 0.235665 |
| Disease_Pe | Down synd | 4/381 | 0.084611 | 0.74408 | 0 | 0 | 2.497332 | 6.167647 |
| Disease_Pe | nephroscle | 4/382 | 0.085235 | 0.74408 | 0 | 0 | 2.490597 | 6.1327 |
| Disease_Pe | polycystic ( | 4/382 | 0.085235 | 0.74408 | 0 | 0 | 2.490597 | 6.1327 |
| GO_Molec | Transcripti | 1/224 | 0.625446 | 0.744579 | 0 | 0 | 1.026697 | 0.481818 |
| Disease_Pe | Gamma-hy | 4/384 | 0.086491 | 0.744977 | 0 | 0 | 2.477235 | 6.063563 |
| PhenGenI_ | Monocytes | 1/217 | 0.613709 | 0.746141 | 0 | 0 | 1.060347 | 0.517698 |
| CellMarker | Ionocyte C | 1/294 | 0.725056 | 0.746862 | 0 | 0 | 0.778633 | 0.250335 |
| Gene_Pert | Map2k3 Se | 5/337 | 0.015835 | 0.74715 | 0 | 0 | 3.596275 | 14.90853 |
| GO_Biologi | Positive Re | 2/594 | 0.734801 | 0.747434 | 0 | 0 | 0.767925 | 0.236641 |
| Rare_Disea | Synovial ca | 1/309 | 0.742718 | 0.748756 | 0 | 0 | 0.740146 | 0.220149 |
| GeneSigDB | 17284527-1 | 2/526 | 0.671018 | 0.749012 | 0 | 0 | 0.870633 | 0.347347 |
| GeneSigDB | 18310505-1 | 5/1294 | 0.670941 | 0.749012 | 0 | 0 | 0.881001 | 0.351584 |
| Gene_Pert | Aldh1a2 KC | 5/355 | 0.019371 | 0.749142 | 0 | 0 | 3.408188 | 13.44184 |
| Gene_Pert | Prkag3 KO | 5/353 | 0.018954 | 0.749142 | 0 | 0 | 3.428126 | 13.59498 |
| Gene_Pert | Gata4 INAC | 5/352 | 0.018748 | 0.749142 | 0 | 0 | 3.438181 | 13.67242 |
| Gene_Pert | CMAHP KO | 4/236 | 0.019637 | 0.749142 | 0 | 0 | 4.088284 | 16.06828 |
| Gene_Pert | Retnlb KO r | 4/233 | 0.018834 | 0.749142 | 0 | 0 | 4.142474 | 16.45435 |
| Gene_Pert | EGR3 KD hu | 4/232 | 0.01857 | 0.749142 | 0 | 0 | 4.160854 | 16.58593 |
| Gene_Pert | Apod OE m | 4/228 | 0.017541 | 0.749142 | 0 | 0 | 4.236015 | 17.12723 |
| Gene_Pert | NET1 KD hu | 4/236 | 0.019637 | 0.749142 | 0 | 0 | 4.088284 | 16.06828 |
| Gene_Pert | HMGA2 KC | 4/232 | 0.01857 | 0.749142 | 0 | 0 | 4.160854 | 16.58593 |
| GTEx_Tissu | GTEX-XQ3S | 8/999 | 0.068875 | 0.749442 | 0 | 0 | 1.933554 | 5.173149 |
| GTEx_Tissu | GTEX-XMK | 13/1909 | 0.068908 | 0.749442 | 0 | 0 | 1.669382 | 4.465562 |
| Gene_Pert | Esr2 KO mc | 5/372 | 0.023156 | 0.750089 | 0 | 0 | 3.247491 | 12.22843 |
| Gene_Pert | IRS3P KO m | 4/248 | 0.023063 | 0.750089 | 0 | 0 | 3.884851 | 14.64409 |
| GTEx_Tissu | GTEX-POM | 6/664 | 0.06933 | 0.751205 | 0 | 0 | 2.167624 | 5.785122 |
| DisGeNET | Tuberculos | 2/602 | 0.74158 | 0.75133 | 0 | 0 | 0.757373 | 0.226434 |
| KEGG_202 | Huntingtor | 1/306 | 0.739278 | 0.751768 | 0 | 0 | 0.747541 | 0.225819 |
| KEGG_202 | MicroRNAs | 1/310 | 0.743855 | 0.751768 | 0 | 0 | 0.737714 | 0.218297 |
| GeneSigDB | 15735024-1 | 13/3256 | 0.676658 | 0.751768 | 0 | 0 | 0.903026 | 0.352712 |
| GeneSigDB | 17894856-1 | 1/257 | 0.676202 | 0.751768 | 0 | 0 | 0.892851 | 0.34934 |
| GeneSigDB | 18757322-1 | 7/1801 | 0.677536 | 0.751768 | 0 | 0 | 0.88373 | 0.344029 |
| DisGeNET | Schizophre | Jul-23 | 0.74297 | 0.752157 | 0 | 0 | 0.821888 | 0.244182 |
| DisGeNET | Peripheral | 1/309 | 0.742718 | 0.752157 | 0 | 0 | 0.740146 | 0.220149 |
| KOMP2_M | Abnormal I | 1/262 | 0.683274 | 0.752996 | 0 | 0 | 0.875523 | 0.333451 |
| KOMP2_M | Decreased | 1/254 | 0.671883 | 0.752996 | 0 | 0 | 0.903576 | 0.359325 |
| KOMP2_M | Male Infert | 1/256 | 0.674769 | 0.752996 | 0 | 0 | 0.896398 | 0.35263 |
| KOMP2_M | Decreased | 1/258 | 0.677628 | 0.752996 | 0 | 0 | 0.889331 | 0.346089 |
| GTEx_Tissu | GTEX-S341 | 8/1002 | 0.069827 | 0.753768 | 0 | 0 | 1.927413 | 5.130257 |
| GTEx_Tissu | GTEX-OXRI | 14/2105 | 0.070166 | 0.754616 | 0 | 0 | 1.634585 | 4.342905 |

| | | | | | | | | |
|---|---|---|---|---|---|---|---|---|
| DisGeNET | Colitis | 2/608 | 0.746568 | 0.755507 | 0 | 0 | 0.749641 | 0.219097 |
| GO_Biologi | Positive Re | 1/310 | 0.743855 | 0.75556 | 0 | 0 | 0.737714 | 0.218297 |
| GTEx_Tissu | GTEX-R55G | 4/316 | 0.049098 | 0.756045 | 0 | 0 | 3.027649 | 9.125164 |
| GTEx_Tissu | GTEX-V1D1 | 17/2592 | 0.053057 | 0.756045 | 0 | 0 | 1.635207 | 4.801608 |
| GTEx_Tissu | GTEX-Q2AI | 43/8060 | 0.052538 | 0.756045 | 0 | 0 | 1.450123 | 4.272384 |
| GTEx_Tissu | GTEX-R53T | 23/3772 | 0.051396 | 0.756045 | 0 | 0 | 1.549463 | 4.599104 |
| GTEx_Tissu | GTEX-S95S | 5/464 | 0.051799 | 0.756045 | 0 | 0 | 2.584356 | 7.650711 |
| GTEx_Tissu | GTEX-U3ZH | Feb-88 | 0.05629 | 0.756045 | 0 | 0 | 5.424624 | 15.60794 |
| GTEx_Tissu | GTEX-N7M | 6/620 | 0.053192 | 0.756045 | 0 | 0 | 2.328266 | 6.830763 |
| GTEx_Tissu | GTEX-X585 | 22/3563 | 0.050478 | 0.756045 | 0 | 0 | 1.564895 | 4.673114 |
| GTEx_Tissu | GTEX-XGQ | 49/9396 | 0.050394 | 0.756045 | 0 | 0 | 1.457642 | 4.355273 |
| GTEx_Tissu | GTEX-XXEK | 44/8262 | 0.05031 | 0.756045 | 0 | 0 | 1.456191 | 4.353347 |
| GTEx_Tissu | GTEX-QCQ | Feb-89 | 0.057425 | 0.756045 | 0 | 0 | 5.362001 | 15.32067 |
| GTEx_Tissu | GTEX-VUSH | 22/3602 | 0.055875 | 0.756045 | 0 | 0 | 1.54416 | 4.454349 |
| GTEx_Tissu | GTEX-X4EC | 28/4838 | 0.055859 | 0.756045 | 0 | 0 | 1.49013 | 4.298901 |
| GTEx_Tissu | GTEX-WHS | 5/473 | 0.055375 | 0.756045 | 0 | 0 | 2.533484 | 7.330969 |
| GTEx_Tissu | GTEX-RM2 | 3/197 | 0.054684 | 0.756045 | 0 | 0 | 3.630155 | 10.54993 |
| GTEx_Tissu | GTEX-X4XY | 4/315 | 0.048632 | 0.756045 | 0 | 0 | 3.037539 | 9.183935 |
| GTEx_Tissu | GTEX-X638 | 10/1302 | 0.055929 | 0.756045 | 0 | 0 | 1.871758 | 5.397539 |
| GTEx_Tissu | GTEX-WVLI | 47/8944 | 0.050788 | 0.756045 | 0 | 0 | 1.45485 | 4.335589 |
| GTEx_Tissu | GTEX-T5JC | 6/616 | 0.051853 | 0.756045 | 0 | 0 | 2.344019 | 6.936777 |
| GTEx_Tissu | GTEX-WRH | 5/466 | 0.052581 | 0.756045 | 0 | 0 | 2.57288 | 7.578156 |
| GTEx_Tissu | GTEX-TKQ1 | 32/5697 | 0.057376 | 0.756045 | 0 | 0 | 1.463327 | 4.182384 |
| GTEx_Tissu | GTEX-WW | 6/622 | 0.05387 | 0.756045 | 0 | 0 | 2.320467 | 6.778498 |
| GTEx_Tissu | GTEX-UPIC | 3/196 | 0.054015 | 0.756045 | 0 | 0 | 3.649149 | 10.65003 |
| GTEx_Tissu | GTEX-PLZ6 | 21/3408 | 0.056996 | 0.756045 | 0 | 0 | 1.552487 | 4.447512 |
| GTEx_Tissu | GTEX-REY6 | 3/193 | 0.052032 | 0.756045 | 0 | 0 | 3.707331 | 10.95845 |
| GTEx_Tissu | GTEX-S4P3 | 3/200 | 0.056714 | 0.756045 | 0 | 0 | 3.574329 | 10.25739 |
| GTEx_Tissu | GTEX-U412 | 51/9918 | 0.056739 | 0.756045 | 0 | 0 | 1.442367 | 4.138579 |
| GWAS_Cat | Blood Prot | 3/890 | 0.749714 | 0.756535 | 0 | 0 | 0.766065 | 0.220675 |
| GWAS_Cat | Mean Plate | 2/613 | 0.750662 | 0.756866 | 0 | 0 | 0.743314 | 0.213182 |
| GTEx_Tissu | GTEX-VUSG | 11/1544 | 0.070992 | 0.757056 | 0 | 0 | 1.735331 | 4.590279 |
| GTEx_Tissu | GTEX-WL4 | 7/835 | 0.071269 | 0.757056 | 0 | 0 | 2.016833 | 5.327045 |
| GTEx_Tissu | GTEX-U8XE | 8/1007 | 0.071432 | 0.757056 | 0 | 0 | 1.917259 | 5.059663 |
| GTEx_Tissu | GTEX-WOF | 9/1183 | 0.07144 | 0.757056 | 0 | 0 | 1.841731 | 4.860136 |
| GeneSigDB | 18535662- | 2/540 | 0.685076 | 0.757118 | 0 | 0 | 0.847365 | 0.320495 |
| GeneSigDB | 20460173- | 2/540 | 0.685076 | 0.757118 | 0 | 0 | 0.847365 | 0.320495 |
| Disease_Pe | pulmonary | 3/261 | 0.105224 | 0.75896 | 0 | 0 | 2.720792 | 6.1263 |
| Disease_Pe | polycystic | 4/414 | 0.106396 | 0.75896 | 0 | 0 | 2.292448 | 5.136423 |
| DisGeNET | Agenesis o | 1/317 | 0.751674 | 0.76038 | 0 | 0 | 0.721114 | 0.205844 |
| GeDiPNet_ | Prostatic N | 2/618 | 0.754701 | 0.761229 | 0 | 0 | 0.737089 | 0.207442 |
| GeDiPNet_ | Dysphagia | 1/321 | 0.756036 | 0.761259 | 0 | 0 | 0.711955 | 0.19911 |
| Disease_Pe | ulcerative | 3/264 | 0.107935 | 0.761472 | 0 | 0 | 2.689108 | 5.986573 |
| PhenGenI_ | Monocyte | 1/230 | 0.635226 | 0.761924 | 0 | 0 | 0.999492 | 0.453544 |
| PhenGenI_ | Migraine D | 1/235 | 0.643183 | 0.761924 | 0 | 0 | 0.977887 | 0.431568 |

| | | | | | | | | |
|---|---|---|---|---|---|---|---|---|
| PhenGenI_ | Body Fat D | 1/233 | 0.640021 | 0.761924 | 0 | 0 | 0.986417 | 0.440193 |
| PhenGenI_ | Immunoglc | 2/490 | 0.632583 | 0.761924 | 0 | 0 | 0.936596 | 0.428908 |
| PhenGenI_ | Diabetes M | 1/235 | 0.643183 | 0.761924 | 0 | 0 | 0.977887 | 0.431568 |
| GTEx_Tissu | GTEX-PLZ6 | 48/9244 | 0.058325 | 0.763375 | 0 | 0 | 1.434336 | 4.07598 |
| PhenGenI_ | Menarche | 1/238 | 0.647874 | 0.763566 | 0 | 0 | 0.965362 | 0.419024 |
| SysMyo_M | MGS69 Mu | 3/905 | 0.759867 | 0.763916 | 0 | 0 | 0.752732 | 0.206709 |
| Gene_Pert | Gys1 OE m | 4/239 | 0.020462 | 0.764336 | 0 | 0 | 4.035478 | 15.69475 |
| Gene_Pert | Kdm2b KD | 4/245 | 0.022174 | 0.764592 | 0 | 0 | 3.93381 | 14.98315 |
| Gene_Pert | ABL1 drugi | 4/244 | 0.021883 | 0.764592 | 0 | 0 | 3.950402 | 15.09859 |
| Gene_Pert | Pitx2 KO m | 4/244 | 0.021883 | 0.764592 | 0 | 0 | 3.950402 | 15.09859 |
| Gene_Pert | Vax2 KO m | 4/245 | 0.022174 | 0.764592 | 0 | 0 | 3.93381 | 14.98315 |
| GO_Biologi | Proteasom | 1/319 | 0.753864 | 0.764634 | 0 | 0 | 0.716506 | 0.202444 |
| Disease_Pe | Cancer of t | 2/148 | 0.135804 | 0.765012 | 0 | 0 | 3.185657 | 6.360306 |
| Disease_Pe | hepatitis C | 3/295 | 0.137457 | 0.765012 | 0 | 0 | 2.399829 | 4.762324 |
| Disease_Pe | Crohn's dis | 2/139 | 0.122685 | 0.765012 | 0 | 0 | 3.396479 | 7.126265 |
| Disease_Pe | Dry eye syr | 3/280 | 0.122839 | 0.765012 | 0 | 0 | 2.531717 | 5.308703 |
| Disease_Pe | Hepatocell | 3/302 | 0.144476 | 0.765012 | 0 | 0 | 2.342809 | 4.532498 |
| Disease_Pe | Alzheimer's | 3/301 | 0.143466 | 0.765012 | 0 | 0 | 2.350791 | 4.564431 |
| Disease_Pe | COPD - Chr | 3/289 | 0.131539 | 0.765012 | 0 | 0 | 2.450924 | 4.971583 |
| Disease_Pe | Huntingtor | 3/288 | 0.130562 | 0.765012 | 0 | 0 | 2.459649 | 5.007625 |
| Disease_Pe | Fracture of | 4/432 | 0.119259 | 0.765012 | 0 | 0 | 2.19401 | 4.665472 |
| Disease_Pe | glaucoma a | 3/255 | 0.099887 | 0.765012 | 0 | 0 | 2.786423 | 6.419114 |
| Disease_Pe | hepatitis C | 4/453 | 0.135078 | 0.765012 | 0 | 0 | 2.089141 | 4.182261 |
| Disease_Pe | hepatitis C | 3/293 | 0.135474 | 0.765012 | 0 | 0 | 2.416626 | 4.830773 |
| Disease_Pe | Uterine leic | 3/258 | 0.102542 | 0.765012 | 0 | 0 | 2.753221 | 6.27042 |
| Disease_Pe | monoclona | 3/300 | 0.142458 | 0.765012 | 0 | 0 | 2.358826 | 4.596658 |
| Disease_Pe | Carcinoma, | 3/297 | 0.13945 | 0.765012 | 0 | 0 | 2.38326 | 4.695137 |
| Disease_Pe | nemaline n | 2/150 | 0.138761 | 0.765012 | 0 | 0 | 3.142289 | 6.206032 |
| Disease_Pe | Alzheimer's | 3/275 | 0.118102 | 0.765012 | 0 | 0 | 2.578913 | 5.509085 |
| Disease_Pe | Retinoschis | 4/404 | 0.099543 | 0.765012 | 0 | 0 | 2.350964 | 5.424064 |
| Disease_Pe | inclusion b | 4/431 | 0.118527 | 0.765012 | 0 | 0 | 2.199261 | 4.690176 |
| Disease_Pe | pulmonary | 3/258 | 0.102542 | 0.765012 | 0 | 0 | 2.753221 | 6.27042 |
| Disease_Pe | Vitamin A I | 3/265 | 0.108844 | 0.765012 | 0 | 0 | 2.678708 | 5.940942 |
| Disease_Pe | Uterine leic | 3/262 | 0.106125 | 0.765012 | 0 | 0 | 2.710149 | 6.079245 |
| Disease_Pe | invasive du | 3/265 | 0.108844 | 0.765012 | 0 | 0 | 2.678708 | 5.940942 |
| Disease_Pe | Papillary Ca | 4/400 | 0.096862 | 0.765012 | 0 | 0 | 2.375198 | 5.544825 |
| Disease_Pe | Leukemia, | 3/265 | 0.108844 | 0.765012 | 0 | 0 | 2.678708 | 5.940942 |
| Disease_Pe | Generalize | 3/282 | 0.124754 | 0.765012 | 0 | 0 | 2.513313 | 5.231246 |
| Disease_Pe | psoriasis D | 3/285 | 0.127646 | 0.765012 | 0 | 0 | 2.486196 | 5.117829 |
| Disease_Pe | NASH C040 | 3/275 | 0.118102 | 0.765012 | 0 | 0 | 2.578913 | 5.509085 |
| Disease_Pe | muscular d | 3/275 | 0.118102 | 0.765012 | 0 | 0 | 2.578913 | 5.509085 |
| Disease_Pe | progressive | 4/415 | 0.107093 | 0.765012 | 0 | 0 | 2.286753 | 5.10873 |
| Disease_Pe | acute myo | 4/434 | 0.120729 | 0.765012 | 0 | 0 | 2.183581 | 4.61655 |
| Disease_Pe | Spinal Corc | 3/249 | 0.094665 | 0.765012 | 0 | 0 | 2.855256 | 6.731013 |
| Disease_Pe | Glaucoma ( | 3/290 | 0.132519 | 0.765012 | 0 | 0 | 2.44226 | 4.935882 |

| | | | | | | | | |
|---|---|---|---|---|---|---|---|---|
| Disease_Pe | Alcoholic H | 3/275 | 0.118102 | 0.765012 | 0 | 0 | 2.578913 | 5.509085 |
| Disease_Pe | colon aden | 4/427 | 0.11562 | 0.765012 | 0 | 0 | 2.220513 | 4.790646 |
| Disease_Pe | Infection b | 3/300 | 0.142458 | 0.765012 | 0 | 0 | 2.358826 | 4.596658 |
| Disease_Pe | Hypertroph | 3/287 | 0.129587 | 0.765012 | 0 | 0 | 2.468436 | 5.04401 |
| Disease_Pe | Eczema DC | 3/288 | 0.130562 | 0.765012 | 0 | 0 | 2.459649 | 5.007625 |
| Disease_Pe | Obesity C0 | 3/291 | 0.133501 | 0.765012 | 0 | 0 | 2.433656 | 4.900515 |
| Disease_Pe | Congestive | 3/272 | 0.115294 | 0.765012 | 0 | 0 | 2.608072 | 5.634137 |
| Disease_Pe | Leukemia, | 3/287 | 0.129587 | 0.765012 | 0 | 0 | 2.468436 | 5.04401 |
| Disease_Pe | amyotroph | 4/423 | 0.112745 | 0.765012 | 0 | 0 | 2.242172 | 4.893832 |
| Disease_Pe | Primary pu | 3/270 | 0.113436 | 0.765012 | 0 | 0 | 2.627876 | 5.719604 |
| Disease_Pe | ulcerative ( | 3/286 | 0.128615 | 0.765012 | 0 | 0 | 2.477284 | 5.080743 |
| Disease_Pe | asthma DO | 3/272 | 0.115294 | 0.765012 | 0 | 0 | 2.608072 | 5.634137 |
| Disease_Pe | progressive | 4/459 | 0.13975 | 0.765012 | 0 | 0 | 2.060956 | 4.055757 |
| Disease_Pe | Thymic Car | 3/249 | 0.094665 | 0.765012 | 0 | 0 | 2.855256 | 6.731013 |
| Disease_Pe | Dengue Fe | 3/266 | 0.109757 | 0.76591 | 0 | 0 | 2.668387 | 5.895774 |
| GO_Biologi | Organelle A | 1/322 | 0.757114 | 0.766835 | 0 | 0 | 0.709701 | 0.197468 |
| PhenGenI_ | Cardiovasc | 1/242 | 0.654034 | 0.766913 | 0 | 0 | 0.949146 | 0.403003 |
| GWAS_Cat | Self-report | 1/326 | 0.761381 | 0.76704 | 0 | 0 | 0.700823 | 0.191059 |
| DisGeNET | Hepatitis, T | 1/325 | 0.760322 | 0.767942 | 0 | 0 | 0.703022 | 0.192638 |
| DisGeNET | Drug-Induc | 1/325 | 0.760322 | 0.767942 | 0 | 0 | 0.703022 | 0.192638 |
| DisGeNET | Chemical a | 1/325 | 0.760322 | 0.767942 | 0 | 0 | 0.703022 | 0.192638 |
| DisGeNET | Chemically | 1/325 | 0.760322 | 0.767942 | 0 | 0 | 0.703022 | 0.192638 |
| CellMarker | Paneth Cel | 1/314 | 0.748352 | 0.767972 | 0 | 0 | 0.728137 | 0.211074 |
| KOMP2_M | Increased S | 1/278 | 0.704895 | 0.768255 | 0 | 0 | 0.82428 | 0.288256 |
| KOMP2_M | Abnormal I | 1/283 | 0.711348 | 0.768255 | 0 | 0 | 0.809459 | 0.275697 |
| GTEx_Tissu | GTEX-RU72 | 20/3269 | 0.066978 | 0.768958 | 0 | 0 | 1.531034 | 4.138976 |
| GTEx_Tissu | GTEX-WHS | 9/1177 | 0.069672 | 0.768958 | 0 | 0 | 1.851785 | 4.933069 |
| GTEx_Tissu | GTEX-Q734 | 14/2119 | 0.073278 | 0.768958 | 0 | 0 | 1.622438 | 4.240241 |
| GTEx_Tissu | GTEX-UPJH | 32/5775 | 0.067531 | 0.768958 | 0 | 0 | 1.43555 | 3.869053 |
| GTEx_Tissu | GTEX-T5JW | Feb-98 | 0.067994 | 0.768958 | 0 | 0 | 4.857108 | 13.05753 |
| GTEx_Tissu | GTEX-SN8C | 20/3276 | 0.068191 | 0.768958 | 0 | 0 | 1.5271 | 4.100934 |
| GTEx_Tissu | GTEX-SUCS | 17/2679 | 0.068331 | 0.768958 | 0 | 0 | 1.573827 | 4.223197 |
| GTEx_Tissu | GTEX-SIU7- | 44/8396 | 0.065074 | 0.768958 | 0 | 0 | 1.41641 | 3.869962 |
| GTEx_Tissu | GTEX-WZT( | 8/988 | 0.065454 | 0.768958 | 0 | 0 | 1.956394 | 5.333936 |
| GTEx_Tissu | GTEX-RU72 | 5/518 | 0.07536 | 0.768958 | 0 | 0 | 2.3059 | 5.961845 |
| GTEx_Tissu | GTEX-TMM | 5/518 | 0.07536 | 0.768958 | 0 | 0 | 2.3059 | 5.961845 |
| GTEx_Tissu | GTEX-WHS | 7/804 | 0.060729 | 0.768958 | 0 | 0 | 2.098683 | 5.879111 |
| GTEx_Tissu | GTEX-U3ZN | 3/206 | 0.060881 | 0.768958 | 0 | 0 | 3.467628 | 9.705325 |
| GTEx_Tissu | GTEX-X4XY | 18/2845 | 0.062633 | 0.768958 | 0 | 0 | 1.57666 | 4.368085 |
| GTEx_Tissu | GTEX-NPJ8 | 3/209 | 0.063017 | 0.768958 | 0 | 0 | 3.416609 | 9.444704 |
| GTEx_Tissu | GTEX-WFO | 20/3268 | 0.066806 | 0.768958 | 0 | 0 | 1.531597 | 4.144437 |
| GTEx_Tissu | GTEX-SN8C | 2/102 | 0.072883 | 0.768958 | 0 | 0 | 4.661882 | 12.20902 |
| GTEx_Tissu | GTEX-T6MI | 22/3658 | 0.064343 | 0.768958 | 0 | 0 | 1.515165 | 4.156887 |
| GTEx_Tissu | GTEX-X62C | 34/6188 | 0.065073 | 0.768958 | 0 | 0 | 1.434275 | 3.918795 |
| GTEx_Tissu | GTEX-X15G | 45/8572 | 0.059307 | 0.768958 | 0 | 0 | 1.430666 | 4.041666 |

| | | | | | | | | |
|---|---|---|---|---|---|---|---|---|
| GTEx_Tissu | GTEX-RWS( | 45/8582 | 0.06044 | 0.768958 | 0 | 0 | 1.427735 | 4.006374 |
| GTEx_Tissu | GTEX-QLQ7 | 3/204 | 0.059476 | 0.768958 | 0 | 0 | 3.502488 | 9.88465 |
| GTEx_Tissu | GTEX-XBED | Feb-96 | 0.065593 | 0.768958 | 0 | 0 | 4.960951 | 13.51507 |
| GTEx_Tissu | GTEX-XXEK | 19/3085 | 0.069959 | 0.768958 | 0 | 0 | 1.535307 | 4.083673 |
| GTEx_Tissu | GTEX-Q734 | 32/5800 | 0.071051 | 0.768958 | 0 | 0 | 1.426806 | 3.772994 |
| GTEx_Tissu | GTEX-U4B1 | 3/220 | 0.071143 | 0.768958 | 0 | 0 | 3.241606 | 8.567754 |
| GTEx_Tissu | GTEX-QCQ( | 3/220 | 0.071143 | 0.768958 | 0 | 0 | 3.241606 | 8.567754 |
| GTEx_Tissu | GTEX-S7SF- | 4/359 | 0.071457 | 0.768958 | 0 | 0 | 2.655082 | 7.005859 |
| GTEx_Tissu | GTEX-T6MI | 11/1546 | 0.071509 | 0.768958 | 0 | 0 | 1.732882 | 4.571233 |
| GTEx_Tissu | GTEX-WWY | 9/1167 | 0.066788 | 0.768958 | 0 | 0 | 1.868772 | 5.057333 |
| GTEx_Tissu | GTEX-PWC | 53/10487 | 0.068861 | 0.768958 | 0 | 0 | 1.416148 | 3.78913 |
| GTEx_Tissu | GTEX-XMD | 3/221 | 0.071905 | 0.768958 | 0 | 0 | 3.226573 | 8.493677 |
| GTEx_Tissu | GTEX-N7M | 26/4537 | 0.072711 | 0.768958 | 0 | 0 | 1.455284 | 3.814673 |
| GTEx_Tissu | GTEX-S7SF- | 36/6628 | 0.065761 | 0.768958 | 0 | 0 | 1.426435 | 3.882372 |
| GTEx_Tissu | GTEX-T6M( | 6/655 | 0.06582 | 0.768958 | 0 | 0 | 2.19871 | 5.982331 |
| GTEx_Tissu | GTEX-RNOI | 35/6414 | 0.066264 | 0.768958 | 0 | 0 | 1.428033 | 3.875829 |
| GTEx_Tissu | GTEX-TSE9 | 18/2867 | 0.066549 | 0.768958 | 0 | 0 | 1.56247 | 4.234015 |
| GTEx_Tissu | GTEX-T6M( | 20/3267 | 0.066635 | 0.768958 | 0 | 0 | 1.532161 | 4.149906 |
| GTEx_Tissu | GTEX-REY6 | Feb-97 | 0.06679 | 0.768958 | 0 | 0 | 4.908483 | 13.28336 |
| GTEx_Tissu | GTEX-S4Z8 | 3/225 | 0.074986 | 0.768958 | 0 | 0 | 3.167793 | 8.206005 |
| GTEx_Tissu | GTEX-XOT4 | 7/846 | 0.075258 | 0.768958 | 0 | 0 | 1.989243 | 5.14583 |
| GTEx_Tissu | GTEX-XOT( | 23/3929 | 0.075326 | 0.768958 | 0 | 0 | 1.472738 | 3.808395 |
| GTEx_Tissu | GTEX-XUYS | 19/3115 | 0.075589 | 0.768958 | 0 | 0 | 1.517722 | 3.919436 |
| GTEx_Tissu | GTEX-RNOI | 3/226 | 0.075766 | 0.768958 | 0 | 0 | 3.153427 | 8.136178 |
| GTEx_Tissu | GTEX-S4Q7 | 17/2702 | 0.072831 | 0.768958 | 0 | 0 | 1.558265 | 4.082044 |
| GTEx_Tissu | GTEX-SNM | 3/222 | 0.07267 | 0.768958 | 0 | 0 | 3.211676 | 8.420478 |
| GTEx_Tissu | GTEX-XQ3S | 3/224 | 0.07421 | 0.768958 | 0 | 0 | 3.182288 | 8.276655 |
| GTEx_Tissu | GTEX-N7M | 43/8256 | 0.076032 | 0.768958 | 0 | 0 | 1.392194 | 3.587133 |
| GTEx_Tissu | GTEX-PWC | 2/101 | 0.07165 | 0.768958 | 0 | 0 | 4.70921 | 12.4133 |
| GTEx_Tissu | GTEX-PVO\ | 48/9362 | 0.072431 | 0.768958 | 0 | 0 | 1.400572 | 3.676662 |
| GTEx_Tissu | GTEX-WFO | 51/10082 | 0.076453 | 0.76972 | 0 | 0 | 1.395624 | 3.588257 |
| CellMarker | Neuron Bra | 2/616 | 0.753092 | 0.769952 | 0 | 0 | 0.739567 | 0.209717 |
| CellMarker | CD1C-CD14 | 1/321 | 0.756036 | 0.770088 | 0 | 0 | 0.711955 | 0.19911 |
| GO_Molecu | DNA-bindir | 1/243 | 0.655558 | 0.771244 | 0 | 0 | 0.945176 | 0.399118 |
| GeneSigDB | 18310505- | 1/274 | 0.699631 | 0.771672 | 0 | 0 | 0.836528 | 0.29881 |
| DisGeNET | Hepatitis, D | 1/329 | 0.764533 | 0.771899 | 0 | 0 | 0.694307 | 0.186414 |
| Reactome_ | Signal Tran | Oct-65 | 0.64212 | 0.771935 | 0 | 0 | 0.923533 | 0.409107 |
| Reactome_ | Chromatin | 1/238 | 0.647874 | 0.771935 | 0 | 0 | 0.965362 | 0.419024 |
| Reactome_ | Neddylatio | 1/237 | 0.646317 | 0.771935 | 0 | 0 | 0.969501 | 0.423154 |
| Disease_Pe | Arrhythmo | 3/308 | 0.150586 | 0.772051 | 0 | 0 | 2.296019 | 4.346874 |
| Disease_Pe | Down synd | 3/306 | 0.14854 | 0.772051 | 0 | 0 | 2.31141 | 4.407632 |
| Disease_Pe | Hypertroph | 3/308 | 0.150586 | 0.772051 | 0 | 0 | 2.296019 | 4.346874 |
| Disease_Pe | asthma DO | 3/306 | 0.14854 | 0.772051 | 0 | 0 | 2.31141 | 4.407632 |
| DisGeNET | Fatigue | 1/330 | 0.765575 | 0.772652 | 0 | 0 | 0.692161 | 0.184896 |
| GWAS_Cat | Neuroticisr | 1/332 | 0.767644 | 0.772711 | 0 | 0 | 0.687908 | 0.181903 |

| | | | | | | | | |
|---|---|---|---|---|---|---|---|---|
| GO_Molecı | Metal Ion E | 2/521 | 0.665878 | 0.774276 | 0 | 0 | 0.879247 | 0.357545 |
| GTEx_Tissu | GTEX-U3ZN | 7/843 | 0.074157 | 0.774503 | 0 | 0 | 1.996696 | 5.194533 |
| GTEx_Tissu | GTEX-QVU! | 14/2122 | 0.073956 | 0.774503 | 0 | 0 | 1.619857 | 4.218562 |
| GTEx_Tissu | GTEX-QMR | 7/842 | 0.073793 | 0.774503 | 0 | 0 | 1.999192 | 5.210886 |
| GTEx_Tissu | GTEX-OIZI- | 13/1929 | 0.073542 | 0.774503 | 0 | 0 | 1.650123 | 4.306663 |
| GeneSigDB | 17910759- | Oct-99 | 0.708772 | 0.775506 | 0 | 0 | 0.869011 | 0.299132 |
| GeneSigDB | 14684422- | 1/282 | 0.710068 | 0.775506 | 0 | 0 | 0.812381 | 0.278154 |
| GeneSigDB | 18440302- | 1/280 | 0.707493 | 0.775506 | 0 | 0 | 0.818288 | 0.28315 |
| GeneSigDB | 16135788- | 1/280 | 0.707493 | 0.775506 | 0 | 0 | 0.818288 | 0.28315 |
| GeneSigDB | 20077526- | 1/279 | 0.706197 | 0.775506 | 0 | 0 | 0.821273 | 0.285689 |
| Reactome_ | Leishmania | 1/247 | 0.661585 | 0.775891 | 0 | 0 | 0.929618 | 0.384041 |
| Reactome_ | Transmissi | 1/246 | 0.660088 | 0.775891 | 0 | 0 | 0.93346 | 0.387742 |
| Reactome_ | SLC-mediat | 1/247 | 0.661585 | 0.775891 | 0 | 0 | 0.929618 | 0.384041 |
| Rare_Disea | Cerebellar | 1/335 | 0.770714 | 0.775928 | 0 | 0 | 0.681625 | 0.177521 |
| Rare_Disea | Congenital | 1/337 | 0.772738 | 0.776915 | 0 | 0 | 0.677499 | 0.174669 |
| Disease_Pe | neurofibro | 3/318 | 0.160951 | 0.778901 | 0 | 0 | 2.221995 | 4.058822 |
| Disease_Pe | multiple sc | 3/290 | 0.132519 | 0.778901 | 0 | 0 | 2.44226 | 4.935882 |
| Disease_Pe | RA (rheum | 3/314 | 0.156778 | 0.778901 | 0 | 0 | 2.251034 | 4.17099 |
| Disease_Pe | Cancer of t | 3/315 | 0.157818 | 0.778901 | 0 | 0 | 2.243704 | 4.142577 |
| Disease_Pe | Senescence | 3/314 | 0.156778 | 0.778901 | 0 | 0 | 2.251034 | 4.17099 |
| Disease_Pe | melanoma | 3/315 | 0.157818 | 0.778901 | 0 | 0 | 2.243704 | 4.142577 |
| Disease_Pe | Waldenstr | 3/307 | 0.149562 | 0.778901 | 0 | 0 | 2.303689 | 4.377116 |
| Disease_Pe | Cardiomyo | 2/150 | 0.138761 | 0.778901 | 0 | 0 | 3.142289 | 6.206032 |
| Disease_Pe | prostate ca | 3/315 | 0.157818 | 0.778901 | 0 | 0 | 2.243704 | 4.142577 |
| Disease_Pe | bronchopu | 3/295 | 0.137457 | 0.778901 | 0 | 0 | 2.399829 | 4.762324 |
| Disease_Pe | Hepatocell | 3/298 | 0.14045 | 0.778901 | 0 | 0 | 2.375061 | 4.662008 |
| Disease_Pe | prostate ca | 3/304 | 0.146503 | 0.778901 | 0 | 0 | 2.327005 | 4.469498 |
| Disease_Pe | mental reta | 2/165 | 0.161361 | 0.778901 | 0 | 0 | 2.850956 | 5.200458 |
| Disease_Pe | multiple sc | 3/291 | 0.133501 | 0.778901 | 0 | 0 | 2.433656 | 4.900515 |
| Disease_Pe | Burn C000 | 3/317 | 0.159904 | 0.778901 | 0 | 0 | 2.229186 | 4.086495 |
| Disease_Pe | hepatocell | 3/317 | 0.159904 | 0.778901 | 0 | 0 | 2.229186 | 4.086495 |
| Disease_Pe | swine influ | 3/315 | 0.157818 | 0.778901 | 0 | 0 | 2.243704 | 4.142577 |
| Disease_Pe | Eczema C0 | 2/163 | 0.158308 | 0.778901 | 0 | 0 | 2.886664 | 5.320729 |
| Disease_Pe | pancreatic | 3/286 | 0.128615 | 0.778901 | 0 | 0 | 2.477284 | 5.080743 |
| Disease_Pe | autistic dis | 2/143 | 0.128476 | 0.778901 | 0 | 0 | 3.299458 | 6.770526 |
| Disease_Pe | COPD - Chr | 4/442 | 0.126686 | 0.778901 | 0 | 0 | 2.142818 | 4.427156 |
| Disease_Pe | Lung trans | 3/307 | 0.149562 | 0.778901 | 0 | 0 | 2.303689 | 4.377116 |
| Disease_Pe | hepatitis C | 3/305 | 0.14752 | 0.778901 | 0 | 0 | 2.319182 | 4.438425 |
| Disease_Pe | cervical car | 3/296 | 0.138452 | 0.778901 | 0 | 0 | 2.391516 | 4.728574 |
| Disease_Pe | Breast Can | 3/301 | 0.143466 | 0.778901 | 0 | 0 | 2.350791 | 4.564431 |
| Disease_Pe | ulcerative | 4/429 | 0.117069 | 0.778901 | 0 | 0 | 2.209837 | 4.740077 |
| Disease_Pe | prolactinor | 4/440 | 0.125185 | 0.778901 | 0 | 0 | 2.152868 | 4.473582 |
| Disease_Pe | Hypertensi | 3/282 | 0.124754 | 0.778901 | 0 | 0 | 2.513313 | 5.231246 |
| Disease_Pe | Synovial sa | 3/273 | 0.116227 | 0.778901 | 0 | 0 | 2.59828 | 5.592039 |
| Disease_Pe | Arrhythmo | 3/292 | 0.134486 | 0.778901 | 0 | 0 | 2.425111 | 4.86548 |

| | | | | | | | | |
|---|---|---|---|---|---|---|---|---|
| Disease_Pe | systemic lu | 3/294 | 0.136464 | 0.778901 | 0 | 0 | 2.408198 | 4.796389 |
| Disease_Pe | Vulvar intra | 3/270 | 0.113436 | 0.778901 | 0 | 0 | 2.627876 | 5.719604 |
| Disease_Pe | Cardiomyo | 3/274 | 0.117163 | 0.778901 | 0 | 0 | 2.588561 | 5.550357 |
| Disease_Pe | polycystic c | 3/275 | 0.118102 | 0.778901 | 0 | 0 | 2.578913 | 5.509085 |
| Disease_Pe | Hypertroph | 3/313 | 0.155741 | 0.778901 | 0 | 0 | 2.25841 | 4.199656 |
| Disease_Pe | Insulin Resi | 3/310 | 0.152641 | 0.778901 | 0 | 0 | 2.280828 | 4.287197 |
| Disease_Pe | cerebral pa | 3/308 | 0.150586 | 0.778901 | 0 | 0 | 2.296019 | 4.346874 |
| Disease_Pe | amyotroph | 3/308 | 0.150586 | 0.778901 | 0 | 0 | 2.296019 | 4.346874 |
| Disease_Pe | Adenocarci | 3/282 | 0.124754 | 0.778901 | 0 | 0 | 2.513313 | 5.231246 |
| Disease_Pe | Obesity C0 | 3/281 | 0.123795 | 0.778901 | 0 | 0 | 2.522482 | 5.269787 |
| Disease_Pe | Huntingtor | 3/289 | 0.131539 | 0.778901 | 0 | 0 | 2.450924 | 4.971583 |
| RNA-Seq_D | miR21 Mar | 5/498 | 0.066046 | 0.778947 | 0 | 0 | 2.40192 | 6.526982 |
| RNA-Seq_D | BCAT2 Gas | 5/499 | 0.066495 | 0.778947 | 0 | 0 | 2.396934 | 6.497181 |
| RNA-Seq_D | Dicer1 Reti | 5/499 | 0.066495 | 0.778947 | 0 | 0 | 2.396934 | 6.497181 |
| RNA-Seq_D | Cushing's S | 5/497 | 0.065598 | 0.778947 | 0 | 0 | 2.406925 | 6.556953 |
| RNA-Seq_D | PS2APP Mi | 5/499 | 0.066495 | 0.778947 | 0 | 0 | 2.396934 | 6.497181 |
| RNA-Seq_D | Asxl1 BM-c | 5/498 | 0.066046 | 0.778947 | 0 | 0 | 2.40192 | 6.526982 |
| RNA-Seq_D | Baf60c Car | 5/498 | 0.066046 | 0.778947 | 0 | 0 | 2.40192 | 6.526982 |
| RNA-Seq_D | Bmal1 Live | 5/498 | 0.066046 | 0.778947 | 0 | 0 | 2.40192 | 6.526982 |
| RNA-Seq_D | Bmal1 Live | 5/498 | 0.066046 | 0.778947 | 0 | 0 | 2.40192 | 6.526982 |
| RNA-Seq_D | Bmal1 Live | 5/498 | 0.066046 | 0.778947 | 0 | 0 | 2.40192 | 6.526982 |
| RNA-Seq_D | Bmal1 Live | 5/498 | 0.066046 | 0.778947 | 0 | 0 | 2.40192 | 6.526982 |
| RNA-Seq_D | SAP130 HC | 5/497 | 0.065598 | 0.778947 | 0 | 0 | 2.406925 | 6.556953 |
| RNA-Seq_D | Huntingtor | 5/496 | 0.065153 | 0.778947 | 0 | 0 | 2.411952 | 6.587097 |
| RNA-Seq_D | Olig2 Brain | 5/495 | 0.064708 | 0.778947 | 0 | 0 | 2.416999 | 6.617415 |
| RNA-Seq_D | WZ4002 NS | 5/497 | 0.065598 | 0.778947 | 0 | 0 | 2.406925 | 6.556953 |
| RNA-Seq_D | miR-18a He | 5/497 | 0.065598 | 0.778947 | 0 | 0 | 2.406925 | 6.556953 |
| RNA-Seq_D | Vemurafen | 5/493 | 0.063825 | 0.778947 | 0 | 0 | 2.427154 | 6.678574 |
| RNA-Seq_D | TAF15 Mot | 5/497 | 0.065598 | 0.778947 | 0 | 0 | 2.406925 | 6.556953 |
| RNA-Seq_D | TGM5 HCT | 5/497 | 0.065598 | 0.778947 | 0 | 0 | 2.406925 | 6.556953 |
| RNA-Seq_D | Vemurafen | 5/497 | 0.065598 | 0.778947 | 0 | 0 | 2.406925 | 6.556953 |
| RNA-Seq_D | Gprc5a col | 5/499 | 0.066495 | 0.778947 | 0 | 0 | 2.396934 | 6.497181 |
| RNA-Seq_D | ABCA4 RPE | 5/498 | 0.066046 | 0.778947 | 0 | 0 | 2.40192 | 6.526982 |
| RNA-Seq_D | Akti-1-2 HC | 5/498 | 0.066046 | 0.778947 | 0 | 0 | 2.40192 | 6.526982 |
| RNA-Seq_D | recombina | 5/496 | 0.065153 | 0.778947 | 0 | 0 | 2.411952 | 6.587097 |
| RNA-Seq_D | DICER1 Fib | 5/496 | 0.065153 | 0.778947 | 0 | 0 | 2.411952 | 6.587097 |
| RNA-Seq_D | DMOG Live | 5/495 | 0.064708 | 0.778947 | 0 | 0 | 2.416999 | 6.617415 |
| RNA-Seq_D | ELAVL2 NP | 5/495 | 0.064708 | 0.778947 | 0 | 0 | 2.416999 | 6.617415 |
| RNA-Seq_D | ALS (Spora | 5/496 | 0.065153 | 0.778947 | 0 | 0 | 2.411952 | 6.587097 |
| RNA-Seq_D | Snord116 E | 5/498 | 0.066046 | 0.778947 | 0 | 0 | 2.40192 | 6.526982 |
| RNA-Seq_D | HDAC3 Hip | 5/498 | 0.066046 | 0.778947 | 0 | 0 | 2.40192 | 6.526982 |
| RNA-Seq_D | BRD9-inhib | 5/497 | 0.065598 | 0.778947 | 0 | 0 | 2.406925 | 6.556953 |
| RNA-Seq_D | Vemurafen | 5/498 | 0.066046 | 0.778947 | 0 | 0 | 2.40192 | 6.526982 |
| RNA-Seq_D | Kawasaki D | 5/497 | 0.065598 | 0.778947 | 0 | 0 | 2.406925 | 6.556953 |
| RNA-Seq_D | MeCP2 Vis | 5/497 | 0.065598 | 0.778947 | 0 | 0 | 2.406925 | 6.556953 |

| | | | | | | | | |
|---|---|---|---|---|---|---|---|---|
| RNA-Seq_[ | Bmal1 Live | 5/497 | 0.065598 | 0.778947 | 0 | 0 | 2.406925 | 6.556953 |
| RNA-Seq_[ | Bmal1 Live | 5/497 | 0.065598 | 0.778947 | 0 | 0 | 2.406925 | 6.556953 |
| RNA-Seq_[ | Bmal1 Live | 5/497 | 0.065598 | 0.778947 | 0 | 0 | 2.406925 | 6.556953 |
| RNA-Seq_[ | Brg1 Heart | 5/497 | 0.065598 | 0.778947 | 0 | 0 | 2.406925 | 6.556953 |
| RNA-Seq_[ | Colorectal | 5/497 | 0.065598 | 0.778947 | 0 | 0 | 2.406925 | 6.556953 |
| RNA-Seq_[ | Ezh2 Embr | 5/497 | 0.065598 | 0.778947 | 0 | 0 | 2.406925 | 6.556953 |
| RNA-Seq_[ | HER2 BT-4 | 5/495 | 0.064708 | 0.778947 | 0 | 0 | 2.416999 | 6.617415 |
| RNA-Seq_[ | HER2 BT47 | 5/495 | 0.064708 | 0.778947 | 0 | 0 | 2.416999 | 6.617415 |
| RNA-Seq_[ | Polycystic I | 5/498 | 0.066046 | 0.778947 | 0 | 0 | 2.40192 | 6.526982 |
| RNA-Seq_[ | Rapamycin | 5/498 | 0.066046 | 0.778947 | 0 | 0 | 2.40192 | 6.526982 |
| RNA-Seq_[ | Ezh2 Embr | 5/497 | 0.065598 | 0.778947 | 0 | 0 | 2.406925 | 6.556953 |
| RNA-Seq_[ | FOXC1 Skir | 5/497 | 0.065598 | 0.778947 | 0 | 0 | 2.406925 | 6.556953 |
| RNA-Seq_[ | Gefitinib N | 5/497 | 0.065598 | 0.778947 | 0 | 0 | 2.406925 | 6.556953 |
| RNA-Seq_[ | I-BET MV4- | 5/497 | 0.065598 | 0.778947 | 0 | 0 | 2.406925 | 6.556953 |
| RNA-Seq_[ | DAPT 3 hES | 5/498 | 0.066046 | 0.778947 | 0 | 0 | 2.40192 | 6.526982 |
| RNA-Seq_[ | miR-18a Ta | 5/498 | 0.066046 | 0.778947 | 0 | 0 | 2.40192 | 6.526982 |
| RNA-Seq_[ | Myc RKO k | 5/497 | 0.065598 | 0.778947 | 0 | 0 | 2.406925 | 6.556953 |
| RNA-Seq_[ | Nova1 mid | 5/497 | 0.065598 | 0.778947 | 0 | 0 | 2.406925 | 6.556953 |
| RNA-Seq_[ | LIN41 hESC | 5/498 | 0.066046 | 0.778947 | 0 | 0 | 2.40192 | 6.526982 |
| RNA-Seq_[ | Mef2c Hea | 5/498 | 0.066046 | 0.778947 | 0 | 0 | 2.40192 | 6.526982 |
| RNA-Seq_[ | N-acetylcys | 5/498 | 0.066046 | 0.778947 | 0 | 0 | 2.40192 | 6.526982 |
| RNA-Seq_[ | Panobinost | 5/498 | 0.066046 | 0.778947 | 0 | 0 | 2.40192 | 6.526982 |
| RNA-Seq_[ | Rapamycin | 5/495 | 0.064708 | 0.778947 | 0 | 0 | 2.416999 | 6.617415 |
| RNA-Seq_[ | dimethylox | 5/495 | 0.064708 | 0.778947 | 0 | 0 | 2.416999 | 6.617415 |
| RNA-Seq_[ | Parkinson's | 5/496 | 0.065153 | 0.778947 | 0 | 0 | 2.411952 | 6.587097 |
| RNA-Seq_[ | TGFB3 Pala | 5/496 | 0.065153 | 0.778947 | 0 | 0 | 2.411952 | 6.587097 |
| RNA-Seq_[ | MSI2 Hema | 5/495 | 0.064708 | 0.778947 | 0 | 0 | 2.416999 | 6.617415 |
| Gene_Pert | Ncoa2 KO r | 4/248 | 0.023063 | 0.780216 | 0 | 0 | 3.884851 | 14.64409 |
| DisGeNET | Pneumonit | 1/339 | 0.774745 | 0.781606 | 0 | 0 | 0.673421 | 0.171872 |
| Gene_Pert | Cav1 Defici | 4/253 | 0.024591 | 0.781735 | 0 | 0 | 3.805874 | 14.1022 |
| Gene_Pert | Src overexp | 4/257 | 0.025857 | 0.781735 | 0 | 0 | 3.74494 | 13.68845 |
| Gene_Pert | RPS24 OE H | 5/383 | 0.025845 | 0.781735 | 0 | 0 | 3.151213 | 11.51975 |
| Gene_Pert | Dmd Defici | 4/256 | 0.025537 | 0.781735 | 0 | 0 | 3.759992 | 13.7903 |
| KOMP2_M | Cataract (M | 1/299 | 0.731073 | 0.781741 | 0 | 0 | 0.765374 | 0.239748 |
| Disease_Pe | prolactinor | 2/160 | 0.153751 | 0.78207 | 0 | 0 | 2.941921 | 5.508522 |
| Gene_Pert | Tlr4 mutati | 3/147 | 0.026333 | 0.783078 | 0 | 0 | 4.903026 | 17.83201 |
| Disease_Pe | hepatocellu | 3/314 | 0.156778 | 0.783387 | 0 | 0 | 2.251034 | 4.17099 |
| Disease_Pe | endometri | 3/315 | 0.157818 | 0.783387 | 0 | 0 | 2.243704 | 4.142577 |
| Disease_Pe | Hepatic lipi | 3/316 | 0.15886 | 0.783387 | 0 | 0 | 2.236422 | 4.114413 |
| Disease_Pe | intracrania | 2/163 | 0.158308 | 0.783387 | 0 | 0 | 2.886664 | 5.320729 |
| Disease_Pe | Fracture of | 2/168 | 0.165961 | 0.783432 | 0 | 0 | 2.799008 | 5.027029 |
| Disease_Pe | Dehydratio | 3/322 | 0.165157 | 0.783432 | 0 | 0 | 2.193686 | 3.950513 |
| Disease_Pe | Muscular D | 3/322 | 0.165157 | 0.783432 | 0 | 0 | 2.193686 | 3.950513 |
| Disease_Pe | inclusion b | 2/169 | 0.167499 | 0.784923 | 0 | 0 | 2.782106 | 4.971004 |
| DisGeNET | Squamous | 3/934 | 0.778568 | 0.785161 | 0 | 0 | 0.728172 | 0.182261 |

| | | | | | | | | |
|---|---|---|---|---|---|---|---|---|
| Disease_Pe | Generalize | 3/321 | 0.164103 | 0.785261 | 0 | 0 | 2.200696 | 3.977238 |
| Disease_Pe | juvenile de | 4/489 | 0.164053 | 0.785261 | 0 | 0 | 1.930493 | 3.489493 |
| Disease_Pe | Obesity C0 | 3/319 | 0.161999 | 0.785261 | 0 | 0 | 2.214851 | 4.03139 |
| Disease_Pe | JRA - Juven | 3/320 | 0.16305 | 0.785261 | 0 | 0 | 2.207751 | 4.004196 |
| GTEx_Tissu | GTEX-PVO\ | 16/2518 | 0.075521 | 0.785907 | 0 | 0 | 1.568188 | 4.05117 |
| Gene_Pert | Taf4b KO n | 4/251 | 0.023972 | 0.787348 | 0 | 0 | 3.837081 | 14.31558 |
| Gene_Pert | Emd KO mc | 4/253 | 0.024591 | 0.787348 | 0 | 0 | 3.805874 | 14.1022 |
| Gene_Pert | Cav1 KO m | 4/252 | 0.02428 | 0.787348 | 0 | 0 | 3.821415 | 14.20834 |
| Disease_Pe | Septic Shoc | 4/494 | 0.168247 | 0.787998 | 0 | 0 | 1.910302 | 3.404772 |
| Disease_Pe | Type 2 diak | 3/323 | 0.166214 | 0.787998 | 0 | 0 | 2.186719 | 3.924018 |
| Disease_Pe | Hypoxia C0 | 3/325 | 0.168334 | 0.787998 | 0 | 0 | 2.172915 | 3.871711 |
| GTEx_Tissu | GTEX-QV31 | 7/848 | 0.075998 | 0.788034 | 0 | 0 | 1.984304 | 5.113655 |
| Disease_Pe | Spinal Corc | 3/326 | 0.169397 | 0.788064 | 0 | 0 | 2.166077 | 3.845893 |
| GeneSigDB | 20208059-: | 1/293 | 0.723837 | 0.788996 | 0 | 0 | 0.78134 | 0.25252 |
| GTEx_Tissu | GTEX-TKQ2 | 28/5005 | 0.080247 | 0.790308 | 0 | 0 | 1.424206 | 3.592759 |
| GTEx_Tissu | GTEX-R3RS | 5/528 | 0.080274 | 0.790308 | 0 | 0 | 2.260644 | 5.702045 |
| GTEx_Tissu | GTEX-U4B1 | 3/231 | 0.079718 | 0.790308 | 0 | 0 | 3.08349 | 7.798958 |
| GTEx_Tissu | GTEX-PX3G | 5/526 | 0.079278 | 0.790308 | 0 | 0 | 2.269557 | 5.752869 |
| GTEx_Tissu | GTEX-T5JC- | 48/9419 | 0.080112 | 0.790308 | 0 | 0 | 1.384566 | 3.495109 |
| Gene_Pert | Aldh1a2 KC | 5/418 | 0.035702 | 0.791328 | 0 | 0 | 2.878994 | 9.594359 |
| Gene_Pert | Pparg OE n | 3/170 | 0.038062 | 0.791328 | 0 | 0 | 4.22284 | 13.80251 |
| Gene_Pert | SPDEF KD h | 4/290 | 0.037792 | 0.791328 | 0 | 0 | 3.307271 | 10.83346 |
| Gene_Pert | Calr KO mc | 4/300 | 0.041941 | 0.791328 | 0 | 0 | 3.193911 | 10.12943 |
| Gene_Pert | Gykl1 KO n | 4/300 | 0.041941 | 0.791328 | 0 | 0 | 3.193911 | 10.12943 |
| Gene_Pert | SALL4 KD h | 4/286 | 0.036202 | 0.791328 | 0 | 0 | 3.354866 | 11.13356 |
| Gene_Pert | Klhl40 KO r | 4/286 | 0.036202 | 0.791328 | 0 | 0 | 3.354866 | 11.13356 |
| Gene_Pert | Dmd Defici | 4/285 | 0.035811 | 0.791328 | 0 | 0 | 3.366977 | 11.21034 |
| Gene_Pert | Apln KO mc | 5/382 | 0.025592 | 0.791328 | 0 | 0 | 3.159733 | 11.5819 |
| Gene_Pert | Atp1a1 KD | 4/279 | 0.033515 | 0.791328 | 0 | 0 | 3.44149 | 11.68643 |
| Gene_Pert | Suz12 KD n | 3/165 | 0.035319 | 0.791328 | 0 | 0 | 4.354277 | 14.55783 |
| Gene_Pert | Vax2 KO m | 4/294 | 0.039422 | 0.791328 | 0 | 0 | 3.260989 | 10.54418 |
| Gene_Pert | Kdm6a KO | 4/293 | 0.039011 | 0.791328 | 0 | 0 | 3.272439 | 10.61551 |
| Gene_Pert | Psap hypor | 4/278 | 0.033142 | 0.791328 | 0 | 0 | 3.454226 | 11.76843 |
| Gene_Pert | CMAHP KO | 4/277 | 0.03277 | 0.791328 | 0 | 0 | 3.467055 | 11.85122 |
| Gene_Pert | Sox2 KD mc | 4/287 | 0.036596 | 0.791328 | 0 | 0 | 3.342841 | 11.05749 |
| Gene_Pert | Cbfb KD mc | 5/420 | 0.036327 | 0.791328 | 0 | 0 | 2.864825 | 9.497447 |
| Gene_Pert | MECOM KI | 4/292 | 0.038602 | 0.791328 | 0 | 0 | 3.283969 | 10.6875 |
| Gene_Pert | MIR148B C | 4/313 | 0.047707 | 0.791328 | 0 | 0 | 3.057512 | 9.302998 |
| Gene_Pert | Map2k7 m | 5/414 | 0.034473 | 0.791328 | 0 | 0 | 2.907746 | 9.792057 |
| Gene_Pert | Pcgf6 KD m | 5/397 | 0.029546 | 0.791328 | 0 | 0 | 3.036492 | 10.69391 |
| Gene_Pert | NRAS DEPL | 4/262 | 0.027493 | 0.791328 | 0 | 0 | 3.67143 | 13.19442 |
| Gene_Pert | STAU1 DEL | 4/272 | 0.03095 | 0.791328 | 0 | 0 | 3.532638 | 12.27729 |
| Gene_Pert | Htt KO mou | 4/281 | 0.034271 | 0.791328 | 0 | 0 | 3.416293 | 11.52474 |
| Gene_Pert | Myd88 KO | 4/281 | 0.034271 | 0.791328 | 0 | 0 | 3.416293 | 11.52474 |
| Gene_Pert | Prmt1 KD r | Feb-66 | 0.033522 | 0.791328 | 0 | 0 | 7.297426 | 24.7787 |

| | | | | | | | | |
|---|---|---|---|---|---|---|---|---|
| Gene_Pert | Nfe2l2 KO | 4/307 | 0.044994 | 0.791328 | 0 | 0 | 3.119011 | 9.672767 |
| Gene_Pert | Ackr3 Defic | 4/323 | 0.052429 | 0.791328 | 0 | 0 | 2.960154 | 8.727434 |
| Gene_Pert | Psap hypor | 4/323 | 0.052429 | 0.791328 | 0 | 0 | 2.960154 | 8.727434 |
| Gene_Pert | Abca1 Hete | 4/322 | 0.051945 | 0.791328 | 0 | 0 | 2.969614 | 8.782826 |
| Gene_Pert | SYK druginl | 4/321 | 0.051464 | 0.791328 | 0 | 0 | 2.979134 | 8.838684 |
| Gene_Pert | CMAHP KO | 4/321 | 0.051464 | 0.791328 | 0 | 0 | 2.979134 | 8.838684 |
| Gene_Pert | Dmd Defici | 3/192 | 0.05138 | 0.791328 | 0 | 0 | 3.727135 | 11.06404 |
| Gene_Pert | Tert OE mc | 4/325 | 0.053403 | 0.791328 | 0 | 0 | 2.941411 | 8.618031 |
| Gene_Pert | Htt KO mou | 4/270 | 0.030239 | 0.791328 | 0 | 0 | 3.559562 | 12.45357 |
| Gene_Pert | VAMP7 KD | 4/310 | 0.046339 | 0.791328 | 0 | 0 | 3.08796 | 9.485478 |
| Gene_Pert | CMAHP KO | 4/324 | 0.052914 | 0.791328 | 0 | 0 | 2.950753 | 8.672504 |
| Gene_Pert | THRA HCC | 4/304 | 0.043671 | 0.791328 | 0 | 0 | 3.150683 | 9.865033 |
| Gene_Pert | Htt OE mou | 4/307 | 0.044994 | 0.791328 | 0 | 0 | 3.119011 | 9.672767 |
| Gene_Pert | Casp1 KO r | 3/182 | 0.045077 | 0.791328 | 0 | 0 | 3.93735 | 12.20334 |
| Gene_Pert | Nos3 KO m | 4/275 | 0.032035 | 0.791328 | 0 | 0 | 3.492998 | 12.01919 |
| Gene_Pert | CXCL8 OE h | 4/282 | 0.034652 | 0.791328 | 0 | 0 | 3.403831 | 11.44503 |
| Gene_Pert | RRM2 OE h | 4/285 | 0.035811 | 0.791328 | 0 | 0 | 3.366977 | 11.21034 |
| Gene_Pert | Egfr drugin | 4/299 | 0.041515 | 0.791328 | 0 | 0 | 3.204901 | 10.19702 |
| Gene_Pert | PDCD10 KD | 4/299 | 0.041515 | 0.791328 | 0 | 0 | 3.204901 | 10.19702 |
| Gene_Pert | E2f2 KD mc | 4/300 | 0.041941 | 0.791328 | 0 | 0 | 3.193911 | 10.12943 |
| Gene_Pert | DNMT1 Inh | 4/325 | 0.053403 | 0.791328 | 0 | 0 | 2.941411 | 8.618031 |
| Gene_Pert | Pdpn NULL | 3/194 | 0.052689 | 0.791328 | 0 | 0 | 3.687734 | 10.85427 |
| Gene_Pert | Itga7 OE m | 5/447 | 0.045426 | 0.791328 | 0 | 0 | 2.6861 | 8.304515 |
| Gene_Pert | Vax2 KO m | 4/308 | 0.04544 | 0.791328 | 0 | 0 | 3.108592 | 9.609793 |
| Gene_Pert | Atm knock | 4/301 | 0.04237 | 0.791328 | 0 | 0 | 3.182995 | 10.06245 |
| Gene_Pert | Eed KO mo | 5/467 | 0.052975 | 0.791328 | 0 | 0 | 2.567179 | 7.542205 |
| Gene_Pert | KDM4C nat | 4/292 | 0.038602 | 0.791328 | 0 | 0 | 3.283969 | 10.6875 |
| Gene_Pert | Nfe2l2 KO | 4/292 | 0.038602 | 0.791328 | 0 | 0 | 3.283969 | 10.6875 |
| Gene_Pert | Aldh5a1 KC | 4/292 | 0.038602 | 0.791328 | 0 | 0 | 3.283969 | 10.6875 |
| Gene_Pert | Hmx1 Defic | 4/303 | 0.043235 | 0.791328 | 0 | 0 | 3.161381 | 9.930257 |
| Gene_Pert | TP63 Muta | 3/173 | 0.039759 | 0.791328 | 0 | 0 | 4.147689 | 13.37596 |
| Gene_Pert | Tgfbr2 kno | 4/320 | 0.050986 | 0.791328 | 0 | 0 | 2.988714 | 8.895015 |
| Gene_Pert | Irf4 KO mo | 4/319 | 0.05051 | 0.791328 | 0 | 0 | 2.998355 | 8.951822 |
| Gene_Pert | Pparg OE n | 4/319 | 0.05051 | 0.791328 | 0 | 0 | 2.998355 | 8.951822 |
| Gene_Pert | Insig1 KO n | 4/313 | 0.047707 | 0.791328 | 0 | 0 | 3.057512 | 9.302998 |
| Gene_Pert | Neurod1 C | 4/311 | 0.046793 | 0.791328 | 0 | 0 | 3.077744 | 9.424125 |
| Gene_Pert | Cdkn1b KO | 4/310 | 0.046339 | 0.791328 | 0 | 0 | 3.08796 | 9.485478 |
| Gene_Pert | TP53BP1 K | 3/174 | 0.040333 | 0.791328 | 0 | 0 | 4.123225 | 13.23795 |
| Gene_Pert | Ighmbp2 m | 5/432 | 0.040218 | 0.791328 | 0 | 0 | 2.782601 | 8.941757 |
| Gene_Pert | Hdac2 Defi | 4/262 | 0.027493 | 0.791636 | 0 | 0 | 3.67143 | 13.19442 |
| Gene_Pert | IRF9 OE hu | 5/389 | 0.027392 | 0.791636 | 0 | 0 | 3.101023 | 11.15592 |
| Disease_Pe | non-systen | 3/327 | 0.170462 | 0.792218 | 0 | 0 | 2.159281 | 3.820295 |
| GeneSigDB | 12734205- | 1/297 | 0.728682 | 0.792482 | 0 | 0 | 0.770624 | 0.243916 |
| GeneSigDB | 17150101- | 1/298 | 0.72988 | 0.792482 | 0 | 0 | 0.76799 | 0.241821 |
| KOMP2_M | Abnormal I | 1/328 | 0.763487 | 0.792852 | 0 | 0 | 0.696465 | 0.187947 |

| | | | | | | | | |
|---|---|---|---|---|---|---|---|---|
| KOMP2_M | Decreased | 1/322 | 0.757114 | 0.792852 | 0 | 0 | 0.709701 | 0.197468 |
| KOMP2_M | Decreased | 1/319 | 0.753864 | 0.792852 | 0 | 0 | 0.716506 | 0.202444 |
| GTEx_Tissu | GTEX-T5JW | 46/8969 | 0.080942 | 0.793373 | 0 | 0 | 1.38185 | 3.474003 |
| GeneSigDB | 15257931-: | 2/591 | 0.732221 | 0.793476 | 0 | 0 | 0.771956 | 0.240598 |
| GeneSigDB | 20081105-: | Sep-06 | 0.733829 | 0.793676 | 0 | 0 | 0.843169 | 0.260943 |
| GTEx_Tissu | GTEX-QDV. | 9/1201 | 0.076913 | 0.794676 | 0 | 0 | 1.812177 | 4.64838 |
| GTEx_Tissu | GTEX-RNOI | 12/1857 | 0.106494 | 0.794766 | 0 | 0 | 1.566873 | 3.50927 |
| GTEx_Tissu | GTEX-QDVI | 2/132 | 0.112716 | 0.794766 | 0 | 0 | 3.580633 | 7.816104 |
| GTEx_Tissu | GTEX-VJWI | 43/8475 | 0.110688 | 0.794766 | 0 | 0 | 1.330653 | 2.928825 |
| GTEx_Tissu | GTEX-SNM | 5/583 | 0.11027 | 0.794766 | 0 | 0 | 2.039729 | 4.497247 |
| GTEx_Tissu | GTEX-OOB. | 3/266 | 0.109757 | 0.794766 | 0 | 0 | 2.668387 | 5.895774 |
| GTEx_Tissu | GTEX-SN8( | 3/266 | 0.109757 | 0.794766 | 0 | 0 | 2.668387 | 5.895774 |
| GTEx_Tissu | GTEX-XBEC | 49/9836 | 0.109685 | 0.794766 | 0 | 0 | 1.334138 | 2.94864 |
| GTEx_Tissu | GTEX-WFG | 2/129 | 0.108512 | 0.794766 | 0 | 0 | 3.665771 | 8.141279 |
| GTEx_Tissu | GTEX-RVPV | 46/9014 | 0.087514 | 0.794766 | 0 | 0 | 1.369286 | 3.335528 |
| GTEx_Tissu | GTEX-SNM | 3/268 | 0.111591 | 0.794766 | 0 | 0 | 2.647978 | 5.806801 |
| GTEx_Tissu | GTEX-XQ8I | 3/268 | 0.111591 | 0.794766 | 0 | 0 | 2.647978 | 5.806801 |
| GTEx_Tissu | GTEX-TMM | 3/268 | 0.111591 | 0.794766 | 0 | 0 | 2.647978 | 5.806801 |
| GTEx_Tissu | GTEX-NPJ7 | 33/6125 | 0.087827 | 0.794766 | 0 | 0 | 1.386436 | 3.372345 |
| GTEx_Tissu | GTEX-WFG | 4/415 | 0.107093 | 0.794766 | 0 | 0 | 2.286753 | 5.10873 |
| GTEx_Tissu | GTEX-UPK5 | 22/3879 | 0.106681 | 0.794766 | 0 | 0 | 1.408955 | 3.153112 |
| GTEx_Tissu | GTEX-OXR( | 19/3283 | 0.112871 | 0.794766 | 0 | 0 | 1.425223 | 3.109142 |
| GTEx_Tissu | GTEX-WFO | 16/2647 | 0.106343 | 0.794766 | 0 | 0 | 1.480249 | 3.317369 |
| GTEx_Tissu | GTEX-NPJ8 | 61/12627 | 0.106142 | 0.794766 | 0 | 0 | 1.371733 | 3.076763 |
| GTEx_Tissu | GTEX-SNM | 3/262 | 0.106125 | 0.794766 | 0 | 0 | 2.710149 | 6.079245 |
| GTEx_Tissu | GTEX-SJXC- | 2/127 | 0.105734 | 0.794766 | 0 | 0 | 3.7248 | 8.368983 |
| GTEx_Tissu | GTEX-SUCS | 2/131 | 0.11131 | 0.794766 | 0 | 0 | 3.608573 | 7.922388 |
| GTEx_Tissu | GTEX-R55F | 2/114 | 0.088189 | 0.794766 | 0 | 0 | 4.159874 | 10.1013 |
| GTEx_Tissu | GTEX-QESD | 45/8794 | 0.088567 | 0.794766 | 0 | 0 | 1.367177 | 3.314032 |
| GTEx_Tissu | GTEX-T6M( | 2/115 | 0.089505 | 0.794766 | 0 | 0 | 4.122853 | 9.950336 |
| GTEx_Tissu | GTEX-X5EB | 4/424 | 0.11346 | 0.794766 | 0 | 0 | 2.236718 | 4.867776 |
| GTEx_Tissu | GTEX-S7PN | 34/6405 | 0.098433 | 0.794766 | 0 | 0 | 1.363573 | 3.161278 |
| GTEx_Tissu | GTEX-X4XX | 3/253 | 0.098134 | 0.794766 | 0 | 0 | 2.809 | 6.520881 |
| GTEx_Tissu | GTEX-S4Z8 | 22/3836 | 0.097282 | 0.794766 | 0 | 0 | 1.428656 | 3.328974 |
| GTEx_Tissu | GTEX-TML8 | 5/571 | 0.103306 | 0.794766 | 0 | 0 | 2.084267 | 4.731403 |
| GTEx_Tissu | GTEX-SE5C | 43/8428 | 0.102448 | 0.794766 | 0 | 0 | 1.34359 | 3.061232 |
| GTEx_Tissu | GTEX-QCQ( | 3/261 | 0.105224 | 0.794766 | 0 | 0 | 2.720792 | 6.1263 |
| GTEx_Tissu | GTEX-QCQ( | 3/261 | 0.105224 | 0.794766 | 0 | 0 | 2.720792 | 6.1263 |
| GTEx_Tissu | GTEX-REY6 | 3/261 | 0.105224 | 0.794766 | 0 | 0 | 2.720792 | 6.1263 |
| GTEx_Tissu | GTEX-SNM | 2/126 | 0.104352 | 0.794766 | 0 | 0 | 3.755028 | 8.486292 |
| GTEx_Tissu | GTEX-U3ZN | 2/126 | 0.104352 | 0.794766 | 0 | 0 | 3.755028 | 8.486292 |
| GTEx_Tissu | GTEX-T6MI | 51/10264 | 0.103991 | 0.794766 | 0 | 0 | 1.345507 | 3.045486 |
| GTEx_Tissu | GTEX-UJHI- | 5/572 | 0.103878 | 0.794766 | 0 | 0 | 2.080484 | 4.711335 |
| GTEx_Tissu | GTEX-WFJ( | 40/7828 | 0.115732 | 0.794766 | 0 | 0 | 1.325006 | 2.857345 |
| GTEx_Tissu | GTEX-WYJK | 24-Jan | 0.099394 | 0.794766 | 0 | 0 | 10.05561 | 23.215 |

| | | | | | | | | |
|---|---|---|---|---|---|---|---|---|
| GTEx_Tissu | GTEX-NPJ7 | 13/2029 | 0.099775 | 0.794766 | 0 | 0 | 1.559557 | 3.594527 |
| GTEx_Tissu | GTEX-TSE9 | 3/255 | 0.099887 | 0.794766 | 0 | 0 | 2.786423 | 6.419114 |
| GTEx_Tissu | GTEX-QDT8 | 12/1836 | 0.100004 | 0.794766 | 0 | 0 | 1.586754 | 3.653571 |
| GTEx_Tissu | GTEX-XYKS | 4/407 | 0.101577 | 0.794766 | 0 | 0 | 2.333104 | 5.335676 |
| GTEx_Tissu | GTEX-OHPI | 12/1831 | 0.098496 | 0.794766 | 0 | 0 | 1.591556 | 3.688815 |
| GTEx_Tissu | GTEX-U4B1 | 3/258 | 0.102542 | 0.794766 | 0 | 0 | 2.753221 | 6.27042 |
| GTEx_Tissu | GTEX-XGQ4 | 2/125 | 0.102976 | 0.794766 | 0 | 0 | 3.785748 | 8.60599 |
| GTEx_Tissu | GTEX-U3ZN | 2/120 | 0.096172 | 0.794766 | 0 | 0 | 3.947159 | 9.242743 |
| GTEx_Tissu | GTEX-R45C | 2/134 | 0.115542 | 0.794766 | 0 | 0 | 3.526025 | 7.609588 |
| GTEx_Tissu | GTEX-U3ZG | 16/2680 | 0.115345 | 0.794766 | 0 | 0 | 1.459121 | 3.151449 |
| GTEx_Tissu | GTEX-U3ZH | 17/2882 | 0.115099 | 0.794766 | 0 | 0 | 1.445106 | 3.124269 |
| GTEx_Tissu | GTEX-XMK | 28-Jan | 0.114983 | 0.794766 | 0 | 0 | 8.564169 | 18.52408 |
| GTEx_Tissu | GTEX-WFG | 36/6931 | 0.114367 | 0.794766 | 0 | 0 | 1.33273 | 2.889814 |
| GTEx_Tissu | GTEX-T2IS- | 3/271 | 0.114364 | 0.794766 | 0 | 0 | 2.617937 | 5.676657 |
| GTEx_Tissu | GTEX-TSE9 | 4/425 | 0.114178 | 0.794766 | 0 | 0 | 2.231291 | 4.841894 |
| GTEx_Tissu | GTEX-S341 | 53/10684 | 0.096765 | 0.794766 | 0 | 0 | 1.36102 | 3.178625 |
| GTEx_Tissu | GTEX-UJM( | 41/7945 | 0.096811 | 0.794766 | 0 | 0 | 1.35421 | 3.162078 |
| GTEx_Tissu | GTEX-WHP | 40/7697 | 0.092799 | 0.794766 | 0 | 0 | 1.362236 | 3.238474 |
| GTEx_Tissu | GTEX-VUS( | 17/2769 | 0.087087 | 0.794766 | 0 | 0 | 1.514415 | 3.696463 |
| GTEx_Tissu | GTEX-SUCS | 5/547 | 0.090075 | 0.794766 | 0 | 0 | 2.179259 | 5.245735 |
| GTEx_Tissu | GTEX-T6MI | 18/2999 | 0.093656 | 0.794766 | 0 | 0 | 1.481732 | 3.508923 |
| GTEx_Tissu | GTEX-NL4V | 8/1070 | 0.093603 | 0.794766 | 0 | 0 | 1.797516 | 4.25777 |
| GTEx_Tissu | GTEX-RWS/ | 27/4867 | 0.093347 | 0.794766 | 0 | 0 | 1.401415 | 3.323366 |
| GTEx_Tissu | GTEX-WW\ | 45/8823 | 0.093061 | 0.794766 | 0 | 0 | 1.35912 | 3.227236 |
| GTEx_Tissu | GTEX-WEY! | 53/10660 | 0.092976 | 0.794766 | 0 | 0 | 1.367626 | 3.24868 |
| GTEx_Tissu | GTEX-QDVI | 4/394 | 0.092906 | 0.794766 | 0 | 0 | 2.412481 | 5.732456 |
| GTEx_Tissu | GTEX-Q2AH | 14/2199 | 0.092823 | 0.794766 | 0 | 0 | 1.556014 | 3.698741 |
| GTEx_Tissu | GTEX-X8H( | 7/939 | 0.114174 | 0.794766 | 0 | 0 | 1.782014 | 3.867024 |
| GTEx_Tissu | GTEX-RU72 | 40/7644 | 0.084518 | 0.794766 | 0 | 0 | 1.377662 | 3.403924 |
| GTEx_Tissu | GTEX-WCD | 34/6333 | 0.086253 | 0.794766 | 0 | 0 | 1.386491 | 3.397562 |
| GTEx_Tissu | GTEX-PLZ6 | 2/115 | 0.089505 | 0.794766 | 0 | 0 | 4.122853 | 9.950336 |
| GTEx_Tissu | GTEX-QCQ( | 2/111 | 0.084277 | 0.794766 | 0 | 0 | 4.275013 | 10.57487 |
| GTEx_Tissu | GTEX-V1D1 | 30/5455 | 0.08412 | 0.794766 | 0 | 0 | 1.405578 | 3.479523 |
| GTEx_Tissu | GTEX-U3ZN | 5/535 | 0.083814 | 0.794766 | 0 | 0 | 2.229982 | 5.528463 |
| GTEx_Tissu | GTEX-WRH | 33/6100 | 0.083766 | 0.794766 | 0 | 0 | 1.394667 | 3.458391 |
| GTEx_Tissu | GTEX-X261 | 2/109 | 0.0817 | 0.794766 | 0 | 0 | 4.35536 | 10.90889 |
| GTEx_Tissu | GTEX-QEG4 | 4/389 | 0.089671 | 0.794766 | 0 | 0 | 2.444437 | 5.895038 |
| GTEx_Tissu | GTEX-PX3G | 22/3830 | 0.096016 | 0.794766 | 0 | 0 | 1.43144 | 3.354212 |
| GTEx_Tissu | GTEX-RNOI | 3/250 | 0.095527 | 0.794766 | 0 | 0 | 2.843551 | 6.677635 |
| GTEx_Tissu | GTEX-T2IS- | 3/250 | 0.095527 | 0.794766 | 0 | 0 | 2.843551 | 6.677635 |
| GTEx_Tissu | GTEX-OXRL | 43/8384 | 0.09514 | 0.794766 | 0 | 0 | 1.355833 | 3.189471 |
| GTEx_Tissu | GTEX-V955 | 17/2789 | 0.091678 | 0.794766 | 0 | 0 | 1.501737 | 3.588355 |
| GTEx_Tissu | GTEX-QDT8 | 4/416 | 0.107792 | 0.794766 | 0 | 0 | 2.281086 | 5.081227 |
| GTEx_Tissu | GTEX-QDT8 | 2/115 | 0.089505 | 0.794766 | 0 | 0 | 4.122853 | 9.950336 |
| GTEx_Tissu | GTEX-XQ3S | 2/129 | 0.108512 | 0.794766 | 0 | 0 | 3.665771 | 8.141279 |

| | | | | | | | | |
|---|---|---|---|---|---|---|---|---|
| GTEx_Tissu | GTEX-XQ8I | 2/117 | 0.092155 | 0.794766 | 0 | 0 | 4.050742 | 9.658129 |
| GTEx_Tissu | GTEX-P78B | 2/133 | 0.114127 | 0.794766 | 0 | 0 | 3.553121 | 7.711855 |
| GTEx_Tissu | GTEX-XPT6 | 2/133 | 0.114127 | 0.794766 | 0 | 0 | 3.553121 | 7.711855 |
| GTEx_Tissu | GTEX-S7SF | 5/554 | 0.093837 | 0.794766 | 0 | 0 | 2.150695 | 5.088975 |
| GTEx_Tissu | GTEX-WVLI | 7/906 | 0.099313 | 0.794766 | 0 | 0 | 1.85064 | 4.27401 |
| GTEx_Tissu | GTEX-R55G | 53/10699 | 0.09919 | 0.794766 | 0 | 0 | 1.356906 | 3.135427 |
| GTEx_Tissu | GTEX-Q734 | 2/122 | 0.098877 | 0.794766 | 0 | 0 | 3.88098 | 8.980101 |
| GTEx_Tissu | GTEX-WFG | 5/548 | 0.090607 | 0.794766 | 0 | 0 | 2.175134 | 5.222983 |
| GTEx_Tissu | GTEX-QMR | 34/6366 | 0.091694 | 0.794766 | 0 | 0 | 1.375922 | 3.287489 |
| GTEx_Tissu | GTEX-WH7 | 53/10653 | 0.091892 | 0.794766 | 0 | 0 | 1.369559 | 3.269333 |
| GTEx_Tissu | GTEX-WYV | 24/4260 | 0.098578 | 0.794766 | 0 | 0 | 1.409866 | 3.266534 |
| GTEx_Tissu | GTEX-QDVI | 3/246 | 0.092098 | 0.794766 | 0 | 0 | 2.890947 | 6.894635 |
| GTEx_Tissu | GTEX-T5JW | 2/117 | 0.092155 | 0.794766 | 0 | 0 | 4.050742 | 9.658129 |
| GeneSigDB | 18614019- | 2/597 | 0.73736 | 0.795949 | 0 | 0 | 0.763935 | 0.232755 |
| DisGeNET | Retinal Dis | 1/355 | 0.79018 | 0.796564 | 0 | 0 | 0.642458 | 0.151296 |
| GTEx_Tissu | GTEX-U8XE | 8/1025 | 0.077398 | 0.79684 | 0 | 0 | 1.881533 | 4.814458 |
| GO_Molec | ATP Bindin | 1/279 | 0.706197 | 0.79783 | 0 | 0 | 0.821273 | 0.285689 |
| GO_Molec | Ubiquitin P | 1/271 | 0.695622 | 0.79783 | 0 | 0 | 0.845952 | 0.307038 |
| GO_Molec | mRNA Binc | 1/282 | 0.710068 | 0.79783 | 0 | 0 | 0.812381 | 0.278154 |
| GTEx_Tissu | GTEX-X88G | 5/523 | 0.077796 | 0.798098 | 0 | 0 | 2.283054 | 5.830158 |
| Gene_Pert | Pten KO m | 4/264 | 0.028165 | 0.798306 | 0 | 0 | 3.642817 | 13.00366 |
| GO_Molec | Ubiquitin-L | 1/289 | 0.718907 | 0.798785 | 0 | 0 | 0.792353 | 0.261495 |
| Rare_Disea | Alzheimer's | 1/361 | 0.795694 | 0.798916 | 0 | 0 | 0.631557 | 0.144336 |
| Gene_Pert | Hipk2 OE n | 5/394 | 0.028726 | 0.799726 | 0 | 0 | 3.06038 | 10.86417 |
| Gene_Pert | Tgfbr2 kno | 4/268 | 0.029538 | 0.799726 | 0 | 0 | 3.586893 | 12.63333 |
| Gene_Pert | Dmd KO m | 4/268 | 0.029538 | 0.799726 | 0 | 0 | 3.586893 | 12.63333 |
| Gene_Pert | Stub1 KO n | 4/328 | 0.054882 | 0.800027 | 0 | 0 | 2.913729 | 8.457304 |
| Gene_Pert | Nr1i3 hNR1 | 4/328 | 0.054882 | 0.800027 | 0 | 0 | 2.913729 | 8.457304 |
| Gene_Pert | Aldh5a1 KC | 4/329 | 0.05538 | 0.800778 | 0 | 0 | 2.904615 | 8.404609 |
| Disease_Pe | Endometri | 3/330 | 0.173668 | 0.801356 | 0 | 0 | 2.139144 | 3.744801 |
| GTEx_Tissu | GTEX-QDT8 | 2/136 | 0.118386 | 0.801588 | 0 | 0 | 3.473047 | 7.410799 |
| GTEx_Tissu | GTEX-QDV. | 4/431 | 0.118527 | 0.801588 | 0 | 0 | 2.199261 | 4.690176 |
| GTEx_Tissu | GTEX-OXR( | 3/274 | 0.117163 | 0.801588 | 0 | 0 | 2.588561 | 5.550357 |
| GTEx_Tissu | GTEX-R55C | 3/275 | 0.118102 | 0.801588 | 0 | 0 | 2.578913 | 5.509085 |
| GTEx_Tissu | GTEX-TMM | 3/275 | 0.118102 | 0.801588 | 0 | 0 | 2.578913 | 5.509085 |
| DisGeNET | Small cell c | 2/673 | 0.795558 | 0.801677 | 0 | 0 | 0.674744 | 0.154322 |
| CellMarker | Mitotic Fet | 2/665 | 0.790008 | 0.801712 | 0 | 0 | 0.683169 | 0.161032 |
| Disease_Pe | Leukemia, | 3/335 | 0.179051 | 0.802354 | 0 | 0 | 2.10639 | 3.623169 |
| Disease_Pe | Neurofibro | 3/335 | 0.179051 | 0.802354 | 0 | 0 | 2.10639 | 3.623169 |
| Disease_Pe | Hutchinsor | 3/335 | 0.179051 | 0.802354 | 0 | 0 | 2.10639 | 3.623169 |
| Disease_Pe | Heart Injur | 3/336 | 0.180133 | 0.802354 | 0 | 0 | 2.099957 | 3.599451 |
| Disease_Pe | schizophre | 3/337 | 0.181217 | 0.802354 | 0 | 0 | 2.093563 | 3.575931 |
| Disease_Pe | mental reta | 3/333 | 0.176892 | 0.802354 | 0 | 0 | 2.119372 | 3.671207 |
| Disease_Pe | Vitamin A I | 3/335 | 0.179051 | 0.802354 | 0 | 0 | 2.10639 | 3.623169 |
| KOMP2_M | Decreased | 1/345 | 0.78066 | 0.802965 | 0 | 0 | 0.661472 | 0.163791 |

| | | | | | | | | |
|---|---|---|---|---|---|---|---|---|
| GTEx_Tissu | GTEX-T6M( | 9/1327 | 0.122349 | 0.803481 | 0 | 0 | 1.627904 | 3.420032 |
| GTEx_Tissu | GTEX-TKQ2 | 2/137 | 0.119815 | 0.803481 | 0 | 0 | 3.447146 | 7.31418 |
| GTEx_Tissu | GTEX-N7M | 7/951 | 0.119858 | 0.803481 | 0 | 0 | 1.758249 | 3.730029 |
| GTEx_Tissu | GTEX-W5X | 17/2903 | 0.120862 | 0.803481 | 0 | 0 | 1.432823 | 3.027711 |
| GTEx_Tissu | GTEX-Q2AI | 5/601 | 0.121135 | 0.803481 | 0 | 0 | 1.976285 | 4.171633 |
| GTEx_Tissu | GTEX-Q2A( | 7/954 | 0.121302 | 0.803481 | 0 | 0 | 1.752402 | 3.696639 |
| GTEx_Tissu | GTEX-T6M( | 4/435 | 0.121466 | 0.803481 | 0 | 0 | 2.178403 | 4.59233 |
| GTEx_Tissu | GTEX-XUZ( | 4/436 | 0.122206 | 0.803481 | 0 | 0 | 2.173249 | 4.568268 |
| GTEx_Tissu | GTEX-OHPI | 39/7639 | 0.122418 | 0.803481 | 0 | 0 | 1.316357 | 2.764762 |
| GTEx_Tissu | GTEX-S4P3 | 4/434 | 0.120729 | 0.803481 | 0 | 0 | 2.183581 | 4.61655 |
| DisGeNET | Cognitive d | 3/966 | 0.797822 | 0.80365 | 0 | 0 | 0.702789 | 0.158739 |
| GWAS_Cat | Cognitive P | 1/365 | 0.799291 | 0.803903 | 0 | 0 | 0.624489 | 0.139904 |
| GTEx_Tissu | GTEX-S7SF- | 17/2914 | 0.12395 | 0.804155 | 0 | 0 | 1.426461 | 2.978271 |
| GTEx_Tissu | GTEX-RM2I | 26/4800 | 0.123784 | 0.804155 | 0 | 0 | 1.351631 | 2.823852 |
| GTEx_Tissu | GTEX-X4EP | 54/11072 | 0.124018 | 0.804155 | 0 | 0 | 1.321061 | 2.757494 |
| GTEx_Tissu | GTEX-P4Q1 | 2/140 | 0.124127 | 0.804155 | 0 | 0 | 3.371697 | 7.034881 |
| GTEx_Tissu | GTEX-XMD | Dec-11 | 0.124328 | 0.804155 | 0 | 0 | 1.517767 | 3.164293 |
| Gene_Pert | BST2 OE hu | 4/271 | 0.030593 | 0.804289 | 0 | 0 | 3.546049 | 12.365 |
| Gene_Pert | Dmd Defici | 4/271 | 0.030593 | 0.804289 | 0 | 0 | 3.546049 | 12.365 |
| GTEx_Tissu | GTEX-RNOI | 20/3538 | 0.125155 | 0.805019 | 0 | 0 | 1.39114 | 2.891071 |
| GTEx_Tissu | GTEX-RTLS- | 4/440 | 0.125185 | 0.805019 | 0 | 0 | 2.152868 | 4.473582 |
| GTEx_Tissu | GTEX-NFK9 | 2/141 | 0.125573 | 0.805184 | 0 | 0 | 3.34727 | 6.945157 |
| GTEx_Tissu | GTEX-RNOI | 7/856 | 0.078998 | 0.805864 | 0 | 0 | 1.964782 | 4.987259 |
| GTEx_Tissu | GTEX-OHPI | 9/1208 | 0.07911 | 0.805864 | 0 | 0 | 1.800924 | 4.568791 |
| GO_Biologi | Positive Re | 4/1243 | 0.797221 | 0.806306 | 0 | 0 | 0.726353 | 0.164609 |
| Disease_Pe | progressive | 3/339 | 0.18339 | 0.806415 | 0 | 0 | 2.080889 | 3.529475 |
| Reactome_ | PIP3 Activa | 1/268 | 0.691559 | 0.806819 | 0 | 0 | 0.855587 | 0.315546 |
| GTEx_Tissu | GTEX-WFO | 7/859 | 0.080142 | 0.808704 | 0 | 0 | 1.957556 | 4.940794 |
| GTEx_Tissu | GTEX-SUCS | 14/2150 | 0.080492 | 0.808704 | 0 | 0 | 1.596108 | 4.021545 |
| GTEx_Tissu | GTEX-OHPI | 6/691 | 0.080507 | 0.808704 | 0 | 0 | 2.079265 | 5.238522 |
| GTEx_Tissu | GTEX-XQ8I | 6/691 | 0.080507 | 0.808704 | 0 | 0 | 2.079265 | 5.238522 |
| Gene_Pert | Dmd Defici | 4/312 | 0.047249 | 0.808796 | 0 | 0 | 3.067595 | 9.3633 |
| Gene_Pert | Dmrta2 OE | 4/276 | 0.032401 | 0.808796 | 0 | 0 | 3.479979 | 11.93481 |
| Gene_Pert | PLK1 drugi | 5/408 | 0.032679 | 0.808796 | 0 | 0 | 2.951946 | 10.09863 |
| Gene_Pert | Ache AChE | 3/185 | 0.046925 | 0.808796 | 0 | 0 | 3.87186 | 11.84483 |
| Gene_Pert | E2F1 OE hu | 4/297 | 0.040671 | 0.808796 | 0 | 0 | 3.227106 | 10.33401 |
| Gene_Pert | IRS3P KO m | 4/289 | 0.037391 | 0.808796 | 0 | 0 | 3.319045 | 10.90745 |
| Gene_Pert | TFAP2C KD | 4/290 | 0.037792 | 0.808796 | 0 | 0 | 3.307271 | 10.83346 |
| Gene_Pert | Ppara KO n | 4/291 | 0.038196 | 0.808796 | 0 | 0 | 3.29558 | 10.76015 |
| Gene_Pert | Pou2af1 NU | 4/297 | 0.040671 | 0.808796 | 0 | 0 | 3.227106 | 10.33401 |
| Gene_Pert | SNORD15B | 5/426 | 0.038242 | 0.808796 | 0 | 0 | 2.823127 | 9.214202 |
| Gene_Pert | Mok knock | 4/298 | 0.041092 | 0.808796 | 0 | 0 | 3.215966 | 10.2652 |
| Gene_Pert | Map3k7 KC | 4/313 | 0.047707 | 0.808796 | 0 | 0 | 3.057512 | 9.302998 |
| Gene_Pert | Fmr1 KD m | 4/306 | 0.04455 | 0.808796 | 0 | 0 | 3.129498 | 9.736294 |
| Gene_Pert | Ndel1 KO n | 4/305 | 0.044109 | 0.808796 | 0 | 0 | 3.140055 | 9.800381 |

| | | | | | | | | |
|---|---|---|---|---|---|---|---|---|
| Gene_Pert | Tada3 Con | 3/184 | 0.046305 | 0.808796 | 0 | 0 | 3.893449 | 11.96266 |
| Gene_Pert | Dmd Defici | 4/311 | 0.046793 | 0.808796 | 0 | 0 | 3.077744 | 9.424125 |
| Gene_Pert | Adora2b D | 4/309 | 0.045888 | 0.808796 | 0 | 0 | 3.098242 | 9.547366 |
| Gene_Pert | CHEK1 KD I | 4/309 | 0.045888 | 0.808796 | 0 | 0 | 3.098242 | 9.547366 |
| Gene_Pert | EGFR drug | 4/302 | 0.042801 | 0.808796 | 0 | 0 | 3.172152 | 9.996061 |
| Gene_Pert | SMC3 KD h | 4/302 | 0.042801 | 0.808796 | 0 | 0 | 3.172152 | 9.996061 |
| Gene_Pert | CTNNB1 Ac | 4/295 | 0.039836 | 0.808796 | 0 | 0 | 3.249617 | 10.47349 |
| Gene_Pert | Zbtb7a KO | 4/305 | 0.044109 | 0.808796 | 0 | 0 | 3.140055 | 9.800381 |
| Gene_Pert | Srf KO mou | 4/305 | 0.044109 | 0.808796 | 0 | 0 | 3.140055 | 9.800381 |
| Gene_Pert | RAP1A KO | 4/288 | 0.036992 | 0.808796 | 0 | 0 | 3.330901 | 10.98213 |
| Gene_Pert | Ache AChE | 4/275 | 0.032035 | 0.808796 | 0 | 0 | 3.492998 | 12.01919 |
| Gene_Pert | Dmd Defici | 4/301 | 0.04237 | 0.808796 | 0 | 0 | 3.182995 | 10.06245 |
| Gene_Pert | C20orf197 | 3/186 | 0.047549 | 0.808796 | 0 | 0 | 3.850507 | 11.72863 |
| Gene_Pert | Dmd Defici | 3/175 | 0.040912 | 0.808796 | 0 | 0 | 4.099045 | 13.10196 |
| Gene_Pert | HDAC2 KD | 4/300 | 0.041941 | 0.808796 | 0 | 0 | 3.193911 | 10.12943 |
| Gene_Pert | Hunk KO m | 4/311 | 0.046793 | 0.808796 | 0 | 0 | 3.077744 | 9.424125 |
| Gene_Pert | Rap1a KO r | 4/278 | 0.033142 | 0.808796 | 0 | 0 | 3.454226 | 11.76843 |
| Gene_Pert | TFAP2C Sil | 4/280 | 0.033892 | 0.808796 | 0 | 0 | 3.428846 | 11.6052 |
| Gene_Pert | SMC3 KD h | 4/275 | 0.032035 | 0.808796 | 0 | 0 | 3.492998 | 12.01919 |
| Gene_Pert | Naca KO m | 5/419 | 0.036014 | 0.808796 | 0 | 0 | 2.871892 | 9.545744 |
| Gene_Pert | Igf1r knock | 4/292 | 0.038602 | 0.808796 | 0 | 0 | 3.283969 | 10.6875 |
| Gene_Pert | Nr2c2 KO r | 4/283 | 0.035036 | 0.808796 | 0 | 0 | 3.391458 | 11.36607 |
| Gene_Pert | Runx1 KO r | 4/281 | 0.034271 | 0.808796 | 0 | 0 | 3.416293 | 11.52474 |
| Gene_Pert | Psmc4 KO | 4/283 | 0.035036 | 0.808796 | 0 | 0 | 3.391458 | 11.36607 |
| Gene_Pert | Klhl40 KO r | 4/314 | 0.048168 | 0.809049 | 0 | 0 | 3.047493 | 9.243211 |
| Gene_Pert | Npc1 KO m | 4/332 | 0.056889 | 0.809541 | 0 | 0 | 2.877608 | 8.249101 |
| Gene_Pert | Gys1 KO m | 4/332 | 0.056889 | 0.809541 | 0 | 0 | 2.877608 | 8.249101 |
| GTEx_Tissu | GTEX-WHV | 37/7216 | 0.126977 | 0.809813 | 0 | 0 | 1.312601 | 2.708881 |
| GTEx_Tissu | GTEX-R55F | 2/142 | 0.127022 | 0.809813 | 0 | 0 | 3.323193 | 6.857052 |
| GeDiPNet_ | Developme | 5/1532 | 0.80576 | 0.809928 | 0 | 0 | 0.734183 | 0.158561 |
| Rare_Disea | Ectropion i | 1/375 | 0.808011 | 0.810189 | 0 | 0 | 0.60748 | 0.129503 |
| Gene_Pert | Pdpn KO m | 4/333 | 0.057397 | 0.810337 | 0 | 0 | 2.868715 | 8.198108 |
| GeneSigDB | 15902281- | 2/616 | 0.753092 | 0.81036 | 0 | 0 | 0.739567 | 0.209717 |
| GeneSigDB | 19218430- | Nov-48 | 0.753621 | 0.81036 | 0 | 0 | 0.836586 | 0.236642 |
| Human_Ph | Cryptorchi | 1/379 | 0.811393 | 0.811393 | 0 | 0 | 0.600929 | 0.125596 |
| GWAS_Cat | Resistance | 1/375 | 0.808011 | 0.812004 | 0 | 0 | 0.60748 | 0.129503 |
| GTEx_Tissu | GTEX-T5JW | 13/2121 | 0.128489 | 0.812399 | 0 | 0 | 1.483826 | 3.044685 |
| GTEx_Tissu | GTEX-VUS( | 2/144 | 0.129934 | 0.812399 | 0 | 0 | 3.276056 | 6.685543 |
| GTEx_Tissu | GTEX-RN6 | 4/446 | 0.129711 | 0.812399 | 0 | 0 | 2.12299 | 4.336092 |
| GTEx_Tissu | GTEX-WOF | 55/11333 | 0.129355 | 0.812399 | 0 | 0 | 1.315961 | 2.691394 |
| GTEx_Tissu | GTEX-TMN | 5/613 | 0.128652 | 0.812399 | 0 | 0 | 1.936076 | 3.970207 |
| GTEx_Tissu | GTEX-P4PC | 44/8796 | 0.128643 | 0.812399 | 0 | 0 | 1.304908 | 2.675989 |
| GTEx_Tissu | GTEX-XUJ4 | 19/3348 | 0.129984 | 0.812399 | 0 | 0 | 1.391939 | 2.840037 |
| Gene_Pert | Rock1 INHI | 3/202 | 0.058087 | 0.813673 | 0 | 0 | 3.538047 | 10.06862 |
| GeneSigDB | 20035825- | 1/323 | 0.758188 | 0.813701 | 0 | 0 | 0.707461 | 0.195842 |

| | | | | | | | | |
|---|---|---|---|---|---|---|---|---|
| Disease_Pe | Type 1 diat | 3/343 | 0.187758 | 0.813722 | 0 | 0 | 2.055987 | 3.438845 |
| Disease_Pe | pulmonary | 3/342 | 0.186664 | 0.813722 | 0 | 0 | 2.062158 | 3.461222 |
| Disease_Pe | prostate ca | 3/344 | 0.188855 | 0.813722 | 0 | 0 | 2.049853 | 3.416651 |
| Jensen_TIS | Erythroid c | 17/4448 | 0.765709 | 0.813724 | 0 | 0 | 0.848548 | 0.226522 |
| GO_Molecu | Tubulin Bin | 1/322 | 0.757114 | 0.814101 | 0 | 0 | 0.709701 | 0.197468 |
| GO_Molecu | Cadherin B | 1/319 | 0.753864 | 0.814101 | 0 | 0 | 0.716506 | 0.202444 |
| GO_Molecu | Adenyl Rib | 1/309 | 0.742718 | 0.814101 | 0 | 0 | 0.740146 | 0.220149 |
| GTEx_Tissu | GTEX-XQ8I | 7/865 | 0.082457 | 0.814156 | 0 | 0 | 1.943255 | 4.849353 |
| GTEx_Tissu | GTEX-U412 | 6/695 | 0.082245 | 0.814156 | 0 | 0 | 2.066763 | 5.162884 |
| GTEx_Tissu | GTEX-U8XE | 7/863 | 0.081681 | 0.814156 | 0 | 0 | 1.947999 | 4.879615 |
| GTEx_Tissu | GTEX-WFO | 6/694 | 0.081809 | 0.814156 | 0 | 0 | 2.069875 | 5.181671 |
| GTEx_Tissu | GTEX-PWO | 9/1218 | 0.082316 | 0.814156 | 0 | 0 | 1.785073 | 4.457677 |
| SysMyo_M | MGS71 Hu | 1/379 | 0.811393 | 0.81427 | 0 | 0 | 0.600929 | 0.125596 |
| Jensen_TIS | Thymus | 19/4940 | 0.768838 | 0.814361 | 0 | 0 | 0.851238 | 0.223769 |
| PhenGenI_ | Gastrointe: | 1/273 | 0.6983 | 0.814683 | 0 | 0 | 0.839646 | 0.301522 |
| Rare_Disea | Acute myel | 1/382 | 0.813891 | 0.814986 | 0 | 0 | 0.596106 | 0.122756 |
| GTEx_Tissu | GTEX-QDT8 | 8/1041 | 0.082948 | 0.815158 | 0 | 0 | 1.850822 | 4.607687 |
| GTEx_Tissu | GTEX-RU1J | 6/697 | 0.083122 | 0.815158 | 0 | 0 | 2.060567 | 5.125552 |
| DisGeNET | Drug-Induc | 1/377 | 0.809709 | 0.815311 | 0 | 0 | 0.604187 | 0.127532 |
| Gene_Pert | Tmod3 KO | 3/204 | 0.059476 | 0.815559 | 0 | 0 | 3.502488 | 9.88465 |
| Gene_Pert | BAP1 KO hu | 3/205 | 0.060177 | 0.815559 | 0 | 0 | 3.484972 | 9.794417 |
| Gene_Pert | Hras mous | 4/339 | 0.060496 | 0.815559 | 0 | 0 | 2.816472 | 7.900705 |
| Gene_Pert | CDK19 kno | 4/337 | 0.059453 | 0.815559 | 0 | 0 | 2.833677 | 7.99824 |
| Gene_Pert | Hras OE mo | 4/338 | 0.059973 | 0.815559 | 0 | 0 | 2.825049 | 7.949276 |
| Reactome_ | SARS-CoV-: | 1/283 | 0.711348 | 0.815874 | 0 | 0 | 0.809459 | 0.275697 |
| Reactome_ | Metabolisr | 1/285 | 0.713889 | 0.815874 | 0 | 0 | 0.803677 | 0.270861 |
| Reactome_ | Deubiquitir | 1/279 | 0.706197 | 0.815874 | 0 | 0 | 0.821273 | 0.285689 |
| Reactome_ | Asparagine | 1/282 | 0.710068 | 0.815874 | 0 | 0 | 0.812381 | 0.278154 |
| DisGeNET | Secondary | 4/1271 | 0.811367 | 0.816666 | 0 | 0 | 0.709236 | 0.148255 |
| Gene_Pert | ZHX2 KD hu | 4/316 | 0.049098 | 0.817093 | 0 | 0 | 3.027649 | 9.125164 |
| GTEx_Tissu | GTEX-SJXC- | 9/1222 | 0.08362 | 0.817271 | 0 | 0 | 1.778807 | 4.414064 |
| Gene_Pert | NANOG OE | 4/318 | 0.050037 | 0.818531 | 0 | 0 | 3.008058 | 9.009113 |
| Gene_Pert | Plagl2 OE n | 3/190 | 0.050087 | 0.818531 | 0 | 0 | 3.76738 | 11.27955 |
| GTEx_Tissu | GTEX-XBEC | 2/111 | 0.084277 | 0.818658 | 0 | 0 | 4.275013 | 10.57487 |
| GTEx_Tissu | GTEX-TKQ2 | 6/700 | 0.084447 | 0.818658 | 0 | 0 | 2.05134 | 5.070152 |
| GTEx_Tissu | GTEX-X261 | 4/381 | 0.084611 | 0.818658 | 0 | 0 | 2.497332 | 6.167647 |
| GTEx_Tissu | GTEX-UTH( | Dec-41 | 0.134941 | 0.818932 | 0 | 0 | 1.491674 | 2.987701 |
| GTEx_Tissu | GTEX-SE5C | 4/453 | 0.135078 | 0.818932 | 0 | 0 | 2.089141 | 4.182261 |
| GTEx_Tissu | GTEX-OHPI | 5/622 | 0.134428 | 0.818932 | 0 | 0 | 1.906945 | 3.826721 |
| GTEx_Tissu | GTEX-SNOS | 3/292 | 0.134486 | 0.818932 | 0 | 0 | 2.425111 | 4.86548 |
| GTEx_Tissu | GTEX-TSE9 | 4/451 | 0.133535 | 0.818932 | 0 | 0 | 2.098704 | 4.225511 |
| GTEx_Tissu | GTEX-R55C | 14/2337 | 0.13374 | 0.818932 | 0 | 0 | 1.452185 | 2.921588 |
| GTEx_Tissu | GTEX-S4P3 | 45/9045 | 0.133039 | 0.818932 | 0 | 0 | 1.299167 | 2.620571 |
| GTEx_Tissu | GTEX-TSE9 | 8/1165 | 0.133781 | 0.818932 | 0 | 0 | 1.641609 | 3.302178 |
| GTEx_Tissu | GTEX-PVO\ | 11/1737 | 0.13228 | 0.818932 | 0 | 0 | 1.525104 | 3.085032 |

| | | | | | | | | |
|---|---|---|---|---|---|---|---|---|
| GTEx_Tissu | GTEX-NPJ8 | 4/450 | 0.132767 | 0.818932 | 0 | 0 | 2.103517 | 4.247345 |
| GTEx_Tissu | GTEX-TKQ2 | 17/2946 | 0.133208 | 0.818932 | 0 | 0 | 1.408223 | 2.838761 |
| GTEx_Tissu | GTEX-RTLS | Dec-43 | 0.135666 | 0.820265 | 0 | 0 | 1.489964 | 2.976288 |
| GO_Molec | RNA Polym | 4/1225 | 0.787693 | 0.820514 | 0 | 0 | 0.737772 | 0.176067 |
| GO_Molec | DNA-bindir | 1/348 | 0.78356 | 0.820514 | 0 | 0 | 0.655653 | 0.159919 |
| GO_Molec | Calcium Ior | 1/346 | 0.781631 | 0.820514 | 0 | 0 | 0.659521 | 0.162488 |
| DisGeNET | Prostatic N | 5/1554 | 0.815713 | 0.820725 | 0 | 0 | 0.72289 | 0.147247 |
| DisGeNET | Invasive ca | 1/385 | 0.816356 | 0.821057 | 0 | 0 | 0.591358 | 0.119989 |
| DisGeNET | Colon Carc | Jul-91 | 0.817328 | 0.821719 | 0 | 0 | 0.748578 | 0.150999 |
| DisGeNET | Finding of I | 1/387 | 0.817981 | 0.821745 | 0 | 0 | 0.588234 | 0.118185 |
| DisGeNET | Erythrocyte | 1/387 | 0.817981 | 0.821745 | 0 | 0 | 0.588234 | 0.118185 |
| Rare_Disea | Bone cance | 1/393 | 0.822773 | 0.822773 | 0 | 0 | 0.579052 | 0.112959 |
| Jensen_TIS | Melanoma | 1/347 | 0.782598 | 0.823518 | 0 | 0 | 0.657582 | 0.161197 |
| Jensen_TIS | Umbilical c | 1/345 | 0.78066 | 0.823518 | 0 | 0 | 0.661472 | 0.163791 |
| Gene_Pert | Il10 KO mo | 3/207 | 0.061589 | 0.824098 | 0 | 0 | 3.450455 | 9.617355 |
| PhenGenI_ | Neuroblast | 1/282 | 0.710068 | 0.82425 | 0 | 0 | 0.812381 | 0.278154 |
| GWAS_Cat | Protein Qu | 3/1008 | 0.820986 | 0.824364 | 0 | 0 | 0.671926 | 0.132537 |
| PhenGenI_ | Intelligence | 1/285 | 0.713889 | 0.824542 | 0 | 0 | 0.803677 | 0.270861 |
| PhenGenI_ | Multiple Sc | 1/288 | 0.717661 | 0.824774 | 0 | 0 | 0.795154 | 0.263799 |
| Jensen_TIS | BTO:00003 | 1/351 | 0.786422 | 0.824846 | 0 | 0 | 0.649934 | 0.156154 |
| Gene_Pert | Gnas KO m | 4/320 | 0.050986 | 0.825794 | 0 | 0 | 2.988714 | 8.895015 |
| Gene_Pert | THRB HCC | 4/327 | 0.054386 | 0.826561 | 0 | 0 | 2.922899 | 8.510436 |
| Gene_Pert | Lmna KO m | 4/334 | 0.057907 | 0.826561 | 0 | 0 | 2.859876 | 8.147531 |
| Gene_Pert | Met active | 4/327 | 0.054386 | 0.826561 | 0 | 0 | 2.922899 | 8.510436 |
| Gene_Pert | Pou5f1 KD | 3/201 | 0.057398 | 0.826561 | 0 | 0 | 3.556097 | 10.16239 |
| Gene_Pert | Krt1 KO mc | 4/332 | 0.056889 | 0.826561 | 0 | 0 | 2.877608 | 8.249101 |
| Gene_Pert | HSF1 KD hu | 4/333 | 0.057397 | 0.826561 | 0 | 0 | 2.868715 | 8.198108 |
| Gene_Pert | Dysf Defici | 4/329 | 0.05538 | 0.826561 | 0 | 0 | 2.904615 | 8.404609 |
| Gene_Pert | Bmpr2 Del | 3/199 | 0.056033 | 0.826561 | 0 | 0 | 3.592748 | 10.35363 |
| Gene_Pert | ESRRA KO | 4/335 | 0.05842 | 0.826561 | 0 | 0 | 2.85109 | 8.097363 |
| Gene_Pert | Prkag3 Mu | 4/334 | 0.057907 | 0.826561 | 0 | 0 | 2.859876 | 8.147531 |
| Gene_Pert | Pcgf6 KD m | 3/203 | 0.05878 | 0.826561 | 0 | 0 | 3.520179 | 9.976043 |
| Gene_Pert | Cdkn1b KO | 3/194 | 0.052689 | 0.826561 | 0 | 0 | 3.687734 | 10.85427 |
| Gene_Pert | Neurog3 O | 4/327 | 0.054386 | 0.826561 | 0 | 0 | 2.922899 | 8.510436 |
| Gene_Pert | Hras mous | 3/197 | 0.054684 | 0.826561 | 0 | 0 | 3.630155 | 10.54993 |
| Gene_Pert | Nos3 KO m | 4/325 | 0.053403 | 0.826561 | 0 | 0 | 2.941411 | 8.618031 |
| Gene_Pert | RNF40 KD I | 4/325 | 0.053403 | 0.826561 | 0 | 0 | 2.941411 | 8.618031 |
| Gene_Pert | Rbl1 Defici | 5/480 | 0.058253 | 0.826561 | 0 | 0 | 2.49525 | 7.093875 |
| KOMP2_M | Abnormal I | 1/379 | 0.811393 | 0.826702 | 0 | 0 | 0.600929 | 0.125596 |
| GeneSigDB | 15902281- | 2/640 | 0.771814 | 0.826732 | 0 | 0 | 0.710861 | 0.184121 |
| Reactome_ | Post-transl | 5/1383 | 0.727277 | 0.826954 | 0 | 0 | 0.820162 | 0.261179 |
| Gene_Pert | Ghr KO mo | Feb-91 | 0.05972 | 0.826966 | 0 | 0 | 5.240978 | 14.76951 |
| Gene_Pert | Lat2 KD mc | 4/337 | 0.059453 | 0.826966 | 0 | 0 | 2.833677 | 7.99824 |
| Disease_Pe | COPD - Chr | 2/188 | 0.197153 | 0.827268 | 0 | 0 | 2.495509 | 4.05215 |
| Disease_Pe | Infertility d | 2/187 | 0.195575 | 0.827268 | 0 | 0 | 2.509126 | 4.094425 |

| | | | | | | | | |
|---|---|---|---|---|---|---|---|---|
| Disease_Pe | schizophre | 3/350 | 0.195467 | 0.827268 | 0 | 0 | 2.013792 | 3.287236 |
| Disease_Pe | nemaline n | 2/188 | 0.197153 | 0.827268 | 0 | 0 | 2.495509 | 4.05215 |
| Jensen_TIS | Globus pall | 14/3792 | 0.791538 | 0.827517 | 0 | 0 | 0.819053 | 0.191476 |
| Disease_Pe | Sjogren's s | 1/308 | 0.741576 | 0.82776 | 0 | 0 | 0.742595 | 0.222019 |
| Disease_Pe | Hyperlipide | 1/323 | 0.758188 | 0.82776 | 0 | 0 | 0.707461 | 0.195842 |
| Disease_Pe | autism spe | 1/357 | 0.792034 | 0.82776 | 0 | 0 | 0.638784 | 0.148933 |
| Disease_Pe | Asthma C0 | 1/357 | 0.792034 | 0.82776 | 0 | 0 | 0.638784 | 0.148933 |
| Disease_Pe | Ulcerative | 1/354 | 0.789247 | 0.82776 | 0 | 0 | 0.644311 | 0.152493 |
| Disease_Pe | oligodendr | 1/313 | 0.747235 | 0.82776 | 0 | 0 | 0.730508 | 0.212852 |
| Disease_Pe | systemic lu | 1/313 | 0.747235 | 0.82776 | 0 | 0 | 0.730508 | 0.212852 |
| Disease_Pe | Down synd | 1/320 | 0.754952 | 0.82776 | 0 | 0 | 0.714223 | 0.200769 |
| Disease_Pe | fetal alcoh | 1/320 | 0.754952 | 0.82776 | 0 | 0 | 0.714223 | 0.200769 |
| Disease_Pe | Huntingtor | 1/319 | 0.753864 | 0.82776 | 0 | 0 | 0.716506 | 0.202444 |
| Disease_Pe | schizophre | 1/304 | 0.736959 | 0.82776 | 0 | 0 | 0.752552 | 0.229696 |
| Disease_Pe | Leber cong | 1/309 | 0.742718 | 0.82776 | 0 | 0 | 0.740146 | 0.220149 |
| Disease_Pe | myelodysp | 1/309 | 0.742718 | 0.82776 | 0 | 0 | 0.740146 | 0.220149 |
| Disease_Pe | autism spe | 1/308 | 0.741576 | 0.82776 | 0 | 0 | 0.742595 | 0.222019 |
| Disease_Pe | adenosqua | 1/308 | 0.741576 | 0.82776 | 0 | 0 | 0.742595 | 0.222019 |
| Disease_Pe | Sjogren's s | 1/377 | 0.809709 | 0.82776 | 0 | 0 | 0.604187 | 0.127532 |
| Disease_Pe | Senescence | 1/307 | 0.740429 | 0.82776 | 0 | 0 | 0.74506 | 0.223909 |
| Disease_Pe | hepatitis C | 1/307 | 0.740429 | 0.82776 | 0 | 0 | 0.74506 | 0.223909 |
| Disease_Pe | Breast Can | 1/302 | 0.73462 | 0.82776 | 0 | 0 | 0.75763 | 0.233655 |
| Disease_Pe | Type 2 diak | 1/306 | 0.739278 | 0.82776 | 0 | 0 | 0.747541 | 0.225819 |
| Disease_Pe | Cancer of p | 1/306 | 0.739278 | 0.82776 | 0 | 0 | 0.747541 | 0.225819 |
| Disease_Pe | polycystic | 1/306 | 0.739278 | 0.82776 | 0 | 0 | 0.747541 | 0.225819 |
| Disease_Pe | schizophre | 1/305 | 0.738121 | 0.82776 | 0 | 0 | 0.750038 | 0.227747 |
| Disease_Pe | Neurofibro | 1/305 | 0.738121 | 0.82776 | 0 | 0 | 0.750038 | 0.227747 |
| Disease_Pe | MDS - Mye | 1/305 | 0.738121 | 0.82776 | 0 | 0 | 0.750038 | 0.227747 |
| Disease_Pe | psoriasis D | 1/305 | 0.738121 | 0.82776 | 0 | 0 | 0.750038 | 0.227747 |
| Disease_Pe | Familial co | 1/302 | 0.73462 | 0.82776 | 0 | 0 | 0.75763 | 0.233655 |
| Disease_Pe | schizophre | 2/291 | 0.36192 | 0.82776 | 0 | 0 | 1.59772 | 1.623813 |
| Disease_Pe | Depression | 2/309 | 0.389926 | 0.82776 | 0 | 0 | 1.502663 | 1.415207 |
| Disease_Pe | Obesity C0 | 2/309 | 0.389926 | 0.82776 | 0 | 0 | 1.502663 | 1.415207 |
| Disease_Pe | amyotroph | 1/325 | 0.760322 | 0.82776 | 0 | 0 | 0.703022 | 0.192638 |
| Disease_Pe | diffuse larg | 1/337 | 0.772738 | 0.82776 | 0 | 0 | 0.677499 | 0.174669 |
| Disease_Pe | invasive du | 1/335 | 0.770714 | 0.82776 | 0 | 0 | 0.681625 | 0.177521 |
| Disease_Pe | Squamous | 1/335 | 0.770714 | 0.82776 | 0 | 0 | 0.681625 | 0.177521 |
| Disease_Pe | APECED - A | 1/334 | 0.769695 | 0.82776 | 0 | 0 | 0.683707 | 0.178968 |
| Disease_Pe | Macular de | 1/337 | 0.772738 | 0.82776 | 0 | 0 | 0.677499 | 0.174669 |
| Disease_Pe | type 2 diab | 1/347 | 0.782598 | 0.82776 | 0 | 0 | 0.657582 | 0.161197 |
| Disease_Pe | Emery-Dre | 1/347 | 0.782598 | 0.82776 | 0 | 0 | 0.657582 | 0.161197 |
| Disease_Pe | autism spe | 1/346 | 0.781631 | 0.82776 | 0 | 0 | 0.659521 | 0.162488 |
| Disease_Pe | Endometri | 1/341 | 0.776734 | 0.82776 | 0 | 0 | 0.669391 | 0.169127 |
| Disease_Pe | ulcerative | 1/329 | 0.764533 | 0.82776 | 0 | 0 | 0.694307 | 0.186414 |
| Disease_Pe | systemic lu | 1/329 | 0.764533 | 0.82776 | 0 | 0 | 0.694307 | 0.186414 |

| | | | | | | | | |
|---|---|---|---|---|---|---|---|---|
| Disease_Pe | glaucoma a | 1/328 | 0.763487 | 0.82776 | 0 | 0 | 0.696465 | 0.187947 |
| Disease_Pe | tuberculosi | 1/325 | 0.760322 | 0.82776 | 0 | 0 | 0.703022 | 0.192638 |
| Disease_Pe | Alzheimer's | 1/325 | 0.760322 | 0.82776 | 0 | 0 | 0.703022 | 0.192638 |
| Disease_Pe | endometria | 2/298 | 0.37287 | 0.82776 | 0 | 0 | 1.55938 | 1.538369 |
| Disease_Pe | Wolfram sy | 1/324 | 0.759257 | 0.82776 | 0 | 0 | 0.705234 | 0.194232 |
| Disease_Pe | autism spe | 1/334 | 0.769695 | 0.82776 | 0 | 0 | 0.683707 | 0.178968 |
| Disease_Pe | autism spe | 1/339 | 0.774745 | 0.82776 | 0 | 0 | 0.673421 | 0.171872 |
| Disease_Pe | Crohn's dis | 1/339 | 0.774745 | 0.82776 | 0 | 0 | 0.673421 | 0.171872 |
| Disease_Pe | acute myoc | 1/339 | 0.774745 | 0.82776 | 0 | 0 | 0.673421 | 0.171872 |
| Disease_Pe | endometric | 1/318 | 0.752771 | 0.82776 | 0 | 0 | 0.718803 | 0.204135 |
| Disease_Pe | Leukemia, . | 1/318 | 0.752771 | 0.82776 | 0 | 0 | 0.718803 | 0.204135 |
| Disease_Pe | Spinal Corc | 1/351 | 0.786422 | 0.82776 | 0 | 0 | 0.649934 | 0.156154 |
| Disease_Pe | Wolfram sy | 1/350 | 0.785472 | 0.82776 | 0 | 0 | 0.651829 | 0.157397 |
| Disease_Pe | nasophary | 1/373 | 0.806297 | 0.82776 | 0 | 0 | 0.610809 | 0.131509 |
| Disease_Pe | atopic derr | 1/319 | 0.753864 | 0.82776 | 0 | 0 | 0.716506 | 0.202444 |
| Disease_Pe | Bacterial In | 1/316 | 0.750571 | 0.82776 | 0 | 0 | 0.72344 | 0.20757 |
| Disease_Pe | MS (Multip | 1/336 | 0.771728 | 0.82776 | 0 | 0 | 0.679556 | 0.176089 |
| Disease_Pe | colorectal c | 1/336 | 0.771728 | 0.82776 | 0 | 0 | 0.679556 | 0.176089 |
| Disease_Pe | Papillary Ca | 1/292 | 0.722612 | 0.82776 | 0 | 0 | 0.784065 | 0.254729 |
| Disease_Pe | autism spe | 1/297 | 0.728682 | 0.82776 | 0 | 0 | 0.770624 | 0.243916 |
| Disease_Pe | colon canc | 1/296 | 0.727479 | 0.82776 | 0 | 0 | 0.773276 | 0.246034 |
| Disease_Pe | schizophre | 1/296 | 0.727479 | 0.82776 | 0 | 0 | 0.773276 | 0.246034 |
| Disease_Pe | anemia DO | 1/294 | 0.725056 | 0.82776 | 0 | 0 | 0.778633 | 0.250335 |
| Disease_Pe | Type 1 diak | 1/294 | 0.725056 | 0.82776 | 0 | 0 | 0.778633 | 0.250335 |
| Disease_Pe | systemic lu | 1/318 | 0.752771 | 0.82776 | 0 | 0 | 0.718803 | 0.204135 |
| Disease_Pe | myelodysp | 1/317 | 0.751674 | 0.82776 | 0 | 0 | 0.721114 | 0.205844 |
| Disease_Pe | Familial hy | 1/324 | 0.759257 | 0.82776 | 0 | 0 | 0.705234 | 0.194232 |
| Disease_Pe | asthma DO | 1/324 | 0.759257 | 0.82776 | 0 | 0 | 0.705234 | 0.194232 |
| Disease_Pe | Sepsis C02 | 1/316 | 0.750571 | 0.82776 | 0 | 0 | 0.72344 | 0.20757 |
| Disease_Pe | psoriasis D | 1/315 | 0.749464 | 0.82776 | 0 | 0 | 0.725781 | 0.209313 |
| Disease_Pe | Asthma DC | 1/314 | 0.748352 | 0.82776 | 0 | 0 | 0.728137 | 0.211074 |
| Disease_Pe | asthma DO | 1/292 | 0.722612 | 0.82776 | 0 | 0 | 0.784065 | 0.254729 |
| Disease_Pe | Hypertroph | 1/292 | 0.722612 | 0.82776 | 0 | 0 | 0.784065 | 0.254729 |
| Disease_Pe | psoriasis D | 1/297 | 0.728682 | 0.82776 | 0 | 0 | 0.770624 | 0.243916 |
| Disease_Pe | Barrett's es | 1/291 | 0.721383 | 0.82776 | 0 | 0 | 0.786808 | 0.25696 |
| Disease_Pe | anemia DO | 1/290 | 0.720147 | 0.82776 | 0 | 0 | 0.789571 | 0.259216 |
| Disease_Pe | Hypertroph | 1/311 | 0.744986 | 0.82776 | 0 | 0 | 0.735296 | 0.216463 |
| Disease_Pe | anemia DO | 1/311 | 0.744986 | 0.82776 | 0 | 0 | 0.735296 | 0.216463 |
| Disease_Pe | cardiomyo | 1/310 | 0.743855 | 0.82776 | 0 | 0 | 0.737714 | 0.218297 |
| Disease_Pe | fragile X sy | 1/288 | 0.717661 | 0.82776 | 0 | 0 | 0.795154 | 0.263799 |
| Disease_Pe | Psoriasis vu | 1/310 | 0.743855 | 0.82776 | 0 | 0 | 0.737714 | 0.218297 |
| Disease_Pe | polycystic c | 2/359 | 0.46465 | 0.82776 | 0 | 0 | 1.288911 | 0.987914 |
| Disease_Pe | Hepatic lipi | 1/284 | 0.712621 | 0.82776 | 0 | 0 | 0.806558 | 0.273266 |
| Disease_Pe | Sendai viru | 1/292 | 0.722612 | 0.82776 | 0 | 0 | 0.784065 | 0.254729 |
| Disease_Pe | Idiopathic t | 1/277 | 0.703588 | 0.82776 | 0 | 0 | 0.827309 | 0.290851 |

| | | | | | | | | |
|---|---|---|---|---|---|---|---|---|
| Disease_Pe | Type 2 diak | 1/277 | 0.703588 | 0.82776 | 0 | 0 | 0.827309 | 0.290851 |
| Disease_Pe | bipolar diso | 1/276 | 0.702274 | 0.82776 | 0 | 0 | 0.830359 | 0.293475 |
| Disease_Pe | myotonic d | 1/284 | 0.712621 | 0.82776 | 0 | 0 | 0.806558 | 0.273266 |
| Disease_Pe | Down synd | 2/247 | 0.29182 | 0.82776 | 0 | 0 | 1.888884 | 2.326383 |
| Disease_Pe | Hyperchole | 2/298 | 0.37287 | 0.82776 | 0 | 0 | 1.55938 | 1.538369 |
| Disease_Pe | ulcerative c | 2/297 | 0.37131 | 0.82776 | 0 | 0 | 1.564746 | 1.550222 |
| Disease_Pe | Septic Shoc | 1/106 | 0.370812 | 0.82776 | 0 | 0 | 2.193577 | 2.176161 |
| Disease_Pe | endometria | 2/295 | 0.368186 | 0.82776 | 0 | 0 | 1.575587 | 1.574275 |
| Disease_Pe | systemic lu | 2/285 | 0.352481 | 0.82776 | 0 | 0 | 1.632093 | 1.70188 |
| Disease_Pe | Down Sync | 2/306 | 0.385293 | 0.82776 | 0 | 0 | 1.517724 | 1.44753 |
| Disease_Pe | Asthma, all | 2/304 | 0.382197 | 0.82776 | 0 | 0 | 1.527931 | 1.469594 |
| Disease_Pe | melanoma | 2/293 | 0.365056 | 0.82776 | 0 | 0 | 1.586578 | 1.598802 |
| Disease_Pe | Huntingtor | 2/293 | 0.365056 | 0.82776 | 0 | 0 | 1.586578 | 1.598802 |
| Disease_Pe | Premature | 2/292 | 0.363489 | 0.82776 | 0 | 0 | 1.59213 | 1.611246 |
| Disease_Pe | Acne C070. | 2/292 | 0.363489 | 0.82776 | 0 | 0 | 1.59213 | 1.611246 |
| Disease_Pe | Muscular D | 2/291 | 0.36192 | 0.82776 | 0 | 0 | 1.59772 | 1.623813 |
| Disease_Pe | Dry eye syr | 2/320 | 0.40678 | 0.82776 | 0 | 0 | 1.449871 | 1.304135 |
| Disease_Pe | pancreatic | 2/248 | 0.29343 | 0.82776 | 0 | 0 | 1.88111 | 2.306459 |
| Disease_Pe | Huntingtor | 1/337 | 0.772738 | 0.82776 | 0 | 0 | 0.677499 | 0.174669 |
| Disease_Pe | Bipolar Dis | 2/288 | 0.357207 | 0.82776 | 0 | 0 | 1.614726 | 1.662266 |
| Disease_Pe | Zaire ebola | 2/287 | 0.355633 | 0.82776 | 0 | 0 | 1.620475 | 1.675339 |
| Disease_Pe | Barrett's es | 2/286 | 0.354057 | 0.82776 | 0 | 0 | 1.626263 | 1.688543 |
| Disease_Pe | schizophre | 2/286 | 0.354057 | 0.82776 | 0 | 0 | 1.626263 | 1.688543 |
| Disease_Pe | hepatocellu | 2/286 | 0.354057 | 0.82776 | 0 | 0 | 1.626263 | 1.688543 |
| Disease_Pe | Overexertic | 2/314 | 0.397613 | 0.82776 | 0 | 0 | 1.478205 | 1.363314 |
| Disease_Pe | nemaline n | 1/128 | 0.428675 | 0.82776 | 0 | 0 | 1.811573 | 1.534505 |
| Disease_Pe | asthma DO | 2/333 | 0.426412 | 0.82776 | 0 | 0 | 1.392003 | 1.186473 |
| Disease_Pe | Nemaline N | 2/324 | 0.412854 | 0.82776 | 0 | 0 | 1.431567 | 1.266451 |
| Disease_Pe | Huntingtor | 2/324 | 0.412854 | 0.82776 | 0 | 0 | 1.431567 | 1.266451 |
| Disease_Pe | endometria | 2/322 | 0.409821 | 0.82776 | 0 | 0 | 1.440662 | 1.285121 |
| Disease_Pe | PTSD - Post | 2/321 | 0.408301 | 0.82776 | 0 | 0 | 1.445252 | 1.294585 |
| Disease_Pe | allergic cor | 2/321 | 0.408301 | 0.82776 | 0 | 0 | 1.445252 | 1.294585 |
| Disease_Pe | pancreatic | 1/288 | 0.717661 | 0.82776 | 0 | 0 | 0.795154 | 0.263799 |
| Disease_Pe | pancreatic | 2/284 | 0.350903 | 0.82776 | 0 | 0 | 1.637964 | 1.71535 |
| Disease_Pe | endometria | 2/380 | 0.494441 | 0.82776 | 0 | 0 | 1.215998 | 0.85646 |
| Disease_Pe | allergic cor | 2/368 | 0.477542 | 0.82776 | 0 | 0 | 1.256638 | 0.928785 |
| Disease_Pe | Breast Can | 2/367 | 0.476119 | 0.82776 | 0 | 0 | 1.260145 | 0.935139 |
| Disease_Pe | diffuse larg | 2/366 | 0.474693 | 0.82776 | 0 | 0 | 1.263672 | 0.941545 |
| Disease_Pe | bronchopu | 2/364 | 0.471835 | 0.82776 | 0 | 0 | 1.270783 | 0.954519 |
| Disease_Pe | Neurofibro | 2/360 | 0.466091 | 0.82776 | 0 | 0 | 1.285245 | 0.981122 |
| Disease_Pe | psoriasis D | 2/319 | 0.405256 | 0.82776 | 0 | 0 | 1.454518 | 1.313773 |
| Disease_Pe | systemic lu | 2/318 | 0.403731 | 0.82776 | 0 | 0 | 1.459196 | 1.323499 |
| Disease_Pe | epidermoly | 3/356 | 0.202138 | 0.82776 | 0 | 0 | 1.978956 | 3.163964 |
| Disease_Pe | lung large c | 3/374 | 0.222461 | 0.82776 | 0 | 0 | 1.881209 | 2.827462 |
| Disease_Pe | myotonic d | 2/252 | 0.299865 | 0.82776 | 0 | 0 | 1.850635 | 2.228949 |

| | | | | | | | | |
|---|---|---|---|---|---|---|---|---|
| Disease_Pe | glaucoma a | 2/268 | 0.325502 | 0.82776 | 0 | 0 | 1.737904 | 1.950602 |
| Disease_Pe | Huntingtor | 2/294 | 0.366622 | 0.82776 | 0 | 0 | 1.581064 | 1.586479 |
| Disease_Pe | large cell n | 2/294 | 0.366622 | 0.82776 | 0 | 0 | 1.581064 | 1.586479 |
| Disease_Pe | acute myoc | 1/157 | 0.496981 | 0.82776 | 0 | 0 | 1.472645 | 1.029678 |
| Disease_Pe | Osteoarthr | 2/283 | 0.349324 | 0.82776 | 0 | 0 | 1.643877 | 1.728956 |
| Disease_Pe | endometria | 2/282 | 0.347744 | 0.82776 | 0 | 0 | 1.649832 | 1.7427 |
| Disease_Pe | Schizophre | 2/281 | 0.346162 | 0.82776 | 0 | 0 | 1.65583 | 1.756583 |
| Disease_Pe | pancreatic | 2/280 | 0.34458 | 0.82776 | 0 | 0 | 1.661871 | 1.770607 |
| Disease_Pe | Lung Injury | 1/346 | 0.781631 | 0.82776 | 0 | 0 | 0.659521 | 0.162488 |
| Disease_Pe | allergic cor | 1/345 | 0.78066 | 0.82776 | 0 | 0 | 0.661472 | 0.163791 |
| Disease_Pe | glaucoma a | 1/345 | 0.78066 | 0.82776 | 0 | 0 | 0.661472 | 0.163791 |
| Disease_Pe | Peripheral | 1/344 | 0.779685 | 0.82776 | 0 | 0 | 0.663435 | 0.165106 |
| Disease_Pe | glomerulos | 1/362 | 0.796599 | 0.82776 | 0 | 0 | 0.629775 | 0.143213 |
| Disease_Pe | Breast canc | 1/358 | 0.792955 | 0.82776 | 0 | 0 | 0.636962 | 0.147768 |
| Disease_Pe | systemic lu | 1/358 | 0.792955 | 0.82776 | 0 | 0 | 0.636962 | 0.147768 |
| Disease_Pe | MS (Multip | 1/340 | 0.775741 | 0.82776 | 0 | 0 | 0.6714 | 0.170493 |
| Disease_Pe | Infantile ne | 2/262 | 0.31591 | 0.82776 | 0 | 0 | 1.778552 | 2.04942 |
| Disease_Pe | Cardiomyo | 2/291 | 0.36192 | 0.82776 | 0 | 0 | 1.59772 | 1.623813 |
| Disease_Pe | Dehydratio | 2/375 | 0.487441 | 0.82776 | 0 | 0 | 1.232613 | 0.885738 |
| Disease_Pe | Primary op | 2/249 | 0.295039 | 0.82776 | 0 | 0 | 1.873398 | 2.286757 |
| Disease_Pe | lung cancer | 3/365 | 0.212244 | 0.82776 | 0 | 0 | 1.928867 | 2.989777 |
| Disease_Pe | breast canc | 2/197 | 0.211424 | 0.82776 | 0 | 0 | 2.379246 | 3.697085 |
| Disease_Pe | breast canc | 1/376 | 0.808862 | 0.82776 | 0 | 0 | 0.605829 | 0.128513 |
| Disease_Pe | Familial cor | 1/374 | 0.807156 | 0.82776 | 0 | 0 | 0.60914 | 0.130501 |
| Disease_Pe | Lewy body | 2/234 | 0.270864 | 0.82776 | 0 | 0 | 1.996045 | 2.607109 |
| Disease_Pe | Huntingtor | 2/231 | 0.266025 | 0.82776 | 0 | 0 | 2.022502 | 2.678127 |
| Disease_Pe | Cancer of p | 3/369 | 0.216772 | 0.82776 | 0 | 0 | 1.907397 | 2.916234 |
| Disease_Pe | asthma DO | 3/369 | 0.216772 | 0.82776 | 0 | 0 | 1.907397 | 2.916234 |
| Disease_Pe | COPD - Chr | 2/200 | 0.216205 | 0.82776 | 0 | 0 | 2.34284 | 3.588123 |
| Disease_Pe | asthma DO | 3/365 | 0.212244 | 0.82776 | 0 | 0 | 1.928867 | 2.989777 |
| Disease_Pe | Down synd | 2/219 | 0.246681 | 0.82776 | 0 | 0 | 2.135647 | 2.989182 |
| Disease_Pe | type 2 diab | 2/380 | 0.494441 | 0.82776 | 0 | 0 | 1.215998 | 0.85646 |
| Disease_Pe | melanoma | 2/379 | 0.493046 | 0.82776 | 0 | 0 | 1.219285 | 0.862221 |
| Disease_Pe | Niemann-P | 2/290 | 0.36035 | 0.82776 | 0 | 0 | 1.60335 | 1.636504 |
| Disease_Pe | nasophary | 2/375 | 0.487441 | 0.82776 | 0 | 0 | 1.232613 | 0.885738 |
| Disease_Pe | type 1 diab | 2/373 | 0.484624 | 0.82776 | 0 | 0 | 1.239385 | 0.897787 |
| Disease_Pe | Sickle Cell | 1/197 | 0.578132 | 0.82776 | 0 | 0 | 1.169732 | 0.640958 |
| Disease_Pe | amyotroph | 2/462 | 0.600376 | 0.82776 | 0 | 0 | 0.995038 | 0.507667 |
| Disease_Pe | Crohn's dis | 2/461 | 0.599188 | 0.82776 | 0 | 0 | 0.997257 | 0.510775 |
| Disease_Pe | colitis DOIE | 2/359 | 0.46465 | 0.82776 | 0 | 0 | 1.288911 | 0.987914 |
| Disease_Pe | Ulcerative | 2/397 | 0.517793 | 0.82776 | 0 | 0 | 1.162651 | 0.765232 |
| Disease_Pe | Epilepsy DO | 2/397 | 0.517793 | 0.82776 | 0 | 0 | 1.162651 | 0.765232 |
| Disease_Pe | breast canc | 2/443 | 0.577361 | 0.82776 | 0 | 0 | 1.038922 | 0.570668 |
| Disease_Pe | idiopathic | 1/192 | 0.568743 | 0.82776 | 0 | 0 | 1.200657 | 0.677563 |
| Disease_Pe | idiopathic | 1/192 | 0.568743 | 0.82776 | 0 | 0 | 1.200657 | 0.677563 |

| | | | | | | | | |
|---|---|---|---|---|---|---|---|---|
| Disease_Pe | nasophary | 2/433 | 0.564871 | 0.82776 | 0 | 0 | 1.063573 | 0.607468 |
| Disease_Pe | Neurologic | 1/189 | 0.563011 | 0.82776 | 0 | 0 | 1.220002 | 0.700838 |
| Disease_Pe | autism spe | 2/347 | 0.44718 | 0.82776 | 0 | 0 | 1.334561 | 1.074048 |
| Disease_Pe | psoriasis D | 1/373 | 0.806297 | 0.82776 | 0 | 0 | 0.610809 | 0.131509 |
| Disease_Pe | Type 1 diak | 1/321 | 0.756036 | 0.82776 | 0 | 0 | 0.711955 | 0.19911 |
| Disease_Pe | Asthma DC | 1/321 | 0.756036 | 0.82776 | 0 | 0 | 0.711955 | 0.19911 |
| Disease_Pe | familial cor | 1/321 | 0.756036 | 0.82776 | 0 | 0 | 0.711955 | 0.19911 |
| Disease_Pe | Ulcerative | 1/328 | 0.763487 | 0.82776 | 0 | 0 | 0.696465 | 0.187947 |
| Disease_Pe | Anaplasmo | 1/373 | 0.806297 | 0.82776 | 0 | 0 | 0.610809 | 0.131509 |
| Disease_Pe | muscular d | 1/325 | 0.760322 | 0.82776 | 0 | 0 | 0.703022 | 0.192638 |
| Disease_Pe | Crohn's dis | 1/370 | 0.803699 | 0.82776 | 0 | 0 | 0.615869 | 0.134586 |
| Disease_Pe | Alzheimer's | 1/369 | 0.802825 | 0.82776 | 0 | 0 | 0.617575 | 0.135631 |
| Disease_Pe | ALS (Amyo | 1/368 | 0.801947 | 0.82776 | 0 | 0 | 0.619289 | 0.136685 |
| Disease_Pe | Cancer of ( | 1/362 | 0.796599 | 0.82776 | 0 | 0 | 0.629775 | 0.143213 |
| Disease_Pe | Alcohol poi | 1/333 | 0.768672 | 0.82776 | 0 | 0 | 0.685801 | 0.180429 |
| Disease_Pe | Acute Lung | 1/330 | 0.765575 | 0.82776 | 0 | 0 | 0.692161 | 0.184896 |
| Disease_Pe | cystic fibro | 1/322 | 0.757114 | 0.82776 | 0 | 0 | 0.709701 | 0.197468 |
| Disease_Pe | Idiopathic | 1/374 | 0.807156 | 0.82776 | 0 | 0 | 0.60914 | 0.130501 |
| Disease_Pe | psoriasis D | 1/322 | 0.757114 | 0.82776 | 0 | 0 | 0.709701 | 0.197468 |
| Disease_Pe | Small cell c | 1/373 | 0.806297 | 0.82776 | 0 | 0 | 0.610809 | 0.131509 |
| Disease_Pe | systemic lu | 1/327 | 0.762437 | 0.82776 | 0 | 0 | 0.698637 | 0.189496 |
| Disease_Pe | Parkinson's | 1/326 | 0.761381 | 0.82776 | 0 | 0 | 0.700823 | 0.191059 |
| Disease_Pe | Dehydratio | 1/370 | 0.803699 | 0.82776 | 0 | 0 | 0.615869 | 0.134586 |
| Disease_Pe | Aplastic an | 1/346 | 0.781631 | 0.82776 | 0 | 0 | 0.659521 | 0.162488 |
| Disease_Pe | Oligodendr | 1/343 | 0.778706 | 0.82776 | 0 | 0 | 0.665409 | 0.166434 |
| Disease_Pe | Uterine lei | 1/342 | 0.777722 | 0.82776 | 0 | 0 | 0.667394 | 0.167774 |
| Disease_Pe | atopic derr | 1/330 | 0.765575 | 0.82776 | 0 | 0 | 0.692161 | 0.184896 |
| Disease_Pe | Nephrolith | 1/330 | 0.765575 | 0.82776 | 0 | 0 | 0.692161 | 0.184896 |
| Disease_Pe | B-cell chro | 1/372 | 0.805435 | 0.82776 | 0 | 0 | 0.612487 | 0.132526 |
| Disease_Pe | Type 1 diak | 1/371 | 0.804569 | 0.82776 | 0 | 0 | 0.614173 | 0.133551 |
| Disease_Pe | colon canc | 1/317 | 0.751674 | 0.82776 | 0 | 0 | 0.721114 | 0.205844 |
| Disease_Pe | Dystonia C | 1/317 | 0.751674 | 0.82776 | 0 | 0 | 0.721114 | 0.205844 |
| Disease_Pe | myotonic d | 1/337 | 0.772738 | 0.82776 | 0 | 0 | 0.677499 | 0.174669 |
| Disease_Pe | ovarian car | 3/364 | 0.211116 | 0.82776 | 0 | 0 | 1.934309 | 3.008525 |
| Disease_Pe | Lewy body | 2/277 | 0.339825 | 0.82776 | 0 | 0 | 1.680257 | 1.813544 |
| Disease_Pe | Hypoxia C0 | 2/275 | 0.336649 | 0.82776 | 0 | 0 | 1.692739 | 1.842909 |
| Disease_Pe | Polycystic ( | 2/275 | 0.336649 | 0.82776 | 0 | 0 | 1.692739 | 1.842909 |
| Disease_Pe | schizophre | 2/274 | 0.335059 | 0.82776 | 0 | 0 | 1.699048 | 1.85782 |
| Disease_Pe | endometri | 2/273 | 0.333469 | 0.82776 | 0 | 0 | 1.705405 | 1.872885 |
| Disease_Pe | ulcerative ( | 2/252 | 0.299865 | 0.82776 | 0 | 0 | 1.850635 | 2.228949 |
| Disease_Pe | Type 2 diak | 2/285 | 0.352481 | 0.82776 | 0 | 0 | 1.632093 | 1.70188 |
| Disease_Pe | chronic lyn | 2/285 | 0.352481 | 0.82776 | 0 | 0 | 1.632093 | 1.70188 |
| Disease_Pe | Emery-Dre | 2/265 | 0.32071 | 0.82776 | 0 | 0 | 1.757996 | 1.999226 |
| Disease_Pe | Down synd | 2/265 | 0.32071 | 0.82776 | 0 | 0 | 1.757996 | 1.999226 |
| Disease_Pe | Lung trans | 2/265 | 0.32071 | 0.82776 | 0 | 0 | 1.757996 | 1.999226 |

| | | | | | | | | |
|---|---|---|---|---|---|---|---|---|
| Disease_Pe | Dehydratio | 2/263 | 0.317511 | 0.82776 | 0 | 0 | 1.771648 | 2.03251 |
| Disease_Pe | psoriasis D | 2/263 | 0.317511 | 0.82776 | 0 | 0 | 1.771648 | 2.03251 |
| Disease_Pe | multiple sc | 2/299 | 0.374428 | 0.82776 | 0 | 0 | 1.55405 | 1.526629 |
| Disease_Pe | systemic lu | 1/294 | 0.725056 | 0.82776 | 0 | 0 | 0.778633 | 0.250335 |
| Disease_Pe | Parkinson's | 3/361 | 0.207738 | 0.82776 | 0 | 0 | 1.950818 | 3.065664 |
| Disease_Pe | Parkinson's | 2/258 | 0.3095 | 0.82776 | 0 | 0 | 1.80671 | 2.118903 |
| Disease_Pe | breast canc | 2/256 | 0.306291 | 0.82776 | 0 | 0 | 1.821121 | 2.154787 |
| Disease_Pe | Systemic lu | 2/256 | 0.306291 | 0.82776 | 0 | 0 | 1.821121 | 2.154787 |
| Disease_Pe | fetal alcoh | 2/255 | 0.304685 | 0.82776 | 0 | 0 | 1.828412 | 2.173024 |
| Disease_Pe | ulcerative c | 2/252 | 0.299865 | 0.82776 | 0 | 0 | 1.850635 | 2.228949 |
| Disease_Pe | skin squam | 2/295 | 0.368186 | 0.82776 | 0 | 0 | 1.575587 | 1.574275 |
| Disease_Pe | Cardiac Hyp | 2/313 | 0.396079 | 0.82776 | 0 | 0 | 1.483034 | 1.3735 |
| Disease_Pe | invasive lob | 2/313 | 0.396079 | 0.82776 | 0 | 0 | 1.483034 | 1.3735 |
| Disease_Pe | Hepatic Cir | 2/303 | 0.380646 | 0.82776 | 0 | 0 | 1.533086 | 1.480784 |
| Disease_Pe | endometria | 2/302 | 0.379094 | 0.82776 | 0 | 0 | 1.538275 | 1.492081 |
| Disease_Pe | fragile X sy | 2/301 | 0.37754 | 0.82776 | 0 | 0 | 1.543498 | 1.503487 |
| Disease_Pe | Granuloma | 2/301 | 0.37754 | 0.82776 | 0 | 0 | 1.543498 | 1.503487 |
| Disease_Pe | Infection b | 2/300 | 0.375985 | 0.82776 | 0 | 0 | 1.548756 | 1.515003 |
| Disease_Pe | RA (rheuma | 2/332 | 0.424913 | 0.82776 | 0 | 0 | 1.396292 | 1.195045 |
| Disease_Pe | HIV - Huma | 1/226 | 0.628735 | 0.82776 | 0 | 0 | 1.017468 | 0.472152 |
| Disease_Pe | systemic lu | 1/240 | 0.650968 | 0.82776 | 0 | 0 | 0.957186 | 0.410916 |
| Disease_Pe | esophagus | 2/329 | 0.420406 | 0.82776 | 0 | 0 | 1.409318 | 1.221224 |
| Disease_Pe | Chronic Lyr | 2/359 | 0.46465 | 0.82776 | 0 | 0 | 1.288911 | 0.987914 |
| Disease_Pe | prostate ca | 2/359 | 0.46465 | 0.82776 | 0 | 0 | 1.288911 | 0.987914 |
| Disease_Pe | Uterine leio | 2/338 | 0.433875 | 0.82776 | 0 | 0 | 1.370938 | 1.144732 |
| Disease_Pe | multiple m | 2/338 | 0.433875 | 0.82776 | 0 | 0 | 1.370938 | 1.144732 |
| Disease_Pe | Neurologic | 2/338 | 0.433875 | 0.82776 | 0 | 0 | 1.370938 | 1.144732 |
| Disease_Pe | amyotroph | 2/337 | 0.432387 | 0.82776 | 0 | 0 | 1.375101 | 1.152933 |
| Disease_Pe | schizophre | 2/337 | 0.432387 | 0.82776 | 0 | 0 | 1.375101 | 1.152933 |
| Disease_Pe | Huntingtor | 2/327 | 0.417391 | 0.82776 | 0 | 0 | 1.418136 | 1.239071 |
| Disease_Pe | Anterior Ho | 2/346 | 0.44571 | 0.82776 | 0 | 0 | 1.338509 | 1.081632 |
| Disease_Pe | Diabetes D | 2/346 | 0.44571 | 0.82776 | 0 | 0 | 1.338509 | 1.081632 |
| Disease_Pe | Diabetic Ne | 2/333 | 0.426412 | 0.82776 | 0 | 0 | 1.392003 | 1.186473 |
| Disease_Pe | polycystic c | 2/492 | 0.634806 | 0.82776 | 0 | 0 | 0.932677 | 0.423842 |
| Disease_Pe | endometria | 2/331 | 0.423413 | 0.82776 | 0 | 0 | 1.400608 | 1.203693 |
| Disease_Pe | Uterine leio | 2/330 | 0.42191 | 0.82776 | 0 | 0 | 1.40495 | 1.212419 |
| Disease_Pe | Primary pu | 2/330 | 0.42191 | 0.82776 | 0 | 0 | 1.40495 | 1.212419 |
| Disease_Pe | Teratosper | 2/372 | 0.483213 | 0.82776 | 0 | 0 | 1.242798 | 0.903885 |
| Disease_Pe | SCID - Seve | 2/299 | 0.374428 | 0.82776 | 0 | 0 | 1.55405 | 1.526629 |
| Disease_Pe | Carcinoma, | 2/298 | 0.37287 | 0.82776 | 0 | 0 | 1.55938 | 1.538369 |
| Disease_Pe | Sendai viru | 2/328 | 0.418899 | 0.82776 | 0 | 0 | 1.413713 | 1.230107 |
| Disease_Pe | Hypoxia C0 | 2/328 | 0.418899 | 0.82776 | 0 | 0 | 1.413713 | 1.230107 |
| Disease_Pe | endometria | 2/328 | 0.418899 | 0.82776 | 0 | 0 | 1.413713 | 1.230107 |
| Disease_Pe | asthma DO | 2/328 | 0.418899 | 0.82776 | 0 | 0 | 1.413713 | 1.230107 |
| Disease_Pe | Androgen i | 2/327 | 0.417391 | 0.82776 | 0 | 0 | 1.418136 | 1.239071 |

| | | | | | | | | |
|---|---|---|---|---|---|---|---|---|
| Disease_Pe | Sclerodern | 1/189 | 0.563011 | 0.82776 | 0 | 0 | 1.220002 | 0.700838 |
| Disease_Pe | Essential Tl | 1/174 | 0.533201 | 0.82776 | 0 | 0 | 1.326791 | 0.834362 |
| Disease_Pe | Pulmonary | 1/173 | 0.531144 | 0.82776 | 0 | 0 | 1.334573 | 0.844414 |
| Disease_Pe | Huntingtor | 1/246 | 0.660088 | 0.82776 | 0 | 0 | 0.93346 | 0.387742 |
| Disease_Pe | lupus eryth | 1/267 | 0.690194 | 0.82776 | 0 | 0 | 0.858848 | 0.318446 |
| Disease_Pe | Urothelial ( | 1/220 | 0.618783 | 0.82776 | 0 | 0 | 1.045662 | 0.501918 |
| Disease_Pe | Breast Can | 2/449 | 0.58473 | 0.82776 | 0 | 0 | 1.024661 | 0.549838 |
| Disease_Pe | nephroscle | 1/218 | 0.615408 | 0.82776 | 0 | 0 | 1.055407 | 0.512369 |
| Disease_Pe | cellular pha | 1/218 | 0.615408 | 0.82776 | 0 | 0 | 1.055407 | 0.512369 |
| Disease_Pe | polycystic ( | 1/218 | 0.615408 | 0.82776 | 0 | 0 | 1.055407 | 0.512369 |
| Disease_Pe | idiopathic | | 1/307 | 0.740429 | 0.82776 | 0 | 0 | 0.74506 | 0.223909 |
| Disease_Pe | Retinitis Pi | 1/262 | 0.683274 | 0.82776 | 0 | 0 | 0.875523 | 0.333451 |
| Disease_Pe | non-systen | 1/262 | 0.683274 | 0.82776 | 0 | 0 | 0.875523 | 0.333451 |
| Disease_Pe | nemaline n | 1/210 | 0.601608 | 0.82776 | 0 | 0 | 1.09625 | 0.557059 |
| Disease_Pe | AML - Acut | 1/261 | 0.681872 | 0.82776 | 0 | 0 | 0.878936 | 0.336556 |
| Disease_Pe | Pauciarticu | 1/270 | 0.694273 | 0.82776 | 0 | 0 | 0.84914 | 0.309842 |
| Disease_Pe | Endometri | 1/270 | 0.694273 | 0.82776 | 0 | 0 | 0.84914 | 0.309842 |
| Disease_Pe | hepatocell | 1/269 | 0.692919 | 0.82776 | 0 | 0 | 0.852352 | 0.312678 |
| Disease_Pe | neurofibro | 1/142 | 0.462724 | 0.82776 | 0 | 0 | 1.630546 | 1.256538 |
| Disease_Pe | Type 2 diat | 1/245 | 0.658585 | 0.82776 | 0 | 0 | 0.937333 | 0.391489 |
| Disease_Pe | non-systen | 1/241 | 0.652504 | 0.82776 | 0 | 0 | 0.953149 | 0.406935 |
| Disease_Pe | nemaline n | 2/450 | 0.585949 | 0.82776 | 0 | 0 | 1.022321 | 0.546454 |
| Disease_Pe | ulcerative ( | 1/255 | 0.673329 | 0.82776 | 0 | 0 | 0.899973 | 0.355958 |
| Disease_Pe | Protozoan | 1/266 | 0.688822 | 0.82776 | 0 | 0 | 0.862133 | 0.32138 |
| Disease_Pe | cervical car | 1/265 | 0.687444 | 0.82776 | 0 | 0 | 0.865442 | 0.324346 |
| Disease_Pe | autism spe | 1/264 | 0.68606 | 0.82776 | 0 | 0 | 0.868777 | 0.327347 |
| Disease_Pe | myotonic d | 1/239 | 0.649424 | 0.82776 | 0 | 0 | 0.961257 | 0.414945 |
| Disease_Pe | Rett Syndr | 1/219 | 0.617099 | 0.82776 | 0 | 0 | 1.050512 | 0.507109 |
| Disease_Pe | head and n | 1/237 | 0.646317 | 0.82776 | 0 | 0 | 0.969501 | 0.423154 |
| Disease_Pe | Cardiac Hy | 1/235 | 0.643183 | 0.82776 | 0 | 0 | 0.977887 | 0.431568 |
| Disease_Pe | Nemaline N | 1/234 | 0.641605 | 0.82776 | 0 | 0 | 0.982134 | 0.435854 |
| Disease_Pe | Neurofibro | 1/231 | 0.636831 | 0.82776 | 0 | 0 | 0.995096 | 0.449038 |
| Disease_Pe | nicotine de | 1/230 | 0.635226 | 0.82776 | 0 | 0 | 0.999492 | 0.453544 |
| Disease_Pe | Hypertensi | 3/418 | 0.273635 | 0.82776 | 0 | 0 | 1.677969 | 2.174583 |
| Disease_Pe | skin squam | 3/373 | 0.221321 | 0.82776 | 0 | 0 | 1.88639 | 2.844943 |
| Disease_Pe | Nutritional | 3/370 | 0.217908 | 0.82776 | 0 | 0 | 1.902102 | 2.898203 |
| Disease_Pe | Duchenne | 2/267 | 0.323905 | 0.82776 | 0 | 0 | 1.74455 | 1.966639 |
| Disease_Pe | Breast Can | 2/250 | 0.296648 | 0.82776 | 0 | 0 | 1.86575 | 2.267274 |
| Disease_Pe | intracrania | 3/437 | 0.296154 | 0.82776 | 0 | 0 | 1.602946 | 1.950585 |
| Disease_Pe | acute myel | 3/437 | 0.296154 | 0.82776 | 0 | 0 | 1.602946 | 1.950585 |
| Disease_Pe | salivary gla | 2/249 | 0.295039 | 0.82776 | 0 | 0 | 1.873398 | 2.286757 |
| Disease_Pe | Erectile dys | 2/249 | 0.295039 | 0.82776 | 0 | 0 | 1.873398 | 2.286757 |
| Disease_Pe | low-grade : | 2/259 | 0.311104 | 0.82776 | 0 | 0 | 1.799588 | 2.10125 |
| Disease_Pe | osteoarthri | 2/245 | 0.288599 | 0.82776 | 0 | 0 | 1.904624 | 2.366908 |
| Disease_Pe | Schizophre | 2/242 | 0.283765 | 0.82776 | 0 | 0 | 1.928725 | 2.429439 |

| | | | | | | | | |
|---|---|---|---|---|---|---|---|---|
| Disease_Pe | nemaline n | 2/239 | 0.278929 | 0.82776 | 0 | 0 | 1.953438 | 2.494149 |
| Disease_Pe | Carcinoma, | 2/237 | 0.275703 | 0.82776 | 0 | 0 | 1.970263 | 2.538546 |
| Disease_Pe | Psoriasis vu | 2/236 | 0.27409 | 0.82776 | 0 | 0 | 1.978783 | 2.561135 |
| Disease_Pe | stomach ca | 3/353 | 0.198796 | 0.82776 | 0 | 0 | 1.996224 | 3.224856 |
| Disease_Pe | schizophre | 2/268 | 0.325502 | 0.82776 | 0 | 0 | 1.737904 | 1.950602 |
| Disease_Pe | diffuse larg | 3/440 | 0.299723 | 0.82776 | 0 | 0 | 1.591697 | 1.917827 |
| Disease_Pe | Down synd | 2/229 | 0.262799 | 0.82776 | 0 | 0 | 2.040529 | 2.726893 |
| Disease_Pe | colon canc | 2/228 | 0.261186 | 0.82776 | 0 | 0 | 2.049662 | 2.751716 |
| Disease_Pe | Primary op | 2/227 | 0.259573 | 0.82776 | 0 | 0 | 2.058876 | 2.776839 |
| Disease_Pe | Hepatitis, A | 2/227 | 0.259573 | 0.82776 | 0 | 0 | 2.058876 | 2.776839 |
| Disease_Pe | Viral cardic | 2/227 | 0.259573 | 0.82776 | 0 | 0 | 2.058876 | 2.776839 |
| Disease_Pe | polycystic c | 3/397 | 0.248995 | 0.82776 | 0 | 0 | 1.769307 | 2.459905 |
| Disease_Pe | Cancer of t | 3/452 | 0.314024 | 0.82776 | 0 | 0 | 1.548202 | 1.793262 |
| Disease_Pe | autism spe | 2/279 | 0.342996 | 0.82776 | 0 | 0 | 1.667955 | 1.784774 |
| Disease_Pe | facioscapul | 2/214 | 0.238636 | 0.82776 | 0 | 0 | 2.18657 | 3.132957 |
| Disease_Pe | Huntingtor | 2/213 | 0.237028 | 0.82776 | 0 | 0 | 2.197045 | 3.162813 |
| Disease_Pe | Parkinson's | 2/207 | 0.227399 | 0.82776 | 0 | 0 | 2.262037 | 3.350186 |
| Disease_Pe | Malignant | 2/205 | 0.224196 | 0.82776 | 0 | 0 | 2.284555 | 3.415945 |
| Disease_Pe | dermatomy | 1/171 | 0.527002 | 0.82776 | 0 | 0 | 1.35041 | 0.865006 |
| Disease_Pe | allergic cor | 2/357 | 0.46176 | 0.82776 | 0 | 0 | 1.296305 | 1.001668 |
| Disease_Pe | Alpha-1 an | 2/356 | 0.460312 | 0.82776 | 0 | 0 | 1.300033 | 1.008633 |
| Disease_Pe | severe asth | 2/353 | 0.455954 | 0.82776 | 0 | 0 | 1.311346 | 1.029883 |
| Disease_Pe | idiopathic | 1/199 | 0.581831 | 0.82776 | 0 | 0 | 1.157799 | 0.627036 |
| Disease_Pe | Diamond-B | 2/218 | 0.245071 | 0.82776 | 0 | 0 | 2.145643 | 3.017223 |
| Disease_Pe | Alzheimer's | 1/181 | 0.547354 | 0.82776 | 0 | 0 | 1.274742 | 0.768234 |
| Disease_Pe | Nephroblas | 2/419 | 0.546954 | 0.82776 | 0 | 0 | 1.100071 | 0.663773 |
| Disease_Pe | schizophre | 2/415 | 0.541742 | 0.82776 | 0 | 0 | 1.110953 | 0.680975 |
| Disease_Pe | Diabetic Ne | 2/414 | 0.540433 | 0.82776 | 0 | 0 | 1.113706 | 0.685357 |
| Disease_Pe | testis semi | 2/414 | 0.540433 | 0.82776 | 0 | 0 | 1.113706 | 0.685357 |
| Disease_Pe | amyotroph | 1/177 | 0.53932 | 0.82776 | 0 | 0 | 1.303977 | 0.805137 |
| Disease_Pe | Psoriasis vu | 1/182 | 0.549341 | 0.82776 | 0 | 0 | 1.267635 | 0.759358 |
| Disease_Pe | Breast Can | 2/382 | 0.497225 | 0.82776 | 0 | 0 | 1.209474 | 0.845075 |
| Disease_Pe | H1N1 DOID | 1/173 | 0.531144 | 0.82776 | 0 | 0 | 1.334573 | 0.844414 |
| Disease_Pe | asthma DO | 1/263 | 0.68467 | 0.82776 | 0 | 0 | 0.872137 | 0.330382 |
| Disease_Pe | prostate ca | 1/170 | 0.524918 | 0.82776 | 0 | 0 | 1.35847 | 0.875552 |
| Disease_Pe | Dental cavi | 2/401 | 0.523185 | 0.82776 | 0 | 0 | 1.150759 | 0.745484 |
| Disease_Pe | Arthropath | 2/401 | 0.523185 | 0.82776 | 0 | 0 | 1.150759 | 0.745484 |
| Disease_Pe | Parkinson's | 1/167 | 0.51861 | 0.82776 | 0 | 0 | 1.383231 | 0.908233 |
| Disease_Pe | precursor E | 1/184 | 0.55329 | 0.82776 | 0 | 0 | 1.253654 | 0.742005 |
| Disease_Pe | Bipolar Dis | 2/341 | 0.438329 | 0.82776 | 0 | 0 | 1.358598 | 1.120553 |
| Disease_Pe | adrenoleuk | 2/338 | 0.433875 | 0.82776 | 0 | 0 | 1.370938 | 1.144732 |
| Disease_Pe | acute myoc | 1/166 | 0.516489 | 0.82776 | 0 | 0 | 1.391684 | 0.919487 |
| Disease_Pe | esophagus | 2/391 | 0.509632 | 0.82776 | 0 | 0 | 1.180947 | 0.796037 |
| Disease_Pe | bipolar disc | 2/388 | 0.505518 | 0.82776 | 0 | 0 | 1.190308 | 0.811994 |
| Disease_Pe | CML - Chro | 1/159 | 0.501382 | 0.82776 | 0 | 0 | 1.453856 | 1.003722 |

| | | | | | | | | |
|---|---|---|---|---|---|---|---|---|
| Disease_Pe | endometri( | 2/384 | 0.499999 | 0.82776 | 0 | 0 | 1.203018 | 0.833871 |
| Disease_Pe | HIV - Huma | 2/204 | 0.222596 | 0.82776 | 0 | 0 | 2.295981 | 3.449476 |
| Disease_Pe | endometri | 1/273 | 0.6983 | 0.82776 | 0 | 0 | 0.839646 | 0.301522 |
| Disease_Pe | Type 2 diak | 2/308 | 0.388383 | 0.82776 | 0 | 0 | 1.507651 | 1.42588 |
| Disease_Pe | Rotavirus i | 2/307 | 0.386839 | 0.82776 | 0 | 0 | 1.512671 | 1.436654 |
| Disease_Pe | asthma DO | 2/307 | 0.386839 | 0.82776 | 0 | 0 | 1.512671 | 1.436654 |
| Disease_Pe | autism spe | 2/307 | 0.386839 | 0.82776 | 0 | 0 | 1.512671 | 1.436654 |
| Disease_Pe | tibial musc | 2/335 | 0.429403 | 0.82776 | 0 | 0 | 1.383501 | 1.169555 |
| Disease_Pe | osteoarthri | 2/371 | 0.481798 | 0.82776 | 0 | 0 | 1.24623 | 0.910034 |
| Disease_Pe | ulcerative ( | 2/370 | 0.480382 | 0.82776 | 0 | 0 | 1.24968 | 0.916233 |
| Disease_Pe | testis semi | 2/345 | 0.444238 | 0.82776 | 0 | 0 | 1.34248 | 1.089282 |
| Disease_Pe | amyotroph | 2/344 | 0.442764 | 0.82776 | 0 | 0 | 1.346474 | 1.096999 |
| Disease_Pe | testis semi | 2/343 | 0.441287 | 0.82776 | 0 | 0 | 1.350492 | 1.104782 |
| Disease_Pe | Treacher C | 2/342 | 0.439809 | 0.82776 | 0 | 0 | 1.354533 | 1.112633 |
| Disease_Pe | peripartum | 2/342 | 0.439809 | 0.82776 | 0 | 0 | 1.354533 | 1.112633 |
| Disease_Pe | Carcinoma | 1/273 | 0.6983 | 0.82776 | 0 | 0 | 0.839646 | 0.301522 |
| Disease_Pe | Depression | 1/273 | 0.6983 | 0.82776 | 0 | 0 | 0.839646 | 0.301522 |
| Disease_Pe | Gamma-hy | 1/273 | 0.6983 | 0.82776 | 0 | 0 | 0.839646 | 0.301522 |
| Disease_Pe | psoriasis D | 2/309 | 0.389926 | 0.82776 | 0 | 0 | 1.502663 | 1.415207 |
| Disease_Pe | Huntingtor | 1/272 | 0.696964 | 0.82776 | 0 | 0 | 0.842787 | 0.304265 |
| Disease_Pe | polycystic ( | 1/280 | 0.707493 | 0.82776 | 0 | 0 | 0.818288 | 0.28315 |
| Disease_Pe | systemic lu | 1/210 | 0.601608 | 0.82776 | 0 | 0 | 1.09625 | 0.557059 |
| Disease_Pe | Escherichia | 1/225 | 0.627094 | 0.82776 | 0 | 0 | 1.022062 | 0.476954 |
| Disease_Pe | Leukemia [ | 1/303 | 0.735792 | 0.82776 | 0 | 0 | 0.755082 | 0.231665 |
| Disease_Pe | dilated car( | 1/303 | 0.735792 | 0.82776 | 0 | 0 | 0.755082 | 0.231665 |
| Disease_Pe | Carcinoma, | 1/303 | 0.735792 | 0.82776 | 0 | 0 | 0.755082 | 0.231665 |
| Disease_Pe | ductal carc | 1/303 | 0.735792 | 0.82776 | 0 | 0 | 0.755082 | 0.231665 |
| Disease_Pe | asthma DO | 1/303 | 0.735792 | 0.82776 | 0 | 0 | 0.755082 | 0.231665 |
| Disease_Pe | Type 2 diak | 1/268 | 0.691559 | 0.82776 | 0 | 0 | 0.855587 | 0.315546 |
| Disease_Pe | CML - Chro | 1/266 | 0.688822 | 0.82776 | 0 | 0 | 0.862133 | 0.32138 |
| Disease_Pe | Cystic Fibr( | 1/275 | 0.700955 | 0.82776 | 0 | 0 | 0.833432 | 0.296128 |
| Disease_Pe | Inflammat( | 1/301 | 0.733443 | 0.82776 | 0 | 0 | 0.760194 | 0.235665 |
| Disease_Pe | Atheroscle | 1/301 | 0.733443 | 0.82776 | 0 | 0 | 0.760194 | 0.235665 |
| Disease_Pe | mytonic dy | 2/317 | 0.402204 | 0.82776 | 0 | 0 | 1.463903 | 1.333316 |
| Disease_Pe | Huntingtor | 1/312 | 0.746113 | 0.82776 | 0 | 0 | 0.732895 | 0.214649 |
| Disease_Pe | Huntingtor | 1/312 | 0.746113 | 0.82776 | 0 | 0 | 0.732895 | 0.214649 |
| Disease_Pe | Eczema DC | 1/312 | 0.746113 | 0.82776 | 0 | 0 | 0.732895 | 0.214649 |
| Disease_Pe | Hypoxia C0 | 1/310 | 0.743855 | 0.82776 | 0 | 0 | 0.737714 | 0.218297 |
| Disease_Pe | idiopathic ( | 1/271 | 0.695622 | 0.82776 | 0 | 0 | 0.845952 | 0.307038 |
| Disease_Pe | endometri( | 1/279 | 0.706197 | 0.82776 | 0 | 0 | 0.821273 | 0.285689 |
| Disease_Pe | Retinal dar | 1/279 | 0.706197 | 0.82776 | 0 | 0 | 0.821273 | 0.285689 |
| Disease_Pe | asthma DO | 1/278 | 0.704895 | 0.82776 | 0 | 0 | 0.82428 | 0.288256 |
| Disease_Pe | ulcerative ( | 1/298 | 0.72988 | 0.82776 | 0 | 0 | 0.76799 | 0.241821 |
| Disease_Pe | Simian Acq | 1/247 | 0.661585 | 0.82776 | 0 | 0 | 0.929618 | 0.384041 |
| Disease_Pe | silicosis DO | 1/260 | 0.680464 | 0.82776 | 0 | 0 | 0.882374 | 0.339697 |

| | | | | | | | | |
|---|---|---|---|---|---|---|---|---|
| Disease_Pe | Schizophre | 2/309 | 0.389926 | 0.82776 | 0 | 0 | 1.502663 | 1.415207 |
| Disease_Pe | Turner Syn | 2/329 | 0.420406 | 0.82776 | 0 | 0 | 1.409318 | 1.221224 |
| Disease_Pe | multiple sc | 2/329 | 0.420406 | 0.82776 | 0 | 0 | 1.409318 | 1.221224 |
| Disease_Pe | Generalize | 2/318 | 0.403731 | 0.82776 | 0 | 0 | 1.459196 | 1.323499 |
| Disease_Pe | myelodysp | 1/276 | 0.702274 | 0.82776 | 0 | 0 | 0.830359 | 0.293475 |
| Disease_Pe | Duchenne | 2/315 | 0.399145 | 0.82776 | 0 | 0 | 1.473407 | 1.353222 |
| Disease_Pe | schizophre | 2/315 | 0.399145 | 0.82776 | 0 | 0 | 1.473407 | 1.353222 |
| Disease_Pe | schizophre | 2/315 | 0.399145 | 0.82776 | 0 | 0 | 1.473407 | 1.353222 |
| Disease_Pe | bipolar dis | 2/306 | 0.385293 | 0.82776 | 0 | 0 | 1.517724 | 1.44753 |
| Disease_Pe | autism spe | 2/325 | 0.414369 | 0.82776 | 0 | 0 | 1.427062 | 1.257241 |
| Disease_Pe | prostate ca | 2/325 | 0.414369 | 0.82776 | 0 | 0 | 1.427062 | 1.257241 |
| Disease_Pe | Schistosom | 1/115 | 0.395152 | 0.82776 | 0 | 0 | 2.019482 | 1.875059 |
| Disease_Pe | bipolar dis | 2/312 | 0.394543 | 0.82776 | 0 | 0 | 1.487894 | 1.383781 |
| Disease_Pe | endometri | 2/311 | 0.393006 | 0.82776 | 0 | 0 | 1.492785 | 1.394158 |
| Disease_Pe | Oligodendr | 2/311 | 0.393006 | 0.82776 | 0 | 0 | 1.492785 | 1.394158 |
| Disease_Pe | Glaucoma | 2/310 | 0.391467 | 0.82776 | 0 | 0 | 1.497708 | 1.404633 |
| Disease_Pe | colorectal | 2/342 | 0.439809 | 0.82776 | 0 | 0 | 1.354533 | 1.112633 |
| Disease_Pe | Generalize | 2/279 | 0.342996 | 0.82776 | 0 | 0 | 1.667955 | 1.784774 |
| Disease_Pe | diabetic re | 2/278 | 0.341411 | 0.82776 | 0 | 0 | 1.674084 | 1.799086 |
| Disease_Pe | Down Synd | 1/224 | 0.625446 | 0.82776 | 0 | 0 | 1.026697 | 0.481818 |
| Disease_Pe | cystic fibro | 1/256 | 0.674769 | 0.82776 | 0 | 0 | 0.896398 | 0.35263 |
| Disease_Pe | Pseudomo | 1/239 | 0.649424 | 0.82776 | 0 | 0 | 0.961257 | 0.414945 |
| Disease_Pe | cystic fibro | 1/255 | 0.673329 | 0.82776 | 0 | 0 | 0.899973 | 0.355958 |
| Disease_Pe | Cushing Sy | 1/284 | 0.712621 | 0.82776 | 0 | 0 | 0.806558 | 0.273266 |
| Disease_Pe | Breast Can | 1/299 | 0.731073 | 0.82776 | 0 | 0 | 0.765374 | 0.239748 |
| Disease_Pe | Kidney disc | 1/282 | 0.710068 | 0.82776 | 0 | 0 | 0.812381 | 0.278154 |
| Disease_Pe | autistic dis | 1/280 | 0.707493 | 0.82776 | 0 | 0 | 0.818288 | 0.28315 |
| Disease_Pe | JRA - Juven | 1/280 | 0.707493 | 0.82776 | 0 | 0 | 0.818288 | 0.28315 |
| Disease_Pe | autism spe | 1/247 | 0.661585 | 0.82776 | 0 | 0 | 0.929618 | 0.384041 |
| Disease_Pe | myotonic d | 1/279 | 0.706197 | 0.82776 | 0 | 0 | 0.821273 | 0.285689 |
| Disease_Pe | cystic fibro | 1/288 | 0.717661 | 0.82776 | 0 | 0 | 0.795154 | 0.263799 |
| Disease_Pe | Preeclamps | 1/225 | 0.627094 | 0.82776 | 0 | 0 | 1.022062 | 0.476954 |
| Disease_Pe | LGLL - Larg | 1/225 | 0.627094 | 0.82776 | 0 | 0 | 1.022062 | 0.476954 |
| Disease_Pe | autism spe | 1/283 | 0.711348 | 0.82776 | 0 | 0 | 0.809459 | 0.275697 |
| Disease_Pe | pancreatic | 1/224 | 0.625446 | 0.82776 | 0 | 0 | 1.026697 | 0.481818 |
| Disease_Pe | chronic my | 1/220 | 0.618783 | 0.82776 | 0 | 0 | 1.045662 | 0.501918 |
| Disease_Pe | ulcerative | 1/239 | 0.649424 | 0.82776 | 0 | 0 | 0.961257 | 0.414945 |
| Disease_Pe | anaplastic | 1/276 | 0.702274 | 0.82776 | 0 | 0 | 0.830359 | 0.293475 |
| Disease_Pe | Infection b | 1/284 | 0.712621 | 0.82776 | 0 | 0 | 0.806558 | 0.273266 |
| Disease_Pe | autism spe | 2/472 | 0.612113 | 0.82776 | 0 | 0 | 0.973367 | 0.477766 |
| Disease_Pe | ulcerative | 1/214 | 0.608568 | 0.82776 | 0 | 0 | 1.075445 | 0.534116 |
| Disease_Pe | Type 1 diab | 1/213 | 0.606839 | 0.82776 | 0 | 0 | 1.080573 | 0.539737 |
| Disease_Pe | Setleis syn | 1/207 | 0.596308 | 0.82776 | 0 | 0 | 1.112384 | 0.575101 |
| Disease_Pe | osteoarthri | 1/206 | 0.594525 | 0.82776 | 0 | 0 | 1.117867 | 0.581282 |
| Disease_Pe | acute myel | 2/455 | 0.592006 | 0.82776 | 0 | 0 | 1.010778 | 0.529889 |

| | | | | | | | | |
|---|---|---|---|---|---|---|---|---|
| Disease_Pe | Malignant | 1/203 | 0.589132 | 0.82776 | 0 | 0 | 1.134642 | 0.600344 |
| Disease_Pe | atopic derr | 1/269 | 0.692919 | 0.82776 | 0 | 0 | 0.852352 | 0.312678 |
| Disease_Pe | autism spe | 1/285 | 0.713889 | 0.82776 | 0 | 0 | 0.803677 | 0.270861 |
| Disease_Pe | hepatitis C | 1/299 | 0.731073 | 0.82776 | 0 | 0 | 0.765374 | 0.239748 |
| Disease_Pe | renal cell c | 1/298 | 0.72988 | 0.82776 | 0 | 0 | 0.76799 | 0.241821 |
| Disease_Pe | allergic ast | 1/271 | 0.695622 | 0.82776 | 0 | 0 | 0.845952 | 0.307038 |
| Disease_Pe | Appendicit | 1/298 | 0.72988 | 0.82776 | 0 | 0 | 0.76799 | 0.241821 |
| Disease_Pe | invasive du | 1/304 | 0.736959 | 0.82776 | 0 | 0 | 0.752552 | 0.229696 |
| Disease_Pe | autism spe | 1/259 | 0.679049 | 0.82776 | 0 | 0 | 0.885839 | 0.342875 |
| Disease_Pe | Alzheimer's | 1/259 | 0.679049 | 0.82776 | 0 | 0 | 0.885839 | 0.342875 |
| Disease_Pe | Malignant | 1/258 | 0.677628 | 0.82776 | 0 | 0 | 0.889331 | 0.346089 |
| Disease_Pe | Rhinovirus | 1/256 | 0.674769 | 0.82776 | 0 | 0 | 0.896398 | 0.35263 |
| Disease_Pe | methylmal | 1/256 | 0.674769 | 0.82776 | 0 | 0 | 0.896398 | 0.35263 |
| Disease_Pe | idiopathic | 1/267 | 0.690194 | 0.82776 | 0 | 0 | 0.858848 | 0.318446 |
| Disease_Pe | Huntingtor | 1/293 | 0.723837 | 0.82776 | 0 | 0 | 0.78134 | 0.25252 |
| Disease_Pe | Essential H | 1/301 | 0.733443 | 0.82776 | 0 | 0 | 0.760194 | 0.235665 |
| Disease_Pe | psoriasis D | 1/255 | 0.673329 | 0.82776 | 0 | 0 | 0.899973 | 0.355958 |
| Disease_Pe | Type 2 diak | 1/254 | 0.671883 | 0.82776 | 0 | 0 | 0.903576 | 0.359325 |
| Disease_Pe | Asthma, all | 1/262 | 0.683274 | 0.82776 | 0 | 0 | 0.875523 | 0.333451 |
| Disease_Pe | Wilson dise | 1/284 | 0.712621 | 0.82776 | 0 | 0 | 0.806558 | 0.273266 |
| Disease_Pe | Huntingtor | 1/285 | 0.713889 | 0.82776 | 0 | 0 | 0.803677 | 0.270861 |
| Disease_Pe | endometri | 1/285 | 0.713889 | 0.82776 | 0 | 0 | 0.803677 | 0.270861 |
| Disease_Pe | facioscapul | 1/286 | 0.715152 | 0.82776 | 0 | 0 | 0.800816 | 0.268482 |
| Disease_Pe | Carcinoma, | 1/247 | 0.661585 | 0.82776 | 0 | 0 | 0.929618 | 0.384041 |
| Disease_Pe | nemaline n | 1/226 | 0.628735 | 0.82776 | 0 | 0 | 1.017468 | 0.472152 |
| Disease_Pe | lung squan | 1/253 | 0.670431 | 0.82776 | 0 | 0 | 0.907207 | 0.362732 |
| Disease_Pe | Malignant | 1/250 | 0.666037 | 0.82776 | 0 | 0 | 0.918278 | 0.373197 |
| Disease_Pe | psoriatic ar | 1/251 | 0.667508 | 0.82776 | 0 | 0 | 0.914558 | 0.369667 |
| Gene_Pert | Esrrg Defic | 4/344 | 0.063146 | 0.828117 | 0 | 0 | 2.774344 | 7.663604 |
| Gene_Pert | Srf NULL M | 4/343 | 0.062611 | 0.828117 | 0 | 0 | 2.782671 | 7.710271 |
| Gene_Pert | UBB KO mc | 4/345 | 0.063683 | 0.828117 | 0 | 0 | 2.766067 | 7.617307 |
| Gene_Pert | Srf KD mou | 3/210 | 0.063737 | 0.828117 | 0 | 0 | 3.399931 | 9.359987 |
| GeneSigDB | 16651414- | 6/1733 | 0.775717 | 0.829317 | 0 | 0 | 0.78003 | 0.198102 |
| GTEx_Tissu | GTEX-W5W | 17/2962 | 0.137988 | 0.829833 | 0 | 0 | 1.399253 | 2.771347 |
| GTEx_Tissu | GTEX-PVO\ | 8/1174 | 0.137995 | 0.829833 | 0 | 0 | 1.628156 | 3.22463 |
| GTEx_Tissu | GTEX-TML8 | 11/1754 | 0.138779 | 0.830064 | 0 | 0 | 1.508817 | 2.979719 |
| GTEx_Tissu | GTEX-OHPI | 47/9529 | 0.138731 | 0.830064 | 0 | 0 | 1.292599 | 2.553162 |
| DisGeNET | Mental anc | 3/1021 | 0.82769 | 0.83118 | 0 | 0 | 0.662889 | 0.125363 |
| GTEx_Tissu | GTEX-NPJ8 | 4/384 | 0.086491 | 0.831334 | 0 | 0 | 2.477235 | 6.063563 |
| GTEx_Tissu | GTEX-WFG | 10/1414 | 0.086495 | 0.831334 | 0 | 0 | 1.712084 | 4.190605 |
| Reactome_ | Membrane | 2/599 | 0.739055 | 0.831489 | 0 | 0 | 0.761297 | 0.230203 |
| Reactome_ | DNA Repai | 1/310 | 0.743855 | 0.831489 | 0 | 0 | 0.737714 | 0.218297 |
| Reactome_ | Intracellula | 1/306 | 0.739278 | 0.831489 | 0 | 0 | 0.747541 | 0.225819 |
| Reactome_ | G Alpha (I) | 1/312 | 0.746113 | 0.831489 | 0 | 0 | 0.732895 | 0.214649 |
| Disease_Pe | ulcerative ( | 1/384 | 0.815538 | 0.832393 | 0 | 0 | 0.592932 | 0.120903 |

| | | | | | | | | |
|---|---|---|---|---|---|---|---|---|
| Disease_Pe | nasophary | 1/386 | 0.81717 | 0.832735 | 0 | 0 | 0.589792 | 0.119083 |
| Jensen_TIS | Amygdala | 20/5279 | 0.799188 | 0.832811 | 0 | 0 | 0.831779 | 0.18645 |
| Gene_Pert | Muc2 &quo | 1/249 | 0.66456 | 0.832891 | 0 | 0 | 0.922027 | 0.376768 |
| Gene_Pert | Ache OE m | 1/239 | 0.649424 | 0.832891 | 0 | 0 | 0.961257 | 0.414945 |
| Gene_Pert | MEG3 KO r | 1/238 | 0.647874 | 0.832891 | 0 | 0 | 0.965362 | 0.419024 |
| Gene_Pert | S1pr2 KO n | 1/255 | 0.673329 | 0.832891 | 0 | 0 | 0.899973 | 0.355958 |
| Gene_Pert | Hoxa10 KO | 1/251 | 0.667508 | 0.832891 | 0 | 0 | 0.914558 | 0.369667 |
| Gene_Pert | IPMK KD hu | 1/251 | 0.667508 | 0.832891 | 0 | 0 | 0.914558 | 0.369667 |
| Gene_Pert | Jak1 drugir | 1/254 | 0.671883 | 0.832891 | 0 | 0 | 0.903576 | 0.359325 |
| Gene_Pert | SERPINI1 M | 1/254 | 0.671883 | 0.832891 | 0 | 0 | 0.903576 | 0.359325 |
| Gene_Pert | SQSTM1 OI | 1/254 | 0.671883 | 0.832891 | 0 | 0 | 0.903576 | 0.359325 |
| Gene_Pert | Trp53inp2 | 1/253 | 0.670431 | 0.832891 | 0 | 0 | 0.907207 | 0.362732 |
| Gene_Pert | Myc knock- | 1/253 | 0.670431 | 0.832891 | 0 | 0 | 0.907207 | 0.362732 |
| Gene_Pert | Itk KO mou | 1/253 | 0.670431 | 0.832891 | 0 | 0 | 0.907207 | 0.362732 |
| Gene_Pert | Lep KO mo | 1/249 | 0.66456 | 0.832891 | 0 | 0 | 0.922027 | 0.376768 |
| Gene_Pert | HIST1H4C I | 1/239 | 0.649424 | 0.832891 | 0 | 0 | 0.961257 | 0.414945 |
| Gene_Pert | Hdac1 KO r | 1/252 | 0.668973 | 0.832891 | 0 | 0 | 0.910868 | 0.366179 |
| Gene_Pert | Il10 KO mo | 2/524 | 0.668969 | 0.832891 | 0 | 0 | 0.874059 | 0.351387 |
| Gene_Pert | Eed KO mo | 1/251 | 0.667508 | 0.832891 | 0 | 0 | 0.914558 | 0.369667 |
| Gene_Pert | Des KO mo | 1/251 | 0.667508 | 0.832891 | 0 | 0 | 0.914558 | 0.369667 |
| Gene_Pert | Ppard KO n | 1/251 | 0.667508 | 0.832891 | 0 | 0 | 0.914558 | 0.369667 |
| Gene_Pert | Foxp3 Knoc | 1/251 | 0.667508 | 0.832891 | 0 | 0 | 0.914558 | 0.369667 |
| Gene_Pert | MYC KD hu | 1/248 | 0.663076 | 0.832891 | 0 | 0 | 0.925807 | 0.380383 |
| Gene_Pert | Ffar4 KO m | 1/248 | 0.663076 | 0.832891 | 0 | 0 | 0.925807 | 0.380383 |
| Gene_Pert | Trim28 knc | 1/251 | 0.667508 | 0.832891 | 0 | 0 | 0.914558 | 0.369667 |
| Gene_Pert | NPTX1 KO I | 1/250 | 0.666037 | 0.832891 | 0 | 0 | 0.918278 | 0.373197 |
| Gene_Pert | Mat1a KO r | 1/250 | 0.666037 | 0.832891 | 0 | 0 | 0.918278 | 0.373197 |
| Gene_Pert | TCF4 DN hu | 1/250 | 0.666037 | 0.832891 | 0 | 0 | 0.918278 | 0.373197 |
| Gene_Pert | SERPINI1 O | 1/244 | 0.657075 | 0.832891 | 0 | 0 | 0.941238 | 0.39528 |
| Gene_Pert | OLR1 OE hu | 1/247 | 0.661585 | 0.832891 | 0 | 0 | 0.929618 | 0.384041 |
| Gene_Pert | Atxn1 Knoc | 1/246 | 0.660088 | 0.832891 | 0 | 0 | 0.93346 | 0.387742 |
| Gene_Pert | OTX2 silen | 1/246 | 0.660088 | 0.832891 | 0 | 0 | 0.93346 | 0.387742 |
| Gene_Pert | KDM3A KD | 1/246 | 0.660088 | 0.832891 | 0 | 0 | 0.93346 | 0.387742 |
| Gene_Pert | Por KO mo | 1/246 | 0.660088 | 0.832891 | 0 | 0 | 0.93346 | 0.387742 |
| Gene_Pert | Iltifb KO m | 1/242 | 0.654034 | 0.832891 | 0 | 0 | 0.949146 | 0.403003 |
| Gene_Pert | TRPM7 def | 1/241 | 0.652504 | 0.832891 | 0 | 0 | 0.953149 | 0.406935 |
| Gene_Pert | Il10 KO mo | 1/246 | 0.660088 | 0.832891 | 0 | 0 | 0.93346 | 0.387742 |
| Gene_Pert | Htt Mutatic | 1/246 | 0.660088 | 0.832891 | 0 | 0 | 0.93346 | 0.387742 |
| Gene_Pert | Ahr KO mo | 1/245 | 0.658585 | 0.832891 | 0 | 0 | 0.937333 | 0.391489 |
| Gene_Pert | Psap hypor | 1/245 | 0.658585 | 0.832891 | 0 | 0 | 0.937333 | 0.391489 |
| Gene_Pert | Irf8 KO mo | 1/244 | 0.657075 | 0.832891 | 0 | 0 | 0.941238 | 0.39528 |
| Gene_Pert | Tfam KD m | 1/250 | 0.666037 | 0.832891 | 0 | 0 | 0.918278 | 0.373197 |
| Gene_Pert | MYC KD hu | 1/237 | 0.646317 | 0.832891 | 0 | 0 | 0.969501 | 0.423154 |
| Gene_Pert | Map3k1 kn | 1/237 | 0.646317 | 0.832891 | 0 | 0 | 0.969501 | 0.423154 |
| Gene_Pert | Rp1 KO mc | 1/243 | 0.655558 | 0.832891 | 0 | 0 | 0.945176 | 0.399118 |

| | | | | | | | | |
|---|---|---|---|---|---|---|---|---|
| Gene_Pert | Neurog3 C | 1/242 | 0.654034 | 0.832891 | 0 | 0 | 0.949146 | 0.403003 |
| Gene_Pert | Srf KO mou | 1/242 | 0.654034 | 0.832891 | 0 | 0 | 0.949146 | 0.403003 |
| Gene_Pert | PIK3CA KD | 1/242 | 0.654034 | 0.832891 | 0 | 0 | 0.949146 | 0.403003 |
| Gene_Pert | Por KO mo | 1/242 | 0.654034 | 0.832891 | 0 | 0 | 0.949146 | 0.403003 |
| Gene_Pert | Myc Myc + | 1/242 | 0.654034 | 0.832891 | 0 | 0 | 0.949146 | 0.403003 |
| Gene_Pert | Vdr KO mo | 1/253 | 0.670431 | 0.832891 | 0 | 0 | 0.907207 | 0.362732 |
| Gene_Pert | Irs2 KO mo | 1/252 | 0.668973 | 0.832891 | 0 | 0 | 0.910868 | 0.366179 |
| Gene_Pert | FGFR3 kno | 1/255 | 0.673329 | 0.832891 | 0 | 0 | 0.899973 | 0.355958 |
| Gene_Pert | Ahr low-aff | 1/255 | 0.673329 | 0.832891 | 0 | 0 | 0.899973 | 0.355958 |
| Gene_Pert | UBB KO mo | 1/255 | 0.673329 | 0.832891 | 0 | 0 | 0.899973 | 0.355958 |
| Gene_Pert | POU1F1 OE | 1/254 | 0.671883 | 0.832891 | 0 | 0 | 0.903576 | 0.359325 |
| Gene_Pert | Cftr Deficie | 1/256 | 0.674769 | 0.832891 | 0 | 0 | 0.896398 | 0.35263 |
| Gene_Pert | CDH1 OE h | 1/256 | 0.674769 | 0.832891 | 0 | 0 | 0.896398 | 0.35263 |
| Gene_Pert | CDK8 knocl | 1/256 | 0.674769 | 0.832891 | 0 | 0 | 0.896398 | 0.35263 |
| Gene_Pert | Emx2 Defic | 1/256 | 0.674769 | 0.832891 | 0 | 0 | 0.896398 | 0.35263 |
| Gene_Pert | FLI1 KD hu | 1/256 | 0.674769 | 0.832891 | 0 | 0 | 0.896398 | 0.35263 |
| Gene_Pert | Zfx KO mou | 1/256 | 0.674769 | 0.832891 | 0 | 0 | 0.896398 | 0.35263 |
| Gene_Pert | BRAF Drug | 1/268 | 0.691559 | 0.832891 | 0 | 0 | 0.855587 | 0.315546 |
| Gene_Pert | ZNF750 KD | 1/268 | 0.691559 | 0.832891 | 0 | 0 | 0.855587 | 0.315546 |
| Gene_Pert | NELFE KD h | 1/271 | 0.695622 | 0.832891 | 0 | 0 | 0.845952 | 0.307038 |
| Gene_Pert | Snca KO m | 1/270 | 0.694273 | 0.832891 | 0 | 0 | 0.84914 | 0.309842 |
| Gene_Pert | Psap KO m | 1/255 | 0.673329 | 0.832891 | 0 | 0 | 0.899973 | 0.355958 |
| Gene_Pert | Cftr mouse | 1/255 | 0.673329 | 0.832891 | 0 | 0 | 0.899973 | 0.355958 |
| Gene_Pert | Satb1 KO m | 1/255 | 0.673329 | 0.832891 | 0 | 0 | 0.899973 | 0.355958 |
| Gene_Pert | PFKL OE hu | 1/270 | 0.694273 | 0.832891 | 0 | 0 | 0.84914 | 0.309842 |
| Gene_Pert | MYC KD hu | 1/267 | 0.690194 | 0.832891 | 0 | 0 | 0.858848 | 0.318446 |
| Gene_Pert | PTEN KD hu | 1/266 | 0.688822 | 0.832891 | 0 | 0 | 0.862133 | 0.32138 |
| Gene_Pert | Pten KO m | 1/269 | 0.692919 | 0.832891 | 0 | 0 | 0.852352 | 0.312678 |
| Gene_Pert | BRAF Drug | 1/269 | 0.692919 | 0.832891 | 0 | 0 | 0.852352 | 0.312678 |
| Gene_Pert | IRF9 OE hu | 1/269 | 0.692919 | 0.832891 | 0 | 0 | 0.852352 | 0.312678 |
| Gene_Pert | CTNNB1 KC | 1/269 | 0.692919 | 0.832891 | 0 | 0 | 0.852352 | 0.312678 |
| Gene_Pert | Erbb2 drug | 1/269 | 0.692919 | 0.832891 | 0 | 0 | 0.852352 | 0.312678 |
| Gene_Pert | ZNF658 KD | 1/269 | 0.692919 | 0.832891 | 0 | 0 | 0.852352 | 0.312678 |
| Gene_Pert | LRRK2 Gly2 | 1/265 | 0.687444 | 0.832891 | 0 | 0 | 0.865442 | 0.324346 |
| Gene_Pert | SNORD14E | 1/264 | 0.68606 | 0.832891 | 0 | 0 | 0.868777 | 0.327347 |
| Gene_Pert | Klf1 KO mo | 1/263 | 0.68467 | 0.832891 | 0 | 0 | 0.872137 | 0.330382 |
| Gene_Pert | Creb1 DN r | 1/249 | 0.66456 | 0.832891 | 0 | 0 | 0.922027 | 0.376768 |
| Gene_Pert | ERG KD hu | 1/261 | 0.681872 | 0.832891 | 0 | 0 | 0.878936 | 0.336556 |
| Gene_Pert | FLI1 KD hu | 1/261 | 0.681872 | 0.832891 | 0 | 0 | 0.878936 | 0.336556 |
| Gene_Pert | SYK KD hur | 1/264 | 0.68606 | 0.832891 | 0 | 0 | 0.868777 | 0.327347 |
| Gene_Pert | RNF185 KD | 1/264 | 0.68606 | 0.832891 | 0 | 0 | 0.868777 | 0.327347 |
| Gene_Pert | Atm KO mc | 1/249 | 0.66456 | 0.832891 | 0 | 0 | 0.922027 | 0.376768 |
| Gene_Pert | CCND1 KD | 1/249 | 0.66456 | 0.832891 | 0 | 0 | 0.922027 | 0.376768 |
| Gene_Pert | Esr1 KO mc | 1/249 | 0.66456 | 0.832891 | 0 | 0 | 0.922027 | 0.376768 |
| Gene_Pert | CDK19 kno | 1/263 | 0.68467 | 0.832891 | 0 | 0 | 0.872137 | 0.330382 |

| | | | | | | | | |
|---|---|---|---|---|---|---|---|---|
| Gene_Pert | TP53 DEPLI | 1/258 | 0.677628 | 0.832891 | 0 | 0 | 0.889331 | 0.346089 |
| Gene_Pert | Stat3 KO m | 1/257 | 0.676202 | 0.832891 | 0 | 0 | 0.892851 | 0.34934 |
| Gene_Pert | Pou1f1 Sne | 1/263 | 0.68467 | 0.832891 | 0 | 0 | 0.872137 | 0.330382 |
| Gene_Pert | Por KO mo | 1/247 | 0.661585 | 0.832891 | 0 | 0 | 0.929618 | 0.384041 |
| Gene_Pert | Rp1 KO mo | 1/262 | 0.683274 | 0.832891 | 0 | 0 | 0.875523 | 0.333451 |
| Gene_Pert | Mbd3 KD n | 1/262 | 0.683274 | 0.832891 | 0 | 0 | 0.875523 | 0.333451 |
| Gene_Pert | Ager KO m | 1/262 | 0.683274 | 0.832891 | 0 | 0 | 0.875523 | 0.333451 |
| Gene_Pert | HNF1B OE | 1/261 | 0.681872 | 0.832891 | 0 | 0 | 0.878936 | 0.336556 |
| Gene_Pert | SYK Deplet | 1/256 | 0.674769 | 0.832891 | 0 | 0 | 0.896398 | 0.35263 |
| Gene_Pert | Mut KO mo | 1/256 | 0.674769 | 0.832891 | 0 | 0 | 0.896398 | 0.35263 |
| Gene_Pert | Kcnip3 KO | 1/261 | 0.681872 | 0.832891 | 0 | 0 | 0.878936 | 0.336556 |
| Gene_Pert | Ovol2 OE n | 1/260 | 0.680464 | 0.832891 | 0 | 0 | 0.882374 | 0.339697 |
| Gene_Pert | Por KO mo | 1/260 | 0.680464 | 0.832891 | 0 | 0 | 0.882374 | 0.339697 |
| Gene_Pert | NR5A2 KD | 1/259 | 0.679049 | 0.832891 | 0 | 0 | 0.885839 | 0.342875 |
| Gene_Pert | KLF13 KD h | 1/258 | 0.677628 | 0.832891 | 0 | 0 | 0.889331 | 0.346089 |
| Gene_Pert | Tgfb1 OE m | 1/258 | 0.677628 | 0.832891 | 0 | 0 | 0.889331 | 0.346089 |
| Gene_Pert | Erbb2 drug | 1/223 | 0.623791 | 0.832891 | 0 | 0 | 1.031374 | 0.486746 |
| Gene_Pert | Twist1 OE | 1/199 | 0.581831 | 0.832891 | 0 | 0 | 1.157799 | 0.627036 |
| Gene_Pert | ARHGDIB T | 1/198 | 0.579985 | 0.832891 | 0 | 0 | 1.163735 | 0.633948 |
| Gene_Pert | Eed VR mo | 1/221 | 0.62046 | 0.832891 | 0 | 0 | 1.040856 | 0.496795 |
| Gene_Pert | Rnf2 VR mo | 1/221 | 0.62046 | 0.832891 | 0 | 0 | 1.040856 | 0.496795 |
| Gene_Pert | Aplp2 KO n | 1/226 | 0.628735 | 0.832891 | 0 | 0 | 1.017468 | 0.472152 |
| Gene_Pert | Il10 KO mo | 1/225 | 0.627094 | 0.832891 | 0 | 0 | 1.022062 | 0.476954 |
| Gene_Pert | FLI1 KD hu | 1/195 | 0.574401 | 0.832891 | 0 | 0 | 1.181911 | 0.655284 |
| Gene_Pert | ERBB3 OE | 1/194 | 0.572523 | 0.832891 | 0 | 0 | 1.188095 | 0.662603 |
| Gene_Pert | Myd88 KO | 1/193 | 0.570637 | 0.832891 | 0 | 0 | 1.194344 | 0.670029 |
| Gene_Pert | FLI1 KD hu | 1/225 | 0.627094 | 0.832891 | 0 | 0 | 1.022062 | 0.476954 |
| Gene_Pert | Nfe2l2 KO | 2/475 | 0.615583 | 0.832891 | 0 | 0 | 0.967044 | 0.469196 |
| Gene_Pert | Nrl Deficie | 1/218 | 0.615408 | 0.832891 | 0 | 0 | 1.055407 | 0.512369 |
| Gene_Pert | MYC KD hu | 2/448 | 0.583508 | 0.832891 | 0 | 0 | 1.027011 | 0.553248 |
| Gene_Pert | Prkcz KO m | 1/223 | 0.623791 | 0.832891 | 0 | 0 | 1.031374 | 0.486746 |
| Gene_Pert | POU3F3 KD | 1/222 | 0.622129 | 0.832891 | 0 | 0 | 1.036094 | 0.491738 |
| Gene_Pert | IRF4 KD hu | 1/222 | 0.622129 | 0.832891 | 0 | 0 | 1.036094 | 0.491738 |
| Gene_Pert | Alk KD mo | 2/480 | 0.621315 | 0.832891 | 0 | 0 | 0.956682 | 0.455302 |
| Gene_Pert | Nfe2l2 KO | 2/480 | 0.621315 | 0.832891 | 0 | 0 | 0.956682 | 0.455302 |
| Gene_Pert | Snai1 KO m | 1/212 | 0.605103 | 0.832891 | 0 | 0 | 1.085749 | 0.545433 |
| Gene_Pert | HNRNPU-A | 1/211 | 0.603359 | 0.832891 | 0 | 0 | 1.090975 | 0.551207 |
| Gene_Pert | TFPI OE hu | 1/221 | 0.62046 | 0.832891 | 0 | 0 | 1.040856 | 0.496795 |
| Gene_Pert | Trp53 KO n | 1/220 | 0.618783 | 0.832891 | 0 | 0 | 1.045662 | 0.501918 |
| Gene_Pert | Stat1 Stat1 | 1/220 | 0.618783 | 0.832891 | 0 | 0 | 1.045662 | 0.501918 |
| Gene_Pert | Abcb4 KO r | 1/219 | 0.617099 | 0.832891 | 0 | 0 | 1.050512 | 0.507109 |
| Gene_Pert | Creb1 DN r | 1/219 | 0.617099 | 0.832891 | 0 | 0 | 1.050512 | 0.507109 |
| Gene_Pert | RB1 KD hu | 1/198 | 0.579985 | 0.832891 | 0 | 0 | 1.163735 | 0.633948 |
| Gene_Pert | Ppargc1a D | 3/339 | 0.18339 | 0.832891 | 0 | 0 | 2.080889 | 3.529475 |
| Gene_Pert | Cstb KO mo | 3/339 | 0.18339 | 0.832891 | 0 | 0 | 2.080889 | 3.529475 |

| | | | | | | | | |
|---|---|---|---|---|---|---|---|---|
| Gene_Pert | Prdm16 DE | 3/340 | 0.18448 | 0.832891 | 0 | 0 | 2.074608 | 3.506535 |
| Gene_Pert | Myc JoMa1 | 1/210 | 0.601608 | 0.832891 | 0 | 0 | 1.09625 | 0.557059 |
| Gene_Pert | Nr2e3 Ecto | 2/454 | 0.5908 | 0.832891 | 0 | 0 | 1.013066 | 0.533155 |
| Gene_Pert | Dmd Defici | 1/203 | 0.589132 | 0.832891 | 0 | 0 | 1.134642 | 0.600344 |
| Gene_Pert | Sod1 G93A | 2/462 | 0.600376 | 0.832891 | 0 | 0 | 0.995038 | 0.507667 |
| Gene_Pert | IRF1 OE hu | 1/209 | 0.599849 | 0.832891 | 0 | 0 | 1.101576 | 0.562991 |
| Gene_Pert | FLI1 KD hu | 1/209 | 0.599849 | 0.832891 | 0 | 0 | 1.101576 | 0.562991 |
| Gene_Pert | Slc6a4 KO r | 1/209 | 0.599849 | 0.832891 | 0 | 0 | 1.101576 | 0.562991 |
| Gene_Pert | Il1rn KO mo | 1/209 | 0.599849 | 0.832891 | 0 | 0 | 1.101576 | 0.562991 |
| Gene_Pert | UPF1 KD hu | 1/208 | 0.598082 | 0.832891 | 0 | 0 | 1.106954 | 0.569005 |
| Gene_Pert | MECP2 het | 1/219 | 0.617099 | 0.832891 | 0 | 0 | 1.050512 | 0.507109 |
| Gene_Pert | RB1 KD hur | 2/444 | 0.578595 | 0.832891 | 0 | 0 | 1.036518 | 0.567133 |
| Gene_Pert | SOX11 DEP | 1/207 | 0.596308 | 0.832891 | 0 | 0 | 1.112384 | 0.575101 |
| Gene_Pert | Myc Activa | 1/207 | 0.596308 | 0.832891 | 0 | 0 | 1.112384 | 0.575101 |
| Gene_Pert | TWIST2 Q1 | 1/207 | 0.596308 | 0.832891 | 0 | 0 | 1.112384 | 0.575101 |
| Gene_Pert | Ifng KO mo | 1/206 | 0.594525 | 0.832891 | 0 | 0 | 1.117867 | 0.581282 |
| Gene_Pert | ATM KD hu | 1/204 | 0.590938 | 0.832891 | 0 | 0 | 1.128995 | 0.593902 |
| Gene_Pert | IL1B Challe | 1/204 | 0.590938 | 0.832891 | 0 | 0 | 1.128995 | 0.593902 |
| Gene_Pert | RB1 KD hur | 1/193 | 0.570637 | 0.832891 | 0 | 0 | 1.194344 | 0.670029 |
| Gene_Pert | RBP2 KD hu | 2/437 | 0.569898 | 0.832891 | 0 | 0 | 1.053577 | 0.592424 |
| Gene_Pert | Lhx1 NULL | 1/202 | 0.587319 | 0.832891 | 0 | 0 | 1.140345 | 0.606877 |
| Gene_Pert | Rb1 KO mo | 2/450 | 0.585949 | 0.832891 | 0 | 0 | 1.022321 | 0.546454 |
| Gene_Pert | Psap KO m | 1/201 | 0.585497 | 0.832891 | 0 | 0 | 1.146105 | 0.613503 |
| Gene_Pert | UPF1 Defic | 1/230 | 0.635226 | 0.832891 | 0 | 0 | 0.999492 | 0.453544 |
| Gene_Pert | IL2 OE hum | 1/227 | 0.630368 | 0.832891 | 0 | 0 | 1.012914 | 0.46741 |
| Gene_Pert | Nfyc KD mo | 1/235 | 0.643183 | 0.832891 | 0 | 0 | 0.977887 | 0.431568 |
| Gene_Pert | SYK KD hur | 1/235 | 0.643183 | 0.832891 | 0 | 0 | 0.977887 | 0.431568 |
| Gene_Pert | Ksr1 knock | 1/234 | 0.641605 | 0.832891 | 0 | 0 | 0.982134 | 0.435854 |
| Gene_Pert | TNF Stimul | 1/234 | 0.641605 | 0.832891 | 0 | 0 | 0.982134 | 0.435854 |
| Gene_Pert | Ddr1 KO m | 1/233 | 0.640021 | 0.832891 | 0 | 0 | 0.986417 | 0.440193 |
| Gene_Pert | CDK19 kno | 1/232 | 0.63843 | 0.832891 | 0 | 0 | 0.990738 | 0.444588 |
| Gene_Pert | Pten KO mo | 1/224 | 0.625446 | 0.832891 | 0 | 0 | 1.026697 | 0.481818 |
| Gene_Pert | Fgf23 Trans | 1/223 | 0.623791 | 0.832891 | 0 | 0 | 1.031374 | 0.486746 |
| Gene_Pert | Nf1 Hetero | 1/231 | 0.636831 | 0.832891 | 0 | 0 | 0.995096 | 0.449038 |
| Gene_Pert | Clpp KO mo | 1/230 | 0.635226 | 0.832891 | 0 | 0 | 0.999492 | 0.453544 |
| Gene_Pert | Hdc KO mo | 1/230 | 0.635226 | 0.832891 | 0 | 0 | 0.999492 | 0.453544 |
| Gene_Pert | NPM1 mut | 1/227 | 0.630368 | 0.832891 | 0 | 0 | 1.012914 | 0.46741 |
| Gene_Pert | Arid3a KD r | 1/228 | 0.631995 | 0.832891 | 0 | 0 | 1.008401 | 0.462729 |
| Gene_Pert | Hdac1 Drug | 1/228 | 0.631995 | 0.832891 | 0 | 0 | 1.008401 | 0.462729 |
| Gene_Pert | Fmr1 KO m | 1/246 | 0.660088 | 0.832891 | 0 | 0 | 0.93346 | 0.387742 |
| Gene_Pert | Crem KO m | 1/246 | 0.660088 | 0.832891 | 0 | 0 | 0.93346 | 0.387742 |
| Gene_Pert | Ppargc1b H | 1/249 | 0.66456 | 0.832891 | 0 | 0 | 0.922027 | 0.376768 |
| Gene_Pert | HSF1 KD hu | 1/249 | 0.66456 | 0.832891 | 0 | 0 | 0.922027 | 0.376768 |
| Gene_Pert | Polg KO mo | 1/225 | 0.627094 | 0.832891 | 0 | 0 | 1.022062 | 0.476954 |
| Gene_Pert | Flnb KO mo | 1/225 | 0.627094 | 0.832891 | 0 | 0 | 1.022062 | 0.476954 |

| | | | | | | | | |
|---|---|---|---|---|---|---|---|---|
| Gene_Pert | KDM1B KD | 1/225 | 0.627094 | 0.832891 | 0 | 0 | 1.022062 | 0.476954 |
| Gene_Pert | Ikzf1 IkL/L | 1/248 | 0.663076 | 0.832891 | 0 | 0 | 0.925807 | 0.380383 |
| Gene_Pert | Esr1 KO mc | 1/244 | 0.657075 | 0.832891 | 0 | 0 | 0.941238 | 0.39528 |
| Gene_Pert | Fosl1 KO m | 1/243 | 0.655558 | 0.832891 | 0 | 0 | 0.945176 | 0.399118 |
| Gene_Pert | Psap KO m | 2/464 | 0.602744 | 0.832891 | 0 | 0 | 0.990629 | 0.501518 |
| Gene_Pert | LMO3 KD h | 2/463 | 0.601562 | 0.832891 | 0 | 0 | 0.992829 | 0.504582 |
| Gene_Pert | RBP2 KD hu | 2/463 | 0.601562 | 0.832891 | 0 | 0 | 0.992829 | 0.504582 |
| Gene_Pert | Retnlb KO | 1/216 | 0.612003 | 0.832891 | 0 | 0 | 1.065333 | 0.523098 |
| Gene_Pert | RB1 KD hur | 1/215 | 0.610289 | 0.832891 | 0 | 0 | 1.070365 | 0.528571 |
| Gene_Pert | USF1 OE hu | 1/214 | 0.608568 | 0.832891 | 0 | 0 | 1.075445 | 0.534116 |
| Gene_Pert | Rad21 KD r | 1/214 | 0.608568 | 0.832891 | 0 | 0 | 1.075445 | 0.534116 |
| Gene_Pert | Cstb KO mc | 1/213 | 0.606839 | 0.832891 | 0 | 0 | 1.080573 | 0.539737 |
| Gene_Pert | ERBB2 over | 1/212 | 0.605103 | 0.832891 | 0 | 0 | 1.085749 | 0.545433 |
| Gene_Pert | Prkag3 KO | 1/235 | 0.643183 | 0.832891 | 0 | 0 | 0.977887 | 0.431568 |
| Gene_Pert | Cdk4 Heter | 1/235 | 0.643183 | 0.832891 | 0 | 0 | 0.977887 | 0.431568 |
| Gene_Pert | Sirt7 KO m | 1/241 | 0.652504 | 0.832891 | 0 | 0 | 0.953149 | 0.406935 |
| Gene_Pert | Rnf2 KO m | 1/240 | 0.650968 | 0.832891 | 0 | 0 | 0.957186 | 0.410916 |
| Gene_Pert | Gys1 KO m | 1/268 | 0.691559 | 0.832891 | 0 | 0 | 0.855587 | 0.315546 |
| Gene_Pert | Pcdh12 De | 1/210 | 0.601608 | 0.832891 | 0 | 0 | 1.09625 | 0.557059 |
| Gene_Pert | Arid3b KO | 1/210 | 0.601608 | 0.832891 | 0 | 0 | 1.09625 | 0.557059 |
| Gene_Pert | Pgr KO mo | 1/238 | 0.647874 | 0.832891 | 0 | 0 | 0.965362 | 0.419024 |
| Gene_Pert | Pten PTEN | 1/232 | 0.63843 | 0.832891 | 0 | 0 | 0.990738 | 0.444588 |
| Gene_Pert | Rfwd2 KO | 1/231 | 0.636831 | 0.832891 | 0 | 0 | 0.995096 | 0.449038 |
| Gene_Pert | Ndel1 KD n | 1/237 | 0.646317 | 0.832891 | 0 | 0 | 0.969501 | 0.423154 |
| Gene_Pert | LITAF KD h | 1/237 | 0.646317 | 0.832891 | 0 | 0 | 0.969501 | 0.423154 |
| Gene_Pert | Fgf23 expr | 1/237 | 0.646317 | 0.832891 | 0 | 0 | 0.969501 | 0.423154 |
| Gene_Pert | L1cam KO | 1/237 | 0.646317 | 0.832891 | 0 | 0 | 0.969501 | 0.423154 |
| Gene_Pert | HNF4A DEF | 1/236 | 0.644753 | 0.832891 | 0 | 0 | 0.973676 | 0.427334 |
| Gene_Pert | Zfp42 KD m | 1/235 | 0.643183 | 0.832891 | 0 | 0 | 0.977887 | 0.431568 |
| Gene_Pert | PTEN KD hu | 1/287 | 0.716409 | 0.832891 | 0 | 0 | 0.797975 | 0.266128 |
| Gene_Pert | Gata4 Dele | 1/296 | 0.727479 | 0.832891 | 0 | 0 | 0.773276 | 0.246034 |
| Gene_Pert | Htra2 KO m | 1/296 | 0.727479 | 0.832891 | 0 | 0 | 0.773276 | 0.246034 |
| Gene_Pert | THRA HCC | 1/296 | 0.727479 | 0.832891 | 0 | 0 | 0.773276 | 0.246034 |
| Gene_Pert | Pmp22 KO | 1/296 | 0.727479 | 0.832891 | 0 | 0 | 0.773276 | 0.246034 |
| Gene_Pert | ZMPSTE24 | 1/298 | 0.72988 | 0.832891 | 0 | 0 | 0.76799 | 0.241821 |
| Gene_Pert | Tia1 KO mc | 1/298 | 0.72988 | 0.832891 | 0 | 0 | 0.76799 | 0.241821 |
| Gene_Pert | Akt1 knock | 1/286 | 0.715152 | 0.832891 | 0 | 0 | 0.800816 | 0.268482 |
| Gene_Pert | Lhx1 KO m | 1/286 | 0.715152 | 0.832891 | 0 | 0 | 0.800816 | 0.268482 |
| Gene_Pert | RNF40 KD | 1/286 | 0.715152 | 0.832891 | 0 | 0 | 0.800816 | 0.268482 |
| Gene_Pert | ARNTL2 KD | 1/285 | 0.713889 | 0.832891 | 0 | 0 | 0.803677 | 0.270861 |
| Gene_Pert | Irf4 KO mo | 1/285 | 0.713889 | 0.832891 | 0 | 0 | 0.803677 | 0.270861 |
| Gene_Pert | Myrf KO m | 1/285 | 0.713889 | 0.832891 | 0 | 0 | 0.803677 | 0.270861 |
| Gene_Pert | RAF1 overe | 1/296 | 0.727479 | 0.832891 | 0 | 0 | 0.773276 | 0.246034 |
| Gene_Pert | Dusp1 KO | 1/287 | 0.716409 | 0.832891 | 0 | 0 | 0.797975 | 0.266128 |
| Gene_Pert | AGR2 OE h | 1/284 | 0.712621 | 0.832891 | 0 | 0 | 0.806558 | 0.273266 |

| | | | | | | | | |
|---|---|---|---|---|---|---|---|---|
| Gene_Pert | Vdr KO mo | 1/284 | 0.712621 | 0.832891 | 0 | 0 | 0.806558 | 0.273266 |
| Gene_Pert | MELK drug | 1/284 | 0.712621 | 0.832891 | 0 | 0 | 0.806558 | 0.273266 |
| Gene_Pert | Lpin1 OE m | 1/284 | 0.712621 | 0.832891 | 0 | 0 | 0.806558 | 0.273266 |
| Gene_Pert | Rif1 KD mo | 1/293 | 0.723837 | 0.832891 | 0 | 0 | 0.78134 | 0.25252 |
| Gene_Pert | RNF40 KD l | 1/293 | 0.723837 | 0.832891 | 0 | 0 | 0.78134 | 0.25252 |
| Gene_Pert | Trim28 KO | 1/295 | 0.72627 | 0.832891 | 0 | 0 | 0.775945 | 0.248173 |
| Gene_Pert | Gast KO mo | 1/295 | 0.72627 | 0.832891 | 0 | 0 | 0.775945 | 0.248173 |
| Gene_Pert | SRF KD hur | 1/293 | 0.723837 | 0.832891 | 0 | 0 | 0.78134 | 0.25252 |
| Gene_Pert | OGT siOGT | 1/293 | 0.723837 | 0.832891 | 0 | 0 | 0.78134 | 0.25252 |
| Gene_Pert | CTNNB1 Ac | 1/293 | 0.723837 | 0.832891 | 0 | 0 | 0.78134 | 0.25252 |
| Gene_Pert | Tcof1 OE m | 1/288 | 0.717661 | 0.832891 | 0 | 0 | 0.795154 | 0.263799 |
| Gene_Pert | Setd2 Defic | 1/293 | 0.723837 | 0.832891 | 0 | 0 | 0.78134 | 0.25252 |
| Gene_Pert | NR5A2 KD | 1/283 | 0.711348 | 0.832891 | 0 | 0 | 0.809459 | 0.275697 |
| Gene_Pert | LRRK2 dom | 1/282 | 0.710068 | 0.832891 | 0 | 0 | 0.812381 | 0.278154 |
| Gene_Pert | Bcl2 OE mo | 1/282 | 0.710068 | 0.832891 | 0 | 0 | 0.812381 | 0.278154 |
| Gene_Pert | Sfrp5 KO m | 1/282 | 0.710068 | 0.832891 | 0 | 0 | 0.812381 | 0.278154 |
| Gene_Pert | Rps6ka3 KO | 1/281 | 0.708784 | 0.832891 | 0 | 0 | 0.815324 | 0.280639 |
| Gene_Pert | Pik3ca kno | 1/281 | 0.708784 | 0.832891 | 0 | 0 | 0.815324 | 0.280639 |
| Gene_Pert | Trp53 KO n | 1/287 | 0.716409 | 0.832891 | 0 | 0 | 0.797975 | 0.266128 |
| Gene_Pert | BRAF KD hu | 1/287 | 0.716409 | 0.832891 | 0 | 0 | 0.797975 | 0.266128 |
| Gene_Pert | SOX2 Defic | 1/287 | 0.716409 | 0.832891 | 0 | 0 | 0.797975 | 0.266128 |
| Gene_Pert | Cldn16 Def | 1/287 | 0.716409 | 0.832891 | 0 | 0 | 0.797975 | 0.266128 |
| Gene_Pert | MAP4K5 kr | 1/287 | 0.716409 | 0.832891 | 0 | 0 | 0.797975 | 0.266128 |
| Gene_Pert | Pde6b RD1 | 1/292 | 0.722612 | 0.832891 | 0 | 0 | 0.784065 | 0.254729 |
| Gene_Pert | Ebf1 OE mo | 1/288 | 0.717661 | 0.832891 | 0 | 0 | 0.795154 | 0.263799 |
| Gene_Pert | FH KD hum | 1/290 | 0.720147 | 0.832891 | 0 | 0 | 0.789571 | 0.259216 |
| Gene_Pert | Foxd3 KO r | 1/290 | 0.720147 | 0.832891 | 0 | 0 | 0.789571 | 0.259216 |
| Gene_Pert | Lmna KO m | 1/288 | 0.717661 | 0.832891 | 0 | 0 | 0.795154 | 0.263799 |
| Gene_Pert | VIM KD hu | 1/288 | 0.717661 | 0.832891 | 0 | 0 | 0.795154 | 0.263799 |
| Gene_Pert | Cited2 Defi | 1/288 | 0.717661 | 0.832891 | 0 | 0 | 0.795154 | 0.263799 |
| Gene_Pert | DOCK4 OE | 1/287 | 0.716409 | 0.832891 | 0 | 0 | 0.797975 | 0.266128 |
| Gene_Pert | Insig1 KO n | 1/287 | 0.716409 | 0.832891 | 0 | 0 | 0.797975 | 0.266128 |
| Gene_Pert | MIR148B C | 1/287 | 0.716409 | 0.832891 | 0 | 0 | 0.797975 | 0.266128 |
| Gene_Pert | Nrl Deficie | 1/289 | 0.718907 | 0.832891 | 0 | 0 | 0.792353 | 0.261495 |
| Gene_Pert | Irf1 OE mo | 1/289 | 0.718907 | 0.832891 | 0 | 0 | 0.792353 | 0.261495 |
| Gene_Pert | SCYL1 knoc | 1/296 | 0.727479 | 0.832891 | 0 | 0 | 0.773276 | 0.246034 |
| Gene_Pert | FAM60A KI | 1/306 | 0.739278 | 0.832891 | 0 | 0 | 0.747541 | 0.225819 |
| Gene_Pert | SYK druginl | 1/298 | 0.72988 | 0.832891 | 0 | 0 | 0.76799 | 0.241821 |
| Gene_Pert | Gfi1b OE m | 1/300 | 0.73226 | 0.832891 | 0 | 0 | 0.762775 | 0.237696 |
| Gene_Pert | Gykl1 KO m | 1/300 | 0.73226 | 0.832891 | 0 | 0 | 0.762775 | 0.237696 |
| Gene_Pert | FLI1 KD hu | 1/298 | 0.72988 | 0.832891 | 0 | 0 | 0.76799 | 0.241821 |
| Gene_Pert | RB1 KD hu | 1/298 | 0.72988 | 0.832891 | 0 | 0 | 0.76799 | 0.241821 |
| Gene_Pert | MCAM KD | 1/297 | 0.728682 | 0.832891 | 0 | 0 | 0.770624 | 0.243916 |
| Gene_Pert | Clpp KO mo | 1/297 | 0.728682 | 0.832891 | 0 | 0 | 0.770624 | 0.243916 |
| Gene_Pert | Nod2 KO m | 1/297 | 0.728682 | 0.832891 | 0 | 0 | 0.770624 | 0.243916 |

| | | | | | | | | |
|---|---|---|---|---|---|---|---|---|
| Gene_Pert | RARA Activ | 1/297 | 0.728682 | 0.832891 | 0 | 0 | 0.770624 | 0.243916 |
| Gene_Pert | Pou2af1 OI | 1/299 | 0.731073 | 0.832891 | 0 | 0 | 0.765374 | 0.239748 |
| Gene_Pert | Bptf KO mc | 1/299 | 0.731073 | 0.832891 | 0 | 0 | 0.765374 | 0.239748 |
| Gene_Pert | PPARGC1A | 1/307 | 0.740429 | 0.832891 | 0 | 0 | 0.74506 | 0.223909 |
| Gene_Pert | Igf1r drugir | 1/298 | 0.72988 | 0.832891 | 0 | 0 | 0.76799 | 0.241821 |
| Gene_Pert | SP3 KD hur | 1/306 | 0.739278 | 0.832891 | 0 | 0 | 0.747541 | 0.225819 |
| Gene_Pert | FLI1 KD hu | 1/306 | 0.739278 | 0.832891 | 0 | 0 | 0.747541 | 0.225819 |
| Gene_Pert | Nf1 Hetero | 1/305 | 0.738121 | 0.832891 | 0 | 0 | 0.750038 | 0.227747 |
| Gene_Pert | AGO1 KD h | 1/305 | 0.738121 | 0.832891 | 0 | 0 | 0.750038 | 0.227747 |
| Gene_Pert | TARBP2 OE | 1/308 | 0.741576 | 0.832891 | 0 | 0 | 0.742595 | 0.222019 |
| Gene_Pert | HDAC9 INH | 1/308 | 0.741576 | 0.832891 | 0 | 0 | 0.742595 | 0.222019 |
| Gene_Pert | FLI1 KD hu | 1/295 | 0.72627 | 0.832891 | 0 | 0 | 0.775945 | 0.248173 |
| Gene_Pert | Socs3 KO n | 1/295 | 0.72627 | 0.832891 | 0 | 0 | 0.775945 | 0.248173 |
| Gene_Pert | JUND KD h | 1/295 | 0.72627 | 0.832891 | 0 | 0 | 0.775945 | 0.248173 |
| Gene_Pert | Socs3 KO n | 1/295 | 0.72627 | 0.832891 | 0 | 0 | 0.775945 | 0.248173 |
| Gene_Pert | Jun KO mo | 1/295 | 0.72627 | 0.832891 | 0 | 0 | 0.775945 | 0.248173 |
| Gene_Pert | Gata4 abla | 1/295 | 0.72627 | 0.832891 | 0 | 0 | 0.775945 | 0.248173 |
| Gene_Pert | Bach2 KO r | 1/290 | 0.720147 | 0.832891 | 0 | 0 | 0.789571 | 0.259216 |
| Gene_Pert | Vax2 KO m | 1/292 | 0.722612 | 0.832891 | 0 | 0 | 0.784065 | 0.254729 |
| Gene_Pert | Akt1 KO m | 1/292 | 0.722612 | 0.832891 | 0 | 0 | 0.784065 | 0.254729 |
| Gene_Pert | Dbi KO mo | 1/294 | 0.725056 | 0.832891 | 0 | 0 | 0.778633 | 0.250335 |
| Gene_Pert | Hsf1 KO m | 1/294 | 0.725056 | 0.832891 | 0 | 0 | 0.778633 | 0.250335 |
| Gene_Pert | SREBF2 KD | 1/301 | 0.733443 | 0.832891 | 0 | 0 | 0.760194 | 0.235665 |
| Gene_Pert | Gfi1b OE m | 1/301 | 0.733443 | 0.832891 | 0 | 0 | 0.760194 | 0.235665 |
| Gene_Pert | Aire NULL I | 1/301 | 0.733443 | 0.832891 | 0 | 0 | 0.760194 | 0.235665 |
| Gene_Pert | Pink1 KO m | 1/301 | 0.733443 | 0.832891 | 0 | 0 | 0.760194 | 0.235665 |
| Gene_Pert | PDCD10 KD | 1/301 | 0.733443 | 0.832891 | 0 | 0 | 0.760194 | 0.235665 |
| Gene_Pert | Zfp36l2 de | 1/301 | 0.733443 | 0.832891 | 0 | 0 | 0.760194 | 0.235665 |
| Gene_Pert | SYK knockc | 1/303 | 0.735792 | 0.832891 | 0 | 0 | 0.755082 | 0.231665 |
| Gene_Pert | GPLD1 OE I | 1/303 | 0.735792 | 0.832891 | 0 | 0 | 0.755082 | 0.231665 |
| Gene_Pert | FLI1 KD hu | 1/293 | 0.723837 | 0.832891 | 0 | 0 | 0.78134 | 0.25252 |
| Gene_Pert | GSK3A KD I | 1/290 | 0.720147 | 0.832891 | 0 | 0 | 0.789571 | 0.259216 |
| Gene_Pert | Myod1 Mu | 1/290 | 0.720147 | 0.832891 | 0 | 0 | 0.789571 | 0.259216 |
| Gene_Pert | ESR1 KD hu | 1/290 | 0.720147 | 0.832891 | 0 | 0 | 0.789571 | 0.259216 |
| Gene_Pert | CMAHP KO | 1/290 | 0.720147 | 0.832891 | 0 | 0 | 0.789571 | 0.259216 |
| Gene_Pert | VAMP7 KD | 1/290 | 0.720147 | 0.832891 | 0 | 0 | 0.789571 | 0.259216 |
| Gene_Pert | SRF KD hur | 1/292 | 0.722612 | 0.832891 | 0 | 0 | 0.784065 | 0.254729 |
| Gene_Pert | Myd88 KO | 1/292 | 0.722612 | 0.832891 | 0 | 0 | 0.784065 | 0.254729 |
| Gene_Pert | Met knock | 1/289 | 0.718907 | 0.832891 | 0 | 0 | 0.792353 | 0.261495 |
| Gene_Pert | Neurod1 C | 1/289 | 0.718907 | 0.832891 | 0 | 0 | 0.792353 | 0.261495 |
| Gene_Pert | Trim24 Def | 1/289 | 0.718907 | 0.832891 | 0 | 0 | 0.792353 | 0.261495 |
| Gene_Pert | Runx1 KO r | 1/289 | 0.718907 | 0.832891 | 0 | 0 | 0.792353 | 0.261495 |
| Gene_Pert | FLI1 KD hu | 1/272 | 0.696964 | 0.832891 | 0 | 0 | 0.842787 | 0.304265 |
| Gene_Pert | Med23 KO | 1/274 | 0.699631 | 0.832891 | 0 | 0 | 0.836528 | 0.29881 |
| Gene_Pert | Il1rn KO m | 1/274 | 0.699631 | 0.832891 | 0 | 0 | 0.836528 | 0.29881 |

| | | | | | | | | |
|---|---|---|---|---|---|---|---|---|
| Gene_Pert | ERO1L OE I | 1/273 | 0.6983 | 0.832891 | 0 | 0 | 0.839646 | 0.301522 |
| Gene_Pert | Grid2 KO m | 1/273 | 0.6983 | 0.832891 | 0 | 0 | 0.839646 | 0.301522 |
| Gene_Pert | PIK3CA act | 1/273 | 0.6983 | 0.832891 | 0 | 0 | 0.839646 | 0.301522 |
| Gene_Pert | Ndel1 KO n | 1/271 | 0.695622 | 0.832891 | 0 | 0 | 0.845952 | 0.307038 |
| Gene_Pert | Tnf KD mo | 1/271 | 0.695622 | 0.832891 | 0 | 0 | 0.845952 | 0.307038 |
| Gene_Pert | MTDH DEP | 1/273 | 0.6983 | 0.832891 | 0 | 0 | 0.839646 | 0.301522 |
| Gene_Pert | CD24 KD hu | 1/273 | 0.6983 | 0.832891 | 0 | 0 | 0.839646 | 0.301522 |
| Gene_Pert | GRN deficie | 1/273 | 0.6983 | 0.832891 | 0 | 0 | 0.839646 | 0.301522 |
| Gene_Pert | Aldh5a1 KO | 1/273 | 0.6983 | 0.832891 | 0 | 0 | 0.839646 | 0.301522 |
| Gene_Pert | IRF5 KD hu | 1/272 | 0.696964 | 0.832891 | 0 | 0 | 0.842787 | 0.304265 |
| Gene_Pert | HMGN5 OE | 1/274 | 0.699631 | 0.832891 | 0 | 0 | 0.836528 | 0.29881 |
| Gene_Pert | NR2F2 KD I | 1/270 | 0.694273 | 0.832891 | 0 | 0 | 0.84914 | 0.309842 |
| Gene_Pert | CMAHP KO | 1/270 | 0.694273 | 0.832891 | 0 | 0 | 0.84914 | 0.309842 |
| Gene_Pert | Akt1 drugir | 1/272 | 0.696964 | 0.832891 | 0 | 0 | 0.842787 | 0.304265 |
| Gene_Pert | Bcl2 OE mo | 1/271 | 0.695622 | 0.832891 | 0 | 0 | 0.845952 | 0.307038 |
| Gene_Pert | Pten KD mo | 1/271 | 0.695622 | 0.832891 | 0 | 0 | 0.845952 | 0.307038 |
| Gene_Pert | ERBB3 kno | 1/271 | 0.695622 | 0.832891 | 0 | 0 | 0.845952 | 0.307038 |
| Gene_Pert | Smarca5 KO | 1/271 | 0.695622 | 0.832891 | 0 | 0 | 0.845952 | 0.307038 |
| Gene_Pert | Ppara agor | 1/271 | 0.695622 | 0.832891 | 0 | 0 | 0.845952 | 0.307038 |
| Gene_Pert | Trp53inp2 | 1/280 | 0.707493 | 0.832891 | 0 | 0 | 0.818288 | 0.28315 |
| Gene_Pert | Klf7 KO mo | 1/280 | 0.707493 | 0.832891 | 0 | 0 | 0.818288 | 0.28315 |
| Gene_Pert | Abca1 OE r | 1/270 | 0.694273 | 0.832891 | 0 | 0 | 0.84914 | 0.309842 |
| Gene_Pert | Adam10 KO | 1/270 | 0.694273 | 0.832891 | 0 | 0 | 0.84914 | 0.309842 |
| Gene_Pert | ETS2 KD hu | 1/265 | 0.687444 | 0.832891 | 0 | 0 | 0.865442 | 0.324346 |
| Gene_Pert | KDM5A KD | 1/268 | 0.691559 | 0.832891 | 0 | 0 | 0.855587 | 0.315546 |
| Gene_Pert | HMGB1 KO | 1/268 | 0.691559 | 0.832891 | 0 | 0 | 0.855587 | 0.315546 |
| Gene_Pert | Npc1 KO m | 1/268 | 0.691559 | 0.832891 | 0 | 0 | 0.855587 | 0.315546 |
| Gene_Pert | IFNL3 muta | 1/267 | 0.690194 | 0.832891 | 0 | 0 | 0.858848 | 0.318446 |
| Gene_Pert | CDH1 OE h | 1/267 | 0.690194 | 0.832891 | 0 | 0 | 0.858848 | 0.318446 |
| Gene_Pert | CMAHP KO | 1/263 | 0.68467 | 0.832891 | 0 | 0 | 0.872137 | 0.330382 |
| Gene_Pert | Lrp5 KO mo | 1/263 | 0.68467 | 0.832891 | 0 | 0 | 0.872137 | 0.330382 |
| Gene_Pert | Mat1a KO r | 1/266 | 0.688822 | 0.832891 | 0 | 0 | 0.862133 | 0.32138 |
| Gene_Pert | Bach2 KO r | 1/266 | 0.688822 | 0.832891 | 0 | 0 | 0.862133 | 0.32138 |
| Gene_Pert | HDAC2 KD | 1/266 | 0.688822 | 0.832891 | 0 | 0 | 0.862133 | 0.32138 |
| Gene_Pert | Mmp8 KO r | 1/266 | 0.688822 | 0.832891 | 0 | 0 | 0.862133 | 0.32138 |
| Gene_Pert | Hprt KO mo | 2/543 | 0.688025 | 0.832891 | 0 | 0 | 0.842536 | 0.31505 |
| Gene_Pert | Tfrc KO mo | 1/270 | 0.694273 | 0.832891 | 0 | 0 | 0.84914 | 0.309842 |
| Gene_Pert | SYK druginl | 1/273 | 0.6983 | 0.832891 | 0 | 0 | 0.839646 | 0.301522 |
| Gene_Pert | SYK druginl | 1/273 | 0.6983 | 0.832891 | 0 | 0 | 0.839646 | 0.301522 |
| Gene_Pert | YWHAZ OE | 1/276 | 0.702274 | 0.832891 | 0 | 0 | 0.830359 | 0.293475 |
| Gene_Pert | Tert OE mo | 1/275 | 0.700955 | 0.832891 | 0 | 0 | 0.833432 | 0.296128 |
| Gene_Pert | RB1 KD hur | 1/263 | 0.68467 | 0.832891 | 0 | 0 | 0.872137 | 0.330382 |
| Gene_Pert | IFITM1 KO | 1/263 | 0.68467 | 0.832891 | 0 | 0 | 0.872137 | 0.330382 |
| Gene_Pert | Kdm1a KD | 1/263 | 0.68467 | 0.832891 | 0 | 0 | 0.872137 | 0.330382 |
| Gene_Pert | Fcgr3 KO m | 1/275 | 0.700955 | 0.832891 | 0 | 0 | 0.833432 | 0.296128 |

| | | | | | | | | |
|---|---|---|---|---|---|---|---|---|
| Gene_Pert | Cftr KO mo | 1/275 | 0.700955 | 0.832891 | 0 | 0 | 0.833432 | 0.296128 |
| Gene_Pert | Dlx5 Defici | 1/275 | 0.700955 | 0.832891 | 0 | 0 | 0.833432 | 0.296128 |
| Gene_Pert | Htt Knock-i | 1/272 | 0.696964 | 0.832891 | 0 | 0 | 0.842787 | 0.304265 |
| Gene_Pert | PIAS1 Depl | 1/272 | 0.696964 | 0.832891 | 0 | 0 | 0.842787 | 0.304265 |
| Gene_Pert | AGR2 KD h | 1/283 | 0.711348 | 0.832891 | 0 | 0 | 0.809459 | 0.275697 |
| Gene_Pert | Myc knock- | 1/281 | 0.708784 | 0.832891 | 0 | 0 | 0.815324 | 0.280639 |
| Gene_Pert | Dmd Defici | 1/284 | 0.712621 | 0.832891 | 0 | 0 | 0.806558 | 0.273266 |
| Gene_Pert | MYC KD hu | 1/284 | 0.712621 | 0.832891 | 0 | 0 | 0.806558 | 0.273266 |
| Gene_Pert | ESRRA Defi | 1/275 | 0.700955 | 0.832891 | 0 | 0 | 0.833432 | 0.296128 |
| Gene_Pert | Tgfbr1 Stim | 1/286 | 0.715152 | 0.832891 | 0 | 0 | 0.800816 | 0.268482 |
| Gene_Pert | Ppt1 KO m | 1/288 | 0.717661 | 0.832891 | 0 | 0 | 0.795154 | 0.263799 |
| Gene_Pert | Srf homozy | 1/288 | 0.717661 | 0.832891 | 0 | 0 | 0.795154 | 0.263799 |
| Gene_Pert | Atp7b KO r | 1/284 | 0.712621 | 0.832891 | 0 | 0 | 0.806558 | 0.273266 |
| Gene_Pert | MYC KD hu | 1/284 | 0.712621 | 0.832891 | 0 | 0 | 0.806558 | 0.273266 |
| Gene_Pert | PTK7 KD hu | 1/286 | 0.715152 | 0.832891 | 0 | 0 | 0.800816 | 0.268482 |
| Gene_Pert | Gata6 KO r | 1/286 | 0.715152 | 0.832891 | 0 | 0 | 0.800816 | 0.268482 |
| Gene_Pert | Trpm7 KO | 1/284 | 0.712621 | 0.832891 | 0 | 0 | 0.806558 | 0.273266 |
| Gene_Pert | EZH2 DEPL | 1/281 | 0.708784 | 0.832891 | 0 | 0 | 0.815324 | 0.280639 |
| Gene_Pert | IRF7 KD hu | 1/283 | 0.711348 | 0.832891 | 0 | 0 | 0.809459 | 0.275697 |
| Gene_Pert | Xrcc5 KO m | 1/283 | 0.711348 | 0.832891 | 0 | 0 | 0.809459 | 0.275697 |
| Gene_Pert | Nr4a2 OE r | 1/283 | 0.711348 | 0.832891 | 0 | 0 | 0.809459 | 0.275697 |
| Gene_Pert | Gata1 Delt | 1/283 | 0.711348 | 0.832891 | 0 | 0 | 0.809459 | 0.275697 |
| Gene_Pert | FTO OE hu | 1/285 | 0.713889 | 0.832891 | 0 | 0 | 0.803677 | 0.270861 |
| Gene_Pert | Pten KO m | 1/284 | 0.712621 | 0.832891 | 0 | 0 | 0.806558 | 0.273266 |
| Gene_Pert | Bdkrb1 KO | 1/291 | 0.721383 | 0.832891 | 0 | 0 | 0.786808 | 0.25696 |
| Gene_Pert | Myc KO mo | 1/291 | 0.721383 | 0.832891 | 0 | 0 | 0.786808 | 0.25696 |
| Gene_Pert | Ovol1 KO n | 1/291 | 0.721383 | 0.832891 | 0 | 0 | 0.786808 | 0.25696 |
| Gene_Pert | Nr1i3 hNR1 | 1/290 | 0.720147 | 0.832891 | 0 | 0 | 0.789571 | 0.259216 |
| Gene_Pert | Ret KO mo | 1/290 | 0.720147 | 0.832891 | 0 | 0 | 0.789571 | 0.259216 |
| Gene_Pert | TERC KD hu | 1/290 | 0.720147 | 0.832891 | 0 | 0 | 0.789571 | 0.259216 |
| Gene_Pert | Cldn16 Def | 1/279 | 0.706197 | 0.832891 | 0 | 0 | 0.821273 | 0.285689 |
| Gene_Pert | Trim33 knc | 1/279 | 0.706197 | 0.832891 | 0 | 0 | 0.821273 | 0.285689 |
| Gene_Pert | Pparg Defic | 1/278 | 0.704895 | 0.832891 | 0 | 0 | 0.82428 | 0.288256 |
| Gene_Pert | RB1 KD hu | 1/281 | 0.708784 | 0.832891 | 0 | 0 | 0.815324 | 0.280639 |
| Gene_Pert | Ppargc1b H | 1/281 | 0.708784 | 0.832891 | 0 | 0 | 0.815324 | 0.280639 |
| Gene_Pert | Ddit3 knoc | 1/280 | 0.707493 | 0.832891 | 0 | 0 | 0.818288 | 0.28315 |
| Gene_Pert | Dusp1 Defi | 1/280 | 0.707493 | 0.832891 | 0 | 0 | 0.818288 | 0.28315 |
| Gene_Pert | CIITA KD hu | 1/280 | 0.707493 | 0.832891 | 0 | 0 | 0.818288 | 0.28315 |
| Gene_Pert | RAF1 active | 1/280 | 0.707493 | 0.832891 | 0 | 0 | 0.818288 | 0.28315 |
| Gene_Pert | Dicer1 KO | 1/276 | 0.702274 | 0.832891 | 0 | 0 | 0.830359 | 0.293475 |
| Gene_Pert | HDAC1 KD | 1/276 | 0.702274 | 0.832891 | 0 | 0 | 0.830359 | 0.293475 |
| Gene_Pert | Tgfbr2 kno | 1/280 | 0.707493 | 0.832891 | 0 | 0 | 0.818288 | 0.28315 |
| Gene_Pert | Gm10480 I | 1/279 | 0.706197 | 0.832891 | 0 | 0 | 0.821273 | 0.285689 |
| Gene_Pert | Bach2 KO r | 1/211 | 0.603359 | 0.832891 | 0 | 0 | 1.090975 | 0.551207 |
| Gene_Pert | SYK druginl | 1/279 | 0.706197 | 0.832891 | 0 | 0 | 0.821273 | 0.285689 |

| | | | | | | | | |
|---|---|---|---|---|---|---|---|---|
| Gene_Pert | Tmem173 | 1/279 | 0.706197 | 0.832891 | 0 | 0 | 0.821273 | 0.285689 |
| Gene_Pert | EGFR druga | 1/279 | 0.706197 | 0.832891 | 0 | 0 | 0.821273 | 0.285689 |
| Gene_Pert | Tgfbr2 kno | 1/274 | 0.699631 | 0.832891 | 0 | 0 | 0.836528 | 0.29881 |
| Gene_Pert | LLGL1 KD h | 1/274 | 0.699631 | 0.832891 | 0 | 0 | 0.836528 | 0.29881 |
| Gene_Pert | Aire KO mc | 1/278 | 0.704895 | 0.832891 | 0 | 0 | 0.82428 | 0.288256 |
| Gene_Pert | GTF2B KD I | 1/278 | 0.704895 | 0.832891 | 0 | 0 | 0.82428 | 0.288256 |
| Gene_Pert | Psap hypor | 1/277 | 0.703588 | 0.832891 | 0 | 0 | 0.827309 | 0.290851 |
| Gene_Pert | NCSTN KD | 1/277 | 0.703588 | 0.832891 | 0 | 0 | 0.827309 | 0.290851 |
| Gene_Pert | Nfkbiz KO r | 1/277 | 0.703588 | 0.832891 | 0 | 0 | 0.827309 | 0.290851 |
| Gene_Pert | SON KD hu | 1/276 | 0.702274 | 0.832891 | 0 | 0 | 0.830359 | 0.293475 |
| Gene_Pert | Hivep2 KO | 3/262 | 0.106125 | 0.832891 | 0 | 0 | 2.710149 | 6.079245 |
| Gene_Pert | ERBB3 drug | 3/300 | 0.142458 | 0.832891 | 0 | 0 | 2.358826 | 4.596658 |
| Gene_Pert | GAPDH KD | 3/300 | 0.142458 | 0.832891 | 0 | 0 | 2.358826 | 4.596658 |
| Gene_Pert | Scd1 Null n | 3/300 | 0.142458 | 0.832891 | 0 | 0 | 2.358826 | 4.596658 |
| Gene_Pert | Dicer1 KO r | 2/152 | 0.141732 | 0.832891 | 0 | 0 | 3.100078 | 6.056983 |
| Gene_Pert | CYR61 dox | 2/152 | 0.141732 | 0.832891 | 0 | 0 | 3.100078 | 6.056983 |
| Gene_Pert | LOXL2 DEP | 3/299 | 0.141453 | 0.832891 | 0 | 0 | 2.366916 | 4.629183 |
| Gene_Pert | CHEK1 KD I | 4/408 | 0.102259 | 0.832891 | 0 | 0 | 2.32721 | 5.30662 |
| Gene_Pert | Atxn1 Knoc | 3/265 | 0.108844 | 0.832891 | 0 | 0 | 2.678708 | 5.940942 |
| Gene_Pert | CMAHP KO | 3/265 | 0.108844 | 0.832891 | 0 | 0 | 2.678708 | 5.940942 |
| Gene_Pert | ATM KD hu | 4/417 | 0.108494 | 0.832891 | 0 | 0 | 2.275446 | 5.053909 |
| Gene_Pert | TOX3 OE hu | 3/263 | 0.107028 | 0.832891 | 0 | 0 | 2.699588 | 6.032672 |
| Gene_Pert | Hras OE mc | 3/262 | 0.106125 | 0.832891 | 0 | 0 | 2.710149 | 6.079245 |
| Gene_Pert | Calr KO mc | 3/300 | 0.142458 | 0.832891 | 0 | 0 | 2.358826 | 4.596658 |
| Gene_Pert | Cstb KO mc | 4/413 | 0.105701 | 0.832891 | 0 | 0 | 2.298171 | 5.164304 |
| Gene_Pert | Ilk activem | 3/252 | 0.097262 | 0.832891 | 0 | 0 | 2.820425 | 6.572577 |
| Gene_Pert | STK19 OE h | 3/261 | 0.105224 | 0.832891 | 0 | 0 | 2.720792 | 6.1263 |
| Gene_Pert | LMNA HGP | 3/261 | 0.105224 | 0.832891 | 0 | 0 | 2.720792 | 6.1263 |
| Gene_Pert | ATM KD hu | 4/412 | 0.105009 | 0.832891 | 0 | 0 | 2.303922 | 5.192378 |
| Gene_Pert | Hey2 KO m | 3/260 | 0.104327 | 0.832891 | 0 | 0 | 2.731518 | 6.173843 |
| Gene_Pert | Mmp7 KO r | 4/411 | 0.104318 | 0.832891 | 0 | 0 | 2.309701 | 5.220644 |
| Gene_Pert | Hdac1 Drug | 3/258 | 0.102542 | 0.832891 | 0 | 0 | 2.753221 | 6.27042 |
| Gene_Pert | Klf15 Defic | 3/258 | 0.102542 | 0.832891 | 0 | 0 | 2.753221 | 6.27042 |
| Gene_Pert | CDK9 knoc | 3/287 | 0.129587 | 0.832891 | 0 | 0 | 2.468436 | 5.04401 |
| Gene_Pert | Emd KO mc | 3/293 | 0.135474 | 0.832891 | 0 | 0 | 2.416626 | 4.830773 |
| Gene_Pert | CMAHP KO | 3/293 | 0.135474 | 0.832891 | 0 | 0 | 2.416626 | 4.830773 |
| Gene_Pert | NTRK3 drug | 3/279 | 0.121886 | 0.832891 | 0 | 0 | 2.54102 | 5.348 |
| Gene_Pert | Esr2 KO mc | 4/433 | 0.119993 | 0.832891 | 0 | 0 | 2.188783 | 4.64093 |
| Gene_Pert | YY1AP1 OE | 3/286 | 0.128615 | 0.832891 | 0 | 0 | 2.477284 | 5.080743 |
| Gene_Pert | Srf KO mou | 2/142 | 0.127022 | 0.832891 | 0 | 0 | 3.323193 | 6.857052 |
| Gene_Pert | Cap1 OE m | 3/284 | 0.126679 | 0.832891 | 0 | 0 | 2.49517 | 5.155272 |
| Gene_Pert | Esrrg Defic | 3/284 | 0.126679 | 0.832891 | 0 | 0 | 2.49517 | 5.155272 |
| Gene_Pert | Runx1 KO r | 3/283 | 0.125715 | 0.832891 | 0 | 0 | 2.504209 | 5.193076 |
| Gene_Pert | PRLH OE hu | 4/440 | 0.125185 | 0.832891 | 0 | 0 | 2.152868 | 4.473582 |
| Gene_Pert | Dmd Defici | 3/282 | 0.124754 | 0.832891 | 0 | 0 | 2.513313 | 5.231246 |

| | | | | | | | | |
|---|---|---|---|---|---|---|---|---|
| Gene_Pert | S1pr3 KO n | 3/275 | 0.118102 | 0.832891 | 0 | 0 | 2.578913 | 5.509085 |
| Gene_Pert | Fshr KO mc | 3/282 | 0.124754 | 0.832891 | 0 | 0 | 2.513313 | 5.231246 |
| Gene_Pert | EWSR1 KD | 3/280 | 0.122839 | 0.832891 | 0 | 0 | 2.531717 | 5.308703 |
| Gene_Pert | Pitx2 KO m | 2/139 | 0.122685 | 0.832891 | 0 | 0 | 3.396479 | 7.126265 |
| Gene_Pert | MIR26B OE | 3/292 | 0.134486 | 0.832891 | 0 | 0 | 2.425111 | 4.86548 |
| Gene_Pert | Nfe2l2 KO | 3/279 | 0.121886 | 0.832891 | 0 | 0 | 2.54102 | 5.348 |
| Gene_Pert | CCND1 KD | 3/278 | 0.120936 | 0.832891 | 0 | 0 | 2.55039 | 5.387681 |
| Gene_Pert | THRB RCCC | 3/278 | 0.120936 | 0.832891 | 0 | 0 | 2.55039 | 5.387681 |
| Gene_Pert | Slc18a3 KD | 3/299 | 0.141453 | 0.832891 | 0 | 0 | 2.366916 | 4.629183 |
| Gene_Pert | Insm1 Defi | 3/303 | 0.145488 | 0.832891 | 0 | 0 | 2.334881 | 4.500855 |
| Gene_Pert | BRAF Drug | 3/301 | 0.143466 | 0.832891 | 0 | 0 | 2.350791 | 4.564431 |
| Gene_Pert | Pgr KO mo | 3/301 | 0.143466 | 0.832891 | 0 | 0 | 2.350791 | 4.564431 |
| Gene_Pert | TGM2 KD h | 3/301 | 0.143466 | 0.832891 | 0 | 0 | 2.350791 | 4.564431 |
| Gene_Pert | Dmd Defici | 3/301 | 0.143466 | 0.832891 | 0 | 0 | 2.350791 | 4.564431 |
| Gene_Pert | Ogt KD mo | 3/301 | 0.143466 | 0.832891 | 0 | 0 | 2.350791 | 4.564431 |
| Gene_Pert | Cbfb KO m | 2/153 | 0.143223 | 0.832891 | 0 | 0 | 3.079392 | 5.984343 |
| Gene_Pert | Cstb KO mc | 4/458 | 0.138967 | 0.832891 | 0 | 0 | 2.065602 | 4.076509 |
| Gene_Pert | HSF1 KD hu | 4/406 | 0.100897 | 0.832891 | 0 | 0 | 2.339028 | 5.364934 |
| Gene_Pert | Igf1r drugir | 4/428 | 0.116343 | 0.832891 | 0 | 0 | 2.215163 | 4.765278 |
| Gene_Pert | Hdac6 KO r | 3/273 | 0.116227 | 0.832891 | 0 | 0 | 2.59828 | 5.592039 |
| Gene_Pert | HSF1 KD hu | 4/427 | 0.11562 | 0.832891 | 0 | 0 | 2.220513 | 4.790646 |
| Gene_Pert | TP53BP1 K | 4/426 | 0.114898 | 0.832891 | 0 | 0 | 2.225889 | 4.816185 |
| Gene_Pert | App KO mc | 3/269 | 0.112512 | 0.832891 | 0 | 0 | 2.637889 | 5.762983 |
| Gene_Pert | CHEK2 KD l | 4/421 | 0.111319 | 0.832891 | 0 | 0 | 2.253157 | 4.946473 |
| Gene_Pert | Ret mutant | 3/267 | 0.110672 | 0.832891 | 0 | 0 | 2.658144 | 5.851062 |
| Gene_Pert | Setd2 Defic | 3/253 | 0.098134 | 0.832891 | 0 | 0 | 2.809 | 6.520881 |
| Gene_Pert | Runx1 KO r | 3/252 | 0.097262 | 0.832891 | 0 | 0 | 2.820425 | 6.572577 |
| Gene_Pert | Mfi2 KO m | 4/395 | 0.09356 | 0.832891 | 0 | 0 | 2.406187 | 5.700628 |
| Gene_Pert | PARK7 KD l | 4/394 | 0.092906 | 0.832891 | 0 | 0 | 2.412481 | 5.732456 |
| Gene_Pert | Ppargc1a D | 3/246 | 0.092098 | 0.832891 | 0 | 0 | 2.890947 | 6.894635 |
| Gene_Pert | Hras mous | 3/261 | 0.105224 | 0.832891 | 0 | 0 | 2.720792 | 6.1263 |
| Gene_Pert | Pink1 KO m | 3/256 | 0.100769 | 0.832891 | 0 | 0 | 2.775268 | 6.369029 |
| Gene_Pert | St6gal1 De | 3/255 | 0.099887 | 0.832891 | 0 | 0 | 2.786423 | 6.419114 |
| Gene_Pert | Acvr1 KO n | 3/255 | 0.099887 | 0.832891 | 0 | 0 | 2.786423 | 6.419114 |
| Gene_Pert | Lhb OE mo | 4/403 | 0.098869 | 0.832891 | 0 | 0 | 2.356977 | 5.453938 |
| Gene_Pert | Vhl KO mo | 2/386 | 0.502763 | 0.832891 | 0 | 0 | 1.19663 | 0.822846 |
| Gene_Pert | Nsun2 KO r | 2/352 | 0.454497 | 0.832891 | 0 | 0 | 1.31516 | 1.037088 |
| Gene_Pert | IRF9 OE hu | 2/352 | 0.454497 | 0.832891 | 0 | 0 | 1.31516 | 1.037088 |
| Gene_Pert | Tcof1 KD m | 2/352 | 0.454497 | 0.832891 | 0 | 0 | 1.31516 | 1.037088 |
| Gene_Pert | PFKL OE hu | 2/356 | 0.460312 | 0.832891 | 0 | 0 | 1.300033 | 1.008633 |
| Gene_Pert | Kdm2b KD | 2/355 | 0.458861 | 0.832891 | 0 | 0 | 1.303783 | 1.015656 |
| Gene_Pert | BST2 OE hu | 2/354 | 0.457409 | 0.832891 | 0 | 0 | 1.307553 | 1.02274 |
| Gene_Pert | FOXP3 FOX | 2/354 | 0.457409 | 0.832891 | 0 | 0 | 1.307553 | 1.02274 |
| Gene_Pert | BST2 OE hu | 4/400 | 0.096862 | 0.832891 | 0 | 0 | 2.375198 | 5.544825 |
| Gene_Pert | Nfe2l2 kno | 2/147 | 0.134331 | 0.832891 | 0 | 0 | 3.207789 | 6.43948 |

| | | | | | | | | |
|---|---|---|---|---|---|---|---|---|
| Gene_Pert | Cstb KO mc | 4/451 | 0.133535 | 0.832891 | 0 | 0 | 2.098704 | 4.225511 |
| Gene_Pert | THRA RCCC | 3/291 | 0.133501 | 0.832891 | 0 | 0 | 2.433656 | 4.900515 |
| Gene_Pert | Pde10a dru | 3/291 | 0.133501 | 0.832891 | 0 | 0 | 2.433656 | 4.900515 |
| Gene_Pert | Met knock | 3/282 | 0.124754 | 0.832891 | 0 | 0 | 2.513313 | 5.231246 |
| Gene_Pert | Dysf Defici | 3/291 | 0.133501 | 0.832891 | 0 | 0 | 2.433656 | 4.900515 |
| Gene_Pert | Cdk4 Heter | 3/290 | 0.132519 | 0.832891 | 0 | 0 | 2.44226 | 4.935882 |
| Gene_Pert | Ebf1 OE mc | 3/290 | 0.132519 | 0.832891 | 0 | 0 | 2.44226 | 4.935882 |
| Gene_Pert | Acvrl1 mo | 3/289 | 0.131539 | 0.832891 | 0 | 0 | 2.450924 | 4.971583 |
| Gene_Pert | GOLGA8A ( | 3/289 | 0.131539 | 0.832891 | 0 | 0 | 2.450924 | 4.971583 |
| Gene_Pert | B4galnt2 O | 3/289 | 0.131539 | 0.832891 | 0 | 0 | 2.450924 | 4.971583 |
| Gene_Pert | Appl2 KO n | 3/289 | 0.131539 | 0.832891 | 0 | 0 | 2.450924 | 4.971583 |
| Gene_Pert | NANOG KD | 3/311 | 0.153672 | 0.832891 | 0 | 0 | 2.273307 | 4.257756 |
| Gene_Pert | EGFR drugi | 3/251 | 0.096393 | 0.832891 | 0 | 0 | 2.831941 | 6.624826 |
| Gene_Pert | Rock1 INHI | 4/398 | 0.095534 | 0.832891 | 0 | 0 | 2.387499 | 5.606488 |
| Gene_Pert | Dmd Defici | 4/397 | 0.094874 | 0.832891 | 0 | 0 | 2.393697 | 5.637647 |
| Gene_Pert | TP53BP1 K | 4/396 | 0.094216 | 0.832891 | 0 | 0 | 2.399926 | 5.669026 |
| Gene_Pert | Pde6b RD1 | 4/419 | 0.109902 | 0.832891 | 0 | 0 | 2.264247 | 4.999827 |
| Gene_Pert | Ackr3 Defic | 3/277 | 0.119989 | 0.832891 | 0 | 0 | 2.559828 | 5.427752 |
| Gene_Pert | Ywhae KO | 3/277 | 0.119989 | 0.832891 | 0 | 0 | 2.559828 | 5.427752 |
| Gene_Pert | Rbm15 KO | 3/277 | 0.119989 | 0.832891 | 0 | 0 | 2.559828 | 5.427752 |
| Gene_Pert | HNF4A KD | 3/276 | 0.119044 | 0.832891 | 0 | 0 | 2.569335 | 5.468219 |
| Gene_Pert | CMAHP KO | 3/276 | 0.119044 | 0.832891 | 0 | 0 | 2.569335 | 5.468219 |
| Gene_Pert | CHEK1 KD I | 3/276 | 0.119044 | 0.832891 | 0 | 0 | 2.569335 | 5.468219 |
| Gene_Pert | Dmd Defici | 3/275 | 0.118102 | 0.832891 | 0 | 0 | 2.578913 | 5.509085 |
| Gene_Pert | Pafah1b1 K | 3/342 | 0.186664 | 0.832891 | 0 | 0 | 2.062158 | 3.461222 |
| Gene_Pert | Clpp KO mc | 3/348 | 0.193257 | 0.832891 | 0 | 0 | 2.025673 | 3.329673 |
| Gene_Pert | Klf5 KO mo | 3/347 | 0.192154 | 0.832891 | 0 | 0 | 2.031665 | 3.351151 |
| Gene_Pert | Hdc KO mo | 3/347 | 0.192154 | 0.832891 | 0 | 0 | 2.031665 | 3.351151 |
| Gene_Pert | Clock muta | 3/346 | 0.191052 | 0.832891 | 0 | 0 | 2.037693 | 3.372806 |
| Gene_Pert | Nfia Deficie | 3/345 | 0.189953 | 0.832891 | 0 | 0 | 2.043755 | 3.394639 |
| Gene_Pert | Des KO mo | 3/337 | 0.181217 | 0.832891 | 0 | 0 | 2.093563 | 3.575931 |
| Gene_Pert | Lmna KO m | 3/344 | 0.188855 | 0.832891 | 0 | 0 | 2.049853 | 3.416651 |
| Gene_Pert | Prkag3 Mu | 3/343 | 0.187758 | 0.832891 | 0 | 0 | 2.055987 | 3.438845 |
| Gene_Pert | Foxo1 KO r | 3/342 | 0.186664 | 0.832891 | 0 | 0 | 2.062158 | 3.461222 |
| Gene_Pert | PARK2 KD I | 3/342 | 0.186664 | 0.832891 | 0 | 0 | 2.062158 | 3.461222 |
| Gene_Pert | Lin28a KD | 3/342 | 0.186664 | 0.832891 | 0 | 0 | 2.062158 | 3.461222 |
| Gene_Pert | SIN3A Siler | 3/342 | 0.186664 | 0.832891 | 0 | 0 | 2.062158 | 3.461222 |
| Gene_Pert | Cav1 KO m | 3/348 | 0.193257 | 0.832891 | 0 | 0 | 2.025673 | 3.329673 |
| Gene_Pert | PPARGC1A | 3/304 | 0.146503 | 0.832891 | 0 | 0 | 2.327005 | 4.469498 |
| Gene_Pert | INSR mutat | 3/310 | 0.152641 | 0.832891 | 0 | 0 | 2.280828 | 4.287197 |
| Gene_Pert | CTNNB1 KD | 3/310 | 0.152641 | 0.832891 | 0 | 0 | 2.280828 | 4.287197 |
| Gene_Pert | Msgn1 OE | 3/309 | 0.151612 | 0.832891 | 0 | 0 | 2.288399 | 4.316902 |
| Gene_Pert | NME2 Indu | 3/309 | 0.151612 | 0.832891 | 0 | 0 | 2.288399 | 4.316902 |
| Gene_Pert | PPARGC1A | 3/308 | 0.150586 | 0.832891 | 0 | 0 | 2.296019 | 4.346874 |
| Gene_Pert | Pparg OE n | 3/308 | 0.150586 | 0.832891 | 0 | 0 | 2.296019 | 4.346874 |

| | | | | | | | | |
|---|---|---|---|---|---|---|---|---|
| Gene_Pert | FGFR3 kno | 3/308 | 0.150586 | 0.832891 | 0 | 0 | 2.296019 | 4.346874 |
| Gene_Pert | Pink1 KO m | 3/300 | 0.142458 | 0.832891 | 0 | 0 | 2.358826 | 4.596658 |
| Gene_Pert | Sost KO mo | 3/307 | 0.149562 | 0.832891 | 0 | 0 | 2.303689 | 4.377116 |
| Gene_Pert | CHRNA9 KO | 2/157 | 0.14922 | 0.832891 | 0 | 0 | 2.999317 | 5.705706 |
| Gene_Pert | Erbb3 drug | 2/157 | 0.14922 | 0.832891 | 0 | 0 | 2.999317 | 5.705706 |
| Gene_Pert | Zxdc Deple | 3/324 | 0.167273 | 0.832891 | 0 | 0 | 2.179795 | 3.897752 |
| Gene_Pert | Mapk14 KD | 2/196 | 0.209833 | 0.832891 | 0 | 0 | 2.391631 | 3.734399 |
| Gene_Pert | DOT1L INH | 3/362 | 0.208863 | 0.832891 | 0 | 0 | 1.945285 | 3.046467 |
| Gene_Pert | MIR140 KO | 3/359 | 0.205494 | 0.832891 | 0 | 0 | 1.961978 | 3.104515 |
| Gene_Pert | Twist1 OE | 2/193 | 0.205067 | 0.832891 | 0 | 0 | 2.429566 | 3.84945 |
| Gene_Pert | ERBB2 KO | 3/358 | 0.204374 | 0.832891 | 0 | 0 | 1.967606 | 3.124174 |
| Gene_Pert | WNT1 OE h | 2/192 | 0.203481 | 0.832891 | 0 | 0 | 2.442477 | 3.888868 |
| Gene_Pert | TFAP2C KD | 3/357 | 0.203255 | 0.832891 | 0 | 0 | 1.973265 | 3.143989 |
| Gene_Pert | CITED1 KD | 3/319 | 0.161999 | 0.832891 | 0 | 0 | 2.214851 | 4.03139 |
| Gene_Pert | PARK7 KD | 3/326 | 0.169397 | 0.832891 | 0 | 0 | 2.166077 | 3.845893 |
| Gene_Pert | Tcf3 KO mo | 3/326 | 0.169397 | 0.832891 | 0 | 0 | 2.166077 | 3.845893 |
| Gene_Pert | Tmem100 | 3/326 | 0.169397 | 0.832891 | 0 | 0 | 2.166077 | 3.845893 |
| Gene_Pert | Cav3 Defici | 3/325 | 0.168334 | 0.832891 | 0 | 0 | 2.172915 | 3.871711 |
| Gene_Pert | Met knock | 3/306 | 0.14854 | 0.832891 | 0 | 0 | 2.31141 | 4.407632 |
| Gene_Pert | Cldn2 OE m | 4/492 | 0.166565 | 0.832891 | 0 | 0 | 1.918329 | 3.438356 |
| Gene_Pert | Dmd Defici | 3/323 | 0.166214 | 0.832891 | 0 | 0 | 2.186719 | 3.924018 |
| Gene_Pert | Prkca KO m | 3/315 | 0.157818 | 0.832891 | 0 | 0 | 2.243704 | 4.142577 |
| Gene_Pert | MS4A1 blo | 3/323 | 0.166214 | 0.832891 | 0 | 0 | 2.186719 | 3.924018 |
| Gene_Pert | POU5F1 KD | 3/322 | 0.165157 | 0.832891 | 0 | 0 | 2.193686 | 3.950513 |
| Gene_Pert | Dmd Defici | 3/322 | 0.165157 | 0.832891 | 0 | 0 | 2.193686 | 3.950513 |
| Gene_Pert | Dysf Defici | 3/322 | 0.165157 | 0.832891 | 0 | 0 | 2.193686 | 3.950513 |
| Gene_Pert | Pou4f1 KO | 2/167 | 0.164425 | 0.832891 | 0 | 0 | 2.816114 | 5.083934 |
| Gene_Pert | Bmpr1a KO | 3/321 | 0.164103 | 0.832891 | 0 | 0 | 2.200696 | 3.977238 |
| Gene_Pert | Myd88 KO | 3/319 | 0.161999 | 0.832891 | 0 | 0 | 2.214851 | 4.03139 |
| Gene_Pert | RB1 KD hu | 3/340 | 0.18448 | 0.832891 | 0 | 0 | 2.074608 | 3.506535 |
| Gene_Pert | Mycn OE m | 3/348 | 0.193257 | 0.832891 | 0 | 0 | 2.025673 | 3.329673 |
| Gene_Pert | CXCL8 OE h | 3/318 | 0.160951 | 0.832891 | 0 | 0 | 2.221995 | 4.058822 |
| Gene_Pert | ANGPT1 St | 3/298 | 0.14045 | 0.832891 | 0 | 0 | 2.375061 | 4.662008 |
| Gene_Pert | Hmx1 Defic | 3/297 | 0.13945 | 0.832891 | 0 | 0 | 2.38326 | 4.695137 |
| Gene_Pert | Ifnb1 Induc | 3/291 | 0.133501 | 0.832891 | 0 | 0 | 2.433656 | 4.900515 |
| Gene_Pert | STAT6 KD h | 3/296 | 0.138452 | 0.832891 | 0 | 0 | 2.391516 | 4.728574 |
| Gene_Pert | SOX2 OE hu | 3/296 | 0.138452 | 0.832891 | 0 | 0 | 2.391516 | 4.728574 |
| Gene_Pert | Rora KO mo | 4/457 | 0.138185 | 0.832891 | 0 | 0 | 2.070268 | 4.097392 |
| Gene_Pert | Satb2 OE m | 3/295 | 0.137457 | 0.832891 | 0 | 0 | 2.399829 | 4.762324 |
| Gene_Pert | Dicer1 KO | 2/149 | 0.13728 | 0.832891 | 0 | 0 | 3.163826 | 6.282501 |
| Gene_Pert | Cd47 KO m | 4/455 | 0.136628 | 0.832891 | 0 | 0 | 2.079662 | 4.139557 |
| Gene_Pert | MYB OE hu | 2/148 | 0.135804 | 0.832891 | 0 | 0 | 3.185657 | 6.360306 |
| Gene_Pert | Runx1 KO r | 3/311 | 0.153672 | 0.832891 | 0 | 0 | 2.273307 | 4.257756 |
| Gene_Pert | Scd1 KO mo | 3/319 | 0.161999 | 0.832891 | 0 | 0 | 2.214851 | 4.03139 |
| Gene_Pert | Mstn DEPL | 3/299 | 0.141453 | 0.832891 | 0 | 0 | 2.366916 | 4.629183 |

| | | | | | | | | |
|---|---|---|---|---|---|---|---|---|
| Gene_Pert | Nf1 mouse | 3/318 | 0.160951 | 0.832891 | 0 | 0 | 2.221995 | 4.058822 |
| Gene_Pert | PCGF2 KD I | 3/317 | 0.159904 | 0.832891 | 0 | 0 | 2.229186 | 4.086495 |
| Gene_Pert | Gfap OE m | 3/315 | 0.157818 | 0.832891 | 0 | 0 | 2.243704 | 4.142577 |
| Gene_Pert | IL6 anti-IL6 | 3/315 | 0.157818 | 0.832891 | 0 | 0 | 2.243704 | 4.142577 |
| Gene_Pert | Crebbp cpb | 3/315 | 0.157818 | 0.832891 | 0 | 0 | 2.243704 | 4.142577 |
| Gene_Pert | VDR OE hu | 3/308 | 0.150586 | 0.832891 | 0 | 0 | 2.296019 | 4.346874 |
| Gene_Pert | Mn1 reotrv | 3/315 | 0.157818 | 0.832891 | 0 | 0 | 2.243704 | 4.142577 |
| Gene_Pert | Hdac3 KO r | 3/314 | 0.156778 | 0.832891 | 0 | 0 | 2.251034 | 4.17099 |
| Gene_Pert | PDGFRA ac | 3/313 | 0.155741 | 0.832891 | 0 | 0 | 2.25841 | 4.199656 |
| Gene_Pert | IL6 anti-IL6 | 3/313 | 0.155741 | 0.832891 | 0 | 0 | 2.25841 | 4.199656 |
| Gene_Pert | Ikzf1 KO m | 2/161 | 0.155267 | 0.832891 | 0 | 0 | 2.92327 | 5.444911 |
| Gene_Pert | Dnmt1 KO | 2/161 | 0.155267 | 0.832891 | 0 | 0 | 2.92327 | 5.444911 |
| Gene_Pert | Pmp22 OE | 3/330 | 0.173668 | 0.832891 | 0 | 0 | 2.139144 | 3.744801 |
| Gene_Pert | Rara KO m | 2/156 | 0.147716 | 0.832891 | 0 | 0 | 3.018946 | 5.773631 |
| Gene_Pert | Hes6 OE m | 3/305 | 0.14752 | 0.832891 | 0 | 0 | 2.319182 | 4.438425 |
| Gene_Pert | GSK3B KD I | 3/305 | 0.14752 | 0.832891 | 0 | 0 | 2.319182 | 4.438425 |
| Gene_Pert | Pmp22 Tre | 3/327 | 0.170462 | 0.832891 | 0 | 0 | 2.159281 | 3.820295 |
| Gene_Pert | Pink1 KO m | 3/333 | 0.176892 | 0.832891 | 0 | 0 | 2.119372 | 3.671207 |
| Gene_Pert | Grhl2 KD m | 3/333 | 0.176892 | 0.832891 | 0 | 0 | 2.119372 | 3.671207 |
| Gene_Pert | NOTCH1 Re | 3/333 | 0.176892 | 0.832891 | 0 | 0 | 2.119372 | 3.671207 |
| Gene_Pert | Msgn1 OE | 3/333 | 0.176892 | 0.832891 | 0 | 0 | 2.119372 | 3.671207 |
| Gene_Pert | Dnajc7 KO | 2/175 | 0.176781 | 0.832891 | 0 | 0 | 2.684801 | 4.652344 |
| Gene_Pert | Insr KO mo | 3/332 | 0.175816 | 0.832891 | 0 | 0 | 2.125923 | 3.695531 |
| Gene_Pert | Pgr KO mo | 3/332 | 0.175816 | 0.832891 | 0 | 0 | 2.125923 | 3.695531 |
| Gene_Pert | CMAHP KO | 3/323 | 0.166214 | 0.832891 | 0 | 0 | 2.186719 | 3.924018 |
| Gene_Pert | PTEN KD hu | 2/354 | 0.457409 | 0.832891 | 0 | 0 | 1.307553 | 1.02274 |
| Gene_Pert | APP Induce | 3/330 | 0.173668 | 0.832891 | 0 | 0 | 2.139144 | 3.744801 |
| Gene_Pert | Snca KO m | 3/330 | 0.173668 | 0.832891 | 0 | 0 | 2.139144 | 3.744801 |
| Gene_Pert | Tnfaip6 KO | 3/329 | 0.172598 | 0.832891 | 0 | 0 | 2.145815 | 3.769752 |
| Gene_Pert | LAMP3 KD | 3/328 | 0.171529 | 0.832891 | 0 | 0 | 2.152527 | 3.794916 |
| Gene_Pert | EZR KD hur | 3/328 | 0.171529 | 0.832891 | 0 | 0 | 2.152527 | 3.794916 |
| Gene_Pert | Ikbkb Cons | 3/327 | 0.170462 | 0.832891 | 0 | 0 | 2.159281 | 3.820295 |
| Gene_Pert | TNKS INHIE | 3/293 | 0.135474 | 0.832891 | 0 | 0 | 2.416626 | 4.830773 |
| Gene_Pert | Atm knock | 3/299 | 0.141453 | 0.832891 | 0 | 0 | 2.366916 | 4.629183 |
| Gene_Pert | PTGS2 INH | 3/299 | 0.141453 | 0.832891 | 0 | 0 | 2.366916 | 4.629183 |
| Gene_Pert | SOX2 KD hu | 3/299 | 0.141453 | 0.832891 | 0 | 0 | 2.366916 | 4.629183 |
| Gene_Pert | Myc KD mc | 3/299 | 0.141453 | 0.832891 | 0 | 0 | 2.366916 | 4.629183 |
| Gene_Pert | Psap hypor | 4/341 | 0.061549 | 0.832891 | 0 | 0 | 2.799471 | 7.804726 |
| Gene_Pert | Lat2 KO mc | 4/353 | 0.068068 | 0.832891 | 0 | 0 | 2.701557 | 7.25977 |
| Gene_Pert | ATR KD hur | 4/352 | 0.067511 | 0.832891 | 0 | 0 | 2.709459 | 7.303248 |
| Gene_Pert | Clpp KO mc | 16-Jan | 0.067399 | 0.832891 | 0 | 0 | 15.42481 | 41.60265 |
| Gene_Pert | Hnf4a KO r | 4/351 | 0.066957 | 0.832891 | 0 | 0 | 2.717406 | 7.347065 |
| Gene_Pert | BRAF overe | 3/212 | 0.065188 | 0.832891 | 0 | 0 | 3.367054 | 9.193681 |
| Gene_Pert | STAG2 KO I | 3/210 | 0.063737 | 0.832891 | 0 | 0 | 3.399931 | 9.359987 |
| Gene_Pert | ATM KD hu | 4/345 | 0.063683 | 0.832891 | 0 | 0 | 2.766067 | 7.617307 |

| | | | | | | | | |
|---|---|---|---|---|---|---|---|---|
| Gene_Pert | Zbtb20 Def | 4/342 | 0.062078 | 0.832891 | 0 | 0 | 2.791046 | 7.75731 |
| Gene_Pert | Rb1 Deficie | 3/207 | 0.061589 | 0.832891 | 0 | 0 | 3.450455 | 9.617355 |
| Gene_Pert | Dnajc7 KO | 4/390 | 0.090313 | 0.832891 | 0 | 0 | 2.43798 | 5.862055 |
| Gene_Pert | Nanog OE | 4/388 | 0.08903 | 0.832891 | 0 | 0 | 2.450929 | 5.928259 |
| Gene_Pert | PFKL OE hu | 3/242 | 0.088721 | 0.832891 | 0 | 0 | 2.939928 | 7.121261 |
| Gene_Pert | Mecp2 KO | 4/353 | 0.068068 | 0.832891 | 0 | 0 | 2.701557 | 7.25977 |
| Gene_Pert | GATA3 Ect | 3/233 | 0.081323 | 0.832891 | 0 | 0 | 3.056366 | 7.669407 |
| Gene_Pert | Akt2 knock | 3/232 | 0.080519 | 0.832891 | 0 | 0 | 3.069869 | 7.733813 |
| Gene_Pert | Dmd Defici | 3/231 | 0.079718 | 0.832891 | 0 | 0 | 3.08349 | 7.798958 |
| Gene_Pert | Tnf siTNF-C | 2/197 | 0.211424 | 0.832891 | 0 | 0 | 2.379246 | 3.697085 |
| Gene_Pert | ARHGDIB K | 2/397 | 0.517793 | 0.832891 | 0 | 0 | 1.162651 | 0.765232 |
| Gene_Pert | Mybl1 KO | 2/396 | 0.516439 | 0.832891 | 0 | 0 | 1.165661 | 0.770266 |
| Gene_Pert | BAP1 KO h | 2/395 | 0.515083 | 0.832891 | 0 | 0 | 1.168687 | 0.77534 |
| Gene_Pert | Dicer1 KO | 2/408 | 0.532525 | 0.832891 | 0 | 0 | 1.130513 | 0.712364 |
| Gene_Pert | Dmd Defici | 2/408 | 0.532525 | 0.832891 | 0 | 0 | 1.130513 | 0.712364 |
| Gene_Pert | Pdpn NULL | 2/406 | 0.529869 | 0.832891 | 0 | 0 | 1.136226 | 0.721646 |
| Gene_Pert | Stk11 KO m | 1/172 | 0.529077 | 0.832891 | 0 | 0 | 1.342445 | 0.854628 |
| Gene_Pert | Ffar4 KO m | 1/171 | 0.527002 | 0.832891 | 0 | 0 | 1.35041 | 0.865006 |
| Gene_Pert | MTDH KD | 3/223 | 0.073438 | 0.832891 | 0 | 0 | 3.196916 | 8.348142 |
| Gene_Pert | Psap KO m | 3/239 | 0.086224 | 0.832891 | 0 | 0 | 2.977754 | 7.297891 |
| Gene_Pert | Htt Mutati | 4/371 | 0.078492 | 0.832891 | 0 | 0 | 2.566692 | 6.531619 |
| Gene_Pert | Htt CAG kn | 4/371 | 0.078492 | 0.832891 | 0 | 0 | 2.566692 | 6.531619 |
| Gene_Pert | MTDH KD | 4/370 | 0.077893 | 0.832891 | 0 | 0 | 2.573836 | 6.569522 |
| Gene_Pert | NPTX1 KD | 3/237 | 0.084577 | 0.832891 | 0 | 0 | 3.00351 | 7.418958 |
| Gene_Pert | Dmrta2 OE | 4/380 | 0.083988 | 0.832891 | 0 | 0 | 2.504102 | 6.202851 |
| Gene_Pert | Dmd Defici | 3/236 | 0.083758 | 0.832891 | 0 | 0 | 3.016554 | 7.480516 |
| Gene_Pert | Dlx5 OE m | 3/236 | 0.083758 | 0.832891 | 0 | 0 | 3.016554 | 7.480516 |
| Gene_Pert | LEF1 KD hu | 3/235 | 0.082943 | 0.832891 | 0 | 0 | 3.029711 | 7.542769 |
| Gene_Pert | Ppara OE n | 4/372 | 0.079093 | 0.832891 | 0 | 0 | 2.559586 | 6.493998 |
| Gene_Pert | Mapk8 KO | 3/230 | 0.07892 | 0.832891 | 0 | 0 | 3.097231 | 7.864854 |
| Gene_Pert | Sirt1 KO m | 4/364 | 0.074347 | 0.832891 | 0 | 0 | 2.617537 | 6.803013 |
| Gene_Pert | Lipe KO mc | 2/391 | 0.509632 | 0.832891 | 0 | 0 | 1.180947 | 0.796037 |
| Gene_Pert | Gys1 OE m | 4/361 | 0.072606 | 0.832891 | 0 | 0 | 2.639938 | 6.923791 |
| Gene_Pert | PIR KD hun | 4/359 | 0.071457 | 0.832891 | 0 | 0 | 2.655082 | 7.005859 |
| Gene_Pert | Rps6ka3 kr | 3/220 | 0.071143 | 0.832891 | 0 | 0 | 3.241606 | 8.567754 |
| Gene_Pert | Tnf siTNF-C | 3/220 | 0.071143 | 0.832891 | 0 | 0 | 3.241606 | 8.567754 |
| Gene_Pert | Mecp2 KD | 4/357 | 0.070318 | 0.832891 | 0 | 0 | 2.670398 | 7.089194 |
| Gene_Pert | Pitx2 KO m | 4/357 | 0.070318 | 0.832891 | 0 | 0 | 2.670398 | 7.089194 |
| Gene_Pert | Htt Mutati | 4/356 | 0.069752 | 0.832891 | 0 | 0 | 2.678122 | 7.131345 |
| Gene_Pert | Apln KO m | 3/218 | 0.069632 | 0.832891 | 0 | 0 | 3.272093 | 8.718593 |
| Gene_Pert | IRS4 KO mc | 3/217 | 0.068882 | 0.832891 | 0 | 0 | 3.28755 | 8.795386 |
| Gene_Pert | E2f4 KO m | 4/354 | 0.068627 | 0.832891 | 0 | 0 | 2.693701 | 7.21663 |
| Gene_Pert | Tardbp KD | 3/216 | 0.068136 | 0.832891 | 0 | 0 | 3.303152 | 8.873113 |
| Gene_Pert | Cdh1 KO m | 3/216 | 0.068136 | 0.832891 | 0 | 0 | 3.303152 | 8.873113 |
| Gene_Pert | Lmx1b KO | 2/415 | 0.541742 | 0.832891 | 0 | 0 | 1.110953 | 0.680975 |

| | | | | | | | | |
|---|---|---|---|---|---|---|---|---|
| Gene_Pert | Notch1 KO | 2/425 | 0.554694 | 0.832891 | 0 | 0 | 1.084133 | 0.638922 |
| Gene_Pert | GRN defici | 2/425 | 0.554694 | 0.832891 | 0 | 0 | 1.084133 | 0.638922 |
| Gene_Pert | Pde6b rd1 | 2/424 | 0.55341 | 0.832891 | 0 | 0 | 1.086758 | 0.642986 |
| Gene_Pert | Erbb3 knoc | 2/424 | 0.55341 | 0.832891 | 0 | 0 | 1.086758 | 0.642986 |
| Gene_Pert | Nfe2l2 KO | 2/423 | 0.552124 | 0.832891 | 0 | 0 | 1.089395 | 0.647082 |
| Gene_Pert | KLF4 OE hu | 1/183 | 0.55132 | 0.832891 | 0 | 0 | 1.260606 | 0.750615 |
| Gene_Pert | PARK2 KD | 3/335 | 0.179051 | 0.832891 | 0 | 0 | 2.10639 | 3.623169 |
| Gene_Pert | SOX2 KO h | 3/335 | 0.179051 | 0.832891 | 0 | 0 | 2.10639 | 3.623169 |
| Gene_Pert | Nf1 Hetero | 3/335 | 0.179051 | 0.832891 | 0 | 0 | 2.10639 | 3.623169 |
| Gene_Pert | Pde10a KO | 3/335 | 0.179051 | 0.832891 | 0 | 0 | 2.10639 | 3.623169 |
| Gene_Pert | Dhcr7 KO n | 2/443 | 0.577361 | 0.832891 | 0 | 0 | 1.038922 | 0.570668 |
| Gene_Pert | NOTCH1 IN | 1/196 | 0.576271 | 0.832891 | 0 | 0 | 1.17579 | 0.64807 |
| Gene_Pert | Irak4 IRAK4 | 2/426 | 0.555975 | 0.832891 | 0 | 0 | 1.081521 | 0.634887 |
| Gene_Pert | Dmd Defici | 1/178 | 0.541341 | 0.832891 | 0 | 0 | 1.296544 | 0.795696 |
| Gene_Pert | KDR KD hu | 3/336 | 0.180133 | 0.832891 | 0 | 0 | 2.099957 | 3.599451 |
| Gene_Pert | Hdac3 Inte | 3/336 | 0.180133 | 0.832891 | 0 | 0 | 2.099957 | 3.599451 |
| Gene_Pert | Rptor KO m | 3/337 | 0.181217 | 0.832891 | 0 | 0 | 2.093563 | 3.575931 |
| Gene_Pert | HIF1A KD h | 3/331 | 0.174741 | 0.832891 | 0 | 0 | 2.132513 | 3.720062 |
| Gene_Pert | Dicer1 KO | 3/338 | 0.182303 | 0.832891 | 0 | 0 | 2.087207 | 3.552606 |
| Gene_Pert | SIRT1 KD h | 3/338 | 0.182303 | 0.832891 | 0 | 0 | 2.087207 | 3.552606 |
| Gene_Pert | CCND1 KD | 3/338 | 0.182303 | 0.832891 | 0 | 0 | 2.087207 | 3.552606 |
| Gene_Pert | Srf homozy | 3/338 | 0.182303 | 0.832891 | 0 | 0 | 2.087207 | 3.552606 |
| Gene_Pert | Zbtb7b KO | 2/459 | 0.596805 | 0.832891 | 0 | 0 | 1.001725 | 0.517056 |
| Gene_Pert | IRF7 stable | 1/207 | 0.596308 | 0.832891 | 0 | 0 | 1.112384 | 0.575101 |
| Gene_Pert | Stat6 KO m | 1/211 | 0.603359 | 0.832891 | 0 | 0 | 1.090975 | 0.551207 |
| Gene_Pert | FTO KD hu | 3/366 | 0.213374 | 0.832891 | 0 | 0 | 1.923455 | 2.971175 |
| Gene_Pert | NCOA1 KO | 1/161 | 0.505746 | 0.832891 | 0 | 0 | 1.435538 | 0.978637 |
| Gene_Pert | RBBP8 DEP | 2/388 | 0.505518 | 0.832891 | 0 | 0 | 1.190308 | 0.811994 |
| Gene_Pert | Dmd Defici | 1/169 | 0.522824 | 0.832891 | 0 | 0 | 1.366625 | 0.886269 |
| Gene_Pert | Foxa2 KO n | 1/168 | 0.520722 | 0.832891 | 0 | 0 | 1.374878 | 0.897162 |
| Gene_Pert | Runx1 KO | 2/399 | 0.520494 | 0.832891 | 0 | 0 | 1.156675 | 0.755281 |
| Gene_Pert | Trim12a OI | 2/398 | 0.519145 | 0.832891 | 0 | 0 | 1.159655 | 0.760237 |
| Gene_Pert | Ahnak KO | 2/397 | 0.517793 | 0.832891 | 0 | 0 | 1.162651 | 0.765232 |
| Gene_Pert | Nrl Deficie | 2/428 | 0.55853 | 0.832891 | 0 | 0 | 1.076333 | 0.626907 |
| Gene_Pert | CDH11 KD | 2/197 | 0.211424 | 0.832891 | 0 | 0 | 2.379246 | 3.697085 |
| Gene_Pert | HMGA2 KD | 2/198 | 0.213017 | 0.832891 | 0 | 0 | 2.366987 | 3.660273 |
| Gene_Pert | TGM2 KD h | 3/366 | 0.213374 | 0.832891 | 0 | 0 | 1.923455 | 2.971175 |
| Gene_Pert | Nrl Deficie | 3/366 | 0.213374 | 0.832891 | 0 | 0 | 1.923455 | 2.971175 |
| Gene_Pert | YAP1 KD hu | 4/384 | 0.086491 | 0.832891 | 0 | 0 | 2.477235 | 6.063563 |
| Gene_Pert | Srf homozy | 3/369 | 0.216772 | 0.832891 | 0 | 0 | 1.907397 | 2.916234 |
| Gene_Pert | TFAP2C Sile | 3/356 | 0.202138 | 0.832891 | 0 | 0 | 1.978956 | 3.163964 |
| Gene_Pert | Nfe2l2 KO | 2/421 | 0.549544 | 0.832891 | 0 | 0 | 1.094707 | 0.655365 |
| Gene_Pert | IKBKB expr | 1/191 | 0.566841 | 0.832891 | 0 | 0 | 1.207038 | 0.685208 |
| Gene_Pert | Srf homozy | 1/190 | 0.56493 | 0.832891 | 0 | 0 | 1.213486 | 0.692965 |
| Gene_Pert | S1pr2 KO n | 2/432 | 0.563608 | 0.832891 | 0 | 0 | 1.066101 | 0.611299 |

| | | | | | | | | |
|---|---|---|---|---|---|---|---|---|
| Gene_Pert | MIR196A2 | 2/431 | 0.562342 | 0.832891 | 0 | 0 | 1.068641 | 0.615158 |
| Gene_Pert | Neurod1 D | 2/430 | 0.561074 | 0.832891 | 0 | 0 | 1.071193 | 0.619045 |
| Gene_Pert | GPLD1 OE | 2/429 | 0.559803 | 0.832891 | 0 | 0 | 1.073757 | 0.622962 |
| Gene_Pert | MIR20A KD | 1/193 | 0.570637 | 0.832891 | 0 | 0 | 1.194344 | 0.670029 |
| Gene_Pert | Rb1 Deficie | 1/178 | 0.541341 | 0.832891 | 0 | 0 | 1.296544 | 0.795696 |
| Gene_Pert | PAX3 KD hu | 2/361 | 0.46753 | 0.832891 | 0 | 0 | 1.281599 | 0.974388 |
| Gene_Pert | NCSTN KD | 2/360 | 0.466091 | 0.832891 | 0 | 0 | 1.285245 | 0.981122 |
| Gene_Pert | RRP1B OE | 2/366 | 0.474693 | 0.832891 | 0 | 0 | 1.263672 | 0.941545 |
| Gene_Pert | Htt Knock-i | 2/365 | 0.473265 | 0.832891 | 0 | 0 | 1.267218 | 0.948005 |
| Gene_Pert | Ghr trunca | 1/146 | 0.472079 | 0.832891 | 0 | 0 | 1.585245 | 1.1899 |
| Gene_Pert | CMAHP KO | 2/364 | 0.471835 | 0.832891 | 0 | 0 | 1.270783 | 0.954519 |
| Gene_Pert | NET1 KD hu | 2/364 | 0.471835 | 0.832891 | 0 | 0 | 1.270783 | 0.954519 |
| Gene_Pert | CTNNB1 KD | 2/358 | 0.463206 | 0.832891 | 0 | 0 | 1.292597 | 0.994762 |
| Gene_Pert | CMAHP KO | 2/358 | 0.463206 | 0.832891 | 0 | 0 | 1.292597 | 0.994762 |
| Gene_Pert | Nf1 KO mo | 1/142 | 0.462724 | 0.832891 | 0 | 0 | 1.630546 | 1.256538 |
| Gene_Pert | Fgf21 KO m | 1/145 | 0.469755 | 0.832891 | 0 | 0 | 1.596334 | 1.2061 |
| Gene_Pert | Rnh1 KO m | 2/362 | 0.468968 | 0.832891 | 0 | 0 | 1.277974 | 0.96771 |
| Gene_Pert | KMT2A ML | 2/361 | 0.46753 | 0.832891 | 0 | 0 | 1.281599 | 0.974388 |
| Gene_Pert | Nf1 Hetero | 2/360 | 0.466091 | 0.832891 | 0 | 0 | 1.285245 | 0.981122 |
| Gene_Pert | Dicer1 KO | 2/361 | 0.46753 | 0.832891 | 0 | 0 | 1.281599 | 0.974388 |
| Gene_Pert | TP63 DEPL | 2/385 | 0.501382 | 0.832891 | 0 | 0 | 1.199816 | 0.828337 |
| Gene_Pert | NOTCH1 Re | 2/385 | 0.501382 | 0.832891 | 0 | 0 | 1.199816 | 0.828337 |
| Gene_Pert | Myoc OE m | 2/385 | 0.501382 | 0.832891 | 0 | 0 | 1.199816 | 0.828337 |
| Gene_Pert | Dmd Defici | 2/395 | 0.515083 | 0.832891 | 0 | 0 | 1.168687 | 0.77534 |
| Gene_Pert | Bmpr1a KC | 1/165 | 0.514359 | 0.832891 | 0 | 0 | 1.400241 | 0.930927 |
| Gene_Pert | Psap hypor | 2/394 | 0.513724 | 0.832891 | 0 | 0 | 1.171729 | 0.780453 |
| Gene_Pert | Myd88 Def | 2/392 | 0.510998 | 0.832891 | 0 | 0 | 1.177858 | 0.790801 |
| Gene_Pert | Tmem87a | 2/392 | 0.510998 | 0.832891 | 0 | 0 | 1.177858 | 0.790801 |
| Gene_Pert | Pde6b RD1 | 2/381 | 0.495834 | 0.832891 | 0 | 0 | 1.212727 | 0.850744 |
| Gene_Pert | HSF1 KD hu | 2/381 | 0.495834 | 0.832891 | 0 | 0 | 1.212727 | 0.850744 |
| Gene_Pert | Cbfb Trunc | 1/156 | 0.494766 | 0.832891 | 0 | 0 | 1.482221 | 1.042994 |
| Gene_Pert | Ppargc1a K | 2/376 | 0.488846 | 0.832891 | 0 | 0 | 1.229254 | 0.879787 |
| Gene_Pert | IRF9 OE hu | 2/349 | 0.450113 | 0.832891 | 0 | 0 | 1.326733 | 1.059074 |
| Gene_Pert | Ndel1 KD n | 2/349 | 0.450113 | 0.832891 | 0 | 0 | 1.326733 | 1.059074 |
| Gene_Pert | Arx KO mo | 2/349 | 0.450113 | 0.832891 | 0 | 0 | 1.326733 | 1.059074 |
| Gene_Pert | SOX7 OE hu | 2/354 | 0.457409 | 0.832891 | 0 | 0 | 1.307553 | 1.02274 |
| Gene_Pert | Sox7 OE m | 2/353 | 0.455954 | 0.832891 | 0 | 0 | 1.311346 | 1.029883 |
| Gene_Pert | Hmgn1 KO | 2/353 | 0.455954 | 0.832891 | 0 | 0 | 1.311346 | 1.029883 |
| Gene_Pert | HMGA2 OE | 2/353 | 0.455954 | 0.832891 | 0 | 0 | 1.311346 | 1.029883 |
| Gene_Pert | TYK2 KD hu | 2/352 | 0.454497 | 0.832891 | 0 | 0 | 1.31516 | 1.037088 |
| Gene_Pert | Elavl1 KO n | 2/368 | 0.477542 | 0.832891 | 0 | 0 | 1.256638 | 0.928785 |
| Gene_Pert | Clpp KO mo | 2/367 | 0.476119 | 0.832891 | 0 | 0 | 1.260145 | 0.935139 |
| Gene_Pert | WEE1 drug | 2/366 | 0.474693 | 0.832891 | 0 | 0 | 1.263672 | 0.941545 |
| Gene_Pert | Mgea5 KO | 2/377 | 0.490248 | 0.832891 | 0 | 0 | 1.225914 | 0.873884 |
| Gene_Pert | Zfp42 KD m | 1/160 | 0.503569 | 0.832891 | 0 | 0 | 1.444639 | 0.991073 |

| | | | | | | | | |
|---|---|---|---|---|---|---|---|---|
| Gene_Pert | Itga7 OE m | 1/153 | 0.488063 | 0.832891 | 0 | 0 | 1.511704 | 1.084362 |
| Gene_Pert | Msgn1 OE | 2/375 | 0.487441 | 0.832891 | 0 | 0 | 1.232613 | 0.885738 |
| Gene_Pert | Otx2 KO m | 2/375 | 0.487441 | 0.832891 | 0 | 0 | 1.232613 | 0.885738 |
| Gene_Pert | Pten KO mo | 2/364 | 0.471835 | 0.832891 | 0 | 0 | 1.270783 | 0.954519 |
| Gene_Pert | Igf1 KO mo | 2/364 | 0.471835 | 0.832891 | 0 | 0 | 1.270783 | 0.954519 |
| Gene_Pert | Crhr2 KO m | 2/363 | 0.470402 | 0.832891 | 0 | 0 | 1.274369 | 0.961087 |
| Gene_Pert | FOXO3 KO | 2/372 | 0.483213 | 0.832891 | 0 | 0 | 1.242798 | 0.903885 |
| Gene_Pert | EPHB2 drug | 2/371 | 0.481798 | 0.832891 | 0 | 0 | 1.24623 | 0.910034 |
| Gene_Pert | Tfap2c KD | 2/370 | 0.480382 | 0.832891 | 0 | 0 | 1.24968 | 0.916233 |
| Gene_Pert | Gata3 OE n | 3/594 | 0.480046 | 0.832891 | 0 | 0 | 1.167634 | 0.856895 |
| Gene_Pert | Foxo1 KD n | 1/175 | 0.535249 | 0.832891 | 0 | 0 | 1.319099 | 0.824467 |
| Gene_Pert | Atf6b KO m | 2/361 | 0.46753 | 0.832891 | 0 | 0 | 1.281599 | 0.974388 |
| Gene_Pert | RB1 KD hur | 4/382 | 0.085235 | 0.832891 | 0 | 0 | 2.490597 | 6.1327 |
| Gene_Pert | Dmd Defici | 1/181 | 0.547354 | 0.832891 | 0 | 0 | 1.274742 | 0.768234 |
| Gene_Pert | Twist1 OE | 1/181 | 0.547354 | 0.832891 | 0 | 0 | 1.274742 | 0.768234 |
| Gene_Pert | Myc myc +, | 2/417 | 0.544353 | 0.832891 | 0 | 0 | 1.105485 | 0.672309 |
| Gene_Pert | Ezh2 Defici | 1/179 | 0.543355 | 0.832891 | 0 | 0 | 1.289195 | 0.7864 |
| Gene_Pert | Bcl11b KO | 1/171 | 0.527002 | 0.832891 | 0 | 0 | 1.35041 | 0.865006 |
| Gene_Pert | Frzb KO mo | 2/403 | 0.525866 | 0.832891 | 0 | 0 | 1.144902 | 0.735838 |
| Gene_Pert | UBB KO mo | 2/402 | 0.524527 | 0.832891 | 0 | 0 | 1.147824 | 0.740643 |
| Gene_Pert | MALAT1 OI | 1/177 | 0.53932 | 0.832891 | 0 | 0 | 1.303977 | 0.805137 |
| Gene_Pert | Sod1 G93A | 1/177 | 0.53932 | 0.832891 | 0 | 0 | 1.303977 | 0.805137 |
| Gene_Pert | PAX5 OE hu | 1/177 | 0.53932 | 0.832891 | 0 | 0 | 1.303977 | 0.805137 |
| Gene_Pert | COMMD5 ( | 2/411 | 0.536491 | 0.832891 | 0 | 0 | 1.122048 | 0.698706 |
| Gene_Pert | Lama2 Defi | 3/238 | 0.085399 | 0.832891 | 0 | 0 | 2.990578 | 7.358087 |
| Gene_Pert | Myc myc +, | 1/181 | 0.547354 | 0.832891 | 0 | 0 | 1.274742 | 0.768234 |
| Gene_Pert | Eed KO mo | 3/237 | 0.084577 | 0.832891 | 0 | 0 | 3.00351 | 7.418958 |
| Gene_Pert | Sirt3 KO m | 3/246 | 0.092098 | 0.832891 | 0 | 0 | 2.890947 | 6.894635 |
| Gene_Pert | ESR1 KD hu | 3/245 | 0.091249 | 0.832891 | 0 | 0 | 2.90304 | 6.950367 |
| Gene_Pert | Tnf siTNF n | 3/245 | 0.091249 | 0.832891 | 0 | 0 | 2.90304 | 6.950367 |
| Gene_Pert | Nfe2l2 KO | 4/391 | 0.090958 | 0.832891 | 0 | 0 | 2.431556 | 5.829307 |
| Gene_Pert | Foxo1 KO r | 3/244 | 0.090403 | 0.832891 | 0 | 0 | 2.915234 | 7.00671 |
| Gene_Pert | CHEK1 KD | 4/377 | 0.082135 | 0.832891 | 0 | 0 | 2.524629 | 6.310023 |
| Gene_Pert | Sod1 G93A | 2/109 | 0.0817 | 0.832891 | 0 | 0 | 4.35536 | 10.90889 |
| Gene_Pert | CHEK2 KD | 4/376 | 0.081522 | 0.832891 | 0 | 0 | 2.531546 | 6.346276 |
| Gene_Pert | Dmd Defici | 3/242 | 0.088721 | 0.832891 | 0 | 0 | 2.939928 | 7.121261 |
| Gene_Pert | Irs1 KO mo | 3/242 | 0.088721 | 0.832891 | 0 | 0 | 2.939928 | 7.121261 |
| Gene_Pert | DDIT4 Repr | 4/384 | 0.086491 | 0.832891 | 0 | 0 | 2.477235 | 6.063563 |
| Gene_Pert | Il17rc KO m | 2/381 | 0.495834 | 0.832891 | 0 | 0 | 1.212727 | 0.850744 |
| Gene_Pert | Dgcr8 KO n | 1/160 | 0.503569 | 0.832891 | 0 | 0 | 1.444639 | 0.991073 |
| Gene_Pert | FLI1 KD hur | 1/160 | 0.503569 | 0.832891 | 0 | 0 | 1.444639 | 0.991073 |
| Gene_Pert | CTNNB1 m | 1/160 | 0.503569 | 0.832891 | 0 | 0 | 1.444639 | 0.991073 |
| Gene_Pert | Nfe2l2 KO | 3/353 | 0.198796 | 0.832891 | 0 | 0 | 1.996224 | 3.224856 |
| Gene_Pert | Akt1 KO mo | 1/187 | 0.559148 | 0.832891 | 0 | 0 | 1.233246 | 0.716937 |
| Gene_Pert | Neurod1 C | 2/379 | 0.493046 | 0.832891 | 0 | 0 | 1.219285 | 0.862221 |

| | | | | | | | | |
|---|---|---|---|---|---|---|---|---|
| Gene_Pert | Stat1 KO m | 2/379 | 0.493046 | 0.832891 | 0 | 0 | 1.219285 | 0.862221 |
| Gene_Pert | Prmt6 OE r | 1/155 | 0.492541 | 0.832891 | 0 | 0 | 1.491921 | 1.056544 |
| Gene_Pert | EWSR1 KD | 1/158 | 0.499187 | 0.832891 | 0 | 0 | 1.463191 | 1.016589 |
| Gene_Pert | Pparg OE m | 2/383 | 0.498613 | 0.832891 | 0 | 0 | 1.206237 | 0.839451 |
| Gene_Pert | Ahr KO mo | 2/382 | 0.497225 | 0.832891 | 0 | 0 | 1.209474 | 0.845075 |
| Gene_Pert | EHMT2 KD | 2/382 | 0.497225 | 0.832891 | 0 | 0 | 1.209474 | 0.845075 |
| Gene_Pert | Insm1 KO r | 1/297 | 0.728682 | 0.832891 | 0 | 0 | 0.770624 | 0.243916 |
| Gene_Pert | TET3 DEPLI | 2/373 | 0.484624 | 0.832891 | 0 | 0 | 1.239385 | 0.897787 |
| Gene_Pert | ERG KD hu | 2/373 | 0.484624 | 0.832891 | 0 | 0 | 1.239385 | 0.897787 |
| Gene_Pert | NFKB1 Inac | 2/372 | 0.483213 | 0.832891 | 0 | 0 | 1.242798 | 0.903885 |
| Gene_Pert | Atxn7 OE n | 1/156 | 0.494766 | 0.832891 | 0 | 0 | 1.482221 | 1.042994 |
| Gene_Pert | Gata5 KO r | 2/380 | 0.494441 | 0.832891 | 0 | 0 | 1.215998 | 0.85646 |
| Gene_Pert | Kdm6a KD | 2/380 | 0.494441 | 0.832891 | 0 | 0 | 1.215998 | 0.85646 |
| Gene_Pert | Rock1 INHI | 2/379 | 0.493046 | 0.832891 | 0 | 0 | 1.219285 | 0.862221 |
| Gene_Pert | Gata5 KO r | 2/379 | 0.493046 | 0.832891 | 0 | 0 | 1.219285 | 0.862221 |
| Gene_Pert | Src overexp | 2/410 | 0.535171 | 0.832891 | 0 | 0 | 1.124856 | 0.703224 |
| Gene_Pert | Dmd Defici | 2/409 | 0.53385 | 0.832891 | 0 | 0 | 1.127677 | 0.707777 |
| Gene_Pert | BST2 OE hu | 2/408 | 0.532525 | 0.832891 | 0 | 0 | 1.130513 | 0.712364 |
| Gene_Pert | Stat3 defici | 2/278 | 0.341411 | 0.832891 | 0 | 0 | 1.674084 | 1.799086 |
| Gene_Pert | S1pr3 KO n | 2/288 | 0.357207 | 0.832891 | 0 | 0 | 1.614726 | 1.662266 |
| Gene_Pert | ZNF148 KD | 2/292 | 0.363489 | 0.832891 | 0 | 0 | 1.59213 | 1.611246 |
| Gene_Pert | E2f3 KD mc | 2/293 | 0.365056 | 0.832891 | 0 | 0 | 1.586578 | 1.598802 |
| Gene_Pert | Ctcf DEPLE | 2/293 | 0.365056 | 0.832891 | 0 | 0 | 1.586578 | 1.598802 |
| Gene_Pert | Akt1 KO m | 2/293 | 0.365056 | 0.832891 | 0 | 0 | 1.586578 | 1.598802 |
| Gene_Pert | Atxn1 Knoc | 2/293 | 0.365056 | 0.832891 | 0 | 0 | 1.586578 | 1.598802 |
| Gene_Pert | Rock1 INHI | 3/490 | 0.359359 | 0.832891 | 0 | 0 | 1.424611 | 1.457995 |
| Gene_Pert | ATM knock | 2/290 | 0.36035 | 0.832891 | 0 | 0 | 1.60335 | 1.636504 |
| Gene_Pert | Cdkn1b KO | 2/290 | 0.36035 | 0.832891 | 0 | 0 | 1.60335 | 1.636504 |
| Gene_Pert | Tfap2c KO | 2/277 | 0.339825 | 0.832891 | 0 | 0 | 1.680257 | 1.813544 |
| Gene_Pert | Gata6 KO r | 2/277 | 0.339825 | 0.832891 | 0 | 0 | 1.680257 | 1.813544 |
| Gene_Pert | Bnc1 KD m | 2/277 | 0.339825 | 0.832891 | 0 | 0 | 1.680257 | 1.813544 |
| Gene_Pert | PPARGC1A | 2/292 | 0.363489 | 0.832891 | 0 | 0 | 1.59213 | 1.611246 |
| Gene_Pert | DIP2B KD h | 2/279 | 0.342996 | 0.832891 | 0 | 0 | 1.667955 | 1.784774 |
| Gene_Pert | CMAHP KO | 2/279 | 0.342996 | 0.832891 | 0 | 0 | 1.667955 | 1.784774 |
| Gene_Pert | Fhl2 Defici | 2/279 | 0.342996 | 0.832891 | 0 | 0 | 1.667955 | 1.784774 |
| Gene_Pert | SYNCRIP OI | 3/469 | 0.334315 | 0.832891 | 0 | 0 | 1.49042 | 1.633009 |
| Gene_Pert | SYK druginl | 2/279 | 0.342996 | 0.832891 | 0 | 0 | 1.667955 | 1.784774 |
| Gene_Pert | Klf3 KO mo | 2/280 | 0.34458 | 0.832891 | 0 | 0 | 1.661871 | 1.770607 |
| Gene_Pert | TFAP2C KD | 2/280 | 0.34458 | 0.832891 | 0 | 0 | 1.661871 | 1.770607 |
| Gene_Pert | Dicer1 KO | 2/280 | 0.34458 | 0.832891 | 0 | 0 | 1.661871 | 1.770607 |
| Gene_Pert | PARK7 KD l | 2/281 | 0.346162 | 0.832891 | 0 | 0 | 1.65583 | 1.756583 |
| Gene_Pert | Aire KO mc | 2/281 | 0.346162 | 0.832891 | 0 | 0 | 1.65583 | 1.756583 |
| Gene_Pert | Rap1a Rap: | 2/281 | 0.346162 | 0.832891 | 0 | 0 | 1.65583 | 1.756583 |
| Gene_Pert | Trim24 knc | 2/275 | 0.336649 | 0.832891 | 0 | 0 | 1.692739 | 1.842909 |
| Gene_Pert | Lmna LMN. | 2/265 | 0.32071 | 0.832891 | 0 | 0 | 1.757996 | 1.999226 |

| | | | | | | | | |
|---|---|---|---|---|---|---|---|---|
| Gene_Pert | TGFB1I1 KD | 3/484 | 0.352211 | 0.832891 | 0 | 0 | 1.442827 | 1.505624 |
| Gene_Pert | Gykl1 KO m | 2/285 | 0.352481 | 0.832891 | 0 | 0 | 1.632093 | 1.70188 |
| Gene_Pert | PSIP1 Defic | 2/285 | 0.352481 | 0.832891 | 0 | 0 | 1.632093 | 1.70188 |
| Gene_Pert | Pycard KO | 2/286 | 0.354057 | 0.832891 | 0 | 0 | 1.626263 | 1.688543 |
| Gene_Pert | EOMES OE | 2/281 | 0.346162 | 0.832891 | 0 | 0 | 1.65583 | 1.756583 |
| Gene_Pert | IRF9 KD hu | 2/269 | 0.327097 | 0.832891 | 0 | 0 | 1.731306 | 1.934732 |
| Gene_Pert | Slc9a3 KO r | 2/269 | 0.327097 | 0.832891 | 0 | 0 | 1.731306 | 1.934732 |
| Gene_Pert | Prkaca KO | 2/269 | 0.327097 | 0.832891 | 0 | 0 | 1.731306 | 1.934732 |
| Gene_Pert | Pdpn NULL | 2/269 | 0.327097 | 0.832891 | 0 | 0 | 1.731306 | 1.934732 |
| Gene_Pert | Ep300 KD r | 2/269 | 0.327097 | 0.832891 | 0 | 0 | 1.731306 | 1.934732 |
| Gene_Pert | TRPM7 ove | 2/269 | 0.327097 | 0.832891 | 0 | 0 | 1.731306 | 1.934732 |
| Gene_Pert | Atm knock | 2/269 | 0.327097 | 0.832891 | 0 | 0 | 1.731306 | 1.934732 |
| Gene_Pert | TREM2 KD | 3/502 | 0.373623 | 0.832891 | 0 | 0 | 1.389493 | 1.367968 |
| Gene_Pert | CBFA2T3 K | 2/270 | 0.328691 | 0.832891 | 0 | 0 | 1.724759 | 1.919028 |
| Gene_Pert | TBK1 KD hu | 2/270 | 0.328691 | 0.832891 | 0 | 0 | 1.724759 | 1.919028 |
| Gene_Pert | Trim71 KD | 2/270 | 0.328691 | 0.832891 | 0 | 0 | 1.724759 | 1.919028 |
| Gene_Pert | MYB KD hu | 2/270 | 0.328691 | 0.832891 | 0 | 0 | 1.724759 | 1.919028 |
| Gene_Pert | Map2k3 OE | 2/271 | 0.330285 | 0.832891 | 0 | 0 | 1.718259 | 1.903487 |
| Gene_Pert | Neurod1 C | 2/271 | 0.330285 | 0.832891 | 0 | 0 | 1.718259 | 1.903487 |
| Gene_Pert | TFAP2C Sile | 2/271 | 0.330285 | 0.832891 | 0 | 0 | 1.718259 | 1.903487 |
| Gene_Pert | ZMPSTE24 | 2/268 | 0.325502 | 0.832891 | 0 | 0 | 1.737904 | 1.950602 |
| Gene_Pert | Apod KO m | 2/290 | 0.36035 | 0.832891 | 0 | 0 | 1.60335 | 1.636504 |
| Gene_Pert | STK33 knoc | 2/290 | 0.36035 | 0.832891 | 0 | 0 | 1.60335 | 1.636504 |
| Gene_Pert | LRRK2 mut | 2/291 | 0.36192 | 0.832891 | 0 | 0 | 1.59772 | 1.623813 |
| Gene_Pert | Tfap2c KO | 2/292 | 0.363489 | 0.832891 | 0 | 0 | 1.59213 | 1.611246 |
| Gene_Pert | Kitl stimula | 2/303 | 0.380646 | 0.832891 | 0 | 0 | 1.533086 | 1.480784 |
| Gene_Pert | PPIB Deple | 2/289 | 0.358779 | 0.832891 | 0 | 0 | 1.609018 | 1.649321 |
| Gene_Pert | FOXM1 KD | 2/289 | 0.358779 | 0.832891 | 0 | 0 | 1.609018 | 1.649321 |
| Gene_Pert | EPAS1 KD h | 2/289 | 0.358779 | 0.832891 | 0 | 0 | 1.609018 | 1.649321 |
| Gene_Pert | Zmpste24 I | 2/289 | 0.358779 | 0.832891 | 0 | 0 | 1.609018 | 1.649321 |
| Gene_Pert | Rybp KO m | 2/289 | 0.358779 | 0.832891 | 0 | 0 | 1.609018 | 1.649321 |
| Gene_Pert | ZMPSTE24 | 2/286 | 0.354057 | 0.832891 | 0 | 0 | 1.626263 | 1.688543 |
| Gene_Pert | Srf homozy | 2/304 | 0.382197 | 0.832891 | 0 | 0 | 1.527931 | 1.469594 |
| Gene_Pert | Pkd1 KO m | 2/304 | 0.382197 | 0.832891 | 0 | 0 | 1.527931 | 1.469594 |
| Gene_Pert | IRF9 OE hu | 2/304 | 0.382197 | 0.832891 | 0 | 0 | 1.527931 | 1.469594 |
| Gene_Pert | Gata4 OE n | 2/305 | 0.383746 | 0.832891 | 0 | 0 | 1.522811 | 1.45851 |
| Gene_Pert | Nfe2l2 KO | 2/305 | 0.383746 | 0.832891 | 0 | 0 | 1.522811 | 1.45851 |
| Gene_Pert | Arntl KO m | 2/303 | 0.380646 | 0.832891 | 0 | 0 | 1.533086 | 1.480784 |
| Gene_Pert | Ppp1r1a KO | 2/288 | 0.357207 | 0.832891 | 0 | 0 | 1.614726 | 1.662266 |
| Gene_Pert | Sirt3 KO m | 2/303 | 0.380646 | 0.832891 | 0 | 0 | 1.533086 | 1.480784 |
| Gene_Pert | MYC KD hu | 2/305 | 0.383746 | 0.832891 | 0 | 0 | 1.522811 | 1.45851 |
| Gene_Pert | BATF KD hu | 2/305 | 0.383746 | 0.832891 | 0 | 0 | 1.522811 | 1.45851 |
| Gene_Pert | MIR126 KD | 2/305 | 0.383746 | 0.832891 | 0 | 0 | 1.522811 | 1.45851 |
| Gene_Pert | H19 Deplet | 2/306 | 0.385293 | 0.832891 | 0 | 0 | 1.517724 | 1.44753 |
| Gene_Pert | Mecp2 KO | 2/307 | 0.386839 | 0.832891 | 0 | 0 | 1.512671 | 1.436654 |

| | | | | | | | | |
|---|---|---|---|---|---|---|---|---|
| Gene_Pert | Plagl2 defic | 2/304 | 0.382197 | 0.832891 | 0 | 0 | 1.527931 | 1.469594 |
| Gene_Pert | Ptrf KO mo | 2/304 | 0.382197 | 0.832891 | 0 | 0 | 1.527931 | 1.469594 |
| Gene_Pert | Tnfsf14 OE | 2/304 | 0.382197 | 0.832891 | 0 | 0 | 1.527931 | 1.469594 |
| Gene_Pert | Sftpc KO m | 2/294 | 0.366622 | 0.832891 | 0 | 0 | 1.581064 | 1.586479 |
| Gene_Pert | Htt knock-i | 2/294 | 0.366622 | 0.832891 | 0 | 0 | 1.581064 | 1.586479 |
| Gene_Pert | NR2C2 KO | 2/295 | 0.368186 | 0.832891 | 0 | 0 | 1.575587 | 1.574275 |
| Gene_Pert | BRAF Drug | 2/297 | 0.37131 | 0.832891 | 0 | 0 | 1.564746 | 1.550222 |
| Gene_Pert | Irf6 KO mo | 2/299 | 0.374428 | 0.832891 | 0 | 0 | 1.55405 | 1.526629 |
| Gene_Pert | Lama2 Defi | 2/299 | 0.374428 | 0.832891 | 0 | 0 | 1.55405 | 1.526629 |
| Gene_Pert | STAT3 KD h | 2/299 | 0.374428 | 0.832891 | 0 | 0 | 1.55405 | 1.526629 |
| Gene_Pert | Snca KO m | 2/299 | 0.374428 | 0.832891 | 0 | 0 | 1.55405 | 1.526629 |
| Gene_Pert | Srf KO mou | 2/295 | 0.368186 | 0.832891 | 0 | 0 | 1.575587 | 1.574275 |
| Gene_Pert | SCARNA13 | 2/296 | 0.369749 | 0.832891 | 0 | 0 | 1.570148 | 1.56219 |
| Gene_Pert | Myod1 Mu | 2/296 | 0.369749 | 0.832891 | 0 | 0 | 1.570148 | 1.56219 |
| Gene_Pert | Nr2f2 KO m | 2/300 | 0.375985 | 0.832891 | 0 | 0 | 1.548756 | 1.515003 |
| Gene_Pert | LMO3 KD h | 2/300 | 0.375985 | 0.832891 | 0 | 0 | 1.548756 | 1.515003 |
| Gene_Pert | Dot1l DELE | 2/300 | 0.375985 | 0.832891 | 0 | 0 | 1.548756 | 1.515003 |
| Gene_Pert | Vax2 KO m | 2/301 | 0.37754 | 0.832891 | 0 | 0 | 1.543498 | 1.503487 |
| Gene_Pert | Htt Huntin | 2/301 | 0.37754 | 0.832891 | 0 | 0 | 1.543498 | 1.503487 |
| Gene_Pert | PPARGC1A | 2/284 | 0.350903 | 0.832891 | 0 | 0 | 1.637964 | 1.71535 |
| Gene_Pert | DUX4 OE h | 2/298 | 0.37287 | 0.832891 | 0 | 0 | 1.55938 | 1.538369 |
| Gene_Pert | Nrl Deficie | 2/298 | 0.37287 | 0.832891 | 0 | 0 | 1.55938 | 1.538369 |
| Gene_Pert | Scd1 KO m | 2/286 | 0.354057 | 0.832891 | 0 | 0 | 1.626263 | 1.688543 |
| Gene_Pert | Dmd Defici | 2/287 | 0.355633 | 0.832891 | 0 | 0 | 1.620475 | 1.675339 |
| Gene_Pert | NFKB1 Inac | 2/287 | 0.355633 | 0.832891 | 0 | 0 | 1.620475 | 1.675339 |
| Gene_Pert | HHLA1 KD | 2/287 | 0.355633 | 0.832891 | 0 | 0 | 1.620475 | 1.675339 |
| Gene_Pert | Nfe2l2 Mut | 2/287 | 0.355633 | 0.832891 | 0 | 0 | 1.620475 | 1.675339 |
| Gene_Pert | FOXL2 KD h | 2/288 | 0.357207 | 0.832891 | 0 | 0 | 1.614726 | 1.662266 |
| Gene_Pert | CREB1 DEP | 2/288 | 0.357207 | 0.832891 | 0 | 0 | 1.614726 | 1.662266 |
| Gene_Pert | Met knock | 2/284 | 0.350903 | 0.832891 | 0 | 0 | 1.637964 | 1.71535 |
| Gene_Pert | Mecom OE | 2/288 | 0.357207 | 0.832891 | 0 | 0 | 1.614726 | 1.662266 |
| Gene_Pert | Mapk1 INH | 2/266 | 0.322308 | 0.832891 | 0 | 0 | 1.751248 | 1.982846 |
| Gene_Pert | FLI1 KD hu | 2/239 | 0.278929 | 0.832891 | 0 | 0 | 1.953438 | 2.494149 |
| Gene_Pert | Des KO mo | 2/233 | 0.269251 | 0.832891 | 0 | 0 | 2.004787 | 2.630503 |
| Gene_Pert | Nfe2l2 kno | 2/263 | 0.317511 | 0.832891 | 0 | 0 | 1.771648 | 2.03251 |
| Gene_Pert | Plin5 OE m | 2/264 | 0.319111 | 0.832891 | 0 | 0 | 1.764796 | 2.015779 |
| Gene_Pert | MED12 KD | 2/264 | 0.319111 | 0.832891 | 0 | 0 | 1.764796 | 2.015779 |
| Gene_Pert | Tal1 KO mc | 2/264 | 0.319111 | 0.832891 | 0 | 0 | 1.764796 | 2.015779 |
| Gene_Pert | IRF9 OE hu | 2/264 | 0.319111 | 0.832891 | 0 | 0 | 1.764796 | 2.015779 |
| Gene_Pert | EWSR1 KD | 2/264 | 0.319111 | 0.832891 | 0 | 0 | 1.764796 | 2.015779 |
| Gene_Pert | Mecp2 KO | 2/265 | 0.32071 | 0.832891 | 0 | 0 | 1.757996 | 1.999226 |
| Gene_Pert | Tgfbr2 KO | 2/260 | 0.312707 | 0.832891 | 0 | 0 | 1.792522 | 2.083787 |
| Gene_Pert | POU5F1 OE | 2/265 | 0.32071 | 0.832891 | 0 | 0 | 1.757996 | 1.999226 |
| Gene_Pert | MAML1 M/ | Jan-89 | 0.322165 | 0.832891 | 0 | 0 | 2.619582 | 2.967177 |
| Gene_Pert | WNT1 OE h | 2/238 | 0.277316 | 0.832891 | 0 | 0 | 1.961815 | 2.516219 |

| | | | | | | | | |
|---|---|---|---|---|---|---|---|---|
| Gene_Pert | Pafah1b1 K | 2/266 | 0.322308 | 0.832891 | 0 | 0 | 1.751248 | 1.982846 |
| Gene_Pert | EGR3 KD hu | 3/460 | 0.323571 | 0.832891 | 0 | 0 | 1.520475 | 1.715609 |
| Gene_Pert | Gmnn KD r | 2/267 | 0.323905 | 0.832891 | 0 | 0 | 1.74455 | 1.966639 |
| Gene_Pert | Pten KO mo | 2/267 | 0.323905 | 0.832891 | 0 | 0 | 1.74455 | 1.966639 |
| Gene_Pert | LRRK2 dom | 2/263 | 0.317511 | 0.832891 | 0 | 0 | 1.771648 | 2.03251 |
| Gene_Pert | Cd53 OE m | 3/431 | 0.289025 | 0.832891 | 0 | 0 | 1.625918 | 2.018155 |
| Gene_Pert | Abca1 Hete | 2/246 | 0.29021 | 0.832891 | 0 | 0 | 1.896721 | 2.346531 |
| Gene_Pert | Aurka drug | 2/247 | 0.29182 | 0.832891 | 0 | 0 | 1.888884 | 2.326383 |
| Gene_Pert | Twist1 OE | 2/247 | 0.29182 | 0.832891 | 0 | 0 | 1.888884 | 2.326383 |
| Gene_Pert | Prkag3 KO | 2/247 | 0.29182 | 0.832891 | 0 | 0 | 1.888884 | 2.326383 |
| Gene_Pert | PTHLH siRN | Jan-79 | 0.291832 | 0.832891 | 0 | 0 | 2.956917 | 3.641673 |
| Gene_Pert | Gimap4 OE | 3/434 | 0.292588 | 0.832891 | 0 | 0 | 1.614352 | 1.984021 |
| Gene_Pert | RAP1A KO | 3/415 | 0.270096 | 0.832891 | 0 | 0 | 1.690447 | 2.212759 |
| Gene_Pert | Brd2 OE m | 3/440 | 0.299723 | 0.832891 | 0 | 0 | 1.591697 | 1.917827 |
| Gene_Pert | NET1 knocl | 3/441 | 0.300914 | 0.832891 | 0 | 0 | 1.587981 | 1.907057 |
| Gene_Pert | Furin Defic | 2/253 | 0.301472 | 0.832891 | 0 | 0 | 1.843169 | 2.210102 |
| Gene_Pert | Zfx KO mou | 2/248 | 0.29343 | 0.832891 | 0 | 0 | 1.88111 | 2.306459 |
| Gene_Pert | SYNCRIP OI | 2/253 | 0.301472 | 0.832891 | 0 | 0 | 1.843169 | 2.210102 |
| Gene_Pert | VGLL4 OE h | 2/254 | 0.303079 | 0.832891 | 0 | 0 | 1.835761 | 2.191461 |
| Gene_Pert | IRF9 OE hu | 3/443 | 0.303295 | 0.832891 | 0 | 0 | 1.580601 | 1.885733 |
| Gene_Pert | Kdm2b OE | 2/255 | 0.304685 | 0.832891 | 0 | 0 | 1.828412 | 2.173024 |
| Gene_Pert | Esrrg Defici | 2/256 | 0.306291 | 0.832891 | 0 | 0 | 1.821121 | 2.154787 |
| Gene_Pert | Dmd Defici | 2/256 | 0.306291 | 0.832891 | 0 | 0 | 1.821121 | 2.154787 |
| Gene_Pert | Tgfbr1 acti | 2/256 | 0.306291 | 0.832891 | 0 | 0 | 1.821121 | 2.154787 |
| Gene_Pert | Pink1 KO m | 2/250 | 0.296648 | 0.832891 | 0 | 0 | 1.86575 | 2.267274 |
| Gene_Pert | Igf1 KO mo | 2/242 | 0.283765 | 0.832891 | 0 | 0 | 1.928725 | 2.429439 |
| Gene_Pert | RBP2 KD hu | 3/415 | 0.270096 | 0.832891 | 0 | 0 | 1.690447 | 2.212759 |
| Gene_Pert | Pten KD mo | 3/415 | 0.270096 | 0.832891 | 0 | 0 | 1.690447 | 2.212759 |
| Gene_Pert | ARID1A KD | 3/416 | 0.271275 | 0.832891 | 0 | 0 | 1.686268 | 2.199943 |
| Gene_Pert | TP63 Over- | 2/235 | 0.272477 | 0.832891 | 0 | 0 | 1.987377 | 2.583987 |
| Gene_Pert | Dgcr8 KO n | 2/235 | 0.272477 | 0.832891 | 0 | 0 | 1.987377 | 2.583987 |
| Gene_Pert | PSMB9 KO | 2/236 | 0.27409 | 0.832891 | 0 | 0 | 1.978783 | 2.561135 |
| Gene_Pert | IRX5 KO mo | 2/228 | 0.261186 | 0.832891 | 0 | 0 | 2.049662 | 2.751716 |
| Gene_Pert | Hoxb4 indu | 2/237 | 0.275703 | 0.832891 | 0 | 0 | 1.970263 | 2.538546 |
| Gene_Pert | Abcb4 KO r | 2/237 | 0.275703 | 0.832891 | 0 | 0 | 1.970263 | 2.538546 |
| Gene_Pert | UBB KO mo | 3/420 | 0.275997 | 0.832891 | 0 | 0 | 1.66975 | 2.14958 |
| Gene_Pert | Hif1an Defi | 3/420 | 0.275997 | 0.832891 | 0 | 0 | 1.66975 | 2.14958 |
| Gene_Pert | Rb1 KO mo | 2/238 | 0.277316 | 0.832891 | 0 | 0 | 1.961815 | 2.516219 |
| Gene_Pert | Dysf Defici | 2/263 | 0.317511 | 0.832891 | 0 | 0 | 1.771648 | 2.03251 |
| Gene_Pert | ZBTB38 KD | 2/257 | 0.307896 | 0.832891 | 0 | 0 | 1.813887 | 2.136747 |
| Gene_Pert | Irs2 KO mo | 2/257 | 0.307896 | 0.832891 | 0 | 0 | 1.813887 | 2.136747 |
| Gene_Pert | Eed KO mo | 2/257 | 0.307896 | 0.832891 | 0 | 0 | 1.813887 | 2.136747 |
| Gene_Pert | Trim24 knc | 2/258 | 0.3095 | 0.832891 | 0 | 0 | 1.80671 | 2.118903 |
| Gene_Pert | Map2k3 m | 2/259 | 0.311104 | 0.832891 | 0 | 0 | 1.799588 | 2.10125 |
| Gene_Pert | Shh KO mo | 2/259 | 0.311104 | 0.832891 | 0 | 0 | 1.799588 | 2.10125 |

| | | | | | | | | |
|---|---|---|---|---|---|---|---|---|
| Gene_Pert | Des deplet | 2/253 | 0.301472 | 0.832891 | 0 | 0 | 1.843169 | 2.210102 |
| Gene_Pert | Snca KO m | 2/261 | 0.314309 | 0.832891 | 0 | 0 | 1.78551 | 2.066511 |
| Gene_Pert | HSF1 KD hu | 2/261 | 0.314309 | 0.832891 | 0 | 0 | 1.78551 | 2.066511 |
| Gene_Pert | Map2k3 Se | 2/263 | 0.317511 | 0.832891 | 0 | 0 | 1.771648 | 2.03251 |
| Gene_Pert | FLI1 KD hu | 2/263 | 0.317511 | 0.832891 | 0 | 0 | 1.771648 | 2.03251 |
| Gene_Pert | Socs3 KO n | 2/263 | 0.317511 | 0.832891 | 0 | 0 | 1.771648 | 2.03251 |
| Gene_Pert | FLI1 KD hu | 2/257 | 0.307896 | 0.832891 | 0 | 0 | 1.813887 | 2.136747 |
| Gene_Pert | HSF1 KD hu | 2/263 | 0.317511 | 0.832891 | 0 | 0 | 1.771648 | 2.03251 |
| Gene_Pert | Aplp2 KO n | 2/256 | 0.306291 | 0.832891 | 0 | 0 | 1.821121 | 2.154787 |
| Gene_Pert | FLI1 KD hu | 2/281 | 0.346162 | 0.832891 | 0 | 0 | 1.65583 | 1.756583 |
| Gene_Pert | GAPDH KD | 2/282 | 0.347744 | 0.832891 | 0 | 0 | 1.649832 | 1.7427 |
| Gene_Pert | BRMS1 KD | 2/282 | 0.347744 | 0.832891 | 0 | 0 | 1.649832 | 1.7427 |
| Gene_Pert | Kdm1a KD | 2/282 | 0.347744 | 0.832891 | 0 | 0 | 1.649832 | 1.7427 |
| Gene_Pert | SNRK overe | 2/282 | 0.347744 | 0.832891 | 0 | 0 | 1.649832 | 1.7427 |
| Gene_Pert | Wt1 KO mc | 2/283 | 0.349324 | 0.832891 | 0 | 0 | 1.643877 | 1.728956 |
| Gene_Pert | Srf KO mou | 2/283 | 0.349324 | 0.832891 | 0 | 0 | 1.643877 | 1.728956 |
| Gene_Pert | HNRNPU-A | 2/279 | 0.342996 | 0.832891 | 0 | 0 | 1.667955 | 1.784774 |
| Gene_Pert | Akt1 KO m | 2/284 | 0.350903 | 0.832891 | 0 | 0 | 1.637964 | 1.71535 |
| Gene_Pert | Mok knock | 2/284 | 0.350903 | 0.832891 | 0 | 0 | 1.637964 | 1.71535 |
| Gene_Pert | Npc1 KD m | 2/272 | 0.331878 | 0.832891 | 0 | 0 | 1.711808 | 1.888106 |
| Gene_Pert | CTNNB1 Ac | 2/248 | 0.29343 | 0.832891 | 0 | 0 | 1.88111 | 2.306459 |
| Gene_Pert | Gata4 INAC | 2/248 | 0.29343 | 0.832891 | 0 | 0 | 1.88111 | 2.306459 |
| Gene_Pert | Tcf12 KO n | 2/249 | 0.295039 | 0.832891 | 0 | 0 | 1.873398 | 2.286757 |
| Gene_Pert | COMMD3-I | 2/249 | 0.295039 | 0.832891 | 0 | 0 | 1.873398 | 2.286757 |
| Gene_Pert | Pparg V29C | 2/250 | 0.296648 | 0.832891 | 0 | 0 | 1.86575 | 2.267274 |
| Gene_Pert | Scd1 KO m | 2/250 | 0.296648 | 0.832891 | 0 | 0 | 1.86575 | 2.267274 |
| Gene_Pert | CEBPA VR I | 2/250 | 0.296648 | 0.832891 | 0 | 0 | 1.86575 | 2.267274 |
| Gene_Pert | Aldh1a2 KC | 2/245 | 0.288599 | 0.832891 | 0 | 0 | 1.904624 | 2.366908 |
| Gene_Pert | B4galnt2 O | 2/271 | 0.330285 | 0.832891 | 0 | 0 | 1.718259 | 1.903487 |
| Gene_Pert | AMER1 OE | 2/271 | 0.330285 | 0.832891 | 0 | 0 | 1.718259 | 1.903487 |
| Gene_Pert | Psap KO m | 2/272 | 0.331878 | 0.832891 | 0 | 0 | 1.711808 | 1.888106 |
| Gene_Pert | Elane Inhib | 2/272 | 0.331878 | 0.832891 | 0 | 0 | 1.711808 | 1.888106 |
| Gene_Pert | JAK1 knock | 2/295 | 0.368186 | 0.832891 | 0 | 0 | 1.575587 | 1.574275 |
| Gene_Pert | Syncrip OE | 2/272 | 0.331878 | 0.832891 | 0 | 0 | 1.711808 | 1.888106 |
| Gene_Pert | Rif1 KD mo | 2/273 | 0.333469 | 0.832891 | 0 | 0 | 1.705405 | 1.872885 |
| Gene_Pert | Atf6b KO n | 2/270 | 0.328691 | 0.832891 | 0 | 0 | 1.724759 | 1.919028 |
| Gene_Pert | XBP1 OE hu | 2/274 | 0.335059 | 0.832891 | 0 | 0 | 1.699048 | 1.85782 |
| Gene_Pert | JAG1 OE hu | 2/274 | 0.335059 | 0.832891 | 0 | 0 | 1.699048 | 1.85782 |
| Gene_Pert | Syncrip OE | 2/274 | 0.335059 | 0.832891 | 0 | 0 | 1.699048 | 1.85782 |
| Gene_Pert | Smchd1 KC | 2/274 | 0.335059 | 0.832891 | 0 | 0 | 1.699048 | 1.85782 |
| Gene_Pert | Smad4 KO | 2/275 | 0.336649 | 0.832891 | 0 | 0 | 1.692739 | 1.842909 |
| Gene_Pert | DNMT1 Inh | 2/275 | 0.336649 | 0.832891 | 0 | 0 | 1.692739 | 1.842909 |
| Gene_Pert | Ulk1 knock | 2/275 | 0.336649 | 0.832891 | 0 | 0 | 1.692739 | 1.842909 |
| Gene_Pert | TBK1 knocl | 2/271 | 0.330285 | 0.832891 | 0 | 0 | 1.718259 | 1.903487 |
| Gene_Pert | AR KD hum | 2/346 | 0.44571 | 0.832891 | 0 | 0 | 1.338509 | 1.081632 |

| | | | | | | | | |
|---|---|---|---|---|---|---|---|---|
| Gene_Pert | Elavl4 OE n | 2/324 | 0.412854 | 0.832891 | 0 | 0 | 1.431567 | 1.266451 |
| Gene_Pert | Por DELETI | 2/324 | 0.412854 | 0.832891 | 0 | 0 | 1.431567 | 1.266451 |
| Gene_Pert | RAD21 KD | 2/324 | 0.412854 | 0.832891 | 0 | 0 | 1.431567 | 1.266451 |
| Gene_Pert | Cftr OE mo | 2/327 | 0.417391 | 0.832891 | 0 | 0 | 1.418136 | 1.239071 |
| Gene_Pert | Jun KO mo | 2/327 | 0.417391 | 0.832891 | 0 | 0 | 1.418136 | 1.239071 |
| Gene_Pert | OGT small | 2/327 | 0.417391 | 0.832891 | 0 | 0 | 1.418136 | 1.239071 |
| Gene_Pert | Srf homozy | 2/328 | 0.418899 | 0.832891 | 0 | 0 | 1.413713 | 1.230107 |
| Gene_Pert | STAU1 DEL | 2/328 | 0.418899 | 0.832891 | 0 | 0 | 1.413713 | 1.230107 |
| Gene_Pert | Prkaca KO | 2/326 | 0.415881 | 0.832891 | 0 | 0 | 1.422585 | 1.248115 |
| Gene_Pert | Adam17 Co | 2/326 | 0.415881 | 0.832891 | 0 | 0 | 1.422585 | 1.248115 |
| Gene_Pert | Htt Knock-i | 2/326 | 0.415881 | 0.832891 | 0 | 0 | 1.422585 | 1.248115 |
| Gene_Pert | Fgfr1 active | 2/346 | 0.44571 | 0.832891 | 0 | 0 | 1.338509 | 1.081632 |
| Gene_Pert | REST Drug | 2/327 | 0.417391 | 0.832891 | 0 | 0 | 1.418136 | 1.239071 |
| Gene_Pert | Tlr4 mutati | 2/347 | 0.44718 | 0.832891 | 0 | 0 | 1.334561 | 1.074048 |
| Gene_Pert | Bhlha15 KO | 1/136 | 0.448384 | 0.832891 | 0 | 0 | 1.703531 | 1.36641 |
| Gene_Pert | Trp53inp2 | 2/348 | 0.448647 | 0.832891 | 0 | 0 | 1.330636 | 1.066529 |
| Gene_Pert | Pafah1b1 K | 2/343 | 0.441287 | 0.832891 | 0 | 0 | 1.350492 | 1.104782 |
| Gene_Pert | HNF1A OE | 2/343 | 0.441287 | 0.832891 | 0 | 0 | 1.350492 | 1.104782 |
| Gene_Pert | Kmt2a KO | 2/343 | 0.441287 | 0.832891 | 0 | 0 | 1.350492 | 1.104782 |
| Gene_Pert | Myb KO mo | 2/349 | 0.450113 | 0.832891 | 0 | 0 | 1.326733 | 1.059074 |
| Gene_Pert | Taf4b KO m | 2/349 | 0.450113 | 0.832891 | 0 | 0 | 1.326733 | 1.059074 |
| Gene_Pert | Nrl KO mou | 2/350 | 0.451576 | 0.832891 | 0 | 0 | 1.322853 | 1.051682 |
| Gene_Pert | IRF9 OE hu | 2/350 | 0.451576 | 0.832891 | 0 | 0 | 1.322853 | 1.051682 |
| Gene_Pert | VPRBP KD | 2/351 | 0.453038 | 0.832891 | 0 | 0 | 1.318995 | 1.044354 |
| Gene_Pert | Pebp1 KO | 2/346 | 0.44571 | 0.832891 | 0 | 0 | 1.338509 | 1.081632 |
| Gene_Pert | Akt1 KO mo | 2/336 | 0.430896 | 0.832891 | 0 | 0 | 1.379288 | 1.161207 |
| Gene_Pert | Psap hypor | 2/322 | 0.409821 | 0.832891 | 0 | 0 | 1.440662 | 1.285121 |
| Gene_Pert | MYB KD hu | 2/322 | 0.409821 | 0.832891 | 0 | 0 | 1.440662 | 1.285121 |
| Gene_Pert | SYK knocko | 2/317 | 0.402204 | 0.832891 | 0 | 0 | 1.463903 | 1.333316 |
| Gene_Pert | MYC KD hu | 2/317 | 0.402204 | 0.832891 | 0 | 0 | 1.463903 | 1.333316 |
| Gene_Pert | Tet1 KD mo | 2/317 | 0.402204 | 0.832891 | 0 | 0 | 1.463903 | 1.333316 |
| Gene_Pert | FH Heteroz | 2/338 | 0.433875 | 0.832891 | 0 | 0 | 1.370938 | 1.144732 |
| Gene_Pert | Ssfa2 Defic | 2/338 | 0.433875 | 0.832891 | 0 | 0 | 1.370938 | 1.144732 |
| Gene_Pert | Jak2 active | 2/338 | 0.433875 | 0.832891 | 0 | 0 | 1.370938 | 1.144732 |
| Gene_Pert | EIF1AY OE | 2/338 | 0.433875 | 0.832891 | 0 | 0 | 1.370938 | 1.144732 |
| Gene_Pert | Apod OE m | 2/338 | 0.433875 | 0.832891 | 0 | 0 | 1.370938 | 1.144732 |
| Gene_Pert | Twist1 OE | 2/336 | 0.430896 | 0.832891 | 0 | 0 | 1.379288 | 1.161207 |
| Gene_Pert | Akt1 knock | 2/336 | 0.430896 | 0.832891 | 0 | 0 | 1.379288 | 1.161207 |
| Gene_Pert | SCYL1 KD h | 2/346 | 0.44571 | 0.832891 | 0 | 0 | 1.338509 | 1.081632 |
| Gene_Pert | Hsp90b1 O | 2/339 | 0.435362 | 0.832891 | 0 | 0 | 1.3668 | 1.136601 |
| Gene_Pert | BMI1 OE m | 1/131 | 0.436146 | 0.832891 | 0 | 0 | 1.769499 | 1.468291 |
| Gene_Pert | TGFB1I1 KD | 2/340 | 0.436846 | 0.832891 | 0 | 0 | 1.362687 | 1.128542 |
| Gene_Pert | SOX17 OE | 2/340 | 0.436846 | 0.832891 | 0 | 0 | 1.362687 | 1.128542 |
| Gene_Pert | Pmp22 KO | 2/341 | 0.438329 | 0.832891 | 0 | 0 | 1.358598 | 1.120553 |
| Gene_Pert | Fgfr1 active | 2/337 | 0.432387 | 0.832891 | 0 | 0 | 1.375101 | 1.152933 |

| | | | | | | | | |
|---|---|---|---|---|---|---|---|---|
| Gene_Pert | MTDH KD I | 1/130 | 0.433666 | 0.832891 | 0 | 0 | 1.783306 | 1.489916 |
| Gene_Pert | Neurog3 O | 2/338 | 0.433875 | 0.832891 | 0 | 0 | 1.370938 | 1.144732 |
| Gene_Pert | TET1 DEPLI | 2/326 | 0.415881 | 0.832891 | 0 | 0 | 1.422585 | 1.248115 |
| Gene_Pert | Pink1 KO m | 2/326 | 0.415881 | 0.832891 | 0 | 0 | 1.422585 | 1.248115 |
| Gene_Pert | Pten KO mo | 2/326 | 0.415881 | 0.832891 | 0 | 0 | 1.422585 | 1.248115 |
| Gene_Pert | Crem KO m | 3/539 | 0.417184 | 0.832891 | 0 | 0 | 1.291111 | 1.128725 |
| Gene_Pert | Hsd17b4 K | 2/343 | 0.441287 | 0.832891 | 0 | 0 | 1.350492 | 1.104782 |
| Gene_Pert | Slc39a13 D | 2/358 | 0.463206 | 0.832891 | 0 | 0 | 1.292597 | 0.994762 |
| Gene_Pert | Stk11 knoc | 2/359 | 0.46465 | 0.832891 | 0 | 0 | 1.288911 | 0.987914 |
| Gene_Pert | MAPK1 KD | 2/359 | 0.46465 | 0.832891 | 0 | 0 | 1.288911 | 0.987914 |
| Gene_Pert | TYROBP KD | 2/359 | 0.46465 | 0.832891 | 0 | 0 | 1.288911 | 0.987914 |
| Gene_Pert | RELA KD hu | 2/356 | 0.460312 | 0.832891 | 0 | 0 | 1.300033 | 1.008633 |
| Gene_Pert | Nfe2l2 Defi | 2/356 | 0.460312 | 0.832891 | 0 | 0 | 1.300033 | 1.008633 |
| Gene_Pert | Il17rc KO m | 2/356 | 0.460312 | 0.832891 | 0 | 0 | 1.300033 | 1.008633 |
| Gene_Pert | Adnp Defic | 1/132 | 0.438615 | 0.832891 | 0 | 0 | 1.755903 | 1.447098 |
| Gene_Pert | Tcof1 KD m | 2/342 | 0.439809 | 0.832891 | 0 | 0 | 1.354533 | 1.112633 |
| Gene_Pert | Nfe2l2 KO | 2/335 | 0.429403 | 0.832891 | 0 | 0 | 1.383501 | 1.169555 |
| Gene_Pert | HSF1 KD hu | 2/342 | 0.439809 | 0.832891 | 0 | 0 | 1.354533 | 1.112633 |
| Gene_Pert | SOX2 Defic | 2/342 | 0.439809 | 0.832891 | 0 | 0 | 1.354533 | 1.112633 |
| Gene_Pert | MIR22 OE I | 2/354 | 0.457409 | 0.832891 | 0 | 0 | 1.307553 | 1.02274 |
| Gene_Pert | Pmp22 Tre | 2/338 | 0.433875 | 0.832891 | 0 | 0 | 1.370938 | 1.144732 |
| Gene_Pert | Fhl2 KO mo | 2/339 | 0.435362 | 0.832891 | 0 | 0 | 1.3668 | 1.136601 |
| Gene_Pert | THOC5 KD | 2/339 | 0.435362 | 0.832891 | 0 | 0 | 1.3668 | 1.136601 |
| Gene_Pert | Sarm1 KO r | 2/344 | 0.442764 | 0.832891 | 0 | 0 | 1.346474 | 1.096999 |
| Gene_Pert | Pafah1b1 K | 2/344 | 0.442764 | 0.832891 | 0 | 0 | 1.346474 | 1.096999 |
| Gene_Pert | TARDBP KD | 2/344 | 0.442764 | 0.832891 | 0 | 0 | 1.346474 | 1.096999 |
| Gene_Pert | Dhcr7 KO n | 2/345 | 0.444238 | 0.832891 | 0 | 0 | 1.34248 | 1.089282 |
| Gene_Pert | Tcf3 Deficie | 2/345 | 0.444238 | 0.832891 | 0 | 0 | 1.34248 | 1.089282 |
| Gene_Pert | Jak1 drugin | 2/341 | 0.438329 | 0.832891 | 0 | 0 | 1.358598 | 1.120553 |
| Gene_Pert | Ywhae KO | 2/341 | 0.438329 | 0.832891 | 0 | 0 | 1.358598 | 1.120553 |
| Gene_Pert | Psap hypor | 2/341 | 0.438329 | 0.832891 | 0 | 0 | 1.358598 | 1.120553 |
| Gene_Pert | BCOR VR h | 2/369 | 0.478963 | 0.832891 | 0 | 0 | 1.25315 | 0.922483 |
| Gene_Pert | Zfx KO mou | 2/337 | 0.432387 | 0.832891 | 0 | 0 | 1.375101 | 1.152933 |
| Gene_Pert | Neurog3 O | 2/346 | 0.44571 | 0.832891 | 0 | 0 | 1.338509 | 1.081632 |
| Gene_Pert | ERBB2 OE I | 2/334 | 0.427909 | 0.832891 | 0 | 0 | 1.387739 | 1.177976 |
| Gene_Pert | SYK DEPLET | 2/335 | 0.429403 | 0.832891 | 0 | 0 | 1.383501 | 1.169555 |
| Gene_Pert | BRAF knocl | 2/359 | 0.46465 | 0.832891 | 0 | 0 | 1.288911 | 0.987914 |
| Gene_Pert | Ppargc1a K | 2/335 | 0.429403 | 0.832891 | 0 | 0 | 1.383501 | 1.169555 |
| Gene_Pert | MYC KD hu | 2/335 | 0.429403 | 0.832891 | 0 | 0 | 1.383501 | 1.169555 |
| Gene_Pert | Ezh2 defici | 2/331 | 0.423413 | 0.832891 | 0 | 0 | 1.400608 | 1.203693 |
| Gene_Pert | Tet2 KO mo | 2/331 | 0.423413 | 0.832891 | 0 | 0 | 1.400608 | 1.203693 |
| Gene_Pert | CDC73 KD I | 2/331 | 0.423413 | 0.832891 | 0 | 0 | 1.400608 | 1.203693 |
| Gene_Pert | Pax3 Knock | 2/336 | 0.430896 | 0.832891 | 0 | 0 | 1.379288 | 1.161207 |
| Gene_Pert | Sirt1 KO mo | 2/337 | 0.432387 | 0.832891 | 0 | 0 | 1.375101 | 1.152933 |
| Gene_Pert | FGFR3 KD I | 2/337 | 0.432387 | 0.832891 | 0 | 0 | 1.375101 | 1.152933 |

| | | | | | | | | |
|---|---|---|---|---|---|---|---|---|
| Gene_Pert | GSK3B KD l | 2/322 | 0.409821 | 0.832891 | 0 | 0 | 1.440662 | 1.285121 |
| Gene_Pert | Sod1 G93A | 2/337 | 0.432387 | 0.832891 | 0 | 0 | 1.375101 | 1.152933 |
| Gene_Pert | Ddb1 KD m | 2/333 | 0.426412 | 0.832891 | 0 | 0 | 1.392003 | 1.186473 |
| Gene_Pert | SYNCRIP KI | 2/333 | 0.426412 | 0.832891 | 0 | 0 | 1.392003 | 1.186473 |
| Gene_Pert | Apc KO mo | 2/334 | 0.427909 | 0.832891 | 0 | 0 | 1.387739 | 1.177976 |
| Gene_Pert | IL6 anti-IL6 | 2/357 | 0.46176 | 0.832891 | 0 | 0 | 1.296305 | 1.001668 |
| Gene_Pert | Atf6 KO mc | 2/357 | 0.46176 | 0.832891 | 0 | 0 | 1.296305 | 1.001668 |
| Gene_Pert | Cxcr4 KO m | 2/357 | 0.46176 | 0.832891 | 0 | 0 | 1.296305 | 1.001668 |
| Gene_Pert | Prkaca KO | 2/357 | 0.46176 | 0.832891 | 0 | 0 | 1.296305 | 1.001668 |
| Gene_Pert | CTNNB1 In | 2/357 | 0.46176 | 0.832891 | 0 | 0 | 1.296305 | 1.001668 |
| Gene_Pert | SOX17 OE l | 2/354 | 0.457409 | 0.832891 | 0 | 0 | 1.307553 | 1.02274 |
| Gene_Pert | Prdm14 KC | 2/354 | 0.457409 | 0.832891 | 0 | 0 | 1.307553 | 1.02274 |
| Gene_Pert | Dusp5 KO r | 2/303 | 0.380646 | 0.832891 | 0 | 0 | 1.533086 | 1.480784 |
| Gene_Pert | FOXA1 KD l | 2/313 | 0.396079 | 0.832891 | 0 | 0 | 1.483034 | 1.3735 |
| Gene_Pert | SALL4 KD h | 2/314 | 0.397613 | 0.832891 | 0 | 0 | 1.478205 | 1.363314 |
| Gene_Pert | CLOCK KD l | 2/301 | 0.37754 | 0.832891 | 0 | 0 | 1.543498 | 1.503487 |
| Gene_Pert | ARF1 KD hu | 2/301 | 0.37754 | 0.832891 | 0 | 0 | 1.543498 | 1.503487 |
| Gene_Pert | Aplp2 KO n | 2/302 | 0.379094 | 0.832891 | 0 | 0 | 1.538275 | 1.492081 |
| Gene_Pert | PARK7 KD l | 2/302 | 0.379094 | 0.832891 | 0 | 0 | 1.538275 | 1.492081 |
| Gene_Pert | Traf3 OE m | 2/303 | 0.380646 | 0.832891 | 0 | 0 | 1.533086 | 1.480784 |
| Gene_Pert | Map2k3 Se | 2/299 | 0.374428 | 0.832891 | 0 | 0 | 1.55405 | 1.526629 |
| Gene_Pert | Socs3 KO n | 2/299 | 0.374428 | 0.832891 | 0 | 0 | 1.55405 | 1.526629 |
| Gene_Pert | Nfatc3 OE | 2/300 | 0.375985 | 0.832891 | 0 | 0 | 1.548756 | 1.515003 |
| Gene_Pert | Ikbkb KO m | 2/303 | 0.380646 | 0.832891 | 0 | 0 | 1.533086 | 1.480784 |
| Gene_Pert | Pfkl OE mo | 2/303 | 0.380646 | 0.832891 | 0 | 0 | 1.533086 | 1.480784 |
| Gene_Pert | IRF9 OE hu | 2/312 | 0.394543 | 0.832891 | 0 | 0 | 1.487894 | 1.383781 |
| Gene_Pert | Ppargc1b K | 2/303 | 0.380646 | 0.832891 | 0 | 0 | 1.533086 | 1.480784 |
| Gene_Pert | MAPK1 KD | 2/303 | 0.380646 | 0.832891 | 0 | 0 | 1.533086 | 1.480784 |
| Gene_Pert | MAPK7 cor | 2/301 | 0.37754 | 0.832891 | 0 | 0 | 1.543498 | 1.503487 |
| Gene_Pert | Irak4 Defici | 2/301 | 0.37754 | 0.832891 | 0 | 0 | 1.543498 | 1.503487 |
| Gene_Pert | Gata1 KO r | 2/301 | 0.37754 | 0.832891 | 0 | 0 | 1.543498 | 1.503487 |
| Gene_Pert | Dnmt1 KO | 2/323 | 0.411339 | 0.832891 | 0 | 0 | 1.4361 | 1.275744 |
| Gene_Pert | Met knock | 2/323 | 0.411339 | 0.832891 | 0 | 0 | 1.4361 | 1.275744 |
| Gene_Pert | CXCR4 Stim | 2/324 | 0.412854 | 0.832891 | 0 | 0 | 1.431567 | 1.266451 |
| Gene_Pert | Nrl Deficier | 2/324 | 0.412854 | 0.832891 | 0 | 0 | 1.431567 | 1.266451 |
| Gene_Pert | Htt Mutati | 2/324 | 0.412854 | 0.832891 | 0 | 0 | 1.431567 | 1.266451 |
| Gene_Pert | Pde10a KO | 2/320 | 0.40678 | 0.832891 | 0 | 0 | 1.449871 | 1.304135 |
| Gene_Pert | IGF1 OE hu | 2/320 | 0.40678 | 0.832891 | 0 | 0 | 1.449871 | 1.304135 |
| Gene_Pert | RET drugin | 2/314 | 0.397613 | 0.832891 | 0 | 0 | 1.478205 | 1.363314 |
| Gene_Pert | Cftr OE mo | 2/295 | 0.368186 | 0.832891 | 0 | 0 | 1.575587 | 1.574275 |
| Gene_Pert | Fgfr1 active | 2/292 | 0.363489 | 0.832891 | 0 | 0 | 1.59213 | 1.611246 |
| Gene_Pert | GAPDH KD | 2/292 | 0.363489 | 0.832891 | 0 | 0 | 1.59213 | 1.611246 |
| Gene_Pert | OGT small | 2/292 | 0.363489 | 0.832891 | 0 | 0 | 1.59213 | 1.611246 |
| Gene_Pert | Pink1 KO m | 2/296 | 0.369749 | 0.832891 | 0 | 0 | 1.570148 | 1.56219 |
| Gene_Pert | GAPDH KD | 2/296 | 0.369749 | 0.832891 | 0 | 0 | 1.570148 | 1.56219 |

| | | | | | | | | |
|---|---|---|---|---|---|---|---|---|
| Gene_Pert | Mecp2 KO | 2/296 | 0.369749 | 0.832891 | 0 | 0 | 1.570148 | 1.56219 |
| Gene_Pert | Notch3 KO | 2/296 | 0.369749 | 0.832891 | 0 | 0 | 1.570148 | 1.56219 |
| Gene_Pert | Nrl Deficier | 2/297 | 0.37131 | 0.832891 | 0 | 0 | 1.564746 | 1.550222 |
| Gene_Pert | CDH1 OE h | 2/293 | 0.365056 | 0.832891 | 0 | 0 | 1.586578 | 1.598802 |
| Gene_Pert | Tet2 KO mo | 2/293 | 0.365056 | 0.832891 | 0 | 0 | 1.586578 | 1.598802 |
| Gene_Pert | Htt OE mou | 2/293 | 0.365056 | 0.832891 | 0 | 0 | 1.586578 | 1.598802 |
| Gene_Pert | Hif1a NULL | 1/120 | 0.408269 | 0.832891 | 0 | 0 | 1.934141 | 1.732658 |
| Gene_Pert | Yy1 KO mo | 2/314 | 0.397613 | 0.832891 | 0 | 0 | 1.478205 | 1.363314 |
| Gene_Pert | KIT activem | 2/314 | 0.397613 | 0.832891 | 0 | 0 | 1.478205 | 1.363314 |
| Gene_Pert | Socs3 DELE | 2/315 | 0.399145 | 0.832891 | 0 | 0 | 1.473407 | 1.353222 |
| Gene_Pert | RRM2 OE h | 2/315 | 0.399145 | 0.832891 | 0 | 0 | 1.473407 | 1.353222 |
| Gene_Pert | BST1 OE hu | 2/309 | 0.389926 | 0.832891 | 0 | 0 | 1.502663 | 1.415207 |
| Gene_Pert | Sox2 KO m | 2/310 | 0.391467 | 0.832891 | 0 | 0 | 1.497708 | 1.404633 |
| Gene_Pert | Dusp1 KO r | 2/310 | 0.391467 | 0.832891 | 0 | 0 | 1.497708 | 1.404633 |
| Gene_Pert | Rb1 KO mo | 2/316 | 0.400676 | 0.832891 | 0 | 0 | 1.46864 | 1.343223 |
| Gene_Pert | Alkbh1 KO | 2/316 | 0.400676 | 0.832891 | 0 | 0 | 1.46864 | 1.343223 |
| Gene_Pert | Pgr KO mou | 2/316 | 0.400676 | 0.832891 | 0 | 0 | 1.46864 | 1.343223 |
| Gene_Pert | JAG1 OE hu | 2/317 | 0.402204 | 0.832891 | 0 | 0 | 1.463903 | 1.333316 |
| Gene_Pert | MYC KD hu | 2/317 | 0.402204 | 0.832891 | 0 | 0 | 1.463903 | 1.333316 |
| Gene_Pert | FGFR1 mut | 2/328 | 0.418899 | 0.832891 | 0 | 0 | 1.413713 | 1.230107 |
| Gene_Pert | Kmt2a KO r | 2/329 | 0.420406 | 0.832891 | 0 | 0 | 1.409318 | 1.221224 |
| Gene_Pert | Gata6 KO r | 2/329 | 0.420406 | 0.832891 | 0 | 0 | 1.409318 | 1.221224 |
| Gene_Pert | YAP1 KD hu | 2/330 | 0.42191 | 0.832891 | 0 | 0 | 1.40495 | 1.212419 |
| Gene_Pert | PARK7 KD h | 2/330 | 0.42191 | 0.832891 | 0 | 0 | 1.40495 | 1.212419 |
| Gene_Pert | Gfap OE m | 2/327 | 0.417391 | 0.832891 | 0 | 0 | 1.418136 | 1.239071 |
| Gene_Pert | PIAS1 KD h | 2/327 | 0.417391 | 0.832891 | 0 | 0 | 1.418136 | 1.239071 |
| Gene_Pert | Dot1l DELE | 2/327 | 0.417391 | 0.832891 | 0 | 0 | 1.418136 | 1.239071 |
| Gene_Pert | Pmp22 KO | 2/332 | 0.424913 | 0.832891 | 0 | 0 | 1.396292 | 1.195045 |
| Gene_Pert | Bcl2 OE mo | 2/332 | 0.424913 | 0.832891 | 0 | 0 | 1.396292 | 1.195045 |
| Gene_Pert | Egr1 KO mo | 2/332 | 0.424913 | 0.832891 | 0 | 0 | 1.396292 | 1.195045 |
| Gene_Pert | FOXP1 KD h | 2/333 | 0.426412 | 0.832891 | 0 | 0 | 1.392003 | 1.186473 |
| Gene_Pert | Pde10a KO | 2/333 | 0.426412 | 0.832891 | 0 | 0 | 1.392003 | 1.186473 |
| Gene_Pert | H3F3A Mut | 2/328 | 0.418899 | 0.832891 | 0 | 0 | 1.413713 | 1.230107 |
| Gene_Pert | SYK knocko | 2/328 | 0.418899 | 0.832891 | 0 | 0 | 1.413713 | 1.230107 |
| Gene_Pert | Shank3 KD | 2/328 | 0.418899 | 0.832891 | 0 | 0 | 1.413713 | 1.230107 |
| Gene_Pert | PTEN Delet | 2/318 | 0.403731 | 0.832891 | 0 | 0 | 1.459196 | 1.323499 |
| Gene_Pert | Cstf2t OE n | 2/319 | 0.405256 | 0.832891 | 0 | 0 | 1.454518 | 1.313773 |
| Gene_Pert | TFPI OE hu | 2/320 | 0.40678 | 0.832891 | 0 | 0 | 1.449871 | 1.304135 |
| Gene_Pert | Irs1 KO mo | 2/320 | 0.40678 | 0.832891 | 0 | 0 | 1.449871 | 1.304135 |
| Gene_Pert | FGFR3 kno | 2/320 | 0.40678 | 0.832891 | 0 | 0 | 1.449871 | 1.304135 |
| Gene_Pert | Gata4 OE n | 2/315 | 0.399145 | 0.832891 | 0 | 0 | 1.473407 | 1.353222 |
| Gene_Pert | Dmd Defici | 2/315 | 0.399145 | 0.832891 | 0 | 0 | 1.473407 | 1.353222 |
| Gene_Pert | Pten KO mo | 2/316 | 0.400676 | 0.832891 | 0 | 0 | 1.46864 | 1.343223 |
| Gene_Pert | Adrb1 mut | 2/321 | 0.408301 | 0.832891 | 0 | 0 | 1.445252 | 1.294585 |
| Gene_Pert | Bcl2 OE mo | 2/321 | 0.408301 | 0.832891 | 0 | 0 | 1.445252 | 1.294585 |

| | | | | | | | | |
|---|---|---|---|---|---|---|---|---|
| Gene_Pert | Insm1 KO r | 2/309 | 0.389926 | 0.832891 | 0 | 0 | 1.502663 | 1.415207 |
| Gene_Pert | SYK druginl | 2/325 | 0.414369 | 0.832891 | 0 | 0 | 1.427062 | 1.257241 |
| Gene_Pert | OGT small | 2/325 | 0.414369 | 0.832891 | 0 | 0 | 1.427062 | 1.257241 |
| Gene_Pert | DLX4 OE hu | 2/325 | 0.414369 | 0.832891 | 0 | 0 | 1.427062 | 1.257241 |
| Gene_Pert | NR2F2 KD l | 2/325 | 0.414369 | 0.832891 | 0 | 0 | 1.427062 | 1.257241 |
| Gene_Pert | Mef2a KD r | 2/325 | 0.414369 | 0.832891 | 0 | 0 | 1.427062 | 1.257241 |
| Gene_Pert | Gys1 KO m | 2/322 | 0.409821 | 0.832891 | 0 | 0 | 1.440662 | 1.285121 |
| Gene_Pert | ZXDC deple | 2/322 | 0.409821 | 0.832891 | 0 | 0 | 1.440662 | 1.285121 |
| Gene_Pert | TRPM2-AS | 2/322 | 0.409821 | 0.832891 | 0 | 0 | 1.440662 | 1.285121 |
| Gene_Pert | MECOM KD | 2/308 | 0.388383 | 0.832891 | 0 | 0 | 1.507651 | 1.42588 |
| Gene_Pert | Ssfa2 Defic | 2/308 | 0.388383 | 0.832891 | 0 | 0 | 1.507651 | 1.42588 |
| Gene_Pert | Dcx KO mo | 2/309 | 0.389926 | 0.832891 | 0 | 0 | 1.502663 | 1.415207 |
| Gene_Pert | Pten KO m | 2/309 | 0.389926 | 0.832891 | 0 | 0 | 1.502663 | 1.415207 |
| Gene_Pert | Ghr Mutati | 2/251 | 0.298257 | 0.832891 | 0 | 0 | 1.858162 | 2.248005 |
| Gene_Pert | Il10 KO mo | 2/305 | 0.383746 | 0.832891 | 0 | 0 | 1.522811 | 1.45851 |
| Gene_Pert | PTPRC C77 | 2/305 | 0.383746 | 0.832891 | 0 | 0 | 1.522811 | 1.45851 |
| Gene_Pert | Fgfr2 active | 2/305 | 0.383746 | 0.832891 | 0 | 0 | 1.522811 | 1.45851 |
| Gene_Pert | Ovol1 KO n | 2/310 | 0.391467 | 0.832891 | 0 | 0 | 1.497708 | 1.404633 |
| Gene_Pert | MIR221 KD | 2/311 | 0.393006 | 0.832891 | 0 | 0 | 1.492785 | 1.394158 |
| Gene_Pert | PPARGC1A | 2/312 | 0.394543 | 0.832891 | 0 | 0 | 1.487894 | 1.383781 |
| Gene_Pert | Rif1 KD mo | 2/312 | 0.394543 | 0.832891 | 0 | 0 | 1.487894 | 1.383781 |
| Gene_Pert | MAP3K7 KI | 2/312 | 0.394543 | 0.832891 | 0 | 0 | 1.487894 | 1.383781 |
| Gene_Pert | App OE mc | 2/308 | 0.388383 | 0.832891 | 0 | 0 | 1.507651 | 1.42588 |
| Gene_Pert | Trim28 knc | 2/308 | 0.388383 | 0.832891 | 0 | 0 | 1.507651 | 1.42588 |
| Gene_Pert | Kdm1a KD | 2/308 | 0.388383 | 0.832891 | 0 | 0 | 1.507651 | 1.42588 |
| Gene_Pert | Pkd1 KO m | 1/328 | 0.763487 | 0.832891 | 0 | 0 | 0.696465 | 0.187947 |
| Gene_Pert | H3F3A Mut | 1/315 | 0.749464 | 0.832891 | 0 | 0 | 0.725781 | 0.209313 |
| Gene_Pert | Ebf1 OE mc | 1/315 | 0.749464 | 0.832891 | 0 | 0 | 0.725781 | 0.209313 |
| Gene_Pert | Ret knocko | 1/315 | 0.749464 | 0.832891 | 0 | 0 | 0.725781 | 0.209313 |
| Gene_Pert | Cd70 trans | 1/314 | 0.748352 | 0.832891 | 0 | 0 | 0.728137 | 0.211074 |
| Gene_Pert | TGFB1I1 KD | 1/314 | 0.748352 | 0.832891 | 0 | 0 | 0.728137 | 0.211074 |
| Gene_Pert | EGR4 KD hu | 1/314 | 0.748352 | 0.832891 | 0 | 0 | 0.728137 | 0.211074 |
| Gene_Pert | Pctp KO mc | 1/317 | 0.751674 | 0.832891 | 0 | 0 | 0.721114 | 0.205844 |
| Gene_Pert | Cebpa KO r | 1/317 | 0.751674 | 0.832891 | 0 | 0 | 0.721114 | 0.205844 |
| Gene_Pert | GSK3A KD l | 1/313 | 0.747235 | 0.832891 | 0 | 0 | 0.730508 | 0.212852 |
| Gene_Pert | ALAS2 OE h | 1/313 | 0.747235 | 0.832891 | 0 | 0 | 0.730508 | 0.212852 |
| Gene_Pert | Aire KO mc | 1/313 | 0.747235 | 0.832891 | 0 | 0 | 0.730508 | 0.212852 |
| Gene_Pert | Dusp1 KO r | 1/313 | 0.747235 | 0.832891 | 0 | 0 | 0.730508 | 0.212852 |
| Gene_Pert | PDCD10 KD | 1/333 | 0.768672 | 0.832891 | 0 | 0 | 0.685801 | 0.180429 |
| Gene_Pert | Met KO mc | 1/328 | 0.763487 | 0.832891 | 0 | 0 | 0.696465 | 0.187947 |
| Gene_Pert | Xbp1 KO m | 1/330 | 0.765575 | 0.832891 | 0 | 0 | 0.692161 | 0.184896 |
| Gene_Pert | TRIM33 KD | 1/330 | 0.765575 | 0.832891 | 0 | 0 | 0.692161 | 0.184896 |
| Gene_Pert | Hdac6 KO r | 1/326 | 0.761381 | 0.832891 | 0 | 0 | 0.700823 | 0.191059 |
| Gene_Pert | Commd1 K | 1/326 | 0.761381 | 0.832891 | 0 | 0 | 0.700823 | 0.191059 |
| Gene_Pert | Clpp KO mc | 1/326 | 0.761381 | 0.832891 | 0 | 0 | 0.700823 | 0.191059 |

| | | | | | | | | |
|---|---|---|---|---|---|---|---|---|
| Gene_Pert | NPM1 mut | 1/326 | 0.761381 | 0.832891 | 0 | 0 | 0.700823 | 0.191059 |
| Gene_Pert | Nfe2l2 KO | 1/326 | 0.761381 | 0.832891 | 0 | 0 | 0.700823 | 0.191059 |
| Gene_Pert | MAP3K1 IN | 1/325 | 0.760322 | 0.832891 | 0 | 0 | 0.703022 | 0.192638 |
| Gene_Pert | Trpv4 DEPL | 1/330 | 0.765575 | 0.832891 | 0 | 0 | 0.692161 | 0.184896 |
| Gene_Pert | SNCA SNCA | 1/329 | 0.764533 | 0.832891 | 0 | 0 | 0.694307 | 0.186414 |
| Gene_Pert | TOP2B Inhi | 1/342 | 0.777722 | 0.832891 | 0 | 0 | 0.667394 | 0.167774 |
| Gene_Pert | GSK3B KD | 1/318 | 0.752771 | 0.832891 | 0 | 0 | 0.718803 | 0.204135 |
| Gene_Pert | Emd KO mo | 1/311 | 0.744986 | 0.832891 | 0 | 0 | 0.735296 | 0.216463 |
| Gene_Pert | HIF1A KD h | 1/309 | 0.742718 | 0.832891 | 0 | 0 | 0.740146 | 0.220149 |
| Gene_Pert | Psmd14 KD | 1/308 | 0.741576 | 0.832891 | 0 | 0 | 0.742595 | 0.222019 |
| Gene_Pert | FMR1 KO h | 1/308 | 0.741576 | 0.832891 | 0 | 0 | 0.742595 | 0.222019 |
| Gene_Pert | KDM4C nat | 1/308 | 0.741576 | 0.832891 | 0 | 0 | 0.742595 | 0.222019 |
| Gene_Pert | LRRK2 dom | 1/320 | 0.754952 | 0.832891 | 0 | 0 | 0.714223 | 0.200769 |
| Gene_Pert | Nfe2l2 KO | 1/320 | 0.754952 | 0.832891 | 0 | 0 | 0.714223 | 0.200769 |
| Gene_Pert | SON KD hu | 1/324 | 0.759257 | 0.832891 | 0 | 0 | 0.705234 | 0.194232 |
| Gene_Pert | Wfs1 KO m | 1/324 | 0.759257 | 0.832891 | 0 | 0 | 0.705234 | 0.194232 |
| Gene_Pert | BST2 OE hu | 1/319 | 0.753864 | 0.832891 | 0 | 0 | 0.716506 | 0.202444 |
| Gene_Pert | Htt KO mou | 1/319 | 0.753864 | 0.832891 | 0 | 0 | 0.716506 | 0.202444 |
| Gene_Pert | Trim28 knc | 1/318 | 0.752771 | 0.832891 | 0 | 0 | 0.718803 | 0.204135 |
| Gene_Pert | Dnajc7 KO | 1/341 | 0.776734 | 0.832891 | 0 | 0 | 0.669391 | 0.169127 |
| Gene_Pert | FOXP1 KD | 1/317 | 0.751674 | 0.832891 | 0 | 0 | 0.721114 | 0.205844 |
| Gene_Pert | Ddit3 knoc | 1/317 | 0.751674 | 0.832891 | 0 | 0 | 0.721114 | 0.205844 |
| Gene_Pert | Nfyb KD m | 1/322 | 0.757114 | 0.832891 | 0 | 0 | 0.709701 | 0.197468 |
| Gene_Pert | Adra2b KO | 1/322 | 0.757114 | 0.832891 | 0 | 0 | 0.709701 | 0.197468 |
| Gene_Pert | TP63 hum | 1/331 | 0.766611 | 0.832891 | 0 | 0 | 0.690028 | 0.183392 |
| Gene_Pert | Nfe2l2 KO | 1/331 | 0.766611 | 0.832891 | 0 | 0 | 0.690028 | 0.183392 |
| Gene_Pert | Lrp5 KO m | 1/331 | 0.766611 | 0.832891 | 0 | 0 | 0.690028 | 0.183392 |
| Gene_Pert | ZNF750 KD | 1/331 | 0.766611 | 0.832891 | 0 | 0 | 0.690028 | 0.183392 |
| Gene_Pert | GRN defici | 1/331 | 0.766611 | 0.832891 | 0 | 0 | 0.690028 | 0.183392 |
| Gene_Pert | MIR142 OE | 1/331 | 0.766611 | 0.832891 | 0 | 0 | 0.690028 | 0.183392 |
| Gene_Pert | Pparg OE n | 1/333 | 0.768672 | 0.832891 | 0 | 0 | 0.685801 | 0.180429 |
| Gene_Pert | HIPK2 KD h | 1/336 | 0.771728 | 0.832891 | 0 | 0 | 0.679556 | 0.176089 |
| Gene_Pert | Lamtor2 K | 3/384 | 0.233929 | 0.832891 | 0 | 0 | 1.830896 | 2.659811 |
| Gene_Pert | Pink1 KO m | 1/334 | 0.769695 | 0.832891 | 0 | 0 | 0.683707 | 0.178968 |
| Gene_Pert | Nr3c1 KO r | 1/334 | 0.769695 | 0.832891 | 0 | 0 | 0.683707 | 0.178968 |
| Gene_Pert | NRAS DEPL | 1/338 | 0.773744 | 0.832891 | 0 | 0 | 0.675454 | 0.173264 |
| Gene_Pert | YY1 KD hur | 1/338 | 0.773744 | 0.832891 | 0 | 0 | 0.675454 | 0.173264 |
| Gene_Pert | Satb1 OE n | 1/332 | 0.767644 | 0.832891 | 0 | 0 | 0.687908 | 0.181903 |
| Gene_Pert | Il10 KO mo | 1/332 | 0.767644 | 0.832891 | 0 | 0 | 0.687908 | 0.181903 |
| Gene_Pert | LMO3 KD h | 1/332 | 0.767644 | 0.832891 | 0 | 0 | 0.687908 | 0.181903 |
| Gene_Pert | MAPK7 KD | 1/332 | 0.767644 | 0.832891 | 0 | 0 | 0.687908 | 0.181903 |
| Gene_Pert | Cyp27b1 K | 1/332 | 0.767644 | 0.832891 | 0 | 0 | 0.687908 | 0.181903 |
| Gene_Pert | Spi1 KD mc | 1/331 | 0.766611 | 0.832891 | 0 | 0 | 0.690028 | 0.183392 |
| Gene_Pert | ETS2 KD hu | 1/336 | 0.771728 | 0.832891 | 0 | 0 | 0.679556 | 0.176089 |
| Gene_Pert | Hoxc13 OE | 3/384 | 0.233929 | 0.832891 | 0 | 0 | 1.830896 | 2.659811 |

| | | | | | | | | |
|---|---|---|---|---|---|---|---|---|
| Gene_Pert | UBR5 DEPL | 1/350 | 0.785472 | 0.832891 | 0 | 0 | 0.651829 | 0.157397 |
| Gene_Pert | SDC1 KD hu | 1/349 | 0.784518 | 0.832891 | 0 | 0 | 0.653736 | 0.158652 |
| Gene_Pert | ZNF217 OE | 1/349 | 0.784518 | 0.832891 | 0 | 0 | 0.653736 | 0.158652 |
| Gene_Pert | Traf3 KO m | 1/349 | 0.784518 | 0.832891 | 0 | 0 | 0.653736 | 0.158652 |
| Gene_Pert | SOX4 RNAi | 1/349 | 0.784518 | 0.832891 | 0 | 0 | 0.653736 | 0.158652 |
| Gene_Pert | Pla2g4a KC | 2/251 | 0.298257 | 0.832891 | 0 | 0 | 1.858162 | 2.248005 |
| Gene_Pert | Elavl1 KO n | 1/352 | 0.787368 | 0.832891 | 0 | 0 | 0.648049 | 0.154923 |
| Gene_Pert | Wfs1 KO m | 1/352 | 0.787368 | 0.832891 | 0 | 0 | 0.648049 | 0.154923 |
| Gene_Pert | Ncoa3 KD r | 1/330 | 0.765575 | 0.832891 | 0 | 0 | 0.692161 | 0.184896 |
| Gene_Pert | Pmp22 OE | 1/330 | 0.765575 | 0.832891 | 0 | 0 | 0.692161 | 0.184896 |
| Gene_Pert | Htt KO mou | 1/330 | 0.765575 | 0.832891 | 0 | 0 | 0.692161 | 0.184896 |
| Gene_Pert | Inha KO mc | 1/325 | 0.760322 | 0.832891 | 0 | 0 | 0.703022 | 0.192638 |
| Gene_Pert | Il10 KO mo | 1/341 | 0.776734 | 0.832891 | 0 | 0 | 0.669391 | 0.169127 |
| Gene_Pert | TNKS INHIE | 1/340 | 0.775741 | 0.832891 | 0 | 0 | 0.6714 | 0.170493 |
| Gene_Pert | Hey2 KO m | 1/340 | 0.775741 | 0.832891 | 0 | 0 | 0.6714 | 0.170493 |
| Gene_Pert | Il10 KO mo | 1/338 | 0.773744 | 0.832891 | 0 | 0 | 0.675454 | 0.173264 |
| Gene_Pert | Ikzf3 NULL | 1/345 | 0.78066 | 0.832891 | 0 | 0 | 0.661472 | 0.163791 |
| Gene_Pert | CCL3 KO m | 1/345 | 0.78066 | 0.832891 | 0 | 0 | 0.661472 | 0.163791 |
| Gene_Pert | Rpe65 KD r | 1/324 | 0.759257 | 0.832891 | 0 | 0 | 0.705234 | 0.194232 |
| Gene_Pert | MAPK1 KD | 1/324 | 0.759257 | 0.832891 | 0 | 0 | 0.705234 | 0.194232 |
| Gene_Pert | PBRM1 KO | 1/324 | 0.759257 | 0.832891 | 0 | 0 | 0.705234 | 0.194232 |
| Gene_Pert | Trip12 KO r | 1/323 | 0.758188 | 0.832891 | 0 | 0 | 0.707461 | 0.195842 |
| Gene_Pert | Gtf2i KO m | 1/323 | 0.758188 | 0.832891 | 0 | 0 | 0.707461 | 0.195842 |
| Gene_Pert | CFTR D508 | 1/322 | 0.757114 | 0.832891 | 0 | 0 | 0.709701 | 0.197468 |
| Gene_Pert | Foxd3 OE n | 1/311 | 0.744986 | 0.832891 | 0 | 0 | 0.735296 | 0.216463 |
| Gene_Pert | Fgfr1 druga | 1/325 | 0.760322 | 0.832891 | 0 | 0 | 0.703022 | 0.192638 |
| Gene_Pert | Uqcr10 OE | 1/321 | 0.756036 | 0.832891 | 0 | 0 | 0.711955 | 0.19911 |
| Gene_Pert | GSK3B KD l | 1/321 | 0.756036 | 0.832891 | 0 | 0 | 0.711955 | 0.19911 |
| Gene_Pert | NFKB1 Acti | 1/321 | 0.756036 | 0.832891 | 0 | 0 | 0.711955 | 0.19911 |
| Gene_Pert | Pparg OE n | 1/321 | 0.756036 | 0.832891 | 0 | 0 | 0.711955 | 0.19911 |
| Gene_Pert | TYROBP KD | 2/208 | 0.229002 | 0.832891 | 0 | 0 | 2.250942 | 3.317943 |
| Gene_Pert | Foxp3 Indu | 3/380 | 0.229328 | 0.832891 | 0 | 0 | 1.850701 | 2.725344 |
| Gene_Pert | Rtn4 KO m | 3/382 | 0.231626 | 0.832891 | 0 | 0 | 1.840746 | 2.692329 |
| Gene_Pert | S1pr3 KO n | 2/210 | 0.23221 | 0.832891 | 0 | 0 | 2.229072 | 3.254694 |
| Gene_Pert | YAP1 OE hu | 2/204 | 0.222596 | 0.832891 | 0 | 0 | 2.295981 | 3.449476 |
| Gene_Pert | Rora gene | 3/384 | 0.233929 | 0.832891 | 0 | 0 | 1.830896 | 2.659811 |
| Gene_Pert | Lmna Lmna | 3/335 | 0.179051 | 0.832891 | 0 | 0 | 2.10639 | 3.623169 |
| Gene_Pert | Gfi1b KO m | 1/300 | 0.73226 | 0.832891 | 0 | 0 | 0.762775 | 0.237696 |
| Gene_Pert | E2f2 KD mc | 1/300 | 0.73226 | 0.832891 | 0 | 0 | 0.762775 | 0.237696 |
| Gene_Pert | PRKCDBP K | 1/300 | 0.73226 | 0.832891 | 0 | 0 | 0.762775 | 0.237696 |
| Gene_Pert | SYK KD hur | 1/300 | 0.73226 | 0.832891 | 0 | 0 | 0.762775 | 0.237696 |
| Gene_Pert | Cbx3 KD m | 1/299 | 0.731073 | 0.832891 | 0 | 0 | 0.765374 | 0.239748 |
| Gene_Pert | Stat1 KO m | 1/299 | 0.731073 | 0.832891 | 0 | 0 | 0.765374 | 0.239748 |
| Gene_Pert | FLI1 KD hui | 1/302 | 0.73462 | 0.832891 | 0 | 0 | 0.75763 | 0.233655 |
| Gene_Pert | BRD4 drugi | 1/302 | 0.73462 | 0.832891 | 0 | 0 | 0.75763 | 0.233655 |

| | | | | | | | | |
|---|---|---|---|---|---|---|---|---|
| Gene_Pert | TGFB1I1 KD | 1/299 | 0.731073 | 0.832891 | 0 | 0 | 0.765374 | 0.239748 |
| Gene_Pert | Epha4 knoc | 1/299 | 0.731073 | 0.832891 | 0 | 0 | 0.765374 | 0.239748 |
| Gene_Pert | Tet2 KO mo | 1/299 | 0.731073 | 0.832891 | 0 | 0 | 0.765374 | 0.239748 |
| Gene_Pert | ETS1 OE hu | 1/299 | 0.731073 | 0.832891 | 0 | 0 | 0.765374 | 0.239748 |
| Gene_Pert | Mecp2 KO | 1/312 | 0.746113 | 0.832891 | 0 | 0 | 0.732895 | 0.214649 |
| Gene_Pert | Map2k1 dr | 1/308 | 0.741576 | 0.832891 | 0 | 0 | 0.742595 | 0.222019 |
| Gene_Pert | Cbx2 KO m | 1/308 | 0.741576 | 0.832891 | 0 | 0 | 0.742595 | 0.222019 |
| Gene_Pert | Adora2a KO | 1/309 | 0.742718 | 0.832891 | 0 | 0 | 0.740146 | 0.220149 |
| Gene_Pert | LAMC2 KD | 1/309 | 0.742718 | 0.832891 | 0 | 0 | 0.740146 | 0.220149 |
| Gene_Pert | Vdr KO mo | 1/308 | 0.741576 | 0.832891 | 0 | 0 | 0.742595 | 0.222019 |
| Gene_Pert | PPARGC1A | 1/307 | 0.740429 | 0.832891 | 0 | 0 | 0.74506 | 0.223909 |
| Gene_Pert | SYNCRIP OI | 1/307 | 0.740429 | 0.832891 | 0 | 0 | 0.74506 | 0.223909 |
| Gene_Pert | USF1 KD hu | 1/307 | 0.740429 | 0.832891 | 0 | 0 | 0.74506 | 0.223909 |
| Gene_Pert | NRAS DEPL | 1/307 | 0.740429 | 0.832891 | 0 | 0 | 0.74506 | 0.223909 |
| Gene_Pert | Snca KO mo | 1/307 | 0.740429 | 0.832891 | 0 | 0 | 0.74506 | 0.223909 |
| Gene_Pert | IL13RA1 Sti | 1/309 | 0.742718 | 0.832891 | 0 | 0 | 0.740146 | 0.220149 |
| Gene_Pert | MIR210 OE | 2/594 | 0.734801 | 0.832891 | 0 | 0 | 0.767925 | 0.236641 |
| Gene_Pert | Pmp22 OE | 1/297 | 0.728682 | 0.832891 | 0 | 0 | 0.770624 | 0.243916 |
| Gene_Pert | ZNF395 KD | 1/294 | 0.725056 | 0.832891 | 0 | 0 | 0.778633 | 0.250335 |
| Gene_Pert | Pten KO mo | 1/293 | 0.723837 | 0.832891 | 0 | 0 | 0.78134 | 0.25252 |
| Gene_Pert | Setd2 KO n | 1/293 | 0.723837 | 0.832891 | 0 | 0 | 0.78134 | 0.25252 |
| Gene_Pert | Pbx1 Expre | 1/293 | 0.723837 | 0.832891 | 0 | 0 | 0.78134 | 0.25252 |
| Gene_Pert | Ring1 KO m | 1/303 | 0.735792 | 0.832891 | 0 | 0 | 0.755082 | 0.231665 |
| Gene_Pert | Sh2b3 KO r | 1/303 | 0.735792 | 0.832891 | 0 | 0 | 0.755082 | 0.231665 |
| Gene_Pert | Npc1 KO m | 1/305 | 0.738121 | 0.832891 | 0 | 0 | 0.750038 | 0.227747 |
| Gene_Pert | Dgcr8 KO n | 1/305 | 0.738121 | 0.832891 | 0 | 0 | 0.750038 | 0.227747 |
| Gene_Pert | TGFB1I1 KD | 1/303 | 0.735792 | 0.832891 | 0 | 0 | 0.755082 | 0.231665 |
| Gene_Pert | TCF12 KD h | 1/303 | 0.735792 | 0.832891 | 0 | 0 | 0.755082 | 0.231665 |
| Gene_Pert | Srf KO mou | 1/303 | 0.735792 | 0.832891 | 0 | 0 | 0.755082 | 0.231665 |
| Gene_Pert | Clpp KO mo | 1/309 | 0.742718 | 0.832891 | 0 | 0 | 0.740146 | 0.220149 |
| Gene_Pert | Dot1l DELE | 1/302 | 0.73462 | 0.832891 | 0 | 0 | 0.75763 | 0.233655 |
| Gene_Pert | MIF DEPLE | 1/302 | 0.73462 | 0.832891 | 0 | 0 | 0.75763 | 0.233655 |
| Gene_Pert | JAK2 knock | 1/304 | 0.736959 | 0.832891 | 0 | 0 | 0.752552 | 0.229696 |
| Gene_Pert | FOXA1 OE I | 1/303 | 0.735792 | 0.832891 | 0 | 0 | 0.755082 | 0.231665 |
| Gene_Pert | SPDEF KD h | 1/310 | 0.743855 | 0.832891 | 0 | 0 | 0.737714 | 0.218297 |
| Gene_Pert | DENND4B ( | 1/310 | 0.743855 | 0.832891 | 0 | 0 | 0.737714 | 0.218297 |
| Gene_Pert | LAMP3 KD | 1/310 | 0.743855 | 0.832891 | 0 | 0 | 0.737714 | 0.218297 |
| Gene_Pert | Egln3 KO m | 1/310 | 0.743855 | 0.832891 | 0 | 0 | 0.737714 | 0.218297 |
| Gene_Pert | Notch1 OE | 1/310 | 0.743855 | 0.832891 | 0 | 0 | 0.737714 | 0.218297 |
| Gene_Pert | Ctla4 KO m | 1/310 | 0.743855 | 0.832891 | 0 | 0 | 0.737714 | 0.218297 |
| Gene_Pert | Fosl1 KO m | 1/312 | 0.746113 | 0.832891 | 0 | 0 | 0.732895 | 0.214649 |
| Gene_Pert | Sod1 G93A | 1/325 | 0.760322 | 0.832891 | 0 | 0 | 0.703022 | 0.192638 |
| Gene_Pert | AKAP12 OE | 1/315 | 0.749464 | 0.832891 | 0 | 0 | 0.725781 | 0.209313 |
| Gene_Pert | RNF185 KD | 1/315 | 0.749464 | 0.832891 | 0 | 0 | 0.725781 | 0.209313 |
| Gene_Pert | Htt CAG kn | 1/312 | 0.746113 | 0.832891 | 0 | 0 | 0.732895 | 0.214649 |

| | | | | | | | | |
|---|---|---|---|---|---|---|---|---|
| Gene_Pert | NRAS DEPL | 1/312 | 0.746113 | 0.832891 | 0 | 0 | 0.732895 | 0.214649 |
| Gene_Pert | Htt Mutati | 1/312 | 0.746113 | 0.832891 | 0 | 0 | 0.732895 | 0.214649 |
| Gene_Pert | Pde10a KO | 1/312 | 0.746113 | 0.832891 | 0 | 0 | 0.732895 | 0.214649 |
| Gene_Pert | MIR126 OE | 1/312 | 0.746113 | 0.832891 | 0 | 0 | 0.732895 | 0.214649 |
| Gene_Pert | Acadm KO | 1/311 | 0.744986 | 0.832891 | 0 | 0 | 0.735296 | 0.216463 |
| Gene_Pert | MALAT1 OI | 1/314 | 0.748352 | 0.832891 | 0 | 0 | 0.728137 | 0.211074 |
| Gene_Pert | Il10 KO mo | 1/314 | 0.748352 | 0.832891 | 0 | 0 | 0.728137 | 0.211074 |
| Gene_Pert | PAGE5 KD | 1/325 | 0.760322 | 0.832891 | 0 | 0 | 0.703022 | 0.192638 |
| Gene_Pert | TTN TMD n | 1/325 | 0.760322 | 0.832891 | 0 | 0 | 0.703022 | 0.192638 |
| Gene_Pert | PTEN KD hu | 1/312 | 0.746113 | 0.832891 | 0 | 0 | 0.732895 | 0.214649 |
| Gene_Pert | Neurod1 C | 1/324 | 0.759257 | 0.832891 | 0 | 0 | 0.705234 | 0.194232 |
| Gene_Pert | Cftr KO mo | 1/324 | 0.759257 | 0.832891 | 0 | 0 | 0.705234 | 0.194232 |
| Gene_Pert | FH mutatio | 1/324 | 0.759257 | 0.832891 | 0 | 0 | 0.705234 | 0.194232 |
| Gene_Pert | Ppara KO n | 1/328 | 0.763487 | 0.832891 | 0 | 0 | 0.696465 | 0.187947 |
| Gene_Pert | Gnb5 KO m | 1/327 | 0.762437 | 0.832891 | 0 | 0 | 0.698637 | 0.189496 |
| Gene_Pert | WHSC1 KD | 1/309 | 0.742718 | 0.832891 | 0 | 0 | 0.740146 | 0.220149 |
| Gene_Pert | Calcr KO m | 1/309 | 0.742718 | 0.832891 | 0 | 0 | 0.740146 | 0.220149 |
| Gene_Pert | Pten KO mo | 1/309 | 0.742718 | 0.832891 | 0 | 0 | 0.740146 | 0.220149 |
| Gene_Pert | SYK druginl | 1/309 | 0.742718 | 0.832891 | 0 | 0 | 0.740146 | 0.220149 |
| Gene_Pert | Rpe65 KO r | 1/309 | 0.742718 | 0.832891 | 0 | 0 | 0.740146 | 0.220149 |
| Gene_Pert | Dbh KO mo | 1/309 | 0.742718 | 0.832891 | 0 | 0 | 0.740146 | 0.220149 |
| Gene_Pert | HNRNPU-A | 1/304 | 0.736959 | 0.832891 | 0 | 0 | 0.752552 | 0.229696 |
| Gene_Pert | Tet2 KO mo | 1/317 | 0.751674 | 0.832891 | 0 | 0 | 0.721114 | 0.205844 |
| Gene_Pert | H3f3b KO r | 1/316 | 0.750571 | 0.832891 | 0 | 0 | 0.72344 | 0.20757 |
| Gene_Pert | Uqcr10 OE | 1/316 | 0.750571 | 0.832891 | 0 | 0 | 0.72344 | 0.20757 |
| Gene_Pert | RASGRF1 K | 1/316 | 0.750571 | 0.832891 | 0 | 0 | 0.72344 | 0.20757 |
| Gene_Pert | NFKB1 Inac | 1/316 | 0.750571 | 0.832891 | 0 | 0 | 0.72344 | 0.20757 |
| Gene_Pert | RNF2 KD hu | 1/315 | 0.749464 | 0.832891 | 0 | 0 | 0.725781 | 0.209313 |
| Gene_Pert | Ppargc1a N | 1/320 | 0.754952 | 0.832891 | 0 | 0 | 0.714223 | 0.200769 |
| Gene_Pert | Ptbp2 KO n | 1/320 | 0.754952 | 0.832891 | 0 | 0 | 0.714223 | 0.200769 |
| Gene_Pert | Jun KO mo | 1/304 | 0.736959 | 0.832891 | 0 | 0 | 0.752552 | 0.229696 |
| Gene_Pert | IGF1 OE hu | 1/304 | 0.736959 | 0.832891 | 0 | 0 | 0.752552 | 0.229696 |
| Gene_Pert | Batf KO mo | 1/304 | 0.736959 | 0.832891 | 0 | 0 | 0.752552 | 0.229696 |
| Gene_Pert | EWSR1 KD | 1/304 | 0.736959 | 0.832891 | 0 | 0 | 0.752552 | 0.229696 |
| Gene_Pert | CLDN1 KD | 1/349 | 0.784518 | 0.832891 | 0 | 0 | 0.653736 | 0.158652 |
| Gene_Pert | Atf7ip2 De | 1/304 | 0.736959 | 0.832891 | 0 | 0 | 0.752552 | 0.229696 |
| Gene_Pert | Dicer1 Defi | 1/307 | 0.740429 | 0.832891 | 0 | 0 | 0.74506 | 0.223909 |
| Gene_Pert | FGFR3 KD h | 1/307 | 0.740429 | 0.832891 | 0 | 0 | 0.74506 | 0.223909 |
| Gene_Pert | Sox2 KD mo | 1/303 | 0.735792 | 0.832891 | 0 | 0 | 0.755082 | 0.231665 |
| Gene_Pert | SDC1 OE hu | 1/303 | 0.735792 | 0.832891 | 0 | 0 | 0.755082 | 0.231665 |
| Gene_Pert | OPTN KD h | 1/303 | 0.735792 | 0.832891 | 0 | 0 | 0.755082 | 0.231665 |
| Gene_Pert | HIF1A KD h | 1/303 | 0.735792 | 0.832891 | 0 | 0 | 0.755082 | 0.231665 |
| Gene_Pert | Notch1 De | 3/378 | 0.227035 | 0.832891 | 0 | 0 | 1.860762 | 2.758864 |
| Gene_Pert | Rp1 KO mo | 2/207 | 0.227399 | 0.832891 | 0 | 0 | 2.262037 | 3.350186 |
| Gene_Pert | PDE1C KD | 3/379 | 0.228181 | 0.832891 | 0 | 0 | 1.855718 | 2.74204 |

| | | | | | | | | |
|---|---|---|---|---|---|---|---|---|
| Gene_Pert | IGF1 OE hu | 1/312 | 0.746113 | 0.832891 | 0 | 0 | 0.732895 | 0.214649 |
| Gene_Pert | Pde6b RD1 | 1/372 | 0.805435 | 0.832891 | 0 | 0 | 0.612487 | 0.132526 |
| Gene_Pert | NOTCH1 Re | 1/380 | 0.812229 | 0.832891 | 0 | 0 | 0.599313 | 0.124641 |
| Gene_Pert | Il1rn KO mo | 1/380 | 0.812229 | 0.832891 | 0 | 0 | 0.599313 | 0.124641 |
| Gene_Pert | KDM4C nat | 1/380 | 0.812229 | 0.832891 | 0 | 0 | 0.599313 | 0.124641 |
| Gene_Pert | Nfe2l2 KO | 1/374 | 0.807156 | 0.832891 | 0 | 0 | 0.60914 | 0.130501 |
| Gene_Pert | S1pr3 KO n | 1/373 | 0.806297 | 0.832891 | 0 | 0 | 0.610809 | 0.131509 |
| Gene_Pert | Kdm2b KD | 1/373 | 0.806297 | 0.832891 | 0 | 0 | 0.610809 | 0.131509 |
| Gene_Pert | Arhgap5 KI | 1/373 | 0.806297 | 0.832891 | 0 | 0 | 0.610809 | 0.131509 |
| Gene_Pert | ZNF207 OE | 1/373 | 0.806297 | 0.832891 | 0 | 0 | 0.610809 | 0.131509 |
| Gene_Pert | NOTCH1 Re | 1/372 | 0.805435 | 0.832891 | 0 | 0 | 0.612487 | 0.132526 |
| Gene_Pert | CPSF6 KD h | 1/378 | 0.810553 | 0.832891 | 0 | 0 | 0.602554 | 0.12656 |
| Gene_Pert | Pou1f1 Sne | 1/378 | 0.810553 | 0.832891 | 0 | 0 | 0.602554 | 0.12656 |
| Gene_Pert | Hey2 KO m | 1/372 | 0.805435 | 0.832891 | 0 | 0 | 0.612487 | 0.132526 |
| Gene_Pert | Znrf2 KO m | 2/227 | 0.259573 | 0.832891 | 0 | 0 | 2.058876 | 2.776839 |
| Gene_Pert | Taf7l KO m | 1/372 | 0.805435 | 0.832891 | 0 | 0 | 0.612487 | 0.132526 |
| Gene_Pert | Pml KO mo | 1/371 | 0.804569 | 0.832891 | 0 | 0 | 0.614173 | 0.133551 |
| Gene_Pert | Dicer1 Defi | 2/227 | 0.259573 | 0.832891 | 0 | 0 | 2.058876 | 2.776839 |
| Gene_Pert | Myc myc +, | 3/407 | 0.260688 | 0.832891 | 0 | 0 | 1.724629 | 2.318644 |
| Gene_Pert | RB1 KD hur | 2/217 | 0.243461 | 0.832891 | 0 | 0 | 2.155732 | 3.045615 |
| Gene_Pert | Nrf1 KO mo | 2/230 | 0.264412 | 0.832891 | 0 | 0 | 2.031476 | 2.702365 |
| Gene_Pert | SP3 KD hur | 3/411 | 0.265386 | 0.832891 | 0 | 0 | 1.70737 | 2.264943 |
| Gene_Pert | Ppargc1a K | 2/231 | 0.266025 | 0.832891 | 0 | 0 | 2.022502 | 2.678127 |
| Gene_Pert | HSPA1A tra | 2/232 | 0.267638 | 0.832891 | 0 | 0 | 2.013606 | 2.654174 |
| Gene_Pert | Psap hypor | 2/232 | 0.267638 | 0.832891 | 0 | 0 | 2.013606 | 2.654174 |
| Gene_Pert | Hif1a KO m | 3/414 | 0.268918 | 0.832891 | 0 | 0 | 1.694647 | 2.225665 |
| Gene_Pert | PIK3CA OE | 2/233 | 0.269251 | 0.832891 | 0 | 0 | 2.004787 | 2.630503 |
| Gene_Pert | FGFR1 mut | 3/355 | 0.201022 | 0.832891 | 0 | 0 | 1.984679 | 3.184098 |
| Gene_Pert | MIR128-1 I | 1/354 | 0.789247 | 0.832891 | 0 | 0 | 0.644311 | 0.152493 |
| Gene_Pert | Polg Mutat | 1/354 | 0.789247 | 0.832891 | 0 | 0 | 0.644311 | 0.152493 |
| Gene_Pert | Dmd Defici | 3/377 | 0.225889 | 0.832891 | 0 | 0 | 1.865833 | 2.775817 |
| Gene_Pert | Map2k7 m | 2/186 | 0.193998 | 0.832891 | 0 | 0 | 2.52289 | 4.137304 |
| Gene_Pert | TNF OE hur | 3/349 | 0.194361 | 0.832891 | 0 | 0 | 2.019715 | 3.308368 |
| Gene_Pert | CDK4 knocl | 3/349 | 0.194361 | 0.832891 | 0 | 0 | 2.019715 | 3.308368 |
| Gene_Pert | RNF40 KD I | 3/350 | 0.195467 | 0.832891 | 0 | 0 | 2.013792 | 3.287236 |
| Gene_Pert | Cadm1 KO | 2/187 | 0.195575 | 0.832891 | 0 | 0 | 2.509126 | 4.094425 |
| Gene_Pert | Mtf2 KD m | 3/351 | 0.196575 | 0.832891 | 0 | 0 | 2.007902 | 3.266274 |
| Gene_Pert | Lat2 KO mo | 3/352 | 0.197685 | 0.832891 | 0 | 0 | 2.002047 | 3.245481 |
| Gene_Pert | Eif2ak3 KO | 2/183 | 0.189279 | 0.832891 | 0 | 0 | 2.565096 | 4.269683 |
| Gene_Pert | CTNNB1 OI | 3/355 | 0.201022 | 0.832891 | 0 | 0 | 1.984679 | 3.184098 |
| Gene_Pert | GSK3B kno | 2/223 | 0.253125 | 0.832891 | 0 | 0 | 2.096566 | 2.880418 |
| Gene_Pert | Vax2 KO m | 3/355 | 0.201022 | 0.832891 | 0 | 0 | 1.984679 | 3.184098 |
| Gene_Pert | Akt1 knock | 2/191 | 0.201897 | 0.832891 | 0 | 0 | 2.455524 | 3.928837 |
| Gene_Pert | ABL1 drugi | 3/356 | 0.202138 | 0.832891 | 0 | 0 | 1.978956 | 3.163964 |
| Gene_Pert | BRD4 knoc | 3/356 | 0.202138 | 0.832891 | 0 | 0 | 1.978956 | 3.163964 |

| | | | | | | | | |
|---|---|---|---|---|---|---|---|---|
| Gene_Pert | TNF Stimul | 3/356 | 0.202138 | 0.832891 | 0 | 0 | 1.978956 | 3.163964 |
| Gene_Pert | Prmt1 KD r | 1/379 | 0.811393 | 0.832891 | 0 | 0 | 0.600929 | 0.125596 |
| Gene_Pert | Chsy1 KO n | 1/379 | 0.811393 | 0.832891 | 0 | 0 | 0.600929 | 0.125596 |
| Gene_Pert | Chrna4 KD | 3/348 | 0.193257 | 0.832891 | 0 | 0 | 2.025673 | 3.329673 |
| Gene_Pert | Suz12 KO n | 3/402 | 0.254832 | 0.832891 | 0 | 0 | 1.746688 | 2.387989 |
| Gene_Pert | Hmgn1 KO | 2/225 | 0.256348 | 0.832891 | 0 | 0 | 2.077552 | 2.828 |
| Gene_Pert | Scd1 Defici | 2/225 | 0.256348 | 0.832891 | 0 | 0 | 2.077552 | 2.828 |
| Gene_Pert | Top2b KO r | 2/226 | 0.257961 | 0.832891 | 0 | 0 | 2.068172 | 2.802265 |
| Gene_Pert | Dgcr8 KO n | 2/215 | 0.240243 | 0.832891 | 0 | 0 | 2.176194 | 3.103476 |
| Gene_Pert | TP63 Muta | 3/427 | 0.284281 | 0.832891 | 0 | 0 | 1.641594 | 2.064784 |
| Gene_Pert | Runx1 KO r | 2/243 | 0.285377 | 0.832891 | 0 | 0 | 1.920625 | 2.408358 |
| Gene_Pert | Rest KD mc | 2/244 | 0.286988 | 0.832891 | 0 | 0 | 1.912591 | 2.387516 |
| Gene_Pert | Abcb4 KO r | 2/244 | 0.286988 | 0.832891 | 0 | 0 | 1.912591 | 2.387516 |
| Gene_Pert | Zmpste24 I | 2/245 | 0.288599 | 0.832891 | 0 | 0 | 1.904624 | 2.366908 |
| Gene_Pert | Twist1 OE r | 2/245 | 0.288599 | 0.832891 | 0 | 0 | 1.904624 | 2.366908 |
| Gene_Pert | Chrna4 KO | 2/240 | 0.280541 | 0.832891 | 0 | 0 | 1.945131 | 2.472331 |
| Gene_Pert | BRCA1 DEF | 3/387 | 0.237391 | 0.832891 | 0 | 0 | 1.816313 | 2.611943 |
| Gene_Pert | Foxp3 Defi | 3/388 | 0.238547 | 0.832891 | 0 | 0 | 1.811503 | 2.596225 |
| Gene_Pert | Pdk1 knock | 2/214 | 0.238636 | 0.832891 | 0 | 0 | 2.18657 | 3.132957 |
| Gene_Pert | Cebpg KO r | 2/214 | 0.238636 | 0.832891 | 0 | 0 | 2.18657 | 3.132957 |
| Gene_Pert | BST2 OE hu | 2/214 | 0.238636 | 0.832891 | 0 | 0 | 2.18657 | 3.132957 |
| Gene_Pert | Nfe2l2 KO r | 2/242 | 0.283765 | 0.832891 | 0 | 0 | 1.928725 | 2.429439 |
| Gene_Pert | MYC KD hu | 2/216 | 0.241852 | 0.832891 | 0 | 0 | 2.165915 | 3.074364 |
| Gene_Pert | Psap KO m | 2/210 | 0.23221 | 0.832891 | 0 | 0 | 2.229072 | 3.254694 |
| Gene_Pert | Prkaca kno | 2/218 | 0.245071 | 0.832891 | 0 | 0 | 2.145643 | 3.017223 |
| Gene_Pert | WTAP KD h | 2/219 | 0.246681 | 0.832891 | 0 | 0 | 2.135647 | 2.989182 |
| Gene_Pert | BMI1 OE m | 2/219 | 0.246681 | 0.832891 | 0 | 0 | 2.135647 | 2.989182 |
| Gene_Pert | Dmd Defici | 2/221 | 0.249902 | 0.832891 | 0 | 0 | 2.115928 | 2.934131 |
| Gene_Pert | RNF40 KD I | 3/400 | 0.252495 | 0.832891 | 0 | 0 | 1.755668 | 2.41644 |
| Gene_Pert | Nfix KD mo | 2/223 | 0.253125 | 0.832891 | 0 | 0 | 2.096566 | 2.880418 |
| Gene_Pert | Nfe2l1 KO r | 2/223 | 0.253125 | 0.832891 | 0 | 0 | 2.096566 | 2.880418 |
| Gene_Pert | Sp3 Deficie | 3/387 | 0.237391 | 0.832891 | 0 | 0 | 1.816313 | 2.611943 |
| Gene_Pert | Clpp KO mc | 2/251 | 0.298257 | 0.832891 | 0 | 0 | 1.858162 | 2.248005 |
| Gene_Pert | Ulk1 knock | 2/251 | 0.298257 | 0.832891 | 0 | 0 | 1.858162 | 2.248005 |
| Gene_Pert | Sox3 KO m | 2/206 | 0.225797 | 0.832891 | 0 | 0 | 2.273241 | 3.382851 |
| Gene_Pert | Akt1 active | 3/370 | 0.217908 | 0.832891 | 0 | 0 | 1.902102 | 2.898203 |
| Gene_Pert | Fgfr1 active | 3/371 | 0.219044 | 0.832891 | 0 | 0 | 1.896836 | 2.880312 |
| Gene_Pert | Zfp335 KD | 2/202 | 0.219398 | 0.832891 | 0 | 0 | 2.319176 | 3.517879 |
| Gene_Pert | HSF1 KD hu | 3/372 | 0.220182 | 0.832891 | 0 | 0 | 1.891599 | 2.862559 |
| Gene_Pert | Apod OE m | 3/372 | 0.220182 | 0.832891 | 0 | 0 | 1.891599 | 2.862559 |
| Gene_Pert | Nrl Deficier | 3/373 | 0.221321 | 0.832891 | 0 | 0 | 1.88639 | 2.844943 |
| Gene_Pert | PADI4 KD h | 3/374 | 0.222461 | 0.832891 | 0 | 0 | 1.881209 | 2.827462 |
| Gene_Pert | SMARCB1 ( | 3/364 | 0.211116 | 0.832891 | 0 | 0 | 1.934309 | 3.008525 |
| Gene_Pert | FLI1 EWS-F | 2/205 | 0.224196 | 0.832891 | 0 | 0 | 2.284555 | 3.415945 |
| Gene_Pert | Zfp322a KD | 2/205 | 0.224196 | 0.832891 | 0 | 0 | 2.284555 | 3.415945 |

| | | | | | | | | |
|---|---|---|---|---|---|---|---|---|
| Gene_Pert | NFKB1 Inac | 3/376 | 0.224746 | 0.832891 | 0 | 0 | 1.870931 | 2.7929 |
| Gene_Pert | IGH mutati | 3/376 | 0.224746 | 0.832891 | 0 | 0 | 1.870931 | 2.7929 |
| Gene_Pert | Abca1 Hete | 1/330 | 0.765575 | 0.832891 | 0 | 0 | 0.692161 | 0.184896 |
| Gene_Pert | Kcnip3 KO | 2/206 | 0.225797 | 0.832891 | 0 | 0 | 2.273241 | 3.382851 |
| Gene_Pert | Dmd mutat | 2/206 | 0.225797 | 0.832891 | 0 | 0 | 2.273241 | 3.382851 |
| Gene_Pert | YAP1 KD hu | 3/369 | 0.216772 | 0.832891 | 0 | 0 | 1.907397 | 2.916234 |
| Gene_Pert | Chrnb4 Def | 2/240 | 0.280541 | 0.832891 | 0 | 0 | 1.945131 | 2.472331 |
| Gene_Pert | EWSR1 KD | 2/240 | 0.280541 | 0.832891 | 0 | 0 | 1.945131 | 2.472331 |
| Gene_Pert | E2f1 Null m | 2/241 | 0.282153 | 0.832891 | 0 | 0 | 1.936894 | 2.450762 |
| Gene_Pert | Pik3cb OE | 2/241 | 0.282153 | 0.832891 | 0 | 0 | 1.936894 | 2.450762 |
| Gene_Pert | Nr2c2 KO r | 2/241 | 0.282153 | 0.832891 | 0 | 0 | 1.936894 | 2.450762 |
| Gene_Pert | MIR122 An | 3/426 | 0.283096 | 0.832891 | 0 | 0 | 1.645559 | 2.076645 |
| Gene_Pert | SPDEF KD h | 2/242 | 0.283765 | 0.832891 | 0 | 0 | 1.928725 | 2.429439 |
| Gene_Pert | Tsc1 Defici | 2/236 | 0.27409 | 0.832891 | 0 | 0 | 1.978783 | 2.561135 |
| Gene_Pert | Gtf2i KO m | 1/335 | 0.770714 | 0.832891 | 0 | 0 | 0.681625 | 0.177521 |
| Gene_Pert | Nod2 KO m | 1/362 | 0.796599 | 0.832891 | 0 | 0 | 0.629775 | 0.143213 |
| Gene_Pert | Sod1 SOD1 | 1/368 | 0.801947 | 0.832891 | 0 | 0 | 0.619289 | 0.136685 |
| Gene_Pert | EGR3 KD hu | 1/368 | 0.801947 | 0.832891 | 0 | 0 | 0.619289 | 0.136685 |
| Gene_Pert | SYK drugin | 1/338 | 0.773744 | 0.832891 | 0 | 0 | 0.675454 | 0.173264 |
| Gene_Pert | Smarca4 KI | 1/338 | 0.773744 | 0.832891 | 0 | 0 | 0.675454 | 0.173264 |
| Gene_Pert | SIRT1 OE h | 1/337 | 0.772738 | 0.832891 | 0 | 0 | 0.677499 | 0.174669 |
| Gene_Pert | Htt Mutati | 1/337 | 0.772738 | 0.832891 | 0 | 0 | 0.677499 | 0.174669 |
| Gene_Pert | SYK drugin | 1/336 | 0.771728 | 0.832891 | 0 | 0 | 0.679556 | 0.176089 |
| Gene_Pert | Rora KO m | 1/336 | 0.771728 | 0.832891 | 0 | 0 | 0.679556 | 0.176089 |
| Gene_Pert | GSK3A KD | 1/342 | 0.777722 | 0.832891 | 0 | 0 | 0.667394 | 0.167774 |
| Gene_Pert | Nfya KD m | 1/342 | 0.777722 | 0.832891 | 0 | 0 | 0.667394 | 0.167774 |
| Gene_Pert | NFKB1 Inac | 1/335 | 0.770714 | 0.832891 | 0 | 0 | 0.681625 | 0.177521 |
| Gene_Pert | Ndel1 KD n | 1/362 | 0.796599 | 0.832891 | 0 | 0 | 0.629775 | 0.143213 |
| Gene_Pert | Irf8 KO mo | 1/334 | 0.769695 | 0.832891 | 0 | 0 | 0.683707 | 0.178968 |
| Gene_Pert | Aire KO mc | 1/334 | 0.769695 | 0.832891 | 0 | 0 | 0.683707 | 0.178968 |
| Gene_Pert | Uqcr10 OE | 1/353 | 0.788309 | 0.832891 | 0 | 0 | 0.646175 | 0.153702 |
| Gene_Pert | RPS4X KO r | 1/353 | 0.788309 | 0.832891 | 0 | 0 | 0.646175 | 0.153702 |
| Gene_Pert | Atxn1l KO | 1/360 | 0.794785 | 0.832891 | 0 | 0 | 0.633348 | 0.14547 |
| Gene_Pert | PTEN KD hu | 1/359 | 0.793872 | 0.832891 | 0 | 0 | 0.63515 | 0.146613 |
| Gene_Pert | ABL1 muta | 1/352 | 0.787368 | 0.832891 | 0 | 0 | 0.648049 | 0.154923 |
| Gene_Pert | Myod1 Mu | 1/352 | 0.787368 | 0.832891 | 0 | 0 | 0.648049 | 0.154923 |
| Gene_Pert | GPLD1 OE | 1/351 | 0.786422 | 0.832891 | 0 | 0 | 0.649934 | 0.156154 |
| Gene_Pert | Tcf3 KO mc | 1/351 | 0.786422 | 0.832891 | 0 | 0 | 0.649934 | 0.156154 |
| Gene_Pert | Vhl Deficie | 1/351 | 0.786422 | 0.832891 | 0 | 0 | 0.649934 | 0.156154 |
| Gene_Pert | Ikbkb delet | 1/345 | 0.78066 | 0.832891 | 0 | 0 | 0.661472 | 0.163791 |
| Gene_Pert | Gata4 KO r | 1/330 | 0.765575 | 0.832891 | 0 | 0 | 0.692161 | 0.184896 |
| Gene_Pert | Clcn5 KO m | 1/330 | 0.765575 | 0.832891 | 0 | 0 | 0.692161 | 0.184896 |
| Gene_Pert | Rpe65 KD r | 1/331 | 0.766611 | 0.832891 | 0 | 0 | 0.690028 | 0.183392 |
| Gene_Pert | Rb1 Deficie | 1/331 | 0.766611 | 0.832891 | 0 | 0 | 0.690028 | 0.183392 |
| Gene_Pert | Bhlha15 KC | 1/329 | 0.764533 | 0.832891 | 0 | 0 | 0.694307 | 0.186414 |

| | | | | | | | | |
|---|---|---|---|---|---|---|---|---|
| Gene_Pert | FOXM1 INA | 1/329 | 0.764533 | 0.832891 | 0 | 0 | 0.694307 | 0.186414 |
| Gene_Pert | Ep300 KD r | 1/328 | 0.763487 | 0.832891 | 0 | 0 | 0.696465 | 0.187947 |
| Gene_Pert | PIN1 DEPLE | 1/328 | 0.763487 | 0.832891 | 0 | 0 | 0.696465 | 0.187947 |
| Gene_Pert | Aurka KD n | 1/346 | 0.781631 | 0.832891 | 0 | 0 | 0.659521 | 0.162488 |
| Gene_Pert | Nlgn3 KD n | 1/345 | 0.78066 | 0.832891 | 0 | 0 | 0.661472 | 0.163791 |
| Gene_Pert | Hnf1a KO r | 1/349 | 0.784518 | 0.832891 | 0 | 0 | 0.653736 | 0.158652 |
| Gene_Pert | MYB KD hu | 1/349 | 0.784518 | 0.832891 | 0 | 0 | 0.653736 | 0.158652 |
| Gene_Pert | Cldn18 KO | 1/354 | 0.789247 | 0.832891 | 0 | 0 | 0.644311 | 0.152493 |
| Gene_Pert | Gata1 Trun | 1/344 | 0.779685 | 0.832891 | 0 | 0 | 0.663435 | 0.165106 |
| Gene_Pert | E2F1 OE hu | 1/343 | 0.778706 | 0.832891 | 0 | 0 | 0.665409 | 0.166434 |
| Gene_Pert | Trps1 Muta | 1/343 | 0.778706 | 0.832891 | 0 | 0 | 0.665409 | 0.166434 |
| Gene_Pert | Bmpr2 don | 1/343 | 0.778706 | 0.832891 | 0 | 0 | 0.665409 | 0.166434 |
| Gene_Pert | Sarm1 KO r | 1/342 | 0.777722 | 0.832891 | 0 | 0 | 0.667394 | 0.167774 |
| Gene_Pert | EPHA4 drug | 1/347 | 0.782598 | 0.832891 | 0 | 0 | 0.657582 | 0.161197 |
| Gene_Pert | ERBB2 OE h | 1/346 | 0.781631 | 0.832891 | 0 | 0 | 0.659521 | 0.162488 |
| Gene_Pert | HNF4A KD | 1/364 | 0.798398 | 0.832891 | 0 | 0 | 0.626241 | 0.140997 |
| Gene_Pert | Ikzf4 KO m | 1/364 | 0.798398 | 0.832891 | 0 | 0 | 0.626241 | 0.140997 |
| Gene_Pert | Hdac2 KO r | 1/363 | 0.797501 | 0.832891 | 0 | 0 | 0.628003 | 0.1421 |
| Gene_Pert | RUNX1 KD | 1/362 | 0.796599 | 0.832891 | 0 | 0 | 0.629775 | 0.143213 |
| Gene_Pert | Xbp1 KO m | 1/365 | 0.799291 | 0.832891 | 0 | 0 | 0.624489 | 0.139904 |
| Gene_Pert | Kdm2b OE | 1/346 | 0.781631 | 0.832891 | 0 | 0 | 0.659521 | 0.162488 |
| Gene_Pert | NRAS KD h | 1/350 | 0.785472 | 0.832891 | 0 | 0 | 0.651829 | 0.157397 |
| Gene_Pert | Atxn1 Knoc | 1/370 | 0.803699 | 0.832891 | 0 | 0 | 0.615869 | 0.134586 |
| Gene_Pert | Dcx KO mo | 1/354 | 0.789247 | 0.832891 | 0 | 0 | 0.644311 | 0.152493 |
| Gene_Pert | Cdk5rap1 C | 1/370 | 0.803699 | 0.832891 | 0 | 0 | 0.615869 | 0.134586 |
| Gene_Pert | Rnf2 KD mo | 1/374 | 0.807156 | 0.832891 | 0 | 0 | 0.60914 | 0.130501 |
| Gene_Pert | ALK drugin | 1/374 | 0.807156 | 0.832891 | 0 | 0 | 0.60914 | 0.130501 |
| Gene_Pert | Ache AChE | 1/368 | 0.801947 | 0.832891 | 0 | 0 | 0.619289 | 0.136685 |
| Gene_Pert | EP300 KD h | 1/367 | 0.801066 | 0.832891 | 0 | 0 | 0.621013 | 0.137748 |
| Gene_Pert | Retnlb KO r | 1/367 | 0.801066 | 0.832891 | 0 | 0 | 0.621013 | 0.137748 |
| Gene_Pert | Esrrb Muta | 1/366 | 0.80018 | 0.832891 | 0 | 0 | 0.622746 | 0.138822 |
| Gene_Pert | TCF7 KO m | 1/365 | 0.799291 | 0.832891 | 0 | 0 | 0.624489 | 0.139904 |
| Gene_Pert | KDM4C nat | 1/356 | 0.791109 | 0.832891 | 0 | 0 | 0.640616 | 0.150109 |
| Gene_Pert | Prdm14 KC | 1/372 | 0.805435 | 0.832891 | 0 | 0 | 0.612487 | 0.132526 |
| Gene_Pert | Gata1 Delta | 1/372 | 0.805435 | 0.832891 | 0 | 0 | 0.612487 | 0.132526 |
| Gene_Pert | S1pr2 KO n | 2/212 | 0.235422 | 0.832891 | 0 | 0 | 2.207619 | 3.19305 |
| Gene_Pert | Pparg OE n | 2/213 | 0.237028 | 0.832891 | 0 | 0 | 2.197045 | 3.162813 |
| Gene_Pert | E2f1 KD mo | 1/358 | 0.792955 | 0.832891 | 0 | 0 | 0.636962 | 0.147768 |
| Gene_Pert | CTNNB1 KD | 1/357 | 0.792034 | 0.832891 | 0 | 0 | 0.638784 | 0.148933 |
| Gene_Pert | CREBBP KD | 1/357 | 0.792034 | 0.832891 | 0 | 0 | 0.638784 | 0.148933 |
| Gene_Pert | Fosl1 KO m | 1/357 | 0.792034 | 0.832891 | 0 | 0 | 0.638784 | 0.148933 |
| Gene_Pert | HMGN5 Ov | 1/357 | 0.792034 | 0.832891 | 0 | 0 | 0.638784 | 0.148933 |
| Gene_Pert | Map3k8 KC | 1/356 | 0.791109 | 0.832891 | 0 | 0 | 0.640616 | 0.150109 |
| Gene_Pert | MIR140 OE | 1/365 | 0.799291 | 0.832891 | 0 | 0 | 0.624489 | 0.139904 |
| Gene_Pert | Cbx2 KO m | 1/365 | 0.799291 | 0.832891 | 0 | 0 | 0.624489 | 0.139904 |

| | | | | | | | | |
|---|---|---|---|---|---|---|---|---|
| Gene_Pert | MIR210 KD | 1/346 | 0.781631 | 0.832891 | 0 | 0 | 0.659521 | 0.162488 |
| Gene_Pert | Pink1 KO m | 1/348 | 0.78356 | 0.832891 | 0 | 0 | 0.655653 | 0.159919 |
| Gene_Pert | TGFB1I1 KD | 1/348 | 0.78356 | 0.832891 | 0 | 0 | 0.655653 | 0.159919 |
| Gene_Pert | Neurog3 C | 1/347 | 0.782598 | 0.832891 | 0 | 0 | 0.657582 | 0.161197 |
| Gene_Pert | Emd KO m | 1/347 | 0.782598 | 0.832891 | 0 | 0 | 0.657582 | 0.161197 |
| Gene_Pert | Dicer1 KO | 1/347 | 0.782598 | 0.832891 | 0 | 0 | 0.657582 | 0.161197 |
| Gene_Pert | Mylk knock | 1/348 | 0.78356 | 0.832891 | 0 | 0 | 0.655653 | 0.159919 |
| Gene_Pert | Srf homozy | 2/212 | 0.235422 | 0.832891 | 0 | 0 | 2.207619 | 3.19305 |
| Gene_Pert | Bmpr2 R89 | 3/385 | 0.235082 | 0.832891 | 0 | 0 | 1.82601 | 2.643735 |
| Gene_Pert | Dlx5 KO m | 1/350 | 0.785472 | 0.832891 | 0 | 0 | 0.651829 | 0.157397 |
| Gene_Pert | Wfs1 KO m | 1/350 | 0.785472 | 0.832891 | 0 | 0 | 0.651829 | 0.157397 |
| Gene_Pert | Apod KO m | 1/346 | 0.781631 | 0.832891 | 0 | 0 | 0.659521 | 0.162488 |
| Gene_Pert | Irak2 knock | 1/375 | 0.808011 | 0.832891 | 0 | 0 | 0.60748 | 0.129503 |
| Gene_Pert | STAT1 KD | 1/378 | 0.810553 | 0.832891 | 0 | 0 | 0.602554 | 0.12656 |
| Gene_Pert | FOXM1 KD | 1/378 | 0.810553 | 0.832891 | 0 | 0 | 0.602554 | 0.12656 |
| Gene_Pert | Prkcz KO m | 1/378 | 0.810553 | 0.832891 | 0 | 0 | 0.602554 | 0.12656 |
| Gene_Pert | S1pr2 KO n | 1/378 | 0.810553 | 0.832891 | 0 | 0 | 0.602554 | 0.12656 |
| Gene_Pert | Pdss2 KO n | 1/378 | 0.810553 | 0.832891 | 0 | 0 | 0.602554 | 0.12656 |
| Gene_Pert | BCAR1 OE | 1/354 | 0.789247 | 0.832891 | 0 | 0 | 0.644311 | 0.152493 |
| Gene_Pert | GSK3A KD | 1/346 | 0.781631 | 0.832891 | 0 | 0 | 0.659521 | 0.162488 |
| GTEx_Tissu | GTEX-P78B | Jan-35 | 0.141623 | 0.833627 | 0 | 0 | 6.798564 | 13.28839 |
| GTEx_Tissu | GTEX-U4B1 | 2/151 | 0.140245 | 0.833627 | 0 | 0 | 3.121042 | 6.130869 |
| GTEx_Tissu | GTEX-R3RS | 46/9309 | 0.140504 | 0.833627 | 0 | 0 | 1.289947 | 2.531546 |
| GTEx_Tissu | GTEX-NPJ8 | 4/460 | 0.140535 | 0.833627 | 0 | 0 | 2.056331 | 4.035136 |
| GTEx_Tissu | GTEX-WHS | 10/1564 | 0.14099 | 0.833627 | 0 | 0 | 1.534289 | 3.005779 |
| GTEx_Tissu | GTEX-P4PP | 3/299 | 0.141453 | 0.833627 | 0 | 0 | 2.366916 | 4.629183 |
| GTEx_Tissu | GTEX-OIZH | 7/878 | 0.087607 | 0.833703 | 0 | 0 | 1.912945 | 4.65783 |
| GTEx_Tissu | GTEX-U3ZN | 10/1417 | 0.087433 | 0.833703 | 0 | 0 | 1.708157 | 4.162577 |
| GTEx_Tissu | GTEX-WWY | 4/385 | 0.087122 | 0.833703 | 0 | 0 | 2.470607 | 6.029369 |
| Gene_Pert | Rorc KO m | 1/382 | 0.813891 | 0.834123 | 0 | 0 | 0.596106 | 0.122756 |
| PhenGenI_ | Stroke | 4/1127 | 0.729647 | 0.834398 | 0 | 0 | 0.80636 | 0.254161 |
| PhenGenI_ | Lipoprotein | 1/301 | 0.733443 | 0.834607 | 0 | 0 | 0.760194 | 0.235665 |
| Disease_Pe | Bipolar Dis | 1/395 | 0.824342 | 0.835475 | 0 | 0 | 0.576054 | 0.111276 |
| Disease_Pe | sporadic br | 1/400 | 0.828206 | 0.835475 | 0 | 0 | 0.568689 | 0.107194 |
| Disease_Pe | Sjogren's s | 1/393 | 0.822773 | 0.835475 | 0 | 0 | 0.579052 | 0.112959 |
| Disease_Pe | Streptococ | 1/400 | 0.828206 | 0.835475 | 0 | 0 | 0.568689 | 0.107194 |
| Disease_Pe | diffuse larg | 1/397 | 0.825898 | 0.835475 | 0 | 0 | 0.573086 | 0.109622 |
| Disease_Pe | bipolar dis | 1/401 | 0.828968 | 0.835475 | 0 | 0 | 0.567238 | 0.106399 |
| Disease_Pe | nasophary | 1/392 | 0.821983 | 0.835475 | 0 | 0 | 0.580563 | 0.113811 |
| Disease_Pe | Severe acu | 1/405 | 0.831985 | 0.835891 | 0 | 0 | 0.561507 | 0.103284 |
| Disease_Pe | melanoma | 1/404 | 0.831236 | 0.835891 | 0 | 0 | 0.562929 | 0.104053 |
| Gene_Pert | Adnp Defic | 1/385 | 0.816356 | 0.836177 | 0 | 0 | 0.591358 | 0.119989 |
| GeneSigDB | 18537972- | 2/657 | 0.784327 | 0.836667 | 0 | 0 | 0.691801 | 0.168058 |
| GeneSigDB | 15489324- | 6/1758 | 0.787477 | 0.836667 | 0 | 0 | 0.767842 | 0.183454 |
| GeneSigDB | 16081686- | 2/663 | 0.7886 | 0.836667 | 0 | 0 | 0.685307 | 0.162758 |

| | | | | | | | | |
|---|---|---|---|---|---|---|---|---|
| GeneSigDB | 19139136-1 | 1/352 | 0.787368 | 0.836667 | 0 | 0 | 0.648049 | 0.154923 |
| Disease_Pe | esophagus | 1/408 | 0.834213 | 0.83682 | 0 | 0 | 0.557282 | 0.101017 |
| Gene_Pert | TERC KO m | 1/387 | 0.817981 | 0.837369 | 0 | 0 | 0.588234 | 0.118185 |
| GTEx_Tissu | GTEX-XOTC | Dec-64 | 0.143414 | 0.837523 | 0 | 0 | 1.472213 | 2.859067 |
| GTEx_Tissu | GTEX-X585 | 6/814 | 0.143273 | 0.837523 | 0 | 0 | 1.751467 | 3.403105 |
| GTEx_Tissu | GTEX-OHPI | 4/463 | 0.142901 | 0.837523 | 0 | 0 | 2.042576 | 3.974044 |
| Gene_Pert | NR2C2 KD | 1/388 | 0.818789 | 0.837723 | 0 | 0 | 0.586683 | 0.117295 |
| Gene_Pert | Epha4 knoc | 1/389 | 0.819593 | 0.838073 | 0 | 0 | 0.585141 | 0.116413 |
| Disease_Pe | low-grade : | 3/341 | 0.185571 | 0.838313 | 0 | 0 | 2.068364 | 3.483785 |
| Disease_Pe | autism spe | 3/341 | 0.185571 | 0.838313 | 0 | 0 | 2.068364 | 3.483785 |
| Disease_Pe | pulmonary | 3/339 | 0.18339 | 0.838313 | 0 | 0 | 2.080889 | 3.529475 |
| GTEx_Tissu | GTEX-T2IS- | 2/154 | 0.144717 | 0.839228 | 0 | 0 | 3.058978 | 5.912923 |
| GTEx_Tissu | GTEX-X4LF- | 33/6410 | 0.144206 | 0.839228 | 0 | 0 | 1.297162 | 2.511969 |
| GTEx_Tissu | GTEX-NFK9 | 13/2168 | 0.144838 | 0.839228 | 0 | 0 | 1.447633 | 2.797033 |
| Gene_Pert | Hspa9 hete | 1/391 | 0.82119 | 0.839233 | 0 | 0 | 0.582081 | 0.114671 |
| GTEx_Tissu | GTEX-RTLS- | 5/544 | 0.088487 | 0.839321 | 0 | 0 | 2.191728 | 5.314722 |
| Gene_Pert | SYK drugin | 1/392 | 0.821983 | 0.839571 | 0 | 0 | 0.580563 | 0.113811 |
| Gene_Pert | SYK drugin | 1/393 | 0.822773 | 0.839904 | 0 | 0 | 0.579052 | 0.112959 |
| Gene_Pert | ZXDC KD hu | 1/394 | 0.823559 | 0.840234 | 0 | 0 | 0.577549 | 0.112114 |
| GTEx_Tissu | GTEX-SUCS | 3/303 | 0.145488 | 0.840809 | 0 | 0 | 2.334881 | 4.500855 |
| Gene_Pert | Atf2 KO mc | 1/397 | 0.825898 | 0.841201 | 0 | 0 | 0.573086 | 0.109622 |
| Gene_Pert | Tcf3 KO mc | 1/397 | 0.825898 | 0.841201 | 0 | 0 | 0.573086 | 0.109622 |
| Gene_Pert | Tmod3 KO | 1/396 | 0.825122 | 0.841201 | 0 | 0 | 0.574566 | 0.110446 |
| Human_Ge | Testis | 3/384 | 0.233929 | 0.841246 | 0 | 0 | 1.830896 | 2.659811 |
| Human_Ge | TestisGerm | 2/314 | 0.397613 | 0.841246 | 0 | 0 | 1.478205 | 1.363314 |
| Human_Ge | Cerebellum | 1/108 | 0.376303 | 0.841246 | 0 | 0 | 2.152358 | 2.10363 |
| Human_Ge | Spinalcord | 1/105 | 0.368048 | 0.841246 | 0 | 0 | 2.214781 | 2.213764 |
| Human_Ge | pineal day | 4/694 | 0.357045 | 0.841246 | 0 | 0 | 1.342623 | 1.382756 |
| Human_Ge | Hypothalar | Jan-97 | 0.345504 | 0.841246 | 0 | 0 | 2.400315 | 2.550937 |
| Human_Ge | Fetalliver | 1/126 | 0.42364 | 0.841246 | 0 | 0 | 1.840744 | 1.580964 |
| Human_Ge | CD71+ Earl | 3/553 | 0.433437 | 0.841246 | 0 | 0 | 1.257338 | 1.051146 |
| Human_Ge | AdrenalCor | 2/106 | 0.077881 | 0.841246 | 0 | 0 | 4.481674 | 11.43977 |
| Human_Ge | CD105+ En | 4/370 | 0.077893 | 0.841246 | 0 | 0 | 2.573836 | 6.569522 |
| Human_Ge | Liver | 5/618 | 0.131846 | 0.841246 | 0 | 0 | 1.919787 | 3.889717 |
| Human_Ge | Uterus | 2/201 | 0.217801 | 0.841246 | 0 | 0 | 2.330949 | 3.552767 |
| Human_Ge | Skin | 1/111 | 0.384451 | 0.841246 | 0 | 0 | 2.09334 | 2.001105 |
| Human_Ge | SkeletalMu | 3/441 | 0.300914 | 0.841246 | 0 | 0 | 1.587981 | 1.907057 |
| Human_Ge | Thalamus | Jan-79 | 0.291832 | 0.841246 | 0 | 0 | 2.956917 | 3.641673 |
| Human_Ge | DorsalRoot | Jan-69 | 0.260157 | 0.841246 | 0 | 0 | 3.393468 | 4.569198 |
| Human_Ge | TestisInter: | 3/568 | 0.450673 | 0.841246 | 0 | 0 | 1.223009 | 0.974755 |
| Human_Ge | FetalThyroi | Jan-66 | 0.250385 | 0.841246 | 0 | 0 | 3.550626 | 4.916752 |
| Human_Ge | Adrenalgla | Jan-65 | 0.247099 | 0.841246 | 0 | 0 | 3.606286 | 5.041467 |
| Human_Ge | Leukemia p | Jan-62 | 0.237156 | 0.841246 | 0 | 0 | 3.784217 | 5.445629 |
| Human_Ge | Cerebellum | 1/137 | 0.4508 | 0.841246 | 0 | 0 | 1.69092 | 1.347208 |
| Human_Ge | Lymphoma | 2/355 | 0.458861 | 0.841246 | 0 | 0 | 1.303783 | 1.015656 |

| | | | | | | | | |
|---|---|---|---|---|---|---|---|---|
| Human_Ge | Placenta | 3/405 | 0.258343 | 0.841246 | 0 | 0 | 1.733387 | 2.346081 |
| Gene_Pert | LMO3 KD h | 1/399 | 0.82744 | 0.842133 | 0 | 0 | 0.570147 | 0.107997 |
| Gene_Pert | Il1rn KO m | 1/400 | 0.828206 | 0.842133 | 0 | 0 | 0.568689 | 0.107194 |
| Gene_Pert | Dmrt1 KO r | 1/400 | 0.828206 | 0.842133 | 0 | 0 | 0.568689 | 0.107194 |
| GTEx_Tissu | GTEX-QDVI | 8/1058 | 0.089101 | 0.842386 | 0 | 0 | 1.819216 | 4.398832 |
| Gene_Pert | Ilk activem | 4/348 | 0.065309 | 0.842439 | 0 | 0 | 2.741524 | 7.480592 |
| Disease_Pe | Type 2 diak | 2/332 | 0.424913 | 0.842782 | 0 | 0 | 1.396292 | 1.195045 |
| Disease_Pe | Lung trans | 2/335 | 0.429403 | 0.842782 | 0 | 0 | 1.383501 | 1.169555 |
| Disease_Pe | Status Epile | 2/336 | 0.430896 | 0.842782 | 0 | 0 | 1.379288 | 1.161207 |
| Disease_Pe | Myocardial | 2/336 | 0.430896 | 0.842782 | 0 | 0 | 1.379288 | 1.161207 |
| Disease_Pe | psoriasis D | 2/337 | 0.432387 | 0.842782 | 0 | 0 | 1.375101 | 1.152933 |
| Disease_Pe | bipolar disc | 2/324 | 0.412854 | 0.842782 | 0 | 0 | 1.431567 | 1.266451 |
| Disease_Pe | Polycystic ( | 2/325 | 0.414369 | 0.842782 | 0 | 0 | 1.427062 | 1.257241 |
| Disease_Pe | Cardiomyo | 2/326 | 0.415881 | 0.842782 | 0 | 0 | 1.422585 | 1.248115 |
| Disease_Pe | amyotroph | 2/341 | 0.438329 | 0.842782 | 0 | 0 | 1.358598 | 1.120553 |
| Disease_Pe | atheroscler | 2/341 | 0.438329 | 0.842782 | 0 | 0 | 1.358598 | 1.120553 |
| Disease_Pe | Parkinson's | 2/342 | 0.439809 | 0.842782 | 0 | 0 | 1.354533 | 1.112633 |
| Disease_Pe | Cardiac Hyp | 1/133 | 0.441073 | 0.842782 | 0 | 0 | 1.742512 | 1.426323 |
| Disease_Pe | methylmal | 2/344 | 0.442764 | 0.842782 | 0 | 0 | 1.346474 | 1.096999 |
| Disease_Pe | Cardiomyo | 2/330 | 0.42191 | 0.842782 | 0 | 0 | 1.40495 | 1.212419 |
| Disease_Pe | Chronic ph | 2/330 | 0.42191 | 0.842782 | 0 | 0 | 1.40495 | 1.212419 |
| Disease_Pe | Dengue Fe | 2/334 | 0.427909 | 0.842782 | 0 | 0 | 1.387739 | 1.177976 |
| Disease_Pe | schizophre | 2/295 | 0.368186 | 0.842782 | 0 | 0 | 1.575587 | 1.574275 |
| Disease_Pe | invasive du | 2/296 | 0.369749 | 0.842782 | 0 | 0 | 1.570148 | 1.56219 |
| Disease_Pe | ductal carc | 2/297 | 0.37131 | 0.842782 | 0 | 0 | 1.564746 | 1.550222 |
| Disease_Pe | Leukemia D | 2/297 | 0.37131 | 0.842782 | 0 | 0 | 1.564746 | 1.550222 |
| Disease_Pe | Huntingtor | 2/354 | 0.457409 | 0.842782 | 0 | 0 | 1.307553 | 1.02274 |
| Disease_Pe | inflammato | 2/337 | 0.432387 | 0.842782 | 0 | 0 | 1.375101 | 1.152933 |
| Disease_Pe | multiple sc | 2/337 | 0.432387 | 0.842782 | 0 | 0 | 1.375101 | 1.152933 |
| Disease_Pe | Alzheimer's | 2/341 | 0.438329 | 0.842782 | 0 | 0 | 1.358598 | 1.120553 |
| Disease_Pe | Pseudomo | 2/361 | 0.46753 | 0.842782 | 0 | 0 | 1.281599 | 0.974388 |
| Disease_Pe | ulcerative ( | 2/361 | 0.46753 | 0.842782 | 0 | 0 | 1.281599 | 0.974388 |
| Disease_Pe | acute myel | 1/145 | 0.469755 | 0.842782 | 0 | 0 | 1.596334 | 1.2061 |
| Disease_Pe | Nemaline N | 2/366 | 0.474693 | 0.842782 | 0 | 0 | 1.263672 | 0.941545 |
| Disease_Pe | systemic ju | 2/367 | 0.476119 | 0.842782 | 0 | 0 | 1.260145 | 0.935139 |
| Disease_Pe | psoriasis D | 2/345 | 0.444238 | 0.842782 | 0 | 0 | 1.34248 | 1.089282 |
| Disease_Pe | Muscular D | 2/271 | 0.330285 | 0.842782 | 0 | 0 | 1.718259 | 1.903487 |
| Disease_Pe | Down synd | 3/516 | 0.390193 | 0.842782 | 0 | 0 | 1.350599 | 1.271068 |
| Disease_Pe | Granuloma | 2/299 | 0.374428 | 0.842782 | 0 | 0 | 1.55405 | 1.526629 |
| Disease_Pe | Breast Can | 2/299 | 0.374428 | 0.842782 | 0 | 0 | 1.55405 | 1.526629 |
| Disease_Pe | juvenile de | 3/503 | 0.374809 | 0.842782 | 0 | 0 | 1.386643 | 1.360766 |
| Disease_Pe | fragile X sy | 2/312 | 0.394543 | 0.842782 | 0 | 0 | 1.487894 | 1.383781 |
| Disease_Pe | cerebral pa | 2/312 | 0.394543 | 0.842782 | 0 | 0 | 1.487894 | 1.383781 |
| Disease_Pe | Duchenne | 2/313 | 0.396079 | 0.842782 | 0 | 0 | 1.483034 | 1.3735 |
| Disease_Pe | chronic lyn | 2/315 | 0.399145 | 0.842782 | 0 | 0 | 1.473407 | 1.353222 |

| | | | | | | | | |
|---|---|---|---|---|---|---|---|---|
| Disease_Pe | Wilson dise | 2/316 | 0.400676 | 0.842782 | 0 | 0 | 1.46864 | 1.343223 |
| Disease_Pe | hepatitis C | 2/306 | 0.385293 | 0.842782 | 0 | 0 | 1.517724 | 1.44753 |
| Disease_Pe | anemia DO | 2/306 | 0.385293 | 0.842782 | 0 | 0 | 1.517724 | 1.44753 |
| Disease_Pe | melanoma | 2/307 | 0.386839 | 0.842782 | 0 | 0 | 1.512671 | 1.436654 |
| Disease_Pe | colorectal a | 2/270 | 0.328691 | 0.842782 | 0 | 0 | 1.724759 | 1.919028 |
| Disease_Pe | multiple sc | 2/271 | 0.330285 | 0.842782 | 0 | 0 | 1.718259 | 1.903487 |
| Disease_Pe | ulcerative c | 2/345 | 0.444238 | 0.842782 | 0 | 0 | 1.34248 | 1.089282 |
| Disease_Pe | Nemaline M | 3/466 | 0.330734 | 0.842782 | 0 | 0 | 1.500309 | 1.660003 |
| Disease_Pe | myelodysp | 2/324 | 0.412854 | 0.842782 | 0 | 0 | 1.431567 | 1.266451 |
| Disease_Pe | anemia DO | 2/310 | 0.391467 | 0.842782 | 0 | 0 | 1.497708 | 1.404633 |
| Disease_Pe | Huntingtor | 2/312 | 0.394543 | 0.842782 | 0 | 0 | 1.487894 | 1.383781 |
| Disease_Pe | pancreatic | 2/312 | 0.394543 | 0.842782 | 0 | 0 | 1.487894 | 1.383781 |
| Disease_Pe | colon aden | 2/326 | 0.415881 | 0.842782 | 0 | 0 | 1.422585 | 1.248115 |
| Disease_Pe | Wolfram Sy | 2/327 | 0.417391 | 0.842782 | 0 | 0 | 1.418136 | 1.239071 |
| Disease_Pe | Synovial sa | 2/327 | 0.417391 | 0.842782 | 0 | 0 | 1.418136 | 1.239071 |
| Disease_Pe | Carcinoma | 2/327 | 0.417391 | 0.842782 | 0 | 0 | 1.418136 | 1.239071 |
| Disease_Pe | Ventilator- | 2/328 | 0.418899 | 0.842782 | 0 | 0 | 1.413713 | 1.230107 |
| Disease_Pe | Diabetes ar | 2/318 | 0.403731 | 0.842782 | 0 | 0 | 1.459196 | 1.323499 |
| Disease_Pe | Adenocarci | 2/318 | 0.403731 | 0.842782 | 0 | 0 | 1.459196 | 1.323499 |
| Disease_Pe | Kidney disc | 2/318 | 0.403731 | 0.842782 | 0 | 0 | 1.459196 | 1.323499 |
| Disease_Pe | nasophary | 1/167 | 0.51861 | 0.842782 | 0 | 0 | 1.383231 | 0.908233 |
| Disease_Pe | ulcerative c | 2/424 | 0.55341 | 0.842782 | 0 | 0 | 1.086758 | 0.642986 |
| Disease_Pe | schizophre | 1/185 | 0.555251 | 0.842782 | 0 | 0 | 1.246777 | 0.733523 |
| Disease_Pe | schizophre | 2/384 | 0.499999 | 0.842782 | 0 | 0 | 1.203018 | 0.833871 |
| Disease_Pe | diffuse larg | 1/160 | 0.503569 | 0.842782 | 0 | 0 | 1.444639 | 0.991073 |
| Disease_Pe | Type 1 diak | 2/387 | 0.504142 | 0.842782 | 0 | 0 | 1.193461 | 0.817399 |
| Disease_Pe | melanoma | 1/196 | 0.576271 | 0.842782 | 0 | 0 | 1.17579 | 0.64807 |
| Disease_Pe | prostate ca | 1/241 | 0.652504 | 0.842782 | 0 | 0 | 0.953149 | 0.406935 |
| Disease_Pe | melanoma | 1/221 | 0.62046 | 0.842782 | 0 | 0 | 1.040856 | 0.496795 |
| Disease_Pe | type 2 diab | 1/220 | 0.618783 | 0.842782 | 0 | 0 | 1.045662 | 0.501918 |
| Disease_Pe | hepatocellu | 1/214 | 0.608568 | 0.842782 | 0 | 0 | 1.075445 | 0.534116 |
| Disease_Pe | Cardiomyo | 2/450 | 0.585949 | 0.842782 | 0 | 0 | 1.022321 | 0.546454 |
| Disease_Pe | diffuse larg | 1/203 | 0.589132 | 0.842782 | 0 | 0 | 1.134642 | 0.600344 |
| Disease_Pe | multiple m | 2/454 | 0.5908 | 0.842782 | 0 | 0 | 1.013066 | 0.533155 |
| Disease_Pe | autistic dis | 2/457 | 0.59441 | 0.842782 | 0 | 0 | 1.006231 | 0.523427 |
| Disease_Pe | pulmonary | 2/423 | 0.552124 | 0.842782 | 0 | 0 | 1.089395 | 0.647082 |
| Disease_Pe | B-cell chror | 1/228 | 0.631995 | 0.842782 | 0 | 0 | 1.008401 | 0.462729 |
| Disease_Pe | actinic kera | 1/170 | 0.524918 | 0.842782 | 0 | 0 | 1.35847 | 0.875552 |
| Disease_Pe | Actinic kera | 1/171 | 0.527002 | 0.842782 | 0 | 0 | 1.35041 | 0.865006 |
| Disease_Pe | Sjogren's sy | 1/223 | 0.623791 | 0.842782 | 0 | 0 | 1.031374 | 0.486746 |
| Disease_Pe | Dehydratio | 1/225 | 0.627094 | 0.842782 | 0 | 0 | 1.022062 | 0.476954 |
| Disease_Pe | pulmonary | 2/486 | 0.628107 | 0.842782 | 0 | 0 | 0.944531 | 0.439249 |
| Disease_Pe | Idiopathic f | 1/226 | 0.628735 | 0.842782 | 0 | 0 | 1.017468 | 0.472152 |
| Disease_Pe | Sepsis C02 | 1/226 | 0.628735 | 0.842782 | 0 | 0 | 1.017468 | 0.472152 |
| Disease_Pe | Huntingtor | 2/436 | 0.568645 | 0.842782 | 0 | 0 | 1.056059 | 0.596144 |

| | | | | | | | | |
|---|---|---|---|---|---|---|---|---|
| Disease_Pe | esophagus | 1/192 | 0.568743 | 0.842782 | 0 | 0 | 1.200657 | 0.677563 |
| Disease_Pe | Severe acu | 1/195 | 0.574401 | 0.842782 | 0 | 0 | 1.181911 | 0.655284 |
| Disease_Pe | Barrett's es | 2/354 | 0.457409 | 0.842782 | 0 | 0 | 1.307553 | 1.02274 |
| Disease_Pe | Down synd | 2/353 | 0.455954 | 0.842782 | 0 | 0 | 1.311346 | 1.029883 |
| Disease_Pe | amyotroph | 1/138 | 0.453206 | 0.842782 | 0 | 0 | 1.678493 | 1.328374 |
| Disease_Pe | Parkinson's | 1/158 | 0.499187 | 0.842782 | 0 | 0 | 1.463191 | 1.016589 |
| Disease_Pe | HIV enceph | 2/351 | 0.453038 | 0.842782 | 0 | 0 | 1.318995 | 1.044354 |
| Disease_Pe | Huntingtor | 2/369 | 0.478963 | 0.842782 | 0 | 0 | 1.25315 | 0.922483 |
| Disease_Pe | nicotine de | 2/370 | 0.480382 | 0.842782 | 0 | 0 | 1.24968 | 0.916233 |
| Disease_Pe | sickle cell a | 2/371 | 0.481798 | 0.842782 | 0 | 0 | 1.24623 | 0.910034 |
| Disease_Pe | Obesity C0 | 2/371 | 0.481798 | 0.842782 | 0 | 0 | 1.24623 | 0.910034 |
| Disease_Pe | Down synd | 2/371 | 0.481798 | 0.842782 | 0 | 0 | 1.24623 | 0.910034 |
| Disease_Pe | in situ carc | 2/354 | 0.457409 | 0.842782 | 0 | 0 | 1.307553 | 1.02274 |
| Disease_Pe | Schizophre | 2/358 | 0.463206 | 0.842782 | 0 | 0 | 1.292597 | 0.994762 |
| Disease_Pe | Pompe dise | 2/358 | 0.463206 | 0.842782 | 0 | 0 | 1.292597 | 0.994762 |
| Disease_Pe | Preeclamp: | 2/375 | 0.487441 | 0.842782 | 0 | 0 | 1.232613 | 0.885738 |
| Disease_Pe | Rett Syndro | 2/381 | 0.495834 | 0.842782 | 0 | 0 | 1.212727 | 0.850744 |
| Disease_Pe | breast canc | 1/157 | 0.496981 | 0.842782 | 0 | 0 | 1.472645 | 1.029678 |
| Disease_Pe | Diamond-B | 2/382 | 0.497225 | 0.842782 | 0 | 0 | 1.209474 | 0.845075 |
| Disease_Pe | Neurologic | 1/230 | 0.635226 | 0.842782 | 0 | 0 | 0.999492 | 0.453544 |
| Disease_Pe | ALS (Amyo | 2/345 | 0.444238 | 0.842782 | 0 | 0 | 1.34248 | 1.089282 |
| Disease_Pe | multiple sc | 1/148 | 0.476695 | 0.842782 | 0 | 0 | 1.563518 | 1.158376 |
| Disease_Pe | Dermatom | 2/368 | 0.477542 | 0.842782 | 0 | 0 | 1.256638 | 0.928785 |
| Disease_Pe | Neurofibro | 2/369 | 0.478963 | 0.842782 | 0 | 0 | 1.25315 | 0.922483 |
| Disease_Pe | Dermatom | 2/320 | 0.40678 | 0.842782 | 0 | 0 | 1.449871 | 1.304135 |
| Disease_Pe | endometri | 2/321 | 0.408301 | 0.842782 | 0 | 0 | 1.445252 | 1.294585 |
| Disease_Pe | Tendonopa | 2/322 | 0.409821 | 0.842782 | 0 | 0 | 1.440662 | 1.285121 |
| Disease_Pe | Osteoporo: | 2/323 | 0.411339 | 0.842782 | 0 | 0 | 1.4361 | 1.275744 |
| Disease_Pe | Malignant | 2/397 | 0.517793 | 0.842782 | 0 | 0 | 1.162651 | 0.765232 |
| Disease_Pe | Breast Can | 1/151 | 0.483545 | 0.842782 | 0 | 0 | 1.532016 | 1.113178 |
| Disease_Pe | dengue hei | 2/374 | 0.486034 | 0.842782 | 0 | 0 | 1.23599 | 0.891738 |
| Disease_Pe | dengue dis | 2/375 | 0.487441 | 0.842782 | 0 | 0 | 1.232613 | 0.885738 |
| Disease_Pe | COPD - Chr | 2/412 | 0.537807 | 0.842782 | 0 | 0 | 1.119254 | 0.694223 |
| Disease_Pe | Psoriasis vu | 2/418 | 0.545655 | 0.842782 | 0 | 0 | 1.102771 | 0.668025 |
| Disease_Pe | testis semi | 2/255 | 0.304685 | 0.842782 | 0 | 0 | 1.828412 | 2.173024 |
| Disease_Pe | endometri | 2/216 | 0.241852 | 0.842782 | 0 | 0 | 2.165915 | 3.074364 |
| Disease_Pe | Parkinson's | 2/235 | 0.272477 | 0.842782 | 0 | 0 | 1.987377 | 2.583987 |
| Disease_Pe | bronchopu | 2/236 | 0.27409 | 0.842782 | 0 | 0 | 1.978783 | 2.561135 |
| Disease_Pe | Alzheimer's | 3/419 | 0.274815 | 0.842782 | 0 | 0 | 1.67385 | 2.162037 |
| Disease_Pe | Rift Valley f | 2/237 | 0.275703 | 0.842782 | 0 | 0 | 1.970263 | 2.538546 |
| Disease_Pe | colon carci | 3/421 | 0.277179 | 0.842782 | 0 | 0 | 1.66567 | 2.13721 |
| Disease_Pe | Thrombocy | 2/226 | 0.257961 | 0.842782 | 0 | 0 | 2.068172 | 2.802265 |
| Disease_Pe | Anaplasmo | 2/227 | 0.259573 | 0.842782 | 0 | 0 | 2.058876 | 2.776839 |
| Disease_Pe | Carcinoma | 3/408 | 0.261862 | 0.842782 | 0 | 0 | 1.720282 | 2.305074 |
| Disease_Pe | schizophre | 2/250 | 0.296648 | 0.842782 | 0 | 0 | 1.86575 | 2.267274 |

| | | | | | | | | |
|---|---|---|---|---|---|---|---|---|
| Disease_Pe | Breast Can | 2/233 | 0.269251 | 0.842782 | 0 | 0 | 2.004787 | 2.630503 |
| Disease_Pe | amyotroph | 3/416 | 0.271275 | 0.842782 | 0 | 0 | 1.686268 | 2.199943 |
| Disease_Pe | lung cance | 2/235 | 0.272477 | 0.842782 | 0 | 0 | 1.987377 | 2.583987 |
| Disease_Pe | acute myo | 3/443 | 0.303295 | 0.842782 | 0 | 0 | 1.580601 | 1.885733 |
| Disease_Pe | nasophary | 2/214 | 0.238636 | 0.842782 | 0 | 0 | 2.18657 | 3.132957 |
| Disease_Pe | testis semi | 2/257 | 0.307896 | 0.842782 | 0 | 0 | 1.813887 | 2.136747 |
| Disease_Pe | Endometri | 2/259 | 0.311104 | 0.842782 | 0 | 0 | 1.799588 | 2.10125 |
| Disease_Pe | Rift Valley | 2/240 | 0.280541 | 0.842782 | 0 | 0 | 1.945131 | 2.472331 |
| Disease_Pe | polycystic | 2/241 | 0.282153 | 0.842782 | 0 | 0 | 1.936894 | 2.450762 |
| Disease_Pe | Breast can | 2/242 | 0.283765 | 0.842782 | 0 | 0 | 1.928725 | 2.429439 |
| Disease_Pe | multiple m | 2/262 | 0.31591 | 0.842782 | 0 | 0 | 1.778552 | 2.04942 |
| Disease_Pe | adrenoleuk | 2/262 | 0.31591 | 0.842782 | 0 | 0 | 1.778552 | 2.04942 |
| Disease_Pe | diffuse larg | 2/263 | 0.317511 | 0.842782 | 0 | 0 | 1.771648 | 2.03251 |
| Disease_Pe | hepatocell | 3/455 | 0.317603 | 0.842782 | 0 | 0 | 1.53769 | 1.763658 |
| Disease_Pe | ulcerative | 2/264 | 0.319111 | 0.842782 | 0 | 0 | 1.764796 | 2.015779 |
| Disease_Pe | Wolfram sy | 2/250 | 0.296648 | 0.842782 | 0 | 0 | 1.86575 | 2.267274 |
| Disease_Pe | ulcerative | 2/250 | 0.296648 | 0.842782 | 0 | 0 | 1.86575 | 2.267274 |
| Disease_Pe | CML - Chro | 3/441 | 0.300914 | 0.842782 | 0 | 0 | 1.587981 | 1.907057 |
| Disease_Pe | autism spe | 2/266 | 0.322308 | 0.842782 | 0 | 0 | 1.751248 | 1.982846 |
| Disease_Pe | Sickle Cell | 3/403 | 0.256001 | 0.842782 | 0 | 0 | 1.742232 | 2.373918 |
| Disease_Pe | psoriasis D | 3/346 | 0.191052 | 0.842782 | 0 | 0 | 2.037693 | 3.372806 |
| Disease_Pe | lung squan | 3/347 | 0.192154 | 0.842782 | 0 | 0 | 2.031665 | 3.351151 |
| Disease_Pe | testis semi | 2/186 | 0.193998 | 0.842782 | 0 | 0 | 2.52289 | 4.137304 |
| Disease_Pe | Diabetic Ne | 2/186 | 0.193998 | 0.842782 | 0 | 0 | 2.52289 | 4.137304 |
| Disease_Pe | Psoriasis vu | 3/364 | 0.211116 | 0.842782 | 0 | 0 | 1.934309 | 3.008525 |
| Disease_Pe | adrenoleuk | 3/365 | 0.212244 | 0.842782 | 0 | 0 | 1.928867 | 2.989777 |
| Disease_Pe | Dental cavi | 2/199 | 0.21461 | 0.842782 | 0 | 0 | 2.354852 | 3.623955 |
| Disease_Pe | sporadic br | 2/200 | 0.216205 | 0.842782 | 0 | 0 | 2.34284 | 3.588123 |
| Disease_Pe | facioscapul | 3/372 | 0.220182 | 0.842782 | 0 | 0 | 1.891599 | 2.862559 |
| Disease_Pe | endometri | 2/209 | 0.230606 | 0.842782 | 0 | 0 | 2.239955 | 3.286115 |
| Disease_Pe | Erectile dys | 3/351 | 0.196575 | 0.842782 | 0 | 0 | 2.007902 | 3.266274 |
| Disease_Pe | Bone fractu | 3/359 | 0.205494 | 0.842782 | 0 | 0 | 1.961978 | 3.104515 |
| Disease_Pe | Type 1 diab | 2/257 | 0.307896 | 0.842782 | 0 | 0 | 1.813887 | 2.136747 |
| Disease_Pe | Duchenne | 3/363 | 0.209989 | 0.842782 | 0 | 0 | 1.939782 | 3.027421 |
| Disease_Pe | Bipolar Dis | 2/266 | 0.322308 | 0.842782 | 0 | 0 | 1.751248 | 1.982846 |
| Disease_Pe | ulcerative | 2/216 | 0.241852 | 0.842782 | 0 | 0 | 2.165915 | 3.074364 |
| Disease_Pe | retinoblast | 3/393 | 0.244342 | 0.842782 | 0 | 0 | 1.787821 | 2.51937 |
| Disease_Pe | COPD - Chr | 3/400 | 0.252495 | 0.842782 | 0 | 0 | 1.755668 | 2.41644 |
| Disease_Pe | breast canc | 3/403 | 0.256001 | 0.842782 | 0 | 0 | 1.742232 | 2.373918 |
| Disease_Pe | Bipolar Dis | 2/205 | 0.224196 | 0.842782 | 0 | 0 | 2.284555 | 3.415945 |
| Disease_Pe | Squamous | 3/376 | 0.224746 | 0.842782 | 0 | 0 | 1.870931 | 2.7929 |
| Disease_Pe | nemaline n | 3/379 | 0.228181 | 0.842782 | 0 | 0 | 1.855718 | 2.74204 |
| Disease_Pe | idiopathic | 3/408 | 0.261862 | 0.842782 | 0 | 0 | 1.720282 | 2.305074 |
| Disease_Pe | Type 1 diab | 2/229 | 0.262799 | 0.842782 | 0 | 0 | 2.040529 | 2.726893 |
| Disease_Pe | Crohn's dis | 2/230 | 0.264412 | 0.842782 | 0 | 0 | 2.031476 | 2.702365 |

| | | | | | | | | |
|---|---|---|---|---|---|---|---|---|
| Disease_Pe | nemaline n | 3/412 | 0.266563 | 0.842782 | 0 | 0 | 1.703109 | 2.251757 |
| Disease_Pe | ulcerative ( | 3/413 | 0.26774 | 0.842782 | 0 | 0 | 1.698868 | 2.238665 |
| Disease_Pe | cellular pha | 3/382 | 0.231626 | 0.842782 | 0 | 0 | 1.840746 | 2.692329 |
| Disease_Pe | Hypertroph | 2/289 | 0.358779 | 0.842782 | 0 | 0 | 1.609018 | 1.649321 |
| Disease_Pe | Huntingtor | 2/276 | 0.338237 | 0.842782 | 0 | 0 | 1.686475 | 1.828152 |
| Disease_Pe | amyotroph | 3/491 | 0.360549 | 0.842782 | 0 | 0 | 1.421619 | 1.450231 |
| Disease_Pe | cerebral pa | 2/291 | 0.36192 | 0.842782 | 0 | 0 | 1.59772 | 1.623813 |
| Disease_Pe | cerebral pa | 2/291 | 0.36192 | 0.842782 | 0 | 0 | 1.59772 | 1.623813 |
| Disease_Pe | nemaline n | 3/493 | 0.362929 | 0.842782 | 0 | 0 | 1.415671 | 1.434849 |
| Disease_Pe | Sjogren's s | 2/292 | 0.363489 | 0.842782 | 0 | 0 | 1.59213 | 1.611246 |
| Disease_Pe | endometri | 2/282 | 0.347744 | 0.842782 | 0 | 0 | 1.649832 | 1.7427 |
| Disease_Pe | myelodysp | 2/283 | 0.349324 | 0.842782 | 0 | 0 | 1.643877 | 1.728956 |
| Disease_Pe | hepatocell | 2/283 | 0.349324 | 0.842782 | 0 | 0 | 1.643877 | 1.728956 |
| Disease_Pe | H1N1 DOID | 3/427 | 0.284281 | 0.842782 | 0 | 0 | 1.641594 | 2.064784 |
| Disease_Pe | allergic cor | 2/243 | 0.285377 | 0.842782 | 0 | 0 | 1.920625 | 2.408358 |
| Disease_Pe | sickle-cell a | 3/429 | 0.286652 | 0.842782 | 0 | 0 | 1.633719 | 2.041308 |
| Disease_Pe | stomach ca | 2/247 | 0.29182 | 0.842782 | 0 | 0 | 1.888884 | 2.326383 |
| Disease_Pe | hepatitis C | 2/298 | 0.37287 | 0.842782 | 0 | 0 | 1.55938 | 1.538369 |
| Disease_Pe | asthma DO | 2/276 | 0.338237 | 0.842782 | 0 | 0 | 1.686475 | 1.828152 |
| Disease_Pe | Psoriasis vu | 2/290 | 0.36035 | 0.842782 | 0 | 0 | 1.60335 | 1.636504 |
| Disease_Pe | oligodendr | 2/290 | 0.36035 | 0.842782 | 0 | 0 | 1.60335 | 1.636504 |
| Disease_Pe | hepatitis C | 2/301 | 0.37754 | 0.842782 | 0 | 0 | 1.543498 | 1.503487 |
| Disease_Pe | multiple sc | 2/301 | 0.37754 | 0.842782 | 0 | 0 | 1.543498 | 1.503487 |
| Disease_Pe | acute myo | 2/301 | 0.37754 | 0.842782 | 0 | 0 | 1.543498 | 1.503487 |
| Disease_Pe | psoriasis D | 2/303 | 0.380646 | 0.842782 | 0 | 0 | 1.533086 | 1.480784 |
| Disease_Pe | Polycystic ( | 2/306 | 0.385293 | 0.842782 | 0 | 0 | 1.517724 | 1.44753 |
| Disease_Pe | amyotroph | 2/292 | 0.363489 | 0.842782 | 0 | 0 | 1.59213 | 1.611246 |
| Disease_Pe | Waldenstr | 2/293 | 0.365056 | 0.842782 | 0 | 0 | 1.586578 | 1.598802 |
| Disease_Pe | psoriasis D | 2/295 | 0.368186 | 0.842782 | 0 | 0 | 1.575587 | 1.574275 |
| Disease_Pe | Alzheimer's | 2/308 | 0.388383 | 0.842782 | 0 | 0 | 1.507651 | 1.42588 |
| Disease_Pe | Senescence | 2/308 | 0.388383 | 0.842782 | 0 | 0 | 1.507651 | 1.42588 |
| Disease_Pe | schizophre | 2/309 | 0.389926 | 0.842782 | 0 | 0 | 1.502663 | 1.415207 |
| Disease_Pe | Familial hy | 2/276 | 0.338237 | 0.842782 | 0 | 0 | 1.686475 | 1.828152 |
| Disease_Pe | Down Sync | 3/460 | 0.323571 | 0.842782 | 0 | 0 | 1.520475 | 1.715609 |
| Disease_Pe | Diabetic Ne | 2/267 | 0.323905 | 0.842782 | 0 | 0 | 1.74455 | 1.966639 |
| Disease_Pe | systemic lu | 2/267 | 0.323905 | 0.842782 | 0 | 0 | 1.74455 | 1.966639 |
| Disease_Pe | MS (Multip | 2/260 | 0.312707 | 0.842782 | 0 | 0 | 1.792522 | 2.083787 |
| Disease_Pe | sickle-cell a | 3/452 | 0.314024 | 0.842782 | 0 | 0 | 1.548202 | 1.793262 |
| Disease_Pe | thymic car | 2/261 | 0.314309 | 0.842782 | 0 | 0 | 1.78551 | 2.066511 |
| Disease_Pe | nasophary | 2/208 | 0.229002 | 0.842782 | 0 | 0 | 2.250942 | 3.317943 |
| Disease_Pe | Neurogeni | 2/208 | 0.229002 | 0.842782 | 0 | 0 | 2.250942 | 3.317943 |
| Disease_Pe | bipolar dis | 3/380 | 0.229328 | 0.842782 | 0 | 0 | 1.850701 | 2.725344 |
| Disease_Pe | Gastrointe: | 3/380 | 0.229328 | 0.842782 | 0 | 0 | 1.850701 | 2.725344 |
| Disease_Pe | glaucoma a | 2/272 | 0.331878 | 0.842782 | 0 | 0 | 1.711808 | 1.888106 |
| Disease_Pe | tibial musc | 2/265 | 0.32071 | 0.842782 | 0 | 0 | 1.757996 | 1.999226 |

| | | | | | | | | |
|---|---|---|---|---|---|---|---|---|
| Disease_Pe | neurofibro | 3/458 | 0.321183 | 0.842782 | 0 | 0 | 1.527316 | 1.734638 |
| Disease_Pe | systemic lu | 2/266 | 0.322308 | 0.842782 | 0 | 0 | 1.751248 | 1.982846 |
| Disease_Pe | Muscular D | 2/309 | 0.389926 | 0.842782 | 0 | 0 | 1.502663 | 1.415207 |
| Disease_Pe | Hyperlipide | 2/277 | 0.339825 | 0.842782 | 0 | 0 | 1.680257 | 1.813544 |
| Disease_Pe | Muscular D | 2/278 | 0.341411 | 0.842782 | 0 | 0 | 1.674084 | 1.799086 |
| Disease_Pe | Hereditary | 2/279 | 0.342996 | 0.842782 | 0 | 0 | 1.667955 | 1.784774 |
| Disease_Pe | Adenocarci | 2/281 | 0.346162 | 0.842782 | 0 | 0 | 1.65583 | 1.756583 |
| Disease_Pe | asthma DO | 2/267 | 0.323905 | 0.842782 | 0 | 0 | 1.74455 | 1.966639 |
| Disease_Pe | Type 2 diab | 2/269 | 0.327097 | 0.842782 | 0 | 0 | 1.731306 | 1.934732 |
| Disease_Pe | Acute Lung | 2/270 | 0.328691 | 0.842782 | 0 | 0 | 1.724759 | 1.919028 |
| Disease_Pe | schizophre | 2/285 | 0.352481 | 0.842782 | 0 | 0 | 1.632093 | 1.70188 |
| Disease_Pe | Alexander | 2/286 | 0.354057 | 0.842782 | 0 | 0 | 1.626263 | 1.688543 |
| Disease_Pe | pancreatic | 3/487 | 0.355786 | 0.842782 | 0 | 0 | 1.433663 | 1.481583 |
| Disease_Pe | Huntingtor | 2/288 | 0.357207 | 0.842782 | 0 | 0 | 1.614726 | 1.662266 |
| Disease_Pe | Oligodendr | 2/289 | 0.358779 | 0.842782 | 0 | 0 | 1.609018 | 1.649321 |
| Disease_Pe | systemic lu | 2/273 | 0.333469 | 0.842782 | 0 | 0 | 1.705405 | 1.872885 |
| Disease_Pe | myelodysp | 1/327 | 0.762437 | 0.842782 | 0 | 0 | 0.698637 | 0.189496 |
| Disease_Pe | Schizophre | 1/291 | 0.721383 | 0.842782 | 0 | 0 | 0.786808 | 0.25696 |
| Disease_Pe | adenosqua | 1/292 | 0.722612 | 0.842782 | 0 | 0 | 0.784065 | 0.254729 |
| Disease_Pe | Lung transp | 1/293 | 0.723837 | 0.842782 | 0 | 0 | 0.78134 | 0.25252 |
| Disease_Pe | myotonic d | 1/321 | 0.756036 | 0.842782 | 0 | 0 | 0.711955 | 0.19911 |
| Disease_Pe | asthma DO | 1/314 | 0.748352 | 0.842782 | 0 | 0 | 0.728137 | 0.211074 |
| Disease_Pe | Cystic Fibro | 1/314 | 0.748352 | 0.842782 | 0 | 0 | 0.728137 | 0.211074 |
| Disease_Pe | facioscapul | 1/314 | 0.748352 | 0.842782 | 0 | 0 | 0.728137 | 0.211074 |
| Disease_Pe | asthma DO | 1/322 | 0.757114 | 0.842782 | 0 | 0 | 0.709701 | 0.197468 |
| Disease_Pe | Lewy body | 1/323 | 0.758188 | 0.842782 | 0 | 0 | 0.707461 | 0.195842 |
| Disease_Pe | anaplastic | 1/324 | 0.759257 | 0.842782 | 0 | 0 | 0.705234 | 0.194232 |
| Disease_Pe | Duchenne | 1/324 | 0.759257 | 0.842782 | 0 | 0 | 0.705234 | 0.194232 |
| Disease_Pe | schizophre | 1/326 | 0.761381 | 0.842782 | 0 | 0 | 0.700823 | 0.191059 |
| Disease_Pe | pancreatic | 1/316 | 0.750571 | 0.842782 | 0 | 0 | 0.72344 | 0.20757 |
| Disease_Pe | autism spe | 1/317 | 0.751674 | 0.842782 | 0 | 0 | 0.721114 | 0.205844 |
| Disease_Pe | autism spe | 1/317 | 0.751674 | 0.842782 | 0 | 0 | 0.721114 | 0.205844 |
| Disease_Pe | psoriasis D | 1/291 | 0.721383 | 0.842782 | 0 | 0 | 0.786808 | 0.25696 |
| Disease_Pe | endometri | 1/327 | 0.762437 | 0.842782 | 0 | 0 | 0.698637 | 0.189496 |
| Disease_Pe | Turner syn | 1/328 | 0.763487 | 0.842782 | 0 | 0 | 0.696465 | 0.187947 |
| Disease_Pe | Huntingtor | 1/328 | 0.763487 | 0.842782 | 0 | 0 | 0.696465 | 0.187947 |
| Disease_Pe | idiopathic | 1/329 | 0.764533 | 0.842782 | 0 | 0 | 0.694307 | 0.186414 |
| Disease_Pe | Polymyosit | 1/321 | 0.756036 | 0.842782 | 0 | 0 | 0.711955 | 0.19911 |
| Disease_Pe | diabetic re | 1/322 | 0.757114 | 0.842782 | 0 | 0 | 0.709701 | 0.197468 |
| Disease_Pe | esophagus | 1/322 | 0.757114 | 0.842782 | 0 | 0 | 0.709701 | 0.197468 |
| Disease_Pe | Pauciarticu | 1/330 | 0.765575 | 0.842782 | 0 | 0 | 0.692161 | 0.184896 |
| Disease_Pe | atopic derr | 1/331 | 0.766611 | 0.842782 | 0 | 0 | 0.690028 | 0.183392 |
| Disease_Pe | systemic lu | 1/331 | 0.766611 | 0.842782 | 0 | 0 | 0.690028 | 0.183392 |
| Disease_Pe | schizophre | 1/332 | 0.767644 | 0.842782 | 0 | 0 | 0.687908 | 0.181903 |
| Disease_Pe | Type 2 diab | 1/332 | 0.767644 | 0.842782 | 0 | 0 | 0.687908 | 0.181903 |

| | | | | | | | | |
|---|---|---|---|---|---|---|---|---|
| Disease_Pe | Atheroscle | 1/326 | 0.761381 | 0.842782 | 0 | 0 | 0.700823 | 0.191059 |
| Disease_Pe | endometri | 1/327 | 0.762437 | 0.842782 | 0 | 0 | 0.698637 | 0.189496 |
| Disease_Pe | Barrett's e | 1/311 | 0.744986 | 0.842782 | 0 | 0 | 0.735296 | 0.216463 |
| Disease_Pe | idiopathic | 1/277 | 0.703588 | 0.842782 | 0 | 0 | 0.827309 | 0.290851 |
| Disease_Pe | Type 1 dial | 1/306 | 0.739278 | 0.842782 | 0 | 0 | 0.747541 | 0.225819 |
| Disease_Pe | idiopathic | 1/300 | 0.73226 | 0.842782 | 0 | 0 | 0.762775 | 0.237696 |
| Disease_Pe | Breast Can | 1/301 | 0.733443 | 0.842782 | 0 | 0 | 0.760194 | 0.235665 |
| Disease_Pe | SCID - Seve | 1/301 | 0.733443 | 0.842782 | 0 | 0 | 0.760194 | 0.235665 |
| Disease_Pe | nephronop | 1/307 | 0.740429 | 0.842782 | 0 | 0 | 0.74506 | 0.223909 |
| Disease_Pe | Papillary C | 1/308 | 0.741576 | 0.842782 | 0 | 0 | 0.742595 | 0.222019 |
| Disease_Pe | acute myo | 1/308 | 0.741576 | 0.842782 | 0 | 0 | 0.742595 | 0.222019 |
| Disease_Pe | asthma DO | 1/308 | 0.741576 | 0.842782 | 0 | 0 | 0.742595 | 0.222019 |
| Disease_Pe | Premature | 1/308 | 0.741576 | 0.842782 | 0 | 0 | 0.742595 | 0.222019 |
| Disease_Pe | schizophre | 1/304 | 0.736959 | 0.842782 | 0 | 0 | 0.752552 | 0.229696 |
| Disease_Pe | colon canc | 1/304 | 0.736959 | 0.842782 | 0 | 0 | 0.752552 | 0.229696 |
| Disease_Pe | bronchopu | 1/305 | 0.738121 | 0.842782 | 0 | 0 | 0.750038 | 0.227747 |
| Disease_Pe | multiple sc | 1/310 | 0.743855 | 0.842782 | 0 | 0 | 0.737714 | 0.218297 |
| Disease_Pe | Gamma-hy | 1/327 | 0.762437 | 0.842782 | 0 | 0 | 0.698637 | 0.189496 |
| Disease_Pe | Huntingtor | 1/311 | 0.744986 | 0.842782 | 0 | 0 | 0.735296 | 0.216463 |
| Disease_Pe | Bipolar Dis | 1/312 | 0.746113 | 0.842782 | 0 | 0 | 0.732895 | 0.214649 |
| Disease_Pe | pancreatic | 1/314 | 0.748352 | 0.842782 | 0 | 0 | 0.728137 | 0.211074 |
| Disease_Pe | Lymphatic | 1/306 | 0.739278 | 0.842782 | 0 | 0 | 0.747541 | 0.225819 |
| Disease_Pe | Pulmonary | 1/306 | 0.739278 | 0.842782 | 0 | 0 | 0.747541 | 0.225819 |
| Disease_Pe | Huntingtor | 1/307 | 0.740429 | 0.842782 | 0 | 0 | 0.74506 | 0.223909 |
| Disease_Pe | schizophre | 1/314 | 0.748352 | 0.842782 | 0 | 0 | 0.728137 | 0.211074 |
| Disease_Pe | Barrett's e | 1/314 | 0.748352 | 0.842782 | 0 | 0 | 0.728137 | 0.211074 |
| Disease_Pe | autism spe | 1/315 | 0.749464 | 0.842782 | 0 | 0 | 0.725781 | 0.209313 |
| Disease_Pe | familial hyp | 1/315 | 0.749464 | 0.842782 | 0 | 0 | 0.725781 | 0.209313 |
| Disease_Pe | myotonic d | 1/316 | 0.750571 | 0.842782 | 0 | 0 | 0.72344 | 0.20757 |
| Disease_Pe | Sendai viru | 1/308 | 0.741576 | 0.842782 | 0 | 0 | 0.742595 | 0.222019 |
| Disease_Pe | Barrett's e | 1/309 | 0.742718 | 0.842782 | 0 | 0 | 0.740146 | 0.220149 |
| Disease_Pe | multiple sc | 1/309 | 0.742718 | 0.842782 | 0 | 0 | 0.740146 | 0.220149 |
| Disease_Pe | myotonic d | 1/361 | 0.795694 | 0.842782 | 0 | 0 | 0.631557 | 0.144336 |
| Disease_Pe | ulcerative | 1/348 | 0.78356 | 0.842782 | 0 | 0 | 0.655653 | 0.159919 |
| Disease_Pe | ulcerative | 1/348 | 0.78356 | 0.842782 | 0 | 0 | 0.655653 | 0.159919 |
| Disease_Pe | endometri | 1/318 | 0.752771 | 0.842782 | 0 | 0 | 0.718803 | 0.204135 |
| Disease_Pe | cystic fibro | 1/319 | 0.753864 | 0.842782 | 0 | 0 | 0.716506 | 0.202444 |
| Disease_Pe | Retinal dar | 1/321 | 0.756036 | 0.842782 | 0 | 0 | 0.711955 | 0.19911 |
| Disease_Pe | familial hyp | 1/321 | 0.756036 | 0.842782 | 0 | 0 | 0.711955 | 0.19911 |
| Disease_Pe | Huntingtor | 1/372 | 0.805435 | 0.842782 | 0 | 0 | 0.612487 | 0.132526 |
| Disease_Pe | acute myo | 1/352 | 0.787368 | 0.842782 | 0 | 0 | 0.648049 | 0.154923 |
| Disease_Pe | Carcinoma | 1/353 | 0.788309 | 0.842782 | 0 | 0 | 0.646175 | 0.153702 |
| Disease_Pe | Simian Acq | 1/353 | 0.788309 | 0.842782 | 0 | 0 | 0.646175 | 0.153702 |
| Disease_Pe | HIV - Huma | 1/374 | 0.807156 | 0.842782 | 0 | 0 | 0.60914 | 0.130501 |
| Disease_Pe | LGLL - Larg | 1/375 | 0.808011 | 0.842782 | 0 | 0 | 0.60748 | 0.129503 |

| | | | | | | | | |
|---|---|---|---|---|---|---|---|---|
| Disease_Pe | Escherichia | 1/375 | 0.808011 | 0.842782 | 0 | 0 | 0.60748 | 0.129503 |
| Disease_Pe | pancreatic | 1/376 | 0.808862 | 0.842782 | 0 | 0 | 0.605829 | 0.128513 |
| Disease_Pe | discoid lup | 1/360 | 0.794785 | 0.842782 | 0 | 0 | 0.633348 | 0.14547 |
| Disease_Pe | non-systen | 1/348 | 0.78356 | 0.842782 | 0 | 0 | 0.655653 | 0.159919 |
| Disease_Pe | nemaline n | 1/361 | 0.795694 | 0.842782 | 0 | 0 | 0.631557 | 0.144336 |
| Disease_Pe | pancreatiti | 1/362 | 0.796599 | 0.842782 | 0 | 0 | 0.629775 | 0.143213 |
| Disease_Pe | colon canc | 1/372 | 0.805435 | 0.842782 | 0 | 0 | 0.612487 | 0.132526 |
| Disease_Pe | Acute Lung | 1/372 | 0.805435 | 0.842782 | 0 | 0 | 0.612487 | 0.132526 |
| Disease_Pe | Primary op | 1/373 | 0.806297 | 0.842782 | 0 | 0 | 0.610809 | 0.131509 |
| Disease_Pe | osteoarthri | 1/374 | 0.807156 | 0.842782 | 0 | 0 | 0.60914 | 0.130501 |
| Disease_Pe | Gastrointe: | 1/376 | 0.808862 | 0.842782 | 0 | 0 | 0.605829 | 0.128513 |
| Disease_Pe | lupus eryth | 1/333 | 0.768672 | 0.842782 | 0 | 0 | 0.685801 | 0.180429 |
| Disease_Pe | Down synd | 1/333 | 0.768672 | 0.842782 | 0 | 0 | 0.685801 | 0.180429 |
| Disease_Pe | Protozoan | 1/334 | 0.769695 | 0.842782 | 0 | 0 | 0.683707 | 0.178968 |
| Disease_Pe | cervical car | 1/335 | 0.770714 | 0.842782 | 0 | 0 | 0.681625 | 0.177521 |
| Disease_Pe | head and n | 1/363 | 0.797501 | 0.842782 | 0 | 0 | 0.628003 | 0.1421 |
| Disease_Pe | Carcinoma, | 1/363 | 0.797501 | 0.842782 | 0 | 0 | 0.628003 | 0.1421 |
| Disease_Pe | Spinal Mus | 1/368 | 0.801947 | 0.842782 | 0 | 0 | 0.619289 | 0.136685 |
| Disease_Pe | Duchenne | 1/332 | 0.767644 | 0.842782 | 0 | 0 | 0.687908 | 0.181903 |
| Disease_Pe | skin squam | 1/305 | 0.738121 | 0.842782 | 0 | 0 | 0.750038 | 0.227747 |
| Disease_Pe | endometri: | 1/305 | 0.738121 | 0.842782 | 0 | 0 | 0.750038 | 0.227747 |
| Disease_Pe | large cell n | 1/306 | 0.739278 | 0.842782 | 0 | 0 | 0.747541 | 0.225819 |
| Disease_Pe | systemic lu | 1/306 | 0.739278 | 0.842782 | 0 | 0 | 0.747541 | 0.225819 |
| Disease_Pe | Crohn's dis | 1/337 | 0.772738 | 0.842782 | 0 | 0 | 0.677499 | 0.174669 |
| Disease_Pe | allergic ast | 1/329 | 0.764533 | 0.842782 | 0 | 0 | 0.694307 | 0.186414 |
| Disease_Pe | atheroscler | 1/329 | 0.764533 | 0.842782 | 0 | 0 | 0.694307 | 0.186414 |
| Disease_Pe | Duchenne | 1/329 | 0.764533 | 0.842782 | 0 | 0 | 0.694307 | 0.186414 |
| Disease_Pe | Hypotensiv | 1/339 | 0.774745 | 0.842782 | 0 | 0 | 0.673421 | 0.171872 |
| Disease_Pe | HIV - Huma | 1/340 | 0.775741 | 0.842782 | 0 | 0 | 0.6714 | 0.170493 |
| Disease_Pe | Bacterial Ir | 1/342 | 0.777722 | 0.842782 | 0 | 0 | 0.667394 | 0.167774 |
| Disease_Pe | breast canc | 1/344 | 0.779685 | 0.842782 | 0 | 0 | 0.663435 | 0.165106 |
| Disease_Pe | GERD - Gas | 1/346 | 0.781631 | 0.842782 | 0 | 0 | 0.659521 | 0.162488 |
| Disease_Pe | sickle-cell a | 1/332 | 0.767644 | 0.842782 | 0 | 0 | 0.687908 | 0.181903 |
| Disease_Pe | rheumatoid | 1/232 | 0.63843 | 0.842782 | 0 | 0 | 0.990738 | 0.444588 |
| Disease_Pe | idiopathic p | 1/333 | 0.768672 | 0.842782 | 0 | 0 | 0.685801 | 0.180429 |
| Disease_Pe | psoriatic ar | 1/349 | 0.784518 | 0.842782 | 0 | 0 | 0.653736 | 0.158652 |
| Disease_Pe | glaucoma a | 1/350 | 0.785472 | 0.842782 | 0 | 0 | 0.651829 | 0.157397 |
| Disease_Pe | Breast Can | 1/350 | 0.785472 | 0.842782 | 0 | 0 | 0.651829 | 0.157397 |
| Disease_Pe | Fracture of | 1/351 | 0.786422 | 0.842782 | 0 | 0 | 0.649934 | 0.156154 |
| Disease_Pe | Primary op | 1/351 | 0.786422 | 0.842782 | 0 | 0 | 0.649934 | 0.156154 |
| Disease_Pe | Dehydratio | 1/337 | 0.772738 | 0.842782 | 0 | 0 | 0.677499 | 0.174669 |
| Disease_Pe | non-systen | 1/338 | 0.773744 | 0.842782 | 0 | 0 | 0.675454 | 0.173264 |
| Disease_Pe | Retinitis Pi | 1/338 | 0.773744 | 0.842782 | 0 | 0 | 0.675454 | 0.173264 |
| Disease_Pe | autism spe | 1/353 | 0.788309 | 0.842782 | 0 | 0 | 0.646175 | 0.153702 |
| Disease_Pe | osteoarthri | 1/355 | 0.79018 | 0.842782 | 0 | 0 | 0.642458 | 0.151296 |

| | | | | | | | | |
|---|---|---|---|---|---|---|---|---|
| Disease_Pe | systemic lu | 1/357 | 0.792034 | 0.842782 | 0 | 0 | 0.638784 | 0.148933 |
| Disease_Pe | Influenza C | 1/359 | 0.793872 | 0.842782 | 0 | 0 | 0.63515 | 0.146613 |
| Disease_Pe | systemic ju | 1/360 | 0.794785 | 0.842782 | 0 | 0 | 0.633348 | 0.14547 |
| Disease_Pe | Neurologic | 1/262 | 0.683274 | 0.842782 | 0 | 0 | 0.875523 | 0.333451 |
| Disease_Pe | Huntingtor | 1/263 | 0.68467 | 0.842782 | 0 | 0 | 0.872137 | 0.330382 |
| Disease_Pe | myotonic d | 1/263 | 0.68467 | 0.842782 | 0 | 0 | 0.872137 | 0.330382 |
| Disease_Pe | Macular de | 1/263 | 0.68467 | 0.842782 | 0 | 0 | 0.872137 | 0.330382 |
| Disease_Pe | melanoma | 1/263 | 0.68467 | 0.842782 | 0 | 0 | 0.872137 | 0.330382 |
| Disease_Pe | Peripheral | 1/256 | 0.674769 | 0.842782 | 0 | 0 | 0.896398 | 0.35263 |
| Disease_Pe | amyotroph | 1/256 | 0.674769 | 0.842782 | 0 | 0 | 0.896398 | 0.35263 |
| Disease_Pe | type 2 diab | 1/256 | 0.674769 | 0.842782 | 0 | 0 | 0.896398 | 0.35263 |
| Disease_Pe | RA (rheum | 1/268 | 0.691559 | 0.842782 | 0 | 0 | 0.855587 | 0.315546 |
| Disease_Pe | Nephrolith | 1/270 | 0.694273 | 0.842782 | 0 | 0 | 0.84914 | 0.309842 |
| Disease_Pe | atopic derr | 1/270 | 0.694273 | 0.842782 | 0 | 0 | 0.84914 | 0.309842 |
| Disease_Pe | autism spe | 1/270 | 0.694273 | 0.842782 | 0 | 0 | 0.84914 | 0.309842 |
| Disease_Pe | systemic lu | 1/271 | 0.695622 | 0.842782 | 0 | 0 | 0.845952 | 0.307038 |
| Disease_Pe | Crohn's dis | 1/261 | 0.681872 | 0.842782 | 0 | 0 | 0.878936 | 0.336556 |
| Disease_Pe | autism spe | 1/261 | 0.681872 | 0.842782 | 0 | 0 | 0.878936 | 0.336556 |
| Disease_Pe | schizophre | 1/263 | 0.68467 | 0.842782 | 0 | 0 | 0.872137 | 0.330382 |
| Disease_Pe | Hypoxia C0 | 1/272 | 0.696964 | 0.842782 | 0 | 0 | 0.842787 | 0.304265 |
| Disease_Pe | multiple m | 1/273 | 0.6983 | 0.842782 | 0 | 0 | 0.839646 | 0.301522 |
| Disease_Pe | Huntingtor | 1/273 | 0.6983 | 0.842782 | 0 | 0 | 0.839646 | 0.301522 |
| Disease_Pe | Parkinson's | 1/274 | 0.699631 | 0.842782 | 0 | 0 | 0.836528 | 0.29881 |
| Disease_Pe | Ankylosing | 1/274 | 0.699631 | 0.842782 | 0 | 0 | 0.836528 | 0.29881 |
| Disease_Pe | MS (Multip | 1/264 | 0.68606 | 0.842782 | 0 | 0 | 0.868777 | 0.327347 |
| Disease_Pe | breast adei | 1/266 | 0.688822 | 0.842782 | 0 | 0 | 0.862133 | 0.32138 |
| Disease_Pe | Alcohol poi | 1/267 | 0.690194 | 0.842782 | 0 | 0 | 0.858848 | 0.318446 |
| Disease_Pe | Small cell c | 1/227 | 0.630368 | 0.842782 | 0 | 0 | 1.012914 | 0.46741 |
| Disease_Pe | psoriasis D | 1/227 | 0.630368 | 0.842782 | 0 | 0 | 1.012914 | 0.46741 |
| Disease_Pe | skin squam | 1/227 | 0.630368 | 0.842782 | 0 | 0 | 1.012914 | 0.46741 |
| Disease_Pe | Teratosper | 1/228 | 0.631995 | 0.842782 | 0 | 0 | 1.008401 | 0.462729 |
| Disease_Pe | psoriasis D | 1/278 | 0.704895 | 0.842782 | 0 | 0 | 0.82428 | 0.288256 |
| Disease_Pe | esophagus | 1/271 | 0.695622 | 0.842782 | 0 | 0 | 0.845952 | 0.307038 |
| Disease_Pe | Lung Injury | 1/254 | 0.671883 | 0.842782 | 0 | 0 | 0.903576 | 0.359325 |
| Disease_Pe | ALS (Amyo | 1/232 | 0.63843 | 0.842782 | 0 | 0 | 0.990738 | 0.444588 |
| Disease_Pe | diffuse larg | 1/234 | 0.641605 | 0.842782 | 0 | 0 | 0.982134 | 0.435854 |
| Disease_Pe | pancreatic | 2/499 | 0.642504 | 0.842782 | 0 | 0 | 0.919209 | 0.406642 |
| Disease_Pe | lung large ( | 1/226 | 0.628735 | 0.842782 | 0 | 0 | 1.017468 | 0.472152 |
| Disease_Pe | Familial coi | 1/226 | 0.628735 | 0.842782 | 0 | 0 | 1.017468 | 0.472152 |
| Disease_Pe | type 1 diab | 1/227 | 0.630368 | 0.842782 | 0 | 0 | 1.012914 | 0.46741 |
| Disease_Pe | colitis DOID | 1/241 | 0.652504 | 0.842782 | 0 | 0 | 0.953149 | 0.406935 |
| Disease_Pe | relapsing-r | 1/241 | 0.652504 | 0.842782 | 0 | 0 | 0.953149 | 0.406935 |
| Disease_Pe | Fibrosis of | 1/242 | 0.654034 | 0.842782 | 0 | 0 | 0.949146 | 0.403003 |
| Disease_Pe | Niemann-P | 1/243 | 0.655558 | 0.842782 | 0 | 0 | 0.945176 | 0.399118 |
| Disease_Pe | Huntingtor | 1/229 | 0.633614 | 0.842782 | 0 | 0 | 1.003927 | 0.458108 |

| | | | | | | | | |
|---|---|---|---|---|---|---|---|---|
| Disease_Pe | Huntingtor | 1/229 | 0.633614 | 0.842782 | 0 | 0 | 1.003927 | 0.458108 |
| Disease_Pe | Nutritional | 1/230 | 0.635226 | 0.842782 | 0 | 0 | 0.999492 | 0.453544 |
| Disease_Pe | Ulcerative | 1/246 | 0.660088 | 0.842782 | 0 | 0 | 0.93346 | 0.387742 |
| Disease_Pe | Obstructive | 1/275 | 0.700955 | 0.842782 | 0 | 0 | 0.833432 | 0.296128 |
| Disease_Pe | diabetes m | 1/254 | 0.671883 | 0.842782 | 0 | 0 | 0.903576 | 0.359325 |
| Disease_Pe | Aplastic an | 1/254 | 0.671883 | 0.842782 | 0 | 0 | 0.903576 | 0.359325 |
| Disease_Pe | Autism DO | 1/255 | 0.673329 | 0.842782 | 0 | 0 | 0.899973 | 0.355958 |
| Disease_Pe | asthma DO | 1/235 | 0.643183 | 0.842782 | 0 | 0 | 0.977887 | 0.431568 |
| Disease_Pe | Parkinson's | 1/239 | 0.649424 | 0.842782 | 0 | 0 | 0.961257 | 0.414945 |
| Disease_Pe | Neurofibro | 1/240 | 0.650968 | 0.842782 | 0 | 0 | 0.957186 | 0.410916 |
| Disease_Pe | Huntingtor | 2/387 | 0.504142 | 0.842782 | 0 | 0 | 1.193461 | 0.817399 |
| Disease_Pe | Smoldering | 2/388 | 0.505518 | 0.842782 | 0 | 0 | 1.190308 | 0.811994 |
| Disease_Pe | nemaline n | 2/390 | 0.508263 | 0.842782 | 0 | 0 | 1.184051 | 0.801314 |
| Disease_Pe | Setleis sync | 2/393 | 0.512362 | 0.842782 | 0 | 0 | 1.174786 | 0.785607 |
| Disease_Pe | acute myo | 1/261 | 0.681872 | 0.842782 | 0 | 0 | 0.878936 | 0.336556 |
| Disease_Pe | Asthma C0 | 1/243 | 0.655558 | 0.842782 | 0 | 0 | 0.945176 | 0.399118 |
| Disease_Pe | Niemann-P | 1/244 | 0.657075 | 0.842782 | 0 | 0 | 0.941238 | 0.39528 |
| Disease_Pe | Huntingtor | 1/244 | 0.657075 | 0.842782 | 0 | 0 | 0.941238 | 0.39528 |
| Disease_Pe | Essential H | 1/299 | 0.731073 | 0.842782 | 0 | 0 | 0.765374 | 0.239748 |
| Disease_Pe | Senescence | 1/286 | 0.715152 | 0.842782 | 0 | 0 | 0.800816 | 0.268482 |
| Disease_Pe | Senescence | 1/286 | 0.715152 | 0.842782 | 0 | 0 | 0.800816 | 0.268482 |
| Disease_Pe | RA (rheum | 1/286 | 0.715152 | 0.842782 | 0 | 0 | 0.800816 | 0.268482 |
| Disease_Pe | bipolar diso | 1/294 | 0.725056 | 0.842782 | 0 | 0 | 0.778633 | 0.250335 |
| Disease_Pe | MDS - Mye | 1/295 | 0.72627 | 0.842782 | 0 | 0 | 0.775945 | 0.248173 |
| Disease_Pe | Neurofibro | 1/295 | 0.72627 | 0.842782 | 0 | 0 | 0.775945 | 0.248173 |
| Disease_Pe | prostate ca | 1/296 | 0.727479 | 0.842782 | 0 | 0 | 0.773276 | 0.246034 |
| Disease_Pe | type 2 diab | 1/296 | 0.727479 | 0.842782 | 0 | 0 | 0.773276 | 0.246034 |
| Disease_Pe | Williams-B | 1/290 | 0.720147 | 0.842782 | 0 | 0 | 0.789571 | 0.259216 |
| Disease_Pe | myelodysp | 1/291 | 0.721383 | 0.842782 | 0 | 0 | 0.786808 | 0.25696 |
| Disease_Pe | Leber cong | 1/291 | 0.721383 | 0.842782 | 0 | 0 | 0.786808 | 0.25696 |
| Disease_Pe | Familial co | 1/298 | 0.72988 | 0.842782 | 0 | 0 | 0.76799 | 0.241821 |
| Disease_Pe | Atheroscle | 1/299 | 0.731073 | 0.842782 | 0 | 0 | 0.765374 | 0.239748 |
| Disease_Pe | autism spe | 1/299 | 0.731073 | 0.842782 | 0 | 0 | 0.765374 | 0.239748 |
| Disease_Pe | endometri | 1/272 | 0.696964 | 0.842782 | 0 | 0 | 0.842787 | 0.304265 |
| Disease_Pe | myelodysp | 1/300 | 0.73226 | 0.842782 | 0 | 0 | 0.762775 | 0.237696 |
| Disease_Pe | Senescence | 1/293 | 0.723837 | 0.842782 | 0 | 0 | 0.78134 | 0.25252 |
| Disease_Pe | Cancer of p | 1/294 | 0.725056 | 0.842782 | 0 | 0 | 0.778633 | 0.250335 |
| Disease_Pe | Type 2 diab | 1/294 | 0.725056 | 0.842782 | 0 | 0 | 0.778633 | 0.250335 |
| Disease_Pe | Appendicit | 1/302 | 0.73462 | 0.842782 | 0 | 0 | 0.75763 | 0.233655 |
| Disease_Pe | ulcerative c | 1/302 | 0.73462 | 0.842782 | 0 | 0 | 0.75763 | 0.233655 |
| Disease_Pe | Hyperchole | 1/302 | 0.73462 | 0.842782 | 0 | 0 | 0.75763 | 0.233655 |
| Disease_Pe | autism spe | 1/303 | 0.735792 | 0.842782 | 0 | 0 | 0.755082 | 0.231665 |
| Disease_Pe | acute myo | 1/303 | 0.735792 | 0.842782 | 0 | 0 | 0.755082 | 0.231665 |
| Disease_Pe | Asthma, all | 1/296 | 0.727479 | 0.842782 | 0 | 0 | 0.773276 | 0.246034 |
| Disease_Pe | schizophre | 1/296 | 0.727479 | 0.842782 | 0 | 0 | 0.773276 | 0.246034 |

| | | | | | | | | |
|---|---|---|---|---|---|---|---|---|
| Disease_Pe | asthma DO | 1/297 | 0.728682 | 0.842782 | 0 | 0 | 0.770624 | 0.243916 |
| Disease_Pe | autism spe | 1/275 | 0.700955 | 0.842782 | 0 | 0 | 0.833432 | 0.296128 |
| Disease_Pe | Wolfram sy | 1/276 | 0.702274 | 0.842782 | 0 | 0 | 0.830359 | 0.293475 |
| Disease_Pe | Bipolar Dis | 1/259 | 0.679049 | 0.842782 | 0 | 0 | 0.885839 | 0.342875 |
| Disease_Pe | pulmonary | 2/552 | 0.696737 | 0.842782 | 0 | 0 | 0.828364 | 0.299327 |
| Disease_Pe | familial cor | 1/279 | 0.706197 | 0.842782 | 0 | 0 | 0.821273 | 0.285689 |
| Disease_Pe | Asthma DC | 1/279 | 0.706197 | 0.842782 | 0 | 0 | 0.821273 | 0.285689 |
| Disease_Pe | Type 1 diat | 1/279 | 0.706197 | 0.842782 | 0 | 0 | 0.821273 | 0.285689 |
| Disease_Pe | atopic derr | 1/281 | 0.708784 | 0.842782 | 0 | 0 | 0.815324 | 0.280639 |
| Disease_Pe | neurofibro | 1/282 | 0.710068 | 0.842782 | 0 | 0 | 0.812381 | 0.278154 |
| Disease_Pe | Spinal Corc | 1/274 | 0.699631 | 0.842782 | 0 | 0 | 0.836528 | 0.29881 |
| Disease_Pe | amyotroph | 1/275 | 0.700955 | 0.842782 | 0 | 0 | 0.833432 | 0.296128 |
| Disease_Pe | prostate ca | 1/275 | 0.700955 | 0.842782 | 0 | 0 | 0.833432 | 0.296128 |
| Disease_Pe | mytonic dy | 1/283 | 0.711348 | 0.842782 | 0 | 0 | 0.809459 | 0.275697 |
| Disease_Pe | schizophre | 1/284 | 0.712621 | 0.842782 | 0 | 0 | 0.806558 | 0.273266 |
| Disease_Pe | Sepsis C02 | 1/284 | 0.712621 | 0.842782 | 0 | 0 | 0.806558 | 0.273266 |
| Disease_Pe | melanoma | 1/285 | 0.713889 | 0.842782 | 0 | 0 | 0.803677 | 0.270861 |
| Disease_Pe | schizophre | 1/285 | 0.713889 | 0.842782 | 0 | 0 | 0.803677 | 0.270861 |
| Disease_Pe | Dehydratio | 1/278 | 0.704895 | 0.842782 | 0 | 0 | 0.82428 | 0.288256 |
| Disease_Pe | allergic cor | 1/279 | 0.706197 | 0.842782 | 0 | 0 | 0.821273 | 0.285689 |
| Disease_Pe | Asthma DC | 1/286 | 0.715152 | 0.842782 | 0 | 0 | 0.800816 | 0.268482 |
| Disease_Pe | Overexerti | 1/286 | 0.715152 | 0.842782 | 0 | 0 | 0.800816 | 0.268482 |
| Disease_Pe | Huntingtor | 1/288 | 0.717661 | 0.842782 | 0 | 0 | 0.795154 | 0.263799 |
| Disease_Pe | schizophre | 1/289 | 0.718907 | 0.842782 | 0 | 0 | 0.792353 | 0.261495 |
| Disease_Pe | cardiomyo | 1/290 | 0.720147 | 0.842782 | 0 | 0 | 0.789571 | 0.259216 |
| Disease_Pe | Leukemia, | 1/282 | 0.710068 | 0.842782 | 0 | 0 | 0.812381 | 0.278154 |
| Disease_Pe | cystic fibro | 1/278 | 0.704895 | 0.842782 | 0 | 0 | 0.82428 | 0.288256 |
| Disease_Pe | idiopathic | 1/293 | 0.723837 | 0.842782 | 0 | 0 | 0.78134 | 0.25252 |
| Disease_Pe | glaucoma a | 1/259 | 0.679049 | 0.842782 | 0 | 0 | 0.885839 | 0.342875 |
| Disease_Pe | multiple m | 1/282 | 0.710068 | 0.842782 | 0 | 0 | 0.812381 | 0.278154 |
| Disease_Pe | sarcoidosis | 1/283 | 0.711348 | 0.842782 | 0 | 0 | 0.809459 | 0.275697 |
| Disease_Pe | Purpura, Id | 2/531 | 0.676095 | 0.842782 | 0 | 0 | 0.862182 | 0.337476 |
| Disease_Pe | colorectal a | 1/258 | 0.677628 | 0.842782 | 0 | 0 | 0.889331 | 0.346089 |
| Jensen_TIS | Bronchial e | 18/4837 | 0.811668 | 0.843088 | 0 | 0 | 0.817092 | 0.170497 |
| Disease_Pe | chronic my | 1/380 | 0.812229 | 0.843569 | 0 | 0 | 0.599313 | 0.124641 |
| Disease_Pe | Urothelial | 1/380 | 0.812229 | 0.843569 | 0 | 0 | 0.599313 | 0.124641 |
| GTEx_Tissu | GTEX-QV44 | 6/712 | 0.089866 | 0.8441 | 0 | 0 | 2.015214 | 4.855524 |
| GTEx_Tissu | GTEX-RTLS | 9/1240 | 0.089645 | 0.8441 | 0 | 0 | 1.751109 | 4.223495 |
| GTEx_Tissu | GTEX-SN8C | 3/304 | 0.146503 | 0.844481 | 0 | 0 | 2.327005 | 4.469498 |
| Gene_Pert | TTN TMD n | 3/275 | 0.118102 | 0.844507 | 0 | 0 | 2.578913 | 5.509085 |
| Gene_Pert | Apc KD mo | 4/427 | 0.11562 | 0.844507 | 0 | 0 | 2.220513 | 4.790646 |
| Gene_Pert | FH Heteroz | 3/262 | 0.106125 | 0.844507 | 0 | 0 | 2.710149 | 6.079245 |
| Gene_Pert | SIRT1 KD h | 3/262 | 0.106125 | 0.844507 | 0 | 0 | 2.710149 | 6.079245 |
| Gene_Pert | Foxo1 KO r | 3/271 | 0.114364 | 0.844507 | 0 | 0 | 2.617937 | 5.676657 |
| Gene_Pert | Dnajc7 KO | 4/425 | 0.114178 | 0.844507 | 0 | 0 | 2.231291 | 4.841894 |

| | | | | | | | | |
|---|---|---|---|---|---|---|---|---|
| Gene_Pert | Cidec KO m | 3/281 | 0.123795 | 0.844507 | 0 | 0 | 2.522482 | 5.269787 |
| Gene_Pert | Uqcr10 OE | 3/279 | 0.121886 | 0.844507 | 0 | 0 | 2.54102 | 5.348 |
| Gene_Pert | Nfyb KD m | 3/278 | 0.120936 | 0.844507 | 0 | 0 | 2.55039 | 5.387681 |
| Gene_Pert | Il10 KO mo | 3/278 | 0.120936 | 0.844507 | 0 | 0 | 2.55039 | 5.387681 |
| Gene_Pert | GRN deficie | 3/269 | 0.112512 | 0.844507 | 0 | 0 | 2.637889 | 5.762983 |
| Gene_Pert | Ddb1 KD m | 3/267 | 0.110672 | 0.844507 | 0 | 0 | 2.658144 | 5.851062 |
| Gene_Pert | FH mutatio | 3/276 | 0.119044 | 0.844507 | 0 | 0 | 2.569335 | 5.468219 |
| Gene_Pert | Nfe2l2 KO | 3/274 | 0.117163 | 0.844507 | 0 | 0 | 2.588561 | 5.550357 |
| Gene_Pert | Otx2 KO m | 3/225 | 0.074986 | 0.844507 | 0 | 0 | 3.167793 | 8.206005 |
| Gene_Pert | Nfyc KD m | 4/365 | 0.074932 | 0.844507 | 0 | 0 | 2.610153 | 6.763361 |
| Gene_Pert | PARK2 KD | 4/386 | 0.087756 | 0.844507 | 0 | 0 | 2.464013 | 5.995422 |
| Gene_Pert | Rad21 KD r | 4/386 | 0.087756 | 0.844507 | 0 | 0 | 2.464013 | 5.995422 |
| Gene_Pert | RB1 KD hu | 4/385 | 0.087122 | 0.844507 | 0 | 0 | 2.470607 | 6.029369 |
| Gene_Pert | Aldh5a1 K | 4/384 | 0.086491 | 0.844507 | 0 | 0 | 2.477235 | 6.063563 |
| Gene_Pert | CTNNB1 K | 4/383 | 0.085862 | 0.844507 | 0 | 0 | 2.483899 | 6.098006 |
| Gene_Pert | Prkaca kno | 4/382 | 0.085235 | 0.844507 | 0 | 0 | 2.490597 | 6.1327 |
| Gene_Pert | Erbb2 drug | 4/377 | 0.082135 | 0.844507 | 0 | 0 | 2.524629 | 6.310023 |
| Gene_Pert | Mn1 OE m | 4/373 | 0.079697 | 0.844507 | 0 | 0 | 2.552519 | 6.456655 |
| Gene_Pert | Onecut2 K | 3/246 | 0.092098 | 0.844507 | 0 | 0 | 2.890947 | 6.894635 |
| Gene_Pert | SOX17 OE | 3/260 | 0.104327 | 0.844507 | 0 | 0 | 2.731518 | 6.173843 |
| Gene_Pert | Cstb KO m | 4/459 | 0.13975 | 0.844507 | 0 | 0 | 2.060956 | 4.055757 |
| Gene_Pert | MAPK1 KD | 3/297 | 0.13945 | 0.844507 | 0 | 0 | 2.38326 | 4.695137 |
| Gene_Pert | Adora2b D | 3/291 | 0.133501 | 0.844507 | 0 | 0 | 2.433656 | 4.900515 |
| Gene_Pert | DENND4B | 3/290 | 0.132519 | 0.844507 | 0 | 0 | 2.44226 | 4.935882 |
| Gene_Pert | Ing1 KO m | 3/297 | 0.13945 | 0.844507 | 0 | 0 | 2.38326 | 4.695137 |
| Gene_Pert | GPLD1 OE | 3/297 | 0.13945 | 0.844507 | 0 | 0 | 2.38326 | 4.695137 |
| Gene_Pert | Twist1 OE | 3/264 | 0.107935 | 0.844507 | 0 | 0 | 2.689108 | 5.986573 |
| Gene_Pert | Cstb KO m | 4/415 | 0.107093 | 0.844507 | 0 | 0 | 2.286753 | 5.10873 |
| Gene_Pert | Des KO mo | 3/263 | 0.107028 | 0.844507 | 0 | 0 | 2.699588 | 6.032672 |
| Gene_Pert | Zfx KO mo | 3/263 | 0.107028 | 0.844507 | 0 | 0 | 2.699588 | 6.032672 |
| Gene_Pert | Twist1 OE | 4/401 | 0.097529 | 0.844507 | 0 | 0 | 2.369094 | 5.514317 |
| Gene_Pert | Arx KO mo | 3/251 | 0.096393 | 0.844507 | 0 | 0 | 2.831941 | 6.624826 |
| Gene_Pert | SOX17 OE | 3/246 | 0.092098 | 0.844507 | 0 | 0 | 2.890947 | 6.894635 |
| Gene_Pert | Hey2 KO m | 3/260 | 0.104327 | 0.844507 | 0 | 0 | 2.731518 | 6.173843 |
| Gene_Pert | Fgf23 expr | 4/363 | 0.073764 | 0.844507 | 0 | 0 | 2.624962 | 6.842967 |
| Gene_Pert | Sox9 OE m | 4/361 | 0.072606 | 0.844507 | 0 | 0 | 2.639938 | 6.923791 |
| Gene_Pert | Prmt1 KD r | 3/221 | 0.071905 | 0.844507 | 0 | 0 | 3.226573 | 8.493677 |
| Gene_Pert | Igf1 KO mo | 4/358 | 0.070886 | 0.844507 | 0 | 0 | 2.662719 | 7.047366 |
| Gene_Pert | PARK2 KD | 3/258 | 0.102542 | 0.844507 | 0 | 0 | 2.753221 | 6.27042 |
| Gene_Pert | Thrb d337T | 3/257 | 0.101654 | 0.844507 | 0 | 0 | 2.764201 | 6.319467 |
| Gene_Pert | Pde10a KO | 3/265 | 0.108844 | 0.844507 | 0 | 0 | 2.678708 | 5.940942 |
| Gene_Pert | Eed KO mo | 4/417 | 0.108494 | 0.844507 | 0 | 0 | 2.275446 | 5.053909 |
| Gene_Pert | TET1 DEPLI | 3/274 | 0.117163 | 0.844507 | 0 | 0 | 2.588561 | 5.550357 |
| Gene_Pert | Hdac6 KO | 3/274 | 0.117163 | 0.844507 | 0 | 0 | 2.588561 | 5.550357 |
| Gene_Pert | Chrna4 KO | 1/360 | 0.794785 | 0.844507 | 0 | 0 | 0.633348 | 0.14547 |

| | | | | | | | | |
|---|---|---|---|---|---|---|---|---|
| Gene_Pert | Ppargc1b H | 4/351 | 0.066957 | 0.844507 | 0 | 0 | 2.717406 | 7.347065 |
| Gene_Pert | Pparg OE m | 3/217 | 0.068882 | 0.844507 | 0 | 0 | 3.28755 | 8.795386 |
| Gene_Pert | HIRA KD hu | 4/353 | 0.068068 | 0.844507 | 0 | 0 | 2.701557 | 7.25977 |
| Gene_Pert | MTDH KD h | 3/230 | 0.07892 | 0.844507 | 0 | 0 | 3.097231 | 7.864854 |
| Gene_Pert | Arid3a OE | 4/370 | 0.077893 | 0.844507 | 0 | 0 | 2.573836 | 6.569522 |
| Gene_Pert | TERT OE hu | 4/370 | 0.077893 | 0.844507 | 0 | 0 | 2.573836 | 6.569522 |
| Gene_Pert | Psap hypor | 4/368 | 0.076701 | 0.844507 | 0 | 0 | 2.588243 | 6.646184 |
| Gene_Pert | Kdm2b KD | 3/227 | 0.076549 | 0.844507 | 0 | 0 | 3.13919 | 8.067162 |
| Gene_Pert | Ksr1 knock | 4/366 | 0.075519 | 0.844507 | 0 | 0 | 2.602809 | 6.724007 |
| Gene_Pert | EWSR1 KD | 1/360 | 0.794785 | 0.844507 | 0 | 0 | 0.633348 | 0.14547 |
| Gene_Pert | Myc Activa | 1/360 | 0.794785 | 0.844507 | 0 | 0 | 0.633348 | 0.14547 |
| Gene_Pert | Chrnb4 Def | 1/360 | 0.794785 | 0.844507 | 0 | 0 | 0.633348 | 0.14547 |
| Gene_Pert | Clpp KO mo | 16-Jan | 0.067399 | 0.844507 | 0 | 0 | 15.42481 | 41.60265 |
| Gene_Pert | TRPM7 def | 1/359 | 0.793872 | 0.844507 | 0 | 0 | 0.63515 | 0.146613 |
| Gene_Pert | Nr2c2 KO r | 1/359 | 0.793872 | 0.844507 | 0 | 0 | 0.63515 | 0.146613 |
| Gene_Pert | E2f1 Null m | 1/359 | 0.793872 | 0.844507 | 0 | 0 | 0.63515 | 0.146613 |
| Gene_Pert | TRAF6 shRI | 1/359 | 0.793872 | 0.844507 | 0 | 0 | 0.63515 | 0.146613 |
| Gene_Pert | HNF4A DEF | 1/364 | 0.798398 | 0.844507 | 0 | 0 | 0.626241 | 0.140997 |
| Gene_Pert | Abcb4 KO r | 1/363 | 0.797501 | 0.844507 | 0 | 0 | 0.628003 | 0.1421 |
| Gene_Pert | Map3k1 kn | 1/363 | 0.797501 | 0.844507 | 0 | 0 | 0.628003 | 0.1421 |
| Gene_Pert | Dmd Defici | 1/363 | 0.797501 | 0.844507 | 0 | 0 | 0.628003 | 0.1421 |
| Gene_Pert | Foxo1 KD n | 1/363 | 0.797501 | 0.844507 | 0 | 0 | 0.628003 | 0.1421 |
| Gene_Pert | Aire Ectopi | 1/361 | 0.795694 | 0.844507 | 0 | 0 | 0.631557 | 0.144336 |
| Gene_Pert | FLI1 KD hu | 1/361 | 0.795694 | 0.844507 | 0 | 0 | 0.631557 | 0.144336 |
| Gene_Pert | Dmd Defici | 4/397 | 0.094874 | 0.844507 | 0 | 0 | 2.393697 | 5.637647 |
| Gene_Pert | TYROBP KD | 4/392 | 0.091605 | 0.844507 | 0 | 0 | 2.425165 | 5.796793 |
| Gene_Pert | Gata4 abla | 3/245 | 0.091249 | 0.844507 | 0 | 0 | 2.90304 | 6.950367 |
| Gene_Pert | CTNNB1 OI | 3/245 | 0.091249 | 0.844507 | 0 | 0 | 2.90304 | 6.950367 |
| Gene_Pert | PFKL OE hu | 3/244 | 0.090403 | 0.844507 | 0 | 0 | 2.915234 | 7.00671 |
| Gene_Pert | Lmna KO m | 3/256 | 0.100769 | 0.844507 | 0 | 0 | 2.775268 | 6.369029 |
| Gene_Pert | Sarm1 KO r | 3/256 | 0.100769 | 0.844507 | 0 | 0 | 2.775268 | 6.369029 |
| Gene_Pert | Rs1 KO mo | 4/404 | 0.099543 | 0.844507 | 0 | 0 | 2.350964 | 5.424064 |
| Gene_Pert | Ebf1 OE mo | 3/254 | 0.099009 | 0.844507 | 0 | 0 | 2.797666 | 6.469729 |
| Gene_Pert | Hnf4a KO r | 3/243 | 0.08956 | 0.844507 | 0 | 0 | 2.92753 | 7.063672 |
| Gene_Pert | Snai1 KO m | 4/388 | 0.08903 | 0.844507 | 0 | 0 | 2.450929 | 5.928259 |
| Gene_Pert | Psap KO m | 4/399 | 0.096197 | 0.844507 | 0 | 0 | 2.381333 | 5.575548 |
| Gene_Pert | Pitx2 KO m | 4/461 | 0.141322 | 0.844507 | 0 | 0 | 2.051726 | 4.014644 |
| Gene_Pert | NET1 KD hu | 4/410 | 0.103629 | 0.844507 | 0 | 0 | 2.315508 | 5.249106 |
| Gene_Pert | Psap hypor | 3/259 | 0.103433 | 0.844507 | 0 | 0 | 2.742327 | 6.221881 |
| Gene_Pert | Dnajc7 KO | 3/259 | 0.103433 | 0.844507 | 0 | 0 | 2.742327 | 6.221881 |
| Gene_Pert | Foxo1 KO r | 3/258 | 0.102542 | 0.844507 | 0 | 0 | 2.753221 | 6.27042 |
| Gene_Pert | Uqcr10 OE | 3/247 | 0.09295 | 0.844507 | 0 | 0 | 2.878952 | 6.839504 |
| Gene_Pert | MIR122 KC | 4/393 | 0.092254 | 0.844507 | 0 | 0 | 2.418806 | 5.76451 |
| Gene_Pert | Rbp2 defici | 4/407 | 0.101577 | 0.844507 | 0 | 0 | 2.333104 | 5.335676 |
| Gene_Pert | Rap1a KO r | 4/407 | 0.101577 | 0.844507 | 0 | 0 | 2.333104 | 5.335676 |

| | | | | | | | | |
|---|---|---|---|---|---|---|---|---|
| Gene_Pert | Tfam KD m | 4/350 | 0.066405 | 0.844507 | 0 | 0 | 2.725399 | 7.391226 |
| Gene_Pert | Dlx5 OE mc | 4/364 | 0.074347 | 0.844507 | 0 | 0 | 2.617537 | 6.803013 |
| Gene_Pert | Eed KO mo | 4/363 | 0.073764 | 0.844507 | 0 | 0 | 2.624962 | 6.842967 |
| Gene_Pert | Pink1 KO m | 2/312 | 0.394543 | 0.844507 | 0 | 0 | 1.487894 | 1.383781 |
| Gene_Pert | MYC KD hu | 2/316 | 0.400676 | 0.844507 | 0 | 0 | 1.46864 | 1.343223 |
| Gene_Pert | Myrf KO m | 2/315 | 0.399145 | 0.844507 | 0 | 0 | 1.473407 | 1.353222 |
| Gene_Pert | Gadd45a K | 2/315 | 0.399145 | 0.844507 | 0 | 0 | 1.473407 | 1.353222 |
| Gene_Pert | Scd1 KO m | 2/314 | 0.397613 | 0.844507 | 0 | 0 | 1.478205 | 1.363314 |
| Gene_Pert | Srf KO mou | 3/317 | 0.159904 | 0.844507 | 0 | 0 | 2.229186 | 4.086495 |
| Gene_Pert | Ywhae KO | 3/323 | 0.166214 | 0.844507 | 0 | 0 | 2.186719 | 3.924018 |
| Gene_Pert | Ifnar2 KO n | 3/323 | 0.166214 | 0.844507 | 0 | 0 | 2.186719 | 3.924018 |
| Gene_Pert | BRMS1 KD | 2/318 | 0.403731 | 0.844507 | 0 | 0 | 1.459196 | 1.323499 |
| Gene_Pert | FGFR3 mut | 2/317 | 0.402204 | 0.844507 | 0 | 0 | 1.463903 | 1.333316 |
| Gene_Pert | Dmd Defici | 2/317 | 0.402204 | 0.844507 | 0 | 0 | 1.463903 | 1.333316 |
| Gene_Pert | Cdkn1a DN | 2/317 | 0.402204 | 0.844507 | 0 | 0 | 1.463903 | 1.333316 |
| Gene_Pert | NR5A2 KD | 2/317 | 0.402204 | 0.844507 | 0 | 0 | 1.463903 | 1.333316 |
| Gene_Pert | Fmr1 KO m | 3/319 | 0.161999 | 0.844507 | 0 | 0 | 2.214851 | 4.03139 |
| Gene_Pert | Htt Mutati | 2/312 | 0.394543 | 0.844507 | 0 | 0 | 1.487894 | 1.383781 |
| Gene_Pert | Lmna KO m | 2/312 | 0.394543 | 0.844507 | 0 | 0 | 1.487894 | 1.383781 |
| Gene_Pert | TET1 OE hu | 2/312 | 0.394543 | 0.844507 | 0 | 0 | 1.487894 | 1.383781 |
| Gene_Pert | E2f3 KD mc | 3/307 | 0.149562 | 0.844507 | 0 | 0 | 2.303689 | 4.377116 |
| Gene_Pert | CTNNB1 Ac | 3/307 | 0.149562 | 0.844507 | 0 | 0 | 2.303689 | 4.377116 |
| Gene_Pert | RB1 KD hur | 2/156 | 0.147716 | 0.844507 | 0 | 0 | 3.018946 | 5.773631 |
| Gene_Pert | ADRB1 Bet | 4/469 | 0.14768 | 0.844507 | 0 | 0 | 2.015598 | 3.855244 |
| Gene_Pert | ABL1 muta | 2/149 | 0.13728 | 0.844507 | 0 | 0 | 3.163826 | 6.282501 |
| Gene_Pert | Ghr trunca | 4/454 | 0.135852 | 0.844507 | 0 | 0 | 2.084391 | 4.160841 |
| Gene_Pert | Insm1 KO r | 3/303 | 0.145488 | 0.844507 | 0 | 0 | 2.334881 | 4.500855 |
| Gene_Pert | PTGS2 INH | 3/301 | 0.143466 | 0.844507 | 0 | 0 | 2.350791 | 4.564431 |
| Gene_Pert | Sox10 KO r | 3/317 | 0.159904 | 0.844507 | 0 | 0 | 2.229186 | 4.086495 |
| Gene_Pert | Runx1 KO r | 3/311 | 0.153672 | 0.844507 | 0 | 0 | 2.273307 | 4.257756 |
| Gene_Pert | FH KD hum | 2/310 | 0.391467 | 0.844507 | 0 | 0 | 1.497708 | 1.404633 |
| Gene_Pert | GSK3A KD | 2/310 | 0.391467 | 0.844507 | 0 | 0 | 1.497708 | 1.404633 |
| Gene_Pert | Myod1 Mu | 2/310 | 0.391467 | 0.844507 | 0 | 0 | 1.497708 | 1.404633 |
| Gene_Pert | ESR1 KD hu | 2/310 | 0.391467 | 0.844507 | 0 | 0 | 1.497708 | 1.404633 |
| Gene_Pert | Srf KO mou | 2/305 | 0.383746 | 0.844507 | 0 | 0 | 1.522811 | 1.45851 |
| Gene_Pert | CTNNB1 Ac | 2/305 | 0.383746 | 0.844507 | 0 | 0 | 1.522811 | 1.45851 |
| Gene_Pert | JAK1 knock | 2/305 | 0.383746 | 0.844507 | 0 | 0 | 1.522811 | 1.45851 |
| Gene_Pert | Jak2 active | 2/305 | 0.383746 | 0.844507 | 0 | 0 | 1.522811 | 1.45851 |
| Gene_Pert | RB1 KD hur | 3/319 | 0.161999 | 0.844507 | 0 | 0 | 2.214851 | 4.03139 |
| Gene_Pert | LRRK2 dom | 3/318 | 0.160951 | 0.844507 | 0 | 0 | 2.221995 | 4.058822 |
| Gene_Pert | Dmd Defici | 3/318 | 0.160951 | 0.844507 | 0 | 0 | 2.221995 | 4.058822 |
| Gene_Pert | Abca1 Hete | 3/270 | 0.113436 | 0.844507 | 0 | 0 | 2.627876 | 5.719604 |
| Gene_Pert | PRLH OE hu | 2/160 | 0.153751 | 0.844507 | 0 | 0 | 2.941921 | 5.508522 |
| Gene_Pert | Psmc4 KO | 3/317 | 0.159904 | 0.844507 | 0 | 0 | 2.229186 | 4.086495 |
| Gene_Pert | Gata1 Delt | 3/317 | 0.159904 | 0.844507 | 0 | 0 | 2.229186 | 4.086495 |

| | | | | | | | | |
|---|---|---|---|---|---|---|---|---|
| Gene_Pert | Pten KO m | 3/316 | 0.15886 | 0.844507 | 0 | 0 | 2.236422 | 4.114413 |
| Gene_Pert | Gys1 KO m | 3/292 | 0.134486 | 0.844507 | 0 | 0 | 2.425111 | 4.86548 |
| Gene_Pert | Pparg OE n | 3/292 | 0.134486 | 0.844507 | 0 | 0 | 2.425111 | 4.86548 |
| Gene_Pert | Dicer1 KO | 4/451 | 0.133535 | 0.844507 | 0 | 0 | 2.098704 | 4.225511 |
| Gene_Pert | Hes1 KO m | 3/291 | 0.133501 | 0.844507 | 0 | 0 | 2.433656 | 4.900515 |
| Gene_Pert | RBP2 KD hu | 2/163 | 0.158308 | 0.844507 | 0 | 0 | 2.886664 | 5.320729 |
| Gene_Pert | Klf3 KO mo | 3/320 | 0.16305 | 0.844507 | 0 | 0 | 2.207751 | 4.004196 |
| Gene_Pert | Pax6 KO m | 3/319 | 0.161999 | 0.844507 | 0 | 0 | 2.214851 | 4.03139 |
| Gene_Pert | RBP2 KD hu | 2/137 | 0.119815 | 0.844507 | 0 | 0 | 3.447146 | 7.31418 |
| Gene_Pert | Yy1 KO mo | 3/286 | 0.128615 | 0.844507 | 0 | 0 | 2.477284 | 5.080743 |
| Gene_Pert | Uqcr10 OE | 3/284 | 0.126679 | 0.844507 | 0 | 0 | 2.49517 | 5.155272 |
| Gene_Pert | FOXP1 KD l | 3/283 | 0.125715 | 0.844507 | 0 | 0 | 2.504209 | 5.193076 |
| Gene_Pert | Sod2 KO m | 3/272 | 0.115294 | 0.844507 | 0 | 0 | 2.608072 | 5.634137 |
| Gene_Pert | Met KO mc | 3/272 | 0.115294 | 0.844507 | 0 | 0 | 2.608072 | 5.634137 |
| Gene_Pert | Prmt8 KD r | 2/140 | 0.124127 | 0.844507 | 0 | 0 | 3.371697 | 7.034881 |
| Gene_Pert | CITED1 KD | 3/281 | 0.123795 | 0.844507 | 0 | 0 | 2.522482 | 5.269787 |
| Gene_Pert | ELK1 KD hu | 3/249 | 0.094665 | 0.844507 | 0 | 0 | 2.855256 | 6.731013 |
| Gene_Pert | Eed KO mo | 4/396 | 0.094216 | 0.844507 | 0 | 0 | 2.399926 | 5.669026 |
| Gene_Pert | IRF9 OE hu | 3/248 | 0.093806 | 0.844507 | 0 | 0 | 2.867055 | 6.784966 |
| Gene_Pert | Sox7 OE m | 3/247 | 0.09295 | 0.844507 | 0 | 0 | 2.878952 | 6.839504 |
| Gene_Pert | Nfe2l2 defi | 3/277 | 0.119989 | 0.844507 | 0 | 0 | 2.559828 | 5.427752 |
| Gene_Pert | Klf15 KO m | 3/286 | 0.128615 | 0.844507 | 0 | 0 | 2.477284 | 5.080743 |
| Gene_Pert | CXCL8 hum | 3/288 | 0.130562 | 0.844507 | 0 | 0 | 2.459649 | 5.007625 |
| Gene_Pert | Htt CAG kn | 3/288 | 0.130562 | 0.844507 | 0 | 0 | 2.459649 | 5.007625 |
| Gene_Pert | Tnfsf14 OE | 3/296 | 0.138452 | 0.844507 | 0 | 0 | 2.391516 | 4.728574 |
| Gene_Pert | Ptrf KO mo | 3/296 | 0.138452 | 0.844507 | 0 | 0 | 2.391516 | 4.728574 |
| Gene_Pert | Nfe2l2 KO | 3/295 | 0.137457 | 0.844507 | 0 | 0 | 2.399829 | 4.762324 |
| Gene_Pert | Ahr KO mo | 2/149 | 0.13728 | 0.844507 | 0 | 0 | 3.163826 | 6.282501 |
| Gene_Pert | Tet2 KO m | 3/283 | 0.125715 | 0.844507 | 0 | 0 | 2.504209 | 5.193076 |
| Gene_Pert | Dgcr8 KO n | 4/440 | 0.125185 | 0.844507 | 0 | 0 | 2.152868 | 4.473582 |
| Gene_Pert | Usp22 DEP | 3/293 | 0.135474 | 0.844507 | 0 | 0 | 2.416626 | 4.830773 |
| Gene_Pert | Cstb KO m | 4/453 | 0.135078 | 0.844507 | 0 | 0 | 2.089141 | 4.182261 |
| Gene_Pert | ARF1 KD hu | 3/299 | 0.141453 | 0.844507 | 0 | 0 | 2.366916 | 4.629183 |
| Gene_Pert | BMI1 OE m | 4/469 | 0.14768 | 0.844507 | 0 | 0 | 2.015598 | 3.855244 |
| Gene_Pert | Sod1 G93A | 4/423 | 0.112745 | 0.844507 | 0 | 0 | 2.242172 | 4.893832 |
| Gene_Pert | Map2k7 Se | 4/423 | 0.112745 | 0.844507 | 0 | 0 | 2.242172 | 4.893832 |
| Gene_Pert | Lrp5 KO m | 3/269 | 0.112512 | 0.844507 | 0 | 0 | 2.637889 | 5.762983 |
| Gene_Pert | Ikbkb KO n | 3/297 | 0.13945 | 0.844507 | 0 | 0 | 2.38326 | 4.695137 |
| Gene_Pert | Kitl stimula | 3/297 | 0.13945 | 0.844507 | 0 | 0 | 2.38326 | 4.695137 |
| Gene_Pert | PARK7 KD l | 3/308 | 0.150586 | 0.844507 | 0 | 0 | 2.296019 | 4.346874 |
| Gene_Pert | Emd KO m | 3/307 | 0.149562 | 0.844507 | 0 | 0 | 2.303689 | 4.377116 |
| Gene_Pert | NFKB1 Inac | 3/313 | 0.155741 | 0.844507 | 0 | 0 | 2.25841 | 4.199656 |
| Gene_Pert | Ppp1r1a KC | 3/312 | 0.154705 | 0.844507 | 0 | 0 | 2.265834 | 4.228577 |
| Gene_Pert | S1pr3 KO n | 3/312 | 0.154705 | 0.844507 | 0 | 0 | 2.265834 | 4.228577 |
| Gene_Pert | Srf homozy | 3/312 | 0.154705 | 0.844507 | 0 | 0 | 2.265834 | 4.228577 |

| | | | | | | | | |
|---|---|---|---|---|---|---|---|---|
| Gene_Pert | Ache OE m | 1/361 | 0.795694 | 0.844507 | 0 | 0 | 0.631557 | 0.144336 |
| Gene_Pert | MECOM KI | 3/304 | 0.146503 | 0.844507 | 0 | 0 | 2.327005 | 4.469498 |
| Gene_Pert | B4galnt2 O | 3/311 | 0.153672 | 0.844507 | 0 | 0 | 2.273307 | 4.257756 |
| Gene_Pert | Irf1 OE mo | 3/311 | 0.153672 | 0.844507 | 0 | 0 | 2.273307 | 4.257756 |
| Gene_Pert | AGR2 OE h | 3/316 | 0.15886 | 0.844507 | 0 | 0 | 2.236422 | 4.114413 |
| Gene_Pert | Lpin1 OE m | 3/316 | 0.15886 | 0.844507 | 0 | 0 | 2.236422 | 4.114413 |
| Gene_Pert | Dmd Defici | 3/316 | 0.15886 | 0.844507 | 0 | 0 | 2.236422 | 4.114413 |
| Gene_Pert | Esrrg Defic | 3/316 | 0.15886 | 0.844507 | 0 | 0 | 2.236422 | 4.114413 |
| Gene_Pert | ESRRA KO r | 3/308 | 0.150586 | 0.844507 | 0 | 0 | 2.296019 | 4.346874 |
| Gene_Pert | IRS4 KO mc | 3/315 | 0.157818 | 0.844507 | 0 | 0 | 2.243704 | 4.142577 |
| Gene_Pert | ERBB2 OE I | 3/315 | 0.157818 | 0.844507 | 0 | 0 | 2.243704 | 4.142577 |
| Gene_Pert | Zfp335 KD | 3/313 | 0.155741 | 0.844507 | 0 | 0 | 2.25841 | 4.199656 |
| Gene_Pert | Tfrc KO mo | 2/330 | 0.42191 | 0.844507 | 0 | 0 | 1.40495 | 1.212419 |
| Gene_Pert | Hdac1 KO r | 2/348 | 0.448647 | 0.844507 | 0 | 0 | 1.330636 | 1.066529 |
| Gene_Pert | Pink1 KO m | 2/344 | 0.442764 | 0.844507 | 0 | 0 | 1.346474 | 1.096999 |
| Gene_Pert | Zfx KO mou | 2/344 | 0.442764 | 0.844507 | 0 | 0 | 1.346474 | 1.096999 |
| Gene_Pert | NELFE KD h | 2/329 | 0.420406 | 0.844507 | 0 | 0 | 1.409318 | 1.221224 |
| Gene_Pert | Mef2d KD r | 2/329 | 0.420406 | 0.844507 | 0 | 0 | 1.409318 | 1.221224 |
| Gene_Pert | Nlgn3 KD n | 2/328 | 0.418899 | 0.844507 | 0 | 0 | 1.413713 | 1.230107 |
| Gene_Pert | FGFR3 KD I | 2/332 | 0.424913 | 0.844507 | 0 | 0 | 1.396292 | 1.195045 |
| Gene_Pert | KDM5A KD | 2/332 | 0.424913 | 0.844507 | 0 | 0 | 1.396292 | 1.195045 |
| Gene_Pert | UBB KO mc | 1/126 | 0.42364 | 0.844507 | 0 | 0 | 1.840744 | 1.580964 |
| Gene_Pert | Pebp1 KO r | 2/346 | 0.44571 | 0.844507 | 0 | 0 | 1.338509 | 1.081632 |
| Gene_Pert | SERPINI1 M | 2/346 | 0.44571 | 0.844507 | 0 | 0 | 1.338509 | 1.081632 |
| Gene_Pert | Abca1 OE r | 2/330 | 0.42191 | 0.844507 | 0 | 0 | 1.40495 | 1.212419 |
| Gene_Pert | EGFR druga | 2/348 | 0.448647 | 0.844507 | 0 | 0 | 1.330636 | 1.066529 |
| Gene_Pert | Snca KO mi | 2/330 | 0.42191 | 0.844507 | 0 | 0 | 1.40495 | 1.212419 |
| Gene_Pert | CMAHP KO | 2/335 | 0.429403 | 0.844507 | 0 | 0 | 1.383501 | 1.169555 |
| Gene_Pert | Mecp2 KO | 2/335 | 0.429403 | 0.844507 | 0 | 0 | 1.383501 | 1.169555 |
| Gene_Pert | SOX2 Defic | 2/335 | 0.429403 | 0.844507 | 0 | 0 | 1.383501 | 1.169555 |
| Gene_Pert | Neurod1 C | 2/329 | 0.420406 | 0.844507 | 0 | 0 | 1.409318 | 1.221224 |
| Gene_Pert | Ppara agor | 2/329 | 0.420406 | 0.844507 | 0 | 0 | 1.409318 | 1.221224 |
| Gene_Pert | Mapk1 INH | 2/334 | 0.427909 | 0.844507 | 0 | 0 | 1.387739 | 1.177976 |
| Gene_Pert | KLF2 KD hu | 2/334 | 0.427909 | 0.844507 | 0 | 0 | 1.387739 | 1.177976 |
| Gene_Pert | YY1 KD hur | 1/127 | 0.426163 | 0.844507 | 0 | 0 | 1.826043 | 1.557494 |
| Gene_Pert | HOXA9 OE | 2/336 | 0.430896 | 0.844507 | 0 | 0 | 1.379288 | 1.161207 |
| Gene_Pert | SYK KD hur | 2/336 | 0.430896 | 0.844507 | 0 | 0 | 1.379288 | 1.161207 |
| Gene_Pert | CHRNA9 KC | 2/344 | 0.442764 | 0.844507 | 0 | 0 | 1.346474 | 1.096999 |
| Gene_Pert | Shh KO mo | 2/341 | 0.438329 | 0.844507 | 0 | 0 | 1.358598 | 1.120553 |
| Gene_Pert | Atm Deficie | 2/341 | 0.438329 | 0.844507 | 0 | 0 | 1.358598 | 1.120553 |
| Gene_Pert | Mkl2 KO m | 1/143 | 0.465078 | 0.844507 | 0 | 0 | 1.618981 | 1.239412 |
| Gene_Pert | Rora KO mi | 1/143 | 0.465078 | 0.844507 | 0 | 0 | 1.618981 | 1.239412 |
| Gene_Pert | HNF1B OE | 2/339 | 0.435362 | 0.844507 | 0 | 0 | 1.3668 | 1.136601 |
| Gene_Pert | ERG KD hui | 2/339 | 0.435362 | 0.844507 | 0 | 0 | 1.3668 | 1.136601 |
| Gene_Pert | Ksr2 knock | 2/339 | 0.435362 | 0.844507 | 0 | 0 | 1.3668 | 1.136601 |

| | | | | | | | | |
|---|---|---|---|---|---|---|---|---|
| Gene_Pert | Stap2 KO m | 2/344 | 0.442764 | 0.844507 | 0 | 0 | 1.346474 | 1.096999 |
| Gene_Pert | Dmd Defici | 2/344 | 0.442764 | 0.844507 | 0 | 0 | 1.346474 | 1.096999 |
| Gene_Pert | Deaf1 OE n | 2/344 | 0.442764 | 0.844507 | 0 | 0 | 1.346474 | 1.096999 |
| Gene_Pert | FGFR3 kno | 2/338 | 0.433875 | 0.844507 | 0 | 0 | 1.370938 | 1.144732 |
| Gene_Pert | TP63 knock | 2/337 | 0.432387 | 0.844507 | 0 | 0 | 1.375101 | 1.152933 |
| Gene_Pert | RNF185 KD | 2/336 | 0.430896 | 0.844507 | 0 | 0 | 1.379288 | 1.161207 |
| Gene_Pert | Mut KO mc | 2/344 | 0.442764 | 0.844507 | 0 | 0 | 1.346474 | 1.096999 |
| Gene_Pert | Eed KO mo | 2/343 | 0.441287 | 0.844507 | 0 | 0 | 1.350492 | 1.104782 |
| Gene_Pert | Setd2 Defic | 2/347 | 0.44718 | 0.844507 | 0 | 0 | 1.334561 | 1.074048 |
| Gene_Pert | YY2 KD hur | 1/135 | 0.445958 | 0.844507 | 0 | 0 | 1.716331 | 1.38599 |
| Gene_Pert | TP53 Muta | 1/135 | 0.445958 | 0.844507 | 0 | 0 | 1.716331 | 1.38599 |
| Gene_Pert | Tgfbr2 KO | 2/340 | 0.436846 | 0.844507 | 0 | 0 | 1.362687 | 1.128542 |
| Gene_Pert | Por KO mo | 2/340 | 0.436846 | 0.844507 | 0 | 0 | 1.362687 | 1.128542 |
| Gene_Pert | VGLL4 OE h | 2/346 | 0.44571 | 0.844507 | 0 | 0 | 1.338509 | 1.081632 |
| Gene_Pert | FGFR3 kno | 2/345 | 0.444238 | 0.844507 | 0 | 0 | 1.34248 | 1.089282 |
| Gene_Pert | Aplp2 KO n | 2/344 | 0.442764 | 0.844507 | 0 | 0 | 1.346474 | 1.096999 |
| Gene_Pert | Hoxa10 KO | 2/349 | 0.450113 | 0.844507 | 0 | 0 | 1.326733 | 1.059074 |
| Gene_Pert | Rif1 KD mo | 2/327 | 0.417391 | 0.844507 | 0 | 0 | 1.418136 | 1.239071 |
| Gene_Pert | GTF2B KD h | 2/322 | 0.409821 | 0.844507 | 0 | 0 | 1.440662 | 1.285121 |
| Gene_Pert | MIR1-2 KO | 2/321 | 0.408301 | 0.844507 | 0 | 0 | 1.445252 | 1.294585 |
| Gene_Pert | JAG1 OE hu | 2/326 | 0.415881 | 0.844507 | 0 | 0 | 1.422585 | 1.248115 |
| Gene_Pert | XBP1 OE hu | 2/326 | 0.415881 | 0.844507 | 0 | 0 | 1.422585 | 1.248115 |
| Gene_Pert | Mecp2 KD | 2/325 | 0.414369 | 0.844507 | 0 | 0 | 1.427062 | 1.257241 |
| Gene_Pert | Npc1 KD m | 2/328 | 0.418899 | 0.844507 | 0 | 0 | 1.413713 | 1.230107 |
| Gene_Pert | Htt Knock-i | 2/328 | 0.418899 | 0.844507 | 0 | 0 | 1.413713 | 1.230107 |
| Gene_Pert | PIAS1 Depl | 2/328 | 0.418899 | 0.844507 | 0 | 0 | 1.413713 | 1.230107 |
| Gene_Pert | Ache AChE | 2/325 | 0.414369 | 0.844507 | 0 | 0 | 1.427062 | 1.257241 |
| Gene_Pert | CMAHP KO | 2/324 | 0.412854 | 0.844507 | 0 | 0 | 1.431567 | 1.266451 |
| Gene_Pert | LAMC2 KD | 2/328 | 0.418899 | 0.844507 | 0 | 0 | 1.413713 | 1.230107 |
| Gene_Pert | Gnmt KO m | 1/124 | 0.418561 | 0.844507 | 0 | 0 | 1.870864 | 1.629399 |
| Gene_Pert | MIR34A OE | 2/326 | 0.415881 | 0.844507 | 0 | 0 | 1.422585 | 1.248115 |
| Gene_Pert | CMAHP KO | 2/330 | 0.42191 | 0.844507 | 0 | 0 | 1.40495 | 1.212419 |
| Gene_Pert | Pten KD mc | 2/329 | 0.420406 | 0.844507 | 0 | 0 | 1.409318 | 1.221224 |
| Gene_Pert | Smarca5 KC | 2/329 | 0.420406 | 0.844507 | 0 | 0 | 1.409318 | 1.221224 |
| Gene_Pert | Mef2a KD r | 2/326 | 0.415881 | 0.844507 | 0 | 0 | 1.422585 | 1.248115 |
| Gene_Pert | Syncrip OE | 2/326 | 0.415881 | 0.844507 | 0 | 0 | 1.422585 | 1.248115 |
| Gene_Pert | Psap KO m | 2/328 | 0.418899 | 0.844507 | 0 | 0 | 1.413713 | 1.230107 |
| Gene_Pert | Akt1 drugir | 2/328 | 0.418899 | 0.844507 | 0 | 0 | 1.413713 | 1.230107 |
| Gene_Pert | XIAP Deple | 2/380 | 0.494441 | 0.844507 | 0 | 0 | 1.215998 | 0.85646 |
| Gene_Pert | Arid3a OE | 2/380 | 0.494441 | 0.844507 | 0 | 0 | 1.215998 | 0.85646 |
| Gene_Pert | Tnf siTNF-C | 2/380 | 0.494441 | 0.844507 | 0 | 0 | 1.215998 | 0.85646 |
| Gene_Pert | POU3F3 KD | 2/378 | 0.491648 | 0.844507 | 0 | 0 | 1.222591 | 0.868029 |
| Gene_Pert | Pink1 KO m | 2/321 | 0.408301 | 0.844507 | 0 | 0 | 1.445252 | 1.294585 |
| Gene_Pert | Ep300 KD r | 2/331 | 0.423413 | 0.844507 | 0 | 0 | 1.400608 | 1.203693 |
| Gene_Pert | AKAP12 OE | 2/330 | 0.42191 | 0.844507 | 0 | 0 | 1.40495 | 1.212419 |

| | | | | | | | | |
|---|---|---|---|---|---|---|---|---|
| Gene_Pert | CMAHP KO | 2/335 | 0.429403 | 0.844507 | 0 | 0 | 1.383501 | 1.169555 |
| Gene_Pert | CDK8 knocl | 2/335 | 0.429403 | 0.844507 | 0 | 0 | 1.383501 | 1.169555 |
| Gene_Pert | NR2F2 KD l | 3/330 | 0.173668 | 0.844507 | 0 | 0 | 2.139144 | 3.744801 |
| Gene_Pert | MYB KD hu | 2/339 | 0.435362 | 0.844507 | 0 | 0 | 1.3668 | 1.136601 |
| Gene_Pert | Ager KO m | 2/338 | 0.433875 | 0.844507 | 0 | 0 | 1.370938 | 1.144732 |
| Gene_Pert | NOTCH3 KI | 2/338 | 0.433875 | 0.844507 | 0 | 0 | 1.370938 | 1.144732 |
| Gene_Pert | PTEN KD hu | 2/334 | 0.427909 | 0.844507 | 0 | 0 | 1.387739 | 1.177976 |
| Gene_Pert | Bach2 KO r | 2/334 | 0.427909 | 0.844507 | 0 | 0 | 1.387739 | 1.177976 |
| Gene_Pert | Tmem173 | 2/321 | 0.408301 | 0.844507 | 0 | 0 | 1.445252 | 1.294585 |
| Gene_Pert | PPARA ago | 2/343 | 0.441287 | 0.844507 | 0 | 0 | 1.350492 | 1.104782 |
| Gene_Pert | Fhl2 Deficie | 2/321 | 0.408301 | 0.844507 | 0 | 0 | 1.445252 | 1.294585 |
| Gene_Pert | Dmd Defici | 2/325 | 0.414369 | 0.844507 | 0 | 0 | 1.427062 | 1.257241 |
| Gene_Pert | Fcgr3 KO m | 2/325 | 0.414369 | 0.844507 | 0 | 0 | 1.427062 | 1.257241 |
| Gene_Pert | Smad4 KO | 2/325 | 0.414369 | 0.844507 | 0 | 0 | 1.427062 | 1.257241 |
| Gene_Pert | Plin5 OE m | 2/336 | 0.430896 | 0.844507 | 0 | 0 | 1.379288 | 1.161207 |
| Gene_Pert | POU5F1 OI | 2/335 | 0.429403 | 0.844507 | 0 | 0 | 1.383501 | 1.169555 |
| Gene_Pert | Shank3 KD | 2/324 | 0.412854 | 0.844507 | 0 | 0 | 1.431567 | 1.266451 |
| Gene_Pert | HNF4A KD | 2/324 | 0.412854 | 0.844507 | 0 | 0 | 1.431567 | 1.266451 |
| Gene_Pert | Tfap2c KO | 2/323 | 0.411339 | 0.844507 | 0 | 0 | 1.4361 | 1.275744 |
| Gene_Pert | Nlgn1 KD n | 2/327 | 0.417391 | 0.844507 | 0 | 0 | 1.418136 | 1.239071 |
| Gene_Pert | Wfs1 KO m | 2/327 | 0.417391 | 0.844507 | 0 | 0 | 1.418136 | 1.239071 |
| Gene_Pert | Pgr KO mo | 2/362 | 0.468968 | 0.844507 | 0 | 0 | 1.277974 | 0.96771 |
| Gene_Pert | Smarca4 K | 1/357 | 0.792034 | 0.844507 | 0 | 0 | 0.638784 | 0.148933 |
| Gene_Pert | Esr1 KO mc | 1/356 | 0.791109 | 0.844507 | 0 | 0 | 0.640616 | 0.150109 |
| Gene_Pert | ESR1 KD hu | 1/355 | 0.79018 | 0.844507 | 0 | 0 | 0.642458 | 0.151296 |
| Gene_Pert | Sirt7 KO m | 1/359 | 0.793872 | 0.844507 | 0 | 0 | 0.63515 | 0.146613 |
| Gene_Pert | Rag1 KO m | 1/358 | 0.792955 | 0.844507 | 0 | 0 | 0.636962 | 0.147768 |
| Gene_Pert | Neurog3 C | 1/358 | 0.792955 | 0.844507 | 0 | 0 | 0.636962 | 0.147768 |
| Gene_Pert | Srf KO mou | 1/358 | 0.792955 | 0.844507 | 0 | 0 | 0.636962 | 0.147768 |
| Gene_Pert | PIK3CA KD | 1/358 | 0.792955 | 0.844507 | 0 | 0 | 0.636962 | 0.147768 |
| Gene_Pert | Por KO mo | 1/358 | 0.792955 | 0.844507 | 0 | 0 | 0.636962 | 0.147768 |
| Gene_Pert | Myc Myc + | 1/358 | 0.792955 | 0.844507 | 0 | 0 | 0.636962 | 0.147768 |
| Gene_Pert | SPDEF KD h | 1/358 | 0.792955 | 0.844507 | 0 | 0 | 0.636962 | 0.147768 |
| Gene_Pert | Rb1 KO mo | 2/362 | 0.468968 | 0.844507 | 0 | 0 | 1.277974 | 0.96771 |
| Gene_Pert | Rp1 KO mo | 1/357 | 0.792034 | 0.844507 | 0 | 0 | 0.638784 | 0.148933 |
| Gene_Pert | Psap KO m | 2/361 | 0.46753 | 0.844507 | 0 | 0 | 1.281599 | 0.974388 |
| Gene_Pert | GATA3 Ect | 2/367 | 0.476119 | 0.844507 | 0 | 0 | 1.260145 | 0.935139 |
| Gene_Pert | CDK8 knocl | 2/367 | 0.476119 | 0.844507 | 0 | 0 | 1.260145 | 0.935139 |
| Gene_Pert | Nfe2l2 KO | 1/147 | 0.474392 | 0.844507 | 0 | 0 | 1.574307 | 1.173995 |
| Gene_Pert | LEF1 KD hu | 2/365 | 0.473265 | 0.844507 | 0 | 0 | 1.267218 | 0.948005 |
| Gene_Pert | Cdk4 Heter | 2/365 | 0.473265 | 0.844507 | 0 | 0 | 1.267218 | 0.948005 |
| Gene_Pert | Klf9 Deficie | 2/365 | 0.473265 | 0.844507 | 0 | 0 | 1.267218 | 0.948005 |
| Gene_Pert | Rfwd2 KO r | 2/369 | 0.478963 | 0.844507 | 0 | 0 | 1.25315 | 0.922483 |
| Gene_Pert | Dmd Defici | 2/369 | 0.478963 | 0.844507 | 0 | 0 | 1.25315 | 0.922483 |
| Gene_Pert | Nf1 Hetero | 2/369 | 0.478963 | 0.844507 | 0 | 0 | 1.25315 | 0.922483 |

| | | | | | | | | |
|---|---|---|---|---|---|---|---|---|
| Gene_Pert | MEG3 KO r | 2/362 | 0.468968 | 0.844507 | 0 | 0 | 1.277974 | 0.96771 |
| Gene_Pert | Psap hypor | 1/355 | 0.79018 | 0.844507 | 0 | 0 | 0.642458 | 0.151296 |
| Gene_Pert | Stat3 Actva | 1/365 | 0.799291 | 0.844507 | 0 | 0 | 0.624489 | 0.139904 |
| Gene_Pert | TP63 Over- | 1/365 | 0.799291 | 0.844507 | 0 | 0 | 0.624489 | 0.139904 |
| Gene_Pert | SYK KD hur | 1/365 | 0.799291 | 0.844507 | 0 | 0 | 0.624489 | 0.139904 |
| Gene_Pert | Aurka drug | 1/353 | 0.788309 | 0.844507 | 0 | 0 | 0.646175 | 0.153702 |
| Gene_Pert | Ikzf1 IkL/L | 1/352 | 0.787368 | 0.844507 | 0 | 0 | 0.648049 | 0.154923 |
| Gene_Pert | IRS3P KO m | 1/352 | 0.787368 | 0.844507 | 0 | 0 | 0.648049 | 0.154923 |
| Gene_Pert | CTNNB1 Ac | 1/352 | 0.787368 | 0.844507 | 0 | 0 | 0.648049 | 0.154923 |
| Gene_Pert | Zfx KO mou | 1/352 | 0.787368 | 0.844507 | 0 | 0 | 0.648049 | 0.154923 |
| Gene_Pert | TIAM1 KD l | 1/352 | 0.787368 | 0.844507 | 0 | 0 | 0.648049 | 0.154923 |
| Gene_Pert | Creb1 DN r | 1/351 | 0.786422 | 0.844507 | 0 | 0 | 0.649934 | 0.156154 |
| Gene_Pert | Muc2 &quo | 1/351 | 0.786422 | 0.844507 | 0 | 0 | 0.649934 | 0.156154 |
| Gene_Pert | WNT1 OE h | 2/362 | 0.468968 | 0.844507 | 0 | 0 | 1.277974 | 0.96771 |
| Gene_Pert | Tnf siTNF n | 1/355 | 0.79018 | 0.844507 | 0 | 0 | 0.642458 | 0.151296 |
| Gene_Pert | KDM3A KD | 1/354 | 0.789247 | 0.844507 | 0 | 0 | 0.644311 | 0.152493 |
| Gene_Pert | Crem KO m | 1/354 | 0.789247 | 0.844507 | 0 | 0 | 0.644311 | 0.152493 |
| Gene_Pert | MIR517A C | 1/353 | 0.788309 | 0.844507 | 0 | 0 | 0.646175 | 0.153702 |
| Gene_Pert | Mmp10 KC | 1/353 | 0.788309 | 0.844507 | 0 | 0 | 0.646175 | 0.153702 |
| Gene_Pert | Twist1 OE | 1/353 | 0.788309 | 0.844507 | 0 | 0 | 0.646175 | 0.153702 |
| Gene_Pert | Por KO mo | 1/353 | 0.788309 | 0.844507 | 0 | 0 | 0.646175 | 0.153702 |
| Gene_Pert | Iltifb KO m | 1/358 | 0.792955 | 0.844507 | 0 | 0 | 0.636962 | 0.147768 |
| Gene_Pert | Fosl1 KO m | 1/357 | 0.792034 | 0.844507 | 0 | 0 | 0.638784 | 0.148933 |
| Gene_Pert | CHRNA9 K( | 1/357 | 0.792034 | 0.844507 | 0 | 0 | 0.638784 | 0.148933 |
| Gene_Pert | PTPN1 KD l | 1/357 | 0.792034 | 0.844507 | 0 | 0 | 0.638784 | 0.148933 |
| Gene_Pert | COMMD3-I | 2/351 | 0.453038 | 0.844507 | 0 | 0 | 1.318995 | 1.044354 |
| Gene_Pert | Runx1 KO r | 2/357 | 0.46176 | 0.844507 | 0 | 0 | 1.296305 | 1.001668 |
| Gene_Pert | Ovol2 KO n | 2/356 | 0.460312 | 0.844507 | 0 | 0 | 1.300033 | 1.008633 |
| Gene_Pert | Abcb4 KO r | 2/356 | 0.460312 | 0.844507 | 0 | 0 | 1.300033 | 1.008633 |
| Gene_Pert | Trp53inp2 | 2/347 | 0.44718 | 0.844507 | 0 | 0 | 1.334561 | 1.074048 |
| Gene_Pert | CXCL5 KO r | 2/347 | 0.44718 | 0.844507 | 0 | 0 | 1.334561 | 1.074048 |
| Gene_Pert | Twist1 OE | 2/355 | 0.458861 | 0.844507 | 0 | 0 | 1.303783 | 1.015656 |
| Gene_Pert | Gfi1 KO mc | 2/355 | 0.458861 | 0.844507 | 0 | 0 | 1.303783 | 1.015656 |
| Gene_Pert | Met KO mc | 2/354 | 0.457409 | 0.844507 | 0 | 0 | 1.307553 | 1.02274 |
| Gene_Pert | Nfia Deficie | 2/361 | 0.46753 | 0.844507 | 0 | 0 | 1.281599 | 0.974388 |
| Gene_Pert | Dicer1 Defi | 2/361 | 0.46753 | 0.844507 | 0 | 0 | 1.281599 | 0.974388 |
| Gene_Pert | Rnf2 KO m | 2/360 | 0.466091 | 0.844507 | 0 | 0 | 1.285245 | 0.981122 |
| Gene_Pert | Lep KO mo | 2/351 | 0.453038 | 0.844507 | 0 | 0 | 1.318995 | 1.044354 |
| Gene_Pert | Des KO mo | 2/349 | 0.450113 | 0.844507 | 0 | 0 | 1.326733 | 1.059074 |
| Gene_Pert | PFKL OE hu | 2/358 | 0.463206 | 0.844507 | 0 | 0 | 1.292597 | 0.994762 |
| Gene_Pert | Dmd Defici | 2/358 | 0.463206 | 0.844507 | 0 | 0 | 1.292597 | 0.994762 |
| Gene_Pert | Klf15 Defic | 1/142 | 0.462724 | 0.844507 | 0 | 0 | 1.630546 | 1.256538 |
| Gene_Pert | Gnmt KO n | 2/364 | 0.471835 | 0.844507 | 0 | 0 | 1.270783 | 0.954519 |
| Gene_Pert | Dmd Defici | 2/364 | 0.471835 | 0.844507 | 0 | 0 | 1.270783 | 0.954519 |
| Gene_Pert | MTDH KD h | 2/364 | 0.471835 | 0.844507 | 0 | 0 | 1.270783 | 0.954519 |

| | | | | | | | | |
|---|---|---|---|---|---|---|---|---|
| Gene_Pert | Rest KD mc | 2/356 | 0.460312 | 0.844507 | 0 | 0 | 1.300033 | 1.008633 |
| Gene_Pert | NPTX1 KD l | 2/356 | 0.460312 | 0.844507 | 0 | 0 | 1.300033 | 1.008633 |
| Gene_Pert | Sirt1 OE mc | 2/337 | 0.432387 | 0.844507 | 0 | 0 | 1.375101 | 1.152933 |
| Gene_Pert | Dysf Defici | 2/337 | 0.432387 | 0.844507 | 0 | 0 | 1.375101 | 1.152933 |
| Gene_Pert | CABIN1 KD | 2/337 | 0.432387 | 0.844507 | 0 | 0 | 1.375101 | 1.152933 |
| Gene_Pert | WT1 OE hu | 2/368 | 0.477542 | 0.844507 | 0 | 0 | 1.256638 | 0.928785 |
| Gene_Pert | XIAP DEPLE | 2/368 | 0.477542 | 0.844507 | 0 | 0 | 1.256638 | 0.928785 |
| Gene_Pert | HSPA1A tra | 2/368 | 0.477542 | 0.844507 | 0 | 0 | 1.256638 | 0.928785 |
| Gene_Pert | Des KO mo | 2/367 | 0.476119 | 0.844507 | 0 | 0 | 1.260145 | 0.935139 |
| Gene_Pert | Top2b KO r | 2/374 | 0.486034 | 0.844507 | 0 | 0 | 1.23599 | 0.891738 |
| Gene_Pert | Alk KD mou | 2/373 | 0.484624 | 0.844507 | 0 | 0 | 1.239385 | 0.897787 |
| Gene_Pert | NPM1 mut | 2/373 | 0.484624 | 0.844507 | 0 | 0 | 1.239385 | 0.897787 |
| Gene_Pert | Htt Knock-i | 2/365 | 0.473265 | 0.844507 | 0 | 0 | 1.267218 | 0.948005 |
| Gene_Pert | Zfp42 KD m | 2/365 | 0.473265 | 0.844507 | 0 | 0 | 1.267218 | 0.948005 |
| Gene_Pert | Cdx2 KO m | 3/594 | 0.480046 | 0.844507 | 0 | 0 | 1.167634 | 0.856895 |
| Gene_Pert | Ahnak KO r | 1/149 | 0.478989 | 0.844507 | 0 | 0 | 1.552876 | 1.143039 |
| Gene_Pert | Ppargc1a K | 2/369 | 0.478963 | 0.844507 | 0 | 0 | 1.25315 | 0.922483 |
| Gene_Pert | PTEN DELE | 2/318 | 0.403731 | 0.844507 | 0 | 0 | 1.459196 | 1.323499 |
| Gene_Pert | Mat1a KO r | 2/368 | 0.477542 | 0.844507 | 0 | 0 | 1.256638 | 0.928785 |
| Gene_Pert | TGM2 KD h | 2/347 | 0.44718 | 0.844507 | 0 | 0 | 1.334561 | 1.074048 |
| Gene_Pert | BAHD1 KD | 2/347 | 0.44718 | 0.844507 | 0 | 0 | 1.334561 | 1.074048 |
| Gene_Pert | Klf13 KO m | 2/347 | 0.44718 | 0.844507 | 0 | 0 | 1.334561 | 1.074048 |
| Gene_Pert | OTX2 silen | 2/354 | 0.457409 | 0.844507 | 0 | 0 | 1.307553 | 1.02274 |
| Gene_Pert | Htt Mutati | 2/354 | 0.457409 | 0.844507 | 0 | 0 | 1.307553 | 1.02274 |
| Gene_Pert | Sod1 G93A | 1/138 | 0.453206 | 0.844507 | 0 | 0 | 1.678493 | 1.328374 |
| Gene_Pert | Clpp KO mc | 2/370 | 0.480382 | 0.844507 | 0 | 0 | 1.24968 | 0.916233 |
| Gene_Pert | UPF1 Defic | 2/370 | 0.480382 | 0.844507 | 0 | 0 | 1.24968 | 0.916233 |
| Gene_Pert | Pparg V29C | 2/350 | 0.451576 | 0.844507 | 0 | 0 | 1.322853 | 1.051682 |
| Gene_Pert | Eed KO mo | 2/349 | 0.450113 | 0.844507 | 0 | 0 | 1.326733 | 1.059074 |
| Gene_Pert | Wfs1 KO m | 2/250 | 0.296648 | 0.844507 | 0 | 0 | 1.86575 | 2.267274 |
| Gene_Pert | Kdm2b OE | 2/254 | 0.303079 | 0.844507 | 0 | 0 | 1.835761 | 2.191461 |
| Gene_Pert | Neurog3 C | 2/253 | 0.301472 | 0.844507 | 0 | 0 | 1.843169 | 2.210102 |
| Gene_Pert | Dicer1 KO r | 2/253 | 0.301472 | 0.844507 | 0 | 0 | 1.843169 | 2.210102 |
| Gene_Pert | Mylk knock | 2/252 | 0.299865 | 0.844507 | 0 | 0 | 1.850635 | 2.228949 |
| Gene_Pert | BST2 OE hu | 2/246 | 0.29021 | 0.844507 | 0 | 0 | 1.896721 | 2.346531 |
| Gene_Pert | Cldn18 KO | 2/246 | 0.29021 | 0.844507 | 0 | 0 | 1.896721 | 2.346531 |
| Gene_Pert | Dcx KO mo | 2/246 | 0.29021 | 0.844507 | 0 | 0 | 1.896721 | 2.346531 |
| Gene_Pert | Polg Mutat | 2/246 | 0.29021 | 0.844507 | 0 | 0 | 1.896721 | 2.346531 |
| Gene_Pert | YAP1 KD hu | 2/231 | 0.266025 | 0.844507 | 0 | 0 | 2.022502 | 2.678127 |
| Gene_Pert | Atxn1 Knoc | 2/230 | 0.264412 | 0.844507 | 0 | 0 | 2.031476 | 2.702365 |
| Gene_Pert | Srf homozy | 3/410 | 0.264211 | 0.844507 | 0 | 0 | 1.711653 | 2.278224 |
| Gene_Pert | Ppara OE n | 2/229 | 0.262799 | 0.844507 | 0 | 0 | 2.040529 | 2.726893 |
| Gene_Pert | E2f1 KD mc | 2/242 | 0.283765 | 0.844507 | 0 | 0 | 1.928725 | 2.429439 |
| Gene_Pert | GPLD1 OE l | 2/249 | 0.295039 | 0.844507 | 0 | 0 | 1.873398 | 2.286757 |
| Gene_Pert | Vhl Deficie | 2/249 | 0.295039 | 0.844507 | 0 | 0 | 1.873398 | 2.286757 |

| | | | | | | | | |
|---|---|---|---|---|---|---|---|---|
| Gene_Pert | Elavl1 KO n | 2/248 | 0.29343 | 0.844507 | 0 | 0 | 1.88111 | 2.306459 |
| Gene_Pert | Eed KO mo | 3/418 | 0.273635 | 0.844507 | 0 | 0 | 1.677969 | 2.174583 |
| Gene_Pert | Tada3 Con | 3/416 | 0.271275 | 0.844507 | 0 | 0 | 1.686268 | 2.199943 |
| Gene_Pert | TGM2 KD h | 2/234 | 0.270864 | 0.844507 | 0 | 0 | 1.996045 | 2.607109 |
| Gene_Pert | C20orf197 | 3/414 | 0.268918 | 0.844507 | 0 | 0 | 1.694647 | 2.225665 |
| Gene_Pert | CDH11 KD | 3/403 | 0.256001 | 0.844507 | 0 | 0 | 1.742232 | 2.373918 |
| Gene_Pert | Mgea5 KO | 2/223 | 0.253125 | 0.844507 | 0 | 0 | 2.096566 | 2.880418 |
| Gene_Pert | CTNNB1 be | 3/400 | 0.252495 | 0.844507 | 0 | 0 | 1.755668 | 2.41644 |
| Gene_Pert | CPSF6 KD h | 2/222 | 0.251513 | 0.844507 | 0 | 0 | 2.106203 | 2.90711 |
| Gene_Pert | Rtn4 KO m | 2/218 | 0.245071 | 0.844507 | 0 | 0 | 2.145643 | 3.017223 |
| Gene_Pert | NCOA1 KD | 3/412 | 0.266563 | 0.844507 | 0 | 0 | 1.703109 | 2.251757 |
| Gene_Pert | Psap KO m | 3/390 | 0.240862 | 0.844507 | 0 | 0 | 1.801956 | 2.565139 |
| Gene_Pert | STAG2 KO | 3/390 | 0.240862 | 0.844507 | 0 | 0 | 1.801956 | 2.565139 |
| Gene_Pert | Myc JoMa1 | 3/390 | 0.240862 | 0.844507 | 0 | 0 | 1.801956 | 2.565139 |
| Gene_Pert | Myoc OE m | 2/215 | 0.240243 | 0.844507 | 0 | 0 | 2.176194 | 3.103476 |
| Gene_Pert | Ezh2 Defici | 3/421 | 0.277179 | 0.844507 | 0 | 0 | 1.66567 | 2.13721 |
| Gene_Pert | Mef2d KD | 2/236 | 0.27409 | 0.844507 | 0 | 0 | 1.978783 | 2.561135 |
| Gene_Pert | Pten KO m | 2/236 | 0.27409 | 0.844507 | 0 | 0 | 1.978783 | 2.561135 |
| Gene_Pert | SMARCB1 ( | 2/236 | 0.27409 | 0.844507 | 0 | 0 | 1.978783 | 2.561135 |
| Gene_Pert | YAP1 OE hu | 3/396 | 0.247831 | 0.844507 | 0 | 0 | 1.7739 | 2.474607 |
| Gene_Pert | Pde6b RD1 | 2/219 | 0.246681 | 0.844507 | 0 | 0 | 2.135647 | 2.989182 |
| Gene_Pert | Akt1 knock | 3/395 | 0.246667 | 0.844507 | 0 | 0 | 1.778517 | 2.489418 |
| Gene_Pert | SYK druginl | 2/262 | 0.31591 | 0.844507 | 0 | 0 | 1.778552 | 2.04942 |
| Gene_Pert | IRS4 KO mo | 3/383 | 0.232777 | 0.844507 | 0 | 0 | 1.835808 | 2.676009 |
| Gene_Pert | Nrl Deficie | 3/382 | 0.231626 | 0.844507 | 0 | 0 | 1.840746 | 2.692329 |
| Gene_Pert | Tlr4 KO mo | 2/209 | 0.230606 | 0.844507 | 0 | 0 | 2.239955 | 3.286115 |
| Gene_Pert | Hspa9 hete | 2/209 | 0.230606 | 0.844507 | 0 | 0 | 2.239955 | 3.286115 |
| Gene_Pert | Ikzf1 KO m | 3/439 | 0.298533 | 0.844507 | 0 | 0 | 1.595429 | 1.928672 |
| Gene_Pert | NRAS KD h | 2/250 | 0.296648 | 0.844507 | 0 | 0 | 1.86575 | 2.267274 |
| Gene_Pert | IRF9 OE hu | 2/250 | 0.296648 | 0.844507 | 0 | 0 | 1.86575 | 2.267274 |
| Gene_Pert | CDK9 drugi | 2/250 | 0.296648 | 0.844507 | 0 | 0 | 1.86575 | 2.267274 |
| Gene_Pert | Stk11 KO m | 3/428 | 0.285466 | 0.844507 | 0 | 0 | 1.637647 | 2.053005 |
| Gene_Pert | Selenbp2 K | 2/243 | 0.285377 | 0.844507 | 0 | 0 | 1.920625 | 2.408358 |
| Gene_Pert | IL6 anti-IL6 | 2/243 | 0.285377 | 0.844507 | 0 | 0 | 1.920625 | 2.408358 |
| Gene_Pert | Srf KO mou | 3/458 | 0.321183 | 0.844507 | 0 | 0 | 1.527316 | 1.734638 |
| Gene_Pert | ABL1 muta | 2/248 | 0.29343 | 0.844507 | 0 | 0 | 1.88111 | 2.306459 |
| Gene_Pert | Myod1 Mu | 2/248 | 0.29343 | 0.844507 | 0 | 0 | 1.88111 | 2.306459 |
| Gene_Pert | Klf5 KO mo | 2/247 | 0.29182 | 0.844507 | 0 | 0 | 1.888884 | 2.326383 |
| Gene_Pert | Gsk3b KO r | 2/241 | 0.282153 | 0.844507 | 0 | 0 | 1.936894 | 2.450762 |
| Gene_Pert | Cyfip1 OE r | 3/424 | 0.280728 | 0.844507 | 0 | 0 | 1.653546 | 2.100616 |
| Gene_Pert | IRF9 OE hu | 2/240 | 0.280541 | 0.844507 | 0 | 0 | 1.945131 | 2.472331 |
| Gene_Pert | Atxn1l KO r | 2/240 | 0.280541 | 0.844507 | 0 | 0 | 1.945131 | 2.472331 |
| Gene_Pert | Ubr5 KD m | 2/270 | 0.328691 | 0.844507 | 0 | 0 | 1.724759 | 1.919028 |
| Gene_Pert | TRIM33 KD | 2/270 | 0.328691 | 0.844507 | 0 | 0 | 1.724759 | 1.919028 |
| Gene_Pert | Trpv4 DEPL | 2/270 | 0.328691 | 0.844507 | 0 | 0 | 1.724759 | 1.919028 |

| | | | | | | | | |
|---|---|---|---|---|---|---|---|---|
| Gene_Pert | Bhlha15 KC | 3/464 | 0.328346 | 0.844507 | 0 | 0 | 1.506972 | 1.678296 |
| Gene_Pert | Nf1 KO mo | 3/458 | 0.321183 | 0.844507 | 0 | 0 | 1.527316 | 1.734638 |
| Gene_Pert | ATR KD hu | 2/248 | 0.29343 | 0.844507 | 0 | 0 | 1.88111 | 2.306459 |
| Gene_Pert | MYC KD hu | 2/265 | 0.32071 | 0.844507 | 0 | 0 | 1.757996 | 1.999226 |
| Gene_Pert | ESRRA KO | 2/265 | 0.32071 | 0.844507 | 0 | 0 | 1.757996 | 1.999226 |
| Gene_Pert | Gata6 KO r | 2/271 | 0.330285 | 0.844507 | 0 | 0 | 1.718259 | 1.903487 |
| Gene_Pert | Tnfaip6 KO | 2/271 | 0.330285 | 0.844507 | 0 | 0 | 1.718259 | 1.903487 |
| Gene_Pert | RB1 KD hu | 2/271 | 0.330285 | 0.844507 | 0 | 0 | 1.718259 | 1.903487 |
| Gene_Pert | Met knock | 2/271 | 0.330285 | 0.844507 | 0 | 0 | 1.718259 | 1.903487 |
| Gene_Pert | LMO3 KD h | 2/268 | 0.325502 | 0.844507 | 0 | 0 | 1.737904 | 1.950602 |
| Gene_Pert | SIN3A knoc | 2/268 | 0.325502 | 0.844507 | 0 | 0 | 1.737904 | 1.950602 |
| Gene_Pert | NPAP1 OE | 2/268 | 0.325502 | 0.844507 | 0 | 0 | 1.737904 | 1.950602 |
| Gene_Pert | Pmp22 KO | 2/268 | 0.325502 | 0.844507 | 0 | 0 | 1.737904 | 1.950602 |
| Gene_Pert | ATM KD hu | 2/255 | 0.304685 | 0.844507 | 0 | 0 | 1.828412 | 2.173024 |
| Gene_Pert | Cbfb KO m | 3/447 | 0.308061 | 0.844507 | 0 | 0 | 1.566039 | 1.843943 |
| Gene_Pert | Smarca4 KI | 2/262 | 0.31591 | 0.844507 | 0 | 0 | 1.778552 | 2.04942 |
| Gene_Pert | Snca KO m | 2/261 | 0.314309 | 0.844507 | 0 | 0 | 1.78551 | 2.066511 |
| Gene_Pert | Hsp90b1 O | 2/261 | 0.314309 | 0.844507 | 0 | 0 | 1.78551 | 2.066511 |
| Gene_Pert | Fgfr1 active | 2/254 | 0.303079 | 0.844507 | 0 | 0 | 1.835761 | 2.191461 |
| Gene_Pert | GSK3A KD | 2/254 | 0.303079 | 0.844507 | 0 | 0 | 1.835761 | 2.191461 |
| Gene_Pert | Neurog3 O | 2/254 | 0.303079 | 0.844507 | 0 | 0 | 1.835761 | 2.191461 |
| Gene_Pert | ESR1 L540 | 2/254 | 0.303079 | 0.844507 | 0 | 0 | 1.835761 | 2.191461 |
| Gene_Pert | SOX2 KO h | 2/265 | 0.32071 | 0.844507 | 0 | 0 | 1.757996 | 1.999226 |
| Gene_Pert | NFKB1 Inac | 2/265 | 0.32071 | 0.844507 | 0 | 0 | 1.757996 | 1.999226 |
| Gene_Pert | PARK2 KD | 2/265 | 0.32071 | 0.844507 | 0 | 0 | 1.757996 | 1.999226 |
| Gene_Pert | NRAS DEPL | 2/263 | 0.317511 | 0.844507 | 0 | 0 | 1.771648 | 2.03251 |
| Gene_Pert | Ache AChE | 2/232 | 0.267638 | 0.844507 | 0 | 0 | 2.013606 | 2.654174 |
| Gene_Pert | Kmt2a KO | 2/257 | 0.307896 | 0.844507 | 0 | 0 | 1.813887 | 2.136747 |
| Gene_Pert | Prkag3 Mu | 2/257 | 0.307896 | 0.844507 | 0 | 0 | 1.813887 | 2.136747 |
| Gene_Pert | Bmpr2 don | 2/257 | 0.307896 | 0.844507 | 0 | 0 | 1.813887 | 2.136747 |
| Gene_Pert | Dmd Defici | 3/431 | 0.289025 | 0.844507 | 0 | 0 | 1.625918 | 2.018155 |
| Gene_Pert | FGFR1 mut | 2/245 | 0.288599 | 0.844507 | 0 | 0 | 1.904624 | 2.366908 |
| Gene_Pert | KDM4C nat | 2/244 | 0.286988 | 0.844507 | 0 | 0 | 1.912591 | 2.387516 |
| Gene_Pert | BRD4 knoc | 2/244 | 0.286988 | 0.844507 | 0 | 0 | 1.912591 | 2.387516 |
| Gene_Pert | MYB OE hu | 3/452 | 0.314024 | 0.844507 | 0 | 0 | 1.548202 | 1.793262 |
| Gene_Pert | RB1 KD hu | 2/260 | 0.312707 | 0.844507 | 0 | 0 | 1.792522 | 2.083787 |
| Gene_Pert | TGFB1I1 KI | 2/260 | 0.312707 | 0.844507 | 0 | 0 | 1.792522 | 2.083787 |
| Gene_Pert | Smad3 KO | 2/259 | 0.311104 | 0.844507 | 0 | 0 | 1.799588 | 2.10125 |
| Gene_Pert | KDM6B KD | 3/348 | 0.193257 | 0.844507 | 0 | 0 | 2.025673 | 3.329673 |
| Gene_Pert | HNRNPU-A | 3/367 | 0.214506 | 0.844507 | 0 | 0 | 1.918073 | 2.952718 |
| Gene_Pert | Dgcr8 KO n | 3/365 | 0.212244 | 0.844507 | 0 | 0 | 1.928867 | 2.989777 |
| Gene_Pert | Tsc1 Defici | 3/364 | 0.211116 | 0.844507 | 0 | 0 | 1.934309 | 3.008525 |
| Gene_Pert | BRCA1 DEF | 2/213 | 0.237028 | 0.844507 | 0 | 0 | 2.197045 | 3.162813 |
| Gene_Pert | USF1 OE hu | 3/386 | 0.236236 | 0.844507 | 0 | 0 | 1.821149 | 2.627779 |
| Gene_Pert | Nanog OE | 2/212 | 0.235422 | 0.844507 | 0 | 0 | 2.207619 | 3.19305 |

| | | | | | | | | |
|---|---|---|---|---|---|---|---|---|
| Gene_Pert | Trp53inp2 | 3/384 | 0.233929 | 0.844507 | 0 | 0 | 1.830896 | 2.659811 |
| Gene_Pert | CD24 Redu | 2/204 | 0.222596 | 0.844507 | 0 | 0 | 2.295981 | 3.449476 |
| Gene_Pert | Mybl1 KO r | 2/204 | 0.222596 | 0.844507 | 0 | 0 | 2.295981 | 3.449476 |
| Gene_Pert | Aplp2 KO n | 3/374 | 0.222461 | 0.844507 | 0 | 0 | 1.881209 | 2.827462 |
| Gene_Pert | Ahnak KO r | 2/203 | 0.220997 | 0.844507 | 0 | 0 | 2.307521 | 3.483452 |
| Gene_Pert | Clpp KO mo | 3/349 | 0.194361 | 0.844507 | 0 | 0 | 2.019715 | 3.308368 |
| Gene_Pert | CDK19 kno | 3/368 | 0.215638 | 0.844507 | 0 | 0 | 1.91272 | 2.934405 |
| Gene_Pert | Gata3 KO r | 3/348 | 0.193257 | 0.844507 | 0 | 0 | 2.025673 | 3.329673 |
| Gene_Pert | Pten KD mo | 2/185 | 0.192423 | 0.844507 | 0 | 0 | 2.536805 | 4.180799 |
| Gene_Pert | MTDH KD h | 3/377 | 0.225889 | 0.844507 | 0 | 0 | 1.865833 | 2.775817 |
| Gene_Pert | Fgf23 Trans | 3/377 | 0.225889 | 0.844507 | 0 | 0 | 1.865833 | 2.775817 |
| Gene_Pert | PAX5 OE hu | 4/558 | 0.224989 | 0.844507 | 0 | 0 | 1.68405 | 2.512102 |
| Gene_Pert | Pten KO mo | 3/376 | 0.224746 | 0.844507 | 0 | 0 | 1.870931 | 2.7929 |
| Gene_Pert | Ppargc1a D | 3/354 | 0.199908 | 0.844507 | 0 | 0 | 1.990435 | 3.204395 |
| Gene_Pert | Tlr4 KO mo | 2/189 | 0.198732 | 0.844507 | 0 | 0 | 2.482038 | 4.010467 |
| Gene_Pert | ATM KD hu | 2/188 | 0.197153 | 0.844507 | 0 | 0 | 2.495509 | 4.05215 |
| Gene_Pert | Esr1 KO mo | 3/351 | 0.196575 | 0.844507 | 0 | 0 | 2.007902 | 3.266274 |
| Gene_Pert | Psap KO m | 3/345 | 0.189953 | 0.844507 | 0 | 0 | 2.043755 | 3.394639 |
| Gene_Pert | SNORD14E | 3/336 | 0.180133 | 0.844507 | 0 | 0 | 2.099957 | 3.599451 |
| Gene_Pert | S1pr2 KO n | 2/168 | 0.165961 | 0.844507 | 0 | 0 | 2.799008 | 5.027029 |
| Gene_Pert | MED12 KD | 3/342 | 0.186664 | 0.844507 | 0 | 0 | 2.062158 | 3.461222 |
| Gene_Pert | Pde6b RD1 | 2/181 | 0.186143 | 0.844507 | 0 | 0 | 2.594019 | 4.361174 |
| Gene_Pert | NFKB2 KD l | 3/341 | 0.185571 | 0.844507 | 0 | 0 | 2.068364 | 3.483785 |
| Gene_Pert | Cstb KO mo | 3/340 | 0.18448 | 0.844507 | 0 | 0 | 2.074608 | 3.506535 |
| Gene_Pert | SQSTM1 OI | 3/346 | 0.191052 | 0.844507 | 0 | 0 | 2.037693 | 3.372806 |
| Gene_Pert | Agtr1a KO | 4/520 | 0.190656 | 0.844507 | 0 | 0 | 1.811619 | 3.002372 |
| Gene_Pert | Ahr low-afl | 3/345 | 0.189953 | 0.844507 | 0 | 0 | 2.043755 | 3.394639 |
| Gene_Pert | BRCA1 stak | 3/345 | 0.189953 | 0.844507 | 0 | 0 | 2.043755 | 3.394639 |
| Gene_Pert | Hras OE mo | 3/338 | 0.182303 | 0.844507 | 0 | 0 | 2.087207 | 3.552606 |
| Gene_Pert | LRRK2 dom | 3/337 | 0.181217 | 0.844507 | 0 | 0 | 2.093563 | 3.575931 |
| Gene_Pert | Pou1f1 Sne | 3/337 | 0.181217 | 0.844507 | 0 | 0 | 2.093563 | 3.575931 |
| Gene_Pert | Dicer1 KO r | 3/345 | 0.189953 | 0.844507 | 0 | 0 | 2.043755 | 3.394639 |
| Gene_Pert | PRSS2 OE h | 3/344 | 0.188855 | 0.844507 | 0 | 0 | 2.049853 | 3.416651 |
| Gene_Pert | Emx2 Defic | 3/344 | 0.188855 | 0.844507 | 0 | 0 | 2.049853 | 3.416651 |
| Gene_Pert | SYK Deplet | 3/344 | 0.188855 | 0.844507 | 0 | 0 | 2.049853 | 3.416651 |
| Gene_Pert | ZBTB38 KD | 3/343 | 0.187758 | 0.844507 | 0 | 0 | 2.055987 | 3.438845 |
| Gene_Pert | FGFR3 KD h | 3/333 | 0.176892 | 0.844507 | 0 | 0 | 2.119372 | 3.671207 |
| Gene_Pert | Gmnn KD r | 3/333 | 0.176892 | 0.844507 | 0 | 0 | 2.119372 | 3.671207 |
| Gene_Pert | DPP3 OE hu | 3/332 | 0.175816 | 0.844507 | 0 | 0 | 2.125923 | 3.695531 |
| Gene_Pert | Lipe KO mo | 2/209 | 0.230606 | 0.844507 | 0 | 0 | 2.239955 | 3.286115 |
| Gene_Pert | BMI1 OE m | 3/381 | 0.230477 | 0.844507 | 0 | 0 | 1.845711 | 2.708774 |
| Gene_Pert | Rps6ka3 kr | 3/380 | 0.229328 | 0.844507 | 0 | 0 | 1.850701 | 2.725344 |
| Gene_Pert | Eed VR mo | 3/379 | 0.228181 | 0.844507 | 0 | 0 | 1.855718 | 2.74204 |
| Gene_Pert | Keap1 OE r | 2/238 | 0.277316 | 0.844507 | 0 | 0 | 1.961815 | 2.516219 |
| Gene_Pert | KDM4C nat | 2/220 | 0.248291 | 0.844507 | 0 | 0 | 2.125742 | 2.961486 |

| | | | | | | | | |
|---|---|---|---|---|---|---|---|---|
| Gene_Pert | Foxp3 Indu | 2/220 | 0.248291 | 0.844507 | 0 | 0 | 2.125742 | 2.961486 |
| Gene_Pert | Dmd Defici | 2/205 | 0.224196 | 0.844507 | 0 | 0 | 2.284555 | 3.415945 |
| Gene_Pert | Flnb KO mc | 3/375 | 0.223603 | 0.844507 | 0 | 0 | 1.876056 | 2.810115 |
| Gene_Pert | KDM1B KD | 3/375 | 0.223603 | 0.844507 | 0 | 0 | 1.876056 | 2.810115 |
| Gene_Pert | TP53BP1 K | 2/204 | 0.222596 | 0.844507 | 0 | 0 | 2.295981 | 3.449476 |
| Gene_Pert | DNMT1 KO | 3/362 | 0.208863 | 0.844507 | 0 | 0 | 1.945285 | 3.046467 |
| Gene_Pert | NRAS KD h | 2/195 | 0.208243 | 0.844507 | 0 | 0 | 2.404145 | 3.772225 |
| Gene_Pert | Dicer1 KO | 2/194 | 0.206654 | 0.844507 | 0 | 0 | 2.416789 | 3.810572 |
| Gene_Pert | Irs1 KO mo | 3/358 | 0.204374 | 0.844507 | 0 | 0 | 1.967606 | 3.124174 |
| Gene_Pert | PAX3 KD hu | 2/239 | 0.278929 | 0.844507 | 0 | 0 | 1.953438 | 2.494149 |
| Gene_Pert | Dmd Defici | 3/422 | 0.278361 | 0.844507 | 0 | 0 | 1.661609 | 2.124926 |
| Gene_Pert | NOTCH1 Re | 2/220 | 0.248291 | 0.844507 | 0 | 0 | 2.125742 | 2.961486 |
| Gene_Pert | Apc KD mo | 3/421 | 0.277179 | 0.844507 | 0 | 0 | 1.66567 | 2.13721 |
| Gene_Pert | NOTCH1 Re | 2/228 | 0.261186 | 0.844507 | 0 | 0 | 2.049662 | 2.751716 |
| Gene_Pert | Prdm14 KC | 2/228 | 0.261186 | 0.844507 | 0 | 0 | 2.049662 | 2.751716 |
| Gene_Pert | Pde6b RD1 | 2/228 | 0.261186 | 0.844507 | 0 | 0 | 2.049662 | 2.751716 |
| Gene_Pert | S1pr3 KO n | 2/227 | 0.259573 | 0.844507 | 0 | 0 | 2.058876 | 2.776839 |
| Gene_Pert | CMAHP KO | 2/242 | 0.283765 | 0.844507 | 0 | 0 | 1.928725 | 2.429439 |
| Gene_Pert | MIR140 KC | 2/241 | 0.282153 | 0.844507 | 0 | 0 | 1.936894 | 2.450762 |
| Gene_Pert | PIR KD hun | 2/241 | 0.282153 | 0.844507 | 0 | 0 | 1.936894 | 2.450762 |
| Gene_Pert | BRAF knocl | 2/241 | 0.282153 | 0.844507 | 0 | 0 | 1.936894 | 2.450762 |
| Gene_Pert | Apc APC(M | 3/413 | 0.26774 | 0.844507 | 0 | 0 | 1.698868 | 2.238665 |
| Gene_Pert | Elavl1 KO n | 2/232 | 0.267638 | 0.844507 | 0 | 0 | 2.013606 | 2.654174 |
| Gene_Pert | Rnf2 KD mo | 2/226 | 0.257961 | 0.844507 | 0 | 0 | 2.068172 | 2.802265 |
| Gene_Pert | ATM KD hu | 2/183 | 0.189279 | 0.844507 | 0 | 0 | 2.565096 | 4.269683 |
| Gene_Pert | Tgfbr1 acti | 3/344 | 0.188855 | 0.844507 | 0 | 0 | 2.049853 | 3.416651 |
| Gene_Pert | Pou1f1 Sne | 2/222 | 0.251513 | 0.844507 | 0 | 0 | 2.106203 | 2.90711 |
| Gene_Pert | S1pr2 KO n | 2/222 | 0.251513 | 0.844507 | 0 | 0 | 2.106203 | 2.90711 |
| Gene_Pert | GRN deficie | 2/222 | 0.251513 | 0.844507 | 0 | 0 | 2.106203 | 2.90711 |
| Gene_Pert | Cnr1 KO m | 3/399 | 0.251327 | 0.844507 | 0 | 0 | 1.760191 | 2.430822 |
| Gene_Pert | Cav3 KO m | 3/389 | 0.239704 | 0.844507 | 0 | 0 | 1.806717 | 2.580624 |
| Gene_Pert | Bach2 KO r | 3/389 | 0.239704 | 0.844507 | 0 | 0 | 1.806717 | 2.580624 |
| Gene_Pert | Vhl KO mo | 2/214 | 0.238636 | 0.844507 | 0 | 0 | 2.18657 | 3.132957 |
| Gene_Pert | Pparg OE n | 3/387 | 0.237391 | 0.844507 | 0 | 0 | 1.816313 | 2.611943 |
| Gene_Pert | ZNF207 OE | 2/227 | 0.259573 | 0.844507 | 0 | 0 | 2.058876 | 2.776839 |
| Gene_Pert | Ikzf3 NULL | 2/255 | 0.304685 | 0.844507 | 0 | 0 | 1.828412 | 2.173024 |
| Gene_Pert | Gata1 KD n | 2/226 | 0.257961 | 0.844507 | 0 | 0 | 2.068172 | 2.802265 |
| Gene_Pert | Nfe2l2 KO | 2/226 | 0.257961 | 0.844507 | 0 | 0 | 2.068172 | 2.802265 |
| Gene_Pert | Rb1 Deficie | 3/393 | 0.244342 | 0.844507 | 0 | 0 | 1.787821 | 2.51937 |
| Gene_Pert | Phgdh KO r | 3/393 | 0.244342 | 0.844507 | 0 | 0 | 1.787821 | 2.51937 |
| Gene_Pert | DDIT4 Repr | 2/216 | 0.241852 | 0.844507 | 0 | 0 | 2.165915 | 3.074364 |
| Gene_Pert | Pcdh12 De | 3/390 | 0.240862 | 0.844507 | 0 | 0 | 1.801956 | 2.565139 |
| Gene_Pert | Arid3a KD | 3/372 | 0.220182 | 0.844507 | 0 | 0 | 1.891599 | 2.862559 |
| Gene_Pert | MIR30A KD | 3/372 | 0.220182 | 0.844507 | 0 | 0 | 1.891599 | 2.862559 |
| Gene_Pert | IRX5 KO mc | 3/372 | 0.220182 | 0.844507 | 0 | 0 | 1.891599 | 2.862559 |

| | | | | | | | | |
|---|---|---|---|---|---|---|---|---|
| Gene_Pert | CDK19 kno | 3/370 | 0.217908 | 0.844507 | 0 | 0 | 1.902102 | 2.898203 |
| Gene_Pert | PDE1C KD I | 2/221 | 0.249902 | 0.844507 | 0 | 0 | 2.115928 | 2.934131 |
| Gene_Pert | JAK2 knock | 2/296 | 0.369749 | 0.844507 | 0 | 0 | 1.570148 | 1.56219 |
| Gene_Pert | Epha4 knoc | 2/301 | 0.37754 | 0.844507 | 0 | 0 | 1.543498 | 1.503487 |
| Gene_Pert | Irf6 KO mo | 2/301 | 0.37754 | 0.844507 | 0 | 0 | 1.543498 | 1.503487 |
| Gene_Pert | ERBB3 drug | 2/300 | 0.375985 | 0.844507 | 0 | 0 | 1.548756 | 1.515003 |
| Gene_Pert | Ikbkg KO m | 3/496 | 0.366497 | 0.844507 | 0 | 0 | 1.406839 | 1.412136 |
| Gene_Pert | H6pd KO m | 2/293 | 0.365056 | 0.844507 | 0 | 0 | 1.586578 | 1.598802 |
| Gene_Pert | Emd KO mo | 2/293 | 0.365056 | 0.844507 | 0 | 0 | 1.586578 | 1.598802 |
| Gene_Pert | CDH1 OE h | 2/293 | 0.365056 | 0.844507 | 0 | 0 | 1.586578 | 1.598802 |
| Gene_Pert | Ghr KO mo | 3/509 | 0.381919 | 0.844507 | 0 | 0 | 1.369777 | 1.318475 |
| Gene_Pert | Mylk knock | 2/303 | 0.380646 | 0.844507 | 0 | 0 | 1.533086 | 1.480784 |
| Gene_Pert | Rp1 KO mo | 2/303 | 0.380646 | 0.844507 | 0 | 0 | 1.533086 | 1.480784 |
| Gene_Pert | Pmp22 OE | 2/303 | 0.380646 | 0.844507 | 0 | 0 | 1.533086 | 1.480784 |
| Gene_Pert | Atf7ip2 De | 2/296 | 0.369749 | 0.844507 | 0 | 0 | 1.570148 | 1.56219 |
| Gene_Pert | Bptf KO mo | 2/301 | 0.37754 | 0.844507 | 0 | 0 | 1.543498 | 1.503487 |
| Gene_Pert | MIR126 KD | 2/295 | 0.368186 | 0.844507 | 0 | 0 | 1.575587 | 1.574275 |
| Gene_Pert | Hes6 OE m | 2/295 | 0.368186 | 0.844507 | 0 | 0 | 1.575587 | 1.574275 |
| Gene_Pert | Sod1 G93A | 3/491 | 0.360549 | 0.844507 | 0 | 0 | 1.421619 | 1.450231 |
| Gene_Pert | LAMP3 KD | 2/290 | 0.36035 | 0.844507 | 0 | 0 | 1.60335 | 1.636504 |
| Gene_Pert | Notch1 OE | 2/290 | 0.36035 | 0.844507 | 0 | 0 | 1.60335 | 1.636504 |
| Gene_Pert | Gata4 KO r | 2/290 | 0.36035 | 0.844507 | 0 | 0 | 1.60335 | 1.636504 |
| Gene_Pert | FOXL2 KD h | 2/312 | 0.394543 | 0.844507 | 0 | 0 | 1.487894 | 1.383781 |
| Gene_Pert | Mnat1 Defi | 2/311 | 0.393006 | 0.844507 | 0 | 0 | 1.492785 | 1.394158 |
| Gene_Pert | GOLGA8A ( | 2/311 | 0.393006 | 0.844507 | 0 | 0 | 1.492785 | 1.394158 |
| Gene_Pert | FOXA1 OE I | 2/311 | 0.393006 | 0.844507 | 0 | 0 | 1.492785 | 1.394158 |
| Gene_Pert | Pten KO mo | 2/307 | 0.386839 | 0.844507 | 0 | 0 | 1.512671 | 1.436654 |
| Gene_Pert | Gata4 abla | 2/305 | 0.383746 | 0.844507 | 0 | 0 | 1.522811 | 1.45851 |
| Gene_Pert | Traf3 OE m | 2/297 | 0.37131 | 0.844507 | 0 | 0 | 1.564746 | 1.550222 |
| Gene_Pert | IRF9 OE hu | 2/296 | 0.369749 | 0.844507 | 0 | 0 | 1.570148 | 1.56219 |
| Gene_Pert | Pou2af1 OI | 2/296 | 0.369749 | 0.844507 | 0 | 0 | 1.570148 | 1.56219 |
| Gene_Pert | HNRNPU-A | 2/296 | 0.369749 | 0.844507 | 0 | 0 | 1.570148 | 1.56219 |
| Gene_Pert | Hdac3 KO r | 2/286 | 0.354057 | 0.844507 | 0 | 0 | 1.626263 | 1.688543 |
| Gene_Pert | RET drugin | 2/286 | 0.354057 | 0.844507 | 0 | 0 | 1.626263 | 1.688543 |
| Gene_Pert | Gata4 OE n | 2/285 | 0.352481 | 0.844507 | 0 | 0 | 1.632093 | 1.70188 |
| Gene_Pert | ESRRA KD h | 2/285 | 0.352481 | 0.844507 | 0 | 0 | 1.632093 | 1.70188 |
| Gene_Pert | Pparg OE n | 2/279 | 0.342996 | 0.844507 | 0 | 0 | 1.667955 | 1.784774 |
| Gene_Pert | Pou4f1 KO | 3/476 | 0.34267 | 0.844507 | 0 | 0 | 1.467834 | 1.572031 |
| Gene_Pert | Dmd Defici | 2/278 | 0.341411 | 0.844507 | 0 | 0 | 1.674084 | 1.799086 |
| Gene_Pert | MYB KD hu | 2/278 | 0.341411 | 0.844507 | 0 | 0 | 1.674084 | 1.799086 |
| Gene_Pert | Kdm1a KD | 2/307 | 0.386839 | 0.844507 | 0 | 0 | 1.512671 | 1.436654 |
| Gene_Pert | SCYL1 knoc | 2/304 | 0.382197 | 0.844507 | 0 | 0 | 1.527931 | 1.469594 |
| Gene_Pert | PAX5 KD hu | 2/304 | 0.382197 | 0.844507 | 0 | 0 | 1.527931 | 1.469594 |
| Gene_Pert | SCARNA13 | 2/304 | 0.382197 | 0.844507 | 0 | 0 | 1.527931 | 1.469594 |
| Gene_Pert | EGFR OE hu | 2/299 | 0.374428 | 0.844507 | 0 | 0 | 1.55405 | 1.526629 |

| | | | | | | | | |
|---|---|---|---|---|---|---|---|---|
| Gene_Pert | TGM2 KD h | 2/299 | 0.374428 | 0.844507 | 0 | 0 | 1.55405 | 1.526629 |
| Gene_Pert | Irak4 Defici | 2/299 | 0.374428 | 0.844507 | 0 | 0 | 1.55405 | 1.526629 |
| Gene_Pert | Zfp36l2 def | 2/299 | 0.374428 | 0.844507 | 0 | 0 | 1.55405 | 1.526629 |
| Gene_Pert | CMAHP KO | 2/307 | 0.386839 | 0.844507 | 0 | 0 | 1.512671 | 1.436654 |
| Gene_Pert | Setd2 Defic | 2/307 | 0.386839 | 0.844507 | 0 | 0 | 1.512671 | 1.436654 |
| Gene_Pert | HDAC1 KD | 2/306 | 0.385293 | 0.844507 | 0 | 0 | 1.517724 | 1.44753 |
| Gene_Pert | MAML1 M | 3/511 | 0.384285 | 0.844507 | 0 | 0 | 1.364244 | 1.304722 |
| Gene_Pert | THRA RCC | 3/309 | 0.151612 | 0.844507 | 0 | 0 | 2.288399 | 4.316902 |
| Gene_Pert | OPTN KD h | 2/297 | 0.37131 | 0.844507 | 0 | 0 | 1.564746 | 1.550222 |
| Gene_Pert | Zfx KO mo | 2/297 | 0.37131 | 0.844507 | 0 | 0 | 1.564746 | 1.550222 |
| Gene_Pert | LMNA OE h | 2/321 | 0.408301 | 0.844507 | 0 | 0 | 1.445252 | 1.294585 |
| Gene_Pert | Nfe2l2 KO | 1/120 | 0.408269 | 0.844507 | 0 | 0 | 1.934141 | 1.732658 |
| Gene_Pert | BMI1 DEPL | 2/320 | 0.40678 | 0.844507 | 0 | 0 | 1.449871 | 1.304135 |
| Gene_Pert | EOMES OE | 2/319 | 0.405256 | 0.844507 | 0 | 0 | 1.454518 | 1.313773 |
| Gene_Pert | Gata6 KO r | 2/314 | 0.397613 | 0.844507 | 0 | 0 | 1.478205 | 1.363314 |
| Gene_Pert | Nfe2l2 Mut | 2/313 | 0.396079 | 0.844507 | 0 | 0 | 1.483034 | 1.3735 |
| Gene_Pert | HHLA1 KD | 2/313 | 0.396079 | 0.844507 | 0 | 0 | 1.483034 | 1.3735 |
| Gene_Pert | Cldn16 Def | 2/313 | 0.396079 | 0.844507 | 0 | 0 | 1.483034 | 1.3735 |
| Gene_Pert | Acadm KO | 3/310 | 0.152641 | 0.844507 | 0 | 0 | 2.280828 | 4.287197 |
| Gene_Pert | TFAP2C KD | 3/310 | 0.152641 | 0.844507 | 0 | 0 | 2.280828 | 4.287197 |
| Gene_Pert | Sox2 KD m | 2/297 | 0.37131 | 0.844507 | 0 | 0 | 1.564746 | 1.550222 |
| Gene_Pert | Acadm KO | 3/308 | 0.150586 | 0.844507 | 0 | 0 | 2.296019 | 4.346874 |
| Gene_Pert | Vdr KO mo | 2/316 | 0.400676 | 0.844507 | 0 | 0 | 1.46864 | 1.343223 |
| Gene_Pert | Mok knock | 2/316 | 0.400676 | 0.844507 | 0 | 0 | 1.46864 | 1.343223 |
| Gene_Pert | Atp7b KO r | 2/316 | 0.400676 | 0.844507 | 0 | 0 | 1.46864 | 1.343223 |
| Gene_Pert | RARRES1 K | 2/316 | 0.400676 | 0.844507 | 0 | 0 | 1.46864 | 1.343223 |
| Gene_Pert | Tusc2 KD m | 2/308 | 0.388383 | 0.844507 | 0 | 0 | 1.507651 | 1.42588 |
| Gene_Pert | Fgfr1 activ | 2/308 | 0.388383 | 0.844507 | 0 | 0 | 1.507651 | 1.42588 |
| Gene_Pert | Tfap2c KO | 2/308 | 0.388383 | 0.844507 | 0 | 0 | 1.507651 | 1.42588 |
| Gene_Pert | Igf1r knock | 2/308 | 0.388383 | 0.844507 | 0 | 0 | 1.507651 | 1.42588 |
| Gene_Pert | Rap1a Rap | 2/319 | 0.405256 | 0.844507 | 0 | 0 | 1.454518 | 1.313773 |
| Gene_Pert | GSK3A KD | 2/319 | 0.405256 | 0.844507 | 0 | 0 | 1.454518 | 1.313773 |
| Gene_Pert | Gfi1b KO m | 2/300 | 0.375985 | 0.844507 | 0 | 0 | 1.548756 | 1.515003 |
| Gene_Pert | Tet2 KO m | 2/307 | 0.386839 | 0.844507 | 0 | 0 | 1.512671 | 1.436654 |
| Gene_Pert | RNF40 KD | 2/307 | 0.386839 | 0.844507 | 0 | 0 | 1.512671 | 1.436654 |
| Gene_Pert | MAP4K5 kr | 2/313 | 0.396079 | 0.844507 | 0 | 0 | 1.483034 | 1.3735 |
| Gene_Pert | Parp2 KO n | 1/115 | 0.395152 | 0.844507 | 0 | 0 | 2.019482 | 1.875059 |
| Gene_Pert | RAP1A KO | 2/312 | 0.394543 | 0.844507 | 0 | 0 | 1.487894 | 1.383781 |
| Gene_Pert | Ebf1 OE m | 2/312 | 0.394543 | 0.844507 | 0 | 0 | 1.487894 | 1.383781 |
| Gene_Pert | CMAHP KO | 2/310 | 0.391467 | 0.844507 | 0 | 0 | 1.497708 | 1.404633 |
| Gene_Pert | Bdkrb1 KO | 2/309 | 0.389926 | 0.844507 | 0 | 0 | 1.502663 | 1.415207 |
| Gene_Pert | LRRK2 mut | 2/309 | 0.389926 | 0.844507 | 0 | 0 | 1.502663 | 1.415207 |
| Gene_Pert | Myc KO m | 2/309 | 0.389926 | 0.844507 | 0 | 0 | 1.502663 | 1.415207 |
| Gene_Pert | Nr2f2 KO n | 2/300 | 0.375985 | 0.844507 | 0 | 0 | 1.548756 | 1.515003 |
| Gene_Pert | Yy1 KD mo | 2/282 | 0.347744 | 0.844507 | 0 | 0 | 1.649832 | 1.7427 |

| | | | | | | | | |
|---|---|---|---|---|---|---|---|---|
| Gene_Pert | HDAC2 KD | 2/300 | 0.375985 | 0.844507 | 0 | 0 | 1.548756 | 1.515003 |
| Gene_Pert | Nfatc3 OE | 2/300 | 0.375985 | 0.844507 | 0 | 0 | 1.548756 | 1.515003 |
| Gene_Pert | Appl2 KO n | 2/311 | 0.393006 | 0.844507 | 0 | 0 | 1.492785 | 1.394158 |
| Gene_Pert | Trim24 Def | 2/311 | 0.393006 | 0.844507 | 0 | 0 | 1.492785 | 1.394158 |
| Gene_Pert | Nr1i3 hNR1 | 2/310 | 0.391467 | 0.844507 | 0 | 0 | 1.497708 | 1.404633 |
| Gene_Pert | Dusp1 KO r | 2/310 | 0.391467 | 0.844507 | 0 | 0 | 1.497708 | 1.404633 |
| Gene_Pert | Igf1r drugin | 2/302 | 0.379094 | 0.844507 | 0 | 0 | 1.538275 | 1.492081 |
| Gene_Pert | Neurog3 O | 2/302 | 0.379094 | 0.844507 | 0 | 0 | 1.538275 | 1.492081 |
| Gene_Pert | ANGPT1 St | 2/302 | 0.379094 | 0.844507 | 0 | 0 | 1.538275 | 1.492081 |
| Gene_Pert | Cbx3 KD m | 2/301 | 0.37754 | 0.844507 | 0 | 0 | 1.543498 | 1.503487 |
| Gene_Pert | Ppargc1b K | 2/297 | 0.37131 | 0.844507 | 0 | 0 | 1.564746 | 1.550222 |
| Gene_Pert | Nrl Deficier | 2/276 | 0.338237 | 0.844507 | 0 | 0 | 1.686475 | 1.828152 |
| Gene_Pert | Bmpr1a KC | 2/279 | 0.342996 | 0.844507 | 0 | 0 | 1.667955 | 1.784774 |
| Gene_Pert | Bcl2 OE mc | 2/279 | 0.342996 | 0.844507 | 0 | 0 | 1.667955 | 1.784774 |
| Gene_Pert | MTDH KD h | 3/470 | 0.335509 | 0.844507 | 0 | 0 | 1.487152 | 1.624127 |
| Gene_Pert | Ptpn2 KD n | 2/274 | 0.335059 | 0.844507 | 0 | 0 | 1.699048 | 1.85782 |
| Gene_Pert | Prkaca KO | 2/274 | 0.335059 | 0.844507 | 0 | 0 | 1.699048 | 1.85782 |
| Gene_Pert | Jun KO mo | 2/273 | 0.333469 | 0.844507 | 0 | 0 | 1.705405 | 1.872885 |
| Gene_Pert | Pparg OE n | 2/267 | 0.323905 | 0.844507 | 0 | 0 | 1.74455 | 1.966639 |
| Gene_Pert | Pde10a KO | 2/267 | 0.323905 | 0.844507 | 0 | 0 | 1.74455 | 1.966639 |
| Gene_Pert | PDCD10 KD | 2/267 | 0.323905 | 0.844507 | 0 | 0 | 1.74455 | 1.966639 |
| Gene_Pert | Prkag3 Mu | 2/266 | 0.322308 | 0.844507 | 0 | 0 | 1.751248 | 1.982846 |
| Gene_Pert | Htt Mutati | 2/276 | 0.338237 | 0.844507 | 0 | 0 | 1.686475 | 1.828152 |
| Gene_Pert | MAPK1 KD | 2/276 | 0.338237 | 0.844507 | 0 | 0 | 1.686475 | 1.828152 |
| Gene_Pert | Irs1 KO mo | 2/280 | 0.34458 | 0.844507 | 0 | 0 | 1.661871 | 1.770607 |
| Gene_Pert | Bcl6 KO mc | 3/471 | 0.336703 | 0.844507 | 0 | 0 | 1.483898 | 1.615303 |
| Gene_Pert | Onecut1 K( | 2/269 | 0.327097 | 0.844507 | 0 | 0 | 1.731306 | 1.934732 |
| Gene_Pert | NEUROG1 | 2/269 | 0.327097 | 0.844507 | 0 | 0 | 1.731306 | 1.934732 |
| Gene_Pert | MIR142 OE | 2/269 | 0.327097 | 0.844507 | 0 | 0 | 1.731306 | 1.934732 |
| Gene_Pert | Egr1 KO mc | 2/268 | 0.325502 | 0.844507 | 0 | 0 | 1.737904 | 1.950602 |
| Gene_Pert | ERBB2 OE I | 2/263 | 0.317511 | 0.844507 | 0 | 0 | 1.771648 | 2.03251 |
| Gene_Pert | Sirt1 KO m | 2/263 | 0.317511 | 0.844507 | 0 | 0 | 1.771648 | 2.03251 |
| Gene_Pert | Id4 KO mou | 2/262 | 0.31591 | 0.844507 | 0 | 0 | 1.778552 | 2.04942 |
| Gene_Pert | Apod OE m | 2/262 | 0.31591 | 0.844507 | 0 | 0 | 1.778552 | 2.04942 |
| Gene_Pert | Foxd3 OE n | 2/289 | 0.358779 | 0.844507 | 0 | 0 | 1.609018 | 1.649321 |
| Gene_Pert | Mecp2 KO | 2/288 | 0.357207 | 0.844507 | 0 | 0 | 1.614726 | 1.662266 |
| Gene_Pert | Clpp KO mc | 2/252 | 0.299865 | 0.844507 | 0 | 0 | 1.850635 | 2.228949 |
| Gene_Pert | Cbfb Trunc | 3/444 | 0.304486 | 0.844507 | 0 | 0 | 1.576936 | 1.875179 |
| Gene_Pert | Erbb3 drug | 3/443 | 0.303295 | 0.844507 | 0 | 0 | 1.580601 | 1.885733 |
| Gene_Pert | Spi1 KD mc | 2/269 | 0.327097 | 0.844507 | 0 | 0 | 1.731306 | 1.934732 |
| Gene_Pert | CDC73 KD I | 2/269 | 0.327097 | 0.844507 | 0 | 0 | 1.731306 | 1.934732 |
| Gene_Pert | Rb1 Deficie | 2/269 | 0.327097 | 0.844507 | 0 | 0 | 1.731306 | 1.934732 |
| Gene_Pert | Nfe2l2 KO | 2/269 | 0.327097 | 0.844507 | 0 | 0 | 1.731306 | 1.934732 |
| Gene_Pert | Zbtb20 Def | 2/258 | 0.3095 | 0.844507 | 0 | 0 | 1.80671 | 2.118903 |
| Gene_Pert | Cdh1 KO m | 2/258 | 0.3095 | 0.844507 | 0 | 0 | 1.80671 | 2.118903 |

| | | | | | | | | |
|---|---|---|---|---|---|---|---|---|
| Gene_Pert | MYC KD hu | 2/258 | 0.3095 | 0.844507 | 0 | 0 | 1.80671 | 2.118903 |
| Gene_Pert | Pafah1b1 K | 2/258 | 0.3095 | 0.844507 | 0 | 0 | 1.80671 | 2.118903 |
| Gene_Pert | Pink1 KO m | 2/252 | 0.299865 | 0.844507 | 0 | 0 | 1.850635 | 2.228949 |
| Gene_Pert | Htt Mutati | 2/288 | 0.357207 | 0.844507 | 0 | 0 | 1.614726 | 1.662266 |
| Gene_Pert | TGFB1I1 KD | 2/252 | 0.299865 | 0.844507 | 0 | 0 | 1.850635 | 2.228949 |
| Gene_Pert | Zfp42 KD m | 3/440 | 0.299723 | 0.844507 | 0 | 0 | 1.591697 | 1.917827 |
| Gene_Pert | Egr2 KD mo | 2/278 | 0.341411 | 0.844507 | 0 | 0 | 1.674084 | 1.799086 |
| Gene_Pert | Dysf Defici | 2/278 | 0.341411 | 0.844507 | 0 | 0 | 1.674084 | 1.799086 |
| Gene_Pert | Neurod1 C | 2/276 | 0.338237 | 0.844507 | 0 | 0 | 1.686475 | 1.828152 |
| Gene_Pert | Por DELETI | 2/276 | 0.338237 | 0.844507 | 0 | 0 | 1.686475 | 1.828152 |
| Gene_Pert | PIN1 DEPLE | 2/272 | 0.331878 | 0.844507 | 0 | 0 | 1.711808 | 1.888106 |
| Gene_Pert | Pkd1 KO m | 2/272 | 0.331878 | 0.844507 | 0 | 0 | 1.711808 | 1.888106 |
| Gene_Pert | EZR KD hur | 2/272 | 0.331878 | 0.844507 | 0 | 0 | 1.711808 | 1.888106 |
| Gene_Pert | Srf homozy | 2/272 | 0.331878 | 0.844507 | 0 | 0 | 1.711808 | 1.888106 |
| Gene_Pert | Nr1i3 KO m | 2/280 | 0.34458 | 0.844507 | 0 | 0 | 1.661871 | 1.770607 |
| Gene_Pert | SYK druginl | 2/291 | 0.36192 | 0.844507 | 0 | 0 | 1.59772 | 1.623813 |
| Gene_Pert | SYK druginl | 2/275 | 0.336649 | 0.844507 | 0 | 0 | 1.692739 | 1.842909 |
| Gene_Pert | Kmt2a KO r | 2/271 | 0.330285 | 0.844507 | 0 | 0 | 1.718259 | 1.903487 |
| Gene_Pert | Dysf Defici | 2/271 | 0.330285 | 0.844507 | 0 | 0 | 1.718259 | 1.903487 |
| Gene_Pert | FOXM1 INA | 2/271 | 0.330285 | 0.844507 | 0 | 0 | 1.718259 | 1.903487 |
| Gene_Pert | Zfp36 Defic | 2/270 | 0.328691 | 0.844507 | 0 | 0 | 1.724759 | 1.919028 |
| Gene_Pert | Sirt3 KO mo | 2/297 | 0.37131 | 0.844507 | 0 | 0 | 1.564746 | 1.550222 |
| Gene_Pert | NRAS DEPL | 2/297 | 0.37131 | 0.844507 | 0 | 0 | 1.564746 | 1.550222 |
| Gene_Pert | Sh2b3 KO r | 2/297 | 0.37131 | 0.844507 | 0 | 0 | 1.564746 | 1.550222 |
| Gene_Pert | RAP1A KO | 2/297 | 0.37131 | 0.844507 | 0 | 0 | 1.564746 | 1.550222 |
| Gene_Pert | Ssfa2 Defic | 2/292 | 0.363489 | 0.844507 | 0 | 0 | 1.59213 | 1.611246 |
| Gene_Pert | Egr1 KO mo | 2/292 | 0.363489 | 0.844507 | 0 | 0 | 1.59213 | 1.611246 |
| Gene_Pert | MIR140 Sil | 2/291 | 0.36192 | 0.844507 | 0 | 0 | 1.59772 | 1.623813 |
| Gene_Pert | PAGE5 KD I | 2/275 | 0.336649 | 0.844507 | 0 | 0 | 1.692739 | 1.842909 |
| Gene_Pert | Ogt KD mo | 2/299 | 0.374428 | 0.844507 | 0 | 0 | 1.55405 | 1.526629 |
| Gene_Pert | Vax2 KO m | 2/299 | 0.374428 | 0.844507 | 0 | 0 | 1.55405 | 1.526629 |
| Gene_Pert | Xbp1 OE m | 2/298 | 0.37287 | 0.844507 | 0 | 0 | 1.55938 | 1.538369 |
| Gene_Pert | Aplp2 KO n | 2/298 | 0.37287 | 0.844507 | 0 | 0 | 1.55938 | 1.538369 |
| Gene_Pert | Fgfr2 active | 2/295 | 0.368186 | 0.844507 | 0 | 0 | 1.575587 | 1.574275 |
| Gene_Pert | Idh2 R140C | 2/294 | 0.366622 | 0.844507 | 0 | 0 | 1.581064 | 1.586479 |
| Gene_Pert | SYK druginl | 2/294 | 0.366622 | 0.844507 | 0 | 0 | 1.581064 | 1.586479 |
| Gene_Pert | SP3 KD hur | 2/294 | 0.366622 | 0.844507 | 0 | 0 | 1.581064 | 1.586479 |
| Gene_Pert | RASGRF1 K | 2/284 | 0.350903 | 0.844507 | 0 | 0 | 1.637964 | 1.71535 |
| Gene_Pert | ZHX2 KD hu | 2/284 | 0.350903 | 0.844507 | 0 | 0 | 1.637964 | 1.71535 |
| Gene_Pert | IKBKB trans | 3/482 | 0.349827 | 0.844507 | 0 | 0 | 1.449001 | 1.521909 |
| Gene_Pert | H3f3b KO r | 2/284 | 0.350903 | 0.844507 | 0 | 0 | 1.637964 | 1.71535 |
| Gene_Pert | NRAS DEPL | 2/288 | 0.357207 | 0.844507 | 0 | 0 | 1.614726 | 1.662266 |
| Gene_Pert | Mut KD mo | 2/282 | 0.347744 | 0.844507 | 0 | 0 | 1.649832 | 1.7427 |
| Gene_Pert | Scd1 KO mo | 2/281 | 0.346162 | 0.844507 | 0 | 0 | 1.65583 | 1.756583 |
| Gene_Pert | Nfe2l2 KO r | 2/280 | 0.34458 | 0.844507 | 0 | 0 | 1.661871 | 1.770607 |

| | | | | | | | | |
|---|---|---|---|---|---|---|---|---|
| Gene_Pert | Ppargc1a N | 2/280 | 0.34458 | 0.844507 | 0 | 0 | 1.661871 | 1.770607 |
| Gene_Pert | FGFR3 KD h | 2/291 | 0.36192 | 0.844507 | 0 | 0 | 1.59772 | 1.623813 |
| Gene_Pert | Pten KO mo | 2/291 | 0.36192 | 0.844507 | 0 | 0 | 1.59772 | 1.623813 |
| Gene_Pert | CHEK1 KD h | 2/291 | 0.36192 | 0.844507 | 0 | 0 | 1.59772 | 1.623813 |
| Gene_Pert | Dcx KO mo | 2/291 | 0.36192 | 0.844507 | 0 | 0 | 1.59772 | 1.623813 |
| Gene_Pert | H3F3A Mut | 2/285 | 0.352481 | 0.844507 | 0 | 0 | 1.632093 | 1.70188 |
| Gene_Pert | Socs3 DELE | 2/285 | 0.352481 | 0.844507 | 0 | 0 | 1.632093 | 1.70188 |
| Gene_Pert | Atxn1 Knoc | 2/335 | 0.429403 | 0.844507 | 0 | 0 | 1.383501 | 1.169555 |
| Gene_Pert | CLDN1 OE | 2/284 | 0.350903 | 0.844507 | 0 | 0 | 1.637964 | 1.71535 |
| Gene_Pert | Pafah1b1 K | 2/273 | 0.333469 | 0.844507 | 0 | 0 | 1.705405 | 1.872885 |
| Gene_Pert | Gfap OE mo | 2/273 | 0.333469 | 0.844507 | 0 | 0 | 1.705405 | 1.872885 |
| Gene_Pert | AMER1 OE | 2/273 | 0.333469 | 0.844507 | 0 | 0 | 1.705405 | 1.872885 |
| Gene_Pert | Ep300 KD r | 2/272 | 0.331878 | 0.844507 | 0 | 0 | 1.711808 | 1.888106 |
| Gene_Pert | Pde10a KO | 2/288 | 0.357207 | 0.844507 | 0 | 0 | 1.614726 | 1.662266 |
| Gene_Pert | FOXA1 KD | 2/287 | 0.355633 | 0.844507 | 0 | 0 | 1.620475 | 1.675339 |
| Gene_Pert | Map3k7 KO | 2/287 | 0.355633 | 0.844507 | 0 | 0 | 1.620475 | 1.675339 |
| Gene_Pert | TGFB1I1 KD | 2/286 | 0.354057 | 0.844507 | 0 | 0 | 1.626263 | 1.688543 |
| Gene_Pert | Mef2a KD r | 2/275 | 0.336649 | 0.844507 | 0 | 0 | 1.692739 | 1.842909 |
| Gene_Pert | Fgfr1 druga | 2/275 | 0.336649 | 0.844507 | 0 | 0 | 1.692739 | 1.842909 |
| Gene_Pert | BRAF Drug | 1/299 | 0.731073 | 0.844507 | 0 | 0 | 0.765374 | 0.239748 |
| Gene_Pert | TCF12 KD h | 1/297 | 0.728682 | 0.844507 | 0 | 0 | 0.770624 | 0.243916 |
| Gene_Pert | Srf KO mou | 1/297 | 0.728682 | 0.844507 | 0 | 0 | 0.770624 | 0.243916 |
| Gene_Pert | RFWD2 KD | 1/297 | 0.728682 | 0.844507 | 0 | 0 | 0.770624 | 0.243916 |
| Gene_Pert | Dot1l DELE | 1/298 | 0.72988 | 0.844507 | 0 | 0 | 0.76799 | 0.241821 |
| Gene_Pert | Ikbkb OE m | 1/298 | 0.72988 | 0.844507 | 0 | 0 | 0.76799 | 0.241821 |
| Gene_Pert | MIF DEPLET | 1/298 | 0.72988 | 0.844507 | 0 | 0 | 0.76799 | 0.241821 |
| Gene_Pert | HSF1 KD hu | 1/298 | 0.72988 | 0.844507 | 0 | 0 | 0.76799 | 0.241821 |
| Gene_Pert | Pfkl OE mo | 1/297 | 0.728682 | 0.844507 | 0 | 0 | 0.770624 | 0.243916 |
| Gene_Pert | SDC1 OE hu | 1/297 | 0.728682 | 0.844507 | 0 | 0 | 0.770624 | 0.243916 |
| Gene_Pert | HIF1A KD h | 1/297 | 0.728682 | 0.844507 | 0 | 0 | 0.770624 | 0.243916 |
| Gene_Pert | Ring1 KO m | 1/297 | 0.728682 | 0.844507 | 0 | 0 | 0.770624 | 0.243916 |
| Gene_Pert | Gfi1b OE m | 1/300 | 0.73226 | 0.844507 | 0 | 0 | 0.762775 | 0.237696 |
| Gene_Pert | SYK KD hur | 1/300 | 0.73226 | 0.844507 | 0 | 0 | 0.762775 | 0.237696 |
| Gene_Pert | Clpp KO mo | 1/297 | 0.728682 | 0.844507 | 0 | 0 | 0.770624 | 0.243916 |
| Gene_Pert | Dmd Defici | 1/299 | 0.731073 | 0.844507 | 0 | 0 | 0.765374 | 0.239748 |
| Gene_Pert | Gfi1b OE m | 1/299 | 0.731073 | 0.844507 | 0 | 0 | 0.765374 | 0.239748 |
| Gene_Pert | CLOCK KD | 1/299 | 0.731073 | 0.844507 | 0 | 0 | 0.765374 | 0.239748 |
| Gene_Pert | MAPK7 cor | 1/299 | 0.731073 | 0.844507 | 0 | 0 | 0.765374 | 0.239748 |
| Gene_Pert | Dmd Defici | 1/299 | 0.731073 | 0.844507 | 0 | 0 | 0.765374 | 0.239748 |
| Gene_Pert | Socs3 KO m | 1/301 | 0.733443 | 0.844507 | 0 | 0 | 0.760194 | 0.235665 |
| Gene_Pert | STAT3 KD h | 1/301 | 0.733443 | 0.844507 | 0 | 0 | 0.760194 | 0.235665 |
| Gene_Pert | OLR1 OE hu | 1/301 | 0.733443 | 0.844507 | 0 | 0 | 0.760194 | 0.235665 |
| Gene_Pert | Pou2af1 OE | 1/301 | 0.733443 | 0.844507 | 0 | 0 | 0.760194 | 0.235665 |
| Gene_Pert | TGFB1I1 KD | 1/301 | 0.733443 | 0.844507 | 0 | 0 | 0.760194 | 0.235665 |
| Gene_Pert | ALK KD hur | 1/301 | 0.733443 | 0.844507 | 0 | 0 | 0.760194 | 0.235665 |

| | | | | | | | | |
|---|---|---|---|---|---|---|---|---|
| Gene_Pert | Dot1l DELE | 1/300 | 0.73226 | 0.844507 | 0 | 0 | 0.762775 | 0.237696 |
| Gene_Pert | LMO3 KD h | 1/300 | 0.73226 | 0.844507 | 0 | 0 | 0.762775 | 0.237696 |
| Gene_Pert | Gata4 OE n | 1/295 | 0.72627 | 0.844507 | 0 | 0 | 0.775945 | 0.248173 |
| Gene_Pert | Dgcr8 KO n | 1/295 | 0.72627 | 0.844507 | 0 | 0 | 0.775945 | 0.248173 |
| Gene_Pert | Xbp1 OE m | 1/294 | 0.725056 | 0.844507 | 0 | 0 | 0.778633 | 0.250335 |
| Gene_Pert | Fmr1 KD m | 1/294 | 0.725056 | 0.844507 | 0 | 0 | 0.778633 | 0.250335 |
| Gene_Pert | H19 Deplet | 1/294 | 0.725056 | 0.844507 | 0 | 0 | 0.778633 | 0.250335 |
| Gene_Pert | FAM60A KI | 1/294 | 0.725056 | 0.844507 | 0 | 0 | 0.778633 | 0.250335 |
| Gene_Pert | Mecp2 KO | 1/293 | 0.723837 | 0.844507 | 0 | 0 | 0.78134 | 0.25252 |
| Gene_Pert | PPARGC1A | 1/293 | 0.723837 | 0.844507 | 0 | 0 | 0.78134 | 0.25252 |
| Gene_Pert | NRAS DEPL | 1/293 | 0.723837 | 0.844507 | 0 | 0 | 0.78134 | 0.25252 |
| Gene_Pert | Nf1 Hetero | 1/295 | 0.72627 | 0.844507 | 0 | 0 | 0.775945 | 0.248173 |
| Gene_Pert | Pparg KO n | 1/295 | 0.72627 | 0.844507 | 0 | 0 | 0.775945 | 0.248173 |
| Gene_Pert | BATF KD hu | 1/295 | 0.72627 | 0.844507 | 0 | 0 | 0.775945 | 0.248173 |
| Gene_Pert | PTPRC C77 | 1/295 | 0.72627 | 0.844507 | 0 | 0 | 0.775945 | 0.248173 |
| Gene_Pert | Il10 KO mo | 1/295 | 0.72627 | 0.844507 | 0 | 0 | 0.775945 | 0.248173 |
| Gene_Pert | RARA Activ | 1/303 | 0.735792 | 0.844507 | 0 | 0 | 0.755082 | 0.231665 |
| Gene_Pert | PPARGC1A | 1/296 | 0.727479 | 0.844507 | 0 | 0 | 0.773276 | 0.246034 |
| Gene_Pert | Pkd1 KO m | 1/296 | 0.727479 | 0.844507 | 0 | 0 | 0.773276 | 0.246034 |
| Gene_Pert | GLI1 INHIB | 1/296 | 0.727479 | 0.844507 | 0 | 0 | 0.773276 | 0.246034 |
| Gene_Pert | Rbm15 KO | 1/296 | 0.727479 | 0.844507 | 0 | 0 | 0.773276 | 0.246034 |
| Gene_Pert | Jun KO mo | 1/296 | 0.727479 | 0.844507 | 0 | 0 | 0.773276 | 0.246034 |
| Gene_Pert | Srf homozy | 1/296 | 0.727479 | 0.844507 | 0 | 0 | 0.773276 | 0.246034 |
| Gene_Pert | Batf KO mc | 1/296 | 0.727479 | 0.844507 | 0 | 0 | 0.773276 | 0.246034 |
| Gene_Pert | Trp53inp2 | 1/296 | 0.727479 | 0.844507 | 0 | 0 | 0.773276 | 0.246034 |
| Gene_Pert | TET2 DEPLI | 1/297 | 0.728682 | 0.844507 | 0 | 0 | 0.770624 | 0.243916 |
| Gene_Pert | Insm1 Defi | 1/297 | 0.728682 | 0.844507 | 0 | 0 | 0.770624 | 0.243916 |
| Gene_Pert | Arntl KO m | 1/297 | 0.728682 | 0.844507 | 0 | 0 | 0.770624 | 0.243916 |
| Gene_Pert | SYK knockc | 1/297 | 0.728682 | 0.844507 | 0 | 0 | 0.770624 | 0.243916 |
| Gene_Pert | ZNF148 KD | 1/308 | 0.741576 | 0.844507 | 0 | 0 | 0.742595 | 0.222019 |
| Gene_Pert | Trim28 KO | 1/305 | 0.738121 | 0.844507 | 0 | 0 | 0.750038 | 0.227747 |
| Gene_Pert | Cftr OE mo | 1/305 | 0.738121 | 0.844507 | 0 | 0 | 0.750038 | 0.227747 |
| Gene_Pert | IKZF1 IkL/L | 1/305 | 0.738121 | 0.844507 | 0 | 0 | 0.750038 | 0.227747 |
| Gene_Pert | FLI1 KD hu | 1/305 | 0.738121 | 0.844507 | 0 | 0 | 0.750038 | 0.227747 |
| Gene_Pert | Glis2 KO m | 1/307 | 0.740429 | 0.844507 | 0 | 0 | 0.74506 | 0.223909 |
| Gene_Pert | SNAI1 OE h | 1/307 | 0.740429 | 0.844507 | 0 | 0 | 0.74506 | 0.223909 |
| Gene_Pert | Atxn1 Knoc | 1/307 | 0.740429 | 0.844507 | 0 | 0 | 0.74506 | 0.223909 |
| Gene_Pert | Akt1 KO m | 1/307 | 0.740429 | 0.844507 | 0 | 0 | 0.74506 | 0.223909 |
| Gene_Pert | Ctcf DEPLE | 1/307 | 0.740429 | 0.844507 | 0 | 0 | 0.74506 | 0.223909 |
| Gene_Pert | Dicer1 KO | 1/307 | 0.740429 | 0.844507 | 0 | 0 | 0.74506 | 0.223909 |
| Gene_Pert | OGT siOGT | 1/307 | 0.740429 | 0.844507 | 0 | 0 | 0.74506 | 0.223909 |
| Gene_Pert | Gnmt KO m | 1/306 | 0.739278 | 0.844507 | 0 | 0 | 0.747541 | 0.225819 |
| Gene_Pert | Pde6b RD1 | 1/308 | 0.741576 | 0.844507 | 0 | 0 | 0.742595 | 0.222019 |
| Gene_Pert | Tsc2 inacti | 1/305 | 0.738121 | 0.844507 | 0 | 0 | 0.750038 | 0.227747 |
| Gene_Pert | Vax2 KO m | 1/308 | 0.741576 | 0.844507 | 0 | 0 | 0.742595 | 0.222019 |

| | | | | | | | | |
|---|---|---|---|---|---|---|---|---|
| Gene_Pert | OGT small | 1/308 | 0.741576 | 0.844507 | 0 | 0 | 0.742595 | 0.222019 |
| Gene_Pert | Notch1 OE | 1/308 | 0.741576 | 0.844507 | 0 | 0 | 0.742595 | 0.222019 |
| Gene_Pert | SRF KD hur | 1/308 | 0.741576 | 0.844507 | 0 | 0 | 0.742595 | 0.222019 |
| Gene_Pert | Myd88 KO | 1/308 | 0.741576 | 0.844507 | 0 | 0 | 0.742595 | 0.222019 |
| Gene_Pert | Snca KO m | 1/307 | 0.740429 | 0.844507 | 0 | 0 | 0.74506 | 0.223909 |
| Gene_Pert | Foxd3 KO r | 1/310 | 0.743855 | 0.844507 | 0 | 0 | 0.737714 | 0.218297 |
| Gene_Pert | Bach2 KO r | 1/310 | 0.743855 | 0.844507 | 0 | 0 | 0.737714 | 0.218297 |
| Gene_Pert | Ebf1 OE m | 1/310 | 0.743855 | 0.844507 | 0 | 0 | 0.737714 | 0.218297 |
| Gene_Pert | STK33 knoc | 1/310 | 0.743855 | 0.844507 | 0 | 0 | 0.737714 | 0.218297 |
| Gene_Pert | MIR222 KD | 1/309 | 0.742718 | 0.844507 | 0 | 0 | 0.740146 | 0.220149 |
| Gene_Pert | Pde10a dru | 1/309 | 0.742718 | 0.844507 | 0 | 0 | 0.740146 | 0.220149 |
| Gene_Pert | MCAM KD | 1/303 | 0.735792 | 0.844507 | 0 | 0 | 0.755082 | 0.231665 |
| Gene_Pert | SYK druginl | 1/302 | 0.73462 | 0.844507 | 0 | 0 | 0.75763 | 0.233655 |
| Gene_Pert | Nrl Deficier | 1/302 | 0.73462 | 0.844507 | 0 | 0 | 0.75763 | 0.233655 |
| Gene_Pert | Pafah1b1 K | 1/302 | 0.73462 | 0.844507 | 0 | 0 | 0.75763 | 0.233655 |
| Gene_Pert | FLI1 KD hui | 1/302 | 0.73462 | 0.844507 | 0 | 0 | 0.75763 | 0.233655 |
| Gene_Pert | RB1 KD hur | 1/302 | 0.73462 | 0.844507 | 0 | 0 | 0.75763 | 0.233655 |
| Gene_Pert | LOXL2 DEP | 1/301 | 0.733443 | 0.844507 | 0 | 0 | 0.760194 | 0.235665 |
| Gene_Pert | Stat1 KO m | 1/301 | 0.733443 | 0.844507 | 0 | 0 | 0.760194 | 0.235665 |
| Gene_Pert | Gata4 Dele | 1/304 | 0.736959 | 0.844507 | 0 | 0 | 0.752552 | 0.229696 |
| Gene_Pert | Pink1 KO m | 1/304 | 0.736959 | 0.844507 | 0 | 0 | 0.752552 | 0.229696 |
| Gene_Pert | Htra2 KO m | 1/304 | 0.736959 | 0.844507 | 0 | 0 | 0.752552 | 0.229696 |
| Gene_Pert | Pmp22 KO | 1/304 | 0.736959 | 0.844507 | 0 | 0 | 0.752552 | 0.229696 |
| Gene_Pert | FGFR3 KD h | 1/304 | 0.736959 | 0.844507 | 0 | 0 | 0.752552 | 0.229696 |
| Gene_Pert | Pou2af1 NU | 1/303 | 0.735792 | 0.844507 | 0 | 0 | 0.755082 | 0.231665 |
| Gene_Pert | IRF8 KD hu | 1/292 | 0.722612 | 0.844507 | 0 | 0 | 0.784065 | 0.254729 |
| Gene_Pert | BRAF Drug | 1/303 | 0.735792 | 0.844507 | 0 | 0 | 0.755082 | 0.231665 |
| Gene_Pert | JUND KD h | 1/305 | 0.738121 | 0.844507 | 0 | 0 | 0.750038 | 0.227747 |
| Gene_Pert | NR2C2 KO | 1/305 | 0.738121 | 0.844507 | 0 | 0 | 0.750038 | 0.227747 |
| Gene_Pert | Socs3 KO m | 1/305 | 0.738121 | 0.844507 | 0 | 0 | 0.750038 | 0.227747 |
| Gene_Pert | Jun KO mo | 1/305 | 0.738121 | 0.844507 | 0 | 0 | 0.750038 | 0.227747 |
| Gene_Pert | STAT6 KD h | 1/304 | 0.736959 | 0.844507 | 0 | 0 | 0.752552 | 0.229696 |
| Gene_Pert | GAPDH KD | 1/304 | 0.736959 | 0.844507 | 0 | 0 | 0.752552 | 0.229696 |
| Gene_Pert | RAF1 overe | 1/304 | 0.736959 | 0.844507 | 0 | 0 | 0.752552 | 0.229696 |
| Gene_Pert | NFE2L1 KD | 1/304 | 0.736959 | 0.844507 | 0 | 0 | 0.752552 | 0.229696 |
| Gene_Pert | ZNF395 KD | 1/306 | 0.739278 | 0.844507 | 0 | 0 | 0.747541 | 0.225819 |
| Gene_Pert | Sftpc KO m | 1/306 | 0.739278 | 0.844507 | 0 | 0 | 0.747541 | 0.225819 |
| Gene_Pert | Dock8 KO r | 1/306 | 0.739278 | 0.844507 | 0 | 0 | 0.747541 | 0.225819 |
| Gene_Pert | NFKB1 Acti | 1/279 | 0.706197 | 0.844507 | 0 | 0 | 0.821273 | 0.285689 |
| Gene_Pert | MYC KD hu | 1/275 | 0.700955 | 0.844507 | 0 | 0 | 0.833432 | 0.296128 |
| Gene_Pert | Gys1 KO m | 1/278 | 0.704895 | 0.844507 | 0 | 0 | 0.82428 | 0.288256 |
| Gene_Pert | CFTR D508 | 1/278 | 0.704895 | 0.844507 | 0 | 0 | 0.82428 | 0.288256 |
| Gene_Pert | GSK3B KD I | 1/278 | 0.704895 | 0.844507 | 0 | 0 | 0.82428 | 0.288256 |
| Gene_Pert | ZFX KD hur | 1/278 | 0.704895 | 0.844507 | 0 | 0 | 0.82428 | 0.288256 |
| Gene_Pert | GRN deficie | 1/277 | 0.703588 | 0.844507 | 0 | 0 | 0.827309 | 0.290851 |

| | | | | | | | | |
|---|---|---|---|---|---|---|---|---|
| Gene_Pert | Dmd Defici | 1/277 | 0.703588 | 0.844507 | 0 | 0 | 0.827309 | 0.290851 |
| Gene_Pert | Trip12 KO r | 1/277 | 0.703588 | 0.844507 | 0 | 0 | 0.827309 | 0.290851 |
| Gene_Pert | Gtf2i KO m | 1/277 | 0.703588 | 0.844507 | 0 | 0 | 0.827309 | 0.290851 |
| Gene_Pert | ELK1 KD hu | 1/279 | 0.706197 | 0.844507 | 0 | 0 | 0.821273 | 0.285689 |
| Gene_Pert | Nr1i3 hNR1 | 1/279 | 0.706197 | 0.844507 | 0 | 0 | 0.821273 | 0.285689 |
| Gene_Pert | CTNNB1 KC | 1/279 | 0.706197 | 0.844507 | 0 | 0 | 0.821273 | 0.285689 |
| Gene_Pert | GSK3B KD h | 1/279 | 0.706197 | 0.844507 | 0 | 0 | 0.821273 | 0.285689 |
| Gene_Pert | CXCR4 Stim | 1/276 | 0.702274 | 0.844507 | 0 | 0 | 0.830359 | 0.293475 |
| Gene_Pert | NRAS DEPL | 1/278 | 0.704895 | 0.844507 | 0 | 0 | 0.82428 | 0.288256 |
| Gene_Pert | POU5F1 KD | 1/278 | 0.704895 | 0.844507 | 0 | 0 | 0.82428 | 0.288256 |
| Gene_Pert | ZXDC deple | 1/278 | 0.704895 | 0.844507 | 0 | 0 | 0.82428 | 0.288256 |
| Gene_Pert | Cebpa KO r | 1/283 | 0.711348 | 0.844507 | 0 | 0 | 0.809459 | 0.275697 |
| Gene_Pert | Pdk1 knock | 1/283 | 0.711348 | 0.844507 | 0 | 0 | 0.809459 | 0.275697 |
| Gene_Pert | Nf1 mouse | 1/282 | 0.710068 | 0.844507 | 0 | 0 | 0.812381 | 0.278154 |
| Gene_Pert | Nr4a2 OE r | 1/282 | 0.710068 | 0.844507 | 0 | 0 | 0.812381 | 0.278154 |
| Gene_Pert | Cstf2t OE n | 1/281 | 0.708784 | 0.844507 | 0 | 0 | 0.815324 | 0.280639 |
| Gene_Pert | IGF1 OE hu | 1/280 | 0.707493 | 0.844507 | 0 | 0 | 0.818288 | 0.28315 |
| Gene_Pert | Pde10a KO | 1/280 | 0.707493 | 0.844507 | 0 | 0 | 0.818288 | 0.28315 |
| Gene_Pert | TFPI OE hu | 1/280 | 0.707493 | 0.844507 | 0 | 0 | 0.818288 | 0.28315 |
| Gene_Pert | Ebf1 OE mo | 1/285 | 0.713889 | 0.844507 | 0 | 0 | 0.803677 | 0.270861 |
| Gene_Pert | Sod1 G93A | 1/275 | 0.700955 | 0.844507 | 0 | 0 | 0.833432 | 0.296128 |
| Gene_Pert | LRRK2 dom | 1/271 | 0.695622 | 0.844507 | 0 | 0 | 0.845952 | 0.307038 |
| Gene_Pert | PARK7 KD h | 1/270 | 0.694273 | 0.844507 | 0 | 0 | 0.84914 | 0.309842 |
| Gene_Pert | YAP1 KD hu | 1/270 | 0.694273 | 0.844507 | 0 | 0 | 0.84914 | 0.309842 |
| Gene_Pert | Pmp22 OE | 1/270 | 0.694273 | 0.844507 | 0 | 0 | 0.84914 | 0.309842 |
| Gene_Pert | GSK3B kno | 2/377 | 0.490248 | 0.844507 | 0 | 0 | 1.225914 | 0.873884 |
| Gene_Pert | THRB HCC | 1/273 | 0.6983 | 0.844507 | 0 | 0 | 0.839646 | 0.301522 |
| Gene_Pert | Pink1 KO m | 1/273 | 0.6983 | 0.844507 | 0 | 0 | 0.839646 | 0.301522 |
| Gene_Pert | H3F3A Mut | 1/272 | 0.696964 | 0.844507 | 0 | 0 | 0.842787 | 0.304265 |
| Gene_Pert | Shank3 KD | 1/272 | 0.696964 | 0.844507 | 0 | 0 | 0.842787 | 0.304265 |
| Gene_Pert | MYC OE hu | 1/272 | 0.696964 | 0.844507 | 0 | 0 | 0.842787 | 0.304265 |
| Gene_Pert | SYK knocko | 1/272 | 0.696964 | 0.844507 | 0 | 0 | 0.842787 | 0.304265 |
| Gene_Pert | Foxp3 ABL/ | 1/272 | 0.696964 | 0.844507 | 0 | 0 | 0.842787 | 0.304265 |
| Gene_Pert | DLX4 OE hu | 1/275 | 0.700955 | 0.844507 | 0 | 0 | 0.833432 | 0.296128 |
| Gene_Pert | Stat1 KO m | 1/285 | 0.713889 | 0.844507 | 0 | 0 | 0.803677 | 0.270861 |
| Gene_Pert | Hnf4a NUL | 1/274 | 0.699631 | 0.844507 | 0 | 0 | 0.836528 | 0.29881 |
| Gene_Pert | Clpp KO mo | 1/274 | 0.699631 | 0.844507 | 0 | 0 | 0.836528 | 0.29881 |
| Gene_Pert | NPM1 mut | 1/274 | 0.699631 | 0.844507 | 0 | 0 | 0.836528 | 0.29881 |
| Gene_Pert | Neurog3 O | 1/273 | 0.6983 | 0.844507 | 0 | 0 | 0.839646 | 0.301522 |
| Gene_Pert | Dot1l DELE | 1/273 | 0.6983 | 0.844507 | 0 | 0 | 0.839646 | 0.301522 |
| Gene_Pert | PIAS1 KD h | 1/273 | 0.6983 | 0.844507 | 0 | 0 | 0.839646 | 0.301522 |
| Gene_Pert | RPS24 OE h | 1/277 | 0.703588 | 0.844507 | 0 | 0 | 0.827309 | 0.290851 |
| Gene_Pert | RAD21 KD | 1/276 | 0.702274 | 0.844507 | 0 | 0 | 0.830359 | 0.293475 |
| Gene_Pert | SON KD hu | 1/276 | 0.702274 | 0.844507 | 0 | 0 | 0.830359 | 0.293475 |
| Gene_Pert | Wfs1 KO m | 1/276 | 0.702274 | 0.844507 | 0 | 0 | 0.830359 | 0.293475 |

| | | | | | | | | |
|---|---|---|---|---|---|---|---|---|
| Gene_Pert | Rpe65 KD r | 1/276 | 0.702274 | 0.844507 | 0 | 0 | 0.830359 | 0.293475 |
| Gene_Pert | Elavl4 OE n | 1/276 | 0.702274 | 0.844507 | 0 | 0 | 0.830359 | 0.293475 |
| Gene_Pert | Trim28 knc | 1/292 | 0.722612 | 0.844507 | 0 | 0 | 0.784065 | 0.254729 |
| Gene_Pert | MAP3K7 KI | 1/288 | 0.717661 | 0.844507 | 0 | 0 | 0.795154 | 0.263799 |
| Gene_Pert | PPARGC1A | 1/288 | 0.717661 | 0.844507 | 0 | 0 | 0.795154 | 0.263799 |
| Gene_Pert | BCL6 KD hu | 1/288 | 0.717661 | 0.844507 | 0 | 0 | 0.795154 | 0.263799 |
| Gene_Pert | Msgn1 OE | 1/291 | 0.721383 | 0.844507 | 0 | 0 | 0.786808 | 0.25696 |
| Gene_Pert | LAMC2 KD | 1/291 | 0.721383 | 0.844507 | 0 | 0 | 0.786808 | 0.25696 |
| Gene_Pert | WHSC1 KD | 1/291 | 0.721383 | 0.844507 | 0 | 0 | 0.786808 | 0.25696 |
| Gene_Pert | Calcr KO m | 1/291 | 0.721383 | 0.844507 | 0 | 0 | 0.786808 | 0.25696 |
| Gene_Pert | Pten KO m | 1/291 | 0.721383 | 0.844507 | 0 | 0 | 0.786808 | 0.25696 |
| Gene_Pert | Rpe65 KO r | 1/291 | 0.721383 | 0.844507 | 0 | 0 | 0.786808 | 0.25696 |
| Gene_Pert | IL13RA1 St | 1/291 | 0.721383 | 0.844507 | 0 | 0 | 0.786808 | 0.25696 |
| Gene_Pert | Clpp KO m | 1/291 | 0.721383 | 0.844507 | 0 | 0 | 0.786808 | 0.25696 |
| Gene_Pert | Cbx2 KO m | 1/292 | 0.722612 | 0.844507 | 0 | 0 | 0.784065 | 0.254729 |
| Gene_Pert | TARBP2 OE | 1/292 | 0.722612 | 0.844507 | 0 | 0 | 0.784065 | 0.254729 |
| Gene_Pert | PTEN KD hu | 1/288 | 0.717661 | 0.844507 | 0 | 0 | 0.795154 | 0.263799 |
| Gene_Pert | FGFR3 kno | 1/292 | 0.722612 | 0.844507 | 0 | 0 | 0.784065 | 0.254729 |
| Gene_Pert | App OE mc | 1/292 | 0.722612 | 0.844507 | 0 | 0 | 0.784065 | 0.254729 |
| Gene_Pert | HDAC9 INH | 1/292 | 0.722612 | 0.844507 | 0 | 0 | 0.784065 | 0.254729 |
| Gene_Pert | Vdr KO mo | 1/292 | 0.722612 | 0.844507 | 0 | 0 | 0.784065 | 0.254729 |
| Gene_Pert | BST1 OE hu | 1/291 | 0.721383 | 0.844507 | 0 | 0 | 0.786808 | 0.25696 |
| Gene_Pert | FGFR3 KD h | 1/293 | 0.723837 | 0.844507 | 0 | 0 | 0.78134 | 0.25252 |
| Gene_Pert | PPARGC1A | 1/293 | 0.723837 | 0.844507 | 0 | 0 | 0.78134 | 0.25252 |
| Gene_Pert | Psmd14 KD | 1/292 | 0.722612 | 0.844507 | 0 | 0 | 0.784065 | 0.254729 |
| Gene_Pert | PPARGC1A | 1/292 | 0.722612 | 0.844507 | 0 | 0 | 0.784065 | 0.254729 |
| Gene_Pert | PTEN KD hu | 1/292 | 0.722612 | 0.844507 | 0 | 0 | 0.784065 | 0.254729 |
| Gene_Pert | SOX2 Defic | 1/292 | 0.722612 | 0.844507 | 0 | 0 | 0.784065 | 0.254729 |
| Gene_Pert | Map2k1 dr | 1/292 | 0.722612 | 0.844507 | 0 | 0 | 0.784065 | 0.254729 |
| Gene_Pert | IL6 anti-IL6 | 1/285 | 0.713889 | 0.844507 | 0 | 0 | 0.803677 | 0.270861 |
| Gene_Pert | Pgr KO mou | 1/284 | 0.712621 | 0.844507 | 0 | 0 | 0.806558 | 0.273266 |
| Gene_Pert | Pten KD m | 1/284 | 0.712621 | 0.844507 | 0 | 0 | 0.806558 | 0.273266 |
| Gene_Pert | Rb1 KO mc | 1/284 | 0.712621 | 0.844507 | 0 | 0 | 0.806558 | 0.273266 |
| Gene_Pert | Batf KO mc | 1/284 | 0.712621 | 0.844507 | 0 | 0 | 0.806558 | 0.273266 |
| Gene_Pert | SYK knockc | 1/283 | 0.711348 | 0.844507 | 0 | 0 | 0.809459 | 0.275697 |
| Gene_Pert | PCGF2 KD h | 1/283 | 0.711348 | 0.844507 | 0 | 0 | 0.809459 | 0.275697 |
| Gene_Pert | EGR4 KD hu | 1/286 | 0.715152 | 0.844507 | 0 | 0 | 0.800816 | 0.268482 |
| Gene_Pert | MALAT1 OI | 1/286 | 0.715152 | 0.844507 | 0 | 0 | 0.800816 | 0.268482 |
| Gene_Pert | BCL3 KD hu | 1/286 | 0.715152 | 0.844507 | 0 | 0 | 0.800816 | 0.268482 |
| Gene_Pert | RNF2 KD hu | 1/285 | 0.713889 | 0.844507 | 0 | 0 | 0.803677 | 0.270861 |
| Gene_Pert | RNF185 KD | 1/285 | 0.713889 | 0.844507 | 0 | 0 | 0.803677 | 0.270861 |
| Gene_Pert | TNIK knock | 1/285 | 0.713889 | 0.844507 | 0 | 0 | 0.803677 | 0.270861 |
| Gene_Pert | Gfap OE m | 1/285 | 0.713889 | 0.844507 | 0 | 0 | 0.803677 | 0.270861 |
| Gene_Pert | Ifnb1 Induc | 1/309 | 0.742718 | 0.844507 | 0 | 0 | 0.740146 | 0.220149 |
| Gene_Pert | Htt Mutati | 1/288 | 0.717661 | 0.844507 | 0 | 0 | 0.795154 | 0.263799 |

| | | | | | | | | |
|---|---|---|---|---|---|---|---|---|
| Gene_Pert | Dmd Defici | 1/288 | 0.717661 | 0.844507 | 0 | 0 | 0.795154 | 0.263799 |
| Gene_Pert | Ezh2 KO m | 1/287 | 0.716409 | 0.844507 | 0 | 0 | 0.797975 | 0.266128 |
| Gene_Pert | STAT3 KD h | 1/287 | 0.716409 | 0.844507 | 0 | 0 | 0.797975 | 0.266128 |
| Gene_Pert | RXRA KD hu | 1/287 | 0.716409 | 0.844507 | 0 | 0 | 0.797975 | 0.266128 |
| Gene_Pert | FOXP1 OE l | 1/287 | 0.716409 | 0.844507 | 0 | 0 | 0.797975 | 0.266128 |
| Gene_Pert | Dusp1 KO r | 1/287 | 0.716409 | 0.844507 | 0 | 0 | 0.797975 | 0.266128 |
| Gene_Pert | KIT activen | 1/286 | 0.715152 | 0.844507 | 0 | 0 | 0.800816 | 0.268482 |
| Gene_Pert | Insm1 KO r | 1/291 | 0.721383 | 0.844507 | 0 | 0 | 0.786808 | 0.25696 |
| Gene_Pert | Acadm KO | 1/289 | 0.718907 | 0.844507 | 0 | 0 | 0.792353 | 0.261495 |
| Gene_Pert | Runx1 KO r | 1/289 | 0.718907 | 0.844507 | 0 | 0 | 0.792353 | 0.261495 |
| Gene_Pert | MIR221 KD | 1/289 | 0.718907 | 0.844507 | 0 | 0 | 0.792353 | 0.261495 |
| Gene_Pert | Myc myc +, | 1/339 | 0.774745 | 0.844507 | 0 | 0 | 0.673421 | 0.171872 |
| Gene_Pert | Mbd3 KD n | 1/338 | 0.773744 | 0.844507 | 0 | 0 | 0.675454 | 0.173264 |
| Gene_Pert | SOX2 KO hu | 1/338 | 0.773744 | 0.844507 | 0 | 0 | 0.675454 | 0.173264 |
| Gene_Pert | PCDH12 OE | 1/338 | 0.773744 | 0.844507 | 0 | 0 | 0.675454 | 0.173264 |
| Gene_Pert | Hdac2 Defi | 1/338 | 0.773744 | 0.844507 | 0 | 0 | 0.675454 | 0.173264 |
| Gene_Pert | RB1 KD hur | 1/337 | 0.772738 | 0.844507 | 0 | 0 | 0.677499 | 0.174669 |
| Gene_Pert | SCARA5 Dc | 1/337 | 0.772738 | 0.844507 | 0 | 0 | 0.677499 | 0.174669 |
| Gene_Pert | Nfe2l2 kno | 1/337 | 0.772738 | 0.844507 | 0 | 0 | 0.677499 | 0.174669 |
| Gene_Pert | HSF1 KD hu | 1/337 | 0.772738 | 0.844507 | 0 | 0 | 0.677499 | 0.174669 |
| Gene_Pert | NR5A2 KD | 1/341 | 0.776734 | 0.844507 | 0 | 0 | 0.669391 | 0.169127 |
| Gene_Pert | Hey2 KO m | 1/340 | 0.775741 | 0.844507 | 0 | 0 | 0.6714 | 0.170493 |
| Gene_Pert | ERO1L over | 1/340 | 0.775741 | 0.844507 | 0 | 0 | 0.6714 | 0.170493 |
| Gene_Pert | HSF1 KD hu | 1/339 | 0.774745 | 0.844507 | 0 | 0 | 0.673421 | 0.171872 |
| Gene_Pert | KRAS onco | 1/339 | 0.774745 | 0.844507 | 0 | 0 | 0.673421 | 0.171872 |
| Gene_Pert | CFTR F508 | 1/335 | 0.770714 | 0.844507 | 0 | 0 | 0.681625 | 0.177521 |
| Gene_Pert | FLI1 KD hur | 1/339 | 0.774745 | 0.844507 | 0 | 0 | 0.673421 | 0.171872 |
| Gene_Pert | Rp1 KO mo | 1/338 | 0.773744 | 0.844507 | 0 | 0 | 0.675454 | 0.173264 |
| Gene_Pert | Src overexp | 1/343 | 0.778706 | 0.844507 | 0 | 0 | 0.665409 | 0.166434 |
| Gene_Pert | Stat3 KO m | 1/343 | 0.778706 | 0.844507 | 0 | 0 | 0.665409 | 0.166434 |
| Gene_Pert | Trim24 knc | 1/342 | 0.777722 | 0.844507 | 0 | 0 | 0.667394 | 0.167774 |
| Gene_Pert | Tgfb1 OE n | 1/342 | 0.777722 | 0.844507 | 0 | 0 | 0.667394 | 0.167774 |
| Gene_Pert | TP53 DEPLI | 1/342 | 0.777722 | 0.844507 | 0 | 0 | 0.667394 | 0.167774 |
| Gene_Pert | Itk KO mou | 1/342 | 0.777722 | 0.844507 | 0 | 0 | 0.667394 | 0.167774 |
| Gene_Pert | Hdac1 Drug | 1/342 | 0.777722 | 0.844507 | 0 | 0 | 0.667394 | 0.167774 |
| Gene_Pert | ERBB2 OE I | 1/342 | 0.777722 | 0.844507 | 0 | 0 | 0.667394 | 0.167774 |
| Gene_Pert | Dicer1 KO r | 3/320 | 0.16305 | 0.844507 | 0 | 0 | 2.207751 | 4.004196 |
| Gene_Pert | Ikbkb KO n | 3/320 | 0.16305 | 0.844507 | 0 | 0 | 2.207751 | 4.004196 |
| Gene_Pert | Tgfbr2 kno | 3/326 | 0.169397 | 0.844507 | 0 | 0 | 2.166077 | 3.845893 |
| Gene_Pert | HDAC2 KD | 1/334 | 0.769695 | 0.844507 | 0 | 0 | 0.683707 | 0.178968 |
| Gene_Pert | CBFA2T3 K | 1/330 | 0.765575 | 0.844507 | 0 | 0 | 0.692161 | 0.184896 |
| Gene_Pert | Rpe65 KD r | 1/330 | 0.765575 | 0.844507 | 0 | 0 | 0.692161 | 0.184896 |
| Gene_Pert | Adrb1 mut | 1/330 | 0.765575 | 0.844507 | 0 | 0 | 0.692161 | 0.184896 |
| Gene_Pert | Bcl2 OE mc | 1/329 | 0.764533 | 0.844507 | 0 | 0 | 0.694307 | 0.186414 |
| Gene_Pert | ZNF750 KD | 1/332 | 0.767644 | 0.844507 | 0 | 0 | 0.687908 | 0.181903 |

| | | | | | | | | |
|---|---|---|---|---|---|---|---|---|
| Gene_Pert | Rb1 KO mo | 1/332 | 0.767644 | 0.844507 | 0 | 0 | 0.687908 | 0.181903 |
| Gene_Pert | Dmd KO m | 1/332 | 0.767644 | 0.844507 | 0 | 0 | 0.687908 | 0.181903 |
| Gene_Pert | Dmd Defici | 1/331 | 0.766611 | 0.844507 | 0 | 0 | 0.690028 | 0.183392 |
| Gene_Pert | Atm knock | 1/331 | 0.766611 | 0.844507 | 0 | 0 | 0.690028 | 0.183392 |
| Gene_Pert | App KO mo | 1/331 | 0.766611 | 0.844507 | 0 | 0 | 0.690028 | 0.183392 |
| Gene_Pert | Pten KO m | 1/331 | 0.766611 | 0.844507 | 0 | 0 | 0.690028 | 0.183392 |
| Gene_Pert | BRAF Drug | 1/331 | 0.766611 | 0.844507 | 0 | 0 | 0.690028 | 0.183392 |
| Gene_Pert | Ppp2r2d KI | 1/334 | 0.769695 | 0.844507 | 0 | 0 | 0.683707 | 0.178968 |
| Gene_Pert | LLGL1 KD h | 3/326 | 0.169397 | 0.844507 | 0 | 0 | 2.166077 | 3.845893 |
| Gene_Pert | Pten KO m | 1/333 | 0.768672 | 0.844507 | 0 | 0 | 0.685801 | 0.180429 |
| Gene_Pert | Cbfb Defici | 1/333 | 0.768672 | 0.844507 | 0 | 0 | 0.685801 | 0.180429 |
| Gene_Pert | PIK3CA dru | 1/333 | 0.768672 | 0.844507 | 0 | 0 | 0.685801 | 0.180429 |
| Gene_Pert | MYC KD hu | 1/333 | 0.768672 | 0.844507 | 0 | 0 | 0.685801 | 0.180429 |
| Gene_Pert | BRAF Drug | 1/332 | 0.767644 | 0.844507 | 0 | 0 | 0.687908 | 0.181903 |
| Gene_Pert | Lrp5 KO m | 1/337 | 0.772738 | 0.844507 | 0 | 0 | 0.677499 | 0.174669 |
| Gene_Pert | Klf1 KO mo | 1/337 | 0.772738 | 0.844507 | 0 | 0 | 0.677499 | 0.174669 |
| Gene_Pert | Pten KO m | 1/336 | 0.771728 | 0.844507 | 0 | 0 | 0.679556 | 0.176089 |
| Gene_Pert | Tal1 KO mo | 1/336 | 0.771728 | 0.844507 | 0 | 0 | 0.679556 | 0.176089 |
| Gene_Pert | Ppif KO mo | 1/335 | 0.770714 | 0.844507 | 0 | 0 | 0.681625 | 0.177521 |
| Gene_Pert | Ghrh KO m | 1/335 | 0.770714 | 0.844507 | 0 | 0 | 0.681625 | 0.177521 |
| Gene_Pert | ETS2 KD hu | 1/335 | 0.770714 | 0.844507 | 0 | 0 | 0.681625 | 0.177521 |
| Gene_Pert | LRRK2 Gly2 | 3/335 | 0.179051 | 0.844507 | 0 | 0 | 2.10639 | 3.623169 |
| Gene_Pert | Des deplet | 3/347 | 0.192154 | 0.844507 | 0 | 0 | 2.031665 | 3.351151 |
| Gene_Pert | Cav1 Defici | 3/347 | 0.192154 | 0.844507 | 0 | 0 | 2.031665 | 3.351151 |
| Gene_Pert | Prmt5 KO r | 3/346 | 0.191052 | 0.844507 | 0 | 0 | 2.037693 | 3.372806 |
| Gene_Pert | PSMB9 KO | 3/364 | 0.211116 | 0.844507 | 0 | 0 | 1.934309 | 3.008525 |
| Gene_Pert | Ndel1 KD n | 3/363 | 0.209989 | 0.844507 | 0 | 0 | 1.939782 | 3.027421 |
| Gene_Pert | Hoxb4 indu | 3/363 | 0.209989 | 0.844507 | 0 | 0 | 1.939782 | 3.027421 |
| Gene_Pert | L1cam KO | 3/363 | 0.209989 | 0.844507 | 0 | 0 | 1.939782 | 3.027421 |
| Gene_Pert | Jun Overex | 2/187 | 0.195575 | 0.844507 | 0 | 0 | 2.509126 | 4.094425 |
| Gene_Pert | NPTX1 KO | 3/350 | 0.195467 | 0.844507 | 0 | 0 | 2.013792 | 3.287236 |
| Gene_Pert | Ghr Mutati | 3/349 | 0.194361 | 0.844507 | 0 | 0 | 2.019715 | 3.308368 |
| Gene_Pert | Ulk1 knock | 3/349 | 0.194361 | 0.844507 | 0 | 0 | 2.019715 | 3.308368 |
| Gene_Pert | MED12 KD | 3/336 | 0.180133 | 0.844507 | 0 | 0 | 2.099957 | 3.599451 |
| Gene_Pert | Hras Select | 3/335 | 0.179051 | 0.844507 | 0 | 0 | 2.10639 | 3.623169 |
| Gene_Pert | RAP1A KO | 2/185 | 0.192423 | 0.844507 | 0 | 0 | 2.536805 | 4.180799 |
| Gene_Pert | PARK2 KD | 3/334 | 0.177971 | 0.844507 | 0 | 0 | 2.112861 | 3.647087 |
| Gene_Pert | Ahr KO mo | 3/355 | 0.201022 | 0.844507 | 0 | 0 | 1.984679 | 3.184098 |
| Gene_Pert | Zmpste24 | 3/355 | 0.201022 | 0.844507 | 0 | 0 | 1.984679 | 3.184098 |
| Gene_Pert | Src overexp | 2/190 | 0.200314 | 0.844507 | 0 | 0 | 2.468711 | 3.969367 |
| Gene_Pert | Atxn1 Knoc | 3/354 | 0.199908 | 0.844507 | 0 | 0 | 1.990435 | 3.204395 |
| Gene_Pert | Akt1 knock | 3/340 | 0.18448 | 0.844507 | 0 | 0 | 2.074608 | 3.506535 |
| Gene_Pert | Dicer1 KO | 3/339 | 0.18339 | 0.844507 | 0 | 0 | 2.080889 | 3.529475 |
| Gene_Pert | Kcnip3 KO | 3/339 | 0.18339 | 0.844507 | 0 | 0 | 2.080889 | 3.529475 |
| Gene_Pert | BAP1 KO h | 2/179 | 0.183014 | 0.844507 | 0 | 0 | 2.623596 | 4.455375 |

| | | | | | | | | |
|---|---|---|---|---|---|---|---|---|
| Gene_Pert | Snca KO m | 3/330 | 0.173668 | 0.844507 | 0 | 0 | 2.139144 | 3.744801 |
| Gene_Pert | Trim71 KD | 3/330 | 0.173668 | 0.844507 | 0 | 0 | 2.139144 | 3.744801 |
| Gene_Pert | PFKL OE hu | 3/330 | 0.173668 | 0.844507 | 0 | 0 | 2.139144 | 3.744801 |
| Gene_Pert | GRN defici | 3/327 | 0.170462 | 0.844507 | 0 | 0 | 2.159281 | 3.820295 |
| Gene_Pert | Med23 KO | 3/326 | 0.169397 | 0.844507 | 0 | 0 | 2.166077 | 3.845893 |
| Gene_Pert | PARK7 KO | 3/326 | 0.169397 | 0.844507 | 0 | 0 | 2.166077 | 3.845893 |
| Gene_Pert | Rbm15 KO | 3/323 | 0.166214 | 0.844507 | 0 | 0 | 2.186719 | 3.924018 |
| Gene_Pert | HMGB1 KC | 3/332 | 0.175816 | 0.844507 | 0 | 0 | 2.125923 | 3.695531 |
| Gene_Pert | Dicer1 KO | 3/324 | 0.167273 | 0.844507 | 0 | 0 | 2.179795 | 3.897752 |
| Gene_Pert | DDR1 knoc | 3/324 | 0.167273 | 0.844507 | 0 | 0 | 2.179795 | 3.897752 |
| Gene_Pert | BST2 OE hu | 3/329 | 0.172598 | 0.844507 | 0 | 0 | 2.145815 | 3.769752 |
| Gene_Pert | PTEN KD hu | 3/329 | 0.172598 | 0.844507 | 0 | 0 | 2.145815 | 3.769752 |
| Gene_Pert | Ndel1 KO n | 3/329 | 0.172598 | 0.844507 | 0 | 0 | 2.145815 | 3.769752 |
| Gene_Pert | NR2F2 KD | 3/328 | 0.171529 | 0.844507 | 0 | 0 | 2.152527 | 3.794916 |
| Gene_Pert | Runx1 KO r | 3/321 | 0.164103 | 0.844507 | 0 | 0 | 2.200696 | 3.977238 |
| Gene_Pert | TFAP2C Sile | 3/320 | 0.16305 | 0.844507 | 0 | 0 | 2.207751 | 4.004196 |
| Gene_Pert | CD24 KD hu | 3/327 | 0.170462 | 0.844507 | 0 | 0 | 2.159281 | 3.820295 |
| Gene_Pert | TBK1 KD hu | 1/330 | 0.765575 | 0.844507 | 0 | 0 | 0.692161 | 0.184896 |
| Gene_Pert | Zfpm2 KO r | 3/332 | 0.175816 | 0.844507 | 0 | 0 | 2.125923 | 3.695531 |
| Gene_Pert | TRPM7 ove | 3/331 | 0.174741 | 0.844507 | 0 | 0 | 2.132513 | 3.720062 |
| Gene_Pert | Pdpn NULL | 3/331 | 0.174741 | 0.844507 | 0 | 0 | 2.132513 | 3.720062 |
| Gene_Pert | CTNNB1 KC | 3/331 | 0.174741 | 0.844507 | 0 | 0 | 2.132513 | 3.720062 |
| Gene_Pert | Neurod1 D | 2/170 | 0.16904 | 0.844507 | 0 | 0 | 2.765406 | 4.91584 |
| Gene_Pert | Ulk1 knock | 3/325 | 0.168334 | 0.844507 | 0 | 0 | 2.172915 | 3.871711 |
| Gene_Pert | Grid2 KO m | 3/327 | 0.170462 | 0.844507 | 0 | 0 | 2.159281 | 3.820295 |
| Gene_Pert | SYK druginl | 3/327 | 0.170462 | 0.844507 | 0 | 0 | 2.159281 | 3.820295 |
| Gene_Pert | BST2 OE hu | 2/192 | 0.203481 | 0.844507 | 0 | 0 | 2.442477 | 3.888868 |
| Gene_Pert | CHEK1 KD | 2/192 | 0.203481 | 0.844507 | 0 | 0 | 2.442477 | 3.888868 |
| Gene_Pert | Atf6 tamox | 3/357 | 0.203255 | 0.844507 | 0 | 0 | 1.973265 | 3.143989 |
| Gene_Pert | Foxo1 KO r | 3/356 | 0.202138 | 0.844507 | 0 | 0 | 1.978956 | 3.163964 |
| Gene_Pert | BRAF Drug | 1/315 | 0.749464 | 0.844507 | 0 | 0 | 0.725781 | 0.209313 |
| Gene_Pert | Aqp11 Defi | 1/315 | 0.749464 | 0.844507 | 0 | 0 | 0.725781 | 0.209313 |
| Gene_Pert | Cftr KO mo | 1/314 | 0.748352 | 0.844507 | 0 | 0 | 0.728137 | 0.211074 |
| Gene_Pert | PTK7 KD hu | 1/314 | 0.748352 | 0.844507 | 0 | 0 | 0.728137 | 0.211074 |
| Gene_Pert | Pycard KO | 1/314 | 0.748352 | 0.844507 | 0 | 0 | 0.728137 | 0.211074 |
| Gene_Pert | RNF40 KD | 1/314 | 0.748352 | 0.844507 | 0 | 0 | 0.728137 | 0.211074 |
| Gene_Pert | DOCK4 OE | 1/313 | 0.747235 | 0.844507 | 0 | 0 | 0.730508 | 0.212852 |
| Gene_Pert | Wt1 KO mc | 1/317 | 0.751674 | 0.844507 | 0 | 0 | 0.721114 | 0.205844 |
| Gene_Pert | Runx1 KO r | 1/317 | 0.751674 | 0.844507 | 0 | 0 | 0.721114 | 0.205844 |
| Gene_Pert | Cap1 OE m | 1/316 | 0.750571 | 0.844507 | 0 | 0 | 0.72344 | 0.20757 |
| Gene_Pert | Trpm7 KO | 1/316 | 0.750571 | 0.844507 | 0 | 0 | 0.72344 | 0.20757 |
| Gene_Pert | Met knock | 1/316 | 0.750571 | 0.844507 | 0 | 0 | 0.72344 | 0.20757 |
| Gene_Pert | Mapk14 KC | 1/316 | 0.750571 | 0.844507 | 0 | 0 | 0.72344 | 0.20757 |
| Gene_Pert | Nr1i3 hNR1 | 1/315 | 0.749464 | 0.844507 | 0 | 0 | 0.725781 | 0.209313 |
| Gene_Pert | Gykl1 KO m | 1/315 | 0.749464 | 0.844507 | 0 | 0 | 0.725781 | 0.209313 |

| | | | | | | | | |
|---|---|---|---|---|---|---|---|---|
| Gene_Pert | Met knock | 1/318 | 0.752771 | 0.844507 | 0 | 0 | 0.718803 | 0.204135 |
| Gene_Pert | SNRK overe | 1/318 | 0.752771 | 0.844507 | 0 | 0 | 0.718803 | 0.204135 |
| Gene_Pert | GAPDH KD | 1/318 | 0.752771 | 0.844507 | 0 | 0 | 0.718803 | 0.204135 |
| Gene_Pert | Fshr KO mo | 1/318 | 0.752771 | 0.844507 | 0 | 0 | 0.718803 | 0.204135 |
| Gene_Pert | IRF7 KD hu | 1/317 | 0.751674 | 0.844507 | 0 | 0 | 0.721114 | 0.205844 |
| Gene_Pert | Xrcc5 KO m | 1/317 | 0.751674 | 0.844507 | 0 | 0 | 0.721114 | 0.205844 |
| Gene_Pert | Tgfb2 KD m | 1/317 | 0.751674 | 0.844507 | 0 | 0 | 0.721114 | 0.205844 |
| Gene_Pert | Nr1i3 KO m | 1/317 | 0.751674 | 0.844507 | 0 | 0 | 0.721114 | 0.205844 |
| Gene_Pert | Sirt4 KO mo | 1/319 | 0.753864 | 0.844507 | 0 | 0 | 0.716506 | 0.202444 |
| Gene_Pert | Cftr Deficie | 1/319 | 0.753864 | 0.844507 | 0 | 0 | 0.716506 | 0.202444 |
| Gene_Pert | THOC1 KD | 1/319 | 0.753864 | 0.844507 | 0 | 0 | 0.716506 | 0.202444 |
| Gene_Pert | EZH2 DEPL | 1/319 | 0.753864 | 0.844507 | 0 | 0 | 0.716506 | 0.202444 |
| Gene_Pert | Acvrl1 mo | 1/311 | 0.744986 | 0.844507 | 0 | 0 | 0.735296 | 0.216463 |
| Gene_Pert | Ovol1 KO n | 1/309 | 0.742718 | 0.844507 | 0 | 0 | 0.740146 | 0.220149 |
| Gene_Pert | Eomes KO | 1/311 | 0.744986 | 0.844507 | 0 | 0 | 0.735296 | 0.216463 |
| Gene_Pert | SMARCB1 I | 1/311 | 0.744986 | 0.844507 | 0 | 0 | 0.735296 | 0.216463 |
| Gene_Pert | FOXM1 KD | 1/311 | 0.744986 | 0.844507 | 0 | 0 | 0.735296 | 0.216463 |
| Gene_Pert | PPIB Deple | 1/311 | 0.744986 | 0.844507 | 0 | 0 | 0.735296 | 0.216463 |
| Gene_Pert | Vldlr KO mo | 1/310 | 0.743855 | 0.844507 | 0 | 0 | 0.737714 | 0.218297 |
| Gene_Pert | TERC KD hu | 1/310 | 0.743855 | 0.844507 | 0 | 0 | 0.737714 | 0.218297 |
| Gene_Pert | Cdk4 Heter | 1/310 | 0.743855 | 0.844507 | 0 | 0 | 0.737714 | 0.218297 |
| Gene_Pert | Nxnl1 KO n | 1/310 | 0.743855 | 0.844507 | 0 | 0 | 0.737714 | 0.218297 |
| Gene_Pert | RRM2B KD | 1/312 | 0.746113 | 0.844507 | 0 | 0 | 0.732895 | 0.214649 |
| Gene_Pert | Mecom OE | 1/312 | 0.746113 | 0.844507 | 0 | 0 | 0.732895 | 0.214649 |
| Gene_Pert | CREB1 DEP | 1/312 | 0.746113 | 0.844507 | 0 | 0 | 0.732895 | 0.214649 |
| Gene_Pert | Cited2 Defi | 1/312 | 0.746113 | 0.844507 | 0 | 0 | 0.732895 | 0.214649 |
| Gene_Pert | Runx1 KO r | 1/319 | 0.753864 | 0.844507 | 0 | 0 | 0.716506 | 0.202444 |
| Gene_Pert | Rybp KO m | 1/311 | 0.744986 | 0.844507 | 0 | 0 | 0.735296 | 0.216463 |
| Gene_Pert | IRS3P KO m | 1/311 | 0.744986 | 0.844507 | 0 | 0 | 0.735296 | 0.216463 |
| Gene_Pert | Nrl Deficier | 1/311 | 0.744986 | 0.844507 | 0 | 0 | 0.735296 | 0.216463 |
| Gene_Pert | PTEN KD hu | 1/313 | 0.747235 | 0.844507 | 0 | 0 | 0.730508 | 0.212852 |
| Gene_Pert | Dmd Defici | 1/313 | 0.747235 | 0.844507 | 0 | 0 | 0.730508 | 0.212852 |
| Gene_Pert | Dusp1 KO r | 1/313 | 0.747235 | 0.844507 | 0 | 0 | 0.730508 | 0.212852 |
| Gene_Pert | Trp53 KO n | 1/313 | 0.747235 | 0.844507 | 0 | 0 | 0.730508 | 0.212852 |
| Gene_Pert | Fosl1 KO m | 1/313 | 0.747235 | 0.844507 | 0 | 0 | 0.730508 | 0.212852 |
| Gene_Pert | SOX2 Defic | 1/313 | 0.747235 | 0.844507 | 0 | 0 | 0.730508 | 0.212852 |
| Gene_Pert | CDK9 knocl | 1/313 | 0.747235 | 0.844507 | 0 | 0 | 0.730508 | 0.212852 |
| Gene_Pert | Ppt1 KO mo | 1/312 | 0.746113 | 0.844507 | 0 | 0 | 0.732895 | 0.214649 |
| Gene_Pert | FTO OE hur | 1/315 | 0.749464 | 0.844507 | 0 | 0 | 0.725781 | 0.209313 |
| Gene_Pert | Hdac6 KO r | 1/327 | 0.762437 | 0.844507 | 0 | 0 | 0.698637 | 0.189496 |
| Gene_Pert | Il1rn KO mo | 1/326 | 0.761381 | 0.844507 | 0 | 0 | 0.700823 | 0.191059 |
| Gene_Pert | SMC3 KD h | 1/325 | 0.760322 | 0.844507 | 0 | 0 | 0.703022 | 0.192638 |
| Gene_Pert | Ache AChE | 1/325 | 0.760322 | 0.844507 | 0 | 0 | 0.703022 | 0.192638 |
| Gene_Pert | Dlx5 Defici | 1/325 | 0.760322 | 0.844507 | 0 | 0 | 0.703022 | 0.192638 |
| Gene_Pert | FGFR3 kno | 1/325 | 0.760322 | 0.844507 | 0 | 0 | 0.703022 | 0.192638 |

| | | | | | | | | |
|---|---|---|---|---|---|---|---|---|
| Gene_Pert | SON KD hu | 1/324 | 0.759257 | 0.844507 | 0 | 0 | 0.705234 | 0.194232 |
| Gene_Pert | HDAC1 KD | 1/324 | 0.759257 | 0.844507 | 0 | 0 | 0.705234 | 0.194232 |
| Gene_Pert | Syncrip OE | 1/328 | 0.763487 | 0.844507 | 0 | 0 | 0.696465 | 0.187947 |
| Gene_Pert | FLI1 KD hu | 1/328 | 0.763487 | 0.844507 | 0 | 0 | 0.696465 | 0.187947 |
| Gene_Pert | Htt Knock-i | 1/328 | 0.763487 | 0.844507 | 0 | 0 | 0.696465 | 0.187947 |
| Gene_Pert | Elane Inhib | 1/328 | 0.763487 | 0.844507 | 0 | 0 | 0.696465 | 0.187947 |
| Gene_Pert | Tcf3 KO mo | 1/327 | 0.762437 | 0.844507 | 0 | 0 | 0.698637 | 0.189496 |
| Gene_Pert | SYK druginl | 1/327 | 0.762437 | 0.844507 | 0 | 0 | 0.698637 | 0.189496 |
| Gene_Pert | HNF1B Ove | 1/326 | 0.761381 | 0.844507 | 0 | 0 | 0.700823 | 0.191059 |
| Gene_Pert | Aldh5a1 KO | 1/327 | 0.762437 | 0.844507 | 0 | 0 | 0.698637 | 0.189496 |
| Gene_Pert | Dmd Defici | 1/329 | 0.764533 | 0.844507 | 0 | 0 | 0.694307 | 0.186414 |
| Gene_Pert | ERBB3 kno | 1/329 | 0.764533 | 0.844507 | 0 | 0 | 0.694307 | 0.186414 |
| Gene_Pert | TBK1 knock | 1/329 | 0.764533 | 0.844507 | 0 | 0 | 0.694307 | 0.186414 |
| Gene_Pert | TFAP2C Sile | 1/329 | 0.764533 | 0.844507 | 0 | 0 | 0.694307 | 0.186414 |
| Gene_Pert | Tnf KD mou | 1/329 | 0.764533 | 0.844507 | 0 | 0 | 0.694307 | 0.186414 |
| Gene_Pert | Ppara KO m | 1/329 | 0.764533 | 0.844507 | 0 | 0 | 0.694307 | 0.186414 |
| Gene_Pert | IRF5 KD hu | 1/328 | 0.763487 | 0.844507 | 0 | 0 | 0.696465 | 0.187947 |
| Gene_Pert | Fmr1 KD m | 1/328 | 0.763487 | 0.844507 | 0 | 0 | 0.696465 | 0.187947 |
| Gene_Pert | Slc9a3 KO r | 1/331 | 0.766611 | 0.844507 | 0 | 0 | 0.690028 | 0.183392 |
| Gene_Pert | Erbb2 drug | 1/331 | 0.766611 | 0.844507 | 0 | 0 | 0.690028 | 0.183392 |
| Gene_Pert | ZNF658 KD | 1/331 | 0.766611 | 0.844507 | 0 | 0 | 0.690028 | 0.183392 |
| Gene_Pert | Rictor KO n | 1/321 | 0.756036 | 0.844507 | 0 | 0 | 0.711955 | 0.19911 |
| Gene_Pert | Myc knock- | 1/319 | 0.753864 | 0.844507 | 0 | 0 | 0.716506 | 0.202444 |
| Gene_Pert | Ppargc1b H | 1/319 | 0.753864 | 0.844507 | 0 | 0 | 0.716506 | 0.202444 |
| Gene_Pert | Bcl2 OE mo | 1/318 | 0.752771 | 0.844507 | 0 | 0 | 0.718803 | 0.204135 |
| Gene_Pert | RAF1 active | 1/320 | 0.754952 | 0.844507 | 0 | 0 | 0.714223 | 0.200769 |
| Gene_Pert | Trp53inp2 | 1/320 | 0.754952 | 0.844507 | 0 | 0 | 0.714223 | 0.200769 |
| Gene_Pert | TFAP2C KD | 1/320 | 0.754952 | 0.844507 | 0 | 0 | 0.714223 | 0.200769 |
| Gene_Pert | Nrg4 KO m | 1/320 | 0.754952 | 0.844507 | 0 | 0 | 0.714223 | 0.200769 |
| Gene_Pert | Klf7 KO mo | 1/320 | 0.754952 | 0.844507 | 0 | 0 | 0.714223 | 0.200769 |
| Gene_Pert | H6pd KO m | 1/320 | 0.754952 | 0.844507 | 0 | 0 | 0.714223 | 0.200769 |
| Gene_Pert | EWSR1 KD | 1/320 | 0.754952 | 0.844507 | 0 | 0 | 0.714223 | 0.200769 |
| Gene_Pert | SYK druginl | 1/321 | 0.756036 | 0.844507 | 0 | 0 | 0.711955 | 0.19911 |
| Gene_Pert | HNRNPU-A | 1/321 | 0.756036 | 0.844507 | 0 | 0 | 0.711955 | 0.19911 |
| Gene_Pert | NTRK3 drug | 1/321 | 0.756036 | 0.844507 | 0 | 0 | 0.711955 | 0.19911 |
| Gene_Pert | Arhgap5 KD | 1/273 | 0.6983 | 0.844507 | 0 | 0 | 0.839646 | 0.301522 |
| Gene_Pert | DIP2B KD h | 1/321 | 0.756036 | 0.844507 | 0 | 0 | 0.711955 | 0.19911 |
| Gene_Pert | Trim33 knc | 1/321 | 0.756036 | 0.844507 | 0 | 0 | 0.711955 | 0.19911 |
| Gene_Pert | Dusp1 Defi | 1/320 | 0.754952 | 0.844507 | 0 | 0 | 0.714223 | 0.200769 |
| Gene_Pert | CIITA KD hu | 1/320 | 0.754952 | 0.844507 | 0 | 0 | 0.714223 | 0.200769 |
| Gene_Pert | YWHAZ OE | 1/324 | 0.759257 | 0.844507 | 0 | 0 | 0.705234 | 0.194232 |
| Gene_Pert | CHEK1 KD | 1/324 | 0.759257 | 0.844507 | 0 | 0 | 0.705234 | 0.194232 |
| Gene_Pert | Bnc1 KD m | 1/323 | 0.758188 | 0.844507 | 0 | 0 | 0.707461 | 0.195842 |
| Gene_Pert | IPMK KD hu | 1/323 | 0.758188 | 0.844507 | 0 | 0 | 0.707461 | 0.195842 |
| Gene_Pert | Il2 KO mou | 1/323 | 0.758188 | 0.844507 | 0 | 0 | 0.707461 | 0.195842 |

| | | | | | | | | |
|---|---|---|---|---|---|---|---|---|
| Gene_Pert | NCSTN KD | 1/323 | 0.758188 | 0.844507 | 0 | 0 | 0.707461 | 0.195842 |
| Gene_Pert | CCND1 KD | 1/322 | 0.757114 | 0.844507 | 0 | 0 | 0.709701 | 0.197468 |
| Gene_Pert | Ahr KO mo | 1/322 | 0.757114 | 0.844507 | 0 | 0 | 0.709701 | 0.197468 |
| Gene_Pert | CHEK2 KD I | 1/224 | 0.625446 | 0.844507 | 0 | 0 | 1.026697 | 0.481818 |
| Gene_Pert | Ep400 KD r | 2/473 | 0.613272 | 0.844507 | 0 | 0 | 0.97125 | 0.47489 |
| Gene_Pert | Msr1 KO m | 1/204 | 0.590938 | 0.844507 | 0 | 0 | 1.128995 | 0.593902 |
| Gene_Pert | Prkcz KO m | 1/222 | 0.622129 | 0.844507 | 0 | 0 | 1.036094 | 0.491738 |
| Gene_Pert | Notch1 Def | 1/222 | 0.622129 | 0.844507 | 0 | 0 | 1.036094 | 0.491738 |
| Gene_Pert | FOXM1 KD | 1/222 | 0.622129 | 0.844507 | 0 | 0 | 1.036094 | 0.491738 |
| Gene_Pert | STAT1 KD h | 1/222 | 0.622129 | 0.844507 | 0 | 0 | 1.036094 | 0.491738 |
| Gene_Pert | Hif1a NULL | 2/480 | 0.621315 | 0.844507 | 0 | 0 | 0.956682 | 0.455302 |
| Gene_Pert | Satb1 KO n | 1/221 | 0.62046 | 0.844507 | 0 | 0 | 1.040856 | 0.496795 |
| Gene_Pert | Trnp1 KD n | 2/466 | 0.605102 | 0.844507 | 0 | 0 | 0.986258 | 0.495455 |
| Gene_Pert | Stat1 KO m | 1/221 | 0.62046 | 0.844507 | 0 | 0 | 1.040856 | 0.496795 |
| Gene_Pert | NFKB1 Inac | 1/224 | 0.625446 | 0.844507 | 0 | 0 | 1.026697 | 0.481818 |
| Gene_Pert | Rp1 KO mo | 1/224 | 0.625446 | 0.844507 | 0 | 0 | 1.026697 | 0.481818 |
| Gene_Pert | RPS24 OE h | 1/217 | 0.613709 | 0.844507 | 0 | 0 | 1.060347 | 0.517698 |
| Gene_Pert | Dmd Defici | 1/223 | 0.623791 | 0.844507 | 0 | 0 | 1.031374 | 0.486746 |
| Gene_Pert | CHEK1 KD I | 1/223 | 0.623791 | 0.844507 | 0 | 0 | 1.031374 | 0.486746 |
| Gene_Pert | Nfe2l2 KO r | 1/223 | 0.623791 | 0.844507 | 0 | 0 | 1.031374 | 0.486746 |
| Gene_Pert | Lamtor2 KC | 1/216 | 0.612003 | 0.844507 | 0 | 0 | 1.065333 | 0.523098 |
| Gene_Pert | Pdss2 KO n | 1/222 | 0.622129 | 0.844507 | 0 | 0 | 1.036094 | 0.491738 |
| Gene_Pert | ZXDC KD hu | 1/226 | 0.628735 | 0.844507 | 0 | 0 | 1.017468 | 0.472152 |
| Gene_Pert | PTK7 KD hu | 1/226 | 0.628735 | 0.844507 | 0 | 0 | 1.017468 | 0.472152 |
| Gene_Pert | Irak2 knock | 1/225 | 0.627094 | 0.844507 | 0 | 0 | 1.022062 | 0.476954 |
| Gene_Pert | AATF KD hu | 1/225 | 0.627094 | 0.844507 | 0 | 0 | 1.022062 | 0.476954 |
| Gene_Pert | Irf8 KO mo | 1/225 | 0.627094 | 0.844507 | 0 | 0 | 1.022062 | 0.476954 |
| Gene_Pert | IGH mutati | 1/224 | 0.625446 | 0.844507 | 0 | 0 | 1.026697 | 0.481818 |
| Gene_Pert | CBFB KD hu | 1/216 | 0.612003 | 0.844507 | 0 | 0 | 1.065333 | 0.523098 |
| Gene_Pert | BST2 OE hu | 1/200 | 0.583668 | 0.844507 | 0 | 0 | 1.151922 | 0.620221 |
| Gene_Pert | Dmrt1 KO r | 1/200 | 0.583668 | 0.844507 | 0 | 0 | 1.151922 | 0.620221 |
| Gene_Pert | Htt Knock-i | 2/436 | 0.568645 | 0.844507 | 0 | 0 | 1.056059 | 0.596144 |
| Gene_Pert | IRF9 OE hu | 1/211 | 0.603359 | 0.844507 | 0 | 0 | 1.090975 | 0.551207 |
| Gene_Pert | NRAS KD h | 1/209 | 0.599849 | 0.844507 | 0 | 0 | 1.101576 | 0.562991 |
| Gene_Pert | Atf6 KO mc | 1/208 | 0.598082 | 0.844507 | 0 | 0 | 1.106954 | 0.569005 |
| Gene_Pert | SYK drugin | 1/208 | 0.598082 | 0.844507 | 0 | 0 | 1.106954 | 0.569005 |
| Gene_Pert | Myd88 Def | 1/208 | 0.598082 | 0.844507 | 0 | 0 | 1.106954 | 0.569005 |
| Gene_Pert | Hipk2 OE n | 1/206 | 0.594525 | 0.844507 | 0 | 0 | 1.117867 | 0.581282 |
| Gene_Pert | Mfi2 KO m | 1/205 | 0.592736 | 0.844507 | 0 | 0 | 1.123404 | 0.587548 |
| Gene_Pert | CHRNA9 KC | 2/443 | 0.577361 | 0.844507 | 0 | 0 | 1.038922 | 0.570668 |
| Gene_Pert | Gata5 KO r | 1/221 | 0.62046 | 0.844507 | 0 | 0 | 1.040856 | 0.496795 |
| Gene_Pert | Rora gene | 1/216 | 0.612003 | 0.844507 | 0 | 0 | 1.065333 | 0.523098 |
| Gene_Pert | Adnp Defic | 1/215 | 0.610289 | 0.844507 | 0 | 0 | 1.070365 | 0.528571 |
| Gene_Pert | ADRB1 Bet | 2/470 | 0.609786 | 0.844507 | 0 | 0 | 0.977627 | 0.48358 |
| Gene_Pert | SIRT1 KD h | 1/214 | 0.608568 | 0.844507 | 0 | 0 | 1.075445 | 0.534116 |

| | | | | | | | | |
|---|---|---|---|---|---|---|---|---|
| Gene_Pert | Sp3 Deficie | 1/213 | 0.606839 | 0.844507 | 0 | 0 | 1.080573 | 0.539737 |
| Gene_Pert | NR2C2 KD | 1/212 | 0.605103 | 0.844507 | 0 | 0 | 1.085749 | 0.545433 |
| Gene_Pert | Dmd Defici | 1/198 | 0.579985 | 0.844507 | 0 | 0 | 1.163735 | 0.633948 |
| Gene_Pert | Gata5 KO r | 1/220 | 0.618783 | 0.844507 | 0 | 0 | 1.045662 | 0.501918 |
| Gene_Pert | HSF1 KD hu | 1/219 | 0.617099 | 0.844507 | 0 | 0 | 1.050512 | 0.507109 |
| Gene_Pert | Rorc KO mo | 1/218 | 0.615408 | 0.844507 | 0 | 0 | 1.055407 | 0.512369 |
| Gene_Pert | Ahr KO mo | 1/218 | 0.615408 | 0.844507 | 0 | 0 | 1.055407 | 0.512369 |
| Gene_Pert | SOX7 OE hu | 1/246 | 0.660088 | 0.844507 | 0 | 0 | 0.93346 | 0.387742 |
| Gene_Pert | Htt CAG kn | 1/229 | 0.633614 | 0.844507 | 0 | 0 | 1.003927 | 0.458108 |
| Gene_Pert | Hmgn1 KO | 1/229 | 0.633614 | 0.844507 | 0 | 0 | 1.003927 | 0.458108 |
| Gene_Pert | RELA KD hu | 1/244 | 0.657075 | 0.844507 | 0 | 0 | 0.941238 | 0.39528 |
| Gene_Pert | Prkaca KO | 1/243 | 0.655558 | 0.844507 | 0 | 0 | 0.945176 | 0.399118 |
| Gene_Pert | Fosl1 KO m | 1/243 | 0.655558 | 0.844507 | 0 | 0 | 0.945176 | 0.399118 |
| Gene_Pert | WEE1 drug | 1/234 | 0.641605 | 0.844507 | 0 | 0 | 0.982134 | 0.435854 |
| Gene_Pert | RRP1B OE | 1/234 | 0.641605 | 0.844507 | 0 | 0 | 0.982134 | 0.435854 |
| Gene_Pert | PPRC1 Dep | 1/234 | 0.641605 | 0.844507 | 0 | 0 | 0.982134 | 0.435854 |
| Gene_Pert | PPRC1 DEP | 1/233 | 0.640021 | 0.844507 | 0 | 0 | 0.986417 | 0.440193 |
| Gene_Pert | Clpp KO mo | 1/233 | 0.640021 | 0.844507 | 0 | 0 | 0.986417 | 0.440193 |
| Gene_Pert | Prdm14 KO | 1/246 | 0.660088 | 0.844507 | 0 | 0 | 0.93346 | 0.387742 |
| Gene_Pert | Htt Mutatio | 1/229 | 0.633614 | 0.844507 | 0 | 0 | 1.003927 | 0.458108 |
| Gene_Pert | MIR128-1 | 1/246 | 0.660088 | 0.844507 | 0 | 0 | 0.93346 | 0.387742 |
| Gene_Pert | Atf6b KO m | 1/239 | 0.649424 | 0.844507 | 0 | 0 | 0.961257 | 0.414945 |
| Gene_Pert | HIST1H4C | 1/239 | 0.649424 | 0.844507 | 0 | 0 | 0.961257 | 0.414945 |
| Gene_Pert | Rnh1 KO m | 1/238 | 0.647874 | 0.844507 | 0 | 0 | 0.965362 | 0.419024 |
| Gene_Pert | Ndel1 KD n | 1/238 | 0.647874 | 0.844507 | 0 | 0 | 0.965362 | 0.419024 |
| Gene_Pert | Hdac2 KO r | 1/237 | 0.646317 | 0.844507 | 0 | 0 | 0.969501 | 0.423154 |
| Gene_Pert | Nsun2 KO r | 1/248 | 0.663076 | 0.844507 | 0 | 0 | 0.925807 | 0.380383 |
| Gene_Pert | Wfs1 KO m | 1/248 | 0.663076 | 0.844507 | 0 | 0 | 0.925807 | 0.380383 |
| Gene_Pert | HMGN5 Ov | 1/243 | 0.655558 | 0.844507 | 0 | 0 | 0.945176 | 0.399118 |
| Gene_Pert | CTNNB1 KD | 1/242 | 0.654034 | 0.844507 | 0 | 0 | 0.949146 | 0.403003 |
| Gene_Pert | Fosl1 KO m | 1/242 | 0.654034 | 0.844507 | 0 | 0 | 0.949146 | 0.403003 |
| Gene_Pert | BCOR VR h | 1/231 | 0.636831 | 0.844507 | 0 | 0 | 0.995096 | 0.449038 |
| Gene_Pert | Elf4 KO mo | 1/224 | 0.625446 | 0.844507 | 0 | 0 | 1.026697 | 0.481818 |
| Gene_Pert | Ppara OE n | 1/228 | 0.631995 | 0.844507 | 0 | 0 | 1.008401 | 0.462729 |
| Gene_Pert | FOXO3 KO | 1/228 | 0.631995 | 0.844507 | 0 | 0 | 1.008401 | 0.462729 |
| Gene_Pert | Esr2 KO mo | 1/228 | 0.631995 | 0.844507 | 0 | 0 | 1.008401 | 0.462729 |
| Gene_Pert | CFAP45 OE | 1/227 | 0.630368 | 0.844507 | 0 | 0 | 1.012914 | 0.46741 |
| Gene_Pert | Nrl Deficier | 1/227 | 0.630368 | 0.844507 | 0 | 0 | 1.012914 | 0.46741 |
| Gene_Pert | TET3 DEPLI | 1/227 | 0.630368 | 0.844507 | 0 | 0 | 1.012914 | 0.46741 |
| Gene_Pert | DTX1 OE hu | 2/482 | 0.623589 | 0.844507 | 0 | 0 | 0.952598 | 0.449878 |
| Gene_Pert | ALK drugin | 1/226 | 0.628735 | 0.844507 | 0 | 0 | 1.017468 | 0.472152 |
| Gene_Pert | Sod1 SOD1 | 1/232 | 0.63843 | 0.844507 | 0 | 0 | 0.990738 | 0.444588 |
| Gene_Pert | Srf homozy | 1/231 | 0.636831 | 0.844507 | 0 | 0 | 0.995096 | 0.449038 |
| Gene_Pert | Runx1 KO r | 1/201 | 0.585497 | 0.844507 | 0 | 0 | 1.146105 | 0.613503 |
| Gene_Pert | PADI4 KD h | 1/226 | 0.628735 | 0.844507 | 0 | 0 | 1.017468 | 0.472152 |

| | | | | | | | | |
|---|---|---|---|---|---|---|---|---|
| Gene_Pert | Ppargc1a K | 1/224 | 0.625446 | 0.844507 | 0 | 0 | 1.026697 | 0.481818 |
| Gene_Pert | NFKB1 Inac | 1/228 | 0.631995 | 0.844507 | 0 | 0 | 1.008401 | 0.462729 |
| Gene_Pert | Crhr2 KO m | 1/237 | 0.646317 | 0.844507 | 0 | 0 | 0.969501 | 0.423154 |
| Gene_Pert | HNF4A KD | 1/236 | 0.644753 | 0.844507 | 0 | 0 | 0.973676 | 0.427334 |
| Gene_Pert | MYC KD hu | 1/235 | 0.643183 | 0.844507 | 0 | 0 | 0.977887 | 0.431568 |
| Gene_Pert | Stk11 knoc | 1/241 | 0.652504 | 0.844507 | 0 | 0 | 0.953149 | 0.406935 |
| Gene_Pert | Nf1 Hetero | 1/240 | 0.650968 | 0.844507 | 0 | 0 | 0.957186 | 0.410916 |
| Gene_Pert | KMT2A ML | 1/239 | 0.649424 | 0.844507 | 0 | 0 | 0.961257 | 0.414945 |
| Gene_Pert | Akt1 active | 1/230 | 0.635226 | 0.844507 | 0 | 0 | 0.999492 | 0.453544 |
| Gene_Pert | MECP2 OE | 1/230 | 0.635226 | 0.844507 | 0 | 0 | 0.999492 | 0.453544 |
| Gene_Pert | Usp2 KO m | 2/405 | 0.528537 | 0.844507 | 0 | 0 | 1.139104 | 0.72634 |
| Gene_Pert | NR4A2 OE | 1/161 | 0.505746 | 0.844507 | 0 | 0 | 1.435538 | 0.978637 |
| Gene_Pert | Creb1 DN r | 2/381 | 0.495834 | 0.844507 | 0 | 0 | 1.212727 | 0.850744 |
| Gene_Pert | DNMT3B K | 2/388 | 0.505518 | 0.844507 | 0 | 0 | 1.190308 | 0.811994 |
| Gene_Pert | TRAC Stimu | 2/395 | 0.515083 | 0.844507 | 0 | 0 | 1.168687 | 0.77534 |
| Gene_Pert | Dgcr8 KO n | 2/394 | 0.513724 | 0.844507 | 0 | 0 | 1.171729 | 0.780453 |
| Gene_Pert | Dmd mutat | 2/394 | 0.513724 | 0.844507 | 0 | 0 | 1.171729 | 0.780453 |
| Gene_Pert | Htt Gain of | 2/394 | 0.513724 | 0.844507 | 0 | 0 | 1.171729 | 0.780453 |
| Gene_Pert | Ifng KO mo | 2/394 | 0.513724 | 0.844507 | 0 | 0 | 1.171729 | 0.780453 |
| Gene_Pert | Sox3 KO m | 2/394 | 0.513724 | 0.844507 | 0 | 0 | 1.171729 | 0.780453 |
| Gene_Pert | Cebpg KO r | 2/386 | 0.502763 | 0.844507 | 0 | 0 | 1.19663 | 0.822846 |
| Gene_Pert | TWIST2 Q1 | 2/393 | 0.512362 | 0.844507 | 0 | 0 | 1.174786 | 0.785607 |
| Gene_Pert | Fosl1 KO m | 2/405 | 0.528537 | 0.844507 | 0 | 0 | 1.139104 | 0.72634 |
| Gene_Pert | Mecom KO | 1/162 | 0.507913 | 0.844507 | 0 | 0 | 1.426549 | 0.966408 |
| Gene_Pert | NOTCH1 IN | 2/404 | 0.527203 | 0.844507 | 0 | 0 | 1.141996 | 0.731071 |
| Gene_Pert | PIK3CA mu | 2/404 | 0.527203 | 0.844507 | 0 | 0 | 1.141996 | 0.731071 |
| Gene_Pert | Htt Knock-i | 2/403 | 0.525866 | 0.844507 | 0 | 0 | 1.144902 | 0.735838 |
| Gene_Pert | Tnf siTNF-C | 2/403 | 0.525866 | 0.844507 | 0 | 0 | 1.144902 | 0.735838 |
| Gene_Pert | FGFR3 drug | 1/161 | 0.505746 | 0.844507 | 0 | 0 | 1.435538 | 0.978637 |
| Gene_Pert | Lhx1 NULL | 2/398 | 0.519145 | 0.844507 | 0 | 0 | 1.159655 | 0.760237 |
| Gene_Pert | GRN deficie | 1/175 | 0.535249 | 0.844507 | 0 | 0 | 1.319099 | 0.824467 |
| Gene_Pert | Plagl2 OE n | 2/410 | 0.535171 | 0.844507 | 0 | 0 | 1.124856 | 0.703224 |
| Gene_Pert | Akt1 knock | 2/409 | 0.53385 | 0.844507 | 0 | 0 | 1.127677 | 0.707777 |
| Gene_Pert | Twist1 OE r | 2/407 | 0.531199 | 0.844507 | 0 | 0 | 1.133362 | 0.716987 |
| Gene_Pert | PSIP1 KD h | 2/406 | 0.529869 | 0.844507 | 0 | 0 | 1.136226 | 0.721646 |
| Gene_Pert | S1pr2 KO n | 2/388 | 0.505518 | 0.844507 | 0 | 0 | 1.190308 | 0.811994 |
| Gene_Pert | Pparg Defic | 2/322 | 0.409821 | 0.844507 | 0 | 0 | 1.440662 | 1.285121 |
| Gene_Pert | Aire KO mc | 2/322 | 0.409821 | 0.844507 | 0 | 0 | 1.440662 | 1.285121 |
| Gene_Pert | THRB RCCC | 2/322 | 0.409821 | 0.844507 | 0 | 0 | 1.440662 | 1.285121 |
| Gene_Pert | Dgcr8 KO n | 2/385 | 0.501382 | 0.844507 | 0 | 0 | 1.199816 | 0.828337 |
| Gene_Pert | Tardbp KD | 2/384 | 0.499999 | 0.844507 | 0 | 0 | 1.203018 | 0.833871 |
| Gene_Pert | NOTCH1 Re | 2/384 | 0.499999 | 0.844507 | 0 | 0 | 1.203018 | 0.833871 |
| Gene_Pert | IRF9 OE hu | 1/157 | 0.496981 | 0.844507 | 0 | 0 | 1.472645 | 1.029678 |
| Gene_Pert | WTAP KD h | 2/381 | 0.495834 | 0.844507 | 0 | 0 | 1.212727 | 0.850744 |
| Gene_Pert | Xbp1 expre | 2/381 | 0.495834 | 0.844507 | 0 | 0 | 1.212727 | 0.850744 |

| | | | | | | | | |
|---|---|---|---|---|---|---|---|---|
| Gene_Pert | MECP2 het | 2/381 | 0.495834 | 0.844507 | 0 | 0 | 1.212727 | 0.850744 |
| Gene_Pert | Stat1 Stat1 | 2/380 | 0.494441 | 0.844507 | 0 | 0 | 1.215998 | 0.85646 |
| Gene_Pert | ERBB3 OE I | 2/406 | 0.529869 | 0.844507 | 0 | 0 | 1.136226 | 0.721646 |
| Gene_Pert | Panct1 KD | 2/387 | 0.504142 | 0.844507 | 0 | 0 | 1.193461 | 0.817399 |
| Gene_Pert | Klf1 OE mo | 2/387 | 0.504142 | 0.844507 | 0 | 0 | 1.193461 | 0.817399 |
| Gene_Pert | Met knock | 1/160 | 0.503569 | 0.844507 | 0 | 0 | 1.444639 | 0.991073 |
| Gene_Pert | Brd2 OE m | 1/160 | 0.503569 | 0.844507 | 0 | 0 | 1.444639 | 0.991073 |
| Gene_Pert | Tet1 KD mo | 2/386 | 0.502763 | 0.844507 | 0 | 0 | 1.19663 | 0.822846 |
| Gene_Pert | Hmgn1 KO | 2/375 | 0.487441 | 0.844507 | 0 | 0 | 1.232613 | 0.885738 |
| Gene_Pert | Pdk1 knock | 2/386 | 0.502763 | 0.844507 | 0 | 0 | 1.19663 | 0.822846 |
| Gene_Pert | Klf3 KO mo | 2/393 | 0.512362 | 0.844507 | 0 | 0 | 1.174786 | 0.785607 |
| Gene_Pert | DROSHA KI | 1/164 | 0.51222 | 0.844507 | 0 | 0 | 1.408903 | 0.942558 |
| Gene_Pert | CTNNB1 VF | 2/391 | 0.509632 | 0.844507 | 0 | 0 | 1.180947 | 0.796037 |
| Gene_Pert | IRF1 OE hu | 2/391 | 0.509632 | 0.844507 | 0 | 0 | 1.180947 | 0.796037 |
| Gene_Pert | Ikbkb KO m | 1/196 | 0.576271 | 0.844507 | 0 | 0 | 1.17579 | 0.64807 |
| Gene_Pert | Wt1 KO mo | 2/411 | 0.536491 | 0.844507 | 0 | 0 | 1.122048 | 0.698706 |
| Gene_Pert | ARID1A KD | 1/184 | 0.55329 | 0.844507 | 0 | 0 | 1.253654 | 0.742005 |
| Gene_Pert | IFNAR1 Stir | 2/442 | 0.576123 | 0.844507 | 0 | 0 | 1.041337 | 0.574228 |
| Gene_Pert | EWSR1 KD | 2/442 | 0.576123 | 0.844507 | 0 | 0 | 1.041337 | 0.574228 |
| Gene_Pert | SOX7 OE hu | 1/195 | 0.574401 | 0.844507 | 0 | 0 | 1.181911 | 0.655284 |
| Gene_Pert | FLI1 KD hu | 2/440 | 0.573641 | 0.844507 | 0 | 0 | 1.046199 | 0.581427 |
| Gene_Pert | Kdm1a KD | 1/194 | 0.572523 | 0.844507 | 0 | 0 | 1.188095 | 0.662603 |
| Gene_Pert | Hras Select | 2/439 | 0.572396 | 0.844507 | 0 | 0 | 1.048647 | 0.585066 |
| Gene_Pert | CHEK2 KD I | 1/179 | 0.543355 | 0.844507 | 0 | 0 | 1.289195 | 0.7864 |
| Gene_Pert | Bmpr1a KC | 2/435 | 0.56739 | 0.844507 | 0 | 0 | 1.058552 | 0.599891 |
| Gene_Pert | Smarca4 D | 1/184 | 0.55329 | 0.844507 | 0 | 0 | 1.253654 | 0.742005 |
| Gene_Pert | FTO OE hu | 1/184 | 0.55329 | 0.844507 | 0 | 0 | 1.253654 | 0.742005 |
| Gene_Pert | Suz12 KO n | 1/198 | 0.579985 | 0.844507 | 0 | 0 | 1.163735 | 0.633948 |
| Gene_Pert | UBB KO mo | 1/198 | 0.579985 | 0.844507 | 0 | 0 | 1.163735 | 0.633948 |
| Gene_Pert | Rara KO m | 2/444 | 0.578595 | 0.844507 | 0 | 0 | 1.036518 | 0.567133 |
| Gene_Pert | Atxn7 OE n | 2/444 | 0.578595 | 0.844507 | 0 | 0 | 1.036518 | 0.567133 |
| Gene_Pert | Lhb OE mo | 1/197 | 0.578132 | 0.844507 | 0 | 0 | 1.169732 | 0.640958 |
| Gene_Pert | NRAS DEPL | 1/197 | 0.578132 | 0.844507 | 0 | 0 | 1.169732 | 0.640958 |
| Gene_Pert | Frzb KO mo | 1/197 | 0.578132 | 0.844507 | 0 | 0 | 1.169732 | 0.640958 |
| Gene_Pert | Atf2 KO mo | 1/203 | 0.589132 | 0.844507 | 0 | 0 | 1.134642 | 0.600344 |
| Gene_Pert | Dmd Defici | 1/203 | 0.589132 | 0.844507 | 0 | 0 | 1.134642 | 0.600344 |
| Gene_Pert | Ncoa2 KO r | 1/202 | 0.587319 | 0.844507 | 0 | 0 | 1.140345 | 0.606877 |
| Gene_Pert | LMO3 KD h | 1/201 | 0.585497 | 0.844507 | 0 | 0 | 1.146105 | 0.613503 |
| Gene_Pert | Dmd Defici | 2/419 | 0.546954 | 0.844507 | 0 | 0 | 1.100071 | 0.663773 |
| Gene_Pert | Rp1 KO mo | 2/393 | 0.512362 | 0.844507 | 0 | 0 | 1.174786 | 0.785607 |
| Gene_Pert | Igf1r drugir | 1/172 | 0.529077 | 0.844507 | 0 | 0 | 1.342445 | 0.854628 |
| Gene_Pert | Dicer1 KD r | 2/414 | 0.540433 | 0.844507 | 0 | 0 | 1.113706 | 0.685357 |
| Gene_Pert | Akt1 KO m | 2/413 | 0.539122 | 0.844507 | 0 | 0 | 1.116473 | 0.689773 |
| Gene_Pert | Ffar4 KO m | 2/412 | 0.537807 | 0.844507 | 0 | 0 | 1.119254 | 0.694223 |
| Gene_Pert | Pde6b rd1 | 1/176 | 0.537289 | 0.844507 | 0 | 0 | 1.311495 | 0.814726 |

| | | | | | | | | |
|---|---|---|---|---|---|---|---|---|
| Gene_Pert | RNF2 KD hu | 1/176 | 0.537289 | 0.844507 | 0 | 0 | 1.311495 | 0.814726 |
| Gene_Pert | NFE2L2 KD | 2/411 | 0.536491 | 0.844507 | 0 | 0 | 1.122048 | 0.698706 |
| Gene_Pert | Pou5f1 KD | 2/399 | 0.520494 | 0.844507 | 0 | 0 | 1.156675 | 0.755281 |
| Gene_Pert | Notch1 KO | 1/175 | 0.535249 | 0.844507 | 0 | 0 | 1.319099 | 0.824467 |
| Gene_Pert | YY2 KD hur | 1/183 | 0.55132 | 0.844507 | 0 | 0 | 1.260606 | 0.750615 |
| Gene_Pert | Bhlha15 KC | 1/271 | 0.695622 | 0.844507 | 0 | 0 | 0.845952 | 0.307038 |
| Gene_Pert | Twist1 OE | 2/419 | 0.546954 | 0.844507 | 0 | 0 | 1.100071 | 0.663773 |
| Gene_Pert | MIR122 KC | 2/419 | 0.546954 | 0.844507 | 0 | 0 | 1.100071 | 0.663773 |
| Gene_Pert | Hif1an Defi | 1/180 | 0.545359 | 0.844507 | 0 | 0 | 1.281928 | 0.777247 |
| Gene_Pert | Eif2ak3 KO | 2/417 | 0.544353 | 0.844507 | 0 | 0 | 1.105485 | 0.672309 |
| Gene_Pert | Tsc1 KO mc | 2/406 | 0.529869 | 0.844507 | 0 | 0 | 1.136226 | 0.721646 |
| Gene_Pert | Ing2 KO mc | 1/178 | 0.541341 | 0.844507 | 0 | 0 | 1.296544 | 0.795696 |
| Gene_Pert | NRAS DEPL | 1/190 | 0.56493 | 0.844507 | 0 | 0 | 1.213486 | 0.692965 |
| Gene_Pert | Pou4f1 KO | 2/433 | 0.564871 | 0.844507 | 0 | 0 | 1.063573 | 0.607468 |
| Gene_Pert | PTEN KD hu | 2/431 | 0.562342 | 0.844507 | 0 | 0 | 1.068641 | 0.615158 |
| Gene_Pert | Ffar4 KO m | 2/429 | 0.559803 | 0.844507 | 0 | 0 | 1.073757 | 0.622962 |
| Gene_Pert | Lmx1b KO | 1/185 | 0.555251 | 0.844507 | 0 | 0 | 1.246777 | 0.733523 |
| Gene_Pert | Kdm6a KD | 1/345 | 0.78066 | 0.844507 | 0 | 0 | 0.661472 | 0.163791 |
| Gene_Pert | SOX2 Defic | 1/258 | 0.677628 | 0.844507 | 0 | 0 | 0.889331 | 0.346089 |
| Gene_Pert | Nfe2l2 KO | 1/260 | 0.680464 | 0.844507 | 0 | 0 | 0.882374 | 0.339697 |
| Gene_Pert | TNKS INHIE | 1/260 | 0.680464 | 0.844507 | 0 | 0 | 0.882374 | 0.339697 |
| Gene_Pert | PDE1C KD I | 2/535 | 0.680112 | 0.844507 | 0 | 0 | 0.855535 | 0.329807 |
| Gene_Pert | Jak1 drugir | 1/259 | 0.679049 | 0.844507 | 0 | 0 | 0.885839 | 0.342875 |
| Gene_Pert | Lin28a KD | 1/258 | 0.677628 | 0.844507 | 0 | 0 | 0.889331 | 0.346089 |
| Gene_Pert | GSK3A KD I | 1/258 | 0.677628 | 0.844507 | 0 | 0 | 0.889331 | 0.346089 |
| Gene_Pert | Nfya KD mc | 1/258 | 0.677628 | 0.844507 | 0 | 0 | 0.889331 | 0.346089 |
| Gene_Pert | TAL1 KD hu | 1/258 | 0.677628 | 0.844507 | 0 | 0 | 0.889331 | 0.346089 |
| Gene_Pert | Ppara Defic | 1/346 | 0.781631 | 0.844507 | 0 | 0 | 0.659521 | 0.162488 |
| Gene_Pert | St6gal1 De | 1/345 | 0.78066 | 0.844507 | 0 | 0 | 0.661472 | 0.163791 |
| Gene_Pert | FOXA1 OE I | 1/345 | 0.78066 | 0.844507 | 0 | 0 | 0.661472 | 0.163791 |
| Gene_Pert | Satb1 KO n | 1/345 | 0.78066 | 0.844507 | 0 | 0 | 0.661472 | 0.163791 |
| Gene_Pert | Kdm2b OE | 1/345 | 0.78066 | 0.844507 | 0 | 0 | 0.661472 | 0.163791 |
| Gene_Pert | Sarm1 KO | 1/258 | 0.677628 | 0.844507 | 0 | 0 | 0.889331 | 0.346089 |
| Gene_Pert | CDH1 OE h | 1/344 | 0.779685 | 0.844507 | 0 | 0 | 0.663435 | 0.165106 |
| Gene_Pert | CDK8 knocl | 1/344 | 0.779685 | 0.844507 | 0 | 0 | 0.663435 | 0.165106 |
| Gene_Pert | SYNCRIP OI | 1/347 | 0.782598 | 0.844507 | 0 | 0 | 0.657582 | 0.161197 |
| Gene_Pert | Itk KO mou | 1/347 | 0.782598 | 0.844507 | 0 | 0 | 0.657582 | 0.161197 |
| Gene_Pert | Furin Defic | 1/347 | 0.782598 | 0.844507 | 0 | 0 | 0.657582 | 0.161197 |
| Gene_Pert | Myc knock | 1/347 | 0.782598 | 0.844507 | 0 | 0 | 0.657582 | 0.161197 |
| Gene_Pert | Itk KO mou | 1/347 | 0.782598 | 0.844507 | 0 | 0 | 0.657582 | 0.161197 |
| Gene_Pert | Vdr KO mo | 1/347 | 0.782598 | 0.844507 | 0 | 0 | 0.657582 | 0.161197 |
| Gene_Pert | POU1F1 OE | 1/346 | 0.781631 | 0.844507 | 0 | 0 | 0.659521 | 0.162488 |
| Gene_Pert | Bcl11a KO | 1/346 | 0.781631 | 0.844507 | 0 | 0 | 0.659521 | 0.162488 |
| Gene_Pert | CEBPA VR I | 1/350 | 0.785472 | 0.844507 | 0 | 0 | 0.651829 | 0.157397 |
| Gene_Pert | BRCA1 DEP | 1/349 | 0.784518 | 0.844507 | 0 | 0 | 0.653736 | 0.158652 |

| | | | | | | | | |
|---|---|---|---|---|---|---|---|---|
| Gene_Pert | Ppard KO n | 1/349 | 0.784518 | 0.844507 | 0 | 0 | 0.653736 | 0.158652 |
| Gene_Pert | AR KD hum | 1/254 | 0.671883 | 0.844507 | 0 | 0 | 0.903576 | 0.359325 |
| Gene_Pert | ESR1 KD hu | 1/253 | 0.670431 | 0.844507 | 0 | 0 | 0.907207 | 0.362732 |
| Gene_Pert | Vdr KO mo | 1/251 | 0.667508 | 0.844507 | 0 | 0 | 0.914558 | 0.369667 |
| Gene_Pert | Myb KO mo | 1/251 | 0.667508 | 0.844507 | 0 | 0 | 0.914558 | 0.369667 |
| Gene_Pert | Traf3 KO m | 1/251 | 0.667508 | 0.844507 | 0 | 0 | 0.914558 | 0.369667 |
| Gene_Pert | FOXP1 KD l | 1/241 | 0.652504 | 0.844507 | 0 | 0 | 0.953149 | 0.406935 |
| Gene_Pert | BST2 OE hu | 1/253 | 0.670431 | 0.844507 | 0 | 0 | 0.907207 | 0.362732 |
| Gene_Pert | Tlr4 mutati | 1/253 | 0.670431 | 0.844507 | 0 | 0 | 0.907207 | 0.362732 |
| Gene_Pert | Chrna4 KD | 1/252 | 0.668973 | 0.844507 | 0 | 0 | 0.910868 | 0.366179 |
| Gene_Pert | TAF1 KD hu | 1/252 | 0.668973 | 0.844507 | 0 | 0 | 0.910868 | 0.366179 |
| Gene_Pert | CCL3 KO m | 1/255 | 0.673329 | 0.844507 | 0 | 0 | 0.899973 | 0.355958 |
| Gene_Pert | Tcf3 Deficie | 1/255 | 0.673329 | 0.844507 | 0 | 0 | 0.899973 | 0.355958 |
| Gene_Pert | Nfia Deficie | 1/255 | 0.673329 | 0.844507 | 0 | 0 | 0.899973 | 0.355958 |
| Gene_Pert | Dhcr7 KO n | 1/255 | 0.673329 | 0.844507 | 0 | 0 | 0.899973 | 0.355958 |
| Gene_Pert | EGFR drugi | 1/349 | 0.784518 | 0.844507 | 0 | 0 | 0.653736 | 0.158652 |
| Gene_Pert | ERBB2 OE h | 1/254 | 0.671883 | 0.844507 | 0 | 0 | 0.903576 | 0.359325 |
| Gene_Pert | MIR210 KD | 1/254 | 0.671883 | 0.844507 | 0 | 0 | 0.903576 | 0.359325 |
| Gene_Pert | Apod KO m | 1/254 | 0.671883 | 0.844507 | 0 | 0 | 0.903576 | 0.359325 |
| Gene_Pert | SIN3A Siler | 1/258 | 0.677628 | 0.844507 | 0 | 0 | 0.889331 | 0.346089 |
| Gene_Pert | TOP2B Inhi | 1/258 | 0.677628 | 0.844507 | 0 | 0 | 0.889331 | 0.346089 |
| Gene_Pert | E2F1 OE hu | 1/257 | 0.676202 | 0.844507 | 0 | 0 | 0.892851 | 0.34934 |
| Gene_Pert | Trps1 Muta | 1/257 | 0.676202 | 0.844507 | 0 | 0 | 0.892851 | 0.34934 |
| Gene_Pert | Nat6 KD m | 1/257 | 0.676202 | 0.844507 | 0 | 0 | 0.892851 | 0.34934 |
| Gene_Pert | TARDBP KD | 1/256 | 0.674769 | 0.844507 | 0 | 0 | 0.896398 | 0.35263 |
| Gene_Pert | Pafah1b1 K | 1/256 | 0.674769 | 0.844507 | 0 | 0 | 0.896398 | 0.35263 |
| Gene_Pert | Por KO mo | 1/256 | 0.674769 | 0.844507 | 0 | 0 | 0.896398 | 0.35263 |
| Gene_Pert | Pmp22 KO | 1/259 | 0.679049 | 0.844507 | 0 | 0 | 0.885839 | 0.342875 |
| Gene_Pert | Il10 KO mo | 1/259 | 0.679049 | 0.844507 | 0 | 0 | 0.885839 | 0.342875 |
| Gene_Pert | Nxnl1 Defic | 1/268 | 0.691559 | 0.844507 | 0 | 0 | 0.855587 | 0.315546 |
| Gene_Pert | MAPK7 KD | 1/268 | 0.691559 | 0.844507 | 0 | 0 | 0.855587 | 0.315546 |
| Gene_Pert | Grhl2 KD m | 1/267 | 0.690194 | 0.844507 | 0 | 0 | 0.858848 | 0.318446 |
| Gene_Pert | FOXP1 KD l | 1/267 | 0.690194 | 0.844507 | 0 | 0 | 0.858848 | 0.318446 |
| Gene_Pert | NFXL1 KD h | 1/267 | 0.690194 | 0.844507 | 0 | 0 | 0.858848 | 0.318446 |
| Gene_Pert | Mut KO mo | 1/267 | 0.690194 | 0.844507 | 0 | 0 | 0.858848 | 0.318446 |
| Gene_Pert | Nr3c1 KO r | 1/266 | 0.688822 | 0.844507 | 0 | 0 | 0.862133 | 0.32138 |
| Gene_Pert | Ppargc1a K | 1/265 | 0.687444 | 0.844507 | 0 | 0 | 0.865442 | 0.324346 |
| Gene_Pert | Nfe2l2 KO | 1/265 | 0.687444 | 0.844507 | 0 | 0 | 0.865442 | 0.324346 |
| Gene_Pert | Tet2 KO mo | 1/269 | 0.692919 | 0.844507 | 0 | 0 | 0.852352 | 0.312678 |
| Gene_Pert | Ezh2 defici | 1/269 | 0.692919 | 0.844507 | 0 | 0 | 0.852352 | 0.312678 |
| Gene_Pert | ZNF750 KD | 1/269 | 0.692919 | 0.844507 | 0 | 0 | 0.852352 | 0.312678 |
| Gene_Pert | Rarb KO mo | 1/269 | 0.692919 | 0.844507 | 0 | 0 | 0.852352 | 0.312678 |
| Gene_Pert | Krt1 KO mo | 1/268 | 0.691559 | 0.844507 | 0 | 0 | 0.855587 | 0.315546 |
| Gene_Pert | SIRT1 OE h | 1/263 | 0.68467 | 0.844507 | 0 | 0 | 0.872137 | 0.330382 |
| Gene_Pert | Il10 KO mo | 1/268 | 0.691559 | 0.844507 | 0 | 0 | 0.855587 | 0.315546 |

| | | | | | | | | |
|---|---|---|---|---|---|---|---|---|
| Gene_Pert | Mnat1 Def | 1/268 | 0.691559 | 0.844507 | 0 | 0 | 0.855587 | 0.315546 |
| Gene_Pert | Gata4 KO r | 1/270 | 0.694273 | 0.844507 | 0 | 0 | 0.84914 | 0.309842 |
| Gene_Pert | Pmp22 OE | 1/270 | 0.694273 | 0.844507 | 0 | 0 | 0.84914 | 0.309842 |
| Gene_Pert | Clcn5 KO m | 1/270 | 0.694273 | 0.844507 | 0 | 0 | 0.84914 | 0.309842 |
| Gene_Pert | Snca KO m | 1/270 | 0.694273 | 0.844507 | 0 | 0 | 0.84914 | 0.309842 |
| Gene_Pert | HIF1A KD h | 1/269 | 0.692919 | 0.844507 | 0 | 0 | 0.852352 | 0.312678 |
| Gene_Pert | Rpe65 KD r | 1/269 | 0.692919 | 0.844507 | 0 | 0 | 0.852352 | 0.312678 |
| Gene_Pert | JAG1 OE hu | 1/269 | 0.692919 | 0.844507 | 0 | 0 | 0.852352 | 0.312678 |
| Gene_Pert | TP63 huma | 1/269 | 0.692919 | 0.844507 | 0 | 0 | 0.852352 | 0.312678 |
| Gene_Pert | FGFR1 mut | 1/272 | 0.696964 | 0.844507 | 0 | 0 | 0.842787 | 0.304265 |
| Gene_Pert | SNCA SNCA | 1/271 | 0.695622 | 0.844507 | 0 | 0 | 0.845952 | 0.307038 |
| Gene_Pert | SUZ12 DEP | 1/271 | 0.695622 | 0.844507 | 0 | 0 | 0.845952 | 0.307038 |
| Gene_Pert | Fgfr1 active | 1/263 | 0.68467 | 0.844507 | 0 | 0 | 0.872137 | 0.330382 |
| Gene_Pert | Pla2g4a KC | 1/349 | 0.784518 | 0.844507 | 0 | 0 | 0.653736 | 0.158652 |
| Gene_Pert | ZNF217 OE | 1/349 | 0.784518 | 0.844507 | 0 | 0 | 0.653736 | 0.158652 |
| Gene_Pert | Pink1 KO m | 1/349 | 0.784518 | 0.844507 | 0 | 0 | 0.653736 | 0.158652 |
| Gene_Pert | Irs2 KO mo | 1/348 | 0.78356 | 0.844507 | 0 | 0 | 0.655653 | 0.159919 |
| Gene_Pert | HSF1 KD hu | 1/351 | 0.786422 | 0.844507 | 0 | 0 | 0.649934 | 0.156154 |
| Gene_Pert | Aldh1a2 KC | 1/351 | 0.786422 | 0.844507 | 0 | 0 | 0.649934 | 0.156154 |
| Gene_Pert | Tcf12 KO m | 1/351 | 0.786422 | 0.844507 | 0 | 0 | 0.649934 | 0.156154 |
| Gene_Pert | Pink1 KO m | 1/350 | 0.785472 | 0.844507 | 0 | 0 | 0.651829 | 0.157397 |
| Gene_Pert | TCF4 DN hu | 1/350 | 0.785472 | 0.844507 | 0 | 0 | 0.651829 | 0.157397 |
| Gene_Pert | FLI1 KD hur | 1/344 | 0.779685 | 0.844507 | 0 | 0 | 0.663435 | 0.165106 |
| Gene_Pert | Dmd Defici | 1/344 | 0.779685 | 0.844507 | 0 | 0 | 0.663435 | 0.165106 |
| Gene_Pert | Irs2 KO mo | 1/343 | 0.778706 | 0.844507 | 0 | 0 | 0.665409 | 0.166434 |
| Gene_Pert | Htt Mutatio | 1/263 | 0.68467 | 0.844507 | 0 | 0 | 0.872137 | 0.330382 |
| Gene_Pert | Hnf1a KO r | 1/251 | 0.667508 | 0.844507 | 0 | 0 | 0.914558 | 0.369667 |
| Gene_Pert | Erbb2 drug | 1/263 | 0.68467 | 0.844507 | 0 | 0 | 0.872137 | 0.330382 |
| Gene_Pert | Lat2 KD mc | 1/263 | 0.68467 | 0.844507 | 0 | 0 | 0.872137 | 0.330382 |
| Gene_Pert | Kdm1a KD | 1/262 | 0.683274 | 0.844507 | 0 | 0 | 0.875523 | 0.333451 |
| Gene_Pert | CCND1 KD | 1/262 | 0.683274 | 0.844507 | 0 | 0 | 0.875523 | 0.333451 |
| Gene_Pert | Neurog3 O | 1/262 | 0.683274 | 0.844507 | 0 | 0 | 0.875523 | 0.333451 |
| Gene_Pert | Ppargc1a D | 1/261 | 0.681872 | 0.844507 | 0 | 0 | 0.878936 | 0.336556 |
| Gene_Pert | Gtf2i KO m | 1/265 | 0.687444 | 0.844507 | 0 | 0 | 0.865442 | 0.324346 |
| Gene_Pert | SYK DEPLET | 1/265 | 0.687444 | 0.844507 | 0 | 0 | 0.865442 | 0.324346 |
| Gene_Pert | SYK drugin | 1/264 | 0.68606 | 0.844507 | 0 | 0 | 0.868777 | 0.327347 |
| Gene_Pert | Pax3 Knock | 1/264 | 0.68606 | 0.844507 | 0 | 0 | 0.868777 | 0.327347 |
| Gene_Pert | Rora KO m | 1/264 | 0.68606 | 0.844507 | 0 | 0 | 0.868777 | 0.327347 |
| Gene_Pert | MED1 OE h | 1/264 | 0.68606 | 0.844507 | 0 | 0 | 0.868777 | 0.327347 |
| Gene_Pert | ETS2 KD hu | 1/264 | 0.68606 | 0.844507 | 0 | 0 | 0.868777 | 0.327347 |
| Gene_Pert | Lat2 KO mc | 1/248 | 0.663076 | 0.844507 | 0 | 0 | 0.925807 | 0.380383 |
| Gene_Pert | Ndel1 KD n | 1/251 | 0.667508 | 0.844507 | 0 | 0 | 0.914558 | 0.369667 |
| Gene_Pert | Dlx5 KO mc | 1/250 | 0.666037 | 0.844507 | 0 | 0 | 0.918278 | 0.373197 |
| Gene_Pert | MIR22 OE l | 1/246 | 0.660088 | 0.844507 | 0 | 0 | 0.93346 | 0.387742 |
| Gene_Pert | Lat2 KO mc | 1/247 | 0.661585 | 0.844507 | 0 | 0 | 0.929618 | 0.384041 |

| | | | | | | | | |
|---|---|---|---|---|---|---|---|---|
| Gene_Pert | Nfe2l2 KO | 1/247 | 0.661585 | 0.844507 | 0 | 0 | 0.929618 | 0.384041 |
| Gene_Pert | Hmgn1 KO | 1/247 | 0.661585 | 0.844507 | 0 | 0 | 0.929618 | 0.384041 |
| Gene_Pert | Mecp2 KO | 1/247 | 0.661585 | 0.844507 | 0 | 0 | 0.929618 | 0.384041 |
| Gene_Pert | CDK4 knocl | 1/251 | 0.667508 | 0.844507 | 0 | 0 | 0.914558 | 0.369667 |
| Gene_Pert | SDC1 KD hu | 1/251 | 0.667508 | 0.844507 | 0 | 0 | 0.914558 | 0.369667 |
| Gene_Pert | Map3k8 KC | 1/244 | 0.657075 | 0.844507 | 0 | 0 | 0.941238 | 0.39528 |
| Gene_Pert | Htt Mutati | 1/244 | 0.657075 | 0.844507 | 0 | 0 | 0.941238 | 0.39528 |
| Gene_Pert | Npc1 KO m | 1/244 | 0.657075 | 0.844507 | 0 | 0 | 0.941238 | 0.39528 |
| Gene_Pert | Hdc KO mo | 1/253 | 0.670431 | 0.844507 | 0 | 0 | 0.907207 | 0.362732 |
| Gene_Pert | Sod2 KO m | 1/245 | 0.658585 | 0.844507 | 0 | 0 | 0.937333 | 0.391489 |
| Gene_Pert | Lat2 KD mc | 1/245 | 0.658585 | 0.844507 | 0 | 0 | 0.937333 | 0.391489 |
| Gene_Pert | Nrl KO mou | 1/250 | 0.666037 | 0.844507 | 0 | 0 | 0.918278 | 0.373197 |
| Gene_Pert | Ptpn2 KD n | 1/250 | 0.666037 | 0.844507 | 0 | 0 | 0.918278 | 0.373197 |
| Gene_Pert | PTEN KD hu | 1/241 | 0.652504 | 0.844507 | 0 | 0 | 0.953149 | 0.406935 |
| Gene_Pert | UBR5 DEPL | 1/250 | 0.666037 | 0.844507 | 0 | 0 | 0.918278 | 0.373197 |
| Gene_Pert | Klf5 KO mo | 1/253 | 0.670431 | 0.844507 | 0 | 0 | 0.907207 | 0.362732 |
| Gene_Pert | Gsk3a KO r | 1/254 | 0.671883 | 0.844507 | 0 | 0 | 0.903576 | 0.359325 |
| Gene_Pert | Trp53inp2 | 1/252 | 0.668973 | 0.844507 | 0 | 0 | 0.910868 | 0.366179 |
| Gene_Pert | TYK2 KD hu | 1/248 | 0.663076 | 0.844507 | 0 | 0 | 0.925807 | 0.380383 |
| Gene_Pert | Tcf3 KO mc | 1/249 | 0.66456 | 0.844507 | 0 | 0 | 0.922027 | 0.376768 |
| Gene_Pert | Mtf2 KD m | 1/249 | 0.66456 | 0.844507 | 0 | 0 | 0.922027 | 0.376768 |
| Gene_Pert | PTHLH siRN | 2/521 | 0.665878 | 0.844507 | 0 | 0 | 0.879247 | 0.357545 |
| GO_Biologi | Vesicle-Me | 1/411 | 0.836412 | 0.84474 | 0 | 0 | 0.55312 | 0.098806 |
| Gene_Pert | Il4 KO mou | 1/404 | 0.831236 | 0.84474 | 0 | 0 | 0.562929 | 0.104053 |
| Gene_Pert | Dicer1 KO r | 1/406 | 0.832731 | 0.84484 | 0 | 0 | 0.560092 | 0.102522 |
| Gene_Pert | SOX7 OE hu | 1/405 | 0.831985 | 0.84484 | 0 | 0 | 0.561507 | 0.103284 |
| Gene_Pert | Kdm1a KD | 1/406 | 0.832731 | 0.84484 | 0 | 0 | 0.560092 | 0.102522 |
| Gene_Pert | TNF Stimul | 1/366 | 0.80018 | 0.844949 | 0 | 0 | 0.622746 | 0.138822 |
| GTEx_Tissu | GTEX-TKQ2 | 29/5280 | 0.090876 | 0.845349 | 0 | 0 | 1.396115 | 3.348249 |
| GTEx_Tissu | GTEX-VUSC | 7/886 | 0.090866 | 0.845349 | 0 | 0 | 1.894738 | 4.544287 |
| GTEx_Tissu | GTEX-OXRF | 6/714 | 0.090788 | 0.845349 | 0 | 0 | 2.009312 | 4.820806 |
| GeneSigDB | 17676974-1 | 1/364 | 0.798398 | 0.845452 | 0 | 0 | 0.626241 | 0.140997 |
| Gene_Pert | Pten PTEN | 1/368 | 0.801947 | 0.845818 | 0 | 0 | 0.619289 | 0.136685 |
| Gene_Pert | SYK druginl | 1/368 | 0.801947 | 0.845818 | 0 | 0 | 0.619289 | 0.136685 |
| Gene_Pert | Nsun2 KO r | 1/408 | 0.834213 | 0.845871 | 0 | 0 | 0.557282 | 0.101017 |
| Jensen_TIS | Thalamus | 18/4856 | 0.817117 | 0.846018 | 0 | 0 | 0.812858 | 0.164175 |
| Gene_Pert | Hey2 KO m | 1/372 | 0.805435 | 0.84624 | 0 | 0 | 0.612487 | 0.132526 |
| Gene_Pert | Il10 KO mo | 1/375 | 0.808011 | 0.84624 | 0 | 0 | 0.60748 | 0.129503 |
| Gene_Pert | Polg KO mc | 1/375 | 0.808011 | 0.84624 | 0 | 0 | 0.60748 | 0.129503 |
| Gene_Pert | HNRNPU-A | 1/372 | 0.805435 | 0.84624 | 0 | 0 | 0.612487 | 0.132526 |
| Gene_Pert | Htt Mutati | 1/372 | 0.805435 | 0.84624 | 0 | 0 | 0.612487 | 0.132526 |
| Gene_Pert | OLR1 OE hu | 1/372 | 0.805435 | 0.84624 | 0 | 0 | 0.612487 | 0.132526 |
| Gene_Pert | Kras Knock | 1/373 | 0.806297 | 0.84624 | 0 | 0 | 0.610809 | 0.131509 |
| Gene_Pert | Met knock | 1/373 | 0.806297 | 0.84624 | 0 | 0 | 0.610809 | 0.131509 |
| Gene_Pert | FOXL2 OE h | 1/374 | 0.807156 | 0.84624 | 0 | 0 | 0.60914 | 0.130501 |

| | | | | | | | | |
|---|---|---|---|---|---|---|---|---|
| Gene_Pert | IL2 OE hum | 1/373 | 0.806297 | 0.84624 | 0 | 0 | 0.610809 | 0.131509 |
| Gene_Pert | FOXP1 OE l | 1/370 | 0.803699 | 0.84624 | 0 | 0 | 0.615869 | 0.134586 |
| Gene_Pert | FLI1 KD hu | 1/375 | 0.808011 | 0.84624 | 0 | 0 | 0.60748 | 0.129503 |
| Disease_Pe | Bacterial In | 1/385 | 0.816356 | 0.846494 | 0 | 0 | 0.591358 | 0.119989 |
| Gene_Pert | FOXL2 OE h | 1/376 | 0.808862 | 0.846637 | 0 | 0 | 0.605829 | 0.128513 |
| DisGeNET | Malignant | 1/421 | 0.843535 | 0.846767 | 0 | 0 | 0.539673 | 0.091827 |
| Disease_Pe | Meningocc | 1/387 | 0.817981 | 0.84682 | 0 | 0 | 0.588234 | 0.118185 |
| GTEx_Tissu | GTEX-WFG | 5/551 | 0.092214 | 0.846909 | 0 | 0 | 2.162847 | 5.155447 |
| GTEx_Tissu | GTEX-XUZC | 6/717 | 0.09218 | 0.846909 | 0 | 0 | 2.000521 | 4.769276 |
| GTEx_Tissu | GTEX-OOB. | 8/1065 | 0.091711 | 0.846909 | 0 | 0 | 1.806498 | 4.315924 |
| GTEx_Tissu | GTEX-PWO | 5/550 | 0.091677 | 0.846909 | 0 | 0 | 2.166928 | 5.17784 |
| Gene_Pert | Pycard KO | 1/378 | 0.810553 | 0.846924 | 0 | 0 | 0.602554 | 0.12656 |
| Gene_Pert | IRF4 KD hu | 1/378 | 0.810553 | 0.846924 | 0 | 0 | 0.602554 | 0.12656 |
| Gene_Pert | Prkcz KO m | 1/377 | 0.809709 | 0.846924 | 0 | 0 | 0.604187 | 0.127532 |
| GWAS_Cat | Platelet Dis | 1/422 | 0.844231 | 0.847008 | 0 | 0 | 0.538364 | 0.091161 |
| Disease_Pe | osteoarthri | 1/394 | 0.823559 | 0.847164 | 0 | 0 | 0.577549 | 0.112114 |
| Disease_Pe | Waldenstro | 1/393 | 0.822773 | 0.847164 | 0 | 0 | 0.579052 | 0.112959 |
| Disease_Pe | Parkinson's | 1/393 | 0.822773 | 0.847164 | 0 | 0 | 0.579052 | 0.112959 |
| Disease_Pe | systemic lu | 1/390 | 0.820393 | 0.847164 | 0 | 0 | 0.583607 | 0.115538 |
| RNA-Seq_D | Erlotinib 11 | 7/840 | 0.073066 | 0.847273 | 0 | 0 | 2.004202 | 5.243772 |
| Gene_Pert | Dmd Defici | 1/379 | 0.811393 | 0.847308 | 0 | 0 | 0.600929 | 0.125596 |
| Gene_Pert | HMGA2 OE | 1/382 | 0.813891 | 0.847686 | 0 | 0 | 0.596106 | 0.122756 |
| Gene_Pert | Pink1 KO m | 1/382 | 0.813891 | 0.847686 | 0 | 0 | 0.596106 | 0.122756 |
| Gene_Pert | Retnlb KO | 1/384 | 0.815538 | 0.847686 | 0 | 0 | 0.592932 | 0.120903 |
| Gene_Pert | Btk KO mo | 1/384 | 0.815538 | 0.847686 | 0 | 0 | 0.592932 | 0.120903 |
| Gene_Pert | Btk Missen | 1/381 | 0.813062 | 0.847686 | 0 | 0 | 0.597705 | 0.123694 |
| Gene_Pert | MYC KD hu | 1/384 | 0.815538 | 0.847686 | 0 | 0 | 0.592932 | 0.120903 |
| Gene_Pert | RB1 KD hu | 1/383 | 0.814716 | 0.847686 | 0 | 0 | 0.594515 | 0.121825 |
| Gene_Pert | Nr0b2 KO r | 1/382 | 0.813891 | 0.847686 | 0 | 0 | 0.596106 | 0.122756 |
| Jensen_TIS | Subthalam | 16/4383 | 0.822035 | 0.848382 | 0 | 0 | 0.802227 | 0.157215 |
| DisGeNET | Alcoholic In | 1/424 | 0.845612 | 0.848527 | 0 | 0 | 0.535763 | 0.089845 |
| PhenGenI_ | Insulin Resi | 1/315 | 0.749464 | 0.848658 | 0 | 0 | 0.725781 | 0.209313 |
| Gene_Pert | BST2 OE hu | 1/386 | 0.81717 | 0.848891 | 0 | 0 | 0.589792 | 0.119083 |
| Gene_Pert | MIR124-1 ( | 1/415 | 0.839299 | 0.849378 | 0 | 0 | 0.547663 | 0.095944 |
| Gene_Pert | Smarca4 D | 1/416 | 0.840013 | 0.849378 | 0 | 0 | 0.546315 | 0.095244 |
| Gene_Pert | Smarca4 K( | 1/415 | 0.839299 | 0.849378 | 0 | 0 | 0.547663 | 0.095944 |
| Gene_Pert | FTO OE hu | 1/416 | 0.840013 | 0.849378 | 0 | 0 | 0.546315 | 0.095244 |
| Gene_Pert | Ncoa3 KD r | 1/416 | 0.840013 | 0.849378 | 0 | 0 | 0.546315 | 0.095244 |
| Gene_Pert | BRAF overe | 1/388 | 0.818789 | 0.849588 | 0 | 0 | 0.586683 | 0.117295 |
| Gene_Pert | ERBB2 over | 1/388 | 0.818789 | 0.849588 | 0 | 0 | 0.586683 | 0.117295 |
| Gene_Pert | Il1rn KO m | 1/391 | 0.82119 | 0.849951 | 0 | 0 | 0.582081 | 0.114671 |
| Gene_Pert | FLI1 KD hu | 1/391 | 0.82119 | 0.849951 | 0 | 0 | 0.582081 | 0.114671 |
| Gene_Pert | Ifng KO mo | 1/392 | 0.821983 | 0.849951 | 0 | 0 | 0.580563 | 0.113811 |
| Gene_Pert | Arid3b KO | 1/390 | 0.820393 | 0.849951 | 0 | 0 | 0.583607 | 0.115538 |
| Gene_Pert | UPF1 KD hu | 1/392 | 0.821983 | 0.849951 | 0 | 0 | 0.580563 | 0.113811 |

| | | | | | | | | |
|---|---|---|---|---|---|---|---|---|
| Gene_Pert | Ash2l KD m | 1/392 | 0.821983 | 0.849951 | 0 | 0 | 0.580563 | 0.113811 |
| GTEx_Tissu | GTEX-SNOS | 8/1068 | 0.092843 | 0.849987 | 0 | 0 | 1.801099 | 4.280926 |
| Disease_Pe | idiopathic | 1/401 | 0.828968 | 0.850021 | 0 | 0 | 0.567238 | 0.106399 |
| Disease_Pe | Type 1 diak | 1/400 | 0.828206 | 0.850021 | 0 | 0 | 0.568689 | 0.107194 |
| Disease_Pe | Atheroscle | 1/403 | 0.830483 | 0.850225 | 0 | 0 | 0.564358 | 0.104828 |
| Gene_Pert | CDK8 knocl | 1/393 | 0.822773 | 0.850278 | 0 | 0 | 0.579052 | 0.112959 |
| Disease_Pe | Acute panc | 1/412 | 0.837139 | 0.851036 | 0 | 0 | 0.551746 | 0.098081 |
| Disease_Pe | Infertility d | 1/413 | 0.837862 | 0.851036 | 0 | 0 | 0.550378 | 0.097363 |
| Disease_Pe | Neurologic | 1/411 | 0.836412 | 0.851036 | 0 | 0 | 0.55312 | 0.098806 |
| Disease_Pe | Sarcoidosis | 1/413 | 0.837862 | 0.851036 | 0 | 0 | 0.550378 | 0.097363 |
| Disease_Pe | idiopathic | 1/408 | 0.834213 | 0.851036 | 0 | 0 | 0.557282 | 0.101017 |
| Gene_Pert | Tsc1 Defici | 1/396 | 0.825122 | 0.851233 | 0 | 0 | 0.574566 | 0.110446 |
| Gene_Pert | IL1B Challe | 1/396 | 0.825122 | 0.851233 | 0 | 0 | 0.574566 | 0.110446 |
| Gene_Pert | ATM KD hu | 1/396 | 0.825122 | 0.851233 | 0 | 0 | 0.574566 | 0.110446 |
| Gene_Pert | BRCA1 DEF | 1/398 | 0.82667 | 0.851361 | 0 | 0 | 0.571613 | 0.108806 |
| Gene_Pert | Nfkbia KO | 1/398 | 0.82667 | 0.851361 | 0 | 0 | 0.571613 | 0.108806 |
| Gene_Pert | Zfp335 KD | 1/398 | 0.82667 | 0.851361 | 0 | 0 | 0.571613 | 0.108806 |
| GeneSigDB | 18614019- | 1/374 | 0.807156 | 0.851488 | 0 | 0 | 0.60914 | 0.130501 |
| GeneSigDB | 17899371- | 1/373 | 0.806297 | 0.851488 | 0 | 0 | 0.610809 | 0.131509 |
| Gene_Pert | Cdkn1b KO | 1/406 | 0.832731 | 0.85152 | 0 | 0 | 0.560092 | 0.102522 |
| Gene_Pert | RB1 KD hur | 1/402 | 0.829727 | 0.85152 | 0 | 0 | 0.565795 | 0.10561 |
| Gene_Pert | Pten KO m | 1/406 | 0.832731 | 0.85152 | 0 | 0 | 0.560092 | 0.102522 |
| Gene_Pert | Bmpr2 Del | 1/401 | 0.828968 | 0.85152 | 0 | 0 | 0.567238 | 0.106399 |
| Gene_Pert | FLI1 KD hur | 1/405 | 0.831985 | 0.85152 | 0 | 0 | 0.561507 | 0.103284 |
| Gene_Pert | Il13 OE mo | 1/405 | 0.831985 | 0.85152 | 0 | 0 | 0.561507 | 0.103284 |
| Gene_Pert | Il2 KO mou | 1/402 | 0.829727 | 0.85152 | 0 | 0 | 0.565795 | 0.10561 |
| Gene_Pert | Map3k1 KC | 1/406 | 0.832731 | 0.85152 | 0 | 0 | 0.560092 | 0.102522 |
| Gene_Pert | RB1 KD hur | 1/407 | 0.833474 | 0.85152 | 0 | 0 | 0.558684 | 0.101766 |
| Gene_Pert | MIR20A KD | 1/407 | 0.833474 | 0.85152 | 0 | 0 | 0.558684 | 0.101766 |
| Gene_Pert | Myd88 KO | 1/407 | 0.833474 | 0.85152 | 0 | 0 | 0.558684 | 0.101766 |
| Gene_Pert | HMGA2 KD | 1/402 | 0.829727 | 0.85152 | 0 | 0 | 0.565795 | 0.10561 |
| Gene_Pert | Muc2 &qu | 1/407 | 0.833474 | 0.85152 | 0 | 0 | 0.558684 | 0.101766 |
| Gene_Pert | ARHGDIB T | 1/402 | 0.829727 | 0.85152 | 0 | 0 | 0.565795 | 0.10561 |
| Gene_Pert | WNT1 OE h | 1/408 | 0.834213 | 0.851791 | 0 | 0 | 0.557282 | 0.101017 |
| Reactome_ | Generic Tra | 4/1190 | 0.768175 | 0.851837 | 0 | 0 | 0.760966 | 0.200696 |
| PhenGenI_ | Muscle, Ske | 1/332 | 0.767644 | 0.851841 | 0 | 0 | 0.687908 | 0.181903 |
| PhenGenI_ | Schizophre | 5/1453 | 0.766595 | 0.851841 | 0 | 0 | 0.777565 | 0.206675 |
| PhenGenI_ | Colitis, Ulc | 1/321 | 0.756036 | 0.851841 | 0 | 0 | 0.711955 | 0.19911 |
| PhenGenI_ | 3-Hydroxy- | 1/335 | 0.770714 | 0.851841 | 0 | 0 | 0.681625 | 0.177521 |
| PhenGenI_ | Child Devel | Oct-26 | 0.76451 | 0.851841 | 0 | 0 | 0.822304 | 0.220805 |
| GTEx_Tissu | GTEX-P44H | 3/344 | 0.188855 | 0.852032 | 0 | 0 | 2.049853 | 3.416651 |
| GTEx_Tissu | GTEX-U8XE | 18/3275 | 0.171215 | 0.852032 | 0 | 0 | 1.334063 | 2.354398 |
| GTEx_Tissu | GTEX-QDVI | 2/171 | 0.170583 | 0.852032 | 0 | 0 | 2.748904 | 4.86152 |
| GTEx_Tissu | GTEX-T5JC- | 18/3273 | 0.170552 | 0.852032 | 0 | 0 | 1.335043 | 2.361307 |
| GTEx_Tissu | GTEX-QCQ | 3/327 | 0.170462 | 0.852032 | 0 | 0 | 2.159281 | 3.820295 |

| | | | | | | | | |
|---|---|---|---|---|---|---|---|---|
| GTEx_Tissu | GTEX-UTH( | 5/697 | 0.186799 | 0.852032 | 0 | 0 | 1.693659 | 2.841486 |
| GTEx_Tissu | GTEX-UPIC | 3/341 | 0.185571 | 0.852032 | 0 | 0 | 2.068364 | 3.483785 |
| GTEx_Tissu | GTEX-PLZ4 | 4/521 | 0.191536 | 0.852032 | 0 | 0 | 1.808021 | 2.988077 |
| GTEx_Tissu | GTEX-XLM4 | 3/346 | 0.191052 | 0.852032 | 0 | 0 | 2.037693 | 3.372806 |
| GTEx_Tissu | GTEX-RTLS | 2/184 | 0.19085 | 0.852032 | 0 | 0 | 2.550873 | 4.224921 |
| GTEx_Tissu | GTEX-X4XX | 6/890 | 0.190787 | 0.852032 | 0 | 0 | 1.59452 | 2.64148 |
| GTEx_Tissu | GTEX-TMM | 27/5271 | 0.190597 | 0.852032 | 0 | 0 | 1.258781 | 2.086547 |
| GTEx_Tissu | GTEX-QDVI | 4/519 | 0.189776 | 0.852032 | 0 | 0 | 1.81523 | 3.016749 |
| GTEx_Tissu | GTEX-R55E | 2/180 | 0.184577 | 0.852032 | 0 | 0 | 2.608724 | 4.407929 |
| GTEx_Tissu | GTEX-NL3H | 44/9044 | 0.184522 | 0.852032 | 0 | 0 | 1.240755 | 2.096859 |
| GTEx_Tissu | GTEX-XOT( | 7/1049 | 0.171549 | 0.852032 | 0 | 0 | 1.584657 | 2.793571 |
| GTEx_Tissu | GTEX-WZT( | 42/8606 | 0.188656 | 0.852032 | 0 | 0 | 1.236852 | 2.062859 |
| GTEx_Tissu | GTEX-OXRI | 9/1469 | 0.187846 | 0.852032 | 0 | 0 | 1.458351 | 2.438553 |
| GTEx_Tissu | GTEX-Q734 | 3/343 | 0.187758 | 0.852032 | 0 | 0 | 2.055987 | 3.438845 |
| GTEx_Tissu | GTEX-XMD | 2/182 | 0.18771 | 0.852032 | 0 | 0 | 2.579477 | 4.315096 |
| GTEx_Tissu | GTEX-WH7 | 8/1271 | 0.187534 | 0.852032 | 0 | 0 | 1.495335 | 2.50288 |
| GTEx_Tissu | GTEX-RVPL | 25/4806 | 0.181923 | 0.852032 | 0 | 0 | 1.276221 | 2.174901 |
| GTEx_Tissu | GTEX-QDVI | 10/1656 | 0.181782 | 0.852032 | 0 | 0 | 1.441274 | 2.457297 |
| GTEx_Tissu | GTEX-S4Q7 | 36/7164 | 0.165404 | 0.852032 | 0 | 0 | 1.266092 | 2.278161 |
| GTEx_Tissu | GTEX-S33H | 13/2222 | 0.164983 | 0.852032 | 0 | 0 | 1.40795 | 2.537 |
| GTEx_Tissu | GTEX-XXEK | 4/490 | 0.164889 | 0.852032 | 0 | 0 | 1.926422 | 3.472345 |
| GTEx_Tissu | GTEX-RVPL | 14/2387 | 0.150804 | 0.852032 | 0 | 0 | 1.417546 | 2.681676 |
| GTEx_Tissu | GTEX-XUJ4 | 2/167 | 0.164425 | 0.852032 | 0 | 0 | 2.816114 | 5.083934 |
| GTEx_Tissu | GTEX-T5JC | 3/321 | 0.164103 | 0.852032 | 0 | 0 | 2.200696 | 3.977238 |
| GTEx_Tissu | GTEX-S3XE | 3/313 | 0.155741 | 0.852032 | 0 | 0 | 2.25841 | 4.199656 |
| GTEx_Tissu | GTEX-X261 | 22/4192 | 0.192503 | 0.852032 | 0 | 0 | 1.277794 | 2.105351 |
| GTEx_Tissu | GTEX-UTH( | 10/1617 | 0.163846 | 0.852032 | 0 | 0 | 1.479404 | 2.675992 |
| GTEx_Tissu | GTEX-XBED | Jan-41 | 0.163825 | 0.852032 | 0 | 0 | 5.777035 | 10.45039 |
| GTEx_Tissu | GTEX-U3ZH | 10/1629 | 0.169266 | 0.852032 | 0 | 0 | 1.467476 | 2.606651 |
| GTEx_Tissu | GTEX-Q2AI | 4/494 | 0.168247 | 0.852032 | 0 | 0 | 1.910302 | 3.404772 |
| GTEx_Tissu | GTEX-TML8 | 13/2229 | 0.167699 | 0.852032 | 0 | 0 | 1.402948 | 2.505083 |
| GTEx_Tissu | GTEX-WL4( | Dec-26 | 0.167671 | 0.852032 | 0 | 0 | 1.421966 | 2.539281 |
| GTEx_Tissu | GTEX-POM | 40/8069 | 0.167491 | 0.852032 | 0 | 0 | 1.259689 | 2.250844 |
| GTEx_Tissu | GTEX-RM2 | 3/323 | 0.166214 | 0.852032 | 0 | 0 | 2.186719 | 3.924018 |
| GTEx_Tissu | GTEX-XMD | 47/9585 | 0.150695 | 0.852032 | 0 | 0 | 1.278111 | 2.418818 |
| GTEx_Tissu | GTEX-QV31 | 3/339 | 0.18339 | 0.852032 | 0 | 0 | 2.080889 | 3.529475 |
| GTEx_Tissu | GTEX-XBEV | 2/179 | 0.183014 | 0.852032 | 0 | 0 | 2.623596 | 4.455375 |
| GTEx_Tissu | GTEX-PWO | 38/7680 | 0.182607 | 0.852032 | 0 | 0 | 1.245261 | 2.117468 |
| GTEx_Tissu | GTEX-TKQ1 | 3/338 | 0.182303 | 0.852032 | 0 | 0 | 2.087207 | 3.552606 |
| GTEx_Tissu | GTEX-X5EB | 52/10913 | 0.192617 | 0.852032 | 0 | 0 | 1.238255 | 2.039471 |
| GTEx_Tissu | GTEX-WZT( | 10/1619 | 0.164743 | 0.852032 | 0 | 0 | 1.477404 | 2.664304 |
| GTEx_Tissu | GTEX-TSE9 | 13/2265 | 0.18203 | 0.852032 | 0 | 0 | 1.377712 | 2.347048 |
| GTEx_Tissu | GTEX-XMD | Dec-12 | 0.162016 | 0.852032 | 0 | 0 | 1.43304 | 2.608217 |
| GTEx_Tissu | GTEX-QDV. | 6/845 | 0.161931 | 0.852032 | 0 | 0 | 1.684015 | 3.065892 |
| GTEx_Tissu | GTEX-R55E | 13/2213 | 0.161527 | 0.852032 | 0 | 0 | 1.414429 | 2.578623 |

| | | | | | | | | |
|---|---|---|---|---|---|---|---|---|
| GTEx_Tissu | GTEX-PX3G | 2/176 | 0.178336 | 0.852032 | 0 | 0 | 2.669236 | 4.601995 |
| GTEx_Tissu | GTEX-WRH | 18/3296 | 0.178262 | 0.852032 | 0 | 0 | 1.323845 | 2.28297 |
| GTEx_Tissu | GTEX-T8EM | 3/318 | 0.160951 | 0.852032 | 0 | 0 | 2.221995 | 4.058822 |
| GTEx_Tissu | GTEX-VUSG | 2/185 | 0.192423 | 0.852032 | 0 | 0 | 2.536805 | 4.180799 |
| GTEx_Tissu | GTEX-SNOS | 17/3123 | 0.191558 | 0.852032 | 0 | 0 | 1.314134 | 2.171694 |
| GTEx_Tissu | GTEX-OXRF | 53/10989 | 0.155141 | 0.852032 | 0 | 0 | 1.279587 | 2.384406 |
| GTEx_Tissu | GTEX-W5W | 4/478 | 0.154966 | 0.852032 | 0 | 0 | 1.976412 | 3.685118 |
| GTEx_Tissu | GTEX-X3Y1 | 41/8237 | 0.154064 | 0.852032 | 0 | 0 | 1.274209 | 2.383262 |
| GTEx_Tissu | GTEX-QLQV | 41/8233 | 0.153157 | 0.852032 | 0 | 0 | 1.275266 | 2.392773 |
| GTEx_Tissu | GTEX-PX3G | 2/159 | 0.152237 | 0.852032 | 0 | 0 | 2.960809 | 5.573176 |
| GTEx_Tissu | GTEX-T2YK | 3/323 | 0.166214 | 0.852032 | 0 | 0 | 2.186719 | 3.924018 |
| GTEx_Tissu | GTEX-UPK5 | 35/6961 | 0.170312 | 0.852032 | 0 | 0 | 1.262092 | 2.234062 |
| GTEx_Tissu | GTEX-QCQG | 4/496 | 0.169936 | 0.852032 | 0 | 0 | 1.902341 | 3.371586 |
| GTEx_Tissu | GTEX-X585 | 23/4351 | 0.175057 | 0.852032 | 0 | 0 | 1.294099 | 2.255151 |
| GTEx_Tissu | GTEX-T2IS- | 4/502 | 0.175038 | 0.852032 | 0 | 0 | 1.878841 | 3.274358 |
| GTEx_Tissu | GTEX-TKQ2 | 11/1795 | 0.15516 | 0.852032 | 0 | 0 | 1.470815 | 2.740564 |
| GTEx_Tissu | GTEX-SN8G | 41/8323 | 0.174408 | 0.852032 | 0 | 0 | 1.251722 | 2.185955 |
| GTEx_Tissu | GTEX-WFO | 6/864 | 0.17387 | 0.852032 | 0 | 0 | 1.645083 | 2.877985 |
| GTEx_Tissu | GTEX-R55C | 2/173 | 0.173677 | 0.852032 | 0 | 0 | 2.716477 | 4.755345 |
| GTEx_Tissu | GTEX-SNM | 25/4830 | 0.189089 | 0.852032 | 0 | 0 | 1.267833 | 2.11162 |
| GTEx_Tissu | GTEX-XLM4 | 4/518 | 0.188898 | 0.852032 | 0 | 0 | 1.818855 | 3.03121 |
| GTEx_Tissu | GTEX-XMD | 14/2444 | 0.171661 | 0.852032 | 0 | 0 | 1.379796 | 2.431525 |
| GTEx_Tissu | GTEX-NPJ8 | 6/828 | 0.151569 | 0.852032 | 0 | 0 | 1.720375 | 3.245852 |
| GTEx_Tissu | GTEX-NL4V | Dec-84 | 0.151016 | 0.852032 | 0 | 0 | 1.455659 | 2.751739 |
| GTEx_Tissu | GTEX-X5EB | 16/2795 | 0.150284 | 0.852032 | 0 | 0 | 1.389415 | 2.633262 |
| GTEx_Tissu | GTEX-T6MI | 8/1247 | 0.174604 | 0.852032 | 0 | 0 | 1.526261 | 2.663682 |
| GTEx_Tissu | GTEX-UTHO | 33/6499 | 0.165717 | 0.852032 | 0 | 0 | 1.270896 | 2.284399 |
| GTEx_Tissu | GTEX-U3ZN | 17/3025 | 0.15778 | 0.852032 | 0 | 0 | 1.36486 | 2.520286 |
| GTEx_Tissu | GTEX-TSE9 | 8/1213 | 0.157027 | 0.852032 | 0 | 0 | 1.572183 | 2.910645 |
| GTEx_Tissu | GTEX-XMD | 5/655 | 0.156579 | 0.852032 | 0 | 0 | 1.807036 | 3.350599 |
| GTEx_Tissu | GTEX-T5JW | 6/835 | 0.155798 | 0.852032 | 0 | 0 | 1.705223 | 3.170337 |
| GTEx_Tissu | GTEX-Q734 | 10/1595 | 0.15414 | 0.852032 | 0 | 0 | 1.501741 | 2.808092 |
| GTEx_Tissu | GTEX-U3ZN | 2/165 | 0.161361 | 0.852032 | 0 | 0 | 2.850956 | 5.200458 |
| GTEx_Tissu | GTEX-R55F | 2/165 | 0.161361 | 0.852032 | 0 | 0 | 2.850956 | 5.200458 |
| GTEx_Tissu | GTEX-T6MO | 2/166 | 0.162892 | 0.852032 | 0 | 0 | 2.833429 | 5.141737 |
| GTEx_Tissu | GTEX-NPJ8 | 5/664 | 0.162872 | 0.852032 | 0 | 0 | 1.781524 | 3.233092 |
| GTEx_Tissu | GTEX-QEG5 | 35/6929 | 0.16237 | 0.852032 | 0 | 0 | 1.271075 | 2.310658 |
| GTEx_Tissu | GTEX-Q2AH | 6/833 | 0.154585 | 0.852032 | 0 | 0 | 1.709526 | 3.191707 |
| GTEx_Tissu | GTEX-T8EM | 2/159 | 0.152237 | 0.852032 | 0 | 0 | 2.960809 | 5.573176 |
| GTEx_Tissu | GTEX-RVPU | 23/4295 | 0.158746 | 0.852032 | 0 | 0 | 1.315773 | 2.421613 |
| GTEx_Tissu | GTEX-X4XX | 8/1267 | 0.18535 | 0.852032 | 0 | 0 | 1.500407 | 2.528947 |
| GTEx_Tissu | GTEX-WVLI | 8/1267 | 0.18535 | 0.852032 | 0 | 0 | 1.500407 | 2.528947 |
| GTEx_Tissu | GTEX-P78B | 36/7240 | 0.184697 | 0.852032 | 0 | 0 | 1.245289 | 2.103342 |
| GTEx_Tissu | GTEX-Q734 | 2/180 | 0.184577 | 0.852032 | 0 | 0 | 2.608724 | 4.407929 |
| GTEx_Tissu | GTEX-T8EM | 2/180 | 0.184577 | 0.852032 | 0 | 0 | 2.608724 | 4.407929 |

| | | | | | | | | |
|---|---|---|---|---|---|---|---|---|
| GTEx_Tissu | GTEX-U412 | 2/180 | 0.184577 | 0.852032 | 0 | 0 | 2.608724 | 4.407929 |
| GTEx_Tissu | GTEX-TSE9 | 4/512 | 0.183658 | 0.852032 | 0 | 0 | 1.840907 | 3.119747 |
| GTEx_Tissu | GTEX-XV7C | 2/164 | 0.159833 | 0.852032 | 0 | 0 | 2.8687 | 5.260115 |
| GTEx_Tissu | GTEX-R55D | 19/3449 | 0.159557 | 0.852032 | 0 | 0 | 1.342724 | 2.464372 |
| GTEx_Tissu | GTEX-TSE9 | 49/10082 | 0.159184 | 0.852032 | 0 | 0 | 1.26981 | 2.333524 |
| GTEx_Tissu | GTEX-TMM | 17/3029 | 0.159089 | 0.852032 | 0 | 0 | 1.362725 | 2.50509 |
| GTEx_Tissu | GTEX-XQ3S | 42/8557 | 0.176329 | 0.852032 | 0 | 0 | 1.24934 | 2.168107 |
| GTEx_Tissu | GTEX-OIZF | 26/5000 | 0.175245 | 0.852032 | 0 | 0 | 1.280145 | 2.22946 |
| GTEx_Tissu | GTEX-R55D | 2/174 | 0.175228 | 0.852032 | 0 | 0 | 2.700547 | 4.703455 |
| GTEx_Tissu | GTEX-NFK9 | 2/173 | 0.173677 | 0.852032 | 0 | 0 | 2.716477 | 4.755345 |
| GTEx_Tissu | GTEX-PWO | 33/6541 | 0.176514 | 0.852032 | 0 | 0 | 1.25875 | 2.183119 |
| GTEx_Tissu | GTEX-PLZ4 | 31/6099 | 0.176534 | 0.852032 | 0 | 0 | 1.263051 | 2.190436 |
| GTEx_Tissu | GTEX-P4PC | 14/2457 | 0.176621 | 0.852032 | 0 | 0 | 1.371433 | 2.37772 |
| GTEx_Tissu | GTEX-V1D1 | 3/333 | 0.176892 | 0.852032 | 0 | 0 | 2.119372 | 3.671207 |
| GTEx_Tissu | GTEX-QV31 | 23/4360 | 0.177764 | 0.852032 | 0 | 0 | 1.290668 | 2.229365 |
| GTEx_Tissu | GTEX-WL4 | 7/1052 | 0.17327 | 0.852032 | 0 | 0 | 1.579856 | 2.769333 |
| GTEx_Tissu | GTEX-OHPI | 2/178 | 0.181452 | 0.852032 | 0 | 0 | 2.638636 | 4.503525 |
| GTEx_Tissu | GTEX-SJXC | 10/1649 | 0.178495 | 0.852032 | 0 | 0 | 1.447985 | 2.495156 |
| GTEx_Tissu | GTEX-T5JC | 6/872 | 0.179006 | 0.852032 | 0 | 0 | 1.629202 | 2.802775 |
| GTEx_Tissu | GTEX-SSA3 | 30/5888 | 0.17902 | 0.852032 | 0 | 0 | 1.262781 | 2.172311 |
| GTEx_Tissu | GTEX-SIU7 | 16/2880 | 0.179583 | 0.852032 | 0 | 0 | 1.34149 | 2.303499 |
| GTEx_Tissu | GTEX-TML8 | 2/177 | 0.179893 | 0.852032 | 0 | 0 | 2.653849 | 4.552394 |
| GTEx_Tissu | GTEX-TKQ1 | 4/524 | 0.194187 | 0.852056 | 0 | 0 | 1.797312 | 2.94568 |
| GTEx_Tissu | GTEX-U3ZN | 2/186 | 0.193998 | 0.852056 | 0 | 0 | 2.52289 | 4.137304 |
| GTEx_Tissu | GTEX-NL4V | 57/12083 | 0.193981 | 0.852056 | 0 | 0 | 1.246075 | 2.043558 |
| GTEx_Tissu | GTEX-TMM | 3/349 | 0.194361 | 0.852056 | 0 | 0 | 2.019715 | 3.308368 |
| GTEx_Tissu | GTEX-X8HC | 37/7501 | 0.194537 | 0.852056 | 0 | 0 | 1.234226 | 2.020592 |
| Gene_Pert | Sst KO mou | 1/409 | 0.834949 | 0.852057 | 0 | 0 | 0.555888 | 0.100273 |
| Gene_Pert | Ing2 KO mo | 1/422 | 0.844231 | 0.852692 | 0 | 0 | 0.538364 | 0.091161 |
| Gene_Pert | CCAR2 KD I | 1/422 | 0.844231 | 0.852692 | 0 | 0 | 0.538364 | 0.091161 |
| GTEx_Tissu | GTEX-O5YV | 12/1816 | 0.094056 | 0.852988 | 0 | 0 | 1.60612 | 3.796655 |
| GTEx_Tissu | GTEX-OXRL | 8/1071 | 0.093984 | 0.852988 | 0 | 0 | 1.79573 | 4.246244 |
| GTEx_Tissu | GTEX-XPVG | 10/1436 | 0.093516 | 0.852988 | 0 | 0 | 1.683667 | 3.989652 |
| Gene_Pert | RB1 KD hur | 1/411 | 0.836412 | 0.853064 | 0 | 0 | 0.55312 | 0.098806 |
| GTEx_Tissu | GTEX-U3ZN | 8/1072 | 0.094366 | 0.853124 | 0 | 0 | 1.793947 | 4.234753 |
| GTEx_Tissu | GTEX-RWS | 17/3137 | 0.196668 | 0.853433 | 0 | 0 | 1.307147 | 2.125732 |
| GTEx_Tissu | GTEX-XGQ | 3/350 | 0.195467 | 0.853433 | 0 | 0 | 2.013792 | 3.287236 |
| GTEx_Tissu | GTEX-T6M | 2/188 | 0.197153 | 0.853433 | 0 | 0 | 2.495509 | 4.05215 |
| GTEx_Tissu | GTEX-UPK5 | 16/2924 | 0.195854 | 0.853433 | 0 | 0 | 1.317783 | 2.148495 |
| GTEx_Tissu | GTEX-WFG | 32/6394 | 0.196823 | 0.853433 | 0 | 0 | 1.239267 | 2.014366 |
| GTEx_Tissu | GTEX-XPVG | 49/10235 | 0.196397 | 0.853433 | 0 | 0 | 1.231368 | 2.004196 |
| Gene_Pert | Cadm1 KO | 1/413 | 0.837862 | 0.854057 | 0 | 0 | 0.550378 | 0.097363 |
| dbGaP | Body Mass | 1/437 | 0.8543 | 0.8543 | 0 | 0 | 0.519442 | 0.081798 |
| Gene_Pert | MIR124-1 ( | 1/415 | 0.839299 | 0.85455 | 0 | 0 | 0.547663 | 0.095944 |
| Gene_Pert | Ache AChE | 1/415 | 0.839299 | 0.85455 | 0 | 0 | 0.547663 | 0.095944 |

| | | | | | | | | |
|---|---|---|---|---|---|---|---|---|
| Reactome_ | Signaling B | 2/644 | 0.774813 | 0.854967 | 0 | 0 | 0.706286 | 0.180197 |
| Jensen_TIS | Cerebral pe | 18/4907 | 0.831207 | 0.855108 | 0 | 0 | 0.801658 | 0.148207 |
| GTEx_Tissu | GTEX-VJYA | 42/8644 | 0.198563 | 0.855377 | 0 | 0 | 1.227265 | 1.984057 |
| GTEx_Tissu | GTEX-SJXC- | 23/4425 | 0.198004 | 0.855377 | 0 | 0 | 1.266303 | 2.050736 |
| GTEx_Tissu | GTEX-NFK9 | 3/354 | 0.199908 | 0.855377 | 0 | 0 | 1.990435 | 3.204395 |
| GTEx_Tissu | GTEX-P4QF | 36/7296 | 0.199729 | 0.855377 | 0 | 0 | 1.230238 | 1.981659 |
| GTEx_Tissu | GTEX-O5YV | 30/5962 | 0.199861 | 0.855377 | 0 | 0 | 1.240462 | 1.997309 |
| GTEx_Tissu | GTEX-NPJ8 | 16/2933 | 0.199271 | 0.855377 | 0 | 0 | 1.313022 | 2.11802 |
| GTEx_Tissu | GTEX-PSDG | 2/190 | 0.200314 | 0.855467 | 0 | 0 | 2.468711 | 3.969367 |
| Gene_Pert | KLF4 OE hu | 1/417 | 0.840724 | 0.855495 | 0 | 0 | 0.544974 | 0.094549 |
| Gene_Pert | Htt Gain of | 1/419 | 0.842136 | 0.855495 | 0 | 0 | 0.542311 | 0.093177 |
| Gene_Pert | PIK3CA Mu | 1/419 | 0.842136 | 0.855495 | 0 | 0 | 0.542311 | 0.093177 |
| Gene_Pert | CD44 KD hu | 1/419 | 0.842136 | 0.855495 | 0 | 0 | 0.542311 | 0.093177 |
| Gene_Pert | Prkcz KO m | 1/427 | 0.847661 | 0.85568 | 0 | 0 | 0.531909 | 0.087911 |
| GTEx_Tissu | GTEX-XUZC | 2/191 | 0.201897 | 0.855808 | 0 | 0 | 2.455524 | 3.928837 |
| GTEx_Tissu | GTEX-NPJ8 | 3/355 | 0.201022 | 0.855808 | 0 | 0 | 1.984679 | 3.184098 |
| GTEx_Tissu | GTEX-XBEV | 36/7304 | 0.201932 | 0.855808 | 0 | 0 | 1.228107 | 1.964754 |
| GTEx_Tissu | GTEX-T2IS- | 2/191 | 0.201897 | 0.855808 | 0 | 0 | 2.455524 | 3.928837 |
| GeneSigDB | 18362358-T | 1/381 | 0.813062 | 0.856097 | 0 | 0 | 0.597705 | 0.123694 |
| Human_Ge | Wholebrain | 1/240 | 0.650968 | 0.856607 | 0 | 0 | 0.957186 | 0.410916 |
| Human_Ge | Lung | 1/299 | 0.731073 | 0.856607 | 0 | 0 | 0.765374 | 0.239748 |
| Human_Ge | WholeBloo | 2/514 | 0.658575 | 0.856607 | 0 | 0 | 0.89159 | 0.372397 |
| Human_Ge | 721 B lymp | 7/1543 | 0.511207 | 0.856607 | 0 | 0 | 1.046867 | 0.702427 |
| Human_Ge | Prostate | 1/249 | 0.66456 | 0.856607 | 0 | 0 | 0.922027 | 0.376768 |
| Human_Ge | CD33+ Mye | 2/679 | 0.799636 | 0.856607 | 0 | 0 | 0.668555 | 0.149488 |
| Human_Ge | SmoothMu | 1/363 | 0.797501 | 0.856607 | 0 | 0 | 0.628003 | 0.1421 |
| Human_Ge | pineal nigh | 2/674 | 0.796242 | 0.856607 | 0 | 0 | 0.673704 | 0.153505 |
| Human_Ge | CD4+ Tcells | 2/533 | 0.678108 | 0.856607 | 0 | 0 | 0.858846 | 0.333617 |
| Human_Ge | Thyroid | 1/388 | 0.818789 | 0.856607 | 0 | 0 | 0.586683 | 0.117295 |
| Human_Ge | PrefrontalC | 2/521 | 0.665878 | 0.856607 | 0 | 0 | 0.879247 | 0.357545 |
| Human_Ge | colon | 1/258 | 0.677628 | 0.856607 | 0 | 0 | 0.889331 | 0.346089 |
| Human_Ge | CD34+ | 2/645 | 0.775558 | 0.856607 | 0 | 0 | 0.70515 | 0.17923 |
| Human_Ge | CiliaryGang | 1/278 | 0.704895 | 0.856607 | 0 | 0 | 0.82428 | 0.288256 |
| Human_Ge | CardiacMyo | 1/273 | 0.6983 | 0.856607 | 0 | 0 | 0.839646 | 0.301522 |
| Human_Ge | CD19+ BCe | 1/412 | 0.837139 | 0.856607 | 0 | 0 | 0.551746 | 0.098081 |
| Human_Ge | Amygdala | 1/345 | 0.78066 | 0.856607 | 0 | 0 | 0.661472 | 0.163791 |
| Human_Ge | TrigeminalG | 1/196 | 0.576271 | 0.856607 | 0 | 0 | 1.17579 | 0.64807 |
| Human_Ge | CD56+ NKC | 3/752 | 0.640261 | 0.856607 | 0 | 0 | 0.91379 | 0.40744 |
| Gene_Pert | Kat5 KD mo | 1/424 | 0.845612 | 0.856811 | 0 | 0 | 0.535763 | 0.089845 |
| Gene_Pert | POU5F1 KD | 1/423 | 0.844923 | 0.856811 | 0 | 0 | 0.537061 | 0.0905 |
| Gene_Pert | Dmd Defici | 1/425 | 0.846298 | 0.856811 | 0 | 0 | 0.534472 | 0.089195 |
| Gene_Pert | Foxo1 KD n | 1/425 | 0.846298 | 0.856811 | 0 | 0 | 0.534472 | 0.089195 |
| Gene_Pert | Rb1 Deficie | 1/422 | 0.844231 | 0.856811 | 0 | 0 | 0.538364 | 0.091161 |
| Gene_Pert | MALAT1 OE | 1/423 | 0.844923 | 0.856811 | 0 | 0 | 0.537061 | 0.0905 |
| GTEx_Tissu | GTEX-P4QS | 4/534 | 0.203104 | 0.856901 | 0 | 0 | 1.762491 | 2.809477 |

| | | | | | | | | |
|---|---|---|---|---|---|---|---|---|
| GTEx_Tissu | GTEX-S7PN | 3/357 | 0.203255 | 0.856901 | 0 | 0 | 1.973265 | 3.143989 |
| GTEx_Tissu | GTEX-UTH( | 13/2316 | 0.203345 | 0.856901 | 0 | 0 | 1.343312 | 2.139694 |
| GO_Biologi | Negative R | 2/763 | 0.849669 | 0.85691 | 0 | 0 | 0.592162 | 0.096468 |
| Gene_Pert | Pparg OE n | 1/430 | 0.849684 | 0.857245 | 0 | 0 | 0.528108 | 0.086024 |
| GTEx_Tissu | GTEX-S32V | 8/1074 | 0.095132 | 0.857375 | 0 | 0 | 1.790391 | 4.211874 |
| Gene_Pert | Prkcz KO m | 1/427 | 0.847661 | 0.857707 | 0 | 0 | 0.531909 | 0.087911 |
| Gene_Pert | Myc Activa | 1/428 | 0.848339 | 0.857908 | 0 | 0 | 0.530636 | 0.087277 |
| GeneSigDB | 19204204-: | 1/385 | 0.816356 | 0.857944 | 0 | 0 | 0.591358 | 0.119989 |
| Gene_Pert | Bcl11b KO | 1/429 | 0.849013 | 0.858106 | 0 | 0 | 0.529369 | 0.086648 |
| Disease_Pe | Pulmonary | 1/427 | 0.847661 | 0.858324 | 0 | 0 | 0.531909 | 0.087911 |
| Disease_Pe | Huntingtor | 1/429 | 0.849013 | 0.858324 | 0 | 0 | 0.529369 | 0.086648 |
| Disease_Pe | prostate ca | 1/430 | 0.849684 | 0.858324 | 0 | 0 | 0.528108 | 0.086024 |
| Disease_Pe | MELAS - M | 1/431 | 0.850352 | 0.858324 | 0 | 0 | 0.526852 | 0.085405 |
| GWAS_Cat | Self-report | 1/440 | 0.856236 | 0.858346 | 0 | 0 | 0.515813 | 0.080059 |
| GO_Biologi | Regulation | 1/434 | 0.852339 | 0.858384 | 0 | 0 | 0.523122 | 0.083579 |
| Reactome_ | Cell Cycle F | 2/654 | 0.782163 | 0.858846 | 0 | 0 | 0.695092 | 0.170778 |
| Disease_Pe | Parkinson's | 1/442 | 0.857512 | 0.858849 | 0 | 0 | 0.513421 | 0.078923 |
| Disease_Pe | mental reta | 1/435 | 0.852996 | 0.859623 | 0 | 0 | 0.521889 | 0.082981 |
| Disease_Pe | Eczema C0 | 1/437 | 0.8543 | 0.859623 | 0 | 0 | 0.519442 | 0.081798 |
| Gene_Pert | Foxa2 KO n | 1/432 | 0.851017 | 0.859647 | 0 | 0 | 0.525603 | 0.084792 |
| Reactome_ | Metabolisr | 2/666 | 0.790709 | 0.8598 | 0 | 0 | 0.682105 | 0.160176 |
| Reactome_ | Transcripti | 1/354 | 0.789247 | 0.8598 | 0 | 0 | 0.644311 | 0.152493 |
| SysMyo_M | MGS70 Hu | 3/1087 | 0.858549 | 0.860069 | 0 | 0 | 0.620355 | 0.094611 |
| Gene_Pert | Suz12 KD n | 1/435 | 0.852996 | 0.860108 | 0 | 0 | 0.521889 | 0.082981 |
| Jensen_TIS | Embryonic | 18/4936 | 0.838872 | 0.860245 | 0 | 0 | 0.795392 | 0.139748 |
| Gene_Pert | DROSHA KI | 1/436 | 0.853649 | 0.860289 | 0 | 0 | 0.520663 | 0.082387 |
| Gene_Pert | RELA KD hu | 1/434 | 0.852339 | 0.860498 | 0 | 0 | 0.523122 | 0.083579 |
| Gene_Pert | Mecom KO | 1/438 | 0.854948 | 0.861119 | 0 | 0 | 0.518227 | 0.081214 |
| Gene_Pert | FGFR3 drug | 1/439 | 0.855593 | 0.861291 | 0 | 0 | 0.517017 | 0.080634 |
| GeneSigDB | 19341462-: | 72/17122 | 0.821136 | 0.861342 | 0 | 0 | 0.806006 | 0.158837 |
| GTEx_Tissu | GTEX-OHPI | 5/558 | 0.096023 | 0.862599 | 0 | 0 | 2.134698 | 5.001963 |
| GTEx_Tissu | GTEX-WFG | 7/899 | 0.096308 | 0.862599 | 0 | 0 | 1.865849 | 4.366465 |
| Gene_Pert | NCOA1 KO | 1/439 | 0.855593 | 0.862811 | 0 | 0 | 0.517017 | 0.080634 |
| Gene_Pert | Dnmt1 KO | 1/439 | 0.855593 | 0.862811 | 0 | 0 | 0.517017 | 0.080634 |
| Gene_Pert | CTNNB1 m | 1/440 | 0.856236 | 0.862974 | 0 | 0 | 0.515813 | 0.080059 |
| GTEx_Tissu | GTEX-R55C | 8/1078 | 0.096676 | 0.863222 | 0 | 0 | 1.78332 | 4.166528 |
| Reactome_ | Metabolisr | Jul-49 | 0.800439 | 0.864581 | 0 | 0 | 0.765775 | 0.170458 |
| Reactome_ | SARS-CoV I | 1/369 | 0.802825 | 0.864581 | 0 | 0 | 0.617575 | 0.135631 |
| GTEx_Tissu | GTEX-NL4V | 6/912 | 0.205588 | 0.864713 | 0 | 0 | 1.554002 | 2.458247 |
| KOMP2_M | Abnormal S | 1/441 | 0.856875 | 0.864883 | 0 | 0 | 0.514614 | 0.079489 |
| GTEx_Tissu | GTEX-Q2AI | 3/377 | 0.225889 | 0.86526 | 0 | 0 | 1.865833 | 2.775817 |
| GTEx_Tissu | GTEX-X5EB | 7/1139 | 0.226315 | 0.86526 | 0 | 0 | 1.451712 | 2.156994 |
| GTEx_Tissu | GTEX-T8EN | 7/1139 | 0.226315 | 0.86526 | 0 | 0 | 1.451712 | 2.156994 |
| GTEx_Tissu | GTEX-T6MI | 7/1139 | 0.226315 | 0.86526 | 0 | 0 | 1.451712 | 2.156994 |
| GTEx_Tissu | GTEX-XV7C | 3/362 | 0.208863 | 0.86526 | 0 | 0 | 1.945285 | 3.046467 |

| | | | | | | | | |
|---|---|---|---|---|---|---|---|---|
| GTEx_Tissu | GTEX-P4QS | Nov-17 | 0.209463 | 0.86526 | 0 | 0 | 1.367406 | 2.137539 |
| GTEx_Tissu | GTEX-R53T | 26/5115 | 0.209694 | 0.86526 | 0 | 0 | 1.241585 | 1.939485 |
| GTEx_Tissu | GTEX-WCD | 14/2544 | 0.211665 | 0.86526 | 0 | 0 | 1.317678 | 2.046027 |
| GTEx_Tissu | GTEX-QVJC | 4/558 | 0.224989 | 0.86526 | 0 | 0 | 1.68405 | 2.512102 |
| GTEx_Tissu | GTEX-PX3G | Jan-58 | 0.223696 | 0.86526 | 0 | 0 | 4.050592 | 6.065622 |
| GTEx_Tissu | GTEX-T5JC- | 8/1335 | 0.223949 | 0.86526 | 0 | 0 | 1.418332 | 2.122302 |
| GTEx_Tissu | GTEX-T5JC- | 10/1743 | 0.224917 | 0.86526 | 0 | 0 | 1.3624 | 2.032736 |
| GTEx_Tissu | GTEX-O5YT | 4/549 | 0.216707 | 0.86526 | 0 | 0 | 1.712656 | 2.61901 |
| GTEx_Tissu | GTEX-UTH( | 6/927 | 0.215918 | 0.86526 | 0 | 0 | 1.527486 | 2.341421 |
| GTEx_Tissu | GTEX-WZT( | 4/549 | 0.216707 | 0.86526 | 0 | 0 | 1.712656 | 2.61901 |
| GTEx_Tissu | GTEX-NPJ7 | 9/1534 | 0.222266 | 0.86526 | 0 | 0 | 1.391274 | 2.092307 |
| GTEx_Tissu | GTEX-PWO | 14/2547 | 0.212928 | 0.86526 | 0 | 0 | 1.315891 | 2.03542 |
| GTEx_Tissu | GTEX-U3ZN | 3/378 | 0.227035 | 0.86526 | 0 | 0 | 1.860762 | 2.758864 |
| GTEx_Tissu | GTEX-S4UY | Jan-59 | 0.227083 | 0.86526 | 0 | 0 | 3.980553 | 5.900928 |
| GTEx_Tissu | GTEX-U3ZH | 37/7617 | 0.227106 | 0.86526 | 0 | 0 | 1.204013 | 1.784753 |
| GTEx_Tissu | GTEX-WFG | 18/3426 | 0.225216 | 0.86526 | 0 | 0 | 1.263396 | 1.883339 |
| GTEx_Tissu | GTEX-T2IS- | 2/206 | 0.225797 | 0.86526 | 0 | 0 | 2.273241 | 3.382851 |
| GTEx_Tissu | GTEX-XLM4 | 4/560 | 0.226841 | 0.86526 | 0 | 0 | 1.677819 | 2.489052 |
| GTEx_Tissu | GTEX-SN8G | 8/1314 | 0.211707 | 0.86526 | 0 | 0 | 1.442767 | 2.239968 |
| GTEx_Tissu | GTEX-SNM | 13/2339 | 0.213329 | 0.86526 | 0 | 0 | 1.328292 | 2.052106 |
| GTEx_Tissu | GTEX-OXRI | 2/198 | 0.213017 | 0.86526 | 0 | 0 | 2.366987 | 3.660273 |
| GTEx_Tissu | GTEX-R55G | 2/198 | 0.213017 | 0.86526 | 0 | 0 | 2.366987 | 3.660273 |
| GTEx_Tissu | GTEX-UJHI- | 4/545 | 0.213054 | 0.86526 | 0 | 0 | 1.725675 | 2.668255 |
| GTEx_Tissu | GTEX-UPIC | 14/2533 | 0.207063 | 0.86526 | 0 | 0 | 1.32427 | 2.085373 |
| GTEx_Tissu | GTEX-Q2A( | 2/194 | 0.206654 | 0.86526 | 0 | 0 | 2.416789 | 3.810572 |
| GTEx_Tissu | GTEX-OHPI | 44/9180 | 0.220647 | 0.86526 | 0 | 0 | 1.207052 | 1.824085 |
| GTEx_Tissu | GTEX-VUSI | 33/6701 | 0.221328 | 0.86526 | 0 | 0 | 1.213882 | 1.830669 |
| GTEx_Tissu | GTEX-PWO | 9/1542 | 0.22667 | 0.86526 | 0 | 0 | 1.383411 | 2.053343 |
| GTEx_Tissu | GTEX-XOT4 | 34/6925 | 0.221523 | 0.86526 | 0 | 0 | 1.212268 | 1.827167 |
| GTEx_Tissu | GTEX-R45C | 58/12392 | 0.214158 | 0.86526 | 0 | 0 | 1.228961 | 1.893877 |
| GTEx_Tissu | GTEX-O5YV | 3/361 | 0.207738 | 0.86526 | 0 | 0 | 1.950818 | 3.065664 |
| GTEx_Tissu | GTEX-U8XE | 2/195 | 0.208243 | 0.86526 | 0 | 0 | 2.404145 | 3.772225 |
| GTEx_Tissu | GTEX-PLZ4 | Nov-29 | 0.21522 | 0.86526 | 0 | 0 | 1.357946 | 2.085935 |
| GTEx_Tissu | GTEX-OXR( | 27/5354 | 0.215858 | 0.86526 | 0 | 0 | 1.232157 | 1.889061 |
| GTEx_Tissu | GTEX-S95S | 40/8275 | 0.22163 | 0.86526 | 0 | 0 | 1.206888 | 1.818474 |
| GTEx_Tissu | GTEX-WH7 | 5/733 | 0.21428 | 0.86526 | 0 | 0 | 1.606892 | 2.475371 |
| GTEx_Tissu | GTEX-QVJC | 4/547 | 0.214878 | 0.86526 | 0 | 0 | 1.719142 | 2.643496 |
| GTEx_Tissu | GTEX-WH7 | Jan-56 | 0.216879 | 0.86526 | 0 | 0 | 4.198309 | 6.416764 |
| GTEx_Tissu | GTEX-PLZ6 | 6/922 | 0.212454 | 0.86526 | 0 | 0 | 1.536228 | 2.379665 |
| GTEx_Tissu | GTEX-WL4( | 10/1719 | 0.212613 | 0.86526 | 0 | 0 | 1.383356 | 2.141825 |
| GTEx_Tissu | GTEX-NFK9 | 2/202 | 0.219398 | 0.86526 | 0 | 0 | 2.319176 | 3.517879 |
| GTEx_Tissu | GTEX-XPT6 | 39/8045 | 0.220503 | 0.86526 | 0 | 0 | 1.208398 | 1.826912 |
| GTEx_Tissu | GTEX-Q2A( | 25/4948 | 0.226518 | 0.86526 | 0 | 0 | 1.227779 | 1.823167 |
| GTEx_Tissu | GTEX-S95S | 16/2984 | 0.219186 | 0.86526 | 0 | 0 | 1.286587 | 1.952826 |
| GTEx_Tissu | GTEX-WEY! | 5/743 | 0.22214 | 0.86526 | 0 | 0 | 1.584292 | 2.383483 |

| | | | | | | | | |
|---|---|---|---|---|---|---|---|---|
| GTEx_Tissu | GTEX-XPVC | 3/375 | 0.223603 | 0.86526 | 0 | 0 | 1.876056 | 2.810115 |
| GTEx_Tissu | GTEX-P4PP | 2/201 | 0.217801 | 0.86526 | 0 | 0 | 2.330949 | 3.552767 |
| GTEx_Tissu | GTEX-XUJ4 | 2/195 | 0.208243 | 0.86526 | 0 | 0 | 2.404145 | 3.772225 |
| GTEx_Tissu | GTEX-UPIC | 15/2791 | 0.226943 | 0.86526 | 0 | 0 | 1.286098 | 1.907356 |
| GTEx_Tissu | GTEX-N7M | 9/1535 | 0.222815 | 0.86526 | 0 | 0 | 1.390286 | 2.087395 |
| Gene_Pert | Prmt6 OE r | 1/445 | 0.859405 | 0.865681 | 0 | 0 | 0.509873 | 0.077254 |
| Gene_Pert | CYR61 dox | 1/448 | 0.861273 | 0.867076 | 0 | 0 | 0.506373 | 0.075624 |
| GTEx_Tissu | GTEX-S4Z8 | 37/7621 | 0.228279 | 0.868241 | 0 | 0 | 1.202988 | 1.777037 |
| Gene_Pert | Nfe2l2 kno | 1/453 | 0.864332 | 0.868326 | 0 | 0 | 0.500643 | 0.072993 |
| Gene_Pert | Htt Gain of | 1/452 | 0.863726 | 0.868326 | 0 | 0 | 0.501779 | 0.073511 |
| Gene_Pert | Elf4 KO mo | 1/454 | 0.864936 | 0.868326 | 0 | 0 | 0.499512 | 0.072479 |
| Gene_Pert | Mbd2 KO n | 1/454 | 0.864936 | 0.868326 | 0 | 0 | 0.499512 | 0.072479 |
| Gene_Pert | Htt Knock-i | 1/454 | 0.864936 | 0.868326 | 0 | 0 | 0.499512 | 0.072479 |
| Gene_Pert | ABL1 muta | 1/451 | 0.863117 | 0.868383 | 0 | 0 | 0.50292 | 0.074033 |
| Gene_Pert | Fgf21 KO m | 1/455 | 0.865537 | 0.868443 | 0 | 0 | 0.498386 | 0.071969 |
| Gene_Pert | Tsc2 inacti | 1/452 | 0.863726 | 0.868514 | 0 | 0 | 0.501779 | 0.073511 |
| Gene_Pert | Nfe2l2 KO | 1/453 | 0.864332 | 0.868642 | 0 | 0 | 0.500643 | 0.072993 |
| KOMP2_M | Abnormal I | 1/462 | 0.869672 | 0.869672 | 0 | 0 | 0.490642 | 0.068513 |
| GTEx_Tissu | GTEX-S95S | 14/2587 | 0.230103 | 0.870469 | 0 | 0 | 1.292452 | 1.898908 |
| GTEx_Tissu | GTEX-WEY! | 3/380 | 0.229328 | 0.870469 | 0 | 0 | 1.850701 | 2.725344 |
| GTEx_Tissu | GTEX-SIU8 | 28/5618 | 0.229705 | 0.870469 | 0 | 0 | 1.215985 | 1.788666 |
| GTEx_Tissu | GTEX-V1D1 | 6/949 | 0.23139 | 0.870469 | 0 | 0 | 1.490122 | 2.181017 |
| GTEx_Tissu | GTEX-RM2 | Nov-62 | 0.231397 | 0.870469 | 0 | 0 | 1.332529 | 1.950316 |
| GTEx_Tissu | GTEX-X3Y1 | 22/4309 | 0.232091 | 0.870469 | 0 | 0 | 1.233683 | 1.801951 |
| GTEx_Tissu | GTEX-RUS | 3/383 | 0.232777 | 0.870469 | 0 | 0 | 1.835808 | 2.676009 |
| GTEx_Tissu | GTEX-PWN | 3/383 | 0.232777 | 0.870469 | 0 | 0 | 1.835808 | 2.676009 |
| GTEx_Tissu | GTEX-S7PN | 3/383 | 0.232777 | 0.870469 | 0 | 0 | 1.835808 | 2.676009 |
| GTEx_Tissu | GTEX-XQ3S | 14/2588 | 0.23054 | 0.870469 | 0 | 0 | 1.291876 | 1.895609 |
| Reactome_ | M Phase R | 1/380 | 0.812229 | 0.870523 | 0 | 0 | 0.599313 | 0.124641 |
| Gene_Pert | Mkl2 KO m | 1/457 | 0.866732 | 0.870571 | 0 | 0 | 0.49615 | 0.070962 |
| Gene_Pert | Hes1 OE m | 1/460 | 0.868504 | 0.870932 | 0 | 0 | 0.492831 | 0.069481 |
| DisGeNET | Liver Cirrho | 2/801 | 0.868381 | 0.87104 | 0 | 0 | 0.56288 | 0.079437 |
| GO_Biologi | Negative R | 1/456 | 0.866136 | 0.871043 | 0 | 0 | 0.497266 | 0.071464 |
| CellMarker | Sertoli Cell | 1/449 | 0.86189 | 0.871431 | 0 | 0 | 0.505217 | 0.075089 |
| Reactome_ | Neuronal S | 1/386 | 0.81717 | 0.871648 | 0 | 0 | 0.589792 | 0.119083 |
| Gene_Pert | Prmt8 KD r | 1/460 | 0.868504 | 0.871868 | 0 | 0 | 0.492831 | 0.069481 |
| GTEx_Tissu | GTEX-S4P3 | 7/903 | 0.098019 | 0.872519 | 0 | 0 | 1.857129 | 4.313358 |
| GTEx_Tissu | GTEX-U3ZN | 2/211 | 0.233816 | 0.872885 | 0 | 0 | 2.218294 | 3.223675 |
| Gene_Pert | TP53 Muta | 1/465 | 0.871405 | 0.874297 | 0 | 0 | 0.487395 | 0.067089 |
| Gene_Pert | DUX4 OE h | 1/468 | 0.873116 | 0.874579 | 0 | 0 | 0.484189 | 0.065698 |
| Gene_Pert | Adnp Defic | 1/468 | 0.873116 | 0.874579 | 0 | 0 | 0.484189 | 0.065698 |
| UK_Bioban | Lymphocyt | 1/460 | 0.868504 | 0.874642 | 0 | 0 | 0.492831 | 0.069481 |
| GeneSigDB | 20003503- | 2/738 | 0.836089 | 0.875379 | 0 | 0 | 0.613075 | 0.109753 |
| DisGeNET | Septicemia | 1/468 | 0.873116 | 0.875455 | 0 | 0 | 0.484189 | 0.065698 |
| Jensen_TIS | Caudate nu | 16/4512 | 0.856562 | 0.875597 | 0 | 0 | 0.772743 | 0.119643 |

| | | | | | | | | |
|---|---|---|---|---|---|---|---|---|
| UK_Bioban | Pulse wave | 2/811 | 0.872941 | 0.876015 | 0 | 0 | 0.555631 | 0.075503 |
| GTEx_Tissu | GTEX-NL4V | 4/569 | 0.235225 | 0.876676 | 0 | 0 | 1.650325 | 2.388372 |
| Jensen_TIS | Helper T-ly | 19/5270 | 0.86063 | 0.876971 | 0 | 0 | 0.780182 | 0.117098 |
| GeDiPNet_ | Prostate Ca | 2/814 | 0.874282 | 0.877291 | 0 | 0 | 0.553492 | 0.074363 |
| GO_Molecu | Cis-Regulat | 3/1098 | 0.863202 | 0.877568 | 0 | 0 | 0.613764 | 0.090289 |
| GO_Molecu | Purine Ribo | 1/476 | 0.877568 | 0.877568 | 0 | 0 | 0.475838 | 0.062145 |
| GO_Molecu | RNA Bindin | 4/1411 | 0.870628 | 0.877568 | 0 | 0 | 0.63387 | 0.087817 |
| GO_Molecu | RNA Polym | 3/1122 | 0.872896 | 0.877568 | 0 | 0 | 0.599834 | 0.081541 |
| GTEx_Tissu | GTEX-XQ3S | Nov-71 | 0.235894 | 0.877699 | 0 | 0 | 1.325745 | 1.91487 |
| Jensen_TIS | Medulla ob | 16/4544 | 0.864312 | 0.877945 | 0 | 0 | 0.76569 | 0.111654 |
| CellMarker | Gonadal Er | 1/466 | 0.871978 | 0.87839 | 0 | 0 | 0.486322 | 0.066622 |
| GeneSigDB | 18801183-: | Jul-55 | 0.84096 | 0.878827 | 0 | 0 | 0.723667 | 0.125347 |
| Gene_Pert | UBB KO mo | 1/474 | 0.87647 | 0.878892 | 0 | 0 | 0.4779 | 0.063013 |
| GTEx_Tissu | GTEX-RNOI | 7/906 | 0.099313 | 0.879335 | 0 | 0 | 1.85064 | 4.27401 |
| GTEx_Tissu | GTEX-VJYA | 9/1272 | 0.100987 | 0.879335 | 0 | 0 | 1.703819 | 3.906457 |
| GTEx_Tissu | GTEX-QVUS | 10/1459 | 0.101216 | 0.879335 | 0 | 0 | 1.654881 | 3.790497 |
| GTEx_Tissu | GTEX-SN8G | 12/1837 | 0.100308 | 0.879335 | 0 | 0 | 1.585797 | 3.646564 |
| GTEx_Tissu | GTEX-OXRF | 6/734 | 0.100287 | 0.879335 | 0 | 0 | 1.952076 | 4.489219 |
| GTEx_Tissu | GTEX-S4Z8 | 7/910 | 0.101054 | 0.879335 | 0 | 0 | 1.842054 | 4.22218 |
| GTEx_Tissu | GTEX-XYKS | 5/566 | 0.100473 | 0.879335 | 0 | 0 | 2.103387 | 4.833309 |
| GTEx_Tissu | GTEX-XUW | 7/907 | 0.099747 | 0.879335 | 0 | 0 | 1.848486 | 4.260985 |
| Gene_Pert | Gnmt KO m | 1/476 | 0.877568 | 0.879508 | 0 | 0 | 0.475838 | 0.062145 |
| Gene_Pert | MYB OE hu | 1/477 | 0.878114 | 0.879568 | 0 | 0 | 0.474814 | 0.061716 |
| GeneSigDB | 18081427-T | 2/751 | 0.843281 | 0.879603 | 0 | 0 | 0.602026 | 0.102618 |
| GeneSigDB | 16622258-: | 1/423 | 0.844923 | 0.879667 | 0 | 0 | 0.537061 | 0.0905 |
| Reactome_ | Metabolism | 2/732 | 0.832672 | 0.879804 | 0 | 0 | 0.618308 | 0.113222 |
| Reactome_ | RNA Polym | 4/1312 | 0.830647 | 0.879804 | 0 | 0 | 0.685494 | 0.127194 |
| RNA-Seq_D | OPC Cereb | 4/495 | 0.169091 | 0.880031 | 0 | 0 | 1.906314 | 3.388129 |
| RNA-Seq_D | Bcl9/9l Col | 4/495 | 0.169091 | 0.880031 | 0 | 0 | 1.906314 | 3.388129 |
| RNA-Seq_D | Cardiomyo | 4/496 | 0.169936 | 0.880031 | 0 | 0 | 1.902341 | 3.371586 |
| RNA-Seq_D | PS2APP Ast | 4/498 | 0.17163 | 0.880031 | 0 | 0 | 1.894444 | 3.338792 |
| RNA-Seq_D | Panobinost | 4/498 | 0.17163 | 0.880031 | 0 | 0 | 1.894444 | 3.338792 |
| RNA-Seq_D | Psoriasis Sk | 4/498 | 0.17163 | 0.880031 | 0 | 0 | 1.894444 | 3.338792 |
| RNA-Seq_D | Rpl11 Hem | 4/498 | 0.17163 | 0.880031 | 0 | 0 | 1.894444 | 3.338792 |
| RNA-Seq_D | UTX Spleer | 4/496 | 0.169936 | 0.880031 | 0 | 0 | 1.902341 | 3.371586 |
| RNA-Seq_D | Myelodysp | 4/495 | 0.169091 | 0.880031 | 0 | 0 | 1.906314 | 3.388129 |
| RNA-Seq_D | Asxl1 BM-c | 4/495 | 0.169091 | 0.880031 | 0 | 0 | 1.906314 | 3.388129 |
| RNA-Seq_D | RGS18 HCT | 4/495 | 0.169091 | 0.880031 | 0 | 0 | 1.906314 | 3.388129 |
| RNA-Seq_D | SMCX Emb | 4/495 | 0.169091 | 0.880031 | 0 | 0 | 1.906314 | 3.388129 |
| RNA-Seq_D | Smc3 Embr | 4/495 | 0.169091 | 0.880031 | 0 | 0 | 1.906314 | 3.388129 |
| RNA-Seq_D | ZRSR2 Bor | 4/495 | 0.169091 | 0.880031 | 0 | 0 | 1.906314 | 3.388129 |
| RNA-Seq_D | CITCO Hepa | 4/496 | 0.169936 | 0.880031 | 0 | 0 | 1.902341 | 3.371586 |
| RNA-Seq_D | UTX Bone r | 4/499 | 0.17248 | 0.880031 | 0 | 0 | 1.89052 | 3.32254 |
| RNA-Seq_D | Msi2 MIA F | 4/496 | 0.169936 | 0.880031 | 0 | 0 | 1.902341 | 3.371586 |
| RNA-Seq_D | NMP JJN3 4 | 4/496 | 0.169936 | 0.880031 | 0 | 0 | 1.902341 | 3.371586 |

| | | | | | | | | |
|---|---|---|---|---|---|---|---|---|
| RNA-Seq_[ | Miconazole | 4/494 | 0.168247 | 0.880031 | 0 | 0 | 1.910302 | 3.404772 |
| RNA-Seq_[ | Down Sydr | 4/494 | 0.168247 | 0.880031 | 0 | 0 | 1.910302 | 3.404772 |
| RNA-Seq_[ | Dexametha | 4/494 | 0.168247 | 0.880031 | 0 | 0 | 1.910302 | 3.404772 |
| RNA-Seq_[ | BRG1 501 I | 4/494 | 0.168247 | 0.880031 | 0 | 0 | 1.910302 | 3.404772 |
| RNA-Seq_[ | Myelodysp | 4/493 | 0.167405 | 0.880031 | 0 | 0 | 1.914308 | 3.421514 |
| RNA-Seq_[ | Dioxin Live | 4/496 | 0.169936 | 0.880031 | 0 | 0 | 1.902341 | 3.371586 |
| RNA-Seq_[ | Dioxin Live | 4/496 | 0.169936 | 0.880031 | 0 | 0 | 1.902341 | 3.371586 |
| RNA-Seq_[ | DDPA5 HCT | 4/498 | 0.17163 | 0.880031 | 0 | 0 | 1.894444 | 3.338792 |
| RNA-Seq_[ | Cyclophosp | 4/498 | 0.17163 | 0.880031 | 0 | 0 | 1.894444 | 3.338792 |
| RNA-Seq_[ | Cardiomyo | 4/498 | 0.17163 | 0.880031 | 0 | 0 | 1.894444 | 3.338792 |
| RNA-Seq_[ | Cardiomyo | 4/498 | 0.17163 | 0.880031 | 0 | 0 | 1.894444 | 3.338792 |
| RNA-Seq_[ | CI-994 Hipp | 4/498 | 0.17163 | 0.880031 | 0 | 0 | 1.894444 | 3.338792 |
| RNA-Seq_[ | Schizophre | 4/496 | 0.169936 | 0.880031 | 0 | 0 | 1.902341 | 3.371586 |
| RNA-Seq_[ | Autism iPS( | 4/497 | 0.170782 | 0.880031 | 0 | 0 | 1.898385 | 3.35514 |
| RNA-Seq_[ | Albuterol 8 | 4/497 | 0.170782 | 0.880031 | 0 | 0 | 1.898385 | 3.35514 |
| RNA-Seq_[ | ZRSR2 Bor | 4/498 | 0.17163 | 0.880031 | 0 | 0 | 1.894444 | 3.338792 |
| RNA-Seq_[ | WZ4002 N! | 4/498 | 0.17163 | 0.880031 | 0 | 0 | 1.894444 | 3.338792 |
| RNA-Seq_[ | WZ4002 N! | 4/498 | 0.17163 | 0.880031 | 0 | 0 | 1.894444 | 3.338792 |
| RNA-Seq_[ | UTX Muscl | 4/499 | 0.17248 | 0.880031 | 0 | 0 | 1.89052 | 3.32254 |
| RNA-Seq_[ | Zfp57 Emb | 4/499 | 0.17248 | 0.880031 | 0 | 0 | 1.89052 | 3.32254 |
| RNA-Seq_[ | Myotonic [ | 4/497 | 0.170782 | 0.880031 | 0 | 0 | 1.898385 | 3.35514 |
| RNA-Seq_[ | SMCX Adul | 4/497 | 0.170782 | 0.880031 | 0 | 0 | 1.898385 | 3.35514 |
| RNA-Seq_[ | TAF15 Mot | 4/497 | 0.170782 | 0.880031 | 0 | 0 | 1.898385 | 3.35514 |
| RNA-Seq_[ | Tamoxifen | 4/497 | 0.170782 | 0.880031 | 0 | 0 | 1.898385 | 3.35514 |
| RNA-Seq_[ | Vemurafen | 4/498 | 0.17163 | 0.880031 | 0 | 0 | 1.894444 | 3.338792 |
| RNA-Seq_[ | dimethylox | 4/498 | 0.17163 | 0.880031 | 0 | 0 | 1.894444 | 3.338792 |
| RNA-Seq_[ | LIN41 hESC | 4/497 | 0.170782 | 0.880031 | 0 | 0 | 1.898385 | 3.35514 |
| RNA-Seq_[ | NFYA HCT1 | 4/497 | 0.170782 | 0.880031 | 0 | 0 | 1.898385 | 3.35514 |
| RNA-Seq_[ | Ezh2 Embr | 4/495 | 0.169091 | 0.880031 | 0 | 0 | 1.906314 | 3.388129 |
| RNA-Seq_[ | ZNA MCF7 | 4/498 | 0.17163 | 0.880031 | 0 | 0 | 1.894444 | 3.338792 |
| RNA-Seq_[ | Lsd1 Oocyt | 4/497 | 0.170782 | 0.880031 | 0 | 0 | 1.898385 | 3.35514 |
| RNA-Seq_[ | Mef2c Hea | 4/497 | 0.170782 | 0.880031 | 0 | 0 | 1.898385 | 3.35514 |
| RNA-Seq_[ | ZNF2 NPC I | 4/496 | 0.169936 | 0.880031 | 0 | 0 | 1.902341 | 3.371586 |
| RNA-Seq_[ | Salmonella | 4/498 | 0.17163 | 0.880031 | 0 | 0 | 1.894444 | 3.338792 |
| RNA-Seq_[ | SMCX Emb | 4/498 | 0.17163 | 0.880031 | 0 | 0 | 1.894444 | 3.338792 |
| RNA-Seq_[ | Trex2 Skin | 4/499 | 0.17248 | 0.880031 | 0 | 0 | 1.89052 | 3.32254 |
| RNA-Seq_[ | Tbx1 P19Cl | 4/499 | 0.17248 | 0.880031 | 0 | 0 | 1.89052 | 3.32254 |
| RNA-Seq_[ | ILK (Epider | 4/499 | 0.17248 | 0.880031 | 0 | 0 | 1.89052 | 3.32254 |
| RNA-Seq_[ | GPR65 BMI | 4/499 | 0.17248 | 0.880031 | 0 | 0 | 1.89052 | 3.32254 |
| RNA-Seq_[ | FFAR3 Pan | 4/499 | 0.17248 | 0.880031 | 0 | 0 | 1.89052 | 3.32254 |
| RNA-Seq_[ | Ezh2 Mami | 4/499 | 0.17248 | 0.880031 | 0 | 0 | 1.89052 | 3.32254 |
| RNA-Seq_[ | Ezh2 Heart | 4/499 | 0.17248 | 0.880031 | 0 | 0 | 1.89052 | 3.32254 |
| RNA-Seq_[ | hnRNPL Fe | 4/499 | 0.17248 | 0.880031 | 0 | 0 | 1.89052 | 3.32254 |
| RNA-Seq_[ | Satb1 Midb | 4/496 | 0.169936 | 0.880031 | 0 | 0 | 1.902341 | 3.371586 |
| RNA-Seq_[ | Retinoic Ac | 4/496 | 0.169936 | 0.880031 | 0 | 0 | 1.902341 | 3.371586 |

| | | | | | | | | |
|---|---|---|---|---|---|---|---|---|
| RNA-Seq_[ | Parkinson's | 4/496 | 0.169936 | 0.880031 | 0 | 0 | 1.902341 | 3.371586 |
| RNA-Seq_[ | PRMT5 MV | 4/496 | 0.169936 | 0.880031 | 0 | 0 | 1.902341 | 3.371586 |
| RNA-Seq_[ | NMP U266 | 4/496 | 0.169936 | 0.880031 | 0 | 0 | 1.902341 | 3.371586 |
| RNA-Seq_[ | PAX2 REP k | 4/498 | 0.17163 | 0.880031 | 0 | 0 | 1.894444 | 3.338792 |
| RNA-Seq_[ | BRD9-inhib | 4/498 | 0.17163 | 0.880031 | 0 | 0 | 1.894444 | 3.338792 |
| RNA-Seq_[ | PRDM14 KI | 4/498 | 0.17163 | 0.880031 | 0 | 0 | 1.894444 | 3.338792 |
| RNA-Seq_[ | Mettl3 Emb | 4/497 | 0.170782 | 0.880031 | 0 | 0 | 1.898385 | 3.35514 |
| RNA-Seq_[ | PKC412 ZR | 4/498 | 0.17163 | 0.880031 | 0 | 0 | 1.894444 | 3.338792 |
| RNA-Seq_[ | Dioxin Live | 4/496 | 0.169936 | 0.880031 | 0 | 0 | 1.902341 | 3.371586 |
| RNA-Seq_[ | Imatinib CN | 4/498 | 0.17163 | 0.880031 | 0 | 0 | 1.894444 | 3.338792 |
| RNA-Seq_[ | Idiopathic I | 4/498 | 0.17163 | 0.880031 | 0 | 0 | 1.894444 | 3.338792 |
| RNA-Seq_[ | Hira Oocyte | 4/498 | 0.17163 | 0.880031 | 0 | 0 | 1.894444 | 3.338792 |
| RNA-Seq_[ | Gefitinib N | 4/498 | 0.17163 | 0.880031 | 0 | 0 | 1.894444 | 3.338792 |
| RNA-Seq_[ | Gcn5 Embr | 4/498 | 0.17163 | 0.880031 | 0 | 0 | 1.894444 | 3.338792 |
| RNA-Seq_[ | FOXA2 End | 4/498 | 0.17163 | 0.880031 | 0 | 0 | 1.894444 | 3.338792 |
| RNA-Seq_[ | FBXL10 Ma | 4/498 | 0.17163 | 0.880031 | 0 | 0 | 1.894444 | 3.338792 |
| RNA-Seq_[ | Dnmt1 Cor | 4/498 | 0.17163 | 0.880031 | 0 | 0 | 1.894444 | 3.338792 |
| RNA-Seq_[ | Ezh1 Mamı | 4/499 | 0.17248 | 0.880031 | 0 | 0 | 1.89052 | 3.32254 |
| RNA-Seq_[ | miR-146a E | 4/498 | 0.17163 | 0.880031 | 0 | 0 | 1.894444 | 3.338792 |
| RNA-Seq_[ | POLR2D HC | 4/498 | 0.17163 | 0.880031 | 0 | 0 | 1.894444 | 3.338792 |
| RNA-Seq_[ | Nod2 Smal | 4/498 | 0.17163 | 0.880031 | 0 | 0 | 1.894444 | 3.338792 |
| RNA-Seq_[ | Myelodysp | 4/498 | 0.17163 | 0.880031 | 0 | 0 | 1.894444 | 3.338792 |
| RNA-Seq_[ | MLL1 Comı | 4/498 | 0.17163 | 0.880031 | 0 | 0 | 1.894444 | 3.338792 |
| RNA-Seq_[ | KH-CB19 Eı | 4/498 | 0.17163 | 0.880031 | 0 | 0 | 1.894444 | 3.338792 |
| RNA-Seq_[ | JAK2 HCT1 | 4/498 | 0.17163 | 0.880031 | 0 | 0 | 1.894444 | 3.338792 |
| RNA-Seq_[ | CBFA2T2 K | 4/498 | 0.17163 | 0.880031 | 0 | 0 | 1.894444 | 3.338792 |
| RNA-Seq_[ | SGC0946 N | 4/498 | 0.17163 | 0.880031 | 0 | 0 | 1.894444 | 3.338792 |
| RNA-Seq_[ | SMCX Adul | 4/497 | 0.170782 | 0.880031 | 0 | 0 | 1.898385 | 3.35514 |
| RNA-Seq_[ | SAP130 HC | 4/497 | 0.170782 | 0.880031 | 0 | 0 | 1.898385 | 3.35514 |
| RNA-Seq_[ | RFWD2 Lur | 4/497 | 0.170782 | 0.880031 | 0 | 0 | 1.898385 | 3.35514 |
| RNA-Seq_[ | RBM47 H3 | 4/497 | 0.170782 | 0.880031 | 0 | 0 | 1.898385 | 3.35514 |
| RNA-Seq_[ | Prmt5 Neu | 4/497 | 0.170782 | 0.880031 | 0 | 0 | 1.898385 | 3.35514 |
| RNA-Seq_[ | PRMT5 A54 | 4/497 | 0.170782 | 0.880031 | 0 | 0 | 1.898385 | 3.35514 |
| RNA-Seq_[ | PBS-1086 8 | 4/497 | 0.170782 | 0.880031 | 0 | 0 | 1.898385 | 3.35514 |
| RNA-Seq_[ | ALS (Spora | 4/498 | 0.17163 | 0.880031 | 0 | 0 | 1.894444 | 3.338792 |
| RNA-Seq_[ | Trichostati | 4/496 | 0.169936 | 0.880031 | 0 | 0 | 1.902341 | 3.371586 |
| RNA-Seq_[ | Tetradotox | 4/496 | 0.169936 | 0.880031 | 0 | 0 | 1.902341 | 3.371586 |
| RNA-Seq_[ | TOX4 HCT1 | 4/496 | 0.169936 | 0.880031 | 0 | 0 | 1.902341 | 3.371586 |
| RNA-Seq_[ | TGFB3 Pala | 4/496 | 0.169936 | 0.880031 | 0 | 0 | 1.902341 | 3.371586 |
| RNA-Seq_[ | TGFB3 Pala | 4/496 | 0.169936 | 0.880031 | 0 | 0 | 1.902341 | 3.371586 |
| RNA-Seq_[ | FBXL10 Ma | 4/497 | 0.170782 | 0.880031 | 0 | 0 | 1.898385 | 3.35514 |
| RNA-Seq_[ | Desipramir | 4/497 | 0.170782 | 0.880031 | 0 | 0 | 1.898385 | 3.35514 |
| RNA-Seq_[ | Ch25h BMI | 4/497 | 0.170782 | 0.880031 | 0 | 0 | 1.898385 | 3.35514 |
| RNA-Seq_[ | Calcific aor | 4/497 | 0.170782 | 0.880031 | 0 | 0 | 1.898385 | 3.35514 |
| RNA-Seq_[ | Calcific aor | 4/497 | 0.170782 | 0.880031 | 0 | 0 | 1.898385 | 3.35514 |

| | | | | | | | | |
|---|---|---|---|---|---|---|---|---|
| RNA-Seq_D | CITCO Hep | 4/497 | 0.170782 | 0.880031 | 0 | 0 | 1.898385 | 3.35514 |
| RNA-Seq_D | CDK9 HCT1 | 4/497 | 0.170782 | 0.880031 | 0 | 0 | 1.898385 | 3.35514 |
| RNA-Seq_D | BCAT2 Gas | 4/497 | 0.170782 | 0.880031 | 0 | 0 | 1.898385 | 3.35514 |
| RNA-Seq_D | Bmal1 Live | 4/498 | 0.17163 | 0.880031 | 0 | 0 | 1.894444 | 3.338792 |
| RNA-Seq_D | heat-inacti | 4/497 | 0.170782 | 0.880031 | 0 | 0 | 1.898385 | 3.35514 |
| RNA-Seq_D | DAPT Coch | 7/937 | 0.113241 | 0.880031 | 0 | 0 | 1.786035 | 3.890403 |
| RNA-Seq_D | Brg1 Heart | 4/498 | 0.17163 | 0.880031 | 0 | 0 | 1.894444 | 3.338792 |
| RNA-Seq_D | DEANR1 Er | 4/498 | 0.17163 | 0.880031 | 0 | 0 | 1.894444 | 3.338792 |
| RNA-Seq_D | DMR1-U2 S | 4/498 | 0.17163 | 0.880031 | 0 | 0 | 1.894444 | 3.338792 |
| RNA-Seq_D | Acromegal | 4/498 | 0.17163 | 0.880031 | 0 | 0 | 1.894444 | 3.338792 |
| RNA-Seq_D | All-trans re | 4/498 | 0.17163 | 0.880031 | 0 | 0 | 1.894444 | 3.338792 |
| RNA-Seq_D | All-trans re | 4/498 | 0.17163 | 0.880031 | 0 | 0 | 1.894444 | 3.338792 |
| RNA-Seq_D | BMAL1 Ren | 4/498 | 0.17163 | 0.880031 | 0 | 0 | 1.894444 | 3.338792 |
| RNA-Seq_D | GAB2 Bone | 4/497 | 0.170782 | 0.880031 | 0 | 0 | 1.898385 | 3.35514 |
| RNA-Seq_D | GFP Retina | 4/497 | 0.170782 | 0.880031 | 0 | 0 | 1.898385 | 3.35514 |
| RNA-Seq_D | Imatinib BN | 4/497 | 0.170782 | 0.880031 | 0 | 0 | 1.898385 | 3.35514 |
| RNA-Seq_D | Klf6 Oligod | 4/497 | 0.170782 | 0.880031 | 0 | 0 | 1.898385 | 3.35514 |
| RNA-Seq_D | Topotecan | 4/497 | 0.170782 | 0.880031 | 0 | 0 | 1.898385 | 3.35514 |
| GTEx_Tissu | GTEX-XGQ4 | 10/1769 | 0.238565 | 0.880538 | 0 | 0 | 1.340342 | 1.920861 |
| GTEx_Tissu | GTEX-T6MI | Nov-75 | 0.237904 | 0.880538 | 0 | 0 | 1.32275 | 1.899321 |
| GTEx_Tissu | GTEX-QDVI | Jan-62 | 0.237156 | 0.880538 | 0 | 0 | 3.784217 | 5.445629 |
| GTEx_Tissu | GTEX-T5JC- | 2/214 | 0.238636 | 0.880538 | 0 | 0 | 2.18657 | 3.132957 |
| GTEx_Tissu | GTEX-RU1J | 25/4984 | 0.23863 | 0.880538 | 0 | 0 | 1.215938 | 1.742248 |
| DisGeNET | Liver carcir | Dec-93 | 0.878729 | 0.880746 | 0 | 0 | 0.729718 | 0.094337 |
| GTEx_Tissu | GTEX-N7M | 10/1770 | 0.239097 | 0.880778 | 0 | 0 | 1.339507 | 1.916684 |
| RNA-Seq_D | CNBP HEK2 | 2/498 | 0.641412 | 0.881236 | 0 | 0 | 0.92111 | 0.40905 |
| RNA-Seq_D | PNN HCET | 2/497 | 0.640317 | 0.881236 | 0 | 0 | 0.923018 | 0.411474 |
| RNA-Seq_D | PRDM11 U | 2/497 | 0.640317 | 0.881236 | 0 | 0 | 0.923018 | 0.411474 |
| RNA-Seq_D | PRMT4 CD3 | 2/497 | 0.640317 | 0.881236 | 0 | 0 | 0.923018 | 0.411474 |
| RNA-Seq_D | PRMT5 A54 | 2/497 | 0.640317 | 0.881236 | 0 | 0 | 0.923018 | 0.411474 |
| RNA-Seq_D | Panobinost | 2/497 | 0.640317 | 0.881236 | 0 | 0 | 0.923018 | 0.411474 |
| RNA-Seq_D | Panobinost | 2/497 | 0.640317 | 0.881236 | 0 | 0 | 0.923018 | 0.411474 |
| RNA-Seq_D | Perhexiline | 2/497 | 0.640317 | 0.881236 | 0 | 0 | 0.923018 | 0.411474 |
| RNA-Seq_D | Ptbp1 Neur | 2/497 | 0.640317 | 0.881236 | 0 | 0 | 0.923018 | 0.411474 |
| RNA-Seq_D | C9orf72 Sp | 2/498 | 0.641412 | 0.881236 | 0 | 0 | 0.92111 | 0.40905 |
| RNA-Seq_D | CFTR Duod | 2/498 | 0.641412 | 0.881236 | 0 | 0 | 0.92111 | 0.40905 |
| RNA-Seq_D | Silvestrol M | 3/494 | 0.364119 | 0.881236 | 0 | 0 | 1.412715 | 1.427231 |
| RNA-Seq_D | Clobetasol | 2/498 | 0.641412 | 0.881236 | 0 | 0 | 0.92111 | 0.40905 |
| RNA-Seq_D | Clobetasol | 2/498 | 0.641412 | 0.881236 | 0 | 0 | 0.92111 | 0.40905 |
| RNA-Seq_D | Cnot3 ESCs | 2/498 | 0.641412 | 0.881236 | 0 | 0 | 0.92111 | 0.40905 |
| RNA-Seq_D | Cyp26b1 Sl | 2/498 | 0.641412 | 0.881236 | 0 | 0 | 0.92111 | 0.40905 |
| RNA-Seq_D | PDPN Macr | 2/497 | 0.640317 | 0.881236 | 0 | 0 | 0.923018 | 0.411474 |
| RNA-Seq_D | Parathyroi | 2/496 | 0.63922 | 0.881236 | 0 | 0 | 0.924935 | 0.413914 |
| RNA-Seq_D | Pcbp2 Feta | 2/496 | 0.63922 | 0.881236 | 0 | 0 | 0.924935 | 0.413914 |
| RNA-Seq_D | Prmt5 Mou | 2/496 | 0.63922 | 0.881236 | 0 | 0 | 0.924935 | 0.413914 |

| | | | | | | | | |
|---|---|---|---|---|---|---|---|---|
| RNA-Seq_[ | Retinoic Ac | 2/496 | 0.63922 | 0.881236 | 0 | 0 | 0.924935 | 0.413914 |
| RNA-Seq_[ | BRD9-inhib | 2/496 | 0.63922 | 0.881236 | 0 | 0 | 0.924935 | 0.413914 |
| RNA-Seq_[ | Ezh2 Mami | 2/497 | 0.640317 | 0.881236 | 0 | 0 | 0.923018 | 0.411474 |
| RNA-Seq_[ | Colorectal | 2/496 | 0.63922 | 0.881236 | 0 | 0 | 0.924935 | 0.413914 |
| RNA-Seq_[ | Crizotinib H | 2/496 | 0.63922 | 0.881236 | 0 | 0 | 0.924935 | 0.413914 |
| RNA-Seq_[ | Cyclophosp | 2/496 | 0.63922 | 0.881236 | 0 | 0 | 0.924935 | 0.413914 |
| RNA-Seq_[ | Cyclophosp | 2/496 | 0.63922 | 0.881236 | 0 | 0 | 0.924935 | 0.413914 |
| RNA-Seq_[ | Cyp26b1 Sl | 2/496 | 0.63922 | 0.881236 | 0 | 0 | 0.924935 | 0.413914 |
| RNA-Seq_[ | DMOG Ret | 2/496 | 0.63922 | 0.881236 | 0 | 0 | 0.924935 | 0.413914 |
| RNA-Seq_[ | Dicer1 Non | 2/496 | 0.63922 | 0.881236 | 0 | 0 | 0.924935 | 0.413914 |
| RNA-Seq_[ | Dioxin Live | 2/496 | 0.63922 | 0.881236 | 0 | 0 | 0.924935 | 0.413914 |
| RNA-Seq_[ | AUF1 Skele | 2/496 | 0.63922 | 0.881236 | 0 | 0 | 0.924935 | 0.413914 |
| RNA-Seq_[ | Rosiglitazo | 2/496 | 0.63922 | 0.881236 | 0 | 0 | 0.924935 | 0.413914 |
| RNA-Seq_[ | CCNK HCT1 | 2/496 | 0.63922 | 0.881236 | 0 | 0 | 0.924935 | 0.413914 |
| RNA-Seq_[ | Cardiomyo | 2/496 | 0.63922 | 0.881236 | 0 | 0 | 0.924935 | 0.413914 |
| RNA-Seq_[ | Cardiomyo | 2/496 | 0.63922 | 0.881236 | 0 | 0 | 0.924935 | 0.413914 |
| RNA-Seq_[ | Chronic Ob | 2/496 | 0.63922 | 0.881236 | 0 | 0 | 0.924935 | 0.413914 |
| RNA-Seq_[ | Clobetasol | 2/496 | 0.63922 | 0.881236 | 0 | 0 | 0.924935 | 0.413914 |
| RNA-Seq_[ | Clobetasol | 2/496 | 0.63922 | 0.881236 | 0 | 0 | 0.924935 | 0.413914 |
| RNA-Seq_[ | Dnmt1 Cor | 3/494 | 0.364119 | 0.881236 | 0 | 0 | 1.412715 | 1.427231 |
| RNA-Seq_[ | Li-Fraumer | 3/494 | 0.364119 | 0.881236 | 0 | 0 | 1.412715 | 1.427231 |
| RNA-Seq_[ | Lin28 Neur | 3/494 | 0.364119 | 0.881236 | 0 | 0 | 1.412715 | 1.427231 |
| RNA-Seq_[ | CDK10 HCT | 2/497 | 0.640317 | 0.881236 | 0 | 0 | 0.923018 | 0.411474 |
| RNA-Seq_[ | LKB1 RBE k | 2/498 | 0.641412 | 0.881236 | 0 | 0 | 0.92111 | 0.40905 |
| RNA-Seq_[ | LMNB1 Em | 2/498 | 0.641412 | 0.881236 | 0 | 0 | 0.92111 | 0.40905 |
| RNA-Seq_[ | Li-Fraumer | 2/498 | 0.641412 | 0.881236 | 0 | 0 | 0.92111 | 0.40905 |
| RNA-Seq_[ | Lsd1 Oocyt | 2/498 | 0.641412 | 0.881236 | 0 | 0 | 0.92111 | 0.40905 |
| RNA-Seq_[ | MeCP2 Hyp | 2/498 | 0.641412 | 0.881236 | 0 | 0 | 0.92111 | 0.40905 |
| RNA-Seq_[ | Mef2c/d pr | 2/498 | 0.641412 | 0.881236 | 0 | 0 | 0.92111 | 0.40905 |
| RNA-Seq_[ | Vemurafen | 2/497 | 0.640317 | 0.881236 | 0 | 0 | 0.923018 | 0.411474 |
| RNA-Seq_[ | Brpf1 Fetal | 2/497 | 0.640317 | 0.881236 | 0 | 0 | 0.923018 | 0.411474 |
| RNA-Seq_[ | C9ORF72 B | 2/497 | 0.640317 | 0.881236 | 0 | 0 | 0.923018 | 0.411474 |
| RNA-Seq_[ | C9orf72 Sp | 2/497 | 0.640317 | 0.881236 | 0 | 0 | 0.923018 | 0.411474 |
| RNA-Seq_[ | LKB1 HuH-: | 2/498 | 0.641412 | 0.881236 | 0 | 0 | 0.92111 | 0.40905 |
| RNA-Seq_[ | CITCO Hep | 2/497 | 0.640317 | 0.881236 | 0 | 0 | 0.923018 | 0.411474 |
| RNA-Seq_[ | CNBP HEK2 | 2/497 | 0.640317 | 0.881236 | 0 | 0 | 0.923018 | 0.411474 |
| RNA-Seq_[ | Casz1 Hear | 2/497 | 0.640317 | 0.881236 | 0 | 0 | 0.923018 | 0.411474 |
| RNA-Seq_[ | Colorectal | 2/497 | 0.640317 | 0.881236 | 0 | 0 | 0.923018 | 0.411474 |
| RNA-Seq_[ | Albuterol H | 2/497 | 0.640317 | 0.881236 | 0 | 0 | 0.923018 | 0.411474 |
| RNA-Seq_[ | BEGAIN HC | 2/497 | 0.640317 | 0.881236 | 0 | 0 | 0.923018 | 0.411474 |
| RNA-Seq_[ | BRD4 HT29 | 2/497 | 0.640317 | 0.881236 | 0 | 0 | 0.923018 | 0.411474 |
| RNA-Seq_[ | BZLT Akata | 2/497 | 0.640317 | 0.881236 | 0 | 0 | 0.923018 | 0.411474 |
| RNA-Seq_[ | Bacillus Cal | 2/497 | 0.640317 | 0.881236 | 0 | 0 | 0.923018 | 0.411474 |
| RNA-Seq_[ | Panobinost | 2/496 | 0.63922 | 0.881236 | 0 | 0 | 0.924935 | 0.413914 |
| RNA-Seq_[ | Rosiglitazo | 2/496 | 0.63922 | 0.881236 | 0 | 0 | 0.924935 | 0.413914 |

| | | | | | | | | |
|---|---|---|---|---|---|---|---|---|
| RNA-Seq_[ | SMCX Adul | 2/496 | 0.63922 | 0.881236 | 0 | 0 | 0.924935 | 0.413914 |
| RNA-Seq_[ | Sarcoma Sa | 2/496 | 0.63922 | 0.881236 | 0 | 0 | 0.924935 | 0.413914 |
| RNA-Seq_[ | NMP JJN3 | 2/496 | 0.63922 | 0.881236 | 0 | 0 | 0.924935 | 0.413914 |
| RNA-Seq_[ | Nova2 Brai | 2/496 | 0.63922 | 0.881236 | 0 | 0 | 0.924935 | 0.413914 |
| RNA-Seq_[ | OVOL1 Tro | 2/496 | 0.63922 | 0.881236 | 0 | 0 | 0.924935 | 0.413914 |
| RNA-Seq_[ | Obesity On | 2/496 | 0.63922 | 0.881236 | 0 | 0 | 0.924935 | 0.413914 |
| RNA-Seq_[ | PATZ1 Emb | 2/496 | 0.63922 | 0.881236 | 0 | 0 | 0.924935 | 0.413914 |
| RNA-Seq_[ | PBS-1086 8 | 2/496 | 0.63922 | 0.881236 | 0 | 0 | 0.924935 | 0.413914 |
| RNA-Seq_[ | Utx Spleen | 2/497 | 0.640317 | 0.881236 | 0 | 0 | 0.923018 | 0.411474 |
| RNA-Seq_[ | Panobinost | 2/496 | 0.63922 | 0.881236 | 0 | 0 | 0.924935 | 0.413914 |
| RNA-Seq_[ | Vitamin E k | 2/497 | 0.640317 | 0.881236 | 0 | 0 | 0.923018 | 0.411474 |
| RNA-Seq_[ | Yersinia ps | 2/497 | 0.640317 | 0.881236 | 0 | 0 | 0.923018 | 0.411474 |
| RNA-Seq_[ | Yersinia ps | 2/497 | 0.640317 | 0.881236 | 0 | 0 | 0.923018 | 0.411474 |
| RNA-Seq_[ | ZFP36L3 Yo | 2/497 | 0.640317 | 0.881236 | 0 | 0 | 0.923018 | 0.411474 |
| RNA-Seq_[ | ZnF1 Thym | 2/497 | 0.640317 | 0.881236 | 0 | 0 | 0.923018 | 0.411474 |
| RNA-Seq_[ | heat-inacti | 2/497 | 0.640317 | 0.881236 | 0 | 0 | 0.923018 | 0.411474 |
| RNA-Seq_[ | heat-inacti | 2/497 | 0.640317 | 0.881236 | 0 | 0 | 0.923018 | 0.411474 |
| RNA-Seq_[ | pterosin B | 2/497 | 0.640317 | 0.881236 | 0 | 0 | 0.923018 | 0.411474 |
| RNA-Seq_[ | STC1 Embr | 3/499 | 0.370061 | 0.881236 | 0 | 0 | 1.398113 | 1.389846 |
| RNA-Seq_[ | miR-18a W | 3/499 | 0.370061 | 0.881236 | 0 | 0 | 1.398113 | 1.389846 |
| RNA-Seq_[ | KCl Embryo | 4/709 | 0.371956 | 0.881236 | 0 | 0 | 1.313031 | 1.298561 |
| RNA-Seq_[ | Xbp1 Liver | 4/711 | 0.373943 | 0.881236 | 0 | 0 | 1.30918 | 1.287776 |
| RNA-Seq_[ | Doxycyclin | 5/925 | 0.375867 | 0.881236 | 0 | 0 | 1.258815 | 1.231777 |
| RNA-Seq_[ | MITF 501 M | 4/727 | 0.389826 | 0.881236 | 0 | 0 | 1.279141 | 1.205022 |
| RNA-Seq_[ | Xbp1 Liver | 4/733 | 0.395771 | 0.881236 | 0 | 0 | 1.268217 | 1.175534 |
| RNA-Seq_[ | SAHA Hipp | 4/790 | 0.451702 | 0.881236 | 0 | 0 | 1.172752 | 0.932024 |
| RNA-Seq_[ | Rb1 Colon | 3/499 | 0.370061 | 0.881236 | 0 | 0 | 1.398113 | 1.389846 |
| RNA-Seq_[ | Rfx2 Testes | 3/499 | 0.370061 | 0.881236 | 0 | 0 | 1.398113 | 1.389846 |
| RNA-Seq_[ | Runx1 Bon | 3/499 | 0.370061 | 0.881236 | 0 | 0 | 1.398113 | 1.389846 |
| RNA-Seq_[ | miR-18a He | 3/499 | 0.370061 | 0.881236 | 0 | 0 | 1.398113 | 1.389846 |
| RNA-Seq_[ | Satb1 Midb | 3/499 | 0.370061 | 0.881236 | 0 | 0 | 1.398113 | 1.389846 |
| RNA-Seq_[ | Shh Forelin | 3/499 | 0.370061 | 0.881236 | 0 | 0 | 1.398113 | 1.389846 |
| RNA-Seq_[ | TGFB3 Pala | 3/499 | 0.370061 | 0.881236 | 0 | 0 | 1.398113 | 1.389846 |
| RNA-Seq_[ | Tet1 Bone | 3/499 | 0.370061 | 0.881236 | 0 | 0 | 1.398113 | 1.389846 |
| RNA-Seq_[ | 2,3,7,8-tetr | 2/497 | 0.640317 | 0.881236 | 0 | 0 | 0.923018 | 0.411474 |
| RNA-Seq_[ | 4-hydroxyt | 2/497 | 0.640317 | 0.881236 | 0 | 0 | 0.923018 | 0.411474 |
| RNA-Seq_[ | 4-hydroxyt | 2/497 | 0.640317 | 0.881236 | 0 | 0 | 0.923018 | 0.411474 |
| RNA-Seq_[ | 4 Skin Biop | 2/497 | 0.640317 | 0.881236 | 0 | 0 | 0.923018 | 0.411474 |
| RNA-Seq_[ | 5-Azacytidi | 2/497 | 0.640317 | 0.881236 | 0 | 0 | 0.923018 | 0.411474 |
| RNA-Seq_[ | Myelodysp | 2/496 | 0.63922 | 0.881236 | 0 | 0 | 0.924935 | 0.413914 |
| RNA-Seq_[ | Salmonella | 3/498 | 0.368874 | 0.881236 | 0 | 0 | 1.40101 | 1.397229 |
| RNA-Seq_[ | Setdb1 C2C | 3/498 | 0.368874 | 0.881236 | 0 | 0 | 1.40101 | 1.397229 |
| RNA-Seq_[ | Setdb1 Ske | 3/498 | 0.368874 | 0.881236 | 0 | 0 | 1.40101 | 1.397229 |
| RNA-Seq_[ | Smchd1 Ne | 3/498 | 0.368874 | 0.881236 | 0 | 0 | 1.40101 | 1.397229 |
| RNA-Seq_[ | Miconazole | 2/496 | 0.63922 | 0.881236 | 0 | 0 | 0.924935 | 0.413914 |

| | | | | | | | | |
|---|---|---|---|---|---|---|---|---|
| RNA-Seq_D | Miconazole | 2/496 | 0.63922 | 0.881236 | 0 | 0 | 0.924935 | 0.413914 |
| RNA-Seq_D | Msi1 MIA F | 2/496 | 0.63922 | 0.881236 | 0 | 0 | 0.924935 | 0.413914 |
| RNA-Seq_D | Msi1 MIA F | 2/496 | 0.63922 | 0.881236 | 0 | 0 | 0.924935 | 0.413914 |
| RNA-Seq_D | Mycobacte | 2/496 | 0.63922 | 0.881236 | 0 | 0 | 0.924935 | 0.413914 |
| RNA-Seq_D | Mycobacte | 2/496 | 0.63922 | 0.881236 | 0 | 0 | 0.924935 | 0.413914 |
| RNA-Seq_D | 5-Azacytidi | 2/497 | 0.640317 | 0.881236 | 0 | 0 | 0.923018 | 0.411474 |
| RNA-Seq_D | Myelodysp | 2/496 | 0.63922 | 0.881236 | 0 | 0 | 0.924935 | 0.413914 |
| RNA-Seq_D | Crizotinib H | 2/497 | 0.640317 | 0.881236 | 0 | 0 | 0.923018 | 0.411474 |
| RNA-Seq_D | Ctcfl Trans | 2/497 | 0.640317 | 0.881236 | 0 | 0 | 0.923018 | 0.411474 |
| RNA-Seq_D | Dexametha | 2/497 | 0.640317 | 0.881236 | 0 | 0 | 0.923018 | 0.411474 |
| RNA-Seq_D | Dexametha | 3/496 | 0.366497 | 0.881236 | 0 | 0 | 1.406839 | 1.412136 |
| RNA-Seq_D | Duchenne | 2/497 | 0.640317 | 0.881236 | 0 | 0 | 0.923018 | 0.411474 |
| RNA-Seq_D | EPAS1 Yolk | 2/497 | 0.640317 | 0.881236 | 0 | 0 | 0.923018 | 0.411474 |
| RNA-Seq_D | EVX1 CJ9 st | 2/497 | 0.640317 | 0.881236 | 0 | 0 | 0.923018 | 0.411474 |
| RNA-Seq_D | MOV10 HE | 2/496 | 0.63922 | 0.881236 | 0 | 0 | 0.924935 | 0.413914 |
| RNA-Seq_D | GLI1 HCT11 | 2/497 | 0.640317 | 0.881236 | 0 | 0 | 0.923018 | 0.411474 |
| RNA-Seq_D | Tunicamyci | 2/494 | 0.637018 | 0.881236 | 0 | 0 | 0.92879 | 0.418845 |
| RNA-Seq_D | Tunicamyci | 2/494 | 0.637018 | 0.881236 | 0 | 0 | 0.92879 | 0.418845 |
| RNA-Seq_D | UTX Muscle | 2/494 | 0.637018 | 0.881236 | 0 | 0 | 0.92879 | 0.418845 |
| RNA-Seq_D | recombina | 2/494 | 0.637018 | 0.881236 | 0 | 0 | 0.92879 | 0.418845 |
| RNA-Seq_D | ALS IPSCs-c | 2/495 | 0.63812 | 0.881236 | 0 | 0 | 0.926858 | 0.416371 |
| RNA-Seq_D | All-trans re | 2/495 | 0.63812 | 0.881236 | 0 | 0 | 0.926858 | 0.416371 |
| RNA-Seq_D | FLT3 Thym | 2/497 | 0.640317 | 0.881236 | 0 | 0 | 0.923018 | 0.411474 |
| RNA-Seq_D | FUS Motor | 2/497 | 0.640317 | 0.881236 | 0 | 0 | 0.923018 | 0.411474 |
| RNA-Seq_D | GATA2 Ch- | 2/497 | 0.640317 | 0.881236 | 0 | 0 | 0.923018 | 0.411474 |
| RNA-Seq_D | GLI1 HCT11 | 2/497 | 0.640317 | 0.881236 | 0 | 0 | 0.923018 | 0.411474 |
| RNA-Seq_D | Tunicamyci | 2/494 | 0.637018 | 0.881236 | 0 | 0 | 0.92879 | 0.418845 |
| RNA-Seq_D | GW3965 Tu | 2/497 | 0.640317 | 0.881236 | 0 | 0 | 0.923018 | 0.411474 |
| RNA-Seq_D | Gcn5 Embr | 2/497 | 0.640317 | 0.881236 | 0 | 0 | 0.923018 | 0.411474 |
| RNA-Seq_D | HNRNPA1 / | 2/497 | 0.640317 | 0.881236 | 0 | 0 | 0.923018 | 0.411474 |
| RNA-Seq_D | Tunicamyci | 2/497 | 0.640317 | 0.881236 | 0 | 0 | 0.923018 | 0.411474 |
| RNA-Seq_D | Tunicamyci | 2/497 | 0.640317 | 0.881236 | 0 | 0 | 0.923018 | 0.411474 |
| RNA-Seq_D | Tunicamyci | 2/497 | 0.640317 | 0.881236 | 0 | 0 | 0.923018 | 0.411474 |
| RNA-Seq_D | Tunicamyci | 2/497 | 0.640317 | 0.881236 | 0 | 0 | 0.923018 | 0.411474 |
| RNA-Seq_D | Tunicamyci | 2/497 | 0.640317 | 0.881236 | 0 | 0 | 0.923018 | 0.411474 |
| RNA-Seq_D | Tunicamyci | 2/497 | 0.640317 | 0.881236 | 0 | 0 | 0.923018 | 0.411474 |
| RNA-Seq_D | 17-betaest | 2/497 | 0.640317 | 0.881236 | 0 | 0 | 0.923018 | 0.411474 |
| RNA-Seq_D | ADAR Fetal | 2/497 | 0.640317 | 0.881236 | 0 | 0 | 0.923018 | 0.411474 |
| RNA-Seq_D | ALS (Famili | 2/497 | 0.640317 | 0.881236 | 0 | 0 | 0.923018 | 0.411474 |
| RNA-Seq_D | Mycobacte | 2/497 | 0.640317 | 0.881236 | 0 | 0 | 0.923018 | 0.411474 |
| RNA-Seq_D | Myelodysp | 2/497 | 0.640317 | 0.881236 | 0 | 0 | 0.923018 | 0.411474 |
| RNA-Seq_D | Myelodysp | 2/497 | 0.640317 | 0.881236 | 0 | 0 | 0.923018 | 0.411474 |
| RNA-Seq_D | N-acetylcys | 2/497 | 0.640317 | 0.881236 | 0 | 0 | 0.923018 | 0.411474 |
| RNA-Seq_D | NHERF2 HC | 2/497 | 0.640317 | 0.881236 | 0 | 0 | 0.923018 | 0.411474 |
| RNA-Seq_D | NHERF2 HC | 2/497 | 0.640317 | 0.881236 | 0 | 0 | 0.923018 | 0.411474 |

| | | | | | | | | |
|---|---|---|---|---|---|---|---|---|
| RNA-Seq_[ | PATZ1 Emb | 2/497 | 0.640317 | 0.881236 | 0 | 0 | 0.923018 | 0.411474 |
| RNA-Seq_[ | Bicuculin H | 2/497 | 0.640317 | 0.881236 | 0 | 0 | 0.923018 | 0.411474 |
| RNA-Seq_[ | Autism iPS( | 2/495 | 0.63812 | 0.881236 | 0 | 0 | 0.926858 | 0.416371 |
| RNA-Seq_[ | Bexarotene | 2/495 | 0.63812 | 0.881236 | 0 | 0 | 0.926858 | 0.416371 |
| RNA-Seq_[ | CPSF2 HEK | 2/495 | 0.63812 | 0.881236 | 0 | 0 | 0.926858 | 0.416371 |
| RNA-Seq_[ | Cyclophosp | 2/495 | 0.63812 | 0.881236 | 0 | 0 | 0.926858 | 0.416371 |
| RNA-Seq_[ | Dioxin Live | 2/495 | 0.63812 | 0.881236 | 0 | 0 | 0.926858 | 0.416371 |
| RNA-Seq_[ | EIF3L HCT1 | 2/495 | 0.63812 | 0.881236 | 0 | 0 | 0.926858 | 0.416371 |
| RNA-Seq_[ | Hira Oocyt | 2/495 | 0.63812 | 0.881236 | 0 | 0 | 0.926858 | 0.416371 |
| RNA-Seq_[ | IL-4 BMDM | 2/495 | 0.63812 | 0.881236 | 0 | 0 | 0.926858 | 0.416371 |
| RNA-Seq_[ | SUZ12 HEK | 2/494 | 0.637018 | 0.881236 | 0 | 0 | 0.92879 | 0.418845 |
| RNA-Seq_[ | Ginkgolic A | 2/498 | 0.641412 | 0.881236 | 0 | 0 | 0.92111 | 0.40905 |
| RNA-Seq_[ | INK128 MD | 2/498 | 0.641412 | 0.881236 | 0 | 0 | 0.92111 | 0.40905 |
| RNA-Seq_[ | IRF8 Spleer | 2/498 | 0.641412 | 0.881236 | 0 | 0 | 0.92111 | 0.40905 |
| RNA-Seq_[ | Idiopathic I | 2/498 | 0.641412 | 0.881236 | 0 | 0 | 0.92111 | 0.40905 |
| RNA-Seq_[ | Idiopathic ı | 2/498 | 0.641412 | 0.881236 | 0 | 0 | 0.92111 | 0.40905 |
| RNA-Seq_[ | Irf4 CD8+ T | 2/498 | 0.641412 | 0.881236 | 0 | 0 | 0.92111 | 0.40905 |
| RNA-Seq_[ | Itch Skin Kr | 2/498 | 0.641412 | 0.881236 | 0 | 0 | 0.92111 | 0.40905 |
| RNA-Seq_[ | FICZ Skin B | 2/498 | 0.641412 | 0.881236 | 0 | 0 | 0.92111 | 0.40905 |
| RNA-Seq_[ | G9a Embry | 2/498 | 0.641412 | 0.881236 | 0 | 0 | 0.92111 | 0.40905 |
| RNA-Seq_[ | GW3965 Tu | 2/498 | 0.641412 | 0.881236 | 0 | 0 | 0.92111 | 0.40905 |
| RNA-Seq_[ | Gcn2 Cerek | 2/498 | 0.641412 | 0.881236 | 0 | 0 | 0.92111 | 0.40905 |
| RNA-Seq_[ | ILK (Epider | 2/498 | 0.641412 | 0.881236 | 0 | 0 | 0.92111 | 0.40905 |
| RNA-Seq_[ | Gtf2ird1 Lip | 2/498 | 0.641412 | 0.881236 | 0 | 0 | 0.92111 | 0.40905 |
| RNA-Seq_[ | HNRNPA2E | 2/498 | 0.641412 | 0.881236 | 0 | 0 | 0.92111 | 0.40905 |
| RNA-Seq_[ | HNRNPA2E | 2/498 | 0.641412 | 0.881236 | 0 | 0 | 0.92111 | 0.40905 |
| RNA-Seq_[ | Phenobarb | 2/498 | 0.641412 | 0.881236 | 0 | 0 | 0.92111 | 0.40905 |
| RNA-Seq_[ | Ptbp2 Neur | 2/498 | 0.641412 | 0.881236 | 0 | 0 | 0.92111 | 0.40905 |
| RNA-Seq_[ | Ptf1a 14 da | 2/498 | 0.641412 | 0.881236 | 0 | 0 | 0.92111 | 0.40905 |
| RNA-Seq_[ | RBM47 H3! | 2/498 | 0.641412 | 0.881236 | 0 | 0 | 0.92111 | 0.40905 |
| RNA-Seq_[ | Renal Cell ( | 2/498 | 0.641412 | 0.881236 | 0 | 0 | 0.92111 | 0.40905 |
| RNA-Seq_[ | Retinoic Ac | 2/498 | 0.641412 | 0.881236 | 0 | 0 | 0.92111 | 0.40905 |
| RNA-Seq_[ | Dlx3(Dlx3-I | 2/499 | 0.642504 | 0.881236 | 0 | 0 | 0.919209 | 0.406642 |
| RNA-Seq_[ | Brg1 Embry | 2/498 | 0.641412 | 0.881236 | 0 | 0 | 0.92111 | 0.40905 |
| RNA-Seq_[ | C9ORF72 B | 2/498 | 0.641412 | 0.881236 | 0 | 0 | 0.92111 | 0.40905 |
| RNA-Seq_[ | 4-hydroxyt | 2/498 | 0.641412 | 0.881236 | 0 | 0 | 0.92111 | 0.40905 |
| RNA-Seq_[ | 5-Azacytidi | 2/498 | 0.641412 | 0.881236 | 0 | 0 | 0.92111 | 0.40905 |
| RNA-Seq_[ | AEMeis2 B | 2/498 | 0.641412 | 0.881236 | 0 | 0 | 0.92111 | 0.40905 |
| RNA-Seq_[ | AMD3100 | 2/498 | 0.641412 | 0.881236 | 0 | 0 | 0.92111 | 0.40905 |
| RNA-Seq_[ | All-trans re | 2/498 | 0.641412 | 0.881236 | 0 | 0 | 0.92111 | 0.40905 |
| RNA-Seq_[ | BRD4 HCT1 | 2/498 | 0.641412 | 0.881236 | 0 | 0 | 0.92111 | 0.40905 |
| RNA-Seq_[ | BRD4 HT29 | 2/498 | 0.641412 | 0.881236 | 0 | 0 | 0.92111 | 0.40905 |
| RNA-Seq_[ | BRD9-inhib | 2/498 | 0.641412 | 0.881236 | 0 | 0 | 0.92111 | 0.40905 |
| RNA-Seq_[ | Rfx2 Testes | 2/498 | 0.641412 | 0.881236 | 0 | 0 | 0.92111 | 0.40905 |
| RNA-Seq_[ | EPAS1 Yolk | 2/499 | 0.642504 | 0.881236 | 0 | 0 | 0.919209 | 0.406642 |

| | | | | | | | | |
|---|---|---|---|---|---|---|---|---|
| RNA-Seq_[ | Ezh1 Mamı | 2/499 | 0.642504 | 0.881236 | 0 | 0 | 0.919209 | 0.406642 |
| RNA-Seq_[ | Ezh2 Splee | 2/499 | 0.642504 | 0.881236 | 0 | 0 | 0.919209 | 0.406642 |
| RNA-Seq_[ | Ezh2 mESC | 2/499 | 0.642504 | 0.881236 | 0 | 0 | 0.919209 | 0.406642 |
| RNA-Seq_[ | FOXC1 Skir | 2/499 | 0.642504 | 0.881236 | 0 | 0 | 0.919209 | 0.406642 |
| RNA-Seq_[ | FancC matı | 2/499 | 0.642504 | 0.881236 | 0 | 0 | 0.919209 | 0.406642 |
| RNA-Seq_[ | Gprc5a col | 2/499 | 0.642504 | 0.881236 | 0 | 0 | 0.919209 | 0.406642 |
| RNA-Seq_[ | Dgcr8 Cone | 2/499 | 0.642504 | 0.881236 | 0 | 0 | 0.919209 | 0.406642 |
| RNA-Seq_[ | Hnrnpk TT2 | 2/498 | 0.641412 | 0.881236 | 0 | 0 | 0.92111 | 0.40905 |
| RNA-Seq_[ | Bicuculin H | 3/495 | 0.365308 | 0.881236 | 0 | 0 | 1.409771 | 1.41966 |
| RNA-Seq_[ | HER2 BT47 | 3/497 | 0.367685 | 0.881236 | 0 | 0 | 1.403918 | 1.404659 |
| RNA-Seq_[ | IL-13 Small | 3/497 | 0.367685 | 0.881236 | 0 | 0 | 1.403918 | 1.404659 |
| RNA-Seq_[ | ISRIB & Tur | 3/497 | 0.367685 | 0.881236 | 0 | 0 | 1.403918 | 1.404659 |
| RNA-Seq_[ | Dioxin Live | 3/496 | 0.366497 | 0.881236 | 0 | 0 | 1.406839 | 1.412136 |
| RNA-Seq_[ | JUND PC3 I | 3/497 | 0.367685 | 0.881236 | 0 | 0 | 1.403918 | 1.404659 |
| RNA-Seq_[ | KCNK1 RKC | 3/497 | 0.367685 | 0.881236 | 0 | 0 | 1.403918 | 1.404659 |
| RNA-Seq_[ | KH-CB19 Eı | 3/497 | 0.367685 | 0.881236 | 0 | 0 | 1.403918 | 1.404659 |
| RNA-Seq_[ | Ezh2 Embr | 3/496 | 0.366497 | 0.881236 | 0 | 0 | 1.406839 | 1.412136 |
| RNA-Seq_[ | 5-Azacytidi | 3/495 | 0.365308 | 0.881236 | 0 | 0 | 1.409771 | 1.41966 |
| RNA-Seq_[ | Acromegal | 3/495 | 0.365308 | 0.881236 | 0 | 0 | 1.409771 | 1.41966 |
| RNA-Seq_[ | Huntingtor | 3/496 | 0.366497 | 0.881236 | 0 | 0 | 1.406839 | 1.412136 |
| RNA-Seq_[ | Trim33 B ly | 2/499 | 0.642504 | 0.881236 | 0 | 0 | 0.919209 | 0.406642 |
| RNA-Seq_[ | Panobinost | 2/498 | 0.641412 | 0.881236 | 0 | 0 | 0.92111 | 0.40905 |
| RNA-Seq_[ | Idiopathic | 3/497 | 0.367685 | 0.881236 | 0 | 0 | 1.403918 | 1.404659 |
| RNA-Seq_[ | Mettl3 Eml | 2/499 | 0.642504 | 0.881236 | 0 | 0 | 0.919209 | 0.406642 |
| RNA-Seq_[ | 2,3,7,8-tetı | 3/494 | 0.364119 | 0.881236 | 0 | 0 | 1.412715 | 1.427231 |
| RNA-Seq_[ | ALS (Famili | 3/494 | 0.364119 | 0.881236 | 0 | 0 | 1.412715 | 1.427231 |
| RNA-Seq_[ | APP/PS1-2 | 3/494 | 0.364119 | 0.881236 | 0 | 0 | 1.412715 | 1.427231 |
| RNA-Seq_[ | Desipramir | 3/496 | 0.366497 | 0.881236 | 0 | 0 | 1.406839 | 1.412136 |
| RNA-Seq_[ | Dexametha | 3/496 | 0.366497 | 0.881236 | 0 | 0 | 1.406839 | 1.412136 |
| RNA-Seq_[ | CFTR Color | 6/888 | 0.189463 | 0.881236 | 0 | 0 | 1.598304 | 2.658878 |
| RNA-Seq_[ | Roxadust L | 2/498 | 0.641412 | 0.881236 | 0 | 0 | 0.92111 | 0.40905 |
| RNA-Seq_[ | Nova1 Brai | 2/498 | 0.641412 | 0.881236 | 0 | 0 | 0.92111 | 0.40905 |
| RNA-Seq_[ | Nrp1neg C | 2/498 | 0.641412 | 0.881236 | 0 | 0 | 0.92111 | 0.40905 |
| RNA-Seq_[ | PER2 Endc | 2/498 | 0.641412 | 0.881236 | 0 | 0 | 0.92111 | 0.40905 |
| RNA-Seq_[ | PRMT9 Hel | 2/498 | 0.641412 | 0.881236 | 0 | 0 | 0.92111 | 0.40905 |
| RNA-Seq_[ | PS2APP As | 2/498 | 0.641412 | 0.881236 | 0 | 0 | 0.92111 | 0.40905 |
| RNA-Seq_[ | PS2APP Mi | 2/498 | 0.641412 | 0.881236 | 0 | 0 | 0.92111 | 0.40905 |
| RNA-Seq_[ | Panobinost | 2/498 | 0.641412 | 0.881236 | 0 | 0 | 0.92111 | 0.40905 |
| RNA-Seq_[ | EPZ6438 K | 3/496 | 0.366497 | 0.881236 | 0 | 0 | 1.406839 | 1.412136 |
| RNA-Seq_[ | Bmpr1a Ha | 2/498 | 0.641412 | 0.881236 | 0 | 0 | 0.92111 | 0.40905 |
| RNA-Seq_[ | Desipramir | 3/493 | 0.362929 | 0.881236 | 0 | 0 | 1.415671 | 1.434849 |
| RNA-Seq_[ | NNK Lung | 3/493 | 0.362929 | 0.881236 | 0 | 0 | 1.415671 | 1.434849 |
| RNA-Seq_[ | Ezh2 mESC | 3/496 | 0.366497 | 0.881236 | 0 | 0 | 1.406839 | 1.412136 |
| RNA-Seq_[ | FBXL10 Fer | 3/496 | 0.366497 | 0.881236 | 0 | 0 | 1.406839 | 1.412136 |
| RNA-Seq_[ | FOXA2 End | 3/496 | 0.366497 | 0.881236 | 0 | 0 | 1.406839 | 1.412136 |

| | | | | | | | | |
|---|---|---|---|---|---|---|---|---|
| RNA-Seq_[ | FUS Motor | 3/496 | 0.366497 | 0.881236 | 0 | 0 | 1.406839 | 1.412136 |
| RNA-Seq_[ | G9a Embry | 3/496 | 0.366497 | 0.881236 | 0 | 0 | 1.406839 | 1.412136 |
| RNA-Seq_[ | Gcn2 Cerel | 3/496 | 0.366497 | 0.881236 | 0 | 0 | 1.406839 | 1.412136 |
| RNA-Seq_[ | Ginkgolic A | 3/496 | 0.366497 | 0.881236 | 0 | 0 | 1.406839 | 1.412136 |
| RNA-Seq_[ | IPTKB Hem | 2/497 | 0.640317 | 0.881236 | 0 | 0 | 0.923018 | 0.411474 |
| RNA-Seq_[ | Klf6 Immat | 2/497 | 0.640317 | 0.881236 | 0 | 0 | 0.923018 | 0.411474 |
| RNA-Seq_[ | Kras(G12D) | 2/497 | 0.640317 | 0.881236 | 0 | 0 | 0.923018 | 0.411474 |
| RNA-Seq_[ | LPS Lung 5 | 2/497 | 0.640317 | 0.881236 | 0 | 0 | 0.923018 | 0.411474 |
| RNA-Seq_[ | Lap2alpha | 2/497 | 0.640317 | 0.881236 | 0 | 0 | 0.923018 | 0.411474 |
| RNA-Seq_[ | Lapatinib H | 2/497 | 0.640317 | 0.881236 | 0 | 0 | 0.923018 | 0.411474 |
| RNA-Seq_[ | Mycobacte | 2/497 | 0.640317 | 0.881236 | 0 | 0 | 0.923018 | 0.411474 |
| RNA-Seq_[ | HNRNPA2E | 2/497 | 0.640317 | 0.881236 | 0 | 0 | 0.923018 | 0.411474 |
| RNA-Seq_[ | Haloperido | 2/497 | 0.640317 | 0.881236 | 0 | 0 | 0.923018 | 0.411474 |
| RNA-Seq_[ | Huntingtor | 2/497 | 0.640317 | 0.881236 | 0 | 0 | 0.923018 | 0.411474 |
| RNA-Seq_[ | I-BET MV4- | 2/497 | 0.640317 | 0.881236 | 0 | 0 | 0.923018 | 0.411474 |
| RNA-Seq_[ | Kawasaki D | 2/497 | 0.640317 | 0.881236 | 0 | 0 | 0.923018 | 0.411474 |
| RNA-Seq_[ | ISRIB HEK2 | 2/497 | 0.640317 | 0.881236 | 0 | 0 | 0.923018 | 0.411474 |
| RNA-Seq_[ | Imatinib BN | 2/497 | 0.640317 | 0.881236 | 0 | 0 | 0.923018 | 0.411474 |
| RNA-Seq_[ | Imatinib CN | 2/497 | 0.640317 | 0.881236 | 0 | 0 | 0.923018 | 0.411474 |
| RNA-Seq_[ | Msi2 MIA F | 2/498 | 0.641412 | 0.881236 | 0 | 0 | 0.92111 | 0.40905 |
| RNA-Seq_[ | Mycobacte | 2/498 | 0.641412 | 0.881236 | 0 | 0 | 0.92111 | 0.40905 |
| RNA-Seq_[ | Mycobacte | 2/498 | 0.641412 | 0.881236 | 0 | 0 | 0.92111 | 0.40905 |
| RNA-Seq_[ | Mycobacte | 2/498 | 0.641412 | 0.881236 | 0 | 0 | 0.92111 | 0.40905 |
| RNA-Seq_[ | Mycobacte | 2/498 | 0.641412 | 0.881236 | 0 | 0 | 0.92111 | 0.40905 |
| RNA-Seq_[ | Mycobacte | 2/498 | 0.641412 | 0.881236 | 0 | 0 | 0.92111 | 0.40905 |
| RNA-Seq_[ | FICZ Skin B | 2/498 | 0.641412 | 0.881236 | 0 | 0 | 0.92111 | 0.40905 |
| RNA-Seq_[ | Blinc1 Panc | 2/497 | 0.640317 | 0.881236 | 0 | 0 | 0.923018 | 0.411474 |
| RNA-Seq_[ | Bmal1 Live | 2/497 | 0.640317 | 0.881236 | 0 | 0 | 0.923018 | 0.411474 |
| RNA-Seq_[ | DGCR8 Cor | 2/498 | 0.641412 | 0.881236 | 0 | 0 | 0.92111 | 0.40905 |
| RNA-Seq_[ | Dgcr8 Cone | 2/498 | 0.641412 | 0.881236 | 0 | 0 | 0.92111 | 0.40905 |
| RNA-Seq_[ | Dioxin Live | 2/498 | 0.641412 | 0.881236 | 0 | 0 | 0.92111 | 0.40905 |
| RNA-Seq_[ | Dnmt3a Co | 2/498 | 0.641412 | 0.881236 | 0 | 0 | 0.92111 | 0.40905 |
| RNA-Seq_[ | Doxycyclin | 2/498 | 0.641412 | 0.881236 | 0 | 0 | 0.92111 | 0.40905 |
| RNA-Seq_[ | ETS2 Skin C | 2/498 | 0.641412 | 0.881236 | 0 | 0 | 0.92111 | 0.40905 |
| RNA-Seq_[ | Epac1-PLN | 2/498 | 0.641412 | 0.881236 | 0 | 0 | 0.92111 | 0.40905 |
| RNA-Seq_[ | Mycobacte | 2/498 | 0.641412 | 0.881236 | 0 | 0 | 0.92111 | 0.40905 |
| RNA-Seq_[ | Roxadust R | 2/498 | 0.641412 | 0.881236 | 0 | 0 | 0.92111 | 0.40905 |
| RNA-Seq_[ | SGC0946 & | 2/498 | 0.641412 | 0.881236 | 0 | 0 | 0.92111 | 0.40905 |
| RNA-Seq_[ | SGC0946 & | 2/498 | 0.641412 | 0.881236 | 0 | 0 | 0.92111 | 0.40905 |
| RNA-Seq_[ | SGC0946 & | 2/498 | 0.641412 | 0.881236 | 0 | 0 | 0.92111 | 0.40905 |
| RNA-Seq_[ | SGC0946 & | 2/498 | 0.641412 | 0.881236 | 0 | 0 | 0.92111 | 0.40905 |
| RNA-Seq_[ | SMYD1 Tib | 2/498 | 0.641412 | 0.881236 | 0 | 0 | 0.92111 | 0.40905 |
| RNA-Seq_[ | SNTA1 RKC | 2/498 | 0.641412 | 0.881236 | 0 | 0 | 0.92111 | 0.40905 |
| RNA-Seq_[ | DAXX PC3 I | 2/498 | 0.641412 | 0.881236 | 0 | 0 | 0.92111 | 0.40905 |
| RNA-Seq_[ | JAK3i (CP-6 | 2/497 | 0.640317 | 0.881236 | 0 | 0 | 0.923018 | 0.411474 |

| | | | | | | | | |
|---|---|---|---|---|---|---|---|---|
| RNA-Seq_[ | Salmonella | 2/498 | 0.641412 | 0.881236 | 0 | 0 | 0.92111 | 0.40905 |
| RNA-Seq_[ | SMCX Adul | 2/497 | 0.640317 | 0.881236 | 0 | 0 | 0.923018 | 0.411474 |
| RNA-Seq_[ | SNTA1 RKC | 2/497 | 0.640317 | 0.881236 | 0 | 0 | 0.923018 | 0.411474 |
| RNA-Seq_[ | SOD1(G93/ | 2/497 | 0.640317 | 0.881236 | 0 | 0 | 0.923018 | 0.411474 |
| RNA-Seq_[ | SUZ12 HEK | 2/497 | 0.640317 | 0.881236 | 0 | 0 | 0.923018 | 0.411474 |
| RNA-Seq_[ | SYKi (R406) | 2/497 | 0.640317 | 0.881236 | 0 | 0 | 0.923018 | 0.411474 |
| RNA-Seq_[ | STAU1 HEK | 2/498 | 0.641412 | 0.881236 | 0 | 0 | 0.92111 | 0.40905 |
| RNA-Seq_[ | STAU1 HEK | 2/498 | 0.641412 | 0.881236 | 0 | 0 | 0.92111 | 0.40905 |
| RNA-Seq_[ | STC1 Embr | 2/498 | 0.641412 | 0.881236 | 0 | 0 | 0.92111 | 0.40905 |
| RNA-Seq_[ | Salmonella | 2/498 | 0.641412 | 0.881236 | 0 | 0 | 0.92111 | 0.40905 |
| RNA-Seq_[ | Salmonella | 2/498 | 0.641412 | 0.881236 | 0 | 0 | 0.92111 | 0.40905 |
| RNA-Seq_[ | Roxadust L | 2/497 | 0.640317 | 0.881236 | 0 | 0 | 0.923018 | 0.411474 |
| RNA-Seq_[ | Shh Forelin | 2/498 | 0.641412 | 0.881236 | 0 | 0 | 0.92111 | 0.40905 |
| RNA-Seq_[ | Sox2 Stoma | 2/498 | 0.641412 | 0.881236 | 0 | 0 | 0.92111 | 0.40905 |
| RNA-Seq_[ | YY1 Bone n | 2/499 | 0.642504 | 0.881236 | 0 | 0 | 0.919209 | 0.406642 |
| RNA-Seq_[ | SOD1(G93/ | 2/498 | 0.641412 | 0.881236 | 0 | 0 | 0.92111 | 0.40905 |
| RNA-Seq_[ | Bacillus Cal | 2/498 | 0.641412 | 0.881236 | 0 | 0 | 0.92111 | 0.40905 |
| RNA-Seq_[ | Bacillus Cal | 2/498 | 0.641412 | 0.881236 | 0 | 0 | 0.92111 | 0.40905 |
| RNA-Seq_[ | Baf60c Car | 2/498 | 0.641412 | 0.881236 | 0 | 0 | 0.92111 | 0.40905 |
| RNA-Seq_[ | Blinc1 Panc | 2/498 | 0.641412 | 0.881236 | 0 | 0 | 0.92111 | 0.40905 |
| RNA-Seq_[ | Bmal1 Live | 2/498 | 0.641412 | 0.881236 | 0 | 0 | 0.92111 | 0.40905 |
| RNA-Seq_[ | SYKi (R406) | 2/497 | 0.640317 | 0.881236 | 0 | 0 | 0.923018 | 0.411474 |
| RNA-Seq_[ | NMP JJN3 4 | 2/498 | 0.641412 | 0.881236 | 0 | 0 | 0.92111 | 0.40905 |
| RNA-Seq_[ | Baf60c Car | 2/499 | 0.642504 | 0.881236 | 0 | 0 | 0.919209 | 0.406642 |
| RNA-Seq_[ | Bmal1 Live | 2/499 | 0.642504 | 0.881236 | 0 | 0 | 0.919209 | 0.406642 |
| RNA-Seq_[ | Bmal1 Live | 2/499 | 0.642504 | 0.881236 | 0 | 0 | 0.919209 | 0.406642 |
| RNA-Seq_[ | Bmal1 Live | 2/499 | 0.642504 | 0.881236 | 0 | 0 | 0.919209 | 0.406642 |
| RNA-Seq_[ | Brg1 Embry | 2/499 | 0.642504 | 0.881236 | 0 | 0 | 0.919209 | 0.406642 |
| RNA-Seq_[ | C9orf72 Sp | 2/499 | 0.642504 | 0.881236 | 0 | 0 | 0.919209 | 0.406642 |
| RNA-Seq_[ | DGCR8 Cor | 2/499 | 0.642504 | 0.881236 | 0 | 0 | 0.919209 | 0.406642 |
| RNA-Seq_[ | Mettl3 Eml | 2/498 | 0.641412 | 0.881236 | 0 | 0 | 0.92111 | 0.40905 |
| RNA-Seq_[ | Dioxin Live | 2/497 | 0.640317 | 0.881236 | 0 | 0 | 0.923018 | 0.411474 |
| RNA-Seq_[ | Salmonella | 2/497 | 0.640317 | 0.881236 | 0 | 0 | 0.923018 | 0.411474 |
| RNA-Seq_[ | Sim2 Embr | 2/497 | 0.640317 | 0.881236 | 0 | 0 | 0.923018 | 0.411474 |
| RNA-Seq_[ | Sim2 Embr | 2/497 | 0.640317 | 0.881236 | 0 | 0 | 0.923018 | 0.411474 |
| RNA-Seq_[ | TCF7L2 Pa | 2/497 | 0.640317 | 0.881236 | 0 | 0 | 0.923018 | 0.411474 |
| RNA-Seq_[ | TDP43 Mot | 2/497 | 0.640317 | 0.881236 | 0 | 0 | 0.923018 | 0.411474 |
| RNA-Seq_[ | TGFB3 Pala | 2/497 | 0.640317 | 0.881236 | 0 | 0 | 0.923018 | 0.411474 |
| RNA-Seq_[ | Trim24 Live | 2/497 | 0.640317 | 0.881236 | 0 | 0 | 0.923018 | 0.411474 |
| RNA-Seq_[ | Rcor1 Fetal | 2/497 | 0.640317 | 0.881236 | 0 | 0 | 0.923018 | 0.411474 |
| RNA-Seq_[ | Rocaglamic | 2/497 | 0.640317 | 0.881236 | 0 | 0 | 0.923018 | 0.411474 |
| RNA-Seq_[ | Nurr1 Subs | 3/498 | 0.368874 | 0.881236 | 0 | 0 | 1.40101 | 1.397229 |
| RNA-Seq_[ | Zeb2 Sciati | 3/497 | 0.367685 | 0.881236 | 0 | 0 | 1.403918 | 1.404659 |
| RNA-Seq_[ | ZnF4 Thym | 3/497 | 0.367685 | 0.881236 | 0 | 0 | 1.403918 | 1.404659 |
| RNA-Seq_[ | heat-inacti | 3/497 | 0.367685 | 0.881236 | 0 | 0 | 1.403918 | 1.404659 |

| | | | | | | | | |
|---|---|---|---|---|---|---|---|---|
| RNA-Seq_D | hnRNPL Fe | 3/497 | 0.367685 | 0.881236 | 0 | 0 | 1.403918 | 1.404659 |
| RNA-Seq_D | miR-30c Liv | 3/497 | 0.367685 | 0.881236 | 0 | 0 | 1.403918 | 1.404659 |
| RNA-Seq_D | nmf205 Ce | 3/497 | 0.367685 | 0.881236 | 0 | 0 | 1.403918 | 1.404659 |
| RNA-Seq_D | NNK Lung 1 | 3/498 | 0.368874 | 0.881236 | 0 | 0 | 1.40101 | 1.397229 |
| RNA-Seq_D | Nfib Primar | 3/498 | 0.368874 | 0.881236 | 0 | 0 | 1.40101 | 1.397229 |
| RNA-Seq_D | Nfix Brain H | 3/498 | 0.368874 | 0.881236 | 0 | 0 | 1.40101 | 1.397229 |
| RNA-Seq_D | Zeb2 Sciati | 3/497 | 0.367685 | 0.881236 | 0 | 0 | 1.403918 | 1.404659 |
| RNA-Seq_D | PKC412 MD | 3/498 | 0.368874 | 0.881236 | 0 | 0 | 1.40101 | 1.397229 |
| RNA-Seq_D | PQBP1 Cor | 3/498 | 0.368874 | 0.881236 | 0 | 0 | 1.40101 | 1.397229 |
| RNA-Seq_D | PS2APP Ne | 3/498 | 0.368874 | 0.881236 | 0 | 0 | 1.40101 | 1.397229 |
| RNA-Seq_D | Vemurafen | 3/497 | 0.367685 | 0.881236 | 0 | 0 | 1.403918 | 1.404659 |
| RNA-Seq_D | MPTP-HCl I | 3/496 | 0.366497 | 0.881236 | 0 | 0 | 1.406839 | 1.412136 |
| RNA-Seq_D | MeCP2 Olfa | 3/496 | 0.366497 | 0.881236 | 0 | 0 | 1.406839 | 1.412136 |
| RNA-Seq_D | Mycobacte | 3/496 | 0.366497 | 0.881236 | 0 | 0 | 1.406839 | 1.412136 |
| RNA-Seq_D | PU.1 K562 | 3/496 | 0.366497 | 0.881236 | 0 | 0 | 1.406839 | 1.412136 |
| RNA-Seq_D | Parkinson's | 3/496 | 0.366497 | 0.881236 | 0 | 0 | 1.406839 | 1.412136 |
| RNA-Seq_D | LNA 30bcd | 3/495 | 0.365308 | 0.881236 | 0 | 0 | 1.409771 | 1.41966 |
| RNA-Seq_D | Kap1 Skele | 3/497 | 0.367685 | 0.881236 | 0 | 0 | 1.403918 | 1.404659 |
| RNA-Seq_D | Autism iPSC | 3/496 | 0.366497 | 0.881236 | 0 | 0 | 1.406839 | 1.412136 |
| RNA-Seq_D | Autism iPSC | 3/496 | 0.366497 | 0.881236 | 0 | 0 | 1.406839 | 1.412136 |
| RNA-Seq_D | Bcl9/9l APC | 3/496 | 0.366497 | 0.881236 | 0 | 0 | 1.406839 | 1.412136 |
| RNA-Seq_D | CCNK HCT1 | 3/496 | 0.366497 | 0.881236 | 0 | 0 | 1.406839 | 1.412136 |
| RNA-Seq_D | CDK13 HCT | 3/496 | 0.366497 | 0.881236 | 0 | 0 | 1.406839 | 1.412136 |
| RNA-Seq_D | CFIm25 He | 3/496 | 0.366497 | 0.881236 | 0 | 0 | 1.406839 | 1.412136 |
| RNA-Seq_D | CPI360 KAR | 3/496 | 0.366497 | 0.881236 | 0 | 0 | 1.406839 | 1.412136 |
| RNA-Seq_D | Cortistatin | 3/496 | 0.366497 | 0.881236 | 0 | 0 | 1.406839 | 1.412136 |
| RNA-Seq_D | RBFOX2 HE | 3/496 | 0.366497 | 0.881236 | 0 | 0 | 1.406839 | 1.412136 |
| RNA-Seq_D | MLL1 Comm | 3/495 | 0.365308 | 0.881236 | 0 | 0 | 1.409771 | 1.41966 |
| RNA-Seq_D | Nfix Brain H | 3/495 | 0.365308 | 0.881236 | 0 | 0 | 1.409771 | 1.41966 |
| RNA-Seq_D | Panobinost | 3/495 | 0.365308 | 0.881236 | 0 | 0 | 1.409771 | 1.41966 |
| RNA-Seq_D | Rcor1 Fetal | 3/495 | 0.365308 | 0.881236 | 0 | 0 | 1.409771 | 1.41966 |
| RNA-Seq_D | SYKi (R406) | 3/495 | 0.365308 | 0.881236 | 0 | 0 | 1.409771 | 1.41966 |
| RNA-Seq_D | beta-caten | 3/495 | 0.365308 | 0.881236 | 0 | 0 | 1.409771 | 1.41966 |
| RNA-Seq_D | Albuterol H | 3/496 | 0.366497 | 0.881236 | 0 | 0 | 1.406839 | 1.412136 |
| RNA-Seq_D | Yersinia ps | 3/497 | 0.367685 | 0.881236 | 0 | 0 | 1.403918 | 1.404659 |
| RNA-Seq_D | Desipramir | 3/497 | 0.367685 | 0.881236 | 0 | 0 | 1.403918 | 1.404659 |
| RNA-Seq_D | Dgcr8 Cond | 3/499 | 0.370061 | 0.881236 | 0 | 0 | 1.398113 | 1.389846 |
| RNA-Seq_D | Dgcr8 Cond | 3/499 | 0.370061 | 0.881236 | 0 | 0 | 1.398113 | 1.389846 |
| RNA-Seq_D | Dlx3 Femur | 3/499 | 0.370061 | 0.881236 | 0 | 0 | 1.398113 | 1.389846 |
| RNA-Seq_D | EVX1AS CJS | 3/499 | 0.370061 | 0.881236 | 0 | 0 | 1.398113 | 1.389846 |
| RNA-Seq_D | FBXL10 Fer | 3/499 | 0.370061 | 0.881236 | 0 | 0 | 1.398113 | 1.389846 |
| RNA-Seq_D | GAB2 Bone | 3/499 | 0.370061 | 0.881236 | 0 | 0 | 1.398113 | 1.389846 |
| RNA-Seq_D | Dlx3 Femur | 3/498 | 0.368874 | 0.881236 | 0 | 0 | 1.40101 | 1.397229 |
| RNA-Seq_D | DRB (dichlo | 3/497 | 0.367685 | 0.881236 | 0 | 0 | 1.403918 | 1.404659 |
| RNA-Seq_D | DRB (dichlo | 3/497 | 0.367685 | 0.881236 | 0 | 0 | 1.403918 | 1.404659 |

| | | | | | | | | |
|---|---|---|---|---|---|---|---|---|
| RNA-Seq_D | Bmpr1a Ha | 3/499 | 0.370061 | 0.881236 | 0 | 0 | 1.398113 | 1.389846 |
| RNA-Seq_D | Dexametha | 3/497 | 0.367685 | 0.881236 | 0 | 0 | 1.403918 | 1.404659 |
| RNA-Seq_D | Dlx3 Femu | 3/497 | 0.367685 | 0.881236 | 0 | 0 | 1.403918 | 1.404659 |
| RNA-Seq_D | Dot1l Com | 3/497 | 0.367685 | 0.881236 | 0 | 0 | 1.403918 | 1.404659 |
| RNA-Seq_D | GLI1 HCT1 | 3/497 | 0.367685 | 0.881236 | 0 | 0 | 1.403918 | 1.404659 |
| RNA-Seq_D | HER2 BT-4 | 3/497 | 0.367685 | 0.881236 | 0 | 0 | 1.403918 | 1.404659 |
| RNA-Seq_D | CDK9 HCT1 | 3/497 | 0.367685 | 0.881236 | 0 | 0 | 1.403918 | 1.404659 |
| RNA-Seq_D | CFIm25 He | 3/497 | 0.367685 | 0.881236 | 0 | 0 | 1.403918 | 1.404659 |
| RNA-Seq_D | CI-994 Hip | 3/497 | 0.367685 | 0.881236 | 0 | 0 | 1.403918 | 1.404659 |
| RNA-Seq_D | LPA2 HCT1 | 3/496 | 0.366497 | 0.881236 | 0 | 0 | 1.406839 | 1.412136 |
| RNA-Seq_D | Renal Cell | 3/496 | 0.366497 | 0.881236 | 0 | 0 | 1.406839 | 1.412136 |
| RNA-Seq_D | Salmonella | 3/496 | 0.366497 | 0.881236 | 0 | 0 | 1.406839 | 1.412136 |
| RNA-Seq_D | Huntingtor | 3/496 | 0.366497 | 0.881236 | 0 | 0 | 1.406839 | 1.412136 |
| RNA-Seq_D | Idiopathic | 3/496 | 0.366497 | 0.881236 | 0 | 0 | 1.406839 | 1.412136 |
| RNA-Seq_D | Itch Skin K | 3/496 | 0.366497 | 0.881236 | 0 | 0 | 1.406839 | 1.412136 |
| RNA-Seq_D | JAK2 HCT1 | 3/496 | 0.366497 | 0.881236 | 0 | 0 | 1.406839 | 1.412136 |
| RNA-Seq_D | Salmonella | 3/498 | 0.368874 | 0.881236 | 0 | 0 | 1.40101 | 1.397229 |
| RNA-Seq_D | JUN PC3 kr | 3/496 | 0.366497 | 0.881236 | 0 | 0 | 1.406839 | 1.412136 |
| RNA-Seq_D | L254F mut | 3/496 | 0.366497 | 0.881236 | 0 | 0 | 1.406839 | 1.412136 |
| RNA-Seq_D | Dioxin Live | 3/498 | 0.368874 | 0.881236 | 0 | 0 | 1.40101 | 1.397229 |
| RNA-Seq_D | Dlx3 Femu | 3/498 | 0.368874 | 0.881236 | 0 | 0 | 1.40101 | 1.397229 |
| RNA-Seq_D | Down Sydr | 3/498 | 0.368874 | 0.881236 | 0 | 0 | 1.40101 | 1.397229 |
| RNA-Seq_D | Doxycyclin | 3/498 | 0.368874 | 0.881236 | 0 | 0 | 1.40101 | 1.397229 |
| RNA-Seq_D | Drc Embry | 3/498 | 0.368874 | 0.881236 | 0 | 0 | 1.40101 | 1.397229 |
| RNA-Seq_D | EHF Calu-3 | 3/498 | 0.368874 | 0.881236 | 0 | 0 | 1.40101 | 1.397229 |
| RNA-Seq_D | FUS Motor | 3/498 | 0.368874 | 0.881236 | 0 | 0 | 1.40101 | 1.397229 |
| RNA-Seq_D | Fanconi An | 3/498 | 0.368874 | 0.881236 | 0 | 0 | 1.40101 | 1.397229 |
| RNA-Seq_D | Fanconi An | 3/498 | 0.368874 | 0.881236 | 0 | 0 | 1.40101 | 1.397229 |
| RNA-Seq_D | MeCP2 Hyp | 2/499 | 0.642504 | 0.881236 | 0 | 0 | 0.919209 | 0.406642 |
| RNA-Seq_D | Nrp1neg C | 2/499 | 0.642504 | 0.881236 | 0 | 0 | 0.919209 | 0.406642 |
| RNA-Seq_D | Nurr1 Vent | 2/499 | 0.642504 | 0.881236 | 0 | 0 | 0.919209 | 0.406642 |
| RNA-Seq_D | TAF15 Stria | 2/499 | 0.642504 | 0.881236 | 0 | 0 | 0.919209 | 0.406642 |
| RNA-Seq_D | Tet3 Blastc | 2/499 | 0.642504 | 0.881236 | 0 | 0 | 0.919209 | 0.406642 |
| RNA-Seq_D | EPAS1 HUV | 3/496 | 0.366497 | 0.881236 | 0 | 0 | 1.406839 | 1.412136 |
| RNA-Seq_D | HnRNP U H | 2/499 | 0.642504 | 0.881236 | 0 | 0 | 0.919209 | 0.406642 |
| RNA-Seq_D | Jmjd1c Bor | 2/499 | 0.642504 | 0.881236 | 0 | 0 | 0.919209 | 0.406642 |
| RNA-Seq_D | Lap2alpha | 2/499 | 0.642504 | 0.881236 | 0 | 0 | 0.919209 | 0.406642 |
| RNA-Seq_D | Maf1 Epidi | 2/499 | 0.642504 | 0.881236 | 0 | 0 | 0.919209 | 0.406642 |
| RNA-Seq_D | Nova2 Brai | 2/499 | 0.642504 | 0.881236 | 0 | 0 | 0.919209 | 0.406642 |
| RNA-Seq_D | Mecp2 Mic | 2/499 | 0.642504 | 0.881236 | 0 | 0 | 0.919209 | 0.406642 |
| RNA-Seq_D | Mecp2 Mic | 2/499 | 0.642504 | 0.881236 | 0 | 0 | 0.919209 | 0.406642 |
| RNA-Seq_D | Dioxin Live | 3/496 | 0.366497 | 0.881236 | 0 | 0 | 1.406839 | 1.412136 |
| RNA-Seq_D | Clobetasol | 3/495 | 0.365308 | 0.881236 | 0 | 0 | 1.409771 | 1.41966 |
| RNA-Seq_D | Desipramir | 3/495 | 0.365308 | 0.881236 | 0 | 0 | 1.409771 | 1.41966 |
| RNA-Seq_D | GW3965 Tı | 3/495 | 0.365308 | 0.881236 | 0 | 0 | 1.409771 | 1.41966 |

| | | | | | | | | |
|---|---|---|---|---|---|---|---|---|
| RNA-Seq_[ | H2A.Z Hipp | 3/495 | 0.365308 | 0.881236 | 0 | 0 | 1.409771 | 1.41966 |
| RNA-Seq_[ | Juvenile Idi | 3/495 | 0.365308 | 0.881236 | 0 | 0 | 1.409771 | 1.41966 |
| RNA-Seq_[ | T0901317 | 2/498 | 0.641412 | 0.881236 | 0 | 0 | 0.92111 | 0.40905 |
| RNA-Seq_[ | WZ4002 N | 2/498 | 0.641412 | 0.881236 | 0 | 0 | 0.92111 | 0.40905 |
| RNA-Seq_[ | YAP Oocyte | 2/498 | 0.641412 | 0.881236 | 0 | 0 | 0.92111 | 0.40905 |
| RNA-Seq_[ | ZNA MCF7 | 2/498 | 0.641412 | 0.881236 | 0 | 0 | 0.92111 | 0.40905 |
| RNA-Seq_[ | ZnF1 Thym | 2/498 | 0.641412 | 0.881236 | 0 | 0 | 0.92111 | 0.40905 |
| RNA-Seq_[ | heat-inacti | 2/498 | 0.641412 | 0.881236 | 0 | 0 | 0.92111 | 0.40905 |
| RNA-Seq_[ | heat-inacti | 2/498 | 0.641412 | 0.881236 | 0 | 0 | 0.92111 | 0.40905 |
| RNA-Seq_[ | miR-92a-1 | 2/498 | 0.641412 | 0.881236 | 0 | 0 | 0.92111 | 0.40905 |
| RNA-Seq_[ | pterosin B | 2/498 | 0.641412 | 0.881236 | 0 | 0 | 0.92111 | 0.40905 |
| RNA-Seq_[ | Staphyloco | 2/498 | 0.641412 | 0.881236 | 0 | 0 | 0.92111 | 0.40905 |
| RNA-Seq_[ | Smchd1 Ne | 3/496 | 0.366497 | 0.881236 | 0 | 0 | 1.406839 | 1.412136 |
| RNA-Seq_[ | Tet1 Bone | 2/498 | 0.641412 | 0.881236 | 0 | 0 | 0.92111 | 0.40905 |
| RNA-Seq_[ | Tet3 Oocyt | 2/498 | 0.641412 | 0.881236 | 0 | 0 | 0.92111 | 0.40905 |
| RNA-Seq_[ | Tex10 Emb | 2/498 | 0.641412 | 0.881236 | 0 | 0 | 0.92111 | 0.40905 |
| RNA-Seq_[ | Trisomy 21 | 2/498 | 0.641412 | 0.881236 | 0 | 0 | 0.92111 | 0.40905 |
| RNA-Seq_[ | Vemurafen | 2/498 | 0.641412 | 0.881236 | 0 | 0 | 0.92111 | 0.40905 |
| RNA-Seq_[ | Vemurafen | 2/498 | 0.641412 | 0.881236 | 0 | 0 | 0.92111 | 0.40905 |
| RNA-Seq_[ | Msi2 Leuke | 2/499 | 0.642504 | 0.881236 | 0 | 0 | 0.919209 | 0.406642 |
| RNA-Seq_[ | Nck2 Epidi | 2/499 | 0.642504 | 0.881236 | 0 | 0 | 0.919209 | 0.406642 |
| RNA-Seq_[ | Mycobacte | 3/497 | 0.367685 | 0.881236 | 0 | 0 | 1.403918 | 1.404659 |
| RNA-Seq_[ | Trex2 Skin | 3/497 | 0.367685 | 0.881236 | 0 | 0 | 1.403918 | 1.404659 |
| RNA-Seq_[ | Tunicamyc | 3/497 | 0.367685 | 0.881236 | 0 | 0 | 1.403918 | 1.404659 |
| RNA-Seq_[ | Tunicamyc | 3/497 | 0.367685 | 0.881236 | 0 | 0 | 1.403918 | 1.404659 |
| RNA-Seq_[ | miR-148a E | 3/496 | 0.366497 | 0.881236 | 0 | 0 | 1.406839 | 1.412136 |
| RNA-Seq_[ | Klf6 Immat | 3/497 | 0.367685 | 0.881236 | 0 | 0 | 1.403918 | 1.404659 |
| RNA-Seq_[ | MEP50 A5 | 3/497 | 0.367685 | 0.881236 | 0 | 0 | 1.403918 | 1.404659 |
| RNA-Seq_[ | Mycobacte | 3/497 | 0.367685 | 0.881236 | 0 | 0 | 1.403918 | 1.404659 |
| RNA-Seq_[ | Mycobacte | 3/497 | 0.367685 | 0.881236 | 0 | 0 | 1.403918 | 1.404659 |
| RNA-Seq_[ | Mycobacte | 3/497 | 0.367685 | 0.881236 | 0 | 0 | 1.403918 | 1.404659 |
| RNA-Seq_[ | TGFB3 Pala | 3/497 | 0.367685 | 0.881236 | 0 | 0 | 1.403918 | 1.404659 |
| RNA-Seq_[ | Myelodysp | 3/497 | 0.367685 | 0.881236 | 0 | 0 | 1.403918 | 1.404659 |
| RNA-Seq_[ | Myotonic [ | 3/497 | 0.367685 | 0.881236 | 0 | 0 | 1.403918 | 1.404659 |
| RNA-Seq_[ | DDX5 Th17 | 3/498 | 0.368874 | 0.881236 | 0 | 0 | 1.40101 | 1.397229 |
| RNA-Seq_[ | DMOG Ret | 3/498 | 0.368874 | 0.881236 | 0 | 0 | 1.40101 | 1.397229 |
| RNA-Seq_[ | DMR1-U2 | 3/498 | 0.368874 | 0.881236 | 0 | 0 | 1.40101 | 1.397229 |
| RNA-Seq_[ | Desipramir | 3/498 | 0.368874 | 0.881236 | 0 | 0 | 1.40101 | 1.397229 |
| RNA-Seq_[ | Dexametha | 3/498 | 0.368874 | 0.881236 | 0 | 0 | 1.40101 | 1.397229 |
| RNA-Seq_[ | Dgcr8 Cone | 3/498 | 0.368874 | 0.881236 | 0 | 0 | 1.40101 | 1.397229 |
| RNA-Seq_[ | ALS IPSCs-c | 3/497 | 0.367685 | 0.881236 | 0 | 0 | 1.403918 | 1.404659 |
| RNA-Seq_[ | TAF15 Mot | 3/496 | 0.366497 | 0.881236 | 0 | 0 | 1.406839 | 1.412136 |
| RNA-Seq_[ | TDP43 Mot | 3/496 | 0.366497 | 0.881236 | 0 | 0 | 1.406839 | 1.412136 |
| RNA-Seq_[ | TGFB3 Pala | 3/496 | 0.366497 | 0.881236 | 0 | 0 | 1.406839 | 1.412136 |
| RNA-Seq_[ | Yersinia ps | 3/496 | 0.366497 | 0.881236 | 0 | 0 | 1.406839 | 1.412136 |

| | | | | | | | | |
|---|---|---|---|---|---|---|---|---|
| RNA-Seq_[ | Zmiz1 T lyn | 3/496 | 0.366497 | 0.881236 | 0 | 0 | 1.406839 | 1.412136 |
| RNA-Seq_[ | hnRNPU He | 3/496 | 0.366497 | 0.881236 | 0 | 0 | 1.406839 | 1.412136 |
| RNA-Seq_[ | Tunicamyci | 3/494 | 0.364119 | 0.881236 | 0 | 0 | 1.412715 | 1.427231 |
| RNA-Seq_[ | nSR100/Srr | 3/496 | 0.366497 | 0.881236 | 0 | 0 | 1.406839 | 1.412136 |
| RNA-Seq_[ | 5-Azacytidi | 3/497 | 0.367685 | 0.881236 | 0 | 0 | 1.403918 | 1.404659 |
| RNA-Seq_[ | JAK3i (CP-6 | 3/496 | 0.366497 | 0.881236 | 0 | 0 | 1.406839 | 1.412136 |
| RNA-Seq_[ | Autism iPS( | 3/497 | 0.367685 | 0.881236 | 0 | 0 | 1.403918 | 1.404659 |
| RNA-Seq_[ | Bacillus Cal | 3/497 | 0.367685 | 0.881236 | 0 | 0 | 1.403918 | 1.404659 |
| RNA-Seq_[ | Bacillus Cal | 3/497 | 0.367685 | 0.881236 | 0 | 0 | 1.403918 | 1.404659 |
| RNA-Seq_[ | Bcl9/9l Col | 3/497 | 0.367685 | 0.881236 | 0 | 0 | 1.403918 | 1.404659 |
| RNA-Seq_[ | Bmal1 U2C | 3/497 | 0.367685 | 0.881236 | 0 | 0 | 1.403918 | 1.404659 |
| RNA-Seq_[ | Slfn5 786-C | 3/497 | 0.367685 | 0.881236 | 0 | 0 | 1.403918 | 1.404659 |
| RNA-Seq_[ | T0901317 ˉ | 3/497 | 0.367685 | 0.881236 | 0 | 0 | 1.403918 | 1.404659 |
| RNA-Seq_[ | TDP43 Mot | 3/497 | 0.367685 | 0.881236 | 0 | 0 | 1.403918 | 1.404659 |
| RNA-Seq_[ | Jmjd1c Bor | 3/498 | 0.368874 | 0.881236 | 0 | 0 | 1.40101 | 1.397229 |
| RNA-Seq_[ | Dnmt3a Cc | 2/496 | 0.63922 | 0.881236 | 0 | 0 | 0.924935 | 0.413914 |
| RNA-Seq_[ | EPAS1 HUV | 2/496 | 0.63922 | 0.881236 | 0 | 0 | 0.924935 | 0.413914 |
| RNA-Seq_[ | GLI1 HCT1: | 2/496 | 0.63922 | 0.881236 | 0 | 0 | 0.924935 | 0.413914 |
| RNA-Seq_[ | GSK126 KA | 2/496 | 0.63922 | 0.881236 | 0 | 0 | 0.924935 | 0.413914 |
| RNA-Seq_[ | Haloperido | 2/496 | 0.63922 | 0.881236 | 0 | 0 | 0.924935 | 0.413914 |
| RNA-Seq_[ | JAK3i (CP-6 | 2/496 | 0.63922 | 0.881236 | 0 | 0 | 0.924935 | 0.413914 |
| RNA-Seq_[ | L254F muta | 2/496 | 0.63922 | 0.881236 | 0 | 0 | 0.924935 | 0.413914 |
| RNA-Seq_[ | MSI2 Hema | 2/491 | 0.633696 | 0.881236 | 0 | 0 | 0.934633 | 0.426367 |
| RNA-Seq_[ | LNA 30bcd | 2/494 | 0.637018 | 0.881236 | 0 | 0 | 0.92879 | 0.418845 |
| RNA-Seq_[ | KDM2B HS | 3/498 | 0.368874 | 0.881236 | 0 | 0 | 1.40101 | 1.397229 |
| RNA-Seq_[ | Kap1 Skele | 3/498 | 0.368874 | 0.881236 | 0 | 0 | 1.40101 | 1.397229 |
| RNA-Seq_[ | LPA2 HCT1 | 3/498 | 0.368874 | 0.881236 | 0 | 0 | 1.40101 | 1.397229 |
| RNA-Seq_[ | LPS Neuror | 3/498 | 0.368874 | 0.881236 | 0 | 0 | 1.40101 | 1.397229 |
| RNA-Seq_[ | METTL3 MI | 3/498 | 0.368874 | 0.881236 | 0 | 0 | 1.40101 | 1.397229 |
| RNA-Seq_[ | MeCP2 Hyp | 3/498 | 0.368874 | 0.881236 | 0 | 0 | 1.40101 | 1.397229 |
| RNA-Seq_[ | Miconazole | 3/498 | 0.368874 | 0.881236 | 0 | 0 | 1.40101 | 1.397229 |
| RNA-Seq_[ | GW3965 Tu | 3/498 | 0.368874 | 0.881236 | 0 | 0 | 1.40101 | 1.397229 |
| RNA-Seq_[ | Asthma Air | 3/498 | 0.368874 | 0.881236 | 0 | 0 | 1.40101 | 1.397229 |
| RNA-Seq_[ | Cyclophosp | 3/498 | 0.368874 | 0.881236 | 0 | 0 | 1.40101 | 1.397229 |
| RNA-Seq_[ | 5-Azacytidi | 3/498 | 0.368874 | 0.881236 | 0 | 0 | 1.40101 | 1.397229 |
| RNA-Seq_[ | ABCA4 Ret | 3/498 | 0.368874 | 0.881236 | 0 | 0 | 1.40101 | 1.397229 |
| RNA-Seq_[ | APP/PS1-2: | 3/498 | 0.368874 | 0.881236 | 0 | 0 | 1.40101 | 1.397229 |
| RNA-Seq_[ | Activin A Er | 3/498 | 0.368874 | 0.881236 | 0 | 0 | 1.40101 | 1.397229 |
| RNA-Seq_[ | Alkbh1 Eml | 3/498 | 0.368874 | 0.881236 | 0 | 0 | 1.40101 | 1.397229 |
| RNA-Seq_[ | All-trans re | 3/498 | 0.368874 | 0.881236 | 0 | 0 | 1.40101 | 1.397229 |
| RNA-Seq_[ | Anankinra | 3/498 | 0.368874 | 0.881236 | 0 | 0 | 1.40101 | 1.397229 |
| RNA-Seq_[ | H2A.Z Hipp | 3/498 | 0.368874 | 0.881236 | 0 | 0 | 1.40101 | 1.397229 |
| RNA-Seq_[ | Mycobacte | 2/492 | 0.634806 | 0.881236 | 0 | 0 | 0.932677 | 0.423842 |
| RNA-Seq_[ | Salmonella | 2/492 | 0.634806 | 0.881236 | 0 | 0 | 0.932677 | 0.423842 |
| RNA-Seq_[ | CPSF2 HEK: | 2/493 | 0.635913 | 0.881236 | 0 | 0 | 0.93073 | 0.421335 |

| | | | | | | | | |
|---|---|---|---|---|---|---|---|---|
| RNA-Seq_[ | Doxycyclin | 2/493 | 0.635913 | 0.881236 | 0 | 0 | 0.93073 | 0.421335 |
| RNA-Seq_[ | RNPK1 A67 | 2/493 | 0.635913 | 0.881236 | 0 | 0 | 0.93073 | 0.421335 |
| RNA-Seq_[ | BRG1 MCF- | 2/494 | 0.637018 | 0.881236 | 0 | 0 | 0.92879 | 0.418845 |
| RNA-Seq_[ | LNA 30bcd | 2/494 | 0.637018 | 0.881236 | 0 | 0 | 0.92879 | 0.418845 |
| RNA-Seq_[ | ZnF1 Thym | 3/498 | 0.368874 | 0.881236 | 0 | 0 | 1.40101 | 1.397229 |
| RNA-Seq_[ | T0901317 | 2/496 | 0.63922 | 0.881236 | 0 | 0 | 0.924935 | 0.413914 |
| RNA-Seq_[ | Thap11 Re | 2/496 | 0.63922 | 0.881236 | 0 | 0 | 0.924935 | 0.413914 |
| RNA-Seq_[ | Trisomy 21 | 2/496 | 0.63922 | 0.881236 | 0 | 0 | 0.924935 | 0.413914 |
| RNA-Seq_[ | ZNA MCF7 | 2/496 | 0.63922 | 0.881236 | 0 | 0 | 0.924935 | 0.413914 |
| RNA-Seq_[ | Retinoic Ac | 2/495 | 0.63812 | 0.881236 | 0 | 0 | 0.926858 | 0.416371 |
| RNA-Seq_[ | Tet3 Blastc | 3/498 | 0.368874 | 0.881236 | 0 | 0 | 1.40101 | 1.397229 |
| RNA-Seq_[ | U2AF1(S34 | 3/498 | 0.368874 | 0.881236 | 0 | 0 | 1.40101 | 1.397229 |
| RNA-Seq_[ | Yersinia ps | 3/498 | 0.368874 | 0.881236 | 0 | 0 | 1.40101 | 1.397229 |
| RNA-Seq_[ | Staphyloco | 2/496 | 0.63922 | 0.881236 | 0 | 0 | 0.924935 | 0.413914 |
| RNA-Seq_[ | miR-92a-1 | 3/498 | 0.368874 | 0.881236 | 0 | 0 | 1.40101 | 1.397229 |
| RNA-Seq_[ | miR-92a-1 | 3/498 | 0.368874 | 0.881236 | 0 | 0 | 1.40101 | 1.397229 |
| RNA-Seq_[ | Bcl9/9l Col | 3/499 | 0.370061 | 0.881236 | 0 | 0 | 1.398113 | 1.389846 |
| RNA-Seq_[ | Blinc1 Panc | 3/499 | 0.370061 | 0.881236 | 0 | 0 | 1.398113 | 1.389846 |
| RNA-Seq_[ | SRA MCF7 | 3/498 | 0.368874 | 0.881236 | 0 | 0 | 1.40101 | 1.397229 |
| RNA-Seq_[ | Salmonella | 3/498 | 0.368874 | 0.881236 | 0 | 0 | 1.40101 | 1.397229 |
| RNA-Seq_[ | Salmonella | 3/498 | 0.368874 | 0.881236 | 0 | 0 | 1.40101 | 1.397229 |
| RNA-Seq_[ | Tunicamyci | 2/495 | 0.63812 | 0.881236 | 0 | 0 | 0.926858 | 0.416371 |
| RNA-Seq_[ | H2AZ Embr | 3/498 | 0.368874 | 0.881236 | 0 | 0 | 1.40101 | 1.397229 |
| RNA-Seq_[ | Hnrnpk TT | 3/498 | 0.368874 | 0.881236 | 0 | 0 | 1.40101 | 1.397229 |
| RNA-Seq_[ | Huntingtor | 3/498 | 0.368874 | 0.881236 | 0 | 0 | 1.40101 | 1.397229 |
| RNA-Seq_[ | Huntingtor | 3/498 | 0.368874 | 0.881236 | 0 | 0 | 1.40101 | 1.397229 |
| RNA-Seq_[ | I-BET MV4- | 3/498 | 0.368874 | 0.881236 | 0 | 0 | 1.40101 | 1.397229 |
| RNA-Seq_[ | Idiopathic | 3/498 | 0.368874 | 0.881236 | 0 | 0 | 1.40101 | 1.397229 |
| RNA-Seq_[ | TAF15 Stria | 2/495 | 0.63812 | 0.881236 | 0 | 0 | 0.926858 | 0.416371 |
| RNA-Seq_[ | TCF21 HCA | 2/495 | 0.63812 | 0.881236 | 0 | 0 | 0.926858 | 0.416371 |
| RNA-Seq_[ | Cnot3 ESCs | 3/497 | 0.367685 | 0.881236 | 0 | 0 | 1.403918 | 1.404659 |
| RNA-Seq_[ | Tunicamyci | 2/495 | 0.63812 | 0.881236 | 0 | 0 | 0.926858 | 0.416371 |
| RNA-Seq_[ | nSR100/Sri | 2/495 | 0.63812 | 0.881236 | 0 | 0 | 0.926858 | 0.416371 |
| RNA-Seq_[ | 17-betaest | 2/496 | 0.63922 | 0.881236 | 0 | 0 | 0.924935 | 0.413914 |
| RNA-Seq_[ | 4-hydroxyt | 2/496 | 0.63922 | 0.881236 | 0 | 0 | 0.924935 | 0.413914 |
| RNA-Seq_[ | AE9aMeis2 | 2/496 | 0.63922 | 0.881236 | 0 | 0 | 0.924935 | 0.413914 |
| RNA-Seq_[ | Silvestrol N | 2/496 | 0.63922 | 0.881236 | 0 | 0 | 0.924935 | 0.413914 |
| RNA-Seq_[ | Staphyloco | 2/496 | 0.63922 | 0.881236 | 0 | 0 | 0.924935 | 0.413914 |
| RNA-Seq_[ | PU.1 K562 | 3/497 | 0.367685 | 0.881236 | 0 | 0 | 1.403918 | 1.404659 |
| RNA-Seq_[ | SENCR HCA | 3/497 | 0.367685 | 0.881236 | 0 | 0 | 1.403918 | 1.404659 |
| RNA-Seq_[ | SGC0946 M | 3/497 | 0.367685 | 0.881236 | 0 | 0 | 1.403918 | 1.404659 |
| RNA-Seq_[ | Salmonella | 3/497 | 0.367685 | 0.881236 | 0 | 0 | 1.403918 | 1.404659 |
| RNA-Seq_[ | Salmonella | 3/497 | 0.367685 | 0.881236 | 0 | 0 | 1.403918 | 1.404659 |
| RNA-Seq_[ | Silvestrol K | 3/497 | 0.367685 | 0.881236 | 0 | 0 | 1.403918 | 1.404659 |
| RNA-Seq_[ | Slfn5 786-C | 3/497 | 0.367685 | 0.881236 | 0 | 0 | 1.403918 | 1.404659 |

| | | | | | | | | |
|---|---|---|---|---|---|---|---|---|
| RNA-Seq_[ | NHD13 My | 3/497 | 0.367685 | 0.881236 | 0 | 0 | 1.403918 | 1.404659 |
| RNA-Seq_[ | Notch1 T ly | 3/497 | 0.367685 | 0.881236 | 0 | 0 | 1.403918 | 1.404659 |
| RNA-Seq_[ | Nurr1 Subs | 3/497 | 0.367685 | 0.881236 | 0 | 0 | 1.403918 | 1.404659 |
| RNA-Seq_[ | Nutlin-3a N | 3/497 | 0.367685 | 0.881236 | 0 | 0 | 1.403918 | 1.404659 |
| RNA-Seq_[ | Nutlin-3a N | 3/497 | 0.367685 | 0.881236 | 0 | 0 | 1.403918 | 1.404659 |
| RNA-Seq_[ | SAHA Liver | 3/497 | 0.367685 | 0.881236 | 0 | 0 | 1.403918 | 1.404659 |
| RNA-Seq_[ | Panobinost | 3/497 | 0.367685 | 0.881236 | 0 | 0 | 1.403918 | 1.404659 |
| RNA-Seq_[ | Parkin Hea | 3/497 | 0.367685 | 0.881236 | 0 | 0 | 1.403918 | 1.404659 |
| RNA-Seq_[ | LMNB1 Em | 3/499 | 0.370061 | 0.881236 | 0 | 0 | 1.398113 | 1.389846 |
| RNA-Seq_[ | Maf1 Epidi | 3/499 | 0.370061 | 0.881236 | 0 | 0 | 1.398113 | 1.389846 |
| RNA-Seq_[ | MeCP2 Hyp | 3/499 | 0.370061 | 0.881236 | 0 | 0 | 1.398113 | 1.389846 |
| RNA-Seq_[ | Mef2d Reti | 3/499 | 0.370061 | 0.881236 | 0 | 0 | 1.398113 | 1.389846 |
| RNA-Seq_[ | Nck2 Epidi | 3/499 | 0.370061 | 0.881236 | 0 | 0 | 1.398113 | 1.389846 |
| RNA-Seq_[ | Nurr1 Vent | 3/499 | 0.370061 | 0.881236 | 0 | 0 | 1.398113 | 1.389846 |
| RNA-Seq_[ | PS2APP Mi | 3/499 | 0.370061 | 0.881236 | 0 | 0 | 1.398113 | 1.389846 |
| RNA-Seq_[ | RFWD2 Lur | 3/499 | 0.370061 | 0.881236 | 0 | 0 | 1.398113 | 1.389846 |
| RNA-Seq_[ | Rpl11 Hem | 3/498 | 0.368874 | 0.881236 | 0 | 0 | 1.40101 | 1.397229 |
| RNA-Seq_[ | Chd7 Spina | 3/497 | 0.367685 | 0.881236 | 0 | 0 | 1.403918 | 1.404659 |
| RNA-Seq_[ | Cortistatin | 3/497 | 0.367685 | 0.881236 | 0 | 0 | 1.403918 | 1.404659 |
| RNA-Seq_[ | CrX Retina | 3/497 | 0.367685 | 0.881236 | 0 | 0 | 1.403918 | 1.404659 |
| RNA-Seq_[ | DICER1 Hip | 3/497 | 0.367685 | 0.881236 | 0 | 0 | 1.403918 | 1.404659 |
| RNA-Seq_[ | Parathyroi | 3/498 | 0.368874 | 0.881236 | 0 | 0 | 1.40101 | 1.397229 |
| RNA-Seq_[ | Phenobarb | 3/498 | 0.368874 | 0.881236 | 0 | 0 | 1.40101 | 1.397229 |
| RNA-Seq_[ | Phenobarb | 3/498 | 0.368874 | 0.881236 | 0 | 0 | 1.40101 | 1.397229 |
| RNA-Seq_[ | Prmt5 Neu | 3/498 | 0.368874 | 0.881236 | 0 | 0 | 1.40101 | 1.397229 |
| RNA-Seq_[ | Ptbp1 Neu | 3/498 | 0.368874 | 0.881236 | 0 | 0 | 1.40101 | 1.397229 |
| RNA-Seq_[ | Rhinovirus | 3/498 | 0.368874 | 0.881236 | 0 | 0 | 1.40101 | 1.397229 |
| RNA-Seq_[ | Cardiomyo | 3/498 | 0.368874 | 0.881236 | 0 | 0 | 1.40101 | 1.397229 |
| RNA-Seq_[ | SOD1(G93/ | 3/498 | 0.368874 | 0.881236 | 0 | 0 | 1.40101 | 1.397229 |
| RNA-Seq_[ | SAHA Hipp | 4/835 | 0.494742 | 0.881236 | 0 | 0 | 1.106636 | 0.77876 |
| RNA-Seq_[ | KCl Embryc | 4/880 | 0.536375 | 0.881236 | 0 | 0 | 1.047313 | 0.652394 |
| RNA-Seq_[ | DAPT Coch | 4/893 | 0.548095 | 0.881236 | 0 | 0 | 1.031293 | 0.620123 |
| RNA-Seq_[ | DAPT Coch | 4/923 | 0.574561 | 0.881236 | 0 | 0 | 0.996054 | 0.551963 |
| RNA-Seq_[ | Vitamin D / | 3/712 | 0.603052 | 0.881236 | 0 | 0 | 0.967358 | 0.489243 |
| RNA-Seq_[ | 2,3,7,8-tetı | 2/488 | 0.63035 | 0.881236 | 0 | 0 | 0.940547 | 0.434043 |
| RNA-Seq_[ | 2,3,7,8-tetı | 2/491 | 0.633696 | 0.881236 | 0 | 0 | 0.934633 | 0.426367 |
| RNA-Seq_[ | PSMD13 H | 3/498 | 0.368874 | 0.881236 | 0 | 0 | 1.40101 | 1.397229 |
| RNA-Seq_[ | RNPK1 A67 | 3/497 | 0.367685 | 0.881236 | 0 | 0 | 1.403918 | 1.404659 |
| RNA-Seq_[ | Myelodysp | 2/495 | 0.63812 | 0.881236 | 0 | 0 | 0.926858 | 0.416371 |
| RNA-Seq_[ | Cocaine Nu | 3/498 | 0.368874 | 0.881236 | 0 | 0 | 1.40101 | 1.397229 |
| RNA-Seq_[ | C9orf72 Sp | 3/498 | 0.368874 | 0.881236 | 0 | 0 | 1.40101 | 1.397229 |
| RNA-Seq_[ | Bmal1 Live | 3/498 | 0.368874 | 0.881236 | 0 | 0 | 1.40101 | 1.397229 |
| RNA-Seq_[ | BS69 Hela | 3/498 | 0.368874 | 0.881236 | 0 | 0 | 1.40101 | 1.397229 |
| RNA-Seq_[ | BMAL1 Rer | 3/498 | 0.368874 | 0.881236 | 0 | 0 | 1.40101 | 1.397229 |
| RNA-Seq_[ | Autism iPS | 3/498 | 0.368874 | 0.881236 | 0 | 0 | 1.40101 | 1.397229 |

| | | | | | | | | |
|---|---|---|---|---|---|---|---|---|
| RNA-Seq_[ | Jmjd1c Bor | 3/499 | 0.370061 | 0.881236 | 0 | 0 | 1.398113 | 1.389846 |
| RNA-Seq_[ | Relapsing-r | 2/495 | 0.63812 | 0.881236 | 0 | 0 | 0.926858 | 0.416371 |
| RNA-Seq_[ | Ptbp2 Neu | 2/495 | 0.63812 | 0.881236 | 0 | 0 | 0.926858 | 0.416371 |
| RNA-Seq_[ | PRDM11 U | 2/495 | 0.63812 | 0.881236 | 0 | 0 | 0.926858 | 0.416371 |
| RNA-Seq_[ | KCNK1 RKC | 2/495 | 0.63812 | 0.881236 | 0 | 0 | 0.926858 | 0.416371 |
| RNA-Seq_[ | PER2 Endc | 2/495 | 0.63812 | 0.881236 | 0 | 0 | 0.926858 | 0.416371 |
| RNA-Seq_[ | PKC412 MI | 2/495 | 0.63812 | 0.881236 | 0 | 0 | 0.926858 | 0.416371 |
| Gene_Pert | Utrn OE m | 1/481 | 0.880272 | 0.881244 | 0 | 0 | 0.470761 | 0.060033 |
| Gene_Pert | Tcf4 Deleti | 1/484 | 0.881866 | 0.882851 | 0 | 0 | 0.467764 | 0.058805 |
| GTEx_Tissu | GTEX-NL3G | 13/2402 | 0.241772 | 0.885067 | 0 | 0 | 1.288631 | 1.829547 |
| GTEx_Tissu | GTEX-Q2A( | Dec-90 | 0.240949 | 0.885067 | 0 | 0 | 1.302847 | 1.854172 |
| GTEx_Tissu | GTEX-SSA3 | 2/216 | 0.241852 | 0.885067 | 0 | 0 | 2.165915 | 3.074364 |
| GTEx_Tissu | GTEX-Q2AI | 46/9709 | 0.241252 | 0.885067 | 0 | 0 | 1.190107 | 1.69223 |
| Jensen_TIS | Pons | 15/4336 | 0.874171 | 0.885167 | 0 | 0 | 0.751755 | 0.101095 |
| GTEx_Tissu | GTEX-X8H( | 3/395 | 0.246667 | 0.885506 | 0 | 0 | 1.778517 | 2.489418 |
| GTEx_Tissu | GTEX-RU72 | 27/5453 | 0.248137 | 0.885506 | 0 | 0 | 1.201465 | 1.67457 |
| GTEx_Tissu | GTEX-RU72 | 13/2415 | 0.24783 | 0.885506 | 0 | 0 | 1.280706 | 1.786603 |
| GTEx_Tissu | GTEX-XBED | 2/217 | 0.243461 | 0.885506 | 0 | 0 | 2.155732 | 3.045615 |
| GTEx_Tissu | GTEX-NPJ7 | 10/1778 | 0.243363 | 0.885506 | 0 | 0 | 1.332858 | 1.883594 |
| GTEx_Tissu | GTEX-RM2 | Jan-65 | 0.247099 | 0.885506 | 0 | 0 | 3.606286 | 5.041467 |
| GTEx_Tissu | GTEX-QDV. | 13/2413 | 0.246894 | 0.885506 | 0 | 0 | 1.28192 | 1.793146 |
| GTEx_Tissu | GTEX-RM2 | 3/392 | 0.243181 | 0.885506 | 0 | 0 | 1.792508 | 2.534513 |
| GTEx_Tissu | GTEX-X261 | 7/1167 | 0.244526 | 0.885506 | 0 | 0 | 1.414558 | 1.992314 |
| GTEx_Tissu | GTEX-XAJ8 | 42/8807 | 0.244393 | 0.885506 | 0 | 0 | 1.187085 | 1.672578 |
| GTEx_Tissu | GTEX-T2IS- | 8/1372 | 0.246149 | 0.885506 | 0 | 0 | 1.377111 | 1.930459 |
| GTEx_Tissu | GTEX-R55E | 4/580 | 0.245574 | 0.885506 | 0 | 0 | 1.617888 | 2.271767 |
| GTEx_Tissu | GTEX-RU1J | 5/774 | 0.247052 | 0.885506 | 0 | 0 | 1.517968 | 2.122354 |
| GTEx_Tissu | GTEX-U412 | 10/1788 | 0.248738 | 0.885506 | 0 | 0 | 1.324631 | 1.843035 |
| GTEx_Tissu | GTEX-TML8 | 2/220 | 0.248291 | 0.885506 | 0 | 0 | 2.125742 | 2.961486 |
| GTEx_Tissu | GTEX-N7M | 8/1376 | 0.248594 | 0.885506 | 0 | 0 | 1.372789 | 1.91083 |
| GTEx_Tissu | GTEX-N7M | 6/973 | 0.248669 | 0.885506 | 0 | 0 | 1.4513 | 2.019674 |
| Disease_Pe | Schistosom | 1/485 | 0.882393 | 0.886509 | 0 | 0 | 0.466774 | 0.058402 |
| GTEx_Tissu | GTEX-SUCS | 7/914 | 0.102811 | 0.887857 | 0 | 0 | 1.833545 | 4.171063 |
| GTEx_Tissu | GTEX-X5EB | 6/739 | 0.102743 | 0.887857 | 0 | 0 | 1.938255 | 4.410553 |
| Disease_Pe | Nicotine ac | 1/497 | 0.888534 | 0.888534 | 0 | 0 | 0.4552 | 0.053797 |
| GTEx_Tissu | GTEX-POM | 19/3708 | 0.250922 | 0.888796 | 0 | 0 | 1.228836 | 1.699005 |
| GTEx_Tissu | GTEX-U4B1 | 25/5026 | 0.253144 | 0.888796 | 0 | 0 | 1.20234 | 1.651773 |
| GTEx_Tissu | GTEX-TSE9 | 8/1384 | 0.253509 | 0.888796 | 0 | 0 | 1.364218 | 1.872192 |
| GTEx_Tissu | GTEX-QDV. | 2/223 | 0.253125 | 0.888796 | 0 | 0 | 2.096566 | 2.880418 |
| GTEx_Tissu | GTEX-NPJ7 | 5/782 | 0.253603 | 0.888796 | 0 | 0 | 1.501711 | 2.060326 |
| GTEx_Tissu | GTEX-WEY! | Jan-67 | 0.253656 | 0.888796 | 0 | 0 | 3.496653 | 4.79662 |
| GTEx_Tissu | GTEX-NPJ7 | 5/779 | 0.251141 | 0.888796 | 0 | 0 | 1.507768 | 2.083344 |
| GTEx_Tissu | GTEX-TKQ2 | 4/587 | 0.252215 | 0.888796 | 0 | 0 | 1.597884 | 2.201043 |
| GTEx_Tissu | GTEX-XXEK | 4/586 | 0.251264 | 0.888796 | 0 | 0 | 1.600712 | 2.210987 |
| GTEx_Tissu | GTEX-QVU! | 6/978 | 0.252318 | 0.888796 | 0 | 0 | 1.443454 | 1.987732 |

| | | | | | | | | |
|---|---|---|---|---|---|---|---|---|
| KEGG_202: | Herpes sim | 1/498 | 0.889031 | 0.889031 | 0 | 0 | 0.45426 | 0.053432 |
| ARCHS4_Ti | SUBCUTAN | 15/2316 | 0.074101 | 0.889216 | 0 | 0 | 1.594597 | 4.149652 |
| ARCHS4_Ti | ADIPOSE (E | 15/2316 | 0.074101 | 0.889216 | 0 | 0 | 1.594597 | 4.149652 |
| ARCHS4_Ti | RESPIRATO | 15/2316 | 0.074101 | 0.889216 | 0 | 0 | 1.594597 | 4.149652 |
| Reactome_ | Diseases O | 1/424 | 0.845612 | 0.889282 | 0 | 0 | 0.535763 | 0.089845 |
| GTEx_Tissu | GTEX-QMR | 11/1657 | 0.104067 | 0.8895 | 0 | 0 | 1.606262 | 3.634528 |
| GTEx_Tissu | GTEX-XLM4 | 7/917 | 0.10414 | 0.8895 | 0 | 0 | 1.827212 | 4.133186 |
| GTEx_Tissu | GTEX-WFO | 22/3864 | 0.103337 | 0.8895 | 0 | 0 | 1.415777 | 3.213474 |
| GTEx_Tissu | GTEX-N7M | 6/742 | 0.104231 | 0.8895 | 0 | 0 | 1.930052 | 4.364128 |
| GTEx_Tissu | GTEX-PWN | 2/224 | 0.254736 | 0.889778 | 0 | 0 | 2.087016 | 2.85405 |
| GTEx_Tissu | GTEX-TMM | 2/224 | 0.254736 | 0.889778 | 0 | 0 | 2.087016 | 2.85405 |
| GTEx_Tissu | GTEX-OXRI | 9/1282 | 0.104696 | 0.890145 | 0 | 0 | 1.689528 | 3.81275 |
| GTEx_Tissu | GTEX-WW\ | 7/919 | 0.105031 | 0.890145 | 0 | 0 | 1.823013 | 4.108151 |
| GTEx_Tissu | GTEX-XUJ4 | 6/744 | 0.10523 | 0.890145 | 0 | 0 | 1.924621 | 4.333495 |
| GO_Biologi | Regulation | Jun-28 | 0.886658 | 0.89042 | 0 | 0 | 0.65542 | 0.078844 |
| GWAS_Cat | Obesity-rel | 1/498 | 0.889031 | 0.890491 | 0 | 0 | 0.45426 | 0.053432 |
| GTEx_Tissu | GTEX-N7M | 13/2432 | 0.255842 | 0.890679 | 0 | 0 | 1.270471 | 1.731901 |
| GTEx_Tissu | GTEX-RVPL | 45/9530 | 0.256054 | 0.890679 | 0 | 0 | 1.17795 | 1.604801 |
| GTEx_Tissu | GTEX-UJHI | 2/225 | 0.256348 | 0.890679 | 0 | 0 | 2.077552 | 2.828 |
| GTEx_Tissu | GTEX-TSE9 | Dec-22 | 0.256596 | 0.890679 | 0 | 0 | 1.281665 | 1.743391 |
| PhenGenI_ | Urine | 1/377 | 0.809709 | 0.89068 | 0 | 0 | 0.604187 | 0.127532 |
| GTEx_Tissu | GTEX-WFG | Jan-69 | 0.260157 | 0.891318 | 0 | 0 | 3.393468 | 4.569198 |
| GTEx_Tissu | GTEX-T5JC | 10/1808 | 0.259615 | 0.891318 | 0 | 0 | 1.308452 | 1.764519 |
| GTEx_Tissu | GTEX-WL46 | Jan-69 | 0.260157 | 0.891318 | 0 | 0 | 3.393468 | 4.569198 |
| GTEx_Tissu | GTEX-REY6 | 2/228 | 0.261186 | 0.891318 | 0 | 0 | 2.049662 | 2.751716 |
| GTEx_Tissu | GTEX-U3ZN | Jan-69 | 0.260157 | 0.891318 | 0 | 0 | 3.393468 | 4.569198 |
| GTEx_Tissu | GTEX-WHP | Jan-69 | 0.260157 | 0.891318 | 0 | 0 | 3.393468 | 4.569198 |
| GTEx_Tissu | GTEX-T2YK | 13/2440 | 0.259647 | 0.891318 | 0 | 0 | 1.265705 | 1.706717 |
| GTEx_Tissu | GTEX-R55C | 2/226 | 0.257961 | 0.891318 | 0 | 0 | 2.068172 | 2.802265 |
| GTEx_Tissu | GTEX-WZT( | 24/4821 | 0.258556 | 0.891318 | 0 | 0 | 1.200433 | 1.623757 |
| GTEx_Tissu | GTEX-XYKS | 2/228 | 0.261186 | 0.891318 | 0 | 0 | 2.049662 | 2.751716 |
| GTEx_Tissu | GTEX-T8EN | 2/228 | 0.261186 | 0.891318 | 0 | 0 | 2.049662 | 2.751716 |
| GTEx_Tissu | GTEX-RM2 | 20/3953 | 0.261702 | 0.891711 | 0 | 0 | 1.212853 | 1.625888 |
| DisGeNET | Renal carci | 1/500 | 0.890019 | 0.891721 | 0 | 0 | 0.452393 | 0.052709 |
| GTEx_Tissu | GTEX-PLZ6 | Jan-70 | 0.263387 | 0.891962 | 0 | 0 | 3.344119 | 4.461494 |
| GTEx_Tissu | GTEX-SNOS | 20/3955 | 0.262482 | 0.891962 | 0 | 0 | 1.212084 | 1.621251 |
| GTEx_Tissu | GTEX-XYKS | 4/599 | 0.263688 | 0.891962 | 0 | 0 | 1.564686 | 2.085708 |
| GTEx_Tissu | GTEX-XLM4 | Nov-25 | 0.263584 | 0.891962 | 0 | 0 | 1.286318 | 1.715153 |
| GTEx_Tissu | GTEX-SSA3 | 15/2876 | 0.26378 | 0.891962 | 0 | 0 | 1.241699 | 1.654736 |
| GeDiPNet_ | Rheumatoi | 1/501 | 0.89051 | 0.89204 | 0 | 0 | 0.451465 | 0.052353 |
| GTEx_Tissu | GTEX-TMZS | 9/1608 | 0.264232 | 0.892132 | 0 | 0 | 1.321547 | 1.758886 |
| DisGeNET | Glioblastor | 2/854 | 0.890961 | 0.892323 | 0 | 0 | 0.526402 | 0.060776 |
| GO_Biologi | Protein Pho | 1/500 | 0.890019 | 0.892533 | 0 | 0 | 0.452393 | 0.052709 |
| GTEx_Tissu | GTEX-PX3G | 5/576 | 0.10618 | 0.893411 | 0 | 0 | 2.065482 | 4.632088 |
| GTEx_Tissu | GTEX-R55C | 6/746 | 0.106233 | 0.893411 | 0 | 0 | 1.919219 | 4.303113 |

| | | | | | | | | |
|---|---|---|---|---|---|---|---|---|
| Jensen_TIS | Spinal gang | Dec-23 | 0.885685 | 0.894014 | 0 | 0 | 0.722326 | 0.087686 |
| CellMarker | Oogenesis | 2/855 | 0.891351 | 0.894616 | 0 | 0 | 0.525757 | 0.060471 |
| GWAS_Cat | Coronary A | 1/508 | 0.893884 | 0.894618 | 0 | 0 | 0.445071 | 0.049928 |
| RNA-Seq_D | ABCA4 RPE | 1/499 | 0.889526 | 0.894983 | 0 | 0 | 0.453325 | 0.053069 |
| RNA-Seq_D | Cushing's S | 1/498 | 0.889031 | 0.894983 | 0 | 0 | 0.45426 | 0.053432 |
| RNA-Seq_D | Colorectal | 1/498 | 0.889031 | 0.894983 | 0 | 0 | 0.45426 | 0.053432 |
| RNA-Seq_D | Col4a3 Kidi | 1/498 | 0.889031 | 0.894983 | 0 | 0 | 0.45426 | 0.053432 |
| RNA-Seq_D | Ch25h BMI | 1/498 | 0.889031 | 0.894983 | 0 | 0 | 0.45426 | 0.053432 |
| RNA-Seq_D | CH-223191 | 1/498 | 0.889031 | 0.894983 | 0 | 0 | 0.45426 | 0.053432 |
| RNA-Seq_D | CFTR Ileum | 1/498 | 0.889031 | 0.894983 | 0 | 0 | 0.45426 | 0.053432 |
| RNA-Seq_D | CFTR Color | 1/498 | 0.889031 | 0.894983 | 0 | 0 | 0.45426 | 0.053432 |
| RNA-Seq_D | AE9aMeis2 | 1/499 | 0.889526 | 0.894983 | 0 | 0 | 0.453325 | 0.053069 |
| RNA-Seq_D | ABCA4 Ret | 1/499 | 0.889526 | 0.894983 | 0 | 0 | 0.453325 | 0.053069 |
| RNA-Seq_D | Cxcr4 T-cel | 1/498 | 0.889031 | 0.894983 | 0 | 0 | 0.45426 | 0.053432 |
| RNA-Seq_D | nmf205 Ce | 1/498 | 0.889031 | 0.894983 | 0 | 0 | 0.45426 | 0.053432 |
| RNA-Seq_D | miR-18a W | 1/498 | 0.889031 | 0.894983 | 0 | 0 | 0.45426 | 0.053432 |
| RNA-Seq_D | DICER1 Fib | 1/498 | 0.889031 | 0.894983 | 0 | 0 | 0.45426 | 0.053432 |
| RNA-Seq_D | Cyclophosp | 1/498 | 0.889031 | 0.894983 | 0 | 0 | 0.45426 | 0.053432 |
| RNA-Seq_D | Cyclophosp | 1/498 | 0.889031 | 0.894983 | 0 | 0 | 0.45426 | 0.053432 |
| RNA-Seq_D | INK128 MD | 1/498 | 0.889031 | 0.894983 | 0 | 0 | 0.45426 | 0.053432 |
| RNA-Seq_D | IL-4 BMDM | 1/498 | 0.889031 | 0.894983 | 0 | 0 | 0.45426 | 0.053432 |
| RNA-Seq_D | HDAC3 Hip | 1/498 | 0.889031 | 0.894983 | 0 | 0 | 0.45426 | 0.053432 |
| RNA-Seq_D | Gefitinib N | 1/498 | 0.889031 | 0.894983 | 0 | 0 | 0.45426 | 0.053432 |
| RNA-Seq_D | Phenobarb | 1/498 | 0.889031 | 0.894983 | 0 | 0 | 0.45426 | 0.053432 |
| RNA-Seq_D | MeCP2 Visi | 1/498 | 0.889031 | 0.894983 | 0 | 0 | 0.45426 | 0.053432 |
| RNA-Seq_D | TGFB3 Pala | 1/499 | 0.889526 | 0.894983 | 0 | 0 | 0.453325 | 0.053069 |
| RNA-Seq_D | Sip1 Lens K | 1/499 | 0.889526 | 0.894983 | 0 | 0 | 0.453325 | 0.053069 |
| RNA-Seq_D | Sip1 Lens K | 1/499 | 0.889526 | 0.894983 | 0 | 0 | 0.453325 | 0.053069 |
| RNA-Seq_D | Satb1 Canc | 1/499 | 0.889526 | 0.894983 | 0 | 0 | 0.453325 | 0.053069 |
| RNA-Seq_D | SOD1 Astro | 1/499 | 0.889526 | 0.894983 | 0 | 0 | 0.453325 | 0.053069 |
| RNA-Seq_D | Nod2 Smal | 1/498 | 0.889031 | 0.894983 | 0 | 0 | 0.45426 | 0.053432 |
| RNA-Seq_D | NMP U266 | 1/498 | 0.889031 | 0.894983 | 0 | 0 | 0.45426 | 0.053432 |
| RNA-Seq_D | NMP U266 | 1/498 | 0.889031 | 0.894983 | 0 | 0 | 0.45426 | 0.053432 |
| RNA-Seq_D | Firre Embry | 1/498 | 0.889031 | 0.894983 | 0 | 0 | 0.45426 | 0.053432 |
| RNA-Seq_D | Panobinost | 1/498 | 0.889031 | 0.894983 | 0 | 0 | 0.45426 | 0.053432 |
| RNA-Seq_D | Panobinost | 1/498 | 0.889031 | 0.894983 | 0 | 0 | 0.45426 | 0.053432 |
| RNA-Seq_D | PTP1B Mac | 1/498 | 0.889031 | 0.894983 | 0 | 0 | 0.45426 | 0.053432 |
| RNA-Seq_D | PSMD13 H | 1/498 | 0.889031 | 0.894983 | 0 | 0 | 0.45426 | 0.053432 |
| RNA-Seq_D | PSMD13 H | 1/498 | 0.889031 | 0.894983 | 0 | 0 | 0.45426 | 0.053432 |
| RNA-Seq_D | TTP BMDM | 1/499 | 0.889526 | 0.894983 | 0 | 0 | 0.453325 | 0.053069 |
| RNA-Seq_D | TNFR2 Colo | 1/499 | 0.889526 | 0.894983 | 0 | 0 | 0.453325 | 0.053069 |
| RNA-Seq_D | BZLT Akata | 1/498 | 0.889031 | 0.894983 | 0 | 0 | 0.45426 | 0.053432 |
| RNA-Seq_D | Nfib Primai | 1/497 | 0.888534 | 0.894983 | 0 | 0 | 0.4552 | 0.053797 |
| RNA-Seq_D | EIF3L HCT1 | 1/498 | 0.889031 | 0.894983 | 0 | 0 | 0.45426 | 0.053432 |
| RNA-Seq_D | Duchenne | 1/498 | 0.889031 | 0.894983 | 0 | 0 | 0.45426 | 0.053432 |

| | | | | | | | | |
|---|---|---|---|---|---|---|---|---|
| RNA-Seq_[ | Ptbp1 Emb | 1/497 | 0.888534 | 0.894983 | 0 | 0 | 0.4552 | 0.053797 |
| RNA-Seq_[ | Prox1 Lens | 1/497 | 0.888534 | 0.894983 | 0 | 0 | 0.4552 | 0.053797 |
| RNA-Seq_[ | PS2APP Ne | 1/498 | 0.889031 | 0.894983 | 0 | 0 | 0.45426 | 0.053432 |
| RNA-Seq_[ | PS2APP Ast | 1/498 | 0.889031 | 0.894983 | 0 | 0 | 0.45426 | 0.053432 |
| RNA-Seq_[ | PRMT9 Hel | 1/498 | 0.889031 | 0.894983 | 0 | 0 | 0.45426 | 0.053432 |
| RNA-Seq_[ | PKC412 ZR | 1/497 | 0.888534 | 0.894983 | 0 | 0 | 0.4552 | 0.053797 |
| RNA-Seq_[ | PBS-1086 1 | 1/497 | 0.888534 | 0.894983 | 0 | 0 | 0.4552 | 0.053797 |
| RNA-Seq_[ | Doxycyclin | 1/497 | 0.888534 | 0.894983 | 0 | 0 | 0.4552 | 0.053797 |
| RNA-Seq_[ | TGM5 HCT | 1/497 | 0.888534 | 0.894983 | 0 | 0 | 0.4552 | 0.053797 |
| RNA-Seq_[ | T0901317 | 1/497 | 0.888534 | 0.894983 | 0 | 0 | 0.4552 | 0.053797 |
| RNA-Seq_[ | Staphyloco | 1/497 | 0.888534 | 0.894983 | 0 | 0 | 0.4552 | 0.053797 |
| RNA-Seq_[ | Notch1 T ly | 1/498 | 0.889031 | 0.894983 | 0 | 0 | 0.45426 | 0.053432 |
| RNA-Seq_[ | RGS18 HCT | 1/497 | 0.888534 | 0.894983 | 0 | 0 | 0.4552 | 0.053797 |
| RNA-Seq_[ | RBM38 NA | 1/497 | 0.888534 | 0.894983 | 0 | 0 | 0.4552 | 0.053797 |
| RNA-Seq_[ | miR-18a Ta | 1/497 | 0.888534 | 0.894983 | 0 | 0 | 0.4552 | 0.053797 |
| RNA-Seq_[ | miR-148a E | 1/497 | 0.888534 | 0.894983 | 0 | 0 | 0.4552 | 0.053797 |
| RNA-Seq_[ | PDPN Mac | 1/496 | 0.888034 | 0.894983 | 0 | 0 | 0.456143 | 0.054165 |
| RNA-Seq_[ | IL-10 Macr | 2/679 | 0.799636 | 0.894983 | 0 | 0 | 0.668555 | 0.149488 |
| RNA-Seq_[ | Ezh2 Small | 1/498 | 0.889031 | 0.894983 | 0 | 0 | 0.45426 | 0.053432 |
| RNA-Seq_[ | Blimp1 Bor | 1/499 | 0.889526 | 0.894983 | 0 | 0 | 0.453325 | 0.053069 |
| RNA-Seq_[ | AEMeis2 B | 1/499 | 0.889526 | 0.894983 | 0 | 0 | 0.453325 | 0.053069 |
| RNA-Seq_[ | Tunicamyc | 1/497 | 0.888534 | 0.894983 | 0 | 0 | 0.4552 | 0.053797 |
| RNA-Seq_[ | Topotecan | 1/497 | 0.888534 | 0.894983 | 0 | 0 | 0.4552 | 0.053797 |
| RNA-Seq_[ | Drc Embry | 1/498 | 0.889031 | 0.894983 | 0 | 0 | 0.45426 | 0.053432 |
| RNA-Seq_[ | Panobinost | 1/497 | 0.888534 | 0.894983 | 0 | 0 | 0.4552 | 0.053797 |
| RNA-Seq_[ | PSMD13 H | 1/497 | 0.888534 | 0.894983 | 0 | 0 | 0.4552 | 0.053797 |
| RNA-Seq_[ | PRMT4 CD | 1/497 | 0.888534 | 0.894983 | 0 | 0 | 0.4552 | 0.053797 |
| RNA-Seq_[ | Meis1-Jmjc | 1/498 | 0.889031 | 0.894983 | 0 | 0 | 0.45426 | 0.053432 |
| RNA-Seq_[ | Vitamin D / | 2/663 | 0.7886 | 0.894983 | 0 | 0 | 0.685307 | 0.162758 |
| RNA-Seq_[ | Doxycyclin | 3/924 | 0.772256 | 0.894983 | 0 | 0 | 0.736467 | 0.190332 |
| RNA-Seq_[ | DAPT Coch | 3/871 | 0.736374 | 0.894983 | 0 | 0 | 0.783616 | 0.2398 |
| RNA-Seq_[ | NMP JJN3 | 1/497 | 0.888534 | 0.894983 | 0 | 0 | 0.4552 | 0.053797 |
| RNA-Seq_[ | Mycobacte | 1/497 | 0.888534 | 0.894983 | 0 | 0 | 0.4552 | 0.053797 |
| RNA-Seq_[ | PRMT5 MV | 1/498 | 0.889031 | 0.894983 | 0 | 0 | 0.45426 | 0.053432 |
| RNA-Seq_[ | PRDM14 KI | 1/498 | 0.889031 | 0.894983 | 0 | 0 | 0.45426 | 0.053432 |
| RNA-Seq_[ | PNN HCET | 1/498 | 0.889031 | 0.894983 | 0 | 0 | 0.45426 | 0.053432 |
| RNA-Seq_[ | TGFB3 Pala | 1/498 | 0.889031 | 0.894983 | 0 | 0 | 0.45426 | 0.053432 |
| RNA-Seq_[ | Ptf1a 14 da | 1/499 | 0.889526 | 0.894983 | 0 | 0 | 0.453325 | 0.053069 |
| RNA-Seq_[ | Prox1 Lens | 1/499 | 0.889526 | 0.894983 | 0 | 0 | 0.453325 | 0.053069 |
| RNA-Seq_[ | PS2APP Mi | 1/499 | 0.889526 | 0.894983 | 0 | 0 | 0.453325 | 0.053069 |
| RNA-Seq_[ | TGFB3 Pala | 1/498 | 0.889031 | 0.894983 | 0 | 0 | 0.45426 | 0.053432 |
| RNA-Seq_[ | Irf4 CD8+ T | 1/498 | 0.889031 | 0.894983 | 0 | 0 | 0.45426 | 0.053432 |
| RNA-Seq_[ | Dot1l Com | 1/499 | 0.889526 | 0.894983 | 0 | 0 | 0.453325 | 0.053069 |
| RNA-Seq_[ | Dgcr8 Con | 1/499 | 0.889526 | 0.894983 | 0 | 0 | 0.453325 | 0.053069 |
| RNA-Seq_[ | Ctcfl Trans | 1/499 | 0.889526 | 0.894983 | 0 | 0 | 0.453325 | 0.053069 |

| | | | | | | | | |
|---|---|---|---|---|---|---|---|---|
| RNA-Seq_D | Tamoxifen | 1/498 | 0.889031 | 0.894983 | 0 | 0 | 0.45426 | 0.053432 |
| RNA-Seq_D | MeCP2 Olfa | 1/499 | 0.889526 | 0.894983 | 0 | 0 | 0.453325 | 0.053069 |
| RNA-Seq_D | Tex10 Emb | 1/499 | 0.889526 | 0.894983 | 0 | 0 | 0.453325 | 0.053069 |
| RNA-Seq_D | Tet1 Bone | 1/499 | 0.889526 | 0.894983 | 0 | 0 | 0.453325 | 0.053069 |
| RNA-Seq_D | Ezh2 Embr | 1/499 | 0.889526 | 0.894983 | 0 | 0 | 0.453325 | 0.053069 |
| RNA-Seq_D | Asthma Tra | 1/498 | 0.889031 | 0.894983 | 0 | 0 | 0.45426 | 0.053432 |
| RNA-Seq_D | AUF1 Skele | 1/498 | 0.889031 | 0.894983 | 0 | 0 | 0.45426 | 0.053432 |
| RNA-Seq_D | ADAR E861 | 1/498 | 0.889031 | 0.894983 | 0 | 0 | 0.45426 | 0.053432 |
| RNA-Seq_D | 5-Azacytidi | 1/498 | 0.889031 | 0.894983 | 0 | 0 | 0.45426 | 0.053432 |
| RNA-Seq_D | Runx1 Bon | 1/498 | 0.889031 | 0.894983 | 0 | 0 | 0.45426 | 0.053432 |
| RNA-Seq_D | Ptbp1 Emb | 1/498 | 0.889031 | 0.894983 | 0 | 0 | 0.45426 | 0.053432 |
| RNA-Seq_D | TCF21 HCA | 1/498 | 0.889031 | 0.894983 | 0 | 0 | 0.45426 | 0.053432 |
| RNA-Seq_D | KH-CB19 Er | 1/495 | 0.887533 | 0.894983 | 0 | 0 | 0.45709 | 0.054535 |
| RNA-Seq_D | nSR100/Srr | 1/499 | 0.889526 | 0.894983 | 0 | 0 | 0.453325 | 0.053069 |
| RNA-Seq_D | miR-146a E | 1/499 | 0.889526 | 0.894983 | 0 | 0 | 0.453325 | 0.053069 |
| RNA-Seq_D | beta-caten | 1/499 | 0.889526 | 0.894983 | 0 | 0 | 0.453325 | 0.053069 |
| RNA-Seq_D | UTX Spleer | 1/499 | 0.889526 | 0.894983 | 0 | 0 | 0.453325 | 0.053069 |
| RNA-Seq_D | Trex1 Splee | 1/499 | 0.889526 | 0.894983 | 0 | 0 | 0.453325 | 0.053069 |
| RNA-Seq_D | Lin28 Neur | 1/499 | 0.889526 | 0.894983 | 0 | 0 | 0.453325 | 0.053069 |
| RNA-Seq_D | Kras(G12D) | 1/499 | 0.889526 | 0.894983 | 0 | 0 | 0.453325 | 0.053069 |
| RNA-Seq_D | Tet1 Bone | 1/499 | 0.889526 | 0.894983 | 0 | 0 | 0.453325 | 0.053069 |
| RNA-Seq_D | Psoriasis Sk | 1/498 | 0.889031 | 0.894983 | 0 | 0 | 0.45426 | 0.053432 |
| RNA-Seq_D | SRA MCF7 | 1/498 | 0.889031 | 0.894983 | 0 | 0 | 0.45426 | 0.053432 |
| RNA-Seq_D | SENCR HCA | 1/498 | 0.889031 | 0.894983 | 0 | 0 | 0.45426 | 0.053432 |
| RNA-Seq_D | SAHA Liver | 1/498 | 0.889031 | 0.894983 | 0 | 0 | 0.45426 | 0.053432 |
| RNA-Seq_D | nSR100/Srr | 1/499 | 0.889526 | 0.894983 | 0 | 0 | 0.453325 | 0.053069 |
| RNA-Seq_D | ZnF4 Thym | 1/499 | 0.889526 | 0.894983 | 0 | 0 | 0.453325 | 0.053069 |
| RNA-Seq_D | Oct4 Ather | 1/499 | 0.889526 | 0.894983 | 0 | 0 | 0.453325 | 0.053069 |
| RNA-Seq_D | Nova1 mid | 1/499 | 0.889526 | 0.894983 | 0 | 0 | 0.453325 | 0.053069 |
| RNA-Seq_D | Mrpp3 Hea | 1/499 | 0.889526 | 0.894983 | 0 | 0 | 0.453325 | 0.053069 |
| RNA-Seq_D | LPS Astrocy | 1/497 | 0.888534 | 0.894983 | 0 | 0 | 0.4552 | 0.053797 |
| RNA-Seq_D | KDM2B HS | 1/499 | 0.889526 | 0.894983 | 0 | 0 | 0.453325 | 0.053069 |
| RNA-Seq_D | IRF8 Spleer | 1/499 | 0.889526 | 0.894983 | 0 | 0 | 0.453325 | 0.053069 |
| RNA-Seq_D | IRF1 Prima | 1/499 | 0.889526 | 0.894983 | 0 | 0 | 0.453325 | 0.053069 |
| RNA-Seq_D | LPS Lung 5 | 1/498 | 0.889031 | 0.894983 | 0 | 0 | 0.45426 | 0.053432 |
| RNA-Seq_D | LNC277 A6 | 1/498 | 0.889031 | 0.894983 | 0 | 0 | 0.45426 | 0.053432 |
| RNA-Seq_D | LPS Microg | 1/498 | 0.889031 | 0.894983 | 0 | 0 | 0.45426 | 0.053432 |
| RNA-Seq_D | Mycobacte | 1/497 | 0.888534 | 0.894983 | 0 | 0 | 0.4552 | 0.053797 |
| RNA-Seq_D | MSI2 Hema | 1/497 | 0.888534 | 0.894983 | 0 | 0 | 0.4552 | 0.053797 |
| RNA-Seq_D | METTL3 He | 1/497 | 0.888534 | 0.894983 | 0 | 0 | 0.4552 | 0.053797 |
| RNA-Seq_D | Blimp1 Bor | 1/499 | 0.889526 | 0.894983 | 0 | 0 | 0.453325 | 0.053069 |
| RNA-Seq_D | Influenza A | 1/498 | 0.889031 | 0.894983 | 0 | 0 | 0.45426 | 0.053432 |
| RNA-Seq_D | IRF1 Prima | 1/498 | 0.889031 | 0.894983 | 0 | 0 | 0.45426 | 0.053432 |
| RNA-Seq_D | IPTKB Hem | 1/498 | 0.889031 | 0.894983 | 0 | 0 | 0.45426 | 0.053432 |
| RNA-Seq_D | NHD13 My | 1/498 | 0.889031 | 0.894983 | 0 | 0 | 0.45426 | 0.053432 |

| | | | | | | | | |
|---|---|---|---|---|---|---|---|---|
| RNA-Seq_[ | Myc RKO k | 1/498 | 0.889031 | 0.894983 | 0 | 0 | 0.45426 | 0.053432 |
| RNA-Seq_[ | Multiple Sc | 1/498 | 0.889031 | 0.894983 | 0 | 0 | 0.45426 | 0.053432 |
| RNA-Seq_[ | Multiple Sc | 1/498 | 0.889031 | 0.894983 | 0 | 0 | 0.45426 | 0.053432 |
| RNA-Seq_[ | Msi2 Leuke | 1/498 | 0.889031 | 0.894983 | 0 | 0 | 0.45426 | 0.053432 |
| RNA-Seq_[ | Mettl3 Eml | 1/498 | 0.889031 | 0.894983 | 0 | 0 | 0.45426 | 0.053432 |
| RNA-Seq_[ | Ezh2 Embr | 1/499 | 0.889526 | 0.894983 | 0 | 0 | 0.453325 | 0.053069 |
| RNA-Seq_[ | CFTR Duod | 1/499 | 0.889526 | 0.894983 | 0 | 0 | 0.453325 | 0.053069 |
| RNA-Seq_[ | Vemurafen | 1/498 | 0.889031 | 0.894983 | 0 | 0 | 0.45426 | 0.053432 |
| RNA-Seq_[ | Utx Spleen | 1/498 | 0.889031 | 0.894983 | 0 | 0 | 0.45426 | 0.053432 |
| RNA-Seq_[ | UTX Bone r | 1/498 | 0.889031 | 0.894983 | 0 | 0 | 0.45426 | 0.053432 |
| RNA-Seq_[ | Trim33 B ly | 1/498 | 0.889031 | 0.894983 | 0 | 0 | 0.45426 | 0.053432 |
| RNA-Seq_[ | Trim24 Live | 1/498 | 0.889031 | 0.894983 | 0 | 0 | 0.45426 | 0.053432 |
| RNA-Seq_[ | GFP Retina | 1/499 | 0.889526 | 0.894983 | 0 | 0 | 0.453325 | 0.053069 |
| RNA-Seq_[ | FancC matu | 1/499 | 0.889526 | 0.894983 | 0 | 0 | 0.453325 | 0.053069 |
| RNA-Seq_[ | Jmjd1c Bor | 1/498 | 0.889031 | 0.894983 | 0 | 0 | 0.45426 | 0.053432 |
| RNA-Seq_[ | LKB1 RBE k | 1/495 | 0.887533 | 0.894983 | 0 | 0 | 0.45709 | 0.054535 |
| RNA-Seq_[ | ZnF1 Thym | 1/498 | 0.889031 | 0.894983 | 0 | 0 | 0.45426 | 0.053432 |
| RNA-Seq_[ | Yersinia ps | 1/498 | 0.889031 | 0.894983 | 0 | 0 | 0.45426 | 0.053432 |
| RNA-Seq_[ | YY1 Bone n | 1/498 | 0.889031 | 0.894983 | 0 | 0 | 0.45426 | 0.053432 |
| RNA-Seq_[ | WZ4002 N | 1/498 | 0.889031 | 0.894983 | 0 | 0 | 0.45426 | 0.053432 |
| RNA-Seq_[ | Col4a3 Kid | 1/499 | 0.889526 | 0.894983 | 0 | 0 | 0.453325 | 0.053069 |
| RNA-Seq_[ | CFTR Ileum | 1/499 | 0.889526 | 0.894983 | 0 | 0 | 0.453325 | 0.053069 |
| RNA-Seq_[ | CrX Retina | 1/499 | 0.889526 | 0.894983 | 0 | 0 | 0.453325 | 0.053069 |
| RNA-Seq_[ | Bmal1 Live | 1/499 | 0.889526 | 0.894983 | 0 | 0 | 0.453325 | 0.053069 |
| RNA-Seq_[ | BRD4 HCT1 | 1/496 | 0.888034 | 0.894983 | 0 | 0 | 0.456143 | 0.054165 |
| RNA-Seq_[ | Albuterol A | 2/757 | 0.846505 | 0.894983 | 0 | 0 | 0.597055 | 0.099493 |
| RNA-Seq_[ | CNS2 Regu | 2/750 | 0.842738 | 0.894983 | 0 | 0 | 0.602863 | 0.103149 |
| RNA-Seq_[ | CNS2 Regu | 2/729 | 0.830939 | 0.894983 | 0 | 0 | 0.620956 | 0.115 |
| RNA-Seq_[ | Gsa Cerebe | 2/719 | 0.825049 | 0.894983 | 0 | 0 | 0.629945 | 0.121146 |
| RNA-Seq_[ | Estradiol G | 1/496 | 0.888034 | 0.894983 | 0 | 0 | 0.456143 | 0.054165 |
| RNA-Seq_[ | EHF A549 ( | 1/496 | 0.888034 | 0.894983 | 0 | 0 | 0.456143 | 0.054165 |
| RNA-Seq_[ | Dexametha | 1/496 | 0.888034 | 0.894983 | 0 | 0 | 0.456143 | 0.054165 |
| RNA-Seq_[ | DMOG Sple | 1/496 | 0.888034 | 0.894983 | 0 | 0 | 0.456143 | 0.054165 |
| RNA-Seq_[ | DEANR1 Er | 1/496 | 0.888034 | 0.894983 | 0 | 0 | 0.456143 | 0.054165 |
| RNA-Seq_[ | Bardet-Bie | 1/496 | 0.888034 | 0.894983 | 0 | 0 | 0.456143 | 0.054165 |
| RNA-Seq_[ | Bacillus Cal | 1/496 | 0.888034 | 0.894983 | 0 | 0 | 0.456143 | 0.054165 |
| RNA-Seq_[ | Relapsing-r | 1/490 | 0.884991 | 0.894983 | 0 | 0 | 0.461882 | 0.056432 |
| RNA-Seq_[ | BEGAIN HC | 1/493 | 0.886523 | 0.894983 | 0 | 0 | 0.458995 | 0.055285 |
| RNA-Seq_[ | 4-hydroxyt | 1/496 | 0.888034 | 0.894983 | 0 | 0 | 0.456143 | 0.054165 |
| RNA-Seq_[ | 2,3,7,8-tet | 1/496 | 0.888034 | 0.894983 | 0 | 0 | 0.456143 | 0.054165 |
| RNA-Seq_[ | Phenobarb | 1/495 | 0.887533 | 0.894983 | 0 | 0 | 0.45709 | 0.054535 |
| RNA-Seq_[ | Phenobarb | 1/495 | 0.887533 | 0.894983 | 0 | 0 | 0.45709 | 0.054535 |
| RNA-Seq_[ | Perhexiline | 1/495 | 0.887533 | 0.894983 | 0 | 0 | 0.45709 | 0.054535 |
| RNA-Seq_[ | Nova1 Brai | 1/495 | 0.887533 | 0.894983 | 0 | 0 | 0.45709 | 0.054535 |
| RNA-Seq_[ | Desipramir | 1/494 | 0.887029 | 0.894983 | 0 | 0 | 0.45804 | 0.054909 |

| | | | | | | | | |
|---|---|---|---|---|---|---|---|---|
| RNA-Seq_[ | DAXX PC3 I | 1/494 | 0.887029 | 0.894983 | 0 | 0 | 0.45804 | 0.054909 |
| RNA-Seq_[ | DAPT 3 hES | 1/494 | 0.887029 | 0.894983 | 0 | 0 | 0.45804 | 0.054909 |
| RNA-Seq_[ | Albuterol A | 2/712 | 0.820818 | 0.894983 | 0 | 0 | 0.636388 | 0.125657 |
| RNA-Seq_[ | ELAVL2 NP | 1/495 | 0.887533 | 0.894983 | 0 | 0 | 0.45709 | 0.054535 |
| RNA-Seq_[ | DRB (dichlo | 1/495 | 0.887533 | 0.894983 | 0 | 0 | 0.45709 | 0.054535 |
| RNA-Seq_[ | Hobit Bone | 1/496 | 0.888034 | 0.894983 | 0 | 0 | 0.456143 | 0.054165 |
| RNA-Seq_[ | BRG1 MCF- | 1/497 | 0.888534 | 0.894983 | 0 | 0 | 0.4552 | 0.053797 |
| RNA-Seq_[ | Albuterol 8 | 1/497 | 0.888534 | 0.894983 | 0 | 0 | 0.4552 | 0.053797 |
| RNA-Seq_[ | Akti-1-2 M | 1/497 | 0.888534 | 0.894983 | 0 | 0 | 0.4552 | 0.053797 |
| RNA-Seq_[ | AMD3100 | 1/497 | 0.888534 | 0.894983 | 0 | 0 | 0.4552 | 0.053797 |
| RNA-Seq_[ | ADAR Fetal | 1/497 | 0.888534 | 0.894983 | 0 | 0 | 0.4552 | 0.053797 |
| RNA-Seq_[ | ADAR E861 | 1/497 | 0.888534 | 0.894983 | 0 | 0 | 0.4552 | 0.053797 |
| RNA-Seq_[ | 5-Azacytidi | 1/497 | 0.888534 | 0.894983 | 0 | 0 | 0.4552 | 0.053797 |
| RNA-Seq_[ | Anankinra | 1/494 | 0.887029 | 0.894983 | 0 | 0 | 0.45804 | 0.054909 |
| RNA-Seq_[ | Gsa Cerebe | 2/703 | 0.815246 | 0.894983 | 0 | 0 | 0.64486 | 0.131722 |
| RNA-Seq_[ | MEP50 A54 | 1/496 | 0.888034 | 0.894983 | 0 | 0 | 0.456143 | 0.054165 |
| RNA-Seq_[ | MEF2 Skele | 1/496 | 0.888034 | 0.894983 | 0 | 0 | 0.456143 | 0.054165 |
| RNA-Seq_[ | LKB1 HuH-: | 1/496 | 0.888034 | 0.894983 | 0 | 0 | 0.456143 | 0.054165 |
| RNA-Seq_[ | Klf3 Fetal L | 1/496 | 0.888034 | 0.894983 | 0 | 0 | 0.456143 | 0.054165 |
| RNA-Seq_[ | JAK3i (CP-6 | 1/496 | 0.888034 | 0.894983 | 0 | 0 | 0.456143 | 0.054165 |
| RNA-Seq_[ | Huntingtor | 1/496 | 0.888034 | 0.894983 | 0 | 0 | 0.456143 | 0.054165 |
| RNA-Seq_[ | Asthma Air | 1/496 | 0.888034 | 0.894983 | 0 | 0 | 0.456143 | 0.054165 |
| RNA-Seq_[ | Vemurafen | 1/491 | 0.885504 | 0.894983 | 0 | 0 | 0.460916 | 0.056047 |
| RNA-Seq_[ | Juvenile Idi | 1/491 | 0.885504 | 0.894983 | 0 | 0 | 0.460916 | 0.056047 |
| RNA-Seq_[ | MOV10 HE | 1/496 | 0.888034 | 0.894983 | 0 | 0 | 0.456143 | 0.054165 |
| RNA-Seq_[ | GW3965 Tu | 1/497 | 0.888534 | 0.894983 | 0 | 0 | 0.4552 | 0.053797 |
| RNA-Seq_[ | GPR65 BM | 1/497 | 0.888534 | 0.894983 | 0 | 0 | 0.4552 | 0.053797 |
| RNA-Seq_[ | GATA2 Ch- | 1/497 | 0.888534 | 0.894983 | 0 | 0 | 0.4552 | 0.053797 |
| RNA-Seq_[ | Firre Embry | 1/497 | 0.888534 | 0.894983 | 0 | 0 | 0.4552 | 0.053797 |
| RNA-Seq_[ | Rosiglitazo | 1/497 | 0.888534 | 0.894983 | 0 | 0 | 0.4552 | 0.053797 |
| RNA-Seq_[ | OVOL1 Tro | 1/496 | 0.888034 | 0.894983 | 0 | 0 | 0.456143 | 0.054165 |
| RNA-Seq_[ | OPC Cerebr | 1/496 | 0.888034 | 0.894983 | 0 | 0 | 0.456143 | 0.054165 |
| RNA-Seq_[ | NAF1 MCF- | 1/496 | 0.888034 | 0.894983 | 0 | 0 | 0.456143 | 0.054165 |
| RNA-Seq_[ | Myelodysp | 1/496 | 0.888034 | 0.894983 | 0 | 0 | 0.456143 | 0.054165 |
| RNA-Seq_[ | Miconazole | 1/496 | 0.888034 | 0.894983 | 0 | 0 | 0.456143 | 0.054165 |
| RNA-Seq_[ | Mef2c/d pr | 1/496 | 0.888034 | 0.894983 | 0 | 0 | 0.456143 | 0.054165 |
| RNA-Seq_[ | GW3965 Tu | 1/497 | 0.888534 | 0.894983 | 0 | 0 | 0.4552 | 0.053797 |
| RNA-Seq_[ | MOV10 HE | 1/496 | 0.888034 | 0.894983 | 0 | 0 | 0.456143 | 0.054165 |
| RNA-Seq_[ | Id3 Spleen | 2/847 | 0.888196 | 0.894983 | 0 | 0 | 0.530957 | 0.062952 |
| RNA-Seq_[ | mir-93 Kidr | 1/496 | 0.888034 | 0.894983 | 0 | 0 | 0.456143 | 0.054165 |
| RNA-Seq_[ | miR21 Mar | 1/496 | 0.888034 | 0.894983 | 0 | 0 | 0.456143 | 0.054165 |
| RNA-Seq_[ | miR-92a-1 | 1/496 | 0.888034 | 0.894983 | 0 | 0 | 0.456143 | 0.054165 |
| RNA-Seq_[ | miR-92a-1 | 1/496 | 0.888034 | 0.894983 | 0 | 0 | 0.456143 | 0.054165 |
| RNA-Seq_[ | ZnF4 Thym | 1/496 | 0.888034 | 0.894983 | 0 | 0 | 0.456143 | 0.054165 |
| RNA-Seq_[ | ZnF4 Thym | 1/496 | 0.888034 | 0.894983 | 0 | 0 | 0.456143 | 0.054165 |

| | | | | | | | | |
|---|---|---|---|---|---|---|---|---|
| RNA-Seq_D | PAX2 REP k | 1/496 | 0.888034 | 0.894983 | 0 | 0 | 0.456143 | 0.054165 |
| RNA-Seq_D | NFYA HCT1 | 1/495 | 0.887533 | 0.894983 | 0 | 0 | 0.45709 | 0.054535 |
| RNA-Seq_D | TDP43 Mot | 1/494 | 0.887029 | 0.894983 | 0 | 0 | 0.45804 | 0.054909 |
| RNA-Seq_D | All-trans re | 1/494 | 0.887029 | 0.894983 | 0 | 0 | 0.45804 | 0.054909 |
| RNA-Seq_D | Cocaine Nu | 1/493 | 0.886523 | 0.894983 | 0 | 0 | 0.458995 | 0.055285 |
| RNA-Seq_D | BS69 Hela I | 1/493 | 0.886523 | 0.894983 | 0 | 0 | 0.458995 | 0.055285 |
| RNA-Seq_D | BRD9-inhib | 1/493 | 0.886523 | 0.894983 | 0 | 0 | 0.458995 | 0.055285 |
| RNA-Seq_D | Rhinovirus | 1/497 | 0.888534 | 0.894983 | 0 | 0 | 0.4552 | 0.053797 |
| RNA-Seq_D | CDK12 HCT | 1/495 | 0.887533 | 0.894983 | 0 | 0 | 0.45709 | 0.054535 |
| RNA-Seq_D | Bmal1 U2C | 1/495 | 0.887533 | 0.894983 | 0 | 0 | 0.45709 | 0.054535 |
| RNA-Seq_D | Blinc1 Panc | 1/495 | 0.887533 | 0.894983 | 0 | 0 | 0.45709 | 0.054535 |
| RNA-Seq_D | All-trans re | 1/495 | 0.887533 | 0.894983 | 0 | 0 | 0.45709 | 0.054535 |
| RNA-Seq_D | Activin A Er | 1/495 | 0.887533 | 0.894983 | 0 | 0 | 0.45709 | 0.054535 |
| RNA-Seq_D | Trisomy 21 | 1/496 | 0.888034 | 0.894983 | 0 | 0 | 0.456143 | 0.054165 |
| RNA-Seq_D | SYKi (R406) | 1/494 | 0.887029 | 0.894983 | 0 | 0 | 0.45804 | 0.054909 |
| RNA-Seq_D | 4 Skin Biop | 1/497 | 0.888534 | 0.894983 | 0 | 0 | 0.4552 | 0.053797 |
| RNA-Seq_D | 4-hydroxyt | 1/497 | 0.888534 | 0.894983 | 0 | 0 | 0.4552 | 0.053797 |
| RNA-Seq_D | Silvestrol M | 1/497 | 0.888534 | 0.894983 | 0 | 0 | 0.4552 | 0.053797 |
| RNA-Seq_D | Salmonella | 1/497 | 0.888534 | 0.894983 | 0 | 0 | 0.4552 | 0.053797 |
| RNA-Seq_D | STAU1 HEK | 1/497 | 0.888534 | 0.894983 | 0 | 0 | 0.4552 | 0.053797 |
| RNA-Seq_D | SOD1 Astro | 1/497 | 0.888534 | 0.894983 | 0 | 0 | 0.4552 | 0.053797 |
| RNA-Seq_D | Rosiglitazo | 1/497 | 0.888534 | 0.894983 | 0 | 0 | 0.4552 | 0.053797 |
| RNA-Seq_D | H2AZ Embr | 1/497 | 0.888534 | 0.894983 | 0 | 0 | 0.4552 | 0.053797 |
| RNA-Seq_D | Ginkgolic A | 1/497 | 0.888534 | 0.894983 | 0 | 0 | 0.4552 | 0.053797 |
| RNA-Seq_D | Dgcr8 Cone | 1/498 | 0.889031 | 0.894983 | 0 | 0 | 0.45426 | 0.053432 |
| RNA-Seq_D | DRB (dichlo | 1/497 | 0.888534 | 0.894983 | 0 | 0 | 0.4552 | 0.053797 |
| RNA-Seq_D | DDX5 Th17 | 1/497 | 0.888534 | 0.894983 | 0 | 0 | 0.4552 | 0.053797 |
| RNA-Seq_D | CFTR Color | 3/845 | 0.717237 | 0.894983 | 0 | 0 | 0.808916 | 0.268842 |
| RNA-Seq_D | MITF 501 M | 3/816 | 0.694671 | 0.894983 | 0 | 0 | 0.839044 | 0.305678 |
| RNA-Seq_D | CBFA2T2 K | 1/497 | 0.888534 | 0.894983 | 0 | 0 | 0.4552 | 0.053797 |
| RNA-Seq_D | Dlx3(Dlx3-K | 1/498 | 0.889031 | 0.894983 | 0 | 0 | 0.45426 | 0.053432 |
| RNA-Seq_D | Dicer1 Reti | 1/498 | 0.889031 | 0.894983 | 0 | 0 | 0.45426 | 0.053432 |
| RNA-Seq_D | Dicer1 Non | 1/498 | 0.889031 | 0.894983 | 0 | 0 | 0.45426 | 0.053432 |
| RNA-Seq_D | Desipramir | 1/497 | 0.888534 | 0.894983 | 0 | 0 | 0.4552 | 0.053797 |
| RNA-Seq_D | FFAR3 Panc | 1/497 | 0.888534 | 0.894983 | 0 | 0 | 0.4552 | 0.053797 |
| RNA-Seq_D | I-BET MV4- | 1/497 | 0.888534 | 0.894983 | 0 | 0 | 0.4552 | 0.053797 |
| RNA-Seq_D | HNRNPA2E | 1/496 | 0.888034 | 0.894983 | 0 | 0 | 0.456143 | 0.054165 |
| RNA-Seq_D | GLI1 HCT1: | 1/496 | 0.888034 | 0.894983 | 0 | 0 | 0.456143 | 0.054165 |
| RNA-Seq_D | GLI1 HCT1: | 1/496 | 0.888034 | 0.894983 | 0 | 0 | 0.456143 | 0.054165 |
| RNA-Seq_D | GATA2 Ch- | 1/496 | 0.888034 | 0.894983 | 0 | 0 | 0.456143 | 0.054165 |
| RNA-Seq_D | GATA2 Ch- | 1/496 | 0.888034 | 0.894983 | 0 | 0 | 0.456143 | 0.054165 |
| RNA-Seq_D | GATA2 Ch- | 1/496 | 0.888034 | 0.894983 | 0 | 0 | 0.456143 | 0.054165 |
| RNA-Seq_D | Bmal1 Live | 1/497 | 0.888534 | 0.894983 | 0 | 0 | 0.4552 | 0.053797 |
| RNA-Seq_D | Zmiz1 T lyn | 1/496 | 0.888034 | 0.894983 | 0 | 0 | 0.456143 | 0.054165 |
| RNA-Seq_D | Trex1 Splee | 1/496 | 0.888034 | 0.894983 | 0 | 0 | 0.456143 | 0.054165 |

| | | | | | | | | |
|---|---|---|---|---|---|---|---|---|
| RNA-Seq_[ | Tetradotox | 1/496 | 0.888034 | 0.894983 | 0 | 0 | 0.456143 | 0.054165 |
| RNA-Seq_[ | Tbx1 P19Cl | 1/496 | 0.888034 | 0.894983 | 0 | 0 | 0.456143 | 0.054165 |
| RNA-Seq_[ | DDPA5 HCT | 1/497 | 0.888534 | 0.894983 | 0 | 0 | 0.4552 | 0.053797 |
| RNA-Seq_[ | CDK13 HCT | 1/497 | 0.888534 | 0.894983 | 0 | 0 | 0.4552 | 0.053797 |
| RNA-Seq_[ | Schizophre | 1/496 | 0.888034 | 0.894983 | 0 | 0 | 0.456143 | 0.054165 |
| RNA-Seq_[ | Brpf1 Fetal | 1/497 | 0.888534 | 0.894983 | 0 | 0 | 0.4552 | 0.053797 |
| RNA-Seq_[ | Estradiol G | 1/495 | 0.887533 | 0.894983 | 0 | 0 | 0.45709 | 0.054535 |
| RNA-Seq_[ | Bmal1 Live | 1/497 | 0.888534 | 0.894983 | 0 | 0 | 0.4552 | 0.053797 |
| RNA-Seq_[ | Blimp1 Bor | 1/497 | 0.888534 | 0.894983 | 0 | 0 | 0.4552 | 0.053797 |
| RNA-Seq_[ | 4-hydroxyt | 1/498 | 0.889031 | 0.894983 | 0 | 0 | 0.45426 | 0.053432 |
| RNA-Seq_[ | Ezh2 Splee | 1/497 | 0.888534 | 0.894983 | 0 | 0 | 0.4552 | 0.053797 |
| RNA-Seq_[ | ETS2 Skin C | 1/497 | 0.888534 | 0.894983 | 0 | 0 | 0.4552 | 0.053797 |
| RNA-Seq_[ | Vemurafen | 1/497 | 0.888534 | 0.894983 | 0 | 0 | 0.4552 | 0.053797 |
| RNA-Seq_[ | Dioxin Live | 1/497 | 0.888534 | 0.894983 | 0 | 0 | 0.4552 | 0.053797 |
| RNA-Seq_[ | Dioxin Live | 1/497 | 0.888534 | 0.894983 | 0 | 0 | 0.4552 | 0.053797 |
| RNA-Seq_[ | TOX4 HCT1 | 1/496 | 0.888034 | 0.894983 | 0 | 0 | 0.456143 | 0.054165 |
| RNA-Seq_[ | HD (R6/1; t | 1/497 | 0.888534 | 0.894983 | 0 | 0 | 0.4552 | 0.053797 |
| RNA-Seq_[ | Satb1 Canc | 1/496 | 0.888034 | 0.894983 | 0 | 0 | 0.456143 | 0.054165 |
| RNA-Seq_[ | SOD1 Astro | 1/496 | 0.888034 | 0.894983 | 0 | 0 | 0.456143 | 0.054165 |
| RNA-Seq_[ | SOD1(G93/ | 1/496 | 0.888034 | 0.894983 | 0 | 0 | 0.456143 | 0.054165 |
| RNA-Seq_[ | EWSFLI1 A( | 1/495 | 0.887533 | 0.894983 | 0 | 0 | 0.45709 | 0.054535 |
| RNA-Seq_[ | LKB1 SSP2! | 1/497 | 0.888534 | 0.894983 | 0 | 0 | 0.4552 | 0.053797 |
| RNA-Seq_[ | Cyclophosp | 1/495 | 0.887533 | 0.894983 | 0 | 0 | 0.45709 | 0.054535 |
| RNA-Seq_[ | RBM38 NA | 1/496 | 0.888034 | 0.894983 | 0 | 0 | 0.456143 | 0.054165 |
| RNA-Seq_[ | Influenza A | 1/497 | 0.888534 | 0.894983 | 0 | 0 | 0.4552 | 0.053797 |
| RNA-Seq_[ | LNA 30bcd | 1/497 | 0.888534 | 0.894983 | 0 | 0 | 0.4552 | 0.053797 |
| RNA-Seq_[ | RBFOX2 HE | 1/496 | 0.888034 | 0.894983 | 0 | 0 | 0.456143 | 0.054165 |
| RNA-Seq_[ | PU.1 K562 | 1/496 | 0.888034 | 0.894983 | 0 | 0 | 0.456143 | 0.054165 |
| RNA-Seq_[ | LNC277 A6 | 1/497 | 0.888534 | 0.894983 | 0 | 0 | 0.4552 | 0.053797 |
| RNA-Seq_[ | HD (R6/1; t | 1/497 | 0.888534 | 0.894983 | 0 | 0 | 0.4552 | 0.053797 |
| RNA-Seq_[ | Klf3 Fetal L | 1/497 | 0.888534 | 0.894983 | 0 | 0 | 0.4552 | 0.053797 |
| RNA-Seq_[ | Smc3 Embr | 1/496 | 0.888034 | 0.894983 | 0 | 0 | 0.456143 | 0.054165 |
| RNA-Seq_[ | Huntingtor | 1/495 | 0.887533 | 0.894983 | 0 | 0 | 0.45709 | 0.054535 |
| RNA-Seq_[ | FICZ Skin B | 1/495 | 0.887533 | 0.894983 | 0 | 0 | 0.45709 | 0.054535 |
| RNA-Seq_[ | KH-CB19 Er | 1/497 | 0.888534 | 0.894983 | 0 | 0 | 0.4552 | 0.053797 |
| RNA-Seq_[ | PQBP1 Cor | 1/496 | 0.888034 | 0.894983 | 0 | 0 | 0.456143 | 0.054165 |
| RNA-Seq_[ | FLT3 Thym | 1/496 | 0.888034 | 0.894983 | 0 | 0 | 0.456143 | 0.054165 |
| GTEx_Tissu | GTEX-XAJ8 | 6/747 | 0.106737 | 0.895046 | 0 | 0 | 1.916529 | 4.288015 |
| Gene_Pert | Kat5 KD mc | 1/510 | 0.894829 | 0.895329 | 0 | 0 | 0.443277 | 0.049258 |
| GTEx_Tissu | GTEX-PLZ4 | 4/601 | 0.26561 | 0.895429 | 0 | 0 | 1.559282 | 2.067179 |
| GTEx_Tissu | GTEX-OXRI | 42/8919 | 0.278829 | 0.895882 | 0 | 0 | 1.160332 | 1.481928 |
| GTEx_Tissu | GTEX-SNOS | 33/6966 | 0.307172 | 0.895882 | 0 | 0 | 1.144126 | 1.350465 |
| GTEx_Tissu | GTEX-R53T | Jan-77 | 0.285606 | 0.895882 | 0 | 0 | 3.035037 | 3.803331 |
| GTEx_Tissu | GTEX-X8H( | 14/2758 | 0.309556 | 0.895882 | 0 | 0 | 1.199958 | 1.407091 |
| GTEx_Tissu | GTEX-RTLS | 7/1224 | 0.28297 | 0.895882 | 0 | 0 | 1.344207 | 1.696947 |

| | | | | | | | | |
|---|---|---|---|---|---|---|---|---|
| GTEx_Tissu | GTEX-XK95 | 4/620 | 0.284002 | 0.895882 | 0 | 0 | 1.509701 | 1.900371 |
| GTEx_Tissu | GTEX-S32W | Jan-82 | 0.30107 | 0.895882 | 0 | 0 | 2.846971 | 3.417539 |
| GTEx_Tissu | GTEX-T5JW | 16/3165 | 0.29657 | 0.895882 | 0 | 0 | 1.199683 | 1.458183 |
| GTEx_Tissu | GTEX-P4PP | Nov-86 | 0.296163 | 0.895882 | 0 | 0 | 1.244249 | 1.514056 |
| GTEx_Tissu | GTEX-U3ZN | 38/8062 | 0.295623 | 0.895882 | 0 | 0 | 1.149058 | 1.400324 |
| GTEx_Tissu | GTEX-QMR | 10/1872 | 0.295474 | 0.895882 | 0 | 0 | 1.259015 | 1.53496 |
| GTEx_Tissu | GTEX-NPJ8 | 14/2754 | 0.307603 | 0.895882 | 0 | 0 | 1.20199 | 1.41708 |
| GTEx_Tissu | GTEX-Q734 | 2/257 | 0.307896 | 0.895882 | 0 | 0 | 1.813887 | 2.136747 |
| GTEx_Tissu | GTEX-RU72 | 2/257 | 0.307896 | 0.895882 | 0 | 0 | 1.813887 | 2.136747 |
| GTEx_Tissu | GTEX-OXRL | Jan-75 | 0.279326 | 0.895882 | 0 | 0 | 3.117379 | 3.975827 |
| GTEx_Tissu | GTEX-WWY | 15/2912 | 0.280053 | 0.895882 | 0 | 0 | 1.22368 | 1.55747 |
| GTEx_Tissu | GTEX-RU72 | 2/240 | 0.280541 | 0.895882 | 0 | 0 | 1.945131 | 2.472331 |
| GTEx_Tissu | GTEX-X5EB | 2/240 | 0.280541 | 0.895882 | 0 | 0 | 1.945131 | 2.472331 |
| GTEx_Tissu | GTEX-XQ8I | 4/617 | 0.281084 | 0.895882 | 0 | 0 | 1.517325 | 1.92564 |
| GTEx_Tissu | GTEX-XMK | Jan-80 | 0.294925 | 0.895882 | 0 | 0 | 2.919341 | 3.564619 |
| GTEx_Tissu | GTEX-U4B1 | Jan-73 | 0.272992 | 0.895882 | 0 | 0 | 3.204296 | 4.160181 |
| GTEx_Tissu | GTEX-XMD | 6/1034 | 0.294148 | 0.895882 | 0 | 0 | 1.360787 | 1.665157 |
| GTEx_Tissu | GTEX-S4Q7 | 9/1619 | 0.270687 | 0.895882 | 0 | 0 | 1.31173 | 1.714159 |
| GTEx_Tissu | GTEX-P4PC | Jan-85 | 0.310189 | 0.895882 | 0 | 0 | 2.744878 | 3.21308 |
| GTEx_Tissu | GTEX-TMM | Jan-84 | 0.307163 | 0.895882 | 0 | 0 | 2.778089 | 3.279195 |
| GTEx_Tissu | GTEX-R55G | Nov-95 | 0.301072 | 0.895882 | 0 | 0 | 1.23825 | 1.486404 |
| GTEx_Tissu | GTEX-QVUS | 4/603 | 0.267536 | 0.895882 | 0 | 0 | 1.553915 | 2.048841 |
| GTEx_Tissu | GTEX-QLQ7 | 3/413 | 0.26774 | 0.895882 | 0 | 0 | 1.698868 | 2.238665 |
| GTEx_Tissu | GTEX-T2IS- | 2/233 | 0.269251 | 0.895882 | 0 | 0 | 2.004787 | 2.630503 |
| GTEx_Tissu | GTEX-PWN | 3/442 | 0.302104 | 0.895882 | 0 | 0 | 1.584282 | 1.896359 |
| GTEx_Tissu | GTEX-TMZS | 3/442 | 0.302104 | 0.895882 | 0 | 0 | 1.584282 | 1.896359 |
| GTEx_Tissu | GTEX-X4EC | 2/251 | 0.298257 | 0.895882 | 0 | 0 | 1.858162 | 2.248005 |
| GTEx_Tissu | GTEX-P4PC | 3/441 | 0.300914 | 0.895882 | 0 | 0 | 1.587981 | 1.907057 |
| GTEx_Tissu | GTEX-SNM | Jan-85 | 0.310189 | 0.895882 | 0 | 0 | 2.744878 | 3.21308 |
| GTEx_Tissu | GTEX-P4QT | 3/445 | 0.305678 | 0.895882 | 0 | 0 | 1.573287 | 1.864697 |
| GTEx_Tissu | GTEX-OXRL | Jan-81 | 0.298004 | 0.895882 | 0 | 0 | 2.882703 | 3.48994 |
| GTEx_Tissu | GTEX-PLZ6- | Jan-81 | 0.298004 | 0.895882 | 0 | 0 | 2.882703 | 3.48994 |
| GTEx_Tissu | GTEX-Q2A( | 9/1665 | 0.298197 | 0.895882 | 0 | 0 | 1.272088 | 1.539225 |
| GTEx_Tissu | GTEX-SNOS | 2/251 | 0.298257 | 0.895882 | 0 | 0 | 1.858162 | 2.248005 |
| GTEx_Tissu | GTEX-WOF | Jan-81 | 0.298004 | 0.895882 | 0 | 0 | 2.882703 | 3.48994 |
| GTEx_Tissu | GTEX-T2IS- | 13/2532 | 0.304845 | 0.895882 | 0 | 0 | 1.213062 | 1.441057 |
| GTEx_Tissu | GTEX-S33H | 26/5388 | 0.303982 | 0.895882 | 0 | 0 | 1.15667 | 1.377347 |
| GTEx_Tissu | GTEX-RVPU | 23/4675 | 0.286159 | 0.895882 | 0 | 0 | 1.178938 | 1.475098 |
| GTEx_Tissu | GTEX-S4P3 | 2/254 | 0.303079 | 0.895882 | 0 | 0 | 1.835761 | 2.191461 |
| GTEx_Tissu | GTEX-RM2I | 34/7196 | 0.308721 | 0.895882 | 0 | 0 | 1.142123 | 1.342355 |
| GTEx_Tissu | GTEX-T6MI | Jan-83 | 0.304123 | 0.895882 | 0 | 0 | 2.81211 | 3.34732 |
| GTEx_Tissu | GTEX-PSDG | Jan-76 | 0.282473 | 0.895882 | 0 | 0 | 3.075659 | 3.888164 |
| GTEx_Tissu | GTEX-Q734 | Dec-10 | 0.301446 | 0.895882 | 0 | 0 | 1.226458 | 1.470723 |
| GTEx_Tissu | GTEX-Q2AI | 2/253 | 0.301472 | 0.895882 | 0 | 0 | 1.843169 | 2.210102 |
| GTEx_Tissu | GTEX-RWS/ | 35/7401 | 0.301598 | 0.895882 | 0 | 0 | 1.146497 | 1.37426 |

| | | | | | | | | |
|---|---|---|---|---|---|---|---|---|
| GTEx_Tissu | GTEX-R3RS | 10/1855 | 0.285805 | 0.895882 | 0 | 0 | 1.271812 | 1.592875 |
| GTEx_Tissu | GTEX-TMM | Jan-83 | 0.304123 | 0.895882 | 0 | 0 | 2.81211 | 3.34732 |
| GTEx_Tissu | GTEX-RNOI | 7/1247 | 0.298918 | 0.895882 | 0 | 0 | 1.317651 | 1.591178 |
| GTEx_Tissu | GTEX-R55G | 3/443 | 0.303295 | 0.895882 | 0 | 0 | 1.580601 | 1.885733 |
| GTEx_Tissu | GTEX-WFG | 2/252 | 0.299865 | 0.895882 | 0 | 0 | 1.850635 | 2.228949 |
| GTEx_Tissu | GTEX-T5JW | 2/252 | 0.299865 | 0.895882 | 0 | 0 | 1.850635 | 2.228949 |
| GTEx_Tissu | GTEX-XBEC | 10/1880 | 0.300056 | 0.895882 | 0 | 0 | 1.253073 | 1.508432 |
| GTEx_Tissu | GTEX-QDVI | Jan-83 | 0.304123 | 0.895882 | 0 | 0 | 2.81211 | 3.34732 |
| GTEx_Tissu | GTEX-XQ3S | 2/255 | 0.304685 | 0.895882 | 0 | 0 | 1.828412 | 2.173024 |
| GTEx_Tissu | GTEX-NL3H | 27/5528 | 0.274024 | 0.895882 | 0 | 0 | 1.178949 | 1.526197 |
| GTEx_Tissu | GTEX-XMD | 24/4915 | 0.293283 | 0.895882 | 0 | 0 | 1.17004 | 1.43519 |
| GTEx_Tissu | GTEX-X4XX | 8/1447 | 0.293259 | 0.895882 | 0 | 0 | 1.300059 | 1.594781 |
| GTEx_Tissu | GTEX-R55F | 45/9567 | 0.26736 | 0.895882 | 0 | 0 | 1.16921 | 1.542373 |
| GTEx_Tissu | GTEX-UTH( | 7/1238 | 0.292651 | 0.895882 | 0 | 0 | 1.327924 | 1.631718 |
| GTEx_Tissu | GTEX-PX3G | 7/1206 | 0.270651 | 0.895882 | 0 | 0 | 1.365701 | 1.784869 |
| GTEx_Tissu | GTEX-P78B | Jan-74 | 0.276166 | 0.895882 | 0 | 0 | 3.160242 | 4.066453 |
| GTEx_Tissu | GTEX-RNOI | 2/238 | 0.277316 | 0.895882 | 0 | 0 | 1.961815 | 2.516219 |
| GTEx_Tissu | GTEX-NFK9 | 6/1012 | 0.277519 | 0.895882 | 0 | 0 | 1.392166 | 1.784569 |
| GTEx_Tissu | GTEX-OXRI | 6/1013 | 0.27827 | 0.895882 | 0 | 0 | 1.390709 | 1.778945 |
| GTEx_Tissu | GTEX-P4QT | 3/422 | 0.278361 | 0.895882 | 0 | 0 | 1.661609 | 2.124926 |
| GTEx_Tissu | GTEX-U4B1 | 3/416 | 0.271275 | 0.895882 | 0 | 0 | 1.686268 | 2.199943 |
| GTEx_Tissu | GTEX-NPJ8 | 3/417 | 0.272454 | 0.895882 | 0 | 0 | 1.682108 | 2.187218 |
| GTEx_Tissu | GTEX-S3XE | 2/239 | 0.278929 | 0.895882 | 0 | 0 | 1.953438 | 2.494149 |
| GTEx_Tissu | GTEX-WHP | Jan-81 | 0.298004 | 0.895882 | 0 | 0 | 2.882703 | 3.48994 |
| GTEx_Tissu | GTEX-RU1J | 2/236 | 0.27409 | 0.895882 | 0 | 0 | 1.978783 | 2.561135 |
| GTEx_Tissu | GTEX-U3ZH | 2/236 | 0.27409 | 0.895882 | 0 | 0 | 1.978783 | 2.561135 |
| GTEx_Tissu | GTEX-U3ZN | 3/419 | 0.274815 | 0.895882 | 0 | 0 | 1.67385 | 2.162037 |
| GTEx_Tissu | GTEX-RWS( | 9/1627 | 0.275413 | 0.895882 | 0 | 0 | 1.304673 | 1.682355 |
| GTEx_Tissu | GTEX-UPJH | 17/3413 | 0.309914 | 0.895882 | 0 | 0 | 1.181175 | 1.383702 |
| GTEx_Tissu | GTEX-S4Q7 | Jan-71 | 0.266602 | 0.895882 | 0 | 0 | 3.296179 | 4.35754 |
| GTEx_Tissu | GTEX-QDT8 | Jan-74 | 0.276166 | 0.895882 | 0 | 0 | 3.160242 | 4.066453 |
| GTEx_Tissu | GTEX-OIZH | 5/847 | 0.308262 | 0.895882 | 0 | 0 | 1.381076 | 1.625257 |
| GTEx_Tissu | GTEX-P78B | Dec-23 | 0.308269 | 0.895882 | 0 | 0 | 1.218659 | 1.434094 |
| GTEx_Tissu | GTEX-WFO | Jan-81 | 0.298004 | 0.895882 | 0 | 0 | 2.882703 | 3.48994 |
| GTEx_Tissu | GTEX-Q2A( | 3/445 | 0.305678 | 0.895882 | 0 | 0 | 1.573287 | 1.864697 |
| GTEx_Tissu | GTEX-X4XY | 10/1898 | 0.310438 | 0.895882 | 0 | 0 | 1.239888 | 1.450385 |
| GTEx_Tissu | GTEX-QMR | 7/1240 | 0.294041 | 0.895882 | 0 | 0 | 1.325629 | 1.622617 |
| GTEx_Tissu | GTEX-U3ZH | 2/237 | 0.275703 | 0.895882 | 0 | 0 | 1.970263 | 2.538546 |
| GTEx_Tissu | GTEX-WHS | 7/1214 | 0.276107 | 0.895882 | 0 | 0 | 1.356069 | 1.745215 |
| GTEx_Tissu | GTEX-T6M( | Jan-81 | 0.298004 | 0.895882 | 0 | 0 | 2.882703 | 3.48994 |
| GTEx_Tissu | GTEX-X4XX | 4/637 | 0.300626 | 0.895882 | 0 | 0 | 1.467862 | 1.764205 |
| GTEx_Tissu | GTEX-WI4N | 9/1669 | 0.300626 | 0.895882 | 0 | 0 | 1.268744 | 1.524891 |
| GTEx_Tissu | GTEX-T6MI | 14/2738 | 0.29983 | 0.895882 | 0 | 0 | 1.210176 | 1.457705 |
| GTEx_Tissu | GTEX-OIZI-( | 4/624 | 0.287901 | 0.895882 | 0 | 0 | 1.49965 | 1.867272 |
| GTEx_Tissu | GTEX-T8EN | 7/1253 | 0.303113 | 0.895882 | 0 | 0 | 1.310885 | 1.564739 |

| | | | | | | | | |
|---|---|---|---|---|---|---|---|---|
| GTEx_Tissu | GTEX-PX3G | 16/3125 | 0.278669 | 0.895882 | 0 | 0 | 1.218018 | 1.5563 |
| GTEx_Tissu | GTEX-T8EM | 26/5340 | 0.286282 | 0.895882 | 0 | 0 | 1.170968 | 1.464621 |
| GTEx_Tissu | GTEX-WH7 | 2/248 | 0.29343 | 0.895882 | 0 | 0 | 1.88111 | 2.306459 |
| GTEx_Tissu | GTEX-OXRI | 15/2940 | 0.292956 | 0.895882 | 0 | 0 | 1.209972 | 1.485522 |
| GTEx_Tissu | GTEX-QXCU | 5/816 | 0.281911 | 0.895882 | 0 | 0 | 1.436197 | 1.818461 |
| GTEx_Tissu | GTEX-XUZC | 4/617 | 0.281084 | 0.895882 | 0 | 0 | 1.517325 | 1.92564 |
| GTEx_Tissu | GTEX-XUYS | 16/3153 | 0.291158 | 0.895882 | 0 | 0 | 1.205135 | 1.487004 |
| GTEx_Tissu | GTEX-S33H | 4/630 | 0.293764 | 0.895882 | 0 | 0 | 1.484815 | 1.818864 |
| GTEx_Tissu | GTEX-X585 | 3/429 | 0.286652 | 0.895882 | 0 | 0 | 1.633719 | 2.041308 |
| GTEx_Tissu | GTEX-PX3G | 2/249 | 0.295039 | 0.895882 | 0 | 0 | 1.873398 | 2.286757 |
| GTEx_Tissu | GTEX-X88G | 5/824 | 0.288668 | 0.895882 | 0 | 0 | 1.421573 | 1.766271 |
| GTEx_Tissu | GTEX-WHS | 6/1028 | 0.289591 | 0.895882 | 0 | 0 | 1.369211 | 1.696845 |
| GTEx_Tissu | GTEX-WL4 | 7/1234 | 0.289877 | 0.895882 | 0 | 0 | 1.332539 | 1.650082 |
| GTEx_Tissu | GTEX-QXCU | Dec-89 | 0.290524 | 0.895882 | 0 | 0 | 1.239245 | 1.531793 |
| GTEx_Tissu | GTEX-VUSG | 2/250 | 0.296648 | 0.895882 | 0 | 0 | 1.86575 | 2.267274 |
| GTEx_Tissu | GTEX-P4PP | 48/10330 | 0.291093 | 0.895882 | 0 | 0 | 1.152844 | 1.42274 |
| GTEx_Tissu | GTEX-XQ8I | 3/451 | 0.312831 | 0.89688 | 0 | 0 | 1.551738 | 1.803263 |
| GTEx_Tissu | GTEX-QDVI | Jan-86 | 0.313203 | 0.89688 | 0 | 0 | 2.712449 | 3.148895 |
| GTEx_Tissu | GTEX-S7SF | Jan-86 | 0.313203 | 0.89688 | 0 | 0 | 2.712449 | 3.148895 |
| GTEx_Tissu | GTEX-VJYA | 3/450 | 0.311638 | 0.89688 | 0 | 0 | 1.555289 | 1.813331 |
| GTEx_Tissu | GTEX-XLM | 5/851 | 0.311692 | 0.89688 | 0 | 0 | 1.374258 | 1.602028 |
| GTEx_Tissu | GTEX-VJYA | 2/260 | 0.312707 | 0.89688 | 0 | 0 | 1.792522 | 2.083787 |
| GTEx_Tissu | GTEX-QVUS | 3/452 | 0.314024 | 0.896923 | 0 | 0 | 1.548202 | 1.793262 |
| GTEx_Tissu | GTEX-WFG | 3/452 | 0.314024 | 0.896923 | 0 | 0 | 1.548202 | 1.793262 |
| GTEx_Tissu | GTEX-T5JC | 9/1692 | 0.314686 | 0.897662 | 0 | 0 | 1.249829 | 1.445027 |
| GTEx_Tissu | GTEX-XMD | 5/855 | 0.315127 | 0.89777 | 0 | 0 | 1.367504 | 1.579164 |
| Gene_Pert | SIRT6 OE h | 1/518 | 0.898528 | 0.898528 | 0 | 0 | 0.436238 | 0.046677 |
| GTEx_Tissu | GTEX-WFG | 2/262 | 0.31591 | 0.898534 | 0 | 0 | 1.778552 | 2.04942 |
| GTEx_Tissu | GTEX-TKQ1 | Jan-87 | 0.316203 | 0.898534 | 0 | 0 | 2.680773 | 3.086563 |
| GeneSigDB | 20081105- | 3/1105 | 0.866093 | 0.900026 | 0 | 0 | 0.609638 | 0.087643 |
| GTEx_Tissu | GTEX-Q2A | 6/1064 | 0.317153 | 0.900084 | 0 | 0 | 1.320101 | 1.515964 |
| Disease_Pe | autistic dis | 1/517 | 0.898072 | 0.900861 | 0 | 0 | 0.437106 | 0.046991 |
| GTEx_Tissu | GTEX-OOBI | Nov-26 | 0.318152 | 0.901207 | 0 | 0 | 1.217979 | 1.394862 |
| GTEx_Tissu | GTEX-S341 | 4/655 | 0.318359 | 0.901207 | 0 | 0 | 1.425943 | 1.632101 |
| GTEx_Tissu | GTEX-O5YV | 8/1106 | 0.107887 | 0.902076 | 0 | 0 | 1.735261 | 3.863848 |
| GTEx_Tissu | GTEX-SIU8 | Jan-88 | 0.319191 | 0.902413 | 0 | 0 | 2.649826 | 3.026014 |
| GTEx_Tissu | GTEX-QESD | 11/1670 | 0.108382 | 0.9036 | 0 | 0 | 1.592541 | 3.538776 |
| GTEx_Tissu | GTEX-R55G | 6/751 | 0.108765 | 0.904185 | 0 | 0 | 1.905841 | 4.228238 |
| GeneSigDB | 15297395- | 1/472 | 0.875362 | 0.904595 | 0 | 0 | 0.479978 | 0.063894 |
| GeneSigDB | 12917485- | 1/469 | 0.873681 | 0.904595 | 0 | 0 | 0.48313 | 0.065242 |
| GeneSigDB | 19204204- | 1/472 | 0.875362 | 0.904595 | 0 | 0 | 0.479978 | 0.063894 |
| GTEx_Tissu | GTEX-XOT4 | 6/752 | 0.109275 | 0.905479 | 0 | 0 | 1.903187 | 4.213446 |
| GTEx_Tissu | GTEX-VJWI | 8/1110 | 0.109546 | 0.905479 | 0 | 0 | 1.728595 | 3.822627 |
| GTEx_Tissu | GTEX-S7SE | 7/1278 | 0.320727 | 0.905606 | 0 | 0 | 1.283379 | 1.459414 |
| Gene_Pert | Prmt1 KD r | 1/534 | 0.905542 | 0.906042 | 0 | 0 | 0.422793 | 0.04195 |

| | | | | | | | | |
|---|---|---|---|---|---|---|---|---|
| Disease_Pe | West Nile f | 1/532 | 0.904693 | 0.906095 | 0 | 0 | 0.42443 | 0.042511 |
| GeneSigDB | 18381933-: | 2/825 | 0.879086 | 0.906761 | 0 | 0 | 0.545779 | 0.070336 |
| GTEx_Tissu | GTEX-S7SF- | 28/5885 | 0.322561 | 0.907072 | 0 | 0 | 1.138918 | 1.288643 |
| GTEx_Tissu | GTEX-PSDG | 5/864 | 0.322877 | 0.907072 | 0 | 0 | 1.352537 | 1.529022 |
| GTEx_Tissu | GTEX-WOF | Jan-89 | 0.322165 | 0.907072 | 0 | 0 | 2.619582 | 2.967177 |
| GTEx_Tissu | GTEX-SIU8- | 2/266 | 0.322308 | 0.907072 | 0 | 0 | 1.751248 | 1.982846 |
| GTEx_Tissu | GTEX-X638 | 22/3894 | 0.110096 | 0.90743 | 0 | 0 | 1.402185 | 3.093784 |
| GeneSigDB | 20460173- | 1/484 | 0.881866 | 0.907947 | 0 | 0 | 0.467764 | 0.058805 |
| PhenGenI_ | Uric Acid | 1/403 | 0.830483 | 0.909202 | 0 | 0 | 0.564358 | 0.104828 |
| GTEx_Tissu | GTEX-S7SF- | 8/1495 | 0.324571 | 0.909947 | 0 | 0 | 1.254825 | 1.411994 |
| GTEx_Tissu | GTEX-WI4N | Jan-90 | 0.325127 | 0.909947 | 0 | 0 | 2.590018 | 2.909988 |
| GTEx_Tissu | GTEX-X4EP | Jan-90 | 0.325127 | 0.909947 | 0 | 0 | 2.590018 | 2.909988 |
| GTEx_Tissu | GTEX-QMR | 7/1285 | 0.325694 | 0.91039 | 0 | 0 | 1.275871 | 1.431266 |
| RNA-Seq_D | Id3 Spleen | 2/899 | 0.907287 | 0.912054 | 0 | 0 | 0.498813 | 0.048533 |
| RNA-Seq_D | Erlotinib 11 | 2/902 | 0.908291 | 0.912264 | 0 | 0 | 0.497072 | 0.047814 |
| GTEx_Tissu | GTEX-XPVG | Dec-58 | 0.326851 | 0.912476 | 0 | 0 | 1.19809 | 1.339765 |
| GeneSigDB | 18535662-1 | 1/500 | 0.890019 | 0.912966 | 0 | 0 | 0.452393 | 0.052709 |
| GeneSigDB | 20368555-1 | 2/851 | 0.889784 | 0.912966 | 0 | 0 | 0.528345 | 0.061698 |
| GTEx_Tissu | GTEX-POYV | 17/3453 | 0.327938 | 0.913032 | 0 | 0 | 1.164598 | 1.298444 |
| GTEx_Tissu | GTEX-XUJ4 | 4/665 | 0.328253 | 0.913032 | 0 | 0 | 1.403642 | 1.563615 |
| GTEx_Tissu | GTEX-XOT( | 7/1289 | 0.328539 | 0.913032 | 0 | 0 | 1.271617 | 1.415435 |
| GTEx_Tissu | GTEX-WXY( | 2/270 | 0.328691 | 0.913032 | 0 | 0 | 1.724759 | 1.919028 |
| GTEx_Tissu | GTEX-QEG4 | 13/2580 | 0.329328 | 0.91366 | 0 | 0 | 1.187094 | 1.318506 |
| GTEx_Tissu | GTEX-PVO\ | 2/271 | 0.330285 | 0.914391 | 0 | 0 | 1.718259 | 1.903487 |
| GTEx_Tissu | GTEX-OIZH | Jan-92 | 0.331012 | 0.914391 | 0 | 0 | 2.532839 | 2.800307 |
| GTEx_Tissu | GTEX-U4B1 | 9/1722 | 0.333252 | 0.914391 | 0 | 0 | 1.225919 | 1.347108 |
| GTEx_Tissu | GTEX-QXCU | 17/3463 | 0.332492 | 0.914391 | 0 | 0 | 1.160513 | 1.277886 |
| GTEx_Tissu | GTEX-U4B1 | 2/273 | 0.333469 | 0.914391 | 0 | 0 | 1.705405 | 1.872885 |
| GTEx_Tissu | GTEX-T5JW | 5/877 | 0.334112 | 0.914391 | 0 | 0 | 1.331464 | 1.459655 |
| GTEx_Tissu | GTEX-QDT8 | 8/1509 | 0.333832 | 0.914391 | 0 | 0 | 1.242176 | 1.362811 |
| GTEx_Tissu | GTEX-U3ZN | Nov-53 | 0.333227 | 0.914391 | 0 | 0 | 1.200802 | 1.319599 |
| GTEx_Tissu | GTEX-T2IS- | 30/6360 | 0.332059 | 0.914391 | 0 | 0 | 1.129376 | 1.24507 |
| GTEx_Tissu | GTEX-P4QT | Dec-71 | 0.333825 | 0.914391 | 0 | 0 | 1.190606 | 1.306261 |
| GTEx_Tissu | GTEX-Q734 | 2/273 | 0.333469 | 0.914391 | 0 | 0 | 1.705405 | 1.872885 |
| GTEx_Tissu | GTEX-XOT( | 43/9317 | 0.334973 | 0.914496 | 0 | 0 | 1.121113 | 1.226169 |
| GTEx_Tissu | GTEX-OHPI | 29/6142 | 0.33464 | 0.914496 | 0 | 0 | 1.128742 | 1.235634 |
| GTEx_Tissu | GTEX-PLZ4- | 3/477 | 0.343864 | 0.915484 | 0 | 0 | 1.464662 | 1.563542 |
| GTEx_Tissu | GTEX-U8T8 | Oct-55 | 0.343881 | 0.915484 | 0 | 0 | 1.199746 | 1.28068 |
| GTEx_Tissu | GTEX-U3ZN | 5/888 | 0.343651 | 0.915484 | 0 | 0 | 1.314118 | 1.403647 |
| GTEx_Tissu | GTEX-T5JC- | 6/1098 | 0.343577 | 0.915484 | 0 | 0 | 1.276692 | 1.363947 |
| GTEx_Tissu | GTEX-UPK5 | Jan-95 | 0.339745 | 0.915484 | 0 | 0 | 2.451633 | 2.646685 |
| GTEx_Tissu | GTEX-OXRN | 2/276 | 0.338237 | 0.915484 | 0 | 0 | 1.686475 | 1.828152 |
| GTEx_Tissu | GTEX-T6M( | Oct-53 | 0.342695 | 0.915484 | 0 | 0 | 1.201115 | 1.286291 |
| GTEx_Tissu | GTEX-XQ8I | 55/12110 | 0.34693 | 0.915484 | 0 | 0 | 1.12036 | 1.186049 |
| GTEx_Tissu | GTEX-P4PP | 4/683 | 0.346113 | 0.915484 | 0 | 0 | 1.365154 | 1.448414 |

| | | | | | | | | |
|---|---|---|---|---|---|---|---|---|
| GTEx_Tissu | GTEX-PWO | Jan-97 | 0.345504 | 0.915484 | 0 | 0 | 2.400315 | 2.550937 |
| GTEx_Tissu | GTEX-TKQ2 | 31/6604 | 0.33892 | 0.915484 | 0 | 0 | 1.123481 | 1.215597 |
| GTEx_Tissu | GTEX-QV31 | 3/481 | 0.348635 | 0.915484 | 0 | 0 | 1.452107 | 1.530129 |
| GTEx_Tissu | GTEX-QV31 | 9/1728 | 0.336993 | 0.915484 | 0 | 0 | 1.221238 | 1.328334 |
| GTEx_Tissu | GTEX-RTLS | 2/275 | 0.336649 | 0.915484 | 0 | 0 | 1.692739 | 1.842909 |
| GTEx_Tissu | GTEX-OHPI | Dec-76 | 0.336516 | 0.915484 | 0 | 0 | 1.18775 | 1.293588 |
| GTEx_Tissu | GTEX-WL4( | 8/1521 | 0.341809 | 0.915484 | 0 | 0 | 1.231521 | 1.32204 |
| GTEx_Tissu | GTEX-U8XE | 4/677 | 0.340155 | 0.915484 | 0 | 0 | 1.377755 | 1.485709 |
| GTEx_Tissu | GTEX-WFJC | 7/1307 | 0.341392 | 0.915484 | 0 | 0 | 1.252798 | 1.346412 |
| GTEx_Tissu | GTEX-OHPI | 20/4147 | 0.341417 | 0.915484 | 0 | 0 | 1.141807 | 1.227045 |
| GTEx_Tissu | GTEX-NFK9 | 2/283 | 0.349324 | 0.915484 | 0 | 0 | 1.643877 | 1.728956 |
| GTEx_Tissu | GTEX-WVLI | 8/1532 | 0.349149 | 0.915484 | 0 | 0 | 1.221901 | 1.285752 |
| GTEx_Tissu | GTEX-U3ZN | 2/279 | 0.342996 | 0.915484 | 0 | 0 | 1.667955 | 1.784774 |
| GTEx_Tissu | GTEX-N7M | 5/886 | 0.341915 | 0.915484 | 0 | 0 | 1.317239 | 1.413653 |
| GTEx_Tissu | GTEX-PLZ6 | Jan-96 | 0.342631 | 0.915484 | 0 | 0 | 2.425704 | 2.598176 |
| GTEx_Tissu | GTEX-QDT8 | 7/1305 | 0.33996 | 0.915484 | 0 | 0 | 1.254863 | 1.353906 |
| GTEx_Tissu | GTEX-WFO | Jan-98 | 0.348365 | 0.915484 | 0 | 0 | 2.37545 | 2.504922 |
| GTEx_Tissu | GTEX-SSA3 | 9/1747 | 0.348889 | 0.915484 | 0 | 0 | 1.206626 | 1.27058 |
| GTEx_Tissu | GTEX-WL4( | 9/1741 | 0.345124 | 0.915484 | 0 | 0 | 1.211205 | 1.288543 |
| GTEx_Tissu | GTEX-S3XE | Jan-97 | 0.345504 | 0.915484 | 0 | 0 | 2.400315 | 2.550937 |
| GTEx_Tissu | GTEX-S4UY | Jan-97 | 0.345504 | 0.915484 | 0 | 0 | 2.400315 | 2.550937 |
| GTEx_Tissu | GTEX-P78B | Jan-98 | 0.348365 | 0.915484 | 0 | 0 | 2.37545 | 2.504922 |
| GTEx_Tissu | GTEX-XUW | 8/1513 | 0.336488 | 0.915484 | 0 | 0 | 1.238605 | 1.349081 |
| GTEx_Tissu | GTEX-W5X | Nov-78 | 0.347327 | 0.915484 | 0 | 0 | 1.185279 | 1.253418 |
| GTEx_Tissu | GTEX-VUS( | 26/5501 | 0.347165 | 0.915484 | 0 | 0 | 1.124 | 1.189141 |
| GTEx_Tissu | GTEX-T5JW | 8/1533 | 0.349818 | 0.9157 | 0 | 0 | 1.221033 | 1.282503 |
| GTEx_Tissu | GTEX-POM | 5/585 | 0.111452 | 0.915999 | 0 | 0 | 2.032485 | 4.459594 |
| GeneSigDB | 18614019- | 2/864 | 0.894802 | 0.916185 | 0 | 0 | 0.520022 | 0.057802 |
| GTEx_Tissu | GTEX-QVJC | 47/10278 | 0.350566 | 0.916579 | 0 | 0 | 1.111949 | 1.165553 |
| GTEx_Tissu | GTEX-TKQ2 | 7/934 | 0.11185 | 0.91666 | 0 | 0 | 1.792098 | 3.92577 |
| GTEx_Tissu | GTEX-NPJ8 | 2/285 | 0.352481 | 0.917994 | 0 | 0 | 1.632093 | 1.70188 |
| GTEx_Tissu | GTEX-WI4N | 4/689 | 0.352075 | 0.917994 | 0 | 0 | 1.352775 | 1.412175 |
| GTEx_Tissu | GTEX-X261 | 2/285 | 0.352481 | 0.917994 | 0 | 0 | 1.632093 | 1.70188 |
| GTEx_Tissu | GTEX-XLM4 | 27/5740 | 0.352757 | 0.917994 | 0 | 0 | 1.118502 | 1.165451 |
| GeneSigDB | 12917485- | 1/521 | 0.899881 | 0.91801 | 0 | 0 | 0.433654 | 0.045747 |
| GeneSigDB | 18593951- | 1/520 | 0.899432 | 0.91801 | 0 | 0 | 0.434512 | 0.046055 |
| GTEx_Tissu | GTEX-SNO9 | 1/100 | 0.35405 | 0.918171 | 0 | 0 | 2.327226 | 2.416399 |
| GTEx_Tissu | GTEX-QDT8 | 6/1111 | 0.353752 | 0.918171 | 0 | 0 | 1.260801 | 1.310174 |
| GTEx_Tissu | GTEX-QXCU | 4/691 | 0.354063 | 0.918171 | 0 | 0 | 1.348696 | 1.400323 |
| GTEx_Tissu | GTEX-U8XE | 8/1541 | 0.355173 | 0.919976 | 0 | 0 | 1.214133 | 1.256811 |
| GTEx_Tissu | GTEX-X5EB | 2/287 | 0.355633 | 0.920096 | 0 | 0 | 1.620475 | 1.675339 |
| Disease_Pe | acute myo | 1/572 | 0.920339 | 0.920339 | 0 | 0 | 0.393883 | 0.032697 |
| GeneSigDB | 18387200- | 3/1213 | 0.904419 | 0.920954 | 0 | 0 | 0.552037 | 0.055459 |
| GTEx_Tissu | GTEX-PWO | 4/713 | 0.37593 | 0.921329 | 0 | 0 | 1.305351 | 1.277092 |
| GTEx_Tissu | GTEX-X261 | 7/1349 | 0.371638 | 0.921329 | 0 | 0 | 1.210851 | 1.198544 |

| | | | | | | | | |
|---|---|---|---|---|---|---|---|---|
| GTEx_Tissu | GTEX-Q734 | 29/6240 | 0.371752 | 0.921329 | 0 | 0 | 1.103043 | 1.091493 |
| GTEx_Tissu | GTEX-WWY | 4/705 | 0.367981 | 0.921329 | 0 | 0 | 1.320798 | 1.320435 |
| GTEx_Tissu | GTEX-NL3H | 15/3109 | 0.37444 | 0.921329 | 0 | 0 | 1.132501 | 1.112482 |
| GTEx_Tissu | GTEX-OHPI | 17/3553 | 0.374196 | 0.921329 | 0 | 0 | 1.124794 | 1.105646 |
| GTEx_Tissu | GTEX-PVOV | Oct-96 | 0.368355 | 0.921329 | 0 | 0 | 1.172297 | 1.170782 |
| GTEx_Tissu | GTEX-WI4N | 8/1571 | 0.375345 | 0.921329 | 0 | 0 | 1.188885 | 1.165 |
| GTEx_Tissu | GTEX-N7M | 5/919 | 0.370634 | 0.921329 | 0 | 0 | 1.267479 | 1.258022 |
| GTEx_Tissu | GTEX-TKQ2 | 1/106 | 0.370812 | 0.921329 | 0 | 0 | 2.193577 | 2.176161 |
| GTEx_Tissu | GTEX-WXY | 2/297 | 0.37131 | 0.921329 | 0 | 0 | 1.564746 | 1.550222 |
| GTEx_Tissu | GTEX-NPJ7 | 7/1345 | 0.368745 | 0.921329 | 0 | 0 | 1.214733 | 1.211877 |
| GTEx_Tissu | GTEX-XQ8I | 2/292 | 0.363489 | 0.921329 | 0 | 0 | 1.59213 | 1.611246 |
| GTEx_Tissu | GTEX-R55G | 2/300 | 0.375985 | 0.921329 | 0 | 0 | 1.548756 | 1.515003 |
| GTEx_Tissu | GTEX-WFO | 2/292 | 0.363489 | 0.921329 | 0 | 0 | 1.59213 | 1.611246 |
| GTEx_Tissu | GTEX-WWY | 5/904 | 0.357564 | 0.921329 | 0 | 0 | 1.289644 | 1.326323 |
| GTEx_Tissu | GTEX-P44H | 4/709 | 0.371956 | 0.921329 | 0 | 0 | 1.313031 | 1.298561 |
| GTEx_Tissu | GTEX-QVU | Oct-84 | 0.361163 | 0.921329 | 0 | 0 | 1.180213 | 1.201959 |
| GTEx_Tissu | GTEX-XLM4 | 4/700 | 0.36301 | 0.921329 | 0 | 0 | 1.330633 | 1.348362 |
| GTEx_Tissu | GTEX-T2IS- | 1/107 | 0.373563 | 0.921329 | 0 | 0 | 2.172773 | 2.139459 |
| GTEx_Tissu | GTEX-R55G | 2/289 | 0.358779 | 0.921329 | 0 | 0 | 1.609018 | 1.649321 |
| GTEx_Tissu | GTEX-XAJ8 | 2/294 | 0.366622 | 0.921329 | 0 | 0 | 1.581064 | 1.586479 |
| GTEx_Tissu | GTEX-OHPI | 9/1763 | 0.358961 | 0.921329 | 0 | 0 | 1.194566 | 1.223882 |
| GTEx_Tissu | GTEX-SN8G | Oct-81 | 0.359369 | 0.921329 | 0 | 0 | 1.182207 | 1.209878 |
| GTEx_Tissu | GTEX-WWY | 4/701 | 0.364004 | 0.921329 | 0 | 0 | 1.328655 | 1.342724 |
| GTEx_Tissu | GTEX-SUCS | Dec-48 | 0.375768 | 0.921329 | 0 | 0 | 1.147915 | 1.123559 |
| GTEx_Tissu | GTEX-XYKS | 34/7358 | 0.366557 | 0.921329 | 0 | 0 | 1.102671 | 1.10664 |
| GTEx_Tissu | GTEX-OXRI | 2/299 | 0.374428 | 0.921329 | 0 | 0 | 1.55405 | 1.526629 |
| GTEx_Tissu | GTEX-XGQ4 | 1/105 | 0.368048 | 0.921329 | 0 | 0 | 2.214781 | 2.213764 |
| GTEx_Tissu | GTEX-S4UY | 1/107 | 0.373563 | 0.921329 | 0 | 0 | 2.172773 | 2.139459 |
| GTEx_Tissu | GTEX-QMR | Nov-23 | 0.372988 | 0.921329 | 0 | 0 | 1.158222 | 1.14225 |
| GTEx_Tissu | GTEX-WHS | 42/9197 | 0.372899 | 0.921329 | 0 | 0 | 1.096756 | 1.081892 |
| GTEx_Tissu | GTEX-R55E | 6/1118 | 0.359243 | 0.921329 | 0 | 0 | 1.252398 | 1.282152 |
| GTEx_Tissu | GTEX-SNOS | 1/107 | 0.373563 | 0.921329 | 0 | 0 | 2.172773 | 2.139459 |
| GTEx_Tissu | GTEX-S32W | 3/502 | 0.373623 | 0.921329 | 0 | 0 | 1.389493 | 1.367968 |
| GTEx_Tissu | GTEX-T5JC- | 8/1566 | 0.371974 | 0.921329 | 0 | 0 | 1.193026 | 1.17982 |
| GTEx_Tissu | GTEX-TML8 | 13/2649 | 0.365337 | 0.921329 | 0 | 0 | 1.151422 | 1.159408 |
| GTEx_Tissu | GTEX-PVOV | 9/1773 | 0.365278 | 0.921329 | 0 | 0 | 1.18714 | 1.195564 |
| GTEx_Tissu | GTEX-XOT4 | 3/494 | 0.364119 | 0.921329 | 0 | 0 | 1.412715 | 1.427231 |
| GTEx_Tissu | GTEX-TMM | 9/1772 | 0.364646 | 0.921329 | 0 | 0 | 1.187879 | 1.198366 |
| GTEx_Tissu | GTEX-WFG | 1/102 | 0.359686 | 0.921329 | 0 | 0 | 2.280912 | 2.332289 |
| GTEx_Tissu | GTEX-XBEC | 34/7340 | 0.359968 | 0.921329 | 0 | 0 | 1.106968 | 1.131034 |
| GTEx_Tissu | GTEX-NL3H | 8/1557 | 0.365915 | 0.921329 | 0 | 0 | 1.200546 | 1.206974 |
| GTEx_Tissu | GTEX-XUW | 3/489 | 0.358168 | 0.921329 | 0 | 0 | 1.427616 | 1.465808 |
| GTEx_Tissu | GTEX-VUSG | 24/5095 | 0.36422 | 0.921329 | 0 | 0 | 1.114986 | 1.126134 |
| GTEx_Tissu | GTEX-R53T | 1/101 | 0.356874 | 0.921329 | 0 | 0 | 2.303837 | 2.373811 |
| GTEx_Tissu | GTEX-NPJ8 | 7/1338 | 0.363689 | 0.921329 | 0 | 0 | 1.221582 | 1.235577 |

| | | | | | | | | |
|---|---|---|---|---|---|---|---|---|
| GTEx_Tissu | GTEX-R55D | Nov-03 | 0.361544 | 0.921329 | 0 | 0 | 1.17011 | 1.190437 |
| DisGeNET | Autism Spe | 1/572 | 0.920339 | 0.921394 | 0 | 0 | 0.393883 | 0.032697 |
| GTEx_Tissu | GTEX-WH7 | 20/4228 | 0.376617 | 0.921863 | 0 | 0 | 1.114083 | 1.08793 |
| GTEx_Tissu | GTEX-S341 | 10/1492 | 0.112905 | 0.922697 | 0 | 0 | 1.615139 | 3.522952 |
| GTEx_Tissu | GTEX-Q2A( | 5/927 | 0.377611 | 0.923279 | 0 | 0 | 1.255952 | 1.223161 |
| RNA-Seq_D | LPS BMDM | 1/573 | 0.920696 | 0.923915 | 0 | 0 | 0.393174 | 0.032486 |
| GeneSigDB | 18450603-' | 1/543 | 0.909275 | 0.924209 | 0 | 0 | 0.41558 | 0.039525 |
| GTEx_Tissu | GTEX-V1D1 | 1/109 | 0.379031 | 0.924719 | 0 | 0 | 2.132321 | 2.068645 |
| GTEx_Tissu | GTEX-XUZ( | 1/109 | 0.379031 | 0.924719 | 0 | 0 | 2.132321 | 2.068645 |
| Reactome_ | Gene Expre | 4/1449 | 0.883706 | 0.925001 | 0 | 0 | 0.615934 | 0.076148 |
| Reactome_ | Signaling B | 1/496 | 0.888034 | 0.925208 | 0 | 0 | 0.456143 | 0.054165 |
| DisGeNET | Influenza | 1/589 | 0.926193 | 0.926901 | 0 | 0 | 0.382159 | 0.029301 |
| Jensen_TIS | Cingulate c | 16/4839 | 0.921697 | 0.927457 | 0 | 0 | 0.705072 | 0.057491 |
| GTEx_Tissu | GTEX-PWO | Oct-17 | 0.380988 | 0.92746 | 0 | 0 | 1.158672 | 1.118103 |
| GTEx_Tissu | GTEX-WZT( | 4/718 | 0.380896 | 0.92746 | 0 | 0 | 1.295873 | 1.250813 |
| Gene_Pert | Rock1 INHI | 1/593 | 0.927508 | 0.927508 | 0 | 0 | 0.379498 | 0.028559 |
| GTEx_Tissu | GTEX-TMM | 22/4694 | 0.38421 | 0.928614 | 0 | 0 | 1.104129 | 1.056174 |
| GTEx_Tissu | GTEX-OOBI | 34/7428 | 0.392499 | 0.928614 | 0 | 0 | 1.086159 | 1.015799 |
| GTEx_Tissu | GTEX-VJYA | 1/114 | 0.392494 | 0.928614 | 0 | 0 | 2.037456 | 1.905499 |
| GTEx_Tissu | GTEX-OXRI | 8/1607 | 0.399683 | 0.928614 | 0 | 0 | 1.159839 | 1.063669 |
| GTEx_Tissu | GTEX-O5Y\ | 9/1815 | 0.391952 | 0.928614 | 0 | 0 | 1.156849 | 1.083524 |
| GTEx_Tissu | GTEX-SNM | 2/305 | 0.383746 | 0.928614 | 0 | 0 | 1.522811 | 1.45851 |
| GTEx_Tissu | GTEX-S4Q7 | 1/118 | 0.403056 | 0.928614 | 0 | 0 | 1.967402 | 1.787737 |
| GTEx_Tissu | GTEX-NPJ8 | 9/1826 | 0.398965 | 0.928614 | 0 | 0 | 1.149147 | 1.05593 |
| GTEx_Tissu | GTEX-T6MI | 7/1387 | 0.399183 | 0.928614 | 0 | 0 | 1.1751 | 1.079135 |
| GTEx_Tissu | GTEX-S3XE | 17/3608 | 0.400178 | 0.928614 | 0 | 0 | 1.103847 | 1.010955 |
| GTEx_Tissu | GTEX-WFG | 2/316 | 0.400676 | 0.928614 | 0 | 0 | 1.46864 | 1.343223 |
| GTEx_Tissu | GTEX-NFK9 | 4/733 | 0.395771 | 0.928614 | 0 | 0 | 1.268217 | 1.175534 |
| GTEx_Tissu | GTEX-OXR( | 3/524 | 0.399617 | 0.928614 | 0 | 0 | 1.329312 | 1.219308 |
| GTEx_Tissu | GTEX-WFO | 4/743 | 0.405663 | 0.928614 | 0 | 0 | 1.250404 | 1.128156 |
| GTEx_Tissu | GTEX-P4PC | Dec-80 | 0.393441 | 0.928614 | 0 | 0 | 1.130956 | 1.054982 |
| GTEx_Tissu | GTEX-XV7C | 2/316 | 0.400676 | 0.928614 | 0 | 0 | 1.46864 | 1.343223 |
| GTEx_Tissu | GTEX-T6MI | 8/1609 | 0.401038 | 0.928614 | 0 | 0 | 1.158263 | 1.058305 |
| GTEx_Tissu | GTEX-O5YT | 15/3166 | 0.402888 | 0.928614 | 0 | 0 | 1.108246 | 1.007502 |
| GTEx_Tissu | GTEX-RVPU | 3/515 | 0.389012 | 0.928614 | 0 | 0 | 1.353306 | 1.277717 |
| GTEx_Tissu | GTEX-XV7C | 1/116 | 0.397798 | 0.928614 | 0 | 0 | 2.00182 | 1.845299 |
| GTEx_Tissu | GTEX-X585 | 1/116 | 0.397798 | 0.928614 | 0 | 0 | 2.00182 | 1.845299 |
| GTEx_Tissu | GTEX-PSD( | 9/1825 | 0.398327 | 0.928614 | 0 | 0 | 1.149843 | 1.05841 |
| GTEx_Tissu | GTEX-RUS( | 2/310 | 0.391467 | 0.928614 | 0 | 0 | 1.497708 | 1.404633 |
| GTEx_Tissu | GTEX-S95S | 2/310 | 0.391467 | 0.928614 | 0 | 0 | 1.497708 | 1.404633 |
| GTEx_Tissu | GTEX-OIZI- | 1/114 | 0.392494 | 0.928614 | 0 | 0 | 2.037456 | 1.905499 |
| GTEx_Tissu | GTEX-T6M( | 2/312 | 0.394543 | 0.928614 | 0 | 0 | 1.487894 | 1.383781 |
| GTEx_Tissu | GTEX-S4UY | 1/118 | 0.403056 | 0.928614 | 0 | 0 | 1.967402 | 1.787737 |
| GTEx_Tissu | GTEX-R55E | 3/523 | 0.398441 | 0.928614 | 0 | 0 | 1.331937 | 1.225642 |
| GTEx_Tissu | GTEX-T2IS- | 8/1585 | 0.384797 | 0.928614 | 0 | 0 | 1.177432 | 1.124493 |

| | | | | | | | | |
|---|---|---|---|---|---|---|---|---|
| GTEx_Tissu | GTEX-S7SF· | 4/736 | 0.398741 | 0.928614 | 0 | 0 | 1.262822 | 1.161092 |
| GTEx_Tissu | GTEX-T6MI | 2/318 | 0.403731 | 0.928614 | 0 | 0 | 1.459196 | 1.323499 |
| GTEx_Tissu | GTEX-QESD | 7/1395 | 0.40499 | 0.928614 | 0 | 0 | 1.167822 | 1.055585 |
| GTEx_Tissu | GTEX-WFJC | 3/528 | 0.404316 | 0.928614 | 0 | 0 | 1.318912 | 1.194352 |
| GTEx_Tissu | GTEX-S32W | 1/113 | 0.389825 | 0.928614 | 0 | 0 | 2.055752 | 1.936638 |
| GTEx_Tissu | GTEX-XQ3S | 2/309 | 0.389926 | 0.928614 | 0 | 0 | 1.502663 | 1.415207 |
| GTEx_Tissu | GTEX-SJXC· | 36/7898 | 0.398071 | 0.928614 | 0 | 0 | 1.081988 | 0.996647 |
| GTEx_Tissu | GTEX-S7SE· | 7/1370 | 0.386848 | 0.928614 | 0 | 0 | 1.190847 | 1.130974 |
| GTEx_Tissu | GTEX-V955 | 14/2915 | 0.388625 | 0.928614 | 0 | 0 | 1.124638 | 1.062941 |
| GTEx_Tissu | GTEX-XBEC | 4/726 | 0.388834 | 0.928614 | 0 | 0 | 1.28098 | 1.210016 |
| GTEx_Tissu | GTEX-T6MI | 6/1163 | 0.394661 | 0.928614 | 0 | 0 | 1.200807 | 1.116422 |
| GTEx_Tissu | GTEX-S7SE· | 2/306 | 0.385293 | 0.928614 | 0 | 0 | 1.517724 | 1.44753 |
| GTEx_Tissu | GTEX-R53T | 2/306 | 0.385293 | 0.928614 | 0 | 0 | 1.517724 | 1.44753 |
| GTEx_Tissu | GTEX-OXRI | 9/1833 | 0.403432 | 0.928614 | 0 | 0 | 1.144294 | 1.038731 |
| GTEx_Tissu | GTEX-PVO\ | 2/318 | 0.403731 | 0.928614 | 0 | 0 | 1.459196 | 1.323499 |
| GTEx_Tissu | GTEX-NPJ7 | 2/311 | 0.393006 | 0.928614 | 0 | 0 | 1.492785 | 1.394158 |
| GTEx_Tissu | GTEX-TSE9 | 3/511 | 0.384285 | 0.928614 | 0 | 0 | 1.364244 | 1.304722 |
| GTEx_Tissu | GTEX-T2IS- | 9/1834 | 0.40407 | 0.928614 | 0 | 0 | 1.143604 | 1.036296 |
| GTEx_Tissu | GTEX-QV44 | 16/3361 | 0.388924 | 0.928614 | 0 | 0 | 1.116183 | 1.054091 |
| GTEx_Tissu | GTEX-OIZG | Dec-79 | 0.392888 | 0.928614 | 0 | 0 | 1.13148 | 1.057064 |
| GTEx_Tissu | GTEX-QDV. | 1/119 | 0.405668 | 0.928614 | 0 | 0 | 1.950631 | 1.759896 |
| GTEx_Tissu | GTEX-OXRI | 1/119 | 0.405668 | 0.928614 | 0 | 0 | 1.950631 | 1.759896 |
| GTEx_Tissu | GTEX-Q2AI | 2/316 | 0.400676 | 0.928614 | 0 | 0 | 1.46864 | 1.343223 |
| GTEx_Tissu | GTEX-T6M( | 2/308 | 0.388383 | 0.928614 | 0 | 0 | 1.507651 | 1.42588 |
| GTEx_Tissu | GTEX-WYJI | 22/4693 | 0.383784 | 0.928614 | 0 | 0 | 1.104438 | 1.057693 |
| GTEx_Tissu | GTEX-XUJ4 | 2/316 | 0.400676 | 0.928614 | 0 | 0 | 1.46864 | 1.343223 |
| GTEx_Tissu | GTEX-XUZC | 7/1375 | 0.390475 | 0.928614 | 0 | 0 | 1.186175 | 1.115469 |
| GTEx_Tissu | GTEX-N7M | 6/1171 | 0.400966 | 0.928614 | 0 | 0 | 1.192052 | 1.08939 |
| GTEx_Tissu | GTEX-WFO | Dec-75 | 0.390673 | 0.928614 | 0 | 0 | 1.133577 | 1.06543 |
| PhenGenI_ | Obesity | 1/434 | 0.852339 | 0.928728 | 0 | 0 | 0.523122 | 0.083579 |
| GTEx_Tissu | GTEX-XBED | 2/320 | 0.40678 | 0.929245 | 0 | 0 | 1.449871 | 1.304135 |
| GTEx_Tissu | GTEX-S95S | 2/320 | 0.40678 | 0.929245 | 0 | 0 | 1.449871 | 1.304135 |
| GTEx_Tissu | GTEX-XMD | 1/120 | 0.408269 | 0.929383 | 0 | 0 | 1.934141 | 1.732658 |
| GTEx_Tissu | GTEX-TMN | 2/321 | 0.408301 | 0.929383 | 0 | 0 | 1.445252 | 1.294585 |
| GTEx_Tissu | GTEX-QVJC | 3/532 | 0.409005 | 0.929383 | 0 | 0 | 1.308669 | 1.169987 |
| GTEx_Tissu | GTEX-TML8 | 3/532 | 0.409005 | 0.929383 | 0 | 0 | 1.308669 | 1.169987 |
| GTEx_Tissu | GTEX-V955 | 5/962 | 0.408114 | 0.929383 | 0 | 0 | 1.207789 | 1.08243 |
| GTEx_Tissu | GTEX-WRH | 7/1401 | 0.409346 | 0.929383 | 0 | 0 | 1.162419 | 1.038266 |
| GTEx_Tissu | GTEX-S3XE | 8/1622 | 0.409845 | 0.929567 | 0 | 0 | 1.148119 | 1.024094 |
| GTEx_Tissu | GTEX-OHPI | 20/4304 | 0.410297 | 0.929645 | 0 | 0 | 1.089023 | 0.970181 |
| GTEx_Tissu | GTEX-XUW | 4/749 | 0.411585 | 0.929871 | 0 | 0 | 1.239945 | 1.100749 |
| GTEx_Tissu | GTEX-S4Z8 | 13/2736 | 0.411651 | 0.929871 | 0 | 0 | 1.109021 | 0.984345 |
| GTEx_Tissu | GTEX-QVJC | 5/966 | 0.411593 | 0.929871 | 0 | 0 | 1.202508 | 1.067489 |
| Human_Ge | SuperiorCe | 1/602 | 0.930381 | 0.930381 | 0 | 0 | 0.373641 | 0.026962 |
| GTEx_Tissu | GTEX-U3ZN | 21/4537 | 0.413643 | 0.930591 | 0 | 0 | 1.08482 | 0.957628 |

| | | | | | | | | |
|---|---|---|---|---|---|---|---|---|
| GTEx_Tissu | GTEX-T5JC- | 2/324 | 0.412854 | 0.930591 | 0 | 0 | 1.431567 | 1.266451 |
| GTEx_Tissu | GTEX-NFK9 | 1/122 | 0.413437 | 0.930591 | 0 | 0 | 1.90198 | 1.679923 |
| GTEx_Tissu | GTEX-T2YK | 1/122 | 0.413437 | 0.930591 | 0 | 0 | 1.90198 | 1.679923 |
| GTEx_Tissu | GTEX-Q2AI | 2/326 | 0.415881 | 0.931197 | 0 | 0 | 1.422585 | 1.248115 |
| GTEx_Tissu | GTEX-V955 | 1/123 | 0.416004 | 0.931197 | 0 | 0 | 1.886294 | 1.654392 |
| GTEx_Tissu | GTEX-OHPI | 1/123 | 0.416004 | 0.931197 | 0 | 0 | 1.886294 | 1.654392 |
| GTEx_Tissu | GTEX-OHPI | 2/325 | 0.414369 | 0.931197 | 0 | 0 | 1.427062 | 1.257241 |
| GTEx_Tissu | GTEX-R55G | 2/326 | 0.415881 | 0.931197 | 0 | 0 | 1.422585 | 1.248115 |
| GTEx_Tissu | GTEX-XK95 | 9/1854 | 0.416843 | 0.9312 | 0 | 0 | 1.129956 | 0.988764 |
| GTEx_Tissu | GTEX-TKQ2 | 4/754 | 0.416512 | 0.9312 | 0 | 0 | 1.231357 | 1.078473 |
| GTEx_Tissu | GTEX-RM2I | 3/540 | 0.41835 | 0.933163 | 0 | 0 | 1.288641 | 1.122969 |
| GTEx_Tissu | GTEX-R55E | 1/124 | 0.418561 | 0.933163 | 0 | 0 | 1.870864 | 1.629399 |
| UK_Bioban | No surgery | 1/612 | 0.933442 | 0.933442 | 0 | 0 | 0.367335 | 0.025301 |
| PhenGenI_ | Parkinson I | 1/448 | 0.861273 | 0.934057 | 0 | 0 | 0.506373 | 0.075624 |
| GTEx_Tissu | GTEX-X4EP | 1/125 | 0.421106 | 0.93408 | 0 | 0 | 1.855683 | 1.604928 |
| GTEx_Tissu | GTEX-XGQ4 | 1/125 | 0.421106 | 0.93408 | 0 | 0 | 1.855683 | 1.604928 |
| GTEx_Tissu | GTEX-TKQ1 | 21/4554 | 0.421104 | 0.93408 | 0 | 0 | 1.079558 | 0.933682 |
| GTEx_Tissu | GTEX-N7M | 2/330 | 0.42191 | 0.93408 | 0 | 0 | 1.40495 | 1.212419 |
| GTEx_Tissu | GTEX-RU72 | 3/541 | 0.419515 | 0.93408 | 0 | 0 | 1.286179 | 1.117247 |
| GTEx_Tissu | GTEX-S3XE | 2/329 | 0.420406 | 0.93408 | 0 | 0 | 1.409318 | 1.221224 |
| GTEx_Tissu | GTEX-Q2AI | 2/330 | 0.42191 | 0.93408 | 0 | 0 | 1.40495 | 1.212419 |
| Reactome_ | Cell Cycle, I | 1/523 | 0.900773 | 0.934135 | 0 | 0 | 0.431948 | 0.045139 |
| GTEx_Tissu | GTEX-NL4V | 7/1419 | 0.422408 | 0.934252 | 0 | 0 | 1.146485 | 0.988023 |
| PhenGenI_ | Asthma | 2/818 | 0.876048 | 0.934827 | 0 | 0 | 0.550663 | 0.072872 |
| PhenGenI_ | Coronary A | 1/471 | 0.874804 | 0.934827 | 0 | 0 | 0.481024 | 0.06434 |
| PhenGenI_ | Respiratory | 2/859 | 0.892897 | 0.934827 | 0 | 0 | 0.523193 | 0.059269 |
| PhenGenI_ | Autistic Dis | 1/505 | 0.892451 | 0.934827 | 0 | 0 | 0.44779 | 0.050951 |
| PhenGenI_ | Creatinine | 1/493 | 0.886523 | 0.934827 | 0 | 0 | 0.458995 | 0.055285 |
| PhenGenI_ | Insulin | 1/480 | 0.879736 | 0.934827 | 0 | 0 | 0.471768 | 0.060449 |
| PhenGenI_ | Sleep | 1/463 | 0.870252 | 0.934827 | 0 | 0 | 0.489555 | 0.068034 |
| PhenGenI_ | Depressive | 1/509 | 0.894358 | 0.934827 | 0 | 0 | 0.444172 | 0.049592 |
| GTEx_Tissu | GTEX-XGQ4 | 20/3497 | 0.114714 | 0.934836 | 0 | 0 | 1.411064 | 3.055399 |
| PhenGenI_ | Prostatic N | 1/529 | 0.903403 | 0.935813 | 0 | 0 | 0.426907 | 0.043368 |
| PhenGenI_ | Myocardial | 2/885 | 0.902466 | 0.935813 | 0 | 0 | 0.507095 | 0.05204 |
| GTEx_Tissu | GTEX-WHS | 3/549 | 0.428809 | 0.936478 | 0 | 0 | 1.266811 | 1.072665 |
| GTEx_Tissu | GTEX-R45C | 9/1867 | 0.425148 | 0.936478 | 0 | 0 | 1.121243 | 0.959019 |
| GTEx_Tissu | GTEX-W5X | 1/128 | 0.428675 | 0.936478 | 0 | 0 | 1.811573 | 1.534505 |
| GTEx_Tissu | GTEX-VUSC | 1/128 | 0.428675 | 0.936478 | 0 | 0 | 1.811573 | 1.534505 |
| GTEx_Tissu | GTEX-V1D1 | 4/766 | 0.428301 | 0.936478 | 0 | 0 | 1.211207 | 1.027017 |
| GTEx_Tissu | GTEX-X4XX | 2/333 | 0.426412 | 0.936478 | 0 | 0 | 1.392003 | 1.186473 |
| GTEx_Tissu | GTEX-XYKS | 1/128 | 0.428675 | 0.936478 | 0 | 0 | 1.811573 | 1.534505 |
| GTEx_Tissu | GTEX-QEL4 | 16/3436 | 0.425538 | 0.936478 | 0 | 0 | 1.086764 | 0.928532 |
| GTEx_Tissu | GTEX-XUW | 1/127 | 0.426163 | 0.936478 | 0 | 0 | 1.826043 | 1.557494 |
| GTEx_Tissu | GTEX-R55G | 2/334 | 0.427909 | 0.936478 | 0 | 0 | 1.387739 | 1.177976 |
| GTEx_Tissu | GTEX-S341 | 1/127 | 0.426163 | 0.936478 | 0 | 0 | 1.826043 | 1.557494 |

| | | | | | | | | |
|---|---|---|---|---|---|---|---|---|
| GTEx_Tissu | GTEX-XQ8I | 5/983 | 0.426351 | 0.936478 | 0 | 0 | 1.180545 | 1.006406 |
| GTEx_Tissu | GTEX-PLZ5 | 22/4798 | 0.428886 | 0.936478 | 0 | 0 | 1.072716 | 0.908124 |
| RNA-Seq_D | IFNg Prima | 1/614 | 0.934038 | 0.936485 | 0 | 0 | 0.366099 | 0.024982 |
| GeneSigDB | 19636063- | 1/583 | 0.924177 | 0.937645 | 0 | 0 | 0.386218 | 0.030454 |
| PhenGenI_ | Echocardio | 2/907 | 0.909942 | 0.938378 | 0 | 0 | 0.494196 | 0.046639 |
| RNA-Seq_D | IFNg Prima | 1/628 | 0.938065 | 0.939702 | 0 | 0 | 0.357665 | 0.022868 |
| GTEx_Tissu | GTEX-PWC | 2/336 | 0.430896 | 0.939944 | 0 | 0 | 1.379288 | 1.161207 |
| GTEx_Tissu | GTEX-S4UY | Dec-49 | 0.431801 | 0.940997 | 0 | 0 | 1.095845 | 0.92028 |
| GTEx_Tissu | GTEX-QLQ | 2/337 | 0.432387 | 0.94135 | 0 | 0 | 1.375101 | 1.152933 |
| GTEx_Tissu | GTEX-T8EM | 1/130 | 0.433666 | 0.941421 | 0 | 0 | 1.783306 | 1.489916 |
| GTEx_Tissu | GTEX-RM2 | 14/3002 | 0.433688 | 0.941421 | 0 | 0 | 1.086309 | 0.907534 |
| GTEx_Tissu | GTEX-N7M | 1/130 | 0.433666 | 0.941421 | 0 | 0 | 1.783306 | 1.489916 |
| DisGeNET | Squamous | 2/1023 | 0.941313 | 0.941672 | 0 | 0 | 0.435375 | 0.026331 |
| GTEx_Tissu | GTEX-R55E | 16/3454 | 0.43437 | 0.941982 | 0 | 0 | 1.079894 | 0.900479 |
| GTEx_Tissu | GTEX-X5EB | 2/339 | 0.435362 | 0.942411 | 0 | 0 | 1.3668 | 1.136601 |
| GTEx_Tissu | GTEX-NPJ8 | 8/1661 | 0.436262 | 0.942411 | 0 | 0 | 1.118641 | 0.927926 |
| GTEx_Tissu | GTEX-XMK | Dec-57 | 0.436259 | 0.942411 | 0 | 0 | 1.091898 | 0.905751 |
| GTEx_Tissu | GTEX-XPT6 | 8/1660 | 0.435586 | 0.942411 | 0 | 0 | 1.11938 | 0.930276 |
| GTEx_Tissu | GTEX-UTH( | 5/995 | 0.436732 | 0.942511 | 0 | 0 | 1.165496 | 0.965538 |
| GTEx_Tissu | GTEX-XMK | 27/5953 | 0.437476 | 0.942617 | 0 | 0 | 1.062125 | 0.878093 |
| GTEx_Tissu | GTEX-TKQ2 | 15/3235 | 0.437629 | 0.942617 | 0 | 0 | 1.080034 | 0.892523 |
| GTEx_Tissu | GTEX-XUZC | 6/765 | 0.116019 | 0.942815 | 0 | 0 | 1.869321 | 4.026525 |
| GTEx_Tissu | GTEX-VUS( | 1/132 | 0.438615 | 0.942916 | 0 | 0 | 1.755903 | 1.447098 |
| GTEx_Tissu | GTEX-POM | 2/341 | 0.438329 | 0.942916 | 0 | 0 | 1.358598 | 1.120553 |
| GO_Biologi | Negative R | 2/1025 | 0.941751 | 0.943079 | 0 | 0 | 0.434478 | 0.026075 |
| GTEx_Tissu | GTEX-P44H | 3/558 | 0.439204 | 0.943271 | 0 | 0 | 1.245689 | 1.024943 |
| GTEx_Tissu | GTEX-UJM( | 1/144 | 0.467422 | 0.943336 | 0 | 0 | 1.607578 | 1.222601 |
| GTEx_Tissu | GTEX-WFO | 2/364 | 0.471835 | 0.943336 | 0 | 0 | 1.270783 | 0.954519 |
| GTEx_Tissu | GTEX-PW2 | 8/1703 | 0.464601 | 0.943336 | 0 | 0 | 1.088413 | 0.834352 |
| GTEx_Tissu | GTEX-V955 | 2/358 | 0.463206 | 0.943336 | 0 | 0 | 1.292597 | 0.994762 |
| GTEx_Tissu | GTEX-X585 | 3/579 | 0.463182 | 0.943336 | 0 | 0 | 1.198971 | 0.92277 |
| GTEx_Tissu | GTEX-X4EP | 1/142 | 0.462724 | 0.943336 | 0 | 0 | 1.630546 | 1.256538 |
| GTEx_Tissu | GTEX-RU1J | 2/362 | 0.468968 | 0.943336 | 0 | 0 | 1.277974 | 0.96771 |
| GTEx_Tissu | GTEX-WFJ( | 2/361 | 0.46753 | 0.943336 | 0 | 0 | 1.281599 | 0.974388 |
| GTEx_Tissu | GTEX-PWO | 1/144 | 0.467422 | 0.943336 | 0 | 0 | 1.607578 | 1.222601 |
| GTEx_Tissu | GTEX-XPVC | 1/147 | 0.474392 | 0.943336 | 0 | 0 | 1.574307 | 1.173995 |
| GTEx_Tissu | GTEX-QMR | Oct-57 | 0.465796 | 0.943336 | 0 | 0 | 1.07465 | 0.82104 |
| GTEx_Tissu | GTEX-NL4V | Sep-29 | 0.464673 | 0.943336 | 0 | 0 | 1.08131 | 0.828738 |
| GTEx_Tissu | GTEX-X585 | 7/1481 | 0.467177 | 0.943336 | 0 | 0 | 1.094581 | 0.833028 |
| GTEx_Tissu | GTEX-PWO | Oct-58 | 0.4664 | 0.943336 | 0 | 0 | 1.074089 | 0.819219 |
| GTEx_Tissu | GTEX-PSDC | 1/141 | 0.46036 | 0.943336 | 0 | 0 | 1.642276 | 1.27399 |
| GTEx_Tissu | GTEX-WRH | 5/1034 | 0.470186 | 0.943336 | 0 | 0 | 1.119012 | 0.844436 |
| GTEx_Tissu | GTEX-X585 | 3/589 | 0.474451 | 0.943336 | 0 | 0 | 1.177901 | 0.87824 |
| GTEx_Tissu | GTEX-XMD | 6/1259 | 0.469881 | 0.943336 | 0 | 0 | 1.10313 | 0.833168 |
| GTEx_Tissu | GTEX-XOT( | 8/1715 | 0.472658 | 0.943336 | 0 | 0 | 1.08005 | 0.80937 |

| | | | | | | | | |
|---|---|---|---|---|---|---|---|---|
| GTEx_Tissu | GTEX-S7SE | Oct-72 | 0.474848 | 0.943336 | 0 | 0 | 1.066293 | 0.794133 |
| GTEx_Tissu | GTEX-QCQ( | 2/366 | 0.474693 | 0.943336 | 0 | 0 | 1.263672 | 0.941545 |
| GTEx_Tissu | GTEX-NL3H | Oct-45 | 0.458541 | 0.943336 | 0 | 0 | 1.08142 | 0.84319 |
| GTEx_Tissu | GTEX-X3Y1 | Oct-51 | 0.46217 | 0.943336 | 0 | 0 | 1.078025 | 0.832044 |
| GTEx_Tissu | GTEX-P4QF | Nov-77 | 0.462073 | 0.943336 | 0 | 0 | 1.073414 | 0.82871 |
| GTEx_Tissu | GTEX-RTLS | 2/357 | 0.46176 | 0.943336 | 0 | 0 | 1.296305 | 1.001668 |
| GTEx_Tissu | GTEX-UPIC | 1/141 | 0.46036 | 0.943336 | 0 | 0 | 1.642276 | 1.27399 |
| GTEx_Tissu | GTEX-PVO\ | Oct-51 | 0.46217 | 0.943336 | 0 | 0 | 1.078025 | 0.832044 |
| GTEx_Tissu | GTEX-PSDG | 3/588 | 0.473328 | 0.943336 | 0 | 0 | 1.179976 | 0.882581 |
| GTEx_Tissu | GTEX-T5JC | 4/801 | 0.462334 | 0.943336 | 0 | 0 | 1.155901 | 0.891741 |
| GTEx_Tissu | GTEX-XUZ( | 3/568 | 0.450673 | 0.943336 | 0 | 0 | 1.223009 | 0.974755 |
| GTEx_Tissu | GTEX-Q2AI | Dec-99 | 0.459648 | 0.943336 | 0 | 0 | 1.071573 | 0.832927 |
| GTEx_Tissu | GTEX-OIZH | 3/575 | 0.458647 | 0.943336 | 0 | 0 | 1.207605 | 0.941298 |
| GTEx_Tissu | GTEX-S32W | 2/366 | 0.474693 | 0.943336 | 0 | 0 | 1.263672 | 0.941545 |
| GTEx_Tissu | GTEX-P78B | 1/140 | 0.457986 | 0.943336 | 0 | 0 | 1.654174 | 1.291774 |
| GTEx_Tissu | GTEX-WK1 | 24/5320 | 0.4579 | 0.943336 | 0 | 0 | 1.051431 | 0.821279 |
| GTEx_Tissu | GTEX-T6MI | 5/1019 | 0.457378 | 0.943336 | 0 | 0 | 1.136468 | 0.888995 |
| GTEx_Tissu | GTEX-PWN | 5/1009 | 0.448797 | 0.943336 | 0 | 0 | 1.148394 | 0.920076 |
| GTEx_Tissu | GTEX-X4XX | 7/1465 | 0.455674 | 0.943336 | 0 | 0 | 1.107553 | 0.870512 |
| GTEx_Tissu | GTEX-T6M( | 1/137 | 0.4508 | 0.943336 | 0 | 0 | 1.69092 | 1.347208 |
| GTEx_Tissu | GTEX-XV7C | 2/356 | 0.460312 | 0.943336 | 0 | 0 | 1.300033 | 1.008633 |
| GTEx_Tissu | GTEX-U8T8 | 3/581 | 0.465444 | 0.943336 | 0 | 0 | 1.194698 | 0.913662 |
| GTEx_Tissu | GTEX-RVPV | Dec-09 | 0.465209 | 0.943336 | 0 | 0 | 1.066831 | 0.816412 |
| GTEx_Tissu | GTEX-OIZI- | Nov-47 | 0.444695 | 0.943336 | 0 | 0 | 1.089058 | 0.882535 |
| GTEx_Tissu | GTEX-U3ZN | 1/136 | 0.448384 | 0.943336 | 0 | 0 | 1.703531 | 1.36641 |
| GTEx_Tissu | GTEX-OHPI | Dec-78 | 0.447958 | 0.943336 | 0 | 0 | 1.081652 | 0.868627 |
| GTEx_Tissu | GTEX-SUCS | 3/565 | 0.447241 | 0.943336 | 0 | 0 | 1.229728 | 0.989509 |
| GTEx_Tissu | GTEX-N7M | 3/565 | 0.447241 | 0.943336 | 0 | 0 | 1.229728 | 0.989509 |
| GTEx_Tissu | GTEX-POY\ | 3/564 | 0.446096 | 0.943336 | 0 | 0 | 1.231984 | 0.994484 |
| GTEx_Tissu | GTEX-P4PC | Nov-48 | 0.445275 | 0.943336 | 0 | 0 | 1.08853 | 0.880689 |
| GTEx_Tissu | GTEX-REY6 | 2/360 | 0.466091 | 0.943336 | 0 | 0 | 1.285245 | 0.981122 |
| GTEx_Tissu | GTEX-QEG4 | 6/1239 | 0.454338 | 0.943336 | 0 | 0 | 1.122225 | 0.885339 |
| GTEx_Tissu | GTEX-S341 | 3/569 | 0.451815 | 0.943336 | 0 | 0 | 1.220785 | 0.969893 |
| GTEx_Tissu | GTEX-RWS. | 4/790 | 0.451702 | 0.943336 | 0 | 0 | 1.172752 | 0.932024 |
| GTEx_Tissu | GTEX-RNOI | 7/1484 | 0.469328 | 0.943336 | 0 | 0 | 1.09218 | 0.826183 |
| GTEx_Tissu | GTEX-XOT( | Sep-40 | 0.471653 | 0.943336 | 0 | 0 | 1.074493 | 0.807494 |
| GTEx_Tissu | GTEX-WYV. | 21/4665 | 0.470077 | 0.943336 | 0 | 0 | 1.046149 | 0.789696 |
| GTEx_Tissu | GTEX-S4P3 | 7/1490 | 0.473626 | 0.943336 | 0 | 0 | 1.087407 | 0.81266 |
| GTEx_Tissu | GTEX-Q734 | 1/145 | 0.469755 | 0.943336 | 0 | 0 | 1.596334 | 1.2061 |
| GTEx_Tissu | GTEX-S32W | 1/145 | 0.469755 | 0.943336 | 0 | 0 | 1.596334 | 1.2061 |
| GTEx_Tissu | GTEX-QVU! | 1/145 | 0.469755 | 0.943336 | 0 | 0 | 1.596334 | 1.2061 |
| GTEx_Tissu | GTEX-QEG4 | 1/145 | 0.469755 | 0.943336 | 0 | 0 | 1.596334 | 1.2061 |
| GTEx_Tissu | GTEX-RN64 | 3/579 | 0.463182 | 0.943336 | 0 | 0 | 1.198971 | 0.92277 |
| GTEx_Tissu | GTEX-UJHI | 5/1036 | 0.471888 | 0.943336 | 0 | 0 | 1.116723 | 0.838675 |
| GTEx_Tissu | GTEX-OHPI | 1/146 | 0.472079 | 0.943336 | 0 | 0 | 1.585245 | 1.1899 |

| | | | | | | | | |
|---|---|---|---|---|---|---|---|---|
| GTEx_Tissu | GTEX-S32V | 5/1016 | 0.454807 | 0.943336 | 0 | 0 | 1.140021 | 0.898201 |
| GTEx_Tissu | GTEX-RNOI | 25/5545 | 0.456779 | 0.943336 | 0 | 0 | 1.051382 | 0.823817 |
| GTEx_Tissu | GTEX-QMR | 3/560 | 0.441505 | 0.943336 | 0 | 0 | 1.241087 | 1.014672 |
| GTEx_Tissu | GTEX-R53T | 1/133 | 0.441073 | 0.943336 | 0 | 0 | 1.742512 | 1.426323 |
| GTEx_Tissu | GTEX-WQL | 1/133 | 0.441073 | 0.943336 | 0 | 0 | 1.742512 | 1.426323 |
| GTEx_Tissu | GTEX-RN64 | 1/134 | 0.443521 | 0.943336 | 0 | 0 | 1.729323 | 1.405958 |
| GTEx_Tissu | GTEX-WH7 | 2/344 | 0.442764 | 0.943336 | 0 | 0 | 1.346474 | 1.096999 |
| GTEx_Tissu | GTEX-U4B1 | 17/3695 | 0.441714 | 0.943336 | 0 | 0 | 1.071992 | 0.875917 |
| GTEx_Tissu | GTEX-N7M | 2/345 | 0.444238 | 0.943336 | 0 | 0 | 1.34248 | 1.089282 |
| GTEx_Tissu | GTEX-X261 | 20/4394 | 0.45065 | 0.943336 | 0 | 0 | 1.060473 | 0.845265 |
| GTEx_Tissu | GTEX-NFK9 | Dec-82 | 0.450186 | 0.943336 | 0 | 0 | 1.07972 | 0.861719 |
| GTEx_Tissu | GTEX-WWY | 2/349 | 0.450113 | 0.943336 | 0 | 0 | 1.326733 | 1.059074 |
| GTEx_Tissu | GTEX-PSDG | 14/3033 | 0.44981 | 0.943336 | 0 | 0 | 1.073185 | 0.857399 |
| GTEx_Tissu | GTEX-UPIC | 14/3033 | 0.44981 | 0.943336 | 0 | 0 | 1.073185 | 0.857399 |
| GTEx_Tissu | GTEX-R55E | 4/788 | 0.449762 | 0.943336 | 0 | 0 | 1.175867 | 0.939559 |
| GTEx_Tissu | GTEX-SE5C | 1/145 | 0.469755 | 0.943336 | 0 | 0 | 1.596334 | 1.2061 |
| GTEx_Tissu | GTEX-WZT( | 4/787 | 0.448792 | 0.943336 | 0 | 0 | 1.17743 | 0.943352 |
| GTEx_Tissu | GTEX-XQ8I | 1/139 | 0.455601 | 0.943336 | 0 | 0 | 1.666245 | 1.309899 |
| GTEx_Tissu | GTEX-X261 | 21/4632 | 0.455494 | 0.943336 | 0 | 0 | 1.055914 | 0.830342 |
| GTEx_Tissu | GTEX-RUS( | 2/342 | 0.439809 | 0.943336 | 0 | 0 | 1.354533 | 1.112633 |
| PhenGenI_ | Iron | 1/568 | 0.918896 | 0.9434 | 0 | 0 | 0.396743 | 0.033557 |
| GTEx_Tissu | GTEX-PX3G | 6/1267 | 0.476068 | 0.943636 | 0 | 0 | 1.095662 | 0.813194 |
| GTEx_Tissu | GTEX-X261 | 1/148 | 0.476695 | 0.943636 | 0 | 0 | 1.563518 | 1.158376 |
| GTEx_Tissu | GTEX-S4Q7 | 1/148 | 0.476695 | 0.943636 | 0 | 0 | 1.563518 | 1.158376 |
| GTEx_Tissu | GTEX-P4QS | 2/367 | 0.476119 | 0.943636 | 0 | 0 | 1.260145 | 0.935139 |
| GTEx_Tissu | GTEX-UJM( | 2/368 | 0.477542 | 0.943723 | 0 | 0 | 1.256638 | 0.928785 |
| GTEx_Tissu | GTEX-P4PP | 21/4682 | 0.477587 | 0.943723 | 0 | 0 | 1.041173 | 0.769436 |
| GeneSigDB | 19096012- | 1/608 | 0.932234 | 0.944099 | 0 | 0 | 0.369833 | 0.025952 |
| GTEx_Tissu | GTEX-XLM4 | 1/149 | 0.478989 | 0.945652 | 0 | 0 | 1.552876 | 1.143039 |
| GTEx_Tissu | GTEX-WOF | 13/2865 | 0.480883 | 0.946169 | 0 | 0 | 1.050913 | 0.769406 |
| GTEx_Tissu | GTEX-XAJ8 | 2/377 | 0.490248 | 0.946169 | 0 | 0 | 1.225914 | 0.873884 |
| GTEx_Tissu | GTEX-XBED | 2/377 | 0.490248 | 0.946169 | 0 | 0 | 1.225914 | 0.873884 |
| GTEx_Tissu | GTEX-OXRK | 43/9731 | 0.485041 | 0.946169 | 0 | 0 | 1.031442 | 0.746272 |
| GTEx_Tissu | GTEX-QLQ\ | 4/824 | 0.484338 | 0.946169 | 0 | 0 | 1.122128 | 0.813512 |
| GTEx_Tissu | GTEX-XQ8I | 21/4696 | 0.483769 | 0.946169 | 0 | 0 | 1.037103 | 0.75309 |
| GTEx_Tissu | GTEX-XOT( | 5/1050 | 0.483751 | 0.946169 | 0 | 0 | 1.100945 | 0.79949 |
| GTEx_Tissu | GTEX-T5JC- | 5/1050 | 0.483751 | 0.946169 | 0 | 0 | 1.100945 | 0.79949 |
| GTEx_Tissu | GTEX-TMM | 3/594 | 0.480046 | 0.946169 | 0 | 0 | 1.167634 | 0.856895 |
| GTEx_Tissu | GTEX-WFG | 4/825 | 0.485287 | 0.946169 | 0 | 0 | 1.120702 | 0.810284 |
| GTEx_Tissu | GTEX-PWC | Sep-56 | 0.481778 | 0.946169 | 0 | 0 | 1.064715 | 0.777531 |
| GTEx_Tissu | GTEX-WHS | 8/1729 | 0.482029 | 0.946169 | 0 | 0 | 1.07044 | 0.781155 |
| GTEx_Tissu | GTEX-R55E | 6/1275 | 0.482237 | 0.946169 | 0 | 0 | 1.088288 | 0.793709 |
| GTEx_Tissu | GTEX-WHS | 7/1512 | 0.489315 | 0.946169 | 0 | 0 | 1.070233 | 0.764948 |
| GTEx_Tissu | GTEX-W5X | 19/4244 | 0.485914 | 0.946169 | 0 | 0 | 1.037494 | 0.748784 |
| GTEx_Tissu | GTEX-OOBI | 1/151 | 0.483545 | 0.946169 | 0 | 0 | 1.532016 | 1.113178 |

| | | | | | | | | |
|---|---|---|---|---|---|---|---|---|
| GTEx_Tissu | GTEX-RU72 | 6/1283 | 0.488386 | 0.946169 | 0 | 0 | 1.081006 | 0.774702 |
| GTEx_Tissu | GTEX-Q2A( | 37/8370 | 0.489792 | 0.946169 | 0 | 0 | 1.028345 | 0.734007 |
| GTEx_Tissu | GTEX-R55D | 1/154 | 0.490307 | 0.946169 | 0 | 0 | 1.501748 | 1.070331 |
| GTEx_Tissu | GTEX-WK1 | 1/152 | 0.485809 | 0.946169 | 0 | 0 | 1.521793 | 1.098642 |
| GTEx_Tissu | GTEX-S95S | 1/153 | 0.488063 | 0.946169 | 0 | 0 | 1.511704 | 1.084362 |
| GTEx_Tissu | GTEX-XXEK | 1/153 | 0.488063 | 0.946169 | 0 | 0 | 1.511704 | 1.084362 |
| GTEx_Tissu | GTEX-OOB. | 1/153 | 0.488063 | 0.946169 | 0 | 0 | 1.511704 | 1.084362 |
| GTEx_Tissu | GTEX-SE5C | 2/375 | 0.487441 | 0.946169 | 0 | 0 | 1.232613 | 0.885738 |
| GTEx_Tissu | GTEX-S32V | 8/1736 | 0.486701 | 0.946169 | 0 | 0 | 1.065694 | 0.767411 |
| GTEx_Tissu | GTEX-PW2 | Sep-63 | 0.486196 | 0.946169 | 0 | 0 | 1.060487 | 0.764763 |
| GTEx_Tissu | GTEX-QCQ | 7/1514 | 0.490735 | 0.946175 | 0 | 0 | 1.068696 | 0.760752 |
| GTEx_Tissu | GTEX-XUW | Dec-56 | 0.491255 | 0.946358 | 0 | 0 | 1.045023 | 0.742793 |
| GTEx_Tissu | GTEX-S3XE | 37/8375 | 0.491687 | 0.946369 | 0 | 0 | 1.027285 | 0.729283 |
| GTEx_Tissu | GTEX-WHS | 8/1128 | 0.117187 | 0.947603 | 0 | 0 | 1.699186 | 3.643027 |
| GTEx_Tissu | GTEX-OHPI | 8/1130 | 0.118054 | 0.947603 | 0 | 0 | 1.695977 | 3.62365 |
| GTEx_Tissu | GTEX-OIZH | 5/596 | 0.118067 | 0.947603 | 0 | 0 | 1.993521 | 4.259156 |
| GTEx_Tissu | GTEX-XXEK | 6/769 | 0.118136 | 0.947603 | 0 | 0 | 1.859133 | 3.97096 |
| GTEx_Tissu | GTEX-XAJ8 | 6/770 | 0.118668 | 0.947603 | 0 | 0 | 1.856603 | 3.957208 |
| GTEx_Tissu | GTEX-RVPV | 7/949 | 0.118901 | 0.947603 | 0 | 0 | 1.762168 | 3.752478 |
| GTEx_Tissu | GTEX-QLQV | 7/949 | 0.118901 | 0.947603 | 0 | 0 | 1.762168 | 3.752478 |
| GTEx_Tissu | GTEX-OOBI | 5/598 | 0.11929 | 0.948094 | 0 | 0 | 1.986592 | 4.223885 |
| GTEx_Tissu | GTEX-WFG | 8/1752 | 0.497343 | 0.948386 | 0 | 0 | 1.054988 | 0.736883 |
| GTEx_Tissu | GTEX-VUSI | 7/1524 | 0.497822 | 0.948386 | 0 | 0 | 1.061074 | 0.740114 |
| GTEx_Tissu | GTEX-QMR | 5/1066 | 0.497204 | 0.948386 | 0 | 0 | 1.083423 | 0.757048 |
| GTEx_Tissu | GTEX-OHPI | 2/386 | 0.502763 | 0.948386 | 0 | 0 | 1.19663 | 0.822846 |
| GTEx_Tissu | GTEX-QEG! | 1/159 | 0.501382 | 0.948386 | 0 | 0 | 1.453856 | 1.003722 |
| GTEx_Tissu | GTEX-T5JW | 7/1518 | 0.493573 | 0.948386 | 0 | 0 | 1.065635 | 0.752428 |
| GTEx_Tissu | GTEX-RN64 | 8/1765 | 0.505948 | 0.948386 | 0 | 0 | 1.046433 | 0.712957 |
| GTEx_Tissu | GTEX-U3ZH | 3/612 | 0.499961 | 0.948386 | 0 | 0 | 1.132067 | 0.784777 |
| GTEx_Tissu | GTEX-QV44 | 8/1758 | 0.50132 | 0.948386 | 0 | 0 | 1.051024 | 0.725744 |
| GTEx_Tissu | GTEX-T5JC | 2/385 | 0.501382 | 0.948386 | 0 | 0 | 1.199816 | 0.828337 |
| GTEx_Tissu | GTEX-UJHI | 17/3828 | 0.505322 | 0.948386 | 0 | 0 | 1.026105 | 0.700377 |
| GTEx_Tissu | GTEX-SNO! | Oct-23 | 0.505419 | 0.948386 | 0 | 0 | 1.038726 | 0.708793 |
| GTEx_Tissu | GTEX-Q2AI | 1/161 | 0.505746 | 0.948386 | 0 | 0 | 1.435538 | 0.978637 |
| GTEx_Tissu | GTEX-S4P3 | 1/161 | 0.505746 | 0.948386 | 0 | 0 | 1.435538 | 0.978637 |
| GTEx_Tissu | GTEX-RWS | 4/840 | 0.499444 | 0.948386 | 0 | 0 | 1.099729 | 0.763498 |
| GTEx_Tissu | GTEX-SUCS | 14/3123 | 0.496477 | 0.948386 | 0 | 0 | 1.036566 | 0.725822 |
| GTEx_Tissu | GTEX-O5YV | 1/160 | 0.503569 | 0.948386 | 0 | 0 | 1.444639 | 0.991073 |
| GTEx_Tissu | GTEX-XUZ( | 2/387 | 0.504142 | 0.948386 | 0 | 0 | 1.193461 | 0.817399 |
| GTEx_Tissu | GTEX-QMR | 6/1306 | 0.505942 | 0.948386 | 0 | 0 | 1.06057 | 0.722602 |
| GTEx_Tissu | GTEX-XOT( | 2/381 | 0.495834 | 0.948386 | 0 | 0 | 1.212727 | 0.850744 |
| GTEx_Tissu | GTEX-PSD( | 2/386 | 0.502763 | 0.948386 | 0 | 0 | 1.19663 | 0.822846 |
| GTEx_Tissu | GTEX-X5EB | 2/386 | 0.502763 | 0.948386 | 0 | 0 | 1.19663 | 0.822846 |
| GTEx_Tissu | GTEX-WHS | 3/615 | 0.503244 | 0.948386 | 0 | 0 | 1.126342 | 0.773436 |
| GTEx_Tissu | GTEX-QMR | 7/1532 | 0.503471 | 0.948386 | 0 | 0 | 1.055049 | 0.724006 |

| | | | | | | | | |
|---|---|---|---|---|---|---|---|---|
| GTEx_Tissu | GTEX-WFJ( | 1/160 | 0.503569 | 0.948386 | 0 | 0 | 1.444639 | 0.991073 |
| GTEx_Tissu | GTEX-XV7( | 1/160 | 0.503569 | 0.948386 | 0 | 0 | 1.444639 | 0.991073 |
| GTEx_Tissu | GTEX-WL4( | 2/380 | 0.494441 | 0.948386 | 0 | 0 | 1.215998 | 0.85646 |
| GTEx_Tissu | GTEX-T6MI | 5/1063 | 0.494691 | 0.948386 | 0 | 0 | 1.086668 | 0.764822 |
| GTEx_Tissu | GTEX-QMR | 1/156 | 0.494766 | 0.948386 | 0 | 0 | 1.482221 | 1.042994 |
| GTEx_Tissu | GTEX-WOF | 37/8385 | 0.495477 | 0.948386 | 0 | 0 | 1.025168 | 0.719909 |
| GTEx_Tissu | GTEX-S4P3 | 2/381 | 0.495834 | 0.948386 | 0 | 0 | 1.212727 | 0.850744 |
| SysMyo_M | MGS137 M | 1/669 | 0.948511 | 0.948511 | 0 | 0 | 0.334999 | 0.017709 |
| GTEx_Tissu | GTEX-N7M | 3/619 | 0.507605 | 0.948872 | 0 | 0 | 1.118796 | 0.758602 |
| GTEx_Tissu | GTEX-QLQ7 | 1/162 | 0.507913 | 0.948872 | 0 | 0 | 1.426549 | 0.966408 |
| GTEx_Tissu | GTEX-S7SF- | 2/389 | 0.506892 | 0.948872 | 0 | 0 | 1.187171 | 0.806633 |
| GTEx_Tissu | GTEX-XUJ4 | 21/4750 | 0.507556 | 0.948872 | 0 | 0 | 1.021627 | 0.692814 |
| GTEx_Tissu | GTEX-WZT( | 6/1315 | 0.512758 | 0.948963 | 0 | 0 | 1.052769 | 0.703199 |
| GTEx_Tissu | GTEX-U8XE | 3/625 | 0.51411 | 0.948963 | 0 | 0 | 1.10766 | 0.736946 |
| GTEx_Tissu | GTEX-VUS( | 7/1541 | 0.509803 | 0.948963 | 0 | 0 | 1.048346 | 0.706302 |
| GTEx_Tissu | GTEX-TSE9- | 7/1544 | 0.511909 | 0.948963 | 0 | 0 | 1.046129 | 0.700497 |
| GTEx_Tissu | GTEX-WRH | 1/164 | 0.51222 | 0.948963 | 0 | 0 | 1.408903 | 0.942558 |
| GTEx_Tissu | GTEX-NFK9 | 22/4991 | 0.512619 | 0.948963 | 0 | 0 | 1.017905 | 0.680186 |
| GTEx_Tissu | GTEX-UTH( | Oct-35 | 0.512554 | 0.948963 | 0 | 0 | 1.032424 | 0.690019 |
| GTEx_Tissu | GTEX-P78B | 21/4761 | 0.512386 | 0.948963 | 0 | 0 | 1.018517 | 0.681059 |
| GTEx_Tissu | GTEX-T5JC- | 4/850 | 0.508793 | 0.948963 | 0 | 0 | 1.08616 | 0.733934 |
| GTEx_Tissu | GTEX-S4Q7 | 3/625 | 0.51411 | 0.948963 | 0 | 0 | 1.10766 | 0.736946 |
| GTEx_Tissu | GTEX-XV7( | Sep-04 | 0.511903 | 0.948963 | 0 | 0 | 1.036322 | 0.693942 |
| GTEx_Tissu | GTEX-WVLI | Oct-38 | 0.514334 | 0.948963 | 0 | 0 | 1.030859 | 0.6854 |
| GTEx_Tissu | GTEX-T2IS- | Oct-31 | 0.510179 | 0.948963 | 0 | 0 | 1.034517 | 0.696224 |
| GTEx_Tissu | GTEX-OIZG | 1/165 | 0.514359 | 0.948963 | 0 | 0 | 1.400241 | 0.930927 |
| GTEx_Tissu | GTEX-OOB. | 16/3608 | 0.509829 | 0.948963 | 0 | 0 | 1.023934 | 0.689804 |
| GTEx_Tissu | GTEX-U8XE | 4/857 | 0.515292 | 0.949111 | 0 | 0 | 1.076851 | 0.713975 |
| GTEx_Tissu | GTEX-W5X | 8/1779 | 0.515169 | 0.949111 | 0 | 0 | 1.03736 | 0.68804 |
| CellMarker | Type I Spira | 2/1061 | 0.949128 | 0.949128 | 0 | 0 | 0.418908 | 0.021872 |
| GTEx_Tissu | GTEX-U3ZN | 1/166 | 0.516489 | 0.949743 | 0 | 0 | 1.391684 | 0.919487 |
| GTEx_Tissu | GTEX-RN64 | 2/396 | 0.516439 | 0.949743 | 0 | 0 | 1.165661 | 0.770266 |
| GTEx_Tissu | GTEX-WHS | 9/1321 | 0.119905 | 0.950368 | 0 | 0 | 1.635876 | 3.469787 |
| GTEx_Tissu | GTEX-PWO | 1/171 | 0.527002 | 0.950861 | 0 | 0 | 1.35041 | 0.865006 |
| GTEx_Tissu | GTEX-UJM( | 2/404 | 0.527203 | 0.950861 | 0 | 0 | 1.141996 | 0.731071 |
| GTEx_Tissu | GTEX-T5JC- | 6/1326 | 0.521043 | 0.950861 | 0 | 0 | 1.043378 | 0.680201 |
| GTEx_Tissu | GTEX-Q734 | 2/400 | 0.521841 | 0.950861 | 0 | 0 | 1.15371 | 0.750364 |
| GTEx_Tissu | GTEX-QLQ\ | 16/3633 | 0.521956 | 0.950861 | 0 | 0 | 1.015299 | 0.660119 |
| GTEx_Tissu | GTEX-OIZI- | 3/635 | 0.524853 | 0.950861 | 0 | 0 | 1.089568 | 0.702376 |
| GTEx_Tissu | GTEX-S4Q7 | 1/170 | 0.524918 | 0.950861 | 0 | 0 | 1.35847 | 0.875552 |
| GTEx_Tissu | GTEX-XUJ4 | 2/412 | 0.537807 | 0.950861 | 0 | 0 | 1.119254 | 0.694223 |
| GTEx_Tissu | GTEX-U4B1 | 4/882 | 0.538187 | 0.950861 | 0 | 0 | 1.044817 | 0.647315 |
| GTEx_Tissu | GTEX-S4P3 | 8/1801 | 0.529553 | 0.950861 | 0 | 0 | 1.023389 | 0.650591 |
| GTEx_Tissu | GTEX-R55C | 19/4340 | 0.529615 | 0.950861 | 0 | 0 | 1.008236 | 0.640839 |
| GTEx_Tissu | GTEX-XQ8I | 16/3652 | 0.531131 | 0.950861 | 0 | 0 | 1.008816 | 0.638326 |

| | | | | | | | | |
|---|---|---|---|---|---|---|---|---|
| GTEx_Tissu | GTEX-OXRK | Sep-25 | 0.524936 | 0.950861 | 0 | 0 | 1.024325 | 0.660155 |
| GTEx_Tissu | GTEX-OHPI | 8/1794 | 0.524991 | 0.950861 | 0 | 0 | 1.027797 | 0.662286 |
| GTEx_Tissu | GTEX-N7M | 7/1563 | 0.525175 | 0.950861 | 0 | 0 | 1.032286 | 0.664818 |
| GTEx_Tissu | GTEX-S95S | 7/1553 | 0.518208 | 0.950861 | 0 | 0 | 1.039529 | 0.683365 |
| GTEx_Tissu | GTEX-SE5C | 1/177 | 0.53932 | 0.950861 | 0 | 0 | 1.303977 | 0.805137 |
| GTEx_Tissu | GTEX-SJXC | 1/177 | 0.53932 | 0.950861 | 0 | 0 | 1.303977 | 0.805137 |
| GTEx_Tissu | GTEX-UJM( | 1/167 | 0.51861 | 0.950861 | 0 | 0 | 1.383231 | 0.908233 |
| GTEx_Tissu | GTEX-NFK9 | 1/173 | 0.531144 | 0.950861 | 0 | 0 | 1.334573 | 0.844414 |
| GTEx_Tissu | GTEX-XBED | 1/173 | 0.531144 | 0.950861 | 0 | 0 | 1.334573 | 0.844414 |
| GTEx_Tissu | GTEX-WK1 | 21/4784 | 0.522461 | 0.950861 | 0 | 0 | 1.012063 | 0.657036 |
| GTEx_Tissu | GTEX-QEL4 | 2/413 | 0.539122 | 0.950861 | 0 | 0 | 1.116473 | 0.689773 |
| GTEx_Tissu | GTEX-QXCU | 2/413 | 0.539122 | 0.950861 | 0 | 0 | 1.116473 | 0.689773 |
| GTEx_Tissu | GTEX-POYV | 2/413 | 0.539122 | 0.950861 | 0 | 0 | 1.116473 | 0.689773 |
| GTEx_Tissu | GTEX-P44H | 8/1816 | 0.539279 | 0.950861 | 0 | 0 | 1.014058 | 0.626204 |
| GTEx_Tissu | GTEX-TSE9 | 7/1581 | 0.537626 | 0.950861 | 0 | 0 | 1.019481 | 0.632681 |
| GTEx_Tissu | GTEX-TSE9 | 2/412 | 0.537807 | 0.950861 | 0 | 0 | 1.119254 | 0.694223 |
| GTEx_Tissu | GTEX-S7SF | 6/1332 | 0.525541 | 0.950861 | 0 | 0 | 1.038322 | 0.66798 |
| GTEx_Tissu | GTEX-OHPI | 1/167 | 0.51861 | 0.950861 | 0 | 0 | 1.383231 | 0.908233 |
| GTEx_Tissu | GTEX-XQ8I | 2/402 | 0.524527 | 0.950861 | 0 | 0 | 1.147824 | 0.740643 |
| GTEx_Tissu | GTEX-XPVG | 14/3196 | 0.533858 | 0.950861 | 0 | 0 | 1.008386 | 0.632889 |
| GTEx_Tissu | GTEX-SNM | 15/3427 | 0.533913 | 0.950861 | 0 | 0 | 1.007535 | 0.63225 |
| GTEx_Tissu | GTEX-XUW | 17/3891 | 0.535109 | 0.950861 | 0 | 0 | 1.005469 | 0.628704 |
| GTEx_Tissu | GTEX-WYB | 1/175 | 0.535249 | 0.950861 | 0 | 0 | 1.319099 | 0.824467 |
| GTEx_Tissu | GTEX-UTH( | 6/1346 | 0.535974 | 0.950861 | 0 | 0 | 1.0267 | 0.640321 |
| GTEx_Tissu | GTEX-XUYS | Oct-75 | 0.536137 | 0.950861 | 0 | 0 | 1.011898 | 0.630782 |
| GTEx_Tissu | GTEX-WFG | 1/172 | 0.529077 | 0.950861 | 0 | 0 | 1.342445 | 0.854628 |
| GTEx_Tissu | GTEX-XUZ( | 1/176 | 0.537289 | 0.950861 | 0 | 0 | 1.311495 | 0.814726 |
| GTEx_Tissu | GTEX-P4QS | 1/176 | 0.537289 | 0.950861 | 0 | 0 | 1.311495 | 0.814726 |
| GTEx_Tissu | GTEX-RNOI | Sep-36 | 0.531722 | 0.950861 | 0 | 0 | 1.01814 | 0.643093 |
| GTEx_Tissu | GTEX-U3ZN | 2/409 | 0.53385 | 0.950861 | 0 | 0 | 1.127677 | 0.707777 |
| GTEx_Tissu | GTEX-S32V | 4/883 | 0.539092 | 0.950861 | 0 | 0 | 1.043574 | 0.644791 |
| GTEx_Tissu | GTEX-QVJC | 4/883 | 0.539092 | 0.950861 | 0 | 0 | 1.043574 | 0.644791 |
| GTEx_Tissu | GTEX-RU1J | 1/177 | 0.53932 | 0.950861 | 0 | 0 | 1.303977 | 0.805137 |
| GTEx_Tissu | GTEX-Q2A( | 3/630 | 0.519497 | 0.950861 | 0 | 0 | 1.098542 | 0.719429 |
| GTEx_Tissu | GTEX-QEG4 | 7/1557 | 0.520998 | 0.950861 | 0 | 0 | 1.036621 | 0.675885 |
| GTEx_Tissu | GTEX-UPIC | 1/176 | 0.537289 | 0.950861 | 0 | 0 | 1.311495 | 0.814726 |
| GTEx_Tissu | GTEX-WFG | 1/176 | 0.537289 | 0.950861 | 0 | 0 | 1.311495 | 0.814726 |
| GTEx_Tissu | GTEX-VUS( | 1/176 | 0.537289 | 0.950861 | 0 | 0 | 1.311495 | 0.814726 |
| GTEx_Tissu | GTEX-S4Z8 | 1/176 | 0.537289 | 0.950861 | 0 | 0 | 1.311495 | 0.814726 |
| GTEx_Tissu | GTEX-XUJ4 | 3/634 | 0.523784 | 0.950861 | 0 | 0 | 1.091352 | 0.70575 |
| GTEx_Tissu | GTEX-U3ZN | 5/1126 | 0.546453 | 0.95119 | 0 | 0 | 1.022171 | 0.617706 |
| GTEx_Tissu | GTEX-X261 | 6/1472 | 0.625183 | 0.95119 | 0 | 0 | 0.93209 | 0.437813 |
| GTEx_Tissu | GTEX-RWS( | 3/736 | 0.62567 | 0.95119 | 0 | 0 | 0.934516 | 0.438225 |
| GTEx_Tissu | GTEX-R55D | Nov-58 | 0.618295 | 0.95119 | 0 | 0 | 0.944098 | 0.453912 |
| GTEx_Tissu | GTEX-PWC | 2/478 | 0.61903 | 0.95119 | 0 | 0 | 0.960801 | 0.460802 |

| | | | | | | | | |
|---|---|---|---|---|---|---|---|---|
| GTEx_Tissu | GTEX-POYV | 14/3384 | 0.62603 | 0.95119 | 0 | 0 | 0.941433 | 0.440926 |
| GTEx_Tissu | GTEX-TML8 | 1/225 | 0.627094 | 0.95119 | 0 | 0 | 1.022062 | 0.476954 |
| GTEx_Tissu | GTEX-R55D | 1/225 | 0.627094 | 0.95119 | 0 | 0 | 1.022062 | 0.476954 |
| GTEx_Tissu | GTEX-RU1J | 1/225 | 0.627094 | 0.95119 | 0 | 0 | 1.022062 | 0.476954 |
| GTEx_Tissu | GTEX-QDVI | 1/225 | 0.627094 | 0.95119 | 0 | 0 | 1.022062 | 0.476954 |
| GTEx_Tissu | GTEX-OHPI | 7/1717 | 0.627144 | 0.95119 | 0 | 0 | 0.93144 | 0.43459 |
| GTEx_Tissu | GTEX-TKQ1 | 1/195 | 0.574401 | 0.95119 | 0 | 0 | 1.181911 | 0.655284 |
| GTEx_Tissu | GTEX-QMR | 2/436 | 0.568645 | 0.95119 | 0 | 0 | 1.056059 | 0.596144 |
| GTEx_Tissu | GTEX-OOB. | 1/192 | 0.568743 | 0.95119 | 0 | 0 | 1.200657 | 0.677563 |
| GTEx_Tissu | GTEX-XXEK | 2/480 | 0.621315 | 0.95119 | 0 | 0 | 0.956682 | 0.455302 |
| GTEx_Tissu | GTEX-PWC | 2/419 | 0.546954 | 0.95119 | 0 | 0 | 1.100071 | 0.663773 |
| GTEx_Tissu | GTEX-XUJ4 | 2/415 | 0.541742 | 0.95119 | 0 | 0 | 1.110953 | 0.680975 |
| GTEx_Tissu | GTEX-N7M | 2/415 | 0.541742 | 0.95119 | 0 | 0 | 1.110953 | 0.680975 |
| GTEx_Tissu | GTEX-XLM4 | 4/887 | 0.542704 | 0.95119 | 0 | 0 | 1.038628 | 0.6348 |
| GTEx_Tissu | GTEX-PWC | 1/185 | 0.555251 | 0.95119 | 0 | 0 | 1.246777 | 0.733523 |
| GTEx_Tissu | GTEX-WZT( | 4/902 | 0.556122 | 0.95119 | 0 | 0 | 1.020474 | 0.598782 |
| GTEx_Tissu | GTEX-QMR | 4/884 | 0.539997 | 0.95119 | 0 | 0 | 1.042333 | 0.642278 |
| GTEx_Tissu | GTEX-QVJC | 2/439 | 0.572396 | 0.95119 | 0 | 0 | 1.048647 | 0.585066 |
| GTEx_Tissu | GTEX-T6MI | 5/1159 | 0.572594 | 0.95119 | 0 | 0 | 0.991197 | 0.55267 |
| GTEx_Tissu | GTEX-Q2A( | 5/1160 | 0.573374 | 0.95119 | 0 | 0 | 0.990286 | 0.550813 |
| GTEx_Tissu | GTEX-XLM4 | 1/195 | 0.574401 | 0.95119 | 0 | 0 | 1.181911 | 0.655284 |
| GTEx_Tissu | GTEX-UJM( | Dec-77 | 0.557046 | 0.95119 | 0 | 0 | 0.992289 | 0.580596 |
| GTEx_Tissu | GTEX-WWY | 5/1224 | 0.621779 | 0.95119 | 0 | 0 | 0.935093 | 0.444328 |
| GTEx_Tissu | GTEX-U412 | 1/215 | 0.610289 | 0.95119 | 0 | 0 | 1.070365 | 0.528571 |
| GTEx_Tissu | GTEX-X4EC | 1/216 | 0.612003 | 0.95119 | 0 | 0 | 1.065333 | 0.523098 |
| GTEx_Tissu | GTEX-RNOI | Sep-71 | 0.612018 | 0.95119 | 0 | 0 | 0.94736 | 0.465147 |
| GTEx_Tissu | GTEX-NFK9 | 42/9831 | 0.606818 | 0.95119 | 0 | 0 | 0.965274 | 0.48218 |
| GTEx_Tissu | GTEX-N7M | 2/481 | 0.622453 | 0.95119 | 0 | 0 | 0.954636 | 0.452581 |
| GTEx_Tissu | GTEX-PVO\ | 7/1710 | 0.622762 | 0.95119 | 0 | 0 | 0.935628 | 0.443105 |
| GTEx_Tissu | GTEX-WEY! | 1/213 | 0.606839 | 0.95119 | 0 | 0 | 1.080573 | 0.539737 |
| GTEx_Tissu | GTEX-RM2 | 1/213 | 0.606839 | 0.95119 | 0 | 0 | 1.080573 | 0.539737 |
| GTEx_Tissu | GTEX-P44H | 3/718 | 0.608788 | 0.95119 | 0 | 0 | 0.958941 | 0.475908 |
| GTEx_Tissu | GTEX-OXRI | 14/3348 | 0.608974 | 0.95119 | 0 | 0 | 0.95367 | 0.473001 |
| GTEx_Tissu | GTEX-Q2A( | 4/964 | 0.609326 | 0.95119 | 0 | 0 | 0.951456 | 0.471353 |
| GTEx_Tissu | GTEX-XQ8I | 2/481 | 0.622453 | 0.95119 | 0 | 0 | 0.954636 | 0.452581 |
| GTEx_Tissu | GTEX-T6MI | 4/979 | 0.621616 | 0.95119 | 0 | 0 | 0.936077 | 0.445042 |
| GTEx_Tissu | GTEX-XMD | 1/186 | 0.557204 | 0.95119 | 0 | 0 | 1.239975 | 0.725168 |
| GTEx_Tissu | GTEX-PLZ6 | 1/222 | 0.622129 | 0.95119 | 0 | 0 | 1.036094 | 0.491738 |
| GTEx_Tissu | GTEX-UPK5 | 7/1709 | 0.622134 | 0.95119 | 0 | 0 | 0.936229 | 0.444334 |
| GTEx_Tissu | GTEX-NPJ8 | 3/669 | 0.560403 | 0.95119 | 0 | 0 | 1.032121 | 0.5977 |
| GTEx_Tissu | GTEX-N7M | 2/430 | 0.561074 | 0.95119 | 0 | 0 | 1.071193 | 0.619045 |
| GTEx_Tissu | GTEX-WHP | 16/3705 | 0.556492 | 0.95119 | 0 | 0 | 0.991085 | 0.580878 |
| GTEx_Tissu | GTEX-U3ZI | 4/976 | 0.619177 | 0.95119 | 0 | 0 | 0.939114 | 0.450178 |
| GTEx_Tissu | GTEX-RWS. | 2/479 | 0.620173 | 0.95119 | 0 | 0 | 0.958737 | 0.458043 |
| GTEx_Tissu | GTEX-U4B1 | 2/479 | 0.620173 | 0.95119 | 0 | 0 | 0.958737 | 0.458043 |

| | | | | | | | | |
|---|---|---|---|---|---|---|---|---|
| GTEx_Tissu | GTEX-R53T | Oct-24 | 0.620267 | 0.95119 | 0 | 0 | 0.941424 | 0.449629 |
| GTEx_Tissu | GTEX-S7SF- | 1/221 | 0.62046 | 0.95119 | 0 | 0 | 1.040856 | 0.496795 |
| GTEx_Tissu | GTEX-SIU8- | 1/218 | 0.615408 | 0.95119 | 0 | 0 | 1.055407 | 0.512369 |
| GTEx_Tissu | GTEX-WFG | 1/218 | 0.615408 | 0.95119 | 0 | 0 | 1.055407 | 0.512369 |
| GTEx_Tissu | GTEX-QEL4 | 6/1367 | 0.55145 | 0.95119 | 0 | 0 | 1.009715 | 0.600987 |
| GTEx_Tissu | GTEX-WFO | 2/443 | 0.577361 | 0.95119 | 0 | 0 | 1.038922 | 0.570668 |
| GTEx_Tissu | GTEX-O5YV | 8/1838 | 0.553412 | 0.95119 | 0 | 0 | 1.00065 | 0.592037 |
| GTEx_Tissu | GTEX-T6M( | 2/425 | 0.554694 | 0.95119 | 0 | 0 | 1.084133 | 0.638922 |
| GTEx_Tissu | GTEX-XXEK | 8/1857 | 0.565484 | 0.95119 | 0 | 0 | 0.989327 | 0.563989 |
| GTEx_Tissu | GTEX-QMR | Dec-28 | 0.583941 | 0.95119 | 0 | 0 | 0.97142 | 0.52258 |
| GTEx_Tissu | GTEX-NPJ7 | 3/688 | 0.579577 | 0.95119 | 0 | 0 | 1.002503 | 0.546821 |
| GTEx_Tissu | GTEX-XPVG | 6/1414 | 0.585254 | 0.95119 | 0 | 0 | 0.973537 | 0.521534 |
| GTEx_Tissu | GTEX-UJM( | 1/201 | 0.585497 | 0.95119 | 0 | 0 | 1.146105 | 0.613503 |
| GTEx_Tissu | GTEX-RWS( | 1/201 | 0.585497 | 0.95119 | 0 | 0 | 1.146105 | 0.613503 |
| GTEx_Tissu | GTEX-UPIC- | 35/8067 | 0.548744 | 0.95119 | 0 | 0 | 0.995621 | 0.597495 |
| GTEx_Tissu | GTEX-POYV | 20/4619 | 0.551143 | 0.95119 | 0 | 0 | 0.993986 | 0.592178 |
| GTEx_Tissu | GTEX-PWO | 1/202 | 0.587319 | 0.95119 | 0 | 0 | 1.140345 | 0.606877 |
| GTEx_Tissu | GTEX-R55C | 4/931 | 0.581475 | 0.95119 | 0 | 0 | 0.987042 | 0.535161 |
| GTEx_Tissu | GTEX-N7M | 5/1164 | 0.576489 | 0.95119 | 0 | 0 | 0.986658 | 0.54345 |
| GTEx_Tissu | GTEX-WEY! | 1/179 | 0.543355 | 0.95119 | 0 | 0 | 1.289195 | 0.7864 |
| GTEx_Tissu | GTEX-RVPL | 4/889 | 0.544504 | 0.95119 | 0 | 0 | 1.036172 | 0.629867 |
| GTEx_Tissu | GTEX-O5YV | 1/184 | 0.55329 | 0.95119 | 0 | 0 | 1.253654 | 0.742005 |
| GTEx_Tissu | GTEX-SJXC- | 3/683 | 0.574581 | 0.95119 | 0 | 0 | 1.010137 | 0.55973 |
| GTEx_Tissu | GTEX-UJM( | 1/197 | 0.578132 | 0.95119 | 0 | 0 | 1.169732 | 0.640958 |
| GTEx_Tissu | GTEX-UJHI- | 8/1878 | 0.578671 | 0.95119 | 0 | 0 | 0.97708 | 0.534484 |
| GTEx_Tissu | GTEX-WOF | Sep-60 | 0.546417 | 0.95119 | 0 | 0 | 1.004876 | 0.60732 |
| GTEx_Tissu | GTEX-RVPL | 22/5127 | 0.57065 | 0.95119 | 0 | 0 | 0.981771 | 0.550754 |
| GTEx_Tissu | GTEX-NL3H | 20/4667 | 0.572082 | 0.95119 | 0 | 0 | 0.980636 | 0.547659 |
| GTEx_Tissu | GTEX-WK1 | 2/420 | 0.54825 | 0.95119 | 0 | 0 | 1.097382 | 0.659553 |
| GTEx_Tissu | GTEX-PWN | 8/1830 | 0.548292 | 0.95119 | 0 | 0 | 1.005488 | 0.604246 |
| GTEx_Tissu | GTEX-RWS( | 13/3020 | 0.562391 | 0.95119 | 0 | 0 | 0.987686 | 0.56847 |
| GTEx_Tissu | GTEX-RU72 | 1/179 | 0.543355 | 0.95119 | 0 | 0 | 1.289195 | 0.7864 |
| GTEx_Tissu | GTEX-R53T | 1/180 | 0.545359 | 0.95119 | 0 | 0 | 1.281928 | 0.777247 |
| GTEx_Tissu | GTEX-XUJ4 | 2/418 | 0.545655 | 0.95119 | 0 | 0 | 1.102771 | 0.668025 |
| GTEx_Tissu | GTEX-WHS | 3/655 | 0.545952 | 0.95119 | 0 | 0 | 1.05505 | 0.638541 |
| GTEx_Tissu | GTEX-OIZI- | 6/1360 | 0.546315 | 0.95119 | 0 | 0 | 1.015318 | 0.61382 |
| GTEx_Tissu | GTEX-XMD | 8/1827 | 0.546366 | 0.95119 | 0 | 0 | 1.007314 | 0.608888 |
| GTEx_Tissu | GTEX-S7PN | 19/4417 | 0.564104 | 0.95119 | 0 | 0 | 0.985692 | 0.564326 |
| GTEx_Tissu | GTEX-XQ8I | 1/190 | 0.56493 | 0.95119 | 0 | 0 | 1.213486 | 0.692965 |
| GTEx_Tissu | GTEX-T8EN | Dec-92 | 0.565018 | 0.95119 | 0 | 0 | 0.986072 | 0.562946 |
| GTEx_Tissu | GTEX-U412 | 4/913 | 0.565832 | 0.95119 | 0 | 0 | 1.007542 | 0.573753 |
| GTEx_Tissu | GTEX-WYJK | 17/3957 | 0.565845 | 0.95119 | 0 | 0 | 0.984558 | 0.560642 |
| GTEx_Tissu | GTEX-XOT4 | 2/423 | 0.552124 | 0.95119 | 0 | 0 | 1.089395 | 0.647082 |
| GTEx_Tissu | GTEX-QEG! | 2/423 | 0.552124 | 0.95119 | 0 | 0 | 1.089395 | 0.647082 |
| GTEx_Tissu | GTEX-QVU! | 7/1635 | 0.574209 | 0.95119 | 0 | 0 | 0.982763 | 0.545199 |

| | | | | | | | | |
|---|---|---|---|---|---|---|---|---|
| GTEx_Tissu | GTEX-UJM( | 1/204 | 0.590938 | 0.95119 | 0 | 0 | 1.128995 | 0.593902 |
| GTEx_Tissu | GTEX-QDV. | 1/186 | 0.557204 | 0.95119 | 0 | 0 | 1.239975 | 0.725168 |
| GTEx_Tissu | GTEX-WY7( | 1/186 | 0.557204 | 0.95119 | 0 | 0 | 1.239975 | 0.725168 |
| GTEx_Tissu | GTEX-POM | Sep-81 | 0.559141 | 0.95119 | 0 | 0 | 0.993522 | 0.577587 |
| GTEx_Tissu | GTEX-O5YV | 3/669 | 0.560403 | 0.95119 | 0 | 0 | 1.032121 | 0.5977 |
| GTEx_Tissu | GTEX-SNM | 1/189 | 0.563011 | 0.95119 | 0 | 0 | 1.220002 | 0.700838 |
| GTEx_Tissu | GTEX-W5X | 1/189 | 0.563011 | 0.95119 | 0 | 0 | 1.220002 | 0.700838 |
| GTEx_Tissu | GTEX-U8XE | 3/722 | 0.612582 | 0.95119 | 0 | 0 | 0.953408 | 0.467239 |
| GTEx_Tissu | GTEX-SE5C | 7/1654 | 0.586774 | 0.95119 | 0 | 0 | 0.970416 | 0.517344 |
| GTEx_Tissu | GTEX-QEL4 | 7/1601 | 0.551317 | 0.95119 | 0 | 0 | 1.005591 | 0.598774 |
| GTEx_Tissu | GTEX-PWO | 1/206 | 0.594525 | 0.95119 | 0 | 0 | 1.117867 | 0.581282 |
| GTEx_Tissu | GTEX-U8XE | 1/196 | 0.576271 | 0.95119 | 0 | 0 | 1.17579 | 0.64807 |
| GTEx_Tissu | GTEX-NL3H | Oct-45 | 0.576489 | 0.95119 | 0 | 0 | 0.977669 | 0.538499 |
| GTEx_Tissu | GTEX-PVO\ | 2/447 | 0.582284 | 0.95119 | 0 | 0 | 1.029372 | 0.556682 |
| GTEx_Tissu | GTEX-T8EN | 1/198 | 0.579985 | 0.95119 | 0 | 0 | 1.163735 | 0.633948 |
| GTEx_Tissu | GTEX-WHS | 4/929 | 0.579752 | 0.95119 | 0 | 0 | 0.98928 | 0.53931 |
| GTEx_Tissu | GTEX-V955 | 5/1168 | 0.579593 | 0.95119 | 0 | 0 | 0.983055 | 0.536187 |
| GTEx_Tissu | GTEX-R55D | 1/208 | 0.598082 | 0.95119 | 0 | 0 | 1.106954 | 0.569005 |
| GTEx_Tissu | GTEX-OOBI | 1/213 | 0.606839 | 0.95119 | 0 | 0 | 1.080573 | 0.539737 |
| GTEx_Tissu | GTEX-PLZ6 | 1/200 | 0.583668 | 0.95119 | 0 | 0 | 1.151922 | 0.620221 |
| GTEx_Tissu | GTEX-XUZC | 2/448 | 0.583508 | 0.95119 | 0 | 0 | 1.027011 | 0.553248 |
| GTEx_Tissu | GTEX-POY\ | 4/933 | 0.583194 | 0.95119 | 0 | 0 | 0.984813 | 0.531046 |
| GTEx_Tissu | GTEX-S33H | 3/689 | 0.580572 | 0.95119 | 0 | 0 | 1.000989 | 0.544279 |
| GTEx_Tissu | GTEX-PLZ5 | 1/199 | 0.581831 | 0.95119 | 0 | 0 | 1.157799 | 0.627036 |
| GTEx_Tissu | GTEX-R55C | 3/690 | 0.581566 | 0.95119 | 0 | 0 | 0.99948 | 0.541749 |
| GTEx_Tissu | GTEX-NL3H | Oct-43 | 0.575355 | 0.95119 | 0 | 0 | 0.978618 | 0.540949 |
| GTEx_Tissu | GTEX-U8XE | 27/6382 | 0.609424 | 0.95119 | 0 | 0 | 0.960047 | 0.475454 |
| GTEx_Tissu | GTEX-RVPU | 5/1215 | 0.615164 | 0.95119 | 0 | 0 | 0.942502 | 0.45793 |
| GTEx_Tissu | GTEX-S7SE | 7/1697 | 0.614553 | 0.95119 | 0 | 0 | 0.943499 | 0.459351 |
| GTEx_Tissu | GTEX-WZT( | 5/1214 | 0.614425 | 0.95119 | 0 | 0 | 0.943332 | 0.459467 |
| GTEx_Tissu | GTEX-P4PP | 21/4998 | 0.613691 | 0.95119 | 0 | 0 | 0.954865 | 0.466226 |
| GTEx_Tissu | GTEX-RU72 | 17/4040 | 0.603564 | 0.95119 | 0 | 0 | 0.959234 | 0.484321 |
| GTEx_Tissu | GTEX-S7SF | 1/223 | 0.623791 | 0.95119 | 0 | 0 | 1.031374 | 0.486746 |
| GTEx_Tissu | GTEX-REY6 | 2/482 | 0.623589 | 0.95119 | 0 | 0 | 0.952598 | 0.449878 |
| GTEx_Tissu | GTEX-QESD | 4/981 | 0.623237 | 0.95119 | 0 | 0 | 0.934062 | 0.441652 |
| GTEx_Tissu | GTEX-WFJC | 4/981 | 0.623237 | 0.95119 | 0 | 0 | 0.934062 | 0.441652 |
| GTEx_Tissu | GTEX-R55F | Aug-44 | 0.618931 | 0.95119 | 0 | 0 | 0.940318 | 0.451128 |
| GTEx_Tissu | GTEX-PWC | 1/220 | 0.618783 | 0.95119 | 0 | 0 | 1.045662 | 0.501918 |
| GTEx_Tissu | GTEX-VJWI | 15/3575 | 0.605207 | 0.95119 | 0 | 0 | 0.956987 | 0.480584 |
| GTEx_Tissu | GTEX-XBED | 1/212 | 0.605103 | 0.95119 | 0 | 0 | 1.085749 | 0.545433 |
| GTEx_Tissu | GTEX-XPT6 | 2/473 | 0.613272 | 0.95119 | 0 | 0 | 0.97125 | 0.47489 |
| GTEx_Tissu | GTEX-PWN | 1/211 | 0.603359 | 0.95119 | 0 | 0 | 1.090975 | 0.551207 |
| GTEx_Tissu | GTEX-WEY! | 7/1678 | 0.602395 | 0.95119 | 0 | 0 | 0.955221 | 0.484146 |
| GTEx_Tissu | GTEX-QVU! | 1/221 | 0.62046 | 0.95119 | 0 | 0 | 1.040856 | 0.496795 |
| GTEx_Tissu | GTEX-R55D | 1/221 | 0.62046 | 0.95119 | 0 | 0 | 1.040856 | 0.496795 |

| | | | | | | | | |
|---|---|---|---|---|---|---|---|---|
| GTEx_Tissu | GTEX-XBEV | 6/1472 | 0.625183 | 0.95119 | 0 | 0 | 0.93209 | 0.437813 |
| GTEx_Tissu | GTEX-UPK5 | 1/197 | 0.578132 | 0.95119 | 0 | 0 | 1.169732 | 0.640958 |
| GTEx_Tissu | GTEX-PLZ6 | Sep-18 | 0.581224 | 0.95119 | 0 | 0 | 0.974067 | 0.528547 |
| GTEx_Tissu | GTEX-QMR | 8/1882 | 0.581163 | 0.95119 | 0 | 0 | 0.974778 | 0.529036 |
| GTEx_Tissu | GTEX-R55D | 3/671 | 0.562446 | 0.95119 | 0 | 0 | 1.028924 | 0.592106 |
| GTEx_Tissu | GTEX-XBEC | 2/449 | 0.58473 | 0.95119 | 0 | 0 | 1.024661 | 0.549838 |
| GTEx_Tissu | GTEX-PWC | 2/454 | 0.5908 | 0.95119 | 0 | 0 | 1.013066 | 0.533155 |
| GTEx_Tissu | GTEX-P78B | 1/193 | 0.570637 | 0.95119 | 0 | 0 | 1.194344 | 0.670029 |
| GTEx_Tissu | GTEX-SUCS | 4/918 | 0.570208 | 0.95119 | 0 | 0 | 1.001766 | 0.562746 |
| GTEx_Tissu | GTEX-W5W | 4/917 | 0.569335 | 0.95119 | 0 | 0 | 1.002916 | 0.56493 |
| GTEx_Tissu | GTEX-WRH | 37/8581 | 0.569254 | 0.95119 | 0 | 0 | 0.984675 | 0.554793 |
| GTEx_Tissu | GTEX-WL4 | 6/1391 | 0.568862 | 0.95119 | 0 | 0 | 0.990935 | 0.559003 |
| GTEx_Tissu | GTEX-O5YT | 1/192 | 0.568743 | 0.95119 | 0 | 0 | 1.200657 | 0.677563 |
| GTEx_Tissu | GTEX-NL3H | Oct-28 | 0.566811 | 0.95119 | 0 | 0 | 0.985792 | 0.559663 |
| GTEx_Tissu | GTEX-WY7 | 14/3317 | 0.594035 | 0.95119 | 0 | 0 | 0.96442 | 0.502286 |
| GTEx_Tissu | GTEX-XAJ8 | 17/4034 | 0.600878 | 0.95119 | 0 | 0 | 0.96103 | 0.489513 |
| GTEx_Tissu | GTEX-XBED | 6/1436 | 0.600644 | 0.95119 | 0 | 0 | 0.95742 | 0.488047 |
| GTEx_Tissu | GTEX-PLZ6 | 2/462 | 0.600376 | 0.95119 | 0 | 0 | 0.995038 | 0.507667 |
| GTEx_Tissu | GTEX-QEG | 22/5194 | 0.598521 | 0.95119 | 0 | 0 | 0.964668 | 0.495157 |
| GTEx_Tissu | GTEX-PWC | 7/1672 | 0.598517 | 0.95119 | 0 | 0 | 0.958979 | 0.492245 |
| GTEx_Tissu | GTEX-XUYS | 1/208 | 0.598082 | 0.95119 | 0 | 0 | 1.106954 | 0.569005 |
| GTEx_Tissu | GTEX-XXEK | 1/201 | 0.585497 | 0.95119 | 0 | 0 | 1.146105 | 0.613503 |
| GTEx_Tissu | GTEX-WFJ | 8/1892 | 0.587365 | 0.95119 | 0 | 0 | 0.969067 | 0.515649 |
| GTEx_Tissu | GTEX-UTH | 6/1454 | 0.613017 | 0.95119 | 0 | 0 | 0.944598 | 0.462251 |
| GTEx_Tissu | GTEX-S32V | 1/213 | 0.606839 | 0.95119 | 0 | 0 | 1.080573 | 0.539737 |
| GTEx_Tissu | GTEX-QCQ | 3/706 | 0.597263 | 0.95119 | 0 | 0 | 0.97592 | 0.502987 |
| GTEx_Tissu | GTEX-T2IS- | 4/941 | 0.59003 | 0.95119 | 0 | 0 | 0.975994 | 0.514916 |
| GTEx_Tissu | GTEX-V1D1 | 1/203 | 0.589132 | 0.95119 | 0 | 0 | 1.134642 | 0.600344 |
| GTEx_Tissu | GTEX-XQ8I | 1/203 | 0.589132 | 0.95119 | 0 | 0 | 1.134642 | 0.600344 |
| GTEx_Tissu | GTEX-QDV. | 1/203 | 0.589132 | 0.95119 | 0 | 0 | 1.134642 | 0.600344 |
| GTEx_Tissu | GTEX-XOT | 8/1893 | 0.587983 | 0.95119 | 0 | 0 | 0.968499 | 0.514328 |
| GTEx_Tissu | GTEX-S7SE | 4/965 | 0.610152 | 0.95119 | 0 | 0 | 0.950416 | 0.469549 |
| GTEx_Tissu | GTEX-XQ3S | 3/719 | 0.609739 | 0.95119 | 0 | 0 | 0.957552 | 0.473725 |
| GTEx_Tissu | GTEX-UJHI | 3/690 | 0.581566 | 0.95119 | 0 | 0 | 0.99948 | 0.541749 |
| GTEx_Tissu | GTEX-OIZH | 25/5813 | 0.567489 | 0.95119 | 0 | 0 | 0.98403 | 0.557485 |
| GTEx_Tissu | GTEX-U8XE | 2/435 | 0.56739 | 0.95119 | 0 | 0 | 1.058552 | 0.599891 |
| GTEx_Tissu | GTEX-OIZG | 14/3262 | 0.567017 | 0.95119 | 0 | 0 | 0.983999 | 0.558287 |
| GTEx_Tissu | GTEX-P4QT | 1/191 | 0.566841 | 0.95119 | 0 | 0 | 1.207038 | 0.685208 |
| GTEx_Tissu | GTEX-RWS | 1/191 | 0.566841 | 0.95119 | 0 | 0 | 1.207038 | 0.685208 |
| GTEx_Tissu | GTEX-U4B1 | 4/944 | 0.592577 | 0.95119 | 0 | 0 | 0.972725 | 0.509001 |
| GTEx_Tissu | GTEX-X4EC | 1/207 | 0.596308 | 0.95119 | 0 | 0 | 1.112384 | 0.575101 |
| GTEx_Tissu | GTEX-Q2AI | 4/949 | 0.596803 | 0.95119 | 0 | 0 | 0.967323 | 0.499302 |
| GTEx_Tissu | GTEX-NL4V | 13/3085 | 0.595326 | 0.95119 | 0 | 0 | 0.963071 | 0.499494 |
| GTEx_Tissu | GTEX-QVJC | 4/947 | 0.595116 | 0.95119 | 0 | 0 | 0.969477 | 0.503158 |
| GTEx_Tissu | GTEX-N7M | 1/202 | 0.587319 | 0.95119 | 0 | 0 | 1.140345 | 0.606877 |

| | | | | | | | | |
|---|---|---|---|---|---|---|---|---|
| GTEx_Tissu | GTEX-OOBI | 2/459 | 0.596805 | 0.95119 | 0 | 0 | 1.001725 | 0.517056 |
| GTEx_Tissu | GTEX-WHS | 19/4492 | 0.596967 | 0.95119 | 0 | 0 | 0.96448 | 0.497568 |
| GTEx_Tissu | GTEX-UJHI- | Sep-72 | 0.612589 | 0.95119 | 0 | 0 | 0.946869 | 0.464023 |
| GTEx_Tissu | GTEX-QDVI | 4/983 | 0.624853 | 0.95119 | 0 | 0 | 0.932055 | 0.438288 |
| GTEx_Tissu | GTEX-XQ8I | 1/204 | 0.590938 | 0.95119 | 0 | 0 | 1.128995 | 0.593902 |
| GTEx_Tissu | GTEX-TKQ2 | 22/5178 | 0.591919 | 0.95119 | 0 | 0 | 0.968712 | 0.507978 |
| GTEx_Tissu | GTEX-OXRI | 15/3548 | 0.592504 | 0.95119 | 0 | 0 | 0.965893 | 0.505546 |
| GTEx_Tissu | GTEX-OHPI | 6/1425 | 0.592985 | 0.95119 | 0 | 0 | 0.965416 | 0.504513 |
| GTEx_Tissu | GTEX-T2IS- | 1/202 | 0.587319 | 0.95119 | 0 | 0 | 1.140345 | 0.606877 |
| GTEx_Tissu | GTEX-OOB. | 1/215 | 0.610289 | 0.95119 | 0 | 0 | 1.070365 | 0.528571 |
| GTEx_Tissu | GTEX-XXEK | 1/210 | 0.601608 | 0.95119 | 0 | 0 | 1.09625 | 0.557059 |
| GTEx_Tissu | GTEX-RWS( | 1/210 | 0.601608 | 0.95119 | 0 | 0 | 1.09625 | 0.557059 |
| GTEx_Tissu | GTEX-WQL | 17/4053 | 0.60936 | 0.95119 | 0 | 0 | 0.955362 | 0.473235 |
| GTEx_Tissu | GTEX-O5YL | Dec-83 | 0.612215 | 0.95119 | 0 | 0 | 0.949746 | 0.466013 |
| GTEx_Tissu | GTEX-P4QS | 3/727 | 0.61729 | 0.95119 | 0 | 0 | 0.946577 | 0.456644 |
| GTEx_Tissu | GTEX-S4P3 | 1/219 | 0.617099 | 0.95119 | 0 | 0 | 1.050512 | 0.507109 |
| GTEx_Tissu | GTEX-RTLS | 1/219 | 0.617099 | 0.95119 | 0 | 0 | 1.050512 | 0.507109 |
| GTEx_Tissu | GTEX-T5JC- | 22/5238 | 0.616483 | 0.95119 | 0 | 0 | 0.953675 | 0.461316 |
| GTEx_Tissu | GTEX-P4QT | Aug-39 | 0.615948 | 0.95119 | 0 | 0 | 0.943015 | 0.456978 |
| GTEx_Tissu | GTEX-P4QT | 4/945 | 0.593424 | 0.95119 | 0 | 0 | 0.97164 | 0.507046 |
| GTEx_Tissu | GTEX-QCQ( | 34/8051 | 0.628093 | 0.951981 | 0 | 0 | 0.951902 | 0.442698 |
| GTEx_Tissu | GTEX-X8H( | 4/989 | 0.629677 | 0.952772 | 0 | 0 | 0.926084 | 0.428358 |
| GTEx_Tissu | GTEX-XXEK | 1/227 | 0.630368 | 0.952772 | 0 | 0 | 1.012914 | 0.46741 |
| GTEx_Tissu | GTEX-QMR | 5/1234 | 0.629053 | 0.952772 | 0 | 0 | 0.926988 | 0.429695 |
| GTEx_Tissu | GTEX-WRH | 13/3157 | 0.630603 | 0.952772 | 0 | 0 | 0.936993 | 0.432028 |
| GTEx_Tissu | GTEX-REY6 | 25/5973 | 0.630756 | 0.952772 | 0 | 0 | 0.946713 | 0.436279 |
| GTEx_Tissu | GTEX-WL4( | Aug-66 | 0.631917 | 0.953877 | 0 | 0 | 0.928615 | 0.426232 |
| GTEx_Tissu | GTEX-XGQ4 | Sep-08 | 0.632866 | 0.954662 | 0 | 0 | 0.929478 | 0.425233 |
| GTEx_Tissu | GTEX-XMD | Dec-11 | 0.124328 | 0.954721 | 0 | 0 | 1.517767 | 3.164293 |
| GTEx_Tissu | GTEX-WYB. | 8/1142 | 0.123327 | 0.954721 | 0 | 0 | 1.676958 | 3.509736 |
| GTEx_Tissu | GTEX-S4P3 | 5/611 | 0.127384 | 0.954721 | 0 | 0 | 1.942667 | 4.002958 |
| GTEx_Tissu | GTEX-X4EC | 6/786 | 0.127351 | 0.954721 | 0 | 0 | 1.816999 | 3.744493 |
| GTEx_Tissu | GTEX-RVPL | 8/1149 | 0.12646 | 0.954721 | 0 | 0 | 1.666049 | 3.445099 |
| GTEx_Tissu | GTEX-PLZ5 | 6/786 | 0.127351 | 0.954721 | 0 | 0 | 1.816999 | 3.744493 |
| GTEx_Tissu | GTEX-W5W | 7/954 | 0.121302 | 0.954721 | 0 | 0 | 1.752402 | 3.696639 |
| GTEx_Tissu | GTEX-X261 | 5/610 | 0.126753 | 0.954721 | 0 | 0 | 1.945979 | 4.019454 |
| GTEx_Tissu | GTEX-U3ZH | 8/1150 | 0.126912 | 0.954721 | 0 | 0 | 1.664502 | 3.435971 |
| GTEx_Tissu | GTEX-U3ZG | 20/3545 | 0.126995 | 0.954721 | 0 | 0 | 1.387784 | 2.863843 |
| GTEx_Tissu | GTEX-SJXC- | 18/3112 | 0.121971 | 0.954721 | 0 | 0 | 1.418088 | 2.983615 |
| GTEx_Tissu | GTEX-RU72 | 7/957 | 0.122755 | 0.954721 | 0 | 0 | 1.746592 | 3.663584 |
| GTEx_Tissu | GTEX-VUS( | 5/609 | 0.126123 | 0.954721 | 0 | 0 | 1.949301 | 4.036031 |
| GTEx_Tissu | GTEX-OHPI | 5/607 | 0.124867 | 0.954721 | 0 | 0 | 1.95598 | 4.069431 |
| GTEx_Tissu | GTEX-W5W | Dec-02 | 0.121242 | 0.954721 | 0 | 0 | 1.525757 | 3.219302 |
| GTEx_Tissu | GTEX-OHPI | 9/1328 | 0.122759 | 0.954721 | 0 | 0 | 1.626582 | 3.411814 |
| GTEx_Tissu | GTEX-XUJ4 | 8/1143 | 0.123772 | 0.954721 | 0 | 0 | 1.675392 | 3.500422 |

| | | | | | | | | |
|---|---|---|---|---|---|---|---|---|
| GTEx_Tissu | GTEX-REY6 | 4/439 | 0.124437 | 0.954721 | 0 | 0 | 2.157928 | 4.497022 |
| GTEx_Tissu | GTEX-PSDG | 8/1142 | 0.123327 | 0.954721 | 0 | 0 | 1.676958 | 3.509736 |
| GTEx_Tissu | GTEX-P4QT | 7/964 | 0.126181 | 0.954721 | 0 | 0 | 1.733177 | 3.587737 |
| GTEx_Tissu | GTEX-PSDG | 7/965 | 0.126675 | 0.954721 | 0 | 0 | 1.731276 | 3.577046 |
| GTEx_Tissu | GTEX-P44H | 16/3873 | 0.633752 | 0.955351 | 0 | 0 | 0.9381 | 0.427865 |
| GTEx_Tissu | GTEX-OHPI | 8/1152 | 0.127816 | 0.955486 | 0 | 0 | 1.661415 | 3.417796 |
| GTEx_Tissu | GTEX-OOBI | 7/1740 | 0.64135 | 0.955613 | 0 | 0 | 0.917917 | 0.40772 |
| GTEx_Tissu | GTEX-UJHI- | 17/4133 | 0.644256 | 0.955613 | 0 | 0 | 0.932073 | 0.409795 |
| GTEx_Tissu | GTEX-NPJ8 | 7/1741 | 0.641961 | 0.955613 | 0 | 0 | 0.917337 | 0.406589 |
| GTEx_Tissu | GTEX-U3ZN | 1/239 | 0.649424 | 0.955613 | 0 | 0 | 0.961257 | 0.414945 |
| GTEx_Tissu | GTEX-SSA3 | 2/500 | 0.643593 | 0.955613 | 0 | 0 | 0.917316 | 0.404251 |
| GTEx_Tissu | GTEX-SUCS | 7/1729 | 0.634593 | 0.955613 | 0 | 0 | 0.924339 | 0.420363 |
| GTEx_Tissu | GTEX-XPVG | 15/3653 | 0.64097 | 0.955613 | 0 | 0 | 0.932002 | 0.41453 |
| GTEx_Tissu | GTEX-OHPI | Sep-30 | 0.644989 | 0.955613 | 0 | 0 | 0.919129 | 0.403059 |
| GTEx_Tissu | GTEX-X4EC | 19/4604 | 0.644274 | 0.955613 | 0 | 0 | 0.934095 | 0.410657 |
| GTEx_Tissu | GTEX-RU1J | 2/503 | 0.646846 | 0.955613 | 0 | 0 | 0.911683 | 0.397172 |
| GTEx_Tissu | GTEX-S7SE- | 7/1752 | 0.648642 | 0.955613 | 0 | 0 | 0.911003 | 0.394349 |
| GTEx_Tissu | GTEX-S341 | 13/3190 | 0.646289 | 0.955613 | 0 | 0 | 0.925435 | 0.40396 |
| GTEx_Tissu | GTEX-QDT8 | 4/1009 | 0.645478 | 0.955613 | 0 | 0 | 0.906695 | 0.396919 |
| GTEx_Tissu | GTEX-XOTC | 6/1492 | 0.638452 | 0.955613 | 0 | 0 | 0.918548 | 0.412161 |
| GTEx_Tissu | GTEX-R53T | 19/4591 | 0.638908 | 0.955613 | 0 | 0 | 0.937545 | 0.420016 |
| GTEx_Tissu | GTEX-QVUS | 6/1493 | 0.639108 | 0.955613 | 0 | 0 | 0.917881 | 0.410918 |
| GTEx_Tissu | GTEX-WZTC | 5/1249 | 0.63981 | 0.955613 | 0 | 0 | 0.915075 | 0.408658 |
| GTEx_Tissu | GTEX-TML8 | 1/233 | 0.640021 | 0.955613 | 0 | 0 | 0.986417 | 0.440193 |
| GTEx_Tissu | GTEX-QESD | 2/504 | 0.647925 | 0.955613 | 0 | 0 | 0.90982 | 0.394844 |
| GTEx_Tissu | GTEX-P44H | 23/5550 | 0.647543 | 0.955613 | 0 | 0 | 0.935402 | 0.406498 |
| GTEx_Tissu | GTEX-R55C | 26/6250 | 0.647483 | 0.955613 | 0 | 0 | 0.937445 | 0.407473 |
| GTEx_Tissu | GTEX-S4P3 | Aug-97 | 0.649817 | 0.955613 | 0 | 0 | 0.912563 | 0.393373 |
| GTEx_Tissu | GTEX-OXRK | 19/4616 | 0.649196 | 0.955613 | 0 | 0 | 0.930927 | 0.402179 |
| GTEx_Tissu | GTEX-S32W | 3/762 | 0.649181 | 0.955613 | 0 | 0 | 0.90128 | 0.389392 |
| GTEx_Tissu | GTEX-XOT4 | 17/4135 | 0.64511 | 0.955613 | 0 | 0 | 0.931503 | 0.40831 |
| GTEx_Tissu | GTEX-WYJK | 1/237 | 0.646317 | 0.955613 | 0 | 0 | 0.969501 | 0.423154 |
| GTEx_Tissu | GTEX-R55E | 3/755 | 0.642953 | 0.955613 | 0 | 0 | 0.910002 | 0.401932 |
| GTEx_Tissu | GTEX-TSE9 | 1/232 | 0.63843 | 0.955613 | 0 | 0 | 0.990738 | 0.444588 |
| GTEx_Tissu | GTEX-XXEK | 13/3171 | 0.637297 | 0.955613 | 0 | 0 | 0.93206 | 0.419912 |
| GTEx_Tissu | GTEX-REY6 | 2/494 | 0.637018 | 0.955613 | 0 | 0 | 0.92879 | 0.418845 |
| GTEx_Tissu | GTEX-RN64 | 24/5754 | 0.636142 | 0.955613 | 0 | 0 | 0.94294 | 0.426524 |
| GTEx_Tissu | GTEX-XBED | 1/232 | 0.63843 | 0.955613 | 0 | 0 | 0.990738 | 0.444588 |
| GTEx_Tissu | GTEX-XQ8I | 1/232 | 0.63843 | 0.955613 | 0 | 0 | 0.990738 | 0.444588 |
| GTEx_Tissu | GTEX-UPIC | 1/239 | 0.649424 | 0.955613 | 0 | 0 | 0.961257 | 0.414945 |
| GTEx_Tissu | GTEX-XPVG | 1/239 | 0.649424 | 0.955613 | 0 | 0 | 0.961257 | 0.414945 |
| GTEx_Tissu | GTEX-S95S | 3/747 | 0.635743 | 0.955613 | 0 | 0 | 0.920171 | 0.416801 |
| GTEx_Tissu | GTEX-U3ZN | Aug-73 | 0.636 | 0.955613 | 0 | 0 | 0.924946 | 0.418591 |
| Reactome_ | Infectious I | 2/961 | 0.926103 | 0.955977 | 0 | 0 | 0.465043 | 0.035701 |
| GTEx_Tissu | GTEX-X4XY | 1/240 | 0.650968 | 0.956042 | 0 | 0 | 0.957186 | 0.410916 |

| | | | | | | | | |
|---|---|---|---|---|---|---|---|---|
| GTEx_Tissu | GTEX-X585 | 1/240 | 0.650968 | 0.956042 | 0 | 0 | 0.957186 | 0.410916 |
| GTEx_Tissu | GTEX-RU72 | Sep-44 | 0.652593 | 0.956113 | 0 | 0 | 0.912648 | 0.38952 |
| GTEx_Tissu | GTEX-X4XX | 6/1514 | 0.652735 | 0.956113 | 0 | 0 | 0.904067 | 0.38566 |
| GTEx_Tissu | GTEX-WL4( | 7/1757 | 0.651656 | 0.956113 | 0 | 0 | 0.90815 | 0.388905 |
| GTEx_Tissu | GTEX-NL3H | 7/1758 | 0.652257 | 0.956113 | 0 | 0 | 0.907581 | 0.387824 |
| GTEx_Tissu | GTEX-NPJ7 | 2/509 | 0.653282 | 0.956284 | 0 | 0 | 0.900615 | 0.383434 |
| GTEx_Tissu | GTEX-PWO | 1/242 | 0.654034 | 0.956582 | 0 | 0 | 0.949146 | 0.403003 |
| GTEx_Tissu | GTEX-Q734 | 2/510 | 0.654345 | 0.956582 | 0 | 0 | 0.898796 | 0.381197 |
| GTEx_Tissu | GTEX-POM | Aug-10 | 0.657181 | 0.956955 | 0 | 0 | 0.90598 | 0.380327 |
| GTEx_Tissu | GTEX-U8XE | 3/771 | 0.657077 | 0.956955 | 0 | 0 | 0.890299 | 0.373885 |
| GTEx_Tissu | GTEX-SIU8- | Aug-09 | 0.656617 | 0.956955 | 0 | 0 | 0.906483 | 0.381316 |
| GTEx_Tissu | GTEX-PWO | 3/771 | 0.657077 | 0.956955 | 0 | 0 | 0.890299 | 0.373885 |
| GTEx_Tissu | GTEX-QDT8 | 1/244 | 0.657075 | 0.956955 | 0 | 0 | 0.941238 | 0.39528 |
| GTEx_Tissu | GTEX-S341 | 1/244 | 0.657075 | 0.956955 | 0 | 0 | 0.941238 | 0.39528 |
| GTEx_Tissu | GTEX-TKQ1 | 28/5250 | 0.12838 | 0.957228 | 0 | 0 | 1.335121 | 2.740679 |
| GTEx_Tissu | GTEX-P44H | 6/1523 | 0.658482 | 0.957731 | 0 | 0 | 0.898264 | 0.375311 |
| GTEx_Tissu | GTEX-T6MI | 2/514 | 0.658575 | 0.957731 | 0 | 0 | 0.89159 | 0.372397 |
| GTEx_Tissu | GTEX-QESD | 28/6793 | 0.67548 | 0.957813 | 0 | 0 | 0.922355 | 0.361869 |
| GTEx_Tissu | GTEX-OIZH | 3/859 | 0.727669 | 0.957813 | 0 | 0 | 0.795102 | 0.25277 |
| GTEx_Tissu | GTEX-WVLI | 6/1641 | 0.728408 | 0.957813 | 0 | 0 | 0.828089 | 0.262416 |
| GTEx_Tissu | GTEX-NPJ8 | 1/297 | 0.728682 | 0.957813 | 0 | 0 | 0.770624 | 0.243916 |
| GTEx_Tissu | GTEX-QVU! | 3/813 | 0.692263 | 0.957813 | 0 | 0 | 0.842284 | 0.309783 |
| GTEx_Tissu | GTEX-S4UY | 4/1082 | 0.699398 | 0.957813 | 0 | 0 | 0.842032 | 0.301057 |
| GTEx_Tissu | GTEX-XBEV | 1/274 | 0.699631 | 0.957813 | 0 | 0 | 0.836528 | 0.29881 |
| GTEx_Tissu | GTEX-W5W | 6/1591 | 0.70003 | 0.957813 | 0 | 0 | 0.856549 | 0.305473 |
| GTEx_Tissu | GTEX-U4B1 | 6/1592 | 0.700616 | 0.957813 | 0 | 0 | 0.855962 | 0.304548 |
| GTEx_Tissu | GTEX-UJM( | 2/549 | 0.693855 | 0.957813 | 0 | 0 | 0.833036 | 0.304468 |
| GTEx_Tissu | GTEX-QLQ7 | 1/270 | 0.694273 | 0.957813 | 0 | 0 | 0.84914 | 0.309842 |
| GTEx_Tissu | GTEX-PW2( | 1/256 | 0.674769 | 0.957813 | 0 | 0 | 0.896398 | 0.35263 |
| GTEx_Tissu | GTEX-V955 | 1/270 | 0.694273 | 0.957813 | 0 | 0 | 0.84914 | 0.309842 |
| GTEx_Tissu | GTEX-UTH( | 7/1795 | 0.674076 | 0.957813 | 0 | 0 | 0.88699 | 0.34984 |
| GTEx_Tissu | GTEX-WWY | 1/256 | 0.674769 | 0.957813 | 0 | 0 | 0.896398 | 0.35263 |
| GTEx_Tissu | GTEX-X4EP | 1/261 | 0.681872 | 0.957813 | 0 | 0 | 0.878936 | 0.336556 |
| GTEx_Tissu | GTEX-XV7C | 1/271 | 0.695622 | 0.957813 | 0 | 0 | 0.845952 | 0.307038 |
| GTEx_Tissu | GTEX-U8T8 | 2/531 | 0.676095 | 0.957813 | 0 | 0 | 0.862182 | 0.337476 |
| GTEx_Tissu | GTEX-X5EB | 25/6096 | 0.676775 | 0.957813 | 0 | 0 | 0.919363 | 0.358934 |
| GTEx_Tissu | GTEX-RWS/ | Nov-76 | 0.677349 | 0.957813 | 0 | 0 | 0.89763 | 0.349688 |
| GTEx_Tissu | GTEX-S7SF- | Sep-91 | 0.677468 | 0.957813 | 0 | 0 | 0.891475 | 0.347133 |
| GTEx_Tissu | GTEX-S33H | 1/262 | 0.683274 | 0.957813 | 0 | 0 | 0.875523 | 0.333451 |
| GTEx_Tissu | GTEX-S341 | 3/802 | 0.683312 | 0.957813 | 0 | 0 | 0.854372 | 0.325348 |
| GTEx_Tissu | GTEX-X4XX | 1/285 | 0.713889 | 0.957813 | 0 | 0 | 0.803677 | 0.270861 |
| GTEx_Tissu | GTEX-SIU7- | 2/534 | 0.679111 | 0.957813 | 0 | 0 | 0.857187 | 0.331706 |
| GTEx_Tissu | GTEX-PLZ5- | 23/5635 | 0.679684 | 0.957813 | 0 | 0 | 0.915791 | 0.353612 |
| GTEx_Tissu | GTEX-W5W | 17/4218 | 0.679705 | 0.957813 | 0 | 0 | 0.908301 | 0.350691 |
| GTEx_Tissu | GTEX-X5EB | 7/1806 | 0.680403 | 0.957813 | 0 | 0 | 0.881031 | 0.339259 |

| | | | | | | | | |
|---|---|---|---|---|---|---|---|---|
| GTEx_Tissu | GTEX-QLQ\ | 2/536 | 0.68111 | 0.957813 | 0 | 0 | 0.853889 | 0.327921 |
| GTEx_Tissu | GTEX-U8XE | Sep-07 | 0.6857 | 0.957813 | 0 | 0 | 0.884465 | 0.333722 |
| GTEx_Tissu | GTEX-X4XX | 6/1567 | 0.685749 | 0.957813 | 0 | 0 | 0.870857 | 0.328525 |
| GTEx_Tissu | GTEX-WL4( | 3/809 | 0.68903 | 0.957813 | 0 | 0 | 0.846641 | 0.315349 |
| GTEx_Tissu | GTEX-XQ3S | 6/1621 | 0.717281 | 0.957813 | 0 | 0 | 0.839262 | 0.278876 |
| GTEx_Tissu | GTEX-OXR( | 5/1353 | 0.709078 | 0.957813 | 0 | 0 | 0.839772 | 0.288705 |
| GTEx_Tissu | GTEX-WFJ( | 1/282 | 0.710068 | 0.957813 | 0 | 0 | 0.812381 | 0.278154 |
| GTEx_Tissu | GTEX-N7M | 1/265 | 0.687444 | 0.957813 | 0 | 0 | 0.865442 | 0.324346 |
| GTEx_Tissu | GTEX-WI4N | 1/284 | 0.712621 | 0.957813 | 0 | 0 | 0.806558 | 0.273266 |
| GTEx_Tissu | GTEX-WEY! | 1/272 | 0.696964 | 0.957813 | 0 | 0 | 0.842787 | 0.304265 |
| GTEx_Tissu | GTEX-X585 | 1/259 | 0.679049 | 0.957813 | 0 | 0 | 0.885839 | 0.342875 |
| GTEx_Tissu | GTEX-R53T | 6/1589 | 0.698856 | 0.957813 | 0 | 0 | 0.857724 | 0.307332 |
| GTEx_Tissu | GTEX-OOB. | 20/5046 | 0.723318 | 0.957813 | 0 | 0 | 0.884178 | 0.286391 |
| GTEx_Tissu | GTEX-TMZ! | 18/4569 | 0.723331 | 0.957813 | 0 | 0 | 0.880571 | 0.285207 |
| GTEx_Tissu | GTEX-QDT8 | 6/1632 | 0.723438 | 0.957813 | 0 | 0 | 0.833083 | 0.269702 |
| GTEx_Tissu | GTEX-WEY! | 7/1874 | 0.717834 | 0.957813 | 0 | 0 | 0.845755 | 0.280382 |
| GTEx_Tissu | GTEX-XLM4 | 6/1625 | 0.719531 | 0.957813 | 0 | 0 | 0.837005 | 0.275505 |
| GTEx_Tissu | GTEX-OOB. | 25/6233 | 0.724634 | 0.957813 | 0 | 0 | 0.890176 | 0.286715 |
| GTEx_Tissu | GTEX-XYKS | 1/266 | 0.688822 | 0.957813 | 0 | 0 | 0.862133 | 0.32138 |
| GTEx_Tissu | GTEX-QEL4 | 1/264 | 0.68606 | 0.957813 | 0 | 0 | 0.868777 | 0.327347 |
| GTEx_Tissu | GTEX-WEY! | Sep-14 | 0.689263 | 0.957813 | 0 | 0 | 0.881428 | 0.328008 |
| GTEx_Tissu | GTEX-PWN | 3/810 | 0.68984 | 0.957813 | 0 | 0 | 0.845548 | 0.313948 |
| GTEx_Tissu | GTEX-UTH( | Sep-62 | 0.713045 | 0.957813 | 0 | 0 | 0.861094 | 0.291231 |
| GTEx_Tissu | GTEX-QDT8 | 4/1072 | 0.692365 | 0.957813 | 0 | 0 | 0.850368 | 0.312631 |
| GTEx_Tissu | GTEX-XGQ4 | 1/269 | 0.692919 | 0.957813 | 0 | 0 | 0.852352 | 0.312678 |
| GTEx_Tissu | GTEX-XQ8I | 1/269 | 0.692919 | 0.957813 | 0 | 0 | 0.852352 | 0.312678 |
| GTEx_Tissu | GTEX-QV44 | 3/814 | 0.693067 | 0.957813 | 0 | 0 | 0.841201 | 0.308408 |
| GTEx_Tissu | GTEX-OOBI | 2/541 | 0.686061 | 0.957813 | 0 | 0 | 0.845749 | 0.318668 |
| GTEx_Tissu | GTEX-QDV. | 1/265 | 0.687444 | 0.957813 | 0 | 0 | 0.865442 | 0.324346 |
| GTEx_Tissu | GTEX-XUW | 1/270 | 0.694273 | 0.957813 | 0 | 0 | 0.84914 | 0.309842 |
| GTEx_Tissu | GTEX-TKQ1 | 1/266 | 0.688822 | 0.957813 | 0 | 0 | 0.862133 | 0.32138 |
| GTEx_Tissu | GTEX-WFG | 6/1582 | 0.694724 | 0.957813 | 0 | 0 | 0.861863 | 0.313926 |
| GTEx_Tissu | GTEX-QMR | Sep-25 | 0.694813 | 0.957813 | 0 | 0 | 0.876694 | 0.319215 |
| GTEx_Tissu | GTEX-P78B | 4/1076 | 0.695191 | 0.957813 | 0 | 0 | 0.847015 | 0.307948 |
| GTEx_Tissu | GTEX-PWO | 1/310 | 0.743855 | 0.957813 | 0 | 0 | 0.737714 | 0.218297 |
| GTEx_Tissu | GTEX-WI4N | 5/1287 | 0.666213 | 0.957813 | 0 | 0 | 0.886144 | 0.359904 |
| GTEx_Tissu | GTEX-P44H | Aug-27 | 0.666679 | 0.957813 | 0 | 0 | 0.897499 | 0.363888 |
| GTEx_Tissu | GTEX-PWC | 4/1037 | 0.666863 | 0.957813 | 0 | 0 | 0.880813 | 0.35688 |
| GTEx_Tissu | GTEX-R55E | 4/1037 | 0.666863 | 0.957813 | 0 | 0 | 0.880813 | 0.35688 |
| GTEx_Tissu | GTEX-TML8 | 2/552 | 0.696737 | 0.957813 | 0 | 0 | 0.828364 | 0.299327 |
| GTEx_Tissu | GTEX-WFO | 2/524 | 0.668969 | 0.957813 | 0 | 0 | 0.874059 | 0.351387 |
| GTEx_Tissu | GTEX-PWN | 2/525 | 0.669995 | 0.957813 | 0 | 0 | 0.872343 | 0.34936 |
| GTEx_Tissu | GTEX-TSE9 | 5/1294 | 0.670941 | 0.957813 | 0 | 0 | 0.881001 | 0.351584 |
| GTEx_Tissu | GTEX-PX3G | 13/3226 | 0.663026 | 0.957813 | 0 | 0 | 0.913098 | 0.375229 |
| GTEx_Tissu | GTEX-XBED | 1/248 | 0.663076 | 0.957813 | 0 | 0 | 0.925807 | 0.380383 |

| | | | | | | | | |
|---|---|---|---|---|---|---|---|---|
| GTEx_Tissu | GTEX-QEL4 | Aug-34 | 0.670546 | 0.957813 | 0 | 0 | 0.894048 | 0.357318 |
| GTEx_Tissu | GTEX-X261 | 1/304 | 0.736959 | 0.957813 | 0 | 0 | 0.752552 | 0.229696 |
| GTEx_Tissu | GTEX-NPJ8 | 2/598 | 0.738209 | 0.957813 | 0 | 0 | 0.762614 | 0.231475 |
| GTEx_Tissu | GTEX-WYJK | 1/300 | 0.73226 | 0.957813 | 0 | 0 | 0.762775 | 0.237696 |
| GTEx_Tissu | GTEX-PWO | 1/300 | 0.73226 | 0.957813 | 0 | 0 | 0.762775 | 0.237696 |
| GTEx_Tissu | GTEX-NPJ8 | 1/255 | 0.673329 | 0.957813 | 0 | 0 | 0.899973 | 0.355958 |
| GTEx_Tissu | GTEX-T6MI | Jul-25 | 0.743963 | 0.957813 | 0 | 0 | 0.82094 | 0.242804 |
| GTEx_Tissu | GTEX-X8HC | 13/3422 | 0.746465 | 0.957813 | 0 | 0 | 0.850499 | 0.248692 |
| GTEx_Tissu | GTEX-S7SE | 1/313 | 0.747235 | 0.957813 | 0 | 0 | 0.730508 | 0.212852 |
| GTEx_Tissu | GTEX-WFG | 16/4137 | 0.741211 | 0.957813 | 0 | 0 | 0.863567 | 0.258612 |
| GTEx_Tissu | GTEX-UTHC | 1/309 | 0.742718 | 0.957813 | 0 | 0 | 0.740146 | 0.220149 |
| GTEx_Tissu | GTEX-T5JC | Jul-24 | 0.743467 | 0.957813 | 0 | 0 | 0.821414 | 0.243492 |
| GTEx_Tissu | GTEX-N7M | 6/1669 | 0.743481 | 0.957813 | 0 | 0 | 0.812899 | 0.240953 |
| GTEx_Tissu | GTEX-U8T8 | 1/310 | 0.743855 | 0.957813 | 0 | 0 | 0.737714 | 0.218297 |
| GTEx_Tissu | GTEX-OXRC | 1/311 | 0.744986 | 0.957813 | 0 | 0 | 0.735296 | 0.216463 |
| GTEx_Tissu | GTEX-WFG | 1/311 | 0.744986 | 0.957813 | 0 | 0 | 0.735296 | 0.216463 |
| GTEx_Tissu | GTEX-PW2 | 1/311 | 0.744986 | 0.957813 | 0 | 0 | 0.735296 | 0.216463 |
| GTEx_Tissu | GTEX-V1D1 | 3/886 | 0.74695 | 0.957813 | 0 | 0 | 0.769697 | 0.224564 |
| GTEx_Tissu | GTEX-XK95 | 1/302 | 0.73462 | 0.957813 | 0 | 0 | 0.75763 | 0.233655 |
| GTEx_Tissu | GTEX-P44H | 1/313 | 0.747235 | 0.957813 | 0 | 0 | 0.730508 | 0.212852 |
| GTEx_Tissu | GTEX-SJXC | 6/1536 | 0.666682 | 0.957813 | 0 | 0 | 0.890002 | 0.360844 |
| GTEx_Tissu | GTEX-N7M | 3/800 | 0.681665 | 0.957813 | 0 | 0 | 0.856605 | 0.328266 |
| GTEx_Tissu | GTEX-OXRF | 1/261 | 0.681872 | 0.957813 | 0 | 0 | 0.878936 | 0.336556 |
| GTEx_Tissu | GTEX-R55D | 25/6111 | 0.6822 | 0.957813 | 0 | 0 | 0.916103 | 0.350347 |
| GTEx_Tissu | GTEX-QDT8 | 4/1058 | 0.682329 | 0.957813 | 0 | 0 | 0.862303 | 0.32961 |
| GTEx_Tissu | GTEX-OHPI | 18/4462 | 0.682352 | 0.957813 | 0 | 0 | 0.908054 | 0.347067 |
| GTEx_Tissu | GTEX-XMK | 2/518 | 0.662763 | 0.957813 | 0 | 0 | 0.884496 | 0.363827 |
| GTEx_Tissu | GTEX-RU1J | 7/1813 | 0.68439 | 0.957813 | 0 | 0 | 0.877277 | 0.332687 |
| GTEx_Tissu | GTEX-SJXC | 1/263 | 0.68467 | 0.957813 | 0 | 0 | 0.872137 | 0.330382 |
| GTEx_Tissu | GTEX-XMK | 2/519 | 0.663804 | 0.957813 | 0 | 0 | 0.88274 | 0.361719 |
| GTEx_Tissu | GTEX-QVJC | 2/540 | 0.685076 | 0.957813 | 0 | 0 | 0.847365 | 0.320495 |
| GTEx_Tissu | GTEX-U3ZN | 6/1566 | 0.685145 | 0.957813 | 0 | 0 | 0.871462 | 0.329522 |
| GTEx_Tissu | GTEX-UJHI | 1/249 | 0.66456 | 0.957813 | 0 | 0 | 0.922027 | 0.376768 |
| GTEx_Tissu | GTEX-X8HC | 4/1035 | 0.665364 | 0.957813 | 0 | 0 | 0.882615 | 0.359596 |
| GTEx_Tissu | GTEX-WWY | 5/1286 | 0.665534 | 0.957813 | 0 | 0 | 0.886883 | 0.361108 |
| GTEx_Tissu | GTEX-S3XE | 1/308 | 0.741576 | 0.957813 | 0 | 0 | 0.742595 | 0.222019 |
| GTEx_Tissu | GTEX-VUSC | 7/1803 | 0.678685 | 0.957813 | 0 | 0 | 0.882649 | 0.342114 |
| GTEx_Tissu | GTEX-XYKS | 1/259 | 0.679049 | 0.957813 | 0 | 0 | 0.885839 | 0.342875 |
| GTEx_Tissu | GTEX-UJMC | 1/263 | 0.68467 | 0.957813 | 0 | 0 | 0.872137 | 0.330382 |
| GTEx_Tissu | GTEX-X8HC | 1/251 | 0.667508 | 0.957813 | 0 | 0 | 0.914558 | 0.369667 |
| GTEx_Tissu | GTEX-X4XY | 29/7004 | 0.667753 | 0.957813 | 0 | 0 | 0.927455 | 0.37454 |
| GTEx_Tissu | GTEX-RWS. | 4/1039 | 0.668357 | 0.957813 | 0 | 0 | 0.879018 | 0.354185 |
| GTEx_Tissu | GTEX-N7M | Aug-72 | 0.691082 | 0.957813 | 0 | 0 | 0.875724 | 0.323577 |
| GTEx_Tissu | GTEX-WCD | 1/268 | 0.691559 | 0.957813 | 0 | 0 | 0.855587 | 0.315546 |
| GTEx_Tissu | GTEX-XGQ | 3/821 | 0.698655 | 0.957813 | 0 | 0 | 0.833697 | 0.298963 |

| | | | | | | | | |
|---|---|---|---|---|---|---|---|---|
| GTEx_Tissu | GTEX-R55E | 3/779 | 0.66399 | 0.957813 | 0 | 0 | 0.880753 | 0.360658 |
| GTEx_Tissu | GTEX-X4XY | 21/5124 | 0.664162 | 0.957813 | 0 | 0 | 0.923432 | 0.377896 |
| GTEx_Tissu | GTEX-WL4( | Aug-35 | 0.671096 | 0.957813 | 0 | 0 | 0.893557 | 0.356389 |
| GTEx_Tissu | GTEX-WL4( | 19/4671 | 0.671354 | 0.957813 | 0 | 0 | 0.916617 | 0.365235 |
| GTEx_Tissu | GTEX-XV7C | 5/1295 | 0.671613 | 0.957813 | 0 | 0 | 0.88027 | 0.350412 |
| GTEx_Tissu | GTEX-R55E | 24/6042 | 0.739519 | 0.957813 | 0 | 0 | 0.879583 | 0.26542 |
| GTEx_Tissu | GTEX-WHP | 1/299 | 0.731073 | 0.957813 | 0 | 0 | 0.765374 | 0.239748 |
| GTEx_Tissu | GTEX-WHP | 1/295 | 0.72627 | 0.957813 | 0 | 0 | 0.775945 | 0.248173 |
| GTEx_Tissu | GTEX-OHPI | Oct-38 | 0.726687 | 0.957813 | 0 | 0 | 0.854188 | 0.272707 |
| GTEx_Tissu | GTEX-V955 | 1/291 | 0.721383 | 0.957813 | 0 | 0 | 0.786808 | 0.25696 |
| GTEx_Tissu | GTEX-R55F | 2/580 | 0.72258 | 0.957813 | 0 | 0 | 0.787095 | 0.255749 |
| GTEx_Tissu | GTEX-XV7C | 5/1376 | 0.723103 | 0.957813 | 0 | 0 | 0.824661 | 0.267358 |
| GTEx_Tissu | GTEX-RVPV | 3/840 | 0.713439 | 0.957813 | 0 | 0 | 0.813961 | 0.274841 |
| GTEx_Tissu | GTEX-XUW | 1/268 | 0.691559 | 0.957813 | 0 | 0 | 0.855587 | 0.315546 |
| GTEx_Tissu | GTEX-XMD | Nov-57 | 0.714806 | 0.957813 | 0 | 0 | 0.867963 | 0.291413 |
| GTEx_Tissu | GTEX-U8XE | 5/1396 | 0.734911 | 0.957813 | 0 | 0 | 0.811927 | 0.250078 |
| GTEx_Tissu | GTEX-NL3H | 15/3880 | 0.735492 | 0.957813 | 0 | 0 | 0.865028 | 0.26575 |
| GTEx_Tissu | GTEX-OIZI- | 17/4321 | 0.720114 | 0.957813 | 0 | 0 | 0.880752 | 0.289191 |
| GTEx_Tissu | GTEX-OIZI- | 1/290 | 0.720147 | 0.957813 | 0 | 0 | 0.789571 | 0.259216 |
| GTEx_Tissu | GTEX-XXEK | 1/306 | 0.739278 | 0.957813 | 0 | 0 | 0.747541 | 0.225819 |
| GTEx_Tissu | GTEX-P4QS | 1/299 | 0.731073 | 0.957813 | 0 | 0 | 0.765374 | 0.239748 |
| GTEx_Tissu | GTEX-PW2( | 1/294 | 0.725056 | 0.957813 | 0 | 0 | 0.778633 | 0.250335 |
| GTEx_Tissu | GTEX-U8XE | 1/294 | 0.725056 | 0.957813 | 0 | 0 | 0.778633 | 0.250335 |
| GTEx_Tissu | GTEX-NPJ7 | 4/1141 | 0.738591 | 0.957813 | 0 | 0 | 0.795838 | 0.241147 |
| GTEx_Tissu | GTEX-V1D1 | Nov-13 | 0.739112 | 0.957813 | 0 | 0 | 0.848421 | 0.256483 |
| GTEx_Tissu | GTEX-R55C | 1/292 | 0.722612 | 0.957813 | 0 | 0 | 0.784065 | 0.254729 |
| GTEx_Tissu | GTEX-SSA3 | 1/292 | 0.722612 | 0.957813 | 0 | 0 | 0.784065 | 0.254729 |
| GTEx_Tissu | GTEX-PWO | 1/307 | 0.740429 | 0.957813 | 0 | 0 | 0.74506 | 0.223909 |
| GTEx_Tissu | GTEX-T5JW | 1/307 | 0.740429 | 0.957813 | 0 | 0 | 0.74506 | 0.223909 |
| GTEx_Tissu | GTEX-OIZI- | 1/307 | 0.740429 | 0.957813 | 0 | 0 | 0.74506 | 0.223909 |
| GTEx_Tissu | GTEX-XGQ4 | 5/1406 | 0.74068 | 0.957813 | 0 | 0 | 0.805696 | 0.241859 |
| GTEx_Tissu | GTEX-REY6 | 5/1378 | 0.7243 | 0.957813 | 0 | 0 | 0.823371 | 0.265578 |
| GTEx_Tissu | GTEX-WFG | 14/3554 | 0.701668 | 0.957813 | 0 | 0 | 0.887013 | 0.314265 |
| GTEx_Tissu | GTEX-NPJ7 | 1/276 | 0.702274 | 0.957813 | 0 | 0 | 0.830359 | 0.293475 |
| GTEx_Tissu | GTEX-VJYA | 1/304 | 0.736959 | 0.957813 | 0 | 0 | 0.752552 | 0.229696 |
| GTEx_Tissu | GTEX-PLZ6 | 1/314 | 0.748352 | 0.957813 | 0 | 0 | 0.728137 | 0.211074 |
| GTEx_Tissu | GTEX-U3ZN | 1/301 | 0.733443 | 0.957813 | 0 | 0 | 0.760194 | 0.235665 |
| GTEx_Tissu | GTEX-QV44 | 1/301 | 0.733443 | 0.957813 | 0 | 0 | 0.760194 | 0.235665 |
| GTEx_Tissu | GTEX-OHPI | 1/303 | 0.735792 | 0.957813 | 0 | 0 | 0.755082 | 0.231665 |
| GTEx_Tissu | GTEX-R55G | 1/303 | 0.735792 | 0.957813 | 0 | 0 | 0.755082 | 0.231665 |
| GTEx_Tissu | GTEX-X585 | 2/596 | 0.736509 | 0.957813 | 0 | 0 | 0.76526 | 0.234042 |
| GTEx_Tissu | GTEX-XUYS | 26/6494 | 0.733119 | 0.957813 | 0 | 0 | 0.886001 | 0.275056 |
| GTEx_Tissu | GTEX-RUSC | 13/3427 | 0.748412 | 0.957813 | 0 | 0 | 0.848996 | 0.246041 |
| GTEx_Tissu | GTEX-O5YV | 18/4639 | 0.748438 | 0.957813 | 0 | 0 | 0.86328 | 0.25015 |
| GTEx_Tissu | GTEX-XXEK | 4/1157 | 0.748545 | 0.957813 | 0 | 0 | 0.784125 | 0.227102 |

| | | | | | | | | |
|---|---|---|---|---|---|---|---|---|
| GTEx_Tissu | GTEX-WYV. | 3/889 | 0.749025 | 0.957813 | 0 | 0 | 0.76697 | 0.221641 |
| GTEx_Tissu | GTEX-T2IS- | 2/588 | 0.729619 | 0.957813 | 0 | 0 | 0.776029 | 0.244629 |
| GTEx_Tissu | GTEX-WFG | 4/1127 | 0.729647 | 0.957813 | 0 | 0 | 0.80636 | 0.254161 |
| GTEx_Tissu | GTEX-UPK5 | 1/293 | 0.723837 | 0.957813 | 0 | 0 | 0.78134 | 0.25252 |
| GTEx_Tissu | GTEX-XLM4 | 5/1366 | 0.717063 | 0.957813 | 0 | 0 | 0.831168 | 0.276439 |
| GTEx_Tissu | GTEX-WFO | 1/299 | 0.731073 | 0.957813 | 0 | 0 | 0.765374 | 0.239748 |
| GTEx_Tissu | GTEX-UPJH | 24/6017 | 0.731244 | 0.957813 | 0 | 0 | 0.884842 | 0.276962 |
| GTEx_Tissu | GTEX-OHPI | 2/590 | 0.731356 | 0.957813 | 0 | 0 | 0.773309 | 0.241934 |
| GTEx_Tissu | GTEX-O5YV | 2/611 | 0.749031 | 0.957813 | 0 | 0 | 0.745832 | 0.215526 |
| GTEx_Tissu | GTEX-OOBI | 25/6293 | 0.744327 | 0.957813 | 0 | 0 | 0.877795 | 0.259191 |
| GTEx_Tissu | GTEX-Q2AI | 3/889 | 0.749025 | 0.957813 | 0 | 0 | 0.76697 | 0.221641 |
| GTEx_Tissu | GTEX-P4PP | 5/1412 | 0.744099 | 0.957813 | 0 | 0 | 0.802 | 0.237056 |
| GTEx_Tissu | GTEX-S4UY | Nov-92 | 0.730153 | 0.957813 | 0 | 0 | 0.85566 | 0.269106 |
| GTEx_Tissu | GTEX-T2IS- | 4/1135 | 0.734785 | 0.957813 | 0 | 0 | 0.800315 | 0.246639 |
| GTEx_Tissu | GTEX-QEL4 | 2/594 | 0.734801 | 0.957813 | 0 | 0 | 0.767925 | 0.236641 |
| GTEx_Tissu | GTEX-QCQ( | 16/4107 | 0.730024 | 0.957813 | 0 | 0 | 0.871552 | 0.274258 |
| GTEx_Tissu | GTEX-XUZC | 14/3670 | 0.747961 | 0.957813 | 0 | 0 | 0.852785 | 0.247653 |
| GTEx_Tissu | GTEX-P78B | 1/299 | 0.731073 | 0.957813 | 0 | 0 | 0.765374 | 0.239748 |
| GTEx_Tissu | GTEX-SSA3 | Aug-42 | 0.72683 | 0.957813 | 0 | 0 | 0.843676 | 0.269186 |
| GTEx_Tissu | GTEX-PWC | 1/286 | 0.715152 | 0.957813 | 0 | 0 | 0.800816 | 0.268482 |
| GTEx_Tissu | GTEX-OXR( | 1/275 | 0.700955 | 0.957813 | 0 | 0 | 0.833432 | 0.296128 |
| GTEx_Tissu | GTEX-WHS | 2/557 | 0.701491 | 0.957813 | 0 | 0 | 0.820689 | 0.290973 |
| GTEx_Tissu | GTEX-N7M | Nov-49 | 0.711226 | 0.957813 | 0 | 0 | 0.870818 | 0.296744 |
| GTEx_Tissu | GTEX-RU72 | 6/1611 | 0.711605 | 0.957813 | 0 | 0 | 0.844952 | 0.28748 |
| GTEx_Tissu | GTEX-OIZG | 20/5015 | 0.712135 | 0.957813 | 0 | 0 | 0.891518 | 0.302659 |
| GTEx_Tissu | GTEX-UPJH | 4/1088 | 0.703563 | 0.957813 | 0 | 0 | 0.837105 | 0.294324 |
| GTEx_Tissu | GTEX-U3ZN | 1/277 | 0.703588 | 0.957813 | 0 | 0 | 0.827309 | 0.290851 |
| GTEx_Tissu | GTEX-S7SE- | 1/278 | 0.704895 | 0.957813 | 0 | 0 | 0.82428 | 0.288256 |
| GTEx_Tissu | GTEX-TSE9 | 6/1627 | 0.720651 | 0.957813 | 0 | 0 | 0.835881 | 0.273835 |
| GTEx_Tissu | GTEX-OHPI | 2/569 | 0.712653 | 0.957813 | 0 | 0 | 0.802822 | 0.271965 |
| GTEx_Tissu | GTEX-RU72 | 2/569 | 0.712653 | 0.957813 | 0 | 0 | 0.802822 | 0.271965 |
| GTEx_Tissu | GTEX-RTLS- | 4/1091 | 0.705631 | 0.957813 | 0 | 0 | 0.834662 | 0.291016 |
| GTEx_Tissu | GTEX-RWS( | 1/276 | 0.702274 | 0.957813 | 0 | 0 | 0.830359 | 0.293475 |
| GTEx_Tissu | GTEX-RVPV | 4/1091 | 0.705631 | 0.957813 | 0 | 0 | 0.834662 | 0.291016 |
| GTEx_Tissu | GTEX-UJM( | 1/280 | 0.707493 | 0.957813 | 0 | 0 | 0.818288 | 0.28315 |
| GTEx_Tissu | GTEX-S3XE | 1/280 | 0.707493 | 0.957813 | 0 | 0 | 0.818288 | 0.28315 |
| GTEx_Tissu | GTEX-U8T8 | 3/833 | 0.708057 | 0.957813 | 0 | 0 | 0.821127 | 0.283479 |
| GTEx_Tissu | GTEX-Q2A( | 17/4285 | 0.706326 | 0.957813 | 0 | 0 | 0.89023 | 0.309514 |
| GTEx_Tissu | GTEX-RWS. | 3/850 | 0.720997 | 0.957813 | 0 | 0 | 0.80393 | 0.262982 |
| GTEx_Tissu | GTEX-QDT8 | 4/1114 | 0.721143 | 0.957813 | 0 | 0 | 0.816368 | 0.266885 |
| GTEx_Tissu | GTEX-WWY | 3/845 | 0.717237 | 0.957813 | 0 | 0 | 0.808916 | 0.268842 |
| GTEx_Tissu | GTEX-OIZG | 22/5515 | 0.721317 | 0.957813 | 0 | 0 | 0.888515 | 0.290258 |
| GTEx_Tissu | GTEX-X585 | 3/855 | 0.724719 | 0.957813 | 0 | 0 | 0.799002 | 0.257256 |
| GTEx_Tissu | GTEX-UPK5 | 1/297 | 0.728682 | 0.957813 | 0 | 0 | 0.770624 | 0.243916 |
| GTEx_Tissu | GTEX-NPJ8 | 6/1602 | 0.706433 | 0.957813 | 0 | 0 | 0.850135 | 0.295445 |

| | | | | | | | | |
|---|---|---|---|---|---|---|---|---|
| GTEx_Tissu | GTEX-T2IS- | 4/1092 | 0.706317 | 0.957813 | 0 | 0 | 0.83385 | 0.289922 |
| GTEx_Tissu | GTEX-SNM | Dec-31 | 0.72762 | 0.957813 | 0 | 0 | 0.861507 | 0.273939 |
| GTEx_Tissu | GTEX-PW2 | 18/4645 | 0.750524 | 0.958071 | 0 | 0 | 0.861822 | 0.247328 |
| GTEx_Tissu | GTEX-XUJ4 | 6/1682 | 0.750278 | 0.958071 | 0 | 0 | 0.80602 | 0.231578 |
| GTEx_Tissu | GTEX-SE5C | Sep-42 | 0.750097 | 0.958071 | 0 | 0 | 0.828986 | 0.238377 |
| GTEx_Tissu | GTEX-U3ZM | 1/317 | 0.751674 | 0.958105 | 0 | 0 | 0.721114 | 0.205844 |
| GTEx_Tissu | GTEX-S4UY | 3/892 | 0.751087 | 0.958105 | 0 | 0 | 0.764262 | 0.218757 |
| GTEx_Tissu | GTEX-PWN | 3/895 | 0.753135 | 0.958105 | 0 | 0 | 0.761571 | 0.215913 |
| GTEx_Tissu | GTEX-WZT | 2/616 | 0.753092 | 0.958105 | 0 | 0 | 0.739567 | 0.209717 |
| GTEx_Tissu | GTEX-UTH | Jul-42 | 0.752297 | 0.958105 | 0 | 0 | 0.812959 | 0.231387 |
| GTEx_Tissu | GTEX-R55F | 4/1164 | 0.752809 | 0.958105 | 0 | 0 | 0.779103 | 0.221221 |
| GTEx_Tissu | GTEX-T2IS- | Jul-45 | 0.753748 | 0.958225 | 0 | 0 | 0.811565 | 0.229427 |
| GTEx_Tissu | GTEX-POYV | 6/1690 | 0.754398 | 0.958225 | 0 | 0 | 0.801839 | 0.225986 |
| GTEx_Tissu | GTEX-QLQ\ | 1/320 | 0.754952 | 0.958225 | 0 | 0 | 0.714223 | 0.200769 |
| GTEx_Tissu | GTEX-XBEC | 1/320 | 0.754952 | 0.958225 | 0 | 0 | 0.714223 | 0.200769 |
| GTEx_Tissu | GTEX-XPT6 | Nov-54 | 0.756054 | 0.959077 | 0 | 0 | 0.834585 | 0.233385 |
| Jensen_TIS | Prefrontal | 18/5632 | 0.956343 | 0.959322 | 0 | 0 | 0.664441 | 0.02966 |
| GeneSigDB | 19549311- | 1/672 | 0.949203 | 0.95954 | 0 | 0 | 0.333449 | 0.017383 |
| GeneSigDB | 17683518- | 1/680 | 0.951004 | 0.959618 | 0 | 0 | 0.329383 | 0.016547 |
| Reactome_ | Sensory Pe | 1/616 | 0.934629 | 0.960352 | 0 | 0 | 0.36487 | 0.024667 |
| DisGeNET | B-Cell Lym | 1/730 | 0.960913 | 0.960913 | 0 | 0 | 0.305994 | 0.0122 |
| GTEx_Tissu | GTEX-XPT6 | Dec-07 | 0.758379 | 0.960926 | 0 | 0 | 0.837208 | 0.231548 |
| GTEx_Tissu | GTEX-RUS | 4/1174 | 0.758807 | 0.960926 | 0 | 0 | 0.772032 | 0.213086 |
| GTEx_Tissu | GTEX-XUZ | 1/323 | 0.758188 | 0.960926 | 0 | 0 | 0.707461 | 0.195842 |
| GTEx_Tissu | GTEX-UPJH | 1/324 | 0.759257 | 0.960948 | 0 | 0 | 0.705234 | 0.194232 |
| GTEx_Tissu | GTEX-S7SE | 5/1441 | 0.760166 | 0.961468 | 0 | 0 | 0.784573 | 0.215144 |
| GTEx_Tissu | GTEX-VJWI | 3/906 | 0.760532 | 0.961468 | 0 | 0 | 0.751859 | 0.205811 |
| GTEx_Tissu | GTEX-RU72 | 3/907 | 0.761196 | 0.961761 | 0 | 0 | 0.750988 | 0.204918 |
| GTEx_Tissu | GTEX-QMR | 1/328 | 0.763487 | 0.961925 | 0 | 0 | 0.696465 | 0.187947 |
| GTEx_Tissu | GTEX-XV7C | 1/328 | 0.763487 | 0.961925 | 0 | 0 | 0.696465 | 0.187947 |
| GTEx_Tissu | GTEX-R53T | Oct-21 | 0.762459 | 0.961925 | 0 | 0 | 0.82406 | 0.22349 |
| GTEx_Tissu | GTEX-XUJ4 | 2/628 | 0.762611 | 0.961925 | 0 | 0 | 0.724939 | 0.196464 |
| GTEx_Tissu | GTEX-WHV | 2/629 | 0.76339 | 0.961925 | 0 | 0 | 0.723745 | 0.195402 |
| RNA-Seq_D | Eed Small i | 1/732 | 0.961265 | 0.962103 | 0 | 0 | 0.305125 | 0.012054 |
| PhenGenI_ | Personality | 1/641 | 0.941586 | 0.962417 | 0 | 0 | 0.350164 | 0.021076 |
| GTEx_Tissu | GTEX-X585 | 6/1711 | 0.764983 | 0.963264 | 0 | 0 | 0.79105 | 0.211924 |
| GTEx_Tissu | GTEX-XUJ4 | 8/1160 | 0.13147 | 0.963428 | 0 | 0 | 1.649174 | 3.346135 |
| GTEx_Tissu | GTEX-XOT | 9/1349 | 0.131543 | 0.963428 | 0 | 0 | 1.599282 | 3.244017 |
| GTEx_Tissu | GTEX-QXCU | 5/616 | 0.130564 | 0.963428 | 0 | 0 | 1.92627 | 3.921678 |
| GTEx_Tissu | GTEX-SUCS | 7/974 | 0.13116 | 0.963428 | 0 | 0 | 1.714349 | 3.482414 |
| GTEx_Tissu | GTEX-V1D1 | 4/447 | 0.130472 | 0.963428 | 0 | 0 | 2.118089 | 4.313691 |
| GTEx_Tissu | GTEX-NPJ7 | 9/1346 | 0.130268 | 0.963428 | 0 | 0 | 1.60313 | 3.26744 |
| GTEx_Tissu | GTEX-P4PP | 6/792 | 0.130685 | 0.963428 | 0 | 0 | 1.802563 | 3.668148 |
| GTEx_Tissu | GTEX-RTLS | 1/330 | 0.765575 | 0.963463 | 0 | 0 | 0.692161 | 0.184896 |
| GTEx_Tissu | GTEX-VUS | 1/331 | 0.766611 | 0.963574 | 0 | 0 | 0.690028 | 0.183392 |

| | | | | | | | | |
|---|---|---|---|---|---|---|---|---|
| GTEx_Tissu | GTEX-S3XE | 1/331 | 0.766611 | 0.963574 | 0 | 0 | 0.690028 | 0.183392 |
| GTEx_Tissu | GTEX-XUZ( | 6/1715 | 0.766961 | 0.963574 | 0 | 0 | 0.789025 | 0.209343 |
| GTEx_Tissu | GTEX-OHPI | 15/3969 | 0.768101 | 0.963917 | 0 | 0 | 0.840868 | 0.22185 |
| GTEx_Tissu | GTEX-S32V | 2/635 | 0.768017 | 0.963917 | 0 | 0 | 0.716662 | 0.189158 |
| GTEx_Tissu | GTEX-QMR | 9/1350 | 0.13197 | 0.964112 | 0 | 0 | 1.598004 | 3.23625 |
| GTEx_Tissu | GTEX-S7SE | 2/636 | 0.768781 | 0.964226 | 0 | 0 | 0.715495 | 0.188139 |
| GTEx_Tissu | GTEX-PVO\ | 6/1764 | 0.79023 | 0.964449 | 0 | 0 | 0.764969 | 0.180097 |
| GTEx_Tissu | GTEX-RWS( | 4/1210 | 0.77949 | 0.964449 | 0 | 0 | 0.747547 | 0.186225 |
| GTEx_Tissu | GTEX-S7PN | 19/4945 | 0.770463 | 0.964449 | 0 | 0 | 0.85009 | 0.221672 |
| GTEx_Tissu | GTEX-X585 | 2/663 | 0.7886 | 0.964449 | 0 | 0 | 0.685307 | 0.162758 |
| GTEx_Tissu | GTEX-XUJ4 | 2/646 | 0.7763 | 0.964449 | 0 | 0 | 0.704019 | 0.178269 |
| GTEx_Tissu | GTEX-P44H | 6/1757 | 0.787015 | 0.964449 | 0 | 0 | 0.768323 | 0.184019 |
| GTEx_Tissu | GTEX-PLZ6 | 3/929 | 0.77543 | 0.964449 | 0 | 0 | 0.732297 | 0.186251 |
| GTEx_Tissu | GTEX-T2YK | 19/4980 | 0.78164 | 0.964449 | 0 | 0 | 0.842121 | 0.207466 |
| GTEx_Tissu | GTEX-OIZG | 4/1224 | 0.787154 | 0.964449 | 0 | 0 | 0.738416 | 0.176726 |
| GTEx_Tissu | GTEX-SIU8 | 1/343 | 0.778706 | 0.964449 | 0 | 0 | 0.665409 | 0.166434 |
| GTEx_Tissu | GTEX-XUW | 1/337 | 0.772738 | 0.964449 | 0 | 0 | 0.677499 | 0.174669 |
| GTEx_Tissu | GTEX-U3ZN | 1/344 | 0.779685 | 0.964449 | 0 | 0 | 0.663435 | 0.165106 |
| GTEx_Tissu | GTEX-OXRI | 29/7353 | 0.780096 | 0.964449 | 0 | 0 | 0.859435 | 0.213431 |
| GTEx_Tissu | GTEX-T6M( | 1/346 | 0.781631 | 0.964449 | 0 | 0 | 0.659521 | 0.162488 |
| GTEx_Tissu | GTEX-RN64 | Jul-26 | 0.790715 | 0.964449 | 0 | 0 | 0.775495 | 0.1821 |
| GTEx_Tissu | GTEX-WHP | 1/348 | 0.78356 | 0.964449 | 0 | 0 | 0.655653 | 0.159919 |
| GTEx_Tissu | GTEX-XYKS | 6/1756 | 0.786553 | 0.964449 | 0 | 0 | 0.768804 | 0.184586 |
| GTEx_Tissu | GTEX-W5W | 3/940 | 0.782287 | 0.964449 | 0 | 0 | 0.723281 | 0.17759 |
| GTEx_Tissu | GTEX-S7SE | 2/657 | 0.784327 | 0.964449 | 0 | 0 | 0.691801 | 0.168058 |
| GTEx_Tissu | GTEX-X62C | Aug-72 | 0.785797 | 0.964449 | 0 | 0 | 0.789417 | 0.190294 |
| GTEx_Tissu | GTEX-VUS( | 3/921 | 0.770335 | 0.964449 | 0 | 0 | 0.73899 | 0.192825 |
| GTEx_Tissu | GTEX-OIZI- | 17/4463 | 0.770771 | 0.964449 | 0 | 0 | 0.844865 | 0.219973 |
| GTEx_Tissu | GTEX-R55E | 3/923 | 0.771617 | 0.964449 | 0 | 0 | 0.737306 | 0.191159 |
| GTEx_Tissu | GTEX-PLZ6 | 3/928 | 0.774798 | 0.964449 | 0 | 0 | 0.733127 | 0.18706 |
| GTEx_Tissu | GTEX-WYJI | 3/927 | 0.774165 | 0.964449 | 0 | 0 | 0.733959 | 0.187872 |
| GTEx_Tissu | GTEX-Q2A( | 5/1467 | 0.773933 | 0.964449 | 0 | 0 | 0.769536 | 0.197209 |
| GTEx_Tissu | GTEX-W5W | 1/337 | 0.772738 | 0.964449 | 0 | 0 | 0.677499 | 0.174669 |
| GTEx_Tissu | GTEX-V955 | 2/669 | 0.792799 | 0.964449 | 0 | 0 | 0.678931 | 0.157638 |
| GTEx_Tissu | GTEX-PWO | 1/337 | 0.772738 | 0.964449 | 0 | 0 | 0.677499 | 0.174669 |
| GTEx_Tissu | GTEX-U3ZI | 3/934 | 0.778568 | 0.964449 | 0 | 0 | 0.728172 | 0.182261 |
| GTEx_Tissu | GTEX-R53T | 3/933 | 0.777943 | 0.964449 | 0 | 0 | 0.728994 | 0.183052 |
| GTEx_Tissu | GTEX-XUW | 15/3997 | 0.777806 | 0.964449 | 0 | 0 | 0.83349 | 0.209438 |
| GTEx_Tissu | GTEX-WZT( | 1/347 | 0.782598 | 0.964449 | 0 | 0 | 0.657582 | 0.161197 |
| GTEx_Tissu | GTEX-OHPI | Dec-70 | 0.782108 | 0.964449 | 0 | 0 | 0.817925 | 0.201016 |
| GTEx_Tissu | GTEX-WYB: | Sep-99 | 0.774481 | 0.964449 | 0 | 0 | 0.807368 | 0.206333 |
| GTEx_Tissu | GTEX-NPJ8 | 1/354 | 0.789247 | 0.964449 | 0 | 0 | 0.644311 | 0.152493 |
| GTEx_Tissu | GTEX-UPK5 | 1/353 | 0.788309 | 0.964449 | 0 | 0 | 0.646175 | 0.153702 |
| GTEx_Tissu | GTEX-R55D | 1/353 | 0.788309 | 0.964449 | 0 | 0 | 0.646175 | 0.153702 |
| GTEx_Tissu | GTEX-UJHI- | Aug-76 | 0.787457 | 0.964449 | 0 | 0 | 0.787846 | 0.188253 |

| | | | | | | | | |
|---|---|---|---|---|---|---|---|---|
| GTEx_Tissu | GTEX-UPK5 | 6/1769 | 0.792505 | 0.964449 | 0 | 0 | 0.762589 | 0.177345 |
| GTEx_Tissu | GTEX-U3ZH | 5/1504 | 0.792499 | 0.964449 | 0 | 0 | 0.749036 | 0.174199 |
| GTEx_Tissu | GTEX-POYW | 1/356 | 0.791109 | 0.964449 | 0 | 0 | 0.640616 | 0.150109 |
| GTEx_Tissu | GTEX-X4EP | 1/356 | 0.791109 | 0.964449 | 0 | 0 | 0.640616 | 0.150109 |
| GTEx_Tissu | GTEX-X4XY | Oct-53 | 0.775381 | 0.964449 | 0 | 0 | 0.812931 | 0.20681 |
| GTEx_Tissu | GTEX-VUSC | 3/920 | 0.769691 | 0.964449 | 0 | 0 | 0.739835 | 0.193663 |
| GTEx_Tissu | GTEX-VJYA | 1/355 | 0.79018 | 0.964449 | 0 | 0 | 0.642458 | 0.151296 |
| GTEx_Tissu | GTEX-P4QT | 1/354 | 0.789247 | 0.964449 | 0 | 0 | 0.644311 | 0.152493 |
| GTEx_Tissu | GTEX-XQ3S | 1/355 | 0.79018 | 0.964449 | 0 | 0 | 0.642458 | 0.151296 |
| GTEx_Tissu | GTEX-P4QS | 3/942 | 0.783515 | 0.964449 | 0 | 0 | 0.721664 | 0.176061 |
| GTEx_Tissu | GTEX-XMK | 6/1759 | 0.787937 | 0.964449 | 0 | 0 | 0.767362 | 0.18289 |
| GTEx_Tissu | GTEX-XOTC | 18/4729 | 0.778629 | 0.964449 | 0 | 0 | 0.841804 | 0.210636 |
| GTEx_Tissu | GTEX-RU1J | 1/342 | 0.777722 | 0.964449 | 0 | 0 | 0.667394 | 0.167774 |
| GTEx_Tissu | GTEX-XAJ8 | 1/342 | 0.777722 | 0.964449 | 0 | 0 | 0.667394 | 0.167774 |
| GTEx_Tissu | GTEX-R53T | 1/355 | 0.79018 | 0.964449 | 0 | 0 | 0.642458 | 0.151296 |
| GTEx_Tissu | GTEX-P44H | 3/957 | 0.79255 | 0.964449 | 0 | 0 | 0.709756 | 0.165018 |
| Jensen_TIS | MOLT-4 ce | 13/4430 | 0.965122 | 0.965122 | 0 | 0 | 0.616317 | 0.021879 |
| Reactome_ | Innate Imm | 2/1035 | 0.943895 | 0.965445 | 0 | 0 | 0.430044 | 0.024831 |
| GTEx_Tissu | GTEX-P4PC | 1/361 | 0.795694 | 0.967443 | 0 | 0 | 0.631557 | 0.144336 |
| GTEx_Tissu | GTEX-WFO | 1/362 | 0.796599 | 0.968014 | 0 | 0 | 0.629775 | 0.143213 |
| GeneSigDB | 18787218- | 1/742 | 0.962979 | 0.968194 | 0 | 0 | 0.300851 | 0.011349 |
| GeneSigDB | 18310505- | 1/732 | 0.961265 | 0.968194 | 0 | 0 | 0.305125 | 0.012054 |
| GTEx_Tissu | GTEX-PLZ4 | 9/1353 | 0.133254 | 0.968603 | 0 | 0 | 1.594179 | 3.213066 |
| GTEx_Tissu | GTEX-QV44 | 9/1353 | 0.133254 | 0.968603 | 0 | 0 | 1.594179 | 3.213066 |
| GTEx_Tissu | GTEX-QV31 | 17/4550 | 0.798748 | 0.969037 | 0 | 0 | 0.823989 | 0.185158 |
| GTEx_Tissu | GTEX-WYJK | 1/364 | 0.798398 | 0.969037 | 0 | 0 | 0.626241 | 0.140997 |
| GTEx_Tissu | GTEX-WH7 | 1/364 | 0.798398 | 0.969037 | 0 | 0 | 0.626241 | 0.140997 |
| GTEx_Tissu | GTEX-XOTC | 1/365 | 0.799291 | 0.969167 | 0 | 0 | 0.624489 | 0.139904 |
| GTEx_Tissu | GTEX-PX3G | Sep-63 | 0.799846 | 0.969312 | 0 | 0 | 0.784245 | 0.175151 |
| GTEx_Tissu | GTEX-QVJC | 3/981 | 0.806363 | 0.970054 | 0 | 0 | 0.691462 | 0.148817 |
| GTEx_Tissu | GTEX-T6MI | 2/691 | 0.80758 | 0.970054 | 0 | 0 | 0.656501 | 0.140303 |
| GTEx_Tissu | GTEX-SIU7 | 3/985 | 0.80859 | 0.970054 | 0 | 0 | 0.6885 | 0.146281 |
| GTEx_Tissu | GTEX-XQ3S | 2/689 | 0.806275 | 0.970054 | 0 | 0 | 0.658481 | 0.141791 |
| GTEx_Tissu | GTEX-S3XE | 2/685 | 0.803643 | 0.970054 | 0 | 0 | 0.662475 | 0.144817 |
| GTEx_Tissu | GTEX-W5W | 2/689 | 0.806275 | 0.970054 | 0 | 0 | 0.658481 | 0.141791 |
| GTEx_Tissu | GTEX-QDT8 | 1/380 | 0.812229 | 0.970054 | 0 | 0 | 0.599313 | 0.124641 |
| GTEx_Tissu | GTEX-OXRC | 13/3597 | 0.808948 | 0.970054 | 0 | 0 | 0.800393 | 0.1697 |
| GTEx_Tissu | GTEX-PWN | 3/985 | 0.80859 | 0.970054 | 0 | 0 | 0.6885 | 0.146281 |
| GTEx_Tissu | GTEX-SE5C | 5/1539 | 0.808971 | 0.970054 | 0 | 0 | 0.730555 | 0.154872 |
| GTEx_Tissu | GTEX-POM | 5/1533 | 0.806221 | 0.970054 | 0 | 0 | 0.733663 | 0.158029 |
| GTEx_Tissu | GTEX-PWO | 3/991 | 0.81189 | 0.970054 | 0 | 0 | 0.684102 | 0.14256 |
| GTEx_Tissu | GTEX-WL46 | 1/369 | 0.802825 | 0.970054 | 0 | 0 | 0.617575 | 0.135631 |
| GTEx_Tissu | GTEX-XPT6 | Nov-86 | 0.805612 | 0.970054 | 0 | 0 | 0.792546 | 0.171312 |
| GTEx_Tissu | GTEX-SNM | 5/1545 | 0.811691 | 0.970054 | 0 | 0 | 0.727471 | 0.151777 |
| GTEx_Tissu | GTEX-T8EM | 1/379 | 0.811393 | 0.970054 | 0 | 0 | 0.600929 | 0.125596 |

| | | | | | | | | |
|---|---|---|---|---|---|---|---|---|
| GTEx_Tissu | GTEX-NFK9 | 13/3603 | 0.810883 | 0.970054 | 0 | 0 | 0.798762 | 0.167446 |
| GTEx_Tissu | GTEX-QESD | 2/686 | 0.804304 | 0.970054 | 0 | 0 | 0.661472 | 0.144054 |
| GTEx_Tissu | GTEX-S341 | 2/695 | 0.810166 | 0.970054 | 0 | 0 | 0.652576 | 0.137378 |
| GTEx_Tissu | GTEX-T6MI | 1/378 | 0.810553 | 0.970054 | 0 | 0 | 0.602554 | 0.12656 |
| GTEx_Tissu | GTEX-WH7 | 1/371 | 0.804569 | 0.970054 | 0 | 0 | 0.614173 | 0.133551 |
| GTEx_Tissu | GTEX-QDT8 | 5/1530 | 0.804835 | 0.970054 | 0 | 0 | 0.735226 | 0.159631 |
| GTEx_Tissu | GTEX-P4QS | 19/5053 | 0.803813 | 0.970054 | 0 | 0 | 0.825858 | 0.180357 |
| GTEx_Tissu | GTEX-RM2 | 14/3833 | 0.804977 | 0.970054 | 0 | 0 | 0.808201 | 0.175332 |
| GTEx_Tissu | GTEX-WH7 | 6/1813 | 0.811732 | 0.970054 | 0 | 0 | 0.742216 | 0.154816 |
| GTEx_Tissu | GTEX-RWS | 1/373 | 0.806297 | 0.970054 | 0 | 0 | 0.610809 | 0.131509 |
| GTEx_Tissu | GTEX-QDT8 | 3/991 | 0.81189 | 0.970054 | 0 | 0 | 0.684102 | 0.14256 |
| Reactome_ | Cellular Re | 1/722 | 0.959472 | 0.970626 | 0 | 0 | 0.309518 | 0.012805 |
| Reactome_ | Cellular Re | 1/736 | 0.96196 | 0.970626 | 0 | 0 | 0.303401 | 0.011767 |
| Reactome_ | Adaptive Ir | 1/733 | 0.96144 | 0.970626 | 0 | 0 | 0.304692 | 0.011981 |
| GTEx_Tissu | GTEX-RU72 | 1/382 | 0.813891 | 0.970996 | 0 | 0 | 0.596106 | 0.122756 |
| GTEx_Tissu | GTEX-XBED | 4/1276 | 0.813809 | 0.970996 | 0 | 0 | 0.706259 | 0.14551 |
| GTEx_Tissu | GTEX-XBEV | 3/997 | 0.815143 | 0.971969 | 0 | 0 | 0.679757 | 0.138936 |
| GTEx_Tissu | GTEX-R3RS | Aug-51 | 0.816892 | 0.972746 | 0 | 0 | 0.759386 | 0.153585 |
| GTEx_Tissu | GTEX-RM2 | 1/387 | 0.817981 | 0.972746 | 0 | 0 | 0.588234 | 0.118185 |
| GTEx_Tissu | GTEX-W5X | 1/386 | 0.81717 | 0.972746 | 0 | 0 | 0.589792 | 0.119083 |
| GTEx_Tissu | GTEX-SN8C | 2/707 | 0.817741 | 0.972746 | 0 | 0 | 0.641068 | 0.128989 |
| GTEx_Tissu | GTEX-O5YT | 2/705 | 0.816498 | 0.972746 | 0 | 0 | 0.642959 | 0.130348 |
| GTEx_Tissu | GTEX-PW2 | 1/388 | 0.818789 | 0.973186 | 0 | 0 | 0.586683 | 0.117295 |
| GTEx_Tissu | GTEX-XBED | 1/389 | 0.819593 | 0.973622 | 0 | 0 | 0.585141 | 0.116413 |
| GTEx_Tissu | GTEX-S32W | 1/390 | 0.820393 | 0.974052 | 0 | 0 | 0.583607 | 0.115538 |
| GeneSigDB | 15902281- | 1/793 | 0.970621 | 0.974119 | 0 | 0 | 0.280729 | 0.008371 |
| GTEx_Tissu | GTEX-X638 | 4/1291 | 0.820981 | 0.974231 | 0 | 0 | 0.697466 | 0.137579 |
| GTEx_Tissu | GTEX-SJXC- | 7/981 | 0.134705 | 0.974252 | 0 | 0 | 1.701399 | 3.410743 |
| GTEx_Tissu | GTEX-TSE9 | 9/1356 | 0.134545 | 0.974252 | 0 | 0 | 1.590372 | 3.190058 |
| GTEx_Tissu | GTEX-OHPI | 9/1358 | 0.135409 | 0.974333 | 0 | 0 | 1.587843 | 3.174817 |
| GTEx_Tissu | GTEX-PLZ5 | 7/982 | 0.135215 | 0.974333 | 0 | 0 | 1.699564 | 3.400639 |
| GTEx_Tissu | GTEX-TKQ1 | 7/983 | 0.135726 | 0.974333 | 0 | 0 | 1.697733 | 3.390568 |
| GTEx_Tissu | GTEX-X261 | 3/294 | 0.136464 | 0.974794 | 0 | 0 | 2.408198 | 4.796389 |
| GTEx_Tissu | GTEX-QLQ | 5/625 | 0.136379 | 0.974794 | 0 | 0 | 1.897423 | 3.780268 |
| GTEx_Tissu | GTEX-RVPU | Aug-66 | 0.822399 | 0.975393 | 0 | 0 | 0.753911 | 0.147412 |
| GTEx_Tissu | GTEX-P4QT | 20/5373 | 0.825537 | 0.976418 | 0 | 0 | 0.811931 | 0.155664 |
| GTEx_Tissu | GTEX-NFK9 | 2/720 | 0.825646 | 0.976418 | 0 | 0 | 0.629035 | 0.120516 |
| GTEx_Tissu | GTEX-QDVI | 1/398 | 0.82667 | 0.976418 | 0 | 0 | 0.571613 | 0.108806 |
| GTEx_Tissu | GTEX-VUSC | 1/398 | 0.82667 | 0.976418 | 0 | 0 | 0.571613 | 0.108806 |
| GTEx_Tissu | GTEX-QEG4 | 1/395 | 0.824342 | 0.976418 | 0 | 0 | 0.576054 | 0.111276 |
| GTEx_Tissu | GTEX-O5YV | 15/4152 | 0.826672 | 0.976418 | 0 | 0 | 0.794457 | 0.151223 |
| GTEx_Tissu | GTEX-WYB | 3/1021 | 0.82769 | 0.976418 | 0 | 0 | 0.662889 | 0.125363 |
| GTEx_Tissu | GTEX-XXEK | 1/400 | 0.828206 | 0.976418 | 0 | 0 | 0.568689 | 0.107194 |
| GTEx_Tissu | GTEX-XOTC | 5/1583 | 0.828219 | 0.976418 | 0 | 0 | 0.708484 | 0.133533 |
| GTEx_Tissu | GTEX-O5YV | 1/396 | 0.825122 | 0.976418 | 0 | 0 | 0.574566 | 0.110446 |

| | | | | | | | | |
|---|---|---|---|---|---|---|---|---|
| GTEx_Tissu | GTEX-WHV | 1/401 | 0.828968 | 0.976418 | 0 | 0 | 0.567238 | 0.106399 |
| GTEx_Tissu | GTEX-WL4( | 1/401 | 0.828968 | 0.976418 | 0 | 0 | 0.567238 | 0.106399 |
| GTEx_Tissu | GTEX-QCQ( | 3/1023 | 0.828703 | 0.976418 | 0 | 0 | 0.66152 | 0.124295 |
| GTEx_Tissu | GTEX-WH7 | 1/403 | 0.830483 | 0.977685 | 0 | 0 | 0.564358 | 0.104828 |
| GTEx_Tissu | GTEX-QDVI | 3/1029 | 0.83171 | 0.978095 | 0 | 0 | 0.657442 | 0.121148 |
| GTEx_Tissu | GTEX-QDT8 | 5/1591 | 0.831548 | 0.978095 | 0 | 0 | 0.704603 | 0.129975 |
| RNA-Seq_E | Eed Small i | 1/858 | 0.978139 | 0.978139 | 0 | 0 | 0.258555 | 0.005715 |
| GTEx_Tissu | GTEX-QVJC | Jul-31 | 0.832391 | 0.97819 | 0 | 0 | 0.732833 | 0.13444 |
| GTEx_Tissu | GTEX-NPJ7 | 6/1865 | 0.83267 | 0.97819 | 0 | 0 | 0.719383 | 0.131732 |
| GTEx_Tissu | GTEX-NL4V | 1/408 | 0.834213 | 0.978658 | 0 | 0 | 0.557282 | 0.101017 |
| GTEx_Tissu | GTEX-XXEK | 2/735 | 0.834388 | 0.978658 | 0 | 0 | 0.615681 | 0.111473 |
| GTEx_Tissu | GTEX-RVPV | 2/735 | 0.834388 | 0.978658 | 0 | 0 | 0.615681 | 0.111473 |
| GTEx_Tissu | GTEX-T6MI | 4/1322 | 0.835098 | 0.978974 | 0 | 0 | 0.679928 | 0.122527 |
| GTEx_Tissu | GTEX-OHPI | 6/804 | 0.137482 | 0.979362 | 0 | 0 | 1.774343 | 3.52076 |
| GTEx_Tissu | GTEX-R53T | 7/987 | 0.137781 | 0.979362 | 0 | 0 | 1.690446 | 3.350617 |
| GTEx_Tissu | GTEX-UJHI- | 4/457 | 0.138185 | 0.979828 | 0 | 0 | 2.070268 | 4.097392 |
| GTEx_Tissu | GTEX-T2IS- | 7/1002 | 0.145623 | 0.980133 | 0 | 0 | 1.663643 | 3.205394 |
| GTEx_Tissu | GTEX-T6MI | 22/4038 | 0.146477 | 0.980133 | 0 | 0 | 1.339772 | 2.573546 |
| GTEx_Tissu | GTEX-XXEK | 7/1004 | 0.146685 | 0.980133 | 0 | 0 | 1.66013 | 3.186564 |
| GTEx_Tissu | GTEX-X4EC | Dec-69 | 0.145294 | 0.980133 | 0 | 0 | 1.468043 | 2.831847 |
| GTEx_Tissu | GTEX-SSA3 | 8/1180 | 0.140841 | 0.980133 | 0 | 0 | 1.619303 | 3.17403 |
| GTEx_Tissu | GTEX-WHP | 6/820 | 0.146802 | 0.980133 | 0 | 0 | 1.738011 | 3.334671 |
| GTEx_Tissu | GTEX-RWS( | 10/1565 | 0.141404 | 0.980133 | 0 | 0 | 1.533219 | 2.999182 |
| GTEx_Tissu | GTEX-X638 | Dec-58 | 0.141176 | 0.980133 | 0 | 0 | 1.477246 | 2.892077 |
| GTEx_Tissu | GTEX-U3ZN | 7/997 | 0.142985 | 0.980133 | 0 | 0 | 1.672487 | 3.25301 |
| GTEx_Tissu | GTEX-R55E | 6/823 | 0.148581 | 0.980133 | 0 | 0 | 1.731357 | 3.301043 |
| GTEx_Tissu | GTEX-XQ3S | 20/3598 | 0.141469 | 0.980133 | 0 | 0 | 1.362806 | 2.6652 |
| GTEx_Tissu | GTEX-N7M | 13/2161 | 0.142332 | 0.980133 | 0 | 0 | 1.452923 | 2.83261 |
| GTEx_Tissu | GTEX-R45C | 3/306 | 0.14854 | 0.980133 | 0 | 0 | 2.31141 | 4.407632 |
| GTEx_Tissu | GTEX-PWC | 6/824 | 0.149177 | 0.980133 | 0 | 0 | 1.72915 | 3.28992 |
| GTEx_Tissu | GTEX-X261 | 8/1198 | 0.14956 | 0.980133 | 0 | 0 | 1.593277 | 3.027318 |
| GTEx_Tissu | GTEX-T6M( | 4/471 | 0.149287 | 0.980133 | 0 | 0 | 2.006759 | 3.816618 |
| GTEx_Tissu | GTEX-XMD | 10/1579 | 0.147275 | 0.980133 | 0 | 0 | 1.51838 | 2.908383 |
| GTEx_Tissu | GTEX-RM2I | 7/990 | 0.139332 | 0.980133 | 0 | 0 | 1.68502 | 3.320998 |
| GTEx_Tissu | GTEX-PX3G | 14/2384 | 0.149747 | 0.980133 | 0 | 0 | 1.419583 | 2.695512 |
| GTEx_Tissu | GTEX-XMD | Dec-72 | 0.146429 | 0.980133 | 0 | 0 | 1.465551 | 2.815641 |
| GTEx_Tissu | GTEX-SNM | 10/1583 | 0.148976 | 0.980133 | 0 | 0 | 1.514188 | 2.882968 |
| GTEx_Tissu | GTEX-X4XY | 6/824 | 0.149177 | 0.980133 | 0 | 0 | 1.72915 | 3.28992 |
| GTEx_Tissu | GTEX-X5EB | 6/820 | 0.146802 | 0.980133 | 0 | 0 | 1.738011 | 3.334671 |
| GTEx_Tissu | GTEX-WYJK | 7/990 | 0.139332 | 0.980133 | 0 | 0 | 1.68502 | 3.320998 |
| GTEx_Tissu | GTEX-UJHI- | 6/821 | 0.147394 | 0.980133 | 0 | 0 | 1.735787 | 3.323419 |
| GTEx_Tissu | GTEX-XBEC | 7/1006 | 0.147751 | 0.980133 | 0 | 0 | 1.656632 | 3.167857 |
| GTEx_Tissu | GTEX-WEY! | 21/3803 | 0.140008 | 0.980133 | 0 | 0 | 1.35711 | 2.668151 |
| GTEx_Tissu | GTEX-RTLS- | 10/1574 | 0.145164 | 0.980133 | 0 | 0 | 1.523649 | 2.940479 |
| GTEx_Tissu | GTEX-PWO | 7/997 | 0.142985 | 0.980133 | 0 | 0 | 1.672487 | 3.25301 |

| | | | | | | | | |
|---|---|---|---|---|---|---|---|---|
| GTEx_Tissu | GTEX-U8XE | 9/1378 | 0.144216 | 0.980133 | 0 | 0 | 1.56296 | 3.026587 |
| GTEx_Tissu | GTEX-QEG4 | 6/817 | 0.145032 | 0.980133 | 0 | 0 | 1.744714 | 3.36869 |
| GTEx_Tissu | GTEX-POYW | 11/1769 | 0.144657 | 0.980133 | 0 | 0 | 1.494708 | 2.889849 |
| GTEx_Tissu | GTEX-SNOS | 5/631 | 0.14032 | 0.980133 | 0 | 0 | 1.878653 | 3.689354 |
| GTEx_Tissu | GTEX-W5X | 14/2358 | 0.140764 | 0.980133 | 0 | 0 | 1.437456 | 2.818375 |
| GTEx_Tissu | GTEX-S3XE | 3/1039 | 0.836624 | 0.980247 | 0 | 0 | 0.650752 | 0.116081 |
| GeneSigDB | 16288205- | 1/866 | 0.978921 | 0.980682 | 0 | 0 | 0.256056 | 0.005455 |
| GTEx_Tissu | GTEX-WFJC | 1/413 | 0.837862 | 0.98096 | 0 | 0 | 0.550378 | 0.097363 |
| GTEx_Tissu | GTEX-S4Z8 | 2/742 | 0.838333 | 0.98096 | 0 | 0 | 0.609634 | 0.107503 |
| GTEx_Tissu | GTEX-WZT( | 3/1043 | 0.838555 | 0.98096 | 0 | 0 | 0.648111 | 0.114116 |
| GTEx_Tissu | GTEX-O5YV | 1/415 | 0.839299 | 0.981314 | 0 | 0 | 0.547663 | 0.095944 |
| GTEx_Tissu | GTEX-TML8 | 10/1587 | 0.150687 | 0.981472 | 0 | 0 | 1.510018 | 2.857782 |
| GTEx_Tissu | GTEX-T5JC- | 6/827 | 0.15097 | 0.981472 | 0 | 0 | 1.722561 | 3.256805 |
| GTEx_Tissu | GTEX-QDV. | 6/827 | 0.15097 | 0.981472 | 0 | 0 | 1.722561 | 3.256805 |
| GTEx_Tissu | GTEX-V955 | Jul-57 | 0.841659 | 0.981494 | 0 | 0 | 0.722913 | 0.124616 |
| GTEx_Tissu | GTEX-XOT( | 2/747 | 0.841099 | 0.981494 | 0 | 0 | 0.605385 | 0.104759 |
| GTEx_Tissu | GTEX-X4EP | 3/1048 | 0.840942 | 0.981494 | 0 | 0 | 0.644839 | 0.111707 |
| GTEx_Tissu | GTEX-REY6 | Aug-21 | 0.841531 | 0.981494 | 0 | 0 | 0.734419 | 0.126711 |
| GTEx_Tissu | GTEX-UPIC | 1/416 | 0.840013 | 0.981494 | 0 | 0 | 0.546315 | 0.095244 |
| GTEx_Tissu | GTEX-TKQ1 | Jun-25 | 0.854522 | 0.981575 | 0 | 0 | 0.694575 | 0.109196 |
| GTEx_Tissu | GTEX-XXEK | 2/765 | 0.85071 | 0.981575 | 0 | 0 | 0.590548 | 0.095482 |
| GTEx_Tissu | GTEX-TSE9 | Oct-55 | 0.845818 | 0.981575 | 0 | 0 | 0.748264 | 0.125297 |
| GTEx_Tissu | GTEX-WRH | Oct-59 | 0.847023 | 0.981575 | 0 | 0 | 0.747073 | 0.124034 |
| GTEx_Tissu | GTEX-XQ8I | 13/3716 | 0.844775 | 0.981575 | 0 | 0 | 0.769026 | 0.129723 |
| GTEx_Tissu | GTEX-QDT8 | 3/1055 | 0.844233 | 0.981575 | 0 | 0 | 0.640311 | 0.108422 |
| GTEx_Tissu | GTEX-XK95 | 1/422 | 0.844231 | 0.981575 | 0 | 0 | 0.538364 | 0.091161 |
| GTEx_Tissu | GTEX-X4XX | 2/752 | 0.843823 | 0.981575 | 0 | 0 | 0.601192 | 0.10209 |
| GTEx_Tissu | GTEX-QV31 | 3/1071 | 0.851539 | 0.981575 | 0 | 0 | 0.630183 | 0.101277 |
| GTEx_Tissu | GTEX-QXCU | 4/1367 | 0.853957 | 0.981575 | 0 | 0 | 0.655888 | 0.103548 |
| GTEx_Tissu | GTEX-RU1J | 18/4997 | 0.854184 | 0.981575 | 0 | 0 | 0.782452 | 0.123321 |
| GTEx_Tissu | GTEX-S7SE | 4/1340 | 0.842868 | 0.981575 | 0 | 0 | 0.670118 | 0.114553 |
| GTEx_Tissu | GTEX-X62C | 3/1074 | 0.852875 | 0.981575 | 0 | 0 | 0.628318 | 0.099992 |
| GTEx_Tissu | GTEX-WHP | Jun-20 | 0.852792 | 0.981575 | 0 | 0 | 0.696583 | 0.110923 |
| GTEx_Tissu | GTEX-WXY( | 18/4991 | 0.852726 | 0.981575 | 0 | 0 | 0.78371 | 0.124858 |
| GTEx_Tissu | GTEX-WVLI | 1/425 | 0.846298 | 0.981575 | 0 | 0 | 0.534472 | 0.089195 |
| GTEx_Tissu | GTEX-Q2A( | 3/1077 | 0.854202 | 0.981575 | 0 | 0 | 0.626463 | 0.098723 |
| GTEx_Tissu | GTEX-Q2AI | 1/437 | 0.8543 | 0.981575 | 0 | 0 | 0.519442 | 0.081798 |
| GTEx_Tissu | GTEX-XUZC | 1/437 | 0.8543 | 0.981575 | 0 | 0 | 0.519442 | 0.081798 |
| GTEx_Tissu | GTEX-OHPI | Jun-05 | 0.847505 | 0.981575 | 0 | 0 | 0.70267 | 0.116263 |
| GTEx_Tissu | GTEX-O5YT | 2/766 | 0.851228 | 0.981575 | 0 | 0 | 0.589744 | 0.094993 |
| GTEx_Tissu | GTEX-RNOI | 3/1076 | 0.853761 | 0.981575 | 0 | 0 | 0.62708 | 0.099144 |
| GTEx_Tissu | GTEX-S3XE | 1/429 | 0.849013 | 0.981575 | 0 | 0 | 0.529369 | 0.086648 |
| GTEx_Tissu | GTEX-PWN | 1/431 | 0.850352 | 0.981575 | 0 | 0 | 0.526852 | 0.085405 |
| GTEx_Tissu | GTEX-WVLI | 3/1076 | 0.853761 | 0.981575 | 0 | 0 | 0.62708 | 0.099144 |
| GTEx_Tissu | GTEX-XOT( | 1/428 | 0.848339 | 0.981575 | 0 | 0 | 0.530636 | 0.087277 |

| | | | | | | | | |
|---|---|---|---|---|---|---|---|---|
| GTEx_Tissu | GTEX-TKQ1 | 1/427 | 0.847661 | 0.981575 | 0 | 0 | 0.531909 | 0.087911 |
| GTEx_Tissu | GTEX-QV44 | 5/1633 | 0.848192 | 0.981575 | 0 | 0 | 0.684852 | 0.11276 |
| GTEx_Tissu | GTEX-VUSG | 1/428 | 0.848339 | 0.981575 | 0 | 0 | 0.530636 | 0.087277 |
| PhenGenI_ | Erythrocyte | 1/752 | 0.964618 | 0.981616 | 0 | 0 | 0.29669 | 0.010688 |
| PhenGenI_ | Body Weig | 1/788 | 0.969947 | 0.982709 | 0 | 0 | 0.282586 | 0.008623 |
| GTEx_Tissu | GTEX-QEG4 | 7/1013 | 0.15151 | 0.982778 | 0 | 0 | 1.644496 | 3.103332 |
| GTEx_Tissu | GTEX-RU72 | Nov-48 | 0.856128 | 0.982913 | 0 | 0 | 0.745638 | 0.115824 |
| GTEx_Tissu | GTEX-S7PM | 3/312 | 0.154705 | 0.983653 | 0 | 0 | 2.265834 | 4.228577 |
| GTEx_Tissu | GTEX-QMR | 5/651 | 0.153815 | 0.983653 | 0 | 0 | 1.818602 | 3.404429 |
| GTEx_Tissu | GTEX-XMD | 4/477 | 0.15415 | 0.983653 | 0 | 0 | 1.980692 | 3.70356 |
| GTEx_Tissu | GTEX-WHW | 5/652 | 0.154504 | 0.983653 | 0 | 0 | 1.815697 | 3.390877 |
| GTEx_Tissu | GTEX-QESD | 10/1592 | 0.152841 | 0.983653 | 0 | 0 | 1.504835 | 2.82662 |
| GTEx_Tissu | GTEX-T5JC | 6/830 | 0.152772 | 0.983653 | 0 | 0 | 1.716019 | 3.22407 |
| GTEx_Tissu | GTEX-XQ8I | 3/311 | 0.153672 | 0.983653 | 0 | 0 | 2.273307 | 4.257756 |
| GTEx_Tissu | GTEX-OIZI- | 7/1014 | 0.152051 | 0.983653 | 0 | 0 | 1.642776 | 3.094233 |
| GTEx_Tissu | GTEX-UPJH | 9/1399 | 0.153775 | 0.983653 | 0 | 0 | 1.537604 | 2.8788 |
| PhenGenI_ | Erythrocyte | 1/830 | 0.975167 | 0.983684 | 0 | 0 | 0.26768 | 0.006731 |
| GTEx_Tissu | GTEX-WFG | 5/1660 | 0.858172 | 0.984281 | 0 | 0 | 0.672684 | 0.102888 |
| GTEx_Tissu | GTEX-WZT( | 2/780 | 0.858317 | 0.984281 | 0 | 0 | 0.578709 | 0.088416 |
| GTEx_Tissu | GTEX-U8T8 | 14/4020 | 0.858647 | 0.984281 | 0 | 0 | 0.761522 | 0.116054 |
| GTEx_Tissu | GTEX-V1D1 | 1/445 | 0.859405 | 0.984642 | 0 | 0 | 0.509873 | 0.077254 |
| GO_Biologi | Positive Re | 1/938 | 0.984826 | 0.984826 | 0 | 0 | 0.235487 | 0.003601 |
| GTEx_Tissu | GTEX-O5YV | 3/1092 | 0.860681 | 0.985597 | 0 | 0 | 0.617342 | 0.092621 |
| GTEx_Tissu | GTEX-PWO | 3/1095 | 0.861946 | 0.986538 | 0 | 0 | 0.615548 | 0.091447 |
| GTEx_Tissu | GTEX-S4Q7 | 6/835 | 0.155798 | 0.9868 | 0 | 0 | 1.705223 | 3.170337 |
| GTEx_Tissu | GTEX-R55E | 10/1599 | 0.155882 | 0.9868 | 0 | 0 | 1.497634 | 2.783583 |
| GTEx_Tissu | GTEX-WL4( | 2/798 | 0.866984 | 0.987273 | 0 | 0 | 0.56509 | 0.080658 |
| GTEx_Tissu | GTEX-QEL4 | 1/463 | 0.870252 | 0.987273 | 0 | 0 | 0.489555 | 0.068034 |
| GTEx_Tissu | GTEX-QVU! | Nov-96 | 0.869005 | 0.987273 | 0 | 0 | 0.732628 | 0.102866 |
| GTEx_Tissu | GTEX-NPJ8 | 14/4043 | 0.864406 | 0.987273 | 0 | 0 | 0.75608 | 0.11017 |
| GTEx_Tissu | GTEX-S32W | 4/1394 | 0.864387 | 0.987273 | 0 | 0 | 0.642212 | 0.093593 |
| GTEx_Tissu | GTEX-R55F | Aug-25 | 0.873339 | 0.987273 | 0 | 0 | 0.699889 | 0.094787 |
| GTEx_Tissu | GTEX-OXRK | 1/469 | 0.873681 | 0.987273 | 0 | 0 | 0.48313 | 0.065242 |
| GTEx_Tissu | GTEX-XPVG | 1/469 | 0.873681 | 0.987273 | 0 | 0 | 0.48313 | 0.065242 |
| GTEx_Tissu | GTEX-T5JW | Dec-32 | 0.86358 | 0.987273 | 0 | 0 | 0.745136 | 0.109288 |
| GTEx_Tissu | GTEX-QDT8 | 3/1122 | 0.872896 | 0.987273 | 0 | 0 | 0.599834 | 0.081541 |
| GTEx_Tissu | GTEX-SE5C | 1/454 | 0.864936 | 0.987273 | 0 | 0 | 0.499512 | 0.072479 |
| GTEx_Tissu | GTEX-XBED | 17/4815 | 0.869463 | 0.987273 | 0 | 0 | 0.765066 | 0.107017 |
| GTEx_Tissu | GTEX-RVPU | 24/6541 | 0.872636 | 0.987273 | 0 | 0 | 0.783066 | 0.106683 |
| GTEx_Tissu | GTEX-OXRI | 5/1694 | 0.86997 | 0.987273 | 0 | 0 | 0.657916 | 0.091645 |
| GTEx_Tissu | GTEX-S33H | 5/1692 | 0.8693 | 0.987273 | 0 | 0 | 0.658768 | 0.092272 |
| GTEx_Tissu | GTEX-OXRF | 1/466 | 0.871978 | 0.987273 | 0 | 0 | 0.486322 | 0.066622 |
| GTEx_Tissu | GTEX-WEY! | 1/466 | 0.871978 | 0.987273 | 0 | 0 | 0.486322 | 0.066622 |
| GTEx_Tissu | GTEX-OHPI | 2/799 | 0.867451 | 0.987273 | 0 | 0 | 0.564352 | 0.080249 |
| GTEx_Tissu | GTEX-S7SE | 2/802 | 0.868843 | 0.987273 | 0 | 0 | 0.562147 | 0.079034 |

| | | | | | | | | |
|---|---|---|---|---|---|---|---|---|
| GTEx_Tissu | GTEX-QVJC | 1/459 | 0.867916 | 0.987273 | 0 | 0 | 0.493932 | 0.069971 |
| GTEx_Tissu | GTEX-WHS | 4/1405 | 0.868454 | 0.987273 | 0 | 0 | 0.636791 | 0.089814 |
| GTEx_Tissu | GTEX-NFK9 | 1/461 | 0.869089 | 0.987273 | 0 | 0 | 0.491734 | 0.068995 |
| GTEx_Tissu | GTEX-SE5C | 1/468 | 0.873116 | 0.987273 | 0 | 0 | 0.484189 | 0.065698 |
| GTEx_Tissu | GTEX-SN8C | Oct-53 | 0.873333 | 0.987273 | 0 | 0 | 0.719983 | 0.097513 |
| GTEx_Tissu | GTEX-RNOI | 1/454 | 0.864936 | 0.987273 | 0 | 0 | 0.499512 | 0.072479 |
| PhenGenI_ | Smoking | 1/938 | 0.984826 | 0.98764 | 0 | 0 | 0.235487 | 0.003601 |
| PhenGenI_ | Platelet Fu | 23/7446 | 0.98764 | 0.98764 | 0 | 0 | 0.604687 | 0.007521 |
| GTEx_Tissu | GTEX-UPK5 | 6/836 | 0.156407 | 0.987959 | 0 | 0 | 1.703079 | 3.159713 |
| GTEx_Tissu | GTEX-WFG | 4/1423 | 0.874886 | 0.988133 | 0 | 0 | 0.628102 | 0.083953 |
| GWAS_Cat | Schizophre | 1/996 | 0.988366 | 0.988366 | 0 | 0 | 0.221082 | 0.002587 |
| GTEx_Tissu | GTEX-XV7C | 7/1024 | 0.157509 | 0.988448 | 0 | 0 | 1.625762 | 3.004849 |
| GTEx_Tissu | GTEX-PX3G | 8/1213 | 0.157027 | 0.988448 | 0 | 0 | 1.572183 | 2.910645 |
| GTEx_Tissu | GTEX-NPJ7 | 5/656 | 0.157273 | 0.988448 | 0 | 0 | 1.804166 | 3.337298 |
| Reactome_ | Disease R-I | 3/1736 | 0.984183 | 0.988597 | 0 | 0 | 0.37466 | 0.005973 |
| GTEx_Tissu | GTEX-T6MI | 2/818 | 0.876048 | 0.988943 | 0 | 0 | 0.550663 | 0.072872 |
| GTEx_Tissu | GTEX-XXEK | 5/657 | 0.157968 | 0.989182 | 0 | 0 | 1.801306 | 3.324059 |
| GTEx_Tissu | GTEX-QDT8 | 5/1715 | 0.876845 | 0.98934 | 0 | 0 | 0.649087 | 0.085307 |
| GTEx_Tissu | GTEX-QLQ7 | 5/1720 | 0.878436 | 0.989364 | 0 | 0 | 0.647017 | 0.083861 |
| GTEx_Tissu | GTEX-QDT8 | 14/4105 | 0.879051 | 0.989364 | 0 | 0 | 0.741715 | 0.095616 |
| GTEx_Tissu | GTEX-Q2AC | 3/1135 | 0.877894 | 0.989364 | 0 | 0 | 0.592535 | 0.077166 |
| GTEx_Tissu | GTEX-SIU8- | 16/4609 | 0.879089 | 0.989364 | 0 | 0 | 0.751664 | 0.096866 |
| GTEx_Tissu | GTEX-XGQ4 | 1/476 | 0.877568 | 0.989364 | 0 | 0 | 0.475838 | 0.062145 |
| GTEx_Tissu | GTEX-RWS( | 6/841 | 0.159465 | 0.989432 | 0 | 0 | 1.692437 | 3.107195 |
| GTEx_Tissu | GTEX-QDVI | 7/1026 | 0.158612 | 0.989432 | 0 | 0 | 1.622399 | 2.987316 |
| GTEx_Tissu | GTEX-Q2AC | 10/1607 | 0.159397 | 0.989432 | 0 | 0 | 1.489481 | 2.735224 |
| GTEx_Tissu | GTEX-T5JC- | 5/659 | 0.159363 | 0.989432 | 0 | 0 | 1.795611 | 3.297766 |
| GTEx_Tissu | GTEX-UPK5 | 7/1028 | 0.159718 | 0.989432 | 0 | 0 | 1.61905 | 2.969895 |
| GTEx_Tissu | GTEX-VJYA | 1/480 | 0.879736 | 0.989592 | 0 | 0 | 0.471768 | 0.060449 |
| GTEx_Tissu | GTEX-Q2AI | 5/1727 | 0.880635 | 0.989794 | 0 | 0 | 0.644139 | 0.081878 |
| GTEx_Tissu | GTEX-XUJ4 | 1/482 | 0.880806 | 0.989794 | 0 | 0 | 0.469758 | 0.059621 |
| GTEx_Tissu | GTEX-S32V | 13/3863 | 0.881887 | 0.99001 | 0 | 0 | 0.732955 | 0.092126 |
| GTEx_Tissu | GTEX-S4Q7 | Sep-22 | 0.881588 | 0.99001 | 0 | 0 | 0.701414 | 0.0884 |
| GTEx_Tissu | GTEX-QVU! | 1/486 | 0.882917 | 0.990666 | 0 | 0 | 0.465788 | 0.058002 |
| GTEx_Tissu | GTEX-PSDC | 1/487 | 0.883439 | 0.990752 | 0 | 0 | 0.464805 | 0.057605 |
| GeneSigDB | 16166618-: | 2/1495 | 0.990785 | 0.990785 | 0 | 0 | 0.290296 | 0.002687 |
| GTEx_Tissu | GTEX-PLZ6- | 1/490 | 0.884991 | 0.991493 | 0 | 0 | 0.461882 | 0.056432 |
| GTEx_Tissu | GTEX-WFO | 1/490 | 0.884991 | 0.991493 | 0 | 0 | 0.461882 | 0.056432 |
| GTEx_Tissu | GTEX-QVU! | 5/1867 | 0.918083 | 0.99168 | 0 | 0 | 0.591123 | 0.050522 |
| GTEx_Tissu | GTEX-S341 | 4/1496 | 0.898306 | 0.99168 | 0 | 0 | 0.595013 | 0.063812 |
| GTEx_Tissu | GTEX-S341 | 1/519 | 0.898981 | 0.99168 | 0 | 0 | 0.435373 | 0.046365 |
| GTEx_Tissu | GTEX-WEY! | 1/520 | 0.899432 | 0.99168 | 0 | 0 | 0.434512 | 0.046055 |
| GTEx_Tissu | GTEX-OHPI | 1/522 | 0.900328 | 0.99168 | 0 | 0 | 0.432799 | 0.045442 |
| GTEx_Tissu | GTEX-O5YT | 1/522 | 0.900328 | 0.99168 | 0 | 0 | 0.432799 | 0.045442 |
| GTEx_Tissu | GTEX-QEG! | 1/571 | 0.919981 | 0.99168 | 0 | 0 | 0.394594 | 0.03291 |

| | | | | | | | | |
|---|---|---|---|---|---|---|---|---|
| GTEx_Tissu | GTEX-Q2AI | 15/4575 | 0.920282 | 0.99168 | 0 | 0 | 0.701435 | 0.058272 |
| GTEx_Tissu | GTEX-P78B | 4/1695 | 0.943894 | 0.99168 | 0 | 0 | 0.519319 | 0.029986 |
| GTEx_Tissu | GTEX-P44H | 4/1495 | 0.898013 | 0.99168 | 0 | 0 | 0.595444 | 0.064053 |
| GTEx_Tissu | GTEX-QXCU | 1/654 | 0.944909 | 0.99168 | 0 | 0 | 0.342961 | 0.019435 |
| GTEx_Tissu | GTEX-P44H | 1/656 | 0.945403 | 0.99168 | 0 | 0 | 0.341878 | 0.019194 |
| GTEx_Tissu | GTEX-OHPI | 20/6029 | 0.945457 | 0.99168 | 0 | 0 | 0.690705 | 0.038739 |
| GTEx_Tissu | GTEX-WWY | 15/4758 | 0.945469 | 0.99168 | 0 | 0 | 0.666333 | 0.037364 |
| GTEx_Tissu | GTEX-WY7( | 4/1706 | 0.945773 | 0.99168 | 0 | 0 | 0.515651 | 0.028749 |
| GTEx_Tissu | GTEX-T8EM | Nov-63 | 0.940165 | 0.99168 | 0 | 0 | 0.644459 | 0.039763 |
| GTEx_Tissu | GTEX-S95S | 15/4719 | 0.940744 | 0.99168 | 0 | 0 | 0.673585 | 0.041145 |
| GTEx_Tissu | GTEX-QEG5 | 2/973 | 0.929307 | 0.99168 | 0 | 0 | 0.459005 | 0.033653 |
| GTEx_Tissu | GTEX-N7M | 1/645 | 0.942629 | 0.99168 | 0 | 0 | 0.347916 | 0.020556 |
| GTEx_Tissu | GTEX-XUJ4 | 2/955 | 0.92445 | 0.99168 | 0 | 0 | 0.468119 | 0.036774 |
| GTEx_Tissu | GTEX-UJM( | 15/4639 | 0.92999 | 0.99168 | 0 | 0 | 0.688843 | 0.049998 |
| GTEx_Tissu | GTEX-OIZG | 1/596 | 0.928478 | 0.99168 | 0 | 0 | 0.377526 | 0.028016 |
| GTEx_Tissu | GTEX-R55E | 1/597 | 0.928799 | 0.99168 | 0 | 0 | 0.376873 | 0.027837 |
| GTEx_Tissu | GTEX-XPVG | 3/1305 | 0.929027 | 0.99168 | 0 | 0 | 0.510506 | 0.037582 |
| GTEx_Tissu | GTEX-XLM4 | 1/598 | 0.929118 | 0.99168 | 0 | 0 | 0.376222 | 0.02766 |
| GTEx_Tissu | GTEX-PSDG | 17/5143 | 0.929125 | 0.99168 | 0 | 0 | 0.700571 | 0.051501 |
| GTEx_Tissu | GTEX-QCQ( | 3/1251 | 0.915383 | 0.99168 | 0 | 0 | 0.53414 | 0.047225 |
| GTEx_Tissu | GTEX-TSE9 | 1/583 | 0.924177 | 0.99168 | 0 | 0 | 0.386218 | 0.030454 |
| GTEx_Tissu | GTEX-OIZH | 3/1289 | 0.925207 | 0.99168 | 0 | 0 | 0.517302 | 0.040214 |
| GTEx_Tissu | GTEX-QVUS | 3/1290 | 0.925451 | 0.99168 | 0 | 0 | 0.516872 | 0.040044 |
| GTEx_Tissu | GTEX-O5YT | 20/5872 | 0.925857 | 0.99168 | 0 | 0 | 0.717244 | 0.055253 |
| GTEx_Tissu | GTEX-QLQ7 | Jun-95 | 0.926163 | 0.99168 | 0 | 0 | 0.599767 | 0.046005 |
| GTEx_Tissu | GTEX-UJM( | 3/1298 | 0.927378 | 0.99168 | 0 | 0 | 0.513458 | 0.038712 |
| GTEx_Tissu | GTEX-S3XE | 18/5384 | 0.927524 | 0.99168 | 0 | 0 | 0.707206 | 0.053208 |
| GTEx_Tissu | GTEX-Q2AI | 1/647 | 0.943143 | 0.99168 | 0 | 0 | 0.346803 | 0.020301 |
| GTEx_Tissu | GTEX-S341 | 5/1823 | 0.907583 | 0.99168 | 0 | 0 | 0.606905 | 0.058852 |
| GTEx_Tissu | GTEX-NPJ8 | Jun-14 | 0.908779 | 0.99168 | 0 | 0 | 0.625659 | 0.059846 |
| GTEx_Tissu | GTEX-QV31 | 1/543 | 0.909275 | 0.99168 | 0 | 0 | 0.41558 | 0.039525 |
| GTEx_Tissu | GTEX-T6MI | 2/905 | 0.909285 | 0.99168 | 0 | 0 | 0.495342 | 0.047105 |
| GTEx_Tissu | GTEX-Q2AH | Dec-66 | 0.944864 | 0.99168 | 0 | 0 | 0.645787 | 0.036625 |
| GTEx_Tissu | GTEX-O5YT | May-83 | 0.940958 | 0.99168 | 0 | 0 | 0.55288 | 0.033646 |
| GTEx_Tissu | GTEX-REY6 | 1/641 | 0.941586 | 0.99168 | 0 | 0 | 0.350164 | 0.021076 |
| GTEx_Tissu | GTEX-OXRL | May-95 | 0.942968 | 0.99168 | 0 | 0 | 0.549179 | 0.032249 |
| GTEx_Tissu | GTEX-X62C | 3/1223 | 0.907421 | 0.99168 | 0 | 0 | 0.547219 | 0.053162 |
| GTEx_Tissu | GTEX-XAJ8 | 1/625 | 0.937223 | 0.99168 | 0 | 0 | 0.35944 | 0.023304 |
| GTEx_Tissu | GTEX-XYKS | May-64 | 0.937647 | 0.99168 | 0 | 0 | 0.558834 | 0.035978 |
| GTEx_Tissu | GTEX-R55G | Sep-04 | 0.937798 | 0.99168 | 0 | 0 | 0.626991 | 0.040266 |
| GTEx_Tissu | GTEX-RVPV | 2/1014 | 0.939303 | 0.99168 | 0 | 0 | 0.439456 | 0.027517 |
| GTEx_Tissu | GTEX-S33H | May-75 | 0.939584 | 0.99168 | 0 | 0 | 0.555373 | 0.03461 |
| GTEx_Tissu | GTEX-T6MI | 17/4997 | 0.906036 | 0.99168 | 0 | 0 | 0.72823 | 0.071859 |
| GTEx_Tissu | GTEX-QESD | Dec-92 | 0.900497 | 0.99168 | 0 | 0 | 0.705783 | 0.073972 |
| GTEx_Tissu | GTEX-QXCU | 20/5783 | 0.912466 | 0.99168 | 0 | 0 | 0.732931 | 0.06714 |

| | | | | | | | | |
|---|---|---|---|---|---|---|---|---|
| GTEx_Tissu | GTEX-XV7C | 1/559 | 0.915555 | 0.99168 | 0 | 0 | 0.40333 | 0.035584 |
| GTEx_Tissu | GTEX-QMR | Jun-45 | 0.915804 | 0.99168 | 0 | 0 | 0.615518 | 0.054137 |
| GTEx_Tissu | GTEX-X4XX | 2/926 | 0.915969 | 0.99168 | 0 | 0 | 0.48355 | 0.042443 |
| GTEx_Tissu | GTEX-XUZC | 5/1858 | 0.916023 | 0.99168 | 0 | 0 | 0.59429 | 0.052128 |
| GTEx_Tissu | GTEX-UPJH | 1/564 | 0.917428 | 0.99168 | 0 | 0 | 0.399645 | 0.034442 |
| GTEx_Tissu | GTEX-XOT4 | 3/1231 | 0.909762 | 0.99168 | 0 | 0 | 0.543421 | 0.051393 |
| GTEx_Tissu | GTEX-S7SF- | 2/1005 | 0.937229 | 0.99168 | 0 | 0 | 0.44361 | 0.028758 |
| GTEx_Tissu | GTEX-XQ3S | 24/6755 | 0.911201 | 0.99168 | 0 | 0 | 0.746058 | 0.069377 |
| GTEx_Tissu | GTEX-OOB. | Jul-48 | 0.89801 | 0.99168 | 0 | 0 | 0.656792 | 0.070654 |
| GTEx_Tissu | GTEX-QEL4 | 16/4783 | 0.91263 | 0.99168 | 0 | 0 | 0.716002 | 0.06546 |
| GTEx_Tissu | GTEX-V1D1 | Aug-90 | 0.913158 | 0.99168 | 0 | 0 | 0.650601 | 0.059105 |
| GTEx_Tissu | GTEX-OXRL | 3/1244 | 0.913452 | 0.99168 | 0 | 0 | 0.537355 | 0.048643 |
| GTEx_Tissu | GTEX-QEG5 | 1/554 | 0.91364 | 0.99168 | 0 | 0 | 0.407082 | 0.036767 |
| GTEx_Tissu | GTEX-XXEK | 13/4160 | 0.935618 | 0.99168 | 0 | 0 | 0.667881 | 0.044446 |
| GTEx_Tissu | GTEX-WHS | 2/999 | 0.93581 | 0.99168 | 0 | 0 | 0.446422 | 0.029617 |
| GTEx_Tissu | GTEX-TSE9 | 4/1674 | 0.940145 | 0.99168 | 0 | 0 | 0.526456 | 0.032494 |
| GTEx_Tissu | GTEX-RWS | Jun-28 | 0.886658 | 0.99168 | 0 | 0 | 0.65542 | 0.078844 |
| GTEx_Tissu | GTEX-QEG4 | 21/5914 | 0.89253 | 0.99168 | 0 | 0 | 0.756984 | 0.086065 |
| GTEx_Tissu | GTEX-PSDC | 14/4322 | 0.920898 | 0.99168 | 0 | 0 | 0.694694 | 0.057247 |
| GTEx_Tissu | GTEX-XUJ4 | 1/575 | 0.921405 | 0.99168 | 0 | 0 | 0.391763 | 0.032068 |
| GTEx_Tissu | GTEX-TKQ1 | 1/577 | 0.922107 | 0.99168 | 0 | 0 | 0.390363 | 0.031656 |
| GTEx_Tissu | GTEX-T6M( | 1/578 | 0.922456 | 0.99168 | 0 | 0 | 0.389666 | 0.031452 |
| GTEx_Tissu | GTEX-P4QF | Jul-64 | 0.923318 | 0.99168 | 0 | 0 | 0.621652 | 0.049597 |
| GTEx_Tissu | GTEX-XBED | Jun-83 | 0.923783 | 0.99168 | 0 | 0 | 0.603481 | 0.047843 |
| GTEx_Tissu | GTEX-P4PC | 1/564 | 0.917428 | 0.99168 | 0 | 0 | 0.399645 | 0.034442 |
| GTEx_Tissu | GTEX-QLQ7 | 14/4167 | 0.892453 | 0.99168 | 0 | 0 | 0.727779 | 0.082808 |
| GTEx_Tissu | GTEX-WFO | Nov-26 | 0.918742 | 0.99168 | 0 | 0 | 0.675219 | 0.057225 |
| GTEx_Tissu | GTEX-QDV. | Dec-94 | 0.919547 | 0.99168 | 0 | 0 | 0.682433 | 0.057238 |
| GTEx_Tissu | GTEX-X4XX | 2/938 | 0.91958 | 0.99168 | 0 | 0 | 0.477049 | 0.039995 |
| GTEx_Tissu | GTEX-R55E | 1/570 | 0.919621 | 0.99168 | 0 | 0 | 0.395308 | 0.033124 |
| GTEx_Tissu | GTEX-WFG | 3/1267 | 0.919653 | 0.99168 | 0 | 0 | 0.526927 | 0.044135 |
| GTEx_Tissu | GTEX-X88G | 1/571 | 0.919981 | 0.99168 | 0 | 0 | 0.394594 | 0.03291 |
| GTEx_Tissu | GTEX-Q2A( | 4/1554 | 0.914118 | 0.99168 | 0 | 0 | 0.570944 | 0.051269 |
| GTEx_Tissu | GTEX-X5EB | 3/1247 | 0.914285 | 0.99168 | 0 | 0 | 0.535973 | 0.04803 |
| GTEx_Tissu | GTEX-XPT6 | 2/1025 | 0.941751 | 0.99168 | 0 | 0 | 0.434478 | 0.026075 |
| GTEx_Tissu | GTEX-U3ZN | 13/3997 | 0.909368 | 0.99168 | 0 | 0 | 0.702394 | 0.066731 |
| GTEx_Tissu | GTEX-OIZF- | Jul-03 | 0.886487 | 0.99168 | 0 | 0 | 0.67138 | 0.080894 |
| GTEx_Tissu | GTEX-S341 | 1/494 | 0.887029 | 0.99168 | 0 | 0 | 0.45804 | 0.054909 |
| GTEx_Tissu | GTEX-SNOS | Jun-35 | 0.888611 | 0.99168 | 0 | 0 | 0.652903 | 0.077105 |
| GTEx_Tissu | GTEX-WQL | Jun-35 | 0.888611 | 0.99168 | 0 | 0 | 0.652903 | 0.077105 |
| GTEx_Tissu | GTEX-SIU7- | 5/1762 | 0.891139 | 0.99168 | 0 | 0 | 0.630093 | 0.072621 |
| GTEx_Tissu | GTEX-RTLS- | 3/1173 | 0.891531 | 0.99168 | 0 | 0 | 0.572131 | 0.065689 |
| GTEx_Tissu | GTEX-Q2A( | 3/1356 | 0.940053 | 0.99168 | 0 | 0 | 0.489917 | 0.030286 |
| GTEx_Tissu | GTEX-WFG | 1/524 | 0.901217 | 0.99168 | 0 | 0 | 0.4311 | 0.044839 |
| GTEx_Tissu | GTEX-WVLI | 1/530 | 0.903835 | 0.99168 | 0 | 0 | 0.426078 | 0.04308 |

| | | | | | | | | |
|---|---|---|---|---|---|---|---|---|
| GTEx_Tissu | GTEX-TSE9 | 4/1519 | 0.904855 | 0.99168 | 0 | 0 | 0.585248 | 0.058513 |
| GTEx_Tissu | GTEX-S7PM | Jun-98 | 0.904963 | 0.99168 | 0 | 0 | 0.631011 | 0.063013 |
| GTEx_Tissu | GTEX-SNOS | 1/533 | 0.905118 | 0.99168 | 0 | 0 | 0.42361 | 0.042229 |
| GTEx_Tissu | GTEX-PVOV | 3/1216 | 0.905329 | 0.99168 | 0 | 0 | 0.550583 | 0.054759 |
| GTEx_Tissu | GTEX-T6M( | 1/534 | 0.905542 | 0.99168 | 0 | 0 | 0.422793 | 0.04195 |
| GTEx_Tissu | GTEX-PLZ6 | Oct-31 | 0.892364 | 0.99168 | 0 | 0 | 0.698744 | 0.079574 |
| GTEx_Tissu | GTEX-RVPV | May-51 | 0.935289 | 0.99168 | 0 | 0 | 0.562975 | 0.037663 |
| GTEx_Tissu | GTEX-OXRL | Oct-70 | 0.936814 | 0.99168 | 0 | 0 | 0.639804 | 0.041761 |
| GTEx_Tissu | GTEX-WZT( | 1/599 | 0.929436 | 0.99168 | 0 | 0 | 0.375574 | 0.027483 |
| GTEx_Tissu | GTEX-S7SE | 3/1214 | 0.904723 | 0.99168 | 0 | 0 | 0.551551 | 0.055225 |
| GTEx_Tissu | GTEX-PVOV | 1/606 | 0.931622 | 0.99168 | 0 | 0 | 0.371094 | 0.026284 |
| GTEx_Tissu | GTEX-PLZ4 | 1/614 | 0.934038 | 0.99168 | 0 | 0 | 0.366099 | 0.024982 |
| GTEx_Tissu | GTEX-UJHI | 1/616 | 0.934629 | 0.99168 | 0 | 0 | 0.36487 | 0.024667 |
| GTEx_Tissu | GTEX-R55F | 14/4415 | 0.934816 | 0.99168 | 0 | 0 | 0.675961 | 0.045564 |
| GTEx_Tissu | GTEX-WFG | 1/617 | 0.934922 | 0.99168 | 0 | 0 | 0.364259 | 0.024512 |
| GTEx_Tissu | GTEX-XQ3S | May-58 | 0.936569 | 0.99168 | 0 | 0 | 0.560738 | 0.036747 |
| GTEx_Tissu | GTEX-NPJ8 | 1/594 | 0.927833 | 0.99168 | 0 | 0 | 0.378838 | 0.028376 |
| GTEx_Tissu | GTEX-SIU7 | Oct-13 | 0.927928 | 0.99168 | 0 | 0 | 0.653086 | 0.048851 |
| GTEx_Tissu | GTEX-VJYA | 3/1204 | 0.901644 | 0.99168 | 0 | 0 | 0.556441 | 0.057611 |
| GTEx_Tissu | GTEX-OHPI | 1/525 | 0.901658 | 0.99168 | 0 | 0 | 0.430255 | 0.04454 |
| GTEx_Tissu | GTEX-Q2A( | 3/1205 | 0.901956 | 0.99168 | 0 | 0 | 0.555948 | 0.057368 |
| GTEx_Tissu | GTEX-P4PP | 2/885 | 0.902466 | 0.99168 | 0 | 0 | 0.507095 | 0.05204 |
| GTEx_Tissu | GTEX-WEY! | 1/528 | 0.90297 | 0.99168 | 0 | 0 | 0.427739 | 0.043658 |
| GTEx_Tissu | GTEX-PLZ4 | Dec-26 | 0.886363 | 0.99168 | 0 | 0 | 0.721594 | 0.087045 |
| GTEx_Tissu | GTEX-N7M | 4/1495 | 0.898013 | 0.99168 | 0 | 0 | 0.595444 | 0.064053 |
| GTEx_Tissu | GTEX-X88G | Jul-32 | 0.894028 | 0.99168 | 0 | 0 | 0.661914 | 0.074146 |
| GTEx_Tissu | GTEX-U8XE | 2/863 | 0.894423 | 0.99168 | 0 | 0 | 0.520653 | 0.058093 |
| GTEx_Tissu | GTEX-NPJ8 | 1/510 | 0.894829 | 0.99168 | 0 | 0 | 0.443277 | 0.049258 |
| GTEx_Tissu | GTEX-S4Z8 | Oct-45 | 0.895524 | 0.99168 | 0 | 0 | 0.695043 | 0.076695 |
| GTEx_Tissu | GTEX-U412 | 17/4945 | 0.896515 | 0.99168 | 0 | 0 | 0.738477 | 0.080672 |
| GTEx_Tissu | GTEX-P4PP | 1/516 | 0.897615 | 0.99168 | 0 | 0 | 0.437977 | 0.047308 |
| GTEx_Tissu | GTEX-U8XE | Jul-72 | 0.903749 | 0.99168 | 0 | 0 | 0.649239 | 0.065705 |
| GTEx_Tissu | GTEX-SNM | 1/547 | 0.910887 | 0.99168 | 0 | 0 | 0.41245 | 0.038496 |
| GTEx_Tissu | GTEX-N7M | 5/1820 | 0.906827 | 0.99168 | 0 | 0 | 0.608009 | 0.059465 |
| GTEx_Tissu | GTEX-WVLI | 5/1821 | 0.90708 | 0.99168 | 0 | 0 | 0.607641 | 0.05926 |
| GTEx_Tissu | GTEX-PWC | 1/621 | 0.936083 | 0.99168 | 0 | 0 | 0.361834 | 0.0239 |
| GTEx_Tissu | GTEX-WZT( | 4/1653 | 0.936172 | 0.99168 | 0 | 0 | 0.533774 | 0.035205 |
| GTEx_Tissu | GTEX-WY7( | 1/622 | 0.93637 | 0.99168 | 0 | 0 | 0.361233 | 0.023749 |
| GTEx_Tissu | GTEX-WEY! | 1/622 | 0.93637 | 0.99168 | 0 | 0 | 0.361233 | 0.023749 |
| GTEx_Tissu | GTEX-XOT( | 4/1710 | 0.946442 | 0.991914 | 0 | 0 | 0.514329 | 0.028312 |
| GTEx_Tissu | GTEX-PW2 | 4/1713 | 0.946939 | 0.991968 | 0 | 0 | 0.513342 | 0.027988 |
| GTEx_Tissu | GTEX-U4B1 | 2/1159 | 0.965004 | 0.992005 | 0 | 0 | 0.381433 | 0.013588 |
| GTEx_Tissu | GTEX-R55E | 4/1799 | 0.959529 | 0.992005 | 0 | 0 | 0.486438 | 0.020096 |
| GTEx_Tissu | GTEX-V1D1 | 1/738 | 0.962303 | 0.992005 | 0 | 0 | 0.302546 | 0.011626 |
| GTEx_Tissu | GTEX-P44H | 2/1076 | 0.951935 | 0.992005 | 0 | 0 | 0.412729 | 0.02033 |

| | | | | | | | | |
|---|---|---|---|---|---|---|---|---|
| GTEx_Tissu | GTEX-U3ZG | Jul-80 | 0.956415 | 0.992005 | 0 | 0 | 0.564347 | 0.025149 |
| GTEx_Tissu | GTEX-WHS | Nov-40 | 0.960621 | 0.992005 | 0 | 0 | 0.607978 | 0.024426 |
| GTEx_Tissu | GTEX-RWS( | 1/733 | 0.96144 | 0.992005 | 0 | 0 | 0.304692 | 0.011981 |
| GTEx_Tissu | GTEX-XQ3S | 2/1142 | 0.962637 | 0.992005 | 0 | 0 | 0.387472 | 0.014754 |
| GTEx_Tissu | GTEX-NPJ7 | 3/1496 | 0.962771 | 0.992005 | 0 | 0 | 0.440628 | 0.016717 |
| GTEx_Tissu | GTEX-RU72 | 2/1147 | 0.963349 | 0.992005 | 0 | 0 | 0.385677 | 0.014401 |
| GTEx_Tissu | GTEX-REY6 | 3/1501 | 0.963411 | 0.992005 | 0 | 0 | 0.439038 | 0.016365 |
| GTEx_Tissu | GTEX-T5JW | 1/752 | 0.964618 | 0.992005 | 0 | 0 | 0.29669 | 0.010688 |
| GTEx_Tissu | GTEX-RU72 | 1/753 | 0.964778 | 0.992005 | 0 | 0 | 0.29628 | 0.010624 |
| GTEx_Tissu | GTEX-R53T | 3/1444 | 0.95547 | 0.992005 | 0 | 0 | 0.457817 | 0.020854 |
| GTEx_Tissu | GTEX-PVO\ | 4/1795 | 0.95901 | 0.992005 | 0 | 0 | 0.487632 | 0.020409 |
| GTEx_Tissu | GTEX-PWO | 2/1058 | 0.948549 | 0.992005 | 0 | 0 | 0.420165 | 0.022194 |
| GTEx_Tissu | GTEX-O5Y\ | 3/1402 | 0.948634 | 0.992005 | 0 | 0 | 0.472634 | 0.024923 |
| GTEx_Tissu | GTEX-XV7C | 1/672 | 0.949203 | 0.992005 | 0 | 0 | 0.333449 | 0.017383 |
| GTEx_Tissu | GTEX-XLM4 | 1/674 | 0.94966 | 0.992005 | 0 | 0 | 0.332423 | 0.01717 |
| GTEx_Tissu | GTEX-Q2A( | 1/675 | 0.949886 | 0.992005 | 0 | 0 | 0.331913 | 0.017065 |
| GTEx_Tissu | GTEX-XMK | 1/678 | 0.95056 | 0.992005 | 0 | 0 | 0.330391 | 0.016752 |
| GTEx_Tissu | GTEX-OOB. | 1/669 | 0.948511 | 0.992005 | 0 | 0 | 0.334999 | 0.017709 |
| GTEx_Tissu | GTEX-VJYA | 2/1097 | 0.95562 | 0.992005 | 0 | 0 | 0.404362 | 0.018356 |
| GTEx_Tissu | GTEX-WYJK | 3/1434 | 0.953923 | 0.992005 | 0 | 0 | 0.461266 | 0.021759 |
| GTEx_Tissu | GTEX-U412 | 1/694 | 0.954005 | 0.992005 | 0 | 0 | 0.322494 | 0.015185 |
| GTEx_Tissu | GTEX-QDV. | 1/698 | 0.954829 | 0.992005 | 0 | 0 | 0.320577 | 0.014818 |
| GTEx_Tissu | GTEX-T2IS- | 15/4876 | 0.957891 | 0.992005 | 0 | 0 | 0.6451 | 0.027753 |
| GTEx_Tissu | GTEX-WVLI | 2/1114 | 0.958406 | 0.992005 | 0 | 0 | 0.397821 | 0.016901 |
| GTEx_Tissu | GTEX-QEL4 | 3/1421 | 0.951838 | 0.992005 | 0 | 0 | 0.465822 | 0.022993 |
| GTEx_Tissu | GTEX-R45C | Oct-76 | 0.961613 | 0.992005 | 0 | 0 | 0.595341 | 0.023304 |
| GTEx_Tissu | GTEX-QDVI | 3/1488 | 0.961725 | 0.992005 | 0 | 0 | 0.443194 | 0.017297 |
| GTEx_Tissu | GTEX-QDVI | 4/1755 | 0.953473 | 0.992005 | 0 | 0 | 0.499873 | 0.023816 |
| GTEx_Tissu | GTEX-V1D1 | Aug-52 | 0.965255 | 0.992005 | 0 | 0 | 0.561187 | 0.019845 |
| GTEx_Tissu | GTEX-R55G | 2/1160 | 0.965139 | 0.992005 | 0 | 0 | 0.381083 | 0.013522 |
| GTEx_Tissu | GTEX-XV7C | 1/736 | 0.96196 | 0.992005 | 0 | 0 | 0.303401 | 0.011767 |
| GTEx_Tissu | GTEX-XGQ4 | Aug-21 | 0.96225 | 0.992005 | 0 | 0 | 0.568003 | 0.021857 |
| GTEx_Tissu | GTEX-WHW | 3/1447 | 0.955925 | 0.992005 | 0 | 0 | 0.456792 | 0.02059 |
| GTEx_Tissu | GTEX-N7M | 13/4335 | 0.956444 | 0.992005 | 0 | 0 | 0.633725 | 0.028221 |
| GTEx_Tissu | GTEX-X4XX | 1/708 | 0.956824 | 0.992005 | 0 | 0 | 0.315878 | 0.013941 |
| GTEx_Tissu | GTEX-X4XY | 2/1114 | 0.958406 | 0.992005 | 0 | 0 | 0.397821 | 0.016901 |
| GTEx_Tissu | GTEX-S4P3 | 14/4611 | 0.957598 | 0.992005 | 0 | 0 | 0.638963 | 0.027685 |
| GTEx_Tissu | GTEX-PWO | Oct-40 | 0.967352 | 0.992167 | 0 | 0 | 0.582555 | 0.019336 |
| GTEx_Tissu | GTEX-SUCS | 3/1522 | 0.96599 | 0.992167 | 0 | 0 | 0.432474 | 0.014965 |
| GTEx_Tissu | GTEX-WEY! | Nov-66 | 0.971209 | 0.992167 | 0 | 0 | 0.583998 | 0.017061 |
| GTEx_Tissu | GTEX-OOBI | 1/797 | 0.97115 | 0.992167 | 0 | 0 | 0.27926 | 0.008175 |
| GTEx_Tissu | GTEX-SNM | 1/791 | 0.970353 | 0.992167 | 0 | 0 | 0.281469 | 0.008471 |
| GTEx_Tissu | GTEX-UJHI- | 1/780 | 0.968836 | 0.992167 | 0 | 0 | 0.285608 | 0.009042 |
| GTEx_Tissu | GTEX-QVJC | 4/1895 | 0.970328 | 0.992167 | 0 | 0 | 0.459297 | 0.013835 |
| GTEx_Tissu | GTEX-QLQ\ | 13/4489 | 0.969724 | 0.992167 | 0 | 0 | 0.605877 | 0.018627 |

| | | | | | | | | |
|---|---|---|---|---|---|---|---|---|
| GTEx_Tissu | GTEX-O5YT | 2/1196 | 0.969672 | 0.992167 | 0 | 0 | 0.368884 | 0.011361 |
| GTEx_Tissu | GTEX-WFG | Jul-99 | 0.968637 | 0.992167 | 0 | 0 | 0.536564 | 0.017098 |
| GTEx_Tissu | GTEX-PWC | May-97 | 0.968804 | 0.992167 | 0 | 0 | 0.492951 | 0.015623 |
| GTEx_Tissu | GTEX-RWS | Jun-99 | 0.968419 | 0.992167 | 0 | 0 | 0.517597 | 0.01661 |
| GTEx_Tissu | GTEX-PSDG | 2/1171 | 0.966588 | 0.992167 | 0 | 0 | 0.377276 | 0.012821 |
| GTEx_Tissu | GTEX-RUSC | 3/1579 | 0.972163 | 0.992686 | 0 | 0 | 0.415541 | 0.011732 |
| Reactome_ | Immune Sy | Mar-43 | 0.992701 | 0.992701 | 0 | 0 | 0.330873 | 0.002424 |
| GTEx_Tissu | GTEX-OHPI | 1/820 | 0.974012 | 0.993207 | 0 | 0 | 0.271091 | 0.007138 |
| GTEx_Tissu | GTEX-P44H | 1/818 | 0.973775 | 0.993207 | 0 | 0 | 0.271783 | 0.007223 |
| GTEx_Tissu | GTEX-U3ZH | 3/1595 | 0.973697 | 0.993207 | 0 | 0 | 0.411006 | 0.010955 |
| GTEx_Tissu | GTEX-RWS | 18/5881 | 0.974924 | 0.993435 | 0 | 0 | 0.625144 | 0.015876 |
| GTEx_Tissu | GTEX-WFG | 1/843 | 0.976594 | 0.993435 | 0 | 0 | 0.263368 | 0.006238 |
| GTEx_Tissu | GTEX-XV7C | 1/852 | 0.977533 | 0.993435 | 0 | 0 | 0.26046 | 0.005918 |
| GTEx_Tissu | GTEX-PSDG | 2/1251 | 0.975531 | 0.993435 | 0 | 0 | 0.351604 | 0.00871 |
| GTEx_Tissu | GTEX-WFG | 2/1281 | 0.978254 | 0.993435 | 0 | 0 | 0.342805 | 0.007537 |
| GTEx_Tissu | GTEX-NPJ8 | 1/858 | 0.978139 | 0.993435 | 0 | 0 | 0.258555 | 0.005715 |
| GTEx_Tissu | GTEX-OHPI | Oct-54 | 0.975731 | 0.993435 | 0 | 0 | 0.560863 | 0.013779 |
| GTEx_Tissu | GTEX-S4UY | May-01 | 0.977455 | 0.993435 | 0 | 0 | 0.46786 | 0.010669 |
| GTEx_Tissu | GTEX-Q2AH | 2/1274 | 0.977646 | 0.993435 | 0 | 0 | 0.344821 | 0.007796 |
| GTEx_Tissu | GTEX-S7PM | 14/4905 | 0.978946 | 0.993684 | 0 | 0 | 0.589027 | 0.012534 |
| GTEx_Tissu | GTEX-PWO | 7/1030 | 0.160828 | 0.994179 | 0 | 0 | 1.615714 | 2.952586 |
| GTEx_Tissu | GTEX-U8XE | 1/883 | 0.980493 | 0.9948 | 0 | 0 | 0.250896 | 0.004943 |
| GTEx_Tissu | GTEX-XQ3S | 1/893 | 0.981363 | 0.995229 | 0 | 0 | 0.247953 | 0.004665 |
| GTEx_Tissu | GTEX-UTHO | Apr-51 | 0.982381 | 0.995807 | 0 | 0 | 0.420621 | 0.007477 |
| GTEx_Tissu | GTEX-S7PM | 1/911 | 0.982833 | 0.995812 | 0 | 0 | 0.242819 | 0.004205 |
| GTEx_Tissu | GTEX-XYKS | 1/921 | 0.9836 | 0.995983 | 0 | 0 | 0.240053 | 0.00397 |
| GTEx_Tissu | GTEX-V955 | 1/925 | 0.983897 | 0.995983 | 0 | 0 | 0.238964 | 0.003879 |
| GTEx_Tissu | GTEX-XUJ4 | Apr-27 | 0.986429 | 0.997359 | 0 | 0 | 0.403839 | 0.005518 |
| GTEx_Tissu | GTEX-PWN | 14/5072 | 0.986254 | 0.997359 | 0 | 0 | 0.563247 | 0.007796 |
| GTEx_Tissu | GTEX-Q2AH | May-62 | 0.986601 | 0.997359 | 0 | 0 | 0.433207 | 0.005844 |
| GTEx_Tissu | GTEX-WFG | May-73 | 0.987079 | 0.997389 | 0 | 0 | 0.431005 | 0.005605 |
| GTEx_Tissu | GTEX-RNOI | 1/990 | 0.988042 | 0.997767 | 0 | 0 | 0.222494 | 0.002677 |
| GTEx_Tissu | GTEX-P4PC | Jun-85 | 0.990415 | 0.997767 | 0 | 0 | 0.438269 | 0.004221 |
| GTEx_Tissu | GTEX-RWS | 1/1064 | 0.991488 | 0.997767 | 0 | 0 | 0.206196 | 0.001763 |
| GTEx_Tissu | GTEX-OXRK | Apr-57 | 0.991405 | 0.997767 | 0 | 0 | 0.377756 | 0.003261 |
| GTEx_Tissu | GTEX-Q2AH | 1/1060 | 0.99133 | 0.997767 | 0 | 0 | 0.207018 | 0.001803 |
| GTEx_Tissu | GTEX-S3XE | Oct-30 | 0.988648 | 0.997767 | 0 | 0 | 0.513439 | 0.005862 |
| GTEx_Tissu | GTEX-XUJ4 | 3/1845 | 0.989434 | 0.997767 | 0 | 0 | 0.350376 | 0.003722 |
| GTEx_Tissu | GTEX-W5W | 2/1482 | 0.990284 | 0.997767 | 0 | 0 | 0.293052 | 0.002861 |
| GTEx_Tissu | GTEX-RTLS | May-55 | 0.990171 | 0.997767 | 0 | 0 | 0.415184 | 0.004101 |
| GTEx_Tissu | GTEX-RWS | Sep-70 | 0.992278 | 0.998109 | 0 | 0 | 0.479708 | 0.003719 |
| GTEx_Tissu | GTEX-WWY | 1/1115 | 0.993272 | 0.998207 | 0 | 0 | 0.196224 | 0.001325 |
| GTEx_Tissu | GTEX-PWO | 2/1562 | 0.992997 | 0.998207 | 0 | 0 | 0.276817 | 0.001945 |
| GTEx_Tissu | GTEX-OOB | May-61 | 0.996607 | 0.998452 | 0 | 0 | 0.364167 | 0.001238 |
| GTEx_Tissu | GTEX-SIU8 | 1/1266 | 0.996658 | 0.998452 | 0 | 0 | 0.171413 | 0.000574 |

| | | | | | | | | |
|---|---|---|---|---|---|---|---|---|
| GTEx_Tissu | GTEX-TSE9 | 1/1165 | 0.99466 | 0.998452 | 0 | 0 | 0.187295 | 0.001003 |
| GTEx_Tissu | GTEX-QVJC | 1/1222 | 0.995899 | 0.998452 | 0 | 0 | 0.178009 | 0.000731 |
| GTEx_Tissu | GTEX-TSE9 | 1/1241 | 0.996246 | 0.998452 | 0 | 0 | 0.175103 | 0.000659 |
| GTEx_Tissu | GTEX-PLZ5 | May-17 | 0.996031 | 0.998452 | 0 | 0 | 0.370819 | 0.001475 |
| GTEx_Tissu | GTEX-OXRI | 1/1230 | 0.996049 | 0.998452 | 0 | 0 | 0.176774 | 0.0007 |
| GeDiPNet_ | Schizophre | 1/1471 | 0.998718 | 0.998718 | 0 | 0 | 0.145887 | 0.000187 |
| GTEx_Tissu | GTEX-WH7 | Feb-80 | 0.998801 | 0.999341 | 0 | 0 | 0.213347 | 0.000256 |
| GTEx_Tissu | GTEX-QDV. | Mar-94 | 0.998768 | 0.999341 | 0 | 0 | 0.261726 | 0.000323 |
| GTEx_Tissu | GTEX-VUSC | 1/1502 | 0.998892 | 0.999341 | 0 | 0 | 0.142634 | 0.000158 |
| GTEx_Tissu | GTEX-OIZH | 1/1664 | 0.999484 | 0.999484 | 0 | 0 | 0.127606 | 6.59E-05 |
| GTEx_Tissu | GTEX-U8XE | 3/965 | 0.797241 | 0.999971 | 0 | 0 | 0.703557 | 0.159424 |
| GTEx_Tissu | GTEX-O5YV | 4/1261 | 0.806408 | 0.999971 | 0 | 0 | 0.715262 | 0.1539 |
| GTEx_Tissu | GTEX-UTH( | 4/1238 | 0.794608 | 0.999971 | 0 | 0 | 0.729492 | 0.167714 |
| GTEx_Tissu | GTEX-U3ZN | 4/1263 | 0.807408 | 0.999971 | 0 | 0 | 0.714049 | 0.152754 |
| GTEx_Tissu | GTEX-QMR | 4/1264 | 0.807907 | 0.999971 | 0 | 0 | 0.713444 | 0.152184 |
| GTEx_Tissu | GTEX-T6M( | 6/1804 | 0.807913 | 0.999971 | 0 | 0 | 0.746302 | 0.159187 |
| GTEx_Tissu | GTEX-PWO | Oct-40 | 0.808056 | 0.999971 | 0 | 0 | 0.783948 | 0.167078 |
| GTEx_Tissu | GTEX-OHPI | 1/376 | 0.808862 | 0.999971 | 0 | 0 | 0.605829 | 0.128513 |
| GTEx_Tissu | GTEX-XLM4 | Jul-70 | 0.809023 | 0.999971 | 0 | 0 | 0.757089 | 0.160449 |
| GTEx_Tissu | GTEX-PLZ5 | 3/986 | 0.809143 | 0.999971 | 0 | 0 | 0.687763 | 0.145654 |
| GTEx_Tissu | GTEX-XQ8I | 1/373 | 0.806297 | 0.999971 | 0 | 0 | 0.610809 | 0.131509 |
| GTEx_Tissu | GTEX-P4PP | 1/357 | 0.792034 | 0.999971 | 0 | 0 | 0.638784 | 0.148933 |
| GTEx_Tissu | GTEX-XAJ8 | 6/1768 | 0.792051 | 0.999971 | 0 | 0 | 0.763064 | 0.177893 |
| GTEx_Tissu | GTEX-WZT( | Sep-45 | 0.792926 | 0.999971 | 0 | 0 | 0.79063 | 0.183446 |
| GTEx_Tissu | GTEX-PLZ6 | Nov-52 | 0.793577 | 0.999971 | 0 | 0 | 0.803025 | 0.185663 |
| GTEx_Tissu | GTEX-PSDG | Sep-48 | 0.794091 | 0.999971 | 0 | 0 | 0.78956 | 0.182039 |
| GTEx_Tissu | GTEX-XXEK | 2/671 | 0.794182 | 0.999971 | 0 | 0 | 0.676831 | 0.15597 |
| GTEx_Tissu | GTEX-T5JC | 2/671 | 0.794182 | 0.999971 | 0 | 0 | 0.676831 | 0.15597 |
| GTEx_Tissu | GTEX-U4B1 | 2/705 | 0.816498 | 0.999971 | 0 | 0 | 0.642959 | 0.130348 |
| GTEx_Tissu | GTEX-NFK9 | 4/1257 | 0.804395 | 0.999971 | 0 | 0 | 0.717699 | 0.156218 |
| GTEx_Tissu | GTEX-QCQ( | Aug-66 | 0.78329 | 0.999971 | 0 | 0 | 0.791784 | 0.193395 |
| GTEx_Tissu | GTEX-WFG | 6/1749 | 0.783297 | 0.999971 | 0 | 0 | 0.772189 | 0.188602 |
| GTEx_Tissu | GTEX-XV7C | 3/942 | 0.783515 | 0.999971 | 0 | 0 | 0.721664 | 0.176061 |
| GTEx_Tissu | GTEX-XQ8I | 1/348 | 0.78356 | 0.999971 | 0 | 0 | 0.655653 | 0.159919 |
| GTEx_Tissu | GTEX-XBED | 2/690 | 0.806929 | 0.999971 | 0 | 0 | 0.65749 | 0.141045 |
| GTEx_Tissu | GTEX-T6MI | Aug-17 | 0.803943 | 0.999971 | 0 | 0 | 0.772059 | 0.168484 |
| GTEx_Tissu | GTEX-Q2AI | 3/977 | 0.804115 | 0.999971 | 0 | 0 | 0.694449 | 0.151399 |
| GTEx_Tissu | GTEX-S341 | Jul-58 | 0.804152 | 0.999971 | 0 | 0 | 0.762031 | 0.166098 |
| GTEx_Tissu | GTEX-POM | Nov-82 | 0.804222 | 0.999971 | 0 | 0 | 0.793767 | 0.172946 |
| GTEx_Tissu | GTEX-U4B1 | Aug-38 | 0.812018 | 0.999971 | 0 | 0 | 0.764188 | 0.159129 |
| GTEx_Tissu | GTEX-WFG | 2/687 | 0.804963 | 0.999971 | 0 | 0 | 0.660472 | 0.143295 |
| GTEx_Tissu | GTEX-Q2AI | 2/687 | 0.804963 | 0.999971 | 0 | 0 | 0.660472 | 0.143295 |
| GTEx_Tissu | GTEX-XOT( | 6/1807 | 0.809192 | 0.999971 | 0 | 0 | 0.744936 | 0.157717 |
| GTEx_Tissu | GTEX-P4Q1 | 3/979 | 0.805242 | 0.999971 | 0 | 0 | 0.692952 | 0.150102 |
| GTEx_Tissu | GTEX-PLZ4 | 6/1798 | 0.805335 | 0.999971 | 0 | 0 | 0.749049 | 0.162167 |

| | | | | | | | | |
|---|---|---|---|---|---|---|---|---|
| GTEx_Tissu | GTEX-R3RS | 6/1799 | 0.805766 | 0.999971 | 0 | 0 | 0.74859 | 0.161667 |
| GTEx_Tissu | GTEX-PWN | 3/980 | 0.805803 | 0.999971 | 0 | 0 | 0.692206 | 0.149458 |
| GTEx_Tissu | GTEX-X4XY | 2/689 | 0.806275 | 0.999971 | 0 | 0 | 0.658481 | 0.141791 |
| GTEx_Tissu | GTEX-PVO\ | 3/974 | 0.802415 | 0.999971 | 0 | 0 | 0.696704 | 0.153365 |
| GTEx_Tissu | GTEX-RM2 | 1/388 | 0.818789 | 0.999971 | 0 | 0 | 0.586683 | 0.117295 |
| GTEx_Tissu | GTEX-T5JW | 1/388 | 0.818789 | 0.999971 | 0 | 0 | 0.586683 | 0.117295 |
| GTEx_Tissu | GTEX-RN64 | Oct-72 | 0.819176 | 0.999971 | 0 | 0 | 0.77373 | 0.154325 |
| GTEx_Tissu | GTEX-SIU7 | Sep-77 | 0.805112 | 0.999971 | 0 | 0 | 0.77934 | 0.168941 |
| GTEx_Tissu | GTEX-S4UY | 2/682 | 0.801648 | 0.999971 | 0 | 0 | 0.665502 | 0.147132 |
| GTEx_Tissu | GTEX-XBED | 2/683 | 0.802315 | 0.999971 | 0 | 0 | 0.66449 | 0.146356 |
| GTEx_Tissu | GTEX-S3XE | 2/683 | 0.802315 | 0.999971 | 0 | 0 | 0.66449 | 0.146356 |
| GTEx_Tissu | GTEX-N7M | 4/1253 | 0.802366 | 0.999971 | 0 | 0 | 0.720152 | 0.15857 |
| GTEx_Tissu | GTEX-O5Y\ | 3/974 | 0.802415 | 0.999971 | 0 | 0 | 0.696704 | 0.153365 |
| GTEx_Tissu | GTEX-OXRI | 4/1286 | 0.818616 | 0.999971 | 0 | 0 | 0.700374 | 0.140173 |
| GTEx_Tissu | GTEX-WFG | 1/369 | 0.802825 | 0.999971 | 0 | 0 | 0.617575 | 0.135631 |
| GTEx_Tissu | GTEX-QV44 | 4/1303 | 0.826557 | 0.999971 | 0 | 0 | 0.690578 | 0.131545 |
| GTEx_Tissu | GTEX-XYKS | 2/714 | 0.822036 | 0.999971 | 0 | 0 | 0.634534 | 0.12435 |
| GTEx_Tissu | GTEX-OXRI | 3/1011 | 0.822552 | 0.999971 | 0 | 0 | 0.66982 | 0.130845 |
| GTEx_Tissu | GTEX-WL46 | Sep-26 | 0.822755 | 0.999971 | 0 | 0 | 0.762588 | 0.148779 |
| GTEx_Tissu | GTEX-XXEK | 2/716 | 0.823247 | 0.999971 | 0 | 0 | 0.632691 | 0.123058 |
| GTEx_Tissu | GTEX-WWY | Oct-85 | 0.823558 | 0.999971 | 0 | 0 | 0.769644 | 0.149405 |
| GTEx_Tissu | GTEX-SE5C | 3/1014 | 0.824107 | 0.999971 | 0 | 0 | 0.667726 | 0.129175 |
| GTEx_Tissu | GTEX-PSDG | 2/702 | 0.814618 | 0.999971 | 0 | 0 | 0.645815 | 0.132415 |
| GTEx_Tissu | GTEX-WZTC | Aug-38 | 0.812018 | 0.999971 | 0 | 0 | 0.764188 | 0.159129 |
| GTEx_Tissu | GTEX-Q734 | 2/698 | 0.812085 | 0.999971 | 0 | 0 | 0.649662 | 0.135227 |
| GTEx_Tissu | GTEX-SNOS | 1/380 | 0.812229 | 0.999971 | 0 | 0 | 0.599313 | 0.124641 |
| GTEx_Tissu | GTEX-XLM4 | Jul-79 | 0.812616 | 0.999971 | 0 | 0 | 0.753421 | 0.156332 |
| GTEx_Tissu | GTEX-QV31 | 1/381 | 0.813062 | 0.999971 | 0 | 0 | 0.597705 | 0.123694 |
| GTEx_Tissu | GTEX-T5JW | 3/994 | 0.813523 | 0.999971 | 0 | 0 | 0.681923 | 0.140736 |
| GTEx_Tissu | GTEX-S4Z8 | 5/1563 | 0.819668 | 0.999971 | 0 | 0 | 0.718362 | 0.14285 |
| GTEx_Tissu | GTEX-WFG | 3/995 | 0.814064 | 0.999971 | 0 | 0 | 0.6812 | 0.140134 |
| GTEx_Tissu | GTEX-N7M | Jul-83 | 0.814197 | 0.999971 | 0 | 0 | 0.7518 | 0.154535 |
| GTEx_Tissu | GTEX-PWC | 3/940 | 0.782287 | 0.999971 | 0 | 0 | 0.723281 | 0.17759 |
| GTEx_Tissu | GTEX-Q734 | 2/702 | 0.814618 | 0.999971 | 0 | 0 | 0.645815 | 0.132415 |
| GTEx_Tissu | GTEX-XPVG | 5/1552 | 0.814825 | 0.999971 | 0 | 0 | 0.723903 | 0.148242 |
| GTEx_Tissu | GTEX-Q2AI | Jul-85 | 0.814984 | 0.999971 | 0 | 0 | 0.750993 | 0.153644 |
| GTEx_Tissu | GTEX-X261 | 5/1553 | 0.81527 | 0.999971 | 0 | 0 | 0.723396 | 0.147744 |
| GTEx_Tissu | GTEX-RWS | 5/1526 | 0.802975 | 0.999971 | 0 | 0 | 0.73732 | 0.161792 |
| GTEx_Tissu | GTEX-QDVI | 2/706 | 0.81712 | 0.999971 | 0 | 0 | 0.642012 | 0.129666 |
| GTEx_Tissu | GTEX-P4PC | 2/707 | 0.817741 | 0.999971 | 0 | 0 | 0.641068 | 0.128989 |
| GTEx_Tissu | GTEX-WFJC | Sep-12 | 0.817838 | 0.999971 | 0 | 0 | 0.76731 | 0.154299 |
| GTEx_Tissu | GTEX-U8T8 | 5/1494 | 0.787599 | 0.999971 | 0 | 0 | 0.754476 | 0.180144 |
| GTEx_Tissu | GTEX-R55G | 3/943 | 0.784127 | 0.999971 | 0 | 0 | 0.720859 | 0.175301 |
| GTEx_Tissu | GTEX-T6MI | Nov-28 | 0.784777 | 0.999971 | 0 | 0 | 0.810565 | 0.196445 |
| GTEx_Tissu | GTEX-OIZH | 2/658 | 0.785045 | 0.999971 | 0 | 0 | 0.69071 | 0.167162 |

| | | | | | | | | |
|---|---|---|---|---|---|---|---|---|
| GTEx_Tissu | GTEX-POM | Sep-27 | 0.785839 | 0.999971 | 0 | 0 | 0.797107 | 0.192105 |
| GTEx_Tissu | GTEX-R55F | Oct-80 | 0.785906 | 0.999971 | 0 | 0 | 0.803741 | 0.193636 |
| GTEx_Tissu | GTEX-R55E | Jul-28 | 0.791574 | 0.999971 | 0 | 0 | 0.774641 | 0.181058 |
| GTEx_Tissu | GTEX-SSA3 | 4/1223 | 0.786613 | 0.999971 | 0 | 0 | 0.739061 | 0.177388 |
| GTEx_Tissu | GTEX-WWY | Aug-75 | 0.787043 | 0.999971 | 0 | 0 | 0.788239 | 0.188762 |
| GTEx_Tissu | GTEX-S7SF- | 2/661 | 0.787184 | 0.999971 | 0 | 0 | 0.687459 | 0.164504 |
| GTEx_Tissu | GTEX-XMD | 5/1487 | 0.784117 | 0.999971 | 0 | 0 | 0.758328 | 0.184423 |
| GTEx_Tissu | GTEX-WEY! | 3/949 | 0.787771 | 0.999971 | 0 | 0 | 0.71606 | 0.170815 |
| GTEx_Tissu | GTEX-SNO! | 2/664 | 0.789305 | 0.999971 | 0 | 0 | 0.684237 | 0.161892 |
| GTEx_Tissu | GTEX-T5JC- | 6/1764 | 0.79023 | 0.999971 | 0 | 0 | 0.764969 | 0.180097 |
| GTEx_Tissu | GTEX-RM2I | 3/927 | 0.774165 | 0.999971 | 0 | 0 | 0.733959 | 0.187872 |
| GTEx_Tissu | GTEX-QV44 | 6/1765 | 0.790687 | 0.999971 | 0 | 0 | 0.764492 | 0.179544 |
| GTEx_Tissu | GTEX-V1D1 | Jul-26 | 0.790715 | 0.999971 | 0 | 0 | 0.775495 | 0.1821 |
| GTEx_Tissu | GTEX-WZT( | Aug-84 | 0.790749 | 0.999971 | 0 | 0 | 0.784721 | 0.184232 |
| GTEx_Tissu | GTEX-X4XX | 4/1231 | 0.790907 | 0.999971 | 0 | 0 | 0.733928 | 0.172161 |
| GTEx_Tissu | GTEX-XXEK | 4/1182 | 0.763526 | 0.999971 | 0 | 0 | 0.766461 | 0.206797 |
| GTEx_Tissu | GTEX-R55D | 2/633 | 0.766483 | 0.999971 | 0 | 0 | 0.719008 | 0.191215 |
| GTEx_Tissu | GTEX-QLQ\ | 1/344 | 0.779685 | 0.999971 | 0 | 0 | 0.663435 | 0.165106 |
| GTEx_Tissu | GTEX-T6MI | Oct-64 | 0.779711 | 0.999971 | 0 | 0 | 0.809165 | 0.201346 |
| GTEx_Tissu | GTEX-WZT( | Aug-58 | 0.779915 | 0.999971 | 0 | 0 | 0.794959 | 0.197603 |
| GTEx_Tissu | GTEX-PW2( | 3/937 | 0.780434 | 0.999971 | 0 | 0 | 0.725719 | 0.17991 |
| GTEx_Tissu | GTEX-O5YV | 3/937 | 0.780434 | 0.999971 | 0 | 0 | 0.725719 | 0.17991 |
| GTEx_Tissu | GTEX-S7SE- | Aug-61 | 0.781185 | 0.999971 | 0 | 0 | 0.793766 | 0.196015 |
| GTEx_Tissu | GTEX-O5YT | 6/1745 | 0.78142 | 0.999971 | 0 | 0 | 0.774136 | 0.190935 |
| GTEx_Tissu | GTEX-X5EB | 2/636 | 0.768781 | 0.999971 | 0 | 0 | 0.715495 | 0.188139 |
| GTEx_Tissu | GTEX-SIU7- | 3/955 | 0.791364 | 0.999971 | 0 | 0 | 0.711322 | 0.166448 |
| GTEx_Tissu | GTEX-T6M( | 3/911 | 0.763836 | 0.999971 | 0 | 0 | 0.747522 | 0.201384 |
| GTEx_Tissu | GTEX-PWC | 3/912 | 0.764492 | 0.999971 | 0 | 0 | 0.74666 | 0.200511 |
| GTEx_Tissu | GTEX-O5YV | 3/912 | 0.764492 | 0.999971 | 0 | 0 | 0.74666 | 0.200511 |
| GTEx_Tissu | GTEX-U3ZN | 4/1187 | 0.76644 | 0.999971 | 0 | 0 | 0.763018 | 0.202962 |
| GTEx_Tissu | GTEX-PSDG | 6/1714 | 0.766468 | 0.999971 | 0 | 0 | 0.789531 | 0.209986 |
| GTEx_Tissu | GTEX-XUJ4 | 2/633 | 0.766483 | 0.999971 | 0 | 0 | 0.719008 | 0.191215 |
| GTEx_Tissu | GTEX-R55E | 5/1500 | 0.790549 | 0.999971 | 0 | 0 | 0.751203 | 0.176553 |
| GTEx_Tissu | GTEX-RM2I | 4/1218 | 0.783895 | 0.999971 | 0 | 0 | 0.742304 | 0.180736 |
| GTEx_Tissu | GTEX-NPJ7 | Aug-06 | 0.799614 | 0.999971 | 0 | 0 | 0.776239 | 0.173587 |
| GTEx_Tissu | GTEX-QDVI | 5/1515 | 0.797789 | 0.999971 | 0 | 0 | 0.743135 | 0.167883 |
| GTEx_Tissu | GTEX-S341 | 6/1781 | 0.797888 | 0.999971 | 0 | 0 | 0.756933 | 0.170906 |
| GTEx_Tissu | GTEX-P4QT | 3/967 | 0.798401 | 0.999971 | 0 | 0 | 0.702023 | 0.158057 |
| GTEx_Tissu | GTEX-S7SF- | 3/967 | 0.798401 | 0.999971 | 0 | 0 | 0.702023 | 0.158057 |
| GTEx_Tissu | GTEX-QVJC | Oct-14 | 0.798666 | 0.999971 | 0 | 0 | 0.792421 | 0.178146 |
| GTEx_Tissu | GTEX-U4B1 | Aug-04 | 0.79882 | 0.999971 | 0 | 0 | 0.777003 | 0.17453 |
| GTEx_Tissu | GTEX-XPT6 | 2/678 | 0.798961 | 0.999971 | 0 | 0 | 0.669579 | 0.150282 |
| GTEx_Tissu | GTEX-SNO! | 4/1218 | 0.783895 | 0.999971 | 0 | 0 | 0.742304 | 0.180736 |
| GTEx_Tissu | GTEX-X4EC | Jul-47 | 0.799607 | 0.999971 | 0 | 0 | 0.766612 | 0.171441 |
| GTEx_Tissu | GTEX-S4UY | 2/681 | 0.80098 | 0.999971 | 0 | 0 | 0.666517 | 0.147913 |

| | | | | | | | | |
|---|---|---|---|---|---|---|---|---|
| GTEx_Tissu | GTEX-T2IS- | 2/679 | 0.799636 | 0.999971 | 0 | 0 | 0.668555 | 0.149488 |
| GTEx_Tissu | GTEX-SJXC- | 5/1519 | 0.799686 | 0.999971 | 0 | 0 | 0.741011 | 0.165642 |
| GTEx_Tissu | GTEX-OOBI | Sep-65 | 0.800604 | 0.999971 | 0 | 0 | 0.783541 | 0.174251 |
| GTEx_Tissu | GTEX-WFG | 3/971 | 0.800703 | 0.999971 | 0 | 0 | 0.698974 | 0.155358 |
| GTEx_Tissu | GTEX-R55G | 2/681 | 0.80098 | 0.999971 | 0 | 0 | 0.666517 | 0.147913 |
| GTEx_Tissu | GTEX-QLQV | 3/946 | 0.785955 | 0.999971 | 0 | 0 | 0.718452 | 0.173043 |
| GTEx_Tissu | GTEX-NPJ8 | 4/1214 | 0.781701 | 0.999971 | 0 | 0 | 0.744917 | 0.18346 |
| GTEx_Tissu | GTEX-NPJ8 | 2/654 | 0.782163 | 0.999971 | 0 | 0 | 0.695092 | 0.170778 |
| GTEx_Tissu | GTEX-U8XE | 3/927 | 0.774165 | 0.999971 | 0 | 0 | 0.733959 | 0.187872 |
| GTEx_Tissu | GTEX-OXRK | 3/955 | 0.791364 | 0.999971 | 0 | 0 | 0.711322 | 0.166448 |
| GTEx_Tissu | GTEX-QVUS | 2/667 | 0.791407 | 0.999971 | 0 | 0 | 0.681044 | 0.159325 |
| GTEx_Tissu | GTEX-RVPL | 5/1471 | 0.775998 | 0.999971 | 0 | 0 | 0.767269 | 0.194583 |
| GTEx_Tissu | GTEX-TSE9 | Dec-44 | 0.772511 | 0.999971 | 0 | 0 | 0.825792 | 0.213144 |
| GTEx_Tissu | GTEX-S4P3 | 6/1727 | 0.772825 | 0.999971 | 0 | 0 | 0.783007 | 0.201783 |
| GTEx_Tissu | GTEX-T2IS- | Sep-96 | 0.773239 | 0.999971 | 0 | 0 | 0.808481 | 0.207914 |
| GTEx_Tissu | GTEX-X638 | 2/642 | 0.773318 | 0.999971 | 0 | 0 | 0.708566 | 0.182147 |
| GTEx_Tissu | GTEX-PLZ5 | 2/642 | 0.773318 | 0.999971 | 0 | 0 | 0.708566 | 0.182147 |
| GTEx_Tissu | GTEX-P4QT | 3/926 | 0.77353 | 0.999971 | 0 | 0 | 0.734793 | 0.188688 |
| GTEx_Tissu | GTEX-QMR | Sep-30 | 0.824141 | 0.999971 | 0 | 0 | 0.761248 | 0.147236 |
| GTEx_Tissu | GTEX-S33H | 4/1247 | 0.799291 | 0.999971 | 0 | 0 | 0.723861 | 0.162166 |
| GTEx_Tissu | GTEX-POM | 3/961 | 0.794907 | 0.999971 | 0 | 0 | 0.706643 | 0.162196 |
| GTEx_Tissu | GTEX-T2IS- | Nov-56 | 0.795019 | 0.999971 | 0 | 0 | 0.80178 | 0.18392 |
| GTEx_Tissu | GTEX-QESD | 4/1241 | 0.796179 | 0.999971 | 0 | 0 | 0.727606 | 0.165844 |
| GTEx_Tissu | GTEX-Q734 | 5/1512 | 0.796356 | 0.999971 | 0 | 0 | 0.744736 | 0.169583 |
| GTEx_Tissu | GTEX-WK1 | Jul-40 | 0.796674 | 0.999971 | 0 | 0 | 0.769552 | 0.174926 |
| GTEx_Tissu | GTEX-QVJC | Oct-10 | 0.797194 | 0.999971 | 0 | 0 | 0.793738 | 0.179907 |
| GTEx_Tissu | GTEX-RN64 | 4/1243 | 0.797221 | 0.999971 | 0 | 0 | 0.726353 | 0.164609 |
| GTEx_Tissu | GTEX-NL3H | Sep-84 | 0.842079 | 0.999971 | 0 | 0 | 0.743551 | 0.127803 |
| GTEx_Tissu | GTEX-RTLS | 4/1359 | 0.850741 | 0.999971 | 0 | 0 | 0.660045 | 0.106695 |
| GTEx_Tissu | GTEX-REY6 | 5/1651 | 0.854906 | 0.999971 | 0 | 0 | 0.676696 | 0.106081 |
| GTEx_Tissu | GTEX-WZT( | Oct-72 | 0.850891 | 0.999971 | 0 | 0 | 0.743224 | 0.120009 |
| GTEx_Tissu | GTEX-T2IS- | 13/3754 | 0.855107 | 0.999971 | 0 | 0 | 0.75943 | 0.118873 |
| GTEx_Tissu | GTEX-XGQ4 | 2/774 | 0.855317 | 0.999971 | 0 | 0 | 0.583389 | 0.091174 |
| GTEx_Tissu | GTEX-WXY( | 3/1080 | 0.855518 | 0.999971 | 0 | 0 | 0.624619 | 0.097471 |
| GTEx_Tissu | GTEX-OIZH | 3/1080 | 0.855518 | 0.999971 | 0 | 0 | 0.624619 | 0.097471 |
| GTEx_Tissu | GTEX-N7M | Jun-18 | 0.852096 | 0.999971 | 0 | 0 | 0.697389 | 0.111621 |
| GTEx_Tissu | GTEX-WEY5 | 2/768 | 0.85226 | 0.999971 | 0 | 0 | 0.588143 | 0.094022 |
| GTEx_Tissu | GTEX-U3ZH | 1/431 | 0.850352 | 0.999971 | 0 | 0 | 0.526852 | 0.085405 |
| GTEx_Tissu | GTEX-OXRL | 1/438 | 0.854948 | 0.999971 | 0 | 0 | 0.518227 | 0.081214 |
| GTEx_Tissu | GTEX-PLZ6 | 1/439 | 0.855593 | 0.999971 | 0 | 0 | 0.517017 | 0.080634 |
| GTEx_Tissu | GTEX-QDT8 | 14/4009 | 0.855829 | 0.999971 | 0 | 0 | 0.764147 | 0.118966 |
| GTEx_Tissu | GTEX-XUZC | Jun-29 | 0.855894 | 0.999971 | 0 | 0 | 0.692976 | 0.107833 |
| GTEx_Tissu | GTEX-WFO | 3/1073 | 0.852431 | 0.999971 | 0 | 0 | 0.628939 | 0.100418 |
| GTEx_Tissu | GTEX-S4Z8 | 3/1076 | 0.853761 | 0.999971 | 0 | 0 | 0.62708 | 0.099144 |
| GTEx_Tissu | GTEX-X4XY | 2/771 | 0.853796 | 0.999971 | 0 | 0 | 0.585757 | 0.092587 |

| | | | | | | | | |
|---|---|---|---|---|---|---|---|---|
| GTEx_Tissu | GTEX-PSDG | 2/753 | 0.844363 | 0.999971 | 0 | 0 | 0.60036 | 0.101565 |
| GTEx_Tissu | GTEX-OIZG | 5/1588 | 0.830306 | 0.999971 | 0 | 0 | 0.706054 | 0.131299 |
| GTEx_Tissu | GTEX-UPK5 | 4/1351 | 0.847467 | 0.999971 | 0 | 0 | 0.664252 | 0.109936 |
| GTEx_Tissu | GTEX-S7SF- | Jun-05 | 0.847505 | 0.999971 | 0 | 0 | 0.70267 | 0.116263 |
| GTEx_Tissu | GTEX-QVJC | Jun-06 | 0.847862 | 0.999971 | 0 | 0 | 0.702261 | 0.115899 |
| GTEx_Tissu | GTEX-XV7C | 2/760 | 0.848094 | 0.999971 | 0 | 0 | 0.594599 | 0.097968 |
| GTEx_Tissu | GTEX-RUSC | 2/735 | 0.834388 | 0.999971 | 0 | 0 | 0.615681 | 0.111473 |
| GTEx_Tissu | GTEX-SUCS | 3/1024 | 0.829207 | 0.999971 | 0 | 0 | 0.660837 | 0.123765 |
| GTEx_Tissu | GTEX-T2IS- | Oct-03 | 0.829495 | 0.999971 | 0 | 0 | 0.764048 | 0.142829 |
| GTEx_Tissu | GTEX-WHS | 6/1857 | 0.829572 | 0.999971 | 0 | 0 | 0.722812 | 0.135054 |
| GTEx_Tissu | GTEX-QLQ7 | 4/1311 | 0.830196 | 0.999971 | 0 | 0 | 0.686056 | 0.12767 |
| GTEx_Tissu | GTEX-NPJ7 | 6/1883 | 0.83948 | 0.999971 | 0 | 0 | 0.711774 | 0.124541 |
| GTEx_Tissu | GTEX-XUJ4 | 2/728 | 0.830358 | 0.999971 | 0 | 0 | 0.621844 | 0.1156 |
| GTEx_Tissu | GTEX-P4PC | 3/1027 | 0.830713 | 0.999971 | 0 | 0 | 0.658796 | 0.122188 |
| GTEx_Tissu | GTEX-QEL4 | 1/434 | 0.852339 | 0.999971 | 0 | 0 | 0.523122 | 0.083579 |
| GTEx_Tissu | GTEX-OIZH | 5/1634 | 0.848571 | 0.999971 | 0 | 0 | 0.684394 | 0.112378 |
| GTEx_Tissu | GTEX-S7PM | 3/1073 | 0.852431 | 0.999971 | 0 | 0 | 0.628939 | 0.100418 |
| GTEx_Tissu | GTEX-WOF | 5/1646 | 0.853066 | 0.999971 | 0 | 0 | 0.678944 | 0.107897 |
| GTEx_Tissu | GTEX-OHPI | 2/770 | 0.853286 | 0.999971 | 0 | 0 | 0.58655 | 0.093063 |
| GTEx_Tissu | GTEX-Q2A( | Sep-21 | 0.853546 | 0.999971 | 0 | 0 | 0.731833 | 0.11589 |
| GTEx_Tissu | GTEX-U3ZN | Sep-47 | 0.861213 | 0.999971 | 0 | 0 | 0.723788 | 0.108144 |
| GTEx_Tissu | GTEX-XMK | Aug-91 | 0.863549 | 0.999971 | 0 | 0 | 0.710859 | 0.104287 |
| GTEx_Tissu | GTEX-XK95 | 3/1091 | 0.860256 | 0.999971 | 0 | 0 | 0.617942 | 0.093016 |
| GTEx_Tissu | GTEX-PWO | 5/1666 | 0.860315 | 0.999971 | 0 | 0 | 0.670034 | 0.100811 |
| GTEx_Tissu | GTEX-WWY | Jul-24 | 0.863707 | 0.999971 | 0 | 0 | 0.698421 | 0.102334 |
| GTEx_Tissu | GTEX-TSE9 | 4/1384 | 0.860599 | 0.999971 | 0 | 0 | 0.647215 | 0.097164 |
| GTEx_Tissu | GTEX-WYV | 5/1635 | 0.84895 | 0.999971 | 0 | 0 | 0.683937 | 0.111998 |
| GTEx_Tissu | GTEX-WYJK | 4/1357 | 0.849928 | 0.999971 | 0 | 0 | 0.661092 | 0.107496 |
| GTEx_Tissu | GTEX-QVJC | 3/1068 | 0.850192 | 0.999971 | 0 | 0 | 0.632059 | 0.102579 |
| GTEx_Tissu | GTEX-WI4N | 4/1358 | 0.850335 | 0.999971 | 0 | 0 | 0.660568 | 0.107094 |
| GTEx_Tissu | GTEX-U3ZN | Aug-91 | 0.863549 | 0.999971 | 0 | 0 | 0.710859 | 0.104287 |
| GTEx_Tissu | GTEX-OHPI | 1/449 | 0.86189 | 0.999971 | 0 | 0 | 0.505217 | 0.075089 |
| GTEx_Tissu | GTEX-TMM | 4/1359 | 0.850741 | 0.999971 | 0 | 0 | 0.660045 | 0.106695 |
| GTEx_Tissu | GTEX-SNOS | 5/1673 | 0.862783 | 0.999971 | 0 | 0 | 0.666966 | 0.098439 |
| GTEx_Tissu | GTEX-XPT6 | 1/432 | 0.851017 | 0.999971 | 0 | 0 | 0.525603 | 0.084792 |
| GTEx_Tissu | GTEX-OHPI | 1/432 | 0.851017 | 0.999971 | 0 | 0 | 0.525603 | 0.084792 |
| GTEx_Tissu | GTEX-TML8 | 3/1071 | 0.851539 | 0.999971 | 0 | 0 | 0.630183 | 0.101277 |
| GTEx_Tissu | GTEX-WWY | Jun-18 | 0.852096 | 0.999971 | 0 | 0 | 0.697389 | 0.111621 |
| GTEx_Tissu | GTEX-N7M | Jun-52 | 0.863582 | 0.999971 | 0 | 0 | 0.68391 | 0.100307 |
| GTEx_Tissu | GTEX-W5W | 1/447 | 0.860653 | 0.999971 | 0 | 0 | 0.507535 | 0.076163 |
| GTEx_Tissu | GTEX-Q734 | 6/1883 | 0.83948 | 0.999971 | 0 | 0 | 0.711774 | 0.124541 |
| GTEx_Tissu | GTEX-UPK5 | Dec-48 | 0.840425 | 0.999971 | 0 | 0 | 0.767264 | 0.133387 |
| GTEx_Tissu | GTEX-TSE9 | Jul-54 | 0.84061 | 0.999971 | 0 | 0 | 0.724045 | 0.125714 |
| GTEx_Tissu | GTEX-Q2A( | Sep-80 | 0.840799 | 0.999971 | 0 | 0 | 0.744838 | 0.129157 |
| GTEx_Tissu | GTEX-REY6 | 5/1616 | 0.841622 | 0.999971 | 0 | 0 | 0.692722 | 0.119442 |

| | | | | | | | | |
|---|---|---|---|---|---|---|---|---|
| GTEx_Tissu | GTEX-U3ZN | 3/1050 | 0.841889 | 0.999971 | 0 | 0 | 0.643539 | 0.110758 |
| GTEx_Tissu | GTEX-XYKS | 5/1666 | 0.860315 | 0.999971 | 0 | 0 | 0.670034 | 0.100811 |
| GTEx_Tissu | GTEX-N7M | Sep-29 | 0.855939 | 0.999971 | 0 | 0 | 0.729341 | 0.113454 |
| GTEx_Tissu | GTEX-QLQV | 4/1384 | 0.860599 | 0.999971 | 0 | 0 | 0.647215 | 0.097164 |
| GTEx_Tissu | GTEX-T5JC- | 4/1350 | 0.847054 | 0.999971 | 0 | 0 | 0.664781 | 0.110348 |
| GTEx_Tissu | GTEX-XK95 | 3/1092 | 0.860681 | 0.999971 | 0 | 0 | 0.617342 | 0.092621 |
| GTEx_Tissu | GTEX-S3XE | 2/785 | 0.860774 | 0.999971 | 0 | 0 | 0.574863 | 0.086185 |
| GTEx_Tissu | GTEX-VUSC | 2/779 | 0.857821 | 0.999971 | 0 | 0 | 0.579484 | 0.088869 |
| GTEx_Tissu | GTEX-S3XE | 3/1086 | 0.858119 | 0.999971 | 0 | 0 | 0.62096 | 0.095014 |
| GTEx_Tissu | GTEX-R55E | Sep-50 | 0.862077 | 0.999971 | 0 | 0 | 0.722869 | 0.107281 |
| GTEx_Tissu | GTEX-T5JC- | Oct-97 | 0.858119 | 0.999971 | 0 | 0 | 0.735916 | 0.112604 |
| GTEx_Tissu | GTEX-P4PP | 2/790 | 0.863193 | 0.999971 | 0 | 0 | 0.571066 | 0.084014 |
| GTEx_Tissu | GTEX-XV7C | 4/1391 | 0.86326 | 0.999971 | 0 | 0 | 0.643705 | 0.09465 |
| GTEx_Tissu | GTEX-XUJ4 | 3/1033 | 0.833691 | 0.999971 | 0 | 0 | 0.65475 | 0.119094 |
| GTEx_Tissu | GTEX-XBEC | 2/696 | 0.810808 | 0.999971 | 0 | 0 | 0.651602 | 0.136657 |
| GTEx_Tissu | GTEX-P4PC | 2/696 | 0.810808 | 0.999971 | 0 | 0 | 0.651602 | 0.136657 |
| GTEx_Tissu | GTEX-RTLS- | 2/696 | 0.810808 | 0.999971 | 0 | 0 | 0.651602 | 0.136657 |
| GTEx_Tissu | GTEX-UPJH | 4/1270 | 0.810876 | 0.999971 | 0 | 0 | 0.709835 | 0.14881 |
| GTEx_Tissu | GTEX-W5W | 3/990 | 0.811344 | 0.999971 | 0 | 0 | 0.684831 | 0.143173 |
| GTEx_Tissu | GTEX-U3ZN | 6/1875 | 0.836481 | 0.999971 | 0 | 0 | 0.715138 | 0.127689 |
| GTEx_Tissu | GTEX-U3ZN | Oct-09 | 0.831442 | 0.999971 | 0 | 0 | 0.762197 | 0.140697 |
| GTEx_Tissu | GTEX-UTH( | 3/1030 | 0.832207 | 0.999971 | 0 | 0 | 0.656767 | 0.120631 |
| GTEx_Tissu | GTEX-S7SF- | 4/1318 | 0.833329 | 0.999971 | 0 | 0 | 0.682144 | 0.124373 |
| GTEx_Tissu | GTEX-OIZG | 3/989 | 0.810795 | 0.999971 | 0 | 0 | 0.685562 | 0.143789 |
| GTEx_Tissu | GTEX-WH7 | 2/734 | 0.833818 | 0.999971 | 0 | 0 | 0.616554 | 0.112053 |
| GTEx_Tissu | GTEX-T5JC- | 3/1034 | 0.834183 | 0.999971 | 0 | 0 | 0.654081 | 0.118587 |
| GTEx_Tissu | GTEX-N7M | 6/1869 | 0.834203 | 0.999971 | 0 | 0 | 0.71768 | 0.1301 |
| GTEx_Tissu | GTEX-P4QS | 2/743 | 0.83889 | 0.999971 | 0 | 0 | 0.60878 | 0.106948 |
| GTEx_Tissu | GTEX-RU1J | Jul-37 | 0.834566 | 0.999971 | 0 | 0 | 0.730522 | 0.13211 |
| GTEx_Tissu | GTEX-RM2 | 5/1599 | 0.834826 | 0.999971 | 0 | 0 | 0.70076 | 0.12651 |
| GTEx_Tissu | GTEX-R53T | 2/736 | 0.834957 | 0.999971 | 0 | 0 | 0.61481 | 0.110896 |
| GTEx_Tissu | GTEX-S4Q7 | 2/737 | 0.835524 | 0.999971 | 0 | 0 | 0.613942 | 0.110323 |
| GTEx_Tissu | GTEX-R55F | Aug-78 | 0.826714 | 0.999971 | 0 | 0 | 0.749581 | 0.142642 |
| GTEx_Tissu | GTEX-N7M | 3/1023 | 0.828703 | 0.999971 | 0 | 0 | 0.66152 | 0.124295 |
| GTEx_Tissu | GTEX-P78B | Oct-89 | 0.82489 | 0.999971 | 0 | 0 | 0.768395 | 0.14792 |
| GTEx_Tissu | GTEX-WHS | 6/1847 | 0.825637 | 0.999971 | 0 | 0 | 0.727141 | 0.13932 |
| GTEx_Tissu | GTEX-W5W | 1/397 | 0.825898 | 0.999971 | 0 | 0 | 0.573086 | 0.109622 |
| GTEx_Tissu | GTEX-WYJK | 4/1302 | 0.826098 | 0.999971 | 0 | 0 | 0.691147 | 0.132038 |
| GTEx_Tissu | GTEX-N7M | 4/1302 | 0.826098 | 0.999971 | 0 | 0 | 0.691147 | 0.132038 |
| GTEx_Tissu | GTEX-XUZC | 5/1578 | 0.826112 | 0.999971 | 0 | 0 | 0.71093 | 0.135805 |
| GTEx_Tissu | GTEX-R55E | 5/1579 | 0.826535 | 0.999971 | 0 | 0 | 0.710439 | 0.135348 |
| GTEx_Tissu | GTEX-T6MI | Nov-03 | 0.81144 | 0.999971 | 0 | 0 | 0.787393 | 0.164522 |
| GTEx_Tissu | GTEX-U3ZN | 4/1323 | 0.835537 | 0.999971 | 0 | 0 | 0.679376 | 0.12207 |
| GTEx_Tissu | GTEX-O5YT | 2/722 | 0.826835 | 0.999971 | 0 | 0 | 0.627222 | 0.119266 |
| GTEx_Tissu | GTEX-U412 | 2/722 | 0.826835 | 0.999971 | 0 | 0 | 0.627222 | 0.119266 |

| | | | | | | | | |
|---|---|---|---|---|---|---|---|---|
| GTEx_Tissu | GTEX-RU72 | 5/1581 | 0.827379 | 0.999971 | 0 | 0 | 0.709461 | 0.134438 |
| GTEx_Tissu | GTEX-U412 | 2/723 | 0.827427 | 0.999971 | 0 | 0 | 0.62632 | 0.118647 |
| GTEx_Tissu | GTEX-WH7 | 1/400 | 0.828206 | 0.999971 | 0 | 0 | 0.568689 | 0.107194 |
| GTEx_Tissu | GTEX-X4EP | 4/1307 | 0.828385 | 0.999971 | 0 | 0 | 0.68831 | 0.129593 |
| GTEx_Tissu | GTEX-WZT( | Oct-56 | 0.813676 | 0.999971 | 0 | 0 | 0.77881 | 0.160585 |
| GTEx_Tissu | GTEX-PW2( | 3/988 | 0.810246 | 0.999971 | 0 | 0 | 0.686294 | 0.144408 |
| GTEx_Tissu | GTEX-U8T8 | 4/1344 | 0.844554 | 0.999971 | 0 | 0 | 0.667973 | 0.112852 |
| GTEx_Tissu | GTEX-R55E | Aug-23 | 0.842196 | 0.999971 | 0 | 0 | 0.733726 | 0.126012 |
| GTEx_Tissu | GTEX-PX3G | Aug-24 | 0.842528 | 0.999971 | 0 | 0 | 0.733381 | 0.125664 |
| GTEx_Tissu | GTEX-RU72 | Aug-24 | 0.842528 | 0.999971 | 0 | 0 | 0.733381 | 0.125664 |
| GTEx_Tissu | GTEX-UJHI- | Sep-86 | 0.842715 | 0.999971 | 0 | 0 | 0.74291 | 0.127131 |
| GTEx_Tissu | GTEX-T6MI | 2/752 | 0.843823 | 0.999971 | 0 | 0 | 0.601192 | 0.10209 |
| GTEx_Tissu | GTEX-X4EP | 4/1343 | 0.844134 | 0.999971 | 0 | 0 | 0.668508 | 0.113275 |
| GTEx_Tissu | GTEX-QLQ7 | 3/1055 | 0.844233 | 0.999971 | 0 | 0 | 0.640311 | 0.108422 |
| GTEx_Tissu | GTEX-R55C | Jul-76 | 0.848176 | 0.999971 | 0 | 0 | 0.715814 | 0.117871 |
| GTEx_Tissu | GTEX-SIU8- | Aug-30 | 0.844506 | 0.999971 | 0 | 0 | 0.731313 | 0.123594 |
| GTEx_Tissu | GTEX-QEG4 | 3/1060 | 0.846549 | 0.999971 | 0 | 0 | 0.637113 | 0.106135 |
| GTEx_Tissu | GTEX-NPJ8 | 4/1345 | 0.844973 | 0.999971 | 0 | 0 | 0.667439 | 0.112431 |
| GTEx_Tissu | GTEX-OXRL | Sep-94 | 0.845243 | 0.999971 | 0 | 0 | 0.740352 | 0.124476 |
| GTEx_Tissu | GTEX-XAJ8 | 5/1626 | 0.845514 | 0.999971 | 0 | 0 | 0.688073 | 0.115466 |
| GTEx_Tissu | GTEX-RWS( | 1/425 | 0.846298 | 0.999971 | 0 | 0 | 0.534472 | 0.089195 |
| GTEx_Tissu | GTEX-RWS( | 3/1060 | 0.846549 | 0.999971 | 0 | 0 | 0.637113 | 0.106135 |
| GTEx_Tissu | GTEX-WVLI | Jul-28 | 0.831295 | 0.999971 | 0 | 0 | 0.733993 | 0.13562 |
| GTEx_Tissu | GTEX-OXRK | 1/426 | 0.846981 | 0.999971 | 0 | 0 | 0.533187 | 0.08855 |
| GTEx_Tissu | GTEX-WH7 | 2/758 | 0.847036 | 0.999971 | 0 | 0 | 0.596234 | 0.098982 |
| GTEx_Tissu | GTEX-QCQ( | 4/1330 | 0.838589 | 0.999971 | 0 | 0 | 0.675535 | 0.118917 |
| GTEx_Tissu | GTEX-WHS | 2/738 | 0.836089 | 0.999971 | 0 | 0 | 0.613075 | 0.109753 |
| GTEx_Tissu | GTEX-RNOI | Aug-06 | 0.836476 | 0.999971 | 0 | 0 | 0.739646 | 0.132069 |
| GTEx_Tissu | GTEX-WVLI | 6/1838 | 0.822036 | 0.999971 | 0 | 0 | 0.731077 | 0.14327 |
| GTEx_Tissu | GTEX-OOB. | Sep-67 | 0.836587 | 0.999971 | 0 | 0 | 0.749045 | 0.133648 |
| GTEx_Tissu | GTEX-PW2( | 2/739 | 0.836653 | 0.999971 | 0 | 0 | 0.612212 | 0.109186 |
| GTEx_Tissu | GTEX-QVU! | 3/1040 | 0.837109 | 0.999971 | 0 | 0 | 0.65009 | 0.115587 |
| GTEx_Tissu | GTEX-P4QS | 3/1042 | 0.838074 | 0.999971 | 0 | 0 | 0.648769 | 0.114604 |
| GTEx_Tissu | GTEX-XLM4 | 6/1880 | 0.83836 | 0.999971 | 0 | 0 | 0.713032 | 0.125713 |
| GTEx_Tissu | GTEX-R53T | 1/414 | 0.838582 | 0.999971 | 0 | 0 | 0.549017 | 0.096651 |
| GTEx_Tissu | GTEX-PLZ4 | 6/1741 | 0.779531 | 0.999971 | 0 | 0 | 0.776091 | 0.193296 |
| GTEx_Tissu | GTEX-T5JW | 4/1299 | 0.824715 | 0.999971 | 0 | 0 | 0.692859 | 0.133526 |
| GTEx_Tissu | GTEX-RUSC | Jul-99 | 0.82042 | 0.999971 | 0 | 0 | 0.745381 | 0.14754 |
| GTEx_Tissu | GTEX-Q734 | Oct-77 | 0.820871 | 0.999971 | 0 | 0 | 0.772154 | 0.152415 |
| GTEx_Tissu | GTEX-QDVI | Jul-01 | 0.821187 | 0.999971 | 0 | 0 | 0.744586 | 0.146687 |
| GTEx_Tissu | GTEX-R53T | 4/1293 | 0.821921 | 0.999971 | 0 | 0 | 0.696309 | 0.136554 |
| GTEx_Tissu | GTEX-WXY( | 4/1293 | 0.821921 | 0.999971 | 0 | 0 | 0.696309 | 0.136554 |
| GTEx_Tissu | GTEX-XBED | 4/1293 | 0.821921 | 0.999971 | 0 | 0 | 0.696309 | 0.136554 |
| GTEx_Tissu | GTEX-S7SF- | 6/1838 | 0.822036 | 0.999971 | 0 | 0 | 0.731077 | 0.14327 |
| GTEx_Tissu | GTEX-V1D1 | 2/576 | 0.719003 | 0.999971 | 0 | 0 | 0.792744 | 0.261518 |

| | | | | | | | | |
|---|---|---|---|---|---|---|---|---|
| GTEx_Tissu | GTEX-S7SF· | 6/1609 | 0.710461 | 0.999971 | 0 | 0 | 0.846099 | 0.289231 |
| GTEx_Tissu | GTEX-WHP | 16/4072 | 0.716621 | 0.999971 | 0 | 0 | 0.881018 | 0.293562 |
| GTEx_Tissu | GTEX-XGQ4 | 3/837 | 0.711141 | 0.999971 | 0 | 0 | 0.817018 | 0.278508 |
| GTEx_Tissu | GTEX-S32V | 5/1366 | 0.717063 | 0.999971 | 0 | 0 | 0.831168 | 0.276439 |
| GTEx_Tissu | GTEX-S3XE | Aug-23 | 0.717399 | 0.999971 | 0 | 0 | 0.852165 | 0.283024 |
| GTEx_Tissu | GTEX-PWO | 4/1109 | 0.717822 | 0.999971 | 0 | 0 | 0.82028 | 0.271951 |
| GTEx_Tissu | GTEX-PX3G | 3/846 | 0.717992 | 0.999971 | 0 | 0 | 0.807914 | 0.267659 |
| GTEx_Tissu | GTEX-OXRK | 2/569 | 0.712653 | 0.999971 | 0 | 0 | 0.802822 | 0.271965 |
| GTEx_Tissu | GTEX-WYV. | 5/1360 | 0.713397 | 0.999971 | 0 | 0 | 0.835118 | 0.282034 |
| GTEx_Tissu | GTEX-RWS. | 6/1609 | 0.710461 | 0.999971 | 0 | 0 | 0.846099 | 0.289231 |
| GTEx_Tissu | GTEX-RU72 | Aug-17 | 0.714378 | 0.999971 | 0 | 0 | 0.854878 | 0.287532 |
| GTEx_Tissu | GTEX-X261 | 7/1878 | 0.719944 | 0.999971 | 0 | 0 | 0.84376 | 0.277244 |
| GTEx_Tissu | GTEX-QMR | 3/849 | 0.720248 | 0.999971 | 0 | 0 | 0.804922 | 0.264143 |
| GTEx_Tissu | GTEX-WFG | 7/1880 | 0.720995 | 0.999971 | 0 | 0 | 0.842766 | 0.275688 |
| GTEx_Tissu | GTEX-S4P3 | 2/579 | 0.721689 | 0.999971 | 0 | 0 | 0.7885 | 0.257178 |
| GTEx_Tissu | GTEX-QVU: | 5/1364 | 0.715845 | 0.999971 | 0 | 0 | 0.832481 | 0.278292 |
| GTEx_Tissu | GTEX-PWC' | 2/573 | 0.716296 | 0.999971 | 0 | 0 | 0.797033 | 0.26594 |
| GTEx_Tissu | GTEX-UJM( | 3/851 | 0.721745 | 0.999971 | 0 | 0 | 0.80294 | 0.261826 |
| GTEx_Tissu | GTEX-POYV | 4/1064 | 0.686657 | 0.999971 | 0 | 0 | 0.857149 | 0.32222 |
| GTEx_Tissu | GTEX-SNM | 7/1860 | 0.710367 | 0.999971 | 0 | 0 | 0.852806 | 0.291637 |
| GTEx_Tissu | GTEX-RVPU | 3/836 | 0.710373 | 0.999971 | 0 | 0 | 0.818042 | 0.279742 |
| GTEx_Tissu | GTEX-XBED | 4/1087 | 0.702872 | 0.999971 | 0 | 0 | 0.837922 | 0.295436 |
| GTEx_Tissu | GTEX-WI4N | 3/827 | 0.703383 | 0.999971 | 0 | 0 | 0.827367 | 0.291112 |
| GTEx_Tissu | GTEX-TSE9 | 6/1609 | 0.710461 | 0.999971 | 0 | 0 | 0.846099 | 0.289231 |
| GTEx_Tissu | GTEX-RM2I | 4/1089 | 0.704253 | 0.999971 | 0 | 0 | 0.836289 | 0.293217 |
| GTEx_Tissu | GTEX-VJYA | Sep-08 | 0.686211 | 0.999971 | 0 | 0 | 0.88403 | 0.3329 |
| GTEx_Tissu | GTEX-XOT4 | 7/1817 | 0.686655 | 0.999971 | 0 | 0 | 0.875145 | 0.328987 |
| GTEx_Tissu | GTEX-XV7C | 4/1064 | 0.686657 | 0.999971 | 0 | 0 | 0.857149 | 0.32222 |
| GTEx_Tissu | GTEX-RWS( | 7/1872 | 0.716775 | 0.999971 | 0 | 0 | 0.846756 | 0.281964 |
| GTEx_Tissu | GTEX-XBEV | 6/1604 | 0.707588 | 0.999971 | 0 | 0 | 0.848978 | 0.293656 |
| GTEx_Tissu | GTEX-SNM | 2/564 | 0.708044 | 0.999971 | 0 | 0 | 0.810174 | 0.279711 |
| GTEx_Tissu | GTEX-PWO | Oct-55 | 0.687736 | 0.999971 | 0 | 0 | 0.886281 | 0.331779 |
| GTEx_Tissu | GTEX-XV7C | 6/1607 | 0.709314 | 0.999971 | 0 | 0 | 0.847248 | 0.290993 |
| GTEx_Tissu | GTEX-UTH( | 5/1321 | 0.688777 | 0.999971 | 0 | 0 | 0.861674 | 0.321265 |
| GTEx_Tissu | GTEX-T6MI | 2/544 | 0.689003 | 0.999971 | 0 | 0 | 0.840938 | 0.313258 |
| GTEx_Tissu | GTEX-WFJ( | 3/809 | 0.68903 | 0.999971 | 0 | 0 | 0.846641 | 0.315349 |
| GTEx_Tissu | GTEX-N7M | 4/1068 | 0.68952 | 0.999971 | 0 | 0 | 0.853746 | 0.317388 |
| GTEx_Tissu | GTEX-W5X | Nov-81 | 0.725386 | 0.999971 | 0 | 0 | 0.859495 | 0.275942 |
| GTEx_Tissu | GTEX-S4Q7 | Aug-43 | 0.727321 | 0.999971 | 0 | 0 | 0.843234 | 0.268475 |
| GTEx_Tissu | GTEX-XV7C | 6/1633 | 0.723994 | 0.999971 | 0 | 0 | 0.832525 | 0.268883 |
| GTEx_Tissu | GTEX-U8XE | 5/1378 | 0.7243 | 0.999971 | 0 | 0 | 0.823371 | 0.265578 |
| GTEx_Tissu | GTEX-U412 | 5/1384 | 0.727869 | 0.999971 | 0 | 0 | 0.819523 | 0.260308 |
| GTEx_Tissu | GTEX-T6MI | 3/855 | 0.724719 | 0.999971 | 0 | 0 | 0.799002 | 0.257256 |
| GTEx_Tissu | GTEX-RU72 | Nov-90 | 0.72929 | 0.999971 | 0 | 0 | 0.856355 | 0.270337 |
| GTEx_Tissu | GTEX-WYJK | 3/862 | 0.729866 | 0.999971 | 0 | 0 | 0.7922 | 0.24946 |

| | | | | | | | | |
|---|---|---|---|---|---|---|---|---|
| GTEx_Tissu | GTEX-WL46 | Sep-99 | 0.730589 | 0.999971 | 0 | 0 | 0.845977 | 0.265556 |
| GTEx_Tissu | GTEX-P4QF | 4/1129 | 0.730938 | 0.999971 | 0 | 0 | 0.804841 | 0.252259 |
| GTEx_Tissu | GTEX-XUJ4 | 5/1383 | 0.727277 | 0.999971 | 0 | 0 | 0.820162 | 0.261179 |
| GTEx_Tissu | GTEX-XUW | 7/1889 | 0.725694 | 0.999971 | 0 | 0 | 0.838317 | 0.268787 |
| GTEx_Tissu | GTEX-U412 | 5/1391 | 0.731993 | 0.999971 | 0 | 0 | 0.815076 | 0.254291 |
| GTEx_Tissu | GTEX-SNM | 7/1890 | 0.726213 | 0.999971 | 0 | 0 | 0.837825 | 0.268031 |
| GTEx_Tissu | GTEX-REY6 | 5/1357 | 0.711551 | 0.999971 | 0 | 0 | 0.837107 | 0.284874 |
| GTEx_Tissu | GTEX-WFJC | 2/568 | 0.711736 | 0.999971 | 0 | 0 | 0.804282 | 0.273495 |
| GTEx_Tissu | GTEX-XMK | 4/1100 | 0.711772 | 0.999971 | 0 | 0 | 0.827412 | 0.281318 |
| GTEx_Tissu | GTEX-QV31 | 3/838 | 0.711909 | 0.999971 | 0 | 0 | 0.815997 | 0.27728 |
| GTEx_Tissu | GTEX-PLZ6 | 3/859 | 0.727669 | 0.999971 | 0 | 0 | 0.795102 | 0.25277 |
| GTEx_Tissu | GTEX-OOBI | 4/1086 | 0.702179 | 0.999971 | 0 | 0 | 0.838741 | 0.296551 |
| GTEx_Tissu | GTEX-PWO | 3/820 | 0.697861 | 0.999971 | 0 | 0 | 0.834761 | 0.300293 |
| GTEx_Tissu | GTEX-S32V | 4/1081 | 0.698699 | 0.999971 | 0 | 0 | 0.842859 | 0.302194 |
| GTEx_Tissu | GTEX-R55F | 7/1839 | 0.698933 | 0.999971 | 0 | 0 | 0.863585 | 0.309337 |
| GTEx_Tissu | GTEX-XYKS | 5/1337 | 0.699042 | 0.999971 | 0 | 0 | 0.850591 | 0.30455 |
| GTEx_Tissu | GTEX-QDVI | 3/847 | 0.718746 | 0.999971 | 0 | 0 | 0.806914 | 0.266482 |
| GTEx_Tissu | GTEX-RWS | Oct-21 | 0.718964 | 0.999971 | 0 | 0 | 0.860595 | 0.283948 |
| GTEx_Tissu | GTEX-Q2AC | 3/824 | 0.701026 | 0.999971 | 0 | 0 | 0.83052 | 0.295009 |
| GTEx_Tissu | GTEX-QLQV | 3/848 | 0.719498 | 0.999971 | 0 | 0 | 0.805917 | 0.265309 |
| GTEx_Tissu | GTEX-XXEK | 5/1341 | 0.701572 | 0.999971 | 0 | 0 | 0.847862 | 0.300509 |
| GTEx_Tissu | GTEX-XXEK | Nov-47 | 0.710327 | 0.999971 | 0 | 0 | 0.871534 | 0.298091 |
| GTEx_Tissu | GTEX-XPVG | 21/5225 | 0.702271 | 0.999971 | 0 | 0 | 0.899334 | 0.317857 |
| GTEx_Tissu | GTEX-PWN | 3/826 | 0.702599 | 0.999971 | 0 | 0 | 0.828415 | 0.292405 |
| GTEx_Tissu | GTEX-UJMC | 2/579 | 0.721689 | 0.999971 | 0 | 0 | 0.7885 | 0.257178 |
| GTEx_Tissu | GTEX-XLM4 | Oct-27 | 0.721705 | 0.999971 | 0 | 0 | 0.858324 | 0.279933 |
| GTEx_Tissu | GTEX-XMK | 5/1381 | 0.726089 | 0.999971 | 0 | 0 | 0.821442 | 0.26293 |
| GTEx_Tissu | GTEX-WHS | 7/1882 | 0.722044 | 0.999971 | 0 | 0 | 0.841773 | 0.27414 |
| GTEx_Tissu | GTEX-T5JW | Aug-41 | 0.726339 | 0.999971 | 0 | 0 | 0.844119 | 0.269898 |
| GTEx_Tissu | GTEX-V955 | Aug-42 | 0.72683 | 0.999971 | 0 | 0 | 0.843676 | 0.269186 |
| GTEx_Tissu | GTEX-SJXC | 13/3235 | 0.667146 | 0.999971 | 0 | 0 | 0.910057 | 0.368342 |
| GTEx_Tissu | GTEX-XGQ4 | Nov-49 | 0.664286 | 0.999971 | 0 | 0 | 0.907909 | 0.371373 |
| GTEx_Tissu | GTEX-QDVJ | Dec-91 | 0.665113 | 0.999971 | 0 | 0 | 0.909513 | 0.370898 |
| GTEx_Tissu | GTEX-RVPV | Oct-10 | 0.665349 | 0.999971 | 0 | 0 | 0.904571 | 0.368562 |
| GTEx_Tissu | GTEX-TML8 | 7/1807 | 0.680974 | 0.999971 | 0 | 0 | 0.880493 | 0.338312 |
| GTEx_Tissu | GTEX-TSE9 | 7/1807 | 0.680974 | 0.999971 | 0 | 0 | 0.880493 | 0.338312 |
| GTEx_Tissu | GTEX-UJHI | 6/1535 | 0.666056 | 0.999971 | 0 | 0 | 0.890633 | 0.361937 |
| GTEx_Tissu | GTEX-O5YT | 2/537 | 0.682105 | 0.999971 | 0 | 0 | 0.852248 | 0.326046 |
| GTEx_Tissu | GTEX-S33H | 6/1536 | 0.666682 | 0.999971 | 0 | 0 | 0.890002 | 0.360844 |
| GTEx_Tissu | GTEX-NPJ7 | 4/1037 | 0.666863 | 0.999971 | 0 | 0 | 0.880813 | 0.35688 |
| GTEx_Tissu | GTEX-XYKS | 2/519 | 0.663804 | 0.999971 | 0 | 0 | 0.88274 | 0.361719 |
| GTEx_Tissu | GTEX-RUSC | Aug-29 | 0.667786 | 0.999971 | 0 | 0 | 0.896511 | 0.362 |
| GTEx_Tissu | GTEX-QDVI | 4/1060 | 0.683776 | 0.999971 | 0 | 0 | 0.860579 | 0.327127 |
| GTEx_Tissu | GTEX-SNOS | 7/1813 | 0.68439 | 0.999971 | 0 | 0 | 0.877277 | 0.332687 |
| GTEx_Tissu | GTEX-PWC | 2/546 | 0.690951 | 0.999971 | 0 | 0 | 0.83776 | 0.309708 |

| | | | | | | | | |
|---|---|---|---|---|---|---|---|---|
| GTEx_Tissu | GTEX-VUS( | 7/1816 | 0.68609 | 0.999971 | 0 | 0 | 0.875677 | 0.329909 |
| GTEx_Tissu | GTEX-XUJ4 | 2/547 | 0.691922 | 0.999971 | 0 | 0 | 0.836179 | 0.30795 |
| GTEx_Tissu | GTEX-WHS | Aug-75 | 0.69267 | 0.999971 | 0 | 0 | 0.874306 | 0.321047 |
| GTEx_Tissu | GTEX-REY6 | 4/1074 | 0.69378 | 0.999971 | 0 | 0 | 0.848688 | 0.31028 |
| GTEx_Tissu | GTEX-RWS( | 4/1051 | 0.677228 | 0.999971 | 0 | 0 | 0.86839 | 0.338452 |
| GTEx_Tissu | GTEX-RU1J | 5/1307 | 0.679609 | 0.999971 | 0 | 0 | 0.871595 | 0.336643 |
| GTEx_Tissu | GTEX-OHPI | 2/530 | 0.675085 | 0.999971 | 0 | 0 | 0.863859 | 0.339425 |
| GTEx_Tissu | GTEX-TMZ! | 3/798 | 0.680011 | 0.999971 | 0 | 0 | 0.85885 | 0.331212 |
| GTEx_Tissu | GTEX-QV31 | Aug-52 | 0.68037 | 0.999971 | 0 | 0 | 0.885283 | 0.340939 |
| GTEx_Tissu | GTEX-N7M | 7/1806 | 0.680403 | 0.999971 | 0 | 0 | 0.881031 | 0.339259 |
| GTEx_Tissu | GTEX-N7M | 3/799 | 0.680839 | 0.999971 | 0 | 0 | 0.857726 | 0.329736 |
| GTEx_Tissu | GTEX-T6M( | 4/1050 | 0.676495 | 0.999971 | 0 | 0 | 0.869267 | 0.339735 |
| GTEx_Tissu | GTEX-PWO | 3/794 | 0.676685 | 0.999971 | 0 | 0 | 0.863374 | 0.33719 |
| GTEx_Tissu | GTEX-RM2I | 4/1057 | 0.681604 | 0.999971 | 0 | 0 | 0.863168 | 0.330858 |
| GTEx_Tissu | GTEX-RVPU | 1/270 | 0.694273 | 0.999971 | 0 | 0 | 0.84914 | 0.309842 |
| GTEx_Tissu | GTEX-SNM | 7/1810 | 0.682685 | 0.999971 | 0 | 0 | 0.878882 | 0.335488 |
| GTEx_Tissu | GTEX-QDVI | 2/538 | 0.683098 | 0.999971 | 0 | 0 | 0.850615 | 0.324184 |
| GTEx_Tissu | GTEX-RU72 | 6/1563 | 0.683328 | 0.999971 | 0 | 0 | 0.873284 | 0.33253 |
| GTEx_Tissu | GTEX-S7SE- | Sep-03 | 0.683654 | 0.999971 | 0 | 0 | 0.886208 | 0.337028 |
| GTEx_Tissu | GTEX-QDT8 | Aug-49 | 0.678745 | 0.999971 | 0 | 0 | 0.886733 | 0.343618 |
| GTEx_Tissu | GTEX-U3ZN | 5/1306 | 0.678947 | 0.999971 | 0 | 0 | 0.872312 | 0.337769 |
| GTEx_Tissu | GTEX-U4B1 | 3/805 | 0.685772 | 0.999971 | 0 | 0 | 0.851042 | 0.321022 |
| GTEx_Tissu | GTEX-PVO\ | 3/798 | 0.680011 | 0.999971 | 0 | 0 | 0.85885 | 0.331212 |
| GTEx_Tissu | GTEX-P78B | 3/829 | 0.704947 | 0.999971 | 0 | 0 | 0.825277 | 0.288543 |
| GTEx_Tissu | GTEX-UJM( | 2/551 | 0.695779 | 0.999971 | 0 | 0 | 0.829915 | 0.30103 |
| GTEx_Tissu | GTEX-XUW | 7/1801 | 0.677536 | 0.999971 | 0 | 0 | 0.88373 | 0.344029 |
| GTEx_Tissu | GTEX-XOT( | 7/1801 | 0.677536 | 0.999971 | 0 | 0 | 0.88373 | 0.344029 |
| GTEx_Tissu | GTEX-WOF | 5/1305 | 0.678285 | 0.999971 | 0 | 0 | 0.87303 | 0.3389 |
| GTEx_Tissu | GTEX-QVU! | 5/1305 | 0.678285 | 0.999971 | 0 | 0 | 0.87303 | 0.3389 |
| GTEx_Tissu | GTEX-T6M( | 4/1079 | 0.6973 | 0.999971 | 0 | 0 | 0.844517 | 0.304482 |
| GTEx_Tissu | GTEX-U3ZN | 2/553 | 0.697693 | 0.999971 | 0 | 0 | 0.826818 | 0.297635 |
| GTEx_Tissu | GTEX-WHV | 3/828 | 0.704166 | 0.999971 | 0 | 0 | 0.82632 | 0.289824 |
| GTEx_Tissu | GTEX-QCQ( | 3/820 | 0.697861 | 0.999971 | 0 | 0 | 0.834761 | 0.300293 |
| GTEx_Tissu | GTEX-P4PC | 2/532 | 0.677103 | 0.999971 | 0 | 0 | 0.860511 | 0.33554 |
| GTEx_Tissu | GTEX-R3RS | 6/1600 | 0.705276 | 0.999971 | 0 | 0 | 0.851294 | 0.297244 |
| GTEx_Tissu | GTEX-WCD | 5/1347 | 0.705341 | 0.999971 | 0 | 0 | 0.843799 | 0.294548 |
| GTEx_Tissu | GTEX-QVJC | Sep-50 | 0.707206 | 0.999971 | 0 | 0 | 0.866099 | 0.300045 |
| GTEx_Tissu | GTEX-WFG | 3/822 | 0.699447 | 0.999971 | 0 | 0 | 0.832636 | 0.297639 |
| GTEx_Tissu | GTEX-XMK | 5/1338 | 0.699676 | 0.999971 | 0 | 0 | 0.849908 | 0.303535 |
| GTEx_Tissu | GTEX-UTH( | 6/1607 | 0.709314 | 0.999971 | 0 | 0 | 0.847248 | 0.290993 |
| GTEx_Tissu | GTEX-XOT4 | Aug-91 | 0.701055 | 0.999971 | 0 | 0 | 0.866812 | 0.307864 |
| GTEx_Tissu | GTEX-SN8G | Dec-90 | 0.710078 | 0.999971 | 0 | 0 | 0.875114 | 0.299621 |
| GTEx_Tissu | GTEX-QESD | 5/1323 | 0.690072 | 0.999971 | 0 | 0 | 0.860274 | 0.319126 |
| GTEx_Tissu | GTEX-X8H( | 4/1064 | 0.686657 | 0.999971 | 0 | 0 | 0.857149 | 0.32222 |
| GTEx_Tissu | GTEX-S4Z8 | 2/542 | 0.687044 | 0.999971 | 0 | 0 | 0.844139 | 0.316853 |

| | | | | | | | | |
|---|---|---|---|---|---|---|---|---|
| GTEx_Tissu | GTEX-QVJC | 2/550 | 0.694818 | 0.999971 | 0 | 0 | 0.831473 | 0.302743 |
| GTEx_Tissu | GTEX-X261 | 4/1066 | 0.688091 | 0.999971 | 0 | 0 | 0.855444 | 0.319795 |
| GTEx_Tissu | GTEX-U3ZN | Aug-81 | 0.695831 | 0.999971 | 0 | 0 | 0.871482 | 0.316042 |
| GTEx_Tissu | GTEX-XQ3S | 4/1077 | 0.695895 | 0.999971 | 0 | 0 | 0.846181 | 0.306788 |
| GTEx_Tissu | GTEX-X585 | 5/1333 | 0.696497 | 0.999971 | 0 | 0 | 0.853337 | 0.308645 |
| GTEx_Tissu | GTEX-S341 | 6/1586 | 0.69709 | 0.999971 | 0 | 0 | 0.859494 | 0.310141 |
| GTEx_Tissu | GTEX-S341 | 5/1323 | 0.690072 | 0.999971 | 0 | 0 | 0.860274 | 0.319126 |
| GTEx_Tissu | GTEX-XYKS | 4/1124 | 0.727701 | 0.999971 | 0 | 0 | 0.808649 | 0.257041 |
| GTEx_Tissu | GTEX-XGQ4 | 15/3785 | 0.697824 | 0.999971 | 0 | 0 | 0.892076 | 0.320958 |
| GTEx_Tissu | GTEX-WHP | 6/1576 | 0.691154 | 0.999971 | 0 | 0 | 0.86544 | 0.319687 |
| GTEx_Tissu | GTEX-W5X | 4/1049 | 0.675761 | 0.999971 | 0 | 0 | 0.870145 | 0.341024 |
| GTEx_Tissu | GTEX-RWS | 3/793 | 0.675849 | 0.999971 | 0 | 0 | 0.864512 | 0.338703 |
| GTEx_Tissu | GTEX-WFG | 16/3972 | 0.676329 | 0.999971 | 0 | 0 | 0.908985 | 0.355482 |
| GTEx_Tissu | GTEX-UPK5 | 7/1799 | 0.676385 | 0.999971 | 0 | 0 | 0.884814 | 0.345956 |
| GTEx_Tissu | GTEX-REY6 | 3/816 | 0.694671 | 0.999971 | 0 | 0 | 0.839044 | 0.305678 |
| GTEx_Tissu | GTEX-OHPI | Sep-25 | 0.694813 | 0.999971 | 0 | 0 | 0.876694 | 0.319215 |
| GTEx_Tissu | GTEX-R53T | 6/1721 | 0.769907 | 0.999971 | 0 | 0 | 0.786006 | 0.205529 |
| GTEx_Tissu | GTEX-XLM4 | Sep-64 | 0.759709 | 0.999971 | 0 | 0 | 0.820523 | 0.225496 |
| GTEx_Tissu | GTEX-XQ8I | 13/3427 | 0.748412 | 0.999971 | 0 | 0 | 0.848996 | 0.246041 |
| GTEx_Tissu | GTEX-PX3G | Oct-18 | 0.761223 | 0.999971 | 0 | 0 | 0.825117 | 0.225116 |
| GTEx_Tissu | GTEX-XGQ4 | Aug-89 | 0.749273 | 0.999971 | 0 | 0 | 0.823313 | 0.237651 |
| GTEx_Tissu | GTEX-VUSC | 3/890 | 0.749714 | 0.999971 | 0 | 0 | 0.766065 | 0.220675 |
| GTEx_Tissu | GTEX-WYJK | 4/1159 | 0.749769 | 0.999971 | 0 | 0 | 0.782684 | 0.225406 |
| GTEx_Tissu | GTEX-X5EB | 2/612 | 0.749848 | 0.999971 | 0 | 0 | 0.744571 | 0.214351 |
| GTEx_Tissu | GTEX-PLZ5 | 2/606 | 0.744914 | 0.999971 | 0 | 0 | 0.752201 | 0.221513 |
| GTEx_Tissu | GTEX-RWS | Oct-82 | 0.746054 | 0.999971 | 0 | 0 | 0.837983 | 0.245493 |
| GTEx_Tissu | GTEX-OIZH | 2/629 | 0.76339 | 0.999971 | 0 | 0 | 0.723745 | 0.195402 |
| GTEx_Tissu | GTEX-T2IS- | 6/1715 | 0.766961 | 0.999971 | 0 | 0 | 0.789025 | 0.209343 |
| GTEx_Tissu | GTEX-WWY | 6/1715 | 0.766961 | 0.999971 | 0 | 0 | 0.789025 | 0.209343 |
| GTEx_Tissu | GTEX-XYKS | 3/916 | 0.767103 | 0.999971 | 0 | 0 | 0.743233 | 0.197056 |
| GTEx_Tissu | GTEX-UPK5 | Aug-29 | 0.767371 | 0.999971 | 0 | 0 | 0.806661 | 0.213591 |
| GTEx_Tissu | GTEX-SIU7- | 6/1716 | 0.767454 | 0.999971 | 0 | 0 | 0.788521 | 0.208703 |
| GTEx_Tissu | GTEX-XMK | 5/1455 | 0.767654 | 0.999971 | 0 | 0 | 0.776409 | 0.205295 |
| GTEx_Tissu | GTEX-QCQ | 3/918 | 0.7684 | 0.999971 | 0 | 0 | 0.74153 | 0.195352 |
| GTEx_Tissu | GTEX-X4LF- | Jul-84 | 0.772079 | 0.999971 | 0 | 0 | 0.793829 | 0.205338 |
| GTEx_Tissu | GTEX-XOTC | 2/622 | 0.757891 | 0.999971 | 0 | 0 | 0.732182 | 0.202972 |
| GTEx_Tissu | GTEX-NL4V | 4/1175 | 0.759401 | 0.999971 | 0 | 0 | 0.771331 | 0.21229 |
| GTEx_Tissu | GTEX-XBEV | 3/898 | 0.75517 | 0.999971 | 0 | 0 | 0.758899 | 0.213108 |
| GTEx_Tissu | GTEX-NPJ8 | Sep-64 | 0.759709 | 0.999971 | 0 | 0 | 0.820523 | 0.225496 |
| GTEx_Tissu | GTEX-S341 | 3/905 | 0.759867 | 0.999971 | 0 | 0 | 0.752732 | 0.206709 |
| GTEx_Tissu | GTEX-X88G | 6/1701 | 0.759984 | 0.999971 | 0 | 0 | 0.796154 | 0.218511 |
| GTEx_Tissu | GTEX-WL46 | 5/1441 | 0.760166 | 0.999971 | 0 | 0 | 0.784573 | 0.215144 |
| GTEx_Tissu | GTEX-POM | Aug-13 | 0.760243 | 0.999971 | 0 | 0 | 0.81325 | 0.222926 |
| GTEx_Tissu | GTEX-NPJ8 | Jul-59 | 0.760443 | 0.999971 | 0 | 0 | 0.805117 | 0.220485 |
| GTEx_Tissu | GTEX-XMK | 3/907 | 0.761196 | 0.999971 | 0 | 0 | 0.750988 | 0.204918 |

| | | | | | | | | |
|---|---|---|---|---|---|---|---|---|
| GTEx_Tissu | GTEX-XBED | 2/636 | 0.768781 | 0.999971 | 0 | 0 | 0.715495 | 0.188139 |
| GTEx_Tissu | GTEX-WCD | 14/3706 | 0.761375 | 0.999971 | 0 | 0 | 0.842599 | 0.229717 |
| GTEx_Tissu | GTEX-OXRI | 3/908 | 0.761858 | 0.999971 | 0 | 0 | 0.750118 | 0.204028 |
| GTEx_Tissu | GTEX-P4PC | 2/609 | 0.747391 | 0.999971 | 0 | 0 | 0.748367 | 0.217899 |
| GTEx_Tissu | GTEX-S7SF | 4/1156 | 0.747931 | 0.999971 | 0 | 0 | 0.784848 | 0.227955 |
| GTEx_Tissu | GTEX-QCQ | Jul-63 | 0.762332 | 0.999971 | 0 | 0 | 0.803291 | 0.217992 |
| GTEx_Tissu | GTEX-SSA3 | Oct-21 | 0.762459 | 0.999971 | 0 | 0 | 0.82406 | 0.22349 |
| GTEx_Tissu | GTEX-OIZG | 3/909 | 0.762519 | 0.999971 | 0 | 0 | 0.749251 | 0.203143 |
| GTEx_Tissu | GTEX-RM2 | 4/1181 | 0.76294 | 0.999971 | 0 | 0 | 0.767154 | 0.207573 |
| GTEx_Tissu | GTEX-QXCI | 2/645 | 0.775558 | 0.999971 | 0 | 0 | 0.70515 | 0.17923 |
| GTEx_Tissu | GTEX-RN6 | 21/5424 | 0.770177 | 0.999971 | 0 | 0 | 0.854492 | 0.223137 |
| GTEx_Tissu | GTEX-S4Z8 | 2/622 | 0.757891 | 0.999971 | 0 | 0 | 0.732182 | 0.202972 |
| GTEx_Tissu | GTEX-UJM | 2/623 | 0.758683 | 0.999971 | 0 | 0 | 0.730965 | 0.201871 |
| GTEx_Tissu | GTEX-P4QT | 5/1478 | 0.779578 | 0.999971 | 0 | 0 | 0.763333 | 0.190072 |
| GTEx_Tissu | GTEX-WY7 | 4/1201 | 0.774452 | 0.999971 | 0 | 0 | 0.75353 | 0.192602 |
| GTEx_Tissu | GTEX-PX3G | Aug-46 | 0.774783 | 0.999971 | 0 | 0 | 0.799765 | 0.204077 |
| GTEx_Tissu | GTEX-XBED | 2/644 | 0.774813 | 0.999971 | 0 | 0 | 0.706286 | 0.180197 |
| GTEx_Tissu | GTEX-U3ZH | Oct-52 | 0.774985 | 0.999971 | 0 | 0 | 0.813275 | 0.207313 |
| GTEx_Tissu | GTEX-S33H | 3/929 | 0.77543 | 0.999971 | 0 | 0 | 0.732297 | 0.186251 |
| GTEx_Tissu | GTEX-T6MI | Sep-56 | 0.756243 | 0.999971 | 0 | 0 | 0.823583 | 0.230103 |
| GTEx_Tissu | GTEX-X4XY | 4/1203 | 0.775579 | 0.999971 | 0 | 0 | 0.752193 | 0.191166 |
| GTEx_Tissu | GTEX-XQ8I | 3/939 | 0.781671 | 0.999971 | 0 | 0 | 0.724092 | 0.17836 |
| GTEx_Tissu | GTEX-N7M | Jul-95 | 0.77707 | 0.999971 | 0 | 0 | 0.788952 | 0.198993 |
| GTEx_Tissu | GTEX-OXRL | 3/932 | 0.777317 | 0.999971 | 0 | 0 | 0.729817 | 0.183846 |
| GTEx_Tissu | GTEX-UTH | Sep-06 | 0.777359 | 0.999971 | 0 | 0 | 0.804781 | 0.202687 |
| GTEx_Tissu | GTEX-Q2A | Aug-54 | 0.778214 | 0.999971 | 0 | 0 | 0.796555 | 0.19974 |
| GTEx_Tissu | GTEX-U3ZH | Aug-56 | 0.779065 | 0.999971 | 0 | 0 | 0.795757 | 0.198669 |
| GTEx_Tissu | GTEX-XLM | Aug-56 | 0.779065 | 0.999971 | 0 | 0 | 0.795757 | 0.198669 |
| GTEx_Tissu | GTEX-WW | Aug-37 | 0.77088 | 0.999971 | 0 | 0 | 0.803403 | 0.209063 |
| GTEx_Tissu | GTEX-OXRI | 3/919 | 0.769047 | 0.999971 | 0 | 0 | 0.740682 | 0.194506 |
| GTEx_Tissu | GTEX-N7M | Aug-33 | 0.76913 | 0.999971 | 0 | 0 | 0.805029 | 0.211316 |
| GTEx_Tissu | GTEX-TKQ2 | 5/1458 | 0.769236 | 0.999971 | 0 | 0 | 0.77468 | 0.203244 |
| GTEx_Tissu | GTEX-T2YK | 6/1720 | 0.769418 | 0.999971 | 0 | 0 | 0.786508 | 0.20616 |
| GTEx_Tissu | GTEX-PSDG | 2/637 | 0.769543 | 0.999971 | 0 | 0 | 0.714331 | 0.187125 |
| GTEx_Tissu | GTEX-X15G | 5/1459 | 0.769761 | 0.999971 | 0 | 0 | 0.774105 | 0.202564 |
| GTEx_Tissu | GTEX-NPJ7 | 4/1168 | 0.755222 | 0.999971 | 0 | 0 | 0.77626 | 0.21793 |
| GTEx_Tissu | GTEX-QMR | Oct-04 | 0.755397 | 0.999971 | 0 | 0 | 0.830079 | 0.232847 |
| GTEx_Tissu | GTEX-WL4 | Aug-36 | 0.770444 | 0.999971 | 0 | 0 | 0.803809 | 0.209624 |
| GTEx_Tissu | GTEX-Q2AI | Sep-63 | 0.759277 | 0.999971 | 0 | 0 | 0.820905 | 0.226068 |
| GTEx_Tissu | GTEX-UJM | 4/1195 | 0.771044 | 0.999971 | 0 | 0 | 0.757569 | 0.196975 |
| GTEx_Tissu | GTEX-OXRF | 4/1195 | 0.771044 | 0.999971 | 0 | 0 | 0.757569 | 0.196975 |
| GTEx_Tissu | GTEX-SIU8 | 4/1196 | 0.771615 | 0.999971 | 0 | 0 | 0.756893 | 0.196239 |
| GTEx_Tissu | GTEX-WL4 | Oct-44 | 0.771796 | 0.999971 | 0 | 0 | 0.816035 | 0.211381 |
| GTEx_Tissu | GTEX-R3RS | 6/1692 | 0.75542 | 0.999971 | 0 | 0 | 0.8008 | 0.224609 |
| GTEx_Tissu | GTEX-OXRL | 3/899 | 0.755846 | 0.999971 | 0 | 0 | 0.758012 | 0.212181 |

| | | | | | | | | |
|---|---|---|---|---|---|---|---|---|
| GTEx_Tissu | GTEX-OXRI | 3/899 | 0.755846 | 0.999971 | 0 | 0 | 0.758012 | 0.212181 |
| GTEx_Tissu | GTEX-QLQ7 | 6/1693 | 0.75593 | 0.999971 | 0 | 0 | 0.800281 | 0.223923 |
| GTEx_Tissu | GTEX-O5YT | 5/1377 | 0.723702 | 0.999971 | 0 | 0 | 0.824015 | 0.266467 |
| GTEx_Tissu | GTEX-S3XE | 2/604 | 0.743252 | 0.999971 | 0 | 0 | 0.754778 | 0.223958 |
| GTEx_Tissu | GTEX-XMD | Sep-27 | 0.7434 | 0.999971 | 0 | 0 | 0.834844 | 0.247549 |
| GTEx_Tissu | GTEX-V955 | 4/1149 | 0.743604 | 0.999971 | 0 | 0 | 0.789941 | 0.234017 |
| GTEx_Tissu | GTEX-WL4( | Jul-10 | 0.736453 | 0.999971 | 0 | 0 | 0.8281 | 0.253324 |
| GTEx_Tissu | GTEX-PWN | 3/872 | 0.73709 | 0.999971 | 0 | 0 | 0.782673 | 0.238751 |
| GTEx_Tissu | GTEX-S32V | Sep-30 | 0.744749 | 0.999971 | 0 | 0 | 0.833667 | 0.245688 |
| GTEx_Tissu | GTEX-T6MI | Sep-16 | 0.738415 | 0.999971 | 0 | 0 | 0.839187 | 0.254483 |
| GTEx_Tissu | GTEX-O5YU | Sep-84 | 0.72356 | 0.999971 | 0 | 0 | 0.852049 | 0.275699 |
| GTEx_Tissu | GTEX-WHP | 5/1377 | 0.723702 | 0.999971 | 0 | 0 | 0.824015 | 0.266467 |
| GTEx_Tissu | GTEX-SIU8 | 4/1148 | 0.742981 | 0.999971 | 0 | 0 | 0.790673 | 0.234897 |
| GTEx_Tissu | GTEX-XUJ4 | 3/854 | 0.723978 | 0.999971 | 0 | 0 | 0.799983 | 0.25839 |
| GTEx_Tissu | GTEX-OXRI | Jul-19 | 0.740977 | 0.999971 | 0 | 0 | 0.823791 | 0.246961 |
| GTEx_Tissu | GTEX-U3ZI | 3/878 | 0.741352 | 0.999971 | 0 | 0 | 0.777061 | 0.232559 |
| GTEx_Tissu | GTEX-UPIC | 4/1119 | 0.724436 | 0.999971 | 0 | 0 | 0.812491 | 0.261916 |
| GTEx_Tissu | GTEX-S4Z8 | 6/1667 | 0.742424 | 0.999971 | 0 | 0 | 0.813967 | 0.242428 |
| GTEx_Tissu | GTEX-S7SE | Aug-38 | 0.724861 | 0.999971 | 0 | 0 | 0.845451 | 0.272045 |
| GTEx_Tissu | GTEX-WYV | Oct-34 | 0.724882 | 0.999971 | 0 | 0 | 0.855688 | 0.275314 |
| GTEx_Tissu | GTEX-NPJ8 | 5/1379 | 0.724897 | 0.999971 | 0 | 0 | 0.822727 | 0.264693 |
| GTEx_Tissu | GTEX-PWC | 3/865 | 0.732049 | 0.999971 | 0 | 0 | 0.789319 | 0.246195 |
| GTEx_Tissu | GTEX-WFO | 5/1360 | 0.713397 | 0.999971 | 0 | 0 | 0.835118 | 0.282034 |
| GTEx_Tissu | GTEX-XUW | 2/586 | 0.727874 | 0.999971 | 0 | 0 | 0.778767 | 0.247358 |
| GTEx_Tissu | GTEX-WVLI | Dec-00 | 0.714417 | 0.999971 | 0 | 0 | 0.871762 | 0.293164 |
| GTEx_Tissu | GTEX-NFK9 | 2/572 | 0.715389 | 0.999971 | 0 | 0 | 0.798473 | 0.267432 |
| GTEx_Tissu | GTEX-S95S | 3/843 | 0.715722 | 0.999971 | 0 | 0 | 0.810927 | 0.271225 |
| GTEx_Tissu | GTEX-U412 | Oct-14 | 0.715745 | 0.999971 | 0 | 0 | 0.863257 | 0.2887 |
| GTEx_Tissu | GTEX-XQ3S | 5/1390 | 0.731406 | 0.999971 | 0 | 0 | 0.815708 | 0.255142 |
| GTEx_Tissu | GTEX-QEG5 | 5/1391 | 0.731993 | 0.999971 | 0 | 0 | 0.815076 | 0.254291 |
| GTEx_Tissu | GTEX-OHPI | 3/873 | 0.737804 | 0.999971 | 0 | 0 | 0.781732 | 0.237707 |
| GTEx_Tissu | GTEX-VUSI | 4/1120 | 0.725091 | 0.999971 | 0 | 0 | 0.81172 | 0.260933 |
| GTEx_Tissu | GTEX-X3Y1 | 1/306 | 0.739278 | 0.999971 | 0 | 0 | 0.747541 | 0.225819 |
| GTEx_Tissu | GTEX-WFG | 1/306 | 0.739278 | 0.999971 | 0 | 0 | 0.747541 | 0.225819 |
| GTEx_Tissu | GTEX-S7SF | 4/1144 | 0.740479 | 0.999971 | 0 | 0 | 0.793617 | 0.238448 |
| GTEx_Tissu | GTEX-NFK9 | 2/601 | 0.740741 | 0.999971 | 0 | 0 | 0.758676 | 0.227682 |
| GTEx_Tissu | GTEX-X261 | 4/1133 | 0.733507 | 0.999971 | 0 | 0 | 0.801818 | 0.248498 |
| GTEx_Tissu | GTEX-XLM4 | 2/593 | 0.733943 | 0.999971 | 0 | 0 | 0.769264 | 0.237952 |
| GTEx_Tissu | GTEX-WXY( | 2/602 | 0.74158 | 0.999971 | 0 | 0 | 0.757373 | 0.226434 |
| GTEx_Tissu | GTEX-XUZ( | 6/1652 | 0.7344 | 0.999971 | 0 | 0 | 0.82206 | 0.253771 |
| GTEx_Tissu | GTEX-WHS | Jul-06 | 0.734426 | 0.999971 | 0 | 0 | 0.830029 | 0.256202 |
| GTEx_Tissu | GTEX-NPJ7 | 4/1161 | 0.750988 | 0.999971 | 0 | 0 | 0.781248 | 0.223722 |
| GTEx_Tissu | GTEX-XUYS | 4/1131 | 0.732225 | 0.999971 | 0 | 0 | 0.803327 | 0.250371 |
| GTEx_Tissu | GTEX-TMM | 6/1649 | 0.732775 | 0.999971 | 0 | 0 | 0.823696 | 0.256101 |
| GTEx_Tissu | GTEX-Q2A( | 2/592 | 0.733083 | 0.999971 | 0 | 0 | 0.770608 | 0.239271 |

| | | | | | | | | |
|---|---|---|---|---|---|---|---|---|
| GTEx_Tissu | GTEX-P4QF | 4/1133 | 0.733507 | 0.999971 | 0 | 0 | 0.801818 | 0.248498 |
| GTEx_Tissu | GTEX-XMK: | 3/894 | 0.752454 | 0.999971 | 0 | 0 | 0.762466 | 0.216857 |
| GTEx_Tissu | GTEX-S4P3 | Oct-98 | 0.752871 | 0.999971 | 0 | 0 | 0.832222 | 0.236235 |
| GTEx_Tissu | GTEX-SE5C | 3/868 | 0.734218 | 0.999971 | 0 | 0 | 0.786457 | 0.242975 |
| GTEx_Tissu | GTEX-S7PM | Nov-47 | 0.753213 | 0.999971 | 0 | 0 | 0.83692 | 0.237189 |
| GTEx_Tissu | GTEX-WHP | 2/618 | 0.754701 | 0.999971 | 0 | 0 | 0.737089 | 0.207442 |
| GTEx_Tissu | GTEX-X5EB | 4/1135 | 0.734785 | 0.999971 | 0 | 0 | 0.800315 | 0.246639 |
| GTEx_Tissu | GTEX-S33H | Jul-09 | 0.735947 | 0.999971 | 0 | 0 | 0.828582 | 0.25404 |
| GTEx_Tissu | GTEX-OOB. | 5/1398 | 0.736072 | 0.999971 | 0 | 0 | 0.810674 | 0.248412 |
| GTEx_Tissu | GTEX-RVPL | 4/1166 | 0.754018 | 0.999971 | 0 | 0 | 0.777679 | 0.219569 |
| GTEx_Tissu | GTEX-VJYA | Oct-01 | 0.754136 | 0.999971 | 0 | 0 | 0.83115 | 0.234536 |
| GTEx_Tissu | GTEX-UPIC | 2/623 | 0.758683 | 0.999971 | 0 | 0 | 0.730965 | 0.201871 |
| GTEx_Tissu | GTEX-NPJ8 | 3/904 | 0.759201 | 0.999971 | 0 | 0 | 0.753607 | 0.207611 |
| GTEx_Tissu | GTEX-T6MI | 3/904 | 0.759201 | 0.999971 | 0 | 0 | 0.753607 | 0.207611 |
| GTEx_Tissu | GTEX-WFG | 3/888 | 0.748335 | 0.999971 | 0 | 0 | 0.767877 | 0.222611 |
| GTEx_Tissu | GTEX-P78B | 2/605 | 0.744084 | 0.999971 | 0 | 0 | 0.753487 | 0.222732 |
| GTEx_Tissu | GTEX-XV7C | 2/605 | 0.744084 | 0.999971 | 0 | 0 | 0.753487 | 0.222732 |
| GTEx_Tissu | GTEX-WHP | 4/1161 | 0.750988 | 0.999971 | 0 | 0 | 0.781248 | 0.223722 |
| GTEx_Tissu | GTEX-WI4N | 2/606 | 0.744914 | 0.999971 | 0 | 0 | 0.752201 | 0.221513 |
| GTEx_Tissu | GTEX-XUW | 2/614 | 0.751474 | 0.999971 | 0 | 0 | 0.742061 | 0.21202 |
| GTEx_Tissu | GTEX-WOF | 3/893 | 0.751771 | 0.999971 | 0 | 0 | 0.763363 | 0.217805 |
| GTEx_Tissu | GTEX-OIZG | 6/1685 | 0.751829 | 0.999971 | 0 | 0 | 0.804447 | 0.229466 |
| GTEx_Tissu | GTEX-S7SF | 3/894 | 0.752454 | 0.999971 | 0 | 0 | 0.762466 | 0.216857 |
| GTEx_Tissu | GTEX-WOF | 2/610 | 0.748212 | 0.999971 | 0 | 0 | 0.747098 | 0.216709 |
| GTEx_Tissu | GTEX-XQ8I | 2/791 | 0.863672 | 0.999971 | 0 | 0 | 0.570312 | 0.083586 |
| GTEx_Tissu | GTEX-TMM | 2/616 | 0.753092 | 0.999971 | 0 | 0 | 0.739567 | 0.209717 |
| GTEx_Tissu | GTEX-PWO | 2/611 | 0.749031 | 0.999971 | 0 | 0 | 0.745832 | 0.215526 |
| GTEx_Tissu | GTEX-QCQ( | 4/1165 | 0.753414 | 0.999971 | 0 | 0 | 0.77839 | 0.220393 |
| GTEx_Tissu | GTEX-XPT6 | 4/1165 | 0.753414 | 0.999971 | 0 | 0 | 0.77839 | 0.220393 |
| GTEx_Tissu | GTEX-RU1J | Aug-99 | 0.753884 | 0.999971 | 0 | 0 | 0.819093 | 0.231407 |
| GTEx_Tissu | GTEX-XPVG | 2/617 | 0.753897 | 0.999971 | 0 | 0 | 0.738326 | 0.208576 |
| GTEx_Tissu | GTEX-X585 | Oct-92 | 0.750329 | 0.999971 | 0 | 0 | 0.834374 | 0.239669 |
| GTEx_Tissu | GTEX-U3ZN | 6/1683 | 0.750796 | 0.999971 | 0 | 0 | 0.805495 | 0.230872 |
| GTEx_Tissu | GTEX-X585 | Jul-11 | 0.947552 | 0.999971 | 0 | 0 | 0.58162 | 0.031334 |
| GTEx_Tissu | GTEX-WZT( | 2/1113 | 0.958247 | 0.999971 | 0 | 0 | 0.3982 | 0.016983 |
| GTEx_Tissu | GTEX-VUS( | 2/1051 | 0.947172 | 0.999971 | 0 | 0 | 0.423126 | 0.022965 |
| GTEx_Tissu | GTEX-NPJ8 | 3/1395 | 0.947405 | 0.999971 | 0 | 0 | 0.47519 | 0.025674 |
| GTEx_Tissu | GTEX-P78B | 4/1796 | 0.95914 | 0.999971 | 0 | 0 | 0.487333 | 0.020331 |
| GTEx_Tissu | GTEX-NPJ8 | May-09 | 0.959239 | 0.999971 | 0 | 0 | 0.516119 | 0.021478 |
| GTEx_Tissu | GTEX-UJHI- | 3/1470 | 0.959271 | 0.999971 | 0 | 0 | 0.44907 | 0.018673 |
| GTEx_Tissu | GTEX-QDV. | 2/1120 | 0.959349 | 0.999971 | 0 | 0 | 0.395559 | 0.016416 |
| GTEx_Tissu | GTEX-XGQ4 | 1/722 | 0.959472 | 0.999971 | 0 | 0 | 0.309518 | 0.012805 |
| GTEx_Tissu | GTEX-X4XX | May-14 | 0.959847 | 0.999971 | 0 | 0 | 0.514751 | 0.021095 |
| GTEx_Tissu | GTEX-WFO | 1/715 | 0.958169 | 0.999971 | 0 | 0 | 0.312667 | 0.013361 |
| GTEx_Tissu | GTEX-QDT8 | 3/1466 | 0.958707 | 0.999971 | 0 | 0 | 0.450395 | 0.018993 |

| | | | | | | | | |
|---|---|---|---|---|---|---|---|---|
| GTEx_Tissu | GTEX-UJHI- | 4/1722 | 0.948405 | 0.999971 | 0 | 0 | 0.5104 | 0.027038 |
| GTEx_Tissu | GTEX-SUCS | 1/671 | 0.948974 | 0.999971 | 0 | 0 | 0.333964 | 0.017491 |
| GTEx_Tissu | GTEX-U8XE | 2/1061 | 0.949128 | 0.999971 | 0 | 0 | 0.418908 | 0.021872 |
| GTEx_Tissu | GTEX-UJHI- | 2/1067 | 0.950269 | 0.999971 | 0 | 0 | 0.416415 | 0.021241 |
| GTEx_Tissu | GTEX-S95S- | Jun-17 | 0.946948 | 0.999971 | 0 | 0 | 0.564194 | 0.030755 |
| GTEx_Tissu | GTEX-R55G | Jul-08 | 0.947134 | 0.999971 | 0 | 0 | 0.582391 | 0.031633 |
| GTEx_Tissu | GTEX-SN8G | 2/1170 | 0.966459 | 0.999971 | 0 | 0 | 0.377619 | 0.012883 |
| GTEx_Tissu | GTEX-P4PP | 1/707 | 0.956629 | 0.999971 | 0 | 0 | 0.316342 | 0.014027 |
| GTEx_Tissu | GTEX-T6MI | 3/1364 | 0.941633 | 0.999971 | 0 | 0 | 0.486827 | 0.029278 |
| GTEx_Tissu | GTEX-X4XX | Aug-55 | 0.941967 | 0.999971 | 0 | 0 | 0.607026 | 0.036291 |
| GTEx_Tissu | GTEX-P4PP | 1/690 | 0.953167 | 0.999971 | 0 | 0 | 0.324434 | 0.015562 |
| GTEx_Tissu | GTEX-WEY! | 4/1755 | 0.953473 | 0.999971 | 0 | 0 | 0.499873 | 0.023816 |
| GTEx_Tissu | GTEX-WFJC | 1/717 | 0.958546 | 0.999971 | 0 | 0 | 0.311761 | 0.013199 |
| GTEx_Tissu | GTEX-UPIC- | 1/704 | 0.956037 | 0.999971 | 0 | 0 | 0.317741 | 0.014285 |
| GTEx_Tissu | GTEX-XMD | 4/1773 | 0.956043 | 0.999971 | 0 | 0 | 0.494296 | 0.02222 |
| GTEx_Tissu | GTEX-NPJ7 | Aug-67 | 0.956468 | 0.999971 | 0 | 0 | 0.580217 | 0.025824 |
| GTEx_Tissu | GTEX-XOTC | May-88 | 0.956598 | 0.999971 | 0 | 0 | 0.521937 | 0.023159 |
| GTEx_Tissu | GTEX-S7SE- | Oct-86 | 0.962564 | 0.999971 | 0 | 0 | 0.593313 | 0.022638 |
| GTEx_Tissu | GTEX-UJM( | 1/707 | 0.956629 | 0.999971 | 0 | 0 | 0.316342 | 0.014027 |
| GTEx_Tissu | GTEX-PWO | 2/1104 | 0.956788 | 0.999971 | 0 | 0 | 0.401644 | 0.017742 |
| GTEx_Tissu | GTEX-NL4V | 3/1477 | 0.960242 | 0.999971 | 0 | 0 | 0.446768 | 0.018125 |
| GTEx_Tissu | GTEX-XQ3S | May-91 | 0.956985 | 0.999971 | 0 | 0 | 0.521099 | 0.022912 |
| GTEx_Tissu | GTEX-QVJC | Aug-72 | 0.957035 | 0.999971 | 0 | 0 | 0.579067 | 0.02543 |
| GTEx_Tissu | GTEX-RTLS- | 2/1108 | 0.957442 | 0.999971 | 0 | 0 | 0.400106 | 0.017401 |
| GTEx_Tissu | GTEX-U3ZN | 2/1111 | 0.957927 | 0.999971 | 0 | 0 | 0.39896 | 0.017149 |
| GTEx_Tissu | GTEX-T5JC- | Aug-80 | 0.957927 | 0.999971 | 0 | 0 | 0.577236 | 0.024812 |
| GTEx_Tissu | GTEX-QDT8 | Aug-59 | 0.955549 | 0.999971 | 0 | 0 | 0.582064 | 0.026466 |
| GTEx_Tissu | GTEX-WOF | 3/1530 | 0.966927 | 0.999971 | 0 | 0 | 0.430022 | 0.014463 |
| GTEx_Tissu | GTEX-O5YT | Jun-84 | 0.967003 | 0.999971 | 0 | 0 | 0.521179 | 0.017488 |
| GTEx_Tissu | GTEX-V955 | 1/768 | 0.967093 | 0.999971 | 0 | 0 | 0.290258 | 0.009712 |
| GTEx_Tissu | GTEX-PVO\ | Aug-70 | 0.956809 | 0.999971 | 0 | 0 | 0.579526 | 0.025587 |
| GTEx_Tissu | GTEX-UTH( | 3/1433 | 0.953766 | 0.999971 | 0 | 0 | 0.461613 | 0.021852 |
| GTEx_Tissu | GTEX-SN8G | 3/1436 | 0.954236 | 0.999971 | 0 | 0 | 0.460572 | 0.021575 |
| GTEx_Tissu | GTEX-U8XE | 3/1440 | 0.954857 | 0.999971 | 0 | 0 | 0.459191 | 0.021212 |
| GTEx_Tissu | GTEX-QEL4 | 4/1768 | 0.955342 | 0.999971 | 0 | 0 | 0.495834 | 0.022652 |
| GTEx_Tissu | GTEX-U3ZN | 3/1444 | 0.95547 | 0.999971 | 0 | 0 | 0.457817 | 0.020854 |
| GTEx_Tissu | GTEX-NPJ8 | 3/1530 | 0.966927 | 0.999971 | 0 | 0 | 0.430022 | 0.014463 |
| GTEx_Tissu | GTEX-XUJ4 | Jun-81 | 0.955643 | 0.999971 | 0 | 0 | 0.546994 | 0.024818 |
| GTEx_Tissu | GTEX-UTH( | 2/1214 | 0.971723 | 0.999971 | 0 | 0 | 0.363056 | 0.010414 |
| GTEx_Tissu | GTEX-T5JW | May-94 | 0.968515 | 0.999971 | 0 | 0 | 0.49371 | 0.015794 |
| GTEx_Tissu | GTEX-VJYA | 2/1189 | 0.968837 | 0.999971 | 0 | 0 | 0.371198 | 0.011752 |
| GTEx_Tissu | GTEX-NFK9 | May-99 | 0.968996 | 0.999971 | 0 | 0 | 0.492446 | 0.01551 |
| GTEx_Tissu | GTEX-RNOI | Sep-85 | 0.969443 | 0.999971 | 0 | 0 | 0.5652 | 0.01754 |
| GTEx_Tissu | GTEX-N7M | 3/1553 | 0.969489 | 0.999971 | 0 | 0 | 0.423111 | 0.01311 |
| GTEx_Tissu | GTEX-SUCS | 4/1889 | 0.969739 | 0.999971 | 0 | 0 | 0.460912 | 0.014163 |

| | | | | | | | | |
|---|---|---|---|---|---|---|---|---|
| GTEx_Tissu | GTEX-X585 | May-65 | 0.965591 | 0.999971 | 0 | 0 | 0.501157 | 0.017548 |
| GTEx_Tissu | GTEX-XOT( | 2/1144 | 0.962923 | 0.999971 | 0 | 0 | 0.386752 | 0.014612 |
| GTEx_Tissu | GTEX-WVLI | Dec-37 | 0.963017 | 0.999971 | 0 | 0 | 0.612383 | 0.023077 |
| GTEx_Tissu | GTEX-WHS | May-46 | 0.963544 | 0.999971 | 0 | 0 | 0.506146 | 0.018797 |
| GTEx_Tissu | GTEX-XUJ4 | 2/1149 | 0.96363 | 0.999971 | 0 | 0 | 0.384963 | 0.014262 |
| GTEx_Tissu | GTEX-XLM4 | Jul-47 | 0.963732 | 0.999971 | 0 | 0 | 0.548408 | 0.020259 |
| GTEx_Tissu | GTEX-XBED | 4/1838 | 0.964288 | 0.999971 | 0 | 0 | 0.475069 | 0.017276 |
| GTEx_Tissu | GTEX-XQ8I | 2/1175 | 0.967101 | 0.999971 | 0 | 0 | 0.375909 | 0.012575 |
| GTEx_Tissu | GTEX-O5Y\ | 2/1157 | 0.964733 | 0.999971 | 0 | 0 | 0.382134 | 0.01372 |
| GTEx_Tissu | GTEX-WVLI | 1/753 | 0.964778 | 0.999971 | 0 | 0 | 0.29628 | 0.010624 |
| GTEx_Tissu | GTEX-T2IS- | 2/1023 | 0.941313 | 0.999971 | 0 | 0 | 0.435375 | 0.026331 |
| GTEx_Tissu | GTEX-PLZ4- | 2/1164 | 0.965672 | 0.999971 | 0 | 0 | 0.37969 | 0.013263 |
| GTEx_Tissu | GTEX-WRH | 2/1167 | 0.966068 | 0.999971 | 0 | 0 | 0.378652 | 0.013072 |
| GTEx_Tissu | GTEX-X5EB | 2/1167 | 0.966068 | 0.999971 | 0 | 0 | 0.378652 | 0.013072 |
| GTEx_Tissu | GTEX-QVJC | 2/1168 | 0.966198 | 0.999971 | 0 | 0 | 0.378307 | 0.013009 |
| GTEx_Tissu | GTEX-XMD | 4/1772 | 0.955904 | 0.999971 | 0 | 0 | 0.494603 | 0.022306 |
| GTEx_Tissu | GTEX-OXR( | 1/764 | 0.966491 | 0.999971 | 0 | 0 | 0.291841 | 0.009947 |
| GTEx_Tissu | GTEX-T5JC- | 2/1171 | 0.966588 | 0.999971 | 0 | 0 | 0.377276 | 0.012821 |
| GTEx_Tissu | GTEX-P4QS | 2/1173 | 0.966845 | 0.999971 | 0 | 0 | 0.376591 | 0.012697 |
| GTEx_Tissu | GTEX-RN64 | 2/1038 | 0.944524 | 0.999971 | 0 | 0 | 0.42873 | 0.02447 |
| GTEx_Tissu | GTEX-X4XX | Jul-81 | 0.943233 | 0.999971 | 0 | 0 | 0.589418 | 0.034447 |
| GTEx_Tissu | GTEX-TKQ1 | 1/648 | 0.943399 | 0.999971 | 0 | 0 | 0.346249 | 0.020175 |
| GTEx_Tissu | GTEX-S341 | 1/649 | 0.943653 | 0.999971 | 0 | 0 | 0.345697 | 0.020049 |
| GTEx_Tissu | GTEX-S341 | 2/1021 | 0.940871 | 0.999971 | 0 | 0 | 0.436275 | 0.02659 |
| GTEx_Tissu | GTEX-PWN | 2/1021 | 0.940871 | 0.999971 | 0 | 0 | 0.436275 | 0.02659 |
| GTEx_Tissu | GTEX-WL4( | 1/650 | 0.943907 | 0.999971 | 0 | 0 | 0.345146 | 0.019925 |
| GTEx_Tissu | GTEX-UPIC- | 4/1680 | 0.941238 | 0.999971 | 0 | 0 | 0.524398 | 0.031757 |
| GTEx_Tissu | GTEX-TMZS | 2/1036 | 0.944105 | 0.999971 | 0 | 0 | 0.429605 | 0.02471 |
| GTEx_Tissu | GTEX-WFG | May-04 | 0.944436 | 0.999971 | 0 | 0 | 0.546432 | 0.031238 |
| GTEx_Tissu | GTEX-PLZ6- | 4/1691 | 0.943197 | 0.999971 | 0 | 0 | 0.520665 | 0.030449 |
| GTEx_Tissu | GTEX-WQL | May-10 | 0.945396 | 0.999971 | 0 | 0 | 0.544614 | 0.030581 |
| GTEx_Tissu | GTEX-S7SE- | Jul-73 | 0.94203 | 0.999971 | 0 | 0 | 0.591529 | 0.035325 |
| GTEx_Tissu | GTEX-SUCS | 3/1368 | 0.942409 | 0.999971 | 0 | 0 | 0.485296 | 0.028786 |
| GTEx_Tissu | GTEX-VUS( | 1/662 | 0.94686 | 0.999971 | 0 | 0 | 0.338669 | 0.018493 |
| GTEx_Tissu | GTEX-QLQ\ | 1/646 | 0.942887 | 0.999971 | 0 | 0 | 0.347359 | 0.020428 |
| GTEx_Tissu | GTEX-X4EC | 2/991 | 0.933871 | 0.999971 | 0 | 0 | 0.450223 | 0.030803 |
| GTEx_Tissu | GTEX-S4UY | 3/1327 | 0.933991 | 0.999971 | 0 | 0 | 0.50143 | 0.034242 |
| GTEx_Tissu | GTEX-RU72 | 2/992 | 0.934117 | 0.999971 | 0 | 0 | 0.449745 | 0.030652 |
| GTEx_Tissu | GTEX-Q2AI | 3/1349 | 0.938638 | 0.999971 | 0 | 0 | 0.49265 | 0.031197 |
| GTEx_Tissu | GTEX-NL4V | Aug-41 | 0.93989 | 0.999971 | 0 | 0 | 0.610526 | 0.037848 |
| GTEx_Tissu | GTEX-OHPI | 2/1006 | 0.937463 | 0.999971 | 0 | 0 | 0.443145 | 0.028617 |
| GTEx_Tissu | GTEX-R55E | May-01 | 0.925475 | 0.999971 | 0 | 0 | 0.579429 | 0.044876 |
| GTEx_Tissu | GTEX-RTLS- | Sep-28 | 0.925487 | 0.999971 | 0 | 0 | 0.64568 | 0.049998 |
| GTEx_Tissu | GTEX-NFK9 | 1/589 | 0.926193 | 0.999971 | 0 | 0 | 0.382159 | 0.029301 |
| GTEx_Tissu | GTEX-UTH( | 3/1294 | 0.92642 | 0.999971 | 0 | 0 | 0.51516 | 0.039372 |

| | | | | | | | | |
|---|---|---|---|---|---|---|---|---|
| GTEx_Tissu | GTEX-W5W | 2/1010 | 0.938389 | 0.999971 | 0 | 0 | 0.441293 | 0.028062 |
| GTEx_Tissu | GTEX-PWN | 3/1348 | 0.938433 | 0.999971 | 0 | 0 | 0.493043 | 0.03133 |
| GTEx_Tissu | GTEX-PLZ5 | 4/1611 | 0.927517 | 0.999971 | 0 | 0 | 0.548984 | 0.041308 |
| GTEx_Tissu | GTEX-TMZ | 4/1644 | 0.934399 | 0.999971 | 0 | 0 | 0.536967 | 0.036434 |
| GTEx_Tissu | GTEX-T6M | 2/968 | 0.927988 | 0.999971 | 0 | 0 | 0.461503 | 0.034491 |
| GTEx_Tissu | GTEX-WRH | Jul-90 | 0.928195 | 0.999971 | 0 | 0 | 0.614227 | 0.045768 |
| GTEx_Tissu | GTEX-PWO | 2/970 | 0.928518 | 0.999971 | 0 | 0 | 0.460501 | 0.034153 |
| GTEx_Tissu | GTEX-WYJK | 2/972 | 0.929045 | 0.999971 | 0 | 0 | 0.459503 | 0.033819 |
| GTEx_Tissu | GTEX-T6MI | Jun-71 | 0.939805 | 0.999971 | 0 | 0 | 0.577156 | 0.035832 |
| GTEx_Tissu | GTEX-TML8 | 3/1355 | 0.939852 | 0.999971 | 0 | 0 | 0.490305 | 0.030415 |
| GTEx_Tissu | GTEX-V1D1 | 3/1369 | 0.942602 | 0.999971 | 0 | 0 | 0.484914 | 0.028664 |
| GTEx_Tissu | GTEX-TML8 | 2/1018 | 0.940204 | 0.999971 | 0 | 0 | 0.437633 | 0.026984 |
| GTEx_Tissu | GTEX-PSDG | 1/637 | 0.940524 | 0.999971 | 0 | 0 | 0.352439 | 0.021611 |
| GTEx_Tissu | GTEX-REY6 | 1/665 | 0.947574 | 0.999971 | 0 | 0 | 0.337087 | 0.018152 |
| GTEx_Tissu | GTEX-QLQ7 | 3/1422 | 0.952001 | 0.999971 | 0 | 0 | 0.465469 | 0.022896 |
| GTEx_Tissu | GTEX-OHPI | 1/685 | 0.952098 | 0.999971 | 0 | 0 | 0.32689 | 0.016046 |
| GTEx_Tissu | GTEX-WL4 | 3/1425 | 0.952488 | 0.999971 | 0 | 0 | 0.464411 | 0.022606 |
| GTEx_Tissu | GTEX-QLQ7 | 4/1750 | 0.952735 | 0.999971 | 0 | 0 | 0.501442 | 0.024279 |
| GTEx_Tissu | GTEX-TSE9 | Jun-41 | 0.95037 | 0.999971 | 0 | 0 | 0.557633 | 0.028386 |
| GTEx_Tissu | GTEX-PWO | 3/1414 | 0.950679 | 0.999971 | 0 | 0 | 0.46831 | 0.023686 |
| GTEx_Tissu | GTEX-TSE9 | 4/1757 | 0.953765 | 0.999971 | 0 | 0 | 0.499247 | 0.023633 |
| GTEx_Tissu | GTEX-XPVG | 1/680 | 0.951004 | 0.999971 | 0 | 0 | 0.329383 | 0.016547 |
| GTEx_Tissu | GTEX-S7SE | May-53 | 0.951854 | 0.999971 | 0 | 0 | 0.531899 | 0.026246 |
| GTEx_Tissu | GTEX-QMR | Nov-66 | 0.940575 | 0.999971 | 0 | 0 | 0.643812 | 0.039443 |
| GTEx_Tissu | GTEX-RWS | May-81 | 0.940617 | 0.999971 | 0 | 0 | 0.553502 | 0.033885 |
| GTEx_Tissu | GTEX-X4XX | Aug-46 | 0.940639 | 0.999971 | 0 | 0 | 0.609272 | 0.037285 |
| GTEx_Tissu | GTEX-WEY | 2/1072 | 0.951201 | 0.999971 | 0 | 0 | 0.41436 | 0.02073 |
| GTEx_Tissu | GTEX-S7SF | Jul-40 | 0.951453 | 0.999971 | 0 | 0 | 0.57425 | 0.028578 |
| GTEx_Tissu | GTEX-OHPI | 2/1022 | 0.941092 | 0.999971 | 0 | 0 | 0.435825 | 0.026461 |
| GTEx_Tissu | GTEX-R53T | 1/684 | 0.951881 | 0.999971 | 0 | 0 | 0.327386 | 0.016145 |
| GTEx_Tissu | GTEX-OHPI | 3/1362 | 0.941242 | 0.999971 | 0 | 0 | 0.487596 | 0.029527 |
| GTEx_Tissu | GTEX-XXEK | 2/1047 | 0.94637 | 0.999971 | 0 | 0 | 0.424835 | 0.023417 |
| GTEx_Tissu | GTEX-X4XX | 2/1034 | 0.943684 | 0.999971 | 0 | 0 | 0.430483 | 0.024953 |
| GTEx_Tissu | GTEX-S4UY | May-00 | 0.943788 | 0.999971 | 0 | 0 | 0.54765 | 0.031684 |
| GTEx_Tissu | GTEX-VUSG | Sep-84 | 0.934739 | 0.999971 | 0 | 0 | 0.63182 | 0.04264 |
| GTEx_Tissu | GTEX-PWN | 3/1376 | 0.943933 | 0.999971 | 0 | 0 | 0.48226 | 0.027827 |
| GTEx_Tissu | GTEX-PX3G | 3/1334 | 0.935503 | 0.999971 | 0 | 0 | 0.498605 | 0.033242 |
| GTEx_Tissu | GTEX-QDV | 1/620 | 0.935794 | 0.999971 | 0 | 0 | 0.362438 | 0.024051 |
| GTEx_Tissu | GTEX-T5JW | 1/621 | 0.936083 | 0.999971 | 0 | 0 | 0.361834 | 0.0239 |
| GTEx_Tissu | GTEX-XYKS | 1/623 | 0.936655 | 0.999971 | 0 | 0 | 0.360633 | 0.0236 |
| GTEx_Tissu | GTEX-XGQ | 1/656 | 0.945403 | 0.999971 | 0 | 0 | 0.341878 | 0.019194 |
| GTEx_Tissu | GTEX-N7M | 3/1557 | 0.969916 | 0.999971 | 0 | 0 | 0.42193 | 0.012888 |
| GTEx_Tissu | GTEX-N7M | 4/1738 | 0.950921 | 0.999971 | 0 | 0 | 0.505246 | 0.025426 |
| GTEx_Tissu | GTEX-T6MI | Jun-17 | 0.946948 | 0.999971 | 0 | 0 | 0.564194 | 0.030755 |
| GTEx_Tissu | GTEX-POM | 3/1416 | 0.951013 | 0.999971 | 0 | 0 | 0.467597 | 0.023486 |

| | | | | | | | | |
|---|---|---|---|---|---|---|---|---|
| GTEx_Tissu | GTEX-V1D1 | 2/1071 | 0.951016 | 0.999971 | 0 | 0 | 0.414769 | 0.020831 |
| GTEx_Tissu | GTEX-QV31 | Jul-37 | 0.951062 | 0.999971 | 0 | 0 | 0.575005 | 0.028852 |
| GTEx_Tissu | GTEX-WL46 | Oct-78 | 0.951127 | 0.999971 | 0 | 0 | 0.615835 | 0.030858 |
| GTEx_Tissu | GTEX-S32W | Jul-10 | 0.947413 | 0.999971 | 0 | 0 | 0.581877 | 0.031433 |
| GTEx_Tissu | GTEX-TKQ1 | May-23 | 0.947425 | 0.999971 | 0 | 0 | 0.540713 | 0.029203 |
| GTEx_Tissu | GTEX-PVOV | Oct-66 | 0.982045 | 0.999971 | 0 | 0 | 0.5408 | 0.009798 |
| GTEx_Tissu | GTEX-XXEK | 3/1813 | 0.988095 | 0.999971 | 0 | 0 | 0.357202 | 0.004278 |
| GTEx_Tissu | GTEX-WHS | Apr-13 | 0.989954 | 0.999971 | 0 | 0 | 0.38624 | 0.0039 |
| GTEx_Tissu | GTEX-S4Q7 | 2/1437 | 0.988338 | 0.999971 | 0 | 0 | 0.30298 | 0.003554 |
| GTEx_Tissu | GTEX-PWO | Jun-26 | 0.988411 | 0.999971 | 0 | 0 | 0.448989 | 0.005234 |
| GTEx_Tissu | GTEX-PSDG | 1/997 | 0.988419 | 0.999971 | 0 | 0 | 0.220849 | 0.002572 |
| GTEx_Tissu | GTEX-V955 | 1/998 | 0.988472 | 0.999971 | 0 | 0 | 0.220615 | 0.002558 |
| GTEx_Tissu | GTEX-OIZI- | 1/1001 | 0.98863 | 0.999971 | 0 | 0 | 0.219919 | 0.002515 |
| GTEx_Tissu | GTEX-XUW | 2/1444 | 0.988663 | 0.999971 | 0 | 0 | 0.301395 | 0.003436 |
| GTEx_Tissu | GTEX-UTH( | Jun-33 | 0.988668 | 0.999971 | 0 | 0 | 0.447694 | 0.005102 |
| GTEx_Tissu | GTEX-Q2A( | Aug-22 | 0.987843 | 0.999971 | 0 | 0 | 0.489392 | 0.005986 |
| GTEx_Tissu | GTEX-T2IS- | Aug-52 | 0.988887 | 0.999971 | 0 | 0 | 0.484247 | 0.005411 |
| GTEx_Tissu | GTEX-N7M | 1/1008 | 0.98899 | 0.999971 | 0 | 0 | 0.218309 | 0.002417 |
| GTEx_Tissu | GTEX-T6M( | 2/1460 | 0.989375 | 0.999971 | 0 | 0 | 0.297829 | 0.003181 |
| GTEx_Tissu | GTEX-XPVG | 2/1460 | 0.989375 | 0.999971 | 0 | 0 | 0.297829 | 0.003181 |
| GTEx_Tissu | GTEX-WFO | 2/1469 | 0.989756 | 0.999971 | 0 | 0 | 0.295858 | 0.003046 |
| GTEx_Tissu | GTEX-QMR | 3/1854 | 0.989784 | 0.999971 | 0 | 0 | 0.348499 | 0.003578 |
| GTEx_Tissu | GTEX-X4EC | 3/1857 | 0.989899 | 0.999971 | 0 | 0 | 0.347877 | 0.003532 |
| GTEx_Tissu | GTEX-XBED | 1/915 | 0.983144 | 0.999971 | 0 | 0 | 0.241705 | 0.004109 |
| GTEx_Tissu | GTEX-RTLS | 1/834 | 0.975615 | 0.999971 | 0 | 0 | 0.266339 | 0.006575 |
| GTEx_Tissu | GTEX-QVUS | May-33 | 0.985262 | 0.999971 | 0 | 0 | 0.43911 | 0.00652 |
| GTEx_Tissu | GTEX-XPVG | 3/1757 | 0.985356 | 0.999971 | 0 | 0 | 0.369747 | 0.005454 |
| GTEx_Tissu | GTEX-WWY | Jul-60 | 0.985377 | 0.999971 | 0 | 0 | 0.483213 | 0.007118 |
| GTEx_Tissu | GTEX-UTH( | Apr-10 | 0.985607 | 0.999971 | 0 | 0 | 0.407487 | 0.005907 |
| GTEx_Tissu | GTEX-Q2A( | May-04 | 0.977668 | 0.999971 | 0 | 0 | 0.46717 | 0.010551 |
| GTEx_Tissu | GTEX-X585 | Jun-80 | 0.975161 | 0.999971 | 0 | 0 | 0.498978 | 0.01255 |
| GTEx_Tissu | GTEX-PLZ5 | 1/831 | 0.97528 | 0.999971 | 0 | 0 | 0.267344 | 0.006692 |
| GTEx_Tissu | GTEX-QMR | Jul-84 | 0.975388 | 0.999971 | 0 | 0 | 0.518127 | 0.012912 |
| GTEx_Tissu | GTEX-XLM4 | 1/833 | 0.975504 | 0.999971 | 0 | 0 | 0.266673 | 0.006614 |
| GTEx_Tissu | GTEX-NL3H | Sep-56 | 0.980792 | 0.999971 | 0 | 0 | 0.532389 | 0.010326 |
| GTEx_Tissu | GTEX-UPIC | 3/1617 | 0.975679 | 0.999971 | 0 | 0 | 0.404917 | 0.00997 |
| GTEx_Tissu | GTEX-WFG | Apr-58 | 0.975901 | 0.999971 | 0 | 0 | 0.442934 | 0.010805 |
| GTEx_Tissu | GTEX-WHS | May-18 | 0.988873 | 0.999971 | 0 | 0 | 0.422195 | 0.004724 |
| GTEx_Tissu | GTEX-PSDG | May-72 | 0.987036 | 0.999971 | 0 | 0 | 0.431204 | 0.005627 |
| GTEx_Tissu | GTEX-NL4V | 2/1411 | 0.987048 | 0.999971 | 0 | 0 | 0.309005 | 0.004028 |
| GTEx_Tissu | GTEX-N7M | Jul-09 | 0.987405 | 0.999971 | 0 | 0 | 0.474198 | 0.00601 |
| GTEx_Tissu | GTEX-WHS | 2/1421 | 0.987559 | 0.999971 | 0 | 0 | 0.306662 | 0.003839 |
| GTEx_Tissu | GTEX-XUYS | 3/1804 | 0.987691 | 0.999971 | 0 | 0 | 0.359166 | 0.004449 |
| GTEx_Tissu | GTEX-N7M | 1/1204 | 0.995542 | 0.999971 | 0 | 0 | 0.180846 | 0.000808 |
| GTEx_Tissu | GTEX-P44H | 2/1577 | 0.993417 | 0.999971 | 0 | 0 | 0.273957 | 0.00181 |

| | | | | | | | | |
|---|---|---|---|---|---|---|---|---|
| GTEx_Tissu | GTEX-S7SF | 1/1138 | 0.99395 | 0.999971 | 0 | 0 | 0.192019 | 0.001165 |
| GTEx_Tissu | GTEX-T6MI | Jul-51 | 0.994132 | 0.999971 | 0 | 0 | 0.433549 | 0.002552 |
| GTEx_Tissu | GTEX-WFG | 3/1788 | 0.986939 | 0.999971 | 0 | 0 | 0.362705 | 0.004769 |
| GTEx_Tissu | GTEX-WZT( | Jul-88 | 0.994798 | 0.999971 | 0 | 0 | 0.427847 | 0.002231 |
| GTEx_Tissu | GTEX-NFKS | 1/1186 | 0.995155 | 0.999971 | 0 | 0 | 0.18377 | 0.000893 |
| GTEx_Tissu | GTEX-WYV. | Mar-50 | 0.995161 | 0.999971 | 0 | 0 | 0.311711 | 0.001512 |
| GTEx_Tissu | GTEX-PSDG | 1/1194 | 0.995331 | 0.999971 | 0 | 0 | 0.18246 | 0.000854 |
| GTEx_Tissu | GTEX-NPJ7 | Jun-03 | 0.995352 | 0.999971 | 0 | 0 | 0.402205 | 0.001874 |
| GTEx_Tissu | GTEX-OOBI | 1/1114 | 0.99324 | 0.999971 | 0 | 0 | 0.19641 | 0.001332 |
| GTEx_Tissu | GTEX-T5JC | Jul-40 | 0.995616 | 0.999971 | 0 | 0 | 0.420041 | 0.001845 |
| GTEx_Tissu | GTEX-QV31 | May-03 | 0.988302 | 0.999971 | 0 | 0 | 0.425096 | 0.005002 |
| GTEx_Tissu | GTEX-OXR( | 2/1388 | 0.985793 | 0.999971 | 0 | 0 | 0.314523 | 0.0045 |
| GTEx_Tissu | GTEX-X8H( | May-47 | 0.985923 | 0.999971 | 0 | 0 | 0.436243 | 0.006185 |
| GTEx_Tissu | GTEX-R55G | 1/956 | 0.986026 | 0.999971 | 0 | 0 | 0.230829 | 0.003248 |
| GTEx_Tissu | GTEX-XUJ4 | Apr-20 | 0.986096 | 0.999971 | 0 | 0 | 0.405334 | 0.005675 |
| GTEx_Tissu | GTEX-P4QS | 2/1395 | 0.986187 | 0.999971 | 0 | 0 | 0.312825 | 0.004351 |
| GTEx_Tissu | GTEX-Q2AI | 1/963 | 0.986467 | 0.999971 | 0 | 0 | 0.229065 | 0.003121 |
| GTEx_Tissu | GTEX-QV44 | Jul-71 | 0.99241 | 0.999971 | 0 | 0 | 0.44632 | 0.0034 |
| GTEx_Tissu | GTEX-W5W | 2/1315 | 0.98099 | 0.999971 | 0 | 0 | 0.333318 | 0.006397 |
| GTEx_Tissu | GTEX-P4QT | 1/889 | 0.98102 | 0.999971 | 0 | 0 | 0.249123 | 0.004774 |
| GTEx_Tissu | GTEX-X4XX | 3/1690 | 0.981297 | 0.999971 | 0 | 0 | 0.38585 | 0.007285 |
| GTEx_Tissu | GTEX-WRH | Apr-34 | 0.981333 | 0.999971 | 0 | 0 | 0.424547 | 0.008 |
| GTEx_Tissu | GTEX-SNM | 1/896 | 0.981616 | 0.999971 | 0 | 0 | 0.247083 | 0.004585 |
| GTEx_Tissu | GTEX-WL4( | Aug-88 | 0.981993 | 0.999971 | 0 | 0 | 0.513523 | 0.009331 |
| GTEx_Tissu | GTEX-NPJ8 | 2/1623 | 0.994557 | 0.999971 | 0 | 0 | 0.265515 | 0.001449 |
| GTEx_Tissu | GTEX-WY7( | 1/1077 | 0.991983 | 0.999971 | 0 | 0 | 0.203564 | 0.001639 |
| GTEx_Tissu | GTEX-WHS | 2/1530 | 0.992013 | 0.999971 | 0 | 0 | 0.283107 | 0.00227 |
| GTEx_Tissu | GTEX-RUS( | 3/1753 | 0.98514 | 0.999971 | 0 | 0 | 0.370673 | 0.00555 |
| GTEx_Tissu | GTEX-QMR | Aug-77 | 0.992412 | 0.999971 | 0 | 0 | 0.46374 | 0.003532 |
| GTEx_Tissu | GTEX-P4PC | 1/1089 | 0.992414 | 0.999971 | 0 | 0 | 0.201191 | 0.001532 |
| GTEx_Tissu | GTEX-XUZ( | Mar-34 | 0.992447 | 0.999971 | 0 | 0 | 0.332581 | 0.002522 |
| GTEx_Tissu | GTEX-NPJ8 | Jun-77 | 0.992908 | 0.999971 | 0 | 0 | 0.422404 | 0.003006 |
| GTEx_Tissu | GTEX-XQ8I | 1/1237 | 0.996175 | 0.999971 | 0 | 0 | 0.175707 | 0.000673 |
| GTEx_Tissu | GTEX-T8EN | 1/1109 | 0.993083 | 0.999971 | 0 | 0 | 0.197349 | 0.00137 |
| GTEx_Tissu | GTEX-QVJC | Aug-07 | 0.993088 | 0.999971 | 0 | 0 | 0.45903 | 0.003184 |
| GTEx_Tissu | GTEX-UPIC | 2/1569 | 0.993196 | 0.999971 | 0 | 0 | 0.275476 | 0.001881 |
| GTEx_Tissu | GTEX-R55G | 3/1627 | 0.976533 | 0.999971 | 0 | 0 | 0.402204 | 0.009551 |
| GTEx_Tissu | GTEX-PWO | 1/733 | 0.96144 | 0.999971 | 0 | 0 | 0.304692 | 0.011981 |
| GTEx_Tissu | GTEX-S95S | 3/1487 | 0.961592 | 0.999971 | 0 | 0 | 0.443517 | 0.01737 |
| GTEx_Tissu | GTEX-O5Y\ | 1/734 | 0.961614 | 0.999971 | 0 | 0 | 0.304261 | 0.011909 |
| GTEx_Tissu | GTEX-XBEV | Jun-33 | 0.961752 | 0.999971 | 0 | 0 | 0.533687 | 0.020813 |
| GTEx_Tissu | GTEX-UTH( | Jun-37 | 0.962189 | 0.999971 | 0 | 0 | 0.532687 | 0.020532 |
| GTEx_Tissu | GTEX-R55E | Oct-15 | 0.979379 | 0.999971 | 0 | 0 | 0.549789 | 0.011455 |
| GTEx_Tissu | GTEX-X4XX | May-82 | 0.97606 | 0.999971 | 0 | 0 | 0.472273 | 0.011444 |
| GTEx_Tissu | GTEX-X5EB | 1/840 | 0.976272 | 0.999971 | 0 | 0 | 0.264351 | 0.006348 |

| | | | | | | | | |
|---|---|---|---|---|---|---|---|---|
| GTEx_Tissu | GTEX-QEL4 | Jul-97 | 0.976296 | 0.999971 | 0 | 0 | 0.515402 | 0.012364 |
| GTEx_Tissu | GTEX-X4EC | 3/1484 | 0.961192 | 0.999971 | 0 | 0 | 0.444487 | 0.017593 |
| GTEx_Tissu | GTEX-QXCU | Jun-03 | 0.976822 | 0.999971 | 0 | 0 | 0.493903 | 0.011582 |
| GTEx_Tissu | GTEX-WFO | May-94 | 0.97695 | 0.999971 | 0 | 0 | 0.469478 | 0.010948 |
| GTEx_Tissu | GTEX-NPJ8 | 1/848 | 0.97712 | 0.999971 | 0 | 0 | 0.261745 | 0.006058 |
| GTEx_Tissu | GTEX-OXRI | 1/886 | 0.980758 | 0.999971 | 0 | 0 | 0.250007 | 0.004857 |
| GTEx_Tissu | GTEX-WFO | 1/856 | 0.977939 | 0.999971 | 0 | 0 | 0.259187 | 0.005782 |
| GTEx_Tissu | GTEX-P44H | Nov-70 | 0.977974 | 0.999971 | 0 | 0 | 0.565326 | 0.012591 |
| GTEx_Tissu | GTEX-WFJC | 1/857 | 0.978039 | 0.999971 | 0 | 0 | 0.25887 | 0.005748 |
| GTEx_Tissu | GTEX-PWC | 2/1284 | 0.97851 | 0.999971 | 0 | 0 | 0.341947 | 0.007429 |
| GTEx_Tissu | GTEX-PLZ5 | 2/1214 | 0.971723 | 0.999971 | 0 | 0 | 0.363056 | 0.010414 |
| GTEx_Tissu | GTEX-OOB. | Jun-73 | 0.974635 | 0.999971 | 0 | 0 | 0.500541 | 0.01286 |
| GTEx_Tissu | GTEX-V1D1 | Aug-11 | 0.970394 | 0.999971 | 0 | 0 | 0.548591 | 0.016487 |
| GTEx_Tissu | GTEX-WL4( | 4/1896 | 0.970425 | 0.999971 | 0 | 0 | 0.459029 | 0.013781 |
| GTEx_Tissu | GTEX-S32V | 3/1564 | 0.970648 | 0.999971 | 0 | 0 | 0.419877 | 0.012509 |
| GTEx_Tissu | GTEX-PLZ5 | 2/1207 | 0.970941 | 0.999971 | 0 | 0 | 0.365301 | 0.010772 |
| GTEx_Tissu | GTEX-WHS | Aug-18 | 0.970956 | 0.999971 | 0 | 0 | 0.547128 | 0.016126 |
| GTEx_Tissu | GTEX-XV7C | 1/797 | 0.97115 | 0.999971 | 0 | 0 | 0.27926 | 0.008175 |
| GTEx_Tissu | GTEX-SNOS | 1/797 | 0.97115 | 0.999971 | 0 | 0 | 0.27926 | 0.008175 |
| GTEx_Tissu | GTEX-N7M | Jul-32 | 0.962196 | 0.999971 | 0 | 0 | 0.551908 | 0.021269 |
| GTEx_Tissu | GTEX-NPJ7 | Jun-30 | 0.978643 | 0.999971 | 0 | 0 | 0.488059 | 0.010537 |
| GTEx_Tissu | GTEX-P44H | 3/1577 | 0.971965 | 0.999971 | 0 | 0 | 0.416115 | 0.011832 |
| GTEx_Tissu | GTEX-QEG5 | 1/808 | 0.972556 | 0.999971 | 0 | 0 | 0.275295 | 0.007661 |
| GTEx_Tissu | GTEX-TMM | Apr-23 | 0.972937 | 0.999971 | 0 | 0 | 0.451892 | 0.012398 |
| GTEx_Tissu | GTEX-SE5C | Jun-55 | 0.973234 | 0.999971 | 0 | 0 | 0.504599 | 0.01369 |
| GTEx_Tissu | GTEX-RNOI | Sep-41 | 0.973686 | 0.999971 | 0 | 0 | 0.554095 | 0.014776 |
| GTEx_Tissu | GTEX-SN8G | May-57 | 0.974105 | 0.999971 | 0 | 0 | 0.478193 | 0.012546 |
| GTEx_Tissu | GTEX-XUZC | 1/751 | 0.964458 | 0.999971 | 0 | 0 | 0.297101 | 0.010752 |
| GTEx_Tissu | GTEX-NFK9 | 3/1480 | 0.960652 | 0.999971 | 0 | 0 | 0.445788 | 0.017895 |
| GTEx_Tissu | GTEX-NPJ8 | 1/924 | 0.983823 | 0.999971 | 0 | 0 | 0.239235 | 0.003902 |
| GTEx_Tissu | GTEX-OXRI | 1/902 | 0.982113 | 0.999971 | 0 | 0 | 0.24536 | 0.004429 |
| GTEx_Tissu | GTEX-WFJC | 2/1332 | 0.982231 | 0.999971 | 0 | 0 | 0.328757 | 0.005894 |
| GTEx_Tissu | GTEX-QDV. | 1/904 | 0.982275 | 0.999971 | 0 | 0 | 0.244791 | 0.004378 |
| GTEx_Tissu | GTEX-REY6 | 2/1337 | 0.982581 | 0.999971 | 0 | 0 | 0.327438 | 0.005754 |
| GTEx_Tissu | GTEX-NPJ7 | Apr-57 | 0.982737 | 0.999971 | 0 | 0 | 0.419251 | 0.007301 |
| GTEx_Tissu | GTEX-X4XY | 3/1716 | 0.982984 | 0.999971 | 0 | 0 | 0.379451 | 0.006512 |
| GTEx_Tissu | GTEX-XQ8I | 2/1345 | 0.983127 | 0.999971 | 0 | 0 | 0.325347 | 0.005536 |
| GTEx_Tissu | GTEX-WFG | 1/952 | 0.985768 | 0.999971 | 0 | 0 | 0.231849 | 0.003323 |
| GTEx_Tissu | GTEX-S341 | 1/922 | 0.983674 | 0.999971 | 0 | 0 | 0.23978 | 0.003947 |
| GTEx_Tissu | GTEX-N7M | Aug-51 | 0.985004 | 0.999971 | 0 | 0 | 0.501937 | 0.007584 |
| GTEx_Tissu | GTEX-T8EM | 1/926 | 0.98397 | 0.999971 | 0 | 0 | 0.238693 | 0.003857 |
| GTEx_Tissu | GTEX-X585 | May-08 | 0.984009 | 0.999971 | 0 | 0 | 0.444312 | 0.007162 |
| GTEx_Tissu | GTEX-X4LF | 3/1734 | 0.984067 | 0.999971 | 0 | 0 | 0.375134 | 0.006025 |
| GTEx_Tissu | GTEX-Q2AI | 1/929 | 0.984189 | 0.999971 | 0 | 0 | 0.237883 | 0.003791 |
| GTEx_Tissu | GTEX-S4UY | 2/1371 | 0.984791 | 0.999971 | 0 | 0 | 0.318721 | 0.004885 |

| | | | | | | | | |
|---|---|---|---|---|---|---|---|---|
| GTEx_Tissu | GTEX-RTLS· | Aug-87 | 0.976011 | 0.999971 | 0 | 0 | 0.533055 | 0.012943 |
| GTEx_Tissu | GTEX-Q2A( | 1/942 | 0.985101 | 0.999971 | 0 | 0 | 0.234436 | 0.003519 |
| GTEx_Tissu | GTEX-WZT( | Aug-54 | 0.985135 | 0.999971 | 0 | 0 | 0.501396 | 0.007509 |
| GTEx_Tissu | GTEX-Q2A( | Jun-61 | 0.980572 | 0.999971 | 0 | 0 | 0.481495 | 0.009447 |
| GTEx_Tissu | GTEX-RM2 | Apr-00 | 0.979061 | 0.999971 | 0 | 0 | 0.4326 | 0.009154 |
| GTEx_Tissu | GTEX-RU72 | 1/868 | 0.979112 | 0.999971 | 0 | 0 | 0.255438 | 0.005392 |
| GTEx_Tissu | GTEX-WQL | 2/1186 | 0.968472 | 0.999971 | 0 | 0 | 0.372198 | 0.011924 |
| GTEx_Tissu | GTEX-U3ZN | 2/1295 | 0.979423 | 0.999971 | 0 | 0 | 0.338838 | 0.007045 |
| GTEx_Tissu | GTEX-X5EB | 3/1664 | 0.979453 | 0.999971 | 0 | 0 | 0.392449 | 0.008148 |
| GTEx_Tissu | GTEX-QCQ( | 1/875 | 0.979768 | 0.999971 | 0 | 0 | 0.253299 | 0.005177 |
| GTEx_Tissu | GTEX-UTH( | Sep-39 | 0.979864 | 0.999971 | 0 | 0 | 0.535509 | 0.010893 |
| GTEx_Tissu | GTEX-RN64 | Jul-53 | 0.979873 | 0.999971 | 0 | 0 | 0.503942 | 0.010246 |
| GTEx_Tissu | GTEX-PLZ6· | 2/1306 | 0.9803 | 0.999971 | 0 | 0 | 0.335781 | 0.006681 |
| GTEx_Tissu | GTEX-R55D | 4/1657 | 0.936947 | 0.999971 | 0 | 0 | 0.532365 | 0.034672 |
| GTEx_Tissu | GTEX-T6MI | Dec-23 | 0.970008 | 0.999971 | 0 | 0 | 0.596609 | 0.018167 |
| GTEx_Tissu | GTEX-QEG5 | 4/1864 | 0.96717 | 0.999971 | 0 | 0 | 0.467755 | 0.015614 |
| GTEx_Tissu | GTEX-S4P3 | 1/772 | 0.967684 | 0.999971 | 0 | 0 | 0.288692 | 0.009483 |
| GTEx_Tissu | GTEX-UJM( | Jul-89 | 0.967742 | 0.999971 | 0 | 0 | 0.538808 | 0.017667 |
| GTEx_Tissu | GTEX-RU72 | Jul-90 | 0.967833 | 0.999971 | 0 | 0 | 0.538582 | 0.01761 |
| GTEx_Tissu | GTEX-QXCU | Jun-97 | 0.968233 | 0.999971 | 0 | 0 | 0.518072 | 0.016725 |
| GTEx_Tissu | GTEX-S4Q7 | 1/777 | 0.968409 | 0.999971 | 0 | 0 | 0.286757 | 0.009205 |
| GTEx_Tissu | GTEX-X585 | May-93 | 0.968418 | 0.999971 | 0 | 0 | 0.493964 | 0.015852 |
| GTEx_Tissu | GTEX-VUS( | 2/863 | 0.894423 | 0.999971 | 0 | 0 | 0.520653 | 0.058093 |
| GTEx_Tissu | GTEX-WH7 | Jul-14 | 0.889398 | 0.999971 | 0 | 0 | 0.667761 | 0.078269 |
| GTEx_Tissu | GTEX-Q734 | Jul-28 | 0.893013 | 0.999971 | 0 | 0 | 0.663206 | 0.075044 |
| GTEx_Tissu | GTEX-OIZI- | 1/499 | 0.889526 | 0.999971 | 0 | 0 | 0.453325 | 0.053069 |
| GTEx_Tissu | GTEX-WHP | 3/1178 | 0.893222 | 0.999971 | 0 | 0 | 0.569544 | 0.064313 |
| GTEx_Tissu | GTEX-T5JC· | Jun-52 | 0.893238 | 0.999971 | 0 | 0 | 0.646863 | 0.073032 |
| GTEx_Tissu | GTEX-PWO | 1/507 | 0.893409 | 0.999971 | 0 | 0 | 0.445974 | 0.050266 |
| GTEx_Tissu | GTEX-WFG | 1/507 | 0.893409 | 0.999971 | 0 | 0 | 0.445974 | 0.050266 |
| GTEx_Tissu | GTEX-WCD | 1/501 | 0.89051 | 0.999971 | 0 | 0 | 0.451465 | 0.052353 |
| GTEx_Tissu | GTEX-U4B1 | 1/501 | 0.89051 | 0.999971 | 0 | 0 | 0.451465 | 0.052353 |
| GTEx_Tissu | GTEX-S3XE | 2/850 | 0.889389 | 0.999971 | 0 | 0 | 0.528996 | 0.062009 |
| GTEx_Tissu | GTEX-U3ZN | 1/502 | 0.890998 | 0.999971 | 0 | 0 | 0.450541 | 0.051998 |
| GTEx_Tissu | GTEX-X4XX | Jul-35 | 0.894785 | 0.999971 | 0 | 0 | 0.660948 | 0.073479 |
| GTEx_Tissu | GTEX-RU72 | 2/864 | 0.894802 | 0.999971 | 0 | 0 | 0.520022 | 0.057802 |
| GTEx_Tissu | GTEX-RUS( | 1/511 | 0.895299 | 0.999971 | 0 | 0 | 0.442385 | 0.048927 |
| GTEx_Tissu | GTEX-X3Y1 | 2/866 | 0.895555 | 0.999971 | 0 | 0 | 0.518764 | 0.057226 |
| GTEx_Tissu | GTEX-XV7C | Jun-50 | 0.892702 | 0.999971 | 0 | 0 | 0.647568 | 0.0735 |
| GTEx_Tissu | GTEX-Q2AI | Oct-33 | 0.89282 | 0.999971 | 0 | 0 | 0.698213 | 0.079157 |
| GTEx_Tissu | GTEX-RVPL | 3/1187 | 0.896207 | 0.999971 | 0 | 0 | 0.564943 | 0.061909 |
| GTEx_Tissu | GTEX-S4P3 | 3/1132 | 0.876756 | 0.999971 | 0 | 0 | 0.594205 | 0.078154 |
| GTEx_Tissu | GTEX-P4PP | 4/1466 | 0.889182 | 0.999971 | 0 | 0 | 0.608211 | 0.071437 |
| GTEx_Tissu | GTEX-Q734 | 4/1466 | 0.889182 | 0.999971 | 0 | 0 | 0.608211 | 0.071437 |
| GTEx_Tissu | GTEX-S7SE· | Sep-43 | 0.886846 | 0.999971 | 0 | 0 | 0.695361 | 0.083502 |

| | | | | | | | | |
|---|---|---|---|---|---|---|---|---|
| GTEx_Tissu | GTEX-PVO\ | Jul-05 | 0.887021 | 0.999971 | 0 | 0 | 0.670719 | 0.08041 |
| GTEx_Tissu | GTEX-POY\ | Dec-40 | 0.889485 | 0.999971 | 0 | 0 | 0.718192 | 0.084109 |
| GTEx_Tissu | GTEX-S341 | 4/1462 | 0.887914 | 0.999971 | 0 | 0 | 0.610012 | 0.072519 |
| GTEx_Tissu | GTEX-RN6 | 4/1426 | 0.875932 | 0.999971 | 0 | 0 | 0.626675 | 0.083014 |
| GTEx_Tissu | GTEX-PVO\ | Jul-65 | 0.875948 | 0.999971 | 0 | 0 | 0.684151 | 0.090615 |
| GTEx_Tissu | GTEX-U8T8 | Sep-02 | 0.876402 | 0.999971 | 0 | 0 | 0.707263 | 0.09331 |
| GTEx_Tissu | GTEX-U4B1 | Jul-28 | 0.893013 | 0.999971 | 0 | 0 | 0.663206 | 0.075044 |
| GTEx_Tissu | GTEX-X585 | Oct-16 | 0.888893 | 0.999971 | 0 | 0 | 0.702745 | 0.082769 |
| GTEx_Tissu | GTEX-XOT4 | 2/849 | 0.888993 | 0.999971 | 0 | 0 | 0.529648 | 0.062322 |
| GTEx_Tissu | GTEX-XMK | Jun-98 | 0.877965 | 0.999971 | 0 | 0 | 0.666406 | 0.086732 |
| GTEx_Tissu | GTEX-T2IS- | Jul-13 | 0.889136 | 0.999971 | 0 | 0 | 0.668089 | 0.078504 |
| GTEx_Tissu | GTEX-PWN | Jun-00 | 0.878561 | 0.999971 | 0 | 0 | 0.665664 | 0.086184 |
| GTEx_Tissu | GTEX-R55G | 3/1137 | 0.878647 | 0.999971 | 0 | 0 | 0.591427 | 0.076514 |
| GTEx_Tissu | GTEX-XBEV | 5/1721 | 0.878752 | 0.999971 | 0 | 0 | 0.646604 | 0.083575 |
| GTEx_Tissu | GTEX-SUCS | 1/479 | 0.879198 | 0.999971 | 0 | 0 | 0.472779 | 0.060868 |
| GTEx_Tissu | GTEX-QCQ | Jul-54 | 0.899471 | 0.999971 | 0 | 0 | 0.654889 | 0.069385 |
| GTEx_Tissu | GTEX-P44H | Aug-35 | 0.901227 | 0.999971 | 0 | 0 | 0.666342 | 0.069298 |
| GTEx_Tissu | GTEX-QVJC | Jun-69 | 0.897705 | 0.999971 | 0 | 0 | 0.640922 | 0.069164 |
| GTEx_Tissu | GTEX-X88G | 2/872 | 0.897784 | 0.999971 | 0 | 0 | 0.515024 | 0.055533 |
| GTEx_Tissu | GTEX-V1D1 | Jul-63 | 0.901629 | 0.999971 | 0 | 0 | 0.652053 | 0.067521 |
| GTEx_Tissu | GTEX-REY6 | 1/519 | 0.898981 | 0.999971 | 0 | 0 | 0.435373 | 0.046365 |
| GTEx_Tissu | GTEX-TSE9 | Oct-75 | 0.902047 | 0.999971 | 0 | 0 | 0.687224 | 0.070845 |
| GTEx_Tissu | GTEX-SE5C | 2/887 | 0.903169 | 0.999971 | 0 | 0 | 0.505896 | 0.051523 |
| GTEx_Tissu | GTEX-P4PC | Jul-70 | 0.903282 | 0.999971 | 0 | 0 | 0.649862 | 0.066105 |
| GTEx_Tissu | GTEX-OXR( | Jul-70 | 0.903282 | 0.999971 | 0 | 0 | 0.649862 | 0.066105 |
| GTEx_Tissu | GTEX-RVPL | 1/524 | 0.901217 | 0.999971 | 0 | 0 | 0.4311 | 0.044839 |
| GTEx_Tissu | GTEX-U8T8 | 4/1500 | 0.899472 | 0.999971 | 0 | 0 | 0.593293 | 0.062858 |
| GTEx_Tissu | GTEX-Q2A( | 4/1516 | 0.904022 | 0.999971 | 0 | 0 | 0.586505 | 0.059179 |
| GTEx_Tissu | GTEX-S32V | 2/878 | 0.89997 | 0.999971 | 0 | 0 | 0.511335 | 0.053891 |
| GTEx_Tissu | GTEX-T5JC- | Aug-87 | 0.889745 | 0.999971 | 0 | 0 | 0.680629 | 0.079511 |
| GTEx_Tissu | GTEX-WYV | 5/1758 | 0.889979 | 0.999971 | 0 | 0 | 0.63167 | 0.073626 |
| GTEx_Tissu | GTEX-X261 | 2/852 | 0.890177 | 0.999971 | 0 | 0 | 0.527696 | 0.061389 |
| GTEx_Tissu | GTEX-Q2A( | Jul-17 | 0.890181 | 0.999971 | 0 | 0 | 0.66678 | 0.077567 |
| GTEx_Tissu | GTEX-QMR | 3/1203 | 0.901332 | 0.999971 | 0 | 0 | 0.556935 | 0.057856 |
| GTEx_Tissu | GTEX-S95S | 1/492 | 0.886014 | 0.999971 | 0 | 0 | 0.459954 | 0.055665 |
| GTEx_Tissu | GTEX-P44H | Jul-95 | 0.884331 | 0.999971 | 0 | 0 | 0.674033 | 0.082855 |
| GTEx_Tissu | GTEX-S341 | Jun-20 | 0.884391 | 0.999971 | 0 | 0 | 0.658318 | 0.080878 |
| GTEx_Tissu | GTEX-N7M | 3/1153 | 0.88453 | 0.999971 | 0 | 0 | 0.582702 | 0.071497 |
| GTEx_Tissu | GTEX-XXEK | 3/1154 | 0.88489 | 0.999971 | 0 | 0 | 0.582165 | 0.071194 |
| GTEx_Tissu | GTEX-XUW | 5/1770 | 0.893427 | 0.999971 | 0 | 0 | 0.626961 | 0.070652 |
| GTEx_Tissu | GTEX-R55G | 1/508 | 0.893884 | 0.999971 | 0 | 0 | 0.445071 | 0.049928 |
| GTEx_Tissu | GTEX-WEY! | 3/1157 | 0.885961 | 0.999971 | 0 | 0 | 0.580558 | 0.070295 |
| GTEx_Tissu | GTEX-R55C | 2/863 | 0.894423 | 0.999971 | 0 | 0 | 0.520653 | 0.058093 |
| GTEx_Tissu | GTEX-WYB: | 4/1456 | 0.885988 | 0.999971 | 0 | 0 | 0.612732 | 0.074172 |
| GTEx_Tissu | GTEX-WYJK | 4/1466 | 0.889182 | 0.999971 | 0 | 0 | 0.608211 | 0.071437 |

| | | | | | | | | |
|---|---|---|---|---|---|---|---|---|
| GTEx_Tissu | GTEX-Q2A( | 4/1457 | 0.886311 | 0.999971 | 0 | 0 | 0.612277 | 0.073894 |
| GTEx_Tissu | GTEX-OHPI | 3/1159 | 0.886671 | 0.999971 | 0 | 0 | 0.579492 | 0.069702 |
| GTEx_Tissu | GTEX-R53T | Sep-81 | 0.895883 | 0.999971 | 0 | 0 | 0.684634 | 0.075272 |
| GTEx_Tissu | GTEX-QMR | Jun-62 | 0.895885 | 0.999971 | 0 | 0 | 0.643356 | 0.070733 |
| GTEx_Tissu | GTEX-WFO | 1/521 | 0.899881 | 0.999971 | 0 | 0 | 0.433654 | 0.045747 |
| GTEx_Tissu | GTEX-VUS( | 5/1780 | 0.896231 | 0.999971 | 0 | 0 | 0.623085 | 0.068264 |
| GTEx_Tissu | GTEX-X8H( | 3/1202 | 0.901018 | 0.999971 | 0 | 0 | 0.557429 | 0.058101 |
| GTEx_Tissu | GTEX-XQ3S | 4/1506 | 0.9012 | 0.999971 | 0 | 0 | 0.59073 | 0.061453 |
| GTEx_Tissu | GTEX-T6M( | Aug-77 | 0.859349 | 0.999971 | 0 | 0 | 0.715464 | 0.108451 |
| GTEx_Tissu | GTEX-U3ZH | 5/1656 | 0.856728 | 0.999971 | 0 | 0 | 0.674462 | 0.104295 |
| GTEx_Tissu | GTEX-XAJ8 | 3/1083 | 0.856824 | 0.999971 | 0 | 0 | 0.622784 | 0.096235 |
| GTEx_Tissu | GTEX-T8EN | 2/779 | 0.857821 | 0.999971 | 0 | 0 | 0.579484 | 0.088869 |
| GTEx_Tissu | GTEX-RU72 | 4/1413 | 0.871346 | 0.999971 | 0 | 0 | 0.632902 | 0.087161 |
| GTEx_Tissu | GTEX-X585 | Dec-63 | 0.87145 | 0.999971 | 0 | 0 | 0.737235 | 0.101441 |
| GTEx_Tissu | GTEX-Q734 | 3/1086 | 0.858119 | 0.999971 | 0 | 0 | 0.62096 | 0.095014 |
| GTEx_Tissu | GTEX-OXRI | 5/1699 | 0.871635 | 0.999971 | 0 | 0 | 0.655794 | 0.090096 |
| GTEx_Tissu | GTEX-PLZ6 | Jul-07 | 0.858358 | 0.999971 | 0 | 0 | 0.704494 | 0.107601 |
| GTEx_Tissu | GTEX-W5W | 4/1380 | 0.859059 | 0.999971 | 0 | 0 | 0.649236 | 0.09863 |
| GTEx_Tissu | GTEX-PLZ6 | 3/1082 | 0.85639 | 0.999971 | 0 | 0 | 0.623395 | 0.096645 |
| GTEx_Tissu | GTEX-WRH | 2/783 | 0.859796 | 0.999971 | 0 | 0 | 0.576395 | 0.08707 |
| GTEx_Tissu | GTEX-T2IS- | Sep-92 | 0.873742 | 0.999971 | 0 | 0 | 0.710219 | 0.095858 |
| GTEx_Tissu | GTEX-P4QI | 2/813 | 0.873836 | 0.999971 | 0 | 0 | 0.554203 | 0.074741 |
| GTEx_Tissu | GTEX-T6M( | 2/827 | 0.879941 | 0.999971 | 0 | 0 | 0.544399 | 0.069629 |
| GTEx_Tissu | GTEX-S3XE | Jul-62 | 0.875084 | 0.999971 | 0 | 0 | 0.685177 | 0.091427 |
| GTEx_Tissu | GTEX-QMR | Jul-80 | 0.880199 | 0.999971 | 0 | 0 | 0.679059 | 0.086653 |
| GTEx_Tissu | GTEX-R55D | 5/1726 | 0.880323 | 0.999971 | 0 | 0 | 0.644549 | 0.082158 |
| GTEx_Tissu | GTEX-Q2AI | 2/828 | 0.880367 | 0.999971 | 0 | 0 | 0.543712 | 0.069278 |
| GTEx_Tissu | GTEX-UPIC | 3/1105 | 0.866093 | 0.999971 | 0 | 0 | 0.609638 | 0.087643 |
| GTEx_Tissu | GTEX-T2IS- | Dec-50 | 0.868193 | 0.999971 | 0 | 0 | 0.740531 | 0.104668 |
| GTEx_Tissu | GTEX-S7SE | 4/1394 | 0.864387 | 0.999971 | 0 | 0 | 0.642212 | 0.093593 |
| GTEx_Tissu | GTEX-XUJ4 | 2/805 | 0.870222 | 0.999971 | 0 | 0 | 0.559959 | 0.077838 |
| GTEx_Tissu | GTEX-XPV( | 2/806 | 0.870679 | 0.999971 | 0 | 0 | 0.559233 | 0.077444 |
| GTEx_Tissu | GTEX-T5JC- | Sep-81 | 0.870765 | 0.999971 | 0 | 0 | 0.713495 | 0.098736 |
| GTEx_Tissu | GTEX-QDVI | 1/464 | 0.87083 | 0.999971 | 0 | 0 | 0.488473 | 0.06756 |
| GTEx_Tissu | GTEX-WYB: | 5/1681 | 0.865558 | 0.999971 | 0 | 0 | 0.663492 | 0.095795 |
| GTEx_Tissu | GTEX-QDT8 | 3/1104 | 0.865683 | 0.999971 | 0 | 0 | 0.610224 | 0.088016 |
| GTEx_Tissu | GTEX-XV7( | 2/808 | 0.871589 | 0.999971 | 0 | 0 | 0.557787 | 0.076661 |
| GTEx_Tissu | GTEX-TSE9 | Jul-81 | 0.880478 | 0.999971 | 0 | 0 | 0.678721 | 0.086394 |
| GTEx_Tissu | GTEX-XUYS | 2/810 | 0.872492 | 0.999971 | 0 | 0 | 0.556348 | 0.075887 |
| GTEx_Tissu | GTEX-O5Y\ | 2/810 | 0.872492 | 0.999971 | 0 | 0 | 0.556348 | 0.075887 |
| GTEx_Tissu | GTEX-VUS( | Aug-22 | 0.872498 | 0.999971 | 0 | 0 | 0.700845 | 0.095592 |
| GTEx_Tissu | GTEX-OXRI | 5/1704 | 0.873282 | 0.999971 | 0 | 0 | 0.653684 | 0.088572 |
| GTEx_Tissu | GTEX-PWC | Jun-64 | 0.86746 | 0.999971 | 0 | 0 | 0.679265 | 0.096582 |
| GTEx_Tissu | GTEX-R55E | 5/1687 | 0.86761 | 0.999971 | 0 | 0 | 0.660907 | 0.093858 |
| GTEx_Tissu | GTEX-T5JW | Oct-58 | 0.874627 | 0.999971 | 0 | 0 | 0.718589 | 0.09626 |

| | | | | | | | | |
|---|---|---|---|---|---|---|---|---|
| GTEx_Tissu | GTEX-U8XE | 4/1406 | 0.868818 | 0.999971 | 0 | 0 | 0.636303 | 0.089478 |
| GTEx_Tissu | GTEX-PWN | 4/1462 | 0.887914 | 0.999971 | 0 | 0 | 0.610012 | 0.072519 |
| GTEx_Tissu | GTEX-PVO\ | Nov-44 | 0.880975 | 0.999971 | 0 | 0 | 0.719993 | 0.091242 |
| GTEx_Tissu | GTEX-WFG | Jun-61 | 0.866499 | 0.999971 | 0 | 0 | 0.680421 | 0.0975 |
| GTEx_Tissu | GTEX-WFG | 1/457 | 0.866732 | 0.999971 | 0 | 0 | 0.49615 | 0.070962 |
| GTEx_Tissu | GTEX-SE5C | 2/798 | 0.866984 | 0.999971 | 0 | 0 | 0.56509 | 0.080658 |
| GTEx_Tissu | GTEX-W5W | 4/1401 | 0.866987 | 0.999971 | 0 | 0 | 0.638753 | 0.09117 |
| GTEx_Tissu | GTEX-XUW | 3/1150 | 0.883447 | 0.999971 | 0 | 0 | 0.584319 | 0.072411 |
| GTEx_Tissu | GTEX-Q2A( | Jul-92 | 0.883514 | 0.999971 | 0 | 0 | 0.675033 | 0.083601 |
| GTEx_Tissu | GTEX-S4Q7 | 5/1749 | 0.887332 | 0.999971 | 0 | 0 | 0.635244 | 0.075935 |
| GTEx_Tissu | GTEX-S7SF- | 3/1152 | 0.88417 | 0.999971 | 0 | 0 | 0.58324 | 0.0718 |
| GTEx_Tissu | GTEX-P4QS | 2/796 | 0.866045 | 0.999971 | 0 | 0 | 0.566573 | 0.081484 |
| GTEx_Tissu | GTEX-X4XX | 4/1463 | 0.888232 | 0.999971 | 0 | 0 | 0.609561 | 0.072247 |
| GTEx_Tissu | GTEX-XLM4 | Jun-34 | 0.888334 | 0.999971 | 0 | 0 | 0.653262 | 0.077351 |
| GTEx_Tissu | GTEX-RNOI | 3/1164 | 0.888428 | 0.999971 | 0 | 0 | 0.576843 | 0.068241 |
| GTEx_Tissu | GTEX-QXCl | 2/839 | 0.884959 | 0.999971 | 0 | 0 | 0.536257 | 0.065538 |
| GTEx_Tissu | GTEX-PWC | Aug-72 | 0.885942 | 0.999971 | 0 | 0 | 0.685203 | 0.082981 |
| GTEx_Tissu | GTEX-RU72 | 1/498 | 0.889031 | 0.999971 | 0 | 0 | 0.45426 | 0.053432 |
| GTEx_Tissu | GTEX-QXCl | 4/1456 | 0.885988 | 0.999971 | 0 | 0 | 0.612732 | 0.074172 |
| GTEx_Tissu | GTEX-U4B1 | Jun-37 | 0.889164 | 0.999971 | 0 | 0 | 0.652187 | 0.076615 |
| GTEx_Tissu | GTEX-WHS | Jul-79 | 0.879919 | 0.999971 | 0 | 0 | 0.679396 | 0.086912 |
| GTEx_Tissu | GTEX-QDT8 | Jul-69 | 0.877094 | 0.999971 | 0 | 0 | 0.682786 | 0.089542 |
| GTEx_Tissu | GTEX-XPT6 | 3/1133 | 0.877136 | 0.999971 | 0 | 0 | 0.593647 | 0.077823 |
| GTEx_Tissu | GTEX-QDT8 | 15/4366 | 0.880881 | 0.999971 | 0 | 0 | 0.745135 | 0.094507 |
| GTEx_Tissu | GTEX-QDVI | 4/1433 | 0.878343 | 0.999971 | 0 | 0 | 0.62337 | 0.080862 |
| GTEx_Tissu | GTEX-RWS( | 2/831 | 0.881635 | 0.999971 | 0 | 0 | 0.541659 | 0.068236 |
| GTEx_Tissu | GTEX-SNOS | Sep-25 | 0.882351 | 0.999971 | 0 | 0 | 0.700544 | 0.087684 |
| GTEx_Tissu | GTEX-WWY | 2/833 | 0.882474 | 0.999971 | 0 | 0 | 0.540299 | 0.067551 |
| GTEx_Tissu | GTEX-TSE9 | Nov-54 | 0.883357 | 0.999971 | 0 | 0 | 0.717406 | 0.088977 |
| GTEx_Tissu | GTEX-S4P3 | 2/826 | 0.879514 | 0.999971 | 0 | 0 | 0.545089 | 0.069982 |
| GTEx_Tissu | GTEX-U3ZN | Jun-84 | 0.901515 | 0.999971 | 0 | 0 | 0.635761 | 0.065915 |
| GTEx_Tissu | GTEX-WFG | Jun-18 | 0.883819 | 0.999971 | 0 | 0 | 0.659046 | 0.081394 |
| GTEx_Tissu | GTEX-RU72 | Sep-16 | 0.880051 | 0.999971 | 0 | 0 | 0.70316 | 0.089847 |
| GTEx_Tissu | GTEX-Q2AI | 5/1679 | 0.864869 | 0.999971 | 0 | 0 | 0.664357 | 0.09645 |
| GTEx_Tissu | GTEX-O5Y\ | 1/454 | 0.864936 | 0.999971 | 0 | 0 | 0.499512 | 0.072479 |
| GTEx_Tissu | GTEX-RTLS- | Sep-62 | 0.865493 | 0.999971 | 0 | 0 | 0.719215 | 0.103895 |
| GTEx_Tissu | GTEX-S3XE | 1/455 | 0.865537 | 0.999971 | 0 | 0 | 0.498386 | 0.071969 |
| GTEx_Tissu | GTEX-X62C | 5/1727 | 0.880635 | 0.999971 | 0 | 0 | 0.644139 | 0.081878 |
| GTEx_Tissu | GTEX-WZT( | Nov-43 | 0.880734 | 0.999971 | 0 | 0 | 0.720253 | 0.091472 |
| GTEx_Tissu | GTEX-PX3G | 1/600 | 0.929752 | 0.999971 | 0 | 0 | 0.374927 | 0.027309 |
| GTEx_Tissu | GTEX-X5EB | 1/583 | 0.924177 | 0.999971 | 0 | 0 | 0.386218 | 0.030454 |
| GTEx_Tissu | GTEX-P44H | 5/1848 | 0.913682 | 0.999971 | 0 | 0 | 0.597846 | 0.053969 |
| GTEx_Tissu | GTEX-OXRI | 4/1596 | 0.924183 | 0.999971 | 0 | 0 | 0.55461 | 0.043728 |
| GTEx_Tissu | GTEX-XXEK | 3/1246 | 0.914008 | 0.999971 | 0 | 0 | 0.536433 | 0.048234 |
| GTEx_Tissu | GTEX-RU1J | Jun-37 | 0.914037 | 0.999971 | 0 | 0 | 0.618107 | 0.055558 |

| | | | | | | | | |
|---|---|---|---|---|---|---|---|---|
| GTEx_Tissu | GTEX-QV31 | 2/920 | 0.914107 | 0.999971 | 0 | 0 | 0.486864 | 0.043724 |
| GTEx_Tissu | GTEX-Q734 | 2/921 | 0.91442 | 0.999971 | 0 | 0 | 0.486309 | 0.043508 |
| GTEx_Tissu | GTEX-QMR | 5/1898 | 0.924847 | 0.999971 | 0 | 0 | 0.580444 | 0.045349 |
| GTEx_Tissu | GTEX-XBED | 2/958 | 0.925281 | 0.999971 | 0 | 0 | 0.466576 | 0.036233 |
| GTEx_Tissu | GTEX-XV7C | 4/1595 | 0.923956 | 0.999971 | 0 | 0 | 0.554989 | 0.043895 |
| GTEx_Tissu | GTEX-TKQ2 | 3/1290 | 0.925451 | 0.999971 | 0 | 0 | 0.516872 | 0.040044 |
| GTEx_Tissu | GTEX-Q734 | May-25 | 0.930337 | 0.999971 | 0 | 0 | 0.571424 | 0.041262 |
| GTEx_Tissu | GTEX-R45C | May-25 | 0.930337 | 0.999971 | 0 | 0 | 0.571424 | 0.041262 |
| GTEx_Tissu | GTEX-UPIC | 3/1313 | 0.93087 | 0.999971 | 0 | 0 | 0.50717 | 0.036332 |
| GTEx_Tissu | GTEX-NPJ8 | Jul-07 | 0.931238 | 0.999971 | 0 | 0 | 0.609455 | 0.043418 |
| GTEx_Tissu | GTEX-X5EB | May-08 | 0.926922 | 0.999971 | 0 | 0 | 0.577073 | 0.043791 |
| GTEx_Tissu | GTEX-XUW | 1/592 | 0.927181 | 0.999971 | 0 | 0 | 0.38016 | 0.028742 |
| GTEx_Tissu | GTEX-WI4N | 3/1319 | 0.932223 | 0.999971 | 0 | 0 | 0.504695 | 0.035421 |
| GTEx_Tissu | GTEX-S4Q7 | 2/956 | 0.924728 | 0.999971 | 0 | 0 | 0.467604 | 0.036593 |
| GTEx_Tissu | GTEX-PW2 | 1/582 | 0.923836 | 0.999971 | 0 | 0 | 0.386903 | 0.030651 |
| GTEx_Tissu | GTEX-QLQ7 | 1/582 | 0.923836 | 0.999971 | 0 | 0 | 0.386903 | 0.030651 |
| GTEx_Tissu | GTEX-XBED | 3/1272 | 0.920947 | 0.999971 | 0 | 0 | 0.52471 | 0.043211 |
| GTEx_Tissu | GTEX-WYJK | 2/943 | 0.921042 | 0.999971 | 0 | 0 | 0.474389 | 0.039018 |
| GTEx_Tissu | GTEX-XAJ8 | 4/1596 | 0.924183 | 0.999971 | 0 | 0 | 0.55461 | 0.043728 |
| GTEx_Tissu | GTEX-OXRL | 4/1585 | 0.921653 | 0.999971 | 0 | 0 | 0.558804 | 0.045591 |
| GTEx_Tissu | GTEX-WYJK | 5/1895 | 0.924214 | 0.999971 | 0 | 0 | 0.581462 | 0.045826 |
| GTEx_Tissu | GTEX-X585 | 3/1287 | 0.924717 | 0.999971 | 0 | 0 | 0.518163 | 0.040555 |
| GTEx_Tissu | GTEX-SJXC | 2/956 | 0.924728 | 0.999971 | 0 | 0 | 0.467604 | 0.036593 |
| GTEx_Tissu | GTEX-N7M | 4/1612 | 0.927735 | 0.999971 | 0 | 0 | 0.548612 | 0.041151 |
| GTEx_Tissu | GTEX-XUJ4 | 2/949 | 0.922764 | 0.999971 | 0 | 0 | 0.471234 | 0.037879 |
| GTEx_Tissu | GTEX-OIZH | 4/1590 | 0.922812 | 0.999971 | 0 | 0 | 0.556891 | 0.044735 |
| GTEx_Tissu | GTEX-X5EB | 3/1290 | 0.925451 | 0.999971 | 0 | 0 | 0.516872 | 0.040044 |
| GTEx_Tissu | GTEX-QVJC | Jun-80 | 0.923177 | 0.999971 | 0 | 0 | 0.604416 | 0.048313 |
| GTEx_Tissu | GTEX-U4B1 | 2/916 | 0.912845 | 0.999971 | 0 | 0 | 0.489098 | 0.0446 |
| GTEx_Tissu | GTEX-V955 | 1/552 | 0.912862 | 0.999971 | 0 | 0 | 0.408602 | 0.037252 |
| GTEx_Tissu | GTEX-WFJC | 3/1243 | 0.913173 | 0.999971 | 0 | 0 | 0.537817 | 0.04885 |
| GTEx_Tissu | GTEX-UPK5 | Sep-62 | 0.9132 | 0.999971 | 0 | 0 | 0.66269 | 0.060173 |
| GTEx_Tissu | GTEX-O5YV | 1/616 | 0.934629 | 0.999971 | 0 | 0 | 0.36487 | 0.024667 |
| GTEx_Tissu | GTEX-P4PC | 2/948 | 0.922479 | 0.999971 | 0 | 0 | 0.471757 | 0.038066 |
| GTEx_Tissu | GTEX-S4UY | 2/988 | 0.93313 | 0.999971 | 0 | 0 | 0.451664 | 0.03126 |
| GTEx_Tissu | GTEX-Q2AI | 1/611 | 0.933142 | 0.999971 | 0 | 0 | 0.367957 | 0.025462 |
| GTEx_Tissu | GTEX-XV7C | 2/1005 | 0.937229 | 0.999971 | 0 | 0 | 0.44361 | 0.028758 |
| GTEx_Tissu | GTEX-OIZH | 2/990 | 0.933625 | 0.999971 | 0 | 0 | 0.450703 | 0.030954 |
| GTEx_Tissu | GTEX-XV7C | 1/626 | 0.937505 | 0.999971 | 0 | 0 | 0.358847 | 0.023158 |
| GTEx_Tissu | GTEX-XMD | 4/1661 | 0.937713 | 0.999971 | 0 | 0 | 0.530964 | 0.034147 |
| GTEx_Tissu | GTEX-X4EP | 3/1346 | 0.938022 | 0.999971 | 0 | 0 | 0.49383 | 0.031596 |
| GTEx_Tissu | GTEX-RVPL | 3/1347 | 0.938228 | 0.999971 | 0 | 0 | 0.493436 | 0.031463 |
| GTEx_Tissu | GTEX-WH7 | 4/1588 | 0.92235 | 0.999971 | 0 | 0 | 0.557655 | 0.045075 |
| GTEx_Tissu | GTEX-Q2AI | 1/616 | 0.934629 | 0.999971 | 0 | 0 | 0.36487 | 0.024667 |
| GTEx_Tissu | GTEX-XUW | May-69 | 0.938534 | 0.999971 | 0 | 0 | 0.557256 | 0.03535 |

| | | | | | | | | |
|---|---|---|---|---|---|---|---|---|
| GTEx_Tissu | GTEX-R45C | 3/1332 | 0.935074 | 0.999971 | 0 | 0 | 0.499409 | 0.033525 |
| GTEx_Tissu | GTEX-T5JC- | Jun-64 | 0.938647 | 0.999971 | 0 | 0 | 0.579175 | 0.036671 |
| GTEx_Tissu | GTEX-V955 | 2/1013 | 0.939076 | 0.999971 | 0 | 0 | 0.439914 | 0.027653 |
| GTEx_Tissu | GTEX-X5EB | 3/1353 | 0.93945 | 0.999971 | 0 | 0 | 0.491085 | 0.030674 |
| GTEx_Tissu | GTEX-XOTC | Jun-69 | 0.939476 | 0.999971 | 0 | 0 | 0.577732 | 0.03607 |
| GTEx_Tissu | GTEX-R53T | 1/624 | 0.93694 | 0.999971 | 0 | 0 | 0.360036 | 0.023451 |
| GTEx_Tissu | GTEX-XPVG | Jul-48 | 0.920179 | 0.999971 | 0 | 0 | 0.626301 | 0.0521 |
| GTEx_Tissu | GTEX-QDT8 | 4/1635 | 0.932582 | 0.999971 | 0 | 0 | 0.540196 | 0.037704 |
| GTEx_Tissu | GTEX-P4QT | 3/1321 | 0.932669 | 0.999971 | 0 | 0 | 0.503875 | 0.035122 |
| GTEx_Tissu | GTEX-U3ZN | May-38 | 0.932853 | 0.999971 | 0 | 0 | 0.567171 | 0.039423 |
| GTEx_Tissu | GTEX-X62C | 2/987 | 0.932881 | 0.999971 | 0 | 0 | 0.452147 | 0.031414 |
| GTEx_Tissu | GTEX-XQ3S | 4/1621 | 0.929667 | 0.999971 | 0 | 0 | 0.545291 | 0.039767 |
| GTEx_Tissu | GTEX-T8EN | 1/600 | 0.929752 | 0.999971 | 0 | 0 | 0.374927 | 0.027309 |
| GTEx_Tissu | GTEX-OOBJ | Jul-19 | 0.93332 | 0.999971 | 0 | 0 | 0.606126 | 0.041827 |
| GTEx_Tissu | GTEX-X638 | 3/1310 | 0.930184 | 0.999971 | 0 | 0 | 0.508416 | 0.036796 |
| GTEx_Tissu | GTEX-S32W | 1/571 | 0.919981 | 0.999971 | 0 | 0 | 0.394594 | 0.03291 |
| GTEx_Tissu | GTEX-RVPV | 5/1892 | 0.923577 | 0.999971 | 0 | 0 | 0.582483 | 0.046308 |
| GTEx_Tissu | GTEX-TKQ2 | Jul-49 | 0.920378 | 0.999971 | 0 | 0 | 0.626008 | 0.05194 |
| GTEx_Tissu | GTEX-QLQV | Aug-27 | 0.92049 | 0.999971 | 0 | 0 | 0.640369 | 0.053054 |
| GTEx_Tissu | GTEX-W5X | May-33 | 0.931895 | 0.999971 | 0 | 0 | 0.5688 | 0.04012 |
| GTEx_Tissu | GTEX-QDVI | 1/607 | 0.931929 | 0.999971 | 0 | 0 | 0.370462 | 0.026117 |
| GTEx_Tissu | GTEX-P4QT | 1/574 | 0.921051 | 0.999971 | 0 | 0 | 0.392467 | 0.032276 |
| GTEx_Tissu | GTEX-OOBJ | 1/608 | 0.932234 | 0.999971 | 0 | 0 | 0.369833 | 0.025952 |
| GTEx_Tissu | GTEX-U3ZH | Jun-74 | 0.921954 | 0.999971 | 0 | 0 | 0.606294 | 0.049267 |
| GTEx_Tissu | GTEX-WWY | Oct-78 | 0.921972 | 0.999971 | 0 | 0 | 0.661471 | 0.053738 |
| GTEx_Tissu | GTEX-QVUS | 4/1493 | 0.897424 | 0.999971 | 0 | 0 | 0.596309 | 0.064537 |
| GTEx_Tissu | GTEX-P4QS | 2/911 | 0.911243 | 0.999971 | 0 | 0 | 0.491917 | 0.045722 |
| GTEx_Tissu | GTEX-OIZI- | Sep-53 | 0.911401 | 0.999971 | 0 | 0 | 0.665068 | 0.0617 |
| GTEx_Tissu | GTEX-N7M | Aug-82 | 0.911501 | 0.999971 | 0 | 0 | 0.65285 | 0.060495 |
| GTEx_Tissu | GTEX-R55C | 1/537 | 0.906803 | 0.999971 | 0 | 0 | 0.420362 | 0.041124 |
| GTEx_Tissu | GTEX-T6M | 4/1527 | 0.907046 | 0.999971 | 0 | 0 | 0.581921 | 0.056774 |
| GTEx_Tissu | GTEX-RNOI | Jun-27 | 0.911783 | 0.999971 | 0 | 0 | 0.62137 | 0.057385 |
| GTEx_Tissu | GTEX-SNOS | 3/1222 | 0.907125 | 0.999971 | 0 | 0 | 0.547697 | 0.053387 |
| GTEx_Tissu | GTEX-WEYS | 1/514 | 0.896695 | 0.999971 | 0 | 0 | 0.43973 | 0.047948 |
| GTEx_Tissu | GTEX-QDT8 | 2/870 | 0.897046 | 0.999971 | 0 | 0 | 0.516265 | 0.056091 |
| GTEx_Tissu | GTEX-TML8 | 2/909 | 0.910595 | 0.999971 | 0 | 0 | 0.493054 | 0.046178 |
| GTEx_Tissu | GTEX-OOBJ | Oct-54 | 0.897517 | 0.999971 | 0 | 0 | 0.692682 | 0.074895 |
| GTEx_Tissu | GTEX-PWN | 1/542 | 0.908868 | 0.999971 | 0 | 0 | 0.416369 | 0.039786 |
| GTEx_Tissu | GTEX-R55E | Oct-09 | 0.909037 | 0.999971 | 0 | 0 | 0.67854 | 0.064712 |
| GTEx_Tissu | GTEX-U3ZN | 1/517 | 0.898072 | 0.999971 | 0 | 0 | 0.437106 | 0.046991 |
| GTEx_Tissu | GTEX-R55D | 3/1229 | 0.909181 | 0.999971 | 0 | 0 | 0.544366 | 0.051829 |
| GTEx_Tissu | GTEX-WY7 | 3/1196 | 0.899118 | 0.999971 | 0 | 0 | 0.560412 | 0.059595 |
| GTEx_Tissu | GTEX-WH7 | 4/1499 | 0.899182 | 0.999971 | 0 | 0 | 0.593722 | 0.063095 |
| GTEx_Tissu | GTEX-PX3G | 5/1791 | 0.899242 | 0.999971 | 0 | 0 | 0.618872 | 0.065726 |
| GTEx_Tissu | GTEX-RUSC | 1/531 | 0.904265 | 0.999971 | 0 | 0 | 0.425252 | 0.042794 |

| | | | | | | | | |
|---|---|---|---|---|---|---|---|---|
| GTEx_Tissu | GTEX-TKQ1 | Sep-60 | 0.890963 | 0.999971 | 0 | 0 | 0.690527 | 0.079723 |
| GTEx_Tissu | GTEX-SJXC- | 5/1800 | 0.90165 | 0.999971 | 0 | 0 | 0.615463 | 0.063718 |
| GTEx_Tissu | GTEX-RVPV | 2/855 | 0.891351 | 0.999971 | 0 | 0 | 0.525757 | 0.060471 |
| GTEx_Tissu | GTEX-QDT8 | Nov-93 | 0.892293 | 0.999971 | 0 | 0 | 0.707464 | 0.080623 |
| GTEx_Tissu | GTEX-RWS. | 2/858 | 0.892512 | 0.999971 | 0 | 0 | 0.523832 | 0.059568 |
| GTEx_Tissu | GTEX-SN8G | 2/858 | 0.892512 | 0.999971 | 0 | 0 | 0.523832 | 0.059568 |
| GTEx_Tissu | GTEX-S7SF- | Oct-83 | 0.90373 | 0.999971 | 0 | 0 | 0.685164 | 0.069356 |
| GTEx_Tissu | GTEX-S32W | 2/889 | 0.903867 | 0.999971 | 0 | 0 | 0.504702 | 0.051012 |
| GTEx_Tissu | GTEX-UJM( | 4/1527 | 0.907046 | 0.999971 | 0 | 0 | 0.581921 | 0.056774 |
| GTEx_Tissu | GTEX-Q2AI | 1/520 | 0.899432 | 0.999971 | 0 | 0 | 0.434512 | 0.046055 |
| GTEx_Tissu | GTEX-R3RS | 1/539 | 0.907635 | 0.999971 | 0 | 0 | 0.418756 | 0.040583 |
| GTEx_Tissu | GTEX-N7M | 4/1531 | 0.908124 | 0.999971 | 0 | 0 | 0.58027 | 0.055923 |
| GTEx_Tissu | GTEX-WY7( | 2/902 | 0.908291 | 0.999971 | 0 | 0 | 0.497072 | 0.047814 |
| GTEx_Tissu | GTEX-OIZF- | Jun-14 | 0.908779 | 0.999971 | 0 | 0 | 0.625659 | 0.059846 |
| GTEx_Tissu | GTEX-WK1 | 4/1520 | 0.905131 | 0.999971 | 0 | 0 | 0.58483 | 0.058293 |
| GTEx_Tissu | GTEX-X261 | Jul-78 | 0.905142 | 0.999971 | 0 | 0 | 0.647375 | 0.06452 |
| GTEx_Tissu | GTEX-PVO\ | Aug-71 | 0.909179 | 0.999971 | 0 | 0 | 0.655965 | 0.062456 |
| GTEx_Tissu | GTEX-TMM | 3/1218 | 0.905931 | 0.999971 | 0 | 0 | 0.549618 | 0.054298 |
| GTEx_Tissu | GTEX-SNM | Aug-56 | 0.905933 | 0.999971 | 0 | 0 | 0.660255 | 0.065227 |
| GTEx_Tissu | GTEX-T2IS- | 4/1557 | 0.914874 | 0.999971 | 0 | 0 | 0.569748 | 0.05069 |
| GTEx_Tissu | GTEX-U3ZH | 2/891 | 0.90456 | 0.999971 | 0 | 0 | 0.503514 | 0.050506 |
| GTEx_Tissu | GTEX-X8HC | 3/1215 | 0.905027 | 0.999971 | 0 | 0 | 0.551067 | 0.054992 |
| GTEx_Tissu | GTEX-XYKS | 1/533 | 0.905118 | 0.999971 | 0 | 0 | 0.42361 | 0.042229 |
| GTEx_Tissu | GTEX-R55G | 1/533 | 0.905118 | 0.999971 | 0 | 0 | 0.42361 | 0.042229 |
| GTEx_Tissu | GTEX-WVLI | Jul-26 | 0.915686 | 0.999971 | 0 | 0 | 0.632792 | 0.055738 |
| GTEx_Tissu | GTEX-WVLI | 1/560 | 0.915933 | 0.999971 | 0 | 0 | 0.402588 | 0.035352 |
| GTEx_Tissu | GTEX-XOT4 | 3/1216 | 0.905329 | 0.999971 | 0 | 0 | 0.550583 | 0.054759 |
| GTEx_Tissu | GTEX-P78B | 3/1256 | 0.916739 | 0.999971 | 0 | 0 | 0.531866 | 0.046236 |
| GTEx_Tissu | GTEX-X4EP | 3/1267 | 0.919653 | 0.999971 | 0 | 0 | 0.526927 | 0.044135 |
| GTEx_Tissu | GTEX-OHPI | 2/896 | 0.906273 | 0.999971 | 0 | 0 | 0.500566 | 0.049263 |
| GTEx_Tissu | GTEX-R55C | 2/896 | 0.906273 | 0.999971 | 0 | 0 | 0.500566 | 0.049263 |
| GTEx_Tissu | GTEX-T5JW | Jul-85 | 0.906745 | 0.999971 | 0 | 0 | 0.645211 | 0.063163 |
| GTEx_Tissu | GTEX-PWO | 1/567 | 0.918532 | 0.999971 | 0 | 0 | 0.397465 | 0.033776 |
| GTEx_Tissu | GTEX-TML8 | 2/938 | 0.91958 | 0.999971 | 0 | 0 | 0.477049 | 0.039995 |
| GTEx_Tissu | GTEX-TKQ1 | 1/580 | 0.923149 | 0.999971 | 0 | 0 | 0.38828 | 0.031049 |
| GTEx_Tissu | GTEX-SE5C | 1/571 | 0.919981 | 0.999971 | 0 | 0 | 0.394594 | 0.03291 |
| GTEx_Tissu | GTEX-XBED | 2/951 | 0.92333 | 0.999971 | 0 | 0 | 0.470192 | 0.037507 |
| GTEx_Tissu | GTEX-P4PC | 1/554 | 0.91364 | 0.999971 | 0 | 0 | 0.407082 | 0.036767 |
| GTEx_Tissu | GTEX-S4Q7 | 1/549 | 0.911683 | 0.999971 | 0 | 0 | 0.410902 | 0.037993 |
| GTEx_Tissu | GTEX-QDV. | 5/1840 | 0.911769 | 0.999971 | 0 | 0 | 0.600718 | 0.055487 |
| GTEx_Tissu | GTEX-T8EM | Jun-40 | 0.914703 | 0.999971 | 0 | 0 | 0.617134 | 0.055021 |
| GTEx_Tissu | GTEX-T5JW | 2/914 | 0.912207 | 0.999971 | 0 | 0 | 0.490222 | 0.045045 |
| GTEx_Tissu | GTEX-TKQ1 | 2/923 | 0.915043 | 0.999971 | 0 | 0 | 0.485202 | 0.043078 |
| GTEx_Tissu | GTEX-XBED | 1/558 | 0.915175 | 0.999971 | 0 | 0 | 0.404075 | 0.035817 |
| GTEx_Tissu | GTEX-PSDG | 4/1559 | 0.915374 | 0.999971 | 0 | 0 | 0.568954 | 0.050308 |

| | | | | | | | | |
|---|---|---|---|---|---|---|---|---|
| GTEx_Tissu | GTEX-T5JC- | 3/1252 | 0.915656 | 0.999971 | 0 | 0 | 0.533684 | 0.047025 |
| GTEx_Tissu | GTEX-RWS( | 5/1847 | 0.913445 | 0.999971 | 0 | 0 | 0.598203 | 0.054157 |
| GTEx_Tissu | GTEX-XUZ( | 6/1547 | 0.673527 | 0.999971 | 0 | 0 | 0.883121 | 0.349033 |
| GTEx_Tissu | GTEX-OHPI | 3/1254 | 0.916199 | 0.999971 | 0 | 0 | 0.532774 | 0.046629 |
| GTEx_Tissu | GTEX-T6MI | Jul-18 | 0.914 | 0.999971 | 0 | 0 | 0.635182 | 0.057119 |
| GTEx_Tissu | GTEX-QEG4 | 3/1257 | 0.917008 | 0.999971 | 0 | 0 | 0.531414 | 0.046041 |
| GTEx_Tissu | GTEX-R55C | 3/1259 | 0.917543 | 0.999971 | 0 | 0 | 0.530511 | 0.045654 |
| GTEx_Tissu | GTEX-WFG | 4/1569 | 0.91784 | 0.999971 | 0 | 0 | 0.56501 | 0.04844 |
| GTEx_Tissu | GTEX-U3ZN | 2/933 | 0.918093 | 0.999971 | 0 | 0 | 0.479737 | 0.040997 |
| GTEx_Tissu | GTEX-R55F | 3/1248 | 0.914561 | 0.999971 | 0 | 0 | 0.535513 | 0.047828 |
| GTEx_Tissu | GTEX-T5JW | 5/1852 | 0.914625 | 0.999971 | 0 | 0 | 0.596419 | 0.053225 |
| GTEx_Tissu | GTEX-O5YT | 8/1481 | 0.315363 | 0.999971 | 0 | 0 | 1.267713 | 1.462982 |
| GTEx_Tissu | GTEX-VUS( | 5/886 | 0.341915 | 0.999971 | 0 | 0 | 1.317239 | 1.413653 |
| GTEx_Tissu | GTEX-P44H | 4/679 | 0.34214 | 0.999971 | 0 | 0 | 1.37353 | 1.473158 |
| GTEx_Tissu | GTEX-VUS( | 13/2605 | 0.342277 | 0.999971 | 0 | 0 | 1.17395 | 1.258633 |
| GTEx_Tissu | GTEX-RWS. | 3/476 | 0.34267 | 0.999971 | 0 | 0 | 1.467834 | 1.572031 |
| GTEx_Tissu | GTEX-R55C | 6/1090 | 0.337333 | 0.999971 | 0 | 0 | 1.286661 | 1.398196 |
| GTEx_Tissu | GTEX-P4QS | 5/861 | 0.320291 | 0.999971 | 0 | 0 | 1.357491 | 1.545539 |
| GTEx_Tissu | GTEX-R55E | 6/1060 | 0.314067 | 0.999971 | 0 | 0 | 1.325392 | 1.535002 |
| GTEx_Tissu | GTEX-QLQ\ | 5/855 | 0.315127 | 0.999971 | 0 | 0 | 1.367504 | 1.579164 |
| GTEx_Tissu | GTEX-SSA3 | 5/855 | 0.315127 | 0.999971 | 0 | 0 | 1.367504 | 1.579164 |
| GTEx_Tissu | GTEX-QDV. | 4/677 | 0.340155 | 0.999971 | 0 | 0 | 1.377755 | 1.485709 |
| GTEx_Tissu | GTEX-X4LF- | 4/652 | 0.315396 | 0.999971 | 0 | 0 | 1.432768 | 1.65331 |
| GTEx_Tissu | GTEX-V1D1 | 4/652 | 0.315396 | 0.999971 | 0 | 0 | 1.432768 | 1.65331 |
| GTEx_Tissu | GTEX-S3XE | 5/883 | 0.339312 | 0.999971 | 0 | 0 | 1.321948 | 1.428809 |
| GTEx_Tissu | GTEX-OXRI | 5/894 | 0.348864 | 0.999971 | 0 | 0 | 1.304837 | 1.37409 |
| GTEx_Tissu | GTEX-OIZH | 6/1105 | 0.349052 | 0.999971 | 0 | 0 | 1.268089 | 1.334708 |
| GTEx_Tissu | GTEX-XGQ4 | 6/1105 | 0.349052 | 0.999971 | 0 | 0 | 1.268089 | 1.334708 |
| GTEx_Tissu | GTEX-RU72 | 5/895 | 0.349733 | 0.999971 | 0 | 0 | 1.303302 | 1.36923 |
| GTEx_Tissu | GTEX-WFO | 6/1107 | 0.350618 | 0.999971 | 0 | 0 | 1.265651 | 1.326477 |
| GTEx_Tissu | GTEX-NL3H | 3/472 | 0.337897 | 0.999971 | 0 | 0 | 1.480658 | 1.606536 |
| GTEx_Tissu | GTEX-NL3G | Nov-64 | 0.339416 | 0.999971 | 0 | 0 | 1.193928 | 1.290075 |
| GTEx_Tissu | GTEX-SIU7- | 6/1086 | 0.334216 | 0.999971 | 0 | 0 | 1.291701 | 1.415662 |
| GTEx_Tissu | GTEX-XGQ4 | 8/1510 | 0.334496 | 0.999971 | 0 | 0 | 1.241282 | 1.359366 |
| GTEx_Tissu | GTEX-XAJ8 | 5/878 | 0.334978 | 0.999971 | 0 | 0 | 1.329869 | 1.454464 |
| GTEx_Tissu | GTEX-PVO\ | 9/1727 | 0.336369 | 0.999971 | 0 | 0 | 1.222016 | 1.331445 |
| GTEx_Tissu | GTEX-XBEV | 18/3693 | 0.336693 | 0.999971 | 0 | 0 | 1.152653 | 1.254759 |
| GTEx_Tissu | GTEX-RU72 | Nov-60 | 0.337162 | 0.999971 | 0 | 0 | 1.196419 | 1.300737 |
| GTEx_Tissu | GTEX-U4B1 | 26/5454 | 0.328967 | 0.999971 | 0 | 0 | 1.137423 | 1.264584 |
| GTEx_Tissu | GTEX-P78B | 3/452 | 0.314024 | 0.999971 | 0 | 0 | 1.548202 | 1.793262 |
| GTEx_Tissu | GTEX-UPK5 | 5/887 | 0.342783 | 0.999971 | 0 | 0 | 1.315677 | 1.40864 |
| GTEx_Tissu | GTEX-UTH( | 6/1092 | 0.338892 | 0.999971 | 0 | 0 | 1.284155 | 1.389549 |
| GTEx_Tissu | GTEX-QDV. | 3/473 | 0.33909 | 0.999971 | 0 | 0 | 1.477432 | 1.597826 |
| GTEx_Tissu | GTEX-QCQ( | 4/676 | 0.339162 | 0.999971 | 0 | 0 | 1.379877 | 1.49203 |
| GTEx_Tissu | GTEX-UJHI- | 4/676 | 0.339162 | 0.999971 | 0 | 0 | 1.379877 | 1.49203 |

| | | | | | | | | |
|---|---|---|---|---|---|---|---|---|
| GTEx_Tissu | GTEX-QESD | 7/1304 | 0.339244 | 0.999971 | 0 | 0 | 1.255898 | 1.357669 |
| GTEx_Tissu | GTEX-U8XE | 4/670 | 0.333209 | 0.999971 | 0 | 0 | 1.392742 | 1.530603 |
| GTEx_Tissu | GTEX-Q2AI | 6/1062 | 0.315609 | 0.999971 | 0 | 0 | 1.322741 | 1.525451 |
| GTEx_Tissu | GTEX-X3Y1 | 8/1518 | 0.339812 | 0.999971 | 0 | 0 | 1.234169 | 1.332115 |
| GTEx_Tissu | GTEX-T8EM | 26/5547 | 0.365259 | 0.999971 | 0 | 0 | 1.111084 | 1.119026 |
| GTEx_Tissu | GTEX-QESD | 5/872 | 0.329786 | 0.999971 | 0 | 0 | 1.339494 | 1.485918 |
| GTEx_Tissu | GTEX-R55F | 7/1292 | 0.330676 | 0.999971 | 0 | 0 | 1.268444 | 1.403681 |
| GTEx_Tissu | GTEX-N7M | 26/5459 | 0.330888 | 0.999971 | 0 | 0 | 1.135984 | 1.25637 |
| GTEx_Tissu | GTEX-PX3G | 4/668 | 0.331226 | 0.999971 | 0 | 0 | 1.397082 | 1.543712 |
| GTEx_Tissu | GTEX-XGQ4 | 4/668 | 0.331226 | 0.999971 | 0 | 0 | 1.397082 | 1.543712 |
| GTEx_Tissu | GTEX-QDVI | 8/1507 | 0.332506 | 0.999971 | 0 | 0 | 1.243969 | 1.369729 |
| GTEx_Tissu | GTEX-XYKS | 6/1114 | 0.356104 | 0.999971 | 0 | 0 | 1.257187 | 1.298086 |
| GTEx_Tissu | GTEX-U4B1 | Oct-90 | 0.364757 | 0.999971 | 0 | 0 | 1.176243 | 1.186271 |
| GTEx_Tissu | GTEX-WRH | 4/702 | 0.364998 | 0.999971 | 0 | 0 | 1.326682 | 1.337113 |
| GTEx_Tissu | GTEX-QEL4 | 5/881 | 0.337578 | 0.999971 | 0 | 0 | 1.325106 | 1.439011 |
| GTEx_Tissu | GTEX-XV7C | Dec-30 | 0.36588 | 0.999971 | 0 | 0 | 1.157651 | 1.163961 |
| GTEx_Tissu | GTEX-XMK | 4/703 | 0.365993 | 0.999971 | 0 | 0 | 1.324715 | 1.331527 |
| GTEx_Tissu | GTEX-WYJK | 6/1119 | 0.360028 | 0.999971 | 0 | 0 | 1.251206 | 1.2782 |
| GTEx_Tissu | GTEX-REY6 | 25/5327 | 0.36721 | 0.999971 | 0 | 0 | 1.111191 | 1.113214 |
| GTEx_Tissu | GTEX-T6M( | 20/4207 | 0.367411 | 0.999971 | 0 | 0 | 1.121168 | 1.122596 |
| GTEx_Tissu | GTEX-S33H | 7/1345 | 0.368745 | 0.999971 | 0 | 0 | 1.214733 | 1.211877 |
| GTEx_Tissu | GTEX-VJYA | 4/706 | 0.368975 | 0.999971 | 0 | 0 | 1.318848 | 1.314928 |
| GTEx_Tissu | GTEX-X638 | 4/706 | 0.368975 | 0.999971 | 0 | 0 | 1.318848 | 1.314928 |
| GTEx_Tissu | GTEX-W5W | 20/4183 | 0.356955 | 0.999971 | 0 | 0 | 1.129352 | 1.163398 |
| GTEx_Tissu | GTEX-T6MI | 25/5289 | 0.352019 | 0.999971 | 0 | 0 | 1.122123 | 1.171574 |
| GTEx_Tissu | GTEX-XYKS | 7/1322 | 0.352159 | 0.999971 | 0 | 0 | 1.23751 | 1.291556 |
| GTEx_Tissu | GTEX-WRH | 3/485 | 0.353403 | 0.999971 | 0 | 0 | 1.43976 | 1.49756 |
| GTEx_Tissu | GTEX-T2IS- | 7/1324 | 0.353598 | 0.999971 | 0 | 0 | 1.235497 | 1.284418 |
| GTEx_Tissu | GTEX-T6MI | 7/1326 | 0.355037 | 0.999971 | 0 | 0 | 1.233491 | 1.277321 |
| GTEx_Tissu | GTEX-S33H | 6/1113 | 0.35532 | 0.999971 | 0 | 0 | 1.258389 | 1.302102 |
| GTEx_Tissu | GTEX-XMD | 5/902 | 0.355823 | 0.999971 | 0 | 0 | 1.292656 | 1.33573 |
| GTEx_Tissu | GTEX-WW | 6/1103 | 0.347486 | 0.999971 | 0 | 0 | 1.270536 | 1.342994 |
| GTEx_Tissu | GTEX-XAJ8 | 25/5300 | 0.356398 | 0.999971 | 0 | 0 | 1.118942 | 1.154419 |
| GTEx_Tissu | GTEX-PX3G | 6/1072 | 0.323342 | 0.999971 | 0 | 0 | 1.309638 | 1.478642 |
| GTEx_Tissu | GTEX-OHPI | 5/877 | 0.334112 | 0.999971 | 0 | 0 | 1.331464 | 1.459655 |
| GTEx_Tissu | GTEX-OOBI | 3/488 | 0.356977 | 0.999971 | 0 | 0 | 1.430633 | 1.473671 |
| GTEx_Tissu | GTEX-P78B | Dec-16 | 0.35822 | 0.999971 | 0 | 0 | 1.165324 | 1.196331 |
| GTEx_Tissu | GTEX-RNOI | 5/905 | 0.358435 | 0.999971 | 0 | 0 | 1.288144 | 1.321647 |
| GTEx_Tissu | GTEX-RVPV | 7/1331 | 0.358639 | 0.999971 | 0 | 0 | 1.228503 | 1.259754 |
| GTEx_Tissu | GTEX-Q734 | 4/696 | 0.359034 | 0.999971 | 0 | 0 | 1.338603 | 1.371183 |
| GTEx_Tissu | GTEX-POM | 5/897 | 0.351472 | 0.999971 | 0 | 0 | 1.300243 | 1.359566 |
| GTEx_Tissu | GTEX-NL4V | Nov-46 | 0.329302 | 0.999971 | 0 | 0 | 1.205214 | 1.338726 |
| GTEx_Tissu | GTEX-S4Z8 | 3/428 | 0.285466 | 0.999971 | 0 | 0 | 1.637647 | 2.053005 |
| GTEx_Tissu | GTEX-XAJ8 | 4/634 | 0.297683 | 0.999971 | 0 | 0 | 1.475081 | 1.787395 |
| GTEx_Tissu | GTEX-S7SE | Nov-04 | 0.306003 | 0.999971 | 0 | 0 | 1.232303 | 1.459243 |

| | | | | | | | | |
|---|---|---|---|---|---|---|---|---|
| GTEx_Tissu | GTEX-POYV | 6/1051 | 0.307143 | 0.999971 | 0 | 0 | 1.337445 | 1.578777 |
| GTEx_Tissu | GTEX-XUW | 4/645 | 0.308493 | 0.999971 | 0 | 0 | 1.448941 | 1.704035 |
| GTEx_Tissu | GTEX-X585 | 10/1895 | 0.308701 | 0.999971 | 0 | 0 | 1.242068 | 1.459904 |
| GTEx_Tissu | GTEX-V1D1 | 13/2540 | 0.308888 | 0.999971 | 0 | 0 | 1.208665 | 1.419912 |
| GTEx_Tissu | GTEX-NFK9 | 7/1262 | 0.30943 | 0.999971 | 0 | 0 | 1.300857 | 1.525937 |
| GTEx_Tissu | GTEX-NPJ8 | 7/1262 | 0.30943 | 0.999971 | 0 | 0 | 1.300857 | 1.525937 |
| GTEx_Tissu | GTEX-XLM4 | 25/5157 | 0.30084 | 0.999971 | 0 | 0 | 1.161356 | 1.394995 |
| GTEx_Tissu | GTEX-WH7 | 5/844 | 0.305694 | 0.999971 | 0 | 0 | 1.386232 | 1.642923 |
| GTEx_Tissu | GTEX-WFO | 3/428 | 0.285466 | 0.999971 | 0 | 0 | 1.637647 | 2.053005 |
| GTEx_Tissu | GTEX-XPVC | 4/647 | 0.310464 | 0.999971 | 0 | 0 | 1.444284 | 1.689362 |
| GTEx_Tissu | GTEX-S4Q7 | 4/647 | 0.310464 | 0.999971 | 0 | 0 | 1.444284 | 1.689362 |
| GTEx_Tissu | GTEX-XPT6 | 19/3855 | 0.311131 | 0.999971 | 0 | 0 | 1.171038 | 1.367236 |
| GTEx_Tissu | GTEX-XUZC | 4/648 | 0.311449 | 0.999971 | 0 | 0 | 1.441967 | 1.68208 |
| GTEx_Tissu | GTEX-WXY | 5/851 | 0.311692 | 0.999971 | 0 | 0 | 1.374258 | 1.602028 |
| GTEx_Tissu | GTEX-XXEK | 4/649 | 0.312436 | 0.999971 | 0 | 0 | 1.439656 | 1.674834 |
| GTEx_Tissu | GTEX-R55C | 26/5346 | 0.288471 | 0.999971 | 0 | 0 | 1.169167 | 1.453461 |
| GTEx_Tissu | GTEX-OXRF | 6/993 | 0.263355 | 0.999971 | 0 | 0 | 1.420391 | 1.895162 |
| GTEx_Tissu | GTEX-WHS | 5/832 | 0.295457 | 0.999971 | 0 | 0 | 1.407232 | 1.715741 |
| GTEx_Tissu | GTEX-P4QT | Nov-85 | 0.29562 | 0.999971 | 0 | 0 | 1.244918 | 1.517159 |
| GTEx_Tissu | GTEX-RM2 | 6/1036 | 0.295671 | 0.999971 | 0 | 0 | 1.358001 | 1.654736 |
| GTEx_Tissu | GTEX-RU1J | 4/632 | 0.295723 | 0.999971 | 0 | 0 | 1.479932 | 1.80305 |
| GTEx_Tissu | GTEX-QCQ | 6/1005 | 0.272278 | 0.999971 | 0 | 0 | 1.402439 | 1.824478 |
| GTEx_Tissu | GTEX-V955 | 3/417 | 0.272454 | 0.999971 | 0 | 0 | 1.682108 | 2.187218 |
| GTEx_Tissu | GTEX-XQ8I | 7/1209 | 0.272693 | 0.999971 | 0 | 0 | 1.362074 | 1.769888 |
| GTEx_Tissu | GTEX-SE5C | 3/431 | 0.289025 | 0.999971 | 0 | 0 | 1.625918 | 2.018155 |
| GTEx_Tissu | GTEX-OHPI | 6/993 | 0.263355 | 0.999971 | 0 | 0 | 1.420391 | 1.895162 |
| GTEx_Tissu | GTEX-N7M | 6/1027 | 0.288833 | 0.999971 | 0 | 0 | 1.370624 | 1.70219 |
| GTEx_Tissu | GTEX-NPJ8 | 4/599 | 0.263688 | 0.999971 | 0 | 0 | 1.564686 | 2.085708 |
| GTEx_Tissu | GTEX-Q2AI | 7/1203 | 0.268613 | 0.999971 | 0 | 0 | 1.369346 | 1.799984 |
| GTEx_Tissu | GTEX-XYKS | 6/1055 | 0.310216 | 0.999971 | 0 | 0 | 1.332062 | 1.559159 |
| GTEx_Tissu | GTEX-U4B1 | 7/1253 | 0.303113 | 0.999971 | 0 | 0 | 1.310885 | 1.564739 |
| GTEx_Tissu | GTEX-S3XE | 4/640 | 0.303573 | 0.999971 | 0 | 0 | 1.460711 | 1.741359 |
| GTEx_Tissu | GTEX-S7SE | 27/5610 | 0.303592 | 0.999971 | 0 | 0 | 1.155024 | 1.37687 |
| GTEx_Tissu | GTEX-S95S | 8/1464 | 0.304262 | 0.999971 | 0 | 0 | 1.283697 | 1.527429 |
| GTEx_Tissu | GTEX-U8XE | 25/5169 | 0.305376 | 0.999971 | 0 | 0 | 1.157706 | 1.373286 |
| GTEx_Tissu | GTEX-POYV | Dec-61 | 0.328457 | 0.999971 | 0 | 0 | 1.196356 | 1.331962 |
| GTEx_Tissu | GTEX-V955 | 24/4992 | 0.323003 | 0.999971 | 0 | 0 | 1.146001 | 1.295088 |
| GTEx_Tissu | GTEX-Q2AC | 4/660 | 0.323303 | 0.999971 | 0 | 0 | 1.414708 | 1.597439 |
| GTEx_Tissu | GTEX-PWC | 6/1062 | 0.315609 | 0.999971 | 0 | 0 | 1.322741 | 1.525451 |
| GTEx_Tissu | GTEX-P78B | 8/1459 | 0.301015 | 0.999971 | 0 | 0 | 1.28847 | 1.546931 |
| GTEx_Tissu | GTEX-S4UY | 7/1282 | 0.323564 | 0.999971 | 0 | 0 | 1.279078 | 1.44326 |
| GTEx_Tissu | GTEX-UPK5 | 4/661 | 0.324292 | 0.999971 | 0 | 0 | 1.412481 | 1.590608 |
| GTEx_Tissu | GTEX-XOT4 | 13/2572 | 0.325212 | 0.999971 | 0 | 0 | 1.191354 | 1.338224 |
| GTEx_Tissu | GTEX-T8EN | 6/1076 | 0.326443 | 0.999971 | 0 | 0 | 1.304465 | 1.460349 |
| GTEx_Tissu | GTEX-QDVI | 5/869 | 0.327193 | 0.999971 | 0 | 0 | 1.344357 | 1.501924 |

| | | | | | | | | |
|---|---|---|---|---|---|---|---|---|
| GTEx_Tissu | GTEX-Q2AI | 4/658 | 0.321324 | 0.999971 | 0 | 0 | 1.419181 | 1.611202 |
| GTEx_Tissu | GTEX-QDT8 | 3/457 | 0.31999 | 0.999971 | 0 | 0 | 1.530758 | 1.744247 |
| GTEx_Tissu | GTEX-T2IS- | 7/1258 | 0.306619 | 0.999971 | 0 | 0 | 1.305296 | 1.543057 |
| GTEx_Tissu | GTEX-XUW | 5/835 | 0.29801 | 0.999971 | 0 | 0 | 1.401925 | 1.697208 |
| GTEx_Tissu | GTEX-WEY! | 8/1455 | 0.298424 | 0.999971 | 0 | 0 | 1.292311 | 1.562715 |
| GTEx_Tissu | GTEX-R55G | 3/439 | 0.298533 | 0.999971 | 0 | 0 | 1.595429 | 1.928672 |
| GTEx_Tissu | GTEX-R55E | 10/1879 | 0.299482 | 0.999971 | 0 | 0 | 1.253813 | 1.511723 |
| GTEx_Tissu | GTEX-VUS( | 8/1458 | 0.300367 | 0.999971 | 0 | 0 | 1.289428 | 1.550861 |
| GTEx_Tissu | GTEX-U3Z( | 5/838 | 0.300568 | 0.999971 | 0 | 0 | 1.396656 | 1.678895 |
| GTEx_Tissu | GTEX-WL4( | 5/857 | 0.316847 | 0.999971 | 0 | 0 | 1.36415 | 1.567867 |
| GTEx_Tissu | GTEX-WFG | 3/431 | 0.289025 | 0.999971 | 0 | 0 | 1.625918 | 2.018155 |
| GTEx_Tissu | GTEX-UPK5 | 20/4003 | 0.281491 | 0.999971 | 0 | 0 | 1.19388 | 1.513427 |
| GTEx_Tissu | GTEX-WOF | 3/425 | 0.281912 | 0.999971 | 0 | 0 | 1.649543 | 2.088589 |
| GTEx_Tissu | GTEX-RN64 | 5/817 | 0.282754 | 0.999971 | 0 | 0 | 1.434354 | 1.811844 |
| GTEx_Tissu | GTEX-WFG | 8/1431 | 0.283002 | 0.999971 | 0 | 0 | 1.315815 | 1.660954 |
| GTEx_Tissu | GTEX-X261 | 5/820 | 0.285286 | 0.999971 | 0 | 0 | 1.428849 | 1.792155 |
| GTEx_Tissu | GTEX-WH7 | 6/1072 | 0.323342 | 0.999971 | 0 | 0 | 1.309638 | 1.478642 |
| GTEx_Tissu | GTEX-R55C | 7/1271 | 0.315774 | 0.999971 | 0 | 0 | 1.290971 | 1.488138 |
| GTEx_Tissu | GTEX-PW2( | 5/856 | 0.315987 | 0.999971 | 0 | 0 | 1.365825 | 1.573504 |
| GTEx_Tissu | GTEX-S33H | 26/5557 | 0.369226 | 0.999971 | 0 | 0 | 1.108305 | 1.104255 |
| GTEx_Tissu | GTEX-OOBI | 8/1485 | 0.317988 | 0.999971 | 0 | 0 | 1.264006 | 1.448226 |
| GTEx_Tissu | GTEX-TKQ1 | 20/4094 | 0.31891 | 0.999971 | 0 | 0 | 1.160545 | 1.326326 |
| GTEx_Tissu | GTEX-RWS( | 4/656 | 0.319347 | 0.999971 | 0 | 0 | 1.423682 | 1.6251 |
| GTEx_Tissu | GTEX-UPIC | 7/1268 | 0.313656 | 0.999971 | 0 | 0 | 1.294251 | 1.500628 |
| GTEx_Tissu | GTEX-XUYS | 4/634 | 0.297683 | 0.999971 | 0 | 0 | 1.475081 | 1.787395 |
| GTEx_Tissu | GTEX-PW2( | 6/1069 | 0.321019 | 0.999971 | 0 | 0 | 1.313543 | 1.492521 |
| GTEx_Tissu | GTEX-S3XE | 5/862 | 0.321152 | 0.999971 | 0 | 0 | 1.355836 | 1.540011 |
| GTEx_Tissu | GTEX-XPV( | 3/458 | 0.321183 | 0.999971 | 0 | 0 | 1.527316 | 1.734638 |
| GTEx_Tissu | GTEX-XLM4 | 5/940 | 0.388948 | 0.999971 | 0 | 0 | 1.237642 | 1.168716 |
| GTEx_Tissu | GTEX-XUZ( | 3/536 | 0.413683 | 0.999971 | 0 | 0 | 1.298579 | 1.146199 |
| GTEx_Tissu | GTEX-XUW | 7/1408 | 0.414427 | 0.999971 | 0 | 0 | 1.156174 | 1.018426 |
| GTEx_Tissu | GTEX-XGQ4 | 2/320 | 0.40678 | 0.999971 | 0 | 0 | 1.449871 | 1.304135 |
| GTEx_Tissu | GTEX-XBED | 26/5640 | 0.402535 | 0.999971 | 0 | 0 | 1.085617 | 0.987883 |
| GTEx_Tissu | GTEX-R55E | 9/1839 | 0.407262 | 0.999971 | 0 | 0 | 1.140164 | 1.024208 |
| GTEx_Tissu | GTEX-OOBI | 4/745 | 0.407638 | 0.999971 | 0 | 0 | 1.246899 | 1.118937 |
| GTEx_Tissu | GTEX-QEG4 | 4/725 | 0.387843 | 0.999971 | 0 | 0 | 1.282823 | 1.215034 |
| GTEx_Tissu | GTEX-WI4N | 4/726 | 0.388834 | 0.999971 | 0 | 0 | 1.28098 | 1.210016 |
| GTEx_Tissu | GTEX-X8H( | 5/940 | 0.388948 | 0.999971 | 0 | 0 | 1.237642 | 1.168716 |
| GTEx_Tissu | GTEX-T6MI | 4/751 | 0.413557 | 0.999971 | 0 | 0 | 1.236496 | 1.091778 |
| GTEx_Tissu | GTEX-UJM( | 5/967 | 0.412463 | 0.999971 | 0 | 0 | 1.201194 | 1.063789 |
| GTEx_Tissu | GTEX-NPJ8 | 7/1395 | 0.40499 | 0.999971 | 0 | 0 | 1.167822 | 1.055585 |
| GTEx_Tissu | GTEX-QVU! | 6/1186 | 0.412782 | 0.999971 | 0 | 0 | 1.175957 | 1.040528 |
| GTEx_Tissu | GTEX-WY7( | 4/751 | 0.413557 | 0.999971 | 0 | 0 | 1.236496 | 1.091778 |
| GTEx_Tissu | GTEX-QEL4 | 6/1160 | 0.392297 | 0.999971 | 0 | 0 | 1.204121 | 1.126739 |
| GTEx_Tissu | GTEX-WFJ( | 5/945 | 0.393307 | 0.999971 | 0 | 0 | 1.230734 | 1.148477 |

| | | | | | | | | |
|---|---|---|---|---|---|---|---|---|
| GTEx_Tissu | GTEX-WFJ( | 8/1600 | 0.394943 | 0.999971 | 0 | 0 | 1.165384 | 1.082656 |
| GTEx_Tissu | GTEX-T5JW | 17/3597 | 0.394959 | 0.999971 | 0 | 0 | 1.107985 | 1.029289 |
| GTEx_Tissu | GTEX-OXRI | 4/745 | 0.407638 | 0.999971 | 0 | 0 | 1.246899 | 1.118937 |
| GTEx_Tissu | GTEX-T8EN | 6/1158 | 0.390721 | 0.999971 | 0 | 0 | 1.20634 | 1.133672 |
| GTEx_Tissu | GTEX-X8H( | 3/504 | 0.375995 | 0.999971 | 0 | 0 | 1.383804 | 1.353608 |
| GTEx_Tissu | GTEX-P78B | 6/1140 | 0.376541 | 0.999971 | 0 | 0 | 1.226664 | 1.198119 |
| GTEx_Tissu | GTEX-T8EN | 8/1573 | 0.376694 | 0.999971 | 0 | 0 | 1.187237 | 1.159125 |
| GTEx_Tissu | GTEX-TSE9 | 8/1574 | 0.377369 | 0.999971 | 0 | 0 | 1.186414 | 1.156199 |
| GTEx_Tissu | GTEX-OXRL | 8/1575 | 0.378044 | 0.999971 | 0 | 0 | 1.185592 | 1.15328 |
| GTEx_Tissu | GTEX-T5JW | 4/737 | 0.399731 | 0.999971 | 0 | 0 | 1.261033 | 1.156322 |
| GTEx_Tissu | GTEX-WEY! | 3/506 | 0.378366 | 0.999971 | 0 | 0 | 1.37816 | 1.339424 |
| GTEx_Tissu | GTEX-XBEV | 3/506 | 0.378366 | 0.999971 | 0 | 0 | 1.37816 | 1.339424 |
| GTEx_Tissu | GTEX-RVPL | 5/947 | 0.395051 | 0.999971 | 0 | 0 | 1.227992 | 1.140487 |
| GTEx_Tissu | GTEX-S4P3 | 3/531 | 0.407833 | 0.999971 | 0 | 0 | 1.311215 | 1.176024 |
| GTEx_Tissu | GTEX-OXRI | 6/1183 | 0.41042 | 0.999971 | 0 | 0 | 1.179143 | 1.050115 |
| GTEx_Tissu | GTEX-QDV. | 9/1844 | 0.410455 | 0.999971 | 0 | 0 | 1.136743 | 1.012257 |
| GTEx_Tissu | GTEX-SIU8 | 4/742 | 0.404675 | 0.999971 | 0 | 0 | 1.252163 | 1.132797 |
| GTEx_Tissu | GTEX-T5JW | 8/1651 | 0.429493 | 0.999971 | 0 | 0 | 1.126066 | 0.951693 |
| GTEx_Tissu | GTEX-OHPI | 3/529 | 0.405489 | 0.999971 | 0 | 0 | 1.316336 | 1.188206 |
| GTEx_Tissu | GTEX-QCQ( | 7/1396 | 0.405716 | 0.999971 | 0 | 0 | 1.166919 | 1.052678 |
| GTEx_Tissu | GTEX-OHPI | 4/751 | 0.413557 | 0.999971 | 0 | 0 | 1.236496 | 1.091778 |
| GTEx_Tissu | GTEX-OHPI | 5/960 | 0.406374 | 0.999971 | 0 | 0 | 1.210446 | 1.089985 |
| GTEx_Tissu | GTEX-S4P3 | 3/528 | 0.404316 | 0.999971 | 0 | 0 | 1.318912 | 1.194352 |
| GTEx_Tissu | GTEX-TMM | 4/742 | 0.404675 | 0.999971 | 0 | 0 | 1.252163 | 1.132797 |
| GTEx_Tissu | GTEX-XUW | 4/770 | 0.432219 | 0.999971 | 0 | 0 | 1.204631 | 1.010472 |
| GTEx_Tissu | GTEX-WFJ( | 18/3901 | 0.432254 | 0.999971 | 0 | 0 | 1.076935 | 0.90327 |
| GTEx_Tissu | GTEX-XUZ( | 3/552 | 0.432281 | 0.999971 | 0 | 0 | 1.259693 | 1.056478 |
| GTEx_Tissu | GTEX-POYV | Oct-90 | 0.425191 | 0.999971 | 0 | 0 | 1.113449 | 0.95224 |
| GTEx_Tissu | GTEX-QEG4 | 24/5250 | 0.428432 | 0.999971 | 0 | 0 | 1.070618 | 0.90748 |
| GTEx_Tissu | GTEX-WH7 | 3/549 | 0.428809 | 0.999971 | 0 | 0 | 1.266811 | 1.072665 |
| GTEx_Tissu | GTEX-QVU! | Nov-80 | 0.40585 | 0.999971 | 0 | 0 | 1.12549 | 1.014935 |
| GTEx_Tissu | GTEX-P44H | Oct-87 | 0.42337 | 0.999971 | 0 | 0 | 1.115245 | 0.958562 |
| GTEx_Tissu | GTEX-SNM | 7/1397 | 0.406442 | 0.999971 | 0 | 0 | 1.166016 | 1.04978 |
| GTEx_Tissu | GTEX-U3ZN | 19/4071 | 0.406724 | 0.999971 | 0 | 0 | 1.093719 | 0.983933 |
| GTEx_Tissu | GTEX-PX3G | 6/1173 | 0.402542 | 0.999971 | 0 | 0 | 1.189882 | 1.08274 |
| GTEx_Tissu | GTEX-U8XE | 6/1174 | 0.40333 | 0.999971 | 0 | 0 | 1.1888 | 1.07943 |
| GTEx_Tissu | GTEX-P78B | 4/741 | 0.403686 | 0.999971 | 0 | 0 | 1.253928 | 1.137459 |
| GTEx_Tissu | GTEX-XGQ4 | 4/741 | 0.403686 | 0.999971 | 0 | 0 | 1.253928 | 1.137459 |
| GTEx_Tissu | GTEX-S4Z8 | 3/558 | 0.439204 | 0.999971 | 0 | 0 | 1.245689 | 1.024943 |
| GTEx_Tissu | GTEX-T8EN | 4/771 | 0.433197 | 0.999971 | 0 | 0 | 1.202997 | 1.006381 |
| GTEx_Tissu | GTEX-T2YK | 9/1858 | 0.419398 | 0.999971 | 0 | 0 | 1.127262 | 0.979517 |
| GTEx_Tissu | GTEX-RTLS | 4/752 | 0.414542 | 0.999971 | 0 | 0 | 1.234779 | 1.087323 |
| GTEx_Tissu | GTEX-TMM | 9/1839 | 0.407262 | 0.999971 | 0 | 0 | 1.140164 | 1.024208 |
| GTEx_Tissu | GTEX-QMR | 6/1154 | 0.387569 | 0.999971 | 0 | 0 | 1.210801 | 1.147673 |
| GTEx_Tissu | GTEX-SNM | 27/5919 | 0.42373 | 0.999971 | 0 | 0 | 1.07085 | 0.919494 |

| | | | | | | | | |
|---|---|---|---|---|---|---|---|---|
| GTEx_Tissu | GTEX-P44H | 6/1189 | 0.415144 | 0.999971 | 0 | 0 | 1.172787 | 1.031033 |
| GTEx_Tissu | GTEX-SIU7- | 3/546 | 0.425329 | 0.999971 | 0 | 0 | 1.274007 | 1.089138 |
| GTEx_Tissu | GTEX-N7M | 6/1203 | 0.426154 | 0.999971 | 0 | 0 | 1.158204 | 0.987896 |
| GTEx_Tissu | GTEX-R55D | 23/5022 | 0.427836 | 0.999971 | 0 | 0 | 1.072158 | 0.910279 |
| GTEx_Tissu | GTEX-X4XX | 7/1427 | 0.428208 | 0.999971 | 0 | 0 | 1.139533 | 0.966491 |
| GTEx_Tissu | GTEX-XQ8I | 4/737 | 0.399731 | 0.999971 | 0 | 0 | 1.261033 | 1.156322 |
| GTEx_Tissu | GTEX-WEY! | 6/1195 | 0.419865 | 0.999971 | 0 | 0 | 1.166495 | 1.012311 |
| GTEx_Tissu | GTEX-S7SF- | 4/767 | 0.429281 | 0.999971 | 0 | 0 | 1.209556 | 1.022853 |
| GTEx_Tissu | GTEX-Q2A( | 5/978 | 0.422016 | 0.999971 | 0 | 0 | 1.186925 | 1.023975 |
| GTEx_Tissu | GTEX-PLZ6- | 3/551 | 0.431124 | 0.999971 | 0 | 0 | 1.262057 | 1.061842 |
| GTEx_Tissu | GTEX-OOB. | 4/769 | 0.43124 | 0.999971 | 0 | 0 | 1.206268 | 1.01458 |
| GTEx_Tissu | GTEX-WFG | 23/5030 | 0.431244 | 0.999971 | 0 | 0 | 1.069871 | 0.899848 |
| GTEx_Tissu | GTEX-QDT8 | 7/1432 | 0.431831 | 0.999971 | 0 | 0 | 1.135228 | 0.953275 |
| GTEx_Tissu | GTEX-XBED | 7/1420 | 0.423133 | 0.999971 | 0 | 0 | 1.145612 | 0.985305 |
| GTEx_Tissu | GTEX-SSA3 | 6/1148 | 0.382841 | 0.999971 | 0 | 0 | 1.217552 | 1.169015 |
| GTEx_Tissu | GTEX-OIZI- | 4/685 | 0.3481 | 0.999971 | 0 | 0 | 1.361003 | 1.436219 |
| GTEx_Tissu | GTEX-NPJ7 | 3/506 | 0.378366 | 0.999971 | 0 | 0 | 1.37816 | 1.339424 |
| GTEx_Tissu | GTEX-S32V | 5/928 | 0.378483 | 0.999971 | 0 | 0 | 1.254525 | 1.218877 |
| GTEx_Tissu | GTEX-S7SE- | 8/1576 | 0.378719 | 0.999971 | 0 | 0 | 1.184771 | 1.150368 |
| GTEx_Tissu | GTEX-TSE9- | 5/930 | 0.380227 | 0.999971 | 0 | 0 | 1.251681 | 1.210358 |
| GTEx_Tissu | GTEX-NL4V | 4/718 | 0.380896 | 0.999971 | 0 | 0 | 1.295873 | 1.250813 |
| GTEx_Tissu | GTEX-XQ8I | 5/923 | 0.374122 | 0.999971 | 0 | 0 | 1.26169 | 1.240459 |
| GTEx_Tissu | GTEX-QESD | Oct-19 | 0.382194 | 0.999971 | 0 | 0 | 1.157389 | 1.113208 |
| GTEx_Tissu | GTEX-T6MI | 9/1800 | 0.382404 | 0.999971 | 0 | 0 | 1.167504 | 1.122297 |
| GTEx_Tissu | GTEX-RU72 | 6/1103 | 0.347486 | 0.999971 | 0 | 0 | 1.270536 | 1.342994 |
| GTEx_Tissu | GTEX-P4PP | 6/1149 | 0.383629 | 0.999971 | 0 | 0 | 1.216422 | 1.165429 |
| GTEx_Tissu | GTEX-NFK9 | 5/934 | 0.383716 | 0.999971 | 0 | 0 | 1.246029 | 1.193513 |
| GTEx_Tissu | GTEX-QLQ7 | 5/934 | 0.383716 | 0.999971 | 0 | 0 | 1.246029 | 1.193513 |
| GTEx_Tissu | GTEX-XMD | 5/935 | 0.384588 | 0.999971 | 0 | 0 | 1.244624 | 1.189341 |
| GTEx_Tissu | GTEX-T5JC- | 3/506 | 0.378366 | 0.999971 | 0 | 0 | 1.37816 | 1.339424 |
| GTEx_Tissu | GTEX-R55D | 7/1367 | 0.384673 | 0.999971 | 0 | 0 | 1.193667 | 1.140383 |
| GTEx_Tissu | GTEX-S4Z8- | 4/723 | 0.385859 | 0.999971 | 0 | 0 | 1.286526 | 1.225138 |
| GTEx_Tissu | GTEX-SN8( | 6/1152 | 0.385993 | 0.999971 | 0 | 0 | 1.213044 | 1.154741 |
| GTEx_Tissu | GTEX-RN64 | 6/1128 | 0.367099 | 0.999971 | 0 | 0 | 1.240576 | 1.243211 |
| GTEx_Tissu | GTEX-NPJ8 | 6/1131 | 0.369458 | 0.999971 | 0 | 0 | 1.23707 | 1.231773 |
| GTEx_Tissu | GTEX-PVO\ | 13/2645 | 0.363228 | 0.999971 | 0 | 0 | 1.153439 | 1.168116 |
| GTEx_Tissu | GTEX-XOT4 | 5/919 | 0.370634 | 0.999971 | 0 | 0 | 1.267479 | 1.258022 |
| GTEx_Tissu | GTEX-WZT( | 7/1348 | 0.370914 | 0.999971 | 0 | 0 | 1.21182 | 1.201863 |
| GTEx_Tissu | GTEX-OXRI | 3/500 | 0.371249 | 0.999971 | 0 | 0 | 1.395229 | 1.382508 |
| GTEx_Tissu | GTEX-WYV. | 9/1783 | 0.37161 | 0.999971 | 0 | 0 | 1.179798 | 1.167893 |
| GTEx_Tissu | GTEX-T6MI | 4/709 | 0.371956 | 0.999971 | 0 | 0 | 1.313031 | 1.298561 |
| GTEx_Tissu | GTEX-TSE9- | 4/709 | 0.371956 | 0.999971 | 0 | 0 | 1.313031 | 1.298561 |
| GTEx_Tissu | GTEX-OXR( | 5/922 | 0.37325 | 0.999971 | 0 | 0 | 1.263133 | 1.244825 |
| GTEx_Tissu | GTEX-QLQ\ | 25/5375 | 0.386629 | 0.999971 | 0 | 0 | 1.097603 | 1.043042 |
| GTEx_Tissu | GTEX-RM2I | Nov-70 | 0.342802 | 0.999971 | 0 | 0 | 1.190207 | 1.27424 |

| | | | | | | | | |
|---|---|---|---|---|---|---|---|---|
| GTEx_Tissu | GTEX-RNOI | 7/1309 | 0.342825 | 0.999971 | 0 | 0 | 1.250739 | 1.338961 |
| GTEx_Tissu | GTEX-SJXC- | 8/1523 | 0.343142 | 0.999971 | 0 | 0 | 1.229761 | 1.315366 |
| GTEx_Tissu | GTEX-RU72 | 8/1526 | 0.345143 | 0.999971 | 0 | 0 | 1.227131 | 1.305419 |
| GTEx_Tissu | GTEX-OIZI- | 24/5049 | 0.34563 | 0.999971 | 0 | 0 | 1.12868 | 1.199095 |
| GTEx_Tissu | GTEX-TML8 | 8/1527 | 0.34581 | 0.999971 | 0 | 0 | 1.226256 | 1.30212 |
| GTEx_Tissu | GTEX-S32W | 3/479 | 0.34625 | 0.999971 | 0 | 0 | 1.458358 | 1.546729 |
| GTEx_Tissu | GTEX-S32W | 7/1346 | 0.369468 | 0.999971 | 0 | 0 | 1.21376 | 1.20853 |
| GTEx_Tissu | GTEX-V1D1 | 25/5396 | 0.395194 | 0.999971 | 0 | 0 | 1.091735 | 1.013543 |
| GTEx_Tissu | GTEX-XUYS | 5/952 | 0.399408 | 0.999971 | 0 | 0 | 1.221186 | 1.120772 |
| GTEx_Tissu | GTEX-W5X | 5/941 | 0.38982 | 0.999971 | 0 | 0 | 1.236254 | 1.164638 |
| GTEx_Tissu | GTEX-OHPI | 25/5384 | 0.390295 | 0.999971 | 0 | 0 | 1.095083 | 1.030312 |
| GTEx_Tissu | GTEX-WH7 | 5/967 | 0.412463 | 0.999971 | 0 | 0 | 1.201194 | 1.063789 |
| GTEx_Tissu | GTEX-RWS. | 4/738 | 0.40072 | 0.999971 | 0 | 0 | 1.25925 | 1.151574 |
| GTEx_Tissu | GTEX-OOB. | 4/739 | 0.401709 | 0.999971 | 0 | 0 | 1.257471 | 1.146848 |
| GTEx_Tissu | GTEX-WFG | 8/1610 | 0.401715 | 0.999971 | 0 | 0 | 1.157477 | 1.055634 |
| GTEx_Tissu | GTEX-Q734 | 6/1172 | 0.401754 | 0.999971 | 0 | 0 | 1.190966 | 1.08606 |
| GTEx_Tissu | GTEX-SNOS | 5/955 | 0.402021 | 0.999971 | 0 | 0 | 1.217137 | 1.109119 |
| GTEx_Tissu | GTEX-V1D1 | 7/1384 | 0.397006 | 0.999971 | 0 | 0 | 1.17785 | 1.088104 |
| GTEx_Tissu | GTEX-V955 | 6/1164 | 0.39545 | 0.999971 | 0 | 0 | 1.199706 | 1.113005 |
| GTEx_Tissu | GTEX-U4B1 | 9/1808 | 0.387494 | 0.999971 | 0 | 0 | 1.161799 | 1.101451 |
| GTEx_Tissu | GTEX-UPK5 | 7/1391 | 0.402087 | 0.999971 | 0 | 0 | 1.171451 | 1.067294 |
| GTEx_Tissu | GTEX-S95S- | 24/5187 | 0.402101 | 0.999971 | 0 | 0 | 1.08833 | 0.991525 |
| GTEx_Tissu | GTEX-SUCS | 4/733 | 0.395771 | 0.999971 | 0 | 0 | 1.268217 | 1.175534 |
| GTEx_Tissu | GTEX-WFJC | 3/503 | 0.374809 | 0.999971 | 0 | 0 | 1.386643 | 1.360766 |
| GTEx_Tissu | GTEX-R53T | 4/713 | 0.37593 | 0.999971 | 0 | 0 | 1.305351 | 1.277092 |
| GTEx_Tissu | GTEX-POYV | 8/1607 | 0.399683 | 0.999971 | 0 | 0 | 1.159839 | 1.063669 |
| GTEx_Tissu | GTEX-S7SE- | 4/700 | 0.36301 | 0.999971 | 0 | 0 | 1.330633 | 1.348362 |
| GTEx_Tissu | GTEX-PVOV | 6/1153 | 0.386781 | 0.999971 | 0 | 0 | 1.211922 | 1.151202 |
| GTEx_Tissu | GTEX-TML8 | 8/1588 | 0.386825 | 0.999971 | 0 | 0 | 1.175004 | 1.115998 |
| GTEx_Tissu | GTEX-XXEK | 7/1370 | 0.386848 | 0.999971 | 0 | 0 | 1.190847 | 1.130974 |
| GTEx_Tissu | GTEX-PLZ4- | 5/932 | 0.381972 | 0.999971 | 0 | 0 | 1.248849 | 1.201904 |
| GTEx_Tissu | GTEX-UJHI- | 4/697 | 0.360028 | 0.999971 | 0 | 0 | 1.336602 | 1.365437 |
| GTEx_Tissu | GTEX-NPJ7 | 5/907 | 0.360177 | 0.999971 | 0 | 0 | 1.285152 | 1.312347 |
| GTEx_Tissu | GTEX-X4EP | 3/492 | 0.361739 | 0.999971 | 0 | 0 | 1.418639 | 1.442516 |
| GTEx_Tissu | GTEX-TKQ1 | 5/909 | 0.361919 | 0.999971 | 0 | 0 | 1.282174 | 1.303119 |
| GTEx_Tissu | GTEX-WFO | 3/428 | 0.285466 | 0.999971 | 0 | 0 | 1.637647 | 2.053005 |
| GTEx_Tissu | GTEX-OHPI | 4/700 | 0.36301 | 0.999971 | 0 | 0 | 1.330633 | 1.348362 |
| GTEx_Tissu | GTEX-OXRI | 4/700 | 0.36301 | 0.999971 | 0 | 0 | 1.330633 | 1.348362 |
| GTEx_Tissu | GTEX-T6M( | 5/935 | 0.384588 | 0.999971 | 0 | 0 | 1.244624 | 1.189341 |
| GTEx_Tissu | GTEX-UPJH | 26/5545 | 0.364467 | 0.999971 | 0 | 0 | 1.111641 | 1.122 |
| GTEx_Tissu | GTEX-PWO | 9/1772 | 0.364646 | 0.999971 | 0 | 0 | 1.187879 | 1.198366 |
| GTEx_Tissu | GTEX-WOF | 7/1383 | 0.39628 | 0.999971 | 0 | 0 | 1.17877 | 1.09111 |
| GTEx_Tissu | GTEX-OHPI | Oct-43 | 0.396694 | 0.999971 | 0 | 0 | 1.142193 | 1.056059 |
| GTEx_Tissu | GTEX-VUS( | 4/734 | 0.396761 | 0.999971 | 0 | 0 | 1.266414 | 1.170698 |
| GTEx_Tissu | GTEX-S7PN | 3/352 | 0.197685 | 0.999971 | 0 | 0 | 2.002047 | 3.245481 |

| | | | | | | | | |
|---|---|---|---|---|---|---|---|---|
| GTEx_Tissu | GTEX-UJHI- | 5/704 | 0.192036 | 0.999971 | 0 | 0 | 1.676088 | 2.765662 |
| GTEx_Tissu | GTEX-P4QT | 5/704 | 0.192036 | 0.999971 | 0 | 0 | 1.676088 | 2.765662 |
| GTEx_Tissu | GTEX-WOF | 6/892 | 0.192114 | 0.999971 | 0 | 0 | 1.590753 | 2.624208 |
| GTEx_Tissu | GTEX-P4QT | 7/1078 | 0.188508 | 0.999971 | 0 | 0 | 1.539379 | 2.568635 |
| GTEx_Tissu | GTEX-Q2AI | 27/5281 | 0.19355 | 0.999971 | 0 | 0 | 1.255529 | 2.061855 |
| GTEx_Tissu | GTEX-T6M( | 7/1088 | 0.194514 | 0.999971 | 0 | 0 | 1.524329 | 2.495707 |
| GTEx_Tissu | GTEX-N7M | 6/899 | 0.19679 | 0.999971 | 0 | 0 | 1.577703 | 2.564743 |
| GTEx_Tissu | GTEX-R55F | 10/1672 | 0.189401 | 0.999971 | 0 | 0 | 1.426149 | 2.37295 |
| GTEx_Tissu | GTEX-S7SE- | 7/1093 | 0.197547 | 0.999971 | 0 | 0 | 1.516908 | 2.460088 |
| GTEx_Tissu | GTEX-OHPI | 4/516 | 0.187146 | 0.999971 | 0 | 0 | 1.826148 | 3.060381 |
| GTEx_Tissu | GTEX-QV44 | 8/1290 | 0.198061 | 0.999971 | 0 | 0 | 1.471672 | 2.382899 |
| GTEx_Tissu | GTEX-UPIC | 5/703 | 0.191285 | 0.999971 | 0 | 0 | 1.678576 | 2.77635 |
| GTEx_Tissu | GTEX-RVPV | 4/530 | 0.199522 | 0.999971 | 0 | 0 | 1.776261 | 2.863033 |
| GTEx_Tissu | GTEX-T5JC- | 4/531 | 0.200416 | 0.999971 | 0 | 0 | 1.772799 | 2.84953 |
| GTEx_Tissu | GTEX-WVLI | 4/531 | 0.200416 | 0.999971 | 0 | 0 | 1.772799 | 2.84953 |
| GTEx_Tissu | GTEX-QCQ( | 9/1477 | 0.191945 | 0.999971 | 0 | 0 | 1.449775 | 2.392917 |
| GTEx_Tissu | GTEX-S33H | 8/1241 | 0.171438 | 0.999971 | 0 | 0 | 1.534181 | 2.705579 |
| GTEx_Tissu | GTEX-S7SF- | 7/1049 | 0.171549 | 0.999971 | 0 | 0 | 1.584657 | 2.793571 |
| GTEx_Tissu | GTEX-U3ZN | 11/1822 | 0.16648 | 0.999971 | 0 | 0 | 1.446729 | 2.59381 |
| GTEx_Tissu | GTEX-O5YT | 5/694 | 0.184571 | 0.999971 | 0 | 0 | 1.701299 | 2.874717 |
| GTEx_Tissu | GTEX-VUS( | 22/4173 | 0.186439 | 0.999971 | 0 | 0 | 1.285192 | 2.158674 |
| GTEx_Tissu | GTEX-TKQ2 | 5/697 | 0.186799 | 0.999971 | 0 | 0 | 1.693659 | 2.841486 |
| GTEx_Tissu | GTEX-SE5C | 5/697 | 0.186799 | 0.999971 | 0 | 0 | 1.693659 | 2.841486 |
| GTEx_Tissu | GTEX-VJYA | 8/1270 | 0.186987 | 0.999971 | 0 | 0 | 1.4966 | 2.50937 |
| GTEx_Tissu | GTEX-SJXC- | 8/1256 | 0.179404 | 0.999971 | 0 | 0 | 1.514525 | 2.602128 |
| GTEx_Tissu | GTEX-WYV | 5/691 | 0.182354 | 0.999971 | 0 | 0 | 1.709006 | 2.908397 |
| GTEx_Tissu | GTEX-SNOS | 5/666 | 0.164285 | 0.999971 | 0 | 0 | 1.775949 | 3.207638 |
| GTEx_Tissu | GTEX-X5EB | 9/1425 | 0.166043 | 0.999971 | 0 | 0 | 1.507252 | 2.706284 |
| GTEx_Tissu | GTEX-U3ZN | 6/861 | 0.171961 | 0.999971 | 0 | 0 | 1.651115 | 2.906771 |
| GTEx_Tissu | GTEX-P4QS | 10/1623 | 0.166545 | 0.999971 | 0 | 0 | 1.473418 | 2.641088 |
| GTEx_Tissu | GTEX-U3ZN | 4/493 | 0.167405 | 0.999971 | 0 | 0 | 1.914308 | 3.421514 |
| GTEx_Tissu | GTEX-U3ZN | 6/854 | 0.16754 | 0.999971 | 0 | 0 | 1.665356 | 2.97521 |
| GTEx_Tissu | GTEX-R55E | 5/673 | 0.169268 | 0.999971 | 0 | 0 | 1.7567 | 3.120378 |
| GTEx_Tissu | GTEX-PWO | 6/846 | 0.16255 | 0.999971 | 0 | 0 | 1.681922 | 3.055663 |
| GTEx_Tissu | GTEX-OOBI | 4/521 | 0.191536 | 0.999971 | 0 | 0 | 1.808021 | 2.988077 |
| GTEx_Tissu | GTEX-R53T | 13/2289 | 0.191916 | 0.999971 | 0 | 0 | 1.361332 | 2.247144 |
| GTEx_Tissu | GTEX-PWN | 8/1279 | 0.191936 | 0.999971 | 0 | 0 | 1.485285 | 2.451599 |
| GTEx_Tissu | GTEX-XOT( | 6/918 | 0.209697 | 0.999971 | 0 | 0 | 1.543291 | 2.410759 |
| GTEx_Tissu | GTEX-WH7 | 20/3823 | 0.213263 | 0.999971 | 0 | 0 | 1.264516 | 1.953966 |
| GTEx_Tissu | GTEX-QDT8 | 6/913 | 0.206271 | 0.999971 | 0 | 0 | 1.552207 | 2.45026 |
| GTEx_Tissu | GTEX-SNM | 6/925 | 0.21453 | 0.999971 | 0 | 0 | 1.530972 | 2.356636 |
| GTEx_Tissu | GTEX-XBED | 20/3827 | 0.214683 | 0.999971 | 0 | 0 | 1.262874 | 1.943048 |
| GTEx_Tissu | GTEX-Q2AH | 27/5351 | 0.214916 | 0.999971 | 0 | 0 | 1.233105 | 1.89591 |
| GTEx_Tissu | GTEX-SNOS | 7/1109 | 0.20738 | 0.999971 | 0 | 0 | 1.493614 | 2.349756 |
| GTEx_Tissu | GTEX-S3XE | 27/5355 | 0.216173 | 0.999971 | 0 | 0 | 1.231841 | 1.886783 |

| | | | | | | | | |
|---|---|---|---|---|---|---|---|---|
| GTEx_Tissu | GTEX-X4EC | 5/736 | 0.216628 | 0.999971 | 0 | 0 | 1.600047 | 2.447387 |
| GTEx_Tissu | GTEX-P4PP | 7/1126 | 0.218033 | 0.999971 | 0 | 0 | 1.469593 | 2.238351 |
| GTEx_Tissu | GTEX-U3ZN | 4/545 | 0.213054 | 0.999971 | 0 | 0 | 1.725675 | 2.668255 |
| GTEx_Tissu | GTEX-WY7( | 7/1076 | 0.187316 | 0.999971 | 0 | 0 | 1.542423 | 2.583496 |
| GTEx_Tissu | GTEX-NPJ7 | 7/1076 | 0.187316 | 0.999971 | 0 | 0 | 1.542423 | 2.583496 |
| GTEx_Tissu | GTEX-RWS. | 6/885 | 0.187484 | 0.999971 | 0 | 0 | 1.604011 | 2.685216 |
| GTEx_Tissu | GTEX-XBEC | 7/1116 | 0.211742 | 0.999971 | 0 | 0 | 1.483634 | 2.303177 |
| GTEx_Tissu | GTEX-V955 | 20/3751 | 0.188529 | 0.999971 | 0 | 0 | 1.294679 | 2.160174 |
| GTEx_Tissu | GTEX-XUYS | 5/700 | 0.189037 | 0.999971 | 0 | 0 | 1.686085 | 2.8087 |
| GTEx_Tissu | GTEX-XMD | 7/1079 | 0.189105 | 0.999971 | 0 | 0 | 1.537861 | 2.561239 |
| GTEx_Tissu | GTEX-P78B | 9/1519 | 0.214105 | 0.999971 | 0 | 0 | 1.40624 | 2.167426 |
| GTEx_Tissu | GTEX-WFG | 4/539 | 0.207609 | 0.999971 | 0 | 0 | 1.745569 | 2.744206 |
| GTEx_Tissu | GTEX-N7M | 11/1882 | 0.193079 | 0.999971 | 0 | 0 | 1.395693 | 2.295437 |
| GTEx_Tissu | GTEX-XQ3S | 5/678 | 0.172864 | 0.999971 | 0 | 0 | 1.743196 | 3.059742 |
| GTEx_Tissu | GTEX-WH7 | 5/680 | 0.174311 | 0.999971 | 0 | 0 | 1.73785 | 3.035873 |
| GTEx_Tissu | GTEX-SNOS | 28/5433 | 0.174489 | 0.999971 | 0 | 0 | 1.273849 | 2.224007 |
| GTEx_Tissu | GTEX-OOBI | 6/900 | 0.197461 | 0.999971 | 0 | 0 | 1.575855 | 2.556371 |
| GTEx_Tissu | GTEX-RN64 | 9/1445 | 0.175795 | 0.999971 | 0 | 0 | 1.484653 | 2.580973 |
| GTEx_Tissu | GTEX-S341 | 6/868 | 0.17643 | 0.999971 | 0 | 0 | 1.637106 | 2.8401 |
| GTEx_Tissu | GTEX-V955 | 8/1251 | 0.17673 | 0.999971 | 0 | 0 | 1.521024 | 2.636135 |
| GTEx_Tissu | GTEX-OHPI | 4/529 | 0.198629 | 0.999971 | 0 | 0 | 1.779736 | 2.876612 |
| GTEx_Tissu | GTEX-S33H | 8/1227 | 0.164157 | 0.999971 | 0 | 0 | 1.552964 | 2.806105 |
| GTEx_Tissu | GTEX-XOT( | 7/1110 | 0.208001 | 0.999971 | 0 | 0 | 1.49218 | 2.34304 |
| GTEx_Tissu | GTEX-NL4V | 4/541 | 0.209419 | 0.999971 | 0 | 0 | 1.738889 | 2.718606 |
| GTEx_Tissu | GTEX-TSE9 | 5/716 | 0.201137 | 0.999971 | 0 | 0 | 1.64677 | 2.641039 |
| GTEx_Tissu | GTEX-WH7 | 6/920 | 0.211074 | 0.999971 | 0 | 0 | 1.539752 | 2.395156 |
| GTEx_Tissu | GTEX-S95S | 3/364 | 0.211116 | 0.999971 | 0 | 0 | 1.934309 | 3.008525 |
| GTEx_Tissu | GTEX-RNOI | 8/1314 | 0.211707 | 0.999971 | 0 | 0 | 1.442767 | 2.239968 |
| GTEx_Tissu | GTEX-U3ZH | 21/4012 | 0.204173 | 0.999971 | 0 | 0 | 1.269379 | 2.016772 |
| GTEx_Tissu | GTEX-UPIC | 5/731 | 0.212719 | 0.999971 | 0 | 0 | 1.611486 | 2.49423 |
| GTEx_Tissu | GTEX-X3Y1 | 2/1496 | 0.990822 | 0.999971 | 0 | 0 | 0.290086 | 0.002675 |
| GTEx_Tissu | GTEX-XAJ8 | Apr-82 | 0.99621 | 0.999971 | 0 | 0 | 0.33908 | 0.001288 |
| GTEx_Tissu | GTEX-XLM4 | Apr-76 | 0.999833 | 0.999971 | 0 | 0 | 0.245103 | 4.09E-05 |
| GTEx_Tissu | GTEX-NPJ8 | Jun-64 | 0.996227 | 0.999971 | 0 | 0 | 0.393005 | 0.001486 |
| GTEx_Tissu | GTEX-P44H | Sep-10 | 0.996334 | 0.999971 | 0 | 0 | 0.444882 | 0.001634 |
| GTEx_Tissu | GTEX-OHPI | 2/1719 | 0.996354 | 0.999971 | 0 | 0 | 0.249354 | 0.000911 |
| GTEx_Tissu | GTEX-RNOI | Mar-26 | 0.996403 | 0.999971 | 0 | 0 | 0.299273 | 0.001079 |
| GTEx_Tissu | GTEX-S7SE | Mar-31 | 0.996472 | 0.999971 | 0 | 0 | 0.298486 | 0.001055 |
| GTEx_Tissu | GTEX-U8XE | 1/1051 | 0.990964 | 0.999971 | 0 | 0 | 0.208893 | 0.001896 |
| GTEx_Tissu | GTEX-T5JC | Aug-18 | 0.990902 | 0.999971 | 0 | 0 | 0.473237 | 0.004325 |
| GTEx_Tissu | GTEX-TKQ1 | 2/1519 | 0.991645 | 0.999971 | 0 | 0 | 0.285331 | 0.002394 |
| GTEx_Tissu | GTEX-N7M | May-69 | 0.990624 | 0.999971 | 0 | 0 | 0.412584 | 0.003887 |
| GTEx_Tissu | GTEX-N7M | 2/1475 | 0.990003 | 0.999971 | 0 | 0 | 0.294557 | 0.002959 |
| GTEx_Tissu | GTEX-NL3H | Apr-15 | 0.993017 | 0.999971 | 0 | 0 | 0.367066 | 0.002572 |
| GTEx_Tissu | GTEX-RVPV | Jun-08 | 0.998888 | 0.999971 | 0 | 0 | 0.347124 | 0.000386 |

| | | | | | | | | |
|---|---|---|---|---|---|---|---|---|
| GTEx_Tissu | GTEX-X4XX | Jul-09 | 0.998764 | 0.999971 | 0 | 0 | 0.370782 | 0.000459 |
| GTEx_Tissu | GTEX-X4XX | Jul-60 | 0.997917 | 0.999971 | 0 | 0 | 0.389474 | 0.000812 |
| GTEx_Tissu | GTEX-S7SE | Jun-59 | 0.998671 | 0.999971 | 0 | 0 | 0.353101 | 0.00047 |
| GTEx_Tissu | GTEX-X261 | May-13 | 0.998652 | 0.999971 | 0 | 0 | 0.329696 | 0.000445 |
| GTEx_Tissu | GTEX-U4B1 | Feb-96 | 0.999276 | 0.999971 | 0 | 0 | 0.200225 | 0.000145 |
| GTEx_Tissu | GTEX-OXRI | Apr-14 | 0.999517 | 0.999971 | 0 | 0 | 0.270632 | 0.000131 |
| GTEx_Tissu | GTEX-XMD | Apr-95 | 0.999652 | 0.999971 | 0 | 0 | 0.262277 | 9.14E-05 |
| GTEx_Tissu | GTEX-PWO | 3/327 | 0.170462 | 0.999971 | 0 | 0 | 2.159281 | 3.820295 |
| GTEx_Tissu | GTEX-QEL4 | 7/1047 | 0.170405 | 0.999971 | 0 | 0 | 1.587873 | 2.809859 |
| GTEx_Tissu | GTEX-QCQ | 5/674 | 0.169985 | 0.999971 | 0 | 0 | 1.753983 | 3.108139 |
| GTEx_Tissu | GTEX-WVLI | May-98 | 0.999021 | 0.999971 | 0 | 0 | 0.319296 | 0.000313 |
| GTEx_Tissu | GTEX-WL4 | May-16 | 0.999086 | 0.999971 | 0 | 0 | 0.317164 | 0.00029 |
| GTEx_Tissu | GTEX-TKQ1 | 1/1562 | 0.999164 | 0.999971 | 0 | 0 | 0.136704 | 0.000114 |
| GTEx_Tissu | GTEX-UTH | Feb-95 | 0.999273 | 0.999971 | 0 | 0 | 0.200332 | 0.000146 |
| GTEx_Tissu | GTEX-Q2A | May-11 | 0.998642 | 0.999971 | 0 | 0 | 0.329948 | 0.000448 |
| GTEx_Tissu | GTEX-XQ3 | 5/673 | 0.169268 | 0.999971 | 0 | 0 | 1.7567 | 3.120378 |
| GTEx_Tissu | GTEX-S4Q7 | 8/1232 | 0.16674 | 0.999971 | 0 | 0 | 1.546207 | 2.769751 |
| GTEx_Tissu | GTEX-WK1 | 2/1736 | 0.996606 | 0.999971 | 0 | 0 | 0.246679 | 0.000839 |
| GTEx_Tissu | GTEX-X4XY | 1/1272 | 0.99675 | 0.999971 | 0 | 0 | 0.170549 | 0.000555 |
| GTEx_Tissu | GTEX-REY6 | Apr-69 | 0.991765 | 0.999971 | 0 | 0 | 0.375499 | 0.003105 |
| GTEx_Tissu | GTEX-X585 | Apr-92 | 0.999954 | 0.999971 | 0 | 0 | 0.219272 | 1.01E-05 |
| GTEx_Tissu | GTEX-NPJ8 | Mar-92 | 0.999971 | 0.999971 | 0 | 0 | 0.180515 | 5.25E-06 |
| GTEx_Tissu | GTEX-P44H | Feb-23 | 0.999005 | 0.999971 | 0 | 0 | 0.208307 | 0.000207 |
| GTEx_Tissu | GTEX-XOT4 | 5/678 | 0.172864 | 0.999971 | 0 | 0 | 1.743196 | 3.059742 |
| GTEx_Tissu | GTEX-UTH | Jun-34 | 0.998545 | 0.999971 | 0 | 0 | 0.356217 | 0.000519 |
| GTEx_Tissu | GTEX-QDVI | Apr-28 | 0.986476 | 0.999971 | 0 | 0 | 0.403626 | 0.005496 |
| GTEx_Tissu | GTEX-XQ8I | 9/1447 | 0.176785 | 0.999971 | 0 | 0 | 1.482428 | 2.56878 |
| GTEx_Tissu | GTEX-OIZG | Dec-50 | 0.177598 | 0.999971 | 0 | 0 | 1.403337 | 2.425293 |
| GTEx_Tissu | GTEX-UPK5 | 5/686 | 0.178681 | 0.999971 | 0 | 0 | 1.722001 | 2.965554 |
| GTEx_Tissu | GTEX-Q2AI | 5/675 | 0.170703 | 0.999971 | 0 | 0 | 1.751274 | 3.095957 |
| GTEx_Tissu | GTEX-P4QS | 27/5235 | 0.180179 | 0.999971 | 0 | 0 | 1.270593 | 2.17755 |
| GTEx_Tissu | GTEX-WFG | 6/874 | 0.1803 | 0.999971 | 0 | 0 | 1.625277 | 2.784319 |
| GTEx_Tissu | GTEX-TMM | 9/1457 | 0.181774 | 0.999971 | 0 | 0 | 1.471393 | 2.50871 |
| GTEx_Tissu | GTEX-PLZ4 | Apr-42 | 0.996971 | 0.999971 | 0 | 0 | 0.329925 | 0.001001 |
| GTEx_Tissu | GTEX-XMK | 21/3947 | 0.182763 | 0.999971 | 0 | 0 | 1.295663 | 2.202065 |
| GTEx_Tissu | GTEX-XXEK | 9/1459 | 0.18278 | 0.999971 | 0 | 0 | 1.469204 | 2.496873 |
| GTEx_Tissu | GTEX-RWS | 20/3734 | 0.182924 | 0.999971 | 0 | 0 | 1.301972 | 2.21164 |
| GTEx_Tissu | GTEX-TSE9 | 8/1248 | 0.175135 | 0.999971 | 0 | 0 | 1.524949 | 2.656766 |
| GTEx_Tissu | GTEX-NFK9 | 9/1463 | 0.184799 | 0.999971 | 0 | 0 | 1.464845 | 2.473373 |
| GTEx_Tissu | GTEX-OXRI | 5/695 | 0.185313 | 0.999971 | 0 | 0 | 1.698745 | 2.86359 |
| GTEx_Tissu | GTEX-XQ3 | 6/882 | 0.185513 | 0.999971 | 0 | 0 | 1.609758 | 2.711847 |
| GTEx_Tissu | GTEX-NPJ8 | 4/504 | 0.17675 | 0.999971 | 0 | 0 | 1.871133 | 3.242707 |
| GTEx_Tissu | GTEX-XOT | May-10 | 0.998025 | 0.999971 | 0 | 0 | 0.343087 | 0.000678 |
| GTEx_Tissu | GTEX-SE5C | Mar-59 | 0.99858 | 0.999971 | 0 | 0 | 0.266144 | 0.000378 |
| GTEx_Tissu | GTEX-RU72 | Jun-38 | 0.998565 | 0.999971 | 0 | 0 | 0.355715 | 0.000511 |

| | | | | | | | | |
|---|---|---|---|---|---|---|---|---|
| GTEx_Tissu | GTEX-X62C | 1/1075 | 0.991909 | 0.999971 | 0 | 0 | 0.203965 | 0.001657 |
| GTEx_Tissu | GTEX-XMD | Jun-17 | 0.998452 | 0.999971 | 0 | 0 | 0.358361 | 0.000555 |
| GTEx_Tissu | GTEX-Q2A( | Jul-37 | 0.998407 | 0.999971 | 0 | 0 | 0.379628 | 0.000605 |
| GTEx_Tissu | GTEX-N7M | Jun-62 | 0.998115 | 0.999971 | 0 | 0 | 0.365448 | 0.00069 |
| GTEx_Tissu | GTEX-XBEV | 1/1386 | 0.99809 | 0.999971 | 0 | 0 | 0.155554 | 0.000297 |
| GTEx_Tissu | GTEX-N7M | Jun-49 | 0.998026 | 0.999971 | 0 | 0 | 0.367157 | 0.000726 |
| GTEx_Tissu | GTEX-SIU8- | 6/888 | 0.189463 | 0.999971 | 0 | 0 | 1.598304 | 2.658878 |
| GTEx_Tissu | GTEX-UPIC | Mar-77 | 0.998024 | 0.999971 | 0 | 0 | 0.277029 | 0.000548 |
| GTEx_Tissu | GTEX-RU72 | Feb-03 | 0.998915 | 0.999971 | 0 | 0 | 0.210624 | 0.000229 |
| GTEx_Tissu | GTEX-XOT( | Apr-24 | 0.997778 | 0.999971 | 0 | 0 | 0.318091 | 0.000708 |
| GTEx_Tissu | GTEX-R55F | Jun-85 | 0.997523 | 0.999971 | 0 | 0 | 0.375769 | 0.000932 |
| GTEx_Tissu | GTEX-XLM4 | May-30 | 0.997354 | 0.999971 | 0 | 0 | 0.354138 | 0.000938 |
| GTEx_Tissu | GTEX-WVLI | May-25 | 0.997306 | 0.999971 | 0 | 0 | 0.354849 | 0.000957 |
| GTEx_Tissu | GTEX-OXRI | 1/1307 | 0.997238 | 0.999971 | 0 | 0 | 0.165667 | 0.000458 |
| GTEx_Tissu | GTEX-QVU! | Aug-78 | 0.997108 | 0.999971 | 0 | 0 | 0.419795 | 0.001216 |
| GTEx_Tissu | GTEX-OHPI | 6/1002 | 0.27004 | 0.999971 | 0 | 0 | 1.406887 | 1.841877 |
| GTEx_Tissu | GTEX-S95S- | 7/1190 | 0.259834 | 0.999971 | 0 | 0 | 1.385355 | 1.867062 |
| GTEx_Tissu | GTEX-WWY | 6/956 | 0.236389 | 0.999971 | 0 | 0 | 1.478596 | 2.132546 |
| GTEx_Tissu | GTEX-OOBI | 4/572 | 0.238037 | 0.999971 | 0 | 0 | 1.641354 | 2.355886 |
| GTEx_Tissu | GTEX-SIU8- | 15/2818 | 0.238387 | 0.999971 | 0 | 0 | 1.271703 | 1.823443 |
| GTEx_Tissu | GTEX-RU72 | 7/1192 | 0.261179 | 0.999971 | 0 | 0 | 1.382869 | 1.856572 |
| GTEx_Tissu | GTEX-TSE9- | 9/1628 | 0.276006 | 0.999971 | 0 | 0 | 1.303796 | 1.678421 |
| GTEx_Tissu | GTEX-RU72 | 6/1010 | 0.276019 | 0.999971 | 0 | 0 | 1.395086 | 1.795874 |
| GTEx_Tissu | GTEX-Q2A( | 7/1214 | 0.276107 | 0.999971 | 0 | 0 | 1.356069 | 1.745215 |
| GTEx_Tissu | GTEX-WHS | 9/1660 | 0.29517 | 0.999971 | 0 | 0 | 1.276289 | 1.557334 |
| GTEx_Tissu | GTEX-W5X | 6/955 | 0.235673 | 0.999971 | 0 | 0 | 1.480233 | 2.139398 |
| GTEx_Tissu | GTEX-R55E | 27/5520 | 0.271208 | 0.999971 | 0 | 0 | 1.181322 | 1.541472 |
| GTEx_Tissu | GTEX-SIU7- | 4/607 | 0.271394 | 0.999971 | 0 | 0 | 1.543288 | 2.01273 |
| GTEx_Tissu | GTEX-S4UY | 5/804 | 0.27184 | 0.999971 | 0 | 0 | 1.458683 | 1.899997 |
| GTEx_Tissu | GTEX-W5X | 20/3999 | 0.279887 | 0.999971 | 0 | 0 | 1.19538 | 1.522162 |
| GTEx_Tissu | GTEX-U8XE | 5/815 | 0.281069 | 0.999971 | 0 | 0 | 1.438046 | 1.825104 |
| GTEx_Tissu | GTEX-U3ZH | 8/1428 | 0.281091 | 0.999971 | 0 | 0 | 1.318809 | 1.673672 |
| GTEx_Tissu | GTEX-XV7C | 5/793 | 0.262682 | 0.999971 | 0 | 0 | 1.479897 | 1.978341 |
| GTEx_Tissu | GTEX-S7PM | 5/806 | 0.273513 | 0.999971 | 0 | 0 | 1.454889 | 1.886128 |
| GTEx_Tissu | GTEX-OXR( | 5/792 | 0.261853 | 0.999971 | 0 | 0 | 1.481855 | 1.985641 |
| GTEx_Tissu | GTEX-U3ZN | 27/5485 | 0.259039 | 0.999971 | 0 | 0 | 1.191783 | 1.609833 |
| GTEx_Tissu | GTEX-U3ZN | 18/3512 | 0.25917 | 0.999971 | 0 | 0 | 1.225878 | 1.655266 |
| GTEx_Tissu | GTEX-X88G | 5/789 | 0.259371 | 0.999971 | 0 | 0 | 1.487758 | 2.007722 |
| GTEx_Tissu | GTEX-PW2( | 25/5025 | 0.252793 | 0.999971 | 0 | 0 | 1.202661 | 1.653879 |
| GTEx_Tissu | GTEX-OHPI | 3/407 | 0.260688 | 0.999971 | 0 | 0 | 1.724629 | 2.318644 |
| GTEx_Tissu | GTEX-X5EB | Dec-31 | 0.261065 | 0.999971 | 0 | 0 | 1.275818 | 1.713408 |
| GTEx_Tissu | GTEX-XPVG | 6/990 | 0.261136 | 0.999971 | 0 | 0 | 1.424947 | 1.913294 |
| GTEx_Tissu | GTEX-T6MI | 8/1388 | 0.255979 | 0.999971 | 0 | 0 | 1.359971 | 1.853178 |
| GTEx_Tissu | GTEX-XAJ8- | 9/1603 | 0.261314 | 0.999971 | 0 | 0 | 1.326054 | 1.779606 |
| GTEx_Tissu | GTEX-NL3H | 5/806 | 0.273513 | 0.999971 | 0 | 0 | 1.454889 | 1.886128 |

| | | | | | | | | |
|---|---|---|---|---|---|---|---|---|
| GTEx_Tissu | GTEX-UPIC | 20/3954 | 0.262092 | 0.999971 | 0 | 0 | 1.212468 | 1.623568 |
| GTEx_Tissu | GTEX-WYB: | 4/593 | 0.257938 | 0.999971 | 0 | 0 | 1.581116 | 2.142469 |
| GTEx_Tissu | GTEX-QEG! | 9/1550 | 0.231108 | 0.999971 | 0 | 0 | 1.37563 | 2.015122 |
| GTEx_Tissu | GTEX-OIZG | 4/565 | 0.231489 | 0.999971 | 0 | 0 | 1.662436 | 2.432513 |
| GTEx_Tissu | GTEX-SNM | 4/565 | 0.231489 | 0.999971 | 0 | 0 | 1.662436 | 2.432513 |
| GTEx_Tissu | GTEX-X638 | 9/1599 | 0.258989 | 0.999971 | 0 | 0 | 1.329681 | 1.79636 |
| GTEx_Tissu | GTEX-PLZ6 | 5/758 | 0.234096 | 0.999971 | 0 | 0 | 1.551518 | 2.252839 |
| GTEx_Tissu | GTEX-S341 | 7/1152 | 0.234709 | 0.999971 | 0 | 0 | 1.434236 | 2.078796 |
| GTEx_Tissu | GTEX-S341 | 4/634 | 0.297683 | 0.999971 | 0 | 0 | 1.475081 | 1.787395 |
| GTEx_Tissu | GTEX-R55C | 5/830 | 0.293757 | 0.999971 | 0 | 0 | 1.410791 | 1.728222 |
| GTEx_Tissu | GTEX-NPJ8 | 3/435 | 0.293777 | 0.999971 | 0 | 0 | 1.610532 | 1.972799 |
| GTEx_Tissu | GTEX-TMM | 6/1055 | 0.310216 | 0.999971 | 0 | 0 | 1.332062 | 1.559159 |
| GTEx_Tissu | GTEX-X638 | 7/1228 | 0.285727 | 0.999971 | 0 | 0 | 1.339517 | 1.678036 |
| GTEx_Tissu | GTEX-V955 | 5/821 | 0.286131 | 0.999971 | 0 | 0 | 1.427024 | 1.785645 |
| GTEx_Tissu | GTEX-S32V | 8/1437 | 0.286837 | 0.999971 | 0 | 0 | 1.309865 | 1.635815 |
| GTEx_Tissu | GTEX-OIZG | 4/624 | 0.287901 | 0.999971 | 0 | 0 | 1.49965 | 1.867272 |
| GTEx_Tissu | GTEX-PWO | 5/833 | 0.296308 | 0.999971 | 0 | 0 | 1.405458 | 1.709539 |
| GTEx_Tissu | GTEX-WZT | 5/833 | 0.296308 | 0.999971 | 0 | 0 | 1.405458 | 1.709539 |
| GTEx_Tissu | GTEX-VUSH | 9/1657 | 0.293357 | 0.999971 | 0 | 0 | 1.278823 | 1.568302 |
| GTEx_Tissu | GTEX-X62C | 18/3565 | 0.281101 | 0.999971 | 0 | 0 | 1.203663 | 1.527496 |
| GTEx_Tissu | GTEX-Q2AI | 8/1441 | 0.289401 | 0.999971 | 0 | 0 | 1.305926 | 1.619273 |
| GTEx_Tissu | GTEX-R55C | 6/1030 | 0.291108 | 0.999971 | 0 | 0 | 1.366392 | 1.686211 |
| GTEx_Tissu | GTEX-XMD | 7/1236 | 0.291263 | 0.999971 | 0 | 0 | 1.330228 | 1.640873 |
| GTEx_Tissu | GTEX-XYKS | 7/1239 | 0.293346 | 0.999971 | 0 | 0 | 1.326776 | 1.627161 |
| GTEx_Tissu | GTEX-T8EM | 8/1409 | 0.269072 | 0.999971 | 0 | 0 | 1.338068 | 1.756585 |
| GTEx_Tissu | GTEX-QDV. | 6/1001 | 0.269295 | 0.999971 | 0 | 0 | 1.408375 | 1.847716 |
| GTEx_Tissu | GTEX-RVPL | 5/801 | 0.269335 | 0.999971 | 0 | 0 | 1.46441 | 1.921013 |
| GTEx_Tissu | GTEX-VJYA | 19/3780 | 0.279781 | 0.999971 | 0 | 0 | 1.199962 | 1.528448 |
| GTEx_Tissu | GTEX-XPVC | 4/611 | 0.275263 | 0.999971 | 0 | 0 | 1.5328 | 1.977357 |
| GTEx_Tissu | GTEX-X88C | 19/3770 | 0.275696 | 0.999971 | 0 | 0 | 1.203906 | 1.551179 |
| GTEx_Tissu | GTEX-T5JC | 7/1175 | 0.249816 | 0.999971 | 0 | 0 | 1.40427 | 1.947765 |
| GTEx_Tissu | GTEX-WHP | 5/778 | 0.250322 | 0.999971 | 0 | 0 | 1.509797 | 2.091081 |
| GTEx_Tissu | GTEX-R55C | 4/609 | 0.273327 | 0.999971 | 0 | 0 | 1.538026 | 1.994952 |
| GTEx_Tissu | GTEX-QDV. | 4/605 | 0.269464 | 0.999971 | 0 | 0 | 1.548584 | 2.030692 |
| GTEx_Tissu | GTEX-QVJC | Nov-54 | 0.278916 | 0.999971 | 0 | 0 | 1.266005 | 1.616492 |
| GTEx_Tissu | GTEX-P4QS | 6/1014 | 0.279021 | 0.999971 | 0 | 0 | 1.389256 | 1.773341 |
| GTEx_Tissu | GTEX-R53T | 8/1425 | 0.279182 | 0.999971 | 0 | 0 | 1.321816 | 1.686491 |
| GTEx_Tissu | GTEX-WY7 | 5/779 | 0.251141 | 0.999971 | 0 | 0 | 1.507768 | 2.083344 |
| GTEx_Tissu | GTEX-P4PP | 10/1793 | 0.251441 | 0.999971 | 0 | 0 | 1.320553 | 1.823085 |
| GTEx_Tissu | GTEX-SIU7 | 19/3711 | 0.252098 | 0.999971 | 0 | 0 | 1.227611 | 1.691571 |
| GTEx_Tissu | GTEX-XOT | 6/978 | 0.252318 | 0.999971 | 0 | 0 | 1.443454 | 1.987732 |
| GTEx_Tissu | GTEX-XXEK | 4/612 | 0.276231 | 0.999971 | 0 | 0 | 1.5302 | 1.968627 |
| GTEx_Tissu | GTEX-OOB. | 4/589 | 0.254119 | 0.999971 | 0 | 0 | 1.592256 | 2.181313 |
| GTEx_Tissu | GTEX-S4Q7 | 26/5253 | 0.255343 | 0.999971 | 0 | 0 | 1.197552 | 1.634836 |
| GTEx_Tissu | GTEX-TMM | 8/1387 | 0.255361 | 0.999971 | 0 | 0 | 1.36103 | 1.857913 |

| | | | | | | | | |
|---|---|---|---|---|---|---|---|---|
| GTEx_Tissu | GTEX-RWS. | 7/1175 | 0.249816 | 0.999971 | 0 | 0 | 1.40427 | 1.947765 |
| GTEx_Tissu | GTEX-XUZC | 6/910 | 0.204225 | 0.999971 | 0 | 0 | 1.557604 | 2.474306 |
| GTEx_Tissu | GTEX-SNM | 4/563 | 0.229627 | 0.999971 | 0 | 0 | 1.668556 | 2.454945 |
| GTEx_Tissu | GTEX-XBEC | 4/563 | 0.229627 | 0.999971 | 0 | 0 | 1.668556 | 2.454945 |
| GTEx_Tissu | GTEX-X4EP | Nov-60 | 0.230402 | 0.999971 | 0 | 0 | 1.334044 | 1.958281 |
| GTEx_Tissu | GTEX-T5JC- | Nov-48 | 0.224473 | 0.999971 | 0 | 0 | 1.343206 | 2.006749 |
| GTEx_Tissu | GTEX-O5YV | 6/906 | 0.201509 | 0.999971 | 0 | 0 | 1.564856 | 2.506775 |
| GTEx_Tissu | GTEX-WQL | 4/534 | 0.203104 | 0.999971 | 0 | 0 | 1.762491 | 2.809477 |
| GTEx_Tissu | GTEX-VJYA | 5/719 | 0.203435 | 0.999971 | 0 | 0 | 1.639595 | 2.610901 |
| GTEx_Tissu | GTEX-OOB. | 7/1141 | 0.227599 | 0.999971 | 0 | 0 | 1.448997 | 2.144761 |
| GTEx_Tissu | GTEX-X8HC | 5/720 | 0.204204 | 0.999971 | 0 | 0 | 1.637216 | 2.600943 |
| GTEx_Tissu | GTEX-OIZI- | 6/938 | 0.223608 | 0.999971 | 0 | 0 | 1.508584 | 2.259651 |
| GTEx_Tissu | GTEX-R53T | 8/1302 | 0.204837 | 0.999971 | 0 | 0 | 1.457085 | 2.310268 |
| GTEx_Tissu | GTEX-S4Z8 | 6/945 | 0.22855 | 0.999971 | 0 | 0 | 1.496785 | 2.209257 |
| GTEx_Tissu | GTEX-R55G | 5/723 | 0.206514 | 0.999971 | 0 | 0 | 1.630121 | 2.57133 |
| GTEx_Tissu | GTEX-R55E | 6/914 | 0.206954 | 0.999971 | 0 | 0 | 1.550416 | 2.442303 |
| GTEx_Tissu | GTEX-QXCL | 5/724 | 0.207286 | 0.999971 | 0 | 0 | 1.627769 | 2.561544 |
| GTEx_Tissu | GTEX-T2IS- | 5/752 | 0.229291 | 0.999971 | 0 | 0 | 1.564469 | 2.304095 |
| GTEx_Tissu | GTEX-TSE9 | 9/1564 | 0.238954 | 0.999971 | 0 | 0 | 1.362206 | 1.94998 |
| GTEx_Tissu | GTEX-QXCL | Nov-79 | 0.239921 | 0.999971 | 0 | 0 | 1.319768 | 1.883897 |
| GTEx_Tissu | GTEX-WY7 | 4/559 | 0.225915 | 0.999971 | 0 | 0 | 1.680929 | 2.500545 |
| GTEx_Tissu | GTEX-T5JC- | 6/890 | 0.190787 | 0.999971 | 0 | 0 | 1.59452 | 2.64148 |
| GTEx_Tissu | GTEX-UTH | 8/1277 | 0.190832 | 0.999971 | 0 | 0 | 1.487786 | 2.464314 |
| GTEx_Tissu | GTEX-RU72 | 5/735 | 0.215845 | 0.999971 | 0 | 0 | 1.602322 | 2.456674 |
| GTEx_Tissu | GTEX-XLM4 | 7/1135 | 0.223754 | 0.999971 | 0 | 0 | 1.45717 | 2.181683 |
| GTEx_Tissu | GTEX-XPVG | 4/557 | 0.224065 | 0.999971 | 0 | 0 | 1.687183 | 2.523721 |
| GTEx_Tissu | GTEX-XUYS | 20/3854 | 0.22439 | 0.999971 | 0 | 0 | 1.251878 | 1.870772 |
| GTEx_Tissu | GTEX-QCQ | Nov-35 | 0.218124 | 0.999971 | 0 | 0 | 1.353259 | 2.060599 |
| GTEx_Tissu | GTEX-RTLS | 3/376 | 0.224746 | 0.999971 | 0 | 0 | 1.870931 | 2.7929 |
| GTEx_Tissu | GTEX-RUSC | 26/5163 | 0.225063 | 0.999971 | 0 | 0 | 1.226001 | 1.828429 |
| GTEx_Tissu | GTEX-T5JC- | 20/3898 | 0.240649 | 0.999971 | 0 | 0 | 1.234288 | 1.758138 |
| GTEx_Tissu | GTEX-WEY! | 5/740 | 0.219772 | 0.999971 | 0 | 0 | 1.591007 | 2.410634 |
| GTEx_Tissu | GTEX-RWS. | 26/5171 | 0.227678 | 0.999971 | 0 | 0 | 1.223432 | 1.810461 |
| GTEx_Tissu | GTEX-XXEK | 3/379 | 0.228181 | 0.999971 | 0 | 0 | 1.855718 | 2.74204 |
| GTEx_Tissu | GTEX-V955 | 3/379 | 0.228181 | 0.999971 | 0 | 0 | 1.855718 | 2.74204 |
| GTEx_Tissu | GTEX-SSA3 | Nov-41 | 0.221044 | 0.999971 | 0 | 0 | 1.348602 | 2.035569 |
| GTEx_Tissu | GTEX-PX3G | 7/1143 | 0.228886 | 0.999971 | 0 | 0 | 1.446292 | 2.132603 |
| GTEx_Tissu | GTEX-OXRI | 6/946 | 0.229259 | 0.999971 | 0 | 0 | 1.495114 | 2.20216 |
| GTEx_Tissu | GTEX-WFG | 8/1344 | 0.229277 | 0.999971 | 0 | 0 | 1.408095 | 2.073875 |
| GTEx_Tissu | GTEX-QMR | 4/556 | 0.223141 | 0.999971 | 0 | 0 | 1.690327 | 2.535405 |
| GTEx_Tissu | GTEX-RWS | 5/739 | 0.218985 | 0.999971 | 0 | 0 | 1.593258 | 2.419763 |
| GTEx_Tissu | GTEX-QDT8 | 6/932 | 0.219401 | 0.999971 | 0 | 0 | 1.518838 | 2.303855 |
| GTEx_Tissu | GTEX-SE5C | 4/578 | 0.243685 | 0.999971 | 0 | 0 | 1.623693 | 2.292461 |
| GTEx_Tissu | GTEX-UPIC | 8/1328 | 0.219838 | 0.999971 | 0 | 0 | 1.42639 | 2.160786 |
| GTEx_Tissu | GTEX-TKQ1 | 6/933 | 0.2201 | 0.999971 | 0 | 0 | 1.51712 | 2.296422 |

| | | | | | | | | |
|---|---|---|---|---|---|---|---|---|
| GTEx_Tissu | GTEX-S4Q7 | 20/3844 | 0.22077 | 0.999971 | 0 | 0 | 1.255933 | 1.897254 |
| GTEx_Tissu | GTEX-Q734 | 4/580 | 0.245574 | 0.999971 | 0 | 0 | 1.617888 | 2.271767 |
| GTEx_Tissu | GTEX-XGQ4 | 6/936 | 0.222202 | 0.999971 | 0 | 0 | 1.511987 | 2.274281 |
| GTEx_Tissu | GTEX-QCQ( | 3/374 | 0.222461 | 0.999971 | 0 | 0 | 1.881209 | 2.827462 |
| GTEx_Tissu | GTEX-P78B | 7/1127 | 0.218666 | 0.999971 | 0 | 0 | 1.468203 | 2.231977 |
| GTEx_Tissu | GTEX-VJYA | 8/1374 | 0.247371 | 0.999971 | 0 | 0 | 1.374947 | 1.920618 |
| GTEx_Tissu | GTEX-T6MI | 9/1594 | 0.256091 | 0.999971 | 0 | 0 | 1.334239 | 1.817528 |
| GTEx_Tissu | GTEX-S95S | 5/786 | 0.256895 | 0.999971 | 0 | 0 | 1.493707 | 2.03008 |
| GTEx_Tissu | GTEX-U3ZH | 4/592 | 0.256982 | 0.999971 | 0 | 0 | 1.583887 | 2.152104 |
| GTEx_Tissu | GTEX-P4QS | 4/584 | 0.249364 | 0.999971 | 0 | 0 | 1.606398 | 2.231033 |
| GTEx_Tissu | GTEX-T5JC | 10/1805 | 0.257973 | 0.999971 | 0 | 0 | 1.310856 | 1.776079 |
| GTEx_Tissu | GTEX-SNOS | 18/3511 | 0.258764 | 0.999971 | 0 | 0 | 1.226304 | 1.657767 |
| GTEx_Tissu | GTEX-RU1J | 6/987 | 0.258924 | 0.999971 | 0 | 0 | 1.429531 | 1.931614 |
| GTEx_Tissu | GTEX-OOBI | 7/1154 | 0.23601 | 0.999971 | 0 | 0 | 1.431582 | 2.067037 |
| GTEx_Tissu | GTEX-X88G | 9/1550 | 0.231108 | 0.999971 | 0 | 0 | 1.37563 | 2.015122 |
| GTEx_Tissu | GTEX-QXCU | 9/1569 | 0.24178 | 0.999971 | 0 | 0 | 1.35747 | 1.927239 |
| GTEx_Tissu | GTEX-WHS | 5/768 | 0.242171 | 0.999971 | 0 | 0 | 1.530384 | 2.170258 |
| GTEx_Tissu | GTEX-OXRK | 7/1164 | 0.242551 | 0.999971 | 0 | 0 | 1.418453 | 2.009298 |
| GTEx_Tissu | GTEX-XV7C | 10/1758 | 0.232751 | 0.999971 | 0 | 0 | 1.349594 | 1.967417 |
| GTEx_Tissu | GTEX-TKQ2 | 6/967 | 0.244313 | 0.999971 | 0 | 0 | 1.460824 | 2.058747 |
| GTEx_Tissu | GTEX-S341 | 4/579 | 0.244629 | 0.999971 | 0 | 0 | 1.620786 | 2.282086 |
| GTEx_Tissu | GTEX-XMK | 27/5444 | 0.245111 | 0.999971 | 0 | 0 | 1.204209 | 1.69317 |
| GTEx_Tissu | GTEX-XOTC | 3/790 | 0.673334 | 0.999971 | 0 | 0 | 0.867943 | 0.343284 |
| GTEx_Tissu | GTEX-OIZH | 26/5226 | 0.246066 | 0.999971 | 0 | 0 | 1.205984 | 1.690978 |
| GTEx_Tissu | GTEX-XBED | Dec-02 | 0.246771 | 0.999971 | 0 | 0 | 1.294831 | 1.811851 |
| GTEx_Tissu | GTEX-Q2A( | 6/971 | 0.247215 | 0.999971 | 0 | 0 | 1.454462 | 2.032608 |
| GTEx_Tissu | GTEX-T5JC | 6/959 | 0.238542 | 0.999971 | 0 | 0 | 1.473709 | 2.112136 |
| GTEx_Tissu | GTEX-XLM4 | 4/582 | 0.247467 | 0.999971 | 0 | 0 | 1.612123 | 2.251291 |
| GTEx_Tissu | GTEX-NL4V | 3/397 | 0.248995 | 0.999971 | 0 | 0 | 1.769307 | 2.459905 |
| GTEx_Tissu | GTEX-TMM | 9/1582 | 0.249185 | 0.999971 | 0 | 0 | 1.345298 | 1.869373 |
| GTEx_Tissu | GTEX-XQ3S | 6/962 | 0.240701 | 0.999971 | 0 | 0 | 1.468852 | 2.091939 |
| GTEx_Tissu | GTEX-O5YT | 3/694 | 0.585525 | 0.999971 | 0 | 0 | 0.993488 | 0.53176 |
| GTEx_Tissu | GTEX-U3ZN | 2/461 | 0.599188 | 0.999971 | 0 | 0 | 0.997257 | 0.510775 |
| GTEx_Tissu | GTEX-S7SE | Sep-60 | 0.605714 | 0.999971 | 0 | 0 | 0.952795 | 0.477681 |
| GTEx_Tissu | GTEX-QEL4 | 7/1674 | 0.599812 | 0.999971 | 0 | 0 | 0.957723 | 0.489531 |
| GTEx_Tissu | GTEX-O5YV | 6/1412 | 0.58384 | 0.999971 | 0 | 0 | 0.975028 | 0.524689 |
| GTEx_Tissu | GTEX-Q2AI | 4/934 | 0.584052 | 0.999971 | 0 | 0 | 0.983703 | 0.529001 |
| GTEx_Tissu | GTEX-QVJC | 3/693 | 0.584538 | 0.999971 | 0 | 0 | 0.994979 | 0.534238 |
| GTEx_Tissu | GTEX-NFK9 | 3/693 | 0.584538 | 0.999971 | 0 | 0 | 0.994979 | 0.534238 |
| GTEx_Tissu | GTEX-P44H | Aug-15 | 0.601473 | 0.999971 | 0 | 0 | 0.956158 | 0.486085 |
| GTEx_Tissu | GTEX-X4XY | Sep-53 | 0.601678 | 0.999971 | 0 | 0 | 0.956282 | 0.485823 |
| GTEx_Tissu | GTEX-NL4V | 5/1193 | 0.598723 | 0.999971 | 0 | 0 | 0.961084 | 0.492994 |
| GTEx_Tissu | GTEX-XMK | 3/712 | 0.603052 | 0.999971 | 0 | 0 | 0.967358 | 0.489243 |
| GTEx_Tissu | GTEX-S3XE | 4/937 | 0.58662 | 0.999971 | 0 | 0 | 0.980385 | 0.522915 |
| GTEx_Tissu | GTEX-P4QF | 6/1416 | 0.586665 | 0.999971 | 0 | 0 | 0.972051 | 0.518397 |

| | | | | | | | | |
|---|---|---|---|---|---|---|---|---|
| GTEx_Tissu | GTEX-WK1 | 6/1417 | 0.587369 | 0.999971 | 0 | 0 | 0.97131 | 0.516835 |
| GTEx_Tissu | GTEX-RWS( | 4/938 | 0.587474 | 0.999971 | 0 | 0 | 0.979283 | 0.520903 |
| GTEx_Tissu | GTEX-OHPI | 7/1682 | 0.60497 | 0.999971 | 0 | 0 | 0.952731 | 0.47882 |
| GTEx_Tissu | GTEX-WFG | 16/3810 | 0.605433 | 0.999971 | 0 | 0 | 0.95742 | 0.480443 |
| GTEx_Tissu | GTEX-QCQ( | 2/470 | 0.609786 | 0.999971 | 0 | 0 | 0.977627 | 0.48358 |
| GTEx_Tissu | GTEX-XUJ4 | 17/4035 | 0.601326 | 0.999971 | 0 | 0 | 0.96073 | 0.488645 |
| GTEx_Tissu | GTEX-U3ZN | 5/1164 | 0.576489 | 0.999971 | 0 | 0 | 0.986658 | 0.54345 |
| GTEx_Tissu | GTEX-XQ8I | 5/1164 | 0.576489 | 0.999971 | 0 | 0 | 0.986658 | 0.54345 |
| GTEx_Tissu | GTEX-XQ3S | 23/5413 | 0.593089 | 0.999971 | 0 | 0 | 0.968312 | 0.505857 |
| GTEx_Tissu | GTEX-P4QS | Oct-75 | 0.593356 | 0.999971 | 0 | 0 | 0.96362 | 0.502972 |
| GTEx_Tissu | GTEX-T2IS- | 5/1194 | 0.599479 | 0.999971 | 0 | 0 | 0.960225 | 0.491342 |
| GTEx_Tissu | GTEX-U3ZH | 6/1426 | 0.593685 | 0.999971 | 0 | 0 | 0.964684 | 0.502993 |
| GTEx_Tissu | GTEX-WWY | Sep-51 | 0.600521 | 0.999971 | 0 | 0 | 0.957283 | 0.488173 |
| GTEx_Tissu | GTEX-XV7C | 5/1196 | 0.600987 | 0.999971 | 0 | 0 | 0.95851 | 0.488056 |
| GTEx_Tissu | GTEX-U8XE | 4/954 | 0.601003 | 0.999971 | 0 | 0 | 0.961978 | 0.489797 |
| GTEx_Tissu | GTEX-OOBI | 23/5445 | 0.606065 | 0.999971 | 0 | 0 | 0.960476 | 0.480975 |
| GTEx_Tissu | GTEX-XGQ4 | Sep-43 | 0.59588 | 0.999971 | 0 | 0 | 0.961304 | 0.497683 |
| GTEx_Tissu | GTEX-X4LF- | 4/949 | 0.596803 | 0.999971 | 0 | 0 | 0.967323 | 0.499302 |
| GTEx_Tissu | GTEX-S4P3 | Aug-17 | 0.602689 | 0.999971 | 0 | 0 | 0.95505 | 0.483593 |
| GTEx_Tissu | GTEX-RNOI | Sep-46 | 0.597623 | 0.999971 | 0 | 0 | 0.959793 | 0.494096 |
| GTEx_Tissu | GTEX-TSE9 | 6/1440 | 0.603411 | 0.999971 | 0 | 0 | 0.954543 | 0.482194 |
| GTEx_Tissu | GTEX-Q2A( | 3/713 | 0.604012 | 0.999971 | 0 | 0 | 0.965946 | 0.486992 |
| GTEx_Tissu | GTEX-QEL4 | 6/1441 | 0.604101 | 0.999971 | 0 | 0 | 0.953826 | 0.480741 |
| GTEx_Tissu | GTEX-Q2AI | 5/1201 | 0.604745 | 0.999971 | 0 | 0 | 0.954248 | 0.479937 |
| GTEx_Tissu | GTEX-WWY | 5/1215 | 0.615164 | 0.999971 | 0 | 0 | 0.942502 | 0.45793 |
| GTEx_Tissu | GTEX-XAJ8 | 3/703 | 0.594348 | 0.999971 | 0 | 0 | 0.980255 | 0.510016 |
| GTEx_Tissu | GTEX-WFO | 4/966 | 0.610978 | 0.999971 | 0 | 0 | 0.949378 | 0.467753 |
| GTEx_Tissu | GTEX-WFJ( | Aug-31 | 0.611152 | 0.999971 | 0 | 0 | 0.947359 | 0.466489 |
| GTEx_Tissu | GTEX-REY6 | 2/459 | 0.596805 | 0.999971 | 0 | 0 | 1.001725 | 0.517056 |
| GTEx_Tissu | GTEX-V955 | 6/1452 | 0.611652 | 0.999971 | 0 | 0 | 0.946007 | 0.465049 |
| GTEx_Tissu | GTEX-T6MI | Dec-55 | 0.597923 | 0.999971 | 0 | 0 | 0.960675 | 0.494069 |
| GTEx_Tissu | GTEX-PLZ4 | Sep-47 | 0.598204 | 0.999971 | 0 | 0 | 0.95929 | 0.492906 |
| GTEx_Tissu | GTEX-XBED | 4/951 | 0.598486 | 0.999971 | 0 | 0 | 0.965179 | 0.495477 |
| GTEx_Tissu | GTEX-WRH | 5/1193 | 0.598723 | 0.999971 | 0 | 0 | 0.961084 | 0.492994 |
| GTEx_Tissu | GTEX-S32W | 3/703 | 0.594348 | 0.999971 | 0 | 0 | 0.980255 | 0.510016 |
| GTEx_Tissu | GTEX-QEG4 | 6/1458 | 0.615738 | 0.999971 | 0 | 0 | 0.941792 | 0.456706 |
| GTEx_Tissu | GTEX-POM | 3/732 | 0.62196 | 0.999971 | 0 | 0 | 0.939839 | 0.44631 |
| GTEx_Tissu | GTEX-XV7C | 3/726 | 0.616351 | 0.999971 | 0 | 0 | 0.947935 | 0.458742 |
| GTEx_Tissu | GTEX-WOF | 2/482 | 0.623589 | 0.999971 | 0 | 0 | 0.952598 | 0.449878 |
| GTEx_Tissu | GTEX-XBED | 2/482 | 0.623589 | 0.999971 | 0 | 0 | 0.952598 | 0.449878 |
| GTEx_Tissu | GTEX-XXEK | 5/1227 | 0.62397 | 0.999971 | 0 | 0 | 0.932647 | 0.439886 |
| GTEx_Tissu | GTEX-WYB | 7/1713 | 0.624643 | 0.999971 | 0 | 0 | 0.933829 | 0.439436 |
| GTEx_Tissu | GTEX-PLZ5 | 20/4775 | 0.618024 | 0.999971 | 0 | 0 | 0.951583 | 0.457928 |
| GTEx_Tissu | GTEX-XOT( | 3/723 | 0.613526 | 0.999971 | 0 | 0 | 0.952034 | 0.465099 |
| GTEx_Tissu | GTEX-TML8 | Sep-68 | 0.610304 | 0.999971 | 0 | 0 | 0.948837 | 0.468534 |

| | | | | | | | | |
|---|---|---|---|---|---|---|---|---|
| GTEx_Tissu | GTEX-R55C | 3/720 | 0.610688 | 0.999971 | 0 | 0 | 0.956167 | 0.471552 |
| GTEx_Tissu | GTEX-U4B1 | 3/720 | 0.610688 | 0.999971 | 0 | 0 | 0.956167 | 0.471552 |
| GTEx_Tissu | GTEX-TMM | 2/471 | 0.610951 | 0.999971 | 0 | 0 | 0.975492 | 0.480663 |
| GTEx_Tissu | GTEX-S7SE | 5/1204 | 0.606991 | 0.999971 | 0 | 0 | 0.951708 | 0.475132 |
| GTEx_Tissu | GTEX-Q2A( | Nov-37 | 0.607283 | 0.999971 | 0 | 0 | 0.952805 | 0.475221 |
| GTEx_Tissu | GTEX-V955 | 3/721 | 0.611636 | 0.999971 | 0 | 0 | 0.954785 | 0.46939 |
| GTEx_Tissu | GTEX-Q2A( | 5/1206 | 0.608484 | 0.999971 | 0 | 0 | 0.950021 | 0.471956 |
| GTEx_Tissu | GTEX-S33H | 7/1693 | 0.61201 | 0.999971 | 0 | 0 | 0.945945 | 0.464466 |
| GTEx_Tissu | GTEX-XUJ4 | 4/925 | 0.576295 | 0.999971 | 0 | 0 | 0.993786 | 0.547711 |
| GTEx_Tissu | GTEX-WFO | Aug-37 | 0.614752 | 0.999971 | 0 | 0 | 0.944098 | 0.459338 |
| GTEx_Tissu | GTEX-S4P3 | 4/971 | 0.61509 | 0.999971 | 0 | 0 | 0.944219 | 0.458877 |
| GTEx_Tissu | GTEX-XXEK | 4/964 | 0.609326 | 0.999971 | 0 | 0 | 0.951456 | 0.471353 |
| GTEx_Tissu | GTEX-S95S | 6/1449 | 0.6096 | 0.999971 | 0 | 0 | 0.948128 | 0.469278 |
| GTEx_Tissu | GTEX-QCQ( | 3/726 | 0.616351 | 0.999971 | 0 | 0 | 0.947935 | 0.458742 |
| GTEx_Tissu | GTEX-S32W | 5/1208 | 0.609974 | 0.999971 | 0 | 0 | 0.948341 | 0.468802 |
| GTEx_Tissu | GTEX-TMZS | 6/1426 | 0.593685 | 0.999971 | 0 | 0 | 0.964684 | 0.502993 |
| GTEx_Tissu | GTEX-O5YV | 7/1665 | 0.593969 | 0.999971 | 0 | 0 | 0.963397 | 0.50186 |
| GTEx_Tissu | GTEX-S3XE | 3/690 | 0.581566 | 0.999971 | 0 | 0 | 0.99948 | 0.541749 |
| GTEx_Tissu | GTEX-X3Y1 | 3/687 | 0.578581 | 0.999971 | 0 | 0 | 1.00402 | 0.549376 |
| GTEx_Tissu | GTEX-XMK | 8/1878 | 0.578671 | 0.999971 | 0 | 0 | 0.97708 | 0.534484 |
| GTEx_Tissu | GTEX-RNOI | 7/1636 | 0.574874 | 0.999971 | 0 | 0 | 0.982106 | 0.543697 |
| GTEx_Tissu | GTEX-OIZH | 7/1636 | 0.574874 | 0.999971 | 0 | 0 | 0.982106 | 0.543697 |
| GTEx_Tissu | GTEX-OHPI | 4/929 | 0.579752 | 0.999971 | 0 | 0 | 0.98928 | 0.53931 |
| GTEx_Tissu | GTEX-QVUS | 13/3047 | 0.576185 | 0.999971 | 0 | 0 | 0.977333 | 0.53883 |
| GTEx_Tissu | GTEX-SNM | 14/3289 | 0.580358 | 0.999971 | 0 | 0 | 0.974305 | 0.530129 |
| GTEx_Tissu | GTEX-XGQ4 | 2/446 | 0.581057 | 0.999971 | 0 | 0 | 1.031744 | 0.56014 |
| GTEx_Tissu | GTEX-NL3H | 8/1882 | 0.581163 | 0.999971 | 0 | 0 | 0.974778 | 0.529036 |
| GTEx_Tissu | GTEX-WH7 | 3/687 | 0.578581 | 0.999971 | 0 | 0 | 1.00402 | 0.549376 |
| GTEx_Tissu | GTEX-W5X | 5/1164 | 0.576489 | 0.999971 | 0 | 0 | 0.986658 | 0.54345 |
| GTEx_Tissu | GTEX-UPK5 | 6/1403 | 0.577452 | 0.999971 | 0 | 0 | 0.981786 | 0.539129 |
| GTEx_Tissu | GTEX-TSE9 | Oct-55 | 0.582142 | 0.999971 | 0 | 0 | 0.972946 | 0.526403 |
| GTEx_Tissu | GTEX-PLZ6 | 4/927 | 0.578026 | 0.999971 | 0 | 0 | 0.991528 | 0.543493 |
| GTEx_Tissu | GTEX-QMR | 3/670 | 0.561425 | 0.999971 | 0 | 0 | 1.03052 | 0.594896 |
| GTEx_Tissu | GTEX-UPJH | 4/909 | 0.562314 | 0.999971 | 0 | 0 | 1.012208 | 0.582724 |
| GTEx_Tissu | GTEX-P44H | Nov-54 | 0.56249 | 0.999971 | 0 | 0 | 0.988627 | 0.568837 |
| GTEx_Tissu | GTEX-QEG5 | 4/910 | 0.563195 | 0.999971 | 0 | 0 | 1.011038 | 0.580467 |
| GTEx_Tissu | GTEX-QEG5 | 6/1369 | 0.552913 | 0.999971 | 0 | 0 | 1.008125 | 0.59737 |
| GTEx_Tissu | GTEX-UPK5 | 4/912 | 0.564954 | 0.999971 | 0 | 0 | 1.008704 | 0.575982 |
| GTEx_Tissu | GTEX-X4EP | 13/2997 | 0.550527 | 0.999971 | 0 | 0 | 0.996653 | 0.594882 |
| GTEx_Tissu | GTEX-WFG | 5/1132 | 0.55126 | 0.999971 | 0 | 0 | 1.016404 | 0.605319 |
| GTEx_Tissu | GTEX-UPJH | 3/661 | 0.552178 | 0.999971 | 0 | 0 | 1.045104 | 0.620671 |
| GTEx_Tissu | GTEX-UJHI | 3/661 | 0.552178 | 0.999971 | 0 | 0 | 1.045104 | 0.620671 |
| GTEx_Tissu | GTEX-S4Q7 | 6/1389 | 0.567423 | 0.999971 | 0 | 0 | 0.992475 | 0.562386 |
| GTEx_Tissu | GTEX-Q734 | 3/676 | 0.567527 | 0.999971 | 0 | 0 | 1.021015 | 0.578371 |
| GTEx_Tissu | GTEX-W5X | 8/1837 | 0.552773 | 0.999971 | 0 | 0 | 1.001253 | 0.59355 |

| | | | | | | | | |
|---|---|---|---|---|---|---|---|---|
| GTEx_Tissu | GTEX-RUS( | 5/1153 | 0.567896 | 0.999971 | 0 | 0 | 0.996696 | 0.563947 |
| GTEx_Tissu | GTEX-OHPI | 5/1167 | 0.578818 | 0.999971 | 0 | 0 | 0.983953 | 0.537993 |
| GTEx_Tissu | GTEX-U8T8 | 3/663 | 0.554243 | 0.999971 | 0 | 0 | 1.041829 | 0.614838 |
| GTEx_Tissu | GTEX-Q2AI | 4/900 | 0.554344 | 0.999971 | 0 | 0 | 1.022859 | 0.603456 |
| GTEx_Tissu | GTEX-Q734 | 7/1606 | 0.554715 | 0.999971 | 0 | 0 | 1.002173 | 0.590582 |
| GTEx_Tissu | GTEX-QDT8 | 7/1629 | 0.570206 | 0.999971 | 0 | 0 | 0.986722 | 0.554298 |
| GTEx_Tissu | GTEX-XOT4 | 4/918 | 0.570208 | 0.999971 | 0 | 0 | 1.001766 | 0.562746 |
| GTEx_Tissu | GTEX-R55E | 6/1403 | 0.577452 | 0.999971 | 0 | 0 | 0.981786 | 0.539129 |
| GTEx_Tissu | GTEX-XUJ4 | 3/680 | 0.571567 | 0.999971 | 0 | 0 | 1.014771 | 0.567637 |
| GTEx_Tissu | GTEX-XBEC | 3/687 | 0.578581 | 0.999971 | 0 | 0 | 1.00402 | 0.549376 |
| GTEx_Tissu | GTEX-XUW | 3/680 | 0.571567 | 0.999971 | 0 | 0 | 1.014771 | 0.567637 |
| GTEx_Tissu | GTEX-VJYA | 6/1415 | 0.58596 | 0.999971 | 0 | 0 | 0.972794 | 0.519963 |
| GTEx_Tissu | GTEX-S4Z8 | 5/1185 | 0.592652 | 0.999971 | 0 | 0 | 0.968014 | 0.506414 |
| GTEx_Tissu | GTEX-U3ZN | 6/1425 | 0.592985 | 0.999971 | 0 | 0 | 0.965416 | 0.504513 |
| GTEx_Tissu | GTEX-QDT8 | 6/1425 | 0.592985 | 0.999971 | 0 | 0 | 0.965416 | 0.504513 |
| GTEx_Tissu | GTEX-OHPI | 6/1425 | 0.592985 | 0.999971 | 0 | 0 | 0.965416 | 0.504513 |
| GTEx_Tissu | GTEX-WXY( | 7/1656 | 0.588087 | 0.999971 | 0 | 0 | 0.969133 | 0.514494 |
| GTEx_Tissu | GTEX-V1D1 | 3/697 | 0.588479 | 0.999971 | 0 | 0 | 0.989039 | 0.524401 |
| GTEx_Tissu | GTEX-XMD | Aug-02 | 0.593527 | 0.999971 | 0 | 0 | 0.963415 | 0.502588 |
| GTEx_Tissu | GTEX-P4PP | 3/698 | 0.589461 | 0.999971 | 0 | 0 | 0.987564 | 0.521973 |
| GTEx_Tissu | GTEX-XMK | 4/943 | 0.591729 | 0.999971 | 0 | 0 | 0.973812 | 0.510965 |
| GTEx_Tissu | GTEX-RM2 | 5/1158 | 0.571813 | 0.999971 | 0 | 0 | 0.99211 | 0.554534 |
| GTEx_Tissu | GTEX-O5YT | 4/921 | 0.572823 | 0.999971 | 0 | 0 | 0.998331 | 0.55625 |
| GTEx_Tissu | GTEX-SE5C | 7/1636 | 0.574874 | 0.999971 | 0 | 0 | 0.982106 | 0.543697 |
| GTEx_Tissu | GTEX-W5W | 7/1661 | 0.59136 | 0.999971 | 0 | 0 | 0.965939 | 0.507438 |
| GTEx_Tissu | GTEX-POY\ | 6/1423 | 0.591585 | 0.999971 | 0 | 0 | 0.966884 | 0.507566 |
| GTEx_Tissu | GTEX-QDT8 | 7/1636 | 0.574874 | 0.999971 | 0 | 0 | 0.982106 | 0.543697 |
| GTEx_Tissu | GTEX-X5EB | 23/5410 | 0.591865 | 0.999971 | 0 | 0 | 0.969052 | 0.508244 |
| GTEx_Tissu | GTEX-PLZ6 | 4/925 | 0.576295 | 0.999971 | 0 | 0 | 0.993786 | 0.547711 |
| GTEx_Tissu | GTEX-XQ8I | 6/1409 | 0.581716 | 0.999971 | 0 | 0 | 0.977271 | 0.529459 |
| GTEx_Tissu | GTEX-WHV | 7/1642 | 0.578858 | 0.999971 | 0 | 0 | 0.97818 | 0.53477 |
| GTEx_Tissu | GTEX-XK95 | Dec-89 | 0.563427 | 0.999971 | 0 | 0 | 0.98731 | 0.566436 |
| GTEx_Tissu | GTEX-T8EN | 6/1407 | 0.580297 | 0.999971 | 0 | 0 | 0.978772 | 0.532663 |
| GTEx_Tissu | GTEX-WVLI | 8/1854 | 0.563587 | 0.999971 | 0 | 0 | 0.991099 | 0.56833 |
| GTEx_Tissu | GTEX-WFO | 23/5342 | 0.563833 | 0.999971 | 0 | 0 | 0.986035 | 0.564995 |
| GTEx_Tissu | GTEX-N7M | 5/1148 | 0.563962 | 0.999971 | 0 | 0 | 1.001323 | 0.573526 |
| GTEx_Tissu | GTEX-O5Y\ | 4/911 | 0.564074 | 0.999971 | 0 | 0 | 1.00987 | 0.57822 |
| GTEx_Tissu | GTEX-UJM( | 3/690 | 0.581566 | 0.999971 | 0 | 0 | 0.99948 | 0.541749 |
| GTEx_Tissu | GTEX-RUS( | 3/728 | 0.618227 | 0.999971 | 0 | 0 | 0.945222 | 0.454557 |
| GTEx_Tissu | GTEX-XOT( | 3/697 | 0.588479 | 0.999971 | 0 | 0 | 0.989039 | 0.524401 |
| GTEx_Tissu | GTEX-PWC | 8/1886 | 0.583649 | 0.999971 | 0 | 0 | 0.972486 | 0.523641 |
| GTEx_Tissu | GTEX-WZT( | 8/1896 | 0.589835 | 0.999971 | 0 | 0 | 0.966799 | 0.510386 |
| GTEx_Tissu | GTEX-VJYA | 21/4942 | 0.590379 | 0.999971 | 0 | 0 | 0.969352 | 0.51084 |
| GTEx_Tissu | GTEX-QDT8 | 7/1660 | 0.590706 | 0.999971 | 0 | 0 | 0.966576 | 0.508841 |
| GTEx_Tissu | GTEX-O5YT | Sep-35 | 0.591215 | 0.999971 | 0 | 0 | 0.965356 | 0.507368 |

| | | | | | | | | |
|---|---|---|---|---|---|---|---|---|
| GTEx_Tissu | GTEX-N7M | 4/935 | 0.584909 | 0.999971 | 0 | 0 | 0.982594 | 0.526964 |
| GTEx_Tissu | GTEX-S7PM | 6/1414 | 0.585254 | 0.999971 | 0 | 0 | 0.973537 | 0.521534 |
| GTEx_Tissu | GTEX-OOB. | 6/1521 | 0.65721 | 0.999971 | 0 | 0 | 0.899548 | 0.377587 |
| GTEx_Tissu | GTEX-R3RS | Oct-91 | 0.655641 | 0.999971 | 0 | 0 | 0.912493 | 0.385202 |
| GTEx_Tissu | GTEX-QDVI | 5/1272 | 0.655938 | 0.999971 | 0 | 0 | 0.897357 | 0.378405 |
| GTEx_Tissu | GTEX-QLQ\ | 4/1028 | 0.660083 | 0.999971 | 0 | 0 | 0.888978 | 0.369272 |
| GTEx_Tissu | GTEX-NPJ8 | 5/1273 | 0.656629 | 0.999971 | 0 | 0 | 0.896601 | 0.377143 |
| GTEx_Tissu | GTEX-XOT( | 4/1028 | 0.660083 | 0.999971 | 0 | 0 | 0.888978 | 0.369272 |
| GTEx_Tissu | GTEX-RM2I | 3/775 | 0.660546 | 0.999971 | 0 | 0 | 0.885501 | 0.367207 |
| GTEx_Tissu | GTEX-OHPI | 2/516 | 0.660674 | 0.999971 | 0 | 0 | 0.888029 | 0.368084 |
| GTEx_Tissu | GTEX-T2IS- | 5/1279 | 0.660757 | 0.999971 | 0 | 0 | 0.892091 | 0.369655 |
| GTEx_Tissu | GTEX-UJHI- | Aug-10 | 0.657181 | 0.999971 | 0 | 0 | 0.90598 | 0.380327 |
| GTEx_Tissu | GTEX-OXR( | 6/1524 | 0.659117 | 0.999971 | 0 | 0 | 0.897624 | 0.374178 |
| GTEx_Tissu | GTEX-QMR | 6/1528 | 0.66165 | 0.999971 | 0 | 0 | 0.89507 | 0.369681 |
| GTEx_Tissu | GTEX-WFJ( | 2/513 | 0.657521 | 0.999971 | 0 | 0 | 0.893381 | 0.374575 |
| GTEx_Tissu | GTEX-PX3G | 3/752 | 0.640261 | 0.999971 | 0 | 0 | 0.91379 | 0.40744 |
| GTEx_Tissu | GTEX-P78B | 3/753 | 0.64116 | 0.999971 | 0 | 0 | 0.912524 | 0.405595 |
| GTEx_Tissu | GTEX-SNO! | 2/498 | 0.641412 | 0.999971 | 0 | 0 | 0.92111 | 0.40905 |
| GTEx_Tissu | GTEX-P4PP | Dec-43 | 0.642064 | 0.999971 | 0 | 0 | 0.927028 | 0.410735 |
| GTEx_Tissu | GTEX-QDV. | 5/1277 | 0.659385 | 0.999971 | 0 | 0 | 0.89359 | 0.372134 |
| GTEx_Tissu | GTEX-UTH( | 7/1771 | 0.660017 | 0.999971 | 0 | 0 | 0.900248 | 0.374044 |
| GTEx_Tissu | GTEX-TSE9 | 4/991 | 0.631277 | 0.999971 | 0 | 0 | 0.92411 | 0.425101 |
| GTEx_Tissu | GTEX-RWS. | 6/1477 | 0.628525 | 0.999971 | 0 | 0 | 0.92867 | 0.431255 |
| GTEx_Tissu | GTEX-POM | 6/1477 | 0.628525 | 0.999971 | 0 | 0 | 0.92867 | 0.431255 |
| GTEx_Tissu | GTEX-WRH | 6/1478 | 0.629192 | 0.999971 | 0 | 0 | 0.927989 | 0.429955 |
| GTEx_Tissu | GTEX-S4P3 | 2/505 | 0.649001 | 0.999971 | 0 | 0 | 0.907964 | 0.392531 |
| GTEx_Tissu | GTEX-SUCS | 6/1509 | 0.649518 | 0.999971 | 0 | 0 | 0.907321 | 0.391531 |
| GTEx_Tissu | GTEX-P4PP | 5/1236 | 0.630498 | 0.999971 | 0 | 0 | 0.925383 | 0.426828 |
| GTEx_Tissu | GTEX-OIZI- | Sep-42 | 0.651512 | 0.999971 | 0 | 0 | 0.913569 | 0.391428 |
| GTEx_Tissu | GTEX-QVU! | Aug-64 | 0.630746 | 0.999971 | 0 | 0 | 0.929668 | 0.42844 |
| GTEx_Tissu | GTEX-XUZ( | 6/1481 | 0.631187 | 0.999971 | 0 | 0 | 0.925951 | 0.426079 |
| GTEx_Tissu | GTEX-WFG | 3/756 | 0.643848 | 0.999971 | 0 | 0 | 0.908746 | 0.400115 |
| GTEx_Tissu | GTEX-TMM | Aug-65 | 0.631331 | 0.999971 | 0 | 0 | 0.929141 | 0.427335 |
| GTEx_Tissu | GTEX-WL4( | Sep-47 | 0.654211 | 0.999971 | 0 | 0 | 0.91127 | 0.386675 |
| GTEx_Tissu | GTEX-WEY! | 2/510 | 0.654345 | 0.999971 | 0 | 0 | 0.898796 | 0.381197 |
| GTEx_Tissu | GTEX-WL4( | 2/513 | 0.657521 | 0.999971 | 0 | 0 | 0.893381 | 0.374575 |
| GTEx_Tissu | GTEX-S7SE | 5/1270 | 0.654554 | 0.999971 | 0 | 0 | 0.898872 | 0.380944 |
| GTEx_Tissu | GTEX-QMR | 4/1025 | 0.657804 | 0.999971 | 0 | 0 | 0.891731 | 0.373501 |
| GTEx_Tissu | GTEX-P4QS | 3/772 | 0.657947 | 0.999971 | 0 | 0 | 0.889095 | 0.372203 |
| GTEx_Tissu | GTEX-PWO | 7/1768 | 0.658235 | 0.999971 | 0 | 0 | 0.901931 | 0.377182 |
| GTEx_Tissu | GTEX-X4XY | 1/243 | 0.655558 | 0.999971 | 0 | 0 | 0.945176 | 0.399118 |
| GTEx_Tissu | GTEX-S3XE | 2/528 | 0.673056 | 0.999971 | 0 | 0 | 0.867233 | 0.34336 |
| GTEx_Tissu | GTEX-SUCS | 2/528 | 0.673056 | 0.999971 | 0 | 0 | 0.867233 | 0.34336 |
| GTEx_Tissu | GTEX-U412 | 4/1040 | 0.669102 | 0.999971 | 0 | 0 | 0.878123 | 0.352846 |
| GTEx_Tissu | GTEX-S7SF | 6/1540 | 0.669181 | 0.999971 | 0 | 0 | 0.887489 | 0.356505 |

| | | | | | | | | |
|---|---|---|---|---|---|---|---|---|
| GTEx_Tissu | GTEX-VJYA | 4/1048 | 0.675026 | 0.999971 | 0 | 0 | 0.871024 | 0.342317 |
| GTEx_Tissu | GTEX-WHV | 5/1293 | 0.670268 | 0.999971 | 0 | 0 | 0.881732 | 0.352761 |
| GTEx_Tissu | GTEX-PSDG | 6/1517 | 0.654657 | 0.999971 | 0 | 0 | 0.902125 | 0.38218 |
| GTEx_Tissu | GTEX-UJHI- | Aug-06 | 0.654924 | 0.999971 | 0 | 0 | 0.907997 | 0.384297 |
| GTEx_Tissu | GTEX-XMD | 4/1022 | 0.655514 | 0.999971 | 0 | 0 | 0.894501 | 0.37778 |
| GTEx_Tissu | GTEX-Y8E5 | 2/528 | 0.673056 | 0.999971 | 0 | 0 | 0.867233 | 0.34336 |
| GTEx_Tissu | GTEX-R55F | 3/788 | 0.671649 | 0.999971 | 0 | 0 | 0.870246 | 0.346375 |
| GTEx_Tissu | GTEX-QV31 | 2/527 | 0.672038 | 0.999971 | 0 | 0 | 0.86893 | 0.345347 |
| GTEx_Tissu | GTEX-S3XE | 3/770 | 0.656206 | 0.999971 | 0 | 0 | 0.891507 | 0.375574 |
| GTEx_Tissu | GTEX-S95S- | 3/789 | 0.672492 | 0.999971 | 0 | 0 | 0.869093 | 0.344826 |
| GTEx_Tissu | GTEX-TML8 | 17/4163 | 0.65697 | 0.999971 | 0 | 0 | 0.923572 | 0.388008 |
| GTEx_Tissu | GTEX-OIZI- | 7/1766 | 0.657044 | 0.999971 | 0 | 0 | 0.903056 | 0.379287 |
| GTEx_Tissu | GTEX-XYKS | 3/771 | 0.657077 | 0.999971 | 0 | 0 | 0.890299 | 0.373885 |
| GTEx_Tissu | GTEX-OIZH | 3/771 | 0.657077 | 0.999971 | 0 | 0 | 0.890299 | 0.373885 |
| GTEx_Tissu | GTEX-S95S- | 16/3958 | 0.670463 | 0.999971 | 0 | 0 | 0.913013 | 0.365011 |
| GTEx_Tissu | GTEX-SUCS | 4/1028 | 0.660083 | 0.999971 | 0 | 0 | 0.888978 | 0.369272 |
| GTEx_Tissu | GTEX-R55D | 2/501 | 0.64468 | 0.999971 | 0 | 0 | 0.915431 | 0.401876 |
| GTEx_Tissu | GTEX-WH7 | 3/757 | 0.644741 | 0.999971 | 0 | 0 | 0.907493 | 0.398306 |
| GTEx_Tissu | GTEX-REY6 | 6/1502 | 0.644985 | 0.999971 | 0 | 0 | 0.911913 | 0.399899 |
| GTEx_Tissu | GTEX-OOB. | 5/1257 | 0.645471 | 0.999971 | 0 | 0 | 0.908839 | 0.397867 |
| GTEx_Tissu | GTEX-OOBI | 6/1527 | 0.661018 | 0.999971 | 0 | 0 | 0.895707 | 0.3708 |
| GTEx_Tissu | GTEX-TML8 | 3/776 | 0.661409 | 0.999971 | 0 | 0 | 0.88431 | 0.365558 |
| GTEx_Tissu | GTEX-OHPI | 4/1041 | 0.669847 | 0.999971 | 0 | 0 | 0.877229 | 0.351512 |
| GTEx_Tissu | GTEX-PWC | 4/1032 | 0.663107 | 0.999971 | 0 | 0 | 0.885331 | 0.36371 |
| GTEx_Tissu | GTEX-O5YT | 5/1233 | 0.62833 | 0.999971 | 0 | 0 | 0.927793 | 0.431136 |
| GTEx_Tissu | GTEX-VJWI | 4/1042 | 0.67059 | 0.999971 | 0 | 0 | 0.876338 | 0.350183 |
| GTEx_Tissu | GTEX-S7SE- | Oct-21 | 0.6709 | 0.999971 | 0 | 0 | 0.90004 | 0.359237 |
| GTEx_Tissu | GTEX-TML8 | Oct-22 | 0.671403 | 0.999971 | 0 | 0 | 0.89963 | 0.3584 |
| GTEx_Tissu | GTEX-PSDG | 3/781 | 0.665703 | 0.999971 | 0 | 0 | 0.878397 | 0.35743 |
| GTEx_Tissu | GTEX-Q2AI | 6/1535 | 0.666056 | 0.999971 | 0 | 0 | 0.890633 | 0.361937 |
| GTEx_Tissu | GTEX-Q2AG | Aug-37 | 0.672195 | 0.999971 | 0 | 0 | 0.892577 | 0.354537 |
| GTEx_Tissu | GTEX-QLQ7 | 3/782 | 0.666557 | 0.999971 | 0 | 0 | 0.877224 | 0.355828 |
| GTEx_Tissu | GTEX-WRH | Aug-38 | 0.672744 | 0.999971 | 0 | 0 | 0.892087 | 0.353615 |
| GTEx_Tissu | GTEX-QDT8 | 4/993 | 0.632872 | 0.999971 | 0 | 0 | 0.922144 | 0.421869 |
| GTEx_Tissu | GTEX-R55D | 3/718 | 0.608788 | 0.999971 | 0 | 0 | 0.958941 | 0.475908 |
| GTEx_Tissu | GTEX-WFG | 6/1448 | 0.608915 | 0.999971 | 0 | 0 | 0.948836 | 0.470695 |
| GTEx_Tissu | GTEX-XYKS | 4/964 | 0.609326 | 0.999971 | 0 | 0 | 0.951456 | 0.471353 |
| GTEx_Tissu | GTEX-WK1 | 4/985 | 0.626465 | 0.999971 | 0 | 0 | 0.930057 | 0.434952 |
| GTEx_Tissu | GTEX-OIZH | 4/985 | 0.626465 | 0.999971 | 0 | 0 | 0.930057 | 0.434952 |
| GTEx_Tissu | GTEX-RTLS- | 5/1238 | 0.63194 | 0.999971 | 0 | 0 | 0.923783 | 0.42398 |
| GTEx_Tissu | GTEX-XUZC | 2/485 | 0.626981 | 0.999971 | 0 | 0 | 0.946535 | 0.441879 |
| GTEx_Tissu | GTEX-U3ZH | 5/1239 | 0.63266 | 0.999971 | 0 | 0 | 0.922985 | 0.422564 |
| GTEx_Tissu | GTEX-S33H | 4/993 | 0.632872 | 0.999971 | 0 | 0 | 0.922144 | 0.421869 |
| GTEx_Tissu | GTEX-WH7 | 2/469 | 0.608619 | 0.999971 | 0 | 0 | 0.979771 | 0.486518 |
| GTEx_Tissu | GTEX-WCD | Oct-49 | 0.633667 | 0.999971 | 0 | 0 | 0.930443 | 0.424497 |

| | | | | | | | | |
|---|---|---|---|---|---|---|---|---|
| GTEx_Tissu | GTEX-N7M | Aug-62 | 0.629573 | 0.999971 | 0 | 0 | 0.930723 | 0.430658 |
| GTEx_Tissu | GTEX-P4QS | 2/488 | 0.63035 | 0.999971 | 0 | 0 | 0.940547 | 0.434043 |
| GTEx_Tissu | GTEX-XBEV | 2/492 | 0.634806 | 0.999971 | 0 | 0 | 0.932677 | 0.423842 |
| GTEx_Tissu | GTEX-RVPV | Aug-64 | 0.630746 | 0.999971 | 0 | 0 | 0.929668 | 0.42844 |
| GTEx_Tissu | GTEX-NFKS | 2/494 | 0.637018 | 0.999971 | 0 | 0 | 0.92879 | 0.418845 |
| GTEx_Tissu | GTEX-T5JC- | 7/1734 | 0.637673 | 0.999971 | 0 | 0 | 0.92141 | 0.41457 |
| GTEx_Tissu | GTEX-S32W | Aug-77 | 0.638323 | 0.999971 | 0 | 0 | 0.922861 | 0.414283 |
| GTEx_Tissu | GTEX-P4PP | 2/481 | 0.622453 | 0.999971 | 0 | 0 | 0.954636 | 0.452581 |
| GTEx_Tissu | GTEX-TSE9- | 4/983 | 0.624853 | 0.999971 | 0 | 0 | 0.932055 | 0.438288 |
| GTEx_Tissu | GTEX-P4QT | 6/1462 | 0.61845 | 0.999971 | 0 | 0 | 0.939001 | 0.451227 |
| GTEx_Tissu | GTEX-XQ3S | Aug-55 | 0.625452 | 0.999971 | 0 | 0 | 0.934433 | 0.438511 |
| GTEx_Tissu | GTEX-WRH | 4/984 | 0.62566 | 0.999971 | 0 | 0 | 0.931055 | 0.436617 |
| GTEx_Tissu | GTEX-XOT4 | 3/736 | 0.62567 | 0.999971 | 0 | 0 | 0.934516 | 0.438225 |
| GTEx_Tissu | GTEX-P44H | Sep-96 | 0.626166 | 0.999971 | 0 | 0 | 0.935212 | 0.437809 |
| GTEx_Tissu | GTEX-PX3G | 4/977 | 0.619991 | 0.999971 | 0 | 0 | 0.9381 | 0.448459 |
| GTEx_Tissu | GTEX-WFO | 2/480 | 0.621315 | 0.999971 | 0 | 0 | 0.956682 | 0.455302 |
| GTEx_Tissu | GTEX-WVLI | Aug-57 | 0.626632 | 0.999971 | 0 | 0 | 0.933371 | 0.436253 |
| GTEx_Tissu | GTEX-VJYA | 5/1247 | 0.638387 | 0.999971 | 0 | 0 | 0.916647 | 0.411401 |
| GTEx_Tissu | GTEX-XPVG | 2/467 | 0.606277 | 0.999971 | 0 | 0 | 0.984086 | 0.492455 |
| GTEx_Tissu | GTEX-OXRL | 3/716 | 0.606882 | 0.999971 | 0 | 0 | 0.961731 | 0.480309 |
| GTEx_Tissu | GTEX-UJHI- | 3/716 | 0.606882 | 0.999971 | 0 | 0 | 0.961731 | 0.480309 |
| GTEx_Tissu | GTEX-QCQ( | 5/1204 | 0.606991 | 0.999971 | 0 | 0 | 0.951708 | 0.475132 |
| GTEx_Tissu | GTEX-XUW | 3/735 | 0.624745 | 0.999971 | 0 | 0 | 0.935841 | 0.440231 |
| GTEx_Tissu | GTEX-SUCS | 23/5492 | 0.624853 | 0.999971 | 0 | 0 | 0.949134 | 0.44632 |
| GTEx_Tissu | GTEX-PWN | Aug-25 | 0.607535 | 0.999971 | 0 | 0 | 0.950641 | 0.473747 |
| GTEx_Tissu | GTEX-RM2I | Aug-55 | 0.625452 | 0.999971 | 0 | 0 | 0.934433 | 0.438511 |
| GTEx_Tissu | GTEX-PLZ5- | Aug-88 | 0.644669 | 0.999971 | 0 | 0 | 0.917172 | 0.402655 |
| GTEx_Tissu | GTEX-O5YV | 2/503 | 0.646846 | 0.999971 | 0 | 0 | 0.911683 | 0.397172 |
| GTEx_Tissu | GTEX-P4PP | 7/1738 | 0.640127 | 0.999971 | 0 | 0 | 0.919079 | 0.409991 |
| GTEx_Tissu | GTEX-QLQ7 | 3/761 | 0.648296 | 0.999971 | 0 | 0 | 0.902516 | 0.391157 |
| GTEx_Tissu | GTEX-RU1J | 3/761 | 0.648296 | 0.999971 | 0 | 0 | 0.902516 | 0.391157 |
| GTEx_Tissu | GTEX-PVOV | Nov-17 | 0.648449 | 0.999971 | 0 | 0 | 0.920357 | 0.398674 |
| GTEx_Tissu | GTEX-SNOS | Aug-95 | 0.648677 | 0.999971 | 0 | 0 | 0.913584 | 0.395418 |
| GTEx_Tissu | GTEX-UPJH | 6/1498 | 0.64238 | 0.999971 | 0 | 0 | 0.914557 | 0.40476 |
| GTEx_Tissu | GTEX-P4QS | 2/500 | 0.643593 | 0.999971 | 0 | 0 | 0.917316 | 0.404251 |
| GTEx_Tissu | GTEX-WFG | 4/1015 | 0.650133 | 0.999971 | 0 | 0 | 0.901028 | 0.387964 |
| GTEx_Tissu | GTEX-PWO | 6/1493 | 0.639108 | 0.999971 | 0 | 0 | 0.917881 | 0.410918 |
| GTEx_Tissu | GTEX-V1D1 | 7/1757 | 0.651656 | 0.999971 | 0 | 0 | 0.90815 | 0.388905 |
| GTEx_Tissu | GTEX-W5W | 2/509 | 0.653282 | 0.999971 | 0 | 0 | 0.900615 | 0.383434 |
| GTEx_Tissu | GTEX-XUJ4 | 2/509 | 0.653282 | 0.999971 | 0 | 0 | 0.900615 | 0.383434 |
| GTEx_Tissu | GTEX-Q2AI | 7/1760 | 0.653457 | 0.999971 | 0 | 0 | 0.906446 | 0.385673 |
| GTEx_Tissu | GTEX-WEY! | 3/759 | 0.646521 | 0.999971 | 0 | 0 | 0.904998 | 0.394714 |
| GTEx_Tissu | GTEX-V1D1 | 3/759 | 0.646521 | 0.999971 | 0 | 0 | 0.904998 | 0.394714 |
| GTEx_Tissu | GTEX-XYKS | 3/768 | 0.654459 | 0.999971 | 0 | 0 | 0.893931 | 0.378979 |
| GTEx_Tissu | GTEX-TSE9- | 7/1750 | 0.647433 | 0.999971 | 0 | 0 | 0.912149 | 0.396548 |

| | | | | | | | | |
|---|---|---|---|---|---|---|---|---|
| GTEx_Tissu | GTEX-Q2AI | 2/491 | 0.633696 | 0.999971 | 0 | 0 | 0.934633 | 0.426367 |
| GTEx_Tissu | GTEX-S32V | 2/489 | 0.631468 | 0.999971 | 0 | 0 | 0.938567 | 0.431467 |
| GTEx_Tissu | GTEX-XPVC | 6/1482 | 0.631851 | 0.999971 | 0 | 0 | 0.925274 | 0.424795 |
| GTEx_Tissu | GTEX-OXRI | 2/496 | 0.63922 | 0.999971 | 0 | 0 | 0.924935 | 0.413914 |
| GTEx_Tissu | GTEX-RWS | 3/743 | 0.632102 | 0.999971 | 0 | 0 | 0.925338 | 0.424457 |
| GTEx_Tissu | GTEX-WRH | 6/1459 | 0.616417 | 0.999971 | 0 | 0 | 0.941093 | 0.45533 |
| GTEx_Tissu | GTEX-T5JC- | 5/1217 | 0.61664 | 0.999971 | 0 | 0 | 0.940846 | 0.454871 |
| GTEx_Tissu | GTEX-R53T | 4/973 | 0.616728 | 0.999971 | 0 | 0 | 0.942171 | 0.455377 |
| GTEx_Tissu | GTEX-WVLI | Nov-57 | 0.617774 | 0.999971 | 0 | 0 | 0.944509 | 0.454906 |
| GTEx_Tissu | GTEX-QDT8 | 9/1891 | 0.440474 | 0.999971 | 0 | 0 | 1.105473 | 0.906382 |
| GTEx_Tissu | GTEX-WY7 | 5/1241 | 0.634096 | 0.999971 | 0 | 0 | 0.921393 | 0.419744 |
| GTEx_Tissu | GTEX-U8T8 | 7/1703 | 0.618353 | 0.999971 | 0 | 0 | 0.939851 | 0.451782 |
| GTEx_Tissu | GTEX-PWO | Aug-74 | 0.636581 | 0.999971 | 0 | 0 | 0.924424 | 0.41751 |
| GTEx_Tissu | GTEX-XPT6 | 23/5477 | 0.618894 | 0.999971 | 0 | 0 | 0.952733 | 0.457142 |
| GTEx_Tissu | GTEX-WZT | 6/1463 | 0.619126 | 0.999971 | 0 | 0 | 0.938305 | 0.449867 |
| GTEx_Tissu | GTEX-U8XE | 6/1463 | 0.619126 | 0.999971 | 0 | 0 | 0.938305 | 0.449867 |
| GTEx_Tissu | GTEX-R55C | 5/1221 | 0.619581 | 0.999971 | 0 | 0 | 0.93755 | 0.448816 |
| GTEx_Tissu | GTEX-WFO | 3/750 | 0.638459 | 0.999971 | 0 | 0 | 0.916332 | 0.411157 |
| GTEx_Tissu | GTEX-U8T8 | 4/825 | 0.485287 | 0.999971 | 0 | 0 | 1.120702 | 0.810284 |
| GTEx_Tissu | GTEX-XMK | 3/589 | 0.474451 | 0.999971 | 0 | 0 | 1.177901 | 0.87824 |
| GTEx_Tissu | GTEX-T6M | 4/821 | 0.481486 | 0.999971 | 0 | 0 | 1.126425 | 0.823278 |
| GTEx_Tissu | GTEX-XQ8I | 3/596 | 0.482277 | 0.999971 | 0 | 0 | 1.163575 | 0.848523 |
| GTEx_Tissu | GTEX-WHV | 4/822 | 0.482438 | 0.999971 | 0 | 0 | 1.124989 | 0.820009 |
| GTEx_Tissu | GTEX-WHS | 8/1730 | 0.482697 | 0.999971 | 0 | 0 | 1.06976 | 0.779177 |
| GTEx_Tissu | GTEX-XV7C | 7/1494 | 0.476487 | 0.999971 | 0 | 0 | 1.084247 | 0.803769 |
| GTEx_Tissu | GTEX-NPJ7 | 4/817 | 0.477675 | 0.999971 | 0 | 0 | 1.132204 | 0.836499 |
| GTEx_Tissu | GTEX-OHPI | 3/598 | 0.484503 | 0.999971 | 0 | 0 | 1.159544 | 0.840243 |
| GTEx_Tissu | GTEX-VJWI | 7/1496 | 0.477916 | 0.999971 | 0 | 0 | 1.082673 | 0.79936 |
| GTEx_Tissu | GTEX-T6M | 3/595 | 0.481162 | 0.999971 | 0 | 0 | 1.165601 | 0.852697 |
| GTEx_Tissu | GTEX-PW2 | 3/599 | 0.485614 | 0.999971 | 0 | 0 | 1.157538 | 0.836137 |
| GTEx_Tissu | GTEX-WRH | 4/826 | 0.486236 | 0.999971 | 0 | 0 | 1.11928 | 0.80707 |
| GTEx_Tissu | GTEX-S341 | Sep-67 | 0.488717 | 0.999971 | 0 | 0 | 1.058085 | 0.757558 |
| GTEx_Tissu | GTEX-XUZC | 3/594 | 0.480046 | 0.999971 | 0 | 0 | 1.167634 | 0.856895 |
| GTEx_Tissu | GTEX-OIZI- | Nov-10 | 0.481124 | 0.999971 | 0 | 0 | 1.056657 | 0.773083 |
| GTEx_Tissu | GTEX-WFG | 4/829 | 0.489077 | 0.999971 | 0 | 0 | 1.115035 | 0.797512 |
| GTEx_Tissu | GTEX-RU72 | 3/595 | 0.481162 | 0.999971 | 0 | 0 | 1.165601 | 0.852697 |
| GTEx_Tissu | GTEX-RVPL | 3/579 | 0.463182 | 0.999971 | 0 | 0 | 1.198971 | 0.92277 |
| GTEx_Tissu | GTEX-XQ8I | 4/810 | 0.470982 | 0.999971 | 0 | 0 | 1.142456 | 0.860195 |
| GTEx_Tissu | GTEX-XXEK | 18/3979 | 0.468779 | 0.999971 | 0 | 0 | 1.050591 | 0.795952 |
| GTEx_Tissu | GTEX-OXRL | 3/584 | 0.468829 | 0.999971 | 0 | 0 | 1.188345 | 0.900192 |
| GTEx_Tissu | GTEX-OXRI | 4/813 | 0.473854 | 0.999971 | 0 | 0 | 1.13804 | 0.849952 |
| GTEx_Tissu | GTEX-TMM | 6/1258 | 0.469106 | 0.999971 | 0 | 0 | 1.104071 | 0.8357 |
| GTEx_Tissu | GTEX-R3RS | 4/789 | 0.450733 | 0.999971 | 0 | 0 | 1.174307 | 0.935783 |
| GTEx_Tissu | GTEX-P78B | 6/1235 | 0.451218 | 0.999971 | 0 | 0 | 1.126119 | 0.89617 |
| GTEx_Tissu | GTEX-RNOI | 8/1684 | 0.451804 | 0.999971 | 0 | 0 | 1.1019 | 0.875468 |

| | | | | | | | | |
|---|---|---|---|---|---|---|---|---|
| GTEx_Tissu | GTEX-XPT6 | 6/1236 | 0.451999 | 0.999971 | 0 | 0 | 1.125143 | 0.893449 |
| GTEx_Tissu | GTEX-QV44 | 4/810 | 0.470982 | 0.999971 | 0 | 0 | 1.142456 | 0.860195 |
| GTEx_Tissu | GTEX-XV7C | 7/1476 | 0.463587 | 0.999971 | 0 | 0 | 1.098604 | 0.844564 |
| GTEx_Tissu | GTEX-U8XE | 5/1014 | 0.453092 | 0.999971 | 0 | 0 | 1.142401 | 0.904394 |
| GTEx_Tissu | GTEX-X15G | Sep-40 | 0.471653 | 0.999971 | 0 | 0 | 1.074493 | 0.807494 |
| GTEx_Tissu | GTEX-R55C | 3/571 | 0.454096 | 0.999971 | 0 | 0 | 1.216361 | 0.960253 |
| GTEx_Tissu | GTEX-R53T | 5/1016 | 0.454807 | 0.999971 | 0 | 0 | 1.140021 | 0.898201 |
| GTEx_Tissu | GTEX-RWS( | 7/1464 | 0.454954 | 0.999971 | 0 | 0 | 1.108373 | 0.87291 |
| GTEx_Tissu | GTEX-PLZ6 | 6/1240 | 0.455118 | 0.999971 | 0 | 0 | 1.121256 | 0.882652 |
| GTEx_Tissu | GTEX-RU72 | 4/812 | 0.472898 | 0.999971 | 0 | 0 | 1.139509 | 0.853351 |
| GTEx_Tissu | GTEX-PWC | 3/588 | 0.473328 | 0.999971 | 0 | 0 | 1.179976 | 0.882581 |
| GTEx_Tissu | GTEX-NPJ8 | 15/3365 | 0.503091 | 0.999971 | 0 | 0 | 1.030037 | 0.70762 |
| GTEx_Tissu | GTEX-P78B | 6/1287 | 0.491453 | 0.999971 | 0 | 0 | 1.077399 | 0.765373 |
| GTEx_Tissu | GTEX-S7SF | 15/3359 | 0.500088 | 0.999971 | 0 | 0 | 1.032259 | 0.715326 |
| GTEx_Tissu | GTEX-X5EB | 17/3802 | 0.492938 | 0.999971 | 0 | 0 | 1.034822 | 0.732004 |
| GTEx_Tissu | GTEX-XUJ4 | 8/1757 | 0.500657 | 0.999971 | 0 | 0 | 1.051682 | 0.727589 |
| GTEx_Tissu | GTEX-S95S | 3/613 | 0.501057 | 0.999971 | 0 | 0 | 1.130152 | 0.780976 |
| GTEx_Tissu | GTEX-SJXC | 4/842 | 0.501319 | 0.999971 | 0 | 0 | 1.096989 | 0.757484 |
| GTEx_Tissu | GTEX-NFK9 | 5/1071 | 0.501383 | 0.999971 | 0 | 0 | 1.078056 | 0.744274 |
| GTEx_Tissu | GTEX-S4P3 | 5/1063 | 0.494691 | 0.999971 | 0 | 0 | 1.086668 | 0.764822 |
| GTEx_Tissu | GTEX-RWS( | 6/1293 | 0.496042 | 0.999971 | 0 | 0 | 1.072031 | 0.751594 |
| GTEx_Tissu | GTEX-S4P3 | Sep-71 | 0.491236 | 0.999971 | 0 | 0 | 1.055693 | 0.750419 |
| GTEx_Tissu | GTEX-W5W | 3/609 | 0.496667 | 0.999971 | 0 | 0 | 1.137848 | 0.796305 |
| GTEx_Tissu | GTEX-WFO | 5/1040 | 0.475285 | 0.999971 | 0 | 0 | 1.112172 | 0.827277 |
| GTEx_Tissu | GTEX-OXR( | Sep-46 | 0.475454 | 0.999971 | 0 | 0 | 1.070807 | 0.79613 |
| GTEx_Tissu | GTEX-PW2( | Oct-74 | 0.476053 | 0.999971 | 0 | 0 | 1.065187 | 0.79061 |
| GTEx_Tissu | GTEX-RN64 | 5/1041 | 0.476134 | 0.999971 | 0 | 0 | 1.111039 | 0.824454 |
| GTEx_Tissu | GTEX-SN8( | 5/1068 | 0.498877 | 0.999971 | 0 | 0 | 1.08127 | 0.751911 |
| GTEx_Tissu | GTEX-QEG! | 7/1527 | 0.499942 | 0.999971 | 0 | 0 | 1.058808 | 0.734032 |
| GTEx_Tissu | GTEX-S341 | 3/592 | 0.477811 | 0.999971 | 0 | 0 | 1.17172 | 0.865362 |
| GTEx_Tissu | GTEX-XQ8I | 4/807 | 0.468105 | 0.999971 | 0 | 0 | 1.146904 | 0.870573 |
| GTEx_Tissu | GTEX-RVPL | 3/580 | 0.464313 | 0.999971 | 0 | 0 | 1.196831 | 0.918203 |
| GTEx_Tissu | GTEX-T5JW | 6/1252 | 0.464452 | 0.999971 | 0 | 0 | 1.109744 | 0.851059 |
| GTEx_Tissu | GTEX-WZT( | Sep-60 | 0.484304 | 0.999971 | 0 | 0 | 1.062295 | 0.77021 |
| GTEx_Tissu | GTEX-XGQ4 | 3/598 | 0.484503 | 0.999971 | 0 | 0 | 1.159544 | 0.840243 |
| GTEx_Tissu | GTEX-T5JW | 5/1028 | 0.465073 | 0.999971 | 0 | 0 | 1.125933 | 0.86197 |
| GTEx_Tissu | GTEX-WHS | 7/1506 | 0.485047 | 0.999971 | 0 | 0 | 1.074867 | 0.777675 |
| GTEx_Tissu | GTEX-VUS( | 7/1480 | 0.466459 | 0.999971 | 0 | 0 | 1.095384 | 0.835323 |
| GTEx_Tissu | GTEX-X4EP | 3/582 | 0.466573 | 0.999971 | 0 | 0 | 1.192573 | 0.909147 |
| GTEx_Tissu | GTEX-QDVI | 5/1031 | 0.467631 | 0.999971 | 0 | 0 | 1.122462 | 0.853156 |
| GTEx_Tissu | GTEX-O5YT | 7/1488 | 0.472194 | 0.999971 | 0 | 0 | 1.088994 | 0.817143 |
| GTEx_Tissu | GTEX-RU1J | Sep-67 | 0.488717 | 0.999971 | 0 | 0 | 1.058085 | 0.757558 |
| GTEx_Tissu | GTEX-XOT( | 5/1056 | 0.48881 | 0.999971 | 0 | 0 | 1.094312 | 0.783289 |
| GTEx_Tissu | GTEX-RWS( | 4/837 | 0.496625 | 0.999971 | 0 | 0 | 1.103863 | 0.772616 |
| GTEx_Tissu | GTEX-XV7C | 4/829 | 0.489077 | 0.999971 | 0 | 0 | 1.115035 | 0.797512 |

| | | | | | | | | |
|---|---|---|---|---|---|---|---|---|
| GTEx_Tissu | GTEX-XPVG | 3/609 | 0.496667 | 0.999971 | 0 | 0 | 1.137848 | 0.796305 |
| GTEx_Tissu | GTEX-U8XE | Oct-09 | 0.497064 | 0.999971 | 0 | 0 | 1.046166 | 0.731308 |
| GTEx_Tissu | GTEX-XPVG | 3/610 | 0.497767 | 0.999971 | 0 | 0 | 1.135914 | 0.792441 |
| GTEx_Tissu | GTEX-WH7 | 23/5187 | 0.498367 | 0.999971 | 0 | 0 | 1.026418 | 0.714816 |
| GTEx_Tissu | GTEX-QVJC | Sep-03 | 0.448128 | 0.999971 | 0 | 0 | 1.097738 | 0.881129 |
| GTEx_Tissu | GTEX-T5JW | 4/780 | 0.441984 | 0.999971 | 0 | 0 | 1.188486 | 0.970378 |
| GTEx_Tissu | GTEX-T8EM | 3/561 | 0.442654 | 0.999971 | 0 | 0 | 1.238799 | 1.009581 |
| GTEx_Tissu | GTEX-NL4V | 6/1197 | 0.421438 | 0.999971 | 0 | 0 | 1.164412 | 1.006149 |
| GTEx_Tissu | GTEX-T6M( | 6/1225 | 0.443405 | 0.999971 | 0 | 0 | 1.135965 | 0.923849 |
| GTEx_Tissu | GTEX-X5EB | 5/979 | 0.422883 | 0.999971 | 0 | 0 | 1.185644 | 1.020435 |
| GTEx_Tissu | GTEX-NFK9 | 3/544 | 0.423006 | 0.999971 | 0 | 0 | 1.278849 | 1.100282 |
| GTEx_Tissu | GTEX-WEY! | 3/544 | 0.423006 | 0.999971 | 0 | 0 | 1.278849 | 1.100282 |
| GTEx_Tissu | GTEX-NL3H | 3/544 | 0.423006 | 0.999971 | 0 | 0 | 1.278849 | 1.100282 |
| GTEx_Tissu | GTEX-SE5C | 6/1231 | 0.448095 | 0.999971 | 0 | 0 | 1.130038 | 0.907137 |
| GTEx_Tissu | GTEX-SSA3 | 16/3420 | 0.417698 | 0.999971 | 0 | 0 | 1.092931 | 0.954125 |
| GTEx_Tissu | GTEX-OOB. | 24/5316 | 0.456213 | 0.999971 | 0 | 0 | 1.052514 | 0.826009 |
| GTEx_Tissu | GTEX-PLZ6 | 3/567 | 0.44953 | 0.999971 | 0 | 0 | 1.225241 | 0.979645 |
| GTEx_Tissu | GTEX-S3XE | 7/1466 | 0.456394 | 0.999971 | 0 | 0 | 1.106734 | 0.86812 |
| GTEx_Tissu | GTEX-UPK5 | 5/1018 | 0.456522 | 0.999971 | 0 | 0 | 1.13765 | 0.892053 |
| GTEx_Tissu | GTEX-XV7C | 4/795 | 0.456543 | 0.999971 | 0 | 0 | 1.165034 | 0.913472 |
| GTEx_Tissu | GTEX-PLZ6 | 7/1467 | 0.457114 | 0.999971 | 0 | 0 | 1.105916 | 0.865735 |
| GTEx_Tissu | GTEX-TSE9 | Oct-35 | 0.452487 | 0.999971 | 0 | 0 | 1.08712 | 0.862083 |
| GTEx_Tissu | GTEX-X4XX | 6/1237 | 0.452779 | 0.999971 | 0 | 0 | 1.124169 | 0.890737 |
| GTEx_Tissu | GTEX-OXRL | 25/5518 | 0.445549 | 0.999971 | 0 | 0 | 1.058532 | 0.855769 |
| GTEx_Tissu | GTEX-P4QS | 5/999 | 0.440185 | 0.999971 | 0 | 0 | 1.160561 | 0.952311 |
| GTEx_Tissu | GTEX-R55F | 6/1222 | 0.441057 | 0.999971 | 0 | 0 | 1.13895 | 0.932324 |
| GTEx_Tissu | GTEX-QDT8 | 7/1445 | 0.441239 | 0.999971 | 0 | 0 | 1.124174 | 0.919763 |
| GTEx_Tissu | GTEX-VJYA | 5/992 | 0.43414 | 0.999971 | 0 | 0 | 1.169224 | 0.975587 |
| GTEx_Tissu | GTEX-RTLS | 5/992 | 0.43414 | 0.999971 | 0 | 0 | 1.169224 | 0.975587 |
| GTEx_Tissu | GTEX-P4QF | 7/1447 | 0.442685 | 0.999971 | 0 | 0 | 1.122491 | 0.914714 |
| GTEx_Tissu | GTEX-X4XY | 6/1216 | 0.436356 | 0.999971 | 0 | 0 | 1.144965 | 0.949515 |
| GTEx_Tissu | GTEX-XUZC | 6/1227 | 0.444969 | 0.999971 | 0 | 0 | 1.133982 | 0.918243 |
| GTEx_Tissu | GTEX-S4Q7 | 6/1227 | 0.444969 | 0.999971 | 0 | 0 | 1.133982 | 0.918243 |
| GTEx_Tissu | GTEX-XGQ4 | 3/574 | 0.45751 | 0.999971 | 0 | 0 | 1.209782 | 0.945996 |
| GTEx_Tissu | GTEX-OHPI | 5/1008 | 0.447937 | 0.999971 | 0 | 0 | 1.1496 | 0.923247 |
| GTEx_Tissu | GTEX-T8EM | 5/997 | 0.438459 | 0.999971 | 0 | 0 | 1.163024 | 0.9589 |
| GTEx_Tissu | GTEX-XUJ4 | 8/1665 | 0.438968 | 0.999971 | 0 | 0 | 1.115696 | 0.918584 |
| GTEx_Tissu | GTEX-SE5C | 3/566 | 0.448386 | 0.999971 | 0 | 0 | 1.22748 | 0.984563 |
| GTEx_Tissu | GTEX-P4PC | 3/558 | 0.439204 | 0.999971 | 0 | 0 | 1.245689 | 1.024943 |
| GTEx_Tissu | GTEX-S3XE | 5/971 | 0.415939 | 0.999971 | 0 | 0 | 1.195968 | 1.049123 |
| GTEx_Tissu | GTEX-XPT6 | 3/539 | 0.417184 | 0.999971 | 0 | 0 | 1.291111 | 1.128725 |
| GTEx_Tissu | GTEX-XBEC | 5/973 | 0.417676 | 0.999971 | 0 | 0 | 1.193371 | 1.041871 |
| GTEx_Tissu | GTEX-SIU7 | 4/809 | 0.470024 | 0.999971 | 0 | 0 | 1.143935 | 0.863639 |
| GTEx_Tissu | GTEX-RU1J | 5/995 | 0.436732 | 0.999971 | 0 | 0 | 1.165496 | 0.965538 |
| GTEx_Tissu | GTEX-P78B | 4/775 | 0.437107 | 0.999971 | 0 | 0 | 1.196506 | 0.990202 |

| | | | | | | | | |
|---|---|---|---|---|---|---|---|---|
| GTEx_Tissu | GTEX-QDT8 | Oct-10 | 0.437329 | 0.999971 | 0 | 0 | 1.101608 | 0.911107 |
| GTEx_Tissu | GTEX-PLZ4 | 3/577 | 0.460916 | 0.999971 | 0 | 0 | 1.203273 | 0.931981 |
| GTEx_Tissu | GTEX-XPVC | 4/801 | 0.462334 | 0.999971 | 0 | 0 | 1.155901 | 0.891741 |
| GTEx_Tissu | GTEX-NPJ8 | 4/808 | 0.469064 | 0.999971 | 0 | 0 | 1.145417 | 0.867099 |
| GTEx_Tissu | GTEX-WFJC | Dec-05 | 0.462985 | 0.999971 | 0 | 0 | 1.068723 | 0.822981 |
| GTEx_Tissu | GTEX-QEL4 | 6/1258 | 0.469106 | 0.999971 | 0 | 0 | 1.104071 | 0.8357 |
| GTEx_Tissu | GTEX-T2IS- | Oct-64 | 0.470023 | 0.999971 | 0 | 0 | 1.070735 | 0.808377 |
| GTEx_Tissu | GTEX-P4PP | 5/995 | 0.436732 | 0.999971 | 0 | 0 | 1.165496 | 0.965538 |
| GTEx_Tissu | GTEX-V1D1 | 6/1260 | 0.470655 | 0.999971 | 0 | 0 | 1.102192 | 0.830644 |
| GTEx_Tissu | GTEX-XOTC | 6/1252 | 0.464452 | 0.999971 | 0 | 0 | 1.109744 | 0.851059 |
| GTEx_Tissu | GTEX-PLZ6 | 5/1028 | 0.465073 | 0.999971 | 0 | 0 | 1.125933 | 0.86197 |
| GTEx_Tissu | GTEX-RTLS | 3/586 | 0.471081 | 0.999971 | 0 | 0 | 1.184146 | 0.891337 |
| GTEx_Tissu | GTEX-T6MC | 6/1254 | 0.466004 | 0.999971 | 0 | 0 | 1.107847 | 0.845907 |
| GTEx_Tissu | GTEX-REY6 | 4/811 | 0.47194 | 0.999971 | 0 | 0 | 1.14098 | 0.856766 |
| GTEx_Tissu | GTEX-OHPI | 4/811 | 0.47194 | 0.999971 | 0 | 0 | 1.14098 | 0.856766 |
| GTEx_Tissu | GTEX-TSE9 | 8/1714 | 0.471988 | 0.999971 | 0 | 0 | 1.080743 | 0.811424 |
| GTEx_Tissu | GTEX-P44H | Nov-67 | 0.456285 | 0.999971 | 0 | 0 | 1.078584 | 0.846298 |
| GTEx_Tissu | GTEX-WFO | 3/571 | 0.454096 | 0.999971 | 0 | 0 | 1.216361 | 0.960253 |
| GTEx_Tissu | GTEX-XUJ4 | 2/355 | 0.458861 | 0.999971 | 0 | 0 | 1.303783 | 1.015656 |
| GTEx_Tissu | GTEX-XYKS | 18/3959 | 0.459411 | 0.999971 | 0 | 0 | 1.057247 | 0.822336 |
| GTEx_Tissu | GTEX-NL3H | 4/798 | 0.459441 | 0.999971 | 0 | 0 | 1.16045 | 0.902535 |
| GTEx_Tissu | GTEX-QDVI | 5/1022 | 0.459946 | 0.999971 | 0 | 0 | 1.132935 | 0.879889 |
| GTEx_Tissu | GTEX-RWS | 3/572 | 0.455235 | 0.999971 | 0 | 0 | 1.21416 | 0.955473 |
| GTEx_Tissu | GTEX-SIU7 | 8/1690 | 0.45585 | 0.999971 | 0 | 0 | 1.097608 | 0.862272 |
| GTEx_Tissu | GTEX-OIZH | Nov-78 | 0.462651 | 0.999971 | 0 | 0 | 1.072899 | 0.826971 |
| GTEx_Tissu | GTEX-Q2AI | 7/1621 | 0.564844 | 0.999971 | 0 | 0 | 0.992046 | 0.566662 |
| GTEx_Tissu | GTEX-R53T | 5/991 | 0.433275 | 0.999971 | 0 | 0 | 1.170472 | 0.978961 |
| GTEx_Tissu | GTEX-XAJ8 | 7/1434 | 0.433279 | 0.999971 | 0 | 0 | 1.133514 | 0.94804 |
| GTEx_Tissu | GTEX-XYKS | 7/1434 | 0.433279 | 0.999971 | 0 | 0 | 1.133514 | 0.94804 |
| GTEx_Tissu | GTEX-UPIC | 7/1435 | 0.434004 | 0.999971 | 0 | 0 | 1.132659 | 0.945433 |
| GTEx_Tissu | GTEX-W5W | Dec-95 | 0.457423 | 0.999971 | 0 | 0 | 1.07348 | 0.839619 |
| GTEx_Tissu | GTEX-S4UY | 3/574 | 0.45751 | 0.999971 | 0 | 0 | 1.209782 | 0.945996 |
| GTEx_Tissu | GTEX-X585 | 6/1215 | 0.435572 | 0.999971 | 0 | 0 | 1.145973 | 0.952411 |
| GTEx_Tissu | GTEX-TMM | 3/574 | 0.45751 | 0.999971 | 0 | 0 | 1.209782 | 0.945996 |
| GTEx_Tissu | GTEX-R55D | 7/1598 | 0.549274 | 0.999971 | 0 | 0 | 1.007652 | 0.603743 |
| GTEx_Tissu | GTEX-XUJ4 | 8/1790 | 0.522378 | 0.999971 | 0 | 0 | 1.030331 | 0.66906 |
| GTEx_Tissu | GTEX-WFO | 2/418 | 0.545655 | 0.999971 | 0 | 0 | 1.102771 | 0.668025 |
| GTEx_Tissu | GTEX-W5X | Dec-16 | 0.524168 | 0.999971 | 0 | 0 | 1.018284 | 0.657754 |
| GTEx_Tissu | GTEX-R55E | 8/1793 | 0.524338 | 0.999971 | 0 | 0 | 1.02843 | 0.663973 |
| GTEx_Tissu | GTEX-Q2AC | Sep-25 | 0.524936 | 0.999971 | 0 | 0 | 1.024325 | 0.660155 |
| GTEx_Tissu | GTEX-P4PP | Oct-58 | 0.526155 | 0.999971 | 0 | 0 | 1.020532 | 0.655345 |
| GTEx_Tissu | GTEX-XUZC | 3/657 | 0.548033 | 0.999971 | 0 | 0 | 1.051715 | 0.632522 |
| GTEx_Tissu | GTEX-WFG | 6/1364 | 0.549252 | 0.999971 | 0 | 0 | 1.012109 | 0.606453 |
| GTEx_Tissu | GTEX-RM2 | 2/426 | 0.555975 | 0.999971 | 0 | 0 | 1.081521 | 0.634887 |
| GTEx_Tissu | GTEX-OHPI | 7/1589 | 0.543122 | 0.999971 | 0 | 0 | 1.013883 | 0.618896 |

| | | | | | | | | |
|---|---|---|---|---|---|---|---|---|
| GTEx_Tissu | GTEX-UJM( | 7/1608 | 0.556071 | 0.999971 | 0 | 0 | 1.000812 | 0.587336 |
| GTEx_Tissu | GTEX-S32W | 3/665 | 0.556302 | 0.999971 | 0 | 0 | 1.038574 | 0.609065 |
| GTEx_Tissu | GTEX-SNOS | 3/665 | 0.556302 | 0.999971 | 0 | 0 | 1.038574 | 0.609065 |
| GTEx_Tissu | GTEX-NL3H | 8/1843 | 0.556602 | 0.999971 | 0 | 0 | 0.997648 | 0.584527 |
| GTEx_Tissu | GTEX-X5EB | 6/1368 | 0.552182 | 0.999971 | 0 | 0 | 1.008919 | 0.599175 |
| GTEx_Tissu | GTEX-WVLI | 4/898 | 0.552563 | 0.999971 | 0 | 0 | 1.025255 | 0.608169 |
| GTEx_Tissu | GTEX-WYV. | 4/904 | 0.557895 | 0.999971 | 0 | 0 | 1.018099 | 0.594146 |
| GTEx_Tissu | GTEX-S4Q7 | 5/1134 | 0.552857 | 0.999971 | 0 | 0 | 1.014496 | 0.601247 |
| GTEx_Tissu | GTEX-NPJ7 | Oct-80 | 0.53906 | 0.999971 | 0 | 0 | 1.009383 | 0.623725 |
| GTEx_Tissu | GTEX-PWN | 4/887 | 0.542704 | 0.999971 | 0 | 0 | 1.038628 | 0.6348 |
| GTEx_Tissu | GTEX-QV44 | 3/646 | 0.536523 | 0.999971 | 0 | 0 | 1.070318 | 0.66643 |
| GTEx_Tissu | GTEX-R3RS | Dec-39 | 0.536644 | 0.999971 | 0 | 0 | 1.008346 | 0.627615 |
| GTEx_Tissu | GTEX-Q2AI | 5/1123 | 0.544041 | 0.999971 | 0 | 0 | 1.025077 | 0.623997 |
| GTEx_Tissu | GTEX-S4UY | 15/3433 | 0.536873 | 0.999971 | 0 | 0 | 1.0054 | 0.625353 |
| GTEx_Tissu | GTEX-XGQ4 | 23/5302 | 0.547112 | 0.999971 | 0 | 0 | 0.996229 | 0.600828 |
| GTEx_Tissu | GTEX-SSA3 | 5/1127 | 0.547255 | 0.999971 | 0 | 0 | 1.021206 | 0.615623 |
| GTEx_Tissu | GTEX-QVUS | Dec-59 | 0.547417 | 0.999971 | 0 | 0 | 0.99984 | 0.602448 |
| GTEx_Tissu | GTEX-QEG4 | 13/2992 | 0.547935 | 0.999971 | 0 | 0 | 0.998621 | 0.600769 |
| GTEx_Tissu | GTEX-TMZS | 3/667 | 0.558355 | 0.999971 | 0 | 0 | 1.035338 | 0.603353 |
| GTEx_Tissu | GTEX-XPVG | 3/649 | 0.539678 | 0.999971 | 0 | 0 | 1.065181 | 0.656986 |
| GTEx_Tissu | GTEX-UPJH | 7/1598 | 0.549274 | 0.999971 | 0 | 0 | 1.007652 | 0.603743 |
| GTEx_Tissu | GTEX-V1D1 | 3/651 | 0.541775 | 0.999971 | 0 | 0 | 1.061784 | 0.650772 |
| GTEx_Tissu | GTEX-S4UY | 4/859 | 0.517143 | 0.999971 | 0 | 0 | 1.07422 | 0.70838 |
| GTEx_Tissu | GTEX-S95S | 3/628 | 0.517346 | 0.999971 | 0 | 0 | 1.102171 | 0.726379 |
| GTEx_Tissu | GTEX-OXR( | 3/630 | 0.519497 | 0.999971 | 0 | 0 | 1.098542 | 0.719429 |
| GTEx_Tissu | GTEX-WXY( | 7/1555 | 0.519604 | 0.999971 | 0 | 0 | 1.038073 | 0.679615 |
| GTEx_Tissu | GTEX-PLZ4 | 4/887 | 0.542704 | 0.999971 | 0 | 0 | 1.038628 | 0.6348 |
| GTEx_Tissu | GTEX-U412 | Sep-88 | 0.563353 | 0.999971 | 0 | 0 | 0.989788 | 0.567988 |
| GTEx_Tissu | GTEX-XOT( | 5/1143 | 0.560011 | 0.999971 | 0 | 0 | 1.00599 | 0.583272 |
| GTEx_Tissu | GTEX-QEL4 | 5/1144 | 0.560803 | 0.999971 | 0 | 0 | 1.005054 | 0.581309 |
| GTEx_Tissu | GTEX-XUW | 2/433 | 0.564871 | 0.999971 | 0 | 0 | 1.063573 | 0.607468 |
| GTEx_Tissu | GTEX-S4Q7 | Sep-84 | 0.560948 | 0.999971 | 0 | 0 | 0.991918 | 0.573455 |
| GTEx_Tissu | GTEX-TKQ2 | 16/3723 | 0.565016 | 0.999971 | 0 | 0 | 0.985178 | 0.56244 |
| GTEx_Tissu | GTEX-X3Y1 | 6/1387 | 0.565982 | 0.999971 | 0 | 0 | 0.994019 | 0.56579 |
| GTEx_Tissu | GTEX-P4PP | 2/434 | 0.566132 | 0.999971 | 0 | 0 | 1.061057 | 0.603666 |
| GTEx_Tissu | GTEX-XUJ4 | 3/675 | 0.566513 | 0.999971 | 0 | 0 | 1.022587 | 0.58109 |
| GTEx_Tissu | GTEX-TKQ1 | 4/910 | 0.563195 | 0.999971 | 0 | 0 | 1.011038 | 0.580467 |
| GTEx_Tissu | GTEX-VUSH | 18/4129 | 0.538493 | 0.999971 | 0 | 0 | 1.002739 | 0.620676 |
| GTEx_Tissu | GTEX-T6M( | 4/915 | 0.567585 | 0.999971 | 0 | 0 | 1.005224 | 0.569323 |
| GTEx_Tissu | GTEX-S7PN | 3/672 | 0.563465 | 0.999971 | 0 | 0 | 1.027333 | 0.58933 |
| GTEx_Tissu | GTEX-XBEC | 24/5584 | 0.568323 | 0.999971 | 0 | 0 | 0.983419 | 0.555697 |
| GTEx_Tissu | GTEX-TMM | 3/677 | 0.568539 | 0.999971 | 0 | 0 | 1.019447 | 0.575667 |
| GTEx_Tissu | GTEX-OHPI | 5/1154 | 0.568681 | 0.999971 | 0 | 0 | 0.995776 | 0.562052 |
| GTEx_Tissu | GTEX-X585 | 7/1627 | 0.568868 | 0.999971 | 0 | 0 | 0.988048 | 0.557364 |
| GTEx_Tissu | GTEX-X15G | 6/1385 | 0.564538 | 0.999971 | 0 | 0 | 0.995568 | 0.569214 |

| | | | | | | | | |
|---|---|---|---|---|---|---|---|---|
| GTEx_Tissu | GTEX-U3ZH | 3/679 | 0.570559 | 0.999971 | 0 | 0 | 1.016325 | 0.5703 |
| GTEx_Tissu | GTEX-X4XY | 4/879 | 0.535467 | 0.999971 | 0 | 0 | 1.048565 | 0.65495 |
| GTEx_Tissu | GTEX-REY6 | 8/1846 | 0.558511 | 0.999971 | 0 | 0 | 0.995854 | 0.580066 |
| GTEx_Tissu | GTEX-RM2 | 25/5794 | 0.55978 | 0.999971 | 0 | 0 | 0.988599 | 0.573597 |
| GTEx_Tissu | GTEX-XV7C | Oct-16 | 0.55993 | 0.999971 | 0 | 0 | 0.991597 | 0.57507 |
| GTEx_Tissu | GTEX-X4XX | 6/1372 | 0.555102 | 0.999971 | 0 | 0 | 1.005748 | 0.591986 |
| GTEx_Tissu | GTEX-RUSC | 4/901 | 0.555233 | 0.999971 | 0 | 0 | 1.021665 | 0.601114 |
| GTEx_Tissu | GTEX-WYB | 5/1144 | 0.560803 | 0.999971 | 0 | 0 | 1.005054 | 0.581309 |
| GTEx_Tissu | GTEX-XK95 | 5/1138 | 0.556043 | 0.999971 | 0 | 0 | 1.010699 | 0.593189 |
| GTEx_Tissu | GTEX-UPJH | 17/3891 | 0.535109 | 0.999971 | 0 | 0 | 1.005469 | 0.628704 |
| GTEx_Tissu | GTEX-XUW | 6/1345 | 0.535232 | 0.999971 | 0 | 0 | 1.027522 | 0.642258 |
| GTEx_Tissu | GTEX-OXRI | 4/818 | 0.478629 | 0.999971 | 0 | 0 | 1.130754 | 0.833173 |
| GTEx_Tissu | GTEX-UJM( | Nov-07 | 0.536351 | 0.999971 | 0 | 0 | 1.009969 | 0.629176 |
| GTEx_Tissu | GTEX-T5JW | 7/1609 | 0.556748 | 0.999971 | 0 | 0 | 1.000133 | 0.58572 |
| GTEx_Tissu | GTEX-P4PC | 7/1609 | 0.556748 | 0.999971 | 0 | 0 | 1.000133 | 0.58572 |
| GTEx_Tissu | GTEX-QDT8 | Oct-76 | 0.536722 | 0.999971 | 0 | 0 | 1.011394 | 0.629365 |
| GTEx_Tissu | GTEX-RWS | 4/904 | 0.557895 | 0.999971 | 0 | 0 | 1.018099 | 0.594146 |
| GTEx_Tissu | GTEX-QMR | 16/3664 | 0.536904 | 0.999971 | 0 | 0 | 1.004757 | 0.624894 |
| GTEx_Tissu | GTEX-OHPI | 6/1348 | 0.537457 | 0.999971 | 0 | 0 | 1.025059 | 0.636466 |
| GTEx_Tissu | GTEX-OIZH | 3/647 | 0.537576 | 0.999971 | 0 | 0 | 1.0686 | 0.663265 |
| GTEx_Tissu | GTEX-QVJC | Oct-35 | 0.512554 | 0.999971 | 0 | 0 | 1.032424 | 0.690019 |
| GTEx_Tissu | GTEX-V1D1 | 16/3617 | 0.514201 | 0.999971 | 0 | 0 | 1.020812 | 0.678983 |
| GTEx_Tissu | GTEX-X4XY | 5/1078 | 0.507212 | 0.999971 | 0 | 0 | 1.070625 | 0.726768 |
| GTEx_Tissu | GTEX-XBED | 6/1310 | 0.508975 | 0.999971 | 0 | 0 | 1.057089 | 0.713912 |
| GTEx_Tissu | GTEX-S7PN | 23/5226 | 0.514983 | 0.999971 | 0 | 0 | 1.01603 | 0.674259 |
| GTEx_Tissu | GTEX-TKQ1 | 3/621 | 0.509778 | 0.999971 | 0 | 0 | 1.11506 | 0.751305 |
| GTEx_Tissu | GTEX-QMR | 8/1740 | 0.489367 | 0.999971 | 0 | 0 | 1.062999 | 0.759665 |
| GTEx_Tissu | GTEX-R55D | 3/603 | 0.490049 | 0.999971 | 0 | 0 | 1.149583 | 0.81994 |
| GTEx_Tissu | GTEX-QV31 | 3/603 | 0.490049 | 0.999971 | 0 | 0 | 1.149583 | 0.81994 |
| GTEx_Tissu | GTEX-Q2AI | 3/604 | 0.491155 | 0.999971 | 0 | 0 | 1.147611 | 0.815946 |
| GTEx_Tissu | GTEX-WFO | 3/625 | 0.51411 | 0.999971 | 0 | 0 | 1.10766 | 0.736946 |
| GTEx_Tissu | GTEX-WH7 | 3/624 | 0.513029 | 0.999971 | 0 | 0 | 1.109501 | 0.740506 |
| GTEx_Tissu | GTEX-TMZ | 16/3571 | 0.491787 | 0.999971 | 0 | 0 | 1.036937 | 0.735925 |
| GTEx_Tissu | GTEX-P4PC | 4/855 | 0.513439 | 0.999971 | 0 | 0 | 1.079495 | 0.719617 |
| GTEx_Tissu | GTEX-U3ZH | 25/5632 | 0.493 | 0.999971 | 0 | 0 | 1.028812 | 0.727622 |
| GTEx_Tissu | GTEX-WL4( | Sep-76 | 0.49438 | 0.999971 | 0 | 0 | 1.052716 | 0.741586 |
| GTEx_Tissu | GTEX-T8EN | 3/607 | 0.494466 | 0.999971 | 0 | 0 | 1.141734 | 0.804097 |
| GTEx_Tissu | GTEX-T2YK | 6/1291 | 0.494514 | 0.999971 | 0 | 0 | 1.073815 | 0.756159 |
| GTEx_Tissu | GTEX-Q2AI | 6/1317 | 0.514268 | 0.999971 | 0 | 0 | 1.05105 | 0.698959 |
| GTEx_Tissu | GTEX-X585 | 8/1772 | 0.510565 | 0.999971 | 0 | 0 | 1.041878 | 0.70039 |
| GTEx_Tissu | GTEX-U8T8 | 22/4963 | 0.50051 | 0.999971 | 0 | 0 | 1.025591 | 0.709839 |
| GTEx_Tissu | GTEX-RU1J | 5/1044 | 0.478677 | 0.999971 | 0 | 0 | 1.107655 | 0.816042 |
| GTEx_Tissu | GTEX-OHPI | 3/593 | 0.478929 | 0.999971 | 0 | 0 | 1.169673 | 0.861117 |
| GTEx_Tissu | GTEX-XUZC | 4/819 | 0.479582 | 0.999971 | 0 | 0 | 1.129307 | 0.82986 |
| GTEx_Tissu | GTEX-W5W | 3/614 | 0.502151 | 0.999971 | 0 | 0 | 1.128244 | 0.777196 |

| | | | | | | | | |
|---|---|---|---|---|---|---|---|---|
| GTEx_Tissu | GTEX-QESD | 5/1072 | 0.502217 | 0.999971 | 0 | 0 | 1.076988 | 0.741746 |
| GTEx_Tissu | GTEX-QEG5 | 4/851 | 0.509723 | 0.999971 | 0 | 0 | 1.084821 | 0.731047 |
| GTEx_Tissu | GTEX-XLM4 | 18/4053 | 0.503346 | 0.999971 | 0 | 0 | 1.02654 | 0.704696 |
| GTEx_Tissu | GTEX-XGQ4 | 6/1312 | 0.510489 | 0.999971 | 0 | 0 | 1.055357 | 0.709607 |
| GTEx_Tissu | GTEX-RUSQ | 5/1121 | 0.542429 | 0.999971 | 0 | 0 | 1.027024 | 0.628228 |
| GTEx_Tissu | GTEX-Q2AG | 6/1314 | 0.512002 | 0.999971 | 0 | 0 | 1.05363 | 0.705328 |
| GTEx_Tissu | GTEX-QEG5 | 4/854 | 0.512511 | 0.999971 | 0 | 0 | 1.080822 | 0.722456 |
| GTEx_Tissu | GTEX-NL3H | 2/387 | 0.504142 | 0.999971 | 0 | 0 | 1.193461 | 0.817399 |
| GTEx_Tissu | GTEX-POYW | Sep-92 | 0.504412 | 0.999971 | 0 | 0 | 1.043291 | 0.713988 |
| GTEx_Tissu | GTEX-UJHI | 3/624 | 0.513029 | 0.999971 | 0 | 0 | 1.109501 | 0.740506 |
| GTEx_Tissu | GTEX-VJYA | 4/846 | 0.505062 | 0.999971 | 0 | 0 | 1.091549 | 0.745609 |
| GTEx_Tissu | GTEX-X88G | 6/1316 | 0.513513 | 0.999971 | 0 | 0 | 1.051908 | 0.701076 |
| GTEx_Tissu | GTEX-S95S | 3/625 | 0.51411 | 0.999971 | 0 | 0 | 1.10766 | 0.736946 |
| GTEx_Tissu | GTEX-PWC | 3/617 | 0.505427 | 0.999971 | 0 | 0 | 1.122557 | 0.765979 |
| GTEx_Tissu | GTEX-X585 | 17/3824 | 0.50342 | 0.999971 | 0 | 0 | 1.027438 | 0.705162 |
| GTEx_Tissu | GTEX-O5YT | 24/5428 | 0.503437 | 0.999971 | 0 | 0 | 1.022805 | 0.701949 |
| GTEx_Tissu | GTEX-Q2AG | 6/1303 | 0.503663 | 0.999971 | 0 | 0 | 1.063194 | 0.72919 |
| GTEx_Tissu | GTEX-SN8G | 17/3825 | 0.503896 | 0.999971 | 0 | 0 | 1.027105 | 0.703963 |
| GTEx_Tissu | GTEX-SNM | 7/1570 | 0.530031 | 0.999971 | 0 | 0 | 1.027271 | 0.652132 |
| GTEx_Tissu | GTEX-SUCS | 16/3650 | 0.530167 | 0.999971 | 0 | 0 | 1.009496 | 0.640589 |
| GTEx_Tissu | GTEX-XOTC | 6/1304 | 0.504423 | 0.999971 | 0 | 0 | 1.062318 | 0.726987 |
| GTEx_Tissu | GTEX-QLQ7 | 4/875 | 0.531829 | 0.999971 | 0 | 0 | 1.053601 | 0.66528 |
| GTEx_Tissu | GTEX-P4QT | 6/1305 | 0.505183 | 0.999971 | 0 | 0 | 1.061443 | 0.724791 |
| GTEx_Tissu | GTEX-X8HC | Oct-60 | 0.527332 | 0.999971 | 0 | 0 | 1.019509 | 0.652409 |
| GTEx_Tissu | GTEX-WOF | 4/847 | 0.505996 | 0.999971 | 0 | 0 | 1.090197 | 0.742672 |
| GTEx_Tissu | GTEX-S32W | 4/848 | 0.506929 | 0.999971 | 0 | 0 | 1.088848 | 0.739747 |
| GTEx_Tissu | GTEX-WFG | 2/409 | 0.53385 | 0.999971 | 0 | 0 | 1.127677 | 0.707777 |
| GTEx_Tissu | GTEX-WZTC | 3/644 | 0.534413 | 0.999971 | 0 | 0 | 1.073769 | 0.67281 |
| GTEx_Tissu | GTEX-PSDG | 3/650 | 0.540727 | 0.999971 | 0 | 0 | 1.06348 | 0.653871 |
| GTEx_Tissu | GTEX-X62C | 4/878 | 0.534559 | 0.999971 | 0 | 0 | 1.049819 | 0.657516 |
| GTEx_Tissu | GTEX-OXRK | 6/1354 | 0.541895 | 0.999971 | 0 | 0 | 1.020167 | 0.625039 |
| GTEx_Tissu | GTEX-R55D | 5/1121 | 0.542429 | 0.999971 | 0 | 0 | 1.027024 | 0.628228 |
| GTEx_Tissu | GTEX-X4XX | 6/1344 | 0.534489 | 0.999971 | 0 | 0 | 1.028345 | 0.6442 |
| GTEx_Tissu | GTEX-VJWI | Sep-08 | 0.514393 | 0.999971 | 0 | 0 | 1.034017 | 0.687381 |
| GTEx_Tissu | GTEX-OIZI- | 7/1550 | 0.516111 | 0.999971 | 0 | 0 | 1.041721 | 0.689029 |
| GTEx_Tissu | GTEX-U3ZG | 2/405 | 0.528537 | 0.999971 | 0 | 0 | 1.139104 | 0.72634 |
| GTEx_Tissu | GTEX-OHPI | 5/1104 | 0.528634 | 0.999971 | 0 | 0 | 1.043854 | 0.665413 |
| GTEx_Tissu | GTEX-NL3H | 4/873 | 0.530004 | 0.999971 | 0 | 0 | 1.056137 | 0.67051 |
| GTEx_Tissu | GTEX-RU72 | 6/1338 | 0.530023 | 0.999971 | 0 | 0 | 1.033311 | 0.655981 |
| GTEx_Tissu | GTEX-OIZG | 6/1326 | 0.521043 | 0.999971 | 0 | 0 | 1.043378 | 0.680201 |
| GTEx_Tissu | GTEX-U8T8 | 3/632 | 0.521643 | 0.999971 | 0 | 0 | 1.094935 | 0.712553 |
| GTEx_Tissu | GTEX-U4B1 | 3/640 | 0.530177 | 0.999971 | 0 | 0 | 1.080736 | 0.685775 |
| GTEx_Tissu | GTEX-S4Q7 | 3/634 | 0.523784 | 0.999971 | 0 | 0 | 1.091352 | 0.70575 |
| GTEx_Tissu | GTEX-T2YK | Sep-37 | 0.532337 | 0.999971 | 0 | 0 | 1.017581 | 0.641563 |
| GTEx_Tissu | GTEX-XLM4 | Sep-37 | 0.532337 | 0.999971 | 0 | 0 | 1.017581 | 0.641563 |

| | | | | | | | | |
|---|---|---|---|---|---|---|---|---|
| GTEx_Tissu | GTEX-RWS( | 7/1574 | 0.532798 | 0.999971 | 0 | 0 | 1.024426 | 0.644991 |
| GTEx_Tissu | GTEX-X8H( | 4/877 | 0.53365 | 0.999971 | 0 | 0 | 1.051077 | 0.660093 |
| GTEx_Tissu | GTEX-XQ3S | 4/869 | 0.526346 | 0.999971 | 0 | 0 | 1.061244 | 0.681103 |
| GTEx_Tissu | GTEX-O5Y\ | 6/1334 | 0.527037 | 0.999971 | 0 | 0 | 1.036647 | 0.663956 |
| GTEx_Tissu | GTEX-S4Z8 | Sep-30 | 0.528024 | 0.999971 | 0 | 0 | 1.021505 | 0.652346 |
| ARCHS4_Ti | THYMUS (E | Sep-16 | 0.690276 | 0.999975 | 0 | 0 | 0.880564 | 0.326393 |
| ARCHS4_Ti | CHONDRO( | 13/2316 | 0.203345 | 0.999975 | 0 | 0 | 1.343312 | 2.139694 |
| ARCHS4_Ti | DORSAL ST | Aug-16 | 0.803553 | 0.999975 | 0 | 0 | 0.772437 | 0.168942 |
| ARCHS4_Ti | DENTATE G | Jul-16 | 0.88992 | 0.999975 | 0 | 0 | 0.667107 | 0.0778 |
| ARCHS4_Ti | OLIGODEN | Sep-16 | 0.690276 | 0.999975 | 0 | 0 | 0.880564 | 0.326393 |
| ARCHS4_Ti | MACROPH/ | Sep-16 | 0.690276 | 0.999975 | 0 | 0 | 0.880564 | 0.326393 |
| ARCHS4_Ti | NEURONAI | 13/2316 | 0.203345 | 0.999975 | 0 | 0 | 1.343312 | 2.139694 |
| ARCHS4_Ti | PANCREAT | Jul-16 | 0.88992 | 0.999975 | 0 | 0 | 0.667107 | 0.0778 |
| ARCHS4_Ti | ENDOMETI | Aug-16 | 0.803553 | 0.999975 | 0 | 0 | 0.772437 | 0.168942 |
| ARCHS4_Ti | MORULA | Dec-16 | 0.30459 | 0.999975 | 0 | 0 | 1.222847 | 1.453707 |
| ARCHS4_Ti | SUPERIOR I | Dec-16 | 0.30459 | 0.999975 | 0 | 0 | 1.222847 | 1.453707 |
| ARCHS4_Ti | BREAST (BU | Nov-16 | 0.426714 | 0.999975 | 0 | 0 | 1.105651 | 0.941617 |
| ARCHS4_Ti | ADRENAL ( | 13/2316 | 0.203345 | 0.999975 | 0 | 0 | 1.343312 | 2.139694 |
| ARCHS4_Ti | COLON (BU | Sep-16 | 0.690276 | 0.999975 | 0 | 0 | 0.880564 | 0.326393 |
| ARCHS4_Ti | FETAL BRAI | 13/2316 | 0.203345 | 0.999975 | 0 | 0 | 1.343312 | 2.139694 |
| ARCHS4_Ti | FIBROBLAS | 13/2316 | 0.203345 | 0.999975 | 0 | 0 | 1.343312 | 2.139694 |
| ARCHS4_Ti | HUMAN EN | 13/2316 | 0.203345 | 0.999975 | 0 | 0 | 1.343312 | 2.139694 |
| ARCHS4_Ti | VASCULAR | 13/2316 | 0.203345 | 0.999975 | 0 | 0 | 1.343312 | 2.139694 |
| ARCHS4_Ti | PERICARDI | 13/2316 | 0.203345 | 0.999975 | 0 | 0 | 1.343312 | 2.139694 |
| ARCHS4_Ti | STROMAL ( | 13/2316 | 0.203345 | 0.999975 | 0 | 0 | 1.343312 | 2.139694 |
| ARCHS4_Ti | CEREBRAL | Oct-16 | 0.55993 | 0.999975 | 0 | 0 | 0.991597 | 0.57507 |
| ARCHS4_Ti | HAIR FOLLI | Oct-16 | 0.55993 | 0.999975 | 0 | 0 | 0.991597 | 0.57507 |
| ARCHS4_Ti | NEURONAI | Sep-16 | 0.690276 | 0.999975 | 0 | 0 | 0.880564 | 0.326393 |
| ARCHS4_Ti | VALVE | 14/2316 | 0.126925 | 0.999975 | 0 | 0 | 1.467182 | 3.0285 |
| ARCHS4_Ti | OMENTUM | 14/2316 | 0.126925 | 0.999975 | 0 | 0 | 1.467182 | 3.0285 |
| ARCHS4_Ti | CEREBELLU | 14/2316 | 0.126925 | 0.999975 | 0 | 0 | 1.467182 | 3.0285 |
| ARCHS4_Ti | MYOFIBRO | Sep-16 | 0.690276 | 0.999975 | 0 | 0 | 0.880564 | 0.326393 |
| ARCHS4_Ti | LUNG (BUL | Sep-16 | 0.690276 | 0.999975 | 0 | 0 | 0.880564 | 0.326393 |
| ARCHS4_Ti | FORESKIN I | Nov-16 | 0.426714 | 0.999975 | 0 | 0 | 1.105651 | 0.941617 |
| ARCHS4_Ti | BETA CELL | Jun-16 | 0.946801 | 0.999975 | 0 | 0 | 0.56447 | 0.030858 |
| ARCHS4_Ti | GASTRIC EI | Jun-16 | 0.946801 | 0.999975 | 0 | 0 | 0.56447 | 0.030858 |
| ARCHS4_Ti | GASTRIC TI | Jun-16 | 0.946801 | 0.999975 | 0 | 0 | 0.56447 | 0.030858 |
| ARCHS4_Ti | KUPFFER C | Aug-16 | 0.803553 | 0.999975 | 0 | 0 | 0.772437 | 0.168942 |
| ARCHS4_Ti | MELANOCY | Aug-16 | 0.803553 | 0.999975 | 0 | 0 | 0.772437 | 0.168942 |
| ARCHS4_Ti | NEUTROPH | Aug-16 | 0.803553 | 0.999975 | 0 | 0 | 0.772437 | 0.168942 |
| ARCHS4_Ti | ERYTHROB | Sep-16 | 0.690276 | 0.999975 | 0 | 0 | 0.880564 | 0.326393 |
| ARCHS4_Ti | SPLEEN (BU | Oct-16 | 0.55993 | 0.999975 | 0 | 0 | 0.991597 | 0.57507 |
| ARCHS4_Ti | ASTROCYTI | Dec-16 | 0.30459 | 0.999975 | 0 | 0 | 1.222847 | 1.453707 |
| ARCHS4_Ti | CINGULATE | Nov-16 | 0.426714 | 0.999975 | 0 | 0 | 1.105651 | 0.941617 |
| ARCHS4_Ti | SKIN (BULK | Jul-16 | 0.88992 | 0.999975 | 0 | 0 | 0.667107 | 0.0778 |

| | | | | | | | | |
|---|---|---|---|---|---|---|---|---|
| ARCHS4_Ti | FETAL BRAI | Jul-16 | 0.88992 | 0.999975 | 0 | 0 | 0.667107 | 0.0778 |
| ARCHS4_Ti | INTESTINAI | Jul-16 | 0.88992 | 0.999975 | 0 | 0 | 0.667107 | 0.0778 |
| ARCHS4_Ti | LUNG EPITI | Jul-16 | 0.88992 | 0.999975 | 0 | 0 | 0.667107 | 0.0778 |
| ARCHS4_Ti | BLASTOCYS | Aug-16 | 0.803553 | 0.999975 | 0 | 0 | 0.772437 | 0.168942 |
| ARCHS4_Ti | MIDBRAIN | Oct-16 | 0.55993 | 0.999975 | 0 | 0 | 0.991597 | 0.57507 |
| ARCHS4_Ti | KIDNEY (BU | Oct-16 | 0.55993 | 0.999975 | 0 | 0 | 0.991597 | 0.57507 |
| ARCHS4_Ti | LIVER (BUL | Sep-16 | 0.690276 | 0.999975 | 0 | 0 | 0.880564 | 0.326393 |
| ARCHS4_Ti | TLYMPHOC | Feb-16 | 0.999725 | 0.999975 | 0 | 0 | 0.178952 | 4.92E-05 |
| ARCHS4_Ti | PODOCYTE | Nov-16 | 0.426714 | 0.999975 | 0 | 0 | 1.105651 | 0.941617 |
| ARCHS4_Ti | SPINAL COI | Oct-16 | 0.55993 | 0.999975 | 0 | 0 | 0.991597 | 0.57507 |
| ARCHS4_Ti | CORD BLOC | Apr-16 | 0.993042 | 0.999975 | 0 | 0 | 0.366886 | 0.002562 |
| ARCHS4_Ti | CD4+ T CEL | Apr-16 | 0.993042 | 0.999975 | 0 | 0 | 0.366886 | 0.002562 |
| ARCHS4_Ti | CD34+ CELI | Apr-16 | 0.993042 | 0.999975 | 0 | 0 | 0.366886 | 0.002562 |
| ARCHS4_Ti | TRACHEA ( | Jun-16 | 0.946801 | 0.999975 | 0 | 0 | 0.56447 | 0.030858 |
| ARCHS4_Ti | PLACENTA | Aug-16 | 0.803553 | 0.999975 | 0 | 0 | 0.772437 | 0.168942 |
| ARCHS4_Ti | TESTIS (BU | Jun-16 | 0.946801 | 0.999975 | 0 | 0 | 0.56447 | 0.030858 |
| ARCHS4_Ti | MAMMARY | Jun-16 | 0.946801 | 0.999975 | 0 | 0 | 0.56447 | 0.030858 |
| ARCHS4_Ti | GRANULOC | Jun-16 | 0.946801 | 0.999975 | 0 | 0 | 0.56447 | 0.030858 |
| ARCHS4_Ti | KERATINOC | May-16 | 0.978503 | 0.999975 | 0 | 0 | 0.464428 | 0.010093 |
| ARCHS4_Ti | ESOPHAGU | May-16 | 0.978503 | 0.999975 | 0 | 0 | 0.464428 | 0.010093 |
| ARCHS4_Ti | DENDRITIC | May-16 | 0.978503 | 0.999975 | 0 | 0 | 0.464428 | 0.010093 |
| ARCHS4_Ti | SPINAL COI | Oct-16 | 0.55993 | 0.999975 | 0 | 0 | 0.991597 | 0.57507 |
| ARCHS4_Ti | AMNIOTIC | May-16 | 0.978503 | 0.999975 | 0 | 0 | 0.464428 | 0.010093 |
| ARCHS4_Ti | ALVEOLAR | May-16 | 0.978503 | 0.999975 | 0 | 0 | 0.464428 | 0.010093 |
| ARCHS4_Ti | ALPHA CEL | May-16 | 0.978503 | 0.999975 | 0 | 0 | 0.464428 | 0.010093 |
| ARCHS4_Ti | WHARTON | Jun-16 | 0.946801 | 0.999975 | 0 | 0 | 0.56447 | 0.030858 |
| ARCHS4_Ti | MYOBLAST | Nov-16 | 0.426714 | 0.999975 | 0 | 0 | 1.105651 | 0.941617 |
| ARCHS4_Ti | HUMAN ZY | Nov-16 | 0.426714 | 0.999975 | 0 | 0 | 1.105651 | 0.941617 |
| ARCHS4_Ti | HEPATOCY | Nov-16 | 0.426714 | 0.999975 | 0 | 0 | 1.105651 | 0.941617 |
| ARCHS4_Ti | GRANULOS | Dec-16 | 0.30459 | 0.999975 | 0 | 0 | 1.222847 | 1.453707 |
| ARCHS4_Ti | FETAL LUN | Dec-16 | 0.30459 | 0.999975 | 0 | 0 | 1.222847 | 1.453707 |
| ARCHS4_Ti | BRAIN (BUI | Dec-16 | 0.30459 | 0.999975 | 0 | 0 | 1.222847 | 1.453707 |
| ARCHS4_Ti | SPERM | Nov-16 | 0.426714 | 0.999975 | 0 | 0 | 1.105651 | 0.941617 |
| ARCHS4_Ti | SMALL INT | Nov-16 | 0.426714 | 0.999975 | 0 | 0 | 1.105651 | 0.941617 |
| ARCHS4_Ti | MOTOR NE | Oct-16 | 0.55993 | 0.999975 | 0 | 0 | 0.991597 | 0.57507 |
| ARCHS4_Ti | THYROID (E | Aug-16 | 0.803553 | 0.999975 | 0 | 0 | 0.772437 | 0.168942 |
| ARCHS4_Ti | SKELETAL N | Aug-16 | 0.803553 | 0.999975 | 0 | 0 | 0.772437 | 0.168942 |
| ARCHS4_Ti | ILEUM (BU | Aug-16 | 0.803553 | 0.999975 | 0 | 0 | 0.772437 | 0.168942 |
| ARCHS4_Ti | HEPATIC ST | Aug-16 | 0.803553 | 0.999975 | 0 | 0 | 0.772437 | 0.168942 |
| ARCHS4_Ti | OOCYTE | Oct-16 | 0.55993 | 0.999975 | 0 | 0 | 0.991597 | 0.57507 |
| ARCHS4_Ti | REGULATO | Apr-16 | 0.993042 | 0.999975 | 0 | 0 | 0.366886 | 0.002562 |
| ARCHS4_Ti | PLASMACY | Apr-16 | 0.993042 | 0.999975 | 0 | 0 | 0.366886 | 0.002562 |
| ARCHS4_Ti | PLASMA CE | Apr-16 | 0.993042 | 0.999975 | 0 | 0 | 0.366886 | 0.002562 |
| ARCHS4_Ti | NATURAL K | Apr-16 | 0.993042 | 0.999975 | 0 | 0 | 0.366886 | 0.002562 |
| ARCHS4_Ti | RENAL COF | May-16 | 0.978503 | 0.999975 | 0 | 0 | 0.464428 | 0.010093 |

| | | | | | | | |
|---|---|---|---|---|---|---|---|
| ARCHS4_Ti RETINA | Jan-16 | 0.999975 | 0.999975 | 0 | 0 | 0.088392 | 2.22E-06 |
| ARCHS4_Ti COLONIC M | Jan-16 | 0.999975 | 0.999975 | 0 | 0 | 0.088392 | 2.22E-06 |
| ARCHS4_Ti ALVEOLAR | Jul-16 | 0.88992 | 0.999975 | 0 | 0 | 0.667107 | 0.0778 |
| ARCHS4_Ti PREFRONT. | May-16 | 0.978503 | 0.999975 | 0 | 0 | 0.464428 | 0.010093 |
| ARCHS4_Ti PREADIPOC | May-16 | 0.978503 | 0.999975 | 0 | 0 | 0.464428 | 0.010093 |
| ARCHS4_Ti PERIPHERA | May-16 | 0.978503 | 0.999975 | 0 | 0 | 0.464428 | 0.010093 |
| ARCHS4_Ti OVARY (BU | May-16 | 0.978503 | 0.999975 | 0 | 0 | 0.464428 | 0.010093 |
| ARCHS4_Ti BONE MAR | Feb-16 | 0.999725 | 0.999975 | 0 | 0 | 0.178952 | 4.92E-05 |
| ARCHS4_Ti BLOOD DEI | Mar-16 | 0.998311 | 0.999975 | 0 | 0 | 0.271756 | 0.000459 |
| ARCHS4_Ti SENSORY N | Apr-16 | 0.993042 | 0.999975 | 0 | 0 | 0.366886 | 0.002562 |
| ARCHS4_Ti CD19+ B CI | May-16 | 0.978503 | 0.999975 | 0 | 0 | 0.464428 | 0.010093 |
| ARCHS4_Ti BLYMPHOC | May-16 | 0.978503 | 0.999975 | 0 | 0 | 0.464428 | 0.010093 |
| ARCHS4_Ti BLOOD PBI | May-16 | 0.978503 | 0.999975 | 0 | 0 | 0.464428 | 0.010093 |
| ARCHS4_Ti BASAL CELI | May-16 | 0.978503 | 0.999975 | 0 | 0 | 0.464428 | 0.010093 |

Genes
GCKR;CEP85L;SLC35F1;KIAA1755;GJA1;B3GNT7;APOC1;ZNF219;SPEG;ARHGEF40;CCDC141;CD34;MYH6
KIAA1755;GJA1;SLC35F1;CCDC141;MYH6
SLC5A6;GCKR;SLC4A1AP;SNX17;ALX4;ZNF513;C2ORF16;ZNF512;APOE;GPN1;NRBP1;PPM1G
GCKR;CEP85L;SLC35F1;KIAA1755;GJA1;CCDC91;B3GNT7;APOC1;GNB4;SPEG;ARHGEF40;CCDC141;CD34
SLC5A6;GCKR;SLC4A1AP;SNX17;ALX4;ZNF513;C2ORF16;ZNF512;GPN1;NRBP1;PPM1G
CCDC121;GCKR;TOMM40;C2ORF16;ZNF512;GPN1
KIAA1755;GJA1;GCKR;APOC1;SLC35F1;ARHGEF40;CCDC141;MYH6
KIAA1755;GCKR;GJA1;B3GNT7;GNB4;SPEG;SLC35F1;APOE;ARHGEF40;CCDC141;CD34;MYH6
CENPW;SERPINA1;SHOX2;SSC5D;CCDC91;NOV;ZNF628;RSRC1;APOC1;ZNF219;GPR126;HORMAD2;LCOI
SLC4A1AP;CR1;TOMM40;APOC1;APOE;PVRL2;DGKH
SLC5A6;EIF2B4;GCKR;SLC4A1AP;SNX17;ZNF512;GPN1;NRBP1;PPM1G
CENPW;SERPINA1;SHOX2;AFAP1;MLF1;CCDC91;ZNF628;RSRC1;ZNF219;HORMAD2;APOE;LCORL;CD34
GJA1;GCKR;APOC1;SLC35F1;ARHGEF40;CCDC141;MYH6
PLN;DES;MYH6;TTN;MYH7
PLN;DES;MYH6;TTN;MYH7
GCKR;C2ORF16;ZNF512;GPN1
GCKR;CENPW;SERPINA1;MTMR3;SHOX2;SSC5D;WARS2;TBX15;CCDC91;NOV;ZNF628;GPR126;HORMAD
PLN;DES;MYH6;TTN;MYH7
PLN;MYH6;TTN;MYH7
PLN;DES;MYH6;TTN;MYH7
DES;MYH6;TTN;MYH7
CCDC121;GCKR;TOMM40;C2ORF16;ZNF512;GPN1
DES;MYH6;MYH7
EIF2B4;TBX15;CENPW;SERPINA1;CCDC91;NOV;ZNF628;DLEU1;AFAP1;GPR126;HORMAD2;LCORL
PLN;DES;MYH6;TTN;MYH7
MLLT10;CENPW;SERPINA1;CCDC91;ZNF628;RSRC1;SHOX2;ZNF219;GPR126;SSC5D;HORMAD2;LCORL
KIAA1755;GJA1;CCDC141
EIF2B4;GCKR;ZCCHC10;SLC4A1AP;TOMM40;C2ORF16;GPN1;NIPSNAP1;NRBP1;PPM1G;SNX17;APOC1;[illegible]
HSPA4;AFAP1;CCDC141;ARHGEF40;MYH7
SLC5A6;GCKR;C2ORF16;GPN1
GCKR;CENPW;MTMR3;PTEN;MLLT10;DNAJC1;CCDC91;NOV;ZNF628;DLEU1;GPR126;LCORL;PVRL2;CD3[illegible]
TOMM40;APOC1;APOE;PVRL2
TOMM40;APOC1;APOE;WNT16
PLN;DES;TTN;MYH7
GCKR;TOMM40;SNX17;APOC1;C2ORF16;ZNF512;APOE;PVRL2;GPN1
PLN;DES;SPEG;MYH6;MYH7;TTN
KIAA1755;CEP85L;FAM3C;CCDC141;MYH6
KIAA1755;CCDC141;MYH6
APOC1;APOE;PVRL2
GCKR;DES;PLN;CEP85L;GNB4;SLC35F1;MYH6;MYH7;TTN
PLN;DES;MYH6;TTN;MYH7
GJA1;DES;PLN;PTEN;MYH6;MYH7;TTN
GCKR;SERPINA1;MTMR3;ZNF628;DLEU1;ARHGEF40;LCORL
GCKR;C2ORF16;GPN1

MLLT10;KIAA1755;SERPINA1;MTMR3;CCDC91;RSPO2;ALX4;FAM3C;APOE;ARHGEF40;WARS2
CENPW;SERPINA1;CCDC91;ZNF628;RSRC1;ZNF219;AFAP1;SSC5D;HORMAD2;LCORL
GCKR;CENPW;MTMR3;PTEN;MLLT10;DNAJC1;CCDC91;NOV;DLEU1;GPR126;LCORL;PVRL2;CD34;DGKH
PLN;DES;CEP85L;TTN
GCKR;SERPINA1;DLEU1;APOE;LCORL
MYH6;TTN;MYH7
GCKR;CENPW;SERPINA1;MTMR3;PTEN;MLLT10;CCDC91;NOV;ZNF628;GPR126;LCORL;PVRL2;CD34;DGK
GJA1;PLN;DES;HSPA4;TTN
MTMR3;DES;PTEN;SPEG;MYH7;TTN
TOMM40;SERPINA1;APOC1;APOE
GJA1;DES;PLN;PTEN;MYH6;MYH7;TTN
TBX15;GCKR;SERPINA1;CCDC91;SNX17
GCKR;CENPW;SERPINA1;MTMR3;PTEN;MLLT10;CCDC91;NOV;ZNF628;GPR126;ARHGEF40;LCORL;CD34;
GCKR;CENPW;MTMR3;PTEN;MLLT10;DNAJC1;CCDC91;NOV;DLEU1;GPR126;LCORL;PVRL2;DGKH
GCKR;CENPW;MTMR3;SLC35F1;SSC5D;CWC27;MLLT10;CCDC91;GPR126;ZNF512;ARHGEF40;LCORL;DG
PLN;DES;CEP85L;MYH6;MYH7;TTN
GCKR;CENPW;SERPINA1;MTMR3;PTEN;MLLT10;CCDC91;NOV;ZNF628;GPR126;ARHGEF40;LCORL;CD34;
GCKR;CENPW;SERPINA1;PTEN;AFAP1;SSC5D;TTN;CCDC91;NOV;ZNF628;RSRC1;GPR126;HORMAD2;ARH
PLN;DES;MYH6;MYH7;TTN
PLN;DES;MYH6;MYH7;TTN
GCKR;CENPW;SERPINA1;MTMR3;PTEN;MLLT10;CCDC91;NOV;ZNF628;GPR126;ARHGEF40;LCORL;CD34;
DES;PLN;GNB4;SLC35F1;MYH6;MYH7;TTN
GCKR;CENPW;SERPINA1;MTMR3;SHOX2;PTEN;AFAP1;MLF1;TBX15;CCDC91;NOV;ZNF628;RSRC1;GPR12
PLN;DES;MYH6;MYH7;TTN
GJA1;PLN;APOE;CD34;MYH7;TTN
GJA1;DES;PLN;CD34;MYH6;MYH7;TTN
GJA1;DES;PLN;CD34;MYH6;MYH7;TTN
TOMM40;SERPINA1;APOE
SLC5A6;GCKR;TOMM40;APOC1;DLEU1;ZNF512;LCORL;PPM1G
TBX15;GCKR;SERPINA1;SNX17
MLLT10;KIAA1755;SERPINA1;CCDC91;RSPO2;ALX4;APOE;WNT16;ARHGEF40;WARS2
GFM1;LXN;HSPA4;CEP85L;SLC35F1;MLF1;TTN;KIAA1755;RSRC1;GNB4;DLEU1;CCDC141;LCORL;MYH6
TOMM40;CR1;APOE
SERPINA1;RSPO2;ALX4;WNT16;CWC27;LCORL
DNAJC1;GJA1;PLN;NOV;RSRC1;SLC35F1;CCDC141;LCORL
GCKR;CENPW;MTMR3;PTEN;MLLT10;DNAJC1;CCDC91;NOV;DLEU1;GPR126;LCORL;PVRL2;CD34;DGKH
MLLT10;GCKR;CENPW;MTMR3;CCDC91;NOV;GPR126;SLC35F1;SSC5D;ZNF512;LCORL;DGKH
MLLT10;DNAJC1;GCKR;CENPW
GCKR;CENPW;SERPINA1;MTMR3;PTEN;MLLT10;CCDC91;NOV;GPR126;ARHGEF40;LCORL;CD34;DGKH
SERPINA1;PTEN;AFAP1;SLC35F1;CWC27;GPN1;MLF1;TTN;MLLT10;RSRC1;FAM174B;DLEU1;APOE;LCOR
MLLT10;GJA1;PTEN;NDRG2
TBX15;GCKR;TOMM40;CCDC91;CWC27;LCORL
GCKR;APOE;ZNF512;GPN1
TBX15;GCKR;CENPW;CCDC91;RSRC1;FAM174B;DLEU1;GPR126;LCORL
MLLT10;GCKR;CENPW;MTMR3;CCDC91;GPR126;SSC5D;ZNF512;CWC27;ARHGEF40;LCORL;DGKH

TBX15;MLLT10;DCAF16;DNAJC1;GCKR;CENPW;GPR126;APOE;LCORL;WARS2
KIAA1755;CEP85L;GNB4;SLC35F1;MYH6;MYH7;TTN
PLN;DES;MYH6;MYH7;TTN
GJA1;MYH6;TTN;MYH7
SHOX2;MYH6;MYH7
DES;PTEN;SPEG;MYH7;TTN
DES;MYH7;TTN
GCKR;GJA1;DES;PLN;HSPA4;SHOX2;SLC35F1;MYH7;TTN
DES;TTN;MYH7
GCKR;TOMM40;C2ORF16;ZNF512;GPN1
PLN;DES;MYH6;MYH7;TTN
CR1;TOMM40;CR1L;APOC1;APOE
DES;SPEG;TTN;MYH7
TOMM40;CR1;APOC1;APOE
GCKR;APOE;PVRL2
DES;PLN;CCDC141;MYH6;MYH7;TTN
MLLT10;DNAJC1;SNX17;DLEU1;CWC27
DNAJC1;APOE;PVRL2
AFAP1;CCDC141;ARHGEF40;MYH6
KIAA1755;CEP85L;MYH6
MLLT10;HSPA4;PTEN;SLC35F1;APOE;LCORL;MLF1
TOMM40;APOC1;APOE
MLLT10;SKIDA1
PLN;TTN;MYH7
GJA1;PLN;FAM174B;MFN1;MLF1;MYH6;TTN
GJA1;DES;PLN;APOC1;PTEN;MYH6;MYH7;TTN
SPEG;TTN;MYH7
EIF2B4;MLLT10;GCKR;SERPINA1;TOMM40;APOC1;C2ORF16;ZNF512;PPM1G
KIAA1755;GJA1;MYH6
TOMM40;APOC1;APOE;PVRL2
TOMM40;APOC1;APOE;PVRL2
MLLT10;GCKR;CENPW;MTMR3;CCDC91;NOV;GPR126;SSC5D;ZNF512;LCORL;DGKH
DES;PTEN;TTN
DES;TTN;MYH7
DES;MYH6;MYH7
GCKR;MTMR3;ZCCHC10;TOMM40;SNX17;ZMAT5;DLEU1;SPEG;APOE;DGKH
GCKR;ZCCHC10;TOMM40;SNX17;DLEU1;SPEG;APOE;DGKH
GCKR;SERPINA1;SNX17;RSRC1;RSPO2;C2ORF16;HORMAD2;NRBP1;PPM1G
GCKR;APOE;ZNF512
GCKR;C2ORF16;ZNF513
MLLT10;MFN1;DLEU1
CR1;CR1L
GNB4;ALX4;ZNF512;CD34
GCKR;SERPINA1;CR1;TOMM40;PTEN;APOE;TTN
SERPINA1;RSPO2;ALX4;CWC27;LCORL

KIAA1755;CCDC141;MYH6
DES;PLN;MYH6;MYH7;TTN
MLLT10;DNAJC1;APOE
PLN;DES;MYH6;MYH7;TTN
KIAA1755;CCDC91;RSPO2;AFAP1;ALX4;APOE
GCKR;TOMM40;APOE
GCKR;APOE
GCKR;APOE
CCDC141;TTN
EIF2B4;MLLT10;GCKR;SERPINA1;SLC4A1AP;TOMM40;SNX17;APOC1;APOE
KIAA1755;SERPINA1;CCDC91;RSPO2;AFAP1;ALX4;APOE;LCORL;WARS2
GCKR;TOMM40;HSPA4;SNX17;APOC1;DLEU1;APOE;LCORL
KIAA1755;DLEU1;SLC35F1;CCDC141;MYH6;WARS2;TTN
MLLT10;GCKR;SERPINA1;APOE
CR1;CR1L
GCKR;APOE
SNX17;APOE
GCKR;TOMM40
GJA1;MYH6;TTN
MLLT10;DNAJC1;APOE
SERPINA1;RSPO2;ALX4;CWC27;LCORL
GCKR;SERPINA1;MTMR3;CCDC91;HSPA4;HORMAD2;APOE;LCORL
MLLT10;DNAJC1;GCKR;CENPW;MTMR3;CCDC91;GPR126;LCORL;PVRL2;DGKH
SERPINA1;RSPO2;ALX4;CWC27;LCORL
PLN;DES;MYH6;MYH7;TTN
TOMM40;APOE
GCKR;SERPINA1;NDRG2
TOMM40;APOE
GCKR;APOE
GCKR;APOE
CR1;CR1L
EIF2B4;GCKR;SERPINA1;ZCCHC10;SNX17;APOE;PPM1G
GCKR;SERPINA1;CCDC91;ZNF628;SNX17;SHOX2;DLEU1;HORMAD2;ZNF512;NDRG2;LCORL;DGKH
MLLT10;CCDC91;AFAP1;ALX4;ASCC2
ZNF219;TMEM253
SNX17;APOE
GCKR;APOC1
GCKR;APOE
APOC1;APOE
EIF2B4;GCKR;SERPINA1;SLC4A1AP;TOMM40;SNX17;APOC1;DLEU1;HORMAD2;ZNF512;APOE;NDRG2
SNX17;APOE
TBX15;ALX4
GJA1;DES;PLN;PTEN;APOE;MYH6;MYH7;TTN
GJA1;PLN;APOE;MYH6
GJA1;PLN;DES;TTN

SERPINA1;ZCCHC10;TOMM40;APOC1;APOE
TBX15;MTMR3;CCDC91;ZNF628;DLEU1;AFAP1;NDRG2;LCORL
GCKR;TOMM40;CR1L;APOC1;PTEN;APOE;DGKH
DES;PLN;MYH6;MYH7;TTN
DES;TTN;MYH7
DES;PLN
PLN;MYH7
MYH6;TTN
PLN;MYH7
DES;PLN
GJA1;NDRG2
DES;PLN
TTN;MYH7
GJA1;APOE
GCKR;DNAJC1;APOE
TBX15;CENPW;DLEU1;WNT16;WARS2
TBX15;CENPW;DLEU1;WNT16;WARS2
SERPINA1;RSPO2;WNT16;CWC27;LCORL
TBX15;CENPW;CCDC91;HSPA4;FAM3C;WNT16;LCORL
APOC1;PVRL2
MTMR3;SKIDA1
SERPINA1;RSPO2;EIF3E;CWC27;LCORL
GCKR;TOMM40;APOC1;C2ORF16;CCDC141;TTN
TOMM40;CR1;CR1L
GCKR;TOMM40;APOC1;APOE
CCDC141;ARHGEF40;MYH6
GCKR;APOE
TOMM40;APOE
GCKR;SERPINA1;APOE
GCKR;APOE
KIAA1755;CCDC141
CCDC141;TTN
GCKR;APOC1
GCKR;TOMM40;C2ORF16
MLLT10;DNAJC1;SKIDA1
DES;TTN;MYH7
GJA1;DES;SPEG;MYH7;TTN
GJA1;DES;PLN;HSPA4;PTEN;CD34;MYH6;MYH7;TTN
GJA1;DES;SPEG;MYH7;TTN
GJA1;DES;SPEG;MYH7;TTN
MYH7;TTN
MYH7;TTN
CR1;SLC4A1AP;TOMM40;APOC1;APOE;PVRL2
TBX15;GCKR;PTEN;ALX4
TBX15;CENPW;DLEU1;WNT16;WARS2

GCKR;APOC1;C2ORF16
EIF2B4;GCKR;SERPINA1;SLC4A1AP;TOMM40;SNX17;APOC1;APOE
DES;PLN;PTEN;SPEG;MYH6;MYH7;TTN
MLLT10;GCKR;SERPINA1;TOMM40;APOE
SPEG;TTN;MYH7
TOMM40;APOC1;APOE
GCKR;SNX17
KIAA1755;CCDC141
SERPINA1;APOE
GCKR;SERPINA1
CEP85L;ASCC2
EIF2B4;GCKR;SERPINA1;MTMR3;DLEU1;HORMAD2;CD34
GCKR;SERPINA1;MTMR3;TOMM40;CR1L;APOE
IFI27L2;SNX17;FAM3C;CCDC141;NRBP1;DGKH;PPM1G
ALX4;LCORL;WARS2
GCKR;TOMM40;APOC1;APOE
DES;SPEG;MYH7
MTMR3;HORMAD2
MLLT10;KIAA1755;AFAP1;ALX4
GNB4;SLC35F1;MYH7;TTN
EIF2B4;GCKR;SERPINA1;ZCCHC10;SLC4A1AP;TOMM40;SNX17;APOC1;APOE
GCKR;CCDC121;TOMM40;SLC35F1;C2ORF16;GPN1;NRBP1;TTN;SNX17;APOC1;ZNF512;APOE;CCDC141;
GCKR;NRBP1
DLEU1;CCDC141
GCKR;MTMR3;CCDC91;HSPA4;SNX17;DLEU1;APOE;LCORL
APOC1;APOE
GCKR;SERPINA1;NDRG2
APOC1;APOE
MTMR3;PTEN;MYH6;PPM1G
GCKR;SLC4A1AP;ZNF512;GPN1
GCKR;SLC4A1AP;ZNF512;GPN1
GCKR;APOC1;APOE
MTMR3;ZNF628;DLEU1;NDRG2
TBX15;SERPINA1;PTEN;HORMAD2;LCORL
SERPINA1;CCDC91;GPR126;HORMAD2;LCORL
TOMM40;APOE
DES;TTN;MYH7
MLLT10;CENPW;RSRC1;APOC1;DLEU1;APOE;LCORL;TRIB2
APOC1;APOE
DES;TTN;MYH7
MYH6;TTN;MYH7
DES;TTN;MYH7
DES;TTN;MYH7
GCKR;APOC1;APOE
CEP85L;RSPO2;FAM3C

GCKR;SERPINA1;RSRC1;GPN1
CCDC121;CMTM5;SHOX2;AFAP1;VSTM2L;SSC5D;MLF1;TTN;TBX15;KIAA1755;GJA1;DES;PLN;NOV;ZNF6[illegible]
MYH6;MYH7
MYH6;MYH7
MYH7;TTN
TBX15;CCDC91;RSRC1;CWC27;TTN
GCKR;APOC1;APOE
CEP85L;SLC35F1;MYH6;TTN
GCKR;APOC1;PTEN
GCKR;ZNF512
CCDC141;TTN
APOE;LCORL
MLLT10;DNAJC1;MTMR3;HSPA4;SKIDA1;DLEU1;HORMAD2;CD34
APOC1;APOE
APOC1;PVRL2
GCKR;TOMM40;APOE
MLLT10;MLF1
TBX15;CCDC91;LCORL
EIF2B4;GCKR;SERPINA1;ZCCHC10;SNX17;DGKH
MTMR3;ZNF219;NCAPG;LCORL;TTN
TBX15;CENPW;SERPINA1;MTMR3;PTEN;ARHGEF40;LCORL
MLLT10;CENPW;RSRC1;APOC1;DLEU1;APOE;LCORL;TRIB2
EIF2B4;GCKR;SERPINA1;SLC4A1AP;ARHGEF40
DES;PLN
GFM1;SNX17;FAM174B;APOC1;MFN1;CD34;MLF1;MYH6;TTN
GCKR;APOE;PPM1G
TOMM40;APOC1;APOE
DES;SPEG;MYH7
GJA1;DES;PLN;PTEN;MYH7
DES;TTN;MYH7
DES;TTN
MYH7;TTN
MLLT10;PTEN
MYH6;MYH7
MLLT10;PTEN
MLLT10;PTEN
MLLT10;PTEN
MLLT10;PTEN
MLLT10;PTEN
MLLT10;PTEN
MLLT10;PTEN
MLLT10;PTEN
SPEG;NRBP1;TTN
MYH6;TTN;MYH7
PLN;SPEG;MYH7;TTN

GJA1;DES;SPEG;MYH7;TTN
GJA1;SERPINA1;MTMR3;PTEN;ALX4;HORMAD2;APOE
MLLT10;PTEN
GFM1;LXN;WARS2
PTEN;GPN1
MLLT10;PTEN
MLLT10;PTEN
MLLT10;PTEN
CR1;TOMM40;APOC1;APOE
GJA1;PTEN;RSPO2;SPEG;TTN
DES;PLN;CEP85L;SPEG;MYH6;MYH7;TTN
MLLT10;PTEN
MLLT10;PTEN
MLLT10;PTEN
MLLT10;PTEN
MLLT10;PTEN
MLLT10;PTEN
MLLT10;PTEN
MLLT10;PTEN
TOMM40;APOE
CCDC141;TTN
TOMM40;APOE
TBX15;GJA1;RSPO2;ALX4;EIF3E;WARS2
EIF2B4;GCKR;SERPINA1;SLC4A1AP;SNX17;APOC1;PTEN;DLEU1;LCORL;PPM1G
CEP85L;MYH6;TTN
GCKR;SERPINA1;APOC1
GJA1;DES;PLN;MYH6;MYH7;TTN
ARHGEF40;CCDC141
MLLT10;HSPA4
SNX17;CWC27
GCKR;TOMM40
GCKR;APOC1;MYH6;MYH7;TTN
TOMM40;APOC1;PVRL2
MLLT10;DNAJC1;CCDC91;GPR126;APOE;LCORL;PVRL2;PPM1G
GCKR;SERPINA1;NDRG2
CR1L;CD34
CR1;APOE;CD34;MYH7
EIF2B4;GCKR;SERPINA1;TOMM40;SNX17;APOC1;DLEU1;ASCC2;APOE;NDRG2
TBX15;CCDC91;LCORL
TBX15;RSPO2;EIF3E
SLC5A6;EIF2B4;GCKR;SLC4A1AP;SNX17;TRIB2;PPM1G
TBX15;MLLT10;ZCCHC10;TOMM40;HSPA4;RSRC1;DLEU1;APOE;WARS2
TBX15;CENPW;WNT16;WARS2
TTN;MYH7
GCKR;RSRC1;LCORL

SERPINA1;TOMM40;HSPA4;APOC1;PTEN;APOE
SERPINA1;TOMM40;HSPA4;APOC1;PTEN;APOE
DES;TOMM40;HSPA4;APOE;MLF1
GCKR;CCDC91;HSPA4;SNX17;DLEU1;APOE;LCORL
GJA1;DES;TTN
NCAPG;LCORL
TBX15;GCKR;CENPW;SERPINA1;CCDC91;GPR126;SPEG;CD34
MLLT10;PTEN
GCKR;DGKH
GCKR;SERPINA1;TOMM40;SNX17;APOC1;ZNF512;APOE;GPN1
CENPW;APOE;LCORL
TBX15;CCDC91;LCORL
DLEU1;CWC27;TTN
TBX15;WARS2
MLLT10;GJA1;PTEN
TBX15;SSC5D;TRIB2
GCKR;MTMR3;CABP7;HORMAD2;TRIB2
GCKR;TOMM40;APOC1
GCKR;TOMM40;APOE
GCKR;HSPA4;PTEN;WARS2;TTN;GJA1;DES;PLN;APOC1;APOE;CD34;MYH6;MYH7
CR1;CR1L
GJA1;DES;MYH6;MYH7;TTN
MYH6;TTN
GCKR;SERPINA1
GFM1;DES;MTMR3;PLN;UQCR10;SUPT7L;MYH6;MYH7
MLLT10;RSPO2;AFAP1;APOE
TBX15;WARS2
SERPINA1;APOE
RSRC1;SHOX2
MYH6;MYH7
MTMR3;PTEN
TBX15;RSPO2;EIF3E
CCDC91;HSPA4;RSRC1;DLEU1;EIF3E
TBX15;MTMR3;HORMAD2;NIPSNAP1
EIF2B4;GCKR;TOMM40;PTEN;ASCC2;APOE
TBX15;MLLT10;GCKR;CENPW;SERPINA1;VSTM2L;GPR126;SPEG;ARHGEF40
TBX15;CENPW;CCDC91;HSPA4;WNT16;LCORL
TBX15;CENPW;CCDC91;HSPA4;WNT16;LCORL
GCKR;SERPINA1;APOE
CABP7;ZMAT5
SERPINA1;APOC1
GCKR;MTMR3;TOMM40;APOC1;APOE;NDRG2
DES;PLN;MFN1;GNB4;MYH6;MYH7;TTN
MLLT10;PTEN
MYH6;MYH7

GJA1;DES;PLN;NDRG2;MYH6;MYH7;TTN
SERPINA1;APOC1;APOE
SLC5A6;GCKR;APOC1;C2ORF16;PPM1G
APOC1;APOE;PVRL2
MLLT10;PTEN;TTN
MTMR3;APOE
GCKR;APOC1;APOE
TBX15;CCDC91;SKIDA1;AFAP1
PTEN;HORMAD2;TRIB2
MYH7;TTN
SPEG;TTN
MYH7;TTN
DES;PLN
TOMM40;APOE
GNB4;TTN
GCKR;CWC27
CR1;TOMM40;APOE
GCKR;APOC1;APOE
EIF2B4;GCKR;SNX17;ZNF513;NRBP1
APOC1;CCDC141;PVRL2;TTN
DES;TTN;MYH7
CR1;TOMM40;HSPA4;APOC1;APOE
DES;TTN;MYH7
SERPINA1;RSPO2;CWC27;LCORL
GCKR;TOMM40;APOE
GCKR;TOMM40
GCKR;DGKH
MLLT10;CCDC91;RSPO2
SERPINA1;RSPO2;CWC27;LCORL
GJA1;TTN;MYH7
CCDC91;RSPO2
GCKR;APOE
GCKR;APOE
TBX15;CCDC91;HORMAD2
GCKR;RSPO2
DNAJC1;CD34
MYH6;MYH7
HSPA4;CD34;TTN
SPEG;NRBP1;TTN
DES;PLN;CEP85L;SPEG;CD34;MYH6;MYH7;TTN
TBX15;SERPINA1
MLLT10;CENPW;CCDC91;APOC1;GPR126;APOE;LCORL
GJA1;PLN;APOE;MYH6
GJA1;DES;PLN;APOC1;APOE;MYH7;TTN
GCKR;TOMM40;APOE

SERPINA1;PTEN;APOE
HSPA4;RSRC1;PTEN;SLC35F1;LCORL
GCKR;APOC1;APOE
GCKR;SERPINA1;DLEU1;APOE;DGKH
TOMM40;APOE
CCDC141;TTN
GCKR;APOE
DES;GNB4;TTN
KIAA1755;GCKR;MTMR3;APOE;DGKH;TTN
PLN;DES;MYH6
MYH6;MYH7
CR1;CR1L
SHOX2;MYH6;TTN
MYH6;TTN;MYH7
SNX17;APOE
MYH6;TTN;MYH7
CR1;CR1L
MFN1;MYH6;MYH7
CR1;CR1L
CR1;CR1L
MYH6;TTN;MYH7
SNX17;APOE
APOC1;APOE;DGKH
SNX17;APOE
CR1;CR1L
MYH6;TTN;MYH7
DES;MYH6;MYH7;TTN
MTMR3;PTEN
MTMR3;PTEN
DES;MYH6;MYH7;TTN
MYH6;MYH7
GCKR;TOMM40;SNX17;APOC1;ZNF512;APOE;GPN1
PTEN;CD34
MYH7;TTN
MYH6;MYH7
PLN;DES;TTN
GCKR;SERPINA1
MTMR3;HORMAD2
GCKR;CENPW;MTMR3;HSPA4;PTEN;DLEU1;CD34;TRIB2
GCKR;APOE
DNAJC1;GCKR;CENPW;CCDC91;GPR126;LCORL;PVRL2;DGKH
GCKR;GJA1;CENPW;PTEN;CD34;TRIB2
DES;MTMR3;MYH7;TTN
GJA1;APOE;MYH6
CR1;APOC1;APOE

GJA1;PLN;MYH6;MYH7
MLLT10;CCDC91;RSRC1;DLEU1;LCORL;WARS2;TTN
MTMR3;HORMAD2
SERPINA1;APOE
PLN;TTN
GJA1;PTEN;MYH7;TTN
SPEG;MYH7;TTN
PTEN;CWC27;TTN
SHOX2;MYH6
SHOX2;MYH6
GCKR;CR1
GCKR;SERPINA1;APOE
TOMM40;APOE
HSPA4;PTEN;SLC35F1;LCORL;MLF1
SKIDA1;APOE
FAM3C;WNT16
GCKR;MTMR3;HSPA4;ZMAT5;DLEU1;SPEG;APOE;DGKH;TTN
GJA1;CCDC91;AFAP1;EIF3E;FAM3C
GCKR;TOMM40
AFAP1;MYH7
TBX15;RSRC1
CR1;SLC4A1AP;TOMM40;APOE
HORMAD2;TRIB2
TTN;MYH7
CENPW;RSPO2;ALX4;WARS2
GJA1;PLN;FAM174B;MFN1;SPEG;NDRG2;CCDC141;MLF1;MYH6;TTN
GFM1;GJA1;DES;PLN;TRIB2;NRBP1;MYH6;MYH7;TTN;PPM1G
GJA1;HSPA4
DES;TTN;MYH7
MYH6;MYH7
GCKR;GJA1;DES;PLN;HSPA4;APOC1;PTEN;APOE;MYH6;MYH7;TTN
GJA1;APOE
MYH6;MYH7
APOC1;APOE
CR1;CR1L
MYH6;TTN;MYH7
SHOX2;MYH6
MLLT10;CCDC91;WARS2
TBX15;APOC1;DLEU1;UQCR10;WARS2
TBX15;SERPINA1;CCDC91;HSPA4;DLEU1;RSPO2;SLC35F1;EIF3E;FAM3C;WNT16;LCORL
GPR126;ARHGEF40;LCORL
TBX15;SERPINA1;HORMAD2;LCORL
APOC1;APOE
GJA1;APOE
DNAJC1;ASCC2

PLN;TTN
DES
GJA1
NDRG2
MYH7
APOE
GJA1
LXN
RSPO2
PVRL2
AC109829.1
APOC1
APOE
PLN
CENPW
SERPINA1
CD34
SERPINA1
CMTM5
MYH7
SERPINA1
CMTM5
MYH7
CABP7
APOC1
CMTM5
NDRG2
APOE
CD34
APOC1
NDRG2
RSPO2
APOE
LXN
APOE
APOE
APOC1
TTN
B3GNT7
CD34
CD34
WNT16
APOC1
SERPINA1
CENPW

RSPO2
CD34
PLN
APOC1
TTN
TTN
APOE
MYH7
APOC1
GJA1
GJA1
APOE
GJA1
SERPINA1
NDRG2
NDRG2
SERPINA1
APOC1
RSPO2
APOE
SERPINA1
VSTM2L
MYH7
WNT16
NCAPG
CD34
PLN
CR1
TTN
TOMM40;HSPA4
TOMM40;HSPA4
HSPA4;PTEN;APOE
DES;PTEN;CD34
DES;TTN;MYH7
DES;MYH6;MYH7;TTN
DES;SPEG;APOE;TTN
DES;MYH6;MYH7;TTN
MLLT10;CCDC91;APOE
TOMM40;APOE
DES;PLN;MLF1;MYH6;TTN
TBX15;DES;MFN1;MLF1;MYH7;TTN
DES;MYH6;MYH7;TTN
GJA1;NOV;HSPA4
DES;MYH6;MYH7;TTN
TOMM40;APOE

TBX15;TOMM40;HSPA4;APOC1;PTEN;DLEU1;APOE;WARS2
TBX15;RSRC1;WARS2
DES;MYH6;MYH7;TTN
GCKR;SERPINA1;NDRG2
DES;PTEN;CD34
SERPINA1;CR1;HSPA4;PTEN;APOE;CD34
GJA1;MYH6
GJA1;DES;PLN;CEP85L;CD34;MYH6;MYH7;TTN
MLLT10;GCKR;TOMM40
GCKR;APOE
GFM1;RSRC1;PTEN;SPEG;WARS2;TTN
GJA1;PTEN;MYH6;MYH7;TTN
TTN;MYH7
MLLT10;PTEN
APOC1;APOE
MLLT10;PTEN
MLLT10;PTEN
MLLT10;PTEN
MLLT10;PTEN
MLLT10;PTEN
MYH6;MYH7
DES;SPEG;MYH7;TTN
MLLT10;PTEN
MLLT10;PTEN
MLLT10;PTEN
MLLT10;PTEN
MLLT10;PTEN
MLLT10;PTEN
GCKR;B3GNT7;PTEN;DLEU1;TRIB2
DNAJC1;AFAP1;LCORL
GFM1;NOV;HSPA4;SHOX2;PTEN;RSPO2;CWC27
GCKR;MTMR3;TOMM40;CR1L;APOC1;APOE
MYH6;TTN
MLLT10;PLN;CMTM5;PTEN;CD34;TRIB2;MLF1;NRBP1
SERPINA1;CCDC91;HSPA4;RSRC1;MYH7
KIAA1755;CENPW;SERPINA1;RSPO2;AFAP1;ALX4;WARS2
GCKR;APOE
GCKR;APOE
IFI27L2;CCDC91;AFAP1
CENPW;RSPO2;ALX4;WARS2
GJA1;NOV;RSRC1;LCORL;MYH6
CR1;TOMM40;APOE
GJA1;APOE;WARS2
SERPINA1;TOMM40;APOC1;DLEU1;APOE;LCORL
GCKR;TOMM40;APOC1;PTEN;DGKH

CEP85L;AFAP1;WARS2
MLLT10;PTEN
MLLT10;PTEN
DES;PTEN;APOE;MYH7;TTN
GJA1;PLN;MYH6
CMTM5;VSTM2L;SSC5D;UQCR10;MLF1;TTN;KIAA1755;DES;PLN;CCDC141;CD34;MYH6;MYH7
NCAPG;TTN
APOC1;APOE
GCKR;ALX4
DES;SPEG;MYH6
MYH6;MYH7;TTN
MTMR3;PTEN
CR1;CR1L
GJA1;PLN;FAM174B;CCDC141;MYH6
AFAP1
CR1;TOMM40;CR1L;APOC1;EIF3E;APOE
EIF2B4;MLLT10;GCKR;SERPINA1;SNX17
SLC4A1AP;PLN;UQCR10;NDRG2;MYH6;MYH7;TTN
DES;UQCR10;NDRG2;MLF1;MYH6;MYH7
GCKR;CCDC91;DLEU1;ASCC2;APOE
MLLT10;PTEN
MLLT10;PTEN
MLLT10;PTEN
IFI27L2;GCKR;LXN;TOMM40;CABP7;C2ORF16;UQCR10;NIPSNAP1;WARS2;GJA1;NOV;SNX17;APOC1;ZNF
PLN;CEP85L;MYH6;MYH7;TTN
HORMAD2;C2ORF16
MLLT10;APOC1;PTEN;CD34
TTN;MYH7
SERPINA1;APOC1;PTEN;APOE
MLLT10;PTEN
MLLT10;PTEN
MLLT10;PTEN
CCDC91;RSRC1
MLLT10;PTEN
SPEG;MYH7
GJA1;PLN;APOE;TTN
TOMM40;APOE
RSPO2;EIF3E
RSRC1;NAT14
APOC1;APOE
SHOX2;LCORL
MYH7
PTEN
DES
EIF2B4

MYH6
MLLT10;DCAF16;CENPW;APOC1;LCORL;PVRL2
GCKR;PTEN;ZNF512
TBX15;RSPO2;CD34
LCORL
APOE
DNAJC1
MTMR3;ZNF219;LCORL;TTN
CWC27
SERPINA1;ASCC2;HORMAD2;UQCR10;APOE
APOE
GCKR;DGKH
GCKR;ZNF512
GCKR;SERPINA1;APOC1;PTEN;LCORL;CD34;TRIB2
GCKR;GJA1;MTMR3;HSPA4;PTEN;APOE
SHOX2;MYH6
SPEG;MYH7
GJA1;HSPA4;PTEN;ALX4
GCKR;SERPINA1;TOMM40;APOC1;ZNF512;APOE;CD34;GPN1
TTN;MYH7
PTEN;CD34
PTEN;APOE
SERPINA1;TOMM40;HSPA4;APOE
SLC5A6;GJA1;HSPA4;PTEN
CD34;MLF1
HSPA4;APOE
GJA1;LXN
SERPINA1;APOE
GFM1;WARS2
GCKR;CCDC91;RSRC1;DLEU1;NCAPG;LCORL
TBX15;GJA1;PTEN;ALX4;CWC27
HORMAD2;APOE
GCKR;DNAJC1
GJA1;PTEN
GJA1;SERPINA1;PTEN;ALX4;APOE
DES;HSPA4;PTEN;CD34
GCKR;APOE
GCKR;APOC1
MLLT10;LCORL;NIPSNAP1
APOE
SERPINA1
GJA1;SERPINA1
PTEN;HORMAD2;TRIB2
WNT16
DGKH

APOE
CR1;HSPA4;CD34;TRIB2;MLF1
DES;PTEN;APOE;MYH7;TTN
GJA1;APOE;CD34
MLLT10;CD34
GJA1;PTEN
MYH6;MYH7
GJA1;DES;PLN;MYH6;MYH7;TTN
GCKR;SERPINA1;APOC1
GCKR;TOMM40;APOE
GCKR;SERPINA1;NDRG2
TOMM40;APOC1;APOE
TOMM40;APOE
SLC5A6;GCKR;SERPINA1;APOC1;APOE;NDRG2
TTN;MYH7
GPR126;LCORL
GCKR;GJA1;PTEN;HORMAD2;APOE;CD34
CENPW;ALX4
SNX17;CWC27
GCKR;SERPINA1;SNX17;APOC1;PTEN;DLEU1;LCORL
SPEG;TTN
DES;SERPINA1;PTEN;SPEG;MLF1;TTN
KIAA1755;WARS2
GCKR;SERPINA1
GCKR;APOE
GCKR;APOC1
GCKR;TOMM40;APOE
KIAA1755;SERPINA1;RSPO2;AFAP1;ALX4;WARS2
GCKR;SERPINA1;TOMM40;APOE
CEP85L
GCKR
TOMM40
TBX15;GJA1;ALX4
MLLT10;DNAJC1
GCKR;MTMR3;HSPA4;DLEU1;APOE;LCORL
GCKR;APOC1
GCKR;APOE
KIAA1755;TTN
GCKR;APOE
GCKR;FAM3C;WNT16
APOE
GCKR;APOC1
GCKR;APOC1
GCKR;SERPINA1;APOE
GCKR;APOC1

SERPINA1;APOE
GCKR;APOC1
TBX15;WARS2
MLLT10;KIAA1755;RSPO2;APOE
GJA1;ALX4;CWC27
PTEN;CD34
CR1;CR1L
PLN;DES;MYH6
WNT16
SKIDA1
CWC27
APOE
SNX17;FAM174B;APOE;NDRG2;PVRL2
FAM3C;WNT16
GCKR;MTMR3;DLEU1;ARHGEF40;LCORL
SERPINA1;APOC1
AFAP1;APOE
APOC1;APOE
GCKR;APOE;DGKH
GCKR;B3GNT7;ASCC2;HORMAD2;NIPSNAP1
PTEN
MLLT10;KIAA1755;SERPINA1;CCDC91;AFAP1;ALX4;ARHGEF40
DES;MYH6;TTN
FAM3C;WNT16
GCKR;APOC1
GJA1;SERPINA1;PTEN;WARS2
APOE
SERPINA1;APOE
SERPINA1;APOE
MLLT10;PTEN
TTN;MYH7
TTN;MYH7
ASCC2;HORMAD2;ZNF512
GCKR;APOC1
GCKR;APOC1
GCKR;APOC1
MLLT10;GCKR;MTMR3;CCDC91;DLEU1;LCORL;DGKH
APOE
APOE
APOE
APOE
APOE
GCKR
APOE
APOE

GCKR;APOC1
GCKR;APOE
GCKR;APOC1
SERPINA1
SNX17
GCKR
TTN
APOC1
CENPW
GCKR
GCKR;APOC1
MLLT10;GCKR;RSRC1;DLEU1;APOE;CWC27;LCORL;GPN1
GCKR;DGKH
GCKR;SERPINA1;DLEU1
SERPINA1;APOE
RSPO2;APOE
GCKR;APOE
MLLT10
RSPO2
APOE
GCKR
GCKR
SERPINA1
APOE
CCDC141
DLEU1
TTN
DLEU1
APOE
MLLT10;GCKR;SERPINA1
DLEU1
APOE
APOE
APOE
APOE
GCKR
GCKR
APOE
TTN
APOE
CCDC141
SNX17
APOE
CCDC141
TOMM40

APOE
MYH6
PPM1G
SERPINA1
GCKR
GCKR
SNX17
MLLT10
APOE
GCKR;DLEU1;APOE;LCORL
APOE
APOE
APOE
APOC1
SNX17
CMTM5
CCDC91
GCKR
FAM3C
GCKR
APOE
GCKR;TOMM40;VSTM2L;C2ORF16;CWC27;WARS2;TBX15;MLLT10;CCDC91;RSRC1;APOC1;DLEU1;SPEG;
APOC1;APOE
APOC1;APOE
GCKR;APOC1
GCKR;APOC1
SERPINA1
GCKR;SERPINA1
GCKR;APOC1
GCKR;APOE
GCKR;SERPINA1
GCKR;ALX4
GCKR;APOC1
GCKR
GCKR;SERPINA1
GCKR;PPM1G
AFAP1;CWC27;CD34
MLLT10;SKIDA1
CCDC91;LCORL
GCKR;APOE
MLLT10;DLEU1
GCKR;PTEN
GCKR;APOC1
GCKR;APOC1
GCKR;APOE

APOE
APOE
WARS2
APOE
GCKR
APOC1;APOE
GCKR;APOE
FAM3C;TTN
GCKR;APOC1
GCKR;APOE
GCKR;APOC1
GCKR;SERPINA1
RSPO2
SERPINA1
GCKR
APOE
APOE
APOE
APOE
APOE
DLEU1
APOE
APOE
APOE
APOE
APOE
GCKR
APOE
APOE
APOE
GCKR
GCKR
APOC1
GCKR
APOE
GCKR
DLEU1
DLEU1
ZCCHC10
GCKR
APOE
SERPINA1
GCKR
GCKR;APOC1
GCKR;APOC1

GCKR
CCDC141
GCKR
APOC1
GCKR;APOE
GCKR;APOC1
GCKR;APOC1
GCKR;APOE
GCKR;APOE
GCKR;APOC1
APOE
GCKR
CWC27
CCDC141
MLLT10;RSRC1;APOE;CWC27;LCORL
GCKR;APOC1
GCKR;APOC1
GCKR;SERPINA1;RSRC1
CCDC91;HSPA4;AFAP1
GCKR;APOE
CCDC91;RSRC1;DLEU1
GCKR
APOE
TBX15
GCKR
APOE
APOE
GCKR
CWC27
APOE
GCKR
APOE
APOE
APOE
APOE
APOE
GCKR
TBX15;SERPINA1;CCDC91;ZNF628;DLEU1;RSPO2;NCAPG;NDRG2;LCORL
TBX15;TOMM40;HSPA4;CABP7;APOC1;PTEN;DLEU1;APOE;WARS2
GCKR;APOE
APOE
MLLT10
SERPINA1
APOE
GCKR

APOE
SERPINA1
APOE
GCKR;APOE
CCDC141;TTN
APOC1;APOE
DLEU1;SLC35F1;CCDC141
GCKR;SERPINA1;AFAP1;C2ORF16
GCKR
GCKR
CEP85L
TOMM40
GCKR
SERPINA1;RSRC1;C2ORF16;LCORL
APOE
APOE
APOC1
TTN
GCKR
CCDC91
CCDC141
APOE
SERPINA1
APOE
GCKR
GCKR
APOE
APOC1
FAM3C
GCKR
GCKR;APOC1
GCKR;SERPINA1;ZMAT5;APOE;LCORL
GCKR;APOC1
APOC1
APOE
MYH6;TTN
GCKR;SERPINA1
GCKR;SERPINA1
APOE
TTN
TTN
GCKR
GCKR
SKIDA1
SERPINA1

GCKR
DGKH
GCKR;APOC1
GCKR;APOE
TBX15;CCDC91
GCKR;APOE
GCKR;APOC1
GCKR;APOC1
FAM3C
GCKR;SERPINA1;HSPA4;LCORL
TBX15;CCDC91
CCDC141
GCKR
GCKR;APOC1
APOE
TOMM40;APOC1
APOE
APOE
GCKR;APOE
GCKR;SNX17
GCKR
CCDC141
GCKR
LCORL
APOE
APOE
GCKR
SKIDA1
HSPA4;PTEN;MYH7
SERPINA1;PTEN;APOE
DLEU1;APOE
GCKR;APOC1
GCKR;APOE
GCKR;APOC1
MLLT10;LCORL
MYH6;MYH7
FAM174B;ALX4
MYH6;MYH7
FAM3C;DGKH
TOMM40;NDRG2
SERPINA1;HSPA4
MLLT10;PTEN
GCKR;APOC1
GCKR;APOC1
GCKR;SERPINA1;MFN1;PTEN;APOE;ARHGEF40

GCKR
GCKR
GCKR
GCKR;APOC1
APOE
GCKR;APOE
GCKR
CEP85L
GPN1
RSRC1
DGKH
CR1L
APOE
GCKR
GCKR;ASCC2
APOE
GCKR
FAM3C
LCORL
GCKR;APOE
GCKR
GCKR
GCKR
TOMM40
SERPINA1
TBX15
GCKR
GCKR;APOC1
APOE
SERPINA1
GCKR;APOC1
GCKR;APOE
GCKR;APOE
TBX15;WARS2
GCKR
APOC1
SERPINA1
SERPINA1
GCKR
GJA1;DES;LXN;PTEN;WNT16;CD34
GJA1;SERPINA1;MFN1;APOE
GJA1;PTEN;APOE
GCKR;MTMR3;CR1;DLEU1;C2ORF16;ARHGEF40
RSRC1;DLEU1
GCKR;SERPINA1

CCDC91;LCORL
GCKR;APOC1
SERPINA1;PTEN;APOE;CD34
TBX15;GJA1;MYH6;TTN
TTN;MYH7
GCKR;HSPA4;SNX17;DLEU1;APOE
GCKR;APOC1
GCKR;SERPINA1
SNX17;APOE
PTEN
HORMAD2
GCKR
APOE
DLEU1
CMTM5
DGKH
GCKR
APOE
DLEU1
DLEU1
KIAA1755
GCKR
RSRC1
CCDC141
MLLT10
APOE
GCKR
GCKR
APOE
GCKR
GCKR
GCKR
CCDC141
WNT16
DES;FAM174B;UQCR10;APOE;NDRG2;MYH6;MYH7
DES;SNX17;EIF3E;APOE;NDRG2;MYH6;MYH7;TTN
CCDC141
APOE
TTN
GJA1;CD34;TRIB2
PTEN;TTN
GCKR;TOMM40
DES;TTN
CCDC121;GFM1;HSPA4;CEP85L;UQCR10;MLF1;TTN;PLN;CCDC91;MFN1;DLEU1;CCDC141;MYH6;MYH7
APOE

GCKR;CENPW;MTMR3;ARHGEF40;TRIB2
APOC1;APOE
CR1;CR1L
CR1;PTEN;APOE;NDRG2
SERPINA1;CR1;APOE
MLLT10;PTEN;WNT16;CD34;TRIB2
APOE;CD34
EIF2B4;GCKR
MYH6;MYH7;TTN
EIF2B4;GCKR
GCKR;APOE
KIAA1755;IFI27L2;DES;CMTM5;ZNF219;VSTM2L;SSC5D;UQCR10;CCDC141;MLF1;MYH6;MYH7
TBX15;GJA1;PTEN;RSPO2;SPEG;ALX4;CWC27
GCKR;TOMM40;CR1L;APOE
TBX15;GJA1;PTEN;ALX4;CWC27
EIF2B4;GCKR;TOMM40;APOC1;APOE
SERPINA1;PTEN;APOE
GJA1;HSPA4;APOE
SHOX2;DLEU1;APOE
HSPA4;APOE
TOMM40;APOE
GCKR
AFAP1
FAM3C
APOE
LCORL
APOE
APOE
DLEU1
GCKR
TOMM40
SERPINA1
APOE
APOE
SERPINA1
GCKR
GCKR
GCKR;ALX4
LCORL
KIAA1755;FAM174B;AFAP1;APOE;CCDC141;LCORL;MYH6
GCKR
GCKR
MYH6
DLEU1
TOMM40

APOE
TOMM40
RSRC1
MLLT10
APOE
GJA1;PTEN;NDRG2
SERPINA1;PLN;CD34
GJA1;DES;PLN;HSPA4;APOC1;PTEN;APOE;MYH6;MYH7;TTN
GJA1;SHOX2
MYH6;MYH7
GJA1;SKIDA1;DLEU1;AFAP1;EIF3E;WNT16
MLLT10;PVRL2
AFAP1
GCKR
APOE
WNT16
KIAA1755;ARHGEF40;TTN
CR1
TBX15;SPEG;MYH6;TTN
TBX15;DES;MFN1;MLF1;MYH7;TTN
GJA1;PLN;SNX17;APOE;TTN
TTN;MYH7
GFM1;MFN1;NDRG2
AFAP1;CWC27;CD34
LXN;APOE
APOE;CD34
GCKR;MTMR3;PTEN;DLEU1;ASCC2;HORMAD2;APOE;TTN
MLLT10;PLN;CD34
APOE
SERPINA1
CCDC91
GCKR
GCKR
CCDC91;LCORL
MLLT10;GCKR;SERPINA1;C2ORF16;TTN
EIF2B4;GCKR;SERPINA1;LCORL
GCKR;SERPINA1;HSPA4
DGKH
DLEU1
TTN
GCKR
DLEU1
APOE
NDRG2
TTN

WNT16
APOE
APOE
TTN
CWC27
TTN
GJA1;MTMR3;GNB4;PTEN
GJA1;MTMR3;GNB4;PTEN
SERPINA1;PTEN;RSPO2
PLN;APOE;MYH7;TTN
SERPINA1;MTMR3;CEP85L;PTEN;SLC35F1;CWC27;NDRG2;GJA1;CCDC91;RSRC1;DLEU1;RSPO2;ALX4;FAI
TBX15;CENPW;HSPA4;WNT16
RSPO2;ALX4;NRBP1
MYH6;TTN
MLLT10;DLEU1
TBX15;GCKR;TOMM40;CCDC91;ZNF512;LCORL
TBX15;GCKR;TOMM40;CCDC91;ZNF512;LCORL
DES;NOV;HSPA4;PTEN;CD34;TTN
MFN1;MYH6;MYH7
GCKR;CENPW
AFAP1
GCKR;MTMR3;TOMM40;APOC1;DLEU1;APOE;LCORL
SERPINA1;APOE
MLLT10;DNAJC1;GCKR;CCDC91;NOV;SNX17;RSPO2
GJA1;CR1;APOE;MYH7
TBX15;GJA1
PTEN;DLEU1;ALX4;HORMAD2;APOE
GJA1;PTEN;CCDC141
TTN;MYH7
DES;MYH7
PTEN;MYH6;MYH7;TTN
DES;MYH7;TTN
SERPINA1
GCKR
SERPINA1
AFAP1
SERPINA1
APOE
LCORL
TBX15
APOE
TOMM40
GCKR
PTEN;ALX4
DES;MYH7

GJA1;PTEN;CCDC141
TBX15;GJA1;RSPO2;ALX4
EIF2B4;TBX15;GJA1;PTEN;CWC27
PTEN;APOE
TOMM40;HSPA4
GJA1;PTEN;MYH6;MYH7
CENPW;LCORL
DES;MYH7
DES;APOE
DES;MYH7
DES;MYH7;TTN
GJA1;PLN;PTEN
GCKR;TOMM40;HSPA4;CR1L;CMTM5;APOC1;APOE;WNT16;CD34;MYH6
PTEN;APOE;CD34
APOC1
APOE
APOE
EIF3E
PTEN;MYH7
GNB4;TTN
DES;MYH7
TBX15;GJA1
MLLT10;MLF1
MTMR3;PTEN
PTEN
PTEN
MTMR3
DES;APOE;TTN
GJA1;HSPA4;PTEN;APOE;CD34
MLLT10;CD34
NOV;HSPA4;PTEN
HSPA4;PTEN
GJA1;DES;SERPINA1;MYH6
WNT16
APOE
GCKR
APOE
GCKR
TOMM40
APOE
TTN
SNX17
TBX15;RSPO2;EIF3E;WARS2
GCKR
WNT16

TOMM40
GCKR
ZNF219
GCKR;SERPINA1;PTEN;DLEU1;AFAP1;APOE;LCORL;TRIB2
GCKR
GCKR
APOE
ARHGEF40
DLEU1
TTN;MYH7
MLLT10;HSPA4;PTEN;CD34;MLF1;NRBP1
SNX17;APOE
PLN;CD34;MYH7;TTN
DES;PLN;ALX4;TTN
HSPA4;PTEN
SPEG;MYH7
GCKR;TOMM40
PTEN;ALX4
DES;MYH7
PTEN;APOE
MLLT10;CD34
TBX15;GCKR;CENPW;SERPINA1;CCDC91;SPEG
PLN;CCDC141;MYH7
PLN;CCDC141;MYH7
PLN;CCDC141;MYH7
PLN;CCDC141;MYH7
PLN;CCDC141;MYH7
PLN;CCDC141;MYH7
GCKR;SERPINA1;APOC1
DES;ZNF219;SSC5D
PLN;CCDC141;MYH7
SLC5A6;SERPINA1;APOC1
IFI27L2;CMTM5;NDRG2
PLN;CCDC141;MYH7
PLN;CCDC141;MYH7
PLN;CCDC141;MYH7
APOC1
TBX15
SKIDA1
SKIDA1
GCKR
DLEU1
APOE
GCKR
DGKH

GCKR
LCORL
TBX15;CCDC91;RSRC1;DLEU1;HORMAD2;CWC27;LCORL;WARS2
TTN
GCKR
GCKR
DLEU1
DLEU1
ALX4
WNT16
GCKR
LCORL
MLLT10;DNAJC1
TBX15;GJA1;RSPO2
CR1;APOE
DES;MTMR3;HORMAD2
HSPA4;RSPO2;FAM3C;CD34
GCKR;APOE
TOMM40;MFN1
TBX15;GJA1;PTEN;SPEG;ALX4;CWC27
TBX15;GJA1;PTEN;SPEG;ALX4;CWC27
PLN;CD34
TBX15;GJA1;PTEN;SPEG;ALX4;CWC27
TBX15;GJA1;PTEN;SPEG;ALX4;CWC27
TTN;MYH7
SERPINA1;APOE
SERPINA1;PTEN;APOE
DES;PLN;PTEN;TTN
CCDC121;CMTM5;APOC1;GNB4;ALX4;WNT16;TTN
PLN;EIF3E;UQCR10;MYH6;MYH7;TTN
MTMR3;PTEN
CR1;SSC5D;CD34;TRIB2
TBX15;GJA1;PTEN;SPEG;ALX4;CWC27
DCAF16;NCAPG
GJA1;APOE;CD34;MYH6;MYH7;TTN
CR1;HSPA4;SPEG
SERPINA1;MTMR3;PTEN;NCAPG;CWC27;NDRG2;TTN;DCAF16;CCDC91;ZNF628;RSRC1;FAM174B;MFN1;
EIF2B4
PTEN
APOE
GJA1;DES;PLN;ZNF513;CCDC141;MYH6;MYH7;TTN
PTEN
PTEN
APOE
EIF2B4

APOE
GJA1
PTEN
GJA1
GJA1
PTEN
APOE
APOE
APOC1
WNT16
APOC1
TBX15;DES;SPEG;TTN
SERPINA1;APOE
TBX15;GJA1;PTEN;SPEG;ALX4;CWC27
TOMM40;APOE
PTEN;CD34
ALX4
RSRC1
APOE
TTN
MLLT10
APOE
CWC27
RSRC1;APOE
ALX4
APOE
DLEU1
AFAP1
SERPINA1
CR1;CD34
IFI27L2;CCDC121;GFM1;SERPINA1;ZCCHC10;SLC4A1AP;PTEN;SLC35F1;CWC27;NAT14;GJA1;CCDC91;RS
SERPINA1;CR1;CD34
MLLT10;HSPA4;CD34
CR1;CD34
GCKR;ZNF513
IFI27L2;CCDC121;GFM1;SERPINA1;ZCCHC10;SLC4A1AP;PTEN;SLC35F1;CWC27;NAT14;GJA1;CCDC91;RS
GJA1;CD34
SERPINA1;GPR126
GPN1
SLC5A6
FAM3C
CD34
NDRG2
GJA1
GPR126;MYH6

GCKR;APOC1
PTEN
DGKH
ZNF512
DES
MLF1
APOE
SERPINA1;CD34
HSPA4;APOE
SPEG
SERPINA1;CD34
CR1;PTEN;TTN
PTEN
FAM174B
TBX15;PTEN;CCDC141;MYH6;MYH7;TTN
MLLT10;DNAJC1;SNX17;PTEN;DGKH;TTN
PTEN
PTEN
DLEU1
MYH6
WARS2
FAM3C
GPR126
TOMM40;APOC1
SLC5A6
DLEU1
KIAA1755;MTMR3;PTEN;CCDC141;TTN
CENPW;MTMR3;HORMAD2
ZNF512
GJA1;ALX4
TBX15;PTEN;CCDC141;MYH6;MYH7;TTN
GCKR;MTMR3;HORMAD2;PPM1G
DES
EIF2B4;MFN1
GCKR;APOE
SLC5A6;GJA1
MYH6;TTN
NCAPG;TTN
SPEG
DLEU1
SERPINA1
MTMR3
EIF2B4
GJA1
SERPINA1

GFM1
SERPINA1
PTEN
TTN
MLLT10;PTEN;FAM3C;NDRG2
DLEU1
GFM1;PTEN;ARHGEF40;MYH6;MYH7;TTN
HSPA4;APOE
MLF1
CD34
GJA1
APOE
CD34
TOMM40;APOE
IFI27L2;CCDC121;GFM1;SERPINA1;ZCCHC10;SLC4A1AP;PTEN;CWC27;GJA1;CCDC91;RSRC1;FAM174B;D
MLLT10;CENPW;WNT16
MYH6;MYH7
PTEN;APOE
MLLT10
GCKR
GCKR
DLEU1
AFAP1
APOC1
TTN
EIF3E
TOMM40;HSPA4;DLEU1;APOE
CCDC91
AFAP1
MYH6
DLEU1
GCKR
DES;CD34
DES;CD34
GJA1;ZNF628;ZNF219;ZNF513;ASCC2;APOE;WNT16;CD34;MLF1;SUPT7L;TTN
MYH6;MYH7
GJA1;SERPINA1;PTEN;ALX4;CD34
GJA1;SERPINA1;CR1;HSPA4;PTEN;APOE;CD34;MYH6;MYH7;TTN
HSPA4;APOE
SERPINA1;TOMM40;HSPA4
IFI27L2;CCDC121;GFM1;SERPINA1;ZCCHC10;SLC4A1AP;PTEN;SLC35F1;CWC27;NAT14;GJA1;CCDC91;RS
IFI27L2;CCDC121;GFM1;SERPINA1;SLC4A1AP;PTEN;SLC35F1;CWC27;GJA1;CCDC91;RSRC1;FAM174B;DL
CCDC121;GFM1;SERPINA1;ZCCHC10;SLC4A1AP;PTEN;CWC27;CCDC91;RSRC1;FAM174B;DLEU1;RSPO2;S
GCKR;ZNF219;PTEN;DLEU1;C2ORF16;ZNF512;APOE;ARHGEF40
TOMM40

DNAJC1
GCKR
CR1
CCDC91
RSRC1
DLEU1
GCKR
RSRC1
WARS2
MLLT10
DLEU1
GCKR
GCKR
DLEU1
APOE
TBX15
PTEN
GJA1;PTEN;ALX4;TTN
MYH7
SHOX2
MYH7
DNAJC1;APOE;TTN
EIF2B4;APOE
CCDC91;PTEN
DES;MYH7
GJA1
HSPA4
PTEN
CD34
CD34
MLLT10;TTN
TTN
GFM1
GJA1
PTEN
EIF2B4
SLC35F1;MYH6;TTN
APOC1
GJA1;SERPINA1;CEP85L;PTEN;ASCC2;APOE
GCKR;DES;PLN;APOC1;RSPO2;APOE
PLN;EIF3E;UQCR10;NDRG2;MYH6;TTN
GJA1;UQCR10;APOE;NDRG2;MYH6;MYH7
TBX15;DES;NOV;MLF1;MYH6;MYH7;TTN
PLN;UQCR10;NDRG2;MLF1;MYH6;MYH7
GJA1;MFN1;UQCR10;NDRG2;MLF1;MYH7

PLN;EIF3E;UQCR10;NDRG2;MLF1;MYH6
CENPW;LXN;PLN;SHOX2;APOC1;GPR126
GJA1;DES;PTEN
NOV;SLC35F1
MTMR3;TOMM40;APOE
GCKR;SERPINA1;PTEN;APOE;NDRG2
CR1;HSPA4;PTEN
GCKR
CD34
CD34
MYH7
MYH7
GJA1
CD34
CD34
PLN
DES
MLLT10;DNAJC1;CCDC121;SLC4A1AP;SKIDA1;MFN1;HORMAD2;LCORL;MLF1
LXN;B3GNT7;NAT14;MLF1
CD34
CD34
CD34
APOE
CD34
CD34
APOE
CD34
MYH7
CD34
CD34
NDRG2;CD34
LXN;APOE;PPM1G
DES;MTMR3;ZCCHC10;CCDC91;SKIDA1;SHOX2;CEP85L;NDRG2;MLF1
DES
CD34
CD34
SHOX2
PLN
CD34
CD34
GJA1;GNB4;SPEG;APOE;NDRG2;CD34;TRIB2
GJA1
SERPINA1;APOE
SHOX2
CD34

GJA1;PTEN;CD34
CD34
CD34
DES
CD34
CD34
CD34
NCAPG
CD34
CCDC141;TTN
CR1;CD34
APOC1;APOE
PTEN;CD34
APOE
CD34
CD34
CD34
SHOX2
DES
PLN
APOC1;APOE
PTEN;CD34
APOE
APOE
GJA1
TTN
PTEN;CD34
CD34
CD34
CD34
CD34
CR1
TTN
CD34
CD34
CD34
CD34
CR1
CR1
CCDC141
SERPINA1
DNAJC1
DLEU1
GPN1
GCKR

KIAA1755
GJA1;APOC1;DLEU1;APOE;LCORL
DES;PTEN;APOE;MYH6;MYH7;TTN
DES;MYH7
TTN;MYH7
GFM1;CD34
MLLT10;RSRC1;DLEU1
GJA1;PTEN;APOE;NDRG2
NRBP1
CD34
PTEN
SERPINA1
GCKR;HORMAD2
CD34
AFAP1
DNAJC1;DLEU1
SHOX2;PVRL2
MYH7
SERPINA1
CR1
LXN
CD34
CD34
GFM1
ALX4
APOC1;APOE;CD34
PTEN
CD34
MYH6
PTEN
SERPINA1
NIPSNAP1
TBX15;APOC1
CD34
PTEN
TTN
ZNF513
PTEN
PTEN
MLLT10;NDRG2;CD34;MLF1
TBX15;GJA1;CWC27
GJA1;SERPINA1
HSPA4;PTEN;APOE
SERPINA1;APOE
EIF2B4;GFM1;PTEN

GJA1;PVRL2
IFI27L2;CCDC121;GFM1;SERPINA1;ZCCHC10;SLC4A1AP;PTEN;SLC35F1;CWC27;NAT14;GJA1;CCDC91;RS
GJA1;MYH6;TTN
IFI27L2;SERPINA1;SLC4A1AP;PTEN;CWC27;NAT14;GJA1;CCDC91;RSRC1;FAM174B;SPEG;FAM3C;ARHGE
MLLT10
GCKR
SERPINA1
GCKR;SERPINA1;APOE
AFAP1;CWC27
GCKR
SERPINA1
GCKR
PTEN;CD34
GNB4;TTN
SERPINA1;PTEN
DES
TTN
CD34
MFN1;CD34
CD34
CD34
CD34
GJA1
CD34
CD34
CD34
MLF1
CD34
DGKH
DES;TTN
SKIDA1
GCKR
DES;PLN;MYH7;TTN
PTEN;APOE
GJA1;PTEN
PLN
GCKR;DLEU1;APOE;LCORL
DLEU1
GJA1
PTEN
PTEN
TTN
ALX4
PTEN
RSRC1;WARS2

MYH7
PTEN
PTEN
CR1
APOE
APOE
SERPINA1
GCKR
RSRC1
APOE
CCDC141
CWC27
SKIDA1
HORMAD2
APOE
GCKR
DLEU1
GCKR
GCKR
DLEU1
GCKR
SHOX2;APOE;NDRG2
MTMR3;PTEN;DGKH
GCKR;MTMR3;DLEU1;ARHGEF40;GPN1
CR1L;LCORL
CR1;TOMM40;CD34
GJA1;HSPA4;APOE
APOC1
APOE
GJA1
APOC1
ALX4
APOE
SERPINA1
SERPINA1;APOE
SERPINA1;HSPA4;APOE;CD34
HSPA4;APOE
CD34
CD34
PLN;DES
GJA1;AFAP1;APOE
GJA1;PLN;TTN
GCKR;HSPA4
MLLT10;PTEN;CD34;MLF1;NRBP1
DES;MYH7;TTN

DES;CD34
GJA1;PLN;PTEN
DES;CD34
GJA1;PLN;PTEN
SERPINA1;APOE
GJA1;PTEN;APOE
HSPA4;PTEN;AFAP1;APOE
SERPINA1;APOE
GJA1;APOE
DES;TOMM40;APOE
LCORL
DLEU1
GJA1
AC022431.2
CR1
AC022431.2
FAM174B
DLEU1
DGKH
AC022431.2
APOC1
FAM174B
EIF3E
CR1L
PTEN
PTEN
VSTM2L
AFAP1
DGKH
CWC27
RSRC1
CR1
AC022431.2
DLEU1
DGKH
SSC5D
GFM1
RSRC1
AC074091.13
CEP85L
SSC5D
CWC27
FAM3C
CCDC141
DLEU1

HSPA4
LCORL
SERPINA1
SHOX2
TRIB2
GPR126
RSRC1
TTN
FAM174B
GJA1
AFAP1
TRIB2
SERPINA1
TBX15
TRIB2
NOV
HSPA4
IFI27L2
DLEU1
TRIB2
EIF3E
SERPINA1
LCORL
EIF3E
DGKH
FAM174B
GJA1
CEP85L
CEP85L
DLEU1
GJA1
DLEU1
TRIB2
FAM3C
CWC27
RSPO2
HSPA4
DNAJC1
MTMR3
NDRG2
TRIB2
AFAP1
LCORL
SERPINA1
CWC27

CEP85L
TRIB2
SERPINA1
DLEU1
RSRC1
TRIB2
GJA1
TRIB2
CEP85L
TRIB2
CCDC91
FAM3C
LCORL
SERPINA1
EIF3E
SERPINA1
GCKR
APOE
APOE
GCKR
HORMAD2
DNAJC1
GCKR
GCKR;TOMM40;APOC1;PTEN;APOE
APOE
GJA1;PLN;FAM174B;UQCR10;MLF1;MYH6
DES;PLN;EIF3E;UQCR10;NDRG2;MLF1;MYH7
EIF3E;UQCR10;MYH6;MYH7;TTN
GCKR;MTMR3;HSPA4;PTEN;APOE
PTEN
GJA1;PTEN;SPEG
GJA1;DES;PTEN;WNT16;CD34
SPEG;MYH7
GJA1;APOE
SERPINA1;HSPA4;MFN1;PTEN;APOE
CCDC91
PTEN;DLEU1
PTEN
TBX15
MYH7
GJA1
PTEN
PTEN
PTEN
MLLT10;DNAJC1;GJA1;DES;HSPA4;PTEN;HORMAD2;CD34

APOE
ZNF628;ZNF512;WARS2
PLN
CD34
SHOX2
CD34
GJA1;PTEN
PTEN;APOE
FAM3C;WNT16
HSPA4;APOE
PTEN;TTN
GJA1;PTEN
GCKR;SNX17;ASCC2;APOE
PTEN;APOE
SLC5A6;SNX17;APOE
VSTM2L
CENPW
EIF3E
DLEU1
WNT16
LCORL
GCKR
ZNF512
TBX15
GCKR;APOC1;NRBP1
MTMR3;HORMAD2
PTEN;EIF3E;NDRG2
GCKR;SERPINA1;NDRG2
TOMM40;APOC1;APOE;PVRL2;CD34
SERPINA1;CR1;CR1L;TRIB2
GCKR;APOC1;NRBP1
SERPINA1
CD34
CD34
CD34
APOE
CD34
DES
GJA1;APOE;NDRG2;TRIB2
CD34
DNAJC1;PTEN;CD34
APOE
GJA1;APOE;NDRG2
CD34
CD34

TRIB2
SERPINA1
TTN;MYH7
CEP85L;SLC35F1;TTN
GPN1
PTEN
GPN1
GPN1
PTEN
GPN1
PTEN
GFM1;SERPINA1;WARS2
APOE
MYH7
APOE
RSRC1;PTEN;WARS2
PTEN
ALX4;TTN
GCKR;FAM3C;WNT16
APOE;CCDC141
APOE;CCDC141
GCKR;CCDC91
GCKR
APOE
APOE
GCKR
DLEU1
APOE
GCKR
TOMM40
CCDC91
ALX4
SERPINA1;CR1;CR1L
MTMR3;PTEN;DGKH
UQCR10;MYH6;MYH7
CR1;CR1L;CD34
GCKR;LXN;TOMM40;HSPA4;CABP7;NIPSNAP1;WARS2;GJA1;B3GNT7;NOV;CR1L;SNX17;APOC1;ZNF219;
IFI27L2;GCKR;LXN;TOMM40;CABP7;SUPT7L;TMEM253;WARS2;GJA1;NOV;SNX17;APOC1;ZNF219;ZMAT
IFI27L2;GCKR;LXN;TOMM40;CABP7;C2ORF16;NIPSNAP1;TMEM253;WARS2;GJA1;NOV;SNX17;APOC1;Z
TBX15;CCDC91;WARS2
SHOX2
HSPA4
LXN
ALX4
CD34

GJA1
CD34
SERPINA1
CD34
EIF3E
GCKR
APOE
CR1;CD34
CENPW
NIPSNAP1
GPR126
GJA1
DNAJC1
DES;NDRG2;TTN
CR1L
IFI27L2;CCDC121;GFM1;SERPINA1;ZCCHC10;SLC4A1AP;PTEN;SLC35F1;CWC27;NAT14;GJA1;CCDC91;RS
APOE;CD34
ALX4
CCDC121;GFM1;SERPINA1;SLC4A1AP;PTEN;SLC35F1;CWC27;GJA1;CCDC91;RSRC1;FAM174B;DLEU1;RS
CD34
IFI27L2;CCDC121;GFM1;SERPINA1;ZCCHC10;SLC4A1AP;PTEN;CWC27;NAT14;GJA1;CCDC91;RSRC1;FAM
IFI27L2;CCDC121;GFM1;SERPINA1;ZCCHC10;SLC4A1AP;PTEN;CWC27;GJA1;CCDC91;RSRC1;FAM174B;D
GJA1;HSPA4;APOE
GJA1;CD34
GCKR
PVRL2
MLF1
SERPINA1;CD34
GFM1;GJA1;DES;PLN;HSPA4;ZNF513;ASCC2;UQCR10;NDRG2;SUPT7L
SERPINA1;APOE
CR1;ZMAT5
PTEN;CD34
IFI27L2;CCDC121;GFM1;SERPINA1;ZCCHC10;SLC4A1AP;PTEN;CWC27;NAT14;GJA1;CCDC91;RSRC1;FAM
WNT16
RSPO2
CD34
TTN
AC074091.13
HSPA4
TRIB2
NDRG2
LCORL
CD34
APOE
CD34

GJA1
CD34
IFI27L2;GCKR;LXN;CABP7;C2ORF16;UQCR10;NIPSNAP1;TMEM253;GJA1;SNX17;APOC1;ZNF219;ZMAT5;
SERPINA1;HSPA4;TTN
GJA1;APOC1;APOE
RSRC1;CCDC141
EIF2B4
AFAP1
CD34
HSPA4;APOE
PTEN;APOE
PTEN;APOE
CR1;APOE;TTN
GCKR;ALX4
GCKR;SERPINA1;PTEN;APOE
B3GNT7;NDRG2
CR1L
DNAJC1
CCDC141
FAM174B
GCKR
APOE
CEP85L
APOC1
APOC1
SERPINA1
SKIDA1
IFI27L2;GFM1;MTMR3;CR1;SHOX2;PTEN;TTN;TBX15;DNAJC1;GJA1;CCDC91;B3GNT7;NOV;CR1L;RSRC1;
APOE
MLLT10;DNAJC1;GCKR;LCORL;PVRL2
DES;SERPINA1;CR1;NOV
RSPO2
TTN
GCKR;GJA1;DES;HSPA4;APOE;TTN
PTEN
PTEN
GJA1;PTEN;ALX4
TBX15
PTEN
PTEN
TBX15;GJA1;PTEN;CWC27
WARS2
CCDC91;CD34
MLLT10;CENPW;RSRC1;APOC1;LCORL
SNX17

APOE
APOE
SERPINA1
GCKR;TOMM40;APOC1;PTEN;APOE
CD34
CD34
CR1
PTEN
APOE
SERPINA1;APOE
MLLT10;PTEN
SERPINA1;PTEN;APOE
SERPINA1;PTEN;APOE
SLC35F1;TRIB2
DES
CD34
CENPW;CD34
APOE
CD34
CD34
CD34
CD34
CD34
GJA1
CD34
CD34
CR1
EIF2B4;TBX15;GJA1;PTEN
EIF2B4;TBX15;GJA1;PTEN
FAM3C;WNT16
TTN
APOE
APOE
APOE
TTN
GCKR
SHOX2
EIF2B4
TTN
SERPINA1;PTEN;APOE
GCKR;SLC35F1;CCDC141
DES;HSPA4;APOE
PTEN;CCDC141
PLN;APOE;MYH7
EIF2B4;TBX15;GJA1;PTEN

TOMM40;HSPA4;PTEN;APOE
PTEN
TTN
PTEN
GJA1
ALX4
DES
APOE
SERPINA1;HSPA4
MYH7;TTN
PTEN
PTEN
APOE
RSRC1;WARS2
MLLT10
APOE
APOE
CR1
GFM1;MYH6;TTN
MTMR3
GCKR
GCKR
GCKR;APOC1;NRBP1
SERPINA1;HSPA4;MYH7
DES;MYH7;TTN
CCDC91;GPR126
GFM1;GJA1
TBX15;GJA1;ALX4
GJA1;PTEN
PLN;MYH7
PTEN;ALX4
GNB4;MYH7;TTN
DES;MYH7
PTEN;ALX4
MLLT10;GJA1;DES;PLN;GNB4;PTEN;ALX4;MLF1;MYH6;MYH7;TTN
MLLT10;SERPINA1
GNB4;NDRG2
MYH6;MYH7
GFM1;ASCC2
CD34;MYH6
SPEG
GJA1;DES
WNT16;TTN
APOE;NDRG2
SERPINA1;APOE

PTEN;CD34
PTEN;TTN
DGKH
CD34
TTN
GCKR;SERPINA1;HORMAD2;ARHGEF40
DLEU1;CWC27
DLEU1
GCKR
CR1L
APOE
APOE
RSRC1
CD34
APOE
EIF3E
TRIB2
NOV
APOE;CD34
ZNF512
IFI27L2;CCDC121;GFM1;SERPINA1;ZCCHC10;SLC4A1AP;PTEN;SLC35F1;CWC27;NAT14;GJA1;CCDC91;RS
GPN1
SHOX2
APOE;CD34
CCDC121;GFM1;SERPINA1;ZCCHC10;SLC4A1AP;PTEN;SLC35F1;CWC27;GJA1;CCDC91;RSRC1;FAM174B;I
SLC4A1AP
IFI27L2;CCDC121;GFM1;SERPINA1;ZCCHC10;SLC4A1AP;PTEN;SLC35F1;CWC27;NAT14;GJA1;CCDC91;FA
DLEU1
LXN
DNAJC1
SHOX2
SERPINA1
WARS2
CD34
GFM1;NRBP1
IFI27L2;GCKR;GFM1;LXN;TOMM40;HSPA4;CABP7;C2ORF16;UQCR10;GPN1;NIPSNAP1;TMEM253;GJA1;
PTEN
APOE
CWC27
SERPINA1
TOMM40
APOE
GJA1
PTEN
SERPINA1

GFM1;MTMR3;LXN;AFAP1;SLC35F1;NRBP1;MLLT10;DNAJC1;RSRC1;DLEU1;ASCC2;LCORL;DGKH
GJA1
SERPINA1
SSC5D
CWC27
GFM1
IFI27L2;CCDC121;GFM1;SERPINA1;SLC4A1AP;PTEN;CWC27;NAT14;GJA1;CCDC91;RSRC1;FAM174B;SPE
HSPA4;CD34
CD34
CCDC121;GFM1;SERPINA1;ZCCHC10;SLC4A1AP;PTEN;CWC27;GJA1;CCDC91;RSRC1;DLEU1;RSPO2;SPEG
GJA1;CD34
GJA1;PVRL2
PTEN
CD34
WNT16
UQCR10
LXN
GFM1
MYH6
GFM1
PTEN
GCKR;HSPA4;PTEN;APOE
GFM1;WARS2
SERPINA1;PTEN
GJA1
GJA1
CD34
TTN
MYH7
PTEN
SLC5A6
PTEN
DES;CD34
ALX4;CD34
CR1;HSPA4;PTEN
RSRC1;WARS2
TOMM40;SHOX2;APOE;CD34
NCAPG;UQCR10
CD34
CD34
CCDC91;APOE
APOE
IFI27L2;CCDC121;GFM1;SERPINA1;ZCCHC10;SLC4A1AP;PTEN;SLC35F1;CWC27;NAT14;GJA1;CCDC91;RS
DLEU1;LCORL
APOE;CD34

PTEN
SHOX2
DES
AFAP1
PTEN;CD34
SHOX2
SLC5A6
CCDC141
WARS2
KIAA1755
RSRC1
DLEU1
GCKR;MTMR3
SERPINA1;APOE
PTEN;EIF3E;CD34
HSPA4
MLLT10;GCKR;RSRC1;LCORL;PVRL2
GJA1;APOE;CD34
TBX15;GJA1;DES;ALX4
TTN
EIF2B4;TOMM40;EIF3E;TTN
PTEN
HSPA4
PTEN
APOE
EIF2B4
PTEN
APOE
PTEN
SERPINA1
MYH7
PTEN
PTEN
PTEN
GJA1
EIF2B4;GJA1;SPEG;CCDC141;TTN
PTEN
MYH7
MYH7
RSPO2
SERPINA1
CD34
APOC1
CD34
CR1

EIF3E
APOE
CD34
GJA1
CD34
SERPINA1;CD34
DES;MYH6;MYH7
NDRG2;MLF1;MYH6;MYH7
DES;TOMM40;MLF1;MYH7
SERPINA1;PTEN;APOE
CWC27
APOE
PTEN;CCDC141
APOE;CD34
PLN;CD34
PTEN;CD34
APOE
PTEN
APOE
PTEN
PTEN;APOE
PTEN
NCAPG
TTN
PLN
TTN
MTMR3;PTEN;PPM1G
PLN
CR1;CD34
MTMR3;PTEN;PPM1G
PTEN;APOE
APOC1
TTN
CD34
EIF3E
SHOX2
WNT16
APOE
APOE
PTEN
GJA1
APOE
PTEN
PLN
APOC1;APOE

PTEN
CR1
GFM1
GJA1;SHOX2;ALX4
CD34
APOE
APOE
GJA1
SSC5D
PLN
GJA1;TTN
PTEN
TTN
APOE
CENPW
PLN
EIF3E
CD34
PTEN
EIF3E
SHOX2
WARS2
TOMM40;HSPA4
APOC1;APOE
NCAPG
CENPW
APOC1;APOE
ASCC2
PLN
APOE
ASCC2
PTEN
PLN
WNT16
RSPO2;ALX4;CCDC141
GJA1;PTEN;CD34
GCKR
GCKR
GCKR
APOE
AFAP1
TTN
ASCC2
GCKR
TOMM40

GCKR
PTEN;C2ORF16;TTN
SERPINA1;APOE;CD34
HSPA4;PTEN;CD34
GJA1;PTEN;CD34;TTN
GJA1;PLN;APOE;CWC27
PTEN;NAT14
SERPINA1;APOE
DES;TTN
APOE
APOE
CD34
CD34
IFI27L2;CCDC121;GFM1;SERPINA1;ZCCHC10;SLC4A1AP;PTEN;CWC27;GJA1;CCDC91;RSRC1;FAM174B;D
SKIDA1
CD34
PTEN
GFM1;SERPINA1;MTMR3;ZCCHC10;LXN;SLC4A1AP;TOMM40;PTEN;NCAPG;UQCR10;CWC27;NDRG2;TTI
SERPINA1;TOMM40;HSPA4
CD34
HSPA4
TRIB2
ZMAT5
SLC5A6
GJA1
CD34
IFI27L2;CCDC121;GFM1;SERPINA1;ZCCHC10;SLC4A1AP;PTEN;SLC35F1;CWC27;NAT14;GJA1;CCDC91;RS
CCDC121;GFM1;SERPINA1;ZCCHC10;SLC4A1AP;PTEN;CWC27;GJA1;CCDC91;RSRC1;FAM174B;DLEU1;RS
CCDC121;GFM1;SERPINA1;SLC4A1AP;PTEN;SLC35F1;CWC27;GJA1;CCDC91;RSRC1;FAM174B;DLEU1;RSI
CCDC121;GFM1;SERPINA1;ZCCHC10;SLC4A1AP;PTEN;CWC27;GJA1;CCDC91;RSRC1;FAM174B;DLEU1;RS
GFM1;MFN1;UQCR10;NDRG2;TRIB2
GJA1;LXN;CD34;MLF1
GJA1;LXN;NCAPG;CD34
DES;APOC1;APOE
MTMR3;PTEN
PTEN;APOE;TTN
CR1
PTEN
APOE
APOE
TTN
PTEN
CR1L
APOC1
SNX17;APOE

MTMR3;ZNF513;APOE
PTEN;WNT16
SERPINA1;CR1
GJA1
PTEN
SNX17;APOE
TBX15
PTEN
SERPINA1
MYH6
DES
PTEN
RSPO2
GCKR
SERPINA1
SLC35F1
CR1
SERPINA1
APOE
SNX17
GCKR
SNX17
GCKR;UQCR10
WARS2
CD34
CD34
APOE
APOE
TTN
IFI27L2;CCDC121;GFM1;SERPINA1;ZCCHC10;SLC4A1AP;PTEN;SLC35F1;CWC27;NAT14;GJA1;CCDC91;RS
WARS2
HSPA4
PTEN
GJA1;APOE
RSRC1
CD34
IFI27L2;GFM1;SERPINA1;SLC4A1AP;PTEN;SLC35F1;CWC27;NAT14;GJA1;CCDC91;RSRC1;FAM174B;RSPC
GFM1;MTMR3;SLC4A1AP;CEP85L;NCAPG;CWC27;NDRG2;PPM1G;CCDC91;SPEG;ALX4;ASCC2;ARHGEF4
LXN
IFI27L2;CCDC121;GFM1;SERPINA1;ZCCHC10;SLC4A1AP;PTEN;SLC35F1;CWC27;NAT14;GJA1;CCDC91;RS
MTMR3
IFI27L2;CCDC121;GFM1;SERPINA1;ZCCHC10;SLC4A1AP;PTEN;SLC35F1;CWC27;NAT14;GJA1;CCDC91;RS
CR1;CD34
GCKR;TOMM40;APOC1;APOE;TTN
MTMR3;HORMAD2

GCKR;C2ORF16
IFI27L2;MTMR3;SLC4A1AP;TOMM40;PTEN;NDRG2;NAT14;TTN;PPM1G;DCAF16;GJA1;CCDC91;RSRC1;F
PTEN;CD34
SHOX2
MLLT10;PTEN;NDRG2
GJA1;PTEN;EIF3E
CR1;PTEN
APOE
PTEN
APOE
GJA1
PTEN;CD34
GJA1;DES;TTN
GJA1
GFM1
CCDC141;TTN
EIF2B4;TOMM40;APOE
CR1;PTEN
PTEN;APOE
DES;TTN
GCKR;ZNF512
IFI27L2;CCDC121;GFM1;SERPINA1;ZCCHC10;SLC4A1AP;PTEN;SLC35F1;CWC27;NAT14;GJA1;CCDC91;RS
SERPINA1;PTEN;APOE
PTEN;APOE
GJA1;TTN
CD34
GJA1
MLLT10;DNAJC1;LXN;PTEN
TBX15;GJA1
GJA1
GFM1;RSRC1;ALX4;WARS2
ALX4
SERPINA1;PTEN;APOE
PTEN
MYH6
DLEU1;HORMAD2
CENPW
CR1
GCKR;TOMM40;APOC1;PTEN;C2ORF16;APOE;TRIB2;NRBP1;DGKH
CCDC91
CCDC141
GCKR;SERPINA1;APOC1
MLLT10;DNAJC1
CR1;CR1L
IFI27L2;CCDC121;GFM1;SERPINA1;ZCCHC10;SLC4A1AP;PTEN;SLC35F1;CWC27;CCDC91;RSRC1;FAM174

DES;RSPO2
MLLT10
GCKR
APOE
GCKR
APOE
APOE
DES;LXN;PLN;VSTM2L;UQCR10;CCDC141;MLF1;MYH6;MYH7
IFI27L2;GCKR;LXN;C2ORF16;UQCR10;NIPSNAP1;TMEM253;WARS2;GJA1;NOV;SNX17;APOC1;ZNF219;ZI
TTN;MYH7
PTEN
PTEN
PLN
GJA1
PTEN
MTMR3
APOE
CENPW
PTEN
PTEN
DGKH
CD34
PTEN;TTN
IFI27L2;CCDC121;GFM1;SERPINA1;ZCCHC10;SLC4A1AP;PTEN;SLC35F1;CWC27;NAT14;GJA1;CCDC91;RS
MYH6;TTN
CCDC121;GFM1;SERPINA1;ZCCHC10;SLC4A1AP;PTEN;CWC27;GJA1;CCDC91;RSRC1;FAM174B;RSPO2;AI
MYH6
ALX4
GJA1;PTEN
PTEN
PTEN
PTEN
GJA1
GJA1
TBX15
TBX15
PTEN
PTEN
PTEN
SPEG
PTEN
PTEN
PTEN
EIF2B4
GJA1

GJA1
GJA1
GFM1
ALX4
EIF2B4
PTEN
APOE
PTEN
EIF2B4;GJA1;PTEN;ZNF513
APOE
GJA1
PTEN
PTEN;SPEG
GJA1
GJA1
PTEN
TBX15
PTEN
GJA1
PTEN
GJA1
PTEN
GJA1
APOE
GJA1
GJA1;ALX4;ZNF513
EIF2B4
ZNF513
APOE
GJA1;PTEN
SPEG
TTN
GFM1
GJA1
APOE
PTEN
TBX15
MYH7
APOE
GJA1
TBX15
TTN
PTEN
APOE
TBX15

DES
PTEN
MYH7
ALX4
PTEN
GJA1
TBX15
GJA1
TTN
GJA1
MYH7
PTEN
TBX15;GJA1
PTEN
GJA1;ALX4
PTEN;ALX4
SERPINA1
PTEN
GJA1
GFM1;ALX4
TBX15
APOE
GFM1
MYH7
TBX15
PTEN
SPEG
MYH7
GJA1
PTEN
MYH7
EIF2B4
GJA1
GFM1
TBX15;GJA1;PTEN
TBX15
PTEN
GJA1
ZNF513
GJA1
ALX4
MYH7
TBX15
PTEN
TBX15

GJA1;PTEN;ALX4
PTEN
GJA1
DES
PTEN
TTN
PTEN;SPEG;MYH7
EIF2B4
PTEN
PTEN
GJA1
PTEN
APOE
DES
PTEN
TBX15;GJA1;PTEN
MYH7
GJA1;ALX4
TBX15
GJA1
GJA1
PTEN;MYH7
EIF2B4;PTEN
GJA1
GJA1
GJA1
TBX15
PTEN
TBX15
GFM1
ZNF513
ALX4
TBX15
MYH7
GJA1
TBX15
PTEN
GJA1
TBX15
MYH7
PTEN
MYH7
GJA1
GJA1
TBX15

GJA1
GJA1
TBX15
GFM1
GJA1
TBX15
GJA1
GJA1
GJA1
GJA1
MYH7
GJA1
PTEN
GFM1
DES
PTEN
APOE
EIF2B4
SERPINA1
PTEN
GJA1
MYH7
GJA1;ALX4
GJA1
EIF2B4
PTEN
APOE
GJA1
ZNF513
PTEN
SERPINA1
PTEN
APOE
DES;PLN;CD34
GJA1;APOE;CD34
KIAA1755;PLN;CD34
GJA1;DES;CD34
GJA1;DES;CD34
GJA1;DES;CD34
DES;APOE;CD34
SERPINA1;APOC1;APOE
CR1;CR1L;APOC1
GJA1;NOV;CD34
DES;PLN;CD34
SERPINA1;APOC1;APOE

KIAA1755;CMTM5;NDRG2
GJA1;APOE;CD34
DES;PLN;CD34
PLN;MYH7;TTN
CENPW;APOE;NAT14
GJA1;APOE;CD34
GJA1;LXN;APOE;NDRG2;CCDC141
PLN;APOC1;APOE;MYH7
APOE
ASCC2
APOE
SHOX2
SERPINA1
PTEN
SERPINA1
GJA1
APOE
PTEN
NOV
MYH7
GJA1
PTEN
APOC1
APOE
APOE
APOE
APOE
CD34
PLN
PTEN
GNB4
APOE
GJA1
PTEN
HSPA4
ZNF513
PTEN
DES;MYH6
SERPINA1;CR1;TOMM40;APOC1;APOE
APOE
PTEN
EIF2B4;GFM1;GJA1;TOMM40;HSPA4;PTEN;ALX4;SLC35F1;APOE;CD34
TTN
TTN
SERPINA1;LXN;APOE;NDRG2;TTN

PTEN;APOE;MYH6;MYH7
PTEN;AFAP1
SLC5A6;PTEN
GJA1;CCDC91;PTEN;APOE
PTEN
APOE
GJA1
APOE
APOE
HSPA4
GJA1
ALX4
GJA1
GJA1
PTEN
APOE
TBX15
ALX4
SERPINA1
PTEN
GJA1
GJA1
GJA1
NDRG2
PTEN
MYH7;TTN
GJA1
SPEG;TTN
TBX15;GJA1
CR1;CD34
EIF2B4;SERPINA1;TOMM40;HSPA4;PTEN;EIF3E;APOE;CD34;SUPT7L;TTN
PTEN;APOE
HSPA4
APOE
TBX15;DES
GJA1
APOC1;CD34
APOE
CD34
EIF2B4
PTEN
CENPW;APOE;CD34
HSPA4;APOE;CD34
HSPA4;APOE;CD34
PLN;APOE;MYH7

HSPA4;PTEN;APOE
EIF2B4;TOMM40;PTEN;APOE
GJA1;PTEN
TBX15
APOE
PTEN
GJA1;PLN
APOE
HSPA4
APOE
GFM1;DES;CD34
APOE
GJA1
GJA1;DES;SERPINA1;HSPA4;PTEN
PTEN;APOE
GJA1;PTEN
ALX4
TTN
PTEN
PLN
DES
MYH7
GJA1;TOMM40;HSPA4;APOE;CD34;DGKH
TBX15;GJA1;ALX4
GJA1;NOV;HSPA4;PTEN;CD34
ALX4
HSPA4;CEP85L;CD34;MYH7
DES
TTN
PTEN
TTN
CD34
DES
MLLT10;PTEN;NDRG2;CD34
GJA1;HSPA4;APOE
GCKR;APOC1;APOE
APOE
PTEN
APOE
DES;PTEN;APOE
SPEG
DES;MYH7
TOMM40;APOE
CR1;LXN;HSPA4;CMTM5;PTEN;WNT16;MLF1;TTN;MLLT10;GJA1;PLN;DLEU1;APOE;CD34;TRIB2
MTMR3;PTEN;HORMAD2;APOE

DLEU1;CD34
APOE;PPM1G
HSPA4;PTEN
LXN;APOE
PTEN
APOE;DGKH
MLLT10;GFM1;GJA1;HSPA4;PTEN;ZNF513;APOE
GJA1;PLN
HSPA4;PTEN
HSPA4;APOE
GJA1
CR1;TOMM40;APOC1;APOE
ALX4
MLLT10;CD34
DES;LXN;HSPA4;MFN1;PTEN;APOE;MYH6
TBX15;DES
SERPINA1;SHOX2
GCKR;APOC1;APOE
TBX15;GJA1;ALX4
GJA1;HSPA4;PTEN;ZNF513;APOE;NDRG2
CCDC91
APOE
APOE
APOE
EIF2B4;GFM1;PTEN;APOE
AFAP1;ALX4;APOE;NDRG2
APOE
MYH7
MYH7;TTN
GJA1;APOE
PTEN
EIF2B4
APOE
PTEN
TBX15
GJA1
EIF2B4;SERPINA1;HSPA4;SHOX2;CEP85L;ASCC2;APOE;DGKH
PTEN;APOE;NDRG2
APOE
PTEN
APOE;MYH7
APOE;PPM1G
GJA1;APOE
EIF2B4;TOMM40
PTEN

GJA1
TOMM40;APOC1;APOE
HSPA4;PTEN;ALX4;NDRG2
TOMM40;APOE
EIF2B4;GFM1;PTEN;APOE
GJA1;ALX4
PTEN
GJA1;PTEN;APOE
PTEN
SERPINA1;APOE
HSPA4
APOE
CD34
PTEN
NOV;HSPA4;PTEN;APOE;TRIB2
GJA1;PTEN;APOE
GJA1;PTEN
PLN;CEP85L
PTEN;APOE
APOE
SERPINA1
EIF2B4
MYH7
MYH7
GJA1
PTEN
HSPA4
TTN
GJA1;DES
APOE
TOMM40;APOE
GJA1;PTEN
GJA1
APOE
PTEN;MYH6;MYH7
PTEN
APOE
APOE
DES
PTEN
DES;TTN
GCKR;SERPINA1;RSPO2;APOE;NDRG2;CD34
CD34
GJA1
MYH7

GJA1
MYH7
PTEN
DES
PTEN
PTEN
PTEN
TTN
DES
DES
PTEN
PTEN
TTN
PTEN
PTEN
PTEN;NDRG2;CD34
PTEN
APOE
PTEN
CD34
ALX4
PTEN
HORMAD2
APOE
DGKH
APOE
CD34
APOC1
GJA1
APOE
CD34
PTEN;MLF1
CENPW
APOC1
VSTM2L;APOE
APOE
APOE
RSRC1;CEP85L;PTEN;ASCC2
PTEN
ALX4
APOE
TBX15
APOE
PTEN
SERPINA1;APOE

APOE
ALX4
PTEN
PTEN
TRIB2
GJA1;MFN1
HSPA4;CD34
APOE
PLN;MYH6;TTN
EIF2B4;TBX15;UQCR10;MYH7
NOV;CD34;MLF1;MYH6
NOV;CCDC141;MYH7;TTN
TBX15;UQCR10;MLF1
GJA1;SERPINA1;PTEN
GJA1;ALX4
GCKR
SERPINA1
CD34
CD34
CD34
CD34
RSRC1
FAM3C
RSRC1
CR1
RSRC1
GCKR
GCKR
LCORL
RSRC1
PTEN;WNT16
DES;PTEN;APOE;NDRG2;CCDC141
HSPA4;CD34
APOE;CD34
NCAPG
GJA1
HSPA4
HSPA4
CD34
GJA1
GNB4
ALX4
PTEN
GJA1;MYH7
TBX15

GCKR;APOE
APOE
CR1;CD34
MYH7;TTN
LXN
TOMM40
GJA1
CR1
SERPINA1;NOV;PTEN;APOE
AFAP1;APOE
CD34
APOE
GJA1
GFM1;CENPW;SERPINA1;MTMR3;TOMM40;HSPA4;SHOX2;PTEN;AFAP1;GJA1;ALX4;EIF3E;HORMAD2;F[illegible]
SERPINA1;APOE;CD34
TOMM40
TOMM40
GJA1
PTEN
TBX15
DES
GJA1;HSPA4;APOC1;PTEN;APOE;DGKH
TBX15
GCKR
ALX4
APOE
GCKR;DES;SERPINA1;APOC1;PTEN;APOE
GJA1;NOV;PTEN;TRIB2;TTN
ALX4
PTEN
SERPINA1
GCKR;ALX4
ALX4;CWC27
PTEN
APOE
APOE
MYH7;TTN
GJA1
SERPINA1;APOE
DES
GJA1
DES
APOE
CD34
GJA1

PTEN
GJA1
GJA1
PTEN
GJA1
APOE;CD34
PTEN;CD34
SERPINA1;HSPA4
HSPA4
APOE
TBX15
TBX15
PTEN
PTEN
TBX15
CR1
PTEN
APOE
PTEN
TTN
TBX15;GJA1;ALX4;CWC27
TOMM40;APOE
GCKR;APOE
MLLT10;PTEN;DLEU1;WNT16;CD34;TRIB2;MLF1
APOE
CD34
PTEN
PTEN
CD34
APOE
SERPINA1
TBX15;GJA1
MLLT10;GJA1;PTEN;NDRG2;CD34
DES;PTEN
CD34
TBX15
TBX15
PTEN
APOE
PTEN
SERPINA1
ALX4
SERPINA1
PTEN
PTEN

PTEN
CD34
PTEN
APOE
APOE
APOE
AFAP1
APOE
EIF2B4
ZNF513
SERPINA1
HSPA4
DES
GCKR
GJA1;PTEN
PTEN;CD34
SERPINA1;PTEN;EIF3E;TTN
GCKR
PTEN
APOE
SERPINA1
DGKH
APOE
GCKR
DLEU1
DLEU1;SLC35F1
MTMR3
HORMAD2
MTMR3
GNB4
TBX15;GJA1
HSPA4;APOE
GJA1
APOE
EIF2B4;GJA1;RSPO2;ZNF513;WARS2
MYH7
PTEN
APOE
PTEN
PTEN
PTEN
PTEN
CCDC91
APOE
PLN

APOE
CR1
GJA1;SERPINA1
MTMR3;CR1;HSPA4;CEP85L;PTEN;AFAP1;NCAPG;C2ORF16;TTN;RSPO2;GPR126;CCDC141;MYH6;MYH7;
GPR126
MLLT10
CD34
HSPA4
APOE
GCKR
GFM1;DES;ALX4;APOE
PTEN;APOE
MLLT10;GJA1;SERPINA1;CR1;HSPA4;PTEN;APOE;CD34
PTEN
PTEN
GCKR;SERPINA1;APOC1;PTEN;APOE
TBX15
APOE
GJA1;FAM3C;APOE
NDRG2
NDRG2
APOE
APOE
FAM3C
PTEN
APOE
APOE
MLLT10
SERPINA1
APOE
TBX15
NRBP1
NRBP1
GCKR;SERPINA1;PTEN
GCKR;SERPINA1;MTMR3;HSPA4;RSPO2;HORMAD2;UQCR10;APOE
PTEN
PTEN
SERPINA1;APOE
DES
DES
NDRG2
NDRG2
APOE
GJA1
MYH7

APOE
EIF2B4
GJA1
APOE
MYH6
NDRG2
NDRG2
TBX15
HSPA4;CD34
GJA1;PTEN
MYH7
DES;PTEN;TTN
TBX15;DES
GNB4;MYH7
GCKR
LCORL
GCKR
GCKR;APOC1;ASCC2
CR1L
ALX4
APOE
GCKR;HSPA4;PTEN;APOE
MLLT10;GCKR;RSRC1;LCORL;PVRL2
PTEN;TRIB2
CD34
UQCR10
PLN
SSC5D
APOE
ASCC2
GJA1
NCAPG
KIAA1755;PTEN;ARHGEF40
PTEN
HSPA4;APOE
HSPA4;APOE
MLLT10;GCKR;LCORL;PVRL2
GFM1;SERPINA1;ZCCHC10;SLC4A1AP;PTEN;SLC35F1;CWC27;NAT14;GJA1;CCDC91;RSRC1;RSPO2;SPEG;[illegible]
APOE;CD34
WNT16
TBX15
PTEN
GNB4
APOE
GCKR;APOE

GCKR;SERPINA1;APOE
PTEN;NDRG2
MFN1
EIF2B4;GJA1;SERPINA1;NOV;HSPA4;DLEU1;HORMAD2;APOE;CD34
CD34
SERPINA1
APOE
APOE
APOE
PTEN
TBX15;PTEN;ALX4
PTEN
GNB4;APOE
APOE;CD34
GJA1
GJA1
ZNF513
PTEN
PTEN
HSPA4
HSPA4
SERPINA1
PTEN
GCKR
PTEN
PTEN
APOE
GJA1
DES
GJA1
SERPINA1
HSPA4
APOE
PTEN
PTEN
SHOX2
SERPINA1
PTEN
GFM1;SERPINA1;CR1;PTEN;APOE;CD34
MFN1
PTEN
APOE
TOMM40;SHOX2;APOE;CD34
CR1;TOMM40;APOE
APOE;CD34

IFI27L2;GCKR;LXN;TOMM40;CABP7;C2ORF16;UQCR10;NIPSNAP1;GJA1;NOV;SNX17;APOC1;ZNF219;ZM
MLLT10;MTMR3;RSRC1;SPEG;ASCC2;LCORL
CWC27
GCKR
APOE
PTEN
PTEN;CD34
PTEN;CD34
PTEN;CCDC141
GJA1
HSPA4
APOE
MYH7;TTN
MYH7
APOE
GJA1
APOE
GCKR
MYH7
SERPINA1
GJA1
CD34
APOE
GFM1
GJA1;DES;PTEN;CD34
CR1;APOE
CR1;APOE
HSPA4;APOE
PTEN
APOE
CD34
APOE
APOE
GJA1;PTEN
GJA1;PTEN
GJA1;PTEN
GJA1;PTEN
MLLT10
GJA1
SERPINA1
GJA1;PTEN
SERPINA1;APOE
PTEN;APOE
GJA1;PTEN
PTEN

PTEN
DES
PTEN
GJA1
APOE
HSPA4
HSPA4
APOE
MFN1
SERPINA1
MTMR3;PTEN;TTN
PTEN;APOE
EIF2B4;APOE
GJA1;CR1;HSPA4;PTEN;DLEU1;APOE;WNT16;CD34;TRIB2
DES
RSPO2
DES
GJA1
ALX4
EIF2B4
PTEN
ALX4
TBX15
PTEN
PTEN
PTEN
SLC5A6;GJA1;TOMM40;RSRC1;PTEN;SPEG;ALX4;ZNF513;CWC27;MYH7;WARS2;TTN
PTEN
GJA1
SERPINA1;HSPA4;CEP85L;ASCC2;APOE;DGKH
SERPINA1
EIF3E
MYH7
PTEN
DES;SERPINA1;TOMM40;PTEN;APOE
TTN
MYH7
DLEU1
APOE
GJA1
MYH7
APOE
PTEN;SPEG;APOE;MYH7
APOE
ZNF513

ALX4
TTN
ALX4
GJA1;PTEN;MYH7
GJA1;PTEN
RSPO2
SERPINA1
PTEN
TBX15
PTEN
PTEN
SERPINA1
APOE
GCKR
GJA1;NOV;PTEN;APOE
SERPINA1;PTEN
PTEN
PTEN
TBX15
APOE
PTEN
GJA1;DES
GJA1;PTEN
TBX15;GJA1
PTEN
GJA1;NOV;PTEN
TBX15
DES
ASCC2
ALX4
CCDC91;LCORL
GFM1;MYH6
PLN;CEP85L;APOE
APOE
CD34
PTEN
CD34
APOE
DES;PLN
PLN;CD34;MYH7
SERPINA1;HSPA4;APOE;NDRG2
APOE
DES
GJA1
CD34

CD34
SERPINA1
PTEN
PTEN
PTEN
APOE
APOE
GJA1
PTEN
APOE
APOE
HSPA4
HSPA4;APOE
PTEN
GJA1
APOE
CR1
CR1L
CR1L
EIF3E
MFN1
NCAPG
PLN
GJA1
CD34
PLN;CMTM5;MFN1;VSTM2L;UQCR10;CCDC141;MLF1;MYH6;TTN
LXN;CMTM5;VSTM2L;SSC5D;CCDC141;CD34;MLF1;MYH6;MYH7
GCKR;LXN;CABP7;C2ORF16;NIPSNAP1;WARS2;GJA1;NOV;SNX17;APOC1;ZNF219;ZMAT5;APOE;ARHGEF
DES;PLN;UQCR10;CCDC141;MLF1;MYH6;MYH7;TTN
IFI27L2;GCKR;GFM1;TOMM40;CABP7;C2ORF16;UQCR10;NIPSNAP1;GJA1;B3GNT7;NOV;SNX17;APOC1;
GFM1;DES;PLN;DLEU1;UQCR10;CCDC141;MLF1;MYH6;MYH7;TTN
GCKR;GFM1;LXN;SLC4A1AP;TOMM40;HSPA4;CABP7;C2ORF16;GPN1;NIPSNAP1;NRBP1;SUPT7L;TMEM
IFI27L2;GCKR;GFM1;TOMM40;CABP7;C2ORF16;UQCR10;GPN1;NIPSNAP1;PPM1G;DNAJC1;GJA1;NOV;
DES;PLN;HSPA4;UQCR10;CCDC141;MYH6;MYH7;TTN
PTEN;ALX4
PTEN;ALX4
PTEN;APOE;CD34
PTEN;ALX4
EIF2B4;GFM1;GJA1
PTEN;ALX4
KIAA1755;GCKR
PTEN;ALX4
CR1
CD34
TTN

TTN
DES;TTN
APOE
TBX15
APOE
GJA1
APOE
APOE
TTN
TBX15
DES;PLN
HSPA4;CD34
GJA1;SERPINA1;PTEN;ALX4;NDRG2;CD34;TRIB2
GCKR;SERPINA1;NOV;PTEN;APOE
GJA1
TBX15
PTEN
SERPINA1
TTN
CWC27
DES;PTEN;TTN
ALX4
ALX4
SERPINA1;HSPA4
TBX15;CCDC91;LCORL
HSPA4;AFAP1;APOE
PTEN;ALX4
CD34;MLF1
GCKR;GJA1;SERPINA1;TOMM40;PLN;NOV;HSPA4;PTEN;APOE;CD34;TTN
PTEN
PTEN
PTEN
TBX15
TOMM40;APOE
HSPA4
APOE
GJA1
GJA1;PTEN;ZNF513;NDRG2
GJA1;DES;MYH7;TTN
CD34
APOE
TTN
SERPINA1
SERPINA1;CCDC91
RSPO2

TOMM40
EIF3E
TOMM40
GCKR
GCKR
APOE
MLLT10
CD34
APOE
MYH7;TTN
GJA1;APOE;CD34
GJA1;PTEN;CD34
RSRC1;WARS2
PTEN
PTEN
GJA1;PTEN;ALX4;APOE;MYH7
VSTM2L;APOE
APOE
SERPINA1
APOC1
MYH6
EIF3E
APOE
APOE
TTN
CENPW
PTEN
PTEN;WNT16
GJA1;EIF3E;WNT16;CD34;TTN
DES;SERPINA1
PPM1G
LXN;CD34
CR1
EIF2B4;EIF3E
PTEN
PTEN
GJA1
GJA1
TBX15
PTEN
GFM1;TTN
ALX4
PTEN
PTEN
PTEN

PTEN
PTEN
TOMM40;APOE
EIF2B4;CCDC141
GCKR;APOE
APOE
APOC1
APOE
SLC35F1
PVRL2
MTMR3;ZCCHC10;CR1;SLC4A1AP;TOMM40;APOC1;APOE;PVRL2;DGKH
B3GNT7
CCDC91;NOV;RSRC1
APOC1;APOE
GCKR
SHOX2;ALX4
GCKR
GCKR;APOC1;ALX4;NRBP1
PTEN
PTEN
PTEN
CD34
GJA1;ALX4
SERPINA1;CD34
GJA1
PTEN
PTEN
PTEN
TTN
GJA1
CD34
DES
PTEN
GJA1
PTEN
CD34
PTEN
CD34
GFM1;ALX4;CWC27
MLLT10;TTN
PTEN;DLEU1;EIF3E;NDRG2;CD34
CD34
CD34
HSPA4
TTN

PTEN
EIF2B4
PTEN
SERPINA1
GJA1;NOV;HSPA4;GNB4;PTEN;WNT16;CWC27;NDRG2;TTN
PTEN;CD34;MYH6
PTEN
GCKR;SERPINA1
PTEN
TBX15
PLN
PTEN
PTEN
CR1
PTEN
B3GNT7
PTEN
APOE
APOE
APOE
MTMR3
APOC1
PTEN
PTEN
TTN
CD34
CR1
RSPO2
PTEN
APOE
GJA1
RSPO2
PTEN
ALX4
MTMR3;PTEN;LCORL;TTN
GJA1;TTN
TRIB2
PTEN
SERPINA1;PTEN;TTN
PTEN
PTEN
TTN
GCKR;NOV;APOE
GJA1;APOC1;ALX4
EIF2B4;GFM1;GJA1;APOE

EIF3E
APOE
PPM1G
TRIB2
HSPA4;PTEN;DLEU1;ZNF512
GJA1
CCDC91
PTEN
PPM1G
PTEN
MTMR3
PTEN
TTN
PLN
EIF2B4;APOE
APOE
TRIB2
RSRC1
GJA1
B3GNT7
TTN
APOE
CR1
APOE
PTEN
EIF2B4
GJA1
PLN
APOE
SSC5D
APOE
RSRC1;ZMAT5
MTMR3;PTEN
GJA1;SERPINA1;HSPA4;APOE;CD34
MLLT10;GCKR;LCORL;PVRL2
CWC27
DCAF16;GJA1;CCDC91;HSPA4;FAM3C;PVRL2;TRIB2;DGKH
MLLT10;GCKR;LCORL;PVRL2
EIF3E
GJA1
PTEN
ALX4
PTEN
TBX15
TTN

PTEN
PTEN
GJA1;PTEN
DES
PTEN
GJA1
DLEU1
APOE
GCKR
RSPO2
MLLT10
DLEU1
WNT16
GCKR
MTMR3
TOMM40
DES;PLN;UQCR10;CCDC141;MLF1;MYH6;MYH7
IFI27L2;GCKR;GFM1;LXN;C2ORF16;UQCR10;NIPSNAP1;TMEM253;WARS2;GJA1;NOV;APOC1;ZNF219;ZN
IFI27L2;GFM1;HSPA4;CABP7;CEP85L;C2ORF16;UQCR10;GPN1;NIPSNAP1;TMEM253;WARS2;GJA1;NOV;
PLN;CMTM5;MFN1;VSTM2L;CCDC141;MLF1;MYH6;MYH7;TTN
CMTM5;VSTM2L;CCDC141;CD34;MLF1;MYH6;MYH7
TBX15;GFM1;DES;SLC4A1AP;CCDC91;HSPA4;UQCR10;CWC27;NRBP1;MYH6;MYH7;TTN
DES;PLN;VSTM2L;UQCR10;CCDC141;MLF1;MYH6;MYH7;TTN
IFI27L2;GCKR;TOMM40;CABP7;TMEM253;B3GNT7;NOV;SNX17;APOC1;ZNF219;ZMAT5;ZNF513;APOE;A
EIF2B4;GFM1;SLC4A1AP;TOMM40;HSPA4;UQCR10;NDRG2;NRBP1;TTN;PPM1G;TBX15;DES;ZNF628;ZNF
EIF2B4;DES;TOMM40;B3GNT7;CMTM5;ZMAT5;UQCR10;CCDC141;MLF1;MYH6;MYH7
EIF2B4;IFI27L2;GCKR;LXN;TOMM40;HSPA4;UQCR10;NIPSNAP1;TMEM253;GJA1;NOV;SNX17;APOC1;ZN
DES;PLN;UQCR10;CCDC141;MLF1;MYH6;MYH7;TTN
EIF2B4;GFM1;GJA1;SERPINA1;TOMM40;HSPA4;PTEN;ALX4;APOE
MLLT10;PTEN;CD34
GJA1
PTEN
PLN
PTEN
GJA1
PTEN
GCKR;SERPINA1;APOE
PLN
SUPT7L
APOC1
MFN1
TOMM40
APOE
SUPT7L
GJA1;WNT16;CD34;TTN

NCAPG
APOE
TTN
CD34
MLF1
GJA1;DES;HSPA4;PTEN;ALX4;APOE;NDRG2;CD34
APOE;CD34
IFI27L2;CCDC121;GFM1;SERPINA1;ZCCHC10;SLC4A1AP;PTEN;CWC27;NAT14;GJA1;CCDC91;RSRC1;FAM
GJA1
APOE
CCDC91;RSRC1
PTEN;CD34
PTEN
APOE
APOE
RSRC1;MLF1
PTEN
PTEN
PTEN
PTEN
HSPA4
GCKR;ARHGEF40
PTEN
ALX4
DES;SERPINA1;PTEN;APOE;NDRG2;CD34;MLF1
GPN1;NRBP1
CEP85L;PTEN;CD34
GJA1
GJA1
APOE
GJA1
MLLT10
GJA1
GJA1
PTEN
APOE
APOE
SERPINA1
TOMM40;DLEU1
VSTM2L;APOE
PTEN;APOE
APOC1
APOE
NDRG2
GJA1;LXN;TOMM40;APOC1;PTEN;APOE

TBX15
PTEN
TBX15
APOE
SERPINA1;APOC1;LCORL
LXN
APOE
PTEN
GJA1
CD34
DES;SERPINA1;PLN;AFAP1;EIF3E;UQCR10;NDRG2;MLF1;NRBP1;MYH7;TTN;PPM1G
MYH7
GJA1;PTEN;CD34
SERPINA1;PTEN;APOE
APOE
GCKR;GJA1;MTMR3;DLEU1;ZNF513;ASCC2;HORMAD2;APOE
CD34
APOC1;ALX4;APOE
CD34
GJA1;PTEN
GJA1;PTEN
PTEN;WNT16
GCKR;DES;SERPINA1;HSPA4;PTEN;APOE;CD34
GJA1;PTEN
GJA1;PTEN;CWC27
SHOX2;PTEN
GJA1;TOMM40;HSPA4;APOE;CD34
MLLT10;GJA1;DES;NOV;HSPA4;PTEN;DLEU1
GCKR
APOE
LCORL
SNX17;HORMAD2
TRIB2
PTEN
PLN
TBX15
GJA1
PLN
GJA1
PTEN
PTEN
GJA1
PTEN
CD34;MLF1;TTN
DES;APOE;NDRG2;CD34;MYH6;MYH7;TTN

APOE
GJA1;PTEN
TBX15
DES
DLEU1
PTEN
DES
DES
TTN
DES;PTEN
SERPINA1;APOE
GJA1
APOE
PTEN
GJA1
CD34
TTN
APOE
APOE
APOE
CD34
APOE
TBX15
PTEN
APOE
DLEU1
HSPA4;APOE
CD34
CD34
GFM1;SERPINA1;SLC4A1AP;PTEN;CWC27;GJA1;CCDC91;RSRC1;FAM174B;DLEU1;RSPO2;SPEG;FAM3C;A
TTN
SERPINA1
GCKR
GFM1;SERPINA1;ZCCHC10;SLC4A1AP;PTEN;SLC35F1;CWC27;GJA1;CCDC91;RSRC1;FAM174B;DLEU1;RSI
GJA1;HSPA4
EIF3E
APOE
GJA1
SERPINA1;CD34
GFM1;SERPINA1;MTMR3;LXN;SLC4A1AP;TOMM40;PTEN;NCAPG;UQCR10;CWC27;NDRG2;NAT14;TTN;I
EIF2B4
APOE
APOC1
PLN
PTEN

MYH7
PTEN
CENPW;MTMR3;ARHGEF40;TRIB2
EIF3E
TBX15;GCKR;VSTM2L;ARHGEF40;LCORL
GCKR;HORMAD2;UQCR10
KIAA1755
PTEN
PTEN
RSPO2
SERPINA1;SPEG
MLLT10;CD34
APOC1;APOE
TBX15;GJA1;PTEN;ALX4
GFM1
PTEN
GJA1
PTEN
SERPINA1
PTEN
EIF2B4
PTEN
PTEN
GJA1;TOMM40;RSRC1;PTEN;ALX4;CD34;DGKH;WARS2
EIF2B4
TBX15
PTEN
GJA1
MLLT10;PTEN
CENPW;SERPINA1;PTEN;NCAPG;CD34
MYH6
APOE
APOE
APOE
GJA1
GJA1
GJA1
APOE
HSPA4
GJA1
HSPA4
GJA1
HSPA4
GJA1
GJA1

GJA1
APOE
APOE
PTEN
PTEN
WARS2
WARS2
APOE
PTEN
PTEN
HSPA4
CD34
HSPA4
HSPA4
PTEN
PTEN
PTEN
HSPA4
PTEN
HSPA4
HSPA4;PTEN
APOE
PTEN
PTEN
GJA1
PTEN
PTEN
PTEN
WARS2
PTEN
PTEN
PTEN
PTEN
PTEN
APOE
TRIB2
TRIB2
TRIB2
PTEN
HSPA4;PTEN
HSPA4;PTEN
HSPA4
PTEN
PTEN
TRIB2

HSPA4;PTEN
PTEN
APOE
APOE
PTEN
HSPA4
PTEN
APOE
APOE
HSPA4
PTEN
PTEN
PTEN
HSPA4
HSPA4
HSPA4
HSPA4
APOE
HSPA4
APOE
TRIB2
CD34
CD34
HSPA4
CD34
GJA1
GJA1
APOE
PTEN
CD34
APOE
GCKR
GFM1;HSPA4;PTEN;CD34;TRIB2;MLF1
APOE
APOE
GFM1;SPEG;TTN
CD34
CD34
APOE
APOE
ALX4
ALX4
APOE
APOE
PTEN

PTEN
ALX4
PTEN
ALX4
APOE
APOE
APOE
TBX15
HSPA4
PTEN
APOE
SERPINA1;CD34
PTEN
APOE
GFM1
CD34
GJA1
PTEN
GFM1;DES;MFN1;UQCR10;NDRG2
GFM1;TOMM40;MFN1;UQCR10;NDRG2
GJA1;LXN;RSPO2
DES;PLN;VSTM2L;MLF1;MYH7
PLN;CCDC141;MYH6;MYH7;TTN
GJA1;TOMM40;NOV;FAM3C;TRIB2
GJA1;PLN;FAM174B
ALX4
APOE
MLLT10;DNAJC1;MTMR3;PTEN;ALX4;MLF1;TTN
SERPINA1;PTEN;APOE
NOV;APOC1;APOE
GFM1;SPEG;TTN
GCKR;APOE
GCKR;APOE
GFM1;CD34
GJA1;PTEN
DES;PLN;HSPA4;MFN1;DLEU1;CCDC141;MLF1;MYH6;MYH7;TTN
HSPA4;PTEN
SERPINA1;PTEN
KIAA1755
ZNF513
PTEN
TTN
ALX4
APOE
PTEN

PTEN
TBX15
PTEN
APOE
PTEN
PTEN
APOE
GJA1
SERPINA1
APOE
ALX4
PTEN
APOE
PLN
GFM1;PTEN
CD34
SERPINA1;LXN;APOE
APOC1;PTEN;ZNF513;APOE
PTEN
DES
PLN;HSPA4;PTEN;CD34
GJA1
CD34
SERPINA1
MYH6
SERPINA1;LXN;HSPA4;SHOX2;PTEN;AFAP1;NDRG2;NIPSNAP1;NRBP1;TTN;SLC5A6;TBX15;GJA1;NOV;MF
SERPINA1;PTEN;CD34
LXN;PLN;VSTM2L;UQCR10;CCDC141;MLF1;MYH6;MYH7
GCKR;GJA1;HSPA4;APOE
PTEN
PTEN
GFM1
GJA1
PLN
GJA1
GJA1
NIPSNAP1
PTEN
PLN;HSPA4;CMTM5;CCDC141;MLF1;MYH6;MYH7;TTN
DES;APOE
NCAPG;TTN
APOE
GJA1
GCKR
GCKR

APOE
GCKR
APOC1
APOE
APOE
PLN;MYH7
TBX15;MLLT10;GCKR;ARHGEF40;LCORL
APOE
CR1;SLC35F1;TMEM253;TTN;TBX15;CCDC91;NOV;GNB4;RSPO2;HORMAD2;FAM3C;LCORL;CD34;TRIB2;
CR1;CR1L
APOC1;APOE
PLN;MYH6;MYH7
RSRC1;RSPO2;CD34
PTEN
PTEN
PTEN
GFM1
EIF2B4;KIAA1755;ARHGEF40
CR1
APOC1
DGKH
MTMR3;PTEN
LXN
B3GNT7
LCORL
MTMR3
MTMR3
B3GNT7
TTN
CR1
APOC1
PLN;CMTM5;VSTM2L;CCDC141;CD34;MLF1;MYH6;MYH7;TTN
IFI27L2;GCKR;LXN;CABP7;NIPSNAP1;TMEM253;WARS2;GJA1;B3GNT7;NOV;APOC1;ZNF219;ZMAT5;APC
DES;PLN;UQCR10;CCDC141;MLF1;MYH6;MYH7;TTN
GCKR;LXN;CABP7;C2ORF16;NIPSNAP1;GJA1;SNX17;APOC1;ZNF219;ZMAT5;APOE;ARHGEF40;PVRL2;TRI
GCKR;APOC1;CEP85L;SLC35F1;CWC27;CCDC141;NRBP1;TTN
GCKR;ALX4
GJA1
MYH7
MYH7
CD34
APOE
PTEN
GJA1
GJA1

SERPINA1
GJA1
PTEN
PTEN
APOE
MYH6
HSPA4;PTEN
SERPINA1;CR1;NOV;HSPA4;PTEN;CD34;TRIB2
SERPINA1;CR1;TOMM40;HSPA4;PTEN;NDRG2;WARS2;GJA1;ZNF628;APOC1;APOE;CD34;MYH6
DES;MYH7
MYH7
GFM1;CWC27;MYH7
MYH7
APOE
APOE
GJA1
GJA1
HSPA4;PTEN;SPEG;TTN
GCKR
PTEN
ALX4
APOE
PTEN
RSRC1;WARS2
MLLT10;PTEN
CD34
DES;MYH7;TTN
CD34
TBX15;NOV;CCDC141;TTN
TBX15;RSRC1
TBX15
ALX4
PTEN
PLN
RSRC1
ZNF512
APOC1
GCKR
DES;PTEN
SERPINA1;APOE
CD34
CR1
APOE
HSPA4
APOE

SLC5A6
CCDC141
APOE
EIF2B4
IFI27L2;GJA1;CENPW;APOC1;NCAPG;APOE;ARHGEF40
DCAF16;GJA1;DES;PLN;MLF1;TTN
DES;SERPINA1;APOC1;APOE;NDRG2;MYH6;MYH7
DES;SNX17;APOC1;APOE;CD34;MYH6;MYH7
DNAJC1;LXN;APOC1;ZNF513;NAT14;NIPSNAP1
KIAA1755;LCORL
APOC1;PTEN;APOE
APOE;TRIB2
SERPINA1
APOE
MLLT10;DNAJC1;LCORL;PVRL2
APOE
MFN1
GJA1;ALX4
EIF2B4
GJA1;PTEN;ALX4
PTEN
SERPINA1;APOC1;GNB4;APOE
CD34
CD34
MFN1;UQCR10;MLF1;MYH7
TBX15;DES;TTN
TBX15;SHOX2;APOC1;APOE
GCKR;UQCR10;APOE
GJA1;PTEN;RSPO2;ZNF513;NDRG2
APOE
APOE
MLLT10
MTMR3;DLEU1;C2ORF16;ARHGEF40
GCKR
APOE
SNX17
GFM1;SERPINA1;APOE
DES;PLN;CCDC91;DLEU1;UQCR10;CCDC141;MLF1;MYH6;MYH7;TTN
IFI27L2;GCKR;LXN;CABP7;C2ORF16;UQCR10;NIPSNAP1;WARS2;GJA1;NOV;APOC1;ZNF219;ZMAT5;APOI
GFM1;PLN;CCDC91;MFN1;UQCR10;CCDC141;MLF1;MYH6;MYH7;TTN
GCKR;GFM1;LXN;TOMM40;HSPA4;CABP7;C2ORF16;UQCR10;NIPSNAP1;WARS2;GJA1;NOV;SNX17;APO(
PTEN
APOE
CD34
TBX15;WNT16

MTMR3;RSPO2;APOE
APOE
CCDC141
KIAA1755
GJA1
GJA1
SERPINA1
TBX15
TTN
SPEG
CR1;APOE
SERPINA1
GJA1
APOE
MYH7
APOE
DLEU1
APOC1;APOE
PTEN
PTEN
APOE
PTEN
CR1
WARS2
PTEN
CEP85L;GPR126;LCORL;DGKH
GCKR
CCDC91
MTMR3
APOC1;NCAPG;APOE
CCDC91
GCKR
DLEU1
PTEN;CD34
WNT16
APOE
GCKR
RSRC1;SHOX2
MLLT10
GCKR
ALX4;CD34
APOE
DGKH
CR1
ASCC2

APOE
APOC1
GJA1
EIF2B4
TRIB2;TTN
APOC1
MYH6;MYH7
CR1;CR1L
PLN;MYH6;MYH7
PTEN
HSPA4;APOE;CD34
APOE
APOE
CD34
APOE
APOE
APOE
PTEN
HSPA4;APOE
CD34
PTEN
GCKR;CCDC141
PTEN
APOE
APOE
ALX4
CD34
SERPINA1
HSPA4
APOE
APOE
DES;NOV;PTEN;TTN
PLN;APOE
PTEN
GJA1
NDRG2
TBX15
GJA1
HSPA4;APOE
GJA1;NIPSNAP1;MYH7
CEP85L
MLLT10;DNAJC1;RSRC1;CEP85L;DLEU1;RSPO2;ALX4;TRIB2;WARS2
GJA1;PTEN;HORMAD2;PVRL2
IFI27L2;GCKR;GJA1;LXN;NOV;CABP7;APOC1;ZNF219;C2ORF16;APOE;ARHGEF40
IFI27L2;GCKR;LXN;CABP7;C2ORF16;NIPSNAP1;GJA1;NOV;APOC1;ZNF219;ZMAT5;APOE;ARHGEF40;TRIB

HSPA4;PTEN
APOE
GCKR
TOMM40
GCKR
APOC1
DES
ALX4
HSPA4
PTEN
PTEN
GJA1;ALX4
PTEN;CD34
CD34
GFM1;SERPINA1
CR1
PTEN
TBX15;TOMM40
CD34
PTEN;CD34
GJA1
PTEN
CCDC141
CD34
SERPINA1
GJA1;ALX4
CD34
GJA1
APOE
APOE
GFM1
CEP85L
APOE
CD34
MYH6
GJA1
GJA1
PTEN
GJA1
PTEN
CD34
GCKR;SERPINA1;TOMM40;PLN;NOV;PTEN;APOE;CD34;TTN
DES
PTEN
GPR126

UQCR10
CD34
SERPINA1;GPR126;CWC27
MLLT10;CR1;LXN;HSPA4;PTEN;DLEU1;CD34
GCKR;SUPT7L
GJA1
PTEN
PLN
PTEN
APOE
APOE
GJA1;SERPINA1;CR1;NOV;HSPA4;SPEG;APOE;CD34
CCDC91;PTEN
SHOX2
GCKR;SERPINA1;APOE
SERPINA1
HSPA4;MFN1;APOE
HSPA4
HSPA4;APOE
PTEN;APOE;CD34
APOE
APOE
ASCC2
PTEN
PTEN
PTEN
PTEN
PTEN
PTEN
HSPA4
CR1;MFN1;FAM3C;WNT16;TTN
APOE
CD34
IFI27L2;CCDC121;GFM1;SERPINA1;ZCCHC10;SLC4A1AP;PTEN;CWC27;NAT14;GJA1;CCDC91;RSRC1;FAM
HSPA4
GCKR
APOE
RSRC1
GCKR
APOC1
GCKR
APOC1
SNX17
TRIB2
SERPINA1;LXN;HSPA4;SHOX2;PTEN;AFAP1;NDRG2;NIPSNAP1;NRBP1;TTN;SLC5A6;TBX15;GJA1;NOV;MF

APOE
APOE;CD34
GCKR;GJA1;SERPINA1;NOV;HSPA4;APOC1;PTEN;RSPO2;FAM3C;NDRG2;CD34;WARS2
GFM1
DLEU1
PTEN
APOE
TTN
PTEN
APOE
APOE
PTEN
PTEN
GNB4
GJA1
CD34
CR1;HSPA4
PTEN
PTEN
HSPA4
PTEN
ALX4
GCKR;MTMR3;HSPA4;RSPO2;HORMAD2;APOE;TRIB2
GJA1
APOE
HSPA4
PTEN
PTEN;DLEU1
TBX15;GJA1;PTEN
PTEN
GJA1;DES;NOV;PTEN;APOE;NDRG2
CD34
PTEN
CCDC141
APOC1;APOE
GJA1;PTEN
HSPA4
HSPA4
HSPA4
APOC1
MLLT10
SNX17
GCKR
GCKR;APOE;CWC27
APOC1

GCKR
KIAA1755;ARHGEF40
SERPINA1
GFM1
PTEN
GJA1
GFM1;DES;PLN;CCDC91;MFN1;UQCR10;CCDC141;MLF1;MYH6;MYH7;TTN
GCKR;CABP7;C2ORF16;UQCR10;NIPSNAP1;WARS2;GJA1;B3GNT7;NOV;SNX17;APOC1;ZNF219;ZMAT5;A
SERPINA1;PTEN
PTEN
DES
MLLT10;PTEN
APOE
DES;SERPINA1
GJA1
PTEN
SERPINA1
PTEN
GFM1;CENPW;SERPINA1;HSPA4;SHOX2;PTEN;AFAP1;GJA1;ALX4;EIF3E;HORMAD2;FAM3C;APOE;TRIB2;
PTEN
CD34
PTEN
EIF2B4
SERPINA1;PTEN
GCKR
APOE
CD34
APOE
AFAP1
APOE
GJA1
PTEN
MLF1
APOC1
APOE
ZNF628;LCORL
GFM1;GJA1;SERPINA1;HSPA4;PTEN;ASCC2;NDRG2;CD34
MLLT10;CEP85L;CD34
GJA1;EIF3E;APOE
EIF2B4
GJA1;CD34
HSPA4;APOE
GJA1
CD34
APOE

APOE;CD34
SERPINA1;PTEN;APOE;WARS2
GJA1;TTN
SERPINA1;PTEN;MYH7;TTN
PTEN;APOE
PTEN
SERPINA1;APOE
PTEN
APOE;MYH7
HSPA4
CD34
SERPINA1;TRIB2
HSPA4
NOV
PTEN;APOE
PTEN;CD34
PTEN
GJA1
B3GNT7;PTEN;NDRG2
HSPA4;APOE;CD34
PTEN;AFAP1;NDRG2;NRBP1
GCKR;GJA1;SERPINA1;NOV;HSPA4;APOC1;PTEN;RSPO2;FAM3C;NDRG2;CD34;WARS2
GNB4;MYH7
GJA1;ALX4
MYH7
APOE
ALX4
SERPINA1
GJA1
CD34
ZNF513
PTEN
PTEN
DES;NDRG2;MYH6;TTN
IFI27L2;CCDC121;GFM1;SERPINA1;ZCCHC10;SLC4A1AP;PTEN;SLC35F1;CWC27;NAT14;GJA1;CCDC91;RS
CABP7
MLLT10;RSRC1
APOE
APOC1
APOE
APOE
GCKR
GCKR;SNX17;APOE
GFM1;CENPW;SERPINA1;HSPA4;SHOX2;PTEN;AFAP1;GJA1;ALX4;EIF3E;HORMAD2;FAM3C;APOE;MYH6
PLN

PTEN
APOC1
APOC1
MFN1
PLN
APOC1
APOE
GCKR;PTEN;HORMAD2
DES
GNB4
ALX4
PTEN
DLEU1;GPN1
HSPA4;APOE
MLLT10;CENPW;APOC1;LCORL
DLEU1;HORMAD2;APOE
GCKR;HORMAD2;UQCR10
HSPA4;CMTM5;APOC1;APOE;CD34;TTN
APOC1
GCKR
SNX17
APOC1
APOE
DES;PLN;SNX17;APOE;CD34;MYH7;TTN
TOMM40;APOC1;HORMAD2;APOE
TBX15;TOMM40;CCDC91;PVRL2;WARS2
CEP85L
MYH7
PTEN
CR1
APOE
PTEN
PTEN
TBX15;WNT16
TBX15;WNT16
DLEU1
GJA1
GJA1
CD34
HSPA4
CD34
GCKR
PTEN
SERPINA1
APOE

APOE
PTEN
PTEN;CWC27
PTEN;CWC27
PTEN;CWC27
PTEN;NDRG2
PTEN
CR1
SUPT7L
MYH6
PTEN
SUPT7L
CENPW;DES;FAM174B;APOE
KIAA1755;GCKR
PTEN
GJA1
EIF3E
NCAPG
MLF1
PTEN
DNAJC1
APOE
RSPO2
APOE
CR1
GJA1;PTEN
PTEN
PTEN
APOE
DES
HSPA4
SERPINA1
PTEN
PTEN
DES
APOE
TOMM40
GJA1
GJA1
ALX4
APOE
DES
MYH7
GJA1;PTEN
SERPINA1

CCDC141
PTEN
APOE
PTEN
EIF3E
PTEN
GJA1;PTEN;ALX4
HSPA4;PTEN
MYH7
GJA1;PTEN;ALX4
GJA1;HSPA4;PTEN
GCKR
APOE
GCKR
GCKR
WARS2
GCKR
HSPA4;PTEN
TOMM40;APOE
PTEN
SERPINA1
APOE
MLLT10;HSPA4;PTEN;CD34
SERPINA1;PTEN;APOE
APOE
APOE
PTEN
PTEN
PTEN
HSPA4
APOE
CD34
CR1
SERPINA1
WARS2
APOE
GFM1
GCKR
PTEN
CR1
CD34
NAT14
APOE
DGKH
SLC5A6

GJA1
CR1
APOE
GJA1
CENPW;MYH6
SUPT7L
APOE
GJA1;PTEN;ALX4
PTEN
PTEN
HSPA4
GJA1
APOE
ALX4
ALX4
MLLT10
GCKR
GCKR
GCKR
GCKR
PTEN
DES
APOE
PTEN
SERPINA1
APOE
PTEN
CWC27
PTEN
PTEN
TTN
MLLT10;PTEN
EIF2B4;TOMM40;PTEN;APOE
SERPINA1;TOMM40;APOC1;APOE
PTEN
PTEN
SHOX2
DES
GJA1;PTEN
GCKR;SERPINA1;MFN1;APOE
PTEN;SPEG;MYH7
GJA1;DES;PTEN;RSPO2;EIF3E;NDRG2
PTEN
APOE
EIF3E

GJA1
ASCC2
PTEN
PLN
TTN
PTEN
GJA1
PTEN
SERPINA1
GJA1
APOE
GCKR;UQCR10
APOE
PTEN
PTEN
PTEN
PTEN
DES
GJA1;APOE;CD34
APOE
SERPINA1;PTEN;NDRG2;CD34;TRIB2
CD34
GJA1;ALX4;ZNF513
SERPINA1;CCDC91
CD34
APOE
APOE
CD34
GJA1
SERPINA1;TOMM40;HSPA4;APOC1;PTEN;APOE
PTEN;CD34
PTEN
CD34
EIF2B4
DES;SERPINA1;PTEN
GCKR
MLLT10
GCKR
GCKR
HSPA4
GCKR
PTEN
GFM1;GJA1;ALX4;ZNF513;CCDC141;WARS2
PTEN;ALX4
GJA1

GFM1
SLC5A6;TBX15;CR1;CCDC91;PTEN;RSPO2;AFAP1;FAM3C;WNT16;TRIB2;DGKH
ZCCHC10
PTEN;ALX4;CD34
ALX4;ZNF513;CCDC141
APOE
GJA1
GJA1
TBX15;TTN
CR1
PTEN
SERPINA1
PTEN
PTEN
PTEN
PTEN
APOE
APOE
TBX15;WNT16
GCKR;HSPA4;CABP7;CEP85L;NIPSNAP1;WARS2;GJA1;B3GNT7;NOV;APOC1;ZNF219;ZMAT5;APOE;ARHG
PLN;CCDC91;MFN1;CCDC141;MLF1;MYH6;MYH7;TTN
DES;PLN;MFN1;VSTM2L;UQCR10;CCDC141;MLF1;MYH6;MYH7;TTN
GFM1;PLN;MFN1;VSTM2L;CCDC141;MLF1;MYH6;TTN
DES;PLN;UQCR10;CCDC141;MLF1;MYH6;MYH7;TTN
PTEN
GJA1;PTEN
TBX15;PTEN
APOE
SERPINA1
HSPA4
PTEN
APOE
DES
PTEN
SERPINA1
TRIB2
APOE
GJA1
HSPA4
CD34
APOE
SPEG;ZNF513
DES
PTEN
APOE

PTEN
SERPINA1
GCKR
GCKR
GCKR
GCKR
GCKR
RSPO2
TTN
FAM174B
HORMAD2
DGKH
LCORL
GCKR
TOMM40
DGKH
APOC1
CCDC91;GPR126
SLC35F1
GCKR;HORMAD2
TRIB2
TBX15;MLLT10;ZNF628;SHOX2;ZNF219;ALX4;ASCC2;APOE;LCORL;NAT14;MLF1;SUPT7L
PLN
TOMM40
APOE
MLLT10
PTEN
PTEN
WNT16
GCKR
GCKR
APOC1
TOMM40
DLEU1
GCKR
SNX17
CCDC91
GCKR
AFAP1;HORMAD2
MTMR3;TOMM40
GCKR
GCKR
PTEN
PTEN
CCDC141

GJA1
TBX15;PTEN;RSPO2
HSPA4;CD34
APOC1;APOE
APOE
APOC1
DNAJC1;APOE
PTEN;APOE
DGKH
APOE
NCAPG
ZNF628;ZNF219;ASCC2;MLF1
SUPT7L
APOE
IFI27L2;CCDC121;GFM1;ZCCHC10;SLC4A1AP;PTEN;CWC27;GJA1;CCDC91;RSRC1;FAM174B;DLEU1;RSPC
ZNF219
SERPINA1
GJA1;CD34
MLLT10;LXN;HSPA4;PTEN;CD34
ZNF513;CWC27
GFM1
GCKR
TOMM40
GCKR
DLEU1;HORMAD2
GCKR
GCKR;HORMAD2
GCKR
MLLT10
GCKR
CD34
CD34
HSPA4
PTEN;ALX4
CD34
ZNF513
APOE;CD34
PTEN
APOE
CR1
PTEN;CD34
TOMM40
NOV
ALX4
CD34

APOE
CD34
DES
SERPINA1
PTEN;ALX4
AFAP1;APOE
KIAA1755;ARHGEF40
GJA1
APOE
GJA1
PTEN;TRIB2
PLN
SUPT7L
CCDC121;GFM1;SERPINA1;LXN;TOMM40;PTEN;UQCR10;TTN;CCDC91;MFN1;FAM3C;APOE;CCDC141;CI
AFAP1
MYH6
PTEN;DLEU1;HORMAD2
APOE
PTEN;ARHGEF40;MLF1
GJA1
PTEN
PLN
WARS2
PTEN
PTEN
DES
GCKR
GCKR
GCKR
GCKR
GCKR
GCKR
GCKR
CCDC141
SLC5A6;ALX4
GCKR;APOC1
CCDC141;MYH6
CCDC141;MYH6
RSPO2;SSC5D
GCKR;APOC1
VSTM2L;MYH6
CCDC141;MYH6
GCKR;APOC1
DES;SSC5D
TBX15;TTN

SLC5A6;ALX4
DES;SSC5D
CCDC141;MYH6
CCDC141;MYH6
TBX15;TTN
SHOX2;SPEG
ZNF219;SSC5D
TBX15;TTN
CCDC141;MYH6
CCDC141;MYH6
CABP7;VSTM2L
TBX15;TTN
CCDC141;MYH6
CCDC141;MYH6
GCKR;APOC1
CCDC141;MYH6
CABP7;VSTM2L
APOE;NDRG2
GCKR
CCDC91
GCKR
GCKR
GCKR
GCKR
GCKR
GCKR
SNX17
GCKR
GCKR
GCKR;ZMAT5;DLEU1;HORMAD2;LCORL
DES;PLN;UQCR10;CCDC141;MLF1;MYH6;MYH7;TTN
DES;PLN;UQCR10;CCDC141;MLF1;MYH6;MYH7;TTN
KIAA1755;LXN;VSTM2L;CCDC141;CD34;MLF1;MYH6
PLN;B3GNT7;CEP85L;NCAPG;TMEM253
TBX15;PTEN;APOE
PTEN;NDRG2;CD34
SNX17;APOE
PTEN;ALX4;APOE;CD34
PTEN
PTEN
ALX4
GJA1
MLLT10;PTEN
SERPINA1
TTN

APOE
GJA1
HSPA4
SERPINA1
IFI27L2;CCDC121;GFM1;SERPINA1;ZCCHC10;SLC4A1AP;PTEN;CWC27;NAT14;GJA1;CCDC91;RSRC1;FAM
ZNF628
DES;PLN;B3GNT7;UQCR10;CCDC141;MYH6;MYH7;TTN
GFM1;PLN;CCDC91;HSPA4;MFN1;UQCR10;CCDC141;MLF1;MYH6;MYH7;TTN
GCKR
GCKR
APOE
GCKR
SLC35F1
GCKR
PTEN
CR1
SUPT7L
CR1
SERPINA1
GJA1
PTEN;APOE;NDRG2
PTEN
SERPINA1;LXN
PTEN
DGKH
B3GNT7
TTN
IFI27L2;GCKR;LXN;CABP7;UQCR10;NIPSNAP1;GJA1;NOV;SNX17;APOC1;ZNF219;ZMAT5;APOE;ARHGEF4
SERPINA1
SERPINA1
GJA1;SERPINA1;HSPA4;PTEN;ALX4;EIF3E;APOE;TRIB2
NAT14
MTMR3
TTN
GPN1
GCKR;APOC1;CWC27;NRBP1
APOE;TTN
HSPA4
CD34
EIF2B4;TBX15;GFM1;GJA1;DES;SERPINA1;PTEN;ZNF513;APOE;TTN
ALX4
SERPINA1;NDRG2
PTEN;CD34
SERPINA1;PTEN
PTEN

GJA1
GCKR;APOC1
PTEN
GJA1
PTEN
APOE
APOE
GJA1
HSPA4
PTEN;CD34
APOE;CD34
PTEN
GJA1
SERPINA1
TOMM40;APOE;CD34
PTEN;CD34
GJA1;PTEN
PTEN;APOE
GJA1;APOE
PTEN;NDRG2
PTEN;CD34
SERPINA1;CR1;SHOX2
GCKR;APOC1
APOE;NDRG2
SERPINA1
GJA1;PTEN;CD34
MLF1
DES;GNB4;WARS2;TTN
EIF2B4;APOE
PTEN;CD34
PTEN
GJA1
PTEN
GJA1
PTEN;CD34
SERPINA1
CD34
PTEN
PTEN
GJA1;HSPA4;PTEN
APOE
PTEN
PTEN
CCDC141
GJA1

GJA1
GJA1;PTEN
GCKR
HORMAD2
GCKR
GCKR
DES;SERPINA1;MLF1
SLC5A6;HSPA4;CD34
TBX15;GCKR;GJA1;SERPINA1;NOV;RSRC1;APOC1;PTEN;ZNF513;FAM3C;APOE;CD34
DES;PTEN
APOE
APOE
APOE
APOE
SPEG
AFAP1
TBX15
ZNF513
UQCR10
SPEG
MFN1
GFM1
AFAP1
GCKR
GCKR
GCKR
GCKR
APOE
GCKR
LXN;APOE;NDRG2;TTN
LXN
DES
CD34
GJA1
APOE
APOE
TOMM40;APOE
PTEN
GJA1
APOE
GNB4
ALX4
PTEN;CD34
GCKR;GJA1;NOV;HORMAD2;APOE
GJA1;DES

CR1;CR1L
MTMR3;PTEN
PLN;MYH6;MYH7
GCKR
ASCC2
GCKR
GCKR
GCKR
MLLT10;RSRC1;SPEG;ASCC2;LCORL
GCKR
APOE
PTEN
APOE
SLC5A6
EIF2B4
EIF2B4;MFN1;NDRG2;MYH7
EIF2B4;DES;NRBP1;TTN
GJA1;CENPW;GNB4
MYH6;MYH7
APOE
CD34
GJA1
RSRC1
TOMM40;MFN1
APOE
SLC5A6
SLC5A6
APOC1
PTEN
CR1
APOE
MLLT10;GJA1;NOV;PTEN;CD34
GJA1;PTEN
MLF1
APOE
MTMR3
APOE
PLN
PTEN
APOE
APOE;NDRG2
CD34
GCKR;ZNF512
GCKR
PLN

GJA1;ALX4;CD34
GJA1
APOE
IFI27L2;GCKR;LXN;HSPA4;CABP7;C2ORF16;NIPSNAP1;GJA1;SNX17;APOC1;ZNF219;ZMAT5;APOE;ARHG[illegible]
IFI27L2;GCKR;TOMM40;HSPA4;CABP7;UQCR10;TMEM253;NOV;APOC1;ZNF219;ZMAT5;APOE;ARHGEF[illegible]
MYH7;TTN
WNT16
PTEN
SERPINA1
GJA1
PTEN
SERPINA1
SERPINA1
GJA1
CCDC141
GJA1
CD34
GJA1
TBX15
TBX15;GJA1
PTEN
GJA1
PTEN
PTEN
GCKR
GCKR
GCKR
DES;MYH7;TTN
GJA1;PTEN;CD34
PTEN
ALX4
PTEN
GJA1;PTEN
GJA1;APOE
APOE;CWC27
GJA1;APOE
GJA1;PTEN
SERPINA1;PTEN
PTEN
PTEN
PTEN;APOE
HSPA4;PTEN;APOE
SERPINA1;PTEN
SERPINA1
PTEN;UQCR10

SERPINA1;APOE
PTEN;UQCR10
PTEN;UQCR10
GJA1;APOE
SERPINA1;PTEN;MYH7
TOMM40;APOE
PTEN;APOE;CCDC141
HSPA4;PTEN
PLN
GJA1
APOE
GFM1;SERPINA1;PTEN
CR1;CD34
PTEN
PTEN;MLF1
GJA1;CD34
RSRC1;WARS2
PTEN
PTEN;CD34
GJA1;LXN;RSPO2
GFM1;DES;PLN;UQCR10;CCDC141;MLF1;MYH6;MYH7;TTN
GJA1;HSPA4;NDRG2
GCKR;UQCR10
HSPA4;PTEN;NRBP1;WARS2
SLC5A6;RSRC1;WARS2
PTEN
PTEN
DES;PLN;UQCR10;NDRG2;MYH6
GJA1;NOV;MFN1;UQCR10;MYH7
SERPINA1;NDRG2;MYH6;MYH7;TTN
DNAJC1
EIF2B4
B3GNT7
ASCC2
TBX15
LCORL
MLLT10
GCKR
DLEU1
CD34
GJA1
PTEN
HSPA4
SERPINA1
NOV

APOE
APOE
PTEN
TBX15;GJA1
SERPINA1;APOE;CD34;TTN
GJA1
HSPA4
PTEN
MYH7;TTN
APOE
EIF2B4;GJA1
PTEN;SPEG;MYH7;TTN
APOE
TOMM40
MLLT10;PTEN
SLC5A6;EIF2B4;PTEN
PTEN
MYH7
CR1;CD34
GJA1;PTEN;APOE
PTEN
GJA1;PTEN
SERPINA1
APOE
GJA1
NAT14
GFM1;MFN1;GPN1
GFM1;MFN1;GPN1
SERPINA1;PTEN
SERPINA1;CD34
TTN
PTEN
APOE
SERPINA1
GCKR;UQCR10
GJA1;PTEN
SERPINA1;PTEN
GJA1;PTEN
GJA1;PTEN
GJA1;PTEN
APOE
PTEN
CD34;WARS2
PTEN
CR1;APOE

APOE
GCKR;CCDC91;SNX17;PTEN;AFAP1
GCKR;SERPINA1;NOV;PTEN;TRIB2
APOE
PTEN
HSPA4
PTEN
SERPINA1;PTEN
ZNF513;CWC27
GJA1;PTEN;TRIB2
GCKR
GCKR
GCKR;C2ORF16
TOMM40
GCKR
MTMR3
DGKH
PTEN
PTEN
WNT16
PTEN
DES;SERPINA1;CR1;TOMM40;HSPA4;APOE
SERPINA1;DLEU1;CD34
PTEN;APOE
ALX4
CD34
GCKR;MTMR3;CR1;TOMM40;CR1L;APOC1;PTEN;APOE;DGKH
GJA1;WARS2;TTN
PVRL2
GJA1
GJA1
APOE
GJA1
SERPINA1
APOE
EIF2B4
TBX15
TTN
CR1
GCKR
GJA1
NOV;CD34
APOE
NRBP1
DES;SERPINA1;MLF1

LXN;GNB4;SPEG;DGKH
PTEN
GJA1;PTEN
GJA1;PTEN
PTEN
PTEN
GCKR
DLEU1
DES;NDRG2
GJA1
PTEN
APOE
UQCR10
ALX4
APOE
PTEN
MLLT10;MLF1
TOMM40
GJA1;PTEN;ALX4
GJA1;HSPA4;APOE
CEP85L;WNT16
PTEN
GFM1
DGKH
PTEN;APOE;TRIB2
PTEN;APOE
GCKR
TTN
SERPINA1
APOE
GJA1
CCDC141
SERPINA1;CR1;APOE
PTEN
CR1
TBX15
SERPINA1
TBX15
GCKR
GCKR
GCKR
DLEU1
TOMM40
HSPA4
APOE

GJA1
PTEN;APOE
DNAJC1;FAM174B
SLC35F1
GCKR;CENPW;MTMR3;HSPA4;CR1L;APOE
IFI27L2;GCKR;LXN;TOMM40;CABP7;NIPSNAP1;GJA1;NOV;APOC1;ZNF219;ZMAT5;APOE;ARHGEF40;TRIB
ALX4
GJA1;PTEN
APOE
PTEN
PTEN;ALX4
GNB4
APOE
PTEN
GJA1
GJA1
ZNF513
MYH6;MYH7;TTN
PLN;EIF3E;UQCR10;NDRG2;TTN
CCDC121;GFM1;SERPINA1;SLC4A1AP;PTEN;SLC35F1;CWC27;GJA1;CCDC91;RSRC1;RSPO2;ARHGEF40;CR
HSPA4
APOE
APOE
HSPA4
APOE
APOE
MTMR3
APOE
CWC27
CCDC121;GFM1;SERPINA1;ZCCHC10;SLC4A1AP;PTEN;CWC27;GJA1;CCDC91;RSRC1;FAM174B;RSPO2;SP
IFI27L2;CCDC121;GFM1;SERPINA1;ZCCHC10;SLC4A1AP;PTEN;SLC35F1;CWC27;NAT14;GJA1;CCDC91;RS
APOE
HSPA4;PTEN
PTEN
PTEN;NDRG2
CD34
CD34
APOC1;PTEN;APOE;NDRG2;CD34
PTEN
HSPA4
PTEN
SERPINA1
SLC5A6;GFM1;GJA1;RSRC1;ALX4;WARS2
CENPW;NCAPG
HSPA4;PTEN;DLEU1;APOE;DGKH

MYH7
CENPW
APOC1;APOE
MYH6;TTN
GPR126;APOE
MYH6;TTN
GCKR
GCKR
PTEN
GCKR
GJA1;CD34
EIF3E
GCKR
IFI27L2;CENPW;HSPA4;NCAPG;SSC5D;C2ORF16;CWC27;GPN1;WARS2;PPM1G;SNX17;GNB4;DLEU1;EIF3
IFI27L2;GCKR;TOMM40;CABP7;UQCR10;NIPSNAP1;TMEM253;NOV;SNX17;APOC1;ZNF219;ZMAT5;APO
MLLT10;KIAA1755;DES;ZNF219;RSPO2;AFAP1
DES;PLN;CMTM5;VSTM2L;UQCR10;CCDC141;MLF1;MYH6
GFM1;PLN;CCDC91;MFN1;DLEU1;UQCR10;CCDC141;MLF1;MYH6;MYH7;TTN
APOE
APOE
PTEN
MYH6
RSPO2
CD34
PTEN
SERPINA1
MYH7
APOE
HSPA4
MLLT10;HSPA4;PTEN;CD34
GJA1;PTEN
HSPA4;PTEN;GPN1;WARS2
PTEN
HSPA4;PTEN;NDRG2
APOE
GJA1
MYH7
GJA1
PTEN;CD34
MLLT10;PTEN
APOE
GJA1;MTMR3
GJA1;ALX4
GJA1;PTEN
PTEN

TRIB2
PTEN
EIF2B4
TOMM40
CENPW
GFM1;SERPINA1
DLEU1;GPR126
GCKR
GCKR
SPEG;MYH7
EIF2B4;PTEN
GJA1;PLN;APOE;CD34;MYH7;TTN
GJA1;SNX17;APOE;NDRG2;CD34;MYH7
GJA1;PLN;APOE;MYH6;TTN
GJA1;PLN;FAM174B;NDRG2;MYH6;MYH7;TTN
DES;SNX17;APOC1;APOE;CD34;MYH6;MYH7
SLC5A6;EIF2B4;GPN1;NRBP1;PPM1G
GJA1;DES;APOE;CD34;MLF1;MYH6;MYH7
SLC5A6;DCAF16;TOMM40;RSRC1;DLEU1;PPM1G
DCAF16;GJA1;DES;PLN;MLF1;TTN
GJA1;LXN;CMTM5;NCAPG;APOE;NDRG2;CD34
APOE
GJA1
GJA1;SERPINA1
SERPINA1;APOE
EIF2B4
GJA1
APOC1
APOE
GJA1
CR1
PTEN
PTEN
GJA1
PTEN
GJA1;AFAP1;ZNF513
EIF2B4
PTEN
RSRC1;APOE;WARS2
CCDC91;RSPO2
MLF1
APOE
MLF1
RSRC1;CWC27
GJA1

RSRC1
DLEU1
DLEU1
PTEN;CWC27
APOE
TOMM40
LCORL
PTEN
CD34
HSPA4
PTEN
PTEN
APOE
APOE
GJA1;NOV;APOE
PTEN
GJA1;ALX4;ZNF513
PTEN
TTN
PTEN
HSPA4
SERPINA1
APOE
GFM1;CENPW;SERPINA1;MTMR3;CR1;LXN;HSPA4;PTEN;AFAP1;NDRG2;WARS2;TTN;GJA1;DES;B3GNT7;
HSPA4
CR1;APOE
GJA1;PTEN
APOE
GJA1;PLN;NOV;EIF3E;CD34;PPM1G
NOV;SLC35F1
TBX15;MLF1;MYH6;MYH7;TTN
TBX15;DES;MLF1;MYH7;TTN
CR1;LXN;HSPA4;PTEN;NCAPG;TTN;MLLT10;DNAJC1;GJA1;NOV;FAM3C;APOE;CD34;TRIB2
APOE
GJA1;DES;SERPINA1;PTEN
GJA1;ALX4;ZNF513
APOE
GJA1
DES;PTEN
APOE
CD34
PTEN
PTEN
APOE
HSPA4

PPM1G
DES;PTEN
GJA1;PTEN
DES
APOE
GFM1;SERPINA1
PTEN;EIF3E
APOE
GCKR;SERPINA1;APOE
SERPINA1;CR1;HSPA4;PTEN;APOE;CD34;TTN
TBX15;SERPINA1;PTEN;NDRG2
SERPINA1;CR1;PTEN;APOE;CD34
GJA1
RSRC1;WARS2
GJA1;PTEN
MYH6
MLLT10;SLC4A1AP;HSPA4;GPN1;PPM1G
HSPA4;CD34;WARS2
PTEN;DLEU1
FAM3C;WNT16;TRIB2
SERPINA1;PTEN;ALX4;TTN
GCKR
PTEN
RSRC1;ZMAT5
EIF2B4
PTEN
ALX4;APOE
GJA1;PTEN
PTEN;CD34
TBX15;DLEU1;WARS2
B3GNT7;PTEN;ALX4;CD34
CD34
APOE
PTEN
PTEN
PTEN
APOE
PTEN
CCDC141
GJA1
CD34
PTEN
APOE
SERPINA1
PTEN;APOE

GJA1
GCKR;APOE
GJA1;PTEN
TBX15
ALX4
PTEN
AFAP1
ASCC2
TBX15;GJA1;B3GNT7;SHOX2;DLEU1;RSPO2;SLC35F1;CWC27;LCORL
SERPINA1
CD34
GCKR
APOE
CEP85L
KIAA1755;SNX17;ZNF219;ZMAT5;ALX4;SSC5D;ZNF512;ARHGEF40;CD34;TRIB2;NAT14;NRBP1
IFI27L2;GCKR;GFM1;TOMM40;HSPA4;CABP7;GPN1;NIPSNAP1;WARS2;PPM1G;NOV;SNX17;APOC1;ZMA
IFI27L2;GCKR;CABP7;UQCR10;WARS2;B3GNT7;NOV;SNX17;APOC1;ZMAT5;APOE;ARHGEF40;TRIB2
CD34
GJA1;PTEN
HORMAD2;TRIB2
GJA1;SLC35F1
APOE;CD34
RSRC1;WARS2
PTEN
PTEN
SERPINA1
SERPINA1
PTEN
HSPA4
TOMM40
SERPINA1
SLC5A6
MYH6;MYH7
PTEN
GCKR
CD34
EIF2B4;MYH6;TTN;PPM1G
PTEN
SERPINA1;APOE
PTEN
SERPINA1
CD34
CENPW;HORMAD2
HSPA4;APOE
PTEN;CD34

SERPINA1
SERPINA1;APOE
PTEN;CD34
CR1
GJA1
CD34
GJA1
APOE
HSPA4
GJA1
MYH7
CD34
APOE
DLEU1
TOMM40;CWC27;WARS2
VSTM2L
GCKR
MLLT10;DNAJC1;TOMM40;RSRC1;CEP85L;DLEU1;EIF3E
IFI27L2;GCKR;LXN;TOMM40;C2ORF16;UQCR10;NIPSNAP1;TMEM253;WARS2;GJA1;NOV;APOC1;ZNF21
MLLT10;IFI27L2;PLN;DLEU1;VSTM2L;UQCR10;CCDC141;CD34;MLF1;MYH6;TTN
GJA1
WNT16
GJA1;ZNF513
SERPINA1
EIF2B4;GFM1;EIF3E;WARS2
APOE
MFN1
TOMM40;MFN1
TOMM40;MFN1
TOMM40;MFN1
APOC1
MTMR3;PTEN
APOC1;APOE
APOC1;APOE
MTMR3;TOMM40;MFN1
APOC1;APOE
DES;MYH6
ASCC2
EIF2B4
NCAPG
MFN1
APOC1;APOE
CR1
SERPINA1;GNB4;FAM3C;DGKH
GJA1

APOE
GJA1
APOE
ASCC2
APOE
PTEN
GJA1
APOC1;APOE
MTMR3;TOMM40;MFN1
SSC5D
SSC5D;APOE
APOE;CD34
PTEN
CR1;PTEN;CD34
GCKR
CD34
SERPINA1
CR1;PTEN;CD34
SLC5A6
SLC5A6
GNB4;ARHGEF40
PTEN
RSRC1
RSRC1
GCKR
APOE
PLN
RSRC1
TBX15
MYH7
PTEN
NDRG2
TBX15;GJA1
TTN
APOE
DES;PTEN
EIF3E
SERPINA1;CR1
PTEN
APOE
DES
SERPINA1
TBX15;PTEN
PTEN
APOE

AFAP1
APOE
GFM1
SPEG;MYH7
GJA1;MFN1
CR1;PTEN;CD34
MYH7
PLN
GJA1
PTEN;APOE
APOE
APOE
ASCC2
MLF1
CD34
DES;PLN;ASCC2;CCDC141;MLF1;MYH6;MYH7;TTN
SERPINA1;APOE
PTEN
EIF2B4
GCKR
LCORL
IFI27L2;MTMR3;SLC4A1AP;TOMM40;CEP85L;PTEN;UQCR10;CWC27;NDRG2;NAT14;TTN;PPM1G;DCAF1
SERPINA1;APOE
HSPA4
HSPA4
GJA1;FAM174B;EIF3E;APOE;CD34;MLF1;TTN
DES;SNX17;APOE;NDRG2;CD34;MYH7;TTN
PLN;FAM174B;MFN1;NDRG2;MYH6;TTN
GJA1;DES;FAM174B;APOE;NDRG2;MLF1;MYH7
DES;SNX17;FAM174B;MYH6;MYH7;TTN
APOE
GJA1
MLLT10;CD34
HSPA4;APOE
SHOX2;PTEN;NDRG2
MYH7
MYH7
PTEN
EIF2B4
SERPINA1
GJA1
CD34
EIF2B4;GCKR;SNX17
APOE
PTEN

APOE
NIPSNAP1
HSPA4;APOE;TTN
GCKR;SERPINA1;UQCR10
GFM1;ALX4
IFI27L2;GFM1;SERPINA1;ZCCHC10;SLC4A1AP;PTEN;CWC27;NAT14;GJA1;CCDC91;RSRC1;FAM174B;DLEU
APOE
APOE
APOE
PTEN
CD34
GJA1
GJA1
MLLT10
HSPA4;MFN1
HSPA4
PTEN
APOE
ALX4
PTEN
PTEN
GJA1;PTEN
PTEN
GJA1
PTEN
APOE
DES
APOE
HSPA4;PTEN;CD34
HSPA4
GFM1;ALX4
PTEN
TBX15;DES;NOV;APOC1;RSPO2;APOE
LXN;NOV;APOC1;EIF3E;APOE;TRIB2
GJA1;DES;SNX17;APOE;MYH6;MYH7
CD34
PTEN;RSPO2
APOE;CD34
DES;PTEN
GFM1
GJA1
DNAJC1
PTEN;APOE
CR1
PTEN

NOV
APOE
APOE
GJA1;PTEN
APOE
SERPINA1
ZNF513
PTEN
APOE
PTEN
TOMM40
DCAF16;GCKR;CENPW;CCDC91;RSRC1;SHOX2;CEP85L;DLEU1;NCAPG;LCORL;DGKH
APOE
GCKR
CCDC91
MLLT10;DNAJC1;TOMM40;APOE;LCORL
PTEN
APOE
ASCC2
APOE
APOC1
CR1;FAM174B
WNT16
APOE
TOMM40
GJA1;PTEN;SPEG;WNT16
APOE
EIF2B4
APOC1
EIF3E;APOE
PTEN
SLC5A6
PTEN
PTEN
APOE
GPN1
PTEN;ZNF513;APOE;CCDC141
TOMM40
GJA1;TTN
GJA1;PTEN
MTMR3;PTEN
DLEU1;SPEG;CWC27;LCORL
HSPA4;PTEN
PLN
APOE

HSPA4
CR1;APOE
APOE
CR1;PTEN
PTEN
GJA1
PTEN;CD34
PTEN
PTEN
NAT14
WARS2
PTEN
GJA1
CD34
APOE
PTEN
EIF2B4
PTEN
APOE
PTEN
CR1
EIF2B4
PTEN
GCKR;HORMAD2
CCDC141
GJA1
GCKR;PTEN
MFN1;GNB4;PTEN;NCAPG;WNT16;TTN
PTEN
LXN;DLEU1
GJA1;PTEN
PTEN;APOE
CEP85L
EIF2B4
CD34
PTEN
GJA1
GJA1
GCKR;GJA1;PTEN;ARHGEF40
NAT14;MLF1
APOE
GCKR
MLLT10
RSRC1
MLLT10;GJA1;CR1;PLN;NOV;HSPA4;PTEN;CD34;TRIB2;MLF1

CWC27
PTEN
GFM1
GJA1
LCORL
SERPINA1;APOE
HSPA4;PTEN
MYH7
MYH7
GJA1
PTEN
ALX4
NOV;APOE
CD34
PTEN
PTEN
CD34
TOMM40
SERPINA1
APOE
MYH7
LXN;SHOX2;CD34
DCAF16;SLC4A1AP
TBX15;NDRG2;CCDC141;TTN
GJA1;APOE;NDRG2;MYH6
TBX15;DES;MFN1;NDRG2
TBX15;DES;NDRG2;MYH7
NDRG2;CCDC141;MYH7;TTN
DES;SPEG;MYH7
PTEN;CCDC141;MYH6
LXN;NOV;FAM3C;CD34
NOV;CD34;MLF1
PLN;TRIB2;MLF1
TBX15;DES;NOV;MYH7
CENPW;RSRC1;SHOX2
NDRG2;MLF1
NOV;MYH6
APOE;MYH7
GFM1;MFN1;UQCR10
TOMM40;UQCR10;TRIB2
LXN;SSC5D;APOE;CCDC141
LXN;FAM174B;CD34
TBX15;PLN;NDRG2;TTN
DES;PLN;MLF1;MYH7
GJA1;LXN;GNB4

TBX15
MLF1
APOE;MYH7
DES;NRBP1;TTN
TBX15;DES;MYH7
LXN;SSC5D;CCDC141;CD34
PLN;CD34;MYH7;TTN
TBX15;MFN1
TBX15;MYH7;TTN
TBX15;CCDC141;MYH7
TBX15;NOV;MYH7
EIF2B4;DES;MYH7
DNAJC1;CWC27;MYH7
UQCR10
MFN1;ALX4;UQCR10
SLC35F1;CD34;TRIB2
PLN;NDRG2;MLF1
APOE
PTEN
EIF2B4
DCAF16;NIPSNAP1;MYH7
GJA1
GJA1;NOV
GCKR;SERPINA1;HSPA4;UQCR10;APOE;CD34
HSPA4;PTEN;DLEU1;CD34
MLLT10;SERPINA1;LXN;HSPA4;PTEN;DLEU1;APOE;CD34
GJA1
DNAJC1;EIF3E
GJA1;MYH7
SERPINA1;B3GNT7
DES;PLN
DES;PLN
DES;PLN
CD34;MLF1
CENPW;NCAPG
CENPW;NCAPG
CMTM5;CD34
B3GNT7;NDRG2
DES;PLN
DES;CMTM5
CR1;WNT16
DES;SPEG
SERPINA1;CD34
SERPINA1;CD34
DES;ARHGEF40

SLC35F1;TRIB2
DES;MYH7
SERPINA1;CMTM5
PLN;CD34
PLN;CD34
TBX15;SSC5D
SERPINA1;APOE
DES;APOE
SERPINA1;CR1
APOC1;APOE
SERPINA1;CR1
APOE;CD34
SERPINA1;ARHGEF40
APOC1;CD34
DES;RSPO2
SERPINA1;CD34
TOMM40;NAT14
RSRC1;FAM174B
DES;CD34
APOC1;APOE
SERPINA1;CR1
SPEG;CD34
CR1;APOE
CMTM5;NDRG2
PLN;CD34
KIAA1755;FAM174B
NCAPG;CD34
DES;CD34
SERPINA1;CR1
PTEN;CD34
DES;SPEG
GNB4;ARHGEF40
SSC5D;CD34
DES;CD34
CD34
CCDC121;GFM1;SERPINA1;ZCCHC10;SLC4A1AP;PTEN;CWC27;GJA1;CCDC91;RSRC1;FAM174B;DLEU1;RS
MLLT10
CCDC91;PTEN
GJA1
IFI27L2;CCDC121;GFM1;SERPINA1;ZCCHC10;SLC4A1AP;PTEN;SLC35F1;CWC27;NAT14;GJA1;CCDC91;RS
PTEN
PTEN
GJA1
APOE
EIF2B4;GJA1;ZNF513

PTEN
GJA1;SERPINA1;PLN;HSPA4;APOC1;PTEN;APOE;MYH7
SERPINA1;PTEN
HSPA4;CD34
PTEN
MYH6
EIF2B4
EIF2B4;EIF3E
PLN
GCKR
GJA1
GCKR
SERPINA1
PTEN
RSPO2
NOV;SHOX2;NDRG2
SLC35F1;APOE
GJA1;NDRG2
APOE
GJA1;PTEN
HSPA4;CD34
GCKR;TOMM40;HSPA4;CABP7;C2ORF16;NIPSNAP1;WARS2;GJA1;NOV;SNX17;APOC1;ZNF219;ZMAT5;A
NOV
PVRL2
FAM174B
GJA1
B3GNT7
APOE
PTEN
PTEN
PTEN
PTEN;CD34
SERPINA1;RSRC1;PTEN
MYH7
PTEN
APOE
TOMM40
CD34
PTEN
TBX15;DES
PTEN
SERPINA1
GJA1;PTEN;TTN
ALX4
PTEN;CCDC141;MYH7;TTN

TOMM40;APOC1;PVRL2
PTEN;WNT16;CD34
MTMR3
APOE
B3GNT7
GJA1;MTMR3;PTEN;HORMAD2
TTN
APOE
MYH6
WNT16
PTEN;TRIB2
APOE
NAT14
GJA1
PTEN;SPEG;APOE
RSPO2;ALX4
MLLT10;PTEN
GJA1
PTEN
EIF2B4
PTEN;CWC27
APOE
PTEN
HSPA4
PTEN
PTEN
APOE
SERPINA1
SUPT7L
GCKR;PTEN
GCKR;PTEN
HSPA4;EIF3E;CWC27;PPM1G
GCKR
NOV
APOE
GCKR;GJA1
APOE
CCDC141
APOE;MYH7
SPEG;UQCR10;TTN
PTEN
CR1;PTEN
AFAP1
DES;PLN;MYH6
IFI27L2;CCDC121;GFM1;SERPINA1;ZCCHC10;SLC4A1AP;PTEN;CWC27;NAT14;GJA1;CCDC91;RSRC1;FAM

HSPA4;TTN
IFI27L2;CCDC121;GFM1;SERPINA1;ZCCHC10;SLC4A1AP;PTEN;CWC27;NAT14;GJA1;CCDC91;RSRC1;FAM
GJA1;PTEN;APOE
APOE
PTEN
HSPA4
HSPA4
APOE
SERPINA1
ALX4
CCDC141
SERPINA1
APOE
CR1;CR1L
PTEN
CD34
PTEN
HSPA4
HSPA4
CD34
HSPA4
CCDC141;TTN;PPM1G
CD34
APOC1
APOC1;APOE
GJA1
PTEN
PTEN
DES;PTEN
PTEN;CD34
GFM1;SERPINA1;APOE
DES;TTN
TBX15;CWC27
SERPINA1
EIF2B4;SERPINA1;HSPA4;APOE;NDRG2
SERPINA1
DES
PTEN
PTEN
GJA1
GJA1
APOE
GJA1;CR1;NOV;HSPA4;APOC1;APOE;CD34
ALX4
CD34

APOE
TTN
ALX4
GJA1
EIF2B4;PTEN;APOE
EIF2B4;PTEN;APOE
TTN
DES;PTEN
LXN;HSPA4;PTEN;CD34
ALX4;ZNF513
DES;UQCR10;NDRG2
CCDC141;MLF1
PLN;PTEN;UQCR10
MYH7;TTN
CD34
PTEN
RSRC1;CWC27
TBX15;DES;MYH7
GFM1;TOMM40;MFN1
APOE;NDRG2;CD34
PTEN
SERPINA1
APOE
DES
PTEN
PTEN;CWC27
APOC1
ALX4
APOE
PTEN
GJA1
SERPINA1
APOE
APOE
GJA1;APOE
CD34
SERPINA1
GNB4
APOE
IFI27L2;GCKR;GFM1;TOMM40;CABP7;C2ORF16;UQCR10;NIPSNAP1;WARS2;GJA1;NOV;ZMAT5;APOE;AI
GFM1;DES;PLN;UQCR10;CCDC141;MLF1;MYH6;MYH7;TTN
GCKR
SERPINA1;HSPA4;APOE;TTN
EIF2B4;HSPA4
PTEN;MYH6

DGKH
PTEN
TRIB2
DLEU1;RSPO2
GJA1
DGKH
GJA1;NOV;HSPA4;GNB4;PTEN;WNT16;NDRG2
DGKH
PTEN
GJA1
GJA1
GJA1;ZNF513;APOE
PTEN
APOE
GCKR;APOE
SERPINA1
MLLT10;GCKR;HSPA4;DLEU1
GJA1
PTEN;TTN
APOE
GJA1;PTEN;APOE
APOE
SERPINA1;TOMM40;APOE
MLLT10;DES;NOV;HSPA4;PTEN;APOE;NDRG2;CD34
EIF2B4;GJA1;HSPA4;PTEN
PVRL2
RSRC1;CWC27
MTMR3;PLN;MFN1;UQCR10;SUPT7L
MTMR3;CD34
NOV;PTEN
MLLT10
GJA1;HSPA4;PTEN
CD34
APOE
SERPINA1
TTN
GFM1;SERPINA1;WARS2
PTEN
DES
GFM1
DGKH
LXN;AFAP1;ARHGEF40;PVRL2;CD34;TRIB2
GJA1;DES;SNX17;APOE;MYH6;MYH7
DES;APOC1;APOE;NDRG2;CD34;TTN
GJA1;DES;NDRG2;CD34;MYH6;MYH7

NOV;SHOX2;APOC1;APOE;NDRG2;CD34
DES;APOE;NDRG2;CD34;MYH6;MYH7
DES;PLN;UQCR10;NDRG2;MYH6
PTEN
NDRG2
SLC5A6
MTMR3
TBX15;NOV;TTN
APOE
PTEN;NDRG2
GJA1;PTEN;CD34
HSPA4;CD34
APOE
CD34
ZNF513
DES;PLN;HSPA4;UQCR10;CCDC141;MLF1;MYH6;MYH7;TTN
PLN;CCDC91;UQCR10;CCDC141;MLF1;MYH6;MYH7;TTN
PLN;UQCR10;CCDC141;MLF1;MYH6;MYH7;TTN
DES;TOMM40;PLN;VSTM2L;UQCR10;CCDC141;MLF1;MYH6;MYH7
GCKR;LXN;C2ORF16;NIPSNAP1;WARS2;GJA1;NOV;APOC1;ZNF219;ZMAT5;APOE;ARHGEF40;PVRL2;TRIB
PLN;UQCR10;CCDC141;MLF1;MYH6;MYH7;TTN
DES;PLN;UQCR10;CCDC141;MLF1;MYH6;MYH7;TTN
TBX15;DES;ZNF628;SPEG;NDRG2;NRBP1;MYH6;MYH7;TTN
PLN;CMTM5;VSTM2L;UQCR10;CCDC141;MLF1;MYH6
GCKR;CABP7;NIPSNAP1;GJA1;B3GNT7;NOV;SNX17;APOC1;ZNF219;ZMAT5;APOE;ARHGEF40;TRIB2
DES;PLN;UQCR10;CCDC141;MLF1;MYH6;MYH7;TTN
WNT16
SNX17
DNAJC1
CR1
TTN
TOMM40
SERPINA1;CCDC91;RSRC1;SHOX2;DLEU1;DGKH
SERPINA1
DES;SHOX2;RSPO2;CCDC141;TTN
GJA1;LXN;EIF3E;MYH6;MYH7
APOE
NDRG2
PTEN;CWC27
DES;PTEN
PTEN
TBX15
GJA1
DES;SERPINA1;CR1;HSPA4;PTEN;APOE
PTEN

KIAA1755;PLN;VSTM2L;SSC5D;CCDC141;CD34;MLF1;MYH6
MLLT10;PTEN
GFM1;PLN;CCDC91;UQCR10;CCDC141;MLF1;MYH6;MYH7;TTN
GCKR
ALX4
GCKR
AFAP1
GJA1
MYH7
PTEN;NDRG2
ALX4
CCDC141
DGKH
MLLT10
SERPINA1;MTMR3;LXN;HSPA4;PTEN;AFAP1;NDRG2;MLF1;GJA1;DES;CCDC91;APOE;CD34
ZNF513
HSPA4
APOE
HSPA4;CD34
PTEN
GJA1
MYH7
TBX15
PTEN;APOE
APOE
GCKR;SERPINA1;APOE
PTEN
APOE
CD34
GJA1;TOMM40;APOE;NDRG2
HSPA4;PTEN;EIF3E;NDRG2;CCDC141;MYH6;MYH7;TTN
GFM1;DES;FAM174B;NDRG2;MLF1;MYH6;MYH7;TTN
GJA1;DES;PLN;PTEN;EIF3E;FAM3C;APOE;CD34
CD34
TBX15
GJA1
CD34
CWC27
GCKR;GJA1;HSPA4;APOE;CD34
PTEN
PTEN
PTEN
ALX4
SHOX2;PTEN;AFAP1
PTEN

PTEN
GJA1
PTEN
GJA1;MFN1;PTEN
APOE
APOE
DES;PTEN
EIF3E
APOE
APOE
SERPINA1
SLC4A1AP
MYH7
GJA1;PTEN
PTEN
GJA1;PTEN
GJA1;APOE;CD34
PLN;HSPA4
HSPA4
GJA1
PTEN;GPN1
APOE
APOE
PTEN
APOE;TTN
PTEN;APOE
SERPINA1;SHOX2;PTEN
AFAP1
APOC1;CD34
DES;APOC1;CD34;MYH7;TTN
MYH6;MYH7;TTN
CD34
GNB4
UQCR10
FAM3C;WNT16
CEP85L;EIF3E
DLEU1
SERPINA1
PTEN
APOE
GCKR
GJA1
APOE
CCDC141;MYH6;MYH7
DES;PLN;UQCR10;CCDC141;MLF1;MYH6;MYH7;TTN

SERPINA1
APOE
PTEN;NDRG2
CD34
CR1
APOE
CD34
RSPO2
MLLT10;CCDC121;ZNF219;GNB4;RSPO2;AFAP1;SPEG;SSC5D;DGKH
PTEN;CWC27
DES
DES
APOE
HSPA4
GJA1
GJA1
PTEN
PTEN
EIF3E
NDRG2
PTEN
AFAP1
CR1;APOE
AFAP1
GCKR
LCORL
PTEN
GJA1
GJA1
GCKR
CD34
GJA1
APOE
SERPINA1
PTEN
DES
TTN
APOE
APOE
PTEN
GJA1
APOE
PTEN
CWC27
APOE

PPM1G
CD34
HSPA4;PTEN
APOE
PTEN
SHOX2
IFI27L2;CCDC121;GFM1;SERPINA1;ZCCHC10;SLC4A1AP;PTEN;SLC35F1;CWC27;NAT14;GJA1;CCDC91;RS
FAM174B;NCAPG;ASCC2;ARHGEF40
PTEN
SLC5A6;DNAJC1;GCKR;SERPINA1;TOMM40;APOC1;GPR126;ZNF513;C2ORF16;FAM3C;APOE;PVRL2
APOC1
DLEU1
APOE
GNB4
GCKR;PTEN
MTMR3;SNX17;PTEN;EIF3E;HORMAD2
GFM1;MTMR3;SLC4A1AP;HSPA4;SHOX2;CEP85L;PTEN;VSTM2L;NCAPG;CWC27;GPN1;TTN;PPM1G;ZMA
APOE
GJA1
GJA1
MLLT10;SERPINA1
NOV
GJA1
HSPA4
PTEN;ASCC2;APOE;CD34
APOE
TBX15
PTEN;CD34
SERPINA1;APOE
PTEN
CD34
CD34
TBX15;PTEN;ALX4
TBX15;LCORL
SLC35F1;APOE
HSPA4;CD34
APOC1;APOE;WNT16;CD34;MYH7
MYH7
TBX15
PTEN
UQCR10
DES
WNT16
MYH6
HSPA4;APOE

GJA1
GJA1
PTEN
PTEN
APOE
GJA1
CD34
SERPINA1;HSPA4;PTEN;AFAP1;NCAPG;WNT16;NDRG2;MLLT10;GJA1;DES;NOV;MFN1;GNB4;RSPO2;ALX
APOE
GCKR
SERPINA1
APOE
CCDC141
DES;PLN;MFN1;UQCR10;CCDC141;MLF1;MYH6;MYH7;TTN
KIAA1755;RSRC1;CMTM5;CEP85L;AFAP1;ALX4;SLC35F1;CWC27;CD34;TRIB2
EIF2B4;TBX15;DES;ZNF628;SPEG;NDRG2;NRBP1;MYH6;MYH7;TTN
IFI27L2;TOMM40;HSPA4;C2ORF16;UQCR10;NIPSNAP1;TMEM253;GJA1;NOV;SNX17;APOC1;ZMAT5;APO
EIF2B4;DES;B3GNT7;UQCR10;CCDC141;MYH6;MYH7
GCKR;APOC1
MTMR3;PTEN
APOC1;APOE
B3GNT7;ALX4
EIF2B4;EIF3E
PTEN
PLN;PTEN
APOE
APOE
GJA1;DES
APOC1;APOE
APOC1;APOE
GJA1;PLN;GNB4
SERPINA1;CR1
PTEN
HSPA4;PTEN
GJA1;PTEN
TOMM40;APOE
PTEN
SERPINA1
SNX17
CR1
GJA1;PTEN
PLN
PTEN;APOE
HSPA4
ALX4

PTEN
SERPINA1;PTEN
MLLT10;PTEN;CD34
PTEN;MYH7;TTN
CD34
GFM1
TBX15
PTEN
TBX15
TBX15
CD34
GJA1;PTEN
GJA1
APOE
CD34
WNT16
SERPINA1
PTEN;ZNF513;CWC27
GNB4;SLC35F1;LCORL
B3GNT7
UQCR10
APOE
TTN
EIF2B4
WARS2
APOE
PTEN
GJA1
SLC5A6
ALX4
VSTM2L
WNT16
CABP7
VSTM2L
SLC35F1
RSPO2
TMEM253
DNAJC1
APOC1
TTN
TMEM253
SSC5D
CMTM5
WNT16
DES

SLC35F1
CABP7
SLC35F1
CD34
APOE
RSPO2
TMEM253
LXN
MYH6
DGKH
CD34
SPEG
RSPO2
ZNF219
WNT16
DES
TMEM253
LXN
RSPO2
TRIB2
WNT16
CABP7
SSC5D
TMEM253
APOE
CMTM5
LXN
APOE
CABP7
LXN
RSPO2
ALX4
LXN
WNT16
PLN
SPEG
TTN
RSPO2
TTN
HORMAD2
SLC35F1
VSTM2L
CD34
ALX4
TRIB2

SSC5D
TTN
CMTM5
CMTM5
CMTM5
TMEM253
WNT16
HORMAD2
TMEM253
TMEM253
SLC35F1
CMTM5
SSC5D
TBX15
CMTM5
SLC35F1
SPEG
TTN
SHOX2
LXN
SSC5D
LXN
DES
CMTM5
VSTM2L
CR1L
CABP7
SPEG
RSPO2
TRIB2
TMEM253
WNT16
WNT16
SLC35F1
CMTM5
GCKR
ALX4
CMTM5
VSTM2L
CABP7
CD34
APOE
SHOX2
NCAPG
SHOX2

ZNF219
RSPO2
RSPO2
TMEM253
SERPINA1
GJA1;PTEN
PTEN
APOE
GJA1
PTEN
ALX4
RSRC1
CR1L;DLEU1
AFAP1
LCORL
DES
CD34
GJA1
IFI27L2;DES;PLN;UQCR10;CCDC141;MYH6;MYH7
PLN;CMTM5;VSTM2L;UQCR10;CCDC141;MLF1;MYH6
DES;B3GNT7;UQCR10;CCDC141;MYH6;MYH7;TTN
GCKR;CCDC91
PVRL2
HORMAD2
SLC35F1
CWC27
PVRL2
DGKH
CWC27
RSPO2
CWC27
TBX15;GCKR;CENPW;TOMM40;CCDC91;B3GNT7;NOV;APOC1;SLC35F1;LCORL;NRBP1;WARS2
CCDC91;DLEU1
ASCC2
TOMM40;APOE
DNAJC1;APOC1;PVRL2
APOC1
GJA1;APOE
APOE
CR1
APOE
EIF2B4;NDRG2;GPN1;NIPSNAP1;PPM1G
GJA1;CMTM5;VSTM2L;NDRG2;MYH7
GFM1;NOV;MFN1;UQCR10;TTN
CCDC121;CMTM5;SKIDA1;CEP85L;SLC35F1;KIAA1755;ZMAT5;GPR126;ALX4;ZNF512;ARHGEF40;CD34;T

SLC5A6
APOE
TTN
APOE
CD34
MLLT10
MLLT10
EIF3E;TRIB2
DNAJC1
HSPA4
APOE
SERPINA1;APOE
SLC35F1
ALX4;CCDC141;MYH7
DGKH
CD34
HSPA4
CR1;NDRG2
CD34
DGKH
CD34
SLC4A1AP;NDRG2;MLF1
NOV;MLF1
TBX15;NOV;MLF1
GFM1;MFN1;UQCR10
LXN;APOE;CD34
LXN;TRIB2
DES;SNX17;APOE;MYH6;MYH7
GJA1;SNX17;EIF3E;APOE;MYH6
EIF2B4;DES;UQCR10;MLF1;NRBP1;MYH7
GNB4;WNT16
GJA1;CD34
CCDC121;CMTM5;SKIDA1;AFAP1;VSTM2L;NAT14;MLF1;TTN;GJA1;DES;PLN;ZNF219;SPEG;ALX4;APOE;C
CCDC121;CMTM5;VSTM2L;SSC5D;NDRG2;MLF1;TTN;KIAA1755;GJA1;DES;PLN;NOV;ZNF219;SPEG;CCDC
HORMAD2
CD34;TTN
PTEN
HSPA4;APOE
GJA1;PTEN;CD34;MYH6
PTEN
PTEN
APOE
IFI27L2;CCDC121;GFM1;SERPINA1;ZCCHC10;SLC4A1AP;PTEN;CWC27;NAT14;GJA1;CCDC91;RSRC1;FAM
GCKR
HSPA4

SERPINA1
PTEN
GJA1
APOE
CD34
PTEN
DES;SERPINA1;CR1;HSPA4;PTEN;RSPO2;APOE;CD34;WARS2;TTN
GJA1;SERPINA1;HSPA4;APOE;CD34
PTEN
DGKH
TBX15;DES;SPEG;NDRG2;NRBP1;MYH6;MYH7;TTN;PPM1G
CR1;HSPA4
CD34
PTEN
MYH7
PTEN
CD34
PTEN
GJA1;PTEN
GJA1;PTEN
SHOX2;TRIB2
HSPA4;APOE;MLF1
HSPA4;APOE;MLF1
PTEN
SERPINA1;APOE
GJA1
PTEN
PTEN
CD34
GJA1
KIAA1755
RSPO2;ALX4
PTEN
RSRC1
PTEN
PTEN;ALX4
TRIB2
PTEN
UQCR10
SPEG;CWC27
SLC5A6;FAM174B;NIPSNAP1
DES;PTEN
PTEN
DGKH
PTEN

HSPA4
PLN;MFN1;SSC5D
PTEN
TTN
MYH7
GCKR;APOE
HORMAD2
RSRC1
PLN;CMTM5;UQCR10;CCDC141;MLF1;MYH6;MYH7;TTN
PTEN
PTEN
PTEN
PTEN
PTEN
PTEN
PTEN
MLLT10;CD34
ALX4
GPR126
GJA1;NOV;HSPA4;GNB4;PTEN;WNT16;NDRG2
APOE
APOE
CD34
PTEN
HSPA4;NDRG2
SERPINA1
CD34
DNAJC1
LCORL
PTEN
APOE;NRBP1
TTN
PLN;MFN1;DLEU1;VSTM2L;UQCR10;CCDC141;MLF1;MYH6
GFM1;SLC4A1AP;HSPA4;CEP85L;UQCR10;GPN1;NRBP1;TTN;TBX15;CCDC91;CCDC141;MYH6;MYH7
PLN;NDRG2;MLF1;MYH6;MYH7
TTN
DLEU1
SERPINA1
DES
APOE
GCKR
APOE
GCKR
MYH7
SERPINA1

GCKR
CD34
APOE
WNT16
CR1
GCKR;SERPINA1;UQCR10;APOE;CD34
SPEG;TRIB2
CCDC91;MFN1;DLEU1;ASCC2
ZNF513
APOE
PTEN
HSPA4;APOE
PTEN
PTEN
APOE
PTEN
ZNF513
GJA1
PTEN;APOE
TBX15
PTEN
PTEN
PTEN
WARS2
SERPINA1;HSPA4;PTEN
AFAP1
PTEN
APOE
APOE
PTEN
SERPINA1
PTEN
SERPINA1
MLLT10
APOE
APOE
GJA1
AFAP1
APOE
GCKR
PTEN
PTEN
TOMM40
SERPINA1
PTEN

TBX15;GJA1;PTEN;CWC27;MYH7;TTN
DGKH
GJA1
GFM1;PTEN;CWC27;WARS2;TTN;TBX15;GJA1;DES;SPEG;ALX4;ZNF513;APOE;DGKH
DES;SERPINA1;HSPA4
SERPINA1
ASCC2
UQCR10
PLN
MLLT10;CD34
GJA1
HSPA4
GNB4
APOE;CD34
PLN;NDRG2;CCDC141;MLF1;MYH6;TTN
DCAF16;GFM1;NCAPG;FAM3C;WNT16
TOMM40;PLN;PTEN;APOE;CD34;MYH6
GJA1;SNX17;APOE;NDRG2;CD34;MYH7
DES;HSPA4;RSRC1;PTEN;APOE;NDRG2;CD34;MLF1;WARS2
GJA1
APOE
MTMR3
MTMR3
GFM1;SERPINA1;MTMR3;ZCCHC10;LXN;SLC4A1AP;TOMM40;PTEN;NCAPG;UQCR10;CWC27;NDRG2;NA
DES;SERPINA1;HSPA4;PTEN
SERPINA1
GJA1
ALX4
GJA1
PTEN
PTEN
GJA1
MLF1
DLEU1
APOE
GNB4;APOE
CD34
HORMAD2
WARS2
GJA1;CD34
SSC5D
EIF2B4;MLLT10;KIAA1755;DES;ZNF628;ZNF219;RSPO2;SPEG;SSC5D;NRBP1
PTEN
MYH7
NDRG2

GJA1
GCKR
GJA1;MYH7
GJA1;RSRC1;PTEN;WARS2
GJA1;LXN
KIAA1755;CENPW;ZCCHC10;SNX17;ALX4;SSC5D;ARHGEF40;PVRL2;NRBP1
GCKR;APOE
TBX15;DES;ZNF628;SPEG;NDRG2;CCDC141;MLF1;MYH6;MYH7;TTN
GNB4
RSPO2
GNB4
EIF2B4;EIF3E
GNB4
WARS2
APOE
SERPINA1;FAM3C
GJA1
GNB4
GNB4
SERPINA1;FAM3C
GNB4
SERPINA1;APOE
SERPINA1;APOE
GJA1
PTEN
WNT16
GNB4
GNB4
SLC5A6
DGKH
APOC1;GNB4;APOE
SERPINA1;MFN1;GNB4;FAM3C;DGKH
GNB4
GNB4
GJA1
CCDC121;CMTM5;SKIDA1;SHOX2;CEP85L;SLC35F1;KIAA1755;DCAF16;GPR126;ZNF512;ARHGEF40;CD3[illegible]
PTEN
GJA1
PTEN
APOE
PTEN
GCKR
GCKR
TBX15;PTEN
TOMM40

SERPINA1
B3GNT7
GNB4
SNX17
PTEN
SLC5A6
TRIB2
GNB4
SERPINA1
GNB4
APOE
B3GNT7
GNB4
APOE
GNB4
GNB4
GNB4
GNB4
FAM3C
SERPINA1
GCKR
DLEU1
GCKR;APOC1;NRBP1
GCKR
GJA1;CEP85L;DLEU1;ASCC2;DGKH
ALX4
B3GNT7
AFAP1;ALX4
GCKR;TOMM40;APOC1;APOE
CCDC91
SHOX2
DGKH
CCDC91
GCKR
DGKH
KIAA1755
SLC5A6
SERPINA1
PTEN
GCKR
SERPINA1
PTEN
DNAJC1;GJA1;APOC1;EIF3E;WARS2
PTEN
ZCCHC10;PTEN;DLEU1

GNB4;RSPO2;WNT16
GJA1;ALX4
GCKR;TOMM40
CCDC141
GCKR
PTEN
TTN
SLC5A6
EIF3E
UQCR10
EIF3E
GCKR
GCKR
MLLT10
APOE
SERPINA1
CD34
CD34
DLEU1
CR1
CD34
APOE
AFAP1
AFAP1
CD34
TRIB2
SSC5D
CD34
CD34
DES
DGKH
CEP85L
SPEG
SERPINA1
SUPT7L
CD34
CD34
APOC1
NAT14
AFAP1
NCAPG
SLC5A6
MFN1
SLC35F1
EIF3E

APOE
TMEM253
SERPINA1
CR1
GJA1
CD34
DES
CD34
CD34
CD34
CD34
RSPO2
GJA1
GNB4
IFI27L2
NCAPG
CEP85L
APOE
SLC35F1
KIAA1755
APOE
GPN1
DES
KIAA1755
CD34
CD34
NCAPG
ZNF512
SERPINA1
CD34
SERPINA1
CR1
DES
KIAA1755
TBX15
DES
APOC1
GCKR
CD34
SERPINA1
CD34
NDRG2
NDRG2
CR1
DLEU1

SLC5A6
TRIB2
SLC35F1
CEP85L
VSTM2L
CD34
CD34
TMEM253
CR1L
CR1
CMTM5
CD34
CD34
TRIB2
CD34
SERPINA1
CD34
CR1
CD34
CR1
APOC1
CD34
CD34
HSPA4
MFN1
NOV
SERPINA1
CD34
SSC5D
CD34
SERPINA1
APOE
CMTM5
FAM174B
SLC5A6
CD34
CCDC141
FAM174B
PLN
CR1L
TTN
KIAA1755
CD34
CD34
AFAP1

AFAP1
CCDC91
VSTM2L
APOE
SLC35F1
CD34
CR1
CD34
CD34
WNT16
CD34
FAM174B
ASCC2
SLC35F1
GJA1
NCAPG
DES
ZNF513
NAT14
SERPINA1
SERPINA1
SERPINA1
TTN
CR1
CD34
CR1
CD34
CR1L
ALX4
ZNF219
MLF1
CMTM5
SPEG
SERPINA1
DES
PLN
CD34
DES
CR1
ZNF513
CMTM5
CR1
CD34
ZNF628
GNB4

CD34
CMTM5
SERPINA1
GJA1
DES
SPEG
SERPINA1
SERPINA1
SSC5D
DLEU1
APOE
APOE
CD34
CD34
NDRG2
SERPINA1
CR1
CD34
SERPINA1
CD34
APOE
APOC1
CR1
CD34
CD34
ZNF512
PLN
CD34
CD34
APOE
NOV
CD34
MLF1
PLN
CD34
SERPINA1
TOMM40
TOMM40
TMEM253
CR1
CD34
APOE
PTEN
B3GNT7
AFAP1

SLC5A6
MLLT10
GJA1;CD34
ALX4
CD34
GJA1
MYH7
NDRG2;CCDC141
APOC1
GCKR
HSPA4;PTEN
PTEN;CD34
DES;PLN;DLEU1;UQCR10;CCDC141;MLF1;MYH6;MYH7;TTN
CD34
CR1
CR1
GCKR
GJA1
ALX4
ALX4
CCDC141
ALX4
DES
ZNF513
TBX15
CR1
PTEN
PLN
PTEN
PTEN
MYH7
CCDC91;EIF3E;MLF1
GCKR;SERPINA1;CCDC91;NOV;SHOX2;AFAP1;C2ORF16
DLEU1;PVRL2
GNB4
TBX15
PTEN
CR1
PTEN
APOE
GJA1
HSPA4
PTEN
PTEN;DLEU1
TBX15;PTEN

GFM1
SERPINA1
EIF3E
TRIB2
DES;PLN;UQCR10;CCDC141;MLF1;MYH6;MYH7
SLC5A6;DNAJC1;GCKR;GFM1;SERPINA1;TOMM40;APOC1;GPR126;FAM3C;APOE
GFM1;PLN;UQCR10;CCDC141;MLF1;MYH6;MYH7;TTN
DES;PLN;UQCR10;CCDC141;MYH6;MYH7;TTN
DES;PLN;UQCR10;CCDC141;MLF1;MYH6;MYH7;TTN
GCKR;LXN;CABP7;C2ORF16;NIPSNAP1;WARS2;GJA1;NOV;APOC1;ZNF219;ZMAT5;APOE;ARHGEF40;TRIE
IFI27L2;GCKR;LXN;C2ORF16;NIPSNAP1;WARS2;GJA1;NOV;APOC1;ZNF219;ZMAT5;APOE;ARHGEF40;TRII
SLC5A6;DNAJC1;GCKR;SERPINA1;APOC1;GPR126;ZNF513;C2ORF16;FAM3C;APOE;PVRL2;NIPSNAP1
DES;PLN;UQCR10;CCDC141;MLF1;MYH6;MYH7;TTN
CR1
GCKR
SERPINA1;HSPA4
ALX4;ZNF513
GCKR
CR1
APOE
NDRG2
TTN
PTEN
PTEN
TBX15
SERPINA1
PTEN;APOE
APOE
TRIB2
PTEN;NDRG2
RSPO2
IFI27L2;MTMR3;SLC4A1AP;TOMM40;CEP85L;PTEN;UQCR10;CWC27;NDRG2;NAT14;TTN;PPM1G;DCAF1
IFI27L2
EIF2B4;DNAJC1;TOMM40;HSPA4;PTEN;APOE
GJA1
EIF2B4
HSPA4;APOE
GNB4
PTEN;TTN
PTEN;RSPO2
SNX17
SLC5A6
DES;HSPA4;RSRC1;PTEN;APOE;NDRG2;CD34;MLF1;WARS2
GJA1
PPM1G

ZNF513
APOE
PTEN
FAM174B
ALX4
TRIB2
APOE
ALX4
GCKR;SERPINA1;HSPA4;SLC35F1;NDRG2;NRBP1;TTN;DNAJC1;GJA1;CCDC91;NOV;APOC1;ALX4;CCDC14[illegible]
DLEU1
TTN
PTEN
SERPINA1;MFN1
GJA1
TRIB2
PTEN
SERPINA1
NIPSNAP1
APOE;CD34
GJA1
DES;NOV;PTEN;CD34
HSPA4
GJA1
CR1
APOE
TBX15;CENPW;MTMR3;RSRC1;AFAP1;SLC35F1;TRIB2
APOE
PTEN
PTEN
SUPT7L
APOE
DES;PLN;UQCR10;CCDC141;MLF1;MYH6;MYH7
SLC5A6;DNAJC1;GCKR;GFM1;SERPINA1;TOMM40;APOC1;GPR126;APOE;PVRL2;NIPSNAP1
APOE
CD34
CWC27
SERPINA1
GJA1
GCKR
CWC27
SLC35F1
CEP85L
TTN
LXN;CD34;MYH7
NCAPG;APOE;CCDC141

DES;PLN;MYH7
GJA1;LXN;CD34
RSRC1
APOE
APOE
B3GNT7
APOE
PTEN;APOE
APOE
PTEN
APOE
RSRC1
EIF3E
UQCR10
PTEN;WNT16
APOE
PTEN
PTEN;WNT16
DES
GCKR
PTEN
EIF2B4;MLLT10;DNAJC1;MTMR3;SLC4A1AP;HSPA4;SNX17;NCAPG;NDRG2;NRBP1;SUPT7L;PPM1G
PTEN
TTN
GJA1;HSPA4
APOE
PTEN
GJA1
RSPO2
TBX15;TOMM40;CCDC91;CWC27;LCORL
NIPSNAP1
PLN
DES;PLN;CCDC91;MFN1;UQCR10;CCDC141;MLF1;MYH6;MYH7;TTN
APOC1;APOE
APOE;TTN
CD34
CMTM5
PTEN
GNB4
ZNF513
SERPINA1
PTEN
PTEN;CD34
APOE
CCDC141;MLF1

TBX15
TBX15;CCDC141
GFM1;PLN
CCDC121;SHOX2;NCAPG;GPN1
SLC4A1AP;MFN1;PTEN;NIPSNAP1;SUPT7L
ZCCHC10;SNX17;GPR126;CWC27
SNX17;EIF3E;UQCR10;GPN1
EIF2B4;DES;SHOX2;ARHGEF40
GFM1;MTMR3;MYH6;PPM1G
TOMM40;SHOX2;APOC1;NCAPG
EIF3E;ZNF512;PVRL2;TTN
EIF2B4;MLLT10;ZNF512;NRBP1
GFM1;NCAPG;MLF1;NIPSNAP1
GJA1;NOV;GNB4;RSPO2;FAM3C
DCAF16;GCKR;SERPINA1;DLEU1;ZNF512
SERPINA1
SERPINA1
APOE
RSRC1
EIF2B4
PTEN
TBX15;GJA1;PTEN;ALX4
GPR126
TBX15
IFI27L2;CCDC121;GFM1;SERPINA1;MTMR3;ZCCHC10;LXN;SLC4A1AP;TOMM40;CEP85L;PTEN;NCAPG;U[illegible]
GNB4
APOE
GFM1;RSRC1;PVRL2
GJA1;HSPA4;PTEN;APOE;CD34
PTEN
CD34
HSPA4
GJA1;CENPW
ASCC2;UQCR10;MYH6
HSPA4
APOE
APOE
GFM1;GJA1;DES;NOV;HSPA4;PTEN;EIF3E;APOE;NDRG2;CD34
APOE
APOE
TBX15;PTEN
EIF2B4;GFM1;PTEN
PTEN
CR1;HSPA4;APOE
TBX15;GCKR;CENPW;CR1L;GNB4;LCORL;PVRL2

APOE
TBX15;GJA1
HSPA4;PTEN;CD34
DLEU1
APOE
TTN
GJA1;PTEN;AFAP1;CD34
HSPA4;APOE
GFM1;ALX4
PTEN
MYH7
VSTM2L
GJA1;SERPINA1;MTMR3;LXN;CCDC91;HSPA4;PTEN;AFAP1;APOE;NDRG2;CD34;MLF1
HSPA4;APOE
PTEN
MLLT10
APOE
PTEN
PTEN
CCDC141
PTEN;APOE
APOE
APOE
APOE
DES
APOE
GJA1
CWC27
PTEN
GCKR
GJA1;LXN;PTEN;APOE
CEP85L;LCORL
PLN
GJA1;DES;PTEN
DGKH
PTEN
GJA1
GJA1
GJA1
APOE
LXN
PTEN
HSPA4;APOE
GFM1;ALX4
GJA1;DES;SERPINA1;NOV;HSPA4;PTEN;RSPO2;AFAP1;EIF3E;APOE;WNT16;NDRG2

APOE
PTEN
GJA1
SERPINA1;PTEN
PTEN
PTEN
GFM1
GJA1
IFI27L2;CCDC121;GFM1;SERPINA1;ZCCHC10;SLC4A1AP;PTEN;CWC27;GJA1;CCDC91;RSRC1;FAM174B;D
GJA1;CD34
APOC1;NIPSNAP1;MYH7
FAM174B;MLF1
CD34;TRIB2
GNB4;CD34
LXN;CD34;MLF1
PTEN
SERPINA1
APOE
PTEN
PTEN
CMTM5;SKIDA1;SHOX2;CEP85L;SLC35F1;CWC27;KIAA1755;GPR126;ALX4;ZNF512;ARHGEF40;LCORL;TR
GJA1;CR1;PTEN
PTEN
PTEN
GJA1;PTEN
GJA1
TOMM40
GJA1;GNB4
TTN
PTEN
PLN
PTEN
PTEN;WNT16
APOE
WNT16
PTEN
SLC5A6;SERPINA1
PTEN
GJA1;PTEN;FAM3C;CD34
RSRC1;HSPA4
PTEN
MLLT10;LCORL;PVRL2
GJA1
GJA1
NDRG2

APOE
EIF2B4;GJA1
CD34
PTEN
HSPA4
PTEN
HSPA4;APOE
PTEN;TTN
CD34;TRIB2
TBX15
PTEN
GJA1;PTEN
GJA1;PTEN
HSPA4;PTEN
PTEN
IFI27L2;CCDC121;CMTM5;SKIDA1;CEP85L;SLC35F1;NDRG2;KIAA1755;ZMAT5;GPR126;ZNF512;ARHGEF
GFM1
DGKH
GJA1;SERPINA1;HSPA4;PTEN;APOE;NDRG2;TRIB2
GJA1;CENPW;CCDC91;DLEU1;LCORL;TTN
GJA1;PLN;FAM174B;UQCR10;MLF1;MYH6
GJA1;CMTM5;APOE;NDRG2
GJA1
HSPA4;CD34
NDRG2
HORMAD2
CEP85L
CD34
MFN1
APOE
EIF2B4
MYH7
APOE
APOE
APOE
APOE
APOE
APOE
APOE
MTMR3;CR1;HSPA4;PTEN;APOE;CD34
PTEN;CD34
APOE
APOE
APOE
PTEN

PTEN
APOE
APOE
APOE
PTEN
DGKH
RSRC1
DGKH
CENPW;DES;PLN;VSTM2L
PLN;B3GNT7;TMEM253;TTN
DES;SERPINA1;LXN;APOC1
CENPW;B3GNT7;APOC1;ALX4
GJA1;UQCR10;TMEM253;MYH7
PLN;SLC35F1;MYH7;TTN
ZCCHC10;APOC1;APOE;CCDC141;MLF1
MLF1;MYH6;MYH7;TTN
SKIDA1;ALX4;SLC35F1;NAT14
PLN
SUPT7L
APOE
SLC5A6
APOE
GNB4
GNB4
GNB4
GNB4
WARS2
PTEN
PTEN
TRIB2
ALX4
KIAA1755
EIF2B4;DES;SNX17;MFN1;EIF3E;NDRG2;MYH7
DES;SNX17;NDRG2;MYH6;MYH7
GJA1;FAM174B;EIF3E;APOE;CD34;MLF1
DCAF16;HSPA4;VSTM2L;FAM3C;ARHGEF40
GJA1;DES;NDRG2;CD34;MLF1;MYH7
PLN;FAM174B;MYH6;TTN
TOMM40;NOV;MYH6;MYH7;TTN
PLN;FAM174B;NDRG2;MYH6;TTN
SLC5A6;TBX15;LXN;NOV;EIF3E
LXN;APOC1;GPN1;NIPSNAP1;WARS2
GJA1;NOV;SHOX2;PTEN;RSPO2
GJA1;MFN1;NDRG2;MLF1;MYH7
GJA1;MFN1;UQCR10;NDRG2;MYH6

CENPW;NCAPG;MYH6;MYH7;TTN
MFN1;UQCR10;NDRG2;MLF1;MYH7
DES;SNX17;NDRG2;MYH6;MYH7
GJA1;SNX17;EIF3E;FAM3C;APOE
FAM174B;NDRG2;MLF1;MYH6;TTN;PPM1G
ZCCHC10;SLC4A1AP;TRIB2;NRBP1
GJA1;PLN;SNX17;TTN
MFN1;NDRG2;MLF1;MYH7;TTN
PLN;GNB4;MYH6;MYH7;TTN
GJA1;MFN1;NDRG2;MLF1;MYH6
TBX15;PTEN
ZCCHC10;GNB4;RSPO2;LCORL;MYH6
CD34
APOC1
PTEN
CD34
APOE
GJA1
MFN1;APOE
TBX15
MYH6
APOE;NDRG2
GJA1
GCKR
CR1
PTEN
CR1
SERPINA1;APOC1;APOE
GJA1
SERPINA1
APOE
PTEN
TBX15
EIF2B4;UQCR10;NRBP1
MYH7
APOE
HSPA4;PTEN
PTEN
PTEN
PTEN
APOE
APOE
APOE
CD34
MTMR3

CR1
APOE
TBX15
GJA1
CCDC91
WNT16
CENPW;ASCC2
MLLT10
WNT16;TRIB2
RSPO2
PLN
DES
GJA1
TTN
SERPINA1
APOE
PTEN
SERPINA1
MLLT10;PTEN;CD34
PTEN
TTN
SLC4A1AP;EIF3E;MYH6
PLN;SLC35F1
MYH6;MYH7
MYH6;MYH7
HSPA4;PTEN
NIPSNAP1
NCAPG;CCDC141
FAM174B;MLF1
NOV;SKIDA1
GJA1;LXN;GNB4
TBX15;FAM174B;NRBP1
PTEN;CCDC141;TTN
TBX15;DES;NDRG2
GJA1;LXN;GNB4
TBX15;DES;NDRG2
GJA1;MYH7
NDRG2;MYH7
SKIDA1;AFAP1
EIF3E;MYH7
EIF2B4;TBX15;SLC4A1AP;APOC1;EIF3E;MLF1
DNAJC1;NDRG2
GJA1;FAM174B
NOV;APOC1;MYH7
TBX15;TTN

APOE;NDRG2
CCDC91;UQCR10
MYH6
NOV;VSTM2L
TTN
DES;TOMM40;SPEG
APOE;MLF1
PLN;MYH7
MLF1;MYH6
GJA1;MYH7
SHOX2
APOE
TTN
GJA1;HSPA4;GNB4
LXN;SNX17;APOE
GJA1;NCAPG;CD34
GJA1;LXN;SHOX2
UQCR10
TBX15;MYH7
NOV;NCAPG
APOC1;APOE
TBX15;MYH7
LXN;CD34
APOC1;UQCR10
SLC5A6;CCDC141
GFM1;TOMM40
EIF2B4;TBX15;DES
NOV;MYH7
NOV;MYH7
GJA1;GNB4;NCAPG
GJA1;NOV;CD34
NOV;TRIB2;TTN
NDRG2;MYH7;TTN
GJA1
CD34;NIPSNAP1;TTN
EIF3E
MTMR3;PTEN;TRIB2;PPM1G
SUPT7L
GCKR;APOC1;NRBP1
GCKR;HORMAD2;NRBP1
GCKR;CCDC91;APOC1;SLC35F1;NRBP1
CWC27
GJA1
EIF3E
APOE

AFAP1
GFM1;ALX4;ZNF513
GJA1;PLN
PTEN;TTN
HSPA4;APOE
APOE
APOE
APOC1
SERPINA1
TRIB2
RSPO2
CD34
RSPO2
APOE
PTEN
GJA1;DES;PTEN;APOE
TBX15;MYH7
GJA1;SNX17;EIF3E;APOE;MYH6
DES;SNX17;EIF3E;APOE;NDRG2;TTN
PTEN
APOE
SHOX2
APOE
GCKR
GFM1;DES;ZNF628;SPEG;NDRG2;MLF1;MYH6;MYH7;TTN
GFM1;DES;PLN;UQCR10;CCDC141;MLF1;MYH6;MYH7;TTN
EIF2B4;KIAA1755;ARHGEF40
CR1
MLLT10
PLN
SLC5A6;APOE
ZMAT5
GNB4
SLC5A6;PLN;APOC1;GNB4;APOE
GNB4
APOE
EIF3E
PTEN
WNT16
DGKH
CD34
TOMM40;FAM174B;ASCC2;EIF3E;PPM1G
AFAP1;NCAPG
GJA1;NOV;SNX17;RSPO2;CD34
SERPINA1

ZMAT5;CWC27
APOE
GJA1
MYH7
APOC1
IFI27L2
CCDC141
APOE
PTEN
ZNF513
APOE
APOE;NDRG2
GFM1;GJA1
SLC5A6;CENPW
PTEN
APOE
TBX15
GJA1
CENPW
ARHGEF40
NRBP1
GJA1
GNB4
PLN
EIF3E
EIF3E
B3GNT7
EIF3E
PTEN
GNB4
GNB4;DGKH
MTMR3;PTEN
PTEN
APOE
CR1
TRIB2
APOE
APOE
GJA1
APOC1;APOE
MTMR3;TOMM40;PTEN;UQCR10;NDRG2;NAT14;PPM1G;GJA1;FAM174B;ZMAT5;ASCC2;APOE;EIF2B4;H
APOE
MYH6
GJA1
APOE

EIF2B4
GCKR
DGKH
APOE
CCDC141
PTEN
SERPINA1;APOC1;EIF3E;UQCR10;APOE;NIPSNAP1
APOE
DNAJC1
GCKR;ALX4
GCKR;ALX4
GCKR;ALX4
APOE
GCKR;ALX4
GJA1
NOV;GNB4;NRBP1;DGKH;TTN
APOC1
CR1;APOC1;MFN1
APOE;CD34
PTEN;WNT16
SLC5A6;MLLT10;GJA1;HSPA4;SHOX2;ALX4;WNT16;CD34;TTN
LXN;PLN;CMTM5;PTEN;SPEG;FAM3C
CENPW;LXN;NDRG2;NAT14;SLC5A6;GJA1;B3GNT7;RSRC1;FAM174B;ZNF219;GNB4;GPR126;SPEG;PVRL[illegible]
PLN;DES
DES;SHOX2
SHOX2
CR1
MLLT10;RSRC1;CD34
SNX17
TOMM40
NCAPG
IFI27L2
NOV;TTN
LXN
GPR126
PLN
HSPA4
SPEG;TRIB2;NRBP1;TTN
DGKH
LCORL
GJA1;APOE
MYH6
PTEN
SLC5A6;TRIB2
TRIB2

SLC5A6;EIF2B4;GFM1;SPEG;UQCR10;FAM3C;NIPSNAP1;TTN
GJA1
GJA1;SNX17;APOE;NIPSNAP1
APOC1;TRIB2
CD34
CENPW;CD34
MYH7
SERPINA1
GPR126
SLC5A6;TTN
SLC5A6;TBX15;GJA1;SHOX2;FAM3C;NDRG2;CD34;MLF1
CD34
MLLT10
APOC1
SHOX2
TRIB2
GJA1;APOC1;AFAP1;APOE
CD34
GJA1;APOC1;APOE
DNAJC1;TOMM40;APOE;MLF1
MTMR3
ZCCHC10
DES
GPR126;TRIB2
CENPW;TOMM40;NCAPG;CD34
NOV
MLF1
TRIB2
SLC5A6;GJA1;APOE;CD34;GPN1
SERPINA1
CCDC121;ZCCHC10;TOMM40;HSPA4;GPN1;NIPSNAP1;WARS2;SLC5A6;GJA1;B3GNT7;ZNF219;GNB4;DLE
GFM1;SLC4A1AP;HSPA4;PTEN;NCAPG;SLC35F1;MLF1;SUPT7L;WARS2;TTN;MLLT10;DCAF16;GJA1;MFN1
SERPINA1;TOMM40;GPR126;GPN1
APOE
APOC1
PLN
FAM3C;WNT16
LXN;NCAPG;CD34;TRIB2
SHOX2
APOE
SLC4A1AP;HSPA4;SPEG;APOE
PPM1G
PLN
SERPINA1;GPR126;FAM3C;LCORL;MLF1;TTN
EIF2B4;TOMM40;HSPA4;UQCR10;GPN1;NIPSNAP1;PPM1G;SLC5A6;DNAJC1;GJA1;CCDC91;B3GNT7;FAM

ALX4;CD34
WNT16;MYH7
APOC1;APOE
ALX4;CD34
PTEN
SHOX2
CD34
DNAJC1;CD34;TRIB2
SERPINA1;PTEN
SERPINA1;CR1;PTEN
CCDC121;LXN
IFI27L2;CR1;SLC35F1;WNT16;NDRG2
NCAPG;APOE
GJA1;APOC1
SHOX2;GPR126
GJA1;FAM174B;APOE
PTEN
HSPA4
DLEU1;TRIB2
NOV;CD34
DCAF16
SERPINA1;HSPA4;NDRG2
APOC1
APOC1
DNAJC1
PPM1G
SERPINA1
NCAPG
WNT16
SLC4A1AP;HSPA4;SPEG;APOE
MTMR3;ZCCHC10;CR1;TOMM40;PVRL2;NIPSNAP1;SUPT7L;DGKH
PTEN
NCAPG
CMTM5
APOC1;APOE
SERPINA1
SERPINA1;ZCCHC10;LXN;HSPA4;SHOX2;NCAPG;NDRG2;MLF1;SLC5A6;DNAJC1;GJA1;PLN;CCDC91;NOV;[illegible]
GJA1;FAM3C;WNT16;TRIB2
UQCR10;TTN
PTEN
TBX15;SERPINA1;PTEN;NDRG2
GJA1;PTEN;CD34
SERPINA1
TBX15
PTEN

HSPA4
GJA1
CR1
PTEN
CD34
APOE
IFI27L2
PTEN
WNT16
PTEN
PTEN
APOE;WNT16
PTEN
CD34
APOE
MYH6
PTEN
PTEN;CD34
APOE;WNT16
SERPINA1
WNT16
PTEN
PTEN
PTEN
PTEN
GCKR
PTEN
CR1
PTEN
PTEN
EIF2B4;PTEN
PTEN
PTEN
PTEN
MYH6
PTEN
PTEN
WNT16
DES
GCKR;ALX4
PTEN;TTN
APOE
SERPINA1
GJA1
SHOX2;CD34

MLLT10;GJA1;LXN;NOV;HSPA4;PTEN;ALX4;APOE;CD34;MYH6
APOE
MYH7
SERPINA1
HSPA4
SERPINA1
WNT16
CR1
PTEN
PTEN
GJA1
GJA1
NRBP1
PLN;EIF3E;MLF1;MYH7
SNX17;APOE
MLLT10;MLF1
DNAJC1
DNAJC1;ZNF219
NCAPG
SLC5A6;DNAJC1;SERPINA1;MTMR3;SNX17;PTEN;ZNF513;EIF3E;PVRL2;NRBP1
GJA1
PVRL2
FAM3C
SLC5A6
MYH6
CD34
SLC5A6;DNAJC1;LXN;CCDC91;GNB4;PTEN;SPEG;FAM3C;NDRG2;LCORL;MLF1
SNX17;APOE;GPN1;NIPSNAP1
SERPINA1;TOMM40;GPR126;GPN1
DES
PLN
GPR126
PTEN
PTEN
FAM3C
GJA1
DCAF16;MTMR3;APOC1;DLEU1;APOE;MLF1
GPR126
MLLT10;NOV
PPM1G
MYH6
NCAPG
NOV;GPR126
CENPW;NCAPG
SLC5A6;FAM174B;PVRL2

NOV;CD34
DES;PLN;GPR126;NCAPG;CCDC141;NIPSNAP1
GJA1
WARS2
MYH6
MLLT10
DLEU1
PPM1G
TBX15
MYH7
CR1
GJA1
GJA1
GJA1
HSPA4
TOMM40
SNX17
GJA1
CD34
APOC1;GPR126;UQCR10;TRIB2;MLF1
MFN1
PTEN
CABP7
CD34
CD34
GJA1
APOC1
GPR126;CD34
PLN;SHOX2
HSPA4
CR1
PTEN
DES
SERPINA1
MLLT10
PPM1G
NDRG2
WNT16
TOMM40
SLC5A6;DNAJC1;FAM174B;GPR126
NOV
FAM174B
TRIB2
SERPINA1
MLF1

APOE
APOE
DES
DLEU1
CD34
NDRG2
MLF1
GPR126
SLC5A6;LXN;SPEG;MLF1
CENPW
ZNF512
WNT16
HSPA4
APOE
PTEN;NCAPG;PPM1G
CENPW;NCAPG
DGKH
GJA1;LXN;AFAP1
CR1;PVRL2;CD34
TRIB2
APOE
MTMR3;FAM3C;CD34;TRIB2
APOC1
SNX17
PTEN
APOC1
GJA1
EIF2B4;HSPA4
GJA1;NOV;CR1L;NRBP1
NCAPG
ZCCHC10;LXN;SNX17;FAM174B;GPR126;EIF3E;CD34;TRIB2
SLC5A6;DES;HSPA4;PTEN;ASCC2
LXN;HSPA4
ASCC2;UQCR10
GFM1;TOMM40;APOC1;LCORL
GJA1;NOV;CR1L;NRBP1
DES
TRIB2
DLEU1;AFAP1;SPEG
NCAPG
SNX17
GFM1;NCAPG;HORMAD2;GPN1;NIPSNAP1
SLC5A6;PTEN
MFN1
GJA1

PTEN
GJA1
APOE
PTEN
DES;PLN;DLEU1;UQCR10;CCDC141;MLF1;MYH6;MYH7;TTN
NOV;PTEN;RSPO2
TOMM40;APOE
GJA1;RSPO2
PTEN
GJA1;DES;PTEN;ZNF513;APOE;CWC27
DNAJC1;RSRC1
GCKR;HORMAD2
APOE
PTEN
FAM174B;EIF3E
MYH7;DGKH
TTN;PPM1G
GJA1
FAM174B;NDRG2
APOE
PTEN
SERPINA1
PTEN
TTN
GCKR
APOE
NDRG2;MYH7
PTEN
ALX4;DGKH
KIAA1755
TRIB2
NOV
DNAJC1
TRIB2
APOE
CR1
SHOX2;PVRL2
WNT16
DES;MYH6
ARHGEF40
NCAPG
APOE
RSRC1
TTN
APOE

CD34
APOE
DES;APOC1;APOE
SERPINA1;APOC1;APOE
HSPA4;NRBP1
MLLT10;GPR126
DES;PLN;UQCR10;CCDC141;MLF1;MYH6;MYH7;TTN
PLN;MFN1;VSTM2L;CCDC141;MLF1;MYH6;TTN
DES;PLN;CEP85L;UQCR10;CCDC141;MLF1;MYH6;MYH7;TTN
DES;PLN;UQCR10;CCDC141;MLF1;MYH6;MYH7
IFI27L2;GCKR;TOMM40;HSPA4;C2ORF16;UQCR10;GPN1;NIPSNAP1;TMEM253;SNX17;APOC1;ZNF219;Z
KIAA1755;DES;PLN;UQCR10;CCDC141;MLF1;MYH6;MYH7;TTN
KIAA1755;CMTM5;SKIDA1;SHOX2;CEP85L;GPR126;SLC35F1;ZNF512;ARHGEF40;CD34;TRIB2;TMEM253
EIF2B4;GJA1;DES;ZNF219;SSC5D;CWC27
GCKR;GJA1;B3GNT7;NOV;APOC1;ZNF219;CEP85L;APOE;ARHGEF40;TRIB2;WARS2
SLC5A6;DNAJC1;GCKR;GFM1;SERPINA1;TOMM40;APOC1;APOE;NIPSNAP1
RSRC1
MTMR3
PTEN
PTEN
GJA1
CCDC91
CCDC91
APOE
CD34
RSRC1;TRIB2;TTN
PLN;MYH6;MYH7;TTN
ALX4
DES;PLN;DLEU1;UQCR10;CCDC141;MLF1;MYH6;MYH7;TTN
LXN
TTN
PTEN;APOE
CD34
PTEN;APOE
GFM1;SERPINA1
PTEN
TOMM40
CCDC141
PTEN
NOV
NCAPG
GPR126
CD34
CABP7
NIPSNAP1

LCORL
TOMM40
GCKR
GFM1;SERPINA1;MTMR3;CR1;LXN;HSPA4;SHOX2;PTEN;AFAP1;NDRG2;TTN;SLC5A6;MLLT10;DNAJC1;GJ
SERPINA1
SERPINA1
GJA1
SERPINA1
SERPINA1
SERPINA1
SERPINA1
TBX15
APOE
GJA1;TOMM40;APOE;NDRG2
CR1;PTEN
HSPA4
MLLT10;PTEN;MLF1
SLC5A6;LXN;CR1L;FAM3C;MYH6
APOE
TRIB2
SERPINA1
PTEN
MYH7
GFM1
SPEG
CCDC91
APOE
EIF3E
APOE
SLC5A6;DNAJC1;CCDC91;FAM174B;GNB4;NCAPG;FAM3C;NDRG2
GCKR;SNX17
SERPINA1
APOE
PTEN
TRIB2
MYH6
SUPT7L
GJA1
CENPW;HSPA4
SERPINA1
DGKH
GFM1
PTEN
WNT16
PLN;UQCR10;CCDC141;MLF1;MYH6;MYH7;TTN

MLLT10;CCDC121;PLN;NOV;RSRC1;GNB4;AFAP1;ZNF512
GJA1
SPEG;DGKH
CD34
GJA1;DES;PTEN
CCDC141
ALX4
SLC5A6;TRIB2
GJA1
MTMR3
PTEN
PTEN
UQCR10
GNB4;PTEN
CR1
HSPA4;CD34
SERPINA1;MTMR3;TOMM40;PTEN;UQCR10;NDRG2;NAT14;PPM1G;GJA1;ASCC2;APOE;ARHGEF40;CD3[illegible]
GJA1;HSPA4;PTEN;APOE
RSPO2
GCKR
TBX15;GJA1;PTEN;CWC27
CD34
APOE
GJA1
PTEN
PTEN
CCDC91
GJA1;APOE
TOMM40;PPM1G
TBX15;CCDC91
MLF1
PLN;MFN1;VSTM2L;CCDC141;MLF1;MYH6;TTN
PLN;UQCR10;CCDC141;MLF1;MYH6;MYH7;TTN
PTEN
PTEN
SLC5A6;DNAJC1;CCDC91;GNB4;FAM3C;NDRG2;LCORL
ZCCHC10;NOV;GNB4;LCORL
GPR126
GJA1
APOE
TOMM40
APOE
GCKR;APOE
TOMM40
RSPO2

APOE
SUPT7L
APOE;CD34
TOMM40
APOE;AC097381.1
GJA1
NOV;CR1L;SPEG;UQCR10
PTEN
APOE
GJA1;PLN;CMTM5;NDRG2
DES;SNX17;APOE;MYH6;MYH7
PTEN
ALX4
PTEN
TOMM40
SERPINA1;APOC1;RSPO2;AFAP1;ALX4;SLC35F1;LCORL;TRIB2;TTN
FAM174B
PTEN;APOE
DES;APOE
DLEU1;APOE
CENPW;SERPINA1;CR1
WNT16
MFN1
APOE
PTEN
MLLT10
GCKR;CWC27
APOE
CCDC141
MYH6
TBX15
HSPA4
ZNF513
GPR126
APOE;NAT14;SUPT7L
APOE
APOE
GJA1
TOMM40;CMTM5;APOC1;APOE;CD34
APOE
PTEN
GJA1;CENPW;DES;SERPINA1;LXN;NOV;HSPA4;GNB4;PTEN;NDRG2
GJA1;DES;PTEN
WNT16
MFN1;APOE

APOE
PTEN
PTEN;EIF3E
APOC1
APOE
WNT16
SLC5A6;SERPINA1
GJA1
PTEN
DES
NOV;GNB4;GPR126
GCKR
PTEN
APOE
GFM1
EIF2B4
APOC1
SLC35F1
CENPW;SLC35F1;WARS2;TBX15;DNAJC1;CCDC91;FAM174B;RSPO2;GPR126;ALX4;ASCC2;HORMAD2;FA[illegible]
EIF2B4
PTEN
DLEU1
PTEN
SERPINA1;PTEN
CENPW;RSRC1;DLEU1;NCAPG;DGKH
CMTM5;APOC1;SLC35F1;APOE;NDRG2
GJA1;NOV;APOC1;EIF3E;APOE
UQCR10;MLF1;MYH6;MYH7
DES;NDRG2;MYH6;MYH7
TOMM40
ZNF513
MYH7
ALX4
APOE
PTEN
PTEN
PTEN
NOV;SLC35F1;EIF3E
MYH7;TTN
SERPINA1
MTMR3;DLEU1;MLF1
GCKR
CCDC91
NOV;MYH7
CENPW;NCAPG

GJA1;TOMM40;PPM1G
MYH6
NOV;NDRG2
TBX15
GJA1;APOE
LXN;APOC1
TBX15;MYH7
PTEN
FAM174B;MYH7
GJA1;APOC1;APOE
TBX15;MYH7
APOE
NCAPG
SLC4A1AP
TBX15;DNAJC1;LXN;AFAP1;GPR126
CCDC141
TOMM40;APOE
PTEN
PTEN;CD34
RSPO2
TBX15;TTN
PTEN
CR1
PTEN;DLEU1
PPM1G
APOE
PTEN
HSPA4
APOE
PTEN;CD34
APOE
PTEN
PTEN
SERPINA1
PTEN
APOE
SERPINA1;PTEN
PTEN
PTEN;CD34
PTEN
APOE
PTEN
GFM1
APOE
APOE

APOE
SERPINA1
SERPINA1
PTEN
PTEN
APOC1
CCDC141
TBX15;MYH7
LXN;MYH7
LXN;CD34
VSTM2L;CCDC141;MLF1;MYH6;MYH7
TBX15;GFM1;PLN;UQCR10;CCDC141;MYH6;MYH7;TTN
GFM1;PLN;CCDC141;MLF1;MYH6;MYH7;TTN
GCKR;DLEU1
PLN;B3GNT7;TTN
CMTM5;APOE;NDRG2
APOC1;SPEG;APOE
CENPW;APOC1;MLF1
MLF1;MYH7;TTN
PLN;APOC1;ARHGEF40
LXN;APOC1;NDRG2
SLC5A6;APOC1;GNB4
APOC1;APOE;NDRG2
APOC1;RSPO2;MYH7
APOC1;CCDC141;MLF1
CENPW;APOC1;MLF1
NCAPG;MYH6;MYH7
VSTM2L;SLC35F1;CD34
SLC5A6;GJA1;MLF1
GJA1;MLF1;TTN
WNT16;MLF1;TTN
TBX15;HORMAD2;TTN
GCKR;MYH6;MYH7
SLC35F1;MYH7;TTN
SERPINA1;CMTM5;VSTM2L
DES;RSPO2;SPEG
LXN;APOC1;MLF1
CCDC91;MYH6;MYH7
DES;ALX4;SLC35F1
RSPO2;WNT16;MYH6
TBX15;HORMAD2;MLF1
SERPINA1;APOC1;NAT14
GJA1;SNX17
EIF2B4
GJA1

APOE
PTEN
APOE;CD34
ZNF513
PTEN
RSRC1;ZNF219
PTEN
ASCC2;ARHGEF40
PLN
ARHGEF40
GNB4
MLF1
MTMR3;SNX17;TTN
TBX15;NOV
SERPINA1
SERPINA1
HORMAD2
HSPA4;APOE
NOV;MYH7
TOMM40
TBX15;NDRG2
NDRG2
DES;MYH7
CCDC91;SNX17
LXN;CCDC141
PTEN
GNB4
PTEN
ARHGEF40
APOE
PVRL2
APOE
HSPA4
PTEN
SERPINA1;APOC1;APOE
CCDC121;GJA1;WNT16
SERPINA1;APOC1;APOE
SPEG
GCKR;GJA1;TOMM40;PLN;HSPA4;PTEN;ALX4;APOE
PTEN
GJA1
GJA1;HSPA4;RSRC1;PTEN
APOE
SERPINA1
APOE

TTN
APOE
CD34
GFM1;MFN1;UQCR10;MLF1
DES;SHOX2;NCAPG;TTN
GFM1;SERPINA1;HSPA4;CR1L
MLLT10;DNAJC1;SPEG;NDRG2
PTEN;EIF3E;PVRL2;TRIB2
SERPINA1;SNX17;ZMAT5;ZNF512
GNB4;PTEN
PTEN
PTEN
LXN
APOE
SERPINA1
HSPA4
TTN
DNAJC1
DLEU1
APOE
CR1
APOE
SERPINA1
NDRG2
APOE
APOE
SPEG
SPEG
APOC1
NCAPG
CENPW;TOMM40;DLEU1;UQCR10
APOE
DNAJC1
LXN;PTEN
NCAPG
SERPINA1;APOC1;APOE
DES;PLN;NOV;MYH7;TTN
CCDC91
NOV
APOE
PTEN
APOE
GFM1;GJA1;ALX4
HSPA4
GCKR;GJA1;PTEN;DLEU1;LCORL

GJA1;LXN;NOV;SNX17;APOE
DLEU1;CCDC141;TTN
TTN
GNB4
ZMAT5;CWC27
CCDC141
LXN;CD34
UQCR10;MYH7
PTEN
ALX4
PTEN
TTN
PTEN
APOE
PTEN
PTEN
HSPA4
PTEN
PTEN
PTEN
GFM1
PTEN
SERPINA1;APOE
PTEN
SERPINA1
HSPA4
PTEN
GCKR
PTEN
APOE
GJA1
CCDC91
TTN
PTEN
TTN
B3GNT7
IFI27L2;GCKR;CABP7;C2ORF16;NIPSNAP1;DNAJC1;GJA1;NOV;CR1L;SNX17;APOC1;APOE;ARHGEF40
PLN;CCDC91;VSTM2L;UQCR10;CCDC141;MLF1;MYH6;MYH7;TTN
MLLT10;PTEN;ASCC2
DNAJC1
TTN
CD34
APOE
CD34
LXN

CD34;TRIB2
TTN
B3GNT7
PTEN;NDRG2
NOV
APOE
PTEN
ZNF513
APOE;MYH7
APOE
IFI27L2;GCKR;CCDC121;TOMM40;C2ORF16;NAT14;NRBP1;SLC5A6;DCAF16;SNX17;ZMAT5;ZNF512;APO
LCORL
CCDC91
APOE;CD34
GFM1
HSPA4
PTEN
GJA1;SERPINA1;SSC5D;APOE
GNB4
EIF2B4;DCAF16;GFM1;SERPINA1;B3GNT7;GNB4;PTEN;EIF3E;APOE;WARS2
DES;PLN;UQCR10;CCDC141;MLF1;MYH6;MYH7;TTN
PLN;CCDC141;CD34;MLF1;MYH6;MYH7
PLN;CCDC141;MLF1;MYH6;MYH7;TTN
APOE
APOC1
FAM174B
DLEU1
DLEU1
GJA1;NOV
APOE
PTEN
PTEN;WARS2
CD34
PTEN
LXN;NOV;PTEN
NCAPG
GCKR;DLEU1
SERPINA1
ASCC2;AC097381.1
NOV;CD34
GFM1;CD34
SNX17;MYH7
ZNF219
CR1
MLF1

SERPINA1
CR1
CR1
DES
NOV
CR1
WNT16
SNX17;APOE
MLLT10
SKIDA1
EIF2B4;TOMM40;ASCC2;WARS2
TRIB2
MLLT10
MLLT10;CEP85L
GCKR;MTMR3
MLLT10;SKIDA1;CEP85L
TOMM40;CEP85L
SLC4A1AP
APOE
ASCC2
EIF2B4;MLLT10;TOMM40;CR1L;SHOX2;ZMAT5;MFN1;ASCC2;UQCR10;NRBP1;WARS2
MLLT10;CEP85L;LCORL
NRBP1
MLLT10
MLLT10;B3GNT7;GNB4;ASCC2
EIF2B4;CR1L;WARS2
TOMM40;CEP85L;GNB4;LCORL;TRIB2
MLLT10
SKIDA1
ZMAT5
ASCC2
MLLT10
MLLT10
ZMAT5;GNB4;NRBP1
MLLT10
SLC5A6;APOE
WARS2
MTMR3;TOMM40
GFM1
APOE
CEP85L;GNB4
NRBP1
ASCC2
GFM1;APOE
LCORL

MLLT10
NIPSNAP1
RSPO2
ZNF513
APOE
EIF3E
PTEN;CD34
CCDC91
DES;MYH7
GNB4;NDRG2
CENPW;TRIB2
CD34;TTN
CD34
SHOX2;ALX4
GPR126
SERPINA1
MLF1
APOC1;MYH6
NOV
CD34
SLC4A1AP;TOMM40;CEP85L;AFAP1;NCAPG;UQCR10;NDRG2;NIPSNAP1;TTN;GJA1;NOV;RSPO2;EIF3E;FA
SLC5A6;EIF2B4
MLLT10;DNAJC1;SNX17;DLEU1
CENPW;NCAPG
WNT16
APOE
LXN;NIPSNAP1
EIF2B4;NCAPG;EIF3E;GPN1;WARS2
PTEN
IFI27L2;GCKR;LXN;TOMM40;SNX17;APOC1;ZNF219;ZMAT5;APOE;ARHGEF40;NIPSNAP1
DES
DES
APOE
GJA1;SERPINA1;PTEN;ALX4;EIF3E;APOE;CD34;TRIB2
APOE
PTEN
CD34
PTEN;CD34
MFN1;EIF3E
AFAP1;NCAPG
NOV
SERPINA1;NDRG2
APOE
PTEN
PTEN;DLEU1

DES
SERPINA1
GJA1
NOV;CMTM5;FAM3C
GCKR;SERPINA1;NOV;AFAP1;C2ORF16
SNX17;MYH7
CD34
PTEN
APOE
CD34
PTEN
PTEN
APOE
SERPINA1
PTEN
SERPINA1;APOE
PTEN
PTEN
PTEN
HSPA4
CD34
APOE;TTN
PTEN
PTEN;CD34
LXN;PTEN
PTEN
PTEN
PTEN
PTEN
APOE
APOE
MYH7
B3GNT7;NCAPG
NDRG2
LXN;MYH6
LXN;SHOX2
MFN1
TBX15;CCDC141
MLF1
EIF2B4;TTN
ZNF628;CWC27
GJA1;PTEN;ALX4
CR1
PTEN
PTEN

PTEN
PTEN
SLC5A6
GJA1
PTEN
LXN
DCAF16
GJA1;PTEN
APOE
GNB4
GFM1
DLEU1
APOE;MYH7
CD34
RSPO2
UQCR10
GNB4
EIF2B4
EIF2B4
EIF2B4
EIF2B4
SERPINA1;HSPA4;APOE
CD34
EIF2B4
EIF2B4
SLC5A6;SPEG
PTEN
TOMM40
CWC27
APOE
APOE
HORMAD2
SNX17
APOE
HSPA4
GJA1
TRIB2;MYH6;TTN
MTMR3
TBX15;SHOX2;ALX4;ZNF513
APOE
DES
PTEN
PTEN
SERPINA1
PLN;NOV;APOC1;MLF1;MYH6;TTN

LXN;NOV;PTEN;VSTM2L;NDRG2
NOV
SERPINA1
APOE
GJA1
GJA1;GNB4
LCORL
SNX17;APOC1;APOE;MYH7
PTEN
APOE
APOE
PTEN
PTEN
CR1
GJA1
DLEU1
ALX4;TTN
PTEN
PTEN
EIF2B4
PTEN
PLN;VSTM2L;UQCR10;CCDC141;MLF1;MYH6
DES;PLN;UQCR10;CCDC141;MLF1;MYH6;MYH7;TTN
EIF2B4;DNAJC1;GJA1;DES;CABP7;ZNF219;AFAP1;ZNF513
APOE
MTMR3
EIF2B4
PTEN
APOE
SERPINA1
EIF2B4
LXN;SNX17;PTEN;EIF3E;APOE
B3GNT7;ZMAT5;VSTM2L;TMEM253
ZCCHC10;SLC4A1AP;TRIB2;NRBP1
GJA1;PLN;UQCR10;MYH7;TTN
PLN;CR1L;SHOX2;CWC27
DES;CCDC91;SPEG;NDRG2;MYH7
TBX15;DES;SHOX2;CCDC141;CD34;MYH7;TTN
DES;HSPA4;APOE;NDRG2;MYH6;MYH7;TTN
PLN;NDRG2;CD34;TRIB2;MLF1;MYH7;TTN
LXN;NOV;PTEN;AFAP1;SPEG;SSC5D;ARHGEF40
DES;SPEG;UQCR10;APOE;NDRG2;MYH6;MYH7
GJA1;DES;HSPA4;APOE;NDRG2;CD34;TTN
TBX15;DES;SPEG;CCDC141;MYH6;MYH7;TTN
DES;PLN;EIF3E;APOE;NDRG2;MYH6;TTN

PTEN
PTEN;NDRG2
MLLT10;GPR126;MLF1;TTN
EIF2B4;CD34;NIPSNAP1
CD34
CCDC91
APOE
GCKR
RSRC1;CWC27
GJA1;APOE
MLLT10;GJA1;NOV;HSPA4;PTEN;DLEU1;ALX4;APOE;CD34;MYH6
DES;PLN;UQCR10;CCDC141;MLF1;MYH6;MYH7;TTN
TBX15;NDRG2
APOE
APOC1
IFI27L2;GCKR;GJA1;LXN;NOV;APOC1;ZNF219;ZMAT5;C2ORF16;APOE;ARHGEF40
LXN;CD34
APOE
SERPINA1;APOE
SERPINA1;LXN
APOC1
PTEN
NDRG2
FAM3C;WNT16
DLEU1
PPM1G
UQCR10
PTEN
GJA1
PTEN
WNT16
GJA1
APOC1
APOC1;APOE
ZNF513
GJA1
FAM3C
LXN;NOV;PTEN;APOE
PTEN
PTEN
GJA1
NOV;GNB4;AFAP1;GPR126;NCAPG;SPEG;APOE;LCORL;MLF1
PTEN
NOV;GPR126;PVRL2
RSPO2;ALX4;CCDC141

ARHGEF40
HSPA4
GNB4
GNB4;WNT16;ARHGEF40;DGKH
EIF3E
APOE
PTEN
DES;CABP7;SHOX2
EIF2B4;SLC4A1AP;TOMM40
ZCCHC10;ZNF512;SUPT7L
FAM174B;FAM3C;NDRG2
GFM1;ZNF219;SUPT7L
SERPINA1;ZCCHC10;WARS2
LXN;DLEU1;NCAPG
LXN;CR1L;DGKH
APOE;GPN1;TTN
SERPINA1;SLC4A1AP;TOMM40
PLN;PTEN;WARS2
DNAJC1;ZMAT5;FAM3C
LXN;LCORL;SUPT7L
SNX17;UQCR10;DGKH
TOMM40;EIF3E;CD34
SLC4A1AP;TOMM40;EIF3E
SLC4A1AP;TOMM40;NRBP1
MLLT10;RSRC1;NRBP1
SERPINA1;HSPA4;RSPO2
LXN
FAM174B
SHOX2
CR1;APOE
DCAF16;DNAJC1;FAM174B
GFM1;MFN1
PLN
NCAPG;LCORL
GJA1;APOC1;APOE;NDRG2
GJA1;SNX17;FAM3C;APOE;NDRG2
DES
SERPINA1
MYH7
FAM174B;ALX4
TBX15;GCKR;SHOX2
CCDC91;CEP85L;ASCC2
TBX15
PLN;MFN1;VSTM2L;CCDC141;MLF1;MYH6;TTN
APOE

SLC5A6;LXN;NRBP1
APOC1
MTMR3;LCORL;NIPSNAP1
ALX4
GJA1
APOE
AFAP1
CEP85L
GNB4
PTEN
APOC1
CMTM5
MTMR3
PTEN;CD34
DES;UQCR10
PTEN
GJA1
CR1
SERPINA1
SPEG;NIPSNAP1
PTEN
PTEN
GNB4
APOE
APOE
TRIB2
SHOX2;CEP85L
MLLT10
MTMR3
DGKH
TRIB2
MLLT10
MLLT10
MLLT10
MTMR3;SUPT7L
GJA1;CENPW;HSPA4;PTEN;ASCC2;CD34
CENPW
SERPINA1;CR1;HSPA4;PTEN;APOE;CD34
DES;PLN;UQCR10;CCDC141;MLF1;MYH6;MYH7;TTN
ZNF513
NDRG2
GJA1;NDRG2;TRIB2
CD34
PLN
DCAF16;FAM174B;PVRL2

MLLT10
PLN;UQCR10;CCDC141;MLF1;MYH6;MYH7;TTN
HSPA4;APOE
PTEN
MLF1;MYH7;TTN
SERPINA1;VSTM2L;CCDC141
CD34;MYH7;TTN
PTEN
SERPINA1
PTEN
MYH6
PTEN
PTEN
SERPINA1
APOE
SHOX2
APOE
FAM174B
APOE;MYH7
GJA1
GJA1;PLN
MLF1
MYH7
ZNF513
HSPA4
NDRG2
TOMM40;HSPA4;APOE
SERPINA1;PTEN
TTN
SERPINA1
PTEN
GFM1;SERPINA1;MTMR3;LXN;SLC4A1AP;TOMM40;CEP85L;PTEN;NCAPG;UQCR10;NDRG2;TTN;PPM1G;
MTMR3
PTEN
PTEN
ZNF513
EIF2B4
APOE
SERPINA1
ZNF513
AFAP1
GNB4
PTEN
HSPA4
RSPO2

SUPT7L
ALX4
TTN
EIF3E
APOE
B3GNT7
MFN1;NDRG2;MLF1;MYH7
UQCR10;NDRG2;MLF1;MYH7
GJA1;DES;PLN;NDRG2
GJA1;NOV;APOC1;APOE
PLN;EIF3E;MYH6;MYH7
EIF2B4;CCDC91;C2ORF16;PPM1G
GFM1;SNX17;MLF1;MYH7
MLLT10;PTEN;NDRG2
PTEN;CD34
PTEN
PTEN
EIF3E
SLC5A6
EIF3E
HSPA4;PTEN;WARS2
TTN
EIF2B4;PTEN;SPEG
CR1
APOE
GFM1;PLN;UQCR10;CCDC141;MLF1;MYH6;MYH7;TTN
APOE
CD34
CCDC121;CMTM5;SHOX2;SSC5D;NDRG2;MLF1;TTN;TBX15;DES;PLN;NOV;SPEG;ALX4;CCDC141;CD34;M[illegible]
CD34
GJA1
WNT16
PTEN
GJA1;PTEN;CD34
TTN
APOE
EIF3E
MTMR3
MLLT10;DNAJC1;DLEU1;CD34
DES
DNAJC1
DNAJC1
EIF2B4;TBX15;MLLT10;TOMM40;PLN;CD34
PTEN
APOC1;EIF3E;UQCR10;FAM3C;TTN

GJA1;SERPINA1;NCAPG;NIPSNAP1
PLN;EIF3E;MYH6;MYH7
APOC1;EIF3E;UQCR10;FAM3C;TTN
EIF2B4;UQCR10;NDRG2;MYH6
GJA1;DLEU1
LXN;PLN;VSTM2L;CCDC141;MLF1;MYH6
EIF2B4;GPN1;MYH6
FAM174B;RSPO2;ALX4
GNB4;LCORL;WARS2
EIF2B4;NCAPG;NIPSNAP1
FAM174B;SPEG;ASCC2
MTMR3;EIF3E;SUPT7L
SLC5A6;LXN;EIF3E
CMTM5;WNT16;MYH7
APOC1;ALX4;TTN
SERPINA1;AFAP1;FAM3C
APOC1;ASCC2;ARHGEF40
CCDC91;GNB4;GPN1
DNAJC1;TOMM40;GPN1
LXN;CD34;MYH7
DNAJC1;PTEN;CWC27
SHOX2;PTEN;DLEU1
GJA1
MTMR3
FAM3C;LCORL
PVRL2
PTEN
PTEN
TOMM40;B3GNT7;PVRL2;MYH7
MLLT10;RSRC1;CD34
TTN
MLF1
KIAA1755;CCDC121;CMTM5;SHOX2;SSC5D;ARHGEF40;CD34
UQCR10
HSPA4;DLEU1
APOE
GCKR;APOC1;APOE;NDRG2;NIPSNAP1
TBX15;DES;EIF3E;NDRG2;MLF1
PTEN
GFM1;GJA1;MLF1;MYH6
UQCR10;NDRG2;MLF1;MYH6
GJA1;ALX4
PTEN
GJA1
SERPINA1

CCDC91
APOE
CENPW
CD34
PTEN
CCDC121;TRIB2;MLF1
CD34
PTEN
WNT16
PTEN
PTEN
APOE
PTEN
PTEN
APOE
DES;PLN;UQCR10;CCDC141;MLF1;MYH6;MYH7;TTN
MLLT10;KIAA1755;CENPW;RSPO2;SSC5D;ZNF512;APOE;ARHGEF40;NRBP1
DES;PLN;UQCR10;CCDC141;MLF1;MYH6;MYH7
TBX15;GFM1;DES;ZNF628;UQCR10;NDRG2;MYH6;MYH7;TTN;PPM1G
SNX17
HSPA4
PTEN
LXN;NOV;SNX17;APOE;NDRG2
HSPA4;PTEN;NDRG2;CD34
PTEN
PTEN
PTEN
CMTM5;SHOX2;MYH6;MYH7
CD34
DES
PTEN
GJA1;MYH6
DLEU1
TBX15;DES;ZNF628;SPEG;UQCR10;NDRG2;MYH6;MYH7;TTN
TBX15;GFM1;UQCR10;CCDC141;MLF1;MYH6;MYH7;TTN
DES;PLN;UQCR10;CCDC141;MLF1;MYH6;MYH7;TTN
PLN;UQCR10;CCDC141;MLF1;MYH6;MYH7;TTN
DES;PLN;UQCR10;CCDC141;MLF1;MYH6;MYH7;TTN
GFM1;SERPINA1;MTMR3;ZCCHC10;LXN;SLC4A1AP;TOMM40;CEP85L;PTEN;NCAPG;UQCR10;CWC27;ND
SLC5A6;DNAJC1;GCKR;SERPINA1;TOMM40;APOC1;GPR126;APOE;PVRL2
DES;PLN;UQCR10;CCDC141;MLF1;MYH6;MYH7;TTN
KIAA1755;DCAF16;CMTM5;SKIDA1;SHOX2;CEP85L;ZMAT5;GPR126;ZNF512;ARHGEF40;CD34;TRIB2
MLLT10;DNAJC1;DES;LXN;ZNF628;ZNF219;AFAP1
EIF2B4;MLLT10;DES;ZNF628;ZNF219;AFAP1;CD34;NRBP1
DES;PLN;UQCR10;CCDC141;MLF1;MYH6;MYH7;TTN

TBX15;GFM1;PLN;UQCR10;CCDC141;MYH6;MYH7;TTN
IFI27L2;CCDC121;SLC4A1AP;CCDC91;CMTM5;APOC1;RSPO2;VSTM2L;SLC35F1;APOE;CWC27;NDRG2
PTEN
WNT16;MLF1
TTN
TTN
ALX4
NCAPG
SNX17;APOE;NIPSNAP1
DNAJC1;GJA1;FAM174B;SHOX2;NCAPG
NCAPG
ZCCHC10
EIF2B4;LXN;TOMM40;HSPA4;PTEN;EIF3E;UQCR10;FAM3C;APOE;GPN1;PPM1G
TBX15;B3GNT7;PVRL2
RSRC1
PTEN
GJA1
CD34
GNB4
GNB4
NAT14
NRBP1;TTN
DGKH
CENPW
DNAJC1;SNX17;APOE
FAM3C
EIF3E
APOE
SPEG;TTN
CMTM5;HORMAD2
PTEN;CCDC141
PLN;MYH6
MYH6;MYH7
MYH7;TTN
PLN;SHOX2
CENPW;APOC1
CEP85L;MYH7
APOC1;MLF1
CENPW;ALX4
CR1;APOC1
APOC1;WNT16
PLN;NCAPG
IFI27L2;CMTM5
CEP85L;CCDC141
RSPO2;WNT16

SLC35F1;HORMAD2
VSTM2L;MLF1
CR1;APOC1
MLF1;TTN
CR1;APOC1
CENPW;MLF1
APOC1;SSC5D
GCKR;APOC1
CABP7;ALX4
CCDC91;MYH6
DES;VSTM2L
CEP85L;LCORL
CMTM5;MYH6
VSTM2L;MYH7
PLN;CEP85L
SHOX2;NCAPG
CENPW;B3GNT7
RSPO2;WNT16
APOC1;TMEM253
TBX15;PLN
SHOX2;CCDC141
PLN;TMEM253
APOC1;GNB4
TBX15;ALX4
CENPW;SHOX2
PLN;NDRG2
GJA1;NDRG2
CENPW;MLF1
GCKR;PLN
APOC1;NDRG2
CCDC121;SLC35F1
B3GNT7;SPEG
ZNF513;MYH7
ALX4;CCDC141
SERPINA1;APOC1
APOC1;NDRG2
TBX15;PLN
TBX15;MYH7
SERPINA1;HORMAD2
VSTM2L;APOE
DES;MYH7
SHOX2;WNT16
CENPW;APOC1
CENPW;CEP85L
DES;NCAPG

KIAA1755;DES;PLN
TBX15;SHOX2
HORMAD2;LCORL
TBX15;TTN
DES;PLN
WNT16;CCDC141
PLN;SHOX2
CEP85L;DGKH
NCAPG;HORMAD2
VSTM2L;MYH7
CMTM5;MYH6
MYH6;MYH7
MYH6;TMEM253
CABP7;APOC1
RSPO2;WNT16
B3GNT7;TMEM253
TBX15;RSPO2
CCDC141;MLF1
B3GNT7;APOC1
CCDC91;APOC1
TBX15;HORMAD2
CEP85L;WNT16
NCAPG;DGKH
DES;PLN
APOC1;MLF1
SLC35F1;MYH7
DGKH
MLLT10
APOE
MLLT10;B3GNT7;GNB4
KIAA1755;DGKH
SERPINA1;LXN;PTEN
TTN
ZCCHC10;SLC4A1AP;TOMM40;CEP85L;NCAPG;UQCR10;NDRG2;TTN;PPM1G;GJA1;CCDC91;MFN1;RSPO
PTEN
ZNF513
MYH7
GJA1;PPM1G
MLF1
MLF1
SERPINA1;ZCCHC10;LXN;SLC4A1AP;TOMM40;CEP85L;PTEN;UQCR10;NDRG2;TTN;PPM1G;CCDC91;MFN
GJA1
APOE
PTEN
APOE

CD34
NOV;APOC1
B3GNT7
PTEN
APOE
TBX15;GFM1;UQCR10;CCDC141;MLF1;MYH6;MYH7;TTN
NDRG2;MYH7
FAM3C;WNT16;NDRG2
GCKR;PTEN;DGKH
CD34
GJA1
PTEN
PTEN;WNT16
SLC5A6
GFM1;PLN;CCDC141;MLF1;MYH6;MYH7;TTN
PTEN
GJA1
TRIB2;DGKH
B3GNT7;CABP7
NCAPG;TRIB2
CENPW;APOC1
CABP7;VSTM2L
DES;CCDC141
LCORL
GJA1;UQCR10;NDRG2;MLF1;MYH6
EIF2B4;KIAA1755;DCAF16;SHOX2;CEP85L;MFN1;GPR126;ZNF512;ARHGEF40;LCORL;TRIB2;WARS2
PLN;VSTM2L;UQCR10;CCDC141;MLF1;MYH6
EIF2B4;TBX15;GFM1;DES;SLC4A1AP;CCDC91;UQCR10;NDRG2;GPN1;NRBP1;TTN
APOE
PTEN
CEP85L
SHOX2;PTEN
PLN
EIF3E
NDRG2
CR1
GJA1;NOV;NCAPG;ZNF512;FAM3C
PTEN
PTEN
SERPINA1
RSRC1;APOC1
ALX4
MTMR3;PTEN;HORMAD2;APOE
DES;PLN;DLEU1;UQCR10;CCDC141;MLF1;MYH6;MYH7
IFI27L2;GCKR;LXN;TOMM40;APOC1;ZNF219;ZNF513;APOE;ARHGEF40;TRIB2;SUPT7L

TBX15;GFM1;DES;UQCR10;NDRG2;MYH6;MYH7;TTN;PPM1G
TBX15;GFM1;DES;ZNF628;UQCR10;NDRG2;MYH6;MYH7;TTN
DES;UQCR10;CCDC141;MLF1;MYH6;MYH7
APOE
APOE
PTEN
SERPINA1;HSPA4;PTEN
ZNF513
SERPINA1
DGKH
PTEN
CD34
GJA1
FAM174B
TBX15
DES
HSPA4
LCORL
SPEG;NAT14
MYH6;MYH7
PLN;CR1L
DES;CR1L
CABP7;VSTM2L
SHOX2;MYH6
CENPW;NCAPG
WNT16
GCKR;TOMM40;NOV;SNX17;APOC1;ZNF219;ZMAT5;C2ORF16;APOE;ARHGEF40;TRIB2
DES;PLN;UQCR10;CCDC141;MLF1;MYH6;MYH7;TTN
KIAA1755;CMTM5;SKIDA1;SHOX2;CEP85L;ZMAT5;GPR126;SLC35F1;ARHGEF40;CD34;TRIB2
KIAA1755;IFI27L2;CMTM5;SHOX2;ZNF219;ZMAT5;GPR126;SLC35F1;ARHGEF40;TRIB2
MLLT10;DES;B3GNT7;ZNF219;RSPO2;SPEG;SSC5D;TMEM253
MLLT10;GJA1;DES;ZNF219;RSPO2;C2ORF16
PTEN
TBX15
TBX15;GFM1;DES;ZNF628;SPEG;MYH6;MYH7;TTN
DES;PLN;UQCR10;CCDC141;MLF1;MYH6;MYH7
SLC5A6;GCKR;SERPINA1;APOC1;GPR126;C2ORF16;APOE;PVRL2;NIPSNAP1
PLN;MFN1;VSTM2L;CCDC141;MLF1;MYH6
SLC5A6;GCKR;SERPINA1;SKIDA1;APOC1;GPR126;ZNF513;C2ORF16;APOE;NIPSNAP1
EIF2B4;KIAA1755;IFI27L2;CMTM5;ZNF219;CEP85L;ZMAT5;GPR126;SLC35F1;NDRG2;ARHGEF40
MLLT10;NOV;RSRC1;SHOX2;SPEG;SSC5D;ZNF512
SLC5A6;GCKR;SERPINA1;APOC1;GPR126;APOE;PVRL2;NIPSNAP1
DES;PLN;UQCR10;CCDC141;MLF1;MYH6;MYH7;TTN
KIAA1755;DCAF16;CCDC121;CMTM5;SHOX2;CEP85L;GPR126;ARHGEF40;CD34;TRIB2
PLN;DLEU1;UQCR10;CCDC141;MLF1;MYH6;MYH7;TTN

IFI27L2;CENPW;SHOX2;NCAPG;CWC27;GPN1;NIPSNAP1;WARS2;DNAJC1;GNB4;DLEU1;EIF3E;LCORL
TBX15;ZCCHC10;CEP85L;NDRG2;MLF1;MYH6;MYH7;TTN
SERPINA1;APOE
PLN
SLC5A6
KIAA1755;DCAF16;CCDC121;CMTM5;SKIDA1;CEP85L;GPR126;SLC35F1;ZNF512;ARHGEF40;CD34;TRIB2
FAM3C
RSRC1
IFI27L2
LXN
PTEN
DNAJC1;CENPW
SLC35F1;APOE
GJA1;SERPINA1
DES;APOE
MFN1;NDRG2
APOC1;APOE
HSPA4;FAM174B;NCAPG
GCKR
GJA1;DES;MTMR3;CEP85L;PTEN
MYH7
HSPA4
PTEN
MTMR3;PTEN
CENPW
CENPW;MFN1;DLEU1;SLC35F1;ZNF512;LCORL;DGKH;TTN
KIAA1755;CMTM5;CCDC141;CD34;MLF1;MYH6
MYH6;MYH7
APOE
NRBP1
PLN;MFN1;CCDC141;MLF1;MYH6
IFI27L2;GCKR;TOMM40;B3GNT7;NOV;APOC1;ZNF219;ZMAT5;ZNF513;UQCR10;APOE;ARHGEF40
ZCCHC10
TTN
LXN
KIAA1755;CCDC121;CMTM5;SKIDA1;ZNF219;CEP85L;GPR126;SLC35F1;ARHGEF40;LCORL;TRIB2
MTMR3
DES
MYH7
DES;CMTM5
TBX15;GFM1;DES;CCDC91;UQCR10;NDRG2;NRBP1;MYH6;MYH7;TTN
PLN;VSTM2L;CCDC141;MLF1;MYH6;MYH7;TTN
DES;PLN;UQCR10;CCDC141;MLF1;MYH6;MYH7
DES;UQCR10;CCDC141;MYH6;MYH7
TBX15;CENPW;RSRC1;SHOX2;AFAP1;SSC5D;DGKH

SLC5A6;DNAJC1;GCKR;SERPINA1;APOC1;GPR126;APOE;PVRL2;NIPSNAP1
SLC35F1
AFAP1;NCAPG
CD34;TRIB2
GNB4
APOE;WNT16
SERPINA1;MTMR3
APOE;MYH7
CCDC141;CD34
GJA1;GPR126
APOC1;NIPSNAP1
GJA1
MLF1
APOE;MYH6
GJA1;LXN
GJA1;LXN
GJA1;LXN
GJA1;LXN
GJA1;LXN
MYH7
SLC35F1
MLF1
TTN
UQCR10;MYH7
MLF1
CCDC141
UQCR10
LXN;MYH7
GJA1;NCAPG
GPR126;TTN
DLEU1
TBX15
APOC1
NCAPG;APOE
SNX17
SLC35F1
DES;MYH7
NCAPG;CD34
LXN;APOE
PLN
DES;CD34
GPN1;MYH7
NCAPG
ZCCHC10
LXN;RSRC1

MLF1
MYH7
GJA1;GNB4
GJA1;GNB4
MYH7
MYH7
SLC35F1;MYH7
GJA1;LCORL
LXN;NCAPG
MLF1
APOC1;APOE
MLF1
SPEG
ARHGEF40
NOV;TRIB2
CEP85L;CD34
MLLT10
TOMM40;CR1L;ASCC2;FAM3C;MYH6
PTEN
PTEN
GJA1;PTEN;CD34
GJA1;DES;HSPA4;PTEN;APOE;NDRG2;CD34;DGKH
PTEN;CD34
APOE
NCAPG
CCDC91
CCDC141
LXN;SLC4A1AP;TOMM40;HSPA4;CEP85L;AFAP1;UQCR10;NDRG2;GPN1;NIPSNAP1;TTN;PPM1G;GJA1;N[illegible]
GJA1;UQCR10;APOE;NDRG2;MYH7
GNB4
PTEN;EIF3E
NDRG2
SNX17;NIPSNAP1;NRBP1;PPM1G
GFM1;SNX17;MFN1;NDRG2
GCKR
LCORL
EIF2B4;MLLT10;GFM1;LXN;TOMM40;UQCR10
SERPINA1
PTEN
GJA1
DNAJC1;HSPA4;FAM174B;GPR126;SLC35F1;CD34;MLF1;MYH7
APOC1
APOE
NCAPG;WNT16
SERPINA1

EIF2B4
SPEG;TRIB2;NRBP1;TTN
SNX17
CD34
SLC5A6
PLN;TRIB2
CD34
GPN1
GNB4
DNAJC1
PTEN
MLLT10
UQCR10
TTN
FAM174B
MLF1
APOE
NDRG2
GJA1;PTEN;APOE;CD34
SERPINA1;APOE
SERPINA1;APOE
NOV;SLC35F1
TTN
APOE
TRIB2
APOE
CD34
MFN1
GFM1;TOMM40;SHOX2;PTEN;GPR126;APOE;LCORL;TRIB2;TTN
NIPSNAP1;MYH7
MLLT10;DNAJC1;ZNF219;RSPO2;AFAP1;GPR126;NRBP1
MTMR3
SHOX2
HSPA4;APOC1
TBX15
PTEN
NOV
CCDC141
MYH7
GJA1
TTN
CD34
TBX15
MLF1
NIPSNAP1

EIF3E
GJA1;APOC1;PTEN
PTEN
PTEN
ARHGEF40
EIF2B4;TBX15;GFM1;DES;UQCR10;MYH6;MYH7;TTN
CD34
DGKH
PTEN
GJA1;PTEN;ZNF513;CWC27
GJA1;PTEN;ZNF513;CWC27
HSPA4;FAM174B;NCAPG;UQCR10
SERPINA1
CD34
CD34
CD34
MYH6
LXN
CMTM5
UQCR10
SLC5A6
PLN
APOE
MLF1
TBX15
GJA1
DES
FAM174B
TRIB2
NOV
GJA1
SLC5A6
MLF1
CWC27
TBX15
MLF1
NOV
PTEN
PTEN
PTEN
DES
PTEN
SERPINA1;HSPA4
AFAP1;NCAPG
TBX15;TOMM40;NOV;PTEN;APOE;CD34

NOV
HSPA4
NDRG2;CD34
GJA1
ZMAT5
MLF1
TBX15
GJA1
MYH7
CCDC141
SLC35F1
CCDC121;GFM1;SERPINA1;MTMR3;LXN;SLC4A1AP;TOMM40;CEP85L;PTEN;CWC27;NDRG2;TTN;PPM1G
GJA1;NCAPG;EIF3E;APOE;PVRL2;SUPT7L
LXN
NOV
CD34
LCORL
MLF1
CCDC141
LXN;NOV;CABP7;SHOX2;NCAPG
GJA1;ZCCHC10;CD34;TRIB2
TOMM40;PLN;MFN1;NDRG2;MYH6
NDRG2
RSRC1
SLC5A6
GNB4
TTN
GJA1
SNX17
HSPA4
GJA1
TBX15
NCAPG
APOE
FAM3C
FAM174B
MLF1
MYH6
NDRG2
LCORL
TTN
MYH7
APOC1
CD34
MYH7

GFM1
MYH7
LXN
APOE
CD34
MLF1
PPM1G
MYH6
SLC35F1
HSPA4
PTEN
PTEN
HSPA4
SERPINA1
APOE
PTEN
PTEN
PTEN
PTEN
APOE
PTEN
GPN1
GNB4
GCKR;NOV;HSPA4;PTEN;NDRG2;GPN1
EIF2B4;MTMR3;SLC4A1AP;HSPA4;AFAP1;NCAPG;NDRG2;GPN1;NRBP1;SUPT7L;PPM1G;MLLT10;DNAJC[illegible]
ZNF513
HSPA4
NDRG2
MLF1
SHOX2
NCAPG
PPM1G
PLN
MYH6
GJA1;SHOX2;GNB4;LCORL;WARS2
LXN;PLN;NOV;GPR126
AFAP1;WNT16;MLF1;MYH7
GJA1;CD34
MLLT10
SLC5A6;GJA1
GPR126;CD34
GFM1
CMTM5;VSTM2L;UQCR10;CCDC141;MLF1;MYH6
SLC5A6;EIF2B4;DCAF16;TOMM40;CMTM5;SHOX2;CEP85L;SLC35F1;ARHGEF40;PVRL2;TRIB2;SUPT7L
CCDC121;CMTM5;SKIDA1;SHOX2;CEP85L;SLC35F1;KIAA1755;DCAF16;RSRC1;GPR126;ALX4;ZNF512;LC[illegible]

MLF1
GNB4
NDRG2
B3GNT7
PLN
DES
TBX15
NAT14
LXN
NOV
ZCCHC10
NDRG2
NOV
GJA1;FAM174B;MFN1;UQCR10;MYH6
TTN
GCKR
MLLT10
NCAPG
DGKH
NOV
HSPA4;PTEN;NDRG2;CD34
MLF1
DES
GNB4;PTEN
GJA1
ZNF513;MYH6
RSRC1
PTEN;WNT16
TOMM40;MFN1;UQCR10;WNT16
GNB4
RSPO2;WNT16
PTEN
EIF3E
PTEN;WNT16
HSPA4
CR1;CR1L
DGKH
CR1;CR1L
EIF2B4;EIF3E
GNB4;PTEN;WNT16
PTEN;WNT16
MFN1
WARS2
GNB4
WNT16

PTEN
DGKH
UQCR10;APOE;WNT16
PTEN
PTEN
WNT16
PTEN;WNT16
MFN1;UQCR10
UQCR10
WNT16
GJA1
PTEN
B3GNT7
GNB4
TOMM40;UQCR10
CD34
GNB4
PTEN
GNB4
PTEN
GNB4
DGKH
DGKH
WNT16
PTEN
PTEN
PTEN
PTEN
WNT16;MLF1
GNB4
PTEN
GNB4;PTEN
GNB4
UQCR10
GNB4
GNB4
WNT16
PTEN
SERPINA1
MYH6
ZNF219;ZNF512
PVRL2;SUPT7L
EIF2B4;DLEU1
MLLT10;SHOX2
ZMAT5;NIPSNAP1

HSPA4;CD34
EIF2B4;ZNF219
CCDC91;WARS2
MLLT10;SHOX2
EIF3E;APOE
ZNF628;TTN
PLN;TTN
EIF2B4;NIPSNAP1
MFN1;DLEU1
SLC4A1AP;GPR126
HSPA4;PTEN
MFN1;MLF1
PVRL2;MLF1
DNAJC1;PLN
PTEN;EIF3E
CCDC121;GJA1
ZNF513;SLC35F1
IFI27L2;GFM1
PTEN;WARS2
SSC5D;APOE
CCDC121;RSPO2
CEP85L;HORMAD2
AFAP1;NCAPG
CD34;GPN1
DLEU1;ZNF512
SLC4A1AP;PPM1G
EIF2B4;GCKR
SLC4A1AP;MYH7
CCDC91;NOV
NOV;CD34
SNX17
UQCR10
EIF2B4
TBX15
DES
SLC35F1
TTN
MYH6
MYH7
TBX15;DES;TOMM40;ZNF628;SPEG;UQCR10;MYH6;MYH7;TTN
KIAA1755;IFI27L2;CCDC91;CMTM5;APOC1;ZNF219;RSPO2;SLC35F1;APOE;NDRG2;NAT14
MYH6;MYH7
CD34
MYH7
CR1

RSPO2
ARHGEF40
CENPW;SERPINA1;MTMR3;LXN;HSPA4;SHOX2;PTEN;NDRG2;TBX15;GJA1;NOV;RSPO2;ALX4;EIF3E;APOE
LCORL
HORMAD2
MYH7
MYH7
VSTM2L
SHOX2
SHOX2
TBX15
DES
B3GNT7
LCORL
CR1L
HORMAD2
DES
TMEM253
MLF1
UQCR10
TBX15
ZNF628
TMEM253
CABP7
VSTM2L
SLC35F1
NCAPG
NDRG2
GJA1
TTN
PTEN
SKIDA1
ZCCHC10
SHOX2
CENPW
CWC27
SERPINA1
CCDC141
CR1
UQCR10
CD34
GCKR
TBX15
APOC1
APOC1

RSPO2
NAT14
TBX15
HORMAD2
TTN
SPEG
GCKR
APOE
ALX4
SERPINA1
RSPO2
PTEN
SHOX2
CEP85L
WNT16
HORMAD2
CR1
SKIDA1
APOC1
TBX15
SHOX2
GCKR
NAT14
TBX15
SHOX2
VSTM2L
GCKR
CENPW
APOC1
DES
CR1L
MFN1
CR1L
PLN
SSC5D
SLC35F1
NDRG2
CCDC141
HORMAD2
DES
SERPINA1
MLF1
TTN
CABP7
NCAPG

SHOX2
CR1
GJA1
MYH6
CCDC141
TTN
VSTM2L
PTEN
DGKH
SLC35F1
SHOX2
APOC1
APOC1
SLC35F1
SPEG
MLF1
CMTM5
GCKR
TMEM253
RSPO2
SKIDA1
CABP7
RSPO2
SHOX2
SLC35F1
PTEN
CEP85L
NCAPG
MYH7
MYH7
NCAPG
SLC35F1
CCDC141
TMEM253
NCAPG
MYH6
B3GNT7
TRIB2
FAM3C
CR1
SLC35F1
CR1L
PTEN
CABP7
DES

APOE
SHOX2
GCKR
SLC5A6
CENPW
CMTM5
CABP7
APOC1
PLN
GFM1
WARS2
CCDC141
APOC1
CABP7
SERPINA1
CCDC121
APOC1
SERPINA1
CABP7
CCDC121
GJA1
DES
WNT16
SSC5D
RSPO2
MYH7
NDRG2
SSC5D
CCDC141
SPEG
APOC1
TRIB2
RSPO2
MLF1
SSC5D
APOE
CABP7
CMTM5
TTN
B3GNT7
VSTM2L
B3GNT7
CEP85L
MLF1
CMTM5

SHOX2
SERPINA1
SHOX2
DGKH
CENPW
SLC35F1
APOC1
CABP7
APOE
ZNF628
CCDC121
APOC1
GJA1
APOC1
DGKH
RSPO2
TBX15
RSPO2
HORMAD2
VSTM2L
TMEM253
TTN
CEP85L
FAM174B
TBX15
APOC1
PTEN
MFN1
CD34;MLF1
GJA1
PTEN;NDRG2
TRIB2
MLLT10
LCORL
LCORL
PTEN
PPM1G
CR1
GJA1;HSPA4;PTEN;APOE
PLN
MLLT10
SLC5A6
MTMR3
MLLT10;CCDC121;SKIDA1;RSPO2;AFAP1;SSC5D
MFN1

WNT16
PTEN
SERPINA1
VSTM2L;APOE
EIF3E
NDRG2
ZNF219;ALX4;ZNF513;MLF1
TBX15;MYH7
APOE
UQCR10
SERPINA1
TOMM40;EIF3E
MLF1;NIPSNAP1
SLC4A1AP;SHOX2
LXN;PPM1G
KIAA1755;SKIDA1
PLN;PVRL2
GCKR;PVRL2
SERPINA1;HSPA4
EIF3E;PPM1G
SLC4A1AP;ASCC2
NCAPG;NRBP1
HSPA4;GPN1
HSPA4;SUPT7L
SLC4A1AP;NRBP1
SLC5A6;AFAP1
SLC4A1AP;PVRL2
CCDC91;ARHGEF40
GNB4;CD34
PLN;ZMAT5
GNB4;VSTM2L
GJA1;B3GNT7
PTEN;GPN1
GJA1;SUPT7L
TOMM40;MFN1
MLLT10;MTMR3
SLC4A1AP;TOMM40
NDRG2;WARS2
DLEU1;EIF3E
SLC5A6;SLC4A1AP
CR1;PVRL2
SNX17;EIF3E
PVRL2;GPN1
MTMR3;EIF3E
DES;PVRL2

SERPINA1;HORMAD2
ZNF219
DES;NDRG2;MYH7;TTN
PLN;EIF3E;NDRG2;TTN
PLN;APOE;MYH6;MYH7
APOC1;EIF3E;APOE;NDRG2
GJA1;GNB4;EIF3E;APOE
GJA1;APOC1;EIF3E;APOE
GJA1;PTEN;UQCR10;NDRG2
GJA1;APOC1;EIF3E;APOE
EIF2B4;MLLT10;APOC1;APOE;NDRG2
APOC1;SLC35F1;EIF3E;APOE
GJA1;CENPW;APOC1;NCAPG
APOC1;UQCR10;APOE;NDRG2
GJA1;DES;PLN;APOC1;TTN
LXN;APOE;NDRG2;DGKH
CR1;CR1L;CMTM5;ARHGEF40;DGKH
GJA1;APOC1;EIF3E;NDRG2
EIF2B4;DES;UQCR10;NRBP1;MYH7
DES;EIF3E;APOE;CCDC141;MYH7
DES;UQCR10;APOE;MYH7
DNAJC1;CABP7;VSTM2L;WNT16
PLN;EIF3E;NDRG2;MLF1;TTN
LXN;SNX17;GNB4;APOE;TRIB2
DES;UQCR10;CD34;MYH7
GJA1;NDRG2;MYH7;TTN
GJA1;APOC1;EIF3E;APOE
GJA1;APOC1;EIF3E;APOE
GJA1;SERPINA1;EIF3E;UQCR10
DLEU1
APOE
WARS2
HSPA4
DNAJC1
APOE
MLLT10;RSRC1
HSPA4;APOC1
NDRG2
GJA1;PTEN;ZNF513;CWC27
SERPINA1;NIPSNAP1
HSPA4;NCAPG
SERPINA1;PVRL2
NOV;MLF1
SERPINA1;ZCCHC10;CD34;MLF1
GJA1

PTEN;EIF3E;FAM3C;APOE
SLC5A6;FAM174B
IFI27L2
TBX15;GFM1;UQCR10;CCDC141;MYH6;MYH7;TTN
DES;PLN;UQCR10;CCDC141;MLF1;MYH6;MYH7
WARS2
APOE
DES
MFN1
APOE
MYH7
GNB4
DLEU1
GJA1;HSPA4;PTEN;FAM3C;NDRG2
MLF1
RSRC1;CD34
GJA1
UQCR10
APOE
MLF1
PTEN;CD34
GJA1
MLF1
APOC1
PPM1G
ZCCHC10
CD34
TBX15;GFM1;DES;SLC4A1AP;ZNF628;UQCR10;NDRG2;MYH6;MYH7;TTN
TBX15;GFM1;TOMM40;ZNF628;CMTM5;DLEU1;UQCR10;NDRG2;MYH6;MYH7;PPM1G
CCDC91
PTEN
PTEN
TOMM40
SKIDA1
MLLT10;GFM1;GJA1;NOV;HSPA4;PTEN;EIF3E;NDRG2;CD34
GJA1
PTEN
SLC5A6;IFI27L2;GJA1;EIF3E
TBX15;GJA1;NOV;FAM3C
DLEU1;ZNF513;ASCC2;TRIB2
GJA1;APOC1;EIF3E;APOE
GJA1;CENPW;NOV;GNB4
FAM174B;APOE;CD34;MYH6
GJA1;APOC1;EIF3E;APOE
MLLT10;GJA1;DES;SERPINA1;NOV;HSPA4;GNB4;PTEN;RSPO2;APOE;NDRG2;CD34

GFM1;SERPINA1;MTMR3;CR1;LXN;HSPA4;SHOX2;CEP85L;PTEN;AFAP1;NDRG2;TTN;SLC5A6;GJA1;DES;N
CD34
DES;RSPO2;MLF1;TTN
PLN;EIF3E;MYH6;MYH7;TTN
PLN;SNX17;FAM174B;MLF1
SLC5A6;TOMM40;HSPA4;VSTM2L
WARS2
PTEN
AFAP1
PTEN
EIF2B4;DCAF16;CMTM5;SKIDA1;SHOX2;CEP85L;MFN1;GPR126;NDRG2;LCORL;TRIB2;WARS2
SLC5A6;GCKR;GFM1;SERPINA1;APOC1;GPR126;APOE;NIPSNAP1
DGKH
HSPA4;APOE
APOC1;GPR126;CD34;MLF1
LXN;APOC1;EIF3E;TRIB2
HSPA4;UQCR10;NDRG2
EIF2B4;DES;APOE;MLF1;MYH7
NOV;CABP7;APOC1;NDRG2
SERPINA1;GPR126;EIF3E;CD34
DES;NDRG2;MLF1;TTN
GJA1;LXN;EIF3E;APOE
GJA1;LXN;EIF3E
DLEU1
PTEN
PTEN
CD34
APOE
ZNF513
MLF1
GPR126;PVRL2
CCDC121;GFM1;ZCCHC10;SLC35F1;CWC27;NAT14;GJA1;CCDC91;RSRC1;FAM174B;DLEU1;SPEG;FAM3C
DLEU1;ALX4;MYH6;MYH7
TBX15;MYH6;MYH7
VSTM2L;WNT16;MYH7
GCKR;CCDC121;CMTM5;SKIDA1;AFAP1;SLC35F1;SSC5D;NDRG2;KIAA1755;GJA1;PLN;NOV;ZNF219;GPR1
CCDC121;GFM1;ZCCHC10;SLC4A1AP;CWC27;NAT14;GJA1;CCDC91;RSRC1;FAM174B;SPEG;FAM3C;ARH
PLN;GPR126;ALX4;CCDC141
CENPW;PTEN;NRBP1;DGKH;TTN;PPM1G
CCDC121;GFM1;ZCCHC10;SLC4A1AP;SLC35F1;CWC27;NAT14;GJA1;CCDC91;RSRC1;FAM174B;DLEU1;SP
GCKR;SERPINA1;CMTM5;AFAP1;VSTM2L;SSC5D;NDRG2;MLF1;KIAA1755;GJA1;NOV;ZNF219;GPR126;SP
SLC5A6;CCDC121;GFM1;SLC4A1AP;TOMM40;ZMAT5;ASCC2;UQCR10;GPN1
MTMR3;PTEN;NRBP1;TMEM253;DGKH;TTN
SHOX2;DLEU1;VSTM2L;NCAPG;MYH6;MYH7
MYH6;MYH7

HSPA4;ALX4;CCDC141;GPN1
CMTM5;SHOX2;DLEU1
CCDC121;GFM1;ZCCHC10;SLC4A1AP;SLC35F1;CWC27;NAT14;GJA1;CCDC91;RSRC1;FAM174B;DLEU1;SP
CCDC121;GFM1;ZCCHC10;LXN;SLC4A1AP;TOMM40;SLC35F1;UQCR10;CWC27;NDRG2;NAT14;DCAF16;G
TBX15;SHOX2;ALX4;MYH6;MYH7
CCDC121;ZMAT5;ALX4
CCDC121;GFM1;ZCCHC10;SLC4A1AP;TOMM40;SLC35F1;UQCR10;CWC27;NAT14;GJA1;CCDC91;RSRC1;F
IFI27L2;CCDC121;MTMR3;ZCCHC10;SLC4A1AP;CEP85L;NCAPG;CWC27;TTN;DCAF16;PLN;RSRC1;ZMAT5
CCDC121;GFM1;ZCCHC10;SLC4A1AP;CCDC91;HSPA4;ALX4
CENPW;MTMR3;ZNF628;PTEN;EIF3E;NRBP1;TMEM253;WARS2;PPM1G
GFM1;ZCCHC10;SLC4A1AP;TOMM40;SLC35F1;CWC27;NDRG2;NAT14;PPM1G;DCAF16;GJA1;CCDC91;RS
GFM1;ZCCHC10;TOMM40;CEP85L;NCAPG;UQCR10;CWC27;NAT14;DCAF16;GJA1;CCDC91;ZNF628;RSRC
GCKR;SERPINA1;CMTM5;SKIDA1;AFAP1;VSTM2L;SSC5D;NDRG2;TMEM253;TTN;KIAA1755;GJA1;PLN;NC
IFI27L2;MTMR3;CEP85L;SLC35F1;CWC27;MLF1;TTN;MLLT10;DCAF16;DES;PLN;ZMAT5;DLEU1;SPEG;ALX
GCKR;SERPINA1;MTMR3;CMTM5;SKIDA1;AFAP1;SSC5D;NDRG2;KIAA1755;GJA1;PLN;NOV;ZNF219;GPR1
CCDC121;GFM1;ZCCHC10;SLC4A1AP;PTEN;SLC35F1;CWC27;GJA1;CCDC91;RSRC1;FAM174B;DLEU1;SPE
CCDC121;SLC4A1AP;ZNF628;ZMAT5;ALX4;C2ORF16;NIPSNAP1
GCKR;CCDC121;SERPINA1;MTMR3;CMTM5;SKIDA1;AFAP1;SSC5D;NDRG2;KIAA1755;GJA1;PLN;NOV;ZN
IFI27L2;CCDC121;MTMR3;ZCCHC10;SLC4A1AP;CEP85L;PTEN;SLC35F1;CWC27;NAT14;TTN;DCAF16;PLN;
EIF2B4;MTMR3;ZCCHC10;SLC4A1AP;TOMM40;HSPA4;PTEN;NCAPG;GPN1;NRBP1;SUPT7L;WARS2;TTN;
GCKR;CMTM5;SKIDA1;SSC5D;NDRG2;TMEM253;KIAA1755;GJA1;PLN;NOV;ZNF219;GPR126;SPEG;ALX4;
GCKR;SERPINA1;MTMR3;CMTM5;AFAP1;SSC5D;NDRG2;KIAA1755;GJA1;DES;NOV;ZNF219;GPR126;SPE
CCDC121;GFM1;ZCCHC10;TOMM40;SHOX2;SLC35F1;UQCR10;CWC27;NAT14;DCAF16;GJA1;PLN;CCDC9
GCKR;MTMR3;CMTM5;AFAP1;SLC35F1;SSC5D;NDRG2;MLF1;KIAA1755;GJA1;PLN;NOV;APOC1;ZNF219;
IFI27L2;MTMR3;LXN;SLC4A1AP;CEP85L;CWC27;TTN;PPM1G;GJA1;CCDC91;RSRC1;MFN1;SPEG;ALX4;EIF
HSPA4;MYH6;MYH7
IFI27L2;MTMR3;ZCCHC10;SLC4A1AP;SHOX2;CEP85L;PTEN;CWC27;TTN;PPM1G;DCAF16;CCDC91;RSRC1
IFI27L2;MTMR3;SLC4A1AP;SHOX2;CEP85L;PTEN;SLC35F1;CWC27;TTN;PPM1G;DCAF16;PLN;CCDC91;RS
GCKR;SERPINA1;SKIDA1;AFAP1;SSC5D;NDRG2;TMEM253;KIAA1755;GJA1;PLN;NOV;ZNF219;GPR126;SP
ALX4;MYH6;MYH7
GCKR;SERPINA1;MTMR3;CMTM5;AFAP1;SSC5D;NDRG2;KIAA1755;GJA1;NOV;APOC1;GNB4;GPR126;SPI
TBX15;ZMAT5;ALX4;MYH6;MYH7;TTN
IFI27L2;LXN;SLC4A1AP;SHOX2;CEP85L;PTEN;SLC35F1;CWC27;TTN;PPM1G;DCAF16;CCDC91;RSRC1;MFN
GCKR;MTMR3;CMTM5;SKIDA1;AFAP1;NDRG2;MLF1;KIAA1755;GJA1;PLN;NOV;ZNF219;GPR126;SPEG;A
IFI27L2;MTMR3;LXN;SHOX2;CEP85L;PTEN;AFAP1;SLC35F1;CWC27;GPN1;MLF1;TTN;DCAF16;GJA1;DES;
ALX4;MYH6;MYH7
GCKR;SERPINA1;MTMR3;CMTM5;SKIDA1;AFAP1;VSTM2L;SSC5D;NDRG2;MLF1;KIAA1755;GJA1;DES;PLN
TBX15;VSTM2L;MYH6;MYH7;TTN
HSPA4;C2ORF16;WNT16;TMEM253
GFM1;HSPA4;SHOX2;ALX4
IFI27L2;CCDC121;MTMR3;SLC4A1AP;CMTM5;CEP85L;PTEN;AFAP1;SSC5D;CWC27;GPN1;MLF1;TTN;MLL
IFI27L2;CCDC121;GFM1;ZCCHC10;SLC4A1AP;TOMM40;SHOX2;SLC35F1;UQCR10;CWC27;NAT14;GJA1;C
DLEU1;NCAPG
MTMR3;PTEN;C2ORF16;GPN1;NRBP1;TMEM253;WARS2;TTN;PPM1G;TBX15;MLLT10;ZNF628;MFN1;TR
IFI27L2;MTMR3;LXN;SLC4A1AP;HSPA4;AFAP1;CWC27;GPN1;MLF1;NRBP1;PPM1G;DCAF16;CCDC91;NO

IFI27L2;MTMR3;ZCCHC10;LXN;SLC4A1AP;SHOX2;CEP85L;CWC27;TTN;PPM1G;PLN;CCDC91;RSRC1;MFN
GFM1;ZCCHC10;SLC4A1AP;TOMM40;SLC35F1;CWC27;NDRG2;PPM1G;DCAF16;GJA1;PLN;CCDC91;RSRC
MTMR3;NCAPG;MYH6;MYH7
TBX15;MYH6;MYH7
TBX15;SHOX2;ALX4;MYH6;MYH7
ZMAT5;VSTM2L;MYH6
NIPSNAP1;MYH6;MYH7
GCKR;CCDC121;SERPINA1;MTMR3;CMTM5;AFAP1;SSC5D;NDRG2;MLF1;KIAA1755;GJA1;PLN;NOV;ZNF2
CMTM5;CEP85L;CCDC141;MYH6;MYH7
ALX4;CCDC141;MYH6;MYH7
GCKR;SERPINA1;CMTM5;SKIDA1;AFAP1;SLC35F1;SSC5D;NDRG2;MLF1;TMEM253;TBX15;KIAA1755;GJA
SHOX2;ALX4;MYH6
FAM174B;SHOX2;NCAPG;MYH6;MYH7
IFI27L2;CCDC121;MTMR3;SLC4A1AP;SHOX2;NCAPG;SLC35F1;CWC27;TTN;DCAF16;GJA1;CCDC91;SPEG;
WNT16;NIPSNAP1;MYH6;TMEM253;MYH7
SLC4A1AP;ZNF628;SNX17;SHOX2;PTEN;GPN1;NRBP1;DGKH;WARS2;TTN;PPM1G
TBX15;SHOX2;CEP85L;ALX4;ARHGEF40;MYH6;MYH7;TTN
CCDC121;GFM1;ZCCHC10;SLC4A1AP;SLC35F1;CWC27;NAT14;GJA1;CCDC91;RSRC1;FAM174B;DLEU1;SP
GCKR;CCDC121;SKIDA1;SHOX2;VSTM2L;SLC35F1;SSC5D;C2ORF16;UQCR10;MLF1;TMEM253;KIAA1755;
TBX15;CCDC141
DLEU1;VSTM2L;NCAPG;MYH6;MYH7
SLC4A1AP;GNB4;NCAPG;GPN1
GCKR;HSPA4;ALX4
IFI27L2;CCDC121;GFM1;ZCCHC10;SLC4A1AP;TOMM40;SHOX2;SLC35F1;UQCR10;CWC27;NDRG2;NAT14
EIF2B4;CENPW;MTMR3;LXN;TOMM40;SHOX2;PTEN;SSC5D;GPN1;NRBP1;TMEM253;WARS2;PPM1G;SL
CCDC121;GFM1;ZCCHC10;TOMM40;SHOX2;SLC35F1;UQCR10;CWC27;NDRG2;NAT14;PPM1G;DCAF16;C
LXN;ZNF628;PTEN;NRBP1;TMEM253;DGKH;WARS2;TTN;PPM1G
IFI27L2;MTMR3;LXN;SLC4A1AP;SHOX2;CEP85L;NCAPG;SLC35F1;CWC27;TTN;PPM1G;DCAF16;GJA1;CCD
SHOX2;ALX4;MYH6;MYH7
CCDC121;GFM1;ZCCHC10;SLC4A1AP;TOMM40;SLC35F1;CWC27;NDRG2;NAT14;PPM1G;DCAF16;GJA1;P
IFI27L2;APOC1;ZMAT5;VSTM2L;UQCR10;WNT16;NAT14;NIPSNAP1
CENPW;MTMR3;ZNF628;PTEN;NRBP1;TMEM253;DGKH;TTN;PPM1G
TBX15;SHOX2;CEP85L;ALX4;ARHGEF40;MYH6;MYH7
GCKR;MTMR3;CMTM5;SKIDA1;AFAP1;SSC5D;NDRG2;TMEM253;KIAA1755;GJA1;NOV;ZNF219;GPR126;
CCDC121;GFM1;ZCCHC10;SLC4A1AP;SLC35F1;CWC27;GJA1;CCDC91;RSRC1;FAM174B;DLEU1;SPEG;FAM
MYH6;MYH7
TBX15;SHOX2;CEP85L;ARHGEF40;MYH7
NOV
ZNF513
PTEN
MFN1
SHOX2;FAM3C
APOE
APOE
CCDC121;SLC4A1AP;TOMM40;HSPA4;CEP85L;AFAP1;NCAPG;UQCR10;NDRG2;NIPSNAP1;TTN;GJA1;DES

EIF3E
SSC5D
LXN
TTN
NIPSNAP1
GNB4
CD34
PTEN
ZCCHC10;SLC4A1AP;TOMM40;HSPA4;CEP85L;PTEN;UQCR10;GPN1;NIPSNAP1;NRBP1;SUPT7L;TTN;PPM
NOV
CR1;PTEN;SPEG
APOE
MTMR3
GNB4
GNB4
GNB4
CENPW;GNB4;NCAPG;MLF1
ZCCHC10;PLN;NOV;HSPA4;AFAP1;SPEG;SSC5D
SHOX2
SPEG
GNB4;ARHGEF40;DGKH
GNB4
TOMM40
SERPINA1
UQCR10
MYH6
PTEN
APOE
DES;UQCR10;NDRG2;MYH7
PTEN
GJA1;SERPINA1;HSPA4;PTEN;AFAP1;EIF3E;HORMAD2;CD34;TRIB2
GJA1;B3GNT7;HSPA4;PTEN;ALX4;ASCC2;APOE;NDRG2
NOV
GJA1;NOV;APOC1;TTN
ZCCHC10;LXN;AFAP1
SERPINA1;APOC1;EIF3E;APOE
HSPA4;NCAPG;FAM3C;WNT16;NIPSNAP1;PPM1G
SLC4A1AP;HSPA4;EIF3E;APOE;NDRG2;NRBP1
TOMM40;HSPA4;AFAP1;NCAPG;TRIB2;PPM1G
CCDC91;SPEG;EIF3E;NDRG2;MYH7;TTN
DES;APOC1;PTEN;UQCR10;APOE;NDRG2
TOMM40;HSPA4;EIF3E;FAM3C;WNT16;PPM1G
DCAF16;ZMAT5;NCAPG;FAM3C;WNT16;NIPSNAP1
MLLT10;GFM1;ZCCHC10;HSPA4;RSPO2;NCAPG
GJA1;DES;NOV;MYH6;MYH7;TTN

TOMM40;HSPA4;NCAPG;FAM3C;TRIB2;PPM1G
GJA1;DES;HSPA4;EIF3E;UQCR10;MYH6
DES;PLN;CMTM5;MLF1;MYH6;MYH7
TBX15;GJA1;PTEN;EIF3E;CD34;TTN
GCKR;APOC1;PTEN;APOE;NDRG2;NIPSNAP1
GJA1;CMTM5;MFN1;APOE;NDRG2;MYH6
DES;B3GNT7;NOV;EIF3E;APOE;NDRG2
DES;HSPA4;APOC1;APOE;NDRG2;TTN
DNAJC1;FAM3C;APOE;CCDC141;CD34;TTN
TBX15;GJA1;DES;NDRG2;CD34;TTN
TBX15;DES;UQCR10;NDRG2;CCDC141;TTN
GCKR;APOC1;EIF3E;APOE;NDRG2;NIPSNAP1
GFM1;DES;HSPA4;SPEG;CCDC141;MYH6
TOMM40;HSPA4;NCAPG;EIF3E;NRBP1;PPM1G
GJA1;MTMR3;AFAP1;ARHGEF40;GPN1;NRBP1
IFI27L2;HSPA4;GNB4;AFAP1;ALX4;EIF3E;CWC27;NIPSNAP1;WARS2
RSPO2
NOV
NDRG2
LXN
DES;APOC1
DES
GJA1
B3GNT7
APOE
LXN
GJA1
CD34
DES;SSC5D;CCDC141;MYH7;TTN
PVRL2;TTN
CR1
SLC5A6
CR1
SERPINA1
DES;PLN;MFN1;UQCR10;CCDC141;MYH6;MYH7;TTN
MLLT10;DCAF16;CEP85L;GNB4
NIPSNAP1
PTEN
WNT16
PLN
PTEN
HSPA4
GNB4
UQCR10
GNB4

GNB4
GJA1;FAM174B;NCAPG;EIF3E;FAM3C;TRIB2
SLC5A6
DES;NDRG2
SHOX2
HSPA4
PTEN;ALX4
MLLT10
MLLT10
GCKR
TOMM40
SKIDA1
SKIDA1;CWC27;LCORL;TRIB2
SKIDA1
TRIB2
CEP85L
SSC5D
MLLT10
CEP85L
SKIDA1
SLC35F1
SKIDA1
SUPT7L
GNB4;APOE
MLLT10
SKIDA1
SKIDA1
GCKR
MTMR3
MLF1
TRIB2
NCAPG
MFN1
GJA1
NCAPG
DNAJC1
TRIB2
TOMM40
PTEN
NCAPG;APOE;CD34;MLF1;NIPSNAP1
DES;TOMM40;HSPA4;DLEU1;VSTM2L;ASCC2;GPN1;MYH6
PTEN;MYH7;TTN
GFM1;ZCCHC10;SLC4A1AP;TOMM40;SLC35F1;CWC27;DCAF16;GJA1;PLN;CCDC91;RSRC1;FAM174B;MF
PTEN
HSPA4

CD34
PTEN
APOC1;EIF3E;APOE;MYH6;MYH7
APOE
PLN
ZNF513
NOV;HSPA4;PTEN;GPN1
GJA1;PTEN;ZNF513;CWC27
SLC5A6;PPM1G
ZNF628;ZMAT5;DLEU1;UQCR10
EIF3E;UQCR10;APOE;TRIB2
SLC5A6;TOMM40;MFN1;DLEU1;EIF3E;TRIB2;NIPSNAP1
TTN
DNAJC1;SERPINA1;NOV;CMTM5;GPR126;NCAPG;TRIB2
MFN1;NDRG2;MLF1;MYH7
CD34
EIF2B4;TBX15;DES;SLC4A1AP;TOMM40;UQCR10;MYH7;TTN;PPM1G
TBX15;GFM1;ZNF628;CEP85L;SPEG;MYH7;TTN
APOE
GCKR;MTMR3;SKIDA1;AFAP1;NDRG2;MLF1;TMEM253;KIAA1755;GJA1;NOV;ZNF219;GPR126;SPEG;APC
HSPA4
PTEN
DES;UQCR10;CCDC141;MLF1;MYH6;MYH7;TTN
HSPA4;CMTM5;APOC1;APOE
LXN
GJA1
APOE
CD34
NOV;APOE;MYH6;MYH7;TTN
KIAA1755;CCDC121;CMTM5;CEP85L;ZMAT5;GPR126;SLC35F1;ARHGEF40;LCORL;CD34;TRIB2
APOE
GJA1;WNT16
TBX15;SHOX2;ALX4
GCKR;CCDC91;APOE
DLEU1
IFI27L2;CCDC121;GFM1;ZCCHC10;LXN;SLC4A1AP;TOMM40;SHOX2;CEP85L;SLC35F1;UQCR10;CWC27;N
GCKR;ALX4;TMEM253
CMTM5;ZMAT5;ALX4;ARHGEF40;MYH6;MYH7
CCDC121;GFM1;ZCCHC10;SLC4A1AP;TOMM40;CEP85L;SLC35F1;CWC27;NDRG2;DCAF16;GJA1;PLN;CCD
GCKR;SERPINA1;MTMR3;CMTM5;AFAP1;SSC5D;NDRG2;TMEM253;KIAA1755;GJA1;NOV;ZNF219;GPR12
CCDC121;GFM1;ZCCHC10;SLC4A1AP;PTEN;SLC35F1;CWC27;NAT14;GJA1;CCDC91;RSRC1;FAM174B;SPE
RSRC1;SHOX2;ZMAT5;ALX4;NIPSNAP1
DLEU1;WNT16;MYH7
RSPO2
MTMR3;PTEN;LCORL

DES;NOV;HSPA4;PTEN
PTEN;APOE
AFAP1;MLF1
TBX15;GFM1;DES;ZNF628;UQCR10;MYH6;MYH7;TTN
GCKR;CENPW;B3GNT7;FAM174B;NCAPG;ASCC2;C2ORF16;UQCR10;PVRL2
SLC5A6;CCDC121;ZCCHC10;RSRC1;PTEN;ALX4;LCORL;CD34
EIF2B4;TBX15;DES;ZNF628;ZMAT5;MYH6;MYH7;TTN
SLC5A6;TOMM40;MFN1;NIPSNAP1;TTN
GJA1;HSPA4
GCKR
CCDC91
DLEU1
NOV
LXN
PLN;MFN1;UQCR10;MLF1
MYH6
APOE
GJA1;MYH7
HSPA4;APOC1
DCAF16;ZCCHC10;NCAPG;NIPSNAP1
EIF2B4;UQCR10;PPM1G
LXN;VSTM2L;SLC35F1;UQCR10
PLN;NDRG2;MYH6
PTEN
TOMM40;APOC1;APOE;PPM1G
GPR126;EIF3E;GPN1
GJA1;DES;SERPINA1
GJA1;LXN;EIF3E;FAM3C
SERPINA1;APOC1;APOE
NOV;ASCC2;ARHGEF40
NOV;GNB4;VSTM2L;CD34
GJA1;VSTM2L;APOE;NDRG2
TBX15;SKIDA1;EIF3E
GCKR;NDRG2;MLF1
UQCR10;MYH6;MYH7
GJA1;LXN;APOE;NDRG2
GJA1;NOV;APOC1;TTN
FAM174B;APOE;MYH6
NOV;ASCC2;ARHGEF40
TBX15;RSRC1;SHOX2;AFAP1;SSC5D
TBX15
NCAPG
SHOX2;NCAPG;MYH6;MYH7
CCDC121;GFM1;ZCCHC10;TOMM40;SHOX2;CEP85L;SLC35F1;CWC27;NDRG2;DCAF16;GJA1;CCDC91;RS[illegible]
TBX15;SHOX2;CEP85L;ALX4;ARHGEF40;MYH7

CCDC121;GFM1;SLC4A1AP;TOMM40;CWC27;DCAF16;GJA1;PLN;CCDC91;FAM174B;MFN1;SPEG;FAM3C
IFI27L2;MTMR3;SLC4A1AP;HSPA4;SHOX2;AFAP1;CWC27;GPN1;MLF1;NRBP1;TTN;PPM1G;DCAF16;CCD
PLN;APOE;NRBP1
TRIB2
HSPA4
PLN
GCKR;PVRL2
SHOX2
CENPW;RSRC1;DLEU1;NCAPG;DGKH
PLN;MYH6;MYH7
SNX17;MYH7
GFM1;CENPW;GNB4;UQCR10
UQCR10;MLF1;TTN
LXN;NOV;DLEU1;RSPO2
GCKR;APOC1;UQCR10;APOE
CEP85L;NDRG2;MLF1
NDRG2;MYH6;MYH7
APOC1;NCAPG;NAT14
APOE
SERPINA1
GCKR;CMTM5;SKIDA1;AFAP1;SSC5D;NDRG2;KIAA1755;GJA1;PLN;NOV;ZNF219;GPR126;SPEG;APOE;ARI
SERPINA1
GJA1
CR1
GJA1;LXN;CCDC91
DCAF16;NCAPG;NDRG2;MLF1;NRBP1
HSPA4;APOE
SNX17
APOE
GJA1;MYH7
HSPA4;PTEN
SERPINA1;ZCCHC10;SLC4A1AP;TOMM40;CEP85L;PTEN;NCAPG;UQCR10;CWC27;TTN;PPM1G;RSRC1;MF
DGKH
TBX15;DES;NDRG2;MYH7;TTN
CCDC91
HSPA4;PTEN
PPM1G
GFM1
GJA1;MYH7
KIAA1755;DCAF16;CENPW;ZNF628;ZNF219;AFAP1;ALX4;SSC5D;EIF3E;ZNF512;CD34;NRBP1
SKIDA1
DES;SERPINA1;SLC4A1AP
GNB4
GJA1;MFN1;ARHGEF40
CWC27

TRIB2
SERPINA1;PTEN
TRIB2
PTEN;DGKH
CCDC121;GFM1;ZCCHC10;SLC4A1AP;TOMM40;CEP85L;CWC27;NDRG2;DCAF16;GJA1;CCDC91;RSRC1;FA
EIF2B4;GFM1;TOMM40;AFAP1;SSC5D;UQCR10;NAT14;MLF1;WARS2;SLC5A6;GJA1;CCDC91;FAM174B;A
GFM1;ZCCHC10;SLC4A1AP;SLC35F1;CWC27;NAT14;GJA1;CCDC91;RSRC1;FAM174B;SPEG;FAM3C;CD34;
SERPINA1;APOC1;NCAPG;GPN1
SERPINA1;DLEU1;VSTM2L;MYH6;MYH7
TBX15;RSRC1;ALX4
CCDC121;GFM1;ZCCHC10;SLC4A1AP;CEP85L;SLC35F1;CWC27;DCAF16;GJA1;CCDC91;RSRC1;FAM174B;
CMTM5;SKIDA1;AFAP1;VSTM2L;SLC35F1;SSC5D;NDRG2;MLF1;KIAA1755;GJA1;NOV;ZNF219;GPR126;SP
SERPINA1;NCAPG;LCORL;MYH7
CCDC121;GFM1;ZCCHC10;SLC35F1;CWC27;GJA1;CCDC91;RSRC1;FAM174B;DLEU1;SPEG;FAM3C;CD34;S
GFM1;TOMM40;HSPA4
SLC5A6;GFM1;HSPA4;NCAPG;UQCR10;MYH6;MYH7
TBX15;MYH6;MYH7
GCKR;CMTM5;AFAP1;SSC5D;NDRG2;MLF1;KIAA1755;GJA1;PLN;NOV;ZNF219;GPR126;SPEG;ALX4;ARHG
CCDC121;GFM1;ZCCHC10;TOMM40;SLC35F1;CWC27;NDRG2;NAT14;PPM1G;DCAF16;GJA1;PLN;CCDC9
CCDC121;GFM1;CENPW;SLC4A1AP;SHOX2;ZMAT5;GPN1;NIPSNAP1
MYH6;MYH7
SLC5A6;APOC1;NCAPG;MYH7
GFM1;SERPINA1;MTMR3;ZCCHC10;LXN;SLC4A1AP;TOMM40;CEP85L;PTEN;UQCR10;TTN;PPM1G;RSPO
PTEN
MLLT10;CEP85L
CR1L
NCAPG
APOE
NCAPG
SERPINA1
SERPINA1
GJA1;GNB4;PTEN;APOE
WARS2
APOE
MLLT10;IFI27L2;LXN;MLF1;PPM1G
MLLT10
MLLT10;NOV;PTEN;DLEU1;CD34
GNB4
GJA1
LXN
CCDC121;CD34;TTN
GJA1;APOC1;EIF3E;UQCR10
SLC5A6;LXN
MLLT10
GPN1

EIF3E;MYH6;MYH7;TTN
APOE
TBX15;GFM1;DES;ZNF628;SPEG;NDRG2;MYH6;MYH7;TTN
TBX15;DES;UQCR10;MLF1;MYH6;MYH7;TTN
CD34
SHOX2
GJA1
IFI27L2
NOV;EIF3E
NCAPG
NCAPG
UQCR10
TOMM40;HSPA4;TRIB2
UQCR10;MLF1;MYH6
IFI27L2;SERPINA1;CR1;CMTM5;PTEN;MLF1;MLLT10;GJA1;SNX17;APOC1;ZNF219;DLEU1;ALX4;FAM3C;A
SHOX2
GJA1;PLN;APOC1;GPN1
MYH7
TBX15
GJA1
GJA1;ARHGEF40
RSPO2
MYH6;MYH7
CCDC91
NIPSNAP1
ZMAT5
GNB4
CCDC121;GFM1;ZCCHC10;SLC4A1AP;TOMM40;SHOX2;CEP85L;UQCR10;CWC27;DCAF16;GJA1;CCDC91;
PTEN
UQCR10
GJA1;SHOX2
PTEN
DNAJC1
DNAJC1
LXN
SSC5D
SHOX2;FAM3C
CR1
KIAA1755;DCAF16;CMTM5;SKIDA1;SHOX2;CEP85L;ZMAT5;GPR126;ARHGEF40;TRIB2;WARS2
CCDC121;GFM1;ZCCHC10;SLC4A1AP;CWC27;NAT14;GJA1;CCDC91;RSRC1;FAM174B;SPEG;FAM3C;CD3
DNAJC1;CENPW;SHOX2;MFN1;NCAPG;ASCC2;NRBP1
APOE
TBX15
HSPA4;PTEN;ALX4;CWC27
GNB4;WNT16

DES;NOV;HSPA4;PTEN
PTEN
MLLT10
SERPINA1;PTEN
PTEN
MLLT10
RSPO2
ZCCHC10;APOC1
FAM3C
CENPW;ZCCHC10;CR1;TOMM40;PTEN;WNT16;TTN;PPM1G;SLC5A6;DCAF16;CCDC91;SNX17;DLEU1;EIF[illegible]
ASCC2
CD34
SNX17;EIF3E;NIPSNAP1
PLN;PTEN;SPEG;FAM3C
PTEN;NCAPG;EIF3E;UQCR10
APOE
APOC1;APOE;NDRG2
DES;B3GNT7;NOV
GCKR;GJA1;LXN;B3GNT7;NOV;HSPA4;APOC1;APOE;ARHGEF40;TRIB2;NIPSNAP1
CMTM5;VSTM2L;SSC5D;NDRG2;MLF1;TTN;KIAA1755;GJA1;DES;PLN;ZNF219;SPEG;CCDC141;CD34;MY[illegible]
SHOX2;AFAP1;VSTM2L;SSC5D;WNT16;NAT14;TBX15;KIAA1755;GJA1;DES;NOV;ZNF628;RSPO2;SPEG;AL[illegible]
DES;ZNF219;AFAP1;TRIB2
FAM3C;MYH7
DES;ZNF219;TRIB2
PLN
MLLT10;RSPO2;FAM3C;DGKH;TTN
PLN
DNAJC1
CD34
NOV;PTEN
DNAJC1
MTMR3
SERPINA1
HSPA4
PVRL2
TBX15
CD34
NRBP1
SNX17
CR1L
CWC27
PTEN
NRBP1
CCDC121
ZCCHC10

MTMR3
NDRG2
MLF1
MLLT10
SERPINA1
EIF2B4
NDRG2
ZNF219
CR1L
WARS2
MFN1
GPR126
NIPSNAP1
MYH7
SERPINA1
DES
PPM1G
SNX17
SPEG
EIF3E
FAM174B
NDRG2
NAT14
CCDC121
GCKR
SPEG
GPR126
ARHGEF40
SLC35F1
WARS2
MLLT10
LXN
EIF2B4
NCAPG
NCAPG
ASCC2
FAM3C
DLEU1
GFM1
CMTM5
CWC27
NIPSNAP1
SHOX2
WARS2
SHOX2

MYH7
GNB4
MYH7
SLC4A1AP
PVRL2
SLC5A6
NOV
ARHGEF40
LXN
SLC4A1AP
NIPSNAP1
MLLT10
SLC4A1AP
SPEG
SERPINA1
AFAP1
EIF3E
MYH7
PLN
LXN
SLC4A1AP
DES
MTMR3
EIF3E
ARHGEF40
LXN
PTEN
SNX17
NRBP1
LXN;CABP7;NCAPG;MYH7
GJA1;EIF3E;MYH6;MYH7
GJA1;SERPINA1;NOV
CABP7;VSTM2L;WNT16
TOMM40;APOC1;VSTM2L
EIF3E;UQCR10;NDRG2
FAM3C
DLEU1
FAM3C
ASCC2
SERPINA1;CR1
TBX15
GCKR;TOMM40;APOC1;APOE;PVRL2;NRBP1
TTN
GCKR;HORMAD2
LXN;SLC4A1AP;TOMM40;HSPA4;CEP85L;AFAP1;NCAPG;UQCR10;GPN1;NIPSNAP1;NRBP1;TTN;PPM1G;[illegible]

TBX15;PLN;RSRC1;SHOX2;GNB4;AFAP1;EIF3E;DGKH
LXN;SHOX2;ALX4;ASCC2;EIF3E;GPN1;TTN
CCDC121;SKIDA1;GNB4;ALX4;MYH6;MYH7;TTN
CCDC121;GFM1;ZCCHC10;LXN;SLC4A1AP;CEP85L;SLC35F1;CWC27;NDRG2;DCAF16;GJA1;CCDC91;RSRC
GFM1;ZCCHC10;TOMM40;CEP85L;SLC35F1;CWC27;DCAF16;GJA1;CCDC91;ZNF628;RSRC1;FAM174B;DL
SERPINA1;MTMR3;CR1;CR1L;PTEN;ASCC2;ARHGEF40;TMEM253
SSC5D
ASCC2
TBX15
GNB4
MYH7
NCAPG
NCAPG
SNX17
NOV
DES
APOE
TBX15;LXN;LCORL;NRBP1
SLC35F1
GJA1
GNB4
GNB4
PPM1G
GNB4
TTN
CCDC141
CD34
DNAJC1
UQCR10
SPEG
PPM1G
PTEN
TBX15;GFM1;DES;UQCR10;NDRG2;MYH6;MYH7;TTN
MYH6;TTN
SLC5A6;GCKR;SERPINA1;LXN;APOC1;GPR126;CD34
GFM1;UQCR10;CCDC141;MLF1;MYH6;MYH7;TTN
TBX15;DES;UQCR10;CCDC141;MYH6;MYH7;TTN
TBX15;GFM1;DES;MLF1;MYH6;MYH7;TTN
KIAA1755;CMTM5;SHOX2;CEP85L;AFAP1;GPR126;SLC35F1;ARHGEF40;TRIB2
ALX4;ZNF513
TBX15;SHOX2;ALX4
TBX15;ALX4;CCDC141;MYH6;MYH7
PTEN
PTEN
SLC4A1AP;TOMM40;HSPA4;CEP85L;AFAP1;UQCR10;NDRG2;GPN1;NIPSNAP1;TTN;GJA1;DES;SNX17;RSF

EIF3E;TTN
TOMM40;CEP85L;UQCR10;NDRG2;NIPSNAP1;TTN;PPM1G;GJA1;SNX17;ZNF219;RSPO2;GPR126;EIF3E;F
SERPINA1;LXN;SLC4A1AP;TOMM40;HSPA4;CEP85L;UQCR10;NDRG2;GPN1;NIPSNAP1;TTN;PPM1G;DNA
SERPINA1;MTMR3;TOMM40;CABP7;CEP85L;NCAPG;UQCR10;CWC27;NDRG2;TTN;PPM1G;GJA1;PLN;ZN
PTEN
LXN;APOC1;EIF3E;TRIB2
DNAJC1;ZCCHC10;LXN;CD34
DNAJC1;SERPINA1;CR1;PTEN
ZNF513
GJA1;NOV;APOE;NDRG2
DGKH
FAM174B
GJA1;DES;NDRG2;CD34;MYH7
CR1;PTEN
PTEN
MLF1;TTN
ZCCHC10;HSPA4;RSRC1;SHOX2;MFN1
DES;APOE;NDRG2;CCDC141;LCORL
TBX15;CCDC91;APOE;NDRG2;MYH7
PLN;CD34;MYH6;MYH7;TTN
SNX17;FAM174B;MYH6;MYH7
PLN;NDRG2;MYH6;MYH7
LXN;NOV;VSTM2L;TRIB2
MTMR3;CABP7;APOC1;TRIB2
SLC5A6;EIF2B4;NRBP1;MYH6
PLN;NCAPG;SLC35F1;MYH6
DES;PLN;UQCR10;CCDC141;MLF1;MYH6;MYH7;TTN
CMTM5;SKIDA1;SHOX2;CEP85L;SLC35F1;SUPT7L;KIAA1755;DCAF16;GPR126;ALX4;ZNF512;ARHGEF40;T
PLN;FAM174B;NIPSNAP1;MYH6;TTN
GJA1;NOV;RSPO2;APOE
CENPW;AFAP1;ALX4;SSC5D;CD34;DGKH
DES;HSPA4
UQCR10
PTEN
EIF2B4;MTMR3;ZCCHC10;LXN;HSPA4;PTEN;NAT14;NIPSNAP1;PPM1G;SLC5A6;RSRC1;MFN1;ASCC2
NCAPG
SLC5A6;SERPINA1;TOMM40;HSPA4;NCAPG;APOE;WARS2
DES;SERPINA1;HSPA4;RSRC1;SHOX2;APOE;DGKH
APOE
CEP85L
SUPT7L
CWC27
MLLT10
SERPINA1;MTMR3;CR1;CR1L;CMTM5;ZMAT5;ARHGEF40;TMEM253
CENPW;HSPA4;AFAP1;NCAPG;C2ORF16;CWC27;GPN1;NIPSNAP1;WARS2;SNX17;MFN1;GNB4;DLEU1;EI

PTEN;RSPO2
APOE
NCAPG;CCDC141;MYH6;MYH7
GCKR;CMTM5;AFAP1;VSTM2L;SSC5D;NDRG2;KIAA1755;GJA1;ZNF219;GPR126;SPEG;ALX4;APOE;ARHG
GFM1;ZCCHC10;TOMM40;CEP85L;SLC35F1;CWC27;NDRG2;NAT14;PPM1G;DCAF16;GJA1;PLN;CCDC91;
EIF2B4;IFI27L2;CENPW;MTMR3;LXN;SLC4A1AP;CMTM5;CEP85L;AFAP1;SLC35F1;CWC27;MLF1;TTN;DC
GFM1;CCDC91;RSRC1;CEP85L;ALX4
TBX15;MYH7
LXN;ZNF628;GPN1;NRBP1;TTN;PPM1G
GCKR;CCDC121;SERPINA1;CMTM5;AFAP1;SSC5D;NDRG2;MLF1;KIAA1755;GJA1;DES;PLN;NOV;ZNF219;
CCDC121;GFM1;ZCCHC10;CWC27;NAT14;GJA1;CCDC91;RSRC1;FAM174B;DLEU1;SPEG;FAM3C;CD34;SK
CCDC121;GFM1;ZCCHC10;SLC4A1AP;TOMM40;CEP85L;SLC35F1;CWC27;PPM1G;DCAF16;GJA1;PLN;CC
MYH6;MYH7
GCKR;SERPINA1;MTMR3;CMTM5;SKIDA1;AFAP1;VSTM2L;NDRG2;MLF1;TMEM253;KIAA1755;GJA1;PLN
CCDC121;GFM1;ZCCHC10;LXN;SLC35F1;UQCR10;CWC27;NAT14;GJA1;CCDC91;FAM174B;MFN1;FAM3
PLN;SHOX2;ZMAT5;EIF3E;LCORL
NCAPG;MYH6;MYH7
ZMAT5;UQCR10;NIPSNAP1;TMEM253
SERPINA1;CR1;PLN;B3GNT7;FAM174B;CMTM5;SLC35F1;NDRG2;CCDC141;MYH6
IFI27L2;CCDC121;GFM1;ZCCHC10;SLC4A1AP;TOMM40;UQCR10;CWC27;NAT14;PPM1G;DCAF16;GJA1;
CENPW;PLN;ZMAT5;GPR126;WNT16;CCDC141
SLC5A6;CCDC121;SLC4A1AP;ZMAT5;C2ORF16
GFM1;TOMM40;UQCR10;CWC27;NAT14;GJA1;CCDC91;ZNF628;FAM174B;DLEU1;SPEG;ALX4;FAM3C;A
CENPW;LXN;ZNF628;MFN1;NRBP1;TTN
SHOX2;CEP85L;PTEN
IFI27L2;GFM1;MTMR3;ZCCHC10;SLC4A1AP;TOMM40;HSPA4;CEP85L;PTEN;UQCR10;CWC27;GPN1;NRB
NCAPG;MYH6;MYH7
GCKR;APOC1;NCAPG
IFI27L2;CCDC121;GFM1;ZCCHC10;SLC4A1AP;SLC35F1;CWC27;NAT14;GJA1;CCDC91;RSRC1;FAM174B;D
RSPO2;EIF3E;APOE
GCKR;AFAP1
EIF2B4;DNAJC1;CENPW;SNX17;RSRC1;FAM174B;ALX4;C2ORF16;PVRL2;NRBP1;WARS2
TBX15;GFM1;UQCR10;CCDC141;MYH6;MYH7;TTN
SLC5A6;DNAJC1;GCKR;SERPINA1;APOC1;GPR126;APOE;PVRL2
TBX15;GFM1;DES;ZCCHC10;SLC4A1AP;CCDC91;NDRG2;MYH7;TTN
MLF1;PPM1G
SPEG;NIPSNAP1
DES;PLN;TTN
GJA1;SERPINA1;ZNF512;NDRG2
ALX4
GJA1;PTEN
MYH7
EIF2B4;DLEU1;NDRG2
AFAP1
GJA1

PVRL2
EIF2B4;CCDC91
CENPW
IFI27L2;GFM1;ZCCHC10;SLC4A1AP;TOMM40;SLC35F1;UQCR10;CWC27;NDRG2;NAT14;PPM1G;DCAF16
CENPW
CCDC91;MFN1;NDRG2
EIF2B4;DES;NDRG2;MLF1
SKIDA1;GPR126;HORMAD2;NDRG2
LXN;APOC1;EIF3E;TRIB2
PLN;APOE;MYH6;TTN
TBX15;CABP7;LCORL;TRIB2
ASCC2
SERPINA1;APOC1
APOC1;EIF3E;APOE
ZNF628;ZNF513
CR1L;APOE;NDRG2
EIF3E;UQCR10;NIPSNAP1
SLC5A6;GJA1;CD34
PLN;UQCR10;TTN
NOV;SHOX2;RSPO2
EIF2B4;MLLT10;APOE;MYH6
GCKR;NCAPG;MLF1
CMTM5;ASCC2;NRBP1;PPM1G
GJA1;EIF3E;APOE
DES;PLN;SPEG
SERPINA1;CR1;MLF1
SERPINA1;APOC1;APOE
UQCR10;MYH7
GJA1;CR1;TTN
SNX17;NDRG2;TRIB2;PPM1G
IFI27L2;APOC1;ZNF512;CCDC141
DES;PLN;TTN
APOE;NDRG2;NRBP1
DES;PLN;UQCR10
DNAJC1;GJA1;EIF3E
SERPINA1;APOC1;APOE;MYH6
FAM174B;EIF3E;TRIB2
LXN;HSPA4;APOE
IFI27L2;UQCR10;TRIB2
SERPINA1;APOC1;NIPSNAP1
NOV;SNX17;DLEU1
LXN;SHOX2;APOE;CD34
GJA1;APOE;CD34;MYH7
GJA1;APOC1;APOE
ZCCHC10;TRIB2;MLF1

CENPW;LXN;NCAPG
GJA1;HSPA4;CR1L;GPN1
NOV;SNX17;NDRG2
PLN;APOE;NDRG2
NCAPG;ZNF513;NAT14
APOE;NDRG2;CD34
EIF3E;APOE;MYH7
EIF3E;UQCR10;TRIB2
GJA1;SNX17;NDRG2;MYH7
APOC1;APOE;MYH6
DES;SERPINA1;GPR126
LXN;CCDC91;ASCC2
GJA1;NCAPG;NDRG2;MLF1
UQCR10;APOE;GPN1
NOV;ASCC2;ARHGEF40
CD34
SLC35F1
LCORL
APOE
APOE
APOE
APOE
CR1
NIPSNAP1
EIF2B4
IFI27L2;MTMR3;LXN;SLC4A1AP;CEP85L;AFAP1;NCAPG;CWC27;GPN1;MLF1;TTN;DCAF16;NOV;MFN1;G[illegible]
MLLT10;CR1;LXN;DLEU1;NCAPG;ASCC2;EIF3E;LCORL;TTN
EIF2B4;MTMR3;SLC4A1AP;CMTM5;CWC27;MLF1;TTN;PPM1G;DCAF16;MFN1;SPEG;MYH6;MYH7;DGKH
CCDC121;GFM1;ZCCHC10;LXN;SLC4A1AP;TOMM40;SLC35F1;UQCR10;CWC27;GJA1;CCDC91;RSRC1;FA[illegible]
WNT16;MYH7
GCKR;SERPINA1;CMTM5;SKIDA1;AFAP1;VSTM2L;SSC5D;NDRG2;TMEM253;KIAA1755;GJA1;NOV;ZNF21[illegible]
GCKR;CMTM5;SKIDA1;AFAP1;SSC5D;NDRG2;KIAA1755;GJA1;PLN;NOV;ZNF219;GPR126;SPEG;APOE;AR[illegible]
CCDC121;GFM1;ZCCHC10;TOMM40;CEP85L;CWC27;NDRG2;NAT14;DCAF16;GJA1;PLN;CCDC91;RSRC1;[illegible]
CENPW;DES;ZNF628;PTEN;ASCC2;EIF3E;NRBP1;TTN
NOV;DLEU1;VSTM2L;ALX4;SLC35F1
DLEU1;VSTM2L;NCAPG;MYH6;MYH7
MFN1;PTEN;NCAPG;ASCC2;LCORL;NRBP1;TTN
TBX15;MYH6;MYH7
IFI27L2;MTMR3;SLC4A1AP;CMTM5;SKIDA1;AFAP1;NCAPG;CWC27;GPN1;MLF1;TTN;GJA1;NOV;ZMAT5;
ZNF628;NRBP1;WARS2
IFI27L2;SERPINA1;LXN;SKIDA1;PTEN;AFAP1;C2ORF16;NAT14;NIPSNAP1;SUPT7L;TMEM253;DCAF16;B3[illegible]
HSPA4;MYH6
EIF2B4;GCKR;MTMR3;ZCCHC10;TOMM40;SHOX2;PTEN;NCAPG;GPN1;NRBP1;SUPT7L;TTN;PPM1G;TBX1[illegible]
CCDC121;GFM1;ZCCHC10;SLC4A1AP;TOMM40;SHOX2;UQCR10;CWC27;NAT14;DCAF16;GJA1;CCDC91;[illegible]
CCDC121;GFM1;ZCCHC10;SLC4A1AP;TOMM40;CEP85L;SLC35F1;CWC27;TTN;DCAF16;GJA1;PLN;CCDC9[illegible]

IFI27L2;CCDC121;GFM1;ZCCHC10;TOMM40;CEP85L;NCAPG;UQCR10;CWC27;NAT14;PPM1G;DCAF16;G
CEP85L;MYH6;MYH7
ALX4;MYH6
GCKR;SERPINA1;CMTM5;AFAP1;VSTM2L;NDRG2;TMEM253;KIAA1755;GJA1;PLN;NOV;ZNF219;GPR126;
CCDC121;GFM1;ZCCHC10;SLC4A1AP;TOMM40;DCAF16;GJA1;CCDC91;RSRC1;FAM174B;MFN1;SPEG;AL
WNT16;MYH6;MYH7
ALX4;MYH6;MYH7
SHOX2;CEP85L;ALX4;ARHGEF40
DNAJC1;DES;SERPINA1;CR1;SHOX2;DLEU1;NCAPG;ASCC2;EIF3E;ARHGEF40;PVRL2
CENPW;LXN;ZNF628;SNX17;MFN1;CCDC141;NRBP1;TTN;PPM1G
CCDC121;GFM1;ZCCHC10;SLC4A1AP;SLC35F1;CWC27;NAT14;GJA1;CCDC91;RSRC1;DLEU1;SPEG;FAM3C
CENPW;SHOX2;PPM1G
CCDC121;GFM1;TOMM40;SLC35F1;NAT14;DCAF16;CCDC91;FAM174B;MFN1;SPEG;ALX4;FAM3C;EIF2B4
GFM1;ZCCHC10;TOMM40;SLC35F1;UQCR10;CWC27;NAT14;DCAF16;GJA1;CCDC91;RSRC1;DLEU1;SPEG;
CEP85L;ALX4;WNT16;ARHGEF40;MYH6;MYH7
CCDC121;GFM1;ZCCHC10;SLC4A1AP;SHOX2;CEP85L;SLC35F1;UQCR10;CWC27;NAT14;GJA1;CCDC91;RS
EIF2B4;CENPW;MTMR3;LXN;TOMM40;PTEN;GPN1;NRBP1;SUPT7L;WARS2;TTN;PPM1G;SLC5A6;ZNF628
GCKR;MTMR3;SKIDA1;AFAP1;VSTM2L;SSC5D;NDRG2;MLF1;KIAA1755;GJA1;NOV;ZNF219;GNB4;GPR126
TBX15;MYH7
SHOX2;ALX4;WNT16
CCDC121;GFM1;SLC4A1AP;ZMAT5;ALX4;GPN1;TMEM253
IFI27L2;CCDC121;ZCCHC10;LXN;SLC4A1AP;CEP85L;PTEN;AFAP1;CWC27;GPN1;MLF1;NRBP1;TTN;DCAF1
GFM1;SKIDA1;SHOX2;AFAP1;VSTM2L;SLC35F1;SSC5D;MLF1;TBX15;MLLT10;KIAA1755;GJA1;PLN;FAM17
TBX15;MYH6;MYH7
GCKR;SERPINA1;AFAP1;VSTM2L;NDRG2;GJA1;PLN;NOV;ZNF219;GPR126;SPEG;ALX4;APOE;ARHGEF40;F
TBX15;ALX4;MYH6
TBX15;CR1;SHOX2
CCDC121;GFM1;ZCCHC10;LXN;SLC4A1AP;TOMM40;SLC35F1;UQCR10;CWC27;NAT14;DCAF16;GJA1;PLN
WNT16;MYH7
CCDC121;GFM1;ZCCHC10;LXN;SLC4A1AP;TOMM40;SHOX2;CEP85L;SLC35F1;UQCR10;CWC27;TTN;DCAI
CCDC121;GFM1;ZCCHC10;SLC4A1AP;SLC35F1;CWC27;NAT14;GJA1;CCDC91;RSRC1;DLEU1;SPEG;FAM3C
AFAP1;NCAPG
SERPINA1
ZNF219
PTEN
APOE
GJA1;APOC1;MFN1;GNB4;PTEN;RSPO2;APOE;WNT16;ARHGEF40;DGKH
SUPT7L
DCAF16
PLN;NDRG2;MYH7
CABP7;RSPO2;WARS2
DES;PLN;APOE
GJA1;AFAP1;MYH6
MYH7
RSRC1

APOE;TTN
GCKR;B3GNT7;FAM174B;CEP85L;C2ORF16;PVRL2;TRIB2
IFI27L2;CCDC121;CABP7;CMTM5;WNT16;NDRG2;MLF1;KIAA1755;GJA1;CCDC91;RSRC1;RSPO2;ALX4;LC
PLN;UQCR10;CCDC141;MLF1;MYH6;MYH7;TTN
CCDC121;SLC4A1AP;CMTM5;CEP85L;PTEN;SLC35F1;C2ORF16;KIAA1755;DCAF16;GPR126;FAM3C;ARHG
ZCCHC10;LXN;PLN;RSRC1;ZMAT5;MFN1;PTEN;NCAPG;MLF1;SUPT7L
TRIB2
DCAF16
TRIB2
NOV
GNB4
GNB4
SLC5A6
GJA1
APOE
GJA1;APOE;NDRG2
CMTM5;APOE;NDRG2
APOC1;APOE;CD34
SNX17;EIF3E;APOE
EIF2B4;LXN;NDRG2
DCAF16;RSRC1;APOC1
ZCCHC10;FAM3C;TRIB2
KIAA1755;ZCCHC10
GJA1;DES;EIF3E
DES;PLN;NOV
GJA1;SERPINA1;APOE
GJA1;DES;NDRG2
CMTM5;ASCC2
CMTM5;APOE;NDRG2
APOC1;APOE;NDRG2
GCKR;GPR126;MYH7
IFI27L2;GJA1;EIF3E
APOC1;APOE
LXN;CABP7;SKIDA1
CMTM5;ASCC2
EIF3E;UQCR10;FAM3C;MLF1
FAM174B;MLF1;TTN
GJA1;SERPINA1;NIPSNAP1
GJA1;DES;EIF3E
GJA1;UQCR10;APOE
CENPW;PLN;MFN1;PVRL2
GJA1;DLEU1;EIF3E;MLF1
GJA1;APOC1;UQCR10
UQCR10;NDRG2;MLF1
GJA1;SNX17;APOE

GJA1;LXN;EIF3E
GJA1;APOC1;APOE
SERPINA1;DLEU1;CD34
CENPW;LXN;UQCR10
DES;MYH6;MYH7
EIF3E;MYH6;MYH7
UQCR10;MYH6;MYH7
HSPA4;UQCR10;NIPSNAP1
GJA1;DES;PLN
EIF3E;UQCR10;APOE
GJA1;APOE;NDRG2
HSPA4;APOE;NDRG2;CD34;TTN
MFN1;SPEG;NDRG2;MLF1;TTN
SLC4A1AP;SKIDA1;PTEN;EIF3E;TTN
DES;PTEN;NDRG2;ARHGEF40;CD34
MLLT10;HSPA4;SPEG;APOE;NDRG2
TBX15;NOV;SHOX2;FAM3C;CD34
HSPA4;NCAPG;EIF3E;MYH6;MYH7
GCKR;APOC1;APOE;NDRG2;NIPSNAP1
GCKR;APOC1;EIF3E;NDRG2;NIPSNAP1
GCKR;APOC1;EIF3E;APOE;NDRG2
GCKR;APOC1;APOE;NDRG2;NIPSNAP1
TOMM40;MFN1;ASCC2;FAM3C;NRBP1
DNAJC1;SPEG;EIF3E;APOE;NIPSNAP1
GNB4;AFAP1;SLC35F1;APOE;NDRG2
TOMM40;HSPA4;EIF3E;TRIB2;PPM1G
DES;HSPA4;CCDC141;MYH7;TTN
NOV;ZNF219;PTEN;SLC35F1;EIF3E
GJA1;GNB4;RSPO2;EIF3E;ARHGEF40
HSPA4;EIF3E;FAM3C;NIPSNAP1;PPM1G
GNB4;PTEN;APOE;NDRG2;TRIB2
DES;UQCR10;NDRG2;MLF1;TTN
GJA1;DES;PTEN;APOE;TTN
NCAPG;EIF3E;FAM3C;DGKH;TTN
GFM1;CR1L;PTEN;EIF3E;TRIB2
GJA1;NOV;PTEN;RSPO2;EIF3E
APOC1;PTEN;APOE;NDRG2;NIPSNAP1
GJA1;PTEN;SLC35F1;FAM3C;NDRG2
GJA1;HSPA4;CMTM5;NDRG2;CD34
LXN;VSTM2L;UQCR10;APOE;NDRG2
NOV;CABP7;APOE;CD34;DGKH
SLC5A6;B3GNT7;EIF3E;TTN;PPM1G
B3GNT7;AFAP1;FAM3C;TRIB2;PPM1G
DES;HSPA4;PTEN;MYH6;MYH7
NOV;SPEG;LCORL;CD34;MYH7

GCKR;APOC1;APOE;NDRG2;NIPSNAP1
GCKR;APOC1;APOE;NDRG2;NIPSNAP1
GCKR;APOC1;APOE;NDRG2;NIPSNAP1
PLN;NDRG2;MLF1;MYH6;TTN
SERPINA1;LXN;B3GNT7;UQCR10;NDRG2
HSPA4;MFN1;AFAP1;EIF3E;TTN
GFM1;HSPA4;PTEN;NCAPG;EIF3E
GFM1;HSPA4;PTEN;NCAPG;EIF3E
GJA1;SERPINA1;FAM174B;VSTM2L;TRIB2
FAM174B;APOC1;MLF1;MYH6;MYH7
HSPA4;MFN1;AFAP1;EIF3E;TTN
GJA1;APOC1;PTEN;EIF3E;ARHGEF40
TOMM40;HSPA4;EIF3E;FAM3C;PPM1G
SLC5A6;TOMM40;B3GNT7;EIF3E;TTN
GJA1;HSPA4;APOC1;EIF3E;APOE
EIF2B4;TOMM40;HSPA4;ALX4;PPM1G
SLC5A6;TOMM40;HSPA4;EIF3E;PPM1G
HSPA4;PTEN;APOE;LCORL;TRIB2
SLC5A6;MLLT10;GJA1;EIF3E;PPM1G
DES;UQCR10;NDRG2;MYH7;TTN
GJA1;APOC1;CCDC141;MYH6;MYH7
IFI27L2;DNAJC1;PTEN;EIF3E;NIPSNAP1
GJA1;DES;APOE;CD34;TTN
DES;PLN;HSPA4;APOE;NIPSNAP1
GJA1;HSPA4;VSTM2L;APOE;MLF1
TBX15;GJA1;DES;EIF3E;CD34
HSPA4;GNB4;PTEN;EIF3E;CD34
DES;PLN;NDRG2;MYH6
HSPA4
FAM174B;UQCR10;MLF1;MYH6
LXN;RSPO2;EIF3E;CD34
DNAJC1;DLEU1;TRIB2;NRBP1;PPM1G
MFN1;UQCR10;MLF1;MYH7
SUPT7L
NOV;SHOX2
UQCR10;APOE;MYH6
SLC5A6;EIF3E;APOE
LXN;APOC1;APOE
APOC1;APOE;NDRG2
SERPINA1;APOC1
NDRG2;MYH7
GJA1;CMTM5;APOE
EIF3E;MLF1;MYH7
CEP85L;MLF1
HSPA4;PTEN;CD34

LXN;NOV;NDRG2
DES;EIF3E;UQCR10;NDRG2
DES;NOV;NDRG2
SERPINA1;CR1;TRIB2
ZCCHC10;CMTM5;SKIDA1;SHOX2;CEP85L;SLC35F1;CWC27;KIAA1755;DCAF16;MFN1;GPR126;ZNF512;A
APOE;NDRG2;CD34;TRIB2
GJA1;NDRG2;MYH7;TTN
FAM174B;UQCR10;MLF1;MYH6
CENPW;CR1;CR1L;ARHGEF40
DES;TOMM40;MYH7
DNAJC1;GJA1;EIF3E
TBX15;GFM1;DES;UQCR10;MYH6;MYH7;TTN
TBX15;HSPA4;MLF1
HORMAD2
IFI27L2;MTMR3;LXN;SLC4A1AP;SHOX2;CEP85L;SLC35F1;CWC27;TTN;DCAF16;GJA1;PLN;CCDC91;RSRC1
TBX15;CEP85L;CCDC141;MYH6;MYH7
IFI27L2;ALX4;TTN
GJA1;NOV;SHOX2;MYH7;DGKH
CCDC121;GFM1;ZCCHC10;LXN;SLC4A1AP;TOMM40;SHOX2;CEP85L;SLC35F1;UQCR10;CWC27;NAT14;PI
DES;PLN;ALX4;NDRG2;MYH7
NOV;RSPO2;NIPSNAP1
LXN;FAM3C;APOE;ARHGEF40
APOC1;EIF3E;FAM3C;APOE
SNX17;UQCR10;APOE;NDRG2
GJA1;SLC4A1AP;PTEN;FAM3C
EIF2B4;DES;UQCR10;NDRG2
DES;UQCR10;APOE;MYH7
SNX17;FAM174B;MFN1;MLF1;TTN
GJA1;UQCR10;NDRG2;MYH6
B3GNT7;CABP7;CCDC141
NOV;RSPO2;MYH7;TTN
GJA1;APOC1;FAM3C;APOE
GJA1;LXN;EIF3E;APOE
PLN;SNX17;FAM174B;MYH6
LXN;SNX17;APOC1;APOE
GJA1;NCAPG;APOE;TRIB2;MLF1
CMTM5;UQCR10;APOE;CD34
HSPA4;NCAPG;EIF3E;PVRL2
DES;FAM174B;APOE;MLF1;MYH7
FAM174B;AFAP1;HORMAD2;CD34;MLF1
SNX17;EIF3E;UQCR10;NIPSNAP1
GJA1;CENPW;HSPA4;EIF3E
LXN;APOE;NDRG2;DGKH
APOC1;UQCR10;APOE;MYH7
GJA1;SKIDA1

PLN;NOV;MYH6;TTN
GJA1;EIF3E;FAM3C;APOE
EIF2B4;SNX17;PTEN;EIF3E
UQCR10;APOE;GPN1;PPM1G
CCDC121;LXN;SHOX2;FAM3C
TBX15;SNX17;APOC1;MYH6
DES;UQCR10;APOE
GJA1;LXN;SNX17;EIF3E
LXN;APOE;NDRG2;DGKH
SLC5A6;GJA1;EIF3E;NIPSNAP1
SNX17;FAM174B;MFN1;MYH6
SLC5A6;SLC4A1AP;SNX17;PPM1G
TBX15;GJA1;NOV;FAM3C
NOV;APOC1;NDRG2
DES;NDRG2;MYH7;TTN
DCAF16;NOV;APOC1;NCAPG
DCAF16;NOV;APOC1;NCAPG
APOC1;UQCR10;APOE;NIPSNAP1
GCKR;GPR126;CD34;WARS2
GJA1;SNX17;EIF3E;APOE
CENPW;LXN;CCDC91;FAM174B
APOE;MYH6;MYH7
SNX17;UQCR10;MLF1;MYH6;MYH7
GCKR;TOMM40;APOC1;WNT16
GCKR;RSPO2;SLC35F1;HORMAD2
TBX15;SKIDA1;HORMAD2;CD34;MLF1
GJA1;LXN;AFAP1;EIF3E
PLN;MYH6;MYH7;TTN
GJA1;PTEN;UQCR10;NDRG2
GCKR;GJA1;CMTM5;NDRG2
SERPINA1;DLEU1;AFAP1
GJA1;DES;PLN;CD34
GJA1;PTEN;FAM3C;TRIB2
NOV;RSPO2;GPR126;NIPSNAP1
GJA1;SNX17;APOC1;APOE
GCKR;GJA1;APOE;MLF1
GNB4;WNT16;NDRG2;CD34
GFM1;TOMM40;VSTM2L
LXN;SNX17;GNB4;APOE;TRIB2
DES;TOMM40;MYH6;MYH7
SHOX2;NCAPG;NDRG2;LCORL;WARS2
CR1L;CMTM5;DGKH
LXN
NCAPG
EIF2B4

CEP85L
SLC4A1AP
CCDC121;GFM1;ZCCHC10;TOMM40;CEP85L;SLC35F1;UQCR10;CWC27;NAT14;DCAF16;GJA1;CCDC91;RS
FAM174B;TRIB2
SLC5A6;IFI27L2;GJA1;LXN;TOMM40;AFAP1;SPEG;NDRG2;NIPSNAP1
TBX15;GFM1;DES;ZNF628;CCDC141;NRBP1;MYH7;TTN;PPM1G
DNAJC1;MTMR3;SHOX2;ZMAT5;GNB4;NCAPG;ASCC2;ARHGEF40;CCDC141;PVRL2;NRBP1;TMEM253
MYH6;MYH7
CCDC121;GFM1;ZCCHC10;SLC4A1AP;TOMM40;CEP85L;SLC35F1;UQCR10;CWC27;NDRG2;DCAF16;GJA1
TBX15;CEP85L;ARHGEF40;CCDC141;MYH6
SHOX2;ALX4;MLF1
UQCR10;MYH6;MYH7
CCDC121;GFM1;ZCCHC10;SLC4A1AP;CWC27;NAT14;GJA1;CCDC91;RSRC1;FAM174B;SPEG;FAM3C;CD3
LCORL;TMEM253
CCDC121;GFM1;ZCCHC10;LXN;SLC4A1AP;TOMM40;SHOX2;SLC35F1;UQCR10;CWC27;NAT14;PPM1G;D
APOC1;MYH6;MYH7
GCKR;ZCCHC10;ALX4
CCDC141;MYH6;MYH7
CCDC121;GFM1;ZCCHC10;LXN;SLC4A1AP;SHOX2;SLC35F1;CWC27;DCAF16;GJA1;CCDC91;RSRC1;FAM17
CMTM5;CEP85L;ALX4;ARHGEF40
IFI27L2;MTMR3;LXN;SLC4A1AP;SHOX2;AFAP1;SLC35F1;CWC27;GPN1;MLF1;NRBP1;WARS2;PPM1G;DC
EIF2B4;CENPW;MTMR3;SLC4A1AP;TOMM40;PTEN;NIPSNAP1;NRBP1;TMEM253;WARS2;PPM1G;DCAF1
CMTM5;SKIDA1;AFAP1;NDRG2;KIAA1755;GJA1;PLN;NOV;ZNF219;GPR126;SPEG;APOE;ARHGEF40;PVRL
CCDC121;GFM1;ZCCHC10;SLC4A1AP;SLC35F1;CWC27;NAT14;GJA1;CCDC91;RSRC1;FAM174B;DLEU1;SP
TBX15;SHOX2;MYH7
SHOX2;MYH7
TMEM253;MYH7
MYH6;MYH7
IFI27L2;CCDC121;GFM1;ZCCHC10;LXN;TOMM40;SHOX2;CEP85L;UQCR10;CWC27;PPM1G;DCAF16;GJA1
DLEU1;MYH7
ZMAT5;GNB4;ALX4;MLF1
CCDC121;GFM1;ZCCHC10;LXN;SLC4A1AP;TOMM40;SLC35F1;UQCR10;CWC27;NAT14;GJA1;CCDC91;RS
CCDC91;C2ORF16;CCDC141
IFI27L2;CENPW;MTMR3;ZCCHC10;LXN;SLC4A1AP;CMTM5;CEP85L;AFAP1;SLC35F1;CWC27;GPN1;MLF1;
GFM1;ALX4;WNT16;MYH6;MYH7
CCDC121;GFM1;ZCCHC10;TOMM40;SLC35F1;CWC27;NDRG2;DCAF16;GJA1;CCDC91;RSRC1;MFN1;DLE
SHOX2;WNT16;MYH7
TBX15;MYH6;MYH7
ZMAT5;ALX4;UQCR10
MYH6;MYH7
MYH6;MYH7
IFI27L2;CCDC121;GFM1;ZCCHC10;SLC4A1AP;PTEN;SLC35F1;CWC27;NAT14;GJA1;CCDC91;RSRC1;FAM1
TBX15;CEP85L;ALX4;ARHGEF40;MLF1
GFM1;ZCCHC10;SLC4A1AP;TOMM40;CWC27;NDRG2;NAT14;DCAF16;GJA1;CCDC91;RSRC1;FAM174B;M
ZMAT5

SERPINA1;MTMR3;CR1;CMTM5;SLC35F1;NDRG2;TTN;DES;PLN;B3GNT7;FAM174B;CCDC141;CD34
CEP85L;ALX4;MYH7
DES;CR1;PLN;B3GNT7;FAM174B;CMTM5;SLC35F1;NDRG2;CCDC141;CD34;MYH6;MYH7
CEP85L;ALX4;ARHGEF40;TTN
MLLT10;TOMM40;ZNF628;SNX17;MFN1;PTEN;ASCC2;WNT16;GPN1;NRBP1;SUPT7L;PPM1G
DLEU1;VSTM2L;NIPSNAP1
DLEU1;VSTM2L
CCDC141;MYH7
MYH6;MYH7
GCKR;CMTM5;SKIDA1;SSC5D;NDRG2;TMEM253;KIAA1755;GJA1;PLN;ZNF219;GPR126;SPEG;APOE;ARH
GCKR;MTMR3;SKIDA1;SSC5D;NDRG2;KIAA1755;GJA1;PLN;NOV;ZNF219;GPR126;SPEG;APOE;ARHGEF4
SHOX2
GFM1;TOMM40;SLC35F1;CWC27;NDRG2;NAT14;PPM1G;DCAF16;GJA1;CCDC91;ZNF628;FAM174B;DLE
SERPINA1;MYH6;MYH7
HSPA4;UQCR10;GPN1;MYH7
CCDC121;GFM1;ZCCHC10;SLC4A1AP;SLC35F1;CWC27;NAT14;GJA1;CCDC91;RSRC1;FAM174B;DLEU1;SP
IFI27L2;GFM1;ZCCHC10;TOMM40;SLC35F1;UQCR10;CWC27;NAT14;DCAF16;GJA1;CCDC91;RSRC1;FAM
CCDC121;GFM1;ZCCHC10;TOMM40;CWC27;NDRG2;PPM1G;DCAF16;GJA1;CCDC91;RSRC1;FAM174B;M
GCKR;SERPINA1;CMTM5;AFAP1;SSC5D;NDRG2;KIAA1755;GJA1;NOV;ZNF219;GPR126;SPEG;ALX4;ARHG
SPEG;ALX4;WNT16;MYH6;MYH7
GCKR;SERPINA1;MTMR3;CMTM5;NDRG2;KIAA1755;GJA1;NOV;APOC1;ZNF219;GPR126;SPEG;ALX4;APC
LXN;SHOX2;MFN1;EIF3E;CCDC141;NRBP1;DGKH;TTN
IFI27L2;CCDC121;ZCCHC10;LXN;SLC4A1AP;CEP85L;CWC27;TTN;DCAF16;CCDC91;RSRC1;MFN1;DLEU1;S
CCDC121;GFM1;ZCCHC10;LXN;SLC4A1AP;TOMM40;CEP85L;SLC35F1;UQCR10;CWC27;NAT14;DCAF16;G
IFI27L2;GFM1;ZCCHC10;SLC4A1AP;SLC35F1;CWC27;NAT14;GJA1;CCDC91;RSRC1;DLEU1;SPEG;FAM3C;C
FAM174B;ALX4;MYH6;MYH7
MTMR3;CR1;CMTM5;SLC35F1;NDRG2;TTN;DES;PLN;B3GNT7;FAM174B;CCDC141;CD34;MYH6;MYH7
ZCCHC10;CCDC91;CEP85L;C2ORF16;ZNF512;SUPT7L;TTN
CCDC121;GFM1;ZCCHC10;LXN;SLC4A1AP;TOMM40;SLC35F1;UQCR10;CWC27;GJA1;CCDC91;RSRC1;FAN
CCDC121;GFM1;ZCCHC10;TOMM40;SLC35F1;CWC27;DCAF16;GJA1;CCDC91;RSRC1;FAM174B;MFN1;DL
DLEU1;LCORL
MYH6;MYH7
GFM1;TOMM40;SLC35F1;CWC27;DCAF16;GJA1;CCDC91;FAM174B;MFN1;SPEG;ALX4;FAM3C;APOE;CD
CMTM5;CEP85L;ARHGEF40;MYH6;MYH7
GFM1;ZCCHC10;TOMM40;CWC27;NDRG2;PPM1G;DCAF16;GJA1;CCDC91;RSRC1;DLEU1;SPEG;FAM3C;A
VSTM2L;UQCR10
ZCCHC10;SLC4A1AP;HSPA4;GPN1
GFM1;ZCCHC10;SLC4A1AP;TOMM40;HSPA4;CEP85L;PTEN;UQCR10;CWC27;GPN1;NRBP1;SUPT7L;PPM
NCAPG;GPN1;MYH7
TBX15;MYH6;MYH7
GFM1;ZCCHC10;SLC4A1AP;TOMM40;SLC35F1;UQCR10;CWC27;NDRG2;PPM1G;DCAF16;GJA1;PLN;CCD
GCKR;CMTM5;AFAP1;VSTM2L;SSC5D;NDRG2;KIAA1755;GJA1;NOV;ZNF219;GPR126;SPEG;ALX4;ARHGE
NCAPG;ALX4;MYH6;MYH7
MYH6;MYH7
VSTM2L;WNT16

SHOX2;ALX4
GCKR;APOC1
DLEU1;LCORL
PLN;CEP85L;VSTM2L;CCDC141;TTN
DNAJC1;ZNF628;NCAPG;ASCC2;NRBP1;TTN;PPM1G
CCDC121;GFM1;ZCCHC10;SLC4A1AP;SLC35F1;CWC27;NAT14;GJA1;CCDC91;RSRC1;FAM174B;DLEU1;SP
MYH6;MYH7
CENPW;SHOX2;APOC1;ALX4;APOE
CCDC121;GFM1;ZCCHC10;TOMM40;SLC35F1;UQCR10;CWC27;DCAF16;GJA1;CCDC91;RSRC1;FAM174B;
CCDC121;GFM1;ZCCHC10;SLC4A1AP;SLC35F1;CWC27;NAT14;GJA1;CCDC91;RSRC1;FAM174B;DLEU1;SP
CCDC121;GFM1;ZCCHC10;HSPA4;SKIDA1;AFAP1;VSTM2L;SLC35F1;SSC5D;C2ORF16;CWC27;MLF1;SLC5
ARHGEF40;CCDC141;MYH7
DLEU1;MYH7
APOC1;GNB4
APOE
GFM1;DES;NDRG2;CCDC141;MLF1;MYH6;MYH7;TTN
HSPA4
TRIB2
SLC4A1AP
PLN;RSRC1;SHOX2;SPEG;EIF3E
DES;NCAPG;SLC35F1;APOE
TRIB2
APOE
LXN;FAM3C;NDRG2;NIPSNAP1;PPM1G
TBX15;WNT16;CCDC141;TTN
UQCR10;NDRG2;MLF1;MYH7
GJA1;UQCR10;CD34;MLF1
SLC5A6;CCDC91;CABP7;FAM3C
GJA1;NOV;APOE;NDRG2
SERPINA1;LXN;EIF3E
MYH6;MYH7
ALX4;LCORL;MYH6;MYH7
ZMAT5;UQCR10;WNT16
MYH6;TMEM253;MYH7
NCAPG;MYH6;MYH7
PTEN;TTN
APOC1;APOE
GJA1;LCORL;GPN1
GJA1;UQCR10;APOE
APOC1;APOE;NIPSNAP1
FAM174B;NDRG2;TTN
APOC1;DLEU1;TRIB2
CR1;LXN;FAM3C
GCKR;SERPINA1;APOC1
CCDC141

CR1;APOC1;ZMAT5;DLEU1;UQCR10;APOE;WNT16;NAT14;NIPSNAP1
APOC1;MYH6
EIF2B4;ZNF628;SNX17;GPN1;NRBP1;TTN;PPM1G
GCKR;MTMR3;AFAP1;VSTM2L;NDRG2;KIAA1755;GJA1;PLN;NOV;ZNF219;SPEG;ALX4;APOE;ARHGEF40;F
DES;WNT16;MYH6;MYH7;DGKH
CENPW;MTMR3;ZNF628;EIF3E;NRBP1;DGKH;PPM1G
GCKR;MYH6;TMEM253;MYH7
SERPINA1;DLEU1;VSTM2L;WNT16
CCDC121;GFM1;ZCCHC10;TOMM40;UQCR10;CWC27;NDRG2;NAT14;PPM1G;DCAF16;GJA1;CCDC91;ZN
HSPA4;VSTM2L;NCAPG;ALX4
PTEN;ALX4;CWC27
LCORL
GCKR;CMTM5;SKIDA1;AFAP1;SSC5D;NDRG2;TMEM253;KIAA1755;GJA1;NOV;ZNF219;GPR126;SPEG;AR
IFI27L2;CCDC121;MTMR3;LXN;SLC4A1AP;CEP85L;PTEN;SLC35F1;CWC27;TTN;DCAF16;CCDC91;SPEG;CC
CCDC121;GFM1;ZCCHC10;SLC4A1AP;SLC35F1;CWC27;NAT14;GJA1;CCDC91;RSRC1;FAM174B;DLEU1;SP
APOC1;UQCR10
DNAJC1;DES;LXN;PLN;SNX17;SHOX2;GPR126;ASCC2;GPN1;WARS2;TTN;PPM1G
GJA1;CENPW;NCAPG;MLF1
MFN1;NDRG2;MLF1;MYH7
IFI27L2;MTMR3;SLC4A1AP;HSPA4;AFAP1;NCAPG;CWC27;GPN1;MLF1;NRBP1;PPM1G;CCDC91;NOV;RSF
ZMAT5;DLEU1;VSTM2L;UQCR10
TBX15;ALX4
TBX15;GFM1;UQCR10;CCDC141;MYH6;MYH7;TTN
KIAA1755;IFI27L2;CMTM5;SHOX2;CEP85L;ZMAT5;ARHGEF40;CD34;TRIB2
MLLT10;APOE;NAT14;SUPT7L
ZCCHC10;HSPA4;MLF1
PTEN
SLC5A6;GCKR;SERPINA1;APOC1;GPR126;APOE;NIPSNAP1
IFI27L2;CENPW;TOMM40;HSPA4;AFAP1;NCAPG;CWC27;GPN1;PPM1G;GNB4;DLEU1;ALX4;EIF3E;LCORL
TBX15;GFM1;DES;SPEG;MYH7;TTN
VSTM2L;UQCR10;CCDC141;MLF1;MYH6;MYH7
MFN1;UQCR10;NDRG2;MLF1
SKIDA1;SHOX2;VSTM2L;GPR126
LXN;ZNF219;SPEG;ARHGEF40;NAT14
NOV;RSPO2;CD34
NOV;GPR126;EIF3E;TRIB2
GJA1;NOV;RSPO2;APOE
CENPW;LXN;AFAP1;NCAPG
APOC1;APOE;NDRG2;NIPSNAP1
LXN;PTEN;EIF3E;TRIB2
GJA1;DES;PLN;APOC1;TTN
FAM174B;APOE;MYH7;TTN
APOC1;APOE;NDRG2;NIPSNAP1
SNX17;APOE;NDRG2;PPM1G
CMTM5;SHOX2;VSTM2L;APOE

GJA1;NCAPG;CD34
DES;APOC1;MFN1;UQCR10
GJA1;PLN;APOE;TTN
DNAJC1;NOV;CABP7;NCAPG
SLC5A6;TBX15;NOV;EIF3E
CENPW;LXN;TOMM40;NCAPG
DES;NDRG2;CD34;TTN
NOV;APOE;CD34;MLF1
GJA1;SNX17;UQCR10;APOE
FAM174B;APOC1;ZNF219;NDRG2
NOV;RSPO2;CD34;MLF1
MFN1;NDRG2;MLF1;MYH7
GJA1;DLEU1;NCAPG
DES;APOC1;MFN1
GJA1;TOMM40;ASCC2;EIF3E
FAM174B;PTEN;FAM3C;NIPSNAP1
SLC5A6;SHOX2;APOC1;MYH7
CENPW;LXN;AFAP1;NCAPG
CENPW;LXN;TOMM40;NCAPG
GJA1;PLN;MYH6;MYH7;TTN
KIAA1755;CABP7;SHOX2;NCAPG
GCKR;APOE;NDRG2;NIPSNAP1
SNX17;APOE;MYH7;PPM1G
TBX15;NDRG2;MLF1;TTN
SLC5A6;APOC1;MLF1;TTN
LXN;NOV;APOE;NDRG2
SPEG;MLF1;MYH7;TTN
GFM1;ZCCHC10;TOMM40;CWC27;NDRG2;NAT14;DCAF16;GJA1;PLN;CCDC91;RSRC1;FAM174B;DLEU1;S
ZMAT5;UQCR10
SLC5A6;RSRC1;PTEN;ALX4;WARS2
GJA1
TBX15;GJA1;LXN;CD34
ZCCHC10;HSPA4
MLLT10;DNAJC1;LXN;RSRC1;FAM174B;MFN1;PTEN;GPR126;FAM3C;NIPSNAP1;WARS2
ALX4
GPN1
SERPINA1;CR1;CMTM5;SLC35F1;WNT16;NDRG2;KIAA1755;PLN;B3GNT7;FAM174B;CCDC141;MYH6;MY
SLC4A1AP;NCAPG
DLEU1;NCAPG;MYH6;MYH7
CCDC121;GFM1;ZCCHC10;SLC4A1AP;CWC27;NAT14;GJA1;CCDC91;RSRC1;FAM174B;SPEG;FAM3C;ARH
VSTM2L;UQCR10;LCORL;MYH6;MYH7
GFM1;ZCCHC10;TOMM40;SLC35F1;UQCR10;CWC27;NDRG2;NAT14;PPM1G;DCAF16;GJA1;CCDC91;ZNF
GCKR;CCDC121;MTMR3;AFAP1;SSC5D;NDRG2;MLF1;KIAA1755;GJA1;NOV;APOC1;ZNF219;GPR126;SPE
HSPA4;AFAP1;TRIB2
VSTM2L

GJA1;MLF1;MYH6
SERPINA1;CR1;APOC1
GJA1;LXN;EIF3E
MTMR3;LXN;TOMM40;HSPA4;CEP85L;NCAPG;UQCR10;GPN1;NIPSNAP1;NRBP1;TTN;PPM1G;SNX17;RS
GJA1
EIF3E
HSPA4
MLLT10;DES;ZNF219;RSPO2;SPEG;SSC5D;NRBP1
TBX15;DES;UQCR10;CCDC141;MYH7;TTN
DES;PLN;UQCR10;CCDC141;MYH6;MYH7;TTN
DNAJC1;GCKR;SERPINA1;B3GNT7;C2ORF16;TRIB2
DNAJC1;GCKR;SERPINA1;B3GNT7;FAM174B;NCAPG;PVRL2;TRIB2;PPM1G
TTN
SLC4A1AP;TOMM40;CEP85L;NCAPG;UQCR10;NDRG2;GPN1;NIPSNAP1;SUPT7L;TTN;PPM1G;GJA1;SNX1
TOMM40
MLLT10
PLN;CCDC91;NOV;RSRC1;MFN1;GNB4;PTEN;EIF3E
ZNF219;AFAP1;GPR126;SPEG;SSC5D;DGKH
APOE
GJA1;PLN;APOE
CR1L;APOC1;APOE
FAM174B;APOE;NDRG2;MYH6
SLC5A6;SERPINA1;NOV;TRIB2
APOE;NDRG2;MYH6;MYH7
GJA1
B3GNT7
PTEN
SERPINA1
PTEN;APOE;NDRG2;CD34
DCAF16;CENPW;NCAPG;ARHGEF40
MLLT10;PLN;NOV;RSRC1;MFN1;AFAP1;SPEG;ZNF512;DGKH
GJA1;TOMM40;EIF3E;APOE
GJA1;LXN;NDRG2
MYH6;MYH7
CCDC121;ZNF219;GNB4;RSPO2;AFAP1;SSC5D
GJA1;APOC1;APOE;NDRG2
CENPW;DES;MTMR3;TOMM40;ZNF628;SNX17;SHOX2;ASCC2;GPN1;NRBP1;TTN;PPM1G
TBX15;CEP85L;CCDC141;TTN
GCKR;SERPINA1;APOC1;APOE;WNT16
ALX4;MYH6;TTN
ZMAT5;VSTM2L;C2ORF16;UQCR10
CCDC121;MTMR3;CR1;CMTM5;NDRG2;TTN;DES;PLN;B3GNT7;ZNF219;CCDC141;CD34;MYH6;MYH7
IFI27L2;CCDC121;GFM1;ZCCHC10;SLC4A1AP;TOMM40;CEP85L;UQCR10;CWC27;NAT14;PPM1G;DCAF1
NOV;HSPA4;CMTM5;GNB4;NCAPG;ALX4;SLC35F1;MYH7
SLC5A6;GCKR;CENPW;MTMR3;ZNF628;SNX17;ZMAT5;ASCC2;GPN1;NIPSNAP1;PPM1G

CENPW;ZNF628;SHOX2;NRBP1
EIF2B4;SERPINA1;MTMR3;CMTM5;AFAP1;SSC5D;NDRG2;KIAA1755;GJA1;NOV;ZNF219;GPR126;SPEG;A
KIAA1755;GCKR;CR1;LXN;PLN;B3GNT7;FAM174B;CMTM5;SLC35F1;CCDC141;MYH6;MYH7
TBX15;ZNF628;ZNF219;ALX4
ALX4
TTN
GJA1;SERPINA1;PTEN;APOE;TRIB2
PTEN
GJA1;B3GNT7;HSPA4;PTEN;ALX4;APOE;NDRG2
HORMAD2
HORMAD2
PTEN
GNB4
UQCR10
PLN;NOV;VSTM2L
RSRC1
DLEU1;CWC27;LCORL
ZNF628
DLEU1
DNAJC1
DES;RSPO2;MLF1;TTN
SLC5A6;SNX17;NRBP1;PPM1G
PLN;MFN1;UQCR10;MLF1
GJA1;LXN;RSPO2;NDRG2
ZCCHC10;APOE;NDRG2
DES;LXN;SNX17;APOE
CCDC121;GJA1;LXN;NOV
DES;UQCR10;NDRG2;MYH7
GPR126;APOE;TRIB2
GJA1;MFN1;UQCR10;MLF1
DES;EIF3E;NDRG2;TTN
CENPW;PTEN;NDRG2
CENPW;NOV;NCAPG
GJA1;CENPW;EIF3E;APOE
PLN;UQCR10;TTN
PLN;SNX17;FAM174B;MLF1
GJA1;ZNF512;ARHGEF40;NAT14
ZCCHC10;NOV;GNB4;APOE;CD34
CEP85L
FAM3C;WNT16
DCAF16;SERPINA1;B3GNT7;PTEN;APOE
SNX17;APOE
APOE;MLF1;NIPSNAP1;WARS2
APOC1;APOE
MLF1;PPM1G

CMTM5;SLC35F1;NDRG2
UQCR10;MLF1
SLC4A1AP;TOMM40;CEP85L;UQCR10;NDRG2;NIPSNAP1;TTN;PPM1G;GJA1;SNX17;RSPO2;EIF3E;FAM3C
NDRG2
SERPINA1
EIF3E
LXN
FAM3C
GJA1
CCDC121
EIF3E
ZMAT5
CENPW
DNAJC1
GCKR
EIF3E
TBX15
RSPO2
NDRG2
NOV
NIPSNAP1
B3GNT7
UQCR10
APOE
CR1L
DLEU1
NOV
EIF3E
APOC1
CR1
UQCR10;TRIB2
TRIB2;NIPSNAP1
APOC1;UQCR10
CD34
HSPA4
SERPINA1
GPR126
GJA1
AFAP1
DLEU1
SNX17
EIF3E
TRIB2
UQCR10
LXN

CMTM5
SHOX2
FAM3C
GJA1;EIF3E
APOC1
SERPINA1
SERPINA1
UQCR10
CR1L
TRIB2
CD34
NDRG2
MLF1
B3GNT7
APOE
UQCR10
NIPSNAP1
B3GNT7
GJA1
SERPINA1
NIPSNAP1
AFAP1
EIF3E
FAM3C
ZNF219
EIF3E
UQCR10
CD34
HSPA4
GNB4
CD34
SERPINA1
MYH6
CD34
GJA1
EIF3E
MYH6
EIF3E
EIF3E
TBX15
EIF3E
CENPW;GNB4
EIF3E
GJA1
CR1L

UQCR10
NIPSNAP1
EIF3E
SKIDA1;APOE
APOC1;NDRG2
APOC1;APOE
ASCC2
GJA1;EIF3E
EIF3E;UQCR10
GJA1;PTEN
FAM174B;APOE
APOC1;TTN
SHOX2;CD34
SLC4A1AP;FAM3C
EIF3E;MYH6
NDRG2;MLF1
EIF2B4;SERPINA1
GJA1;EIF3E
APOE
CENPW;PTEN
APOC1;APOE
DES;UQCR10
UQCR10;APOE
SERPINA1;NDRG2
EIF3E;UQCR10
MFN1
APOC1;APOE
APOC1;APOE
GJA1;NDRG2
GJA1;EIF3E
SERPINA1;EIF3E
WNT16;CD34
CR1L
GJA1;EIF3E
GJA1;EIF3E
WNT16;LCORL
APOE;NDRG2
HSPA4;SNX17
DES;NOV
NOV;NDRG2
NOV;CD34
CCDC121;CABP7
DES;PLN;ZNF512
CMTM5;RSPO2;NDRG2
GJA1;EIF3E

SLC5A6;SLC35F1
NOV;CD34
RSPO2;GPR126
MYH6
LXN;EIF3E
GJA1;EIF3E
DNAJC1;DLEU1
GJA1;EIF3E
EIF3E
NOV
PLN
NDRG2
NIPSNAP1
FAM3C
CWC27
GJA1
SNX17;NDRG2
ALX4;NDRG2
LXN;NDRG2
LXN;EIF3E
LXN;GPR126;CD34
NOV;TRIB2
CD34
UQCR10
TBX15;SLC35F1
UQCR10;APOE
GJA1;LXN;EIF3E
DES;APOC1;APOE
APOC1;APOE
CMTM5;APOC1;APOE
DNAJC1;CD34
PLN;NOV
APOC1;TTN
HSPA4;CMTM5
LXN;B3GNT7
FAM3C;APOE
EIF3E
TOMM40;NDRG2
GCKR;ZCCHC10
DLEU1;MLF1
GCKR;ZCCHC10
LXN;APOE
NDRG2;CD34
CCDC141
CD34

B3GNT7;MLF1
GFM1
EIF3E;UQCR10
MLF1
ASCC2
NCAPG
CR1
UQCR10
UQCR10
MLLT10
GJA1
GJA1
GJA1
UQCR10
EIF3E
ARHGEF40
EIF3E
UQCR10
APOC1
CD34
LXN
EIF3E
FAM3C
SERPINA1
CMTM5
GJA1
PLN
UQCR10
SERPINA1
GPN1
TBX15
NDRG2
GJA1
GJA1;LXN;EIF3E
APOC1;NCAPG
SLC35F1;UQCR10
GJA1;LXN
NDRG2;CD34
GJA1;EIF3E
GFM1;CABP7
EIF2B4;GFM1
PTEN;TRIB2
UQCR10;MLF1
DNAJC1;CD34
SERPINA1;APOC1

GJA1;EIF3E
PLN;SPEG
SERPINA1;UQCR10
ASCC2
GJA1;EIF3E;UQCR10
CMTM5;APOC1
EIF3E;FAM3C
EIF2B4;GNB4
DNAJC1;ZCCHC10
CABP7;PPM1G
APOC1;ARHGEF40
MLF1;MYH6
SERPINA1;EIF3E
APOC1;APOE
GJA1;EIF3E
TBX15;ARHGEF40
APOC1;DLEU1
APOC1;APOE
GCKR;MFN1
SERPINA1
EIF3E
PLN;GPR126
CR1;TRIB2
GJA1;LXN
GJA1;EIF3E
CENPW;NCAPG
APOE;NRBP1
HSPA4;SNX17
GJA1;NDRG2
SERPINA1;ASCC2
GJA1;EIF3E
TOMM40;FAM3C
NOV;APOE
CENPW;GPR126
GJA1;EIF3E
DES;PLN
UQCR10;MYH7
EIF3E;MLF1
APOC1;MYH6
APOC1;NDRG2
GJA1;APOE
SNX17;PPM1G
GJA1;EIF3E
SERPINA1;SSC5D
SHOX2;ARHGEF40

GJA1
UQCR10
SNX17
NDRG2
MTMR3
DES
NDRG2;CD34
GPR126
UQCR10
GJA1
CCDC141
APOE
CCDC121
NDRG2
SLC5A6
SHOX2
GJA1
FAM3C
MYH7
NRBP1
CCDC121
APOC1;APOE
CABP7
EIF3E
GJA1
FAM174B
EIF3E
SERPINA1
MYH7
APOC1
DES
APOE
UQCR10
GJA1;APOE;NDRG2
APOC1;EIF3E;TTN
DES;SNX17;TTN
GFM1;MLF1
TOMM40;APOE
DES;PLN;CCDC91
SERPINA1;LXN;TRIB2
DNAJC1;NDRG2
SLC4A1AP;UQCR10
CEP85L;WARS2
MLF1;MYH6
UQCR10;TRIB2

APOC1;APOE
EIF2B4;NDRG2
APOC1;APOE
CR1L;CMTM5;CD34
CMTM5;NDRG2
SNX17;EIF3E;PPM1G
CR1L;CD34
CCDC141;MLF1
GJA1;LXN
APOC1;APOE
PLN;UQCR10
GNB4;ZNF512;MLF1
CCDC121;MLF1;PPM1G
TBX15;ARHGEF40
SERPINA1;MLF1
SERPINA1;ASCC2
GJA1;EIF3E
SLC4A1AP;DLEU1
CEP85L
PLN;ZNF512
APOC1;APOE
APOC1;APOE
CCDC141
DLEU1;GPR126
NOV
UQCR10;TRIB2
DNAJC1;DLEU1
PVRL2;TRIB2
MLF1;PPM1G
LXN
CD34
GJA1;MLF1
ASCC2
PVRL2
ASCC2
GJA1;EIF3E
TBX15;NOV
GJA1
CCDC121
GJA1;TRIB2
SHOX2;NIPSNAP1
MYH6
GJA1;LXN
GJA1;FAM3C
MLF1

NOV;NDRG2
UQCR10;FAM3C
GJA1
GJA1;APOC1
EIF3E;TRIB2
CR1L;ASCC2
SNX17;TRIB2
MLLT10;DNAJC1
EIF3E;UQCR10
GCKR;ZCCHC10
MLF1;PPM1G
CCDC121;GNB4
MLF1;PPM1G
NCAPG;PPM1G
DLEU1;MYH6
UQCR10
MYH7
GJA1
WNT16;CD34
CD34
TRIB2
EIF3E
APOE
GNB4
MYH6
NDRG2
EIF3E
SERPINA1
NIPSNAP1
EIF3E
APOE
EIF3E
APOE
HSPA4;EIF3E
GNB4
NDRG2
PLN
PVRL2
CR1
GJA1
SPEG
PVRL2
RSPO2
EIF3E
EIF3E

UQCR10;NDRG2
GJA1;APOE
GNB4;EIF3E
GJA1;NDRG2
EIF3E
TBX15;MLF1
DNAJC1;GNB4
LXN;AFAP1
LXN;APOC1
TBX15;TRIB2
GJA1;LXN
GJA1
LXN;PVRL2
GJA1;EIF3E
NOV;FAM174B
CWC27;PPM1G
DES;PLN
UQCR10;APOE
SNX17;EIF3E
GJA1
ZCCHC10
EIF3E
HSPA4
NOV
GJA1
EIF3E
FAM3C
EIF3E
SERPINA1
GJA1
APOE
CABP7
EIF3E
CD34
GJA1
GFM1
SLC5A6
GJA1
APOE
CCDC91;CWC27
CABP7
TRIB2
GJA1
LXN
NOV;ARHGEF40

EIF3E
NCAPG
EIF3E
GJA1
CWC27
GPR126
UQCR10
GJA1
CD34
NOV
GJA1
LXN
SLC5A6
CCDC141
EIF3E
NDRG2
C2ORF16
DES
APOE
NDRG2
GPN1
DES
CCDC141
NIPSNAP1
APOE
RSPO2
GJA1
SKIDA1
GJA1;SNX17;MLF1;MYH6
GJA1;PLN;UQCR10;APOE
SNX17;EIF3E;FAM3C;CD34
APOC1;UQCR10;APOE
DCAF16;FAM174B;APOC1;EIF3E;APOE;PPM1G
GCKR;MTMR3;CMTM5;AFAP1;SSC5D;NDRG2;KIAA1755;GJA1;NOV;ZNF219;GPR126;SPEG;APOE;ARHGE
DES;SERPINA1;ZNF628;ZNF219;ASCC2;PVRL2;NRBP1;PPM1G
KIAA1755;CR1;PLN;B3GNT7;FAM174B;CMTM5;SLC35F1;NDRG2;CCDC141;MYH6;MYH7
CCDC121;GFM1;ZCCHC10;SLC4A1AP;TOMM40;CEP85L;SLC35F1;UQCR10;CWC27;NDRG2;DCAF16;GJA1
PTEN;ALX4;CWC27
SERPINA1;CR1;CR1L;ASCC2
TBX15;GFM1;ZCCHC10;SLC4A1AP;CCDC91;HSPA4;MFN1;NDRG2;MYH6;TTN
GJA1;SERPINA1
ASCC2
PTEN
GNB4
FAM3C

B3GNT7
SLC4A1AP;TOMM40;HSPA4;CEP85L;VSTM2L;UQCR10;NDRG2;GPN1;NIPSNAP1;SUPT7L;TTN;PPM1G;GJ[illegible]
DES
APOE
SNX17
UQCR10
GJA1
APOC1
SLC5A6
TOMM40
PPM1G
APOC1
APOE
EIF3E
APOE
EIF3E
GJA1
APOC1;APOE
NDRG2
DES
CABP7
PPM1G
IFI27L2
MLF1
TRIB2
NDRG2
UQCR10
EIF3E
NOV
GJA1
LXN
DLEU1
EIF3E
SNX17
B3GNT7
DNAJC1
UQCR10
NDRG2
CD34
EIF3E
DNAJC1
APOE
NAT14
SHOX2
APOE

GJA1
APOE
DLEU1
NOV
APOE
UQCR10
RSPO2
IFI27L2
CD34
NRBP1
EIF3E
ZCCHC10
AFAP1
PPM1G
NCAPG
SLC5A6
UQCR10
TOMM40
GJA1
CWC27
WARS2
APOE
HSPA4
CCDC91
SERPINA1
MYH7
PTEN
APOC1
TRIB2
NDRG2
GJA1
GJA1
APOE
EIF3E
GPR126
PPM1G
NOV
CENPW
SLC5A6
LXN
NCAPG
APOE
NDRG2
CD34
WNT16

GPR126
EIF3E
NDRG2
APOC1
APOE
GJA1
ZCCHC10
PPM1G
LXN
SLC5A6
LXN
GJA1
CD34
FAM3C
DNAJC1
APOE
NCAPG
LXN
ASCC2
NIPSNAP1
NIPSNAP1
MLF1
NOV
SLC5A6
RSPO2
GJA1
SLC5A6
B3GNT7;PVRL2
MLF1
SLC5A6;MYH7
MYH6
SSC5D
EIF3E
SLC5A6;NCAPG
B3GNT7;PVRL2
NOV
NCAPG
SERPINA1
WNT16
GJA1
NDRG2
CCDC141
SERPINA1
GFM1;MFN1;UQCR10
ZCCHC10;HSPA4;MLF1

APOC1;UQCR10;APOE
GPR126
GCKR;RSPO2
UQCR10
TOMM40;NDRG2
NIPSNAP1
SLC5A6
GPR126
APOE
EIF3E
SERPINA1
GFM1;MFN1
UQCR10
ZCCHC10
GJA1
APOE
EIF3E
DCAF16
SERPINA1
DLEU1;NCAPG
EIF3E
PLN;APOE
CMTM5
EIF3E
EIF3E
NDRG2
LXN
GJA1
UQCR10
GPN1
WNT16
NOV
SLC5A6
APOE
VSTM2L
NCAPG
CD34
TTN
GJA1
MTMR3
TRIB2
UQCR10
GPN1
LXN
NDRG2

EIF3E
UQCR10
ZCCHC10
MYH7
SNX17;APOE
DNAJC1;NCAPG
GJA1;DLEU1
APOC1
SERPINA1
TRIB2
APOC1
DGKH
EIF3E
UQCR10
GPN1
FAM174B
DES
APOE
SNX17
RSPO2
DES
NCAPG
PLN
MYH7
UQCR10
SLC5A6
SLC4A1AP
CENPW
B3GNT7
SLC5A6
APOC1
MYH7
SERPINA1
UQCR10
EIF3E
VSTM2L
MYH7
SNX17
GFM1
NIPSNAP1
PTEN
SNX17
GPN1
APOE
LXN

LXN
NCAPG
EIF3E
HSPA4
SPEG
TRIB2
UQCR10
MFN1
TOMM40
DES
SNX17
APOC1
GPR126
NCAPG
SNX17
PLN
ALX4
PPM1G
UQCR10
NIPSNAP1
GJA1
SLC5A6
EIF3E
GCKR
CD34
GJA1
NIPSNAP1
TTN
TOMM40
TMEM253
PPM1G
MLF1
IFI27L2
FAM3C
LXN
PTEN
CENPW
GFM1
GNB4
TOMM40
SLC5A6
SERPINA1
GJA1
PVRL2
TOMM40

NDRG2
ZCCHC10
GJA1
UQCR10
NOV
DCAF16
SLC5A6
NOV
FAM174B
TOMM40
LXN
SLC5A6
APOE
DCAF16
UQCR10
GJA1
MYH7
GNB4
RSPO2
NOV
MYH6
TOMM40
PTEN
SSC5D
GJA1
SLC5A6
GJA1
SNX17
NIPSNAP1
EIF3E
SLC5A6
PTEN
LXN
LXN
SLC5A6
APOC1
MFN1
MYH7
LXN
FAM3C
NDRG2
SNX17
SLC5A6
NDRG2
APOE

CR1L
MYH7
EIF3E
HORMAD2
APOC1
DCAF16
EIF3E
LXN
GJA1
SUPT7L
UQCR10
GJA1
GJA1
MLF1
SNX17
NDRG2
GNB4
LXN
GCKR
TBX15
NOV
APOE
NDRG2
TRIB2
NCAPG
GJA1
CMTM5
CD34
GFM1
SLC5A6
GJA1
UQCR10
NOV
LXN
TTN
NOV;RSPO2
EIF3E
B3GNT7
PTEN
DNAJC1
NOV
FAM3C
APOC1
LXN
UQCR10

APOE
CMTM5
CD34
GJA1
SNX17
FAM3C
MLF1
CCDC141
DES
UQCR10
CD34
UQCR10
NDRG2
ZNF628
FAM3C
SLC35F1
GJA1
EIF3E
DCAF16
MLF1
GJA1
CD34
EIF3E
PTEN
APOE
ZCCHC10
HSPA4
NIPSNAP1
NCAPG
LXN
NIPSNAP1
AFAP1
SLC5A6
GCKR
APOE
UQCR10
EIF3E
UQCR10
TOMM40
NIPSNAP1
TRIB2
GNB4
APOE
MLF1
SLC5A6

GNB4
LXN
ZCCHC10
DNAJC1
EIF3E
NIPSNAP1
GJA1
NCAPG
APOE
EIF3E
LXN;NOV;DGKH
LXN;FAM3C;NIPSNAP1
TOMM40;NCAPG;EIF3E
APOC1;NDRG2;NIPSNAP1
NDRG2;SUPT7L
CMTM5;TRIB2
GCKR;DGKH;WARS2
LXN;TRIB2;NAT14;NIPSNAP1
LXN;APOE;NDRG2
NDRG2;MLF1;MYH7
LXN;TRIB2;NAT14;NIPSNAP1
LXN;SNX17;FAM174B
GJA1;PLN;TTN
EIF2B4;HSPA4;PPM1G
LXN;SHOX2;APOE;CD34
DES;TOMM40;MYH6
SERPINA1;CR1;CR1L
LXN;FAM174B;GPR126
LXN;TRIB2;NAT14;NIPSNAP1
LXN;GPR126;EIF3E
ZCCHC10;SLC4A1AP;APOE;PVRL2
MLLT10;ZCCHC10;HORMAD2
APOC1;APOE;NDRG2
GJA1;NOV;TRIB2
NDRG2;MYH7;TTN
MFN1;UQCR10;TTN
HSPA4;APOC1;NDRG2
GFM1;APOE;NDRG2;MYH7
HSPA4;EIF3E;PPM1G
DES;WNT16
EIF3E;FAM3C;APOE
LXN;PLN;MYH7
SNX17;MYH7;PPM1G
GJA1;DLEU1;EIF3E;MLF1
MFN1;UQCR10;NDRG2

NOV;UQCR10;APOE
EIF3E;APOE;NDRG2
SLC5A6;TOMM40;NCAPG
PLN;TTN
LXN;MYH6;PPM1G
GCKR;APOC1;APOE
SKIDA1;NDRG2;TTN
SLC5A6;SERPINA1;SNX17
PLN;MYH6;MYH7
GJA1;VSTM2L;TRIB2
TOMM40;EIF3E;PPM1G
DES;APOE;NDRG2
SERPINA1;CR1;CR1L
APOC1;MFN1;NDRG2
APOC1;HORMAD2;UQCR10
APOE;NDRG2
GJA1;NCAPG;NDRG2;MLF1
RSRC1;GPR126;ARHGEF40;GPN1
CENPW;GNB4;NCAPG;MLF1
SLC5A6;GNB4;APOE
LXN;NOV;RSRC1;GPR126
IFI27L2;LXN;TRIB2;NIPSNAP1
HSPA4;MFN1;MLF1
LXN;TRIB2;NAT14;NIPSNAP1
PLN;FAM174B;APOC1
GCKR;APOC1;GPR126
NOV;APOC1;MYH7
GJA1;LXN;SNX17;NDRG2
GJA1;ZMAT5;MFN1;TRIB2
GFM1;MFN1;UQCR10
GJA1;PLN;MYH7
GCKR;ZCCHC10;SNX17
SNX17;FAM3C;APOE
GJA1;DES;UQCR10
GJA1;APOC1;UQCR10;APOE
SNX17;EIF3E
HORMAD2;PPM1G
CENPW;NCAPG
HSPA4;SNX17
CENPW;LXN
GJA1;LXN
CENPW;PVRL2
CENPW;FAM3C
CENPW;SLC4A1AP;SHOX2;LCORL
APOC1;APOE

GJA1;NCAPG;NDRG2;MLF1
SLC5A6;SERPINA1;SNX17
LXN;NOV;RSPO2
ZCCHC10;LXN;TOMM40
UQCR10;NDRG2;MLF1
TOMM40;APOE;PPM1G
LXN;FAM3C;CD34
GJA1;APOE;NDRG2
SNX17;DLEU1;NRBP1
MFN1;NDRG2;MLF1
HSPA4;TRIB2;MLF1
GJA1;EIF3E;APOE
SERPINA1;APOC1;APOE
CMTM5;PVRL2;NAT14;MLF1
MFN1;NDRG2;MLF1;MYH7
IFI27L2;LXN;TRIB2;NIPSNAP1
SLC5A6;ZMAT5;NDRG2;NRBP1
PLN;MYH6;MYH7
HSPA4;SKIDA1;SHOX2
SNX17;CD34;GPN1
SERPINA1;APOC1;APOE
EIF3E;CD34;MYH7
DNAJC1;NOV;CABP7
GJA1;NOV;MYH7
SHOX2;MLF1;MYH7
NOV;APOC1;APOE
SNX17;APOE;PPM1G
LXN;ZMAT5;UQCR10
APOC1;UQCR10;NIPSNAP1
GJA1;APOE;NDRG2
DES;EIF3E;MLF1
MLF1;MYH7;TTN
DES;EIF3E;NDRG2
TOMM40;SNX17;PPM1G
PTEN;PVRL2;TRIB2
LXN;GPR126;HORMAD2
GJA1;SERPINA1;GNB4
APOE;CD34;MYH7
NOV;UQCR10;TRIB2
EIF3E;MYH6;MYH7
GJA1;TOMM40;EIF3E
GJA1;HSPA4;APOE
GJA1;GPR126;EIF3E
NOV;UQCR10;TRIB2
LXN;FAM3C;CD34

IFI27L2;MYH6;MYH7
CABP7;MLF1;MYH7
LXN;TOMM40;PPM1G
MLF1;MYH7
RSPO2;APOE
GJA1;NCAPG;ZNF512
MLF1;MYH6
DNAJC1;CENPW;HSPA4
HSPA4;CR1L;EIF3E
GJA1;GNB4
DNAJC1;GJA1;FAM174B
B3GNT7;DLEU1
CENPW;LXN;NCAPG
CENPW;GNB4;FAM3C
GJA1;ZMAT5;MFN1
LXN;PTEN;EIF3E
LXN;GNB4;TTN
FAM174B;MLF1;MYH6
LXN;HSPA4;MFN1
GJA1;DES;PLN;APOE
UQCR10;NDRG2;MYH7
SNX17;EIF3E;APOE
NOV;APOC1;EIF3E
EIF3E;APOE;NIPSNAP1
TBX15;MFN1;MLF1
EIF3E;MLF1;MYH7
B3GNT7;NDRG2
B3GNT7;FAM3C;PVRL2
NDRG2;CD34;MLF1
LXN;NOV;FAM174B
SNX17;APOE;CD34
CCDC121;LXN;DGKH
GJA1;CENPW;UQCR10
CD34;MLF1;TTN
GJA1;LXN;NOV
TBX15;DNAJC1;CENPW
GJA1;EIF3E;APOE
GCKR;UQCR10;NDRG2;NIPSNAP1
PLN;RSPO2;MYH6
GJA1;B3GNT7
GNB4;NCAPG;APOE;TRIB2
SNX17;UQCR10
GJA1;GPR126;TRIB2
DES;UQCR10;CD34
CD34;MYH7;TTN

GJA1;APOE;NDRG2
EIF3E;UQCR10;TRIB2
GJA1;TOMM40;UQCR10
SERPINA1;LXN;VSTM2L
GJA1;CMTM5;NDRG2
LXN;GNB4;TRIB2
FAM174B;GNB4;GPR126
PLN;CCDC141;MYH6
CENPW;SERPINA1;FAM174B
SERPINA1;B3GNT7;TRIB2
ZMAT5;TRIB2
FAM174B;APOE
GJA1;UQCR10;NDRG2
TBX15;APOE
IFI27L2;FAM174B;APOE
EIF3E;UQCR10;FAM3C
NOV;UQCR10;APOE
CABP7;APOC1;PVRL2
GPR126;MYH6;MYH7
GJA1;DES;SLC35F1
GJA1;APOE;NIPSNAP1
APOC1;APOE
APOC1;UQCR10;APOE
DES;APOE;NDRG2
APOE;NDRG2;MYH7
CENPW;EIF3E
GJA1;EIF3E;UQCR10
NOV;RSPO2;CD34
SLC35F1;PVRL2;CD34
LXN;NOV;NCAPG
SERPINA1;AFAP1;APOE
DES;RSPO2;NDRG2
SKIDA1;NCAPG;TRIB2
DES;PLN;NOV
SLC5A6;SERPINA1;GPR126
GJA1;EIF3E;NIPSNAP1
NOV;GNB4;NCAPG
SNX17;CMTM5;APOE;NDRG2
CENPW;NCAPG;MLF1;NIPSNAP1
LXN;TRIB2;NAT14;NIPSNAP1
APOC1
APOC1;APOE;NDRG2;NIPSNAP1
CENPW;FAM174B;NCAPG
SLC5A6;GNB4;WNT16
LXN;TRIB2;NAT14;NIPSNAP1

LXN;APOC1;VSTM2L;APOE
EIF2B4;UQCR10;PPM1G
GCKR;APOC1;APOE;NIPSNAP1
PLN;FAM174B;MYH6;MYH7
CEP85L;NDRG2;MLF1
GCKR;WNT16;NDRG2;PVRL2
GJA1;GNB4;TRIB2
DES;APOE;MLF1
MFN1;NDRG2;MLF1
NCAPG;NIPSNAP1
APOC1;EIF3E
EIF3E;MLF1
SERPINA1;PLN
EIF2B4;GJA1
GFM1;MLF1
NOV;UQCR10
SNX17
APOE
GFM1;CENPW;CWC27
GJA1;EIF3E;APOE
GJA1;VSTM2L;APOE;NDRG2
NOV;GNB4;VSTM2L;CD34
DNAJC1;SERPINA1;APOC1;GPR126
SKIDA1;DLEU1;RSPO2
SKIDA1;SHOX2;RSPO2;GPR126
MFN1;NDRG2;MYH7
NAT14;NRBP1;PPM1G
CENPW;DLEU1;PPM1G
SNX17;MLF1;MYH7;TTN
APOC1;APOE;NDRG2
TBX15;NOV;CABP7;RSPO2
EIF2B4;NDRG2
APOC1;APOE;MYH7;TTN
CENPW;PLN;APOC1;EIF3E
SNX17;APOE;NDRG2
CENPW;NCAPG;NIPSNAP1
NOV;VSTM2L;APOE;NDRG2
GJA1;SNX17;EIF3E;MLF1
GJA1;LXN;APOE;NDRG2
PLN;MYH6;MYH7
GJA1;NOV;RSPO2
GJA1;LXN;SNX17;PTEN
LXN;RSPO2;GPN1
WNT16;CD34;NIPSNAP1
SHOX2;LCORL

GPR126;NIPSNAP1
LXN;NOV
SLC5A6;GCKR
NCAPG;TRIB2
TOMM40;APOC1
UQCR10
NOV;GPR126;PVRL2
CCDC141;TRIB2;TTN
GJA1;UQCR10;APOE
CABP7;CD34;DGKH
HSPA4;ZNF219
EIF3E
GJA1;SNX17
MLF1
TOMM40;EIF3E;PPM1G
NOV;NDRG2;DGKH
TBX15;HSPA4;CD34
SLC5A6;PTEN;ARHGEF40
LXN;EIF3E;FAM3C
SKIDA1;PTEN;WARS2
CENPW;SNX17;NCAPG
SNX17;APOE;PPM1G
SNX17;EIF3E
UQCR10
TBX15
GJA1;NCAPG;EIF3E
DLEU1
HSPA4;NCAPG
MFN1
MYH7
SNX17;CD34
TOMM40;PPM1G
GNB4;PTEN
SLC35F1;NIPSNAP1
NOV;TRIB2
NOV;TRIB2
SERPINA1;CR1;CR1L
UQCR10;APOE;NRBP1
CENPW;RSRC1;FAM174B;ZNF512
GJA1;UQCR10;APOE
CENPW;LXN;NCAPG
SLC5A6;MLF1
NCAPG
APOE
NOV;NDRG2

UQCR10;NIPSNAP1
SNX17;EIF3E
SERPINA1;UQCR10
PPM1G
NRBP1
LXN;GNB4
APOE;NIPSNAP1
GJA1;NOV
VSTM2L;MYH7
SNX17
NDRG2;MLF1
MFN1;ARHGEF40
CENPW;UQCR10
DES;NDRG2
MYH7
CCDC141
GCKR;GPR126
GPR126;UQCR10
NOV;NDRG2
EIF3E;FAM3C
GJA1;UQCR10
SLC35F1;ZNF512
GJA1;SNX17
GJA1;APOE
SKIDA1
EIF3E;NDRG2
MYH6;PPM1G
TOMM40;PPM1G
GCKR;ZCCHC10
LXN;NOV
NDRG2
GFM1;UQCR10
NDRG2;PVRL2
SHOX2;MYH7
EIF3E;PPM1G
CENPW;EIF3E
CENPW;EIF3E
APOE;NIPSNAP1
CD34;MLF1
EIF3E;GPN1
ALX4;CD34
RSPO2;DGKH
LXN;UQCR10
NOV;APOE
APOE

APOE
TOMM40;APOE
SLC5A6;MLF1
RSPO2;CD34
MYH7;TTN
NDRG2;MLF1
APOE;NDRG2
LXN;CCDC91
GJA1;LXN
GJA1;SNX17;EIF3E
SNX17
PLN;NOV
GJA1;CENPW;APOC1;MLF1
MFN1
GJA1
APOC1;APOE
TTN
GNB4
DES;NDRG2
MLF1;PPM1G
DLEU1
LXN
WARS2
CENPW;SNX17
UQCR10;MLF1;TTN
MLF1
ZCCHC10;ZMAT5;NIPSNAP1
MLF1;MYH7;TTN
LXN;EIF3E;UQCR10
CENPW;NCAPG;NIPSNAP1
PLN;APOC1;MYH6;TTN
FAM3C;NDRG2;TRIB2
LXN;TRIB2;NAT14;NIPSNAP1
SNX17;MYH7
LXN;TRIB2;NAT14;NIPSNAP1
UQCR10;MLF1;MYH7
GJA1;PTEN;RSPO2
SLC5A6;TOMM40;HSPA4;UQCR10
GJA1;CD34
GJA1
SLC5A6
GPR126
TOMM40;B3GNT7;APOC1
MLF1
GJA1;GPR126

NDRG2;CD34
PTEN
SLC5A6
NCAPG;MLF1
GJA1;FAM3C
GJA1;MLF1
UQCR10
LXN;APOE
LXN;EIF3E
CENPW;UQCR10
FAM3C
APOE;CD34
CENPW;MLF1
NAT14;TMEM253
APOE;CD34
GPR126;PVRL2
TOMM40;UQCR10
NDRG2;PVRL2
GJA1;SNX17
NOV;NRBP1
LXN;DLEU1
APOC1;FAM3C
ZCCHC10;NCAPG
NOV;APOE
PTEN;APOE
SLC5A6;SLC4A1AP;SKIDA1
UQCR10;NIPSNAP1
WARS2;PPM1G
NCAPG;NDRG2
APOC1;APOE
SNX17;NCAPG
NOV;UQCR10
LXN;NRBP1
MLF1;MYH7
UQCR10;DGKH
CCDC91;NCAPG;WARS2
ZNF628;DLEU1
APOC1;APOE
CENPW;NCAPG
GJA1;APOE
ZMAT5;MFN1
GJA1;APOE
LXN;NOV
APOE;NDRG2
UQCR10;MLF1

CENPW;LXN;NCAPG
SNX17;APOC1
CENPW;EIF3E
NDRG2;DGKH
SLC35F1;MYH6
DCAF16;SUPT7L
NOV;EIF3E
GNB4;TRIB2
MYH6;MYH7
NCAPG;APOE
DNAJC1;PTEN
DES;CABP7
APOC1;UQCR10;APOE
CCDC121;ASCC2
GJA1;EIF3E
GNB4;SLC35F1
DLEU1;NCAPG
FAM174B;MLF1
EIF3E;FAM3C
CENPW;NCAPG
CENPW;EIF3E
SHOX2;FAM3C
GNB4;EIF3E
LXN;DLEU1
LXN;GPR126
NOV;FAM3C
TOMM40;NIPSNAP1
DLEU1;GPN1
SLC5A6;ARHGEF40
CENPW;NCAPG
GJA1;APOE
PTEN;EIF3E
APOE;GPN1
SLC5A6;UQCR10
CENPW;FAM3C
GJA1;UQCR10
APOC1;APOE
PTEN;TMEM253
CABP7;APOE
GJA1;LXN
UQCR10;TRIB2
DCAF16;NCAPG
SLC5A6;SUPT7L
NOV;TRIB2
NOV;NDRG2

UQCR10;NIPSNAP1
GJA1;UQCR10
GCKR;NDRG2
GJA1;SNX17
NOV;CD34
LXN;TRIB2
TOMM40;PPM1G
CD34;TTN
UQCR10;MLF1
TOMM40;EIF3E
CABP7;CD34
UQCR10;APOE
NDRG2;PVRL2
LXN;SNX17
APOC1;MLF1
DNAJC1;ARHGEF40
SPEG;EIF3E
PLN;FAM174B
APOE;NDRG2
NOV;UQCR10
GFM1;NCAPG
UQCR10;FAM3C
APOC1;NRBP1
APOC1;APOE
DLEU1;UQCR10
TOMM40;EIF3E
GJA1;LXN
CD34;DGKH
EIF3E;APOE
LXN;SLC4A1AP
NOV;CD34
EIF3E;CD34
SLC5A6;NCAPG
DES;TTN
APOC1;APOE
CCDC141;TTN
LXN;TOMM40
CD34;PPM1G
NCAPG;NDRG2
SLC5A6;NCAPG
NOV;MYH7
GJA1;CD34
GJA1;APOE
CENPW
NOV;GPR126

CENPW;MYH7
IFI27L2;GJA1;APOC1
SNX17;EIF3E
GJA1;EIF3E
LXN;DLEU1
TOMM40;SNX17;PPM1G
APOC1;UQCR10
CENPW;MLF1
CENPW;NCAPG
UQCR10;TTN
TOMM40
TOMM40;SNX17;PPM1G
NOV;SKIDA1;GPR126
SNX17;EIF3E;PPM1G
CCDC91;MFN1;NCAPG
FAM3C;PPM1G
APOE;PPM1G
SHOX2;EIF3E
GJA1;APOE
SHOX2;NDRG2;LCORL
LCORL;PVRL2
PLN;MYH7
MFN1;NDRG2
SNX17;FAM3C
NOV;CD34
SHOX2;HORMAD2
DCAF16;DLEU1;NCAPG
GNB4;RSPO2;CD34
GJA1;PTEN;GPR126
CCDC91;NCAPG
APOE;CCDC141
GJA1;PLN
LXN;GPR126
GJA1;EIF3E
EIF2B4;NDRG2
GJA1;MLF1;PPM1G
GJA1;LXN;GPR126
TOMM40;NRBP1
NDRG2;MLF1
DCAF16;SHOX2
RSPO2;APOE
APOE;CD34
APOC1;APOE
FAM174B;UQCR10
RSPO2;WNT16

DES;MLF1
NOV;RSPO2
GPR126;GPN1
PLN;FAM174B
SLC5A6;NCAPG
UQCR10;APOE
SLC5A6;NCAPG
ARHGEF40;GPN1
MLF1;MYH7
SLC5A6;TOMM40
EIF3E;ZNF512
GJA1;TOMM40
LXN;SNX17
TOMM40;ASCC2
ZCCHC10;NCAPG
APOE;GPN1
GJA1;NCAPG
NOV;CD34
ZCCHC10;GNB4
GJA1;APOE
SNX17;APOE
GPR126;EIF3E
SNX17;PTEN
EIF3E;UQCR10
GJA1;NDRG2
APOC1;APOE
SERPINA1;SKIDA1
SHOX2;SLC35F1
EIF3E;MLF1
EIF3E;UQCR10
APOE;NDRG2
LXN;PLN
MFN1;PTEN
PTEN;FAM3C
APOE;NIPSNAP1
CENPW;NOV
UQCR10;NIPSNAP1
GJA1;NAT14
SNX17;GNB4
APOC1;APOE
NDRG2;MLF1
GPR126;TRIB2
SHOX2;CD34
GJA1;EIF3E
GPR126;EIF3E

GJA1;PTEN
NDRG2;NIPSNAP1
SERPINA1;PTEN
CR1L;FAM174B
CCDC91;MYH7
TOMM40;EIF3E
UQCR10;GPN1
DNAJC1;UQCR10
GNB4;TRIB2
APOE;MYH7
GNB4;CD34
PTEN;NDRG2
GJA1;CENPW
GJA1;UQCR10
FAM174B
NOV;APOE
SHOX2;UQCR10
EIF3E;UQCR10
PTEN;RSPO2
LXN;PTEN
GJA1;NCAPG
MLF1;NIPSNAP1
NDRG2;PVRL2
GPR126;EIF3E
TOMM40;FAM3C
NOV;APOE
SNX17;DGKH
NOV;NCAPG
DLEU1;UQCR10
IFI27L2;NAT14
GJA1;PPM1G
GJA1;EIF3E
APOE;NDRG2
NOV;HORMAD2
WNT16;TTN
NOV;RSRC1
NDRG2;PVRL2
GCKR;MYH7
TBX15;TRIB2
TOMM40;SNX17
MLLT10
GNB4;ARHGEF40
CENPW;EIF3E
GJA1;NDRG2
UQCR10;NDRG2

CWC27
FAM3C;APOE
LXN;APOE
CABP7;SHOX2
PTEN;EIF3E
PLN;CCDC91;DGKH
CENPW;NCAPG
GJA1;SNX17
NOV;SPEG
SLC5A6;NCAPG
APOC1;APOE
SERPINA1;ARHGEF40
GJA1;SNX17
GJA1;CD34
APOE
NCAPG;PPM1G
APOC1;NDRG2
LXN;NOV
DCAF16;GJA1
LXN;EIF3E
APOE;NDRG2
GNB4;APOE
LXN;EIF3E
SNX17;NDRG2
UQCR10;MLF1
CCDC121;GNB4
APOE;NIPSNAP1
SNX17;EIF3E
SLC5A6;APOE
APOE;MLF1
NDRG2;DGKH
SKIDA1;MLF1
SNX17;PPM1G
TOMM40;HSPA4
ZNF219;EIF3E
GJA1;NCAPG
SLC5A6;TRIB2
GJA1;NDRG2
IFI27L2;NAT14
SNX17;PPM1G
SNX17;AFAP1
PTEN;FAM3C
SHOX2;PPM1G
APOC1;NDRG2
NCAPG;FAM3C

PTEN;NCAPG
HSPA4;SNX17
TOMM40;SNX17
CENPW;NCAPG
DES;TMEM253
GJA1;SERPINA1
TBX15;CENPW
GJA1;SLC35F1
GNB4;TRIB2
EIF3E;PPM1G
CENPW;EIF3E
CABP7;HORMAD2
GJA1;DES
CENPW;NCAPG
HSPA4;DGKH
DNAJC1;ARHGEF40
TOMM40;NCAPG
MFN1;MYH7
ZMAT5;MFN1
GPR126;CD34
FAM174B;MLF1
UQCR10;APOE
HSPA4;NIPSNAP1
B3GNT7;NCAPG
UQCR10;PPM1G
NDRG2;PVRL2
UQCR10;NDRG2
DCAF16;ARHGEF40
GJA1;EIF3E
GJA1;NOV
MTMR3;TRIB2
HSPA4;NCAPG
SLC4A1AP;MFN1
DNAJC1;SNX17
FAM3C;MLF1
GJA1;NDRG2
NOV;VSTM2L
DNAJC1;EIF3E
SLC5A6;CENPW
CR1L;FAM174B
UQCR10;NDRG2
TOMM40;EIF3E
TOMM40;NCAPG
CABP7;ASCC2
TOMM40;EIF3E

LXN;NDRG2
PLN;FAM174B
GJA1;MLF1
AFAP1;NIPSNAP1
ZCCHC10;SNX17
SHOX2;CD34
APOE
MFN1;MLF1
CENPW;FAM174B
EIF3E;NIPSNAP1
LXN;DGKH
IFI27L2;MLF1
GPR126;WNT16
APOC1;APOE
GJA1;UQCR10
GJA1;FAM3C
DES;GNB4
GJA1;NAT14
TOMM40;EIF3E
B3GNT7;CD34
PTEN;RSPO2
GJA1;CD34
GNB4;GPN1
EIF3E;MLF1
PTEN;NDRG2
UQCR10;NIPSNAP1
EIF3E;NDRG2
UQCR10;NDRG2
SNX17;APOE
SNX17;NDRG2
SSC5D;C2ORF16
CABP7;SHOX2
EIF3E;TRIB2
RSRC1;EIF3E
GJA1;APOE
HSPA4;EIF3E
SNX17;PPM1G
SERPINA1;GPR126
GJA1;RSPO2
CR1L;SSC5D
DES;UQCR10
TBX15;MLF1
MTMR3;PTEN
GFM1;MYH7
GJA1;SNX17

GJA1;VSTM2L
SLC5A6;GFM1
TOMM40;PPM1G
EIF3E;UQCR10
CENPW;EIF3E
GJA1;APOE
NDRG2;MLF1
GJA1;APOC1
EIF3E;PPM1G
LXN;EIF3E
GJA1;SNX17
SHOX2;UQCR10
NOV;MLF1
SNX17;UQCR10
VSTM2L;NCAPG
SLC35F1;NDRG2
CCDC121;PTEN
APOE;NDRG2
CR1L;RSPO2
NOV;UQCR10
APOE;NIPSNAP1
LXN;TRIB2
APOE;NDRG2
EIF3E;TRIB2
LXN;SNX17
UQCR10
EIF3E
CD34
NCAPG
APOE
NCAPG
FAM174B
NDRG2
GJA1
SLC5A6
EIF3E
APOE
TOMM40
GJA1
NCAPG
CD34
GPN1
TTN
MLF1
FAM3C

CD34
FAM174B
SKIDA1
FAM3C
APOC1
EIF3E
EIF3E
TTN
PTEN
TOMM40
TBX15
CENPW
CCDC91
HORMAD2
UQCR10
APOC1
NDRG2
CENPW
UQCR10
GCKR
UQCR10
GJA1
NDRG2
GJA1
NCAPG
B3GNT7
PPM1G
LXN
NIPSNAP1
UQCR10
FAM3C
EIF3E
MLF1;MYH7;TTN
NOV
MLF1
FAM3C
NIPSNAP1
ZCCHC10
CWC27
GJA1
EIF3E
DES
CD34
ZCCHC10
EIF2B4;APOC1;APOE

GJA1
GNB4
EIF3E
TRIB2
UQCR10
NDRG2;CD34
CD34
MLF1
GPR126
NDRG2
CENPW
CD34
CWC27
SKIDA1
GJA1
TTN
LXN
APOE
SSC5D
EIF3E
NCAPG
MLF1
NOV
EIF3E
APOE
MLF1
PPM1G
FAM3C
SLC35F1
FAM3C
SLC5A6;DLEU1
GNB4;MLF1;DGKH
GJA1;UQCR10;NDRG2
UQCR10;APOE
LXN;CABP7
APOC1;APOE;NAT14
APOC1;APOE;NIPSNAP1
CENPW
FAM3C
TOMM40
LXN
NDRG2
FAM174B
SLC5A6
GJA1

CABP7
CABP7
SNX17
HSPA4
NOV
APOE
CD34
APOC1
TOMM40
UQCR10
UQCR10
TRIB2
GPN1
FAM3C
SUPT7L
GJA1
MFN1;NCAPG
NOV
TOMM40
MYH6
WNT16
CABP7
GJA1
GNB4
NDRG2
CCDC141
DNAJC1
DCAF16
SNX17
DGKH
NCAPG
TOMM40
EIF3E
DLEU1
CENPW
LXN
NOV
TBX15
NAT14
FAM3C
MYH7
CD34
TRIB2
NCAPG
GNB4

NCAPG
NDRG2
GNB4
APOC1
APOE
CR1L
APOE
PPM1G
MLLT10
EIF3E
GCKR
SNX17
GJA1
SNX17
NOV
NCAPG
SNX17
APOC1
TTN
GCKR
MYH6
GJA1
AFAP1
MLF1
PPM1G
NDRG2
UQCR10
DCAF16
NDRG2
MLF1
SSC5D
LXN
EIF3E
SLC5A6
LXN
GJA1
CD34
NCAPG
CD34
GJA1
LXN
SERPINA1
GJA1;SNX17;APOE
FAM3C;APOE
CENPW;CCDC91;NCAPG

EIF3E
ASCC2
FAM174B
SNX17
CENPW
APOC1
NOV
LCORL
NCAPG
AFAP1
FAM174B
LXN
NDRG2
LXN
APOC1;APOE
MLF1
DGKH
FAM174B;UQCR10
SKIDA1;APOC1;TTN
IFI27L2;CENPW
APOC1;NDRG2
EIF3E;UQCR10;PPM1G
UQCR10;APOE
B3GNT7;NIPSNAP1
PTEN;EIF3E
LXN;NOV;HORMAD2
UQCR10;NIPSNAP1
B3GNT7;CD34;TRIB2
CWC27
TTN
APOC1;MFN1;NDRG2
PLN;SNX17
GJA1;GPR126;EIF3E
SLC5A6;CENPW;GPN1
GJA1;EIF3E;MLF1
MLF1;PPM1G
GJA1;SNX17;PPM1G
CENPW;NCAPG;MLF1
CR1L;PPM1G
GJA1;TOMM40;PPM1G
SKIDA1;VSTM2L
GCKR;APOC1;CD34
SHOX2;MYH6
GJA1;CENPW;TOMM40
GJA1;FAM3C;CD34

GJA1;GPR126;EIF3E
CD34
NOV
GJA1;UQCR10;GPN1
GJA1;SNX17;PPM1G
APOC1;APOE
APOC1;APOE
GJA1;APOE
APOE;CCDC141
CCDC121;APOC1;APOE
NOV;APOC1
CCDC121;TRIB2
EIF2B4;NDRG2
TOMM40;SNX17
GJA1;GNB4
UQCR10;APOE
CENPW;HSPA4;EIF3E
LXN;FAM174B;GNB4
SLC5A6;NCAPG
NOV;SHOX2
CENPW;NCAPG
APOC1;UQCR10
IFI27L2;NAT14
CMTM5;NDRG2
GJA1;TTN
LXN;EIF3E
NOV;APOE
UQCR10;MYH7
GJA1;NOV;EIF3E
GJA1;B3GNT7
UQCR10;APOE
EIF2B4;APOE;NIPSNAP1
DNAJC1;DGKH
SHOX2;CD34
APOE;NDRG2
APOC1;NDRG2;NIPSNAP1
APOC1;UQCR10;APOE
SNX17;NDRG2
GPR126;ARHGEF40;GPN1
NOV;RSRC1;CWC27
GJA1;SKIDA1;MLF1
LXN;NOV;MYH7
GJA1;TOMM40;PPM1G
NOV;EIF3E
APOC1;NDRG2

CENPW;DLEU1;UQCR10
SERPINA1;NCAPG;TRIB2
GJA1
RSRC1;TRIB2
UQCR10;MYH7
CENPW;GNB4;FAM3C
UQCR10;APOE
SLC5A6;NCAPG
SNX17;APOE
APOE;CD34
SNX17;APOE
APOC1;APOE;NDRG2
CENPW;NCAPG
MLLT10;RSRC1
RSPO2
NCAPG
GJA1
GJA1
NCAPG
DES
NIPSNAP1
APOE
PTEN
CD34
UQCR10
NDRG2
EIF3E
ZCCHC10
APOE
GJA1
PPM1G
SNX17
NDRG2
SLC5A6
CD34
LXN
EIF3E
NDRG2
LXN
APOE
SNX17
UQCR10
GCKR
SNX17
DNAJC1

GPN1
LCORL
NCAPG
APOE
APOE
NIPSNAP1
LXN
DES
APOE
NCAPG
SNX17
MYH6
SNX17
TRIB2
NCAPG
GPN1
MTMR3
APOC1
FAM3C
MTMR3
LXN
PPM1G
NOV
NOV
EIF3E
CWC27
TOMM40
TRIB2
TBX15
NCAPG
APOE
EIF3E
CENPW
ZMAT5
APOE
NOV;UQCR10
FAM3C;CD34
FAM3C
GJA1
FAM174B
APOE
PTEN
SPEG
MLF1
CD34

IFI27L2
CABP7
CENPW
TOMM40
SNX17
NDRG2
MYH7
UQCR10;APOE
DES;APOE;TRIB2
ZCCHC10
MLF1
FAM3C
CD34
CENPW
UQCR10
TTN
NRBP1
CWC27
EIF3E
NCAPG
TBX15
WNT16;MYH7
CCDC121;GFM1;ZCCHC10;LXN;SLC4A1AP;TOMM40;SHOX2;CEP85L;PTEN;SLC35F1;UQCR10;CWC27;GJA
TBX15;CEP85L;ALX4;ARHGEF40
MTMR3;LXN;SHOX2;MFN1;PTEN;NCAPG;EIF3E;LCORL;NRBP1;TTN
TBX15;SHOX2;CCDC141
TBX15;DES;ZNF628;SPEG;MYH6;MYH7;TTN
DCAF16;CCDC121;CENPW;ZCCHC10;ZNF219;AFAP1;SSC5D;EIF3E;ZNF512;NRBP1
CCDC141;MLF1;MYH6;MYH7
NCAPG
DNAJC1;GCKR;APOC1;NRBP1
TBX15
FAM3C
CD34
UQCR10
SNX17
HSPA4
CMTM5
B3GNT7
EIF3E
EIF3E
SNX17
ZCCHC10;SHOX2
TBX15;MLLT10;SHOX2;ZNF219;ALX4;ZNF513
EIF3E;PPM1G

CMTM5
LXN
EIF2B4
EIF2B4;DNAJC1;LXN;MFN1;GNB4;PTEN;NCAPG;ASCC2;EIF3E;LCORL;GPN1;NRBP1
SNX17;ASCC2;GPN1;WARS2;TTN;PPM1G
FAM174B;APOC1;NCAPG;UQCR10
HSPA4
DES
DLEU1;GPR126;TRIB2
APOC1;APOE;NDRG2
SERPINA1;CR1;APOC1
MYH6;MYH7
GFM1;ZCCHC10;TOMM40;SLC35F1;UQCR10;CWC27;DCAF16;GJA1;CCDC91;RSRC1;SPEG;ALX4;FAM3C;[illegible]
SERPINA1;MTMR3;CR1;CMTM5;SLC35F1;NDRG2;DES;PLN;B3GNT7;FAM174B;APOE;CCDC141;CD34
TRIB2
NOV;GNB4;AFAP1;SSC5D;MYH6
SLC5A6
DGKH
GJA1
ZCCHC10;CEP85L;ARHGEF40
PPM1G
PPM1G
GFM1
EIF2B4;ZNF628;PPM1G
MLF1;PPM1G
NDRG2
GJA1
CCDC91;HSPA4;WNT16;SUPT7L
SUPT7L
SERPINA1
MTMR3;NCAPG;ASCC2
NOV;APOC1
LXN;RSRC1;DLEU1;NCAPG
GCKR;SERPINA1;APOC1;ZMAT5;APOE
PLN;SPEG
TBX15
DES;MYH7;TTN
SHOX2
MLLT10
C2ORF16;HORMAD2;MLF1
FAM174B
NOV
NIPSNAP1
NDRG2
MFN1;NIPSNAP1

SLC5A6;GPR126;CD34
APOE
SNX17
MLF1
EIF2B4;TBX15;DES;TOMM40;MYH6;MYH7;TTN;PPM1G
SNX17;FAM174B;UQCR10;APOE
UQCR10;NDRG2
EIF3E;APOE
GJA1;APOE
EIF3E;APOE
CD34;NRBP1
GJA1;EIF3E
UQCR10;CD34
NOV;TRIB2
APOC1;APOE
PLN;MYH6
GJA1;APOE
UQCR10
EIF3E;APOE
GJA1;HSPA4
LXN;DLEU1
CENPW;NCAPG
IFI27L2;GJA1
DNAJC1;APOC1
GJA1;APOC1
NDRG2;DGKH
MFN1;APOE
DNAJC1;SERPINA1
SERPINA1;APOC1
GJA1;APOC1
UQCR10;FAM3C
SERPINA1;APOC1
CD34
NDRG2;MYH7
CR1;CR1L
KIAA1755;WNT16
GJA1;MYH6
DNAJC1;CCDC141;LCORL
PLN;TRIB2
APOC1;FAM3C
DES;EIF3E;UQCR10
PLN;APOC1
DES;MYH7
DES;APOE
FAM3C;TTN

GCKR;APOE
EIF3E;APOE
DNAJC1;CD34
MLLT10;ZMAT5
NOV;APOE
SERPINA1;PTEN
APOC1;APOE
UQCR10;MLF1;MYH7
DNAJC1;APOC1
APOC1;PTEN
GJA1;APOE
CMTM5;CD34
SERPINA1;EIF3E
ZCCHC10;NOV
SNX17;APOE
SERPINA1;EIF3E
LXN;PTEN
APOC1;APOE
SERPINA1;APOC1
LXN;UQCR10
APOC1
CENPW;SERPINA1
GJA1
IFI27L2;GJA1
APOE
GNB4;FAM3C
APOE
APOC1
ZMAT5
GJA1
CD34
GJA1;CCDC141
APOE
CCDC91;EIF3E
ZCCHC10;ZNF513
LXN;LCORL
UQCR10
GJA1
GJA1
APOC1
APOE
APOC1;NCAPG
TRIB2
SERPINA1
GJA1;ZMAT5

EIF3E
SERPINA1
SERPINA1;LXN
GJA1;FAM174B
APOE
GJA1
GJA1;APOE
GJA1;LXN
HSPA4;NCAPG
LXN;CMTM5
PLN;NDRG2
GJA1;DGKH
NCAPG;APOE
EIF3E;FAM3C
CENPW;LXN
FAM3C;MYH7
GJA1;APOE
NCAPG
LXN;NOV
SERPINA1
APOC1;APOE
EIF3E
DES;UQCR10
GJA1;NOV
MYH7;TTN
LXN;EIF3E
GJA1;EIF3E
ZCCHC10;FAM3C
GJA1;APOC1
NCAPG
CENPW;NCAPG
CENPW;NCAPG
SERPINA1;UQCR10
CENPW;NCAPG
APOC1;APOE
GJA1;EIF3E
GJA1;SHOX2
GJA1;PLN
GJA1;CR1;APOC1
LXN;NOV
GJA1;HSPA4;GPN1
EIF3E;APOE
GJA1;EIF3E
GJA1;HSPA4;GPN1
GJA1;EIF3E

GJA1;LXN
SERPINA1;APOC1;APOE
APOE;MYH7
GFM1;UQCR10;NDRG2
GJA1;NCAPG
APOC1;APOE
GJA1;EIF3E
LXN;NOV
CCDC141;TTN
SNX17;APOE
LXN;NOV
LXN;PLN
EIF3E;APOE
CMTM5;ASCC2;NRBP1
TBX15;CD34
APOC1;SUPT7L
CENPW;NCAPG
GCKR;RSPO2;SPEG
EIF3E;UQCR10
GJA1;UQCR10;PVRL2
DES;MLF1;TTN
CENPW;SHOX2;NCAPG
APOC1;APOE
APOE;CD34
CENPW;PLN;NCAPG
DNAJC1;DLEU1;TRIB2
SERPINA1;APOE
CENPW;NCAPG
GJA1;MYH6;TTN
GJA1;EIF3E
DES;APOE;NDRG2
DES;LXN;APOE
NDRG2;MYH7
GJA1;APOC1;FAM3C
APOE;NIPSNAP1
CR1;TOMM40
GJA1;APOC1;APOE
SERPINA1;SNX17;EIF3E
GJA1;LXN;EIF3E
UQCR10;APOE
SERPINA1;APOE;TTN
UQCR10;NDRG2;MYH7
SLC5A6;GFM1;HSPA4
APOC1;NDRG2
SERPINA1;CR1

APOE;NDRG2;MYH7
GJA1;CENPW;SERPINA1
GPR126;EIF3E;CD34
NDRG2;MYH7
UQCR10;APOE
GJA1;EIF3E;NDRG2
DES;MYH7
DES;TTN
NDRG2;MLF1;MYH7
APOC1;APOE
GJA1;EIF3E
GJA1;APOC1
GJA1;NCAPG
CR1;CMTM5;NCAPG
TOMM40;APOC1
GJA1;CR1L;RSRC1
LXN;ZNF512
GJA1;EIF3E
NCAPG;WNT16
GJA1;UQCR10
SLC35F1;EIF3E
TOMM40;EIF3E
DNAJC1;EIF3E
APOE;MYH7
SERPINA1;TRIB2
NDRG2;MLF1
FAM3C;PPM1G
LXN;RSRC1
NOV;TRIB2
GJA1;APOC1
APOE;NDRG2
GJA1;NDRG2
APOC1;APOE
GJA1;SERPINA1;NDRG2
LXN;APOC1
LXN;MYH6
APOE;NDRG2
CR1L;RSRC1;ASCC2
UQCR10;APOE
NOV;APOC1
DES;NDRG2
IFI27L2;GJA1;EIF3E
LXN;APOC1;WNT16
GCKR;MLF1
NOV;SNX17

PLN;APOE;TTN
SLC5A6;GFM1
GJA1;NOV
APOC1;APOE
GJA1;MYH6
GJA1;EIF3E
APOE;NIPSNAP1
DES;WNT16
SERPINA1;UQCR10
MFN1;ASCC2
CENPW;CR1L
NOV;APOE
SERPINA1;LXN;NOV
APOE;NDRG2
GNB4;ARHGEF40
ZCCHC10;NIPSNAP1
DNAJC1
EIF3E
SHOX2
MYH7
EIF3E
GCKR
UQCR10
CR1
B3GNT7
NDRG2
LXN
APOE
SHOX2
DNAJC1
EIF3E
APOC1
PLN
SERPINA1
ALX4
NOV
APOC1
TTN
FAM3C
SERPINA1
PLN
PLN
EIF3E
GJA1
NDRG2

DES
CENPW
SERPINA1
APOC1
GJA1
AFAP1
GNB4
CD34
APOE
SERPINA1
GJA1
LXN
EIF3E
CENPW
MLF1
EIF3E
FAM3C
NOV
SNX17
EIF3E
GJA1
SLC5A6
APOE
SERPINA1
EIF3E
SERPINA1
SERPINA1
EIF3E
GJA1
PVRL2
SERPINA1
FAM3C
GJA1
APOC1
TBX15
SERPINA1
NCAPG
APOE
EIF3E
APOE
GJA1
NRBP1
HSPA4
UQCR10
UQCR10

NDRG2
SERPINA1
SERPINA1
CR1L
MYH7
SNX17
MYH7
CD34
RSPO2
SERPINA1
CD34
FAM3C
EIF3E
TOMM40
GPR126
PPM1G
APOE
GJA1
APOE
GPR126
SERPINA1
NCAPG
APOE
SERPINA1
CD34
GPR126
APOE
MFN1
UQCR10
SLC5A6
UQCR10
PVRL2
GJA1
SERPINA1
PLN
WNT16
MLF1
GJA1
APOE
ARHGEF40
NDRG2
DLEU1
GJA1
EIF3E
SERPINA1

EIF3E
TOMM40
CMTM5
NDRG2
UQCR10
LXN
EIF3E
NCAPG
APOE
SLC5A6
VSTM2L
NRBP1
NIPSNAP1
NCAPG
EIF3E
ZNF219
EIF3E
EIF3E
SLC35F1
NDRG2
APOE
EIF3E
SERPINA1
NDRG2
EIF3E
NCAPG
UQCR10
NIPSNAP1
WNT16
GJA1
NDRG2
PLN
APOC1
UQCR10
APOE
APOE
SNX17;APOE
NCAPG
EIF3E
UQCR10
SERPINA1
EIF3E
MYH7
APOE
FAM174B

SNX17
MYH7
DES
CCDC91
DES
UQCR10
SERPINA1
VSTM2L
APOC1
UQCR10
PTEN;EIF3E
DNAJC1;EIF3E
UQCR10;MYH7
NOV;EIF3E
CR1
GJA1
APOE
SNX17
APOE
GJA1
APOE
SERPINA1
DGKH
DNAJC1
APOE
EIF3E
TBX15
LXN
LXN
MLF1
PVRL2
APOC1
WNT16
SERPINA1
DNAJC1
GJA1
SNX17
APOE
SERPINA1
IFI27L2
APOE
EIF3E
MYH7
GJA1
PTEN

EIF3E
GJA1
HORMAD2
EIF3E
NCAPG;APOE
FAM3C
CD34
GJA1
IFI27L2
NOV
APOE
APOE
EIF2B4
GJA1
APOE
TRIB2
ALX4
PTEN
NDRG2
TOMM40
NCAPG
ARHGEF40
APOE
GPR126
ASCC2
EIF3E
SERPINA1
APOC1
MLF1
MLF1
CENPW
EIF3E;TRIB2
SLC5A6
SLC4A1AP;TOMM40;HSPA4;CEP85L;PTEN;UQCR10;GPN1;NIPSNAP1;NRBP1;TTN;PPM1G;GJA1;SNX17;M
MLF1
GJA1
PLN;NOV;RSRC1;SHOX2;SPEG;DGKH
SLC5A6;GCKR;SERPINA1;APOC1;GPR126;C2ORF16;APOE;PVRL2;NIPSNAP1
ALX4;MYH6;MYH7
NOV;SNX17;DLEU1
GJA1;HSPA4;CR1L;GPN1
DES;PLN;UQCR10
GJA1;GPR126;FAM3C
APOC1;ZMAT5;NAT14
GCKR;UQCR10;NDRG2;NIPSNAP1

APOC1;UQCR10;APOE
EIF3E;FAM3C;APOE
SLC35F1;CD34;MLF1
MTMR3;PLN;MYH6
TOMM40;RSRC1;CD34
GJA1;HSPA4;APOE
DES;PLN;UQCR10
PLN;NCAPG;MYH6
GJA1;NOV;SHOX2
LXN;SLC35F1;CD34;MLF1
GJA1;AFAP1;SPEG;TRIB2
GJA1;NOV;MLF1;MYH7
TBX15;CENPW;DLEU1;WARS2
GJA1;NOV;APOE;NDRG2
NOV;GNB4;APOE;CD34
DES;MFN1;UQCR10;NDRG2
NOV;APOE;CD34;MLF1
CCDC91;GPR126;APOE;CD34
GCKR;APOE;MLF1
GJA1;APOC1;APOE
GJA1;NCAPG;NDRG2;MLF1
SHOX2;ZMAT5;NRBP1
FAM174B;NDRG2;MLF1
TBX15;DCAF16;NCAPG
GJA1;SNX17;UQCR10
SERPINA1;UQCR10;APOE
GJA1;GNB4;MLF1
LXN;SHOX2;APOE;CD34
CCDC91;NDRG2;MYH7
LXN;GNB4;CD34
CENPW;SLC4A1AP;SHOX2;LCORL
LXN;SNX17;TRIB2
GJA1;APOC1;APOE
LXN;GPR126;EIF3E
LXN;APOC1;APOE;NDRG2
GJA1;APOC1;GPR126;APOE
DES;APOC1;LCORL
DES;MLF1;MYH7;TTN
GJA1;PLN;DLEU1
GJA1;SNX17;MYH7
ZCCHC10;NOV;MLF1
TBX15;NOV;CD34;MLF1
TBX15;CR1L;MLF1
SLC5A6;GNB4;APOE
NDRG2

DES;SNX17;EIF3E;TTN
GJA1;RSPO2;NIPSNAP1
SKIDA1;APOC1;SSC5D;TRIB2
DCAF16;GJA1;CCDC91
LCORL;MLF1;MYH6;TTN
GJA1;CENPW;SERPINA1;LXN
LXN;SNX17;VSTM2L;APOE
TBX15;SKIDA1;CD34
LXN;NOV;RSPO2;UQCR10
LXN
PPM1G
NDRG2
APOC1
EIF3E
FAM3C
FAM3C
NRBP1
GPN1
APOE
SERPINA1
APOE
CD34
SNX17
LXN
GJA1;MFN1;NDRG2;MYH7
TBX15;CENPW;SKIDA1;SHOX2
UQCR10;MLF1;TTN
SNX17;EIF3E;APOE
CEP85L;NDRG2;MLF1
DES;EIF3E;NDRG2
LXN;NOV;APOE
SNX17;NDRG2;TRIB2;PPM1G
LXN;CD34;NRBP1
SNX17;UQCR10;APOE
TOMM40;APOC1;ZMAT5;APOE
GJA1;LXN;EIF3E;APOE
FAM174B;APOE;MLF1;MYH6
DCAF16;RSRC1;NCAPG;FAM3C
LXN;SNX17;EIF3E
APOC1;UQCR10;APOE
LXN;APOE;NDRG2
EIF3E;FAM3C;APOE
SLC5A6;GJA1;SNX17;CD34
GJA1;SNX17;RSPO2;EIF3E
NOV;RSPO2;GPR126;APOE

CENPW;SHOX2;NCAPG;CD34
ZCCHC10;B3GNT7;NOV;MLF1
TBX15;MLLT10;NOV;CD34
GCKR;NOV
CCDC121;DES
LXN;NDRG2
GJA1;PPM1G
UQCR10;NDRG2
GJA1;HSPA4;APOC1
NOV;UQCR10;APOE
GJA1;UQCR10;APOE
CENPW;EIF3E
EIF2B4;CR1L
UQCR10;APOE
APOE;MYH7
FAM174B;APOE
GJA1;APOE;NDRG2
GJA1;APOE
DES;APOE
GJA1;SERPINA1
GJA1;SNX17;APOE
SNX17;APOC1;TTN
IFI27L2;CENPW
GJA1;PTEN;UQCR10;FAM3C
LXN;APOC1
APOC1;UQCR10;APOE;NIPSNAP1
GJA1;VSTM2L;TRIB2
DES;PLN;TRIB2
PTEN;UQCR10;FAM3C
GJA1;EIF3E;APOE
PLN;EIF3E
SLC5A6;EIF3E
GJA1;PPM1G
GJA1;NOV
HSPA4;APOC1
GJA1;APOE
SLC5A6;NCAPG
UQCR10;TRIB2
APOC1;APOE;TRIB2
LXN;NOV;GPR126
DES;APOE;MYH7
UQCR10;GPN1;PPM1G
NOV;SHOX2
EIF2B4;DES;APOE
APOC1;EIF3E;APOE

FAM174B;NCAPG;MLF1
GCKR;APOC1;NDRG2
SLC5A6;APOC1;UQCR10
LXN;SKIDA1;GPR126;CCDC141
GCKR;APOC1;UQCR10
SKIDA1;PVRL2
CENPW;PVRL2;GPN1
GJA1;LXN;APOE
SKIDA1;PVRL2
DES;MYH6;MYH7
EIF3E;FAM3C;APOE
HSPA4;NCAPG;EIF3E
EIF3E;APOE;MYH7
GJA1;SNX17;GNB4
GJA1;SKIDA1
NOV;FAM174B;SLC35F1
LXN;HSPA4;UQCR10
TBX15;NOV;APOC1;MLF1
GJA1;LXN;GNB4
GJA1;APOC1;APOE
GCKR;APOC1;APOE
SNX17;SPEG;MYH7
DES;B3GNT7;NDRG2
NOV;SHOX2;RSPO2
EIF2B4;APOC1;APOE
PLN;APOE;NDRG2
GCKR;UQCR10;NDRG2
APOC1;APOE
GJA1;SNX17;CD34
LXN;SKIDA1;GPR126;CCDC141
GJA1;APOC1;UQCR10
GJA1;NCAPG;NDRG2;MLF1
CENPW;EIF3E;PPM1G
SLC35F1;HORMAD2;CD34;MLF1
GJA1;SNX17;NDRG2;MYH7
DES;MLF1;MYH6;MYH7
GJA1;RSPO2;NDRG2
NDRG2;CCDC141;MYH7
LXN;SNX17;EIF3E
EIF3E;UQCR10;FAM3C
DES;SNX17;APOE
CENPW;NCAPG;EIF3E
LXN;NOV;UQCR10
UQCR10;APOE;PPM1G
GJA1;HSPA4;APOE

PTEN
GJA1;CENPW;UQCR10
DES;SNX17;MYH7
TOMM40;UQCR10;PPM1G
GJA1;FAM3C;TRIB2
APOC1;APOE;NDRG2
UQCR10;APOE;MYH7
GJA1;EIF3E;MYH6
APOC1;APOE;NDRG2
LXN;UQCR10;APOE
GJA1;MLF1;NIPSNAP1
GJA1;LXN;CD34
NOV;SNX17
UQCR10;APOE
CABP7;VSTM2L
HSPA4;CD34
SLC5A6;ZNF512
SNX17;APOE
SNX17;APOE
CCDC121;GJA1
GJA1;NOV
SNX17
TOMM40;FAM3C
DCAF16;NIPSNAP1
TOMM40;APOC1
GJA1;TRIB2
CABP7;CD34
APOE;CD34
ZCCHC10;GPR126
APOE;DGKH
APOE;MLF1
ZCCHC10;DGKH
GJA1;TOMM40
HSPA4;MFN1
LXN
GJA1;SERPINA1
GJA1;NCAPG
MYH7;DGKH
GCKR;APOC1
UQCR10;APOE
CD34
APOE
EIF3E;UQCR10
LXN;EIF3E
SNX17;APOE

APOE;NDRG2
UQCR10;APOE
ALX4;WNT16
GJA1;ZNF513
DCAF16;LXN
EIF3E;NIPSNAP1
EIF3E;APOE
GJA1;NOV
FAM174B;MLF1
LXN
SERPINA1
APOC1;APOE
GJA1;APOE
TOMM40;EIF3E
CCDC121;GJA1
AFAP1;CD34
NOV;GNB4
EIF2B4;LXN
CENPW;RSPO2
SLC35F1;MYH7
AFAP1;EIF3E
SNX17;GPR126
SNX17;APOE
NOV;NDRG2
GNB4;CD34
UQCR10;NIPSNAP1
PTEN;LCORL
SNX17;APOE
LXN;EIF3E
WARS2
CENPW;NCAPG
GFM1;APOC1
SNX17;APOE
APOE;NDRG2
SNX17;APOE
APOE;PVRL2
SNX17;EIF3E
MTMR3;TRIB2
CENPW;LCORL
GJA1;LXN
GJA1;APOE
CENPW;DES
NOV;WNT16
CR1L;TTN
DLEU1;NCAPG

APOE;NDRG2
WNT16;TRIB2
ZNF219;APOE;NIPSNAP1
GFM1;GPR126
FAM3C;PVRL2
APOC1;NCAPG
NCAPG;UQCR10
AFAP1;SSC5D
SNX17;APOE
TBX15;VSTM2L
NOV;GPR126
EIF2B4;MLF1
SNX17;APOE
CD34;MYH6
TBX15;MYH7
TOMM40;PPM1G
SNX17;APOE
GJA1;EIF3E
LXN;GPR126
SNX17;APOE
ZCCHC10;NOV
GJA1;APOE
DES
TOMM40
PPM1G
NCAPG
GJA1
APOE
MTMR3
IFI27L2
APOE
CWC27
TRIB2
UQCR10;PPM1G
NRBP1
LXN;VSTM2L
NOV;FAM174B
SKIDA1;TRIB2
PLN
MFN1;NIPSNAP1
GJA1;APOE
DES;SNX17
NDRG2;MYH7
DES;APOE
GJA1;NOV

APOE;NIPSNAP1
APOE
LXN
SERPINA1
NCAPG
NOV
APOE
LXN
NDRG2
GJA1
CABP7
APOC1
NOV
SLC35F1;TRIB2
APOE
UQCR10
APOC1
GJA1
MLF1
LXN
APOE
APOE
APOE
TBX15
TRIB2
SLC5A6;GJA1
PLN;TRIB2
APOE;NDRG2
GCKR;APOE
NOV;APOE
DES;APOE
CENPW;PVRL2
SNX17;PPM1G
PLN;GPR126
APOE;PPM1G
EIF3E;FAM3C
GJA1;HORMAD2
APOC1;NIPSNAP1
EIF3E;MLF1
CENPW;LXN
APOE;NDRG2
TTN
LXN;CD34
UQCR10;MLF1
SKIDA1;ARHGEF40

VSTM2L;HORMAD2
CEP85L;GPR126
RSRC1;SLC35F1
GJA1;APOE
SKIDA1;APOC1
HSPA4;UQCR10
CENPW;NCAPG
CMTM5;UQCR10
SNX17;MLF1
SHOX2;EIF3E
APOE;NDRG2
SERPINA1;EIF3E
VSTM2L;MYH7
EIF3E;UQCR10
GJA1;SNX17;EIF3E
SLC5A6
GJA1;LXN
HSPA4;EIF3E
LXN;CD34
GJA1;APOC1
EIF3E;UQCR10
APOE;CD34
HSPA4;UQCR10
MFN1;APOE
APOE
RSPO2;DGKH
UQCR10;FAM3C
LXN;NOV
TBX15;NOV
APOC1;SUPT7L
LCORL;PVRL2
GJA1;APOE
SNX17;AFAP1
PLN;NOV
GJA1;GNB4
GJA1;APOC1
SHOX2;UQCR10
SNX17;APOE
GJA1;NIPSNAP1
SNX17;APOE
GJA1;HSPA4;APOC1
SNX17;MYH6
SNX17;APOE
SERPINA1;APOE
EIF3E;NDRG2

SHOX2;RSPO2
TBX15;GPR126;CD34
FAM174B;APOE;NDRG2
GJA1;MLF1
DNAJC1;CR1L;AFAP1
GJA1;LXN;EIF3E
ZCCHC10;FAM174B
GJA1;EIF3E;FAM3C
NCAPG;EIF3E
SNX17;APOE
GJA1;NOV;GNB4
GJA1;SNX17;APOE
NOV;GPR126;NIPSNAP1
SHOX2;CD34;MLF1
EIF3E;APOE
TBX15;NCAPG;MYH7
SNX17;NDRG2
NOV;NDRG2
APOC1;APOE
IFI27L2;FAM174B;VSTM2L
NCAPG;MYH6
WNT16;MYH6;MYH7
LXN;SNX17
LXN;UQCR10;APOE
GCKR;GJA1;APOE
NOV;CD34
GNB4;SSC5D
LXN;APOE;CCDC141
EIF3E;UQCR10
GJA1;GNB4
EIF3E;FAM3C
HSPA4;EIF3E;PPM1G
ZNF628;APOE
DLEU1;ALX4
APOE;CD34;MLF1
GJA1;CMTM5
SNX17;PPM1G
GJA1;DGKH
GJA1;SNX17
GJA1;RSPO2;APOE
GJA1;NDRG2
GJA1;RSRC1
GJA1;EIF3E
GJA1;EIF3E
LXN;APOC1

SNX17;EIF3E;NDRG2
PLN;APOE;TTN
GJA1;ZNF219
NCAPG;MYH7
NDRG2;MYH6
SNX17;APOE
APOE;WNT16
IFI27L2;CENPW
SLC5A6;NOV
CENPW;GNB4
CCDC121;CR1L
ZNF219;APOE
GNB4;APOE
GJA1;ZNF219
GJA1;TOMM40;NIPSNAP1
NOV;MYH7
NOV;RSPO2
EIF3E;MYH7
PLN;APOC1
PTEN;EIF3E
GJA1;APOE
EIF3E;UQCR10
SKIDA1;NDRG2
CENPW;NCAPG
GJA1;SERPINA1
UQCR10;NIPSNAP1
LXN;LCORL
NDRG2;CD34
UQCR10;MLF1
APOC1;APOE
GJA1;UQCR10;APOE
SLC5A6;GJA1
GJA1;EIF3E
SNX17;UQCR10
LXN;EIF3E;GPN1
TBX15;WARS2
IFI27L2;ZNF219
GJA1;FAM3C
GJA1;NOV;CABP7
DNAJC1;CR1L;DLEU1
GCKR;CABP7;GNB4
CENPW;LXN;NCAPG
SERPINA1;FAM3C
APOC1;GPR126;APOE
APOC1;EIF3E

FAM3C;CWC27;MYH7
APOC1;UQCR10
GJA1;SNX17
CENPW;AFAP1;CD34
SLC5A6;NOV
NOV;APOC1;APOE
SLC5A6;SERPINA1;ZNF512
NCAPG;UQCR10;TTN
NOV;NDRG2
NOV;APOE;GPN1
LXN;APOC1;APOE
SLC5A6;LXN;SLC35F1;NDRG2
GPR126;SLC35F1;APOE
APOC1;APOE;MYH7
SNX17;APOE
GFM1;GJA1
DES;SNX17;APOE
EIF2B4;SNX17;AFAP1
GJA1;CR1;LXN
UQCR10;APOE
CCDC91;PTEN;APOE
GJA1;ZCCHC10
SLC5A6;SERPINA1;FAM174B
ZCCHC10;HSPA4;MLF1
CENPW;EIF3E;UQCR10
GJA1;LXN;FAM174B;APOE
FAM3C;APOE;TRIB2
SHOX2;GNB4;MLF1
DES;CD34;MYH7
NOV;FAM3C;WNT16
APOC1;APOE;NIPSNAP1
LXN;EIF3E;FAM3C
GJA1;EIF3E;UQCR10
GJA1;LXN;PPM1G
GJA1;ZNF219;NCAPG
SLC5A6;SERPINA1;FAM174B
TOMM40;ZNF513;FAM3C
TOMM40;APOE;PVRL2
CENPW;EIF3E;UQCR10
MFN1;UQCR10
PLN;MYH6;MYH7
NOV;PTEN;EIF3E
SNX17;APOC1;FAM3C
APOC1;APOE
GJA1;EIF3E

GJA1;APOE
UQCR10;MYH7
LXN;GPR126;APOE
DNAJC1;GJA1;GNB4
TOMM40;VSTM2L
SLC5A6;CCDC121;APOC1
NOV;EIF3E
CENPW;GPR126
LXN;NOV;UQCR10
NOV;TRIB2
GJA1;NDRG2;MYH7
CMTM5;C2ORF16
GJA1;HSPA4;GPN1
CMTM5;C2ORF16
GJA1;LXN
ZMAT5;MLF1
UQCR10;APOE
PLN;MLF1
SNX17;SPEG
TRIB2;NAT14
NOV;CMTM5
GJA1;HSPA4;GPN1
SHOX2;RSPO2
SHOX2;NRBP1
GJA1;CENPW
LXN;EIF3E;APOE
APOC1;APOE
UQCR10;APOE
DCAF16;RSRC1
APOC1;APOE;NDRG2
LXN;NCAPG;FAM3C
SHOX2;GNB4;GPR126
APOE;NDRG2
SLC5A6;LXN;UQCR10
DLEU1;NCAPG
FAM3C;TRIB2
EIF3E;APOE
MYH7;DGKH
GJA1;APOC1;APOE
GJA1;SNX17;EIF3E
CR1L;EIF3E
GJA1;DES;PVRL2
APOC1;NDRG2;MLF1
EIF2B4;TOMM40;EIF3E
DES;MLF1;MYH6

SNX17;EIF3E;UQCR10
APOE;NDRG2
ZMAT5;UQCR10
FAM174B;CD34
GJA1;APOC1
SERPINA1;VSTM2L
GJA1;NCAPG;CD34
PLN;NAT14
SNX17;APOE
AFAP1;NIPSNAP1
APOC1;UQCR10;NDRG2
CCDC141;MYH7
GJA1;APOE
UQCR10;APOE
EIF3E;TTN
DES;UQCR10
CR1L;RSRC1
TOMM40;VSTM2L
GJA1;EIF3E;NDRG2
ASCC2;NIPSNAP1
FAM3C;NDRG2
TBX15;MYH7
LCORL;TRIB2
DES;UQCR10
LXN;AFAP1
B3GNT7;GPR126
GJA1;FAM174B
MLF1;TTN
GJA1;APOC1
GJA1;NDRG2
LXN;TRIB2
DNAJC1;CR1L
DES;APOE
LXN;ZNF512
GJA1;LXN
SLC5A6;PPM1G
UQCR10;NRBP1
NCAPG;APOE;MLF1
UQCR10;APOE
NAT14;NIPSNAP1
LXN;SNX17
DES;TRIB2
TBX15;SNX17
CENPW;MLF1
GJA1;GPR126

GJA1;DLEU1
RSPO2;EIF3E
APOE;NIPSNAP1
SNX17;MYH7
FAM174B;MLF1
APOC1;UQCR10
GJA1;RSRC1;EIF3E
HSPA4;GPR126;EIF3E
EIF3E;UQCR10
NOV;CD34
GJA1;NOV
PLN
UQCR10;FAM3C
GJA1;APOE
CCDC91;MYH7
EIF3E;APOE
GJA1;APOE
UQCR10;NIPSNAP1
APOE;NDRG2;NIPSNAP1
GJA1;SERPINA1;TRIB2
SPEG;NIPSNAP1
APOE;NDRG2;NIPSNAP1
SNX17;APOE
UQCR10;TTN
GCKR;APOE
CENPW;LXN
DGKH;WARS2
GJA1;APOE
NDRG2;DGKH
GCKR;B3GNT7
ZCCHC10;CD34
EIF3E;UQCR10
GNB4;APOE
GJA1;FAM3C
CENPW;NCAPG
NOV;UQCR10
GNB4
GJA1;NOV
TOMM40;UQCR10
APOC1;NDRG2
GJA1;SNX17
FAM3C;WNT16
MTMR3;LXN
PLN;MYH7
LXN;GPN1

APOE;NIPSNAP1
GJA1;FAM3C
CENPW;NOV
APOC1;APOE
GJA1;SNX17
APOC1;APOE
SLC5A6;ZCCHC10
GJA1;EIF3E
LXN;SNX17
LXN;MLF1
APOC1;APOE
RSPO2;SPEG
GPR126;HORMAD2
CR1L;PPM1G
APOC1;ZNF513;APOE
NAT14;MLF1
TOMM40;RSRC1
ALX4;HORMAD2
LXN;NRBP1
ALX4;WNT16
GPR126;CD34
UQCR10;MLF1
UQCR10;APOE
DNAJC1;UQCR10
LXN;NOV
DNAJC1;LXN;NCAPG
GJA1;SNX17
RSPO2;GPR126
CENPW;NCAPG
EIF3E;FAM3C
SERPINA1;TRIB2
GCKR;APOE
GJA1;APOE
MTMR3;APOC1
GJA1;SNX17
GJA1;APOE
DNAJC1;DGKH
GJA1;CD34;MLF1
GJA1;NCAPG;TRIB2
LXN;PPM1G
GJA1;CENPW
MTMR3;SHOX2
PLN;MYH6
NOV;SNX17
DES;NOV

FAM174B;GPN1
NOV;APOE
MYH6;MYH7
APOE;NDRG2
LXN;ZNF512
ZCCHC10;EIF3E;UQCR10
APOE;NDRG2
GJA1;MYH6
HSPA4;SNX17
APOC1;UQCR10
GJA1;NIPSNAP1
TRIB2;NIPSNAP1
RSRC1;TRIB2
SLC5A6;GJA1
EIF2B4;SNX17
CCDC121;NCAPG
CCDC121;SHOX2
NDRG2;CD34
GJA1;MYH6
EIF3E;UQCR10
PVRL2;NIPSNAP1
GPR126;HORMAD2
NOV;EIF3E
NDRG2;DGKH
LXN;DLEU1
APOE;NDRG2
SNX17;APOE
ZCCHC10;CABP7
EIF3E;UQCR10
TBX15;LXN
GCKR;TRIB2
EIF2B4;SNX17
GNB4;CD34
PLN;APOC1
DES;CD34
LXN;TRIB2
SERPINA1;FAM174B
NOV;EIF3E
SERPINA1;NAT14
GJA1;SHOX2;AFAP1
GJA1;UQCR10
EIF3E;UQCR10
PLN;APOE
GJA1;APOE
PLN;NOV

APOC1;APOE
GNB4;FAM3C
FAM174B;APOC1
SLC5A6;TOMM40
ZCCHC10;MYH7
GJA1;TRIB2
GJA1;APOE
CR1L;SNX17
GPR126;NIPSNAP1
UQCR10;WNT16
NOV;APOE
EIF3E;UQCR10
CABP7;MLF1
NOV;CABP7
LXN;SKIDA1
GJA1;UQCR10
ZNF219;SLC35F1
NOV;SNX17
APOC1;MYH7
LXN
SERPINA1
CD34
DGKH
SNX17
NOV
EIF3E
TTN
GJA1
GPR126
RSRC1
HORMAD2
SSC5D
GJA1
PVRL2
APOE
GNB4
UQCR10
NOV
APOE
GJA1
GPN1
UQCR10
FAM174B
ZNF219
GJA1

NOV
TRIB2
DES
CABP7
SNX17
FAM3C
PLN
EIF2B4
LXN
FAM3C
EIF3E
APOE
APOE
SUPT7L
DES
NDRG2
APOC1
FAM3C
GJA1
B3GNT7
TRIB2
GJA1
APOC1
SNX17
TBX15
CR1L
UQCR10
CENPW
UQCR10
UQCR10
UQCR10
SNX17
APOE
LXN
APOE
TRIB2
SNX17
LXN
EIF2B4
CD34
EIF3E
SLC5A6
NCAPG
APOE
GCKR

EIF3E
EIF3E
DGKH
NOV
SUPT7L
APOE
APOE
UQCR10
GJA1
GJA1
GNB4
UQCR10
TRIB2
APOE
APOC1
LXN
TRIB2
FAM3C
MYH7
NCAPG
APOC1
APOC1
APOE
GJA1
SNX17
CENPW
PTEN
UQCR10
ZNF219
SNX17
SSC5D
RSPO2
NCAPG
EIF3E
NAT14
IFI27L2
APOC1
CENPW
CENPW
TRIB2
TTN
SERPINA1
EIF3E
PTEN
RSRC1

MLF1
LXN
HORMAD2
UQCR10
SNX17
EIF3E
UQCR10
APOC1
NOV
GJA1
NCAPG
GNB4
APOE
NOV
GJA1
NDRG2
TOMM40
GNB4
APOE
PTEN
APOE
TTN
FAM3C
SERPINA1
APOE
CD34;TRIB2
GPR126
LXN
PLN
SNX17
NOV
UQCR10
EIF3E
LXN
FAM3C
NDRG2
MYH7
SERPINA1
LXN
NOV
GJA1
NCAPG
SLC5A6
EIF3E
HORMAD2

MLF1
APOE
UQCR10
CENPW
FAM3C
CCDC141
SNX17
NOV
SERPINA1
APOE
FAM174B
MLF1
EIF3E
APOC1
NDRG2
EIF3E
SLC5A6
ASCC2
UQCR10
GJA1
APOE
NOV
GJA1
FAM3C
EIF3E
FAM3C
CENPW
HORMAD2
NDRG2
NDRG2
GJA1
EIF3E
PPM1G
SNX17
EIF3E
GJA1
C2ORF16
PPM1G
TOMM40
TRIB2
EIF3E
SKIDA1
NOV
GNB4
APOE

APOE
LXN
DLEU1
SNX17
UQCR10
APOE
CD34
UQCR10
NDRG2
FAM3C
GJA1
NOV
VSTM2L
NDRG2
APOE
EIF3E
SLC35F1
B3GNT7
UQCR10
DCAF16
FAM174B
GJA1
CR1L
DCAF16
EIF3E
SERPINA1
LXN
GJA1
GJA1
SNX17
GCKR
MYH7
TRIB2
APOE
NOV
SERPINA1
LXN;SNX17;NIPSNAP1
TBX15;NOV;GPR126
TBX15;MYH7;TTN
UQCR10
CR1L
MLF1
MYH7
APOE
LXN

PPM1G
APOE
APOE
SLC35F1
GNB4
FAM174B
NOV
MLF1
CENPW;TOMM40;GNB4
PPM1G
GJA1
EIF3E
RSPO2
EIF3E
SNX17
NDRG2
UQCR10
GJA1
CCDC91
FAM174B
ZCCHC10
GJA1;GPR126;UQCR10
CCDC91;NDRG2;MYH7
APOE;CD34;MYH7
GJA1;VSTM2L;UQCR10
SERPINA1;CR1;CR1L
NOV;UQCR10;APOE
TOMM40;APOE;PPM1G
GNB4;EIF3E;NIPSNAP1
APOC1;APOE
RSPO2;CCDC141;TRIB2
APOC1;APOE;NDRG2
LXN;SLC4A1AP;PVRL2
NOV;PTEN;APOE
DES;CD34;MYH7
FAM3C;WNT16
GJA1;SPEG;TRIB2
LXN;APOE;TRIB2
LXN;NOV;EIF3E
GJA1;CD34
SNX17;UQCR10;APOE
NCAPG;MYH6;MYH7
LXN;EIF3E;FAM3C
GJA1;AFAP1;UQCR10
APOC1;NCAPG

NOV;RSPO2;CD34
LXN;CABP7;GPR126
CCDC91;PTEN;TRIB2
GJA1;APOC1;ARHGEF40
GNB4;ARHGEF40;TTN
GJA1;ZMAT5;MFN1
CR1L;FAM174B;APOE
LXN;SNX17;UQCR10
GJA1;DES;PPM1G
CABP7;VSTM2L;DGKH
LXN;NDRG2;PVRL2
NCAPG;EIF3E;UQCR10
SHOX2;UQCR10;APOE
LXN;EIF3E;FAM3C
NOV;FAM174B;APOE
GJA1;SERPINA1;TRIB2
SERPINA1;UQCR10;NIPSNAP1
UQCR10
CABP7;SHOX2;MYH7
CENPW;EIF3E;UQCR10
APOC1;APOE;PPM1G
SNX17;EIF3E;APOE
NOV;GPR126
LXN;SLC4A1AP;PVRL2
FAM174B;APOC1;MLF1
CCDC121;LXN;TRIB2
GJA1;GNB4
GJA1;CENPW
GJA1;PLN;EIF3E
CENPW;NCAPG;MLF1
EIF3E
APOE
UQCR10
PTEN
GPR126
SNX17
CR1L
CD34
EIF3E
PPM1G
FAM3C
GCKR
FAM174B
SNX17
UQCR10

GCKR
EIF3E
TOMM40
CD34
ZMAT5
PPM1G
MLF1
FAM3C
APOE
NCAPG
DGKH
UQCR10
SNX17
NDRG2
RSPO2
GJA1
UQCR10
CENPW
NOV
LCORL
GJA1
GCKR
SLC35F1
APOE
TRIB2
APOC1
EIF3E
UQCR10
LXN
APOE
EIF3E
MYH7
TOMM40
GJA1
MYH7
APOE
GPR126
GJA1
UQCR10
TTN
PPM1G
FAM174B
CWC27
MYH7
FAM3C

UQCR10
LXN
MLF1
LXN
NOV
NOV
EIF3E
FAM3C
FAM3C
NOV
APOE
SLC5A6
UQCR10
GJA1
NOV
SNX17
SERPINA1
APOE
B3GNT7
APOC1
CENPW
EIF3E
PPM1G
UQCR10
APOE
FAM174B
CD34
GJA1
FAM3C
EIF3E
PLN
LXN
LXN
DNAJC1
GJA1
APOE
SLC5A6
GPR126
APOC1
EIF3E
MFN1
SERPINA1
ZCCHC10
TRIB2
SNX17

VSTM2L
CEP85L
SLC5A6
GJA1
SLC35F1;MLF1
UQCR10
MYH6
EIF2B4
EIF3E
EIF3E
UQCR10;NDRG2
GPR126
APOE;CD34
GJA1
EIF3E
GJA1
LXN
UQCR10
GJA1
PLN
FAM174B
GCKR
DGKH
GPR126
UQCR10
NIPSNAP1
LCORL
WNT16
DNAJC1
LXN
SNX17
GJA1;ZMAT5
LXN
NOV
NCAPG
FAM3C
SNX17
SNX17
APOE
TOMM40;APOE
MYH7
DES
APOE
UQCR10;FAM3C
CENPW

SNX17
UQCR10
APOE
MYH7
FAM3C
GJA1
APOE
GJA1
FAM174B
APOC1
SLC5A6
TOMM40
MYH7
EIF3E
SERPINA1
GJA1
GJA1
VSTM2L
GJA1
SNX17
APOE
NCAPG
EIF3E
DES
UQCR10
PLN
NDRG2
CABP7
ZCCHC10
EIF3E
MYH7
CMTM5
EIF3E
DES
SLC35F1
ZCCHC10
CENPW
RSPO2
CR1L
GJA1;EIF3E
NCAPG
APOE
NRBP1
GJA1
GJA1

MYH7
EIF3E
MYH7
CENPW
GPN1
TBX15
UQCR10
CD34
APOE
NOV
CCDC141;TRIB2
GJA1
CD34;MYH7
CCDC121;GJA1
GNB4;UQCR10
SNX17;GNB4
LXN;APOE
GJA1;CD34
APOC1;UQCR10
TBX15;LXN
APOE;CD34
NOV;EIF3E
PLN;MYH6
EIF3E
CENPW;NCAPG
GJA1;NOV
GJA1;CD34
GJA1;APOE
LXN
SNX17;PPM1G
APOC1
APOE;PPM1G
CMTM5;VSTM2L
NDRG2;PVRL2
GJA1;TOMM40
APOE;NDRG2
B3GNT7;UQCR10
SNX17;GNB4
HSPA4;APOE
CABP7;GNB4
NOV;GPR126
CCDC121;DLEU1
LXN
GJA1;GPR126
EIF3E;APOE

GJA1;APOE
RSRC1;CD34
LXN;FAM3C
SLC35F1;MLF1
TOMM40;PPM1G
NCAPG
APOE
LXN;APOE
NDRG2;NIPSNAP1
GJA1;NOV
CENPW;CCDC141
EIF3E
NOV;UQCR10
GJA1;GNB4
APOE
APOE;TRIB2
NCAPG
SERPINA1;UQCR10
LXN;TRIB2
GJA1
LXN;TRIB2
SLC5A6
CD34;MYH7
GJA1
NOV;HORMAD2
GJA1
UQCR10
APOE
GJA1
NOV;GPR126
SNX17;APOE
FAM3C
NOV
RSPO2
APOC1
APOE
FAM3C
VSTM2L
UQCR10;APOE
GJA1;NRBP1
LXN
APOE;NDRG2
APOE;MYH7
GCKR;NIPSNAP1
MLF1

NRBP1
LXN;NOV
LXN;MYH7
NCAPG
LXN
NDRG2
NDRG2;PVRL2
SNX17;CD34
CD34
SNX17;APOE
CCDC141;TTN
APOC1
NOV
NOV;PVRL2
GJA1;EIF3E
APOC1;NIPSNAP1
ALX4
SLC35F1
APOE
NOV
SERPINA1
NCAPG;LCORL
NCAPG
TTN
EIF3E
MLF1
GJA1
SNX17
GJA1
GPR126
NDRG2
LXN
NOV
APOE
EIF3E
UQCR10
FAM3C
SNX17
UQCR10
LXN
APOE
PVRL2
NDRG2
GJA1
UQCR10

GPR126
DNAJC1
CENPW
APOE
EIF3E
APOE
HSPA4
LXN
APOE
APOE
SUPT7L
UQCR10
CD34
NOV
SERPINA1
CR1L
FAM174B
LXN
CABP7
MLF1
UQCR10
UQCR10
RSPO2
DGKH
SLC5A6
VSTM2L
SLC5A6
GNB4
TRIB2
GCKR
UQCR10
DGKH
PLN
EIF3E
APOE
APOE
APOC1
GJA1
GJA1
FAM3C
GJA1
CD34
GJA1
APOC1
PLN

DES
DES
NOV
NIPSNAP1
WARS2
FAM3C
MLF1
CENPW
LXN
GJA1
CR1L
UQCR10
APOE
GJA1
EIF3E
CABP7
UQCR10
DCAF16
NDRG2
CD34
HORMAD2
UQCR10
LXN
APOE
UQCR10
UQCR10
APOC1
GJA1
NOV
LXN
EIF3E
GJA1
NOV
CD34
LXN
LXN
EIF3E
APOE
UQCR10
TRIB2
NOV
NOV
MYH7
DNAJC1
NOV

MYH7
NDRG2
LXN
EIF3E
TRIB2
NDRG2
SNX17
APOE
APOE
MTMR3
NOV
GCKR
NDRG2
CENPW
UQCR10
EIF3E
PPM1G
APOC1
CENPW
APOE
ZCCHC10
EIF3E;UQCR10
SNX17
UQCR10
NIPSNAP1
CENPW
PPM1G
GJA1
CCDC121;MTMR3;ZCCHC10;SLC4A1AP;CEP85L;NCAPG;CWC27;PPM1G;CCDC91;ZNF628;DLEU1;ALX4;A[illegible]
LXN;PLN;CCDC91;DLEU1;CCDC141;MLF1;MYH6
TBX15;DES;CCDC141;MYH6;MYH7;TTN
GNB4
FAM174B
TRIB2
AFAP1
SLC4A1AP;TOMM40;SHOX2;CEP85L;VSTM2L;UQCR10;NDRG2;GPN1;NIPSNAP1;TTN;PPM1G;GJA1;SNX1[illegible]
LXN
SSC5D
EIF3E
PPM1G
APOE
CENPW
GPR126
RSPO2
NRBP1

SLC5A6
NCAPG
LXN
TRIB2
NRBP1
PTEN
GJA1
MLLT10;ZNF219;RSPO2;AFAP1;SSC5D
MLLT10;ZNF219;RSPO2;AFAP1;SPEG;SSC5D
SLC5A6;GCKR;SERPINA1;SKIDA1;APOC1;GPR126;APOE;NIPSNAP1
IFI27L2;CABP7;CMTM5;APOE;NDRG2
MLF1
SERPINA1
TTN
DLEU1
CENPW
EIF3E
APOC1
SERPINA1
SLC5A6;GFM1;TOMM40;HSPA4;EIF3E;FAM3C;PPM1G
APOE
CENPW
APOC1
PLN
LXN
APOE
GPN1
ZNF512
MTMR3
SLC4A1AP;TOMM40;CEP85L;UQCR10;NDRG2;GPN1;NIPSNAP1;TTN;PPM1G;GJA1;SNX17;RSPO2;EIF3E;[illegible]
APOE
GCKR
NDRG2
DNAJC1
EIF2B4
DES
EIF3E
ZCCHC10
TBX15
HSPA4
PPM1G
LXN
APOE
NDRG2
ASCC2

SHOX2
TBX15;GFM1;DES;ZNF628;MYH6;MYH7;TTN;PPM1G
PLN
APOE
DGKH
TRIB2
SNX17
UQCR10
GNB4
SLC5A6
APOC1
LXN
NCAPG
PPM1G
UQCR10
MLF1
NRBP1
GNB4
GPR126
CD34
TRIB2
APOC1
NCAPG
LXN
B3GNT7
SNX17
TTN
TBX15
GJA1
PPM1G
SLC35F1
NOV
GJA1
TRIB2
CR1;PTEN;APOE;NRBP1
DGKH
SERPINA1;CR1;CR1L;CWC27;TRIB2
HORMAD2
DNAJC1
TBX15;ZNF628;FAM174B;SHOX2;DLEU1;CWC27;NDRG2;CCDC141;TRIB2;TTN
VSTM2L;ALX4;UQCR10
CCDC121;GFM1;SKIDA1;AFAP1;VSTM2L;SLC35F1;SSC5D;C2ORF16;MLF1;KIAA1755;GJA1;PLN;NOV;FAM
ALX4;MYH7
GCKR;SERPINA1;SKIDA1;AFAP1;VSTM2L;SSC5D;NDRG2;KIAA1755;GJA1;NOV;ZNF219;GPR126;SPEG;APC
CMTM5;ALX4;MYH6

ZNF628;ZMAT5;DLEU1;VSTM2L;NAT14
CMTM5;NCAPG;WNT16
VSTM2L;C2ORF16;WNT16;MYH6
PLN;ZMAT5;ALX4
HSPA4;APOC1
ZNF628;PTEN;NRBP1;DGKH;WARS2;TTN
CCDC121;GFM1;ZCCHC10;SLC4A1AP;TOMM40;SLC35F1;UQCR10;CWC27;NAT14;DCAF16;CCDC91;RSRC
ALX4;MYH6;TMEM253;MYH7
GCKR;WNT16
CCDC121;GFM1;ZCCHC10;SLC4A1AP;TOMM40;SHOX2;CEP85L;UQCR10;CWC27;NAT14;PPM1G;DCAF16
TBX15;CCDC121;SLC4A1AP;SKIDA1;ZMAT5;PTEN;ARHGEF40
CCDC121;GFM1;ZCCHC10;LXN;SLC4A1AP;TOMM40;CEP85L;PTEN;SLC35F1;UQCR10;CWC27;NAT14;DCA
TBX15;CCDC121;GFM1;SLC4A1AP;ALX4;ARHGEF40;CCDC141;GPN1;MYH6
ALX4;MYH6;MYH7
CENPW;ZNF628
SLC5A6;EIF2B4;MTMR3;ALX4;ZNF513;WNT16;NRBP1;PPM1G
MTMR3;ZCCHC10;LXN;TOMM40;PTEN;PPM1G;ZNF628;MFN1;ALX4;ASCC2;ARHGEF40;EIF2B4;C2ORF16
KIAA1755;DES;CR1;PLN;B3GNT7;CMTM5;SLC35F1;NDRG2;MYH6;MYH7
GFM1;TOMM40;SLC35F1;CWC27;NDRG2;NAT14;PPM1G;DCAF16;GJA1;PLN;CCDC91;ZNF628;FAM174B
IFI27L2;MTMR3;AFAP1;CWC27;GPN1;MLF1;TTN;PPM1G;DCAF16;NOV;GNB4;EIF3E;DGKH
TBX15;NOV;SHOX2;TMEM253
LXN;SHOX2;NCAPG;GPN1;TTN;MLLT10;GNB4;DLEU1;GPR126;ASCC2;EIF3E;ARHGEF40;LCORL;DGKH
GCKR;ALX4
ZNF628;VSTM2L;WNT16
WNT16;LCORL;TMEM253
IFI27L2;CCDC121;CENPW;SERPINA1;CR1;LXN;SKIDA1;PTEN;SSC5D;C2ORF16;NAT14;NIPSNAP1;TMEM2
DES;ZNF628;SNX17;ASCC2;CCDC141;GPN1;NRBP1;DGKH;WARS2;PPM1G
NCAPG
CR1;PLN;B3GNT7;FAM174B;CMTM5;SLC35F1;NDRG2;CCDC141;MYH6;MYH7
ALX4;MYH6;MYH7;DGKH
IFI27L2;CENPW;MTMR3;SLC4A1AP;TOMM40;NCAPG;UQCR10;WNT16;CWC27;TBX15;DES;GNB4;DGKH
DNAJC1;MTMR3;CR1;LXN;DLEU1;NCAPG;ASCC2;EIF3E;ARHGEF40;PVRL2;TMEM253;TTN
GFM1;ZCCHC10;TOMM40;UQCR10;CWC27;NAT14;PPM1G;DCAF16;CCDC91;ZNF628;FAM174B;MFN1;S
CEP85L;MYH6;MYH7
CCDC121;GFM1;ZCCHC10;LXN;SLC4A1AP;TOMM40;SHOX2;CEP85L;SLC35F1;CWC27;NAT14;TTN;PPM1
ZMAT5;DLEU1;VSTM2L
NCAPG;LCORL
CCDC121;GFM1;ZCCHC10;SLC4A1AP;TOMM40;CWC27;NAT14;DCAF16;GJA1;CCDC91;RSRC1;FAM174B;
SHOX2;DLEU1;MYH7
CCDC121;GFM1;ZCCHC10;SLC4A1AP;SLC35F1;CWC27;NAT14;GJA1;CCDC91;RSRC1;FAM174B;DLEU1;SP
DNAJC1;DES;CR1;LXN;PLN;ASCC2;EIF3E;ARHGEF40;PVRL2;TTN
EIF2B4;MTMR3;ZCCHC10;NCAPG;GPN1;NRBP1;SUPT7L;MLLT10;SNX17;MFN1;ASCC2;LCORL;DGKH
CCDC121;SERPINA1;MTMR3;CR1;PLN;B3GNT7;FAM174B;CMTM5;SLC35F1;NDRG2;CCDC141;CD34
CEP85L;ALX4;ARHGEF40;MYH6;MYH7;TTN
MTMR3;CR1;CMTM5;SLC35F1;NDRG2;TTN;PLN;B3GNT7;FAM174B;CCDC141;CD34;MYH6;MYH7

DLEU1;MYH7
GCKR;CCDC121;GFM1;HSPA4;SLC35F1;SSC5D;C2ORF16;MLF1;TMEM253;TBX15;KIAA1755;GJA1;FAM17
MYH6;TMEM253;MYH7
GCKR;SLC4A1AP
GCKR;CMTM5;AFAP1;VSTM2L;SSC5D;NDRG2;MLF1;KIAA1755;GJA1;NOV;ZNF219;GPR126;SPEG;ALX4;A
CCDC121;GFM1;ZCCHC10;SLC4A1AP;SLC35F1;CWC27;NAT14;GJA1;CCDC91;RSRC1;FAM174B;SPEG;FAM
CMTM5;SHOX2;MYH6;DGKH
CCDC121;GFM1;ZCCHC10;SLC4A1AP;TOMM40;CEP85L;SLC35F1;UQCR10;CWC27;NDRG2;DCAF16;GJA1
CCDC121;GFM1;ZCCHC10;SLC4A1AP;TOMM40;SLC35F1;UQCR10;CWC27;NAT14;PPM1G;GJA1;CCDC91;
MYH6;MYH7
B3GNT7;ZMAT5;WNT16
CCDC121;GFM1;ZCCHC10;SLC4A1AP;TOMM40;UQCR10;CWC27;NAT14;DCAF16;GJA1;CCDC91;FAM174
TBX15;WNT16;MYH6;MYH7
CCDC121;GFM1;ZCCHC10;HSPA4;SKIDA1;SLC35F1;SSC5D;UQCR10;CWC27;MLF1;TBX15;MLLT10;KIAA17
GFM1;SERPINA1;MYH6;MYH7
KIAA1755;SERPINA1;CR1;PLN;B3GNT7;CMTM5;SLC35F1;NDRG2;CCDC141;MYH6;MYH7
CCDC121;GFM1;ZCCHC10;TOMM40;CWC27;NAT14;PPM1G;DCAF16;GJA1;CCDC91;ZNF628;RSRC1;FAM
TBX15;SHOX2;ALX4;CCDC141;NRBP1;DGKH
MYH6;MYH7
GFM1;ZCCHC10;TOMM40;NCAPG;NAT14;ZNF628;ALX4;FAM3C;APOE;EIF2B4;GCKR;CENPW;SKIDA1;VST
SHOX2;MFN1;ASCC2;TTN
GFM1;CENPW;MTMR3;SLC4A1AP;HSPA4;SHOX2;PTEN;NRBP1;WARS2;TTN;PPM1G;ZNF628;SNX17;MFN
TBX15;GFM1;SLC4A1AP;HSPA4;CWC27;PPM1G
MLLT10;MTMR3;LXN;ZNF628;SHOX2;MFN1;PTEN;EIF3E;NRBP1;SUPT7L;WARS2;PPM1G
GCKR;CMTM5;AFAP1;SSC5D;NDRG2;KIAA1755;GJA1;PLN;NOV;ZNF219;GPR126;SPEG;ARHGEF40;PVRL2
DES;MTMR3;ZNF628;NCAPG;ASCC2;ARHGEF40;PVRL2;TTN
CCDC121;GFM1;ZCCHC10;SLC4A1AP;TOMM40;SHOX2;SLC35F1;CWC27;NAT14;DCAF16;GJA1;CCDC91;F
SERPINA1;AFAP1;VSTM2L;SSC5D;NDRG2;MLF1;KIAA1755;GJA1;NOV;ZNF219;GPR126;SPEG;APOE;ARHG
CCDC121;SLC4A1AP;ZMAT5;ALX4;C2ORF16;UQCR10;GPN1;NIPSNAP1
SHOX2;MFN1;WNT16;NRBP1;PPM1G
SERPINA1;ZCCHC10;CR1;CEP85L;C2ORF16;CCDC141
GCKR;DES;CR1;B3GNT7;CMTM5;SLC35F1;NDRG2;CCDC141;MYH6;MYH7
TMEM253;MYH7
HSPA4;MYH7
MYH6;MYH7
MTMR3;ZNF628;PTEN;NRBP1;WARS2
CCDC121;GFM1;ZCCHC10;CEP85L;CWC27;NAT14;DCAF16;GJA1;CCDC91;FAM174B;MFN1;SPEG;ALX4;FA
TBX15;CEP85L;ARHGEF40;CCDC141;MYH6;MYH7
SHOX2;GPN1
MTMR3;ZCCHC10;LXN;SLC4A1AP;HSPA4;SHOX2;PTEN;GPN1;NRBP1;WARS2;TTN;PPM1G;MLLT10;DNAJ
ZNF628;SNX17;ASCC2;EIF3E;GPN1;NRBP1;TTN;PPM1G
DNAJC1;LXN;DLEU1;NCAPG;ASCC2;EIF3E;ARHGEF40;PVRL2
CCDC121;GFM1;SLC4A1AP;TOMM40;SLC35F1;UQCR10;CWC27;NAT14;GJA1;CCDC91;RSRC1;FAM174B;
MYH6;MYH7
MYH6;MYH7

ALX4;CCDC141
ZMAT5;GPR126;NCAPG;ALX4
TBX15;ALX4
IFI27L2;GFM1;CENPW;MTMR3;CR1;SLC4A1AP;TOMM40;HSPA4;CMTM5;PTEN;NCAPG;UQCR10;CWC27
CCDC121;GFM1;ZCCHC10;SLC4A1AP;SLC35F1;CWC27;NAT14;GJA1;CCDC91;RSRC1;FAM174B;SPEG;FAM
IFI27L2;GFM1;MTMR3;ZCCHC10;CR1;SLC4A1AP;HSPA4;CMTM5;UQCR10;CWC27;GPN1;TBX15;RSRC1;Z
CCDC121;GFM1;ZCCHC10;LXN;SLC4A1AP;CEP85L;PTEN;CWC27;TTN;DCAF16;GJA1;PLN;CCDC91;RSRC1;
GFM1;ZCCHC10;TOMM40;SLC35F1;GJA1;CCDC91;SPEG;ALX4;FAM3C;APOE;CD34;SKIDA1;VSTM2L;SSC5
MYH6;MYH7
ZMAT5;DLEU1
GFM1;TOMM40;NDRG2;NAT14;DCAF16;GJA1;PLN;CCDC91;ZNF628;FAM174B;DLEU1;SPEG;FAM3C;APC
CCDC121;GFM1;ZCCHC10;SHOX2;SLC35F1;UQCR10;CWC27;NAT14;DCAF16;CCDC91;RSRC1;FAM174B;N
GFM1;MTMR3;ZCCHC10;SLC4A1AP;TOMM40;HSPA4;CWC27;PPM1G;SLC5A6;ZNF628;SNX17;GNB4;SPE
CEP85L;GNB4;CCDC141
IFI27L2;MTMR3;ZCCHC10;SLC4A1AP;TOMM40;HSPA4;PTEN;NCAPG;UQCR10;WNT16;CWC27;NRBP1;PF
DNAJC1;LXN;NCAPG;ASCC2;PVRL2;DGKH;TTN
GCKR;ALX4
SERPINA1;CR1;PLN;B3GNT7;SLC35F1;WNT16;NDRG2;CCDC141;MYH6;MYH7
ZNF628;PTEN;NRBP1;TMEM253;WARS2;PPM1G
GFM1;SLC4A1AP;TOMM40;UQCR10;CWC27;GJA1;PLN;CCDC91;ZNF628;FAM174B;SPEG;ALX4;FAM3C;A
SERPINA1;MTMR3;CMTM5;SSC5D;NDRG2;KIAA1755;GJA1;NOV;ZNF219;GPR126;SPEG;APOE;ARHGEF4
DES;DLEU1
ZCCHC10;CR1;CMTM5;CEP85L
ALX4;MYH6
CCDC121;GFM1;ZCCHC10;SLC4A1AP;CWC27;NAT14;GJA1;CCDC91;RSRC1;FAM174B;SPEG;FAM3C;ARH
WNT16;MYH6;MYH7
CCDC121;GFM1;ZCCHC10;SLC4A1AP;SLC35F1;CWC27;DCAF16;GJA1;PLN;CCDC91;RSRC1;FAM174B;MF
PVRL2
LXN
TRIB2
EIF2B4;IFI27L2;GCKR;LXN;TOMM40;SNX17;APOC1;ZNF219;C2ORF16;UQCR10;LCORL;SUPT7L
TBX15;GFM1;DES;SPEG;NDRG2;MYH6;MYH7;TTN
KIAA1755;CMTM5;SKIDA1;SHOX2;CEP85L;SLC35F1;NDRG2;ARHGEF40;TRIB2;SUPT7L
APOC1
TBX15;GFM1;DES;ZNF628;SPEG;MYH6;MYH7;TTN
IFI27L2;GFM1;MTMR3;ZCCHC10;CR1;SLC4A1AP;TOMM40;HSPA4;CMTM5;CWC27;NRBP1;TBX15;SNX17
TBX15;GJA1;ALX4
DLEU1;MYH7
SERPINA1;CMTM5;AFAP1;VSTM2L;SSC5D;NDRG2;KIAA1755;GJA1;NOV;ZNF219;GPR126;SPEG;APOE;AR
GFM1;ZCCHC10;TOMM40;SLC35F1;CWC27;NDRG2;DCAF16;GJA1;CCDC91;ZNF628;FAM174B;SPEG;ALX
CCDC121;GFM1;ZCCHC10;SLC4A1AP;SLC35F1;CWC27;GJA1;CCDC91;RSRC1;FAM174B;SPEG;FAM3C;CD
UQCR10
SLC35F1
DNAJC1
CABP7

GJA1;ARHGEF40
SLC4A1AP;TOMM40;CEP85L;UQCR10;NDRG2;GPN1;NIPSNAP1;NRBP1;TTN;PPM1G;GJA1;SNX17;ZNF21
CCDC121;GFM1;ZCCHC10;SLC4A1AP;TOMM40;CEP85L;SLC35F1;CWC27;TTN;DCAF16;GJA1;PLN;CCDC9
EIF2B4;IFI27L2;GCKR;CENPW;SERPINA1;CR1;TOMM40;PTEN;VSTM2L;NAT14;NIPSNAP1;TMEM253;PPM
GFM1;CEP85L;GNB4
CCDC121;GFM1;ZCCHC10;TOMM40;SLC35F1;CWC27;NAT14;DCAF16;GJA1;PLN;CCDC91;FAM174B;MFN
CCDC121;GFM1;TOMM40;SLC35F1;UQCR10;CWC27;NAT14;DCAF16;GJA1;CCDC91;FAM174B;MFN1;SP
GFM1;MTMR3;SLC4A1AP;TOMM40;HSPA4;SHOX2;CEP85L;NRBP1;TTN;PPM1G;MLLT10;DCAF16;ZNF62
TBX15;ALX4
GPR126
ZMAT5
GJA1
DES
ZCCHC10
CEP85L;NCAPG
ZNF628;MFN1;ASCC2
CCDC121;GFM1;ZCCHC10;SLC4A1AP;TOMM40;SLC35F1;CWC27;DCAF16;GJA1;CCDC91;RSRC1;FAM174
VSTM2L;UQCR10
GFM1
NDRG2
CABP7
CR1;PTEN
ZCCHC10;TOMM40;HSPA4;RSRC1;NCAPG;NIPSNAP1;WARS2
FAM174B
MLLT10;CR1
PVRL2
WNT16;SUPT7L
ZNF512;TRIB2
ZNF219
NDRG2;SUPT7L
PLN
DLEU1;ZNF512
TTN
AFAP1
RSRC1
CCDC91
MLLT10
DNAJC1;GPN1;NRBP1
MLF1
FAM174B
MLF1
CD34
UQCR10
AFAP1
SHOX2;CEP85L;PTEN;EIF3E

ZCCHC10;CCDC91;SUPT7L
IFI27L2;GCKR;CR1;LXN;CMTM5;NAT14;NIPSNAP1;TMEM253;B3GNT7;SNX17;ZNF513;ZNF512;PVRL2
SHOX2;ZNF219
TRIB2
TBX15;GFM1;ZCCHC10;CEP85L;ALX4;MYH6;MYH7;TTN
ZCCHC10
PPM1G
SLC5A6
MLF1
EIF3E
CMTM5
EIF3E
TTN
LCORL
PTEN
CENPW;NCAPG
UQCR10
LCORL
GJA1
B3GNT7
ZMAT5;CWC27
PTEN
APOC1;AFAP1;DGKH
NDRG2
SLC4A1AP;TOMM40;CEP85L;VSTM2L;UQCR10;NDRG2;GPN1;NIPSNAP1;NRBP1;TTN;PPM1G;GJA1;RSPC
CENPW
UQCR10
CD34
CENPW
IFI27L2;CCDC121;GFM1;SERPINA1;ZCCHC10;SLC4A1AP;PTEN;SLC35F1;NAT14;GJA1;CCDC91;RSRC1;FAM
TBX15;SHOX2;AFAP1;CD34;WARS2
GCKR;CENPW;SERPINA1;B3GNT7;FAM174B;NCAPG;PVRL2
NOV
TTN
NDRG2
IFI27L2;SHOX2;GNB4;AFAP1;ALX4;SSC5D;EIF3E;NIPSNAP1
SLC5A6;MTMR3;B3GNT7;GNB4;PTEN;UQCR10;APOE
GJA1
CENPW;DES;ZNF628;SHOX2;MFN1;NRBP1
MLLT10
CEP85L;CCDC141;MYH6
IFI27L2;ZCCHC10;SLC4A1AP;ZMAT5;ALX4;UQCR10;CWC27
TOMM40;APOC1;ZMAT5;DLEU1;UQCR10;NAT14;TMEM253
GCKR;SERPINA1;ZCCHC10;CR1;CEP85L;PTEN;SUPT7L
NOV;CMTM5;VSTM2L

DES;SERPINA1;CR1;PLN;B3GNT7;FAM174B;SLC35F1;NDRG2;CCDC141;CD34;MYH6
GFM1;TOMM40;SLC35F1;NAT14;DCAF16;GJA1;CCDC91;FAM174B;ALX4;FAM3C;APOE;CD34;GCKR;SKID
CCDC121;SERPINA1;MTMR3;CR1;CMTM5;PTEN;NDRG2;PLN;B3GNT7;FAM174B;APOC1;APOE;CCDC141
DES;NCAPG;ASCC2;TMEM253
MYH7
EIF2B4;DNAJC1;LXN;SHOX2;MFN1;ASCC2;WNT16;NRBP1
SERPINA1;CR1;PLN;B3GNT7;FAM174B;CMTM5;SLC35F1;CCDC141;MYH6;MYH7
CEP85L;ARHGEF40;MYH6;TTN
DCAF16;HSPA4;ALX4;CCDC141;GPN1;PPM1G
ASCC2;EIF3E;TMEM253;TTN
MTMR3;LXN;SHOX2;PTEN;ALX4;NRBP1;TMEM253;WARS2;PPM1G
LXN;TOMM40;C2ORF16;WNT16;GPN1;NRBP1;PPM1G;MLLT10;ZNF628;SNX17;MFN1;ALX4;CCDC141;D
TBX15;MYH6;MYH7
DLEU1
CCDC121;GFM1;ZCCHC10;SLC4A1AP;TOMM40;CEP85L;CWC27;NAT14;DCAF16;GJA1;CCDC91;RSRC1;FA
MTMR3;CMTM5;SKIDA1;AFAP1;SSC5D;NDRG2;MLF1;KIAA1755;GJA1;PLN;NOV;ZNF219;GPR126;SPEG;
GCKR;WNT16
NCAPG;ASCC2;EIF3E;TTN
TOMM40;ZNF628;APOC1;ZMAT5;DLEU1;UQCR10;APOE;NAT14
SERPINA1;CR1;CMTM5;WNT16;NDRG2;TTN;KIAA1755;PLN;B3GNT7;FAM174B;CCDC141;MYH6;MYH7
DLEU1;VSTM2L
MYH6;MYH7
TOMM40;VSTM2L;ALX4;WNT16
IFI27L2;CENPW;ZCCHC10;SLC4A1AP;TOMM40;HSPA4;CMTM5;PTEN;UQCR10;CWC27;ZNF628;GNB4;DL
SERPINA1;NCAPG
CCDC121;GFM1;ZCCHC10;SLC4A1AP;TOMM40;CEP85L;SLC35F1;UQCR10;CWC27;NAT14;DCAF16;GJA1;
CCDC121;GFM1;ZCCHC10;TOMM40;CEP85L;SLC35F1;DCAF16;GJA1;CCDC91;FAM174B;DLEU1;SPEG;AL
CENPW;ZNF628;SHOX2;ASCC2;EIF3E;WNT16;NRBP1;DGKH;PPM1G
CCDC121;GFM1;ZCCHC10;LXN;SLC4A1AP;UQCR10;CWC27;DCAF16;GJA1;CCDC91;RSRC1;FAM174B;MF
IFI27L2;CCDC121;GFM1;ZCCHC10;SLC4A1AP;PTEN;SLC35F1;CWC27;NAT14;GJA1;CCDC91;RSRC1;FAM1
CMTM5;NIPSNAP1;MYH6
CCDC121;ZMAT5
DES;CR1;PLN;B3GNT7;CMTM5;ALX4;SLC35F1;NDRG2;CCDC141;MYH6;TTN
GFM1;MTMR3;SLC4A1AP;TOMM40;PTEN;TTN;PPM1G;DCAF16;ZNF628;RSRC1;MFN1;ASCC2;ARHGEF4
CCDC121;GFM1;ZCCHC10;TOMM40;CEP85L;UQCR10;CWC27;NAT14;PPM1G;DCAF16;GJA1;CCDC91;RS
CCDC121;CEP85L;ARHGEF40;CCDC141;TTN
ZNF628;NCAPG;ASCC2;NRBP1
ALX4
APOC1;ZMAT5;DLEU1;VSTM2L;UQCR10;NAT14
EIF2B4;MTMR3;SHOX2;CEP85L;MFN1;ASCC2;EIF3E;GPN1;NRBP1;TTN
SLC4A1AP;ZMAT5
GFM1;ZCCHC10;TOMM40;CWC27;PPM1G;DCAF16;GJA1;PLN;CCDC91;RSRC1;FAM174B;MFN1;DLEU1;S
MTMR3;ZCCHC10;SLC4A1AP;TOMM40;SHOX2;PTEN;NCAPG;PPM1G;ZNF628;MFN1;ASCC2;ARHGEF40;
GCKR;CMTM5;AFAP1;SSC5D;NDRG2;MLF1;KIAA1755;GJA1;NOV;ZNF219;GPR126;SPEG;ARHGEF40;PVR
GCKR;SERPINA1;NCAPG;LCORL;WARS2

SERPINA1;C2ORF16;ZNF512
GCKR;NCAPG
DLEU1;ALX4
IFI27L2;MTMR3;CMTM5;CEP85L;PTEN;AFAP1;SLC35F1;CWC27;MLF1;GJA1;NOV;MFN1;GNB4;SPEG;DGI
DNAJC1;MTMR3;ZNF628;SHOX2;MFN1;NCAPG;ASCC2;GPN1;NRBP1
LXN
ASCC2
CCDC121;GFM1;ZCCHC10;TOMM40;UQCR10;CWC27;DCAF16;GJA1;CCDC91;RSRC1;FAM174B;MFN1;DL
NDRG2
ASCC2
SNX17
APOE
GJA1
SERPINA1
CCDC91
CABP7
PVRL2
MLLT10
IFI27L2;GFM1;CENPW;MTMR3;ZCCHC10;SLC4A1AP;HSPA4;NCAPG;UQCR10;CWC27;RSRC1;ZMAT5;GNE
SHOX2;NCAPG;ALX4
CCDC121;GFM1;ZCCHC10;LXN;SLC4A1AP;CWC27;GJA1;CCDC91;RSRC1;FAM174B;MFN1;ALX4;FAM3C;A
TBX15;GFM1;HSPA4;DLEU1;UQCR10;GPN1
IFI27L2;GCKR;SERPINA1;TOMM40;APOC1;ZMAT5;DLEU1;VSTM2L;UQCR10;APOE;NAT14
IFI27L2;GFM1;CENPW;MTMR3;ZCCHC10;SLC4A1AP;TOMM40;HSPA4;PTEN;UQCR10;CWC27;GPN1;NRB
CMTM5;SPEG;WNT16
NCAPG;MYH6;TMEM253
APOC1;ZMAT5;UQCR10
SSC5D;NDRG2;TMEM253;KIAA1755;GJA1;PLN;ZNF219;GPR126;SPEG;APOE;ARHGEF40;PVRL2;CD34;TRI
NCAPG
HORMAD2
MLF1
GJA1;NDRG2
ZNF219
MLF1
GNB4
HORMAD2
TBX15;DES;NDRG2;MLF1;MYH6;MYH7;TTN
GCKR;UQCR10
EIF3E
GJA1
PPM1G
HORMAD2
PTEN;PPM1G
HSPA4
SLC4A1AP;TOMM40;CEP85L;UQCR10;NDRG2;GPN1;NIPSNAP1;TTN;PPM1G;GJA1;SNX17;RSPO2;EIF3E;F

SERPINA1;CR1
ZNF628;MFN1;NRBP1;TTN
SLC4A1AP;TOMM40;HSPA4;CEP85L;UQCR10;NDRG2;GPN1;NIPSNAP1;NRBP1;SUPT7L;TTN;PPM1G;DNA
GJA1;PTEN
TBX15;ZNF628;ZNF219
HSPA4
GFM1;SLC4A1AP;PTEN;EIF3E
TBX15;ZNF628;ZNF219
TBX15;SERPINA1;CCDC91;RSRC1;CEP85L;PTEN;ARHGEF40;CCDC141;LCORL;TRIB2;TTN
SLC4A1AP;TOMM40;CEP85L;UQCR10;NDRG2;GPN1;NIPSNAP1;TTN;PPM1G;GJA1;SNX17;RSPO2;EIF3E;F
PPM1G
DES;SERPINA1;CR1;PLN;NOV;APOE;NIPSNAP1
PPM1G
MLLT10;KIAA1755;DES;ZNF219;SPEG;SSC5D;CD34
SLC5A6;CCDC121;RSRC1;PTEN;SSC5D;EIF3E;CD34;GPN1;WARS2
CCDC121;GJA1;CABP7;CMTM5;APOC1;ZMAT5;SLC35F1;UQCR10;APOE;NDRG2
MLLT10;DCAF16;PLN;NOV;RSRC1;CEP85L;MFN1;GNB4;PTEN;SPEG;EIF3E;ZNF512
SERPINA1;MTMR3;CR1;CR1L;PTEN;ASCC2
KIAA1755;ZMAT5;GPR126;SLC35F1;CWC27;ARHGEF40;TRIB2
MLLT10;ZNF219;DLEU1;SPEG;SSC5D
TBX15;DES;ZNF628;UQCR10;MYH6;MYH7;TTN
APOC1
SERPINA1
CR1L;SPEG
GNB4
MTMR3;PTEN
CR1;PTEN;APOE;NRBP1
GJA1;LXN;SLC35F1;TRIB2
B3GNT7;PTEN;APOE;CCDC141
PLN;CCDC141;MYH7;TTN
GJA1;SLC4A1AP;TRIB2;WARS2
HSPA4;EIF3E;FAM3C;PPM1G
DES;EIF3E;APOE;CD34
NCAPG;UQCR10;TRIB2;PPM1G
HSPA4;PTEN;EIF3E;TRIB2
UQCR10;CD34;GPN1;MLF1
GJA1;HSPA4;RSPO2;AFAP1
HSPA4;FAM3C;NRBP1;PPM1G
PTEN;SLC35F1;EIF3E;LCORL
NOV;PTEN;RSPO2;EIF3E
CR1;APOC1;EIF3E;TRIB2
SERPINA1;APOC1;VSTM2L;EIF3E
GJA1;PTEN;SLC35F1;APOE
SHOX2;PTEN;EIF3E;NRBP1
GJA1;TOMM40;HSPA4;EIF3E

PTEN;SLC35F1;EIF3E;LCORL
GJA1;GNB4;ARHGEF40;TRIB2
EIF3E;APOE;NDRG2;CCDC141
HSPA4;APOE;TRIB2;NIPSNAP1
CR1;APOC1;PTEN;APOE
APOC1;PTEN;APOE;NDRG2
APOC1;UQCR10;APOE;NDRG2
UQCR10;FAM3C;WNT16;NIPSNAP1
GJA1;APOE;CD34;TTN
DES;UQCR10;MYH6;TTN
DES;UQCR10;MYH6;TTN
MFN1;PTEN;SLC35F1;NDRG2
CABP7;SHOX2;FAM3C;ARHGEF40
GJA1;SERPINA1;RSPO2;EIF3E
NOV;SPEG;SSC5D;ARHGEF40
TOMM40;HSPA4;APOE;CD34
TOMM40;HSPA4;TRIB2;PPM1G
TOMM40;HSPA4;EIF3E;PPM1G
TBX15;DES;SLC35F1;CCDC141
HSPA4;EIF3E;TTN;PPM1G
GJA1;PLN;HSPA4;CCDC141
NOV;SPEG;UQCR10;APOE
HSPA4;PTEN;AFAP1;TRIB2
HSPA4;PTEN;EIF3E;APOE
HSPA4;GNB4;EIF3E;TRIB2
MTMR3;ZNF219;CCDC141;TTN
DCAF16;B3GNT7;HSPA4;TRIB2
TOMM40;EIF3E;NRBP1;PPM1G
PTEN;SLC35F1;LCORL;TRIB2
TOMM40;HSPA4;NCAPG;PPM1G
MLLT10;HSPA4;RSPO2;TTN
HSPA4;CCDC141;CD34;MYH6
GNB4;PTEN;APOE;TRIB2
SLC5A6;TOMM40;HSPA4;PPM1G
NOV;SPEG;APOE;NDRG2
GJA1;NDRG2;NRBP1;TTN
GJA1;HSPA4;PTEN;APOE
GJA1;HSPA4;PTEN;NDRG2
PTEN;FAM3C;ARHGEF40;SUPT7L
CABP7;FAM174B;VSTM2L;APOE
DES;APOE;NDRG2;TTN
DES;UQCR10;APOE;MYH7
GFM1;TOMM40;HSPA4;PPM1G
PTEN;UQCR10;NIPSNAP1;NRBP1
GJA1;HSPA4;NCAPG;PPM1G

GJA1;RSPO2;EIF3E;MLF1
SHOX2;PTEN;NCAPG;EIF3E
SNX17;PTEN;EIF3E;NRBP1
HSPA4;EIF3E;ARHGEF40;PPM1G
SLC5A6;GFM1;TOMM40;PPM1G
TOMM40;HSPA4;PTEN;PPM1G
HSPA4;NCAPG;EIF3E;APOE
DNAJC1;HSPA4;EIF3E;UQCR10
GCKR;APOC1;APOE;NDRG2
SLC5A6;TOMM40;CD34;PPM1G
DES;APOC1;SSC5D;APOE
ZCCHC10;RSPO2;FAM3C;TRIB2
TOMM40;HSPA4;TRIB2;PPM1G
GJA1;HSPA4;PTEN;APOE
GJA1;APOE;CWC27;NIPSNAP1
TOMM40;CD34;MYH6;PPM1G
MTMR3;NOV;NDRG2;CCDC141
DES;PTEN;NDRG2;TTN
PTEN;APOE;TRIB2;PPM1G
NCAPG;EIF3E;FAM3C;WNT16
HSPA4;PTEN;EIF3E;APOE
GFM1;HSPA4;PTEN;EIF3E
GFM1;MTMR3;TOMM40;HSPA4
TOMM40;UQCR10;APOE;PPM1G
EIF2B4;MTMR3;TOMM40;NRBP1
TOMM40;HSPA4;PTEN;PPM1G
B3GNT7;PTEN;EIF3E;TTN
VSTM2L;SPEG;APOE;MYH7
HSPA4;NCAPG;EIF3E;NIPSNAP1
GJA1;APOC1;APOE;MYH6
NOV;VSTM2L;FAM3C;LCORL
GJA1;MFN1;PTEN;APOE
GJA1;HSPA4;SNX17;PTEN
GFM1;TOMM40;HSPA4;PPM1G
CR1;APOC1;APOE;TTN
SSC5D;APOE;CD34;TTN
HSPA4;PTEN;EIF3E;NDRG2
HSPA4;NCAPG;FAM3C;NRBP1
GJA1;EIF3E;APOE;MLF1
TBX15;GJA1;HSPA4;ALX4
HSPA4;NCAPG;EIF3E;MYH7
GJA1;HSPA4;APOE;NDRG2
DNAJC1;HSPA4;PTEN;APOE
SERPINA1;NOV;AFAP1;SSC5D
APOE;NDRG2;CD34;MYH7

SERPINA1;VSTM2L;APOE;NIPSNAP1
PTEN;NCAPG;EIF3E;FAM3C
DES;EIF3E;APOE;MYH7
APOC1;APOE;NDRG2;NIPSNAP1
SERPINA1;APOC1;APOE;ARHGEF40
FAM174B;GNB4;SLC35F1;FAM3C;NDRG2;ARHGEF40;TRIB2
GJA1;HSPA4;APOE;MYH7
GJA1;APOE;CWC27;NIPSNAP1
DES;EIF3E;NDRG2;MYH7
DES;NDRG2;ARHGEF40;CD34
TOMM40;HSPA4;PTEN;PPM1G
TOMM40;HSPA4;PTEN;PPM1G
SLC5A6;GFM1;NDRG2;DGKH
HSPA4;NCAPG;EIF3E;PPM1G
GCKR;HSPA4;FAM3C;APOE
SLC5A6;HSPA4;CD34;PPM1G
HSPA4;PTEN;ARHGEF40;TRIB2
GJA1;SLC35F1;APOE;NDRG2
KIAA1755;DES;SERPINA1;CR1;PLN;B3GNT7;CMTM5;SLC35F1;NDRG2;NIPSNAP1
KIAA1755;SERPINA1;CR1;PLN;B3GNT7;CMTM5;SLC35F1;NDRG2;CCDC141;MYH6;MYH7
ALX4
HSPA4;MYH6
GFM1;TOMM40;SLC35F1;DCAF16;GJA1;CCDC91;FAM174B;DLEU1;ALX4;FAM3C;APOE;CD34;EIF2B4;GC[illegible]
GJA1;SERPINA1;NOV;HSPA4;SHOX2;PTEN;RSPO2;FAM3C;APOE;NDRG2;CD34;MYH6
DNAJC1;MTMR3;ZNF628;SHOX2;DLEU1;NCAPG;ASCC2;EIF3E;ARHGEF40;PVRL2
MLLT10;SKIDA1
HSPA4;EIF3E
HSPA4;EIF3E
SNX17;PPM1G
AFAP1;PPM1G
PTEN;EIF3E
GJA1;TOMM40
NCAPG;PPM1G
AFAP1;ARHGEF40
CABP7;APOE
DES;B3GNT7
PTEN;EIF3E;FAM3C
CEP85L;LCORL
NDRG2;CCDC141
EIF2B4;HSPA4
NOV;ARHGEF40
GNB4;FAM3C
GJA1;HSPA4
HSPA4;EIF3E
GJA1;EIF3E

HSPA4;NCAPG
TOMM40;EIF3E
EIF3E;FAM3C
SLC4A1AP;TTN
HSPA4;PPM1G
HSPA4;EIF3E
GJA1;APOE
GJA1;HSPA4
PTEN;TRIB2
GJA1;HSPA4
APOC1;NDRG2
GJA1;CD34
TOMM40;PPM1G
NCAPG;NRBP1
CCDC141;MYH7
CCDC141;MYH7
EIF3E;FAM3C
UQCR10;NDRG2
HSPA4;NDRG2
GJA1;HSPA4;APOE
GJA1;AFAP1;SSC5D
LXN;APOE;TRIB2
AFAP1;WNT16
SERPINA1;EIF3E
TOMM40;HSPA4
TBX15;FAM3C
GJA1;APOE
APOE;NDRG2
TRIB2;PPM1G
NOV;PTEN
PTEN;APOE
APOE;NDRG2
CABP7;NDRG2
GJA1;TRIB2
GJA1;APOE
HSPA4;NCAPG
SHOX2;CCDC141
PTEN;LCORL
LXN;GNB4
SLC5A6;NRBP1
SERPINA1;NIPSNAP1
HSPA4;EIF3E
APOC1;APOE
TOMM40;PPM1G
SERPINA1;EIF3E

HSPA4;PTEN
CR1;APOE
TOMM40;PPM1G
HSPA4;PTEN
HSPA4;AFAP1
NDRG2;ARHGEF40
DES;EIF3E
EIF3E;NIPSNAP1
HSPA4;APOE
TOMM40;EIF3E
APOE;CCDC141
SERPINA1;MTMR3
TOMM40;APOE
APOC1;APOE
PTEN;AFAP1
APOC1;APOE
PTEN;NDRG2
TBX15;GJA1
GJA1;HSPA4;CD34
HSPA4;EIF3E;APOE
HSPA4;PTEN;APOE;LCORL
APOC1;APOE;NDRG2;NIPSNAP1
GJA1;TOMM40;HSPA4;APOE;PPM1G
NOV;PTEN;EIF3E;TRIB2
APOC1;UQCR10;APOE;NDRG2
GJA1;HSPA4;CABP7;PTEN
DES;UQCR10;APOE
EIF2B4;CCDC91;MLF1
FAM174B;GNB4;APOE
CMTM5;CWC27;MYH6
HSPA4;EIF3E;NDRG2
ALX4;EIF3E;ARHGEF40
DES;MYH7;TTN
HSPA4;PTEN;CD34
HSPA4;EIF3E
EIF2B4;EIF3E
LXN;EIF3E
HSPA4;EIF3E
HSPA4;PPM1G
EIF3E;ZNF512
SERPINA1;GNB4;NRBP1
HSPA4;EIF3E;PPM1G
HSPA4;EIF3E;PPM1G
GJA1;CCDC141;NRBP1
APOE;NDRG2

PTEN;LCORL
RSRC1;NRBP1
HSPA4;TRIB2
APOC1;APOE
APOC1;APOE
SNX17;EIF3E
CWC27;CD34
AFAP1;EIF3E
PTEN;NIPSNAP1
TRIB2;PPM1G
UQCR10;NDRG2;CCDC141
DES;PTEN
GJA1;PTEN
B3GNT7;APOE
TOMM40;HSPA4
HSPA4;FAM3C
GJA1;EIF3E
GJA1;EIF3E
MYH7;TTN
HSPA4;PPM1G
ZNF512;TRIB2
GJA1;APOE
NCAPG;APOE
HSPA4;PTEN
SLC5A6;PPM1G
HSPA4;FAM3C
HSPA4;EIF3E
TOMM40;ZNF628
MLLT10;FAM3C
TOMM40;HSPA4
SPEG;ARHGEF40
SPEG;ARHGEF40
SPEG;ARHGEF40
EIF3E;UQCR10
SPEG;ARHGEF40
SPEG;APOE
GJA1;AFAP1
TOMM40;PPM1G
APOC1;APOE
APOC1;APOE
EIF3E;APOE
B3GNT7;CD34
PTEN;APOE
GJA1;NIPSNAP1
DES;FAM3C

GJA1;ARHGEF40
APOE;NDRG2
VSTM2L;TRIB2
NOV;PTEN
EIF3E;NIPSNAP1
HSPA4;EIF3E
HSPA4;APOE
PTEN;NRBP1
MLLT10;TTN
HSPA4;GNB4
HSPA4;EIF3E
HSPA4;NCAPG
NCAPG;EIF3E
HSPA4;PTEN
SERPINA1;TTN
GJA1;SERPINA1
HSPA4;UQCR10
APOE;CD34
GJA1;GNB4
GJA1;HSPA4
SERPINA1;NIPSNAP1
PTEN;TRIB2
EIF3E;CD34
HSPA4;APOE
HSPA4;EIF3E
SERPINA1;TTN
APOC1;NDRG2
GJA1;ALX4
FAM174B;APOE
AFAP1;EIF3E
GFM1;MLF1
RSPO2;CCDC141
APOE;TRIB2
TRIB2;PPM1G
HSPA4;PTEN
GJA1;AFAP1
NIPSNAP1;WARS2
GNB4;FAM3C
HSPA4;PPM1G
PTEN;APOE
FAM3C;WNT16
TOMM40;HSPA4
SERPINA1;MTMR3
HSPA4;PPM1G
DES;FAM3C

APOE;CD34
NOV;CD34
HSPA4;APOE
APOE;CD34
B3GNT7;EIF3E
GJA1;B3GNT7
GCKR;PTEN
SNX17;APOE
HSPA4;AFAP1;TRIB2
SERPINA1;LXN;TTN
DES;B3GNT7;APOE
TOMM40;EIF3E;UQCR10
GCKR;APOC1;NDRG2
SLC5A6;TRIB2;NIPSNAP1
SLC5A6;HSPA4;EIF3E
TOMM40;B3GNT7;SNX17
RSRC1;SHOX2;NDRG2
HSPA4;NCAPG;EIF3E
GJA1;PTEN;EIF3E
GJA1;APOE;NDRG2
GJA1;EIF3E
HSPA4;PPM1G
DES;APOC1;APOE
TTN;PPM1G
HSPA4;GNB4;EIF3E
GJA1;HSPA4;NDRG2
NOV;HSPA4;CABP7
NOV;CABP7;SPEG
HSPA4;APOE;TRIB2
SLC5A6;DES;B3GNT7;NOV;APOE;NDRG2
APOC1;NDRG2
EIF3E;APOE
CCDC141;TRIB2
HSPA4;NRBP1
EIF3E;PPM1G
APOE;NDRG2
PTEN;LCORL
EIF3E;FAM3C
PTEN;NCAPG;TRIB2
GJA1;APOE
GNB4;EIF3E;LCORL
CD34;MYH6;TTN
GJA1;PTEN;NCAPG
HSPA4;EIF3E;PPM1G
B3GNT7;GNB4;NCAPG

HSPA4;PTEN;TTN
B3GNT7;APOE;PPM1G
SPEG;APOE;NDRG2
GFM1;HSPA4;EIF3E
HSPA4;PPM1G
APOE;NDRG2
GJA1;HSPA4
MYH6;TTN
PTEN;EIF3E
NCAPG;UQCR10
IFI27L2;APOC1
GJA1;EIF3E
HSPA4;PTEN
CMTM5;EIF3E
TOMM40;B3GNT7
NOV;NIPSNAP1
EIF3E;TTN
APOE;NDRG2
EIF3E;NDRG2
HSPA4;AFAP1
SERPINA1;NDRG2
SERPINA1;NDRG2
SERPINA1;NDRG2
SERPINA1;NDRG2
SERPINA1;NDRG2
GJA1;PTEN
APOE;CD34
APOE;NDRG2
GCKR;PTEN
CD34;TTN
HSPA4;APOE
APOE;CD34
HSPA4;EIF3E
GJA1;APOE
GJA1;PLN
SERPINA1;NDRG2
HSPA4;FAM3C
TOMM40;B3GNT7
EIF3E;NRBP1
TOMM40;B3GNT7
MTMR3;NRBP1
CEP85L;CD34
GFM1;NRBP1
SERPINA1;UQCR10
KIAA1755;APOC1

SERPINA1;MTMR3
SHOX2;NDRG2
SLC5A6;HSPA4
HSPA4;TRIB2
TOMM40;SNX17
GJA1;NDRG2
TOMM40;UQCR10
TOMM40;SNX17
DES;APOE
EIF3E;NIPSNAP1
EIF3E;NIPSNAP1
APOE;NIPSNAP1
HSPA4;NCAPG
HSPA4;FAM3C
PTEN;APOE
TOMM40;PPM1G
SERPINA1;NDRG2
SERPINA1;NDRG2
CCDC141;TTN
VSTM2L;NIPSNAP1
APOC1;PTEN
APOC1;APOE
NCAPG;NRBP1
PLN;AFAP1
APOE;NDRG2
GFM1;APOE
GCKR;APOC1
GJA1;HSPA4
APOE;CCDC141
SPEG;CCDC141
GJA1;HSPA4
APOE;NIPSNAP1
EIF3E;NIPSNAP1
HSPA4;EIF3E
B3GNT7;APOE
APOE;TRIB2
HSPA4;PTEN
APOE;NDRG2
APOC1;APOE
TOMM40;PPM1G
HSPA4;NCAPG
APOE;NDRG2;ARHGEF40
APOE;NDRG2;CD34
PTEN;EIF3E;APOE
APOC1;EIF3E;APOE

MTMR3;APOE;CD34
APOC1;APOE;NDRG2
HSPA4;APOE;NDRG2
APOE;CCDC141;MYH7
TBX15;NOV;SLC35F1
NDRG2;ARHGEF40;CD34
NOV;CMTM5;LCORL
TOMM40;UQCR10;PPM1G
HSPA4;PTEN;NDRG2
NOV;LCORL;CD34
NOV;PTEN;NAT14
HSPA4;SLC35F1;UQCR10
EIF3E;LCORL;MLF1
GNB4;APOE;ARHGEF40
EIF3E;APOE;NDRG2
FAM174B;GNB4;AFAP1
GJA1;FAM3C;WNT16
DES;NOV;CD34
GJA1;HSPA4;SLC35F1
GNB4;AFAP1;APOE
HSPA4;PTEN;EIF3E
SNX17;PTEN;NIPSNAP1
EIF3E;FAM3C;WNT16
PTEN;AFAP1;EIF3E
PTEN;NCAPG;TRIB2
HSPA4;NCAPG;NIPSNAP1
NOV;UQCR10;NIPSNAP1
PTEN;EIF3E;CD34
GJA1;LXN;TRIB2
HSPA4;NCAPG;EIF3E
NCAPG;EIF3E;APOE
APOC1;AFAP1;APOE
TBX15;HSPA4;PTEN
NOV;APOE;ARHGEF40
APOC1;EIF3E;APOE
SPEG;APOE;NDRG2
GCKR;HSPA4;PTEN
GCKR;HSPA4;PTEN
NOV;RSPO2;MYH7
GJA1;B3GNT7;APOE
GJA1;APOE;MYH7
GJA1;MTMR3;PTEN
GJA1;DES;TTN
GFM1;GJA1;NIPSNAP1
HSPA4;EIF3E;DGKH

EIF2B4;TOMM40;UQCR10
B3GNT7;FAM174B;NDRG2
CD34;MYH6;MYH7
GJA1;EIF3E;CD34
TOMM40;HSPA4;FAM3C
SERPINA1;LXN;TTN
TOMM40;NIPSNAP1;TTN
HSPA4;NRBP1;PPM1G
GJA1;CABP7;APOE
DES;MYH7;TTN
EIF3E;CD34;TTN
TOMM40;APOC1;APOE
GJA1;GNB4;EIF3E
GJA1;NDRG2;CD34
DES;NDRG2;TTN
HSPA4;FAM3C;NIPSNAP1
APOC1;EIF3E;APOE
LXN;HSPA4;EIF3E
PTEN;EIF3E;NIPSNAP1
GCKR;HSPA4;APOE
GJA1;DES;TTN
B3GNT7;HSPA4;APOE
PTEN;APOE;TRIB2
GJA1;TOMM40;UQCR10
TOMM40;EIF3E;PPM1G
GJA1;GNB4;EIF3E
GJA1;APOE;TTN
TOMM40;B3GNT7;PPM1G
GJA1;CABP7
NIPSNAP1;PPM1G
UQCR10;APOE
GNB4;SPEG
EIF2B4;HSPA4
GJA1;HSPA4;EIF3E
SPEG;CCDC141
CD34;PPM1G
LXN;SHOX2
DES;UQCR10
UQCR10;CCDC141
ARHGEF40;CD34
APOE;CCDC141
APOC1;UQCR10;NDRG2
MTMR3;HSPA4;EIF3E
GJA1;APOE;NDRG2
PLN;EIF3E;NDRG2

NOV;HSPA4;CABP7
CR1;EIF3E;TRIB2
TOMM40;ZNF628
B3GNT7;FAM174B
MFN1;GNB4
EIF3E;NIPSNAP1
APOE;TRIB2
SERPINA1;NDRG2
SERPINA1;NDRG2
GJA1;PTEN
GNB4;APOE
SERPINA1;NDRG2
HSPA4;PTEN;TRIB2
ALX4;TTN
GJA1;PTEN
GJA1;AFAP1
GJA1;SNX17
PTEN;EIF3E
FAM3C;TRIB2
HSPA4;NDRG2
APOE;NDRG2
APOC1;EIF3E;APOE
DES;APOE;CD34
SPEG;ARHGEF40;PPM1G
TOMM40;SPEG;ARHGEF40
HSPA4;EIF3E;PPM1G
HSPA4;PTEN;TRIB2
GJA1;HSPA4;NCAPG
APOC1;EIF3E;APOE
APOC1;EIF3E;APOE
SERPINA1;APOE;ARHGEF40
GJA1;ALX4;ARHGEF40
SERPINA1;CR1;PTEN
DES;APOE;MYH7
HSPA4;NCAPG;EIF3E
GCKR;GJA1;TTN
NOV;MLF1;TTN
GJA1;APOE;NDRG2
HSPA4;EIF3E;PPM1G
GCKR;HSPA4;PTEN
GJA1;GNB4;EIF3E
HSPA4;PTEN;TTN
GJA1;GNB4;TRIB2
GJA1;APOE;NDRG2
TOMM40;APOC1;APOE

GNB4;PTEN;EIF3E
HSPA4;PTEN;EIF3E
GJA1;HSPA4;EIF3E
GJA1;HSPA4;PPM1G
HSPA4;NCAPG;EIF3E
KIAA1755;PTEN;NDRG2
SKIDA1;ZNF219;NCAPG
APOC1;EIF3E;APOE
SERPINA1;APOE;ARHGEF40
NOV;PTEN;NCAPG
SLC5A6;GPN1;NRBP1
SERPINA1;FAM3C;NIPSNAP1
SERPINA1;EIF3E;NIPSNAP1
HSPA4;PTEN;TTN
MTMR3;PTEN;CD34
NOV;HSPA4
AFAP1;CD34
EIF3E;NRBP1
PTEN;NCAPG
VSTM2L;APOE
GJA1;APOE
HSPA4;PPM1G
EIF3E;CD34
GJA1;NRBP1
NOV;APOE;CD34
GJA1;HSPA4;NCAPG
EIF3E;FAM3C;WNT16
SLC4A1AP;CWC27;DGKH
FAM3C;NRBP1;PPM1G
GJA1;APOE;NDRG2
HSPA4;NDRG2;PPM1G
APOC1;APOE;TRIB2
PTEN;AFAP1;EIF3E
FAM174B;APOE;NDRG2
DNAJC1;GJA1;NIPSNAP1
GJA1;EIF3E;APOE
GJA1;APOE;NDRG2
HSPA4;APOE;NDRG2
ZCCHC10;HSPA4;EIF3E
TOMM40;PTEN;PPM1G
SERPINA1;CR1;PTEN
GJA1;APOE;NDRG2
IFI27L2;APOC1
EIF3E;NIPSNAP1
GJA1;TOMM40

GNB4;TRIB2
MFN1;GNB4
HSPA4;EIF3E
GJA1;NRBP1
GNB4;NCAPG;EIF3E
TOMM40;PPM1G
GJA1;SNX17
GJA1;AFAP1
HSPA4;EIF3E
EIF3E;LCORL
GJA1;NOV;EIF3E
HSPA4;EIF3E;CD34
SERPINA1;GNB4;NRBP1
APOC1;NDRG2
EIF3E;MYH7;TTN
GJA1;APOE;MYH6
B3GNT7;UQCR10;NDRG2
LXN;VSTM2L;NDRG2
ZNF219;FAM3C;NRBP1
SLC5A6;TOMM40;NRBP1
SLC5A6;TOMM40;HSPA4
HSPA4;EIF3E
GJA1;HSPA4;PTEN
HSPA4;NIPSNAP1;PPM1G
GJA1;HSPA4;PTEN
HSPA4;ZNF219;APOE
MTMR3;NCAPG;NRBP1
SERPINA1;APOC1;APOE
APOE;NDRG2
HSPA4;EIF3E
GJA1;FAM3C;APOE
HSPA4;EIF3E
SLC35F1;LCORL
LXN;LCORL
EIF2B4;APOE
GNB4;FAM3C
HSPA4;NCAPG
APOC1;APOE
DCAF16;ZNF512;PPM1G
GJA1;HSPA4;AFAP1
B3GNT7;EIF3E;ARHGEF40
TOMM40;HSPA4;PPM1G
TOMM40;APOC1;APOE
HSPA4;PTEN;TRIB2
HSPA4;EIF3E;PPM1G

GNB4;NCAPG;CD34
GNB4;PTEN;TRIB2
MFN1;SLC35F1;NIPSNAP1
TOMM40;NCAPG;PPM1G
LXN;FAM174B;ZNF219
GCKR;APOC1;APOE
HSPA4;PTEN;PPM1G
PLN;CR1L;TTN
GJA1;APOE;TTN
PTEN;APOE;NDRG2
CABP7;RSRC1;SHOX2
PTEN;APOE;TRIB2
DES;SPEG;FAM3C
MFN1;ARHGEF40;GPN1
LXN;CCDC91;SPEG
FAM174B;CD34;PPM1G
EIF3E;APOE;CD34
VSTM2L;APOE;NDRG2
SPEG;FAM3C;WNT16
GCKR;SKIDA1;FAM3C
GJA1;NOV;HSPA4
NOV;SHOX2;RSPO2
SERPINA1;APOC1;APOE
GCKR;APOC1;APOE
HSPA4;NCAPG;PPM1G
EIF3E;UQCR10;PPM1G
B3GNT7;GNB4;TRIB2
DES;CMTM5;VSTM2L
GJA1;NOV;PTEN
CABP7;SPEG;APOE;CD34
NOV;APOE;NDRG2;TRIB2
NOV;AFAP1;EIF3E;APOE
HSPA4;GNB4;FAM3C;ARHGEF40
IFI27L2;GJA1;SSC5D
PTEN;FAM3C
GNB4;UQCR10
HSPA4;WNT16;PPM1G
HSPA4;EIF3E;PPM1G
PTEN;EIF3E
VSTM2L;APOE;NDRG2
GJA1;CMTM5;NDRG2
GCKR;APOC1;NIPSNAP1
TOMM40;HSPA4;FAM3C
SLC5A6;NDRG2;DGKH
SKIDA1;ZNF219;TRIB2

MLLT10;EIF3E;PPM1G
CR1;PTEN
TRIB2;NRBP1
DCAF16;NCAPG
NRBP1;TTN
GJA1;SSC5D
LXN;NIPSNAP1
APOE
APOE
MTMR3;CR1;CMTM5;SLC35F1;NDRG2;DES;PLN;B3GNT7;NOV;FAM174B;ZNF219;CCDC141;CD34
KIAA1755;DES;MTMR3;CR1;PLN;B3GNT7;CMTM5;SLC35F1;NDRG2;MYH6;MYH7;TTN
GCKR;DES
CCDC121;GFM1;ZCCHC10;TOMM40;CEP85L;SLC35F1;UQCR10;CWC27;NDRG2;NAT14;DCAF16;GJA1;CC
SLC4A1AP;TOMM40;CEP85L;UQCR10;NDRG2;GPN1;NIPSNAP1;TTN;GJA1;SNX17;RSPO2;EIF3E;FAM3C;A
ZMAT5;WNT16;NIPSNAP1
GFM1;MTMR3;ZCCHC10;LXN;SLC4A1AP;TOMM40;SHOX2;PTEN;NCAPG;PPM1G;ZNF628;ZMAT5;MFN1;
EIF2B4;MTMR3;LXN;GPN1;NRBP1;SUPT7L;PPM1G;MLLT10;ZNF628;SNX17;ZMAT5;ASCC2;EIF3E
NCAPG;MYH6
MLLT10;LXN;ZNF628;SHOX2;MFN1;ASCC2;GPN1;NRBP1;TTN;PPM1G
TBX15
CR1;LXN;CMTM5;VSTM2L;NDRG2;NIPSNAP1;TTN;KIAA1755;DES;PLN;B3GNT7;MYH6;MYH7
CMTM5;SHOX2;ALX4
CENPW;ZNF628;SHOX2;MFN1;EIF3E;NRBP1;TTN
CCDC121;GFM1;ZCCHC10;TOMM40;CEP85L;SLC35F1;CWC27;DCAF16;GJA1;PLN;CCDC91;RSRC1;MFN1;
DNAJC1;CENPW;DES;CR1;SHOX2;GNB4;ASCC2;PVRL2
ZNF628;SHOX2;ASCC2;PPM1G
TBX15;CEP85L;ALX4;WNT16;ARHGEF40
KIAA1755;SERPINA1;CR1;PLN;B3GNT7;CMTM5;NDRG2;CCDC141;MYH6;MYH7
MYH6;MYH7
ZNF628;SHOX2;PTEN;ASCC2;EIF3E;PVRL2;NRBP1;PPM1G
MLLT10;MTMR3;ZNF628;ASCC2;GPN1;NRBP1
NCAPG
TBX15;GFM1;DES;ZNF628;MYH6;MYH7;TTN
TBX15;DES;CEP85L;MYH6;MYH7;TTN
PPM1G
IFI27L2;CENPW;LXN;PTEN;AFAP1;VSTM2L;C2ORF16;NAT14;NIPSNAP1;SUPT7L;TMEM253;WARS2;DCAF
GFM1;TOMM40;SLC35F1;DCAF16;GJA1;CCDC91;FAM174B;SPEG;FAM3C;APOE;CD34;SKIDA1;AFAP1;VS
DNAJC1;LXN;DLEU1;NCAPG;ASCC2;EIF3E;ARHGEF40;TMEM253
TBX15;MYH6
DNAJC1;CENPW;MFN1;ASCC2;NRBP1
SHOX2
LXN;ZNF628;ASCC2;TMEM253;PPM1G
SHOX2;SPEG;MYH6;MYH7
IFI27L2;CCDC141;TMEM253;DGKH
CENPW;LXN;ZNF628;NCAPG;ASCC2;DGKH

EIF2B4
CCDC121;SHOX2;SSC5D;NDRG2;TMEM253;TBX15;PLN;NOV;APOC1;ZNF219;ALX4;HORMAD2;APOE;ARH
CCDC121;SHOX2;SSC5D;NDRG2;TMEM253;TBX15;GJA1;PLN;NOV;ZNF219;ALX4;HORMAD2;APOE;ARHG
LXN;SHOX2;AFAP1;SSC5D;NAT14;TMEM253;KIAA1755;GJA1;DES;NOV;ZNF628;ZNF219;RSPO2;SPEG;AL
PTEN
KIAA1755;CCDC121;GJA1;CCDC91;RSRC1;CMTM5;RSPO2;SLC35F1;APOE;NDRG2;LCORL
IFI27L2;LXN;GPR126;FAM3C;PVRL2;CD34;WARS2
IFI27L2;GFM1;CENPW;ZCCHC10;TOMM40;HSPA4;NCAPG;CWC27;GPN1;NIPSNAP1;PPM1G;SLC5A6;DCA
DES;UQCR10;CCDC141;MYH6;MYH7;TTN
ZMAT5;UQCR10
MYH6;MYH7
TBX15;KIAA1755;DCAF16;CMTM5;CEP85L;SLC35F1;ARHGEF40;TRIB2;DGKH
TBX15;CEP85L;UQCR10;CCDC141;MYH6;MYH7;TTN
ZNF628;ZNF219;RSPO2;SPEG;SSC5D;EIF3E
TBX15;MLLT10;SHOX2;ZNF219;ALX4;LCORL
DLEU1
MTMR3;CR1;CMTM5;PTEN;SLC35F1;NDRG2;TMEM253;DES;PLN;B3GNT7;FAM174B;CCDC141;CD34
CCDC121;GFM1;ZCCHC10;SLC4A1AP;TOMM40;UQCR10;CWC27;NAT14;PPM1G;DCAF16;GJA1;CCDC91;
DLEU1;VSTM2L
MLLT10;CENPW;MTMR3;LXN;SNX17;MFN1;PTEN;NCAPG;ASCC2;EIF3E;GPN1;NRBP1
CD34
ALX4
SLC5A6;GCKR;SERPINA1;CR1;LXN;TOMM40;APOC1;VSTM2L;GPR126;APOE
HSPA4
MYH6;MYH7
MYH7
ALX4
CCDC121;MTMR3;CR1;CMTM5;SLC35F1;NDRG2;TTN;PLN;B3GNT7;CCDC141;CD34;MYH6;MYH7
MYH6;MYH7
EIF2B4;IFI27L2;GCKR;CENPW;SERPINA1;LXN;PTEN;C2ORF16;NAT14;NIPSNAP1;SUPT7L;TMEM253;PPM
TBX15;DES
VSTM2L;UQCR10
IFI27L2;CENPW;MTMR3;CR1;SLC4A1AP;TOMM40;HSPA4;CMTM5;PTEN;NCAPG;UQCR10;CWC27;GPN1
PTEN
SHOX2
IFI27L2;LXN;SKIDA1;PTEN;AFAP1;VSTM2L;SSC5D;C2ORF16;WNT16;NAT14;NIPSNAP1;TMEM253;DCAF1
ZCCHC10;CR1;CCDC91;CEP85L
SLC5A6;GCKR;DES;SERPINA1;CR1;TOMM40;CMTM5;APOC1;SLC35F1;NDRG2;CCDC141
GCKR;AFAP1;SSC5D;NDRG2;MLF1;KIAA1755;GJA1;NOV;ZNF219;GPR126;SPEG;APOE;ARHGEF40;PVRL2;
WNT16
KIAA1755;SERPINA1;CR1;PLN;B3GNT7;FAM174B;CMTM5;SLC35F1;NDRG2
GJA1;PTEN
TRIB2
DES;ZNF219;RSPO2;AFAP1;SSC5D
TBX15;GFM1;UQCR10;MYH6;MYH7;TTN

GJA1;TOMM40;NOV;CEP85L;RSPO2;EIF3E;UQCR10;FAM3C;APOE;NDRG2;NIPSNAP1;TTN
MFN1;GNB4
APOE
FAM174B
APOE
EIF3E
APOE
GNB4
GJA1
APOE
HSPA4
MTMR3
FAM3C
PPM1G
SPEG
ARHGEF40
SPEG
GNB4
APOE
SERPINA1
APOE
PTEN
B3GNT7
APOE
PTEN
ARHGEF40
HSPA4
GJA1
APOE
PTEN
DES
NRBP1
TOMM40
AFAP1
DNAJC1
ZCCHC10
APOE
NRBP1
NRBP1
APOE
B3GNT7
SERPINA1
HSPA4
HSPA4
NCAPG

HSPA4
GJA1
PTEN
PTEN
AFAP1
NIPSNAP1
GFM1
PPM1G
NRBP1
SERPINA1
APOE
EIF3E
AFAP1
NRBP1
GJA1
TTN
EIF3E
DES;APOE
HSPA4
CCDC141
APOE
SPEG
NOV
TTN
TOMM40
NRBP1
LXN
EIF3E
EIF3E;APOE
HSPA4;PTEN;EIF3E
NOV;AFAP1;APOE
PTEN
APOC1
HSPA4
APOE
NAT14
ARHGEF40
VSTM2L
MTMR3
ARHGEF40
TTN
TRIB2
MTMR3
NCAPG
HSPA4

GJA1
APOE
HSPA4
AFAP1
APOE
NDRG2
NCAPG
APOE
HSPA4
PTEN
TTN
GJA1
EIF3E
EIF3E
LXN
LCORL
NIPSNAP1
HSPA4
EIF3E
DES
HSPA4
GJA1
TOMM40
PLN
NDRG2
APOE
PTEN
NOV
AFAP1
PLN
DNAJC1
HSPA4
FAM174B
GJA1
APOE
PPM1G
ARHGEF40
APOE
CMTM5
TTN
EIF3E
TTN
HSPA4
APOE
APOE

NRBP1
GPN1
TTN
MTMR3
PPM1G
APOE
APOE
FAM3C
TRIB2
PPM1G
HSPA4
TTN
TRIB2
APOE
PTEN
NCAPG
TRIB2
SERPINA1
EIF3E
NIPSNAP1
CD34
B3GNT7
APOE
SLC5A6
EIF3E
LXN;EIF3E
MTMR3;TTN
MTMR3;EIF3E
APOE;NDRG2
MTMR3
EIF3E
GNB4
MTMR3
NCAPG
GJA1
PTEN
SERPINA1
FAM3C
EIF2B4
FAM3C
SERPINA1
VSTM2L
EIF3E
GJA1
NOV

AFAP1
SLC5A6
GJA1;FAM3C
APOE
AFAP1
EIF3E
TOMM40
GNB4
EIF3E
PTEN
MTMR3
PPM1G
NCAPG
EIF3E
CABP7;NDRG2
AFAP1
GJA1
NCAPG
MTMR3
APOE
VSTM2L
APOE
PPM1G
SERPINA1
NRBP1
SERPINA1
DNAJC1
PTEN
HSPA4
SERPINA1
SLC5A6
APOE
NIPSNAP1
EIF3E
NDRG2
PTEN
TOMM40
LCORL
MTMR3;APOE
LXN
ALX4
EIF3E
EIF3E
TRIB2
TRIB2

GJA1
FAM3C
TRIB2
B3GNT7
NOV
NIPSNAP1
NAT14
NDRG2
HSPA4
NCAPG
APOE
B3GNT7
GJA1
SSC5D
GJA1
GJA1
PTEN
SNX17
SERPINA1
SLC4A1AP
HSPA4
TOMM40
APOE
GFM1
GCKR
AFAP1
DNAJC1
B3GNT7;NOV;NDRG2
FAM174B;APOE;ARHGEF40
APOE
GJA1
CEP85L
NRBP1
SPEG
MTMR3
NRBP1
CCDC91
NRBP1
NRBP1
NRBP1
DGKH
PPM1G
GFM1
NCAPG
FAM3C

AFAP1
NRBP1
NRBP1
ZMAT5
NCAPG
CD34
HSPA4
APOE
PPM1G
TOMM40
EIF3E
HSPA4
PTEN
APOC1
HSPA4
EIF3E
PTEN
HSPA4
PTEN
PTEN
AFAP1
FAM3C
EIF3E
CR1
HSPA4
HSPA4
HSPA4
SLC5A6
FAM3C
APOE
PPM1G
PPM1G
NDRG2
DES
HSPA4
AFAP1
PTEN
PLN;RSRC1;SHOX2;AFAP1;SPEG;DGKH
CD34
GFM1;APOC1;CEP85L;ARHGEF40
CCDC121;GFM1;ZCCHC10;SLC4A1AP;TOMM40;CEP85L;CWC27;NAT14;DCAF16;CCDC91;RSRC1;FAM174
GFM1;ZCCHC10;TOMM40;SLC35F1;CWC27;NDRG2;DCAF16;GJA1;PLN;CCDC91;FAM174B;DLEU1;SPEG;
MYH7
GFM1;CENPW;SLC4A1AP;HSPA4;PTEN;C2ORF16;CWC27;GPN1;NIPSNAP1;SLC5A6;CCDC91;RSRC1;EIF3E
SLC4A1AP;ZMAT5;ALX4;LCORL;GPN1;NIPSNAP1;TMEM253

SERPINA1;SHOX2;CEP85L;ARHGEF40
NCAPG
SERPINA1;SKIDA1;AFAP1;SSC5D;NDRG2;TMEM253;KIAA1755;GJA1;NOV;ZNF219;GPR126;SPEG;ARHGE
DES;MTMR3;CR1;PLN;B3GNT7;FAM174B;CMTM5;SLC35F1;NDRG2;CCDC141;CD34
GFM1;ZCCHC10;TOMM40;UQCR10;CWC27;DCAF16;GJA1;CCDC91;ZNF628;RSRC1;FAM174B;MFN1;DLE
DNAJC1;DES;SERPINA1;CR1;PLN;SHOX2;GPR126;ASCC2;PVRL2;WARS2
EIF2B4;CENPW;MTMR3;TOMM40;NRBP1;TMEM253;WARS2;PPM1G;ZNF628;SNX17;ZNF513;ASCC2;AR
MYH6;MYH7
TBX15;CEP85L
ALX4
CCDC121;GFM1;CENPW;SLC4A1AP;PTEN;GPN1;NIPSNAP1;NRBP1;PPM1G;CCDC91;SNX17;RSRC1;ZMAT
NCAPG;TMEM253
CCDC91;CEP85L
CENPW;ALX4;WNT16;LCORL
GCKR
MYH7
ZNF628;PTEN;NRBP1;DGKH;TTN;PPM1G
DES;SERPINA1;CR1;PLN;B3GNT7;CMTM5;VSTM2L;SLC35F1;NDRG2
WNT16
MYH7
DES;MTMR3;CR1;PLN;B3GNT7;FAM174B;NDRG2;CCDC141;MYH6;MYH7;TTN
CCDC91;CEP85L;ARHGEF40;CCDC141
TBX15;IFI27L2;SKIDA1
FAM174B;C2ORF16
SLC5A6;NCAPG;LCORL
CCDC91;CEP85L;DGKH
SLC5A6;TTN
SHOX2;CEP85L;ARHGEF40
MYH6
APOC1;DLEU1;LCORL
ARHGEF40
TBX15
DNAJC1;SERPINA1;CR1;SHOX2;DLEU1;ASCC2;EIF3E;ARHGEF40;TMEM253
MYH6;MYH7
VSTM2L
EIF2B4;SERPINA1;CR1;SHOX2;NRBP1;DNAJC1;DES;ZNF628;ZNF219;ZMAT5;ASCC2;ARHGEF40;PVRL2
CCDC121;ZCCHC10;LXN;SHOX2;SLC35F1;CWC27;NAT14;DCAF16;GJA1;CCDC91;RSRC1;FAM174B;MFN1;
EIF2B4;LXN;SLC4A1AP;HSPA4;SHOX2;NCAPG;GPN1;NRBP1;SUPT7L;WARS2;TTN;PPM1G;MLLT10;DNAJC
VSTM2L;LCORL
GFM1;TOMM40;UQCR10;CWC27;NAT14;PPM1G;DCAF16;GJA1;CCDC91;ZNF628;RSRC1;DLEU1;SPEG;FA
DLEU1
APOC1
IFI27L2;DNAJC1;GCKR;CENPW;NOV;SNX17;PTEN;VSTM2L;ZNF513;C2ORF16;ZNF512;NIPSNAP1
ZMAT5;NIPSNAP1
CCDC121;GFM1;ZCCHC10;TOMM40;SLC35F1;UQCR10;CWC27;NAT14;DCAF16;GJA1;CCDC91;RSRC1;FA

KIAA1755;DES;CR1;PLN;B3GNT7;CMTM5;SLC35F1;CCDC141;MYH6;MYH7
UQCR10
CR1;ZNF628;SHOX2;NCAPG;NRBP1;TTN;PPM1G
DLEU1;WNT16;NIPSNAP1
SHOX2;ALX4
MYH6;MYH7
DES;SERPINA1;CR1;PLN;B3GNT7;FAM174B;CMTM5;SLC35F1;NDRG2;CD34
MYH6
SERPINA1;APOC1
GFM1;MTMR3;ZCCHC10;SLC4A1AP;SHOX2;CEP85L;PTEN;NCAPG;TTN;PPM1G;DCAF16;ZNF628;MFN1;A
CCDC121;GFM1;ZCCHC10;SLC4A1AP;HSPA4;SKIDA1;VSTM2L;SLC35F1;C2ORF16;CWC27;MLF1;WARS2;
GCKR;SERPINA1;CR1;PLN;CMTM5;SLC35F1;NDRG2;CCDC141
IFI27L2;CCDC121;GFM1;ZCCHC10;LXN;SLC4A1AP;TOMM40;SLC35F1;UQCR10;CWC27;NAT14;PPM1G;D
DNAJC1;DES;LXN;ZNF628;NCAPG;ASCC2;PVRL2
DES;SERPINA1;ZCCHC10;CR1;CEP85L;SUPT7L;TTN
TBX15
GCKR;SERPINA1
SLC5A6;VSTM2L;ALX4;WNT16;LCORL;TTN
APOC1;ZMAT5;VSTM2L;ALX4;UQCR10;NAT14
DLEU1;VSTM2L;ALX4
TBX15;ALX4;MYH6
GFM1;ALX4;MYH6
ALX4;DGKH
WNT16
DLEU1;WNT16
MYH6;MYH7
ALX4;MYH6;MYH7
LXN;TOMM40;APOC1;ZMAT5;VSTM2L;UQCR10;APOE;WNT16;NAT14
MTMR3;CMTM5;AFAP1;SSC5D;NDRG2;TMEM253;KIAA1755;GJA1;NOV;ZNF219;GPR126;SPEG;APOE;A
NOV
MYH7
CR1;CEP85L;C2ORF16;ZNF512;ARHGEF40
KIAA1755;DES;CR1;PLN;B3GNT7;CMTM5;SLC35F1;NDRG2;CCDC141;NIPSNAP1;MYH6;TTN
CEP85L
PLN;NCAPG;GPN1
DES;SERPINA1;CR1;PLN;B3GNT7;CMTM5;SLC35F1;NDRG2;CCDC141;CD34
DES;ZNF628;SNX17;ASCC2;PVRL2;NRBP1;PPM1G
MYH6;MYH7
MLLT10;ZCCHC10;LXN;DLEU1;ASCC2;EIF3E;TTN
MYH7
EIF3E;DGKH;WARS2;TTN
CCDC121;ZCCHC10;SLC4A1AP;CCDC91;HSPA4;CEP85L;CWC27;LCORL;SUPT7L
IFI27L2;MTMR3;LXN;SLC4A1AP;AFAP1;CWC27;GPN1;MLF1;PPM1G;DCAF16;RSRC1;GNB4;ALX4;DGKH
ZNF628;ZMAT5;DLEU1;WNT16
TBX15;SHOX2;CEP85L;ARHGEF40;MYH6;MYH7;TTN

CCDC121;MTMR3;CR1;CMTM5;NDRG2;TMEM253;TTN;DES;PLN;B3GNT7;FAM174B;CCDC141;CD34;TRI
CCDC121;GFM1;ZCCHC10;SHOX2;SLC35F1;NAT14;DCAF16;CCDC91;RSRC1;FAM174B;MFN1;ALX4;FAM3
CEP85L;WNT16
EIF2B4;CENPW;LXN;TOMM40;SHOX2;GPN1;NRBP1;PPM1G;DNAJC1;ZNF628;SNX17;MFN1;ZNF513;ASC
CCDC121;ALX4;ZNF512;MYH6;MYH7
CMTM5;CEP85L;WNT16;ARHGEF40
GCKR;CCDC121;SERPINA1;CMTM5;AFAP1;NDRG2;KIAA1755;GJA1;ZNF219;GPR126;SPEG;APOE;ARHGEI
TBX15;CEP85L;ARHGEF40;MLF1
PLN;SHOX2;CCDC141
SLC5A6;MYH7
ZCCHC10;CCDC91;CEP85L;LCORL;TTN
ZCCHC10;CCDC91;RSRC1;ZMAT5;C2ORF16;ZNF512
DES;CR1;MFN1;ASCC2;LCORL;NRBP1;TTN
MTMR3;CR1;PLN;B3GNT7;FAM174B;CMTM5;SLC35F1;NDRG2;CCDC141;CD34;MYH6;MYH7
ALX4;CCDC141
CCDC121;GFM1;ZCCHC10;SLC4A1AP;TOMM40;CEP85L;SLC35F1;UQCR10;CWC27;NDRG2;NAT14;PPM1
TBX15;SHOX2;ARHGEF40
WNT16
NCAPG
GCKR;CENPW;WNT16
DES;NCAPG;ASCC2;EIF3E;TTN
NCAPG;TMEM253
LXN;DLEU1;NCAPG
TBX15;SHOX2;ALX4
DNAJC1;CENPW;DES;CR1;ZNF628;ASCC2;PVRL2;WARS2;TTN
GFM1;SERPINA1;VSTM2L;NCAPG;PPM1G
ARHGEF40
APOC1;ALX4
NCAPG
SLC5A6;TOMM40;NIPSNAP1
DNAJC1;CENPW;ZCCHC10;PTEN;ASCC2;EIF3E
CMTM5
CENPW;DES;MTMR3;ZCCHC10;SLC4A1AP;HSPA4;RSRC1;CMTM5;ZMAT5;SPEG;CWC27
TBX15;CEP85L;ZMAT5;ARHGEF40
KIAA1755;CMTM5;SHOX2;ZMAT5;SLC35F1;NDRG2;ARHGEF40;TRIB2
GCKR
DCAF16;SHOX2;ZNF219;GNB4;SSC5D;FAM3C;CWC27;LCORL;GPN1;TMEM253;WARS2
CCDC121;RSRC1;SHOX2;PTEN;CWC27;TRIB2
DNAJC1
DLEU1
SLC5A6
TBX15;UQCR10;CCDC141;MLF1;MYH7;TTN
TBX15;GFM1;DES;CCDC141;MLF1;MYH6;MYH7;TTN
SHOX2;DLEU1;NCAPG;ASCC2;EIF3E;ARHGEF40;TMEM253
LXN

GNB4
PVRL2;MLF1
IFI27L2;CCDC121;LXN;CWC27;NAT14;ZMAT5;DLEU1;FAM3C;ARHGEF40;CENPW;SKIDA1;VSTM2L;SSC5D
SLC5A6;GCKR;CEP85L;ARHGEF40;SUPT7L
DLEU1
CEP85L;CCDC141
GFM1;CENPW;ZCCHC10;TOMM40;HSPA4;NCAPG;C2ORF16;CWC27;GPN1;NIPSNAP1;WARS2;PPM1G;SL
DES
GCKR
SLC5A6;IFI27L2;FAM174B;APOC1;ZMAT5;DLEU1;VSTM2L;UQCR10
ZMAT5
ZMAT5
DNAJC1;CENPW;DES;SERPINA1;SHOX2;ASCC2;PVRL2
EIF3E;FAM3C
EIF3E;NIPSNAP1
SERPINA1;MTMR3;CR1;PLN;B3GNT7;FAM174B;CMTM5;ZMAT5;SLC35F1;NDRG2;CCDC141;CD34
LXN
NCAPG;PVRL2
GFM1;SLC4A1AP;CEP85L;CWC27;GPN1;SUPT7L;WARS2;TTN;TBX15;DCAF16;CCDC91;RSRC1;ALX4;ARHG
SHOX2;CEP85L;ARHGEF40;TTN
DNAJC1;LXN;SHOX2;PTEN;DLEU1;ASCC2;EIF3E
CCDC91;CEP85L
CR1;CMTM5;SLC35F1;NDRG2;NIPSNAP1;TTN;KIAA1755;DES;PLN;B3GNT7;FAM174B;MYH6;MYH7
GCKR;ALX4
ALX4
CR1;PLN;B3GNT7;CMTM5;SLC35F1;NDRG2;CCDC141;MYH6;MYH7
EIF2B4;GCKR;CCDC121;GFM1;SKIDA1;SHOX2;SSC5D;UQCR10;CWC27;MLF1;WARS2;KIAA1755;FAM174
TBX15;WNT16
TBX15;GNB4;ALX4;DGKH;TTN
EIF2B4;DES;SERPINA1;ZNF628;ZMAT5;ASCC2;PVRL2;PPM1G
IFI27L2;GCKR;B3GNT7;NOV;GNB4;PTEN;ZNF513;ZNF512;TRIB2;NAT14;NIPSNAP1
GFM1;SERPINA1;MTMR3;ZCCHC10;SLC4A1AP;TOMM40;PTEN;CWC27;PPM1G;DCAF16;CCDC91;ZNF628
CCDC121;DES;MTMR3;CR1;PLN;CMTM5;SLC35F1;NDRG2;CCDC141;CD34;MYH6;TTN
MYH6;MYH7
CCDC121;GFM1;ZCCHC10;LXN;SLC4A1AP;TOMM40;CEP85L;UQCR10;CWC27;NAT14;PPM1G;DCAF16;G
CCDC121;GFM1;ZCCHC10;SLC35F1;DCAF16;GJA1;CCDC91;FAM174B;MFN1;SPEG;ALX4;FAM3C;ARHGEF
GFM1;HSPA4;VSTM2L
KIAA1755;CR1;PLN;B3GNT7;CMTM5;SLC35F1;NDRG2;CCDC141;MYH6;MYH7
TBX15;FAM174B;CEP85L;CCDC141;TTN
DES;LXN;MFN1;ASCC2;NRBP1;TTN
TBX15
DES;PLN
KIAA1755;SERPINA1;CR1;PLN;CMTM5;SLC35F1;NDRG2;CCDC141;MYH6;MYH7
IFI27L2;CCDC121;GFM1;ZCCHC10;SLC4A1AP;SLC35F1;CWC27;NAT14;GJA1;CCDC91;RSRC1;FAM174B;D
SERPINA1;CEP85L;SUPT7L;TTN

MYH7
GFM1;ZCCHC10;TOMM40;SLC35F1;CWC27;DCAF16;GJA1;CCDC91;FAM174B;MFN1;SPEG;ALX4;FAM3C;
GJA1;ALX4;WARS2
KIAA1755;SERPINA1;CR1;LXN;PLN;B3GNT7;CMTM5;EIF3E;MYH6
SLC4A1AP;HSPA4
DES;MTMR3;CR1;PLN;B3GNT7;FAM174B;CMTM5;APOC1;PTEN;NDRG2;CCDC141;CD34
MTMR3;TOMM40;ZNF628;ZNF219;PTEN;NRBP1;DGKH;PPM1G
CCDC121;ZMAT5;PTEN;TMEM253
ZNF628;FAM174B;ZMAT5;DLEU1;SPEG;NAT14;NIPSNAP1
EIF2B4;GFM1;MTMR3;TOMM40;HSPA4;SHOX2;PTEN;GPN1;NIPSNAP1;NRBP1;SUPT7L;WARS2;PPM1G;
GCKR;SERPINA1
EIF2B4;ZNF628;DLEU1;ASCC2;EIF3E;GPN1;NRBP1;PPM1G
MYH6;MYH7
LXN;ASCC2;EIF3E;TMEM253;TTN
TBX15
CENPW;ZNF628;ALX4;WNT16;TMEM253;DGKH;PPM1G
DLEU1
IFI27L2;DNAJC1;B3GNT7;ZNF628;WNT16;PVRL2;NAT14;NIPSNAP1;TMEM253
DNAJC1;SERPINA1;CR1;LXN;DLEU1;NCAPG;ASCC2;EIF3E;PVRL2
NCAPG
DLEU1
DLEU1
SERPINA1;CR1;PLN;B3GNT7;CMTM5;SLC35F1;NDRG2;CCDC141
SLC5A6;GCKR;CENPW;MTMR3;ZNF628;ALX4;ZNF513;ARHGEF40;NRBP1;DGKH;PPM1G
CCDC121;GFM1;ZCCHC10;SLC35F1;NAT14;DCAF16;GJA1;PLN;FAM174B;MFN1;SPEG;FAM3C;APOE;CD3
TBX15;CCDC121;CEP85L;ZNF512;ARHGEF40;MYH6;MYH7;TTN
DES;ZNF219;RSPO2;AFAP1;SSC5D
CD34;MLF1
CCDC121;GFM1;ZCCHC10;SLC4A1AP;TOMM40;SHOX2;CEP85L;SLC35F1;UQCR10;CWC27;NAT14;PPM1(
SLC5A6;GCKR;SERPINA1;APOC1;GPR126;APOE;NIPSNAP1
GFM1;C2ORF16
CCDC121;SLC4A1AP;LCORL;GPN1
SERPINA1;C2ORF16
CCDC121;GFM1;ZCCHC10;SLC4A1AP;SLC35F1;CWC27;DCAF16;GJA1;CCDC91;RSRC1;MFN1;SPEG;ALX4;F
CD34
CR1
MYH6
EIF2B4;ZNF628;ASCC2;NRBP1;DGKH;PPM1G
SLC5A6;UQCR10;LCORL;MYH7
DES;SERPINA1;CR1;PLN;CMTM5;SLC35F1;NDRG2;CCDC141
APOC1;APOE
APOC1
TOMM40;NCAPG;NRBP1
ZNF628;ASCC2;NRBP1;PPM1G
DCAF16;SNX17;EIF3E;CCDC141;DGKH;WARS2;TTN

GFM1;MTMR3;ZCCHC10;SLC4A1AP;TOMM40;PTEN;UQCR10;CWC27;PPM1G;DCAF16;PLN;CCDC91;ZNF
LXN;ZNF628;MFN1;NRBP1
CENPW;MTMR3;SLC4A1AP;NRBP1;SUPT7L;TMEM253;WARS2;TTN;MLLT10;DCAF16;ZNF628;SNX17;MF
EIF2B4;GFM1;MTMR3;SLC4A1AP;TOMM40;SHOX2;GPN1;NRBP1;SUPT7L;PPM1G;MLLT10;NOV;ZNF628
SLC5A6;IFI27L2;GCKR;TOMM40;ZNF628;APOC1;ZMAT5;UQCR10;APOE;NAT14
CCDC121;ZCCHC10;CCDC91;CEP85L;C2ORF16;ZNF512;CWC27;SUPT7L
DNAJC1;SHOX2;MFN1;ASCC2;PVRL2
MYH6
CCDC91;CEP85L
ZNF628;SHOX2;WNT16;NRBP1;DGKH;WARS2;PPM1G
ZCCHC10;CCDC91
TBX15;MYH6
NCAPG;MYH7
DNAJC1;SHOX2;ASCC2;WARS2;TTN
ZMAT5;DLEU1;UQCR10;NAT14
KIAA1755;DES;SERPINA1;CR1;PLN;B3GNT7;FAM174B;CMTM5;NDRG2;CCDC141
DNAJC1;CENPW;SHOX2;ASCC2
ALX4
MYH6;MYH7
TBX15;MLF1
MTMR3;CR1;PLN;FAM174B;SLC35F1;WNT16;NDRG2;CCDC141;CD34
KIAA1755;SERPINA1;CR1;PLN;B3GNT7;CMTM5;NDRG2;CCDC141;MYH6;MYH7
DNAJC1;ZNF628;ASCC2;TTN
CCDC121;SERPINA1;MTMR3;CR1;PLN;B3GNT7;SLC35F1;NDRG2;CCDC141;CD34;MYH6;MYH7
CCDC121;GFM1;ZCCHC10;SLC4A1AP;TOMM40;SLC35F1;CWC27;DCAF16;GJA1;CCDC91;ZNF628;RSRC1;
SLC35F1;MYH6
MLF1
DLEU1
LXN;ZNF628;SNX17;ZMAT5;ASCC2;PVRL2;NRBP1;TMEM253;DGKH;WARS2;PPM1G
CCDC121;GFM1;ZCCHC10;LXN;SLC4A1AP;TOMM40;CEP85L;SLC35F1;CWC27;NAT14;DCAF16;GJA1;CCD
DNAJC1;ZNF628;SHOX2;ASCC2;NRBP1;PPM1G
MYH7
CMTM5;DLEU1;VSTM2L
DNAJC1;LXN;DLEU1;GPR126;ASCC2;EIF3E;ARHGEF40;CCDC141
IFI27L2;GCKR;SERPINA1;PTEN;VSTM2L;NAT14;NIPSNAP1;B3GNT7;NOV;ZNF513;APOE;PVRL2;TRIB2
EIF2B4;DNAJC1;DES;SERPINA1;MTMR3;ZNF628;DLEU1;ASCC2;ARHGEF40
CENPW;LCORL;PPM1G
KIAA1755;CR1;PLN;B3GNT7;CMTM5;NDRG2;CCDC141;MYH6;MYH7
SHOX2
GFM1;MTMR3;ZCCHC10;SLC4A1AP;TOMM40;PTEN;NCAPG;CWC27;PPM1G;DCAF16;CCDC91;RSRC1;ZN
DCAF16;DES;ZNF628;SNX17;MFN1;ASCC2;NRBP1;TTN
SERPINA1;CMTM5;DLEU1
IFI27L2;CCDC121;MTMR3;ZCCHC10;SLC4A1AP;PTEN;AFAP1;NCAPG;CWC27;GPN1;MLF1;WARS2;MLLT1
MYH7
GFM1;TOMM40;ZNF628;SHOX2;MFN1;NRBP1;WARS2

MTMR3;CR1;PLN;B3GNT7;CMTM5;SLC35F1;NDRG2;CCDC141;CD34;MYH7;TTN
PTEN
IFI27L2;MTMR3;ZCCHC10;CR1;SLC4A1AP;TOMM40;HSPA4;CMTM5;UQCR10;WNT16;CWC27;NRBP1;MI
GCKR;GJA1;NOV;APOC1;ZNF219;C2ORF16;APOE;ARHGEF40;NIPSNAP1;WARS2
DNAJC1;CENPW;SHOX2;ASCC2;NRBP1
APOE
SERPINA1
TMEM253
NCAPG
CR1;PTEN;APOE;NRBP1
APOE
HSPA4
SLC4A1AP;TOMM40;CEP85L;UQCR10;NDRG2;GPN1;NIPSNAP1;TTN;PPM1G;GJA1;SNX17;RSPO2;EIF3E;F
TOMM40;ZNF628;SNX17;MFN1;ASCC2;GPN1;NRBP1;DGKH;TTN;PPM1G
CCDC121;SLC4A1AP;ZMAT5;GPN1
TRIB2
IFI27L2;CCDC121;CENPW;SERPINA1;SKIDA1;PTEN;VSTM2L;C2ORF16;NAT14;NIPSNAP1;SUPT7L;TMEM2
CCDC121;GFM1;ZCCHC10;TOMM40;CEP85L;DCAF16;GJA1;CCDC91;ZNF628;FAM174B;MFN1;DLEU1;SPI
LCORL
DNAJC1;LXN;ZNF628;SHOX2;NCAPG;ASCC2;EIF3E;NRBP1
DES;CR1;PLN;B3GNT7;FAM174B;CMTM5;SLC35F1;NDRG2;CCDC141
MYH6;MYH7
CMTM5
DNAJC1;DES;PLN;ZNF628;ASCC2;GPN1;MYH6;WARS2;TTN
DNAJC1;SERPINA1;PLN;DLEU1;NCAPG;ASCC2;EIF3E
IFI27L2;MTMR3;ZCCHC10;CR1;SLC4A1AP;HSPA4;CMTM5;NCAPG;CWC27;SUPT7L;TBX15;ZMAT5;GNB4;
SHOX2;ALX4
TBX15;CEP85L;ALX4;ARHGEF40
SERPINA1;CCDC91;NIPSNAP1
SERPINA1;CEP85L;SUPT7L;TTN
DES;MTMR3;CR1;PLN;B3GNT7;CMTM5;GPR126;SLC35F1;NDRG2;CCDC141;MYH6;TTN
APOC1;APOE
DNAJC1;DES;SERPINA1;CR1;ZNF628;ZMAT5;PVRL2;TTN
CCDC121;MTMR3;CR1;CMTM5;PTEN;SLC35F1;NDRG2;DES;PLN;B3GNT7;FAM174B;ZNF219;APOE;CCDC
ZMAT5;DLEU1;VSTM2L
TBX15
GFM1
CENPW;DES;ZCCHC10;SLC4A1AP;HSPA4;RSRC1;GNB4;DLEU1;CWC27
SKIDA1;ALX4
MTMR3;ALX4
ALX4
ALX4;MYH6
SHOX2
PTEN;EIF3E;TMEM253
CR1;PLN;SLC35F1;NDRG2;CCDC141;CD34;MYH6;MYH7

IFI27L2;ZMAT5;VSTM2L;NIPSNAP1
CCDC121;ZMAT5
SERPINA1;CR1;B3GNT7;CMTM5;SLC35F1;NDRG2;CCDC141
SHOX2;CEP85L;ARHGEF40
NCAPG
CCDC121;ZMAT5
IFI27L2;CCDC121;GFM1;ZCCHC10;LXN;SLC4A1AP;TOMM40;CEP85L;UQCR10;CWC27;DCAF16;GJA1;CCD
DNAJC1;SERPINA1;SHOX2;ASCC2;CCDC141;DGKH;PPM1G
CENPW;MTMR3;TOMM40;WNT16;NAT14;NRBP1;PPM1G;ZNF628;SNX17;APOC1;ZMAT5;ZNF513;ASCC
CENPW;PTEN;NCAPG;LCORL
DNAJC1;DES;CR1;PLN;SHOX2;NRBP1
GCKR;SERPINA1
TBX15;CR1
CCDC121;GFM1;SLC4A1AP;CCDC91;CEP85L;ALX4;ARHGEF40;GPN1;TTN
VSTM2L;UQCR10
GPR126;CCDC141
HSPA4;NCAPG;MYH7
DNAJC1;DES;CR1;PLN;SHOX2;ASCC2;PVRL2;MYH6;TTN
IFI27L2;GCKR;CENPW;SERPINA1;LXN;NAT14;NIPSNAP1;SUPT7L;B3GNT7;NOV;GNB4;GPR126;ZNF513;Z
KIAA1755;MTMR3;CR1;PLN;B3GNT7;FAM174B;CMTM5;PTEN;SLC35F1;NDRG2;CCDC141;CD34
ZMAT5
C2ORF16
MYH6;MYH7
MYH6;MYH7
IFI27L2;SERPINA1;CR1;LXN;SKIDA1;PTEN;AFAP1;SSC5D;C2ORF16;NIPSNAP1;SUPT7L;DCAF16;GJA1;B3G
APOC1;LCORL
IFI27L2;CR1;APOC1;WNT16;CCDC141;LCORL;TTN
CENPW;ZNF628;PTEN;NRBP1;WARS2;PPM1G
SLC5A6;EIF2B4;GCKR;MTMR3;ZNF628;FAM174B;APOC1;ALX4;ZNF513;WNT16;NRBP1;PPM1G
GCKR
NCAPG;ALX4
RSRC1;DGKH
HSPA4
GCKR;UQCR10
ZNF628;SHOX2;ASCC2
TBX15;SKIDA1;MYH6
TOMM40;ZNF628;VSTM2L;WNT16;NIPSNAP1
APOC1;DLEU1;VSTM2L;NCAPG;UQCR10;APOE;TMEM253
SERPINA1;CR1;PLN;B3GNT7;FAM174B;CMTM5;NDRG2;CCDC141
IFI27L2;LXN;SKIDA1;PTEN;AFAP1;C2ORF16;NAT14;NIPSNAP1;TMEM253;DCAF16;B3GNT7;NOV;SNX17;
APOC1;UQCR10;APOE;TMEM253
MTMR3;CR1;TOMM40;CMTM5;SLC35F1;NDRG2;DES;PLN;B3GNT7;ZNF628;FAM174B;CCDC141;CD34
SHOX2;NCAPG;ASCC2;NRBP1;TTN
PTEN
EIF2B4;IFI27L2;MTMR3;LXN;AFAP1;NCAPG;WNT16;NAT14;MLF1;NRBP1;TMEM253;DCAF16;GJA1;NOV

APOC1;MYH7
NCAPG
MYH7
MYH6;MYH7
SLC4A1AP
DLEU1
SHOX2;NRBP1
MYH6;MYH7
KIAA1755;DES;SERPINA1;CR1;PLN;B3GNT7;CMTM5;NDRG2;CCDC141
SLC5A6;TOMM40;APOC1;APOE
WNT16;MYH6;MYH7
WNT16
DNAJC1
DNAJC1
TTN
DLEU1
IFI27L2;SERPINA1;SKIDA1;VSTM2L;SSC5D;C2ORF16;NAT14;NIPSNAP1;TMEM253;DCAF16;B3GNT7;NOV
CEP85L;MYH6
CENPW;SHOX2;ASCC2
WNT16;NIPSNAP1
MYH6;MYH7
NCAPG
DNAJC1;ZNF628;MFN1;ASCC2;NRBP1;TTN;PPM1G
CCDC91;DGKH
APOC1
CCDC91;DGKH
NDRG2
GCKR
GCKR
LCORL
DLEU1
GFM1;ZCCHC10;SLC4A1AP;TOMM40;HSPA4;SHOX2;C2ORF16;UQCR10;CWC27;GPN1;PPM1G;SLC5A6;D
IFI27L2
APOC1;ALX4
PLN;GPR126;CCDC141
DES;CR1;PLN;B3GNT7;FAM174B;NDRG2;CCDC141;MYH6;MYH7
VSTM2L
VSTM2L
TBX15;CEP85L;ALX4;ARHGEF40
SHOX2;TTN
ALX4
IFI27L2;GFM1;CENPW;ZCCHC10;SLC4A1AP;HSPA4;PTEN;CWC27;GPN1;MLF1;NRBP1;PPM1G;TBX15;ZM
NCAPG
MYH6;MYH7
WNT16

SKIDA1;APOC1;NCAPG;APOE;NAT14
IFI27L2;LXN;SKIDA1;PTEN;AFAP1;SSC5D;C2ORF16;NAT14;NIPSNAP1;SUPT7L;DCAF16;B3GNT7;NOV;SN[illegible]
HSPA4
PPM1G
CCDC91;DLEU1
GJA1
SHOX2;ALX4
KIAA1755;DES;SERPINA1;CR1;PLN;B3GNT7;CMTM5;VSTM2L;SPEG;SLC35F1;NDRG2;CCDC141
NCAPG;WNT16
NCAPG
CCDC121;MTMR3;CR1;CMTM5;TTN;PLN;B3GNT7;FAM174B;ZNF513;APOE;CCDC141;CD34;TRIB2;MYH[illegible]
ZMAT5
PTEN;NDRG2
GCKR;CCDC121;GFM1;ZCCHC10;SHOX2;SLC35F1;C2ORF16;MLLT10;GJA1;CCDC91;FAM174B;ZNF219;M[illegible]
SUPT7L;TTN
DCAF16;GFM1;ZNF628;MFN1;NRBP1;DGKH;WARS2;PPM1G
IFI27L2;GFM1;ZCCHC10;SLC4A1AP;TOMM40;HSPA4;RSRC1;PTEN;DLEU1;UQCR10;CWC27;DGKH
SERPINA1;CR1;B3GNT7;CMTM5;SLC35F1;NDRG2;CCDC141;CD34
ZNF628;ZMAT5;VSTM2L;C2ORF16;WNT16
CCDC121;GFM1;ZCCHC10;SLC35F1;CWC27;DCAF16;CCDC91;RSRC1;FAM174B;MFN1;ALX4;FAM3C;CD3[illegible]
IFI27L2;MTMR3;SLC4A1AP;HSPA4;CMTM5;PTEN;UQCR10;CWC27;TBX15;PLN;SNX17;RSRC1;ZMAT5;GN[illegible]
SLC5A6;SNX17;ZNF219;PTEN;ARHGEF40;CD34
C2ORF16
GJA1;NOV
SHOX2;ZNF219
GFM1;HSPA4;PPM1G
APOC1
TBX15;ALX4
GFM1;SLC4A1AP;HSPA4;ZMAT5;GNB4;CWC27;GPN1;DGKH
ALX4;DGKH
DNAJC1;ASCC2;TTN
ALX4
APOC1;WNT16
APOC1;NCAPG
ZNF628
DLEU1
EIF2B4;CENPW;ZNF628;SHOX2;ZNF219;NCAPG;PVRL2;NRBP1;WARS2;PPM1G
DNAJC1;CR1;SHOX2;MFN1;GNB4;CCDC141;GPN1;WARS2;TTN
GFM1;CR1;CCDC91;CEP85L;MFN1;PTEN;TTN
KIAA1755;DES;CR1;PLN;B3GNT7;CMTM5;SLC35F1;NDRG2;NIPSNAP1;TTN
MYH6
SLC4A1AP;SNX17;ZMAT5;GPN1;PPM1G
ASCC2;CCDC141;TTN
MFN1;GPN1;NRBP1;WARS2;TTN;PPM1G
LXN;PTEN;ASCC2;EIF3E;GPN1;NRBP1;TTN;PPM1G

KIAA1755;DES;SERPINA1;CR1;PLN;B3GNT7;FAM174B;CMTM5;NDRG2;CCDC141
TBX15;MYH6
DNAJC1;DES;CR1;LXN;PLN;NCAPG;ASCC2;EIF3E;ARHGEF40;PVRL2
SLC5A6;DNAJC1;CENPW;RSRC1;ZNF513;WNT16;LCORL;NRBP1;TMEM253;PPM1G
CCDC121;DES;MTMR3;CR1;PLN;FAM174B;CMTM5;SLC35F1;APOE;NDRG2;CCDC141
HSPA4;UQCR10
TBX15
KIAA1755;DES;SERPINA1;CR1;B3GNT7;CMTM5;NDRG2;CCDC141;NIPSNAP1;TTN
ZMAT5;C2ORF16;TMEM253
LXN;NRBP1;DGKH;TTN
IFI27L2;SHOX2;GNB4
KIAA1755;DES;CR1;PLN;B3GNT7;CMTM5;VSTM2L;SPEG;NDRG2;NIPSNAP1;MYH6;MYH7
CMTM5;SPEG;DGKH
ZMAT5;DLEU1
ALX4
EIF2B4;IFI27L2;GCKR;CENPW;SERPINA1;CR1;LXN;PTEN;C2ORF16;NAT14;NIPSNAP1;SUPT7L;PPM1G;SLC
CENPW;DES;ZNF628;TTN;PPM1G
SHOX2;CEP85L;ZMAT5;ZNF512;ARHGEF40
DNAJC1;SNX17;GPR126;ASCC2;CCDC141;PVRL2;TTN
UQCR10
TBX15;TTN
CCDC121;SLC4A1AP;ZMAT5
CCDC121;MTMR3;CR1;PLN;B3GNT7;NOV;FAM174B;CMTM5;APOE;NDRG2;CCDC141;CD34
SERPINA1;CR1;TOMM40;B3GNT7;ZNF628;FAM174B;CMTM5;ZNF219;ZMAT5;SLC35F1;NDRG2
IFI27L2
EIF2B4;MLLT10;MTMR3;TOMM40;ZNF628;SNX17;ASCC2;GPN1;NRBP1;SUPT7L;DGKH;PPM1G
CEP85L;ARHGEF40;MYH7
GCKR;WNT16;GPN1
GCKR;ZNF628;ALX4
IFI27L2;LXN;NOV;GNB4;NCAPG;ARHGEF40;PVRL2;TRIB2;NAT14;NIPSNAP1;TMEM253
NCAPG;MYH7
GCKR;CENPW;ZNF628;APOC1;ZNF513;PPM1G
VSTM2L;WNT16;TMEM253
CCDC121;ZMAT5;ALX4;TMEM253
DNAJC1;DES;SERPINA1;CR1;ASCC2;EIF3E;ARHGEF40
SERPINA1;CR1;B3GNT7;FAM174B;CMTM5;SLC35F1;NDRG2;CCDC141;CD34
IFI27L2;SERPINA1;LXN;PTEN;AFAP1;C2ORF16;NAT14;NIPSNAP1;SUPT7L;SLC5A6;DNAJC1;B3GNT7;NOV;
TBX15;CCDC121;SKIDA1;CEP85L;PTEN;ZNF512;ARHGEF40
MYH7
TMEM253
MYH7
WNT16
GCKR;APOC1;UQCR10
CCDC91;CEP85L;C2ORF16;ZNF512;SUPT7L
SHOX2

SERPINA1;CEP85L;C2ORF16;ZNF512;ARHGEF40
IFI27L2;SERPINA1;LXN;PTEN;NCAPG;NAT14;FAM174B;FAM3C;EIF2B4;GCKR;CENPW;CR1;SKIDA1;AFAP1
ZMAT5;DLEU1;ALX4
MYH7
NCAPG
CR1
DLEU1;NIPSNAP1
IFI27L2;GCKR;LXN;SKIDA1;PTEN;C2ORF16;NAT14;NIPSNAP1;TMEM253;DCAF16;B3GNT7;NOV;SNX17;Z
HSPA4;LCORL
EIF2B4;GFM1;CENPW;SLC4A1AP;HSPA4;PTEN;NCAPG;GPN1;NRBP1;SUPT7L;WARS2;TTN;PPM1G;MLLT
ZCCHC10;CR1;SLC4A1AP;TOMM40;HSPA4;ZMAT5;GNB4;UQCR10;CWC27;NRBP1;DGKH;PPM1G
SHOX2;NRBP1
MTMR3;CR1;CMTM5;SLC35F1;NDRG2;TTN;DES;PLN;B3GNT7;FAM174B;SPEG;CCDC141;CD34;MYH6
MTMR3;AFAP1;SSC5D;NDRG2;KIAA1755;GJA1;NOV;ZNF219;GPR126;SPEG;ARHGEF40;PVRL2;CD34;TRI
SHOX2;ASCC2;NRBP1;PPM1G
CMTM5
ZNF628;ASCC2;NRBP1;PPM1G
DLEU1
EIF2B4;GFM1;CENPW;MTMR3;ZCCHC10;SLC4A1AP;HSPA4;PTEN;GPN1;NRBP1;SUPT7L;WARS2;TTN;PP
ALX4;CCDC141
SHOX2
TBX15;SLC4A1AP;HSPA4;PTEN;MYH6;DGKH
VSTM2L
WNT16
GJA1;ALX4
WNT16;DGKH
EIF2B4;GCKR;CENPW;MTMR3;TOMM40;SHOX2;PTEN;NCAPG;WNT16;NRBP1;TMEM253;PPM1G;SLC5A
ZCCHC10
VSTM2L
IFI27L2;LXN;AFAP1;NCAPG;NAT14;NIPSNAP1;B3GNT7;NOV;GNB4;ZNF513;ARHGEF40;PVRL2;TRIB2
CR1;CEP85L
FAM174B;ARHGEF40
CCDC121;GFM1;ZCCHC10;SLC4A1AP;TOMM40;CEP85L;PTEN;SLC35F1;CWC27;DCAF16;GJA1;CCDC91;R
CR1;SHOX2;ALX4;DGKH
GFM1;CENPW;MTMR3;ZCCHC10;SLC4A1AP;TOMM40;HSPA4;PTEN;UQCR10;CWC27;GPN1;NRBP1;PPM
LXN;ASCC2;EIF3E;DGKH;TTN
DNAJC1;LXN;NCAPG;ASCC2;EIF3E
TOMM40;ZMAT5;UQCR10
ZCCHC10;CMTM5;UQCR10;ARHGEF40
KIAA1755;DES;CR1;PLN;B3GNT7;CMTM5;SLC35F1;NDRG2;CCDC141
SLC5A6;CCDC121;SLC4A1AP;APOC1;ZMAT5;ASCC2;UQCR10;NIPSNAP1
DNAJC1;CR1;SHOX2;ASCC2;PVRL2;TTN
MLLT10;LXN;NCAPG;ASCC2;EIF3E;NRBP1;TTN
AFAP1;SLC35F1;MLF1;TBX15;MLLT10;KIAA1755;GJA1;PLN;CCDC91;FAM174B;APOC1;GNB4;GPR126;AL
SHOX2

LXN;ZNF628;SHOX2;NCAPG;GPN1;NRBP1
CCDC121;GFM1;ZCCHC10;SLC4A1AP;TOMM40;CEP85L;UQCR10;CWC27;NAT14;DCAF16;CCDC91;RSRC1
MYH7
GCKR
DLEU1
TMEM253
SHOX2
CEP85L;ARHGEF40
GJA1;MTMR3;CMTM5;CEP85L;AFAP1;MLF1;DGKH;TTN
DES;MTMR3;B3GNT7;NOV;GPR126;NDRG2;CCDC141;MYH6;TTN
DES;PLN;B3GNT7;CMTM5;NDRG2;MYH6;MYH7
IFI27L2;DCAF16;SERPINA1;B3GNT7;NOV;SNX17;C2ORF16;ZNF512;FAM3C;TRIB2;NIPSNAP1;TMEM253
CCDC121;GFM1;ZCCHC10;TOMM40;CEP85L;UQCR10;CWC27;DCAF16;GJA1;CCDC91;RSRC1;FAM174B;N
EIF2B4;KIAA1755;IFI27L2;CMTM5;ZMAT5;NDRG2;ARHGEF40;TRIB2
TBX15;GFM1;ZNF628;SPEG;UQCR10;NDRG2;MYH6;TTN
KIAA1755;ZNF219;RSPO2;GPR126;DGKH
GJA1;ZCCHC10;NOV;HSPA4;SPEG;SSC5D
KIAA1755;DES;ZNF219;RSPO2;AFAP1;SPEG
SERPINA1;MTMR3;CR1;CR1L;APOC1;PTEN;ASCC2
KIAA1755;CMTM5;SHOX2;SLC35F1;ARHGEF40;TRIB2;WARS2
DES;LXN;ZNF219;RSPO2;ZNF513
KIAA1755;SERPINA1;CR1;PLN;B3GNT7;CMTM5;NDRG2;CCDC141
SERPINA1;CR1;PLN;B3GNT7;NDRG2;CCDC141;CD34
DES;LXN;SHOX2;DLEU1;ASCC2
WNT16;TMEM253
CEP85L
MTMR3;SLC4A1AP;RSRC1;ZMAT5;GNB4;NCAPG;CWC27
TBX15;CCDC91;CEP85L;ARHGEF40;TRIB2;MYH6;MYH7;TTN
VSTM2L;ALX4;MYH7
KIAA1755;SERPINA1;CR1;B3GNT7;FAM174B;CMTM5;NDRG2;CCDC141
ZNF512;SUPT7L
IFI27L2;LXN;PTEN;C2ORF16;NAT14;NIPSNAP1;TMEM253;B3GNT7;NOV;SNX17;APOC1;GNB4;ZNF513;ZN
KIAA1755;CCDC121;SERPINA1;CR1;B3GNT7;CMTM5;VSTM2L;NDRG2;MYH6;MYH7
SPEG
VSTM2L
DLEU1;VSTM2L;NCAPG;MYH6
IFI27L2;CCDC121;PTEN;C2ORF16;NAT14;NIPSNAP1;TMEM253;NOV;SNX17;RSRC1;GNB4;ZNF513;ZNF51
ALX4
GCKR;NCAPG
DES;ZNF628;NRBP1;DGKH;TTN;PPM1G
ZCCHC10;CCDC91
LCORL;MYH6
APOC1;NCAPG
PTEN;EIF3E;TTN
CR1;PLN;B3GNT7;CMTM5;SLC35F1;NDRG2;CCDC141

ALX4
C2ORF16
TOMM40;HSPA4
DNAJC1;DES;SERPINA1;SHOX2;NRBP1
MYH6
GFM1;ZCCHC10;SLC4A1AP;TOMM40;CEP85L;SLC35F1;CWC27;NDRG2;DCAF16;GJA1;CCDC91;RSRC1;MI
VSTM2L;NIPSNAP1
NIPSNAP1
MFN1;GPN1;NRBP1
MYH6
DLEU1;VSTM2L
IFI27L2;CCDC121;LXN;PTEN;SLC35F1;SSC5D;C2ORF16;NAT14;NIPSNAP1;TMEM253;KIAA1755;DCAF16;I
DNAJC1;SERPINA1;DLEU1;ASCC2;EIF3E;TTN
KIAA1755;CCDC141;NIPSNAP1
KIAA1755;CR1;B3GNT7;CMTM5;SLC35F1;NDRG2;CCDC141
MLLT10;SHOX2;MFN1;PTEN;LCORL;GPN1;NRBP1
DLEU1
IFI27L2;GCKR;SERPINA1;LXN;PTEN;AFAP1;SSC5D;C2ORF16;NIPSNAP1;SUPT7L;MLLT10;DCAF16;B3GNT
MTMR3;ZNF628;SNX17;ZMAT5;ASCC2;CCDC141;GPN1;NRBP1;WARS2;PPM1G
IFI27L2;GCKR;LXN;PTEN;AFAP1;SLC35F1;NAT14;NIPSNAP1;TMEM253;KIAA1755;B3GNT7;NOV;SNX17;R
DNAJC1;MTMR3;ASCC2;EIF3E
TBX15;CEP85L;ARHGEF40
KIAA1755;CCDC121;SERPINA1;CR1;B3GNT7;CMTM5;NDRG2;CCDC141;CD34
EIF2B4;ZNF628;SNX17;SHOX2;NCAPG;EIF3E;GPN1;NRBP1;TTN;PPM1G
DNAJC1;DES;MTMR3;CR1;ZNF628;DLEU1;ASCC2;ARHGEF40;PVRL2;MYH6
SHOX2
IFI27L2;MTMR3;SLC4A1AP;AFAP1;CWC27;MLF1;TTN;PPM1G;DCAF16;GJA1;NOV;ZNF219;SPEG;EIF3E;ZI
MTMR3;SKIDA1;ZMAT5;NIPSNAP1
SERPINA1;CR1;B3GNT7;CMTM5;SLC35F1;NDRG2;CCDC141;MYH6
EIF2B4;PTEN
UQCR10
GCKR;ZMAT5
GJA1;TOMM40;GPR126;ALX4;ASCC2;WNT16;PVRL2;DGKH;PPM1G
MYH6
DLEU1;LCORL
DNAJC1;LXN;ZNF628;ASCC2;LCORL;NRBP1
TBX15;MYH7
IFI27L2;GFM1;MTMR3;ZCCHC10;SLC4A1AP;HSPA4;PTEN;UQCR10;CWC27;TBX15;MLLT10;SNX17;RSRC1
TOMM40;ALX4;LCORL
ALX4
CCDC121;CCDC91
CMTM5;CEP85L;ARHGEF40;MYH7
IFI27L2;MTMR3;SLC4A1AP;HSPA4;GNB4;ZNF512;CWC27;DGKH
EIF2B4;GCKR;CENPW;MTMR3;PTEN;AFAP1;WNT16;NRBP1;PPM1G;DNAJC1;ZNF628;SNX17;APOC1;ZM
GCKR;CENPW;SERPINA1;MTMR3;CR1;LXN;PTEN;AFAP1;NCAPG;WNT16;GPN1;NRBP1;PPM1G;DCAF16;I

MTMR3;CR1;PLN;B3GNT7;FAM174B;SLC35F1;NDRG2;CCDC141;CD34
MTMR3;CR1;PLN;B3GNT7;FAM174B;CMTM5;NDRG2;CD34
DNAJC1;SHOX2;DLEU1;EIF3E;PVRL2;GPN1;TTN
SERPINA1;CR1;PLN;B3GNT7;CMTM5;SLC35F1;NDRG2
CENPW
DLEU1
ALX4
WNT16
ZMAT5
EIF2B4;GCKR;CCDC121;GFM1;ZCCHC10;SLC4A1AP;SKIDA1;SSC5D;C2ORF16;CWC27;MLF1;MLLT10;KIAA
GCKR;WNT16
DLEU1;ALX4
ALX4;MYH6
DES;CR1;PLN;B3GNT7;CMTM5;SLC35F1;NDRG2;MYH6
CR1;PLN;B3GNT7;NDRG2;CCDC141;MYH6;MYH7
C2ORF16;UQCR10
CENPW;APOC1;NCAPG;ALX4;WNT16;TMEM253
WNT16
TBX15;GJA1
IFI27L2;SERPINA1;MTMR3;LXN;PTEN;WNT16;NAT14;NIPSNAP1;B3GNT7;NOV;GNB4;ZNF513;ZNF512;F
IFI27L2;LXN;AFAP1;VSTM2L;SSC5D;NAT14;NIPSNAP1;B3GNT7;NOV;SNX17;GNB4;ZNF513;PVRL2;TRIB2;
IFI27L2;GFM1;CENPW;ZCCHC10;SLC4A1AP;TOMM40;HSPA4;PTEN;NCAPG;SSC5D;CWC27;TBX15;SNX17
NCAPG
DNAJC1;PLN;GPR126;ASCC2;CCDC141;TTN
GCKR;SERPINA1;CR1;PLN;B3GNT7;CMTM5;SLC35F1;NDRG2;CCDC141;CD34
WNT16
WNT16
DLEU1
MTMR3;CR1;PLN;FAM174B;SLC35F1;NDRG2;CCDC141;MYH7;TTN
MYH6;MYH7
APOC1;ZMAT5;UQCR10;WNT16
ZNF628;SHOX2;ASCC2;TMEM253
ALX4
ALX4;MYH6;MYH7
SLC5A6;GCKR;SERPINA1;APOC1;APOE;WNT16;TMEM253
APOC1
APOC1
ALX4
DGKH
ZCCHC10;CEP85L;SUPT7L
SLC5A6;GCKR;CENPW;ZNF513;PPM1G
DNAJC1;CR1;PLN;SHOX2;PVRL2;TTN
SERPINA1;CEP85L;ARHGEF40
IFI27L2;GCKR;LXN;B3GNT7;NOV;GNB4;SSC5D;C2ORF16;ZNF512;APOE;NIPSNAP1
DLEU1;NIPSNAP1

CCDC121;MTMR3;CR1;CMTM5;NDRG2;TTN;DES;PLN;B3GNT7;FAM174B;ZNF513;CCDC141;CD34;MYH6
DES
WNT16
GCKR
MYH7
TBX15;MTMR3;ZCCHC10;SLC4A1AP;GNB4;NCAPG;NRBP1
MYH7
DLEU1;WNT16
CMTM5
SHOX2;ALX4
GCKR;MYH6
APOC1;LCORL
SHOX2;NRBP1
LXN;NCAPG;ASCC2;EIF3E
MYH6
DNAJC1;CR1;ASCC2;TTN
DES;ZNF628;DLEU1;ASCC2
ASCC2;TTN
DNAJC1;SERPINA1;SHOX2;GNB4;TTN
DNAJC1;NCAPG;EIF3E;NRBP1;TTN
HSPA4
CCDC121;SERPINA1;MTMR3;CR1;PLN;B3GNT7;FAM174B;CMTM5;PTEN;SLC35F1;NDRG2;CD34
DNAJC1;LXN;ASCC2;ARHGEF40;TMEM253
HSPA4
C2ORF16
GCKR;CENPW;SERPINA1;MTMR3;APOC1;ZNF513;NRBP1;TMEM253;DGKH
CCDC121;GFM1;ZCCHC10;SLC4A1AP;TOMM40;SHOX2;SLC35F1;UQCR10;CWC27;DCAF16;GJA1;CCDC91
SHOX2;NRBP1
DNAJC1;DES;SERPINA1;CR1;ZMAT5;ASCC2;WARS2
MYH6
TBX15
C2ORF16;UQCR10;SUPT7L
IFI27L2;CCDC121;SERPINA1;LXN;PTEN;WNT16;NIPSNAP1;SUPT7L;B3GNT7;NOV;GNB4;ZNF512;FAM3C;
SHOX2;ASCC2;EIF3E;NRBP1
SERPINA1;C2ORF16
DNAJC1;DES;ZNF628;ASCC2
HSPA4
MYH6
KIAA1755;SERPINA1;CR1;B3GNT7;CMTM5;NDRG2;CCDC141
ZNF628;ZMAT5;ASCC2
TOMM40;HSPA4
IFI27L2;LXN;SKIDA1;PTEN;WNT16;NAT14;NIPSNAP1;SUPT7L;NOV;SNX17;GNB4;ZNF513;ZNF512;PVRL2;
CEP85L;ALX4;ARHGEF40;MYH7
DLEU1;VSTM2L
MYH6;MYH7

MTMR3;CR1;PLN;FAM174B;SLC35F1;NDRG2;CCDC141;MYH6;MYH7;TTN
FAM174B
WNT16
SHOX2
ZNF628;APOC1;NCAPG;UQCR10;APOE;WNT16
ARHGEF40;SUPT7L
CR1;PLN;B3GNT7;FAM174B;SLC35F1;NDRG2;CCDC141;CD34
DLEU1;NIPSNAP1
MTMR3;CR1;PLN;B3GNT7;SLC35F1;NDRG2;CCDC141;CD34
DES;LXN;PLN;ZNF628;SHOX2;NCAPG;ASCC2;GPN1;NRBP1;DGKH;TTN;PPM1G
ZNF628;ASCC2;TMEM253
TBX15;IFI27L2;SLC4A1AP;RSRC1;ZMAT5;CWC27
NCAPG
GCKR
GFM1;ZCCHC10;TOMM40;CWC27;NDRG2;DCAF16;GJA1;CCDC91;RSRC1;MFN1;DLEU1;SPEG;ALX4;FAM
EIF2B4;ZCCHC10;SKIDA1;SHOX2;AFAP1;SSC5D;C2ORF16;NAT14;MLF1;SUPT7L;SLC5A6;CCDC91;ZNF628
WNT16
APOC1;UQCR10;NAT14;TMEM253
MTMR3;ZNF628;NRBP1;TTN;PPM1G
NCAPG
GCKR;C2ORF16;SUPT7L;TTN
VSTM2L
GCKR;UQCR10;WNT16
SHOX2
SERPINA1;CR1;B3GNT7;FAM174B;VSTM2L;NDRG2;CCDC141;CD34
IFI27L2;TOMM40;ZMAT5;VSTM2L;UQCR10;APOE;WNT16;NAT14;NIPSNAP1
IFI27L2;CENPW;LXN;PTEN;NCAPG;C2ORF16;WNT16;NAT14;NIPSNAP1;SUPT7L;TMEM253;DNAJC1;B3G
EIF2B4;GFM1;CENPW;LXN;SLC4A1AP;TOMM40;SHOX2;GPN1;NRBP1;SUPT7L;WARS2;TTN;PPM1G;MLL
SLC5A6;TMEM253
DES;CR1;PLN;B3GNT7;CMTM5;SLC35F1;NDRG2;MYH6
CENPW;MTMR3;PTEN;GPN1;NRBP1;TMEM253;ZNF628;SNX17;APOC1;ZNF513;ASCC2;ARHGEF40;LCOR
TBX15
MYH7
ALX4;NIPSNAP1
PLN;PTEN;TTN
ZMAT5;VSTM2L;UQCR10;WNT16;NAT14;NIPSNAP1
GCKR;SERPINA1;CR1;PLN;B3GNT7;SLC35F1;NDRG2;CCDC141
EIF2B4;CENPW;SLC4A1AP;TOMM40;HSPA4;SHOX2;GPN1;NRBP1;SUPT7L;WARS2;PPM1G;DCAF16;ZNF
LCORL
TBX15;IFI27L2;MTMR3;SLC4A1AP;PLN;HSPA4;RSRC1;CMTM5;GNB4;ZNF512;CWC27;DGKH
CCDC91;CEP85L;ZNF512;SUPT7L
EIF2B4;GFM1;ZCCHC10;SLC4A1AP;TOMM40;HSPA4;SKIDA1;SLC35F1;CWC27;SLC5A6;MLLT10;CCDC91;
SLC5A6;TMEM253
CEP85L;ARHGEF40
DNAJC1;DES;SERPINA1;PLN;SHOX2;ALX4;PVRL2

DLEU1
MYH7
CEP85L
SLC5A6;GCKR;CENPW;SNX17;ZMAT5;PTEN;ZNF513;ASCC2;NRBP1
SERPINA1;CCDC91;C2ORF16
MYH7
ALX4
CEP85L;ARHGEF40;TTN
SERPINA1;CR1;B3GNT7;FAM174B;SLC35F1;NDRG2;CCDC141
SERPINA1;CR1;CMTM5;GPR126;SLC35F1;NDRG2;CCDC141
MYH6
TMEM253
DES;MTMR3;CR1;PLN;B3GNT7;FAM174B;CMTM5;NDRG2;CCDC141;CD34
PLN;CCDC141
UQCR10
DNAJC1;CCDC141;CD34;TTN
SLC4A1AP;GNB4;ALX4;CWC27;GPN1
CMTM5
NCAPG
DLEU1
ZMAT5;CCDC141
ZMAT5;VSTM2L;ZNF513;UQCR10
APOC1;APOE;WNT16
ARHGEF40
ARHGEF40;MYH6;MYH7
DNAJC1;MTMR3;LXN;ZNF628;SHOX2;ASCC2;ARHGEF40;PVRL2;NRBP1;PPM1G
IFI27L2;CCDC121;LXN;SLC4A1AP;PTEN;NAT14;FAM3C;ARHGEF40;CENPW;SKIDA1;AFAP1;SSC5D;C2ORF
SHOX2;DLEU1;GPN1;NRBP1;PPM1G
CR1;SNX17;DLEU1;EIF3E;GPN1;NRBP1;PPM1G
DNAJC1;SHOX2;ASCC2;EIF3E;TTN
IFI27L2;MTMR3;ZCCHC10;SLC4A1AP;TOMM40;HSPA4;PTEN;UQCR10;WNT16;CWC27;GPN1;NRBP1;PPM
EIF2B4;CENPW;MTMR3;TOMM40;SHOX2;PTEN;GPN1;NRBP1;SUPT7L;WARS2;PPM1G;MLLT10;ZNF628;
ALX4
ALX4;C2ORF16
GCKR;CEP85L;C2ORF16;MYH6
ZCCHC10;RSRC1;CMTM5;CWC27
CENPW;LXN;ZNF628;MFN1;NCAPG;CCDC141;NRBP1;TMEM253
CCDC91
IFI27L2;CENPW;SERPINA1;LXN;PTEN;C2ORF16;NAT14;NIPSNAP1;TMEM253;NOV;SNX17;GNB4;ZNF513;
DLEU1
MTMR3;PPM1G
NCAPG
SERPINA1;CR1;PLN;B3GNT7;CMTM5;SLC35F1;NDRG2
ZNF628
SHOX2

SERPINA1;CR1;B3GNT7;CMTM5;NDRG2;CCDC141
ALX4
DES;PLN;B3GNT7;FAM174B;CMTM5;VSTM2L;SLC35F1;WNT16;NDRG2
DNAJC1;DES;SERPINA1;CR1;SHOX2;GPR126;ASCC2;PVRL2
GCKR;SHOX2;ZMAT5
SERPINA1;CMTM5
CENPW;SHOX2
TOMM40
MYH6;MYH7;DGKH;TTN
SHOX2;CEP85L;ALX4;ARHGEF40
CCDC121;GFM1;MTMR3;ZCCHC10;SLC4A1AP;TOMM40;CEP85L;PTEN;CWC27;PPM1G;DCAF16;CCDC91;
DNAJC1;CR1;GNB4;ASCC2;PVRL2;TTN
ALX4
DCAF16;DNAJC1;ZNF628;SNX17;MFN1;ASCC2;CCDC141;PVRL2;WARS2;TTN
IFI27L2;LXN;SKIDA1;PTEN;WNT16;NIPSNAP1;NOV;SNX17;GNB4;ZNF513;ZNF512;PVRL2;TRIB2;DGKH
IFI27L2;CCDC121;LXN;SKIDA1;PTEN;C2ORF16;NAT14;NIPSNAP1;DCAF16;CCDC91;B3GNT7;NOV;SNX17;
SERPINA1;CR1;PLN;B3GNT7;NDRG2;CCDC141
ZMAT5;DLEU1
IFI27L2;CCDC121;LXN;SKIDA1;PTEN;AFAP1;NCAPG;NAT14;NIPSNAP1;SUPT7L;DNAJC1;B3GNT7;SNX17;
APOC1;ZMAT5;DLEU1;VSTM2L;UQCR10;APOE;WNT16
CEP85L
CCDC91
KIAA1755;SERPINA1;CR1;B3GNT7;CMTM5;SLC35F1;WNT16;NDRG2
CENPW;DES;ZNF628;NCAPG;ASCC2;NRBP1
NCAPG
GCKR;ZCCHC10;UQCR10
DES;ZNF628;NRBP1;PPM1G
TMEM253
NCAPG
MYH7
LXN;MFN1;GNB4;PTEN;ASCC2;EIF3E;GPN1;NRBP1
EIF2B4;SHOX2;ASCC2;NRBP1
CR1;GNB4;DGKH
CMTM5;DLEU1;VSTM2L
IFI27L2;CCDC121;SERPINA1;LXN;NAT14;DCAF16;RSRC1;FAM3C;APOE;SKIDA1;AFAP1;SSC5D;C2ORF16;
CEP85L;CCDC141
GCKR;GFM1;CENPW;ZCCHC10;SLC4A1AP;PTEN;WNT16;GPN1;NRBP1;PPM1G;DNAJC1;RSRC1;EIF3E;LCC
SHOX2
SERPINA1
APOC1;VSTM2L;ALX4;UQCR10
NCAPG
CEP85L;ARHGEF40;CCDC141;MYH7
MTMR3;CR1;CMTM5;SLC35F1;NDRG2;PLN;B3GNT7;NOV;FAM174B;ZNF219;ZNF513;CCDC141;CD34
GFM1;NCAPG;LCORL;GPN1
NRBP1

CEP85L;ARHGEF40
IFI27L2;LXN;SKIDA1;PTEN;AFAP1;C2ORF16;NAT14;NIPSNAP1;DCAF16;B3GNT7;NOV;SNX17;GNB4;ZNF5
IFI27L2;SLC4A1AP;SNX17;ZMAT5;GNB4;PTEN;UQCR10;CWC27;NRBP1
CEP85L;GNB4;CCDC141;TTN
TBX15
IFI27L2;GCKR;CENPW;PTEN;AFAP1;VSTM2L;NCAPG;C2ORF16;NAT14;NIPSNAP1;SUPT7L;WARS2;DCAF1
IFI27L2;SERPINA1;LXN;NCAPG;NAT14;NIPSNAP1;TMEM253;GJA1;B3GNT7;NOV;SNX17;GNB4;ZNF513;P
IFI27L2;SLC4A1AP;HSPA4;CMTM5;GNB4;ZNF512
MYH7
UQCR10
CEP85L
ALX4
CCDC121;GFM1;SKIDA1;AFAP1;VSTM2L;SSC5D;CWC27;MLF1;KIAA1755;CCDC91;RSRC1;FAM174B;APO
CCDC121;MTMR3;CR1;PLN;B3GNT7;FAM174B;CMTM5;ZNF219;PTEN;NDRG2;CCDC141;CD34
SLC4A1AP;GNB4;CWC27
TOMM40
HSPA4
IFI27L2;GCKR;CENPW;MTMR3;LXN;SKIDA1;PTEN;VSTM2L;C2ORF16;GPN1;NAT14;NIPSNAP1;MLLT10;D
SLC5A6;CCDC91;CEP85L;C2ORF16;ZNF512;ARHGEF40;SUPT7L;TTN
GCKR;NCAPG;ALX4;UQCR10
CCDC121;GFM1;ZCCHC10;TOMM40;UQCR10;CWC27;NAT14;DCAF16;GJA1;CCDC91;RSRC1;MFN1;DLEU
CCDC121;SLC4A1AP;ZMAT5;TTN
APOC1
DES;LXN;ZNF628;ASCC2;DGKH
EIF2B4;IFI27L2;GCKR;TOMM40;PTEN;NAT14;NIPSNAP1;SUPT7L;SLC5A6;DNAJC1;SNX17;ZNF513;ZNF51
IFI27L2;CCDC121;SERPINA1;LXN;NAT14;PPM1G;ZNF628;RSRC1;ZMAT5;ASCC2;ARHGEF40;EIF2B4;SKID
DNAJC1;SERPINA1;CR1;PLN;GNB4;ASCC2;PVRL2;TTN
IFI27L2;CENPW;MTMR3;SLC4A1AP;CMTM5;GNB4;UQCR10;CWC27;DGKH
IFI27L2;CENPW;HSPA4;SHOX2;GNB4;NCAPG;EIF3E;CWC27;LCORL;GPN1;NIPSNAP1;PPM1G
KIAA1755;CMTM5;SHOX2;CEP85L;ZMAT5;SLC35F1;ARHGEF40;TRIB2
TBX15;PLN;NOV;RSRC1;SHOX2
DES;ZNF219;RSPO2;AFAP1;SPEG;SSC5D
HSPA4;GNB4;AFAP1;NCAPG;SSC5D;C2ORF16;EIF3E;NIPSNAP1
TBX15;SHOX2;ZNF219;WNT16;CD34;WARS2
TBX15;GFM1;DES;UQCR10;MYH6;MYH7;TTN
GJA1;APOC1;SLC35F1;APOE;NDRG2
MLLT10;NOV;RSRC1;SHOX2;SPEG;ALX4;EIF3E;DGKH
IFI27L2;GFM1;CENPW;ZCCHC10;CR1;HSPA4;NCAPG;CWC27;GPN1;NIPSNAP1;NRBP1;PPM1G;SLC5A6;C
CMTM5;CEP85L;PTEN;SLC35F1;CWC27;SUPT7L;MLLT10;KIAA1755;DCAF16;CCDC91;RSRC1;MFN1;GPR1
PLN;UQCR10;CCDC141;MLF1;MYH6;MYH7;TTN
NOV;RSRC1;SHOX2;RSPO2;AFAP1
NOV;RSRC1;GNB4;AFAP1;SSC5D
DCAF16;CCDC121;RSRC1;CMTM5;CEP85L;MFN1;PTEN;GPR126;SLC35F1;ZNF512;LCORL;TRIB2
DCAF16;CENPW;ZCCHC10;MFN1;ZNF512;FAM3C;WNT16;TRIB2;NRBP1
KIAA1755;CMTM5;CEP85L;ZMAT5;GPR126;SLC35F1;NDRG2;ARHGEF40

CCDC141;MLF1;MYH6;MYH7
EIF2B4;IFI27L2;TOMM40;SNX17;ZMAT5;ARHGEF40;TRIB2;SUPT7L
KIAA1755;CMTM5;SHOX2;SLC35F1;ARHGEF40;TRIB2;WARS2
TBX15;DES;ZNF628;SPEG;MYH7;TTN;PPM1G
IFI27L2;CENPW;SKIDA1;SSC5D;NAT14;NIPSNAP1;KIAA1755;DNAJC1;B3GNT7;ZNF628;SNX17;ZNF219;ZN
SLC5A6;CENPW;TOMM40;SNX17;ALX4;SSC5D;DGKH;PPM1G
KIAA1755;SERPINA1;CR1;PLN;B3GNT7;CMTM5;NDRG2
CCDC121;GFM1;ZCCHC10;SKIDA1;SLC35F1;SSC5D;MLF1;MLLT10;DCAF16;GJA1;CCDC91;FAM174B;MFN
ZNF628;SHOX2;MFN1;NCAPG;EIF3E;NRBP1;WARS2
ALX4
TOMM40;MYH6
CENPW;SLC4A1AP;HSPA4;RSRC1;CMTM5;GNB4;CWC27
CENPW;MTMR3;PTEN;GPN1;NRBP1;PPM1G;SLC5A6;DNAJC1;SNX17;RSRC1;ZMAT5;ZNF513;ASCC2;ZNF
MTMR3;CR1;PLN;B3GNT7;FAM174B;SLC35F1;NDRG2;CCDC141;CD34
EIF2B4;IFI27L2;CCDC121;LXN;PTEN;C2ORF16;WNT16;NAT14;NIPSNAP1;TMEM253;SLC5A6;DNAJC1;B3G
NCAPG;UQCR10
CR1;SHOX2;GNB4;DLEU1;NCAPG;LCORL;NRBP1
IFI27L2;SERPINA1;LXN;SSC5D;C2ORF16;NIPSNAP1;B3GNT7;NOV;SNX17;ZNF513;ZNF512;PVRL2;DGKH
EIF2B4;ZNF628;ASCC2;PPM1G
DNAJC1;LXN;DLEU1;ASCC2;EIF3E;TTN
IFI27L2;SERPINA1;SKIDA1;PTEN;AFAP1;NAT14;NIPSNAP1;SUPT7L;TMEM253;DCAF16;B3GNT7;NOV;SNX
DNAJC1;LXN;ZNF628;NCAPG;ALX4;ASCC2
DNAJC1;DES;CR1;PVRL2;TTN
MYH7
CCDC141;MYH6
EIF2B4;CCDC121;GFM1;ZCCHC10;LXN;TOMM40;SSC5D;UQCR10;NAT14;MLF1;SUPT7L;WARS2;PPM1G;S
IFI27L2;CCDC121;LXN;PTEN;SLC35F1;NAT14;DCAF16;RSRC1;FAM3C;AFAP1;SSC5D;C2ORF16;NIPSNAP1;
KIAA1755;SERPINA1;CR1;B3GNT7;CMTM5;SLC35F1;NDRG2;NIPSNAP1
IFI27L2;LXN;PTEN;AFAP1;NCAPG;SSC5D;WNT16;NAT14;NIPSNAP1;SUPT7L;DCAF16;B3GNT7;NOV;GNB4
CMTM5;CEP85L;ARHGEF40
IFI27L2;GCKR;LXN;PTEN;WNT16;NAT14;NIPSNAP1;SUPT7L;TMEM253;SLC5A6;DNAJC1;B3GNT7;SNX17;
DLEU1
CENPW;ASCC2;NRBP1
NCAPG
IFI27L2;CENPW;ZCCHC10;SLC4A1AP;HSPA4;CMTM5;PTEN;UQCR10;CWC27;RSRC1;ZMAT5;GNB4;SPEG
ZMAT5;NIPSNAP1
EIF2B4;IFI27L2;SERPINA1;CR1;LXN;PTEN;WNT16;NAT14;NIPSNAP1;SUPT7L;TMEM253;PPM1G;SLC5A6;
CEP85L
SERPINA1
CEP85L
ZMAT5
FAM174B;APOC1;NCAPG
KIAA1755;CR1;PLN;B3GNT7;CMTM5;NDRG2;MYH6;MYH7
GJA1;GNB4
CMTM5

TTN
MTMR3;CR1;PLN;B3GNT7;FAM174B;CMTM5;NDRG2;CCDC141;TTN
DNAJC1;SNX17;ASCC2;CCDC141;PVRL2;TTN
CENPW;MTMR3;LXN;LCORL;GPN1;NRBP1;PPM1G
DNAJC1;DES;CR1;MFN1;ASCC2;TRIB2;NRBP1
ASCC2;TTN
SHOX2
ALX4;MYH6
DES;SERPINA1;CR1;PLN;B3GNT7;CMTM5;NDRG2;CD34
VSTM2L;UQCR10;LCORL
SERPINA1;PLN;B3GNT7;FAM174B;CMTM5;SLC35F1;NDRG2;CCDC141
DES;PLN;GNB4
SHOX2
NCAPG
CCDC121;MTMR3;ZCCHC10;SLC4A1AP;CEP85L;NCAPG;CWC27;PPM1G;CCDC91;ZNF628;DLEU1;ASCC2;[illegible]
EIF2B4;LXN;SHOX2;NCAPG;ASCC2;DGKH
MYH6;MYH7
IFI27L2;CCDC121;SERPINA1;LXN;PTEN;NAT14;RSRC1;ASCC2;FAM3C;ARHGEF40;GCKR;CENPW;SKIDA1;[illegible]
CEP85L;ALX4;ARHGEF40
DNAJC1;SHOX2;ASCC2;EIF3E;ARHGEF40;TTN
MYH7
ZNF628;NCAPG;NRBP1
APOC1;ZMAT5;NCAPG;WNT16
NCAPG
CR1;B3GNT7;CMTM5;SLC35F1;NDRG2;CCDC141
TBX15;IFI27L2;ZCCHC10;CR1;SLC4A1AP;ZNF512
ALX4;DGKH
ZMAT5
TBX15
ARHGEF40
DNAJC1;ZNF628;ZMAT5;NCAPG;ALX4;ASCC2;WNT16
ZMAT5
C2ORF16
ALX4
APOC1;UQCR10
IFI27L2;CCDC121;LXN;PTEN;CWC27;NAT14;ZMAT5;DLEU1;FAM3C;CENPW;SKIDA1;C2ORF16;GPN1;NIP[illegible]
SERPINA1;MTMR3;CR1;PLN;B3GNT7;FAM174B;CMTM5;SLC35F1;NDRG2;CCDC141;CD34
KIAA1755;SERPINA1;CR1;B3GNT7;FAM174B;CMTM5;SLC35F1;NDRG2;CCDC141
WNT16
CEP85L;ARHGEF40;CCDC141
ASCC2
CMTM5;TTN
GFM1;ZCCHC10;SKIDA1;AFAP1;SSC5D;C2ORF16;CWC27;MLF1;SUPT7L;WARS2;MLLT10;DCAF16;GJA1;[illegible]
IFI27L2;LXN;PTEN;AFAP1;NCAPG;C2ORF16;NAT14;NIPSNAP1;TMEM253;DCAF16;SNX17;GNB4;ZNF513;
SERPINA1;CR1;PLN;SLC35F1;NDRG2;CCDC141;CD34

ALX4;MLF1
CCDC121;SLC4A1AP;SKIDA1;APOC1;ZMAT5;UQCR10;APOE;NIPSNAP1;TMEM253
IFI27L2;SNX17;C2ORF16;ZNF512;NAT14;NIPSNAP1
CR1;APOC1;NCAPG
KIAA1755;SERPINA1;CR1;PLN;B3GNT7;NDRG2
SKIDA1;ZMAT5;DLEU1;NCAPG;UQCR10
DLEU1
HSPA4
ZMAT5
WNT16
TTN
GCKR;CENPW;ZNF628;APOC1;ZNF513;ASCC2
IFI27L2;SERPINA1;CR1;LXN;SKIDA1;PTEN;AFAP1;NIPSNAP1;SUPT7L;DCAF16;B3GNT7;NOV;SNX17;GNB4
IFI27L2;CENPW;LXN;PTEN;C2ORF16;WNT16;NAT14;NIPSNAP1;DNAJC1;NOV;SNX17;GNB4;ZNF513;ZNF
CENPW;LXN;ZNF628;NCAPG;NRBP1;PPM1G
EIF2B4;CENPW;MTMR3;ZNF628;ALX4;ZNF513;NRBP1
SLC5A6;CENPW;PTEN;ALX4;LCORL;PPM1G
IFI27L2;GFM1;MTMR3;ZCCHC10;SLC4A1AP;TOMM40;PTEN;UQCR10;CWC27;PPM1G;RSRC1;ZMAT5;AS
NCAPG
NCAPG
CR1;TOMM40;FAM174B;APOC1;NCAPG;ALX4;UQCR10;APOE;NAT14
ZMAT5;UQCR10;NIPSNAP1
DNAJC1;DES;CR1;SNX17;SHOX2;ALX4;ASCC2;CCDC141;PVRL2
EIF2B4;ZNF628;NRBP1;DGKH
CMTM5
LCORL
VSTM2L;WNT16;MYH6
WNT16;MYH6
KIAA1755
NCAPG
CMTM5
SERPINA1;CR1;B3GNT7;CMTM5;SLC35F1;CCDC141
SLC5A6;CENPW;MTMR3;LXN;ZMAT5;ZNF513;ASCC2;WNT16;NRBP1
SKIDA1;APOC1;WNT16;TMEM253
TBX15
SLC4A1AP;PTEN;LCORL;GPN1;NIPSNAP1
SLC5A6;EIF2B4;DNAJC1;ASCC2;PVRL2;NRBP1;DGKH;PPM1G
CEP85L;ALX4;ARHGEF40;MYH6
DNAJC1;ZNF628;ASCC2;EIF3E
CEP85L;DGKH
TOMM40;WNT16
DLEU1;VSTM2L
CR1;SHOX2;MFN1;ASCC2;NRBP1
IFI27L2;GCKR;CENPW;LXN;PTEN;WNT16;NAT14;NIPSNAP1;SUPT7L;TMEM253;DNAJC1;SNX17;TRIB2
ALX4

DCAF16;MTMR3;NOV;CMTM5;SPEG;ALX4;MLF1;TTN
TTN
SHOX2;WARS2
GPN1
MYH6
LCORL
TOMM40;ZNF628;DLEU1;ASCC2;NRBP1;DGKH;PPM1G
IFI27L2;CCDC121;PTEN;VSTM2L;C2ORF16;NAT14;NIPSNAP1;TMEM253;NOV;RSRC1;ZMAT5;DLEU1;ZNF
SERPINA1
IFI27L2;LXN;SKIDA1;AFAP1;SLC35F1;NAT14;NIPSNAP1;TMEM253;B3GNT7;APOC1;ZNF513;APOE;ARHGI
ZMAT5
DNAJC1;CR1;GNB4;GPR126;PVRL2;CD34;WARS2
DNAJC1;SHOX2;NCAPG;ASCC2;EIF3E;ARHGEF40
CMTM5
CMTM5
NCAPG
MLF1
GPN1;DGKH;PPM1G
C2ORF16
UQCR10
GCKR;TOMM40;APOC1;ZMAT5;UQCR10;NAT14
ASCC2;EIF3E;TMEM253
CENPW
CCDC121;GFM1;TOMM40;SLC35F1;UQCR10;NAT14;GJA1;PLN;CCDC91;RSRC1;FAM174B;MFN1;SPEG;FA
EIF2B4;NCAPG;ASCC2;PPM1G
EIF2B4;IFI27L2;CENPW;MTMR3;LXN;PTEN;WNT16;NRBP1;TMEM253;PPM1G;SLC5A6;DCAF16;SNX17;A
HSPA4;VSTM2L
CR1;PLN;B3GNT7;FAM174B;CMTM5;SLC35F1;NDRG2
GCKR
CEP85L;CCDC141
ASCC2;NRBP1
CR1;B3GNT7;CMTM5;NDRG2;MYH6;MYH7
ALX4
HSPA4;NCAPG;ALX4;WNT16
DNAJC1;ZNF628;ASCC2;CCDC141;TTN
DGKH
SNX17;PTEN;ZNF513;C2ORF16;ZNF512;NIPSNAP1;SUPT7L
DGKH
DLEU1
NCAPG
GFM1;MTMR3;ZCCHC10;SLC4A1AP;TOMM40;SHOX2;CEP85L;PTEN;NCAPG;TTN;PPM1G;ZNF628;ZMAT5
SLC5A6;APOC1;NCAPG;APOE
DNAJC1;CR1;SHOX2;GNB4;GPR126;ALX4;PVRL2;TTN
WNT16
CEP85L;ALX4;ARHGEF40

CEP85L;ARHGEF40;MYH7
IFI27L2;CCDC121;CENPW;MTMR3;LXN;SLC4A1AP;SKIDA1;PTEN;C2ORF16;NAT14;NIPSNAP1;SUPT7L;ML
DNAJC1;MTMR3;CR1;LXN;ASCC2;EIF3E;ARHGEF40;PVRL2
EIF2B4;IFI27L2;GCKR;CENPW;SERPINA1;CR1;LXN;PTEN;C2ORF16;TMEM253;SLC5A6;DCAF16;DNAJC1;S
MLLT10;CCDC121;SKIDA1;MFN1;ALX4
EIF2B4;IFI27L2;GCKR;CR1;PTEN;AFAP1;C2ORF16;WNT16;NAT14;NIPSNAP1;SUPT7L;TMEM253;SLC5A6;
WNT16
MTMR3
MLLT10;CENPW;MTMR3;ZNF628;SNX17;MFN1;ASCC2;NRBP1;SUPT7L;PPM1G
ALX4
C2ORF16;UQCR10
IFI27L2;B3GNT7;SLC35F1;NIPSNAP1;TMEM253
SLC4A1AP;CCDC91;CWC27
NOV
DNAJC1;CR1;ZNF628;SNX17;ASCC2;CCDC141;PVRL2;GPN1;WARS2;TTN;PPM1G
MTMR3;ZMAT5;PTEN;LCORL;TMEM253
CR1;TOMM40;SHOX2;NRBP1;SUPT7L;WARS2;TTN;PPM1G;MLLT10;DCAF16;DES;ZNF628;SNX17;MFN1;
CCDC121;SKIDA1;SHOX2;AFAP1;SSC5D;C2ORF16;CWC27;SUPT7L;SLC5A6;MLLT10;KIAA1755;CCDC91;F
ARHGEF40
NIPSNAP1
DGKH
TBX15
NIPSNAP1
DNAJC1;SHOX2;ASCC2;PPM1G
CCDC121;MTMR3;CR1;PLN;B3GNT7;CMTM5;PTEN;SLC35F1;NDRG2;CCDC141;CD34
CMTM5
WNT16
WNT16
NCAPG
TMEM253
MTMR3;CEP85L;PTEN;LCORL;NRBP1
FAM174B;ZMAT5;DLEU1;UQCR10;NAT14
IFI27L2;MTMR3;LXN;PTEN;NAT14;NIPSNAP1;TMEM253;B3GNT7;NOV;SNX17;ZNF513;APOE;TRIB2;DGK
ALX4
TMEM253
DLEU1
VSTM2L
CCDC121
UQCR10
SHOX2
ASCC2;TTN
IFI27L2;SERPINA1;LXN;PTEN;NAT14;PPM1G;RSRC1;MFN1;ASCC2;FAM3C;ARHGEF40;EIF2B4;GCKR;CEN
CCDC121;CENPW;LXN;SLC4A1AP;PTEN;NCAPG;TMEM253;DCAF16;ZNF628;RSRC1;APOC1;ZMAT5;ZNF5
GCKR;CENPW;MTMR3;LXN;PTEN;NCAPG;WNT16;NRBP1;PPM1G;SLC5A6;DCAF16;RSRC1;ALX4;ZNF513;
TBX15;CEP85L;ALX4;ARHGEF40

GCKR;APOC1;APOE
ALX4;CCDC141
SLC4A1AP;GNB4;ALX4;DGKH
TOMM40
DNAJC1;PLN;SHOX2;ALX4;TTN
WNT16
IFI27L2;GCKR;CCDC121;CENPW;SERPINA1;LXN;PTEN;NAT14;NIPSNAP1;SUPT7L;SLC5A6;DNAJC1;B3GNT
NIPSNAP1;TMEM253
FAM174B;MYH6
IFI27L2;CCDC121;LXN;SLC4A1AP;PTEN;NAT14;DCAF16;FAM3C;APOE;CENPW;SKIDA1;SSC5D;C2ORF16;
NCAPG;WNT16;TMEM253
LXN;B3GNT7;NOV;WNT16;NIPSNAP1
KIAA1755;IFI27L2;LXN;B3GNT7;NOV;GNB4;PTEN;SSC5D;C2ORF16;TRIB2;NIPSNAP1
GCKR;APOC1;C2ORF16;WNT16
ZMAT5;VSTM2L
IFI27L2;GCKR;SERPINA1;LXN;SKIDA1;NAT14;NIPSNAP1;TMEM253;DNAJC1;B3GNT7;NOV;SNX17;GNB4;
IFI27L2;LXN;SKIDA1;PTEN;AFAP1;NIPSNAP1;DCAF16;B3GNT7;NOV;GNB4;ZNF512;FAM3C;APOE;DGKH
ALX4
KIAA1755;SERPINA1;CR1;B3GNT7;CMTM5;VSTM2L;NDRG2;CCDC141
MYH6
SERPINA1
PTEN;TTN
SLC5A6;MTMR3;NOV;SHOX2;CEP85L;MFN1;GNB4;PTEN;NCAPG;TTN;PPM1G
DNAJC1;SHOX2;GNB4;ASCC2;CCDC141;PVRL2
IFI27L2;GCKR;CENPW;SERPINA1;LXN;SKIDA1;PTEN;AFAP1;NAT14;NIPSNAP1;SUPT7L;WARS2;MLLT10;B
GFM1;SLC4A1AP;LCORL;GPN1
GNB4
DLEU1
DNAJC1;LXN;NCAPG;ASCC2;TTN;PPM1G
DLEU1;VSTM2L
CR1;ASCC2
NOV;HSPA4;CMTM5;ALX4
APOC1
LXN;NCAPG;ASCC2;EIF3E
DLEU1
TBX15
ZCCHC10;C2ORF16;ZNF512
IFI27L2;GCKR;CENPW;SERPINA1;CR1;LXN;SKIDA1;PTEN;C2ORF16;NAT14;NIPSNAP1;DNAJC1;B3GNT7;S
SLC5A6;APOC1;UQCR10
SLC5A6;TOMM40;ZNF513;WNT16
LXN;ZNF628;ASCC2
EIF2B4;CCDC121;GFM1;ZCCHC10;SKIDA1;SLC35F1;SSC5D;C2ORF16;UQCR10;MLF1;MLLT10;GJA1;CCDC
NCAPG;ASCC2;ARHGEF40
CEP85L
DNAJC1;ZNF628;SNX17;ASCC2;PVRL2;WARS2

DES;ZNF628;SHOX2;ASCC2
SLC5A6;EIF2B4;GCKR;CENPW;MTMR3;ZNF628;SNX17;ZMAT5;ZNF513;ASCC2;NRBP1;PPM1G
IFI27L2;GCKR;LXN;SKIDA1;PTEN;AFAP1;C2ORF16;NAT14;NIPSNAP1;WARS2;DNAJC1;NOV;SNX17;GNB4;
SERPINA1;PLN;B3GNT7;CMTM5;SLC35F1;NDRG2
GCKR;CENPW;LXN;APOC1;ZNF513;C2ORF16;LCORL;NRBP1;TMEM253
MYH7
ZCCHC10;CMTM5;CEP85L
ZMAT5;DLEU1;WNT16
ASCC2;EIF3E
SERPINA1;CR1;B3GNT7;CMTM5;SLC35F1;NDRG2;CCDC141
ZNF628;DLEU1;ASCC2;TMEM253
DNAJC1;DES;SERPINA1;ZNF628;ASCC2;EIF3E;ARHGEF40
IFI27L2;B3GNT7;NOV;APOE;NAT14;NIPSNAP1
WNT16
MYH7
KIAA1755;GJA1;CMTM5;ZNF219;GPR126;SPEG;NDRG2;ARHGEF40;PVRL2;CD34;TRIB2
SLC4A1AP;TOMM40;CEP85L;UQCR10;NDRG2;GPN1;NIPSNAP1;SUPT7L;TTN;PPM1G;GJA1;CCDC91;SNX[illegible]
DNAJC1
FAM3C
APOE
PTEN
SLC5A6;GCKR;CENPW;MTMR3;ZNF628;SNX17;RSRC1;PTEN;ZNF513;WNT16;LCORL;PPM1G
APOC1;SLC35F1;UQCR10;LCORL
WNT16
ZMAT5
DNAJC1;NCAPG;ALX4;CCDC141;PVRL2
IFI27L2;SERPINA1;VSTM2L
NCAPG;UQCR10;WNT16
GCKR
LCORL
TBX15;MTMR3;SNX17;RSRC1;ZMAT5;GNB4;UQCR10;CWC27;NRBP1;DGKH
GNB4;TRIB2
APOC1;NCAPG
EIF3E
TRIB2
DES;CR1;PLN;CMTM5;NDRG2;CCDC141
DNAJC1;GCKR;SERPINA1;B3GNT7;FAM174B;NCAPG;ZNF513;PVRL2
IFI27L2;CABP7;CMTM5;APOC1;VSTM2L;SLC35F1;APOE;NDRG2;NAT14
PLN;NOV;AFAP1;SPEG;SSC5D
EIF2B4;KIAA1755;SNX17;CMTM5;ZMAT5;ARHGEF40;TRIB2
VSTM2L;CCDC141;MLF1;MYH6
IFI27L2;CENPW;HSPA4;GNB4;AFAP1;NCAPG;EIF3E;FAM3C;NIPSNAP1
ZCCHC10;PLN;NOV;MFN1;GNB4;SPEG
WNT16
ALX4

CCDC121
SERPINA1;CR1;SLC35F1;NDRG2;CCDC141;CD34
IFI27L2;CCDC121;SERPINA1;LXN;PTEN;C2ORF16;NAT14;NIPSNAP1;SUPT7L;TMEM253;MLLT10;DCAF16;
TBX15;CWC27
CENPW;ZCCHC10;TOMM40;B3GNT7;ZNF219;NCAPG;UQCR10;TMEM253;PPM1G
APOC1;ALX4
DNAJC1;MTMR3;NCAPG;ASCC2;ARHGEF40;TMEM253
IFI27L2;CMTM5;ALX4;DGKH
EIF2B4;CENPW;SLC4A1AP;TOMM40;HSPA4;WNT16;GPN1;NRBP1;SUPT7L;WARS2;PPM1G;MLLT10;DCA
SHOX2;TTN
VSTM2L;ALX4
DNAJC1;ZMAT5;GNB4;ASCC2;PVRL2;WARS2
TBX15;CEP85L;ARHGEF40
EIF2B4;GFM1;ZCCHC10;TOMM40;AFAP1;C2ORF16;CWC27;NAT14;WARS2;MLLT10;DCAF16;CCDC91;ZN
GFM1;CCDC91;CEP85L;ARHGEF40
APOC1
SERPINA1
SHOX2
IFI27L2;CCDC121;LXN;PTEN;SLC35F1;NAT14;DCAF16;RSRC1;ZMAT5;MFN1;FAM3C;ARHGEF40;CENPW;V
NCAPG
CR1;PLN;CMTM5;SLC35F1;NDRG2;CCDC141;MYH7
CEP85L
CR1;B3GNT7;CMTM5;SLC35F1;NDRG2;CCDC141
TBX15;SHOX2;NRBP1
ASCC2;NRBP1
KIAA1755;CR1;PLN;B3GNT7;CMTM5;NDRG2;CCDC141;NIPSNAP1
CEP85L;ARHGEF40;MLF1
EIF2B4;IFI27L2;SERPINA1;SKIDA1;NCAPG;NAT14;NIPSNAP1;DNAJC1;B3GNT7;NOV;ZNF628;SNX17;ZMAT
SHOX2;EIF3E;NRBP1
GCKR;ZCCHC10;CEP85L
APOC1;APOE;WNT16
DNAJC1;SHOX2;NCAPG;ASCC2;EIF3E
CCDC141
TOMM40;VSTM2L
CCDC121
IFI27L2;UQCR10;WNT16
TOMM40;VSTM2L;NAT14
EIF2B4;GCKR;CCDC121;GFM1;TOMM40;HSPA4;SKIDA1;SLC35F1;CWC27;MLF1;SUPT7L;CCDC91;RSRC1;Z
ZMAT5
IFI27L2;GCKR;CENPW;MTMR3;ZNF628;SNX17;ZMAT5;AFAP1;ZNF513;ASCC2;ARHGEF40;NRBP1
SLC5A6;GFM1;CENPW;SLC4A1AP;HSPA4;LCORL;GPN1;NRBP1;PPM1G
GPR126
GJA1
MYH7
SLC5A6;GCKR;CENPW;PTEN;ALX4;ZNF513;WNT16;TMEM253

GCKR;CENPW;ZNF513;C2ORF16;WNT16;LCORL
IFI27L2;GCKR;LXN;C2ORF16;NIPSNAP1
UQCR10
ZMAT5
TBX15;CCDC121;CCDC91;RSRC1;CEP85L;ALX4;ARHGEF40;CCDC141;GPN1;TTN
VSTM2L;ALX4;NAT14
ALX4
GCKR
SERPINA1
DLEU1;UQCR10;WNT16
SERPINA1;CR1;PLN;CMTM5;APOE;NDRG2
EIF2B4;IFI27L2;CCDC121;SERPINA1;LXN;PTEN;C2ORF16;WNT16;NIPSNAP1;SUPT7L;DCAF16;DNAJC1;SN
CMTM5
UQCR10
MYH7
CENPW;HSPA4;RSRC1
SLC4A1AP;TOMM40;HSPA4;CEP85L;NCAPG;UQCR10;GPN1;NIPSNAP1;TTN;PPM1G;SNX17;RSPO2;EIF3E
SERPINA1;CR1
DLEU1
ALX4
GPR126
NCAPG
KIAA1755;IFI27L2;DNAJC1;LXN;PTEN;SSC5D;EIF3E;WNT16;SUPT7L
IFI27L2;DNAJC1;ZCCHC10;LXN;TOMM40;APOC1;NCAPG;APOE;SUPT7L
IFI27L2;SERPINA1;LXN;PTEN;C2ORF16;NAT14;NIPSNAP1;TMEM253;DCAF16;B3GNT7;SNX17;GNB4;ZNF
CEP85L
APOC1
C2ORF16
SLC5A6;DNAJC1;GCKR;CENPW;LXN;APOC1;PTEN;ZNF513;PPM1G
SHOX2;NCAPG;ASCC2
CENPW;LCORL
SLC4A1AP;GNB4;DGKH
SERPINA1;LXN
ZCCHC10;CCDC91
GCKR;CENPW
DLEU1
EIF2B4;LXN;NRBP1;SUPT7L;TTN;PPM1G;DCAF16;ZNF628;SNX17;MFN1;ASCC2;EIF3E;ARHGEF40
ZCCHC10;CEP85L;SUPT7L
ZCCHC10;RSRC1;GNB4;CWC27;DGKH
SLC5A6;IFI27L2;APOC1;APOE;WNT16
ZNF628;SHOX2;NRBP1
ASCC2
IFI27L2;LXN;B3GNT7;NOV;SKIDA1;GNB4;PTEN;ZNF513;APOE;TRIB2;NIPSNAP1
IFI27L2;TOMM40;APOC1;UQCR10;NAT14
CEP85L

GCKR;CENPW;ZCCHC10;SLC4A1AP;GPN1;NRBP1;PPM1G;SLC5A6;DNAJC1;RSRC1;ZNF513;EIF3E;LCORL
ZMAT5;NIPSNAP1
CMTM5;ZMAT5
PLN
APOC1
DNAJC1;DES;ZMAT5;PVRL2;TTN
IFI27L2;CENPW;LXN;PTEN;WNT16;NAT14;NIPSNAP1;SUPT7L;TMEM253;B3GNT7;NOV;SNX17;GNB4;ZN
IFI27L2;SERPINA1;LXN;VSTM2L;C2ORF16;NAT14;NIPSNAP1;TMEM253;B3GNT7;NOV;SNX17;ZNF513;AP
SERPINA1;CR1;TOMM40;SLC35F1;CCDC141;CD34
DLEU1
LXN;SHOX2;DLEU1
HSPA4
HSPA4
PTEN
WNT16
ZCCHC10;CMTM5;GNB4;CWC27
CEP85L;ARHGEF40;CCDC141
IFI27L2;CENPW;NOV;SNX17;PTEN;APOE;NIPSNAP1;TMEM253
ZMAT5
VSTM2L
C2ORF16;ZNF512
SPEG;WNT16
VSTM2L
ZNF512
ALX4
ALX4
IFI27L2;DNAJC1;GCKR;NIPSNAP1
TBX15;GFM1;CEP85L;CCDC141;MLF1;MYH7;TTN
DCAF16;DNAJC1;MTMR3;CEP85L;MFN1;GPR126;EIF3E;WNT16;TRIB2
SLC5A6;GCKR;TOMM40;ZNF628;SNX17;ZNF513;C2ORF16;TRIB2;NAT14
TBX15;DES;ZNF628;NDRG2;MYH6;MYH7;TTN
KIAA1755;CMTM5;ZMAT5;GPR126;SLC35F1;ARHGEF40;TRIB2
APOC1;APOE;NDRG2
ZNF219;RSPO2;SPEG;SSC5D;C2ORF16
MTMR3;CR1;B3GNT7;FAM174B;SLC35F1;NDRG2;CCDC141;CD34
IFI27L2;GCKR;CCDC121;CENPW;CR1;LXN;SKIDA1;C2ORF16;WNT16;CWC27;NAT14;NIPSNAP1;SUPT7L;T
SLC5A6;UQCR10
ALX4
ALX4
ZCCHC10
IFI27L2;CCDC121;PTEN;C2ORF16;NAT14;NIPSNAP1;TMEM253;DCAF16;NOV;SNX17;ZNF513;ZNF512;FA
SLC5A6;WNT16;TMEM253
RSRC1
DNAJC1;SERPINA1;CR1;GNB4;GPR126
ALX4

CEP85L
TMEM253
IFI27L2;SLC4A1AP;CMTM5
ALX4
NCAPG;LCORL;TMEM253
DNAJC1;SERPINA1;APOC1;ZMAT5;PVRL2
NCAPG
SERPINA1;CR1;B3GNT7;FAM174B;CMTM5;SLC35F1;NDRG2
CENPW;ZNF628;ASCC2;NRBP1;WARS2;PPM1G
TTN
NCAPG;UQCR10
TOMM40;NCAPG
CR1;DGKH;TTN;PPM1G
TBX15;DES;CCDC141;MYH6;MYH7;TTN
IFI27L2;AFAP1;ALX4;SSC5D;EIF3E;NIPSNAP1;WARS2
RSPO2;GPR126;SSC5D;DGKH
KIAA1755;CCDC121;CMTM5;CEP85L;AFAP1;CD34;TRIB2
GFM1;CENPW;ZCCHC10;TOMM40;HSPA4;NCAPG;C2ORF16;CWC27;GPN1;NIPSNAP1;WARS2;PPM1G;Sl
IFI27L2;SHOX2;GNB4;AFAP1;ALX4;EIF3E;FAM3C
KIAA1755;DCAF16;MTMR3;CMTM5;SKIDA1;CEP85L;ZMAT5;GPR126;SLC35F1;ARHGEF40;TRIB2;WARS2
DCAF16;GFM1;CCDC91;CEP85L;CCDC141;MLF1;MYH7;TTN
TBX15;GFM1;DES;MYH6;MYH7;TTN
KIAA1755;CMTM5;CEP85L;ZMAT5;MFN1;GPR126;SLC35F1;ZNF512;NDRG2;ARHGEF40
KIAA1755;DCAF16;CMTM5;SKIDA1;SHOX2;CEP85L;GPR126;ALX4;ZNF512;ARHGEF40;TRIB2;SUPT7L
GCKR;GFM1;SERPINA1;APOC1;GPR126;APOE;NIPSNAP1
GFM1;UQCR10;CCDC141;MYH6;MYH7;TTN
CENPW;ZCCHC10;TOMM40;HSPA4;NCAPG;CWC27;GPN1;NIPSNAP1;WARS2;PPM1G;SLC5A6;MLLT10;D
IFI27L2;CENPW;TOMM40;HSPA4;NCAPG;CWC27;GPN1;NIPSNAP1;PPM1G;SNX17;GNB4;ALX4;FAM3C
CR1;CR1L;ASCC2
DNAJC1;GCKR;B3GNT7;FAM174B;FAM3C;PVRL2
EIF2B4;TBX15;GFM1;ZNF628;LCORL;MYH7;TTN;PPM1G
DES;CCDC91;ZNF219;RSPO2
EIF2B4;IFI27L2;CMTM5;SHOX2;CEP85L;GPR126;SLC35F1;ZNF512;NDRG2;TRIB2
SLC5A6;GCKR;SERPINA1;APOC1;GPR126;APOE;NIPSNAP1
IFI27L2;GCKR;CCDC121;SKIDA1;SSC5D;NRBP1;KIAA1755;DCAF16;B3GNT7;SNX17;EIF3E;ZNF512;APOE;A
KIAA1755;IFI27L2;CCDC121;GJA1;CMTM5;APOC1;ALX4;APOE;WNT16;NDRG2;NAT14;MLF1
TBX15;GFM1;ZCCHC10;PLN;CCDC91;DLEU1;NDRG2;MYH6;MYH7;TTN
DES;CEP85L;CCDC141;MYH6;MYH7;TTN
SERPINA1;MTMR3;CR1;CR1L;ZMAT5;PVRL2
TBX15;DES;ZNF628;NRBP1;MYH6;MYH7;TTN
SLC5A6;DCAF16;ALX4;SSC5D;ZNF512;CD34
TBX15;PTEN;ALX4;SSC5D;FAM3C;CD34;NRBP1
IFI27L2;GFM1;CENPW;ZCCHC10;CR1;TOMM40;HSPA4;PTEN;NCAPG;CWC27;GPN1;NIPSNAP1;PPM1G;S
KIAA1755;DCAF16;CMTM5;CEP85L;ZMAT5;SLC35F1;ZNF512;NDRG2;ARHGEF40;TRIB2
EIF2B4;TBX15;DES;CEP85L;GPN1;MYH7;TTN

SLC5A6;DNAJC1;TOMM40;CEP85L;AFAP1;ZNF513;ARHGEF40;PVRL2;TRIB2
TBX15;DES;CCDC141;MLF1;MYH7;TTN
SLC5A6;TBX15;GFM1;ZCCHC10;SLC4A1AP;HSPA4;CEP85L;UQCR10;LCORL;MYH6;TTN
GCKR;SERPINA1;B3GNT7;FAM174B;PVRL2
CCDC121;CMTM5;SKIDA1;SHOX2;CEP85L;SUPT7L;KIAA1755;DCAF16;MFN1;GPR126;ZNF512;ARHGEF4(
NCAPG;NIPSNAP1;TMEM253
DNAJC1
APOC1
SLC5A6;APOC1
DNAJC1;ASCC2;PVRL2
UQCR10
TBX15;GFM1;ZCCHC10;CCDC91;NDRG2;CCDC141;GPN1;MYH6;MYH7;TTN
KIAA1755;DES;ZNF219;RSPO2;AFAP1;SSC5D
SLC5A6;GCKR;TOMM40;PTEN;CD34;WARS2
IFI27L2;SERPINA1;B3GNT7;ZNF513;PVRL2;NAT14;NIPSNAP1
EIF3E;TTN
PTEN;LCORL;PPM1G
ZNF628;SKIDA1;APOC1;ZMAT5;ALX4;UQCR10;NAT14;NIPSNAP1
DLEU1
CR1;PLN;B3GNT7;FAM174B;CMTM5;MYH7
ZMAT5;PTEN
IFI27L2;B3GNT7;SNX17;DLEU1;ZNF513;APOE;NAT14;NIPSNAP1;SUPT7L;TMEM253
IFI27L2;NOV;SNX17;GNB4;AFAP1;ZNF513;WNT16;TRIB2;NAT14;NIPSNAP1
IFI27L2;CENPW;SERPINA1;LXN;CMTM5;PTEN;NAT14;NIPSNAP1;B3GNT7;ZMAT5;GNB4;ZNF512;APOE
DES;ZNF219;ASCC2
C2ORF16
DCAF16;SHOX2
ZMAT5;NCAPG;CCDC141
CCDC121;ZMAT5;C2ORF16;NIPSNAP1
IFI27L2;CCDC121;LXN;PTEN;AFAP1;VSTM2L;C2ORF16;NAT14;NIPSNAP1;DCAF16;CCDC91;NOV;SNX17;G
DNAJC1;SERPINA1;CR1;GPR126
GFM1;CCDC91;CEP85L
SLC5A6;GCKR;CENPW;MTMR3;PTEN;ZNF513
IFI27L2;LXN;SKIDA1;AFAP1;SSC5D;C2ORF16;NIPSNAP1;SUPT7L;TMEM253;DCAF16;B3GNT7;NOV;SNX17
NRBP1
CENPW;ASCC2;EIF3E
TBX15
GCKR
DNAJC1;SNX17;SHOX2;ASCC2;PVRL2;GPN1
UQCR10;TMEM253
ZNF628;NRBP1;PPM1G
TMEM253
TBX15
DNAJC1;ASCC2;EIF3E
NCAPG

NCAPG
GCKR;CENPW;ZNF628;APOC1;ZNF513
VSTM2L
ALX4
SHOX2
PLN;UQCR10;CCDC141;MLF1;MYH6;MYH7;TTN
IFI27L2;LXN;SNX17;SKIDA1;PTEN;ZNF513;C2ORF16;ZNF512;NAT14;NIPSNAP1;SUPT7L
APOC1;APOE;NDRG2
DNAJC1;GCKR;B3GNT7;FAM174B;PVRL2
IFI27L2;CMTM5;APOC1;APOE
SHOX2;ARHGEF40;PVRL2;CD34;TRIB2
KIAA1755;DCAF16;CMTM5;CEP85L;AFAP1;GPR126;ARHGEF40;PVRL2;TRIB2;MLF1
SERPINA1;MTMR3;CR1;CR1L;PTEN;ASCC2
CR1;CR1L;ASCC2
TBX15;GFM1;CEP85L;NDRG2;MLF1;MYH7;TTN
KIAA1755;CMTM5;SKIDA1;SHOX2;CEP85L;SLC35F1;ARHGEF40;CD34;TRIB2
ALX4
EIF2B4;ZNF628;ZMAT5;ZNF513;PPM1G
APOC1;APOE
IFI27L2;CCDC121;LXN;SKIDA1;PTEN;NAT14;NIPSNAP1;TMEM253;DCAF16;B3GNT7;SNX17;RSRC1;APOC
SHOX2
MLLT10
CEP85L;ARHGEF40;TTN
DNAJC1;ASCC2;C2ORF16
GCKR;SERPINA1;B3GNT7;FAM174B;FAM3C;PVRL2
IFI27L2;HSPA4;GNB4;NCAPG;EIF3E;CWC27;LCORL;GPN1;NIPSNAP1;WARS2
ASCC2;NRBP1
ARHGEF40
IFI27L2;DNAJC1;GCKR;SERPINA1;LXN;SNX17;GNB4;APOE;ARHGEF40;PVRL2;NIPSNAP1
IFI27L2;LXN;CMTM5;WNT16;NAT14;NIPSNAP1;B3GNT7;NOV;SNX17;RSRC1;ZMAT5;GNB4;ZNF513;TRIB
CENPW;ZNF628;APOC1;ZNF513
DNAJC1;CR1;APOC1;GNB4;ASCC2;CCDC141;PVRL2;GPN1
ARHGEF40
ZMAT5
IFI27L2;LXN;B3GNT7;SNX17;SKIDA1;GNB4;VSTM2L;ZNF513;C2ORF16;ZNF512;FAM3C;NIPSNAP1
ZNF628;SHOX2;NCAPG
TMEM253
IFI27L2;LXN;SKIDA1;C2ORF16;WNT16;NAT14;NIPSNAP1;SUPT7L;B3GNT7;NOV;SNX17;GNB4;ZNF513;ZN
EIF2B4;MTMR3;SLC4A1AP;TOMM40;HSPA4;PTEN;GPN1;NRBP1;SUPT7L;TMEM253;WARS2;PPM1G;SLC
DNAJC1;LXN;GPR126;ASCC2;EIF3E
CR1;PLN;FAM174B;NDRG2;CCDC141
UQCR10
DLEU1
SLC5A6;UQCR10
ASCC2;EIF3E

C2ORF16
SERPINA1;CEP85L;AFAP1;ARHGEF40
CEP85L
TBX15
IFI27L2;LXN;B3GNT7;SNX17;ZMAT5;ZNF513;ARHGEF40;NAT14;NIPSNAP1;TMEM253
UQCR10
DNAJC1
GFM1;SERPINA1;LXN;CEP85L;AFAP1;NDRG2;NIPSNAP1;PPM1G;DNAJC1;CCDC91;FAM174B;MFN1;GNB
MLLT10;GJA1;ZCCHC10;PLN;NOV;SPEG
MTMR3;CMTM5;SHOX2;GNB4
CWC27
MLLT10;ZNF219;RSPO2;AFAP1;SPEG;SSC5D;EIF3E
TBX15;GFM1;DES;FAM174B;CCDC141;MYH6;MYH7;TTN
MLLT10;PLN;NOV;EIF3E;ZNF512
GJA1;GNB4;PTEN
SERPINA1;CMTM5
MLLT10;DES;ZNF219;AFAP1;SSC5D
IFI27L2;DNAJC1;NOV;WNT16;NIPSNAP1
SERPINA1;CR1;B3GNT7;CMTM5;NDRG2
EIF2B4;CCDC121;GFM1;ZCCHC10;SKIDA1;CWC27;MLF1;TMEM253;WARS2;MLLT10;FAM174B;ZNF219;
DNAJC1;SERPINA1;ASCC2
IFI27L2;GCKR;PTEN;AFAP1;VSTM2L;WNT16;NAT14;NIPSNAP1;TMEM253;DNAJC1;B3GNT7;SNX17;GNB
APOC1
TBX15;GFM1;CCDC141;MYH6;MYH7;TTN
DNAJC1;GCKR;SERPINA1;B3GNT7;FAM174B;C2ORF16;PVRL2
SLC5A6;EIF2B4;LXN;TOMM40;SNX17;CMTM5;ARHGEF40;PVRL2;TRIB2;SUPT7L
MLLT10;DES;RSPO2;SPEG;SSC5D
SLC5A6;GCKR;SERPINA1;APOC1;GPR126;APOE;NIPSNAP1
GCKR
TBX15;DES;MTMR3;SLC4A1AP;GNB4
APOC1
IFI27L2;CENPW;NCAPG;NAT14;NIPSNAP1;DNAJC1;B3GNT7;SNX17;RSRC1;ZMAT5;ZNF513;APOE;TRIB2
IFI27L2;CENPW;LXN;NOV;SKIDA1;ZNF513;WNT16;NAT14;NIPSNAP1
ASCC2
DLEU1
GPR126;SPEG
GCKR
ALX4
MTMR3;ZNF628;NCAPG;NRBP1;DGKH
CEP85L;ZNF512;SUPT7L;DGKH
GCKR
NIPSNAP1
MYH6
TBX15
LCORL

IFI27L2;LXN;C2ORF16;WNT16;NAT14;NIPSNAP1;TMEM253;KIAA1755;B3GNT7;NOV;SNX17;ZMAT5;ZNF
CENPW;RSRC1;PTEN;GPN1
DNAJC1;SHOX2;ASCC2;PVRL2
LCORL
TTN
IFI27L2;SERPINA1;LXN;PTEN;AFAP1;C2ORF16;NAT14;NIPSNAP1;TMEM253;DCAF16;DNAJC1;B3GNT7;SN
IFI27L2;CENPW;LXN;PTEN;VSTM2L;C2ORF16;NAT14;NIPSNAP1;TMEM253;DCAF16;DNAJC1;SNX17;ZNF
DCAF16;MTMR3;ZNF513;WNT16
IFI27L2;B3GNT7;NOV;SNX17;AFAP1;ZNF513;PVRL2;TRIB2;NAT14;NIPSNAP1;TMEM253
IFI27L2;MTMR3;SKIDA1;C2ORF16;NAT14;NIPSNAP1;DNAJC1;B3GNT7;NOV;SNX17;GNB4;ZNF513;ZNF51
WNT16;NIPSNAP1
UQCR10
LCORL;TMEM253
IFI27L2;GCKR;SKIDA1;C2ORF16;NAT14;NIPSNAP1;B3GNT7;NOV;SNX17;APOC1;GNB4;ZNF513;ZNF512;A
CEP85L
UQCR10
MTMR3;LCORL;PPM1G
APOC1
EIF2B4;CCDC121;GFM1;HSPA4;SKIDA1;VSTM2L;SSC5D;C2ORF16;CWC27;MLF1;SLC5A6;MLLT10;CCDC9
VSTM2L;UQCR10;LCORL
NIPSNAP1
NCAPG;UQCR10;TMEM253
ZNF628;DGKH;TTN
IFI27L2;SERPINA1;MTMR3;LXN;SKIDA1;SSC5D;C2ORF16;NAT14;NIPSNAP1;SUPT7L;TMEM253;WARS2;D
SLC5A6;DNAJC1;CENPW;MTMR3;ZNF513;NRBP1
CENPW;APOC1;ZNF513
IFI27L2;CCDC121;PTEN;VSTM2L;SSC5D;C2ORF16;WNT16;NAT14;NIPSNAP1;SUPT7L;TMEM253;DCAF16
ZMAT5
CR1;B3GNT7;CMTM5;SLC35F1;NDRG2
MLLT10;DCAF16;GFM1;CENPW;PTEN;LCORL
DGKH
APOC1;WNT16
IFI27L2;CENPW;SNX17;ZMAT5;ZNF513;ZNF512;WNT16;ARHGEF40;PVRL2;TRIB2;NIPSNAP1;SUPT7L
CENPW;MTMR3;APOC1;ZNF513;APOE
UQCR10
CR1;B3GNT7;CMTM5;NDRG2;CCDC141
TOMM40;APOC1;NAT14
ZMAT5
IFI27L2;GCKR;B3GNT7;C2ORF16;ZNF512
IFI27L2;NOV;PTEN;ZNF512;WNT16;TRIB2;NAT14;NIPSNAP1;WARS2
WNT16;NRBP1
TBX15;SLC4A1AP;RSRC1;GNB4;CWC27
EIF2B4;IFI27L2;CENPW;PTEN;AFAP1;C2ORF16;NAT14;NIPSNAP1;TMEM253;SLC5A6;DNAJC1;ZNF628;SN
MLLT10;CENPW;MTMR3;SNX17;RSRC1;ZMAT5;PTEN;NCAPG;ZNF512;LCORL;NRBP1;TMEM253
EIF2B4;IFI27L2;CCDC121;CENPW;PTEN;WNT16;NAT14;NIPSNAP1;SUPT7L;PPM1G;DCAF16;DNAJC1;SNX

CENPW
SERPINA1;CR1;PLN;B3GNT7;CMTM5;NDRG2
ASCC2;TTN
SERPINA1;ZCCHC10;CR1;CCDC91;CEP85L
GCKR
MTMR3;CCDC141;CD34
CEP85L;ZNF512
IFI27L2;GCKR;CENPW;SERPINA1;CR1;LXN;PTEN;C2ORF16;NIPSNAP1;SUPT7L;SLC5A6;MLLT10;DCAF16;D
KIAA1755;SERPINA1;CR1;B3GNT7;FAM174B;SLC35F1;NDRG2
IFI27L2;LXN;PTEN;VSTM2L;SSC5D;NAT14;NIPSNAP1;TMEM253;KIAA1755;NOV;SNX17;APOC1;ZMAT5;Z
DNAJC1;CENPW;SNX17;RSRC1;PTEN;ASCC2;GPN1;PPM1G
FAM174B;APOC1;UQCR10
NIPSNAP1
CENPW;SLC4A1AP;PTEN;WNT16;NRBP1;PPM1G;DCAF16;SNX17;RSRC1;ZMAT5;ZNF513;ARHGEF40;LCO
WARS2;TTN
MLLT10;NCAPG;ASCC2;NRBP1
SERPINA1;CR1;B3GNT7;SLC35F1;NDRG2;CCDC141
IFI27L2;GCKR;CENPW;SERPINA1;LXN;PTEN;WNT16;NAT14;NIPSNAP1;SUPT7L;SLC5A6;DNAJC1;B3GNT7;
IFI27L2;GCKR;LXN;C2ORF16;WNT16;NAT14;NIPSNAP1;TMEM253;B3GNT7;SNX17;RSRC1;GNB4;ZNF512
WNT16
NCAPG
MYH6
IFI27L2;NOV;SNX17;ZNF513;ARHGEF40;NAT14;NIPSNAP1
CENPW;MTMR3;APOC1;NCAPG;ZNF513;WNT16
DGKH
IFI27L2;GCKR;LXN;WNT16;NIPSNAP1;TMEM253;B3GNT7;NOV;SNX17;ZMAT5;ZNF513;ZNF512;PVRL2;TI
SLC5A6;MTMR3;ZNF628;APOC1;ZNF513;ASCC2;WNT16;ARHGEF40;NRBP1;SUPT7L;PPM1G
MLLT10;CCDC121;GFM1;GJA1;CCDC91;NOV;ZNF219;MFN1;SPEG;ALX4;SSC5D;MLF1
NCAPG;ASCC2
SHOX2
MTMR3;ZNF513;NRBP1
NIPSNAP1
DNAJC1;SHOX2;GPR126;PVRL2
APOC1;ZMAT5;ALX4
ZMAT5;DLEU1
IFI27L2;LXN;C2ORF16;NAT14;NIPSNAP1;DCAF16;B3GNT7;NOV;GNB4;ZNF513;ZNF512;APOE;TRIB2
SLC5A6;CENPW;ZNF628;PTEN;ZNF513;NRBP1;PPM1G
WNT16
TBX15;MTMR3;SLC4A1AP;GNB4;ZNF512;CWC27
CENPW;SLC4A1AP;RSRC1;EIF3E;LCORL;GPN1
RSRC1;C2ORF16;ZNF512;NIPSNAP1;DGKH
GCKR;TOMM40;UQCR10
DNAJC1;SHOX2;ASCC2
WNT16
NRBP1

CR1;GNB4;ASCC2;PVRL2
SERPINA1;CR1;B3GNT7;CMTM5;VSTM2L;SPEG
WNT16
SLC4A1AP;CCDC91;TRIB2
IFI27L2
IFI27L2;CENPW;B3GNT7;SNX17;ZNF513;APOE;PVRL2;TRIB2;NAT14;NIPSNAP1
DNAJC1;CR1;SHOX2;PVRL2;TTN
DNAJC1;CENPW;MTMR3;LXN;APOC1;PTEN;NCAPG;ZNF513;LCORL;NRBP1
ASCC2
ZNF628;TMEM253;PPM1G
GCKR
APOC1
TBX15
IFI27L2;CCDC121;CENPW;LXN;SKIDA1;NAT14;NIPSNAP1;TMEM253;B3GNT7;NOV;SNX17;ZNF513;ZNF5[illegible]
WNT16
GCKR;CENPW;MTMR3;APOC1;PTEN
CEP85L
GCKR;CENPW;AFAP1;NCAPG;ALX4;ZNF513;WNT16;NRBP1;TMEM253;DGKH
CMTM5;CEP85L;ARHGEF40
SERPINA1
APOC1;APOE;TMEM253
UQCR10;TMEM253
NCAPG
IFI27L2;B3GNT7;NOV;SNX17;PTEN;ZNF513;C2ORF16;ZNF512;FAM3C;NAT14;NIPSNAP1;TMEM253
DNAJC1;DES;PLN;SHOX2
SERPINA1;CR1;B3GNT7;CMTM5;NDRG2;CCDC141;CD34
ZMAT5;NIPSNAP1
ASCC2
IFI27L2;B3GNT7;SNX17;SKIDA1;ZNF513;C2ORF16;ZNF512;PVRL2;NIPSNAP1;DGKH
IFI27L2;GCKR;CCDC121;SERPINA1;PTEN;C2ORF16;WNT16;NAT14;NIPSNAP1;DNAJC1;CCDC91;B3GNT7;[illegible]
VSTM2L
MTMR3;APOC1;ZMAT5;PTEN;LCORL;NIPSNAP1;TMEM253
ZNF628
DNAJC1;DES;PLN;SHOX2;GPR126
DNAJC1;CR1;ASCC2;PVRL2;TTN
WNT16
GCKR;CENPW;MTMR3;APOC1
VSTM2L
ZCCHC10
DNAJC1;GNB4;PVRL2;TTN
APOC1;NCAPG;UQCR10;APOE
CENPW;CEP85L
GCKR;CENPW;MTMR3;LCORL
ZMAT5
ZNF628;ASCC2

IFI27L2;DNAJC1;GCKR;GNB4;PTEN;FAM3C;NIPSNAP1
CCDC121;CENPW;MTMR3;SLC4A1AP;SNX17;RSRC1;ZMAT5;PTEN;NCAPG;ZNF512;LCORL
MTMR3
SLC5A6;NCAPG
DNAJC1;PVRL2;TTN
LXN;NRBP1
SLC4A1AP;RSRC1;GPN1
CMTM5
CENPW
TOMM40;UQCR10;TMEM253
ZNF628;ASCC2;NRBP1;TMEM253
APOC1;WNT16
SLC4A1AP;EIF3E;GPN1
TBX15
TTN
GCKR
ZMAT5
APOC1
RSRC1;ZMAT5
CENPW;ZNF513;GPN1
C2ORF16
ALX4
EIF2B4;IFI27L2;GCKR;CENPW;SERPINA1;SKIDA1;PTEN;NAT14;NIPSNAP1;TMEM253;DNAJC1;B3GNT7;SN
DNAJC1;TTN
GCKR;CENPW;ALX4
SLC5A6;DCAF16;GFM1;MTMR3;SLC4A1AP;HSPA4;LCORL;GPN1;NRBP1;PPM1G
CENPW;ZNF513;TMEM253
IFI27L2;APOE;NIPSNAP1;TMEM253
IFI27L2;CENPW;B3GNT7;NOV;SKIDA1;APOE;NIPSNAP1;TMEM253
IFI27L2;SLC4A1AP
TTN
CENPW;SERPINA1;MTMR3;PTEN;ZNF513;ASCC2;WNT16;TMEM253
CENPW;ZNF513;WNT16
IFI27L2;SERPINA1;PTEN;VSTM2L;NIPSNAP1;SUPT7L;MLLT10;DNAJC1;SNX17;RSRC1;ZNF513;ZNF512;FA
ASCC2
CCDC121;APOC1
IFI27L2;LXN;AFAP1;C2ORF16;NAT14;NIPSNAP1;B3GNT7;NOV;SNX17;GNB4;ZNF513;ZNF512;PVRL2;DGK
IFI27L2;DNAJC1;SKIDA1;ZMAT5;WNT16;TRIB2;NAT14;NIPSNAP1;SUPT7L;TMEM253
CENPW;ZNF513;WNT16
SLC5A6;EIF2B4;CENPW;MTMR3;SNX17;APOC1;ZNF513;ASCC2;LCORL;NRBP1;PPM1G
SERPINA1
ARHGEF40
APOC1
IFI27L2;B3GNT7;NAT14;NIPSNAP1
IFI27L2;GCKR;CENPW;VSTM2L;NAT14;NIPSNAP1;TMEM253;B3GNT7;SNX17;ZNF513;ZNF512;FAM3C;TF

CENPW;CWC27
GCKR;MTMR3;ZNF628;APOC1;ZMAT5;ZNF513;TMEM253
GCKR;CENPW;CEP85L;ZNF513;WNT16
GCKR;SNX17;GPN1;NIPSNAP1;TMEM253;PPM1G
VSTM2L;UQCR10
APOC1;NCAPG;NAT14
SERPINA1;CR1;PTEN
LCORL
ASCC2
CENPW;ZNF513;PPM1G
EIF2B4;IFI27L2;GCKR;CENPW;LXN;PTEN;VSTM2L;C2ORF16;NAT14;NIPSNAP1;DCAF16;DNAJC1;SNX17;R[illegible]
CMTM5
NCAPG
CENPW;LCORL
APOC1;VSTM2L
VSTM2L
SLC5A6;GCKR;MTMR3;SNX17;RSRC1;CEP85L;ZNF513;GPN1;NRBP1;PPM1G
EIF2B4;TOMM40;SKIDA1;ZNF219;SLC35F1
CCDC121;MTMR3
IFI27L2;CCDC121;SERPINA1;PTEN;C2ORF16;NIPSNAP1;SUPT7L;SLC5A6;DNAJC1;SNX17;RSRC1;ZNF513;[illegible]
SERPINA1;CR1;LXN;TOMM40;B3GNT7;APOC1;VSTM2L
NIPSNAP1
LCORL
ZNF628;SHOX2;ASCC2;NRBP1
ARHGEF40
WNT16
VSTM2L
CENPW;ZNF513;WNT16;LCORL
IFI27L2;LXN;NAT14;NIPSNAP1;TMEM253;CCDC91;B3GNT7;SNX17;ZMAT5;GNB4;ZNF513;ZNF512;FAM3
CENPW;AFAP1;ZNF513;NRBP1;TMEM253
MTMR3;ZNF628;ZMAT5;ZNF513;WNT16
SERPINA1
SLC5A6;GCKR;CENPW;MTMR3;ZNF513;NRBP1
LCORL
DCAF16;CENPW;ZNF513;NRBP1
ZMAT5
SLC5A6;GCKR;CENPW;SNX17;APOC1;ZMAT5;ZNF513;WNT16;NIPSNAP1;TMEM253
CEP85L;ALX4;WNT16
CR1;APOC1
IFI27L2;LXN;B3GNT7;NOV;NIPSNAP1
IFI27L2;B3GNT7;SNX17;AFAP1;ZNF513;FAM3C;PVRL2;NAT14;NIPSNAP1
CR1
CENPW;TMEM253
SLC5A6;CENPW;PTEN;AFAP1;ZNF513
RSRC1

ASCC2
CR1
ASCC2
DCAF16;CENPW;RSRC1;EIF3E;LCORL
TMEM253
APOE
CENPW;LCORL
IFI27L2;NAT14;NIPSNAP1
ALX4
CENPW
GJA1;NOV;AFAP1
ZNF628;ZNF219;ALX4;WNT16
KIAA1755;CCDC121;CCDC91;CMTM5
MLLT10;CCDC121;ALX4;CD34
KIAA1755;CCDC121;CMTM5;NDRG2
DNAJC1;SNX17;FAM174B;SKIDA1;FAM3C;TMEM253
ZCCHC10;SLC4A1AP;TOMM40;HSPA4;ZNF628;SHOX2;CWC27;GPN1;WARS2;PPM1G
FAM174B
KIAA1755;IFI27L2;CCDC121;CMTM5;RSPO2;NDRG2;MLF1
SHOX2;CEP85L;TRIB2
SPEG
FAM174B
SERPINA1;B3GNT7;ZNF513;LCORL;SUPT7L;TMEM253
SLC4A1AP;CCDC91;RSRC1;RSPO2;VSTM2L;SLC35F1;FAM3C;NAT14;NIPSNAP1
GFM1;ZCCHC10;TOMM40;HSPA4;ZNF628;GNB4;AFAP1;CWC27;GPN1;WARS2;PPM1G
CENPW;TOMM40;HSPA4;SNX17;GNB4;NCAPG;CWC27;GPN1;PPM1G
EIF2B4;DNAJC1
NOV;SPEG
DNAJC1;FAM174B
EIF2B4;GJA1;ZNF628;WNT16
ZCCHC10;CR1;SNX17;PTEN;VSTM2L;ZNF512;TRIB2;SUPT7L
ZCCHC10;LXN;TOMM40;B3GNT7;NIPSNAP1;TMEM253
TOMM40;PVRL2;DGKH
FAM174B
CR1;PTEN
SLC4A1AP;CCDC91;CABP7;RSRC1;CMTM5;VSTM2L;SLC35F1;FAM3C
CENPW;DLEU1;NCAPG
DCAF16;MFN1;ALX4;SSC5D;EIF3E;ZNF512;FAM3C
DCAF16;ZCCHC10;TOMM40;HSPA4;ZNF628;ZNF219;ZMAT5;GNB4;CWC27;GPN1;PPM1G
ZCCHC10;CR1;ZNF628;SNX17;APOC1;ZMAT5;ZNF513;TRIB2
DNAJC1;FAM174B
SLC5A6;CD34
CABP7;CMTM5;VSTM2L;SLC35F1;APOE;NAT14
B3GNT7;GPR126;CCDC141
SERPINA1;MTMR3;CR1;CR1L;PTEN;ASCC2

SLC5A6;CENPW;LXN;RSRC1;ALX4;WARS2
SLC5A6;ZNF219;PTEN
NOV;SPEG
CENPW;TOMM40;ASCC2
FAM174B
FAM174B
CR1L;ZNF628;MFN1;ZNF513;FAM3C;LCORL;CD34;TRIB2;NAT14;SUPT7L
DNAJC1;CCDC121;SNX17;FAM174B;SKIDA1;ZNF513;C2ORF16;ZNF512;SUPT7L
PTEN;CD34
ZCCHC10;NOV
ZNF219;TMEM253
KIAA1755;IFI27L2;CCDC91;CMTM5
DNAJC1;LXN;CD34
GJA1;GNB4;GPR126;WNT16
RSPO2
EIF2B4;DLEU1;ALX4;WNT16
CABP7;FAM174B
SLC5A6;PTEN;CD34
SLC4A1AP;CCDC91;RSRC1;RSPO2;VSTM2L;SLC35F1;FAM3C;NAT14;NIPSNAP1
SLC5A6;PTEN
SLC4A1AP;CCDC91;HSPA4;RSRC1;SKIDA1;RSPO2;VSTM2L;SLC35F1;FAM3C;NAT14
GJA1;GPR126;WNT16
EIF2B4;ZNF219
SLC4A1AP;RSPO2;VSTM2L;ALX4;SLC35F1;FAM3C;NAT14;NIPSNAP1
CR1;LXN
ZNF219
SERPINA1;MTMR3;CR1;CR1L;ZNF628;ZMAT5;ASCC2
FAM174B
PTEN;SSC5D;NAT14
SHOX2;GNB4;EIF3E;LCORL;WARS2
ZCCHC10;PLN;RSRC1
GJA1;CMTM5;SLC35F1;APOE;NDRG2;NAT14;NIPSNAP1
TOMM40;WNT16;NRBP1
GCKR;FAM174B
LXN;TOMM40;B3GNT7;ZNF513;TMEM253
EIF2B4;LXN;FAM174B;ZNF513;PVRL2;TRIB2;SUPT7L
EIF2B4;MLLT10;GJA1;GPR126;WNT16
DNAJC1;B3GNT7;GPR126;PVRL2;DGKH
GCKR;NOV
CEP85L;PTEN
ZCCHC10;HSPA4;SHOX2;GNB4;AFAP1;CWC27;GPN1;WARS2;PPM1G
EIF2B4;KIAA1755;CEP85L;SLC35F1;ARHGEF40
SLC5A6;ZNF219;ARHGEF40
IFI27L2;SNX17;FAM174B;CEP85L;VSTM2L;ZNF513;ASCC2;SSC5D;NAT14;NIPSNAP1;SUPT7L
NOV;RSRC1

DCAF16;GCKR;LXN;C2ORF16;EIF3E;ZNF512;TRIB2;NRBP1;SUPT7L
CCDC91;RSRC1;VSTM2L;ALX4;SLC35F1;ZNF512;LCORL;NAT14;MLF1;SUPT7L
CCDC91;APOC1;SLC35F1;APOE;CWC27;NDRG2;MLF1
SERPINA1;CR1;CR1L;ASCC2
CCDC91;HSPA4;APOC1;SLC35F1;APOE;CWC27;MLF1;DGKH
EIF2B4;ZNF219
KIAA1755;IFI27L2;CCDC121;CCDC91;CMTM5
LXN;PTEN;WARS2
NOV;SPEG
IFI27L2;CABP7;VSTM2L;SLC35F1;NDRG2;NAT14
DNAJC1;B3GNT7;GPR126
SHOX2;GNB4;SSC5D;LCORL;GPN1;WARS2
MLLT10;DCAF16;GJA1;MFN1;GPR126;ALX4;WNT16
SLC4A1AP;CCDC91;RSPO2;VSTM2L;SLC35F1;FAM3C;NAT14;NIPSNAP1
IFI27L2;RSPO2;NDRG2;NAT14
SERPINA1;MTMR3;CR1;PTEN
MLLT10;ZNF219
FAM174B
SLC4A1AP;CCDC91;RSRC1;APOC1;SLC35F1;FAM3C;APOE;NDRG2;NAT14;MLF1
CABP7;APOC1;VSTM2L;SLC35F1;APOE;NDRG2;NAT14;NIPSNAP1
PLN;NOV;SPEG
GNB4;AFAP1;NCAPG
LXN;CR1L;APOC1;CEP85L;RSPO2;GPR126;ZNF512;SUPT7L
CR1;LXN;TOMM40;MFN1;NCAPG;SUPT7L
DES;SPEG
SNX17;APOC1;APOE
SLC5A6;TBX15;PTEN
GCKR;C2ORF16;NRBP1
B3GNT7;SSC5D;TMEM253
EIF2B4;MTMR3;ALX4;WNT16
SERPINA1;MTMR3;CR1;CR1L;ZMAT5;PTEN
NOV;SPEG
SERPINA1;SNX17;ZMAT5;ASCC2;GPN1
MTMR3;MFN1;ALX4;WNT16
SLC4A1AP;CCDC91;RSRC1;SKIDA1;RSPO2;VSTM2L;SLC35F1;FAM3C
SNX17;FAM174B;SKIDA1;ZMAT5;ZNF513
DNAJC1;FAM174B;SKIDA1;ZNF513;C2ORF16;TMEM253
FAM174B;SKIDA1;TMEM253
PLN;NOV;SPEG
SLC4A1AP;CCDC91;RSRC1;SKIDA1;RSPO2;VSTM2L;SLC35F1;FAM3C;CWC27;NIPSNAP1
EIF2B4;DCAF16;ZNF628;FAM174B;MFN1;ZNF513;PVRL2;WARS2
NOV;SPEG
EIF2B4;DNAJC1;LXN;MFN1
DCAF16;CR1L;CEP85L;RSPO2;GPR126;SUPT7L;DGKH
LXN;SSC5D

PLN;NOV
GJA1;MFN1;GPR126;ALX4;WNT16
ZCCHC10;HSPA4;GNB4;DLEU1;AFAP1;CWC27;GPN1;WARS2;PPM1G
PTEN;TRIB2;WARS2
DNAJC1;FAM174B
ALX4;WNT16;CD34
GJA1;CABP7;APOE;NDRG2
CCDC121;CABP7
SLC5A6;TOMM40;ZNF219
ZCCHC10;DLEU1;GPR126
LXN;NOV
CMTM5;VSTM2L;SLC35F1;NDRG2;NAT14
CCDC121;GFM1;ZCCHC10;SLC4A1AP;CCDC91;HSPA4;CABP7;RSRC1;SKIDA1;CEP85L;ZNF512;CWC27
SERPINA1;MTMR3;CR1;CR1L;ZMAT5;PTEN
GJA1;CCDC91;RSRC1;APOC1;SLC35F1;UQCR10;APOE;NDRG2;NIPSNAP1
SHOX2;CD34
NOV;SPEG
RSRC1;SHOX2;AFAP1
GFM1;GJA1;SLC4A1AP;RSRC1;RSPO2;VSTM2L;SLC35F1;C2ORF16;FAM3C
CCDC121;GNB4;ALX4;CD34
PLN;NOV;SPEG
MLLT10;DCAF16;GCKR;LXN;SSC5D;EIF3E;ZNF512;TRIB2;NRBP1;SUPT7L;WARS2
SERPINA1;MTMR3;CR1;CR1L
SERPINA1;MTMR3;CR1;CR1L;ASCC2
DNAJC1;SNX17;ZNF219;ZNF513;FAM3C;ARHGEF40;TMEM253
ZCCHC10;ZNF628;SHOX2;GNB4;AFAP1;CWC27;GPN1;TMEM253;WARS2;PPM1G
IFI27L2;GNB4;EIF3E;WARS2
SLC4A1AP;CCDC91;HSPA4;RSRC1;RSPO2;VSTM2L;SLC35F1;FAM3C;NAT14
DNAJC1;B3GNT7;GPR126;DGKH
DNAJC1;CR1L;RSPO2;GPR126;DGKH
SLC4A1AP;CCDC91;CABP7;RSPO2;VSTM2L;SLC35F1;FAM3C;WNT16;NAT14;NIPSNAP1
GFM1;ZCCHC10;SLC4A1AP;CABP7;CEP85L;PTEN;NAT14;NIPSNAP1;SUPT7L;CCDC91;RSRC1;MFN1;FAM3
LXN;TMEM253
ZNF628;ALX4;WNT16
B3GNT7;ZNF219;TMEM253
RSRC1;RSPO2;VSTM2L;SLC35F1;FAM3C;NAT14
RSPO2;AFAP1
NOV
NOV
FAM174B
GFM1;ZCCHC10;SLC4A1AP;CABP7;SKIDA1;CEP85L;NAT14;SUPT7L;MLLT10;DCAF16;RSRC1;MFN1;ZNF51
SNX17;FAM174B;SKIDA1;ZNF513;NAT14;TMEM253
DNAJC1;GPR126;CCDC141
SKIDA1;GPN1;TMEM253
TOMM40;FAM3C

TBX15;CD34
EIF2B4;GJA1;DLEU1;ALX4;WNT16
DLEU1;ALX4;EIF3E;WNT16
DNAJC1;CCDC121;LXN;SKIDA1;ZNF512;TMEM253
IFI27L2;CCDC91;CMTM5;APOE;NDRG2;NAT14
SLC5A6;LXN
NOV;SPEG
CENPW;ASCC2;NRBP1
SLC4A1AP;CCDC91;CABP7;RSRC1;SKIDA1;APOC1;SLC35F1;FAM3C;CWC27;MLF1
ZNF628;FAM174B;SKIDA1;C2ORF16;NAT14;TMEM253
DCAF16;MFN1;ALX4;WNT16
SERPINA1;MTMR3;CR1;APOC1;ZMAT5;PTEN
APOC1;CCDC141
IFI27L2;EIF3E;FAM3C
FAM174B
EIF2B4;LXN;FAM174B;ZNF513;PVRL2
ZNF628;ALX4;WNT16
ZNF513;ZNF512;CD34;SUPT7L;TMEM253
DNAJC1;APOC1
IFI27L2;DNAJC1;CENPW;SNX17;DLEU1;NCAPG;CWC27;GPN1;WARS2
MLLT10;DNAJC1;CCDC121;GFM1;SKIDA1;ZNF512;FAM3C;SUPT7L;TMEM253
ZCCHC10;CR1;CR1L;SNX17;ZNF513;ZNF512;WNT16;TRIB2
DNAJC1;TRIB2;DGKH
EIF2B4;SKIDA1;ZNF513;SUPT7L;TMEM253
SLC4A1AP;SKIDA1;RSPO2;VSTM2L;SLC35F1;FAM3C;NAT14
TBX15;DCAF16;MFN1;EIF3E
DCAF16;GJA1;MFN1;ALX4;WNT16
EIF2B4;ZNF628;WNT16;NRBP1
NOV;ZNF628;SPEG
MTMR3;GPR126;WNT16;PVRL2
MLLT10;DCAF16;GCKR;LXN;B3GNT7;EIF3E;ZNF512;TRIB2
SLC35F1
EIF2B4;MLLT10;MFN1;WNT16
DNAJC1;SNX17;FAM174B;C2ORF16;TMEM253
NOV
SLC35F1
CCDC121;EIF3E;CD34
DNAJC1;FAM174B;FAM3C;PVRL2;SUPT7L;TMEM253
IFI27L2;CABP7;CMTM5;VSTM2L;UQCR10;NAT14
FAM174B
MLLT10;ALX4;EIF3E;ZNF512;NRBP1;SUPT7L
SLC5A6;ZCCHC10;CR1;TOMM40;CR1L;CEP85L;NCAPG;ZNF513;ZNF512;WNT16;TRIB2;SUPT7L
DCAF16;MTMR3;ZCCHC10;CEP85L;SLC35F1;TMEM253;WARS2
SLC4A1AP;CCDC91;RSRC1;RSPO2;VSTM2L;SLC35F1;FAM3C;NAT14;NIPSNAP1
TBX15;DCAF16;CMTM5;CEP85L;TRIB2

CENPW;NCAPG;NRBP1
DNAJC1;SHOX2;GNB4;AFAP1;NCAPG
SLC4A1AP;CCDC91;RSRC1;RSPO2;VSTM2L;SLC35F1;FAM3C;WNT16;NIPSNAP1
ZCCHC10;B3GNT7;ZNF513;TMEM253
MTMR3;GNB4;ALX4;WNT16
ZCCHC10;DLEU1;ASCC2
ARHGEF40;TRIB2
ZNF219;CD34
ZNF628;ZNF219;WNT16
GFM1;SLC4A1AP;CABP7;SKIDA1;CEP85L;ALX4;WNT16;NAT14;SUPT7L
SLC4A1AP;CCDC91;RSRC1;SKIDA1;RSPO2;VSTM2L;SLC35F1;FAM3C;NAT14;NIPSNAP1
SSC5D;WNT16
EIF2B4;MFN1;GPR126;WNT16
SPEG;SSC5D;SUPT7L
CR1L;ASCC2
IFI27L2;NOV
RSPO2;PVRL2
GJA1;MTMR3;ALX4;WNT16
GPR126;ASCC2;NRBP1
ZCCHC10;LXN;B3GNT7;ZNF513;LCORL;TMEM253
ZCCHC10;ZNF628;SHOX2;GNB4;AFAP1;C2ORF16;CWC27;GPN1;WARS2;PPM1G
CABP7;VSTM2L;SLC35F1
ZCCHC10;LXN;B3GNT7;TMEM253
ASCC2;PVRL2;NRBP1
ALX4;WNT16
SERPINA1;APOE;NIPSNAP1
GJA1;CABP7;SLC35F1;APOE;NDRG2;MLF1
SHOX2;TRIB2
SERPINA1;MTMR3;CR1;CR1L;SNX17;ZMAT5;PTEN
ZCCHC10;B3GNT7;MFN1;ZNF513;TMEM253
FAM174B;RSPO2
SLC5A6;ARHGEF40
DCAF16;CCDC121;ZCCHC10;RSRC1;MFN1;ALX4;PVRL2;WARS2
CMTM5;SLC35F1;NDRG2
ZCCHC10;SLC4A1AP;TOMM40;ZNF628;SHOX2;CWC27;GPN1;NRBP1;WARS2;PPM1G
CABP7;APOC1;SLC35F1;APOE;NAT14;NIPSNAP1
DES
EIF2B4;GJA1;MFN1;ALX4
KIAA1755;IFI27L2;CMTM5;NDRG2
EIF2B4;TOMM40;ZNF628;ZNF219;WNT16
KIAA1755;CMTM5;APOC1;APOE;NDRG2
SLC4A1AP;CCDC91;RSRC1;SKIDA1;RSPO2;VSTM2L;SLC35F1;C2ORF16;FAM3C;NAT14;NIPSNAP1
GJA1;NCAPG;WNT16;TMEM253
TBX15;TTN
SSC5D;CD34

SLC5A6;DCAF16;CENPW;ALX4;PVRL2
NOV;SPEG
FAM174B
SLC5A6;DCAF16;LXN;ALX4
IFI27L2;SLC4A1AP;CCDC91;RSRC1;CMTM5;RSPO2;VSTM2L;SLC35F1;NAT14;NIPSNAP1
B3GNT7;APOC1;DGKH
TOMM40;ZNF628;DLEU1;WNT16
GJA1;SLC4A1AP;CABP7;VSTM2L;SLC35F1;WNT16;NDRG2;NIPSNAP1
MTMR3;FAM174B;SKIDA1;ZMAT5;ALX4;ZNF513;ARHGEF40;TMEM253
IFI27L2;CCDC91;APOC1;VSTM2L;SLC35F1;APOE;LCORL;NAT14
MLLT10;DCAF16;GCKR;SKIDA1;CEP85L;ZNF512;ARHGEF40;TRIB2;SUPT7L
ALX4;CD34
EIF2B4;ZNF628;ALX4;WNT16
CENPW;NCAPG;NRBP1
CCDC121;FAM174B;SKIDA1;ZNF512;FAM3C;SUPT7L;TMEM253
HSPA4;ZNF628;SHOX2;GNB4;AFAP1;CWC27;GPN1;WARS2
GJA1;ALX4;WNT16
SLC5A6;EIF2B4;LXN;SUPT7L
GFM1;CCDC91;HSPA4;GNB4;AFAP1;CWC27;GPN1;WARS2;PPM1G
EIF2B4;GJA1;GNB4;ALX4;WNT16
FAM174B
LXN;RSPO2;GPR126
CCDC91;APOC1;SLC35F1;APOE;NDRG2;NAT14;MLF1
DES
SLC5A6;PTEN
GJA1;ALX4;WNT16;GPN1
CR1L;ASCC2
SLC5A6;SLC4A1AP;SKIDA1;RSPO2;VSTM2L;SLC35F1;FAM3C;NAT14
SKIDA1;ASCC2;FAM3C;PVRL2;SUPT7L;TMEM253
TOMM40;HSPA4;ZNF628;SNX17;ZMAT5;CWC27;GPN1;WARS2;PPM1G
FAM174B;TRIB2
CEP85L;MYH7;TTN
SERPINA1;TOMM40;SNX17
DNAJC1;FAM174B;ZNF513;PVRL2;SUPT7L;TMEM253
FAM174B
IFI27L2;DNAJC1;AFAP1;NCAPG;EIF3E;WARS2
CENPW;TOMM40;PVRL2;PPM1G
GCKR;CR1;SNX17;APOC1;APOE;PVRL2;TRIB2
MLLT10;DCAF16;CCDC121;SSC5D;EIF3E;ZNF512;LCORL;TRIB2;NRBP1;SUPT7L
SHOX2;GNB4;CWC27;LCORL;GPN1;WARS2;PPM1G
SERPINA1;CR1;CR1L;ASCC2
ZNF628;GNB4;WNT16;GPN1
LXN;B3GNT7;ZNF513;TMEM253
SERPINA1;MTMR3;CR1;ZMAT5;PTEN;ARHGEF40
GCKR;CABP7

DNAJC1;SERPINA1;LXN;SNX17;ZMAT5;PPM1G
CENPW;ZCCHC10;PTEN;NCAPG;CWC27;GPN1;NIPSNAP1;WARS2;PPM1G;DCAF16;CR1L;MFN1;DLEU1;EI
SLC5A6;APOE;ARHGEF40
CENPW;TOMM40;DLEU1;NCAPG;GPN1
CR1;ZNF628;SNX17;APOC1;VSTM2L;APOE;PVRL2;TRIB2
GJA1;VSTM2L;SLC35F1;NDRG2
B3GNT7;SSC5D;TMEM253
FAM174B;FAM3C
MLLT10;GJA1;MFN1;ALX4;WNT16
SERPINA1;MTMR3;CR1;CR1L;PTEN;PVRL2
IFI27L2;CENPW;ZCCHC10;TOMM40;DLEU1;GPR126;NCAPG;ASCC2
IFI27L2;CCDC91;APOC1;VSTM2L;SLC35F1;APOE;NAT14
RSRC1;PTEN;CD34
SERPINA1;MTMR3;CR1;CR1L;ZMAT5;PTEN;TMEM253
DNAJC1;FAM174B
CENPW;TOMM40;DLEU1;GPR126;ASCC2
GNB4;DGKH
GCKR;B3GNT7;FAM174B
ASCC2;C2ORF16;PVRL2;NRBP1
DNAJC1;LXN;FAM174B;GPR126;ASCC2;NIPSNAP1;DGKH
KIAA1755;CCDC91;CMTM5
ZNF628;GNB4;ALX4;WNT16
SLC5A6;ALX4;PVRL2
SLC4A1AP;CCDC91;CABP7;VSTM2L;SLC35F1;NDRG2
TBX15;SHOX2;PTEN;CD34
CR1;CR1L;SNX17;APOC1;ZMAT5;VSTM2L;ZNF513;APOE;TRIB2
MTMR3;CR1L;RSPO2;GPR126;WNT16;PVRL2;DGKH
CENPW;ASCC2;PVRL2;DGKH
DCAF16;ZMAT5;SSC5D;EIF3E;ZNF512;NAT14;NRBP1
IFI27L2;SERPINA1;MTMR3;CR1;ZMAT5;PTEN
ZNF219;AFAP1
IFI27L2;GJA1;CCDC91;RSRC1;APOC1;RSPO2;VSTM2L;ALX4;LCORL;MLF1
DNAJC1;SERPINA1;CR1;TOMM40;VSTM2L;PVRL2
KIAA1755;CCDC121;CCDC91;CABP7;CMTM5
CR1L;ASCC2
TBX15;PTEN;CD34
IFI27L2;CMTM5;RSPO2;SLC35F1
MLLT10;DCAF16;CCDC121;FAM174B;SKIDA1;C2ORF16;ZNF512;FAM3C;LCORL;SUPT7L;WARS2
SHOX2;GNB4;SSC5D;FAM3C;CWC27;LCORL;GPN1;WARS2
GFM1;TOMM40;CEP85L;UQCR10;LCORL;TTN
DNAJC1;LXN;TOMM40;ZNF219;ZNF513
DNAJC1;RSPO2;GPR126;PVRL2;DGKH
CCDC121;CCDC91;CMTM5
IFI27L2;DCAF16;DNAJC1;CCDC121;ZNF628;FAM174B;CEP85L;ZNF513;SSC5D;NAT14;SUPT7L
PLN;RSRC1;SPEG

SLC5A6;SLC4A1AP;CABP7;RSPO2;VSTM2L;SLC35F1;FAM3C;NAT14;NIPSNAP1
MYH6;MYH7;TTN;PPM1G
ZNF628;SKIDA1;ZNF513;C2ORF16;TMEM253
SERPINA1;MTMR3;CR1;CR1L;ZMAT5;PTEN;ARHGEF40
EIF2B4;DLEU1;ALX4;WNT16;PVRL2
EIF2B4;ZNF628;FAM174B;ZMAT5;ZNF513;ASCC2;PVRL2
SLC5A6;PTEN;AFAP1;SSC5D;CD34
EIF2B4;RSPO2
CENPW;TOMM40;NCAPG;NRBP1
GCKR;B3GNT7;FAM174B
NOV;SPEG;DGKH
CENPW;DLEU1;PVRL2;NRBP1
IFI27L2;GJA1;NDRG2
LXN;ZNF512;ARHGEF40;NRBP1
APOC1;ZMAT5;SLC35F1;APOE;NDRG2;NAT14;MLF1
SLC5A6;TOMM40;ARHGEF40;TRIB2;WARS2
DNAJC1;GCKR;FAM174B
CENPW;TOMM40;HSPA4;SNX17;GNB4;DLEU1;NCAPG;CWC27;GPN1;PPM1G
PLN;NOV;SPEG
CR1;CR1L;ASCC2
LXN;B3GNT7;FAM3C;LCORL;TMEM253
SLC5A6;GCKR;ZCCHC10;CR1;ZNF628;SNX17;ZNF513;ZNF512;TRIB2;SUPT7L;PPM1G
EIF2B4;CCDC121;CENPW;MTMR3;SLC4A1AP;HSPA4;SKIDA1;CEP85L;NCAPG;C2ORF16;MLF1;NRBP1;PPM
GJA1;SSC5D;WNT16
CENPW;SPEG
IFI27L2;SLC4A1AP;CABP7;RSPO2;VSTM2L;SLC35F1;UQCR10;FAM3C;NAT14;NIPSNAP1
SERPINA1;CR1;CR1L;SNX17;ZMAT5
CABP7;VSTM2L;SLC35F1;WNT16;NDRG2;NAT14;NIPSNAP1
MLLT10;FAM174B;SKIDA1;ZNF513;ZNF512;FAM3C;SUPT7L;TMEM253
DNAJC1;CCDC121;FAM174B;SKIDA1;C2ORF16;ZNF512;FAM3C;TMEM253
CCDC121;GFM1;ZCCHC10;SLC4A1AP;CWC27;TMEM253;MLLT10;DCAF16;DNAJC1;CCDC91;FAM174B;ZN
ZCCHC10;CR1;SNX17;MFN1;VSTM2L;ZNF513;ZNF512;WNT16;ARHGEF40;TRIB2;SUPT7L
ZCCHC10;TOMM40;HSPA4;ZNF628;SHOX2;GNB4;FAM3C;CWC27;GPN1;TRIB2;WARS2;PPM1G
CENPW;TOMM40;HSPA4;GNB4;DLEU1;NCAPG;CWC27;GPN1;NRBP1;PPM1G
CENPW;ZCCHC10;LXN;B3GNT7;NCAPG;LCORL;TMEM253
SLC4A1AP;CCDC91;RSPO2;VSTM2L;SLC35F1;FAM3C;NDRG2
DNAJC1;RSRC1;SHOX2;GNB4;SSC5D;ARHGEF40
EIF2B4;GJA1
CENPW;ZCCHC10;DLEU1;NCAPG;ASCC2;NRBP1
KIAA1755;IFI27L2;CABP7;CMTM5
NOV;SPEG
CENPW;TOMM40;HSPA4;DLEU1;NCAPG;GPN1;DGKH;PPM1G
SERPINA1;MTMR3;CR1;ZMAT5
SERPINA1;MTMR3;CR1;CR1L;SNX17;ZMAT5;PTEN
NOV;SPEG

SERPINA1;MTMR3;CR1;CR1L;SNX17;ZMAT5;PPM1G
ZNF219;RSPO2
EIF2B4;DCAF16;DNAJC1;LXN;ZNF513;PVRL2;NAT14;PPM1G
EIF2B4;TOMM40;ZNF513;NRBP1
MLLT10;MFN1;ALX4;WNT16
NCAPG;ASCC2;PVRL2;TMEM253;PPM1G
PLN;NOV
UQCR10;MLF1;TTN
DNAJC1;CCDC121;GFM1;SKIDA1;C2ORF16;FAM3C;SUPT7L;TMEM253
SERPINA1;MTMR3;CR1;CR1L;PTEN;NCAPG;ASCC2
ALX4;ARHGEF40;CD34
CR1;LXN;B3GNT7;SNX17
GJA1;SLC35F1;NDRG2
KIAA1755;LXN;ZNF219;ZNF513
NDRG2
MLLT10;DCAF16;GJA1;MFN1;GNB4;GPR126;WNT16
EIF2B4;RSPO2
GJA1;APOC1;SLC35F1;APOE;NDRG2;NAT14
SLC4A1AP;CCDC91;CABP7;CMTM5;SLC35F1;APOE;NDRG2;NAT14;MLF1
SLC5A6;SLC4A1AP;CMTM5;SKIDA1;RSPO2;VSTM2L;SLC35F1;NDRG2
DNAJC1;SERPINA1;ZCCHC10;B3GNT7;CCDC141
TBX15;MYH7;TTN
SERPINA1;CR1L;ASCC2
GJA1;SSC5D;WNT16
ZCCHC10;APOC1
EIF2B4;DNAJC1;FAM174B;SKIDA1;ZNF513;SUPT7L;TMEM253
EIF2B4;KIAA1755;LXN;ZNF628;FAM174B;ZNF513;PVRL2
KIAA1755;CMTM5;CEP85L;ZMAT5;GPR126
IFI27L2;CCDC91;CABP7;CMTM5;NDRG2
MLLT10;DCAF16;ZNF628;SSC5D
PLN;NOV
PLN;SHOX2;SPEG
ZNF219;PTEN;CD34
CCDC121;RSPO2
CENPW;ZCCHC10;TOMM40;DLEU1;ASCC2;NRBP1
SERPINA1;CR1;SNX17;ZMAT5;ASCC2
GJA1;SLC4A1AP;CCDC91;RSRC1;SKIDA1;SLC35F1;CWC27;NDRG2;MLF1
DNAJC1;SERPINA1;CCDC141
GJA1;GNB4;AFAP1;NCAPG;SSC5D
SERPINA1;MTMR3;CR1;CR1L;PTEN;ASCC2
SLC5A6;DCAF16;MTMR3;LXN;CCDC91;MFN1;ALX4;EIF3E
ZCCHC10;SLC4A1AP;TOMM40;HSPA4;ZNF628;SHOX2;GNB4;C2ORF16;CWC27;GPN1;WARS2;PPM1G
GJA1;MFN1;GNB4;ALX4;WNT16
MTMR3;ZNF219;WNT16;PVRL2
CCDC121;CD34

CR1L;ASCC2
VSTM2L;SLC35F1;NDRG2;NAT14
GCKR;CR1;CR1L;SNX17;APOC1;APOE;ARHGEF40;TRIB2
TOMM40;ASCC2;PVRL2;PPM1G
SHOX2;GNB4;AFAP1;FAM3C;WARS2
SLC5A6;DCAF16;MFN1;ALX4;SSC5D;WARS2
CCDC121;RSPO2;GPR126;ALX4;CCDC141
TOMM40;DLEU1;ALX4;WNT16
CENPW;PTEN;NCAPG;GPN1;NIPSNAP1;WARS2;PPM1G;DCAF16;B3GNT7;CR1L;MFN1;DLEU1;EIF3E;LCO[illegible]
DNAJC1;FAM174B;SKIDA1;ZMAT5;FAM3C;TMEM253
CR1;PTEN;EIF3E;WARS2
GCKR;B3GNT7;FAM174B
GFM1;CENPW;ZCCHC10;TOMM40;HSPA4;NCAPG;CWC27;GPN1;NIPSNAP1;PPM1G;RSRC1;ZMAT5;MFN[illegible]
LXN;SNX17;NCAPG;CD34;NRBP1;SUPT7L;PPM1G
SLC5A6;GFM1;TOMM40
CCDC121;GFM1;CCDC91;FAM174B;SKIDA1;ZNF512;FAM3C;LCORL;TMEM253
MLLT10;CCDC121;PTEN;ARHGEF40;LCORL;CD34
SLC4A1AP;CCDC91;RSRC1;RSPO2;VSTM2L;SLC35F1;FAM3C;NAT14;NIPSNAP1
CR1;CEP85L;VSTM2L;SSC5D;WNT16;SUPT7L;CR1L;SNX17;APOC1;ZNF513;ZNF512;ARHGEF40;TRIB2
CENPW;TOMM40;HSPA4;SNX17;GNB4;DLEU1;NCAPG;CWC27;GPN1;PPM1G
CENPW;LXN;TOMM40;B3GNT7;NCAPG;FAM3C;LCORL;TMEM253
GJA1;NOV;ZNF219
MTMR3;CR1;CR1L;ASCC2
FAM174B;MFN1
EIF2B4;RSPO2
EIF2B4;DCAF16;MTMR3;LXN;ZNF628;FAM174B;ZNF513;ZNF512;PVRL2;SUPT7L
NOV;GNB4
SLC5A6;CCDC91;ALX4;EIF3E;PVRL2;MLF1
LXN;B3GNT7;CR1L;APOC1;VSTM2L;PVRL2
CENPW;DLEU1;NRBP1
SHOX2;GNB4;AFAP1;ALX4;C2ORF16;CWC27;DGKH;WARS2
SERPINA1;MTMR3;CR1;CR1L;ZMAT5;ARHGEF40
EIF2B4;LXN;FAM174B;ZNF513;PVRL2
B3GNT7;FAM174B;CCDC141
SERPINA1;MTMR3;CR1;ZMAT5;PTEN;ARHGEF40;TMEM253
NOV;SPEG
IFI27L2;CABP7;SLC35F1;NDRG2
ASCC2;C2ORF16;NRBP1
GJA1;SLC4A1AP;SKIDA1;ZMAT5;RSPO2;VSTM2L;SLC35F1;FAM3C;NAT14
TOMM40;HSPA4;NRBP1
SERPINA1;MTMR3;CR1;CR1L;CMTM5;ZMAT5
CABP7;VSTM2L;SLC35F1;WNT16;NAT14
GCKR;CR1;CR1L;ZNF628;SNX17;ZNF513;PVRL2;TRIB2
CENPW;ZCCHC10;HSPA4;DLEU1;NCAPG;EIF3E;DGKH
SLC5A6;ZNF219;ALX4

GJA1;RSPO2
CENPW;ZCCHC10;CR1;PTEN;NCAPG;CWC27;GPN1;NIPSNAP1;PPM1G;MFN1;DLEU1;EIF3E;LCORL;DGKH
B3GNT7;GPR126;CCDC141
PTEN;CD34
GJA1;ASCC2;WNT16;NRBP1
SERPINA1;MTMR3;CR1;ZMAT5;PTEN;ARHGEF40;PVRL2
ZCCHC10;HSPA4;ZNF628;GNB4;AFAP1;CWC27;LCORL;GPN1;WARS2;PPM1G
LXN;B3GNT7;PVRL2
PLN;RSRC1;MFN1;SPEG
GCKR;PVRL2
EIF2B4;CCDC121;CENPW;SLC4A1AP;HSPA4;SKIDA1;CEP85L;NCAPG;C2ORF16;MLF1;NRBP1;PPM1G;SLC
CCDC91;ZNF219
ZCCHC10;NOV
ZCCHC10;B3GNT7;ZNF513;UQCR10;TMEM253
SERPINA1;CR1;SNX17;ZMAT5
GCKR;CR1;CR1L;SNX17;NCAPG;ZNF513;TRIB2;PPM1G
PTEN;CD34
ZCCHC10;LXN;TOMM40;CEP85L;ZNF513;ASCC2;FAM3C;SUPT7L;DGKH;WARS2
KIAA1755;NOV;SPEG
CCDC91;APOC1;SLC35F1;UQCR10;APOE;NDRG2;NAT14;MLF1;NIPSNAP1
GCKR;ASCC2;PVRL2;NRBP1
GPR126;ASCC2;NRBP1
KIAA1755;CMTM5;APOC1;SLC35F1;APOE;NDRG2;NAT14
B3GNT7;GPR126;CCDC141
EIF2B4;DNAJC1;LXN;ZNF219;DLEU1;EIF3E;ZNF512;TRIB2;SUPT7L
SKIDA1;APOC1;SLC35F1;APOE;NDRG2;NAT14;MLF1;NIPSNAP1
MLLT10;DCAF16;MFN1;ALX4;EIF3E;ZNF512;PVRL2;CD34
SLC4A1AP;SKIDA1;RSPO2;VSTM2L;SLC35F1;FAM3C;NDRG2;NAT14
SLC4A1AP;HSPA4;CABP7;RSPO2;VSTM2L;SLC35F1;FAM3C;NAT14
RSRC1;CD34;WARS2
SLC4A1AP;RSRC1;RSPO2;VSTM2L;SLC35F1;FAM3C;NAT14;NIPSNAP1
SERPINA1;MTMR3;CR1;SNX17;ZMAT5
SERPINA1;MTMR3;CR1;CR1L;APOC1;PTEN
NOV;SPEG
SLC5A6;DCAF16;CCDC121;ALX4;ZNF512
KIAA1755;IFI27L2;CMTM5;NDRG2
GFM1;GJA1;SLC4A1AP;CCDC91;RSRC1;RSPO2;VSTM2L;SLC35F1;C2ORF16;FAM3C
KIAA1755;IFI27L2;CABP7;CMTM5;VSTM2L;SLC35F1;APOE;NDRG2
CR1;CR1L;SNX17;APOC1;MFN1;ZNF513;ZNF512;TRIB2;SUPT7L
B3GNT7;APOC1;GPR126;CCDC141
CENPW;ASCC2;PVRL2;NRBP1
MTMR3;B3GNT7;CR1L;ASCC2
GFM1;SLC4A1AP;CCDC91;RSRC1;RSPO2;VSTM2L;SLC35F1;FAM3C;NAT14;NIPSNAP1
MLLT10;DCAF16;GJA1;MFN1;ALX4;WNT16
TOMM40;ZMAT5;ASCC2

ZCCHC10;ASCC2;NRBP1
SERPINA1;MTMR3;CR1;CR1L;ZMAT5;PTEN
CENPW;LXN;B3GNT7;NCAPG;TMEM253
GJA1;GNB4
MLLT10;IFI27L2;DCAF16;GCKR;LXN;B3GNT7;EIF3E;ZNF512;TRIB2
CENPW;ZCCHC10;DLEU1;NCAPG
KIAA1755;CCDC91;CABP7;CMTM5;VSTM2L;WNT16;NAT14
KIAA1755;SLC35F1;ARHGEF40
DCAF16;GCKR;LXN;B3GNT7;C2ORF16;EIF3E;ZNF512;TRIB2;SUPT7L
MLLT10;DNAJC1;FAM174B;SKIDA1;FAM3C;ARHGEF40;CD34;SUPT7L;TMEM253
CENPW;TOMM40;HSPA4;GNB4;DLEU1;NCAPG;CWC27;GPN1;PPM1G
SERPINA1;CR1;CR1L;ZMAT5;PTEN
GJA1;ZNF628;ALX4;WNT16
SLC5A6;ALX4;WARS2
ZCCHC10;LXN;B3GNT7;DLEU1;NCAPG;LCORL;TMEM253
LXN;B3GNT7;TMEM253
LXN;B3GNT7;SNX17;GPR126
SERPINA1;MTMR3;CR1;ZMAT5;PTEN;ARHGEF40
SLC4A1AP;CCDC91;APOC1;SLC35F1;APOE;NAT14;MLF1;NIPSNAP1
SLC5A6;DCAF16;CCDC121;CENPW;CCDC91;MFN1;ALX4;EIF3E;ZNF512;WARS2
EIF2B4;GCKR;CABP7;FAM174B;TRIB2
SLC5A6;TBX15;PTEN
ZCCHC10;LXN;B3GNT7;ZNF513;TMEM253
KIAA1755;SPEG
GFM1;SLC4A1AP;CCDC91;CABP7;RSRC1;RSPO2;VSTM2L;SLC35F1;C2ORF16;FAM3C;NAT14;NIPSNAP1
ZNF513;TMEM253
GCKR;B3GNT7;FAM174B
DNAJC1;LXN;FAM174B;ZNF219;DLEU1;ZNF513;ASCC2;FAM3C;TRIB2;SUPT7L
GJA1;ZNF628;DLEU1;ALX4;WNT16
DNAJC1;SERPINA1;B3GNT7;FAM174B;CCDC141
TBX15;PTEN;CD34
SHOX2;GNB4;EIF3E;WARS2
FAM174B
RSPO2
CENPW;NCAPG;ASCC2;NRBP1
NOV;SNX17
DNAJC1;CR1L;RSPO2;GPR126
NOV;SPEG
PTEN;CD34
ZCCHC10;LXN;TOMM40;B3GNT7;ZNF513;TMEM253
IFI27L2;CABP7;CMTM5;VSTM2L;APOE;NDRG2;NAT14
RSPO2;VSTM2L;WNT16;NDRG2
GJA1;GNB4;ALX4;WNT16
SLC5A6;DCAF16;CCDC121;MFN1;ALX4;EIF3E
CABP7;CMTM5

DNAJC1;MTMR3;LXN;DGKH
CCDC121;ALX4;CD34
ZCCHC10;SLC4A1AP;RSRC1;SHOX2;GNB4;UQCR10;CWC27;LCORL;GPN1;WARS2
B3GNT7;ZNF219;TMEM253
GFM1;ZCCHC10;CCDC91;HSPA4;ZNF628;GNB4;AFAP1;CWC27;GPN1;WARS2;PPM1G
NOV;SPEG
GJA1;ZNF628;ALX4;WNT16
EIF2B4;LXN;ZNF219;ZNF513;PVRL2;TRIB2;SUPT7L
SERPINA1;MTMR3;CR1;ZMAT5;PTEN
TBX15;SHOX2;CD34;WARS2
DNAJC1;CCDC121;GFM1;HSPA4;SNX17;FAM174B;SKIDA1;C2ORF16;FAM3C;TMEM253
KIAA1755;SSC5D
SLC5A6;EIF2B4;SNX17
NOV;GNB4;SPEG
SLC5A6;PTEN;CD34
NOV;SPEG
ZNF219;ZMAT5
GJA1;GPR126;ALX4;WNT16
GNB4;AFAP1
SLC5A6;ALX4
PLN;NOV;EIF3E
DNAJC1;FAM174B;SKIDA1;ZMAT5;FAM3C;TMEM253
CEP85L;MYH7;TTN
MYH6;TTN
SHOX2;CD34
ZNF219;GNB4
NOV;RSRC1
GCKR;B3GNT7;SSC5D;TRIB2
GNB4;NCAPG;EIF3E;FAM3C
EIF2B4;DNAJC1;CCDC121;FAM174B;SKIDA1;ZNF513;SUPT7L;TMEM253
DNAJC1;FAM174B
SLC5A6;SLC4A1AP;CABP7;RSRC1;SKIDA1;RSPO2;VSTM2L;SLC35F1;FAM3C;NAT14
MLLT10;FAM174B;CEP85L;GPR126;ASCC2;DGKH
SLC4A1AP;SKIDA1;RSPO2;VSTM2L;SLC35F1;FAM3C;NAT14
ALX4;WNT16
GPR126;ASCC2
EIF2B4;ALX4;WNT16
CCDC121;FAM174B;SKIDA1;TMEM253
HSPA4;RSPO2;VSTM2L;SLC35F1;NDRG2
EIF2B4;ZNF628;WNT16
GJA1;WNT16
CD34
GJA1;SLC35F1;APOE;NAT14;MLF1
GCKR
MFN1;EIF3E;LCORL

ZCCHC10;ZNF513;UQCR10;TMEM253
FAM174B
AFAP1;DGKH
ZMAT5;ASCC2
SHOX2;GNB4;CWC27;LCORL;GPN1;WARS2
CR1;SNX17;MFN1;NCAPG;ARHGEF40;TRIB2;SUPT7L
SHOX2;CD34
AFAP1
GJA1;ZNF628;WNT16
SLC4A1AP;RSRC1;SKIDA1;RSPO2;VSTM2L;SLC35F1;FAM3C;NAT14
B3GNT7
SHOX2;GNB4;AFAP1;WARS2
FAM174B
FAM174B
SERPINA1;MTMR3;CR1;ASCC2
EIF2B4;IFI27L2;FAM174B;ZNF513;ASCC2;C2ORF16;NAT14;SUPT7L
CABP7;VSTM2L;SLC35F1;NAT14;NIPSNAP1
GFM1;SLC4A1AP;CABP7;RSRC1;ZNF219;CEP85L;MFN1;NDRG2;NAT14;SUPT7L
CENPW
EIF2B4;TRIB2
SLC35F1;APOE;NDRG2
EIF2B4;FAM174B;ZNF513;SSC5D;PVRL2
GJA1;SLC4A1AP;CCDC91;SLC35F1;CWC27;MLF1;NIPSNAP1;DGKH
CD34;DGKH
ASCC2;NRBP1
ZCCHC10;HSPA4;GNB4;AFAP1;CWC27;GPN1;WARS2;PPM1G
CCDC121;SLC4A1AP;CCDC91;RSRC1;SKIDA1;SLC35F1;FAM3C;MLF1
MTMR3;MFN1;WNT16
IFI27L2;CR1;ZNF628;SNX17;ZMAT5;ZNF513
FAM174B
SLC4A1AP;CCDC91;RSRC1;RSPO2;VSTM2L;SLC35F1;FAM3C;NIPSNAP1
DNAJC1;PTEN;EIF3E
MTMR3;ZNF628;WNT16
TOMM40;DLEU1;WNT16
RSPO2;GPR126;ZNF512;SUPT7L
SLC5A6;ALX4;ZNF512
TRIB2;SUPT7L;TMEM253
CR1;SNX17;MFN1;ZNF513;ARHGEF40;TRIB2
DES;TTN
DCAF16;GCKR;ZNF512;TRIB2;SUPT7L
LXN;C2ORF16
EIF2B4;SKIDA1;ZNF513;SUPT7L;TMEM253
GFM1;SLC4A1AP;CABP7;RSRC1;SKIDA1;CEP85L;NDRG2;NAT14;SUPT7L
CMTM5;SLC35F1;NAT14
FAM174B;SKIDA1;SUPT7L;TMEM253

SLC5A6;CABP7;VSTM2L;SLC35F1;NAT14
ZNF628;WNT16
GFM1;ZCCHC10;SLC4A1AP;CABP7;RSRC1;CEP85L;MFN1;PTEN;FAM3C;NAT14;NIPSNAP1;SUPT7L
CABP7;VSTM2L;UQCR10;FAM3C;NAT14
ZNF628;WNT16
IFI27L2;DCAF16;FAM174B;ZNF513;NAT14;NIPSNAP1;SUPT7L
DCAF16;FAM174B;C2ORF16;TMEM253
ALX4;WNT16
GPR126;ASCC2
CD34
ZCCHC10;CD34
DLEU1;ASCC2
MTMR3;WNT16
LXN;MFN1
DLEU1;WNT16
IFI27L2;GJA1;APOE;NAT14
SPEG
IFI27L2;CMTM5
ZNF513;NIPSNAP1
ASCC2;NRBP1
SLC4A1AP;SKIDA1;RSPO2;VSTM2L;SLC35F1;FAM3C;NAT14
RSRC1
SSC5D
PTEN;CD34
C2ORF16;NRBP1
CD34
GJA1;ZNF219;ALX4;WNT16
ALX4;WNT16
SHOX2;GNB4;LCORL;DGKH;WARS2
EIF2B4;FAM174B;RSPO2;ZNF513
SERPINA1;MTMR3;CR1;ZMAT5;PTEN
SLC4A1AP;CCDC91;VSTM2L;SLC35F1;NDRG2;NAT14;NIPSNAP1
EIF2B4;ZNF628;WNT16
NOV
FAM174B
GJA1;WNT16
LXN;B3GNT7;TMEM253
SERPINA1;ASCC2
MFN1;ALX4;WNT16
EIF2B4;IFI27L2;SLC4A1AP;CABP7;RSRC1;UQCR10;CWC27;NAT14
GJA1;WNT16
RSPO2;VSTM2L;SLC35F1;FAM3C;NDRG2
ZCCHC10;SLC4A1AP;HSPA4;ZNF628;GNB4;CWC27;GPN1;WARS2;PPM1G
AFAP1
PTEN;EIF3E;WARS2

DNAJC1;SNX17
ZNF628;ALX4;WNT16
ZCCHC10;B3GNT7;ZNF513;TMEM253
SERPINA1;MTMR3;CR1;ZMAT5
GJA1;WNT16
DCAF16;ZNF219;ZNF512;ARHGEF40;CD34;SUPT7L;TMEM253
TOMM40;WNT16
PTEN;PVRL2
SHOX2;GNB4;CWC27;LCORL;GPN1;WARS2
ALX4;WNT16;GPN1
LXN;APOC1;ZNF513
B3GNT7;RSPO2
SPEG
LXN
FAM174B;FAM3C;PVRL2
CMTM5
KIAA1755;CCDC121;CMTM5
SHOX2;GNB4;AFAP1;WARS2
MLLT10;MFN1;ZNF512;FAM3C;CD34;TMEM253
EIF2B4;MTMR3;WNT16
IFI27L2;GNB4;LCORL;WARS2
CD34
GJA1;CCDC91;VSTM2L;SLC35F1;FAM3C
DCAF16;GFM1;ZCCHC10;SLC4A1AP;CABP7;RSRC1;CEP85L;MFN1;PTEN;FAM3C;LCORL
DCAF16;ZNF512;ARHGEF40;SUPT7L;TMEM253
EIF2B4;SKIDA1;ZNF513;PVRL2;CD34;NAT14;SUPT7L;TMEM253
ALX4;WNT16
MLLT10;DCAF16;GCKR;CCDC121;EIF3E;ZNF512;TRIB2
B3GNT7;TMEM253
NOV
CABP7;APOC1;APOE
GJA1;WNT16
CABP7;SLC35F1
SERPINA1;MTMR3;CR1;CR1L;PTEN
ZCCHC10;CR1L;CEP85L;DLEU1;RSPO2;ZNF512;WNT16;SUPT7L;TMEM253
SKIDA1;ALX4;TMEM253
GJA1;ZNF628;WNT16
NOV
AFAP1
SSC5D
FAM174B
DNAJC1;EIF3E;CD34
GPR126;WNT16;PVRL2;DGKH
DCAF16;FAM174B;MFN1;ALX4;PVRL2;CD34
DNAJC1;B3GNT7;SNX17

ASCC2;NRBP1
ZNF628;SHOX2;GNB4;AFAP1;CWC27;GPN1;PPM1G
B3GNT7;ZNF628;FAM174B;CEP85L;VSTM2L;ZNF513;SSC5D;C2ORF16;NAT14;SUPT7L
DCAF16;CCDC121;LXN;CR1L;ZNF512;NAT14;SUPT7L
SERPINA1;MTMR3;CR1;ZMAT5;PTEN
MLLT10;GFM1;SLC4A1AP;CABP7;RSRC1;CEP85L;MFN1;FAM3C;SUPT7L;DGKH
DNAJC1;FAM3C;TMEM253
CCDC121;ALX4;NAT14;MLF1
ZNF512;SUPT7L
GFM1;SLC4A1AP;CABP7;NDRG2;NAT14;SUPT7L
PTEN
RSRC1
GPR126
MTMR3;WNT16
DNAJC1;FAM174B;ZNF513;C2ORF16;NAT14;SUPT7L
GFM1;SLC4A1AP;CABP7;RSRC1;CEP85L;MFN1;NAT14;SUPT7L
GFM1;SLC4A1AP;CABP7;RSRC1;CEP85L;MFN1;NAT14;SUPT7L
SPEG
EIF3E;ZNF512
ZNF513;NIPSNAP1
ZNF513;ZNF512
SKIDA1;PVRL2;TMEM253
MTMR3;CR1;PTEN
NRBP1
GPR126
ZNF628;GNB4;AFAP1;CWC27;WARS2
MLLT10;ZNF628;WNT16
SLC4A1AP;HSPA4;CABP7;ZNF219;NDRG2;NAT14;SUPT7L
SLC4A1AP;CCDC91;SLC35F1;NDRG2
SLC5A6;RSPO2;VSTM2L;NDRG2;NAT14
IFI27L2;SLC4A1AP;CABP7;VSTM2L;UQCR10;NAT14
DLEU1
SLC4A1AP;CCDC91;RSRC1;SLC35F1;FAM3C;CWC27;MLF1
CD34
ZCCHC10;SLC4A1AP;HSPA4;CABP7;RSRC1;PTEN;DLEU1;NAT14;SUPT7L
ZNF513;PVRL2;NIPSNAP1
ZNF628;FAM174B;ZNF513;PVRL2
SLC4A1AP;CABP7;RSRC1;NDRG2;NAT14
DCAF16;SKIDA1;ZNF512;SUPT7L;TMEM253
IFI27L2;VSTM2L
FAM174B;CEP85L;ZNF513;C2ORF16;NAT14;NIPSNAP1;SUPT7L
GJA1;WNT16
SERPINA1;CR1;ASCC2
ASCC2
DCAF16;CD34

TBX15
SLC4A1AP;CABP7;ZNF219;MFN1;NDRG2;NAT14;SUPT7L
FAM174B;ZNF512;FAM3C
GFM1;SLC4A1AP;CABP7;RSRC1;NAT14;NIPSNAP1;SUPT7L
ASCC2
DNAJC1;ZNF512;FAM3C
B3GNT7
GFM1;CABP7;RSRC1;SKIDA1;NDRG2;NAT14
WNT16
GFM1;SLC4A1AP;CABP7;CEP85L;MFN1;NAT14;SUPT7L
CR1;SNX17;ZNF513;TRIB2;SUPT7L
TOMM40;WNT16
ZNF513;ZNF512;FAM3C;SUPT7L;TMEM253
ASCC2
GNB4;AFAP1;TMEM253;WARS2
FAM174B;SKIDA1
SNX17
DCAF16;CCDC121;SKIDA1;C2ORF16;ZNF512;SUPT7L;TMEM253
ZNF628;WNT16
NRBP1
KIAA1755;CCDC121;CMTM5
MTMR3;CR1;CR1L;ASCC2
ZMAT5
GFM1;SLC4A1AP;CABP7;RSRC1;CEP85L;MFN1;FAM3C;NAT14
LXN;SUPT7L
WNT16
VSTM2L;NAT14
EIF2B4;ZNF513;SUPT7L
GFM1;ZCCHC10;SLC4A1AP;CABP7;RSRC1;CEP85L;MFN1;NDRG2
WNT16
SLC5A6;GPR126;ALX4
CABP7;DLEU1;ZNF513;C2ORF16;NAT14;SUPT7L
TMEM253
LXN
GFM1;SLC4A1AP;CABP7;RSRC1;SKIDA1;NDRG2;NAT14;SUPT7L
B3GNT7;TMEM253
LXN;B3GNT7;TMEM253
SLC5A6
ZNF628;C2ORF16;WNT16
NOV
FAM174B;SKIDA1;ZNF513;ZNF512;SUPT7L;TMEM253
SLC4A1AP;RSRC1;RSPO2;VSTM2L;SLC35F1;FAM3C
GFM1;SLC4A1AP;CABP7;RSRC1;SKIDA1;CEP85L;MFN1;PTEN;NAT14;SUPT7L
GJA1;RSPO2;VSTM2L;SLC35F1;NAT14
SPEG

DNAJC1;SKIDA1;C2ORF16;ZNF512;FAM3C;SUPT7L;TMEM253
GJA1;ZNF628;WNT16
DCAF16;MTMR3;ZNF628;ZNF513;PVRL2;TMEM253
CR1;SNX17;APOC1;ZNF513;TRIB2
ZMAT5
WNT16
NRBP1
DCAF16;GFM1;ZCCHC10;SLC4A1AP;CABP7;RSRC1;CEP85L;MFN1;PTEN;ZNF512;FAM3C
LXN
FAM174B;SKIDA1
GPR126;CCDC141
CCDC121;FAM174B;SKIDA1;ZNF512;SUPT7L;TMEM253
DCAF16;SKIDA1;ZNF513;ASCC2;C2ORF16;ZNF512;SUPT7L;TMEM253
FAM174B;SKIDA1;FAM3C;TMEM253
ZNF512;SUPT7L;TMEM253
MFN1;WNT16
GFM1;SLC4A1AP;CABP7;RSRC1;CEP85L;VSTM2L;NDRG2;NAT14
WARS2
DLEU1
IFI27L2;ZNF628;SNX17;ZNF513;NAT14;NIPSNAP1;SUPT7L
GFM1;SLC4A1AP;CABP7;RSRC1;NDRG2;LCORL
SLC4A1AP;SLC35F1;NDRG2
NOV
SHOX2;GNB4;AFAP1;WARS2
GFM1;SHOX2;GNB4;CWC27;GPN1;WARS2
ZNF628;SNX17;FAM174B;CEP85L;ZNF513;SSC5D;C2ORF16;NAT14;SUPT7L
DNAJC1;ZNF513;ZNF512;SUPT7L;TMEM253
NRBP1
EIF2B4;FAM174B;ASCC2
EIF2B4
ALX4
ZCCHC10;PTEN
ASCC2
NOV;SPEG
CABP7;SLC35F1;NAT14;MLF1
LXN;B3GNT7;ZNF512
ZNF628;WNT16
ALX4
GJA1
GFM1;SLC4A1AP;CABP7;SKIDA1;ZNF219;NAT14;NIPSNAP1;DGKH
NRBP1
SLC4A1AP;SLC35F1;UQCR10;NAT14;MLF1
SKIDA1;ZNF513;TMEM253
WNT16
GJA1;WNT16

MLLT10;KIAA1755;DCAF16;ALX4;SSC5D;EIF3E;ZNF512;SUPT7L
CD34
GFM1;SLC4A1AP;CABP7;RSRC1;CEP85L;MFN1;NAT14;SUPT7L
DNAJC1;CCDC121;FAM174B;SKIDA1;FAM3C;TMEM253
CCDC121;C2ORF16;FAM3C;TMEM253
CD34
ALX4;WNT16
SNX17;ZNF513
FAM174B;ZNF513;SUPT7L
NRBP1
GFM1;SLC4A1AP;CABP7;RSRC1;CEP85L;MFN1;PTEN;DLEU1;FAM3C
DNAJC1;FAM174B;SKIDA1;ASCC2;ZNF512;SUPT7L;TMEM253
DCAF16;WNT16
B3GNT7;ZNF513;LCORL;TMEM253
GFM1;ZCCHC10;SLC4A1AP;CABP7;RSRC1;CEP85L;MFN1;PTEN;FAM3C;NAT14;NIPSNAP1;SUPT7L
FAM174B;ZMAT5;FAM3C;TMEM253
SNX17
DCAF16;CCDC121;FAM174B;SKIDA1;ZMAT5;ZNF512;TMEM253
SLC4A1AP;CABP7;RSRC1;ALX4;APOE;NDRG2;NAT14
MTMR3;CR1;B3GNT7;CR1L;ZNF628;PTEN
CR1
SERPINA1;MTMR3;CR1;ZNF628;PTEN
SLC5A6;PTEN
ZCCHC10;B3GNT7;MFN1;ZNF513;ZNF512;SUPT7L;TMEM253
CCDC121;FAM174B;SKIDA1;ZMAT5;LCORL;SUPT7L;TMEM253
DES
SERPINA1;CR1;B3GNT7
DCAF16;DNAJC1;FAM174B;ZNF513;NAT14;SUPT7L
LXN
ZNF219
NOV
NOV
TBX15;PTEN
NOV
IFI27L2;CCDC91;RSPO2;VSTM2L;SLC35F1;LCORL;NAT14
LXN;SPEG
EIF2B4
ALX4;WNT16
CR1;SNX17;VSTM2L;ARHGEF40;PVRL2;TRIB2
SLC5A6;GFM1;ZCCHC10;TOMM40;HSPA4;ZNF628;GNB4;CWC27;GPN1;PPM1G
APOC1;SLC35F1;NDRG2
CENPW;TOMM40;PVRL2
EIF2B4;ZNF628;ZMAT5;WNT16
ZCCHC10;B3GNT7;DLEU1;TMEM253
SLC4A1AP;CCDC91;RSRC1;RSPO2;VSTM2L;SLC35F1;FAM3C;NAT14;NIPSNAP1

DNAJC1;ZCCHC10;CEP85L;RSPO2;GPR126;LCORL;CD34
EIF2B4;MLLT10;DCAF16;CCDC121;FAM174B;SKIDA1;MFN1;ZNF513;C2ORF16;ZNF512;SUPT7L;TMEM25
GJA1;ZNF628;MFN1;WNT16
DNAJC1;GPR126;PVRL2;DGKH
SLC4A1AP;CCDC91;CABP7;CMTM5;VSTM2L;SLC35F1;FAM3C
ZCCHC10;HSPA4;GNB4;AFAP1;CWC27;GPN1;DGKH;WARS2;PPM1G
SHOX2;GNB4;AFAP1;CWC27;LCORL;GPN1;WARS2
SLC4A1AP;CCDC91;RSRC1;SKIDA1;RSPO2;VSTM2L;SLC35F1;FAM3C;NAT14;NIPSNAP1
CR1L;ASCC2
FAM174B;SKIDA1;ZMAT5;ZNF513;PVRL2;TMEM253
SLC4A1AP;RSPO2;VSTM2L;SLC35F1;FAM3C;NAT14;NIPSNAP1
SHOX2;GNB4;DLEU1;AFAP1;TMEM253;WARS2
MFN1;ALX4;WNT16
SHOX2;GNB4;AFAP1;FAM3C;WARS2
FAM174B
IFI27L2;GNB4;NCAPG;CWC27;GPN1;WARS2;PPM1G
IFI27L2;DNAJC1;GNB4;DLEU1;AFAP1;ALX4;GPN1;WARS2
IFI27L2;CCDC91;CABP7;RSRC1;CMTM5;VSTM2L
LXN;CD34
EIF2B4;FAM174B;ZNF513;ASCC2;PVRL2;TRIB2;SUPT7L
DES
DCAF16;FAM174B;CEP85L;DLEU1;ZNF513;ZNF512;NAT14;MLF1;NRBP1;SUPT7L
DNAJC1;B3GNT7
CCDC121;SKIDA1;ZNF513;C2ORF16;ZNF512;SUPT7L;TMEM253
DNAJC1;HSPA4;ZNF219;ZMAT5;FAM3C;SUPT7L;TMEM253
SPEG
MTMR3;RSPO2;VSTM2L;DGKH
KIAA1755;CCDC91;CABP7;CMTM5
CABP7;FAM174B
SLC4A1AP;SKIDA1;RSPO2;VSTM2L;SLC35F1;FAM3C;NAT14;NIPSNAP1
SERPINA1;MTMR3;CR1;SNX17;ZMAT5
ALX4;CWC27
GJA1;SLC4A1AP;VSTM2L;SLC35F1;FAM3C;NDRG2;NAT14
DNAJC1;RSPO2;GPR126
FAM174B
KIAA1755;SLC4A1AP;CABP7;CMTM5;VSTM2L;SLC35F1;NDRG2
HSPA4;GNB4;AFAP1;SSC5D;GPN1;WARS2
KIAA1755;CMTM5;NDRG2
EIF2B4;ZNF628;WNT16
DCAF16;GCKR;B3GNT7;EIF3E;TRIB2
SPEG
SLC5A6;PTEN;EIF3E
PLN;NOV
MLLT10;GNB4;WNT16;LCORL
DCAF16;MFN1;ALX4;WARS2

SLC5A6;ALX4;PVRL2;CD34
TOMM40;ZNF628;WNT16
DNAJC1;CCDC121;FAM174B;SKIDA1;ZNF512;FAM3C;ARHGEF40;LCORL;SUPT7L
IFI27L2;SHOX2;GNB4;AFAP1;ALX4;WARS2
SPEG
DNAJC1;CR1;B3GNT7;GPR126;FAM3C
WNT16;PVRL2;NRBP1
SERPINA1;MTMR3;ZNF628;ASCC2
SHOX2;GNB4;UQCR10;CWC27;LCORL;GPN1;TMEM253;WARS2
MFN1;RSPO2;GPR126;ZNF512;SUPT7L
TOMM40;ZNF219;WNT16
PTEN;EIF3E
DCAF16;ZNF628;ALX4;WNT16
IFI27L2;SLC4A1AP;CCDC91;HSPA4;RSRC1;RSPO2;VSTM2L;SLC35F1;C2ORF16;LCORL;NAT14;NIPSNAP1
SHOX2;GNB4;WARS2
DNAJC1;FAM174B;FAM3C;PVRL2;TMEM253
GCKR;CR1;SNX17;VSTM2L;ARHGEF40;PVRL2;TRIB2
LXN;B3GNT7;ZNF513;TMEM253
LXN;B3GNT7;TMEM253
PLN;EIF3E
SLC4A1AP;CCDC91;RSRC1;RSPO2;VSTM2L;SLC35F1;FAM3C;WNT16;NIPSNAP1
SHOX2;AFAP1
ASCC2;NRBP1
ZCCHC10;SHOX2;GNB4;CWC27;LCORL;GPN1;WARS2
GNB4;AFAP1;C2ORF16;CWC27;LCORL;GPN1;WARS2
SERPINA1;MTMR3;CR1;ZMAT5;PTEN
PTEN;WARS2
CENPW;ASCC2;NRBP1
DCAF16;DNAJC1;SLC4A1AP;CCDC91;FAM174B;DLEU1;ZNF513;SSC5D;C2ORF16;ZNF512;NAT14;SUPT7L
GJA1;MFN1;ALX4;WNT16
PVRL2;TRIB2
GNB4;AFAP1
SLC4A1AP;CCDC91;SKIDA1;RSPO2;VSTM2L;SLC35F1;FAM3C;NAT14;NIPSNAP1
NOV
SLC5A6;DCAF16;CENPW;LXN;RSRC1
GNB4;EIF3E;WARS2
PLN;NOV
SLC4A1AP;CCDC91;SKIDA1;RSPO2;VSTM2L;SLC35F1;FAM3C
SSC5D;CD34
SLC5A6;TRIB2
DNAJC1;CCDC121;SKIDA1;DLEU1;ZNF512;FAM3C;SUPT7L;TMEM253
SKIDA1;RSPO2;VSTM2L;SLC35F1;NDRG2
DCAF16;GCKR;B3GNT7;EIF3E;ZNF512;TRIB2
KIAA1755;IFI27L2;CCDC91;CABP7;CMTM5
GFM1;ZCCHC10;HSPA4;ZNF628;GNB4;AFAP1;CWC27;GPN1;WARS2;PPM1G

GJA1;TOMM40;DLEU1;WNT16
B3GNT7;ZNF513;NIPSNAP1;TMEM253
EIF2B4;IFI27L2;B3GNT7;ZNF628;SNX17;FAM174B;VSTM2L;ZNF513;SSC5D;NAT14;SUPT7L
SHOX2;GNB4;LCORL;GPN1;TMEM253;WARS2
SPEG
C2ORF16;NRBP1
MLLT10;DCAF16;SSC5D;ZNF512
EIF2B4;WNT16;NRBP1
CCDC91;CABP7;CMTM5;VSTM2L;SLC35F1;NAT14;NIPSNAP1
SERPINA1;MTMR3;CR1;CR1L;PTEN
KIAA1755;ZMAT5;TRIB2
TBX15;CD34
SERPINA1;MTMR3;CR1;ASCC2
CMTM5;SLC35F1;UQCR10;APOE;NAT14;NIPSNAP1
ALX4;WNT16;CD34
NOV;SPEG
ZCCHC10;ZNF628;GNB4;AFAP1;CWC27;GPN1;WARS2;PPM1G
CMTM5
SERPINA1;B3GNT7;PVRL2;DGKH
SERPINA1;MTMR3;CR1;SNX17;ZMAT5;PTEN
SLC4A1AP;CABP7;RSPO2;VSTM2L;SLC35F1;NAT14;NIPSNAP1
DCAF16;MTMR3;CEP85L;MFN1;RSPO2;GPR126;DGKH
KIAA1755;CMTM5;TRIB2
GFM1;ZCCHC10;SLC4A1AP;CABP7;SKIDA1;CEP85L;PTEN;SUPT7L;MLLT10;DCAF16;RSRC1;MFN1;ZNF512
EIF2B4;DLEU1;GPR126;WNT16
SLC5A6;ALX4
GFM1;ZCCHC10;HSPA4;ZNF628;GNB4;AFAP1;CWC27;GPN1;PPM1G
NOV;SHOX2
CCDC121;GFM1;ZCCHC10;SLC4A1AP;HSPA4;CABP7;RSRC1;CEP85L;CWC27;NDRG2;NAT14
GJA1;WNT16
SHOX2;GNB4;CWC27;GPN1;DGKH;WARS2
SERPINA1;MTMR3;CR1;CR1L;ZMAT5;PTEN
KIAA1755;DCAF16;ZCCHC10;SSC5D;EIF3E;ZNF512;NAT14;NRBP1;SUPT7L
CENPW;ZCCHC10;LXN;B3GNT7;TMEM253
SPEG
IFI27L2;DCAF16;GCKR;B3GNT7;CEP85L;EIF3E;ZNF512;TRIB2;SUPT7L
FAM174B
SERPINA1;MTMR3;CR1;CR1L;PTEN
DCAF16;B3GNT7;ZNF628;FAM174B;CEP85L;VSTM2L;ZNF513;SSC5D;C2ORF16;NAT14;SUPT7L
FAM174B
SPEG
SERPINA1;MTMR3;CR1;CR1L;PTEN
RSPO2;VSTM2L;SLC35F1;NDRG2
ZCCHC10;NCAPG;NRBP1
GNB4;AFAP1;SSC5D;EIF3E;LCORL;WARS2

PTEN;EIF3E
ALX4;WNT16
RSPO2;GPR126;EIF3E;GPN1;SUPT7L
GJA1;WNT16
DNAJC1;FAM3C;SUPT7L;TMEM253
ZCCHC10;B3GNT7;TMEM253
DNAJC1;CR1;APOC1;VSTM2L;SUPT7L
IFI27L2;AFAP1;ALX4;EIF3E;WARS2
MFN1;ASCC2;WNT16
GJA1;HSPA4;ALX4;SLC35F1;NAT14;MLF1;DGKH
B3GNT7;VSTM2L;ZNF513;SUPT7L;DGKH
WNT16
MTMR3;WNT16;PVRL2
PVRL2;NRBP1
CCDC141
PLN
KIAA1755;ARHGEF40;TRIB2
FAM174B;GPR126
SERPINA1;MTMR3;CR1;ZMAT5
SNX17;FAM174B;SKIDA1;TMEM253
DNAJC1;ZNF513;FAM3C;SUPT7L;TMEM253
DLEU1;ASCC2;SUPT7L
CR1L;ASCC2
DLEU1;UQCR10;APOE;NIPSNAP1
ZNF628;WNT16
EIF2B4;GJA1;DLEU1;WNT16
ZNF628;WNT16;NRBP1
SLC4A1AP;RSPO2;VSTM2L;SLC35F1;NDRG2;NAT14
GJA1;GNB4
FAM174B
CENPW;ZMAT5;NCAPG
CR1;CR1L;SNX17;APOC1;ZNF513;ZNF512;WNT16;TRIB2;SUPT7L
NOV
HSPA4;NRBP1
B3GNT7;FAM174B
NOV
MFN1;WNT16
GPR126;ASCC2
CABP7
SHOX2;ALX4;EIF3E;TRIB2
EIF2B4;DCAF16;WNT16
DLEU1;WNT16;NRBP1
EIF2B4;LXN;FAM174B;ZNF513
TOMM40
GNB4;AFAP1;ALX4;CWC27;WARS2

CENPW;ZNF219;NCAPG
DCAF16;FAM174B;MFN1;ALX4;PVRL2;DGKH
KIAA1755;RSPO2
CMTM5;CEP85L;SLC35F1
CCDC91;RSPO2;VSTM2L;SLC35F1;FAM3C;NAT14
NOV
CR1;CR1L;SNX17;NCAPG;SSC5D;ARHGEF40;TRIB2
SLC5A6;DCAF16;ALX4;EIF3E
EIF2B4;ZNF628;WNT16
MLLT10;MTMR3;MFN1;ALX4;WNT16
ALX4;WNT16
GJA1;ZNF628;ALX4;WNT16
KIAA1755
MLLT10;DCAF16;LXN;CCDC91;EIF3E;ZNF512;SUPT7L
SERPINA1;SNX17;ASCC2
DNAJC1
DNAJC1;FAM174B;ZMAT5;FAM3C;TMEM253
ZCCHC10;SHOX2;GNB4;CWC27;LCORL;GPN1;WARS2
CR1;CR1L;SNX17;APOC1;ZNF513;ASCC2;TRIB2;SUPT7L
SERPINA1;CR1;SNX17;ZMAT5;GPN1
NOV
RSPO2
PLN
SHOX2;GNB4;AFAP1;CWC27;GPN1;WARS2
SLC4A1AP;HSPA4;CABP7;RSRC1;ZNF219;PTEN;DLEU1;CWC27;NAT14;SUPT7L
CCDC121;GJA1;CMTM5;NDRG2
CENPW;NCAPG
DCAF16;CCDC121;SKIDA1;C2ORF16;ZNF512;FAM3C;SUPT7L;TMEM253;WARS2
MLLT10;KIAA1755;DCAF16;EIF3E;ZNF512;CD34;SUPT7L;DGKH
RSPO2
EIF2B4;MTMR3;MFN1;WNT16
CCDC121;FAM174B;SKIDA1;ASCC2;ZNF512;TMEM253
TOMM40;B3GNT7;TMEM253
FAM3C
PLN;NOV
ASCC2;NRBP1
CR1;CR1L;SNX17;APOC1;MFN1;NCAPG;ZNF512;ARHGEF40;TRIB2;SUPT7L
NOV
DCAF16;CCDC121;ZNF628;SNX17;FAM174B;ZNF513;SSC5D;C2ORF16;NAT14;SUPT7L
NOV
B3GNT7;APOC1;GPR126
GNB4;ALX4;WNT16
FAM174B;SKIDA1;FAM3C;TMEM253
SERPINA1;MTMR3;CR1;CR1L;ASCC2
NOV

GFM1;ZCCHC10;HSPA4;ZNF628;GNB4;CWC27;GPN1;WARS2;PPM1G
EIF2B4;KIAA1755;FAM174B;RSPO2;PVRL2
ASCC2;NRBP1
CCDC121;GFM1;SLC4A1AP;CCDC91;RSRC1;SKIDA1;RSPO2;VSTM2L;SLC35F1;FAM3C;NIPSNAP1
ALX4;CD34
DNAJC1;RSPO2
GFM1;ZCCHC10;HSPA4;ZNF628;GNB4;AFAP1;CWC27;GPN1;WARS2;PPM1G
TBX15;CD34
LXN;B3GNT7;ZNF513;TMEM253
NOV
CR1L
CENPW;LXN;GPR126;DGKH
B3GNT7;GPR126
SERPINA1;MTMR3;CR1;CR1L;ZMAT5;PTEN
MTMR3;B3GNT7;CR1L;ASCC2
DNAJC1;ZNF628;RSPO2;ZNF513;C2ORF16;NAT14;SUPT7L
KIAA1755;CCDC121;SLC4A1AP;CCDC91;RSRC1;CMTM5;SLC35F1;MLF1
MLLT10;B3GNT7;CCDC141
CCDC121;SERPINA1;MTMR3;CR1;MFN1;PTEN;ARHGEF40;TMEM253
DNAJC1;APOC1;RSPO2;GPR126
SERPINA1;CR1
GJA1;MTMR3;WNT16
NOV
GNB4
SLC4A1AP;RSPO2;VSTM2L;SLC35F1;FAM3C;NAT14;NIPSNAP1
IFI27L2
ZNF628;ALX4;WNT16
B3GNT7;APOC1;RSPO2
DNAJC1;CR1L;PVRL2
ALX4;WNT16
B3GNT7;TMEM253
CR1;SNX17;ZMAT5;VSTM2L;ZNF513;ARHGEF40;TRIB2
EIF2B4
GJA1;WNT16
ZNF219
FAM174B
DCAF16;ALX4
FAM174B
RSRC1
SKIDA1;ZNF513;C2ORF16;SUPT7L;TMEM253
DNAJC1;FAM174B;ZNF513;FAM3C;SUPT7L;TMEM253
GJA1;WNT16
SNX17;APOC1
GJA1
GJA1;MFN1;GNB4;WNT16

GCKR;DLEU1;GPR126
SKIDA1;ZNF513;ZNF512;SUPT7L;TMEM253
B3GNT7;CABP7;ZNF513;FAM3C;LCORL;TMEM253
KIAA1755;IFI27L2;CMTM5
SLC4A1AP;RSPO2;VSTM2L;SLC35F1;C2ORF16;NAT14;NIPSNAP1
CR1;CR1L;ASCC2
DNAJC1;GPR126;CCDC141
DNAJC1;MFN1;ZNF512;WNT16
GJA1;WNT16
DLEU1;ALX4;WNT16
EIF2B4;MFN1;GNB4;GPR126;WNT16
SERPINA1;MTMR3;CR1;SLC4A1AP;CR1L;ZMAT5;PTEN;NRBP1
ZNF628;CEP85L;MLF1;NRBP1;TTN
NOV;APOC1;SPEG;APOE
ZCCHC10;HSPA4;SHOX2;CWC27;GPN1;TMEM253;WARS2;PPM1G;ZNF219;GNB4;FAM3C;LCORL;TRIB2
DES;ZNF219;SSC5D
SERPINA1;MTMR3;CR1;CR1L;PTEN;ASCC2
TBX15;GFM1;CEP85L;MYH7;TTN
DCAF16;ZNF628;ALX4;SSC5D;WNT16;CD34
TBX15;NDRG2;NRBP1;MYH7;TTN
ZCCHC10;PLN;SHOX2;SPEG;EIF3E
DNAJC1;GCKR;SERPINA1;FAM174B
TBX15;SHOX2;CD34;WARS2
KIAA1755;NOV;APOC1;APOE
MLLT10;DNAJC1;FAM174B;GPR126;DGKH
SERPINA1;MTMR3;CR1;CR1L;ASCC2
SERPINA1;MTMR3;CR1;CR1L;ZMAT5;ASCC2
KIAA1755;DNAJC1;LXN;SNX17;RSPO2;NRBP1
CABP7;CMTM5;APOC1;FAM3C;APOE
EIF2B4;GJA1;TOMM40;ZNF628;ALX4;WNT16
IFI27L2;RSPO2;ALX4
CENPW;TOMM40;HSPA4;GNB4;DLEU1;NCAPG;EIF3E;CWC27;GPN1;NRBP1;PPM1G
DNAJC1;ZCCHC10;LXN;FAM174B;GPR126;CCDC141
CENPW;TOMM40;ZMAT5;DLEU1;NCAPG;ASCC2;GPN1;PPM1G
SLC5A6;SNX17;ZNF219;PTEN;ARHGEF40
MLLT10;GJA1;MFN1;GNB4;GPR126;ALX4;WNT16;LCORL;GPN1
CENPW;ZCCHC10;CR1;TOMM40;PTEN;NCAPG;CWC27;GPN1;NIPSNAP1;WARS2;PPM1G;DCAF16;CR1L;N
DCAF16;GCKR;CCDC121;LXN;B3GNT7;SKIDA1;CEP85L;C2ORF16;EIF3E;ZNF512;TRIB2
CCDC121;MTMR3;SLC4A1AP;CEP85L;NCAPG;CWC27;PPM1G;CCDC91;ZNF628;DLEU1;ASCC2;EIF2B4;GC
DES;ZNF219;SSC5D
MLLT10;ZNF219;AFAP1;SPEG;SSC5D
DNAJC1;SHOX2;GNB4;EIF3E;FAM3C;LCORL
EIF2B4;GJA1;DES
DNAJC1;FAM174B;EIF3E;FAM3C
MLLT10;PLN;NOV;SPEG

GCKR;SERPINA1;SKIDA1;APOC1;GPR126;APOE;NIPSNAP1
PLN;CCDC141;MLF1;MYH6
DNAJC1;GCKR;SERPINA1;B3GNT7;FAM174B;PVRL2
EIF2B4;KIAA1755;CMTM5;CEP85L;ZMAT5;MFN1;GPR126;ARHGEF40
CCDC121;MTMR3;ZCCHC10;SLC4A1AP;CEP85L;NCAPG;CWC27;PPM1G;CCDC91;ZNF628;DLEU1;EIF2B4;
GFM1;CCDC141;MLF1;MYH7;TTN
ZCCHC10;RSRC1;GNB4;PTEN;FAM3C;CD34;WARS2
CCDC121;SLC4A1AP;CEP85L;NCAPG;UQCR10;CWC27;PPM1G;CCDC91;ZNF628;DLEU1;ASCC2;EIF2B4;GC
GJA1;NOV;SPEG;CD34
DES;B3GNT7;ZNF219;TMEM253
CENPW;LXN;TOMM40;B3GNT7;NCAPG;NIPSNAP1;TMEM253;PPM1G
EIF2B4;TBX15;UQCR10;MYH6;MYH7;TTN
GFM1;CENPW;LXN;TOMM40;B3GNT7;DLEU1;NCAPG;UQCR10;TMEM253;PPM1G
NOV;GNB4;AFAP1;SSC5D
TBX15;GFM1;MYH6;MYH7;TTN
IFI27L2;CENPW;TOMM40;SNX17;ZNF219;ZNF513;ZNF512;CWC27;NAT14;NRBP1;SUPT7L;PPM1G
SLC5A6;LXN;ALX4;CD34
KIAA1755;CMTM5;CEP85L;SLC35F1;NDRG2;TRIB2
CCDC121;SLC4A1AP;CEP85L;NCAPG;CWC27;PPM1G;CCDC91;ZNF628;DLEU1;ASCC2;EIF2B4;GCKR;CENP
GFM1;CENPW;ZCCHC10;TOMM40;HSPA4;PTEN;NCAPG;CWC27;GPN1;NIPSNAP1;WARS2;PPM1G;DCAF
DNAJC1;ZCCHC10;LXN;SNX17;AFAP1;GPR126;FAM3C
NOV;APOC1;SPEG;APOE
DCAF16;ALX4;WNT16;CD34
GFM1;CENPW;ZCCHC10;TOMM40;HSPA4;NCAPG;CWC27;GPN1;NIPSNAP1;PPM1G;SLC5A6;DCAF16;CR
CCDC121;SLC4A1AP;CEP85L;NCAPG;CWC27;PPM1G;CCDC91;ZNF628;DLEU1;ASCC2;EIF2B4;GCKR;CENP
KIAA1755;ZMAT5;SSC5D;ZNF512;ARHGEF40;NAT14;NRBP1
SLC5A6;LXN;CD34
KIAA1755;IFI27L2;CMTM5;APOC1;RSPO2;APOE;NDRG2
LXN;APOC1;GPR126;SSC5D;WNT16;CD34;TRIB2
PLN;NOV;RSRC1;AFAP1;SPEG;ZNF512
KIAA1755;IFI27L2;CABP7;CMTM5;NDRG2
SERPINA1;MTMR3;CR1;CR1L;ASCC2;PVRL2
CCDC121;MTMR3;SLC4A1AP;CEP85L;NCAPG;CWC27;PPM1G;CCDC91;ZNF628;DLEU1;ASCC2;EIF2B4;GC
GCKR;SERPINA1;SKIDA1;APOC1;APOE;NIPSNAP1
IFI27L2;GNB4;AFAP1;SSC5D;EIF3E
KIAA1755;SNX17;ZMAT5
MLLT10;DCAF16;GCKR;CCDC121;LXN;B3GNT7;CEP85L;EIF3E;ZNF512;CWC27;TRIB2;NRBP1
IFI27L2;GNB4;SSC5D;EIF3E;WARS2
KIAA1755;CABP7;CMTM5;APOC1;APOE;NDRG2;NAT14
MLLT10;LXN;SSC5D;ZNF512
SERPINA1;MTMR3;CR1;CR1L;ASCC2
CENPW;TOMM40;HSPA4;GNB4;DLEU1;AFAP1;NCAPG;SSC5D;CWC27;GPN1;PPM1G
PLN;NOV;SPEG
TBX15;SHOX2;ZNF219;CD34
SLC4A1AP;CCDC91;APOC1;VSTM2L;SLC35F1;UQCR10;APOE;CWC27;NDRG2;NAT14;MLF1

EIF2B4;GCKR;LXN;NOV;SNX17;SPEG
EIF2B4;MLLT10;RSPO2;DGKH
SLC5A6;IFI27L2;CCDC91;CABP7;CMTM5;APOC1;RSPO2;VSTM2L;SLC35F1;APOE
CENPW;TOMM40;HSPA4;AFAP1;NCAPG;CWC27;GPN1;PPM1G;DNAJC1;SNX17;GNB4;DLEU1;ALX4
KIAA1755;CMTM5;ZMAT5;SLC35F1;NDRG2;ARHGEF40;TRIB2
KIAA1755;IFI27L2;CCDC121;GJA1;CMTM5;SLC35F1;NDRG2
CCDC121;SLC4A1AP;CEP85L;NCAPG;CWC27;PPM1G;CCDC91;ZNF628;DLEU1;ASCC2;EIF2B4;GCKR;CENP
SERPINA1;MTMR3;CR1;CR1L;ASCC2
DES;ZNF219;SSC5D
KIAA1755;DES;ZNF219;SSC5D
CCDC91;FAM174B;GPR126;DGKH
GFM1;CENPW;ZCCHC10;TOMM40;HSPA4;NCAPG;C2ORF16;CWC27;GPN1;NIPSNAP1;WARS2;PPM1G;C
SLC5A6;SNX17;WNT16;CD34
CCDC121;PTEN;ALX4;EIF3E;WARS2
KIAA1755;ZNF219;RSPO2;SSC5D
CCDC121;MTMR3;ZCCHC10;SLC4A1AP;CEP85L;NCAPG;CWC27;PPM1G;CCDC91;ZNF628;DLEU1;EIF2B4;
DNAJC1;LXN;TOMM40;GPR126;SSC5D;DGKH
B3GNT7;APOC1;RSPO2;GPR126;CCDC141
IFI27L2;DCAF16;GCKR;LXN;CEP85L;EIF3E;ZNF512;APOE;TRIB2;SUPT7L;WARS2
CENPW;MFN1;DLEU1;GPR126;NCAPG;NRBP1
B3GNT7;ZNF219;SSC5D;TMEM253
TBX15;SPEG;NDRG2;MYH6;MYH7;TTN
GJA1;DES;ZNF219
EIF2B4;GFM1;DES;TOMM40;MYH6;TTN;PPM1G
KIAA1755;CCDC121;ALX4
SERPINA1;TOMM40;B3GNT7;GPR126;PVRL2;PPM1G
CMTM5;APOC1;SLC35F1;APOE;NDRG2;NAT14
IFI27L2;GJA1;SLC35F1;NDRG2
SERPINA1;MTMR3;CR1;CR1L;SNX17;ZMAT5;PTEN
SLC5A6;ZCCHC10;APOC1;ALX4;APOE;CD34
TOMM40;HSPA4;MFN1;DLEU1;GPR126;ASCC2;NRBP1
GJA1;ZCCHC10;NOV;SPEG
CCDC121;MTMR3;ZCCHC10;SLC4A1AP;CEP85L;NCAPG;CWC27;PPM1G;CCDC91;ZNF628;DLEU1;ASCC2;
SLC5A6;EIF2B4;KIAA1755;LXN;ZNF628;ZNF513;PVRL2;SUPT7L
CCDC121;SLC4A1AP;CEP85L;NCAPG;CWC27;PPM1G;CCDC91;ZNF628;DLEU1;EIF2B4;GCKR;CENPW;HSP
CENPW;TOMM40;HSPA4;SNX17;GNB4;DLEU1;NCAPG;EIF3E;CWC27;LCORL;GPN1;PPM1G
EIF2B4;GCKR;CCDC121;CENPW;SLC4A1AP;HSPA4;SKIDA1;CEP85L;NCAPG;C2ORF16;CWC27;GPN1;MLF1
MLLT10;DES;SSC5D;CD34
SERPINA1;MTMR3;CR1;CR1L;ZMAT5;TMEM253
IFI27L2;CENPW;TOMM40;DLEU1;NCAPG;ASCC2;SUPT7L;DGKH
MLLT10;FAM174B;CEP85L;GPR126;ASCC2;ZNF512;DGKH
MLLT10;PLN;NOV;SPEG
EIF2B4;CMTM5;SKIDA1;SHOX2;CEP85L;SUPT7L;KIAA1755;DCAF16;MFN1;GPR126;ZNF512;ARHGEF40;T
DNAJC1;GCKR;B3GNT7;ZNF513;C2ORF16;PVRL2
HSPA4;DLEU1;ASCC2;GPN1;NRBP1

EIF2B4;DES;ZNF219;SSC5D
DES;SSC5D;CD34
CENPW;TOMM40;DLEU1;GPR126;NCAPG;ASCC2;NRBP1
MLLT10;CCDC91;ZNF628;GNB4;DGKH
KIAA1755;CMTM5;CEP85L;ZMAT5;SLC35F1;ZNF512;ARHGEF40;TRIB2
PLN;NOV;SPEG
DNAJC1;CCDC121;SNX17;FAM174B;SKIDA1;ZNF219;FAM3C;ARHGEF40;CD34;TMEM253
EIF2B4;MLLT10;GJA1;ZNF628;ZNF219;MFN1;ALX4;WNT16
UQCR10;NDRG2;MLF1;MYH7;TTN
SHOX2;ALX4;SSC5D;ARHGEF40;CD34
PLN;NOV;SPEG
GFM1;CENPW;ZCCHC10;CR1;HSPA4;PTEN;NCAPG;CWC27;GPN1;NIPSNAP1;PPM1G;MLLT10;DCAF16;C
CR1;CR1L;ASCC2
PLN;NOV;RSRC1;GNB4;AFAP1
DNAJC1;GCKR;B3GNT7;FAM174B;ZNF513;ASCC2;C2ORF16;PVRL2
TBX15;ZCCHC10;SHOX2;EIF3E;CD34
SHOX2;GNB4;C2ORF16;EIF3E;FAM3C;LCORL
IFI27L2;GNB4;AFAP1;NCAPG;ALX4;EIF3E;NIPSNAP1
TBX15;DES;NDRG2;MYH7;TTN
CCDC121;GFM1;MTMR3;ZCCHC10;SLC4A1AP;CEP85L;NCAPG;CWC27;PPM1G;CCDC91;ZNF628;DLEU1;
DNAJC1;SNX17;FAM174B;SKIDA1;ZMAT5;PVRL2;TMEM253;PPM1G
IFI27L2;GFM1;CENPW;ZCCHC10;TOMM40;HSPA4;PTEN;NCAPG;CWC27;GPN1;NIPSNAP1;PPM1G;CR1L;
KIAA1755;GCKR;NOV;SPEG
SLC5A6;GCKR;SERPINA1;APOC1;GPR126;APOE;NIPSNAP1
CCDC121;PLN;NOV;SPEG
IFI27L2;LXN;RSRC1;SHOX2;C2ORF16;EIF3E
TBX15;ZNF628;MYH6;MYH7;TTN
RSPO2;SPEG;SSC5D
GJA1;SHOX2;GNB4;AFAP1;FAM3C
KIAA1755;PLN;SPEG
CMTM5;SHOX2;CEP85L;GPR126;SLC35F1;ARHGEF40;TRIB2
NOV;SPEG
CCDC121;MTMR3;ZCCHC10;SLC4A1AP;CEP85L;NCAPG;CWC27;PPM1G;CCDC91;ZNF628;DLEU1;EIF2B4;
KIAA1755;IFI27L2;CCDC91;CMTM5;RSPO2;VSTM2L;SLC35F1;NDRG2;LCORL
DES;SPEG;MYH7;TTN
PLN;NOV;RSRC1;AFAP1
VSTM2L;CCDC141;MLF1;MYH6
SERPINA1;MTMR3;CR1;CR1L;ASCC2
DES;CABP7;ZNF219;SSC5D
TBX15;GFM1;MYH6;MYH7;TTN
EIF2B4;IFI27L2;LXN;SHOX2;ZMAT5;TRIB2;SUPT7L
SERPINA1;MTMR3;CR1;CR1L;PTEN;ASCC2
SLC5A6;GFM1;SNX17;APOC1
SHOX2;GNB4;AFAP1;SSC5D;C2ORF16;WARS2
SERPINA1;B3GNT7;APOC1;GPR126;CCDC141

SERPINA1;MTMR3;CR1;CR1L;CMTM5;ZMAT5;PTEN;TMEM253
CENPW;ZCCHC10;HSPA4;NCAPG;C2ORF16;WNT16;CWC27;GPN1;NIPSNAP1;WARS2;PPM1G;CR1L;MFN
CCDC121;SHOX2;CD34;WARS2
SERPINA1;MTMR3;CR1;CR1L;PTEN;ASCC2
DES;MYH6;MYH7
SERPINA1;MTMR3;CR1;CR1L;PTEN;ASCC2
EIF2B4;KIAA1755;LXN;ZNF628;RSPO2;ZNF513;ZNF512;SUPT7L
IFI27L2;CENPW;TOMM40;ZNF219;DLEU1;NCAPG;TMEM253;PPM1G
SERPINA1;MTMR3;CR1;CR1L;PTEN;ASCC2;ARHGEF40;TMEM253
GCKR;B3GNT7;FAM174B;FAM3C
DES;GNB4;AFAP1
DES;ZNF219;SPEG
SERPINA1;MTMR3;CR1;CR1L;PTEN
PLN;NOV;SPEG
DES;UQCR10;NDRG2;NRBP1;MYH6;TTN
SERPINA1;MTMR3;CR1;CR1L;SNX17;ZMAT5;PTEN;NCAPG;ASCC2
RSRC1;ALX4;CD34;WARS2
LXN;TOMM40;B3GNT7;ZNF513;UQCR10;NIPSNAP1;TMEM253;PPM1G
IFI27L2;CABP7;VSTM2L
LXN;TOMM40;B3GNT7;DLEU1;NCAPG;ZNF513;TMEM253
GJA1;DES;ZNF219;GNB4
CENPW;ZCCHC10;PTEN;WNT16;CWC27
DES;GNB4;AFAP1
CENPW;AFAP1;SSC5D;NRBP1
NOV;SHOX2;SPEG;DGKH
GFM1;CENPW;ZCCHC10;TOMM40;HSPA4;NCAPG;C2ORF16;CWC27;GPN1;NIPSNAP1;NRBP1;PPM1G;D[illegible]
DES;ZNF219;AFAP1
DNAJC1;ZCCHC10;LXN;CR1L;MFN1;RSPO2;GPR126;ZNF512;FAM3C;TRIB2
EIF2B4;GCKR;CCDC121;CENPW;SLC4A1AP;SKIDA1;CEP85L;AFAP1;NCAPG;C2ORF16;CWC27;GPN1;MLF1
DES;ZNF219;AFAP1
GFM1;ZCCHC10;SLC4A1AP;CCDC91;RSRC1;RSPO2;VSTM2L;SLC35F1;C2ORF16;FAM3C;NDRG2
KIAA1755;DCAF16;CMTM5;CEP85L;SLC35F1;ZNF512;ARHGEF40;LCORL;CD34;TRIB2
MLLT10;GFM1;RSRC1;PTEN;EIF3E;LCORL;CD34
IFI27L2;CENPW;ZCCHC10;PTEN;NCAPG;C2ORF16;CWC27;GPN1;NIPSNAP1;PPM1G;DCAF16;CR1L;RSRC1
SNX17;ZNF219;ALX4;ARHGEF40;LCORL;CD34
IFI27L2;SHOX2;GNB4;EIF3E;LCORL;WARS2
ZNF219;EIF3E;CD34;WARS2
LXN;B3GNT7;GPR126;CCDC141
KIAA1755;NOV;SPEG
PLN;RSRC1;SPEG;DGKH
IFI27L2;CENPW;HSPA4;GNB4;DLEU1;NCAPG;EIF3E;CWC27;GPN1
PLN;NOV;RSRC1;SPEG
CCDC121;SHOX2;CEP85L;MFN1;GPR126;SLC35F1;ARHGEF40;LCORL;TRIB2
CCDC121;CCDC91;CMTM5;VSTM2L;SLC35F1;NDRG2
CCDC121;MTMR3;ZCCHC10;SLC4A1AP;CEP85L;NCAPG;CWC27;PPM1G;CCDC91;ZNF628;DLEU1;ALX4;EI

TBX15;DCAF16;GFM1;MYH6;MYH7;TTN
GCKR;B3GNT7;RSPO2
KIAA1755;IFI27L2;CABP7;CMTM5;SLC35F1;NDRG2
EIF2B4;GCKR;CCDC121;CENPW;MTMR3;SLC4A1AP;HSPA4;SKIDA1;CEP85L;NCAPG;C2ORF16;GPN1;MLF
SHOX2;GNB4;AFAP1;EIF3E;LCORL;DGKH;WARS2
PLN;NOV;SHOX2;SPEG
PLN;CCDC91;NOV;RSRC1;SPEG;EIF3E
GCKR;RSPO2;GPR126;SSC5D
SERPINA1;MTMR3;CR1;CR1L;ASCC2
DES;GNB4;SPEG
CCDC121;GJA1;PLN;NOV
EIF2B4;GCKR;CCDC121;CENPW;SLC4A1AP;HSPA4;SKIDA1;CEP85L;NCAPG;C2ORF16;CWC27;GPN1;MLF1
CCDC121;GJA1;CCDC91;CMTM5;SLC35F1;APOE;NDRG2
SERPINA1;LXN;B3GNT7;ZNF513;FAM3C;LCORL;TMEM253
DCAF16;GJA1;MFN1;ALX4;WNT16;CD34
ZCCHC10;PLN;NOV;SPEG
CMTM5;APOE;NDRG2
MLLT10;PLN;NOV;RSRC1;ZNF512
KIAA1755;CCDC121;CCDC91;CABP7;CMTM5;SLC35F1;APOE;NDRG2
TBX15;CEP85L;MYH6;MYH7;TTN
IFI27L2;CABP7;CMTM5;APOC1
B3GNT7;ZNF628;FAM174B;GPR126;CCDC141
SLC5A6;DNAJC1;SERPINA1;LXN;TOMM40;B3GNT7;ZNF513;PVRL2;GPN1;PPM1G
KIAA1755;IFI27L2;CCDC91;CMTM5;VSTM2L;SLC35F1;APOE;NDRG2;NAT14
IFI27L2;GNB4;SSC5D;EIF3E;LCORL;WARS2
SERPINA1;MTMR3;CR1;CR1L;PTEN;ASCC2
RSRC1;SHOX2;MFN1;CD34;DGKH
DNAJC1;GCKR;B3GNT7;FAM174B;WARS2
SERPINA1;MTMR3;CR1;CR1L;APOC1
PTEN;ALX4;CD34
EIF2B4;TBX15;GFM1;DES;HSPA4;ZMAT5;TTN
ZCCHC10;PLN;NOV;RSRC1
SERPINA1;MTMR3;CR1;CR1L;PTEN;ASCC2
MLLT10;NOV;MFN1;SPEG;ZNF512;CCDC141
EIF2B4;TBX15;DES;MYH7;TTN;PPM1G
ZCCHC10;TOMM40;SHOX2;SSC5D;CWC27;GPN1;NRBP1;WARS2;PPM1G;ZNF628;GNB4;ALX4;TRIB2
SHOX2;GNB4;AFAP1;C2ORF16;EIF3E
KIAA1755;CABP7;CMTM5;APOC1;APOE;NDRG2;NAT14
DES;ZNF219;RSPO2
DNAJC1;ZCCHC10;LXN;B3GNT7;APOC1;NCAPG;PVRL2;GPN1;PPM1G
NOV;RSRC1;AFAP1;SPEG
KIAA1755;CCDC91;CMTM5;NDRG2
SERPINA1;MTMR3;CR1;CR1L;ASCC2
CCDC121;MTMR3;ZCCHC10;SLC4A1AP;CEP85L;NCAPG;CWC27;PPM1G;CCDC91;ZNF628;DLEU1;ASCC2;I
IFI27L2;CENPW;TOMM40;HSPA4;SNX17;GNB4;NCAPG;EIF3E;CWC27;GPN1;PPM1G

GCKR;SERPINA1;SKIDA1;APOC1;GPR126;C2ORF16;APOE
IFI27L2;SHOX2;GNB4;AFAP1;NCAPG;EIF3E;LCORL;NIPSNAP1
KIAA1755;DCAF16;CMTM5;CEP85L;SLC35F1;ZNF512;ARHGEF40;TRIB2
EIF2B4;CCDC121;GFM1;CENPW;MTMR3;SLC4A1AP;HSPA4;SKIDA1;CEP85L;NCAPG;C2ORF16;GPN1;ML[illegible]
SERPINA1;MTMR3;CR1;CMTM5;ZMAT5;ASCC2;ARHGEF40;TMEM253
PLN;NOV;SPEG
EIF2B4;DCAF16;CMTM5;ZMAT5;SLC35F1;ZNF512;GPN1
CCDC121;ZCCHC10;SLC4A1AP;CEP85L;NCAPG;CWC27;PPM1G;CCDC91;ZNF628;DLEU1;ASCC2;EIF2B4;G[illegible]
DNAJC1;GNB4;AFAP1;C2ORF16;FAM3C
CCDC121;SHOX2;SSC5D;CD34;WARS2
CCDC121;MTMR3;ZCCHC10;SLC4A1AP;CEP85L;NCAPG;CWC27;PPM1G;CCDC91;ZNF628;ASCC2;EIF2B4;[illegible]
DNAJC1;B3GNT7;APOC1;GPR126;CCDC141
PLN;NOV;RSRC1;AFAP1
PLN;NOV;RSRC1;SPEG
DCAF16;GJA1;MFN1;GNB4;DLEU1;GPR126;ALX4;WNT16
TBX15;DES;NDRG2;CCDC141;MYH7;TTN
EIF2B4;SNX17;ZNF219;ARHGEF40;TRIB2
TBX15;GFM1;CEP85L;NDRG2;CCDC141;NRBP1;TTN
EIF2B4;KIAA1755;SHOX2;CEP85L;TRIB2;SUPT7L
DNAJC1;GCKR;SERPINA1;FAM174B;MFN1;PVRL2;GPN1;WARS2;PPM1G
MLLT10;CCDC121;ZCCHC10;ALX4;LCORL;PVRL2;CD34
EIF2B4;GCKR;CCDC121;CENPW;SLC4A1AP;HSPA4;SKIDA1;CEP85L;NCAPG;C2ORF16;CWC27;GPN1;MLF1
SLC5A6;TOMM40;SNX17;PTEN
GJA1;DES;GNB4
ZNF219;DLEU1;AFAP1;SSC5D
MLLT10;CCDC121;ZCCHC10;HSPA4;AFAP1;EIF3E;FAM3C;TRIB2
PLN;NOV;AFAP1;SPEG
PLN;NOV;RSRC1;GNB4;AFAP1;DGKH
SLC5A6;MLLT10;DCAF16;CCDC121;ALX4;C2ORF16;EIF3E;ZNF512
DNAJC1;SERPINA1;ZCCHC10;LXN;TOMM40;B3GNT7;FAM3C
DES;CEP85L;MYH6;MYH7;TTN
MLLT10;ZNF219;SPEG;SSC5D
CABP7;RSPO2;VSTM2L;SLC35F1;NDRG2
SERPINA1;CR1L;ASCC2
SLC5A6;TBX15;TOMM40;ZNF219;PTEN
ZNF219;GPR126;SSC5D
GJA1;NOV;HSPA4;FAM3C
GJA1;APOC1;APOE;NDRG2
GCKR;B3GNT7;FAM174B;FAM3C;PVRL2
CCDC121;MTMR3;SLC4A1AP;CEP85L;NCAPG;CWC27;PPM1G;CCDC91;ZNF628;DLEU1;ASCC2;EIF2B4;GC
IFI27L2;CCDC121;GJA1;APOC1;ALX4;SLC35F1;APOE;NDRG2;MLF1
SERPINA1;MTMR3;CR1;CR1L;CMTM5;ZMAT5;TMEM253
MLLT10;DCAF16;GCKR;CCDC121;LXN;CEP85L;C2ORF16;EIF3E;ZNF512;TRIB2
SLC5A6;AFAP1;ARHGEF40;CD34
APOC1;APOE;NDRG2

AFAP1;SPEG;SSC5D;ZNF512;WNT16
TBX15;CR1L;SHOX2;CD34;WARS2
LXN;AFAP1;ALX4;PVRL2;CD34;TRIB2
TBX15;DES;ZNF628;CEP85L;NDRG2;MYH6;TTN
CCDC121;SLC4A1AP;CEP85L;NCAPG;CWC27;PPM1G;CCDC91;ZNF628;DLEU1;ASCC2;EIF2B4;GCKR;CENP
SHOX2;CEP85L;MFN1;GPR126;ZNF512;TRIB2;WARS2
NOV;RSPO2;AFAP1;SSC5D;CCDC141;DGKH
EIF2B4;KIAA1755;CMTM5;SHOX2;CEP85L;ARHGEF40;CD34;TRIB2;SUPT7L;WARS2
ZCCHC10;LXN;ALX4;SSC5D;CD34;TRIB2;WARS2
DNAJC1;GCKR;FAM174B;WARS2
TBX15;GFM1;DES;HSPA4;FAM174B;CCDC141;NRBP1;TTN
DES;ZNF219;AFAP1;SPEG;SSC5D
PLN;NOV;SPEG;ALX4
DES;ZNF219;DLEU1;SSC5D
CABP7;APOC1;APOE;NDRG2
DNAJC1;GCKR;CENPW;B3GNT7;NCAPG;ASCC2;C2ORF16;UQCR10;PVRL2
SLC5A6;GCKR;SERPINA1;APOC1;GPR126;APOE;PVRL2;NIPSNAP1
CENPW;AFAP1;SSC5D;EIF3E;CD34;NRBP1;DGKH
GCKR;GFM1;GJA1;LXN;TOMM40;SNX17;APOC1;C2ORF16;APOE;ARHGEF40;NIPSNAP1
TBX15;IFI27L2;SNX17;SHOX2;TRIB2
GFM1;CENPW;ZCCHC10;SLC4A1AP;TOMM40;HSPA4;NCAPG;C2ORF16;CWC27;GPN1;NIPSNAP1;WARS2
GFM1;CCDC141;MYH6;MYH7;TTN
DNAJC1;DES;ZNF219;AFAP1;NRBP1
ZCCHC10;TOMM40;HSPA4;DLEU1;ASCC2;PVRL2;NRBP1;PPM1G
GCKR;SERPINA1;SKIDA1;APOC1;GPR126;C2ORF16;APOE;NIPSNAP1
TBX15;DES;UQCR10;MYH7;TTN
SLC5A6;SNX17;ZNF219;PTEN;CD34
GJA1;SHOX2;GNB4;AFAP1;NCAPG;FAM3C;LCORL;DGKH;WARS2
SERPINA1;ZCCHC10;LXN;B3GNT7;GPR126;CCDC141
EIF2B4;KIAA1755;DNAJC1;LXN;PTEN;RSPO2;ZNF513;ZNF512;NRBP1;SUPT7L
DES;ZNF219;AFAP1;SSC5D
CCDC121;RSRC1;SHOX2;GNB4;ALX4;EIF3E
NOV;SHOX2;AFAP1;SSC5D;CD34
GJA1;CABP7;APOC1;SLC35F1;APOE;NDRG2
EIF2B4;GJA1;ZNF219;RSPO2
GFM1;CENPW;ZCCHC10;LXN;TOMM40;HSPA4;NCAPG;NIPSNAP1;TMEM253;PPM1G;B3GNT7;DLEU1;LC
DNAJC1;GCKR;B3GNT7;FAM174B;NCAPG;C2ORF16;PVRL2;PPM1G
KIAA1755;CMTM5;APOC1;APOE;NDRG2;NAT14
IFI27L2;GFM1;CENPW;ZCCHC10;TOMM40;HSPA4;NCAPG;CWC27;GPN1;NRBP1;WARS2;PPM1G;CR1L;SI
TBX15;DES;UQCR10;MYH6;MYH7;TTN
DNAJC1;GCKR;B3GNT7;FAM174B;C2ORF16;PVRL2
GFM1;CENPW;ZCCHC10;TOMM40;HSPA4;NCAPG;CWC27;GPN1;NIPSNAP1;WARS2;PPM1G;CR1L;RSRC1
CCDC121;MTMR3;ZCCHC10;SLC4A1AP;CEP85L;NCAPG;CWC27;PPM1G;CCDC91;ZNF628;DLEU1;EIF2B4;
DNAJC1;SERPINA1;TOMM40;B3GNT7;CR1L;SNX17;CCDC141
CCDC121;MTMR3;ZCCHC10;SLC4A1AP;CEP85L;NCAPG;CWC27;PPM1G;CCDC91;ZNF628;DLEU1;ASCC2;

TBX15;UQCR10;MYH6;MYH7;TTN
EIF2B4;FAM174B;ZMAT5;NDRG2;MYH6;TTN;PPM1G
MLLT10;DNAJC1;AFAP1;SPEG
KIAA1755;CCDC121;CMTM5;CEP85L;GPR126;SLC35F1;ARHGEF40
EIF2B4;KIAA1755;CMTM5;SHOX2;SLC35F1;ARHGEF40;TRIB2
GFM1;UQCR10;CCDC141;MYH6;MYH7;TTN
SERPINA1;TOMM40;B3GNT7;FAM174B;APOC1;VSTM2L;CCDC141
CENPW;ZCCHC10;CR1;TOMM40;PTEN;NCAPG;C2ORF16;CWC27;GPN1;NIPSNAP1;WARS2;PPM1G;DCAF
UQCR10;CCDC141;MYH6;MYH7;TTN
GJA1;SHOX2;GNB4;AFAP1;SSC5D;EIF3E;FAM3C
MLLT10;KIAA1755;ZNF219;ALX4;SSC5D;EIF3E;ZNF512;NAT14;SUPT7L
ZNF219;RSPO2;SSC5D;NRBP1
KIAA1755;DCAF16;CMTM5;SKIDA1;SHOX2;CEP85L;GPR126;SLC35F1;ZNF512;ARHGEF40;TRIB2
DES;ZNF219;SSC5D;CD34;NAT14
KIAA1755;IFI27L2;CMTM5;ZMAT5;ARHGEF40
CCDC121;MTMR3;ZCCHC10;SLC4A1AP;CEP85L;NCAPG;CWC27;PPM1G;CCDC91;ZNF628;DLEU1;EIF2B4;
SLC5A6;CR1;TOMM40;PTEN;AFAP1;CD34
GCKR;SERPINA1;SKIDA1;APOC1;GPR126;C2ORF16;APOE;ARHGEF40;NIPSNAP1
PTEN;RSPO2;SSC5D;EIF3E;ZNF512;ARHGEF40
TBX15;GFM1;ZCCHC10;HSPA4;NRBP1;MYH6;MYH7;TTN
TBX15;PLN;NOV;SPEG
IFI27L2;LXN;SHOX2;GNB4;EIF3E;FAM3C;APOE;WARS2
SLC5A6;GCKR;SERPINA1;APOC1;GPR126;APOE;NIPSNAP1
IFI27L2;CABP7;CMTM5;APOC1
CCDC121;CMTM5;APOC1;APOE;NDRG2
EIF2B4;GJA1;TOMM40;ZNF628;ALX4;WNT16
GJA1;DES;ZNF219
CENPW;ZCCHC10;TOMM40;DLEU1;GPR126;NCAPG;ASCC2;NRBP1
GFM1;CENPW;ZCCHC10;TOMM40;HSPA4;PTEN;NCAPG;CWC27;GPN1;NIPSNAP1;WARS2;PPM1G;SLC5A
CCDC121;PLN;NOV;SPEG;CCDC141
APOC1;SUPT7L
MTMR3;CR1;CR1L;SNX17
SLC4A1AP;CABP7;NDRG2;NAT14
MLLT10;CABP7;NDRG2;LCORL;NAT14;SUPT7L
DCAF16;GFM1;SLC4A1AP;CABP7;RSRC1;CEP85L;MFN1;ZNF512;NDRG2
CABP7;DLEU1
KIAA1755;CCDC91;CMTM5
MTMR3;ASCC2;GPN1
ASCC2
GFM1;SLC4A1AP;CABP7;RSRC1;MFN1;NAT14;NIPSNAP1;SUPT7L
ZNF628;WNT16
SLC4A1AP;VSTM2L;SLC35F1;NAT14;NIPSNAP1
MTMR3;DGKH
HSPA4;SLC35F1;MLF1;DGKH
GFM1;SLC4A1AP;CABP7;RSRC1;MFN1;NAT14

SLC4A1AP;CABP7;RSRC1;CEP85L;DLEU1;NAT14;SUPT7L
SLC4A1AP;CABP7;RSRC1;CEP85L;DLEU1;NAT14;SUPT7L
GFM1;SLC4A1AP;RSRC1;MFN1;NAT14;SUPT7L
SLC4A1AP;CABP7;RSRC1;NAT14;SUPT7L
SUPT7L;TMEM253
FAM174B;ZNF513;NAT14;SUPT7L
CABP7;NDRG2;NAT14;SUPT7L
GCKR;NOV;SPEG
EIF2B4;MLLT10;DES;ZNF628;ZNF219;SPEG;NRBP1
IFI27L2;CENPW;SSC5D;WNT16;NRBP1
GFM1;SLC4A1AP;CABP7;MFN1;NAT14
GFM1;SLC4A1AP;CABP7;ZNF219;NAT14
DCAF16
CCDC91;SLC35F1
EIF2B4;FAM174B;ZNF513;NAT14;SUPT7L
DES;ZNF219;SSC5D;CD34;NAT14
CMTM5;SKIDA1;SHOX2;CEP85L;ZMAT5;ZNF512;TRIB2;SUPT7L
RSPO2;GPR126
TOMM40
DCAF16;LXN;ZNF512;SUPT7L
MLLT10;CABP7;RSRC1;NAT14
CABP7;NAT14;SUPT7L
MTMR3;CR1
PLN;NOV;SHOX2;AFAP1;SPEG
SLC4A1AP;CABP7;RSRC1;DLEU1;NAT14;SUPT7L
CR1;SNX17;VSTM2L;TRIB2
KIAA1755;CMTM5;SHOX2;CEP85L;ZMAT5;NDRG2;ARHGEF40;TRIB2;SUPT7L
CENPW;TOMM40;HSPA4;SNX17;GNB4;NCAPG;FAM3C;CWC27;GPN1;NIPSNAP1;NRBP1;PPM1G
KIAA1755;CCDC121;LXN;RSPO2;CD34
DNAJC1;GCKR;SERPINA1;CABP7;FAM174B
CCDC121;MTMR3;ZCCHC10;SLC4A1AP;CEP85L;NCAPG;CWC27;PPM1G;CCDC91;ZNF628;DLEU1;EIF2B4;
LXN;RSRC1;SHOX2;PTEN;TRIB2;WARS2
DNAJC1;GCKR;B3GNT7;FAM174B;CEP85L;MFN1;LCORL;PVRL2;TRIB2
MLLT10;FAM174B;ZNF512;SUPT7L
GFM1;CENPW;ZCCHC10;TOMM40;PTEN;NCAPG;CWC27;GPN1;NIPSNAP1;WARS2;PPM1G;DCAF16;CR1
SLC5A6;EIF2B4;DNAJC1;GJA1;LXN;TOMM40;ZNF513;FAM3C;PPM1G
GFM1;CENPW;ZCCHC10;HSPA4;SHOX2;NCAPG;C2ORF16;CWC27;GPN1;NIPSNAP1;PPM1G;DCAF16;DN
MLLT10;KIAA1755;DCAF16;DNAJC1;EIF3E;FAM3C;WNT16;NRBP1
CENPW;HSPA4;SHOX2;GNB4;AFAP1;NCAPG;EIF3E;FAM3C;CWC27
TBX15;NOV;SHOX2;SPEG;DGKH
KIAA1755;PLN;NOV;AFAP1;SPEG;DGKH
IFI27L2;GJA1;APOE;NDRG2
GFM1;SLC4A1AP;CABP7;RSRC1;NAT14
SKIDA1;SUPT7L;TMEM253
GFM1;SLC4A1AP;CABP7;RSRC1;SKIDA1;NAT14

ASCC2
MLLT10;TOMM40;RSRC1;DLEU1;NAT14;SUPT7L
GFM1;SLC4A1AP;CABP7;RSRC1;NAT14;NIPSNAP1;SUPT7L
GFM1;SLC4A1AP;CABP7;NDRG2;NAT14;SUPT7L
VSTM2L
GFM1;SLC4A1AP;CABP7;RSRC1;SKIDA1;NAT14
GFM1;UQCR10;CCDC141;MYH6;MYH7;TTN
CCDC121;ZNF512;SUPT7L
DCAF16;SUPT7L
GFM1;CABP7;ZNF219;NAT14
GFM1;CABP7;RSRC1;SKIDA1;CEP85L;NAT14
SLC4A1AP;SLC35F1;APOE;NAT14;NIPSNAP1
DCAF16;SNX17;ZNF513;NAT14;SUPT7L
MTMR3
GFM1;SLC4A1AP;CABP7;RSRC1;SKIDA1;MFN1;NDRG2;SUPT7L
DNAJC1;GCKR;B3GNT7;FAM174B;C2ORF16;PVRL2
EIF2B4;KIAA1755;CMTM5;SHOX2;ZNF512;ARHGEF40;SUPT7L
IFI27L2;CABP7;CMTM5;APOC1;APOE;NDRG2
DNAJC1;GCKR;B3GNT7;FAM174B
SKIDA1;CEP85L;SSC5D;SUPT7L;TMEM253;WARS2;MLLT10;DCAF16;FAM174B;MFN1;GPR126;ZNF512;F
SLC5A6;GCKR;SERPINA1;APOC1;GPR126;APOE;NIPSNAP1
MLLT10;KIAA1755;IFI27L2;DCAF16;DNAJC1;EIF3E;ZNF512;ARHGEF40;NRBP1
TBX15;GFM1;UQCR10;MYH6;MYH7;TTN
SLC5A6;GCKR;SERPINA1;SKIDA1;APOC1;GPR126;APOE
IFI27L2;CABP7;CMTM5;APOC1;RSPO2;VSTM2L;SLC35F1;APOE;NDRG2
SHOX2;GNB4;SSC5D;EIF3E;FAM3C;WARS2
CCDC121;ZCCHC10;SLC4A1AP;CEP85L;NCAPG;UQCR10;CWC27;PPM1G;CCDC91;ZNF628;DLEU1;ASCC2;
EIF2B4;GJA1;ZNF219;NRBP1
KIAA1755;CENPW;AFAP1;SSC5D;DGKH
GFM1;CENPW;ZCCHC10;TOMM40;HSPA4;SHOX2;PTEN;NCAPG;CWC27;GPN1;NIPSNAP1;WARS2;PPM1
SLC5A6;TOMM40;SNX17;PVRL2;PPM1G
TBX15;GFM1;TOMM40;HSPA4;ZNF628;UQCR10;NRBP1;TTN
SLC5A6;GFM1;SNX17;ALX4;PVRL2
TBX15;GFM1;CCDC141;MYH7;TTN
CCDC121;HSPA4;VSTM2L;SLC35F1;NDRG2
CCDC121;MTMR3;ZCCHC10;SLC4A1AP;CEP85L;NCAPG;CWC27;PPM1G;CCDC91;ZNF628;DLEU1;EIF2B4;
CENPW;ZCCHC10;TOMM40;HSPA4;PTEN;NCAPG;C2ORF16;GPN1;NIPSNAP1;NRBP1;PPM1G;B3GNT7;C
DNAJC1;SHOX2;GNB4;AFAP1;C2ORF16
CCDC121;SLC4A1AP;CEP85L;NCAPG;CWC27;PPM1G;CCDC91;ZNF628;DLEU1;ASCC2;EIF2B4;GCKR;CENP
TOMM40;PTEN;WARS2
EIF2B4;DNAJC1;TOMM40;ZNF628;SNX17;FAM174B;ZMAT5;MFN1;ZNF513;ASCC2;PVRL2;WARS2
TBX15;GFM1;UQCR10;NDRG2;MYH6;TTN
GCKR;SERPINA1;SKIDA1;APOC1;GPR126;C2ORF16;APOE;NIPSNAP1
SLC5A6;EIF2B4;TOMM40;SHOX2;ZNF219;CEP85L;ARHGEF40;TRIB2;SUPT7L
CCDC121;GJA1;HSPA4;APOE;NDRG2

IFI27L2;CENPW;ZCCHC10;HSPA4;PTEN;NCAPG;CWC27;GPN1;NIPSNAP1;WARS2;PPM1G;DCAF16;CR1L;
PLN;NOV;RSRC1;SPEG
EIF2B4;CENPW;TOMM40;ZMAT5;DLEU1;NCAPG;ASCC2;NRBP1;PPM1G
KIAA1755;ZNF219;RSPO2;SSC5D
B3GNT7;ZNF219;SSC5D;TMEM253
DCAF16;SHOX2;GNB4;C2ORF16;FAM3C;LCORL;TMEM253;DGKH;WARS2
MLLT10;KIAA1755;PLN;NOV;SPEG
GCKR;B3GNT7;FAM174B;ZNF513;ASCC2;C2ORF16;PVRL2
DES;ALX4;SSC5D;DGKH
SERPINA1;MTMR3;CR1;CR1L;ASCC2
GJA1;APOC1;APOE
TBX15;ZNF628;NRBP1;MYH6;MYH7;TTN
TBX15;FAM174B;UQCR10;NDRG2;NRBP1;MYH7;TTN
PLN;NOV;RSRC1;SHOX2;SPEG
SLC5A6;DCAF16;SSC5D;ZNF512;NAT14;NRBP1;SUPT7L;PPM1G
KIAA1755;NOV;SPEG;SSC5D
DNAJC1;GCKR;SERPINA1;FAM174B;TRIB2
PLN;NOV;RSRC1;AFAP1;SPEG
SERPINA1;MTMR3;CR1;CR1L;SNX17;ZMAT5;PTEN;ASCC2;ARHGEF40
GFM1;ZCCHC10;SLC4A1AP;TOMM40;HSPA4;SHOX2;CWC27;GPN1;WARS2;PPM1G;DCAF16;ZNF628;ZN
SLC5A6;SERPINA1;CR1;TOMM40;B3GNT7;SNX17;ZMAT5;PPM1G
DNAJC1;B3GNT7;APOC1;GPR126;NCAPG;CCDC141
KIAA1755;IFI27L2;CCDC121;CMTM5;SLC35F1;CWC27;NDRG2
TOMM40;HSPA4;ZNF628;ZMAT5;DLEU1;ALX4;WNT16
DCAF16;CEP85L;GPR126;SLC35F1;ZNF512;LCORL;TRIB2;WARS2
DNAJC1;GCKR;SERPINA1;B3GNT7;FAM174B;PVRL2
TBX15;GFM1;DES;MYH7;TTN
GFM1;CENPW;ZCCHC10;TOMM40;HSPA4;NCAPG;CWC27;GPN1;NIPSNAP1;WARS2;PPM1G;SLC5A6;DC
SERPINA1;CR1;CR1L;ASCC2
CENPW;ZCCHC10;CR1;TOMM40;HSPA4;NCAPG;CWC27;GPN1;NIPSNAP1;WARS2;PPM1G;DCAF16;CR1L
IFI27L2;CABP7;CMTM5;APOC1;RSPO2;APOE;NDRG2
SLC5A6;CR1;LXN;PTEN;ALX4
MLLT10;ZNF219;AFAP1;SPEG
B3GNT7;ZNF219;ZNF513;TMEM253
IFI27L2;CCDC121;GJA1;CCDC91;CABP7;CMTM5;VSTM2L;ALX4;CWC27;NDRG2;MLF1
IFI27L2;DNAJC1;GNB4;SSC5D;C2ORF16;EIF3E
PLN;NOV;RSRC1;SHOX2;AFAP1;EIF3E;ZNF512;CWC27
SLC5A6;GFM1;TOMM40;SNX17;PTEN
DCAF16;GCKR;CCDC121;LXN;B3GNT7;SSC5D;C2ORF16;EIF3E;ZNF512;TRIB2
GFM1;CENPW;ZCCHC10;HSPA4;NCAPG;CWC27;GPN1;NIPSNAP1;NRBP1;WARS2;PPM1G;SLC5A6;CR1L;
KIAA1755;CMTM5;APOC1;APOE;NDRG2;NAT14
SLC5A6;LXN;TOMM40;PTEN
RSRC1;AFAP1;CD34;WARS2
CCDC121;MTMR3;SLC4A1AP;CEP85L;NCAPG;CWC27;PPM1G;CCDC91;ZNF628;DLEU1;EIF2B4;GCKR;CEN
IFI27L2;GJA1;NOV;APOC1;C2ORF16;APOE;ARHGEF40;NIPSNAP1

SLC5A6;GCKR;SERPINA1;APOC1;GPR126;APOE;NIPSNAP1
DNAJC1;SHOX2;GNB4;AFAP1;ALX4;FAM3C
PLN;NOV;ZMAT5;SPEG
B3GNT7;ZNF219;RSPO2;TMEM253
SLC5A6;KIAA1755;IFI27L2;CR1L;CABP7;CMTM5;APOC1;C2ORF16;APOE;CWC27;LCORL
IFI27L2;GJA1;CMTM5;APOC1;VSTM2L;SLC35F1;UQCR10;FAM3C;APOE;NDRG2;NAT14
TBX15;DES;ZNF628;MYH6;MYH7;TTN
VSTM2L;CCDC141;MLF1;MYH6
DES;ZNF628;ZNF219;AFAP1;SSC5D
TBX15;GFM1;ZNF628;SPEG;NDRG2;MYH7;TTN
EIF2B4;GJA1;DES;ZNF219;ZNF513
B3GNT7;FAM174B;APOC1;RSPO2;GPR126;CCDC141
SLC5A6;GCKR;GFM1;SERPINA1;APOC1;GPR126;APOE;NIPSNAP1
TBX15;GFM1;DES;MYH6;MYH7;TTN
SNX17;AFAP1;ALX4;SSC5D;NRBP1
MLLT10;KIAA1755;PLN;NOV;SPEG;SSC5D
UQCR10;CCDC141;MYH6;MYH7;TTN
DES;ZNF219;ZMAT5;SSC5D;CD34
TBX15;MLLT10;SHOX2;MFN1;EIF3E;FAM3C;CD34;NRBP1;WARS2
CENPW;TOMM40;HSPA4;SNX17;GNB4;NCAPG;FAM3C;CWC27;LCORL;GPN1;PPM1G
PLN;NOV;RSRC1;SPEG
MLLT10;KIAA1755;PLN;NOV;SPEG;SSC5D
SLC5A6;GCKR;SERPINA1;APOC1;DLEU1;GPR126;APOE;NIPSNAP1
MLLT10;DES;ZNF219;SPEG;SSC5D
KIAA1755;IFI27L2;CABP7;CMTM5;RSPO2;APOE;NDRG2
RSPO2;GPR126;SPEG;SSC5D
CENPW;ZCCHC10;CR1;TOMM40;HSPA4;NCAPG;CWC27;GPN1;NIPSNAP1;WARS2;PPM1G;SLC5A6;DCAF
SLC5A6;MLLT10;IFI27L2;DCAF16;TOMM40;ZMAT5;ZNF513;SSC5D;ZNF512;NAT14;SUPT7L
CCDC141;MYH6;MYH7
CCDC121;ZCCHC10;SLC4A1AP;CEP85L;NCAPG;CWC27;PPM1G;CCDC91;ZNF628;DLEU1;EIF2B4;GCKR;CE
CENPW;ZCCHC10;CEP85L;PTEN;NCAPG;CWC27;GPN1;NIPSNAP1;PPM1G;MLLT10;DCAF16;CR1L;RSRC1;
CENPW;AFAP1;ALX4;SSC5D;DGKH
CCDC121;MTMR3;SLC4A1AP;CEP85L;NCAPG;CWC27;PPM1G;CCDC91;ZNF628;DLEU1;ASCC2;EIF2B4;GC
CMTM5;CCDC141;MYH6
DES;ZNF219;RSPO2
KIAA1755;CMTM5;SKIDA1;CEP85L;PTEN;GPR126;SLC35F1;ZNF512;ARHGEF40;LCORL;TRIB2
IFI27L2;CENPW;LXN;B3GNT7;SNX17;RSPO2;NCAPG
GJA1;GNB4;DLEU1;ALX4;WNT16;GPN1
SLC5A6;CENPW;TOMM40;B3GNT7;NCAPG;NIPSNAP1;TMEM253;PPM1G
DES;ZNF219;AFAP1;SSC5D
PLN;NOV;RSRC1;MFN1;SPEG
CCDC121;GJA1;RSRC1;CMTM5;APOE;NDRG2
SLC5A6;LXN;ALX4;ARHGEF40
KIAA1755;CCDC121;ALX4;SSC5D;EIF3E;ZNF512;CD34;NRBP1
TBX15;DES;ZNF628;SPEG;MYH7;TTN

CENPW;ZCCHC10;SLC4A1AP;PTEN;NCAPG;CWC27;GPN1;NIPSNAP1;WARS2;TBX15;MLLT10;DCAF16;CR[illegible]
ZNF219;DLEU1;RSPO2;SSC5D
KIAA1755;CMTM5;ZMAT5;GPR126;ARHGEF40;TRIB2
DES;CABP7;ZNF219
SLC5A6;GCKR;SERPINA1;APOC1;GPR126;APOE;NIPSNAP1
DNAJC1;SERPINA1;LXN;B3GNT7;SNX17;APOC1;ZMAT5;RSPO2
CCDC121;GJA1;CCDC91;CMTM5;APOC1;VSTM2L;SLC35F1;APOE;NDRG2
TBX15;UQCR10;MYH6;MYH7;TTN
MLLT10;DES;ZNF219;SSC5D
TBX15;RSRC1;SHOX2;SPEG
CCDC91;CABP7;CMTM5;APOC1;VSTM2L;SLC35F1;UQCR10;APOE;NDRG2;NAT14
CENPW;ZCCHC10;HSPA4;NCAPG;C2ORF16;WNT16;CWC27;GPN1;NIPSNAP1;PPM1G;MLLT10;SNX17;M[illegible]
MLLT10;CENPW;AFAP1;SSC5D;ARHGEF40;DGKH
DNAJC1;SERPINA1;TOMM40;B3GNT7;SNX17;PVRL2;PPM1G
EIF2B4;TBX15;DES;TOMM40;ZMAT5;DLEU1;UQCR10;NRBP1;TTN
KIAA1755;DCAF16;CMTM5;SKIDA1;CEP85L;SLC35F1;ZNF512;ARHGEF40;TRIB2
NOV;SHOX2;AFAP1;SPEG;DGKH
KIAA1755;CMTM5;SHOX2;CEP85L;ZMAT5;NDRG2;TRIB2
DNAJC1;CENPW;LXN;TOMM40;B3GNT7;DLEU1;UQCR10;NIPSNAP1;TMEM253;PPM1G
PLN;CCDC91;NOV;RSRC1;AFAP1;ZNF512
TBX15;NOV;RSRC1;WARS2
CCDC121;GFM1;MTMR3;ZCCHC10;SLC4A1AP;CEP85L;NCAPG;CWC27;PPM1G;CCDC91;ZNF628;DLEU1;[illegible]
MTMR3;CR1L;ASCC2
CCDC121;GFM1;MTMR3;ZCCHC10;SLC4A1AP;CEP85L;NCAPG;CWC27;PPM1G;CCDC91;ZNF628;DLEU1;[illegible]
DCAF16;HSPA4;SHOX2;GNB4;AFAP1;CWC27;GPN1;TRIB2;TMEM253;DGKH;WARS2;PPM1G
TBX15;UQCR10;CCDC141;MYH6;MYH7;TTN
TBX15;UQCR10;CCDC141;MYH6;MYH7;TTN
IFI27L2;CABP7;CMTM5;NDRG2
SERPINA1;CR1L;ASCC2
KIAA1755;DCAF16;GCKR;LXN;B3GNT7;EIF3E;ZNF512;APOE;TRIB2
LXN;B3GNT7;APOC1;SSC5D;PVRL2;CD34
DNAJC1;GCKR;FAM174B
DES;SPEG
DNAJC1;CCDC121;FAM174B;SKIDA1;ZMAT5;ZNF512;FAM3C;CD34;TMEM253
SLC5A6;EIF2B4;ZCCHC10;ALX4;PVRL2;GPN1;SUPT7L
SERPINA1;MTMR3;CR1;ZMAT5;ARHGEF40;TMEM253
SERPINA1;B3GNT7;GPR126;CCDC141
ALX4;CD34;TRIB2
DES;ZNF628;ZNF219
MLLT10;DCAF16;MFN1;GNB4;GPR126;WNT16;LCORL;GPN1
FAM174B;SKIDA1;ZMAT5;ZNF513;C2ORF16;ZNF512;FAM3C;SUPT7L;TMEM253
SLC5A6;APOC1;SLC35F1;APOE;NDRG2
NOV;AFAP1;SPEG
CENPW;NCAPG;NRBP1;PPM1G
IFI27L2;HSPA4;GNB4;NCAPG;EIF3E;FAM3C

GJA1;APOC1;ALX4;FAM3C;WNT16;LCORL
GCKR;ASCC2;C2ORF16;NRBP1
SERPINA1;MTMR3;CR1;CR1L;ZMAT5;PTEN;ARHGEF40
CENPW;ZCCHC10;TOMM40;PTEN;NCAPG;CWC27;GPN1;NIPSNAP1;PPM1G;CR1L;RSRC1;MFN1;DLEU1;[illegible]
APOC1;APOE
GFM1;CENPW;ZCCHC10;TOMM40;HSPA4;NCAPG;CWC27;GPN1;NIPSNAP1;WARS2;PPM1G;RSRC1;MF[illegible]
SERPINA1;MTMR3;CR1;CR1L;ZMAT5
EIF2B4;MLLT10;RSPO2;SPEG;SUPT7L
EIF2B4;GCKR;CCDC121;CENPW;SLC4A1AP;SKIDA1;CEP85L;NCAPG;C2ORF16;UQCR10;CWC27;GPN1;ML[illegible]
CR1;SNX17;APOC1;VSTM2L;ZNF513;APOE;ARHGEF40;PVRL2;TRIB2;SUPT7L
IFI27L2;PTEN;ZNF513;SSC5D;WNT16
DNAJC1;CCDC121;SKIDA1;DLEU1;CD34;TMEM253
CCDC121;HSPA4;CMTM5;APOC1;SLC35F1;APOE;WNT16;NAT14;MLF1
IFI27L2;SNX17;SSC5D;WNT16;NRBP1
NOV;AFAP1;SPEG;DGKH
EIF2B4;GCKR;CCDC121;CENPW;ZCCHC10;SLC4A1AP;HSPA4;SKIDA1;CEP85L;AFAP1;NCAPG;C2ORF16;C[illegible]
DNAJC1;SNX17;FAM174B;SKIDA1;ZNF219;ALX4;ZNF513;C2ORF16;TMEM253
LXN;B3GNT7;APOC1;CCDC141
DCAF16;GCKR;LXN;B3GNT7;C2ORF16;EIF3E;ZNF512;TRIB2
SLC4A1AP;CCDC91;CABP7;VSTM2L;SLC35F1;WNT16;NAT14;MLF1;DGKH
APOC1;SLC35F1;APOE;CWC27;NDRG2;NAT14
NOV;AFAP1;SPEG
MLLT10;PLN;RSRC1;MFN1;ZNF512;WARS2
DNAJC1;GCKR;B3GNT7;FAM174B;PVRL2
CMTM5;APOC1;SLC35F1;APOE;NDRG2
ZNF219;RSPO2;DGKH
RSRC1;PTEN;EIF3E;CD34
DNAJC1;CCDC121;SKIDA1;C2ORF16;ZNF512;FAM3C;CD34;TMEM253
NOV;SPEG
SERPINA1;MTMR3;CR1;CR1L;ZMAT5;ARHGEF40
CCDC121;SLC4A1AP;CCDC91;RSRC1;APOC1;SLC35F1;FAM3C;APOE;CWC27;NDRG2;NAT14;MLF1
MLLT10;IFI27L2;DCAF16;GCKR;LXN;EIF3E;ZNF512;TRIB2;SUPT7L
GCKR;FAM174B;C2ORF16;PVRL2
MLLT10;PLN;NOV;SPEG;ZNF512
GJA1;NOV;SPEG
DNAJC1;FAM174B;RSPO2;GPR126;PVRL2;DGKH
KIAA1755;NOV;SPEG
MLLT10;ZNF219;DLEU1
LXN;CD34
NOV;RSRC1
SLC5A6;TOMM40;CEP85L;ARHGEF40;TRIB2
DNAJC1;SNX17;FAM174B;ZMAT5;ZNF513;FAM3C;TMEM253
EIF2B4;CCDC121;CENPW;MTMR3;SLC4A1AP;HSPA4;SKIDA1;CEP85L;NCAPG;C2ORF16;MLF1;NRBP1;PP[illegible]
SHOX2;AFAP1;CD34
MLLT10;DCAF16;GCKR;B3GNT7;SKIDA1;EIF3E;ZNF512;LCORL;TRIB2

ZNF219;PTEN;CD34
SLC5A6;TBX15;PTEN
DNAJC1;FAM174B
CCDC121;MFN1;ALX4;CD34;TMEM253
SLC4A1AP;CCDC91;CABP7;RSRC1;RSPO2;VSTM2L;SLC35F1;FAM3C;WNT16;NAT14;NIPSNAP1
PLN;NOV;SPEG
GJA1;MFN1;ALX4;WNT16;CD34
SERPINA1;MTMR3;CR1;CR1L;ZMAT5;ASCC2;PPM1G
CENPW;ASCC2;PVRL2;NRBP1
EIF2B4;MLLT10;GJA1;ZNF628;GNB4;DLEU1;GPR126;WNT16
SLC5A6;DCAF16;LXN;ALX4
GCKR;FAM174B;FAM3C;TRIB2
SERPINA1;MTMR3;CR1;ZMAT5;PTEN;ARHGEF40
PLN;NOV;SPEG
DNAJC1;B3GNT7;VSTM2L;GPR126;SUPT7L
SHOX2;GNB4;CWC27;LCORL;TMEM253;WARS2
CENPW;HSPA4;GNB4;AFAP1;NCAPG;EIF3E;NIPSNAP1
GJA1;LXN;C2ORF16
LXN;PTEN;CD34
CENPW;ZCCHC10;LXN;TOMM40;B3GNT7;NCAPG;TMEM253;PPM1G
SERPINA1;MTMR3;CR1;B3GNT7;CR1L;ZNF628;ASCC2
DNAJC1;SNX17;FAM174B;SKIDA1;ZMAT5;FAM3C;TMEM253
KIAA1755;IFI27L2;CMTM5;ZMAT5
IFI27L2;ZCCHC10;SLC4A1AP;VSTM2L;SLC35F1;NAT14;NIPSNAP1;CCDC91;RSRC1;RSPO2;ZNF512;FAM3C
GFM1;ZCCHC10;SLC4A1AP;TOMM40;HSPA4;SHOX2;UQCR10;CWC27;GPN1;NRBP1;WARS2;PPM1G;ZNF
NOV;SPEG
CENPW;HSPA4;GNB4;DLEU1;NCAPG;EIF3E;CWC27;GPN1
SLC5A6;PTEN;CD34
GJA1;DLEU1;GPR126;NCAPG;WNT16
GCKR;CENPW;DLEU1;GPR126;NCAPG;ASCC2
MLLT10;DCAF16;GCKR;LXN;B3GNT7;C2ORF16;EIF3E;ZNF512;TRIB2;WARS2
TBX15;PTEN;EIF3E;CD34
PLN;NOV;SPEG
KIAA1755;PLN;NOV;SPEG
CCDC121;SLC4A1AP;CCDC91;RSRC1;SKIDA1;APOC1;SLC35F1;APOE;CWC27;NDRG2;MLF1
DES;MYH6;MYH7;TTN
EIF2B4;DCAF16;GJA1;DLEU1;ALX4;WNT16
SERPINA1;CR1;CR1L;ASCC2
LXN;CEP85L;VSTM2L;SSC5D;C2ORF16;NAT14;SUPT7L;DCAF16;DNAJC1;B3GNT7;FAM174B;ZNF513;ZNF5
CENPW;DLEU1;GPR126;NCAPG;NRBP1
NOV;RSRC1;SPEG
PLN;NOV;SPEG
LXN;TOMM40;B3GNT7;ZNF513;PVRL2;TMEM253
NOV;RSRC1;AFAP1
ZCCHC10;LXN;B3GNT7;APOC1;RSPO2;GPR126;LCORL;GPN1

SLC5A6;EIF2B4;DNAJC1;TOMM40;TRIB2
SNX17;FAM174B;SKIDA1;ZNF513;TMEM253
DES;SSC5D;DGKH
EIF2B4;CMTM5;ARHGEF40;WARS2
CR1;LXN;B3GNT7;APOC1;RSPO2;LCORL;SUPT7L
SERPINA1;MTMR3;CR1;CR1L;ZMAT5;PTEN;ASCC2
LXN;GPR126;SSC5D;CD34
CENPW;TOMM40;DLEU1;NCAPG;ASCC2;NRBP1
ASCC2;WNT16;NRBP1
DNAJC1;CABP7;FAM174B
B3GNT7;RSPO2;TMEM253
GJA1;GNB4;DLEU1;ALX4;WNT16;GPN1
DNAJC1;B3GNT7;SNX17;FAM174B;CCDC141
EIF2B4;LXN;SNX17;ZNF513;C2ORF16;PVRL2
DCAF16;MTMR3;SSC5D;ZNF512;WNT16;NRBP1
CENPW;HSPA4;SNX17;GNB4;NCAPG;SSC5D
SERPINA1;MTMR3;CR1;ZMAT5;PTEN;ARHGEF40;TMEM253
MLLT10;ZNF219;DGKH
CCDC121;GJA1;ALX4;APOE;WNT16;NDRG2;NAT14;MLF1
SHOX2;TRIB2;WARS2
LXN;ALX4;FAM3C;PVRL2
TBX15;GFM1;MYH6;MYH7;TTN
PLN;NOV;RSRC1;PTEN
SERPINA1;MTMR3;CR1;CR1L;ZMAT5;PTEN;ARHGEF40
SERPINA1;MTMR3;CR1;CR1L;CMTM5;PTEN;ARHGEF40
SERPINA1;MTMR3;CR1;CR1L;PTEN;ASCC2
GJA1;CCDC91;CABP7;RSRC1;VSTM2L;SLC35F1;NDRG2
EIF2B4;GCKR;CCDC121;CENPW;SLC4A1AP;HSPA4;SKIDA1;CEP85L;AFAP1;NCAPG;C2ORF16;CWC27;MLF
LXN;B3GNT7;GPR126;CCDC141
GCKR;B3GNT7;FAM174B;CEP85L;ASCC2;SUPT7L
SERPINA1;MTMR3;CR1;CR1L;ZMAT5;PTEN;ARHGEF40
ZCCHC10;SLC4A1AP;SHOX2;GNB4;ALX4;UQCR10;CWC27;LCORL;GPN1;TRIB2;WARS2;PPM1G
MLLT10;GJA1;MFN1;GNB4;ALX4;WNT16
KIAA1755;CCDC91;CMTM5;APOC1;SLC35F1;APOE;NDRG2;NAT14
EIF2B4;CCDC121;GFM1;CENPW;MTMR3;ZCCHC10;SLC4A1AP;HSPA4;SKIDA1;CEP85L;NCAPG;C2ORF16;[illegible]
EIF2B4;GJA1;ALX4;WNT16;CD34
DNAJC1;B3GNT7;GPR126;CCDC141
NOV;APOC1;APOE
GJA1;NOV;DGKH
CABP7;CMTM5;VSTM2L
KIAA1755;DCAF16;ZNF628;MFN1;SSC5D;EIF3E;ZNF512;CCDC141
KIAA1755;CCDC121;CCDC91;CABP7;CMTM5;VSTM2L;SLC35F1;NDRG2
EIF2B4;CCDC121;CENPW;MTMR3;ZCCHC10;SLC4A1AP;HSPA4;SKIDA1;CEP85L;NCAPG;C2ORF16;MLF1;[illegible]
CCDC121;GJA1;CCDC91;CMTM5;VSTM2L;SLC35F1;NDRG2
CENPW;TOMM40;HSPA4;GNB4;DLEU1;NCAPG;CWC27;GPN1;PPM1G

MTMR3;B3GNT7;CR1L;ASCC2
SERPINA1;MTMR3;CR1;CR1L;PTEN;ASCC2
DNAJC1;B3GNT7;SNX17;FAM174B;ZMAT5;GPR126
DNAJC1;CCDC121;CCDC91;FAM174B;SKIDA1;ASCC2;ZNF512;FAM3C;SUPT7L;TMEM253
SERPINA1;CR1L;UQCR10;APOE;NIPSNAP1
SLC5A6;PTEN;ARHGEF40;CD34
IFI27L2;GCKR;SPEG;SLC35F1;SUPT7L
CENPW;TOMM40;PVRL2;NRBP1
GCKR;FAM174B;MFN1
PTEN;EIF3E
GJA1;ZNF219;DLEU1;ALX4;WNT16
CR1;CR1L;SNX17;APOC1;VSTM2L;SSC5D;ARHGEF40;TRIB2
EIF2B4;TBX15;TOMM40;FAM174B;ZNF513;TTN
CCDC91;CABP7;CMTM5;SLC35F1;CWC27;NDRG2
RSPO2;SSC5D
SSC5D;WNT16;NRBP1
ZNF219;SSC5D;TMEM253
DNAJC1;FAM174B
SLC5A6;MLLT10;KIAA1755;DCAF16;CCDC121;MTMR3;CEP85L;MFN1;SSC5D;ZNF512;SUPT7L;DGKH
DNAJC1;SERPINA1;TOMM40;B3GNT7;VSTM2L
CCDC91;CABP7;RSPO2;VSTM2L;SLC35F1;FAM3C;WNT16
SERPINA1;MTMR3;CR1;CR1L
MLLT10;GJA1;MFN1;GNB4;ALX4;WNT16
DNAJC1;GCKR;CENPW;B3GNT7;NCAPG;PVRL2
DCAF16;GJA1;GPR126;ALX4;WNT16;LCORL
RSPO2;SSC5D
SERPINA1;MTMR3;CR1;ZMAT5;PTEN;ARHGEF40
ZCCHC10;LXN;B3GNT7;RSPO2;TMEM253
SERPINA1;MTMR3;CR1;CR1L;SNX17;ZMAT5;PTEN;GPN1;TMEM253
KIAA1755;SLC4A1AP;CMTM5;SKIDA1;RSPO2;VSTM2L;SLC35F1;NDRG2
KIAA1755;DCAF16;GCKR;ZNF512;APOE;TRIB2
B3GNT7;LCORL;TMEM253
CENPW;TOMM40;HSPA4;MFN1;DLEU1;NCAPG;GPN1;PPM1G
KIAA1755;IFI27L2;CMTM5;RSPO2;VSTM2L;SLC35F1;FAM3C;NDRG2;NAT14
DES;SPEG
NOV;SPEG
CABP7;RSPO2;VSTM2L;SLC35F1;NDRG2
CCDC121;CMTM5;SKIDA1;NDRG2
DNAJC1;FAM174B;CCDC141
SERPINA1;MTMR3;CR1;CR1L;ZMAT5;PTEN;NCAPG
NDRG2
NOV;SPEG
KIAA1755;RSPO2
SERPINA1;MTMR3;ASCC2;PVRL2
GJA1;ZNF628;GNB4;ALX4;WNT16;GPN1

B3GNT7;ZNF219;ZNF513;TMEM253
GJA1;ZNF628;ZNF219;ALX4;WNT16
CENPW;TOMM40;ZMAT5;MFN1;NCAPG;ASCC2
SERPINA1;MTMR3;CR1;CR1L;SNX17;ZMAT5;NRBP1;PPM1G
IFI27L2;CMTM5;SLC35F1;NDRG2
CR1L;ASCC2
DES;MYH7;TTN
NOV;SPEG
PLN;NOV;RSRC1
ASCC2;PVRL2;NRBP1
GFM1;CENPW;ZCCHC10;TOMM40;HSPA4;PTEN;NCAPG;CWC27;GPN1;NIPSNAP1;PPM1G;SNX17;RSRC1
TBX15;PLN;CCDC91;NOV;RSRC1;MFN1;EIF3E
SLC5A6;CR1;PTEN
GJA1;ZNF219;GNB4
CENPW;ZCCHC10;HSPA4;PTEN;NCAPG;CWC27;GPN1;NIPSNAP1;PPM1G;DCAF16;CR1L;MFN1;DLEU1;EIF
B3GNT7;FAM174B;APOC1;CCDC141
DES;SPEG
B3GNT7;ZNF219;TMEM253
SERPINA1;MTMR3;CR1;CR1L;PTEN;ASCC2
DNAJC1;GCKR;FAM174B;CEP85L;FAM3C
SERPINA1;MTMR3;CR1;CR1L;SNX17;ZMAT5
DNAJC1;FAM174B;EIF3E
CR1;CR1L;ZMAT5;ASCC2
TOMM40;B3GNT7;ZNF513;TMEM253
TBX15;CCDC91;RSRC1;PTEN;LCORL
SERPINA1;MTMR3;CR1L;ASCC2
GFM1;ZCCHC10;CCDC91;HSPA4;SHOX2;GNB4;CWC27;LCORL;GPN1;WARS2
HSPA4;SHOX2;GNB4;C2ORF16;FAM3C;CWC27;LCORL;GPN1;DGKH;WARS2
DNAJC1;FAM174B;CCDC141
SERPINA1;MTMR3;CR1;CR1L;ZMAT5;PTEN
CCDC91;APOC1;SLC35F1;APOE;NDRG2;NAT14;MLF1;NIPSNAP1
SLC5A6;PTEN;AFAP1
SERPINA1;MTMR3;CR1;CR1L;ZMAT5;PTEN;ARHGEF40;TMEM253
MFN1;WNT16;CD34;NRBP1
SLC5A6;PTEN;CD34
FAM174B;PTEN;C2ORF16;ZNF512;FAM3C;TMEM253
SERPINA1;CR1;CR1L;ZMAT5
ZCCHC10;LXN;PVRL2;CD34
DNAJC1;B3GNT7;SNX17;CCDC141
MTMR3;CR1;CR1L;PTEN;ASCC2
DES;ZNF219
DCAF16;GJA1;MFN1;ALX4;WNT16
SLC5A6;CCDC121;ALX4;CD34
DES;RSPO2
CENPW;TOMM40;HSPA4;SNX17;GNB4;AFAP1;NCAPG;CWC27;GPN1;PPM1G

KIAA1755;IFI27L2;CCDC91;CABP7;CMTM5;VSTM2L;SLC35F1;NIPSNAP1
EIF2B4;RSPO2
MLLT10;SPEG
CABP7;CMTM5;RSPO2;VSTM2L;SLC35F1;WNT16;NDRG2;NAT14
FAM174B;RSPO2
CABP7;APOC1;UQCR10;APOE;NDRG2;NAT14;NIPSNAP1
MLLT10;GJA1;MTMR3;MFN1;GNB4;ALX4;WNT16;GPN1
DES;RSPO2
CABP7;VSTM2L;SLC35F1;NDRG2
DCAF16;ZCCHC10;AFAP1;EIF3E;SUPT7L;DGKH
SHOX2;GNB4;SSC5D;FAM3C;CWC27;LCORL;TMEM253;WARS2
SLC5A6;DCAF16;ALX4;CD34
SHOX2;SSC5D;CD34
SLC4A1AP;CCDC91;RSRC1;SKIDA1;RSPO2;VSTM2L;SLC35F1;FAM3C;NDRG2
KIAA1755;LXN;SPEG;CD34
DES;ZNF219
IFI27L2;GJA1;CCDC91;VSTM2L;SLC35F1;UQCR10;APOE;NDRG2
DNAJC1;B3GNT7;FAM3C;LCORL;TMEM253
RSPO2;SPEG
PTEN;ALX4;CD34
SSC5D;CD34;WARS2
KIAA1755;CMTM5;CEP85L;GPR126;ARHGEF40
TBX15;PTEN;CD34
EIF2B4;GCKR;CCDC121;CENPW;MTMR3;ZCCHC10;SLC4A1AP;HSPA4;SKIDA1;CEP85L;NCAPG;C2ORF16;C
DCAF16;GCKR;ZNF628;CEP85L;EIF3E;ZNF512;ARHGEF40;TRIB2
SERPINA1;MTMR3;CR1;CR1L;PTEN;ARHGEF40;PVRL2;TMEM253
SERPINA1;MTMR3;CR1;CR1L;CMTM5;ZMAT5;NCAPG;ASCC2
GJA1;NOV
CR1;CR1L;SNX17;APOC1;SSC5D;ARHGEF40;TRIB2
KIAA1755;CMTM5;SHOX2
ZNF219;GNB4;AFAP1
SLC4A1AP;CCDC91;HSPA4;SKIDA1;RSPO2;VSTM2L;SLC35F1;C2ORF16;FAM3C;NAT14;NIPSNAP1
MLLT10;DNAJC1;LXN;FAM174B;CEP85L;GPR126;ZNF513;DGKH
SERPINA1;MTMR3;CR1;CR1L;PTEN;ASCC2
LXN;NOV
DNAJC1;B3GNT7;GPR126;CCDC141
SERPINA1;MTMR3;CR1;CR1L;PTEN;ASCC2
IFI27L2;SERPINA1;MTMR3;CR1;SNX17;ZMAT5;PTEN
DES;CCDC91
DES;GNB4
EIF2B4;FAM174B;SKIDA1;ZNF513;C2ORF16;SUPT7L;TMEM253
SLC5A6;LXN;PTEN
PLN;NOV;SPEG
B3GNT7;ZNF219;TMEM253
EIF2B4;MLLT10;DCAF16;MTMR3;MFN1;DLEU1;WNT16

APOC1;APOE
GJA1;CCDC91
EIF2B4;ZNF628;MFN1;ALX4;ZNF513;WNT16
CR1L;ASCC2
ZCCHC10;NOV;FAM3C
B3GNT7;APOC1;GPR126;PVRL2;CD34;DGKH
DNAJC1;B3GNT7;GPR126;PVRL2;DGKH
SLC5A6;ALX4;ARHGEF40;CD34
SLC4A1AP;CCDC91;CABP7;RSRC1;RSPO2;VSTM2L;SLC35F1;FAM3C;WNT16;NAT14;NIPSNAP1
CCDC121;GJA1;CCDC91;CABP7;RSPO2;VSTM2L;SLC35F1;FAM3C;NDRG2
DNAJC1;FAM174B;SKIDA1;FAM3C;TMEM253
MLLT10;DNAJC1;SNX17;FAM174B;ZNF513;FAM3C;TMEM253
CENPW;LXN;TOMM40;B3GNT7;NCAPG;NIPSNAP1;TMEM253;PPM1G
EIF2B4;GCKR;CCDC121;CENPW;MTMR3;ZCCHC10;SLC4A1AP;HSPA4;SKIDA1;CEP85L;NCAPG;C2ORF16;N
DNAJC1;B3GNT7;GPR126;WNT16;PVRL2;DGKH
DNAJC1;SERPINA1;TOMM40;B3GNT7;PVRL2;PPM1G
SHOX2;CEP85L;MFN1;GPR126;TRIB2
LXN;PVRL2;CD34
SERPINA1;MTMR3;CR1;CR1L
IFI27L2;AFAP1;SSC5D
PLN;NOV;RSRC1;SPEG
KIAA1755;NOV;SPEG
SLC5A6;CCDC121;LXN;CD34
IFI27L2;CMTM5;VSTM2L;APOE;WNT16;NDRG2;NAT14;MLF1
IFI27L2;SERPINA1;CR1;LXN;CR1L;APOC1;ZMAT5
IFI27L2;VSTM2L;SLC35F1;NDRG2
PLN;NOV;SPEG
MLLT10;GJA1;MFN1;DLEU1;WNT16;LCORL;GPN1
AFAP1;ALX4;SSC5D
CR1L;CMTM5;ASCC2
KIAA1755;NOV;SPEG;CD34
DCAF16;GCKR;LXN;B3GNT7;SKIDA1;C2ORF16;EIF3E;ZNF512;TRIB2
PLN;NOV;SPEG
RSRC1;SHOX2;MFN1;GNB4;C2ORF16;EIF3E;FAM3C;LCORL;GPN1;DGKH;WARS2
DNAJC1;GCKR;FAM174B;EIF3E
APOC1;APOE;NDRG2
KIAA1755;CMTM5;NDRG2
EIF2B4;MLLT10;DES;ZNF219
CENPW;ZCCHC10;TOMM40;HSPA4;NCAPG;CWC27;GPN1;NIPSNAP1;WARS2;PPM1G;DCAF16;CR1L;MFN
PLN;NOV;SPEG
ZCCHC10;NOV;HSPA4;SPEG
SERPINA1;MTMR3;CR1;CR1L;PTEN;ASCC2
TBX15;DES;MYH7;TTN
TBX15;GFM1;NDRG2;MYH6;MYH7;TTN
IFI27L2;GJA1;CABP7;CMTM5;VSTM2L;SLC35F1;APOE;NDRG2

SERPINA1;MTMR3;CR1;CR1L;ZMAT5;PTEN
CCDC121;ALX4;ARHGEF40;CD34
DNAJC1;GCKR;B3GNT7;FAM174B;CEP85L;PVRL2;SUPT7L
SLC5A6;TOMM40;APOC1;ZNF219;PVRL2
DNAJC1;CCDC121;SKIDA1;ZMAT5;ZNF513;ASCC2;ZNF512;SUPT7L;TMEM253
DES;ZNF219;AFAP1
GFM1;CEP85L;MLF1;MYH6;TTN
SERPINA1;MTMR3;CR1;ZMAT5;PTEN;ASCC2;ARHGEF40
KIAA1755;DCAF16;CMTM5;GPR126;ARHGEF40;TRIB2
PLN;RSRC1;AFAP1;SPEG
PTEN;TRIB2;WARS2
ZCCHC10;SLC4A1AP;HSPA4;CABP7;VSTM2L;SLC35F1;CWC27;MLF1;GJA1;CCDC91;RSRC1;RSPO2;ZNF51
GCKR;B3GNT7;FAM174B;CEP85L;LCORL;SUPT7L
SLC4A1AP;PTEN;NCAPG;C2ORF16;CWC27;GPN1;NIPSNAP1;WARS2;MLLT10;CR1L;MFN1;DLEU1;EIF3E;L
CENPW;CR1;TOMM40;NCAPG;C2ORF16;CWC27;GPN1;NIPSNAP1;WARS2;PPM1G;DCAF16;CR1L;MFN1;
DCAF16;GCKR;CCDC121;LXN;SKIDA1;EIF3E;ZNF512;TRIB2
NOV;ZMAT5;SPEG
CCDC121;RSRC1;ALX4;CD34
LXN;APOC1;GPR126;CCDC141;DGKH
TOMM40;B3GNT7;ZNF219;ZNF513;TMEM253
DNAJC1;SHOX2;GNB4;EIF3E;LCORL;WARS2
DNAJC1;FAM174B;SKIDA1;ZNF513;C2ORF16;ZNF512;FAM3C;SUPT7L;TMEM253
PTEN;SSC5D;NRBP1
SERPINA1;MTMR3;CR1;CR1L;ZMAT5
DCAF16;DNAJC1;LXN;B3GNT7;FAM174B;CEP85L;GPR126;ZNF512;DGKH
CR1;CR1L;SNX17;APOC1;VSTM2L;NCAPG;APOE;ARHGEF40;TRIB2;SUPT7L
GJA1;MFN1;ALX4;WNT16;CD34
TBX15;GFM1;MYH6;MYH7;TTN
SERPINA1;MTMR3;CR1;CR1L;CMTM5;ZMAT5;PTEN
DES;ZNF219;SPEG
LXN;GPR126;EIF3E;CD34
VSTM2L;SLC35F1;NDRG2
SLC5A6;DCAF16;TOMM40;ALX4;ZNF513;ARHGEF40
KIAA1755;CMTM5;APOC1;VSTM2L;SLC35F1;APOE;WNT16;NDRG2;NAT14
SLC5A6;LXN;CD34
SLC5A6;GFM1;TOMM40;SNX17;PTEN
SERPINA1;MTMR3;CR1;CR1L;ZMAT5;ARHGEF40;TMEM253
EIF2B4;MLLT10;GJA1;MFN1;DLEU1;ALX4;WNT16
NOV;RSRC1;SPEG
DNAJC1;SERPINA1;CR1;TOMM40;B3GNT7
EIF2B4;TOMM40;ZNF628;SNX17;ZNF219;DLEU1;WNT16
MLLT10;DCAF16;GCKR;B3GNT7;CEP85L;ZNF512;ARHGEF40;PVRL2;TRIB2
CABP7;APOE;WNT16;NDRG2;NAT14
SSC5D;EIF3E;ZNF512;WNT16
DNAJC1;GNB4;EIF3E;FAM3C

PLN;NOV;SPEG
DNAJC1;CENPW;SERPINA1;MTMR3;CR1;CR1L;SNX17;ZMAT5;NCAPG;PPM1G
PLN;RSRC1;SPEG
EIF2B4;CCDC121;CENPW;ZCCHC10;SLC4A1AP;HSPA4;SKIDA1;CEP85L;NCAPG;C2ORF16;CWC27;GPN1;M
IFI27L2;GJA1;CCDC91;CABP7;VSTM2L;SLC35F1;UQCR10;CWC27;NDRG2
CENPW;AFAP1;ALX4;SSC5D
DNAJC1;B3GNT7;FAM174B
SLC5A6;IFI27L2;CABP7;VSTM2L;SLC35F1;NDRG2
DCAF16;GCKR;LXN;EIF3E;ZNF512;TRIB2
TBX15;SHOX2;PTEN;CD34;WARS2
CR1;CR1L;ASCC2
PTEN;SSC5D;WNT16
CABP7;VSTM2L;SLC35F1
SLC5A6;TOMM40;NOV;HSPA4;NRBP1;PPM1G
IFI27L2;CENPW;ZCCHC10;CR1;NCAPG;CWC27;GPN1;NIPSNAP1;PPM1G;DCAF16;CR1L;MFN1;EIF3E;CCD
EIF2B4;GCKR;CCDC121;CENPW;MTMR3;ZCCHC10;SLC4A1AP;HSPA4;SKIDA1;CEP85L;NCAPG;C2ORF16;C
MLLT10;RSPO2;SSC5D
SERPINA1;MTMR3;CR1;CMTM5;ZMAT5;PTEN;ARHGEF40
ZCCHC10;CR1;LXN;EIF3E;CD34
GJA1;NOV;HSPA4;SPEG
SERPINA1;MTMR3;CR1;CR1L;PTEN;ASCC2;ARHGEF40
SLC5A6;EIF2B4;KIAA1755;DCAF16;ZNF219;EIF3E;ZNF512;ARHGEF40;NAT14;NRBP1
TBX15;CEP85L;NDRG2;LCORL;MYH7;TTN
CCDC121;ZCCHC10;SLC4A1AP;CEP85L;NCAPG;CWC27;PPM1G;CCDC91;ZNF628;DLEU1;EIF2B4;GCKR;CE
TOMM40;HSPA4;NCAPG;GPN1;MYH6
SERPINA1;MTMR3;CR1;CR1L;PTEN;ASCC2
CCDC121;GJA1;CCDC91;APOC1;SLC35F1;APOE;NDRG2
SERPINA1;CR1;CR1L;ZMAT5;ASCC2
TBX15;GFM1;UQCR10;MYH7;TTN
CENPW;SERPINA1;MTMR3;CR1;CR1L;ZMAT5;PTEN
SERPINA1;MTMR3;CR1;CR1L;PTEN;ASCC2
SERPINA1;MTMR3;CR1;CR1L;ASCC2;ARHGEF40
DNAJC1;SERPINA1;CR1;LXN;B3GNT7;PVRL2
MLLT10;RSPO2;SPEG
B3GNT7;SNX17;APOC1;PTEN;GPR126
TBX15;GFM1;MYH6;MYH7;TTN
SERPINA1;MTMR3;CR1;CR1L;ZMAT5;PTEN;ARHGEF40;PVRL2
CR1;LXN;SSC5D
RSPO2;GPR126;EIF3E
DNAJC1;GCKR;B3GNT7;FAM174B;MFN1
NOV;SPEG;SSC5D
SHOX2;GNB4;AFAP1;C2ORF16;FAM3C
DNAJC1;FAM174B;MFN1;CCDC141
TBX15;RSRC1;SHOX2;AFAP1;SSC5D
PLN;RSRC1;SHOX2;SPEG

DCAF16;CMTM5;CEP85L;MFN1;PTEN;GPR126;ZNF512;ARHGEF40;CD34;TRIB2
RSRC1;SHOX2;SPEG
DNAJC1;B3GNT7;GPR126;CCDC141
IFI27L2;CMTM5;VSTM2L;APOE
SERPINA1;ZCCHC10;CR1;B3GNT7;CR1L;CEP85L;MFN1;ZNF512;FAM3C;LCORL;SUPT7L;TMEM253
CENPW;ZCCHC10;ZMAT5;DLEU1;NCAPG;ASCC2
MLLT10;DNAJC1;CCDC121;FAM174B;SKIDA1;ZNF512;FAM3C;CD34;TMEM253;WARS2
SERPINA1;B3GNT7;APOC1;CCDC141;PVRL2
MLLT10;GJA1;GNB4;ALX4;WNT16;LCORL
HSPA4;GNB4;AFAP1;NCAPG;C2ORF16;FAM3C
TBX15;GFM1;DES;MYH7;TTN
DES;ZNF219;SSC5D
SERPINA1;MTMR3;CR1;ZMAT5;PTEN;ARHGEF40
TBX15;CEP85L;MYH7;TTN
SLC5A6;APOE;ARHGEF40;CD34
CCDC121;SLC4A1AP;CCDC91;APOC1;SLC35F1;APOE;CWC27;NDRG2
IFI27L2;DCAF16;SLC4A1AP;CCDC91;RSRC1;CMTM5;RSPO2;VSTM2L;SLC35F1;APOE;NDRG2
PLN;NOV;SPEG
NOV;SPEG
IFI27L2;CENPW;ZCCHC10;TOMM40;HSPA4;NCAPG;CWC27;GPN1;NIPSNAP1;NRBP1;WARS2;PPM1G;DC
GJA1;VSTM2L;SLC35F1;NDRG2
EIF2B4;DNAJC1;TOMM40;SNX17;TRIB2
DES;ZNF219;GNB4
SLC5A6;DCAF16;LXN;ZNF628;MFN1;ALX4;ZNF513;PVRL2
CR1;ZNF628;SNX17;APOC1;ZMAT5;VSTM2L;ZNF513;ZNF512;APOE;ARHGEF40;TRIB2
LXN;TOMM40;B3GNT7;UQCR10;NIPSNAP1;TMEM253;PPM1G
HSPA4;DLEU1;GPR126;ASCC2;NRBP1
EIF2B4;DNAJC1;LXN;ZNF628;ZNF219;ZNF513;SUPT7L
DNAJC1;TOMM40;B3GNT7;ZNF628;APOC1;VSTM2L;ZNF513
SERPINA1;MTMR3;CR1;CR1L;ZMAT5;PTEN;ARHGEF40
DCAF16;SHOX2;GNB4;ALX4;C2ORF16;FAM3C;CWC27;LCORL;GPN1;NRBP1;WARS2;PPM1G
NOV;RSRC1;SPEG
DNAJC1;LXN;B3GNT7;FAM174B;RSPO2;GPR126
EIF3E;WNT16;NRBP1
CENPW;TOMM40;ZMAT5;NCAPG;ZNF513;ASCC2;PPM1G
CENPW;LXN;TOMM40;B3GNT7;ZNF513;UQCR10;NIPSNAP1;TMEM253
RSPO2;SSC5D
MLLT10;DCAF16;CCDC121;ZCCHC10;RSRC1;CEP85L;MFN1;ALX4;EIF3E;ZNF512;LCORL;WARS2
KIAA1755;CABP7;CMTM5;APOC1;SLC35F1;APOE;NDRG2;NAT14
CEP85L;GNB4;RSPO2;GPR126;WNT16;PVRL2;CD34;TRIB2;DGKH
IFI27L2;CENPW;TOMM40;HSPA4;SNX17;GNB4;NCAPG;CWC27;GPN1;PPM1G
FAM174B;ZNF219;DGKH
TBX15;GFM1;ZCCHC10;SLC4A1AP;CCDC91;TTN
DES;ZNF219
CENPW;HSPA4;SHOX2;GNB4;NCAPG;EIF3E;GPN1

SERPINA1;MTMR3;CR1;ZMAT5;PTEN;ARHGEF40;TMEM253
SLC5A6;APOC1;ALX4
NOV;RSRC1;SPEG
CCDC91;HSPA4;CABP7;RSPO2;VSTM2L;SLC35F1;FAM3C;WNT16
SERPINA1;B3GNT7;ZNF513;FAM3C;LCORL;TMEM253
MTMR3;CR1;CR1L;ASCC2
PLN;SPEG;EIF3E;ZNF512
TBX15;DES;ZNF628;NDRG2;TTN
IFI27L2;SLC4A1AP;CCDC91;CABP7;RSRC1;CMTM5;RSPO2;VSTM2L;SLC35F1;NIPSNAP1
AFAP1;ALX4;SSC5D;DGKH
DES;ZNF219;AFAP1
IFI27L2;DNAJC1;HSPA4;ZNF628;SNX17;GNB4;AFAP1;NCAPG;EIF3E;CWC27;GPN1;WARS2
HSPA4;ASCC2;PVRL2;NRBP1;PPM1G
GFM1;ZCCHC10;SLC4A1AP;TOMM40;HSPA4;AFAP1;CWC27;GPN1;WARS2;PPM1G;DCAF16;CCDC91;ZN[illegible]
EIF2B4;GCKR;CCDC121;CENPW;SLC4A1AP;HSPA4;SKIDA1;CEP85L;NCAPG;C2ORF16;CWC27;GPN1;MLF1
CCDC121;SSC5D;EIF3E;WNT16;CD34
CCDC121;GJA1;SLC4A1AP;CCDC91;RSRC1;SKIDA1;RSPO2;VSTM2L;SLC35F1;FAM3C;NDRG2;NIPSNAP1
ZCCHC10;SLC4A1AP;TOMM40;HSPA4;SHOX2;CWC27;GPN1;WARS2;PPM1G;ZNF628;ZNF219;GNB4;TRI[illegible]
CCDC121;ZNF219;CD34
CENPW;LXN;ZNF513
MLLT10;GJA1;MFN1;GNB4;GPR126;ALX4;WNT16
GCKR;FAM174B;C2ORF16
SLC5A6;DCAF16;ALX4;EIF3E
NOV;RSRC1;AFAP1
CCDC121;HSPA4;CMTM5
ZNF628;SNX17;SKIDA1;ZNF513;NAT14;SUPT7L;TMEM253
DNAJC1;LXN;TRIB2;DGKH
SHOX2;GNB4;AFAP1;SSC5D;C2ORF16;LCORL;GPN1;DGKH;WARS2
GJA1;ZNF628;ALX4;WNT16;NRBP1
MLLT10;SHOX2;CD34;GPN1;WARS2
FAM174B;CCDC141
EIF2B4;DCAF16;MTMR3;LXN;MFN1;ZNF513;C2ORF16;PVRL2;SUPT7L
CENPW;ZCCHC10;CR1;HSPA4;PTEN;NCAPG;CWC27;GPN1;NIPSNAP1;PPM1G;DCAF16;RSRC1;MFN1;DL[illegible]
SERPINA1;MTMR3;CR1;ZMAT5;ASCC2;ARHGEF40
CCDC121;AFAP1
TBX15;SNX17;PTEN
CENPW;HSPA4;NCAPG;NRBP1
IFI27L2;GFM1;CENPW;ZCCHC10;TOMM40;HSPA4;NCAPG;CWC27;GPN1;NIPSNAP1;WARS2;PPM1G;CR1
SLC5A6;PTEN;ALX4;EIF3E
PLN;NOV;SPEG
EIF2B4;GCKR;CCDC121;CENPW;ZCCHC10;SLC4A1AP;HSPA4;SKIDA1;CEP85L;NCAPG;C2ORF16;CWC27;[illegible]
PLN;NOV;SPEG
TBX15;SLC4A1AP;UQCR10;NDRG2;TTN
SLC5A6;KIAA1755;IFI27L2;CABP7;CMTM5;VSTM2L;SLC35F1
SERPINA1;MTMR3;CR1;CR1L;ZMAT5;ARHGEF40

GJA1;NOV;SPEG
SLC5A6;GCKR;TOMM40;APOC1
EIF2B4;DCAF16;LXN;FAM174B;RSPO2;VSTM2L;ZNF512;PVRL2
CCDC121;MTMR3;ZCCHC10;SLC4A1AP;CEP85L;NCAPG;CWC27;PPM1G;CCDC91;ZNF628;DLEU1;EIF2B4;[illegible]
DCAF16;GCKR;LXN;C2ORF16;EIF3E;ZNF512;CWC27;TRIB2;SUPT7L;DGKH
KIAA1755;CCDC121;CMTM5;APOC1;APOE;NDRG2
SLC5A6;GCKR;TOMM40;PTEN
DNAJC1;LXN;B3GNT7;TRIB2;DGKH
EIF2B4;GJA1;DLEU1;ALX4;WNT16
IFI27L2;CENPW;ZCCHC10;TOMM40;PTEN;NCAPG;CWC27;GPN1;NIPSNAP1;PPM1G;CR1L;RSRC1;MFN1;[illegible]
SLC5A6;MLLT10;DCAF16;SSC5D;ZNF512;DGKH
CMTM5;SHOX2;SLC35F1;TRIB2
CENPW;TOMM40;HSPA4;GNB4;DLEU1;AFAP1;NCAPG;ALX4;CWC27;GPN1;PPM1G
SERPINA1;MTMR3;CR1;ZMAT5;PTEN;ARHGEF40;TMEM253
SERPINA1;MTMR3;CR1;CR1L;PTEN;ASCC2;ARHGEF40
CCDC121;GJA1;SLC4A1AP;CCDC91;RSRC1;CMTM5;RSPO2;SLC35F1;C2ORF16;NDRG2
PLN;B3GNT7;NOV;SPEG
GFM1;ZCCHC10;SLC4A1AP;VSTM2L;SLC35F1;C2ORF16;GPN1;NIPSNAP1;DCAF16;GJA1;CCDC91;RSRC1;[illegible]
SERPINA1;MTMR3;CR1;ZMAT5;ASCC2;ARHGEF40
TBX15;AFAP1;CD34
IFI27L2;CCDC121;GJA1;CCDC91;CABP7;VSTM2L;SLC35F1;APOE;CWC27;NDRG2
CENPW;ZCCHC10;PTEN;NCAPG;CWC27;GPN1;NIPSNAP1;WARS2;PPM1G;DCAF16;CR1L;MFN1;DLEU1;[illegible]
SHOX2;GNB4;NCAPG;FAM3C;LCORL
SERPINA1;MTMR3;CR1;CR1L;ZMAT5;PTEN
EIF2B4;GCKR;CCDC121;CENPW;SLC4A1AP;SKIDA1;CEP85L;AFAP1;NCAPG;C2ORF16;CWC27;GPN1;MLF1[illegible]
ZNF628;NRBP1;DGKH
SERPINA1;MTMR3;CR1;CR1L;SNX17;ZMAT5;PTEN;PVRL2
SHOX2;PTEN;CD34
PLN;NOV;SPEG
DNAJC1;FAM174B;CCDC141
TBX15;SHOX2;CD34
GJA1;PLN;NOV
IFI27L2;CENPW;ZCCHC10;TOMM40;HSPA4;NCAPG;C2ORF16;CWC27;GPN1;NIPSNAP1;PPM1G;CR1L;[illegible]
DNAJC1;B3GNT7;GPR126;CCDC141
CCDC121;GFM1;MTMR3;ZCCHC10;SLC4A1AP;CEP85L;NCAPG;PPM1G;DCAF16;CCDC91;ZNF628;DLEU1;[illegible]
IFI27L2;CMTM5;APOC1;VSTM2L;SLC35F1;UQCR10;APOE;NDRG2;NAT14
PLN;NOV;SPEG
IFI27L2;CMTM5;GPR126;ARHGEF40;TRIB2;SUPT7L
TBX15;GFM1;ZCCHC10;LCORL;MYH7;TTN
EIF2B4;ZNF628;SNX17;ZNF219;DLEU1;ZNF513;WNT16;PVRL2
EIF2B4;GCKR;CCDC121;CENPW;SLC4A1AP;SKIDA1;CEP85L;NCAPG;C2ORF16;CWC27;GPN1;MLF1;NRBP1[illegible]
B3GNT7;SNX17;APOC1;GPR126;CCDC141
GJA1;PLN;NOV
GCKR;B3GNT7;FAM174B;PVRL2
PLN;NOV;SPEG

SLC5A6;TOMM40;APOC1;AFAP1;PVRL2
KIAA1755;NOV;SPEG;NRBP1
GFM1;CENPW;ZCCHC10;CR1;TOMM40;HSPA4;PTEN;NCAPG;CWC27;GPN1;NIPSNAP1;PPM1G;CR1L;MF
SERPINA1;MTMR3;CR1;CR1L;ZMAT5;ARHGEF40
DNAJC1;SERPINA1;B3GNT7;FAM174B;DGKH
KIAA1755;CABP7;CMTM5;SLC35F1;NDRG2;NAT14
SLC5A6;TBX15;TOMM40;PTEN
APOE;NDRG2
EIF2B4;MLLT10;CCDC121;ZCCHC10;CMTM5;SHOX2;CEP85L;SLC35F1;ZNF512
PLN;NOV;SPEG
GCKR;B3GNT7;FAM174B;C2ORF16
IFI27L2;GJA1;ZNF628;ALX4;WNT16;NRBP1
ZNF219;PTEN;C2ORF16
PLN;NOV;SPEG
GFM1;CENPW;ZCCHC10;TOMM40;HSPA4;NCAPG;CWC27;GPN1;NIPSNAP1;PPM1G;SLC5A6;SNX17;RSR
EIF2B4;GCKR;CCDC121;GFM1;CENPW;MTMR3;ZCCHC10;SLC4A1AP;HSPA4;SKIDA1;CEP85L;NCAPG;C2O
IFI27L2;CABP7;CMTM5;VSTM2L;SLC35F1;NDRG2
CENPW;ZCCHC10;CR1;PTEN;NCAPG;CWC27;GPN1;NIPSNAP1;WARS2;SLC5A6;DCAF16;MFN1;DLEU1;EIF
ZCCHC10;PLN;CCDC91;RSRC1;MFN1;GNB4;AFAP1
CENPW;ZCCHC10;CR1;HSPA4;NCAPG;C2ORF16;CWC27;GPN1;NIPSNAP1;PPM1G;DCAF16;CR1L;MFN1;D
KIAA1755;CMTM5;CEP85L;SLC35F1;ARHGEF40;TRIB2
TBX15;GFM1;MYH7;TTN
IFI27L2;DNAJC1;HSPA4;GNB4;NCAPG;FAM3C
DCAF16;GCKR;LXN;CEP85L;C2ORF16;EIF3E;ZNF512;APOE;TRIB2;SUPT7L
B3GNT7;APOC1;GPR126;CCDC141
ZCCHC10;LXN;ZNF512;NRBP1
RSPO2;SSC5D
B3GNT7;CR1L;ASCC2
PLN;NOV;SPEG
MLLT10;PLN;NOV;SPEG
DNAJC1;CCDC121;FAM174B;SKIDA1;SUPT7L;TMEM253
IFI27L2;GNB4;ALX4;EIF3E;NIPSNAP1
EIF2B4;ZNF628;ZNF219;DLEU1;ALX4;WNT16
SLC5A6;GFM1;LXN;CCDC91;RSRC1;MFN1;ALX4;EIF3E;WARS2
TBX15;PTEN;C2ORF16;EIF3E;CD34;GPN1;WARS2
CR1L;ASCC2
SERPINA1;MTMR3;CR1;CR1L;ASCC2
SERPINA1;CR1;CR1L;ASCC2
GJA1;CMTM5;APOC1;APOE;NDRG2;NAT14
SERPINA1;MTMR3;CR1;B3GNT7;CR1L;ASCC2
NOV;SPEG;DGKH
NOV;SHOX2;DGKH
EIF2B4;GJA1;CCDC91
MLLT10;ZCCHC10;SHOX2;ALX4;EIF3E;LCORL;PVRL2;SUPT7L;WARS2
IFI27L2;CMTM5;RSPO2;VSTM2L;SLC35F1;FAM3C;APOE;NDRG2;NAT14

DCAF16;GCKR;LXN;B3GNT7;SSC5D;C2ORF16;TRIB2
DES;MYH6;MYH7;TTN
ALX4;SSC5D;ARHGEF40;CD34
SERPINA1;MTMR3;CR1;CR1L;PTEN;ASCC2
CR1;SNX17;APOC1;ZMAT5;VSTM2L;ZNF513;APOE;ARHGEF40;TRIB2
DES;PLN;ZNF219;CEP85L;NCAPG;ZNF512;CCDC141;NAT14;TTN
SERPINA1;SHOX2;CEP85L;NCAPG;SSC5D;WNT16;TBX15;KIAA1755;DNAJC1;GJA1;PLN;NOV;RSPO2
KIAA1755;GJA1;CMTM5;SLC35F1;APOE;NDRG2;MYH7;DGKH
NOV;RSPO2;VSTM2L;SLC35F1;HORMAD2;MLF1;MYH7
GJA1;CMTM5;AFAP1;VSTM2L;NCAPG;SPEG;SLC35F1;TRIB2;NAT14
GCKR;SERPINA1;CR1L;ZNF628;SHOX2;APOC1;ZNF219;ZNF513;APOE
CENPW;SKIDA1;PTEN;NCAPG;SLC35F1;CWC27;MLLT10;DCAF16;GJA1;RSRC1;ARHGEF40;LCORL;TRIB2
IFI27L2;SERPINA1;CABP7;RSRC1;FAM174B;PTEN;DLEU1
KIAA1755;CCDC121;GJA1;PLN;NOV;AFAP1;SSC5D;DGKH
GJA1;CENPW;B3GNT7;CABP7;APOC1;CEP85L;ZMAT5;MFN1;RSPO2;NCAPG;HORMAD2;MLF1
KIAA1755;GJA1;NOV;CMTM5;RSPO2;VSTM2L;SPEG;SLC35F1;APOE;NDRG2;NAT14;MYH7
SLC5A6;TBX15;KIAA1755;ZNF628;SHOX2;ZNF219;SPEG;ALX4;SSC5D;NDRG2;ARHGEF40
GCKR;CEP85L;NDRG2;TTN;GJA1;DES;PLN;NOV;APOC1;ZNF219;APOE;ARHGEF40;CCDC141
DES;SERPINA1;LXN;PLN;B3GNT7;ZNF219;RSPO2;SSC5D;TMEM253
CMTM5;AFAP1;VSTM2L;NCAPG;SLC35F1;NAT14;GJA1;ZNF628;ZNF219;SPEG;HORMAD2;TRIB2;MYH7
SHOX2;AFAP1;NCAPG;SSC5D;WNT16;NAT14;TBX15;KIAA1755;GJA1;NOV;SPEG;ALX4;ARHGEF40
CENPW;HSPA4;SKIDA1;NCAPG;SLC35F1;CWC27;MLF1;NIPSNAP1;SLC5A6;GJA1;B3GNT7;APOC1;CCDC14
GCKR;LXN;SHOX2;AFAP1;VSTM2L;SSC5D;NAT14;TBX15;KIAA1755;GJA1;PLN;ZNF628;SPEG
CCDC121;SHOX2;AFAP1;VSTM2L;SSC5D;TBX15;KIAA1755;GJA1;DES;SPEG;ALX4;APOE;MYH7
CCDC121;SHOX2;AFAP1;VSTM2L;SSC5D;WNT16;NAT14;TMEM253;TBX15;GJA1;ZNF628;ZNF219;SPEG
KIAA1755;CCDC121;CMTM5;CEP85L;RSPO2;VSTM2L;SLC35F1;CWC27;NDRG2;NAT14
TBX15;GJA1;NOV;SHOX2;AFAP1;SPEG;ALX4;SSC5D;WNT16;ARHGEF40
GJA1;CENPW;SKIDA1;NCAPG;SLC35F1;CWC27;LCORL;MLF1;NIPSNAP1
IFI27L2;CCDC121;CMTM5;SSC5D;NDRG2;NAT14;TBX15;KIAA1755;DES;NOV;RSPO2;SPEG;APOE;CD34
LXN;SSC5D;TBX15;KIAA1755;GJA1;DES;PLN;NOV;ZNF628;APOC1;ZNF219;ALX4;APOE;CD34
SKIDA1;CEP85L;AFAP1;VSTM2L;SLC35F1;WNT16;TTN;KIAA1755;PLN;RSPO2;ALX4;LCORL;MYH7;DGKH
KIAA1755;GJA1;ZNF628;VSTM2L;NCAPG;SSC5D;WNT16;NAT14;TMEM253
GJA1;DES;SERPINA1;PLN;APOC1;RSPO2;VSTM2L;SSC5D;APOE
TBX15;KIAA1755;IFI27L2;GJA1;LXN;NOV;SHOX2;PTEN;AFAP1;SPEG;SSC5D
CABP7;FAM174B;SPEG;ALX4;SSC5D;NAT14
GCKR;SERPINA1;B3GNT7;ZNF628;FAM174B;VSTM2L
KIAA1755;DES;SERPINA1;PLN;RSPO2;SSC5D
GJA1;LXN;NOV;AFAP1;VSTM2L;SSC5D;NAT14;TMEM253
NOV;ZNF628;FAM174B;NCAPG;APOE;ARHGEF40;TRIB2;NAT14
CCDC121;SERPINA1;CR1;NOV;CR1L;ZNF628;ARHGEF40;TMEM253
CCDC121;CENPW;LXN;CR1L;ZNF628;CMTM5;APOC1;NCAPG;NAT14
DES;SERPINA1;CR1;CR1L;APOC1;VSTM2L;SSC5D;APOE;WNT16;TRIB2
TBX15;KIAA1755;GJA1;ZNF628;CABP7;AFAP1;VSTM2L;SPEG;SSC5D;HORMAD2;ARHGEF40;NAT14
GJA1;PLN;NOV;CMTM5;RSPO2;VSTM2L;SLC35F1;FAM3C;WNT16;NDRG2;MYH7
TBX15;GJA1;SHOX2;ZNF219;ALX4;WNT16;TMEM253

DCAF16;SKIDA1;MFN1;PTEN;DLEU1;SLC35F1;LCORL
KIAA1755;NOV;ZNF628;SSC5D;WNT16;NAT14;TMEM253
DNAJC1;ZNF628;SHOX2;AFAP1;NCAPG;NAT14;TMEM253
SLC5A6;GJA1;CENPW;B3GNT7;CABP7;APOC1;NCAPG;HORMAD2
HSPA4;SKIDA1;RSPO2;VSTM2L;SLC35F1;UQCR10;APOE;ARHGEF40;TRIB2;MLF1
CENPW;TOMM40;NOV;HSPA4;APOC1;NCAPG;ALX4;UQCR10;WARS2;PPM1G
TBX15;DNAJC1;GCKR;GJA1;SERPINA1;APOC1;HORMAD2;APOE;NDRG2
HSPA4;UQCR10
TBX15;GJA1;SHOX2;ZNF219;AFAP1;VSTM2L;SPEG;SSC5D;ARHGEF40;NAT14;TMEM253
KIAA1755;GJA1;CABP7;CMTM5;SHOX2;RSPO2;VSTM2L;SPEG;ALX4;SLC35F1
CENPW;CR1L;EIF3E;UQCR10
PLN;CEP85L;CCDC141;WARS2
LXN;SNX17;EIF3E;CD34
GJA1;DES;VSTM2L;ALX4;WARS2;TTN
SLC5A6;GJA1;DES;MTMR3;CR1;PTEN;NRBP1;MYH6
ZNF628;VSTM2L;HORMAD2;CCDC141;MLF1;TTN
SLC5A6;LXN;ZNF628;FAM174B;ZNF219;TMEM253
CCDC121;SERPINA1;CR1;NOV;CR1L;ARHGEF40
GJA1;ZNF628;ZNF219;ALX4;TMEM253
GJA1;DES;ZNF219;NDRG2;TMEM253
SERPINA1;B3GNT7;EIF3E;UQCR10;NDRG2
KIAA1755;GJA1;CABP7;CMTM5;SHOX2;RSPO2;VSTM2L;SPEG;ALX4;SLC35F1
CCDC121;SERPINA1;FAM174B;NDRG2;TMEM253
SERPINA1;CR1;CR1L;APOC1;HORMAD2
SERPINA1;PLN;FAM174B;VSTM2L;NAT14
GJA1;LXN;AFAP1;NCAPG;WNT16;NAT14
TBX15;KIAA1755;IFI27L2;DES;MFN1;GNB4;PTEN;AFAP1;MYH7;DGKH;TTN
GCKR;CCDC121;CENPW;MTMR3;CR1L;FAM174B;RSPO2;NCAPG;CCDC141;MLF1;TMEM253
TBX15;GCKR;SERPINA1;ZNF628;APOC1;ZNF219;ZNF513;HORMAD2;APOE;NDRG2;NIPSNAP1
GCKR;GJA1;ZNF628;SHOX2;APOC1;VSTM2L;ZNF513;SSC5D;APOE;WNT16;TRIB2;NAT14
KIAA1755;GJA1;NOV;ZNF628;SHOX2;RSPO2;AFAP1;SPEG;SSC5D;WNT16;ARHGEF40;NAT14
KIAA1755;GJA1;NOV;CABP7;CMTM5;RSPO2;VSTM2L;SPEG;SLC35F1;APOE;NDRG2;NAT14
TBX15;CCDC121;DES;CCDC91;ZNF628;SHOX2;ZNF513;HORMAD2;CCDC141;MLF1;MYH7
KIAA1755;GJA1;DES;SERPINA1;CR1;PLN;B3GNT7;RSPO2;FAM3C;LCORL;TMEM253
GJA1;SKIDA1;SHOX2;RSPO2;AFAP1;VSTM2L;SLC35F1;TRIB2;NAT14;NIPSNAP1
TBX15;GJA1;NOV;ZNF628;FAM174B;ZNF219;AFAP1;ARHGEF40
TBX15;GJA1;DES;SHOX2;NCAPG;SSC5D;WNT16;TTN
KIAA1755;DES;SERPINA1;B3GNT7;ZNF219;RSPO2;APOE;TMEM253
ZNF628;SHOX2;AFAP1;VSTM2L;NCAPG;SSC5D;HORMAD2;NAT14
CCDC121;CENPW;MTMR3;CR1L;FAM174B;RSPO2;NCAPG;ZNF513;CCDC141;MYH7
LXN;CCDC141;GPN1;WARS2
ZNF628;ZNF513;NAT14;MYH7
TBX15;CCDC121;CABP7;HORMAD2
IFI27L2;B3GNT7;ZNF628;ZMAT5
CCDC121;CR1;CD34;TRIB2;NAT14

CCDC121
B3GNT7
GCKR;GJA1;SERPINA1;B3GNT7;ALX4;CCDC141;MYH6
KIAA1755;MFN1;VSTM2L;SLC35F1;TRIB2
TBX15;SHOX2;NCAPG;SSC5D;WNT16
SERPINA1;CR1;CR1L;CMTM5;WNT16
CCDC121;DES;ALX4;HORMAD2;NDRG2
EIF3E;CD34
B3GNT7;EIF3E;UQCR10
SHOX2;RSPO2;SLC35F1;TRIB2
CR1;CEP85L;WNT16;CCDC141;TTN
ZNF628;ZNF513;WNT16;CCDC141;WARS2
SLC5A6;ZNF628;CEP85L;CCDC141;TRIB2
GJA1;ZNF219;VSTM2L;TMEM253;DGKH

;

;MYH6

RL;CD34

2;ARHGEF40;LCORL;DGKH

LEU1;ZNF512;APOE;LCORL

;DGKH

:H

;DGKH

KH

;DGKH
IGEF40;LCORL

;DGKH

26;HORMAD2;ARHGEF40;LCORL;DGKH

L;MYH6;DGKH

CD34

28;ZNF219;SPEG;ALX4;APOE;CCDC141;CD34;MYH6;MYH7

-219;ZMAT5;APOE;ARHGEF40;PVRL2;TRIB2

APOE;LCORL

[illegible]3C;CCDC141;CENPW;CR1;HSPA4;AFAP1;TBX15;MLLT10;DNAJC1;EIF3E;LCORL;PVRL2;TRIB2;MYH6

;DLEU1;RSPO2;ALX4;ASCC2;HORMAD2;FAM3C;ARHGEF40;CCDC141;GCKR;HSPA4;AFAP1;SSC5D;C2ORF1

RC1;FAM174B;DLEU1;RSPO2;SPEG;FAM3C;ARHGEF40;CD34;CR1;SKIDA1;AFAP1;VSTM2L;MLF1;NIPSNAI

RC1;FAM174B;DLEU1;RSPO2;SPEG;FAM3C;ARHGEF40;CD34;CR1;SKIDA1;AFAP1;VSTM2L;C2ORF16;WNT

LEU1;FAM3C;ARHGEF40;CD34;CR1;AFAP1;MLF1;NIPSNAP1;NRBP1;SUPT7L;SLC5A6;DNAJC1;B3GNT7;NC

RC1;FAM174B;DLEU1;SPEG;FAM3C;ARHGEF40;CD34;CR1;SKIDA1;AFAP1;VSTM2L;WNT16;MLF1;NIPSNA
LEU1;RSPO2;SPEG;FAM3C;ARHGEF40;CD34;SKIDA1;AFAP1;WNT16;MLF1;NRBP1;SUPT7L;DNAJC1;B3GNT
SPEG;FAM3C;SKIDA1;AFAP1;C2ORF16;WNT16;MLF1;NRBP1;SUPT7L;DNAJC1;B3GNT7;GPR126;ZNF513;Z

RC1;FAM174B;DLEU1;RSPO2;SPEG;FAM3C;ARHGEF40;CD34;SKIDA1;AFAP1;VSTM2L;WNT16;MLF1;NIPS

F40;CD34;AFAP1;VSTM2L;MLF1;NIPSNAP1;NRBP1;SLC5A6;B3GNT7;NOV;ZNF513;ZNF512;TRIB2;MYH6;

APOE;ARHGEF40;PVRL2;TRIB2
5;ZNF513;APOE;ARHGEF40;TRIB2
NF219;ZMAT5;APOE;ARHGEF40

RC1;FAM174B;DLEU1;RSPO2;SPEG;HORMAD2;FAM3C;ARHGEF40;CD34;CR1;SKIDA1;AFAP1;VSTM2L;C2[illegible]

[illegible]O2;SPEG;FAM3C;ARHGEF40;CD34;CR1;SKIDA1;AFAP1;VSTM2L;WNT16;MLF1;NIPSNAP1;NRBP1;SUPT7[illegible]

174B;DLEU1;RSPO2;SPEG;FAM3C;ARHGEF40;CD34;CR1;SKIDA1;AFAP1;VSTM2L;C2ORF16;WNT16;MLF1

LEU1;RSPO2;SPEG;FAM3C;ARHGEF40;CD34;CR1;SKIDA1;AFAP1;VSTM2L;C2ORF16;WNT16;MLF1;NIPSN[illegible]

174B;DLEU1;RSPO2;SPEG;FAM3C;ARHGEF40;CD34;CR1;SKIDA1;AFAP1;VSTM2L;WNT16;MLF1;NIPSNAP

;APOE;ARHGEF40;PVRL2;TRIB2

FAM174B;DLEU1;GPR126;EIF3E;FAM3C;CCDC141;LCORL;TRIB2

RC1;FAM174B;DLEU1;RSPO2;SPEG;FAM3C;ARHGEF40;CD34;AFAP1;MLF1;NIPSNAP1;NRBP1;SUPT7L;TM

DLEU1;RSPO2;SPEG;FAM3C;ARHGEF40;CD34;CR1;SKIDA1;AFAP1;C2ORF16;WNT16;MLF1;NIPSNAP1;NRI

M174B;DLEU1;RSPO2;SPEG;FAM3C;ARHGEF40;CD34;SKIDA1;AFAP1;VSTM2L;MLF1;NIPSNAP1;NRBP1;SU

NOV;SNX17;APOC1;ZNF219;ZMAT5;ASCC2;APOE;ARHGEF40

G;FAM3C;ARHGEF40;CD34;AFAP1;MLF1;NIPSNAP1;NRBP1;SLC5A6;DNAJC1;B3GNT7;NOV;ZNF513;ZNF5[illegible]

;FAM3C;ARHGEF40;CR1;SKIDA1;AFAP1;VSTM2L;C2ORF16;WNT16;MLF1;NRBP1;SLC5A6;DNAJC1;B3GNT[illegible]

RC1;FAM174B;DLEU1;RSPO2;SPEG;FAM3C;ARHGEF40;CD34;SKIDA1;AFAP1;VSTM2L;C2ORF16;WNT16;[illegible]

LEU1;RSPO2;SPEG;FAM3C;ARHGEF40;CR1;SKIDA1;AFAP1;VSTM2L;C2ORF16;WNT16;MLF1;NIPSNAP1;N

N;PPM1G;CCDC91;RSRC1;FAM174B;MFN1;SPEG;ASCC2;CCDC141;EIF2B4;GCKR;HSPA4;WNT16;GPN1;NI

RC1;FAM174B;DLEU1;RSPO2;SPEG;FAM3C;ARHGEF40;CD34;CR1;SKIDA1;AFAP1;VSTM2L;C2ORF16;WNT
SPO2;SPEG;FAM3C;ARHGEF40;CD34;CR1;SKIDA1;AFAP1;VSTM2L;C2ORF16;WNT16;MLF1;NIPSNAP1;NRB
PO2;FAM3C;ARHGEF40;CR1;SKIDA1;AFAP1;VSTM2L;C2ORF16;WNT16;MLF1;NIPSNAP1;NRBP1;SUPT7L;D
SPO2;SPEG;FAM3C;CD34;CR1;SKIDA1;AFAP1;VSTM2L;WNT16;MLF1;NIPSNAP1;NRBP1;SUPT7L;DNAJC1;E

RC1;FAM174B;DLEU1;RSPO2;SPEG;FAM3C;ARHGEF40;CD34;CR1;SKIDA1;AFAP1;VSTM2L;C2ORF16;WNT

O2;SPEG;FAM3C;ARHGEF40;CD34;AFAP1;VSTM2L;MLF1;NIPSNAP1;NRBP1;SLC5A6;B3GNT7;NOV;ZNF513
0;EIF2B4;HSPA4;GPN1;MLF1;NRBP1;TBX15;MLLT10;DNAJC1;SNX17;ZNF513;EIF3E;PVRL2

RC1;FAM174B;DLEU1;RSPO2;SPEG;FAM3C;ARHGEF40;CD34;CR1;SKIDA1;AFAP1;VSTM2L;C2ORF16;WNT

RC1;FAM174B;DLEU1;RSPO2;SPEG;FAM3C;ARHGEF40;CD34;CR1;SKIDA1;AFAP1;VSTM2L;C2ORF16;WNT

AM174B;MFN1;RSPO2;SPEG;ALX4;ASCC2;FAM3C;APOE;CD34;EIF2B4;HSPA4;CMTM5;AFAP1;VSTM2L;GI

RC1;FAM174B;DLEU1;RSPO2;SPEG;FAM3C;ARHGEF40;CD34;SKIDA1;AFAP1;VSTM2L;C2ORF16;WNT16;N

B;DLEU1;RSPO2;SPEG;FAM3C;ARHGEF40;CD34;CR1;SKIDA1;AFAP1;VSTM2L;C2ORF16;WNT16;MLF1;NRI

MAT5;APOE;ARHGEF40;TRIB2

RC1;FAM174B;DLEU1;RSPO2;SPEG;FAM3C;ARHGEF40;CD34;SKIDA1;AFAP1;VSTM2L;C2ORF16;WNT16;[illegible]

[illegible]HGEF40;CD34;CR1;SKIDA1;AFAP1;C2ORF16;WNT16;MLF1;NRBP1;SUPT7L;SLC5A6;DNAJC1;ZNF512;LC[illegible]

AM3C;APOE;CD34;MYH6

;DGKH

FAM3C;ARHGEF40;SKIDA1;AFAP1;MLF1;NIPSNAP1;NRBP1;SUPT7L;SLC5A6;GPR126;ZNF512;TRIB2;MYH[illegible]

AT5;APOE;ARHGEF40;PVRL2

40;TRIB2

ZMAT5;APOE;ARHGEF40

253;PPM1G;DNAJC1;CCDC91;CR1L;SNX17;ZMAT5;ASCC2;EIF3E;APOE;PVRL2
CR1L;SNX17;APOC1;ZMAT5;ASCC2;APOE;ARHGEF40

MAT5;APOE;ARHGEF40;TRIB2
;SNX17;APOC1;ZMAT5;APOE;ARHGEF40;LCORL

ARHGEF40;TRIB2
F219;ZMAT5;MYH6

F219;APOE;ARHGEF40;PVRL2

174B;DLEU1;RSPO2;SPEG;FAM3C;ARHGEF40;CD34;CR1;SKIDA1;AFAP1;VSTM2L;C2ORF16;MLF1;NIPSNA

ARHGEF40;CD34;CR1;SKIDA1;AFAP1;VSTM2L;C2ORF16;WNT16;NIPSNAP1;NRBP1;SUPT7L;DNAJC1;B3GN

PO2;SPEG;FAM3C;ARHGEF40;CR1;AFAP1;VSTM2L;C2ORF16;WNT16;MLF1;NIPSNAP1;NRBP1;SUPT7L;TM

PPM1G;GJA1;CCDC91;MFN1;FAM3C;APOE;ARHGEF40;CD34;EIF2B4;HSPA4;AFAP1;NIPSNAP1;NRBP1;WA

N1;ASCC2;APOE;TRIB2;MYH6

DGKH

[illegible]E;ARHGEF40

B2

E;ARHGEF40;TRIB2

C1;ZNF219;APOE;ARHGEF40

174B;DLEU1;SPEG;FAM3C;ARHGEF40;CD34;AFAP1;MLF1;NIPSNAP1;NRBP1;SUPT7L;SLC5A6;DNAJC1;B3

N1;ASCC2;APOE;TRIB2;MYH6

APOE;ARHGEF40

MYH6

RC1;FAM174B;DLEU1;RSPO2;SPEG;HORMAD2;FAM3C;ARHGEF40;CD34;CR1;SKIDA1;AFAP1;VSTM2L;C2[illegible]

EF40;TRIB2

[illegible]2;SPEG;FAM3C;ARHGEF40;CD34;CR1;SKIDA1;AFAP1;C2ORF16;WNT16;MLF1;NIPSNAP1;NRBP1;SUPT7L

)34;EIF2B4;HSPA4;CMTM5;WNT16;NIPSNAP1;NRBP1;KIAA1755;DES;SNX17;APOC1;GNB4;EIF3E;PVRL2;

174B;DLEU1;RSPO2;SPEG;FAM3C;ARHGEF40;CD34;CR1;SKIDA1;AFAP1;VSTM2L;C2ORF16;WNT16;MLF1

40

:EF40;PVRL2;TRIB2
40;PVRL2

32

R1;SKIDA1;AFAP1;C2ORF16;MLF1;NRBP1;SUPT7L;SLC5A6;DNAJC1;B3GNT7;NOV;GPR126;ZNF513;ZNF51

EG;FAM3C;CD34;CR1;SKIDA1;AFAP1;VSTM2L;WNT16;MLF1;NRBP1;SUPT7L;DNAJC1;B3GNT7;ZNF513;Z
RC1;FAM174B;DLEU1;RSPO2;SPEG;FAM3C;ARHGEF40;CD34;CR1;SKIDA1;AFAP1;C2ORF16;WNT16;MLF1

3E;LCORL
E;ARHGEF40

;NOV;EIF3E;FAM3C;APOE;CD34;MYH6

[illegible]T5;APOE;ARHGEF40

9;APOE;ARHGEF40

.6;GJA1;CCDC91;RSRC1;FAM174B;MFN1;RSPO2;SPEG;ALX4;ASCC2;FAM3C;APOE;CD34;EIF2B4;HSPA4;Cl

U1;RSPO2;SPEG;FAM3C;ARHGEF40;CD34;CR1;SKIDA1;AFAP1;WNT16;MLF1;NIPSNAP1;NRBP1;SUPT7L;S

PO2;SPEG;FAM3C;ARHGEF40;CD34;CR1;SKIDA1;AFAP1;VSTM2L;C2ORF16;MLF1;NIPSNAP1;NRBP1;SLC5

RC1;FAM174B;DLEU1;RSPO2;SPEG;FAM3C;ARHGEF40;CD34;CR1;SKIDA1;AFAP1;VSTM2L;C2ORF16;WNT

POE;ARHGEF40;TRIB2

174B;DLEU1;RSPO2;SPEG;FAM3C;ARHGEF40;CD34;CR1;SKIDA1;AFAP1;VSTM2L;WNT16;MLF1;NIPSNAP

174B;DLEU1;RSPO2;SPEG;FAM3C;ARHGEF40;CD34;CR1;SKIDA1;AFAP1;VSTM2L;C2ORF16;WNT16;MLF1

RHGEF40;TRIB2

52

RC1;FAM174B;DLEU1;RSPO2;SPEG;FAM3C;ARHGEF40;CD34;CR1;SKIDA1;AFAP1;VSTM2L;C2ORF16;WNT

[illegible]T5;MFN1;GNB4;CCDC141;MYH6

:4;APOE;CD34

OE;ARHGEF40

RIB2

CDC141;MYH6;MYH7
:141;CD34;MYH6;MYH7

174B;DLEU1;RSPO2;SPEG;FAM3C;ARHGEF40;CD34;CR1;AFAP1;VSTM2L;C2ORF16;MLF1;NIPSNAP1;NRB

T14;TTN;PPM1G;CCDC91;RSRC1;MFN1;ASCC2;FAM3C;APOE;CCDC141;EIF2B4;CR1;HSPA4;GPN1;NIPSN [illegible];TRIB2

32
B2

.6;GJA1;CCDC91;RSRC1;FAM174B;MFN1;RSPO2;SPEG;ASCC2;FAM3C;APOE;ARHGEF40;CD34;EIF2B4;HSI

1;MYH6

QCR10;CWC27;NDRG2;TTN;PPM1G;DCAF16;CCDC91;RSRC1;MFN1;ASCC2;FAM3C;CCDC141;EIF2B4;CR1;

LEU1;RSPO2;SPEG;HORMAD2;FAM3C;ARHGEF40;CD34;SKIDA1;AFAP1;VSTM2L;C2ORF16;WNT16;MLF1;

IB2

40;TRIB2

HSPA4;AFAP1;GPN1;NIPSNAP1;NRBP1;SUPT7L;SLC5A6;DES;SNX17;ZNF219;EIF3E;PVRL2;TRIB2

:U1;EIF3E
L;GNB4;GPR126;EIF3E;FAM3C;LCORL;TRIB2

И174B;ZNF219;DLEU1;EIF3E;PVRL2

FAM174B;APOC1;GPR126;ASCC2;FAM3C;APOE;TRIB2

MAT5;APOE;ARHGEF40

A1;DES;NOV;DLEU1;ASCC2;EIF3E;HORMAD2;APOE;CD34

4;HSPA4;AFAP1;NIPSNAP1;NRBP1;SUPT7L;SLC5A6;DNAJC1;DES;SNX17;EIF3E;PVRL2;TRIB2

M3C;CCDC141;LCORL;TRIB2;DGKH

E;ARHGEF40;TRIB2

AM3C;APOE;PVRL2

RSPO2;SPEG;ASCC2;FAM3C;APOE;ARHGEF40;EIF2B4;CR1;HSPA4;SSC5D;GPN1;NIPSNAP1;NRBP1;MLLT1

YH6;MYH7

RG2;TTN;PPM1G;DCAF16;CCDC91;MFN1;RSPO2;SPEG;ASCC2;FAM3C;ARHGEF40;EIF2B4;CR1;HSPA4;CM

2;FAM3C;APOE;HSPA4;GPN1;MLF1;NIPSNAP1;NRBP1;SUPT7L;SNX17;ZNF219;EIF3E;PVRL2;TRIB2

1;DLEU1;RSPO2;ASCC2;FAM3C;HSPA4;GPN1;NIPSNAP1;NRBP1;SUPT7L;DNAJC1;SNX17;EIF3E

ɔV;SNX17;ZNF219;RSPO2;EIF3E;APOE;PVRL2

;GJA1;PLN;CCDC91;ZNF628;RSRC1;FAM174B;ZMAT5;MFN1;DLEU1;RSPO2;FAM3C;ARHGEF40;CCDC141

1;ZNF628;SNX17;ZNF219;GPR126;ASCC2;EIF3E;ARHGEF40;PVRL2

)RL;TRIB2

:;CD34;TRIB2

ЮV;DLEU1;ASCC2;EIF3E;APOE;CD34

;CD34;SKIDA1;AFAP1;VSTM2L;C2ORF16;WNT16;MLF1;SUPT7L;SLC5A6;B3GNT7;NOV;GPR126;LCORL;TR

126;SPEG;ALX4;ARHGEF40;PVRL2;CD34;TRIB2
GEF40;CD34;SKIDA1;AFAP1;VSTM2L;C2ORF16;MLF1;NIPSNAP1;NRBP1;SUPT7L;TMEM253;SLC5A6;NOV;

EG;FAM3C;CD34;SKIDA1;AFAP1;VSTM2L;C2ORF16;MLF1;NIPSNAP1;NRBP1;SUPT7L;TMEM253;SLC5A6;
EG;APOE;ARHGEF40;PVRL2;CD34;TRIB2

'EG;FAM3C;CD34;SKIDA1;AFAP1;C2ORF16;MLF1;NIPSNAP1;NRBP1;SUPT7L;TMEM253;SLC5A6;DNAJC1;I
GJA1;CCDC91;FAM174B;MFN1;SPEG;ALX4;FAM3C;APOE;CD34;EIF2B4;HSPA4;AFAP1;VSTM2L;SSC5D;C2C

FAM174B;MFN1;SPEG;FAM3C;APOE;EIF2B4;GCKR;HSPA4;SKIDA1;SSC5D;MLF1;SUPT7L;WARS2;SLC5A6;T
;DLEU1;SPEG;ALX4;CCDC141;CD34;CENPW;CMTM5;SKIDA1;AFAP1;MLF1;TMEM253;MLLT10;KIAA1755;

SRC1;FAM174B;DLEU1;SPEG;ALX4;FAM3C;APOE;CD34;EIF2B4;HSPA4;SKIDA1;AFAP1;VSTM2L;SSC5D;GPI
C1;FAM174B;MFN1;DLEU1;SPEG;ALX4;FAM3C;APOE;CCDC141;CD34;EIF2B4;CENPW;HSPA4;SKIDA1;AFA
OV;ZNF219;GNB4;GPR126;SPEG;APOE;ARHGEF40;PVRL2;CD34;TRIB2
X4;ZNF512;MYH6;MYH7;DGKH
126;SPEG;ALX4;APOE;ARHGEF40;PVRL2;CD34;TRIB2
G;FAM3C;ARHGEF40;CD34;SKIDA1;AFAP1;C2ORF16;WNT16;MLF1;NRBP1;SUPT7L;SLC5A6;DNAJC1;GPR

F219;GPR126;SPEG;ALX4;APOE;ARHGEF40;PVRL2;CD34;TRIB2
CCDC91;ZNF628;RSRC1;ZMAT5;DLEU1;SPEG;ALX4;ARHGEF40;CCDC141;CD34;AFAP1;SSC5D;GPN1;MLF1
PPM1G;MLLT10;ZNF628;SNX17;ZMAT5;MFN1;ASCC2;PVRL2;DGKH
APOE;ARHGEF40;PVRL2;CD34;TRIB2
G;ALX4;APOE;ARHGEF40;PVRL2;CD34;TRIB2
)1;RSRC1;MFN1;SPEG;FAM3C;APOE;CCDC141;CD34;EIF2B4;HSPA4;SKIDA1;VSTM2L;SSC5D;WNT16;MLF1
GPR126;SPEG;ALX4;APOE;ARHGEF40;PVRL2;CD34
F2B4;CENPW;HSPA4;AFAP1;GPN1;MLF1;NRBP1;NOV;GNB4;MYH7;DGKH

;FAM174B;MFN1;SPEG;ALX4;EIF2B4;CENPW;AFAP1;GPN1;MLF1;NRBP1;SUPT7L;MLLT10;GNB4;EIF3E;ZN
RC1;MFN1;SPEG;CCDC141;CD34;CENPW;CMTM5;GPN1;MLF1;NRBP1;NOV;GNB4;ZNF512;DGKH
EG;APOE;ARHGEF40;PVRL2;CD34;TRIB2

EG;APOE;ARHGEF40;PVRL2;CD34;TRIB2

N1;SPEG;EIF2B4;HSPA4;CMTM5;AFAP1;GPN1;MLF1;SUPT7L;GNB4;EIF3E;ZNF512;TRIB2;MYH6;MYH7;DG
POE;ARHGEF40;PVRL2;CD34;TRIB2
PLN;NOV;RSRC1;MFN1;GNB4;SPEG;ALX4;ZNF512;DGKH

N;NOV;ZNF219;GPR126;SPEG;ALX4;APOE;ARHGEF40;PVRL2;CD34;TRIB2

LT10;DCAF16;NOV;RSRC1;ZMAT5;GNB4;SPEG;ALX4;EIF3E;ZNF512
CDC91;FAM174B;MFN1;SPEG;ALX4;FAM3C;APOE;CD34;EIF2B4;GCKR;HSPA4;AFAP1;VSTM2L;SSC5D;ML

RIB2;DGKH
V;RSRC1;ZMAT5;MFN1;GNB4;SPEG;ALX4;EIF3E;DGKH

1;SPEG;ALX4;CD34;EIF2B4;CENPW;HSPA4;AFAP1;WNT16;GPN1;MLF1;NOV;GNB4;DGKH
C1;MFN1;DLEU1;SPEG;ALX4;FAM3C;APOE;CD34;EIF2B4;CENPW;HSPA4;SKIDA1;AFAP1;SSC5D;C2ORF16;

219;GPR126;SPEG;ALX4;ARHGEF40;PVRL2;CD34;TRIB2

1;PLN;NOV;ZNF219;GPR126;SPEG;ALX4;ARHGEF40;PVRL2;CD34

ALX4;CCDC141;HSPA4;AFAP1;GPN1;MLF1;NOV;GNB4;EIF3E;ZNF512;MYH6;MYH7

EG;FAM3C;CD34;SKIDA1;VSTM2L;MLF1;SUPT7L;SLC5A6;DNAJC1;NOV;GPR126;ZNF513;ZNF512;LCORL;T
GJA1;FAM174B;APOC1;ZNF219;SPEG;ALX4;APOE;PVRL2

4;PPM1G;GJA1;CCDC91;FAM174B;ZMAT5;MFN1;SPEG;FAM3C;APOE;CD34;EIF2B4;GCKR;HSPA4;SKIDA1;
C5A6;MLLT10;ZNF628;SNX17;ZMAT5;ASCC2;EIF3E;PVRL2;DGKH
GJA1;PLN;CCDC91;ZNF628;FAM174B;SPEG;ALX4;FAM3C;APOE;CD34;EIF2B4;HSPA4;SKIDA1;AFAP1;VSTM

DC91;RSRC1;MFN1;SPEG;ALX4;HSPA4;AFAP1;GPN1;MLF1;NRBP1;NOV;GNB4;EIF3E;ZNF512;LCORL;MYH6

PLN;CCDC91;ZNF628;RSRC1;FAM174B;MFN1;DLEU1;SPEG;ALX4;FAM3C;APOE;CCDC141;CD34;EIF2B4;HS

SPEG;APOE;ARHGEF40;PVRL2;CD34;TRIB2
M3C;CD34;SKIDA1;AFAP1;VSTM2L;WNT16;MLF1;NIPSNAP1;SUPT7L;SLC5A6;DNAJC1;GPR126;ZNF512;LC

;NOV;SNX17;ZNF219;RSPO2;EIF3E;FAM3C;PVRL2

I1G;DNAJC1;CCDC91;SNX17;MFN1;RSPO2;ASCC2;EIF3E;FAM3C;TRIB2

N1;DLEU1;SPEG;FAM3C;APOE;CCDC141;CD34;EIF2B4;HSPA4;AFAP1;VSTM2L;SSC5D;MLF1;SUPT7L;WAR

)E;ARHGEF40;PVRL2;CD34;TRIB2

DRG2;NAT14;DCAF16;GJA1;PLN;CCDC91;ZNF628;RSRC1;FAM174B;MFN1;SPEG;ALX4;FAM3C;APOE;CD3

)C91;RSRC1;FAM174B;MFN1;DLEU1;SPEG;ALX4;FAM3C;APOE;CD34;EIF2B4;HSPA4;SKIDA1;VSTM2L;SSC
26;SPEG;APOE;ARHGEF40;PVRL2;CD34;TRIB2
G;FAM3C;ARHGEF40;CD34;SKIDA1;AFAP1;C2ORF16;MLF1;NIPSNAP1;NRBP1;SUPT7L;TMEM253;SLC5A6

RC1;FAM174B;MFN1;FAM3C;APOE;GCKR;SKIDA1;AFAP1;VSTM2L;SSC5D;C2ORF16;MLF1;SUPT7L;WARS2

:;APOE;CCDC141;CD34;EIF2B4;HSPA4;SKIDA1;AFAP1;VSTM2L;SSC5D;MLF1;SUPT7L;WARS2;SLC5A6;TBX
C91;ZMAT5;MFN1;GNB4;EIF3E;MYH6;MYH7

HGEF40;PVRL2;CD34;TRIB2

N1;DLEU1;RSPO2;ASCC2;FAM3C;HSPA4;GPN1;NIPSNAP1;NRBP1;WARS2;DNAJC1;SNX17;EIF3E

\AM174B;MFN1;DLEU1;SPEG;ALX4;FAM3C;APOE;CCDC141;CD34;EIF2B4;HSPA4;SKIDA1;AFAP1;VSTM2L;S
\POC1;ZNF219;DLEU1;GPR126;ALX4;FAM3C;APOE;PVRL2;CD34;DGKH
;SKIDA1;AFAP1;VSTM2L;C2ORF16;WNT16;MLF1;SUPT7L;SLC5A6;DNAJC1;NOV;GPR126;ZNF513;ZNF512;

MFN1;FAM3C;APOE;CD34;GCKR;CENPW;HSPA4;SKIDA1;AFAP1;VSTM2L;SSC5D;MLF1;TMEM253;WARS2
PEG;ALX4;APOE;ARHGEF40;PVRL2;CD34;TRIB2

SKIDA1;AFAP1;C2ORF16;WNT16;MLF1;SUPT7L;SLC5A6;DNAJC1;NOV;GPR126;ZNF513;ZNF512;LCORL;TR

GEF40;PVRL2;CD34
1;ZNF628;RSRC1;FAM174B;MFN1;DLEU1;SPEG;FAM3C;APOE;CD34;EIF2B4;HSPA4;SKIDA1;AFAP1;SSC5D

2;ASCC2;FAM3C;ARHGEF40;CR1;HSPA4;GPN1;NIPSNAP1;NRBP1;SUPT7L;SNX17;ZNF219;EIF3E

\POE;PVRL2;CD34;TRIB2;DGKH

RSRC1;MFN1;DLEU1;SPEG;ALX4;FAM3C;APOE;CCDC141;CD34;EIF2B4;GCKR;CENPW;HSPA4;SKIDA1;SSC

4;SKIDA1;AFAP1;VSTM2L;C2ORF16;MLF1;NIPSNAP1;SUPT7L;SLC5A6;DNAJC1;NOV;GPR126;ZNF513;ZNF[illegible]

3E;FAM3C;CD34;TRIB2

I6;MYH7
X4;ARHGEF40

DNAJC1;GJA1;SNX17;ZNF219;EIF3E;FAM3C;APOE;PVRL2

1;FAM174B;MFN1;SPEG;FAM3C;CCDC141;CD34;EIF2B4;GCKR;SKIDA1;AFAP1;VSTM2L;SSC5D;C2ORF16;[illegible]
[illegible]EU1;SPEG;ALX4;FAM3C;APOE;CCDC141;CD34;EIF2B4;GCKR;CENPW;SKIDA1;AFAP1;SSC5D;C2ORF16;W[illegible]

[illegible]O2;EIF3E;APOE;PVRL2

:AM3C;APOE;PVRL2;TRIB2
JC1;GJA1;SNX17;ZNF219;RSPO2;EIF3E;FAM3C;PVRL2
IF628;RSPO2;FAM3C;CR1;GPN1;NIPSNAP1;SLC5A6;TBX15;DES;SNX17;ZNF219;EIF3E

'RIB2

F3E

:F40;PVRL2;CD34;TRIB2
FAM174B;MFN1;DLEU1;SPEG;ALX4;FAM3C;APOE;CCDC141;CD34;EIF2B4;HSPA4;SKIDA1;SSC5D;WNT16;
AF16;GJA1;MFN1;GNB4;SPEG;ALX4;ZNF512;MYH6;MYH7;DGKH

GPR126;SPEG;ALX4;APOE;ARHGEF40;PVRL2;CD34;TRIB2
IDA1;AFAP1;VSTM2L;C2ORF16;MLF1;SUPT7L;SLC5A6;DNAJC1;B3GNT7;NOV;GPR126;LCORL;TRIB2;DGKI
)C91;RSRC1;FAM174B;MFN1;SPEG;ALX4;FAM3C;APOE;CCDC141;CD34;EIF2B4;HSPA4;SKIDA1;AFAP1;VS

;NOV;ZNF219;GPR126;SPEG;APOE;ARHGEF40;PVRL2;CD34;TRIB2
;EIF2B4;GCKR;HSPA4;SKIDA1;AFAP1;VSTM2L;SSC5D;C2ORF16;MLF1;WARS2;SLC5A6;MLLT10;KIAA1755

CCDC91;ZNF628;RSRC1;FAM174B;ZMAT5;MFN1;DLEU1;SPEG;ALX4;FAM3C;CD34;EIF2B4;GCKR;HSPA4;SI

POE;CD34;EIF2B4;GCKR;SKIDA1;AFAP1;VSTM2L;SSC5D;MLF1;WARS2;SLC5A6;TBX15;MLLT10;KIAA1755;

P1;TBX15;GJA1;SNX17;RSRC1;ZMAT5;GNB4;DLEU1;TRIB2

LEU1;SPEG;FAM3C;ARHGEF40;CD34;SKIDA1;AFAP1;C2ORF16;MLF1;NIPSNAP1;SUPT7L;SLC5A6;NOV;GP

;GJA1;CCDC91;ZNF628;RSRC1;DLEU1;SPEG;ALX4;FAM3C;APOE;CCDC141;CD34;EIF2B4;HSPA4;SKIDA1;A

NB4;SPEG;ALX4;EIF3E;TRIB2;DGKH

M174B;MFN1;SPEG;FAM3C;APOE;CD34;EIF2B4;GCKR;HSPA4;SKIDA1;VSTM2L;SSC5D;C2ORF16;MLF1;WA

9;GPR126;SPEG;ALX4;ARHGEF40;PVRL2;CD34;TRIB2

HGEF40;PVRL2;CD34

FAM174B;MFN1;DLEU1;SPEG;ALX4;FAM3C;APOE;CCDC141;CD34;EIF2B4;HSPA4;SKIDA1;VSTM2L;SSC5D

GNB4;SPEG;ALX4;EIF3E

GNT7;NOV;SNX17;GNB4;ZNF513;ZNF512;FAM3C;TRIB2

L5;DES;PLN;ZNF628;ZNF219;ALX4;ZNF513;PVRL2;DGKH

RSRC1;FAM174B;MFN1;SPEG;ALX4;FAM3C;CD34;EIF2B4;HSPA4;SKIDA1;SSC5D;C2ORF16;MLF1;WARS2;

1;RSRC1;FAM174B;MFN1;DLEU1;SPEG;FAM3C;APOE;CCDC141;CD34;EIF2B4;HSPA4;SKIDA1;AFAP1;VSTM

JA1;CCDC91;RSRC1;MFN1;DLEU1;SPEG;FAM3C;APOE;CCDC141;CD34;EIF2B4;HSPA4;SKIDA1;AFAP1;SSC

;SPEG;APOE;ARHGEF40;PVRL2;CD34;TRIB2
X4;FAM3C;APOE;EIF2B4;SKIDA1;AFAP1;SSC5D;C2ORF16;MLF1;SUPT7L;TMEM253;WARS2;SLC5A6;TBX1

;CD34;SKIDA1;AFAP1;VSTM2L;C2ORF16;WNT16;MLF1;SUPT7L;SLC5A6;DNAJC1;B3GNT7;NOV;GPR126;Z

4;SKIDA1;AFAP1;VSTM2L;SSC5D;C2ORF16;WNT16;MLF1;SLC5A6;TBX15;MLLT10;KIAA1755;ZNF219;ZNF
;ALX4;FAM3C;APOE;CD34;EIF2B4;HSPA4;SKIDA1;AFAP1;SSC5D;C2ORF16;MLF1;SUPT7L;WARS2;SLC5A6;

RC1;FAM174B;MFN1;SPEG;ALX4;FAM3C;ARHGEF40;EIF2B4;SKIDA1;SSC5D;C2ORF16;MLF1;SUPT7L;WA
3;SNX17;ASCC2;PVRL2;DGKH
6;SPEG;APOE;ARHGEF40;PVRL2;CD34;TRIB2

.6;CCDC91;RSRC1;MFN1;GNB4;SPEG;ALX4;EIF3E;ZNF512;DGKH
74B;APOC1;ALX4;FAM3C;APOE;PVRL2

PVRL2;CD34;TRIB2

N;CCDC91;RSRC1;FAM174B;ZMAT5;MFN1;FAM3C;ARHGEF40;CD34;EIF2B4;HSPA4;SKIDA1;SSC5D;C2ORF

F16;GJA1;CCDC91;RSRC1;FAM174B;ZMAT5;MFN1;SPEG;ALX4;FAM3C;APOE;ARHGEF40;EIF2B4;GCKR;HS
;CD34;SKIDA1;AFAP1;VSTM2L;C2ORF16;WNT16;MLF1;SUPT7L;SLC5A6;DNAJC1;NOV;GPR126;ZNF512;L

:ORL

;EF40;TRIB2

RHGEF40;LCORL;CD34;TRIB2

;ZMAT5;MFN1;SPEG;CCDC141;EIF2B4;AFAP1;MLF1;SUPT7L;NOV;EIF3E;ZNF512;TRIB2;MYH7;DGKH

PM1G;DCAF16;GJA1;CCDC91;RSRC1;FAM174B;MFN1;DLEU1;SPEG;FAM3C;EIF2B4;GCKR;HSPA4;SKIDA1;

SRC1;MFN1;DLEU1;SPEG;ALX4;FAM3C;APOE;CD34;EIF2B4;CENPW;HSPA4;SKIDA1;AFAP1;VSTM2L;SSC5D

;CCDC91;RSRC1;FAM174B;MFN1;SPEG;ALX4;FAM3C;ARHGEF40;CD34;EIF2B4;GCKR;HSPA4;SKIDA1;AFA

4;AFAP1;VSTM2L;C2ORF16;MLF1;SUPT7L;SLC5A6;DNAJC1;GPR126;ZNF513;ZNF512;LCORL;TRIB2;DGKH;

CAF16;GJA1;PLN;CCDC91;RSRC1;FAM174B;ZMAT5;MFN1;SPEG;ALX4;FAM3C;EIF2B4;HSPA4;AFAP1;VSTI

74B;MFN1;SPEG;ALX4;FAM3C;CD34;HSPA4;AFAP1;SSC5D;C2ORF16;MLF1;SUPT7L;TMEM253;WARS2;MI

AF16;GJA1;ZMAT5;MFN1;GNB4;SPEG;EIF3E;ZNF512;DGKH

16;ZNF628;SNX17;ZNF219;MFN1;ZNF513;ASCC2;ARHGEF40

2;CD34;TRIB2

EG;FAM3C;ARHGEF40;CD34;SKIDA1;AFAP1;VSTM2L;C2ORF16;WNT16;MLF1;NIPSNAP1;NRBP1;SUPT7L;

.;CCDC91;RSRC1;FAM174B;MFN1;DLEU1;SPEG;ALX4;FAM3C;APOE;EIF2B4;GCKR;CENPW;HSPA4;SKIDA1

RC1;FAM174B;ZMAT5;MFN1;SPEG;FAM3C;EIF2B4;HSPA4;SKIDA1;AFAP1;SSC5D;C2ORF16;MLF1;TMEM2

;TTN;DCAF16;CCDC91;MFN1;GNB4;SPEG;EIF3E;ZNF512;DGKH

J1;SPEG;ALX4;FAM3C;APOE;CD34;EIF2B4;HSPA4;SKIDA1;AFAP1;VSTM2L;SSC5D;C2ORF16;WNT16;GPN1

74B;SPEG;FAM3C;ARHGEF40;CD34;SKIDA1;AFAP1;VSTM2L;C2ORF16;MLF1;SUPT7L;TMEM253;SLC5A6;I

IFN1;SPEG;ALX4;FAM3C;APOE;CCDC141;CD34;EIF2B4;HSPA4;SKIDA1;AFAP1;SSC5D;C2ORF16;GPN1;MLI

GEF40;PVRL2;CD34
);PVRL2;CD34;TRIB2

U1;SPEG;ALX4;FAM3C;APOE;CD34;EIF2B4;SKIDA1;SSC5D;MLF1;SUPT7L;WARS2;SLC5A6;TBX15;MLLT10;

'EG;FAM3C;CD34;SKIDA1;AFAP1;VSTM2L;C2ORF16;WNT16;MLF1;SUPT7L;SLC5A6;DNAJC1;GPR126;ZNF!
174B;MFN1;DLEU1;SPEG;FAM3C;APOE;CD34;EIF2B4;HSPA4;SKIDA1;AFAP1;VSTM2L;SSC5D;WNT16;GPN
IFN1;SPEG;FAM3C;APOE;CCDC141;CD34;EIF2B4;HSPA4;SKIDA1;AFAP1;VSTM2L;SSC5D;GPN1;MLF1;SUPT
iEF40;PVRL2;CD34;TRIB2

)E;ARHGEF40;PVRL2;CD34;TRIB2

PEG;CENPW;HSPA4;CMTM5;AFAP1;GPN1;MLF1;MLLT10;NOV;GNB4;EIF3E;ZNF512;LCORL;DGKH
GJA1;CCDC91;RSRC1;FAM174B;MFN1;DLEU1;SPEG;ALX4;FAM3C;ARHGEF40;CD34;EIF2B4;SKIDA1;SSC5D
:D34;SKIDA1;AFAP1;VSTM2L;WNT16;MLF1;SUPT7L;SLC5A6;DNAJC1;B3GNT7;GPR126;ZNF513;ZNF512;L

M174B;MFN1;SPEG;ALX4;FAM3C;ARHGEF40;CD34;EIF2B4;HSPA4;SKIDA1;AFAP1;VSTM2L;SSC5D;C2ORF:
LEU1;ALX4;FAM3C;APOE;CD34;EIF2B4;HSPA4;SKIDA1;AFAP1;VSTM2L;SSC5D;MLF1;SUPT7L;WARS2;TBX:

34;EIF2B4;SKIDA1;AFAP1;VSTM2L;SSC5D;MLF1;SUPT7L;WARS2;SLC5A6;MLLT10;KIAA1755;APOC1;ZNF2

APOE;CD34;EIF2B4;GCKR;HSPA4;SKIDA1;AFAP1;SSC5D;C2ORF16;MLF1;SUPT7L;WARS2;SLC5A6;TBX15;M

1G;SLC5A6;CCDC91;SNX17;RSRC1;ZMAT5;GNB4;ASCC2;ZNF512;DGKH

C91;RSRC1;FAM174B;MFN1;DLEU1;SPEG;ALX4;FAM3C;APOE;CD34;EIF2B4;HSPA4;SKIDA1;AFAP1;SSC5D
F40;PVRL2;CD34;TRIB2

EG;FAM3C;CD34;SKIDA1;AFAP1;C2ORF16;WNT16;MLF1;SUPT7L;SLC5A6;DNAJC1;B3GNT7;NOV;GPR12

DLEU1;SPEG;ALX4;FAM3C;APOE;CD34;EIF2B4;HSPA4;SKIDA1;VSTM2L;SSC5D;WNT16;MLF1;SUPT7L;WA
EG;FAM3C;CD34;SKIDA1;AFAP1;VSTM2L;C2ORF16;MLF1;SUPT7L;SLC5A6;DNAJC1;B3GNT7;NOV;GPR12
A6;TBX15;MLLT10;DCAF16;GJA1;CCDC91;FAM174B;ZNF219;MFN1;ALX4;FAM3C;DGKH

'VRL2;CD34;TRIB2

F628;RSRC1;FAM174B;DLEU1;SPEG;ALX4;FAM3C;APOE;CD34;EIF2B4;HSPA4;SKIDA1;SSC5D;GPN1;MLF1

HGEF40;PVRL2;CD34;TRIB2
DC141;CMTM5;AFAP1;GPN1;MLF1;NRBP1;SUPT7L;MLLT10;ZNF219;EIF3E;ZNF512;MYH7;DGKH
'EG;FAM3C;ARHGEF40;CD34;SKIDA1;AFAP1;C2ORF16;MLF1;NRBP1;SUPT7L;TMEM253;SLC5A6;NOV;GPI

C1;MFN1;SPEG;ALX4;EIF3E;MYH7;DGKH

;PEG;ALX4;FAM3C;APOE;CD34;EIF2B4;HSPA4;VSTM2L;SSC5D;GPN1;MLF1;SUPT7L;WARS2;SLC5A6;TBX1

'H7

GEF40;CD34;SKIDA1;AFAP1;VSTM2L;C2ORF16;MLF1;NIPSNAP1;SUPT7L;TMEM253;SLC5A6;DNAJC1;NOV

628;RSRC1;DLEU1;SPEG;ALX4;FAM3C;APOE;CCDC141;CD34;EIF2B4;HSPA4;SKIDA1;SSC5D;C2ORF16;WN
G;ALX4;ARHGEF40;PVRL2;CD34;TRIB2

RC1;RSPO2;ASCC2;EIF3E

7;ZNF219;RSPO2;EIF3E;ZNF512;APOE;TRIB2

;GJA1;CCDC91;RSRC1;MFN1;DLEU1;ALX4;FAM3C;APOE;CCDC141;EIF2B4;GCKR;HSPA4;SKIDA1;AFAP1;\

.POE;ARHGEF40;PVRL2;CD34

:;APOE

F40;PVRL2;CD34;TRIB2

;PLN;CCDC91;RSRC1;FAM174B;MFN1;DLEU1;SPEG;ALX4;FAM3C;APOE;CD34;EIF2B4;HSPA4;SKIDA1;AFA

A1;CCDC91;NOV;SNX17;ZNF219;EIF3E;FAM3C;APOE

\1;CCDC91;RSRC1;FAM174B;MFN1;SPEG;ALX4;FAM3C;APOE;ARHGEF40;CD34;EIF2B4;HSPA4;SKIDA1;AF

\POE;CD34;EIF2B4;HSPA4;SKIDA1;SSC5D;GPN1;MLF1;SUPT7L;WARS2;SLC5A6;MLLT10;KIAA1755;DNAJ(

[illegible]FN1;RSPO2;EIF3E;FAM3C;PVRL2

SCC2;EIF2B4;GCKR;CENPW;HSPA4;SKIDA1;AFAP1;C2ORF16;GPN1;MLF1;NRBP1;SLC5A6;MLLT10;ZNF513

.7;RSPO2;EIF3E;FAM3C;APOE;PVRL2

FAM3C;APOE;PVRL2

I174B;MFN1;GPR126;SPEG;APOE

DE;ARHGEF40;PVRL2;CD34;TRIB2

:1;MFN1;FAM3C;EIF2B4;HSPA4;SKIDA1;SSC5D;C2ORF16;MLF1;SUPT7L;WARS2;SLC5A6;MLLT10;ZNF219;

;;GJA1;CCDC91;RSRC1;FAM174B;MFN1;SPEG;FAM3C;ARHGEF40;CD34;EIF2B4;GCKR;SKIDA1;AFAP1;SSC

AF16;GJA1;CCDC91;RSRC1;FAM174B;MFN1;FAM3C;ARHGEF40;EIF2B4;HSPA4;SKIDA1;AFAP1;VSTM2L;S

;;WNT16;GPN1;NRBP1;TMEM253;WARS2;SLC5A6;MLLT10;SNX17;ZNF219;ZNF513;PVRL2;DGKH

;DLEU1;SPEG;ALX4;FAM3C;APOE;CD34;EIF2B4;HSPA4;SKIDA1;SSC5D;GPN1;MLF1;TMEM253;WARS2;SL

53;KIAA1755;B3GNT7;NOV;SNX17;RSRC1;ZMAT5;GNB4;ZNF512;APOE

PEG;ALX4;FAM3C;APOE;EIF2B4;HSPA4;SKIDA1;AFAP1;SSC5D;C2ORF16;GPN1;MLF1;SUPT7L;TMEM253;

G;DCAF16;GJA1;CCDC91;RSRC1;FAM174B;MFN1;SPEG;ALX4;FAM3C;CCDC141;EIF2B4;HSPA4;AFAP1;SSC

MFN1;SPEG;ALX4;FAM3C;APOE;CD34;EIF2B4;HSPA4;AFAP1;VSTM2L;SSC5D;C2ORF16;GPN1;MLF1;SUP

EG;FAM3C;CD34;SKIDA1;AFAP1;C2ORF16;MLF1;SUPT7L;SLC5A6;DNAJC1;GPR126;ZNF512;LCORL;TRIB2

'4B;ZNF219;MFN1;GPR126;SPEG;ALX4

RHGEF40;PVRL2;CD34
/I3C;ARHGEF40;CD34;SKIDA1;AFAP1;VSTM2L;C2ORF16;MLF1;NIPSNAP1;SUPT7L;TMEM253;SLC5A6;NO

;CCDC91;RSRC1;FAM174B;MFN1;DLEU1;SPEG;ALX4;FAM3C;APOE;CD34;EIF2B4;CENPW;HSPA4;SKIDA1;
;ZNF628;RSRC1;FAM174B;MFN1;SPEG;ALX4;FAM3C;APOE;CD34;EIF2B4;HSPA4;SKIDA1;AFAP1;SSC5D;C

B;MFN1;SPEG;ALX4;FAM3C;CD34;EIF2B4;GCKR;HSPA4;SKIDA1;AFAP1;SSC5D;C2ORF16;MLF1;SUPT7L;W

755;GJA1;CCDC91;NOV;MFN1;GPR126;ALX4;EIF3E;FAM3C;LCORL;DGKH

174B;MFN1;SPEG;ALX4;FAM3C;CD34;EIF2B4;HSPA4;SKIDA1;AFAP1;VSTM2L;SSC5D;C2ORF16;WNT16;G

TM2L;SSC5D;WNT16;MLF1;WARS2;SLC5A6;KIAA1755;APOC1;GPR126;ZNF513;LCORL;PVRL2

N1

;CD34;TRIB2

RSRC1;FAM174B;MFN1;SPEG;FAM3C;CD34;EIF2B4;HSPA4;SKIDA1;AFAP1;VSTM2L;SSC5D;C2ORF16;MLF
GEF40;PVRL2;CD34;TRIB2

AM3C;CD34;EIF2B4;HSPA4;SKIDA1;VSTM2L;SSC5D;C2ORF16;GPN1;MLF1;SUPT7L;WARS2;SLC5A6;TBX1

C1;NOV;ZNF628;SNX17;ZNF219;DLEU1;EIF3E;ARHGEF40;CCDC141;DGKH

MFN1;SPEG;ALX4;FAM3C;CD34;EIF2B4;GCKR;HSPA4;SKIDA1;AFAP1;SSC5D;MLF1;SUPT7L;WARS2;SLC5

';NRBP1;TBX15;SNX17;RSRC1;ZMAT5;GNB4
/I3C;CD34;SKIDA1;AFAP1;VSTM2L;C2ORF16;MLF1;NIPSNAP1;SUPT7L;SLC5A6;DNAJC1;GPR126;ZNF513;Z
MAT5;GNB4;ALX4;ZNF512
FAM174B;MFN1;FAM3C;ARHGEF40;CD34;EIF2B4;HSPA4;SKIDA1;VSTM2L;SSC5D;C2ORF16;GPN1;MLF1;
;D;MLF1;WARS2;SLC5A6;TBX15;MLLT10;KIAA1755;APOC1;ZNF219;GPR126;LCORL;PVRL2;DGKH

)E;CD34;EIF2B4;SKIDA1;VSTM2L;SSC5D;MLF1;TMEM253;WARS2;SLC5A6;TBX15;MLLT10;DES;NOV;APO(
MFN1;DLEU1;ALX4;FAM3C;EIF2B4;HSPA4;SKIDA1;VSTM2L;C2ORF16;MLF1;SUPT7L;WARS2;MLLT10;ZNF
:G;DGKH

'M1G;TBX15;PLN;SNX17;RSRC1;ZMAT5;GNB4;DLEU1;LCORL;TRIB2;DGKH

APOE;CD34;EIF2B4;GCKR;AFAP1;VSTM2L;SSC5D;MLF1;SUPT7L;WARS2;SLC5A6;MLLT10;KIAA1755;APOC
);PVRL2;CD34;TRIB2

GEF40;CD34;SKIDA1;AFAP1;VSTM2L;C2ORF16;MLF1;NIPSNAP1;NRBP1;SUPT7L;TMEM253;SLC5A6;DNAJ

N1;SPEG;ALX4;FAM3C;ARHGEF40;EIF2B4;HSPA4;SKIDA1;SSC5D;GPN1;MLF1;SUPT7L;TMEM253;WARS2;

';RSRC1;ZMAT5;GNB4;ALX4

RHGEF40;PVRL2;CD34
4;FAM3C;APOE;CCDC141;CD34;EIF2B4;SKIDA1;SSC5D;MLF1;SUPT7L;WARS2;SLC5A6;TBX15;MLLT10;KIA
34;SKIDA1;AFAP1;C2ORF16;MLF1;NIPSNAP1;SUPT7L;SLC5A6;DNAJC1;GPR126;ZNF513;ZNF512;LCORL;T

9;RSPO2;EIF3E;FAM3C;APOE;PVRL2
1;RSRC1;FAM174B;MFN1;SPEG;FAM3C;APOE;CCDC141;CD34;EIF2B4;HSPA4;SKIDA1;AFAP1;SSC5D;GPN
11G;SLC5A6;DNAJC1;B3GNT7;ZNF628;SNX17;ZNF219;ZNF513;ASCC2;APOE;PVRL2

N1;SPEG;ALX4;FAM3C;CD34;EIF2B4;AFAP1;SSC5D;C2ORF16;MLF1;NIPSNAP1;SUPT7L;WARS2;SLC5A6;TB
EG;ALX4;CD34;EIF2B4;SKIDA1;AFAP1;SSC5D;C2ORF16;MLF1;WARS2;MLLT10;KIAA1755;NOV;ZNF219;GI
8;SNX17;MFN1;GNB4

B;MFN1;SPEG;FAM3C;CCDC141;CD34;EIF2B4;HSPA4;SKIDA1;AFAP1;SSC5D;GPN1;MLF1;NIPSNAP1;SUPT

)2;EIF3E;FAM3C;APOE;PVRL2;TRIB2

/I174B;RSPO2;SPEG;HORMAD2;FAM3C;CD34;CR1;AFAP1;VSTM2L;C2ORF16;WNT16;MLF1;NIPSNAP1;NF

)A1;AFAP1;VSTM2L;SSC5D;MLF1;WARS2;MLLT10;KIAA1755;ZNF219;GPR126;LCORL;PVRL2;DGKH
;CD34

GKH

M174B;MFN1;SPEG;ALX4;FAM3C;APOE;CD34;EIF2B4;HSPA4;SKIDA1;AFAP1;SSC5D;MLF1;SUPT7L;WARS
RHGEF40;PVRL2;CD34;TRIB2

EU1;ALX4

CCDC91;RSRC1;FAM174B;MFN1;DLEU1;SPEG;ALX4;FAM3C;APOE;CCDC141;CD34;EIF2B4;GCKR;HSPA4;S
X4;FAM3C;APOE;CCDC141;CD34;EIF2B4;HSPA4;AFAP1;SSC5D;MLF1;SUPT7L;WARS2;SLC5A6;MLLT10;KI/

N1;FAM3C;APOE;ARHGEF40;CD34;EIF2B4;GCKR;HSPA4;SKIDA1;VSTM2L;SSC5D;C2ORF16;MLF1;WARS2;
74B;SPEG;FAM3C;ARHGEF40;CD34;SKIDA1;AFAP1;C2ORF16;MLF1;NIPSNAP1;NRBP1;SUPT7L;TMEM253

);EIF2B4;CENPW;C2ORF16;GPN1;NRBP1;SUPT7L;WARS2;SLC5A6;MLLT10;SNX17;ZNF513;EIF3E;LCORL;D
RC1;FAM174B;MFN1;DLEU1;SPEG;ALX4;FAM3C;APOE;CD34;EIF2B4;HSPA4;SKIDA1;SSC5D;WNT16;GPN1

PEG;ALX4;FAM3C;CD34;EIF2B4;HSPA4;SKIDA1;AFAP1;SSC5D;WNT16;GPN1;MLF1;SUPT7L;TMEM253;W
EIF2B4;CENPW;GPN1;NRBP1;SUPT7L;WARS2;MLLT10;SNX17;ZNF219;ZNF513;EIF3E;PVRL2;DGKH
L2;CD34;TRIB2

KH

LEU1;SPEG;ALX4;FAM3C;APOE;CCDC141;CD34;EIF2B4;HSPA4;SKIDA1;SSC5D;WNT16;GPN1;MLF1;SUPT7

34;DLEU1

ARHGEF40;EIF2B4;GCKR;CENPW;HSPA4;SKIDA1;SSC5D;C2ORF16;MLF1;TMEM253;MLLT10;ZNF219;GNB4

P1;PPM1G;TBX15;SNX17;RSRC1;ZMAT5;GNB4;DLEU1;TRIB2;DGKH

B2

FAM3C;APOE;PVRL2

JC1;SNX17;MFN1;RSPO2;ASCC2;EIF3E;TRIB2

AM3C;APOE;PVRL2

KR;SKIDA1;SSC5D;WNT16;MLF1;SLC5A6;MLLT10;KIAA1755;NOV;ZNF219;GPR126;PVRL2

DC91;RSRC1;FAM174B;MFN1;DLEU1;SPEG;ALX4;FAM3C;APOE;CCDC141;CD34;EIF2B4;HSPA4;SKIDA1;A
APOE;PVRL2

ASCC2;ARHGEF40;EIF2B4;SSC5D;C2ORF16;GPN1;NRBP1;SUPT7L;WARS2;SLC5A6;MLLT10;SNX17;ZNF51

DLEU1;SPEG;ALX4;FAM3C;APOE;CCDC141;CD34;EIF2B4;HSPA4;SKIDA1;AFAP1;SSC5D;GPN1;MLF1;SUPT

F16;B3GNT7;SNX17;GNB4;ZNF513;ZNF512;FAM3C
TM2L;SSC5D;MLF1;WARS2;SLC5A6;MLLT10;KIAA1755;NOV;APOC1;ZNF219;GPR126;DGKH

HGEF40;CD34
GEF40;CD34
X4

AF16;CR1L;SNX17;FAM174B;ZMAT5;MFN1;DLEU1;EIF3E;LCORL;DGKH

RSRC1;FAM174B;MFN1;DLEU1;SPEG;ALX4;FAM3C;CCDC141;EIF2B4;HSPA4;SKIDA1;AFAP1;SSC5D;C2OR

1G;SLC5A6;DCAF16;DNAJC1;B3GNT7;ZNF628;SNX17;RSRC1;ZNF513;ZNF512;FAM3C;DGKH

;NRBP1;SUPT7L;TBX15;SNX17;RSRC1;ZMAT5;GNB4

.6;B3GNT7;NOV;GNB4;ZNF512;FAM3C;APOE;PVRL2

:CD34

4B;MFN1;SPEG;ALX4;FAM3C;ARHGEF40;CD34;EIF2B4;HSPA4;SKIDA1;SSC5D;C2ORF16;GPN1;MLF1;NIPSI
ALX4;FAM3C;APOE;CD34;EIF2B4;AFAP1;SSC5D;MLF1;TMEM253;WARS2;SLC5A6;TBX15;MLLT10;KIAA17

;LCORL

F40;PVRL2;CD34;TRIB2

U1;SPEG;FAM3C;APOE;EIF2B4;HSPA4;SKIDA1;AFAP1;VSTM2L;SSC5D;C2ORF16;GPN1;MLF1;SUPT7L;WA

HGEF40;DGKH

'5;EIF3E;LCORL

;FAM3C;APOE;CD34;SKIDA1;VSTM2L;SSC5D;MLF1;TMEM253;MLLT10;KIAA1755;NOV;ZNF219;DGKH
C1;ZNF628;SNX17;MFN1;DLEU1;ASCC2;EIF3E;CCDC141;LCORL;DGKH

AM3C;APOE;CD34;EIF2B4;SKIDA1;SSC5D;C2ORF16;GPN1;MLF1;SUPT7L;WARS2;SLC5A6;MLLT10;KIAA17!

M174B;MFN1;DLEU1;SPEG;ALX4;FAM3C;APOE;EIF2B4;HSPA4;SKIDA1;AFAP1;SSC5D;C2ORF16;GPN1;ML

ASCC2;ARHGEF40;EIF2B4;GPN1;NRBP1;SUPT7L;TMEM253;WARS2;MLLT10;DNAJC1;SNX17;EIF3E;LCORL;
MLLT10;GJA1;CCDC91;NOV;APOC1;ZNF219;MFN1;GNB4;ALX4;FAM3C;LCORL;DGKH

CAF16;GJA1;CCDC91;RSRC1;FAM174B;MFN1;DLEU1;ALX4;FAM3C;APOE;EIF2B4;HSPA4;SKIDA1;AFAP1;S

RHGEF40;PVRL2;CD34;TRIB2

B2;MYH6;MYH7
3C;EIF2B4;GCKR;SKIDA1;SSC5D;MLF1;SUPT7L;SLC5A6;MLLT10;KIAA1755;NOV;ZNF219;ZNF512;TRIB2

C2;EIF3E

F40;PVRL2;CD34;TRIB2

G;DCAF16;GJA1;CCDC91;RSRC1;FAM174B;MFN1;DLEU1;SPEG;FAM3C;APOE;CCDC141;CD34;EIF2B4;HSF

);C2ORF16;NIPSNAP1;SUPT7L;TMEM253;WARS2;SLC5A6;B3GNT7;NOV;SNX17;GNB4;ZNF513;ZNF512;PV

LC5A6;DCAF16;CR1L;RSRC1;MFN1;EIF3E;FAM3C;CCDC141;LCORL;DGKH

GEF40;CCDC141;MYH6;DGKH

B;GPR126;SPEG;ALX4;APOE

3;RSRC1;ZMAT5;ASCC2;EIF2B4;GCKR;CENPW;HSPA4;WNT16;GPN1;NIPSNAP1;NRBP1;SUPT7L;SLC5A6;M

JA1;CCDC91;RSRC1;FAM174B;ZMAT5;MFN1;ALX4;FAM3C;CD34;EIF2B4;GCKR;HSPA4;SKIDA1;SSC5D;C2O
F40;CD34;HSPA4;SKIDA1;AFAP1;VSTM2L;SSC5D;SUPT7L;TMEM253;WARS2;SLC5A6;MLLT10;KIAA1755;ZI

LEU1;SPEG;FAM3C;CD34;SKIDA1;AFAP1;C2ORF16;WNT16;MLF1;NIPSNAP1;NRBP1;SUPT7L;SLC5A6;DNA

;APOE;CD34;HSPA4;SKIDA1;VSTM2L;SSC5D;MLF1;SUPT7L;WARS2;MLLT10;KIAA1755;DES;APOC1;ZNF21

MLLT10;ZNF628;SNX17;MFN1;ASCC2;ARHGEF40;DGKH

4;SKIDA1;VSTM2L;SSC5D;C2ORF16;MLF1;TMEM253;TBX15;MLLT10;KIAA1755;DES;ZNF219;GPR126

G;DCAF16;GJA1;PLN;CCDC91;ZNF628;RSRC1;FAM174B;ZMAT5;MFN1;ALX4;FAM3C;ARHGEF40;EIF2B4;H

FAM3C;CD34;HSPA4;SKIDA1;AFAP1;SSC5D;MLF1;WARS2;TBX15;MLLT10;ZNF219;GPR126;EIF3E;TRIB2

628;RSRC1;ZMAT5;MFN1;ASCC2;EIF2B4;HSPA4;GPN1;NRBP1;WARS2;SLC5A6;MLLT10;SNX17;GNB4;ZNI

N1;ASCC2;ARHGEF40
;SNX17;MFN1;ASCC2;DGKH

FAM174B;MFN1;ALX4;FAM3C;APOE;CD34;EIF2B4;HSPA4;AFAP1;SSC5D;MLF1;SUPT7L;WARS2;SLC5A6;T

C91;RSRC1;FAM174B;MFN1;ALX4;FAM3C;ARHGEF40;EIF2B4;HSPA4;SKIDA1;AFAP1;SSC5D;C2ORF16;GP

AT5;MFN1;DLEU1;ASCC2;ARHGEF40;EIF2B4;CENPW;HSPA4;C2ORF16;GPN1;NIPSNAP1;NRBP1;SUPT7L;

.0;DCAF16;CCDC91;RSRC1;ZMAT5;MFN1;GNB4;SPEG;ALX4;EIF3E;ZNF512;DGKH

LLT10;SNX17;RSRC1;ZMAT5;GNB4;ZNF512;LCORL;TRIB2

FAM3C;APOE;PVRL2

253;DCAF16;B3GNT7;NOV;SNX17;ZMAT5;DLEU1;ZNF513;ZNF512;APOE;PVRL2
EG;FAM3C;APOE;CCDC141;CD34;EIF2B4;SKIDA1;AFAP1;VSTM2L;SSC5D;MLF1;SUPT7L;WARS2;SLC5A6;M

SPEG;ALX4;ZNF512;TRIB2

141;CD34

)C91;RSRC1;FAM174B;MFN1;ALX4;FAM3C;APOE;EIF2B4;GCKR;HSPA4;SKIDA1;SSC5D;C2ORF16;GPN1;MI

2;ARHGEF40

NF512;PVRL2;TRIB2

NT7;NOV;GNB4;DLEU1;ZNF513;ZNF512;PVRL2;TRIB2;DGKH

GNB4;ZNF513;ZNF512;FAM3C;APOE;PVRL2;TRIB2

;ZNF628;ZNF219;ZMAT5;SPEG;ALX4;ZNF512;DGKH

';SNX17;GNB4;GPR126;ZNF513;ZNF512;FAM3C;APOE;PVRL2;TRIB2

CAF16;ZNF628;ZNF219;ZMAT5;GNB4;FAM3C;TRIB2;DGKH

AT5;GNB4;DGKH

[illegible]17;GNB4;ZNF513;ZNF512;FAM3C;APOE;PVRL2;TRIB2;DGKH

,

[illegible]FN1;GPR126;FAM3C;APOE

[illegible]4;EIF2B4;HSPA4;SSC5D;MLF1;SUPT7L;WARS2;TBX15;MLLT10;DNAJC1;ZNF219;GPR126;EIF3E;LCORL;DGI
B4;DLEU1

:5A6;DNAJC1;SNX17;RSRC1;GNB4;ZNF513;ZNF512;FAM3C;PVRL2;TRIB2;DGKH

SNX17;GNB4;ZNF513;ZNF512;FAM3C;PVRL2;TRIB2;DGKH

.;VSTM2L;C2ORF16;WNT16;NIPSNAP1;SUPT7L;DNAJC1;B3GNT7;SNX17;GNB4;ZNF513;ZNF512

NF513;ZNF512;FAM3C;PVRL2

10;DCAF16;ZNF628;SNX17;MFN1;EIF3E;LCORL

32

M1G;MLLT10;DCAF16;ZNF628;SNX17;RSRC1;MFN1;EIF3E

.6;DNAJC1;ZNF628;SNX17;APOC1;ZMAT5;ZNF513;ASCC2;ARHGEF40

SRC1;FAM174B;MFN1;ALX4;FAM3C;CD34;EIF2B4;HSPA4;SKIDA1;AFAP1;SSC5D;C2ORF16;GPN1;MLF1;NI

I1G;TBX15;GJA1;SNX17;RSRC1;ZMAT5;GNB4;TRIB2;DGKH

X4;FAM3C;APOE;LCORL;PVRL2;CD34

[illegible];FAM174B;MFN1;SPEG;ALX4;FAM3C;EIF2B4;HSPA4;SKIDA1;AFAP1;SSC5D;C2ORF16;MLF1;SUPT7L;WA[illegible]

[illegible]MFN1;DLEU1;SPEG;FAM3C;APOE;CCDC141;CD34;EIF2B4;HSPA4;SKIDA1;SSC5D;GPN1;MLF1;SUPT7L;WA[illegible]

[illegible]NF512;FAM3C;APOE;PVRL2

[illegible]12;FAM3C

FN1;DLEU1;SPEG;ALX4;FAM3C;APOE;CD34;EIF2B4;HSPA4;SKIDA1;AFAP1;SSC5D;GPN1;MLF1;SUPT7L;W/

33GNT7;NOV;SNX17;GNB4;ZNF513;ZNF512;FAM3C;TRIB2;DGKH

7;NOV;SNX17;GNB4;ZNF513;ZNF512;FAM3C;PVRL2;TRIB2;DGKH

SRC1;GNB4;ALX4;ZNF513;ZNF512;FAM3C;PVRL2;TRIB2

NF512;DGKH

;ZMAT5;GNB4;ZNF512

AT5;ALX4;ZNF513;ASCC2;APOE;ARHGEF40

RSRC1;ZNF513

.1755;GJA1;CCDC91;FAM174B;ZNF219;MFN1;SPEG;ALX4;FAM3C

AM3C
DGKH
';RSRC1;GNB4;ZNF512;TRIB2

.;RSRC1;FAM174B;MFN1;SPEG;ALX4;FAM3C;EIF2B4;HSPA4;SKIDA1;AFAP1;VSTM2L;SSC5D;C2ORF16;WN

TRIB2

;TRIB2;DGKH

3C;APOE;CD34;EIF2B4;HSPA4;SKIDA1;AFAP1;SSC5D;GPN1;MLF1;SUPT7L;WARS2;SLC5A6;MLLT10;KIAA1
;FAM174B;ZNF219;MFN1;SPEG;ALX4;ZNF513;FAM3C

NT7;NOV;SNX17;APOC1;ZNF513;ZNF512;FAM3C;APOE;ARHGEF40;PVRL2
T10;DCAF16;ZNF628;SNX17;MFN1;ASCC2;EIF3E

L

528;SNX17;FAM174B;MFN1;ASCC2;EIF3E;DGKH

RSRC1;FAM174B;ZNF219;ZMAT5;ZNF512

16;NIPSNAP1;SUPT7L;TMEM253;MLLT10;DNAJC1;B3GNT7;NOV;SNX17;GNB4;GPR126;ZNF513;ZNF512;

M1G;SLC5A6;TBX15;SNX17;RSRC1;ZMAT5;GNB4;ZNF512;DGKH
SNX17;MFN1;ASCC2;EIF3E

;APOE;TRIB2

;RSRC1;ZMAT5;MFN1;DLEU1;ASCC2;FAM3C;EIF2B4;GCKR;CENPW;HSPA4;C2ORF16;GPN1;NIPSNAP1;NR

GNB4;ZNF512;FAM3C;PVRL2

RSRC1;ZMAT5;MFN1;GNB4;GPR126;ZNF513;ZNF512;FAM3C;ARHGEF40

NIPSNAP1;SUPT7L;TMEM253;WARS2;B3GNT7;NOV;SNX17;GNB4;ZNF513;ZNF512;PVRL2;TRIB2

ORL

13;ZNF512;FAM3C;APOE;PVRL2;TRIB2

6;DNAJC1;B3GNT7;NOV;SNX17;ZNF513;ZNF512;FAM3C;APOE;PVRL2
VRL2;TRIB2

C1;ZNF219;MFN1;GPR126;SPEG

CAF16;DNAJC1;B3GNT7;SNX17;GNB4;GPR126;ZNF513;ZNF512;FAM3C

1;FAM3C;APOE;CCDC141;CD34;EIF2B4;CENPW;HSPA4;AFAP1;SSC5D;GPN1;MLF1;SUPT7L;WARS2;SLC5

2
A1;C2ORF16;NIPSNAP1;NRBP1;SLC5A6;MLLT10;DNAJC1;B3GNT7;SNX17;APOC1;ZNF513;ZNF512;LCORL

R1L;SNX17;MFN1;EIF3E;FAM3C;LCORL;DGKH
26;ZNF512;FAM3C;ARHGEF40;LCORL;TRIB2

MAT5;ZNF513;APOE;ARHGEF40

1;GPR126;FAM3C;LCORL;DGKH

512;LCORL

GNT7;SNX17;GNB4;DLEU1;ZNF513;ZNF512;FAM3C

17;GNB4;ZNF513;ZNF512;PVRL2;TRIB2;DGKH

SLC5A6;KIAA1755;ZNF628;FAM174B;ZNF219;SPEG;ALX4;ZNF513;FAM3C;TRIB2
TMEM253;WARS2;DNAJC1;B3GNT7;NOV;SNX17;GNB4;GPR126;ZNF513;ZNF512;PVRL2;TRIB2;DGKH

;ZNF513;ZNF512;FAM3C;PVRL2;TRIB2

ZNF513;ZNF512;FAM3C;PVRL2

DNAJC1;B3GNT7;SNX17;RSRC1;ZNF219;GNB4;ZNF513;ZNF512;FAM3C;ARHGEF40;PVRL2

EIF2B4;GCKR;CENPW;HSPA4;SKIDA1;AFAP1;C2ORF16;GPN1;MLF1;NRBP1;SLC5A6;MLLT10;ZNF513;LCOI

C2ORF16;GPN1;NIPSNAP1;SUPT7L;TMEM253;WARS2;SLC5A6;MLLT10;DNAJC1;B3GNT7;SNX17;GNB4;GI

SNAP1;SUPT7L;MLLT10;DNAJC1;B3GNT7;NOV;SNX17;GNB4;ZNF513;ZNF512;PVRL2;TRIB2

CDC91;RSRC1;APOC1;ZNF219;GPR126;EIF3E;FAM3C;APOE;LCORL;CD34
ZNF512;PVRL2;TRIB2;DGKH

;GPR126;ZNF513;ZNF512;PVRL2;TRIB2;DGKH
512;FAM3C;PVRL2;TRIB2;DGKH

CC2;CENPW;HSPA4;AFAP1;GPN1;MLF1;NRBP1;SLC5A6;MLLT10;SNX17;GNB4;ZNF512;DGKH

513

EF40;PVRL2;TRIB2;DGKH

AM3C;CCDC141;EIF2B4;SKIDA1;VSTM2L;SSC5D;MLF1;SLC5A6;MLLT10;KIAA1755;ZNF219;PVRL2

POC1;ZMAT5;ZNF513;ASCC2;ARHGEF40

;MFN1;DLEU1;ASCC2;ARHGEF40;EIF2B4;CENPW;HSPA4;GPN1;NRBP1;SUPT7L;WARS2;MLLT10;SNX17;

LT10;DNAJC1;CCDC91;SNX17;RSRC1;ZMAT5;MFN1;ZNF512;ARHGEF40

NX17;GNB4;ZNF513;ZNF512;ARHGEF40;DGKH

DCAF16;DNAJC1;B3GNT7;SNX17;RSRC1;GNB4;GPR126;ZNF513;ZNF512;FAM3C;TRIB2

ASCC2
AM174B;SPEG;ALX4;FAM3C;CCDC141

H

'W;WNT16;NIPSNAP1;SUPT7L;SLC5A6;MLLT10;DNAJC1;SNX17;GNB4;ZNF513;ZNF512;PVRL2;DGKH
13
ASCC2;EIF3E;ZNF512;LCORL

7;NOV;SNX17;GNB4;DLEU1;ZNF513;ZNF512;FAM3C;ARHGEF40;PVRL2;TRIB2;DGKH

WNT16;NIPSNAP1;SUPT7L;B3GNT7;NOV;SNX17;GNB4;GPR126;ZNF513;ZNF512;TRIB2;DGKH

APOE;PVRL2;TRIB2

3GNT7;NOV;GNB4;ZNF513;ZNF512;FAM3C;TRIB2

NX17;ZNF513;ZNF512;FAM3C

91;NOV;FAM174B;ZNF219;MFN1;GPR126;SPEG;ZNF512;FAM3C;ARHGEF40

:DLEU1;ZNF513;ZNF512;FAM3C

17;ZNF219;RSPO2;EIF3E;FAM3C;APOE

DNAJC1;ZNF512;TRIB2

F16;ZNF628;SNX17;FAM174B;MFN1;ASCC2;EIF3E

F628;ZNF219;MFN1;ALX4;ZNF513;FAM3C;ARHGEF40

STM2L;C2ORF16;WNT16;NIPSNAP1;SUPT7L;TMEM253;SLC5A6;MLLT10;DNAJC1;SNX17;GNB4;GPR126

T5;ZNF513;ARHGEF40;PVRL2;TRIB2

ZNF219;FAM3C

X17;RSRC1;GNB4;ZNF513;ZNF512;FAM3C

513;ZNF512;PVRL2;TRIB2;DGKH

F513;ZNF512;FAM3C;APOE;TRIB2;DGKH
'OE;PVRL2

MEM253;B3GNT7;SNX17;GNB4;ZNF513;ZNF512;FAM3C

.M3C;APOE;TRIB2

LC5A6;CR1L;SNX17;RSRC1;MFN1;DLEU1;EIF3E;ZNF512;LCORL;DGKH

CAF16;CR1L;RSRC1;FAM174B;MFN1;EIF3E;LCORL;DGKH

RHGEF40

LC5A6;CR1L;RSRC1;MFN1;DLEU1;EIF3E;LCORL;DGKH

);LCORL;TRIB2

GNB4;GPR126;ZNF513;ZNF512;FAM3C

7;GNB4;ZNF513;ZNF512;PVRL2;DGKH

1;ZNF512

2

NF512;FAM3C;APOE;ARHGEF40
5A6;MLLT10;DNAJC1;NOV;ZNF628;SNX17;MFN1;DLEU1;ZNF513;ASCC2;EIF3E;DGKH

4;DLEU1;RSPO2;SPEG;ALX4;EIF3E;APOE;ARHGEF40;CCDC141;CD34;DGKH

MFN1;FAM3C

4;ZNF513;ZNF512;FAM3C

513;APOE;PVRL2

NX17;GNB4;ZNF513;ZNF512;FAM3C;APOE;PVRL2;TRIB2
513;ZNF512;FAM3C

L2;FAM3C;DGKH

APOE;PVRL2

1;ZNF219;ALX4;FAM3C;TRIB2

DCAF16;B3GNT7;NOV;SNX17;ZNF513;ZNF512;FAM3C;APOE

;SNX17;GNB4;ZNF513;ZNF512;PVRL2;TRIB2

NX17;ZNF513;ZNF512;FAM3C;PVRL2

17;RSRC1;GNB4;ZNF513;ASCC2;ZNF512;FAM3C;ARHGEF40

)NAJC1;B3GNT7;SNX17;RSRC1;GNB4;GPR126;ZNF513;ZNF512;FAM3C;PVRL2;TRIB2

NF513;ZNF512;APOE

RL

;SNX17;RSRC1;GNB4;ZNF513;ZNF512;FAM3C;ARHGEF40;PVRL2
;FAM3C

RIB2

12;FAM3C

SNX17;MFN1;ZNF513;ZNF512;FAM3C

NX17;ZNF513;ZNF512

M3C

KH

RIB2

SRC1;ZNF513;ZNF512;ARHGEF40;TRIB2

ZNF512;ARHGEF40

C;TRIB2

;C

.2;LCORL

F3E;LCORL;DGKH

M1G;SLC5A6;MLLT10;CCDC91;ZNF628;DLEU1;ZNF513;LCORL;PVRL2

NF512;FAM3C

RL;DGKH

1;DLEU1;LCORL;DGKH

5A6;MLLT10;DCAF16;CCDC91;ZNF628;DLEU1;ZNF513;LCORL;PVRL2

[illegible]3

;FAM3C;LCORL

MFN1;DLEU1;EIF3E;LCORL;DGKH

KR;CENPW;HSPA4;SKIDA1;C2ORF16;GPN1;MLF1;NRBP1;WARS2;SLC5A6;MLLT10;ZNF513;LCORL;DGKH

GCKR;CENPW;HSPA4;SKIDA1;C2ORF16;GPN1;MLF1;NRBP1;SLC5A6;MLLT10;ZNF513;LCORL;PVRL2;DGKH

CKR;CENPW;HSPA4;SKIDA1;AFAP1;C2ORF16;GPN1;MLF1;NRBP1;SLC5A6;MLLT10;ZNF513;LCORL;DGKH

W;HSPA4;SKIDA1;AFAP1;C2ORF16;GPN1;MLF1;NRBP1;SLC5A6;MLLT10;ZNF513;LCORL;DGKH
16;CR1L;SNX17;RSRC1;MFN1;DLEU1;EIF3E;LCORL

1L;SNX17;RSRC1;MFN1;DLEU1;EIF3E;LCORL;DGKH
W;HSPA4;SKIDA1;AFAP1;C2ORF16;GPN1;MLF1;NRBP1;SLC5A6;MLLT10;ZNF513;LCORL;DGKH

KR;CENPW;SKIDA1;AFAP1;C2ORF16;GPN1;MLF1;NRBP1;SLC5A6;MLLT10;ZNF513;LCORL;DGKH

W;SKIDA1;AFAP1;C2ORF16;GPN1;MLF1;NRBP1;SLC5A6;MLLT10;ZNF513;LCORL;PVRL2;DGKH

R1L;FAM174B;MFN1;DLEU1;EIF3E;LCORL;DGKH

GCKR;CENPW;HSPA4;SKIDA1;C2ORF16;GPN1;MLF1;NRBP1;SLC5A6;MLLT10;ZNF513;LCORL;PVRL2;DGKH

EIF2B4;GCKR;CENPW;HSPA4;SKIDA1;C2ORF16;GPN1;MLF1;NRBP1;SLC5A6;MLLT10;ZNF513;LCORL;PVRL

A4;SKIDA1;AFAP1;C2ORF16;GPN1;MLF1;NRBP1;SLC5A6;MLLT10;ZNF513;LCORL;PVRL2;DGKH

.;NRBP1;PPM1G;SLC5A6;MLLT10;CCDC91;ZNF628;DLEU1;ZNF513;ASCC2;LCORL;DGKH

RIB2

R1L;MFN1;DLEU1;EIF3E;FAM3C;LCORL;DGKH

ALX4;EIF2B4;CENPW;HSPA4;SKIDA1;C2ORF16;GPN1;MLF1;NRBP1;SLC5A6;MLLT10;ZNF513;LCORL;PVRL2

SNX17;RSRC1;MFN1;DLEU1;EIF3E;LCORL;DGKH

GCKR;CENPW;HSPA4;SKIDA1;C2ORF16;GPN1;MLF1;NRBP1;SLC5A6;MLLT10;ZNF513;LCORL;PVRL2;DGKH

1;DLEU1;EIF3E;LCORL;DGKH

CAF16;CR1L;MFN1;EIF3E;LCORL;DGKH

;NRBP1;PPM1G;SLC5A6;MLLT10;CCDC91;ZNF628;DLEU1;ZNF513;ASCC2;LCORL;DGKH

;MFN1;DLEU1;EIF3E;FAM3C;LCORL;DGKH

F2B4;GCKR;CENPW;HSPA4;SKIDA1;C2ORF16;GPN1;MLF1;NRBP1;SLC5A6;MLLT10;ZNF513;LCORL;PVRL2

1;NRBP1;PPM1G;SLC5A6;MLLT10;CCDC91;ZNF628;ZNF513;LCORL;PVRL2;DGKH

.;NRBP1;PPM1G;SLC5A6;MLLT10;CCDC91;ZNF628;DLEU1;ZNF513;LCORL;PVRL2

EIF2B4;CENPW;HSPA4;SKIDA1;C2ORF16;GPN1;MLF1;NRBP1;SLC5A6;MLLT10;ZNF513;LCORL;DGKH

1;NRBP1;PPM1G;SLC5A6;MLLT10;DCAF16;CCDC91;ZNF628;ZNF513;LCORL;PVRL2;DGKH

CKR;CENPW;HSPA4;SKIDA1;C2ORF16;GPN1;MLF1;NRBP1;SLC5A6;MLLT10;ZNF513;LCORL;DGKH

GCKR;CENPW;HSPA4;SKIDA1;C2ORF16;GPN1;MLF1;NRBP1;SLC5A6;MLLT10;ZNF513;LCORL;DGKH

;NRBP1;PPM1G;SLC5A6;MLLT10;CCDC91;ZNF628;DLEU1;ZNF513;ASCC2;LCORL;DGKH

KR;CENPW;HSPA4;SKIDA1;C2ORF16;GPN1;MLF1;NRBP1;SLC5A6;MLLT10;ZNF513;LCORL;PVRL2;DGKH

W;HSPA4;SKIDA1;AFAP1;C2ORF16;GPN1;MLF1;NRBP1;WARS2;SLC5A6;MLLT10;ZNF513;LCORL;PVRL2;D

[illegible];PPM1G;SLC5A6;DCAF16;CR1L;RSRC1;MFN1;DLEU1;EIF3E;LCORL;DGKH

[illegible]ORL

NX17;RSRC1;MFN1;DLEU1;EIF3E;FAM3C;LCORL

[illegible];FAM174B;MFN1;DLEU1;EIF3E;CCDC141;LCORL;DGKH
GCKR;CENPW;HSPA4;SKIDA1;AFAP1;C2ORF16;GPN1;MLF1;NRBP1;SLC5A6;MLLT10;ZNF513;LCORL;PVRL

[illegible]EIF2B4;GCKR;CENPW;HSPA4;SKIDA1;C2ORF16;GPN1;MLF1;NRBP1;SLC5A6;MLLT10;ZNF513;LCORL;PVRL

F16;CR1L;RSRC1;MFN1;DLEU1;EIF3E;LCORL;DGKH

GCKR;CENPW;HSPA4;SKIDA1;AFAP1;C2ORF16;GPN1;MLF1;NRBP1;WARS2;SLC5A6;MLLT10;ZNF513;LCO

A6;CR1L;SNX17;RSRC1;MFN1;DLEU1;EIF3E;LCORL;DGKH

GCKR;CENPW;HSPA4;SKIDA1;AFAP1;C2ORF16;GPN1;MLF1;NRBP1;SLC5A6;MLLT10;ZNF513;LCORL;PVRL

L;RSRC1;FAM174B;MFN1;DLEU1;EIF3E;CCDC141;LCORL;DGKH

AJC1;SNX17;RSRC1;MFN1;DLEU1;EIF3E;FAM3C;LCORL

AM3C;ARHGEF40;CD34

EIF2B4;GCKR;CENPW;HSPA4;SKIDA1;C2ORF16;GPN1;MLF1;NRBP1;SLC5A6;MLLT10;ZNF513;LCORL;PVRI

G;SNX17;RSRC1;MFN1;DLEU1;EIF3E;CCDC141;LCORL

GCKR;CENPW;HSPA4;SKIDA1;C2ORF16;GPN1;MLF1;NRBP1;WARS2;SLC5A6;MLLT10;ZNF513;LCORL;PVR
R1L;MFN1;DLEU1;EIF3E;LCORL;DGKH

W;HSPA4;SKIDA1;C2ORF16;GPN1;MLF1;NRBP1;SLC5A6;MLLT10;ZNF513;LCORL;PVRL2;DGKH

RSRC1;ZMAT5;MFN1;DLEU1;EIF3E;LCORL;DGKH

F219;GNB4;FAM3C;LCORL;TRIB2;DGKH

AF16;CR1L;MFN1;DLEU1;EIF3E;LCORL;DGKH

;RSRC1;MFN1;DLEU1;EIF3E;LCORL;DGKH

RSRC1;MFN1;DLEU1;EIF3E;LCORL;DGKH

IPW;HSPA4;SKIDA1;AFAP1;C2ORF16;GPN1;MLF1;NRBP1;SLC5A6;MLLT10;ZNF513;LCORL;PVRL2;DGKH

16;CR1L;FAM174B;MFN1;EIF3E;ZNF512;LCORL;DGKH

NPW;HSPA4;SKIDA1;AFAP1;C2ORF16;GPN1;MLF1;NRBP1;SLC5A6;MLLT10;ZNF513;LCORL;PVRL2;DGKH
FAM174B;MFN1;DLEU1;EIF3E;FAM3C;LCORL;DGKH

KR;CENPW;HSPA4;SKIDA1;C2ORF16;GPN1;MLF1;NRBP1;SLC5A6;MLLT10;ZNF513;LCORL;PVRL2;DGKH

1L;RSRC1;FAM174B;MFN1;DLEU1;EIF3E;LCORL;DGKH

FN1;DLEU1;EIF3E;FAM3C;LCORL;DGKH

ALX4;EIF2B4;CENPW;HSPA4;SKIDA1;C2ORF16;GPN1;MLF1;NRBP1;SLC5A6;MLLT10;ZNF513;LCORL;PVRL2

EIF2B4;GCKR;CENPW;HSPA4;SKIDA1;C2ORF16;GPN1;MLF1;NRBP1;SLC5A6;MLLT10;ZNF513;LCORL;DGKH

IF3E;LCORL;DGKH

1;DLEU1;EIF3E;LCORL;DGKH

F1;NRBP1;PPM1G;SLC5A6;MLLT10;CCDC91;ZNF628;ZNF513;ASCC2;LCORL;DGKH

C27;MLF1;NRBP1;PPM1G;SLC5A6;MLLT10;CCDC91;ZNF628;ZNF513;LCORL;DGKH

1G;SLC5A6;MLLT10;CCDC91;ZNF628;DLEU1;ZNF513;PVRL2

;LCORL
628;GNB4

512

1;NRBP1;PPM1G;SLC5A6;MLLT10;CCDC91;ZNF628;DLEU1;ZNF513;LCORL;DGKH

MLF1;NRBP1;PPM1G;SLC5A6;MLLT10;CCDC91;ZNF628;DLEU1;ZNF513;LCORL;PVRL2

NRBP1;PPM1G;SLC5A6;MLLT10;CCDC91;ZNF628;ZNF513;LCORL;PVRL2

;MFN1;DLEU1;EIF3E;LCORL

F3E;LCORL;DGKH

:WC27;MLF1;NRBP1;PPM1G;SLC5A6;MLLT10;CCDC91;ZNF628;DLEU1;ZNF513;PVRL2

/ILF1;NRBP1;PPM1G;SLC5A6;MLLT10;CCDC91;ZNF628;DLEU1;ZNF513;LCORL;PVRL2

\1;DLEU1;EIF3E;FAM3C;LCORL;DGKH

2;FAM3C;LCORL

CORL;DGKH
DLEU1;EIF3E;LCORL;DGKH

ILF1;NRBP1;PPM1G;SLC5A6;MLLT10;CCDC91;ZNF628;DLEU1;ZNF513;LCORL;DGKH

C141;LCORL;MYH6
:WC27;MLF1;NRBP1;PPM1G;SLC5A6;MLLT10;CCDC91;ZNF628;DLEU1;ZNF513;LCORL;DGKH

NPW;HSPA4;SKIDA1;AFAP1;C2ORF16;GPN1;MLF1;NRBP1;SLC5A6;MLLT10;ZNF513;LCORL;DGKH

AF16;CR1L;MFN1;EIF3E;LCORL;DGKH

F628;GNB4;DLEU1
.;NRBP1;PPM1G;SLC5A6;MLLT10;CCDC91;ZNF628;ZNF513;LCORL;PVRL2;DGKH

32

U1;EIF3E;LCORL

L;SNX17;RSRC1;MFN1;DLEU1;EIF3E

ILF1;NRBP1;PPM1G;SLC5A6;MLLT10;CCDC91;ZNF628;DLEU1;ZNF513;ASCC2;LCORL;DGKH

CENPW;HSPA4;SKIDA1;C2ORF16;GPN1;MLF1;NRBP1;SLC5A6;MLLT10;ZNF513;LCORL;PVRL2;DGKH

DLEU1;EIF3E;FAM3C;LCORL

MFN1;ZNF512;FAM3C;LCORL

IF3E;LCORL;DGKH

;NRBP1;PPM1G;SLC5A6;MLLT10;CCDC91;ZNF628;DLEU1;ZNF513;ASCC2;DGKH

N1;DLEU1;EIF3E;LCORL

ALX4;EIF2B4;CENPW;HSPA4;SKIDA1;C2ORF16;MLF1;NRBP1;SLC5A6;MLLT10;ZNF513;LCORL;PVRL2

L;PPM1G;SLC5A6;MLLT10;CCDC91;ZNF628;DLEU1;ZNF513;PVRL2;DGKH

N1;DLEU1;EIF3E;LCORL;DGKH

C1;MFN1;EIF3E;LCORL;DGKH
RF16;MLF1;NRBP1;PPM1G;SLC5A6;MLLT10;CCDC91;ZNF628;DLEU1;ZNF513;LCORL;PVRL2

F3E;FAM3C;LCORL;DGKH

DLEU1;EIF3E;LCORL

41

L6;WARS2;TBX15;MLLT10;CR1L;ZNF219;LCORL;TRIB2;DGKH

P1;NRBP1;SUPT7L;SLC5A6;DNAJC1;B3GNT7;NOV;GPR126;ZNF513;ZNF512;LCORL;TRIB2;MYH6;MYH7;D[illegible]

T16;MLF1;NIPSNAP1;NRBP1;SUPT7L;TMEM253;SLC5A6;DNAJC1;B3GNT7;NOV;GPR126;ZNF513;ZNF512;

)V;GPR126;ZNF513;ZNF512;TRIB2;MYH6;MYH7;DGKH;MTMR3;LXN;TOMM40;SHOX2;CEP85L;NCAPG;U

P1;NRBP1;SUPT7L;SLC5A6;DNAJC1;B3GNT7;GPR126;ZNF513;ZNF512;LCORL;TRIB2;MYH6;MYH7;DGKH;
T7;ZNF513;ZNF512;LCORL;DGKH;MTMR3;LXN;TOMM40;CABP7;CEP85L;NCAPG;NDRG2;TTN;PPM1G;DC
NF512;LCORL;TRIB2;DGKH;MTMR3;LXN;TOMM40;CEP85L;NCAPG;NDRG2;TTN;PPM1G;PLN;ZNF628;ZM

NAP1;NRBP1;SUPT7L;SLC5A6;DNAJC1;B3GNT7;NOV;ZNF513;LCORL;TRIB2;MYH7;DGKH;MTMR3;LXN;TC

MYH7;MTMR3;TOMM40;CABP7;NDRG2;TTN;PPM1G;DCAF16;ZMAT5;MFN1;ASCC2;APOE;EIF2B4;HSPA

ORF16;WNT16;MLF1;NIPSNAP1;NRBP1;SUPT7L;TMEM253;SLC5A6;DNAJC1;B3GNT7;NOV;GPR126;ZNF5

L;SLC5A6;DNAJC1;B3GNT7;GPR126;ZNF513;ZNF512;LCORL;TRIB2;MYH6;MYH7;DGKH;MTMR3;LXN;TOM

;NIPSNAP1;NRBP1;SUPT7L;SLC5A6;DNAJC1;B3GNT7;NOV;GPR126;ZNF513;ZNF512;LCORL;TRIB2;MYH6;
AP1;NRBP1;SUPT7L;SLC5A6;DNAJC1;B3GNT7;NOV;GPR126;ZNF513;ZNF512;LCORL;TRIB2;MYH6;MYH7;I

1;NRBP1;SUPT7L;SLC5A6;DNAJC1;NOV;GPR126;ZNF513;ZNF512;LCORL;TRIB2;MYH6;MYH7;DGKH;MTM

IEM253;SLC5A6;DNAJC1;NOV;GPR126;ZNF513;ZNF512;LCORL;TRIB2;MYH6;MYH7;MTMR3;LXN;TOMM4

BP1;SUPT7L;SLC5A6;DNAJC1;B3GNT7;ZNF513;ZNF512;LCORL;TRIB2;MYH6;MYH7;DGKH;MTMR3;LXN;TO

UPT7L;SLC5A6;DNAJC1;NOV;GPR126;ZNF513;ZNF512;LCORL;TRIB2;MYH6;MYH7;DGKH;MTMR3;LXN;TO

12;LCORL;TRIB2;MYH6;MYH7;MTMR3;LXN;TOMM40;UQCR10;NDRG2;TTN;PPM1G;DCAF16;ZMAT5;MF[illegible]

[illegible]7;ZNF513;ZNF512;LCORL;TRIB2;DGKH;LXN;TOMM40;CEP85L;NCAPG;NDRG2;TTN;PPM1G;ZNF628;ZMA[illegible]

[illegible]MLF1;NIPSNAP1;NRBP1;SUPT7L;TMEM253;SLC5A6;DNAJC1;B3GNT7;NOV;ZNF513;ZNF512;LCORL;TRIB2;

RBP1;SUPT7L;TMEM253;DNAJC1;B3GNT7;ZNF513;ZNF512;LCORL;TRIB2;MYH6;MYH7;DGKH;MTMR3;LX

PSNAP1;NRBP1;SUPT7L;WARS2;KIAA1755;DNAJC1;DES;SNX17;GNB4;EIF3E;ZNF512;MYH6;MYH7

T16;MLF1;NIPSNAP1;NRBP1;SUPT7L;TMEM253;SLC5A6;DNAJC1;B3GNT7;NOV;GPR126;ZNF513;ZNF512;
3P1;SUPT7L;TMEM253;SLC5A6;DNAJC1;B3GNT7;ZNF513;ZNF512;LCORL;TRIB2;MYH6;MYH7;DGKH;MTM
DNAJC1;NOV;ZNF513;ZNF512;LCORL;TRIB2;DGKH;MTMR3;LXN;TOMM40;CEP85L;NCAPG;NDRG2;TTN;P
33GNT7;ZNF513;ZNF512;LCORL;TRIB2;MYH6;MYH7;DGKH;MTMR3;LXN;TOMM40;CABP7;CEP85L;NCAP

T16;MLF1;NIPSNAP1;NRBP1;SUPT7L;TMEM253;SLC5A6;DNAJC1;B3GNT7;NOV;GPR126;ZNF513;ZNF512;

3;ZNF512;TRIB2;MYH6;MYH7;DGKH;MTMR3;LXN;TOMM40;NCAPG;UQCR10;NDRG2;TTN;PPM1G;DCAF1

T16;MLF1;NIPSNAP1;NRBP1;SUPT7L;TMEM253;SLC5A6;DNAJC1;B3GNT7;NOV;GPR126;ZNF513;ZNF512;

T16;MLF1;NIPSNAP1;NRBP1;SUPT7L;SLC5A6;DNAJC1;B3GNT7;NOV;GPR126;ZNF513;ZNF512;LCORL;TRIE

PN1;NIPSNAP1;NRBP1;SLC5A6;MLLT10;KIAA1755;B3GNT7;NOV;SNX17;APOC1;ZNF219;GNB4;ZNF513;EI

MLF1;NIPSNAP1;NRBP1;SUPT7L;TMEM253;SLC5A6;DNAJC1;B3GNT7;NOV;GPR126;ZNF513;ZNF512;LCOI

BP1;SUPT7L;TMEM253;DNAJC1;ZNF513;ZNF512;LCORL;TRIB2;MYH6;MYH7;DGKH;MTMR3;LXN;TOMM4

MLF1;NIPSNAP1;NRBP1;SUPT7L;SLC5A6;DNAJC1;NOV;GPR126;ZNF513;ZNF512;LCORL;TRIB2;MYH6;MYH

ORL;TRIB2;DGKH;MTMR3;LXN;TOMM40;CEP85L;NCAPG;NDRG2;TTN;PPM1G;ZNF628;ZMAT5;ASCC2;APC

6;MYH7;DGKH;MTMR3;LXN;TOMM40;SHOX2;CEP85L;NCAPG;UQCR10;NDRG2;TTN;PPM1G;DCAF16;MF

AP1;NRBP1;SUPT7L;SLC5A6;DNAJC1;B3GNT7;NOV;GPR126;ZNF513;ZNF512;LCORL;TRIB2;MYH6;MYH7;D

IT7;ZNF513;ZNF512;LCORL;TRIB2;MYH6;MYH7;DGKH;MTMR3;LXN;TOMM40;CEP85L;NCAPG;NDRG2;TT

1EM253;DNAJC1;NOV;ZNF513;ZNF512;LCORL;DGKH;MTMR3;LXN;TOMM40;CEP85L;NCAPG;NDRG2;TTN

ARS2;SLC5A6;KIAA1755;DES;NOV;SNX17;APOC1;GNB4;GPR126;EIF3E;ZNF512;PVRL2;MYH6;MYH7

GNT7;NOV;GPR126;ZNF513;ZNF512;LCORL;TRIB2;DGKH;MTMR3;LXN;TOMM40;CEP85L;UQCR10;NDRG

ORF16;WNT16;MLF1;NIPSNAP1;NRBP1;SUPT7L;SLC5A6;DNAJC1;B3GNT7;NOV;GPR126;ZNF513;ZNF512

;TMEM253;SLC5A6;DNAJC1;B3GNT7;ZNF513;ZNF512;LCORL;DGKH;MTMR3;LXN;TOMM40;CABP7;CEP8

MYH6;MYH7

;NIPSNAP1;NRBP1;SUPT7L;TMEM253;SLC5A6;DNAJC1;B3GNT7;NOV;GPR126;ZNF513;ZNF512;LCORL;T[illegible]

2;LCORL;TRIB2;MYH6;MYH7;DGKH;MTMR3;SHOX2;CEP85L;NCAPG;NDRG2;TTN;MFN1;ALX4;ASCC2;CCD

NF512;LCORL;TRIB2;MYH6;MYH7;DGKH;MTMR3;LXN;TOMM40;CEP85L;NCAPG;NDRG2;TTN;PPM1G;ZN
.;NIPSNAP1;NRBP1;SUPT7L;SLC5A6;DNAJC1;B3GNT7;GPR126;ZNF513;ZNF512;LCORL;TRIB2;MYH6;MYH

MTM5;AFAP1;VSTM2L;GPN1;MLF1;NIPSNAP1;NRBP1;SLC5A6;MLLT10;KIAA1755;B3GNT7;NOV;SNX17;Z

LC5A6;DNAJC1;B3GNT7;GPR126;ZNF513;ZNF512;LCORL;TRIB2;DGKH;MTMR3;LXN;TOMM40;CABP7;CEI

;A6;DNAJC1;B3GNT7;GPR126;ZNF513;ZNF512;LCORL;TRIB2;MYH6;MYH7;DGKH;MTMR3;LXN;TOMM40;

T16;MLF1;NIPSNAP1;NRBP1;SUPT7L;TMEM253;SLC5A6;DNAJC1;B3GNT7;NOV;GPR126;ZNF513;ZNF512;

1;NRBP1;SUPT7L;SLC5A6;DNAJC1;NOV;GPR126;ZNF513;ZNF512;LCORL;TRIB2;MYH6;MYH7;DGKH;MTM

;NIPSNAP1;NRBP1;SUPT7L;TMEM253;SLC5A6;DNAJC1;B3GNT7;NOV;GPR126;ZNF513;ZNF512;LCORL;TF

T16;MLF1;NIPSNAP1;NRBP1;SUPT7L;SLC5A6;DNAJC1;B3GNT7;NOV;GPR126;ZNF513;ZNF512;LCORL;TRI[illegible]

P1;SUPT7L;SLC5A6;DNAJC1;NOV;GPR126;ZNF513;ZNF512;LCORL;TRIB2;MYH6;MYH7;DGKH;MTMR3;LXI

AP1;NRBP1;SUPT7L;WARS2;KIAA1755;DNAJC1;DES;SNX17;GNB4;EIF3E;ZNF512;MYH6;MYH7

PA4;CMTM5;AFAP1;VSTM2L;GPN1;MLF1;NIPSNAP1;NRBP1;SLC5A6;MLLT10;KIAA1755;NOV;SNX17;ZNF

;HSPA4;WNT16;GPN1;NIPSNAP1;NRBP1;SUPT7L;WARS2;MLLT10;KIAA1755;DNAJC1;DES;SNX17;GNB4;E

;NIPSNAP1;NRBP1;SUPT7L;TMEM253;SLC5A6;DNAJC1;GPR126;ZNF513;ZNF512;LCORL;TRIB2;MYH6;MY

.0;DNAJC1;DES;SNX17;APOC1;ZNF219;GNB4;EIF3E

ʌTM5;GPN1;NIPSNAP1;NRBP1;WARS2;MLLT10;DNAJC1;DES;SNX17;GNB4;EIF3E;ZNF512;PVRL2;TRIB2;M

;HSPA4;SKIDA1;AFAP1;VSTM2L;C2ORF16;WNT16;MLF1;NIPSNAP1;NRBP1;SUPT7L;KIAA1755;DNAJC1;SI

.IB2;MYH6;MYH7;TOMM40;CEP85L;NCAPG;NDRG2;PPM1G;DCAF16;PLN;ZNF628;MFN1;ALX4;APOE;CCD

GPR126;ZNF513;ZNF512;LCORL;TRIB2;DGKH;LXN;TOMM40;CEP85L;UQCR10;NDRG2;PPM1G;DCAF16;ZI

DNAJC1;GPR126;ZNF513;ZNF512;LCORL;TRIB2;DGKH;TOMM40;SHOX2;CEP85L;UQCR10;NDRG2;TTN;PP

NOV;GPR126;ZNF513;ZNF512;LCORL;TRIB2;TOMM40;SHOX2;CEP85L;UQCR10;NDRG2;PPM1G;DCAF16;I
ORF16;GPN1;MLF1;TMEM253;WARS2;SLC5A6;MLLT10;KIAA1755;NOV;ZNF219;GPR126;EIF3E;LCORL

TBX15;MLLT10;KIAA1755;DNAJC1;APOC1;ZNF219;GPR126;ZNF513;EIF3E;PVRL2;DGKH
NOV;ZNF219;GNB4;ZNF512

N1;MLF1;SUPT7L;WARS2;SLC5A6;TBX15;MLLT10;KIAA1755;NOV;APOC1;ZNF219;GPR126;EIF3E;LCORL;T
P1;VSTM2L;SSC5D;WNT16;GPN1;MLF1;SUPT7L;WARS2;SLC5A6;TBX15;MLLT10;DNAJC1;APOC1;ZNF219

126;ZNF513;ZNF512;LCORL;TRIB2;DGKH;TOMM40;SHOX2;CEP85L;UQCR10;NDRG2;TTN;PPM1G;DCAF1

1;TMEM253;MLLT10;KIAA1755;NOV;GNB4;EIF3E;ZNF512

L;SUPT7L;WARS2;SLC5A6;TBX15;MLLT10;KIAA1755;DES;APOC1;ZNF219;GPR126;EIF3E;LCORL;PVRL2;TR

NF512;LCORL

KH

F1;TMEM253;SLC5A6;TBX15;MLLT10;KIAA1755;NOV;APOC1;ZNF219;GPR126;ZNF512

GPN1;MLF1;SUPT7L;WARS2;SLC5A6;TBX15;MLLT10;KIAA1755;DNAJC1;NOV;APOC1;ZNF219;GPR126;EIF

TRIB2;TOMM40;NCAPG;UQCR10;NDRG2;PPM1G;DCAF16;PLN;ZNF628;MFN1;ALX4;APOE;CCDC141;EIF2

AFAP1;VSTM2L;SSC5D;C2ORF16;GPN1;MLF1;SUPT7L;WARS2;SLC5A6;TBX15;SNX17;APOC1;ZNF219;GPI

M2L;SSC5D;C2ORF16;WNT16;MLF1;SUPT7L;WARS2;SLC5A6;MLLT10;KIAA1755;APOC1;ZNF219;GPR126;I

;MYH7;DGKH

SPA4;SKIDA1;AFAP1;SSC5D;C2ORF16;GPN1;MLF1;SUPT7L;WARS2;SLC5A6;TBX15;MLLT10;KIAA1755;NO

ORL;TRIB2;TOMM40;SHOX2;CEP85L;UQCR10;NDRG2;PPM1G;DCAF16;ZNF628;MFN1;ALX4;APOE;EIF2B

[illegible]S2;SLC5A6;TBX15;MLLT10;KIAA1755;APOC1;ZNF219;GPR126;EIF3E;LCORL;PVRL2;DGKH

;4;HSPA4;SKIDA1;AFAP1;VSTM2L;SSC5D;C2ORF16;MLF1;WARS2;SLC5A6;MLLT10;KIAA1755;DES;NOV;ZN

5D;GPN1;MLF1;SUPT7L;WARS2;SLC5A6;MLLT10;KIAA1755;APOC1;ZNF219;GPR126;EIF3E;ZNF512;LCOR

;DNAJC1;GPR126;ZNF513;ZNF512;LCORL;TRIB2;DGKH;MTMR3;TOMM40;CEP85L;UQCR10;NDRG2;TTN;

[illegible];MLLT10;KIAA1755;APOC1;ZNF219;GPR126;LCORL;TRIB2;DGKH

15;MLLT10;KIAA1755;DES;APOC1;GPR126;LCORL;PVRL2;DGKH

SSC5D;MLF1;SUPT7L;WARS2;SLC5A6;TBX15;MLLT10;KIAA1755;NOV;APOC1;ZNF219;GPR126;EIF3E;ZNF[illegible]

;LCORL;TRIB2;DGKH;TOMM40;SHOX2;CEP85L;UQCR10;NDRG2;PPM1G;DCAF16;ZNF628;MFN1;ALX4;AP[illegible]

[illegible];MLLT10;KIAA1755;APOC1;ZNF219;GPR126;EIF3E;LCORL;PVRL2;DGKH

[illegible]IB2;MYH6;MYH7;DGKH;LXN;TOMM40;CEP85L;UQCR10;NDRG2;DCAF16;PLN;MFN1;ALX4;APOE;CCDC1[illegible]

[illegible];GPN1;MLF1;SUPT7L;TMEM253;WARS2;SLC5A6;TBX15;MLLT10;KIAA1755;DNAJC1;NOV;APOC1;ZNF21[illegible]

5D;C2ORF16;GPN1;MLF1;SUPT7L;WARS2;SLC5A6;TBX15;MLLT10;KIAA1755;DNAJC1;APOC1;ZNF219;GP

512;LCORL;TRIB2;TOMM40;CEP85L;NCAPG;UQCR10;PPM1G;DCAF16;ZNF628;MFN1;ALX4;APOE;CCDC1

MLF1;TMEM253;WARS2;TBX15;MLLT10;KIAA1755;ZNF219;GNB4;GPR126;EIF3E;ZNF512;LCORL;TRIB2
NT16;GPN1;MLF1;SUPT7L;TMEM253;WARS2;SLC5A6;TBX15;MLLT10;KIAA1755;NOV;APOC1;ZNF219;GPI

;GPN1;MLF1;SUPT7L;WARS2;SLC5A6;TBX15;MLLT10;KIAA1755;DNAJC1;DES;NOV;ZNF219;GPR126;EIF3E

H;TOMM40;CEP85L;NCAPG;UQCR10;NDRG2;DCAF16;PLN;MFN1;ALX4;APOE;EIF2B4;CENPW;HSPA4;SSC
TM2L;SSC5D;GPN1;MLF1;SUPT7L;TMEM253;WARS2;SLC5A6;TBX15;MLLT10;KIAA1755;DES;NOV;APOC1

;EIF3E;ZNF512

KIDA1;AFAP1;VSTM2L;SSC5D;C2ORF16;GPN1;MLF1;NRBP1;SUPT7L;WARS2;SLC5A6;MLLT10;KIAA1755;S

NOV;APOC1;ZNF219;GPR126;PVRL2

R126;ZNF513;ZNF512;LCORL;TRIB2;DGKH;LXN;TOMM40;SHOX2;CEP85L;UQCR10;DCAF16;MFN1;ALX4;C

FAP1;VSTM2L;SSC5D;GPN1;MLF1;SUPT7L;WARS2;SLC5A6;TBX15;MLLT10;KIAA1755;DNAJC1;DES;B3GN

ARS2;SLC5A6;MLLT10;KIAA1755;ZNF219;GPR126

;C2ORF16;GPN1;MLF1;SUPT7L;WARS2;SLC5A6;TBX15;MLLT10;KIAA1755;DES;NOV;APOC1;ZNF219;GPR

SLC5A6;TBX15;MLLT10;KIAA1755;SNX17;ZNF219;ZNF512;PVRL2
M2L;SSC5D;C2ORF16;MLF1;SUPT7L;WARS2;SLC5A6;TBX15;MLLT10;KIAA1755;NOV;APOC1;ZNF219;GNB

5D;C2ORF16;GPN1;MLF1;SUPT7L;WARS2;SLC5A6;TBX15;MLLT10;KIAA1755;DNAJC1;NOV;APOC1;ZNF21

5;MLLT10;APOC1;ZNF219;LCORL;PVRL2;DGKH

ZNF513;ZNF512;LCORL;TRIB2;DGKH;TOMM40;CEP85L;UQCR10;NDRG2;PPM1G;DCAF16;ZNF628;MFN1;

512
MLLT10;KIAA1755;APOC1;ZNF219;GPR126;EIF3E;LCORL;TRIB2;DGKH

RS2;TBX15;MLLT10;KIAA1755;ZNF219;GNB4;GPR126;LCORL;PVRL2;DGKH

F16;GPN1;MLF1;SUPT7L;TMEM253;WARS2;SLC5A6;MLLT10;KIAA1755;DNAJC1;ZNF219;GNB4;GPR126;Z

SPA4;SKIDA1;AFAP1;SSC5D;C2ORF16;GPN1;MLF1;SUPT7L;WARS2;SLC5A6;TBX15;MLLT10;KIAA1755;DN
CORL;TRIB2;DGKH;TOMM40;CEP85L;NCAPG;UQCR10;NDRG2;PPM1G;DCAF16;MFN1;ALX4;APOE;EIF2B

AFAP1;VSTM2L;SSC5D;C2ORF16;WNT16;GPN1;MLF1;SUPT7L;WARS2;SLC5A6;TBX15;MLLT10;KIAA1755;

);C2ORF16;GPN1;MLF1;SUPT7L;WARS2;SLC5A6;TBX15;MLLT10;KIAA1755;DNAJC1;NOV;APOC1;ZNF219;

P1;VSTM2L;SSC5D;C2ORF16;MLF1;SUPT7L;SLC5A6;TBX15;MLLT10;KIAA1755;ZNF219;GPR126;EIF3E;ZN

LXN;TOMM40;SHOX2;CEP85L;UQCR10;TTN;PPM1G;DCAF16;PLN;MFN1;ALX4;EIF2B4;HSPA4;SSC5D;GPN

M2L;SSC5D;C2ORF16;GPN1;MLF1;SUPT7L;TMEM253;WARS2;SLC5A6;TBX15;MLLT10;KIAA1755;SNX17;Z

LLT10;NOV;ZNF219;GNB4;GPR126;LCORL;TRIB2

TMEM253;SLC5A6;DNAJC1;B3GNT7;NOV;GPR126;ZNF513;ZNF512;LCORL;TRIB2;DGKH;LXN;TOMM40;C

;AFAP1;VSTM2L;SSC5D;C2ORF16;MLF1;SUPT7L;WARS2;SLC5A6;TBX15;MLLT10;KIAA1755;ZNF219;GNB

53;WARS2;SLC5A6;MLLT10;KIAA1755;ZNF219;GPR126;EIF3E;ZNF512

;MLF1;SUPT7L;WARS2;SLC5A6;TBX15;MLLT10;KIAA1755;DNAJC1;NOV;APOC1;ZNF219;GPR126;EIF3E;Z

DNAJC1;NOV;GPR126;ZNF513;ZNF512;LCORL;TRIB2;DGKH;LXN;TOMM40;CEP85L;UQCR10;DCAF16;PLN;

F1;SUPT7L;WARS2;SLC5A6;MLLT10;KIAA1755;DNAJC1;NOV;APOC1;ZNF219;GPR126;EIF3E;LCORL;TRIB2

KIAA1755;DES;B3GNT7;NOV;APOC1;ZNF219;GPR126;PVRL2;TRIB2

513;ZNF512;LCORL;TRIB2;TOMM40;SHOX2;CEP85L;UQCR10;NDRG2;TTN;PPM1G;DCAF16;MFN1;ALX4;A
11;MLF1;SUPT7L;WARS2;SLC5A6;TBX15;MLLT10;KIAA1755;DNAJC1;APOC1;ZNF219;GPR126;EIF3E;LCOR
T7L;WARS2;SLC5A6;TBX15;MLLT10;NOV;APOC1;ZNF219;GPR126;EIF3E;LCORL;PVRL2;TRIB2;DGKH

;C2ORF16;GPN1;MLF1;SUPT7L;TMEM253;WARS2;SLC5A6;MLLT10;KIAA1755;NOV;SNX17;ZNF219;GNB
CORL;TRIB2;TOMM40;SHOX2;CEP85L;UQCR10;NDRG2;PPM1G;DCAF16;ZNF628;MFN1;ALX4;APOE;EIF2

16;SUPT7L;TMEM253;WARS2;SLC5A6;MLLT10;KIAA1755;NOV;ZNF219;GNB4;GPR126;ZNF512;LCORL;TR
15;MLLT10;KIAA1755;APOC1;ZNF219;GPR126;LCORL;DGKH

19;GPR126;LCORL;DGKH

LLT10;KIAA1755;ZNF219;GPR126;LCORL;PVRL2

;C2ORF16;GPN1;MLF1;SUPT7L;WARS2;SLC5A6;MLLT10;KIAA1755;DNAJC1;APOC1;ZNF219;GPR126;EIF

;;ZNF513;ZNF512;LCORL;TRIB2;DGKH;TOMM40;CEP85L;NDRG2;PPM1G;DCAF16;ZNF628;MFN1;ALX4;AI

ARS2;SLC5A6;MLLT10;APOC1;GPR126;EIF3E;LCORL;DGKH
6;ZNF512;LCORL;TRIB2;DGKH;TOMM40;CEP85L;UQCR10;NDRG2;TTN;PPM1G;DCAF16;MFN1;ALX4;APO

;SUPT7L;WARS2;SLC5A6;TBX15;MLLT10;KIAA1755;APOC1;ZNF219;GPR126;EIF3E;LCORL;TRIB2

R126;ZNF513;ZNF512;LCORL;TRIB2;DGKH;LXN;TOMM40;SHOX2;CEP85L;UQCR10;NDRG2;TTN;PPM1G;D

.5;MLLT10;KIAA1755;NOV;APOC1;ZNF219;GPR126;EIF3E;LCORL;DGKH

/;GPR126;ZNF513;ZNF512;LCORL;TRIB2;DGKH;LXN;TOMM40;CEP85L;UQCR10;NDRG2;TTN;PPM1G;DCA

IT16;GPN1;MLF1;SUPT7L;WARS2;SLC5A6;TBX15;MLLT10;KIAA1755;DNAJC1;B3GNT7;NOV;APOC1;ZNF2[illegible]

/STM2L;SSC5D;C2ORF16;WNT16;GPN1;MLF1;SUPT7L;WARS2;SLC5A6;MLLT10;DNAJC1;APOC1;ZNF219;[illegible]

\P1;SSC5D;C2ORF16;GPN1;MLF1;SUPT7L;WARS2;SLC5A6;TBX15;MLLT10;KIAA1755;DNAJC1;APOC1;ZNF

AP1;SSC5D;C2ORF16;MLF1;SUPT7L;TMEM253;WARS2;SLC5A6;MLLT10;KIAA1755;ZNF219;GNB4;GPR12 [illegible]1;APOC1;ZNF219;GPR126;EIF3E;LCORL;PVRL2

[illegible];LCORL;PVRL2;DGKH

;ZNF512;LCORL

5D;C2ORF16;MLF1;NIPSNAP1;SUPT7L;WARS2;SLC5A6;MLLT10;KIAA1755;SNX17;ZNF219;GNB4;GPR126

5C5D;C2ORF16;MLF1;SUPT7L;TMEM253;WARS2;SLC5A6;MLLT10;KIAA1755;ZNF219;GNB4;GPR126;EIF3

C5A6;MLLT10;KIAA1755;NOV;APOC1;ZNF219;GPR126;EIF3E;TRIB2

WARS2;SLC5A6;MLLT10;KIAA1755;B3GNT7;APOC1;ZNF219;GPR126;ZNF513;EIF3E;LCORL;PVRL2;TRIB2;

C5D;C2ORF16;GPN1;MLF1;NRBP1;SUPT7L;WARS2;SLC5A6;MLLT10;KIAA1755;DNAJC1;NOV;SNX17;ZNF2

T7L;WARS2;SLC5A6;TBX15;MLLT10;SNX17;ZNF219;GPR126;ZNF513;EIF3E;ZNF512;TRIB2

;TOMM40;CEP85L;NCAPG;UQCR10;NDRG2;TTN;PPM1G;DCAF16;PLN;MFN1;ALX4;APOE;CCDC141;EIF2B

/;GPR126;ZNF513;ZNF512;LCORL;TRIB2;LXN;TOMM40;CEP85L;UQCR10;NDRG2;PPM1G;DCAF16;ZNF62

AFAP1;SSC5D;GPN1;MLF1;SUPT7L;WARS2;SLC5A6;TBX15;MLLT10;KIAA1755;APOC1;ZNF219;GPR126;EI
2ORF16;MLF1;SUPT7L;WARS2;SLC5A6;TBX15;MLLT10;KIAA1755;APOC1;ZNF219;GPR126;EIF3E;LCORL;P

/ARS2;SLC5A6;TBX15;MLLT10;KIAA1755;ZNF219;EIF3E;ZNF512;LCORL

PN1;MLF1;SUPT7L;TMEM253;WARS2;SLC5A6;MLLT10;KIAA1755;SNX17;ZNF219;GPR126;ZNF513;ZNF5

1;WARS2;SLC5A6;MLLT10;KIAA1755;ZNF219;GPR126;ZNF512

;MLLT10;KIAA1755;NOV;ZNF219;GPR126;LCORL;TRIB2;DGKH

A6;MLLT10;KIAA1755;NOV;ZNF219;GPR126;ZNF513;ZNF512;PVRL2;TRIB2

ZNF512;LCORL;TRIB2;LXN;TOMM40;CEP85L;UQCR10;NDRG2;PPM1G;DCAF16;PLN;MFN1;ALX4;EIF2B4;G

SUPT7L;TMEM253;WARS2;SLC5A6;MLLT10;KIAA1755;DNAJC1;ZNF219;GNB4;GPR126;EIF3E;ZNF512;LC

C1;ZNF219;GPR126;LCORL;TRIB2;DGKH
219;GNB4;EIF3E;ZNF512;LCORL;TRIB2

1;ZNF219;LCORL;DGKH

C1;NOV;GPR126;ZNF513;ZNF512;LCORL;TRIB2;DGKH;LXN;TOMM40;SHOX2;CEP85L;UQCR10;PPM1G;D

SLC5A6;TBX15;MLLT10;KIAA1755;NOV;ZNF219;GPR126;EIF3E;LCORL;TRIB2;DGKH

A1755;DES;B3GNT7;APOC1;ZNF219;GPR126
RIB2;DGKH;TOMM40;CEP85L;UQCR10;NDRG2;PPM1G;DCAF16;PLN;MFN1;ALX4;APOE;CCDC141;EIF2B4

1;MLF1;SUPT7L;WARS2;SLC5A6;TBX15;MLLT10;DNAJC1;NOV;APOC1;GPR126;EIF3E;ZNF512;LCORL;PVR

3X15;MLLT10;KIAA1755;NOV;ZNF219;GPR126;ZNF513;EIF3E;ZNF512;TRIB2
PR126;EIF3E;ZNF512;PVRL2

T7L;WARS2;SLC5A6;MLLT10;ZNF219;GNB4;GPR126;ZNF513;EIF3E;ZNF512;TRIB2 [illegible]BP1;SUPT7L;SLC5A6;DNAJC1;B3GNT7;NOV;GPR126;ZNF513;LCORL;TRIB2;MYH6;MYH7;DGKH;MTMR3;

52;SLC5A6;TBX15;MLLT10;KIAA1755;APOC1;ZNF219;LCORL;PVRL2;TRIB2;DGKH

SKIDA1;AFAP1;SSC5D;C2ORF16;GPN1;MLF1;SUPT7L;WARS2;SLC5A6;MLLT10;KIAA1755;APOC1;ZNF219;
AA1755;NOV;ZNF219;GPR126;LCORL;TRIB2;DGKH

SLC5A6;MLLT10;NOV;ZNF219;GNB4;ZNF512;LCORL;PVRL2
;SLC5A6;DNAJC1;NOV;GPR126;ZNF513;ZNF512;LCORL;TRIB2;DGKH;TOMM40;CEP85L;UQCR10;NDRG2;

DGKH
L;MLF1;SUPT7L;WARS2;SLC5A6;MLLT10;KIAA1755;DNAJC1;APOC1;ZNF219;GPR126;EIF3E;LCORL;TRIB2

ARS2;SLC5A6;TBX15;MLLT10;DNAJC1;NOV;ZNF219;GPR126;EIF3E;LCORL;TRIB2;DGKH

'L;WARS2;SLC5A6;MLLT10;KIAA1755;DNAJC1;ZNF219;GPR126;EIF3E;LCORL;TRIB2

4;GPR126;EIF3E

FAP1;SSC5D;WNT16;GPN1;MLF1;SUPT7L;WARS2;SLC5A6;TBX15;MLLT10;KIAA1755;DES;NOV;APOC1;ZN

3;EIF3E

7L;WARS2;SLC5A6;TBX15;MLLT10;KIAA1755;NOV;APOC1;ZNF219;GNB4;GPR126;EIF3E;ZNF512;LCORL;T

F16;GPN1;MLF1;NRBP1;SUPT7L;TMEM253;WARS2;SLC5A6;MLLT10;KIAA1755;DNAJC1;NOV;SNX17;ZNF

NAP1;SUPT7L;TMEM253;WARS2;SLC5A6;MLLT10;KIAA1755;NOV;SNX17;ZNF219;GNB4;GPR126;ZNF513
55;NOV;APOC1;GPR126;EIF3E;TRIB2;DGKH

.RS2;SLC5A6;MLLT10;DNAJC1;B3GNT7;APOC1;ZNF219;GPR126;EIF3E;LCORL;PVRL2;DGKH

55;DNAJC1;APOC1;ZNF219;GPR126;EIF3E;LCORL;TRIB2

.F1;SUPT7L;WARS2;SLC5A6;MLLT10;KIAA1755;ZNF219;EIF3E;ZNF512

MYH6

SSC5D;C2ORF16;WNT16;GPN1;MLF1;SUPT7L;WARS2;SLC5A6;MLLT10;KIAA1755;SNX17;APOC1;ZNF219;

PA4;SKIDA1;AFAP1;SSC5D;WNT16;GPN1;MLF1;SUPT7L;WARS2;SLC5A6;MLLT10;KIAA1755;DNAJC1;APO(

/RL2;TRIB2;DGKH

ILLT10;DNAJC1;SNX17;ZNF513;EIF3E

ORF16;GPN1;MLF1;SUPT7L;WARS2;SLC5A6;MLLT10;DNAJC1;SNX17;ZNF219;GNB4;GPR126;EIF3E;ZNF51
NF219;GPR126;TRIB2;DGKH

JC1;GPR126;ZNF513;ZNF512;LCORL;TRIB2;TOMM40;CEP85L;UQCR10;NDRG2;TTN;PPM1G;DCAF16;PLN

9;GPR126;LCORL;PVRL2;DGKH

SPA4;AFAP1;SSC5D;GPN1;MLF1;NIPSNAP1;NRBP1;SUPT7L;WARS2;SLC5A6;MLLT10;NOV;SNX17;ZNF21[illegible]

F513;ZNF512;DGKH

BX15;MLLT10;KIAA1755;NOV;GPR126;LCORL;PVRL2;DGKH

N1;MLF1;SUPT7L;WARS2;SLC5A6;MLLT10;KIAA1755;NOV;SNX17;GNB4;GPR126;ZNF513;EIF3E;ZNF512;I

TMEM253;WARS2;SLC5A6;MLLT10;DNAJC1;SNX17;GNB4;EIF3E;LCORL

LLT10;APOC1;ZNF219;GPR126;EIF3E;LCORL;TRIB2;DGKH

LF1;SUPT7L;WARS2;MLLT10;KIAA1755;DNAJC1;GNB4;GPR126;EIF3E;LCORL

KH

IPSNAP1;NRBP1;SUPT7L;WARS2;SLC5A6;MLLT10;KIAA1755;DNAJC1;NOV;ZNF219;GNB4;ZNF513;EIF3E;Z

RS2;SLC5A6;MLLT10;SNX17;ZNF219;GNB4;GPR126;ZNF513;EIF3E;ZNF512;PVRL2;TRIB2

RS2;SLC5A6;MLLT10;KIAA1755;DNAJC1;ZNF219;GPR126;EIF3E;LCORL;TRIB2;DGKH

ARS2;SLC5A6;MLLT10;KIAA1755;DNAJC1;ZNF219;GPR126;EIF3E;LCORL;TRIB2

NT16;MLF1;NRBP1;SUPT7L;WARS2;SLC5A6;MLLT10;KIAA1755;NOV;ZNF219;GNB4;ZNF513;EIF3E;ZNF51[illegible]

.755;APOC1;ZNF219;GPR126;EIF3E;LCORL;TRIB2;DGKH

LCORL;TRIB2

BP1;SUPT7L;TMEM253;WARS2;SLC5A6;MLLT10;DNAJC1;SNX17;ZNF513;EIF3E;ZNF512;LCORL

A6;MLLT10;ZNF219;GPR126;ZNF513;EIF3E;LCORL;TRIB2

RL;PVRL2;DGKH

PR126;ZNF513;ZNF512

GNB4;ZNF513;EIF3E

;ZNF513;ZNF512;TRIB2;DGKH

H

.2;DGKH

;DGKH

DGKH

.2;DGKH

.2;DGKH

ⁱRL;PVRL2;DGKH

.2;DGKH

_2;DGKH

L2;DGKH

2;DGKH

H

GKH;MTMR3;LXN;TOMM40;SHOX2;CEP85L;NCAPG;UQCR10;NDRG2;TTN;PPM1G;DCAF16;PLN;ZMAT5;N

LCORL;TRIB2;MYH6;MYH7;DGKH;MTMR3;LXN;TOMM40;CABP7;CEP85L;NCAPG;UQCR10;NDRG2;TTN;P

QCR10;NDRG2;TTN;PPM1G;DCAF16;PLN;ZMAT5;MFN1;ASCC2;APOE;CCDC141;EIF2B4;CENPW;HSPA4;S[illegible]

MTMR3;LXN;TOMM40;CABP7;CEP85L;NCAPG;UQCR10;NDRG2;TTN;PPM1G;DCAF16;PLN;ZNF628;ZMA[illegible]
AF16;ZMAT5;MFN1;ASCC2;APOE;CCDC141;EIF2B4;HSPA4;CMTM5;SSC5D;KIAA1755;SNX17;APOC1;ZNF[illegible]
IAT5;ASCC2;CCDC141;EIF2B4;GCKR;HSPA4;SSC5D;KIAA1755;DES;SNX17;APOC1;ZNF219;GNB4;PVRL2

)MM40;CABP7;CEP85L;UQCR10;NDRG2;TTN;PPM1G;DCAF16;PLN;MFN1;ASCC2;APOE;EIF2B4;HSPA4;CN

4;CMTM5;GPN1;MLLT10;KIAA1755;DES;SNX17;APOC1;ZNF219;GNB4;EIF3E;PVRL2

13;ZNF512;LCORL;TRIB2;MYH7;DGKH;MTMR3;LXN;TOMM40;CABP7;CEP85L;NCAPG;UQCR10;NDRG2;T

M40;CABP7;CEP85L;NCAPG;NDRG2;TTN;PPM1G;ZNF628;ZMAT5;MFN1;ASCC2;APOE;CCDC141;EIF2B4;

MYH7;DGKH;MTMR3;LXN;TOMM40;SHOX2;CEP85L;NCAPG;UQCR10;NDRG2;TTN;PPM1G;DCAF16;PLN;

DGKH;MTMR3;LXN;TOMM40;CABP7;SHOX2;CEP85L;NCAPG;UQCR10;NDRG2;TTN;PPM1G;DCAF16;PLN;

R3;LXN;TOMM40;SHOX2;CEP85L;NCAPG;UQCR10;NDRG2;TTN;PPM1G;DCAF16;PLN;ZMAT5;MFN1;ASC

40;CABP7;CEP85L;NCAPG;UQCR10;NDRG2;TTN;PPM1G;DCAF16;PLN;ZMAT5;MFN1;ASCC2;APOE;EIF2B4

OMM40;CABP7;CEP85L;NCAPG;NDRG2;TTN;PPM1G;DCAF16;ZNF628;ZMAT5;MFN1;ASCC2;APOE;CCDC1

OMM40;CEP85L;NCAPG;UQCR10;NDRG2;TTN;PPM1G;DCAF16;ZMAT5;MFN1;ASCC2;APOE;CCDC141;EIF2

N1;ALX4;ASCC2;APOE;EIF2B4;HSPA4;CMTM5;SSC5D;GPN1;WARS2;MLLT10;KIAA1755;DES;SNX17;APOC

T5;MFN1;ASCC2;APOE;CCDC141;EIF2B4;GCKR;HSPA4;SSC5D;MLLT10;KIAA1755;SNX17;APOC1;ZNF219;

;MYH6;MYH7;DGKH;MTMR3;LXN;TOMM40;CEP85L;NCAPG;UQCR10;NDRG2;TTN;PPM1G;DCAF16;PLN;

:N;TOMM40;CABP7;CEP85L;NCAPG;NDRG2;TTN;PPM1G;DCAF16;ZNF628;ZMAT5;MFN1;ASCC2;CCDC14

LCORL;TRIB2;MYH6;MYH7;DGKH;MTMR3;LXN;TOMM40;CABP7;CEP85L;NCAPG;UQCR10;NDRG2;TTN;P
MR3;LXN;TOMM40;CABP7;CEP85L;NCAPG;UQCR10;NDRG2;TTN;PPM1G;DCAF16;PLN;ZNF628;ZMAT5;M
PM1G;DCAF16;ZMAT5;MFN1;APOE;CCDC141;HSPA4;CMTM5;SSC5D;TBX15;MLLT10;KIAA1755;DES;CR1
G;NDRG2;TTN;PPM1G;ZMAT5;MFN1;ASCC2;APOE;CCDC141;EIF2B4;HSPA4;SSC5D;KIAA1755;SNX17;ZNF

LCORL;TRIB2;DGKH;MTMR3;LXN;TOMM40;CEP85L;NCAPG;UQCR10;NDRG2;TTN;PPM1G;DCAF16;PLN;Z

L6;MFN1;ALX4;ASCC2;APOE;EIF2B4;HSPA4;CMTM5;GPN1;WARS2;MLLT10;KIAA1755;DES;SNX17;APOC1

LCORL;TRIB2;DGKH;MTMR3;LXN;TOMM40;CABP7;CEP85L;NCAPG;UQCR10;NDRG2;TTN;PPM1G;DCAF1

32;MYH6;MYH7;DGKH;MTMR3;LXN;TOMM40;CABP7;CEP85L;NCAPG;UQCR10;NDRG2;TTN;PPM1G;DCA

F3E;ZNF512;PVRL2;TRIB2

RL;TRIB2;MYH6;MYH7;DGKH;MTMR3;LXN;TOMM40;CABP7;CEP85L;NCAPG;UQCR10;NDRG2;TTN;PPM1

40;CABP7;CEP85L;NCAPG;NDRG2;TTN;PPM1G;DCAF16;ZNF628;ZMAT5;MFN1;ASCC2;APOE;CCDC141;EI

H7;DGKH;MTMR3;LXN;TOMM40;CABP7;CEP85L;NCAPG;UQCR10;NDRG2;TTN;PPM1G;DCAF16;PLN;ZMA

DE;CCDC141;EIF2B4;GCKR;HSPA4;SSC5D;KIAA1755;SNX17;ZNF219;GNB4;EIF3E;PVRL2

N1;ALX4;APOE;EIF2B4;HSPA4;CMTM5;SSC5D;GPN1;WARS2;KIAA1755;DES;SNX17;ZNF219;GNB4;EIF3E;

GKH;MTMR3;LXN;TOMM40;CABP7;SHOX2;CEP85L;NCAPG;UQCR10;NDRG2;TTN;PPM1G;DCAF16;PLN;Z

N;PPM1G;ZNF628;ZMAT5;MFN1;ASCC2;APOE;CCDC141;EIF2B4;GCKR;HSPA4;SSC5D;KIAA1755;SNX17;Z

I;PPM1G;DCAF16;ZMAT5;MFN1;APOE;CCDC141;GCKR;HSPA4;CMTM5;SSC5D;TBX15;KIAA1755;DES;SNX

2;TTN;PPM1G;DCAF16;PLN;ZMAT5;MFN1;ASCC2;APOE;EIF2B4;CENPW;HSPA4;SSC5D;WARS2;MLLT10;K

;LCORL;TRIB2;MYH6;MYH7;DGKH;MTMR3;LXN;TOMM40;CABP7;SHOX2;CEP85L;NCAPG;UQCR10;NDRG

5L;NCAPG;NDRG2;TTN;PPM1G;DCAF16;PLN;ZNF628;ZMAT5;MFN1;ASCC2;CCDC141;EIF2B4;HSPA4;SSC5

RIB2;MYH6;MYH7;DGKH;MTMR3;LXN;TOMM40;CABP7;CEP85L;NCAPG;UQCR10;NDRG2;TTN;PPM1G;DC

)C141;GCKR;HSPA4;SSC5D;WARS2;TBX15;MLLT10;KIAA1755;DES;CR1L;ZNF219;GNB4;EIF3E

F628;ZMAT5;ASCC2;APOE;CCDC141;EIF2B4;GCKR;HSPA4;SSC5D;KIAA1755;SNX17;APOC1;ZNF219;GNB4
7;DGKH;MTMR3;LXN;TOMM40;CABP7;CEP85L;NCAPG;UQCR10;NDRG2;TTN;PPM1G;DCAF16;ZNF628;ZI

:NF219;GNB4;ZNF513;EIF3E;ZNF512;PVRL2;TRIB2;DGKH

P85L;NCAPG;UQCR10;NDRG2;TTN;PPM1G;DCAF16;ZNF628;ZMAT5;MFN1;ASCC2;APOE;CCDC141;EIF2B4

;CEP85L;NCAPG;UQCR10;NDRG2;TTN;PPM1G;DCAF16;ZNF628;ZMAT5;MFN1;ASCC2;APOE;CCDC141;EIF

LCORL;TRIB2;MYH6;MYH7;DGKH;MTMR3;LXN;TOMM40;CABP7;CEP85L;NCAPG;UQCR10;NDRG2;TTN;P

IR3;LXN;TOMM40;CABP7;CEP85L;NCAPG;UQCR10;NDRG2;TTN;PPM1G;DCAF16;PLN;ZNF628;ZMAT5;MF

RIB2;DGKH;MTMR3;LXN;TOMM40;CEP85L;NCAPG;UQCR10;NDRG2;TTN;PPM1G;DCAF16;PLN;ZMAT5;M

[illegible]2;MYH6;MYH7;DGKH;MTMR3;LXN;TOMM40;CABP7;CEP85L;NCAPG;UQCR10;NDRG2;TTN;PPM1G;DCA

N;TOMM40;CEP85L;NCAPG;UQCR10;NDRG2;TTN;PPM1G;DCAF16;PLN;ZNF628;ZMAT5;MFN1;ASCC2;AP

219;GNB4;ZNF513;EIF3E;ZNF512;PVRL2;TRIB2

EIF3E;ZNF512;MYH6;MYH7

H7;DGKH;MTMR3;LXN;TOMM40;CABP7;CEP85L;NCAPG;UQCR10;NDRG2;TTN;PPM1G;DCAF16;ZMAT5;I

ɪYH6;MYH7

NX17;ZNF219;GNB4;ZNF513;EIF3E;PVRL2;DGKH

)C141;EIF2B4;HSPA4;SSC5D;GPN1;WARS2;TBX15;MLLT10;KIAA1755;APOC1;ZNF219;EIF3E

NF628;MFN1;APOE;EIF2B4;GCKR;SSC5D;GPN1;WARS2;TBX15;MLLT10;KIAA1755;SNX17;ZNF219;GNB4;E

'M1G;DCAF16;ZNF628;MFN1;ALX4;APOE;EIF2B4;GCKR;CENPW;HSPA4;SSC5D;GPN1;WARS2;TBX15;MLL

MFN1;ALX4;EIF2B4;GCKR;HSPA4;SSC5D;GPN1;WARS2;MLLT10;KIAA1755;SNX17;ZNF219;EIF3E

TRIB2

;GPR126;LCORL;TRIB2

6;PLN;MFN1;ALX4;APOE;CCDC141;EIF2B4;HSPA4;SSC5D;GPN1;WARS2;TBX15;MLLT10;KIAA1755;DES;AI

IB2;MYH6;DGKH

:3E;ZNF512;LCORL;PVRL2;TRIB2

B4;CENPW;HSPA4;SSC5D;GPN1;WARS2;TBX15;MLLT10;KIAA1755;DES;APOC1;ZNF219;EIF3E

R126;ZNF513;EIF3E;ZNF512;LCORL;TRIB2

EIF3E;LCORL;TRIB2;DGKH

V;ZNF219;GPR126;EIF3E;ZNF512;LCORL;TRIB2

4;HSPA4;SSC5D;GPN1;WARS2;TBX15;MLLT10;KIAA1755;APOC1;ZNF219;EIF3E

NF219;GPR126;ZNF513;LCORL;TRIB2

L;PVRL2;TRIB2;DGKH

PPM1G;DCAF16;ZNF628;ZMAT5;MFN1;ALX4;APOE;EIF2B4;HSPA4;SSC5D;GPN1;WARS2;TBX15;MLLT10;

512;LCORL;TRIB2;DGKH

OE;CCDC141;EIF2B4;HSPA4;SSC5D;GPN1;WARS2;TBX15;MLLT10;KIAA1755;APOC1;ZNF219;EIF3E

41;EIF2B4;GCKR;CENPW;HSPA4;SSC5D;GPN1;WARS2;TBX15;MLLT10;KIAA1755;DES;ZNF219;EIF3E

9;GPR126;EIF3E;LCORL;TRIB2

R126;EIF3E;LCORL;PVRL2;TRIB2

41;EIF2B4;HSPA4;SSC5D;GPN1;WARS2;MLLT10;KIAA1755;SNX17;APOC1;ZNF219;EIF3E;PVRL2

R126;EIF3E;LCORL;TRIB2;DGKH

E;LCORL;TRIB2

5D;GPN1;WARS2;TBX15;MLLT10;KIAA1755;APOC1;EIF3E;PVRL2
L;GPR126;EIF3E;LCORL;DGKH

SNX17;ZNF219;GPR126;ZNF513;ZNF512;LCORL;TRIB2;DGKH

CDC141;EIF2B4;GCKR;HSPA4;SSC5D;GPN1;WARS2;MLLT10;KIAA1755;SNX17;GNB4;EIF3E

T7;APOC1;ZNF219;GPR126;ZNF513;EIF3E;LCORL;PVRL2;TRIB2

126;EIF3E;LCORL;TRIB2

4;GPR126;EIF3E;LCORL;TRIB2;DGKH

l9;GNB4;GPR126;EIF3E;LCORL;PVRL2

ALX4;APOE;EIF2B4;CENPW;HSPA4;SSC5D;GPN1;WARS2;MLLT10;KIAA1755;APOC1;ZNF219;GNB4;EIF3E

ZNF513;EIF3E;ZNF512;PVRL2;TRIB2

AJC1;ZNF219;GNB4;GPR126;EIF3E;ZNF512;LCORL;TRIB2;DGKH
4;CENPW;HSPA4;SSC5D;GPN1;WARS2;MLLT10;KIAA1755;ZNF219;GNB4;EIF3E;PVRL2

;NOV;SNX17;ZNF219;GNB4;ZNF513;ZNF512;LCORL;TRIB2;DGKH

;GNB4;GPR126;EIF3E;LCORL;PVRL2;TRIB2

F512;LCORL;PVRL2;TRIB2;DGKH

N1;WARS2;TBX15;MLLT10;KIAA1755;SNX17;ZNF219;GNB4;EIF3E

ZNF219;GNB4;GPR126;ZNF513;ZNF512;LCORL;TRIB2

EP85L;UQCR10;NDRG2;TTN;PPM1G;DCAF16;PLN;ZNF628;MFN1;CCDC141;EIF2B4;HSPA4;SSC5D;GPN1;\

4;GPR126;EIF3E;ZNF512;LCORL;DGKH

NF512;LCORL;TRIB2

ZMAT5;MFN1;EIF2B4;HSPA4;SSC5D;GPN1;WARS2;MLLT10;ZNF219;GNB4;EIF3E;PVRL2

\POE;CCDC141;EIF2B4;HSPA4;SSC5D;GPN1;WARS2;TBX15;MLLT10;KIAA1755;APOC1;ZNF219;GNB4;EIF3
L;TRIB2

4;GPR126;ZNF513;EIF3E;ZNF512;LCORL;TRIB2
B4;CENPW;HSPA4;SSC5D;GPN1;WARS2;TBX15;MLLT10;KIAA1755;DES;SNX17;APOC1;ZNF219;EIF3E

RIB2;DGKH

3E;LCORL;PVRL2;TRIB2

POE;CCDC141;EIF2B4;HSPA4;SSC5D;GPN1;WARS2;TBX15;MLLT10;KIAA1755;APOC1;ZNF219;GNB4;EIF3E

E;CCDC141;EIF2B4;HSPA4;SSC5D;GPN1;WARS2;TBX15;MLLT10;KIAA1755;APOC1;ZNF219;GNB4;EIF3E

DCAF16;MFN1;ALX4;EIF2B4;GCKR;HSPA4;SSC5D;GPN1;WARS2;TBX15;MLLT10;KIAA1755;SNX17;ZNF219;

F16;ZMAT5;MFN1;ALX4;APOE;EIF2B4;HSPA4;SSC5D;GPN1;WARS2;MLLT10;KIAA1755;SNX17;ZNF219;GI

19;GPR126;EIF3E;LCORL;TRIB2

GPR126;ZNF513;EIF3E;ZNF512;LCORL;TRIB2

219;GNB4;GPR126;EIF3E;ZNF512;LCORL;PVRL2;TRIB2

[illegible]6;ZNF513;EIF3E;ZNF512;LCORL;PVRL2;TRIB2;DGKH

;ZNF513;ZNF512;LCORL;PVRL2;TRIB2;DGKH

E;ZNF512;LCORL;PVRL2;DGKH

DGKH

19;GNB4;GPR126;ZNF513;EIF3E;ZNF512;LCORL;TRIB2

[illegible]4;CENPW;HSPA4;SSC5D;GPN1;WARS2;TBX15;MLLT10;KIAA1755;APOC1;ZNF219;GNB4;EIF3E

8;MFN1;ALX4;EIF2B4;HSPA4;SSC5D;GPN1;WARS2;TBX15;MLLT10;KIAA1755;DES;SNX17;ZNF219;GNB4;I

F3E;LCORL
VRL2;DGKH

12;PVRL2;TRIB2

GCKR;HSPA4;SSC5D;GPN1;WARS2;MLLT10;DES;SNX17;GNB4;EIF3E

ORL;TRIB2;DGKH

CAF16;ZNF628;ZMAT5;MFN1;ALX4;CCDC141;EIF2B4;HSPA4;SSC5D;GPN1;WARS2;TBX15;MLLT10;KIAA17

;HSPA4;SSC5D;GPN1;WARS2;TBX15;MLLT10;KIAA1755;APOC1;ZNF219;EIF3E

L2;DGKH

LXN;TOMM40;CABP7;SHOX2;NCAPG;UQCR10;NDRG2;TTN;PPM1G;DCAF16;PLN;ZNF628;ZMAT5;MFN1;[illegible]

GPR126;EIF3E;ZNF512;LCORL;TRIB2

PPM1G;DCAF16;ZNF628;ZMAT5;MFN1;ALX4;APOE;EIF2B4;GCKR;HSPA4;SSC5D;GPN1;WARS2;MLLT10;K

IF219;GPR126;EIF3E;LCORL;TRIB2;DGKH

ΓRIB2

219;GPR126;ZNF513;EIF3E;ZNF512;LCORL;TRIB2

;EIF3E;ZNF512;LCORL;TRIB2

GPR126;ZNF513;EIF3E;ZNF512;TRIB2;DGKH

C1;ZNF219;GPR126;ZNF513;EIF3E;ZNF512;LCORL;PVRL2;TRIB2;DGKH

.2;LCORL;TRIB2;DGKH

I;ZNF628;ZMAT5;MFN1;EIF2B4;GCKR;HSPA4;SSC5D;GPN1;WARS2;MLLT10;KIAA1755;SNX17;ZNF219;GN [illegible];GNB4;GPR126;ZNF513;EIF3E;ZNF512;LCORL;TRIB2;DGKH

LCORL;TRIB2;DGKH

ZNF512;LCORL;PVRL2;TRIB2

2;LCORL;PVRL2;TRIB2

ΛFN1;ALX4;ASCC2;APOE;CCDC141;EIF2B4;HSPA4;CMTM5;SSC5D;GPN1;WARS2;MLLT10;KIAA1755;DES;S

PM1G;DCAF16;PLN;ZNF628;ZMAT5;MFN1;ALX4;ASCC2;APOE;CCDC141;EIF2B4;GCKR;CENPW;HSPA4;CM

SC5D;GPN1;WARS2;TBX15;MLLT10;DES;SNX17;APOC1;ZNF219;GNB4;EIF3E;PVRL2

T5;MFN1;ASCC2;APOE;CCDC141;EIF2B4;GCKR;CENPW;HSPA4;CMTM5;SSC5D;GPN1;WARS2;MLLT10;KIA
219;GNB4;EIF3E

MTM5;SSC5D;GPN1;WARS2;TBX15;MLLT10;KIAA1755;DES;SNX17;APOC1;ZNF219;GNB4;EIF3E;PVRL2

TN;PPM1G;DCAF16;PLN;ZMAT5;MFN1;ASCC2;APOE;CCDC141;EIF2B4;GCKR;CENPW;HSPA4;CMTM5;SS

HSPA4;SSC5D;MLLT10;KIAA1755;SNX17;APOC1;ZNF219;GNB4;EIF3E;PVRL2

ZNF628;ZMAT5;MFN1;ASCC2;APOE;CCDC141;EIF2B4;GCKR;CENPW;HSPA4;CMTM5;SSC5D;WARS2;TBX

ZMAT5;MFN1;ALX4;ASCC2;APOE;CCDC141;EIF2B4;GCKR;HSPA4;SSC5D;GPN1;WARS2;TBX15;MLLT10;KI

C2;APOE;CCDC141;EIF2B4;CENPW;HSPA4;SSC5D;GPN1;WARS2;TBX15;MLLT10;KIAA1755;DES;CR1L;SN

;GCKR;CENPW;HSPA4;CMTM5;SSC5D;GPN1;WARS2;MLLT10;DES;SNX17;APOC1;ZNF219;GNB4;EIF3E;PV

.41;EIF2B4;GCKR;HSPA4;SSC5D;KIAA1755;SNX17;ZNF219;GNB4;EIF3E;PVRL2

2B4;HSPA4;CMTM5;GPN1;WARS2;MLLT10;KIAA1755;DES;SNX17;APOC1;ZNF219;GNB4;EIF3E;PVRL2

1;ZNF219;GNB4;EIF3E;PVRL2

GNB4;EIF3E

ZMAT5;MFN1;ASCC2;APOE;CCDC141;EIF2B4;GCKR;HSPA4;CMTM5;SSC5D;GPN1;TBX15;MLLT10;KIAA17

1;EIF2B4;HSPA4;SSC5D;KIAA1755;SNX17;ZNF219;GNB4;EIF3E;PVRL2

PM1G;DCAF16;PLN;ZNF628;ZMAT5;MFN1;ASCC2;APOE;CCDC141;EIF2B4;GCKR;CENPW;HSPA4;CMTM5
FN1;ASCC2;APOE;CCDC141;EIF2B4;GCKR;HSPA4;SSC5D;WARS2;TBX15;MLLT10;KIAA1755;DES;SNX17;AI
L;SNX17;ZNF219;GNB4;EIF3E
:219;GNB4;EIF3E;PVRL2

:NF628;ZMAT5;MFN1;ASCC2;APOE;CCDC141;EIF2B4;CENPW;HSPA4;SSC5D;GPN1;WARS2;MLLT10;KIAA:

.;ZNF219;GNB4;EIF3E;PVRL2

6;ZNF628;ZMAT5;MFN1;ASCC2;APOE;CCDC141;EIF2B4;GCKR;CENPW;HSPA4;CMTM5;SSC5D;WARS2;MI

F16;PLN;ZNF628;ZMAT5;MFN1;ASCC2;APOE;CCDC141;EIF2B4;GCKR;CENPW;HSPA4;CMTM5;SSC5D;GPI

G;DCAF16;PLN;ZNF628;ZMAT5;MFN1;ASCC2;APOE;CCDC141;EIF2B4;GCKR;CENPW;HSPA4;SSC5D;GPN1

-2B4;GCKR;HSPA4;SSC5D;KIAA1755;SNX17;ZNF219;GNB4;EIF3E;PVRL2

.T5;MFN1;ASCC2;APOE;CCDC141;EIF2B4;GCKR;HSPA4;CMTM5;SSC5D;WARS2;MLLT10;KIAA1755;DES;SN

PVRL2

ZMAT5;MFN1;ALX4;ASCC2;APOE;CCDC141;EIF2B4;GCKR;HSPA4;SSC5D;GPN1;WARS2;TBX15;MLLT10;KIA

:NF219;GNB4;EIF3E;PVRL2

:17;ZNF219;GNB4;EIF3E;PVRL2

:IAA1755;DES;SNX17;APOC1;ZNF219;GNB4;EIF3E;PVRL2

2;TTN;PPM1G;DCAF16;PLN;ZNF628;ZMAT5;MFN1;ALX4;ASCC2;APOE;CCDC141;EIF2B4;GCKR;CENPW;H[illegible]

[illegible]D;KIAA1755;SNX17;ZNF219;GNB4;EIF3E;PVRL2

CAF16;PLN;ZMAT5;MFN1;ASCC2;APOE;CCDC141;EIF2B4;GCKR;HSPA4;SSC5D;GPN1;WARS2;MLLT10;KIA

4;EIF3E;PVRL2
MAT5;MFN1;ASCC2;APOE;CCDC141;EIF2B4;GCKR;CENPW;HSPA4;SSC5D;WARS2;MLLT10;KIAA1755;DES

4;CENPW;HSPA4;CMTM5;SSC5D;GPN1;WARS2;MLLT10;CR1L;SNX17;APOC1;ZNF219;GNB4;EIF3E;PVRL2

F2B4;GCKR;HSPA4;SSC5D;GPN1;KIAA1755;CR1L;SNX17;ZNF219;GNB4;EIF3E;PVRL2

PM1G;DCAF16;PLN;ZMAT5;MFN1;ASCC2;APOE;CCDC141;EIF2B4;GCKR;HSPA4;CMTM5;SSC5D;GPN1;WA

N1;ASCC2;APOE;CCDC141;EIF2B4;CENPW;HSPA4;SSC5D;GPN1;WARS2;MLLT10;KIAA1755;DES;CR1L;SN

FN1;ASCC2;APOE;CCDC141;EIF2B4;GCKR;CENPW;HSPA4;CMTM5;SSC5D;WARS2;MLLT10;KIAA1755;DES

F16;PLN;ZMAT5;MFN1;ASCC2;APOE;CCDC141;EIF2B4;GCKR;CENPW;HSPA4;SSC5D;GPN1;WARS2;MLLT1

'OE;CCDC141;EIF2B4;CENPW;HSPA4;CMTM5;SSC5D;GPN1;WARS2;TBX15;MLLT10;KIAA1755;DES;SNX17

MFN1;ALX4;ASCC2;APOE;CCDC141;EIF2B4;GCKR;CENPW;HSPA4;SSC5D;GPN1;WARS2;TBX15;MLLT10;KI

:IF3E

T10;KIAA1755;SNX17;APOC1;ZNF219;EIF3E;PVRL2

POC1;ZNF219;GNB4;EIF3E;PVRL2

KIAA1755;SNX17;APOC1;ZNF219;EIF3E;PVRL2

WARS2;MLLT10;KIAA1755;DES;SNX17;ZNF219;GNB4;EIF3E;PVRL2

3E

三

;EIF3E

NB4;EIF3E;PVRL2

EIF3E

755;SNX17;ZNF219;GNB4;EIF3E;PVRL2

ALX4;ASCC2;APOE;CCDC141;EIF2B4;GCKR;CENPW;HSPA4;CMTM5;GPN1;TBX15;MLLT10;DES;SNX17;AP[illegible]

:IAA1755;SNX17;ZNF219;GNB4;EIF3E;PVRL2

[illegible]B4;EIF3E;PVRL2

SNX17;APOC1;ZNF219;GNB4;EIF3E;PVRL2

ITM5;SSC5D;GPN1;WARS2;TBX15;MLLT10;KIAA1755;DES;CR1L;SNX17;APOC1;ZNF219;GNB4;EIF3E;PVR

A1755;DES;CR1L;SNX17;APOC1;ZNF219;GNB4;EIF3E;PVRL2

:5D;WARS2;TBX15;MLLT10;KIAA1755;DES;SNX17;APOC1;ZNF219;GNB4;EIF3E;PVRL2

15;MLLT10;KIAA1755;DES;SNX17;APOC1;ZNF219;GNB4;EIF3E;PVRL2
AA1755;DES;SNX17;APOC1;ZNF219;GNB4;EIF3E;PVRL2

(17;APOC1;ZNF219;GNB4;EIF3E;PVRL2

'RL2

55;DES;SNX17;ZNF219;GNB4;EIF3E;PVRL2

;SSC5D;GPN1;WARS2;TBX15;MLLT10;KIAA1755;DES;SNX17;APOC1;ZNF219;GNB4;EIF3E;PVRL2
POC1;ZNF219;GNB4;EIF3E;PVRL2

1755;DES;SNX17;APOC1;ZNF219;GNB4;EIF3E;PVRL2

LLT10;KIAA1755;DES;SNX17;APOC1;ZNF219;GNB4;EIF3E;PVRL2

N1;WARS2;MLLT10;KIAA1755;DES;SNX17;APOC1;ZNF219;GNB4;EIF3E;PVRL2

;WARS2;MLLT10;KIAA1755;DES;SNX17;APOC1;ZNF219;GNB4;EIF3E;PVRL2

\X17;APOC1;ZNF219;GNB4;EIF3E;PVRL2

\A1755;DES;SNX17;APOC1;ZNF219;GNB4;EIF3E;PVRL2

SPA4;SSC5D;GPN1;WARS2;MLLT10;KIAA1755;DES;CR1L;SNX17;APOC1;ZNF219;GNB4;EIF3E;PVRL2

A1755;DES;SNX17;APOC1;ZNF219;GNB4;EIF3E;PVRL2

;CR1L;SNX17;APOC1;ZNF219;GNB4;EIF3E;PVRL2

ARS2;MLLT10;KIAA1755;DES;CR1L;SNX17;APOC1;ZNF219;GNB4;EIF3E;PVRL2

X17;APOC1;ZNF219;GNB4;EIF3E;PVRL2

;;SNX17;APOC1;ZNF219;GNB4;EIF3E;PVRL2

L0;KIAA1755;DES;SNX17;APOC1;ZNF219;GNB4;EIF3E;PVRL2

';APOC1;ZNF219;GNB4;EIF3E;PVRL2

AA1755;DES;SNX17;APOC1;ZNF219;GNB4;EIF3E;PVRL2

OC1;ZNF219;GNB4;EIF3E;PVRL2

L2
