## Supplementary Table 7 for "Hundreds of cardiac MRI traits derived using 3D diffusion autoencoders share a common genetic architecture"

SNP;CHR;BP;EA;EAFREQ;BETA;SE;P;N;Latent;SNP.rep;CHROM.rep;GENPOS.rep;EA.rep;EA.rep;N.rep;BE
rs10802069;1;119517357;C;0.61501;-0.0385983;0.00582362;3.40524e-11;48368.4;S1993_Z97;rs2645
rs11118288;1;207835812;A;0.132778;0.0566408;0.0094198;1.82176e-09;46881.5;S1701_Z82;rs35707
rs11579568;1;208130627;G;0.33753;-0.0365552;0.00592219;6.71839e-10;48197.7;S42_Z10;rs475774;
rs35330522;2;12877060;A;0.47981;0.0393238;0.00615015;1.61646e-10;47592.2;S2023_Z86;rs130201
rs7558413;2;18721662;A;0.583498;0.0405233;0.00602172;1.70221e-11;48319.6;S1701_Z29;rs755841
rs4832605;2;18722424;C;0.583698;0.03699;0.0057724;1.47364e-10;48279.6;S1994_Z5;rs4637120;2;1
rs1260326;2;27730940;C;0.606543;0.0417682;0.00601653;3.85929e-12;48385.1;S1701_Z53;rs141306
rs142556838;2;179747068;T;0.0909455;0.0765574;0.01099;3.25856e-12;47958.9;S1701_Z82;rs14255
rs13386459;2;220299018;C;0.390501;-0.0381575;0.00606589;3.16439e-10;48628.4;S1701_Z29;rs113
rs763302497;2;232252196;A;0.71681;-0.0352841;0.0058624;1.7581e-09;47362.7;S1994_Z111;rs7633
rs6742228;2;232267116;G;0.325574;0.0292572;0.00457815;1.65218e-10;48301.3;S1994_Z31;rs22901
rs1656376;3;158284681;C;0.556862;0.033081;0.00545708;1.34411e-09;48824;S2023_Z77;rs5627924
rs2339798;3;179173620;A;0.139424;0.0387883;0.00644027;1.71451e-09;47672.4;S1994_Z127;rs7611
rs2339798;3;179173620;A;0.139424;-0.0395575;0.00655498;1.59234e-09;47645.9;S1994_Z35;rs7611
rs2339798;3;179173620;A;0.139424;0.0397286;0.00653979;1.24061e-09;47677.1;S1994_Z94;rs76116
rs2339798;3;179173620;A;0.139424;-0.0425265;0.00693605;8.72036e-10;47643.2;S42_Z105;rs76116
rs2339798;3;179173620;A;0.139424;0.0389536;0.00646758;1.71317e-09;47632.2;S42_Z106;rs761167
rs2339798;3;179173620;A;0.139424;-0.0405872;0.00621921;6.75033e-11;47639.7;S42_Z117;rs76116
rs2339798;3;179173620;A;0.139424;-0.0427439;0.00652487;5.71838e-11;47634.4;S42_Z12;rs761167
rs2339798;3;179173620;A;0.139424;-0.0424266;0.0066587;1.87061e-10;47652;S42_Z4;rs7611674;3;1
rs2339798;3;179173620;A;0.139424;-0.0399834;0.00649709;7.55301e-10;47638.5;S42_Z51;rs761167
rs2339798;3;179173620;A;0.139424;-0.043684;0.00718714;1.21631e-09;47637.7;S42_Z89;rs7611674;
rs28568794;4;7852093;C;0.442003;0.0256081;0.00419636;1.04451e-09;48428.8;S2023_Z95;rs284707
4:17956213_TG_T;4;17956213;T;0.738469;0.0371831;0.00618021;1.78252e-09;47842.1;S1993_Z14;rs
4:17956213_TG_T;4;17956213;T;0.738469;0.042988;0.00697231;7.02466e-10;47858.4;S2023_Z86;rs2
rs464605;5;55807370;T;0.745499;0.0415688;0.00680494;1.00496e-09;48877.5;S1701_Z111;rs459193
rs1309546;5;64290004;C;0.446979;0.0313195;0.00490859;1.76454e-10;48090.5;S1701_Z78;rs540834
rs72801474;5;132444128;A;0.0915467;0.0610096;0.00999342;1.02815e-09;49378.7;S1994_Z1;rs5574
rs6920875;6;2478975;A;0.483184;0.0389913;0.00567112;6.18099e-12;48357.4;S1993_Z97;rs879098;
rs10458143;6;2479086;A;0.4828;0.0365618;0.00592435;6.76687e-10;48392.3;S1701_Z118;rs1124277
rs9503212;6;2501535;G;0.519455;0.0368997;0.00525301;2.14854e-12;48143.5;S1993_Z22;rs3966800
rs4959678;6;2503173;A;0.484052;0.0356157;0.0058948;1.52325e-09;48279.1;S1993_Z28;rs1417958;
rs4959678;6;2503173;A;0.484052;0.0358017;0.00542328;4.0706e-11;48259.4;S1994_Z108;rs1079383
rs4959678;6;2503173;A;0.484052;0.0364636;0.00580553;3.3675e-10;48264.3;S2023_Z44;rs11242779
rs11242779;6;2507901;C;0.489813;0.0289993;0.00442518;5.62991e-11;48107;S1993_Z6;rs1610416;6
rs11242779;6;2507901;C;0.489813;0.0280391;0.0041838;2.05831e-11;48076.6;S2023_Z95;rs1010762
rs3951016;6;118559658;A;0.469273;-0.0265563;0.00431029;7.22231e-10;48043.8;S1994_Z31;rs3951
rs11153730;6;118667522;C;0.492803;-0.0399759;0.00629537;2.15248e-10;48352.7;S1701_Z82;rs776
rs7759673;6;121771621;T;0.540029;0.0272367;0.00450771;1.51982e-09;48216.3;S42_Z12;rs347822
rs58730006;6;122089704;AT;0.0986941;-0.0745815;0.0102927;4.29077e-13;48440.4;S1994_Z92;rs93
rs9388001;6;122092897;A;0.099312;0.0830179;0.0105093;2.80092e-15;48461;S1701_Z82;rs9388001;
rs9388001;6;122092897;A;0.099312;0.0619263;0.00933304;3.2413e-11;48506.1;S42_Z10;rs12196004
rs9388487;6;126678268;T;0.469791;0.0399413;0.00593037;1.63881e-11;48608;S1701_Z29;rs1591805
6:126707845_CT_C;6;126707845;C;0.457082;-0.0364688;0.00584112;4.27995e-10;48589.7;S42_Z125;

rs2184968;6;126760994;C;0.453074;-0.0338303;0.00540086;3.75532e-10;48702;S2023_Z87;rs587980
rs263182;6;142862612;C;0.289586;0.0390443;0.00603027;9.49889e-11;48314.1;S2023_Z100;rs96255
rs35164779;7;46560250;C;0.0952039;-0.0627024;0.0103758;1.51144e-09;47428.3;S42_Z127;rs35336
rs35348547;7;120785124;CTG;0.615928;0.0353766;0.00625661;1.56505e-08;48529.4;S2023_Z86;rs10
rs12531355;7;121027434;T;0.265773;0.0509644;0.00688814;1.37356e-13;48515.1;S2023_Z86;rs1253
rs73221948;8;25464670;T;0.292074;0.0418033;0.00684593;1.01967e-09;44311.3;S1701_Z111;rs7322
rs610891;8;109161003;G;0.460489;-0.0393593;0.00613103;1.36554e-10;48112;S2023_Z86;rs1668180
rs2100837;8;120392313;C;0.250743;0.0400673;0.00637153;3.2057e-10;48350.1;S1701_Z80;rs117810
rs10828265;10;22019212;T;0.592916;-0.0331427;0.00551699;1.88536e-09;48038.9;S1993_Z14;rs474
rs806676;10;89787009;T;0.168264;0.0477705;0.00784666;1.14327e-09;47594.5;S1994_Z1;rs2785079
rs4755800;11;44287681;A;0.653418;-0.0397696;0.0062171;1.58658e-10;48295.8;S2023_Z26;rs47557
rs77282531;11;95023373;A;0.0210414;0.123793;0.020129;7.74914e-10;46839.1;S1994_Z5;rs1413629
rs77282531;11;95023373;A;0.0210414;-0.127922;0.0206515;5.85372e-10;46829.1;S42_Z125;rs14136
rs145910347;11;95035065;T;0.0210915;0.126837;0.0209358;1.37501e-09;47061;S1701_Z29;rs141362
rs76895963;12;4384844;G;0.0196553;0.139134;0.0222016;3.68435e-10;41153;S1994_Z5;rs76895963;
rs4963772;12;24758480;A;0.151465;-0.0672216;0.00887219;3.54554e-14;47326.7;S1701_Z82;rs1104
rs11047527;12;24762501;C;0.151465;-0.0474349;0.0078774;1.72654e-09;47395.2;S42_Z10;rs424622
rs11610461;12;24774691;G;0.177265;0.0412598;0.00636129;8.81017e-11;47714.1;S1994_Z85;rs1104
rs11047539;12;24781446;G;0.150412;0.0523152;0.00793713;4.3629e-11;47523.5;S1701_Z122;rs3421
rs11047543;12;24788339;A;0.150913;-0.0570262;0.00868876;5.26598e-11;47493.1;S1701_Z49;rs128
rs2881860;12;28516201;T;0.22905;-0.0387257;0.00645341;1.96333e-09;48787.8;S2023_Z77;rs20050
rs3741760;12;28544464;A;0.757005;0.0492741;0.00706832;3.1443e-12;48882.3;S2023_Z86;rs107714
rs9535455;13;51088547;A;0.213153;-0.043368;0.00720741;1.77525e-09;47987.5;S1701_Z53;rs13276
rs2319625;14;21565146;T;0.174677;0.0354542;0.00588682;1.71594e-09;48712.2;S42_Z12;rs128892
rs422068;14;23864804;C;0.358255;-0.0380976;0.00586422;8.21454e-11;48391.6;S1701_Z122;rs4127
rs422068;14;23864804;C;0.358255;-0.0329689;0.0054491;1.44542e-09;48402.7;S1994_Z111;rs41276
rs2284651;14;23882144;C;0.378645;0.0397303;0.00647605;8.51813e-10;48545.2;S1701_Z82;rs45203
rs28929474;14;94844947;T;0.0205905;-0.125626;0.0208489;1.6854e-09;46631;S1994_Z38;rs1126352
rs28929474;14;94844947;T;0.0205905;-0.145459;0.0218588;2.84299e-11;46624.7;S2023_Z86;rs1126
rs28929474;14;94844947;T;0.0205905;-0.137261;0.0209143;5.27283e-11;46633.1;S42_Z125;rs11263
rs28929474;14;94844947;T;0.0205905;0.136465;0.0213698;1.70413e-10;46624.4;S1701_Z111;rs2892
rs28929474;14;94844947;T;0.0205905;-0.117298;0.0195331;1.91227e-09;46629.3;S2023_Z77;rs2892
rs139583754;15;93331251;G;0.0107545;-0.164908;0.0265246;5.06195e-10;47128.7;S1701_Z32;rs803
rs139583754;15;93331251;G;0.0107545;-0.177189;0.0271119;6.34102e-11;47151.3;S1701_Z80;rs803
rs11664030;18;20261639;T;0.542083;0.0354844;0.00586224;1.42134e-09;48610.6;S1701_Z20;rs1530
rs429358;19;45411941;C;0.15751;-0.0490037;0.0075527;8.68472e-11;46499.8;S2023_Z87;rs429358;1
rs429358;19;45411941;C;0.15751;-0.0531432;0.00820887;9.55208e-11;46511.4;S42_Z28;rs814573;19
rs147110934;19;55993436;T;0.0246819;0.116306;0.0186035;4.0566e-10;49380.5;S42_Z125;rs546748
rs3746471;20;36841914;A;0.463428;-0.0413851;0.00632912;6.1998e-11;48083.1;S1701_Z82;rs48116
rs140120;22;30138548;C;0.513393;0.0394055;0.00623701;2.64951e-10;48227.9;S1993_Z27;rs131289
rs140120;22;30138548;C;0.513393;0.0387455;0.00624356;5.44629e-10;48241.9;S1993_Z41;rs230140
rs131272;22;30153655;G;0.504743;-0.035868;0.00581765;7.03158e-10;48473.5;S1994_Z1;rs1115975
rs131299;22;30180153;AT;0.648191;0.0398552;0.00651926;9.74971e-10;46534.7;S1701_Z93;rs13129
rs5752961;22;30188084;T;0.492402;-0.0368714;0.00589093;3.87425e-10;48344.2;S1993_Z66;rs3958
rs1076135;22;30238317;T;0.478591;-0.0387768;0.00590672;5.20812e-11;48117.1;S1993_Z28;rs1401

TA.rep;SE.rep;P.rep;id;Replicated
294;1;119574587;T;0.55062;11907;-0.0436782;0.0113203;0.000114135304025367;rs10802069-S1993_
912;1;207829220;T;0.130399;11907;0.00596378;0.018998;0.753584574353222;rs11118288-S1701_Z8
;1;208158787;T;0.304527;11907;-0.0227493;0.0122488;0.0632717723562756;rs11579568-S42_Z10;No
78;2;12978463;A;0.507924;11907;0.0395469;0.0121489;0.0011332350541319;rs35330522-S2023_Z86
3;2;18721662;A;0.602645;11907;0.0270758;0.0122167;0.026671043025936;rs7558413-S1701_Z29;Co
8701627;C;0.65419;11907;0.0280422;0.0122079;0.0216152369147784;rs4832605-S1994_Z5;Concorda
206;2;27745764;G;0.547324;11907;0.0355293;0.0121059;0.00333679864130862;rs1260326-S1701_Z5
6838;2;179747068;T;0.0803108;11907;0.0950785;0.0234041;4.85567935098814e-05;rs142556838-S1
37641;2;220297581;CA;0.572259;11907;-0.00967814;0.0125755;0.441535879086463;rs13386459-S17
02497;2;232252196;A;0.699447;11907;-0.0129659;0.0114704;0.258319386157811;rs763302497-S199
30;2;232263127;A;0.237549;11907;0.0288436;0.0100869;0.00424277498863422;rs6742228-S1994_Z3
2;3;158232862;T;0.453849;11907;0.0398599;0.0111736;0.000360620158561837;rs1656376-S2023_Z77
674;3;179169230;G;0.208752;11907;0.0239324;0.0110539;0.0303822546869739;rs2339798-S1994_Z1
674;3;179169230;G;0.208752;11907;-0.0312994;0.0112001;0.00519708715512298;rs2339798-S1994_
74;3;179169230;G;0.208752;11907;0.0257741;0.0112082;0.0214718764722981;rs2339798-S1994_Z94
74;3;179169230;G;0.208752;11907;-0.0277633;0.0118221;0.0188534138152454;rs2339798-S42_Z105;
4;3;179169230;G;0.208752;11907;0.0363522;0.01106;0.0010132579035892;rs2339798-S42_Z106;Con
74;3;179169230;G;0.208752;11907;-0.027823;0.0106609;0.00905899458376675;rs2339798-S42_Z117;
4;3;179169230;G;0.208752;11907;-0.0193136;0.011212;0.084964987919827;rs2339798-S42_Z12;Not s
79169230;G;0.208752;11907;-0.0168508;0.0114163;0.139938108695905;rs2339798-S42_Z4;Not signi
4;3;179169230;G;0.208752;11907;-0.0289672;0.0111132;0.00914576419951297;rs2339798-S42_Z51;C
3;179169230;G;0.208752;11907;-0.0309044;0.0122614;0.0117197945906848;rs2339798-S42_Z89;Con
31;4;7848144;T;0.497567;11907;0.0135936;0.00844203;0.107346774620177;rs28568794-S2023_Z95;N
2724475;4;17946432;C;0.744949;11907;0.0209172;0.012698;0.0995015852637172;4:17956213_TG_T
724472;4;17978892;T;0.745044;11907;0.0246586;0.0142457;0.0834603112498606;4:17956213_TG_T
;5;55806751;G;0.735702;11907;0.0160717;0.0134417;0.231829660917537;rs464605-S1701_Z111;Not
152;5;64256749;GT;0.447285;11907;0.0225976;0.0101306;0.0257057334555197;rs1309546-S1701_Z7
7751;5;132397351;A;0.0708387;11907;0.029314;0.022957;0.201634573057729;rs72801474-S1994_Z1
;2507448;G;0.38966;11907;0.0191762;0.0116582;0.0999976974414163;rs6920875-S1993_Z97;Not sig
9;6;2507901;C;0.488505;11907;0.0174189;0.0118059;0.140093504665011;rs10458143-S1701_Z118;N
;6;2493113;T;0.488808;11907;0.0215331;0.0104505;0.0393522890918596;rs9503212-S1993_Z22;Con
;2510495;G;0.530721;11907;0.0108881;0.0124288;0.381007916985951;rs4959678-S1993_Z28;Not sig
8;6;2474246;G;0.424494;11907;0.0238032;0.0108621;0.0284223511023194;rs4959678-S1994_Z108;C
;6;2507901;C;0.488505;11907;0.0144105;0.0114004;0.206216779419058;rs4959678-S2023_Z44;Not s
;2474933;A;0.541215;11907;0.007723;0.00911647;0.396912704356787;rs11242779-S1993_Z6;Not sig
;6;2476867;T;0.506632;11907;0.00958962;0.00850895;0.25974069604073;rs11242779-S2023_Z95;No
016;6;118559658;A;0.48235;11907;-0.00955239;0.00866018;0.270016311261912;rs3951016-S1994_Z3
4272;6;118692498;C;0.556242;11907;-0.00546883;0.0127715;0.668500905714942;rs11153730-S1701_
9;6;121766728;GA;0.509755;11907;0.0246582;0.00920107;0.00736376634961514;rs7759673-S42_Z12
38001;6;122092897;A;0.109975;11907;-0.0541517;0.019356;0.00514730615164617;rs58730006-S199
6;122092897;A;0.109975;11907;0.0956455;0.019991;1.7147862797488e-06;rs9388001-S1701_Z82;Co
;6;121933718;G;0.110104;11907;0.0378361;0.0180111;0.0356664594673732;rs9388001-S42_Z10;Con
;6;126717064;G;0.514529;11907;0.0402804;0.0119173;0.000724869789537885;rs9388487-S1701_Z2
rs1361109;6;126771143;T;0.480952;11907;-0.028435;0.0117742;0.0157340309455349;6:126707845_

7;6;126875371;CAT;0.459573;11907;-0.0321694;0.0113941;0.00475247674022448;rs2184968-S2023_
4;6;142734204;C;0.30805;11907;0.0313702;0.0119381;0.00859547734514673;rs263182-S2023_Z100;
348;7;46558672;C;0.10468;11907;-0.0616928;0.0201628;0.00221528700545703;rs35164779-S42_Z127
242653;7;120865801;A;0.591642;11907;0.020217;0.0125585;0.107436041920981;rs35348547-S2023_
1355;7;121027434;T;0.26597;11907;0.0630273;0.013979;6.52213824514443e-06;rs12531355-S2023_
1948;8;25464670;T;0.287054;11907;0.0396541;0.0138163;0.00410355255604658;rs73221948-S1701_
;8;109213500;A;0.426957;11907;-0.0537242;0.0128377;2.85305405476176e-05;rs610891-S2023_Z86;
27;8;120450027;A;0.286671;11907;0.0296404;0.0123432;0.0163342801529616;rs2100837-S1701_Z80
3761;10;21989245;G;0.710743;11907;-0.0309796;0.0120639;0.0102301028503723;rs10828265-S1993_
;10;89782386;A;0.186579;11907;0.0155073;0.0151148;0.304906722726869;rs806676-S1994_Z1;Not si
97;11;44285887;A;0.73349;11907;-0.00471242;0.0134368;0.72580507405669;rs4755800-S2023_Z26;N
55;11;95039669;C;0.0186536;11907;-0.0921148;0.0431678;0.0328526192201754;rs77282531-S1994_
2955;11;95039669;C;0.0186536;11907;0.0903485;0.0448084;0.0437653090905688;rs77282531-S42_Z
2955;11;95039669;C;0.0186536;11907;-0.153874;0.0451997;0.000663315277349058;rs145910347-S17
12;4384844;G;0.0189477;11907;0.164036;0.0464243;0.000410241885920653;rs76895963-S1994_Z5;
7539;12;24781446;G;0.155506;11907;-0.0366381;0.0172713;0.0338937795175651;rs4963772-S1701_
4;12;24784139;A;0.155334;11907;-0.0301381;0.0154293;0.050784361904768;rs11047527-S42_Z10;No
7543;12;24788339;A;0.156043;11907;0.0515777;0.0131914;9.23145166270663e-05;rs11610461-S199
9605;12;24760456;G;0.156251;11907;0.044429;0.0153799;0.0038674971292038;rs11047539-S1701_
26024;12;24776799;A;0.155169;11907;-0.0433652;0.0167337;0.00955608491018313;rs11047543-S17
7341;12;28574477;CT;0.210214;11907;-0.04098;0.0137243;0.00282696219042436;rs2881860-S2023_Z
33;12;28674825;T;0.75141;11907;0.0469156;0.0141498;0.000914365855191293;rs3741760-S2023_Z8
46;13;51122365;G;0.166995;11907;-0.0649501;0.016238;6.33752958894704e-05;rs9535455-S1701_Z5
7;14;21542766;G;0.157344;11907;0.028043;0.012357;0.0232439536095743;rs2319625-S42_Z12;Conc
58;14;23866713;G;0.31933;11907;-0.0249527;0.0120516;0.0384060815725812;rs422068-S1701_Z122;
3;14;23866713;G;0.31933;11907;-0.0185833;0.0112629;0.0989509572680665;rs422068-S1994_Z111;N
6;14;23865885;A;0.367431;11907;0.0301702;0.0129871;0.0201739063294114;rs2284651-S1701_Z82;
99;14;94838142;T;0.0165449;11907;-0.0181491;0.0465315;0.696507825127659;rs28929474-S1994_Z
35299;14;94838142;T;0.0165449;11907;-0.00346404;0.0483236;0.942853244758951;rs28929474-S202
5299;14;94838142;T;0.0165449;11907;-0.00923396;0.0465854;0.842876217489032;rs28929474-S42_Z
9474;14;94844947;T;0.015873;11907;-0.0323672;0.0481018;0.501017620475039;rs28929474-S1701_
9474;14;94844947;T;0.015873;11907;0.0355331;0.0442457;0.421924748314925;rs28929474-S2023_Z
9156;15;93351218;A;0.0124972;11907;-0.0488746;0.0494608;0.32308068871421;rs139583754-S1701_
9156;15;93351218;A;0.0124972;11907;-0.0441935;0.0507365;0.383731984810111;rs139583754-S170
715;18;20290616;C;0.519501;11907;0.0336694;0.0118745;0.00457635809308924;rs11664030-S1701_
9;45411941;C;0.134207;11907;-0.0165566;0.0158843;0.297261044869654;rs429358-S2023_Z87;Not s
;45424351;T;0.174077;11907;-0.0143991;0.0158203;0.362733778633291;rs429358-S42_Z28;Not signi
948;19;55995764;GCC;0.0229506;11907;0.0466048;0.0396719;0.240092724563744;rs147110934-S42_
01;20;36849007;T;0.446215;11907;-0.0472205;0.0127826;0.000220648002406029;rs3746471-S1701_Z
;22;30161133;G;0.366293;11907;0.0553166;0.0127219;1.37290346146731e-05;rs140120-S1993_Z27;
2;22;30234045;A;0.359686;11907;0.0626305;0.0128614;1.11794387274424e-06;rs140120-S1993_Z41;
00;22;30123198;TTAAGGATC;0.479357;11907;-0.056869;0.0118933;1.73920189523564e-06;rs131272-
9;22;30180153;AT;0.609228;11907;0.0199847;0.0126446;0.113994264284271;rs131299-S1701_Z93;N
027;22;30244835;T;0.342301;11907;-0.0604777;0.0122445;7.84584998441426e-07;rs5752961-S1993_
47;22;30184599;A;0.346007;11907;-0.0552977;0.0122205;6.03990350110675e-06;rs1076135-S1993_Z

_Z97;Concordant
2;Not significant
t significant
;Concordant
ncordant
nt
3;Concordant
701_Z82;Concordant
1_Z29;Not significant
_Z111;Not significant
1;Concordant
;Concordant
27;Concordant
Z35;Concordant
;Concordant
Concordant
cordant
Concordant
ignificant
icant
oncordant
cordant
ot significant
-S1993_Z14;Not significant
-S2023_Z86;Not significant
significant
8;Concordant
;Not significant
nificant
ot significant
cordant
nificant
oncordant
ignificant
nificant
t significant
1;Not significant
_Z82;Not significant
;Concordant
_Z92;Concordant
ncordant
cordant
;Concordant
CT_C-S42_Z125;Concordant

Z87;Concordant
Concordant
7;Concordant
_Z86;Not significant
Z86;Concordant
Z111;Concordant
;Concordant
0;Concordant
_Z14;Concordant
ignificant
lot significant
Z5;Discordant
125;Discordant
/01_Z29;Discordant
Concordant
Z82;Concordant
ot significant
4_Z85;Concordant
Z122;Concordant
01_Z49;Concordant
Z77;Concordant
6;Concordant
3;Concordant
ordant
Concordant
lot significant
Concordant
38;Not significant
23_Z86;Not significant
Z125;Not significant
Z111;Not significant
77;Not significant
_Z32;Not significant
1_Z80;Not significant
Z20;Concordant
ignificant
ficant
_Z125;Not significant
Z82;Concordant
Concordant
;Concordant
S1994_Z1;Concordant
ot significant
Z66;Concordant
28;Concordant
