## Supplementary Table 6 for "Hundreds of cardiac MRI traits derived using 3D diffusion autoencoders share a common genetic architecture"

| SNP | CHR | BP | EA | EAFREQ | BETA | SE | P | N |
|---|---|---|---|---|---|---|---|---|
| rs74181240 | 2 | 57775069 | G | 0.656341 | -0.07528 | 0.013613 | 3.21E-08 | 41830 |
| rs77135915 | 3 | 13620165 | T | 0.983384 | 0.271511 | 0.045684 | 2.79E-09 | 46039 |
| rs9833758 | 3 | 39902940 | C | 0.386777 | 0.056335 | 0.010244 | 3.81E-08 | 48182 |
| rs62274713 | 3 | 1.02E+08 | G | 0.157192 | 0.097833 | 0.017286 | 1.52E-08 | 44168 |
| rs14276878 | 6 | 1.18E+08 | C | 0.013343 | 0.294797 | 0.05396 | 4.67E-08 | 44486 |
| rs7001763 | 8 | 73236004 | G | 0.576684 | -0.05595 | 0.010159 | 3.64E-08 | 46757 |
| rs7001763 | 8 | 73236004 | G | 0.576684 | 0.055501 | 0.009868 | 1.86E-08 | 46861 |
| rs72922827 | 11 | 64358795 | T | 0.035987 | 0.18157 | 0.030918 | 4.29E-09 | 47722 |
| rs72925197 | 11 | 64416031 | A | 0.036038 | 0.169159 | 0.025006 | 1.34E-11 | 48483 |
| rs608261 | 11 | 64517317 | T | 0.038877 | 0.157675 | 0.026664 | 3.35E-09 | 46072 |
| rs608261 | 11 | 64517317 | T | 0.038877 | 0.175781 | 0.030925 | 1.31E-08 | 45102 |
| rs11160234 | 14 | 95683873 | C | 0.303413 | -0.05732 | 0.010292 | 2.55E-08 | 46577 |
| rs11160234 | 14 | 95683873 | C | 0.303413 | -0.05962 | 0.010858 | 4.01E-08 | 46810 |
| rs14119741 | 18 | 53812729 | T | 0.026686 | -0.16221 | 0.02906 | 2.38E-08 | 43238 |
| rs11251188 | 19 | 21606390 | A | 0.01371 | 0.305823 | 0.055511 | 3.60E-08 | 45778 |

| Latent | Locus |
|---|---|
| Z127_S4 | 1 |
| Z99_S4 | 2 |
| Z39_S4 | 3 |
| Z127_S4 | 4 |
| Z111_S1 | 5 |
| Z17_S1 | 6 |
| Z57_S1 | 6 |
| Z9_S2 | 7 |
| Z57_S2 | 7 |
| Z3_S2 | 7 |
| Z94_S2 | 7 |
| Z51_S1 | 8 |
| Z73_S1 | 8 |
| Z120_S2 | 9 |
| Z30_S2 | 10 |
