## Supplementary Table 5 for "Hundreds of cardiac MRI traits derived using 3D diffusion autoencoders share a common genetic architecture"

| Disease | Model | predefined | L1_penaliz | CV_OR_He | Fold_num | NumCoeffi | NumSignifi | ClassifRprt_ |
|---|---|---|---|---|---|---|---|---|
| angina-pec | LogRegrs | FALSE | | HeldoutTest | | 66 | 38 | 0.662338 |
| angina-pec | LassoCat | FALSE | 0.359381 | HeldoutTest | | 66 | 26 | 0.62987 |
| angina-pec | LassoCat | TRUE | 0.05 | HeldoutTest | | 66 | 5 | 0.62987 |
| angina-pec | LogRegrs | FALSE | | CV | 0 | 66 | 33 | 0.58216 |
| angina-pec | LogRegrs | FALSE | | CV | 1 | 66 | 35 | 0.619718 |
| angina-pec | LogRegrs | FALSE | | CV | 2 | 66 | 39 | 0.57277 |
| angina-pec | LogRegrs | FALSE | | CV | 3 | 66 | 34 | 0.615023 |
| angina-pec | LogRegrs | FALSE | | CV | 4 | 66 | 37 | 0.599057 |
| angina-pec | LassoCat | FALSE | 0.359381 | CV | 0 | 66 | 22 | 0.58216 |
| angina-pec | LassoCat | FALSE | 0.359381 | CV | 1 | 66 | 20 | 0.624413 |
| angina-pec | LassoCat | FALSE | 21.54435 | CV | 2 | 66 | 39 | 0.57277 |
| angina-pec | LassoCat | FALSE | 2.782559 | CV | 3 | 66 | 34 | 0.605634 |
| angina-pec | LassoCat | FALSE | 0.359381 | CV | 4 | 66 | 18 | 0.613208 |
| angina-pec | LassoCat | TRUE | 0.05 | CV | 0 | 66 | 2 | 0.586854 |
| angina-pec | LassoCat | TRUE | 0.05 | CV | 1 | 66 | 3 | 0.605634 |
| angina-pec | LassoCat | TRUE | 0.05 | CV | 2 | 66 | 3 | 0.544601 |
| angina-pec | LassoCat | TRUE | 0.05 | CV | 3 | 66 | 2 | 0.596244 |
| angina-pec | LassoCat | TRUE | 0.05 | CV | 4 | 66 | 3 | 0.636792 |
| atrial-fibrill | LogRegrs | FALSE | | HeldoutTest | | 66 | 29 | 0.639535 |
| atrial-fibrill | LassoCat | FALSE | 0.005995 | HeldoutTest | | 66 | 1 | 0.616279 |
| atrial-fibrill | LassoCat | TRUE | 0.05 | HeldoutTest | | 66 | 1 | 0.639535 |
| atrial-fibrill | LogRegrs | FALSE | | CV | 0 | 66 | 31 | 0.619247 |
| atrial-fibrill | LogRegrs | FALSE | | CV | 1 | 66 | 35 | 0.644351 |
| atrial-fibrill | LogRegrs | FALSE | | CV | 2 | 66 | 29 | 0.710084 |
| atrial-fibrill | LogRegrs | FALSE | | CV | 3 | 66 | 31 | 0.634454 |
| atrial-fibrill | LogRegrs | FALSE | | CV | 4 | 66 | 29 | 0.655462 |
| atrial-fibrill | LassoCat | FALSE | 0.046416 | CV | 0 | 66 | 1 | 0.594142 |
| atrial-fibrill | LassoCat | FALSE | 166.8101 | CV | 1 | 66 | 36 | 0.640167 |
| atrial-fibrill | LassoCat | FALSE | 2.782559 | CV | 2 | 66 | 28 | 0.705882 |
| atrial-fibrill | LassoCat | FALSE | 0.046416 | CV | 3 | 66 | 1 | 0.684874 |
| atrial-fibrill | LassoCat | FALSE | 0.046416 | CV | 4 | 66 | 1 | 0.634454 |
| atrial-fibrill | LassoCat | TRUE | 0.05 | CV | 0 | 66 | 1 | 0.589958 |
| atrial-fibrill | LassoCat | TRUE | 0.05 | CV | 1 | 66 | 1 | 0.661088 |
| atrial-fibrill | LassoCat | TRUE | 0.05 | CV | 2 | 66 | 1 | 0.739496 |
| atrial-fibrill | LassoCat | TRUE | 0.05 | CV | 3 | 66 | 1 | 0.680672 |
| atrial-fibrill | LassoCat | TRUE | 0.05 | CV | 4 | 66 | 3 | 0.62605 |
| coronary-h | LogRegrs | FALSE | | HeldoutTest | | 67 | 33 | 0.637931 |
| coronary-h | LassoCat | FALSE | 0.359381 | HeldoutTest | | 67 | 29 | 0.633621 |
| coronary-h | LassoCat | TRUE | 0.05 | HeldoutTest | | 67 | 12 | 0.646552 |
| coronary-h | LogRegrs | FALSE | | CV | 0 | 67 | 34 | 0.568106 |
| coronary-h | LogRegrs | FALSE | | CV | 1 | 67 | 31 | 0.627907 |
| coronary-h | LogRegrs | FALSE | | CV | 2 | 67 | 32 | 0.676667 |
| coronary-h | LogRegrs | FALSE | | CV | 3 | 66 | 33 | 0.68 |
| coronary-h | LogRegrs | FALSE | | CV | 4 | 67 | 31 | 0.61 |

| | | | | | | | | |
|---|---|---|---|---|---|---|---|---|
| coronary-h | LassoCat | FALSE | 0.359381 | CV | 0 | 67 | 26 | 0.574751 |
| coronary-h | LassoCat | FALSE | 0.359381 | CV | 1 | 67 | 24 | 0.637874 |
| coronary-h | LassoCat | FALSE | 0.359381 | CV | 2 | 67 | 21 | 0.67 |
| coronary-h | LassoCat | FALSE | 0.359381 | CV | 3 | 66 | 26 | 0.676667 |
| coronary-h | LassoCat | FALSE | 0.359381 | CV | 4 | 67 | 23 | 0.62 |
| coronary-h | LassoCat | TRUE | 0.05 | CV | 0 | 67 | 7 | 0.518272 |
| coronary-h | LassoCat | TRUE | 0.05 | CV | 1 | 67 | 7 | 0.591362 |
| coronary-h | LassoCat | TRUE | 0.05 | CV | 2 | 67 | 7 | 0.656667 |
| coronary-h | LassoCat | TRUE | 0.05 | CV | 3 | 66 | 5 | 0.653333 |
| coronary-h | LassoCat | TRUE | 0.05 | CV | 4 | 67 | 8 | 0.613333 |
| high-choles | LogRegrs | FALSE | | HeldoutTest | | 66 | 36 | 0.584906 |
| high-choles | LassoCat | FALSE | 0.046416 | HeldoutTest | | 66 | 11 | 0.606469 |
| high-choles | LassoCat | TRUE | 0.05 | HeldoutTest | | 66 | 12 | 0.602426 |
| high-choles | LogRegrs | FALSE | | CV | 0 | 66 | 34 | 0.573077 |
| high-choles | LogRegrs | FALSE | | CV | 1 | 66 | 28 | 0.604808 |
| high-choles | LogRegrs | FALSE | | CV | 2 | 66 | 36 | 0.576923 |
| high-choles | LogRegrs | FALSE | | CV | 3 | 66 | 27 | 0.580366 |
| high-choles | LogRegrs | FALSE | | CV | 4 | 66 | 32 | 0.585178 |
| high-choles | LassoCat | FALSE | 0.046416 | CV | 0 | 66 | 7 | 0.573077 |
| high-choles | LassoCat | FALSE | 0.046416 | CV | 1 | 66 | 9 | 0.605769 |
| high-choles | LassoCat | FALSE | 0.046416 | CV | 2 | 66 | 11 | 0.578846 |
| high-choles | LassoCat | FALSE | 0.046416 | CV | 3 | 66 | 10 | 0.582291 |
| high-choles | LassoCat | FALSE | 0.046416 | CV | 4 | 66 | 9 | 0.569779 |
| high-choles | LassoCat | TRUE | 0.05 | CV | 0 | 66 | 8 | 0.574038 |
| high-choles | LassoCat | TRUE | 0.05 | CV | 1 | 66 | 10 | 0.600962 |
| high-choles | LassoCat | TRUE | 0.05 | CV | 2 | 66 | 12 | 0.581731 |
| high-choles | LassoCat | TRUE | 0.05 | CV | 3 | 66 | 11 | 0.584216 |
| high-choles | LassoCat | TRUE | 0.05 | CV | 4 | 66 | 9 | 0.571704 |
| hypertensi | LogRegrs | FALSE | | HeldoutTest | | 66 | 37 | 0.66954 |
| hypertensi | LassoCat | FALSE | 0.359381 | HeldoutTest | | 66 | 34 | 0.673372 |
| hypertensi | LassoCat | TRUE | 0.05 | HeldoutTest | | 66 | 24 | 0.67433 |
| hypertensi | LogRegrs | FALSE | | CV | 0 | 66 | 37 | 0.633067 |
| hypertensi | LogRegrs | FALSE | | CV | 1 | 66 | 36 | 0.647571 |
| hypertensi | LogRegrs | FALSE | | CV | 2 | 66 | 39 | 0.64467 |
| hypertensi | LogRegrs | FALSE | | CV | 3 | 66 | 35 | 0.619289 |
| hypertensi | LogRegrs | FALSE | | CV | 4 | 66 | 38 | 0.639332 |
| hypertensi | LassoCat | FALSE | 0.046416 | CV | 0 | 66 | 20 | 0.642495 |
| hypertensi | LassoCat | FALSE | 0.046416 | CV | 1 | 66 | 19 | 0.640319 |
| hypertensi | LassoCat | FALSE | 0.046416 | CV | 2 | 66 | 23 | 0.646846 |
| hypertensi | LassoCat | FALSE | 0.046416 | CV | 3 | 66 | 20 | 0.633793 |
| hypertensi | LassoCat | FALSE | 0.359381 | CV | 4 | 66 | 35 | 0.642961 |
| hypertensi | LassoCat | TRUE | 0.05 | CV | 0 | 66 | 20 | 0.64322 |
| hypertensi | LassoCat | TRUE | 0.05 | CV | 1 | 66 | 19 | 0.639594 |
| hypertensi | LassoCat | TRUE | 0.05 | CV | 2 | 66 | 24 | 0.64467 |
| hypertensi | LassoCat | TRUE | 0.05 | CV | 3 | 66 | 20 | 0.630167 |

| | | | | | | | | |
|---|---|---|---|---|---|---|---|---|
| hypertensi | LassoCat | TRUE | 0.05 | CV | 4 | 66 | 26 | 0.640784 |
| myocardial | LogRegrs | FALSE | | HeldoutTest | | 67 | 35 | 0.683099 |
| myocardial | LassoCat | FALSE | 0.359381 | HeldoutTest | | 67 | 27 | 0.690141 |
| myocardial | LassoCat | TRUE | 0.05 | HeldoutTest | | 67 | 5 | 0.690141 |
| myocardial | LogRegrs | FALSE | | CV | 0 | 67 | 38 | 0.643275 |
| myocardial | LogRegrs | FALSE | | CV | 1 | 67 | 34 | 0.672515 |
| myocardial | LogRegrs | FALSE | | CV | 2 | 67 | 33 | 0.684211 |
| myocardial | LogRegrs | FALSE | | CV | 3 | 67 | 33 | 0.678363 |
| myocardial | LogRegrs | FALSE | | CV | 4 | 67 | 36 | 0.635294 |
| myocardial | LassoCat | FALSE | 2.782559 | CV | 0 | 67 | 36 | 0.649123 |
| myocardial | LassoCat | FALSE | 21.54435 | CV | 1 | 67 | 35 | 0.678363 |
| myocardial | LassoCat | FALSE | 0.359381 | CV | 2 | 67 | 27 | 0.690058 |
| myocardial | LassoCat | FALSE | 0.359381 | CV | 3 | 67 | 23 | 0.666667 |
| myocardial | LassoCat | FALSE | 0.359381 | CV | 4 | 67 | 26 | 0.652941 |
| myocardial | LassoCat | TRUE | 0.05 | CV | 0 | 67 | 4 | 0.54386 |
| myocardial | LassoCat | TRUE | 0.05 | CV | 1 | 67 | 4 | 0.666667 |
| myocardial | LassoCat | TRUE | 0.05 | CV | 2 | 67 | 3 | 0.637427 |
| myocardial | LassoCat | TRUE | 0.05 | CV | 3 | 67 | 4 | 0.608187 |
| myocardial | LassoCat | TRUE | 0.05 | CV | 4 | 67 | 3 | 0.652941 |
| type-2-diak | LogRegrs | FALSE | | HeldoutTest | | 66 | 32 | 0.692857 |
| type-2-diak | LassoCat | FALSE | 0.046416 | HeldoutTest | | 66 | 13 | 0.685714 |
| type-2-diak | LassoCat | TRUE | 0.05 | HeldoutTest | | 66 | 14 | 0.692857 |
| type-2-diak | LogRegrs | FALSE | | CV | 0 | 66 | 37 | 0.669118 |
| type-2-diak | LogRegrs | FALSE | | CV | 1 | 66 | 29 | 0.661765 |
| type-2-diak | LogRegrs | FALSE | | CV | 2 | 66 | 28 | 0.676471 |
| type-2-diak | LogRegrs | FALSE | | CV | 3 | 66 | 40 | 0.650735 |
| type-2-diak | LogRegrs | FALSE | | CV | 4 | 66 | 35 | 0.676471 |
| type-2-diak | LassoCat | FALSE | 0.359381 | CV | 0 | 66 | 25 | 0.683824 |
| type-2-diak | LassoCat | FALSE | 0.359381 | CV | 1 | 66 | 20 | 0.672794 |
| type-2-diak | LassoCat | FALSE | 0.359381 | CV | 2 | 66 | 21 | 0.691176 |
| type-2-diak | LassoCat | FALSE | 0.046416 | CV | 3 | 66 | 12 | 0.713235 |
| type-2-diak | LassoCat | FALSE | 0.046416 | CV | 4 | 66 | 10 | 0.658088 |
| type-2-diak | LassoCat | TRUE | 0.05 | CV | 0 | 66 | 10 | 0.691176 |
| type-2-diak | LassoCat | TRUE | 0.05 | CV | 1 | 66 | 9 | 0.661765 |
| type-2-diak | LassoCat | TRUE | 0.05 | CV | 2 | 66 | 9 | 0.676471 |
| type-2-diak | LassoCat | TRUE | 0.05 | CV | 3 | 66 | 12 | 0.720588 |
| type-2-diak | LassoCat | TRUE | 0.05 | CV | 4 | 66 | 11 | 0.658088 |

| ClassifRprt_ | ClassifRprt_ | ClassifRprt_ | AUC | TrainSize | TestSize |
|---|---|---|---|---|---|
| 0.662338 | 0.662338 | 0.662338 | 0.694552 | 1064 | 154 |
| 0.629892 | 0.62987 | 0.629855 | 0.698263 | 1064 | 154 |
| 0.629892 | 0.62987 | 0.629855 | 0.675999 | 1064 | 154 |
| 0.586614 | 0.58216 | 0.577449 | 0.628901 | 851 | 213 |
| 0.619864 | 0.619718 | 0.619668 | 0.671751 | 851 | 213 |
| 0.573218 | 0.57277 | 0.572374 | 0.602275 | 851 | 213 |
| 0.616707 | 0.615023 | 0.6139 | 0.651472 | 851 | 213 |
| 0.599065 | 0.599057 | 0.599048 | 0.647383 | 852 | 212 |
| 0.584625 | 0.58216 | 0.57964 | 0.628901 | 851 | 213 |
| 0.624413 | 0.624413 | 0.624413 | 0.65976 | 851 | 213 |
| 0.573218 | 0.57277 | 0.572374 | 0.604567 | 851 | 213 |
| 0.606373 | 0.605634 | 0.605147 | 0.651384 | 851 | 213 |
| 0.613571 | 0.613208 | 0.612897 | 0.653969 | 852 | 212 |
| 0.588639 | 0.586854 | 0.585191 | 0.62079 | 851 | 213 |
| 0.606094 | 0.605634 | 0.605007 | 0.644154 | 851 | 213 |
| 0.547262 | 0.544601 | 0.539467 | 0.603774 | 851 | 213 |
| 0.596658 | 0.596244 | 0.595977 | 0.638776 | 851 | 213 |
| 0.636902 | 0.636792 | 0.63672 | 0.653969 | 852 | 212 |
| 0.639837 | 0.639535 | 0.63934 | 0.684965 | 1192 | 172 |
| 0.617294 | 0.616279 | 0.615447 | 0.679557 | 1192 | 172 |
| 0.640217 | 0.639535 | 0.639096 | 0.694565 | 1192 | 172 |
| 0.619523 | 0.619247 | 0.619114 | 0.688936 | 953 | 239 |
| 0.644374 | 0.644351 | 0.644351 | 0.713866 | 953 | 239 |
| 0.710456 | 0.710084 | 0.709956 | 0.777487 | 954 | 238 |
| 0.635613 | 0.634454 | 0.633671 | 0.708566 | 954 | 238 |
| 0.655858 | 0.655462 | 0.655243 | 0.704541 | 954 | 238 |
| 0.594236 | 0.594142 | 0.593929 | 0.667087 | 953 | 239 |
| 0.640232 | 0.640167 | 0.640155 | 0.712185 | 953 | 239 |
| 0.706407 | 0.705882 | 0.705695 | 0.778194 | 954 | 238 |
| 0.685938 | 0.684874 | 0.684423 | 0.732999 | 954 | 238 |
| 0.634692 | 0.634454 | 0.634292 | 0.714992 | 954 | 238 |
| 0.590002 | 0.589958 | 0.589815 | 0.668207 | 953 | 239 |
| 0.66143 | 0.661088 | 0.660969 | 0.740406 | 953 | 239 |
| 0.741956 | 0.739496 | 0.738832 | 0.768943 | 954 | 238 |
| 0.681493 | 0.680672 | 0.680311 | 0.735259 | 954 | 238 |
| 0.626273 | 0.62605 | 0.625885 | 0.714144 | 954 | 238 |
| 0.638964 | 0.637931 | 0.637257 | 0.695898 | 1502 | 232 |
| 0.634109 | 0.633621 | 0.633287 | 0.699093 | 1502 | 232 |
| 0.646552 | 0.646552 | 0.646552 | 0.688243 | 1502 | 232 |
| 0.568566 | 0.568106 | 0.567103 | 0.614658 | 1201 | 301 |
| 0.627907 | 0.627907 | 0.627907 | 0.67117 | 1201 | 301 |
| 0.676737 | 0.676667 | 0.676634 | 0.745422 | 1202 | 300 |
| 0.680513 | 0.68 | 0.679772 | 0.740489 | 1202 | 300 |
| 0.610397 | 0.61 | 0.609649 | 0.667244 | 1202 | 300 |

| | | | | | |
|---|---|---|---|---|---|
| 0.575665 | 0.574751 | 0.573139 | 0.618764 | 1201 | 301 |
| 0.637986 | 0.637874 | 0.637754 | 0.67479 | 1201 | 301 |
| 0.670189 | 0.67 | 0.669908 | 0.750889 | 1202 | 300 |
| 0.676675 | 0.676667 | 0.676663 | 0.745378 | 1202 | 300 |
| 0.620192 | 0.62 | 0.619848 | 0.669911 | 1202 | 300 |
| 0.518518 | 0.518272 | 0.51479 | 0.561943 | 1201 | 301 |
| 0.591497 | 0.591362 | 0.591109 | 0.666358 | 1201 | 301 |
| 0.656729 | 0.656667 | 0.656632 | 0.722222 | 1202 | 300 |
| 0.653333 | 0.653333 | 0.653333 | 0.722133 | 1202 | 300 |
| 0.614063 | 0.613333 | 0.612714 | 0.6612 | 1202 | 300 |
| 0.584945 | 0.584906 | 0.584857 | 0.643297 | 5198 | 742 |
| 0.606581 | 0.606469 | 0.606366 | 0.647787 | 5198 | 742 |
| 0.602462 | 0.602426 | 0.60239 | 0.648019 | 5198 | 742 |
| 0.573104 | 0.573077 | 0.573037 | 0.595178 | 4158 | 1040 |
| 0.605401 | 0.604808 | 0.604251 | 0.642008 | 4158 | 1040 |
| 0.576941 | 0.576923 | 0.576898 | 0.602371 | 4158 | 1040 |
| 0.580368 | 0.580366 | 0.580365 | 0.609341 | 4159 | 1039 |
| 0.585334 | 0.585178 | 0.585017 | 0.611331 | 4159 | 1039 |
| 0.57326 | 0.573077 | 0.57281 | 0.596283 | 4158 | 1040 |
| 0.606532 | 0.605769 | 0.605062 | 0.638732 | 4158 | 1040 |
| 0.578865 | 0.578846 | 0.578821 | 0.61071 | 4158 | 1040 |
| 0.582292 | 0.582291 | 0.582283 | 0.610167 | 4159 | 1039 |
| 0.569961 | 0.569779 | 0.569539 | 0.608159 | 4159 | 1039 |
| 0.57421 | 0.574038 | 0.573792 | 0.596342 | 4158 | 1040 |
| 0.601866 | 0.600962 | 0.600074 | 0.638961 | 4158 | 1040 |
| 0.581782 | 0.581731 | 0.581665 | 0.610396 | 4158 | 1040 |
| 0.584222 | 0.584216 | 0.584199 | 0.610134 | 4159 | 1039 |
| 0.571919 | 0.571704 | 0.571425 | 0.60827 | 4159 | 1039 |
| 0.669546 | 0.66954 | 0.669538 | 0.728997 | 6894 | 1044 |
| 0.673377 | 0.673372 | 0.673369 | 0.729797 | 6894 | 1044 |
| 0.674586 | 0.67433 | 0.67421 | 0.728802 | 6894 | 1044 |
| 0.633367 | 0.633067 | 0.632876 | 0.697451 | 5515 | 1379 |
| 0.647588 | 0.647571 | 0.647557 | 0.698284 | 5515 | 1379 |
| 0.645035 | 0.64467 | 0.64446 | 0.689586 | 5515 | 1379 |
| 0.619372 | 0.619289 | 0.619213 | 0.677836 | 5515 | 1379 |
| 0.6401 | 0.639332 | 0.638838 | 0.698895 | 5516 | 1378 |
| 0.64292 | 0.642495 | 0.642244 | 0.700446 | 5515 | 1379 |
| 0.640321 | 0.640319 | 0.640319 | 0.699937 | 5515 | 1379 |
| 0.647238 | 0.646846 | 0.646624 | 0.692815 | 5515 | 1379 |
| 0.634052 | 0.633793 | 0.633601 | 0.683984 | 5515 | 1379 |
| 0.643658 | 0.642961 | 0.642527 | 0.70038 | 5516 | 1378 |
| 0.643625 | 0.64322 | 0.642983 | 0.70069 | 5515 | 1379 |
| 0.639595 | 0.639594 | 0.639594 | 0.700151 | 5515 | 1379 |
| 0.645079 | 0.64467 | 0.644434 | 0.692724 | 5515 | 1379 |
| 0.630342 | 0.630167 | 0.63003 | 0.683702 | 5515 | 1379 |

| | | | | | |
|---|---|---|---|---|---|
| 0.641499 | 0.640784 | 0.640329 | 0.701838 | 5516 | 1378 |
| 0.684011 | 0.683099 | 0.682705 | 0.751637 | 854 | 142 |
| 0.690292 | 0.690141 | 0.690079 | 0.761754 | 854 | 142 |
| 0.690141 | 0.690141 | 0.690141 | 0.725848 | 854 | 142 |
| 0.643319 | 0.643275 | 0.643275 | 0.721888 | 683 | 171 |
| 0.672803 | 0.672515 | 0.67229 | 0.745144 | 683 | 171 |
| 0.68655 | 0.684211 | 0.683018 | 0.734063 | 683 | 171 |
| 0.678648 | 0.678363 | 0.678297 | 0.743776 | 683 | 171 |
| 0.635294 | 0.635294 | 0.635294 | 0.712388 | 684 | 170 |
| 0.649123 | 0.649123 | 0.649123 | 0.729001 | 683 | 171 |
| 0.678516 | 0.678363 | 0.678231 | 0.743228 | 683 | 171 |
| 0.693025 | 0.690058 | 0.688652 | 0.73461 | 683 | 171 |
| 0.666712 | 0.666667 | 0.666667 | 0.744596 | 683 | 171 |
| 0.652962 | 0.652941 | 0.652929 | 0.730657 | 684 | 170 |
| 0.544498 | 0.54386 | 0.542985 | 0.604241 | 683 | 171 |
| 0.667632 | 0.666667 | 0.666027 | 0.731737 | 683 | 171 |
| 0.639084 | 0.637427 | 0.636057 | 0.666895 | 683 | 171 |
| 0.608229 | 0.608187 | 0.608187 | 0.695622 | 683 | 171 |
| 0.653132 | 0.652941 | 0.652833 | 0.698547 | 684 | 170 |
| 0.692897 | 0.692857 | 0.692841 | 0.740969 | 1360 | 280 |
| 0.685752 | 0.685714 | 0.685698 | 0.759592 | 1360 | 280 |
| 0.692897 | 0.692857 | 0.692841 | 0.759235 | 1360 | 280 |
| 0.669705 | 0.669118 | 0.668831 | 0.71383 | 1088 | 272 |
| 0.661765 | 0.661765 | 0.661765 | 0.734051 | 1088 | 272 |
| 0.676509 | 0.676471 | 0.676453 | 0.736808 | 1088 | 272 |
| 0.650809 | 0.650735 | 0.650693 | 0.733564 | 1088 | 272 |
| 0.678361 | 0.676471 | 0.675611 | 0.707342 | 1088 | 272 |
| 0.684823 | 0.683824 | 0.683396 | 0.72394 | 1088 | 272 |
| 0.672803 | 0.672794 | 0.67279 | 0.73724 | 1088 | 272 |
| 0.691218 | 0.691176 | 0.69116 | 0.740917 | 1088 | 272 |
| 0.71342 | 0.713235 | 0.713173 | 0.760272 | 1088 | 272 |
| 0.660598 | 0.658088 | 0.656747 | 0.715614 | 1088 | 272 |
| 0.691218 | 0.691176 | 0.69116 | 0.717885 | 1088 | 272 |
| 0.661765 | 0.661765 | 0.661765 | 0.735294 | 1088 | 272 |
| 0.676509 | 0.676471 | 0.676453 | 0.72859 | 1088 | 272 |
| 0.720636 | 0.720588 | 0.720573 | 0.761246 | 1088 | 272 |
| 0.660598 | 0.658088 | 0.656747 | 0.717561 | 1088 | 272 |
