## Supplementary Table 4 for "Hundreds of cardiac MRI traits derived using 3D diffusion autoencoders share a common genetic architecture"

| Disease | Model | predefined | L1_penaliz | CV_OR_He | Fold_num | NumCoeffi | NumSignifi | ClassifRprt_ |
|---|---|---|---|---|---|---|---|---|
| angina-pec | LogRegrs | FALSE | | HeldoutTest | | 21 | 4 | 0.675325 |
| angina-pec | LassoCat | FALSE | 0.046416 | HeldoutTest | | 21 | 3 | 0.597403 |
| angina-pec | LassoCat | TRUE | 0.05 | HeldoutTest | | 21 | 3 | 0.597403 |
| angina-pec | LogRegrs | FALSE | | CV | 0 | 23 | 12 | 0.600939 |
| angina-pec | LogRegrs | FALSE | | CV | 1 | 22 | 10 | 0.605634 |
| angina-pec | LogRegrs | FALSE | | CV | 2 | 22 | 10 | 0.610329 |
| angina-pec | LogRegrs | FALSE | | CV | 3 | 22 | 8 | 0.600939 |
| angina-pec | LogRegrs | FALSE | | CV | 4 | 21 | 10 | 0.608491 |
| angina-pec | LassoCat | FALSE | 0.046416 | CV | 0 | 23 | 3 | 0.600939 |
| angina-pec | LassoCat | FALSE | 0.359381 | CV | 1 | 22 | 5 | 0.615023 |
| angina-pec | LassoCat | FALSE | 0.046416 | CV | 2 | 22 | 3 | 0.58216 |
| angina-pec | LassoCat | FALSE | 0.359381 | CV | 3 | 22 | 6 | 0.615023 |
| angina-pec | LassoCat | FALSE | 0.359381 | CV | 4 | 21 | 4 | 0.627358 |
| angina-pec | LassoCat | TRUE | 0.05 | CV | 0 | 23 | 3 | 0.610329 |
| angina-pec | LassoCat | TRUE | 0.05 | CV | 1 | 22 | 2 | 0.666667 |
| angina-pec | LassoCat | TRUE | 0.05 | CV | 2 | 22 | 3 | 0.586854 |
| angina-pec | LassoCat | TRUE | 0.05 | CV | 3 | 22 | 2 | 0.57277 |
| angina-pec | LassoCat | TRUE | 0.05 | CV | 4 | 21 | 2 | 0.622642 |
| atrial-fibrill | LogRegrs | FALSE | | HeldoutTest | | 22 | 12 | 0.639535 |
| atrial-fibrill | LassoCat | FALSE | 0.359381 | HeldoutTest | | 22 | 7 | 0.616279 |
| atrial-fibrill | LassoCat | TRUE | 0.05 | HeldoutTest | | 22 | 1 | 0.55814 |
| atrial-fibrill | LogRegrs | FALSE | | CV | 0 | 21 | 12 | 0.585774 |
| atrial-fibrill | LogRegrs | FALSE | | CV | 1 | 21 | 8 | 0.543933 |
| atrial-fibrill | LogRegrs | FALSE | | CV | 2 | 22 | 8 | 0.659664 |
| atrial-fibrill | LogRegrs | FALSE | | CV | 3 | 21 | 12 | 0.571429 |
| atrial-fibrill | LogRegrs | FALSE | | CV | 4 | 23 | 13 | 0.592437 |
| atrial-fibrill | LassoCat | FALSE | 0.359381 | CV | 0 | 21 | 8 | 0.60251 |
| atrial-fibrill | LassoCat | FALSE | 21.54435 | CV | 1 | 21 | 9 | 0.543933 |
| atrial-fibrill | LassoCat | FALSE | 166.8101 | CV | 2 | 22 | 8 | 0.663866 |
| atrial-fibrill | LassoCat | FALSE | 0.359381 | CV | 3 | 21 | 10 | 0.588235 |
| atrial-fibrill | LassoCat | FALSE | 0.359381 | CV | 4 | 23 | 8 | 0.592437 |
| atrial-fibrill | LassoCat | TRUE | 0.05 | CV | 0 | 21 | 1 | 0.543933 |
| atrial-fibrill | LassoCat | TRUE | 0.05 | CV | 1 | 21 | 2 | 0.518828 |
| atrial-fibrill | LassoCat | TRUE | 0.05 | CV | 2 | 22 | 1 | 0.62605 |
| atrial-fibrill | LassoCat | TRUE | 0.05 | CV | 3 | 21 | 2 | 0.642857 |
| atrial-fibrill | LassoCat | TRUE | 0.05 | CV | 4 | 23 | 2 | 0.592437 |
| coronary-h | LogRegrs | FALSE | | HeldoutTest | | 23 | 14 | 0.633621 |
| coronary-h | LassoCat | FALSE | 0.359381 | HeldoutTest | | 23 | 10 | 0.668103 |
| coronary-h | LassoCat | TRUE | 0.05 | HeldoutTest | | 23 | 3 | 0.642241 |
| coronary-h | LogRegrs | FALSE | | CV | 0 | 21 | 10 | 0.601329 |
| coronary-h | LogRegrs | FALSE | | CV | 1 | 23 | 15 | 0.591362 |
| coronary-h | LogRegrs | FALSE | | CV | 2 | 22 | 16 | 0.566667 |
| coronary-h | LogRegrs | FALSE | | CV | 3 | 22 | 12 | 0.64 |
| coronary-h | LogRegrs | FALSE | | CV | 4 | 22 | 13 | 0.6 |

| | | | | | | | | |
|---|---|---|---|---|---|---|---|---|
| coronary-h | LassoCat | FALSE | 2.782559 | CV | 0 | 21 | 10 | 0.601329 |
| coronary-h | LassoCat | FALSE | 0.359381 | CV | 1 | 23 | 6 | 0.607973 |
| coronary-h | LassoCat | FALSE | 166.8101 | CV | 2 | 22 | 17 | 0.573333 |
| coronary-h | LassoCat | FALSE | 0.359381 | CV | 3 | 22 | 10 | 0.643333 |
| coronary-h | LassoCat | FALSE | 0.359381 | CV | 4 | 22 | 7 | 0.616667 |
| coronary-h | LassoCat | TRUE | 0.05 | CV | 0 | 21 | 3 | 0.627907 |
| coronary-h | LassoCat | TRUE | 0.05 | CV | 1 | 23 | 3 | 0.604651 |
| coronary-h | LassoCat | TRUE | 0.05 | CV | 2 | 22 | 1 | 0.58 |
| coronary-h | LassoCat | TRUE | 0.05 | CV | 3 | 22 | 1 | 0.616667 |
| coronary-h | LassoCat | TRUE | 0.05 | CV | 4 | 22 | 3 | 0.606667 |
| high-choles | LogRegrs | FALSE | | HeldoutTest | | 21 | 14 | 0.59973 |
| high-choles | LassoCat | FALSE | 0.359381 | HeldoutTest | | 21 | 11 | 0.59973 |
| high-choles | LassoCat | TRUE | 0.05 | HeldoutTest | | 21 | 5 | 0.597035 |
| high-choles | LogRegrs | FALSE | | CV | 0 | 21 | 11 | 0.608654 |
| high-choles | LogRegrs | FALSE | | CV | 1 | 22 | 12 | 0.592308 |
| high-choles | LogRegrs | FALSE | | CV | 2 | 21 | 12 | 0.599038 |
| high-choles | LogRegrs | FALSE | | CV | 3 | 21 | 13 | 0.584216 |
| high-choles | LogRegrs | FALSE | | CV | 4 | 21 | 15 | 0.615977 |
| high-choles | LassoCat | FALSE | 2.782559 | CV | 0 | 21 | 11 | 0.609615 |
| high-choles | LassoCat | FALSE | 2.782559 | CV | 1 | 22 | 12 | 0.590385 |
| high-choles | LassoCat | FALSE | 2.782559 | CV | 2 | 21 | 12 | 0.6 |
| high-choles | LassoCat | FALSE | 0.359381 | CV | 3 | 21 | 10 | 0.587103 |
| high-choles | LassoCat | FALSE | 0.046416 | CV | 4 | 21 | 4 | 0.594803 |
| high-choles | LassoCat | TRUE | 0.05 | CV | 0 | 21 | 6 | 0.591346 |
| high-choles | LassoCat | TRUE | 0.05 | CV | 1 | 22 | 5 | 0.594231 |
| high-choles | LassoCat | TRUE | 0.05 | CV | 2 | 21 | 5 | 0.592308 |
| high-choles | LassoCat | TRUE | 0.05 | CV | 3 | 21 | 6 | 0.576516 |
| high-choles | LassoCat | TRUE | 0.05 | CV | 4 | 21 | 5 | 0.596728 |
| hypertensi | LogRegrs | FALSE | | HeldoutTest | | 21 | 16 | 0.619732 |
| hypertensi | LassoCat | FALSE | 10000 | HeldoutTest | | 21 | 16 | 0.621648 |
| hypertensi | LassoCat | TRUE | 0.05 | HeldoutTest | | 21 | 13 | 0.611111 |
| hypertensi | LogRegrs | FALSE | | CV | 0 | 21 | 18 | 0.626541 |
| hypertensi | LogRegrs | FALSE | | CV | 1 | 21 | 16 | 0.610587 |
| hypertensi | LogRegrs | FALSE | | CV | 2 | 21 | 14 | 0.608412 |
| hypertensi | LogRegrs | FALSE | | CV | 3 | 22 | 14 | 0.621465 |
| hypertensi | LogRegrs | FALSE | | CV | 4 | 22 | 17 | 0.645138 |
| hypertensi | LassoCat | FALSE | 0.359381 | CV | 0 | 21 | 16 | 0.626541 |
| hypertensi | LassoCat | FALSE | 0.359381 | CV | 1 | 21 | 15 | 0.610587 |
| hypertensi | LassoCat | FALSE | 0.359381 | CV | 2 | 21 | 15 | 0.606962 |
| hypertensi | LassoCat | FALSE | 0.359381 | CV | 3 | 22 | 13 | 0.619289 |
| hypertensi | LassoCat | FALSE | 0.359381 | CV | 4 | 22 | 16 | 0.640058 |
| hypertensi | LassoCat | TRUE | 0.05 | CV | 0 | 21 | 13 | 0.633793 |
| hypertensi | LassoCat | TRUE | 0.05 | CV | 1 | 21 | 12 | 0.614938 |
| hypertensi | LassoCat | TRUE | 0.05 | CV | 2 | 21 | 10 | 0.596084 |
| hypertensi | LassoCat | TRUE | 0.05 | CV | 3 | 22 | 10 | 0.618564 |

| | | | | | | | | |
|---|---|---|---|---|---|---|---|---|
| hypertensi | LassoCat | TRUE | 0.05 | CV | 4 | 22 | 10 | 0.632075 |
| myocardial | LogRegrs | FALSE | | HeldoutTest | | 23 | 14 | 0.661972 |
| myocardial | LassoCat | FALSE | 2.782559 | HeldoutTest | | 23 | 14 | 0.661972 |
| myocardial | LassoCat | TRUE | 0.05 | HeldoutTest | | 23 | 0 | 0.612676 |
| myocardial | LogRegrs | FALSE | | CV | 0 | 23 | 11 | 0.590643 |
| myocardial | LogRegrs | FALSE | | CV | 1 | 23 | 10 | 0.660819 |
| myocardial | LogRegrs | FALSE | | CV | 2 | 22 | 11 | 0.608187 |
| myocardial | LogRegrs | FALSE | | CV | 3 | 22 | 13 | 0.643275 |
| myocardial | LogRegrs | FALSE | | CV | 4 | 21 | 14 | 0.535294 |
| myocardial | LassoCat | FALSE | 0.359381 | CV | 0 | 23 | 4 | 0.573099 |
| myocardial | LassoCat | FALSE | 2.782559 | CV | 1 | 23 | 10 | 0.649123 |
| myocardial | LassoCat | FALSE | 0.046416 | CV | 2 | 22 | 0 | 0.649123 |
| myocardial | LassoCat | FALSE | 0.359381 | CV | 3 | 22 | 11 | 0.602339 |
| myocardial | LassoCat | FALSE | 21.54435 | CV | 4 | 21 | 14 | 0.541176 |
| myocardial | LassoCat | TRUE | 0.05 | CV | 0 | 23 | 0 | 0.532164 |
| myocardial | LassoCat | TRUE | 0.05 | CV | 1 | 23 | 0 | 0.666667 |
| myocardial | LassoCat | TRUE | 0.05 | CV | 2 | 22 | 0 | 0.660819 |
| myocardial | LassoCat | TRUE | 0.05 | CV | 3 | 22 | 1 | 0.608187 |
| myocardial | LassoCat | TRUE | 0.05 | CV | 4 | 21 | 1 | 0.523529 |
| type-2-diak | LogRegrs | FALSE | | HeldoutTest | | 21 | 17 | 0.678571 |
| type-2-diak | LassoCat | FALSE | 1291.55 | HeldoutTest | | 21 | 17 | 0.675 |
| type-2-diak | LassoCat | TRUE | 0.05 | HeldoutTest | | 21 | 4 | 0.657143 |
| type-2-diak | LogRegrs | FALSE | | CV | 0 | 22 | 12 | 0.709559 |
| type-2-diak | LogRegrs | FALSE | | CV | 1 | 21 | 18 | 0.676471 |
| type-2-diak | LogRegrs | FALSE | | CV | 2 | 21 | 15 | 0.694853 |
| type-2-diak | LogRegrs | FALSE | | CV | 3 | 22 | 17 | 0.709559 |
| type-2-diak | LogRegrs | FALSE | | CV | 4 | 21 | 14 | 0.694853 |
| type-2-diak | LassoCat | FALSE | 21.54435 | CV | 0 | 22 | 12 | 0.713235 |
| type-2-diak | LassoCat | FALSE | 0.359381 | CV | 1 | 21 | 13 | 0.669118 |
| type-2-diak | LassoCat | FALSE | 2.782559 | CV | 2 | 21 | 13 | 0.694853 |
| type-2-diak | LassoCat | FALSE | 166.8101 | CV | 3 | 22 | 17 | 0.713235 |
| type-2-diak | LassoCat | FALSE | 2.782559 | CV | 4 | 21 | 13 | 0.691176 |
| type-2-diak | LassoCat | TRUE | 0.05 | CV | 0 | 22 | 5 | 0.698529 |
| type-2-diak | LassoCat | TRUE | 0.05 | CV | 1 | 21 | 3 | 0.654412 |
| type-2-diak | LassoCat | TRUE | 0.05 | CV | 2 | 21 | 4 | 0.683824 |
| type-2-diak | LassoCat | TRUE | 0.05 | CV | 3 | 22 | 3 | 0.702206 |
| type-2-diak | LassoCat | TRUE | 0.05 | CV | 4 | 21 | 4 | 0.6875 |

| ClassifRprt_ | ClassifRprt_ | ClassifRprt_ | AUC | TrainSize | TestSize |
|---|---|---|---|---|---|
| 0.683236 | 0.675325 | 0.671782 | 0.704166 | 1064 | 154 |
| 0.597666 | 0.597403 | 0.597131 | 0.653399 | 1064 | 154 |
| 0.597666 | 0.597403 | 0.597131 | 0.654073 | 1064 | 154 |
| 0.600966 | 0.600939 | 0.600833 | 0.643714 | 851 | 213 |
| 0.605636 | 0.605634 | 0.605582 | 0.654206 | 851 | 213 |
| 0.610321 | 0.610329 | 0.610311 | 0.650591 | 851 | 213 |
| 0.60125 | 0.600939 | 0.600763 | 0.626345 | 851 | 213 |
| 0.610147 | 0.608491 | 0.607013 | 0.643378 | 852 | 212 |
| 0.600965 | 0.600939 | 0.600939 | 0.628284 | 851 | 213 |
| 0.615254 | 0.615023 | 0.614922 | 0.668401 | 851 | 213 |
| 0.582249 | 0.58216 | 0.581883 | 0.629607 | 851 | 213 |
| 0.615492 | 0.615023 | 0.614769 | 0.632164 | 851 | 213 |
| 0.629303 | 0.627358 | 0.625952 | 0.65922 | 852 | 212 |
| 0.610321 | 0.610329 | 0.610311 | 0.628461 | 851 | 213 |
| 0.667998 | 0.666667 | 0.666137 | 0.696526 | 851 | 213 |
| 0.586904 | 0.586854 | 0.586672 | 0.629519 | 851 | 213 |
| 0.572758 | 0.57277 | 0.572751 | 0.623523 | 851 | 213 |
| 0.623743 | 0.622642 | 0.6218 | 0.652545 | 852 | 212 |
| 0.642306 | 0.639535 | 0.637772 | 0.636696 | 1192 | 172 |
| 0.617873 | 0.616279 | 0.614978 | 0.630476 | 1192 | 172 |
| 0.558647 | 0.55814 | 0.557182 | 0.611276 | 1192 | 172 |
| 0.585872 | 0.585774 | 0.585731 | 0.658683 | 953 | 239 |
| 0.544328 | 0.543933 | 0.543358 | 0.616246 | 953 | 239 |
| 0.66299 | 0.659664 | 0.657919 | 0.707718 | 954 | 238 |
| 0.571753 | 0.571429 | 0.570944 | 0.615281 | 954 | 238 |
| 0.592496 | 0.592437 | 0.592372 | 0.645435 | 954 | 238 |
| 0.60276 | 0.60251 | 0.602371 | 0.665056 | 953 | 239 |
| 0.544328 | 0.543933 | 0.543358 | 0.617927 | 953 | 239 |
| 0.666882 | 0.663866 | 0.66234 | 0.707365 | 954 | 238 |
| 0.588335 | 0.588235 | 0.588119 | 0.626439 | 954 | 238 |
| 0.592758 | 0.592437 | 0.592084 | 0.654544 | 954 | 238 |
| 0.544009 | 0.543933 | 0.543885 | 0.621289 | 953 | 239 |
| 0.519012 | 0.518828 | 0.518474 | 0.537115 | 953 | 239 |
| 0.626131 | 0.62605 | 0.625991 | 0.659417 | 954 | 238 |
| 0.642867 | 0.642857 | 0.642851 | 0.678836 | 954 | 238 |
| 0.593553 | 0.592437 | 0.591217 | 0.636184 | 954 | 238 |
| 0.634109 | 0.633621 | 0.633287 | 0.733279 | 1502 | 232 |
| 0.668416 | 0.668103 | 0.667949 | 0.736846 | 1502 | 232 |
| 0.642337 | 0.642241 | 0.642182 | 0.713511 | 1502 | 232 |
| 0.601329 | 0.601329 | 0.601329 | 0.643267 | 1201 | 301 |
| 0.591371 | 0.591362 | 0.591308 | 0.647241 | 1201 | 301 |
| 0.567873 | 0.566667 | 0.564732 | 0.618044 | 1202 | 300 |
| 0.643078 | 0.64 | 0.638054 | 0.666578 | 1202 | 300 |
| 0.600018 | 0.6 | 0.599982 | 0.659022 | 1202 | 300 |

| | | | | | |
|---|---|---|---|---|---|
| 0.601329 | 0.601329 | 0.601329 | 0.642561 | 1201 | 301 |
| 0.608015 | 0.607973 | 0.607887 | 0.648256 | 1201 | 301 |
| 0.574661 | 0.573333 | 0.571429 | 0.617467 | 1202 | 300 |
| 0.645671 | 0.643333 | 0.641897 | 0.673733 | 1202 | 300 |
| 0.616672 | 0.616667 | 0.616662 | 0.657911 | 1202 | 300 |
| 0.62803 | 0.627907 | 0.627858 | 0.632539 | 1201 | 301 |
| 0.604723 | 0.604651 | 0.60452 | 0.63585 | 1201 | 301 |
| 0.580014 | 0.58 | 0.579981 | 0.640978 | 1202 | 300 |
| 0.617845 | 0.616667 | 0.615706 | 0.652667 | 1202 | 300 |
| 0.606743 | 0.606667 | 0.606597 | 0.662311 | 1202 | 300 |
| 0.600115 | 0.59973 | 0.599345 | 0.634658 | 5198 | 742 |
| 0.600115 | 0.59973 | 0.599345 | 0.634782 | 5198 | 742 |
| 0.597717 | 0.597035 | 0.59633 | 0.628824 | 5198 | 742 |
| 0.608948 | 0.608654 | 0.60839 | 0.644756 | 4158 | 1040 |
| 0.592375 | 0.592308 | 0.592234 | 0.634101 | 4158 | 1040 |
| 0.5991 | 0.599038 | 0.598976 | 0.630155 | 4158 | 1040 |
| 0.584356 | 0.584216 | 0.584069 | 0.621187 | 4159 | 1039 |
| 0.616018 | 0.615977 | 0.615951 | 0.637576 | 4159 | 1039 |
| 0.609934 | 0.609615 | 0.609332 | 0.64456 | 4158 | 1040 |
| 0.59045 | 0.590385 | 0.59031 | 0.633905 | 4158 | 1040 |
| 0.600053 | 0.6 | 0.599947 | 0.630078 | 4158 | 1040 |
| 0.587262 | 0.587103 | 0.586942 | 0.622629 | 4159 | 1039 |
| 0.594808 | 0.594803 | 0.5948 | 0.63446 | 4159 | 1039 |
| 0.591918 | 0.591346 | 0.59071 | 0.636657 | 4158 | 1040 |
| 0.594432 | 0.594231 | 0.594015 | 0.628136 | 4158 | 1040 |
| 0.592444 | 0.592308 | 0.592157 | 0.625222 | 4158 | 1040 |
| 0.576671 | 0.576516 | 0.576335 | 0.624037 | 4159 | 1039 |
| 0.59675 | 0.596728 | 0.596712 | 0.634749 | 4159 | 1039 |
| 0.619965 | 0.619732 | 0.619547 | 0.661734 | 6894 | 1044 |
| 0.621845 | 0.621648 | 0.621494 | 0.661775 | 6894 | 1044 |
| 0.611703 | 0.611111 | 0.610595 | 0.654182 | 6894 | 1044 |
| 0.626557 | 0.626541 | 0.626533 | 0.660945 | 5515 | 1379 |
| 0.610743 | 0.610587 | 0.610464 | 0.657199 | 5515 | 1379 |
| 0.608431 | 0.608412 | 0.608389 | 0.645411 | 5515 | 1379 |
| 0.621859 | 0.621465 | 0.621138 | 0.668208 | 5515 | 1379 |
| 0.645141 | 0.645138 | 0.645136 | 0.689386 | 5516 | 1378 |
| 0.626557 | 0.626541 | 0.626533 | 0.662914 | 5515 | 1379 |
| 0.610699 | 0.610587 | 0.610501 | 0.657889 | 5515 | 1379 |
| 0.606967 | 0.606962 | 0.606953 | 0.645249 | 5515 | 1379 |
| 0.619827 | 0.619289 | 0.618837 | 0.667584 | 5515 | 1379 |
| 0.640058 | 0.640058 | 0.640058 | 0.689266 | 5516 | 1378 |
| 0.633862 | 0.633793 | 0.633752 | 0.663053 | 5515 | 1379 |
| 0.615126 | 0.614938 | 0.614796 | 0.658558 | 5515 | 1379 |
| 0.596085 | 0.596084 | 0.596082 | 0.636575 | 5515 | 1379 |
| 0.618777 | 0.618564 | 0.618378 | 0.661341 | 5515 | 1379 |

| | | | | | |
|---|---|---|---|---|---|
| 0.632089 | 0.632075 | 0.632066 | 0.683003 | 5516 | 1378 |
| 0.6621 | 0.661972 | 0.661905 | 0.72684 | 854 | 142 |
| 0.6621 | 0.661972 | 0.661905 | 0.727038 | 854 | 142 |
| 0.612878 | 0.612676 | 0.612503 | 0.650665 | 854 | 142 |
| 0.592182 | 0.590643 | 0.58938 | 0.632421 | 683 | 171 |
| 0.660953 | 0.660819 | 0.660796 | 0.726949 | 683 | 171 |
| 0.60824 | 0.608187 | 0.608026 | 0.65212 | 683 | 171 |
| 0.6451 | 0.643275 | 0.642395 | 0.653078 | 683 | 171 |
| 0.535338 | 0.535294 | 0.535149 | 0.576471 | 684 | 170 |
| 0.574183 | 0.573099 | 0.572046 | 0.619562 | 683 | 171 |
| 0.649546 | 0.649123 | 0.648979 | 0.727633 | 683 | 171 |
| 0.649123 | 0.649123 | 0.649123 | 0.690971 | 683 | 171 |
| 0.605544 | 0.602339 | 0.59985 | 0.649521 | 683 | 171 |
| 0.541199 | 0.541176 | 0.541113 | 0.577439 | 684 | 170 |
| 0.5332 | 0.532164 | 0.530043 | 0.588646 | 683 | 171 |
| 0.666667 | 0.666667 | 0.666644 | 0.723666 | 683 | 171 |
| 0.660819 | 0.660819 | 0.660819 | 0.690971 | 683 | 171 |
| 0.610305 | 0.608187 | 0.606708 | 0.632969 | 683 | 171 |
| 0.523533 | 0.523529 | 0.523513 | 0.527336 | 684 | 170 |
| 0.681573 | 0.678571 | 0.677238 | 0.755357 | 1360 | 280 |
| 0.678284 | 0.675 | 0.673497 | 0.755051 | 1360 | 280 |
| 0.658306 | 0.657143 | 0.656512 | 0.733776 | 1360 | 280 |
| 0.70957 | 0.709559 | 0.709555 | 0.764327 | 1088 | 272 |
| 0.676623 | 0.676471 | 0.676401 | 0.740052 | 1088 | 272 |
| 0.695371 | 0.694853 | 0.694651 | 0.782115 | 1088 | 272 |
| 0.710115 | 0.709559 | 0.709366 | 0.766544 | 1088 | 272 |
| 0.69665 | 0.694853 | 0.694154 | 0.777952 | 1088 | 272 |
| 0.713281 | 0.713235 | 0.71322 | 0.763949 | 1088 | 272 |
| 0.669264 | 0.669118 | 0.669046 | 0.744053 | 1088 | 272 |
| 0.695371 | 0.694853 | 0.694651 | 0.782223 | 1088 | 272 |
| 0.713651 | 0.713235 | 0.713096 | 0.764598 | 1088 | 272 |
| 0.692677 | 0.691176 | 0.690574 | 0.778385 | 1088 | 272 |
| 0.698917 | 0.698529 | 0.698383 | 0.749027 | 1088 | 272 |
| 0.654412 | 0.654412 | 0.654412 | 0.729833 | 1088 | 272 |
| 0.683863 | 0.683824 | 0.683806 | 0.792063 | 1088 | 272 |
| 0.702304 | 0.702206 | 0.70217 | 0.760651 | 1088 | 272 |
| 0.687591 | 0.6875 | 0.687462 | 0.78298 | 1088 | 272 |
