## Supplementary Table 3 for "Hundreds of cardiac MRI traits derived using 3D diffusion autoencoders share a common genetic architecture"

| Trait | Model | predefined | L1_penaliz | CV_OR_He | Fold_num | NumCoeffi | NumSignifi | Classificatio |
|---|---|---|---|---|---|---|---|---|
| Year_of_bi | LinRegrs | FALSE | | HeldoutTest | | 22 | 21 | Regression |
| Year_of_bi | Lasso | FALSE | 0.000403 | HeldoutTest | | 22 | 21 | Regression |
| Year_of_bi | Lasso | TRUE | 0.05 | HeldoutTest | | 22 | 9 | Regression |
| Year_of_bi | LinRegrs | FALSE | | CV | 0 | 22 | 21 | Regression |
| Year_of_bi | LinRegrs | FALSE | | CV | 1 | 22 | 21 | Regression |
| Year_of_bi | LinRegrs | FALSE | | CV | 2 | 22 | 20 | Regression |
| Year_of_bi | LinRegrs | FALSE | | CV | 3 | 22 | 20 | Regression |
| Year_of_bi | LinRegrs | FALSE | | CV | 4 | 22 | 20 | Regression |
| Year_of_bi | Lasso | FALSE | 0.000531 | CV | 0 | 22 | 22 | Regression |
| Year_of_bi | Lasso | FALSE | 0.000307 | CV | 1 | 22 | 22 | Regression |
| Year_of_bi | Lasso | FALSE | 0.000331 | CV | 2 | 22 | 22 | Regression |
| Year_of_bi | Lasso | FALSE | 0.000492 | CV | 3 | 22 | 20 | Regression |
| Year_of_bi | Lasso | FALSE | 0.000598 | CV | 4 | 22 | 21 | Regression |
| Year_of_bi | Lasso | TRUE | 0.05 | CV | 0 | 22 | 9 | Regression |
| Year_of_bi | Lasso | TRUE | 0.05 | CV | 1 | 22 | 9 | Regression |
| Year_of_bi | Lasso | TRUE | 0.05 | CV | 2 | 22 | 9 | Regression |
| Year_of_bi | Lasso | TRUE | 0.05 | CV | 3 | 22 | 9 | Regression |
| Year_of_bi | Lasso | TRUE | 0.05 | CV | 4 | 22 | 9 | Regression |
| BMI | LinRegrs | FALSE | | HeldoutTest | | 22 | 22 | Regression |
| BMI | Lasso | FALSE | 0.000448 | HeldoutTest | | 22 | 22 | Regression |
| BMI | Lasso | TRUE | 0.05 | HeldoutTest | | 22 | 10 | Regression |
| BMI | LinRegrs | FALSE | | CV | 0 | 22 | 22 | Regression |
| BMI | LinRegrs | FALSE | | CV | 1 | 22 | 20 | Regression |
| BMI | LinRegrs | FALSE | | CV | 2 | 22 | 22 | Regression |
| BMI | LinRegrs | FALSE | | CV | 3 | 22 | 21 | Regression |
| BMI | LinRegrs | FALSE | | CV | 4 | 22 | 20 | Regression |
| BMI | Lasso | FALSE | 0.000537 | CV | 0 | 22 | 22 | Regression |
| BMI | Lasso | FALSE | 0.000446 | CV | 1 | 22 | 21 | Regression |
| BMI | Lasso | FALSE | 0.000448 | CV | 2 | 22 | 22 | Regression |
| BMI | Lasso | FALSE | 0.000448 | CV | 3 | 22 | 22 | Regression |
| BMI | Lasso | FALSE | 0.00045 | CV | 4 | 22 | 22 | Regression |
| BMI | Lasso | TRUE | 0.05 | CV | 0 | 22 | 9 | Regression |
| BMI | Lasso | TRUE | 0.05 | CV | 1 | 22 | 10 | Regression |
| BMI | Lasso | TRUE | 0.05 | CV | 2 | 22 | 10 | Regression |
| BMI | Lasso | TRUE | 0.05 | CV | 3 | 22 | 10 | Regression |
| BMI | Lasso | TRUE | 0.05 | CV | 4 | 22 | 10 | Regression |
| Age_at_rec | LinRegrs | FALSE | | HeldoutTest | | 22 | 19 | Regression |
| Age_at_rec | Lasso | FALSE | 0.000529 | HeldoutTest | | 22 | 20 | Regression |
| Age_at_rec | Lasso | TRUE | 0.05 | HeldoutTest | | 22 | 9 | Regression |
| Age_at_rec | LinRegrs | FALSE | | CV | 0 | 22 | 22 | Regression |
| Age_at_rec | LinRegrs | FALSE | | CV | 1 | 22 | 21 | Regression |
| Age_at_rec | LinRegrs | FALSE | | CV | 2 | 22 | 20 | Regression |
| Age_at_rec | LinRegrs | FALSE | | CV | 3 | 22 | 20 | Regression |
| Age_at_rec | LinRegrs | FALSE | | CV | 4 | 22 | 20 | Regression |

| | | | | | | | | |
|---|---|---|---|---|---|---|---|---|
| Age_at_rec | Lasso | FALSE | 0.000464 | CV | 0 | 22 | 22 | Regression |
| Age_at_rec | Lasso | FALSE | 0.000375 | CV | 1 | 22 | 21 | Regression |
| Age_at_rec | Lasso | FALSE | 0.000351 | CV | 2 | 22 | 21 | Regression |
| Age_at_rec | Lasso | FALSE | 0.000454 | CV | 3 | 22 | 21 | Regression |
| Age_at_rec | Lasso | FALSE | 0.000454 | CV | 4 | 22 | 21 | Regression |
| Age_at_rec | Lasso | TRUE | 0.05 | CV | 0 | 22 | 9 | Regression |
| Age_at_rec | Lasso | TRUE | 0.05 | CV | 1 | 22 | 9 | Regression |
| Age_at_rec | Lasso | TRUE | 0.05 | CV | 2 | 22 | 9 | Regression |
| Age_at_rec | Lasso | TRUE | 0.05 | CV | 3 | 22 | 9 | Regression |
| Age_at_rec | Lasso | TRUE | 0.05 | CV | 4 | 22 | 9 | Regression |
| Age | LinRegrs | FALSE | | HeldoutTest | | 22 | 22 | Regression |
| Age | Lasso | FALSE | 0.000228 | HeldoutTest | | 22 | 22 | Regression |
| Age | Lasso | TRUE | 0.05 | HeldoutTest | | 22 | 9 | Regression |
| Age | LinRegrs | FALSE | | CV | 0 | 22 | 20 | Regression |
| Age | LinRegrs | FALSE | | CV | 1 | 22 | 22 | Regression |
| Age | LinRegrs | FALSE | | CV | 2 | 22 | 21 | Regression |
| Age | LinRegrs | FALSE | | CV | 3 | 22 | 22 | Regression |
| Age | LinRegrs | FALSE | | CV | 4 | 22 | 22 | Regression |
| Age | Lasso | FALSE | 0.000389 | CV | 0 | 22 | 22 | Regression |
| Age | Lasso | FALSE | 0.000214 | CV | 1 | 22 | 22 | Regression |
| Age | Lasso | FALSE | 0.000401 | CV | 2 | 22 | 21 | Regression |
| Age | Lasso | FALSE | 0.000211 | CV | 3 | 22 | 22 | Regression |
| Age | Lasso | FALSE | 0.000257 | CV | 4 | 22 | 22 | Regression |
| Age | Lasso | TRUE | 0.05 | CV | 0 | 22 | 9 | Regression |
| Age | Lasso | TRUE | 0.05 | CV | 1 | 22 | 9 | Regression |
| Age | Lasso | TRUE | 0.05 | CV | 2 | 22 | 8 | Regression |
| Age | Lasso | TRUE | 0.05 | CV | 3 | 22 | 9 | Regression |
| Age | Lasso | TRUE | 0.05 | CV | 4 | 22 | 9 | Regression |
| MRI_Date | LinRegrs | FALSE | | HeldoutTest | | 22 | 21 | Regression |
| MRI_Date | Lasso | FALSE | 0.0002 | HeldoutTest | | 22 | 21 | Regression |
| MRI_Date | Lasso | TRUE | 0.05 | HeldoutTest | | 22 | 5 | Regression |
| MRI_Date | LinRegrs | FALSE | | CV | 0 | 22 | 21 | Regression |
| MRI_Date | LinRegrs | FALSE | | CV | 1 | 22 | 21 | Regression |
| MRI_Date | LinRegrs | FALSE | | CV | 2 | 22 | 21 | Regression |
| MRI_Date | LinRegrs | FALSE | | CV | 3 | 22 | 21 | Regression |
| MRI_Date | LinRegrs | FALSE | | CV | 4 | 22 | 21 | Regression |
| MRI_Date | Lasso | FALSE | 0.000369 | CV | 0 | 22 | 22 | Regression |
| MRI_Date | Lasso | FALSE | 0.000284 | CV | 1 | 22 | 21 | Regression |
| MRI_Date | Lasso | FALSE | 0.000248 | CV | 2 | 22 | 22 | Regression |
| MRI_Date | Lasso | FALSE | 0.000246 | CV | 3 | 22 | 21 | Regression |
| MRI_Date | Lasso | FALSE | 0.000199 | CV | 4 | 22 | 22 | Regression |
| MRI_Date | Lasso | TRUE | 0.05 | CV | 0 | 22 | 5 | Regression |
| MRI_Date | Lasso | TRUE | 0.05 | CV | 1 | 22 | 5 | Regression |
| MRI_Date | Lasso | TRUE | 0.05 | CV | 2 | 22 | 5 | Regression |
| MRI_Date | Lasso | TRUE | 0.05 | CV | 3 | 22 | 5 | Regression |

| | | | | | | | | |
|---|---|---|---|---|---|---|---|---|
| MRI_Date | Lasso | TRUE | 0.05 | CV | 4 | 22 | 5 | Regression |
| BSA | LinRegrs | FALSE | | HeldoutTest | | 22 | 22 | Regression |
| BSA | Lasso | FALSE | 0.00058 | HeldoutTest | | 22 | 22 | Regression |
| BSA | Lasso | TRUE | 0.05 | HeldoutTest | | 22 | 11 | Regression |
| BSA | LinRegrs | FALSE | | CV | 0 | 22 | 22 | Regression |
| BSA | LinRegrs | FALSE | | CV | 1 | 22 | 22 | Regression |
| BSA | LinRegrs | FALSE | | CV | 2 | 22 | 22 | Regression |
| BSA | LinRegrs | FALSE | | CV | 3 | 22 | 22 | Regression |
| BSA | LinRegrs | FALSE | | CV | 4 | 22 | 22 | Regression |
| BSA | Lasso | FALSE | 0.000668 | CV | 0 | 22 | 22 | Regression |
| BSA | Lasso | FALSE | 0.00058 | CV | 1 | 22 | 22 | Regression |
| BSA | Lasso | FALSE | 0.000579 | CV | 2 | 22 | 22 | Regression |
| BSA | Lasso | FALSE | 0.00058 | CV | 3 | 22 | 22 | Regression |
| BSA | Lasso | FALSE | 0.000581 | CV | 4 | 22 | 22 | Regression |
| BSA | Lasso | TRUE | 0.05 | CV | 0 | 22 | 11 | Regression |
| BSA | Lasso | TRUE | 0.05 | CV | 1 | 22 | 11 | Regression |
| BSA | Lasso | TRUE | 0.05 | CV | 2 | 22 | 11 | Regression |
| BSA | Lasso | TRUE | 0.05 | CV | 3 | 22 | 11 | Regression |
| BSA | Lasso | TRUE | 0.05 | CV | 4 | 22 | 11 | Regression |
| Waist_circu | LinRegrs | FALSE | | HeldoutTest | | 22 | 22 | Regression |
| Waist_circu | Lasso | FALSE | 0.00051 | HeldoutTest | | 22 | 22 | Regression |
| Waist_circu | Lasso | TRUE | 0.05 | HeldoutTest | | 22 | 10 | Regression |
| Waist_circu | LinRegrs | FALSE | | CV | 0 | 22 | 22 | Regression |
| Waist_circu | LinRegrs | FALSE | | CV | 1 | 22 | 22 | Regression |
| Waist_circu | LinRegrs | FALSE | | CV | 2 | 22 | 22 | Regression |
| Waist_circu | LinRegrs | FALSE | | CV | 3 | 22 | 21 | Regression |
| Waist_circu | LinRegrs | FALSE | | CV | 4 | 22 | 21 | Regression |
| Waist_circu | Lasso | FALSE | 0.000643 | CV | 0 | 22 | 22 | Regression |
| Waist_circu | Lasso | FALSE | 0.00051 | CV | 1 | 22 | 22 | Regression |
| Waist_circu | Lasso | FALSE | 0.000509 | CV | 2 | 22 | 22 | Regression |
| Waist_circu | Lasso | FALSE | 0.000509 | CV | 3 | 22 | 21 | Regression |
| Waist_circu | Lasso | FALSE | 0.000511 | CV | 4 | 22 | 21 | Regression |
| Waist_circu | Lasso | TRUE | 0.05 | CV | 0 | 22 | 10 | Regression |
| Waist_circu | Lasso | TRUE | 0.05 | CV | 1 | 22 | 10 | Regression |
| Waist_circu | Lasso | TRUE | 0.05 | CV | 2 | 22 | 10 | Regression |
| Waist_circu | Lasso | TRUE | 0.05 | CV | 3 | 22 | 10 | Regression |
| Waist_circu | Lasso | TRUE | 0.05 | CV | 4 | 22 | 10 | Regression |
| Hip_circum | LinRegrs | FALSE | | HeldoutTest | | 22 | 20 | Regression |
| Hip_circum | Lasso | FALSE | 0.000379 | HeldoutTest | | 22 | 21 | Regression |
| Hip_circum | Lasso | TRUE | 0.05 | HeldoutTest | | 22 | 9 | Regression |
| Hip_circum | LinRegrs | FALSE | | CV | 0 | 22 | 22 | Regression |
| Hip_circum | LinRegrs | FALSE | | CV | 1 | 22 | 21 | Regression |
| Hip_circum | LinRegrs | FALSE | | CV | 2 | 22 | 20 | Regression |
| Hip_circum | LinRegrs | FALSE | | CV | 3 | 22 | 21 | Regression |
| Hip_circum | LinRegrs | FALSE | | CV | 4 | 22 | 20 | Regression |

| | | | | | | | | |
|---|---|---|---|---|---|---|---|---|
| Hip_circum | Lasso | FALSE | 0.000429 | CV | 0 | 22 | 22 | Regression |
| Hip_circum | Lasso | FALSE | 0.000378 | CV | 1 | 22 | 22 | Regression |
| Hip_circum | Lasso | FALSE | 0.000381 | CV | 2 | 22 | 20 | Regression |
| Hip_circum | Lasso | FALSE | 0.000378 | CV | 3 | 22 | 22 | Regression |
| Hip_circum | Lasso | FALSE | 0.000381 | CV | 4 | 22 | 22 | Regression |
| Hip_circum | Lasso | TRUE | 0.05 | CV | 0 | 22 | 10 | Regression |
| Hip_circum | Lasso | TRUE | 0.05 | CV | 1 | 22 | 9 | Regression |
| Hip_circum | Lasso | TRUE | 0.05 | CV | 2 | 22 | 8 | Regression |
| Hip_circum | Lasso | TRUE | 0.05 | CV | 3 | 22 | 9 | Regression |
| Hip_circum | Lasso | TRUE | 0.05 | CV | 4 | 22 | 8 | Regression |
| Standing_h | LinRegrs | FALSE | | HeldoutTest | | 22 | 21 | Regression |
| Standing_h | Lasso | FALSE | 0.000461 | HeldoutTest | | 22 | 21 | Regression |
| Standing_h | Lasso | TRUE | 0.05 | HeldoutTest | | 22 | 10 | Regression |
| Standing_h | LinRegrs | FALSE | | CV | 0 | 22 | 22 | Regression |
| Standing_h | LinRegrs | FALSE | | CV | 1 | 22 | 21 | Regression |
| Standing_h | LinRegrs | FALSE | | CV | 2 | 22 | 21 | Regression |
| Standing_h | LinRegrs | FALSE | | CV | 3 | 22 | 21 | Regression |
| Standing_h | LinRegrs | FALSE | | CV | 4 | 22 | 21 | Regression |
| Standing_h | Lasso | FALSE | 0.000458 | CV | 0 | 22 | 22 | Regression |
| Standing_h | Lasso | FALSE | 0.00046 | CV | 1 | 22 | 21 | Regression |
| Standing_h | Lasso | FALSE | 0.000462 | CV | 2 | 22 | 21 | Regression |
| Standing_h | Lasso | FALSE | 0.000462 | CV | 3 | 22 | 21 | Regression |
| Standing_h | Lasso | FALSE | 0.000461 | CV | 4 | 22 | 21 | Regression |
| Standing_h | Lasso | TRUE | 0.05 | CV | 0 | 22 | 9 | Regression |
| Standing_h | Lasso | TRUE | 0.05 | CV | 1 | 22 | 9 | Regression |
| Standing_h | Lasso | TRUE | 0.05 | CV | 2 | 22 | 10 | Regression |
| Standing_h | Lasso | TRUE | 0.05 | CV | 3 | 22 | 10 | Regression |
| Standing_h | Lasso | TRUE | 0.05 | CV | 4 | 22 | 10 | Regression |
| Seated_hei | LinRegrs | FALSE | | HeldoutTest | | 22 | 22 | Regression |
| Seated_hei | Lasso | FALSE | 0.000351 | HeldoutTest | | 22 | 22 | Regression |
| Seated_hei | Lasso | TRUE | 0.05 | HeldoutTest | | 22 | 6 | Regression |
| Seated_hei | LinRegrs | FALSE | | CV | 0 | 22 | 21 | Regression |
| Seated_hei | LinRegrs | FALSE | | CV | 1 | 22 | 21 | Regression |
| Seated_hei | LinRegrs | FALSE | | CV | 2 | 22 | 19 | Regression |
| Seated_hei | LinRegrs | FALSE | | CV | 3 | 22 | 21 | Regression |
| Seated_hei | LinRegrs | FALSE | | CV | 4 | 22 | 20 | Regression |
| Seated_hei | Lasso | FALSE | 0.000353 | CV | 0 | 22 | 22 | Regression |
| Seated_hei | Lasso | FALSE | 0.00035 | CV | 1 | 22 | 22 | Regression |
| Seated_hei | Lasso | FALSE | 0.000351 | CV | 2 | 22 | 21 | Regression |
| Seated_hei | Lasso | FALSE | 0.000351 | CV | 3 | 22 | 22 | Regression |
| Seated_hei | Lasso | FALSE | 0.000352 | CV | 4 | 22 | 22 | Regression |
| Seated_hei | Lasso | TRUE | 0.05 | CV | 0 | 22 | 6 | Regression |
| Seated_hei | Lasso | TRUE | 0.05 | CV | 1 | 22 | 6 | Regression |
| Seated_hei | Lasso | TRUE | 0.05 | CV | 2 | 22 | 6 | Regression |
| Seated_hei | Lasso | TRUE | 0.05 | CV | 3 | 22 | 6 | Regression |

| | | | | | | | | |
|---|---|---|---|---|---|---|---|---|
| Seated_hei | Lasso | TRUE | 0.05 | CV | 4 | 22 | 6 | Regression |
| Sitting_hei | LinRegrs | FALSE | | HeldoutTest | | 22 | 22 | Regression |
| Sitting_hei | Lasso | FALSE | 0.000422 | HeldoutTest | | 22 | 22 | Regression |
| Sitting_hei | Lasso | TRUE | 0.05 | HeldoutTest | | 22 | 7 | Regression |
| Sitting_hei | LinRegrs | FALSE | | CV | 0 | 22 | 21 | Regression |
| Sitting_hei | LinRegrs | FALSE | | CV | 1 | 22 | 22 | Regression |
| Sitting_hei | LinRegrs | FALSE | | CV | 2 | 22 | 22 | Regression |
| Sitting_hei | LinRegrs | FALSE | | CV | 3 | 22 | 22 | Regression |
| Sitting_hei | LinRegrs | FALSE | | CV | 4 | 22 | 22 | Regression |
| Sitting_hei | Lasso | FALSE | 0.00042 | CV | 0 | 22 | 21 | Regression |
| Sitting_hei | Lasso | FALSE | 0.000422 | CV | 1 | 22 | 22 | Regression |
| Sitting_hei | Lasso | FALSE | 0.000421 | CV | 2 | 22 | 22 | Regression |
| Sitting_hei | Lasso | FALSE | 0.000422 | CV | 3 | 22 | 22 | Regression |
| Sitting_hei | Lasso | FALSE | 0.000423 | CV | 4 | 22 | 22 | Regression |
| Sitting_hei | Lasso | TRUE | 0.05 | CV | 0 | 22 | 10 | Regression |
| Sitting_hei | Lasso | TRUE | 0.05 | CV | 1 | 22 | 7 | Regression |
| Sitting_hei | Lasso | TRUE | 0.05 | CV | 2 | 22 | 7 | Regression |
| Sitting_hei | Lasso | TRUE | 0.05 | CV | 3 | 22 | 7 | Regression |
| Sitting_hei | Lasso | TRUE | 0.05 | CV | 4 | 22 | 7 | Regression |
| Weight | LinRegrs | FALSE | | HeldoutTest | | 22 | 22 | Regression |
| Weight | Lasso | FALSE | 0.000559 | HeldoutTest | | 22 | 22 | Regression |
| Weight | Lasso | TRUE | 0.05 | HeldoutTest | | 22 | 11 | Regression |
| Weight | LinRegrs | FALSE | | CV | 0 | 22 | 21 | Regression |
| Weight | LinRegrs | FALSE | | CV | 1 | 22 | 22 | Regression |
| Weight | LinRegrs | FALSE | | CV | 2 | 22 | 22 | Regression |
| Weight | LinRegrs | FALSE | | CV | 3 | 22 | 22 | Regression |
| Weight | LinRegrs | FALSE | | CV | 4 | 22 | 22 | Regression |
| Weight | Lasso | FALSE | 0.000688 | CV | 0 | 22 | 21 | Regression |
| Weight | Lasso | FALSE | 0.000558 | CV | 1 | 22 | 22 | Regression |
| Weight | Lasso | FALSE | 0.000557 | CV | 2 | 22 | 22 | Regression |
| Weight | Lasso | FALSE | 0.000558 | CV | 3 | 22 | 22 | Regression |
| Weight | Lasso | FALSE | 0.00056 | CV | 4 | 22 | 22 | Regression |
| Weight | Lasso | TRUE | 0.05 | CV | 0 | 22 | 11 | Regression |
| Weight | Lasso | TRUE | 0.05 | CV | 1 | 22 | 11 | Regression |
| Weight | Lasso | TRUE | 0.05 | CV | 2 | 22 | 11 | Regression |
| Weight | Lasso | TRUE | 0.05 | CV | 3 | 22 | 11 | Regression |
| Weight | Lasso | TRUE | 0.05 | CV | 4 | 22 | 11 | Regression |
| BinCAT_Se | LogRegrs | FALSE | | HeldoutTest | | 22 | 21 | Classificatic |
| BinCAT_Se | LassoCat | FALSE | 0.359381 | HeldoutTest | | 22 | 21 | Classificatic |
| BinCAT_Se | LassoCat | TRUE | 0.05 | HeldoutTest | | 22 | 21 | Classificatic |
| BinCAT_Se | LogRegrs | FALSE | | CV | 0 | 22 | 6 | Classificatic |
| BinCAT_Se | LogRegrs | FALSE | | CV | 1 | 22 | 6 | Classificatic |
| BinCAT_Se | LogRegrs | FALSE | | CV | 2 | 22 | 8 | Classificatic |
| BinCAT_Se | LogRegrs | FALSE | | CV | 3 | 22 | 8 | Classificatic |
| BinCAT_Se | LogRegrs | FALSE | | CV | 4 | 22 | 8 | Classificatic |

| | | | | | | | | |
|---|---|---|---|---|---|---|---|---|
| BinCAT_Se | LassoCat | FALSE | 0.046416 | CV | 0 | 22 | 0 | Classificatio |
| BinCAT_Se | LassoCat | FALSE | 2.782559 | CV | 1 | 22 | 6 | Classificatio |
| BinCAT_Se | LassoCat | FALSE | 0.0001 | CV | 2 | 22 | 0 | Classificatio |
| BinCAT_Se | LassoCat | FALSE | 0.0001 | CV | 3 | 22 | 0 | Classificatio |
| BinCAT_Se | LassoCat | FALSE | 0.046416 | CV | 4 | 22 | 0 | Classificatio |
| BinCAT_Se | LassoCat | TRUE | 0.05 | CV | 0 | 22 | 1 | Classificatio |
| BinCAT_Se | LassoCat | TRUE | 0.05 | CV | 1 | 22 | 1 | Classificatio |
| BinCAT_Se | LassoCat | TRUE | 0.05 | CV | 2 | 22 | 1 | Classificatio |
| BinCAT_Se | LassoCat | TRUE | 0.05 | CV | 3 | 22 | 1 | Classificatio |
| BinCAT_Se | LassoCat | TRUE | 0.05 | CV | 4 | 22 | 0 | Classificatio |
| BinCAT_MI | LogRegrs | FALSE | | HeldoutTest | | 22 | 22 | Classificatio |
| BinCAT_MI | LassoCat | FALSE | 0.0001 | HeldoutTest | | 22 | 0 | Classificatio |
| BinCAT_MI | LassoCat | TRUE | 0.05 | HeldoutTest | | 22 | 5 | Classificatio |
| BinCAT_MI | LogRegrs | FALSE | | CV | 0 | 22 | 19 | Classificatio |
| BinCAT_MI | LogRegrs | FALSE | | CV | 1 | 22 | 21 | Classificatio |
| BinCAT_MI | LogRegrs | FALSE | | CV | 2 | 22 | 19 | Classificatio |
| BinCAT_MI | LogRegrs | FALSE | | CV | 3 | 22 | 21 | Classificatio |
| BinCAT_MI | LogRegrs | FALSE | | CV | 4 | 22 | 21 | Classificatio |
| BinCAT_MI | LassoCat | FALSE | 0.0001 | CV | 0 | 22 | 0 | Classificatio |
| BinCAT_MI | LassoCat | FALSE | 0.0001 | CV | 1 | 22 | 0 | Classificatio |
| BinCAT_MI | LassoCat | FALSE | 0.0001 | CV | 2 | 22 | 0 | Classificatio |
| BinCAT_MI | LassoCat | FALSE | 0.0001 | CV | 3 | 22 | 0 | Classificatio |
| BinCAT_MI | LassoCat | FALSE | 0.0001 | CV | 4 | 22 | 0 | Classificatio |
| BinCAT_MI | LassoCat | TRUE | 0.05 | CV | 0 | 22 | 1 | Classificatio |
| BinCAT_MI | LassoCat | TRUE | 0.05 | CV | 1 | 22 | 0 | Classificatio |
| BinCAT_MI | LassoCat | TRUE | 0.05 | CV | 2 | 22 | 1 | Classificatio |
| BinCAT_MI | LassoCat | TRUE | 0.05 | CV | 3 | 22 | 0 | Classificatio |
| BinCAT_MI | LassoCat | TRUE | 0.05 | CV | 4 | 22 | 0 | Classificatio |
| CAT_Montl | LogRegrs | FALSE | | HeldoutTest | | 22 | 9 | Classificatio |
| CAT_Montl | LassoCat | FALSE | 0.046416 | HeldoutTest | | 22 | 5 | Classificatio |
| CAT_Montl | LassoCat | TRUE | 0.05 | HeldoutTest | | 22 | 5 | Classificatio |
| CAT_Montl | LogRegrs | FALSE | | CV | 0 | 22 | 14 | Classificatio |
| CAT_Montl | LogRegrs | FALSE | | CV | 1 | 22 | 15 | Classificatio |
| CAT_Montl | LogRegrs | FALSE | | CV | 2 | 22 | 9 | Classificatio |
| CAT_Montl | LogRegrs | FALSE | | CV | 3 | 22 | 14 | Classificatio |
| CAT_Montl | LogRegrs | FALSE | | CV | 4 | 22 | 16 | Classificatio |
| CAT_Montl | LassoCat | FALSE | 0.046416 | CV | 0 | 22 | 1 | Classificatio |
| CAT_Montl | LassoCat | FALSE | 0.0001 | CV | 1 | 22 | 0 | Classificatio |
| CAT_Montl | LassoCat | FALSE | 0.0001 | CV | 2 | 22 | 0 | Classificatio |
| CAT_Montl | LassoCat | FALSE | 0.000774 | CV | 3 | 22 | 0 | Classificatio |
| CAT_Montl | LassoCat | FALSE | 0.000774 | CV | 4 | 22 | 0 | Classificatio |
| CAT_Montl | LassoCat | TRUE | 0.05 | CV | 0 | 22 | 1 | Classificatio |
| CAT_Montl | LassoCat | TRUE | 0.05 | CV | 1 | 22 | 3 | Classificatio |
| CAT_Montl | LassoCat | TRUE | 0.05 | CV | 2 | 22 | 0 | Classificatio |
| CAT_Montl | LassoCat | TRUE | 0.05 | CV | 3 | 22 | 0 | Classificatio |

| | | | | | | | | |
|---|---|---|---|---|---|---|---|---|
| CAT_Mont | LassoCat | TRUE | 0.05 | CV | 4 | 22 | 2 | Classificatio |
| CAT_Smoki | LogRegrs | FALSE | | HeldoutTest | | 22 | 18 | Classificatio |
| CAT_Smoki | LassoCat | FALSE | 0.359381 | HeldoutTest | | 22 | 14 | Classificatio |
| CAT_Smoki | LassoCat | TRUE | 0.05 | HeldoutTest | | 22 | 10 | Classificatio |
| CAT_Smoki | LogRegrs | FALSE | | CV | 0 | 22 | 12 | Classificatio |
| CAT_Smoki | LogRegrs | FALSE | | CV | 1 | 22 | 11 | Classificatio |
| CAT_Smoki | LogRegrs | FALSE | | CV | 2 | 22 | 8 | Classificatio |
| CAT_Smoki | LogRegrs | FALSE | | CV | 3 | 22 | 9 | Classificatio |
| CAT_Smoki | LogRegrs | FALSE | | CV | 4 | 22 | 10 | Classificatio |
| CAT_Smoki | LassoCat | FALSE | 0.0001 | CV | 0 | 22 | 0 | Classificatio |
| CAT_Smoki | LassoCat | FALSE | 0.0001 | CV | 1 | 22 | 0 | Classificatio |
| CAT_Smoki | LassoCat | FALSE | 0.0001 | CV | 2 | 22 | 0 | Classificatio |
| CAT_Smoki | LassoCat | FALSE | 0.0001 | CV | 3 | 22 | 0 | Classificatio |
| CAT_Smoki | LassoCat | FALSE | 0.0001 | CV | 4 | 22 | 0 | Classificatio |
| CAT_Smoki | LassoCat | TRUE | 0.05 | CV | 0 | 22 | 2 | Classificatio |
| CAT_Smoki | LassoCat | TRUE | 0.05 | CV | 1 | 22 | 4 | Classificatio |
| CAT_Smoki | LassoCat | TRUE | 0.05 | CV | 2 | 22 | 0 | Classificatio |
| CAT_Smoki | LassoCat | TRUE | 0.05 | CV | 3 | 22 | 1 | Classificatio |
| CAT_Smoki | LassoCat | TRUE | 0.05 | CV | 4 | 22 | 5 | Classificatio |
| CAT_BMI | LogRegrs | FALSE | | HeldoutTest | | 22 | 22 | Classificatio |
| CAT_BMI | LassoCat | FALSE | 21.54435 | HeldoutTest | | 22 | 21 | Classificatio |
| CAT_BMI | LassoCat | TRUE | 0.05 | HeldoutTest | | 22 | 11 | Classificatio |
| CAT_BMI | LogRegrs | FALSE | | CV | 0 | 22 | 19 | Classificatio |
| CAT_BMI | LogRegrs | FALSE | | CV | 1 | 22 | 20 | Classificatio |
| CAT_BMI | LogRegrs | FALSE | | CV | 2 | 22 | 19 | Classificatio |
| CAT_BMI | LogRegrs | FALSE | | CV | 3 | 22 | 20 | Classificatio |
| CAT_BMI | LogRegrs | FALSE | | CV | 4 | 22 | 19 | Classificatio |
| CAT_BMI | LassoCat | FALSE | 0.0001 | CV | 0 | 22 | 0 | Classificatio |
| CAT_BMI | LassoCat | FALSE | 0.0001 | CV | 1 | 22 | 0 | Classificatio |
| CAT_BMI | LassoCat | FALSE | 0.359381 | CV | 2 | 22 | 8 | Classificatio |
| CAT_BMI | LassoCat | FALSE | 0.0001 | CV | 3 | 22 | 0 | Classificatio |
| CAT_BMI | LassoCat | FALSE | 0.0001 | CV | 4 | 22 | 0 | Classificatio |
| CAT_BMI | LassoCat | TRUE | 0.05 | CV | 0 | 22 | 0 | Classificatio |
| CAT_BMI | LassoCat | TRUE | 0.05 | CV | 1 | 22 | 0 | Classificatio |
| CAT_BMI | LassoCat | TRUE | 0.05 | CV | 2 | 22 | 0 | Classificatio |
| CAT_BMI | LassoCat | TRUE | 0.05 | CV | 3 | 22 | 0 | Classificatio |
| CAT_BMI | LassoCat | TRUE | 0.05 | CV | 4 | 22 | 0 | Classificatio |
| CAT_MRI_( | LogRegrs | FALSE | | HeldoutTest | | 22 | 17 | Classificatio |
| CAT_MRI_( | LassoCat | FALSE | 21.54435 | HeldoutTest | | 22 | 17 | Classificatio |
| CAT_MRI_( | LassoCat | TRUE | 0.05 | HeldoutTest | | 22 | 9 | Classificatio |
| CAT_MRI_( | LogRegrs | FALSE | | CV | 0 | 22 | 10 | Classificatio |
| CAT_MRI_( | LogRegrs | FALSE | | CV | 1 | 22 | 8 | Classificatio |
| CAT_MRI_( | LogRegrs | FALSE | | CV | 2 | 22 | 12 | Classificatio |
| CAT_MRI_( | LogRegrs | FALSE | | CV | 3 | 22 | 12 | Classificatio |
| CAT_MRI_( | LogRegrs | FALSE | | CV | 4 | 22 | 13 | Classificatio |

| | | | | | | | | |
|---|---|---|---|---|---|---|---|---|
| CAT_MRI_( | LassoCat | FALSE | 0.0001 | CV | 0 | 22 | 0 | Classificatic |
| CAT_MRI_( | LassoCat | FALSE | 0.0001 | CV | 1 | 22 | 0 | Classificatic |
| CAT_MRI_( | LassoCat | FALSE | 0.0001 | CV | 2 | 22 | 0 | Classificatic |
| CAT_MRI_( | LassoCat | FALSE | 0.0001 | CV | 3 | 22 | 0 | Classificatic |
| CAT_MRI_( | LassoCat | FALSE | 0.0001 | CV | 4 | 22 | 0 | Classificatic |
| CAT_MRI_( | LassoCat | TRUE | 0.05 | CV | 0 | 22 | 0 | Classificatic |
| CAT_MRI_( | LassoCat | TRUE | 0.05 | CV | 1 | 22 | 1 | Classificatic |
| CAT_MRI_( | LassoCat | TRUE | 0.05 | CV | 2 | 22 | 0 | Classificatic |
| CAT_MRI_( | LassoCat | TRUE | 0.05 | CV | 3 | 22 | 2 | Classificatic |
| CAT_MRI_( | LassoCat | TRUE | 0.05 | CV | 4 | 22 | 4 | Classificatic |
| Ascending_ | LinRegrs | FALSE | | HeldoutTest | | 22 | 18 | Regression |
| Ascending_ | Lasso | FALSE | 0.000343 | HeldoutTest | | 22 | 22 | Regression |
| Ascending_ | Lasso | TRUE | 0.05 | HeldoutTest | | 22 | 9 | Regression |
| Ascending_ | LinRegrs | FALSE | | CV | 0 | 22 | 18 | Regression |
| Ascending_ | LinRegrs | FALSE | | CV | 1 | 22 | 19 | Regression |
| Ascending_ | LinRegrs | FALSE | | CV | 2 | 21 | 18 | Regression |
| Ascending_ | LinRegrs | FALSE | | CV | 3 | 21 | 20 | Regression |
| Ascending_ | LinRegrs | FALSE | | CV | 4 | 22 | 18 | Regression |
| Ascending_ | Lasso | FALSE | 0.000552 | CV | 0 | 22 | 20 | Regression |
| Ascending_ | Lasso | FALSE | 0.000343 | CV | 1 | 22 | 21 | Regression |
| Ascending_ | Lasso | FALSE | 0.000344 | CV | 2 | 21 | 20 | Regression |
| Ascending_ | Lasso | FALSE | 0.000344 | CV | 3 | 21 | 21 | Regression |
| Ascending_ | Lasso | FALSE | 0.000346 | CV | 4 | 22 | 21 | Regression |
| Ascending_ | Lasso | TRUE | 0.05 | CV | 0 | 22 | 9 | Regression |
| Ascending_ | Lasso | TRUE | 0.05 | CV | 1 | 22 | 9 | Regression |
| Ascending_ | Lasso | TRUE | 0.05 | CV | 2 | 21 | 9 | Regression |
| Ascending_ | Lasso | TRUE | 0.05 | CV | 3 | 21 | 9 | Regression |
| Ascending_ | Lasso | TRUE | 0.05 | CV | 4 | 22 | 8 | Regression |
| Ascending_ | LinRegrs | FALSE | | HeldoutTest | | 22 | 21 | Regression |
| Ascending_ | Lasso | FALSE | 0.000343 | HeldoutTest | | 22 | 21 | Regression |
| Ascending_ | Lasso | TRUE | 0.05 | HeldoutTest | | 22 | 8 | Regression |
| Ascending_ | LinRegrs | FALSE | | CV | 0 | 22 | 20 | Regression |
| Ascending_ | LinRegrs | FALSE | | CV | 1 | 22 | 20 | Regression |
| Ascending_ | LinRegrs | FALSE | | CV | 2 | 21 | 21 | Regression |
| Ascending_ | LinRegrs | FALSE | | CV | 3 | 21 | 21 | Regression |
| Ascending_ | LinRegrs | FALSE | | CV | 4 | 22 | 21 | Regression |
| Ascending_ | Lasso | FALSE | 0.000339 | CV | 0 | 22 | 21 | Regression |
| Ascending_ | Lasso | FALSE | 0.000343 | CV | 1 | 22 | 22 | Regression |
| Ascending_ | Lasso | FALSE | 0.000343 | CV | 2 | 21 | 21 | Regression |
| Ascending_ | Lasso | FALSE | 0.000343 | CV | 3 | 21 | 21 | Regression |
| Ascending_ | Lasso | FALSE | 0.000346 | CV | 4 | 22 | 22 | Regression |
| Ascending_ | Lasso | TRUE | 0.05 | CV | 0 | 22 | 9 | Regression |
| Ascending_ | Lasso | TRUE | 0.05 | CV | 1 | 22 | 8 | Regression |
| Ascending_ | Lasso | TRUE | 0.05 | CV | 2 | 21 | 8 | Regression |
| Ascending_ | Lasso | TRUE | 0.05 | CV | 3 | 21 | 8 | Regression |

| | | | | | | | |
|---|---|---|---|---|---|---|---|
| Ascending_ | Lasso | TRUE | 0.05 | CV | 4 | 22 | 8 Regression |
| Ascending_ | LinRegrs | FALSE | | HeldoutTest | | 22 | 18 Regression |
| Ascending_ | Lasso | FALSE | 0.000818 | HeldoutTest | | 22 | 20 Regression |
| Ascending_ | Lasso | TRUE | 0.05 | HeldoutTest | | 22 | 6 Regression |
| Ascending_ | LinRegrs | FALSE | | CV | 0 | 22 | 16 Regression |
| Ascending_ | LinRegrs | FALSE | | CV | 1 | 22 | 15 Regression |
| Ascending_ | LinRegrs | FALSE | | CV | 2 | 21 | 16 Regression |
| Ascending_ | LinRegrs | FALSE | | CV | 3 | 21 | 17 Regression |
| Ascending_ | LinRegrs | FALSE | | CV | 4 | 22 | 17 Regression |
| Ascending_ | Lasso | FALSE | 0.000936 | CV | 0 | 22 | 18 Regression |
| Ascending_ | Lasso | FALSE | 0.001104 | CV | 1 | 22 | 18 Regression |
| Ascending_ | Lasso | FALSE | 0.001167 | CV | 2 | 21 | 17 Regression |
| Ascending_ | Lasso | FALSE | 0.000886 | CV | 3 | 21 | 18 Regression |
| Ascending_ | Lasso | FALSE | 0.00075 | CV | 4 | 22 | 21 Regression |
| Ascending_ | Lasso | TRUE | 0.05 | CV | 0 | 22 | 6 Regression |
| Ascending_ | Lasso | TRUE | 0.05 | CV | 1 | 22 | 6 Regression |
| Ascending_ | Lasso | TRUE | 0.05 | CV | 2 | 21 | 6 Regression |
| Ascending_ | Lasso | TRUE | 0.05 | CV | 3 | 21 | 6 Regression |
| Ascending_ | Lasso | TRUE | 0.05 | CV | 4 | 22 | 6 Regression |
| Descending | LinRegrs | FALSE | | HeldoutTest | | 22 | 20 Regression |
| Descending | Lasso | FALSE | 0.000601 | HeldoutTest | | 22 | 20 Regression |
| Descending | Lasso | TRUE | 0.05 | HeldoutTest | | 22 | 10 Regression |
| Descending | LinRegrs | FALSE | | CV | 0 | 22 | 20 Regression |
| Descending | LinRegrs | FALSE | | CV | 1 | 22 | 20 Regression |
| Descending | LinRegrs | FALSE | | CV | 2 | 21 | 20 Regression |
| Descending | LinRegrs | FALSE | | CV | 3 | 21 | 19 Regression |
| Descending | LinRegrs | FALSE | | CV | 4 | 22 | 20 Regression |
| Descending | Lasso | FALSE | 0.000526 | CV | 0 | 22 | 21 Regression |
| Descending | Lasso | FALSE | 0.000452 | CV | 1 | 22 | 21 Regression |
| Descending | Lasso | FALSE | 0.00045 | CV | 2 | 21 | 20 Regression |
| Descending | Lasso | FALSE | 0.000456 | CV | 3 | 21 | 20 Regression |
| Descending | Lasso | FALSE | 0.000605 | CV | 4 | 22 | 21 Regression |
| Descending | Lasso | TRUE | 0.05 | CV | 0 | 22 | 10 Regression |
| Descending | Lasso | TRUE | 0.05 | CV | 1 | 22 | 10 Regression |
| Descending | Lasso | TRUE | 0.05 | CV | 2 | 21 | 10 Regression |
| Descending | Lasso | TRUE | 0.05 | CV | 3 | 21 | 10 Regression |
| Descending | Lasso | TRUE | 0.05 | CV | 4 | 22 | 10 Regression |
| Descending | LinRegrs | FALSE | | HeldoutTest | | 22 | 20 Regression |
| Descending | Lasso | FALSE | 0.000569 | HeldoutTest | | 22 | 21 Regression |
| Descending | Lasso | TRUE | 0.05 | HeldoutTest | | 22 | 8 Regression |
| Descending | LinRegrs | FALSE | | CV | 0 | 22 | 19 Regression |
| Descending | LinRegrs | FALSE | | CV | 1 | 22 | 20 Regression |
| Descending | LinRegrs | FALSE | | CV | 2 | 21 | 20 Regression |
| Descending | LinRegrs | FALSE | | CV | 3 | 21 | 18 Regression |
| Descending | LinRegrs | FALSE | | CV | 4 | 22 | 21 Regression |

| | | | | | | | | |
|---|---|---|---|---|---|---|---|---|
| Descending | Lasso | FALSE | 0.000529 | CV | 0 | 22 | 21 | Regression |
| Descending | Lasso | FALSE | 0.000494 | CV | 1 | 22 | 20 | Regression |
| Descending | Lasso | FALSE | 0.000431 | CV | 2 | 21 | 21 | Regression |
| Descending | Lasso | FALSE | 0.00043 | CV | 3 | 21 | 21 | Regression |
| Descending | Lasso | FALSE | 0.000812 | CV | 4 | 22 | 21 | Regression |
| Descending | Lasso | TRUE | 0.05 | CV | 0 | 22 | 8 | Regression |
| Descending | Lasso | TRUE | 0.05 | CV | 1 | 22 | 8 | Regression |
| Descending | Lasso | TRUE | 0.05 | CV | 2 | 21 | 8 | Regression |
| Descending | Lasso | TRUE | 0.05 | CV | 3 | 21 | 8 | Regression |
| Descending | Lasso | TRUE | 0.05 | CV | 4 | 22 | 8 | Regression |
| Descending | LinRegrs | FALSE | | HeldoutTest | | 22 | 18 | Regression |
| Descending | Lasso | FALSE | 0.000989 | HeldoutTest | | 22 | 19 | Regression |
| Descending | Lasso | TRUE | 0.05 | HeldoutTest | | 22 | 6 | Regression |
| Descending | LinRegrs | FALSE | | CV | 0 | 22 | 18 | Regression |
| Descending | LinRegrs | FALSE | | CV | 1 | 22 | 18 | Regression |
| Descending | LinRegrs | FALSE | | CV | 2 | 21 | 16 | Regression |
| Descending | LinRegrs | FALSE | | CV | 3 | 21 | 18 | Regression |
| Descending | LinRegrs | FALSE | | CV | 4 | 22 | 18 | Regression |
| Descending | Lasso | FALSE | 0.000847 | CV | 0 | 22 | 19 | Regression |
| Descending | Lasso | FALSE | 0.001075 | CV | 1 | 22 | 19 | Regression |
| Descending | Lasso | FALSE | 0.000868 | CV | 2 | 21 | 18 | Regression |
| Descending | Lasso | FALSE | 0.000531 | CV | 3 | 21 | 21 | Regression |
| Descending | Lasso | FALSE | 0.001382 | CV | 4 | 22 | 20 | Regression |
| Descending | Lasso | TRUE | 0.05 | CV | 0 | 22 | 6 | Regression |
| Descending | Lasso | TRUE | 0.05 | CV | 1 | 22 | 6 | Regression |
| Descending | Lasso | TRUE | 0.05 | CV | 2 | 21 | 6 | Regression |
| Descending | Lasso | TRUE | 0.05 | CV | 3 | 21 | 6 | Regression |
| Descending | Lasso | TRUE | 0.05 | CV | 4 | 22 | 6 | Regression |
| RA_maxim | LinRegrs | FALSE | | HeldoutTest | | 21 | 20 | Regression |
| RA_maxim | Lasso | FALSE | 0.000346 | HeldoutTest | | 21 | 21 | Regression |
| RA_maxim | Lasso | TRUE | 0.05 | HeldoutTest | | 21 | 8 | Regression |
| RA_maxim | LinRegrs | FALSE | | CV | 0 | 21 | 20 | Regression |
| RA_maxim | LinRegrs | FALSE | | CV | 1 | 21 | 20 | Regression |
| RA_maxim | LinRegrs | FALSE | | CV | 2 | 21 | 21 | Regression |
| RA_maxim | LinRegrs | FALSE | | CV | 3 | 21 | 20 | Regression |
| RA_maxim | LinRegrs | FALSE | | CV | 4 | 21 | 20 | Regression |
| RA_maxim | Lasso | FALSE | 0.000348 | CV | 0 | 21 | 21 | Regression |
| RA_maxim | Lasso | FALSE | 0.000344 | CV | 1 | 21 | 21 | Regression |
| RA_maxim | Lasso | FALSE | 0.000345 | CV | 2 | 21 | 21 | Regression |
| RA_maxim | Lasso | FALSE | 0.000346 | CV | 3 | 21 | 21 | Regression |
| RA_maxim | Lasso | FALSE | 0.000345 | CV | 4 | 21 | 21 | Regression |
| RA_maxim | Lasso | TRUE | 0.05 | CV | 0 | 21 | 8 | Regression |
| RA_maxim | Lasso | TRUE | 0.05 | CV | 1 | 21 | 8 | Regression |
| RA_maxim | Lasso | TRUE | 0.05 | CV | 2 | 21 | 8 | Regression |
| RA_maxim | Lasso | TRUE | 0.05 | CV | 3 | 21 | 8 | Regression |

| | | | | | | | | |
|---|---|---|---|---|---|---|---|---|
| RA_maxim | Lasso | TRUE | 0.05 | CV | 4 | 21 | 8 | Regression |
| RA_minimu | LinRegrs | FALSE | | HeldoutTest | | 21 | 21 | Regression |
| RA_minimu | Lasso | FALSE | 0.000388 | HeldoutTest | | 21 | 21 | Regression |
| RA_minimu | Lasso | TRUE | 0.05 | HeldoutTest | | 21 | 9 | Regression |
| RA_minimu | LinRegrs | FALSE | | CV | 0 | 21 | 21 | Regression |
| RA_minimu | LinRegrs | FALSE | | CV | 1 | 21 | 21 | Regression |
| RA_minimu | LinRegrs | FALSE | | CV | 2 | 21 | 21 | Regression |
| RA_minimu | LinRegrs | FALSE | | CV | 3 | 21 | 21 | Regression |
| RA_minimu | LinRegrs | FALSE | | CV | 4 | 21 | 21 | Regression |
| RA_minimu | Lasso | FALSE | 0.000392 | CV | 0 | 21 | 21 | Regression |
| RA_minimu | Lasso | FALSE | 0.000387 | CV | 1 | 21 | 21 | Regression |
| RA_minimu | Lasso | FALSE | 0.000389 | CV | 2 | 21 | 21 | Regression |
| RA_minimu | Lasso | FALSE | 0.000387 | CV | 3 | 21 | 21 | Regression |
| RA_minimu | Lasso | FALSE | 0.000388 | CV | 4 | 21 | 21 | Regression |
| RA_minimu | Lasso | TRUE | 0.05 | CV | 0 | 21 | 9 | Regression |
| RA_minimu | Lasso | TRUE | 0.05 | CV | 1 | 21 | 9 | Regression |
| RA_minimu | Lasso | TRUE | 0.05 | CV | 2 | 21 | 9 | Regression |
| RA_minimu | Lasso | TRUE | 0.05 | CV | 3 | 21 | 9 | Regression |
| RA_minimu | Lasso | TRUE | 0.05 | CV | 4 | 21 | 9 | Regression |
| RA_stroke_ | LinRegrs | FALSE | | HeldoutTest | | 21 | 21 | Regression |
| RA_stroke_ | Lasso | FALSE | 0.000252 | HeldoutTest | | 21 | 21 | Regression |
| RA_stroke_ | Lasso | TRUE | 0.05 | HeldoutTest | | 21 | 8 | Regression |
| RA_stroke_ | LinRegrs | FALSE | | CV | 0 | 21 | 21 | Regression |
| RA_stroke_ | LinRegrs | FALSE | | CV | 1 | 21 | 21 | Regression |
| RA_stroke_ | LinRegrs | FALSE | | CV | 2 | 21 | 20 | Regression |
| RA_stroke_ | LinRegrs | FALSE | | CV | 3 | 21 | 21 | Regression |
| RA_stroke_ | LinRegrs | FALSE | | CV | 4 | 21 | 21 | Regression |
| RA_stroke_ | Lasso | FALSE | 0.000252 | CV | 0 | 21 | 21 | Regression |
| RA_stroke_ | Lasso | FALSE | 0.000252 | CV | 1 | 21 | 21 | Regression |
| RA_stroke_ | Lasso | FALSE | 0.000251 | CV | 2 | 21 | 21 | Regression |
| RA_stroke_ | Lasso | FALSE | 0.000254 | CV | 3 | 21 | 21 | Regression |
| RA_stroke_ | Lasso | FALSE | 0.000252 | CV | 4 | 21 | 21 | Regression |
| RA_stroke_ | Lasso | TRUE | 0.05 | CV | 0 | 21 | 8 | Regression |
| RA_stroke_ | Lasso | TRUE | 0.05 | CV | 1 | 21 | 8 | Regression |
| RA_stroke_ | Lasso | TRUE | 0.05 | CV | 2 | 21 | 8 | Regression |
| RA_stroke_ | Lasso | TRUE | 0.05 | CV | 3 | 21 | 8 | Regression |
| RA_stroke_ | Lasso | TRUE | 0.05 | CV | 4 | 21 | 8 | Regression |
| RA_ejectio | LinRegrs | FALSE | | HeldoutTest | | 21 | 18 | Regression |
| RA_ejectio | Lasso | FALSE | 0.000286 | HeldoutTest | | 21 | 21 | Regression |
| RA_ejectio | Lasso | TRUE | 0.05 | HeldoutTest | | 21 | 7 | Regression |
| RA_ejectio | LinRegrs | FALSE | | CV | 0 | 21 | 19 | Regression |
| RA_ejectio | LinRegrs | FALSE | | CV | 1 | 21 | 18 | Regression |
| RA_ejectio | LinRegrs | FALSE | | CV | 2 | 21 | 19 | Regression |
| RA_ejectio | LinRegrs | FALSE | | CV | 3 | 21 | 18 | Regression |
| RA_ejectio | LinRegrs | FALSE | | CV | 4 | 21 | 18 | Regression |

| | | | | | | | | |
|---|---|---|---|---|---|---|---|---|
| RA_ejectio | Lasso | FALSE | 0.000627 | CV | 0 | 21 | 20 | Regression |
| RA_ejectio | Lasso | FALSE | 0.000248 | CV | 1 | 21 | 21 | Regression |
| RA_ejectio | Lasso | FALSE | 0.000436 | CV | 2 | 21 | 21 | Regression |
| RA_ejectio | Lasso | FALSE | 0.000263 | CV | 3 | 21 | 21 | Regression |
| RA_ejectio | Lasso | FALSE | 0.000377 | CV | 4 | 21 | 21 | Regression |
| RA_ejectio | Lasso | TRUE | 0.05 | CV | 0 | 21 | 7 | Regression |
| RA_ejectio | Lasso | TRUE | 0.05 | CV | 1 | 21 | 7 | Regression |
| RA_ejectio | Lasso | TRUE | 0.05 | CV | 2 | 21 | 7 | Regression |
| RA_ejectio | Lasso | TRUE | 0.05 | CV | 3 | 21 | 7 | Regression |
| RA_ejectio | Lasso | TRUE | 0.05 | CV | 4 | 21 | 7 | Regression |
| LV_myocar | LinRegrs | FALSE | | HeldoutTest | | 21 | 21 | Regression |
| LV_myocar | Lasso | FALSE | 0.000607 | HeldoutTest | | 21 | 21 | Regression |
| LV_myocar | Lasso | TRUE | 0.05 | HeldoutTest | | 21 | 8 | Regression |
| LV_myocar | LinRegrs | FALSE | | CV | 0 | 21 | 21 | Regression |
| LV_myocar | LinRegrs | FALSE | | CV | 1 | 21 | 21 | Regression |
| LV_myocar | LinRegrs | FALSE | | CV | 2 | 21 | 21 | Regression |
| LV_myocar | LinRegrs | FALSE | | CV | 3 | 21 | 21 | Regression |
| LV_myocar | LinRegrs | FALSE | | CV | 4 | 21 | 21 | Regression |
| LV_myocar | Lasso | FALSE | 0.00061 | CV | 0 | 21 | 21 | Regression |
| LV_myocar | Lasso | FALSE | 0.000607 | CV | 1 | 21 | 21 | Regression |
| LV_myocar | Lasso | FALSE | 0.000606 | CV | 2 | 21 | 21 | Regression |
| LV_myocar | Lasso | FALSE | 0.000607 | CV | 3 | 21 | 21 | Regression |
| LV_myocar | Lasso | FALSE | 0.000608 | CV | 4 | 21 | 21 | Regression |
| LV_myocar | Lasso | TRUE | 0.05 | CV | 0 | 21 | 8 | Regression |
| LV_myocar | Lasso | TRUE | 0.05 | CV | 1 | 21 | 8 | Regression |
| LV_myocar | Lasso | TRUE | 0.05 | CV | 2 | 21 | 8 | Regression |
| LV_myocar | Lasso | TRUE | 0.05 | CV | 3 | 21 | 8 | Regression |
| LV_myocar | Lasso | TRUE | 0.05 | CV | 4 | 21 | 8 | Regression |
| LV_mean_ı | LinRegrs | FALSE | | HeldoutTest | | 21 | 19 | Regression |
| LV_mean_ı | Lasso | FALSE | 0.000409 | HeldoutTest | | 21 | 21 | Regression |
| LV_mean_ı | Lasso | TRUE | 0.05 | HeldoutTest | | 21 | 7 | Regression |
| LV_mean_ı | LinRegrs | FALSE | | CV | 0 | 21 | 20 | Regression |
| LV_mean_ı | LinRegrs | FALSE | | CV | 1 | 21 | 19 | Regression |
| LV_mean_ı | LinRegrs | FALSE | | CV | 2 | 21 | 19 | Regression |
| LV_mean_ı | LinRegrs | FALSE | | CV | 3 | 21 | 19 | Regression |
| LV_mean_ı | LinRegrs | FALSE | | CV | 4 | 21 | 19 | Regression |
| LV_mean_ı | Lasso | FALSE | 0.000627 | CV | 0 | 21 | 21 | Regression |
| LV_mean_ı | Lasso | FALSE | 0.000577 | CV | 1 | 21 | 19 | Regression |
| LV_mean_ı | Lasso | FALSE | 0.00041 | CV | 2 | 21 | 21 | Regression |
| LV_mean_ı | Lasso | FALSE | 0.000536 | CV | 3 | 21 | 20 | Regression |
| LV_mean_ı | Lasso | FALSE | 0.000409 | CV | 4 | 21 | 21 | Regression |
| LV_mean_ı | Lasso | TRUE | 0.05 | CV | 0 | 21 | 7 | Regression |
| LV_mean_ı | Lasso | TRUE | 0.05 | CV | 1 | 21 | 7 | Regression |
| LV_mean_ı | Lasso | TRUE | 0.05 | CV | 2 | 21 | 7 | Regression |
| LV_mean_ı | Lasso | TRUE | 0.05 | CV | 3 | 21 | 7 | Regression |

| | | | | | | | | |
|---|---|---|---|---|---|---|---|---|
| LV_mean_ | Lasso | TRUE | 0.05 | CV | 4 | 21 | 7 | Regression |
| LV_mean_ | LinRegrs | FALSE | | HeldoutTest | | 21 | 18 | Regression |
| LV_mean_ | Lasso | FALSE | 0.000377 | HeldoutTest | | 21 | 19 | Regression |
| LV_mean_ | Lasso | TRUE | 0.05 | HeldoutTest | | 21 | 6 | Regression |
| LV_mean_ | LinRegrs | FALSE | | CV | 0 | 21 | 18 | Regression |
| LV_mean_ | LinRegrs | FALSE | | CV | 1 | 21 | 20 | Regression |
| LV_mean_ | LinRegrs | FALSE | | CV | 2 | 21 | 18 | Regression |
| LV_mean_ | LinRegrs | FALSE | | CV | 3 | 21 | 19 | Regression |
| LV_mean_ | LinRegrs | FALSE | | CV | 4 | 21 | 17 | Regression |
| LV_mean_ | Lasso | FALSE | 0.000309 | CV | 0 | 21 | 21 | Regression |
| LV_mean_ | Lasso | FALSE | 0.00038 | CV | 1 | 21 | 21 | Regression |
| LV_mean_ | Lasso | FALSE | 0.000301 | CV | 2 | 21 | 20 | Regression |
| LV_mean_ | Lasso | FALSE | 0.000305 | CV | 3 | 21 | 21 | Regression |
| LV_mean_ | Lasso | FALSE | 0.000375 | CV | 4 | 21 | 21 | Regression |
| LV_mean_ | Lasso | TRUE | 0.05 | CV | 0 | 21 | 6 | Regression |
| LV_mean_ | Lasso | TRUE | 0.05 | CV | 1 | 21 | 6 | Regression |
| LV_mean_ | Lasso | TRUE | 0.05 | CV | 2 | 21 | 6 | Regression |
| LV_mean_ | Lasso | TRUE | 0.05 | CV | 3 | 21 | 7 | Regression |
| LV_mean_ | Lasso | TRUE | 0.05 | CV | 4 | 21 | 6 | Regression |
| LV_mean_ | LinRegrs | FALSE | | HeldoutTest | | 21 | 20 | Regression |
| LV_mean_ | Lasso | FALSE | 0.000233 | HeldoutTest | | 21 | 21 | Regression |
| LV_mean_ | Lasso | TRUE | 0.05 | HeldoutTest | | 21 | 5 | Regression |
| LV_mean_ | LinRegrs | FALSE | | CV | 0 | 21 | 20 | Regression |
| LV_mean_ | LinRegrs | FALSE | | CV | 1 | 21 | 21 | Regression |
| LV_mean_ | LinRegrs | FALSE | | CV | 2 | 21 | 20 | Regression |
| LV_mean_ | LinRegrs | FALSE | | CV | 3 | 21 | 20 | Regression |
| LV_mean_ | LinRegrs | FALSE | | CV | 4 | 21 | 20 | Regression |
| LV_mean_ | Lasso | FALSE | 0.000236 | CV | 0 | 21 | 21 | Regression |
| LV_mean_ | Lasso | FALSE | 0.000235 | CV | 1 | 21 | 21 | Regression |
| LV_mean_ | Lasso | FALSE | 0.000229 | CV | 2 | 21 | 21 | Regression |
| LV_mean_ | Lasso | FALSE | 0.000229 | CV | 3 | 21 | 21 | Regression |
| LV_mean_ | Lasso | FALSE | 0.000233 | CV | 4 | 21 | 20 | Regression |
| LV_mean_ | Lasso | TRUE | 0.05 | CV | 0 | 21 | 5 | Regression |
| LV_mean_ | Lasso | TRUE | 0.05 | CV | 1 | 21 | 5 | Regression |
| LV_mean_ | Lasso | TRUE | 0.05 | CV | 2 | 21 | 5 | Regression |
| LV_mean_ | Lasso | TRUE | 0.05 | CV | 3 | 21 | 5 | Regression |
| LV_mean_ | Lasso | TRUE | 0.05 | CV | 4 | 21 | 5 | Regression |
| LV_mean_ | LinRegrs | FALSE | | HeldoutTest | | 21 | 20 | Regression |
| LV_mean_ | Lasso | FALSE | 0.00038 | HeldoutTest | | 21 | 21 | Regression |
| LV_mean_ | Lasso | TRUE | 0.05 | HeldoutTest | | 21 | 7 | Regression |
| LV_mean_ | LinRegrs | FALSE | | CV | 0 | 21 | 19 | Regression |
| LV_mean_ | LinRegrs | FALSE | | CV | 1 | 21 | 18 | Regression |
| LV_mean_ | LinRegrs | FALSE | | CV | 2 | 21 | 20 | Regression |
| LV_mean_ | LinRegrs | FALSE | | CV | 3 | 21 | 19 | Regression |
| LV_mean_ | LinRegrs | FALSE | | CV | 4 | 21 | 19 | Regression |

| | | | | | | | | |
|---|---|---|---|---|---|---|---|---|
| LV_mean_ | Lasso | FALSE | 0.000384 | CV | 0 | 21 | 21 | Regression |
| LV_mean_ | Lasso | FALSE | 0.00038 | CV | 1 | 21 | 21 | Regression |
| LV_mean_ | Lasso | FALSE | 0.00038 | CV | 2 | 21 | 20 | Regression |
| LV_mean_ | Lasso | FALSE | 0.000377 | CV | 3 | 21 | 20 | Regression |
| LV_mean_ | Lasso | FALSE | 0.00038 | CV | 4 | 21 | 20 | Regression |
| LV_mean_ | Lasso | TRUE | 0.05 | CV | 0 | 21 | 7 | Regression |
| LV_mean_ | Lasso | TRUE | 0.05 | CV | 1 | 21 | 7 | Regression |
| LV_mean_ | Lasso | TRUE | 0.05 | CV | 2 | 21 | 7 | Regression |
| LV_mean_ | Lasso | TRUE | 0.05 | CV | 3 | 21 | 7 | Regression |
| LV_mean_ | Lasso | TRUE | 0.05 | CV | 4 | 21 | 7 | Regression |
| LV_mean_ | LinRegrs | FALSE | | HeldoutTest | | 21 | 21 | Regression |
| LV_mean_ | Lasso | FALSE | 0.000414 | HeldoutTest | | 21 | 21 | Regression |
| LV_mean_ | Lasso | TRUE | 0.05 | HeldoutTest | | 21 | 8 | Regression |
| LV_mean_ | LinRegrs | FALSE | | CV | 0 | 21 | 21 | Regression |
| LV_mean_ | LinRegrs | FALSE | | CV | 1 | 21 | 21 | Regression |
| LV_mean_ | LinRegrs | FALSE | | CV | 2 | 21 | 21 | Regression |
| LV_mean_ | LinRegrs | FALSE | | CV | 3 | 21 | 21 | Regression |
| LV_mean_ | LinRegrs | FALSE | | CV | 4 | 21 | 21 | Regression |
| LV_mean_ | Lasso | FALSE | 0.000416 | CV | 0 | 21 | 21 | Regression |
| LV_mean_ | Lasso | FALSE | 0.000416 | CV | 1 | 21 | 21 | Regression |
| LV_mean_ | Lasso | FALSE | 0.000414 | CV | 2 | 21 | 21 | Regression |
| LV_mean_ | Lasso | FALSE | 0.000409 | CV | 3 | 21 | 21 | Regression |
| LV_mean_ | Lasso | FALSE | 0.000417 | CV | 4 | 21 | 21 | Regression |
| LV_mean_ | Lasso | TRUE | 0.05 | CV | 0 | 21 | 8 | Regression |
| LV_mean_ | Lasso | TRUE | 0.05 | CV | 1 | 21 | 8 | Regression |
| LV_mean_ | Lasso | TRUE | 0.05 | CV | 2 | 21 | 8 | Regression |
| LV_mean_ | Lasso | TRUE | 0.05 | CV | 3 | 21 | 8 | Regression |
| LV_mean_ | Lasso | TRUE | 0.05 | CV | 4 | 21 | 8 | Regression |
| LV_mean_ | LinRegrs | FALSE | | HeldoutTest | | 21 | 19 | Regression |
| LV_mean_ | Lasso | FALSE | 0.00041 | HeldoutTest | | 21 | 21 | Regression |
| LV_mean_ | Lasso | TRUE | 0.05 | HeldoutTest | | 21 | 7 | Regression |
| LV_mean_ | LinRegrs | FALSE | | CV | 0 | 21 | 19 | Regression |
| LV_mean_ | LinRegrs | FALSE | | CV | 1 | 21 | 18 | Regression |
| LV_mean_ | LinRegrs | FALSE | | CV | 2 | 21 | 19 | Regression |
| LV_mean_ | LinRegrs | FALSE | | CV | 3 | 21 | 19 | Regression |
| LV_mean_ | LinRegrs | FALSE | | CV | 4 | 21 | 19 | Regression |
| LV_mean_ | Lasso | FALSE | 0.000409 | CV | 0 | 21 | 20 | Regression |
| LV_mean_ | Lasso | FALSE | 0.000411 | CV | 1 | 21 | 20 | Regression |
| LV_mean_ | Lasso | FALSE | 0.00041 | CV | 2 | 21 | 21 | Regression |
| LV_mean_ | Lasso | FALSE | 0.000407 | CV | 3 | 21 | 20 | Regression |
| LV_mean_ | Lasso | FALSE | 0.000411 | CV | 4 | 21 | 21 | Regression |
| LV_mean_ | Lasso | TRUE | 0.05 | CV | 0 | 21 | 7 | Regression |
| LV_mean_ | Lasso | TRUE | 0.05 | CV | 1 | 21 | 7 | Regression |
| LV_mean_ | Lasso | TRUE | 0.05 | CV | 2 | 21 | 8 | Regression |
| LV_mean_ | Lasso | TRUE | 0.05 | CV | 3 | 21 | 7 | Regression |

| | | | | | | | | |
|---|---|---|---|---|---|---|---|---|
| LV_mean_ | Lasso | TRUE | 0.05 | CV | 4 | 21 | 7 | Regression |
| LV_mean_ | LinRegrs | FALSE | | HeldoutTest | | 21 | 20 | Regression |
| LV_mean_ | Lasso | FALSE | 0.000432 | HeldoutTest | | 21 | 21 | Regression |
| LV_mean_ | Lasso | TRUE | 0.05 | HeldoutTest | | 21 | 8 | Regression |
| LV_mean_ | LinRegrs | FALSE | | CV | 0 | 21 | 19 | Regression |
| LV_mean_ | LinRegrs | FALSE | | CV | 1 | 21 | 19 | Regression |
| LV_mean_ | LinRegrs | FALSE | | CV | 2 | 21 | 20 | Regression |
| LV_mean_ | LinRegrs | FALSE | | CV | 3 | 21 | 20 | Regression |
| LV_mean_ | LinRegrs | FALSE | | CV | 4 | 21 | 20 | Regression |
| LV_mean_ | Lasso | FALSE | 0.000435 | CV | 0 | 21 | 21 | Regression |
| LV_mean_ | Lasso | FALSE | 0.00043 | CV | 1 | 21 | 21 | Regression |
| LV_mean_ | Lasso | FALSE | 0.00043 | CV | 2 | 21 | 21 | Regression |
| LV_mean_ | Lasso | FALSE | 0.000429 | CV | 3 | 21 | 20 | Regression |
| LV_mean_ | Lasso | FALSE | 0.000434 | CV | 4 | 21 | 21 | Regression |
| LV_mean_ | Lasso | TRUE | 0.05 | CV | 0 | 21 | 8 | Regression |
| LV_mean_ | Lasso | TRUE | 0.05 | CV | 1 | 21 | 8 | Regression |
| LV_mean_ | Lasso | TRUE | 0.05 | CV | 2 | 21 | 8 | Regression |
| LV_mean_ | Lasso | TRUE | 0.05 | CV | 3 | 21 | 8 | Regression |
| LV_mean_ | Lasso | TRUE | 0.05 | CV | 4 | 21 | 8 | Regression |
| LV_mean_ | LinRegrs | FALSE | | HeldoutTest | | 21 | 20 | Regression |
| LV_mean_ | Lasso | FALSE | 0.000414 | HeldoutTest | | 21 | 20 | Regression |
| LV_mean_ | Lasso | TRUE | 0.05 | HeldoutTest | | 21 | 7 | Regression |
| LV_mean_ | LinRegrs | FALSE | | CV | 0 | 21 | 20 | Regression |
| LV_mean_ | LinRegrs | FALSE | | CV | 1 | 21 | 20 | Regression |
| LV_mean_ | LinRegrs | FALSE | | CV | 2 | 21 | 20 | Regression |
| LV_mean_ | LinRegrs | FALSE | | CV | 3 | 21 | 20 | Regression |
| LV_mean_ | LinRegrs | FALSE | | CV | 4 | 21 | 20 | Regression |
| LV_mean_ | Lasso | FALSE | 0.000417 | CV | 0 | 21 | 20 | Regression |
| LV_mean_ | Lasso | FALSE | 0.000412 | CV | 1 | 21 | 20 | Regression |
| LV_mean_ | Lasso | FALSE | 0.000411 | CV | 2 | 21 | 20 | Regression |
| LV_mean_ | Lasso | FALSE | 0.000413 | CV | 3 | 21 | 20 | Regression |
| LV_mean_ | Lasso | FALSE | 0.000415 | CV | 4 | 21 | 21 | Regression |
| LV_mean_ | Lasso | TRUE | 0.05 | CV | 0 | 21 | 7 | Regression |
| LV_mean_ | Lasso | TRUE | 0.05 | CV | 1 | 21 | 7 | Regression |
| LV_mean_ | Lasso | TRUE | 0.05 | CV | 2 | 21 | 7 | Regression |
| LV_mean_ | Lasso | TRUE | 0.05 | CV | 3 | 21 | 7 | Regression |
| LV_mean_ | Lasso | TRUE | 0.05 | CV | 4 | 21 | 7 | Regression |
| LV_mean_ | LinRegrs | FALSE | | HeldoutTest | | 21 | 21 | Regression |
| LV_mean_ | Lasso | FALSE | 0.000495 | HeldoutTest | | 21 | 21 | Regression |
| LV_mean_ | Lasso | TRUE | 0.05 | HeldoutTest | | 21 | 7 | Regression |
| LV_mean_ | LinRegrs | FALSE | | CV | 0 | 21 | 21 | Regression |
| LV_mean_ | LinRegrs | FALSE | | CV | 1 | 21 | 21 | Regression |
| LV_mean_ | LinRegrs | FALSE | | CV | 2 | 21 | 21 | Regression |
| LV_mean_ | LinRegrs | FALSE | | CV | 3 | 21 | 21 | Regression |
| LV_mean_ | LinRegrs | FALSE | | CV | 4 | 21 | 21 | Regression |

| | | | | | | | | |
|---|---|---|---|---|---|---|---|---|
| LV_mean_ | Lasso | FALSE | 0.000499 | CV | 0 | 21 | 21 | Regression |
| LV_mean_ | Lasso | FALSE | 0.000492 | CV | 1 | 21 | 21 | Regression |
| LV_mean_ | Lasso | FALSE | 0.000496 | CV | 2 | 21 | 21 | Regression |
| LV_mean_ | Lasso | FALSE | 0.000496 | CV | 3 | 21 | 21 | Regression |
| LV_mean_ | Lasso | FALSE | 0.000494 | CV | 4 | 21 | 21 | Regression |
| LV_mean_ | Lasso | TRUE | 0.05 | CV | 0 | 21 | 6 | Regression |
| LV_mean_ | Lasso | TRUE | 0.05 | CV | 1 | 21 | 7 | Regression |
| LV_mean_ | Lasso | TRUE | 0.05 | CV | 2 | 21 | 7 | Regression |
| LV_mean_ | Lasso | TRUE | 0.05 | CV | 3 | 21 | 7 | Regression |
| LV_mean_ | Lasso | TRUE | 0.05 | CV | 4 | 21 | 7 | Regression |
| LV_mean_ | LinRegrs | FALSE | | HeldoutTest | | 21 | 20 | Regression |
| LV_mean_ | Lasso | FALSE | 0.00053 | HeldoutTest | | 21 | 20 | Regression |
| LV_mean_ | Lasso | TRUE | 0.05 | HeldoutTest | | 21 | 8 | Regression |
| LV_mean_ | LinRegrs | FALSE | | CV | 0 | 21 | 20 | Regression |
| LV_mean_ | LinRegrs | FALSE | | CV | 1 | 21 | 20 | Regression |
| LV_mean_ | LinRegrs | FALSE | | CV | 2 | 21 | 20 | Regression |
| LV_mean_ | LinRegrs | FALSE | | CV | 3 | 21 | 20 | Regression |
| LV_mean_ | LinRegrs | FALSE | | CV | 4 | 21 | 21 | Regression |
| LV_mean_ | Lasso | FALSE | 0.00053 | CV | 0 | 21 | 20 | Regression |
| LV_mean_ | Lasso | FALSE | 0.000526 | CV | 1 | 21 | 20 | Regression |
| LV_mean_ | Lasso | FALSE | 0.000531 | CV | 2 | 21 | 20 | Regression |
| LV_mean_ | Lasso | FALSE | 0.00053 | CV | 3 | 21 | 21 | Regression |
| LV_mean_ | Lasso | FALSE | 0.00053 | CV | 4 | 21 | 21 | Regression |
| LV_mean_ | Lasso | TRUE | 0.05 | CV | 0 | 21 | 8 | Regression |
| LV_mean_ | Lasso | TRUE | 0.05 | CV | 1 | 21 | 8 | Regression |
| LV_mean_ | Lasso | TRUE | 0.05 | CV | 2 | 21 | 8 | Regression |
| LV_mean_ | Lasso | TRUE | 0.05 | CV | 3 | 21 | 8 | Regression |
| LV_mean_ | Lasso | TRUE | 0.05 | CV | 4 | 21 | 7 | Regression |
| LV_mean_ | LinRegrs | FALSE | | HeldoutTest | | 21 | 21 | Regression |
| LV_mean_ | Lasso | FALSE | 0.000547 | HeldoutTest | | 21 | 21 | Regression |
| LV_mean_ | Lasso | TRUE | 0.05 | HeldoutTest | | 21 | 7 | Regression |
| LV_mean_ | LinRegrs | FALSE | | CV | 0 | 21 | 21 | Regression |
| LV_mean_ | LinRegrs | FALSE | | CV | 1 | 21 | 21 | Regression |
| LV_mean_ | LinRegrs | FALSE | | CV | 2 | 21 | 21 | Regression |
| LV_mean_ | LinRegrs | FALSE | | CV | 3 | 21 | 21 | Regression |
| LV_mean_ | LinRegrs | FALSE | | CV | 4 | 21 | 21 | Regression |
| LV_mean_ | Lasso | FALSE | 0.00055 | CV | 0 | 21 | 21 | Regression |
| LV_mean_ | Lasso | FALSE | 0.000548 | CV | 1 | 21 | 21 | Regression |
| LV_mean_ | Lasso | FALSE | 0.000545 | CV | 2 | 21 | 21 | Regression |
| LV_mean_ | Lasso | FALSE | 0.000547 | CV | 3 | 21 | 21 | Regression |
| LV_mean_ | Lasso | FALSE | 0.000546 | CV | 4 | 21 | 21 | Regression |
| LV_mean_ | Lasso | TRUE | 0.05 | CV | 0 | 21 | 7 | Regression |
| LV_mean_ | Lasso | TRUE | 0.05 | CV | 1 | 21 | 7 | Regression |
| LV_mean_ | Lasso | TRUE | 0.05 | CV | 2 | 21 | 7 | Regression |
| LV_mean_ | Lasso | TRUE | 0.05 | CV | 3 | 21 | 7 | Regression |

| | | | | | | | | |
|---|---|---|---|---|---|---|---|---|
| LV_mean_ı | Lasso | TRUE | 0.05 | CV | 4 | 21 | 7 | Regression |
| LV_mean_ı | LinRegrs | FALSE | | HeldoutTest | | 21 | 21 | Regression |
| LV_mean_ı | Lasso | FALSE | 0.000487 | HeldoutTest | | 21 | 21 | Regression |
| LV_mean_ı | Lasso | TRUE | 0.05 | HeldoutTest | | 21 | 7 | Regression |
| LV_mean_ı | LinRegrs | FALSE | | CV | 0 | 21 | 20 | Regression |
| LV_mean_ı | LinRegrs | FALSE | | CV | 1 | 21 | 20 | Regression |
| LV_mean_ı | LinRegrs | FALSE | | CV | 2 | 21 | 21 | Regression |
| LV_mean_ı | LinRegrs | FALSE | | CV | 3 | 21 | 21 | Regression |
| LV_mean_ı | LinRegrs | FALSE | | CV | 4 | 21 | 21 | Regression |
| LV_mean_ı | Lasso | FALSE | 0.000489 | CV | 0 | 21 | 21 | Regression |
| LV_mean_ı | Lasso | FALSE | 0.00049 | CV | 1 | 21 | 21 | Regression |
| LV_mean_ı | Lasso | FALSE | 0.000484 | CV | 2 | 21 | 21 | Regression |
| LV_mean_ı | Lasso | FALSE | 0.000487 | CV | 3 | 21 | 21 | Regression |
| LV_mean_ı | Lasso | FALSE | 0.000487 | CV | 4 | 21 | 21 | Regression |
| LV_mean_ı | Lasso | TRUE | 0.05 | CV | 0 | 21 | 7 | Regression |
| LV_mean_ı | Lasso | TRUE | 0.05 | CV | 1 | 21 | 7 | Regression |
| LV_mean_ı | Lasso | TRUE | 0.05 | CV | 2 | 21 | 7 | Regression |
| LV_mean_ı | Lasso | TRUE | 0.05 | CV | 3 | 21 | 7 | Regression |
| LV_mean_ı | Lasso | TRUE | 0.05 | CV | 4 | 21 | 7 | Regression |
| LV_mean_ı | LinRegrs | FALSE | | HeldoutTest | | 21 | 21 | Regression |
| LV_mean_ı | Lasso | FALSE | 0.000443 | HeldoutTest | | 21 | 21 | Regression |
| LV_mean_ı | Lasso | TRUE | 0.05 | HeldoutTest | | 21 | 8 | Regression |
| LV_mean_ı | LinRegrs | FALSE | | CV | 0 | 21 | 21 | Regression |
| LV_mean_ı | LinRegrs | FALSE | | CV | 1 | 21 | 21 | Regression |
| LV_mean_ı | LinRegrs | FALSE | | CV | 2 | 21 | 21 | Regression |
| LV_mean_ı | LinRegrs | FALSE | | CV | 3 | 21 | 21 | Regression |
| LV_mean_ı | LinRegrs | FALSE | | CV | 4 | 21 | 21 | Regression |
| LV_mean_ı | Lasso | FALSE | 0.000444 | CV | 0 | 21 | 21 | Regression |
| LV_mean_ı | Lasso | FALSE | 0.000441 | CV | 1 | 21 | 21 | Regression |
| LV_mean_ı | Lasso | FALSE | 0.000444 | CV | 2 | 21 | 21 | Regression |
| LV_mean_ı | Lasso | FALSE | 0.000442 | CV | 3 | 21 | 21 | Regression |
| LV_mean_ı | Lasso | FALSE | 0.000444 | CV | 4 | 21 | 21 | Regression |
| LV_mean_ı | Lasso | TRUE | 0.05 | CV | 0 | 21 | 8 | Regression |
| LV_mean_ı | Lasso | TRUE | 0.05 | CV | 1 | 21 | 8 | Regression |
| LV_mean_ı | Lasso | TRUE | 0.05 | CV | 2 | 21 | 8 | Regression |
| LV_mean_ı | Lasso | TRUE | 0.05 | CV | 3 | 21 | 8 | Regression |
| LV_mean_ı | Lasso | TRUE | 0.05 | CV | 4 | 21 | 8 | Regression |
| LV_mean_ı | LinRegrs | FALSE | | HeldoutTest | | 21 | 21 | Regression |
| LV_mean_ı | Lasso | FALSE | 0.000493 | HeldoutTest | | 21 | 21 | Regression |
| LV_mean_ı | Lasso | TRUE | 0.05 | HeldoutTest | | 21 | 7 | Regression |
| LV_mean_ı | LinRegrs | FALSE | | CV | 0 | 21 | 21 | Regression |
| LV_mean_ı | LinRegrs | FALSE | | CV | 1 | 21 | 21 | Regression |
| LV_mean_ı | LinRegrs | FALSE | | CV | 2 | 21 | 21 | Regression |
| LV_mean_ı | LinRegrs | FALSE | | CV | 3 | 21 | 21 | Regression |
| LV_mean_ı | LinRegrs | FALSE | | CV | 4 | 21 | 21 | Regression |

| | | | | | | | | |
|---|---|---|---|---|---|---|---|---|
| LV_mean_ı | Lasso | FALSE | 0.000497 | CV | 0 | 21 | 21 | Regression |
| LV_mean_ı | Lasso | FALSE | 0.000491 | CV | 1 | 21 | 21 | Regression |
| LV_mean_ı | Lasso | FALSE | 0.000494 | CV | 2 | 21 | 21 | Regression |
| LV_mean_ı | Lasso | FALSE | 0.00049 | CV | 3 | 21 | 21 | Regression |
| LV_mean_ı | Lasso | FALSE | 0.000491 | CV | 4 | 21 | 21 | Regression |
| LV_mean_ı | Lasso | TRUE | 0.05 | CV | 0 | 21 | 6 | Regression |
| LV_mean_ı | Lasso | TRUE | 0.05 | CV | 1 | 21 | 7 | Regression |
| LV_mean_ı | Lasso | TRUE | 0.05 | CV | 2 | 21 | 6 | Regression |
| LV_mean_ı | Lasso | TRUE | 0.05 | CV | 3 | 21 | 7 | Regression |
| LV_mean_ı | Lasso | TRUE | 0.05 | CV | 4 | 21 | 6 | Regression |
| LV_mean_ı | LinRegrs | FALSE | | HeldoutTest | | 21 | 20 | Regression |
| LV_mean_ı | Lasso | FALSE | 0.000507 | HeldoutTest | | 21 | 21 | Regression |
| LV_mean_ı | Lasso | TRUE | 0.05 | HeldoutTest | | 21 | 7 | Regression |
| LV_mean_ı | LinRegrs | FALSE | | CV | 0 | 21 | 20 | Regression |
| LV_mean_ı | LinRegrs | FALSE | | CV | 1 | 21 | 20 | Regression |
| LV_mean_ı | LinRegrs | FALSE | | CV | 2 | 21 | 20 | Regression |
| LV_mean_ı | LinRegrs | FALSE | | CV | 3 | 21 | 20 | Regression |
| LV_mean_ı | LinRegrs | FALSE | | CV | 4 | 21 | 19 | Regression |
| LV_mean_ı | Lasso | FALSE | 0.00051 | CV | 0 | 21 | 21 | Regression |
| LV_mean_ı | Lasso | FALSE | 0.000502 | CV | 1 | 21 | 21 | Regression |
| LV_mean_ı | Lasso | FALSE | 0.000508 | CV | 2 | 21 | 21 | Regression |
| LV_mean_ı | Lasso | FALSE | 0.000506 | CV | 3 | 21 | 20 | Regression |
| LV_mean_ı | Lasso | FALSE | 0.000508 | CV | 4 | 21 | 20 | Regression |
| LV_mean_ı | Lasso | TRUE | 0.05 | CV | 0 | 21 | 7 | Regression |
| LV_mean_ı | Lasso | TRUE | 0.05 | CV | 1 | 21 | 7 | Regression |
| LV_mean_ı | Lasso | TRUE | 0.05 | CV | 2 | 21 | 7 | Regression |
| LV_mean_ı | Lasso | TRUE | 0.05 | CV | 3 | 21 | 7 | Regression |
| LV_mean_ı | Lasso | TRUE | 0.05 | CV | 4 | 21 | 7 | Regression |
| LV_mean_ı | LinRegrs | FALSE | | HeldoutTest | | 21 | 20 | Regression |
| LV_mean_ı | Lasso | FALSE | 0.000482 | HeldoutTest | | 21 | 20 | Regression |
| LV_mean_ı | Lasso | TRUE | 0.05 | HeldoutTest | | 21 | 6 | Regression |
| LV_mean_ı | LinRegrs | FALSE | | CV | 0 | 21 | 20 | Regression |
| LV_mean_ı | LinRegrs | FALSE | | CV | 1 | 21 | 20 | Regression |
| LV_mean_ı | LinRegrs | FALSE | | CV | 2 | 21 | 19 | Regression |
| LV_mean_ı | LinRegrs | FALSE | | CV | 3 | 21 | 19 | Regression |
| LV_mean_ı | LinRegrs | FALSE | | CV | 4 | 21 | 20 | Regression |
| LV_mean_ı | Lasso | FALSE | 0.000485 | CV | 0 | 21 | 21 | Regression |
| LV_mean_ı | Lasso | FALSE | 0.000477 | CV | 1 | 21 | 21 | Regression |
| LV_mean_ı | Lasso | FALSE | 0.000482 | CV | 2 | 21 | 20 | Regression |
| LV_mean_ı | Lasso | FALSE | 0.000482 | CV | 3 | 21 | 21 | Regression |
| LV_mean_ı | Lasso | FALSE | 0.000482 | CV | 4 | 21 | 20 | Regression |
| LV_mean_ı | Lasso | TRUE | 0.05 | CV | 0 | 21 | 6 | Regression |
| LV_mean_ı | Lasso | TRUE | 0.05 | CV | 1 | 21 | 6 | Regression |
| LV_mean_ı | Lasso | TRUE | 0.05 | CV | 2 | 21 | 6 | Regression |
| LV_mean_ı | Lasso | TRUE | 0.05 | CV | 3 | 21 | 6 | Regression |

| | | | | | | | | |
|---|---|---|---|---|---|---|---|---|
| LV_mean_ı | Lasso | TRUE | 0.05 | CV | 4 | 21 | 7 | Regression |
| LV_mean_ı | LinRegrs | FALSE | | HeldoutTest | | 21 | 21 | Regression |
| LV_mean_ı | Lasso | FALSE | 0.000514 | HeldoutTest | | 21 | 21 | Regression |
| LV_mean_ı | Lasso | TRUE | 0.05 | HeldoutTest | | 21 | 8 | Regression |
| LV_mean_ı | LinRegrs | FALSE | | CV | 0 | 21 | 21 | Regression |
| LV_mean_ı | LinRegrs | FALSE | | CV | 1 | 21 | 21 | Regression |
| LV_mean_ı | LinRegrs | FALSE | | CV | 2 | 21 | 21 | Regression |
| LV_mean_ı | LinRegrs | FALSE | | CV | 3 | 21 | 21 | Regression |
| LV_mean_ı | LinRegrs | FALSE | | CV | 4 | 21 | 21 | Regression |
| LV_mean_ı | Lasso | FALSE | 0.000517 | CV | 0 | 21 | 21 | Regression |
| LV_mean_ı | Lasso | FALSE | 0.000514 | CV | 1 | 21 | 21 | Regression |
| LV_mean_ı | Lasso | FALSE | 0.000513 | CV | 2 | 21 | 21 | Regression |
| LV_mean_ı | Lasso | FALSE | 0.000511 | CV | 3 | 21 | 21 | Regression |
| LV_mean_ı | Lasso | FALSE | 0.000515 | CV | 4 | 21 | 21 | Regression |
| LV_mean_ı | Lasso | TRUE | 0.05 | CV | 0 | 21 | 8 | Regression |
| LV_mean_ı | Lasso | TRUE | 0.05 | CV | 1 | 21 | 8 | Regression |
| LV_mean_ı | Lasso | TRUE | 0.05 | CV | 2 | 21 | 8 | Regression |
| LV_mean_ı | Lasso | TRUE | 0.05 | CV | 3 | 21 | 8 | Regression |
| LV_mean_ı | Lasso | TRUE | 0.05 | CV | 4 | 21 | 8 | Regression |
| LV_circumf | LinRegrs | FALSE | | HeldoutTest | | 21 | 20 | Regression |
| LV_circumf | Lasso | FALSE | 0.000171 | HeldoutTest | | 21 | 21 | Regression |
| LV_circumf | Lasso | TRUE | 0.05 | HeldoutTest | | 21 | 2 | Regression |
| LV_circumf | LinRegrs | FALSE | | CV | 0 | 21 | 18 | Regression |
| LV_circumf | LinRegrs | FALSE | | CV | 1 | 21 | 19 | Regression |
| LV_circumf | LinRegrs | FALSE | | CV | 2 | 21 | 20 | Regression |
| LV_circumf | LinRegrs | FALSE | | CV | 3 | 21 | 19 | Regression |
| LV_circumf | LinRegrs | FALSE | | CV | 4 | 21 | 18 | Regression |
| LV_circumf | Lasso | FALSE | 0.000432 | CV | 0 | 21 | 21 | Regression |
| LV_circumf | Lasso | FALSE | 0.000224 | CV | 1 | 21 | 20 | Regression |
| LV_circumf | Lasso | FALSE | 0.000366 | CV | 2 | 21 | 20 | Regression |
| LV_circumf | Lasso | FALSE | 0.000449 | CV | 3 | 21 | 20 | Regression |
| LV_circumf | Lasso | FALSE | 0.000318 | CV | 4 | 21 | 21 | Regression |
| LV_circumf | Lasso | TRUE | 0.05 | CV | 0 | 21 | 2 | Regression |
| LV_circumf | Lasso | TRUE | 0.05 | CV | 1 | 21 | 2 | Regression |
| LV_circumf | Lasso | TRUE | 0.05 | CV | 2 | 21 | 2 | Regression |
| LV_circumf | Lasso | TRUE | 0.05 | CV | 3 | 21 | 2 | Regression |
| LV_circumf | Lasso | TRUE | 0.05 | CV | 4 | 21 | 2 | Regression |
| LV_circumf | LinRegrs | FALSE | | HeldoutTest | | 21 | 16 | Regression |
| LV_circumf | Lasso | FALSE | 0.000328 | HeldoutTest | | 21 | 21 | Regression |
| LV_circumf | Lasso | TRUE | 0.05 | HeldoutTest | | 21 | 8 | Regression |
| LV_circumf | LinRegrs | FALSE | | CV | 0 | 21 | 18 | Regression |
| LV_circumf | LinRegrs | FALSE | | CV | 1 | 21 | 19 | Regression |
| LV_circumf | LinRegrs | FALSE | | CV | 2 | 21 | 18 | Regression |
| LV_circumf | LinRegrs | FALSE | | CV | 3 | 21 | 16 | Regression |
| LV_circumf | LinRegrs | FALSE | | CV | 4 | 21 | 17 | Regression |

| | | | | | | | | |
|---|---|---|---|---|---|---|---|---|
| LV_circumf | Lasso | FALSE | 0.000329 | CV | 0 | 21 | 20 | Regression |
| LV_circumf | Lasso | FALSE | 0.000374 | CV | 1 | 21 | 20 | Regression |
| LV_circumf | Lasso | FALSE | 0.000435 | CV | 2 | 21 | 19 | Regression |
| LV_circumf | Lasso | FALSE | 0.000807 | CV | 3 | 21 | 18 | Regression |
| LV_circumf | Lasso | FALSE | 0.000407 | CV | 4 | 21 | 21 | Regression |
| LV_circumf | Lasso | TRUE | 0.05 | CV | 0 | 21 | 9 | Regression |
| LV_circumf | Lasso | TRUE | 0.05 | CV | 1 | 21 | 8 | Regression |
| LV_circumf | Lasso | TRUE | 0.05 | CV | 2 | 21 | 9 | Regression |
| LV_circumf | Lasso | TRUE | 0.05 | CV | 3 | 21 | 7 | Regression |
| LV_circumf | Lasso | TRUE | 0.05 | CV | 4 | 21 | 9 | Regression |
| LV_circumf | LinRegrs | FALSE | | HeldoutTest | | 21 | 20 | Regression |
| LV_circumf | Lasso | FALSE | 0.000119 | HeldoutTest | | 21 | 21 | Regression |
| LV_circumf | Lasso | TRUE | 0.05 | HeldoutTest | | 21 | 6 | Regression |
| LV_circumf | LinRegrs | FALSE | | CV | 0 | 21 | 19 | Regression |
| LV_circumf | LinRegrs | FALSE | | CV | 1 | 21 | 19 | Regression |
| LV_circumf | LinRegrs | FALSE | | CV | 2 | 21 | 19 | Regression |
| LV_circumf | LinRegrs | FALSE | | CV | 3 | 21 | 21 | Regression |
| LV_circumf | LinRegrs | FALSE | | CV | 4 | 21 | 18 | Regression |
| LV_circumf | Lasso | FALSE | 0.00027 | CV | 0 | 21 | 21 | Regression |
| LV_circumf | Lasso | FALSE | 9.65E-05 | CV | 1 | 21 | 21 | Regression |
| LV_circumf | Lasso | FALSE | 0.000449 | CV | 2 | 21 | 21 | Regression |
| LV_circumf | Lasso | FALSE | 0.000136 | CV | 3 | 21 | 21 | Regression |
| LV_circumf | Lasso | FALSE | 0.000305 | CV | 4 | 21 | 21 | Regression |
| LV_circumf | Lasso | TRUE | 0.05 | CV | 0 | 21 | 5 | Regression |
| LV_circumf | Lasso | TRUE | 0.05 | CV | 1 | 21 | 5 | Regression |
| LV_circumf | Lasso | TRUE | 0.05 | CV | 2 | 21 | 5 | Regression |
| LV_circumf | Lasso | TRUE | 0.05 | CV | 3 | 21 | 5 | Regression |
| LV_circumf | Lasso | TRUE | 0.05 | CV | 4 | 21 | 6 | Regression |
| LV_circumf | LinRegrs | FALSE | | HeldoutTest | | 21 | 19 | Regression |
| LV_circumf | Lasso | FALSE | 0.00026 | HeldoutTest | | 21 | 21 | Regression |
| LV_circumf | Lasso | TRUE | 0.05 | HeldoutTest | | 21 | 2 | Regression |
| LV_circumf | LinRegrs | FALSE | | CV | 0 | 21 | 19 | Regression |
| LV_circumf | LinRegrs | FALSE | | CV | 1 | 21 | 19 | Regression |
| LV_circumf | LinRegrs | FALSE | | CV | 2 | 21 | 19 | Regression |
| LV_circumf | LinRegrs | FALSE | | CV | 3 | 21 | 19 | Regression |
| LV_circumf | LinRegrs | FALSE | | CV | 4 | 21 | 19 | Regression |
| LV_circumf | Lasso | FALSE | 0.000277 | CV | 0 | 21 | 21 | Regression |
| LV_circumf | Lasso | FALSE | 0.000378 | CV | 1 | 21 | 20 | Regression |
| LV_circumf | Lasso | FALSE | 0.000326 | CV | 2 | 21 | 21 | Regression |
| LV_circumf | Lasso | FALSE | 7.06E-05 | CV | 3 | 21 | 21 | Regression |
| LV_circumf | Lasso | FALSE | 0.000586 | CV | 4 | 21 | 20 | Regression |
| LV_circumf | Lasso | TRUE | 0.05 | CV | 0 | 21 | 2 | Regression |
| LV_circumf | Lasso | TRUE | 0.05 | CV | 1 | 21 | 2 | Regression |
| LV_circumf | Lasso | TRUE | 0.05 | CV | 2 | 21 | 2 | Regression |
| LV_circumf | Lasso | TRUE | 0.05 | CV | 3 | 21 | 2 | Regression |

| | | | | | | | | |
|---|---|---|---|---|---|---|---|---|
| LV_circumf | Lasso | TRUE | 0.05 | CV | 4 | 21 | 2 | Regression |
| LV_circumf | LinRegrs | FALSE | | HeldoutTest | | 21 | 19 | Regression |
| LV_circumf | Lasso | FALSE | 0.000218 | HeldoutTest | | 21 | 20 | Regression |
| LV_circumf | Lasso | TRUE | 0.05 | HeldoutTest | | 21 | 6 | Regression |
| LV_circumf | LinRegrs | FALSE | | CV | 0 | 21 | 19 | Regression |
| LV_circumf | LinRegrs | FALSE | | CV | 1 | 21 | 17 | Regression |
| LV_circumf | LinRegrs | FALSE | | CV | 2 | 21 | 18 | Regression |
| LV_circumf | LinRegrs | FALSE | | CV | 3 | 21 | 19 | Regression |
| LV_circumf | LinRegrs | FALSE | | CV | 4 | 21 | 17 | Regression |
| LV_circumf | Lasso | FALSE | 0.000219 | CV | 0 | 21 | 21 | Regression |
| LV_circumf | Lasso | FALSE | 0.000581 | CV | 1 | 21 | 19 | Regression |
| LV_circumf | Lasso | FALSE | 0.00077 | CV | 2 | 21 | 19 | Regression |
| LV_circumf | Lasso | FALSE | 0.000304 | CV | 3 | 21 | 21 | Regression |
| LV_circumf | Lasso | FALSE | 0.000829 | CV | 4 | 21 | 19 | Regression |
| LV_circumf | Lasso | TRUE | 0.05 | CV | 0 | 21 | 6 | Regression |
| LV_circumf | Lasso | TRUE | 0.05 | CV | 1 | 21 | 6 | Regression |
| LV_circumf | Lasso | TRUE | 0.05 | CV | 2 | 21 | 6 | Regression |
| LV_circumf | Lasso | TRUE | 0.05 | CV | 3 | 21 | 6 | Regression |
| LV_circumf | Lasso | TRUE | 0.05 | CV | 4 | 21 | 6 | Regression |
| LV_circumf | LinRegrs | FALSE | | HeldoutTest | | 21 | 20 | Regression |
| LV_circumf | Lasso | FALSE | 0.000103 | HeldoutTest | | 21 | 21 | Regression |
| LV_circumf | Lasso | TRUE | 0.05 | HeldoutTest | | 21 | 4 | Regression |
| LV_circumf | LinRegrs | FALSE | | CV | 0 | 21 | 21 | Regression |
| LV_circumf | LinRegrs | FALSE | | CV | 1 | 21 | 20 | Regression |
| LV_circumf | LinRegrs | FALSE | | CV | 2 | 21 | 20 | Regression |
| LV_circumf | LinRegrs | FALSE | | CV | 3 | 21 | 20 | Regression |
| LV_circumf | LinRegrs | FALSE | | CV | 4 | 21 | 20 | Regression |
| LV_circumf | Lasso | FALSE | 6.90E-05 | CV | 0 | 21 | 21 | Regression |
| LV_circumf | Lasso | FALSE | 8.77E-05 | CV | 1 | 21 | 21 | Regression |
| LV_circumf | Lasso | FALSE | 0.000131 | CV | 2 | 21 | 21 | Regression |
| LV_circumf | Lasso | FALSE | 7.02E-05 | CV | 3 | 21 | 21 | Regression |
| LV_circumf | Lasso | FALSE | 0.000135 | CV | 4 | 21 | 21 | Regression |
| LV_circumf | Lasso | TRUE | 0.05 | CV | 0 | 21 | 5 | Regression |
| LV_circumf | Lasso | TRUE | 0.05 | CV | 1 | 21 | 5 | Regression |
| LV_circumf | Lasso | TRUE | 0.05 | CV | 2 | 21 | 4 | Regression |
| LV_circumf | Lasso | TRUE | 0.05 | CV | 3 | 21 | 4 | Regression |
| LV_circumf | Lasso | TRUE | 0.05 | CV | 4 | 21 | 4 | Regression |
| LV_circumf | LinRegrs | FALSE | | HeldoutTest | | 21 | 19 | Regression |
| LV_circumf | Lasso | FALSE | 0.000352 | HeldoutTest | | 21 | 20 | Regression |
| LV_circumf | Lasso | TRUE | 0.05 | HeldoutTest | | 21 | 8 | Regression |
| LV_circumf | LinRegrs | FALSE | | CV | 0 | 21 | 20 | Regression |
| LV_circumf | LinRegrs | FALSE | | CV | 1 | 21 | 19 | Regression |
| LV_circumf | LinRegrs | FALSE | | CV | 2 | 21 | 19 | Regression |
| LV_circumf | LinRegrs | FALSE | | CV | 3 | 21 | 19 | Regression |
| LV_circumf | LinRegrs | FALSE | | CV | 4 | 21 | 20 | Regression |

| | | | | | | | |
|---|---|---|---|---|---|---|---|
| LV_circumf | Lasso | FALSE | 0.000205 | CV | 0 | 21 | 21 | Regression |
| LV_circumf | Lasso | FALSE | 0.000432 | CV | 1 | 21 | 20 | Regression |
| LV_circumf | Lasso | FALSE | 0.000201 | CV | 2 | 21 | 21 | Regression |
| LV_circumf | Lasso | FALSE | 0.000346 | CV | 3 | 21 | 20 | Regression |
| LV_circumf | Lasso | FALSE | 0.000202 | CV | 4 | 21 | 21 | Regression |
| LV_circumf | Lasso | TRUE | 0.05 | CV | 0 | 21 | 8 | Regression |
| LV_circumf | Lasso | TRUE | 0.05 | CV | 1 | 21 | 8 | Regression |
| LV_circumf | Lasso | TRUE | 0.05 | CV | 2 | 21 | 8 | Regression |
| LV_circumf | Lasso | TRUE | 0.05 | CV | 3 | 21 | 8 | Regression |
| LV_circumf | Lasso | TRUE | 0.05 | CV | 4 | 21 | 8 | Regression |
| LV_circumf | LinRegrs | FALSE | | HeldoutTest | | 21 | 20 | Regression |
| LV_circumf | Lasso | FALSE | 0.000195 | HeldoutTest | | 21 | 21 | Regression |
| LV_circumf | Lasso | TRUE | 0.05 | HeldoutTest | | 21 | 8 | Regression |
| LV_circumf | LinRegrs | FALSE | | CV | 0 | 21 | 19 | Regression |
| LV_circumf | LinRegrs | FALSE | | CV | 1 | 21 | 20 | Regression |
| LV_circumf | LinRegrs | FALSE | | CV | 2 | 21 | 20 | Regression |
| LV_circumf | LinRegrs | FALSE | | CV | 3 | 21 | 19 | Regression |
| LV_circumf | LinRegrs | FALSE | | CV | 4 | 21 | 21 | Regression |
| LV_circumf | Lasso | FALSE | 0.000245 | CV | 0 | 21 | 20 | Regression |
| LV_circumf | Lasso | FALSE | 0.000197 | CV | 1 | 21 | 21 | Regression |
| LV_circumf | Lasso | FALSE | 0.000273 | CV | 2 | 21 | 21 | Regression |
| LV_circumf | Lasso | FALSE | 0.000196 | CV | 3 | 21 | 21 | Regression |
| LV_circumf | Lasso | FALSE | 0.000202 | CV | 4 | 21 | 21 | Regression |
| LV_circumf | Lasso | TRUE | 0.05 | CV | 0 | 21 | 9 | Regression |
| LV_circumf | Lasso | TRUE | 0.05 | CV | 1 | 21 | 8 | Regression |
| LV_circumf | Lasso | TRUE | 0.05 | CV | 2 | 21 | 8 | Regression |
| LV_circumf | Lasso | TRUE | 0.05 | CV | 3 | 21 | 9 | Regression |
| LV_circumf | Lasso | TRUE | 0.05 | CV | 4 | 21 | 8 | Regression |
| LV_circumf | LinRegrs | FALSE | | HeldoutTest | | 21 | 20 | Regression |
| LV_circumf | Lasso | FALSE | 0.000232 | HeldoutTest | | 21 | 21 | Regression |
| LV_circumf | Lasso | TRUE | 0.05 | HeldoutTest | | 21 | 5 | Regression |
| LV_circumf | LinRegrs | FALSE | | CV | 0 | 21 | 20 | Regression |
| LV_circumf | LinRegrs | FALSE | | CV | 1 | 21 | 21 | Regression |
| LV_circumf | LinRegrs | FALSE | | CV | 2 | 21 | 21 | Regression |
| LV_circumf | LinRegrs | FALSE | | CV | 3 | 21 | 20 | Regression |
| LV_circumf | LinRegrs | FALSE | | CV | 4 | 21 | 20 | Regression |
| LV_circumf | Lasso | FALSE | 0.000208 | CV | 0 | 21 | 21 | Regression |
| LV_circumf | Lasso | FALSE | 0.000144 | CV | 1 | 21 | 21 | Regression |
| LV_circumf | Lasso | FALSE | 0.000231 | CV | 2 | 21 | 21 | Regression |
| LV_circumf | Lasso | FALSE | 0.000296 | CV | 3 | 21 | 21 | Regression |
| LV_circumf | Lasso | FALSE | 0.000305 | CV | 4 | 21 | 21 | Regression |
| LV_circumf | Lasso | TRUE | 0.05 | CV | 0 | 21 | 5 | Regression |
| LV_circumf | Lasso | TRUE | 0.05 | CV | 1 | 21 | 5 | Regression |
| LV_circumf | Lasso | TRUE | 0.05 | CV | 2 | 21 | 5 | Regression |
| LV_circumf | Lasso | TRUE | 0.05 | CV | 3 | 21 | 5 | Regression |

| | | | | | | | | |
|---|---|---|---|---|---|---|---|---|
| LV_circumf | Lasso | TRUE | 0.05 | CV | 4 | 21 | 5 | Regression |
| LV_circumf | LinRegrs | FALSE | | HeldoutTest | | 21 | 17 | Regression |
| LV_circumf | Lasso | FALSE | 0.000348 | HeldoutTest | | 21 | 19 | Regression |
| LV_circumf | Lasso | TRUE | 0.05 | HeldoutTest | | 21 | 2 | Regression |
| LV_circumf | LinRegrs | FALSE | | CV | 0 | 21 | 18 | Regression |
| LV_circumf | LinRegrs | FALSE | | CV | 1 | 21 | 17 | Regression |
| LV_circumf | LinRegrs | FALSE | | CV | 2 | 21 | 17 | Regression |
| LV_circumf | LinRegrs | FALSE | | CV | 3 | 21 | 17 | Regression |
| LV_circumf | LinRegrs | FALSE | | CV | 4 | 21 | 17 | Regression |
| LV_circumf | Lasso | FALSE | 0.000518 | CV | 0 | 21 | 20 | Regression |
| LV_circumf | Lasso | FALSE | 0.000291 | CV | 1 | 21 | 21 | Regression |
| LV_circumf | Lasso | FALSE | 0.0007 | CV | 2 | 21 | 20 | Regression |
| LV_circumf | Lasso | FALSE | 0.000184 | CV | 3 | 21 | 21 | Regression |
| LV_circumf | Lasso | FALSE | 0.000739 | CV | 4 | 21 | 18 | Regression |
| LV_circumf | Lasso | TRUE | 0.05 | CV | 0 | 21 | 3 | Regression |
| LV_circumf | Lasso | TRUE | 0.05 | CV | 1 | 21 | 2 | Regression |
| LV_circumf | Lasso | TRUE | 0.05 | CV | 2 | 21 | 3 | Regression |
| LV_circumf | Lasso | TRUE | 0.05 | CV | 3 | 21 | 3 | Regression |
| LV_circumf | Lasso | TRUE | 0.05 | CV | 4 | 21 | 1 | Regression |
| LV_circumf | LinRegrs | FALSE | | HeldoutTest | | 21 | 18 | Regression |
| LV_circumf | Lasso | FALSE | 0.000272 | HeldoutTest | | 21 | 21 | Regression |
| LV_circumf | Lasso | TRUE | 0.05 | HeldoutTest | | 21 | 4 | Regression |
| LV_circumf | LinRegrs | FALSE | | CV | 0 | 21 | 17 | Regression |
| LV_circumf | LinRegrs | FALSE | | CV | 1 | 21 | 20 | Regression |
| LV_circumf | LinRegrs | FALSE | | CV | 2 | 21 | 19 | Regression |
| LV_circumf | LinRegrs | FALSE | | CV | 3 | 21 | 17 | Regression |
| LV_circumf | LinRegrs | FALSE | | CV | 4 | 21 | 18 | Regression |
| LV_circumf | Lasso | FALSE | 0.000308 | CV | 0 | 21 | 20 | Regression |
| LV_circumf | Lasso | FALSE | 0.000208 | CV | 1 | 21 | 20 | Regression |
| LV_circumf | Lasso | FALSE | 0.000356 | CV | 2 | 21 | 21 | Regression |
| LV_circumf | Lasso | FALSE | 0.000361 | CV | 3 | 21 | 20 | Regression |
| LV_circumf | Lasso | FALSE | 0.000156 | CV | 4 | 21 | 21 | Regression |
| LV_circumf | Lasso | TRUE | 0.05 | CV | 0 | 21 | 4 | Regression |
| LV_circumf | Lasso | TRUE | 0.05 | CV | 1 | 21 | 5 | Regression |
| LV_circumf | Lasso | TRUE | 0.05 | CV | 2 | 21 | 5 | Regression |
| LV_circumf | Lasso | TRUE | 0.05 | CV | 3 | 21 | 4 | Regression |
| LV_circumf | Lasso | TRUE | 0.05 | CV | 4 | 21 | 4 | Regression |
| LV_circumf | LinRegrs | FALSE | | HeldoutTest | | 21 | 21 | Regression |
| LV_circumf | Lasso | FALSE | 0.000173 | HeldoutTest | | 21 | 21 | Regression |
| LV_circumf | Lasso | TRUE | 0.05 | HeldoutTest | | 21 | 7 | Regression |
| LV_circumf | LinRegrs | FALSE | | CV | 0 | 21 | 20 | Regression |
| LV_circumf | LinRegrs | FALSE | | CV | 1 | 21 | 21 | Regression |
| LV_circumf | LinRegrs | FALSE | | CV | 2 | 21 | 20 | Regression |
| LV_circumf | LinRegrs | FALSE | | CV | 3 | 21 | 21 | Regression |
| LV_circumf | LinRegrs | FALSE | | CV | 4 | 21 | 21 | Regression |

| | | | | | | | | |
|---|---|---|---|---|---|---|---|---|
| LV_circumf | Lasso | FALSE | 0.000172 | CV | 0 | 21 | 21 | Regression |
| LV_circumf | Lasso | FALSE | 0.000178 | CV | 1 | 21 | 21 | Regression |
| LV_circumf | Lasso | FALSE | 0.000171 | CV | 2 | 21 | 21 | Regression |
| LV_circumf | Lasso | FALSE | 0.000169 | CV | 3 | 21 | 21 | Regression |
| LV_circumf | Lasso | FALSE | 0.000174 | CV | 4 | 21 | 21 | Regression |
| LV_circumf | Lasso | TRUE | 0.05 | CV | 0 | 21 | 6 | Regression |
| LV_circumf | Lasso | TRUE | 0.05 | CV | 1 | 21 | 6 | Regression |
| LV_circumf | Lasso | TRUE | 0.05 | CV | 2 | 21 | 7 | Regression |
| LV_circumf | Lasso | TRUE | 0.05 | CV | 3 | 21 | 6 | Regression |
| LV_circumf | Lasso | TRUE | 0.05 | CV | 4 | 21 | 7 | Regression |
| LV_circumf | LinRegrs | FALSE | | HeldoutTest | | 21 | 19 | Regression |
| LV_circumf | Lasso | FALSE | 0.00016 | HeldoutTest | | 21 | 21 | Regression |
| LV_circumf | Lasso | TRUE | 0.05 | HeldoutTest | | 21 | 6 | Regression |
| LV_circumf | LinRegrs | FALSE | | CV | 0 | 21 | 20 | Regression |
| LV_circumf | LinRegrs | FALSE | | CV | 1 | 21 | 19 | Regression |
| LV_circumf | LinRegrs | FALSE | | CV | 2 | 21 | 19 | Regression |
| LV_circumf | LinRegrs | FALSE | | CV | 3 | 21 | 20 | Regression |
| LV_circumf | LinRegrs | FALSE | | CV | 4 | 21 | 19 | Regression |
| LV_circumf | Lasso | FALSE | 0.000185 | CV | 0 | 21 | 21 | Regression |
| LV_circumf | Lasso | FALSE | 0.000234 | CV | 1 | 21 | 21 | Regression |
| LV_circumf | Lasso | FALSE | 0.000607 | CV | 2 | 21 | 20 | Regression |
| LV_circumf | Lasso | FALSE | 0.000163 | CV | 3 | 21 | 21 | Regression |
| LV_circumf | Lasso | FALSE | 0.000159 | CV | 4 | 21 | 21 | Regression |
| LV_circumf | Lasso | TRUE | 0.05 | CV | 0 | 21 | 6 | Regression |
| LV_circumf | Lasso | TRUE | 0.05 | CV | 1 | 21 | 6 | Regression |
| LV_circumf | Lasso | TRUE | 0.05 | CV | 2 | 21 | 7 | Regression |
| LV_circumf | Lasso | TRUE | 0.05 | CV | 3 | 21 | 6 | Regression |
| LV_circumf | Lasso | TRUE | 0.05 | CV | 4 | 21 | 6 | Regression |
| LV_circumf | LinRegrs | FALSE | | HeldoutTest | | 21 | 19 | Regression |
| LV_circumf | Lasso | FALSE | 0.000126 | HeldoutTest | | 21 | 21 | Regression |
| LV_circumf | Lasso | TRUE | 0.05 | HeldoutTest | | 21 | 4 | Regression |
| LV_circumf | LinRegrs | FALSE | | CV | 0 | 21 | 19 | Regression |
| LV_circumf | LinRegrs | FALSE | | CV | 1 | 21 | 19 | Regression |
| LV_circumf | LinRegrs | FALSE | | CV | 2 | 21 | 19 | Regression |
| LV_circumf | LinRegrs | FALSE | | CV | 3 | 21 | 19 | Regression |
| LV_circumf | LinRegrs | FALSE | | CV | 4 | 21 | 19 | Regression |
| LV_circumf | Lasso | FALSE | 0.000133 | CV | 0 | 21 | 21 | Regression |
| LV_circumf | Lasso | FALSE | 0.000129 | CV | 1 | 21 | 21 | Regression |
| LV_circumf | Lasso | FALSE | 0.000175 | CV | 2 | 21 | 21 | Regression |
| LV_circumf | Lasso | FALSE | 0.000161 | CV | 3 | 21 | 20 | Regression |
| LV_circumf | Lasso | FALSE | 0.000185 | CV | 4 | 21 | 21 | Regression |
| LV_circumf | Lasso | TRUE | 0.05 | CV | 0 | 21 | 4 | Regression |
| LV_circumf | Lasso | TRUE | 0.05 | CV | 1 | 21 | 5 | Regression |
| LV_circumf | Lasso | TRUE | 0.05 | CV | 2 | 21 | 5 | Regression |
| LV_circumf | Lasso | TRUE | 0.05 | CV | 3 | 21 | 5 | Regression |

| | | | | | | | | |
|---|---|---|---|---|---|---|---|---|
| LV_circumf | Lasso | TRUE | 0.05 | CV | 4 | 21 | 4 | Regression |
| LV_circumf | LinRegrs | FALSE | | HeldoutTest | | 21 | 21 | Regression |
| LV_circumf | Lasso | FALSE | 0.000109 | HeldoutTest | | 21 | 21 | Regression |
| LV_circumf | Lasso | TRUE | 0.05 | HeldoutTest | | 21 | 3 | Regression |
| LV_circumf | LinRegrs | FALSE | | CV | 0 | 21 | 20 | Regression |
| LV_circumf | LinRegrs | FALSE | | CV | 1 | 21 | 20 | Regression |
| LV_circumf | LinRegrs | FALSE | | CV | 2 | 21 | 21 | Regression |
| LV_circumf | LinRegrs | FALSE | | CV | 3 | 21 | 19 | Regression |
| LV_circumf | LinRegrs | FALSE | | CV | 4 | 21 | 19 | Regression |
| LV_circumf | Lasso | FALSE | 0.000145 | CV | 0 | 21 | 21 | Regression |
| LV_circumf | Lasso | FALSE | 0.000111 | CV | 1 | 21 | 21 | Regression |
| LV_circumf | Lasso | FALSE | 0.000163 | CV | 2 | 21 | 21 | Regression |
| LV_circumf | Lasso | FALSE | 0.000151 | CV | 3 | 21 | 21 | Regression |
| LV_circumf | Lasso | FALSE | 0.000452 | CV | 4 | 21 | 20 | Regression |
| LV_circumf | Lasso | TRUE | 0.05 | CV | 0 | 21 | 3 | Regression |
| LV_circumf | Lasso | TRUE | 0.05 | CV | 1 | 21 | 3 | Regression |
| LV_circumf | Lasso | TRUE | 0.05 | CV | 2 | 21 | 3 | Regression |
| LV_circumf | Lasso | TRUE | 0.05 | CV | 3 | 21 | 3 | Regression |
| LV_circumf | Lasso | TRUE | 0.05 | CV | 4 | 21 | 3 | Regression |
| LV_circumf | LinRegrs | FALSE | | HeldoutTest | | 21 | 20 | Regression |
| LV_circumf | Lasso | FALSE | 9.26E-05 | HeldoutTest | | 21 | 21 | Regression |
| LV_circumf | Lasso | TRUE | 0.05 | HeldoutTest | | 21 | 4 | Regression |
| LV_circumf | LinRegrs | FALSE | | CV | 0 | 21 | 20 | Regression |
| LV_circumf | LinRegrs | FALSE | | CV | 1 | 21 | 20 | Regression |
| LV_circumf | LinRegrs | FALSE | | CV | 2 | 21 | 21 | Regression |
| LV_circumf | LinRegrs | FALSE | | CV | 3 | 21 | 19 | Regression |
| LV_circumf | LinRegrs | FALSE | | CV | 4 | 21 | 21 | Regression |
| LV_circumf | Lasso | FALSE | 9.03E-05 | CV | 0 | 21 | 21 | Regression |
| LV_circumf | Lasso | FALSE | 8.06E-05 | CV | 1 | 21 | 21 | Regression |
| LV_circumf | Lasso | FALSE | 8.14E-05 | CV | 2 | 21 | 21 | Regression |
| LV_circumf | Lasso | FALSE | 0.000191 | CV | 3 | 21 | 21 | Regression |
| LV_circumf | Lasso | FALSE | 8.40E-05 | CV | 4 | 21 | 21 | Regression |
| LV_circumf | Lasso | TRUE | 0.05 | CV | 0 | 21 | 4 | Regression |
| LV_circumf | Lasso | TRUE | 0.05 | CV | 1 | 21 | 5 | Regression |
| LV_circumf | Lasso | TRUE | 0.05 | CV | 2 | 21 | 3 | Regression |
| LV_circumf | Lasso | TRUE | 0.05 | CV | 3 | 21 | 5 | Regression |
| LV_circumf | Lasso | TRUE | 0.05 | CV | 4 | 21 | 5 | Regression |
| LV_circumf | LinRegrs | FALSE | | HeldoutTest | | 21 | 21 | Regression |
| LV_circumf | Lasso | FALSE | 0.000219 | HeldoutTest | | 21 | 21 | Regression |
| LV_circumf | Lasso | TRUE | 0.05 | HeldoutTest | | 21 | 9 | Regression |
| LV_circumf | LinRegrs | FALSE | | CV | 0 | 21 | 21 | Regression |
| LV_circumf | LinRegrs | FALSE | | CV | 1 | 21 | 21 | Regression |
| LV_circumf | LinRegrs | FALSE | | CV | 2 | 21 | 20 | Regression |
| LV_circumf | LinRegrs | FALSE | | CV | 3 | 21 | 21 | Regression |
| LV_circumf | LinRegrs | FALSE | | CV | 4 | 21 | 21 | Regression |

| | | | | | | | | |
|---|---|---|---|---|---|---|---|---|
| LV_circumf | Lasso | FALSE | 0.000221 | CV | 0 | 21 | 21 | Regression |
| LV_circumf | Lasso | FALSE | 0.000222 | CV | 1 | 21 | 21 | Regression |
| LV_circumf | Lasso | FALSE | 0.000217 | CV | 2 | 21 | 21 | Regression |
| LV_circumf | Lasso | FALSE | 0.000215 | CV | 3 | 21 | 21 | Regression |
| LV_circumf | Lasso | FALSE | 0.00022 | CV | 4 | 21 | 21 | Regression |
| LV_circumf | Lasso | TRUE | 0.05 | CV | 0 | 21 | 9 | Regression |
| LV_circumf | Lasso | TRUE | 0.05 | CV | 1 | 21 | 9 | Regression |
| LV_circumf | Lasso | TRUE | 0.05 | CV | 2 | 21 | 9 | Regression |
| LV_circumf | Lasso | TRUE | 0.05 | CV | 3 | 21 | 9 | Regression |
| LV_circumf | Lasso | TRUE | 0.05 | CV | 4 | 21 | 9 | Regression |
| RV_end_di | LinRegrs | FALSE | | HeldoutTest | | 21 | 20 | Regression |
| RV_end_di | Lasso | FALSE | 0.000522 | HeldoutTest | | 21 | 20 | Regression |
| RV_end_di | Lasso | TRUE | 0.05 | HeldoutTest | | 21 | 9 | Regression |
| RV_end_di | LinRegrs | FALSE | | CV | 0 | 21 | 20 | Regression |
| RV_end_di | LinRegrs | FALSE | | CV | 1 | 21 | 20 | Regression |
| RV_end_di | LinRegrs | FALSE | | CV | 2 | 21 | 20 | Regression |
| RV_end_di | LinRegrs | FALSE | | CV | 3 | 21 | 21 | Regression |
| RV_end_di | LinRegrs | FALSE | | CV | 4 | 21 | 20 | Regression |
| RV_end_di | Lasso | FALSE | 0.000524 | CV | 0 | 21 | 20 | Regression |
| RV_end_di | Lasso | FALSE | 0.000522 | CV | 1 | 21 | 20 | Regression |
| RV_end_di | Lasso | FALSE | 0.000523 | CV | 2 | 21 | 20 | Regression |
| RV_end_di | Lasso | FALSE | 0.000521 | CV | 3 | 21 | 21 | Regression |
| RV_end_di | Lasso | FALSE | 0.000523 | CV | 4 | 21 | 20 | Regression |
| RV_end_di | Lasso | TRUE | 0.05 | CV | 0 | 21 | 9 | Regression |
| RV_end_di | Lasso | TRUE | 0.05 | CV | 1 | 21 | 9 | Regression |
| RV_end_di | Lasso | TRUE | 0.05 | CV | 2 | 21 | 9 | Regression |
| RV_end_di | Lasso | TRUE | 0.05 | CV | 3 | 21 | 9 | Regression |
| RV_end_di | Lasso | TRUE | 0.05 | CV | 4 | 21 | 9 | Regression |
| RV_end_sy | LinRegrs | FALSE | | HeldoutTest | | 21 | 21 | Regression |
| RV_end_sy | Lasso | FALSE | 0.000495 | HeldoutTest | | 21 | 21 | Regression |
| RV_end_sy | Lasso | TRUE | 0.05 | HeldoutTest | | 21 | 9 | Regression |
| RV_end_sy | LinRegrs | FALSE | | CV | 0 | 21 | 21 | Regression |
| RV_end_sy | LinRegrs | FALSE | | CV | 1 | 21 | 20 | Regression |
| RV_end_sy | LinRegrs | FALSE | | CV | 2 | 21 | 20 | Regression |
| RV_end_sy | LinRegrs | FALSE | | CV | 3 | 21 | 21 | Regression |
| RV_end_sy | LinRegrs | FALSE | | CV | 4 | 21 | 20 | Regression |
| RV_end_sy | Lasso | FALSE | 0.000497 | CV | 0 | 21 | 21 | Regression |
| RV_end_sy | Lasso | FALSE | 0.000494 | CV | 1 | 21 | 20 | Regression |
| RV_end_sy | Lasso | FALSE | 0.000495 | CV | 2 | 21 | 20 | Regression |
| RV_end_sy | Lasso | FALSE | 0.000493 | CV | 3 | 21 | 21 | Regression |
| RV_end_sy | Lasso | FALSE | 0.000495 | CV | 4 | 21 | 21 | Regression |
| RV_end_sy | Lasso | TRUE | 0.05 | CV | 0 | 21 | 9 | Regression |
| RV_end_sy | Lasso | TRUE | 0.05 | CV | 1 | 21 | 9 | Regression |
| RV_end_sy | Lasso | TRUE | 0.05 | CV | 2 | 21 | 9 | Regression |
| RV_end_sy | Lasso | TRUE | 0.05 | CV | 3 | 21 | 9 | Regression |

| | | | | | | | | |
|---|---|---|---|---|---|---|---|---|
| RV_end_sy | Lasso | TRUE | 0.05 | CV | 4 | 21 | 9 | Regression |
| RV_stroke_ | LinRegrs | FALSE | | HeldoutTest | | 21 | 18 | Regression |
| RV_stroke_ | Lasso | FALSE | 0.000798 | HeldoutTest | | 21 | 19 | Regression |
| RV_stroke_ | Lasso | TRUE | 0.05 | HeldoutTest | | 21 | 8 | Regression |
| RV_stroke_ | LinRegrs | FALSE | | CV | 0 | 21 | 18 | Regression |
| RV_stroke_ | LinRegrs | FALSE | | CV | 1 | 21 | 19 | Regression |
| RV_stroke_ | LinRegrs | FALSE | | CV | 2 | 21 | 18 | Regression |
| RV_stroke_ | LinRegrs | FALSE | | CV | 3 | 21 | 18 | Regression |
| RV_stroke_ | LinRegrs | FALSE | | CV | 4 | 21 | 18 | Regression |
| RV_stroke_ | Lasso | FALSE | 0.000565 | CV | 0 | 21 | 21 | Regression |
| RV_stroke_ | Lasso | FALSE | 0.000746 | CV | 1 | 21 | 19 | Regression |
| RV_stroke_ | Lasso | FALSE | 0.000853 | CV | 2 | 21 | 18 | Regression |
| RV_stroke_ | Lasso | FALSE | 0.000739 | CV | 3 | 21 | 21 | Regression |
| RV_stroke_ | Lasso | FALSE | 0.000605 | CV | 4 | 21 | 20 | Regression |
| RV_stroke_ | Lasso | TRUE | 0.05 | CV | 0 | 21 | 8 | Regression |
| RV_stroke_ | Lasso | TRUE | 0.05 | CV | 1 | 21 | 8 | Regression |
| RV_stroke_ | Lasso | TRUE | 0.05 | CV | 2 | 21 | 8 | Regression |
| RV_stroke_ | Lasso | TRUE | 0.05 | CV | 3 | 21 | 8 | Regression |
| RV_stroke_ | Lasso | TRUE | 0.05 | CV | 4 | 21 | 8 | Regression |
| RV_ejectio | LinRegrs | FALSE | | HeldoutTest | | 21 | 20 | Regression |
| RV_ejectio | Lasso | FALSE | 0.000216 | HeldoutTest | | 21 | 21 | Regression |
| RV_ejectio | Lasso | TRUE | 0.05 | HeldoutTest | | 21 | 7 | Regression |
| RV_ejectio | LinRegrs | FALSE | | CV | 0 | 21 | 19 | Regression |
| RV_ejectio | LinRegrs | FALSE | | CV | 1 | 21 | 19 | Regression |
| RV_ejectio | LinRegrs | FALSE | | CV | 2 | 21 | 19 | Regression |
| RV_ejectio | LinRegrs | FALSE | | CV | 3 | 21 | 20 | Regression |
| RV_ejectio | LinRegrs | FALSE | | CV | 4 | 21 | 21 | Regression |
| RV_ejectio | Lasso | FALSE | 0.000313 | CV | 0 | 21 | 21 | Regression |
| RV_ejectio | Lasso | FALSE | 0.000215 | CV | 1 | 21 | 21 | Regression |
| RV_ejectio | Lasso | FALSE | 0.000216 | CV | 2 | 21 | 21 | Regression |
| RV_ejectio | Lasso | FALSE | 0.000284 | CV | 3 | 21 | 20 | Regression |
| RV_ejectio | Lasso | FALSE | 0.000218 | CV | 4 | 21 | 21 | Regression |
| RV_ejectio | Lasso | TRUE | 0.05 | CV | 0 | 21 | 7 | Regression |
| RV_ejectio | Lasso | TRUE | 0.05 | CV | 1 | 21 | 7 | Regression |
| RV_ejectio | Lasso | TRUE | 0.05 | CV | 2 | 21 | 7 | Regression |
| RV_ejectio | Lasso | TRUE | 0.05 | CV | 3 | 21 | 7 | Regression |
| RV_ejectio | Lasso | TRUE | 0.05 | CV | 4 | 21 | 7 | Regression |
| LV_longitu | LinRegrs | FALSE | | HeldoutTest | | 21 | 21 | Regression |
| LV_longitu | Lasso | FALSE | 0.000133 | HeldoutTest | | 21 | 21 | Regression |
| LV_longitu | Lasso | TRUE | 0.05 | HeldoutTest | | 21 | 4 | Regression |
| LV_longitu | LinRegrs | FALSE | | CV | 0 | 21 | 21 | Regression |
| LV_longitu | LinRegrs | FALSE | | CV | 1 | 21 | 21 | Regression |
| LV_longitu | LinRegrs | FALSE | | CV | 2 | 21 | 21 | Regression |
| LV_longitu | LinRegrs | FALSE | | CV | 3 | 21 | 19 | Regression |
| LV_longitu | LinRegrs | FALSE | | CV | 4 | 21 | 21 | Regression |

| | | | | | | | | |
|---|---|---|---|---|---|---|---|---|
| LV_longitu | Lasso | FALSE | 0.000138 | CV | 0 | 21 | 21 | Regression |
| LV_longitu | Lasso | FALSE | 0.000132 | CV | 1 | 21 | 21 | Regression |
| LV_longitu | Lasso | FALSE | 0.000131 | CV | 2 | 21 | 21 | Regression |
| LV_longitu | Lasso | FALSE | 0.000307 | CV | 3 | 21 | 21 | Regression |
| LV_longitu | Lasso | FALSE | 0.000131 | CV | 4 | 21 | 21 | Regression |
| LV_longitu | Lasso | TRUE | 0.05 | CV | 0 | 21 | 5 | Regression |
| LV_longitu | Lasso | TRUE | 0.05 | CV | 1 | 21 | 4 | Regression |
| LV_longitu | Lasso | TRUE | 0.05 | CV | 2 | 21 | 4 | Regression |
| LV_longitu | Lasso | TRUE | 0.05 | CV | 3 | 21 | 4 | Regression |
| LV_longitu | Lasso | TRUE | 0.05 | CV | 4 | 21 | 5 | Regression |
| LV_longitu | LinRegrs | FALSE | | HeldoutTest | | 21 | 20 | Regression |
| LV_longitu | Lasso | FALSE | 0.000292 | HeldoutTest | | 21 | 21 | Regression |
| LV_longitu | Lasso | TRUE | 0.05 | HeldoutTest | | 21 | 4 | Regression |
| LV_longitu | LinRegrs | FALSE | | CV | 0 | 21 | 20 | Regression |
| LV_longitu | LinRegrs | FALSE | | CV | 1 | 21 | 20 | Regression |
| LV_longitu | LinRegrs | FALSE | | CV | 2 | 21 | 20 | Regression |
| LV_longitu | LinRegrs | FALSE | | CV | 3 | 21 | 20 | Regression |
| LV_longitu | LinRegrs | FALSE | | CV | 4 | 21 | 20 | Regression |
| LV_longitu | Lasso | FALSE | 0.00029 | CV | 0 | 21 | 21 | Regression |
| LV_longitu | Lasso | FALSE | 0.000292 | CV | 1 | 21 | 21 | Regression |
| LV_longitu | Lasso | FALSE | 0.000292 | CV | 2 | 21 | 21 | Regression |
| LV_longitu | Lasso | FALSE | 0.000295 | CV | 3 | 21 | 21 | Regression |
| LV_longitu | Lasso | FALSE | 0.000291 | CV | 4 | 21 | 21 | Regression |
| LV_longitu | Lasso | TRUE | 0.05 | CV | 0 | 21 | 4 | Regression |
| LV_longitu | Lasso | TRUE | 0.05 | CV | 1 | 21 | 4 | Regression |
| LV_longitu | Lasso | TRUE | 0.05 | CV | 2 | 21 | 4 | Regression |
| LV_longitu | Lasso | TRUE | 0.05 | CV | 3 | 21 | 4 | Regression |
| LV_longitu | Lasso | TRUE | 0.05 | CV | 4 | 21 | 4 | Regression |
| LV_longitu | LinRegrs | FALSE | | HeldoutTest | | 21 | 20 | Regression |
| LV_longitu | Lasso | FALSE | 0.000338 | HeldoutTest | | 21 | 21 | Regression |
| LV_longitu | Lasso | TRUE | 0.05 | HeldoutTest | | 21 | 9 | Regression |
| LV_longitu | LinRegrs | FALSE | | CV | 0 | 21 | 20 | Regression |
| LV_longitu | LinRegrs | FALSE | | CV | 1 | 21 | 20 | Regression |
| LV_longitu | LinRegrs | FALSE | | CV | 2 | 21 | 20 | Regression |
| LV_longitu | LinRegrs | FALSE | | CV | 3 | 21 | 20 | Regression |
| LV_longitu | LinRegrs | FALSE | | CV | 4 | 21 | 20 | Regression |
| LV_longitu | Lasso | FALSE | 0.000413 | CV | 0 | 21 | 21 | Regression |
| LV_longitu | Lasso | FALSE | 0.000339 | CV | 1 | 21 | 21 | Regression |
| LV_longitu | Lasso | FALSE | 0.000378 | CV | 2 | 21 | 20 | Regression |
| LV_longitu | Lasso | FALSE | 0.000422 | CV | 3 | 21 | 21 | Regression |
| LV_longitu | Lasso | FALSE | 0.000299 | CV | 4 | 21 | 21 | Regression |
| LV_longitu | Lasso | TRUE | 0.05 | CV | 0 | 21 | 9 | Regression |
| LV_longitu | Lasso | TRUE | 0.05 | CV | 1 | 21 | 9 | Regression |
| LV_longitu | Lasso | TRUE | 0.05 | CV | 2 | 21 | 9 | Regression |
| LV_longitu | Lasso | TRUE | 0.05 | CV | 3 | 21 | 8 | Regression |

| | | | | | | | | |
|---|---|---|---|---|---|---|---|---|
| LV_longitu | Lasso | TRUE | 0.05 | CV | 4 | 21 | 9 | Regression |
| LV_longitu | LinRegrs | FALSE | | HeldoutTest | | 21 | 20 | Regression |
| LV_longitu | Lasso | FALSE | 0.000178 | HeldoutTest | | 21 | 21 | Regression |
| LV_longitu | Lasso | TRUE | 0.05 | HeldoutTest | | 21 | 8 | Regression |
| LV_longitu | LinRegrs | FALSE | | CV | 0 | 21 | 19 | Regression |
| LV_longitu | LinRegrs | FALSE | | CV | 1 | 21 | 20 | Regression |
| LV_longitu | LinRegrs | FALSE | | CV | 2 | 21 | 20 | Regression |
| LV_longitu | LinRegrs | FALSE | | CV | 3 | 21 | 19 | Regression |
| LV_longitu | LinRegrs | FALSE | | CV | 4 | 21 | 19 | Regression |
| LV_longitu | Lasso | FALSE | 0.000308 | CV | 0 | 21 | 21 | Regression |
| LV_longitu | Lasso | FALSE | 0.000179 | CV | 1 | 21 | 21 | Regression |
| LV_longitu | Lasso | FALSE | 0.000182 | CV | 2 | 21 | 21 | Regression |
| LV_longitu | Lasso | FALSE | 0.000175 | CV | 3 | 21 | 21 | Regression |
| LV_longitu | Lasso | FALSE | 0.000178 | CV | 4 | 21 | 21 | Regression |
| LV_longitu | Lasso | TRUE | 0.05 | CV | 0 | 21 | 8 | Regression |
| LV_longitu | Lasso | TRUE | 0.05 | CV | 1 | 21 | 8 | Regression |
| LV_longitu | Lasso | TRUE | 0.05 | CV | 2 | 21 | 8 | Regression |
| LV_longitu | Lasso | TRUE | 0.05 | CV | 3 | 21 | 8 | Regression |
| LV_longitu | Lasso | TRUE | 0.05 | CV | 4 | 21 | 7 | Regression |
| LV_longitu | LinRegrs | FALSE | | HeldoutTest | | 21 | 20 | Regression |
| LV_longitu | Lasso | FALSE | 0.000403 | HeldoutTest | | 21 | 21 | Regression |
| LV_longitu | Lasso | TRUE | 0.05 | HeldoutTest | | 21 | 3 | Regression |
| LV_longitu | LinRegrs | FALSE | | CV | 0 | 21 | 20 | Regression |
| LV_longitu | LinRegrs | FALSE | | CV | 1 | 21 | 20 | Regression |
| LV_longitu | LinRegrs | FALSE | | CV | 2 | 21 | 20 | Regression |
| LV_longitu | LinRegrs | FALSE | | CV | 3 | 21 | 18 | Regression |
| LV_longitu | LinRegrs | FALSE | | CV | 4 | 21 | 19 | Regression |
| LV_longitu | Lasso | FALSE | 0.000136 | CV | 0 | 21 | 21 | Regression |
| LV_longitu | Lasso | FALSE | 0.000467 | CV | 1 | 21 | 20 | Regression |
| LV_longitu | Lasso | FALSE | 0.000722 | CV | 2 | 21 | 21 | Regression |
| LV_longitu | Lasso | FALSE | 0.000542 | CV | 3 | 21 | 20 | Regression |
| LV_longitu | Lasso | FALSE | 0.00043 | CV | 4 | 21 | 21 | Regression |
| LV_longitu | Lasso | TRUE | 0.05 | CV | 0 | 21 | 3 | Regression |
| LV_longitu | Lasso | TRUE | 0.05 | CV | 1 | 21 | 3 | Regression |
| LV_longitu | Lasso | TRUE | 0.05 | CV | 2 | 21 | 3 | Regression |
| LV_longitu | Lasso | TRUE | 0.05 | CV | 3 | 21 | 3 | Regression |
| LV_longitu | Lasso | TRUE | 0.05 | CV | 4 | 21 | 3 | Regression |
| LV_longitu | LinRegrs | FALSE | | HeldoutTest | | 21 | 21 | Regression |
| LV_longitu | Lasso | FALSE | 0.000144 | HeldoutTest | | 21 | 21 | Regression |
| LV_longitu | Lasso | TRUE | 0.05 | HeldoutTest | | 21 | 4 | Regression |
| LV_longitu | LinRegrs | FALSE | | CV | 0 | 21 | 20 | Regression |
| LV_longitu | LinRegrs | FALSE | | CV | 1 | 21 | 21 | Regression |
| LV_longitu | LinRegrs | FALSE | | CV | 2 | 21 | 21 | Regression |
| LV_longitu | LinRegrs | FALSE | | CV | 3 | 21 | 21 | Regression |
| LV_longitu | LinRegrs | FALSE | | CV | 4 | 21 | 21 | Regression |

| | | | | | | | | |
|---|---|---|---|---|---|---|---|---|
| LV_longitu | Lasso | FALSE | 0.000176 | CV | 0 | 21 | 21 | Regression |
| LV_longitu | Lasso | FALSE | 0.000147 | CV | 1 | 21 | 21 | Regression |
| LV_longitu | Lasso | FALSE | 0.000145 | CV | 2 | 21 | 21 | Regression |
| LV_longitu | Lasso | FALSE | 0.000145 | CV | 3 | 21 | 21 | Regression |
| LV_longitu | Lasso | FALSE | 0.000139 | CV | 4 | 21 | 21 | Regression |
| LV_longitu | Lasso | TRUE | 0.05 | CV | 0 | 21 | 4 | Regression |
| LV_longitu | Lasso | TRUE | 0.05 | CV | 1 | 21 | 4 | Regression |
| LV_longitu | Lasso | TRUE | 0.05 | CV | 2 | 21 | 4 | Regression |
| LV_longitu | Lasso | TRUE | 0.05 | CV | 3 | 21 | 4 | Regression |
| LV_longitu | Lasso | TRUE | 0.05 | CV | 4 | 21 | 4 | Regression |
| LV_longitu | LinRegrs | FALSE | | HeldoutTest | | 21 | 21 | Regression |
| LV_longitu | Lasso | FALSE | 0.00023 | HeldoutTest | | 21 | 21 | Regression |
| LV_longitu | Lasso | TRUE | 0.05 | HeldoutTest | | 21 | 8 | Regression |
| LV_longitu | LinRegrs | FALSE | | CV | 0 | 21 | 21 | Regression |
| LV_longitu | LinRegrs | FALSE | | CV | 1 | 21 | 21 | Regression |
| LV_longitu | LinRegrs | FALSE | | CV | 2 | 21 | 21 | Regression |
| LV_longitu | LinRegrs | FALSE | | CV | 3 | 21 | 20 | Regression |
| LV_longitu | LinRegrs | FALSE | | CV | 4 | 21 | 20 | Regression |
| LV_longitu | Lasso | FALSE | 0.000223 | CV | 0 | 21 | 21 | Regression |
| LV_longitu | Lasso | FALSE | 0.000233 | CV | 1 | 21 | 21 | Regression |
| LV_longitu | Lasso | FALSE | 0.00023 | CV | 2 | 21 | 21 | Regression |
| LV_longitu | Lasso | FALSE | 0.000231 | CV | 3 | 21 | 21 | Regression |
| LV_longitu | Lasso | FALSE | 0.000234 | CV | 4 | 21 | 20 | Regression |
| LV_longitu | Lasso | TRUE | 0.05 | CV | 0 | 21 | 9 | Regression |
| LV_longitu | Lasso | TRUE | 0.05 | CV | 1 | 21 | 7 | Regression |
| LV_longitu | Lasso | TRUE | 0.05 | CV | 2 | 21 | 7 | Regression |
| LV_longitu | Lasso | TRUE | 0.05 | CV | 3 | 21 | 8 | Regression |
| LV_longitu | Lasso | TRUE | 0.05 | CV | 4 | 21 | 9 | Regression |
| LV_radial_s | LinRegrs | FALSE | | HeldoutTest | | 21 | 19 | Regression |
| LV_radial_s | Lasso | FALSE | 0.000125 | HeldoutTest | | 21 | 21 | Regression |
| LV_radial_s | Lasso | TRUE | 0.05 | HeldoutTest | | 21 | 5 | Regression |
| LV_radial_s | LinRegrs | FALSE | | CV | 0 | 21 | 19 | Regression |
| LV_radial_s | LinRegrs | FALSE | | CV | 1 | 21 | 18 | Regression |
| LV_radial_s | LinRegrs | FALSE | | CV | 2 | 21 | 20 | Regression |
| LV_radial_s | LinRegrs | FALSE | | CV | 3 | 21 | 17 | Regression |
| LV_radial_s | LinRegrs | FALSE | | CV | 4 | 21 | 19 | Regression |
| LV_radial_s | Lasso | FALSE | 0.000353 | CV | 0 | 21 | 21 | Regression |
| LV_radial_s | Lasso | FALSE | 0.000325 | CV | 1 | 21 | 20 | Regression |
| LV_radial_s | Lasso | FALSE | 0.00014 | CV | 2 | 21 | 21 | Regression |
| LV_radial_s | Lasso | FALSE | 0.00039 | CV | 3 | 21 | 21 | Regression |
| LV_radial_s | Lasso | FALSE | 0.000275 | CV | 4 | 21 | 20 | Regression |
| LV_radial_s | Lasso | TRUE | 0.05 | CV | 0 | 21 | 5 | Regression |
| LV_radial_s | Lasso | TRUE | 0.05 | CV | 1 | 21 | 5 | Regression |
| LV_radial_s | Lasso | TRUE | 0.05 | CV | 2 | 21 | 5 | Regression |
| LV_radial_s | Lasso | TRUE | 0.05 | CV | 3 | 21 | 5 | Regression |

| | | | | | | | | |
|---|---|---|---|---|---|---|---|---|
| LV_radial_s | Lasso | TRUE | 0.05 | CV | 4 | 21 | 5 | Regression |
| LV_radial_s | LinRegrs | FALSE | | HeldoutTest | | 21 | 14 | Regression |
| LV_radial_s | Lasso | FALSE | 0.00064 | HeldoutTest | | 21 | 18 | Regression |
| LV_radial_s | Lasso | TRUE | 0.05 | HeldoutTest | | 21 | 7 | Regression |
| LV_radial_s | LinRegrs | FALSE | | CV | 0 | 21 | 15 | Regression |
| LV_radial_s | LinRegrs | FALSE | | CV | 1 | 21 | 15 | Regression |
| LV_radial_s | LinRegrs | FALSE | | CV | 2 | 21 | 16 | Regression |
| LV_radial_s | LinRegrs | FALSE | | CV | 3 | 21 | 15 | Regression |
| LV_radial_s | LinRegrs | FALSE | | CV | 4 | 21 | 15 | Regression |
| LV_radial_s | Lasso | FALSE | 0.00045 | CV | 0 | 21 | 21 | Regression |
| LV_radial_s | Lasso | FALSE | 0.000678 | CV | 1 | 21 | 18 | Regression |
| LV_radial_s | Lasso | FALSE | 0.000853 | CV | 2 | 21 | 18 | Regression |
| LV_radial_s | Lasso | FALSE | 0.000795 | CV | 3 | 21 | 19 | Regression |
| LV_radial_s | Lasso | FALSE | 0.001044 | CV | 4 | 21 | 18 | Regression |
| LV_radial_s | Lasso | TRUE | 0.05 | CV | 0 | 21 | 7 | Regression |
| LV_radial_s | Lasso | TRUE | 0.05 | CV | 1 | 21 | 7 | Regression |
| LV_radial_s | Lasso | TRUE | 0.05 | CV | 2 | 21 | 8 | Regression |
| LV_radial_s | Lasso | TRUE | 0.05 | CV | 3 | 21 | 8 | Regression |
| LV_radial_s | Lasso | TRUE | 0.05 | CV | 4 | 21 | 8 | Regression |
| LV_radial_s | LinRegrs | FALSE | | HeldoutTest | | 21 | 20 | Regression |
| LV_radial_s | Lasso | FALSE | 0.000319 | HeldoutTest | | 21 | 21 | Regression |
| LV_radial_s | Lasso | TRUE | 0.05 | HeldoutTest | | 21 | 7 | Regression |
| LV_radial_s | LinRegrs | FALSE | | CV | 0 | 21 | 20 | Regression |
| LV_radial_s | LinRegrs | FALSE | | CV | 1 | 21 | 20 | Regression |
| LV_radial_s | LinRegrs | FALSE | | CV | 2 | 21 | 20 | Regression |
| LV_radial_s | LinRegrs | FALSE | | CV | 3 | 21 | 20 | Regression |
| LV_radial_s | LinRegrs | FALSE | | CV | 4 | 21 | 20 | Regression |
| LV_radial_s | Lasso | FALSE | 0.000297 | CV | 0 | 21 | 21 | Regression |
| LV_radial_s | Lasso | FALSE | 0.000293 | CV | 1 | 21 | 21 | Regression |
| LV_radial_s | Lasso | FALSE | 0.000298 | CV | 2 | 21 | 21 | Regression |
| LV_radial_s | Lasso | FALSE | 0.00032 | CV | 3 | 21 | 21 | Regression |
| LV_radial_s | Lasso | FALSE | 0.000298 | CV | 4 | 21 | 21 | Regression |
| LV_radial_s | Lasso | TRUE | 0.05 | CV | 0 | 21 | 7 | Regression |
| LV_radial_s | Lasso | TRUE | 0.05 | CV | 1 | 21 | 7 | Regression |
| LV_radial_s | Lasso | TRUE | 0.05 | CV | 2 | 21 | 7 | Regression |
| LV_radial_s | Lasso | TRUE | 0.05 | CV | 3 | 21 | 7 | Regression |
| LV_radial_s | Lasso | TRUE | 0.05 | CV | 4 | 21 | 8 | Regression |
| LV_radial_s | LinRegrs | FALSE | | HeldoutTest | | 21 | 19 | Regression |
| LV_radial_s | Lasso | FALSE | 0.00035 | HeldoutTest | | 21 | 20 | Regression |
| LV_radial_s | Lasso | TRUE | 0.05 | HeldoutTest | | 21 | 8 | Regression |
| LV_radial_s | LinRegrs | FALSE | | CV | 0 | 21 | 19 | Regression |
| LV_radial_s | LinRegrs | FALSE | | CV | 1 | 21 | 20 | Regression |
| LV_radial_s | LinRegrs | FALSE | | CV | 2 | 21 | 19 | Regression |
| LV_radial_s | LinRegrs | FALSE | | CV | 3 | 21 | 19 | Regression |
| LV_radial_s | LinRegrs | FALSE | | CV | 4 | 21 | 19 | Regression |

| | | | | | | | | |
|---|---|---|---|---|---|---|---|---|
| LV_radial_s | Lasso | FALSE | 0.000352 | CV | 0 | 21 | 21 | Regression |
| LV_radial_s | Lasso | FALSE | 0.00033 | CV | 1 | 21 | 21 | Regression |
| LV_radial_s | Lasso | FALSE | 0.000321 | CV | 2 | 21 | 21 | Regression |
| LV_radial_s | Lasso | FALSE | 0.000374 | CV | 3 | 21 | 20 | Regression |
| LV_radial_s | Lasso | FALSE | 0.000404 | CV | 4 | 21 | 20 | Regression |
| LV_radial_s | Lasso | TRUE | 0.05 | CV | 0 | 21 | 7 | Regression |
| LV_radial_s | Lasso | TRUE | 0.05 | CV | 1 | 21 | 8 | Regression |
| LV_radial_s | Lasso | TRUE | 0.05 | CV | 2 | 21 | 8 | Regression |
| LV_radial_s | Lasso | TRUE | 0.05 | CV | 3 | 21 | 7 | Regression |
| LV_radial_s | Lasso | TRUE | 0.05 | CV | 4 | 21 | 8 | Regression |
| LV_radial_s | LinRegrs | FALSE | | HeldoutTest | | 21 | 20 | Regression |
| LV_radial_s | Lasso | FALSE | 0.000268 | HeldoutTest | | 21 | 21 | Regression |
| LV_radial_s | Lasso | TRUE | 0.05 | HeldoutTest | | 21 | 9 | Regression |
| LV_radial_s | LinRegrs | FALSE | | CV | 0 | 21 | 18 | Regression |
| LV_radial_s | LinRegrs | FALSE | | CV | 1 | 21 | 19 | Regression |
| LV_radial_s | LinRegrs | FALSE | | CV | 2 | 21 | 19 | Regression |
| LV_radial_s | LinRegrs | FALSE | | CV | 3 | 21 | 19 | Regression |
| LV_radial_s | LinRegrs | FALSE | | CV | 4 | 21 | 20 | Regression |
| LV_radial_s | Lasso | FALSE | 0.000385 | CV | 0 | 21 | 21 | Regression |
| LV_radial_s | Lasso | FALSE | 0.000309 | CV | 1 | 21 | 20 | Regression |
| LV_radial_s | Lasso | FALSE | 0.000353 | CV | 2 | 21 | 20 | Regression |
| LV_radial_s | Lasso | FALSE | 0.000361 | CV | 3 | 21 | 21 | Regression |
| LV_radial_s | Lasso | FALSE | 0.000585 | CV | 4 | 21 | 21 | Regression |
| LV_radial_s | Lasso | TRUE | 0.05 | CV | 0 | 21 | 9 | Regression |
| LV_radial_s | Lasso | TRUE | 0.05 | CV | 1 | 21 | 9 | Regression |
| LV_radial_s | Lasso | TRUE | 0.05 | CV | 2 | 21 | 9 | Regression |
| LV_radial_s | Lasso | TRUE | 0.05 | CV | 3 | 21 | 9 | Regression |
| LV_radial_s | Lasso | TRUE | 0.05 | CV | 4 | 21 | 9 | Regression |
| LV_radial_s | LinRegrs | FALSE | | HeldoutTest | | 21 | 19 | Regression |
| LV_radial_s | Lasso | FALSE | 0.000326 | HeldoutTest | | 21 | 20 | Regression |
| LV_radial_s | Lasso | TRUE | 0.05 | HeldoutTest | | 21 | 5 | Regression |
| LV_radial_s | LinRegrs | FALSE | | CV | 0 | 21 | 19 | Regression |
| LV_radial_s | LinRegrs | FALSE | | CV | 1 | 21 | 18 | Regression |
| LV_radial_s | LinRegrs | FALSE | | CV | 2 | 21 | 20 | Regression |
| LV_radial_s | LinRegrs | FALSE | | CV | 3 | 21 | 20 | Regression |
| LV_radial_s | LinRegrs | FALSE | | CV | 4 | 21 | 19 | Regression |
| LV_radial_s | Lasso | FALSE | 0.000276 | CV | 0 | 21 | 21 | Regression |
| LV_radial_s | Lasso | FALSE | 0.000376 | CV | 1 | 21 | 20 | Regression |
| LV_radial_s | Lasso | FALSE | 0.000141 | CV | 2 | 21 | 21 | Regression |
| LV_radial_s | Lasso | FALSE | 0.000144 | CV | 3 | 21 | 20 | Regression |
| LV_radial_s | Lasso | FALSE | 0.000327 | CV | 4 | 21 | 20 | Regression |
| LV_radial_s | Lasso | TRUE | 0.05 | CV | 0 | 21 | 6 | Regression |
| LV_radial_s | Lasso | TRUE | 0.05 | CV | 1 | 21 | 6 | Regression |
| LV_radial_s | Lasso | TRUE | 0.05 | CV | 2 | 21 | 5 | Regression |
| LV_radial_s | Lasso | TRUE | 0.05 | CV | 3 | 21 | 5 | Regression |

| | | | | | | | | |
|---|---|---|---|---|---|---|---|---|
| LV_radial_s | Lasso | TRUE | 0.05 | CV | 4 | 21 | 6 | Regression |
| LV_radial_s | LinRegrs | FALSE | | HeldoutTest | | 21 | 20 | Regression |
| LV_radial_s | Lasso | FALSE | 0.00014 | HeldoutTest | | 21 | 21 | Regression |
| LV_radial_s | Lasso | TRUE | 0.05 | HeldoutTest | | 21 | 5 | Regression |
| LV_radial_s | LinRegrs | FALSE | | CV | 0 | 21 | 20 | Regression |
| LV_radial_s | LinRegrs | FALSE | | CV | 1 | 21 | 20 | Regression |
| LV_radial_s | LinRegrs | FALSE | | CV | 2 | 21 | 20 | Regression |
| LV_radial_s | LinRegrs | FALSE | | CV | 3 | 21 | 21 | Regression |
| LV_radial_s | LinRegrs | FALSE | | CV | 4 | 21 | 20 | Regression |
| LV_radial_s | Lasso | FALSE | 0.000158 | CV | 0 | 21 | 20 | Regression |
| LV_radial_s | Lasso | FALSE | 0.000149 | CV | 1 | 21 | 21 | Regression |
| LV_radial_s | Lasso | FALSE | 0.000284 | CV | 2 | 21 | 21 | Regression |
| LV_radial_s | Lasso | FALSE | 0.00013 | CV | 3 | 21 | 21 | Regression |
| LV_radial_s | Lasso | FALSE | 0.00015 | CV | 4 | 21 | 21 | Regression |
| LV_radial_s | Lasso | TRUE | 0.05 | CV | 0 | 21 | 5 | Regression |
| LV_radial_s | Lasso | TRUE | 0.05 | CV | 1 | 21 | 5 | Regression |
| LV_radial_s | Lasso | TRUE | 0.05 | CV | 2 | 21 | 5 | Regression |
| LV_radial_s | Lasso | TRUE | 0.05 | CV | 3 | 21 | 4 | Regression |
| LV_radial_s | Lasso | TRUE | 0.05 | CV | 4 | 21 | 5 | Regression |
| LV_radial_s | LinRegrs | FALSE | | HeldoutTest | | 21 | 19 | Regression |
| LV_radial_s | Lasso | FALSE | 0.000163 | HeldoutTest | | 21 | 20 | Regression |
| LV_radial_s | Lasso | TRUE | 0.05 | HeldoutTest | | 21 | 5 | Regression |
| LV_radial_s | LinRegrs | FALSE | | CV | 0 | 21 | 19 | Regression |
| LV_radial_s | LinRegrs | FALSE | | CV | 1 | 21 | 18 | Regression |
| LV_radial_s | LinRegrs | FALSE | | CV | 2 | 21 | 18 | Regression |
| LV_radial_s | LinRegrs | FALSE | | CV | 3 | 21 | 19 | Regression |
| LV_radial_s | LinRegrs | FALSE | | CV | 4 | 21 | 19 | Regression |
| LV_radial_s | Lasso | FALSE | 0.000151 | CV | 0 | 21 | 21 | Regression |
| LV_radial_s | Lasso | FALSE | 0.000156 | CV | 1 | 21 | 21 | Regression |
| LV_radial_s | Lasso | FALSE | 0.000264 | CV | 2 | 21 | 21 | Regression |
| LV_radial_s | Lasso | FALSE | 0.000151 | CV | 3 | 21 | 21 | Regression |
| LV_radial_s | Lasso | FALSE | 0.00016 | CV | 4 | 21 | 21 | Regression |
| LV_radial_s | Lasso | TRUE | 0.05 | CV | 0 | 21 | 6 | Regression |
| LV_radial_s | Lasso | TRUE | 0.05 | CV | 1 | 21 | 5 | Regression |
| LV_radial_s | Lasso | TRUE | 0.05 | CV | 2 | 21 | 6 | Regression |
| LV_radial_s | Lasso | TRUE | 0.05 | CV | 3 | 21 | 5 | Regression |
| LV_radial_s | Lasso | TRUE | 0.05 | CV | 4 | 21 | 5 | Regression |
| LV_radial_s | LinRegrs | FALSE | | HeldoutTest | | 21 | 21 | Regression |
| LV_radial_s | Lasso | FALSE | 0.000151 | HeldoutTest | | 21 | 21 | Regression |
| LV_radial_s | Lasso | TRUE | 0.05 | HeldoutTest | | 21 | 7 | Regression |
| LV_radial_s | LinRegrs | FALSE | | CV | 0 | 21 | 21 | Regression |
| LV_radial_s | LinRegrs | FALSE | | CV | 1 | 21 | 20 | Regression |
| LV_radial_s | LinRegrs | FALSE | | CV | 2 | 21 | 21 | Regression |
| LV_radial_s | LinRegrs | FALSE | | CV | 3 | 21 | 19 | Regression |
| LV_radial_s | LinRegrs | FALSE | | CV | 4 | 21 | 20 | Regression |

| | | | | | | | | |
|---|---|---|---|---|---|---|---|---|
| LV_radial_s | Lasso | FALSE | 0.000149 | CV | 0 | 21 | 21 | Regression |
| LV_radial_s | Lasso | FALSE | 0.00015 | CV | 1 | 21 | 21 | Regression |
| LV_radial_s | Lasso | FALSE | 0.000153 | CV | 2 | 21 | 21 | Regression |
| LV_radial_s | Lasso | FALSE | 0.000433 | CV | 3 | 21 | 21 | Regression |
| LV_radial_s | Lasso | FALSE | 0.00015 | CV | 4 | 21 | 21 | Regression |
| LV_radial_s | Lasso | TRUE | 0.05 | CV | 0 | 21 | 6 | Regression |
| LV_radial_s | Lasso | TRUE | 0.05 | CV | 1 | 21 | 7 | Regression |
| LV_radial_s | Lasso | TRUE | 0.05 | CV | 2 | 21 | 6 | Regression |
| LV_radial_s | Lasso | TRUE | 0.05 | CV | 3 | 21 | 6 | Regression |
| LV_radial_s | Lasso | TRUE | 0.05 | CV | 4 | 21 | 7 | Regression |
| LV_radial_s | LinRegrs | FALSE | | HeldoutTest | | 21 | 21 | Regression |
| LV_radial_s | Lasso | FALSE | 0.000158 | HeldoutTest | | 21 | 21 | Regression |
| LV_radial_s | Lasso | TRUE | 0.05 | HeldoutTest | | 21 | 9 | Regression |
| LV_radial_s | LinRegrs | FALSE | | CV | 0 | 21 | 20 | Regression |
| LV_radial_s | LinRegrs | FALSE | | CV | 1 | 21 | 21 | Regression |
| LV_radial_s | LinRegrs | FALSE | | CV | 2 | 21 | 19 | Regression |
| LV_radial_s | LinRegrs | FALSE | | CV | 3 | 21 | 20 | Regression |
| LV_radial_s | LinRegrs | FALSE | | CV | 4 | 21 | 20 | Regression |
| LV_radial_s | Lasso | FALSE | 0.000165 | CV | 0 | 21 | 21 | Regression |
| LV_radial_s | Lasso | FALSE | 0.000153 | CV | 1 | 21 | 21 | Regression |
| LV_radial_s | Lasso | FALSE | 0.000401 | CV | 2 | 21 | 21 | Regression |
| LV_radial_s | Lasso | FALSE | 0.000157 | CV | 3 | 21 | 21 | Regression |
| LV_radial_s | Lasso | FALSE | 0.000155 | CV | 4 | 21 | 21 | Regression |
| LV_radial_s | Lasso | TRUE | 0.05 | CV | 0 | 21 | 10 | Regression |
| LV_radial_s | Lasso | TRUE | 0.05 | CV | 1 | 21 | 9 | Regression |
| LV_radial_s | Lasso | TRUE | 0.05 | CV | 2 | 21 | 10 | Regression |
| LV_radial_s | Lasso | TRUE | 0.05 | CV | 3 | 21 | 8 | Regression |
| LV_radial_s | Lasso | TRUE | 0.05 | CV | 4 | 21 | 8 | Regression |
| LV_radial_s | LinRegrs | FALSE | | HeldoutTest | | 21 | 20 | Regression |
| LV_radial_s | Lasso | FALSE | 0.000263 | HeldoutTest | | 21 | 21 | Regression |
| LV_radial_s | Lasso | TRUE | 0.05 | HeldoutTest | | 21 | 7 | Regression |
| LV_radial_s | LinRegrs | FALSE | | CV | 0 | 21 | 20 | Regression |
| LV_radial_s | LinRegrs | FALSE | | CV | 1 | 21 | 19 | Regression |
| LV_radial_s | LinRegrs | FALSE | | CV | 2 | 21 | 19 | Regression |
| LV_radial_s | LinRegrs | FALSE | | CV | 3 | 21 | 19 | Regression |
| LV_radial_s | LinRegrs | FALSE | | CV | 4 | 21 | 20 | Regression |
| LV_radial_s | Lasso | FALSE | 0.000199 | CV | 0 | 21 | 21 | Regression |
| LV_radial_s | Lasso | FALSE | 0.000246 | CV | 1 | 21 | 21 | Regression |
| LV_radial_s | Lasso | FALSE | 0.000195 | CV | 2 | 21 | 19 | Regression |
| LV_radial_s | Lasso | FALSE | 0.000203 | CV | 3 | 21 | 21 | Regression |
| LV_radial_s | Lasso | FALSE | 0.000345 | CV | 4 | 21 | 21 | Regression |
| LV_radial_s | Lasso | TRUE | 0.05 | CV | 0 | 21 | 8 | Regression |
| LV_radial_s | Lasso | TRUE | 0.05 | CV | 1 | 21 | 7 | Regression |
| LV_radial_s | Lasso | TRUE | 0.05 | CV | 2 | 21 | 6 | Regression |
| LV_radial_s | Lasso | TRUE | 0.05 | CV | 3 | 21 | 7 | Regression |

| | | | | | | | | |
|---|---|---|---|---|---|---|---|---|
| LV_radial_s | Lasso | TRUE | 0.05 | CV | 4 | 21 | 7 | Regression |
| LV_radial_s | LinRegrs | FALSE | | HeldoutTest | | 21 | 18 | Regression |
| LV_radial_s | Lasso | FALSE | 0.000266 | HeldoutTest | | 21 | 20 | Regression |
| LV_radial_s | Lasso | TRUE | 0.05 | HeldoutTest | | 21 | 6 | Regression |
| LV_radial_s | LinRegrs | FALSE | | CV | 0 | 21 | 18 | Regression |
| LV_radial_s | LinRegrs | FALSE | | CV | 1 | 21 | 18 | Regression |
| LV_radial_s | LinRegrs | FALSE | | CV | 2 | 21 | 19 | Regression |
| LV_radial_s | LinRegrs | FALSE | | CV | 3 | 21 | 19 | Regression |
| LV_radial_s | LinRegrs | FALSE | | CV | 4 | 21 | 18 | Regression |
| LV_radial_s | Lasso | FALSE | 0.000217 | CV | 0 | 21 | 21 | Regression |
| LV_radial_s | Lasso | FALSE | 0.000178 | CV | 1 | 21 | 21 | Regression |
| LV_radial_s | Lasso | FALSE | 0.000231 | CV | 2 | 21 | 21 | Regression |
| LV_radial_s | Lasso | FALSE | 0.000172 | CV | 3 | 21 | 21 | Regression |
| LV_radial_s | Lasso | FALSE | 0.000234 | CV | 4 | 21 | 21 | Regression |
| LV_radial_s | Lasso | TRUE | 0.05 | CV | 0 | 21 | 6 | Regression |
| LV_radial_s | Lasso | TRUE | 0.05 | CV | 1 | 21 | 6 | Regression |
| LV_radial_s | Lasso | TRUE | 0.05 | CV | 2 | 21 | 6 | Regression |
| LV_radial_s | Lasso | TRUE | 0.05 | CV | 3 | 21 | 6 | Regression |
| LV_radial_s | Lasso | TRUE | 0.05 | CV | 4 | 21 | 6 | Regression |
| LV_radial_s | LinRegrs | FALSE | | HeldoutTest | | 21 | 21 | Regression |
| LV_radial_s | Lasso | FALSE | 0.000168 | HeldoutTest | | 21 | 21 | Regression |
| LV_radial_s | Lasso | TRUE | 0.05 | HeldoutTest | | 21 | 3 | Regression |
| LV_radial_s | LinRegrs | FALSE | | CV | 0 | 21 | 18 | Regression |
| LV_radial_s | LinRegrs | FALSE | | CV | 1 | 21 | 20 | Regression |
| LV_radial_s | LinRegrs | FALSE | | CV | 2 | 21 | 19 | Regression |
| LV_radial_s | LinRegrs | FALSE | | CV | 3 | 21 | 19 | Regression |
| LV_radial_s | LinRegrs | FALSE | | CV | 4 | 21 | 21 | Regression |
| LV_radial_s | Lasso | FALSE | 0.000479 | CV | 0 | 21 | 20 | Regression |
| LV_radial_s | Lasso | FALSE | 0.000347 | CV | 1 | 21 | 21 | Regression |
| LV_radial_s | Lasso | FALSE | 0.000388 | CV | 2 | 21 | 21 | Regression |
| LV_radial_s | Lasso | FALSE | 0.000257 | CV | 3 | 21 | 21 | Regression |
| LV_radial_s | Lasso | FALSE | 0.000173 | CV | 4 | 21 | 21 | Regression |
| LV_radial_s | Lasso | TRUE | 0.05 | CV | 0 | 21 | 3 | Regression |
| LV_radial_s | Lasso | TRUE | 0.05 | CV | 1 | 21 | 2 | Regression |
| LV_radial_s | Lasso | TRUE | 0.05 | CV | 2 | 21 | 3 | Regression |
| LV_radial_s | Lasso | TRUE | 0.05 | CV | 3 | 21 | 3 | Regression |
| LV_radial_s | Lasso | TRUE | 0.05 | CV | 4 | 21 | 3 | Regression |
| LV_radial_s | LinRegrs | FALSE | | HeldoutTest | | 21 | 19 | Regression |
| LV_radial_s | Lasso | FALSE | 0.000154 | HeldoutTest | | 21 | 21 | Regression |
| LV_radial_s | Lasso | TRUE | 0.05 | HeldoutTest | | 21 | 2 | Regression |
| LV_radial_s | LinRegrs | FALSE | | CV | 0 | 21 | 18 | Regression |
| LV_radial_s | LinRegrs | FALSE | | CV | 1 | 21 | 19 | Regression |
| LV_radial_s | LinRegrs | FALSE | | CV | 2 | 21 | 18 | Regression |
| LV_radial_s | LinRegrs | FALSE | | CV | 3 | 21 | 20 | Regression |
| LV_radial_s | LinRegrs | FALSE | | CV | 4 | 21 | 19 | Regression |

| | | | | | | | | |
|---|---|---|---|---|---|---|---|---|
| LV_radial_s | Lasso | FALSE | 0.000236 | CV | 0 | 21 | 21 | Regression |
| LV_radial_s | Lasso | FALSE | 0.00015 | CV | 1 | 21 | 21 | Regression |
| LV_radial_s | Lasso | FALSE | 0.000403 | CV | 2 | 21 | 21 | Regression |
| LV_radial_s | Lasso | FALSE | 0.000155 | CV | 3 | 21 | 21 | Regression |
| LV_radial_s | Lasso | FALSE | 0.000168 | CV | 4 | 21 | 21 | Regression |
| LV_radial_s | Lasso | TRUE | 0.05 | CV | 0 | 21 | 2 | Regression |
| LV_radial_s | Lasso | TRUE | 0.05 | CV | 1 | 21 | 2 | Regression |
| LV_radial_s | Lasso | TRUE | 0.05 | CV | 2 | 21 | 2 | Regression |
| LV_radial_s | Lasso | TRUE | 0.05 | CV | 3 | 21 | 2 | Regression |
| LV_radial_s | Lasso | TRUE | 0.05 | CV | 4 | 21 | 2 | Regression |
| LV_radial_s | LinRegrs | FALSE | | HeldoutTest | | 21 | 21 | Regression |
| LV_radial_s | Lasso | FALSE | 0.000136 | HeldoutTest | | 21 | 21 | Regression |
| LV_radial_s | Lasso | TRUE | 0.05 | HeldoutTest | | 21 | 6 | Regression |
| LV_radial_s | LinRegrs | FALSE | | CV | 0 | 21 | 20 | Regression |
| LV_radial_s | LinRegrs | FALSE | | CV | 1 | 21 | 20 | Regression |
| LV_radial_s | LinRegrs | FALSE | | CV | 2 | 21 | 20 | Regression |
| LV_radial_s | LinRegrs | FALSE | | CV | 3 | 21 | 21 | Regression |
| LV_radial_s | LinRegrs | FALSE | | CV | 4 | 21 | 21 | Regression |
| LV_radial_s | Lasso | FALSE | 0.000176 | CV | 0 | 21 | 21 | Regression |
| LV_radial_s | Lasso | FALSE | 0.000163 | CV | 1 | 21 | 21 | Regression |
| LV_radial_s | Lasso | FALSE | 0.000137 | CV | 2 | 21 | 21 | Regression |
| LV_radial_s | Lasso | FALSE | 0.000134 | CV | 3 | 21 | 21 | Regression |
| LV_radial_s | Lasso | FALSE | 0.000137 | CV | 4 | 21 | 21 | Regression |
| LV_radial_s | Lasso | TRUE | 0.05 | CV | 0 | 21 | 6 | Regression |
| LV_radial_s | Lasso | TRUE | 0.05 | CV | 1 | 21 | 5 | Regression |
| LV_radial_s | Lasso | TRUE | 0.05 | CV | 2 | 21 | 5 | Regression |
| LV_radial_s | Lasso | TRUE | 0.05 | CV | 3 | 21 | 6 | Regression |
| LV_radial_s | Lasso | TRUE | 0.05 | CV | 4 | 21 | 6 | Regression |
| LV_radial_s | LinRegrs | FALSE | | HeldoutTest | | 21 | 18 | Regression |
| LV_radial_s | Lasso | FALSE | 0.000583 | HeldoutTest | | 21 | 21 | Regression |
| LV_radial_s | Lasso | TRUE | 0.05 | HeldoutTest | | 21 | 4 | Regression |
| LV_radial_s | LinRegrs | FALSE | | CV | 0 | 21 | 18 | Regression |
| LV_radial_s | LinRegrs | FALSE | | CV | 1 | 21 | 17 | Regression |
| LV_radial_s | LinRegrs | FALSE | | CV | 2 | 21 | 17 | Regression |
| LV_radial_s | LinRegrs | FALSE | | CV | 3 | 21 | 18 | Regression |
| LV_radial_s | LinRegrs | FALSE | | CV | 4 | 21 | 19 | Regression |
| LV_radial_s | Lasso | FALSE | 0.000503 | CV | 0 | 21 | 21 | Regression |
| LV_radial_s | Lasso | FALSE | 0.000557 | CV | 1 | 21 | 21 | Regression |
| LV_radial_s | Lasso | FALSE | 0.00073 | CV | 2 | 21 | 19 | Regression |
| LV_radial_s | Lasso | FALSE | 0.000424 | CV | 3 | 21 | 21 | Regression |
| LV_radial_s | Lasso | FALSE | 0.000512 | CV | 4 | 21 | 20 | Regression |
| LV_radial_s | Lasso | TRUE | 0.05 | CV | 0 | 21 | 4 | Regression |
| LV_radial_s | Lasso | TRUE | 0.05 | CV | 1 | 21 | 4 | Regression |
| LV_radial_s | Lasso | TRUE | 0.05 | CV | 2 | 21 | 4 | Regression |
| LV_radial_s | Lasso | TRUE | 0.05 | CV | 3 | 21 | 4 | Regression |

| | | | | | | | | |
|---|---|---|---|---|---|---|---|---|
| LV_radial_s | Lasso | TRUE | 0.05 | CV | 4 | 21 | 4 | Regression |
| LV_radial_s | LinRegrs | FALSE | | HeldoutTest | | 21 | 21 | Regression |
| LV_radial_s | Lasso | FALSE | 0.000212 | HeldoutTest | | 21 | 21 | Regression |
| LV_radial_s | Lasso | TRUE | 0.05 | HeldoutTest | | 21 | 7 | Regression |
| LV_radial_s | LinRegrs | FALSE | | CV | 0 | 21 | 21 | Regression |
| LV_radial_s | LinRegrs | FALSE | | CV | 1 | 21 | 20 | Regression |
| LV_radial_s | LinRegrs | FALSE | | CV | 2 | 21 | 21 | Regression |
| LV_radial_s | LinRegrs | FALSE | | CV | 3 | 21 | 21 | Regression |
| LV_radial_s | LinRegrs | FALSE | | CV | 4 | 21 | 20 | Regression |
| LV_radial_s | Lasso | FALSE | 0.000213 | CV | 0 | 21 | 21 | Regression |
| LV_radial_s | Lasso | FALSE | 0.000209 | CV | 1 | 21 | 21 | Regression |
| LV_radial_s | Lasso | FALSE | 0.000215 | CV | 2 | 21 | 21 | Regression |
| LV_radial_s | Lasso | FALSE | 0.000214 | CV | 3 | 21 | 21 | Regression |
| LV_radial_s | Lasso | FALSE | 0.000214 | CV | 4 | 21 | 21 | Regression |
| LV_radial_s | Lasso | TRUE | 0.05 | CV | 0 | 21 | 7 | Regression |
| LV_radial_s | Lasso | TRUE | 0.05 | CV | 1 | 21 | 7 | Regression |
| LV_radial_s | Lasso | TRUE | 0.05 | CV | 2 | 21 | 7 | Regression |
| LV_radial_s | Lasso | TRUE | 0.05 | CV | 3 | 21 | 7 | Regression |
| LV_radial_s | Lasso | TRUE | 0.05 | CV | 4 | 21 | 7 | Regression |
| LA_maximu | LinRegrs | FALSE | | HeldoutTest | | 21 | 20 | Regression |
| LA_maximu | Lasso | FALSE | 0.000464 | HeldoutTest | | 21 | 21 | Regression |
| LA_maximu | Lasso | TRUE | 0.05 | HeldoutTest | | 21 | 10 | Regression |
| LA_maximu | LinRegrs | FALSE | | CV | 0 | 21 | 20 | Regression |
| LA_maximu | LinRegrs | FALSE | | CV | 1 | 21 | 21 | Regression |
| LA_maximu | LinRegrs | FALSE | | CV | 2 | 21 | 20 | Regression |
| LA_maximu | LinRegrs | FALSE | | CV | 3 | 21 | 19 | Regression |
| LA_maximu | LinRegrs | FALSE | | CV | 4 | 21 | 19 | Regression |
| LA_maximu | Lasso | FALSE | 0.000464 | CV | 0 | 21 | 20 | Regression |
| LA_maximu | Lasso | FALSE | 0.000467 | CV | 1 | 21 | 21 | Regression |
| LA_maximu | Lasso | FALSE | 0.000465 | CV | 2 | 21 | 20 | Regression |
| LA_maximu | Lasso | FALSE | 0.000463 | CV | 3 | 21 | 20 | Regression |
| LA_maximu | Lasso | FALSE | 0.000461 | CV | 4 | 21 | 21 | Regression |
| LA_maximu | Lasso | TRUE | 0.05 | CV | 0 | 21 | 10 | Regression |
| LA_maximu | Lasso | TRUE | 0.05 | CV | 1 | 21 | 10 | Regression |
| LA_maximu | Lasso | TRUE | 0.05 | CV | 2 | 21 | 10 | Regression |
| LA_maximu | Lasso | TRUE | 0.05 | CV | 3 | 21 | 10 | Regression |
| LA_maximu | Lasso | TRUE | 0.05 | CV | 4 | 21 | 10 | Regression |
| LA_minimu | LinRegrs | FALSE | | HeldoutTest | | 21 | 20 | Regression |
| LA_minimu | Lasso | FALSE | 0.0004 | HeldoutTest | | 21 | 21 | Regression |
| LA_minimu | Lasso | TRUE | 0.05 | HeldoutTest | | 21 | 8 | Regression |
| LA_minimu | LinRegrs | FALSE | | CV | 0 | 21 | 21 | Regression |
| LA_minimu | LinRegrs | FALSE | | CV | 1 | 21 | 19 | Regression |
| LA_minimu | LinRegrs | FALSE | | CV | 2 | 21 | 20 | Regression |
| LA_minimu | LinRegrs | FALSE | | CV | 3 | 21 | 21 | Regression |
| LA_minimu | LinRegrs | FALSE | | CV | 4 | 21 | 20 | Regression |

| | | | | | | | | |
|---|---|---|---|---|---|---|---|---|
| LA_minimu | Lasso | FALSE | 0.000401 | CV | 0 | 21 | 21 | Regression |
| LA_minimu | Lasso | FALSE | 0.000402 | CV | 1 | 21 | 20 | Regression |
| LA_minimu | Lasso | FALSE | 0.0004 | CV | 2 | 21 | 21 | Regression |
| LA_minimu | Lasso | FALSE | 0.000404 | CV | 3 | 21 | 21 | Regression |
| LA_minimu | Lasso | FALSE | 0.000395 | CV | 4 | 21 | 20 | Regression |
| LA_minimu | Lasso | TRUE | 0.05 | CV | 0 | 21 | 8 | Regression |
| LA_minimu | Lasso | TRUE | 0.05 | CV | 1 | 21 | 9 | Regression |
| LA_minimu | Lasso | TRUE | 0.05 | CV | 2 | 21 | 8 | Regression |
| LA_minimu | Lasso | TRUE | 0.05 | CV | 3 | 21 | 8 | Regression |
| LA_minimu | Lasso | TRUE | 0.05 | CV | 4 | 21 | 8 | Regression |
| LA_stroke_ | LinRegrs | FALSE | | HeldoutTest | | 21 | 19 | Regression |
| LA_stroke_ | Lasso | FALSE | 0.000424 | HeldoutTest | | 21 | 21 | Regression |
| LA_stroke_ | Lasso | TRUE | 0.05 | HeldoutTest | | 21 | 11 | Regression |
| LA_stroke_ | LinRegrs | FALSE | | CV | 0 | 21 | 20 | Regression |
| LA_stroke_ | LinRegrs | FALSE | | CV | 1 | 21 | 19 | Regression |
| LA_stroke_ | LinRegrs | FALSE | | CV | 2 | 21 | 18 | Regression |
| LA_stroke_ | LinRegrs | FALSE | | CV | 3 | 21 | 19 | Regression |
| LA_stroke_ | LinRegrs | FALSE | | CV | 4 | 21 | 19 | Regression |
| LA_stroke_ | Lasso | FALSE | 0.000426 | CV | 0 | 21 | 21 | Regression |
| LA_stroke_ | Lasso | FALSE | 0.000426 | CV | 1 | 21 | 21 | Regression |
| LA_stroke_ | Lasso | FALSE | 0.000426 | CV | 2 | 21 | 21 | Regression |
| LA_stroke_ | Lasso | FALSE | 0.000419 | CV | 3 | 21 | 21 | Regression |
| LA_stroke_ | Lasso | FALSE | 0.000424 | CV | 4 | 21 | 21 | Regression |
| LA_stroke_ | Lasso | TRUE | 0.05 | CV | 0 | 21 | 11 | Regression |
| LA_stroke_ | Lasso | TRUE | 0.05 | CV | 1 | 21 | 11 | Regression |
| LA_stroke_ | Lasso | TRUE | 0.05 | CV | 2 | 21 | 11 | Regression |
| LA_stroke_ | Lasso | TRUE | 0.05 | CV | 3 | 21 | 11 | Regression |
| LA_stroke_ | Lasso | TRUE | 0.05 | CV | 4 | 21 | 11 | Regression |
| LA_ejectio | LinRegrs | FALSE | | HeldoutTest | | 21 | 21 | Regression |
| LA_ejectio | Lasso | FALSE | 0.000203 | HeldoutTest | | 21 | 21 | Regression |
| LA_ejectio | Lasso | TRUE | 0.05 | HeldoutTest | | 21 | 6 | Regression |
| LA_ejectio | LinRegrs | FALSE | | CV | 0 | 21 | 20 | Regression |
| LA_ejectio | LinRegrs | FALSE | | CV | 1 | 21 | 20 | Regression |
| LA_ejectio | LinRegrs | FALSE | | CV | 2 | 21 | 20 | Regression |
| LA_ejectio | LinRegrs | FALSE | | CV | 3 | 21 | 21 | Regression |
| LA_ejectio | LinRegrs | FALSE | | CV | 4 | 21 | 21 | Regression |
| LA_ejectio | Lasso | FALSE | 0.000202 | CV | 0 | 21 | 21 | Regression |
| LA_ejectio | Lasso | FALSE | 0.000202 | CV | 1 | 21 | 21 | Regression |
| LA_ejectio | Lasso | FALSE | 0.000205 | CV | 2 | 21 | 21 | Regression |
| LA_ejectio | Lasso | FALSE | 0.000208 | CV | 3 | 21 | 21 | Regression |
| LA_ejectio | Lasso | FALSE | 0.000198 | CV | 4 | 21 | 21 | Regression |
| LA_ejectio | Lasso | TRUE | 0.05 | CV | 0 | 21 | 6 | Regression |
| LA_ejectio | Lasso | TRUE | 0.05 | CV | 1 | 21 | 6 | Regression |
| LA_ejectio | Lasso | TRUE | 0.05 | CV | 2 | 21 | 6 | Regression |
| LA_ejectio | Lasso | TRUE | 0.05 | CV | 3 | 21 | 6 | Regression |

| | | | | | | | | |
|---|---|---|---|---|---|---|---|---|
| LA_ejectio | Lasso | TRUE | 0.05 | CV | 4 | 21 | 6 | Regression |
| LV_end_di | LinRegrs | FALSE | | HeldoutTest | | 21 | 21 | Regression |
| LV_end_di | Lasso | FALSE | 0.000524 | HeldoutTest | | 21 | 21 | Regression |
| LV_end_di | Lasso | TRUE | 0.05 | HeldoutTest | | 21 | 10 | Regression |
| LV_end_di | LinRegrs | FALSE | | CV | 0 | 21 | 21 | Regression |
| LV_end_di | LinRegrs | FALSE | | CV | 1 | 21 | 21 | Regression |
| LV_end_di | LinRegrs | FALSE | | CV | 2 | 21 | 21 | Regression |
| LV_end_di | LinRegrs | FALSE | | CV | 3 | 21 | 21 | Regression |
| LV_end_di | LinRegrs | FALSE | | CV | 4 | 21 | 21 | Regression |
| LV_end_di | Lasso | FALSE | 0.000526 | CV | 0 | 21 | 21 | Regression |
| LV_end_di | Lasso | FALSE | 0.000525 | CV | 1 | 21 | 21 | Regression |
| LV_end_di | Lasso | FALSE | 0.000522 | CV | 2 | 21 | 21 | Regression |
| LV_end_di | Lasso | FALSE | 0.000523 | CV | 3 | 21 | 21 | Regression |
| LV_end_di | Lasso | FALSE | 0.000526 | CV | 4 | 21 | 21 | Regression |
| LV_end_di | Lasso | TRUE | 0.05 | CV | 0 | 21 | 10 | Regression |
| LV_end_di | Lasso | TRUE | 0.05 | CV | 1 | 21 | 10 | Regression |
| LV_end_di | Lasso | TRUE | 0.05 | CV | 2 | 21 | 10 | Regression |
| LV_end_di | Lasso | TRUE | 0.05 | CV | 3 | 21 | 10 | Regression |
| LV_end_di | Lasso | TRUE | 0.05 | CV | 4 | 21 | 10 | Regression |
| LV_end_sy | LinRegrs | FALSE | | HeldoutTest | | 21 | 21 | Regression |
| LV_end_sy | Lasso | FALSE | 0.000442 | HeldoutTest | | 21 | 21 | Regression |
| LV_end_sy | Lasso | TRUE | 0.05 | HeldoutTest | | 21 | 10 | Regression |
| LV_end_sy | LinRegrs | FALSE | | CV | 0 | 21 | 21 | Regression |
| LV_end_sy | LinRegrs | FALSE | | CV | 1 | 21 | 20 | Regression |
| LV_end_sy | LinRegrs | FALSE | | CV | 2 | 21 | 20 | Regression |
| LV_end_sy | LinRegrs | FALSE | | CV | 3 | 21 | 21 | Regression |
| LV_end_sy | LinRegrs | FALSE | | CV | 4 | 21 | 21 | Regression |
| LV_end_sy | Lasso | FALSE | 0.000443 | CV | 0 | 21 | 21 | Regression |
| LV_end_sy | Lasso | FALSE | 0.000445 | CV | 1 | 21 | 21 | Regression |
| LV_end_sy | Lasso | FALSE | 0.000441 | CV | 2 | 21 | 21 | Regression |
| LV_end_sy | Lasso | FALSE | 0.000443 | CV | 3 | 21 | 21 | Regression |
| LV_end_sy | Lasso | FALSE | 0.000441 | CV | 4 | 21 | 21 | Regression |
| LV_end_sy | Lasso | TRUE | 0.05 | CV | 0 | 21 | 10 | Regression |
| LV_end_sy | Lasso | TRUE | 0.05 | CV | 1 | 21 | 10 | Regression |
| LV_end_sy | Lasso | TRUE | 0.05 | CV | 2 | 21 | 10 | Regression |
| LV_end_sy | Lasso | TRUE | 0.05 | CV | 3 | 21 | 10 | Regression |
| LV_end_sy | Lasso | TRUE | 0.05 | CV | 4 | 21 | 10 | Regression |
| LV_stroke_ | LinRegrs | FALSE | | HeldoutTest | | 21 | 20 | Regression |
| LV_stroke_ | Lasso | FALSE | 0.000484 | HeldoutTest | | 21 | 21 | Regression |
| LV_stroke_ | Lasso | TRUE | 0.05 | HeldoutTest | | 21 | 8 | Regression |
| LV_stroke_ | LinRegrs | FALSE | | CV | 0 | 21 | 21 | Regression |
| LV_stroke_ | LinRegrs | FALSE | | CV | 1 | 21 | 21 | Regression |
| LV_stroke_ | LinRegrs | FALSE | | CV | 2 | 21 | 20 | Regression |
| LV_stroke_ | LinRegrs | FALSE | | CV | 3 | 21 | 20 | Regression |
| LV_stroke_ | LinRegrs | FALSE | | CV | 4 | 21 | 20 | Regression |

| | | | | | | | | |
|---|---|---|---|---|---|---|---|---|
| LV_stroke_ | Lasso | FALSE | 0.000485 | CV | 0 | 21 | 21 | Regression |
| LV_stroke_ | Lasso | FALSE | 0.000482 | CV | 1 | 21 | 21 | Regression |
| LV_stroke_ | Lasso | FALSE | 0.000482 | CV | 2 | 21 | 21 | Regression |
| LV_stroke_ | Lasso | FALSE | 0.000482 | CV | 3 | 21 | 20 | Regression |
| LV_stroke_ | Lasso | FALSE | 0.000486 | CV | 4 | 21 | 21 | Regression |
| LV_stroke_ | Lasso | TRUE | 0.05 | CV | 0 | 21 | 9 | Regression |
| LV_stroke_ | Lasso | TRUE | 0.05 | CV | 1 | 21 | 9 | Regression |
| LV_stroke_ | Lasso | TRUE | 0.05 | CV | 2 | 21 | 9 | Regression |
| LV_stroke_ | Lasso | TRUE | 0.05 | CV | 3 | 21 | 9 | Regression |
| LV_stroke_ | Lasso | TRUE | 0.05 | CV | 4 | 21 | 8 | Regression |
| LV_ejectior | LinRegrs | FALSE | | HeldoutTest | | 21 | 20 | Regression |
| LV_ejectior | Lasso | FALSE | 0.000158 | HeldoutTest | | 21 | 21 | Regression |
| LV_ejectior | Lasso | TRUE | 0.05 | HeldoutTest | | 21 | 9 | Regression |
| LV_ejectior | LinRegrs | FALSE | | CV | 0 | 21 | 20 | Regression |
| LV_ejectior | LinRegrs | FALSE | | CV | 1 | 21 | 20 | Regression |
| LV_ejectior | LinRegrs | FALSE | | CV | 2 | 21 | 20 | Regression |
| LV_ejectior | LinRegrs | FALSE | | CV | 3 | 21 | 20 | Regression |
| LV_ejectior | LinRegrs | FALSE | | CV | 4 | 21 | 20 | Regression |
| LV_ejectior | Lasso | FALSE | 0.000157 | CV | 0 | 21 | 21 | Regression |
| LV_ejectior | Lasso | FALSE | 0.000159 | CV | 1 | 21 | 21 | Regression |
| LV_ejectior | Lasso | FALSE | 0.000158 | CV | 2 | 21 | 21 | Regression |
| LV_ejectior | Lasso | FALSE | 0.000162 | CV | 3 | 21 | 21 | Regression |
| LV_ejectior | Lasso | FALSE | 0.000164 | CV | 4 | 21 | 20 | Regression |
| LV_ejectior | Lasso | TRUE | 0.05 | CV | 0 | 21 | 9 | Regression |
| LV_ejectior | Lasso | TRUE | 0.05 | CV | 1 | 21 | 9 | Regression |
| LV_ejectior | Lasso | TRUE | 0.05 | CV | 2 | 21 | 9 | Regression |
| LV_ejectior | Lasso | TRUE | 0.05 | CV | 3 | 21 | 9 | Regression |
| LV_ejectior | Lasso | TRUE | 0.05 | CV | 4 | 21 | 9 | Regression |
| LV_cardiac | LinRegrs | FALSE | | HeldoutTest | | 21 | 19 | Regression |
| LV_cardiac | Lasso | FALSE | 0.00031 | HeldoutTest | | 21 | 21 | Regression |
| LV_cardiac | Lasso | TRUE | 0.05 | HeldoutTest | | 21 | 10 | Regression |
| LV_cardiac | LinRegrs | FALSE | | CV | 0 | 21 | 20 | Regression |
| LV_cardiac | LinRegrs | FALSE | | CV | 1 | 21 | 19 | Regression |
| LV_cardiac | LinRegrs | FALSE | | CV | 2 | 21 | 20 | Regression |
| LV_cardiac | LinRegrs | FALSE | | CV | 3 | 21 | 20 | Regression |
| LV_cardiac | LinRegrs | FALSE | | CV | 4 | 21 | 20 | Regression |
| LV_cardiac | Lasso | FALSE | 0.000312 | CV | 0 | 21 | 21 | Regression |
| LV_cardiac | Lasso | FALSE | 0.000411 | CV | 1 | 21 | 20 | Regression |
| LV_cardiac | Lasso | FALSE | 0.000313 | CV | 2 | 21 | 20 | Regression |
| LV_cardiac | Lasso | FALSE | 0.000303 | CV | 3 | 21 | 21 | Regression |
| LV_cardiac | Lasso | FALSE | 0.000309 | CV | 4 | 21 | 21 | Regression |
| LV_cardiac | Lasso | TRUE | 0.05 | CV | 0 | 21 | 10 | Regression |
| LV_cardiac | Lasso | TRUE | 0.05 | CV | 1 | 21 | 11 | Regression |
| LV_cardiac | Lasso | TRUE | 0.05 | CV | 2 | 21 | 11 | Regression |
| LV_cardiac | Lasso | TRUE | 0.05 | CV | 3 | 21 | 11 | Regression |

| | | | | | | | | |
|---|---|---|---|---|---|---|---|---|
| LV_cardiac | Lasso | TRUE | 0.05 | CV | 4 | 21 | 10 | Regression |

| MSE | R-squared | ClassifRprt | ClassifRprt | ClassifRprt | ClassifRprt | TrainSize | TestSize |
|---|---|---|---|---|---|---|---|
| 0.95325 | 0.09325 | | | | | 47679 | 7501 |
| 0.953068 | 0.093423 | | | | | 47679 | 7501 |
| 0.998939 | 0.04979 | | | | | 47679 | 7501 |
| 0.869399 | 0.132703 | | | | | 38143 | 9536 |
| 0.83145 | 0.156683 | | | | | 38143 | 9536 |
| 0.831661 | 0.164057 | | | | | 38143 | 9536 |
| 0.850448 | 0.155678 | | | | | 38143 | 9536 |
| 0.851373 | 0.15617 | | | | | 38144 | 9535 |
| 0.869379 | 0.132723 | | | | | 38143 | 9536 |
| 0.831446 | 0.156688 | | | | | 38143 | 9536 |
| 0.831693 | 0.164026 | | | | | 38143 | 9536 |
| 0.850218 | 0.155906 | | | | | 38143 | 9536 |
| 0.851471 | 0.156072 | | | | | 38144 | 9535 |
| 0.918767 | 0.083454 | | | | | 38143 | 9536 |
| 0.886752 | 0.100592 | | | | | 38143 | 9536 |
| 0.89351 | 0.10189 | | | | | 38143 | 9536 |
| 0.905391 | 0.10113 | | | | | 38143 | 9536 |
| 0.907982 | 0.100062 | | | | | 38144 | 9535 |
| 0.527641 | 0.504794 | | | | | 47679 | 7501 |
| 0.527681 | 0.504756 | | | | | 47679 | 7501 |
| 0.597574 | 0.439159 | | | | | 47679 | 7501 |
| 0.440148 | 0.547159 | | | | | 38143 | 9536 |
| 0.460426 | 0.540314 | | | | | 38143 | 9536 |
| 0.487924 | 0.520388 | | | | | 38143 | 9536 |
| 0.483196 | 0.520023 | | | | | 38143 | 9536 |
| 0.488467 | 0.512639 | | | | | 38144 | 9535 |
| 0.440219 | 0.547086 | | | | | 38143 | 9536 |
| 0.460627 | 0.540114 | | | | | 38143 | 9536 |
| 0.487885 | 0.520427 | | | | | 38143 | 9536 |
| 0.483179 | 0.52004 | | | | | 38143 | 9536 |
| 0.488487 | 0.512619 | | | | | 38144 | 9535 |
| 0.472987 | 0.513374 | | | | | 38143 | 9536 |
| 0.534691 | 0.466169 | | | | | 38143 | 9536 |
| 0.553853 | 0.455582 | | | | | 38143 | 9536 |
| 0.546651 | 0.45699 | | | | | 38143 | 9536 |
| 0.552799 | 0.448452 | | | | | 38144 | 9535 |
| 0.947588 | 0.097059 | | | | | 47679 | 7501 |
| 0.947368 | 0.097269 | | | | | 47679 | 7501 |
| 0.995677 | 0.051236 | | | | | 47679 | 7501 |
| 0.867449 | 0.130891 | | | | | 38143 | 9536 |
| 0.837568 | 0.153773 | | | | | 38143 | 9536 |
| 0.835022 | 0.161473 | | | | | 38143 | 9536 |
| 0.851316 | 0.153463 | | | | | 38143 | 9536 |
| 0.85396 | 0.154635 | | | | | 38144 | 9535 |

| | | | |
|---|---|---|---|
| 0.867393 | 0.130948 | 38143 | 9536 |
| 0.837578 | 0.153762 | 38143 | 9536 |
| 0.835038 | 0.161457 | 38143 | 9536 |
| 0.851088 | 0.153689 | 38143 | 9536 |
| 0.854042 | 0.154555 | 38144 | 9535 |
| 0.916118 | 0.08213 | 38143 | 9536 |
| 0.891995 | 0.098783 | 38143 | 9536 |
| 0.895732 | 0.100509 | 38143 | 9536 |
| 0.905466 | 0.099617 | 38143 | 9536 |
| 0.910731 | 0.098436 | 38144 | 9535 |
| 0.934138 | 0.125821 | 47679 | 7501 |
| 0.934038 | 0.125915 | 47679 | 7501 |
| 0.989527 | 0.073987 | 47679 | 7501 |
| 0.857711 | 0.149634 | 38143 | 9536 |
| 0.810387 | 0.170578 | 38143 | 9536 |
| 0.804146 | 0.185871 | 38143 | 9536 |
| 0.832581 | 0.176248 | 38143 | 9536 |
| 0.835977 | 0.176773 | 38144 | 9535 |
| 0.857711 | 0.149634 | 38143 | 9536 |
| 0.810378 | 0.170587 | 38143 | 9536 |
| 0.804287 | 0.185728 | 38143 | 9536 |
| 0.832467 | 0.176361 | 38143 | 9536 |
| 0.836018 | 0.176733 | 38144 | 9535 |
| 0.916959 | 0.090894 | 38143 | 9536 |
| 0.869511 | 0.110065 | 38143 | 9536 |
| 0.87721 | 0.111899 | 38143 | 9536 |
| 0.897587 | 0.111932 | 38143 | 9536 |
| 0.90239 | 0.111373 | 38144 | 9535 |
| 0.904105 | 0.093779 | 47679 | 7501 |
| 0.904125 | 0.093759 | 47679 | 7501 |
| 0.949058 | 0.048721 | 47679 | 7501 |
| 0.895419 | 0.093937 | 38143 | 9536 |
| 0.914649 | 0.083544 | 38143 | 9536 |
| 0.906061 | 0.095988 | 38143 | 9536 |
| 0.923962 | 0.084053 | 38143 | 9536 |
| 0.911597 | 0.090577 | 38144 | 9535 |
| 0.895548 | 0.093806 | 38143 | 9536 |
| 0.914498 | 0.083696 | 38143 | 9536 |
| 0.906155 | 0.095894 | 38143 | 9536 |
| 0.923903 | 0.084111 | 38143 | 9536 |
| 0.911563 | 0.090612 | 38144 | 9535 |
| 0.935163 | 0.05372 | 38143 | 9536 |
| 0.94676 | 0.05137 | 38143 | 9536 |
| 0.953063 | 0.049092 | 38143 | 9536 |
| 0.957412 | 0.050892 | 38143 | 9536 |

| | | | |
|---|---|---|---|
| 0.951464 | 0.050806 | 38144 | 9535 |
| 0.406574 | 0.610418 | 47679 | 7501 |
| 0.406472 | 0.610516 | 47679 | 7501 |
| 0.446936 | 0.571744 | 47679 | 7501 |
| 0.358889 | 0.640776 | 38143 | 9536 |
| 0.377027 | 0.626759 | 38143 | 9536 |
| 0.370049 | 0.626659 | 38143 | 9536 |
| 0.38286 | 0.619039 | 38143 | 9536 |
| 0.377747 | 0.620101 | 38144 | 9535 |
| 0.359015 | 0.64065 | 38143 | 9536 |
| 0.377031 | 0.626755 | 38143 | 9536 |
| 0.369966 | 0.626742 | 38143 | 9536 |
| 0.383068 | 0.618831 | 38143 | 9536 |
| 0.377773 | 0.620075 | 38144 | 9535 |
| 0.397509 | 0.60212 | 38143 | 9536 |
| 0.418291 | 0.585909 | 38143 | 9536 |
| 0.407756 | 0.588616 | 38143 | 9536 |
| 0.424097 | 0.578006 | 38143 | 9536 |
| 0.41531 | 0.582324 | 38144 | 9535 |
| 0.410962 | 0.594979 | 47679 | 7501 |
| 0.410922 | 0.595018 | 47679 | 7501 |
| 0.454411 | 0.552158 | 47679 | 7501 |
| 0.38156 | 0.619123 | 38143 | 9536 |
| 0.399807 | 0.608565 | 38143 | 9536 |
| 0.389714 | 0.603827 | 38143 | 9536 |
| 0.393868 | 0.604813 | 38143 | 9536 |
| 0.391431 | 0.607052 | 38144 | 9535 |
| 0.381711 | 0.618972 | 38143 | 9536 |
| 0.399866 | 0.608507 | 38143 | 9536 |
| 0.389617 | 0.603926 | 38143 | 9536 |
| 0.393907 | 0.604774 | 38143 | 9536 |
| 0.391519 | 0.606964 | 38144 | 9535 |
| 0.416071 | 0.584674 | 38143 | 9536 |
| 0.450582 | 0.558854 | 38143 | 9536 |
| 0.4317 | 0.561146 | 38143 | 9536 |
| 0.438202 | 0.56033 | 38143 | 9536 |
| 0.439366 | 0.558932 | 38144 | 9535 |
| 0.686377 | 0.370471 | 47679 | 7501 |
| 0.686561 | 0.370302 | 47679 | 7501 |
| 0.759064 | 0.303804 | 47679 | 7501 |
| 0.616139 | 0.358831 | 38143 | 9536 |
| 0.635084 | 0.358971 | 38143 | 9536 |
| 0.656053 | 0.356308 | 38143 | 9536 |
| 0.659896 | 0.346727 | 38143 | 9536 |
| 0.671275 | 0.34143 | 38144 | 9535 |

| | | | |
|---|---|---|---|
| 0.616074 | 0.358899 | 38143 | 9536 |
| 0.635099 | 0.358955 | 38143 | 9536 |
| 0.656135 | 0.356227 | 38143 | 9536 |
| 0.659859 | 0.346763 | 38143 | 9536 |
| 0.671358 | 0.341348 | 38144 | 9535 |
| 0.65572 | 0.317643 | 38143 | 9536 |
| 0.692871 | 0.300642 | 38143 | 9536 |
| 0.718878 | 0.294667 | 38143 | 9536 |
| 0.714729 | 0.292444 | 38143 | 9536 |
| 0.729994 | 0.283822 | 38144 | 9535 |
| 0.517805 | 0.493095 | 47679 | 7501 |
| 0.51773 | 0.493169 | 47679 | 7501 |
| 0.58217 | 0.430086 | 47679 | 7501 |
| 0.488933 | 0.505743 | 38143 | 9536 |
| 0.494122 | 0.509133 | 38143 | 9536 |
| 0.498455 | 0.502175 | 38143 | 9536 |
| 0.505249 | 0.502439 | 38143 | 9536 |
| 0.496674 | 0.496994 | 38144 | 9535 |
| 0.488978 | 0.505698 | 38143 | 9536 |
| 0.494216 | 0.509039 | 38143 | 9536 |
| 0.498453 | 0.502177 | 38143 | 9536 |
| 0.505382 | 0.502308 | 38143 | 9536 |
| 0.496599 | 0.49707 | 38144 | 9535 |
| 0.544552 | 0.449519 | 38143 | 9536 |
| 0.562054 | 0.441648 | 38143 | 9536 |
| 0.562316 | 0.438394 | 38143 | 9536 |
| 0.568328 | 0.44032 | 38143 | 9536 |
| 0.554361 | 0.438571 | 38144 | 9535 |
| 0.742432 | 0.270464 | 47679 | 7501 |
| 0.742425 | 0.27047 | 47679 | 7501 |
| 0.789598 | 0.224116 | 47679 | 7501 |
| 0.706775 | 0.2725 | 38143 | 9536 |
| 0.731077 | 0.288351 | 38143 | 9536 |
| 0.715403 | 0.277965 | 38143 | 9536 |
| 0.738129 | 0.281854 | 38143 | 9536 |
| 0.714815 | 0.272632 | 38144 | 9535 |
| 0.706729 | 0.272547 | 38143 | 9536 |
| 0.731157 | 0.288273 | 38143 | 9536 |
| 0.71537 | 0.277997 | 38143 | 9536 |
| 0.738184 | 0.2818 | 38143 | 9536 |
| 0.714771 | 0.272677 | 38144 | 9535 |
| 0.753176 | 0.224738 | 38143 | 9536 |
| 0.789296 | 0.23168 | 38143 | 9536 |
| 0.766603 | 0.22629 | 38143 | 9536 |
| 0.790683 | 0.230723 | 38143 | 9536 |

| | | | | | | | | |
|---|---|---|---|---|---|---|---|---|
| | 0.761566 | 0.225061 | | | | | 38144 | 9535 |
| | 0.678163 | 0.368791 | | | | | 47679 | 7501 |
| | 0.678026 | 0.368918 | | | | | 47679 | 7501 |
| | 0.738387 | 0.312736 | | | | | 47679 | 7501 |
| | 0.59864 | 0.382462 | | | | | 38143 | 9536 |
| | 0.638484 | 0.385345 | | | | | 38143 | 9536 |
| | 0.601624 | 0.387741 | | | | | 38143 | 9536 |
| | 0.621686 | 0.387632 | | | | | 38143 | 9536 |
| | 0.619023 | 0.377528 | | | | | 38144 | 9535 |
| | 0.598618 | 0.382485 | | | | | 38143 | 9536 |
| | 0.638619 | 0.385215 | | | | | 38143 | 9536 |
| | 0.601534 | 0.387832 | | | | | 38143 | 9536 |
| | 0.621856 | 0.387465 | | | | | 38143 | 9536 |
| | 0.618987 | 0.377564 | | | | | 38144 | 9535 |
| | 0.65098 | 0.328471 | | | | | 38143 | 9536 |
| | 0.706461 | 0.319906 | | | | | 38143 | 9536 |
| | 0.659517 | 0.328824 | | | | | 38143 | 9536 |
| | 0.683342 | 0.3269 | | | | | 38143 | 9536 |
| | 0.675144 | 0.321094 | | | | | 38144 | 9535 |
| | 0.408012 | 0.610194 | | | | | 47679 | 7501 |
| | 0.408144 | 0.610069 | | | | | 47679 | 7501 |
| | 0.450609 | 0.569498 | | | | | 47679 | 7501 |
| | 0.348207 | 0.650217 | | | | | 38143 | 9536 |
| | 0.369781 | 0.633438 | | | | | 38143 | 9536 |
| | 0.370246 | 0.628444 | | | | | 38143 | 9536 |
| | 0.378368 | 0.622592 | | | | | 38143 | 9536 |
| | 0.375688 | 0.622924 | | | | | 38144 | 9535 |
| | 0.348314 | 0.65011 | | | | | 38143 | 9536 |
| | 0.369783 | 0.633437 | | | | | 38143 | 9536 |
| | 0.370095 | 0.628596 | | | | | 38143 | 9536 |
| | 0.378592 | 0.622368 | | | | | 38143 | 9536 |
| | 0.375713 | 0.6229 | | | | | 38144 | 9535 |
| | 0.38291 | 0.615358 | | | | | 38143 | 9536 |
| | 0.412192 | 0.591397 | | | | | 38143 | 9536 |
| | 0.408561 | 0.589994 | | | | | 38143 | 9536 |
| | 0.419982 | 0.581083 | | | | | 38143 | 9536 |
| | 0.415797 | 0.582667 | | | | | 38144 | 9535 |
| on | | | 0.918811 | 0.919515 | 0.918811 | 0.9188 | 47679 | 7501 |
| on | | | 0.918678 | 0.919391 | 0.918678 | 0.918666 | 47679 | 7501 |
| on | | | 0.918011 | 0.91877 | 0.918011 | 0.917998 | 47679 | 7501 |
| on | | | 0.524853 | 0.486733 | 0.524853 | 0.370739 | 38143 | 9536 |
| on | | | 0.526951 | 0.523468 | 0.526951 | 0.382154 | 38143 | 9536 |
| on | | | 0.526216 | 0.514136 | 0.526216 | 0.383191 | 38143 | 9536 |
| on | | | 0.525902 | 0.510005 | 0.525902 | 0.378472 | 38143 | 9536 |
| on | | | 0.524069 | 0.480967 | 0.524069 | 0.373221 | 38144 | 9535 |

| | | | | | | |
|---|---|---|---|---|---|---|
| on | 0.526216 | 0.750711 | 0.526216 | 0.362978 | 38143 | 9536 |
| on | 0.526531 | 0.51884 | 0.526531 | 0.381268 | 38143 | 9536 |
| on | 0.526216 | 0.276904 | 0.526216 | 0.362863 | 38143 | 9536 |
| on | 0.526216 | 0.276904 | 0.526216 | 0.362863 | 38143 | 9536 |
| on | 0.526691 | 0.583612 | 0.526691 | 0.365095 | 38144 | 9535 |
| on | 0.526321 | 0.75074 | 0.526321 | 0.363212 | 38143 | 9536 |
| on | 0.526846 | 0.554445 | 0.526846 | 0.367524 | 38143 | 9536 |
| on | 0.526216 | 0.513842 | 0.526216 | 0.365835 | 38143 | 9536 |
| on | 0.526112 | 0.502518 | 0.526112 | 0.364678 | 38143 | 9536 |
| on | 0.526481 | 0.547727 | 0.526481 | 0.365184 | 38144 | 9535 |
| on | 0.9976 | 0.995206 | 0.9976 | 0.996402 | 47679 | 7501 |
| on | 0.9976 | 0.995206 | 0.9976 | 0.996402 | 47679 | 7501 |
| on | 0.9976 | 0.995206 | 0.9976 | 0.996402 | 47679 | 7501 |
| on | 0.997588 | 0.995182 | 0.997588 | 0.996384 | 38143 | 9536 |
| on | 0.997588 | 0.995182 | 0.997588 | 0.996384 | 38143 | 9536 |
| on | 0.997483 | 0.994973 | 0.997483 | 0.996226 | 38143 | 9536 |
| on | 0.997483 | 0.994973 | 0.997483 | 0.996226 | 38143 | 9536 |
| on | 0.997588 | 0.995181 | 0.997588 | 0.996383 | 38144 | 9535 |
| on | 0.997588 | 0.995182 | 0.997588 | 0.996384 | 38143 | 9536 |
| on | 0.997588 | 0.995182 | 0.997588 | 0.996384 | 38143 | 9536 |
| on | 0.997483 | 0.994973 | 0.997483 | 0.996226 | 38143 | 9536 |
| on | 0.997483 | 0.994973 | 0.997483 | 0.996226 | 38143 | 9536 |
| on | 0.997588 | 0.995181 | 0.997588 | 0.996383 | 38144 | 9535 |
| on | 0.997588 | 0.995182 | 0.997588 | 0.996384 | 38143 | 9536 |
| on | 0.997588 | 0.995182 | 0.997588 | 0.996384 | 38143 | 9536 |
| on | 0.997483 | 0.994973 | 0.997483 | 0.996226 | 38143 | 9536 |
| on | 0.997483 | 0.994973 | 0.997483 | 0.996226 | 38143 | 9536 |
| on | 0.997588 | 0.995181 | 0.997588 | 0.996383 | 38144 | 9535 |
| on | 0.085589 | 0.080121 | 0.085589 | 0.066705 | 47679 | 7501 |
| on | 0.085989 | 0.074126 | 0.085989 | 0.04468 | 47679 | 7501 |
| on | 0.085855 | 0.074199 | 0.085855 | 0.045884 | 47679 | 7501 |
| on | 0.082424 | 0.081004 | 0.082424 | 0.066631 | 38143 | 9536 |
| on | 0.089346 | 0.086803 | 0.089346 | 0.066749 | 38143 | 9536 |
| on | 0.084522 | 0.066584 | 0.084522 | 0.063809 | 38143 | 9536 |
| on | 0.086619 | 0.09447 | 0.086619 | 0.066491 | 38143 | 9536 |
| on | 0.083797 | 0.080172 | 0.083797 | 0.069817 | 38144 | 9535 |
| on | 0.088926 | 0.091744 | 0.088926 | 0.04913 | 38143 | 9536 |
| on | 0.089555 | 0.00802 | 0.089555 | 0.014722 | 38143 | 9536 |
| on | 0.090185 | 0.008133 | 0.090185 | 0.014921 | 38143 | 9536 |
| on | 0.090185 | 0.008133 | 0.090185 | 0.014921 | 38143 | 9536 |
| on | 0.090194 | 0.008135 | 0.090194 | 0.014924 | 38144 | 9535 |
| on | 0.088612 | 0.092313 | 0.088612 | 0.050311 | 38143 | 9536 |
| on | 0.090919 | 0.068339 | 0.090919 | 0.047913 | 38143 | 9536 |
| on | 0.089451 | 0.064286 | 0.089451 | 0.050067 | 38143 | 9536 |
| on | 0.094065 | 0.078835 | 0.094065 | 0.043641 | 38143 | 9536 |

| | | | | | | |
|---|---|---|---|---|---|---|
| on | 0.089984 | 0.064243 | 0.089984 | 0.04746 | 38144 | 9535 |
| on | 0.592054 | 0.55041 | 0.592054 | 0.459098 | 47679 | 7501 |
| on | 0.591388 | 0.545626 | 0.591388 | 0.457701 | 47679 | 7501 |
| on | 0.590988 | 0.545455 | 0.590988 | 0.456385 | 47679 | 7501 |
| on | 0.613255 | 0.376082 | 0.613255 | 0.46624 | 38143 | 9536 |
| on | 0.613255 | 0.376082 | 0.613255 | 0.46624 | 38143 | 9536 |
| on | 0.613255 | 0.376082 | 0.613255 | 0.46624 | 38143 | 9536 |
| on | 0.613255 | 0.376082 | 0.613255 | 0.46624 | 38143 | 9536 |
| on | 0.613214 | 0.376032 | 0.613214 | 0.46619 | 38144 | 9535 |
| on | 0.613255 | 0.376082 | 0.613255 | 0.46624 | 38143 | 9536 |
| on | 0.613255 | 0.376082 | 0.613255 | 0.46624 | 38143 | 9536 |
| on | 0.613255 | 0.376082 | 0.613255 | 0.46624 | 38143 | 9536 |
| on | 0.613255 | 0.376082 | 0.613255 | 0.46624 | 38143 | 9536 |
| on | 0.613214 | 0.376032 | 0.613214 | 0.46619 | 38144 | 9535 |
| on | 0.613255 | 0.376082 | 0.613255 | 0.46624 | 38143 | 9536 |
| on | 0.613255 | 0.376082 | 0.613255 | 0.46624 | 38143 | 9536 |
| on | 0.613255 | 0.376082 | 0.613255 | 0.46624 | 38143 | 9536 |
| on | 0.613255 | 0.376082 | 0.613255 | 0.46624 | 38143 | 9536 |
| on | 0.613214 | 0.376032 | 0.613214 | 0.46619 | 38144 | 9535 |
| on | 0.643248 | 0.648778 | 0.643248 | 0.636697 | 47679 | 7501 |
| on | 0.643381 | 0.648874 | 0.643381 | 0.636815 | 47679 | 7501 |
| on | 0.643248 | 0.64853 | 0.643248 | 0.636394 | 47679 | 7501 |
| on | 0.464765 | 0.216007 | 0.464765 | 0.294937 | 38143 | 9536 |
| on | 0.464765 | 0.216007 | 0.464765 | 0.294937 | 38143 | 9536 |
| on | 0.46487 | 0.216104 | 0.46487 | 0.295049 | 38143 | 9536 |
| on | 0.46487 | 0.216104 | 0.46487 | 0.295049 | 38143 | 9536 |
| on | 0.464814 | 0.216052 | 0.464814 | 0.294989 | 38144 | 9535 |
| on | 0.464765 | 0.216007 | 0.464765 | 0.294937 | 38143 | 9536 |
| on | 0.464765 | 0.216007 | 0.464765 | 0.294937 | 38143 | 9536 |
| on | 0.46487 | 0.216104 | 0.46487 | 0.295049 | 38143 | 9536 |
| on | 0.46487 | 0.216104 | 0.46487 | 0.295049 | 38143 | 9536 |
| on | 0.464814 | 0.216052 | 0.464814 | 0.294989 | 38144 | 9535 |
| on | 0.464765 | 0.216007 | 0.464765 | 0.294937 | 38143 | 9536 |
| on | 0.464765 | 0.216007 | 0.464765 | 0.294937 | 38143 | 9536 |
| on | 0.46487 | 0.216104 | 0.46487 | 0.295049 | 38143 | 9536 |
| on | 0.46487 | 0.216104 | 0.46487 | 0.295049 | 38143 | 9536 |
| on | 0.464814 | 0.216052 | 0.464814 | 0.294989 | 38144 | 9535 |
| on | 0.47207 | 0.446281 | 0.47207 | 0.381655 | 47679 | 7501 |
| on | 0.472204 | 0.446626 | 0.472204 | 0.381891 | 47679 | 7501 |
| on | 0.471137 | 0.446252 | 0.471137 | 0.376938 | 47679 | 7501 |
| on | 0.512689 | 0.26285 | 0.512689 | 0.347527 | 38143 | 9536 |
| on | 0.512689 | 0.26285 | 0.512689 | 0.347527 | 38143 | 9536 |
| on | 0.512689 | 0.26285 | 0.512689 | 0.347527 | 38143 | 9536 |
| on | 0.512689 | 0.26285 | 0.512689 | 0.347527 | 38143 | 9536 |
| on | 0.512743 | 0.262905 | 0.512743 | 0.347587 | 38144 | 9535 |

| | | | | | | | |
|---|---|---|---|---|---|---|---|
| on | | 0.512689 | 0.26285 | 0.512689 | 0.347527 | 38143 | 9536 |
| on | | 0.512689 | 0.26285 | 0.512689 | 0.347527 | 38143 | 9536 |
| on | | 0.512689 | 0.26285 | 0.512689 | 0.347527 | 38143 | 9536 |
| on | | 0.512689 | 0.26285 | 0.512689 | 0.347527 | 38143 | 9536 |
| on | | 0.512743 | 0.262905 | 0.512743 | 0.347587 | 38144 | 9535 |
| on | | 0.512689 | 0.26285 | 0.512689 | 0.347527 | 38143 | 9536 |
| on | | 0.512689 | 0.26285 | 0.512689 | 0.347527 | 38143 | 9536 |
| on | | 0.512689 | 0.26285 | 0.512689 | 0.347527 | 38143 | 9536 |
| on | | 0.512689 | 0.26285 | 0.512689 | 0.347527 | 38143 | 9536 |
| on | | 0.512743 | 0.262905 | 0.512743 | 0.347587 | 38144 | 9535 |
| 0.705634 | 0.245371 | | | | | 24689 | 3571 |
| 0.705644 | 0.24536 | | | | | 24689 | 3571 |
| 0.741759 | 0.206738 | | | | | 24689 | 3571 |
| 0.773099 | 0.250759 | | | | | 19751 | 4938 |
| 0.786874 | 0.248824 | | | | | 19751 | 4938 |
| 0.745721 | 0.246778 | | | | | 19751 | 4938 |
| 0.735432 | 0.253333 | | | | | 19751 | 4938 |
| 0.692923 | 0.268043 | | | | | 19752 | 4937 |
| 0.773396 | 0.250471 | | | | | 19751 | 4938 |
| 0.786636 | 0.249051 | | | | | 19751 | 4938 |
| 0.745611 | 0.246889 | | | | | 19751 | 4938 |
| 0.735358 | 0.253408 | | | | | 19751 | 4938 |
| 0.692946 | 0.268019 | | | | | 19752 | 4937 |
| 0.810402 | 0.214607 | | | | | 19751 | 4938 |
| 0.832821 | 0.204961 | | | | | 19751 | 4938 |
| 0.781939 | 0.210195 | | | | | 19751 | 4938 |
| 0.774071 | 0.214104 | | | | | 19751 | 4938 |
| 0.737347 | 0.221117 | | | | | 19752 | 4937 |
| 0.732798 | 0.22683 | | | | | 24689 | 3571 |
| 0.732859 | 0.226766 | | | | | 24689 | 3571 |
| 0.772051 | 0.185414 | | | | | 24689 | 3571 |
| 0.792504 | 0.234816 | | | | | 19751 | 4938 |
| 0.794907 | 0.231836 | | | | | 19751 | 4938 |
| 0.76414 | 0.232992 | | | | | 19751 | 4938 |
| 0.754433 | 0.235611 | | | | | 19751 | 4938 |
| 0.714065 | 0.245999 | | | | | 19752 | 4937 |
| 0.792676 | 0.23465 | | | | | 19751 | 4938 |
| 0.794822 | 0.231918 | | | | | 19751 | 4938 |
| 0.76405 | 0.233081 | | | | | 19751 | 4938 |
| 0.754335 | 0.23571 | | | | | 19751 | 4938 |
| 0.71407 | 0.245993 | | | | | 19752 | 4937 |
| 0.832585 | 0.196117 | | | | | 19751 | 4938 |
| 0.841644 | 0.186671 | | | | | 19751 | 4938 |
| 0.804149 | 0.192832 | | | | | 19751 | 4938 |
| 0.794527 | 0.194988 | | | | | 19751 | 4938 |

| | | | |
|---|---|---|---|
| 0.758096 | 0.199506 | 19752 | 4937 |
| 1.064317 | 0.088902 | 24689 | 3571 |
| 1.064778 | 0.088507 | 24689 | 3571 |
| 1.103805 | 0.055098 | 24689 | 3571 |
| 0.862985 | 0.124592 | 19751 | 4938 |
| 0.879384 | 0.10899 | 19751 | 4938 |
| 0.901009 | 0.115649 | 19751 | 4938 |
| 0.876403 | 0.108279 | 19751 | 4938 |
| 0.896429 | 0.125642 | 19752 | 4937 |
| 0.862899 | 0.124679 | 19751 | 4938 |
| 0.8793 | 0.109075 | 19751 | 4938 |
| 0.901444 | 0.115222 | 19751 | 4938 |
| 0.875477 | 0.109222 | 19751 | 4938 |
| 0.896362 | 0.125707 | 19752 | 4937 |
| 0.899185 | 0.08787 | 19751 | 4938 |
| 0.910156 | 0.077811 | 19751 | 4938 |
| 0.936706 | 0.080613 | 19751 | 4938 |
| 0.902609 | 0.081615 | 19751 | 4938 |
| 0.936307 | 0.086746 | 19752 | 4937 |
| 0.560887 | 0.415342 | 24689 | 3571 |
| 0.560868 | 0.415363 | 24689 | 3571 |
| 0.603014 | 0.371431 | 24689 | 3571 |
| 0.577678 | 0.419385 | 19751 | 4938 |
| 0.545063 | 0.438957 | 19751 | 4938 |
| 0.55529 | 0.442975 | 19751 | 4938 |
| 0.569162 | 0.42853 | 19751 | 4938 |
| 0.601213 | 0.422398 | 19752 | 4937 |
| 0.577781 | 0.419282 | 19751 | 4938 |
| 0.544892 | 0.439133 | 19751 | 4938 |
| 0.555339 | 0.442926 | 19751 | 4938 |
| 0.569103 | 0.428588 | 19751 | 4938 |
| 0.601152 | 0.422457 | 19752 | 4937 |
| 0.623987 | 0.37284 | 19751 | 4938 |
| 0.594699 | 0.387866 | 19751 | 4938 |
| 0.6078 | 0.390301 | 19751 | 4938 |
| 0.617289 | 0.380207 | 19751 | 4938 |
| 0.659923 | 0.365994 | 19752 | 4937 |
| 0.579949 | 0.38589 | 24689 | 3571 |
| 0.579955 | 0.385884 | 24689 | 3571 |
| 0.619238 | 0.344287 | 24689 | 3571 |
| 0.605481 | 0.392108 | 19751 | 4938 |
| 0.575679 | 0.409348 | 19751 | 4938 |
| 0.585787 | 0.416461 | 19751 | 4938 |
| 0.591231 | 0.405915 | 19751 | 4938 |
| 0.623949 | 0.394502 | 19752 | 4937 |

| | | | |
|---|---|---|---|
| 0.605659 | 0.39193 | 19751 | 4938 |
| 0.575478 | 0.409555 | 19751 | 4938 |
| 0.585818 | 0.41643 | 19751 | 4938 |
| 0.591163 | 0.405984 | 19751 | 4938 |
| 0.623914 | 0.394536 | 19752 | 4937 |
| 0.652458 | 0.344944 | 19751 | 4938 |
| 0.62429 | 0.359473 | 19751 | 4938 |
| 0.641231 | 0.36123 | 19751 | 4938 |
| 0.639396 | 0.357518 | 19751 | 4938 |
| 0.680038 | 0.340072 | 19752 | 4937 |
| 0.949928 | 0.115829 | 24689 | 3571 |
| 0.949704 | 0.116038 | 24689 | 3571 |
| 0.985167 | 0.08303 | 24689 | 3571 |
| 0.857753 | 0.143115 | 19751 | 4938 |
| 0.900534 | 0.134292 | 19751 | 4938 |
| 0.90239 | 0.128182 | 19751 | 4938 |
| 0.825759 | 0.127077 | 19751 | 4938 |
| 0.846248 | 0.13484 | 19752 | 4937 |
| 0.85767 | 0.143199 | 19751 | 4938 |
| 0.900559 | 0.134268 | 19751 | 4938 |
| 0.902796 | 0.12779 | 19751 | 4938 |
| 0.825227 | 0.12764 | 19751 | 4938 |
| 0.846156 | 0.134934 | 19752 | 4937 |
| 0.899587 | 0.101324 | 19751 | 4938 |
| 0.941099 | 0.095295 | 19751 | 4938 |
| 0.940494 | 0.091369 | 19751 | 4938 |
| 0.854662 | 0.096524 | 19751 | 4938 |
| 0.885176 | 0.095042 | 19752 | 4937 |
| 0.51 | 0.501212 | 27542 | 4025 |
| 0.509946 | 0.501265 | 27542 | 4025 |
| 0.565895 | 0.446546 | 27542 | 4025 |
| 0.498181 | 0.478998 | 22033 | 5509 |
| 0.500005 | 0.491481 | 22033 | 5509 |
| 0.527085 | 0.472864 | 22034 | 5508 |
| 0.514709 | 0.506863 | 22034 | 5508 |
| 0.50232 | 0.505944 | 22034 | 5508 |
| 0.498087 | 0.479097 | 22033 | 5509 |
| 0.500037 | 0.491448 | 22033 | 5509 |
| 0.527015 | 0.472935 | 22034 | 5508 |
| 0.514661 | 0.506909 | 22034 | 5508 |
| 0.502571 | 0.505697 | 22034 | 5508 |
| 0.543011 | 0.432115 | 22033 | 5509 |
| 0.546804 | 0.443885 | 22033 | 5509 |
| 0.568583 | 0.431363 | 22034 | 5508 |
| 0.571797 | 0.452167 | 22034 | 5508 |

| | | | |
|---|---|---|---|
| 0.566997 | 0.442331 | 22034 | 5508 |
| 0.50836 | 0.483761 | 27542 | 4025 |
| 0.508288 | 0.483834 | 27542 | 4025 |
| 0.568686 | 0.4225 | 27542 | 4025 |
| 0.517682 | 0.457561 | 22033 | 5509 |
| 0.509994 | 0.473636 | 22033 | 5509 |
| 0.539216 | 0.468433 | 22034 | 5508 |
| 0.523787 | 0.496462 | 22034 | 5508 |
| 0.517313 | 0.493829 | 22034 | 5508 |
| 0.517598 | 0.457649 | 22033 | 5509 |
| 0.509971 | 0.47366 | 22033 | 5509 |
| 0.539177 | 0.468472 | 22034 | 5508 |
| 0.523725 | 0.496521 | 22034 | 5508 |
| 0.517615 | 0.493534 | 22034 | 5508 |
| 0.572972 | 0.399627 | 22033 | 5509 |
| 0.559453 | 0.422589 | 22033 | 5509 |
| 0.598882 | 0.409614 | 22034 | 5508 |
| 0.594488 | 0.428493 | 22034 | 5508 |
| 0.594943 | 0.417872 | 22034 | 5508 |
| 0.70974 | 0.308258 | 27542 | 4025 |
| 0.709823 | 0.308177 | 27542 | 4025 |
| 0.767056 | 0.252395 | 27542 | 4025 |
| 0.701887 | 0.279106 | 22033 | 5509 |
| 0.716146 | 0.279569 | 22033 | 5509 |
| 0.753598 | 0.252706 | 22034 | 5508 |
| 0.712697 | 0.292419 | 22034 | 5508 |
| 0.722141 | 0.289519 | 22034 | 5508 |
| 0.701764 | 0.279232 | 22033 | 5509 |
| 0.716107 | 0.279608 | 22033 | 5509 |
| 0.753655 | 0.252649 | 22034 | 5508 |
| 0.712642 | 0.292474 | 22034 | 5508 |
| 0.72237 | 0.289293 | 22034 | 5508 |
| 0.743588 | 0.236275 | 22033 | 5509 |
| 0.759211 | 0.236246 | 22033 | 5509 |
| 0.784116 | 0.222443 | 22034 | 5508 |
| 0.75887 | 0.246578 | 22034 | 5508 |
| 0.777422 | 0.23513 | 22034 | 5508 |
| 0.835936 | 0.146632 | 27542 | 4025 |
| 0.835999 | 0.146567 | 27542 | 4025 |
| 0.886386 | 0.09513 | 27542 | 4025 |
| 0.854178 | 0.133025 | 22033 | 5509 |
| 0.849184 | 0.127404 | 22033 | 5509 |
| 0.879335 | 0.147398 | 22034 | 5508 |
| 0.829129 | 0.159768 | 22034 | 5508 |
| 0.858418 | 0.161415 | 22034 | 5508 |

| | | | |
|---|---|---|---|
| 0.85412 | 0.133083 | 22033 | 5509 |
| 0.848954 | 0.12764 | 22033 | 5509 |
| 0.879276 | 0.147455 | 22034 | 5508 |
| 0.829153 | 0.159744 | 22034 | 5508 |
| 0.858688 | 0.161152 | 22034 | 5508 |
| 0.898853 | 0.087681 | 22033 | 5509 |
| 0.880164 | 0.095569 | 22033 | 5509 |
| 0.93163 | 0.096692 | 22034 | 5508 |
| 0.88279 | 0.105388 | 22034 | 5508 |
| 0.921223 | 0.100062 | 22034 | 5508 |
| 0.376927 | 0.622592 | 27542 | 4025 |
| 0.376888 | 0.622631 | 27542 | 4025 |
| 0.427381 | 0.572073 | 27542 | 4025 |
| 0.369833 | 0.620848 | 22033 | 5509 |
| 0.360009 | 0.63235 | 22033 | 5509 |
| 0.35424 | 0.6449 | 22034 | 5508 |
| 0.365714 | 0.637365 | 22034 | 5508 |
| 0.380459 | 0.633797 | 22034 | 5508 |
| 0.37007 | 0.620605 | 22033 | 5509 |
| 0.359869 | 0.632493 | 22033 | 5509 |
| 0.354259 | 0.64488 | 22034 | 5508 |
| 0.36571 | 0.637369 | 22034 | 5508 |
| 0.380602 | 0.633659 | 22034 | 5508 |
| 0.426669 | 0.56258 | 22033 | 5509 |
| 0.409884 | 0.581416 | 22033 | 5509 |
| 0.404399 | 0.594619 | 22034 | 5508 |
| 0.418896 | 0.584631 | 22034 | 5508 |
| 0.441121 | 0.575407 | 22034 | 5508 |
| 0.713232 | 0.279338 | 27542 | 4025 |
| 0.713207 | 0.279364 | 27542 | 4025 |
| 0.742221 | 0.250048 | 27542 | 4025 |
| 0.716511 | 0.273477 | 22033 | 5509 |
| 0.707171 | 0.287276 | 22033 | 5509 |
| 0.715624 | 0.292236 | 22034 | 5508 |
| 0.691593 | 0.295943 | 22034 | 5508 |
| 0.722682 | 0.29708 | 22034 | 5508 |
| 0.716789 | 0.273194 | 22033 | 5509 |
| 0.706882 | 0.287568 | 22033 | 5509 |
| 0.715577 | 0.292282 | 22034 | 5508 |
| 0.691657 | 0.295877 | 22034 | 5508 |
| 0.722767 | 0.296998 | 22034 | 5508 |
| 0.754164 | 0.235297 | 22033 | 5509 |
| 0.735384 | 0.258842 | 22033 | 5509 |
| 0.751694 | 0.256561 | 22034 | 5508 |
| 0.725322 | 0.261605 | 22034 | 5508 |

| | | | |
|---|---|---|---|
| 0.763366 | 0.257509 | 22034 | 5508 |
| 0.860852 | 0.143287 | 27542 | 4025 |
| 0.860608 | 0.14353 | 27542 | 4025 |
| 0.881686 | 0.122553 | 27542 | 4025 |
| 0.836073 | 0.1527 | 22033 | 5509 |
| 0.847246 | 0.149039 | 22033 | 5509 |
| 0.836533 | 0.177404 | 22034 | 5508 |
| 0.794462 | 0.155435 | 22034 | 5508 |
| 0.885908 | 0.16507 | 22034 | 5508 |
| 0.835978 | 0.152796 | 22033 | 5509 |
| 0.846858 | 0.149429 | 22033 | 5509 |
| 0.836651 | 0.177288 | 22034 | 5508 |
| 0.794356 | 0.155547 | 22034 | 5508 |
| 0.886093 | 0.164895 | 22034 | 5508 |
| 0.86625 | 0.122117 | 22033 | 5509 |
| 0.871399 | 0.124781 | 22033 | 5509 |
| 0.871954 | 0.142573 | 22034 | 5508 |
| 0.819482 | 0.128836 | 22034 | 5508 |
| 0.925728 | 0.127541 | 22034 | 5508 |
| 0.923361 | 0.111496 | 27542 | 4025 |
| 0.923232 | 0.11162 | 27542 | 4025 |
| 0.966658 | 0.069834 | 27542 | 4025 |
| 0.87463 | 0.120144 | 22033 | 5509 |
| 0.918434 | 0.104681 | 22033 | 5509 |
| 0.855604 | 0.131278 | 22034 | 5508 |
| 0.85491 | 0.126307 | 22034 | 5508 |
| 0.887546 | 0.126101 | 22034 | 5508 |
| 0.874731 | 0.120042 | 22033 | 5509 |
| 0.918271 | 0.104839 | 22033 | 5509 |
| 0.85561 | 0.131272 | 22034 | 5508 |
| 0.854823 | 0.126396 | 22034 | 5508 |
| 0.887708 | 0.125941 | 22034 | 5508 |
| 0.927027 | 0.067434 | 22033 | 5509 |
| 0.957881 | 0.066226 | 22033 | 5509 |
| 0.903509 | 0.082639 | 22034 | 5508 |
| 0.898126 | 0.082142 | 22034 | 5508 |
| 0.941684 | 0.072795 | 22034 | 5508 |
| 0.732418 | 0.273246 | 27542 | 4025 |
| 0.732444 | 0.273219 | 27542 | 4025 |
| 0.787994 | 0.2181 | 27542 | 4025 |
| 0.715856 | 0.279151 | 22033 | 5509 |
| 0.715469 | 0.268488 | 22033 | 5509 |
| 0.687153 | 0.299025 | 22034 | 5508 |
| 0.708676 | 0.298952 | 22034 | 5508 |
| 0.739188 | 0.287512 | 22034 | 5508 |

| | | | |
|---|---|---|---|
| 0.715982 | 0.279024 | 22033 | 5509 |
| 0.715211 | 0.268752 | 22033 | 5509 |
| 0.687099 | 0.29908 | 22034 | 5508 |
| 0.708721 | 0.298907 | 22034 | 5508 |
| 0.739417 | 0.287291 | 22034 | 5508 |
| 0.779359 | 0.215206 | 22033 | 5509 |
| 0.760651 | 0.222293 | 22033 | 5509 |
| 0.74444 | 0.240586 | 22034 | 5508 |
| 0.771308 | 0.236993 | 22034 | 5508 |
| 0.804697 | 0.224369 | 22034 | 5508 |
| 0.703871 | 0.298957 | 27542 | 4025 |
| 0.703966 | 0.298863 | 27542 | 4025 |
| 0.762816 | 0.240249 | 27542 | 4025 |
| 0.682705 | 0.324153 | 22033 | 5509 |
| 0.65394 | 0.326605 | 22033 | 5509 |
| 0.647896 | 0.34133 | 22034 | 5508 |
| 0.660875 | 0.340193 | 22034 | 5508 |
| 0.697353 | 0.325235 | 22034 | 5508 |
| 0.682782 | 0.324077 | 22033 | 5509 |
| 0.653815 | 0.326733 | 22033 | 5509 |
| 0.647884 | 0.341342 | 22034 | 5508 |
| 0.660888 | 0.34018 | 22034 | 5508 |
| 0.697531 | 0.325063 | 22034 | 5508 |
| 0.745792 | 0.2617 | 22033 | 5509 |
| 0.708313 | 0.270613 | 22033 | 5509 |
| 0.706729 | 0.281519 | 22034 | 5508 |
| 0.722164 | 0.279003 | 22034 | 5508 |
| 0.76122 | 0.263437 | 22034 | 5508 |
| 0.720407 | 0.296931 | 27542 | 4025 |
| 0.72038 | 0.296958 | 27542 | 4025 |
| 0.773479 | 0.245137 | 27542 | 4025 |
| 0.653977 | 0.329897 | 22033 | 5509 |
| 0.660988 | 0.322084 | 22033 | 5509 |
| 0.6749 | 0.334081 | 22034 | 5508 |
| 0.68589 | 0.326283 | 22034 | 5508 |
| 0.688536 | 0.323308 | 22034 | 5508 |
| 0.654092 | 0.329779 | 22033 | 5509 |
| 0.660817 | 0.322259 | 22033 | 5509 |
| 0.674864 | 0.334117 | 22034 | 5508 |
| 0.685958 | 0.326216 | 22034 | 5508 |
| 0.688677 | 0.32317 | 22034 | 5508 |
| 0.717983 | 0.264313 | 22033 | 5509 |
| 0.715519 | 0.266156 | 22033 | 5509 |
| 0.736053 | 0.273742 | 22034 | 5508 |
| 0.746364 | 0.266882 | 22034 | 5508 |

| | | | |
|---|---|---|---|
| 0.751651 | 0.261279 | 22034 | 5508 |
| 0.676294 | 0.364352 | 27542 | 4025 |
| 0.676477 | 0.364179 | 27542 | 4025 |
| 0.74107 | 0.303468 | 27542 | 4025 |
| 0.624037 | 0.363823 | 22033 | 5509 |
| 0.618887 | 0.378474 | 22033 | 5509 |
| 0.610909 | 0.387628 | 22034 | 5508 |
| 0.620195 | 0.389807 | 22034 | 5508 |
| 0.632525 | 0.373168 | 22034 | 5508 |
| 0.624108 | 0.36375 | 22033 | 5509 |
| 0.618693 | 0.378668 | 22033 | 5509 |
| 0.61085 | 0.387688 | 22034 | 5508 |
| 0.620457 | 0.38955 | 22034 | 5508 |
| 0.632578 | 0.373115 | 22034 | 5508 |
| 0.683044 | 0.303668 | 22033 | 5509 |
| 0.671917 | 0.325218 | 22033 | 5509 |
| 0.666386 | 0.332019 | 22034 | 5508 |
| 0.690335 | 0.320798 | 22034 | 5508 |
| 0.690334 | 0.315879 | 22034 | 5508 |
| 0.708979 | 0.323293 | 27542 | 4025 |
| 0.708962 | 0.323309 | 27542 | 4025 |
| 0.776832 | 0.258528 | 27542 | 4025 |
| 0.67863 | 0.329472 | 22033 | 5509 |
| 0.639519 | 0.342641 | 22033 | 5509 |
| 0.629305 | 0.360789 | 22034 | 5508 |
| 0.673355 | 0.348438 | 22034 | 5508 |
| 0.656626 | 0.341456 | 22034 | 5508 |
| 0.678706 | 0.329397 | 22033 | 5509 |
| 0.639393 | 0.342771 | 22033 | 5509 |
| 0.62929 | 0.360804 | 22034 | 5508 |
| 0.673561 | 0.348239 | 22034 | 5508 |
| 0.656642 | 0.34144 | 22034 | 5508 |
| 0.745248 | 0.263649 | 22033 | 5509 |
| 0.697666 | 0.282872 | 22033 | 5509 |
| 0.694488 | 0.29458 | 22034 | 5508 |
| 0.747898 | 0.276308 | 22034 | 5508 |
| 0.720841 | 0.277053 | 22034 | 5508 |
| 0.590637 | 0.422465 | 27542 | 4025 |
| 0.590758 | 0.422346 | 27542 | 4025 |
| 0.63689 | 0.377238 | 27542 | 4025 |
| 0.566595 | 0.422566 | 22033 | 5509 |
| 0.53774 | 0.454357 | 22033 | 5509 |
| 0.550892 | 0.456855 | 22034 | 5508 |
| 0.563723 | 0.444739 | 22034 | 5508 |
| 0.548419 | 0.453558 | 22034 | 5508 |

| | | | |
|---|---|---|---|
| 0.566773 | 0.422385 | 22033 | 5509 |
| 0.537672 | 0.454426 | 22033 | 5509 |
| 0.550968 | 0.45678 | 22034 | 5508 |
| 0.563784 | 0.44468 | 22034 | 5508 |
| 0.548403 | 0.453575 | 22034 | 5508 |
| 0.615293 | 0.372937 | 22033 | 5509 |
| 0.580625 | 0.410841 | 22033 | 5509 |
| 0.601275 | 0.40718 | 22034 | 5508 |
| 0.613432 | 0.395776 | 22034 | 5508 |
| 0.596924 | 0.405229 | 22034 | 5508 |
| 0.53791 | 0.459664 | 27542 | 4025 |
| 0.538259 | 0.459314 | 27542 | 4025 |
| 0.57805 | 0.419344 | 27542 | 4025 |
| 0.524805 | 0.458868 | 22033 | 5509 |
| 0.517634 | 0.480071 | 22033 | 5509 |
| 0.528822 | 0.478602 | 22034 | 5508 |
| 0.539359 | 0.471408 | 22034 | 5508 |
| 0.530069 | 0.469777 | 22034 | 5508 |
| 0.525032 | 0.458634 | 22033 | 5509 |
| 0.51745 | 0.480256 | 22033 | 5509 |
| 0.528832 | 0.478592 | 22034 | 5508 |
| 0.539543 | 0.471227 | 22034 | 5508 |
| 0.529975 | 0.469871 | 22034 | 5508 |
| 0.563599 | 0.418868 | 22033 | 5509 |
| 0.551497 | 0.446057 | 22033 | 5509 |
| 0.568092 | 0.439884 | 22034 | 5508 |
| 0.583924 | 0.427731 | 22034 | 5508 |
| 0.569657 | 0.430178 | 22034 | 5508 |
| 0.553065 | 0.447579 | 27542 | 4025 |
| 0.55344 | 0.447204 | 27542 | 4025 |
| 0.592616 | 0.408074 | 27542 | 4025 |
| 0.551718 | 0.444875 | 22033 | 5509 |
| 0.52732 | 0.456314 | 22033 | 5509 |
| 0.538176 | 0.473451 | 22034 | 5508 |
| 0.525153 | 0.462422 | 22034 | 5508 |
| 0.556193 | 0.463865 | 22034 | 5508 |
| 0.551961 | 0.444631 | 22033 | 5509 |
| 0.527055 | 0.456588 | 22033 | 5509 |
| 0.53809 | 0.473534 | 22034 | 5508 |
| 0.525266 | 0.462307 | 22034 | 5508 |
| 0.556424 | 0.463642 | 22034 | 5508 |
| 0.589369 | 0.406991 | 22033 | 5509 |
| 0.557224 | 0.425482 | 22033 | 5509 |
| 0.574652 | 0.437762 | 22034 | 5508 |
| 0.563632 | 0.423034 | 22034 | 5508 |

| | | | |
|---|---|---|---|
| 0.596801 | 0.424722 | 22034 | 5508 |
| 0.625839 | 0.393731 | 27542 | 4025 |
| 0.625997 | 0.393578 | 27542 | 4025 |
| 0.678768 | 0.342457 | 27542 | 4025 |
| 0.591203 | 0.404549 | 22033 | 5509 |
| 0.589887 | 0.403033 | 22033 | 5509 |
| 0.56637 | 0.433451 | 22034 | 5508 |
| 0.566393 | 0.422865 | 22034 | 5508 |
| 0.611317 | 0.411055 | 22034 | 5508 |
| 0.591366 | 0.404385 | 22033 | 5509 |
| 0.58969 | 0.403233 | 22033 | 5509 |
| 0.566194 | 0.433627 | 22034 | 5508 |
| 0.566565 | 0.422689 | 22034 | 5508 |
| 0.611532 | 0.410848 | 22034 | 5508 |
| 0.644335 | 0.351035 | 22033 | 5509 |
| 0.634905 | 0.357475 | 22033 | 5509 |
| 0.61343 | 0.386376 | 22034 | 5508 |
| 0.622636 | 0.365555 | 22034 | 5508 |
| 0.665981 | 0.358391 | 22034 | 5508 |
| 0.685432 | 0.338067 | 27542 | 4025 |
| 0.68563 | 0.337876 | 27542 | 4025 |
| 0.73553 | 0.289687 | 27542 | 4025 |
| 0.650762 | 0.348727 | 22033 | 5509 |
| 0.629497 | 0.36113 | 22033 | 5509 |
| 0.64686 | 0.354208 | 22034 | 5508 |
| 0.649211 | 0.354416 | 22034 | 5508 |
| 0.667096 | 0.337592 | 22034 | 5508 |
| 0.650852 | 0.348636 | 22033 | 5509 |
| 0.629338 | 0.361291 | 22033 | 5509 |
| 0.646879 | 0.354188 | 22034 | 5508 |
| 0.649264 | 0.354363 | 22034 | 5508 |
| 0.667242 | 0.337446 | 22034 | 5508 |
| 0.707448 | 0.291996 | 22033 | 5509 |
| 0.67662 | 0.313305 | 22033 | 5509 |
| 0.698091 | 0.303061 | 22034 | 5508 |
| 0.698754 | 0.305149 | 22034 | 5508 |
| 0.714045 | 0.290973 | 22034 | 5508 |
| 0.623397 | 0.396769 | 27542 | 4025 |
| 0.623393 | 0.396773 | 27542 | 4025 |
| 0.656255 | 0.364974 | 27542 | 4025 |
| 0.596603 | 0.396708 | 22033 | 5509 |
| 0.575548 | 0.416345 | 22033 | 5509 |
| 0.578316 | 0.418172 | 22034 | 5508 |
| 0.594129 | 0.407197 | 22034 | 5508 |
| 0.606327 | 0.410521 | 22034 | 5508 |

| | | | |
|---|---|---|---|
| 0.596655 | 0.396655 | 22033 | 5509 |
| 0.575369 | 0.416526 | 22033 | 5509 |
| 0.578394 | 0.418093 | 22034 | 5508 |
| 0.594138 | 0.407188 | 22034 | 5508 |
| 0.60654 | 0.410313 | 22034 | 5508 |
| 0.638851 | 0.353986 | 22033 | 5509 |
| 0.612546 | 0.378826 | 22033 | 5509 |
| 0.618261 | 0.377984 | 22034 | 5508 |
| 0.631991 | 0.369419 | 22034 | 5508 |
| 0.649474 | 0.368572 | 22034 | 5508 |
| 0.639303 | 0.376707 | 27542 | 4025 |
| 0.639417 | 0.376596 | 27542 | 4025 |
| 0.669955 | 0.346823 | 27542 | 4025 |
| 0.604696 | 0.387817 | 22033 | 5509 |
| 0.568326 | 0.42033 | 22033 | 5509 |
| 0.612909 | 0.397751 | 22034 | 5508 |
| 0.607286 | 0.393655 | 22034 | 5508 |
| 0.614793 | 0.392538 | 22034 | 5508 |
| 0.604754 | 0.387758 | 22033 | 5509 |
| 0.568041 | 0.42062 | 22033 | 5509 |
| 0.612932 | 0.397729 | 22034 | 5508 |
| 0.607491 | 0.39345 | 22034 | 5508 |
| 0.614969 | 0.392364 | 22034 | 5508 |
| 0.637882 | 0.35422 | 22033 | 5509 |
| 0.599472 | 0.388562 | 22033 | 5509 |
| 0.64426 | 0.366945 | 22034 | 5508 |
| 0.639448 | 0.361543 | 22034 | 5508 |
| 0.647473 | 0.360248 | 22034 | 5508 |
| 0.663345 | 0.3654 | 27542 | 4025 |
| 0.663512 | 0.365239 | 27542 | 4025 |
| 0.70244 | 0.327998 | 27542 | 4025 |
| 0.625828 | 0.368444 | 22033 | 5509 |
| 0.621936 | 0.386328 | 22033 | 5509 |
| 0.628195 | 0.376933 | 22034 | 5508 |
| 0.616837 | 0.372703 | 22034 | 5508 |
| 0.629597 | 0.372784 | 22034 | 5508 |
| 0.625979 | 0.368291 | 22033 | 5509 |
| 0.621731 | 0.386529 | 22033 | 5509 |
| 0.628177 | 0.37695 | 22034 | 5508 |
| 0.616836 | 0.372704 | 22034 | 5508 |
| 0.629764 | 0.372619 | 22034 | 5508 |
| 0.670416 | 0.323447 | 22033 | 5509 |
| 0.657892 | 0.350849 | 22033 | 5509 |
| 0.664875 | 0.340552 | 22034 | 5508 |
| 0.650298 | 0.338674 | 22034 | 5508 |

| | | | |
|---|---|---|---|
| 0.668808 | 0.333722 | 22034 | 5508 |
| 0.570005 | 0.444204 | 27542 | 4025 |
| 0.570096 | 0.444115 | 27542 | 4025 |
| 0.626472 | 0.389144 | 27542 | 4025 |
| 0.528765 | 0.460736 | 22033 | 5509 |
| 0.528262 | 0.462633 | 22033 | 5509 |
| 0.519349 | 0.486501 | 22034 | 5508 |
| 0.52315 | 0.47543 | 22034 | 5508 |
| 0.547259 | 0.467428 | 22034 | 5508 |
| 0.528953 | 0.460544 | 22033 | 5509 |
| 0.527987 | 0.462912 | 22033 | 5509 |
| 0.51935 | 0.4865 | 22034 | 5508 |
| 0.523226 | 0.475354 | 22034 | 5508 |
| 0.547468 | 0.467225 | 22034 | 5508 |
| 0.591245 | 0.397016 | 22033 | 5509 |
| 0.576644 | 0.413417 | 22033 | 5509 |
| 0.577217 | 0.429285 | 22034 | 5508 |
| 0.581758 | 0.416664 | 22034 | 5508 |
| 0.609957 | 0.406412 | 22034 | 5508 |
| 0.942994 | 0.048735 | 27542 | 4025 |
| 0.943044 | 0.048684 | 27542 | 4025 |
| 0.989612 | 0.001708 | 27542 | 4025 |
| 0.921445 | 0.054962 | 22033 | 5509 |
| 0.953056 | 0.050937 | 22033 | 5509 |
| 0.927585 | 0.054206 | 22034 | 5508 |
| 0.971747 | 0.048052 | 22034 | 5508 |
| 0.968801 | 0.04901 | 22034 | 5508 |
| 0.921555 | 0.054849 | 22033 | 5509 |
| 0.952988 | 0.051005 | 22033 | 5509 |
| 0.927531 | 0.054261 | 22034 | 5508 |
| 0.971717 | 0.048081 | 22034 | 5508 |
| 0.968963 | 0.048852 | 22034 | 5508 |
| 0.973547 | 0.001525 | 22033 | 5509 |
| 1.002485 | 0.001715 | 22033 | 5509 |
| 0.979319 | 0.001457 | 22034 | 5508 |
| 1.019281 | 0.001486 | 22034 | 5508 |
| 1.016754 | 0.00194 | 22034 | 5508 |
| 0.804999 | 0.19827 | 27542 | 4025 |
| 0.804857 | 0.198411 | 27542 | 4025 |
| 0.841604 | 0.161814 | 27542 | 4025 |
| 0.847378 | 0.191921 | 22033 | 5509 |
| 0.792383 | 0.206305 | 22033 | 5509 |
| 0.788131 | 0.182374 | 22034 | 5508 |
| 0.786282 | 0.205332 | 22034 | 5508 |
| 0.819616 | 0.180374 | 22034 | 5508 |

| | | | |
|---|---|---|---|
| 0.847399 | 0.191901 | 22033 | 5509 |
| 0.792272 | 0.206416 | 22033 | 5509 |
| 0.788025 | 0.182483 | 22034 | 5508 |
| 0.786255 | 0.205359 | 22034 | 5508 |
| 0.819489 | 0.1805 | 22034 | 5508 |
| 0.884882 | 0.156156 | 22033 | 5509 |
| 0.825588 | 0.173046 | 22033 | 5509 |
| 0.812612 | 0.156976 | 22034 | 5508 |
| 0.826078 | 0.165111 | 22034 | 5508 |
| 0.843461 | 0.156529 | 22034 | 5508 |
| 0.927873 | 0.053158 | 27542 | 4025 |
| 0.927895 | 0.053136 | 27542 | 4025 |
| 0.965069 | 0.015202 | 27542 | 4025 |
| 0.930922 | 0.052907 | 22033 | 5509 |
| 0.954869 | 0.061389 | 22033 | 5509 |
| 0.925356 | 0.074349 | 22034 | 5508 |
| 0.925618 | 0.071894 | 22034 | 5508 |
| 0.944686 | 0.057471 | 22034 | 5508 |
| 0.930791 | 0.05304 | 22033 | 5509 |
| 0.954862 | 0.061396 | 22033 | 5509 |
| 0.925724 | 0.073981 | 22034 | 5508 |
| 0.925726 | 0.071786 | 22034 | 5508 |
| 0.944414 | 0.057743 | 22034 | 5508 |
| 0.96579 | 0.017433 | 22033 | 5509 |
| 0.999482 | 0.017536 | 22033 | 5509 |
| 0.982481 | 0.017206 | 22034 | 5508 |
| 0.978372 | 0.018999 | 22034 | 5508 |
| 0.983385 | 0.018861 | 22034 | 5508 |
| 0.9388 | 0.023182 | 27542 | 4025 |
| 0.938658 | 0.02333 | 27542 | 4025 |
| 0.957648 | 0.003572 | 27542 | 4025 |
| 0.981554 | 0.016655 | 22033 | 5509 |
| 0.978075 | 0.030915 | 22033 | 5509 |
| 0.938498 | 0.038381 | 22034 | 5508 |
| 0.952364 | 0.039319 | 22034 | 5508 |
| 0.996666 | 0.027603 | 22034 | 5508 |
| 0.981412 | 0.016797 | 22033 | 5509 |
| 0.978041 | 0.030949 | 22033 | 5509 |
| 0.938691 | 0.038183 | 22034 | 5508 |
| 0.95238 | 0.039303 | 22034 | 5508 |
| 0.996437 | 0.027827 | 22034 | 5508 |
| 0.994246 | 0.003941 | 22033 | 5509 |
| 1.004526 | 0.004707 | 22033 | 5509 |
| 0.971331 | 0.004738 | 22034 | 5508 |
| 0.986495 | 0.00489 | 22034 | 5508 |

| | | | |
|---|---|---|---|
| 1.019832 | 0.005002 | 22034 | 5508 |
| 0.907061 | 0.076681 | 27542 | 4025 |
| 0.907071 | 0.076671 | 27542 | 4025 |
| 0.934067 | 0.049191 | 27542 | 4025 |
| 0.922145 | 0.093483 | 22033 | 5509 |
| 0.897368 | 0.083576 | 22033 | 5509 |
| 0.863891 | 0.091881 | 22034 | 5508 |
| 0.954122 | 0.102358 | 22034 | 5508 |
| 0.903323 | 0.088094 | 22034 | 5508 |
| 0.922087 | 0.093539 | 22033 | 5509 |
| 0.89738 | 0.083564 | 22033 | 5509 |
| 0.863976 | 0.091791 | 22034 | 5508 |
| 0.954207 | 0.102278 | 22034 | 5508 |
| 0.903224 | 0.088194 | 22034 | 5508 |
| 0.953462 | 0.062696 | 22033 | 5509 |
| 0.921958 | 0.058464 | 22033 | 5509 |
| 0.894078 | 0.060148 | 22034 | 5508 |
| 0.99502 | 0.063881 | 22034 | 5508 |
| 0.933808 | 0.057319 | 22034 | 5508 |
| 0.934012 | 0.038051 | 27542 | 4025 |
| 0.933941 | 0.038123 | 27542 | 4025 |
| 0.965983 | 0.005123 | 27542 | 4025 |
| 0.938914 | 0.049298 | 22033 | 5509 |
| 0.954265 | 0.035587 | 22033 | 5509 |
| 0.926636 | 0.044531 | 22034 | 5508 |
| 0.976245 | 0.047511 | 22034 | 5508 |
| 0.987304 | 0.039837 | 22034 | 5508 |
| 0.93889 | 0.049323 | 22033 | 5509 |
| 0.954244 | 0.035607 | 22033 | 5509 |
| 0.926688 | 0.044477 | 22034 | 5508 |
| 0.976286 | 0.047471 | 22034 | 5508 |
| 0.987238 | 0.039901 | 22034 | 5508 |
| 0.981931 | 0.005742 | 22033 | 5509 |
| 0.983825 | 0.005711 | 22033 | 5509 |
| 0.964312 | 0.005682 | 22034 | 5508 |
| 1.01894 | 0.005855 | 22034 | 5508 |
| 1.023196 | 0.004931 | 22034 | 5508 |
| 0.845366 | 0.12193 | 27542 | 4025 |
| 0.84532 | 0.121978 | 27542 | 4025 |
| 0.884483 | 0.0813 | 27542 | 4025 |
| 0.893593 | 0.121458 | 22033 | 5509 |
| 0.851332 | 0.117054 | 22033 | 5509 |
| 0.884273 | 0.116923 | 22034 | 5508 |
| 0.889254 | 0.116479 | 22034 | 5508 |
| 0.894697 | 0.114499 | 22034 | 5508 |

| | | | |
|---|---|---|---|
| 0.893664 | 0.121389 | 22033 | 5509 |
| 0.851115 | 0.117279 | 22033 | 5509 |
| 0.884373 | 0.116823 | 22034 | 5508 |
| 0.889222 | 0.116511 | 22034 | 5508 |
| 0.894526 | 0.114668 | 22034 | 5508 |
| 0.948545 | 0.067432 | 22033 | 5509 |
| 0.895478 | 0.071269 | 22033 | 5509 |
| 0.930316 | 0.070943 | 22034 | 5508 |
| 0.934353 | 0.071671 | 22034 | 5508 |
| 0.938244 | 0.071399 | 22034 | 5508 |
| 0.862348 | 0.135172 | 27542 | 4025 |
| 0.862307 | 0.135213 | 27542 | 4025 |
| 0.909664 | 0.08772 | 27542 | 4025 |
| 0.909884 | 0.127827 | 22033 | 5509 |
| 0.912916 | 0.120212 | 22033 | 5509 |
| 0.845181 | 0.124285 | 22034 | 5508 |
| 0.85241 | 0.128944 | 22034 | 5508 |
| 0.848504 | 0.130603 | 22034 | 5508 |
| 0.910022 | 0.127695 | 22033 | 5509 |
| 0.912746 | 0.120376 | 22033 | 5509 |
| 0.845078 | 0.124392 | 22034 | 5508 |
| 0.852487 | 0.128866 | 22034 | 5508 |
| 0.848498 | 0.130609 | 22034 | 5508 |
| 0.955133 | 0.084453 | 22033 | 5509 |
| 0.951368 | 0.083155 | 22033 | 5509 |
| 0.880141 | 0.088062 | 22034 | 5508 |
| 0.895168 | 0.085251 | 22034 | 5508 |
| 0.895898 | 0.082042 | 22034 | 5508 |
| 0.916814 | 0.08692 | 27542 | 4025 |
| 0.916716 | 0.087017 | 27542 | 4025 |
| 0.977006 | 0.026974 | 27542 | 4025 |
| 0.892272 | 0.095782 | 22033 | 5509 |
| 0.921764 | 0.084177 | 22033 | 5509 |
| 0.895421 | 0.101907 | 22034 | 5508 |
| 0.920189 | 0.096949 | 22034 | 5508 |
| 0.897871 | 0.092767 | 22034 | 5508 |
| 0.892308 | 0.095747 | 22033 | 5509 |
| 0.921643 | 0.084298 | 22033 | 5509 |
| 0.895491 | 0.101837 | 22034 | 5508 |
| 0.920113 | 0.097023 | 22034 | 5508 |
| 0.898002 | 0.092635 | 22034 | 5508 |
| 0.961751 | 0.025374 | 22033 | 5509 |
| 0.97886 | 0.02745 | 22033 | 5509 |
| 0.969509 | 0.027597 | 22034 | 5508 |
| 0.987908 | 0.030492 | 22034 | 5508 |

| | | | |
|---|---|---|---|
| 0.961083 | 0.028896 | 22034 | 5508 |
| 0.932975 | 0.034747 | 27542 | 4025 |
| 0.932888 | 0.034837 | 27542 | 4025 |
| 0.96332 | 0.003352 | 27542 | 4025 |
| 0.96894 | 0.03052 | 22033 | 5509 |
| 0.97655 | 0.028737 | 22033 | 5509 |
| 0.965135 | 0.019197 | 22034 | 5508 |
| 0.980199 | 0.023488 | 22034 | 5508 |
| 0.976958 | 0.029308 | 22034 | 5508 |
| 0.968909 | 0.030551 | 22033 | 5509 |
| 0.976665 | 0.028622 | 22033 | 5509 |
| 0.965009 | 0.019326 | 22034 | 5508 |
| 0.980095 | 0.023591 | 22034 | 5508 |
| 0.977084 | 0.029182 | 22034 | 5508 |
| 0.994689 | 0.004757 | 22033 | 5509 |
| 1.001744 | 0.003678 | 22033 | 5509 |
| 0.980745 | 0.003334 | 22034 | 5508 |
| 0.999947 | 0.003814 | 22034 | 5508 |
| 1.0024 | 0.004029 | 22034 | 5508 |
| 0.937671 | 0.066822 | 27542 | 4025 |
| 0.937821 | 0.066672 | 27542 | 4025 |
| 0.973359 | 0.031304 | 27542 | 4025 |
| 0.940566 | 0.071846 | 22033 | 5509 |
| 0.919527 | 0.077724 | 22033 | 5509 |
| 0.929805 | 0.072156 | 22034 | 5508 |
| 0.940144 | 0.074679 | 22034 | 5508 |
| 0.913805 | 0.059002 | 22034 | 5508 |
| 0.940761 | 0.071653 | 22033 | 5509 |
| 0.919398 | 0.077853 | 22033 | 5509 |
| 0.929789 | 0.072172 | 22034 | 5508 |
| 0.940162 | 0.074661 | 22034 | 5508 |
| 0.91375 | 0.059058 | 22034 | 5508 |
| 0.978678 | 0.034236 | 22033 | 5509 |
| 0.963958 | 0.033161 | 22033 | 5509 |
| 0.967888 | 0.034153 | 22034 | 5508 |
| 0.98144 | 0.034034 | 22034 | 5508 |
| 0.940456 | 0.031557 | 22034 | 5508 |
| 0.929788 | 0.076147 | 27542 | 4025 |
| 0.929907 | 0.076029 | 27542 | 4025 |
| 0.966385 | 0.039783 | 27542 | 4025 |
| 0.912913 | 0.07335 | 22033 | 5509 |
| 0.93538 | 0.076979 | 22033 | 5509 |
| 0.928485 | 0.080282 | 22034 | 5508 |
| 0.911857 | 0.094842 | 22034 | 5508 |
| 0.907157 | 0.078301 | 22034 | 5508 |

| | | | |
|---|---|---|---|
| 0.912843 | 0.073422 | 22033 | 5509 |
| 0.935436 | 0.076925 | 22033 | 5509 |
| 0.928556 | 0.080212 | 22034 | 5508 |
| 0.911822 | 0.094877 | 22034 | 5508 |
| 0.907167 | 0.078291 | 22034 | 5508 |
| 0.940949 | 0.044893 | 22033 | 5509 |
| 0.972833 | 0.040021 | 22033 | 5509 |
| 0.962492 | 0.046596 | 22034 | 5508 |
| 0.958298 | 0.048743 | 22034 | 5508 |
| 0.940175 | 0.044753 | 22034 | 5508 |
| 0.907238 | 0.095557 | 27542 | 4025 |
| 0.907222 | 0.095572 | 27542 | 4025 |
| 0.948404 | 0.054517 | 27542 | 4025 |
| 0.900909 | 0.098625 | 22033 | 5509 |
| 0.887818 | 0.094949 | 22033 | 5509 |
| 0.920553 | 0.10687 | 22034 | 5508 |
| 0.937732 | 0.104272 | 22034 | 5508 |
| 0.845251 | 0.102451 | 22034 | 5508 |
| 0.90086 | 0.098674 | 22033 | 5509 |
| 0.887741 | 0.095027 | 22033 | 5509 |
| 0.921081 | 0.106357 | 22034 | 5508 |
| 0.93774 | 0.104264 | 22034 | 5508 |
| 0.845186 | 0.102521 | 22034 | 5508 |
| 0.946183 | 0.053327 | 22033 | 5509 |
| 0.932448 | 0.049453 | 22033 | 5509 |
| 0.976905 | 0.052196 | 22034 | 5508 |
| 0.989967 | 0.054377 | 22034 | 5508 |
| 0.886587 | 0.058558 | 22034 | 5508 |
| 0.935434 | 0.078625 | 27542 | 4025 |
| 0.935398 | 0.07866 | 27542 | 4025 |
| 0.984871 | 0.02993 | 27542 | 4025 |
| 0.911933 | 0.083326 | 22033 | 5509 |
| 0.915277 | 0.073737 | 22033 | 5509 |
| 0.941337 | 0.098311 | 22034 | 5508 |
| 0.877556 | 0.084444 | 22034 | 5508 |
| 0.939167 | 0.074538 | 22034 | 5508 |
| 0.91188 | 0.08338 | 22033 | 5509 |
| 0.915212 | 0.073804 | 22033 | 5509 |
| 0.941484 | 0.09817 | 22034 | 5508 |
| 0.877488 | 0.084515 | 22034 | 5508 |
| 0.939204 | 0.074501 | 22034 | 5508 |
| 0.964942 | 0.030042 | 22033 | 5509 |
| 0.961487 | 0.026973 | 22033 | 5509 |
| 1.012701 | 0.029953 | 22034 | 5508 |
| 0.928738 | 0.031045 | 22034 | 5508 |

| | | | |
|---|---|---|---|
| 0.988248 | 0.026173 | 22034 | 5508 |
| 0.994711 | 0.032573 | 27542 | 4025 |
| 0.994707 | 0.032577 | 27542 | 4025 |
| 1.018155 | 0.009772 | 27542 | 4025 |
| 0.98937 | 0.032878 | 22033 | 5509 |
| 0.929176 | 0.040312 | 22033 | 5509 |
| 0.958877 | 0.042791 | 22034 | 5508 |
| 0.983055 | 0.038776 | 22034 | 5508 |
| 0.95001 | 0.034394 | 22034 | 5508 |
| 0.989313 | 0.032934 | 22033 | 5509 |
| 0.929214 | 0.040272 | 22033 | 5509 |
| 0.958886 | 0.042782 | 22034 | 5508 |
| 0.983061 | 0.03877 | 22034 | 5508 |
| 0.950018 | 0.034386 | 22034 | 5508 |
| 1.012709 | 0.010065 | 22033 | 5509 |
| 0.958262 | 0.010271 | 22033 | 5509 |
| 0.990326 | 0.011397 | 22034 | 5508 |
| 1.011056 | 0.011396 | 22034 | 5508 |
| 0.974039 | 0.009971 | 22034 | 5508 |
| 0.965146 | 0.055392 | 27542 | 4025 |
| 0.9652 | 0.055339 | 27542 | 4025 |
| 1.004163 | 0.017205 | 27542 | 4025 |
| 0.953358 | 0.068399 | 22033 | 5509 |
| 0.897091 | 0.066813 | 22033 | 5509 |
| 0.954676 | 0.082347 | 22034 | 5508 |
| 0.934183 | 0.069052 | 22034 | 5508 |
| 0.907417 | 0.066186 | 22034 | 5508 |
| 0.953348 | 0.068408 | 22033 | 5509 |
| 0.8971 | 0.066804 | 22033 | 5509 |
| 0.954694 | 0.082329 | 22034 | 5508 |
| 0.934147 | 0.069088 | 22034 | 5508 |
| 0.907403 | 0.0662 | 22034 | 5508 |
| 1.000302 | 0.022526 | 22033 | 5509 |
| 0.938211 | 0.024038 | 22033 | 5509 |
| 1.017553 | 0.021909 | 22034 | 5508 |
| 0.982065 | 0.021336 | 22034 | 5508 |
| 0.950936 | 0.021401 | 22034 | 5508 |
| 0.823659 | 0.165291 | 27542 | 4025 |
| 0.823629 | 0.165321 | 27542 | 4025 |
| 0.879734 | 0.108464 | 27542 | 4025 |
| 0.834891 | 0.183856 | 22033 | 5509 |
| 0.829998 | 0.173316 | 22033 | 5509 |
| 0.803585 | 0.187585 | 22034 | 5508 |
| 0.831638 | 0.184319 | 22034 | 5508 |
| 0.803908 | 0.166121 | 22034 | 5508 |

| | | | |
|---|---|---|---|
| 0.834931 | 0.183817 | 22033 | 5509 |
| 0.829944 | 0.173369 | 22033 | 5509 |
| 0.803703 | 0.187466 | 22034 | 5508 |
| 0.831661 | 0.184297 | 22034 | 5508 |
| 0.803829 | 0.166202 | 22034 | 5508 |
| 0.908367 | 0.11203 | 22033 | 5509 |
| 0.895387 | 0.108187 | 22033 | 5509 |
| 0.877337 | 0.113022 | 22034 | 5508 |
| 0.900545 | 0.116735 | 22034 | 5508 |
| 0.860149 | 0.107783 | 22034 | 5508 |
| 0.370008 | 0.633422 | 27542 | 4025 |
| 0.369976 | 0.633453 | 27542 | 4025 |
| 0.429628 | 0.574354 | 27542 | 4025 |
| 0.373658 | 0.628359 | 22033 | 5509 |
| 0.353644 | 0.636103 | 22033 | 5509 |
| 0.358398 | 0.633471 | 22034 | 5508 |
| 0.37285 | 0.639101 | 22034 | 5508 |
| 0.364934 | 0.639107 | 22034 | 5508 |
| 0.373798 | 0.62822 | 22033 | 5509 |
| 0.353617 | 0.636131 | 22033 | 5509 |
| 0.358459 | 0.633408 | 22034 | 5508 |
| 0.372776 | 0.639172 | 22034 | 5508 |
| 0.364906 | 0.639135 | 22034 | 5508 |
| 0.437755 | 0.564609 | 22033 | 5509 |
| 0.405373 | 0.582874 | 22033 | 5509 |
| 0.409414 | 0.581297 | 22034 | 5508 |
| 0.432043 | 0.581806 | 22034 | 5508 |
| 0.422486 | 0.582192 | 22034 | 5508 |
| 0.457437 | 0.537318 | 27542 | 4025 |
| 0.457402 | 0.537353 | 27542 | 4025 |
| 0.521627 | 0.472391 | 27542 | 4025 |
| 0.452996 | 0.55233 | 22033 | 5509 |
| 0.434918 | 0.555925 | 22033 | 5509 |
| 0.438608 | 0.552722 | 22034 | 5508 |
| 0.446837 | 0.566054 | 22034 | 5508 |
| 0.440215 | 0.558926 | 22034 | 5508 |
| 0.453151 | 0.552177 | 22033 | 5509 |
| 0.434894 | 0.555949 | 22033 | 5509 |
| 0.438747 | 0.55258 | 22034 | 5508 |
| 0.446647 | 0.566239 | 22034 | 5508 |
| 0.440267 | 0.558874 | 22034 | 5508 |
| 0.527853 | 0.478353 | 22033 | 5509 |
| 0.493658 | 0.495948 | 22033 | 5509 |
| 0.49965 | 0.490473 | 22034 | 5508 |
| 0.518361 | 0.496594 | 22034 | 5508 |

| | | | |
|---|---|---|---|
| 0.506895 | 0.492116 | 22034 | 5508 |
| 0.494582 | 0.514009 | 27542 | 4025 |
| 0.494578 | 0.514013 | 27542 | 4025 |
| 0.546147 | 0.46334 | 27542 | 4025 |
| 0.524802 | 0.479364 | 22033 | 5509 |
| 0.481479 | 0.497853 | 22033 | 5509 |
| 0.503258 | 0.490055 | 22034 | 5508 |
| 0.518562 | 0.49626 | 22034 | 5508 |
| 0.508336 | 0.499765 | 22034 | 5508 |
| 0.524727 | 0.479439 | 22033 | 5509 |
| 0.481497 | 0.497835 | 22033 | 5509 |
| 0.50339 | 0.489922 | 22034 | 5508 |
| 0.518409 | 0.496409 | 22034 | 5508 |
| 0.508299 | 0.499801 | 22034 | 5508 |
| 0.574671 | 0.429891 | 22033 | 5509 |
| 0.525931 | 0.451493 | 22033 | 5509 |
| 0.545843 | 0.446904 | 22034 | 5508 |
| 0.565214 | 0.450941 | 22034 | 5508 |
| 0.557061 | 0.451817 | 22034 | 5508 |
| 0.862681 | 0.115219 | 27542 | 4025 |
| 0.862643 | 0.115257 | 27542 | 4025 |
| 0.920726 | 0.055687 | 27542 | 4025 |
| 0.877732 | 0.141843 | 22033 | 5509 |
| 0.827209 | 0.147273 | 22033 | 5509 |
| 0.840286 | 0.143088 | 22034 | 5508 |
| 0.873674 | 0.156918 | 22034 | 5508 |
| 0.850371 | 0.141522 | 22034 | 5508 |
| 0.877758 | 0.141818 | 22033 | 5509 |
| 0.827155 | 0.147328 | 22033 | 5509 |
| 0.840248 | 0.143128 | 22034 | 5508 |
| 0.873743 | 0.156851 | 22034 | 5508 |
| 0.850355 | 0.141538 | 22034 | 5508 |
| 0.94817 | 0.072977 | 22033 | 5509 |
| 0.893413 | 0.079026 | 22033 | 5509 |
| 0.906174 | 0.075897 | 22034 | 5508 |
| 0.954732 | 0.078698 | 22034 | 5508 |
| 0.919091 | 0.072147 | 22034 | 5508 |
| 0.916939 | 0.068225 | 27542 | 4025 |
| 0.91679 | 0.068375 | 27542 | 4025 |
| 0.963391 | 0.021021 | 27542 | 4025 |
| 0.956124 | 0.05655 | 22033 | 5509 |
| 0.931974 | 0.081654 | 22033 | 5509 |
| 0.900488 | 0.066493 | 22034 | 5508 |
| 0.912495 | 0.078825 | 22034 | 5508 |
| 0.943667 | 0.071925 | 22034 | 5508 |

| | | | |
|---|---|---|---|
| 0.955993 | 0.056679 | 22033 | 5509 |
| 0.932008 | 0.081621 | 22033 | 5509 |
| 0.900466 | 0.066516 | 22034 | 5508 |
| 0.912856 | 0.07846 | 22034 | 5508 |
| 0.94366 | 0.071933 | 22034 | 5508 |
| 0.994662 | 0.018523 | 22033 | 5509 |
| 0.991084 | 0.023409 | 22033 | 5509 |
| 0.945069 | 0.020277 | 22034 | 5508 |
| 0.969343 | 0.021436 | 22034 | 5508 |
| 0.993549 | 0.022868 | 22034 | 5508 |
| 0.850968 | 0.135055 | 27542 | 4025 |
| 0.850823 | 0.135202 | 27542 | 4025 |
| 0.879678 | 0.105873 | 27542 | 4025 |
| 0.85796 | 0.158366 | 22033 | 5509 |
| 0.85197 | 0.153834 | 22033 | 5509 |
| 0.845498 | 0.142174 | 22034 | 5508 |
| 0.873094 | 0.138607 | 22034 | 5508 |
| 0.827916 | 0.150148 | 22034 | 5508 |
| 0.858167 | 0.158162 | 22033 | 5509 |
| 0.852085 | 0.153719 | 22033 | 5509 |
| 0.845348 | 0.142326 | 22034 | 5508 |
| 0.872909 | 0.13879 | 22034 | 5508 |
| 0.827965 | 0.150098 | 22034 | 5508 |
| 0.905059 | 0.112163 | 22033 | 5509 |
| 0.898098 | 0.10802 | 22033 | 5509 |
| 0.878133 | 0.109063 | 22034 | 5508 |
| 0.909504 | 0.102685 | 22034 | 5508 |
| 0.869612 | 0.107347 | 22034 | 5508 |
| 0.897135 | 0.125753 | 27542 | 4025 |
| 0.897263 | 0.125628 | 27542 | 4025 |
| 0.965638 | 0.058998 | 27542 | 4025 |
| 0.928619 | 0.120777 | 22033 | 5509 |
| 0.870931 | 0.117945 | 22033 | 5509 |
| 0.837088 | 0.133864 | 22034 | 5508 |
| 0.886413 | 0.105658 | 22034 | 5508 |
| 0.891712 | 0.107581 | 22034 | 5508 |
| 0.928711 | 0.12069 | 22033 | 5509 |
| 0.870976 | 0.1179 | 22033 | 5509 |
| 0.837223 | 0.133725 | 22034 | 5508 |
| 0.886387 | 0.105685 | 22034 | 5508 |
| 0.891565 | 0.107729 | 22034 | 5508 |
| 0.996496 | 0.05651 | 22033 | 5509 |
| 0.929938 | 0.058185 | 22033 | 5509 |
| 0.907727 | 0.060774 | 22034 | 5508 |
| 0.936766 | 0.054855 | 22034 | 5508 |

| | | | |
|---|---|---|---|
| 0.945192 | 0.054059 | 22034 | 5508 |
| 0.891499 | 0.1226 | 27542 | 4025 |
| 0.891487 | 0.122612 | 27542 | 4025 |
| 0.945648 | 0.069307 | 27542 | 4025 |
| 0.864412 | 0.132833 | 22033 | 5509 |
| 0.86266 | 0.119578 | 22033 | 5509 |
| 0.86877 | 0.126448 | 22034 | 5508 |
| 0.907725 | 0.125764 | 22034 | 5508 |
| 0.858541 | 0.132724 | 22034 | 5508 |
| 0.864444 | 0.132801 | 22033 | 5509 |
| 0.862577 | 0.119664 | 22033 | 5509 |
| 0.868762 | 0.126457 | 22034 | 5508 |
| 0.907746 | 0.125744 | 22034 | 5508 |
| 0.858559 | 0.132706 | 22034 | 5508 |
| 0.919835 | 0.077233 | 22033 | 5509 |
| 0.905482 | 0.075875 | 22033 | 5509 |
| 0.92032 | 0.074615 | 22034 | 5508 |
| 0.95827 | 0.077085 | 22034 | 5508 |
| 0.913735 | 0.076969 | 22034 | 5508 |
| 0.901114 | 0.063137 | 27542 | 4025 |
| 0.901158 | 0.063092 | 27542 | 4025 |
| 0.934287 | 0.028649 | 27542 | 4025 |
| 0.923472 | 0.071018 | 22033 | 5509 |
| 0.920746 | 0.060586 | 22033 | 5509 |
| 0.917481 | 0.052582 | 22034 | 5508 |
| 0.992816 | 0.051014 | 22034 | 5508 |
| 0.949691 | 0.059855 | 22034 | 5508 |
| 0.923532 | 0.070958 | 22033 | 5509 |
| 0.920834 | 0.060497 | 22033 | 5509 |
| 0.917617 | 0.052442 | 22034 | 5508 |
| 0.992719 | 0.051107 | 22034 | 5508 |
| 0.949556 | 0.059988 | 22034 | 5508 |
| 0.966902 | 0.027329 | 22033 | 5509 |
| 0.958257 | 0.022315 | 22033 | 5509 |
| 0.947617 | 0.021464 | 22034 | 5508 |
| 1.023865 | 0.021336 | 22034 | 5508 |
| 0.984789 | 0.025109 | 22034 | 5508 |
| 0.952217 | 0.070023 | 27542 | 4025 |
| 0.952323 | 0.069919 | 27542 | 4025 |
| 0.989441 | 0.033668 | 27542 | 4025 |
| 0.906195 | 0.076061 | 22033 | 5509 |
| 0.9146 | 0.059154 | 22033 | 5509 |
| 0.949538 | 0.065034 | 22034 | 5508 |
| 0.924653 | 0.063366 | 22034 | 5508 |
| 0.972202 | 0.068705 | 22034 | 5508 |

| | | | |
|---|---|---|---|
| 0.90631 | 0.075943 | 22033 | 5509 |
| 0.914562 | 0.059193 | 22033 | 5509 |
| 0.949592 | 0.064981 | 22034 | 5508 |
| 0.924594 | 0.063426 | 22034 | 5508 |
| 0.972173 | 0.068733 | 22034 | 5508 |
| 0.945613 | 0.03587 | 22033 | 5509 |
| 0.944732 | 0.028157 | 22033 | 5509 |
| 0.983084 | 0.032003 | 22034 | 5508 |
| 0.95388 | 0.03376 | 22034 | 5508 |
| 1.006552 | 0.035801 | 22034 | 5508 |
| 0.775915 | 0.190702 | 27542 | 4025 |
| 0.775901 | 0.190716 | 27542 | 4025 |
| 0.852527 | 0.110794 | 27542 | 4025 |
| 0.817754 | 0.18782 | 22033 | 5509 |
| 0.800627 | 0.197647 | 22033 | 5509 |
| 0.792684 | 0.186611 | 22034 | 5508 |
| 0.819563 | 0.177467 | 22034 | 5508 |
| 0.836052 | 0.183657 | 22034 | 5508 |
| 0.817722 | 0.187852 | 22033 | 5509 |
| 0.800722 | 0.197551 | 22033 | 5509 |
| 0.792669 | 0.186626 | 22034 | 5508 |
| 0.819571 | 0.177458 | 22034 | 5508 |
| 0.836045 | 0.183663 | 22034 | 5508 |
| 0.893421 | 0.112669 | 22033 | 5509 |
| 0.894 | 0.104071 | 22033 | 5509 |
| 0.870895 | 0.106357 | 22034 | 5508 |
| 0.895569 | 0.101185 | 22034 | 5508 |
| 0.914078 | 0.10747 | 22034 | 5508 |
| 0.917413 | 0.054903 | 27542 | 4025 |
| 0.917399 | 0.054918 | 27542 | 4025 |
| 0.946817 | 0.024612 | 27542 | 4025 |
| 0.973092 | 0.0519 | 22033 | 5509 |
| 0.948663 | 0.051578 | 22033 | 5509 |
| 0.942748 | 0.041111 | 22034 | 5508 |
| 0.923102 | 0.0466 | 22034 | 5508 |
| 0.965213 | 0.05575 | 22034 | 5508 |
| 0.973188 | 0.051807 | 22033 | 5509 |
| 0.948517 | 0.051724 | 22033 | 5509 |
| 0.942766 | 0.041093 | 22034 | 5508 |
| 0.923217 | 0.046482 | 22034 | 5508 |
| 0.965159 | 0.055803 | 22034 | 5508 |
| 0.999156 | 0.026506 | 22033 | 5509 |
| 0.972907 | 0.02734 | 22033 | 5509 |
| 0.961332 | 0.022208 | 22034 | 5508 |
| 0.944718 | 0.024275 | 22034 | 5508 |

| | | | |
|---|---|---|---|
| 0.995877 | 0.025751 | 22034 | 5508 |
| 0.855858 | 0.1103 | 27542 | 4025 |
| 0.856273 | 0.109868 | 27542 | 4025 |
| 0.882888 | 0.082201 | 27542 | 4025 |
| 0.904378 | 0.09836 | 22033 | 5509 |
| 0.892686 | 0.107625 | 22033 | 5509 |
| 0.881445 | 0.097338 | 22034 | 5508 |
| 0.910595 | 0.112708 | 22034 | 5508 |
| 0.892434 | 0.101607 | 22034 | 5508 |
| 0.904097 | 0.09864 | 22033 | 5509 |
| 0.892482 | 0.107829 | 22033 | 5509 |
| 0.881435 | 0.097348 | 22034 | 5508 |
| 0.910745 | 0.112561 | 22034 | 5508 |
| 0.892585 | 0.101454 | 22034 | 5508 |
| 0.924992 | 0.077807 | 22033 | 5509 |
| 0.916688 | 0.083631 | 22033 | 5509 |
| 0.903171 | 0.075088 | 22034 | 5508 |
| 0.94426 | 0.079904 | 22034 | 5508 |
| 0.915685 | 0.0782 | 22034 | 5508 |
| 0.804645 | 0.168096 | 27542 | 4025 |
| 0.804461 | 0.168286 | 27542 | 4025 |
| 0.822156 | 0.149991 | 27542 | 4025 |
| 0.853704 | 0.158136 | 22033 | 5509 |
| 0.849307 | 0.175405 | 22033 | 5509 |
| 0.810069 | 0.162882 | 22034 | 5508 |
| 0.838085 | 0.179295 | 22034 | 5508 |
| 0.807508 | 0.164228 | 22034 | 5508 |
| 0.853599 | 0.158241 | 22033 | 5509 |
| 0.849415 | 0.175299 | 22033 | 5509 |
| 0.809989 | 0.162964 | 22034 | 5508 |
| 0.838378 | 0.179008 | 22034 | 5508 |
| 0.807296 | 0.164447 | 22034 | 5508 |
| 0.873368 | 0.138745 | 22033 | 5509 |
| 0.877837 | 0.147704 | 22033 | 5509 |
| 0.829919 | 0.142369 | 22034 | 5508 |
| 0.874941 | 0.143204 | 22034 | 5508 |
| 0.821338 | 0.149914 | 22034 | 5508 |
| 0.848516 | 0.163615 | 27542 | 4025 |
| 0.848406 | 0.163724 | 27542 | 4025 |
| 0.876804 | 0.135732 | 27542 | 4025 |
| 0.826246 | 0.173328 | 22033 | 5509 |
| 0.84298 | 0.165855 | 22033 | 5509 |
| 0.809562 | 0.176904 | 22034 | 5508 |
| 0.843286 | 0.170297 | 22034 | 5508 |
| 0.830527 | 0.160862 | 22034 | 5508 |

| | | | |
|---|---|---|---|
| 0.82638 | 0.173194 | 22033 | 5509 |
| 0.843008 | 0.165827 | 22033 | 5509 |
| 0.80963 | 0.176834 | 22034 | 5508 |
| 0.843231 | 0.170351 | 22034 | 5508 |
| 0.83026 | 0.161133 | 22034 | 5508 |
| 0.867076 | 0.132477 | 22033 | 5509 |
| 0.877456 | 0.13174 | 22033 | 5509 |
| 0.840834 | 0.145109 | 22034 | 5508 |
| 0.878571 | 0.13558 | 22034 | 5508 |
| 0.856674 | 0.134445 | 22034 | 5508 |
| 0.902917 | 0.113563 | 27542 | 4025 |
| 0.902924 | 0.113556 | 27542 | 4025 |
| 0.946886 | 0.070397 | 27542 | 4025 |
| 0.890414 | 0.123743 | 22033 | 5509 |
| 0.885367 | 0.107229 | 22033 | 5509 |
| 0.867198 | 0.120638 | 22034 | 5508 |
| 0.913153 | 0.106715 | 22034 | 5508 |
| 0.877808 | 0.107661 | 22034 | 5508 |
| 0.890722 | 0.123439 | 22033 | 5509 |
| 0.885376 | 0.107221 | 22033 | 5509 |
| 0.86715 | 0.120686 | 22034 | 5508 |
| 0.913004 | 0.10686 | 22034 | 5508 |
| 0.877735 | 0.107735 | 22034 | 5508 |
| 0.944317 | 0.070697 | 22033 | 5509 |
| 0.926522 | 0.065731 | 22033 | 5509 |
| 0.916052 | 0.071098 | 22034 | 5508 |
| 0.954619 | 0.066151 | 22034 | 5508 |
| 0.916768 | 0.068056 | 22034 | 5508 |
| 0.912804 | 0.094806 | 27542 | 4025 |
| 0.912864 | 0.094746 | 27542 | 4025 |
| 0.97109 | 0.037005 | 27542 | 4025 |
| 0.901852 | 0.087155 | 22033 | 5509 |
| 0.923912 | 0.075247 | 22033 | 5509 |
| 0.911106 | 0.092762 | 22034 | 5508 |
| 0.92737 | 0.076532 | 22034 | 5508 |
| 0.930043 | 0.073553 | 22034 | 5508 |
| 0.901862 | 0.087145 | 22033 | 5509 |
| 0.923667 | 0.075493 | 22033 | 5509 |
| 0.911156 | 0.092711 | 22034 | 5508 |
| 0.927423 | 0.076479 | 22034 | 5508 |
| 0.92985 | 0.073745 | 22034 | 5508 |
| 0.95198 | 0.036416 | 22033 | 5509 |
| 0.963889 | 0.035234 | 22033 | 5509 |
| 0.968017 | 0.036092 | 22034 | 5508 |
| 0.972386 | 0.031706 | 22034 | 5508 |

| | | | |
|---|---|---|---|
| 0.969225 | 0.034522 | 22034 | 5508 |
| 0.973375 | 0.06015 | 27542 | 4025 |
| 0.973243 | 0.060278 | 27542 | 4025 |
| 1.004466 | 0.03013 | 27542 | 4025 |
| 0.912206 | 0.076676 | 22033 | 5509 |
| 0.974268 | 0.065309 | 22033 | 5509 |
| 0.885982 | 0.072844 | 22034 | 5508 |
| 0.946889 | 0.063889 | 22034 | 5508 |
| 0.930952 | 0.071464 | 22034 | 5508 |
| 0.912277 | 0.076603 | 22033 | 5509 |
| 0.974199 | 0.065375 | 22033 | 5509 |
| 0.886001 | 0.072823 | 22034 | 5508 |
| 0.94681 | 0.063968 | 22034 | 5508 |
| 0.931042 | 0.071374 | 22034 | 5508 |
| 0.955977 | 0.032371 | 22033 | 5509 |
| 1.009746 | 0.031272 | 22033 | 5509 |
| 0.927042 | 0.029876 | 22034 | 5508 |
| 0.981299 | 0.029871 | 22034 | 5508 |
| 0.973295 | 0.02923 | 22034 | 5508 |
| 0.960891 | 0.071444 | 27542 | 4025 |
| 0.960838 | 0.071494 | 27542 | 4025 |
| 0.998527 | 0.035074 | 27542 | 4025 |
| 0.940056 | 0.07445 | 22033 | 5509 |
| 0.955932 | 0.063953 | 22033 | 5509 |
| 0.911334 | 0.064092 | 22034 | 5508 |
| 0.951052 | 0.062653 | 22034 | 5508 |
| 0.915993 | 0.060214 | 22034 | 5508 |
| 0.9401 | 0.074407 | 22033 | 5509 |
| 0.955961 | 0.063926 | 22033 | 5509 |
| 0.911248 | 0.06418 | 22034 | 5508 |
| 0.951108 | 0.062598 | 22034 | 5508 |
| 0.915913 | 0.060297 | 22034 | 5508 |
| 0.984446 | 0.030745 | 22033 | 5509 |
| 0.994005 | 0.026673 | 22033 | 5509 |
| 0.944213 | 0.030326 | 22034 | 5508 |
| 0.986968 | 0.027254 | 22034 | 5508 |
| 0.943893 | 0.031589 | 22034 | 5508 |
| 0.935511 | 0.077479 | 27542 | 4025 |
| 0.935508 | 0.077482 | 27542 | 4025 |
| 0.97833 | 0.035255 | 27542 | 4025 |
| 0.901039 | 0.094851 | 22033 | 5509 |
| 0.901511 | 0.086359 | 22033 | 5509 |
| 0.90892 | 0.089545 | 22034 | 5508 |
| 0.928414 | 0.088354 | 22034 | 5508 |
| 0.914419 | 0.086244 | 22034 | 5508 |

| | | | |
|---|---|---|---|
| 0.900997 | 0.094893 | 22033 | 5509 |
| 0.901519 | 0.08635 | 22033 | 5509 |
| 0.908908 | 0.089556 | 22034 | 5508 |
| 0.928802 | 0.087973 | 22034 | 5508 |
| 0.914367 | 0.086296 | 22034 | 5508 |
| 0.953665 | 0.041985 | 22033 | 5509 |
| 0.949217 | 0.038011 | 22033 | 5509 |
| 0.960596 | 0.037781 | 22034 | 5508 |
| 0.980181 | 0.037521 | 22034 | 5508 |
| 0.96001 | 0.040686 | 22034 | 5508 |
| 0.91669 | 0.091029 | 27542 | 4025 |
| 0.916744 | 0.090976 | 27542 | 4025 |
| 0.951824 | 0.056191 | 27542 | 4025 |
| 0.885413 | 0.092635 | 22033 | 5509 |
| 0.917297 | 0.107275 | 22033 | 5509 |
| 0.927244 | 0.090564 | 22034 | 5508 |
| 0.882694 | 0.113254 | 22034 | 5508 |
| 0.885651 | 0.09773 | 22034 | 5508 |
| 0.88545 | 0.092597 | 22033 | 5509 |
| 0.917209 | 0.10736 | 22033 | 5509 |
| 0.927274 | 0.090535 | 22034 | 5508 |
| 0.88279 | 0.113158 | 22034 | 5508 |
| 0.885642 | 0.097739 | 22034 | 5508 |
| 0.920664 | 0.05651 | 22033 | 5509 |
| 0.962247 | 0.063529 | 22033 | 5509 |
| 0.963586 | 0.05492 | 22034 | 5508 |
| 0.934811 | 0.060898 | 22034 | 5508 |
| 0.921495 | 0.061214 | 22034 | 5508 |
| 0.92286 | 0.10118 | 27542 | 4025 |
| 0.922947 | 0.101096 | 27542 | 4025 |
| 0.967401 | 0.0578 | 27542 | 4025 |
| 0.874836 | 0.113414 | 22033 | 5509 |
| 0.914762 | 0.113269 | 22033 | 5509 |
| 0.887361 | 0.106974 | 22034 | 5508 |
| 0.896745 | 0.108684 | 22034 | 5508 |
| 0.873623 | 0.1104 | 22034 | 5508 |
| 0.874865 | 0.113384 | 22033 | 5509 |
| 0.914668 | 0.11336 | 22033 | 5509 |
| 0.887218 | 0.107118 | 22034 | 5508 |
| 0.896746 | 0.108683 | 22034 | 5508 |
| 0.873648 | 0.110375 | 22034 | 5508 |
| 0.922967 | 0.064637 | 22033 | 5509 |
| 0.964539 | 0.065017 | 22033 | 5509 |
| 0.929254 | 0.064814 | 22034 | 5508 |
| 0.943369 | 0.062342 | 22034 | 5508 |

| | | | |
|---|---|---|---|
| 0.917475 | 0.065746 | 22034 | 5508 |
| 0.937644 | 0.103085 | 27542 | 4025 |
| 0.937549 | 0.103177 | 27542 | 4025 |
| 0.975672 | 0.06671 | 27542 | 4025 |
| 0.875982 | 0.104644 | 22033 | 5509 |
| 0.918698 | 0.115166 | 22033 | 5509 |
| 0.864608 | 0.114083 | 22034 | 5508 |
| 0.872118 | 0.107074 | 22034 | 5508 |
| 0.92089 | 0.10704 | 22034 | 5508 |
| 0.875921 | 0.104707 | 22033 | 5509 |
| 0.91864 | 0.115221 | 22033 | 5509 |
| 0.86456 | 0.114132 | 22034 | 5508 |
| 0.872059 | 0.107135 | 22034 | 5508 |
| 0.920922 | 0.107008 | 22034 | 5508 |
| 0.914403 | 0.065374 | 22033 | 5509 |
| 0.969348 | 0.066382 | 22033 | 5509 |
| 0.912883 | 0.064618 | 22034 | 5508 |
| 0.911069 | 0.067195 | 22034 | 5508 |
| 0.96811 | 0.061251 | 22034 | 5508 |
| 0.968504 | 0.05434 | 27542 | 4025 |
| 0.968418 | 0.054423 | 27542 | 4025 |
| 0.994872 | 0.028594 | 27542 | 4025 |
| 0.959063 | 0.054273 | 22033 | 5509 |
| 0.939888 | 0.059108 | 22033 | 5509 |
| 0.927194 | 0.053103 | 22034 | 5508 |
| 0.963195 | 0.044241 | 22034 | 5508 |
| 0.956263 | 0.043326 | 22034 | 5508 |
| 0.95945 | 0.053892 | 22033 | 5509 |
| 0.939814 | 0.059181 | 22033 | 5509 |
| 0.92736 | 0.052932 | 22034 | 5508 |
| 0.963116 | 0.04432 | 22034 | 5508 |
| 0.956186 | 0.043403 | 22034 | 5508 |
| 0.98802 | 0.025719 | 22033 | 5509 |
| 0.967987 | 0.030979 | 22033 | 5509 |
| 0.953674 | 0.02606 | 22034 | 5508 |
| 0.982546 | 0.02504 | 22034 | 5508 |
| 0.977637 | 0.021943 | 22034 | 5508 |
| 0.944656 | 0.070878 | 27542 | 4025 |
| 0.944535 | 0.070998 | 27542 | 4025 |
| 0.990574 | 0.025715 | 27542 | 4025 |
| 0.951938 | 0.074016 | 22033 | 5509 |
| 0.940519 | 0.073266 | 22033 | 5509 |
| 0.902902 | 0.071419 | 22034 | 5508 |
| 0.92437 | 0.057739 | 22034 | 5508 |
| 0.947554 | 0.055168 | 22034 | 5508 |

| | | | |
|---|---|---|---|
| 0.95217 | 0.073791 | 22033 | 5509 |
| 0.940491 | 0.073293 | 22033 | 5509 |
| 0.902963 | 0.071356 | 22034 | 5508 |
| 0.924258 | 0.057853 | 22034 | 5508 |
| 0.947513 | 0.055208 | 22034 | 5508 |
| 1.00755 | 0.019921 | 22033 | 5509 |
| 0.989918 | 0.02459 | 22033 | 5509 |
| 0.949982 | 0.023 | 22034 | 5508 |
| 0.960281 | 0.021133 | 22034 | 5508 |
| 0.983223 | 0.0196 | 22034 | 5508 |
| 0.906512 | 0.061665 | 27542 | 4025 |
| 0.906474 | 0.061705 | 27542 | 4025 |
| 0.937764 | 0.029316 | 27542 | 4025 |
| 1.001049 | 0.066054 | 22033 | 5509 |
| 0.913058 | 0.074959 | 22033 | 5509 |
| 0.881161 | 0.081798 | 22034 | 5508 |
| 0.93739 | 0.067712 | 22034 | 5508 |
| 0.911217 | 0.067345 | 22034 | 5508 |
| 1.001082 | 0.066023 | 22033 | 5509 |
| 0.913076 | 0.074941 | 22033 | 5509 |
| 0.881282 | 0.081672 | 22034 | 5508 |
| 0.937314 | 0.067788 | 22034 | 5508 |
| 0.911152 | 0.067412 | 22034 | 5508 |
| 1.044857 | 0.025183 | 22033 | 5509 |
| 0.956977 | 0.030464 | 22033 | 5509 |
| 0.931506 | 0.029336 | 22034 | 5508 |
| 0.975848 | 0.029464 | 22034 | 5508 |
| 0.948537 | 0.029148 | 22034 | 5508 |
| 0.965049 | 0.039525 | 27542 | 4025 |
| 0.964989 | 0.039584 | 27542 | 4025 |
| 0.988733 | 0.015953 | 27542 | 4025 |
| 0.962339 | 0.050297 | 22033 | 5509 |
| 0.931279 | 0.053605 | 22033 | 5509 |
| 0.922528 | 0.05463 | 22034 | 5508 |
| 0.983147 | 0.041082 | 22034 | 5508 |
| 0.957402 | 0.044126 | 22034 | 5508 |
| 0.962493 | 0.050145 | 22033 | 5509 |
| 0.931422 | 0.053459 | 22033 | 5509 |
| 0.922887 | 0.054262 | 22034 | 5508 |
| 0.982933 | 0.041291 | 22034 | 5508 |
| 0.95706 | 0.044468 | 22034 | 5508 |
| 0.995833 | 0.017243 | 22033 | 5509 |
| 0.965964 | 0.018357 | 22033 | 5509 |
| 0.960144 | 0.016083 | 22034 | 5508 |
| 1.010167 | 0.014729 | 22034 | 5508 |

| | | | |
|---|---|---|---|
| 0.985437 | 0.016135 | 22034 | 5508 |
| 0.844844 | 0.147159 | 27542 | 4025 |
| 0.844816 | 0.147187 | 27542 | 4025 |
| 0.886853 | 0.104751 | 27542 | 4025 |
| 0.837862 | 0.172214 | 22033 | 5509 |
| 0.848573 | 0.17147 | 22033 | 5509 |
| 0.801465 | 0.165437 | 22034 | 5508 |
| 0.86659 | 0.154809 | 22034 | 5508 |
| 0.826711 | 0.154987 | 22034 | 5508 |
| 0.837968 | 0.17211 | 22033 | 5509 |
| 0.848595 | 0.171449 | 22033 | 5509 |
| 0.801564 | 0.165334 | 22034 | 5508 |
| 0.866551 | 0.154848 | 22034 | 5508 |
| 0.826577 | 0.155124 | 22034 | 5508 |
| 0.904065 | 0.106807 | 22033 | 5509 |
| 0.91069 | 0.11082 | 22033 | 5509 |
| 0.861151 | 0.103286 | 22034 | 5508 |
| 0.920591 | 0.102142 | 22034 | 5508 |
| 0.876212 | 0.10439 | 22034 | 5508 |
| 0.591962 | 0.378614 | 27542 | 4025 |
| 0.591678 | 0.378912 | 27542 | 4025 |
| 0.62882 | 0.339924 | 27542 | 4025 |
| 0.582177 | 0.394011 | 22033 | 5509 |
| 0.59731 | 0.397367 | 22033 | 5509 |
| 0.591527 | 0.403003 | 22034 | 5508 |
| 0.613805 | 0.401318 | 22034 | 5508 |
| 0.600755 | 0.417918 | 22034 | 5508 |
| 0.582015 | 0.394179 | 22033 | 5509 |
| 0.597628 | 0.397046 | 22033 | 5509 |
| 0.591611 | 0.402918 | 22034 | 5508 |
| 0.613628 | 0.401491 | 22034 | 5508 |
| 0.600785 | 0.41789 | 22034 | 5508 |
| 0.626823 | 0.347538 | 22033 | 5509 |
| 0.657394 | 0.336747 | 22033 | 5509 |
| 0.645415 | 0.348616 | 22034 | 5508 |
| 0.667098 | 0.349339 | 22034 | 5508 |
| 0.661574 | 0.35899 | 22034 | 5508 |
| 0.66219 | 0.278259 | 27542 | 4025 |
| 0.661955 | 0.278514 | 27542 | 4025 |
| 0.696294 | 0.241088 | 27542 | 4025 |
| 0.679245 | 0.300289 | 22033 | 5509 |
| 0.692642 | 0.314134 | 22033 | 5509 |
| 0.669758 | 0.311657 | 22034 | 5508 |
| 0.72161 | 0.30383 | 22034 | 5508 |
| 0.66182 | 0.344807 | 22034 | 5508 |

| | | | |
|---|---|---|---|
| 0.679145 | 0.300391 | 22033 | 5509 |
| 0.692997 | 0.313782 | 22033 | 5509 |
| 0.669704 | 0.311712 | 22034 | 5508 |
| 0.721423 | 0.30401 | 22034 | 5508 |
| 0.661879 | 0.344749 | 22034 | 5508 |
| 0.723681 | 0.254514 | 22033 | 5509 |
| 0.759179 | 0.248248 | 22033 | 5509 |
| 0.71784 | 0.26224 | 22034 | 5508 |
| 0.77997 | 0.247527 | 22034 | 5508 |
| 0.730182 | 0.27713 | 22034 | 5508 |
| 0.639704 | 0.348866 | 27542 | 4025 |
| 0.639398 | 0.349178 | 27542 | 4025 |
| 0.694554 | 0.293037 | 27542 | 4025 |
| 0.647306 | 0.339293 | 22033 | 5509 |
| 0.618455 | 0.344541 | 22033 | 5509 |
| 0.667898 | 0.341741 | 22034 | 5508 |
| 0.659079 | 0.35496 | 22034 | 5508 |
| 0.68772 | 0.339106 | 22034 | 5508 |
| 0.647217 | 0.339383 | 22033 | 5509 |
| 0.618667 | 0.344317 | 22033 | 5509 |
| 0.668045 | 0.341596 | 22034 | 5508 |
| 0.658854 | 0.35518 | 22034 | 5508 |
| 0.687647 | 0.339175 | 22034 | 5508 |
| 0.707714 | 0.277634 | 22033 | 5509 |
| 0.681646 | 0.27757 | 22033 | 5509 |
| 0.735476 | 0.275138 | 22034 | 5508 |
| 0.720144 | 0.295196 | 22034 | 5508 |
| 0.75079 | 0.278496 | 22034 | 5508 |
| 0.838312 | 0.088939 | 27542 | 4025 |
| 0.838217 | 0.089042 | 27542 | 4025 |
| 0.879248 | 0.04445 | 27542 | 4025 |
| 0.900484 | 0.116136 | 22033 | 5509 |
| 0.844708 | 0.13441 | 22033 | 5509 |
| 0.887545 | 0.110609 | 22034 | 5508 |
| 0.903696 | 0.116213 | 22034 | 5508 |
| 0.843435 | 0.143692 | 22034 | 5508 |
| 0.900562 | 0.11606 | 22033 | 5509 |
| 0.844817 | 0.134299 | 22033 | 5509 |
| 0.887428 | 0.110727 | 22034 | 5508 |
| 0.903628 | 0.116279 | 22034 | 5508 |
| 0.843472 | 0.143654 | 22034 | 5508 |
| 0.95311 | 0.064481 | 22033 | 5509 |
| 0.909875 | 0.067632 | 22033 | 5509 |
| 0.931484 | 0.066578 | 22034 | 5508 |
| 0.961153 | 0.060022 | 22034 | 5508 |

| | | | |
|---|---|---|---|
| 0.912022 | 0.074058 | 22034 | 5508 |
| 0.365892 | 0.636137 | 27542 | 4025 |
| 0.365884 | 0.636144 | 27542 | 4025 |
| 0.408892 | 0.593375 | 27542 | 4025 |
| 0.373892 | 0.624137 | 22033 | 5509 |
| 0.368167 | 0.622798 | 22033 | 5509 |
| 0.37714 | 0.620073 | 22034 | 5508 |
| 0.364326 | 0.638309 | 22034 | 5508 |
| 0.387567 | 0.623153 | 22034 | 5508 |
| 0.374056 | 0.623972 | 22033 | 5509 |
| 0.368133 | 0.622832 | 22033 | 5509 |
| 0.377208 | 0.620005 | 22034 | 5508 |
| 0.364273 | 0.638362 | 22034 | 5508 |
| 0.38755 | 0.62317 | 22034 | 5508 |
| 0.424255 | 0.573508 | 22033 | 5509 |
| 0.407141 | 0.582866 | 22033 | 5509 |
| 0.41497 | 0.581964 | 22034 | 5508 |
| 0.407521 | 0.595426 | 22034 | 5508 |
| 0.434084 | 0.577923 | 22034 | 5508 |
| 0.488307 | 0.495924 | 27542 | 4025 |
| 0.488232 | 0.496002 | 27542 | 4025 |
| 0.526329 | 0.456674 | 27542 | 4025 |
| 0.495208 | 0.505619 | 22033 | 5509 |
| 0.523094 | 0.486209 | 22033 | 5509 |
| 0.490779 | 0.501434 | 22034 | 5508 |
| 0.483167 | 0.508869 | 22034 | 5508 |
| 0.511495 | 0.494195 | 22034 | 5508 |
| 0.495462 | 0.505365 | 22033 | 5509 |
| 0.523047 | 0.486256 | 22033 | 5509 |
| 0.490967 | 0.501243 | 22034 | 5508 |
| 0.482934 | 0.509106 | 22034 | 5508 |
| 0.511423 | 0.494267 | 22034 | 5508 |
| 0.551828 | 0.449094 | 22033 | 5509 |
| 0.569428 | 0.440699 | 22033 | 5509 |
| 0.536918 | 0.454563 | 22034 | 5508 |
| 0.529656 | 0.461614 | 22034 | 5508 |
| 0.560389 | 0.445845 | 22034 | 5508 |
| 0.479688 | 0.533887 | 27542 | 4025 |
| 0.479709 | 0.533867 | 27542 | 4025 |
| 0.530143 | 0.484859 | 27542 | 4025 |
| 0.496626 | 0.499105 | 22033 | 5509 |
| 0.461024 | 0.514511 | 22033 | 5509 |
| 0.49233 | 0.50402 | 22034 | 5508 |
| 0.490728 | 0.523349 | 22034 | 5508 |
| 0.506736 | 0.511211 | 22034 | 5508 |

| | | | |
|---|---|---|---|
| 0.496545 | 0.499186 | 22033 | 5509 |
| 0.461055 | 0.514478 | 22033 | 5509 |
| 0.492349 | 0.504001 | 22034 | 5508 |
| 0.490823 | 0.523257 | 22034 | 5508 |
| 0.506756 | 0.511192 | 22034 | 5508 |
| 0.541074 | 0.454274 | 22033 | 5509 |
| 0.496687 | 0.476955 | 22033 | 5509 |
| 0.525843 | 0.470259 | 22034 | 5508 |
| 0.531806 | 0.483449 | 22034 | 5508 |
| 0.553301 | 0.466295 | 22034 | 5508 |
| 0.865544 | 0.125126 | 27542 | 4025 |
| 0.865552 | 0.125117 | 27542 | 4025 |
| 0.913318 | 0.076837 | 27542 | 4025 |
| 0.870294 | 0.143897 | 22033 | 5509 |
| 0.879886 | 0.132121 | 22033 | 5509 |
| 0.839304 | 0.138516 | 22034 | 5508 |
| 0.862524 | 0.137065 | 22034 | 5508 |
| 0.872614 | 0.123114 | 22034 | 5508 |
| 0.870329 | 0.143862 | 22033 | 5509 |
| 0.879797 | 0.132209 | 22033 | 5509 |
| 0.839364 | 0.138454 | 22034 | 5508 |
| 0.862538 | 0.137051 | 22034 | 5508 |
| 0.872566 | 0.123162 | 22034 | 5508 |
| 0.930846 | 0.084332 | 22033 | 5509 |
| 0.932259 | 0.080463 | 22033 | 5509 |
| 0.897873 | 0.0784 | 22034 | 5508 |
| 0.922393 | 0.077167 | 22034 | 5508 |
| 0.922143 | 0.073342 | 22034 | 5508 |
| 0.682536 | 0.311843 | 27542 | 4025 |
| 0.682548 | 0.31183 | 27542 | 4025 |
| 0.756416 | 0.237354 | 27542 | 4025 |
| 0.712417 | 0.293024 | 22033 | 5509 |
| 0.674986 | 0.289664 | 22033 | 5509 |
| 0.721902 | 0.287507 | 22034 | 5508 |
| 0.703912 | 0.289219 | 22034 | 5508 |
| 0.743808 | 0.28351 | 22034 | 5508 |
| 0.712293 | 0.293147 | 22033 | 5509 |
| 0.675066 | 0.28958 | 22033 | 5509 |
| 0.721892 | 0.287517 | 22034 | 5508 |
| 0.703897 | 0.289235 | 22034 | 5508 |
| 0.743732 | 0.283583 | 22034 | 5508 |
| 0.781057 | 0.224908 | 22033 | 5509 |
| 0.736309 | 0.22513 | 22033 | 5509 |
| 0.788416 | 0.22186 | 22034 | 5508 |
| 0.761492 | 0.231078 | 22034 | 5508 |

| | | | |
|---|---|---|---|
| 0.807257 | 0.222392 | 22034 | 5508 |
