## Supplementary Table 2 for "Hundreds of cardiac MRI traits derived using 3D diffusion autoencoders share a common genetic architecture"

| Phenotype | Latent | Num samp | Effect Size | StdError | P | Log10P | Category |
|---|---|---|---|---|---|---|---|
| LA_maximu | Z31_S5 | 27446 | 0.277506 | 0.003866 | 0 | 1120.907 | Left atrium |
| LV_end_di | Z12_S4 | 27446 | -0.37271 | 0.005385 | 0 | 1041.947 | Left ventricle |
| LV_end_di | Z31_S5 | 27446 | 0.347311 | 0.005082 | 0 | 1016.038 | Left ventricle |
| RV_end_di | Z12_S4 | 27446 | -0.38694 | 0.005682 | 0 | 1009.04 | Right ventricle |
| LV_end_di | Z106_S4 | 27446 | 0.372427 | 0.005576 | 0 | 970.7225 | Left ventricle |
| LA_minimu | Z31_S5 | 27446 | 0.247181 | 0.003803 | 0 | 919.2234 | Left atrium |
| LV_end_di | Z127_S5 | 27446 | 0.352016 | 0.005417 | 0 | 918.7528 | Left ventricle |
| LA_maximu | Z92_S5 | 27446 | 0.36755 | 0.005914 | 0 | 840.6695 | Left atrium |
| LV_end_di | Z94_S5 | 27446 | 0.342684 | 0.00552 | 0 | 838.7068 | Left ventricle |
| LV_stroke_ | Z12_S4 | 27446 | -0.30131 | 0.004854 | 0 | 838.5273 | Left ventricle |
| LV_end_di | Z99_S3 | 27446 | 0.319209 | 0.005166 | 0 | 831.002 | Left ventricle |
| LV_end_di | Z73_S4 | 27446 | 0.452061 | 0.00736 | 0 | 821.0172 | Left ventricle |
| LV_end_di | Z4_S4 | 27446 | -0.3491 | 0.005698 | 0 | 817.0437 | Left ventricle |
| LA_maximu | Z82_S1 | 27446 | -0.36733 | 0.00608 | 0 | 794.4039 | Left atrium |
| LV_stroke_ | Z82_S1 | 27446 | -0.42522 | 0.007048 | 0 | 792.3323 | Left ventricle |
| LV_end_di | Z117_S4 | 27446 | -0.30835 | 0.005245 | 0 | 752.2947 | Left ventricle |
| RV_end_di | Z73_S4 | 27446 | 0.456821 | 0.007789 | 0 | 748.8961 | Right ventricle |
| RV_end_di | Z4_S4 | 27446 | -0.35305 | 0.006029 | 0 | 746.5979 | Right ventricle |
| LV_end_di | Z89_S4 | 27446 | -0.36474 | 0.006229 | 0 | 746.4819 | Left ventricle |
| LV_stroke_ | Z127_S5 | 27446 | 0.285254 | 0.004873 | 0 | 745.9184 | Left ventricle |
| LV_end_di | Z51_S4 | 27446 | -0.32643 | 0.005584 | 0 | 743.8466 | Left ventricle |
| RV_end_di | Z82_S1 | 27446 | -0.48947 | 0.008383 | 0 | 742.0815 | Right ventricle |
| RV_end_di | Z106_S4 | 27446 | 0.34733 | 0.00597 | 0 | 736.8054 | Right ventricle |
| LA_maximu | Z106_S4 | 27446 | 0.25262 | 0.004348 | 0 | 735.0161 | Left atrium |
| LA_maximu | Z127_S5 | 27446 | 0.243704 | 0.00421 | 0 | 729.4996 | Left atrium |
| LA_maximu | Z99_S3 | 27446 | 0.230182 | 0.00399 | 0 | 724.4167 | Left atrium |
| LV_end_di | Z82_S1 | 27446 | -0.45961 | 0.007977 | 0 | 722.8078 | Left ventricle |
| LA_minimu | Z92_S5 | 27446 | 0.332102 | 0.005787 | 0 | 716.9319 | Left atrium |
| LA_stroke_ | Z31_S5 | 27446 | 0.229389 | 0.004001 | 0 | 715.4752 | Left atrium |
| LV_stroke_ | Z31_S5 | 27446 | 0.264169 | 0.004615 | 0 | 713.3413 | Left ventricle |
| RV_end_di | Z127_S5 | 27446 | 0.33083 | 0.00579 | 0 | 710.7341 | Right ventricle |
| LV_stroke_ | Z94_S5 | 27446 | 0.281288 | 0.004953 | 0 | 702.2851 | Left ventricle |
| RV_end_di | Z31_S5 | 27446 | 0.31062 | 0.005474 | 0 | 700.988 | Right ventricle |
| RA_maxim | Z82_S1 | 27446 | -0.36787 | 0.0065 | 0 | 697.3428 | Right atrium |
| RV_stroke_ | Z12_S4 | 27446 | -0.27993 | 0.004981 | 0 | 687.5473 | Right ventricle |
| RV_end_di | Z94_S5 | 27446 | 0.329556 | 0.005877 | 0 | 684.6185 | Right ventricle |
| LV_end_di | Z40_S5 | 27446 | -0.30959 | 0.005541 | 0 | 679.7461 | Left ventricle |
| RV_end_di | Z29_S1 | 27446 | 0.409947 | 0.007387 | 0 | 670.5972 | Right ventricle |
| RV_stroke_ | Z82_S1 | 27446 | -0.39932 | 0.00722 | 0 | 666.1606 | Right ventricle |
| RA_minimu | Z31_S5 | 27446 | 0.236103 | 0.004302 | 0 | 655.8313 | Right atrium |
| LV_end_di | Z85_S5 | 27446 | 0.332907 | 0.006076 | 0 | 653.7213 | Left ventricle |
| LV_end_di | Z29_S1 | 27446 | 0.384873 | 0.007028 | 0 | 653.1127 | Left ventricle |
| RA_minimu | Z82_S1 | 27446 | -0.35908 | 0.006616 | 0 | 641.4872 | Right atrium |
| LV_stroke_ | Z89_S4 | 27446 | -0.30242 | 0.005576 | 0 | 640.5045 | Left ventricle |

| | | | | | | | |
|---|---|---|---|---|---|---|---|
| LV_end_di | Z50_S4 | 27446 | 0.34008 | 0.006285 | 0 | 637.5596 | Left ventricle |
| LA_minimu | Z82_S1 | 27446 | -0.3218 | 0.005967 | 0 | 633.3182 | Left atrium |
| LV_end_di | Z92_S5 | 27446 | 0.421592 | 0.007839 | 0 | 629.8561 | Left ventricle |
| LA_maximu | Z94_S5 | 27446 | 0.231083 | 0.004297 | 0 | 629.8315 | Left atrium |
| LV_stroke_ | Z92_S5 | 27446 | 0.373968 | 0.006963 | 0 | 628.119 | Left ventricle |
| LA_maximu | Z82_S4 | 27446 | -0.23059 | 0.004297 | 0 | 627.0319 | Left atrium |
| LA_maximu | Z89_S4 | 27446 | -0.25775 | 0.004818 | 0 | 623.3309 | Left atrium |
| LV_stroke_ | Z4_S4 | 27446 | -0.27327 | 0.005137 | 0 | 616.2906 | Left ventricle |
| LA_maximu | Z51_S4 | 27446 | -0.22853 | 0.004323 | 0 | 608.5377 | Left atrium |
| LV_end_di | Z24_S1 | 27446 | 0.291472 | 0.005521 | 0 | 607.059 | Left ventricle |
| RV_end_di | Z117_S4 | 27446 | -0.2945 | 0.005582 | 0 | 606.2682 | Right ventricle |
| LV_stroke_ | Z122_S1 | 27446 | 0.333126 | 0.006331 | 0 | 603.0956 | Left ventricle |
| LV_end_di | Z122_S1 | 27446 | 0.375005 | 0.007128 | 0 | 602.8357 | Left ventricle |
| LA_stroke_ | Z89_S4 | 27446 | -0.2547 | 0.00485 | 0 | 600.5912 | Left atrium |
| LV_stroke_ | Z73_S4 | 27446 | 0.347628 | 0.006649 | 0 | 595.4506 | Left ventricle |
| LV_end_sy | Z106_S4 | 27446 | 0.27552 | 0.005293 | 0 | 590.095 | Left ventricle |
| RA_maxim | Z30_S2 | 27446 | -0.33387 | 0.006432 | 0 | 586.8215 | Right atrium |
| LA_maximu | Z85_S5 | 27446 | 0.242868 | 0.00468 | 0 | 586.5685 | Left atrium |
| LV_stroke_ | Z85_S5 | 27446 | 0.280861 | 0.005425 | 0 | 583.8065 | Left ventricle |
| LV_end_di | Z107_S1 | 27446 | -0.27189 | 0.005253 | 0 | 583.5817 | Left ventricle |
| LV_stroke_ | Z29_S1 | 27446 | 0.324466 | 0.006275 | 0 | 582.3395 | Left ventricle |
| LV_stroke_ | Z106_S4 | 27446 | 0.263474 | 0.005097 | 0 | 582.0118 | Left ventricle |
| LA_stroke_ | Z106_S4 | 27446 | 0.227327 | 0.004422 | 0 | 575.6089 | Left atrium |
| RV_end_di | Z92_S5 | 27446 | 0.425732 | 0.008286 | 0 | 575.1006 | Right ventricle |
| LV_stroke_ | Z51_S4 | 27446 | -0.25759 | 0.005024 | 0 | 572.5939 | Left ventricle |
| RV_end_sy | Z12_S4 | 27446 | -0.2867 | 0.005596 | 0 | 571.7551 | Right ventricle |
| LA_maximu | Z12_S4 | 27446 | -0.21788 | 0.004275 | 0 | 565.7935 | Left atrium |
| RV_end_di | Z89_S4 | 27446 | -0.33689 | 0.006649 | 0 | 559.2072 | Right ventricle |
| LA_stroke_ | Z99_S3 | 27446 | 0.205728 | 0.004061 | 0 | 559.1271 | Left atrium |
| LA_maximu | Z117_S4 | 27446 | -0.20658 | 0.004079 | 0 | 558.7349 | Left atrium |
| LV_end_di | Z78_S4 | 27446 | 0.292624 | 0.005794 | 0 | 555.5888 | Left ventricle |
| RA_minimu | Z127_S4 | 27446 | 0.296867 | 0.005915 | 0 | 548.75 | Right atrium |
| RA_maxim | Z31_S5 | 27446 | 0.214639 | 0.00428 | 0 | 547.9569 | Right atrium |
| LA_stroke_ | Z51_S4 | 27446 | -0.21834 | 0.004366 | 0 | 544.7932 | Left atrium |
| LA_minimu | Z127_S5 | 27446 | 0.206622 | 0.00414 | 0 | 542.5551 | Left atrium |
| LA_stroke_ | Z127_S5 | 27446 | 0.214034 | 0.004292 | 0 | 541.886 | Left atrium |
| LA_stroke_ | Z82_S1 | 27446 | -0.31013 | 0.006229 | 0 | 540.0279 | Left atrium |
| RV_end_di | Z99_S3 | 27446 | 0.276194 | 0.005559 | 0 | 537.9103 | Right ventricle |
| RV_stroke_ | Z127_S5 | 27446 | 0.249268 | 0.005024 | 0 | 536.2997 | Right ventricle |
| RV_stroke_ | Z92_S5 | 27446 | 0.352848 | 0.007116 | 0 | 535.6354 | Right ventricle |
| RV_stroke_ | Z53_S1 | 27446 | 0.304394 | 0.00614 | 0 | 535.4335 | Right ventricle |
| RV_end_di | Z50_S4 | 27446 | 0.330506 | 0.006667 | 0 | 535.3996 | Right ventricle |
| RV_end_di | Z122_S1 | 27446 | 0.37278 | 0.007543 | 0 | 532.121 | Right ventricle |
| LV_end_di | Z95_S3 | 27446 | -0.24875 | 0.005044 | 0 | 529.8096 | Left ventricle |
| LA_maximu | Z73_S4 | 27446 | 0.283941 | 0.005767 | 0 | 528.275 | Left atrium |

| | | | | | | | |
|---|---|---|---|---|---|---|---|
| RV_end_di | Z51_S4 | 27446 | -0.294 | 0.005975 | 0 | 527.5798 | Right ventricle |
| LA_stroke_ | Z12_S4 | 27446 | -0.21198 | 0.004309 | 0 | 527.2667 | Left atrium |
| LV_stroke_ | Z40_S5 | 27446 | -0.24477 | 0.004979 | 0 | 526.6099 | Left ventricle |
| LA_stroke_ | Z92_S5 | 27446 | 0.298142 | 0.006087 | 0 | 522.652 | Left atrium |
| LA_minimu | Z106_S4 | 27446 | 0.209645 | 0.004284 | 0 | 521.7079 | Left atrium |
| LA_minimu | Z99_S3 | 27446 | 0.192193 | 0.003929 | 0 | 521.2695 | Left atrium |
| RV_stroke_ | Z31_S5 | 27446 | 0.232378 | 0.004752 | 0 | 521.0172 | Right ventricle |
| RV_end_di | Z40_S5 | 27446 | -0.28814 | 0.005904 | 0 | 519.0328 | Right ventricle |
| LV_end_sy | Z99_S3 | 27446 | 0.237887 | 0.004882 | 0 | 517.4061 | Left ventricle |
| RV_stroke_ | Z94_S5 | 27446 | 0.248101 | 0.005098 | 0 | 516.1652 | Right ventricle |
| LV_stroke_ | Z53_S1 | 27446 | 0.294764 | 0.00606 | 0 | 515.4934 | Left ventricle |
| LV_end_di | Z105_S4 | 27446 | -0.29045 | 0.005978 | 0 | 514.4549 | Left ventricle |
| LV_end_sy | Z31_S5 | 27446 | 0.236359 | 0.004869 | 0 | 513.4297 | Left ventricle |
| RV_end_sy | Z106_S4 | 27446 | 0.278793 | 0.005794 | 0 | 504.5226 | Right ventricle |
| LA_maximu | Z40_S5 | 27446 | -0.20655 | 0.004304 | 0 | 501.7944 | Left atrium |
| LV_end_di | Z111_S5 | 27446 | 0.311395 | 0.00651 | 0 | 498.6876 | Left ventricle |
| RV_stroke_ | Z73_S4 | 27446 | 0.324606 | 0.006798 | 0 | 496.8779 | Right ventricle |
| LV_end_di | Z1_S1 | 27446 | -0.30466 | 0.006393 | 0 | 494.9285 | Left ventricle |
| LV_stroke_ | Z99_S3 | 27446 | 0.224266 | 0.004706 | 0 | 494.9147 | Left ventricle |
| RV_end_di | Z127_S4 | 27446 | 0.361247 | 0.007584 | 0 | 494.4634 | Right ventricle |
| RV_end_di | Z107_S1 | 27446 | -0.26516 | 0.005567 | 0 | 494.4528 | Right ventricle |
| LV_stroke_ | Z117_S4 | 27446 | -0.22621 | 0.004751 | 0 | 494.0801 | Left ventricle |
| LV_end_di | Z7_S1 | 27446 | -0.28684 | 0.006029 | 0 | 493.3818 | Left ventricle |
| RV_stroke_ | Z127_S4 | 27446 | 0.308333 | 0.006496 | 0 | 491.0349 | Right ventricle |
| LV_end_di | Z52_S4 | 27446 | 0.278863 | 0.005876 | 0 | 490.8896 | Left ventricle |
| RV_stroke_ | Z89_S4 | 27446 | -0.27159 | 0.005724 | 0 | 490.6519 | Right ventricle |
| RV_end_di | Z53_S1 | 27446 | 0.341253 | 0.007196 | 0 | 490.1126 | Right ventricle |
| RV_stroke_ | Z4_S4 | 27446 | -0.24913 | 0.005265 | 0 | 488.0433 | Right ventricle |
| RA_maxim | Z127_S4 | 27446 | 0.276251 | 0.005866 | 0 | 483.4102 | Right atrium |
| LA_stroke_ | Z94_S5 | 27446 | 0.205117 | 0.004368 | 0 | 480.5611 | Left atrium |
| LA_stroke_ | Z117_S4 | 27446 | -0.19321 | 0.004125 | 0 | 478.1369 | Left atrium |
| RV_end_di | Z24_S1 | 27446 | 0.274566 | 0.005869 | 0 | 476.9583 | Right ventricle |
| LV_stroke_ | Z111_S5 | 27446 | 0.270685 | 0.005792 | 0 | 476.1073 | Left ventricle |
| LA_stroke_ | Z73_S4 | 27446 | 0.271433 | 0.00582 | 0 | 474.1373 | Left atrium |
| LA_maximu | Z122_S1 | 27446 | 0.257255 | 0.005517 | 0 | 473.8956 | Left atrium |
| RV_stroke_ | Z122_S1 | 27446 | 0.302344 | 0.006489 | 0 | 473.2341 | Right ventricle |
| LV_end_di | Z35_S5 | 27446 | -0.26278 | 0.005641 | 0 | 473.031 | Left ventricle |
| LV_stroke_ | Z127_S4 | 27446 | 0.298466 | 0.00641 | 0 | 472.4891 | Left ventricle |
| RV_end_di | Z85_S5 | 27446 | 0.301122 | 0.006486 | 0 | 469.8161 | Right ventricle |
| LA_maximu | Z86_S3 | 27446 | -0.26477 | 0.005706 | 0 | 469.354 | Left atrium |
| RA_minimu | Z92_S5 | 27446 | 0.303521 | 0.006543 | 0 | 469.0692 | Right atrium |
| LA_minimu | Z82_S4 | 27446 | -0.19554 | 0.004218 | 0 | 468.4495 | Left atrium |
| LA_stroke_ | Z82_S4 | 27446 | -0.20246 | 0.004372 | 0 | 467.4097 | Left atrium |
| LV_end_di | Z59_S3 | 27446 | -0.24664 | 0.005336 | 0 | 465.7658 | Left ventricle |
| RV_stroke_ | Z29_S1 | 27446 | 0.296628 | 0.006426 | 0 | 464.4444 | Right ventricle |

| | | | | | | | |
|---|---|---|---|---|---|---|---|
| RV_end_di | Z111_S5 | 27446 | 0.316892 | 0.00687 | 0 | 463.7767 | Right ventricle |
| LV_end_di: | Z127_S4 | 27446 | 0.332675 | 0.007223 | 0 | 462.3402 | Left ventricle |
| LA_minimu | Z94_S5 | 27446 | 0.194121 | 0.004221 | 0 | 461.0349 | Left atrium |
| LA_minimu | Z85_S5 | 27446 | 0.210067 | 0.004583 | 0 | 457.9017 | Left atrium |
| RV_end_sy | Z4_S4 | 27446 | -0.26912 | 0.005877 | 0 | 457.1789 | Right ventricle |
| LA_maximu | Z38_S5 | 27446 | -0.24352 | 0.005326 | 0 | 455.6898 | Left atrium |
| RV_end_sy | Z73_S4 | 27446 | 0.345528 | 0.007597 | 0 | 450.9443 | Right ventricle |
| RV_end_di | Z95_S3 | 27446 | -0.24249 | 0.005343 | 0 | 449.0435 | Right ventricle |
| LV_end_di: | Z123_S4 | 27446 | 0.31282 | 0.006906 | 0 | 447.3137 | Left ventricle |
| RA_minimu | Z30_S2 | 27446 | -0.2979 | 0.006591 | 0 | 445.3315 | Right atrium |
| LA_maximu | Z105_S4 | 27446 | -0.20771 | 0.004606 | 0 | 443.2443 | Left atrium |
| LV_end_sy | Z12_S4 | 27446 | -0.23378 | 0.005199 | 0 | 440.8728 | Left ventricle |
| RA_maxim | Z92_S5 | 27446 | 0.290162 | 0.006471 | 0 | 438.3585 | Right atrium |
| LA_maximu | Z4_S4 | 27446 | -0.20149 | 0.004494 | 0 | 438.2542 | Left atrium |
| LV_myocar | Z106_S4 | 27446 | 0.319076 | 0.007122 | 0 | 437.6064 | Left ventricle |
| RV_stroke_ | Z106_S4 | 27446 | 0.233404 | 0.005232 | 0 | 433.9079 | Right ventricle |
| LV_end_di: | Z53_S1 | 27446 | 0.306175 | 0.006867 | 0 | 433.4047 | Left ventricle |
| LA_minimu | Z38_S5 | 27446 | -0.22937 | 0.005176 | 0 | 428.2614 | Left atrium |
| LV_end_di: | Z14_S1 | 27446 | 0.274385 | 0.006205 | 0 | 426.3105 | Left ventricle |
| LV_end_sy | Z117_S4 | 27446 | -0.21912 | 0.004964 | 0 | 424.9387 | Left ventricle |
| LA_minimu | Z86_S3 | 27446 | -0.24485 | 0.005552 | 0 | 424.0287 | Left atrium |
| LA_stroke_ | Z105_S4 | 27446 | -0.20405 | 0.004637 | 0 | 422.2006 | Left atrium |
| LA_maximu | Z13_S3 | 27446 | 0.181537 | 0.004127 | 0 | 421.8931 | Left atrium |
| LV_myocar | Z99_S3 | 27446 | 0.287578 | 0.00654 | 0 | 421.6375 | Left ventricle |
| RV_end_di | Z78_S4 | 27446 | 0.270459 | 0.006163 | 0 | 419.8753 | Right ventricle |
| LA_maximu | Z123_S4 | 27446 | 0.231931 | 0.005305 | 0 | 416.7913 | Left atrium |
| LA_stroke_ | Z85_S5 | 27446 | 0.208298 | 0.004766 | 0 | 416.5385 | Left atrium |
| LV_end_di: | Z1_S2 | 27446 | 0.236272 | 0.00541 | 0 | 415.9065 | Left ventricle |
| LA_stroke_ | Z122_S1 | 27446 | 0.243216 | 0.00557 | 0 | 415.7812 | Left atrium |
| RV_stroke_ | Z85_S5 | 27446 | 0.242853 | 0.005578 | 0 | 413.293 | Right ventricle |
| RV_end_di | Z52_S4 | 27446 | 0.269886 | 0.006223 | 0 | 410.1005 | Right ventricle |
| RV_end_di | Z7_S1 | 27446 | -0.27667 | 0.006387 | 0 | 409.1848 | Right ventricle |
| LV_end_sy | Z73_S4 | 27446 | 0.303427 | 0.00701 | 0 | 408.6248 | Left ventricle |
| LA_stroke_ | Z4_S4 | 27446 | -0.1957 | 0.004527 | 0 | 407.48 | Left atrium |
| RV_stroke_ | Z51_S4 | 27446 | -0.22318 | 0.005164 | 0 | 407.3177 | Right ventricle |
| LV_stroke_ | Z78_S4 | 27446 | 0.22483 | 0.005206 | 0 | 406.6988 | Left ventricle |
| LA_maximu | Z78_S4 | 27446 | 0.193918 | 0.004492 | 0 | 406.3418 | Left atrium |
| LV_end_di: | Z82_S4 | 27446 | -0.2459 | 0.005704 | 0 | 405.2368 | Left ventricle |
| LV_stroke_ | Z24_S1 | 27446 | 0.214088 | 0.004981 | 0 | 402.8011 | Left ventricle |
| LA_stroke_ | Z78_S4 | 27446 | 0.19336 | 0.004517 | 0 | 399.5984 | Left atrium |
| LA_stroke_ | Z40_S5 | 27446 | -0.1865 | 0.004361 | 0 | 398.908 | Left atrium |
| LA_ejectio | Z31_S5 | 27446 | -0.16197 | 0.003792 | 0 | 397.9929 | Left atrium |
| RV_end_sy | Z117_S4 | 27446 | -0.23096 | 0.005409 | 0 | 397.623 | Right ventricle |
| RV_end_di | Z20_S1 | 27446 | 0.345817 | 0.008103 | 0 | 397.2599 | Right ventricle |
| LV_end_di: | Z20_S1 | 27446 | 0.32828 | 0.007699 | 0 | 396.5427 | Left ventricle |

| | | | | | | | |
|---|---|---|---|---|---|---|---|
| LA_maximu | Z50_S4 | 27446 | 0.20912 | 0.004907 | 0 | 396.0405 | Left atrium |
| LV_stroke_ | Z105_S4 | 27446 | -0.22809 | 0.005359 | 0 | 395.1267 | Left ventricle |
| LV_stroke_ | Z107_S1 | 27446 | -0.20136 | 0.004734 | 0 | 394.5526 | Left ventricle |
| LA_maximu | Z59_S3 | 27446 | -0.17483 | 0.004112 | 0 | 394.309 | Left atrium |
| LV_myocar | Z12_S4 | 27446 | -0.29479 | 0.006938 | 0 | 393.752 | Left ventricle |
| LV_stroke_ | Z50_S4 | 27446 | 0.241617 | 0.005688 | 0 | 393.4843 | Left ventricle |
| RV_end_di | Z123_S4 | 27446 | 0.309902 | 0.0073 | 0 | 393.1173 | Right ventricle |
| RV_stroke_ | Z40_S5 | 27446 | -0.21666 | 0.005105 | 0 | 392.8126 | Right ventricle |
| LV_myocar | Z4_S4 | 27446 | -0.30639 | 0.007222 | 0 | 392.5408 | Left ventricle |
| LV_end_sy | Z50_S4 | 27446 | 0.250444 | 0.005912 | 0 | 391.3789 | Left ventricle |
| LA_stroke_ | Z123_S4 | 27446 | 0.226084 | 0.005343 | 0 | 390.5952 | Left atrium |
| LV_end_sy | Z127_S5 | 27446 | 0.220049 | 0.005204 | 0 | 389.9872 | Left ventricle |
| RV_end_sy | Z29_S1 | 27446 | 0.303679 | 0.007199 | 0 | 388.1633 | Right ventricle |
| LV_end_sy | Z4_S4 | 27446 | -0.22895 | 0.005434 | 0 | 387.2104 | Left ventricle |
| RA_minimu | Z38_S5 | 27446 | -0.24267 | 0.00576 | 0 | 387.1181 | Right atrium |
| LV_stroke_ | Z1_S1 | 27446 | -0.24097 | 0.005726 | 0 | 386.24 | Left ventricle |
| RV_end_sy | Z107_S1 | 27446 | -0.22503 | 0.005353 | 0 | 385.4285 | Right ventricle |
| LA_minimu | Z51_S4 | 27446 | -0.17911 | 0.004266 | 0 | 384.6057 | Left atrium |
| LA_maximu | Z24_S1 | 27446 | 0.1801 | 0.004306 | 0 | 381.6743 | Left atrium |
| RV_end_di | Z35_S5 | 27446 | -0.25006 | 0.00598 | 0 | 381.4364 | Right ventricle |
| LV_end_di | Z55_S5 | 27446 | 0.235903 | 0.005656 | 0 | 379.4735 | Left ventricle |
| LA_maximu | Z1_S1 | 27446 | -0.2061 | 0.004944 | 0 | 379.0786 | Left atrium |
| LA_stroke_ | Z50_S4 | 27446 | 0.205863 | 0.004939 | 0 | 379.0388 | Left atrium |
| RV_end_sy | Z50_S4 | 27446 | 0.268107 | 0.006435 | 0 | 378.6082 | Right ventricle |
| LA_maximu | Z111_S5 | 27446 | 0.209736 | 0.005036 | 0 | 378.4005 | Left atrium |
| RV_stroke_ | Z117_S4 | 27446 | -0.20243 | 0.004865 | 0 | 377.637 | Right ventricle |
| RV_end_di | Z105_S4 | 27446 | -0.26354 | 0.006361 | 0 | 374.4733 | Right ventricle |
| LV_end_di | Z114_S5 | 27446 | -0.21904 | 0.005292 | 0 | 373.6545 | Left ventricle |
| LV_stroke_ | Z7_S1 | 27446 | -0.22332 | 0.005405 | 0 | 372.4032 | Left ventricle |
| RV_end_sy | Z31_S5 | 27446 | 0.220956 | 0.005354 | 0 | 371.5693 | Right ventricle |
| LV_end_di | Z62_S1 | 27446 | 0.255894 | 0.00621 | 0 | 370.4253 | Left ventricle |
| LA_minimu | Z117_S4 | 27446 | -0.16536 | 0.004014 | 0 | 370.306 | Left atrium |
| RV_end_sy | Z127_S5 | 27446 | 0.233206 | 0.005668 | 0 | 369.3088 | Right ventricle |
| RV_stroke_ | Z111_S5 | 27446 | 0.243609 | 0.005927 | 0 | 368.5719 | Right ventricle |
| LA_maximu | Z127_S4 | 27446 | 0.228381 | 0.005578 | 0 | 365.7429 | Left atrium |
| LA_minimu | Z89_S4 | 27446 | -0.19501 | 0.004766 | 0 | 365.2467 | Left atrium |
| LA_maximu | Z107_S1 | 27446 | -0.16735 | 0.004095 | 0 | 364.4111 | Left atrium |
| LA_minimu | Z40_S5 | 27446 | -0.17089 | 0.004221 | 0 | 357.6096 | Left atrium |
| RV_end_sy | Z94_S5 | 27446 | 0.232558 | 0.005747 | 0 | 357.24 | Right ventricle |
| LV_stroke_ | Z82_S4 | 27446 | -0.20578 | 0.005086 | 0 | 357.2242 | Left ventricle |
| LV_myocar | Z127_S5 | 27446 | 0.279838 | 0.006939 | 0 | 354.8917 | Left ventricle |
| LA_maximu | Z15_S2 | 27446 | 0.218629 | 0.00543 | 0 | 353.7311 | Left atrium |
| LV_end_di | Z20_S2 | 27446 | 0.22598 | 0.005614 | 0 | 353.577 | Left ventricle |
| LV_end_di | Z38_S5 | 27446 | -0.28198 | 0.007006 | 0 | 353.46 | Left ventricle |
| LA_maximu | Z1_S2 | 27446 | 0.167692 | 0.004166 | 0 | 353.4596 | Left atrium |

| | | | | | | | |
|---|---|---|---|---|---|---|---|
| LV_myocar | Z31_S5 | 27446 | 0.2633 | 0.006557 | 0 | 351.8557 | Left ventricle |
| RV_end_sy | Z95_S3 | 27446 | -0.20532 | 0.005135 | 0 | 348.9066 | Right ventricle |
| LA_maximu | Z7_S1 | 27446 | -0.18661 | 0.004673 | 0 | 347.9964 | Left atrium |
| LA_stroke_ | Z24_S1 | 27446 | 0.173193 | 0.004339 | 0 | 347.7206 | Left atrium |
| RV_end_di | Z14_S1 | 27446 | 0.262331 | 0.006572 | 0 | 347.7001 | Right ventricle |
| LV_end_dia | Z46_S1 | 27446 | -0.26822 | 0.006722 | 0 | 347.4036 | Left ventricle |
| LV_myocar | Z51_S4 | 27446 | -0.28166 | 0.007065 | 0 | 346.7741 | Left ventricle |
| LV_end_sy | Z51_S4 | 27446 | -0.21177 | 0.005314 | 0 | 346.5935 | Left ventricle |
| RV_stroke_ | Z99_S3 | 27446 | 0.192232 | 0.004832 | 0 | 345.4043 | Right ventricle |
| RA_stroke_ | Z30_S2 | 27446 | -0.23755 | 0.005971 | 0 | 345.3563 | Right atrium |
| LV_end_sy | Z24_S1 | 27446 | 0.20684 | 0.0052 | 0 | 345.2513 | Left ventricle |
| LA_minimu | Z12_S4 | 27446 | -0.16759 | 0.004218 | 0 | 344.5739 | Left atrium |
| LV_end_sy | Z94_S5 | 27446 | 0.210206 | 0.005291 | 0 | 344.475 | Left ventricle |
| RV_end_sy | Z99_S3 | 27446 | 0.213736 | 0.005381 | 0 | 344.3041 | Right ventricle |
| LV_myocar | Z94_S5 | 27446 | 0.27861 | 0.007036 | 0 | 342.1581 | Left ventricle |
| LV_end_dia | Z29_S5 | 27446 | 0.281274 | 0.007104 | 0 | 342.1315 | Left ventricle |
| LV_myocar | Z114_S5 | 27446 | -0.25778 | 0.006511 | 0 | 342.062 | Left ventricle |
| RV_end_di | Z1_S1 | 27446 | -0.26943 | 0.00681 | 0 | 341.5848 | Right ventricle |
| RA_maxim | Z75_S5 | 27446 | 0.268565 | 0.006791 | 0 | 341.3458 | Right atrium |
| LV_myocar | Z29_S1 | 27446 | 0.348789 | 0.008836 | 0 | 340.0442 | Left ventricle |
| LV_end_dia | Z15_S2 | 27446 | 0.279516 | 0.007094 | 0 | 338.8326 | Left ventricle |
| LV_stroke_ | Z123_S4 | 27446 | 0.243557 | 0.006186 | 0 | 338.2764 | Left ventricle |
| LA_ejection | Z92_S5 | 27446 | -0.22477 | 0.005711 | 0 | 338.0096 | Left atrium |
| LA_stroke_ | Z59_S3 | 27446 | -0.16327 | 0.004151 | 0 | 337.6233 | Left atrium |
| LV_end_sy | Z95_S3 | 27446 | -0.18572 | 0.004726 | 0 | 336.9738 | Left ventricle |
| LV_end_dia | Z69_S5 | 27446 | -0.22205 | 0.005652 | 0 | 336.8481 | Left ventricle |
| RA_minimu | Z86_S3 | 27446 | -0.24317 | 0.006203 | 0 | 335.4433 | Right atrium |
| LA_minimu | Z73_S4 | 27446 | 0.222417 | 0.005677 | 0 | 335.0459 | Left atrium |
| RA_maxim | Z4_S4 | 27446 | -0.18824 | 0.004809 | 0 | 334.4421 | Right atrium |
| LA_minimu | Z13_S3 | 27446 | 0.157434 | 0.00403 | 0 | 333.0947 | Left atrium |
| RA_maxim | Z122_S1 | 27446 | 0.231258 | 0.005921 | 0 | 332.9709 | Right atrium |
| LA_stroke_ | Z1_S1 | 27446 | -0.19457 | 0.004987 | 0 | 332.2162 | Left atrium |
| LV_stroke_ | Z38_S5 | 27446 | -0.24285 | 0.006233 | 0 | 331.2801 | Left ventricle |
| LV_end_dia | Z44_S4 | 27446 | -0.21888 | 0.005633 | 0 | 329.5444 | Left ventricle |
| LA_stroke_ | Z111_S5 | 27446 | 0.196897 | 0.005081 | 0 | 327.7399 | Left atrium |
| LA_stroke_ | Z7_S1 | 27446 | -0.18195 | 0.004704 | 0 | 326.5331 | Left atrium |
| LV_end_sy | Z107_S1 | 27446 | -0.1911 | 0.004946 | 0 | 325.7969 | Left ventricle |
| LA_maximu | Z52_S4 | 27446 | 0.17606 | 0.004562 | 0 | 325.1163 | Left atrium |
| Descending | Z107_S1 | 24599 | -0.19871 | 0.00515 | 0 | 324.9981 | Aorta |
| LA_maximu | Z44_S4 | 27446 | -0.16661 | 0.004318 | 0 | 324.9313 | Left atrium |
| LV_myocar | Z117_S4 | 27446 | -0.25631 | 0.006653 | 0 | 323.9915 | Left ventricle |
| LA_maximu | Z95_S3 | 27446 | -0.15104 | 0.003929 | 2.9643938 | 322.5565 | Left atrium |
| LA_maximu | Z47_S4 | 27446 | -0.19527 | 0.005096 | 2.7717082 | 320.5576 | Left atrium |
| LA_minimu | Z15_S2 | 27446 | 0.202251 | 0.005279 | 3.9080592 | 320.4081 | Left atrium |
| LV_stroke_ | Z95_S3 | 27446 | -0.17446 | 0.004554 | 5.0394695 | 320.2978 | Left ventricle |

| | | | | | | | |
|---|---|---|---|---|---|---|---|
| LA_minimu | Z14_S2 | 27446 | -0.18889 | 0.004933 | 7.2182990 | 320.1416 | Left atrium |
| LV_stroke_ | Z52_S4 | 27446 | 0.202493 | 0.005289 | 1.0899088 | 319.9626 | Left ventricle |
| LA_stroke_ | Z52_S4 | 27446 | 0.17559 | 0.004587 | 1.1659949 | 319.9333 | Left atrium |
| RV_end_di | Z59_S3 | 27446 | -0.21729 | 0.005681 | 4.3803860 | 319.3585 | Right ventricle |
| LV_end_sy | Z40_S5 | 27446 | -0.20032 | 0.00526 | 2.0799437 | 316.6819 | Left ventricle |
| LA_maximu | Z125_S4 | 27446 | -0.20037 | 0.005263 | 4.0466930 | 316.3929 | Left atrium |
| RV_stroke_ | Z123_S4 | 27446 | 0.239049 | 0.006289 | 3.8912947 | 315.4099 | Right ventricle |
| RV_end_sy | Z24_S1 | 27446 | 0.215346 | 0.005668 | 7.8583477 | 315.1047 | Right ventricle |
| LA_maximu | Z14_S4 | 27446 | 0.215569 | 0.005695 | 1.5962666 | 312.7969 | Left atrium |
| RV_stroke_ | Z50_S4 | 27446 | 0.219749 | 0.00581 | 4.4631705 | 312.3503 | Right ventricle |
| LA_maximu | Z35_S5 | 27446 | -0.16531 | 0.004378 | 5.9519184 | 311.2253 | Left atrium |
| LV_myocar | Z107_S1 | 27446 | -0.24845 | 0.006585 | 1.6222548 | 310.7899 | Left ventricle |
| LV_end_sy | Z1_S2 | 27446 | 0.189591 | 0.005035 | 2.9508595 | 309.5301 | Left ventricle |
| LV_end_di | Z98_S3 | 27446 | -0.21928 | 0.005828 | 8.7376615 | 309.0586 | Left ventricle |
| LA_minimu | Z122_S1 | 27446 | 0.20376 | 0.00542 | 0.00E+00 | 308.6051 | Left atrium |
| RV_end_sy | Z20_S1 | 27446 | 0.292208 | 0.007781 | 0.00E+00 | 307.9212 | Right ventricle |
| LA_maximu | Z14_S2 | 27446 | -0.19117 | 0.005092 | 0.00E+00 | 307.7503 | Left atrium |
| Descending | Z12_S4 | 24599 | -0.20511 | 0.005474 | 2.65E-307 | 306.576 | Aorta |
| LA_stroke_ | Z95_S3 | 27446 | -0.14803 | 0.003954 | 8.99E-307 | 306.0464 | Left atrium |
| RA_stroke_ | Z82_S1 | 27446 | -0.22806 | 0.006105 | 1.89E-305 | 304.7246 | Right atrium |
| Descending | Z4_S4 | 24599 | -0.21249 | 0.005695 | 9.62E-305 | 304.0168 | Aorta |
| RV_end_sy | Z82_S1 | 27446 | -0.3084 | 0.008269 | 1.81E-304 | 303.743 | Right ventricle |
| LA_stroke_ | Z127_S4 | 27446 | 0.209901 | 0.005634 | 7.86E-304 | 303.1045 | Left atrium |
| RV_end_di | Z114_S5 | 27446 | -0.20854 | 0.005602 | 2.54E-303 | 302.5945 | Right ventricle |
| RV_stroke_ | Z7_S1 | 27446 | -0.20475 | 0.005516 | 1.43E-301 | 300.8452 | Right ventricle |
| LA_stroke_ | Z13_S3 | 27446 | 0.155197 | 0.004188 | 1.31E-300 | 299.8817 | Left atrium |
| LV_stroke_ | Z35_S5 | 27446 | -0.18819 | 0.005079 | 1.53E-300 | 299.8144 | Left ventricle |
| LV_end_sy | Z89_S4 | 27446 | -0.22037 | 0.00595 | 3.21E-300 | 299.4937 | Left ventricle |
| RV_stroke_ | Z24_S1 | 27446 | 0.188707 | 0.005097 | 4.37E-300 | 299.3593 | Right ventricle |
| Descending | Z106_S4 | 24599 | 0.208252 | 0.005628 | 9.91E-300 | 299.0039 | Aorta |
| LV_end_sy | Z59_S3 | 27446 | -0.18455 | 0.00499 | 1.89E-299 | 298.7234 | Left ventricle |
| LA_maximu | Z62_S1 | 27446 | 0.176969 | 0.004786 | 3.03E-299 | 298.5189 | Left atrium |
| LV_stroke_ | Z14_S4 | 27446 | 0.243917 | 0.006608 | 3.07E-298 | 297.5128 | Left ventricle |
| LA_maximu | Z53_S1 | 27446 | 0.196259 | 0.00532 | 5.95E-298 | 297.2255 | Left atrium |
| RV_stroke_ | Z78_S4 | 27446 | 0.196576 | 0.005329 | 7.43E-298 | 297.1288 | Right ventricle |
| RV_stroke_ | Z105_S4 | 27446 | -0.20181 | 0.005481 | 8.41E-297 | 296.075 | Right ventricle |
| RA_minimu | Z122_S1 | 27446 | 0.221607 | 0.006018 | 8.53E-297 | 296.069 | Right atrium |
| LV_myocar | Z85_S5 | 27446 | 0.281976 | 0.007667 | 4.15E-296 | 295.382 | Left ventricle |
| LV_myocar | Z40_S5 | 27446 | -0.25745 | 0.007006 | 1.20E-295 | 294.9214 | Left ventricle |
| RV_end_di | Z55_S5 | 27446 | 0.220227 | 0.005993 | 1.33E-295 | 294.8756 | Right ventricle |
| LV_end_di | Z109_S1 | 27446 | -0.24513 | 0.006688 | 4.57E-294 | 293.3401 | Left ventricle |
| LV_end_di | Z30_S2 | 27446 | -0.29578 | 0.008093 | 1.92E-292 | 291.7169 | Left ventricle |
| LA_minimu | Z26_S3 | 27446 | -0.20382 | 0.005582 | 6.84E-292 | 291.1651 | Left atrium |
| LV_myocar | Z89_S4 | 27446 | -0.28875 | 0.007918 | 3.47E-291 | 290.4601 | Left ventricle |
| LV_stroke_ | Z46_S1 | 27446 | -0.21863 | 0.005998 | 6.17E-291 | 290.2096 | Left ventricle |

| | | | | | | | |
|---|---|---|---|---|---|---|---|
| RV_end_di | Z46_S1 | 27446 | -0.259 | 0.007108 | 1.03E-290 | 289.9864 | Right ventricle |
| LV_end_di | Z5_S5 | 27446 | 0.250239 | 0.006876 | 5.51E-290 | 289.2589 | Left ventricle |
| LV_end_sy | Z52_S4 | 27446 | 0.200494 | 0.00551 | 6.31E-290 | 289.1998 | Left ventricle |
| RA_minimu | Z4_S4 | 27446 | -0.17785 | 0.004892 | 2.02E-289 | 288.6948 | Right atrium |
| LA_stroke_ | Z107_S1 | 27446 | -0.15046 | 0.00414 | 3.46E-289 | 288.461 | Left atrium |
| LV_end_sy | Z35_S5 | 27446 | -0.19186 | 0.005283 | 7.88E-289 | 288.1033 | Left ventricle |
| LA_maximu | Z83_S2 | 27446 | 0.175162 | 0.004826 | 2.06E-288 | 287.6861 | Left atrium |
| RV_end_sy | Z52_S4 | 27446 | 0.217312 | 0.005992 | 5.63E-288 | 287.2494 | Right ventricle |
| Descending | Z117_S4 | 24599 | -0.1895 | 0.005227 | 9.24E-288 | 287.0342 | Aorta |
| RA_maxim | Z21_S3 | 27446 | -0.24602 | 0.006802 | 1.95E-286 | 285.7096 | Right atrium |
| LV_end_di | Z80_S4 | 27446 | -0.20756 | 0.00576 | 2.55E-284 | 283.5942 | Left ventricle |
| LV_end_sy | Z78_S4 | 27446 | 0.196627 | 0.005463 | 1.22E-283 | 282.9139 | Left ventricle |
| LV_stroke_ | Z59_S3 | 27446 | -0.17261 | 0.004808 | 3.06E-282 | 281.5137 | Left ventricle |
| LV_end_di | Z5_S1 | 27446 | -0.23626 | 0.006586 | 8.51E-282 | 281.07 | Left ventricle |
| LV_end_sy | Z55_S5 | 27446 | 0.188712 | 0.005261 | 9.01E-282 | 281.045 | Left ventricle |
| LV_end_di | Z62_S2 | 27446 | 0.212139 | 0.005916 | 1.43E-281 | 280.8437 | Left ventricle |
| LA_maximu | Z39_S1 | 27446 | 0.169352 | 0.004728 | 5.86E-281 | 280.2321 | Left atrium |
| LV_stroke_ | Z5_S5 | 27446 | 0.218674 | 0.006112 | 2.27E-280 | 279.6431 | Left ventricle |
| LV_myocar | Z82_S4 | 27446 | -0.25283 | 0.007071 | 5.12E-280 | 279.2911 | Left ventricle |
| LV_stroke_ | Z14_S1 | 27446 | 0.199277 | 0.005576 | 9.89E-280 | 279.0049 | Left ventricle |
| LA_stroke_ | Z86_S3 | 27446 | -0.20802 | 0.005821 | 1.15E-279 | 278.9378 | Left atrium |
| LA_stroke_ | Z44_S4 | 27446 | -0.15561 | 0.004356 | 1.72E-279 | 278.7647 | Left atrium |
| LA_stroke_ | Z47_S4 | 27446 | -0.18353 | 0.005138 | 2.01E-279 | 278.6977 | Left atrium |
| LV_end_sy | Z20_S2 | 27446 | 0.18523 | 0.005213 | 1.69E-276 | 275.7722 | Left ventricle |
| LA_stroke_ | Z62_S1 | 27446 | 0.171204 | 0.004819 | 1.92E-276 | 275.7169 | Left atrium |
| LV_end_di | Z86_S3 | 27446 | -0.26838 | 0.00756 | 5.34E-276 | 275.2724 | Left ventricle |
| RV_end_sy | Z40_S5 | 27446 | -0.20364 | 0.005738 | 7.53E-276 | 275.123 | Right ventricle |
| LV_end_di | Z41_S2 | 27446 | -0.27934 | 0.007901 | 7.79E-274 | 273.1084 | Left ventricle |
| RV_end_di | Z62_S1 | 27446 | 0.232824 | 0.006587 | 1.19E-273 | 272.9253 | Right ventricle |
| LV_end_di | Z120_S4 | 27446 | -0.23399 | 0.006623 | 2.10E-273 | 272.6786 | Left ventricle |
| RV_end_sy | Z51_S4 | 27446 | -0.20525 | 0.005812 | 3.90E-273 | 272.4092 | Right ventricle |
| LV_end_di | Z28_S4 | 27446 | -0.25409 | 0.007199 | 6.75E-273 | 272.1707 | Left ventricle |
| RV_end_di | Z28_S4 | 27446 | -0.2674 | 0.007577 | 8.12E-273 | 272.0905 | Right ventricle |
| LV_end_di | Z83_S2 | 27446 | 0.222062 | 0.006306 | 1.25E-271 | 270.9039 | Left ventricle |
| LA_stroke_ | Z14_S4 | 27446 | 0.202134 | 0.005743 | 2.20E-271 | 270.6575 | Left atrium |
| LA_maximu | Z20_S2 | 27446 | 0.152316 | 0.004331 | 5.66E-271 | 270.2469 | Left atrium |
| LA_minimu | Z4_S4 | 27446 | -0.15526 | 0.004416 | 8.36E-271 | 270.0776 | Left atrium |
| Descending | Z51_S4 | 24599 | -0.19538 | 0.005562 | 2.48E-270 | 269.6053 | Aorta |
| RA_minimu | Z12_S4 | 27446 | -0.16503 | 0.004707 | 2.97E-269 | 268.5266 | Right atrium |
| RA_minimu | Z94_S5 | 27446 | 0.166673 | 0.004754 | 3.15E-269 | 268.502 | Right atrium |
| LA_stroke_ | Z1_S2 | 27446 | 0.147681 | 0.004216 | 7.96E-269 | 268.0988 | Left atrium |
| LA_minimu | Z105_S4 | 27446 | -0.15814 | 0.00453 | 4.80E-267 | 266.3186 | Left atrium |
| RA_minimu | Z127_S5 | 27446 | 0.163752 | 0.004694 | 1.35E-266 | 265.8702 | Right atrium |
| Descending | Z95_S3 | 24599 | -0.17279 | 0.004955 | 1.87E-266 | 265.728 | Aorta |
| LA_minimu | Z125_S4 | 27446 | -0.17868 | 0.005124 | 2.08E-266 | 265.6817 | Left atrium |

| | | | | | | | |
|---|---|---|---|---|---|---|---|
| LA_minimu | Z1_S2 | 27446 | 0.141839 | 0.004068 | 2.66E-266 | 265.5757 | Left atrium |
| LA_maximu | Z29_S1 | 27446 | 0.193355 | 0.005551 | 7.90E-266 | 265.1026 | Left atrium |
| LA_maximu | Z29_S5 | 27446 | 0.190698 | 0.005478 | 1.52E-265 | 264.8175 | Left atrium |
| LA_minimu | Z59_S3 | 27446 | -0.14016 | 0.004028 | 3.07E-265 | 264.5125 | Left atrium |
| RV_stroke_ | Z1_S1 | 27446 | -0.20397 | 0.005868 | 9.23E-265 | 264.0349 | Right ventricle |
| LA_minimu | Z107_S1 | 27446 | -0.13899 | 0.004002 | 3.26E-264 | 263.4868 | Left atrium |
| RV_end_sy | Z114_S5 | 27446 | -0.18568 | 0.005359 | 4.61E-263 | 262.3365 | Right ventricle |
| RV_stroke_ | Z107_S1 | 27446 | -0.16802 | 0.004855 | 1.98E-262 | 261.7027 | Right ventricle |
| LA_stroke_ | Z53_S1 | 27446 | 0.185287 | 0.005362 | 1.13E-261 | 260.9473 | Left atrium |
| LA_maximu | Z5_S1 | 27446 | -0.17457 | 0.005056 | 2.88E-261 | 260.5403 | Left atrium |
| RA_minimu | Z85_S5 | 27446 | 0.178311 | 0.005165 | 3.43E-261 | 260.4651 | Right atrium |
| RV_end_sy | Z35_S5 | 27446 | -0.19836 | 0.005757 | 4.22E-260 | 259.375 | Right ventricle |
| RA_stroke_ | Z75_S5 | 27446 | 0.214147 | 0.006228 | 4.04E-259 | 258.394 | Right atrium |
| LV_stroke_ | Z20_S1 | 27446 | 0.237391 | 0.006914 | 2.23E-258 | 257.651 | Left ventricle |
| LA_stroke_ | Z39_S1 | 27446 | 0.16338 | 0.004761 | 3.99E-258 | 257.3994 | Left atrium |
| LV_end_di | Z105_S5 | 27446 | 0.232375 | 0.006772 | 4.46E-258 | 257.3511 | Left ventricle |
| LV_end_di | Z2_S1 | 27446 | -0.20937 | 0.006103 | 6.95E-258 | 257.1579 | Left ventricle |
| LV_myocar | Z24_S1 | 27446 | 0.238815 | 0.006963 | 8.99E-258 | 257.0462 | Left ventricle |
| LA_stroke_ | Z38_S5 | 27446 | -0.18628 | 0.005438 | 3.55E-257 | 256.4496 | Left atrium |
| LV_myocar | Z95_S3 | 27446 | -0.21663 | 0.006325 | 4.69E-257 | 256.3286 | Left ventricle |
| LV_myocar | Z73_S4 | 27446 | 0.322812 | 0.009434 | 1.36E-256 | 255.8676 | Left ventricle |
| Descending | Z99_S3 | 24599 | 0.177207 | 0.005184 | 3.90E-256 | 255.4087 | Aorta |
| LA_maximu | Z26_S3 | 27446 | -0.19733 | 0.005774 | 4.98E-256 | 255.3025 | Left atrium |
| LA_maximu | Z41_S2 | 27446 | -0.20723 | 0.006063 | 5.18E-256 | 255.2859 | Left atrium |
| LA_minimu | Z123_S4 | 27446 | 0.178048 | 0.00521 | 6.41E-256 | 255.1934 | Left atrium |
| RV_end_di | Z1_S2 | 27446 | 0.196896 | 0.005768 | 2.00E-255 | 254.6998 | Right ventricle |
| LV_stroke_ | Z62_S1 | 27446 | 0.189772 | 0.005567 | 1.06E-254 | 253.9759 | Left ventricle |
| RV_stroke_ | Z38_S5 | 27446 | -0.21694 | 0.006365 | 1.24E-254 | 253.9067 | Right ventricle |
| LA_minimu | Z127_S4 | 27446 | 0.185887 | 0.005457 | 2.59E-254 | 253.5871 | Left atrium |
| Descending | Z114_S5 | 24599 | -0.17505 | 0.005147 | 1.68E-253 | 252.7751 | Aorta |
| LA_maximu | Z14_S1 | 27446 | 0.163968 | 0.004821 | 1.69E-253 | 252.771 | Left atrium |
| LV_end_sy | Z14_S1 | 27446 | 0.197233 | 0.005807 | 7.08E-253 | 252.1501 | Left ventricle |
| RV_end_di | Z69_S5 | 27446 | -0.20333 | 0.00599 | 1.33E-252 | 251.8751 | Right ventricle |
| Descending | Z107_S1 | 24599 | -0.1785 | 0.005258 | 1.38E-252 | 251.8598 | Aorta |
| LA_minimu | Z111_S5 | 27446 | 0.16728 | 0.004933 | 4.21E-252 | 251.3762 | Left atrium |
| LA_maximu | Z114_S5 | 27446 | -0.13874 | 0.004096 | 1.61E-251 | 250.7929 | Left atrium |
| LV_end_sy | Z114_S5 | 27446 | -0.16698 | 0.004933 | 3.37E-251 | 250.4718 | Left ventricle |
| LV_end_sy | Z29_S1 | 27446 | 0.226557 | 0.006693 | 3.66E-251 | 250.4367 | Left ventricle |
| LV_end_sy | Z85_S5 | 27446 | 0.195739 | 0.005787 | 8.90E-251 | 250.0508 | Left ventricle |
| LV_end_di | Z125_S4 | 27446 | -0.23352 | 0.006904 | 9.12E-251 | 250.0402 | Left ventricle |
| LA_minimu | Z37_S2 | 27446 | -0.18257 | 0.0054 | 1.42E-250 | 249.8464 | Left atrium |
| LV_end_di | Z8_S3 | 27446 | 0.192276 | 0.00569 | 2.45E-250 | 249.6111 | Left ventricle |
| RV_end_sy | Z14_S1 | 27446 | 0.21328 | 0.006316 | 5.56E-250 | 249.2547 | Right ventricle |
| LA_maximu | Z98_S3 | 27446 | -0.15149 | 0.004488 | 9.31E-250 | 249.0311 | Left atrium |
| LA_minimu | Z1_S1 | 27446 | -0.16347 | 0.004844 | 1.11E-249 | 248.9554 | Left atrium |

| | | | | | | | |
|---|---|---|---|---|---|---|---|
| LA_stroke_ | Z35_S5 | 27446 | -0.14918 | 0.004422 | 1.80E-249 | 248.7449 | Left atrium |
| LA_stroke_ | Z5_S1 | 27446 | -0.17146 | 0.005086 | 3.76E-249 | 248.4245 | Left atrium |
| LV_end_sy | Z105_S4 | 27446 | -0.18968 | 0.005634 | 1.86E-248 | 247.7293 | Left ventricle |
| LA_ejectio | Z38_S5 | 27446 | 0.169311 | 0.005032 | 3.68E-248 | 247.4344 | Left atrium |
| RV_end_di | Z20_S2 | 27446 | 0.200469 | 0.005959 | 4.14E-248 | 247.3831 | Right ventricle |
| LA_maxim | Z80_S4 | 27446 | -0.14889 | 0.004427 | 6.06E-248 | 247.2177 | Left atrium |
| LA_maxim | Z30_S2 | 27446 | -0.20919 | 0.006224 | 1.26E-247 | 246.8981 | Left atrium |
| RV_end_di | Z5_S5 | 27446 | 0.243986 | 0.007262 | 1.80E-247 | 246.7436 | Right ventricle |
| LA_ejectio | Z82_S1 | 27446 | 0.19781 | 0.005895 | 7.86E-247 | 246.1043 | Left atrium |
| RV_end_di | Z30_S2 | 27446 | -0.28685 | 0.008549 | 8.33E-247 | 246.0794 | Right ventricle |
| LA_maxim | Z62_S2 | 27446 | 0.152546 | 0.004547 | 8.64E-247 | 246.0637 | Left atrium |
| RV_end_di | Z82_S4 | 27446 | -0.204 | 0.006081 | 9.26E-247 | 246.0336 | Right ventricle |
| LA_maxim | Z109_S1 | 27446 | -0.17248 | 0.005145 | 2.28E-246 | 245.6427 | Left atrium |
| Descending | Z106_S4 | 24599 | 0.192255 | 0.005736 | 2.87E-246 | 245.5421 | Aorta |
| RV_stroke_ | Z82_S4 | 27446 | -0.1744 | 0.005207 | 6.19E-246 | 245.2085 | Right ventricle |
| RV_stroke_ | Z52_S4 | 27446 | 0.180794 | 0.005399 | 8.29E-246 | 245.0815 | Right ventricle |
| RV_end_sy | Z78_S4 | 27446 | 0.199296 | 0.005959 | 3.31E-245 | 244.4808 | Right ventricle |
| LV_end_di | Z14_S4 | 27446 | 0.249849 | 0.007472 | 3.97E-245 | 244.4013 | Left ventricle |
| LA_maxim | Z37_S2 | 27446 | -0.1862 | 0.005571 | 6.72E-245 | 244.1724 | Left atrium |
| RV_end_di | Z15_S2 | 27446 | 0.251274 | 0.007523 | 1.41E-244 | 243.8496 | Right ventricle |
| LV_end_sy | Z7_S1 | 27446 | -0.18947 | 0.005675 | 2.05E-244 | 243.689 | Left ventricle |
| Descending | Z95_S3 | 24599 | -0.16771 | 0.005037 | 4.27E-243 | 242.3695 | Aorta |
| RV_end_sy | Z89_S4 | 27446 | -0.21617 | 0.006501 | 1.76E-242 | 241.7552 | Right ventricle |
| RV_end_sy | Z55_S5 | 27446 | 0.190821 | 0.005739 | 2.04E-242 | 241.6897 | Right ventricle |
| LA_minim | Z24_S1 | 27446 | 0.140223 | 0.004222 | 6.84E-242 | 241.1649 | Left atrium |
| LV_myocar | Z50_S4 | 27446 | 0.264082 | 0.007956 | 1.39E-241 | 240.8564 | Left ventricle |
| RV_stroke_ | Z95_S3 | 27446 | -0.15405 | 0.004652 | 1.67E-240 | 239.7769 | Right ventricle |
| RV_end_di | Z29_S5 | 27446 | 0.24921 | 0.007539 | 1.26E-239 | 238.901 | Right ventricle |
| RA_maxim | Z38_S5 | 27446 | -0.18993 | 0.005751 | 3.51E-239 | 238.4547 | Right atrium |
| LV_cardiac | Z10_S4 | 27446 | 0.187913 | 0.00569 | 3.76E-239 | 238.4246 | Left ventricle |
| LA_minim | Z50_S4 | 27446 | 0.158863 | 0.004819 | 2.44E-238 | 237.6134 | Left atrium |
| LV_end_sy | Z20_S1 | 27446 | 0.23712 | 0.007196 | 4.08E-238 | 237.3899 | Left ventricle |
| LV_myocar | Z78_S4 | 27446 | 0.239959 | 0.007292 | 1.63E-237 | 236.7866 | Left ventricle |
| LA_minim | Z78_S4 | 27446 | 0.14527 | 0.004415 | 2.08E-237 | 236.6823 | Left atrium |
| LA_ejectio | Z26_S3 | 27446 | 0.176784 | 0.005374 | 2.39E-237 | 236.6209 | Left atrium |
| LA_maxim | Z69_S5 | 27446 | -0.1435 | 0.004367 | 7.55E-237 | 236.122 | Left atrium |
| RV_stroke_ | Z35_S5 | 27446 | -0.17 | 0.00518 | 3.40E-236 | 235.4683 | Right ventricle |
| LV_end_di | Z51_S1 | 27446 | -0.19296 | 0.005884 | 7.49E-236 | 235.1253 | Left ventricle |
| LV_stroke_ | Z29_S5 | 27446 | 0.208561 | 0.006364 | 1.38E-235 | 234.8604 | Left ventricle |
| LV_end_sy | Z1_S1 | 27446 | -0.19704 | 0.006023 | 1.04E-234 | 233.9846 | Left ventricle |
| RV_end_sy | Z111_S5 | 27446 | 0.217571 | 0.006672 | 2.94E-233 | 232.5323 | Right ventricle |
| LV_myocar | Z59_S3 | 27446 | -0.2175 | 0.00667 | 3.11E-233 | 232.5068 | Left ventricle |
| Descending | Z24_S1 | 24599 | 0.177835 | 0.005464 | 2.37E-232 | 231.6255 | Aorta |
| LA_minim | Z41_S2 | 27446 | -0.1915 | 0.005891 | 8.05E-232 | 231.0944 | Left atrium |
| LV_myocar | Z29_S5 | 27446 | 0.28576 | 0.008796 | 1.55E-231 | 230.8094 | Left ventricle |

| | | | | | | | |
|---|---|---|---|---|---|---|---|
| LA_stroke_ | Z109_S1 | 27446 | -0.1679 | 0.005178 | 1.08E-230 | 229.965 | Left atrium |
| RA_minimu | Z53_S1 | 27446 | 0.186554 | 0.005753 | 1.22E-230 | 229.915 | Right atrium |
| RV_end_sy | Z7_S1 | 27446 | -0.19948 | 0.006179 | 1.15E-228 | 227.9403 | Right ventricle |
| LA_ejectio | Z86_S3 | 27446 | 0.174289 | 0.005406 | 4.44E-228 | 227.3529 | Left atrium |
| LA_maximu | Z65_S2 | 27446 | 0.170384 | 0.005286 | 5.89E-228 | 227.2298 | Left atrium |
| Descending | Z114_S5 | 24599 | -0.16868 | 0.005233 | 6.29E-228 | 227.2011 | Aorta |
| RV_end_sy | Z92_S5 | 27446 | 0.261617 | 0.00812 | 9.18E-228 | 227.0371 | Right ventricle |
| LA_maximu | Z68_S2 | 27446 | -0.15054 | 0.004678 | 2.95E-227 | 226.5305 | Left atrium |
| LA_minimu | Z64_S1 | 27446 | -0.17815 | 0.005537 | 3.88E-227 | 226.4112 | Left atrium |
| RV_end_sy | Z122_S1 | 27446 | 0.237017 | 0.007368 | 5.01E-227 | 226.3 | Right ventricle |
| LA_stroke_ | Z14_S1 | 27446 | 0.156112 | 0.004856 | 9.35E-227 | 226.0292 | Left atrium |
| LV_myocar | Z1_S1 | 27446 | -0.25698 | 0.008015 | 1.44E-225 | 224.8415 | Left ventricle |
| Descending | Z127_S5 | 24599 | 0.175788 | 0.005485 | 2.25E-225 | 224.6479 | Aorta |
| LA_minimu | Z29_S5 | 27446 | 0.170748 | 0.005329 | 2.74E-225 | 224.5628 | Left atrium |
| LV_myocar | Z105_S4 | 27446 | -0.24032 | 0.007506 | 6.24E-225 | 224.2046 | Left ventricle |
| LA_minimu | Z35_S5 | 27446 | -0.13683 | 0.004275 | 9.04E-225 | 224.0436 | Left atrium |
| LV_stroke_ | Z69_S5 | 27446 | -0.16192 | 0.005066 | 3.41E-224 | 223.4674 | Left ventricle |
| LA_stroke_ | Z98_S3 | 27446 | -0.14445 | 0.00452 | 3.98E-224 | 223.4 | Left atrium |
| LV_stroke_ | Z15_S2 | 27446 | 0.20304 | 0.006359 | 1.06E-223 | 222.974 | Left ventricle |
| LV_stroke_ | Z28_S4 | 27446 | -0.20471 | 0.006419 | 3.69E-223 | 222.4328 | Left ventricle |
| LV_end_sy | Z123_S4 | 27446 | 0.206619 | 0.006489 | 1.69E-222 | 221.7725 | Left ventricle |
| LA_stroke_ | Z80_S4 | 27446 | -0.14184 | 0.004459 | 4.31E-222 | 221.3652 | Left atrium |
| RV_end_di | Z44_S4 | 27446 | -0.18997 | 0.005981 | 2.02E-221 | 220.6937 | Right ventricle |
| RV_stroke_ | Z46_S1 | 27446 | -0.19434 | 0.00612 | 2.89E-221 | 220.5387 | Right ventricle |
| LV_stroke_ | Z5_S1 | 27446 | -0.18639 | 0.005878 | 1.20E-220 | 219.9226 | Left ventricle |
| LA_maximu | Z46_S1 | 27446 | -0.165 | 0.005205 | 1.43E-220 | 219.8458 | Left atrium |
| RV_end_di | Z109_S1 | 27446 | -0.22447 | 0.007081 | 1.60E-220 | 219.7965 | Right ventricle |
| LA_minimu | Z65_S2 | 27446 | 0.162446 | 0.005129 | 3.50E-220 | 219.4558 | Left atrium |
| LV_stroke_ | Z125_S4 | 27446 | -0.19459 | 0.006147 | 6.58E-220 | 219.1818 | Left ventricle |
| LA_minimu | Z83_S2 | 27446 | 0.14885 | 0.004706 | 1.37E-219 | 218.8645 | Left atrium |
| LV_stroke_ | Z114_S5 | 27446 | -0.15052 | 0.004759 | 1.47E-219 | 218.8334 | Left ventricle |
| LV_longitu | Z49_S1 | 27446 | 0.188737 | 0.005967 | 1.52E-219 | 218.817 | Left ventricle |
| LA_minimu | Z44_S4 | 27446 | -0.13355 | 0.004223 | 1.70E-219 | 218.7701 | Left atrium |
| LV_end_sy | Z62_S2 | 27446 | 0.173468 | 0.005488 | 2.68E-219 | 218.5725 | Left ventricle |
| RA_minimu | Z14_S2 | 27446 | -0.1741 | 0.005517 | 1.47E-218 | 217.8333 | Right atrium |
| LA_minimu | Z77_S3 | 27446 | -0.16042 | 0.005085 | 2.01E-218 | 217.6968 | Left atrium |
| LV_stroke_ | Z1_S2 | 27446 | 0.153874 | 0.004882 | 4.38E-218 | 217.358 | Left ventricle |
| RV_stroke_ | Z14_S4 | 27446 | 0.212576 | 0.006749 | 9.90E-218 | 217.0045 | Right ventricle |
| Descending | Z50_S4 | 24599 | 0.19615 | 0.006235 | 3.23E-217 | 216.4904 | Aorta |
| LA_stroke_ | Z83_S2 | 27446 | 0.153426 | 0.004878 | 4.12E-217 | 216.3846 | Left atrium |
| LV_stroke_ | Z44_S4 | 27446 | -0.15877 | 0.005048 | 4.36E-217 | 216.36 | Left ventricle |
| LA_stroke_ | Z29_S1 | 27446 | 0.176088 | 0.005601 | 5.77E-217 | 216.2386 | Left atrium |
| RA_minimu | Z75_S5 | 27446 | 0.21851 | 0.006952 | 7.47E-217 | 216.1266 | Right atrium |
| LA_maximu | Z77_S3 | 27446 | -0.16481 | 0.005245 | 1.07E-216 | 215.9687 | Left atrium |
| LV_stroke_ | Z41_S2 | 27446 | -0.22109 | 0.00705 | 6.68E-216 | 215.1754 | Left ventricle |

| | | | | | | | |
|---|---|---|---|---|---|---|---|
| LA_stroke_ | Z20_S2 | 27446 | 0.137021 | 0.004372 | 1.29E-215 | 214.8909 | Left atrium |
| LV_myocar | Z122_S1 | 27446 | 0.282596 | 0.009018 | 1.49E-215 | 214.8271 | Left ventricle |
| LV_stroke_ | Z109_S1 | 27446 | -0.18732 | 0.005978 | 1.54E-215 | 214.8138 | Left ventricle |
| Descending | Z4_S4 | 24599 | -0.18233 | 0.005823 | 3.16E-215 | 214.5004 | Aorta |
| RV_stroke_ | Z20_S1 | 27446 | 0.220196 | 0.00704 | 9.97E-215 | 214.0012 | Right ventricle |
| LA_minimu | Z7_S1 | 27446 | -0.14322 | 0.00458 | 1.19E-214 | 213.9248 | Left atrium |
| RV_end_sy | Z59_S3 | 27446 | -0.17079 | 0.005463 | 1.46E-214 | 213.8351 | Right ventricle |
| RA_minimu | Z37_S2 | 27446 | -0.18759 | 0.006008 | 5.22E-214 | 213.2823 | Right atrium |
| LA_minimu | Z47_S4 | 27446 | -0.15556 | 0.004984 | 7.03E-214 | 213.153 | Left atrium |
| LV_end_sy | Z80_S4 | 27446 | -0.16669 | 0.005347 | 2.29E-213 | 212.6405 | Left ventricle |
| RA_maxim | Z82_S4 | 27446 | -0.14677 | 0.004712 | 5.06E-213 | 212.2955 | Right atrium |
| RA_maxim | Z12_S4 | 27446 | -0.14527 | 0.004666 | 7.64E-213 | 212.1168 | Right atrium |
| LA_maximu | Z56_S2 | 27446 | -0.13752 | 0.00442 | 1.49E-212 | 211.8265 | Left atrium |
| LV_cardiac | Z99_S4 | 27446 | 0.189949 | 0.006107 | 2.11E-212 | 211.6766 | Left ventricle |
| LA_stroke_ | Z15_S2 | 27446 | 0.171674 | 0.00552 | 2.20E-212 | 211.6573 | Left atrium |
| Descending | Z117_S4 | 24599 | -0.16604 | 0.005339 | 2.31E-212 | 211.6361 | Aorta |
| RV_end_di | Z38_S5 | 27446 | -0.23192 | 0.007458 | 2.70E-212 | 211.5682 | Right ventricle |
| LV_end_di | Z120_S2 | 27446 | 0.179264 | 0.005765 | 2.91E-212 | 211.5365 | Left ventricle |
| LV_end_sy | Z83_S2 | 27446 | 0.181757 | 0.005848 | 4.57E-212 | 211.3396 | Left ventricle |
| RA_stroke_ | Z21_S3 | 27446 | -0.19366 | 0.006234 | 7.70E-212 | 211.1136 | Right atrium |
| RV_end_di | Z105_S5 | 27446 | 0.222133 | 0.007154 | 1.13E-211 | 210.9478 | Right ventricle |
| LA_stroke_ | Z125_S4 | 27446 | -0.16537 | 0.005335 | 5.51E-211 | 210.259 | Left atrium |
| RA_maxim | Z18_S4 | 27446 | -0.21109 | 0.006822 | 3.27E-210 | 209.4856 | Right atrium |
| LV_end_sy | Z62_S1 | 27446 | 0.179568 | 0.005806 | 4.81E-210 | 209.3175 | Left ventricle |
| LA_minimu | Z14_S4 | 27446 | 0.172131 | 0.005568 | 8.24E-210 | 209.0842 | Left atrium |
| LV_end_di | Z52_S3 | 27446 | 0.177148 | 0.005733 | 1.21E-209 | 208.9183 | Left ventricle |
| LA_stroke_ | Z69_S5 | 27446 | -0.13577 | 0.004398 | 3.13E-209 | 208.5049 | Left atrium |
| Descending | Z94_S5 | 24599 | 0.171805 | 0.005567 | 3.69E-209 | 208.4333 | Aorta |
| RA_minimu | Z99_S3 | 27446 | 0.137813 | 0.004468 | 7.35E-209 | 208.134 | Right atrium |
| RA_maxim | Z94_S5 | 27446 | 0.144689 | 0.004715 | 7.99E-207 | 206.0975 | Right atrium |
| LA_maximu | Z64_S1 | 27446 | -0.17552 | 0.00572 | 8.37E-207 | 206.0773 | Left atrium |
| RV_end_sy | Z85_S5 | 27446 | 0.193102 | 0.006316 | 2.78E-205 | 204.5565 | Right ventricle |
| RV_stroke_ | Z14_S1 | 27446 | 0.174022 | 0.005692 | 2.93E-205 | 204.5325 | Right ventricle |
| LA_maximu | Z55_S5 | 27446 | 0.134302 | 0.004396 | 5.07E-205 | 204.2953 | Left atrium |
| RA_maxim | Z83_S2 | 27446 | 0.157144 | 0.005157 | 6.49E-204 | 203.1879 | Right atrium |
| Descending | Z78_S4 | 24599 | 0.174294 | 0.005723 | 1.02E-203 | 202.9927 | Aorta |
| RV_end_di | Z80_S4 | 27446 | -0.18584 | 0.006102 | 1.03E-203 | 202.9855 | Right ventricle |
| RV_end_di | Z120_S4 | 27446 | -0.21348 | 0.007011 | 1.16E-203 | 202.9342 | Right ventricle |
| LV_myocar | Z1_S2 | 27446 | 0.205352 | 0.006753 | 4.28E-203 | 202.3681 | Left ventricle |
| LV_end_sy | Z92_S5 | 27446 | 0.2275 | 0.007482 | 4.49E-203 | 202.3481 | Left ventricle |
| LA_minimu | Z114_S5 | 27446 | -0.12111 | 0.003987 | 1.15E-202 | 201.9377 | Left atrium |
| LV_end_sy | Z98_S3 | 27446 | -0.1648 | 0.005425 | 1.15E-202 | 201.9376 | Left ventricle |
| RV_end_di | Z98_S3 | 27446 | -0.18786 | 0.006187 | 1.75E-202 | 201.756 | Right ventricle |
| RA_minimu | Z106_S4 | 27446 | 0.147953 | 0.004875 | 2.53E-202 | 201.5963 | Right atrium |
| RV_stroke_ | Z59_S3 | 27446 | -0.14903 | 0.004911 | 2.62E-202 | 201.5814 | Right ventricle |

| | | | | | | | |
|---|---|---|---|---|---|---|---|
| LV_end_sy | Z15_S2 | 27446 | 0.200883 | 0.00662 | 2.78E-202 | 201.5558 | Left ventricle |
| Ascending_ | Z106_S4 | 24599 | 0.150398 | 0.004956 | 2.84E-202 | 201.5462 | Aorta |
| LV_myocar | Z92_S5 | 27446 | 0.301906 | 0.009949 | 2.94E-202 | 201.5315 | Left ventricle |
| LV_end_di | Z47_S4 | 27446 | -0.2033 | 0.006714 | 2.02E-201 | 200.6943 | Left ventricle |
| LV_stroke_ | Z55_S5 | 27446 | 0.154157 | 0.005096 | 4.99E-201 | 200.3017 | Left ventricle |
| LV_end_di | Z23_S3 | 27446 | 0.180433 | 0.005965 | 5.52E-201 | 200.258 | Left ventricle |
| RA_maxim | Z86_S3 | 27446 | -0.18706 | 0.006187 | 8.26E-201 | 200.0831 | Right atrium |
| Descending | Z52_S4 | 24599 | 0.174558 | 0.005776 | 1.34E-200 | 199.8741 | Aorta |
| RV_stroke_ | Z5_S5 | 27446 | 0.188478 | 0.006242 | 2.76E-200 | 199.5598 | Right ventricle |
| LA_minimu | Z66_S2 | 27446 | 0.172782 | 0.005724 | 3.45E-200 | 199.4617 | Left atrium |
| RV_end_di | Z8_S3 | 27446 | 0.181501 | 0.006013 | 3.71E-200 | 199.4305 | Right ventricle |
| LA_ejectio | Z14_S2 | 27446 | 0.144063 | 0.004774 | 4.60E-200 | 199.3375 | Left atrium |
| LV_end_di | Z55_S1 | 27446 | -0.18803 | 0.006235 | 9.09E-200 | 199.0413 | Left ventricle |
| LV_end_sy | Z69_S5 | 27446 | -0.15889 | 0.005275 | 2.44E-199 | 198.6129 | Left ventricle |
| Descending | Z99_S3 | 24599 | 0.159183 | 0.005285 | 2.82E-199 | 198.5499 | Aorta |
| LV_end_di | Z68_S2 | 27446 | -0.18417 | 0.006118 | 4.29E-199 | 198.3676 | Left ventricle |
| Descending | Z51_S1 | 24599 | -0.17067 | 0.005677 | 1.38E-198 | 197.8604 | Aorta |
| RV_end_di | Z5_S1 | 27446 | -0.20955 | 0.006979 | 4.48E-198 | 197.3484 | Right ventricle |
| LV_myocar | Z52_S4 | 27446 | 0.221525 | 0.007381 | 6.38E-198 | 197.1951 | Left ventricle |
| RA_maxim | Z106_S4 | 27446 | 0.144388 | 0.004812 | 7.71E-198 | 197.1131 | Right atrium |
| LV_end_di | Z57_S1 | 27446 | -0.18604 | 0.006204 | 1.42E-197 | 196.8463 | Left ventricle |
| LV_end_sy | Z44_S4 | 27446 | -0.15755 | 0.005255 | 1.61E-197 | 196.7926 | Left ventricle |
| RV_end_sy | Z123_S4 | 27446 | 0.211795 | 0.007071 | 3.81E-197 | 196.4187 | Right ventricle |
| LA_minimu | Z95_S3 | 27446 | -0.11528 | 0.003849 | 4.00E-197 | 196.3979 | Left atrium |
| LA_maximu | Z111_S1 | 27446 | 0.168279 | 0.00562 | 5.32E-197 | 196.274 | Left atrium |
| LA_minimu | Z53_S1 | 27446 | 0.155665 | 0.0052 | 7.18E-197 | 196.144 | Left atrium |
| LA_minimu | Z20_S2 | 27446 | 0.126442 | 0.004225 | 7.94E-197 | 196.0999 | Left atrium |
| LV_end_sy | Z30_S2 | 27446 | -0.22435 | 0.007528 | 3.59E-195 | 194.445 | Left ventricle |
| Ascending_ | Z114_S5 | 24599 | -0.13458 | 0.004517 | 4.80E-195 | 194.319 | Aorta |
| RA_minimu | Z82_S4 | 27446 | -0.14244 | 0.004782 | 6.30E-195 | 194.2003 | Right atrium |
| LV_end_sy | Z82_S1 | 27446 | -0.22848 | 0.007671 | 6.50E-195 | 194.1869 | Left ventricle |
| LA_maximu | Z66_S2 | 27446 | 0.175763 | 0.005905 | 1.18E-194 | 193.9274 | Left atrium |
| LV_end_sy | Z29_S5 | 27446 | 0.197415 | 0.006635 | 1.52E-194 | 193.8182 | Left ventricle |
| LV_end_di | Z66_S3 | 27446 | -0.22667 | 0.007628 | 4.97E-194 | 193.3037 | Left ventricle |
| RA_stroke_ | Z49_S1 | 27446 | -0.18088 | 0.006088 | 5.09E-194 | 193.2935 | Right atrium |
| LV_longitu | Z23_S1 | 27446 | 0.163345 | 0.005498 | 5.38E-194 | 193.2693 | Left ventricle |
| LV_end_sy | Z122_S1 | 27446 | 0.201823 | 0.006794 | 6.52E-194 | 193.1854 | Left ventricle |
| RV_end_di | Z26_S4 | 27446 | 0.260642 | 0.008775 | 6.77E-194 | 193.1692 | Right ventricle |
| RV_end_sy | Z20_S2 | 27446 | 0.169449 | 0.005707 | 9.88E-194 | 193.0051 | Right ventricle |
| LA_stroke_ | Z62_S2 | 27446 | 0.136178 | 0.004589 | 1.65E-193 | 192.7823 | Left atrium |
| RA_ejectio | Z86_S3 | 27446 | 0.164637 | 0.005551 | 2.51E-193 | 192.5997 | Right atrium |
| RV_end_di | Z61_S4 | 27446 | -0.19786 | 0.006672 | 2.74E-193 | 192.5618 | Right ventricle |
| LV_end_di | Z13_S3 | 27446 | 0.162642 | 0.005485 | 3.05E-193 | 192.515 | Left ventricle |
| RV_end_di | Z41_S2 | 27446 | -0.24811 | 0.00837 | 4.16E-193 | 192.3804 | Right ventricle |
| Descending | Z13_S1 | 24599 | -0.17425 | 0.005879 | 4.29E-193 | 192.3674 | Aorta |

| | | | | | | | |
|---|---|---|---|---|---|---|---|
| Descending | Z31_S5 | 24599 | 0.155278 | 0.005241 | 6.40E-193 | 192.1937 | Aorta |
| LV_end_di | Z111_S1 | 27446 | 0.217267 | 0.007336 | 8.86E-193 | 192.0524 | Left ventricle |
| RA_maxim | Z127_S5 | 27446 | 0.137992 | 0.004659 | 9.27E-193 | 192.0331 | Right atrium |
| LA_minimu | Z30_S2 | 27446 | -0.1794 | 0.006062 | 1.93E-192 | 191.7134 | Left atrium |
| LA_maximu | Z2_S1 | 27446 | -0.13911 | 0.004702 | 2.48E-192 | 191.6063 | Left atrium |
| RV_end_sy | Z105_S4 | 27446 | -0.18189 | 0.006155 | 6.07E-192 | 191.2168 | Right ventricle |
| LA_maximu | Z19_S2 | 27446 | 0.150227 | 0.005087 | 1.08E-191 | 190.9679 | Left atrium |
| RA_ejectio | Z38_S5 | 27446 | 0.152864 | 0.005177 | 1.19E-191 | 190.9242 | Right atrium |
| LA_minimu | Z52_S4 | 27446 | 0.131861 | 0.004472 | 4.05E-191 | 190.3931 | Left atrium |
| RA_maxim | Z49_S1 | 27446 | -0.19707 | 0.006684 | 4.36E-191 | 190.3607 | Right atrium |
| LV_longitu | Z74_S4 | 27446 | 0.151377 | 0.005139 | 1.04E-190 | 189.9818 | Left ventricle |
| LV_myocar | Z14_S2 | 27446 | -0.2422 | 0.008232 | 2.85E-190 | 189.5445 | Left ventricle |
| RV_stroke_ | Z62_S1 | 27446 | 0.167013 | 0.005679 | 3.98E-190 | 189.4002 | Right ventricle |
| RA_maxim | Z53_S1 | 27446 | 0.167352 | 0.005695 | 8.89E-190 | 189.0511 | Right atrium |
| Ascending_ | Z95_S3 | 24599 | -0.12791 | 0.004355 | 1.46E-189 | 188.8355 | Aorta |
| RV_end_sy | Z1_S2 | 27446 | 0.162275 | 0.005529 | 2.38E-189 | 188.6234 | Right ventricle |
| LV_end_sy | Z86_S3 | 27446 | -0.20618 | 0.007027 | 2.97E-189 | 188.5275 | Left ventricle |
| RV_end_di | Z2_S1 | 27446 | -0.18954 | 0.00646 | 3.12E-189 | 188.5055 | Right ventricle |
| LA_maximu | Z24_S5 | 27446 | -0.16475 | 0.00562 | 6.28E-189 | 188.2024 | Left atrium |
| LA_minimu | Z29_S1 | 27446 | 0.158724 | 0.005416 | 8.76E-189 | 188.0573 | Left atrium |
| LV_stroke_ | Z111_S1 | 27446 | 0.190681 | 0.006518 | 3.76E-188 | 187.4245 | Left ventricle |
| LA_minimu | Z62_S1 | 27446 | 0.136937 | 0.004683 | 5.65E-188 | 187.2476 | Left atrium |
| LV_stroke_ | Z98_S3 | 27446 | -0.15281 | 0.005227 | 7.35E-188 | 187.1338 | Left ventricle |
| Descending | Z12_S4 | 24599 | -0.16386 | 0.005613 | 2.27E-187 | 186.644 | Aorta |
| Descending | Z51_S1 | 24599 | -0.16824 | 0.005765 | 3.03E-187 | 186.5183 | Aorta |
| LA_minimu | Z56_S2 | 27446 | -0.12529 | 0.004294 | 3.93E-187 | 186.4055 | Left atrium |
| RA_minimu | Z21_S3 | 27446 | -0.20247 | 0.00695 | 1.40E-186 | 185.854 | Right atrium |
| RA_minimu | Z123_S4 | 27446 | 0.169137 | 0.005813 | 4.00E-186 | 185.3983 | Right atrium |
| LV_end_di | Z39_S1 | 27446 | 0.180868 | 0.006217 | 4.54E-186 | 185.343 | Left ventricle |
| LV_end_sy | Z2_S1 | 27446 | -0.16463 | 0.005666 | 1.29E-185 | 184.8888 | Left ventricle |
| LV_end_di | Z43_S2 | 27446 | -0.17095 | 0.005896 | 7.91E-185 | 184.1019 | Left ventricle |
| LA_stroke_ | Z2_S1 | 27446 | -0.13707 | 0.004729 | 9.87E-185 | 184.0055 | Left atrium |
| RA_stroke_ | Z93_S1 | 27446 | 0.165632 | 0.005715 | 1.11E-184 | 183.9565 | Right atrium |
| Descending | Z89_S4 | 24599 | -0.18106 | 0.006257 | 4.19E-184 | 183.3781 | Aorta |
| LA_minimu | Z24_S5 | 27446 | -0.15759 | 0.005451 | 9.13E-184 | 183.0397 | Left atrium |
| Descending | Z120_S4 | 24599 | -0.18559 | 0.006421 | 1.02E-183 | 182.9893 | Aorta |
| LV_longitu | Z120_S5 | 27446 | -0.17095 | 0.005916 | 1.28E-183 | 182.892 | Left ventricle |
| RV_end_di | Z83_S2 | 27446 | 0.192999 | 0.006685 | 2.80E-183 | 182.5524 | Right ventricle |
| Ascending_ | Z107_S1 | 24599 | -0.13127 | 0.004554 | 9.61E-183 | 182.0172 | Aorta |
| LA_stroke_ | Z30_S2 | 27446 | -0.18127 | 0.006288 | 1.00E-182 | 181.9983 | Left atrium |
| RV_end_sy | Z28_S4 | 27446 | -0.20967 | 0.007278 | 1.58E-182 | 181.8018 | Right ventricle |
| RA_minimu | Z40_S5 | 27446 | -0.13666 | 0.004749 | 4.53E-182 | 181.3443 | Right atrium |
| LA_minimu | Z62_S2 | 27446 | 0.127503 | 0.004432 | 5.09E-182 | 181.293 | Left atrium |
| Ascending_ | Z95_S3 | 24599 | -0.12577 | 0.004374 | 8.99E-182 | 181.0462 | Aorta |
| LV_end_di | Z115_S4 | 27446 | 0.177135 | 0.006172 | 3.69E-181 | 180.4324 | Left ventricle |

| | | | | | | | |
|---|---|---|---|---|---|---|---|
| RV_end_sy | Z1_S1 | 27446 | -0.18877 | 0.006578 | 4.22E-181 | 180.3746 | Right ventricle |
| Descending | Z59_S3 | 24599 | -0.15034 | 0.005244 | 8.65E-181 | 180.063 | Aorta |
| LV_end_sy | Z51_S1 | 27446 | -0.15636 | 0.005455 | 1.13E-180 | 179.9486 | Left ventricle |
| RA_maxim | Z111_S5 | 27446 | 0.155599 | 0.00543 | 1.45E-180 | 179.8399 | Right atrium |
| Descending | Z105_S4 | 24599 | -0.16914 | 0.005906 | 2.23E-180 | 179.6512 | Aorta |
| Descending | Z51_S4 | 24599 | -0.16285 | 0.005688 | 2.80E-180 | 179.5534 | Aorta |
| LV_myocar | Z38_S5 | 27446 | -0.24944 | 0.008719 | 5.07E-180 | 179.2953 | Left ventricle |
| LA_minimu | Z68_S2 | 27446 | -0.13006 | 0.004553 | 1.92E-179 | 178.7157 | Left atrium |
| LV_stroke_ | Z120_S4 | 27446 | -0.16891 | 0.005928 | 1.37E-178 | 177.8645 | Left ventricle |
| RV_end_sy | Z127_S4 | 27446 | 0.211273 | 0.007416 | 1.54E-178 | 177.8134 | Right ventricle |
| LV_longitu | Z65_S5 | 27446 | -0.16612 | 0.005839 | 4.61E-178 | 177.3364 | Left ventricle |
| Ascending_ | Z114_S5 | 24599 | -0.12913 | 0.004541 | 6.60E-178 | 177.1801 | Aorta |
| RA_minimu | Z83_S2 | 27446 | 0.148923 | 0.005238 | 8.55E-178 | 177.0682 | Right atrium |
| LA_minimu | Z39_S1 | 27446 | 0.131421 | 0.004623 | 9.24E-178 | 177.0345 | Left atrium |
| LA_stroke_ | Z114_S5 | 27446 | -0.11766 | 0.004141 | 1.42E-177 | 176.8469 | Left atrium |
| LV_stroke_ | Z20_S2 | 27446 | 0.143676 | 0.005057 | 1.50E-177 | 176.8241 | Left ventricle |
| RV_end_di | Z75_S4 | 27446 | -0.23836 | 0.008391 | 1.59E-177 | 176.798 | Right ventricle |
| Ascending_ | Z106_S4 | 24599 | 0.141604 | 0.004985 | 1.82E-177 | 176.7393 | Aorta |
| RA_minimu | Z111_S5 | 27446 | 0.156286 | 0.005505 | 2.82E-177 | 176.5502 | Right atrium |
| LA_minimu | Z46_S1 | 27446 | -0.14373 | 0.005064 | 3.48E-177 | 176.4579 | Left atrium |
| RV_stroke_ | Z41_S2 | 27446 | -0.20321 | 0.007177 | 2.30E-176 | 175.6385 | Right ventricle |
| LV_myocar | Z35_S5 | 27446 | -0.20066 | 0.007088 | 2.60E-176 | 175.5849 | Left ventricle |
| LV_myocar | Z86_S3 | 27446 | -0.26453 | 0.009353 | 5.80E-176 | 175.2368 | Left ventricle |
| Ascending_ | Z99_S3 | 24599 | 0.128906 | 0.004558 | 5.92E-176 | 175.2274 | Aorta |
| LV_end_sy | Z111_S5 | 27446 | 0.174204 | 0.006164 | 1.07E-175 | 174.972 | Left ventricle |
| LA_stroke_ | Z29_S5 | 27446 | 0.156563 | 0.005545 | 2.31E-175 | 174.637 | Left atrium |
| Descending | Z73_S4 | 24599 | 0.210045 | 0.007443 | 3.16E-175 | 174.5008 | Aorta |
| Descending | Z24_S1 | 24599 | 0.157021 | 0.005572 | 9.86E-175 | 174.0062 | Aorta |
| LV_myocar | Z111_S5 | 27446 | 0.230997 | 0.008197 | 1.02E-174 | 173.9914 | Left ventricle |
| LV_stroke_ | Z47_S4 | 27446 | -0.16833 | 0.005976 | 1.37E-174 | 173.8623 | Left ventricle |
| LV_stroke_ | Z30_S2 | 27446 | -0.20428 | 0.007257 | 2.31E-174 | 173.6363 | Left ventricle |
| LV_myocar | Z120_S4 | 27446 | -0.23048 | 0.008194 | 4.33E-174 | 173.3639 | Left ventricle |
| LV_end_di | Z50_S3 | 27446 | -0.17749 | 0.006312 | 5.26E-174 | 173.2787 | Left ventricle |
| LV_end_di | Z14_S2 | 27446 | -0.18885 | 0.006717 | 6.39E-174 | 173.1946 | Left ventricle |
| RV_stroke_ | Z28_S4 | 27446 | -0.18382 | 0.00654 | 8.07E-174 | 173.0929 | Right ventricle |
| LA_ejectio | Z85_S5 | 27446 | -0.12628 | 0.004494 | 9.65E-174 | 173.0157 | Left atrium |
| LA_maximu | Z50_S3 | 27446 | -0.13591 | 0.004837 | 1.06E-173 | 172.9747 | Left atrium |
| LV_cardiac | Z82_S1 | 27446 | 0.190307 | 0.006778 | 1.78E-173 | 172.7489 | Left ventricle |
| Descending | Z13_S1 | 24599 | -0.16273 | 0.005806 | 6.92E-173 | 172.1597 | Aorta |
| LV_end_di | Z61_S4 | 27446 | -0.17797 | 0.006349 | 7.07E-173 | 172.1508 | Left ventricle |
| LV_myocar | Z14_S1 | 27446 | 0.217761 | 0.007771 | 9.04E-173 | 172.044 | Left ventricle |
| LA_stroke_ | Z55_S5 | 27446 | 0.123953 | 0.004429 | 2.52E-172 | 171.5993 | Left atrium |
| RA_minimu | Z117_S4 | 27446 | -0.12659 | 0.004524 | 2.74E-172 | 171.5627 | Right atrium |
| Descending | Z14_S1 | 24599 | 0.170168 | 0.006084 | 3.75E-172 | 171.4258 | Aorta |
| LV_end_sy | Z105_S5 | 27446 | 0.175974 | 0.006293 | 4.70E-172 | 171.3281 | Left ventricle |

| | | | | | | | |
|---|---|---|---|---|---|---|---|
| LV_cardiac | Z85_S5 | 27446 | -0.14352 | 0.005137 | 8.39E-172 | 171.076 | Left ventricle |
| LV_myocar | Z51_S1 | 27446 | -0.20279 | 0.007259 | 9.50E-172 | 171.0223 | Left ventricle |
| LV_longitu | Z111_S5 | 27446 | -0.13567 | 0.004863 | 2.99E-171 | 170.5238 | Left ventricle |
| LV_end_sy | Z8_S3 | 27446 | 0.147319 | 0.005285 | 5.43E-171 | 170.2651 | Left ventricle |
| LA_ejectio | Z127_S5 | 27446 | -0.11375 | 0.004087 | 1.94E-170 | 169.7131 | Left atrium |
| LA_maximu | Z52_S3 | 27446 | 0.122509 | 0.004408 | 5.08E-170 | 169.2945 | Left atrium |
| Descending | Z1_S2 | 24599 | 0.147124 | 0.005299 | 1.15E-169 | 168.9379 | Aorta |
| RV_stroke_ | Z109_S1 | 27446 | -0.16903 | 0.006089 | 1.26E-169 | 168.8998 | Right ventricle |
| RA_stroke_ | Z4_S4 | 27446 | -0.12314 | 0.00444 | 2.66E-169 | 168.5754 | Right atrium |
| RV_stroke_ | Z69_S5 | 27446 | -0.14318 | 0.005163 | 3.11E-169 | 168.5069 | Right ventricle |
| LV_longitu | Z122_S1 | 27446 | -0.14861 | 0.005369 | 1.34E-168 | 167.8719 | Left ventricle |
| LV_end_di | Z58_S2 | 27446 | 0.174878 | 0.00632 | 1.51E-168 | 167.8212 | Left ventricle |
| Descending | Z50_S4 | 24599 | 0.175674 | 0.006353 | 2.70E-168 | 167.5681 | Aorta |
| LA_stroke_ | Z52_S3 | 27446 | 0.122493 | 0.00443 | 2.90E-168 | 167.5373 | Left atrium |
| LV_end_sy | Z120_S2 | 27446 | 0.147532 | 0.00534 | 5.22E-168 | 167.282 | Left ventricle |
| LV_myocar | Z7_S1 | 27446 | -0.2095 | 0.007593 | 1.37E-167 | 166.8631 | Left ventricle |
| LV_myocar | Z87_S3 | 27446 | -0.22234 | 0.008061 | 1.85E-167 | 166.7328 | Left ventricle |
| RA_minimu | Z111_S1 | 27446 | 0.167108 | 0.00606 | 2.15E-167 | 166.6683 | Right atrium |
| RV_stroke_ | Z29_S5 | 27446 | 0.179023 | 0.006493 | 2.55E-167 | 166.5931 | Right ventricle |
| RV_stroke_ | Z30_S2 | 27446 | -0.20269 | 0.007368 | 1.40E-166 | 165.855 | Right ventricle |
| LA_maximu | Z66_S3 | 27446 | -0.16112 | 0.005859 | 1.78E-166 | 165.7505 | Left atrium |
| RV_end_sy | Z62_S1 | 27446 | 0.174214 | 0.006336 | 1.88E-166 | 165.726 | Right ventricle |
| Descending | Z65_S1 | 24599 | -0.15088 | 0.005487 | 1.97E-166 | 165.7049 | Aorta |
| LV_end_sy | Z120_S4 | 27446 | -0.16935 | 0.006166 | 4.56E-166 | 165.3412 | Left ventricle |
| LA_minimu | Z14_S1 | 27446 | 0.128965 | 0.004709 | 3.74E-165 | 164.4276 | Left atrium |
| LV_myocar | Z15_S2 | 27446 | 0.241695 | 0.008829 | 5.26E-165 | 164.2793 | Left ventricle |
| LA_stroke_ | Z68_S2 | 27446 | -0.1293 | 0.004725 | 6.80E-165 | 164.1673 | Left atrium |
| LV_end_sy | Z82_S4 | 27446 | -0.14645 | 0.005368 | 6.61E-164 | 163.1801 | Left ventricle |
| RV_end_sy | Z120_S4 | 27446 | -0.1829 | 0.006706 | 8.75E-164 | 163.0582 | Right ventricle |
| Ascending_ | Z13_S1 | 24599 | -0.13882 | 0.005091 | 1.03E-163 | 162.9883 | Aorta |
| LV_cardiac | Z12_S5 | 27446 | -0.17768 | 0.006517 | 1.20E-163 | 162.9191 | Left ventricle |
| RV_end_sy | Z8_S3 | 27446 | 0.156732 | 0.00575 | 1.43E-163 | 162.8445 | Right ventricle |
| Descending | Z82_S1 | 24599 | -0.19813 | 0.007272 | 1.79E-163 | 162.7461 | Aorta |
| LV_end_di | Z75_S4 | 27446 | -0.21714 | 0.007981 | 5.42E-163 | 162.2661 | Left ventricle |
| RA_maxim | Z85_S5 | 27446 | 0.139678 | 0.005137 | 7.88E-163 | 162.1037 | Right atrium |
| LV_myocar | Z82_S1 | 27446 | -0.27794 | 0.010227 | 1.26E-162 | 161.899 | Left ventricle |
| LA_minimu | Z19_S2 | 27446 | 0.134286 | 0.004944 | 1.88E-162 | 161.7259 | Left atrium |
| RA_maxim | Z123_S4 | 27446 | 0.156007 | 0.005747 | 2.68E-162 | 161.5717 | Right atrium |
| LA_minimu | Z98_S3 | 27446 | -0.11896 | 0.004383 | 2.95E-162 | 161.5305 | Left atrium |
| RA_maxim | Z99_S3 | 27446 | 0.120108 | 0.004426 | 3.37E-162 | 161.472 | Right atrium |
| LA_ejectio | Z82_S4 | 27446 | 0.112273 | 0.004143 | 1.01E-161 | 160.9939 | Left atrium |
| RV_end_di | Z62_S2 | 27446 | 0.170384 | 0.006288 | 1.04E-161 | 160.9839 | Right ventricle |
| LA_minimu | Z80_S4 | 27446 | -0.11703 | 0.004323 | 2.12E-161 | 160.6733 | Left atrium |
| RV_stroke_ | Z5_S1 | 27446 | -0.16216 | 0.005994 | 3.43E-161 | 160.4653 | Right ventricle |
| RV_end_sy | Z15_S2 | 27446 | 0.195388 | 0.007222 | 3.47E-161 | 160.4595 | Right ventricle |

| | | | | | | | |
|---|---|---|---|---|---|---|---|
| LA_stroke_ | Z8_S3 | 27446 | 0.119351 | 0.004414 | 5.10E-161 | 160.2923 | Left atrium |
| RA_stroke_ | Z122_S1 | 27446 | 0.14787 | 0.00547 | 5.76E-161 | 160.2396 | Right atrium |
| LV_myocar | Z55_S5 | 27446 | 0.190884 | 0.007064 | 8.32E-161 | 160.0797 | Left ventricle |
| LV_myocar | Z20_S2 | 27446 | 0.189031 | 0.006997 | 9.83E-161 | 160.0074 | Left ventricle |
| Ascending_ | Z13_S1 | 24599 | -0.13706 | 0.005074 | 1.05E-160 | 159.978 | Aorta |
| Descending | Z1_S1 | 24599 | -0.17044 | 0.006311 | 1.16E-160 | 159.9352 | Aorta |
| LV_stroke_ | Z86_S3 | 27446 | -0.18301 | 0.006778 | 1.42E-160 | 159.8479 | Left ventricle |
| RV_end_sy | Z69_S5 | 27446 | -0.15536 | 0.005754 | 1.47E-160 | 159.8339 | Right ventricle |
| RA_maxim | Z39_S3 | 27446 | 0.185221 | 0.00687 | 4.32E-160 | 159.3648 | Right atrium |
| RV_end_di | Z55_S1 | 27446 | -0.17743 | 0.006584 | 5.93E-160 | 159.2269 | Right ventricle |
| Descending | Z65_S1 | 24599 | -0.1458 | 0.005412 | 7.71E-160 | 159.1131 | Aorta |
| Descending | Z40_S5 | 24599 | -0.14931 | 0.005547 | 1.34E-159 | 158.8744 | Aorta |
| LA_minimu | Z5_S1 | 27446 | -0.13294 | 0.004943 | 2.46E-159 | 158.6085 | Left atrium |
| LA_stroke_ | Z50_S3 | 27446 | -0.13087 | 0.004867 | 2.87E-159 | 158.5422 | Left atrium |
| LV_end_sy | Z115_S4 | 27446 | 0.153421 | 0.005706 | 3.16E-159 | 158.5006 | Left ventricle |
| LA_minimu | Z111_S1 | 27446 | 0.146715 | 0.005466 | 1.11E-158 | 157.9556 | Left atrium |
| RV_stroke_ | Z15_S2 | 27446 | 0.174106 | 0.006487 | 1.15E-158 | 157.9401 | Right ventricle |
| RV_end_sy | Z80_S4 | 27446 | -0.15674 | 0.00584 | 1.15E-158 | 157.9396 | Right ventricle |
| Descending | Z55_S5 | 24599 | 0.148362 | 0.005531 | 1.71E-158 | 157.7663 | Aorta |
| LV_end_di | Z87_S3 | 27446 | -0.17619 | 0.006574 | 3.04E-158 | 157.5171 | Left ventricle |
| RV_end_sy | Z46_S1 | 27446 | -0.18354 | 0.00685 | 3.91E-158 | 157.4084 | Right ventricle |
| LV_myocar | Z44_S4 | 27446 | -0.18774 | 0.007009 | 4.97E-158 | 157.3032 | Left ventricle |
| LV_myocar | Z98_S3 | 27446 | -0.19392 | 0.00724 | 5.07E-158 | 157.2948 | Left ventricle |
| Descending | Z8_S3 | 24599 | 0.147662 | 0.005516 | 7.08E-158 | 157.1501 | Aorta |
| RA_minimu | Z89_S4 | 27446 | -0.14391 | 0.005376 | 7.99E-158 | 157.0974 | Right atrium |
| RV_end_sy | Z51_S1 | 27446 | -0.15897 | 0.005943 | 1.27E-157 | 156.8971 | Right ventricle |
| RA_ejectio | Z31_S5 | 27446 | -0.10588 | 0.003958 | 1.29E-157 | 156.8904 | Right atrium |
| LV_end_sy | Z68_S2 | 27446 | -0.15151 | 0.005665 | 1.43E-157 | 156.8449 | Left ventricle |
| LV_end_di | Z31_S2 | 27446 | 0.181437 | 0.006785 | 1.56E-157 | 156.8066 | Left ventricle |
| RV_end_di | Z51_S1 | 27446 | -0.16658 | 0.006233 | 2.42E-157 | 156.6169 | Right ventricle |
| LV_longitu | Z12_S4 | 27446 | 0.111969 | 0.004194 | 5.35E-157 | 156.2719 | Left ventricle |
| LA_stroke_ | Z46_S1 | 27446 | -0.14029 | 0.005258 | 7.61E-157 | 156.1186 | Left atrium |
| LV_end_di | Z65_S2 | 27446 | 0.18503 | 0.006937 | 1.01E-156 | 155.9955 | Left ventricle |
| LA_minimu | Z69_S5 | 27446 | -0.11357 | 0.004262 | 1.83E-156 | 155.7383 | Left atrium |
| LV_myocar | Z28_S4 | 27446 | -0.23738 | 0.008918 | 4.26E-156 | 155.3706 | Left ventricle |
| LV_myocar | Z20_S1 | 27446 | 0.256347 | 0.009632 | 4.81E-156 | 155.3177 | Left ventricle |
| RA_minimu | Z29_S1 | 27446 | 0.160273 | 0.006025 | 6.27E-156 | 155.2027 | Right atrium |
| RV_end_sy | Z30_S2 | 27446 | -0.21833 | 0.008212 | 9.94E-156 | 155.0028 | Right ventricle |
| LV_end_sy | Z109_S1 | 27446 | -0.16585 | 0.006242 | 1.60E-155 | 154.7971 | Left ventricle |
| LA_maximu | Z105_S5 | 27446 | 0.13895 | 0.005233 | 2.32E-155 | 154.6342 | Left atrium |
| LV_cardiac | Z62_S4 | 27446 | -0.16694 | 0.006292 | 4.24E-155 | 154.3728 | Left ventricle |
| LA_stroke_ | Z41_S2 | 27446 | -0.16297 | 0.006144 | 4.78E-155 | 154.3204 | Left atrium |
| LV_stroke_ | Z105_S5 | 27446 | 0.160748 | 0.006065 | 8.37E-155 | 154.0775 | Left ventricle |
| Descending | Z23_S3 | 24599 | 0.152504 | 0.00576 | 1.87E-154 | 153.7273 | Aorta |
| LV_end_di | Z17_S1 | 27446 | 0.166961 | 0.006308 | 2.13E-154 | 153.6709 | Left ventricle |

| | | | | | | | |
|---|---|---|---|---|---|---|---|
| LA_stroke_ | Z41_S1 | 27446 | 0.148584 | 0.005615 | 2.72E-154 | 153.5662 | Left atrium |
| LA_maximu | Z43_S2 | 27446 | -0.11963 | 0.00453 | 1.10E-153 | 152.9595 | Left atrium |
| LV_end_sy | Z43_S2 | 27446 | -0.14398 | 0.005456 | 1.72E-153 | 152.7633 | Left ventricle |
| LA_minimu | Z109_S1 | 27446 | -0.1326 | 0.005027 | 2.40E-153 | 152.619 | Left atrium |
| LV_end_sy | Z46_S1 | 27446 | -0.16624 | 0.006302 | 2.47E-153 | 152.6076 | Left ventricle |
| Descending | Z44_S4 | 24599 | -0.1451 | 0.005502 | 3.03E-153 | 152.518 | Aorta |
| LA_maximu | Z5_S5 | 27446 | 0.140499 | 0.005328 | 3.05E-153 | 152.515 | Left atrium |
| LV_myocar | Z46_S1 | 27446 | -0.22083 | 0.00838 | 4.96E-153 | 152.3047 | Left ventricle |
| LA_ejection | Z37_S2 | 27446 | 0.137427 | 0.005217 | 6.05E-153 | 152.2182 | Left atrium |
| Descending | Z29_S1 | 24599 | 0.186683 | 0.007088 | 7.02E-153 | 152.1536 | Aorta |
| RV_end_di | Z52_S3 | 27446 | 0.159529 | 0.006062 | 1.28E-152 | 151.893 | Right ventricle |
| RV_stroke_ | Z55_S5 | 27446 | 0.136281 | 0.005192 | 7.29E-152 | 151.1375 | Right ventricle |
| LV_end_dia | Z37_S2 | 27446 | -0.1917 | 0.007326 | 6.03E-151 | 150.2199 | Left ventricle |
| LA_stroke_ | Z14_S2 | 27446 | -0.13561 | 0.005183 | 6.73E-151 | 150.1718 | Left atrium |
| RA_stroke_ | Z92_S5 | 27446 | 0.157721 | 0.006033 | 1.17E-150 | 149.9327 | Right atrium |
| Descending | Z8_S3 | 24599 | 0.146336 | 0.005599 | 1.43E-150 | 149.8439 | Aorta |
| LA_stroke_ | Z20_S1 | 27446 | 0.158035 | 0.006048 | 1.67E-150 | 149.778 | Left atrium |
| LA_maximu | Z6_S2 | 27446 | -0.10512 | 0.004024 | 2.02E-150 | 149.6943 | Left atrium |
| LV_stroke_ | Z80_S4 | 27446 | -0.13505 | 0.005172 | 2.56E-150 | 149.5913 | Left ventricle |
| LV_longitu | Z18_S4 | 27446 | 0.160173 | 0.006135 | 3.00E-150 | 149.5232 | Left ventricle |
| LA_maximu | Z41_S1 | 27446 | 0.145899 | 0.005589 | 3.27E-150 | 149.486 | Left atrium |
| LV_end_sy | Z23_S3 | 27446 | 0.144233 | 0.005528 | 4.70E-150 | 149.328 | Left ventricle |
| Descending | Z52_S4 | 24599 | 0.15351 | 0.005887 | 6.64E-150 | 149.1776 | Aorta |
| RV_end_sy | Z53_S1 | 27446 | 0.183829 | 0.00705 | 7.24E-150 | 149.1401 | Right ventricle |
| Ascending_ | Z107_S1 | 24599 | -0.11933 | 0.004584 | 2.14E-149 | 148.6686 | Aorta |
| RV_stroke_ | Z44_S4 | 27446 | -0.13408 | 0.005151 | 2.34E-149 | 148.6311 | Right ventricle |
| RV_end_di | Z79_S4 | 27446 | -0.21787 | 0.008373 | 2.83E-149 | 148.5478 | Right ventricle |
| RA_minimu | Z18_S4 | 27446 | -0.1807 | 0.006949 | 4.48E-149 | 148.3492 | Right atrium |
| Descending | Z1_S2 | 24599 | 0.140033 | 0.005385 | 4.70E-149 | 148.3284 | Aorta |
| Ascending_ | Z29_S1 | 24599 | 0.160942 | 0.00619 | 4.88E-149 | 148.3113 | Aorta |
| LV_end_sy | Z57_S1 | 27446 | -0.14939 | 0.005749 | 6.74E-149 | 148.1714 | Left ventricle |
| Descending | Z23_S3 | 24599 | 0.151804 | 0.005846 | 1.23E-148 | 147.9103 | Aorta |
| Descending | Z55_S5 | 24599 | 0.145669 | 0.005616 | 2.48E-148 | 147.6062 | Aorta |
| LV_end_dia | Z70_S5 | 27446 | 0.167453 | 0.006457 | 2.68E-148 | 147.572 | Left ventricle |
| RV_end_di | Z23_S3 | 27446 | 0.163291 | 0.006306 | 7.40E-148 | 147.1306 | Right ventricle |
| LV_end_dia | Z26_S4 | 27446 | 0.216573 | 0.008368 | 1.10E-147 | 146.9593 | Left ventricle |
| RV_end_sy | Z105_S5 | 27446 | 0.177311 | 0.006857 | 1.99E-147 | 146.7013 | Right ventricle |
| RV_stroke_ | Z1_S2 | 27446 | 0.128786 | 0.004982 | 2.47E-147 | 146.6067 | Right ventricle |
| RA_stroke_ | Z31_S5 | 27446 | 0.104023 | 0.004025 | 2.94E-147 | 146.5318 | Right atrium |
| LV_end_dia | Z64_S1 | 27446 | -0.19367 | 0.0075 | 4.75E-147 | 146.323 | Left ventricle |
| LV_stroke_ | Z8_S3 | 27446 | 0.131474 | 0.005096 | 8.97E-147 | 146.0473 | Left ventricle |
| RA_stroke_ | Z127_S4 | 27446 | 0.141536 | 0.00549 | 1.39E-146 | 145.8563 | Right atrium |
| LV_stroke_ | Z61_S4 | 27446 | -0.1456 | 0.005651 | 2.25E-146 | 145.6475 | Left ventricle |
| Ascending_ | Z32_S1 | 24599 | -0.15617 | 0.006066 | 3.56E-146 | 145.4487 | Aorta |
| LA_minimu | Z66_S3 | 27446 | -0.14649 | 0.005691 | 4.00E-146 | 145.3978 | Left atrium |

| | | | | | | | |
|---|---|---|---|---|---|---|---|
| Descending | Z120_S4 | 24599 | -0.16822 | 0.006535 | 4.30E-146 | 145.3663 | Aorta |
| Descending | Z59_S3 | 24599 | -0.13733 | 0.005336 | 4.56E-146 | 145.3407 | Aorta |
| RV_stroke_ | Z114_S5 | 27446 | -0.1249 | 0.004858 | 9.44E-146 | 145.0248 | Right ventricle |
| LV_end_sy | Z127_S4 | 27446 | 0.175762 | 0.006838 | 1.04E-145 | 144.9834 | Left ventricle |
| RV_end_sy | Z29_S5 | 27446 | 0.186169 | 0.007244 | 1.13E-145 | 144.9483 | Right ventricle |
| LA_maximu | Z8_S3 | 27446 | 0.113018 | 0.004398 | 1.16E-145 | 144.9344 | Left atrium |
| LV_end_sy | Z52_S3 | 27446 | 0.13661 | 0.005319 | 1.76E-145 | 144.7548 | Left ventricle |
| RA_ejectio | Z14_S2 | 27446 | 0.125998 | 0.00491 | 3.21E-145 | 144.4934 | Right atrium |
| LV_myocar | Z62_S2 | 27446 | 0.188277 | 0.007341 | 4.56E-145 | 144.3406 | Left ventricle |
| RV_end_sy | Z2_S1 | 27446 | -0.15854 | 0.006182 | 4.70E-145 | 144.3275 | Right ventricle |
| Descending | Z85_S5 | 24599 | 0.141113 | 0.005502 | 4.71E-145 | 144.3273 | Aorta |
| RV_end_sy | Z26_S4 | 27446 | 0.215212 | 0.0084 | 9.20E-145 | 144.0364 | Right ventricle |
| LV_myocar | Z62_S1 | 27446 | 0.198719 | 0.007761 | 1.35E-144 | 143.8707 | Left ventricle |
| RA_maxim | Z57_S2 | 27446 | 0.11677 | 0.004561 | 1.42E-144 | 143.849 | Right atrium |
| Ascending_ | Z99_S3 | 24599 | 0.117359 | 0.004588 | 2.54E-144 | 143.5946 | Aorta |
| LV_end_di | Z6_S2 | 27446 | -0.13432 | 0.005254 | 3.59E-144 | 143.4446 | Left ventricle |
| LA_maximu | Z20_S1 | 27446 | 0.153789 | 0.006021 | 6.88E-144 | 143.1622 | Left atrium |
| LA_ejectio | Z99_S3 | 27446 | -0.09896 | 0.003881 | 2.19E-143 | 142.6587 | Left atrium |
| LA_ejectio | Z94_S5 | 27446 | -0.10575 | 0.00415 | 2.97E-143 | 142.5269 | Left atrium |
| Descending | Z78_S4 | 24599 | 0.148736 | 0.005837 | 3.30E-143 | 142.4812 | Aorta |
| LV_stroke_ | Z2_S1 | 27446 | -0.13938 | 0.005472 | 3.86E-143 | 142.4131 | Left ventricle |
| LV_myocar | Z69_S5 | 27446 | -0.17946 | 0.007046 | 4.17E-143 | 142.3794 | Left ventricle |
| LV_end_sy | Z39_S1 | 27446 | 0.146582 | 0.005758 | 5.99E-143 | 142.2223 | Left ventricle |
| LV_stroke_ | Z62_S2 | 27446 | 0.13525 | 0.005314 | 6.68E-143 | 142.1753 | Left ventricle |
| LV_myocar | Z125_S4 | 27446 | -0.21745 | 0.008548 | 8.96E-143 | 142.0476 | Left ventricle |
| LA_minimu | Z55_S5 | 27446 | 0.108882 | 0.004284 | 1.66E-142 | 141.7789 | Left atrium |
| LA_stroke_ | Z120_S4 | 27446 | -0.13096 | 0.005156 | 2.38E-142 | 141.6234 | Left atrium |
| RA_maxim | Z117_S4 | 27446 | -0.11354 | 0.004475 | 5.10E-142 | 141.2927 | Right atrium |
| RA_stroke_ | Z18_S4 | 27446 | -0.15854 | 0.00625 | 5.82E-142 | 141.2348 | Right atrium |
| Ascending_ | Z80_S1 | 24599 | -0.15666 | 0.006179 | 8.32E-142 | 141.0798 | Aorta |
| RV_end_sy | Z83_S2 | 27446 | 0.162071 | 0.006396 | 1.16E-141 | 140.9349 | Right ventricle |
| Descending | Z35_S5 | 24599 | -0.14117 | 0.005573 | 1.42E-141 | 140.8484 | Aorta |
| LA_maximu | Z120_S4 | 27446 | -0.1298 | 0.00513 | 3.17E-141 | 140.4991 | Left atrium |
| RA_minimu | Z14_S4 | 27446 | 0.157118 | 0.006212 | 3.89E-141 | 140.4105 | Right atrium |
| LA_maximu | Z120_S2 | 27446 | 0.11222 | 0.004444 | 1.08E-140 | 139.9649 | Left atrium |
| LV_myocar | Z57_S1 | 27446 | -0.1931 | 0.007649 | 1.27E-140 | 139.8978 | Left ventricle |
| RV_end_sy | Z44_S4 | 27446 | -0.14478 | 0.00574 | 2.34E-140 | 139.6301 | Right ventricle |
| Ascending_ | Z117_S4 | 24599 | -0.1166 | 0.004624 | 2.84E-140 | 139.5464 | Aorta |
| LA_stroke_ | Z111_S1 | 27446 | 0.142925 | 0.005674 | 5.38E-140 | 139.2692 | Left atrium |
| Ascending_ | Z4_S4 | 24599 | -0.12695 | 0.005046 | 1.08E-139 | 138.9681 | Aorta |
| LV_end_sy | Z55_S1 | 27446 | -0.14506 | 0.005783 | 8.00E-139 | 138.0968 | Left ventricle |
| RV_stroke_ | Z125_S4 | 27446 | -0.15747 | 0.006279 | 8.67E-139 | 138.062 | Right ventricle |
| Descending | Z38_S5 | 24599 | -0.15653 | 0.006248 | 1.63E-138 | 137.787 | Aorta |
| RA_ejectio | Z85_S5 | 27446 | -0.11549 | 0.004615 | 3.28E-138 | 137.4839 | Right atrium |
| Descending | Z7_S1 | 24599 | -0.14945 | 0.005972 | 3.33E-138 | 137.4773 | Aorta |

| | | | | | | | |
|---|---|---|---|---|---|---|---|
| Ascending_ | Z51_S1 | 24599 | -0.12465 | 0.004983 | 4.31E-138 | 137.3652 | Aorta |
| LA_maximu | Z91_S1 | 27446 | 0.125179 | 0.005008 | 6.82E-138 | 137.1659 | Left atrium |
| LV_stroke_ | Z83_S2 | 27446 | 0.141419 | 0.005662 | 1.09E-137 | 136.9616 | Left ventricle |
| LV_end_sy | Z58_S2 | 27446 | 0.145965 | 0.005847 | 1.56E-137 | 136.8061 | Left ventricle |
| LV_cardiac | Z38_S5 | 27446 | 0.14482 | 0.005803 | 1.86E-137 | 136.7312 | Left ventricle |
| LV_end_sy | Z5_S1 | 27446 | -0.15332 | 0.00615 | 3.58E-137 | 136.4456 | Left ventricle |
| RA_minimu | Z105_S4 | 27446 | -0.12631 | 0.005079 | 1.52E-136 | 135.8187 | Right atrium |
| Descending | Z57_S1 | 24599 | -0.14918 | 0.005998 | 1.53E-136 | 135.8148 | Aorta |
| Ascending_ | Z12_S4 | 24599 | -0.1205 | 0.004853 | 4.17E-136 | 135.3799 | Aorta |
| RV_stroke_ | Z20_S2 | 27446 | 0.127599 | 0.005149 | 1.50E-135 | 134.824 | Right ventricle |
| Ascending_ | Z55_S5 | 24599 | 0.11979 | 0.004839 | 2.66E-135 | 134.5757 | Aorta |
| Descending | Z20_S2 | 24599 | 0.135968 | 0.005495 | 3.60E-135 | 134.4443 | Aorta |
| RA_minimu | Z35_S5 | 27446 | -0.11806 | 0.004778 | 8.07E-135 | 134.0931 | Right atrium |
| LA_ejection | Z64_S1 | 27446 | 0.132106 | 0.005347 | 8.97E-135 | 134.0471 | Left atrium |
| LV_end_sy | Z64_S1 | 27446 | -0.17118 | 0.006929 | 9.22E-135 | 134.0352 | Left ventricle |
| LV_stroke_ | Z66_S3 | 27446 | -0.16803 | 0.006808 | 1.68E-134 | 133.7745 | Left ventricle |
| Descending | Z111_S5 | 24599 | 0.159164 | 0.00645 | 1.91E-134 | 133.7198 | Aorta |
| LA_stroke_ | Z56_S2 | 27446 | -0.11031 | 0.00447 | 1.94E-134 | 133.7116 | Left atrium |
| RA_maxim | Z40_S5 | 27446 | -0.11607 | 0.004704 | 2.10E-134 | 133.6775 | Right atrium |
| RV_end_sy | Z23_S3 | 27446 | 0.14832 | 0.006019 | 4.63E-134 | 133.3342 | Right ventricle |
| LV_myocar | Z5_S5 | 27446 | 0.210524 | 0.008545 | 5.23E-134 | 133.2817 | Left ventricle |
| RV_end_sy | Z75_S4 | 27446 | -0.19738 | 0.008029 | 1.97E-133 | 132.7048 | Right ventricle |
| LA_stroke_ | Z23_S3 | 27446 | 0.113527 | 0.00462 | 2.35E-133 | 132.6292 | Left atrium |
| RV_end_di | Z31_S2 | 27446 | 0.175739 | 0.007156 | 3.40E-133 | 132.4691 | Right ventricle |
| RV_stroke_ | Z98_S3 | 27446 | -0.13084 | 0.005328 | 3.68E-133 | 132.4346 | Right ventricle |
| RA_minimu | Z51_S4 | 27446 | -0.11853 | 0.004829 | 4.83E-133 | 132.316 | Right atrium |
| LV_end_di | Z75_S1 | 27446 | -0.17088 | 0.006964 | 5.67E-133 | 132.2461 | Left ventricle |
| Descending | Z53_S1 | 24599 | 0.151192 | 0.006165 | 8.46E-133 | 132.0726 | Aorta |
| RA_minimu | Z118_S1 | 27446 | 0.151965 | 0.0062 | 1.09E-132 | 131.9613 | Right atrium |
| RV_end_sy | Z98_S3 | 27446 | -0.14527 | 0.005934 | 2.26E-132 | 131.6457 | Right ventricle |
| LV_stroke_ | Z75_S4 | 27446 | -0.17397 | 0.007106 | 2.38E-132 | 131.6238 | Left ventricle |
| LV_end_sy | Z41_S2 | 27446 | -0.18049 | 0.007376 | 3.03E-132 | 131.5179 | Left ventricle |
| Ascending_ | Z65_S1 | 24599 | -0.11583 | 0.004737 | 4.77E-132 | 131.3215 | Aorta |
| RV_stroke_ | Z111_S1 | 27446 | 0.162417 | 0.006644 | 5.75E-132 | 131.2402 | Right ventricle |
| Ascending_ | Z50_S4 | 24599 | 0.134051 | 0.005486 | 6.93E-132 | 131.159 | Aorta |
| LV_end_sy | Z13_S3 | 27446 | 0.124295 | 0.005087 | 7.88E-132 | 131.1035 | Left ventricle |
| Ascending_ | Z55_S5 | 24599 | 0.11868 | 0.004858 | 8.34E-132 | 131.0787 | Aorta |
| LV_end_di | Z60_S2 | 27446 | -0.14395 | 0.005893 | 8.72E-132 | 131.0593 | Left ventricle |
| RA_ejectio | Z26_S3 | 27446 | 0.135553 | 0.00555 | 9.68E-132 | 131.014 | Right atrium |
| LV_end_sy | Z14_S2 | 27446 | -0.15182 | 0.006221 | 1.46E-131 | 130.8354 | Left ventricle |
| Descending | Z85_S5 | 24599 | 0.14853 | 0.006086 | 1.48E-131 | 130.8286 | Aorta |
| LA_ejection | Z15_S2 | 27446 | -0.12519 | 0.005138 | 3.97E-131 | 130.4014 | Left atrium |
| LV_end_sy | Z38_S5 | 27446 | -0.16027 | 0.006583 | 6.55E-131 | 130.1841 | Left ventricle |
| LV_end_di | Z79_S4 | 27446 | -0.19395 | 0.007967 | 6.72E-131 | 130.1728 | Left ventricle |
| RV_stroke_ | Z105_S5 | 27446 | 0.150097 | 0.006166 | 7.12E-131 | 130.1476 | Right ventricle |

| | | | | | | | |
|---|---|---|---|---|---|---|---|
| Descending | Z14_S1 | 24599 | 0.150679 | 0.006195 | 1.13E-130 | 129.9464 | Aorta |
| LV_end_sy | Z17_S1 | 27446 | 0.141691 | 0.005833 | 2.39E-130 | 129.622 | Left ventricle |
| RV_end_sy | Z55_S1 | 27446 | -0.15283 | 0.006293 | 2.82E-130 | 129.5498 | Right ventricle |
| RA_minimu | Z64_S1 | 27446 | -0.15003 | 0.006191 | 1.01E-129 | 128.9945 | Right atrium |
| Ascending_ | Z51_S1 | 24599 | -0.12121 | 0.005005 | 1.51E-129 | 128.8221 | Aorta |
| LA_stroke_ | Z6_S2 | 27446 | -0.09807 | 0.004051 | 1.86E-129 | 128.7305 | Left atrium |
| LV_myocar | Z13_S1 | 27446 | -0.17977 | 0.007429 | 2.37E-129 | 128.6246 | Left ventricle |
| Ascending_ | Z7_S3 | 24599 | -0.16408 | 0.006787 | 4.22E-129 | 128.3749 | Aorta |
| LV_longitu | Z93_S1 | 27446 | -0.13623 | 0.005639 | 6.46E-129 | 128.1895 | Left ventricle |
| LA_minimu | Z5_S5 | 27446 | 0.125014 | 0.005177 | 7.55E-129 | 128.122 | Left atrium |
| LA_stroke_ | Z43_S2 | 27446 | -0.11014 | 0.004562 | 8.75E-129 | 128.058 | Left atrium |
| RV_stroke_ | Z47_S4 | 27446 | -0.14692 | 0.006087 | 1.00E-128 | 128 | Right ventricle |
| Descending | Z122_S1 | 24599 | 0.155638 | 0.006449 | 1.12E-128 | 127.9515 | Aorta |
| LV_stroke_ | Z79_S4 | 27446 | -0.17078 | 0.007077 | 1.19E-128 | 127.9247 | Left ventricle |
| LV_myocar | Z80_S4 | 27446 | -0.17268 | 0.007159 | 1.46E-128 | 127.8344 | Left ventricle |
| RV_end_di | Z115_S4 | 27446 | 0.157327 | 0.006524 | 1.72E-128 | 127.7645 | Right ventricle |
| RA_minimu | Z41_S2 | 27446 | -0.15858 | 0.006591 | 6.35E-128 | 127.197 | Right atrium |
| LV_myocar | Z66_S3 | 27446 | -0.22643 | 0.009412 | 6.91E-128 | 127.1606 | Left ventricle |
| Descending | Z2_S1 | 24599 | -0.14275 | 0.005935 | 7.58E-128 | 127.1202 | Aorta |
| LA_stroke_ | Z19_S2 | 27446 | 0.123608 | 0.005139 | 7.66E-128 | 127.1157 | Left atrium |
| Descending | Z98_S3 | 24599 | -0.13657 | 0.005681 | 1.08E-127 | 126.9649 | Aorta |
| RA_ejectio | Z65_S2 | 27446 | -0.12191 | 0.005072 | 1.11E-127 | 126.9528 | Right atrium |
| Ascending_ | Z32_S1 | 24599 | -0.14548 | 0.006054 | 1.29E-127 | 126.8905 | Aorta |
| Descending | Z80_S4 | 24599 | -0.13495 | 0.005617 | 1.50E-127 | 126.8252 | Aorta |
| RV_end_sy | Z61_S4 | 27446 | -0.15359 | 0.006396 | 2.01E-127 | 126.6963 | Right ventricle |
| Descending | Z20_S1 | 24599 | 0.181703 | 0.007568 | 2.20E-127 | 126.6578 | Aorta |
| RV_end_di | Z57_S1 | 27446 | -0.15764 | 0.006567 | 2.58E-127 | 126.5878 | Right ventricle |
| LA_ejection | Z65_S5 | 27446 | 0.137655 | 0.005737 | 3.10E-127 | 126.5088 | Left atrium |
| LA_ejection | Z66_S2 | 27446 | -0.13225 | 0.005519 | 6.55E-127 | 126.1835 | Left atrium |
| RA_maxim | Z37_S2 | 27446 | -0.14304 | 0.00597 | 7.54E-127 | 126.1228 | Right atrium |
| LV_cardiac | Z23_S1 | 27446 | 0.148046 | 0.006182 | 9.33E-127 | 126.0303 | Left ventricle |
| LA_maximu | Z70_S5 | 27446 | 0.118698 | 0.004957 | 1.00E-126 | 125.998 | Left atrium |
| Ascending_ | Z8_S3 | 24599 | 0.115499 | 0.004829 | 2.02E-126 | 125.6936 | Aorta |
| RV_stroke_ | Z61_S4 | 27446 | -0.13732 | 0.005744 | 2.60E-126 | 125.5843 | Right ventricle |
| LA_stroke_ | Z65_S2 | 27446 | 0.127979 | 0.005356 | 3.67E-126 | 125.4348 | Left atrium |
| Ascending_ | Z20_S1 | 24599 | 0.157698 | 0.006608 | 6.92E-126 | 125.1602 | Aorta |
| LV_cardiac | Z49_S1 | 27446 | 0.160472 | 0.006724 | 7.10E-126 | 125.1486 | Left ventricle |
| Ascending_ | Z51_S4 | 24599 | -0.11739 | 0.004919 | 7.40E-126 | 125.1308 | Aorta |
| LV_end_di | Z93_S2 | 27446 | 0.198007 | 0.008299 | 8.26E-126 | 125.0831 | Left ventricle |
| LA_ejection | Z77_S3 | 27446 | 0.117069 | 0.004911 | 1.36E-125 | 124.8667 | Left atrium |
| RV_end_sy | Z31_S2 | 27446 | 0.162721 | 0.006827 | 1.47E-125 | 124.8325 | Right ventricle |
| Ascending_ | Z8_S3 | 24599 | 0.115459 | 0.004847 | 2.10E-125 | 124.6783 | Aorta |
| LV_end_sy | Z28_S4 | 27446 | -0.16016 | 0.006724 | 2.11E-125 | 124.6754 | Left ventricle |
| Descending | Z120_S2 | 24599 | 0.132867 | 0.005581 | 2.72E-125 | 124.5651 | Aorta |
| RV_end_sy | Z5_S5 | 27446 | 0.166416 | 0.006994 | 3.71E-125 | 124.4311 | Right ventricle |

| | | | | | | | |
|---|---|---|---|---|---|---|---|
| Ascending_ | Z38_S5 | 24599 | -0.15161 | 0.006372 | 3.97E-125 | 124.4007 | Aorta |
| LV_end_di | Z99_S5 | 27446 | 0.154257 | 0.006494 | 9.82E-125 | 124.0077 | Left ventricle |
| RA_maxim | Z105_S4 | 27446 | -0.11914 | 0.005016 | 1.02E-124 | 123.993 | Right atrium |
| Descending | Z62_S1 | 24599 | 0.144721 | 0.006093 | 1.08E-124 | 123.9672 | Aorta |
| LV_longitu | Z26_S3 | 27446 | -0.13164 | 0.005543 | 1.08E-124 | 123.9649 | Left ventricle |
| Ascending_ | Z65_S1 | 24599 | -0.11262 | 0.004758 | 7.22E-124 | 123.1416 | Aorta |
| LA_stroke_ | Z37_S2 | 27446 | -0.1338 | 0.005655 | 9.11E-124 | 123.0406 | Left atrium |
| RV_end_sy | Z115_S4 | 27446 | 0.147115 | 0.006223 | 1.50E-123 | 122.8244 | Right ventricle |
| LA_maximu | Z51_S1 | 27446 | -0.10751 | 0.004551 | 2.16E-123 | 122.6662 | Left atrium |
| LA_ejectio | Z65_S2 | 27446 | -0.11696 | 0.004955 | 3.33E-123 | 122.4778 | Left atrium |
| Ascending_ | Z20_S1 | 24599 | 0.156574 | 0.006634 | 3.55E-123 | 122.4499 | Aorta |
| LA_stroke_ | Z120_S2 | 27446 | 0.105566 | 0.004473 | 3.63E-123 | 122.4397 | Left atrium |
| LV_end_di | Z74_S2 | 27446 | 0.157698 | 0.006685 | 5.07E-123 | 122.2951 | Left ventricle |
| RA_minimu | Z73_S4 | 27446 | 0.151054 | 0.006407 | 6.59E-123 | 122.1813 | Right atrium |
| LV_stroke_ | Z51_S1 | 27446 | -0.12431 | 0.005275 | 8.30E-123 | 122.0811 | Left ventricle |
| LA_ejectio | Z24_S5 | 27446 | 0.123699 | 0.005251 | 1.06E-122 | 121.9737 | Left atrium |
| LV_stroke_ | Z26_S4 | 27446 | 0.175438 | 0.007447 | 1.06E-122 | 121.9729 | Left ventricle |
| RV_end_di | Z63_S1 | 27446 | -0.19931 | 0.008462 | 1.16E-122 | 121.9341 | Right ventricle |
| LA_ejectio | Z13_S3 | 27446 | -0.09249 | 0.003929 | 1.62E-122 | 121.7901 | Left atrium |
| LV_end_sy | Z31_S2 | 27446 | 0.147806 | 0.00628 | 1.74E-122 | 121.759 | Left ventricle |
| Descending | Z92_S5 | 24599 | 0.167793 | 0.007131 | 1.94E-122 | 121.7123 | Aorta |
| RA_maxim | Z14_S2 | 27446 | -0.12902 | 0.005486 | 2.76E-122 | 121.5585 | Right atrium |
| RV_end_di | Z120_S2 | 27446 | 0.143701 | 0.006113 | 3.36E-122 | 121.4736 | Right ventricle |
| LV_cardiac | Z112_S1 | 27446 | 0.161116 | 0.006856 | 4.20E-122 | 121.3765 | Left ventricle |
| LV_stroke_ | Z52_S3 | 27446 | 0.120338 | 0.005128 | 9.30E-122 | 121.0313 | Left ventricle |
| LV_cardiac | Z18_S4 | 27446 | 0.161331 | 0.006877 | 1.03E-121 | 120.9867 | Left ventricle |
| Descending | Z120_S2 | 24599 | 0.132855 | 0.005663 | 1.04E-121 | 120.9823 | Aorta |
| Descending | Z73_S1 | 24599 | -0.14521 | 0.006194 | 1.56E-121 | 120.8071 | Aorta |
| RV_end_di | Z43_S2 | 27446 | -0.14611 | 0.006238 | 2.56E-121 | 120.5918 | Right ventricle |
| Descending | Z127_S5 | 24599 | 0.131531 | 0.005617 | 2.84E-121 | 120.5465 | Aorta |
| RA_minimu | Z125_S4 | 27446 | -0.13468 | 0.005752 | 3.00E-121 | 120.5225 | Right atrium |
| Descending | Z82_S4 | 24599 | -0.13154 | 0.005622 | 4.38E-121 | 120.3584 | Aorta |
| RV_end_di | Z58_S2 | 27446 | 0.156223 | 0.006677 | 4.76E-121 | 120.3228 | Right ventricle |
| RV_end_sy | Z62_S2 | 27446 | 0.140731 | 0.006015 | 4.78E-121 | 120.3206 | Right ventricle |
| LV_cardiac | Z111_S5 | 27446 | -0.12776 | 0.005461 | 4.82E-121 | 120.3173 | Left ventricle |
| RV_end_di | Z93_S2 | 27446 | 0.204427 | 0.008739 | 4.97E-121 | 120.3038 | Right ventricle |
| Descending | Z122_S1 | 24599 | 0.166759 | 0.007129 | 5.26E-121 | 120.2789 | Aorta |
| RV_end_di | Z50_S3 | 27446 | -0.156 | 0.006672 | 6.74E-121 | 120.1714 | Right ventricle |
| Ascending_ | Z24_S1 | 24599 | 0.112371 | 0.004819 | 2.82E-120 | 119.5493 | Aorta |
| LA_maximu | Z23_S3 | 27446 | 0.107283 | 0.004602 | 3.18E-120 | 119.4978 | Left atrium |
| LV_end_sy | Z60_S2 | 27446 | -0.12678 | 0.005444 | 5.94E-120 | 119.2259 | Left ventricle |
| LV_myocar | Z23_S3 | 27446 | 0.171527 | 0.007369 | 7.59E-120 | 119.1197 | Left ventricle |
| Descending | Z57_S1 | 24599 | -0.14182 | 0.006094 | 8.47E-120 | 119.0723 | Aorta |
| RA_ejectio | Z77_S3 | 27446 | 0.117083 | 0.005031 | 8.74E-120 | 119.0586 | Right atrium |
| RV_end_sy | Z109_S1 | 27446 | -0.15835 | 0.006808 | 1.13E-119 | 118.9484 | Right ventricle |

| | | | | | | | |
|---|---|---|---|---|---|---|---|
| LV_myocar | Z8_S3 | 27446 | 0.164096 | 0.007057 | 1.33E-119 | 118.8763 | Left ventricle |
| LV_end_di | Z63_S1 | 27446 | -0.18668 | 0.008042 | 3.37E-119 | 118.4727 | Left ventricle |
| LV_end_sy | Z50_S3 | 27446 | -0.1358 | 0.005851 | 3.69E-119 | 118.4332 | Left ventricle |
| RV_end_di | Z14_S4 | 27446 | 0.184353 | 0.007946 | 4.42E-119 | 118.3549 | Right ventricle |
| RV_end_di | Z70_S5 | 27446 | 0.158011 | 0.006812 | 5.04E-119 | 118.2976 | Right ventricle |
| LA_minimu | Z105_S5 | 27446 | 0.118043 | 0.005089 | 5.08E-119 | 118.294 | Left atrium |
| RA_maxim | Z29_S1 | 27446 | 0.138275 | 0.005962 | 5.51E-119 | 118.2592 | Right atrium |
| LV_longitu | Z94_S5 | 27446 | -0.09853 | 0.00425 | 6.24E-119 | 118.205 | Left ventricle |
| Descending | Z20_S2 | 24599 | 0.129401 | 0.005583 | 7.33E-119 | 118.1346 | Aorta |
| LV_end_sy | Z6_S2 | 27446 | -0.11256 | 0.004859 | 9.85E-119 | 118.0064 | Left ventricle |
| LV_stroke_ | Z10_S4 | 27446 | -0.14516 | 0.00627 | 1.44E-118 | 117.8415 | Left ventricle |
| RV_end_sy | Z52_S3 | 27446 | 0.134054 | 0.005797 | 2.53E-118 | 117.5973 | Right ventricle |
| LV_myocar | Z2_S1 | 27446 | -0.17508 | 0.007576 | 3.59E-118 | 117.4453 | Left ventricle |
| LV_myocar | Z111_S1 | 27446 | 0.209284 | 0.009057 | 3.99E-118 | 117.3986 | Left ventricle |
| Ascending_ | Z80_S1 | 24599 | -0.14249 | 0.00617 | 5.39E-118 | 117.2683 | Aorta |
| LA_stroke_ | Z105_S5 | 27446 | 0.121751 | 0.005275 | 7.34E-118 | 117.1343 | Left atrium |
| LA_maximu | Z28_S4 | 27446 | -0.1289 | 0.005587 | 8.81E-118 | 117.0548 | Left atrium |
| Descending | Z14_S4 | 24599 | 0.152287 | 0.006603 | 1.04E-117 | 116.9816 | Aorta |
| LA_stroke_ | Z37_S4 | 27446 | 0.148826 | 0.006456 | 1.44E-117 | 116.8406 | Left atrium |
| LV_end_di | Z19_S2 | 27446 | 0.153921 | 0.006678 | 1.49E-117 | 116.8269 | Left ventricle |
| Ascending_ | Z73_S4 | 24599 | 0.150464 | 0.006532 | 2.04E-117 | 116.6914 | Aorta |
| LA_minimu | Z2_S1 | 27446 | -0.10557 | 0.004588 | 3.52E-117 | 116.4539 | Left atrium |
| LA_stroke_ | Z91_S1 | 27446 | 0.115886 | 0.005042 | 6.55E-117 | 116.184 | Left atrium |
| LA_stroke_ | Z51_S1 | 27446 | -0.10518 | 0.004576 | 6.67E-117 | 116.1757 | Left atrium |
| Descending | Z62_S2 | 24599 | 0.132368 | 0.00576 | 6.98E-117 | 116.156 | Aorta |
| RA_maxim | Z89_S4 | 27446 | -0.12225 | 0.005323 | 9.71E-117 | 116.0129 | Right atrium |
| RA_minimu | Z5_S5 | 27446 | 0.132031 | 0.005749 | 9.85E-117 | 116.0064 | Right atrium |
| Ascending_ | Z1_S2 | 24599 | 0.106747 | 0.004648 | 1.05E-116 | 115.9775 | Aorta |
| Descending | Z73_S4 | 24599 | 0.174212 | 0.00759 | 1.43E-116 | 115.8457 | Aorta |
| Ascending_ | Z82_S1 | 24599 | -0.17066 | 0.007439 | 1.75E-116 | 115.7558 | Aorta |
| Ascending_ | Z117_S4 | 24599 | -0.10668 | 0.004652 | 2.13E-116 | 115.6725 | Aorta |
| RA_ejectio | Z37_S2 | 27446 | 0.122829 | 0.005358 | 2.62E-116 | 115.5819 | Right atrium |
| LA_ejection | Z106_S4 | 27446 | -0.09719 | 0.004242 | 3.43E-116 | 115.4647 | Left atrium |
| LA_stroke_ | Z70_S5 | 27446 | 0.114231 | 0.004986 | 3.63E-116 | 115.4406 | Left atrium |
| Descending | Z32_S1 | 24599 | -0.16129 | 0.007042 | 4.16E-116 | 115.3812 | Aorta |
| Descending | Z52_S3 | 24599 | 0.127236 | 0.005556 | 4.68E-116 | 115.3296 | Aorta |
| LV_stroke_ | Z55_S1 | 27446 | -0.12768 | 0.005576 | 4.76E-116 | 115.3222 | Left ventricle |
| LA_maximu | Z60_S2 | 27446 | -0.1035 | 0.004522 | 6.14E-116 | 115.2119 | Left atrium |
| RA_minimu | Z39_S3 | 27446 | 0.159917 | 0.006988 | 6.80E-116 | 115.1678 | Right atrium |
| RA_minimu | Z63_S1 | 27446 | -0.15161 | 0.006632 | 1.12E-115 | 114.9526 | Right atrium |
| RV_end_sy | Z57_S1 | 27446 | -0.1432 | 0.006268 | 1.59E-115 | 114.7999 | Right ventricle |
| Ascending_ | Z50_S4 | 24599 | 0.125903 | 0.005514 | 2.14E-115 | 114.6687 | Aorta |
| LV_myocar | Z13_S3 | 27446 | 0.154632 | 0.006774 | 2.42E-115 | 114.6163 | Left ventricle |
| Descending | Z44_S4 | 24599 | -0.12781 | 0.005601 | 3.00E-115 | 114.5228 | Aorta |
| RA_ejectio | Z24_S5 | 27446 | 0.122746 | 0.00538 | 3.34E-115 | 114.4764 | Right atrium |

| | | | | | | | |
|---|---|---|---|---|---|---|---|
| RA_ejectio | Z127_S4 | 27446 | -0.12357 | 0.005417 | 3.49E-115 | 114.4567 | Right atrium |
| Descending | Z20_S1 | 24599 | 0.175285 | 0.007686 | 3.93E-115 | 114.4053 | Aorta |
| RA_minimu | Z7_S1 | 27446 | -0.11676 | 0.005122 | 5.29E-115 | 114.2762 | Right atrium |
| LV_end_di | Z66_S2 | 27446 | 0.17663 | 0.007756 | 8.62E-115 | 114.0643 | Left ventricle |
| LA_stroke_ | Z74_S4 | 27446 | -0.12456 | 0.005471 | 1.03E-114 | 113.9871 | Left atrium |
| LA_minimu | Z65_S5 | 27446 | -0.13631 | 0.005992 | 1.40E-114 | 113.8534 | Left atrium |
| LV_myocar | Z123_S4 | 27446 | 0.198019 | 0.008704 | 1.45E-114 | 113.8386 | Left ventricle |
| LV_stroke_ | Z13_S3 | 27446 | 0.111496 | 0.004903 | 1.73E-114 | 113.7611 | Left ventricle |
| RA_maxim | Z7_S1 | 27446 | -0.11484 | 0.005054 | 2.72E-114 | 113.5647 | Right atrium |
| Ascending_ | Z7_S3 | 24599 | -0.15379 | 0.006772 | 3.61E-114 | 113.4419 | Aorta |
| Ascending_ | Z28_S4 | 24599 | -0.14017 | 0.006173 | 3.77E-114 | 113.4239 | Aorta |
| LA_maximu | Z55_S1 | 27446 | -0.10923 | 0.004812 | 4.54E-114 | 113.3429 | Left atrium |
| Descending | Z13_S3 | 24599 | 0.120374 | 0.005306 | 6.06E-114 | 113.2178 | Aorta |
| RA_ejectio | Z93_S1 | 27446 | 0.128221 | 0.005657 | 1.01E-113 | 112.9948 | Right atrium |
| RA_maxim | Z63_S1 | 27446 | -0.14814 | 0.006544 | 1.92E-113 | 112.7161 | Right atrium |
| RA_minimu | Z65_S2 | 27446 | 0.129893 | 0.005739 | 2.09E-113 | 112.6797 | Right atrium |
| Ascending_ | Z4_S4 | 24599 | -0.11486 | 0.005076 | 2.39E-113 | 112.6222 | Aorta |
| LA_maximu | Z75_S1 | 27446 | -0.12086 | 0.005345 | 3.38E-113 | 112.471 | Left atrium |
| RA_ejectio | Z125_S4 | 27446 | 0.115109 | 0.005092 | 3.82E-113 | 112.4182 | Right atrium |
| Descending | Z94_S5 | 24599 | 0.114852 | 0.005083 | 4.97E-113 | 112.3033 | Aorta |
| Descending | Z69_S5 | 24599 | -0.12503 | 0.00554 | 8.47E-113 | 112.0723 | Aorta |
| LA_maximu | Z74_S4 | 27446 | -0.12288 | 0.005445 | 9.20E-113 | 112.0364 | Left atrium |
| LV_circumf | Z10_S4 | 27446 | 0.118763 | 0.005264 | 1.02E-112 | 111.9917 | Left ventricle |
| LV_myocar | Z65_S2 | 27446 | 0.192751 | 0.008544 | 1.10E-112 | 111.9602 | Left ventricle |
| LV_circumf | Z53_S1 | 27446 | -0.11843 | 0.005251 | 1.29E-112 | 111.8884 | Left ventricle |
| LV_myocar | Z109_S1 | 27446 | -0.18783 | 0.008329 | 1.34E-112 | 111.8723 | Left ventricle |
| LV_myocar | Z5_S1 | 27446 | -0.18478 | 0.008194 | 1.36E-112 | 111.8662 | Left ventricle |
| RA_maxim | Z118_S1 | 27446 | 0.138143 | 0.006127 | 1.44E-112 | 111.8415 | Right atrium |
| LA_stroke_ | Z77_S3 | 27446 | -0.11982 | 0.005316 | 1.77E-112 | 111.7512 | Left atrium |
| Ascending_ | Z73_S1 | 24599 | -0.12066 | 0.005355 | 1.96E-112 | 111.7084 | Aorta |
| Ascending_ | Z120_S2 | 24599 | 0.109882 | 0.004878 | 2.22E-112 | 111.6529 | Aorta |
| RV_stroke_ | Z80_S4 | 27446 | -0.11858 | 0.005264 | 2.34E-112 | 111.6305 | Right ventricle |
| RV_stroke_ | Z26_S4 | 27446 | 0.170149 | 0.007564 | 4.68E-112 | 111.33 | Right ventricle |
| RA_maxim | Z109_S1 | 27446 | -0.12418 | 0.005521 | 4.87E-112 | 111.3125 | Right atrium |
| LV_end_sy | Z66_S3 | 27446 | -0.15913 | 0.007088 | 1.23E-111 | 110.9091 | Left ventricle |
| Ascending_ | Z59_S3 | 24599 | -0.10342 | 0.004607 | 1.25E-111 | 110.9033 | Aorta |
| LV_stroke_ | Z74_S4 | 27446 | -0.14159 | 0.006311 | 1.82E-111 | 110.7405 | Left ventricle |
| Descending | Z55_S1 | 24599 | -0.13556 | 0.006043 | 1.89E-111 | 110.7228 | Aorta |
| Descending | Z127_S5 | 24599 | 0.112494 | 0.005017 | 2.33E-111 | 110.633 | Aorta |
| LV_myocar | Z30_S2 | 27446 | -0.22596 | 0.010079 | 2.53E-111 | 110.5973 | Left ventricle |
| Ascending_ | Z23_S3 | 24599 | 0.113177 | 0.005049 | 2.64E-111 | 110.5781 | Aorta |
| LA_minimu | Z50_S3 | 27446 | -0.10567 | 0.004714 | 2.83E-111 | 110.5475 | Left atrium |
| LV_myocar | Z14_S4 | 27446 | 0.207694 | 0.00927 | 3.60E-111 | 110.4438 | Left ventricle |
| Descending | Z105_S4 | 24599 | -0.13501 | 0.006029 | 4.71E-111 | 110.3268 | Aorta |
| Ascending_ | Z23_S3 | 24599 | 0.113413 | 0.005067 | 5.94E-111 | 110.2266 | Aorta |

| | | | | | | | |
|---|---|---|---|---|---|---|---|
| RA_minimu | Z24_S1 | 27446 | 0.105866 | 0.004734 | 8.96E-111 | 110.0476 | Right atrium |
| LV_longitu | Z82_S1 | 27446 | 0.136136 | 0.006092 | 1.32E-110 | 109.8798 | Left ventricle |
| Ascending_ | Z120_S2 | 24599 | 0.109373 | 0.004896 | 1.59E-110 | 109.798 | Aorta |
| RA_maxim | Z51_S4 | 27446 | -0.10659 | 0.004773 | 1.86E-110 | 109.7303 | Right atrium |
| LV_longitu | Z64_S1 | 27446 | -0.12203 | 0.005476 | 5.16E-110 | 109.2877 | Left ventricle |
| Ascending_ | Z120_S4 | 24599 | -0.12557 | 0.005643 | 1.08E-109 | 108.9661 | Aorta |
| RV_end_di | Z125_S4 | 27446 | -0.16332 | 0.007351 | 2.31E-109 | 108.6359 | Right ventricle |
| RA_stroke_ | Z57_S2 | 27446 | 0.092535 | 0.004167 | 3.02E-109 | 108.5204 | Right atrium |
| Descending | Z94_S5 | 24599 | 0.126537 | 0.005698 | 3.03E-109 | 108.5188 | Aorta |
| Descending | Z111_S5 | 24599 | 0.129938 | 0.005852 | 3.22E-109 | 108.4917 | Aorta |
| Ascending_ | Z29_S1 | 24599 | 0.138277 | 0.006235 | 5.84E-109 | 108.2339 | Aorta |
| LV_myocar | Z55_S1 | 27446 | -0.17095 | 0.007709 | 5.86E-109 | 108.2318 | Left ventricle |
| LV_myocar | Z120_S2 | 27446 | 0.15801 | 0.007135 | 1.16E-108 | 107.937 | Left ventricle |
| LV_end_sy | Z13_S1 | 27446 | -0.1239 | 0.005597 | 1.36E-108 | 107.8676 | Left ventricle |
| Ascending_ | Z22_S2 | 24599 | -0.12847 | 0.005804 | 1.48E-108 | 107.8289 | Aorta |
| RV_end_sy | Z93_S2 | 27446 | 0.184618 | 0.008341 | 1.49E-108 | 107.8281 | Right ventricle |
| RV_stroke_ | Z120_S4 | 27446 | -0.13383 | 0.00605 | 2.00E-108 | 107.6999 | Right ventricle |
| LV_stroke_ | Z23_S3 | 27446 | 0.118003 | 0.005338 | 2.80E-108 | 107.5529 | Left ventricle |
| Descending | Z2_S1 | 24599 | -0.13328 | 0.006031 | 3.19E-108 | 107.4956 | Aorta |
| LA_stroke_ | Z55_S1 | 27446 | -0.10686 | 0.004839 | 4.29E-108 | 107.368 | Left atrium |
| LV_myocar | Z37_S2 | 27446 | -0.19903 | 0.009022 | 7.53E-108 | 107.1232 | Left ventricle |
| LA_maximu | Z1_S5 | 27446 | 0.116872 | 0.005298 | 7.54E-108 | 107.1228 | Left atrium |
| RA_minimu | Z1_S1 | 27446 | -0.11991 | 0.005436 | 7.88E-108 | 107.1036 | Right atrium |
| LA_minimu | Z43_S2 | 27446 | -0.09721 | 0.004409 | 9.95E-108 | 107.0022 | Left atrium |
| LV_longitu | Z85_S5 | 27446 | -0.10174 | 0.004618 | 1.38E-107 | 106.8609 | Left ventricle |
| LA_stroke_ | Z26_S3 | 27446 | -0.12933 | 0.005873 | 1.78E-107 | 106.7487 | Left atrium |
| RA_maxim | Z5_S1 | 27446 | -0.11952 | 0.005433 | 3.07E-107 | 106.5127 | Right atrium |
| LV_stroke_ | Z120_S2 | 27446 | 0.113441 | 0.005165 | 6.22E-107 | 106.2059 | Left ventricle |
| LV_end_di | Z13_S1 | 27446 | -0.13317 | 0.006065 | 7.36E-107 | 106.1334 | Left ventricle |
| LA_maximu | Z93_S2 | 27446 | 0.139644 | 0.00637 | 1.63E-106 | 105.7882 | Left atrium |
| RV_stroke_ | Z2_S1 | 27446 | -0.12203 | 0.005569 | 1.95E-106 | 105.7102 | Right ventricle |
| LA_stroke_ | Z66_S3 | 27446 | -0.12961 | 0.005917 | 2.47E-106 | 105.6077 | Left atrium |
| Ascending_ | Z1_S2 | 24599 | 0.102244 | 0.00467 | 2.92E-106 | 105.5344 | Aorta |
| RA_maxim | Z80_S1 | 27446 | 0.130776 | 0.005974 | 3.05E-106 | 105.5155 | Right atrium |
| RA_stroke_ | Z39_S3 | 27446 | 0.137626 | 0.006287 | 3.13E-106 | 105.5049 | Right atrium |
| RV_end_sy | Z93_S1 | 27446 | 0.170414 | 0.00779 | 4.41E-106 | 105.3557 | Right ventricle |
| RV_end_sy | Z58_S2 | 27446 | 0.139449 | 0.006375 | 4.52E-106 | 105.3445 | Right ventricle |
| LA_maximu | Z57_S1 | 27446 | -0.10467 | 0.00479 | 7.61E-106 | 105.1184 | Left atrium |
| LV_end_sy | Z74_S2 | 27446 | 0.134982 | 0.006179 | 8.54E-106 | 105.0686 | Left ventricle |
| LA_maximu | Z37_S4 | 27446 | 0.140471 | 0.006431 | 8.77E-106 | 105.0571 | Left atrium |
| LA_ejectio | Z125_S4 | 27446 | 0.108656 | 0.004976 | 1.04E-105 | 104.9828 | Left atrium |
| RV_end_di | Z66_S3 | 27446 | -0.17659 | 0.008087 | 1.06E-105 | 104.9757 | Right ventricle |
| RA_minimu | Z14_S1 | 27446 | 0.114483 | 0.005249 | 1.90E-105 | 104.7215 | Right atrium |
| LV_stroke_ | Z57_S1 | 27446 | -0.12106 | 0.005552 | 2.01E-105 | 104.696 | Left ventricle |
| RA_maxim | Z24_S1 | 27446 | 0.101857 | 0.004673 | 2.44E-105 | 104.6134 | Right atrium |

| | | | | | | | |
|---|---|---|---|---|---|---|---|
| LV_stroke_ | Z118_S1 | 27446 | 0.145906 | 0.006694 | 2.55E-105 | 104.5941 | Left ventricle |
| Ascending_ | Z52_S4 | 24599 | 0.110804 | 0.005087 | 3.38E-105 | 104.4711 | Aorta |
| LV_cardiac | Z122_S1 | 27446 | -0.1314 | 0.006036 | 4.38E-105 | 104.3584 | Left ventricle |
| RA_maxim | Z35_S5 | 27446 | -0.10284 | 0.004725 | 5.22E-105 | 104.2822 | Right atrium |
| LV_stroke_ | Z87_S3 | 27446 | -0.12761 | 0.005864 | 5.36E-105 | 104.2709 | Left ventricle |
| RA_ejectio | Z64_S1 | 27446 | 0.119391 | 0.005489 | 6.55E-105 | 104.1839 | Right atrium |
| Descending | Z80_S4 | 24599 | -0.12418 | 0.00571 | 7.13E-105 | 104.147 | Aorta |
| RV_end_di | Z99_S5 | 27446 | 0.148883 | 0.006846 | 7.31E-105 | 104.1363 | Right ventricle |
| Descending | Z98_S3 | 24599 | -0.12556 | 0.005775 | 8.46E-105 | 104.0724 | Aorta |
| RV_stroke_ | Z10_S4 | 27446 | -0.13845 | 0.00637 | 9.58E-105 | 104.0188 | Right ventricle |
| RV_stroke_ | Z8_S3 | 27446 | 0.112762 | 0.005189 | 1.05E-104 | 103.9773 | Right ventricle |
| Descending | Z35_S5 | 24599 | -0.12326 | 0.005672 | 1.07E-104 | 103.9697 | Aorta |
| RV_end_di | Z47_S4 | 27446 | -0.15476 | 0.007122 | 1.09E-104 | 103.9644 | Right ventricle |
| RA_maxim | Z7_S5 | 27446 | 0.130884 | 0.006024 | 1.11E-104 | 103.9562 | Right atrium |
| RA_minimu | Z59_S3 | 27446 | -0.09834 | 0.004528 | 1.34E-104 | 103.8723 | Right atrium |
| Descending | Z115_S4 | 24599 | 0.131582 | 0.006059 | 1.40E-104 | 103.8537 | Aorta |
| LV_end_di | Z81_S1 | 27446 | -0.14918 | 0.006869 | 1.40E-104 | 103.8531 | Left ventricle |
| LA_ejectio | Z41_S2 | 27446 | 0.123895 | 0.005705 | 1.41E-104 | 103.8502 | Left atrium |
| LV_end_sy | Z125_S4 | 27446 | -0.14001 | 0.006449 | 1.60E-104 | 103.7955 | Left ventricle |
| RV_end_sy | Z12_S5 | 27446 | -0.17517 | 0.008075 | 2.40E-104 | 103.6196 | Right ventricle |
| Descending | Z80_S1 | 24599 | -0.15559 | 0.007179 | 3.58E-104 | 103.4457 | Aorta |
| RV_end_di | Z7_S3 | 27446 | -0.18415 | 0.008497 | 3.84E-104 | 103.4151 | Right ventricle |
| RA_maxim | Z111_S1 | 27446 | 0.130224 | 0.00601 | 4.14E-104 | 103.3827 | Right atrium |
| RV_stroke_ | Z86_S3 | 27446 | -0.14969 | 0.00691 | 4.56E-104 | 103.3413 | Right ventricle |
| RA_minimu | Z26_S3 | 27446 | -0.13618 | 0.006288 | 5.14E-104 | 103.2888 | Right atrium |
| Ascending_ | Z78_S4 | 24599 | 0.109159 | 0.005042 | 5.90E-104 | 103.2295 | Aorta |
| LA_stroke_ | Z24_S5 | 27446 | -0.12312 | 0.005687 | 6.14E-104 | 103.212 | Left atrium |
| LA_ejectio | Z56_S2 | 27446 | 0.089654 | 0.004144 | 8.70E-104 | 103.0607 | Left atrium |
| LV_myocar | Z52_S3 | 27446 | 0.153523 | 0.007097 | 8.71E-104 | 103.0598 | Left ventricle |
| LV_cardiac | Z92_S5 | 27446 | -0.1439 | 0.006652 | 9.14E-104 | 103.039 | Left ventricle |
| LA_stroke_ | Z5_S5 | 27446 | 0.116297 | 0.005376 | 9.24E-104 | 103.0342 | Left atrium |
| RV_end_sy | Z7_S3 | 27446 | -0.17521 | 0.008103 | 1.07E-103 | 102.9709 | Right ventricle |
| LV_longitu | Z89_S4 | 27446 | 0.103097 | 0.004768 | 1.14E-103 | 102.9427 | Left ventricle |
| Ascending_ | Z12_S4 | 24599 | -0.10562 | 0.004885 | 1.15E-103 | 102.9378 | Aorta |
| RA_stroke_ | Z83_S2 | 27446 | 0.102297 | 0.004737 | 2.05E-103 | 102.6879 | Right atrium |
| LV_end_sy | Z75_S1 | 27446 | -0.13912 | 0.006443 | 2.08E-103 | 102.6824 | Left ventricle |
| Descending | Z52_S3 | 24599 | 0.121828 | 0.005643 | 2.26E-103 | 102.6455 | Aorta |
| Ascending_ | Z14_S4 | 24599 | 0.14529 | 0.006734 | 3.06E-103 | 102.5139 | Aorta |
| LV_stroke_ | Z50_S3 | 27446 | -0.12154 | 0.005638 | 4.70E-103 | 102.3282 | Left ventricle |
| Descending | Z31_S5 | 24599 | 0.115516 | 0.00536 | 5.17E-103 | 102.2869 | Aorta |
| LA_ejectio | Z40_S5 | 27446 | 0.088994 | 0.00413 | 5.50E-103 | 102.2596 | Left atrium |
| LA_minimu | Z6_S2 | 27446 | -0.08435 | 0.003917 | 7.50E-103 | 102.1247 | Left atrium |
| LV_end_sy | Z5_S5 | 27446 | 0.13871 | 0.006443 | 8.52E-103 | 102.0697 | Left ventricle |
| RV_end_di | Z17_S1 | 27446 | 0.143444 | 0.006667 | 1.12E-102 | 101.9522 | Right ventricle |
| LA_stroke_ | Z115_S4 | 27446 | 0.102843 | 0.004784 | 1.67E-102 | 101.778 | Left atrium |

| | | | | | | | |
|---|---|---|---|---|---|---|---|
| RV_stroke_ | Z83_S2 | 27446 | 0.12373 | 0.005762 | 2.82E-102 | 101.5491 | Right ventricle |
| RA_maxim | Z14_S1 | 27446 | 0.111196 | 0.005181 | 3.40E-102 | 101.4688 | Right atrium |
| LA_stroke_ | Z92_S1 | 27446 | -0.11137 | 0.005189 | 3.44E-102 | 101.4634 | Left atrium |
| LA_stroke_ | Z17_S1 | 27446 | 0.104714 | 0.004879 | 3.45E-102 | 101.4619 | Left atrium |
| LV_end_di | Z7_S3 | 27446 | -0.17329 | 0.008075 | 3.66E-102 | 101.4365 | Left ventricle |
| RV_stroke_ | Z75_S4 | 27446 | -0.15514 | 0.007229 | 3.70E-102 | 101.432 | Right ventricle |
| Descending | Z127_S4 | 24599 | 0.139474 | 0.006506 | 5.81E-102 | 101.2358 | Aorta |
| RV_end_di | Z93_S1 | 27446 | 0.175188 | 0.008173 | 6.19E-102 | 101.2084 | Right ventricle |
| LV_end_sy | Z53_S1 | 27446 | 0.139388 | 0.006509 | 9.99E-102 | 101.0003 | Left ventricle |
| Ascending_ | Z85_S5 | 24599 | 0.120286 | 0.005627 | 2.21E-101 | 100.6565 | Aorta |
| Descending | Z115_S4 | 24599 | 0.127635 | 0.005974 | 2.91E-101 | 100.5355 | Aorta |
| Ascending_ | Z73_S1 | 24599 | -0.11409 | 0.005341 | 3.01E-101 | 100.5217 | Aorta |
| Descending | Z28_S4 | 24599 | -0.15122 | 0.007079 | 3.08E-101 | 100.5116 | Aorta |
| LA_stroke_ | Z60_S2 | 27446 | -0.09714 | 0.00455 | 3.95E-101 | 100.4034 | Left atrium |
| LA_maximu | Z115_S4 | 27446 | 0.101592 | 0.004761 | 4.93E-101 | 100.3071 | Left atrium |
| LV_end_sy | Z65_S2 | 27446 | 0.137212 | 0.006432 | 5.67E-101 | 100.2462 | Left ventricle |
| LA_minimu | Z1_S5 | 27446 | 0.109623 | 0.00514 | 6.23E-101 | 100.2057 | Left atrium |
| RA_ejectio | Z127_S5 | 27446 | -0.08975 | 0.004209 | 7.25E-101 | 100.1394 | Right atrium |
| RV_stroke_ | Z79_S4 | 27446 | -0.1534 | 0.007198 | 8.98E-101 | 100.0467 | Right ventricle |
| LV_end_di | Z92_S1 | 27446 | -0.14355 | 0.006738 | 1.01E-100 | 99.99429 | Left ventricle |
| RA_stroke_ | Z3_S4 | 27446 | 0.133311 | 0.00626 | 1.20E-100 | 99.91989 | Right atrium |
| LA_minimu | Z52_S3 | 27446 | 0.091495 | 0.004298 | 1.52E-100 | 99.81837 | Left atrium |
| LV_stroke_ | Z68_S2 | 27446 | -0.11659 | 0.005477 | 1.54E-100 | 99.81151 | Left ventricle |
| RV_end_sy | Z5_S1 | 27446 | -0.14259 | 0.006708 | 2.85E-100 | 99.54576 | Right ventricle |
| Ascending_ | Z24_S1 | 24599 | 0.102897 | 0.004845 | 4.44E-100 | 99.35294 | Aorta |
| RA_stroke_ | Z82_S4 | 27446 | -0.09189 | 0.004332 | 7.92E-100 | 99.10152 | Right atrium |
| Descending | Z46_S1 | 24599 | -0.13964 | 0.00659 | 1.14E-99 | 98.94217 | Aorta |
| LV_stroke_ | Z63_S1 | 27446 | -0.15148 | 0.007154 | 1.68E-99 | 98.77341 | Left ventricle |
| Descending | Z89_S4 | 24599 | -0.13542 | 0.006397 | 1.79E-99 | 98.74736 | Aorta |
| LA_maximu | Z17_S1 | 27446 | 0.102758 | 0.004856 | 2.11E-99 | 98.6748 | Left atrium |
| RA_minimu | Z47_S4 | 27446 | -0.11809 | 0.005581 | 2.24E-99 | 98.65043 | Right atrium |
| Ascending_ | Z127_S5 | 24599 | 0.102448 | 0.004843 | 2.66E-99 | 98.57581 | Aorta |
| LV_circumf | Z14_S4 | 27446 | -0.11912 | 0.005635 | 3.60E-99 | 98.44369 | Left ventricle |
| Ascending_ | Z118_S1 | 24599 | 0.141742 | 0.006707 | 3.81E-99 | 98.41938 | Aorta |
| RA_minimu | Z109_S1 | 27446 | -0.11831 | 0.005602 | 5.05E-99 | 98.29665 | Right atrium |
| LA_maximu | Z92_S1 | 27446 | -0.10896 | 0.005164 | 8.16E-99 | 98.08823 | Left atrium |
| LA_maximu | Z62_S3 | 27446 | 0.114783 | 0.00544 | 8.16E-99 | 98.08806 | Left atrium |
| LV_stroke_ | Z65_S2 | 27446 | 0.130606 | 0.006191 | 8.51E-99 | 98.07018 | Left ventricle |
| RA_minimu | Z24_S5 | 27446 | -0.12832 | 0.00609 | 1.46E-98 | 97.83467 | Right atrium |
| RA_minimu | Z7_S5 | 27446 | 0.128646 | 0.006108 | 1.81E-98 | 97.74234 | Right atrium |
| LV_stroke_ | Z37_S2 | 27446 | -0.13743 | 0.006535 | 3.44E-98 | 97.46328 | Left ventricle |
| LA_stroke_ | Z66_S2 | 27446 | 0.125749 | 0.005982 | 4.22E-98 | 97.37516 | Left atrium |
| LV_stroke_ | Z70_S5 | 27446 | 0.121033 | 0.005758 | 4.34E-98 | 97.36214 | Left ventricle |
| Ascending_ | Z22_S2 | 24599 | -0.12167 | 0.005789 | 4.35E-98 | 97.36184 | Aorta |
| Descending | Z62_S2 | 24599 | 0.122945 | 0.005853 | 5.74E-98 | 97.2412 | Aorta |

| | | | | | | | |
|---|---|---|---|---|---|---|---|
| LA_minimu | Z112_S3 | 27446 | -0.1252 | 0.005975 | 1.73E-97 | 96.76136 | Left atrium |
| Descending | Z15_S2 | 24599 | 0.146036 | 0.00697 | 1.74E-97 | 96.7588 | Aorta |
| Descending | Z73_S1 | 24599 | -0.12819 | 0.00612 | 1.97E-97 | 96.70555 | Aorta |
| Descending | Z1_S1 | 24599 | -0.13487 | 0.006439 | 2.01E-97 | 96.69632 | Aorta |
| LV_stroke_ | Z39_S1 | 27446 | 0.116492 | 0.005562 | 2.11E-97 | 96.67572 | Left ventricle |
| Descending | Z31_S2 | 24599 | 0.137334 | 0.00656 | 2.56E-97 | 96.59185 | Aorta |
| Ascending_ | Z115_S4 | 24599 | 0.109599 | 0.005237 | 2.91E-97 | 96.53567 | Aorta |
| LA_minimu | Z93_S2 | 27446 | 0.129297 | 0.006182 | 3.81E-97 | 96.41914 | Left atrium |
| Descending | Z17_S1 | 24599 | 0.127494 | 0.006096 | 3.90E-97 | 96.40945 | Aorta |
| LV_longitu | Z61_S4 | 27446 | 0.097176 | 0.00465 | 5.79E-97 | 96.23716 | Left ventricle |
| Ascending_ | Z57_S1 | 24599 | -0.10981 | 0.005255 | 5.91E-97 | 96.22826 | Aorta |
| RV_end_sy | Z120_S2 | 27446 | 0.121939 | 0.005841 | 8.66E-97 | 96.06225 | Right ventricle |
| RV_end_sy | Z43_S2 | 27446 | -0.12436 | 0.00596 | 1.12E-96 | 95.9506 | Right ventricle |
| LV_end_dia | Z80_S2 | 27446 | 0.125945 | 0.006039 | 1.37E-96 | 95.8623 | Left ventricle |
| LV_end_sy | Z87_S3 | 27446 | -0.12704 | 0.006098 | 2.15E-96 | 95.66758 | Left ventricle |
| Ascending_ | Z28_S4 | 24599 | -0.12907 | 0.006206 | 4.45E-96 | 95.35163 | Aorta |
| Ascending_ | Z7_S5 | 24599 | -0.13013 | 0.006258 | 4.71E-96 | 95.32698 | Aorta |
| RV_end_di | Z50_S2 | 27446 | 0.181515 | 0.00873 | 5.15E-96 | 95.28778 | Right ventricle |
| RA_maxim | Z14_S4 | 27446 | 0.12789 | 0.006152 | 5.52E-96 | 95.2584 | Right atrium |
| RV_stroke_ | Z63_S1 | 27446 | -0.15096 | 0.007262 | 5.58E-96 | 95.25372 | Right ventricle |
| LA_minimu | Z91_S1 | 27446 | 0.101193 | 0.004873 | 8.66E-96 | 95.06227 | Left atrium |
| LV_end_dia | Z11_S5 | 27446 | -0.13542 | 0.006522 | 8.99E-96 | 95.04604 | Left ventricle |
| RV_end_sy | Z79_S4 | 27446 | -0.16649 | 0.008019 | 9.33E-96 | 95.03021 | Right ventricle |
| LV_cardiac | Z50_S5 | 27446 | -0.14228 | 0.006853 | 9.73E-96 | 95.01179 | Left ventricle |
| Ascending_ | Z59_S3 | 24599 | -0.09612 | 0.00463 | 1.02E-95 | 94.99343 | Aorta |
| LV_end_sy | Z19_S2 | 27446 | 0.128039 | 0.006174 | 1.60E-95 | 94.79563 | Left ventricle |
| LA_maximu | Z118_S1 | 27446 | 0.11987 | 0.005781 | 1.66E-95 | 94.78068 | Left atrium |
| LA_minimu | Z28_S4 | 27446 | -0.11246 | 0.005427 | 2.18E-95 | 94.66136 | Left atrium |
| RA_ejectio | Z66_S2 | 27446 | -0.11722 | 0.005666 | 4.47E-95 | 94.34927 | Right atrium |
| Descending | Z31_S2 | 24599 | 0.137524 | 0.006656 | 7.73E-95 | 94.11208 | Aorta |
| LV_myocar | Z41_S2 | 27446 | -0.2032 | 0.009838 | 8.75E-95 | 94.05787 | Left ventricle |
| LA_minimu | Z120_S2 | 27446 | 0.089324 | 0.004325 | 9.61E-95 | 94.0172 | Left atrium |
| LV_myocar | Z83_S2 | 27446 | 0.161978 | 0.007851 | 1.45E-94 | 93.83964 | Left ventricle |
| LV_stroke_ | Z18_S4 | 27446 | -0.15514 | 0.00752 | 1.47E-94 | 93.83227 | Left ventricle |
| RV_end_di | Z39_S1 | 27446 | 0.135947 | 0.006593 | 1.81E-94 | 93.74187 | Right ventricle |
| Ascending_ | Z92_S5 | 24599 | 0.149961 | 0.007282 | 3.11E-94 | 93.50784 | Aorta |
| LV_myocar | Z75_S4 | 27446 | -0.20284 | 0.00985 | 3.14E-94 | 93.50297 | Left ventricle |
| LV_end_sy | Z37_S2 | 27446 | -0.13981 | 0.006793 | 3.93E-94 | 93.4052 | Left ventricle |
| Ascending_ | Z51_S4 | 24599 | -0.10192 | 0.004952 | 3.97E-94 | 93.40089 | Aorta |
| Ascending_ | Z115_S4 | 24599 | 0.107418 | 0.005219 | 4.00E-94 | 93.39762 | Aorta |
| RA_minimu | Z57_S2 | 27446 | 0.095533 | 0.004642 | 4.03E-94 | 93.39436 | Right atrium |
| RA_ejectio | Z1_S5 | 27446 | -0.10371 | 0.005047 | 8.34E-94 | 93.07909 | Right atrium |
| LV_longitu | Z112_S1 | 27446 | 0.126257 | 0.006146 | 8.65E-94 | 93.06298 | Left ventricle |
| RV_stroke_ | Z62_S2 | 27446 | 0.11119 | 0.005414 | 1.01E-93 | 92.99722 | Right ventricle |
| Ascending_ | Z120_S4 | 24599 | -0.11644 | 0.005672 | 1.18E-93 | 92.92929 | Aorta |

| | | | | | | | |
|---|---|---|---|---|---|---|---|
| RA_minimu | Z28_S4 | 27446 | -0.12361 | 0.006022 | 1.22E-93 | 92.91309 | Right atrium |
| LA_stroke_ | Z64_S1 | 27446 | -0.11882 | 0.005802 | 3.26E-93 | 92.48644 | Left atrium |
| RV_end_sy | Z17_S1 | 27446 | 0.130228 | 0.006362 | 4.05E-93 | 92.39291 | Right ventricle |
| LV_end_di | Z50_S2 | 27446 | 0.169751 | 0.008297 | 4.85E-93 | 92.314 | Left ventricle |
| Ascending_ | Z73_S4 | 24599 | 0.134399 | 0.00657 | 5.39E-93 | 92.26869 | Aorta |
| LV_stroke_ | Z14_S2 | 27446 | -0.12273 | 0.006005 | 7.94E-93 | 92.10014 | Left ventricle |
| LV_cardiac | Z94_S5 | 27446 | -0.09726 | 0.004762 | 1.07E-92 | 91.97118 | Left ventricle |
| RV_end_di | Z68_S2 | 27446 | -0.13262 | 0.006495 | 1.18E-92 | 91.92948 | Right ventricle |
| Ascending_ | Z46_S1 | 24599 | -0.11748 | 0.005756 | 1.43E-92 | 91.84567 | Aorta |
| Descending | Z22_S2 | 24599 | -0.13714 | 0.00673 | 2.63E-92 | 91.57936 | Aorta |
| Ascending_ | Z59_S2 | 24599 | 0.133231 | 0.006541 | 3.18E-92 | 91.49748 | Aorta |
| Descending | Z17_S1 | 24599 | 0.125945 | 0.006187 | 3.95E-92 | 91.40346 | Aorta |
| LV_end_di | Z91_S1 | 27446 | 0.133528 | 0.00656 | 4.08E-92 | 91.38977 | Left ventricle |
| LA_stroke_ | Z37_S1 | 27446 | 0.119656 | 0.005882 | 5.32E-92 | 91.27387 | Left atrium |
| RA_maxim | Z59_S3 | 27446 | -0.09095 | 0.004472 | 5.96E-92 | 91.22498 | Right atrium |
| Descending | Z80_S2 | 24599 | 0.11995 | 0.0059 | 7.00E-92 | 91.15488 | Aorta |
| LA_ejection | Z29_S5 | 27446 | -0.10494 | 0.005163 | 7.97E-92 | 91.0983 | Left atrium |
| LV_end_di | Z59_S2 | 27446 | 0.157377 | 0.007745 | 8.51E-92 | 91.07021 | Left ventricle |
| RA_maxim | Z1_S1 | 27446 | -0.10908 | 0.005371 | 1.03E-91 | 90.98783 | Right atrium |
| RA_ejectio | Z65_S5 | 27446 | 0.119641 | 0.005892 | 1.11E-91 | 90.95617 | Right atrium |
| RA_minimu | Z49_S1 | 27446 | -0.1386 | 0.006829 | 1.45E-91 | 90.84008 | Right atrium |
| Descending | Z83_S2 | 24599 | 0.125146 | 0.006167 | 1.46E-91 | 90.83576 | Aorta |
| LV_myocar | Z64_S1 | 27446 | -0.18756 | 0.009246 | 1.72E-91 | 90.76422 | Left ventricle |
| LV_end_di | Z37_S1 | 27446 | 0.154916 | 0.007637 | 1.77E-91 | 90.7528 | Left ventricle |
| LV_myocar | Z68_S2 | 27446 | -0.15353 | 0.007574 | 2.35E-91 | 90.62901 | Left ventricle |
| RA_minimu | Z44_S4 | 27446 | -0.09596 | 0.004734 | 2.46E-91 | 90.60863 | Right atrium |
| RA_minimu | Z15_S2 | 27446 | 0.120916 | 0.005967 | 2.60E-91 | 90.58504 | Right atrium |
| RV_end_sy | Z82_S4 | 27446 | -0.11897 | 0.005872 | 2.82E-91 | 90.54999 | Right ventricle |
| LV_end_di | Z74_S4 | 27446 | -0.14403 | 0.007118 | 4.86E-91 | 90.3138 | Left ventricle |
| LV_longitu | Z37_S4 | 27446 | -0.12373 | 0.006115 | 5.06E-91 | 90.29588 | Left ventricle |
| LV_end_sy | Z80_S2 | 27446 | 0.112772 | 0.005576 | 6.06E-91 | 90.21717 | Left ventricle |
| LA_minimu | Z75_S1 | 27446 | -0.10501 | 0.005193 | 6.09E-91 | 90.21518 | Left atrium |
| RA_maxim | Z95_S3 | 27446 | -0.08592 | 0.004249 | 6.52E-91 | 90.18561 | Right atrium |
| LV_end_sy | Z70_S5 | 27446 | 0.121017 | 0.005987 | 7.56E-91 | 90.12157 | Left ventricle |
| RV_end_di | Z86_S3 | 27446 | -0.16317 | 0.008078 | 1.00E-90 | 90 | Right ventricle |
| Ascending_ | Z14_S1 | 24599 | 0.108047 | 0.005351 | 1.15E-90 | 89.94072 | Aorta |
| RA_ejectio | Z94_S5 | 27446 | -0.08615 | 0.004268 | 1.28E-90 | 89.89446 | Right atrium |
| LA_ejection | Z127_S4 | 27446 | -0.10696 | 0.005301 | 1.51E-90 | 89.8199 | Left atrium |
| Descending | Z40_S5 | 24599 | -0.11425 | 0.005662 | 1.54E-90 | 89.81343 | Aorta |
| LV_circumf | Z32_S1 | 27446 | -0.10825 | 0.005367 | 1.87E-90 | 89.72746 | Left ventricle |
| LA_stroke_ | Z57_S1 | 27446 | -0.0972 | 0.00482 | 1.96E-90 | 89.70758 | Left atrium |
| RA_minimu | Z87_S3 | 27446 | -0.10975 | 0.00545 | 3.31E-90 | 89.48077 | Right atrium |
| Ascending_ | Z94_S5 | 24599 | 0.098867 | 0.004913 | 4.43E-90 | 89.3532 | Aorta |
| Ascending_ | Z59_S2 | 24599 | 0.131112 | 0.006519 | 5.58E-90 | 89.25299 | Aorta |
| Descending | Z55_S1 | 24599 | -0.12346 | 0.006142 | 7.24E-90 | 89.14043 | Aorta |

| | | | | | | | |
|---|---|---|---|---|---|---|---|
| RA_minimu | Z22_S2 | 27446 | 0.113942 | 0.005675 | 1.18E-89 | 88.92738 | Right atrium |
| LV_myocar | Z47_S4 | 27446 | -0.16684 | 0.008315 | 1.48E-89 | 88.83015 | Left ventricle |
| Ascending_ | Z52_S4 | 24599 | 0.102549 | 0.005113 | 1.74E-89 | 88.75926 | Aorta |
| Descending | Z13_S3 | 24599 | 0.108189 | 0.005395 | 1.82E-89 | 88.7393 | Aorta |
| RA_minimu | Z107_S1 | 27446 | -0.0903 | 0.004504 | 2.03E-89 | 88.69146 | Right atrium |
| LV_longitu | Z4_S4 | 27446 | 0.088006 | 0.00439 | 2.15E-89 | 88.66667 | Left ventricle |
| LA_maximu | Z96_S3 | 27446 | 0.101443 | 0.005062 | 2.39E-89 | 88.62122 | Left atrium |
| LV_end_di | Z114_S4 | 27446 | 0.132235 | 0.006601 | 2.81E-89 | 88.55184 | Left ventricle |
| LV_stroke_ | Z37_S4 | 27446 | 0.149497 | 0.007463 | 2.86E-89 | 88.54407 | Left ventricle |
| RV_end_di | Z111_S1 | 27446 | 0.155854 | 0.007787 | 4.08E-89 | 88.38916 | Right ventricle |
| Descending | Z29_S1 | 24599 | 0.144748 | 0.007232 | 4.14E-89 | 88.38337 | Aorta |
| LA_stroke_ | Z58_S2 | 27446 | 0.097992 | 0.004899 | 5.32E-89 | 88.2737 | Left atrium |
| LV_end_sy | Z73_S1 | 27446 | -0.11736 | 0.005868 | 5.47E-89 | 88.26234 | Left ventricle |
| RA_ejectio | Z5_S5 | 27446 | -0.10195 | 0.005097 | 5.48E-89 | 88.26091 | Right atrium |
| Descending | Z58_S2 | 24599 | 0.122223 | 0.006117 | 7.92E-89 | 88.10138 | Aorta |
| RA_maxim | Z44_S4 | 27446 | -0.09333 | 0.004672 | 8.75E-89 | 88.05813 | Right atrium |
| RA_maxim | Z3_S4 | 27446 | 0.137319 | 0.006877 | 1.08E-88 | 87.967 | Right atrium |
| RA_ejectio | Z111_S1 | 27446 | -0.10766 | 0.005396 | 1.44E-88 | 87.84095 | Right atrium |
| RV_end_sy | Z13_S1 | 27446 | -0.12154 | 0.006096 | 1.94E-88 | 87.7126 | Right ventricle |
| LV_stroke_ | Z43_S2 | 27446 | -0.10523 | 0.005278 | 1.97E-88 | 87.70532 | Left ventricle |
| Ascending_ | Z98_S3 | 24599 | -0.09918 | 0.004978 | 2.49E-88 | 87.60411 | Aorta |
| RV_end_di | Z87_S3 | 27446 | -0.13861 | 0.006959 | 2.83E-88 | 87.54769 | Right ventricle |
| RA_stroke_ | Z28_S2 | 27446 | -0.10668 | 0.005356 | 2.87E-88 | 87.54142 | Right atrium |
| LA_maximu | Z18_S4 | 27446 | -0.12925 | 0.006492 | 3.50E-88 | 87.45591 | Left atrium |
| Descending | Z5_S1 | 24599 | -0.12826 | 0.006446 | 4.33E-88 | 87.36319 | Aorta |
| LA_stroke_ | Z96_S3 | 27446 | 0.101134 | 0.005088 | 6.21E-88 | 87.20661 | Left atrium |
| RA_ejectio | Z112_S3 | 27446 | 0.11657 | 0.005869 | 8.58E-88 | 87.06671 | Right atrium |
| RA_maxim | Z73_S4 | 27446 | 0.125827 | 0.00634 | 1.15E-87 | 86.9382 | Right atrium |
| LA_stroke_ | Z61_S4 | 27446 | -0.09771 | 0.004924 | 1.30E-87 | 86.88485 | Left atrium |
| Descending | Z118_S1 | 24599 | 0.130705 | 0.006591 | 1.64E-87 | 86.78425 | Aorta |
| RA_ejectio | Z28_S2 | 27446 | -0.10453 | 0.005272 | 1.70E-87 | 86.77016 | Right atrium |
| Descending | Z80_S2 | 24599 | 0.115164 | 0.005819 | 3.50E-87 | 86.45575 | Aorta |
| Ascending_ | Z29_S5 | 24599 | 0.120708 | 0.006099 | 3.64E-87 | 86.43889 | Aorta |
| LV_radial_ | Z1_S2 | 27446 | -0.08296 | 0.004197 | 5.90E-87 | 86.22882 | Left ventricle |
| Descending | Z5_S5 | 24599 | 0.121391 | 0.006144 | 6.96E-87 | 86.1574 | Aorta |
| LV_myocar | Z61_S4 | 27446 | -0.15478 | 0.007847 | 1.34E-86 | 85.8743 | Left ventricle |
| Ascending_ | Z20_S2 | 24599 | 0.095033 | 0.004819 | 1.38E-86 | 85.85933 | Aorta |
| Descending | Z7_S1 | 24599 | -0.12001 | 0.006087 | 1.61E-86 | 85.79372 | Aorta |
| LV_circumf | Z73_S1 | 27446 | -0.0934 | 0.004739 | 1.82E-86 | 85.73925 | Left ventricle |
| RA_minimu | Z5_S1 | 27446 | -0.1087 | 0.005516 | 1.96E-86 | 85.70849 | Right atrium |
| RV_stroke_ | Z66_S3 | 27446 | -0.13666 | 0.006936 | 2.00E-86 | 85.69889 | Right ventricle |
| LV_end_sy | Z47_S4 | 27446 | -0.12308 | 0.006255 | 3.41E-86 | 85.46754 | Left ventricle |
| Descending | Z62_S1 | 24599 | 0.121868 | 0.006203 | 6.27E-86 | 85.20284 | Aorta |
| LV_myocar | Z66_S2 | 27446 | 0.187395 | 0.009541 | 6.96E-86 | 85.15714 | Left ventricle |
| LA_maximu | Z74_S2 | 27446 | 0.100912 | 0.005139 | 7.59E-86 | 85.11955 | Left atrium |

| | | | | | | | |
|---|---|---|---|---|---|---|---|
| LV_circumf | Z80_S1 | 27446 | -0.10761 | 0.005482 | 8.91E-86 | 85.05029 | Left ventricle |
| LV_stroke_ | Z66_S2 | 27446 | 0.135519 | 0.006905 | 9.40E-86 | 85.02709 | Left ventricle |
| Ascending_ | Z57_S1 | 24599 | -0.10361 | 0.00528 | 9.81E-86 | 85.00854 | Aorta |
| LA_maximu | Z58_S2 | 27446 | 0.09563 | 0.004876 | 1.23E-85 | 84.9115 | Left atrium |
| RV_end_di | Z6_S2 | 27446 | -0.10895 | 0.005556 | 1.31E-85 | 84.88118 | Right ventricle |
| Ascending_ | Z53_S1 | 24599 | 0.123622 | 0.006307 | 1.53E-85 | 84.81393 | Aorta |
| RA_maxim | Z9_S2 | 27446 | 0.11561 | 0.005899 | 1.65E-85 | 84.78312 | Right atrium |
| LA_minimu | Z41_S1 | 27446 | 0.106719 | 0.005448 | 1.98E-85 | 84.70245 | Left atrium |
| RV_stroke_ | Z52_S3 | 27446 | 0.102199 | 0.00522 | 2.32E-85 | 84.63493 | Right ventricle |
| Descending | Z109_S1 | 24599 | -0.12822 | 0.00655 | 2.50E-85 | 84.60193 | Aorta |
| Ascending_ | Z24_S5 | 24599 | -0.12907 | 0.006599 | 3.44E-85 | 84.46349 | Aorta |
| LV_myocar | Z7_S3 | 27446 | -0.19409 | 0.009923 | 3.44E-85 | 84.46287 | Left ventricle |
| RA_maxim | Z107_S1 | 27446 | -0.08692 | 0.004445 | 3.94E-85 | 84.40472 | Right atrium |
| LV_end_sy | Z99_S5 | 27446 | 0.117558 | 0.006013 | 4.03E-85 | 84.39441 | Left ventricle |
| LV_end_sy | Z91_S1 | 27446 | 0.118341 | 0.006058 | 5.40E-85 | 84.26723 | Left ventricle |
| LV_stroke_ | Z93_S2 | 27446 | 0.144461 | 0.007396 | 5.78E-85 | 84.23827 | Left ventricle |
| Ascending_ | Z31_S2 | 24599 | 0.112297 | 0.005754 | 8.20E-85 | 84.08633 | Aorta |
| RV_stroke_ | Z55_S1 | 27446 | -0.11056 | 0.005673 | 1.37E-84 | 83.86265 | Right ventricle |
| Descending | Z58_S2 | 24599 | 0.120967 | 0.006207 | 1.40E-84 | 83.85447 | Aorta |
| LV_cardiac | Z7_S3 | 27446 | -0.12805 | 0.006582 | 2.74E-84 | 83.56206 | Left ventricle |
| LA_stroke_ | Z28_S4 | 27446 | -0.10938 | 0.00563 | 4.59E-84 | 83.33854 | Left atrium |
| Ascending_ | Z35_S5 | 24599 | -0.09495 | 0.004891 | 5.94E-84 | 83.22623 | Aorta |
| LV_circumf | Z78_S1 | 27446 | 0.089537 | 0.004612 | 6.06E-84 | 83.21783 | Left ventricle |
| Ascending_ | Z75_S1 | 24599 | -0.11374 | 0.005863 | 7.85E-84 | 83.10519 | Aorta |
| Ascending_ | Z40_S5 | 24599 | -0.09456 | 0.004876 | 8.88E-84 | 83.05163 | Aorta |
| LV_end_dia | Z77_S3 | 27446 | -0.13417 | 0.006919 | 9.08E-84 | 83.04191 | Left ventricle |
| LV_myocar | Z50_S3 | 27446 | -0.1513 | 0.007803 | 9.42E-84 | 83.0261 | Left ventricle |
| Ascending_ | Z78_S4 | 24599 | 0.098223 | 0.00507 | 1.25E-83 | 82.90167 | Aorta |
| LV_end_dia | Z122_S5 | 27446 | 0.11824 | 0.006104 | 1.38E-83 | 82.86005 | Left ventricle |
| RV_end_di | Z23_S1 | 27446 | -0.15534 | 0.008022 | 1.53E-83 | 82.81488 | Right ventricle |
| LV_longitu | Z63_S1 | 27446 | 0.113688 | 0.005871 | 1.55E-83 | 82.80923 | Left ventricle |
| LV_longitu | Z127_S5 | 27446 | -0.08145 | 0.004207 | 1.70E-83 | 82.77014 | Left ventricle |
| LA_maximu | Z112_S3 | 27446 | -0.11942 | 0.006169 | 1.78E-83 | 82.74986 | Left atrium |
| RA_ejectio | Z46_S1 | 27446 | 0.096923 | 0.005009 | 2.02E-83 | 82.69521 | Right atrium |
| LV_circumf | Z12_S5 | 27446 | -0.11617 | 0.006007 | 2.44E-83 | 82.61172 | Left ventricle |
| Ascending_ | Z7_S5 | 24599 | -0.12071 | 0.006242 | 2.53E-83 | 82.59721 | Aorta |
| RV_end_sy | Z108_S5 | 27446 | -0.1348 | 0.006976 | 3.47E-83 | 82.46004 | Right ventricle |
| LV_myocar | Z65_S1 | 27446 | -0.1335 | 0.00691 | 3.73E-83 | 82.42824 | Left ventricle |
| LA_maximu | Z63_S1 | 27446 | -0.11931 | 0.006182 | 5.33E-83 | 82.27327 | Left atrium |
| Ascending_ | Z31_S2 | 24599 | 0.110669 | 0.005734 | 5.44E-83 | 82.26438 | Aorta |
| RV_end_sy | Z41_S2 | 27446 | -0.15538 | 0.008053 | 6.00E-83 | 82.22208 | Right ventricle |
| LA_maximu | Z65_S5 | 27446 | -0.11947 | 0.006195 | 7.28E-83 | 82.13798 | Left atrium |
| LV_longitu | Z10_S4 | 27446 | 0.099376 | 0.005154 | 7.65E-83 | 82.1162 | Left ventricle |
| RA_minimu | Z62_S1 | 27446 | 0.100988 | 0.00524 | 9.16E-83 | 82.03806 | Right atrium |
| LV_end_dia | Z41_S1 | 27446 | 0.141329 | 0.007334 | 9.35E-83 | 82.02908 | Left ventricle |

| | | | | | | | |
|---|---|---|---|---|---|---|---|
| Ascending_ | Z122_S1 | 24599 | 0.127089 | 0.006597 | 1.04E-82 | 81.98257 | Aorta |
| Descending | Z123_S4 | 24599 | 0.132102 | 0.00686 | 1.24E-82 | 81.90802 | Aorta |
| LV_stroke_ | Z58_S2 | 27446 | 0.108833 | 0.005653 | 1.31E-82 | 81.88314 | Left ventricle |
| LV_myocar | Z75_S1 | 27446 | -0.1652 | 0.008581 | 1.39E-82 | 81.85778 | Left ventricle |
| RV_end_di | Z81_S1 | 27446 | -0.13942 | 0.007243 | 1.44E-82 | 81.84253 | Right ventricle |
| Descending | Z43_S2 | 24599 | -0.11026 | 0.005731 | 1.67E-82 | 81.77694 | Aorta |
| RV_end_sy | Z50_S3 | 27446 | -0.12267 | 0.006382 | 2.45E-82 | 81.61016 | Right ventricle |
| RA_maxim | Z47_S4 | 27446 | -0.10593 | 0.005514 | 3.03E-82 | 81.51804 | Right atrium |
| Ascending_ | Z113_S2 | 24599 | 0.132421 | 0.006896 | 3.61E-82 | 81.44211 | Aorta |
| LA_minimu | Z120_S4 | 27446 | -0.09596 | 0.004999 | 3.96E-82 | 81.40278 | Left atrium |
| Descending | Z82_S1 | 24599 | -0.15514 | 0.008093 | 6.71E-82 | 81.17307 | Aorta |
| RV_end_di | Z108_S5 | 27446 | -0.14024 | 0.007317 | 7.20E-82 | 81.14273 | Right ventricle |
| LV_stroke_ | Z31_S2 | 27446 | 0.116174 | 0.006064 | 8.11E-82 | 81.09116 | Left ventricle |
| Descending | Z7_S3 | 24599 | -0.1512 | 0.007892 | 8.39E-82 | 81.07607 | Aorta |
| RV_end_sy | Z23_S1 | 27446 | -0.14652 | 0.00765 | 9.06E-82 | 81.04305 | Right ventricle |
| RA_maxim | Z62_S1 | 27446 | 0.099002 | 0.005171 | 1.02E-81 | 80.99174 | Right atrium |
| LA_minimu | Z70_S5 | 27446 | 0.092346 | 0.004824 | 1.12E-81 | 80.94942 | Left atrium |
| LA_stroke_ | Z75_S1 | 27446 | -0.10309 | 0.005386 | 1.17E-81 | 80.93346 | Left atrium |
| LA_maximu | Z87_S3 | 27446 | -0.09665 | 0.00507 | 5.06E-81 | 80.29555 | Left atrium |
| Descending | Z29_S5 | 24599 | 0.133218 | 0.006991 | 5.84E-81 | 80.23367 | Aorta |
| Ascending_ | Z75_S1 | 24599 | -0.11212 | 0.005886 | 6.79E-81 | 80.16815 | Aorta |
| LV_stroke_ | Z115_S4 | 27446 | 0.105238 | 0.005527 | 7.67E-81 | 80.11495 | Left ventricle |
| Descending | Z69_S5 | 24599 | -0.10723 | 0.005636 | 1.05E-80 | 79.98025 | Aorta |
| Ascending_ | Z20_S2 | 24599 | 0.092042 | 0.004839 | 1.13E-80 | 79.94798 | Aorta |
| LV_myocar | Z77_S3 | 27446 | -0.16149 | 0.008493 | 1.31E-80 | 79.88147 | Left ventricle |
| Descending | Z6_S2 | 24599 | -0.09689 | 0.005104 | 2.39E-80 | 79.62176 | Aorta |
| RV_end_di | Z37_S1 | 27446 | 0.152686 | 0.008046 | 2.64E-80 | 79.57876 | Right ventricle |
| RA_maxim | Z93_S1 | 27446 | 0.120046 | 0.006327 | 2.81E-80 | 79.55144 | Right atrium |
| LA_minimu | Z20_S1 | 27446 | 0.111314 | 0.005869 | 3.25E-80 | 79.4881 | Left atrium |
| RV_end_di | Z11_S5 | 27446 | -0.1303 | 0.006873 | 3.84E-80 | 79.41516 | Right ventricle |
| RV_end_sy | Z99_S5 | 27446 | 0.123949 | 0.006541 | 4.48E-80 | 79.3485 | Right ventricle |
| RV_stroke_ | Z57_S2 | 27446 | 0.096025 | 0.005081 | 1.14E-79 | 78.9415 | Right ventricle |
| Descending | Z10_S4 | 24599 | -0.11681 | 0.006182 | 1.26E-79 | 78.89911 | Aorta |
| Descending | Z31_S5 | 24599 | 0.090566 | 0.004793 | 1.29E-79 | 78.89028 | Aorta |
| RA_ejectio | Z92_S5 | 27446 | -0.11283 | 0.005972 | 1.33E-79 | 78.87597 | Right atrium |
| LV_stroke_ | Z24_S5 | 27446 | -0.12411 | 0.006571 | 1.47E-79 | 78.83409 | Left ventricle |
| RV_end_sy | Z80_S2 | 27446 | 0.114485 | 0.006069 | 2.31E-79 | 78.63609 | Right ventricle |
| RA_stroke_ | Z9_S2 | 27446 | 0.101416 | 0.005377 | 2.45E-79 | 78.61158 | Right atrium |
| LV_end_di | Z96_S3 | 27446 | 0.124619 | 0.00661 | 2.82E-79 | 78.55001 | Left ventricle |
| Ascending_ | Z15_S2 | 24599 | 0.11488 | 0.006095 | 2.97E-79 | 78.52747 | Aorta |
| RV_end_sy | Z70_S5 | 27446 | 0.122797 | 0.006517 | 3.34E-79 | 78.47674 | Right ventricle |
| RA_minimu | Z50_S4 | 27446 | 0.102046 | 0.005416 | 3.41E-79 | 78.46662 | Right atrium |
| RA_maxim | Z23_S1 | 27446 | -0.11663 | 0.006201 | 6.54E-79 | 78.1843 | Right atrium |
| LV_stroke_ | Z1_S5 | 27446 | 0.115725 | 0.006154 | 7.04E-79 | 78.15224 | Left ventricle |
| Ascending_ | Z113_S2 | 24599 | 0.129228 | 0.006873 | 7.39E-79 | 78.13135 | Aorta |

| | | | | | | | |
|---|---|---|---|---|---|---|---|
| LV_myocar | Z115_S4 | 27446 | 0.143558 | 0.007638 | 8.15E-79 | 78.08865 | Left ventricle |
| LV_end_di | Z108_S5 | 27446 | -0.13069 | 0.006954 | 8.35E-79 | 78.07852 | Left ventricle |
| Descending | Z56_S5 | 24599 | 0.106477 | 0.005666 | 8.69E-79 | 78.061 | Aorta |
| LA_minimu | Z60_S2 | 27446 | -0.08256 | 0.004398 | 1.28E-78 | 77.89431 | Left atrium |
| LA_ejection | Z112_S3 | 27446 | 0.107659 | 0.005736 | 1.33E-78 | 77.87567 | Left atrium |
| LV_longitu | Z123_S4 | 27446 | -0.09716 | 0.005178 | 1.51E-78 | 77.82191 | Left ventricle |
| Ascending_ | Z87_S3 | 24599 | -0.10518 | 0.005612 | 2.27E-78 | 77.6435 | Aorta |
| RV_end_di | Z57_S2 | 27446 | 0.111209 | 0.005934 | 2.31E-78 | 77.63618 | Right ventricle |
| LV_myocar | Z53_S1 | 27446 | 0.162415 | 0.008672 | 2.89E-78 | 77.53875 | Left ventricle |
| LV_end_sy | Z12_S5 | 27446 | -0.13933 | 0.007442 | 3.21E-78 | 77.49397 | Left ventricle |
| LV_cardiac | Z82_S4 | 27446 | 0.089261 | 0.004768 | 3.23E-78 | 77.4902 | Left ventricle |
| Descending | Z28_S4 | 24599 | -0.13473 | 0.007196 | 3.31E-78 | 77.48014 | Aorta |
| RV_stroke_ | Z70_S5 | 27446 | 0.109544 | 0.005853 | 3.62E-78 | 77.44173 | Right ventricle |
| LA_ejection | Z1_S2 | 27446 | -0.07409 | 0.00396 | 3.98E-78 | 77.40061 | Left atrium |
| LA_stroke_ | Z18_S4 | 27446 | -0.12205 | 0.00653 | 5.99E-78 | 77.22284 | Left atrium |
| LA_maximu | Z108_S5 | 27446 | -0.09953 | 0.00533 | 7.94E-78 | 77.10045 | Left atrium |
| RA_minimu | Z95_S3 | 27446 | -0.08051 | 0.004312 | 8.10E-78 | 77.09178 | Right atrium |
| LV_end_sy | Z92_S1 | 27446 | -0.1163 | 0.00623 | 9.09E-78 | 77.04121 | Left ventricle |
| Descending | Z50_S3 | 24599 | -0.11413 | 0.006118 | 1.15E-77 | 76.93777 | Aorta |
| RV_stroke_ | Z50_S3 | 27446 | -0.10695 | 0.005734 | 1.20E-77 | 76.9211 | Right ventricle |
| RA_ejectio | Z105_S5 | 27446 | -0.09336 | 0.005012 | 1.94E-77 | 76.71263 | Right atrium |
| Ascending_ | Z75_S4 | 24599 | -0.12616 | 0.006777 | 2.34E-77 | 76.63023 | Aorta |
| LV_end_sy | Z56_S2 | 27446 | -0.10012 | 0.005382 | 3.04E-77 | 76.51782 | Left ventricle |
| LV_end_sy | Z75_S4 | 27446 | -0.13796 | 0.007418 | 3.37E-77 | 76.47246 | Left ventricle |
| Ascending_ | Z98_S3 | 24599 | -0.09301 | 0.005001 | 3.39E-77 | 76.46935 | Aorta |
| LV_cardiac | Z5_S5 | 27446 | -0.10586 | 0.005695 | 3.85E-77 | 76.4141 | Left ventricle |
| LV_end_sy | Z61_S4 | 27446 | -0.10971 | 0.005906 | 5.16E-77 | 76.28771 | Left ventricle |
| LA_minimu | Z51_S1 | 27446 | -0.08222 | 0.00443 | 6.57E-77 | 76.1825 | Left atrium |
| LV_myocar | Z17_S1 | 27446 | 0.144339 | 0.00779 | 1.22E-76 | 75.91468 | Left ventricle |
| RA_minimu | Z29_S5 | 27446 | 0.110799 | 0.005984 | 1.53E-76 | 75.81673 | Right atrium |
| LV_end_di | Z56_S2 | 27446 | -0.10798 | 0.005832 | 1.54E-76 | 75.81336 | Left ventricle |
| LV_radial_ | Z99_S3 | 27446 | -0.07664 | 0.00414 | 1.67E-76 | 75.77838 | Left ventricle |
| RV_ejectio | Z14_S4 | 27446 | 0.104909 | 0.005671 | 2.15E-76 | 75.66744 | Right ventricle |
| RA_stroke_ | Z106_S4 | 27446 | 0.08185 | 0.004427 | 2.64E-76 | 75.57855 | Right atrium |
| RV_stroke_ | Z118_S1 | 27446 | 0.125859 | 0.006809 | 2.82E-76 | 75.54948 | Right ventricle |
| LA_minimu | Z87_S3 | 27446 | -0.0909 | 0.004918 | 2.86E-76 | 75.54335 | Left atrium |
| LV_longitu | Z82_S4 | 27446 | 0.078809 | 0.004264 | 2.87E-76 | 75.54172 | Left ventricle |
| LA_minimu | Z62_S3 | 27446 | 0.097561 | 0.005285 | 4.35E-76 | 75.36197 | Left atrium |
| LV_end_sy | Z11_S5 | 27446 | -0.11128 | 0.006029 | 4.50E-76 | 75.3469 | Left ventricle |
| LA_minimu | Z8_S3 | 27446 | 0.079136 | 0.004289 | 4.96E-76 | 75.30447 | Left atrium |
| LA_ejection | Z117_S4 | 27446 | 0.072669 | 0.00394 | 5.72E-76 | 75.24239 | Left atrium |
| Descending | Z61_S4 | 24599 | -0.11408 | 0.00619 | 7.43E-76 | 75.12904 | Aorta |
| LV_end_sy | Z93_S2 | 27446 | 0.141603 | 0.007691 | 1.07E-75 | 74.96883 | Left ventricle |
| Descending | Z7_S5 | 24599 | -0.13346 | 0.007258 | 1.65E-75 | 74.78181 | Aorta |
| RA_ejectio | Z12_S4 | 27446 | 0.077774 | 0.004231 | 1.80E-75 | 74.74551 | Right atrium |

| | | | | | | | |
|---|---|---|---|---|---|---|---|
| RA_minimu | Z80_S1 | 27446 | 0.111539 | 0.00607 | 2.04E-75 | 74.69115 | Right atrium |
| LA_ejectio | Z19_S2 | 27446 | -0.08769 | 0.004773 | 2.19E-75 | 74.6592 | Left atrium |
| RA_stroke_ | Z111_S5 | 27446 | 0.091652 | 0.00499 | 2.44E-75 | 74.61198 | Right atrium |
| RV_end_di | Z75_S5 | 27446 | 0.16486 | 0.008979 | 2.75E-75 | 74.56007 | Right ventricle |
| LA_stroke_ | Z114_S4 | 27446 | 0.093418 | 0.00509 | 3.04E-75 | 74.51726 | Left atrium |
| Descending | Z15_S2 | 24599 | 0.130019 | 0.007085 | 3.15E-75 | 74.50101 | Aorta |
| LA_stroke_ | Z62_S3 | 27446 | 0.100516 | 0.005478 | 3.40E-75 | 74.46879 | Left atrium |
| LA_maximu | Z99_S5 | 27446 | 0.091676 | 0.004997 | 3.51E-75 | 74.45484 | Left atrium |
| LA_stroke_ | Z99_S5 | 27446 | 0.092091 | 0.005022 | 4.13E-75 | 74.38428 | Left atrium |
| LA_minimu | Z118_S1 | 27446 | 0.102932 | 0.005615 | 4.61E-75 | 74.33652 | Left atrium |
| RA_stroke_ | Z62_S4 | 27446 | -0.10568 | 0.005766 | 4.95E-75 | 74.30555 | Right atrium |
| LA_stroke_ | Z74_S2 | 27446 | 0.094741 | 0.005169 | 5.01E-75 | 74.29998 | Left atrium |
| RV_end_sy | Z73_S1 | 27446 | -0.11706 | 0.006388 | 5.24E-75 | 74.28088 | Right ventricle |
| RV_end_di | Z59_S2 | 27446 | 0.14949 | 0.008163 | 6.51E-75 | 74.18672 | Right ventricle |
| LV_stroke_ | Z99_S5 | 27446 | 0.106044 | 0.005791 | 6.72E-75 | 74.17249 | Left ventricle |
| RV_end_di | Z12_S5 | 27446 | -0.15531 | 0.008489 | 9.16E-75 | 74.03826 | Right ventricle |
| LV_ejectio | Z10_S4 | 27446 | -0.09503 | 0.005197 | 1.08E-74 | 73.96796 | Left ventricle |
| RA_ejectio | Z41_S2 | 27446 | 0.107004 | 0.005856 | 1.39E-74 | 73.8574 | Right atrium |
| RV_end_di | Z60_S2 | 27446 | -0.11377 | 0.006232 | 1.81E-74 | 73.74158 | Right ventricle |
| Ascending_ | Z14_S1 | 24599 | 0.098176 | 0.005379 | 1.95E-74 | 73.70942 | Aorta |
| Ascending_ | Z80_S2 | 24599 | 0.093159 | 0.005104 | 2.04E-74 | 73.6909 | Aorta |
| LV_longitu | Z53_S1 | 27446 | -0.09388 | 0.005145 | 2.22E-74 | 73.65457 | Left ventricle |
| LV_end_sy | Z108_S5 | 27446 | -0.11699 | 0.00642 | 3.51E-74 | 73.45503 | Left ventricle |
| RA_maxim | Z39_S4 | 27446 | -0.09013 | 0.004947 | 3.69E-74 | 73.43331 | Right atrium |
| RV_end_sy | Z97_S1 | 27446 | -0.15151 | 0.008322 | 4.62E-74 | 73.33521 | Right ventricle |
| LV_longitu | Z105_S4 | 27446 | 0.082009 | 0.004507 | 5.72E-74 | 73.24284 | Left ventricle |
| Ascending_ | Z78_S1 | 24599 | 0.095184 | 0.005236 | 7.68E-74 | 73.11491 | Aorta |
| LV_myocar | Z43_S2 | 27446 | -0.1327 | 0.007302 | 8.50E-74 | 73.07067 | Left ventricle |
| LV_end_di | Z24_S5 | 27446 | -0.13429 | 0.007403 | 1.50E-73 | 72.82294 | Left ventricle |
| LA_ejectio | Z83_S2 | 27446 | -0.08279 | 0.004565 | 1.61E-73 | 72.79343 | Left atrium |
| LA_maximu | Z79_S4 | 27446 | -0.11126 | 0.006134 | 1.61E-73 | 72.79266 | Left atrium |
| LV_cardiac | Z127_S5 | 27446 | -0.08534 | 0.004709 | 2.04E-73 | 72.69053 | Left ventricle |
| LV_circumf | Z118_S1 | 27446 | -0.10208 | 0.005632 | 2.04E-73 | 72.69014 | Left ventricle |
| LA_minimu | Z57_S1 | 27446 | -0.08436 | 0.004657 | 2.47E-73 | 72.60677 | Left atrium |
| Descending | Z6_S2 | 24599 | -0.09377 | 0.005181 | 3.25E-73 | 72.48785 | Aorta |
| Descending | Z29_S1 | 24599 | 0.116893 | 0.006463 | 4.14E-73 | 72.38265 | Aorta |
| LA_ejectio | Z46_S1 | 27446 | 0.088525 | 0.004896 | 4.38E-73 | 72.35895 | Left atrium |
| RA_stroke_ | Z80_S1 | 27446 | 0.098668 | 0.005457 | 4.51E-73 | 72.34622 | Right atrium |
| Descending | Z43_S2 | 24599 | -0.10514 | 0.005818 | 5.40E-73 | 72.26799 | Aorta |
| LV_myocar | Z60_S2 | 27446 | -0.13122 | 0.007267 | 6.83E-73 | 72.16527 | Left ventricle |
| LV_cardiac | Z22_S4 | 27446 | 0.10341 | 0.005728 | 7.49E-73 | 72.12531 | Left ventricle |
| LV_stroke_ | Z17_S1 | 27446 | 0.101812 | 0.00564 | 7.50E-73 | 72.12487 | Left ventricle |
| Descending | Z89_S4 | 24599 | -0.10312 | 0.005722 | 1.32E-72 | 71.87856 | Aorta |
| LV_end_sy | Z122_S5 | 27446 | 0.101537 | 0.005639 | 1.74E-72 | 71.76045 | Left ventricle |
| Descending | Z122_S2 | 24599 | -0.09757 | 0.005419 | 1.76E-72 | 71.75466 | Aorta |

| | | | | | | | |
|---|---|---|---|---|---|---|---|
| Ascending_ | Z52_S3 | 24599 | 0.087653 | 0.00487 | 2.01E-72 | 71.69693 | Aorta |
| LV_cardiac | Z118_S1 | 27446 | -0.1107 | 0.006152 | 2.12E-72 | 71.67333 | Left ventricle |
| RA_maxim | Z22_S2 | 27446 | 0.100848 | 0.005608 | 2.60E-72 | 71.58473 | Right atrium |
| RV_end_di | Z75_S1 | 27446 | -0.13232 | 0.007366 | 3.81E-72 | 71.41886 | Right ventricle |
| LV_myocar | Z105_S5 | 27446 | 0.151478 | 0.008437 | 4.47E-72 | 71.34958 | Left ventricle |
| RA_minimu | Z105_S5 | 27446 | 0.101706 | 0.005668 | 5.24E-72 | 71.28063 | Right atrium |
| Descending | Z32_S1 | 24599 | -0.12505 | 0.00697 | 5.57E-72 | 71.25388 | Aorta |
| RV_stroke_ | Z68_S2 | 27446 | -0.09983 | 0.005571 | 8.47E-72 | 71.07212 | Right ventricle |
| Ascending_ | Z12_S5 | 24599 | 0.128339 | 0.007164 | 8.96E-72 | 71.04756 | Aorta |
| Ascending_ | Z127_S4 | 24599 | 0.118957 | 0.006645 | 1.14E-71 | 70.94184 | Aorta |
| RV_end_sy | Z50_S2 | 27446 | 0.149239 | 0.008341 | 1.36E-71 | 70.86491 | Right ventricle |
| RA_minimu | Z46_S1 | 27446 | -0.10132 | 0.005667 | 1.72E-71 | 70.76477 | Right atrium |
| LV_longitu | Z3_S4 | 27446 | -0.1102 | 0.006164 | 1.72E-71 | 70.76453 | Left ventricle |
| Ascending_ | Z29_S5 | 24599 | 0.10955 | 0.006131 | 2.05E-71 | 70.68764 | Aorta |
| RV_end_di | Z80_S2 | 27446 | 0.113769 | 0.00637 | 2.36E-71 | 70.62704 | Right ventricle |
| RA_minimu | Z20_S2 | 27446 | 0.084563 | 0.004735 | 2.47E-71 | 70.60771 | Right atrium |
| Descending | Z24_S5 | 24599 | -0.11581 | 0.006487 | 2.79E-71 | 70.55484 | Aorta |
| LV_end_dia | Z118_S1 | 27446 | 0.134885 | 0.007559 | 3.14E-71 | 70.50294 | Left ventricle |
| LA_minimu | Z55_S1 | 27446 | -0.08353 | 0.004683 | 3.51E-71 | 70.45473 | Left atrium |
| LV_radial_s | Z78_S1 | 27446 | -0.0835 | 0.004681 | 3.52E-71 | 70.45313 | Left ventricle |
| LV_myocar | Z58_S2 | 27446 | 0.139456 | 0.007818 | 3.55E-71 | 70.45031 | Left ventricle |
| Ascending_ | Z35_S5 | 24599 | -0.08766 | 0.004915 | 3.65E-71 | 70.43821 | Aorta |
| RA_stroke_ | Z23_S1 | 27446 | -0.10083 | 0.005653 | 3.74E-71 | 70.42655 | Right atrium |
| LA_ejection | Z122_S1 | 27446 | -0.09445 | 0.005296 | 3.84E-71 | 70.41556 | Left atrium |
| RV_stroke_ | Z18_S4 | 27446 | -0.13632 | 0.007646 | 4.23E-71 | 70.37349 | Right ventricle |
| LA_stroke_ | Z118_S1 | 27446 | 0.10371 | 0.005822 | 5.44E-71 | 70.26427 | Left atrium |
| LV_end_sy | Z7_S3 | 27446 | -0.13286 | 0.007472 | 9.82E-71 | 70.00793 | Left ventricle |
| LA_ejection | Z111_S1 | 27446 | -0.09378 | 0.005277 | 1.21E-70 | 69.91567 | Left atrium |
| Ascending_ | Z127_S5 | 24599 | 0.09114 | 0.005129 | 1.23E-70 | 69.91171 | Aorta |
| Ascending_ | Z58_S2 | 24599 | 0.095244 | 0.005369 | 2.02E-70 | 69.69517 | Aorta |
| LV_stroke_ | Z6_S2 | 27446 | -0.08318 | 0.004694 | 2.97E-70 | 69.5273 | Left ventricle |
| LV_radial_s | Z20_S2 | 27446 | -0.07688 | 0.00434 | 3.28E-70 | 69.48374 | Left ventricle |
| LA_ejection | Z51_S4 | 27446 | 0.07416 | 0.004194 | 5.65E-70 | 69.24773 | Left atrium |
| LV_stroke_ | Z37_S1 | 27446 | 0.12015 | 0.006795 | 5.77E-70 | 69.23917 | Left ventricle |
| LV_end_sy | Z99_S4 | 27446 | 0.123839 | 0.007006 | 6.28E-70 | 69.20204 | Left ventricle |
| Descending | Z60_S2 | 24599 | -0.10079 | 0.005704 | 7.21E-70 | 69.142 | Aorta |
| Descending | Z40_S5 | 24599 | -0.08945 | 0.005062 | 7.25E-70 | 69.13962 | Aorta |
| Ascending_ | Z46_S1 | 24599 | -0.10223 | 0.00579 | 8.83E-70 | 69.05426 | Aorta |
| Descending | Z75_S1 | 24599 | -0.12027 | 0.006822 | 1.46E-69 | 68.83611 | Aorta |
| RV_end_di | Z74_S2 | 27446 | 0.124577 | 0.007068 | 1.55E-69 | 68.81052 | Right ventricle |
| Descending | Z83_S2 | 24599 | 0.110401 | 0.006268 | 1.98E-69 | 68.70246 | Aorta |
| LV_stroke_ | Z75_S1 | 27446 | -0.1095 | 0.006217 | 2.00E-69 | 68.69881 | Left ventricle |
| Ascending_ | Z58_S2 | 24599 | 0.094159 | 0.005349 | 2.39E-69 | 68.62181 | Aorta |
| LA_minimu | Z79_S4 | 27446 | -0.1047 | 0.00595 | 2.68E-69 | 68.57245 | Left atrium |
| RV_ejectio | Z32_S1 | 27446 | 0.094993 | 0.005401 | 3.05E-69 | 68.51503 | Right ventricle |

| | | | | | | | |
|---|---|---|---|---|---|---|---|
| LA_ejectio | Z68_S2 | 27446 | 0.077328 | 0.004404 | 5.05E-69 | 68.29668 | Left atrium |
| Ascending_ | Z52_S3 | 24599 | 0.085848 | 0.004889 | 5.19E-69 | 68.28467 | Aorta |
| Ascending_ | Z80_S2 | 24599 | 0.089337 | 0.005088 | 5.23E-69 | 68.28185 | Aorta |
| RA_ejectio | Z40_S5 | 27446 | 0.074442 | 0.004241 | 5.62E-69 | 68.25036 | Right atrium |
| RA_maxim | Z74_S4 | 27446 | -0.10178 | 0.0058 | 6.25E-69 | 68.2044 | Right atrium |
| RV_stroke_ | Z23_S3 | 27446 | 0.095354 | 0.005435 | 6.41E-69 | 68.19346 | Right ventricle |
| Descending | Z75_S1 | 24599 | -0.11792 | 0.006725 | 7.87E-69 | 68.10394 | Aorta |
| Descending | Z39_S1 | 24599 | 0.106009 | 0.006048 | 8.97E-69 | 68.0471 | Aorta |
| RA_ejectio | Z99_S3 | 27446 | -0.07008 | 0.003999 | 9.03E-69 | 68.04411 | Right atrium |
| LV_cardiac | Z53_S1 | 27446 | -0.10085 | 0.005757 | 1.01E-68 | 67.99542 | Left ventricle |
| Ascending_ | Z69_S5 | 24599 | -0.08507 | 0.004856 | 1.01E-68 | 67.99509 | Aorta |
| RA_minimu | Z10_S4 | 27446 | -0.10234 | 0.005844 | 1.16E-68 | 67.93581 | Right atrium |
| RA_minimu | Z1_S2 | 27446 | 0.080309 | 0.004587 | 1.22E-68 | 67.91255 | Right atrium |
| RV_ejectio | Z80_S1 | 27446 | 0.096491 | 0.005515 | 1.54E-68 | 67.81131 | Right ventricle |
| LV_stroke_ | Z41_S1 | 27446 | 0.114063 | 0.006521 | 1.66E-68 | 67.77905 | Left ventricle |
| Ascending_ | Z55_S1 | 24599 | -0.09256 | 0.005296 | 2.19E-68 | 67.66 | Aorta |
| LV_end_sy | Z111_S1 | 27446 | 0.119519 | 0.00684 | 2.26E-68 | 67.64634 | Left ventricle |
| Ascending_ | Z94_S5 | 24599 | 0.090829 | 0.005199 | 2.41E-68 | 67.61774 | Aorta |
| LA_maximu | Z27_S2 | 27446 | -0.10583 | 0.006062 | 3.02E-68 | 67.52057 | Left atrium |
| LV_cardiac | Z37_S4 | 27446 | -0.11956 | 0.006851 | 3.43E-68 | 67.46487 | Left ventricle |
| RA_maxim | Z37_S3 | 27446 | -0.10972 | 0.006291 | 4.02E-68 | 67.3955 | Right atrium |
| LA_maximu | Z75_S5 | 27446 | 0.114054 | 0.006542 | 4.51E-68 | 67.34545 | Left atrium |
| LV_end_sy | Z26_S4 | 27446 | 0.135449 | 0.007774 | 5.48E-68 | 67.26085 | Left ventricle |
| RV_end_sy | Z11_S5 | 27446 | -0.11422 | 0.00656 | 6.90E-68 | 67.16135 | Right ventricle |
| LA_ejectio | Z30_S2 | 27446 | 0.102184 | 0.00587 | 7.30E-68 | 67.13654 | Left atrium |
| Ascending_ | Z5_S5 | 24599 | 0.109102 | 0.006269 | 7.87E-68 | 67.10416 | Aorta |
| LA_minimu | Z75_S5 | 27446 | 0.110382 | 0.006343 | 8.10E-68 | 67.09132 | Left atrium |
| Ascending_ | Z75_S4 | 24599 | -0.1184 | 0.006808 | 9.31E-68 | 67.03114 | Aorta |
| Ascending_ | Z89_S4 | 24599 | -0.09599 | 0.005521 | 1.04E-67 | 66.98243 | Aorta |
| LA_ejectio | Z35_S5 | 27446 | 0.072115 | 0.004151 | 1.28E-67 | 66.89127 | Left atrium |
| LV_stroke_ | Z81_S1 | 27446 | -0.10623 | 0.006119 | 1.66E-67 | 66.78029 | Left ventricle |
| LA_stroke_ | Z81_S1 | 27446 | -0.09212 | 0.005307 | 1.68E-67 | 66.77512 | Left atrium |
| RA_minimu | Z78_S4 | 27446 | 0.0862 | 0.004967 | 1.78E-67 | 66.74847 | Right atrium |
| RV_end_di | Z21_S3 | 27446 | -0.15546 | 0.008961 | 2.01E-67 | 66.69603 | Right ventricle |
| Descending | Z7_S3 | 24599 | -0.13512 | 0.00779 | 2.20E-67 | 66.658 | Aorta |
| LV_longitu | Z107_S1 | 27446 | 0.069085 | 0.003984 | 2.32E-67 | 66.63378 | Left ventricle |
| RV_stroke_ | Z74_S4 | 27446 | -0.11141 | 0.006428 | 2.64E-67 | 66.57784 | Right ventricle |
| LV_radial_ | Z55_S5 | 27446 | -0.07593 | 0.004383 | 3.00E-67 | 66.52289 | Left ventricle |
| RA_maxim | Z78_S1 | 27446 | -0.08733 | 0.005041 | 3.07E-67 | 66.51227 | Right atrium |
| LV_radial_ | Z31_S5 | 27446 | -0.07156 | 0.004131 | 3.27E-67 | 66.48512 | Left ventricle |
| RV_stroke_ | Z120_S2 | 27446 | 0.091071 | 0.005258 | 3.31E-67 | 66.48068 | Right ventricle |
| RV_end_sy | Z6_S2 | 27446 | -0.09173 | 0.005306 | 5.83E-67 | 66.23434 | Right ventricle |
| RA_minimu | Z79_S4 | 27446 | -0.11405 | 0.006602 | 7.33E-67 | 66.13518 | Right atrium |
| RA_maxim | Z41_S2 | 27446 | -0.11281 | 0.006535 | 9.11E-67 | 66.04047 | Right atrium |
| LV_circumf | Z37_S4 | 27446 | -0.10827 | 0.006274 | 9.72E-67 | 66.01236 | Left ventricle |

| | | | | | | | |
|---|---|---|---|---|---|---|---|
| LA_stroke_ | Z63_S1 | 27446 | -0.10735 | 0.006221 | 1.01E-66 | 65.9943 | Left atrium |
| LV_end_sy | Z81_S1 | 27446 | -0.10959 | 0.00636 | 1.54E-66 | 65.81183 | Left ventricle |
| LA_maximu | Z81_S1 | 27446 | -0.09095 | 0.005281 | 1.78E-66 | 65.75069 | Left atrium |
| RV_end_sy | Z63_S1 | 27446 | -0.13955 | 0.008106 | 2.03E-66 | 65.69254 | Right ventricle |
| RV_stroke_ | Z1_S5 | 27446 | 0.107612 | 0.006252 | 2.11E-66 | 65.67544 | Right ventricle |
| LA_stroke_ | Z31_S2 | 27446 | 0.090571 | 0.005265 | 2.55E-66 | 65.59429 | Left atrium |
| RV_stroke_ | Z43_S2 | 27446 | -0.09229 | 0.005366 | 2.74E-66 | 65.56239 | Right ventricle |
| LV_end_sy | Z96_S3 | 27446 | 0.105006 | 0.006107 | 2.99E-66 | 65.52482 | Left ventricle |
| RA_ejectio | Z53_S1 | 27446 | -0.0886 | 0.005159 | 4.08E-66 | 65.38903 | Right atrium |
| LA_ejectio | Z47_S4 | 27446 | 0.083004 | 0.004835 | 4.65E-66 | 65.3327 | Left atrium |
| RV_end_di | Z74_S4 | 27446 | -0.12885 | 0.007507 | 4.97E-66 | 65.30379 | Right ventricle |
| RV_end_di | Z13_S3 | 27446 | 0.100075 | 0.005833 | 5.72E-66 | 65.24236 | Right ventricle |
| Descending | Z22_S2 | 24599 | -0.11406 | 0.00665 | 6.12E-66 | 65.21308 | Aorta |
| Descending | Z75_S4 | 24599 | -0.13308 | 0.007771 | 9.62E-66 | 65.01681 | Aorta |
| Descending | Z12_S4 | 24599 | -0.08673 | 0.005065 | 9.71E-66 | 65.013 | Aorta |
| LV_myocar | Z79_S4 | 27446 | -0.16832 | 0.00983 | 9.77E-66 | 65.00991 | Left ventricle |
| LV_cardiac | Z56_S5 | 27446 | 0.080785 | 0.004719 | 1.04E-65 | 64.98156 | Left ventricle |
| RA_minimu | Z62_S2 | 27446 | 0.084857 | 0.004964 | 1.61E-65 | 64.79242 | Right atrium |
| RA_maxim | Z87_S3 | 27446 | -0.09199 | 0.005388 | 2.37E-65 | 64.62491 | Right atrium |
| LV_radial_ | Z117_S4 | 27446 | 0.071396 | 0.004183 | 2.57E-65 | 64.58997 | Left ventricle |
| LA_minimu | Z74_S4 | 27446 | -0.09037 | 0.005301 | 3.61E-65 | 64.44308 | Left atrium |
| LA_stroke_ | Z93_S2 | 27446 | 0.109515 | 0.006424 | 3.63E-65 | 64.44034 | Left atrium |
| RV_end_di | Z10_S4 | 27446 | -0.12715 | 0.007463 | 4.45E-65 | 64.35203 | Right ventricle |
| Ascending_ | Z1_S1 | 24599 | -0.09463 | 0.005558 | 5.16E-65 | 64.28745 | Aorta |
| LV_end_di | Z1_S5 | 27446 | 0.118077 | 0.006937 | 5.80E-65 | 64.23635 | Left ventricle |
| LV_radial_ | Z106_S4 | 27446 | -0.07683 | 0.004518 | 7.84E-65 | 64.10542 | Left ventricle |
| LV_radial_ | Z120_S2 | 27446 | -0.07487 | 0.004409 | 1.11E-64 | 63.95566 | Left ventricle |
| RA_maxim | Z37_S4 | 27446 | 0.116237 | 0.006848 | 1.30E-64 | 63.88766 | Right atrium |
| RV_stroke_ | Z37_S1 | 27446 | 0.117085 | 0.006898 | 1.31E-64 | 63.88369 | Right ventricle |
| LV_end_sy | Z50_S2 | 27446 | 0.130245 | 0.007675 | 1.38E-64 | 63.86105 | Left ventricle |
| LA_maximu | Z76_S3 | 27446 | -0.09309 | 0.005486 | 1.43E-64 | 63.84445 | Left atrium |
| LV_cardiac | Z13_S1 | 27446 | -0.08401 | 0.004954 | 1.66E-64 | 63.77902 | Left ventricle |
| Descending | Z122_S2 | 24599 | -0.09056 | 0.005346 | 2.25E-64 | 63.6469 | Aorta |
| LV_myocar | Z6_S2 | 27446 | -0.10986 | 0.00649 | 2.76E-64 | 63.55912 | Left ventricle |
| LV_radial_ | Z6_S2 | 27446 | 0.067612 | 0.003995 | 3.08E-64 | 63.51159 | Left ventricle |
| LV_longitu | Z5_S1 | 27446 | 0.082523 | 0.00488 | 3.72E-64 | 63.42935 | Left ventricle |
| LV_longitu | Z32_S1 | 27446 | -0.08883 | 0.005254 | 3.91E-64 | 63.40794 | Left ventricle |
| LA_stroke_ | Z11_S5 | 27446 | -0.08513 | 0.005036 | 4.23E-64 | 63.37413 | Left atrium |
| LV_end_sy | Z59_S2 | 27446 | 0.12108 | 0.007164 | 4.47E-64 | 63.34998 | Left ventricle |
| RV_stroke_ | Z51_S1 | 27446 | -0.09088 | 0.005378 | 4.65E-64 | 63.33259 | Right ventricle |
| LV_stroke_ | Z57_S2 | 27446 | 0.084685 | 0.005013 | 5.15E-64 | 63.2878 | Left ventricle |
| Descending | Z82_S4 | 24599 | -0.08636 | 0.005116 | 6.28E-64 | 63.20208 | Aorta |
| RV_end_di | Z122_S5 | 27446 | 0.108628 | 0.006435 | 6.32E-64 | 63.19929 | Right ventricle |
| LV_cardiac | Z63_S1 | 27446 | 0.110979 | 0.006576 | 6.71E-64 | 63.1731 | Left ventricle |
| LV_end_sy | Z77_S3 | 27446 | -0.10788 | 0.006396 | 8.05E-64 | 63.09432 | Left ventricle |

| | | | | | | | |
|---|---|---|---|---|---|---|---|
| LV_radial_s | Z75_S1 | 27446 | 0.089235 | 0.005291 | 8.25E-64 | 63.0834 | Left ventricle |
| LV_cardiac_ | Z74_S4 | 27446 | 0.097878 | 0.005807 | 9.58E-64 | 63.01854 | Left ventricle |
| LV_longitu | Z28_S2 | 27446 | 0.08877 | 0.005272 | 1.29E-63 | 62.88911 | Left ventricle |
| LA_stroke_ | Z1_S5 | 27446 | 0.089957 | 0.005344 | 1.37E-63 | 62.86287 | Left atrium |
| LA_ejectio | Z1_S5 | 27446 | -0.08319 | 0.004942 | 1.38E-63 | 62.86067 | Left atrium |
| RV_end_sy | Z60_S2 | 27446 | -0.10011 | 0.005947 | 1.39E-63 | 62.85689 | Right ventricle |
| Descending | Z60_S2 | 24599 | -0.09745 | 0.00579 | 1.45E-63 | 62.83847 | Aorta |
| Ascending_ | Z62_S2 | 24599 | 0.085016 | 0.005053 | 1.58E-63 | 62.80003 | Aorta |
| Descending | Z111_S1 | 24599 | 0.109141 | 0.006493 | 2.11E-63 | 62.67618 | Aorta |
| RV_stroke_ | Z37_S4 | 27446 | 0.127534 | 0.007589 | 2.24E-63 | 62.64936 | Right ventricle |
| LV_myocar | Z97_S1 | 27446 | -0.17113 | 0.010185 | 2.38E-63 | 62.62363 | Left ventricle |
| LV_myocar | Z74_S2 | 27446 | 0.138479 | 0.008245 | 2.61E-63 | 62.58346 | Left ventricle |
| LV_circum | Z18_S4 | 27446 | 0.106227 | 0.006327 | 2.85E-63 | 62.54535 | Left ventricle |
| RV_end_sy | Z59_S2 | 27446 | 0.130812 | 0.007791 | 2.88E-63 | 62.54041 | Right ventricle |
| LV_cardiac_ | Z12_S4 | 27446 | 0.07935 | 0.004727 | 3.00E-63 | 62.52326 | Left ventricle |
| RV_end_sy | Z39_S1 | 27446 | 0.105745 | 0.006303 | 3.55E-63 | 62.44932 | Right ventricle |
| RA_stroke_ | Z39_S4 | 27446 | -0.07566 | 0.004511 | 3.87E-63 | 62.41217 | Right atrium |
| LV_myocar | Z31_S2 | 27446 | 0.140698 | 0.008391 | 4.25E-63 | 62.37191 | Left ventricle |
| LV_longitu | Z51_S4 | 27446 | 0.071883 | 0.004289 | 4.65E-63 | 62.33222 | Left ventricle |
| RV_end_di | Z64_S1 | 27446 | -0.13318 | 0.007949 | 5.17E-63 | 62.28682 | Right ventricle |
| LA_minimu | Z27_S2 | 27446 | -0.09847 | 0.005881 | 6.31E-63 | 62.20015 | Left atrium |
| RA_maxim | Z50_S4 | 27446 | 0.08956 | 0.005351 | 6.96E-63 | 62.1575 | Right atrium |
| LV_myocar | Z19_S2 | 27446 | 0.137758 | 0.008232 | 7.37E-63 | 62.13239 | Left ventricle |
| LA_ejectio | Z12_S4 | 27446 | 0.069163 | 0.004136 | 9.01E-63 | 62.04521 | Left atrium |
| Descending | Z37_S4 | 24599 | 0.122677 | 0.007336 | 9.09E-63 | 62.04158 | Aorta |
| Ascending_ | Z56_S5 | 24599 | 0.081931 | 0.004901 | 9.87E-63 | 62.00569 | Aorta |
| RV_stroke_ | Z65_S2 | 27446 | 0.10532 | 0.006301 | 1.02E-62 | 61.99099 | Right ventricle |
| LA_minimu | Z23_S3 | 27446 | 0.074928 | 0.004483 | 1.05E-62 | 61.97838 | Left atrium |
| LV_stroke_ | Z64_S1 | 27446 | -0.11198 | 0.006707 | 1.41E-62 | 61.85076 | Left ventricle |
| RV_end_sy | Z122_S5 | 27446 | 0.102403 | 0.006137 | 1.64E-62 | 61.78444 | Right ventricle |
| RV_end_di | Z97_S1 | 27446 | -0.14577 | 0.008736 | 1.66E-62 | 61.78091 | Right ventricle |
| Ascending_ | Z97_S1 | 24599 | -0.11663 | 0.00699 | 1.69E-62 | 61.7718 | Aorta |
| LA_ejectio | Z111_S5 | 27446 | -0.08004 | 0.004801 | 2.20E-62 | 61.65834 | Left atrium |
| Ascending_ | Z122_S2 | 24599 | -0.07808 | 0.004685 | 2.29E-62 | 61.63932 | Aorta |
| RA_stroke_ | Z5_S1 | 27446 | -0.08279 | 0.004968 | 2.42E-62 | 61.61614 | Right atrium |
| LA_ejectio | Z14_S4 | 27446 | -0.09001 | 0.005402 | 2.46E-62 | 61.60974 | Left atrium |
| LV_stroke_ | Z49_S1 | 27446 | -0.12289 | 0.007376 | 2.49E-62 | 61.60377 | Left ventricle |
| Descending | Z78_S1 | 24599 | 0.101059 | 0.006069 | 2.90E-62 | 61.53692 | Aorta |
| LV_longitu | Z50_S5 | 27446 | -0.10233 | 0.006146 | 2.97E-62 | 61.52757 | Left ventricle |
| LV_myocar | Z74_S4 | 27446 | -0.14568 | 0.008756 | 3.68E-62 | 61.43369 | Left ventricle |
| RV_stroke_ | Z39_S1 | 27446 | 0.094169 | 0.005661 | 3.81E-62 | 61.41872 | Right ventricle |
| LV_cardiac_ | Z44_S1 | 27446 | 0.091817 | 0.005519 | 3.84E-62 | 61.41541 | Left ventricle |
| Ascending_ | Z87_S3 | 24599 | -0.09377 | 0.005641 | 4.84E-62 | 61.31545 | Aorta |
| Ascending_ | Z15_S2 | 24599 | 0.101625 | 0.006127 | 8.64E-62 | 61.06331 | Aorta |
| Ascending_ | Z44_S4 | 24599 | -0.08033 | 0.004844 | 9.32E-62 | 61.03041 | Aorta |

| | | | | | | | |
|---|---|---|---|---|---|---|---|
| LA_ejectioı | Z59_S3 | 27446 | 0.06509 | 0.003926 | 9.64E-62 | 61.01601 | Left atrium |
| LV_radial_ | Z56_S2 | 27446 | 0.073135 | 0.004411 | 9.99E-62 | 61.00054 | Left ventricle |
| Ascending_ | Z111_S1 | 24599 | 0.10964 | 0.006615 | 1.05E-61 | 60.97688 | Aorta |
| Descending | Z105_S5 | 24599 | 0.10949 | 0.006609 | 1.20E-61 | 60.92233 | Aorta |
| Descending | Z81_S1 | 24599 | -0.10995 | 0.006643 | 1.56E-61 | 60.80743 | Aorta |
| LV_end_sy | Z66_S2 | 27446 | 0.118898 | 0.00719 | 2.01E-61 | 60.69629 | Left ventricle |
| LV_circum | Z99_S4 | 27446 | 0.093601 | 0.005661 | 2.06E-61 | 60.68573 | Left ventricle |
| LV_cardiac | Z32_S1 | 27446 | -0.09705 | 0.005876 | 2.86E-61 | 60.54437 | Left ventricle |
| LA_ejectioı | Z89_S4 | 27446 | 0.077314 | 0.004682 | 2.97E-61 | 60.52699 | Left atrium |
| RV_end_sy | Z81_S1 | 27446 | -0.11417 | 0.006919 | 3.57E-61 | 60.44759 | Right ventricle |
| Ascending_ | Z111_S5 | 24599 | 0.098785 | 0.005987 | 3.74E-61 | 60.42678 | Aorta |
| Ascending_ | Z122_S2 | 24599 | -0.077 | 0.004668 | 4.09E-61 | 60.38871 | Aorta |
| LA_minimu | Z63_S1 | 27446 | -0.09902 | 0.006005 | 4.27E-61 | 60.36934 | Left atrium |
| Ascending_ | Z80_S4 | 24599 | -0.08133 | 0.004934 | 4.74E-61 | 60.32412 | Aorta |
| LV_end_sy | Z114_S4 | 27446 | 0.100652 | 0.006106 | 4.81E-61 | 60.31809 | Left ventricle |
| LA_stroke_ | Z108_S5 | 27446 | -0.08838 | 0.005364 | 5.27E-61 | 60.27847 | Left atrium |
| Ascending_ | Z17_S1 | 24599 | 0.087955 | 0.005339 | 5.74E-61 | 60.24116 | Aorta |
| LV_radial_ | Z35_S5 | 27446 | 0.07257 | 0.004405 | 5.75E-61 | 60.24064 | Left ventricle |
| Descending | Z87_S3 | 24599 | -0.10605 | 0.006439 | 5.99E-61 | 60.22287 | Aorta |
| LV_stroke_ | Z7_S3 | 27446 | -0.11841 | 0.007196 | 7.79E-61 | 60.10842 | Left ventricle |
| Descending | Z46_S1 | 24599 | -0.11036 | 0.006709 | 8.26E-61 | 60.08313 | Aorta |
| Descending | Z50_S3 | 24599 | -0.10219 | 0.006216 | 9.86E-61 | 60.00617 | Aorta |
| RV_end_sy | Z65_S1 | 27446 | -0.09307 | 0.005662 | 1.01E-60 | 59.99759 | Right ventricle |
| RA_ejectio | Z14_S4 | 27446 | -0.09086 | 0.005532 | 1.29E-60 | 59.88995 | Right atrium |
| LA_maximu | Z11_S5 | 27446 | -0.0823 | 0.005013 | 1.42E-60 | 59.84736 | Left atrium |
| Ascending_ | Z127_S5 | 24599 | 0.080095 | 0.004879 | 1.43E-60 | 59.84526 | Aorta |
| LV_end_di | Z21_S3 | 27446 | -0.1398 | 0.008518 | 1.57E-60 | 59.80354 | Left ventricle |
| Ascending_ | Z31_S5 | 24599 | 0.075988 | 0.004631 | 1.65E-60 | 59.78207 | Aorta |
| Descending | Z29_S5 | 24599 | 0.116546 | 0.007105 | 1.81E-60 | 59.74167 | Aorta |
| RV_stroke_ | Z99_S5 | 27446 | 0.096345 | 0.005884 | 2.90E-60 | 59.5382 | Right ventricle |
| LV_end_di | Z97_S1 | 27446 | -0.1357 | 0.008302 | 4.71E-60 | 59.32742 | Left ventricle |
| Ascending_ | Z31_S5 | 24599 | 0.079948 | 0.004891 | 4.73E-60 | 59.32504 | Aorta |
| Ascending_ | Z7_S1 | 24599 | -0.08575 | 0.005251 | 6.13E-60 | 59.21277 | Aorta |
| LV_radial_ | Z95_S3 | 27446 | 0.064492 | 0.003957 | 1.05E-59 | 58.97928 | Left ventricle |
| RV_stroke_ | Z87_S3 | 27446 | -0.09732 | 0.005973 | 1.11E-59 | 58.95384 | Right ventricle |
| RA_minimu | Z19_S2 | 27446 | 0.090103 | 0.005531 | 1.17E-59 | 58.93005 | Right atrium |
| RV_stroke_ | Z57_S1 | 27446 | -0.09204 | 0.005655 | 1.48E-59 | 58.82888 | Right ventricle |
| LV_circum | Z56_S5 | 27446 | 0.070269 | 0.004322 | 2.00E-59 | 58.69822 | Left ventricle |
| LA_ejectioı | Z123_S4 | 27446 | -0.08244 | 0.005075 | 2.39E-59 | 58.62159 | Left atrium |
| Descending | Z74_S4 | 24599 | -0.11189 | 0.006888 | 2.42E-59 | 58.61559 | Aorta |
| RA_ejectio | Z120_S5 | 27446 | 0.097265 | 0.005988 | 2.48E-59 | 58.60507 | Right atrium |
| LA_minimu | Z18_S4 | 27446 | -0.10249 | 0.00631 | 2.57E-59 | 58.59089 | Left atrium |
| Ascending_ | Z55_S1 | 24599 | -0.08641 | 0.00532 | 2.57E-59 | 58.58995 | Aorta |
| RV_stroke_ | Z58_S2 | 27446 | 0.093269 | 0.005747 | 3.14E-59 | 58.50306 | Right ventricle |
| RV_stroke_ | Z31_S2 | 27446 | 0.100045 | 0.006165 | 3.15E-59 | 58.50129 | Right ventricle |

| | | | | | | | |
|---|---|---|---|---|---|---|---|
| LA_stroke_ | Z80_S1 | 27446 | 0.092118 | 0.005678 | 3.50E-59 | 58.4558 | Left atrium |
| Descending | Z112_S1 | 24599 | -0.11958 | 0.007372 | 3.53E-59 | 58.45257 | Aorta |
| RV_stroke_ | Z66_S2 | 27446 | 0.113874 | 0.007023 | 3.94E-59 | 58.40415 | Right ventricle |
| Descending | Z111_S5 | 24599 | 0.106787 | 0.006588 | 4.41E-59 | 58.35528 | Aorta |
| LV_stroke_ | Z9_S2 | 27446 | 0.104634 | 0.006456 | 4.47E-59 | 58.34924 | Left ventricle |
| RA_ejectio | Z27_S2 | 27446 | 0.093558 | 0.005774 | 4.71E-59 | 58.32679 | Right atrium |
| LA_ejectio | Z120_S5 | 27446 | 0.094755 | 0.005848 | 4.76E-59 | 58.32223 | Left atrium |
| RA_minimu | Z65_S5 | 27446 | -0.10818 | 0.006677 | 4.96E-59 | 58.30429 | Right atrium |
| Descending | Z80_S1 | 24599 | -0.11511 | 0.007106 | 5.12E-59 | 58.29099 | Aorta |
| RA_ejectio | Z89_S4 | 27446 | 0.077673 | 0.004795 | 5.24E-59 | 58.28091 | Right atrium |
| LA_ejectio | Z66_S3 | 27446 | 0.088913 | 0.005494 | 6.44E-59 | 58.19083 | Left atrium |
| LV_longitu | Z7_S1 | 27446 | 0.073531 | 0.004544 | 6.74E-59 | 58.17144 | Left ventricle |
| Ascending_ | Z17_S1 | 24599 | 0.086705 | 0.00536 | 7.39E-59 | 58.13147 | Aorta |
| LV_circumf | Z82_S1 | 27446 | 0.10127 | 0.006264 | 8.67E-59 | 58.0622 | Left ventricle |
| Ascending_ | Z2_S1 | 24599 | -0.08427 | 0.005214 | 9.25E-59 | 58.03393 | Aorta |
| RA_minimu | Z52_S4 | 27446 | 0.081028 | 0.005015 | 1.01E-58 | 57.99699 | Right atrium |
| LA_minimu | Z115_S4 | 27446 | 0.074824 | 0.004633 | 1.10E-58 | 57.95865 | Left atrium |
| LA_minimu | Z108_S5 | 27446 | -0.08358 | 0.005176 | 1.19E-58 | 57.92412 | Left atrium |
| LV_stroke_ | Z19_S2 | 27446 | 0.09618 | 0.00596 | 1.38E-58 | 57.85959 | Left ventricle |
| RV_end_sy | Z75_S1 | 27446 | -0.11342 | 0.007032 | 1.60E-58 | 57.79709 | Right ventricle |
| LA_minimu | Z74_S2 | 27446 | 0.080479 | 0.004994 | 2.04E-58 | 57.69049 | Left atrium |
| LA_ejectio | Z53_S1 | 27446 | -0.08106 | 0.005041 | 3.62E-58 | 57.44116 | Left atrium |
| RA_maxim | Z64_S1 | 27446 | -0.09867 | 0.006145 | 5.04E-58 | 57.2978 | Right atrium |
| RV_stroke_ | Z80_S1 | 27446 | 0.106682 | 0.006645 | 5.42E-58 | 57.26638 | Right ventricle |
| Ascending_ | Z125_S4 | 24599 | -0.10082 | 0.006282 | 5.73E-58 | 57.24174 | Aorta |
| RA_minimu | Z112_S1 | 27446 | -0.11188 | 0.006981 | 8.31E-58 | 57.08027 | Right atrium |
| RA_minimu | Z120_S4 | 27446 | -0.08892 | 0.005557 | 1.24E-57 | 56.90778 | Right atrium |
| RV_end_di | Z13_S1 | 27446 | -0.10248 | 0.00641 | 1.53E-57 | 56.8148 | Right ventricle |
| RV_stroke_ | Z93_S2 | 27446 | 0.120265 | 0.007522 | 1.54E-57 | 56.81294 | Right ventricle |
| Ascending_ | Z37_S4 | 24599 | 0.119531 | 0.007476 | 1.54E-57 | 56.81126 | Aorta |
| RV_end_di | Z113_S2 | 27446 | 0.137743 | 0.008619 | 1.74E-57 | 56.75991 | Right ventricle |
| Descending | Z74_S2 | 24599 | 0.103194 | 0.00646 | 1.93E-57 | 56.71338 | Aorta |
| RV_end_sy | Z87_S3 | 27446 | -0.10625 | 0.006652 | 1.99E-57 | 56.70028 | Right ventricle |
| Ascending_ | Z69_S5 | 24599 | -0.07783 | 0.004879 | 2.67E-57 | 56.57303 | Aorta |
| LV_myocar | Z73_S1 | 27446 | -0.12477 | 0.007823 | 2.88E-57 | 56.54083 | Left ventricle |
| LV_cardiac | Z7_S5 | 27446 | -0.09651 | 0.006052 | 2.93E-57 | 56.53276 | Left ventricle |
| LV_longitu | Z52_S4 | 27446 | -0.07059 | 0.004429 | 3.38E-57 | 56.47152 | Left ventricle |
| LV_stroke_ | Z74_S2 | 27446 | 0.095129 | 0.00597 | 3.64E-57 | 56.43847 | Left ventricle |
| LV_myocar | Z27_S2 | 27446 | -0.15476 | 0.009717 | 4.09E-57 | 56.38824 | Left ventricle |
| RV_end_sy | Z44_S3 | 27446 | -0.12 | 0.007537 | 4.47E-57 | 56.34964 | Right ventricle |
| LA_stroke_ | Z76_S3 | 27446 | -0.08782 | 0.005517 | 4.75E-57 | 56.32367 | Left atrium |
| RV_end_di | Z44_S3 | 27446 | -0.12581 | 0.007904 | 4.86E-57 | 56.31359 | Right ventricle |
| LV_radial_ | Z65_S1 | 27446 | 0.067824 | 0.004263 | 5.57E-57 | 56.25407 | Left ventricle |
| RV_stroke_ | Z41_S1 | 27446 | 0.105361 | 0.006624 | 5.67E-57 | 56.24624 | Right ventricle |
| LV_ejectio | Z78_S1 | 27446 | -0.07235 | 0.004549 | 6.00E-57 | 56.22207 | Left ventricle |

| | | | | | | | |
|---|---|---|---|---|---|---|---|
| LA_minimu | Z17_S1 | 27446 | 0.075109 | 0.004725 | 6.68E-57 | 56.17533 | Left atrium |
| LA_maximu | Z80_S1 | 27446 | 0.089822 | 0.005651 | 6.94E-57 | 56.15841 | Left atrium |
| LV_stroke_ | Z60_S2 | 27446 | -0.08364 | 0.005266 | 8.44E-57 | 56.07389 | Left ventricle |
| Descending | Z92_S5 | 24599 | 0.125855 | 0.007925 | 8.49E-57 | 56.07106 | Aorta |
| RV_stroke_ | Z50_S2 | 27446 | 0.119026 | 0.0075 | 1.02E-56 | 55.99353 | Right ventricle |
| RV_end_di | Z3_S2 | 27446 | 0.101302 | 0.006383 | 1.02E-56 | 55.9925 | Right ventricle |
| Descending | Z39_S1 | 24599 | 0.097482 | 0.006143 | 1.02E-56 | 55.98951 | Aorta |
| Descending | Z99_S5 | 24599 | 0.099839 | 0.006291 | 1.04E-56 | 55.98347 | Aorta |
| LV_longitu | Z78_S4 | 27446 | -0.06965 | 0.004389 | 1.07E-56 | 55.97116 | Left ventricle |
| RA_stroke_ | Z109_S1 | 27446 | -0.08015 | 0.005053 | 1.15E-56 | 55.9401 | Right atrium |
| Descending | Z61_S4 | 24599 | -0.0997 | 0.00629 | 1.43E-56 | 55.84595 | Aorta |
| Ascending_ | Z122_S5 | 24599 | 0.081772 | 0.005163 | 1.70E-56 | 55.76999 | Aorta |
| RV_end_sy | Z74_S2 | 27446 | 0.10681 | 0.006746 | 1.87E-56 | 55.72931 | Right ventricle |
| Descending | Z93_S2 | 24599 | 0.127017 | 0.008026 | 2.05E-56 | 55.68737 | Aorta |
| LV_radial_ | Z49_S1 | 27446 | -0.09933 | 0.006277 | 2.11E-56 | 55.67573 | Left ventricle |
| Ascending_ | Z78_S1 | 24599 | 0.082653 | 0.005225 | 2.36E-56 | 55.62625 | Aorta |
| Descending | Z122_S5 | 24599 | 0.094492 | 0.005978 | 2.76E-56 | 55.55912 | Aorta |
| Ascending_ | Z40_S5 | 24599 | -0.07755 | 0.004906 | 2.78E-56 | 55.55655 | Aorta |
| LA_minimu | Z92_S1 | 27446 | -0.07937 | 0.005025 | 3.43E-56 | 55.46524 | Left atrium |
| Descending | Z50_S5 | 24599 | 0.116194 | 0.007361 | 3.92E-56 | 55.40648 | Aorta |
| LV_radial_ | Z62_S2 | 27446 | -0.07186 | 0.004553 | 4.00E-56 | 55.3975 | Left ventricle |
| Descending | Z56_S5 | 24599 | 0.08829 | 0.005597 | 4.66E-56 | 55.33167 | Aorta |
| LV_end_di | Z9_S2 | 27446 | 0.114643 | 0.007271 | 5.20E-56 | 55.28386 | Left ventricle |
| RV_stroke_ | Z37_S2 | 27446 | -0.10488 | 0.006654 | 5.72E-56 | 55.24261 | Right ventricle |
| LV_radial_ | Z80_S4 | 27446 | 0.069794 | 0.004434 | 7.88E-56 | 55.1033 | Left ventricle |
| RV_stroke_ | Z14_S2 | 27446 | -0.09619 | 0.006113 | 8.56E-56 | 55.06757 | Right ventricle |
| RA_ejectio | Z68_S2 | 27446 | 0.071027 | 0.004514 | 8.92E-56 | 55.04941 | Right atrium |
| RV_end_di | Z92_S1 | 27446 | -0.11198 | 0.007118 | 9.14E-56 | 55.03888 | Right ventricle |
| Ascending_ | Z10_S4 | 24599 | -0.09925 | 0.006311 | 9.81E-56 | 55.00839 | Aorta |
| LA_ejectio | Z73_S4 | 27446 | -0.08753 | 0.005566 | 1.00E-55 | 55.00013 | Left atrium |
| RV_stroke_ | Z115_S4 | 27446 | 0.088362 | 0.00562 | 1.05E-55 | 54.97783 | Right ventricle |
| LV_radial_ | Z50_S4 | 27446 | -0.07814 | 0.004973 | 1.27E-55 | 54.89524 | Left ventricle |
| LA_maximu | Z75_S4 | 27446 | -0.09693 | 0.006171 | 1.32E-55 | 54.87965 | Left atrium |
| Descending | Z70_S5 | 24599 | 0.098449 | 0.006269 | 1.41E-55 | 54.85184 | Aorta |
| LV_end_di | Z113_S2 | 27446 | 0.128624 | 0.00819 | 1.42E-55 | 54.84887 | Left ventricle |
| RA_maxim | Z120_S4 | 27446 | -0.08611 | 0.005484 | 1.46E-55 | 54.8356 | Right atrium |
| LV_end_sy | Z14_S4 | 27446 | 0.109978 | 0.007004 | 1.46E-55 | 54.8343 | Left ventricle |
| RA_minimu | Z80_S4 | 27446 | -0.07596 | 0.004838 | 1.47E-55 | 54.83219 | Right atrium |
| Ascending_ | Z105_S5 | 24599 | 0.090532 | 0.005773 | 1.99E-55 | 54.70192 | Aorta |
| LV_stroke_ | Z50_S2 | 27446 | 0.115907 | 0.007392 | 2.04E-55 | 54.69014 | Left ventricle |
| RV_end_sy | Z38_S5 | 27446 | -0.11286 | 0.007204 | 2.53E-55 | 54.59681 | Right ventricle |
| LA_minimu | Z37_S4 | 27446 | 0.097958 | 0.006261 | 3.61E-55 | 54.44209 | Left atrium |
| Descending | Z123_S4 | 24599 | 0.097385 | 0.006226 | 3.74E-55 | 54.4272 | Aorta |
| RA_ejectio | Z28_S4 | 27446 | 0.083581 | 0.005344 | 3.92E-55 | 54.40707 | Right atrium |
| Ascending_ | Z62_S2 | 24599 | 0.079361 | 0.005075 | 4.08E-55 | 54.38926 | Aorta |

| | | | | | | | |
|---|---|---|---|---|---|---|---|
| LV_myocar | Z39_S1 | 27446 | 0.120532 | 0.007712 | 4.63E-55 | 54.33444 | Left ventricle |
| LV_circumf | Z38_S5 | 27446 | 0.083598 | 0.005349 | 4.71E-55 | 54.3267 | Left ventricle |
| Ascending_ | Z80_S4 | 24599 | -0.07735 | 0.004955 | 6.07E-55 | 54.2171 | Aorta |
| LV_end_sy | Z63_S1 | 27446 | -0.11647 | 0.007462 | 6.31E-55 | 54.19976 | Left ventricle |
| Descending | Z122_S5 | 24599 | 0.091974 | 0.005893 | 6.59E-55 | 54.18082 | Aorta |
| Ascending_ | Z105_S4 | 24599 | -0.08135 | 0.005216 | 7.54E-55 | 54.12247 | Aorta |
| RA_maxim | Z78_S4 | 27446 | 0.076482 | 0.004906 | 8.39E-55 | 54.07624 | Right atrium |
| RV_ejectio | Z65_S1 | 27446 | 0.065867 | 0.004225 | 8.66E-55 | 54.06233 | Right ventricle |
| LA_maximu | Z31_S2 | 27446 | 0.081726 | 0.005244 | 9.19E-55 | 54.03691 | Left atrium |
| RA_maxim | Z112_S1 | 27446 | -0.10736 | 0.006889 | 9.47E-55 | 54.02382 | Right atrium |
| Descending | Z93_S1 | 24599 | 0.117071 | 0.00753 | 1.65E-54 | 53.78339 | Aorta |
| LV_circumf | Z13_S1 | 27446 | -0.07054 | 0.00454 | 1.87E-54 | 53.72718 | Left ventricle |
| LV_stroke_ | Z59_S2 | 27446 | 0.107162 | 0.0069 | 2.14E-54 | 53.66999 | Left ventricle |
| LV_radial_s | Z122_S2 | 27446 | 0.065219 | 0.004202 | 2.47E-54 | 53.6069 | Left ventricle |
| RV_end_sy | Z66_S3 | 27446 | -0.12015 | 0.007744 | 2.73E-54 | 53.5631 | Right ventricle |
| LA_ejection | Z114_S5 | 27446 | 0.060004 | 0.003868 | 2.90E-54 | 53.5371 | Left atrium |
| Ascending_ | Z66_S3 | 24599 | -0.10095 | 0.00651 | 3.10E-54 | 53.50853 | Aorta |
| Ascending_ | Z94_S5 | 24599 | 0.076691 | 0.004947 | 3.32E-54 | 53.47924 | Aorta |
| Descending | Z80_S1 | 24599 | 0.099675 | 0.006436 | 4.17E-54 | 53.37935 | Aorta |
| RA_stroke_ | Z74_S4 | 27446 | -0.08193 | 0.005291 | 4.30E-54 | 53.36681 | Right atrium |
| RA_stroke_ | Z40_S4 | 27446 | -0.09423 | 0.006085 | 4.36E-54 | 53.36086 | Right atrium |
| LV_stroke_ | Z114_S4 | 27446 | 0.091015 | 0.005879 | 4.73E-54 | 53.32538 | Left ventricle |
| RA_maxim | Z5_S5 | 27446 | 0.088236 | 0.005701 | 5.00E-54 | 53.30146 | Right atrium |
| RA_minimu | Z66_S3 | 27446 | -0.0984 | 0.006361 | 5.60E-54 | 53.25165 | Right atrium |
| LV_mean_r | Z114_S5 | 27446 | -0.08578 | 0.00555 | 6.88E-54 | 53.16244 | Left ventricle |
| LV_radial_s | Z40_S4 | 27446 | -0.09582 | 0.006204 | 8.15E-54 | 53.08887 | Left ventricle |
| LA_minimu | Z96_S3 | 27446 | 0.076008 | 0.004922 | 8.66E-54 | 53.06255 | Left atrium |
| LV_myocar | Z127_S4 | 27446 | 0.141453 | 0.009161 | 8.73E-54 | 53.0588 | Left ventricle |
| LV_radial_s | Z43_S2 | 27446 | 0.06951 | 0.004503 | 9.32E-54 | 53.0304 | Left ventricle |
| RA_maxim | Z10_S4 | 27446 | -0.08909 | 0.005773 | 9.96E-54 | 53.00154 | Right atrium |
| RA_minimu | Z68_S2 | 27446 | -0.07874 | 0.005104 | 1.07E-53 | 52.97102 | Right atrium |
| Descending | Z93_S2 | 24599 | 0.12561 | 0.008144 | 1.12E-53 | 52.9492 | Aorta |
| RV_end_di | Z114_S4 | 27446 | 0.107415 | 0.006968 | 1.28E-53 | 52.89177 | Right ventricle |
| LA_ejection | Z20_S2 | 27446 | -0.06313 | 0.004097 | 1.44E-53 | 52.84143 | Left atrium |
| RA_ejectio | Z66_S3 | 27446 | 0.086711 | 0.005628 | 1.46E-53 | 52.83678 | Right atrium |
| RV_end_di | Z96_S3 | 27446 | 0.1074 | 0.006972 | 1.55E-53 | 52.811 | Right ventricle |
| LA_maximu | Z61_S4 | 27446 | -0.07568 | 0.004914 | 1.59E-53 | 52.79837 | Left atrium |
| LV_radial_s | Z64_S1 | 27446 | 0.087945 | 0.00571 | 1.60E-53 | 52.79692 | Left ventricle |
| LV_stroke_ | Z92_S1 | 27446 | -0.09249 | 0.006008 | 1.76E-53 | 52.75464 | Left ventricle |
| LA_stroke_ | Z80_S2 | 27446 | 0.071756 | 0.004668 | 2.51E-53 | 52.60081 | Left atrium |
| Ascending_ | Z2_S1 | 24599 | -0.08046 | 0.005236 | 2.66E-53 | 52.57516 | Aorta |
| Descending | Z18_S4 | 24599 | -0.11381 | 0.007416 | 3.68E-53 | 52.4338 | Aorta |
| LV_circumf | Z1_S5 | 27446 | -0.07941 | 0.005175 | 3.81E-53 | 52.41857 | Left ventricle |
| RA_stroke_ | Z37_S3 | 27446 | -0.08805 | 0.005739 | 3.90E-53 | 52.40939 | Right atrium |
| LV_radial_s | Z13_S3 | 27446 | -0.06431 | 0.004192 | 4.04E-53 | 52.39384 | Left ventricle |

| | | | | | | | |
|---|---|---|---|---|---|---|---|
| LA_ejectior | Z107_S1 | 27446 | 0.059835 | 0.003903 | 4.75E-53 | 52.32291 | Left atrium |
| LV_cardiac | Z46_S1 | 27446 | 0.085913 | 0.005604 | 4.83E-53 | 52.31647 | Left ventricle |
| LV_radial_s | Z68_S2 | 27446 | 0.071707 | 0.004678 | 4.87E-53 | 52.31282 | Left ventricle |
| RA_maxim | Z62_S4 | 27446 | -0.0972 | 0.006341 | 4.89E-53 | 52.31046 | Right atrium |
| RV_end_sy | Z64_S1 | 27446 | -0.11625 | 0.007585 | 5.20E-53 | 52.28421 | Right ventricle |
| RV_stroke_ | Z75_S5 | 27446 | 0.117895 | 0.007703 | 7.10E-53 | 52.14883 | Right ventricle |
| Ascending_ | Z122_S5 | 24599 | 0.078742 | 0.005146 | 7.42E-53 | 52.12934 | Aorta |
| RA_ejectio | Z29_S5 | 27446 | -0.08113 | 0.005304 | 8.25E-53 | 52.08341 | Right atrium |
| LV_end_dia | Z26_S3 | 27446 | -0.11706 | 0.00766 | 1.01E-52 | 51.99699 | Left ventricle |
| Descending | Z109_S1 | 24599 | -0.10181 | 0.006664 | 1.09E-52 | 51.96197 | Aorta |
| Ascending_ | Z97_S1 | 24599 | -0.10726 | 0.007022 | 1.15E-52 | 51.93876 | Aorta |
| LV_end_sy | Z26_S3 | 27446 | -0.1079 | 0.00707 | 1.37E-52 | 51.86334 | Left ventricle |
| LV_end_dia | Z12_S5 | 27446 | -0.12314 | 0.008081 | 1.94E-52 | 51.71288 | Left ventricle |
| Descending | Z5_S1 | 24599 | -0.09991 | 0.006561 | 2.31E-52 | 51.63571 | Aorta |
| LA_ejectior | Z1_S1 | 27446 | 0.071783 | 0.004718 | 2.84E-52 | 51.54666 | Left atrium |
| RA_stroke_ | Z78_S1 | 27446 | -0.06996 | 0.004598 | 2.87E-52 | 51.54216 | Right atrium |
| Descending | Z82_S4 | 24599 | -0.0873 | 0.005739 | 2.98E-52 | 51.52529 | Aorta |
| Ascending_ | Z26_S4 | 24599 | 0.108023 | 0.007102 | 3.05E-52 | 51.51534 | Aorta |
| Ascending_ | Z1_S5 | 24599 | 0.094463 | 0.006211 | 3.10E-52 | 51.50925 | Aorta |
| LV_radial_s | Z73_S1 | 27446 | 0.073215 | 0.004818 | 3.81E-52 | 51.41896 | Left ventricle |
| RV_end_sy | Z113_S2 | 27446 | 0.124909 | 0.008222 | 4.02E-52 | 51.39591 | Right ventricle |
| LV_longitu | Z127_S4 | 27446 | -0.08253 | 0.005435 | 4.33E-52 | 51.36323 | Left ventricle |
| LV_radial_s | Z59_S3 | 27446 | 0.063282 | 0.004168 | 4.53E-52 | 51.34386 | Left ventricle |
| LA_ejectior | Z62_S2 | 27446 | -0.06512 | 0.004293 | 5.88E-52 | 51.23061 | Left atrium |
| RA_ejectio | Z82_S1 | 27446 | 0.093001 | 0.006134 | 6.27E-52 | 51.2026 | Right atrium |
| Descending | Z97_S1 | 24599 | -0.12153 | 0.008015 | 6.31E-52 | 51.19999 | Aorta |
| LA_minimu | Z39_S3 | 27446 | 0.095993 | 0.006333 | 6.68E-52 | 51.17551 | Left atrium |
| RA_maxim | Z20_S2 | 27446 | 0.070888 | 0.004679 | 7.71E-52 | 51.11272 | Right atrium |
| Ascending_ | Z112_S1 | 24599 | -0.11381 | 0.007514 | 7.96E-52 | 51.09905 | Aorta |
| LV_myocar | Z81_S1 | 27446 | -0.12822 | 0.008466 | 8.21E-52 | 51.08584 | Left ventricle |
| Descending | Z75_S4 | 24599 | -0.11949 | 0.007893 | 9.04E-52 | 51.04401 | Aorta |
| Descending | Z74_S2 | 24599 | 0.099224 | 0.006557 | 9.77E-52 | 51.01024 | Aorta |
| LV_cardiac | Z108_S5 | 27446 | -0.08585 | 0.005673 | 9.85E-52 | 51.0066 | Left ventricle |
| Descending | Z85_S5 | 24599 | 0.09411 | 0.006219 | 9.88E-52 | 51.00503 | Aorta |
| LV_myocar | Z26_S4 | 27446 | 0.156622 | 0.010351 | 1.01E-51 | 50.99718 | Left ventricle |
| LV_cardiac | Z24_S5 | 27446 | 0.091299 | 0.006036 | 1.12E-51 | 50.95216 | Left ventricle |
| Ascending_ | Z18_S4 | 24599 | -0.11414 | 0.007555 | 1.42E-51 | 50.84779 | Aorta |
| LV_cardiac | Z20_S1 | 27446 | 0.097341 | 0.006444 | 1.49E-51 | 50.82546 | Left ventricle |
| RA_maxim | Z50_S5 | 27446 | 0.103772 | 0.006873 | 1.67E-51 | 50.77602 | Right atrium |
| RA_maxim | Z15_S2 | 27446 | 0.089113 | 0.005906 | 1.96E-51 | 50.70803 | Right atrium |
| LV_longitu | Z40_S5 | 27446 | 0.063904 | 0.004239 | 2.34E-51 | 50.63018 | Left ventricle |
| RV_end_sy | Z68_S2 | 27446 | -0.09359 | 0.006215 | 2.97E-51 | 50.52732 | Right ventricle |
| RA_maxim | Z28_S4 | 27446 | -0.08977 | 0.005962 | 3.11E-51 | 50.50655 | Right atrium |
| Ascending_ | Z56_S5 | 24599 | 0.073578 | 0.004888 | 3.37E-51 | 50.47208 | Aorta |
| RV_end_di | Z37_S2 | 27446 | -0.11701 | 0.007774 | 3.40E-51 | 50.46795 | Right ventricle |

| | | | | | | | |
|---|---|---|---|---|---|---|---|
| LV_myocar | Z114_S4 | 27446 | 0.122234 | 0.008126 | 3.86E-51 | 50.41296 | Left ventricle |
| RA_stroke_ | Z12_S4 | 27446 | -0.0647 | 0.004307 | 5.42E-51 | 50.26603 | Right atrium |
| RV_stroke_ | Z13_S3 | 27446 | 0.075091 | 0.005001 | 5.90E-51 | 50.22898 | Right ventricle |
| RV_end_di | Z14_S2 | 27446 | -0.10722 | 0.007142 | 6.01E-51 | 50.22093 | Right ventricle |
| RV_stroke_ | Z32_S1 | 27446 | 0.097755 | 0.006512 | 6.21E-51 | 50.20711 | Right ventricle |
| LA_maximu | Z21_S3 | 27446 | -0.09807 | 0.006533 | 6.29E-51 | 50.20107 | Left atrium |
| Descending | Z59_S2 | 24599 | 0.114032 | 0.007602 | 7.28E-51 | 50.13782 | Aorta |
| LV_cardiac | Z127_S4 | 27446 | -0.09108 | 0.006078 | 9.26E-51 | 50.03357 | Left ventricle |
| LV_ejectio | Z73_S1 | 27446 | 0.069981 | 0.004678 | 1.36E-50 | 49.86747 | Left ventricle |
| Descending | Z87_S3 | 24599 | -0.0978 | 0.006538 | 1.38E-50 | 49.85922 | Aorta |
| LV_cardiac | Z79_S4 | 27446 | 0.09762 | 0.006528 | 1.46E-50 | 49.83492 | Left ventricle |
| Ascending_ | Z38_S4 | 24599 | -0.09895 | 0.006617 | 1.47E-50 | 49.83338 | Aorta |
| Ascending_ | Z50_S2 | 24599 | 0.105187 | 0.007035 | 1.53E-50 | 49.81512 | Aorta |
| RA_stroke_ | Z53_S1 | 27446 | 0.078411 | 0.005248 | 1.81E-50 | 49.74115 | Right atrium |
| RV_stroke_ | Z3_S2 | 27446 | 0.081641 | 0.005469 | 2.13E-50 | 49.67095 | Right ventricle |
| LV_end_di | Z73_S1 | 27446 | -0.09512 | 0.006378 | 2.71E-50 | 49.56723 | Left ventricle |
| LV_circumf | Z111_S1 | 27446 | -0.08242 | 0.005531 | 3.28E-50 | 49.48463 | Left ventricle |
| Ascending_ | Z50_S2 | 24599 | 0.104411 | 0.00701 | 3.57E-50 | 49.44765 | Aorta |
| LV_stroke_ | Z75_S5 | 27446 | 0.113085 | 0.007593 | 3.62E-50 | 49.44112 | Left ventricle |
| Ascending_ | Z38_S4 | 24599 | -0.09819 | 0.006593 | 3.71E-50 | 49.4301 | Aorta |
| LV_end_di | Z87_S5 | 27446 | -0.09982 | 0.006716 | 5.72E-50 | 49.2429 | Left ventricle |
| LV_stroke_ | Z21_S3 | 27446 | -0.11249 | 0.007572 | 6.45E-50 | 49.19071 | Left ventricle |
| LV_radial_s | Z105_S5 | 27446 | -0.07727 | 0.005204 | 7.21E-50 | 49.14222 | Left ventricle |
| LV_end_di | Z18_S4 | 27446 | -0.12615 | 0.008499 | 7.68E-50 | 49.11471 | Left ventricle |
| RV_stroke_ | Z24_S5 | 27446 | -0.09919 | 0.006685 | 8.28E-50 | 49.08206 | Right ventricle |
| Descending | Z1_S5 | 24599 | 0.090462 | 0.006099 | 9.14E-50 | 49.0391 | Aorta |
| LA_stroke_ | Z21_S3 | 27446 | -0.09739 | 0.006567 | 9.25E-50 | 49.03404 | Left atrium |
| RA_stroke_ | Z123_S4 | 27446 | 0.078305 | 0.005284 | 1.10E-49 | 48.95686 | Right atrium |
| Descending | Z125_S4 | 24599 | -0.09141 | 0.006172 | 1.27E-49 | 48.89698 | Aorta |
| RV_end_di | Z9_S2 | 27446 | 0.113317 | 0.007657 | 1.48E-49 | 48.83068 | Right ventricle |
| LA_maximu | Z39_S3 | 27446 | 0.096452 | 0.006532 | 2.45E-49 | 48.61029 | Left atrium |
| LV_stroke_ | Z11_S5 | 27446 | -0.08585 | 0.005815 | 2.47E-49 | 48.60683 | Left ventricle |
| RV_stroke_ | Z81_S1 | 27446 | -0.09181 | 0.006219 | 2.56E-49 | 48.59114 | Right ventricle |
| Descending | Z11_S5 | 24599 | -0.09315 | 0.006311 | 2.62E-49 | 48.58128 | Aorta |
| Ascending_ | Z105_S5 | 24599 | 0.08551 | 0.005797 | 3.07E-49 | 48.51326 | Aorta |
| RA_maxim | Z80_S4 | 27446 | -0.07044 | 0.004776 | 3.07E-49 | 48.51265 | Right atrium |
| LV_longitu | Z120_S4 | 27446 | 0.072402 | 0.00491 | 3.28E-49 | 48.4842 | Left ventricle |
| Ascending_ | Z44_S4 | 24599 | -0.07173 | 0.004867 | 3.70E-49 | 48.43137 | Aorta |
| RA_maxim | Z125_S4 | 27446 | -0.08403 | 0.005709 | 4.91E-49 | 48.30857 | Right atrium |
| RA_ejectio | Z56_S2 | 27446 | 0.06273 | 0.004263 | 5.13E-49 | 48.28973 | Right atrium |
| Ascending_ | Z26_S4 | 24599 | 0.104856 | 0.00713 | 5.95E-49 | 48.22523 | Aorta |
| RA_minimu | Z22_S4 | 27446 | -0.08529 | 0.005813 | 9.66E-49 | 48.01492 | Right atrium |
| LA_maximu | Z114_S4 | 27446 | 0.074467 | 0.005075 | 9.73E-49 | 48.01175 | Left atrium |
| Descending | Z105_S5 | 24599 | 0.098475 | 0.006712 | 9.84E-49 | 48.00703 | Aorta |
| RA_stroke_ | Z95_S3 | 27446 | -0.05693 | 0.003885 | 1.26E-48 | 47.90109 | Right atrium |

| | | | | | | | |
|---|---|---|---|---|---|---|---|
| RV_stroke_ | Z21_S3 | 27446 | -0.1126 | 0.007685 | 1.30E-48 | 47.88545 | Right ventricle |
| LA_minimu | Z58_S2 | 27446 | 0.06945 | 0.004743 | 1.48E-48 | 47.82958 | Left atrium |
| RA_minimu | Z69_S5 | 27446 | -0.06984 | 0.00477 | 1.53E-48 | 47.81654 | Right atrium |
| RV_ejectio | Z37_S4 | 27446 | 0.092407 | 0.006311 | 1.54E-48 | 47.81365 | Right ventricle |
| LV_radial_ | Z115_S4 | 27446 | -0.06902 | 0.004714 | 1.54E-48 | 47.81166 | Left ventricle |
| RV_stroke_ | Z17_S1 | 27446 | 0.083819 | 0.005735 | 2.20E-48 | 47.65713 | Right ventricle |
| LV_myocar | Z99_S5 | 27446 | 0.117185 | 0.008019 | 2.34E-48 | 47.63164 | Left ventricle |
| LA_maximu | Z37_S1 | 27446 | 0.085812 | 0.005874 | 2.44E-48 | 47.6121 | Left atrium |
| LV_longitu | Z44_S4 | 27446 | 0.06125 | 0.004195 | 2.81E-48 | 47.55204 | Left ventricle |
| LV_longitu | Z1_S1 | 27446 | 0.070402 | 0.004824 | 3.00E-48 | 47.52282 | Left ventricle |
| Ascending_ | Z61_S4 | 24599 | -0.07906 | 0.005418 | 3.06E-48 | 47.51493 | Aorta |
| RV_stroke_ | Z6_S2 | 27446 | -0.06965 | 0.004772 | 3.10E-48 | 47.50879 | Right ventricle |
| LV_myocar | Z70_S5 | 27446 | 0.116526 | 0.007989 | 3.47E-48 | 47.45954 | Left ventricle |
| Descending | Z7_S5 | 24599 | -0.10462 | 0.007173 | 3.55E-48 | 47.45024 | Aorta |
| LV_stroke_ | Z50_S5 | 27446 | 0.10935 | 0.007507 | 4.65E-48 | 47.33233 | Left ventricle |
| LV_circumf | Z65_S1 | 27446 | -0.06124 | 0.004209 | 5.77E-48 | 47.23914 | Left ventricle |
| LV_myocar | Z113_S2 | 27446 | 0.146342 | 0.010058 | 5.80E-48 | 47.23639 | Left ventricle |
| Descending | Z1_S1 | 24599 | -0.08395 | 0.005772 | 6.38E-48 | 47.19509 | Aorta |
| RA_stroke_ | Z7_S5 | 27446 | 0.080168 | 0.005514 | 6.76E-48 | 47.17023 | Right atrium |
| LV_end_sy | Z37_S1 | 27446 | 0.102841 | 0.007074 | 6.98E-48 | 47.15604 | Left ventricle |
| LV_end_sy | Z65_S1 | 27446 | -0.07571 | 0.005212 | 8.33E-48 | 47.0792 | Left ventricle |
| LA_stroke_ | Z87_S3 | 27446 | -0.07411 | 0.005109 | 1.15E-47 | 46.94072 | Left atrium |
| Ascending_ | Z62_S1 | 24599 | 0.077673 | 0.005357 | 1.24E-47 | 46.90759 | Aorta |
| RV_end_di | Z41_S1 | 27446 | 0.112212 | 0.007741 | 1.30E-47 | 46.88603 | Right ventricle |
| LV_end_di | Z27_S2 | 27446 | -0.1148 | 0.007924 | 1.46E-47 | 46.83453 | Left ventricle |
| LV_longitu | Z81_S1 | 27446 | 0.072726 | 0.005024 | 1.69E-47 | 46.77114 | Left ventricle |
| RV_end_di | Z66_S2 | 27446 | 0.118716 | 0.008209 | 2.14E-47 | 46.67054 | Right ventricle |
| Ascending_ | Z114_S4 | 24599 | 0.080811 | 0.005588 | 2.14E-47 | 46.66883 | Aorta |
| Descending | Z59_S2 | 24599 | 0.10834 | 0.007496 | 2.41E-47 | 46.61768 | Aorta |
| LV_radial_ | Z60_S2 | 27446 | 0.064738 | 0.004483 | 2.91E-47 | 46.5358 | Left ventricle |
| LV_end_di | Z75_S5 | 27446 | 0.123474 | 0.008551 | 2.91E-47 | 46.53572 | Left ventricle |
| RA_ejectio | Z29_S1 | 27446 | -0.07764 | 0.005378 | 3.02E-47 | 46.51983 | Right atrium |
| Descending | Z12_S5 | 24599 | 0.101743 | 0.007049 | 3.18E-47 | 46.49821 | Aorta |
| Ascending_ | Z85_S5 | 24599 | 0.077262 | 0.005354 | 3.35E-47 | 46.47433 | Aorta |
| Ascending_ | Z5_S5 | 24599 | 0.085806 | 0.005947 | 3.43E-47 | 46.46461 | Aorta |
| RV_end_di | Z65_S2 | 27446 | 0.1063 | 0.007368 | 3.48E-47 | 46.45807 | Right ventricle |
| RV_end_sy | Z75_S5 | 27446 | 0.123796 | 0.008582 | 3.59E-47 | 46.44535 | Right ventricle |
| LV_radial_ | Z52_S4 | 27446 | -0.06632 | 0.004601 | 4.08E-47 | 46.38892 | Left ventricle |
| LA_ejectior | Z44_S4 | 27446 | 0.059144 | 0.004105 | 4.70E-47 | 46.32826 | Left atrium |
| LV_cardiac | Z120_S5 | 27446 | -0.09629 | 0.00669 | 5.66E-47 | 46.24721 | Left ventricle |
| RV_end_sy | Z99_S4 | 27446 | 0.109813 | 0.007632 | 6.17E-47 | 46.20977 | Right ventricle |
| LV_end_di | Z37_S4 | 27446 | 0.121238 | 0.008432 | 7.08E-47 | 46.15022 | Left ventricle |
| LA_maximu | Z62_S4 | 27446 | -0.08592 | 0.005977 | 7.52E-47 | 46.12398 | Left atrium |
| RA_maxim | Z62_S2 | 27446 | 0.070464 | 0.004905 | 8.54E-47 | 46.06841 | Right atrium |
| RA_stroke_ | Z37_S4 | 27446 | 0.089748 | 0.006248 | 8.75E-47 | 46.05781 | Right atrium |

| | | | | | | | |
|---|---|---|---|---|---|---|---|
| Descending | Z92_S1 | 24599 | -0.0936 | 0.006517 | 8.77E-47 | 46.05698 | Aorta |
| LV_myocar | Z92_S1 | 27446 | -0.11928 | 0.008306 | 8.99E-47 | 46.04602 | Left ventricle |
| Descending | Z81_S1 | 24599 | -0.09689 | 0.006748 | 9.45E-47 | 46.02439 | Aorta |
| RV_ejectio | Z73_S1 | 27446 | 0.068537 | 0.004776 | 1.08E-46 | 45.96602 | Right ventricle |
| Descending | Z32_S1 | 24599 | 0.090658 | 0.006324 | 1.32E-46 | 45.87981 | Aorta |
| RV_end_sy | Z96_S3 | 27446 | 0.095344 | 0.006652 | 1.36E-46 | 45.86623 | Right ventricle |
| LV_end_sy | Z79_S4 | 27446 | -0.10606 | 0.007404 | 1.53E-46 | 45.8147 | Left ventricle |
| RA_ejectio | Z123_S4 | 27446 | -0.07449 | 0.005202 | 1.63E-46 | 45.78671 | Right atrium |
| LA_stroke_ | Z122_S5 | 27446 | 0.067457 | 0.004716 | 2.06E-46 | 45.68662 | Left atrium |
| RA_ejectio | Z35_S5 | 27446 | 0.060898 | 0.004258 | 2.09E-46 | 45.68004 | Right atrium |
| LV_cardiac | Z75_S5 | 27446 | -0.09956 | 0.006961 | 2.11E-46 | 45.67642 | Left ventricle |
| RA_ejectio | Z15_S2 | 27446 | -0.07577 | 0.005298 | 2.17E-46 | 45.6645 | Right atrium |
| LV_longitu | Z117_S4 | 27446 | 0.057681 | 0.004036 | 2.51E-46 | 45.6001 | Left ventricle |
| RA_stroke_ | Z7_S1 | 27446 | -0.06611 | 0.004631 | 3.08E-46 | 45.51109 | Right atrium |
| RA_minimu | Z9_S2 | 27446 | 0.085609 | 0.005999 | 3.30E-46 | 45.4809 | Right atrium |
| LV_longitu | Z109_S1 | 27446 | 0.070881 | 0.004968 | 3.45E-46 | 45.46199 | Left ventricle |
| LV_radial_ | Z56_S5 | 27446 | -0.06258 | 0.004387 | 3.57E-46 | 45.44755 | Left ventricle |
| LV_end_di | Z100_S3 | 27446 | 0.099745 | 0.006993 | 3.65E-46 | 45.43729 | Left ventricle |
| RA_ejectio | Z19_S2 | 27446 | -0.06981 | 0.004899 | 4.44E-46 | 45.35247 | Right atrium |
| RV_end_di | Z18_S4 | 27446 | -0.12745 | 0.008948 | 4.88E-46 | 45.31124 | Right ventricle |
| RA_maxim | Z52_S4 | 27446 | 0.070495 | 0.004953 | 5.78E-46 | 45.23811 | Right atrium |
| RV_end_di | Z87_S5 | 27446 | -0.10062 | 0.007071 | 6.05E-46 | 45.21796 | Right ventricle |
| Descending | Z68_S2 | 24599 | -0.08473 | 0.005955 | 6.16E-46 | 45.21053 | Aorta |
| RA_stroke_ | Z94_S5 | 27446 | 0.061914 | 0.004352 | 6.37E-46 | 45.19596 | Right atrium |
| Descending | Z96_S3 | 24599 | 0.090595 | 0.006376 | 8.19E-46 | 45.08652 | Aorta |
| LA_minimu | Z75_S4 | 27446 | -0.08506 | 0.005988 | 8.54E-46 | 45.06852 | Left atrium |
| Descending | Z108_S5 | 24599 | -0.09692 | 0.006831 | 1.08E-45 | 44.96559 | Aorta |
| LV_radial_ | Z57_S2 | 27446 | 0.060511 | 0.004271 | 1.46E-45 | 44.83496 | Left ventricle |
| Descending | Z114_S4 | 24599 | 0.090346 | 0.006379 | 1.55E-45 | 44.80936 | Aorta |
| Descending | Z97_S1 | 24599 | -0.1151 | 0.008136 | 1.95E-45 | 44.71005 | Aorta |
| RA_minimu | Z112_S3 | 27446 | -0.09416 | 0.006657 | 2.03E-45 | 44.69175 | Right atrium |
| LV_stroke_ | Z77_S3 | 27446 | -0.08719 | 0.006165 | 2.06E-45 | 44.68599 | Left ventricle |
| LV_radial_ | Z91_S1 | 27446 | -0.07026 | 0.004975 | 2.74E-45 | 44.56273 | Left ventricle |
| LV_end_di | Z106_S3 | 27446 | -0.11342 | 0.008032 | 2.78E-45 | 44.55613 | Left ventricle |
| RV_end_di | Z51_S3 | 27446 | -0.12349 | 0.008752 | 3.31E-45 | 44.48067 | Right ventricle |
| Ascending_ | Z93_S2 | 24599 | 0.098932 | 0.007014 | 3.52E-45 | 44.45305 | Aorta |
| RV_stroke_ | Z50_S5 | 27446 | 0.107477 | 0.00762 | 3.56E-45 | 44.44798 | Right ventricle |
| LV_myocar | Z9_S2 | 27446 | 0.125929 | 0.008931 | 3.76E-45 | 44.4245 | Left ventricle |
| RA_ejectio | Z1_S2 | 27446 | -0.0573 | 0.004066 | 4.10E-45 | 44.38685 | Right atrium |
| RA_ejectio | Z91_S1 | 27446 | -0.06768 | 0.004803 | 4.21E-45 | 44.37534 | Right atrium |
| Descending | Z122_S1 | 24599 | 0.102623 | 0.007283 | 4.29E-45 | 44.36731 | Aorta |
| Descending | Z99_S5 | 24599 | 0.089912 | 0.006389 | 5.59E-45 | 44.25259 | Aorta |
| LV_myocar | Z26_S3 | 27446 | -0.13229 | 0.009406 | 6.27E-45 | 44.20257 | Left ventricle |
| LA_maximu | Z80_S2 | 27446 | 0.065348 | 0.004648 | 6.71E-45 | 44.1734 | Left atrium |
| LV_cardiac | Z78_S1 | 27446 | 0.070981 | 0.005054 | 8.34E-45 | 44.07893 | Left ventricle |

| | | | | | | | |
|---|---|---|---|---|---|---|---|
| RA_maxim | Z56_S5 | 27446 | -0.06633 | 0.004723 | 8.44E-45 | 44.07362 | Right atrium |
| Ascending_ | Z114_S4 | 24599 | 0.07821 | 0.005569 | 8.48E-45 | 44.07176 | Aorta |
| RA_maxim | Z72_S5 | 27446 | 0.095799 | 0.006823 | 8.87E-45 | 44.05223 | Right atrium |
| LA_stroke_ | Z13_S1 | 27446 | -0.06592 | 0.004695 | 8.93E-45 | 44.04907 | Left atrium |
| RA_stroke_ | Z63_S1 | 27446 | -0.08418 | 0.005996 | 9.08E-45 | 44.042 | Right atrium |
| LV_stroke_ | Z97_S1 | 27446 | -0.10361 | 0.007383 | 9.70E-45 | 44.01316 | Left ventricle |
| RV_end_di | Z94_S2 | 27446 | 0.108368 | 0.007726 | 1.06E-44 | 43.97338 | Right ventricle |
| Descending | Z93_S1 | 24599 | 0.107226 | 0.007645 | 1.10E-44 | 43.96053 | Aorta |
| LV_end_di | Z57_S2 | 27446 | 0.07928 | 0.005654 | 1.13E-44 | 43.94563 | Left ventricle |
| LA_minimu | Z99_S5 | 27446 | 0.068114 | 0.004857 | 1.13E-44 | 43.94507 | Left atrium |
| Descending | Z79_S4 | 24599 | -0.10868 | 0.007752 | 1.18E-44 | 43.92883 | Aorta |
| LV_circumf | Z115_S4 | 27446 | 0.065176 | 0.004651 | 1.31E-44 | 43.88237 | Left ventricle |
| Ascending_ | Z61_S4 | 24599 | -0.0762 | 0.005439 | 1.37E-44 | 43.86485 | Aorta |
| LA_ejection | Z105_S4 | 27446 | 0.061883 | 0.004421 | 1.62E-44 | 43.79017 | Left atrium |
| LV_myocar | Z93_S2 | 27446 | 0.143514 | 0.010253 | 1.63E-44 | 43.78817 | Left ventricle |
| RA_minimu | Z37_S4 | 27446 | 0.097262 | 0.006952 | 1.80E-44 | 43.74369 | Right atrium |
| LV_stroke_ | Z3_S4 | 27446 | 0.105384 | 0.007537 | 2.02E-44 | 43.69468 | Left ventricle |
| RA_ejectio | Z117_S4 | 27446 | 0.056543 | 0.004045 | 2.10E-44 | 43.6787 | Right atrium |
| LV_end_sy | Z87_S5 | 27446 | -0.08669 | 0.006201 | 2.10E-44 | 43.67741 | Left ventricle |
| LV_longitu | Z14_S1 | 27446 | -0.06512 | 0.004658 | 2.11E-44 | 43.67526 | Left ventricle |
| LV_circumf | Z49_S1 | 27446 | 0.0865 | 0.006198 | 2.89E-44 | 43.53859 | Left ventricle |
| RA_maxim | Z115_S3 | 27446 | -0.08223 | 0.005893 | 2.93E-44 | 43.53344 | Right atrium |
| Descending | Z114_S4 | 24599 | 0.090223 | 0.006472 | 3.58E-44 | 43.44672 | Aorta |
| LV_cardiac | Z106_S4 | 27446 | 0.067975 | 0.004876 | 3.63E-44 | 43.43953 | Left ventricle |
| RA_minimu | Z74_S4 | 27446 | -0.08209 | 0.005891 | 3.81E-44 | 43.41896 | Right atrium |
| LV_radial_ | Z107_S1 | 27446 | 0.057694 | 0.004143 | 4.48E-44 | 43.34875 | Left ventricle |
| RA_minimu | Z23_S1 | 27446 | -0.08772 | 0.006303 | 4.99E-44 | 43.30154 | Right atrium |
| LV_ejection | Z49_S1 | 27446 | -0.08484 | 0.0061 | 5.70E-44 | 43.24387 | Left ventricle |
| RV_end_di | Z106_S3 | 27446 | -0.11751 | 0.008454 | 6.37E-44 | 43.19584 | Right ventricle |
| RA_minimu | Z39_S4 | 27446 | -0.06985 | 0.005026 | 6.64E-44 | 43.17804 | Right atrium |
| RA_minimu | Z37_S3 | 27446 | -0.08876 | 0.006389 | 7.02E-44 | 43.15341 | Right atrium |
| Ascending_ | Z64_S1 | 24599 | -0.08865 | 0.006385 | 7.80E-44 | 43.10786 | Aorta |
| LV_radial_ | Z39_S4 | 27446 | -0.06393 | 0.004607 | 8.78E-44 | 43.05638 | Left ventricle |
| RA_stroke_ | Z115_S3 | 27446 | -0.07445 | 0.005369 | 9.87E-44 | 43.00557 | Right atrium |
| RV_stroke_ | Z49_S1 | 27446 | -0.10394 | 0.007496 | 1.01E-43 | 42.99506 | Right ventricle |
| LA_minimu | Z76_S3 | 27446 | -0.07388 | 0.005329 | 1.03E-43 | 42.9853 | Left atrium |
| Descending | Z64_S1 | 24599 | -0.10138 | 0.007314 | 1.08E-43 | 42.96686 | Aorta |
| LV_cardiac | Z115_S4 | 27446 | 0.070373 | 0.005081 | 1.25E-43 | 42.90197 | Left ventricle |
| RV_end_sy | Z87_S5 | 27446 | -0.0934 | 0.006744 | 1.27E-43 | 42.89568 | Right ventricle |
| RV_stroke_ | Z9_S2 | 27446 | 0.090837 | 0.00656 | 1.31E-43 | 42.88301 | Right ventricle |
| LA_ejection | Z105_S5 | 27446 | -0.06792 | 0.004908 | 1.49E-43 | 42.82724 | Left atrium |
| Descending | Z51_S4 | 24599 | -0.07111 | 0.00514 | 1.56E-43 | 42.80796 | Aorta |
| Ascending_ | Z7_S1 | 24599 | -0.07299 | 0.005279 | 1.77E-43 | 42.75227 | Aorta |
| RA_minimu | Z78_S1 | 27446 | -0.07076 | 0.005119 | 1.85E-43 | 42.73186 | Right atrium |
| LA_stroke_ | Z3_S4 | 27446 | 0.090309 | 0.006537 | 2.06E-43 | 42.6867 | Left atrium |

| | | | | | | | |
|---|---|---|---|---|---|---|---|
| RV_stroke_ | Z7_S3 | 27446 | -0.10091 | 0.007313 | 2.58E-43 | 42.58901 | Right ventricle |
| LA_stroke_ | Z79_S4 | 27446 | -0.08524 | 0.006181 | 2.88E-43 | 42.54007 | Left atrium |
| LA_ejectio | Z5_S5 | 27446 | -0.06886 | 0.004997 | 3.35E-43 | 42.47553 | Left atrium |
| Ascending_ | Z66_S3 | 24599 | -0.09004 | 0.00654 | 3.97E-43 | 42.40092 | Aorta |
| Descending | Z11_S5 | 24599 | -0.08816 | 0.006406 | 4.28E-43 | 42.36812 | Aorta |
| Ascending_ | Z7_S3 | 24599 | 0.099057 | 0.007199 | 4.49E-43 | 42.3473 | Aorta |
| RA_minimu | Z98_S3 | 27446 | -0.06752 | 0.00491 | 4.95E-43 | 42.30561 | Right atrium |
| LV_longitu | Z62_S1 | 27446 | -0.06383 | 0.004642 | 5.06E-43 | 42.29545 | Left ventricle |
| LV_cardiac | Z31_S5 | 27446 | -0.06129 | 0.00446 | 5.58E-43 | 42.25351 | Left ventricle |
| LV_stroke_ | Z112_S1 | 27446 | -0.10339 | 0.00753 | 6.69E-43 | 42.17453 | Left ventricle |
| Ascending_ | Z89_S4 | 24599 | -0.07612 | 0.005554 | 9.34E-43 | 42.02977 | Aorta |
| RV_stroke_ | Z3_S4 | 27446 | 0.104844 | 0.00765 | 9.39E-43 | 42.02737 | Right ventricle |
| Ascending_ | Z1_S1 | 24599 | -0.0766 | 0.005589 | 9.54E-43 | 42.02048 | Aorta |
| LV_circumf | Z127_S4 | 27446 | -0.0763 | 0.005568 | 9.81E-43 | 42.00813 | Left ventricle |
| LV_cardiac | Z80_S1 | 27446 | -0.08234 | 0.006009 | 9.89E-43 | 42.00488 | Left ventricle |
| LV_longitu | Z70_S5 | 27446 | -0.06496 | 0.004741 | 9.94E-43 | 42.00282 | Left ventricle |
| RA_stroke_ | Z105_S4 | 27446 | -0.06292 | 0.004601 | 1.42E-42 | 41.8483 | Right atrium |
| LV_radial_s | Z46_S1 | 27446 | 0.071173 | 0.005206 | 1.53E-42 | 41.81473 | Left ventricle |
| RV_end_sy | Z37_S1 | 27446 | 0.10518 | 0.007696 | 1.59E-42 | 41.79792 | Right ventricle |
| LV_longitu | Z35_S5 | 27446 | 0.058093 | 0.004251 | 1.61E-42 | 41.79398 | Left ventricle |
| LV_longitu | Z95_S3 | 27446 | 0.052168 | 0.003818 | 1.69E-42 | 41.77324 | Left ventricle |
| RA_maxim | Z2_S1 | 27446 | -0.06902 | 0.005053 | 1.73E-42 | 41.76215 | Right atrium |
| Descending | Z113_S2 | 24599 | 0.109415 | 0.008014 | 1.93E-42 | 41.71504 | Aorta |
| LV_circumf | Z55_S5 | 27446 | 0.05897 | 0.004332 | 3.38E-42 | 41.47126 | Left ventricle |
| LA_minimu | Z120_S5 | 27446 | -0.08314 | 0.00611 | 3.63E-42 | 41.44061 | Left atrium |
| LA_maximu | Z80_S5 | 27446 | -0.08375 | 0.006159 | 4.10E-42 | 41.38752 | Left atrium |
| Ascending_ | Z14_S2 | 24599 | -0.08231 | 0.006054 | 4.23E-42 | 41.37348 | Aorta |
| LA_ejectio | Z24_S1 | 27446 | -0.05591 | 0.004113 | 4.38E-42 | 41.35875 | Left atrium |
| LV_myocar | Z80_S2 | 27446 | 0.101119 | 0.007445 | 5.06E-42 | 41.29594 | Left ventricle |
| RA_maxim | Z65_S2 | 27446 | 0.077366 | 0.005696 | 5.12E-42 | 41.29089 | Right atrium |
| RA_minimu | Z1_S5 | 27446 | 0.077741 | 0.00573 | 6.24E-42 | 41.20515 | Right atrium |
| RA_minimu | Z43_S2 | 27446 | -0.06672 | 0.004918 | 6.35E-42 | 41.19733 | Right atrium |
| RA_ejectio | Z73_S4 | 27446 | -0.07733 | 0.005706 | 7.74E-42 | 41.11145 | Right atrium |
| Ascending_ | Z12_S5 | 24599 | -0.09249 | 0.006826 | 7.99E-42 | 41.09721 | Aorta |
| LA_maximu | Z13_S1 | 27446 | -0.06328 | 0.004673 | 9.03E-42 | 41.04427 | Left atrium |
| LV_radial_s | Z69_S5 | 27446 | 0.059193 | 0.004374 | 1.01E-41 | 40.99634 | Left ventricle |
| LV_mean_r | Z32_S1 | 27446 | -0.09994 | 0.007389 | 1.10E-41 | 40.95831 | Left ventricle |
| LV_end_dia | Z99_S4 | 27446 | 0.102894 | 0.007608 | 1.13E-41 | 40.94614 | Left ventricle |
| LV_myocar | Z122_S5 | 27446 | 0.101636 | 0.007517 | 1.18E-41 | 40.92724 | Left ventricle |
| LV_radial_s | Z24_S1 | 27446 | -0.05897 | 0.004363 | 1.27E-41 | 40.89518 | Left ventricle |
| LV_radial_s | Z26_S3 | 27446 | 0.07828 | 0.005793 | 1.30E-41 | 40.88667 | Left ventricle |
| LV_myocar | Z18_S4 | 27446 | -0.14096 | 0.010437 | 1.43E-41 | 40.84419 | Left ventricle |
| Ascending_ | Z50_S5 | 24599 | 0.101347 | 0.007508 | 1.58E-41 | 40.80041 | Aorta |
| LA_stroke_ | Z27_S2 | 27446 | -0.08243 | 0.006106 | 1.59E-41 | 40.79965 | Left atrium |
| LA_stroke_ | Z100_S3 | 27446 | 0.07272 | 0.005388 | 1.62E-41 | 40.79023 | Left atrium |

| | | | | | | | |
|---|---|---|---|---|---|---|---|
| Ascending_ | Z93_S2 | 24599 | 0.094994 | 0.007042 | 1.80E-41 | 40.74576 | Aorta |
| LA_ejectio | Z27_S2 | 27446 | 0.076138 | 0.005647 | 1.95E-41 | 40.70916 | Left atrium |
| LA_ejectio | Z7_S1 | 27446 | 0.06001 | 0.004453 | 2.12E-41 | 40.67324 | Left atrium |
| RV_end_di | Z49_S1 | 27446 | -0.11795 | 0.008756 | 2.32E-41 | 40.63461 | Right ventricle |
| RV_stroke_ | Z11_S5 | 27446 | -0.07953 | 0.005904 | 2.34E-41 | 40.63116 | Right ventricle |
| LV_stroke_ | Z27_S2 | 27446 | -0.09483 | 0.007042 | 2.45E-41 | 40.61113 | Left ventricle |
| LV_longitu | Z47_S4 | 27446 | 0.066639 | 0.004951 | 2.74E-41 | 40.56161 | Left ventricle |
| LV_ejectio | Z32_S1 | 27446 | 0.071359 | 0.005304 | 2.91E-41 | 40.53585 | Left ventricle |
| LV_radial_ | Z80_S2 | 27446 | -0.06167 | 0.004584 | 2.92E-41 | 40.5342 | Left ventricle |
| Descending | Z64_S1 | 24599 | -0.09977 | 0.007421 | 3.30E-41 | 40.48191 | Aorta |
| LV_longitu | Z32_S5 | 27446 | -0.08021 | 0.005967 | 3.38E-41 | 40.47097 | Left ventricle |
| LA_stroke_ | Z62_S4 | 27446 | -0.08074 | 0.00601 | 3.78E-41 | 40.42296 | Left atrium |
| RV_end_sy | Z21_S3 | 27446 | -0.11502 | 0.008563 | 3.89E-41 | 40.4101 | Right ventricle |
| LV_end_di | Z10_S4 | 27446 | -0.09543 | 0.007105 | 3.96E-41 | 40.40244 | Left ventricle |
| Ascending_ | Z43_S2 | 24599 | -0.06744 | 0.005022 | 4.06E-41 | 40.39143 | Aorta |
| LV_myocar | Z106_S3 | 27446 | -0.13235 | 0.00986 | 4.42E-41 | 40.35456 | Left ventricle |
| LV_ejectio | Z99_S4 | 27446 | -0.07488 | 0.005581 | 4.77E-41 | 40.32159 | Left ventricle |
| Ascending_ | Z6_S2 | 24599 | -0.06 | 0.004472 | 4.85E-41 | 40.31451 | Aorta |
| Descending | Z5_S5 | 24599 | 0.091421 | 0.006816 | 5.11E-41 | 40.29194 | Aorta |
| LV_circumf | Z112_S1 | 27446 | 0.084622 | 0.00632 | 6.90E-41 | 40.16105 | Left ventricle |
| RA_minimu | Z56_S5 | 27446 | -0.06411 | 0.004789 | 7.19E-41 | 40.14301 | Right atrium |
| LA_ejectio | Z75_S5 | 27446 | -0.08155 | 0.006094 | 7.66E-41 | 40.11552 | Left atrium |
| LV_ejectio | Z12_S5 | 27446 | 0.079275 | 0.005932 | 9.95E-41 | 40.00203 | Left ventricle |
| LA_stroke_ | Z32_S1 | 27446 | 0.074426 | 0.00557 | 1.00E-40 | 39.99923 | Left atrium |
| LV_radial_ | Z12_S4 | 27446 | 0.058731 | 0.004395 | 1.01E-40 | 39.99532 | Left ventricle |
| Ascending_ | Z65_S2 | 24599 | 0.0834 | 0.006243 | 1.05E-40 | 39.97751 | Aorta |
| Ascending_ | Z6_S2 | 24599 | -0.05994 | 0.004489 | 1.12E-40 | 39.95031 | Aorta |
| Descending | Z105_S4 | 24599 | -0.07224 | 0.005415 | 1.35E-40 | 39.87077 | Aorta |
| LA_ejectio | Z4_S4 | 27446 | 0.057535 | 0.004313 | 1.35E-40 | 39.87001 | Left atrium |
| RV_end_di | Z100_S3 | 27446 | 0.098209 | 0.007363 | 1.40E-40 | 39.85402 | Right ventricle |
| RV_ejectio | Z78_S1 | 27446 | -0.06198 | 0.00465 | 1.57E-40 | 39.80361 | Right ventricle |
| Descending | Z66_S3 | 24599 | -0.09944 | 0.007466 | 1.79E-40 | 39.74663 | Aorta |
| LA_maximu | Z50_S5 | 27446 | 0.086264 | 0.006482 | 2.07E-40 | 39.68359 | Left atrium |
| Descending | Z26_S4 | 24599 | 0.108306 | 0.008144 | 2.37E-40 | 39.62515 | Aorta |
| RV_ejectio | Z1_S5 | 27446 | 0.069149 | 0.005204 | 2.74E-40 | 39.56248 | Right ventricle |
| LV_radial_ | Z114_S5 | 27446 | 0.054538 | 0.004108 | 3.23E-40 | 39.4908 | Left ventricle |
| LV_mean_ | Z127_S4 | 27446 | -0.10136 | 0.007637 | 3.34E-40 | 39.47598 | Left ventricle |
| LV_stroke_ | Z80_S2 | 27446 | 0.071517 | 0.005389 | 3.38E-40 | 39.4715 | Left ventricle |
| Descending | Z70_S5 | 24599 | 0.084484 | 0.006368 | 3.63E-40 | 39.44049 | Aorta |
| LV_stroke_ | Z93_S1 | 27446 | 0.091873 | 0.006932 | 4.27E-40 | 39.36924 | Left ventricle |
| LV_circumf | Z83_S2 | 27446 | 0.063479 | 0.004789 | 4.28E-40 | 39.36901 | Left ventricle |
| LA_maximu | Z112_S1 | 27446 | -0.08608 | 0.006499 | 4.83E-40 | 39.31561 | Left atrium |
| RV_ejectio | Z12_S5 | 27446 | 0.08016 | 0.006055 | 5.32E-40 | 39.27383 | Right ventricle |
| Ascending_ | Z13_S3 | 24599 | 0.061724 | 0.004664 | 5.51E-40 | 39.25879 | Aorta |
| Ascending_ | Z82_S4 | 24599 | -0.06911 | 0.005222 | 5.55E-40 | 39.25567 | Aorta |

| | | | | | | | |
|---|---|---|---|---|---|---|---|
| LV_longitu | Z77_S3 | 27446 | -0.06688 | 0.005055 | 5.79E-40 | 39.23755 | Left ventricle |
| Ascending_ | Z80_S1 | 24599 | 0.08669 | 0.006564 | 7.95E-40 | 39.09961 | Aorta |
| RV_stroke_ | Z93_S1 | 27446 | 0.092895 | 0.007034 | 8.07E-40 | 39.09291 | Right ventricle |
| Ascending_ | Z43_S2 | 24599 | -0.06654 | 0.005041 | 8.73E-40 | 39.05901 | Aorta |
| LV_radial_ | Z8_S3 | 27446 | -0.0577 | 0.004373 | 9.46E-40 | 39.02408 | Left ventricle |
| LV_longitu | Z75_S5 | 27446 | -0.08216 | 0.006228 | 9.88E-40 | 39.00519 | Left ventricle |
| LV_stroke_ | Z13_S1 | 27446 | -0.07144 | 0.005417 | 1.01E-39 | 38.99467 | Left ventricle |
| LA_stroke_ | Z50_S5 | 27446 | 0.085924 | 0.006515 | 1.01E-39 | 38.99414 | Left atrium |
| LV_ejectio | Z20_S2 | 27446 | -0.05567 | 0.004224 | 1.16E-39 | 38.93511 | Left ventricle |
| LV_circumf | Z80_S2 | 27446 | 0.059553 | 0.004522 | 1.31E-39 | 38.88437 | Left ventricle |
| RA_maxim | Z1_S2 | 27446 | 0.059711 | 0.004536 | 1.44E-39 | 38.84162 | Right atrium |
| RA_stroke_ | Z14_S1 | 27446 | 0.062443 | 0.004744 | 1.46E-39 | 38.83636 | Right atrium |
| LA_stroke_ | Z75_S4 | 27446 | -0.08167 | 0.00621 | 1.68E-39 | 38.7747 | Left atrium |
| LV_circumf | Z92_S5 | 27446 | -0.08051 | 0.006123 | 1.74E-39 | 38.76037 | Left ventricle |
| RA_ejectio | Z79_S4 | 27446 | 0.076965 | 0.005854 | 1.78E-39 | 38.75042 | Right atrium |
| LV_stroke_ | Z91_S1 | 27446 | 0.076918 | 0.005851 | 1.80E-39 | 38.74365 | Left ventricle |
| Ascending_ | Z29_S1 | 24599 | 0.086792 | 0.006604 | 1.88E-39 | 38.72661 | Aorta |
| RV_stroke_ | Z114_S4 | 27446 | 0.078504 | 0.005973 | 1.89E-39 | 38.72348 | Right ventricle |
| LA_ejectio | Z29_S1 | 27446 | -0.06891 | 0.005255 | 2.78E-39 | 38.55569 | Left atrium |
| RA_stroke_ | Z50_S5 | 27446 | 0.082182 | 0.006268 | 2.86E-39 | 38.54437 | Right atrium |
| RV_stroke_ | Z75_S1 | 27446 | -0.08291 | 0.006325 | 2.92E-39 | 38.53403 | Right ventricle |
| RA_stroke_ | Z24_S1 | 27446 | 0.056109 | 0.00428 | 2.96E-39 | 38.52941 | Right atrium |
| RV_end_di | Z49_S3 | 27446 | -0.10896 | 0.008315 | 3.10E-39 | 38.50892 | Right ventricle |
| LA_ejectio | Z91_S1 | 27446 | -0.06146 | 0.004692 | 3.42E-39 | 38.46659 | Left atrium |
| LA_stroke_ | Z112_S1 | 27446 | -0.08553 | 0.006532 | 3.60E-39 | 38.44319 | Left atrium |
| LA_ejectio | Z93_S1 | 27446 | 0.07273 | 0.005559 | 4.11E-39 | 38.38577 | Left atrium |
| LV_end_sy | Z41_S1 | 27446 | 0.08886 | 0.006793 | 4.21E-39 | 38.37586 | Left ventricle |
| Descending | Z47_S4 | 24599 | -0.08598 | 0.006574 | 4.39E-39 | 38.3577 | Aorta |
| Ascending_ | Z40_S5 | 24599 | -0.0676 | 0.005171 | 4.75E-39 | 38.32331 | Aorta |
| LV_radial_ | Z80_S1 | 27446 | 0.072937 | 0.00558 | 4.76E-39 | 38.32253 | Left ventricle |
| LV_longitu | Z29_S1 | 27446 | -0.0702 | 0.005371 | 4.93E-39 | 38.30737 | Left ventricle |
| LV_ejectio | Z1_S2 | 27446 | -0.05344 | 0.004091 | 5.28E-39 | 38.27713 | Left ventricle |
| RA_minimu | Z66_S2 | 27446 | 0.084052 | 0.006434 | 5.34E-39 | 38.27206 | Right atrium |
| RA_minimu | Z2_S1 | 27446 | -0.06687 | 0.005122 | 6.03E-39 | 38.22 | Right atrium |
| LV_stroke_ | Z122_S5 | 27446 | 0.071031 | 0.005442 | 6.07E-39 | 38.21674 | Left ventricle |
| RA_maxim | Z29_S5 | 27446 | 0.077288 | 0.005922 | 6.34E-39 | 38.19782 | Right atrium |
| LV_end_sy | Z122_S2 | 27446 | -0.06706 | 0.005139 | 6.55E-39 | 38.18401 | Left ventricle |
| LA_minimu | Z81_S1 | 27446 | -0.06695 | 0.005132 | 6.82E-39 | 38.16594 | Left atrium |
| LV_myocar | Z91_S1 | 27446 | 0.10546 | 0.008085 | 6.92E-39 | 38.15972 | Left ventricle |
| RV_stroke_ | Z59_S2 | 27446 | 0.091439 | 0.007011 | 6.99E-39 | 38.15531 | Right ventricle |
| LV_radial_ | Z32_S1 | 27446 | 0.071132 | 0.005465 | 9.85E-39 | 38.00664 | Left ventricle |
| LV_stroke_ | Z96_S3 | 27446 | 0.076666 | 0.005891 | 1.01E-38 | 37.99633 | Left ventricle |
| LV_end_di | Z76_S3 | 27446 | -0.09334 | 0.007174 | 1.06E-38 | 37.97609 | Left ventricle |
| LV_end_sy | Z115_S3 | 27446 | -0.08699 | 0.006689 | 1.16E-38 | 37.936 | Left ventricle |
| RV_end_sy | Z51_S3 | 27446 | -0.10854 | 0.008349 | 1.23E-38 | 37.91054 | Right ventricle |

| | | | | | | | |
|---|---|---|---|---|---|---|---|
| LV_myocar | Z11_S5 | 27446 | -0.10453 | 0.008041 | 1.23E-38 | 37.90923 | Left ventricle |
| LA_ejectio | Z69_S5 | 27446 | 0.053595 | 0.004125 | 1.32E-38 | 37.87846 | Left atrium |
| LV_radial_ | Z58_S2 | 27446 | -0.06266 | 0.004826 | 1.50E-38 | 37.82522 | Left ventricle |
| Ascending_ | Z99_S4 | 24599 | 0.083324 | 0.006421 | 1.64E-38 | 37.78426 | Aorta |
| Descending | Z49_S1 | 24599 | -0.09424 | 0.007264 | 1.71E-38 | 37.76769 | Aorta |
| Ascending_ | Z74_S2 | 24599 | 0.073286 | 0.005649 | 1.76E-38 | 37.75448 | Aorta |
| Descending | Z92_S1 | 24599 | -0.0858 | 0.006616 | 1.84E-38 | 37.73631 | Aorta |
| RV_ejectio | Z40_S4 | 27446 | -0.07977 | 0.006155 | 2.07E-38 | 37.68354 | Right ventricle |
| LV_cardiac | Z22_S2 | 27446 | -0.07291 | 0.005628 | 2.17E-38 | 37.66364 | Left ventricle |
| LV_end_di | Z115_S3 | 27446 | -0.09381 | 0.007248 | 2.58E-38 | 37.58892 | Left ventricle |
| LV_radial_ | Z15_S2 | 27446 | -0.07106 | 0.005492 | 2.75E-38 | 37.56006 | Left ventricle |
| Descending | Z78_S1 | 24599 | -0.07021 | 0.005428 | 2.82E-38 | 37.55008 | Aorta |
| RA_stroke_ | Z117_S4 | 27446 | -0.05317 | 0.004112 | 3.05E-38 | 37.51617 | Right atrium |
| RV_stroke_ | Z60_S2 | 27446 | -0.06913 | 0.005352 | 3.65E-38 | 37.43771 | Right ventricle |
| RV_stroke_ | Z92_S1 | 27446 | -0.07883 | 0.006104 | 3.70E-38 | 37.43166 | Right ventricle |
| RA_minimu | Z72_S5 | 27446 | 0.089326 | 0.006919 | 3.98E-38 | 37.3999 | Right atrium |
| Descending | Z46_S1 | 24599 | -0.07739 | 0.005999 | 4.45E-38 | 37.35136 | Aorta |
| Descending | Z96_S3 | 24599 | 0.083444 | 0.006473 | 5.06E-38 | 37.29547 | Aorta |
| Ascending_ | Z112_S1 | 24599 | 0.092143 | 0.007149 | 5.22E-38 | 37.28275 | Aorta |
| LV_myocar | Z56_S2 | 27446 | -0.09236 | 0.00718 | 7.17E-38 | 37.14441 | Left ventricle |
| LV_stroke_ | Z3_S2 | 27446 | 0.069391 | 0.005395 | 7.27E-38 | 37.13842 | Left ventricle |
| RV_stroke_ | Z74_S2 | 27446 | 0.078035 | 0.006068 | 7.51E-38 | 37.12416 | Right ventricle |
| RV_end_di | Z19_S2 | 27446 | 0.090987 | 0.007075 | 7.53E-38 | 37.12305 | Right ventricle |
| Ascending_ | Z26_S3 | 24599 | -0.08764 | 0.006819 | 8.29E-38 | 37.0814 | Aorta |
| LV_myocar | Z50_S2 | 27446 | 0.131446 | 0.010228 | 8.44E-38 | 37.0736 | Left ventricle |
| RA_minimu | Z55_S5 | 27446 | 0.061589 | 0.004794 | 8.84E-38 | 37.05332 | Right atrium |
| RA_minimu | Z3_S4 | 27446 | 0.089917 | 0.007 | 9.14E-38 | 37.03922 | Right atrium |
| Descending | Z113_S2 | 24599 | 0.101509 | 0.007903 | 9.30E-38 | 37.03148 | Aorta |
| RA_maxim | Z22_S4 | 27446 | -0.07373 | 0.005741 | 9.31E-38 | 37.03102 | Right atrium |
| Ascending_ | Z105_S4 | 24599 | -0.06728 | 0.005243 | 1.08E-37 | 36.96809 | Aorta |
| Descending | Z123_S4 | 24599 | 0.089635 | 0.006988 | 1.16E-37 | 36.93709 | Aorta |
| RV_ejectio | Z118_S1 | 27446 | 0.072731 | 0.005674 | 1.30E-37 | 36.88466 | Right ventricle |
| LV_myocar | Z37_S1 | 27446 | 0.120633 | 0.009414 | 1.37E-37 | 36.86345 | Left ventricle |
| LV_cardiac | Z73_S1 | 27446 | -0.06639 | 0.005197 | 2.29E-37 | 36.64085 | Left ventricle |
| RA_stroke_ | Z99_S3 | 27446 | 0.051991 | 0.004074 | 2.64E-37 | 36.57786 | Right atrium |
| LV_longitu | Z80_S1 | 27446 | -0.06857 | 0.005376 | 2.98E-37 | 36.52516 | Left ventricle |
| LV_longitu | Z65_S2 | 27446 | 0.065023 | 0.0051 | 3.10E-37 | 36.50916 | Left ventricle |
| Descending | Z108_S5 | 24599 | -0.0858 | 0.006739 | 3.91E-37 | 36.40777 | Aorta |
| RA_maxim | Z32_S1 | 27446 | 0.074857 | 0.005884 | 4.44E-37 | 36.35281 | Right atrium |
| LV_end_di | Z112_S3 | 27446 | -0.1028 | 0.008081 | 4.49E-37 | 36.34822 | Left ventricle |
| RV_end_sy | Z13_S3 | 27446 | 0.070918 | 0.005576 | 4.62E-37 | 36.33575 | Right ventricle |
| LV_end_sy | Z56_S5 | 27446 | 0.06818 | 0.005363 | 5.00E-37 | 36.30112 | Left ventricle |
| LV_circumf | Z51_S1 | 27446 | -0.05665 | 0.004457 | 5.17E-37 | 36.28665 | Left ventricle |
| RA_minimu | Z75_S4 | 27446 | -0.08448 | 0.006648 | 5.33E-37 | 36.27338 | Right atrium |
| RV_end_sy | Z100_S3 | 27446 | 0.089213 | 0.007023 | 5.71E-37 | 36.24366 | Right ventricle |

| | | | | | | | |
|---|---|---|---|---|---|---|---|
| LA_stroke_ | Z75_S5 | 27446 | 0.083692 | 0.006592 | 6.16E-37 | 36.21059 | Left atrium |
| LV_circumf | Z20_S2 | 27446 | 0.054447 | 0.004293 | 7.33E-37 | 36.13469 | Left ventricle |
| LA_stroke_ | Z87_S5 | 27446 | -0.06565 | 0.005178 | 7.80E-37 | 36.10807 | Left atrium |
| Descending | Z73_S4 | 24599 | 0.086464 | 0.006823 | 8.49E-37 | 36.07092 | Aorta |
| LV_radial_s | Z98_S3 | 27446 | 0.057024 | 0.004502 | 9.18E-37 | 36.03733 | Left ventricle |
| Descending | Z7_S1 | 24599 | -0.06909 | 0.005457 | 9.76E-37 | 36.01064 | Aorta |
| RV_ejectio | Z24_S5 | 27446 | -0.07037 | 0.005559 | 1.00E-36 | 35.99929 | Right ventricle |
| Ascending_ | Z78_S1 | 24599 | -0.06999 | 0.005529 | 1.01E-36 | 35.99737 | Aorta |
| RV_end_sy | Z49_S3 | 27446 | -0.10036 | 0.00793 | 1.03E-36 | 35.98618 | Right ventricle |
| LV_end_dia | Z34_S2 | 27446 | -0.09882 | 0.00781 | 1.07E-36 | 35.97228 | Left ventricle |
| RA_stroke_ | Z77_S3 | 27446 | 0.065105 | 0.005147 | 1.14E-36 | 35.94292 | Right atrium |
| LV_cardiac | Z80_S5 | 27446 | 0.082572 | 0.006545 | 1.72E-36 | 35.76529 | Left ventricle |
| LA_ejection | Z93_S2 | 27446 | -0.07506 | 0.005955 | 1.97E-36 | 35.7056 | Left atrium |
| LV_radial_s | Z86_S3 | 27446 | 0.073419 | 0.005825 | 2.00E-36 | 35.69857 | Left ventricle |
| LV_cardiac | Z114_S4 | 27446 | 0.068011 | 0.005396 | 2.02E-36 | 35.69366 | Left ventricle |
| Descending | Z63_S1 | 24599 | -0.08904 | 0.007066 | 2.07E-36 | 35.68429 | Aorta |
| LA_minimu | Z80_S5 | 27446 | -0.07528 | 0.005975 | 2.14E-36 | 35.66878 | Left atrium |
| Ascending_ | Z51_S3 | 24599 | -0.08842 | 0.007018 | 2.16E-36 | 35.66626 | Aorta |
| RV_end_sy | Z92_S1 | 27446 | -0.08559 | 0.006798 | 2.41E-36 | 35.61768 | Right ventricle |
| LV_stroke_ | Z113_S2 | 27446 | 0.091629 | 0.007286 | 2.86E-36 | 35.54382 | Left ventricle |
| Descending | Z87_S5 | 24599 | -0.08156 | 0.006488 | 3.01E-36 | 35.52211 | Aorta |
| RV_end_di | Z91_S1 | 27446 | 0.087053 | 0.006936 | 3.94E-36 | 35.40405 | Right ventricle |
| LV_ejection | Z80_S1 | 27446 | 0.067994 | 0.005418 | 3.98E-36 | 35.40015 | Left ventricle |
| LV_ejection | Z56_S5 | 27446 | -0.05346 | 0.004262 | 4.31E-36 | 35.36505 | Left ventricle |
| LA_ejection | Z62_S1 | 27446 | -0.057 | 0.004545 | 4.43E-36 | 35.35403 | Left atrium |
| LV_end_sy | Z100_S3 | 27446 | 0.080968 | 0.006459 | 4.78E-36 | 35.32062 | Left ventricle |
| LV_stroke_ | Z80_S1 | 27446 | 0.082234 | 0.006561 | 4.84E-36 | 35.315 | Left ventricle |
| LV_longitu | Z99_S5 | 27446 | -0.05967 | 0.004762 | 5.04E-36 | 35.29783 | Left ventricle |
| RA_stroke_ | Z127_S5 | 27446 | 0.053815 | 0.0043 | 6.15E-36 | 35.21092 | Right atrium |
| RA_maxim | Z43_S2 | 27446 | -0.06075 | 0.004855 | 6.32E-36 | 35.19913 | Right atrium |
| Ascending_ | Z74_S2 | 24599 | 0.070947 | 0.005672 | 6.64E-36 | 35.17765 | Aorta |
| Ascending_ | Z50_S3 | 24599 | -0.06706 | 0.005362 | 6.78E-36 | 35.16872 | Aorta |
| LA_ejection | Z39_S1 | 27446 | -0.05605 | 0.004483 | 7.15E-36 | 35.14552 | Left atrium |
| Ascending_ | Z62_S1 | 24599 | 0.067299 | 0.005383 | 7.21E-36 | 35.14185 | Aorta |
| LA_maximu | Z122_S2 | 27446 | -0.05336 | 0.004269 | 7.28E-36 | 35.13799 | Left atrium |
| RV_stroke_ | Z23_S1 | 27446 | -0.08619 | 0.006896 | 7.62E-36 | 35.11797 | Right ventricle |
| LA_maximu | Z34_S2 | 27446 | -0.07481 | 0.005986 | 7.67E-36 | 35.11523 | Left atrium |
| RA_ejectio | Z118_S1 | 27446 | -0.06906 | 0.005529 | 8.43E-36 | 35.07407 | Right atrium |
| LA_ejection | Z39_S3 | 27446 | -0.07586 | 0.006078 | 9.40E-36 | 35.02664 | Left atrium |
| LV_ejection | Z18_S4 | 27446 | -0.07789 | 0.006241 | 9.54E-36 | 35.02062 | Left ventricle |
| RA_maxim | Z24_S5 | 27446 | -0.07535 | 0.00604 | 1.01E-35 | 34.99555 | Right atrium |
| LV_end_sy | Z113_S2 | 27446 | 0.094449 | 0.007572 | 1.03E-35 | 34.98529 | Left ventricle |
| LV_myocar | Z32_S1 | 27446 | -0.11058 | 0.008878 | 1.32E-35 | 34.88057 | Left ventricle |
| LV_stroke_ | Z106_S3 | 27446 | -0.08886 | 0.007139 | 1.45E-35 | 34.8373 | Left ventricle |
| LV_cardiac | Z14_S4 | 27446 | -0.07698 | 0.006187 | 1.57E-35 | 34.8035 | Left ventricle |

| | | | | | | | |
|---|---|---|---|---|---|---|---|
| LV_radial_s | Z77_S3 | 27446 | 0.065226 | 0.005249 | 1.86E-35 | 34.73066 | Left ventricle |
| RA_stroke_ | Z61_S4 | 27446 | -0.05901 | 0.004758 | 2.58E-35 | 34.58885 | Right atrium |
| Descending | Z37_S2 | 24599 | -0.08855 | 0.007146 | 2.90E-35 | 34.53708 | Aorta |
| Ascending_ | Z72_S5 | 24599 | -0.08773 | 0.007081 | 2.94E-35 | 34.53104 | Aorta |
| RA_stroke_ | Z44_S3 | 27446 | -0.06906 | 0.005575 | 3.06E-35 | 34.5144 | Right atrium |
| RA_maxim | Z79_S4 | 27446 | -0.08082 | 0.006531 | 3.62E-35 | 34.44154 | Right atrium |
| Ascending_ | Z89_S4 | 24599 | -0.07229 | 0.005849 | 4.33E-35 | 34.36368 | Aorta |
| Descending | Z68_S2 | 24599 | -0.0747 | 0.006047 | 4.72E-35 | 34.3261 | Aorta |
| LV_longitu | Z69_S5 | 27446 | 0.052057 | 0.004217 | 5.13E-35 | 34.29023 | Left ventricle |
| RA_ejectio | Z47_S4 | 27446 | 0.061278 | 0.004964 | 5.16E-35 | 34.28693 | Right atrium |
| LV_mean_ | Z65_S1 | 27446 | -0.07129 | 0.005777 | 5.47E-35 | 34.26215 | Left ventricle |
| Descending | Z78_S1 | 24599 | 0.073997 | 0.005997 | 5.66E-35 | 34.24752 | Aorta |
| LV_circumf | Z23_S3 | 27446 | 0.055582 | 0.004507 | 5.96E-35 | 34.22497 | Left ventricle |
| LV_myocar | Z59_S2 | 27446 | 0.117772 | 0.009549 | 6.00E-35 | 34.22155 | Left ventricle |
| LV_circumf | Z31_S2 | 27446 | 0.062967 | 0.005108 | 6.42E-35 | 34.19217 | Left ventricle |
| LV_circumf | Z40_S4 | 27446 | 0.075501 | 0.006129 | 7.25E-35 | 34.13972 | Left ventricle |
| RV_stroke_ | Z112_S1 | 27446 | -0.09418 | 0.007647 | 7.34E-35 | 34.13414 | Right ventricle |
| Ascending_ | Z51_S3 | 24599 | -0.0861 | 0.006994 | 7.92E-35 | 34.10118 | Aorta |
| RA_minimu | Z77_S3 | 27446 | -0.07047 | 0.005727 | 8.65E-35 | 34.0629 | Right atrium |
| LV_radial_s | Z23_S3 | 27446 | -0.0562 | 0.004569 | 8.91E-35 | 34.05009 | Left ventricle |
| RV_end_di | Z120_S5 | 27446 | 0.106517 | 0.008659 | 8.95E-35 | 34.04811 | Right ventricle |
| LV_myocar | Z63_S1 | 27446 | -0.12225 | 0.009938 | 8.96E-35 | 34.04769 | Left ventricle |
| RA_stroke_ | Z44_S4 | 27446 | -0.05256 | 0.004276 | 9.77E-35 | 34.01031 | Right atrium |
| LV_ejectio | Z55_S5 | 27446 | -0.0524 | 0.004266 | 1.13E-34 | 33.94512 | Left ventricle |
| LV_circumf | Z1_S2 | 27446 | 0.051067 | 0.004158 | 1.15E-34 | 33.94075 | Left ventricle |
| LV_radial_s | Z10_S4 | 27446 | -0.06596 | 0.005371 | 1.16E-34 | 33.93531 | Left ventricle |
| LV_longitu | Z1_S5 | 27446 | 0.062163 | 0.005062 | 1.17E-34 | 33.93177 | Left ventricle |
| LV_longitu | Z41_S2 | 27446 | -0.072 | 0.005864 | 1.20E-34 | 33.92182 | Left ventricle |
| Ascending_ | Z60_S2 | 24599 | -0.06133 | 0.004996 | 1.23E-34 | 33.91158 | Aorta |
| LV_longitu | Z37_S1 | 27446 | -0.0685 | 0.005585 | 1.40E-34 | 33.85507 | Left ventricle |
| LV_longitu | Z66_S3 | 27446 | -0.06894 | 0.005626 | 1.59E-34 | 33.79969 | Left ventricle |
| RV_stroke_ | Z19_S2 | 27446 | 0.074227 | 0.00606 | 1.70E-34 | 33.76835 | Right ventricle |
| Descending | Z87_S5 | 24599 | -0.08062 | 0.006582 | 1.71E-34 | 33.76825 | Aorta |
| LV_longitu | Z87_S3 | 27446 | -0.05912 | 0.004834 | 2.16E-34 | 33.66471 | Left ventricle |
| LV_myocar | Z34_S2 | 27446 | -0.11712 | 0.009586 | 2.51E-34 | 33.60088 | Left ventricle |
| Ascending_ | Z95_S5 | 24599 | -0.08442 | 0.00691 | 2.54E-34 | 33.59512 | Aorta |
| LA_stroke_ | Z112_S3 | 27446 | -0.07603 | 0.006225 | 2.63E-34 | 33.58036 | Left atrium |
| LV_radial_s | Z52_S3 | 27446 | -0.05366 | 0.004394 | 2.71E-34 | 33.5671 | Left ventricle |
| LA_ejectio | Z98_S3 | 27446 | 0.051795 | 0.004245 | 3.07E-34 | 33.51236 | Left atrium |
| RV_end_di | Z118_S1 | 27446 | 0.097359 | 0.00798 | 3.09E-34 | 33.50938 | Right ventricle |
| LV_radial_s | Z78_S4 | 27446 | -0.05568 | 0.004564 | 3.19E-34 | 33.49668 | Left ventricle |
| LV_end_di | Z1_S4 | 27446 | -0.08745 | 0.007178 | 3.82E-34 | 33.4185 | Left ventricle |
| LV_radial_s | Z13_S1 | 27446 | 0.056147 | 0.00461 | 4.00E-34 | 33.39847 | Left ventricle |
| RA_minimu | Z50_S5 | 27446 | 0.084947 | 0.006976 | 4.15E-34 | 33.38174 | Right atrium |
| RV_end_di | Z73_S1 | 27446 | -0.08185 | 0.006722 | 4.18E-34 | 33.37885 | Right ventricle |

| | | | | | | | |
|---|---|---|---|---|---|---|---|
| LV_longitu | Z41_S1 | 27446 | -0.06525 | 0.00536 | 4.29E-34 | 33.36799 | Left ventricle |
| RA_minimu | Z50_S3 | 27446 | -0.06405 | 0.005263 | 4.55E-34 | 33.34202 | Right atrium |
| Ascending_ | Z95_S5 | 24599 | -0.08435 | 0.006936 | 4.95E-34 | 33.30529 | Aorta |
| Ascending_ | Z86_S3 | 24599 | -0.0835 | 0.006872 | 5.65E-34 | 33.24824 | Aorta |
| LA_maximu | Z32_S1 | 27446 | 0.067378 | 0.005545 | 5.68E-34 | 33.2454 | Left atrium |
| LV_circumf | Z58_S2 | 27446 | 0.057852 | 0.004762 | 5.86E-34 | 33.232 | Left ventricle |
| LV_radial_s | Z120_S5 | 27446 | 0.075485 | 0.006216 | 6.24E-34 | 33.20471 | Left ventricle |
| LA_ejectio | Z118_S1 | 27446 | -0.06555 | 0.0054 | 6.53E-34 | 33.18478 | Left atrium |
| RV_stroke_ | Z27_S2 | 27446 | -0.08678 | 0.00715 | 6.71E-34 | 33.17299 | Right ventricle |
| LA_minimu | Z11_S5 | 27446 | -0.0591 | 0.004871 | 7.08E-34 | 33.15015 | Left atrium |
| RA_stroke_ | Z120_S5 | 27446 | 0.073894 | 0.006097 | 8.28E-34 | 33.08218 | Right atrium |
| LV_ejectio | Z99_S3 | 27446 | -0.04888 | 0.004033 | 8.30E-34 | 33.0809 | Left ventricle |
| LV_longitu | Z95_S5 | 27446 | -0.07233 | 0.005977 | 1.05E-33 | 32.977 | Left ventricle |
| Ascending_ | Z60_S2 | 24599 | -0.06068 | 0.005015 | 1.06E-33 | 32.97596 | Aorta |
| LV_cardiac | Z95_S3 | 27446 | -0.05169 | 0.004273 | 1.08E-33 | 32.96691 | Left ventricle |
| LV_radial_s | Z29_S5 | 27446 | -0.06654 | 0.005504 | 1.18E-33 | 32.92817 | Left ventricle |
| LV_radial_s | Z44_S4 | 27446 | 0.052713 | 0.00436 | 1.18E-33 | 32.92739 | Left ventricle |
| LA_minimu | Z34_S2 | 27446 | -0.07012 | 0.005805 | 1.37E-33 | 32.86474 | Left atrium |
| RA_maxim | Z122_S2 | 27446 | 0.054711 | 0.004531 | 1.45E-33 | 32.83999 | Right atrium |
| LV_longitu | Z55_S1 | 27446 | 0.055534 | 0.004601 | 1.53E-33 | 32.81623 | Left ventricle |
| LV_circumf | Z24_S5 | 27446 | 0.066781 | 0.005535 | 1.62E-33 | 32.79071 | Left ventricle |
| LV_radial_s | Z57_S1 | 27446 | 0.057301 | 0.004751 | 1.69E-33 | 32.77116 | Left ventricle |
| RA_ejectio | Z51_S4 | 27446 | 0.05193 | 0.004307 | 1.80E-33 | 32.74355 | Right atrium |
| LA_maximu | Z122_S5 | 27446 | 0.056585 | 0.004697 | 2.04E-33 | 32.68993 | Left atrium |
| RV_stroke_ | Z108_S5 | 27446 | -0.07571 | 0.006291 | 2.33E-33 | 32.63258 | Right ventricle |
| RA_minimu | Z44_S1 | 27446 | -0.06743 | 0.005603 | 2.36E-33 | 32.62651 | Right atrium |
| LV_ejectio | Z37_S4 | 27446 | 0.074434 | 0.006191 | 2.72E-33 | 32.56611 | Left ventricle |
| Ascending_ | Z64_S1 | 24599 | -0.07706 | 0.006414 | 3.02E-33 | 32.52007 | Aorta |
| LV_radial_s | Z122_S5 | 27446 | -0.0556 | 0.004631 | 3.32E-33 | 32.47877 | Left ventricle |
| LV_end_sy | Z76_S3 | 27446 | -0.07952 | 0.006624 | 3.39E-33 | 32.4703 | Left ventricle |
| LV_circumf | Z122_S1 | 27446 | -0.06672 | 0.005559 | 3.50E-33 | 32.45546 | Left ventricle |
| LV_myocar | Z24_S5 | 27446 | -0.10932 | 0.009115 | 3.83E-33 | 32.41665 | Left ventricle |
| LV_radial_s | Z127_S5 | 27446 | -0.05257 | 0.004385 | 4.09E-33 | 32.38799 | Left ventricle |
| LV_longitu | Z92_S5 | 27446 | -0.07173 | 0.005984 | 4.16E-33 | 32.38136 | Left ventricle |
| LV_radial_s | Z19_S2 | 27446 | -0.06089 | 0.005081 | 4.30E-33 | 32.3666 | Left ventricle |
| LA_stroke_ | Z115_S3 | 27446 | -0.06692 | 0.005585 | 4.40E-33 | 32.35694 | Left atrium |
| RA_maxim | Z26_S3 | 27446 | -0.07475 | 0.00624 | 4.64E-33 | 32.3333 | Right atrium |
| LV_stroke_ | Z108_S5 | 27446 | -0.07425 | 0.006199 | 4.71E-33 | 32.32732 | Left ventricle |
| RV_stroke_ | Z64_S1 | 27446 | -0.08171 | 0.006823 | 4.78E-33 | 32.32068 | Right ventricle |
| RA_stroke_ | Z62_S1 | 27446 | 0.056623 | 0.00473 | 4.98E-33 | 32.30238 | Right atrium |
| LV_end_sy | Z97_S1 | 27446 | -0.09191 | 0.007679 | 5.21E-33 | 32.28357 | Left ventricle |
| LV_longitu | Z38_S4 | 27446 | -0.06811 | 0.005691 | 5.24E-33 | 32.28098 | Left ventricle |
| LV_circumf | Z95_S3 | 27446 | -0.04681 | 0.003912 | 5.41E-33 | 32.26694 | Left ventricle |
| LV_end_di | Z65_S1 | 27446 | -0.06756 | 0.005654 | 6.60E-33 | 32.18037 | Left ventricle |
| LV_radial_s | Z83_S2 | 27446 | -0.05802 | 0.004858 | 7.07E-33 | 32.15052 | Left ventricle |

| | | | | | | | |
|---|---|---|---|---|---|---|---|
| LA_minimu | Z50_S2 | 27446 | 0.073974 | 0.006196 | 7.40E-33 | 32.13105 | Left atrium |
| Descending | Z56_S2 | 24599 | -0.06759 | 0.005661 | 7.44E-33 | 32.12824 | Aorta |
| Descending | Z1_S4 | 24599 | -0.08281 | 0.00694 | 8.12E-33 | 32.09047 | Aorta |
| LA_ejection | Z75_S1 | 27446 | 0.059612 | 0.005001 | 9.31E-33 | 32.03099 | Left atrium |
| RA_stroke_ | Z118_S1 | 27446 | 0.066941 | 0.005619 | 1.01E-32 | 31.99781 | Right atrium |
| LV_longitu | Z24_S1 | 27446 | -0.05006 | 0.004207 | 1.18E-32 | 31.92728 | Left ventricle |
| Ascending_ | Z108_S5 | 24599 | 0.073949 | 0.006216 | 1.24E-32 | 31.90588 | Aorta |
| LV_longitu | Z112_S3 | 27446 | -0.06993 | 0.005884 | 1.43E-32 | 31.8435 | Left ventricle |
| LV_ejection | Z106_S4 | 27446 | -0.05225 | 0.004398 | 1.52E-32 | 31.81948 | Left ventricle |
| Ascending_ | Z108_S5 | 24599 | -0.07013 | 0.005907 | 1.67E-32 | 31.77673 | Aorta |
| LA_minimu | Z80_S1 | 27446 | 0.065168 | 0.005491 | 1.72E-32 | 31.7634 | Left atrium |
| RA_maxim | Z19_S2 | 27446 | 0.064849 | 0.00547 | 2.01E-32 | 31.6967 | Right atrium |
| RA_maxim | Z61_S4 | 27446 | -0.06188 | 0.005224 | 2.30E-32 | 31.63838 | Right atrium |
| LV_radial_ | Z17_S1 | 27446 | -0.05698 | 0.004814 | 2.50E-32 | 31.60179 | Left ventricle |
| RV_end_di | Z99_S4 | 27446 | 0.094779 | 0.008014 | 2.85E-32 | 31.5447 | Right ventricle |
| RA_minimu | Z106_S3 | 27446 | -0.07833 | 0.006628 | 3.15E-32 | 31.50141 | Right atrium |
| RV_end_sy | Z114_S4 | 27446 | 0.078634 | 0.006656 | 3.29E-32 | 31.48286 | Right ventricle |
| Ascending_ | Z31_S5 | 24599 | 0.055012 | 0.00466 | 3.68E-32 | 31.43409 | Aorta |
| Ascending_ | Z49_S1 | 24599 | -0.08737 | 0.007403 | 3.81E-32 | 31.41908 | Aorta |
| LA_minimu | Z62_S4 | 27446 | -0.06846 | 0.005803 | 4.06E-32 | 31.39188 | Left atrium |
| Descending | Z12_S5 | 24599 | -0.09323 | 0.00791 | 4.62E-32 | 31.33525 | Aorta |
| LV_cardiac | Z23_S3 | 27446 | 0.05802 | 0.004923 | 4.67E-32 | 31.33066 | Left ventricle |
| RA_stroke_ | Z107_S1 | 27446 | -0.04792 | 0.004068 | 4.93E-32 | 31.3072 | Right atrium |
| LA_maximu | Z26_S4 | 27446 | 0.076234 | 0.006474 | 5.27E-32 | 31.27798 | Left atrium |
| RA_maxim | Z106_S3 | 27446 | -0.07698 | 0.00654 | 5.48E-32 | 31.26147 | Right atrium |
| RA_maxim | Z40_S4 | 27446 | -0.07873 | 0.006691 | 5.80E-32 | 31.23682 | Right atrium |
| Ascending_ | Z1_S1 | 24599 | -0.06924 | 0.005888 | 6.36E-32 | 31.19656 | Aorta |
| RA_stroke_ | Z122_S2 | 27446 | 0.048521 | 0.004129 | 6.90E-32 | 31.16144 | Right atrium |
| RV_ejectio | Z13_S1 | 27446 | 0.053667 | 0.004569 | 7.34E-32 | 31.13419 | Right ventricle |
| RA_maxim | Z69_S5 | 27446 | -0.05528 | 0.004713 | 9.07E-32 | 31.04239 | Right atrium |
| RA_maxim | Z50_S3 | 27446 | -0.0609 | 0.005194 | 9.60E-32 | 31.01777 | Right atrium |
| LA_ejection | Z55_S5 | 27446 | -0.04855 | 0.004144 | 1.04E-31 | 30.985 | Left atrium |
| Descending | Z26_S4 | 24599 | 0.096884 | 0.008269 | 1.04E-31 | 30.98249 | Aorta |
| LV_end_sy | Z62_S3 | 27446 | 0.077183 | 0.006588 | 1.07E-31 | 30.97254 | Left ventricle |
| LV_end_sy | Z62_S4 | 27446 | -0.0844 | 0.007208 | 1.13E-31 | 30.94709 | Left ventricle |
| LA_ejection | Z22_S2 | 27446 | -0.05765 | 0.004927 | 1.26E-31 | 30.90077 | Left atrium |
| Descending | Z19_S2 | 24599 | 0.075687 | 0.006471 | 1.33E-31 | 30.87564 | Aorta |
| Ascending_ | Z34_S2 | 24599 | -0.07675 | 0.006565 | 1.44E-31 | 30.84242 | Aorta |
| LV_circumf | Z108_S5 | 27446 | -0.06081 | 0.005203 | 1.45E-31 | 30.83875 | Left ventricle |
| LA_maximu | Z50_S2 | 27446 | 0.074682 | 0.006391 | 1.50E-31 | 30.82463 | Left atrium |
| LV_ejection | Z62_S2 | 27446 | -0.05172 | 0.004429 | 1.66E-31 | 30.77907 | Left ventricle |
| Descending | Z86_S3 | 24599 | -0.08702 | 0.007454 | 1.73E-31 | 30.76224 | Aorta |
| RV_end_di | Z50_S5 | 27446 | 0.104 | 0.008909 | 1.74E-31 | 30.75869 | Right ventricle |
| LA_ejection | Z80_S4 | 27446 | 0.048866 | 0.004188 | 1.86E-31 | 30.7294 | Left atrium |
| Ascending_ | Z99_S4 | 24599 | -0.07879 | 0.006761 | 2.21E-31 | 30.65554 | Aorta |

| | | | | | | | |
|---|---|---|---|---|---|---|---|
| Descending | Z47_S4 | 24599 | -0.06935 | 0.005955 | 2.42E-31 | 30.6167 | Aorta |
| LV_myocar | Z122_S2 | 27446 | -0.07961 | 0.006838 | 2.49E-31 | 30.60416 | Left ventricle |
| LA_stroke_ | Z26_S4 | 27446 | 0.075726 | 0.006507 | 2.66E-31 | 30.57471 | Left atrium |
| LV_circumf | Z106_S4 | 27446 | 0.052012 | 0.004469 | 2.66E-31 | 30.57435 | Left ventricle |
| Descending | Z66_S3 | 24599 | -0.08819 | 0.00758 | 2.74E-31 | 30.56203 | Aorta |
| LA_minimu | Z21_S3 | 27446 | -0.07379 | 0.006345 | 2.92E-31 | 30.53458 | Left atrium |
| RA_maxim | Z100_S3 | 27446 | -0.06619 | 0.005695 | 3.14E-31 | 30.50276 | Right atrium |
| LV_mean_ | Z80_S1 | 27446 | -0.08775 | 0.007551 | 3.22E-31 | 30.49191 | Left ventricle |
| LV_ejectio | Z53_S1 | 27446 | 0.060464 | 0.005203 | 3.24E-31 | 30.48911 | Left ventricle |
| LV_ejectio | Z115_S4 | 27446 | -0.05323 | 0.004583 | 3.44E-31 | 30.46342 | Left ventricle |
| LV_cardiac | Z55_S5 | 27446 | 0.055009 | 0.004736 | 3.44E-31 | 30.46312 | Left ventricle |
| RV_stroke_ | Z91_S1 | 27446 | 0.068997 | 0.005942 | 3.57E-31 | 30.44726 | Right ventricle |
| LV_radial_ | Z18_S4 | 27446 | -0.07465 | 0.006431 | 3.77E-31 | 30.42396 | Left ventricle |
| LV_longitu | Z44_S3 | 27446 | 0.063565 | 0.005478 | 3.96E-31 | 30.40217 | Left ventricle |
| RV_end_sy | Z115_S3 | 27446 | -0.08442 | 0.007279 | 4.21E-31 | 30.37622 | Right ventricle |
| RV_end_sy | Z106_S3 | 27446 | -0.09352 | 0.00807 | 4.72E-31 | 30.32573 | Right ventricle |
| LV_stroke_ | Z34_S2 | 27446 | -0.0804 | 0.00694 | 4.85E-31 | 30.31417 | Left ventricle |
| Ascending_ | Z24_S5 | 24599 | 0.072972 | 0.006302 | 5.29E-31 | 30.27676 | Aorta |
| RA_minimu | Z32_S1 | 27446 | 0.069072 | 0.005966 | 5.40E-31 | 30.26738 | Right atrium |
| RA_maxim | Z98_S3 | 27446 | -0.05601 | 0.004849 | 7.37E-31 | 30.13276 | Right atrium |
| Ascending_ | Z94_S2 | 24599 | 0.071868 | 0.006234 | 9.44E-31 | 30.02505 | Aorta |
| LV_radial_ | Z65_S5 | 27446 | 0.070607 | 0.006134 | 1.17E-30 | 29.93141 | Left ventricle |
| LV_ejectio | Z64_S1 | 27446 | 0.063915 | 0.005554 | 1.20E-30 | 29.92189 | Left ventricle |
| LV_radial_ | Z51_S1 | 27446 | 0.052009 | 0.004521 | 1.27E-30 | 29.89625 | Left ventricle |
| RA_ejectio | Z62_S2 | 27446 | -0.05065 | 0.004404 | 1.33E-30 | 29.87754 | Right atrium |
| RV_end_sy | Z94_S2 | 27446 | 0.08479 | 0.007375 | 1.37E-30 | 29.86398 | Right ventricle |
| RA_ejectio | Z87_S3 | 27446 | 0.055691 | 0.004845 | 1.41E-30 | 29.8518 | Right atrium |
| LV_ejectio | Z82_S1 | 27446 | -0.07101 | 0.006179 | 1.45E-30 | 29.83854 | Left ventricle |
| RA_stroke_ | Z51_S4 | 27446 | -0.05026 | 0.004378 | 1.62E-30 | 29.78918 | Right atrium |
| LV_radial_ | Z40_S5 | 27446 | 0.050585 | 0.004407 | 1.72E-30 | 29.76509 | Left ventricle |
| Descending | Z38_S4 | 24599 | -0.08809 | 0.007675 | 1.73E-30 | 29.76248 | Aorta |
| RV_ejectio | Z53_S1 | 27446 | 0.060954 | 0.005311 | 1.73E-30 | 29.76212 | Right ventricle |
| LV_radial_ | Z3_S2 | 27446 | 0.052698 | 0.004592 | 1.74E-30 | 29.76035 | Left ventricle |
| LA_stroke_ | Z122_S2 | 27446 | -0.04919 | 0.004292 | 2.04E-30 | 29.69057 | Left atrium |
| Descending | Z125_S4 | 24599 | -0.07825 | 0.00683 | 2.19E-30 | 29.65936 | Aorta |
| LA_ejectio | Z78_S4 | 27446 | -0.04929 | 0.004304 | 2.34E-30 | 29.63161 | Left atrium |
| LV_longitu | Z40_S4 | 27446 | 0.068537 | 0.005989 | 2.52E-30 | 29.59853 | Left ventricle |
| Ascending_ | Z50_S3 | 24599 | -0.06161 | 0.005384 | 2.58E-30 | 29.58852 | Aorta |
| RV_ejectio | Z10_S4 | 27446 | -0.06088 | 0.005324 | 2.81E-30 | 29.55158 | Right ventricle |
| LV_ejectio | Z83_S2 | 27446 | -0.05394 | 0.004717 | 2.82E-30 | 29.54984 | Left ventricle |
| LV_end_di | Z51_S3 | 27446 | -0.09504 | 0.008325 | 3.50E-30 | 29.45646 | Left ventricle |
| LV_radial_ | Z2_S1 | 27446 | 0.053631 | 0.004698 | 3.50E-30 | 29.45602 | Left ventricle |
| RV_ejectio | Z114_S5 | 27446 | 0.046496 | 0.004074 | 3.63E-30 | 29.44046 | Right ventricle |
| RA_ejectio | Z20_S2 | 27446 | -0.04787 | 0.004203 | 4.81E-30 | 29.318 | Right atrium |
| RA_minimu | Z100_S3 | 27446 | -0.06573 | 0.005772 | 4.85E-30 | 29.31446 | Right atrium |

| | | | | | | | |
|---|---|---|---|---|---|---|---|
| LV_circumf | Z125_S4 | 27446 | 0.059643 | 0.005239 | 5.03E-30 | 29.29823 | Left ventricle |
| LV_radial_s | Z73_S4 | 27446 | -0.06736 | 0.005917 | 5.03E-30 | 29.29802 | Left ventricle |
| Descending | Z74_S4 | 24599 | -0.07975 | 0.007006 | 5.09E-30 | 29.2937 | Aorta |
| LV_longitu | Z97_S2 | 27446 | 0.059675 | 0.005244 | 5.28E-30 | 29.27697 | Left ventricle |
| LA_ejection | Z62_S3 | 27446 | -0.05785 | 0.005085 | 5.42E-30 | 29.26613 | Left atrium |
| RA_minimu | Z57_S1 | 27446 | -0.05897 | 0.005185 | 5.68E-30 | 29.24568 | Right atrium |
| Descending | Z115_S3 | 24599 | -0.07955 | 0.006995 | 5.74E-30 | 29.2409 | Aorta |
| LV_myocar | Z96_S3 | 27446 | 0.092603 | 0.008145 | 5.97E-30 | 29.22386 | Left ventricle |
| LV_end_dia | Z93_S1 | 27446 | 0.088776 | 0.007811 | 6.23E-30 | 29.20546 | Left ventricle |
| LV_circumf | Z62_S2 | 27446 | 0.051115 | 0.004501 | 6.82E-30 | 29.16607 | Left ventricle |
| LV_circumf | Z80_S4 | 27446 | -0.04975 | 0.004383 | 7.37E-30 | 29.13255 | Left ventricle |
| LA_maximu | Z87_S5 | 27446 | -0.05847 | 0.005156 | 8.27E-30 | 29.08264 | Left atrium |
| LV_myocar | Z112_S3 | 27446 | -0.11251 | 0.009923 | 8.47E-30 | 29.07221 | Left ventricle |
| Ascending_ | Z81_S1 | 24599 | -0.06589 | 0.005816 | 9.39E-30 | 29.02724 | Aorta |
| Descending | Z99_S4 | 24599 | -0.0752 | 0.006639 | 9.62E-30 | 29.01705 | Aorta |
| LA_ejection | Z109_S1 | 27446 | 0.055094 | 0.004867 | 1.06E-29 | 28.97609 | Left atrium |
| LA_ejection | Z5_S1 | 27446 | 0.05419 | 0.004788 | 1.08E-29 | 28.96555 | Left atrium |
| LV_cardiac | Z39_S1 | 27446 | 0.058006 | 0.005126 | 1.09E-29 | 28.96081 | Left ventricle |
| Ascending_ | Z99_S5 | 24599 | 0.062239 | 0.005506 | 1.26E-29 | 28.89912 | Aorta |
| RV_end_di | Z3_S4 | 27446 | 0.101066 | 0.008943 | 1.30E-29 | 28.88535 | Right ventricle |
| Descending | Z50_S2 | 24599 | 0.092153 | 0.008161 | 1.44E-29 | 28.84146 | Aorta |
| Ascending_ | Z32_S1 | 24599 | 0.072774 | 0.006452 | 1.65E-29 | 28.78138 | Aorta |
| Ascending_ | Z13_S3 | 24599 | 0.052842 | 0.004685 | 1.68E-29 | 28.77396 | Aorta |
| RV_stroke_ | Z94_S2 | 27446 | 0.074651 | 0.006624 | 1.84E-29 | 28.73589 | Right ventricle |
| LA_ejection | Z79_S4 | 27446 | 0.064484 | 0.005722 | 1.86E-29 | 28.73162 | Left atrium |
| LV_longitu | Z27_S2 | 27446 | -0.06505 | 0.005777 | 2.06E-29 | 28.68714 | Left ventricle |
| Ascending_ | Z34_S2 | 24599 | -0.07421 | 0.006591 | 2.06E-29 | 28.68552 | Aorta |
| LV_circumf | Z44_S1 | 27446 | 0.057037 | 0.005067 | 2.15E-29 | 28.66765 | Left ventricle |
| LV_cardiac | Z89_S4 | 27446 | 0.060373 | 0.005365 | 2.24E-29 | 28.64961 | Left ventricle |
| RA_maxim | Z3_S2 | 27446 | 0.055619 | 0.004944 | 2.32E-29 | 28.6354 | Right atrium |
| LV_radial_s | Z74_S2 | 27446 | -0.05726 | 0.00509 | 2.36E-29 | 28.62789 | Left ventricle |
| RA_ejectio | Z80_S5 | 27446 | 0.065991 | 0.005868 | 2.40E-29 | 28.61904 | Right atrium |
| RA_maxim | Z105_S5 | 27446 | 0.063098 | 0.005612 | 2.48E-29 | 28.60468 | Right atrium |
| LV_circumf | Z7_S5 | 27446 | -0.06233 | 0.005554 | 3.15E-29 | 28.50129 | Left ventricle |
| RA_stroke_ | Z59_S3 | 27446 | -0.04594 | 0.004095 | 3.35E-29 | 28.4747 | Right atrium |
| RV_end_sy | Z57_S2 | 27446 | 0.063713 | 0.005681 | 3.46E-29 | 28.4608 | Right ventricle |
| LV_circumf | Z43_S2 | 27446 | -0.04989 | 0.004451 | 3.69E-29 | 28.43293 | Left ventricle |
| RA_minimu | Z91_S1 | 27446 | 0.06091 | 0.005436 | 3.83E-29 | 28.41722 | Right atrium |
| LV_cardiac | Z29_S1 | 27446 | -0.06736 | 0.006011 | 3.83E-29 | 28.41674 | Left ventricle |
| LV_radial_s | Z51_S4 | 27446 | 0.049999 | 0.004464 | 4.04E-29 | 28.39338 | Left ventricle |
| RA_minimu | Z97_S1 | 27446 | -0.07685 | 0.006861 | 4.05E-29 | 28.39288 | Right atrium |
| LV_end_sy | Z21_S3 | 27446 | -0.08828 | 0.007882 | 4.06E-29 | 28.39158 | Left ventricle |
| LV_stroke_ | Z56_S2 | 27446 | -0.05823 | 0.0052 | 4.12E-29 | 28.3854 | Left ventricle |
| LV_end_dia | Z97_S2 | 27446 | 0.080634 | 0.007205 | 4.49E-29 | 28.34821 | Left ventricle |
| LA_ejection | Z50_S4 | 27446 | -0.05257 | 0.004699 | 4.62E-29 | 28.33512 | Left atrium |

| | | | | | | | |
|---|---|---|---|---|---|---|---|
| LV_circumf | Z17_S1 | 27446 | 0.053088 | 0.004749 | 5.25E-29 | 28.28013 | Left ventricle |
| LV_end_dia | Z95_S5 | 27446 | -0.09182 | 0.008215 | 5.27E-29 | 28.27823 | Left ventricle |
| RV_ejectio | Z38_S4 | 27446 | 0.065339 | 0.005855 | 6.47E-29 | 28.18919 | Right ventricle |
| Ascending_ | Z106_S3 | 24599 | -0.07553 | 0.006769 | 6.50E-29 | 28.18683 | Aorta |
| Ascending_ | Z75_S5 | 24599 | 0.084949 | 0.007614 | 6.62E-29 | 28.17936 | Aorta |
| RV_ejectio | Z111_S1 | 27446 | 0.062095 | 0.005566 | 6.71E-29 | 28.17341 | Right ventricle |
| LV_radial_s | Z12_S5 | 27446 | 0.068116 | 0.006117 | 8.43E-29 | 28.07427 | Left ventricle |
| LV_cardiac | Z37_S1 | 27446 | 0.069579 | 0.006249 | 8.49E-29 | 28.07096 | Left ventricle |
| LA_stroke_ | Z99_S4 | 27446 | 0.065295 | 0.005866 | 8.88E-29 | 28.05167 | Left atrium |
| LV_radial_s | Z62_S3 | 27446 | -0.05995 | 0.005395 | 1.09E-28 | 27.96416 | Left ventricle |
| LA_ejection | Z43_S2 | 27446 | 0.047237 | 0.004254 | 1.19E-28 | 27.92528 | Left atrium |
| LV_circumf | Z50_S4 | 27446 | 0.054549 | 0.004917 | 1.33E-28 | 27.87609 | Left ventricle |
| Ascending_ | Z68_S2 | 24599 | -0.05771 | 0.005207 | 1.52E-28 | 27.81701 | Aorta |
| Ascending_ | Z79_S4 | 24599 | -0.07506 | 0.006777 | 1.64E-28 | 27.7854 | Aorta |
| Descending | Z65_S2 | 24599 | 0.067955 | 0.006136 | 1.67E-28 | 27.77722 | Aorta |
| LV_radial_s | Z37_S2 | 27446 | 0.061894 | 0.005591 | 1.74E-28 | 27.75839 | Left ventricle |
| LA_ejection | Z14_S1 | 27446 | -0.05051 | 0.004564 | 1.82E-28 | 27.74102 | Left atrium |
| LV_radial_s | Z66_S2 | 27446 | -0.06529 | 0.005902 | 1.90E-28 | 27.72169 | Left ventricle |
| LV_longitu | Z75_S1 | 27446 | -0.05651 | 0.005113 | 2.15E-28 | 27.66823 | Left ventricle |
| LV_myocar | Z37_S4 | 27446 | 0.114494 | 0.010364 | 2.25E-28 | 27.64848 | Left ventricle |
| LV_circumf | Z85_S5 | 27446 | -0.05256 | 0.004759 | 2.30E-28 | 27.63814 | Left ventricle |
| Descending | Z50_S2 | 24599 | 0.088858 | 0.008046 | 2.35E-28 | 27.62935 | Aorta |
| LA_stroke_ | Z80_S5 | 27446 | -0.06842 | 0.006197 | 2.44E-28 | 27.61275 | Left atrium |
| Descending | Z100_S3 | 24599 | 0.074559 | 0.006756 | 2.54E-28 | 27.59451 | Aorta |
| Descending | Z115_S3 | 24599 | -0.07826 | 0.007097 | 2.80E-28 | 27.55264 | Aorta |
| Ascending_ | Z94_S2 | 24599 | 0.068479 | 0.006212 | 2.97E-28 | 27.52791 | Aorta |
| Descending | Z30_S2 | 24599 | -0.08792 | 0.007977 | 2.98E-28 | 27.52648 | Aorta |
| RV_end_di | Z115_S3 | 27446 | -0.08412 | 0.007635 | 3.17E-28 | 27.49846 | Right ventricle |
| RV_stroke_ | Z106_S3 | 27446 | -0.07964 | 0.007249 | 4.43E-28 | 27.35332 | Right ventricle |
| LV_circumf | Z120_S2 | 27446 | 0.047881 | 0.004362 | 4.93E-28 | 27.30699 | Left ventricle |
| Ascending_ | Z106_S3 | 24599 | -0.07446 | 0.006794 | 5.96E-28 | 27.22507 | Aorta |
| LV_end_dia | Z44_S3 | 27446 | -0.08245 | 0.007528 | 6.45E-28 | 27.19061 | Left ventricle |
| RA_ejectio | Z97_S2 | 27446 | -0.0575 | 0.005255 | 7.25E-28 | 27.13974 | Right atrium |
| LV_radial_s | Z39_S1 | 27446 | -0.05199 | 0.004759 | 8.71E-28 | 27.05998 | Left ventricle |
| RV_stroke_ | Z122_S5 | 27446 | 0.060322 | 0.005527 | 9.88E-28 | 27.00521 | Right ventricle |
| RA_ejectio | Z69_S5 | 27446 | 0.046118 | 0.004227 | 1.04E-27 | 26.98365 | Right atrium |
| LV_radial_s | Z3_S4 | 27446 | 0.070007 | 0.006421 | 1.11E-27 | 26.95325 | Left ventricle |
| RV_stroke_ | Z113_S2 | 27446 | 0.080607 | 0.007399 | 1.22E-27 | 26.91252 | Right ventricle |
| LV_radial_s | Z55_S1 | 27446 | 0.052046 | 0.004778 | 1.25E-27 | 26.90275 | Left ventricle |
| Descending | Z7_S5 | 24599 | 0.070862 | 0.006506 | 1.26E-27 | 26.90007 | Aorta |
| LV_end_sy | Z78_S1 | 27446 | 0.062465 | 0.005739 | 1.36E-27 | 26.86554 | Left ventricle |
| RV_end_sy | Z38_S4 | 27446 | -0.08544 | 0.00785 | 1.38E-27 | 26.86102 | Right ventricle |
| RA_ejectio | Z1_S1 | 27446 | 0.052633 | 0.004841 | 1.57E-27 | 26.80442 | Right atrium |
| RA_minimu | Z60_S2 | 27446 | -0.05326 | 0.0049 | 1.61E-27 | 26.79322 | Right atrium |
| RA_ejectio | Z50_S2 | 27446 | -0.06611 | 0.006084 | 1.67E-27 | 26.77677 | Right atrium |

| | | | | | | | |
|---|---|---|---|---|---|---|---|
| LV_stroke_ | Z112_S3 | 27446 | -0.07791 | 0.007183 | 2.06E-27 | 26.68566 | Left ventricle |
| LV_circumf | Z60_S2 | 27446 | -0.04802 | 0.00443 | 2.23E-27 | 26.65149 | Left ventricle |
| Ascending_ | Z72_S5 | 24599 | -0.07646 | 0.00706 | 2.46E-27 | 26.60822 | Aorta |
| RA_ejectio | Z22_S2 | 27446 | -0.05463 | 0.005047 | 2.63E-27 | 26.57974 | Right atrium |
| RA_ejectio | Z105_S4 | 27446 | 0.049054 | 0.004534 | 2.76E-27 | 26.55831 | Right atrium |
| LA_maximu | Z115_S3 | 27446 | -0.06012 | 0.00556 | 2.96E-27 | 26.52934 | Left atrium |
| RA_ejectio | Z106_S4 | 27446 | -0.04727 | 0.004375 | 3.29E-27 | 26.48258 | Right atrium |
| Descending | Z1_S5 | 24599 | -0.07394 | 0.006849 | 3.61E-27 | 26.4424 | Aorta |
| LA_ejection | Z80_S5 | 27446 | 0.061844 | 0.005731 | 3.81E-27 | 26.41924 | Left atrium |
| RV_stroke_ | Z96_S3 | 27446 | 0.064558 | 0.005983 | 3.86E-27 | 26.41391 | Right ventricle |
| RA_stroke_ | Z27_S2 | 27446 | 0.063447 | 0.005882 | 3.97E-27 | 26.40077 | Right atrium |
| Descending | Z99_S4 | 24599 | 0.080211 | 0.007441 | 4.33E-27 | 26.36344 | Aorta |
| LV_end_dia | Z3_S4 | 27446 | 0.091558 | 0.008499 | 4.62E-27 | 26.33552 | Left ventricle |
| Descending | Z1_S4 | 24599 | -0.07588 | 0.007044 | 4.65E-27 | 26.33241 | Aorta |
| Ascending_ | Z99_S4 | 24599 | 0.068952 | 0.006404 | 4.93E-27 | 26.30697 | Aorta |
| RA_stroke_ | Z1_S1 | 27446 | -0.05295 | 0.004919 | 5.07E-27 | 26.29473 | Right atrium |
| Ascending_ | Z12_S5 | 24599 | -0.0733 | 0.006811 | 5.18E-27 | 26.28551 | Aorta |
| LV_ejection | Z43_S2 | 27446 | 0.047124 | 0.004381 | 5.54E-27 | 26.25654 | Left ventricle |
| RA_maxim | Z39_S1 | 27446 | 0.055108 | 0.005123 | 5.55E-27 | 26.25591 | Right atrium |
| LV_myocar | Z108_S5 | 27446 | -0.09215 | 0.00857 | 5.81E-27 | 26.23593 | Left ventricle |
| RV_ejectio | Z107_S1 | 27446 | 0.044192 | 0.004111 | 5.98E-27 | 26.22333 | Right ventricle |
| LA_ejection | Z28_S2 | 27446 | -0.05558 | 0.005174 | 6.44E-27 | 26.19107 | Left atrium |
| RA_stroke_ | Z40_S5 | 27446 | -0.04644 | 0.004324 | 6.63E-27 | 26.17816 | Right atrium |
| LV_radial_s | Z37_S4 | 27446 | 0.068489 | 0.006381 | 7.13E-27 | 26.14699 | Left ventricle |
| LV_mean_r | Z7_S5 | 27446 | -0.08174 | 0.007616 | 7.20E-27 | 26.14252 | Left ventricle |
| Ascending_ | Z111_S5 | 24599 | 0.060978 | 0.005687 | 7.98E-27 | 26.09826 | Aorta |
| LV_ejection | Z40_S4 | 27446 | -0.0647 | 0.006036 | 8.33E-27 | 26.07928 | Left ventricle |
| Ascending_ | Z99_S5 | 24599 | 0.059228 | 0.005528 | 8.70E-27 | 26.06048 | Aorta |
| RV_end_sy | Z86_S3 | 27446 | -0.08293 | 0.007744 | 9.14E-27 | 26.03908 | Right ventricle |
| LV_mean_r | Z13_S1 | 27446 | -0.06673 | 0.006239 | 1.07E-26 | 25.9721 | Left ventricle |
| Ascending_ | Z49_S3 | 24599 | -0.0714 | 0.006676 | 1.08E-26 | 25.96818 | Aorta |
| LA_ejection | Z87_S3 | 27446 | 0.050603 | 0.004733 | 1.12E-26 | 25.95185 | Left atrium |
| LV_ejection | Z120_S5 | 27446 | 0.064545 | 0.006038 | 1.14E-26 | 25.94246 | Left ventricle |
| LV_end_dia | Z62_S3 | 27446 | 0.076321 | 0.007141 | 1.17E-26 | 25.93219 | Left ventricle |
| Ascending_ | Z80_S5 | 24599 | 0.072578 | 0.006793 | 1.21E-26 | 25.91846 | Aorta |
| LV_ejection | Z60_S2 | 27446 | 0.046573 | 0.00436 | 1.23E-26 | 25.90951 | Left ventricle |
| RV_ejectio | Z7_S5 | 27446 | 0.059547 | 0.00558 | 1.38E-26 | 25.85865 | Right ventricle |
| Descending | Z100_S3 | 24599 | 0.073078 | 0.006854 | 1.54E-26 | 25.81371 | Aorta |
| RV_end_sy | Z3_S2 | 27446 | 0.065033 | 0.006102 | 1.59E-26 | 25.79729 | Right ventricle |
| RV_end_sy | Z47_S4 | 27446 | -0.07272 | 0.006835 | 1.96E-26 | 25.7077 | Right ventricle |
| Descending | Z37_S1 | 24599 | 0.078746 | 0.007403 | 2.01E-26 | 25.69738 | Aorta |
| RA_ejectio | Z59_S3 | 27446 | 0.042867 | 0.004032 | 2.10E-26 | 25.67726 | Right atrium |
| LV_longitu | Z115_S3 | 27446 | 0.056136 | 0.005281 | 2.18E-26 | 25.66199 | Left ventricle |
| RA_stroke_ | Z29_S1 | 27446 | 0.058091 | 0.005474 | 2.60E-26 | 25.58577 | Right atrium |
| LV_cardiac_ | Z28_S1 | 27446 | -0.06224 | 0.005864 | 2.60E-26 | 25.58522 | Left ventricle |

| | | | | | | | |
|---|---|---|---|---|---|---|---|
| LV_cardiac | Z40_S5 | 27446 | 0.050239 | 0.00475 | 3.84E-26 | 25.41619 | Left ventricle |
| LV_longitu | Z50_S4 | 27446 | -0.05079 | 0.004803 | 3.88E-26 | 25.41063 | Left ventricle |
| LA_ejection | Z95_S3 | 27446 | 0.039552 | 0.003741 | 3.99E-26 | 25.3986 | Left atrium |
| RA_minimu | Z50_S2 | 27446 | 0.072687 | 0.006877 | 4.15E-26 | 25.38156 | Right atrium |
| LA_ejection | Z6_S2 | 27446 | 0.039916 | 0.003778 | 4.36E-26 | 25.36077 | Left atrium |
| LV_longitu | Z87_S5 | 27446 | 0.051719 | 0.004898 | 4.65E-26 | 25.33248 | Left ventricle |
| LV_circumf | Z20_S1 | 27446 | 0.062406 | 0.005913 | 4.83E-26 | 25.31595 | Left ventricle |
| RV_ejectio | Z95_S3 | 27446 | 0.041502 | 0.003932 | 4.85E-26 | 25.31453 | Right ventricle |
| RV_end_sy | Z120_S5 | 27446 | 0.087187 | 0.008263 | 4.98E-26 | 25.30274 | Right ventricle |
| LV_end_di | Z122_S2 | 27446 | -0.05878 | 0.005574 | 5.39E-26 | 25.26819 | Left ventricle |
| RA_ejectio | Z111_S5 | 27446 | -0.052 | 0.004932 | 5.45E-26 | 25.26398 | Right atrium |
| Ascending_ | Z125_S4 | 24599 | -0.06289 | 0.005965 | 5.48E-26 | 25.26158 | Aorta |
| LV_radial_s | Z31_S2 | 27446 | -0.05462 | 0.005182 | 5.61E-26 | 25.2514 | Left ventricle |
| RV_end_di | Z56_S2 | 27446 | -0.06495 | 0.006164 | 5.83E-26 | 25.23397 | Right ventricle |
| LA_minimu | Z31_S2 | 27446 | 0.053683 | 0.005097 | 6.11E-26 | 25.21369 | Left atrium |
| LA_maximu | Z100_S3 | 27446 | 0.056534 | 0.005368 | 6.15E-26 | 25.21106 | Left atrium |
| Descending | Z79_S4 | 24599 | -0.08286 | 0.007877 | 7.06E-26 | 25.15136 | Aorta |
| LV_end_sy | Z95_S5 | 27446 | -0.07977 | 0.007584 | 7.15E-26 | 25.14548 | Left ventricle |
| LA_stroke_ | Z121_S4 | 27446 | 0.062295 | 0.005924 | 7.37E-26 | 25.1327 | Left atrium |
| LA_ejection | Z50_S3 | 27446 | 0.047831 | 0.00455 | 7.67E-26 | 25.11537 | Left atrium |
| LA_minimu | Z28_S2 | 27446 | 0.056749 | 0.005399 | 7.69E-26 | 25.114 | Left atrium |
| RA_ejectio | Z75_S4 | 27446 | 0.061772 | 0.005887 | 9.31E-26 | 25.03098 | Right atrium |
| LV_circumf | Z53_S2 | 27446 | -0.05901 | 0.005624 | 9.49E-26 | 25.02272 | Left ventricle |
| Ascending_ | Z49_S3 | 24599 | -0.06978 | 0.006652 | 9.72E-26 | 25.01235 | Aorta |
| LV_stroke_ | Z94_S2 | 27446 | 0.068467 | 0.006529 | 1.00E-25 | 24.99978 | Left ventricle |
| LV_cardiac | Z94_S2 | 27446 | 0.062733 | 0.005984 | 1.03E-25 | 24.98718 | Left ventricle |
| LV_end_di | Z50_S5 | 27446 | 0.088758 | 0.008469 | 1.06E-25 | 24.97593 | Left ventricle |
| LV_ejection | Z80_S4 | 27446 | 0.045193 | 0.004315 | 1.15E-25 | 24.94109 | Left ventricle |
| LV_cardiac | Z123_S4 | 27446 | -0.06091 | 0.005816 | 1.16E-25 | 24.93697 | Left ventricle |
| Ascending_ | Z108_S5 | 24599 | -0.06167 | 0.005889 | 1.16E-25 | 24.93379 | Aorta |
| LV_cardiac | Z31_S2 | 27446 | 0.058453 | 0.005583 | 1.19E-25 | 24.92411 | Left ventricle |
| Ascending_ | Z39_S3 | 24599 | -0.07549 | 0.007211 | 1.20E-25 | 24.92064 | Aorta |
| Ascending_ | Z50_S5 | 24599 | -0.07478 | 0.007145 | 1.23E-25 | 24.91164 | Aorta |
| Descending | Z14_S2 | 24599 | -0.06881 | 0.006576 | 1.26E-25 | 24.89822 | Aorta |
| LV_longitu | Z14_S2 | 27446 | -0.05178 | 0.004949 | 1.28E-25 | 24.89411 | Left ventricle |
| RV_end_sy | Z125_S4 | 27446 | -0.07383 | 0.007058 | 1.31E-25 | 24.88353 | Right ventricle |
| LA_minimu | Z122_S2 | 27446 | -0.04331 | 0.004142 | 1.38E-25 | 24.86122 | Left atrium |
| LV_ejection | Z14_S4 | 27446 | 0.058332 | 0.00558 | 1.41E-25 | 24.85146 | Left ventricle |
| LV_stroke_ | Z97_S2 | 27446 | 0.066904 | 0.006401 | 1.43E-25 | 24.84447 | Left ventricle |
| LV_radial_s | Z94_S5 | 27446 | -0.04646 | 0.004445 | 1.44E-25 | 24.84249 | Left ventricle |
| LV_cardiac | Z8_S3 | 27446 | 0.049291 | 0.004717 | 1.48E-25 | 24.82918 | Left ventricle |
| LV_radial_s | Z50_S3 | 27446 | 0.05041 | 0.004827 | 1.58E-25 | 24.80088 | Left ventricle |
| RV_stroke_ | Z80_S2 | 27446 | 0.057146 | 0.005475 | 1.67E-25 | 24.77821 | Right ventricle |
| LA_stroke_ | Z1_S4 | 27446 | -0.05768 | 0.005533 | 1.91E-25 | 24.71882 | Left atrium |
| Ascending_ | Z37_S4 | 24599 | -0.07423 | 0.007125 | 2.06E-25 | 24.68686 | Aorta |

| | | | | | | | |
|---|---|---|---|---|---|---|---|
| LV_ejectior | Z120_S2 | 27446 | -0.04467 | 0.004294 | 2.39E-25 | 24.62086 | Left ventricle |
| LV_longitu | Z12_S5 | 27446 | -0.06133 | 0.005895 | 2.40E-25 | 24.62009 | Left ventricle |
| RV_end_di | Z34_S2 | 27446 | -0.08557 | 0.008228 | 2.47E-25 | 24.60728 | Right ventricle |
| Descending | Z44_S1 | 24599 | 0.069243 | 0.00667 | 3.03E-25 | 24.51852 | Aorta |
| LV_ejectior | Z68_S2 | 27446 | 0.047232 | 0.004552 | 3.15E-25 | 24.50154 | Left ventricle |
| Descending | Z4_S4 | 24599 | -0.05486 | 0.005287 | 3.22E-25 | 24.49192 | Aorta |
| RV_ejectio | Z38_S5 | 27446 | -0.05587 | 0.005387 | 3.37E-25 | 24.47244 | Right ventricle |
| Descending | Z32_S5 | 24599 | -0.08322 | 0.008028 | 3.55E-25 | 24.44938 | Aorta |
| LA_minimu | Z65_S1 | 27446 | -0.04355 | 0.004204 | 3.85E-25 | 24.41435 | Left atrium |
| Descending | Z56_S2 | 24599 | -0.05951 | 0.005747 | 3.96E-25 | 24.40252 | Aorta |
| LV_myocar | Z118_S1 | 27446 | 0.096417 | 0.009311 | 3.98E-25 | 24.40043 | Left ventricle |
| LV_radial_s | Z105_S4 | 27446 | 0.048632 | 0.004697 | 4.06E-25 | 24.3912 | Left ventricle |
| RA_stroke_ | Z38_S5 | 27446 | -0.0552 | 0.005332 | 4.11E-25 | 24.38622 | Right atrium |
| LV_myocar | Z87_S5 | 27446 | -0.08543 | 0.008259 | 4.47E-25 | 24.34992 | Left ventricle |
| RA_ejectio | Z95_S5 | 27446 | 0.06196 | 0.005993 | 4.74E-25 | 24.32417 | Right atrium |
| LV_ejectior | Z56_S2 | 27446 | 0.044381 | 0.004296 | 5.06E-25 | 24.29556 | Left ventricle |
| LV_circumf | Z2_S1 | 27446 | -0.0479 | 0.004636 | 5.10E-25 | 24.2927 | Left ventricle |
| Ascending_ | Z5_S5 | 24599 | 0.061735 | 0.005981 | 5.62E-25 | 24.25042 | Aorta |
| RV_end_di | Z1_S5 | 27446 | 0.0756 | 0.007326 | 5.77E-25 | 24.23911 | Right ventricle |
| RA_ejectio | Z13_S3 | 27446 | -0.04184 | 0.004056 | 5.92E-25 | 24.22799 | Right atrium |
| Ascending_ | Z47_S4 | 24599 | -0.06261 | 0.006069 | 5.95E-25 | 24.22516 | Aorta |
| LV_stroke_ | Z100_S3 | 27446 | 0.064126 | 0.006221 | 6.54E-25 | 24.18449 | Left ventricle |
| Ascending_ | Z18_S4 | 24599 | 0.074147 | 0.007197 | 6.83E-25 | 24.16561 | Aorta |
| RA_stroke_ | Z75_S1 | 27446 | 0.053609 | 0.005207 | 7.31E-25 | 24.13621 | Right atrium |
| Ascending_ | Z81_S1 | 24599 | -0.06012 | 0.00584 | 7.47E-25 | 24.1264 | Aorta |
| LV_circumf | Z114_S5 | 27446 | -0.04175 | 0.004058 | 7.85E-25 | 24.10488 | Left ventricle |
| Ascending_ | Z109_S1 | 24599 | -0.05915 | 0.00575 | 8.04E-25 | 24.09471 | Aorta |
| RA_stroke_ | Z66_S2 | 27446 | -0.05955 | 0.00579 | 8.30E-25 | 24.08087 | Right atrium |
| RA_maxim | Z49_S3 | 27446 | -0.06615 | 0.006433 | 8.41E-25 | 24.07517 | Right atrium |
| LA_minimu | Z50_S5 | 27446 | 0.064699 | 0.006293 | 8.63E-25 | 24.06413 | Left atrium |
| LA_stroke_ | Z39_S3 | 27446 | 0.067588 | 0.006578 | 9.21E-25 | 24.03596 | Left atrium |
| RA_stroke_ | Z72_S5 | 27446 | 0.063959 | 0.006227 | 9.44E-25 | 24.02509 | Right atrium |
| RV_ejectio | Z55_S5 | 27446 | -0.04475 | 0.004358 | 9.68E-25 | 24.01416 | Right ventricle |
| RV_ejectio | Z51_S1 | 27446 | 0.04602 | 0.004482 | 9.85E-25 | 24.00642 | Right ventricle |
| LA_minimu | Z112_S1 | 27446 | -0.06474 | 0.00631 | 1.07E-24 | 23.97176 | Left atrium |
| RA_stroke_ | Z35_S5 | 27446 | -0.04445 | 0.004334 | 1.12E-24 | 23.9521 | Right atrium |
| RV_end_sy | Z74_S4 | 27446 | -0.07364 | 0.007183 | 1.16E-24 | 23.93589 | Right ventricle |
| Descending | Z78_S4 | 24599 | 0.053871 | 0.005266 | 1.46E-24 | 23.83484 | Aorta |
| Descending | Z26_S3 | 24599 | -0.06856 | 0.006703 | 1.49E-24 | 23.82825 | Aorta |
| LA_maximu | Z3_S4 | 27446 | 0.066619 | 0.006514 | 1.51E-24 | 23.82028 | Left atrium |
| LA_ejectior | Z28_S4 | 27446 | 0.053492 | 0.005232 | 1.56E-24 | 23.80791 | Left atrium |
| LV_longitu | Z66_S2 | 27446 | 0.05815 | 0.005688 | 1.56E-24 | 23.80657 | Left ventricle |
| RV_stroke_ | Z39_S4 | 27446 | -0.05623 | 0.005504 | 1.69E-24 | 23.77168 | Right ventricle |
| LV_stroke_ | Z51_S3 | 27446 | -0.0755 | 0.007398 | 1.86E-24 | 23.73066 | Left ventricle |
| Ascending_ | Z77_S3 | 24599 | -0.0632 | 0.006196 | 1.95E-24 | 23.70896 | Aorta |

| | | | | | | | |
|---|---|---|---|---|---|---|---|
| Descending | Z44_S3 | 24599 | -0.0753 | 0.007382 | 1.98E-24 | 23.70375 | Aorta |
| RA_maxim | Z60_S2 | 27446 | -0.04925 | 0.004836 | 2.33E-24 | 23.63303 | Right atrium |
| Descending | Z75_S5 | 24599 | 0.076123 | 0.007478 | 2.46E-24 | 23.60962 | Aorta |
| LV_radial_ | Z14_S1 | 27446 | -0.04929 | 0.004843 | 2.51E-24 | 23.59954 | Left ventricle |
| Ascending_ | Z77_S3 | 24599 | -0.05968 | 0.005866 | 2.60E-24 | 23.58485 | Aorta |
| Descending | Z5_S1 | 24599 | -0.0597 | 0.00587 | 2.67E-24 | 23.5738 | Aorta |
| LV_longitu | Z37_S2 | 27446 | -0.05474 | 0.005388 | 3.00E-24 | 23.52226 | Left ventricle |
| LV_ejectio | Z13_S1 | 27446 | 0.045412 | 0.004479 | 3.71E-24 | 23.43091 | Left ventricle |
| LV_ejectio | Z65_S5 | 27446 | 0.060344 | 0.005958 | 4.16E-24 | 23.38111 | Left ventricle |
| LV_radial_ | Z96_S3 | 27446 | -0.0508 | 0.005017 | 4.23E-24 | 23.37395 | Left ventricle |
| RV_stroke_ | Z44_S3 | 27446 | -0.0687 | 0.006787 | 4.41E-24 | 23.35566 | Right ventricle |
| Ascending_ | Z123_S4 | 24599 | 0.06434 | 0.00636 | 4.65E-24 | 23.33288 | Aorta |
| Descending | Z38_S4 | 24599 | -0.07653 | 0.007571 | 5.05E-24 | 23.29634 | Aorta |
| LA_maxim | Z99_S4 | 27446 | 0.058994 | 0.005839 | 5.36E-24 | 23.2711 | Left atrium |
| RV_ejectio | Z20_S1 | 27446 | -0.05999 | 0.00594 | 5.62E-24 | 23.25041 | Right ventricle |
| Ascending_ | Z72_S5 | 24599 | 0.07525 | 0.007459 | 6.21E-24 | 23.20694 | Aorta |
| Descending | Z63_S1 | 24599 | -0.07881 | 0.007814 | 6.43E-24 | 23.19181 | Aorta |
| LV_end_di | Z49_S3 | 27446 | -0.07976 | 0.00791 | 6.52E-24 | 23.18601 | Left ventricle |
| LV_circum | Z22_S2 | 27446 | -0.05197 | 0.005159 | 7.14E-24 | 23.14637 | Left ventricle |
| Descending | Z56_S5 | 24599 | -0.05113 | 0.005082 | 8.16E-24 | 23.08812 | Aorta |
| LV_cardiac | Z72_S5 | 27446 | -0.06881 | 0.00684 | 8.35E-24 | 23.07812 | Left ventricle |
| LV_radial_ | Z108_S5 | 27446 | 0.053084 | 0.005278 | 8.47E-24 | 23.07207 | Left ventricle |
| RV_stroke_ | Z34_S2 | 27446 | -0.07077 | 0.007046 | 9.79E-24 | 23.00942 | Right ventricle |
| LV_longitu | Z31_S5 | 27446 | -0.04011 | 0.003994 | 9.93E-24 | 23.00315 | Left ventricle |
| LA_stroke_ | Z65_S5 | 27446 | -0.06275 | 0.006257 | 1.13E-23 | 22.94658 | Left atrium |
| LV_radial_ | Z14_S2 | 27446 | 0.051523 | 0.005138 | 1.15E-23 | 22.93987 | Left ventricle |
| LV_longitu | Z11_S5 | 27446 | 0.047887 | 0.004776 | 1.15E-23 | 22.93809 | Left ventricle |
| LV_radial_ | Z120_S4 | 27446 | 0.051205 | 0.005107 | 1.17E-23 | 22.93093 | Left ventricle |
| RV_end_sy | Z9_S2 | 27446 | 0.07333 | 0.007317 | 1.21E-23 | 22.91565 | Right ventricle |
| LV_circum | Z122_S5 | 27446 | 0.045807 | 0.004572 | 1.25E-23 | 22.90208 | Left ventricle |
| RA_stroke_ | Z89_S4 | 27446 | -0.04895 | 0.004887 | 1.29E-23 | 22.88914 | Right atrium |
| RA_maxim | Z44_S3 | 27446 | -0.06134 | 0.006125 | 1.33E-23 | 22.8768 | Right atrium |
| Ascending_ | Z112_S1 | 24599 | 0.071393 | 0.007133 | 1.39E-23 | 22.85824 | Aorta |
| LV_circum | Z5_S5 | 27446 | -0.05242 | 0.005237 | 1.39E-23 | 22.85802 | Left ventricle |
| Ascending_ | Z39_S3 | 24599 | 0.075966 | 0.00759 | 1.39E-23 | 22.85709 | Aorta |
| RV_stroke_ | Z51_S3 | 27446 | -0.07513 | 0.007507 | 1.41E-23 | 22.84958 | Right ventricle |
| LV_end_sy | Z106_S3 | 27446 | -0.0743 | 0.007426 | 1.43E-23 | 22.84352 | Left ventricle |
| LV_circum | Z28_S1 | 27446 | -0.05373 | 0.00537 | 1.45E-23 | 22.83949 | Left ventricle |
| LV_circum | Z8_S3 | 27446 | 0.043213 | 0.00432 | 1.47E-23 | 22.83273 | Left ventricle |
| RV_stroke_ | Z22_S2 | 27446 | 0.062391 | 0.006239 | 1.52E-23 | 22.81958 | Right ventricle |
| LV_radial_ | Z53_S1 | 27446 | 0.053628 | 0.005363 | 1.54E-23 | 22.81347 | Left ventricle |
| RA_minim | Z13_S3 | 27446 | 0.045842 | 0.004585 | 1.55E-23 | 22.8092 | Right atrium |
| RA_maxim | Z46_S1 | 27446 | -0.05611 | 0.005614 | 1.62E-23 | 22.79118 | Right atrium |
| RA_minim | Z39_S1 | 27446 | 0.051871 | 0.005194 | 1.75E-23 | 22.75741 | Right atrium |
| LV_cardiac | Z120_S2 | 27446 | 0.04753 | 0.004766 | 2.01E-23 | 22.6972 | Left ventricle |

| | | | | | | | |
|---|---|---|---|---|---|---|---|
| LV_mean_ | Z7_S3 | 27446 | -0.08277 | 0.008305 | 2.13E-23 | 22.67158 | Left ventricle |
| LV_ejectio | Z58_S2 | 27446 | -0.04674 | 0.004691 | 2.19E-23 | 22.65931 | Left ventricle |
| Ascending_ | Z122_S1 | 24599 | 0.062566 | 0.00628 | 2.22E-23 | 22.65422 | Aorta |
| LA_ejectio | Z63_S1 | 27446 | 0.05751 | 0.005773 | 2.25E-23 | 22.64718 | Left atrium |
| Ascending_ | Z68_S2 | 24599 | -0.05202 | 0.005229 | 2.57E-23 | 22.5897 | Aorta |
| LV_myocar | Z7_S5 | 27446 | -0.091 | 0.00915 | 2.62E-23 | 22.5817 | Left ventricle |
| Ascending_ | Z44_S1 | 24599 | -0.06027 | 0.006064 | 2.79E-23 | 22.55367 | Aorta |
| RA_ejectio | Z122_S1 | 27446 | -0.05406 | 0.005445 | 3.11E-23 | 22.50677 | Right atrium |
| LV_cardiac | Z59_S2 | 27446 | 0.062948 | 0.00634 | 3.12E-23 | 22.50556 | Left ventricle |
| LA_minimu | Z13_S1 | 27446 | -0.04505 | 0.004538 | 3.15E-23 | 22.50157 | Left atrium |
| Ascending_ | Z100_S3 | 24599 | 0.058745 | 0.005922 | 3.42E-23 | 22.46622 | Aorta |
| LV_stroke_ | Z1_S4 | 27446 | -0.06326 | 0.006381 | 3.62E-23 | 22.44093 | Left ventricle |
| Ascending_ | Z80_S5 | 24599 | -0.07085 | 0.007151 | 3.86E-23 | 22.4136 | Aorta |
| RV_stroke_ | Z97_S2 | 27446 | 0.064361 | 0.006497 | 3.89E-23 | 22.40961 | Right ventricle |
| RA_maxim | Z28_S2 | 27446 | -0.05847 | 0.005911 | 4.48E-23 | 22.34847 | Right atrium |
| LV_stroke_ | Z87_S5 | 27446 | -0.0591 | 0.005978 | 4.79E-23 | 22.32008 | Left ventricle |
| RV_ejectio | Z112_S1 | 27446 | -0.06285 | 0.006358 | 4.84E-23 | 22.3152 | Right ventricle |
| RA_minimu | Z3_S2 | 27446 | 0.049518 | 0.005013 | 5.23E-23 | 22.28144 | Right atrium |
| RA_ejectio | Z50_S4 | 27446 | -0.04752 | 0.004814 | 5.55E-23 | 22.25602 | Right atrium |
| Descending | Z41_S2 | 24599 | -0.07692 | 0.007792 | 5.56E-23 | 22.25458 | Aorta |
| RA_ejectio | Z75_S1 | 27446 | 0.050557 | 0.005125 | 5.93E-23 | 22.22673 | Right atrium |
| RA_maxim | Z55_S5 | 27446 | 0.046628 | 0.004736 | 7.11E-23 | 22.14821 | Right atrium |
| LV_circumf | Z122_S2 | 27446 | -0.04091 | 0.004155 | 7.12E-23 | 22.14735 | Left ventricle |
| LV_longitu | Z80_S4 | 27446 | 0.042161 | 0.004283 | 7.35E-23 | 22.13361 | Left ventricle |
| LV_longitu | Z52_S3 | 27446 | -0.04166 | 0.004238 | 8.24E-23 | 22.08422 | Left ventricle |
| RA_stroke_ | Z47_S4 | 27446 | -0.04962 | 0.005048 | 8.40E-23 | 22.07587 | Right atrium |
| RA_minimu | Z115_S3 | 27446 | -0.05877 | 0.005983 | 8.96E-23 | 22.04762 | Right atrium |
| LV_mean_ | Z82_S4 | 27446 | -0.05905 | 0.006013 | 9.10E-23 | 22.04119 | Left ventricle |
| Descending | Z127_S4 | 24599 | 0.071049 | 0.007239 | 9.76E-23 | 22.01039 | Aorta |
| LA_ejectio | Z94_S2 | 27446 | 0.051377 | 0.005237 | 1.02E-22 | 21.99063 | Left atrium |
| RV_end_di | Z37_S4 | 27446 | 0.087232 | 0.008893 | 1.03E-22 | 21.98902 | Right ventricle |
| LV_radial_ | Z99_S5 | 27446 | -0.04853 | 0.004948 | 1.03E-22 | 21.98687 | Left ventricle |
| RA_ejectio | Z55_S5 | 27446 | -0.04164 | 0.004246 | 1.06E-22 | 21.97639 | Right atrium |
| LV_longitu | Z59_S3 | 27446 | 0.039471 | 0.004025 | 1.06E-22 | 21.97603 | Left ventricle |
| LV_mean_ | Z14_S2 | 27446 | -0.06808 | 0.00695 | 1.17E-22 | 21.93285 | Left ventricle |
| Descending | Z109_S1 | 24599 | -0.0583 | 0.005964 | 1.43E-22 | 21.84552 | Aorta |
| RA_maxim | Z68_S2 | 27446 | -0.04932 | 0.005049 | 1.52E-22 | 21.81806 | Right atrium |
| LV_myocar | Z38_S4 | 27446 | -0.09374 | 0.009603 | 1.66E-22 | 21.77928 | Left ventricle |
| LA_stroke_ | Z10_S4 | 27446 | -0.05346 | 0.005481 | 1.76E-22 | 21.75495 | Left atrium |
| LA_ejectio | Z52_S4 | 27446 | -0.04238 | 0.004346 | 1.80E-22 | 21.74533 | Left atrium |
| LV_end_sy | Z24_S5 | 27446 | -0.06691 | 0.006861 | 1.81E-22 | 21.74165 | Left ventricle |
| LV_cardiac | Z106_S3 | 27446 | -0.06387 | 0.00655 | 1.83E-22 | 21.73865 | Left ventricle |
| LV_myocar | Z76_S3 | 27446 | -0.08588 | 0.008816 | 2.02E-22 | 21.69561 | Left ventricle |
| Ascending_ | Z57_S2 | 24599 | -0.04671 | 0.004797 | 2.06E-22 | 21.68586 | Aorta |
| RV_ejectio | Z18_S4 | 27446 | -0.06206 | 0.006377 | 2.20E-22 | 21.65727 | Right ventricle |

| | | | | | | | |
|---|---|---|---|---|---|---|---|
| Ascending_ | Z63_S1 | 24599 | -0.07002 | 0.007206 | 2.57E-22 | 21.59045 | Aorta |
| Descending | Z51_S3 | 24599 | -0.079 | 0.008136 | 2.74E-22 | 21.56294 | Aorta |
| Descending | Z7_S3 | 24599 | 0.068734 | 0.007082 | 2.85E-22 | 21.54537 | Aorta |
| RA_stroke_ | Z56_S5 | 27446 | -0.04182 | 0.004311 | 3.01E-22 | 21.52181 | Right atrium |
| LV_ejection | Z50_S4 | 27446 | -0.04695 | 0.004841 | 3.08E-22 | 21.51093 | Left ventricle |
| Descending | Z80_S5 | 24599 | -0.06801 | 0.007021 | 3.43E-22 | 21.46467 | Aorta |
| RA_ejectio | Z63_S1 | 27446 | 0.057251 | 0.005912 | 3.56E-22 | 21.44888 | Right atrium |
| LV_longitu | Z21_S3 | 27446 | 0.060201 | 0.00622 | 3.73E-22 | 21.42839 | Left ventricle |
| LV_ejection | Z95_S3 | 27446 | 0.037262 | 0.003854 | 4.08E-22 | 21.389 | Left ventricle |
| LV_radial_s | Z59_S2 | 27446 | -0.05686 | 0.005885 | 4.37E-22 | 21.35954 | Left ventricle |
| LV_ejection | Z91_S1 | 27446 | -0.04674 | 0.004839 | 4.54E-22 | 21.34263 | Left ventricle |
| Descending | Z91_S1 | 24599 | 0.061494 | 0.006368 | 4.58E-22 | 21.33898 | Aorta |
| RA_maxim | Z11_S5 | 27446 | -0.05154 | 0.005337 | 4.61E-22 | 21.33668 | Right atrium |
| Ascending_ | Z118_S1 | 24599 | -0.06197 | 0.006418 | 4.68E-22 | 21.32966 | Aorta |
| LV_stroke_ | Z44_S3 | 27446 | -0.06455 | 0.006689 | 4.93E-22 | 21.30697 | Left ventricle |
| LV_circumf | Z6_S2 | 27446 | -0.03813 | 0.003955 | 5.40E-22 | 21.26756 | Left ventricle |
| LV_end_dia | Z3_S2 | 27446 | 0.058615 | 0.006082 | 5.56E-22 | 21.25512 | Left ventricle |
| LV_end_dia | Z94_S2 | 27446 | 0.070864 | 0.007354 | 5.62E-22 | 21.25022 | Left ventricle |
| LA_minimu | Z80_S2 | 27446 | 0.043477 | 0.004515 | 6.05E-22 | 21.21792 | Left atrium |
| LV_radial_s | Z99_S4 | 27446 | -0.05544 | 0.005758 | 6.07E-22 | 21.21687 | Left ventricle |
| LV_longitu | Z8_S3 | 27446 | -0.04061 | 0.00422 | 6.36E-22 | 21.19668 | Left ventricle |
| LV_ejection | Z17_S1 | 27446 | -0.04495 | 0.004677 | 7.25E-22 | 21.1396 | Left ventricle |
| Descending | Z22_S2 | 24599 | 0.058069 | 0.006045 | 7.49E-22 | 21.1253 | Aorta |
| LV_stroke_ | Z32_S1 | 27446 | 0.06178 | 0.006433 | 7.72E-22 | 21.11262 | Left ventricle |
| LA_maximu | Z44_S3 | 27446 | -0.05541 | 0.005772 | 8.02E-22 | 21.09597 | Left atrium |
| Descending | Z37_S2 | 24599 | -0.06963 | 0.007258 | 8.52E-22 | 21.06974 | Aorta |
| RA_stroke_ | Z120_S4 | 27446 | -0.04806 | 0.00501 | 8.58E-22 | 21.06662 | Right atrium |
| RA_maxim | Z114_S5 | 27446 | -0.04246 | 0.00443 | 9.22E-22 | 21.03547 | Right atrium |
| Ascending_ | Z56_S5 | 24599 | -0.04961 | 0.005177 | 9.43E-22 | 21.0254 | Aorta |
| LV_ejection | Z80_S2 | 27446 | -0.04269 | 0.004457 | 9.81E-22 | 21.00815 | Left ventricle |
| LA_minimu | Z95_S5 | 27446 | -0.0585 | 0.006108 | 1.00E-21 | 20.99786 | Left atrium |
| RV_end_sy | Z65_S5 | 27446 | 0.078046 | 0.008154 | 1.06E-21 | 20.97558 | Right ventricle |
| LA_stroke_ | Z34_S2 | 27446 | -0.05764 | 0.006023 | 1.06E-21 | 20.97312 | Left atrium |
| Descending | Z117_S4 | 24599 | -0.04638 | 0.004848 | 1.10E-21 | 20.95901 | Aorta |
| LV_cardiac | Z32_S5 | 27446 | -0.06385 | 0.006683 | 1.25E-21 | 20.90244 | Left ventricle |
| LV_cardiac | Z80_S2 | 27446 | 0.047217 | 0.004946 | 1.35E-21 | 20.87033 | Left ventricle |
| LV_longitu | Z37_S3 | 27446 | -0.05394 | 0.005652 | 1.38E-21 | 20.85909 | Left ventricle |
| RA_minimu | Z114_S5 | 27446 | -0.04279 | 0.00449 | 1.56E-21 | 20.80738 | Right atrium |
| Descending | Z108_S5 | 24599 | 0.058226 | 0.006109 | 1.56E-21 | 20.80675 | Aorta |
| LV_end_sy | Z1_S4 | 27446 | -0.06315 | 0.006632 | 1.71E-21 | 20.76823 | Left ventricle |
| LV_end_sy | Z27_S2 | 27446 | -0.06975 | 0.007329 | 1.78E-21 | 20.74903 | Left ventricle |
| RA_ejectio | Z98_S3 | 27446 | 0.041361 | 0.004352 | 2.00E-21 | 20.69793 | Right atrium |
| LV_myocar | Z10_S4 | 27446 | -0.083 | 0.008734 | 2.02E-21 | 20.69359 | Left ventricle |
| LA_ejection | Z108_S5 | 27446 | 0.047285 | 0.004976 | 2.06E-21 | 20.68663 | Left atrium |
| Descending | Z74_S4 | 24599 | -0.05945 | 0.006257 | 2.08E-21 | 20.68117 | Aorta |

| | | | | | | | |
|---|---|---|---|---|---|---|---|
| LV_mean_ | Z87_S3 | 27446 | -0.06447 | 0.006794 | 2.32E-21 | 20.63499 | Left ventricle |
| Descending | Z37_S1 | 24599 | 0.071258 | 0.007513 | 2.45E-21 | 20.61164 | Aorta |
| RA_minimu | Z55_S1 | 27446 | -0.04945 | 0.005216 | 2.54E-21 | 20.59471 | Right atrium |
| Ascending_ | Z41_S2 | 24599 | -0.06449 | 0.006804 | 2.58E-21 | 20.58831 | Aorta |
| LA_ejection | Z70_S5 | 27446 | -0.04404 | 0.004647 | 2.60E-21 | 20.58454 | Left atrium |
| LA_maximu | Z120_S5 | 27446 | -0.05979 | 0.006312 | 2.74E-21 | 20.56285 | Left atrium |
| Descending | Z14_S2 | 24599 | -0.05639 | 0.005955 | 2.82E-21 | 20.5499 | Aorta |
| RA_stroke_ | Z32_S1 | 27446 | 0.05077 | 0.005367 | 3.11E-21 | 20.50662 | Right atrium |
| RV_stroke_ | Z120_S5 | 27446 | 0.070172 | 0.007423 | 3.29E-21 | 20.48332 | Right ventricle |
| LV_ejection | Z114_S5 | 27446 | 0.037722 | 0.003995 | 3.61E-21 | 20.44196 | Left ventricle |
| LV_end_dia | Z49_S1 | 27446 | -0.0786 | 0.008333 | 4.01E-21 | 20.39685 | Left ventricle |
| Descending | Z57_S2 | 24599 | -0.05239 | 0.005554 | 4.01E-21 | 20.39639 | Aorta |
| LV_end_sy | Z74_S4 | 27446 | -0.06231 | 0.006608 | 4.11E-21 | 20.38642 | Left ventricle |
| LV_ejection | Z59_S3 | 27446 | 0.038191 | 0.004056 | 4.73E-21 | 20.32528 | Left ventricle |
| RA_minimu | Z56_S2 | 27446 | -0.04546 | 0.00483 | 4.85E-21 | 20.31438 | Right atrium |
| Ascending_ | Z11_S5 | 24599 | -0.05199 | 0.005524 | 4.86E-21 | 20.3136 | Aorta |
| LV_end_sy | Z112_S3 | 27446 | -0.07027 | 0.007468 | 4.96E-21 | 20.30467 | Left ventricle |
| RV_stroke_ | Z87_S5 | 27446 | -0.05708 | 0.006068 | 5.08E-21 | 20.29446 | Right ventricle |
| LA_maximu | Z9_S2 | 27446 | 0.052553 | 0.005588 | 5.24E-21 | 20.28074 | Left atrium |
| RA_stroke_ | Z2_S1 | 27446 | -0.04334 | 0.004611 | 5.48E-21 | 20.26125 | Right atrium |
| Ascending_ | Z92_S1 | 24599 | -0.05359 | 0.005703 | 5.57E-21 | 20.25408 | Aorta |
| RA_minimu | Z6_S2 | 27446 | -0.04109 | 0.004375 | 5.90E-21 | 20.22936 | Right atrium |
| LA_maximu | Z10_S4 | 27446 | -0.0512 | 0.005454 | 6.12E-21 | 20.21294 | Left atrium |
| LV_ejection | Z6_S2 | 27446 | 0.036539 | 0.003893 | 6.19E-21 | 20.20819 | Left ventricle |
| LV_radial_ | Z85_S5 | 27446 | -0.04528 | 0.004827 | 6.67E-21 | 20.1759 | Left ventricle |
| LV_ejection | Z122_S2 | 27446 | 0.038356 | 0.00409 | 6.77E-21 | 20.16932 | Left ventricle |
| LV_ejection | Z86_S3 | 27446 | 0.053066 | 0.005662 | 7.09E-21 | 20.14905 | Left ventricle |
| Descending | Z44_S3 | 24599 | -0.06819 | 0.007279 | 7.44E-21 | 20.12868 | Aorta |
| Ascending_ | Z11_S5 | 24599 | -0.05182 | 0.005544 | 9.04E-21 | 20.04369 | Aorta |
| RA_stroke_ | Z112_S1 | 27446 | -0.05882 | 0.006294 | 9.22E-21 | 20.03514 | Right atrium |
| LV_stroke_ | Z26_S3 | 27446 | -0.06371 | 0.006821 | 9.67E-21 | 20.01452 | Left ventricle |
| LV_circumf | Z74_S2 | 27446 | 0.046923 | 0.005025 | 9.75E-21 | 20.01117 | Left ventricle |
| LV_cardiac_ | Z49_S3 | 27446 | -0.06012 | 0.00644 | 1.02E-20 | 19.9929 | Left ventricle |
| Ascending_ | Z51_S4 | 24599 | -0.04897 | 0.005246 | 1.02E-20 | 19.99248 | Aorta |
| RA_maxim | Z57_S1 | 27446 | -0.04779 | 0.00512 | 1.02E-20 | 19.99027 | Right atrium |
| Ascending_ | Z100_S3 | 24599 | 0.055024 | 0.005902 | 1.13E-20 | 19.94805 | Aorta |
| RV_end_sy | Z95_S5 | 27446 | -0.0769 | 0.00825 | 1.15E-20 | 19.9386 | Right ventricle |
| RV_end_di | Z95_S5 | 27446 | -0.08039 | 0.008653 | 1.53E-20 | 19.81601 | Right ventricle |
| RV_ejectio | Z106_S4 | 27446 | -0.04171 | 0.004494 | 1.68E-20 | 19.77474 | Right ventricle |
| Descending | Z41_S2 | 24599 | -0.06544 | 0.007056 | 1.77E-20 | 19.75145 | Aorta |
| LV_cardiac_ | Z51_S1 | 27446 | -0.04521 | 0.004875 | 1.78E-20 | 19.74933 | Left ventricle |
| LV_circumf | Z7_S3 | 27446 | -0.05617 | 0.006058 | 1.84E-20 | 19.73519 | Left ventricle |
| RV_stroke_ | Z40_S4 | 27446 | -0.0688 | 0.007421 | 1.85E-20 | 19.73316 | Right ventricle |
| Ascending_ | Z85_S5 | 24599 | 0.049935 | 0.005387 | 1.88E-20 | 19.72662 | Aorta |
| LV_circumf | Z11_S5 | 27446 | -0.04533 | 0.004891 | 1.89E-20 | 19.72273 | Left ventricle |

| | | | | | | | |
|---|---|---|---|---|---|---|---|
| RV_end_di | Z39_S4 | 27446 | -0.05946 | 0.00643 | 2.30E-20 | 19.63812 | Right ventricle |
| LV_longitu | Z6_S2 | 27446 | 0.035681 | 0.003863 | 2.56E-20 | 19.59164 | Left ventricle |
| RV_ejectio | Z115_S4 | 27446 | -0.04324 | 0.004682 | 2.58E-20 | 19.58756 | Right ventricle |
| Ascending_ | Z80_S5 | 24599 | 0.062517 | 0.006772 | 2.67E-20 | 19.57374 | Aorta |
| LV_circumf | Z38_S4 | 27446 | -0.05384 | 0.005833 | 2.72E-20 | 19.56517 | Left ventricle |
| RV_stroke_ | Z97_S1 | 27446 | -0.0692 | 0.007507 | 3.01E-20 | 19.52148 | Right ventricle |
| LV_stroke_ | Z49_S3 | 27446 | -0.06476 | 0.007027 | 3.12E-20 | 19.50586 | Left ventricle |
| LV_end_di | Z62_S4 | 27446 | -0.07201 | 0.007817 | 3.19E-20 | 19.49683 | Left ventricle |
| Ascending_ | Z32_S5 | 24599 | -0.06386 | 0.006937 | 3.40E-20 | 19.46838 | Aorta |
| RV_end_sy | Z56_S2 | 27446 | -0.05411 | 0.00588 | 3.48E-20 | 19.45781 | Right ventricle |
| RV_ejectio | Z7_S3 | 27446 | 0.055999 | 0.006086 | 3.54E-20 | 19.45152 | Right ventricle |
| Ascending_ | Z32_S5 | 24599 | -0.06357 | 0.006912 | 3.67E-20 | 19.43556 | Aorta |
| LV_longitu | Z73_S4 | 27446 | -0.05246 | 0.005705 | 3.75E-20 | 19.42651 | Left ventricle |
| LV_end_sy | Z65_S5 | 27446 | -0.06891 | 0.0075 | 4.00E-20 | 19.39771 | Left ventricle |
| LA_ejection | Z80_S1 | 27446 | -0.04837 | 0.005268 | 4.21E-20 | 19.37531 | Left atrium |
| LA_stroke_ | Z38_S4 | 27446 | 0.055347 | 0.006028 | 4.27E-20 | 19.36976 | Left atrium |
| LA_stroke_ | Z44_S3 | 27446 | -0.05321 | 0.005802 | 4.67E-20 | 19.33092 | Left atrium |
| LV_longitu | Z2_S1 | 27446 | 0.041542 | 0.00453 | 4.72E-20 | 19.32645 | Left ventricle |
| LV_cardiac | Z1_S2 | 27446 | 0.041683 | 0.004547 | 4.87E-20 | 19.31226 | Left ventricle |
| RV_ejectio | Z31_S2 | 27446 | -0.04708 | 0.005137 | 4.95E-20 | 19.30548 | Right ventricle |
| LV_radial_ | Z62_S1 | 27446 | -0.04424 | 0.004827 | 4.98E-20 | 19.30294 | Left ventricle |
| RA_ejectio | Z44_S1 | 27446 | 0.045456 | 0.004962 | 5.20E-20 | 19.28414 | Right atrium |
| LV_ejection | Z74_S2 | 27446 | -0.04529 | 0.004945 | 5.26E-20 | 19.27923 | Left ventricle |
| RA_minimu | Z13_S1 | 27446 | 0.046029 | 0.005036 | 6.24E-20 | 19.20496 | Right atrium |
| LA_ejection | Z40_S4 | 27446 | -0.05353 | 0.005865 | 7.05E-20 | 19.15208 | Left atrium |
| LA_minimu | Z26_S4 | 27446 | 0.057349 | 0.006284 | 7.12E-20 | 19.14779 | Left atrium |
| Ascending_ | Z37_S2 | 24599 | -0.06017 | 0.006598 | 7.60E-20 | 19.11913 | Aorta |
| RA_ejectio | Z78_S4 | 27446 | -0.04022 | 0.004411 | 7.64E-20 | 19.11664 | Right atrium |
| LV_ejection | Z51_S1 | 27446 | 0.040019 | 0.004393 | 8.26E-20 | 19.0828 | Left ventricle |
| LV_ejection | Z39_S4 | 27446 | -0.04078 | 0.004481 | 8.94E-20 | 19.04845 | Left ventricle |
| LV_end_di | Z112_S1 | 27446 | -0.07726 | 0.008495 | 9.44E-20 | 19.02521 | Left ventricle |
| RV_ejectio | Z26_S3 | 27446 | -0.05221 | 0.00575 | 1.09E-19 | 18.96141 | Right ventricle |
| Ascending_ | Z5_S1 | 24599 | -0.05142 | 0.005663 | 1.10E-19 | 18.9603 | Aorta |
| RV_stroke_ | Z100_S3 | 27446 | 0.057343 | 0.006316 | 1.10E-19 | 18.95998 | Right ventricle |
| LA_maximu | Z28_S2 | 27446 | 0.050562 | 0.005571 | 1.13E-19 | 18.94832 | Left atrium |
| LV_cardiac | Z58_S2 | 27446 | 0.047242 | 0.005207 | 1.17E-19 | 18.93235 | Left ventricle |
| RA_ejectio | Z10_S4 | 27446 | 0.047055 | 0.005191 | 1.25E-19 | 18.903 | Right atrium |
| LV_longitu | Z50_S3 | 27446 | 0.042134 | 0.004653 | 1.36E-19 | 18.8661 | Left ventricle |
| Descending | Z107_S1 | 24599 | -0.0434 | 0.004793 | 1.37E-19 | 18.86444 | Aorta |
| RV_ejectio | Z23_S3 | 27446 | -0.04104 | 0.004533 | 1.39E-19 | 18.85823 | Right ventricle |
| RV_ejectio | Z47_S4 | 27446 | -0.04618 | 0.005102 | 1.40E-19 | 18.85327 | Right ventricle |
| Ascending_ | Z7_S5 | 24599 | 0.060028 | 0.006632 | 1.41E-19 | 18.85175 | Aorta |
| RV_ejectio | Z97_S1 | 27446 | 0.056332 | 0.006236 | 1.66E-19 | 18.77951 | Right ventricle |
| LV_ejection | Z23_S3 | 27446 | -0.04009 | 0.004441 | 1.77E-19 | 18.7529 | Left ventricle |
| LA_minimu | Z22_S2 | 27446 | 0.046359 | 0.005146 | 2.07E-19 | 18.6832 | Left atrium |

| | | | | | | | |
|---|---|---|---|---|---|---|---|
| RA_stroke_ | Z12_S5 | 27446 | -0.05405 | 0.006004 | 2.21E-19 | 18.65464 | Right atrium |
| LV_ejectior | Z118_S1 | 27446 | 0.050107 | 0.005568 | 2.27E-19 | 18.6448 | Left ventricle |
| Descending | Z76_S3 | 24599 | -0.06216 | 0.006915 | 2.50E-19 | 18.60248 | Aorta |
| Descending | Z19_S2 | 24599 | 0.059071 | 0.006572 | 2.51E-19 | 18.60116 | Aorta |
| LV_circumf | Z87_S5 | 27446 | -0.04509 | 0.005018 | 2.55E-19 | 18.59357 | Left ventricle |
| RA_ejectio | Z24_S1 | 27446 | -0.03791 | 0.00422 | 2.61E-19 | 18.58392 | Right atrium |
| LV_circumf | Z74_S4 | 27446 | 0.047937 | 0.005336 | 2.63E-19 | 18.58037 | Left ventricle |
| RA_ejectio | Z120_S2 | 27446 | -0.03834 | 0.004272 | 2.80E-19 | 18.55257 | Right atrium |
| LV_circumf | Z27_S2 | 27446 | 0.053127 | 0.005919 | 2.83E-19 | 18.54804 | Left ventricle |
| RA_maxim | Z66_S3 | 27446 | -0.05648 | 0.006294 | 2.87E-19 | 18.54162 | Right atrium |
| Ascending_ | Z70_S5 | 24599 | 0.049267 | 0.005491 | 2.90E-19 | 18.53711 | Aorta |
| LV_radial_ | Z34_S2 | 27446 | 0.052995 | 0.005909 | 3.01E-19 | 18.52136 | Left ventricle |
| LV_stroke_ | Z23_S1 | 27446 | -0.06102 | 0.006805 | 3.05E-19 | 18.51618 | Left ventricle |
| Ascending_ | Z39_S3 | 24599 | -0.06444 | 0.007189 | 3.13E-19 | 18.50429 | Aorta |
| LV_radial_ | Z95_S5 | 27446 | 0.055682 | 0.006212 | 3.14E-19 | 18.50327 | Left ventricle |
| RA_minimu | Z70_S5 | 27446 | 0.048168 | 0.00538 | 3.44E-19 | 18.4638 | Right atrium |
| Descending | Z32_S5 | 24599 | -0.07089 | 0.007918 | 3.45E-19 | 18.4618 | Aorta |
| RV_ejectio | Z99_S4 | 27446 | -0.05108 | 0.005706 | 3.50E-19 | 18.45612 | Right ventricle |
| LV_circumf | Z57_S1 | 27446 | -0.04198 | 0.004692 | 3.61E-19 | 18.44308 | Left ventricle |
| Ascending_ | Z79_S4 | 24599 | -0.06086 | 0.006808 | 3.88E-19 | 18.41089 | Aorta |
| RV_ejectio | Z125_S4 | 27446 | -0.04705 | 0.005268 | 4.17E-19 | 18.38021 | Right ventricle |
| RA_stroke_ | Z113_S2 | 27446 | -0.05436 | 0.006088 | 4.27E-19 | 18.36918 | Right atrium |
| LV_circumf | Z39_S1 | 27446 | 0.041944 | 0.004697 | 4.29E-19 | 18.36743 | Left ventricle |
| RA_maxim | Z6_S2 | 27446 | -0.03852 | 0.004317 | 4.54E-19 | 18.34318 | Right atrium |
| LA_ejectior | Z60_S2 | 27446 | 0.037784 | 0.004236 | 4.70E-19 | 18.32803 | Left atrium |
| LV_radial_ | Z7_S1 | 27446 | 0.042185 | 0.004732 | 4.89E-19 | 18.31048 | Left ventricle |
| LA_minimu | Z97_S1 | 27446 | -0.05511 | 0.00619 | 5.46E-19 | 18.26252 | Left atrium |
| Descending | Z72_S5 | 24599 | 0.065207 | 0.007326 | 5.57E-19 | 18.25447 | Aorta |
| LA_maximu | Z72_S5 | 27446 | 0.057332 | 0.006443 | 5.65E-19 | 18.24805 | Left atrium |
| LV_cardiac | Z99_S3 | 27446 | 0.039864 | 0.004481 | 5.83E-19 | 18.23458 | Left ventricle |
| RA_minimu | Z49_S3 | 27446 | -0.05801 | 0.006523 | 5.99E-19 | 18.22253 | Right atrium |
| RA_stroke_ | Z22_S2 | 27446 | 0.045588 | 0.005131 | 6.45E-19 | 18.19056 | Right atrium |
| RV_ejectio | Z80_S2 | 27446 | -0.04042 | 0.00455 | 6.49E-19 | 18.1877 | Right ventricle |
| LV_myocar | Z21_S3 | 27446 | -0.09313 | 0.01049 | 6.78E-19 | 18.16851 | Left ventricle |
| LV_stroke_ | Z76_S3 | 27446 | -0.05665 | 0.006382 | 6.91E-19 | 18.16068 | Left ventricle |
| LV_ejectior | Z108_S5 | 27446 | 0.045491 | 0.005126 | 6.98E-19 | 18.15625 | Left ventricle |
| LV_radial_ | Z93_S1 | 27446 | 0.05238 | 0.005907 | 7.48E-19 | 18.12613 | Left ventricle |
| RA_maxim | Z44_S1 | 27446 | -0.04894 | 0.005535 | 9.37E-19 | 18.02842 | Right atrium |
| Ascending_ | Z41_S2 | 24599 | -0.06354 | 0.007188 | 9.62E-19 | 18.01705 | Aorta |
| LV_radial_ | Z20_S1 | 27446 | -0.05299 | 0.005998 | 1.00E-18 | 17.99962 | Left ventricle |
| LV_circumf | Z50_S5 | 27446 | -0.05578 | 0.006315 | 1.02E-18 | 17.98959 | Left ventricle |
| LV_longitu | Z46_S1 | 27446 | 0.044361 | 0.005026 | 1.08E-18 | 17.96637 | Left ventricle |
| LV_stroke_ | Z40_S4 | 27446 | -0.06449 | 0.007315 | 1.18E-18 | 17.9288 | Left ventricle |
| LV_cardiac | Z111_S1 | 27446 | -0.05338 | 0.006057 | 1.21E-18 | 17.91566 | Left ventricle |
| Descending | Z3_S4 | 24599 | 0.065354 | 0.007421 | 1.29E-18 | 17.88799 | Aorta |

| | | | | | | | |
|---|---|---|---|---|---|---|---|
| RA_stroke_ | Z3_S2 | 27446 | 0.039691 | 0.004508 | 1.32E-18 | 17.88085 | Right atrium |
| LV_ejectio | Z65_S1 | 27446 | 0.036471 | 0.004152 | 1.58E-18 | 17.80057 | Left ventricle |
| RV_ejectio | Z56_S5 | 27446 | -0.03825 | 0.004357 | 1.64E-18 | 17.786 | Right ventricle |
| LV_mean_ | Z57_S2 | 27446 | -0.05082 | 0.00579 | 1.67E-18 | 17.77801 | Left ventricle |
| LV_ejectio | Z2_S1 | 27446 | 0.04007 | 0.004565 | 1.68E-18 | 17.77592 | Left ventricle |
| Ascending_ | Z44_S3 | 24599 | 0.058913 | 0.006713 | 1.70E-18 | 17.76837 | Aorta |
| RV_stroke_ | Z49_S3 | 27446 | -0.06247 | 0.007132 | 1.97E-18 | 17.70543 | Right ventricle |
| Ascending_ | Z1_S4 | 24599 | -0.0531 | 0.006067 | 2.09E-18 | 17.68086 | Aorta |
| LV_stroke_ | Z39_S4 | 27446 | -0.04744 | 0.005427 | 2.29E-18 | 17.63973 | Left ventricle |
| Ascending_ | Z46_S1 | 24599 | -0.05351 | 0.006122 | 2.29E-18 | 17.63936 | Aorta |
| LA_stroke_ | Z22_S4 | 27446 | 0.047577 | 0.005445 | 2.36E-18 | 17.62666 | Left atrium |
| RA_ejectio | Z7_S1 | 27446 | 0.039905 | 0.004568 | 2.43E-18 | 17.61499 | Right atrium |
| LV_ejectio | Z14_S2 | 27446 | 0.043589 | 0.00499 | 2.43E-18 | 17.61435 | Left ventricle |
| RV_ejectio | Z93_S1 | 27446 | -0.05112 | 0.005853 | 2.48E-18 | 17.60582 | Right ventricle |
| LV_end_sy | Z28_S1 | 27446 | -0.05805 | 0.006654 | 2.69E-18 | 17.57077 | Left ventricle |
| LV_ejectio | Z35_S5 | 27446 | 0.037423 | 0.004292 | 2.79E-18 | 17.55457 | Left ventricle |
| LV_end_sy | Z32_S1 | 27446 | -0.05829 | 0.006687 | 2.86E-18 | 17.54357 | Left ventricle |
| Descending | Z51_S3 | 24599 | -0.06987 | 0.008023 | 3.07E-18 | 17.51248 | Aorta |
| RV_ejectio | Z50_S4 | 27446 | -0.04304 | 0.004943 | 3.14E-18 | 17.50288 | Right ventricle |
| RA_minimu | Z122_S2 | 27446 | 0.040031 | 0.004598 | 3.16E-18 | 17.5001 | Right atrium |
| RA_stroke_ | Z86_S3 | 27446 | -0.0498 | 0.005722 | 3.21E-18 | 17.49322 | Right atrium |
| LV_end_sy | Z9_S2 | 27446 | 0.058586 | 0.006731 | 3.22E-18 | 17.49171 | Left ventricle |
| Descending | Z79_S4 | 24599 | -0.06122 | 0.007034 | 3.24E-18 | 17.48952 | Aorta |
| Ascending_ | Z92_S1 | 24599 | -0.0498 | 0.005725 | 3.36E-18 | 17.4734 | Aorta |
| LA_maximu | Z57_S2 | 27446 | 0.037764 | 0.004342 | 3.41E-18 | 17.46664 | Left atrium |
| Ascending_ | Z57_S2 | 24599 | -0.04153 | 0.004781 | 3.77E-18 | 17.42398 | Aorta |
| RA_ejectio | Z52_S4 | 27446 | -0.03864 | 0.004452 | 3.94E-18 | 17.4046 | Right atrium |
| LV_myocar | Z115_S3 | 27446 | -0.07715 | 0.00891 | 4.77E-18 | 17.32163 | Left ventricle |
| LA_ejectio | Z34_S2 | 27446 | 0.048206 | 0.005571 | 5.01E-18 | 17.30017 | Left atrium |
| Descending | Z62_S1 | 24599 | 0.048185 | 0.00557 | 5.14E-18 | 17.28896 | Aorta |
| RV_end_sy | Z91_S1 | 27446 | 0.057278 | 0.006624 | 5.26E-18 | 17.27878 | Right ventricle |
| Descending | Z69_S5 | 24599 | -0.04373 | 0.005059 | 5.43E-18 | 17.26523 | Aorta |
| RA_ejectio | Z14_S1 | 27446 | -0.04043 | 0.004678 | 5.46E-18 | 17.26295 | Right atrium |
| RA_stroke_ | Z1_S5 | 27446 | -0.04449 | 0.005161 | 6.68E-18 | 17.17537 | Right atrium |
| LV_radial_ | Z66_S3 | 27446 | 0.050401 | 0.005847 | 6.74E-18 | 17.17157 | Left ventricle |
| RV_ejectio | Z108_S5 | 27446 | 0.045077 | 0.005232 | 6.96E-18 | 17.15743 | Right ventricle |
| RA_stroke_ | Z85_S5 | 27446 | 0.040758 | 0.004736 | 7.55E-18 | 17.12219 | Right atrium |
| LV_mean_ | Z82_S1 | 27446 | 0.074129 | 0.008618 | 7.88E-18 | 17.10369 | Left ventricle |
| LV_radial_ | Z4_S4 | 27446 | 0.039418 | 0.004584 | 8.01E-18 | 17.09648 | Left ventricle |
| LV_circumf | Z28_S2 | 27446 | -0.04658 | 0.005418 | 8.19E-18 | 17.08671 | Left ventricle |
| LV_ejectio | Z57_S1 | 27446 | 0.039698 | 0.004618 | 8.23E-18 | 17.08434 | Left ventricle |
| LV_ejectio | Z31_S2 | 27446 | -0.04327 | 0.005034 | 8.28E-18 | 17.082 | Left ventricle |
| Ascending_ | Z22_S2 | 24599 | 0.0529 | 0.006159 | 8.80E-18 | 17.05546 | Aorta |
| LV_myocar | Z56_S5 | 27446 | 0.061334 | 0.007142 | 8.90E-18 | 17.05078 | Left ventricle |
| LV_ejectio | Z26_S3 | 27446 | 0.04836 | 0.005634 | 9.25E-18 | 17.034 | Left ventricle |

| | | | | | | | |
|---|---|---|---|---|---|---|---|
| RA_minimu | Z93_S2 | 27446 | 0.059227 | 0.006903 | 9.49E-18 | 17.02273 | Right atrium |
| LV_ejectio | Z112_S1 | 27446 | -0.05344 | 0.006232 | 9.83E-18 | 17.00744 | Left ventricle |
| LV_radial_s | Z100_S3 | 27446 | -0.04541 | 0.005296 | 9.89E-18 | 17.00501 | Left ventricle |
| Descending | Z82_S1 | 24599 | -0.07081 | 0.008259 | 9.93E-18 | 17.00293 | Aorta |
| LV_ejectio | Z50_S5 | 27446 | 0.053247 | 0.006215 | 1.06E-17 | 16.9739 | Left ventricle |
| LA_maximu | Z22_S2 | 27446 | 0.04545 | 0.005307 | 1.10E-17 | 16.96036 | Left atrium |
| LA_stroke_ | Z9_S2 | 27446 | 0.048099 | 0.005618 | 1.11E-17 | 16.95499 | Left atrium |
| Descending | Z44_S1 | 24599 | 0.056297 | 0.00658 | 1.17E-17 | 16.93167 | Aorta |
| LV_myocar | Z75_S5 | 27446 | 0.089953 | 0.01052 | 1.22E-17 | 16.91357 | Left ventricle |
| RV_ejectio | Z22_S2 | 27446 | 0.044314 | 0.005185 | 1.26E-17 | 16.89906 | Right ventricle |
| LA_ejectio | Z120_S2 | 27446 | -0.03564 | 0.004172 | 1.33E-17 | 16.8754 | Left atrium |
| LV_circumf | Z100_S3 | 27446 | 0.044591 | 0.005224 | 1.39E-17 | 16.85853 | Left ventricle |
| RV_stroke_ | Z12_S5 | 27446 | -0.06228 | 0.007304 | 1.51E-17 | 16.82177 | Right ventricle |
| LV_ejectio | Z39_S1 | 27446 | -0.03942 | 0.004624 | 1.53E-17 | 16.81494 | Left ventricle |
| LV_cardiac | Z20_S2 | 27446 | 0.040024 | 0.004696 | 1.56E-17 | 16.80665 | Left ventricle |
| LV_circumf | Z52_S3 | 27446 | 0.036977 | 0.00434 | 1.61E-17 | 16.79362 | Left ventricle |
| LA_ejectio | Z2_S1 | 27446 | 0.037768 | 0.004433 | 1.61E-17 | 16.79288 | Left atrium |
| RA_maxim | Z75_S4 | 27446 | -0.05596 | 0.00657 | 1.62E-17 | 16.79032 | Right atrium |
| LV_circumf | Z115_S3 | 27446 | -0.04605 | 0.005411 | 1.73E-17 | 16.76081 | Left ventricle |
| LV_end_sy | Z34_S2 | 27446 | -0.06142 | 0.007219 | 1.78E-17 | 16.74998 | Left ventricle |
| LV_circumf | Z29_S1 | 27446 | -0.04685 | 0.005509 | 1.82E-17 | 16.74002 | Left ventricle |
| Descending | Z34_S2 | 24599 | -0.06403 | 0.00753 | 1.84E-17 | 16.73536 | Aorta |
| LA_ejectio | Z41_S1 | 27446 | -0.04463 | 0.005252 | 1.91E-17 | 16.71785 | Left atrium |
| Ascending_ | Z26_S3 | 24599 | 0.055157 | 0.006492 | 1.96E-17 | 16.70834 | Aorta |
| RV_end_sy | Z78_S1 | 27446 | 0.053043 | 0.006246 | 2.02E-17 | 16.69553 | Right ventricle |
| RA_maxim | Z13_S1 | 27446 | 0.042181 | 0.00497 | 2.11E-17 | 16.67583 | Right atrium |
| RV_end_sy | Z19_S2 | 27446 | 0.057332 | 0.006758 | 2.17E-17 | 16.66291 | Right ventricle |
| Ascending_ | Z96_S3 | 24599 | 0.04728 | 0.005581 | 2.43E-17 | 16.61416 | Aorta |
| Ascending_ | Z83_S2 | 24599 | 0.045916 | 0.005421 | 2.44E-17 | 16.6128 | Aorta |
| LV_circumf | Z96_S3 | 27446 | 0.041937 | 0.004952 | 2.47E-17 | 16.60795 | Left ventricle |
| LV_longitu | Z15_S2 | 27446 | 0.044884 | 0.005301 | 2.51E-17 | 16.60089 | Left ventricle |
| Ascending_ | Z1_S4 | 24599 | -0.05154 | 0.00609 | 2.59E-17 | 16.58611 | Aorta |
| LV_circumf | Z30_S2 | 27446 | -0.0522 | 0.006168 | 2.60E-17 | 16.58564 | Left ventricle |
| LV_radial_s | Z11_S5 | 27446 | 0.041953 | 0.004959 | 2.70E-17 | 16.5694 | Left ventricle |
| Descending | Z24_S1 | 24599 | 0.042596 | 0.005044 | 3.06E-17 | 16.51404 | Aorta |
| Ascending_ | Z105_S4 | 24599 | -0.04667 | 0.005528 | 3.10E-17 | 16.50853 | Aorta |
| LV_radial_s | Z123_S4 | 27446 | -0.04558 | 0.005402 | 3.25E-17 | 16.48875 | Left ventricle |
| Ascending_ | Z74_S4 | 24599 | -0.0537 | 0.006376 | 3.66E-17 | 16.43659 | Aorta |
| LV_radial_s | Z70_S5 | 27446 | -0.04153 | 0.004931 | 3.69E-17 | 16.43256 | Left ventricle |
| LV_cardiac | Z39_S3 | 27446 | -0.05855 | 0.006957 | 3.87E-17 | 16.41278 | Left ventricle |
| LV_myocar | Z12_S5 | 27446 | -0.08364 | 0.009945 | 4.10E-17 | 16.38728 | Left ventricle |
| RV_end_sy | Z37_S2 | 27446 | -0.06242 | 0.007434 | 4.60E-17 | 16.33733 | Right ventricle |
| RA_maxim | Z97_S1 | 27446 | -0.05681 | 0.006777 | 5.16E-17 | 16.28748 | Right atrium |
| LA_ejectio | Z18_S4 | 27446 | 0.05087 | 0.006069 | 5.23E-17 | 16.28131 | Left atrium |
| LV_circumf | Z52_S4 | 27446 | 0.038121 | 0.004549 | 5.32E-17 | 16.27446 | Left ventricle |

| | | | | | | | |
|---|---|---|---|---|---|---|---|
| Descending | Z52_S4 | 24599 | 0.044548 | 0.005318 | 5.42E-17 | 16.26632 | Aorta |
| RV_end_sy | Z14_S2 | 27446 | -0.05709 | 0.006829 | 6.23E-17 | 16.20519 | Right ventricle |
| Descending | Z24_S5 | 24599 | 0.060925 | 0.007306 | 7.49E-17 | 16.12544 | Aorta |
| LV_ejection | Z75_S1 | 27446 | 0.042985 | 0.005157 | 7.71E-17 | 16.11298 | Left ventricle |
| LV_radial_s | Z118_S1 | 27446 | 0.047794 | 0.005736 | 7.97E-17 | 16.09832 | Left ventricle |
| LV_radial_s | Z65_S2 | 27446 | -0.04415 | 0.005302 | 8.26E-17 | 16.08281 | Left ventricle |
| RA_maxim | Z51_S1 | 27446 | -0.04054 | 0.004873 | 8.79E-17 | 16.05591 | Right atrium |
| LV_longitu | Z105_S5 | 27446 | -0.04182 | 0.005027 | 8.83E-17 | 16.05384 | Left ventricle |
| LV_end_sy | Z44_S1 | 27446 | 0.052204 | 0.006282 | 9.52E-17 | 16.02138 | Left ventricle |
| LA_ejection | Z95_S5 | 27446 | 0.048675 | 0.005857 | 9.53E-17 | 16.02078 | Left atrium |
| RV_end_di | Z27_S2 | 27446 | -0.06949 | 0.008362 | 9.55E-17 | 16.01998 | Right ventricle |
| Descending | Z91_S1 | 24599 | 0.053694 | 0.006463 | 9.72E-17 | 16.01235 | Aorta |
| LA_stroke_ | Z59_S2 | 27446 | 0.049863 | 0.006002 | 9.77E-17 | 16.01026 | Left atrium |
| LV_ejection | Z105_S5 | 27446 | -0.04205 | 0.005066 | 1.03E-16 | 15.98544 | Left ventricle |
| RA_minimu | Z20_S1 | 27446 | 0.054285 | 0.006545 | 1.10E-16 | 15.95901 | Right atrium |
| LA_minimu | Z72_S5 | 27446 | 0.051785 | 0.006248 | 1.15E-16 | 15.93824 | Left atrium |
| LA_stroke_ | Z44_S1 | 27446 | 0.043432 | 0.005242 | 1.18E-16 | 15.92866 | Left atrium |
| Ascending_ | Z93_S1 | 24599 | 0.054812 | 0.006621 | 1.24E-16 | 15.90571 | Aorta |
| Ascending_ | Z7_S1 | 24599 | -0.04609 | 0.005569 | 1.26E-16 | 15.89915 | Aorta |
| Descending | Z12_S5 | 24599 | -0.06461 | 0.007809 | 1.29E-16 | 15.88869 | Aorta |
| RV_end_sy | Z32_S1 | 27446 | -0.06013 | 0.007272 | 1.35E-16 | 15.86865 | Right ventricle |
| RA_minimu | Z62_S4 | 27446 | -0.05328 | 0.006446 | 1.39E-16 | 15.85565 | Right atrium |
| LV_radial_s | Z50_S2 | 27446 | -0.05211 | 0.006308 | 1.46E-16 | 15.83709 | Left ventricle |
| LA_minimu | Z32_S1 | 27446 | 0.044461 | 0.005385 | 1.49E-16 | 15.826 | Left atrium |
| LA_stroke_ | Z57_S2 | 27446 | 0.036026 | 0.004365 | 1.53E-16 | 15.81591 | Left atrium |
| Ascending_ | Z10_S4 | 24599 | 0.049648 | 0.006019 | 1.59E-16 | 15.79725 | Aorta |
| Ascending_ | Z93_S1 | 24599 | 0.054379 | 0.006597 | 1.67E-16 | 15.7765 | Aorta |
| Ascending_ | Z66_S2 | 24599 | 0.0542 | 0.006575 | 1.68E-16 | 15.77499 | Aorta |
| LV_myocar | Z41_S1 | 27446 | 0.074582 | 0.009049 | 1.70E-16 | 15.77017 | Left ventricle |
| LA_ejection | Z53_S2 | 27446 | -0.04429 | 0.005379 | 1.83E-16 | 15.73762 | Left atrium |
| LA_maximu | Z121_S4 | 27446 | 0.048563 | 0.0059 | 1.85E-16 | 15.73388 | Left atrium |
| LV_ejection | Z19_S2 | 27446 | -0.04062 | 0.004939 | 1.96E-16 | 15.70814 | Left ventricle |
| Descending | Z86_S3 | 24599 | -0.06227 | 0.007572 | 1.99E-16 | 15.7018 | Aorta |
| LV_radial_s | Z89_S4 | 27446 | 0.040981 | 0.004985 | 2.03E-16 | 15.6929 | Left ventricle |
| RA_stroke_ | Z49_S3 | 27446 | -0.04818 | 0.005865 | 2.13E-16 | 15.67194 | Right atrium |
| RA_stroke_ | Z80_S4 | 27446 | -0.03581 | 0.004363 | 2.27E-16 | 15.6438 | Right atrium |
| LA_stroke_ | Z50_S2 | 27446 | 0.052758 | 0.006431 | 2.32E-16 | 15.63434 | Left atrium |
| LV_ejection | Z30_S2 | 27446 | 0.049783 | 0.006071 | 2.40E-16 | 15.61924 | Left ventricle |
| Ascending_ | Z28_S2 | 24599 | 0.053272 | 0.006502 | 2.54E-16 | 15.59575 | Aorta |
| Ascending_ | Z109_S1 | 24599 | -0.04732 | 0.005776 | 2.54E-16 | 15.59565 | Aorta |
| RV_ejectio | Z127_S4 | 27446 | 0.04587 | 0.005606 | 2.78E-16 | 15.55569 | Right ventricle |
| LV_ejection | Z62_S4 | 27446 | 0.046924 | 0.005735 | 2.80E-16 | 15.55318 | Left ventricle |
| RV_ejectio | Z23_S1 | 27446 | 0.046924 | 0.005737 | 2.87E-16 | 15.54214 | Right ventricle |
| Descending | Z30_S2 | 24599 | -0.05908 | 0.007229 | 3.03E-16 | 15.51915 | Aorta |
| LV_stroke_ | Z78_S1 | 27446 | -0.04516 | 0.005528 | 3.09E-16 | 15.50962 | Left ventricle |

| | | | | | | | |
|---|---|---|---|---|---|---|---|
| LV_cardiac | Z122_S5 | 27446 | 0.040769 | 0.004996 | 3.36E-16 | 15.47335 | Left ventricle |
| LA_maximu | Z95_S5 | 27446 | -0.05133 | 0.006302 | 3.80E-16 | 15.42045 | Left atrium |
| Descending | Z86_S3 | 24599 | -0.05503 | 0.006757 | 3.84E-16 | 15.41574 | Aorta |
| LV_cardiac | Z17_S1 | 27446 | 0.042254 | 0.005193 | 4.06E-16 | 15.39105 | Left ventricle |
| Ascending_ | Z39_S1 | 24599 | 0.043159 | 0.005306 | 4.16E-16 | 15.38122 | Aorta |
| Ascending_ | Z73_S1 | 24599 | 0.046228 | 0.005685 | 4.24E-16 | 15.37221 | Aorta |
| LV_radial_s | Z113_S2 | 27446 | -0.05046 | 0.006208 | 4.37E-16 | 15.35942 | Left ventricle |
| RA_stroke_ | Z73_S4 | 27446 | 0.047172 | 0.00581 | 4.70E-16 | 15.32827 | Right atrium |
| Descending | Z28_S1 | 24599 | -0.05726 | 0.007062 | 5.12E-16 | 15.29053 | Aorta |
| Descending | Z97_S2 | 24599 | 0.051222 | 0.006323 | 5.47E-16 | 15.26229 | Aorta |
| LV_circumf | Z99_S3 | 27446 | 0.033243 | 0.004104 | 5.50E-16 | 15.25937 | Left ventricle |
| Descending | Z39_S3 | 24599 | 0.060377 | 0.007457 | 5.62E-16 | 15.24996 | Aorta |
| Ascending_ | Z96_S3 | 24599 | 0.045347 | 0.005602 | 5.77E-16 | 15.23895 | Aorta |
| RA_stroke_ | Z50_S4 | 27446 | 0.03955 | 0.004894 | 6.40E-16 | 15.19415 | Right atrium |
| Descending | Z34_S2 | 24599 | -0.06173 | 0.00764 | 6.44E-16 | 15.19083 | Aorta |
| LA_minimu | Z114_S4 | 27446 | 0.039871 | 0.004935 | 6.50E-16 | 15.18704 | Left atrium |
| RA_ejectio | Z74_S2 | 27446 | -0.03974 | 0.004919 | 6.59E-16 | 15.1814 | Right atrium |
| LV_cardiac | Z1_S1 | 27446 | 0.043645 | 0.005408 | 7.03E-16 | 15.15274 | Left ventricle |
| LV_cardiac | Z105_S4 | 27446 | 0.04087 | 0.005065 | 7.05E-16 | 15.15195 | Left ventricle |
| RA_stroke_ | Z11_S5 | 27446 | -0.03925 | 0.004865 | 7.14E-16 | 15.14604 | Right atrium |
| RA_minimu | Z11_S5 | 27446 | -0.04362 | 0.005412 | 7.61E-16 | 15.11889 | Right atrium |
| Ascending_ | Z50_S5 | 24599 | -0.05741 | 0.007125 | 7.75E-16 | 15.11063 | Aorta |
| Descending | Z73_S1 | 24599 | 0.044968 | 0.005582 | 7.85E-16 | 15.10518 | Aorta |
| LV_circumf | Z117_S4 | 27446 | -0.03338 | 0.004143 | 7.89E-16 | 15.10286 | Left ventricle |
| Ascending_ | Z70_S5 | 24599 | 0.044349 | 0.005513 | 8.66E-16 | 15.06241 | Aorta |
| RA_ejectio | Z112_S1 | 27446 | 0.049842 | 0.006197 | 8.81E-16 | 15.05506 | Right atrium |
| RV_ejectio | Z27_S2 | 27446 | -0.04783 | 0.005948 | 8.86E-16 | 15.05237 | Right ventricle |
| RA_ejectio | Z44_S4 | 27446 | 0.033879 | 0.004215 | 9.11E-16 | 15.04056 | Right atrium |
| RA_minimu | Z74_S2 | 27446 | 0.044663 | 0.00556 | 9.56E-16 | 15.0197 | Right atrium |
| LV_radial_s | Z114_S4 | 27446 | -0.0403 | 0.005017 | 9.66E-16 | 15.01505 | Left ventricle |
| LA_maximu | Z22_S4 | 27446 | 0.043458 | 0.005419 | 1.06E-15 | 14.97574 | Left atrium |
| RA_ejectio | Z4_S4 | 27446 | 0.035487 | 0.004425 | 1.07E-15 | 14.97143 | Right atrium |
| RV_stroke_ | Z112_S3 | 27446 | -0.05847 | 0.007296 | 1.12E-15 | 14.95234 | Right ventricle |
| LV_end_sy | Z1_S5 | 27446 | 0.051472 | 0.006429 | 1.18E-15 | 14.92719 | Left ventricle |
| RA_maxim | Z41_S1 | 27446 | 0.048018 | 0.005998 | 1.19E-15 | 14.92429 | Right atrium |
| LV_radial_s | Z92_S1 | 27446 | 0.041032 | 0.005127 | 1.21E-15 | 14.91869 | Left ventricle |
| LA_minimu | Z61_S4 | 27446 | -0.03822 | 0.00478 | 1.28E-15 | 14.89428 | Left atrium |
| LV_mean_r | Z22_S2 | 27446 | -0.0566 | 0.007078 | 1.28E-15 | 14.89291 | Left ventricle |
| Ascending_ | Z37_S2 | 24599 | -0.04996 | 0.00625 | 1.30E-15 | 14.88635 | Aorta |
| RV_stroke_ | Z78_S1 | 27446 | -0.04482 | 0.00561 | 1.34E-15 | 14.87221 | Right ventricle |
| Descending | Z99_S3 | 24599 | 0.038314 | 0.004796 | 1.37E-15 | 14.86325 | Aorta |
| RV_ejectio | Z20_S2 | 27446 | -0.03448 | 0.00432 | 1.45E-15 | 14.83974 | Right ventricle |
| RV_ejectio | Z80_S5 | 27446 | -0.04811 | 0.00603 | 1.48E-15 | 14.83113 | Right ventricle |
| RV_ejectio | Z93_S2 | 27446 | -0.04994 | 0.00627 | 1.64E-15 | 14.78451 | Right ventricle |
| RA_ejectio | Z22_S4 | 27446 | 0.041055 | 0.005157 | 1.70E-15 | 14.76917 | Right atrium |

| | | | | | | | |
|---|---|---|---|---|---|---|---|
| RV_stroke_ | Z56_S2 | 27446 | -0.04199 | 0.005283 | 1.87E-15 | 14.7273 | Right ventricle |
| LV_circumf | Z82_S4 | 27446 | 0.03487 | 0.004388 | 1.91E-15 | 14.71887 | Left ventricle |
| LV_end_sy | Z51_S3 | 27446 | -0.06109 | 0.007693 | 2.00E-15 | 14.69938 | Left ventricle |
| RV_ejectio | Z28_S2 | 27446 | 0.043227 | 0.005444 | 2.02E-15 | 14.69555 | Right ventricle |
| LV_myocar | Z94_S2 | 27446 | 0.071646 | 0.00903 | 2.11E-15 | 14.67551 | Left ventricle |
| Ascending_ | Z5_S1 | 24599 | -0.04746 | 0.005984 | 2.16E-15 | 14.66471 | Aorta |
| Descending | Z44_S1 | 24599 | -0.04724 | 0.005958 | 2.19E-15 | 14.65901 | Aorta |
| LA_minimu | Z94_S2 | 27446 | -0.04336 | 0.005468 | 2.20E-15 | 14.65855 | Left atrium |
| RA_maxim | Z12_S5 | 27446 | -0.05225 | 0.006593 | 2.27E-15 | 14.64363 | Right atrium |
| RA_minimu | Z99_S5 | 27446 | 0.042806 | 0.005402 | 2.29E-15 | 14.63946 | Right atrium |
| LV_ejectio | Z98_S3 | 27446 | 0.034693 | 0.004378 | 2.31E-15 | 14.63695 | Left ventricle |
| Ascending_ | Z86_S3 | 24599 | -0.05163 | 0.006517 | 2.32E-15 | 14.63376 | Aorta |
| LA_minimu | Z9_S2 | 27446 | 0.042942 | 0.005421 | 2.35E-15 | 14.62889 | Left atrium |
| LV_end_sy | Z75_S5 | 27446 | 0.062663 | 0.007913 | 2.39E-15 | 14.62124 | Left ventricle |
| LA_ejectio | Z50_S2 | 27446 | -0.04706 | 0.005947 | 2.52E-15 | 14.59811 | Left atrium |
| RA_maxim | Z65_S5 | 27446 | -0.0523 | 0.006612 | 2.57E-15 | 14.59059 | Right atrium |
| Descending | Z47_S4 | 24599 | -0.05287 | 0.006683 | 2.57E-15 | 14.58951 | Aorta |
| RV_end_di | Z65_S5 | 27446 | 0.067676 | 0.008556 | 2.59E-15 | 14.58719 | Right ventricle |
| RA_ejectio | Z40_S4 | 27446 | -0.04751 | 0.006008 | 2.63E-15 | 14.57927 | Right atrium |
| LV_ejectio | Z8_S3 | 27446 | -0.0336 | 0.004254 | 2.85E-15 | 14.54556 | Left ventricle |
| LV_cardiac | Z75_S1 | 27446 | -0.04519 | 0.005724 | 2.88E-15 | 14.54094 | Left ventricle |
| RV_ejectio | Z57_S1 | 27446 | 0.037187 | 0.004715 | 3.08E-15 | 14.51153 | Right ventricle |
| LA_ejectio | Z44_S1 | 27446 | 0.038202 | 0.004848 | 3.28E-15 | 14.48361 | Left atrium |
| LV_ejectio | Z7_S5 | 27446 | 0.043116 | 0.005472 | 3.30E-15 | 14.48188 | Left ventricle |
| RA_ejectio | Z82_S4 | 27446 | 0.033832 | 0.004294 | 3.30E-15 | 14.48114 | Right atrium |
| RV_ejectio | Z8_S3 | 27446 | -0.03419 | 0.004342 | 3.46E-15 | 14.46056 | Right ventricle |
| RA_stroke_ | Z112_S3 | 27446 | 0.047203 | 0.005999 | 3.59E-15 | 14.44523 | Right atrium |
| RV_stroke_ | Z7_S5 | 27446 | 0.052892 | 0.006724 | 3.66E-15 | 14.43619 | Right ventricle |
| RV_end_di | Z65_S1 | 27446 | -0.04688 | 0.00596 | 3.68E-15 | 14.4347 | Right ventricle |
| Descending | Z106_S3 | 24599 | -0.0619 | 0.007875 | 3.83E-15 | 14.4166 | Aorta |
| RA_ejectio | Z62_S4 | 27446 | -0.0448 | 0.005703 | 4.01E-15 | 14.39721 | Right atrium |
| RV_end_di | Z24_S5 | 27446 | -0.06142 | 0.007829 | 4.33E-15 | 14.36382 | Right ventricle |
| LV_stroke_ | Z115_S3 | 27446 | -0.05059 | 0.00645 | 4.38E-15 | 14.35811 | Left ventricle |
| LV_ejectio | Z117_S4 | 27446 | 0.031966 | 0.004078 | 4.53E-15 | 14.34389 | Left ventricle |
| RA_minimu | Z61_S4 | 27446 | -0.04149 | 0.005303 | 5.08E-15 | 14.29412 | Right atrium |
| Ascending_ | Z24_S5 | 24599 | 0.049197 | 0.006289 | 5.15E-15 | 14.28827 | Aorta |
| RA_stroke_ | Z14_S4 | 27446 | 0.044027 | 0.005642 | 6.05E-15 | 14.21815 | Right atrium |
| LV_circumf | Z24_S1 | 27446 | 0.033604 | 0.004314 | 6.68E-15 | 14.17508 | Left ventricle |
| LV_longitu | Z24_S5 | 27446 | -0.04218 | 0.005415 | 6.72E-15 | 14.17271 | Left ventricle |
| LV_mean_ | Z51_S1 | 27446 | -0.04769 | 0.006123 | 6.73E-15 | 14.17228 | Left ventricle |
| RA_maxim | Z55_S1 | 27446 | -0.04005 | 0.005149 | 7.44E-15 | 14.12864 | Right atrium |
| RV_end_sy | Z49_S1 | 27446 | -0.06506 | 0.008367 | 7.52E-15 | 14.12358 | Right ventricle |
| LV_end_di | Z121_S4 | 27446 | 0.059862 | 0.007699 | 7.54E-15 | 14.12268 | Left ventricle |
| LV_end_sy | Z44_S3 | 27446 | -0.05407 | 0.006955 | 7.64E-15 | 14.11695 | Left ventricle |
| LA_maximu | Z65_S1 | 27446 | -0.0337 | 0.00434 | 8.08E-15 | 14.09266 | Left atrium |

| | | | | | | | |
|---|---|---|---|---|---|---|---|
| LA_stroke_ | Z28_S1 | 27446 | -0.04311 | 0.005554 | 8.31E-15 | 14.08019 | Left atrium |
| RA_stroke_ | Z78_S4 | 27446 | 0.034805 | 0.004484 | 8.36E-15 | 14.07804 | Right atrium |
| RA_stroke_ | Z39_S1 | 27446 | 0.03626 | 0.004672 | 8.46E-15 | 14.07246 | Right atrium |
| RA_ejectio | Z83_S2 | 27446 | -0.03644 | 0.004697 | 8.60E-15 | 14.06552 | Right atrium |
| LV_mean_ı | Z53_S1 | 27446 | -0.0563 | 0.007259 | 8.74E-15 | 14.05834 | Left ventricle |
| Descending | Z35_S5 | 24599 | -0.03959 | 0.005104 | 8.79E-15 | 14.05618 | Aorta |
| Ascending_ | Z37_S1 | 24599 | 0.050349 | 0.006494 | 8.92E-15 | 14.04967 | Aorta |
| RA_ejectio | Z80_S4 | 27446 | 0.033235 | 0.004295 | 1.00E-14 | 13.99853 | Right atrium |
| LV_myocar | Z51_S3 | 27446 | -0.07916 | 0.01023 | 1.01E-14 | 13.99745 | Left ventricle |
| LV_mean_ı | Z73_S1 | 27446 | -0.05054 | 0.006537 | 1.06E-14 | 13.97383 | Left ventricle |
| Descending | Z26_S3 | 24599 | 0.058013 | 0.007518 | 1.20E-14 | 13.9225 | Aorta |
| LA_minimu | Z44_S3 | 27446 | -0.0432 | 0.0056 | 1.21E-14 | 13.91607 | Left atrium |
| Ascending_ | Z44_S3 | 24599 | -0.0492 | 0.006382 | 1.27E-14 | 13.8978 | Aorta |
| Descending | Z66_S2 | 24599 | 0.057988 | 0.007533 | 1.38E-14 | 13.85863 | Aorta |
| Descending | Z21_S3 | 24599 | -0.05742 | 0.007459 | 1.39E-14 | 13.85792 | Aorta |
| RA_ejectio | Z62_S1 | 27446 | -0.03586 | 0.004662 | 1.44E-14 | 13.84207 | Right atrium |
| Descending | Z75_S5 | 24599 | 0.063467 | 0.008269 | 1.65E-14 | 13.78369 | Aorta |
| Descending | Z50_S4 | 24599 | 0.044081 | 0.00575 | 1.77E-14 | 13.75195 | Aorta |
| LV_ejectior | Z122_S5 | 27446 | -0.03451 | 0.004503 | 1.79E-14 | 13.74785 | Left ventricle |
| RA_stroke_ | Z100_S3 | 27446 | -0.03981 | 0.005195 | 1.82E-14 | 13.73948 | Right atrium |
| RV_end_sy | Z80_S5 | 27446 | 0.061916 | 0.008084 | 1.88E-14 | 13.72656 | Right ventricle |
| RV_end_di | Z76_S3 | 27446 | -0.05791 | 0.007566 | 1.95E-14 | 13.70958 | Right ventricle |
| LV_longitu | Z118_S1 | 27446 | -0.04227 | 0.005528 | 2.05E-14 | 13.68723 | Left ventricle |
| LA_minimu | Z23_S1 | 27446 | 0.043515 | 0.005696 | 2.18E-14 | 13.66217 | Left atrium |
| LV_radial_ | Z112_S1 | 27446 | -0.04905 | 0.006421 | 2.19E-14 | 13.65964 | Left ventricle |
| LV_radial_ | Z112_S3 | 27446 | 0.046678 | 0.006117 | 2.33E-14 | 13.63289 | Left ventricle |
| LV_myocar | Z80_S1 | 27446 | -0.06905 | 0.009082 | 2.89E-14 | 13.53918 | Left ventricle |
| LV_stroke_ | Z28_S2 | 27446 | 0.049077 | 0.006459 | 3.01E-14 | 13.52175 | Left ventricle |
| LV_ejectior | Z28_S1 | 27446 | 0.040183 | 0.005289 | 3.03E-14 | 13.51804 | Left ventricle |
| LV_ejectior | Z3_S4 | 27446 | 0.047402 | 0.00624 | 3.04E-14 | 13.51649 | Left ventricle |
| LV_myocar | Z22_S2 | 27446 | -0.06456 | 0.008501 | 3.12E-14 | 13.5064 | Left ventricle |
| Descending | Z62_S3 | 24599 | 0.052196 | 0.00688 | 3.27E-14 | 13.48557 | Aorta |
| RV_ejectio | Z58_S2 | 27446 | -0.03633 | 0.004792 | 3.38E-14 | 13.47095 | Right ventricle |
| Descending | Z77_S3 | 24599 | -0.05098 | 0.006726 | 3.48E-14 | 13.45904 | Aorta |
| RA_maxim | Z70_S5 | 27446 | 0.040175 | 0.00531 | 3.85E-14 | 13.41448 | Right atrium |
| Ascending_ | Z37_S1 | 24599 | 0.048905 | 0.00647 | 4.08E-14 | 13.38977 | Aorta |
| Ascending_ | Z30_S2 | 24599 | -0.05566 | 0.007364 | 4.10E-14 | 13.38757 | Aorta |
| Descending | Z49_S3 | 24599 | -0.05847 | 0.007739 | 4.18E-14 | 13.37903 | Aorta |
| LV_circumf | Z22_S4 | 27446 | 0.039798 | 0.00527 | 4.30E-14 | 13.3667 | Left ventricle |
| LV_circumf | Z62_S4 | 27446 | -0.044 | 0.005829 | 4.40E-14 | 13.35684 | Left ventricle |
| Ascending_ | Z12_S4 | 24599 | -0.03909 | 0.005183 | 4.65E-14 | 13.33267 | Aorta |
| LV_longitu | Z92_S1 | 27446 | 0.037241 | 0.00494 | 4.73E-14 | 13.32485 | Left ventricle |
| Ascending_ | Z77_S3 | 24599 | -0.0444 | 0.005893 | 4.91E-14 | 13.30866 | Aorta |
| LA_minimu | Z87_S5 | 27446 | -0.03769 | 0.005006 | 5.07E-14 | 13.29458 | Left atrium |
| LV_ejectior | Z52_S3 | 27446 | -0.03216 | 0.004273 | 5.18E-14 | 13.28538 | Left ventricle |

| | | | | | | | |
|---|---|---|---|---|---|---|---|
| LV_ejectio | Z122_S1 | 27446 | 0.041238 | 0.00548 | 5.25E-14 | 13.27981 | Left ventricle |
| LV_cardiac | Z122_S2 | 27446 | -0.03416 | 0.004542 | 5.40E-14 | 13.26776 | Left ventricle |
| Descending | Z76_S3 | 24599 | -0.05279 | 0.007019 | 5.40E-14 | 13.26726 | Aorta |
| LV_myocar | Z1_S4 | 27446 | -0.06633 | 0.008824 | 5.58E-14 | 13.25317 | Left ventricle |
| RA_maxim | Z81_S1 | 27446 | -0.0423 | 0.005629 | 5.75E-14 | 13.24057 | Right atrium |
| Descending | Z14_S1 | 24599 | 0.041942 | 0.005588 | 6.11E-14 | 13.21422 | Aorta |
| LV_cardiac | Z87_S3 | 27446 | -0.04061 | 0.005415 | 6.39E-14 | 13.19444 | Left ventricle |
| RA_maxim | Z113_S2 | 27446 | -0.05004 | 0.006685 | 7.10E-14 | 13.1486 | Right atrium |
| LV_end_sy | Z121_S4 | 27446 | 0.053196 | 0.007106 | 7.12E-14 | 13.14777 | Left ventricle |
| RA_minimu | Z81_S1 | 27446 | -0.04268 | 0.005705 | 7.35E-14 | 13.13356 | Right atrium |
| RV_end_sy | Z111_S1 | 27446 | 0.055881 | 0.007471 | 7.46E-14 | 13.12714 | Right ventricle |
| LA_ejectio | Z52_S3 | 27446 | -0.03103 | 0.004149 | 7.47E-14 | 13.12651 | Left atrium |
| LA_ejectio | Z7_S3 | 27446 | -0.04328 | 0.005793 | 7.98E-14 | 13.09827 | Left atrium |
| Ascending_ | Z87_S5 | 24599 | -0.04252 | 0.005696 | 8.39E-14 | 13.07628 | Aorta |
| RA_stroke_ | Z59_S2 | 27446 | -0.04304 | 0.005776 | 9.24E-14 | 13.03441 | Right atrium |
| LV_cardiac | Z95_S5 | 27446 | -0.04986 | 0.006695 | 9.54E-14 | 13.02024 | Left ventricle |
| RV_end_sy | Z53_S2 | 27446 | -0.05646 | 0.007582 | 9.56E-14 | 13.01952 | Right ventricle |
| LV_myocar | Z49_S3 | 27446 | -0.07231 | 0.009715 | 9.85E-14 | 13.0066 | Left ventricle |
| LV_stroke_ | Z95_S5 | 27446 | -0.05434 | 0.007306 | 1.02E-13 | 12.99171 | Left ventricle |
| LA_ejectio | Z32_S1 | 27446 | -0.03839 | 0.005162 | 1.03E-13 | 12.98661 | Left atrium |
| RA_ejectio | Z57_S1 | 27446 | 0.034142 | 0.004594 | 1.06E-13 | 12.97266 | Right atrium |
| Ascending_ | Z44_S1 | 24599 | 0.042787 | 0.005767 | 1.18E-13 | 12.92723 | Aorta |
| Descending | Z41_S1 | 24599 | 0.052767 | 0.007113 | 1.18E-13 | 12.92665 | Aorta |
| LV_myocar | Z1_S5 | 27446 | 0.063414 | 0.00855 | 1.20E-13 | 12.92155 | Left ventricle |
| RA_stroke_ | Z106_S3 | 27446 | -0.04424 | 0.005967 | 1.22E-13 | 12.91362 | Right atrium |
| LV_mean_ | Z57_S1 | 27446 | -0.04767 | 0.006436 | 1.30E-13 | 12.88525 | Left ventricle |
| Descending | Z5_S5 | 24599 | 0.051239 | 0.006932 | 1.45E-13 | 12.83894 | Aorta |
| LA_ejectio | Z76_S3 | 27446 | 0.037835 | 0.00512 | 1.48E-13 | 12.83065 | Left atrium |
| LV_ejectio | Z74_S4 | 27446 | -0.03878 | 0.005254 | 1.57E-13 | 12.80331 | Left ventricle |
| RA_maxim | Z93_S2 | 27446 | 0.050273 | 0.006813 | 1.60E-13 | 12.79646 | Right atrium |
| Descending | Z106_S3 | 24599 | -0.05724 | 0.007764 | 1.68E-13 | 12.77583 | Aorta |
| LA_maximu | Z23_S1 | 27446 | 0.043235 | 0.005875 | 1.84E-13 | 12.73414 | Left atrium |
| LV_longitu | Z57_S1 | 27446 | 0.033735 | 0.004585 | 1.86E-13 | 12.72982 | Left ventricle |
| LV_longitu | Z53_S2 | 27446 | 0.04045 | 0.005499 | 1.89E-13 | 12.7225 | Left ventricle |
| LV_end_di | Z28_S2 | 27446 | 0.053483 | 0.007273 | 1.93E-13 | 12.71422 | Left ventricle |
| LV_mean_ | Z29_S1 | 27446 | 0.055523 | 0.007556 | 2.01E-13 | 12.6966 | Left ventricle |
| RA_stroke_ | Z34_S2 | 27446 | 0.042536 | 0.005798 | 2.20E-13 | 12.65694 | Right atrium |
| Descending | Z30_S2 | 24599 | -0.05943 | 0.008103 | 2.22E-13 | 12.653 | Aorta |
| LV_myocar | Z93_S1 | 27446 | 0.070367 | 0.009599 | 2.29E-13 | 12.64006 | Left ventricle |
| Ascending_ | Z109_S1 | 24599 | -0.04447 | 0.00608 | 2.57E-13 | 12.5907 | Aorta |
| LV_myocar | Z49_S1 | 27446 | -0.07484 | 0.010233 | 2.59E-13 | 12.58639 | Left ventricle |
| RV_end_sy | Z41_S1 | 27446 | 0.054124 | 0.007402 | 2.64E-13 | 12.57841 | Right ventricle |
| LV_radial_ | Z1_S1 | 27446 | 0.036698 | 0.005021 | 2.70E-13 | 12.56911 | Left ventricle |
| LV_end_sy | Z97_S2 | 27446 | 0.048663 | 0.006658 | 2.70E-13 | 12.56907 | Left ventricle |
| RA_ejectio | Z34_S2 | 27446 | 0.041702 | 0.005707 | 2.72E-13 | 12.56488 | Right atrium |

| | | | | | | | |
|---|---|---|---|---|---|---|---|
| Descending | Z22_S4 | 24599 | -0.04517 | 0.006182 | 2.73E-13 | 12.564 | Aorta |
| Descending | Z53_S2 | 24599 | -0.05401 | 0.007394 | 2.79E-13 | 12.55424 | Aorta |
| Descending | Z44_S3 | 24599 | 0.048118 | 0.006594 | 2.94E-13 | 12.53115 | Aorta |
| LV_mean_ | Z29_S5 | 27446 | 0.054319 | 0.007456 | 3.22E-13 | 12.49235 | Left ventricle |
| RA_stroke_ | Z10_S4 | 27446 | -0.03844 | 0.005277 | 3.25E-13 | 12.48817 | Right atrium |
| LA_minimu | Z122_S5 | 27446 | 0.033225 | 0.004563 | 3.28E-13 | 12.48361 | Left atrium |
| LA_minimu | Z115_S3 | 27446 | -0.03925 | 0.005397 | 3.53E-13 | 12.45267 | Left atrium |
| Ascending_ | Z39_S1 | 24599 | 0.038735 | 0.005327 | 3.55E-13 | 12.44919 | Aorta |
| Descending | Z65_S2 | 24599 | 0.049296 | 0.006788 | 3.81E-13 | 12.41917 | Aorta |
| Descending | Z28_S2 | 24599 | 0.046333 | 0.006385 | 3.97E-13 | 12.40109 | Aorta |
| LV_radial_ | Z41_S2 | 27446 | 0.044236 | 0.006098 | 4.03E-13 | 12.39454 | Left ventricle |
| Ascending_ | Z14_S2 | 24599 | -0.04165 | 0.005747 | 4.25E-13 | 12.37174 | Aorta |
| LV_circumf | Z55_S1 | 27446 | -0.03414 | 0.004719 | 4.68E-13 | 12.33017 | Left ventricle |
| LV_circumf | Z112_S3 | 27446 | 0.0436 | 0.006034 | 5.00E-13 | 12.30107 | Left ventricle |
| Ascending_ | Z75_S5 | 24599 | -0.05229 | 0.007246 | 5.31E-13 | 12.2745 | Aorta |
| RV_stroke_ | Z1_S4 | 27446 | -0.04674 | 0.006481 | 5.50E-13 | 12.25995 | Right ventricle |
| RA_ejectio | Z49_S1 | 27446 | -0.04386 | 0.006081 | 5.52E-13 | 12.25769 | Right atrium |
| LV_ejection | Z55_S1 | 27446 | 0.033469 | 0.004644 | 5.73E-13 | 12.24217 | Left ventricle |
| LV_radial_ | Z7_S5 | 27446 | 0.040598 | 0.005638 | 5.98E-13 | 12.22301 | Left ventricle |
| LV_cardiac | Z66_S3 | 27446 | -0.04535 | 0.006302 | 6.18E-13 | 12.20915 | Left ventricle |
| LV_cardiac | Z65_S5 | 27446 | -0.04758 | 0.006618 | 6.52E-13 | 12.18551 | Left ventricle |
| RV_end_di | Z112_S1 | 27446 | -0.0643 | 0.008946 | 6.58E-13 | 12.18199 | Right ventricle |
| LV_circumf | Z35_S5 | 27446 | -0.03134 | 0.004363 | 6.77E-13 | 12.16917 | Left ventricle |
| LA_stroke_ | Z72_S5 | 27446 | 0.046526 | 0.006478 | 6.87E-13 | 12.16289 | Left atrium |
| LV_radial_ | Z75_S4 | 27446 | 0.04379 | 0.006105 | 7.36E-13 | 12.13335 | Left ventricle |
| LV_longitu | Z80_S2 | 27446 | -0.03173 | 0.004426 | 7.55E-13 | 12.12204 | Left ventricle |
| RA_maxim | Z20_S1 | 27446 | 0.04629 | 0.00646 | 7.75E-13 | 12.11073 | Right atrium |
| RA_ejectio | Z30_S2 | 27446 | 0.043251 | 0.006039 | 7.92E-13 | 12.10122 | Right atrium |
| LV_circumf | Z59_S3 | 27446 | -0.02953 | 0.004124 | 8.10E-13 | 12.09148 | Left ventricle |
| LV_ejection | Z52_S4 | 27446 | -0.03201 | 0.004479 | 8.90E-13 | 12.05048 | Left ventricle |
| Ascending_ | Z3_S4 | 24599 | 0.054049 | 0.007563 | 8.91E-13 | 12.04992 | Aorta |
| RA_stroke_ | Z87_S3 | 27446 | -0.03522 | 0.00493 | 9.04E-13 | 12.04366 | Right atrium |
| RA_ejectio | Z43_S2 | 27446 | 0.031138 | 0.004361 | 9.38E-13 | 12.02765 | Right atrium |
| LV_circumf | Z3_S4 | 27446 | -0.04525 | 0.006341 | 9.65E-13 | 12.01528 | Left ventricle |
| RV_ejectio | Z77_S3 | 27446 | -0.03717 | 0.00521 | 9.81E-13 | 12.0083 | Right ventricle |
| Descending | Z14_S2 | 24599 | -0.0476 | 0.006679 | 1.03E-12 | 11.98831 | Aorta |
| Descending | Z18_S4 | 24599 | -0.05852 | 0.008223 | 1.10E-12 | 11.95873 | Aorta |
| RV_end_sy | Z80_S1 | 27446 | -0.05284 | 0.007428 | 1.13E-12 | 11.94538 | Right ventricle |
| LV_longitu | Z60_S2 | 27446 | 0.030812 | 0.004332 | 1.14E-12 | 11.94497 | Left ventricle |
| LV_ejection | Z13_S3 | 27446 | -0.02903 | 0.004083 | 1.17E-12 | 11.93235 | Left ventricle |
| LV_longitu | Z50_S2 | 27446 | 0.043165 | 0.006079 | 1.25E-12 | 11.90461 | Left ventricle |
| LV_cardiac | Z9_S2 | 27446 | 0.042166 | 0.00594 | 1.25E-12 | 11.90162 | Left ventricle |
| LV_circumf | Z120_S4 | 27446 | -0.03579 | 0.005042 | 1.27E-12 | 11.89498 | Left ventricle |
| Ascending_ | Z87_S5 | 24599 | -0.04026 | 0.005676 | 1.32E-12 | 11.88067 | Aorta |
| LV_end_sy | Z53_S2 | 27446 | -0.04938 | 0.006973 | 1.42E-12 | 11.84792 | Left ventricle |

| | | | | | | | |
|---|---|---|---|---|---|---|---|
| LV_radial_s | Z50_S5 | 27446 | 0.045346 | 0.006405 | 1.45E-12 | 11.83991 | Left ventricle |
| RA_ejectio | Z7_S5 | 27446 | -0.03853 | 0.005442 | 1.45E-12 | 11.83863 | Right atrium |
| LA_stroke_ | Z56_S5 | 27446 | 0.031739 | 0.004484 | 1.47E-12 | 11.8334 | Left atrium |
| RV_ejectio | Z52_S4 | 27446 | -0.03231 | 0.004572 | 1.57E-12 | 11.80279 | Right ventricle |
| LV_mean_ | Z107_S1 | 27446 | -0.03971 | 0.005618 | 1.58E-12 | 11.80229 | Left ventricle |
| Ascending_ | Z37_S4 | 24599 | -0.05023 | 0.007108 | 1.59E-12 | 11.79877 | Aorta |
| Ascending_ | Z83_S2 | 24599 | 0.038455 | 0.005443 | 1.60E-12 | 11.79529 | Aorta |
| Ascending_ | Z40_S4 | 24599 | 0.049075 | 0.006953 | 1.69E-12 | 11.7728 | Aorta |
| RV_ejectio | Z97_S2 | 27446 | 0.038083 | 0.005401 | 1.78E-12 | 11.75007 | Right ventricle |
| Ascending_ | Z115_S3 | 24599 | -0.04312 | 0.006116 | 1.78E-12 | 11.74957 | Aorta |
| RA_ejectio | Z97_S1 | 27446 | 0.042828 | 0.006079 | 1.84E-12 | 11.7342 | Right atrium |
| RV_ejectio | Z14_S1 | 27446 | -0.03384 | 0.004804 | 1.86E-12 | 11.73151 | Right ventricle |
| LA_stroke_ | Z40_S4 | 27446 | -0.04469 | 0.006346 | 1.89E-12 | 11.72357 | Left atrium |
| LA_minimu | Z57_S2 | 27446 | 0.029642 | 0.004212 | 1.97E-12 | 11.70614 | Left atrium |
| LV_circumf | Z89_S4 | 27446 | 0.034614 | 0.004919 | 1.97E-12 | 11.70585 | Left ventricle |
| RV_ejectio | Z1_S2 | 27446 | -0.02942 | 0.004185 | 2.06E-12 | 11.68612 | Right ventricle |
| RV_ejectio | Z17_S1 | 27446 | -0.03358 | 0.004778 | 2.08E-12 | 11.68129 | Right ventricle |
| LV_stroke_ | Z120_S5 | 27446 | 0.05145 | 0.00732 | 2.09E-12 | 11.6798 | Left ventricle |
| LV_radial_s | Z75_S5 | 27446 | 0.045528 | 0.006479 | 2.11E-12 | 11.67506 | Left ventricle |
| LA_ejection | Z75_S4 | 27446 | 0.040422 | 0.005755 | 2.17E-12 | 11.6642 | Left atrium |
| RA_ejectio | Z107_S1 | 27446 | 0.028156 | 0.00401 | 2.19E-12 | 11.65865 | Right atrium |
| LV_radial_s | Z76_S3 | 27446 | 0.038133 | 0.005432 | 2.22E-12 | 11.65347 | Left ventricle |
| LV_cardiac_ | Z107_S1 | 27446 | 0.031407 | 0.004475 | 2.26E-12 | 11.64629 | Left ventricle |
| RA_stroke_ | Z87_S5 | 27446 | -0.03502 | 0.004992 | 2.29E-12 | 11.63952 | Right atrium |
| LV_end_dia | Z23_S1 | 27446 | -0.05378 | 0.007666 | 2.29E-12 | 11.63923 | Left ventricle |
| RV_ejectio | Z44_S3 | 27446 | 0.039574 | 0.005643 | 2.33E-12 | 11.63248 | Right ventricle |
| LA_maximu | Z94_S2 | 27446 | -0.03955 | 0.00564 | 2.36E-12 | 11.62794 | Left atrium |
| RA_stroke_ | Z51_S1 | 27446 | -0.0311 | 0.004441 | 2.49E-12 | 11.6034 | Right atrium |
| Ascending_ | Z115_S3 | 24599 | -0.04299 | 0.006139 | 2.50E-12 | 11.60282 | Aorta |
| Ascending_ | Z41_S2 | 24599 | -0.04781 | 0.006834 | 2.63E-12 | 11.58006 | Aorta |
| LV_ejection | Z24_S1 | 27446 | -0.02969 | 0.004246 | 2.73E-12 | 11.56386 | Left ventricle |
| LV_ejection | Z3_S2 | 27446 | 0.031184 | 0.004464 | 2.85E-12 | 11.545 | Left ventricle |
| RV_ejectio | Z26_S4 | 27446 | -0.04423 | 0.006335 | 2.92E-12 | 11.53473 | Right ventricle |
| Ascending_ | Z19_S2 | 24599 | 0.041718 | 0.005977 | 2.97E-12 | 11.52792 | Aorta |
| Descending | Z99_S4 | 24599 | 0.051229 | 0.007346 | 3.08E-12 | 11.51155 | Aorta |
| LV_cardiac_ | Z74_S2 | 27446 | 0.038257 | 0.005492 | 3.26E-12 | 11.48744 | Left ventricle |
| LV_circumf | Z14_S1 | 27446 | 0.033304 | 0.004782 | 3.30E-12 | 11.48083 | Left ventricle |
| Ascending_ | Z73_S4 | 24599 | 0.048418 | 0.006966 | 3.64E-12 | 11.43922 | Aorta |
| LV_ejection | Z96_S3 | 27446 | -0.03387 | 0.004875 | 3.71E-12 | 11.43014 | Left ventricle |
| LV_circumf | Z66_S2 | 27446 | -0.04049 | 0.00583 | 3.78E-12 | 11.4227 | Left ventricle |
| LA_ejection | Z72_S5 | 27446 | -0.04159 | 0.005991 | 3.84E-12 | 11.41548 | Left atrium |
| LV_cardiac_ | Z113_S2 | 27446 | 0.04644 | 0.006691 | 3.90E-12 | 11.40934 | Left ventricle |
| LV_radial_s | Z82_S4 | 27446 | 0.030864 | 0.00445 | 4.02E-12 | 11.39607 | Left ventricle |
| Descending | Z1_S5 | 24599 | -0.04688 | 0.006761 | 4.11E-12 | 11.38637 | Aorta |
| LA_ejection | Z74_S2 | 27446 | -0.0333 | 0.004805 | 4.23E-12 | 11.37395 | Left atrium |

| | | | | | | | |
|---|---|---|---|---|---|---|---|
| RV_end_sy | Z66_S2 | 27446 | 0.054374 | 0.007851 | 4.33E-12 | 11.364 | Right ventricle |
| LV_ejectio | Z62_S3 | 27446 | -0.03631 | 0.005244 | 4.37E-12 | 11.35994 | Left ventricle |
| RV_ejectio | Z83_S2 | 27446 | -0.03339 | 0.004822 | 4.38E-12 | 11.35901 | Right ventricle |
| LV_ejectio | Z92_S5 | 27446 | 0.041804 | 0.00604 | 4.48E-12 | 11.34883 | Left ventricle |
| LV_cardiac | Z52_S3 | 27446 | 0.032804 | 0.004743 | 4.63E-12 | 11.33468 | Left ventricle |
| LV_ejectio | Z31_S5 | 27446 | -0.02786 | 0.004029 | 4.67E-12 | 11.33039 | Left ventricle |
| RA_minimu | Z51_S1 | 27446 | -0.03411 | 0.004941 | 5.08E-12 | 11.2939 | Right atrium |
| RA_stroke_ | Z50_S3 | 27446 | -0.03272 | 0.00474 | 5.09E-12 | 11.29317 | Right atrium |
| RV_ejectio | Z112_S3 | 27446 | -0.04178 | 0.006062 | 5.52E-12 | 11.25812 | Right ventricle |
| Descending | Z14_S4 | 24599 | 0.050749 | 0.007365 | 5.56E-12 | 11.25513 | Aorta |
| LV_longitu | Z31_S2 | 27446 | -0.03444 | 0.004998 | 5.57E-12 | 11.25415 | Left ventricle |
| Descending | Z57_S2 | 24599 | -0.03774 | 0.00548 | 5.71E-12 | 11.24322 | Aorta |
| LV_myocar | Z62_S4 | 27446 | -0.06598 | 0.009599 | 6.25E-12 | 11.20388 | Left ventricle |
| LV_cardiac | Z62_S2 | 27446 | 0.033828 | 0.004923 | 6.36E-12 | 11.1968 | Left ventricle |
| LA_minimu | Z10_S4 | 27446 | -0.03636 | 0.005293 | 6.44E-12 | 11.19113 | Left atrium |
| LV_circumf | Z75_S1 | 27446 | -0.036 | 0.005242 | 6.52E-12 | 11.18573 | Left ventricle |
| RA_maxim | Z108_S5 | 27446 | -0.03906 | 0.005687 | 6.53E-12 | 11.18505 | Right atrium |
| RA_ejectio | Z3_S4 | 27446 | 0.042604 | 0.006206 | 6.66E-12 | 11.17674 | Right atrium |
| LV_ejectio | Z11_S5 | 27446 | 0.033047 | 0.004817 | 6.84E-12 | 11.16475 | Left ventricle |
| Ascending_ | Z18_S4 | 24599 | 0.049197 | 0.007179 | 7.23E-12 | 11.1407 | Aorta |
| LA_minimu | Z99_S4 | 27446 | 0.038821 | 0.005668 | 7.41E-12 | 11.13015 | Left atrium |
| LA_maximu | Z59_S2 | 27446 | 0.040889 | 0.005975 | 7.72E-12 | 11.1124 | Left atrium |
| Descending | Z28_S1 | 24599 | -0.04765 | 0.006964 | 7.82E-12 | 11.10687 | Aorta |
| LA_minimu | Z93_S1 | 27446 | -0.03976 | 0.005813 | 7.97E-12 | 11.09879 | Left atrium |
| LV_longitu | Z99_S4 | 27446 | 0.037955 | 0.005552 | 8.11E-12 | 11.09083 | Left ventricle |
| LA_ejectio | Z57_S1 | 27446 | 0.030669 | 0.004487 | 8.16E-12 | 11.08811 | Left atrium |
| LV_cardiac | Z47_S4 | 27446 | 0.03794 | 0.005551 | 8.18E-12 | 11.08723 | Left ventricle |
| RV_ejectio | Z50_S5 | 27446 | 0.04335 | 0.006347 | 8.50E-12 | 11.07073 | Right ventricle |
| RA_ejectio | Z44_S3 | 27446 | -0.03752 | 0.005498 | 8.75E-12 | 11.05776 | Right atrium |
| LV_radial_ | Z22_S2 | 27446 | 0.035727 | 0.005235 | 8.82E-12 | 11.05455 | Left ventricle |
| Descending | Z95_S5 | 24599 | -0.05489 | 0.008047 | 9.03E-12 | 11.04426 | Aorta |
| Ascending_ | Z44_S3 | 24599 | -0.04338 | 0.00636 | 9.04E-12 | 11.04385 | Aorta |
| LV_longitu | Z43_S2 | 27446 | 0.029693 | 0.004353 | 9.06E-12 | 11.04306 | Left ventricle |
| LA_ejectio | Z112_S1 | 27446 | 0.041258 | 0.006054 | 9.45E-12 | 11.02479 | Left atrium |
| RV_ejectio | Z35_S5 | 27446 | 0.029864 | 0.004383 | 9.51E-12 | 11.02189 | Right ventricle |
| RV_ejectio | Z82_S4 | 27446 | -0.03001 | 0.004409 | 9.99E-12 | 11.0003 | Right ventricle |
| RV_stroke_ | Z77_S3 | 27446 | -0.04269 | 0.006273 | 1.01E-11 | 10.99573 | Right ventricle |
| LV_end_sy | Z49_S3 | 27446 | -0.04973 | 0.007308 | 1.01E-11 | 10.99435 | Left ventricle |
| Ascending_ | Z66_S2 | 24599 | 0.047248 | 0.006948 | 1.04E-11 | 10.98169 | Aorta |
| Ascending_ | Z5_S1 | 24599 | -0.03868 | 0.005688 | 1.04E-11 | 10.98123 | Aorta |
| LA_maximu | Z1_S4 | 27446 | -0.03746 | 0.005511 | 1.07E-11 | 10.97088 | Left atrium |
| LV_cardiac | Z51_S3 | 27446 | -0.0461 | 0.006787 | 1.10E-11 | 10.95832 | Left ventricle |
| RV_ejectio | Z3_S4 | 27446 | 0.043272 | 0.00637 | 1.10E-11 | 10.95779 | Right ventricle |
| RV_ejectio | Z117_S4 | 27446 | 0.028252 | 0.004163 | 1.15E-11 | 10.9385 | Right ventricle |
| RA_minimu | Z80_S5 | 27446 | -0.04504 | 0.006642 | 1.20E-11 | 10.92118 | Right atrium |

| | | | | | | | |
|---|---|---|---|---|---|---|---|
| RA_ejectio | Z70_S5 | 27446 | -0.0322 | 0.004762 | 1.37E-11 | 10.86235 | Right atrium |
| Descending | Z38_S5 | 24599 | -0.04718 | 0.006983 | 1.41E-11 | 10.84971 | Aorta |
| RA_stroke_ | Z41_S1 | 27446 | 0.036925 | 0.005466 | 1.43E-11 | 10.84507 | Right atrium |
| Ascending_ | Z107_S1 | 24599 | -0.03297 | 0.004886 | 1.49E-11 | 10.82589 | Aorta |
| LV_cardiac | Z121_S4 | 27446 | 0.042303 | 0.006269 | 1.50E-11 | 10.82525 | Left ventricle |
| LV_circumf | Z47_S4 | 27446 | 0.034281 | 0.005082 | 1.52E-11 | 10.8168 | Left ventricle |
| LV_longitu | Z23_S3 | 27446 | -0.02974 | 0.00441 | 1.54E-11 | 10.81384 | Left ventricle |
| LV_ejectior | Z50_S3 | 27446 | 0.031619 | 0.004692 | 1.59E-11 | 10.79865 | Left ventricle |
| Ascending_ | Z79_S4 | 24599 | -0.0483 | 0.007169 | 1.61E-11 | 10.79191 | Aorta |
| Ascending_ | Z125_S4 | 24599 | -0.04037 | 0.005995 | 1.65E-11 | 10.78264 | Aorta |
| LA_stroke_ | Z97_S2 | 27446 | 0.037383 | 0.005557 | 1.73E-11 | 10.76144 | Left atrium |
| RV_ejectio | Z55_S1 | 27446 | 0.03188 | 0.004741 | 1.76E-11 | 10.75432 | Right ventricle |
| LV_circumf | Z39_S4 | 27446 | 0.030637 | 0.004556 | 1.77E-11 | 10.7532 | Left ventricle |
| RV_end_sy | Z34_S2 | 27446 | -0.05277 | 0.007854 | 1.84E-11 | 10.73515 | Right ventricle |
| Descending | Z19_S2 | 24599 | 0.039415 | 0.005869 | 1.87E-11 | 10.72886 | Aorta |
| RA_stroke_ | Z111_S1 | 27446 | 0.037054 | 0.005518 | 1.87E-11 | 10.72755 | Right atrium |
| Ascending_ | Z111_S5 | 24599 | 0.038372 | 0.005716 | 1.90E-11 | 10.72083 | Aorta |
| RV_end_sy | Z26_S3 | 27446 | 0.051776 | 0.007714 | 1.92E-11 | 10.71707 | Right ventricle |
| RA_stroke_ | Z52_S4 | 27446 | 0.030361 | 0.004525 | 1.96E-11 | 10.70739 | Right atrium |
| LV_ejectior | Z127_S4 | 27446 | 0.036842 | 0.005495 | 2.01E-11 | 10.69632 | Left ventricle |
| LV_myocar | Z100_S3 | 27446 | 0.057594 | 0.008606 | 2.20E-11 | 10.65821 | Left ventricle |
| Descending | Z28_S4 | 24599 | -0.04324 | 0.006462 | 2.20E-11 | 10.65743 | Aorta |
| Ascending_ | Z9_S2 | 24599 | 0.041238 | 0.006166 | 2.27E-11 | 10.64415 | Aorta |
| Ascending_ | Z22_S4 | 24599 | -0.04211 | 0.006298 | 2.28E-11 | 10.64234 | Aorta |
| Ascending_ | Z97_S2 | 24599 | 0.043072 | 0.006443 | 2.31E-11 | 10.63588 | Aorta |
| LV_ejectior | Z44_S1 | 27446 | -0.03336 | 0.004994 | 2.40E-11 | 10.61997 | Left ventricle |
| Descending | Z49_S3 | 24599 | -0.05096 | 0.00763 | 2.41E-11 | 10.61867 | Aorta |
| Descending | Z120_S5 | 24599 | 0.048091 | 0.007201 | 2.42E-11 | 10.61629 | Aorta |
| RV_ejectio | Z80_S4 | 27446 | 0.029407 | 0.004409 | 2.57E-11 | 10.5895 | Right ventricle |
| RA_stroke_ | Z43_S2 | 27446 | -0.02956 | 0.004432 | 2.58E-11 | 10.58856 | Right atrium |
| LV_circumf | Z64_S1 | 27446 | -0.03766 | 0.005652 | 2.69E-11 | 10.56973 | Left ventricle |
| LV_end_dia | Z56_S5 | 27446 | 0.038692 | 0.005823 | 3.04E-11 | 10.51649 | Left ventricle |
| LV_ejectior | Z53_S2 | 27446 | 0.036807 | 0.005542 | 3.10E-11 | 10.50823 | Left ventricle |
| LV_longitu | Z13_S3 | 27446 | -0.0269 | 0.004053 | 3.19E-11 | 10.49627 | Left ventricle |
| RV_end_sy | Z40_S4 | 27446 | 0.054844 | 0.00827 | 3.33E-11 | 10.47806 | Right ventricle |
| RA_minimu | Z41_S1 | 27446 | 0.040316 | 0.006082 | 3.37E-11 | 10.47204 | Right atrium |
| Descending | Z92_S5 | 24599 | 0.053516 | 0.008073 | 3.38E-11 | 10.47143 | Aorta |
| Ascending_ | Z49_S1 | 24599 | 0.046669 | 0.007048 | 3.56E-11 | 10.44864 | Aorta |
| LV_circumf | Z56_S2 | 27446 | -0.02889 | 0.00437 | 3.82E-11 | 10.41792 | Left ventricle |
| RA_stroke_ | Z37_S2 | 27446 | -0.03629 | 0.005491 | 3.90E-11 | 10.40946 | Right atrium |
| RA_stroke_ | Z108_S5 | 27446 | -0.03418 | 0.005182 | 4.23E-11 | 10.37353 | Right atrium |
| RA_maxim | Z13_S3 | 27446 | 0.029853 | 0.004528 | 4.33E-11 | 10.36376 | Right atrium |
| RV_stroke_ | Z76_S3 | 27446 | -0.04271 | 0.006481 | 4.40E-11 | 10.3567 | Right ventricle |
| Descending | Z125_S4 | 24599 | -0.04573 | 0.006941 | 4.45E-11 | 10.35188 | Aorta |
| Ascending_ | Z40_S4 | 24599 | 0.045623 | 0.006929 | 4.56E-11 | 10.34141 | Aorta |

| | | | | | | | |
|---|---|---|---|---|---|---|---|
| RA_maxim | Z122_S5 | 27446 | -0.03289 | 0.004995 | 4.56E-11 | 10.34108 | Right atrium |
| LV_mean_ | Z97_S1 | 27446 | -0.05596 | 0.008518 | 5.04E-11 | 10.29786 | Left ventricle |
| LV_ejectio | Z20_S1 | 27446 | -0.03827 | 0.005826 | 5.08E-11 | 10.29375 | Left ventricle |
| LV_ejectio | Z92_S1 | 27446 | 0.032653 | 0.004979 | 5.43E-11 | 10.26527 | Left ventricle |
| RV_stroke_ | Z13_S1 | 27446 | -0.03613 | 0.00551 | 5.44E-11 | 10.26409 | Right ventricle |
| LV_circum | Z59_S2 | 27446 | 0.03809 | 0.005811 | 5.55E-11 | 10.25585 | Left ventricle |
| Descending | Z72_S5 | 24599 | -0.05379 | 0.008216 | 5.88E-11 | 10.23079 | Aorta |
| RV_end_di | Z1_S4 | 27446 | -0.04955 | 0.00757 | 5.94E-11 | 10.22619 | Right ventricle |
| RV_ejectio | Z65_S2 | 27446 | 0.034394 | 0.005256 | 6.02E-11 | 10.22069 | Right ventricle |
| LV_radial_ | Z28_S1 | 27446 | 0.035648 | 0.00545 | 6.12E-11 | 10.21359 | Left ventricle |
| LV_ejectio | Z14_S1 | 27446 | -0.03072 | 0.004707 | 6.69E-11 | 10.17465 | Left ventricle |
| LV_radial_ | Z93_S2 | 27446 | -0.0413 | 0.00633 | 6.83E-11 | 10.16572 | Left ventricle |
| LV_circum | Z107_S1 | 27446 | -0.02674 | 0.004098 | 6.85E-11 | 10.16462 | Left ventricle |
| LV_ejectio | Z115_S3 | 27446 | 0.034754 | 0.005328 | 6.92E-11 | 10.15986 | Left ventricle |
| LA_maxim | Z97_S1 | 27446 | -0.04164 | 0.006388 | 7.13E-11 | 10.14715 | Left atrium |
| LV_cardiac | Z30_S2 | 27446 | -0.04391 | 0.006741 | 7.32E-11 | 10.13557 | Left ventricle |
| LV_longitu | Z106_S3 | 27446 | -0.03818 | 0.005863 | 7.41E-11 | 10.13001 | Left ventricle |
| RA_maxim | Z99_S5 | 27446 | 0.03461 | 0.005332 | 8.51E-11 | 10.06996 | Right atrium |
| LV_mean_ | Z38_S4 | 27446 | -0.05195 | 0.008004 | 8.56E-11 | 10.06762 | Left ventricle |
| RA_ejectio | Z99_S5 | 27446 | -0.03101 | 0.004781 | 8.83E-11 | 10.05381 | Right atrium |
| RV_stroke_ | Z115_S3 | 27446 | -0.04244 | 0.006548 | 9.05E-11 | 10.04341 | Right ventricle |
| Descending | Z9_S2 | 24599 | 0.045749 | 0.007064 | 9.39E-11 | 10.02753 | Aorta |
| Descending | Z41_S2 | 24599 | -0.05123 | 0.007914 | 9.57E-11 | 10.01931 | Aorta |
| Ascending_ | Z3_S4 | 24599 | -0.04649 | 0.007188 | 1.00E-10 | 10.00009 | Aorta |
| LV_cardiac | Z1_S5 | 27446 | -0.03662 | 0.005672 | 1.07E-10 | 9.969515 | Left ventricle |
| LA_maxim | Z38_S4 | 27446 | 0.038703 | 0.006003 | 1.14E-10 | 9.943547 | Left atrium |
| RA_minim | Z27_S2 | 27446 | -0.04222 | 0.006552 | 1.17E-10 | 9.931605 | Right atrium |
| LV_ejectio | Z22_S4 | 27446 | -0.03341 | 0.005188 | 1.20E-10 | 9.920344 | Left ventricle |
| RV_end_di | Z39_S3 | 27446 | 0.057888 | 0.009 | 1.26E-10 | 9.899012 | Right ventricle |
| Ascending_ | Z64_S1 | 24599 | -0.04349 | 0.006764 | 1.28E-10 | 9.891591 | Aorta |
| LV_radial_ | Z1_S4 | 27446 | 0.034932 | 0.005434 | 1.29E-10 | 9.889956 | Left ventricle |
| LV_ejectio | Z57_S2 | 27446 | 0.026732 | 0.004158 | 1.29E-10 | 9.889396 | Left ventricle |
| RV_ejectio | Z122_S5 | 27446 | -0.02948 | 0.004597 | 1.43E-10 | 9.845976 | Right ventricle |
| RV_end_di | Z97_S2 | 27446 | 0.048695 | 0.007595 | 1.44E-10 | 9.841471 | Right ventricle |
| LV_longitu | Z122_S5 | 27446 | -0.02863 | 0.00447 | 1.51E-10 | 9.822395 | Left ventricle |
| LV_longitu | Z121_S4 | 27446 | -0.03586 | 0.005606 | 1.59E-10 | 9.799617 | Left ventricle |
| LV_circum | Z99_S5 | 27446 | 0.0312 | 0.004885 | 1.70E-10 | 9.770028 | Left ventricle |
| Descending | Z112_S1 | 24599 | 0.052992 | 0.008298 | 1.70E-10 | 9.769129 | Aorta |
| Descending | Z66_S2 | 24599 | 0.043549 | 0.006822 | 1.73E-10 | 9.762435 | Aorta |
| LV_cardiac | Z44_S3 | 27446 | -0.03917 | 0.006136 | 1.74E-10 | 9.759053 | Left ventricle |
| Descending | Z62_S3 | 24599 | 0.044555 | 0.006981 | 1.75E-10 | 9.757963 | Aorta |
| LV_ejectio | Z69_S5 | 27446 | 0.02717 | 0.004257 | 1.75E-10 | 9.757272 | Left ventricle |
| Descending | Z41_S1 | 24599 | 0.041034 | 0.006441 | 1.88E-10 | 9.72629 | Aorta |
| Ascending_ | Z21_S3 | 24599 | -0.04841 | 0.007601 | 1.90E-10 | 9.72135 | Aorta |
| RA_stroke_ | Z62_S3 | 27446 | -0.03374 | 0.005299 | 1.92E-10 | 9.715662 | Right atrium |

| | | | | | | | |
|---|---|---|---|---|---|---|---|
| LV_longitu | Z125_S4 | 27446 | -0.03261 | 0.005126 | 2.00E-10 | 9.699743 | Left ventricle |
| LV_ejectio | Z93_S1 | 27446 | 0.036493 | 0.005738 | 2.03E-10 | 9.693493 | Left ventricle |
| RA_maxim | Z87_S5 | 27446 | -0.03484 | 0.005481 | 2.07E-10 | 9.684026 | Right atrium |
| Descending | Z57_S2 | 24599 | 0.031521 | 0.004961 | 2.10E-10 | 9.676819 | Aorta |
| RA_stroke_ | Z20_S2 | 27446 | 0.027162 | 0.004278 | 2.16E-10 | 9.665738 | Right atrium |
| LV_radial_ | Z94_S2 | 27446 | 0.035306 | 0.005561 | 2.18E-10 | 9.662516 | Left ventricle |
| RV_ejectio | Z120_S4 | 27446 | 0.032138 | 0.005066 | 2.24E-10 | 9.649035 | Right ventricle |
| Ascending_ | Z1_S5 | 24599 | -0.03758 | 0.005925 | 2.25E-10 | 9.6479 | Aorta |
| RA_maxim | Z91_S1 | 27446 | 0.034048 | 0.005371 | 2.32E-10 | 9.635146 | Right atrium |
| RV_end_sy | Z65_S2 | 27446 | 0.044662 | 0.007047 | 2.33E-10 | 9.632491 | Right ventricle |
| LV_radial_ | Z9_S2 | 27446 | 0.034892 | 0.005514 | 2.49E-10 | 9.604019 | Left ventricle |
| LA_ejectio | Z7_S5 | 27446 | -0.03361 | 0.005316 | 2.56E-10 | 9.591968 | Left atrium |
| LV_cardiac | Z93_S2 | 27446 | 0.043116 | 0.00682 | 2.58E-10 | 9.58766 | Left ventricle |
| RA_ejectio | Z55_S1 | 27446 | 0.029174 | 0.004619 | 2.68E-10 | 9.571969 | Right atrium |
| Ascending_ | Z53_S2 | 24599 | -0.04024 | 0.006389 | 3.00E-10 | 9.522199 | Aorta |
| LA_minimu | Z37_S1 | 27446 | 0.035975 | 0.005713 | 3.04E-10 | 9.516888 | Left atrium |
| LV_cardiac | Z97_S1 | 27446 | 0.042703 | 0.006785 | 3.11E-10 | 9.507448 | Left ventricle |
| LV_cardiac | Z83_S2 | 27446 | 0.033 | 0.005244 | 3.11E-10 | 9.506589 | Left ventricle |
| Descending | Z53_S2 | 24599 | -0.04586 | 0.007291 | 3.18E-10 | 9.497094 | Aorta |
| Descending | Z118_S1 | 24599 | -0.04677 | 0.007439 | 3.23E-10 | 9.490211 | Aorta |
| LV_ejectio | Z77_S3 | 27446 | 0.032099 | 0.005106 | 3.24E-10 | 9.489018 | Left ventricle |
| RV_ejectio | Z92_S5 | 27446 | 0.038579 | 0.006166 | 3.93E-10 | 9.405675 | Right ventricle |
| RA_ejectio | Z50_S3 | 27446 | 0.029167 | 0.004666 | 4.06E-10 | 9.390992 | Right atrium |
| RV_ejectio | Z86_S3 | 27446 | -0.03613 | 0.005784 | 4.20E-10 | 9.376884 | Right ventricle |
| LA_ejectio | Z55_S1 | 27446 | 0.02817 | 0.004511 | 4.24E-10 | 9.373146 | Left atrium |
| RV_end_sy | Z122_S2 | 27446 | -0.03494 | 0.005602 | 4.47E-10 | 9.349585 | Right ventricle |
| LV_radial_ | Z38_S4 | 27446 | 0.03687 | 0.005919 | 4.68E-10 | 9.329966 | Left ventricle |
| Descending | Z13_S1 | 24599 | 0.033154 | 0.005335 | 5.13E-10 | 9.289624 | Aorta |
| Descending | Z80_S5 | 24599 | 0.048942 | 0.007877 | 5.18E-10 | 9.285621 | Aorta |
| LV_end_sy | Z118_S1 | 27446 | 0.043519 | 0.007011 | 5.41E-10 | 9.266994 | Left ventricle |
| LV_cardiac | Z53_S2 | 27446 | -0.03816 | 0.006151 | 5.52E-10 | 9.258169 | Left ventricle |
| Descending | Z37_S2 | 24599 | -0.04017 | 0.006484 | 5.79E-10 | 9.237309 | Aorta |
| LA_ejectio | Z96_S3 | 27446 | -0.02934 | 0.004735 | 5.80E-10 | 9.23687 | Left atrium |
| LV_mean_ | Z3_S2 | 27446 | -0.03853 | 0.006221 | 5.87E-10 | 9.231528 | Left ventricle |
| RV_ejectio | Z24_S1 | 27446 | -0.02685 | 0.004335 | 5.90E-10 | 9.228877 | Right ventricle |
| LV_myocar | Z95_S5 | 27446 | -0.06248 | 0.010098 | 6.11E-10 | 9.213635 | Left ventricle |
| Descending | Z49_S1 | 24599 | -0.04975 | 0.008045 | 6.26E-10 | 9.203261 | Aorta |
| RV_ejectio | Z75_S4 | 27446 | 0.0374 | 0.006051 | 6.37E-10 | 9.196191 | Right ventricle |
| Descending | Z44_S4 | 24599 | -0.03119 | 0.005047 | 6.40E-10 | 9.193894 | Aorta |
| LV_cardiac | Z2_S1 | 27446 | -0.03133 | 0.00507 | 6.44E-10 | 9.191348 | Left ventricle |
| LV_radial_ | Z87_S5 | 27446 | 0.031451 | 0.005091 | 6.51E-10 | 9.186269 | Left ventricle |
| LV_radial_ | Z44_S1 | 27446 | -0.03173 | 0.005145 | 7.00E-10 | 9.155138 | Left ventricle |
| LV_radial_ | Z28_S4 | 27446 | 0.034248 | 0.005557 | 7.14E-10 | 9.146137 | Left ventricle |
| Descending | Z23_S1 | 24599 | -0.04149 | 0.006733 | 7.21E-10 | 9.141791 | Aorta |
| Descending | Z120_S4 | 24599 | -0.03634 | 0.005906 | 7.61E-10 | 9.118776 | Aorta |

| | | | | | | | |
|---|---|---|---|---|---|---|---|
| RA_stroke_ | Z114_S5 | 27446 | -0.02485 | 0.00404 | 7.63E-10 | 9.117192 | Right atrium |
| LV_longitu | Z62_S4 | 27446 | -0.03502 | 0.005695 | 7.76E-10 | 9.109933 | Left ventricle |
| RV_stroke_ | Z28_S2 | 27446 | 0.040322 | 0.006557 | 7.77E-10 | 9.109479 | Right ventricle |
| RA_minimu | Z95_S5 | 27446 | -0.04165 | 0.006783 | 8.26E-10 | 9.083016 | Right atrium |
| LV_mean_ | Z39_S3 | 27446 | -0.05356 | 0.008739 | 8.82E-10 | 9.054385 | Left ventricle |
| Ascending_ | Z112_S3 | 24599 | -0.04417 | 0.007212 | 9.09E-10 | 9.041277 | Aorta |
| Ascending_ | Z66_S2 | 24599 | 0.040372 | 0.006603 | 9.73E-10 | 9.011699 | Aorta |
| LV_radial_ | Z115_S3 | 27446 | 0.033555 | 0.005489 | 9.79E-10 | 9.009077 | Left ventricle |
| LV_radial_ | Z121_S4 | 27446 | -0.03553 | 0.005819 | 1.02E-09 | 8.990694 | Left ventricle |
| LV_circumf | Z98_S3 | 27446 | -0.02718 | 0.004451 | 1.02E-09 | 8.989797 | Left ventricle |
| RV_ejectio | Z99_S3 | 27446 | -0.02513 | 0.004125 | 1.12E-09 | 8.95239 | Right ventricle |
| LV_stroke_ | Z12_S5 | 27446 | -0.04384 | 0.007202 | 1.15E-09 | 8.940378 | Left ventricle |
| LV_radial_ | Z127_S4 | 27446 | 0.034447 | 0.005661 | 1.16E-09 | 8.934391 | Left ventricle |
| RV_end_sy | Z56_S5 | 27446 | 0.035551 | 0.005845 | 1.18E-09 | 8.926613 | Right ventricle |
| LA_maximu | Z56_S5 | 27446 | 0.027143 | 0.004463 | 1.19E-09 | 8.923857 | Left atrium |
| RA_stroke_ | Z122_S5 | 27446 | -0.02766 | 0.004551 | 1.22E-09 | 8.915133 | Right atrium |
| Descending | Z77_S3 | 24599 | -0.04145 | 0.006826 | 1.25E-09 | 8.902257 | Aorta |
| Ascending_ | Z123_S4 | 24599 | 0.036606 | 0.006029 | 1.27E-09 | 8.897125 | Aorta |
| RV_ejectio | Z2_S1 | 27446 | 0.028307 | 0.004663 | 1.28E-09 | 8.894422 | Right ventricle |
| Ascending_ | Z26_S3 | 24599 | 0.039264 | 0.006473 | 1.31E-09 | 8.882282 | Aorta |
| LV_longitu | Z98_S3 | 27446 | 0.02633 | 0.004347 | 1.39E-09 | 8.856515 | Left ventricle |
| LV_ejection | Z15_S2 | 27446 | -0.03232 | 0.005345 | 1.47E-09 | 8.833234 | Left ventricle |
| LV_myocar | Z37_S3 | 27446 | -0.05761 | 0.009538 | 1.54E-09 | 8.81255 | Left ventricle |
| LV_cardiac | Z114_S5 | 27446 | -0.02678 | 0.004437 | 1.58E-09 | 8.800533 | Left ventricle |
| LA_maximu | Z97_S2 | 27446 | 0.033291 | 0.00553 | 1.75E-09 | 8.757268 | Left atrium |
| LV_cardiac | Z64_S1 | 27446 | -0.03716 | 0.006174 | 1.77E-09 | 8.752816 | Left ventricle |
| LA_stroke_ | Z49_S1 | 27446 | -0.03859 | 0.006424 | 1.89E-09 | 8.724423 | Left atrium |
| LA_ejection | Z23_S1 | 27446 | -0.0328 | 0.005462 | 1.92E-09 | 8.717614 | Left atrium |
| Descending | Z62_S4 | 24599 | 0.040945 | 0.006823 | 1.96E-09 | 8.708268 | Aorta |
| RV_end_di | Z80_S1 | 27446 | 0.046722 | 0.007792 | 2.02E-09 | 8.694481 | Right ventricle |
| LV_myocar | Z65_S5 | 27446 | -0.05982 | 0.009981 | 2.05E-09 | 8.687828 | Left ventricle |
| LV_cardiac | Z59_S3 | 27446 | -0.02697 | 0.004506 | 2.17E-09 | 8.663142 | Left ventricle |
| Ascending_ | Z113_S2 | 24599 | -0.04357 | 0.007306 | 2.47E-09 | 8.607438 | Aorta |
| LV_cardiac | Z98_S3 | 27446 | -0.02897 | 0.004862 | 2.54E-09 | 8.59589 | Left ventricle |
| Ascending_ | Z65_S2 | 24599 | 0.035324 | 0.005928 | 2.54E-09 | 8.595015 | Aorta |
| RV_ejectio | Z53_S2 | 27446 | 0.033686 | 0.005657 | 2.61E-09 | 8.583236 | Right ventricle |
| RA_stroke_ | Z22_S4 | 27446 | -0.03116 | 0.005242 | 2.78E-09 | 8.556402 | Right atrium |
| Descending | Z28_S2 | 24599 | -0.04242 | 0.007158 | 3.09E-09 | 8.509506 | Aorta |
| Ascending_ | Z91_S1 | 24599 | 0.032968 | 0.005566 | 3.15E-09 | 8.501561 | Aorta |
| LV_ejection | Z107_S1 | 27446 | 0.023879 | 0.004034 | 3.23E-09 | 8.49132 | Left ventricle |
| LV_ejection | Z87_S5 | 27446 | 0.029251 | 0.004943 | 3.26E-09 | 8.487369 | Left ventricle |
| Ascending_ | Z62_S1 | 24599 | 0.033599 | 0.005678 | 3.28E-09 | 8.484097 | Aorta |
| LA_minimu | Z53_S2 | 27446 | 0.033206 | 0.005616 | 3.36E-09 | 8.473835 | Left atrium |
| RA_ejectio | Z26_S4 | 27446 | -0.03649 | 0.006173 | 3.40E-09 | 8.468105 | Right atrium |
| RV_stroke_ | Z95_S5 | 27446 | -0.04371 | 0.007416 | 3.76E-09 | 8.424376 | Right ventricle |

| | | | | | | | |
|---|---|---|---|---|---|---|---|
| LA_ejectior | Z21_S3 | 27446 | 0.035866 | 0.006093 | 3.94E-09 | 8.404163 | Left atrium |
| LA_minimu | Z106_S3 | 27446 | -0.03518 | 0.005986 | 4.16E-09 | 8.380612 | Left atrium |
| RA_stroke_ | Z95_S5 | 27446 | 0.035832 | 0.006098 | 4.19E-09 | 8.377604 | Right atrium |
| RV_ejectio | Z72_S5 | 27446 | 0.036928 | 0.006299 | 4.55E-09 | 8.341738 | Right ventricle |
| RV_end_sy | Z7_S5 | 27446 | -0.04386 | 0.007491 | 4.79E-09 | 8.319713 | Right ventricle |
| RV_ejectio | Z122_S2 | 27446 | 0.024454 | 0.004179 | 4.86E-09 | 8.313068 | Right ventricle |
| LV_end_sy | Z120_S5 | 27446 | -0.04449 | 0.007609 | 5.00E-09 | 8.3008 | Left ventricle |
| RV_ejectio | Z43_S2 | 27446 | 0.026175 | 0.004478 | 5.07E-09 | 8.295 | Right ventricle |
| RA_ejectio | Z109_S1 | 27446 | 0.029178 | 0.004993 | 5.09E-09 | 8.293301 | Right atrium |
| RV_ejectio | Z52_S3 | 27446 | -0.0255 | 0.004363 | 5.09E-09 | 8.293289 | Right ventricle |
| Ascending_ | Z13_S3 | 24599 | 0.028868 | 0.00494 | 5.10E-09 | 8.292631 | Aorta |
| LA_ejectior | Z97_S1 | 27446 | 0.034691 | 0.005938 | 5.15E-09 | 8.288538 | Left atrium |
| Ascending_ | Z80_S2 | 24599 | -0.03154 | 0.005404 | 5.36E-09 | 8.271192 | Aorta |
| Ascending_ | Z99_S3 | 24599 | 0.028527 | 0.004888 | 5.36E-09 | 8.270604 | Aorta |
| RA_stroke_ | Z62_S2 | 27446 | 0.026127 | 0.004483 | 5.60E-09 | 8.251685 | Right atrium |
| LV_circumf | Z111_S5 | 27446 | -0.02932 | 0.005046 | 6.23E-09 | 8.2054 | Left ventricle |
| RV_end_sy | Z76_S3 | 27446 | -0.04193 | 0.007218 | 6.26E-09 | 8.203492 | Right ventricle |
| RV_ejectio | Z66_S2 | 27446 | 0.033954 | 0.005858 | 6.77E-09 | 8.169497 | Right ventricle |
| LV_radial_s | Z81_S1 | 27446 | 0.030295 | 0.005231 | 6.98E-09 | 8.156199 | Left ventricle |
| LV_radial_s | Z109_S1 | 27446 | 0.02995 | 0.005172 | 7.01E-09 | 8.154007 | Left ventricle |
| LV_ejectior | Z99_S5 | 27446 | -0.0278 | 0.004809 | 7.39E-09 | 8.131226 | Left ventricle |
| Ascending_ | Z9_S2 | 24599 | 0.035739 | 0.00619 | 7.78E-09 | 8.10905 | Aorta |
| LA_ejectior | Z92_S1 | 27446 | 0.027856 | 0.004835 | 8.37E-09 | 8.077146 | Left atrium |
| RV_end_sy | Z72_S5 | 27446 | -0.04853 | 0.008444 | 9.10E-09 | 8.041148 | Right ventricle |
| RA_ejectio | Z60_S2 | 27446 | 0.024935 | 0.004342 | 9.28E-09 | 8.032336 | Right atrium |
| Ascending_ | Z51_S3 | 24599 | 0.042525 | 0.007404 | 9.29E-09 | 8.032154 | Aorta |
| LV_cardiac | Z125_S4 | 27446 | 0.032868 | 0.005732 | 9.83E-09 | 8.007561 | Left ventricle |
| LA_maximu | Z49_S1 | 27446 | -0.03661 | 0.006392 | 1.02E-08 | 7.991394 | Left atrium |
| RA_ejectio | Z62_S3 | 27446 | -0.02987 | 0.005216 | 1.03E-08 | 7.989009 | Right atrium |
| RV_ejectio | Z62_S2 | 27446 | -0.02592 | 0.004529 | 1.05E-08 | 7.979383 | Right ventricle |
| RV_ejectio | Z82_S1 | 27446 | -0.03614 | 0.006319 | 1.07E-08 | 7.972282 | Right ventricle |
| RV_ejectio | Z41_S1 | 27446 | 0.031576 | 0.005524 | 1.09E-08 | 7.963215 | Right ventricle |
| LA_stroke_ | Z22_S2 | 27446 | 0.030495 | 0.005338 | 1.11E-08 | 7.954241 | Left atrium |
| RV_ejectio | Z105_S5 | 27446 | -0.02949 | 0.005174 | 1.20E-08 | 7.92243 | Right ventricle |
| RA_stroke_ | Z97_S2 | 27446 | -0.03046 | 0.005348 | 1.22E-08 | 7.912401 | Right atrium |
| LV_myocar | Z28_S2 | 27446 | 0.050854 | 0.008929 | 1.23E-08 | 7.909764 | Left ventricle |
| Descending | Z32_S5 | 24599 | 0.040856 | 0.007175 | 1.24E-08 | 7.906937 | Aorta |
| Descending | Z37_S4 | 24599 | 0.046244 | 0.008145 | 1.37E-08 | 7.864834 | Aorta |
| LV_circumf | Z50_S3 | 27446 | -0.02707 | 0.004768 | 1.37E-08 | 7.863518 | Left ventricle |
| LV_ejectior | Z44_S4 | 27446 | 0.024035 | 0.004241 | 1.45E-08 | 7.837875 | Left ventricle |
| Ascending_ | Z87_S3 | 24599 | -0.03381 | 0.005966 | 1.45E-08 | 7.837426 | Aorta |
| Descending | Z70_S5 | 24599 | 0.032296 | 0.005699 | 1.46E-08 | 7.836557 | Aorta |
| LV_circumf | Z77_S3 | 27446 | 0.029322 | 0.005189 | 1.59E-08 | 7.797555 | Left ventricle |
| LV_cardiac | Z43_S2 | 27446 | -0.02751 | 0.004869 | 1.60E-08 | 7.795781 | Left ventricle |
| RA_maxim | Z50_S2 | 27446 | 0.038396 | 0.006795 | 1.60E-08 | 7.795074 | Right atrium |

| | | | | | | | |
|---|---|---|---|---|---|---|---|
| Ascending_ | Z44_S1 | 24599 | 0.032471 | 0.005749 | 1.62E-08 | 7.78978 | Aorta |
| LV_longitu | Z17_S1 | 27446 | -0.02622 | 0.004647 | 1.68E-08 | 7.775657 | Left ventricle |
| RA_minimu | Z76_S3 | 27446 | -0.03345 | 0.005929 | 1.68E-08 | 7.775165 | Right atrium |
| Ascending_ | Z118_S1 | 24599 | -0.0361 | 0.006403 | 1.72E-08 | 7.764774 | Aorta |
| RA_stroke_ | Z60_S2 | 27446 | -0.02487 | 0.004411 | 1.73E-08 | 7.761896 | Right atrium |
| LA_ejectio | Z37_S4 | 27446 | -0.03391 | 0.006025 | 1.81E-08 | 7.742234 | Left atrium |
| LA_ejectio | Z99_S5 | 27446 | -0.02628 | 0.00467 | 1.83E-08 | 7.738072 | Left atrium |
| RA_minimu | Z120_S5 | 27446 | -0.03825 | 0.006797 | 1.83E-08 | 7.737548 | Right atrium |
| RV_end_sy | Z39_S3 | 27446 | 0.048268 | 0.008584 | 1.87E-08 | 7.72754 | Right ventricle |
| Ascending_ | Z41_S1 | 24599 | 0.036808 | 0.006562 | 2.03E-08 | 7.692964 | Aorta |
| Descending | Z63_S1 | 24599 | -0.04452 | 0.007938 | 2.04E-08 | 7.690052 | Aorta |
| LV_cardiac | Z51_S4 | 27446 | 0.027013 | 0.004817 | 2.05E-08 | 7.687359 | Left ventricle |
| LV_end_sy | Z38_S4 | 27446 | -0.04054 | 0.007231 | 2.06E-08 | 7.686728 | Left ventricle |
| Ascending_ | Z82_S4 | 24599 | -0.02778 | 0.004959 | 2.12E-08 | 7.674609 | Aorta |
| RA_ejectio | Z32_S5 | 27446 | -0.03357 | 0.005995 | 2.15E-08 | 7.668377 | Right atrium |
| LV_end_sy | Z7_S5 | 27446 | -0.03845 | 0.00689 | 2.40E-08 | 7.620582 | Left ventricle |
| LV_end_dia | Z39_S3 | 27446 | 0.047687 | 0.008553 | 2.47E-08 | 7.60771 | Left ventricle |
| LV_stroke_ | Z62_S3 | 27446 | 0.035404 | 0.006352 | 2.50E-08 | 7.602477 | Left ventricle |
| LV_cardiac | Z26_S3 | 27446 | 0.034875 | 0.006258 | 2.50E-08 | 7.601448 | Left ventricle |
| LV_circumf | Z91_S1 | 27446 | 0.027406 | 0.004922 | 2.58E-08 | 7.587711 | Left ventricle |
| RV_ejectio | Z120_S2 | 27446 | -0.02441 | 0.004389 | 2.67E-08 | 7.573153 | Right ventricle |
| LV_mean_ | Z123_S4 | 27446 | -0.04054 | 0.007312 | 2.94E-08 | 7.531776 | Left ventricle |
| RA_stroke_ | Z94_S2 | 27446 | 0.030228 | 0.005455 | 3.01E-08 | 7.521491 | Right atrium |
| LV_mean_ | Z27_S2 | 27446 | -0.04501 | 0.008124 | 3.02E-08 | 7.520025 | Left ventricle |
| LV_circumf | Z105_S5 | 27446 | 0.028502 | 0.005151 | 3.14E-08 | 7.503485 | Left ventricle |
| RA_minimu | Z17_S1 | 27446 | 0.029122 | 0.005263 | 3.14E-08 | 7.503269 | Right atrium |
| RA_stroke_ | Z37_S1 | 27446 | 0.031515 | 0.005698 | 3.19E-08 | 7.495811 | Right atrium |
| LV_ejectio | Z29_S5 | 27446 | -0.02961 | 0.005354 | 3.21E-08 | 7.493656 | Left ventricle |
| LA_ejectio | Z74_S4 | 27446 | 0.028191 | 0.005104 | 3.34E-08 | 7.476586 | Left atrium |
| LA_minimu | Z22_S4 | 27446 | 0.029031 | 0.005257 | 3.35E-08 | 7.474322 | Left atrium |
| LA_minimu | Z100_S3 | 27446 | 0.028775 | 0.005213 | 3.38E-08 | 7.470865 | Left atrium |
| RA_minimu | Z51_S3 | 27446 | -0.03795 | 0.006876 | 3.39E-08 | 7.469236 | Right atrium |
| LA_ejectio | Z65_S1 | 27446 | 0.022268 | 0.004035 | 3.43E-08 | 7.465054 | Left atrium |
| RA_ejectio | Z17_S1 | 27446 | -0.02567 | 0.004656 | 3.50E-08 | 7.455437 | Right atrium |
| Descending | Z95_S5 | 24599 | -0.04374 | 0.007935 | 3.54E-08 | 7.451551 | Aorta |
| LV_circumf | Z44_S3 | 27446 | -0.03095 | 0.005619 | 3.65E-08 | 7.437778 | Left ventricle |
| RA_ejectio | Z13_S1 | 27446 | -0.02454 | 0.00446 | 3.74E-08 | 7.42667 | Right atrium |
| LV_longitu | Z49_S3 | 27446 | -0.03172 | 0.005765 | 3.74E-08 | 7.4266 | Left ventricle |
| RA_ejectio | Z37_S1 | 27446 | 0.030831 | 0.005608 | 3.86E-08 | 7.413671 | Right atrium |
| Ascending_ | Z13_S1 | 24599 | 0.029803 | 0.005435 | 4.16E-08 | 7.380864 | Aorta |
| LV_circumf | Z92_S1 | 27446 | -0.02772 | 0.00506 | 4.28E-08 | 7.368468 | Left ventricle |
| LV_radial_ | Z14_S4 | 27446 | 0.031434 | 0.005756 | 4.74E-08 | 7.324273 | Left ventricle |
| LV_ejectio | Z65_S2 | 27446 | -0.02812 | 0.005151 | 4.77E-08 | 7.321652 | Left ventricle |
| LV_mean_ | Z4_S4 | 27446 | -0.03385 | 0.006205 | 4.88E-08 | 7.311687 | Left ventricle |
| LV_radial_ | Z97_S1 | 27446 | 0.034358 | 0.006299 | 4.92E-08 | 7.308189 | Left ventricle |

| | | | | | | | |
|---|---|---|---|---|---|---|---|
| RA_maxim | Z74_S2 | 27446 | 0.029922 | 0.00549 | 5.02E-08 | 7.299414 | Right atrium |
| Descending | Z97_S2 | 24599 | -0.03859 | 0.007091 | 5.25E-08 | 7.280116 | Aorta |
| RV_end_sy | Z14_S4 | 27446 | 0.041548 | 0.007646 | 5.52E-08 | 7.25827 | Right ventricle |
| RA_stroke_ | Z14_S2 | 27446 | -0.02739 | 0.005046 | 5.70E-08 | 7.244444 | Right atrium |
| LV_cardiac | Z60_S2 | 27446 | -0.0263 | 0.004846 | 5.74E-08 | 7.240776 | Left ventricle |
| LA_ejectio | Z50_S5 | 27446 | -0.03268 | 0.00604 | 6.32E-08 | 7.199536 | Left atrium |
| LV_end_di | Z44_S1 | 27446 | 0.036834 | 0.006811 | 6.38E-08 | 7.195386 | Left ventricle |
| Ascending_ | Z78_S4 | 24599 | 0.029035 | 0.005372 | 6.50E-08 | 7.187195 | Aorta |
| LV_longitu | Z76_S3 | 27446 | 0.028206 | 0.005235 | 7.14E-08 | 7.146235 | Left ventricle |
| RV_end_sy | Z10_S4 | 27446 | -0.03845 | 0.00715 | 7.56E-08 | 7.121482 | Right ventricle |
| LV_end_di | Z28_S1 | 27446 | -0.03875 | 0.007215 | 7.88E-08 | 7.103384 | Left ventricle |
| Descending | Z39_S4 | 24599 | 0.032123 | 0.005988 | 8.09E-08 | 7.09195 | Aorta |
| RA_ejectio | Z59_S2 | 27446 | -0.0305 | 0.005688 | 8.22E-08 | 7.085016 | Right atrium |
| RV_ejectio | Z73_S4 | 27446 | -0.0314 | 0.005874 | 9.02E-08 | 7.044851 | Right ventricle |
| RA_minimu | Z114_S4 | 27446 | -0.02927 | 0.005478 | 9.17E-08 | 7.037721 | Right atrium |
| RV_ejectio | Z89_S4 | 27446 | -0.02638 | 0.004943 | 9.42E-08 | 7.025841 | Right ventricle |
| LV_circumf | Z114_S4 | 27446 | 0.026366 | 0.004952 | 1.02E-07 | 6.993001 | Left ventricle |
| Ascending_ | Z37_S2 | 24599 | -0.03337 | 0.006277 | 1.06E-07 | 6.976559 | Aorta |
| RV_ejectio | Z60_S2 | 27446 | 0.023617 | 0.004457 | 1.17E-07 | 6.933601 | Right ventricle |
| Ascending_ | Z62_S3 | 24599 | 0.033589 | 0.006347 | 1.21E-07 | 6.917238 | Aorta |
| LV_circumf | Z94_S5 | 27446 | -0.0232 | 0.004391 | 1.27E-07 | 6.896119 | Left ventricle |
| RA_ejectio | Z99_S4 | 27446 | 0.029387 | 0.005564 | 1.28E-07 | 6.891407 | Right atrium |
| LV_longitu | Z94_S2 | 27446 | 0.028289 | 0.005359 | 1.30E-07 | 6.885622 | Left ventricle |
| Descending | Z29_S5 | 24599 | 0.033625 | 0.006371 | 1.31E-07 | 6.882968 | Aorta |
| LV_cardiac | Z66_S2 | 27446 | -0.03359 | 0.00637 | 1.34E-07 | 6.872757 | Left ventricle |
| RV_ejectio | Z59_S2 | 27446 | -0.03076 | 0.005838 | 1.37E-07 | 6.863514 | Right ventricle |
| RA_maxim | Z51_S3 | 27446 | -0.03574 | 0.006784 | 1.37E-07 | 6.862211 | Right atrium |
| LV_circumf | Z62_S3 | 27446 | 0.028 | 0.005331 | 1.50E-07 | 6.824481 | Left ventricle |
| LV_cardiac | Z87_S5 | 27446 | -0.02881 | 0.005486 | 1.52E-07 | 6.819388 | Left ventricle |
| RA_stroke_ | Z120_S2 | 27446 | -0.02281 | 0.004345 | 1.53E-07 | 6.816536 | Right atrium |
| Ascending_ | Z122_S1 | 24599 | 0.033099 | 0.006312 | 1.57E-07 | 6.803253 | Aorta |
| Descending | Z51_S3 | 24599 | 0.03811 | 0.00727 | 1.59E-07 | 6.799068 | Aorta |
| LA_maximu | Z106_S3 | 27446 | -0.03234 | 0.006174 | 1.63E-07 | 6.788639 | Left atrium |
| RV_ejectio | Z64_S1 | 27446 | 0.029733 | 0.005679 | 1.65E-07 | 6.782914 | Right ventricle |
| LV_circumf | Z46_S1 | 27446 | 0.026932 | 0.00515 | 1.70E-07 | 6.768614 | Left ventricle |
| LV_circumf | Z3_S2 | 27446 | -0.02372 | 0.004538 | 1.72E-07 | 6.764923 | Left ventricle |
| RV_ejectio | Z11_S5 | 27446 | 0.025706 | 0.004918 | 1.72E-07 | 6.763325 | Right ventricle |
| LV_circumf | Z68_S2 | 27446 | -0.02419 | 0.004632 | 1.77E-07 | 6.752551 | Left ventricle |
| RA_minimu | Z93_S1 | 27446 | 0.033654 | 0.006451 | 1.82E-07 | 6.739376 | Right atrium |
| LA_stroke_ | Z12_S5 | 27446 | -0.03258 | 0.006247 | 1.83E-07 | 6.737635 | Left atrium |
| LV_myocar | Z99_S4 | 27446 | 0.048766 | 0.009363 | 1.91E-07 | 6.719654 | Left ventricle |
| LV_ejectio | Z95_S5 | 27446 | 0.031418 | 0.006036 | 1.94E-07 | 6.712026 | Left ventricle |
| RV_ejectio | Z115_S3 | 27446 | 0.028313 | 0.00544 | 1.95E-07 | 6.710926 | Right ventricle |
| LV_cardiac | Z7_S1 | 27446 | 0.026512 | 0.005103 | 2.04E-07 | 6.689582 | Left ventricle |
| RA_maxim | Z114_S4 | 27446 | -0.02808 | 0.005405 | 2.04E-07 | 6.689522 | Right atrium |

| | | | | | | | |
|---|---|---|---|---|---|---|---|
| RA_maxim | Z76_S3 | 27446 | -0.03039 | 0.00585 | 2.05E-07 | 6.687769 | Right atrium |
| Descending | Z9_S2 | 24599 | 0.037226 | 0.007168 | 2.07E-07 | 6.684874 | Aorta |
| RA_ejectio | Z9_S2 | 27446 | 0.027644 | 0.005324 | 2.08E-07 | 6.682399 | Right atrium |
| RA_minimu | Z40_S4 | 27446 | -0.03526 | 0.006795 | 2.11E-07 | 6.675618 | Right atrium |
| LV_longitu | Z58_S2 | 27446 | -0.02417 | 0.004661 | 2.15E-07 | 6.666832 | Left ventricle |
| RA_ejectio | Z120_S4 | 27446 | 0.02559 | 0.004937 | 2.18E-07 | 6.662514 | Right atrium |
| LV_end_sy | Z80_S1 | 27446 | -0.03541 | 0.006834 | 2.19E-07 | 6.658907 | Left ventricle |
| LV_mean_ | Z106_S3 | 27446 | -0.04266 | 0.008233 | 2.20E-07 | 6.657533 | Left ventricle |
| LV_longitu | Z114_S5 | 27446 | 0.020545 | 0.003969 | 2.26E-07 | 6.645823 | Left ventricle |
| RV_stroke_ | Z99_S4 | 27446 | 0.035576 | 0.006877 | 2.30E-07 | 6.638864 | Right ventricle |
| RV_end_di | Z22_S2 | 27446 | 0.03769 | 0.007296 | 2.39E-07 | 6.621736 | Right ventricle |
| Descending | Z105_S5 | 24599 | 0.031009 | 0.006013 | 2.50E-07 | 6.601276 | Aorta |
| RV_ejectio | Z59_S3 | 27446 | 0.021373 | 0.004145 | 2.52E-07 | 6.599158 | Right ventricle |
| Descending | Z83_S2 | 24599 | 0.028998 | 0.005626 | 2.54E-07 | 6.594377 | Aorta |
| LA_ejectio | Z37_S1 | 27446 | 0.028232 | 0.005477 | 2.55E-07 | 6.59423 | Left atrium |
| RV_ejectio | Z122_S1 | 27446 | 0.028805 | 0.005596 | 2.64E-07 | 6.577709 | Right ventricle |
| LV_radial_ | Z26_S4 | 27446 | -0.03292 | 0.006396 | 2.65E-07 | 6.577021 | Left ventricle |
| LV_cardiac | Z3_S4 | 27446 | -0.03566 | 0.006929 | 2.65E-07 | 6.57636 | Left ventricle |
| RA_stroke_ | Z7_S3 | 27446 | -0.03102 | 0.006031 | 2.69E-07 | 6.570974 | Right atrium |
| LV_longitu | Z80_S5 | 27446 | 0.030167 | 0.005867 | 2.72E-07 | 6.56569 | Left ventricle |
| Ascending_ | Z28_S1 | 24599 | 0.033022 | 0.006424 | 2.74E-07 | 6.561656 | Aorta |
| LV_longitu | Z29_S5 | 27446 | 0.027315 | 0.005314 | 2.74E-07 | 6.561548 | Left ventricle |
| Descending | Z55_S1 | 24599 | -0.02836 | 0.005523 | 2.81E-07 | 6.551117 | Aorta |
| LV_cardiac | Z21_S3 | 27446 | 0.035699 | 0.006964 | 2.96E-07 | 6.528894 | Left ventricle |
| Ascending_ | Z65_S5 | 24599 | 0.035216 | 0.00688 | 3.08E-07 | 6.511519 | Aorta |
| LV_end_sy | Z57_S2 | 27446 | 0.026775 | 0.005234 | 3.13E-07 | 6.504426 | Left ventricle |
| Ascending_ | Z59_S2 | 24599 | -0.03541 | 0.006938 | 3.32E-07 | 6.478808 | Aorta |
| RA_stroke_ | Z6_S2 | 27446 | -0.02008 | 0.003937 | 3.42E-07 | 6.466279 | Right atrium |
| LV_end_sy | Z39_S3 | 27446 | 0.040246 | 0.007894 | 3.43E-07 | 6.464582 | Left ventricle |
| Ascending_ | Z14_S4 | 24599 | -0.03291 | 0.006456 | 3.44E-07 | 6.463947 | Aorta |
| Descending | Z66_S2 | 24599 | 0.038938 | 0.007647 | 3.54E-07 | 6.4505 | Aorta |
| Ascending_ | Z15_S2 | 24599 | 0.032971 | 0.00648 | 3.62E-07 | 6.441731 | Aorta |
| RV_end_di | Z38_S4 | 27446 | -0.04188 | 0.008247 | 3.81E-07 | 6.419402 | Right ventricle |
| Ascending_ | Z53_S2 | 24599 | -0.03232 | 0.006367 | 3.85E-07 | 6.414416 | Aorta |
| LV_radial_ | Z53_S2 | 27446 | 0.02896 | 0.005711 | 3.95E-07 | 6.402968 | Left ventricle |
| RV_ejectio | Z75_S1 | 27446 | 0.026713 | 0.005268 | 3.95E-07 | 6.402959 | Right ventricle |
| Descending | Z15_S2 | 24599 | 0.032222 | 0.006362 | 4.09E-07 | 6.388453 | Aorta |
| LV_ejectio | Z100_S3 | 27446 | -0.02605 | 0.005145 | 4.13E-07 | 6.383833 | Left ventricle |
| RA_minimu | Z44_S3 | 27446 | -0.03145 | 0.006217 | 4.21E-07 | 6.375793 | Right atrium |
| Ascending_ | Z3_S4 | 24599 | -0.03624 | 0.007165 | 4.24E-07 | 6.372726 | Aorta |
| LV_myocar | Z97_S2 | 27446 | 0.044778 | 0.008858 | 4.30E-07 | 6.366505 | Left ventricle |
| Descending | Z23_S1 | 24599 | -0.03758 | 0.00744 | 4.38E-07 | 6.35881 | Aorta |
| LV_longitu | Z9_S2 | 27446 | 0.026837 | 0.005314 | 4.41E-07 | 6.355752 | Left ventricle |
| Ascending_ | Z91_S1 | 24599 | 0.028188 | 0.005587 | 4.53E-07 | 6.343768 | Aorta |
| LV_ejectio | Z111_S5 | 27446 | 0.025055 | 0.004967 | 4.56E-07 | 6.341286 | Left ventricle |

| | | | | | | | |
|---|---|---|---|---|---|---|---|
| LV_cardiac | Z50_S4 | 27446 | 0.027111 | 0.00538 | 4.66E-07 | 6.33132 | Left ventricle |
| LV_ejection | Z62_S1 | 27446 | -0.02362 | 0.004691 | 4.78E-07 | 6.32028 | Left ventricle |
| Descending | Z40_S4 | 24599 | -0.03618 | 0.007187 | 4.81E-07 | 6.317604 | Aorta |
| LA_stroke_ | Z23_S1 | 27446 | 0.029712 | 0.005907 | 4.91E-07 | 6.309125 | Left atrium |
| RA_minimu | Z108_S5 | 27446 | -0.029 | 0.005767 | 4.93E-07 | 6.30736 | Right atrium |
| LA_ejection | Z62_S4 | 27446 | 0.028008 | 0.005573 | 5.03E-07 | 6.298396 | Left atrium |
| RA_minimu | Z122_S5 | 27446 | -0.02544 | 0.005064 | 5.05E-07 | 6.296597 | Right atrium |
| LV_stroke_ | Z121_S4 | 27446 | 0.034355 | 0.006842 | 5.14E-07 | 6.288715 | Left ventricle |
| LV_myocar | Z32_S5 | 27446 | -0.05065 | 0.010088 | 5.14E-07 | 6.288678 | Left ventricle |
| RV_ejectio | Z100_S3 | 27446 | -0.02636 | 0.005252 | 5.17E-07 | 6.286252 | Right ventricle |
| RV_ejectio | Z113_S2 | 27446 | -0.03089 | 0.006156 | 5.25E-07 | 6.280213 | Right ventricle |
| RA_maxim | Z112_S3 | 27446 | -0.03299 | 0.006589 | 5.53E-07 | 6.257586 | Right atrium |
| RV_end_di | Z53_S2 | 27446 | -0.03982 | 0.007956 | 5.58E-07 | 6.253632 | Right ventricle |
| LV_ejection | Z50_S2 | 27446 | -0.03067 | 0.006129 | 5.58E-07 | 6.253273 | Left ventricle |
| LA_stroke_ | Z113_S2 | 27446 | 0.031665 | 0.006334 | 5.75E-07 | 6.240569 | Left atrium |
| LV_ejection | Z1_S5 | 27446 | 0.025557 | 0.005113 | 5.77E-07 | 6.238573 | Left ventricle |
| LV_longitu | Z5_S5 | 27446 | -0.0256 | 0.005122 | 5.81E-07 | 6.236114 | Left ventricle |
| RV_ejectio | Z28_S4 | 27446 | 0.02752 | 0.005508 | 5.83E-07 | 6.234269 | Right ventricle |
| LA_stroke_ | Z73_S1 | 27446 | -0.02462 | 0.00493 | 5.95E-07 | 6.225348 | Left atrium |
| LA_minimu | Z40_S4 | 27446 | 0.030582 | 0.006126 | 5.97E-07 | 6.224318 | Left atrium |
| LV_longitu | Z14_S4 | 27446 | -0.02767 | 0.005546 | 6.09E-07 | 6.215278 | Left ventricle |
| Descending | Z81_S1 | 24599 | -0.03015 | 0.006044 | 6.11E-07 | 6.214307 | Aorta |
| RA_minimu | Z97_S2 | 27446 | 0.029672 | 0.00595 | 6.14E-07 | 6.211525 | Right atrium |
| Ascending_ | Z53_S2 | 24599 | 0.033476 | 0.006725 | 6.43E-07 | 6.191697 | Aorta |
| RV_stroke_ | Z26_S3 | 27446 | -0.03447 | 0.00693 | 6.54E-07 | 6.184091 | Right ventricle |
| LV_end_di | Z80_S1 | 27446 | 0.036803 | 0.007405 | 6.69E-07 | 6.17467 | Left ventricle |
| RA_stroke_ | Z65_S5 | 27446 | 0.029958 | 0.006028 | 6.70E-07 | 6.173613 | Right atrium |
| RV_ejectio | Z49_S3 | 27446 | 0.029442 | 0.00593 | 6.87E-07 | 6.162962 | Right ventricle |
| Descending | Z28_S1 | 24599 | 0.031295 | 0.006307 | 6.99E-07 | 6.155709 | Aorta |
| LV_ejection | Z59_S2 | 27446 | -0.02836 | 0.00572 | 7.12E-07 | 6.147332 | Left ventricle |
| LA_maximu | Z28_S1 | 27446 | -0.02739 | 0.00553 | 7.31E-07 | 6.136287 | Left atrium |
| RA_stroke_ | Z56_S2 | 27446 | 0.021513 | 0.004346 | 7.44E-07 | 6.128584 | Right atrium |
| LV_mean_ | Z23_S3 | 27446 | 0.030616 | 0.006193 | 7.68E-07 | 6.114522 | Left ventricle |
| LA_stroke_ | Z28_S2 | 27446 | 0.02769 | 0.005605 | 7.79E-07 | 6.108388 | Left atrium |
| Descending | Z106_S4 | 24599 | 0.025799 | 0.005232 | 8.18E-07 | 6.087176 | Aorta |
| LV_ejection | Z37_S2 | 27446 | 0.02681 | 0.005437 | 8.19E-07 | 6.086883 | Left ventricle |
| LV_ejection | Z76_S3 | 27446 | 0.026006 | 0.005276 | 8.25E-07 | 6.083298 | Left ventricle |
| Ascending_ | Z94_S2 | 24599 | -0.0324 | 0.006575 | 8.30E-07 | 6.080785 | Aorta |
| LV_radial_ | Z38_S5 | 27446 | 0.026796 | 0.005445 | 8.59E-07 | 6.065783 | Left ventricle |
| LV_mean_ | Z56_S5 | 27446 | 0.029277 | 0.005953 | 8.73E-07 | 6.059102 | Left ventricle |
| RV_ejectio | Z95_S5 | 27446 | 0.030263 | 0.006161 | 9.03E-07 | 6.044434 | Right ventricle |
| LV_stroke_ | Z65_S1 | 27446 | -0.02471 | 0.005033 | 9.07E-07 | 6.042404 | Left ventricle |
| RV_ejectio | Z74_S4 | 27446 | -0.02635 | 0.005366 | 9.08E-07 | 6.041814 | Right ventricle |
| LV_mean_ | Z99_S3 | 27446 | 0.027625 | 0.005632 | 9.33E-07 | 6.030324 | Left ventricle |
| LV_cardiac | Z120_S4 | 27446 | 0.026978 | 0.00551 | 9.77E-07 | 6.009976 | Left ventricle |

| | | | | | | | |
|---|---|---|---|---|---|---|---|
| Ascending_ | Z14_S1 | 24599 | 0.027856 | 0.005696 | 1.00E-06 | 5.998405 | Aorta |
| LV_end_di | Z65_S5 | 27446 | -0.03978 | 0.008135 | 1.01E-06 | 5.995304 | Left ventricle |
| LV_mean_ | Z62_S3 | 27446 | -0.03574 | 0.007309 | 1.01E-06 | 5.994566 | Left ventricle |
| LV_cardiac | Z50_S2 | 27446 | 0.033225 | 0.006802 | 1.03E-06 | 5.985143 | Left ventricle |
| RV_stroke_ | Z56_S5 | 27446 | -0.02564 | 0.00525 | 1.04E-06 | 5.98196 | Right ventricle |
| LV_cardiac | Z100_S3 | 27446 | 0.027827 | 0.00571 | 1.10E-06 | 5.95889 | Left ventricle |
| RA_stroke_ | Z73_S1 | 27446 | 0.023095 | 0.004744 | 1.12E-06 | 5.949465 | Right atrium |
| RA_stroke_ | Z99_S4 | 27446 | 0.02747 | 0.005654 | 1.18E-06 | 5.92675 | Right atrium |
| RV_ejectio | Z87_S5 | 27446 | 0.024512 | 0.005046 | 1.19E-06 | 5.925353 | Right ventricle |
| LV_ejectio | Z7_S3 | 27446 | 0.028995 | 0.005969 | 1.19E-06 | 5.924439 | Left ventricle |
| Ascending_ | Z28_S1 | 24599 | -0.02964 | 0.006105 | 1.20E-06 | 5.92053 | Aorta |
| Ascending_ | Z75_S5 | 24599 | -0.03506 | 0.007224 | 1.22E-06 | 5.914772 | Aorta |
| RV_stroke_ | Z39_S3 | 27446 | 0.037398 | 0.007709 | 1.23E-06 | 5.910521 | Right ventricle |
| RV_end_sy | Z18_S4 | 27446 | -0.0415 | 0.00856 | 1.24E-06 | 5.905026 | Right ventricle |
| LV_longitu | Z83_S2 | 27446 | -0.02271 | 0.004691 | 1.30E-06 | 5.887451 | Left ventricle |
| RV_ejectio | Z65_S5 | 27446 | -0.02945 | 0.00609 | 1.32E-06 | 5.878162 | Right ventricle |
| LV_circumf | Z97_S2 | 27446 | -0.02601 | 0.005379 | 1.32E-06 | 5.877884 | Left ventricle |
| Ascending_ | Z95_S3 | 24599 | 0.022602 | 0.004678 | 1.35E-06 | 5.868436 | Aorta |
| LV_end_sy | Z40_S4 | 27446 | 0.036704 | 0.007608 | 1.41E-06 | 5.852033 | Left ventricle |
| RA_stroke_ | Z81_S1 | 27446 | -0.02469 | 0.005131 | 1.49E-06 | 5.82571 | Right atrium |
| LV_mean_ | Z9_S2 | 27446 | 0.035866 | 0.00746 | 1.53E-06 | 5.816177 | Left ventricle |
| LV_circumf | Z57_S2 | 27446 | -0.02026 | 0.004227 | 1.65E-06 | 5.782725 | Left ventricle |
| RA_stroke_ | Z65_S1 | 27446 | 0.020118 | 0.004199 | 1.66E-06 | 5.780369 | Right atrium |
| Ascending_ | Z49_S3 | 24599 | 0.033698 | 0.007039 | 1.69E-06 | 5.772535 | Aorta |
| Ascending_ | Z19_S2 | 24599 | 0.027069 | 0.005662 | 1.75E-06 | 5.757656 | Aorta |
| LV_circumf | Z120_S5 | 27446 | -0.02938 | 0.006146 | 1.75E-06 | 5.756452 | Left ventricle |
| Descending | Z39_S4 | 24599 | 0.028204 | 0.005903 | 1.77E-06 | 5.751509 | Aorta |
| Descending | Z75_S4 | 24599 | -0.03379 | 0.007073 | 1.78E-06 | 5.750432 | Aorta |
| LV_circumf | Z93_S2 | 27446 | 0.029836 | 0.006246 | 1.78E-06 | 5.749042 | Left ventricle |
| LV_longitu | Z96_S3 | 27446 | -0.02312 | 0.004841 | 1.79E-06 | 5.747832 | Left ventricle |
| Descending | Z65_S5 | 24599 | 0.037493 | 0.007853 | 1.80E-06 | 5.744098 | Aorta |
| LV_end_di | Z37_S3 | 27446 | -0.03711 | 0.007774 | 1.81E-06 | 5.741498 | Left ventricle |
| LV_radial_ | Z7_S3 | 27446 | 0.029314 | 0.006149 | 1.87E-06 | 5.728786 | Left ventricle |
| LV_circumf | Z32_S5 | 27446 | -0.02917 | 0.006127 | 1.92E-06 | 5.715982 | Left ventricle |
| Descending | Z53_S1 | 24599 | 0.032796 | 0.00689 | 1.94E-06 | 5.712531 | Aorta |
| RV_ejectio | Z4_S4 | 27446 | 0.021632 | 0.004546 | 1.95E-06 | 5.709417 | Right ventricle |
| RA_minimu | Z26_S4 | 27446 | 0.033191 | 0.006979 | 1.98E-06 | 5.703722 | Right atrium |
| LA_minimu | Z3_S4 | 27446 | 0.030074 | 0.006326 | 2.00E-06 | 5.700051 | Left atrium |
| LV_cardiac | Z28_S4 | 27446 | 0.028399 | 0.005989 | 2.12E-06 | 5.674623 | Left ventricle |
| Ascending_ | Z115_S4 | 24599 | -0.02635 | 0.005557 | 2.12E-06 | 5.672961 | Aorta |
| LV_radial_ | Z22_S4 | 27446 | -0.02533 | 0.005346 | 2.17E-06 | 5.664342 | Left ventricle |
| LV_ejectio | Z73_S4 | 27446 | -0.02726 | 0.005755 | 2.18E-06 | 5.662045 | Left ventricle |
| Ascending_ | Z83_S2 | 24599 | 0.027084 | 0.005731 | 2.29E-06 | 5.640281 | Aorta |
| Ascending_ | Z57_S2 | 24599 | 0.023829 | 0.005055 | 2.43E-06 | 5.614074 | Aorta |
| LV_end_sy | Z94_S2 | 27446 | 0.031989 | 0.006796 | 2.51E-06 | 5.599861 | Left ventricle |

| | | | | | | | |
|---|---|---|---|---|---|---|---|
| LV_circumf | Z69_S5 | 27446 | -0.02036 | 0.004327 | 2.55E-06 | 5.593406 | Left ventricle |
| LV_longitu | Z39_S4 | 27446 | -0.02092 | 0.004452 | 2.61E-06 | 5.583683 | Left ventricle |
| Descending | Z77_S3 | 24599 | -0.0286 | 0.006093 | 2.67E-06 | 5.573325 | Aorta |
| Ascending_ | Z10_S4 | 24599 | 0.028126 | 0.006002 | 2.79E-06 | 5.55503 | Aorta |
| RV_ejectio | Z99_S5 | 27446 | -0.02296 | 0.004909 | 2.91E-06 | 5.5368 | Right ventricle |
| LV_longitu | Z20_S2 | 27446 | -0.01965 | 0.004203 | 2.93E-06 | 5.532679 | Left ventricle |
| RA_stroke_ | Z98_S3 | 27446 | -0.02069 | 0.004427 | 2.96E-06 | 5.529039 | Right atrium |
| Descending | Z59_S3 | 24599 | -0.02253 | 0.004823 | 2.99E-06 | 5.524849 | Aorta |
| RA_ejectio | Z94_S2 | 27446 | 0.025079 | 0.00537 | 3.01E-06 | 5.521337 | Right atrium |
| Descending | Z121_S4 | 24599 | 0.034623 | 0.007419 | 3.06E-06 | 5.514534 | Aorta |
| RA_minimu | Z12_S5 | 27446 | -0.03117 | 0.006687 | 3.14E-06 | 5.50256 | Right atrium |
| LV_myocar | Z121_S4 | 27446 | 0.044025 | 0.009455 | 3.22E-06 | 5.492037 | Left ventricle |
| RV_end_di | Z32_S1 | 27446 | 0.03553 | 0.007633 | 3.25E-06 | 5.488585 | Right ventricle |
| Ascending_ | Z86_S3 | 24599 | -0.03047 | 0.006546 | 3.25E-06 | 5.488445 | Aorta |
| RA_ejectio | Z92_S1 | 27446 | 0.023021 | 0.004952 | 3.35E-06 | 5.475362 | Right atrium |
| RA_ejectio | Z58_S2 | 27446 | -0.02167 | 0.004671 | 3.48E-06 | 5.458104 | Right atrium |
| LV_ejectio | Z114_S4 | 27446 | -0.02256 | 0.004875 | 3.69E-06 | 5.43251 | Left ventricle |
| RV_ejectio | Z96_S3 | 27446 | -0.02302 | 0.004979 | 3.75E-06 | 5.42582 | Right ventricle |
| LV_ejectio | Z38_S4 | 27446 | 0.026531 | 0.005747 | 3.91E-06 | 5.407884 | Left ventricle |
| LV_longitu | Z51_S1 | 27446 | 0.020136 | 0.004365 | 3.96E-06 | 5.402246 | Left ventricle |
| Descending | Z94_S2 | 24599 | 0.033345 | 0.007234 | 4.03E-06 | 5.394511 | Aorta |
| RA_ejectio | Z12_S5 | 27446 | -0.02727 | 0.005916 | 4.03E-06 | 5.39421 | Right atrium |
| LV_end_sy | Z93_S1 | 27446 | 0.033275 | 0.007223 | 4.09E-06 | 5.388107 | Left ventricle |
| LA_maximu | Z73_S1 | 27446 | -0.02259 | 0.004906 | 4.14E-06 | 5.383214 | Left atrium |
| LA_maximu | Z7_S5 | 27446 | 0.026325 | 0.005722 | 4.21E-06 | 5.375865 | Left atrium |
| RA_stroke_ | Z13_S1 | 27446 | 0.020847 | 0.004532 | 4.22E-06 | 5.374456 | Right atrium |
| Ascending_ | Z27_S2 | 24599 | -0.03262 | 0.007094 | 4.25E-06 | 5.372034 | Aorta |
| RV_end_sy | Z39_S4 | 27446 | -0.02821 | 0.006138 | 4.33E-06 | 5.363989 | Right ventricle |
| Descending | Z26_S3 | 24599 | 0.034078 | 0.007417 | 4.33E-06 | 5.363163 | Aorta |
| LV_radial_ | Z5_S1 | 27446 | 0.023375 | 0.005089 | 4.37E-06 | 5.359449 | Left ventricle |
| RA_maxim | Z75_S1 | 27446 | 0.02625 | 0.005724 | 4.51E-06 | 5.345377 | Right atrium |
| LV_ejectio | Z70_S5 | 27446 | -0.02193 | 0.004791 | 4.70E-06 | 5.327771 | Left ventricle |
| Descending | Z62_S3 | 24599 | 0.028516 | 0.006232 | 4.75E-06 | 5.323311 | Aorta |
| LV_longitu | Z86_S3 | 27446 | -0.02573 | 0.005626 | 4.81E-06 | 5.317667 | Left ventricle |
| Descending | Z13_S3 | 24599 | 0.022157 | 0.004851 | 4.94E-06 | 5.306301 | Aorta |
| LV_circumf | Z44_S4 | 27446 | -0.01969 | 0.00431 | 4.94E-06 | 5.306121 | Left ventricle |
| LV_stroke_ | Z37_S3 | 27446 | -0.03153 | 0.006905 | 4.96E-06 | 5.30432 | Left ventricle |
| Ascending_ | Z56_S2 | 24599 | -0.02258 | 0.004954 | 5.19E-06 | 5.28444 | Aorta |
| Descending | Z50_S3 | 24599 | -0.02539 | 0.005575 | 5.28E-06 | 5.277642 | Aorta |
| LA_maximu | Z39_S4 | 27446 | 0.021343 | 0.004687 | 5.28E-06 | 5.277103 | Left atrium |
| RV_ejectio | Z12_S4 | 27446 | 0.019878 | 0.004368 | 5.34E-06 | 5.272696 | Right ventricle |
| LV_stroke_ | Z22_S2 | 27446 | 0.027996 | 0.006157 | 5.44E-06 | 5.264634 | Left ventricle |
| RV_ejectio | Z6_S2 | 27446 | 0.018055 | 0.003978 | 5.66E-06 | 5.24734 | Right ventricle |
| RA_stroke_ | Z32_S5 | 27446 | -0.02763 | 0.006092 | 5.74E-06 | 5.241087 | Right atrium |
| LV_ejectio | Z120_S4 | 27446 | 0.022465 | 0.004965 | 6.05E-06 | 5.218042 | Left ventricle |

| | | | | | | | |
|---|---|---|---|---|---|---|---|
| RA_minimu | Z38_S4 | 27446 | -0.02923 | 0.00646 | 6.07E-06 | 5.217027 | Right atrium |
| Descending | Z121_S4 | 24599 | 0.034027 | 0.007526 | 6.15E-06 | 5.210923 | Aorta |
| RA_stroke_ | Z121_S4 | 27446 | -0.02576 | 0.005709 | 6.40E-06 | 5.193638 | Right atrium |
| Descending | Z65_S5 | 24599 | 0.035948 | 0.007967 | 6.41E-06 | 5.192999 | Aorta |
| RA_maxim | Z59_S2 | 27446 | -0.02862 | 0.006344 | 6.44E-06 | 5.191094 | Right atrium |
| RV_ejectio | Z50_S2 | 27446 | -0.02822 | 0.006256 | 6.46E-06 | 5.189971 | Right ventricle |
| LV_stroke_ | Z7_S5 | 27446 | 0.029906 | 0.006631 | 6.49E-06 | 5.187727 | Left ventricle |
| RV_end_sy | Z50_S5 | 27446 | 0.038372 | 0.008513 | 6.56E-06 | 5.183103 | Right ventricle |
| LV_radial_ | Z80_S5 | 27446 | 0.027428 | 0.00609 | 6.68E-06 | 5.174945 | Left ventricle |
| RV_ejectio | Z32_S5 | 27446 | 0.027716 | 0.006155 | 6.69E-06 | 5.174451 | Right ventricle |
| Descending | Z21_S3 | 24599 | -0.0371 | 0.008247 | 6.83E-06 | 5.16532 | Aorta |
| Descending | Z41_S1 | 24599 | 0.032459 | 0.007221 | 6.95E-06 | 5.158279 | Aorta |
| LV_longitu | Z28_S1 | 27446 | 0.023584 | 0.005253 | 7.13E-06 | 5.14701 | Left ventricle |
| LV_cardiac | Z6_S2 | 27446 | -0.01942 | 0.004325 | 7.15E-06 | 5.145529 | Left ventricle |
| Descending | Z68_S2 | 24599 | -0.02429 | 0.005411 | 7.16E-06 | 5.145343 | Aorta |
| Ascending_ | Z23_S3 | 24599 | -0.02413 | 0.005384 | 7.39E-06 | 5.131337 | Aorta |
| LA_ejectio | Z3_S2 | 27446 | 0.019438 | 0.004338 | 7.41E-06 | 5.130045 | Left atrium |
| LV_circum | Z19_S2 | 27446 | 0.022508 | 0.005023 | 7.42E-06 | 5.129461 | Left ventricle |
| RV_ejectio | Z15_S2 | 27446 | -0.02436 | 0.005457 | 8.01E-06 | 5.096295 | Right ventricle |
| LV_mean_ | Z65_S2 | 27446 | 0.031865 | 0.007178 | 9.02E-06 | 5.044864 | Left ventricle |
| RA_minimu | Z92_S1 | 27446 | -0.02484 | 0.005598 | 9.08E-06 | 5.041974 | Right atrium |
| RV_ejectio | Z74_S2 | 27446 | -0.02241 | 0.005054 | 9.20E-06 | 5.036148 | Right ventricle |
| Ascending_ | Z69_S5 | 24599 | -0.02288 | 0.005158 | 9.21E-06 | 5.035654 | Aorta |
| Descending | Z3_S4 | 24599 | 0.036347 | 0.008208 | 9.50E-06 | 5.022485 | Aorta |
| LV_myocar | Z22_S4 | 27446 | -0.03844 | 0.008684 | 9.58E-06 | 5.018459 | Left ventricle |
| LV_stroke_ | Z99_S4 | 27446 | 0.029981 | 0.006777 | 9.70E-06 | 5.013072 | Left ventricle |
| LA_stroke_ | Z7_S5 | 27446 | 0.025425 | 0.005751 | 9.81E-06 | 5.008208 | Left atrium |
| LV_stroke_ | Z73_S1 | 27446 | -0.02514 | 0.005686 | 9.84E-06 | 5.006961 | Left ventricle |
| RV_ejectio | Z51_S3 | 27446 | 0.027542 | 0.006245 | 1.03E-05 | 4.986012 | Right ventricle |
| LV_circum | Z26_S3 | 27446 | 0.025229 | 0.005731 | 1.07E-05 | 4.970286 | Left ventricle |
| RV_end_di | Z112_S3 | 27446 | -0.03738 | 0.008527 | 1.17E-05 | 4.93165 | Right ventricle |
| LV_mean_ | Z41_S1 | 27446 | -0.03301 | 0.007542 | 1.20E-05 | 4.920383 | Left ventricle |
| RA_ejectio | Z7_S3 | 27446 | -0.02597 | 0.005936 | 1.21E-05 | 4.915955 | Right atrium |
| Ascending_ | Z76_S3 | 24599 | -0.02643 | 0.006045 | 1.23E-05 | 4.910786 | Aorta |
| LA_ejectio | Z17_S1 | 27446 | -0.01988 | 0.004548 | 1.24E-05 | 4.907991 | Left atrium |
| RV_ejectio | Z29_S1 | 27446 | -0.02421 | 0.005539 | 1.24E-05 | 4.906666 | Right ventricle |
| LV_mean_ | Z55_S1 | 27446 | -0.02825 | 0.006474 | 1.28E-05 | 4.892269 | Left ventricle |
| Ascending_ | Z114_S4 | 24599 | -0.02574 | 0.005903 | 1.30E-05 | 4.886993 | Aorta |
| LV_mean_ | Z26_S3 | 27446 | -0.03414 | 0.007858 | 1.39E-05 | 4.855937 | Left ventricle |
| LA_minimu | Z121_S4 | 27446 | 0.024861 | 0.005726 | 1.41E-05 | 4.850321 | Left atrium |
| RA_maxim | Z73_S1 | 27446 | 0.022591 | 0.005207 | 1.43E-05 | 4.843311 | Right atrium |
| LV_cardiac | Z4_S4 | 27446 | 0.021446 | 0.004943 | 1.44E-05 | 4.84302 | Left ventricle |
| LV_circum | Z70_S5 | 27446 | 0.021109 | 0.004869 | 1.45E-05 | 4.837778 | Left ventricle |
| LV_mean_ | Z95_S3 | 27446 | -0.02331 | 0.005376 | 1.46E-05 | 4.837111 | Left ventricle |
| LV_end_di | Z53_S2 | 27446 | -0.03275 | 0.007559 | 1.48E-05 | 4.83118 | Left ventricle |

| | | | | | | | |
|---|---|---|---|---|---|---|---|
| Descending | Z99_S5 | 24599 | 0.024784 | 0.005722 | 1.48E-05 | 4.829 | Aorta |
| LV_cardiac | Z75_S4 | 27446 | 0.028475 | 0.006581 | 1.51E-05 | 4.819854 | Left ventricle |
| LV_cardiac | Z3_S2 | 27446 | 0.021425 | 0.004957 | 1.55E-05 | 4.8106 | Left ventricle |
| LV_circumf | Z127_S5 | 27446 | -0.01871 | 0.004335 | 1.59E-05 | 4.798324 | Left ventricle |
| Ascending_ | Z20_S1 | 24599 | -0.03042 | 0.007057 | 1.62E-05 | 4.789951 | Aorta |
| Descending | Z98_S3 | 24599 | -0.02242 | 0.005201 | 1.63E-05 | 4.788568 | Aorta |
| RV_end_sy | Z3_S4 | 27446 | 0.03681 | 0.008545 | 1.65E-05 | 4.783303 | Right ventricle |
| Descending | Z49_S3 | 24599 | 0.029765 | 0.006911 | 1.66E-05 | 4.781045 | Aorta |
| Ascending_ | Z117_S4 | 24599 | -0.02129 | 0.004945 | 1.67E-05 | 4.777342 | Aorta |
| Ascending_ | Z65_S5 | 24599 | 0.029457 | 0.006856 | 1.74E-05 | 4.76053 | Aorta |
| Descending | Z111_S1 | 24599 | 0.030982 | 0.007211 | 1.74E-05 | 4.760498 | Aorta |
| Ascending_ | Z44_S4 | 24599 | -0.02209 | 0.005143 | 1.75E-05 | 4.757323 | Aorta |
| LV_longitu | Z114_S4 | 27446 | -0.02072 | 0.004838 | 1.84E-05 | 4.734012 | Left ventricle |
| Ascending_ | Z24_S1 | 24599 | 0.022019 | 0.005144 | 1.86E-05 | 4.729861 | Aorta |
| RA_ejectio | Z32_S1 | 27446 | -0.02264 | 0.00529 | 1.87E-05 | 4.727264 | Right atrium |
| LV_ejection | Z5_S5 | 27446 | 0.022057 | 0.005162 | 1.93E-05 | 4.714935 | Left ventricle |
| Descending | Z106_S3 | 24599 | 0.030027 | 0.007034 | 1.96E-05 | 4.706811 | Aorta |
| LA_stroke_ | Z95_S5 | 27446 | -0.02705 | 0.00634 | 1.98E-05 | 4.703082 | Left atrium |
| RA_maxim | Z121_S4 | 27446 | -0.02667 | 0.006267 | 2.08E-05 | 4.682393 | Right atrium |
| Ascending_ | Z28_S4 | 24599 | -0.02799 | 0.006585 | 2.14E-05 | 4.670201 | Aorta |
| Descending | Z28_S2 | 24599 | -0.02998 | 0.007058 | 2.16E-05 | 4.664567 | Aorta |
| Ascending_ | Z8_S3 | 24599 | -0.02189 | 0.005158 | 2.20E-05 | 4.658286 | Aorta |
| RA_minimu | Z52_S3 | 27446 | 0.020353 | 0.004806 | 2.29E-05 | 4.640839 | Right atrium |
| RA_maxim | Z1_S5 | 27446 | 0.023969 | 0.005671 | 2.37E-05 | 4.625457 | Right atrium |
| LV_mean_ | Z39_S1 | 27446 | -0.02723 | 0.006448 | 2.41E-05 | 4.617543 | Left ventricle |
| LV_mean_ | Z1_S5 | 27446 | -0.02996 | 0.007124 | 2.60E-05 | 4.584461 | Left ventricle |
| LV_mean_ | Z106_S4 | 27446 | 0.025778 | 0.006141 | 2.70E-05 | 4.568758 | Left ventricle |
| LV_circumf | Z86_S3 | 27446 | 0.024161 | 0.00576 | 2.74E-05 | 4.562642 | Left ventricle |
| LV_mean_ | Z18_S4 | 27446 | -0.03655 | 0.008716 | 2.75E-05 | 4.561269 | Left ventricle |
| LV_cardiac | Z40_S4 | 27446 | -0.02813 | 0.006711 | 2.76E-05 | 4.558801 | Left ventricle |
| Ascending_ | Z31_S2 | 24599 | -0.02556 | 0.0061 | 2.78E-05 | 4.555462 | Aorta |
| RV_ejectio | Z85_S5 | 27446 | 0.020045 | 0.004789 | 2.85E-05 | 4.545214 | Right ventricle |
| LA_ejection | Z49_S3 | 27446 | -0.02358 | 0.005642 | 2.92E-05 | 4.535305 | Left atrium |
| LV_mean_ | Z78_S1 | 27446 | 0.026591 | 0.006366 | 2.95E-05 | 4.530339 | Left ventricle |
| LV_mean_ | Z66_S3 | 27446 | -0.03306 | 0.007917 | 2.96E-05 | 4.52805 | Left ventricle |
| Descending | Z120_S5 | 24599 | 0.033238 | 0.007959 | 2.97E-05 | 4.527475 | Aorta |
| LV_longitu | Z7_S5 | 27446 | -0.02267 | 0.005435 | 3.04E-05 | 4.517458 | Left ventricle |
| RV_end_di | Z62_S3 | 27446 | 0.031401 | 0.00753 | 3.04E-05 | 4.516775 | Right ventricle |
| Ascending_ | Z4_S4 | 24599 | -0.02244 | 0.005395 | 3.19E-05 | 4.496307 | Aorta |
| LV_myocar | Z78_S1 | 27446 | 0.031743 | 0.007645 | 3.29E-05 | 4.482521 | Left ventricle |
| RA_maxim | Z52_S3 | 27446 | 0.019673 | 0.004742 | 3.34E-05 | 4.475662 | Right atrium |
| LA_minimu | Z49_S1 | 27446 | -0.0257 | 0.0062 | 3.41E-05 | 4.467594 | Left atrium |
| Ascending_ | Z22_S4 | 24599 | 0.024795 | 0.005988 | 3.46E-05 | 4.460699 | Aorta |
| LV_longitu | Z100_S3 | 27446 | -0.02112 | 0.005107 | 3.55E-05 | 4.449978 | Left ventricle |
| LV_end_sy | Z22_S4 | 27446 | 0.026979 | 0.006531 | 3.62E-05 | 4.441648 | Left ventricle |

| | | | | | | | |
|---|---|---|---|---|---|---|---|
| LV_circumf | Z41_S2 | 27446 | 0.024834 | 0.006019 | 3.69E-05 | 4.433084 | Left ventricle |
| LA_ejectio | Z51_S1 | 27446 | 0.01762 | 0.004271 | 3.70E-05 | 4.431811 | Left atrium |
| Descending | Z3_S2 | 24599 | -0.02463 | 0.005973 | 3.72E-05 | 4.429407 | Aorta |
| LV_stroke_ | Z122_S2 | 27446 | -0.02044 | 0.004959 | 3.78E-05 | 4.42255 | Left ventricle |
| RA_ejectio | Z6_S2 | 27446 | 0.01595 | 0.003876 | 3.86E-05 | 4.413117 | Right atrium |
| LV_mean_ | Z122_S2 | 27446 | -0.02348 | 0.005706 | 3.87E-05 | 4.412302 | Left ventricle |
| Ascending_ | Z122_S5 | 24599 | -0.02245 | 0.005459 | 3.91E-05 | 4.407589 | Aorta |
| LV_circumf | Z62_S1 | 27446 | 0.019581 | 0.004768 | 4.01E-05 | 4.397377 | Left ventricle |
| LV_radial_ | Z87_S3 | 27446 | 0.020611 | 0.00503 | 4.17E-05 | 4.379821 | Left ventricle |
| LV_radial_ | Z125_S4 | 27446 | 0.021787 | 0.005322 | 4.25E-05 | 4.371516 | Left ventricle |
| RA_maxim | Z94_S2 | 27446 | 0.024503 | 0.00599 | 4.30E-05 | 4.366651 | Right atrium |
| LA_ejectio | Z115_S4 | 27446 | -0.01824 | 0.00446 | 4.33E-05 | 4.36344 | Left atrium |
| LV_mean_ | Z120_S4 | 27446 | -0.02827 | 0.006918 | 4.37E-05 | 4.359031 | Left ventricle |
| LA_ejectio | Z81_S1 | 27446 | 0.020131 | 0.004933 | 4.48E-05 | 4.348562 | Left atrium |
| LV_longitu | Z20_S1 | 27446 | 0.02352 | 0.005785 | 4.79E-05 | 4.319746 | Left ventricle |
| RV_end_sy | Z44_S1 | 27446 | 0.027712 | 0.006837 | 5.05E-05 | 4.29645 | Right ventricle |
| RA_minimu | Z113_S2 | 27446 | -0.02748 | 0.00678 | 5.07E-05 | 4.295363 | Right atrium |
| RA_maxim | Z7_S3 | 27446 | -0.02681 | 0.006621 | 5.12E-05 | 4.290618 | Right atrium |
| LV_cardiac | Z92_S1 | 27446 | -0.02236 | 0.005528 | 5.23E-05 | 4.281174 | Left ventricle |
| RA_minimu | Z58_S2 | 27446 | 0.021351 | 0.00528 | 5.27E-05 | 4.278416 | Right atrium |
| RA_minimu | Z120_S2 | 27446 | 0.019534 | 0.004834 | 5.33E-05 | 4.273624 | Right atrium |
| LA_stroke_ | Z94_S2 | 27446 | -0.0229 | 0.005672 | 5.41E-05 | 4.266707 | Left atrium |
| LV_myocar | Z62_S3 | 27446 | 0.035419 | 0.00878 | 5.48E-05 | 4.261509 | Left ventricle |
| Ascending_ | Z14_S2 | 24599 | -0.02326 | 0.005773 | 5.58E-05 | 4.253694 | Aorta |
| RA_ejectio | Z38_S4 | 27446 | 0.023024 | 0.005715 | 5.62E-05 | 4.250414 | Right atrium |
| LV_end_sy | Z28_S2 | 27446 | 0.027038 | 0.006717 | 5.69E-05 | 4.244597 | Left ventricle |
| LA_ejectio | Z122_S2 | 27446 | 0.015963 | 0.003977 | 5.98E-05 | 4.223655 | Left atrium |
| RV_end_sy | Z28_S1 | 27446 | -0.02905 | 0.007243 | 6.04E-05 | 4.218719 | Right ventricle |
| LV_myocar | Z3_S4 | 27446 | 0.041907 | 0.010449 | 6.05E-05 | 4.218113 | Left ventricle |
| RA_maxim | Z62_S3 | 27446 | -0.02333 | 0.005819 | 6.08E-05 | 4.215941 | Right atrium |
| LA_minimu | Z97_S2 | 27446 | 0.021461 | 0.005365 | 6.32E-05 | 4.199389 | Left atrium |
| LV_cardiac | Z81_S1 | 27446 | 0.022539 | 0.005637 | 6.38E-05 | 4.194926 | Left ventricle |
| LV_circumf | Z93_S1 | 27446 | 0.023255 | 0.005833 | 6.70E-05 | 4.173838 | Left ventricle |
| RA_stroke_ | Z26_S3 | 27446 | 0.0227 | 0.005699 | 6.79E-05 | 4.168049 | Right atrium |
| LA_stroke_ | Z39_S4 | 27446 | 0.018765 | 0.004711 | 6.81E-05 | 4.167126 | Left atrium |
| LA_minimu | Z59_S2 | 27446 | 0.02308 | 0.005797 | 6.84E-05 | 4.164852 | Left atrium |
| LV_cardiac | Z11_S5 | 27446 | -0.02128 | 0.005349 | 6.92E-05 | 4.159887 | Left ventricle |
| Descending | Z22_S4 | 24599 | 0.027577 | 0.006933 | 6.96E-05 | 4.157332 | Aorta |
| LA_minimu | Z39_S4 | 27446 | 0.018078 | 0.004545 | 6.97E-05 | 4.156462 | Left atrium |
| RV_ejectio | Z14_S2 | 27446 | -0.02028 | 0.005099 | 6.99E-05 | 4.155393 | Right ventricle |
| LV_cardiac | Z29_S5 | 27446 | 0.023604 | 0.005943 | 7.15E-05 | 4.145959 | Left ventricle |
| LV_ejectio | Z87_S3 | 27446 | 0.019368 | 0.004883 | 7.30E-05 | 4.136936 | Left ventricle |
| LV_ejectio | Z66_S3 | 27446 | 0.022533 | 0.005683 | 7.33E-05 | 4.134706 | Left ventricle |
| RA_ejectio | Z5_S1 | 27446 | 0.019477 | 0.004913 | 7.37E-05 | 4.132791 | Right atrium |
| Ascending_ | Z74_S4 | 24599 | -0.02395 | 0.006043 | 7.40E-05 | 4.13054 | Aorta |

| | | | | | | | |
|---|---|---|---|---|---|---|---|
| RA_ejectio | Z76_S3 | 27446 | 0.020787 | 0.005247 | 7.44E-05 | 4.128569 | Right atrium |
| LA_minimu | Z3_S2 | 27446 | -0.01791 | 0.004526 | 7.56E-05 | 4.121439 | Left atrium |
| RA_maxim | Z80_S2 | 27446 | -0.01958 | 0.004949 | 7.61E-05 | 4.11836 | Right atrium |
| RA_ejectio | Z2_S1 | 27446 | 0.017973 | 0.004544 | 7.66E-05 | 4.116051 | Right atrium |
| RA_ejectio | Z95_S3 | 27446 | 0.015165 | 0.003837 | 7.76E-05 | 4.110134 | Right atrium |
| Descending | Z27_S2 | 24599 | -0.02751 | 0.006965 | 7.81E-05 | 4.107391 | Aorta |
| Ascending_ | Z106_S3 | 24599 | 0.028262 | 0.007165 | 7.99E-05 | 4.097188 | Aorta |
| RV_ejectio | Z30_S2 | 27446 | 0.024419 | 0.006203 | 8.25E-05 | 4.08357 | Right ventricle |
| LV_cardiac | Z5_S1 | 27446 | 0.021566 | 0.005484 | 8.39E-05 | 4.075999 | Left ventricle |
| RA_stroke_ | Z69_S5 | 27446 | -0.01692 | 0.004303 | 8.41E-05 | 4.075109 | Right atrium |
| Descending | Z87_S3 | 24599 | -0.02301 | 0.005859 | 8.58E-05 | 4.06651 | Aorta |
| LV_circumf | Z65_S2 | 27446 | -0.02053 | 0.005235 | 8.77E-05 | 4.056935 | Left ventricle |
| LA_maximu | Z12_S5 | 27446 | -0.02438 | 0.006217 | 8.77E-05 | 4.056759 | Left atrium |
| RA_minimu | Z87_S5 | 27446 | -0.02171 | 0.005557 | 9.38E-05 | 4.027782 | Right atrium |
| Ascending_ | Z37_S3 | 24599 | -0.02556 | 0.006555 | 9.62E-05 | 4.016745 | Aorta |
| Ascending_ | Z62_S4 | 24599 | 0.02711 | 0.006952 | 9.65E-05 | 4.015651 | Aorta |
| LA_ejectior | Z1_S4 | 27446 | -0.01997 | 0.005125 | 9.72E-05 | 4.012377 | Left atrium |
| Descending | Z26_S4 | 24599 | 0.0288 | 0.007397 | 9.88E-05 | 4.005315 | Aorta |
| LA_maximu | Z113_S2 | 27446 | 0.024513 | 0.006303 | 0.000101 | 3.997031 | Left atrium |
| LV_mean_ | Z32_S5 | 27446 | -0.03265 | 0.008402 | 0.000102 | 3.991731 | Left ventricle |
| RV_end_sy | Z28_S2 | 27446 | -0.02837 | 0.007304 | 0.000103 | 3.987719 | Right ventricle |
| RV_ejectio | Z37_S2 | 27446 | -0.02154 | 0.005551 | 0.000104 | 3.982822 | Right ventricle |
| Descending | Z112_S3 | 24599 | -0.02748 | 0.007083 | 0.000105 | 3.979009 | Aorta |
| LV_longitu | Z39_S1 | 27446 | -0.01778 | 0.004593 | 0.000109 | 3.964224 | Left ventricle |
| LA_ejectior | Z58_S2 | 27446 | -0.01766 | 0.004562 | 0.000109 | 3.963348 | Left atrium |
| Ascending_ | Z66_S3 | 24599 | -0.02668 | 0.006907 | 0.000112 | 3.950119 | Aorta |
| LA_ejectior | Z57_S2 | 27446 | -0.01558 | 0.00404 | 0.000115 | 3.939737 | Left atrium |
| Ascending_ | Z50_S2 | 24599 | -0.02865 | 0.007435 | 0.000116 | 3.934239 | Aorta |
| LV_stroke_ | Z39_S3 | 27446 | 0.029279 | 0.007598 | 0.000117 | 3.933496 | Left ventricle |
| Descending | Z76_S3 | 24599 | -0.02415 | 0.006269 | 0.000117 | 3.931213 | Aorta |
| RA_ejectio | Z72_S5 | 27446 | -0.02361 | 0.006138 | 0.00012 | 3.922462 | Right atrium |
| RA_ejectio | Z93_S2 | 27446 | -0.0235 | 0.006114 | 0.000121 | 3.917002 | Right atrium |
| LA_maximu | Z93_S1 | 27446 | -0.02306 | 0.005999 | 0.000121 | 3.916496 | Left atrium |
| LV_ejectior | Z81_S1 | 27446 | 0.019497 | 0.00508 | 0.000124 | 3.906674 | Left ventricle |
| LV_ejectior | Z23_S1 | 27446 | -0.02159 | 0.005626 | 0.000125 | 3.904773 | Left ventricle |
| LV_mean_ | Z66_S2 | 27446 | 0.030667 | 0.007998 | 0.000126 | 3.899661 | Left ventricle |
| LV_longitu | Z44_S1 | 27446 | -0.01901 | 0.004959 | 0.000126 | 3.899303 | Left ventricle |
| Ascending_ | Z49_S1 | 24599 | 0.026938 | 0.007027 | 0.000126 | 3.898621 | Aorta |
| RV_end_di | Z80_S5 | 27446 | 0.032508 | 0.008485 | 0.000128 | 3.894201 | Right ventricle |
| LV_radial_ | Z47_S4 | 27446 | 0.019724 | 0.005155 | 0.00013 | 3.885708 | Left ventricle |
| Ascending_ | Z28_S1 | 24599 | -0.02326 | 0.006084 | 0.000131 | 3.881094 | Aorta |
| LV_longitu | Z59_S2 | 27446 | 0.021674 | 0.005678 | 0.000135 | 3.869494 | Left ventricle |
| LV_radial_ | Z5_S5 | 27446 | -0.02028 | 0.005318 | 0.000137 | 3.863498 | Left ventricle |
| Ascending_ | Z40_S4 | 24599 | -0.02792 | 0.007322 | 0.000137 | 3.862093 | Aorta |
| LV_mean_ | Z74_S4 | 27446 | -0.02788 | 0.007326 | 0.000141 | 3.849476 | Left ventricle |

| | | | | | | | |
|---|---|---|---|---|---|---|---|
| LA_minimu | Z56_S5 | 27446 | 0.016451 | 0.00433 | 0.000145 | 3.839222 | Left atrium |
| Descending | Z122_S2 | 24599 | 0.018476 | 0.004866 | 0.000147 | 3.833986 | Aorta |
| Descending | Z10_S4 | 24599 | -0.02603 | 0.006876 | 0.000153 | 3.813937 | Aorta |
| Ascending_ | Z27_S2 | 24599 | -0.02541 | 0.006717 | 0.000155 | 3.809425 | Aorta |
| LV_cardiac | Z37_S3 | 27446 | -0.02393 | 0.006329 | 0.000156 | 3.806054 | Left ventricle |
| Descending | Z72_S5 | 24599 | -0.03063 | 0.008104 | 0.000157 | 3.803665 | Aorta |
| LV_longitu | Z1_S4 | 27446 | 0.019781 | 0.005238 | 0.000159 | 3.798689 | Left ventricle |
| LV_ejectio | Z121_S4 | 27446 | -0.02134 | 0.005652 | 0.00016 | 3.796558 | Left ventricle |
| RV_ejectio | Z69_S5 | 27446 | 0.016366 | 0.004348 | 0.000167 | 3.777601 | Right ventricle |
| LV_circumf | Z50_S2 | 27446 | 0.023424 | 0.006228 | 0.000169 | 3.771152 | Left ventricle |
| RV_end_sy | Z37_S3 | 27446 | -0.02924 | 0.007803 | 0.000178 | 3.748412 | Right ventricle |
| LV_cardiac | Z68_S2 | 27446 | -0.01895 | 0.00506 | 0.00018 | 3.745286 | Left ventricle |
| LV_mean_ | Z22_S4 | 27446 | -0.02708 | 0.007232 | 0.00018 | 3.743988 | Left ventricle |
| RA_ejectio | Z1_S4 | 27446 | 0.019646 | 0.005248 | 0.000181 | 3.741366 | Right atrium |
| Descending | Z40_S4 | 24599 | 0.030148 | 0.008056 | 0.000182 | 3.739318 | Aorta |
| LA_ejectio | Z8_S3 | 27446 | -0.01545 | 0.004135 | 0.000186 | 3.730729 | Left atrium |
| LV_circumf | Z79_S4 | 27446 | 0.022394 | 0.006 | 0.00019 | 3.722321 | Left ventricle |
| Descending | Z94_S2 | 24599 | 0.026532 | 0.007132 | 0.000199 | 3.701174 | Aorta |
| RA_maxim | Z17_S1 | 27446 | 0.019235 | 0.005194 | 0.000213 | 3.671983 | Right atrium |
| LV_longitu | Z79_S4 | 27446 | 0.02169 | 0.00586 | 0.000214 | 3.668873 | Left ventricle |
| Ascending_ | Z21_S3 | 24599 | 0.026745 | 0.007227 | 0.000215 | 3.667842 | Aorta |
| Descending | Z66_S3 | 24599 | -0.02503 | 0.006781 | 0.000224 | 3.6505 | Aorta |
| LV_cardiac | Z28_S2 | 27446 | -0.02184 | 0.005924 | 0.000228 | 3.642492 | Left ventricle |
| Ascending_ | Z112_S3 | 24599 | -0.02518 | 0.006831 | 0.000228 | 3.641798 | Aorta |
| Ascending_ | Z65_S5 | 24599 | -0.02668 | 0.007242 | 0.000229 | 3.639655 | Aorta |
| Ascending_ | Z91_S1 | 24599 | 0.021665 | 0.005881 | 0.00023 | 3.638859 | Aorta |
| LV_mean_ | Z113_S2 | 27446 | 0.030931 | 0.008405 | 0.000233 | 3.632425 | Left ventricle |
| LA_minimu | Z7_S5 | 27446 | 0.020406 | 0.005549 | 0.000236 | 3.627725 | Left atrium |
| Descending | Z38_S4 | 24599 | 0.025237 | 0.006866 | 0.000237 | 3.624924 | Aorta |
| LV_circumf | Z4_S4 | 27446 | -0.01659 | 0.004526 | 0.000247 | 3.607954 | Left ventricle |
| Descending | Z65_S2 | 24599 | 0.02525 | 0.006892 | 0.000248 | 3.604906 | Aorta |
| RV_ejectio | Z49_S1 | 27446 | -0.02287 | 0.006247 | 0.000251 | 3.600883 | Right ventricle |
| LV_longitu | Z13_S1 | 27446 | -0.01629 | 0.004452 | 0.000252 | 3.598074 | Left ventricle |
| RA_stroke_ | Z80_S2 | 27446 | -0.01649 | 0.004509 | 0.000255 | 3.593406 | Right atrium |
| LA_ejectio | Z11_S5 | 27446 | 0.017066 | 0.00468 | 0.000266 | 3.575136 | Left atrium |
| RV_stroke_ | Z32_S5 | 27446 | 0.02698 | 0.00741 | 0.000272 | 3.565713 | Right ventricle |
| LV_stroke_ | Z22_S4 | 27446 | -0.02288 | 0.006286 | 0.000272 | 3.56473 | Left ventricle |
| RA_ejectio | Z53_S2 | 27446 | -0.01992 | 0.005514 | 0.000303 | 3.518125 | Right atrium |
| Ascending_ | Z76_S3 | 24599 | -0.02191 | 0.006068 | 0.000305 | 3.516007 | Aorta |
| LV_longitu | Z51_S3 | 27446 | 0.021884 | 0.006073 | 0.000314 | 3.503271 | Left ventricle |
| LV_mean_ | Z73_S4 | 27446 | -0.0289 | 0.00802 | 0.000314 | 3.502605 | Left ventricle |
| Ascending_ | Z56_S2 | 24599 | -0.01791 | 0.004973 | 0.000317 | 3.499516 | Aorta |
| LV_end_di | Z39_S4 | 27446 | -0.02202 | 0.006117 | 0.000318 | 3.497444 | Left ventricle |
| Ascending_ | Z127_S4 | 24599 | -0.02285 | 0.006354 | 0.000323 | 3.490658 | Aorta |
| Ascending_ | Z68_S2 | 24599 | -0.01982 | 0.005512 | 0.000324 | 3.488788 | Aorta |

| | | | | | | | |
|---|---|---|---|---|---|---|---|
| LA_stroke_ | Z7_S3 | 27446 | -0.02253 | 0.00627 | 0.000327 | 3.485868 | Left atrium |
| LV_mean_ | Z85_S5 | 27446 | 0.023432 | 0.006538 | 0.000338 | 3.470475 | Left ventricle |
| Ascending_ | Z1_S5 | 24599 | -0.02103 | 0.005907 | 0.00037 | 3.431241 | Aorta |
| Descending | Z75_S5 | 24599 | 0.029876 | 0.008396 | 0.000373 | 3.428359 | Aorta |
| LV_myocar | Z44_S3 | 27446 | -0.03293 | 0.009256 | 0.000374 | 3.426937 | Left ventricle |
| RA_stroke_ | Z80_S5 | 27446 | 0.02116 | 0.005974 | 0.000397 | 3.400884 | Right atrium |
| LV_mean_ | Z111_S1 | 27446 | 0.026931 | 0.007613 | 0.000404 | 3.3935 | Left ventricle |
| Descending | Z61_S4 | 24599 | -0.01991 | 0.00564 | 0.000417 | 3.380195 | Aorta |
| LA_ejectio | Z20_S1 | 27446 | -0.01993 | 0.005661 | 0.00043 | 3.366731 | Left atrium |
| Descending | Z62_S2 | 24599 | 0.018546 | 0.005268 | 0.000431 | 3.365476 | Aorta |
| LV_radial_ | Z27_S2 | 27446 | 0.021122 | 0.006008 | 0.000439 | 3.357372 | Left ventricle |
| Ascending_ | Z123_S4 | 24599 | 0.021278 | 0.006054 | 0.000441 | 3.355876 | Aorta |
| LV_circumf | Z81_S1 | 27446 | -0.01814 | 0.005162 | 0.000442 | 3.354815 | Left ventricle |
| LV_ejectio | Z78_S4 | 27446 | -0.01559 | 0.004442 | 0.000447 | 3.349782 | Left ventricle |
| Ascending_ | Z76_S3 | 24599 | -0.02241 | 0.006385 | 0.000449 | 3.347662 | Aorta |
| RV_stroke_ | Z65_S5 | 27446 | 0.025707 | 0.007333 | 0.000456 | 3.341196 | Right ventricle |
| LV_longitu | Z75_S4 | 27446 | -0.02063 | 0.005886 | 0.000457 | 3.340389 | Left ventricle |
| RA_ejectio | Z56_S5 | 27446 | 0.014891 | 0.004249 | 0.000458 | 3.33932 | Right atrium |
| LV_mean_ | Z49_S1 | 27446 | -0.02983 | 0.008527 | 0.000469 | 3.329226 | Left ventricle |
| RA_maxim | Z56_S2 | 27446 | -0.01669 | 0.004772 | 0.000471 | 3.326972 | Right atrium |
| LV_mean_ | Z75_S1 | 27446 | -0.02514 | 0.007192 | 0.000472 | 3.325849 | Left ventricle |
| LV_mean_ | Z59_S3 | 27446 | -0.01976 | 0.005659 | 0.000479 | 3.3201 | Left ventricle |
| LV_circumf | Z65_S5 | 27446 | -0.02118 | 0.006064 | 0.000479 | 3.319626 | Left ventricle |
| LV_cardiac | Z80_S4 | 27446 | -0.01672 | 0.004797 | 0.000491 | 3.30911 | Left ventricle |
| LV_longitu | Z3_S2 | 27446 | 0.015451 | 0.004434 | 0.000492 | 3.307956 | Left ventricle |
| LV_ejectio | Z93_S2 | 27446 | -0.02143 | 0.006149 | 0.000492 | 3.307771 | Left ventricle |
| RA_ejectio | Z114_S5 | 27446 | 0.013836 | 0.003978 | 0.000504 | 3.297385 | Right atrium |
| LV_ejectio | Z9_S2 | 27446 | 0.018593 | 0.005356 | 0.000517 | 3.2862 | Left ventricle |
| RA_ejectio | Z122_S2 | 27446 | 0.014115 | 0.004073 | 0.000529 | 3.276761 | Right atrium |
| RA_ejectio | Z23_S1 | 27446 | -0.01938 | 0.005595 | 0.000531 | 3.274951 | Right atrium |
| LV_myocar | Z112_S1 | 27446 | -0.03615 | 0.010438 | 0.000533 | 3.273194 | Left ventricle |
| RA_stroke_ | Z57_S1 | 27446 | -0.01616 | 0.004671 | 0.000539 | 3.2683 | Right atrium |
| RV_ejectio | Z41_S2 | 27446 | -0.0209 | 0.006046 | 0.000549 | 3.260739 | Right ventricle |
| Descending | Z96_S3 | 24599 | 0.020023 | 0.005794 | 0.000549 | 3.260274 | Aorta |
| Descending | Z62_S4 | 24599 | -0.02641 | 0.007649 | 0.000556 | 3.255089 | Aorta |
| Descending | Z91_S1 | 24599 | 0.019923 | 0.005774 | 0.000559 | 3.252278 | Aorta |
| RV_ejectio | Z98_S3 | 27446 | 0.015433 | 0.004473 | 0.00056 | 3.251612 | Right ventricle |
| Descending | Z97_S2 | 24599 | -0.02411 | 0.006993 | 0.000565 | 3.247732 | Aorta |
| Descending | Z95_S5 | 24599 | 0.024756 | 0.007186 | 0.000571 | 3.243728 | Aorta |
| LV_circumf | Z113_S2 | 27446 | 0.021095 | 0.00613 | 0.000579 | 3.237387 | Left ventricle |
| RV_end_sy | Z62_S3 | 27446 | 0.024709 | 0.00718 | 0.000579 | 3.237101 | Right ventricle |
| Ascending_ | Z92_S5 | 24599 | -0.02401 | 0.006977 | 0.00058 | 3.236866 | Aorta |
| Ascending_ | Z58_S2 | 24599 | -0.01952 | 0.005684 | 0.000595 | 3.225605 | Aorta |
| LV_longitu | Z22_S2 | 27446 | -0.01731 | 0.005047 | 0.000605 | 3.218586 | Left ventricle |
| RA_minimu | Z53_S2 | 27446 | 0.021375 | 0.006233 | 0.000605 | 3.218554 | Right atrium |

| | | | | | | | |
|---|---|---|---|---|---|---|---|
| LV_circumf | Z13_S3 | 27446 | 0.014228 | 0.004152 | 0.00061 | 3.214685 | Left ventricle |
| LV_end_di | Z80_S5 | 27446 | -0.02757 | 0.008062 | 0.000626 | 3.203208 | Left ventricle |
| LV_mean_ | Z34_S2 | 27446 | -0.02733 | 0.008002 | 0.000638 | 3.195474 | Left ventricle |
| LA_ejectio | Z23_S3 | 27446 | -0.01475 | 0.004318 | 0.000638 | 3.195302 | Left atrium |
| RA_ejectio | Z20_S1 | 27446 | -0.01978 | 0.005797 | 0.000644 | 3.191341 | Right atrium |
| LV_end_sy | Z39_S4 | 27446 | 0.01922 | 0.005646 | 0.000664 | 3.178109 | Left ventricle |
| LV_mean_ | Z77_S3 | 27446 | -0.02419 | 0.007117 | 0.000676 | 3.169971 | Left ventricle |
| RA_minimu | Z28_S1 | 27446 | 0.020216 | 0.00595 | 0.00068 | 3.167551 | Right atrium |
| Ascending_ | Z93_S1 | 24599 | -0.02369 | 0.006975 | 0.000683 | 3.165589 | Aorta |
| RV_ejectio | Z81_S1 | 27446 | 0.017559 | 0.005185 | 0.000708 | 3.149707 | Right ventricle |
| LV_end_di | Z38_S4 | 27446 | -0.02652 | 0.007838 | 0.000716 | 3.144936 | Left ventricle |
| RA_minimu | Z115_S4 | 27446 | 0.017459 | 0.005163 | 0.00072 | 3.142605 | Right atrium |
| LV_mean_ | Z37_S3 | 27446 | -0.02683 | 0.007946 | 0.000733 | 3.13506 | Left ventricle |
| LV_mean_ | Z40_S5 | 27446 | -0.02014 | 0.005974 | 0.00075 | 3.124698 | Left ventricle |
| Ascending_ | Z121_S4 | 24599 | 0.021825 | 0.006478 | 0.000754 | 3.122478 | Aorta |
| RA_stroke_ | Z15_S2 | 27446 | 0.018189 | 0.005402 | 0.00076 | 3.11918 | Right atrium |
| LA_maximu | Z3_S2 | 27446 | -0.01564 | 0.004668 | 0.000806 | 3.093517 | Left atrium |
| LV_radial_ | Z111_S5 | 27446 | -0.01712 | 0.005118 | 0.000821 | 3.085673 | Left ventricle |
| Ascending_ | Z121_S4 | 24599 | 0.021753 | 0.006502 | 0.000821 | 3.085649 | Aorta |
| LV_ejectio | Z94_S2 | 27446 | 0.018036 | 0.005402 | 0.000841 | 3.075057 | Left ventricle |
| RV_stroke_ | Z62_S3 | 27446 | 0.021515 | 0.006448 | 0.000848 | 3.071356 | Right ventricle |
| Ascending_ | Z29_S5 | 24599 | 0.021648 | 0.006492 | 0.000854 | 3.068572 | Aorta |
| Ascending_ | Z52_S4 | 24599 | 0.018032 | 0.005423 | 0.000884 | 3.053615 | Aorta |
| Ascending_ | Z37_S1 | 24599 | -0.02274 | 0.00684 | 0.000885 | 3.053101 | Aorta |
| Descending | Z80_S5 | 24599 | 0.025801 | 0.007769 | 0.000897 | 3.047218 | Aorta |
| LV_stroke_ | Z62_S4 | 27446 | -0.02302 | 0.006952 | 0.000929 | 3.032058 | Left ventricle |
| LV_mean_ | Z51_S4 | 27446 | -0.02003 | 0.00605 | 0.000929 | 3.03202 | Left ventricle |
| LV_ejectio | Z111_S1 | 27446 | 0.018094 | 0.005465 | 0.000929 | 3.031765 | Left ventricle |
| Descending | Z80_S4 | 24599 | -0.01702 | 0.005143 | 0.000936 | 3.028581 | Aorta |
| Ascending_ | Z38_S5 | 24599 | 0.020225 | 0.006123 | 0.000956 | 3.019408 | Aorta |
| LV_mean_ | Z105_S5 | 27446 | -0.02333 | 0.007065 | 0.000957 | 3.018892 | Left ventricle |
| RV_end_di | Z122_S2 | 27446 | -0.01941 | 0.005878 | 0.000959 | 3.018335 | Right ventricle |
| Ascending_ | Z37_S3 | 24599 | -0.0216 | 0.006579 | 0.00103 | 2.987257 | Aorta |
| Ascending_ | Z56_S2 | 24599 | -0.01717 | 0.005234 | 0.001038 | 2.983791 | Aorta |
| Descending | Z10_S4 | 24599 | 0.022864 | 0.006976 | 0.001047 | 2.979856 | Aorta |
| LA_ejectio | Z10_S4 | 27446 | 0.016631 | 0.005076 | 0.001052 | 2.978101 | Left atrium |
| RV_end_sy | Z1_S4 | 27446 | -0.02363 | 0.007222 | 0.001067 | 2.971839 | Right ventricle |
| RV_end_di | Z37_S3 | 27446 | -0.02676 | 0.008184 | 0.001076 | 2.968094 | Right ventricle |
| LA_ejectio | Z120_S4 | 27446 | 0.015698 | 0.004822 | 0.001133 | 2.945794 | Left atrium |
| LV_cardiac | Z15_S2 | 27446 | 0.01928 | 0.005934 | 0.001158 | 2.936355 | Left ventricle |
| Ascending_ | Z32_S5 | 24599 | 0.023707 | 0.007311 | 0.001185 | 2.926244 | Aorta |
| Ascending_ | Z82_S1 | 24599 | 0.023136 | 0.007142 | 0.001199 | 2.921311 | Aorta |
| Ascending_ | Z81_S1 | 24599 | -0.01993 | 0.006158 | 0.001212 | 2.916506 | Aorta |
| Ascending_ | Z35_S5 | 24599 | -0.01683 | 0.005204 | 0.001221 | 2.913378 | Aorta |
| LV_mean_ | Z50_S5 | 27446 | -0.02803 | 0.008669 | 0.001222 | 2.912797 | Left ventricle |

| | | | | | | | |
|---|---|---|---|---|---|---|---|
| RA_stroke_ | Z93_S2 | 27446 | 0.020076 | 0.006212 | 0.001231 | 2.909908 | Right atrium |
| LA_ejectio | Z106_S3 | 27446 | 0.018545 | 0.00574 | 0.001234 | 2.908527 | Left atrium |
| Ascending_ | Z97_S1 | 24599 | -0.02395 | 0.007423 | 0.001254 | 2.901588 | Aorta |
| LV_cardiac_ | Z115_S3 | 27446 | -0.01907 | 0.005917 | 0.001269 | 2.896685 | Left ventricle |
| Ascending_ | Z70_S5 | 24599 | 0.018714 | 0.005808 | 0.001272 | 2.895351 | Aorta |
| Descending | Z3_S2 | 24599 | -0.01894 | 0.005889 | 0.001296 | 2.887366 | Aorta |
| RV_ejectio | Z50_S3 | 27446 | 0.015392 | 0.004792 | 0.001317 | 2.880426 | Right ventricle |
| Descending | Z1_S4 | 24599 | -0.02017 | 0.0063 | 0.001363 | 2.865433 | Aorta |
| RA_maxim | Z65_S1 | 27446 | 0.014762 | 0.00461 | 0.001364 | 2.865161 | Right atrium |
| Ascending_ | Z55_S5 | 24599 | -0.01656 | 0.005173 | 0.001372 | 2.862675 | Aorta |
| LV_mean_ | Z60_S2 | 27446 | -0.01946 | 0.006086 | 0.001382 | 2.859428 | Left ventricle |
| LA_minimu | Z73_S1 | 27446 | -0.01519 | 0.004758 | 0.001416 | 2.849076 | Left atrium |
| RA_maxim | Z92_S1 | 27446 | -0.01762 | 0.005524 | 0.001428 | 2.845327 | Right atrium |
| LV_mean_ | Z28_S4 | 27446 | -0.02398 | 0.00752 | 0.001431 | 2.844485 | Left ventricle |
| Ascending_ | Z120_S4 | 24599 | -0.01918 | 0.006019 | 0.001436 | 2.842752 | Aorta |
| RA_stroke_ | Z20_S1 | 27446 | 0.018758 | 0.00589 | 0.001448 | 2.839095 | Right atrium |
| RV_ejectio | Z28_S1 | 27446 | 0.017185 | 0.005403 | 0.001471 | 2.832344 | Right ventricle |
| LV_mean_ | Z13_S3 | 27446 | 0.018022 | 0.005693 | 0.001546 | 2.810673 | Left ventricle |
| RA_ejectio | Z113_S2 | 27446 | -0.01896 | 0.005999 | 0.001576 | 2.802456 | Right atrium |
| Ascending_ | Z38_S4 | 24599 | 0.02201 | 0.006994 | 0.001649 | 2.782785 | Aorta |
| LV_circum | Z80_S5 | 27446 | 0.01887 | 0.006009 | 0.001687 | 2.773 | Left ventricle |
| RA_ejectio | Z81_S1 | 27446 | 0.015861 | 0.005052 | 0.001691 | 2.771837 | Right atrium |
| RV_stroke_ | Z121_S4 | 27446 | 0.021781 | 0.006945 | 0.001712 | 2.766451 | Right ventricle |
| RA_stroke_ | Z46_S1 | 27446 | 0.016063 | 0.005123 | 0.001715 | 2.765729 | Right atrium |
| Ascending_ | Z19_S2 | 24599 | 0.017824 | 0.005684 | 0.001715 | 2.765636 | Aorta |
| LV_circum | Z76_S3 | 27446 | -0.0168 | 0.005362 | 0.001725 | 2.763285 | Left ventricle |
| RA_stroke_ | Z1_S2 | 27446 | 0.012965 | 0.004145 | 0.001761 | 2.7542 | Right atrium |
| RA_stroke_ | Z70_S5 | 27446 | 0.01513 | 0.004842 | 0.001779 | 2.749839 | Right atrium |
| LV_mean_ | Z62_S2 | 27446 | 0.019318 | 0.006184 | 0.001787 | 2.747976 | Left ventricle |
| RA_stroke_ | Z114_S4 | 27446 | -0.01538 | 0.004926 | 0.001793 | 2.746459 | Right atrium |
| RV_ejectio | Z29_S5 | 27446 | -0.01706 | 0.005467 | 0.0018 | 2.744751 | Right ventricle |
| LV_longitu | Z106_S4 | 27446 | -0.01365 | 0.004375 | 0.001805 | 2.743544 | Left ventricle |
| Descending | Z57_S1 | 24599 | -0.01714 | 0.005497 | 0.00182 | 2.740012 | Aorta |
| LA_ejectio | Z28_S1 | 27446 | -0.01602 | 0.005141 | 0.001831 | 2.737393 | Left atrium |
| LA_ejectio | Z3_S4 | 27446 | 0.018887 | 0.006065 | 0.001844 | 2.734227 | Left atrium |
| RA_stroke_ | Z1_S4 | 27446 | 0.016598 | 0.005333 | 0.001854 | 2.731852 | Right atrium |
| RV_ejectio | Z79_S4 | 27446 | 0.01869 | 0.006027 | 0.001929 | 2.714594 | Right ventricle |
| LV_ejectio | Z79_S4 | 27446 | -0.01826 | 0.005905 | 0.001986 | 2.701985 | Left ventricle |
| Ascending_ | Z100_S3 | 24599 | -0.01924 | 0.006243 | 0.002054 | 2.687419 | Aorta |
| LV_ejectio | Z75_S5 | 27446 | 0.019396 | 0.006294 | 0.00206 | 2.686106 | Left ventricle |
| Ascending_ | Z122_S2 | 24599 | 0.015247 | 0.004957 | 0.002098 | 2.678131 | Aorta |
| LV_ejectio | Z38_S5 | 27446 | -0.01624 | 0.005287 | 0.002123 | 2.672993 | Left ventricle |
| LV_circum | Z95_S5 | 27446 | -0.01883 | 0.006135 | 0.002144 | 2.668711 | Left ventricle |
| LA_stroke_ | Z106_S3 | 27446 | -0.01905 | 0.006207 | 0.002149 | 2.667845 | Left atrium |
| Ascending_ | Z63_S1 | 24599 | -0.0209 | 0.006835 | 0.002226 | 2.652562 | Aorta |

| | | | | | | | |
|---|---|---|---|---|---|---|---|
| Ascending_ | Z6_S2 | 24599 | 0.014482 | 0.00474 | 0.002247 | 2.648486 | Aorta |
| LV_mean_ | Z114_S4 | 27446 | 0.020751 | 0.006793 | 0.002252 | 2.647491 | Left ventricle |
| Ascending_ | Z55_S1 | 24599 | -0.01719 | 0.005628 | 0.002253 | 2.647322 | Aorta |
| Ascending_ | Z47_S4 | 24599 | -0.01759 | 0.005758 | 0.002255 | 2.646929 | Aorta |
| LV_radial_ | Z82_S1 | 27446 | -0.01948 | 0.00638 | 0.002265 | 2.64487 | Left ventricle |
| LV_circumf | Z73_S4 | 27446 | 0.017846 | 0.005849 | 0.002281 | 2.641931 | Left ventricle |
| Descending | Z113_S2 | 24599 | -0.02188 | 0.007177 | 0.002293 | 2.639508 | Aorta |
| LA_maximu | Z44_S1 | 27446 | 0.015899 | 0.005222 | 0.002328 | 2.633067 | Left atrium |
| LV_ejection | Z123_S4 | 27446 | -0.01598 | 0.00525 | 0.002335 | 2.631652 | Left ventricle |
| RA_stroke_ | Z51_S3 | 27446 | -0.01882 | 0.006183 | 0.00234 | 2.630723 | Right atrium |
| LV_mean_ | Z98_S3 | 27446 | -0.01857 | 0.006106 | 0.002362 | 2.626789 | Left ventricle |
| Ascending_ | Z120_S2 | 24599 | -0.0158 | 0.005204 | 0.002396 | 2.620483 | Aorta |
| RV_end_di | Z77_S3 | 27446 | -0.02225 | 0.007331 | 0.0024 | 2.619725 | Right ventricle |
| LV_circumf | Z34_S2 | 27446 | 0.017712 | 0.005836 | 0.002407 | 2.618528 | Left ventricle |
| LA_maximu | Z53_S2 | 27446 | 0.017564 | 0.005794 | 0.002435 | 2.613534 | Left atrium |
| RA_ejectio | Z39_S1 | 27446 | -0.01391 | 0.004603 | 0.002517 | 2.599096 | Right atrium |
| RA_minimu | Z80_S2 | 27446 | -0.01513 | 0.005017 | 0.002567 | 2.590514 | Right atrium |
| LV_longitu | Z57_S2 | 27446 | 0.012451 | 0.004129 | 0.002568 | 2.590395 | Left ventricle |
| Descending | Z53_S1 | 24599 | -0.02107 | 0.006992 | 0.002586 | 2.587294 | Aorta |
| LV_radial_ | Z51_S3 | 27446 | 0.018946 | 0.006304 | 0.002652 | 2.576349 | Left ventricle |
| LV_end_sy | Z37_S3 | 27446 | -0.02157 | 0.007177 | 0.002653 | 2.576283 | Left ventricle |
| RA_maxim | Z66_S2 | 27446 | 0.019122 | 0.006367 | 0.002671 | 2.573398 | Right atrium |
| LV_ejection | Z22_S2 | 27446 | 0.015269 | 0.005086 | 0.002678 | 2.57223 | Left ventricle |
| LV_mean_ | Z1_S2 | 27446 | 0.017158 | 0.005716 | 0.002686 | 2.570907 | Left ventricle |
| RA_stroke_ | Z28_S4 | 27446 | -0.01636 | 0.005453 | 0.002706 | 2.567709 | Right atrium |
| RV_ejectio | Z70_S5 | 27446 | -0.01467 | 0.004892 | 0.00271 | 2.567094 | Right ventricle |
| Descending | Z80_S2 | 24599 | -0.0158 | 0.005308 | 0.002914 | 2.535472 | Aorta |
| RV_stroke_ | Z22_S4 | 27446 | -0.01894 | 0.006379 | 0.002984 | 2.525151 | Right ventricle |
| LA_ejection | Z78_S1 | 27446 | 0.013134 | 0.004437 | 0.003079 | 2.511608 | Left atrium |
| RV_ejectio | Z57_S2 | 27446 | 0.012536 | 0.004247 | 0.003161 | 2.50024 | Right ventricle |
| LV_circumf | Z78_S4 | 27446 | 0.013316 | 0.004514 | 0.003178 | 2.497792 | Left ventricle |
| LV_longitu | Z55_S5 | 27446 | -0.0125 | 0.004245 | 0.003239 | 2.489604 | Left ventricle |
| LV_ejection | Z24_S5 | 27446 | -0.01607 | 0.005461 | 0.00326 | 2.486793 | Left ventricle |
| Ascending_ | Z65_S2 | 24599 | 0.017496 | 0.005953 | 0.003293 | 2.482374 | Aorta |
| LV_ejection | Z7_S1 | 27446 | 0.013434 | 0.0046 | 0.003494 | 2.456632 | Left ventricle |
| RV_end_di | Z72_S5 | 27446 | -0.02586 | 0.00886 | 0.003515 | 2.454055 | Right ventricle |
| RV_ejectio | Z1_S4 | 27446 | -0.01568 | 0.005387 | 0.003615 | 2.441848 | Right ventricle |
| LV_myocar | Z44_S1 | 27446 | 0.024324 | 0.008362 | 0.003627 | 2.440475 | Left ventricle |
| RV_ejectio | Z56_S2 | 27446 | 0.012766 | 0.004392 | 0.003656 | 2.437031 | Right ventricle |
| LV_circumf | Z1_S4 | 27446 | -0.01558 | 0.005363 | 0.003682 | 2.433879 | Left ventricle |
| LV_circumf | Z15_S2 | 27446 | 0.015759 | 0.005433 | 0.003727 | 2.428693 | Left ventricle |
| LV_mean_ | Z94_S5 | 27446 | 0.017448 | 0.006023 | 0.003769 | 2.423795 | Left ventricle |
| LV_radial_ | Z37_S3 | 27446 | -0.01699 | 0.005875 | 0.00382 | 2.417892 | Left ventricle |
| LV_end_sy | Z80_S5 | 27446 | -0.02149 | 0.007441 | 0.00387 | 2.41224 | Left ventricle |
| RV_stroke_ | Z65_S1 | 27446 | 0.014701 | 0.005108 | 0.004005 | 2.397391 | Right ventricle |

| | | | | | | | |
|---|---|---|---|---|---|---|---|
| RA_stroke_ | Z55_S1 | 27446 | -0.01351 | 0.004696 | 0.004011 | 2.396744 | Right atrium |
| LV_circumf | Z41_S1 | 27446 | -0.01579 | 0.005501 | 0.004094 | 2.387897 | Left ventricle |
| LV_cardiac_ | Z57_S2 | 27446 | 0.013246 | 0.004618 | 0.004125 | 2.384543 | Left ventricle |
| RV_end_di | Z44_S1 | 27446 | 0.020542 | 0.007172 | 0.004179 | 2.378906 | Right ventricle |
| LV_end_sy | Z3_S4 | 27446 | 0.022467 | 0.007859 | 0.004255 | 2.371141 | Left ventricle |
| LV_cardiac_ | Z117_S4 | 27446 | 0.012936 | 0.00453 | 0.004294 | 2.367181 | Left ventricle |
| LV_ejectior | Z109_S1 | 27446 | 0.014287 | 0.005023 | 0.004454 | 2.351208 | Left ventricle |
| RA_stroke_ | Z41_S2 | 27446 | -0.01702 | 0.005985 | 0.004466 | 2.350119 | Right atrium |
| RA_maxim | Z120_S5 | 27446 | 0.019055 | 0.006709 | 0.004508 | 2.345979 | Right atrium |
| LA_ejectior | Z61_S4 | 27446 | -0.01301 | 0.004586 | 0.00456 | 2.341045 | Left atrium |
| LV_mean_ | Z1_S1 | 27446 | -0.01925 | 0.006797 | 0.004618 | 2.335583 | Left ventricle |
| LV_mean_ | Z72_S5 | 27446 | -0.02435 | 0.008602 | 0.00465 | 2.332574 | Left ventricle |
| Descending | Z24_S5 | 24599 | 0.020391 | 0.007211 | 0.004688 | 2.329051 | Aorta |
| LA_stroke_ | Z51_S3 | 27446 | -0.01816 | 0.006426 | 0.004725 | 2.325556 | Left atrium |
| LV_myocar | Z50_S5 | 27446 | 0.029382 | 0.010412 | 0.004773 | 2.32119 | Left ventricle |
| LV_longitu | Z97_S1 | 27446 | -0.0171 | 0.006071 | 0.00486 | 2.313391 | Left ventricle |
| RA_ejectio | Z37_S3 | 27446 | -0.01596 | 0.005671 | 0.004888 | 2.310889 | Right atrium |
| LV_mean_ | Z75_S4 | 27446 | -0.02314 | 0.008264 | 0.005099 | 2.292478 | Left ventricle |
| Descending | Z97_S1 | 24599 | -0.02039 | 0.007288 | 0.005144 | 2.288731 | Aorta |
| LA_maximu | Z51_S3 | 27446 | -0.01787 | 0.006394 | 0.005207 | 2.283385 | Left atrium |
| LV_end_dia | Z40_S4 | 27446 | -0.02302 | 0.008246 | 0.00525 | 2.279848 | Left ventricle |
| LV_cardiac_ | Z14_S2 | 27446 | 0.015428 | 0.005545 | 0.005396 | 2.267912 | Left ventricle |
| RA_ejectio | Z106_S3 | 27446 | 0.01634 | 0.005878 | 0.005439 | 2.264465 | Right atrium |
| RA_minimu | Z121_S4 | 27446 | -0.01759 | 0.006353 | 0.005633 | 2.249242 | Right atrium |
| RV_ejectio | Z121_S4 | 27446 | 0.015967 | 0.005769 | 0.005648 | 2.248128 | Right ventricle |
| LV_radial_s | Z79_S4 | 27446 | 0.016781 | 0.006083 | 0.005805 | 2.236201 | Left ventricle |
| RA_stroke_ | Z76_S3 | 27446 | -0.0147 | 0.005332 | 0.005826 | 2.234608 | Right atrium |
| LA_ejectior | Z59_S2 | 27446 | 0.015264 | 0.005557 | 0.006013 | 2.220906 | Left atrium |
| LV_mean_ | Z81_S1 | 27446 | -0.01944 | 0.007078 | 0.006022 | 2.220271 | Left ventricle |
| LA_ejectior | Z44_S3 | 27446 | 0.014734 | 0.005373 | 0.006099 | 2.214754 | Left atrium |
| LV_cardiac_ | Z65_S1 | 27446 | 0.012633 | 0.004614 | 0.00618 | 2.208982 | Left ventricle |
| LV_ejectior | Z113_S2 | 27446 | -0.01652 | 0.006033 | 0.006191 | 2.208222 | Left ventricle |
| LV_ejectior | Z85_S5 | 27446 | 0.012828 | 0.004693 | 0.006271 | 2.202642 | Left ventricle |
| RA_stroke_ | Z26_S4 | 27446 | -0.01714 | 0.006275 | 0.006308 | 2.200079 | Right atrium |
| LV_mean_ | Z80_S5 | 27446 | 0.022502 | 0.008239 | 0.006311 | 2.199886 | Left ventricle |
| LV_circumf | Z72_S5 | 27446 | -0.01712 | 0.006273 | 0.006341 | 2.197841 | Left ventricle |
| LV_radial_s | Z21_S3 | 27446 | -0.01764 | 0.006466 | 0.006372 | 2.195718 | Left ventricle |
| LV_longitu | Z115_S4 | 27446 | -0.01243 | 0.004559 | 0.006391 | 2.194432 | Left ventricle |
| LV_mean_ | Z41_S2 | 27446 | 0.022487 | 0.008254 | 0.006442 | 2.190981 | Left ventricle |
| Descending | Z50_S5 | 24599 | -0.02255 | 0.008289 | 0.006513 | 2.186207 | Aorta |
| Ascending_ | Z39_S4 | 24599 | 0.014054 | 0.005174 | 0.006601 | 2.180368 | Aorta |
| LV_mean_ | Z99_S4 | 27446 | -0.02118 | 0.007799 | 0.006618 | 2.179294 | Left ventricle |
| LV_radial_s | Z72_S5 | 27446 | 0.01725 | 0.00636 | 0.006684 | 2.174976 | Left ventricle |
| Ascending_ | Z57_S1 | 24599 | -0.01518 | 0.005599 | 0.006698 | 2.174034 | Aorta |
| Ascending_ | Z50_S3 | 24599 | -0.01539 | 0.00568 | 0.006754 | 2.170455 | Aorta |

| | | | | | | | |
|---|---|---|---|---|---|---|---|
| LV_longitu | Z65_S1 | 27446 | -0.01117 | 0.004126 | 0.006795 | 2.167837 | Left ventricle |
| LV_cardiac | Z109_S1 | 27446 | 0.015087 | 0.005575 | 0.006805 | 2.167188 | Left ventricle |
| RA_maxim | Z34_S2 | 27446 | 0.01722 | 0.00637 | 0.006865 | 2.16338 | Right atrium |
| RV_end_sy | Z24_S5 | 27446 | 0.020197 | 0.007473 | 0.006877 | 2.162591 | Right ventricle |
| LV_longitu | Z7_S3 | 27446 | -0.01594 | 0.005926 | 0.007157 | 2.145266 | Left ventricle |
| LV_mean_ | Z52_S3 | 27446 | 0.016013 | 0.005959 | 0.0072 | 2.142679 | Left ventricle |
| LV_cardiac | Z69_S5 | 27446 | 0.012684 | 0.004728 | 0.007298 | 2.136807 | Left ventricle |
| RA_ejectio | Z28_S1 | 27446 | -0.01407 | 0.005264 | 0.00753 | 2.123189 | Right atrium |
| RV_ejectio | Z39_S4 | 27446 | -0.01223 | 0.00458 | 0.007594 | 2.119543 | Right ventricle |
| LV_longitu | Z34_S2 | 27446 | -0.01521 | 0.005701 | 0.007608 | 2.118742 | Left ventricle |
| Ascending_ | Z75_S1 | 24599 | 0.016646 | 0.006239 | 0.007628 | 2.117616 | Aorta |
| Descending | Z65_S1 | 24599 | 0.013197 | 0.004971 | 0.007933 | 2.100577 | Aorta |
| LA_stroke_ | Z65_S1 | 27446 | -0.01157 | 0.004366 | 0.00805 | 2.094218 | Left atrium |
| RA_ejectio | Z80_S1 | 27446 | -0.01431 | 0.005402 | 0.008054 | 2.094012 | Right atrium |
| RA_maxim | Z115_S4 | 27446 | 0.01348 | 0.005094 | 0.008139 | 2.089413 | Right atrium |
| Descending | Z114_S5 | 24599 | -0.01261 | 0.004767 | 0.008177 | 2.087425 | Aorta |
| LV_cardiac | Z55_S1 | 27446 | 0.013635 | 0.005158 | 0.008206 | 2.085846 | Left ventricle |
| LV_mean_ | Z93_S1 | 27446 | 0.021099 | 0.008 | 0.008351 | 2.078239 | Left ventricle |
| LA_maximu | Z78_S1 | 27446 | -0.01257 | 0.004775 | 0.008459 | 2.072676 | Left atrium |
| RA_ejectio | Z100_S3 | 27446 | 0.013476 | 0.005118 | 0.008463 | 2.072481 | Right atrium |
| LA_maximu | Z49_S3 | 27446 | 0.015981 | 0.006072 | 0.008493 | 2.070954 | Left atrium |
| RA_maxim | Z32_S5 | 27446 | -0.01758 | 0.006689 | 0.008585 | 2.066254 | Right atrium |
| RA_stroke_ | Z55_S5 | 27446 | 0.011352 | 0.004321 | 0.008616 | 2.064689 | Right atrium |
| LA_ejection | Z37_S3 | 27446 | -0.01455 | 0.005538 | 0.008621 | 2.06442 | Left atrium |
| LV_circumf | Z49_S3 | 27446 | -0.0155 | 0.005905 | 0.00865 | 2.062971 | Left ventricle |
| LA_ejection | Z113_S2 | 27446 | 0.015345 | 0.005859 | 0.008818 | 2.054625 | Left atrium |
| RA_ejectio | Z39_S4 | 27446 | -0.01168 | 0.004462 | 0.008859 | 2.052622 | Right atrium |
| Ascending_ | Z22_S4 | 24599 | 0.0156 | 0.005967 | 0.008942 | 2.048561 | Aorta |
| RV_ejectio | Z44_S4 | 27446 | 0.011293 | 0.004331 | 0.00912 | 2.040021 | Right ventricle |
| Descending | Z50_S5 | 24599 | 0.021303 | 0.008171 | 0.009132 | 2.039451 | Aorta |
| Descending | Z3_S2 | 24599 | 0.013881 | 0.005331 | 0.00922 | 2.035282 | Aorta |
| RA_minimu | Z73_S1 | 27446 | 0.013728 | 0.005279 | 0.009305 | 2.03127 | Right atrium |
| LA_ejection | Z39_S4 | 27446 | -0.01133 | 0.004357 | 0.009321 | 2.030541 | Left atrium |
| RV_ejectio | Z106_S3 | 27446 | 0.015642 | 0.006034 | 0.00953 | 2.020922 | Right ventricle |
| LV_mean_ | Z23_S1 | 27446 | 0.020317 | 0.00784 | 0.009557 | 2.019674 | Left ventricle |
| LV_ejection | Z29_S1 | 27446 | 0.014066 | 0.005428 | 0.009563 | 2.019423 | Left ventricle |
| RA_ejectio | Z115_S4 | 27446 | -0.01183 | 0.004568 | 0.009568 | 2.019176 | Right atrium |
| LA_minimu | Z38_S4 | 27446 | 0.01508 | 0.005825 | 0.009624 | 2.016662 | Left atrium |
| LV_stroke_ | Z80_S5 | 27446 | -0.01852 | 0.007161 | 0.009695 | 2.013433 | Left ventricle |
| Descending | Z127_S4 | 24599 | 0.019014 | 0.007357 | 0.009755 | 2.010784 | Aorta |
| LA_minimu | Z49_S3 | 27446 | 0.015172 | 0.005888 | 0.009974 | 2.001129 | Left atrium |
| Ascending_ | Z87_S5 | 24599 | 0.015406 | 0.006 | 0.01024 | 1.989684 | Aorta |
| Descending | Z53_S2 | 24599 | 0.016914 | 0.006605 | 0.010439 | 1.981328 | Aorta |
| RV_ejectio | Z62_S1 | 27446 | -0.01224 | 0.00479 | 0.010583 | 1.975409 | Right ventricle |
| LV_mean_ | Z3_S4 | 27446 | -0.02223 | 0.008702 | 0.010645 | 1.972858 | Left ventricle |

| | | | | | | | |
|---|---|---|---|---|---|---|---|
| RA_ejectio | Z61_S4 | 27446 | -0.01198 | 0.004696 | 0.010738 | 1.96906 | Right atrium |
| RA_maxim | Z80_S5 | 27446 | -0.01662 | 0.006558 | 0.011264 | 1.94832 | Right atrium |
| RA_stroke_ | Z52_S3 | 27446 | 0.010939 | 0.004321 | 0.011351 | 1.944948 | Right atrium |
| LV_end_sy | Z18_S4 | 27446 | -0.01987 | 0.007874 | 0.011618 | 1.934872 | Left ventricle |
| RV_end_di | Z62_S4 | 27446 | -0.02077 | 0.008239 | 0.011714 | 1.931296 | Right ventricle |
| LV_cardiac | Z93_S1 | 27446 | -0.01605 | 0.006372 | 0.011787 | 1.928586 | Left ventricle |
| Descending | Z56_S2 | 24599 | -0.01289 | 0.005139 | 0.012158 | 1.915135 | Aorta |
| RA_stroke_ | Z23_S3 | 27446 | 0.011266 | 0.004493 | 0.01216 | 1.91508 | Right atrium |
| LV_circumf | Z97_S1 | 27446 | -0.01558 | 0.006216 | 0.012193 | 1.913884 | Left ventricle |
| LA_minimu | Z78_S1 | 27446 | -0.0116 | 0.00463 | 0.012208 | 1.913366 | Left atrium |
| LV_radial_ | Z74_S4 | 27446 | -0.01351 | 0.005417 | 0.012613 | 1.899191 | Left ventricle |
| RV_ejectio | Z44_S1 | 27446 | -0.01269 | 0.005101 | 0.012881 | 1.890065 | Right ventricle |
| LV_end_di | Z22_S2 | 27446 | 0.017231 | 0.006934 | 0.012958 | 1.887454 | Left ventricle |
| RV_ejectio | Z31_S5 | 27446 | -0.01022 | 0.004115 | 0.013001 | 1.886036 | Right ventricle |
| Descending | Z112_S3 | 24599 | -0.0194 | 0.007826 | 0.013179 | 1.880112 | Aorta |
| LV_mean_ | Z38_S5 | 27446 | -0.01824 | 0.007367 | 0.013309 | 1.875847 | Left ventricle |
| Descending | Z94_S2 | 24599 | -0.01596 | 0.006457 | 0.013468 | 1.870683 | Aorta |
| RA_maxim | Z58_S2 | 27446 | 0.012846 | 0.005211 | 0.013693 | 1.863507 | Right atrium |
| Descending | Z39_S3 | 24599 | -0.02059 | 0.008366 | 0.013838 | 1.858926 | Aorta |
| Ascending_ | Z27_S2 | 24599 | -0.01653 | 0.006743 | 0.014239 | 1.846524 | Aorta |
| LA_ejection | Z26_S4 | 27446 | -0.01478 | 0.006032 | 0.014281 | 1.845231 | Left atrium |
| LV_circumf | Z75_S5 | 27446 | -0.01562 | 0.006396 | 0.014612 | 1.835282 | Left ventricle |
| LV_cardiac | Z65_S2 | 27446 | -0.01394 | 0.005719 | 0.014811 | 1.82941 | Left ventricle |
| LV_ejection | Z40_S5 | 27446 | 0.010408 | 0.004288 | 0.015225 | 1.817443 | Left ventricle |
| LV_end_sy | Z10_S4 | 27446 | 0.015944 | 0.006578 | 0.015362 | 1.813564 | Left ventricle |
| Ascending_ | Z39_S4 | 24599 | 0.012466 | 0.005155 | 0.015601 | 1.806847 | Aorta |
| RV_stroke_ | Z62_S4 | 27446 | -0.01705 | 0.007055 | 0.015678 | 1.804722 | Right ventricle |
| LV_circumf | Z23_S1 | 27446 | -0.01378 | 0.005718 | 0.015926 | 1.797906 | Left ventricle |
| LV_circumf | Z63_S1 | 27446 | 0.01454 | 0.006051 | 0.016274 | 1.788512 | Left ventricle |
| Ascending_ | Z53_S1 | 24599 | -0.0145 | 0.006039 | 0.016353 | 1.786394 | Aorta |
| Ascending_ | Z59_S3 | 24599 | -0.01179 | 0.004914 | 0.016412 | 1.784846 | Aorta |
| Ascending_ | Z95_S5 | 24599 | 0.017532 | 0.00732 | 0.016617 | 1.77946 | Aorta |
| LV_mean_ | Z40_S4 | 27446 | 0.020176 | 0.008427 | 0.016654 | 1.778484 | Left ventricle |
| LV_end_sy | Z37_S4 | 27446 | 0.018659 | 0.007811 | 0.016896 | 1.772216 | Left ventricle |
| RA_stroke_ | Z99_S5 | 27446 | 0.011602 | 0.004861 | 0.01699 | 1.769813 | Right atrium |
| Ascending_ | Z62_S2 | 24599 | 0.012796 | 0.005367 | 0.017114 | 1.766648 | Aorta |
| Descending | Z2_S1 | 24599 | -0.01293 | 0.005434 | 0.017325 | 1.761334 | Aorta |
| LV_mean_ | Z92_S5 | 27446 | -0.02001 | 0.008421 | 0.017504 | 1.756852 | Left ventricle |
| LV_myocar | Z28_S1 | 27446 | -0.02102 | 0.008858 | 0.017627 | 1.753826 | Left ventricle |
| RA_stroke_ | Z24_S5 | 27446 | 0.013095 | 0.005518 | 0.017635 | 1.753632 | Right atrium |
| LV_mean_ | Z97_S2 | 27446 | -0.01748 | 0.007378 | 0.017819 | 1.749108 | Left ventricle |
| LV_mean_ | Z94_S2 | 27446 | 0.017827 | 0.007526 | 0.017856 | 1.748227 | Left ventricle |
| RA_stroke_ | Z50_S2 | 27446 | -0.01465 | 0.006194 | 0.018047 | 1.743598 | Right atrium |
| RV_ejectio | Z87_S3 | 27446 | 0.011772 | 0.004985 | 0.018196 | 1.740031 | Right ventricle |
| Descending | Z92_S1 | 24599 | -0.01397 | 0.005923 | 0.018352 | 1.736317 | Aorta |

| | | | | | | | |
|---|---|---|---|---|---|---|---|
| LV_cardiac | Z62_S3 | 27446 | -0.01373 | 0.005824 | 0.018424 | 1.734617 | Left ventricle |
| LV_mean_ | Z44_S3 | 27446 | 0.018124 | 0.007709 | 0.018713 | 1.727861 | Left ventricle |
| LV_mean_ | Z19_S2 | 27446 | 0.016158 | 0.006889 | 0.019 | 1.721241 | Left ventricle |
| LV_cardiac | Z91_S1 | 27446 | 0.012608 | 0.005379 | 0.019084 | 1.719342 | Left ventricle |
| Ascending_ | Z82_S1 | 24599 | -0.01664 | 0.007117 | 0.01936 | 1.713093 | Aorta |
| RV_end_di | Z28_S1 | 27446 | -0.01775 | 0.007598 | 0.019455 | 1.710959 | Right ventricle |
| Ascending_ | Z41_S1 | 24599 | -0.01454 | 0.006239 | 0.019781 | 1.703761 | Aorta |
| LV_cardiac | Z35_S5 | 27446 | 0.011101 | 0.004769 | 0.019928 | 1.700529 | Left ventricle |
| LV_ejection | Z4_S4 | 27446 | 0.01035 | 0.004455 | 0.020183 | 1.695021 | Left ventricle |
| RA_maxim | Z38_S4 | 27446 | -0.01472 | 0.006375 | 0.020978 | 1.678239 | Right atrium |
| RV_ejectio | Z22_S4 | 27446 | -0.0122 | 0.005299 | 0.021345 | 1.670708 | Right ventricle |
| LV_circumf | Z75_S4 | 27446 | -0.01387 | 0.006027 | 0.021412 | 1.669352 | Left ventricle |
| LV_mean_ | Z46_S1 | 27446 | -0.01622 | 0.007065 | 0.021689 | 1.663759 | Left ventricle |
| RV_ejectio | Z61_S4 | 27446 | 0.011052 | 0.00482 | 0.021854 | 1.660476 | Right ventricle |
| Ascending_ | Z21_S3 | 24599 | 0.01643 | 0.007201 | 0.022523 | 1.647374 | Aorta |
| LV_cardiac | Z37_S2 | 27446 | 0.013678 | 0.006036 | 0.023456 | 1.629754 | Left ventricle |
| LV_ejection | Z82_S4 | 27446 | -0.00978 | 0.004323 | 0.023694 | 1.625358 | Left ventricle |
| Ascending_ | Z50_S4 | 24599 | 0.013247 | 0.005863 | 0.023864 | 1.62225 | Aorta |
| LV_longitu | Z56_S2 | 27446 | 0.009619 | 0.004271 | 0.024306 | 1.614281 | Left ventricle |
| LA_ejection | Z114_S4 | 27446 | 0.010648 | 0.004735 | 0.024519 | 1.610495 | Left atrium |
| Descending | Z115_S4 | 24599 | -0.01227 | 0.005458 | 0.024544 | 1.610048 | Aorta |
| LV_mean_ | Z37_S2 | 27446 | -0.01704 | 0.007578 | 0.024549 | 1.609964 | Left ventricle |
| LA_minimu | Z32_S5 | 27446 | -0.01366 | 0.006111 | 0.025343 | 1.596136 | Left atrium |
| LV_radial_ | Z44_S3 | 27446 | 0.012674 | 0.0057 | 0.026174 | 1.582136 | Left ventricle |
| LV_circumf | Z26_S4 | 27446 | 0.014021 | 0.006311 | 0.026312 | 1.579839 | Left ventricle |
| LV_radial_ | Z49_S3 | 27446 | -0.01321 | 0.005987 | 0.027349 | 1.563051 | Left ventricle |
| RV_end_sy | Z77_S3 | 27446 | 0.015418 | 0.006991 | 0.027422 | 1.561901 | Right ventricle |
| LV_mean_ | Z8_S3 | 27446 | 0.013081 | 0.005933 | 0.027476 | 1.561041 | Left ventricle |
| LV_mean_ | Z74_S2 | 27446 | 0.015198 | 0.0069 | 0.027614 | 1.558867 | Left ventricle |
| RA_ejectio | Z52_S3 | 27446 | -0.00937 | 0.004253 | 0.027632 | 1.558593 | Right atrium |
| Descending | Z23_S1 | 24599 | -0.01659 | 0.00755 | 0.02801 | 1.552686 | Aorta |
| RA_stroke_ | Z65_S2 | 27446 | -0.01142 | 0.005206 | 0.028298 | 1.548243 | Right atrium |
| LA_stroke_ | Z32_S5 | 27446 | 0.013881 | 0.006334 | 0.028409 | 1.546551 | Left atrium |
| RA_stroke_ | Z19_S2 | 27446 | 0.010933 | 0.004996 | 0.028637 | 1.543073 | Right atrium |
| LV_end_sy | Z49_S1 | 27446 | 0.016851 | 0.007702 | 0.028683 | 1.542381 | Left ventricle |
| Ascending_ | Z111_S1 | 24599 | 0.01375 | 0.006297 | 0.028995 | 1.53768 | Aorta |
| LV_mean_ | Z62_S4 | 27446 | -0.01743 | 0.007999 | 0.02931 | 1.532981 | Left ventricle |
| LV_end_sy | Z3_S2 | 27446 | 0.012238 | 0.005622 | 0.0295 | 1.530185 | Left ventricle |
| RA_ejectio | Z121_S4 | 27446 | -0.01222 | 0.005621 | 0.029653 | 1.527925 | Right atrium |
| Ascending_ | Z98_S3 | 24599 | -0.01152 | 0.005299 | 0.029755 | 1.526441 | Aorta |
| RA_ejectio | Z115_S3 | 27446 | -0.01152 | 0.005302 | 0.029863 | 1.524868 | Right atrium |
| LA_minimu | Z44_S1 | 27446 | -0.01099 | 0.005064 | 0.029962 | 1.523435 | Left atrium |
| LV_longitu | Z78_S1 | 27446 | 0.00984 | 0.004535 | 0.030046 | 1.522219 | Left ventricle |
| LV_longitu | Z113_S2 | 27446 | 0.012963 | 0.005988 | 0.030404 | 1.517071 | Left ventricle |
| RA_minimu | Z31_S2 | 27446 | 0.012243 | 0.005665 | 0.030684 | 1.513095 | Right atrium |

| | | | | | | | |
|---|---|---|---|---|---|---|---|
| LV_mean_ı | Z61_S4 | 27446 | -0.01417 | 0.00658 | 0.031307 | 1.504365 | Left ventricle |
| Descending | Z27_S2 | 24599 | 0.016797 | 0.007807 | 0.031431 | 1.502644 | Aorta |
| LV_mean_ı | Z117_S4 | 27446 | -0.01222 | 0.005687 | 0.0316 | 1.500317 | Left ventricle |
| LV_radial_ | Z63_S1 | 27446 | 0.013147 | 0.006135 | 0.032111 | 1.493353 | Left ventricle |
| LV_mean_ı | Z44_S4 | 27446 | -0.01266 | 0.005912 | 0.032215 | 1.491935 | Left ventricle |
| LA_ejectio | Z73_S1 | 27446 | -0.00977 | 0.004561 | 0.032263 | 1.491293 | Left atrium |
| LV_cardiac | Z56_S2 | 27446 | 0.010206 | 0.004776 | 0.032606 | 1.486697 | Left ventricle |
| RA_stroke_ | Z66_S3 | 27446 | 0.012237 | 0.005742 | 0.03309 | 1.480304 | Right atrium |
| RV_stroke_ | Z38_S4 | 27446 | 0.015045 | 0.007065 | 0.0332 | 1.478859 | Right ventricle |
| LA_stroke_ | Z120_S5 | 27446 | -0.01344 | 0.006353 | 0.034422 | 1.463165 | Left atrium |
| RV_stroke_ | Z73_S1 | 27446 | -0.0122 | 0.005771 | 0.03454 | 1.461679 | Right ventricle |
| LA_minimu | Z51_S3 | 27446 | -0.0131 | 0.006201 | 0.034631 | 1.460536 | Left atrium |
| LV_mean_ı | Z122_S5 | 27446 | 0.013255 | 0.00628 | 0.034792 | 1.458516 | Left ventricle |
| Descending | Z37_S3 | 24599 | -0.01578 | 0.00751 | 0.035575 | 1.44885 | Aorta |
| LA_minimu | Z1_S4 | 27446 | -0.01118 | 0.005348 | 0.036503 | 1.437671 | Left atrium |
| LV_mean_ı | Z2_S1 | 27446 | -0.0133 | 0.006369 | 0.036698 | 1.435363 | Left ventricle |
| RA_ejectio | Z96_S3 | 27446 | -0.01009 | 0.004852 | 0.03757 | 1.425158 | Right atrium |
| LV_ejectio | Z66_S2 | 27446 | -0.01194 | 0.005742 | 0.03758 | 1.425043 | Left ventricle |
| LA_stroke_ | Z78_S1 | 27446 | -0.00991 | 0.004799 | 0.038989 | 1.409057 | Left atrium |
| LV_ejectio | Z89_S4 | 27446 | -0.00999 | 0.004845 | 0.039176 | 1.406978 | Left ventricle |
| RV_ejectio | Z39_S1 | 27446 | -0.00973 | 0.004725 | 0.039493 | 1.403483 | Right ventricle |
| LV_cardiac | Z70_S5 | 27446 | -0.01094 | 0.005319 | 0.039717 | 1.401027 | Left ventricle |
| LV_mean_ı | Z20_S2 | 27446 | 0.012082 | 0.005903 | 0.040679 | 1.390629 | Left ventricle |
| RV_end_sy | Z22_S2 | 27446 | -0.01422 | 0.006959 | 0.040998 | 1.387241 | Right ventricle |
| LV_cardiac | Z96_S3 | 27446 | 0.011053 | 0.005415 | 0.041224 | 1.384852 | Left ventricle |
| Ascending_ | Z105_S5 | 24599 | 0.012419 | 0.006127 | 0.042667 | 1.36991 | Aorta |
| LA_minimu | Z113_S2 | 27446 | 0.012365 | 0.006113 | 0.043099 | 1.365536 | Left atrium |
| RA_ejectio | Z39_S3 | 27446 | -0.01258 | 0.006241 | 0.043866 | 1.357873 | Right atrium |
| LV_mean_ı | Z24_S1 | 27446 | 0.011929 | 0.005921 | 0.043933 | 1.357209 | Left ventricle |
| LV_mean_ı | Z69_S5 | 27446 | -0.01194 | 0.005935 | 0.044218 | 1.354401 | Left ventricle |
| Descending | Z59_S2 | 24599 | -0.01367 | 0.006814 | 0.044797 | 1.348751 | Aorta |
| Ascending_ | Z112_S3 | 24599 | -0.01376 | 0.006858 | 0.044846 | 1.348277 | Aorta |
| LV_end_sy | Z23_S1 | 27446 | -0.01418 | 0.007081 | 0.045223 | 1.344642 | Left ventricle |
| LV_cardiac | Z39_S4 | 27446 | -0.00991 | 0.00498 | 0.046536 | 1.33221 | Left ventricle |
| LV_longitu | Z26_S4 | 27446 | -0.01226 | 0.006165 | 0.046796 | 1.329796 | Left ventricle |
| RV_end_di | Z40_S4 | 27446 | -0.01722 | 0.00868 | 0.047205 | 1.326009 | Right ventricle |
| LA_stroke_ | Z97_S1 | 27446 | -0.01272 | 0.006425 | 0.04773 | 1.321213 | Left atrium |
| LA_stroke_ | Z49_S3 | 27446 | 0.012082 | 0.006103 | 0.047738 | 1.321134 | Left atrium |
| Ascending_ | Z62_S4 | 24599 | -0.01306 | 0.006608 | 0.048034 | 1.318454 | Aorta |
| Ascending_ | Z120_S5 | 24599 | -0.01378 | 0.006976 | 0.048149 | 1.317415 | Aorta |
| LV_longitu | Z72_S5 | 27446 | -0.0121 | 0.006128 | 0.048369 | 1.315433 | Left ventricle |
| LV_mean_ı | Z87_S5 | 27446 | -0.01358 | 0.00689 | 0.048734 | 1.312165 | Left ventricle |
| RA_maxim | Z28_S1 | 27446 | 0.011553 | 0.005871 | 0.049097 | 1.308942 | Right atrium |
| Descending | Z40_S4 | 24599 | 0.01561 | 0.007943 | 0.049378 | 1.306464 | Aorta |
| Descending | Z1_S2 | 24599 | 0.009566 | 0.004871 | 0.049537 | 1.305068 | Aorta |

| | | | | | | | |
|---|---|---|---|---|---|---|---|
| LV_longitu | Z62_S3 | 27446 | -0.01022 | 0.005209 | 0.04972 | 1.303465 | Left ventricle |
| RA_stroke_ | Z79_S4 | 27446 | -0.0117 | 0.005967 | 0.04987 | 1.302161 | Right atrium |
| LV_mean_ | Z86_S3 | 27446 | -0.01542 | 0.0079 | 0.050958 | 1.292785 | Left ventricle |
| RA_ejectio | Z31_S2 | 27446 | -0.00975 | 0.005012 | 0.051749 | 1.286095 | Right atrium |
| LV_mean_ | Z112_S1 | 27446 | 0.016889 | 0.008693 | 0.052043 | 1.283635 | Left ventricle |
| RA_minimu | Z7_S3 | 27446 | -0.01299 | 0.006712 | 0.053004 | 1.275694 | Right atrium |
| LV_mean_ | Z120_S2 | 27446 | 0.011564 | 0.005994 | 0.053678 | 1.270203 | Left ventricle |
| RV_ejectio | Z37_S3 | 27446 | 0.011208 | 0.005822 | 0.054211 | 1.265913 | Right ventricle |
| LV_mean_ | Z17_S1 | 27446 | 0.012545 | 0.006527 | 0.054588 | 1.262904 | Left ventricle |
| RA_stroke_ | Z97_S1 | 27446 | -0.01185 | 0.006181 | 0.055298 | 1.257288 | Right atrium |
| LV_mean_ | Z95_S5 | 27446 | 0.016122 | 0.008413 | 0.055331 | 1.257032 | Left ventricle |
| RA_minimu | Z34_S2 | 27446 | -0.01237 | 0.006456 | 0.055446 | 1.256127 | Right atrium |
| Ascending_ | Z93_S2 | 24599 | -0.01411 | 0.007437 | 0.057852 | 1.237683 | Aorta |
| LV_circumf | Z37_S1 | 27446 | 0.010851 | 0.005734 | 0.058418 | 1.233455 | Left ventricle |
| LA_minimu | Z12_S5 | 27446 | -0.0114 | 0.00603 | 0.058645 | 1.231767 | Left atrium |
| RV_end_di | Z22_S4 | 27446 | -0.01407 | 0.00745 | 0.058949 | 1.229522 | Right ventricle |
| RV_ejectio | Z78_S4 | 27446 | -0.00856 | 0.004535 | 0.059027 | 1.228951 | Right ventricle |
| RA_ejectio | Z73_S1 | 27446 | 0.008748 | 0.00467 | 0.061043 | 1.214362 | Right atrium |
| LV_radial_ | Z122_S1 | 27446 | 0.010584 | 0.00565 | 0.061047 | 1.214333 | Left ventricle |
| RA_ejectio | Z87_S5 | 27446 | -0.00919 | 0.004918 | 0.061586 | 1.21052 | Right atrium |
| RA_stroke_ | Z91_S1 | 27446 | -0.00912 | 0.004897 | 0.062686 | 1.202827 | Right atrium |
| Descending | Z37_S3 | 24599 | -0.01418 | 0.007618 | 0.06275 | 1.202388 | Aorta |
| LV_ejection | Z51_S4 | 27446 | 0.008074 | 0.004343 | 0.063006 | 1.200616 | Left ventricle |
| Ascending_ | Z34_S2 | 24599 | 0.012914 | 0.006953 | 0.063264 | 1.198846 | Aorta |
| RA_maxim | Z31_S2 | 27446 | 0.010366 | 0.00559 | 0.063671 | 1.196055 | Right atrium |
| Ascending_ | Z61_S4 | 24599 | 0.010651 | 0.005746 | 0.063801 | 1.19517 | Aorta |
| LV_ejection | Z105_S4 | 27446 | 0.008458 | 0.004569 | 0.064124 | 1.192981 | Left ventricle |
| RA_maxim | Z37_S1 | 27446 | 0.011571 | 0.006258 | 0.064454 | 1.19075 | Right atrium |
| Ascending_ | Z38_S5 | 24599 | -0.01124 | 0.006102 | 0.065441 | 1.184147 | Aorta |
| Ascending_ | Z120_S5 | 24599 | -0.0128 | 0.006951 | 0.065547 | 1.183445 | Aorta |
| Descending | Z75_S1 | 24599 | 0.011234 | 0.006125 | 0.06665 | 1.176201 | Aorta |
| LV_mean_ | Z125_S4 | 27446 | -0.01318 | 0.0072 | 0.067176 | 1.172788 | Left ventricle |
| Descending | Z20_S2 | 24599 | 0.009207 | 0.005036 | 0.067483 | 1.170805 | Aorta |
| Descending | Z74_S2 | 24599 | 0.010728 | 0.005878 | 0.067968 | 1.167695 | Aorta |
| LV_circumf | Z40_S5 | 27446 | 0.007936 | 0.004358 | 0.068594 | 1.163714 | Left ventricle |
| LA_minimu | Z7_S3 | 27446 | 0.010926 | 0.00605 | 0.070925 | 1.149204 | Left atrium |
| LV_end_sy | Z32_S5 | 27446 | -0.01352 | 0.00759 | 0.07479 | 1.126156 | Left ventricle |
| LV_mean_ | Z53_S2 | 27446 | 0.013759 | 0.007727 | 0.074954 | 1.125203 | Left ventricle |
| Descending | Z62_S4 | 24599 | -0.01327 | 0.007542 | 0.078467 | 1.105314 | Aorta |
| LV_cardiac | Z27_S2 | 27446 | -0.01139 | 0.006474 | 0.078643 | 1.104341 | Left ventricle |
| RA_stroke_ | Z44_S1 | 27446 | -0.00888 | 0.00505 | 0.078655 | 1.104274 | Right atrium |
| LV_end_sy | Z50_S5 | 27446 | 0.013768 | 0.007831 | 0.078719 | 1.103923 | Left ventricle |
| LV_cardiac | Z50_S3 | 27446 | -0.00911 | 0.005211 | 0.080285 | 1.095367 | Left ventricle |
| LV_radial_ | Z61_S4 | 27446 | 0.008498 | 0.004865 | 0.080677 | 1.09325 | Left ventricle |
| Ascending_ | Z82_S4 | 24599 | -0.00869 | 0.00498 | 0.081079 | 1.091092 | Aorta |

| | | | | | | | |
|---|---|---|---|---|---|---|---|
| RV_ejectio | Z120_S5 | 27446 | -0.01076 | 0.006176 | 0.081507 | 1.088805 | Right ventricle |
| RV_ejectio | Z94_S2 | 27446 | -0.00959 | 0.005514 | 0.081889 | 1.086777 | Right ventricle |
| LV_cardiac | Z13_S3 | 27446 | 0.007865 | 0.004535 | 0.082879 | 1.081556 | Left ventricle |
| RA_minimu | Z37_S1 | 27446 | -0.011 | 0.006343 | 0.082987 | 1.080989 | Right atrium |
| LA_stroke_ | Z3_S2 | 27446 | -0.00813 | 0.004692 | 0.082994 | 1.080952 | Left atrium |
| LV_circumf | Z39_S3 | 27446 | 0.011043 | 0.006377 | 0.083332 | 1.079189 | Left ventricle |
| RV_ejectio | Z75_S5 | 27446 | -0.01107 | 0.006425 | 0.084852 | 1.071338 | Right ventricle |
| Descending | Z93_S2 | 24599 | -0.01257 | 0.007302 | 0.085269 | 1.069208 | Aorta |
| RA_stroke_ | Z29_S5 | 27446 | 0.009302 | 0.005412 | 0.085653 | 1.067259 | Right atrium |
| LV_circumf | Z14_S2 | 27446 | 0.008704 | 0.005077 | 0.086471 | 1.06313 | Left ventricle |
| LV_longitu | Z56_S5 | 27446 | 0.007259 | 0.004242 | 0.087034 | 1.060309 | Left ventricle |
| Ascending_ | Z43_S2 | 24599 | 0.009086 | 0.005323 | 0.087832 | 1.056347 | Aorta |
| RA_maxim | Z23_S3 | 27446 | 0.008417 | 0.004932 | 0.087877 | 1.056123 | Right atrium |
| LV_ejectio | Z112_S3 | 27446 | 0.010114 | 0.005944 | 0.088841 | 1.051386 | Left ventricle |
| LV_mean_ | Z39_S4 | 27446 | 0.010578 | 0.006252 | 0.090679 | 1.042495 | Left ventricle |
| LA_ejectio | Z12_S5 | 27446 | -0.00972 | 0.005779 | 0.092478 | 1.033962 | Left atrium |
| LV_longitu | Z74_S2 | 27446 | -0.00824 | 0.004915 | 0.093596 | 1.028742 | Left ventricle |
| LV_ejectio | Z39_S3 | 27446 | -0.01049 | 0.006276 | 0.094507 | 1.024535 | Left ventricle |
| RA_minimu | Z94_S2 | 27446 | 0.010147 | 0.006072 | 0.094726 | 1.023529 | Right atrium |
| LA_ejectio | Z9_S2 | 27446 | -0.00867 | 0.005202 | 0.095395 | 1.020476 | Left atrium |
| Ascending_ | Z92_S1 | 24599 | -0.01004 | 0.006033 | 0.09616 | 1.017007 | Aorta |
| Descending | Z23_S3 | 24599 | -0.00876 | 0.005288 | 0.097624 | 1.010443 | Aorta |
| LV_myocar | Z39_S3 | 27446 | -0.01735 | 0.010501 | 0.098585 | 1.006191 | Left ventricle |
| Ascending_ | Z30_S2 | 24599 | -0.01152 | 0.00698 | 0.098851 | 1.005021 | Aorta |
| RV_end_sy | Z62_S4 | 27446 | -0.01296 | 0.007857 | 0.099027 | 1.004248 | Right ventricle |
| Ascending_ | Z53_S1 | 24599 | 0.009911 | 0.006018 | 0.099565 | 1.001895 | Aorta |
| RA_maxim | Z26_S4 | 27446 | 0.011333 | 0.006889 | 0.099932 | 1.000296 | Right atrium |
| RV_stroke_ | Z37_S3 | 27446 | -0.01151 | 0.007009 | 0.100472 | 0.997955 | Right ventricle |
| Descending | Z120_S5 | 24599 | 0.013113 | 0.008077 | 0.10447 | 0.981007 | Aorta |
| RV_end_di | Z28_S2 | 27446 | 0.01244 | 0.007662 | 0.10448 | 0.980967 | Right ventricle |
| LV_longitu | Z62_S2 | 27446 | -0.00714 | 0.004406 | 0.105079 | 0.978483 | Left ventricle |
| RV_end_di | Z121_S4 | 27446 | 0.013137 | 0.008112 | 0.105359 | 0.97733 | Right ventricle |
| LV_circumf | Z9_S2 | 27446 | -0.00874 | 0.005443 | 0.108471 | 0.964686 | Left ventricle |
| LV_cardiac | Z52_S4 | 27446 | -0.00798 | 0.004975 | 0.108484 | 0.964634 | Left ventricle |
| RA_minimu | Z99_S4 | 27446 | -0.01008 | 0.006293 | 0.10934 | 0.96122 | Right atrium |
| RV_ejectio | Z7_S1 | 27446 | 0.007507 | 0.004695 | 0.109847 | 0.959211 | Right ventricle |
| RA_stroke_ | Z53_S2 | 27446 | -0.00895 | 0.005603 | 0.110015 | 0.958548 | Right atrium |
| Ascending_ | Z92_S5 | 24599 | 0.011075 | 0.006953 | 0.11119 | 0.953933 | Aorta |
| RA_maxim | Z27_S2 | 27446 | 0.010287 | 0.00647 | 0.111822 | 0.951474 | Right atrium |
| LV_mean_ | Z64_S1 | 27446 | -0.01229 | 0.007756 | 0.113128 | 0.94643 | Left ventricle |
| Ascending_ | Z62_S3 | 24599 | 0.009514 | 0.006013 | 0.113583 | 0.944688 | Aorta |
| LV_circumf | Z51_S4 | 27446 | 0.00695 | 0.004413 | 0.11529 | 0.938207 | Left ventricle |
| RA_maxim | Z99_S4 | 27446 | 0.009768 | 0.006209 | 0.115668 | 0.936787 | Right atrium |
| Ascending_ | Z17_S1 | 24599 | -0.0089 | 0.00567 | 0.116445 | 0.93388 | Aorta |
| Ascending_ | Z52_S3 | 24599 | -0.00812 | 0.005177 | 0.116744 | 0.932766 | Aorta |

| | | | | | | | |
|---|---|---|---|---|---|---|---|
| LV_stroke_ | Z56_S5 | 27446 | -0.00812 | 0.005176 | 0.116891 | 0.932218 | Left ventricle |
| LV_cardiac | Z24_S1 | 27446 | 0.007357 | 0.004716 | 0.11879 | 0.925221 | Left ventricle |
| LV_longitu | Z39_S3 | 27446 | -0.00971 | 0.006229 | 0.119201 | 0.923721 | Left ventricle |
| Ascending_ | Z97_S2 | 24599 | 0.009513 | 0.006106 | 0.119211 | 0.923684 | Aorta |
| LV_cardiac | Z44_S4 | 27446 | 0.007329 | 0.004709 | 0.119616 | 0.922211 | Left ventricle |
| LV_mean_ | Z127_S5 | 27446 | 0.009236 | 0.005946 | 0.120358 | 0.919526 | Left ventricle |
| Ascending_ | Z99_S5 | 24599 | 0.009053 | 0.00583 | 0.120478 | 0.919092 | Aorta |
| RV_end_di | Z7_S5 | 27446 | 0.012188 | 0.007861 | 0.121048 | 0.917043 | Right ventricle |
| Ascending_ | Z74_S4 | 24599 | -0.0094 | 0.006067 | 0.12143 | 0.915675 | Aorta |
| LV_longitu | Z73_S1 | 27446 | -0.00719 | 0.004661 | 0.123169 | 0.909498 | Left ventricle |
| LA_ejectio | Z13_S1 | 27446 | -0.00672 | 0.004357 | 0.123219 | 0.909323 | Left atrium |
| Ascending_ | Z75_S4 | 24599 | -0.0111 | 0.007207 | 0.123465 | 0.908455 | Aorta |
| RA_ejectio | Z108_S5 | 27446 | -0.00786 | 0.005104 | 0.123652 | 0.907798 | Right atrium |
| Descending | Z64_S1 | 24599 | -0.0102 | 0.006646 | 0.124951 | 0.90326 | Aorta |
| LV_mean_ | Z76_S3 | 27446 | -0.01121 | 0.007353 | 0.127286 | 0.895221 | Left ventricle |
| LV_cardiac | Z78_S4 | 27446 | -0.0075 | 0.004931 | 0.127994 | 0.89281 | Left ventricle |
| Ascending_ | Z39_S1 | 24599 | 0.008526 | 0.005611 | 0.128648 | 0.890596 | Aorta |
| Ascending_ | Z60_S2 | 24599 | 0.008011 | 0.005293 | 0.130147 | 0.885564 | Aorta |
| LV_ejectio | Z1_S4 | 27446 | 0.007983 | 0.005279 | 0.130483 | 0.884445 | Left ventricle |
| RV_end_di | Z32_S5 | 27446 | 0.013028 | 0.008656 | 0.13229 | 0.878472 | Right ventricle |
| RA_ejectio | Z8_S3 | 27446 | -0.00637 | 0.004235 | 0.132706 | 0.877108 | Right atrium |
| LV_mean_ | Z55_S5 | 27446 | 0.008946 | 0.00596 | 0.133347 | 0.875016 | Left ventricle |
| LV_myocar | Z72_S5 | 27446 | -0.01548 | 0.010331 | 0.133932 | 0.873117 | Left ventricle |
| LV_mean_ | Z59_S2 | 27446 | -0.01194 | 0.007973 | 0.134293 | 0.871948 | Left ventricle |
| LV_ejectio | Z1_S1 | 27446 | 0.007285 | 0.004879 | 0.135425 | 0.868302 | Left ventricle |
| RV_end_sy | Z37_S4 | 27446 | -0.01268 | 0.008494 | 0.135602 | 0.867733 | Right ventricle |
| RA_minimu | Z75_S1 | 27446 | -0.00857 | 0.005803 | 0.139921 | 0.854118 | Right atrium |
| RA_ejectio | Z114_S4 | 27446 | 0.007153 | 0.004849 | 0.140149 | 0.853411 | Right atrium |
| RV_ejectio | Z3_S2 | 27446 | 0.006724 | 0.004561 | 0.140397 | 0.852642 | Right ventricle |
| LV_mean_ | Z83_S2 | 27446 | -0.00959 | 0.006588 | 0.14533 | 0.837644 | Left ventricle |
| LV_mean_ | Z93_S2 | 27446 | -0.01234 | 0.008568 | 0.149707 | 0.824758 | Left ventricle |
| LA_ejectio | Z100_S3 | 27446 | 0.007199 | 0.004999 | 0.149832 | 0.824396 | Left atrium |
| LV_mean_ | Z112_S3 | 27446 | -0.01189 | 0.008282 | 0.151126 | 0.820662 | Left ventricle |
| Ascending_ | Z96_S3 | 24599 | 0.008457 | 0.005903 | 0.151962 | 0.818265 | Aorta |
| LV_mean_ | Z12_S5 | 27446 | 0.01183 | 0.008292 | 0.153662 | 0.813433 | Left ventricle |
| Descending | Z38_S5 | 24599 | 0.010115 | 0.00709 | 0.153692 | 0.81335 | Aorta |
| LV_cardiac | Z97_S2 | 27446 | 0.008355 | 0.005878 | 0.155201 | 0.809104 | Left ventricle |
| LV_mean_ | Z6_S2 | 27446 | -0.0077 | 0.005432 | 0.156562 | 0.805314 | Left ventricle |
| Ascending_ | Z111_S1 | 24599 | -0.00895 | 0.006321 | 0.156797 | 0.804661 | Aorta |
| LV_mean_ | Z118_S1 | 27446 | -0.01097 | 0.007768 | 0.157945 | 0.801494 | Left ventricle |
| LV_radial_ | Z62_S4 | 27446 | 0.00833 | 0.005915 | 0.159048 | 0.798473 | Left ventricle |
| Descending | Z22_S4 | 24599 | 0.009615 | 0.006836 | 0.159562 | 0.797069 | Aorta |
| LV_cardiac | Z38_S4 | 27446 | 0.008953 | 0.006381 | 0.160551 | 0.794387 | Left ventricle |
| LV_mean_ | Z50_S3 | 27446 | -0.0091 | 0.006542 | 0.164086 | 0.784928 | Left ventricle |
| RA_maxim | Z53_S2 | 27446 | 0.008534 | 0.00615 | 0.165271 | 0.781804 | Right atrium |

| | | | | | | | |
|---|---|---|---|---|---|---|---|
| LV_circumf | Z37_S3 | 27446 | -0.00804 | 0.005796 | 0.165522 | 0.781144 | Left ventricle |
| RV_end_sy | Z32_S5 | 27446 | -0.01141 | 0.008254 | 0.166788 | 0.777836 | Right ventricle |
| LA_minimu | Z28_S1 | 27446 | -0.0074 | 0.005364 | 0.167984 | 0.774732 | Left atrium |
| RV_ejectio | Z34_S2 | 27446 | -0.00807 | 0.005863 | 0.168831 | 0.772548 | Right ventricle |
| RV_ejectio | Z92_S1 | 27446 | 0.00699 | 0.005085 | 0.169312 | 0.771311 | Right ventricle |
| RV_ejectio | Z5_S1 | 27446 | -0.00692 | 0.005045 | 0.170013 | 0.769517 | Right ventricle |
| Ascending_ | Z28_S2 | 24599 | 0.008456 | 0.006164 | 0.170116 | 0.769255 | Aorta |
| RA_ejectio | Z65_S1 | 27446 | 0.005671 | 0.004134 | 0.170135 | 0.769206 | Right atrium |
| LV_cardiac | Z14_S1 | 27446 | -0.00716 | 0.005228 | 0.170901 | 0.767255 | Left ventricle |
| Descending | Z21_S3 | 24599 | -0.01145 | 0.008369 | 0.171438 | 0.765892 | Aorta |
| LV_circumf | Z109_S1 | 27446 | -0.00696 | 0.005105 | 0.172998 | 0.761958 | Left ventricle |
| Descending | Z39_S1 | 24599 | 0.007491 | 0.005509 | 0.173933 | 0.759619 | Aorta |
| LV_circumf | Z1_S1 | 27446 | 0.006695 | 0.004957 | 0.176858 | 0.752376 | Left ventricle |
| RV_ejectio | Z19_S2 | 27446 | 0.0068 | 0.005047 | 0.177888 | 0.749853 | Right ventricle |
| LV_cardiac | Z19_S2 | 27446 | -0.00736 | 0.005488 | 0.179866 | 0.745052 | Left ventricle |
| LV_myocar | Z23_S1 | 27446 | 0.012607 | 0.009416 | 0.18061 | 0.743259 | Left ventricle |
| Ascending_ | Z51_S1 | 24599 | 0.007108 | 0.005329 | 0.182311 | 0.739188 | Aorta |
| LV_mean_ | Z68_S2 | 27446 | -0.00842 | 0.006354 | 0.18507 | 0.732663 | Left ventricle |
| RV_ejectio | Z46_S1 | 27446 | 0.006833 | 0.005176 | 0.186821 | 0.728575 | Right ventricle |
| LA_stroke_ | Z37_S3 | 27446 | 0.007876 | 0.00599 | 0.18856 | 0.724551 | Left atrium |
| LV_circumf | Z7_S1 | 27446 | -0.00613 | 0.004674 | 0.189416 | 0.722584 | Left ventricle |
| LV_ejectio | Z72_S5 | 27446 | 0.008093 | 0.006175 | 0.189997 | 0.721254 | Left ventricle |
| LV_circumf | Z37_S2 | 27446 | 0.007225 | 0.005527 | 0.191112 | 0.718711 | Left ventricle |
| LV_radial_ | Z28_S2 | 27446 | -0.00712 | 0.0055 | 0.195292 | 0.709315 | Left ventricle |
| Ascending_ | Z62_S4 | 24599 | -0.00852 | 0.006584 | 0.19563 | 0.708565 | Aorta |
| Descending | Z50_S2 | 24599 | -0.00937 | 0.007301 | 0.199588 | 0.699865 | Aorta |
| LV_longitu | Z30_S2 | 27446 | -0.00771 | 0.006032 | 0.20111 | 0.696566 | Left ventricle |
| LV_myocar | Z40_S4 | 27446 | 0.012923 | 0.010121 | 0.201632 | 0.69544 | Left ventricle |
| LV_end_di | Z78_S1 | 27446 | 0.007936 | 0.006231 | 0.202808 | 0.692915 | Left ventricle |
| LV_mean_ | Z49_S3 | 27446 | -0.0103 | 0.008098 | 0.203225 | 0.692024 | Left ventricle |
| LV_longitu | Z22_S4 | 27446 | 0.006553 | 0.005153 | 0.203447 | 0.691549 | Left ventricle |
| LV_radial_ | Z92_S5 | 27446 | 0.007889 | 0.006227 | 0.205166 | 0.687894 | Left ventricle |
| LV_radial_ | Z39_S3 | 27446 | 0.008179 | 0.006465 | 0.205867 | 0.686413 | Left ventricle |
| Ascending_ | Z115_S3 | 24599 | 0.00809 | 0.006466 | 0.210908 | 0.675907 | Aorta |
| Descending | Z11_S5 | 24599 | -0.00717 | 0.005738 | 0.211666 | 0.67435 | Aorta |
| LV_ejectio | Z125_S4 | 27446 | -0.00644 | 0.005168 | 0.212588 | 0.672461 | Left ventricle |
| RA_maxim | Z1_S4 | 27446 | 0.00729 | 0.005854 | 0.213039 | 0.67154 | Right atrium |
| Descending | Z39_S4 | 24599 | -0.00664 | 0.005346 | 0.214482 | 0.668608 | Aorta |
| RA_maxim | Z77_S3 | 27446 | -0.00703 | 0.005666 | 0.214933 | 0.667697 | Right atrium |
| Ascending_ | Z106_S4 | 24599 | 0.006598 | 0.005332 | 0.215854 | 0.665839 | Aorta |
| RA_minimu | Z28_S2 | 27446 | 0.007416 | 0.006001 | 0.216539 | 0.664463 | Right atrium |
| Descending | Z9_S2 | 24599 | 0.007907 | 0.0064 | 0.216658 | 0.664226 | Aorta |
| LV_end_di | Z32_S5 | 27446 | -0.01015 | 0.008224 | 0.216963 | 0.663615 | Left ventricle |
| LV_cardiac | Z73_S4 | 27446 | 0.007888 | 0.00639 | 0.216996 | 0.663547 | Left ventricle |
| RV_end_sy | Z112_S3 | 27446 | 0.010016 | 0.008134 | 0.218149 | 0.661246 | Right ventricle |

| | | | | | | | |
|---|---|---|---|---|---|---|---|
| Ascending_ | Z120_S5 | 24599 | 0.008995 | 0.007341 | 0.220475 | 0.65664 | Aorta |
| RA_stroke_ | Z38_S4 | 27446 | 0.007104 | 0.005809 | 0.221351 | 0.654919 | Right atrium |
| LV_cardiac | Z99_S5 | 27446 | -0.00652 | 0.00534 | 0.221767 | 0.654104 | Left ventricle |
| Descending | Z111_S1 | 24599 | -0.00892 | 0.007318 | 0.222711 | 0.652259 | Aorta |
| LV_ejectio | Z41_S2 | 27446 | 0.007215 | 0.005925 | 0.223326 | 0.651061 | Left ventricle |
| LV_ejectio | Z80_S5 | 27446 | 0.007202 | 0.005914 | 0.223348 | 0.651019 | Left ventricle |
| RV_ejectio | Z105_S4 | 27446 | -0.00566 | 0.004663 | 0.224518 | 0.648749 | Right ventricle |
| LV_mean_ | Z115_S4 | 27446 | 0.007746 | 0.0064 | 0.226161 | 0.645582 | Left ventricle |
| LV_circumf | Z66_S3 | 27446 | 0.006966 | 0.005775 | 0.227761 | 0.64252 | Left ventricle |
| LA_ejectio | Z56_S5 | 27446 | 0.004997 | 0.004151 | 0.228601 | 0.640921 | Left atrium |
| LA_ejectio | Z32_S5 | 27446 | 0.007029 | 0.005857 | 0.230131 | 0.638026 | Left atrium |
| RV_ejectio | Z21_S3 | 27446 | 0.007668 | 0.006408 | 0.231442 | 0.635558 | Right ventricle |
| RA_ejectio | Z78_S1 | 27446 | -0.00544 | 0.004545 | 0.231564 | 0.635329 | Right atrium |
| LV_mean_ | Z12_S4 | 27446 | -0.00713 | 0.005964 | 0.231975 | 0.634559 | Left ventricle |
| RA_ejectio | Z18_S4 | 27446 | 0.007419 | 0.006223 | 0.233192 | 0.632286 | Right atrium |
| RA_ejectio | Z51_S3 | 27446 | 0.007253 | 0.006086 | 0.233385 | 0.631927 | Right atrium |
| RA_ejectio | Z75_S5 | 27446 | 0.007453 | 0.00626 | 0.233784 | 0.631185 | Right atrium |
| LV_longitu | Z1_S2 | 27446 | -0.00481 | 0.004072 | 0.237255 | 0.624785 | Left ventricle |
| RA_ejectio | Z11_S5 | 27446 | 0.005603 | 0.004793 | 0.242452 | 0.615375 | Right atrium |
| RA_ejectio | Z122_S5 | 27446 | -0.00523 | 0.004482 | 0.2437 | 0.613144 | Right atrium |
| LV_ejectio | Z47_S4 | 27446 | -0.00583 | 0.005006 | 0.244193 | 0.612267 | Left ventricle |
| RV_ejectio | Z39_S3 | 27446 | -0.00742 | 0.006406 | 0.247034 | 0.607243 | Right ventricle |
| LV_mean_ | Z108_S5 | 27446 | 0.008271 | 0.007151 | 0.247412 | 0.60658 | Left ventricle |
| LV_cardiac | Z41_S1 | 27446 | 0.006888 | 0.006009 | 0.251688 | 0.599137 | Left ventricle |
| LV_mean_ | Z24_S5 | 27446 | 0.008719 | 0.007609 | 0.251849 | 0.59886 | Left ventricle |
| LV_myocar | Z120_S5 | 27446 | -0.0116 | 0.010124 | 0.25204 | 0.59853 | Left ventricle |
| LV_stroke_ | Z72_S5 | 27446 | 0.00856 | 0.007477 | 0.252264 | 0.598144 | Left ventricle |
| LV_mean_ | Z14_S1 | 27446 | 0.007501 | 0.006563 | 0.253058 | 0.596779 | Left ventricle |
| LV_circumf | Z51_S3 | 27446 | -0.00709 | 0.006219 | 0.254535 | 0.594252 | Left ventricle |
| LA_maximu | Z37_S3 | 27446 | 0.006787 | 0.00596 | 0.254792 | 0.593815 | Left atrium |
| LA_stroke_ | Z53_S2 | 27446 | -0.00663 | 0.005824 | 0.255062 | 0.593354 | Left atrium |
| RA_ejectio | Z57_S2 | 27446 | 0.004706 | 0.004138 | 0.255432 | 0.592724 | Right atrium |
| LV_mean_ | Z58_S2 | 27446 | 0.007417 | 0.006547 | 0.257298 | 0.589564 | Left ventricle |
| RA_ejectio | Z41_S1 | 27446 | -0.00605 | 0.005384 | 0.261131 | 0.583141 | Right atrium |
| LV_cardiac | Z112_S3 | 27446 | 0.00738 | 0.006597 | 0.263238 | 0.579651 | Left ventricle |
| RV_end_sy | Z112_S1 | 27446 | 0.009546 | 0.008538 | 0.263554 | 0.579131 | Right ventricle |
| LV_ejectio | Z28_S2 | 27446 | 0.005949 | 0.00534 | 0.265218 | 0.576397 | Left ventricle |
| LV_circumf | Z61_S4 | 27446 | -0.00534 | 0.004799 | 0.266096 | 0.574961 | Left ventricle |
| LV_circumf | Z21_S3 | 27446 | -0.00704 | 0.006379 | 0.269488 | 0.569461 | Left ventricle |
| RV_stroke_ | Z80_S5 | 27446 | -0.00801 | 0.007268 | 0.270483 | 0.56786 | Right ventricle |
| LA_ejectio | Z122_S5 | 27446 | 0.004789 | 0.004377 | 0.273904 | 0.562402 | Left atrium |
| LA_ejectio | Z97_S2 | 27446 | -0.00563 | 0.005143 | 0.274045 | 0.562179 | Left atrium |
| Descending | Z100_S3 | 24599 | 0.006704 | 0.00613 | 0.274132 | 0.56204 | Aorta |
| LV_ejectio | Z37_S3 | 27446 | -0.00623 | 0.005704 | 0.274851 | 0.560902 | Left ventricle |
| LV_cardiac | Z57_S1 | 27446 | -0.0056 | 0.005132 | 0.27559 | 0.559737 | Left ventricle |

| | | | | | | | |
|---|---|---|---|---|---|---|---|
| Descending | Z52_S3 | 24599 | 0.005537 | 0.005083 | 0.275981 | 0.55912 | Aorta |
| LV_mean_ | Z7_S1 | 27446 | 0.006975 | 0.006409 | 0.276458 | 0.558371 | Left ventricle |
| LV_ejection | Z61_S4 | 27446 | -0.00514 | 0.004723 | 0.276578 | 0.558183 | Left ventricle |
| LV_ejection | Z32_S5 | 27446 | 0.006549 | 0.006032 | 0.277643 | 0.556513 | Left ventricle |
| Descending | Z93_S1 | 24599 | 0.007403 | 0.00685 | 0.279776 | 0.553189 | Aorta |
| LV_mean_ | Z31_S5 | 27446 | -0.00605 | 0.005618 | 0.281254 | 0.550901 | Left ventricle |
| LV_mean_ | Z50_S2 | 27446 | -0.00917 | 0.008542 | 0.282885 | 0.548391 | Left ventricle |
| LV_mean_ | Z80_S4 | 27446 | -0.00646 | 0.006024 | 0.283398 | 0.547603 | Left ventricle |
| RV_end_sy | Z1_S5 | 27446 | -0.00749 | 0.006999 | 0.284722 | 0.545579 | Right ventricle |
| LV_mean_ | Z21_S3 | 27446 | 0.009322 | 0.008747 | 0.286539 | 0.542816 | Left ventricle |
| Ascending_ | Z11_S5 | 24599 | 0.006215 | 0.005845 | 0.287648 | 0.541138 | Aorta |
| Descending | Z122_S5 | 24599 | -0.00569 | 0.005361 | 0.288876 | 0.539288 | Aorta |
| Ascending_ | Z23_S1 | 24599 | 0.00689 | 0.006522 | 0.290757 | 0.53647 | Aorta |
| LV_mean_ | Z78_S4 | 27446 | 0.006536 | 0.00619 | 0.291014 | 0.536086 | Left ventricle |
| Descending | Z112_S3 | 24599 | -0.00836 | 0.00794 | 0.292331 | 0.534126 | Aorta |
| RV_ejectio | Z111_S5 | 27446 | 0.005327 | 0.005072 | 0.29362 | 0.532214 | Right ventricle |
| Ascending_ | Z41_S1 | 24599 | -0.00653 | 0.006216 | 0.293713 | 0.532076 | Aorta |
| LV_ejection | Z37_S1 | 27446 | -0.00592 | 0.005643 | 0.294151 | 0.53143 | Left ventricle |
| LV_ejection | Z5_S1 | 27446 | 0.00514 | 0.004943 | 0.298365 | 0.525252 | Left ventricle |
| LV_cardiac | Z77_S3 | 27446 | -0.00589 | 0.00567 | 0.298936 | 0.524422 | Left ventricle |
| RA_stroke_ | Z125_S4 | 27446 | 0.005416 | 0.005222 | 0.299636 | 0.523405 | Right atrium |
| Ascending_ | Z114_S5 | 24599 | -0.005 | 0.004856 | 0.303529 | 0.5178 | Aorta |
| LV_mean_ | Z20_S1 | 27446 | 0.008334 | 0.008123 | 0.304933 | 0.515796 | Left ventricle |
| Descending | Z65_S5 | 24599 | 0.007271 | 0.007112 | 0.306601 | 0.513427 | Aorta |
| LV_end_di | Z120_S5 | 27446 | 0.008412 | 0.00825 | 0.307857 | 0.511652 | Left ventricle |
| RV_ejectio | Z9_S2 | 27446 | 0.005573 | 0.005468 | 0.308078 | 0.511339 | Right ventricle |
| LV_mean_ | Z105_S4 | 27446 | -0.00648 | 0.006366 | 0.308735 | 0.510414 | Left ventricle |
| LV_stroke_ | Z65_S5 | 27446 | 0.007281 | 0.007228 | 0.31377 | 0.503389 | Left ventricle |
| LV_radial_ | Z41_S1 | 27446 | -0.00561 | 0.005578 | 0.31421 | 0.50278 | Left ventricle |
| LA_ejection | Z121_S4 | 27446 | 0.005487 | 0.005489 | 0.317518 | 0.498231 | Left atrium |
| LV_cardiac | Z41_S2 | 27446 | 0.006538 | 0.006576 | 0.320067 | 0.49476 | Left ventricle |
| Ascending_ | Z63_S1 | 24599 | -0.00682 | 0.006861 | 0.32025 | 0.494511 | Aorta |
| Ascending_ | Z23_S1 | 24599 | 0.006453 | 0.006498 | 0.320674 | 0.493936 | Aorta |
| LV_mean_ | Z63_S1 | 27446 | 0.008204 | 0.008298 | 0.322843 | 0.491008 | Left ventricle |
| LV_myocar | Z57_S2 | 27446 | -0.00686 | 0.006963 | 0.324657 | 0.488576 | Left ventricle |
| LV_radial_ | Z29_S1 | 27446 | -0.00543 | 0.005592 | 0.33113 | 0.480001 | Left ventricle |
| LV_ejection | Z97_S1 | 27446 | 0.005944 | 0.006118 | 0.331282 | 0.479802 | Left ventricle |
| RA_ejectio | Z80_S2 | 27446 | -0.0043 | 0.004439 | 0.333176 | 0.477327 | Right atrium |
| RV_end_sy | Z27_S2 | 27446 | -0.00768 | 0.007983 | 0.33612 | 0.473505 | Right ventricle |
| Descending | Z121_S4 | 24599 | 0.006414 | 0.006719 | 0.339787 | 0.468793 | Aorta |
| RV_ejectio | Z123_S4 | 27446 | 0.005114 | 0.00536 | 0.339983 | 0.468543 | Right ventricle |
| LV_end_di | Z32_S1 | 27446 | 0.006919 | 0.007255 | 0.340283 | 0.468159 | Left ventricle |
| LV_circumf | Z28_S4 | 27446 | 0.005207 | 0.005485 | 0.342458 | 0.465393 | Left ventricle |
| LV_radial_ | Z1_S5 | 27446 | 0.005001 | 0.005269 | 0.342579 | 0.46524 | Left ventricle |
| LV_mean_ | Z28_S1 | 27446 | 0.006986 | 0.007377 | 0.3436 | 0.463947 | Left ventricle |

| | | | | | | | |
|---|---|---|---|---|---|---|---|
| RV_ejectio | Z5_S5 | 27446 | 0.004974 | 0.00527 | 0.34529 | 0.461817 | Right ventricle |
| Descending | Z8_S3 | 24599 | -0.00476 | 0.005065 | 0.34772 | 0.45877 | Aorta |
| LV_mean_ | Z92_S1 | 27446 | -0.00649 | 0.006942 | 0.349885 | 0.456075 | Left ventricle |
| RA_minimu | Z65_S1 | 27446 | 0.004367 | 0.004673 | 0.350043 | 0.455879 | Right atrium |
| LV_mean_ | Z100_S3 | 27446 | -0.0067 | 0.007172 | 0.350272 | 0.455595 | Left ventricle |
| Descending | Z43_S2 | 24599 | -0.00487 | 0.005226 | 0.351384 | 0.454219 | Aorta |
| RV_stroke_ | Z53_S2 | 27446 | -0.00635 | 0.006815 | 0.351832 | 0.453664 | Right ventricle |
| RA_stroke_ | Z105_S5 | 27446 | -0.00469 | 0.005124 | 0.359853 | 0.443874 | Right atrium |
| Descending | Z95_S3 | 24599 | 0.004203 | 0.004595 | 0.360312 | 0.443321 | Aorta |
| Descending | Z14_S4 | 24599 | -0.00684 | 0.007478 | 0.360458 | 0.443145 | Aorta |
| Descending | Z3_S4 | 24599 | 0.007606 | 0.008329 | 0.361171 | 0.442288 | Aorta |
| LA_ejectio | Z87_S5 | 27446 | 0.004374 | 0.004803 | 0.362381 | 0.440834 | Left atrium |
| RV_ejectio | Z94_S5 | 27446 | -0.00397 | 0.004413 | 0.368698 | 0.43333 | Right ventricle |
| LV_longitu | Z38_S5 | 27446 | 0.004703 | 0.005248 | 0.370141 | 0.431633 | Left ventricle |
| LV_mean_ | Z37_S4 | 27446 | 0.007751 | 0.008648 | 0.370142 | 0.431631 | Left ventricle |
| LV_cardiac | Z86_S3 | 27446 | 0.005602 | 0.006294 | 0.373436 | 0.427784 | Left ventricle |
| LV_circumf | Z29_S5 | 27446 | 0.004836 | 0.005443 | 0.374248 | 0.426841 | Left ventricle |
| LV_mean_ | Z5_S5 | 27446 | 0.006361 | 0.007194 | 0.376595 | 0.424125 | Left ventricle |
| Ascending_ | Z3_S2 | 24599 | 0.004797 | 0.005431 | 0.377095 | 0.42355 | Aorta |
| RA_stroke_ | Z5_S5 | 27446 | 0.004573 | 0.005217 | 0.380671 | 0.419451 | Right atrium |
| LV_radial_ | Z111_S1 | 27446 | 0.004923 | 0.00563 | 0.381897 | 0.418054 | Left ventricle |
| RA_minimu | Z62_S3 | 27446 | -0.00512 | 0.005899 | 0.385154 | 0.414365 | Right atrium |
| LV_circumf | Z31_S5 | 27446 | 0.003511 | 0.004097 | 0.391468 | 0.407303 | Left ventricle |
| RA_maxim | Z95_S5 | 27446 | -0.00572 | 0.006697 | 0.392788 | 0.405842 | Right atrium |
| RA_ejectio | Z3_S2 | 27446 | -0.00378 | 0.004443 | 0.39526 | 0.403117 | Right atrium |
| LV_ejectio | Z44_S3 | 27446 | 0.004692 | 0.005533 | 0.396432 | 0.401831 | Left ventricle |
| LV_radial_ | Z37_S1 | 27446 | 0.004923 | 0.005813 | 0.397058 | 0.401146 | Left ventricle |
| LV_cardiac | Z1_S4 | 27446 | -0.00493 | 0.005859 | 0.400206 | 0.397717 | Left ventricle |
| LV_radial_ | Z30_S2 | 27446 | 0.005247 | 0.006261 | 0.402015 | 0.395758 | Left ventricle |
| LV_radial_ | Z106_S3 | 27446 | -0.00509 | 0.00609 | 0.403566 | 0.394085 | Left ventricle |
| LV_mean_ | Z31_S2 | 27446 | 0.005864 | 0.007023 | 0.403711 | 0.393929 | Left ventricle |
| LV_ejectio | Z46_S1 | 27446 | 0.004222 | 0.005071 | 0.405146 | 0.392388 | Left ventricle |
| RA_ejectio | Z23_S3 | 27446 | 0.003665 | 0.004422 | 0.407205 | 0.390187 | Right atrium |
| RA_stroke_ | Z8_S3 | 27446 | -0.00356 | 0.004303 | 0.408285 | 0.389036 | Right atrium |
| LA_ejectio | Z115_S3 | 27446 | 0.004275 | 0.005178 | 0.409033 | 0.388242 | Left atrium |
| Descending | Z20_S1 | 24599 | 0.005672 | 0.006931 | 0.413151 | 0.383891 | Aorta |
| Ascending_ | Z62_S3 | 24599 | 0.004934 | 0.006035 | 0.413581 | 0.383439 | Aorta |
| Ascending_ | Z26_S4 | 24599 | -0.00612 | 0.007536 | 0.416635 | 0.380244 | Aorta |
| LV_mean_ | Z79_S4 | 27446 | -0.00666 | 0.008228 | 0.418172 | 0.378645 | Left ventricle |
| RA_stroke_ | Z31_S2 | 27446 | 0.004106 | 0.005093 | 0.420146 | 0.3766 | Right atrium |
| LA_ejectio | Z51_S3 | 27446 | -0.00478 | 0.005944 | 0.420833 | 0.37589 | Left atrium |
| LV_cardiac | Z62_S1 | 27446 | -0.00418 | 0.005209 | 0.42221 | 0.374471 | Left ventricle |
| RV_end_di | Z26_S3 | 27446 | 0.00647 | 0.008096 | 0.424187 | 0.372443 | Right ventricle |
| Ascending_ | Z23_S1 | 24599 | 0.00548 | 0.006863 | 0.424617 | 0.372002 | Aorta |
| LV_longitu | Z99_S3 | 27446 | -0.00319 | 0.004013 | 0.426757 | 0.369819 | Left ventricle |

| | | | | | | | |
|---|---|---|---|---|---|---|---|
| LA_maximu | Z7_S3 | 27446 | -0.00493 | 0.00624 | 0.429136 | 0.367405 | Left atrium |
| LV_radial_s | Z24_S5 | 27446 | 0.004444 | 0.005626 | 0.42957 | 0.366966 | Left ventricle |
| RA_stroke_ | Z115_S4 | 27446 | 0.00366 | 0.004641 | 0.430356 | 0.366172 | Right atrium |
| LV_mean_ | Z91_S1 | 27446 | -0.00526 | 0.006753 | 0.436351 | 0.360164 | Left ventricle |
| LV_mean_ | Z50_S4 | 27446 | -0.00525 | 0.006756 | 0.436834 | 0.359684 | Left ventricle |
| LV_mean_ | Z15_S2 | 27446 | 0.005791 | 0.007451 | 0.437039 | 0.35948 | Left ventricle |
| Descending | Z115_S3 | 24599 | 0.004931 | 0.006349 | 0.437291 | 0.35923 | Aorta |
| Descending | Z114_S4 | 24599 | -0.00449 | 0.005798 | 0.439033 | 0.357503 | Aorta |
| Descending | Z31_S2 | 24599 | -0.00459 | 0.005991 | 0.443869 | 0.352745 | Aorta |
| Descending | Z49_S1 | 24599 | -0.00624 | 0.008167 | 0.445137 | 0.351507 | Aorta |
| LV_mean_ | Z75_S5 | 27446 | 0.006682 | 0.008771 | 0.446138 | 0.350531 | Left ventricle |
| LV_end_sy | Z22_S2 | 27446 | -0.00487 | 0.0064 | 0.446575 | 0.350106 | Left ventricle |
| RV_stroke_ | Z72_S5 | 27446 | 0.005766 | 0.007588 | 0.447326 | 0.349375 | Right ventricle |
| RA_ejectio | Z74_S4 | 27446 | -0.00397 | 0.00523 | 0.448174 | 0.348553 | Right atrium |
| RV_ejectio | Z37_S1 | 27446 | 0.004352 | 0.00576 | 0.449956 | 0.34683 | Right ventricle |
| LV_mean_ | Z43_S2 | 27446 | -0.00461 | 0.006117 | 0.450561 | 0.346247 | Left ventricle |
| RA_ejectio | Z49_S3 | 27446 | -0.00436 | 0.005779 | 0.451016 | 0.345808 | Right atrium |
| LV_radial_s | Z97_S2 | 27446 | 0.004093 | 0.005456 | 0.453108 | 0.343799 | Left ventricle |
| LV_mean_ | Z96_S3 | 27446 | -0.00509 | 0.006798 | 0.454227 | 0.342727 | Left ventricle |
| Descending | Z27_S2 | 24599 | 0.005733 | 0.007697 | 0.456368 | 0.340685 | Aorta |
| RV_ejectio | Z127_S5 | 27446 | -0.00324 | 0.004356 | 0.457163 | 0.339929 | Right ventricle |
| RA_ejectio | Z37_S4 | 27446 | -0.00458 | 0.006173 | 0.457688 | 0.33943 | Right atrium |
| Ascending_ | Z1_S4 | 24599 | -0.00476 | 0.006418 | 0.458026 | 0.33911 | Aorta |
| RA_minimu | Z59_S2 | 27446 | -0.00477 | 0.006432 | 0.458106 | 0.339034 | Right atrium |
| Descending | Z37_S4 | 24599 | -0.00613 | 0.008268 | 0.458373 | 0.338781 | Aorta |
| Ascending_ | Z20_S2 | 24599 | -0.00379 | 0.005129 | 0.459917 | 0.337321 | Aorta |
| RV_end_sy | Z118_S1 | 27446 | 0.005628 | 0.00763 | 0.460683 | 0.336598 | Right ventricle |
| RV_ejectio | Z40_S5 | 27446 | 0.00318 | 0.004378 | 0.46756 | 0.330163 | Right ventricle |
| RV_stroke_ | Z44_S1 | 27446 | 0.00445 | 0.006142 | 0.468696 | 0.329109 | Right ventricle |
| LA_minimu | Z37_S3 | 27446 | 0.004164 | 0.005779 | 0.471162 | 0.32683 | Left atrium |
| LV_longitu | Z93_S2 | 27446 | 0.004286 | 0.006103 | 0.482492 | 0.31651 | Left ventricle |
| LV_mean_ | Z80_S2 | 27446 | 0.004364 | 0.00622 | 0.482937 | 0.316109 | Left ventricle |
| RV_ejectio | Z51_S4 | 27446 | -0.00311 | 0.004433 | 0.483126 | 0.31594 | Right ventricle |
| LV_mean_ | Z89_S4 | 27446 | 0.004712 | 0.006751 | 0.485203 | 0.314076 | Left ventricle |
| LV_circumf | Z121_S4 | 27446 | 0.003957 | 0.005744 | 0.490932 | 0.308979 | Left ventricle |
| RA_stroke_ | Z96_S3 | 27446 | -0.00339 | 0.00493 | 0.49144 | 0.308529 | Right atrium |
| LV_longitu | Z19_S2 | 27446 | 0.003375 | 0.004908 | 0.491657 | 0.308337 | Left ventricle |
| Descending | Z51_S1 | 24599 | 0.003597 | 0.005232 | 0.491842 | 0.308174 | Aorta |
| Ascending_ | Z3_S2 | 24599 | 0.003525 | 0.005142 | 0.492962 | 0.307186 | Aorta |
| Ascending_ | Z121_S4 | 24599 | 0.004668 | 0.006844 | 0.495175 | 0.305241 | Aorta |
| LA_stroke_ | Z93_S1 | 27446 | 0.004086 | 0.00603 | 0.498036 | 0.30274 | Left atrium |
| LV_cardiac_ | Z105_S5 | 27446 | -0.00381 | 0.005629 | 0.498871 | 0.302012 | Left ventricle |
| Descending | Z112_S1 | 24599 | 0.005512 | 0.008187 | 0.5008 | 0.300336 | Aorta |
| LV_mean_ | Z11_S5 | 27446 | -0.00449 | 0.006716 | 0.503838 | 0.297709 | Left ventricle |
| RA_minimu | Z1_S4 | 27446 | -0.00395 | 0.005933 | 0.506079 | 0.295781 | Right atrium |

| | | | | | | | |
|---|---|---|---|---|---|---|---|
| LV_mean_ | Z56_S2 | 27446 | -0.00398 | 0.005996 | 0.507095 | 0.294911 | Left ventricle |
| Ascending_ | Z74_S2 | 24599 | 0.003957 | 0.005987 | 0.508674 | 0.293561 | Aorta |
| Descending | Z37_S3 | 24599 | 0.00447 | 0.006799 | 0.510921 | 0.291646 | Aorta |
| LA_maximu | Z40_S4 | 27446 | -0.00414 | 0.00632 | 0.512299 | 0.290477 | Left atrium |
| RA_ejectio | Z21_S3 | 27446 | 0.004082 | 0.006243 | 0.513201 | 0.289713 | Right atrium |
| LV_mean_ | Z99_S5 | 27446 | 0.004347 | 0.006703 | 0.516726 | 0.28674 | Left ventricle |
| RV_end_sy | Z121_S4 | 27446 | -0.005 | 0.007735 | 0.518115 | 0.285574 | Right ventricle |
| Descending | Z87_S5 | 24599 | 0.003799 | 0.005892 | 0.518995 | 0.284837 | Aorta |
| Descending | Z120_S2 | 24599 | -0.00329 | 0.00511 | 0.519287 | 0.284593 | Aorta |
| LV_mean_ | Z115_S3 | 27446 | -0.00474 | 0.007429 | 0.523093 | 0.281421 | Left ventricle |
| RV_ejectio | Z114_S4 | 27446 | -0.00316 | 0.004977 | 0.525949 | 0.279056 | Right ventricle |
| LV_ejection | Z26_S4 | 27446 | -0.00391 | 0.006212 | 0.528575 | 0.276894 | Left ventricle |
| RA_minimu | Z8_S3 | 27446 | 0.002968 | 0.004787 | 0.535257 | 0.271437 | Right atrium |
| LV_mean_ | Z70_S5 | 27446 | -0.00413 | 0.006678 | 0.53639 | 0.270519 | Left ventricle |
| Descending | Z18_S4 | 24599 | -0.00514 | 0.00835 | 0.538127 | 0.269115 | Aorta |
| RA_stroke_ | Z68_S2 | 27446 | 0.002832 | 0.004607 | 0.538793 | 0.268578 | Right atrium |
| RV_stroke_ | Z122_S2 | 27446 | 0.003086 | 0.005034 | 0.539869 | 0.267712 | Right ventricle |
| RA_minimu | Z96_S3 | 27446 | 0.003361 | 0.005484 | 0.539942 | 0.267653 | Right atrium |
| LV_stroke_ | Z44_S1 | 27446 | 0.00367 | 0.006052 | 0.544314 | 0.264151 | Left ventricle |
| Descending | Z17_S1 | 24599 | -0.00331 | 0.005567 | 0.551926 | 0.258119 | Aorta |
| LV_circumf | Z105_S4 | 27446 | -0.00275 | 0.004643 | 0.553706 | 0.256721 | Left ventricle |
| LV_mean_ | Z35_S5 | 27446 | -0.00354 | 0.005988 | 0.554131 | 0.256387 | Left ventricle |
| LV_circumf | Z123_S4 | 27446 | 0.003152 | 0.005336 | 0.554725 | 0.255923 | Left ventricle |
| LV_ejection | Z106_S3 | 27446 | 0.003482 | 0.005912 | 0.55584 | 0.255051 | Left ventricle |
| LV_ejection | Z94_S5 | 27446 | 0.002504 | 0.004324 | 0.562486 | 0.249888 | Left ventricle |
| LV_mean_ | Z28_S2 | 27446 | -0.00431 | 0.007439 | 0.562593 | 0.249806 | Left ventricle |
| LV_radial_ | Z32_S5 | 27446 | 0.003407 | 0.006214 | 0.583527 | 0.233939 | Left ventricle |
| LV_mean_ | Z47_S4 | 27446 | 0.003797 | 0.006974 | 0.586081 | 0.232042 | Left ventricle |
| RA_minimu | Z23_S3 | 27446 | 0.002678 | 0.004999 | 0.592115 | 0.227594 | Right atrium |
| LV_cardiac | Z76_S3 | 27446 | 0.003117 | 0.005858 | 0.594622 | 0.225759 | Left ventricle |
| LV_mean_ | Z120_S5 | 27446 | 0.004477 | 0.00843 | 0.595337 | 0.225237 | Left ventricle |
| Descending | Z6_S2 | 24599 | 0.002446 | 0.004654 | 0.599138 | 0.222473 | Aorta |
| LV_circumf | Z87_S3 | 27446 | -0.00259 | 0.004963 | 0.601148 | 0.221019 | Left ventricle |
| Ascending_ | Z14_S4 | 24599 | -0.00334 | 0.006436 | 0.603961 | 0.218991 | Aorta |
| LV_cardiac | Z61_S4 | 27446 | -0.00272 | 0.005242 | 0.604456 | 0.218635 | Left ventricle |
| LA_ejection | Z38_S4 | 27446 | -0.00288 | 0.005583 | 0.605816 | 0.217659 | Left atrium |
| LV_ejection | Z63_S1 | 27446 | 0.003056 | 0.005956 | 0.60784 | 0.216211 | Left ventricle |
| LV_longitu | Z122_S2 | 27446 | -0.00208 | 0.004066 | 0.608468 | 0.215762 | Left ventricle |
| RA_stroke_ | Z92_S1 | 27446 | -0.00257 | 0.005034 | 0.609822 | 0.214797 | Right atrium |
| LV_longitu | Z108_S5 | 27446 | 0.002573 | 0.005094 | 0.613542 | 0.212155 | Left ventricle |
| LV_cardiac | Z26_S4 | 27446 | -0.00348 | 0.006894 | 0.613789 | 0.211981 | Left ventricle |
| LV_ejection | Z12_S4 | 27446 | 0.002145 | 0.004281 | 0.61633 | 0.210186 | Left ventricle |
| LV_end_sy | Z72_S5 | 27446 | -0.00386 | 0.00777 | 0.618954 | 0.208342 | Left ventricle |
| LV_mean_ | Z1_S4 | 27446 | 0.003629 | 0.007355 | 0.621703 | 0.206417 | Left ventricle |
| LV_mean_ | Z111_S5 | 27446 | -0.00339 | 0.006924 | 0.624305 | 0.204603 | Left ventricle |

| | | | | | | | |
|---|---|---|---|---|---|---|---|
| LV_mean_ı | Z37_S1 | 27446 | 0.003837 | 0.007862 | 0.625562 | 0.20373 | Left ventricle |
| RV_ejectio | Z62_S4 | 27446 | -0.00285 | 0.005861 | 0.626726 | 0.202922 | Right ventricle |
| RA_stroke_ | Z28_S1 | 27446 | -0.0026 | 0.005349 | 0.627157 | 0.202624 | Right atrium |
| RV_end_di | Z56_S5 | 27446 | 0.002974 | 0.006134 | 0.627793 | 0.202183 | Right ventricle |
| LV_ejectioı | Z51_S3 | 27446 | 0.00293 | 0.006121 | 0.63216 | 0.199173 | Left ventricle |
| Ascending_ | Z9_S2 | 24599 | 0.00293 | 0.006519 | 0.653092 | 0.185026 | Aorta |
| LA_ejectioı | Z80_S2 | 27446 | -0.00194 | 0.004335 | 0.654042 | 0.184394 | Left atrium |
| LV_end_di | Z7_S5 | 27446 | -0.00335 | 0.007469 | 0.654097 | 0.184358 | Left ventricle |
| RA_stroke_ | Z64_S1 | 27446 | -0.00251 | 0.005625 | 0.655063 | 0.183717 | Right atrium |
| RV_ejectio | Z66_S3 | 27446 | -0.00259 | 0.005802 | 0.655181 | 0.183639 | Right ventricle |
| Descending | Z37_S1 | 24599 | 0.002997 | 0.006717 | 0.655411 | 0.183486 | Aorta |
| LV_end_di | Z72_S5 | 27446 | 0.003715 | 0.008419 | 0.659042 | 0.181087 | Left ventricle |
| RA_ejectio | Z51_S1 | 27446 | -0.00191 | 0.004375 | 0.663061 | 0.178447 | Right atrium |
| LV_ejectioı | Z75_S4 | 27446 | 0.002421 | 0.005932 | 0.683241 | 0.165426 | Left ventricle |
| LV_ejectioı | Z127_S5 | 27446 | -0.00173 | 0.004268 | 0.686086 | 0.163621 | Left ventricle |
| LV_mean_ı | Z26_S4 | 27446 | -0.00347 | 0.008655 | 0.688409 | 0.162153 | Left ventricle |
| LV_radial_ | Z23_S1 | 27446 | 0.002261 | 0.005797 | 0.696527 | 0.157062 | Left ventricle |
| LV_mean_ı | Z122_S1 | 27446 | -0.00295 | 0.007642 | 0.699578 | 0.155164 | Left ventricle |
| RV_ejectio | Z91_S1 | 27446 | 0.001892 | 0.004948 | 0.702163 | 0.153562 | Right ventricle |
| LV_circumf | Z94_S2 | 27446 | -0.00209 | 0.00549 | 0.704031 | 0.152408 | Left ventricle |
| Ascending_ | Z47_S4 | 24599 | -0.00212 | 0.00578 | 0.713493 | 0.14661 | Aorta |
| RV_ejectio | Z63_S1 | 27446 | 0.0022 | 0.006079 | 0.71744 | 0.144215 | Right ventricle |
| LV_end_sy | Z112_S1 | 27446 | -0.00284 | 0.007852 | 0.717578 | 0.144131 | Left ventricle |
| LV_mean_ı | Z121_S4 | 27446 | -0.00281 | 0.007876 | 0.720804 | 0.142183 | Left ventricle |
| LV_ejectioı | Z49_S3 | 27446 | -0.00206 | 0.005812 | 0.723015 | 0.140852 | Left ventricle |
| Ascending_ | Z39_S4 | 24599 | -0.00192 | 0.005445 | 0.724289 | 0.140088 | Aorta |
| RA_stroke_ | Z58_S2 | 27446 | -0.00166 | 0.004748 | 0.726737 | 0.138623 | Right atrium |
| RA_stroke_ | Z75_S4 | 27446 | -0.00209 | 0.005993 | 0.727561 | 0.138131 | Right atrium |
| LV_longitu | Z68_S2 | 27446 | 0.001558 | 0.004526 | 0.730604 | 0.136318 | Left ventricle |
| LV_ejectioı | Z27_S2 | 27446 | 0.002 | 0.005834 | 0.731742 | 0.135642 | Left ventricle |
| LV_longitu | Z120_S2 | 27446 | -0.00144 | 0.00427 | 0.735933 | 0.133161 | Left ventricle |
| RA_stroke_ | Z74_S2 | 27446 | 0.001671 | 0.005004 | 0.73843 | 0.131691 | Right atrium |
| LV_longitu | Z91_S1 | 27446 | 0.001572 | 0.004811 | 0.743838 | 0.128522 | Left ventricle |
| Ascending_ | Z97_S2 | 24599 | 0.001947 | 0.006128 | 0.750731 | 0.124516 | Aorta |
| Ascending_ | Z37_S3 | 24599 | -0.00216 | 0.006925 | 0.755393 | 0.121827 | Aorta |
| LV_ejectioı | Z21_S3 | 27446 | 0.001929 | 0.006278 | 0.758654 | 0.119956 | Left ventricle |
| LV_longitu | Z111_S1 | 27446 | 0.001595 | 0.005424 | 0.768661 | 0.114265 | Left ventricle |
| LV_mean_ı | Z14_S4 | 27446 | 0.002267 | 0.007789 | 0.771074 | 0.112904 | Left ventricle |
| LV_myocar | Z3_S2 | 27446 | -0.00216 | 0.007476 | 0.772692 | 0.111994 | Left ventricle |
| LV_mean_ı | Z52_S4 | 27446 | 0.001803 | 0.006246 | 0.772864 | 0.111897 | Left ventricle |
| LV_circumf | Z5_S1 | 27446 | 0.001441 | 0.005022 | 0.774174 | 0.111161 | Left ventricle |
| Ascending_ | Z30_S2 | 24599 | 0.002005 | 0.007006 | 0.774744 | 0.110842 | Aorta |
| Descending | Z34_S2 | 24599 | 0.001944 | 0.006827 | 0.775794 | 0.110253 | Aorta |
| LA_ejectioı | Z49_S1 | 27446 | 0.00169 | 0.005944 | 0.776153 | 0.110053 | Left atrium |
| LV_mean_ı | Z5_S1 | 27446 | -0.00195 | 0.006886 | 0.777176 | 0.109481 | Left ventricle |

| | | | | | | | |
|---|---|---|---|---|---|---|---|
| Ascending_ | Z28_S2 | 24599 | -0.00172 | 0.006187 | 0.780498 | 0.107628 | Aorta |
| Ascending_ | Z65_S1 | 24599 | 0.001401 | 0.005064 | 0.782071 | 0.106754 | Aorta |
| LA_ejectio | Z22_S4 | 27446 | -0.00138 | 0.005042 | 0.784085 | 0.105637 | Left atrium |
| LA_ejectio | Z99_S4 | 27446 | 0.001481 | 0.005437 | 0.785348 | 0.104938 | Left atrium |
| LV_mean_ | Z51_S3 | 27446 | -0.00231 | 0.008527 | 0.786577 | 0.104259 | Left ventricle |
| RA_minimu | Z32_S5 | 27446 | -0.00183 | 0.00678 | 0.786715 | 0.104183 | Right atrium |
| LV_mean_ | Z44_S1 | 27446 | -0.00185 | 0.006964 | 0.790221 | 0.102252 | Left ventricle |
| Ascending_ | Z80_S4 | 24599 | -0.00139 | 0.00524 | 0.791032 | 0.101806 | Aorta |
| Descending | Z60_S2 | 24599 | -0.00136 | 0.005196 | 0.793402 | 0.100507 | Aorta |
| RA_ejectio | Z50_S5 | 27446 | -0.00162 | 0.006189 | 0.793589 | 0.100404 | Right atrium |
| Ascending_ | Z3_S2 | 24599 | 0.001269 | 0.005161 | 0.805746 | 0.093802 | Aorta |
| LV_circumf | Z12_S4 | 27446 | 0.001066 | 0.00435 | 0.806369 | 0.093466 | Left ventricle |
| LV_stroke_ | Z32_S5 | 27446 | -0.00179 | 0.007304 | 0.806397 | 0.093451 | Left ventricle |
| LV_myocar | Z53_S2 | 27446 | -0.00227 | 0.00928 | 0.806612 | 0.093335 | Left ventricle |
| LA_maximu | Z32_S5 | 27446 | -0.00136 | 0.006303 | 0.829717 | 0.08107 | Left atrium |
| LV_ejectio | Z34_S2 | 27446 | 0.001214 | 0.005745 | 0.832614 | 0.079556 | Left ventricle |
| LV_mean_ | Z62_S1 | 27446 | 0.00137 | 0.006539 | 0.834096 | 0.078784 | Left ventricle |
| Descending | Z118_S1 | 24599 | -0.00152 | 0.007339 | 0.83605 | 0.077768 | Aorta |
| Descending | Z55_S5 | 24599 | 0.001042 | 0.00508 | 0.837425 | 0.077054 | Aorta |
| LV_cardiac | Z34_S2 | 27446 | 0.001303 | 0.006375 | 0.838077 | 0.076716 | Left ventricle |
| RV_ejectio | Z68_S2 | 27446 | 0.000939 | 0.004655 | 0.840048 | 0.075696 | Right ventricle |
| RA_maxim | Z97_S2 | 27446 | 0.00115 | 0.005874 | 0.844742 | 0.073276 | Right atrium |
| LV_ejectio | Z28_S4 | 27446 | 0.001056 | 0.005398 | 0.844894 | 0.073198 | Left ventricle |
| RA_maxim | Z120_S2 | 27446 | -0.00087 | 0.004771 | 0.854611 | 0.068231 | Right atrium |
| RV_ejectio | Z1_S1 | 27446 | -0.00085 | 0.00498 | 0.864704 | 0.063132 | Right ventricle |
| LV_stroke_ | Z28_S1 | 27446 | -0.00105 | 0.006412 | 0.869889 | 0.060536 | Left ventricle |
| RV_end_sy | Z97_S2 | 27446 | 0.001123 | 0.007247 | 0.876896 | 0.057052 | Right ventricle |
| Ascending_ | Z2_S1 | 24599 | -0.00082 | 0.005536 | 0.882549 | 0.054261 | Aorta |
| LV_ejectio | Z97_S2 | 27446 | 0.000766 | 0.005296 | 0.884959 | 0.053077 | Left ventricle |
| LV_end_di | Z22_S4 | 27446 | 0.000965 | 0.007079 | 0.891535 | 0.049862 | Left ventricle |
| LV_ejectio | Z41_S1 | 27446 | 0.000732 | 0.005415 | 0.892537 | 0.049374 | Left ventricle |
| RA_stroke_ | Z17_S1 | 27446 | 0.000626 | 0.004733 | 0.894701 | 0.048322 | Right atrium |
| LV_mean_ | Z10_S4 | 27446 | 0.000955 | 0.007284 | 0.895643 | 0.047865 | Left ventricle |
| LV_mean_ | Z65_S5 | 27446 | -0.00108 | 0.008317 | 0.896565 | 0.047418 | Left ventricle |
| LV_myocar | Z80_S5 | 27446 | -0.00113 | 0.009896 | 0.908938 | 0.041466 | Left ventricle |
| RV_ejectio | Z13_S3 | 27446 | -0.00046 | 0.004171 | 0.91144 | 0.040272 | Right ventricle |
| LV_myocar | Z39_S4 | 27446 | -0.00081 | 0.007509 | 0.914231 | 0.038944 | Left ventricle |
| RV_ejectio | Z76_S3 | 27446 | -0.00057 | 0.005387 | 0.915034 | 0.038563 | Right ventricle |
| LV_stroke_ | Z53_S2 | 27446 | -0.0006 | 0.006716 | 0.928594 | 0.032174 | Left ventricle |
| RV_ejectio | Z62_S3 | 27446 | -0.00047 | 0.005357 | 0.929445 | 0.031776 | Right ventricle |
| LV_mean_ | Z30_S2 | 27446 | -0.00075 | 0.008469 | 0.929588 | 0.031709 | Left ventricle |
| Descending | Z39_S3 | 24599 | -0.00069 | 0.008248 | 0.933757 | 0.029766 | Aorta |
| LV_mean_ | Z109_S1 | 27446 | -0.00052 | 0.007 | 0.940314 | 0.026727 | Left ventricle |
| RV_end_di | Z78_S1 | 27446 | -0.00046 | 0.006559 | 0.943769 | 0.025134 | Right ventricle |
| RA_stroke_ | Z13_S3 | 27446 | 0.000268 | 0.004129 | 0.948204 | 0.023098 | Right atrium |

| | | | | | | | |
|---|---|---|---|---|---|---|---|
| RV_stroke_ | Z28_S1 | 27446 | 0.000406 | 0.006506 | 0.950286 | 0.022146 | Right ventricle |
| LV_longitu | Z28_S4 | 27446 | 0.000331 | 0.005358 | 0.950799 | 0.021911 | Left ventricle |
| Ascending_ | Z1_S2 | 24599 | -0.0003 | 0.004962 | 0.951031 | 0.021805 | Aorta |
| LV_circumf | Z106_S3 | 27446 | 0.000348 | 0.006007 | 0.953805 | 0.02054 | Left ventricle |
| Descending | Z58_S2 | 24599 | -0.00032 | 0.005582 | 0.95422 | 0.020351 | Aorta |
| RA_maxim | Z8_S3 | 27446 | -0.00019 | 0.004723 | 0.96824 | 0.014017 | Right atrium |
| LA_ejection | Z31_S2 | 27446 | -0.00017 | 0.004895 | 0.972047 | 0.012313 | Left atrium |
| RA_maxim | Z96_S3 | 27446 | 0.000165 | 0.005411 | 0.975607 | 0.010725 | Right atrium |
| Ascending_ | Z127_S4 | 24599 | 0.000132 | 0.006332 | 0.983379 | 0.007279 | Aorta |
| RV_ejectio | Z109_S1 | 27446 | -6.54E-05 | 0.005128 | 0.989819 | 0.004444 | Right ventricle |
| RV_end_sy | Z22_S4 | 27446 | 8.91E-05 | 0.007104 | 0.989997 | 0.004366 | Right ventricle |
| LV_stroke_ | Z38_S4 | 27446 | 1.57E-05 | 0.006962 | 0.998206 | 0.00078 | Left ventricle |
