## Supplementary Table 1 for "Hundreds of cardiac MRI traits derived using 3D diffusion autoencoders share a common genetic architecture"

| Disease | Latent | Effect Size | StdError | P | Log10P |
|---|---|---|---|---|---|
| atrial fibrill | Z106_S4 | 0.249323 | 0.020353 | 2.15E-34 | 33.66706 |
| mitral prob | Z106_S4 | 0.215924 | 0.040773 | 1.20E-07 | 6.921939 |
| heart failur | Z106_S4 | 0.17602 | 0.033953 | 2.19E-07 | 6.659849 |
| aortic valve | Z106_S4 | 0.217091 | 0.051728 | 2.72E-05 | 4.565784 |
| type 2 diab | Z106_S4 | -0.06698 | 0.022284 | 0.002653 | 2.576258 |
| hypertensi | Z106_S4 | 0.03305 | 0.011288 | 0.003415 | 2.466602 |
| high choles | Z106_S4 | -0.02958 | 0.012402 | 0.017096 | 1.767096 |
| other heart | Z106_S4 | 0.093095 | 0.049731 | 0.061223 | 1.213082 |
| conduction | Z106_S4 | 0.063939 | 0.03571 | 0.073386 | 1.134384 |
| hypotensio | Z106_S4 | 0.062219 | 0.038794 | 0.108767 | 0.963504 |
| arrhythmia | Z106_S4 | 0.050196 | 0.044276 | 0.256936 | 0.590175 |
| myocardial | Z106_S4 | -0.02682 | 0.026054 | 0.30332 | 0.518099 |
| angina pec | Z106_S4 | -0.02152 | 0.024217 | 0.37419 | 0.426908 |
| coronary h | Z106_S4 | 0.00017 | 0.020077 | 0.993232 | 0.002949 |
| atrial fibrill | Z15_S2 | 0.24793 | 0.023854 | 3.02E-25 | 24.52023 |
| mitral prob | Z15_S2 | 0.230135 | 0.047944 | 1.60E-06 | 5.796884 |
| heart failur | Z15_S2 | 0.152413 | 0.039953 | 0.000137 | 3.864453 |
| type 2 diab | Z15_S2 | -0.0988 | 0.026161 | 0.000159 | 3.797592 |
| high choles | Z15_S2 | -0.03395 | 0.014539 | 0.019528 | 1.709337 |
| aortic valve | Z15_S2 | 0.137708 | 0.060865 | 0.023676 | 1.625697 |
| angina pec | Z15_S2 | -0.04956 | 0.028437 | 0.081405 | 1.089351 |
| conduction | Z15_S2 | 0.06522 | 0.042005 | 0.120515 | 0.918958 |
| coronary h | Z15_S2 | -0.03263 | 0.023602 | 0.166847 | 0.777681 |
| myocardial | Z15_S2 | -0.03659 | 0.030652 | 0.232566 | 0.633454 |
| hypotensio | Z15_S2 | 0.030023 | 0.045618 | 0.510458 | 0.29204 |
| other heart | Z15_S2 | 0.029366 | 0.058483 | 0.61558 | 0.210715 |
| hypertensi | Z15_S2 | 0.005013 | 0.013234 | 0.704823 | 0.15192 |
| arrhythmia | Z15_S2 | -0.01017 | 0.0521 | 0.845165 | 0.073059 |
| atrial fibrill | Z32_S1 | -0.24348 | 0.024439 | 2.47E-23 | 22.6077 |
| heart failur | Z32_S1 | -0.25601 | 0.04085 | 3.75E-10 | 9.426157 |
| type 2 diab | Z32_S1 | -0.13114 | 0.026763 | 9.66E-07 | 6.015226 |
| other heart | Z32_S1 | -0.17703 | 0.059867 | 0.00311 | 2.507242 |
| mitral prob | Z32_S1 | -0.13558 | 0.049116 | 0.005779 | 2.238137 |
| angina pec | Z32_S1 | 0.071763 | 0.029113 | 0.013708 | 1.863019 |
| hypotensio | Z32_S1 | -0.10507 | 0.046694 | 0.024448 | 1.611749 |
| coronary h | Z32_S1 | 0.046884 | 0.024147 | 0.052199 | 1.282334 |
| aortic valve | Z32_S1 | -0.12046 | 0.06233 | 0.053297 | 1.273301 |
| myocardial | Z32_S1 | 0.045573 | 0.031365 | 0.146244 | 0.834921 |
| hypertensi | Z32_S1 | -0.01325 | 0.013535 | 0.327624 | 0.484624 |
| conduction | Z32_S1 | -0.02601 | 0.042988 | 0.545152 | 0.263482 |
| arrhythmia | Z32_S1 | -0.01628 | 0.053348 | 0.760208 | 0.119067 |
| high choles | Z32_S1 | -0.00045 | 0.014886 | 0.975773 | 0.010651 |
| atrial fibrill | Z65_S1 | -0.18492 | 0.019497 | 2.67E-21 | 20.574 |
| heart failur | Z65_S1 | -0.22276 | 0.032531 | 7.70E-12 | 11.11373 |

| | | | | | |
|---|---|---|---|---|---|
| type 2 diab | Z65_S1 | -0.11026 | 0.021317 | 2.33E-07 | 6.632333 |
| hypotensio | Z65_S1 | -0.10379 | 0.037138 | 0.005198 | 2.284165 |
| mitral prob | Z65_S1 | -0.10438 | 0.039077 | 0.007565 | 2.121179 |
| aortic valve | Z65_S1 | -0.12183 | 0.049579 | 0.014004 | 1.853754 |
| other heart | Z65_S1 | -0.11373 | 0.047659 | 0.017027 | 1.768855 |
| myocardial | Z65_S1 | -0.03925 | 0.024958 | 0.115846 | 0.936118 |
| arrhythmia | Z65_S1 | -0.05255 | 0.042447 | 0.21574 | 0.66607 |
| coronary h | Z65_S1 | -0.01997 | 0.019242 | 0.299329 | 0.523851 |
| angina pec | Z65_S1 | 0.007575 | 0.023163 | 0.743652 | 0.12863 |
| high choles | Z65_S1 | -0.00382 | 0.011842 | 0.746771 | 0.126813 |
| conduction | Z65_S1 | 0.005677 | 0.034216 | 0.868224 | 0.061368 |
| hypertensi | Z65_S1 | 0.001373 | 0.0108 | 0.898807 | 0.046334 |
| atrial fibrill | Z64_S1 | -0.23655 | 0.025013 | 3.47E-21 | 20.46016 |
| type 2 diab | Z64_S1 | 0.110117 | 0.027355 | 5.70E-05 | 4.243813 |
| mitral prob | Z64_S1 | -0.19739 | 0.050156 | 8.33E-05 | 4.079284 |
| heart failur | Z64_S1 | -0.16362 | 0.041783 | 9.03E-05 | 4.04414 |
| conduction | Z64_S1 | -0.1053 | 0.043907 | 0.016481 | 1.783009 |
| aortic valve | Z64_S1 | -0.13267 | 0.063659 | 0.037169 | 1.429818 |
| angina pec | Z64_S1 | 0.055948 | 0.029715 | 0.059743 | 1.223714 |
| other heart | Z64_S1 | -0.05373 | 0.06116 | 0.379672 | 0.420591 |
| arrhythmia | Z64_S1 | -0.04628 | 0.05449 | 0.395739 | 0.402591 |
| high choles | Z64_S1 | 0.012719 | 0.015217 | 0.403255 | 0.39442 |
| coronary h | Z64_S1 | 0.019936 | 0.024657 | 0.418785 | 0.378009 |
| hypotensio | Z64_S1 | -0.03406 | 0.04771 | 0.475295 | 0.323037 |
| myocardial | Z64_S1 | -0.01264 | 0.032033 | 0.693224 | 0.159126 |
| hypertensi | Z64_S1 | 0.000208 | 0.013842 | 0.988019 | 0.005235 |
| atrial fibrill | Z31_S5 | 0.174668 | 0.01905 | 5.17E-20 | 19.28649 |
| coronary h | Z31_S5 | 0.086225 | 0.018714 | 4.09E-06 | 5.387824 |
| mitral prob | Z31_S5 | 0.160872 | 0.038067 | 2.39E-05 | 4.621978 |
| myocardial | Z31_S5 | 0.102331 | 0.024298 | 2.55E-05 | 4.594097 |
| heart failur | Z31_S5 | 0.128703 | 0.031722 | 4.98E-05 | 4.302428 |
| type 2 diab | Z31_S5 | -0.07677 | 0.020853 | 0.000233 | 3.633308 |
| angina pec | Z31_S5 | 0.081129 | 0.022565 | 0.000325 | 3.488566 |
| aortic valve | Z31_S5 | 0.10754 | 0.048296 | 0.025977 | 1.585415 |
| high choles | Z31_S5 | -0.02472 | 0.011607 | 0.033169 | 1.479269 |
| conduction | Z31_S5 | 0.059476 | 0.033337 | 0.074427 | 1.128269 |
| other heart | Z31_S5 | 0.076003 | 0.046423 | 0.101601 | 0.993104 |
| arrhythmia | Z31_S5 | 0.053085 | 0.041336 | 0.199077 | 0.700979 |
| hypertensi | Z31_S5 | -0.007 | 0.010559 | 0.507542 | 0.294528 |
| hypotensio | Z31_S5 | 0.011636 | 0.03623 | 0.748077 | 0.126054 |
| type 2 diab | Z53_S1 | -0.24138 | 0.026473 | 8.27E-20 | 19.08229 |
| heart failur | Z53_S1 | -0.20616 | 0.040494 | 3.59E-07 | 6.445172 |
| atrial fibrill | Z53_S1 | -0.06408 | 0.024317 | 0.008411 | 2.07515 |
| other heart | Z53_S1 | -0.12714 | 0.059336 | 0.032153 | 1.492783 |
| aortic valve | Z53_S1 | 0.09008 | 0.061759 | 0.144698 | 0.839538 |

| | | | | | |
|---|---|---|---|---|---|
| myocardial | Z53_S1 | 0.036608 | 0.031091 | 0.239027 | 0.621553 |
| hypertensi | Z53_S1 | 0.013572 | 0.013414 | 0.311652 | 0.50633 |
| angina pec | Z53_S1 | -0.02452 | 0.028855 | 0.395374 | 0.402992 |
| high choles | Z53_S1 | -0.00959 | 0.014826 | 0.517664 | 0.285952 |
| arrhythmia | Z53_S1 | 0.0252 | 0.052845 | 0.633459 | 0.198281 |
| conduction | Z53_S1 | -0.01336 | 0.042646 | 0.754154 | 0.12254 |
| hypotensio | Z53_S1 | 0.014359 | 0.046326 | 0.756602 | 0.121132 |
| coronary h | Z53_S1 | 0.006395 | 0.023945 | 0.789406 | 0.1027 |
| mitral prob | Z53_S1 | -0.00349 | 0.048693 | 0.942814 | 0.025574 |
| type 2 diab | Z127_S4 | -0.24342 | 0.027348 | 5.94E-19 | 18.22613 |
| myocardial | Z127_S4 | 0.082359 | 0.032098 | 0.0103 | 1.987176 |
| heart failur | Z127_S4 | -0.08729 | 0.041851 | 0.037022 | 1.431543 |
| arrhythmia | Z127_S4 | 0.093541 | 0.054562 | 0.086469 | 1.063138 |
| high choles | Z127_S4 | -0.02565 | 0.015281 | 0.093243 | 1.030385 |
| coronary h | Z127_S4 | 0.037932 | 0.024707 | 0.124734 | 0.904013 |
| mitral prob | Z127_S4 | 0.062428 | 0.050279 | 0.214389 | 0.668797 |
| atrial fibrill | Z127_S4 | 0.027889 | 0.025122 | 0.266965 | 0.573546 |
| hypertensi | Z127_S4 | 0.010339 | 0.013824 | 0.454492 | 0.342473 |
| angina pec | Z127_S4 | 0.019311 | 0.029781 | 0.516702 | 0.28676 |
| hypotensio | Z127_S4 | -0.02578 | 0.047846 | 0.589983 | 0.229161 |
| other heart | Z127_S4 | -0.01916 | 0.06131 | 0.754648 | 0.122256 |
| aortic valve | Z127_S4 | -0.00771 | 0.063784 | 0.9038 | 0.043928 |
| conduction | Z127_S4 | 0.000285 | 0.044024 | 0.994829 | 0.002251 |
| atrial fibrill | Z93_S2 | 0.239942 | 0.027062 | 8.09E-19 | 18.09184 |
| mitral prob | Z93_S2 | 0.287803 | 0.054309 | 1.17E-07 | 6.930561 |
| heart failur | Z93_S2 | 0.18756 | 0.045212 | 3.36E-05 | 4.47377 |
| type 2 diab | Z93_S2 | -0.08066 | 0.02963 | 0.006492 | 2.187607 |
| high choles | Z93_S2 | -0.03663 | 0.016484 | 0.0263 | 1.580048 |
| aortic valve | Z93_S2 | 0.110435 | 0.068945 | 0.109218 | 0.961708 |
| other heart | Z93_S2 | 0.091173 | 0.066261 | 0.168842 | 0.772518 |
| myocardial | Z93_S2 | -0.04669 | 0.034671 | 0.17812 | 0.749287 |
| hypotensio | Z93_S2 | 0.061417 | 0.051677 | 0.234656 | 0.629568 |
| conduction | Z93_S2 | 0.053292 | 0.047598 | 0.262886 | 0.580233 |
| angina pec | Z93_S2 | -0.03295 | 0.032169 | 0.305746 | 0.514639 |
| coronary h | Z93_S2 | -0.02435 | 0.026684 | 0.361586 | 0.441788 |
| hypertensi | Z93_S2 | 0.010104 | 0.01495 | 0.499147 | 0.301772 |
| arrhythmia | Z93_S2 | 0.016015 | 0.058959 | 0.785914 | 0.104625 |
| type 2 diab | Z82_S1 | 0.263188 | 0.030273 | 3.74E-18 | 17.42766 |
| myocardial | Z82_S1 | -0.23422 | 0.035436 | 3.94E-11 | 10.40494 |
| coronary h | Z82_S1 | -0.17418 | 0.02728 | 1.75E-10 | 9.75748 |
| atrial fibrill | Z82_S1 | -0.17051 | 0.027777 | 8.47E-10 | 9.072319 |
| angina pec | Z82_S1 | -0.15744 | 0.032915 | 1.74E-06 | 5.760112 |
| mitral prob | Z82_S1 | -0.26414 | 0.055556 | 2.00E-06 | 5.698492 |
| arrhythmia | Z82_S1 | -0.18387 | 0.060321 | 0.002305 | 2.637236 |
| heart failur | Z82_S1 | -0.10229 | 0.046289 | 0.027124 | 1.566648 |

| | | | | | |
|---|---|---|---|---|---|
| hypertensi | Z82_S1 | 0.033771 | 0.015311 | 0.027416 | 1.562002 |
| high choles | Z82_S1 | 0.030261 | 0.016864 | 0.072763 | 1.138088 |
| conduction | Z82_S1 | -0.06226 | 0.048691 | 0.201055 | 0.696684 |
| aortic valve | Z82_S1 | -0.03888 | 0.070532 | 0.581461 | 0.23548 |
| hypotensio | Z82_S1 | -0.02679 | 0.052889 | 0.612549 | 0.212859 |
| other heart | Z82_S1 | -0.02563 | 0.067792 | 0.705417 | 0.151554 |
| atrial fibrill | Z30_S2 | -0.23329 | 0.027183 | 9.91E-18 | 17.004 |
| myocardial | Z30_S2 | -0.17988 | 0.0347 | 2.19E-07 | 6.659366 |
| heart failur | Z30_S2 | -0.206 | 0.045306 | 5.47E-06 | 5.261667 |
| coronary h | Z30_S2 | -0.1199 | 0.02671 | 7.19E-06 | 5.14306 |
| other heart | Z30_S2 | -0.24137 | 0.066321 | 0.000274 | 3.562457 |
| angina pec | Z30_S2 | -0.08751 | 0.0322 | 0.006581 | 2.181691 |
| mitral prob | Z30_S2 | -0.14283 | 0.054402 | 0.008659 | 2.062513 |
| high choles | Z30_S2 | 0.037036 | 0.016512 | 0.024914 | 1.603557 |
| conduction | Z30_S2 | -0.10466 | 0.047666 | 0.028131 | 1.550812 |
| arrhythmia | Z30_S2 | -0.12749 | 0.059009 | 0.030747 | 1.512201 |
| hypotensio | Z30_S2 | -0.08159 | 0.051749 | 0.114871 | 0.939791 |
| aortic valve | Z30_S2 | -0.07064 | 0.068999 | 0.305975 | 0.514313 |
| type 2 diab | Z30_S2 | 0.030003 | 0.029693 | 0.312285 | 0.505448 |
| hypertensi | Z30_S2 | 0.00426 | 0.014992 | 0.776311 | 0.109964 |
| atrial fibrill | Z112_S1 | 0.230263 | 0.02769 | 9.63E-17 | 16.01651 |
| myocardial | Z112_S1 | -0.13953 | 0.035435 | 8.26E-05 | 4.083213 |
| coronary h | Z112_S1 | -0.10559 | 0.027266 | 0.000108 | 3.966678 |
| angina pec | Z112_S1 | -0.10824 | 0.032884 | 0.000998 | 3.000721 |
| heart failur | Z112_S1 | 0.150266 | 0.046227 | 0.001153 | 2.938025 |
| mitral prob | Z112_S1 | 0.152974 | 0.055528 | 0.005876 | 2.230942 |
| other heart | Z112_S1 | 0.108994 | 0.067731 | 0.107582 | 0.96826 |
| high choles | Z112_S1 | -0.02354 | 0.016847 | 0.162371 | 0.789491 |
| conduction | Z112_S1 | 0.045676 | 0.048647 | 0.34778 | 0.458695 |
| aortic valve | Z112_S1 | 0.043558 | 0.070472 | 0.536519 | 0.270415 |
| arrhythmia | Z112_S1 | -0.02741 | 0.06028 | 0.649266 | 0.187577 |
| type 2 diab | Z112_S1 | -0.01261 | 0.030295 | 0.677139 | 0.169322 |
| hypotensio | Z112_S1 | 0.020349 | 0.052831 | 0.700114 | 0.154831 |
| hypertensi | Z112_S1 | 0.005476 | 0.015275 | 0.719996 | 0.14267 |
| atrial fibrill | Z99_S3 | 0.150295 | 0.018547 | 5.61E-16 | 15.25141 |
| aortic valve | Z99_S3 | 0.208057 | 0.046955 | 9.43E-06 | 5.025674 |
| mitral prob | Z99_S3 | 0.12238 | 0.037018 | 0.000948 | 3.023107 |
| heart failur | Z99_S3 | 0.099432 | 0.030852 | 0.001271 | 2.895909 |
| hypertensi | Z99_S3 | 0.025094 | 0.01025 | 0.014364 | 1.842732 |
| high choles | Z99_S3 | -0.02142 | 0.011255 | 0.057062 | 1.243653 |
| type 2 diab | Z99_S3 | -0.03408 | 0.020242 | 0.092271 | 1.034934 |
| hypotensio | Z99_S3 | 0.041061 | 0.035238 | 0.243923 | 0.612747 |
| conduction | Z99_S3 | 0.033334 | 0.032423 | 0.30392 | 0.517241 |
| myocardial | Z99_S3 | -0.02108 | 0.023662 | 0.373039 | 0.428246 |
| angina pec | Z99_S3 | -0.01677 | 0.021989 | 0.445634 | 0.351021 |

| | | | | | |
|---|---|---|---|---|---|
| other heart | Z99_S3 | 0.031242 | 0.045163 | 0.4891 | 0.310603 |
| coronary h | Z99_S3 | -0.00834 | 0.018239 | 0.647593 | 0.188698 |
| arrhythmia | Z99_S3 | 0.008244 | 0.040208 | 0.837538 | 0.076995 |
| atrial fibrill | Z73_S4 | 0.209531 | 0.02587 | 5.80E-16 | 15.23655 |
| type 2 diab | Z73_S4 | -0.13313 | 0.028299 | 2.56E-06 | 5.591539 |
| mitral prob | Z73_S4 | 0.23032 | 0.051852 | 8.96E-06 | 5.047687 |
| aortic valve | Z73_S4 | 0.242144 | 0.06578 | 0.000233 | 3.633093 |
| hypertensi | Z73_S4 | 0.038023 | 0.014268 | 0.007706 | 2.113183 |
| heart failur | Z73_S4 | 0.081321 | 0.043182 | 0.059685 | 1.224134 |
| high choles | Z73_S4 | -0.02875 | 0.015726 | 0.067539 | 1.170447 |
| conduction | Z73_S4 | 0.076634 | 0.045436 | 0.091685 | 1.037701 |
| myocardial | Z73_S4 | -0.04369 | 0.033108 | 0.18703 | 0.72809 |
| other heart | Z73_S4 | 0.057474 | 0.063233 | 0.363395 | 0.439621 |
| hypotensio | Z73_S4 | 0.036713 | 0.04933 | 0.456748 | 0.340323 |
| angina pec | Z73_S4 | -0.01478 | 0.030732 | 0.63047 | 0.200336 |
| coronary h | Z73_S4 | 0.00914 | 0.025483 | 0.719831 | 0.142769 |
| arrhythmia | Z73_S4 | 0.00724 | 0.056307 | 0.897695 | 0.046871 |
| atrial fibrill | Z114_S5 | -0.14996 | 0.018526 | 6.02E-16 | 15.22014 |
| aortic valve | Z114_S5 | -0.19422 | 0.046981 | 3.58E-05 | 4.446495 |
| heart failur | Z114_S5 | -0.10588 | 0.030847 | 0.000599 | 3.222297 |
| mitral prob | Z114_S5 | -0.10258 | 0.037034 | 0.005612 | 2.250891 |
| hypertensi | Z114_S5 | -0.02259 | 0.010248 | 0.027473 | 1.561094 |
| angina pec | Z114_S5 | 0.046418 | 0.021978 | 0.034694 | 1.45974 |
| hypotensio | Z114_S5 | -0.05682 | 0.035233 | 0.10683 | 0.971308 |
| other heart | Z114_S5 | -0.0566 | 0.045181 | 0.210325 | 0.677109 |
| conduction | Z114_S5 | 0.031374 | 0.032432 | 0.333367 | 0.477077 |
| myocardial | Z114_S5 | 0.022432 | 0.023658 | 0.34305 | 0.464642 |
| coronary h | Z114_S5 | 0.01044 | 0.018229 | 0.566846 | 0.246535 |
| high choles | Z114_S5 | 0.006352 | 0.011257 | 0.572567 | 0.242173 |
| arrhythmia | Z114_S5 | -0.01439 | 0.04021 | 0.720375 | 0.142442 |
| type 2 diab | Z114_S5 | -0.00332 | 0.020238 | 0.869886 | 0.060538 |
| atrial fibrill | Z95_S3 | -0.14916 | 0.018601 | 1.12E-15 | 14.95192 |
| mitral prob | Z95_S3 | -0.13911 | 0.037219 | 0.000186 | 3.729828 |
| heart failur | Z95_S3 | -0.11383 | 0.031003 | 0.000242 | 3.616748 |
| aortic valve | Z95_S3 | -0.15684 | 0.047216 | 0.000896 | 3.047587 |
| hypertensi | Z95_S3 | -0.02809 | 0.010317 | 0.006479 | 2.188486 |
| type 2 diab | Z95_S3 | 0.038864 | 0.020352 | 0.056202 | 1.250245 |
| myocardial | Z95_S3 | 0.040475 | 0.023797 | 0.088983 | 1.050692 |
| other heart | Z95_S3 | -0.06128 | 0.045399 | 0.177102 | 0.751776 |
| high choles | Z95_S3 | 0.01062 | 0.011338 | 0.348954 | 0.457232 |
| coronary h | Z95_S3 | 0.012821 | 0.018347 | 0.484679 | 0.314546 |
| hypotensio | Z95_S3 | -0.02424 | 0.03541 | 0.49363 | 0.306599 |
| angina pec | Z95_S3 | 0.015014 | 0.022115 | 0.497209 | 0.303461 |
| conduction | Z95_S3 | -0.00151 | 0.032619 | 0.962957 | 0.016393 |
| arrhythmia | Z95_S3 | -0.00127 | 0.040416 | 0.974928 | 0.011027 |

| | | | | | |
|---|---|---|---|---|---|
| atrial fibrill | Z97_S1 | -0.21358 | 0.027432 | 7.23E-15 | 14.14082 |
| mitral prob | Z97_S1 | -0.22008 | 0.054993 | 6.30E-05 | 4.200411 |
| heart failur | Z97_S1 | -0.18244 | 0.045782 | 6.77E-05 | 4.169585 |
| type 2 diab | Z97_S1 | 0.061561 | 0.029983 | 0.040067 | 1.397213 |
| other heart | Z97_S1 | -0.1152 | 0.067057 | 0.08583 | 1.066363 |
| hypotensio | Z97_S1 | -0.08826 | 0.052309 | 0.091563 | 1.038279 |
| myocardial | Z97_S1 | -0.05262 | 0.035105 | 0.133939 | 0.873094 |
| aortic valve | Z97_S1 | -0.09767 | 0.069786 | 0.161655 | 0.791412 |
| arrhythmia | Z97_S1 | -0.07663 | 0.059722 | 0.19945 | 0.700166 |
| conduction | Z97_S1 | -0.05091 | 0.048175 | 0.29059 | 0.536719 |
| angina pec | Z97_S1 | 0.028207 | 0.032577 | 0.38658 | 0.412761 |
| coronary h | Z97_S1 | -0.01772 | 0.027012 | 0.51181 | 0.290891 |
| high choles | Z97_S1 | -0.00458 | 0.01667 | 0.783549 | 0.105934 |
| hypertensi | Z97_S1 | 0.001304 | 0.015141 | 0.931371 | 0.030877 |
| atrial fibrill | Z12_S5 | -0.20577 | 0.02668 | 1.28E-14 | 13.89262 |
| myocardial | Z12_S5 | 0.145403 | 0.034129 | 2.05E-05 | 4.688443 |
| mitral prob | Z12_S5 | -0.20202 | 0.053475 | 0.000159 | 3.799755 |
| type 2 diab | Z12_S5 | 0.086853 | 0.029185 | 0.002924 | 2.534027 |
| coronary h | Z12_S5 | 0.074628 | 0.02626 | 0.004489 | 2.347852 |
| heart failur | Z12_S5 | -0.12555 | 0.044534 | 0.00482 | 2.316962 |
| angina pec | Z12_S5 | 0.086249 | 0.031669 | 0.006465 | 2.189455 |
| high choles | Z12_S5 | 0.035661 | 0.016244 | 0.028148 | 1.550559 |
| other heart | Z12_S5 | -0.12455 | 0.065227 | 0.056204 | 1.250236 |
| arrhythmia | Z12_S5 | 0.086719 | 0.05803 | 0.135089 | 0.869379 |
| conduction | Z12_S5 | -0.06848 | 0.046873 | 0.144066 | 0.841438 |
| hypertensi | Z12_S5 | -0.01678 | 0.014711 | 0.25399 | 0.595183 |
| aortic valve | Z12_S5 | -0.04617 | 0.067862 | 0.496307 | 0.30425 |
| hypotensio | Z12_S5 | 0.031768 | 0.050899 | 0.532545 | 0.273644 |
| atrial fibrill | Z108_S5 | -0.17872 | 0.023623 | 4.01E-14 | 13.39689 |
| mitral prob | Z108_S5 | -0.19104 | 0.047378 | 5.54E-05 | 4.256186 |
| heart failur | Z108_S5 | -0.13123 | 0.039432 | 0.000876 | 3.057465 |
| high choles | Z108_S5 | 0.041221 | 0.01442 | 0.004259 | 2.370686 |
| myocardial | Z108_S5 | 0.084736 | 0.030247 | 0.005091 | 2.293206 |
| type 2 diab | Z108_S5 | 0.053018 | 0.025852 | 0.040299 | 1.394707 |
| other heart | Z108_S5 | -0.10197 | 0.05777 | 0.077567 | 1.110325 |
| conduction | Z108_S5 | -0.05977 | 0.041512 | 0.149956 | 0.824036 |
| hypertensi | Z108_S5 | -0.01705 | 0.013067 | 0.192038 | 0.716612 |
| aortic valve | Z108_S5 | -0.06806 | 0.060094 | 0.257383 | 0.58942 |
| arrhythmia | Z108_S5 | 0.045388 | 0.051403 | 0.377258 | 0.423361 |
| coronary h | Z108_S5 | 0.01909 | 0.023284 | 0.412303 | 0.384783 |
| hypotensio | Z108_S5 | 0.022334 | 0.045075 | 0.620253 | 0.207431 |
| angina pec | Z108_S5 | 0.008866 | 0.028057 | 0.752004 | 0.12378 |
| atrial fibrill | Z83_S2 | 0.165563 | 0.021984 | 5.22E-14 | 13.28255 |
| heart failur | Z83_S2 | 0.140238 | 0.036567 | 0.000126 | 3.900078 |
| myocardial | Z83_S2 | 0.077302 | 0.028039 | 0.005839 | 2.233694 |

| | | | | | |
|---|---|---|---|---|---|
| coronary h | Z83_S2 | 0.057738 | 0.021595 | 0.007507 | 2.124536 |
| mitral prob | Z83_S2 | 0.101985 | 0.04394 | 0.020296 | 1.692582 |
| angina pec | Z83_S2 | 0.056798 | 0.026068 | 0.029351 | 1.53237 |
| other heart | Z83_S2 | 0.109342 | 0.053551 | 0.041181 | 1.385304 |
| aortic valve | Z83_S2 | 0.088355 | 0.055744 | 0.112976 | 0.947015 |
| conduction | Z83_S2 | 0.041948 | 0.038475 | 0.275615 | 0.559697 |
| arrhythmia | Z83_S2 | 0.050811 | 0.047681 | 0.286594 | 0.542734 |
| type 2 diab | Z83_S2 | 0.023556 | 0.023973 | 0.325821 | 0.487021 |
| high choles | Z83_S2 | -0.01064 | 0.013366 | 0.425888 | 0.370705 |
| hypotensio | Z83_S2 | 0.033101 | 0.041781 | 0.42822 | 0.368333 |
| hypertensi | Z83_S2 | -0.00088 | 0.012151 | 0.942012 | 0.025943 |
| atrial fibrill | Z20_S1 | 0.190323 | 0.026177 | 3.69E-13 | 12.43246 |
| mitral prob | Z20_S1 | 0.160649 | 0.052482 | 0.002208 | 2.655909 |
| aortic valve | Z20_S1 | 0.201778 | 0.066575 | 0.002442 | 2.612323 |
| myocardial | Z20_S1 | -0.10108 | 0.0335 | 0.002553 | 2.592869 |
| hypertensi | Z20_S1 | 0.0419 | 0.014435 | 0.003705 | 2.431241 |
| angina pec | Z20_S1 | -0.07322 | 0.031101 | 0.018564 | 1.731336 |
| coronary h | Z20_S1 | -0.04458 | 0.025791 | 0.083908 | 1.076195 |
| type 2 diab | Z20_S1 | -0.04661 | 0.028627 | 0.10351 | 0.985018 |
| conduction | Z20_S1 | 0.065023 | 0.045956 | 0.157108 | 0.803803 |
| heart failur | Z20_S1 | 0.059618 | 0.043705 | 0.172547 | 0.763092 |
| other heart | Z20_S1 | 0.076126 | 0.063992 | 0.234214 | 0.630387 |
| high choles | Z20_S1 | -0.01776 | 0.015909 | 0.264327 | 0.577859 |
| arrhythmia | Z20_S1 | -0.04662 | 0.056987 | 0.413314 | 0.383719 |
| hypotensio | Z20_S1 | 0.02651 | 0.049931 | 0.595465 | 0.225144 |
| atrial fibrill | Z80_S1 | -0.1803 | 0.024846 | 4.09E-13 | 12.38787 |
| heart failur | Z80_S1 | -0.18155 | 0.041507 | 1.23E-05 | 4.911727 |
| type 2 diab | Z80_S1 | -0.09877 | 0.027198 | 0.000282 | 3.549024 |
| angina pec | Z80_S1 | 0.106335 | 0.02957 | 0.000324 | 3.489667 |
| coronary h | Z80_S1 | 0.077825 | 0.024539 | 0.001518 | 2.818594 |
| myocardial | Z80_S1 | 0.097307 | 0.031868 | 0.002265 | 2.644968 |
| mitral prob | Z80_S1 | -0.11933 | 0.0499 | 0.016797 | 1.774779 |
| other heart | Z80_S1 | -0.12861 | 0.060825 | 0.034485 | 1.462373 |
| aortic valve | Z80_S1 | -0.1199 | 0.063321 | 0.05831 | 1.23426 |
| hypertensi | Z80_S1 | -0.02308 | 0.013733 | 0.092789 | 1.032501 |
| hypotensio | Z80_S1 | -0.07517 | 0.04744 | 0.113109 | 0.946502 |
| conduction | Z80_S1 | 0.023945 | 0.043691 | 0.583654 | 0.233845 |
| high choles | Z80_S1 | -0.00592 | 0.015134 | 0.695711 | 0.157571 |
| arrhythmia | Z80_S1 | -0.01484 | 0.054171 | 0.784155 | 0.105598 |
| atrial fibrill | Z44_S3 | -0.17961 | 0.025058 | 7.84E-13 | 12.10587 |
| mitral prob | Z44_S3 | -0.22674 | 0.050179 | 6.26E-06 | 5.203614 |
| heart failur | Z44_S3 | -0.1659 | 0.041768 | 7.15E-05 | 4.145675 |
| type 2 diab | Z44_S3 | 0.086076 | 0.027376 | 0.001668 | 2.777881 |
| high choles | Z44_S3 | 0.024016 | 0.015265 | 0.115667 | 0.936791 |
| other heart | Z44_S3 | -0.09351 | 0.061215 | 0.126631 | 0.89746 |

| | | | | | |
|---|---|---|---|---|---|
| conduction | Z44_S3 | -0.03963 | 0.043979 | 0.367545 | 0.434689 |
| myocardial | Z44_S3 | 0.025353 | 0.032022 | 0.428526 | 0.368023 |
| hypertensi | Z44_S3 | -0.00844 | 0.013844 | 0.542294 | 0.265766 |
| angina pec | Z44_S3 | -0.01496 | 0.029714 | 0.61473 | 0.211316 |
| coronary h | Z44_S3 | -0.01109 | 0.02464 | 0.652563 | 0.185378 |
| arrhythmia | Z44_S3 | -0.02432 | 0.054451 | 0.65518 | 0.183639 |
| hypotensio | Z44_S3 | 0.006234 | 0.047755 | 0.896143 | 0.047623 |
| aortic valve | Z44_S3 | -0.00298 | 0.063667 | 0.962648 | 0.016532 |
| atrial fibrill | Z32_S5 | -0.19365 | 0.027113 | 9.47E-13 | 12.02388 |
| heart failur | Z32_S5 | -0.19847 | 0.045207 | 1.14E-05 | 4.94416 |
| mitral prob | Z32_S5 | -0.18412 | 0.054298 | 0.000698 | 3.156222 |
| other heart | Z32_S5 | -0.10752 | 0.066216 | 0.104431 | 0.981172 |
| conduction | Z32_S5 | -0.06827 | 0.047579 | 0.151333 | 0.820067 |
| myocardial | Z32_S5 | -0.04723 | 0.034668 | 0.173118 | 0.761657 |
| high choles | Z32_S5 | 0.011552 | 0.016454 | 0.482624 | 0.316391 |
| hypotensio | Z32_S5 | -0.03374 | 0.051649 | 0.51358 | 0.289391 |
| hypertensi | Z32_S5 | 0.008553 | 0.014964 | 0.567604 | 0.245955 |
| coronary h | Z32_S5 | -0.01287 | 0.026667 | 0.629498 | 0.201006 |
| arrhythmia | Z32_S5 | -0.02301 | 0.058963 | 0.696403 | 0.157139 |
| angina pec | Z32_S5 | -0.00861 | 0.032158 | 0.788784 | 0.103042 |
| type 2 diab | Z32_S5 | 0.002533 | 0.029605 | 0.931818 | 0.030669 |
| aortic valve | Z32_S5 | 0.00258 | 0.068909 | 0.97013 | 0.01317 |
| atrial fibrill | Z28_S4 | -0.17396 | 0.02449 | 1.25E-12 | 11.90171 |
| type 2 diab | Z28_S4 | 0.174128 | 0.026696 | 7.05E-11 | 10.15159 |
| mitral prob | Z28_S4 | -0.19686 | 0.049061 | 6.02E-05 | 4.220088 |
| angina pec | Z28_S4 | 0.092595 | 0.029071 | 0.001449 | 2.839013 |
| heart failur | Z28_S4 | -0.07545 | 0.040875 | 0.06492 | 1.187623 |
| aortic valve | Z28_S4 | -0.1095 | 0.062231 | 0.078505 | 1.105102 |
| coronary h | Z28_S4 | 0.037129 | 0.024111 | 0.123599 | 0.907984 |
| hypertensi | Z28_S4 | -0.01583 | 0.013514 | 0.241559 | 0.616976 |
| other heart | Z28_S4 | -0.04883 | 0.059787 | 0.414105 | 0.382889 |
| hypotensio | Z28_S4 | -0.02322 | 0.046661 | 0.618822 | 0.208434 |
| high choles | Z28_S4 | 0.006588 | 0.014875 | 0.657845 | 0.181876 |
| arrhythmia | Z28_S4 | -0.01256 | 0.0533 | 0.813671 | 0.089551 |
| myocardial | Z28_S4 | 0.002612 | 0.03135 | 0.933601 | 0.029838 |
| conduction | Z28_S4 | -0.00347 | 0.042963 | 0.935619 | 0.028901 |
| type 2 diab | Z46_S1 | 0.178038 | 0.025136 | 1.45E-12 | 11.83838 |
| aortic valve | Z46_S1 | -0.13743 | 0.058304 | 0.018424 | 1.734607 |
| mitral prob | Z46_S1 | -0.09452 | 0.045979 | 0.039813 | 1.399979 |
| atrial fibrill | Z46_S1 | -0.04579 | 0.02301 | 0.046578 | 1.331816 |
| angina pec | Z46_S1 | 0.051768 | 0.027273 | 0.057688 | 1.238912 |
| heart failur | Z46_S1 | 0.068263 | 0.038327 | 0.074916 | 1.125427 |
| other heart | Z46_S1 | 0.089023 | 0.056064 | 0.11233 | 0.949506 |
| coronary h | Z46_S1 | 0.030675 | 0.022648 | 0.175612 | 0.755446 |
| myocardial | Z46_S1 | 0.033661 | 0.029387 | 0.252047 | 0.598519 |

| | | | | | |
|---|---|---|---|---|---|
| hypertensi | Z46_S1 | -0.01374 | 0.012722 | 0.280122 | 0.552653 |
| high choles | Z46_S1 | -0.01097 | 0.01396 | 0.431964 | 0.364553 |
| conduction | Z46_S1 | 0.015159 | 0.040307 | 0.706858 | 0.150668 |
| hypotensio | Z46_S1 | 0.01168 | 0.043754 | 0.789504 | 0.102646 |
| arrhythmia | Z46_S1 | 0.003887 | 0.049949 | 0.937964 | 0.027814 |
| atrial fibrill | Z1_S2 | 0.135559 | 0.01916 | 1.53E-12 | 11.81397 |
| aortic valve | Z1_S2 | 0.178447 | 0.048548 | 0.000238 | 3.623894 |
| mitral prob | Z1_S2 | 0.107301 | 0.03826 | 0.005043 | 2.297289 |
| heart failur | Z1_S2 | 0.078251 | 0.031887 | 0.014134 | 1.849737 |
| hypertensi | Z1_S2 | 0.020907 | 0.010587 | 0.048307 | 1.315987 |
| angina pec | Z1_S2 | -0.02193 | 0.022723 | 0.334414 | 0.475715 |
| myocardial | Z1_S2 | -0.02288 | 0.024445 | 0.349234 | 0.456883 |
| hypotensio | Z1_S2 | 0.034057 | 0.036397 | 0.349439 | 0.456629 |
| high choles | Z1_S2 | -0.01072 | 0.011652 | 0.357509 | 0.446713 |
| other heart | Z1_S2 | 0.034768 | 0.04666 | 0.456195 | 0.340849 |
| type 2 diab | Z1_S2 | -0.01323 | 0.020911 | 0.526965 | 0.278218 |
| coronary h | Z1_S2 | -0.01033 | 0.018845 | 0.583649 | 0.233848 |
| conduction | Z1_S2 | 0.008844 | 0.033528 | 0.791965 | 0.101294 |
| arrhythmia | Z1_S2 | -0.00435 | 0.041549 | 0.916609 | 0.037816 |
| atrial fibrill | Z106_S3 | -0.18623 | 0.026471 | 2.05E-12 | 11.68881 |
| type 2 diab | Z106_S3 | 0.146135 | 0.028866 | 4.17E-07 | 6.379948 |
| mitral prob | Z106_S3 | -0.21438 | 0.053016 | 5.28E-05 | 4.277401 |
| heart failur | Z106_S3 | -0.1129 | 0.044158 | 0.010576 | 1.975694 |
| angina pec | Z106_S3 | 0.078587 | 0.031411 | 0.01236 | 1.907979 |
| coronary h | Z106_S3 | 0.047246 | 0.026045 | 0.069693 | 1.156813 |
| conduction | Z106_S3 | -0.07952 | 0.046441 | 0.086852 | 1.061222 |
| high choles | Z106_S3 | 0.022654 | 0.016071 | 0.158681 | 0.799476 |
| hypertensi | Z106_S3 | -0.01866 | 0.014604 | 0.201425 | 0.695887 |
| other heart | Z106_S3 | -0.07429 | 0.064655 | 0.25054 | 0.601123 |
| myocardial | Z106_S3 | 0.033501 | 0.033858 | 0.322461 | 0.491523 |
| hypotensio | Z106_S3 | 0.024009 | 0.050435 | 0.634049 | 0.197877 |
| aortic valve | Z106_S3 | 0.022705 | 0.067302 | 0.73585 | 0.133211 |
| arrhythmia | Z106_S3 | 0.013492 | 0.057585 | 0.814758 | 0.088971 |
| atrial fibrill | Z117_S4 | -0.13399 | 0.019187 | 2.96E-12 | 11.52866 |
| mitral prob | Z117_S4 | -0.12658 | 0.038321 | 0.000957 | 3.018966 |
| aortic valve | Z117_S4 | -0.15881 | 0.048618 | 0.001091 | 2.962311 |
| heart failur | Z117_S4 | -0.09701 | 0.031918 | 0.002373 | 2.624625 |
| type 2 diab | Z117_S4 | 0.057607 | 0.020965 | 0.006006 | 2.221448 |
| hypertensi | Z117_S4 | -0.02357 | 0.010633 | 0.026646 | 1.574367 |
| coronary h | Z117_S4 | -0.02246 | 0.018864 | 0.233773 | 0.631205 |
| other heart | Z117_S4 | -0.05428 | 0.046739 | 0.245518 | 0.609916 |
| arrhythmia | Z117_S4 | -0.04027 | 0.041602 | 0.333025 | 0.477524 |
| high choles | Z117_S4 | 0.009556 | 0.011667 | 0.412765 | 0.384298 |
| myocardial | Z117_S4 | -0.01762 | 0.024485 | 0.471766 | 0.326274 |
| hypotensio | Z117_S4 | -0.01963 | 0.036453 | 0.590186 | 0.229011 |

| | | | | | |
|---|---|---|---|---|---|
| angina pec | Z117_S4 | -0.00526 | 0.022772 | 0.817293 | 0.087622 |
| conduction | Z117_S4 | 0.006871 | 0.033583 | 0.83788 | 0.076818 |
| atrial fibrill | Z107_S1 | -0.13124 | 0.019024 | 5.38E-12 | 11.2692 |
| aortic valve | Z107_S1 | -0.1965 | 0.048235 | 4.64E-05 | 4.333549 |
| heart failur | Z107_S1 | -0.11787 | 0.031656 | 0.000197 | 3.705679 |
| mitral prob | Z107_S1 | -0.12246 | 0.03802 | 0.00128 | 2.892863 |
| hypotensio | Z107_S1 | -0.07601 | 0.036134 | 0.035435 | 1.45057 |
| hypertensi | Z107_S1 | -0.01733 | 0.010518 | 0.099524 | 1.002074 |
| arrhythmia | Z107_S1 | -0.06569 | 0.041273 | 0.111513 | 0.952673 |
| coronary h | Z107_S1 | -0.02678 | 0.018724 | 0.152721 | 0.816102 |
| myocardial | Z107_S1 | -0.03024 | 0.024288 | 0.213061 | 0.671495 |
| other heart | Z107_S1 | -0.04467 | 0.046365 | 0.335357 | 0.474493 |
| high choles | Z107_S1 | 0.005198 | 0.011555 | 0.652801 | 0.185219 |
| type 2 diab | Z107_S1 | 0.007548 | 0.020782 | 0.71646 | 0.144808 |
| conduction | Z107_S1 | 0.006721 | 0.033296 | 0.84003 | 0.075705 |
| angina pec | Z107_S1 | -0.00024 | 0.022577 | 0.991528 | 0.003695 |
| myocardial | Z18_S4 | -0.2387 | 0.035369 | 1.53E-11 | 10.81625 |
| coronary h | Z18_S4 | -0.16099 | 0.027219 | 3.37E-09 | 8.472199 |
| angina pec | Z18_S4 | -0.15399 | 0.032852 | 2.78E-06 | 5.555812 |
| heart failur | Z18_S4 | -0.12739 | 0.046204 | 0.005836 | 2.233914 |
| hypertensi | Z18_S4 | 0.025206 | 0.015277 | 0.09897 | 1.004494 |
| arrhythmia | Z18_S4 | -0.08209 | 0.06023 | 0.172892 | 0.762225 |
| mitral prob | Z18_S4 | -0.06599 | 0.055491 | 0.234337 | 0.630159 |
| high choles | Z18_S4 | 0.013731 | 0.016842 | 0.414941 | 0.382014 |
| other heart | Z18_S4 | -0.04 | 0.067673 | 0.554435 | 0.25615 |
| hypotensio | Z18_S4 | -0.01029 | 0.052793 | 0.845481 | 0.072896 |
| type 2 diab | Z18_S4 | -0.00574 | 0.030274 | 0.849511 | 0.070831 |
| aortic valve | Z18_S4 | 0.012368 | 0.070401 | 0.860545 | 0.065226 |
| conduction | Z18_S4 | -0.00844 | 0.048614 | 0.862242 | 0.064371 |
| atrial fibrill | Z18_S4 | 0.003553 | 0.027753 | 0.898143 | 0.046655 |
| type 2 diab | Z92_S5 | -0.19862 | 0.029703 | 2.33E-11 | 10.63256 |
| atrial fibrill | Z92_S5 | 0.180902 | 0.027227 | 3.11E-11 | 10.50657 |
| coronary h | Z92_S5 | 0.141426 | 0.026749 | 1.25E-07 | 6.901981 |
| myocardial | Z92_S5 | 0.156424 | 0.034737 | 6.73E-06 | 5.171837 |
| angina pec | Z92_S5 | 0.139597 | 0.032263 | 1.52E-05 | 4.818386 |
| mitral prob | Z92_S5 | 0.227022 | 0.054447 | 3.06E-05 | 4.513871 |
| heart failur | Z92_S5 | 0.125309 | 0.045363 | 0.005743 | 2.240863 |
| high choles | Z92_S5 | -0.04445 | 0.016526 | 0.007161 | 2.145022 |
| conduction | Z92_S5 | 0.106052 | 0.0477 | 0.026205 | 1.58162 |
| arrhythmia | Z92_S5 | 0.114807 | 0.059121 | 0.052161 | 1.28265 |
| hypertensi | Z92_S5 | -0.02015 | 0.015024 | 0.179982 | 0.744772 |
| aortic valve | Z92_S5 | 0.08428 | 0.069115 | 0.222702 | 0.652276 |
| other heart | Z92_S5 | 0.041376 | 0.066428 | 0.53337 | 0.272971 |
| hypotensio | Z92_S5 | 0.018103 | 0.051824 | 0.726857 | 0.138551 |
| myocardial | Z49_S1 | -0.23101 | 0.034585 | 2.45E-11 | 10.61 |

| | | | | | |
|---|---|---|---|---|---|
| coronary h | Z49_S1 | -0.16833 | 0.026619 | 2.60E-10 | 9.585149 |
| angina pec | Z49_S1 | -0.16347 | 0.032112 | 3.60E-07 | 6.443715 |
| heart failur | Z49_S1 | -0.12485 | 0.045157 | 0.0057 | 2.244106 |
| arrhythmia | Z49_S1 | -0.14337 | 0.058846 | 0.014846 | 1.828395 |
| hypotensio | Z49_S1 | -0.08005 | 0.051566 | 0.12059 | 0.918688 |
| mitral prob | Z49_S1 | -0.07488 | 0.054227 | 0.167332 | 0.776422 |
| other heart | Z49_S1 | -0.07915 | 0.066118 | 0.23131 | 0.635806 |
| atrial fibrill | Z49_S1 | 0.027396 | 0.027104 | 0.312137 | 0.505655 |
| hypertensi | Z49_S1 | 0.015025 | 0.014927 | 0.314176 | 0.502826 |
| aortic valve | Z49_S1 | 0.033375 | 0.068771 | 0.62746 | 0.202414 |
| type 2 diab | Z49_S1 | -0.00953 | 0.029557 | 0.74708 | 0.126633 |
| conduction | Z49_S1 | 0.010648 | 0.047475 | 0.822541 | 0.084843 |
| high choles | Z49_S1 | -0.00033 | 0.016441 | 0.984124 | 0.00695 |
| atrial fibrill | Z40_S4 | 0.178635 | 0.026764 | 2.54E-11 | 10.59552 |
| mitral prob | Z40_S4 | 0.169893 | 0.053689 | 0.001556 | 2.807949 |
| heart failur | Z40_S4 | 0.117475 | 0.044696 | 0.008586 | 2.066188 |
| coronary h | Z40_S4 | -0.05557 | 0.026375 | 0.035122 | 1.454426 |
| myocardial | Z40_S4 | -0.07056 | 0.034275 | 0.039533 | 1.403045 |
| angina pec | Z40_S4 | -0.05943 | 0.031794 | 0.061614 | 1.210319 |
| high choles | Z40_S4 | -0.01509 | 0.016295 | 0.354528 | 0.450349 |
| conduction | Z40_S4 | 0.037796 | 0.047046 | 0.421762 | 0.374932 |
| hypertensi | Z40_S4 | -0.01133 | 0.014771 | 0.443204 | 0.353396 |
| arrhythmia | Z40_S4 | -0.03314 | 0.058254 | 0.569415 | 0.244571 |
| other heart | Z40_S4 | 0.028788 | 0.065483 | 0.660213 | 0.180316 |
| hypotensio | Z40_S4 | -0.01131 | 0.051062 | 0.824671 | 0.083719 |
| type 2 diab | Z40_S4 | -0.00526 | 0.029274 | 0.857316 | 0.066859 |
| aortic valve | Z40_S4 | -0.00413 | 0.068113 | 0.951703 | 0.021499 |
| atrial fibrill | Z7_S3 | -0.17897 | 0.026818 | 2.55E-11 | 10.59268 |
| mitral prob | Z7_S3 | -0.19325 | 0.05378 | 0.000327 | 3.485375 |
| type 2 diab | Z7_S3 | 0.102441 | 0.029356 | 0.000485 | 3.314622 |
| hypertensi | Z7_S3 | -0.04403 | 0.014805 | 0.002941 | 2.53156 |
| angina pec | Z7_S3 | 0.07113 | 0.031881 | 0.025681 | 1.590383 |
| myocardial | Z7_S3 | 0.075376 | 0.034343 | 0.028191 | 1.549895 |
| heart failur | Z7_S3 | -0.09788 | 0.04478 | 0.028842 | 1.53997 |
| other heart | Z7_S3 | -0.12634 | 0.065572 | 0.054024 | 1.267416 |
| coronary h | Z7_S3 | 0.03964 | 0.026431 | 0.133687 | 0.873911 |
| aortic valve | Z7_S3 | -0.08432 | 0.068256 | 0.216708 | 0.664124 |
| conduction | Z7_S3 | -0.04621 | 0.047115 | 0.326712 | 0.485835 |
| high choles | Z7_S3 | 0.010875 | 0.016312 | 0.504975 | 0.29673 |
| arrhythmia | Z7_S3 | 0.020737 | 0.058416 | 0.722603 | 0.1411 |
| hypotensio | Z7_S3 | -0.00306 | 0.051196 | 0.952339 | 0.021208 |
| atrial fibrill | Z41_S2 | -0.17595 | 0.026608 | 3.85E-11 | 10.41427 |
| type 2 diab | Z41_S2 | 0.165665 | 0.029078 | 1.23E-08 | 7.909307 |
| mitral prob | Z41_S2 | -0.16875 | 0.053324 | 0.001555 | 2.808393 |
| conduction | Z41_S2 | -0.09793 | 0.046699 | 0.03601 | 1.443582 |

| | | | | | |
|---|---|---|---|---|---|
| aortic valve | Z41_S2 | -0.11302 | 0.067675 | 0.094913 | 1.022676 |
| high choles | Z41_S2 | 0.021665 | 0.016182 | 0.180647 | 0.74317 |
| heart failur | Z41_S2 | -0.05662 | 0.044417 | 0.202433 | 0.693719 |
| angina pec | Z41_S2 | 0.021931 | 0.031601 | 0.487706 | 0.311842 |
| hypotensio | Z41_S2 | -0.02505 | 0.050716 | 0.621315 | 0.206688 |
| hypertensi | Z41_S2 | -0.00703 | 0.014683 | 0.632004 | 0.19928 |
| other heart | Z41_S2 | -0.02473 | 0.065017 | 0.703719 | 0.152601 |
| arrhythmia | Z41_S2 | -0.01494 | 0.057911 | 0.796449 | 0.098842 |
| coronary h | Z41_S2 | 0.006652 | 0.026207 | 0.799629 | 0.097112 |
| myocardial | Z41_S2 | -0.00073 | 0.034036 | 0.982791 | 0.007539 |
| atrial fibrill | Z99_S4 | 0.162187 | 0.025434 | 1.84E-10 | 9.734556 |
| myocardial | Z99_S4 | -0.14497 | 0.032508 | 8.25E-06 | 5.08341 |
| hypertensi | Z99_S4 | 0.038117 | 0.014031 | 0.006598 | 2.18058 |
| coronary h | Z99_S4 | -0.0673 | 0.02503 | 0.007178 | 2.14401 |
| angina pec | Z99_S4 | -0.0711 | 0.030163 | 0.018426 | 1.73456 |
| aortic valve | Z99_S4 | 0.129547 | 0.064609 | 0.044966 | 1.347116 |
| heart failur | Z99_S4 | 0.081729 | 0.042392 | 0.053873 | 1.268632 |
| other heart | Z99_S4 | 0.090349 | 0.062071 | 0.145523 | 0.837069 |
| mitral prob | Z99_S4 | 0.068175 | 0.050925 | 0.180672 | 0.743109 |
| hypotensio | Z99_S4 | 0.053175 | 0.048402 | 0.271949 | 0.565513 |
| conduction | Z99_S4 | 0.045201 | 0.044635 | 0.311216 | 0.506938 |
| high choles | Z99_S4 | -0.01283 | 0.015456 | 0.406577 | 0.390857 |
| type 2 diab | Z99_S4 | 0.004791 | 0.027744 | 0.862901 | 0.064039 |
| arrhythmia | Z99_S4 | 0.007752 | 0.055276 | 0.888472 | 0.051356 |
| atrial fibrill | Z51_S1 | -0.12998 | 0.020525 | 2.46E-10 | 9.60974 |
| heart failur | Z51_S1 | -0.14323 | 0.034148 | 2.75E-05 | 4.561298 |
| type 2 diab | Z51_S1 | -0.08475 | 0.022394 | 0.000154 | 3.811748 |
| aortic valve | Z51_S1 | -0.1782 | 0.052071 | 0.000622 | 3.205959 |
| hypotensio | Z51_S1 | -0.08446 | 0.039009 | 0.030392 | 1.517234 |
| hypertensi | Z51_S1 | -0.02267 | 0.011355 | 0.045871 | 1.33846 |
| other heart | Z51_S1 | -0.085 | 0.050031 | 0.089344 | 1.048937 |
| mitral prob | Z51_S1 | -0.06922 | 0.04104 | 0.091673 | 1.037759 |
| angina pec | Z51_S1 | 0.015204 | 0.024359 | 0.532537 | 0.27365 |
| arrhythmia | Z51_S1 | -0.02307 | 0.044549 | 0.604502 | 0.218602 |
| coronary h | Z51_S1 | -0.00707 | 0.020185 | 0.726202 | 0.138943 |
| high choles | Z51_S1 | 0.003988 | 0.012466 | 0.749042 | 0.125494 |
| conduction | Z51_S1 | -0.00866 | 0.035922 | 0.809434 | 0.091819 |
| myocardial | Z51_S1 | 0.002491 | 0.026203 | 0.924261 | 0.034205 |
| atrial fibrill | Z66_S3 | -0.16168 | 0.025557 | 2.55E-10 | 9.592611 |
| type 2 diab | Z66_S3 | 0.114582 | 0.027927 | 4.09E-05 | 4.38801 |
| mitral prob | Z66_S3 | -0.16313 | 0.051223 | 0.001451 | 2.838213 |
| angina pec | Z66_S3 | 0.063691 | 0.030362 | 0.035941 | 1.444416 |
| conduction | Z66_S3 | -0.09378 | 0.044844 | 0.036525 | 1.43741 |
| aortic valve | Z66_S3 | -0.11921 | 0.065005 | 0.066678 | 1.17602 |
| coronary h | Z66_S3 | 0.04608 | 0.02518 | 0.06726 | 1.172241 |

| | | | | | |
|---|---|---|---|---|---|
| myocardial | Z66_S3 | 0.054764 | 0.032711 | 0.094109 | 1.026369 |
| heart failur | Z66_S3 | -0.0644 | 0.042671 | 0.131256 | 0.88188 |
| hypertensi | Z66_S3 | -0.01854 | 0.014118 | 0.189178 | 0.72313 |
| high choles | Z66_S3 | 0.019446 | 0.015534 | 0.210646 | 0.676448 |
| arrhythmia | Z66_S3 | 0.038698 | 0.05565 | 0.486816 | 0.312635 |
| hypotensio | Z66_S3 | 0.009217 | 0.048731 | 0.849988 | 0.070587 |
| other heart | Z66_S3 | 0.006286 | 0.062454 | 0.919827 | 0.036294 |
| type 2 diab | Z38_S5 | 0.165445 | 0.026446 | 4.02E-10 | 9.3959 |
| myocardial | Z38_S5 | -0.12166 | 0.030915 | 8.34E-05 | 4.07893 |
| coronary h | Z38_S5 | -0.06435 | 0.023813 | 0.006892 | 2.161652 |
| atrial fibrill | Z38_S5 | -0.05732 | 0.024235 | 0.018021 | 1.744231 |
| angina pec | Z38_S5 | -0.05007 | 0.028687 | 0.080933 | 1.091874 |
| conduction | Z38_S5 | -0.06888 | 0.042456 | 0.104725 | 0.979951 |
| hypertensi | Z38_S5 | 0.017104 | 0.013369 | 0.200759 | 0.697325 |
| arrhythmia | Z38_S5 | -0.05413 | 0.052574 | 0.303178 | 0.518302 |
| mitral prob | Z38_S5 | -0.03705 | 0.048458 | 0.444483 | 0.352145 |
| heart failur | Z38_S5 | 0.027926 | 0.040382 | 0.489229 | 0.310488 |
| aortic valve | Z38_S5 | -0.04134 | 0.061471 | 0.501273 | 0.299925 |
| other heart | Z38_S5 | 0.026821 | 0.059071 | 0.649804 | 0.187218 |
| high choles | Z38_S5 | 0.001963 | 0.014739 | 0.894076 | 0.048626 |
| hypotensio | Z38_S5 | -0.0031 | 0.046083 | 0.946417 | 0.023918 |
| atrial fibrill | Z50_S4 | 0.138552 | 0.022376 | 6.04E-10 | 9.219119 |
| aortic valve | Z50_S4 | 0.204389 | 0.05675 | 0.000317 | 3.498915 |
| mitral prob | Z50_S4 | 0.137837 | 0.044745 | 0.002069 | 2.684234 |
| hypertensi | Z50_S4 | 0.037408 | 0.012355 | 0.002466 | 2.608052 |
| heart failur | Z50_S4 | 0.07657 | 0.037257 | 0.03987 | 1.399354 |
| type 2 diab | Z50_S4 | -0.03718 | 0.024438 | 0.128146 | 0.892293 |
| other heart | Z50_S4 | 0.064406 | 0.054545 | 0.237699 | 0.623972 |
| myocardial | Z50_S4 | -0.02533 | 0.028582 | 0.375474 | 0.42542 |
| hypotensio | Z50_S4 | 0.037585 | 0.042546 | 0.377022 | 0.423634 |
| high choles | Z50_S4 | -0.01079 | 0.013593 | 0.427339 | 0.369228 |
| conduction | Z50_S4 | 0.028881 | 0.039199 | 0.461264 | 0.33605 |
| arrhythmia | Z50_S4 | 0.023963 | 0.048558 | 0.621663 | 0.206445 |
| coronary h | Z50_S4 | 0.010131 | 0.022006 | 0.645246 | 0.190275 |
| angina pec | Z50_S4 | -0.00452 | 0.026546 | 0.864803 | 0.063083 |
| atrial fibrill | Z20_S2 | 0.122793 | 0.019858 | 6.38E-10 | 9.195258 |
| aortic valve | Z20_S2 | 0.1572 | 0.050327 | 0.001789 | 2.747409 |
| mitral prob | Z20_S2 | 0.086344 | 0.039666 | 0.029507 | 1.530075 |
| heart failur | Z20_S2 | 0.067338 | 0.033041 | 0.041558 | 1.381342 |
| hypertensi | Z20_S2 | 0.021576 | 0.010974 | 0.049293 | 1.307216 |
| myocardial | Z20_S2 | -0.03288 | 0.025338 | 0.194449 | 0.711195 |
| angina pec | Z20_S2 | -0.02426 | 0.023549 | 0.302842 | 0.518784 |
| high choles | Z20_S2 | -0.01169 | 0.012078 | 0.333049 | 0.477492 |
| coronary h | Z20_S2 | -0.01477 | 0.019527 | 0.449304 | 0.34746 |
| other heart | Z20_S2 | 0.036356 | 0.048369 | 0.452275 | 0.344598 |

| | | | | | |
|---|---|---|---|---|---|
| hypotensio | Z20_S2 | 0.028253 | 0.037721 | 0.453861 | 0.343078 |
| type 2 diab | Z20_S2 | -0.00845 | 0.021667 | 0.696412 | 0.157134 |
| arrhythmia | Z20_S2 | -0.01026 | 0.043056 | 0.811637 | 0.090638 |
| conduction | Z20_S2 | -0.00196 | 0.034756 | 0.954975 | 0.020008 |
| atrial fibrill | Z55_S5 | 0.123548 | 0.020098 | 8.01E-10 | 9.096311 |
| aortic valve | Z55_S5 | 0.153424 | 0.050959 | 0.002609 | 2.583476 |
| myocardial | Z55_S5 | -0.07149 | 0.025668 | 0.005353 | 2.271384 |
| hypertensi | Z55_S5 | 0.028586 | 0.011118 | 0.010141 | 1.993925 |
| mitral prob | Z55_S5 | 0.098525 | 0.040169 | 0.014185 | 1.848175 |
| angina pec | Z55_S5 | -0.04874 | 0.023846 | 0.040991 | 1.387314 |
| coronary h | Z55_S5 | -0.0329 | 0.019788 | 0.096401 | 1.015917 |
| heart failur | Z55_S5 | 0.043512 | 0.03347 | 0.193603 | 0.713088 |
| type 2 diab | Z55_S5 | -0.02542 | 0.021955 | 0.247023 | 0.607262 |
| other heart | Z55_S5 | 0.050683 | 0.048968 | 0.300673 | 0.521906 |
| hypotensio | Z55_S5 | 0.029236 | 0.038211 | 0.44421 | 0.352411 |
| high choles | Z55_S5 | -0.00721 | 0.01223 | 0.555482 | 0.25533 |
| arrhythmia | Z55_S5 | -0.00996 | 0.043607 | 0.819349 | 0.086531 |
| conduction | Z55_S5 | -0.00598 | 0.035204 | 0.865078 | 0.062945 |
| atrial fibrill | Z61_S4 | 0.136756 | 0.022264 | 8.26E-10 | 9.083262 |
| heart failur | Z61_S4 | 0.104963 | 0.036972 | 0.00453 | 2.343883 |
| conduction | Z61_S4 | 0.104367 | 0.038874 | 0.007263 | 2.138871 |
| hypertensi | Z61_S4 | -0.03228 | 0.012295 | 0.008658 | 2.062586 |
| other heart | Z61_S4 | 0.110398 | 0.054097 | 0.041287 | 1.384186 |
| mitral prob | Z61_S4 | 0.088933 | 0.044352 | 0.044957 | 1.347206 |
| myocardial | Z61_S4 | 0.049257 | 0.028347 | 0.082288 | 1.084665 |
| coronary h | Z61_S4 | 0.025743 | 0.021839 | 0.238514 | 0.622485 |
| angina pec | Z61_S4 | 0.030689 | 0.026336 | 0.243907 | 0.612775 |
| aortic valve | Z61_S4 | -0.05332 | 0.056276 | 0.343444 | 0.464144 |
| type 2 diab | Z61_S4 | -0.01569 | 0.024197 | 0.516788 | 0.286688 |
| high choles | Z61_S4 | -0.0056 | 0.013495 | 0.678202 | 0.168641 |
| arrhythmia | Z61_S4 | 0.017454 | 0.048101 | 0.716714 | 0.144654 |
| hypotensio | Z61_S4 | 0.006995 | 0.042185 | 0.868298 | 0.061331 |
| atrial fibrill | Z37_S2 | -0.14994 | 0.024619 | 1.14E-09 | 8.942178 |
| type 2 diab | Z37_S2 | 0.136104 | 0.026898 | 4.23E-07 | 6.374025 |
| mitral prob | Z37_S2 | -0.15103 | 0.049325 | 0.002202 | 2.657153 |
| heart failur | Z37_S2 | -0.09872 | 0.04108 | 0.016268 | 1.788659 |
| conduction | Z37_S2 | -0.09579 | 0.043184 | 0.026546 | 1.576 |
| myocardial | Z37_S2 | -0.06592 | 0.031493 | 0.036334 | 1.439685 |
| other heart | Z37_S2 | -0.04936 | 0.060127 | 0.4117 | 0.385419 |
| arrhythmia | Z37_S2 | -0.0341 | 0.053561 | 0.524378 | 0.280355 |
| coronary h | Z37_S2 | -0.01347 | 0.024246 | 0.578529 | 0.237675 |
| hypotensio | Z37_S2 | 0.024947 | 0.046928 | 0.595012 | 0.225474 |
| hypertensi | Z37_S2 | 0.004491 | 0.013593 | 0.741107 | 0.130119 |
| angina pec | Z37_S2 | -0.00949 | 0.0292 | 0.745258 | 0.127693 |
| aortic valve | Z37_S2 | -0.01323 | 0.062599 | 0.832564 | 0.079582 |

| | | | | | |
|---|---|---|---|---|---|
| high choles | Z37_S2 | -0.00068 | 0.014956 | 0.963743 | 0.016039 |
| atrial fibrill | Z29_S5 | 0.145746 | 0.024047 | 1.37E-09 | 8.861758 |
| type 2 diab | Z29_S5 | -0.12595 | 0.026289 | 1.67E-06 | 5.777271 |
| mitral prob | Z29_S5 | 0.166831 | 0.048179 | 0.000536 | 3.271119 |
| aortic valve | Z29_S5 | 0.160437 | 0.061143 | 0.008697 | 2.060638 |
| conduction | Z29_S5 | 0.08179 | 0.042203 | 0.052633 | 1.278739 |
| angina pec | Z29_S5 | -0.05445 | 0.028566 | 0.056641 | 1.246867 |
| myocardial | Z29_S5 | -0.04709 | 0.030782 | 0.126088 | 0.899326 |
| high choles | Z29_S5 | -0.02 | 0.014622 | 0.171286 | 0.766278 |
| hypertensi | Z29_S5 | 0.017945 | 0.013292 | 0.177022 | 0.751973 |
| coronary h | Z29_S5 | -0.0303 | 0.023711 | 0.201247 | 0.696271 |
| heart failur | Z29_S5 | 0.04966 | 0.040162 | 0.216292 | 0.66496 |
| arrhythmia | Z29_S5 | -0.03917 | 0.052345 | 0.454309 | 0.342648 |
| other heart | Z29_S5 | 0.007537 | 0.05875 | 0.897918 | 0.046763 |
| hypotensio | Z29_S5 | -0.00509 | 0.04584 | 0.911505 | 0.040241 |
| coronary h | Z82_S4 | -0.12004 | 0.019811 | 1.39E-09 | 8.857389 |
| angina pec | Z82_S4 | -0.1287 | 0.023892 | 7.25E-08 | 7.139798 |
| myocardial | Z82_S4 | -0.13676 | 0.025711 | 1.05E-07 | 6.978324 |
| heart failur | Z82_S4 | -0.14322 | 0.033576 | 2.00E-05 | 4.698423 |
| atrial fibrill | Z82_S4 | -0.07785 | 0.020219 | 0.000118 | 3.927304 |
| mitral prob | Z82_S4 | -0.11471 | 0.040327 | 0.004452 | 2.351491 |
| aortic valve | Z82_S4 | -0.13306 | 0.051126 | 0.009256 | 2.033579 |
| arrhythmia | Z82_S4 | -0.08698 | 0.04377 | 0.046911 | 1.328727 |
| type 2 diab | Z82_S4 | 0.033374 | 0.021985 | 0.129009 | 0.88938 |
| conduction | Z82_S4 | -0.04721 | 0.035288 | 0.180955 | 0.74243 |
| high choles | Z82_S4 | 0.014775 | 0.012258 | 0.228087 | 0.641899 |
| other heart | Z82_S4 | -0.04683 | 0.049177 | 0.340986 | 0.467263 |
| hypotensio | Z82_S4 | -0.02106 | 0.038337 | 0.582827 | 0.234461 |
| hypertensi | Z82_S4 | 0.004417 | 0.011128 | 0.691391 | 0.160276 |
| atrial fibrill | Z37_S4 | -0.16492 | 0.027637 | 2.45E-09 | 8.611193 |
| myocardial | Z37_S4 | 0.187674 | 0.035261 | 1.03E-07 | 6.985478 |
| coronary h | Z37_S4 | 0.140759 | 0.027126 | 2.13E-07 | 6.671586 |
| angina pec | Z37_S4 | 0.155188 | 0.032725 | 2.13E-06 | 5.672343 |
| mitral prob | Z37_S4 | -0.10677 | 0.055293 | 0.053506 | 1.271601 |
| type 2 diab | Z37_S4 | 0.054211 | 0.030165 | 0.072332 | 1.140672 |
| arrhythmia | Z37_S4 | 0.064563 | 0.060006 | 0.281961 | 0.549811 |
| other heart | Z37_S4 | -0.06993 | 0.067434 | 0.299769 | 0.523213 |
| heart failur | Z37_S4 | -0.03942 | 0.04605 | 0.391942 | 0.406778 |
| aortic valve | Z37_S4 | 0.047106 | 0.070161 | 0.501972 | 0.299321 |
| hypertensi | Z37_S4 | -0.0097 | 0.015217 | 0.523781 | 0.28085 |
| hypotensio | Z37_S4 | 0.02825 | 0.052612 | 0.591309 | 0.228186 |
| high choles | Z37_S4 | 0.003243 | 0.016771 | 0.846685 | 0.072278 |
| conduction | Z37_S4 | -0.00665 | 0.048442 | 0.890861 | 0.05019 |
| atrial fibrill | Z9_S2 | 0.144421 | 0.024476 | 3.68E-09 | 8.434718 |
| mitral prob | Z9_S2 | 0.172561 | 0.048952 | 0.000424 | 3.37254 |

| | | | | | |
|---|---|---|---|---|---|
| heart failur | Z9_S2 | 0.131374 | 0.040752 | 0.001267 | 2.897218 |
| type 2 diab | Z9_S2 | -0.0808 | 0.026698 | 0.002477 | 2.606075 |
| myocardial | Z9_S2 | 0.092524 | 0.031262 | 0.003083 | 2.510989 |
| other heart | Z9_S2 | 0.174856 | 0.059657 | 0.003382 | 2.470821 |
| arrhythmia | Z9_S2 | 0.099823 | 0.053104 | 0.060157 | 1.220715 |
| conduction | Z9_S2 | 0.07412 | 0.042842 | 0.08363 | 1.077639 |
| coronary h | Z9_S2 | 0.040496 | 0.024052 | 0.092258 | 1.034996 |
| high choles | Z9_S2 | -0.01586 | 0.014861 | 0.285759 | 0.544 |
| hypotensio | Z9_S2 | 0.043066 | 0.046546 | 0.35485 | 0.449955 |
| aortic valve | Z9_S2 | 0.042217 | 0.062109 | 0.496682 | 0.303922 |
| hypertensi | Z9_S2 | 0.008493 | 0.013515 | 0.529733 | 0.275943 |
| angina pec | Z9_S2 | 0.012575 | 0.029001 | 0.664575 | 0.177456 |
| atrial fibrill | Z13_S1 | -0.12209 | 0.020706 | 3.77E-09 | 8.4236 |
| type 2 diab | Z13_S1 | -0.11358 | 0.02261 | 5.11E-07 | 6.29123 |
| heart failur | Z13_S1 | -0.14099 | 0.034503 | 4.40E-05 | 4.356852 |
| aortic valve | Z13_S1 | -0.20446 | 0.052635 | 0.000103 | 3.987833 |
| myocardial | Z13_S1 | 0.062478 | 0.026478 | 0.018301 | 1.737528 |
| hypertensi | Z13_S1 | -0.02448 | 0.011443 | 0.032413 | 1.489276 |
| angina pec | Z13_S1 | 0.052316 | 0.0246 | 0.033455 | 1.475539 |
| hypotensio | Z13_S1 | -0.07905 | 0.039412 | 0.044901 | 1.347745 |
| mitral prob | Z13_S1 | -0.06808 | 0.041462 | 0.100618 | 0.997325 |
| coronary h | Z13_S1 | 0.028403 | 0.020406 | 0.163969 | 0.785238 |
| other heart | Z13_S1 | -0.06261 | 0.050562 | 0.215613 | 0.666324 |
| conduction | Z13_S1 | -0.01476 | 0.036286 | 0.684196 | 0.16482 |
| high choles | Z13_S1 | 0.003399 | 0.012577 | 0.786976 | 0.104039 |
| arrhythmia | Z13_S1 | -0.01082 | 0.045022 | 0.810128 | 0.091446 |
| type 2 diab | Z12_S4 | 0.129466 | 0.022059 | 4.45E-09 | 8.352019 |
| atrial fibrill | Z12_S4 | -0.08795 | 0.020188 | 1.33E-05 | 4.877626 |
| mitral prob | Z12_S4 | -0.14754 | 0.040314 | 0.000253 | 3.596696 |
| coronary h | Z12_S4 | -0.05058 | 0.019842 | 0.010803 | 1.966446 |
| aortic valve | Z12_S4 | -0.12694 | 0.051125 | 0.013038 | 1.884796 |
| myocardial | Z12_S4 | -0.05677 | 0.025752 | 0.027497 | 1.560708 |
| hypertensi | Z12_S4 | -0.02021 | 0.011187 | 0.070786 | 1.150051 |
| angina pec | Z12_S4 | -0.02915 | 0.023944 | 0.223417 | 0.650884 |
| heart failur | Z12_S4 | -0.03846 | 0.033598 | 0.252362 | 0.597976 |
| conduction | Z12_S4 | 0.030517 | 0.035342 | 0.387898 | 0.411282 |
| other heart | Z12_S4 | -0.03191 | 0.049175 | 0.516447 | 0.286974 |
| high choles | Z12_S4 | 0.006521 | 0.012269 | 0.595093 | 0.225415 |
| arrhythmia | Z12_S4 | -0.01733 | 0.04376 | 0.692051 | 0.159862 |
| hypotensio | Z12_S4 | 0.01321 | 0.038366 | 0.730622 | 0.136307 |
| atrial fibrill | Z37_S3 | -0.14951 | 0.025734 | 6.34E-09 | 8.198151 |
| heart failur | Z37_S3 | -0.15927 | 0.042869 | 0.000203 | 3.691535 |
| myocardial | Z37_S3 | -0.10582 | 0.032851 | 0.001278 | 2.893498 |
| mitral prob | Z37_S3 | -0.12222 | 0.051481 | 0.017597 | 1.754561 |
| other heart | Z37_S3 | -0.12159 | 0.062767 | 0.052737 | 1.277884 |

| | | | | | |
|---|---|---|---|---|---|
| conduction | Z37_S3 | -0.08514 | 0.045099 | 0.059049 | 1.22879 |
| coronary h | Z37_S3 | -0.04617 | 0.025288 | 0.067914 | 1.168038 |
| type 2 diab | Z37_S3 | 0.033498 | 0.028064 | 0.232634 | 0.633327 |
| hypertensi | Z37_S3 | 0.014718 | 0.014213 | 0.30045 | 0.522228 |
| angina pec | Z37_S3 | -0.02922 | 0.030479 | 0.337666 | 0.471513 |
| aortic valv | Z37_S3 | 0.050016 | 0.065309 | 0.443785 | 0.352828 |
| arrhythmia | Z37_S3 | -0.01975 | 0.055874 | 0.723752 | 0.14041 |
| high choles | Z37_S3 | -0.00467 | 0.015609 | 0.764651 | 0.116537 |
| hypotensio | Z37_S3 | -0.00873 | 0.048937 | 0.858378 | 0.066322 |
| atrial fibrill | Z87_S3 | -0.12977 | 0.022386 | 6.84E-09 | 8.164808 |
| angina pec | Z87_S3 | 0.073711 | 0.026538 | 0.005482 | 2.261039 |
| heart failur | Z87_S3 | -0.09735 | 0.037343 | 0.009146 | 2.038769 |
| mitral prob | Z87_S3 | -0.11592 | 0.044819 | 0.009702 | 2.013158 |
| coronary h | Z87_S3 | 0.045405 | 0.022032 | 0.039325 | 1.405329 |
| type 2 diab | Z87_S3 | 0.047876 | 0.024416 | 0.049913 | 1.301785 |
| conduction | Z87_S3 | -0.06156 | 0.039228 | 0.116576 | 0.933392 |
| hypertensi | Z87_S3 | -0.01556 | 0.012374 | 0.208699 | 0.68048 |
| aortic valv | Z87_S3 | -0.05269 | 0.056865 | 0.354135 | 0.450831 |
| high choles | Z87_S3 | 0.010879 | 0.013588 | 0.423347 | 0.373304 |
| other heart | Z87_S3 | -0.02287 | 0.054642 | 0.67552 | 0.170362 |
| arrhythmia | Z87_S3 | 0.01885 | 0.048692 | 0.698662 | 0.155733 |
| myocardial | Z87_S3 | 0.008623 | 0.028636 | 0.763307 | 0.117301 |
| hypotensio | Z87_S3 | -0.00829 | 0.042599 | 0.845729 | 0.072769 |
| atrial fibrill | Z95_S5 | -0.15538 | 0.026923 | 7.96E-09 | 8.099199 |
| type 2 diab | Z95_S5 | 0.117634 | 0.029408 | 6.35E-05 | 4.19719 |
| mitral prob | Z95_S5 | -0.15823 | 0.053954 | 0.003364 | 2.473204 |
| coronary h | Z95_S5 | 0.069135 | 0.026508 | 0.009112 | 2.0404 |
| angina pec | Z95_S5 | 0.080695 | 0.031969 | 0.011603 | 1.935426 |
| myocardial | Z95_S5 | 0.064784 | 0.034453 | 0.060071 | 1.221338 |
| conduction | Z95_S5 | -0.0721 | 0.047258 | 0.127082 | 0.895917 |
| heart failur | Z95_S5 | -0.05465 | 0.044942 | 0.223968 | 0.649814 |
| high choles | Z95_S5 | 0.017964 | 0.016358 | 0.272143 | 0.565203 |
| arrhythmia | Z95_S5 | 0.051909 | 0.058604 | 0.375751 | 0.4251 |
| other heart | Z95_S5 | -0.03645 | 0.065794 | 0.579541 | 0.236916 |
| hypertensi | Z95_S5 | -0.00778 | 0.014848 | 0.600466 | 0.221512 |
| aortic valv | Z95_S5 | -0.03165 | 0.068484 | 0.643986 | 0.191123 |
| hypotensio | Z95_S5 | 0.011351 | 0.051329 | 0.824987 | 0.083553 |
| type 2 diab | Z79_S4 | 0.166716 | 0.028973 | 8.82E-09 | 8.054609 |
| atrial fibrill | Z79_S4 | -0.12509 | 0.026608 | 2.60E-06 | 5.584712 |
| mitral prob | Z79_S4 | -0.20535 | 0.053243 | 0.000115 | 3.938525 |
| myocardial | Z79_S4 | -0.07997 | 0.034 | 0.018672 | 1.728805 |
| heart failur | Z79_S4 | -0.07509 | 0.044338 | 0.090361 | 1.044019 |
| coronary h | Z79_S4 | -0.03361 | 0.026164 | 0.198908 | 0.701348 |
| other heart | Z79_S4 | -0.06576 | 0.064915 | 0.311051 | 0.507168 |
| arrhythmia | Z79_S4 | -0.05015 | 0.057815 | 0.385741 | 0.413704 |

| | | | | | |
|---|---|---|---|---|---|
| aortic valve | Z79_S4 | -0.02895 | 0.067572 | 0.668305 | 0.175026 |
| high choles | Z79_S4 | 0.006726 | 0.016155 | 0.67719 | 0.16929 |
| conduction | Z79_S4 | -0.01878 | 0.046644 | 0.687186 | 0.162926 |
| hypotensio | Z79_S4 | -0.0078 | 0.050654 | 0.877635 | 0.056686 |
| hypertensi | Z79_S4 | -0.00068 | 0.014655 | 0.962728 | 0.016496 |
| angina pec | Z79_S4 | 0.000417 | 0.03155 | 0.989462 | 0.004601 |
| type 2 diab | Z50_S2 | -0.17122 | 0.029761 | 8.88E-09 | 8.051655 |
| atrial fibrill | Z50_S2 | 0.131554 | 0.027249 | 1.39E-06 | 5.857373 |
| coronary h | Z50_S2 | -0.08847 | 0.02681 | 0.000968 | 3.014037 |
| mitral prob | Z50_S2 | 0.168588 | 0.054593 | 0.002017 | 2.695319 |
| angina pec | Z50_S2 | -0.09208 | 0.032352 | 0.004428 | 2.35383 |
| myocardial | Z50_S2 | -0.0958 | 0.034848 | 0.005979 | 2.223339 |
| hypertensi | Z50_S2 | 0.022984 | 0.01501 | 0.125717 | 0.900607 |
| aortic valve | Z50_S2 | 0.040121 | 0.069295 | 0.5626 | 0.2498 |
| high choles | Z50_S2 | -0.00713 | 0.016566 | 0.666835 | 0.175982 |
| conduction | Z50_S2 | -0.00985 | 0.047832 | 0.836827 | 0.077364 |
| other heart | Z50_S2 | 0.008218 | 0.066579 | 0.901761 | 0.044909 |
| hypotensio | Z50_S2 | -0.00506 | 0.05194 | 0.922457 | 0.035054 |
| heart failur | Z50_S2 | -0.0039 | 0.045468 | 0.931655 | 0.030745 |
| arrhythmia | Z50_S2 | -0.00286 | 0.059282 | 0.961466 | 0.017066 |
| atrial fibrill | Z38_S4 | -0.14835 | 0.026051 | 1.25E-08 | 7.902735 |
| heart failur | Z38_S4 | -0.14337 | 0.043428 | 0.000964 | 3.015871 |
| mitral prob | Z38_S4 | -0.1411 | 0.052163 | 0.006837 | 2.165151 |
| angina pec | Z38_S4 | 0.068921 | 0.030889 | 0.025676 | 1.590476 |
| other heart | Z38_S4 | -0.1064 | 0.063578 | 0.094241 | 1.025758 |
| type 2 diab | Z38_S4 | 0.03337 | 0.028404 | 0.240065 | 0.619672 |
| myocardial | Z38_S4 | -0.03543 | 0.033308 | 0.287491 | 0.541376 |
| coronary h | Z38_S4 | 0.021137 | 0.025625 | 0.409457 | 0.387792 |
| hypotensio | Z38_S4 | -0.03904 | 0.049584 | 0.431122 | 0.3654 |
| arrhythmia | Z38_S4 | -0.04158 | 0.056646 | 0.462906 | 0.334507 |
| hypertensi | Z38_S4 | 0.006835 | 0.014365 | 0.634234 | 0.19775 |
| high choles | Z38_S4 | -0.00703 | 0.015799 | 0.65646 | 0.182792 |
| conduction | Z38_S4 | -0.01206 | 0.045697 | 0.791895 | 0.101332 |
| aortic valve | Z38_S4 | -0.00047 | 0.06617 | 0.994382 | 0.002447 |
| myocardial | Z75_S5 | 0.195561 | 0.035378 | 3.28E-08 | 7.484409 |
| atrial fibrill | Z75_S5 | 0.131476 | 0.027736 | 2.15E-06 | 5.668157 |
| coronary h | Z75_S5 | 0.125587 | 0.027231 | 4.01E-06 | 5.396772 |
| heart failur | Z75_S5 | 0.211371 | 0.046184 | 4.75E-06 | 5.323485 |
| angina pec | Z75_S5 | 0.142542 | 0.032859 | 1.44E-05 | 4.84039 |
| arrhythmia | Z75_S5 | 0.171422 | 0.060215 | 0.00442 | 2.354605 |
| mitral prob | Z75_S5 | 0.130045 | 0.055476 | 0.019078 | 1.719472 |
| other heart | Z75_S5 | 0.127844 | 0.067658 | 0.058829 | 1.230412 |
| hypertensi | Z75_S5 | -0.02864 | 0.015283 | 0.060961 | 1.214951 |
| high choles | Z75_S5 | -0.02222 | 0.016838 | 0.186989 | 0.728185 |
| aortic valve | Z75_S5 | -0.0734 | 0.070381 | 0.297004 | 0.527238 |

| | | | | | |
|---|---|---|---|---|---|
| hypotensio | Z75_S5 | 0.047838 | 0.052783 | 0.364783 | 0.437965 |
| type 2 diab | Z75_S5 | -0.01964 | 0.030265 | 0.516346 | 0.287059 |
| conduction | Z75_S5 | 0.019672 | 0.048599 | 0.685652 | 0.163896 |
| atrial fibrill | Z35_S5 | -0.1116 | 0.020212 | 3.40E-08 | 7.468324 |
| type 2 diab | Z35_S5 | 0.062365 | 0.02206 | 0.004701 | 2.327779 |
| aortic valve | Z35_S5 | -0.13153 | 0.051192 | 0.010194 | 1.991663 |
| mitral prob | Z35_S5 | -0.09815 | 0.040364 | 0.015043 | 1.822662 |
| hypertensi | Z35_S5 | -0.02153 | 0.011168 | 0.053943 | 1.268068 |
| heart failur | Z35_S5 | -0.03909 | 0.033634 | 0.24519 | 0.610498 |
| other heart | Z35_S5 | -0.05479 | 0.049206 | 0.265515 | 0.575911 |
| hypotensio | Z35_S5 | -0.0241 | 0.038392 | 0.530192 | 0.275567 |
| myocardial | Z35_S5 | 0.01247 | 0.025782 | 0.628642 | 0.201597 |
| arrhythmia | Z35_S5 | -0.01987 | 0.043808 | 0.650079 | 0.187034 |
| coronary h | Z35_S5 | -0.00797 | 0.019873 | 0.688488 | 0.162104 |
| conduction | Z35_S5 | 0.011543 | 0.035385 | 0.744265 | 0.128272 |
| high choles | Z35_S5 | 0.001548 | 0.012283 | 0.899716 | 0.045894 |
| angina pec | Z35_S5 | 0.001622 | 0.023959 | 0.94602 | 0.0241 |
| atrial fibrill | Z49_S3 | -0.14387 | 0.026116 | 3.65E-08 | 7.43716 |
| type 2 diab | Z49_S3 | 0.117548 | 0.028445 | 3.60E-05 | 4.443506 |
| mitral prob | Z49_S3 | -0.17995 | 0.052249 | 0.000574 | 3.241212 |
| angina pec | Z49_S3 | 0.072395 | 0.030944 | 0.019316 | 1.71409 |
| heart failur | Z49_S3 | -0.08411 | 0.043511 | 0.053244 | 1.273726 |
| coronary h | Z49_S3 | 0.039795 | 0.02566 | 0.120945 | 0.917412 |
| other heart | Z49_S3 | -0.08304 | 0.063706 | 0.192404 | 0.715787 |
| conduction | Z49_S3 | -0.04942 | 0.045755 | 0.280141 | 0.552624 |
| hypertensi | Z49_S3 | -0.01419 | 0.014388 | 0.32405 | 0.489388 |
| myocardial | Z49_S3 | 0.025055 | 0.033356 | 0.452574 | 0.34431 |
| high choles | Z49_S3 | 0.009891 | 0.015832 | 0.532133 | 0.27398 |
| aortic valve | Z49_S3 | 0.037674 | 0.066298 | 0.569871 | 0.244224 |
| hypotensio | Z49_S3 | 0.020538 | 0.049695 | 0.679408 | 0.167869 |
| arrhythmia | Z49_S3 | -0.00284 | 0.056726 | 0.960024 | 0.017718 |
| atrial fibrill | Z26_S4 | 0.151788 | 0.027668 | 4.15E-08 | 7.38174 |
| type 2 diab | Z26_S4 | -0.14455 | 0.030172 | 1.67E-06 | 5.777273 |
| mitral prob | Z26_S4 | 0.209916 | 0.055362 | 0.00015 | 3.823851 |
| angina pec | Z26_S4 | -0.06141 | 0.032802 | 0.061184 | 1.213365 |
| myocardial | Z26_S4 | -0.06344 | 0.035325 | 0.072537 | 1.139441 |
| aortic valve | Z26_S4 | 0.103909 | 0.070263 | 0.139194 | 0.856378 |
| hypertensi | Z26_S4 | 0.020059 | 0.015233 | 0.187923 | 0.72602 |
| coronary h | Z26_S4 | -0.02538 | 0.027183 | 0.350496 | 0.455317 |
| other heart | Z26_S4 | 0.05819 | 0.067533 | 0.388889 | 0.410175 |
| high choles | Z26_S4 | -0.01326 | 0.01679 | 0.429662 | 0.366874 |
| conduction | Z26_S4 | 0.033245 | 0.048505 | 0.493107 | 0.307059 |
| arrhythmia | Z26_S4 | -0.03855 | 0.06012 | 0.521389 | 0.282838 |
| heart failur | Z26_S4 | 0.01653 | 0.046115 | 0.720004 | 0.142665 |
| hypotensio | Z26_S4 | -0.00799 | 0.052686 | 0.87941 | 0.055809 |

| | | | | | |
|---|---|---|---|---|---|
| type 2 diab | Z112_S3 | 0.159219 | 0.029044 | 4.25E-08 | 7.371665 |
| atrial fibrill | Z112_S3 | -0.09951 | 0.026626 | 0.000186 | 3.729574 |
| coronary h | Z112_S3 | 0.05897 | 0.026193 | 0.024369 | 1.613157 |
| mitral prob | Z112_S3 | -0.10097 | 0.053318 | 0.058273 | 1.234533 |
| angina pec | Z112_S3 | 0.058305 | 0.03159 | 0.064952 | 1.187407 |
| myocardial | Z112_S3 | 0.049291 | 0.03403 | 0.147503 | 0.831198 |
| conduction | Z112_S3 | -0.06242 | 0.046683 | 0.181238 | 0.741751 |
| other heart | Z112_S3 | 0.057866 | 0.064997 | 0.373319 | 0.42792 |
| hypotensio | Z112_S3 | 0.040902 | 0.050712 | 0.419925 | 0.376828 |
| arrhythmia | Z112_S3 | 0.039874 | 0.057886 | 0.490934 | 0.308977 |
| high choles | Z112_S3 | 0.007806 | 0.016175 | 0.62939 | 0.20108 |
| hypertensi | Z112_S3 | -0.004 | 0.014695 | 0.785628 | 0.104783 |
| heart failur | Z112_S3 | 0.011872 | 0.044392 | 0.789131 | 0.102851 |
| aortic valve | Z112_S3 | -0.00653 | 0.067649 | 0.923148 | 0.034729 |
| type 2 diab | Z22_S2 | -0.13591 | 0.024891 | 4.81E-08 | 7.317614 |
| heart failur | Z22_S2 | -0.11467 | 0.038078 | 0.002604 | 2.584434 |
| atrial fibrill | Z22_S2 | -0.05111 | 0.022844 | 0.025261 | 1.597548 |
| high choles | Z22_S2 | -0.02723 | 0.01385 | 0.049324 | 1.306941 |
| angina pec | Z22_S2 | 0.048675 | 0.027139 | 0.072906 | 1.137239 |
| aortic valve | Z22_S2 | -0.09794 | 0.058064 | 0.091672 | 1.037764 |
| mitral prob | Z22_S2 | -0.06217 | 0.045756 | 0.174227 | 0.758885 |
| hypotensio | Z22_S2 | -0.04354 | 0.04349 | 0.316815 | 0.499194 |
| conduction | Z22_S2 | 0.036528 | 0.040065 | 0.361931 | 0.441375 |
| coronary h | Z22_S2 | 0.018229 | 0.022501 | 0.41787 | 0.378958 |
| myocardial | Z22_S2 | 0.015708 | 0.029223 | 0.590905 | 0.228482 |
| other heart | Z22_S2 | -0.02811 | 0.055791 | 0.614391 | 0.211555 |
| arrhythmia | Z22_S2 | 0.022313 | 0.04969 | 0.653412 | 0.184813 |
| hypertensi | Z22_S2 | -0.00351 | 0.012605 | 0.780551 | 0.107599 |
| myocardial | Z10_S4 | -0.1666 | 0.030583 | 5.16E-08 | 7.287512 |
| type 2 diab | Z10_S4 | 0.139296 | 0.026036 | 8.88E-08 | 7.051831 |
| atrial fibrill | Z10_S4 | 0.105578 | 0.023933 | 1.03E-05 | 4.986489 |
| coronary h | Z10_S4 | -0.09359 | 0.023561 | 7.15E-05 | 4.145899 |
| angina pec | Z10_S4 | -0.08269 | 0.028375 | 0.003569 | 2.447509 |
| heart failur | Z10_S4 | 0.0658 | 0.039856 | 0.098767 | 1.005387 |
| hypertensi | Z10_S4 | 0.018737 | 0.013209 | 0.156051 | 0.806734 |
| aortic valve | Z10_S4 | 0.057723 | 0.060738 | 0.341944 | 0.466045 |
| mitral prob | Z10_S4 | -0.04431 | 0.047889 | 0.354779 | 0.450042 |
| high choles | Z10_S4 | -0.01268 | 0.014554 | 0.383487 | 0.416249 |
| hypotensio | Z10_S4 | 0.028686 | 0.045498 | 0.528382 | 0.277052 |
| conduction | Z10_S4 | 0.024006 | 0.04193 | 0.566977 | 0.246434 |
| arrhythmia | Z10_S4 | -0.02783 | 0.051956 | 0.592267 | 0.227482 |
| other heart | Z10_S4 | 0.030782 | 0.058341 | 0.597773 | 0.223464 |
| atrial fibrill | Z59_S3 | -0.10266 | 0.01901 | 6.72E-08 | 7.172582 |
| aortic valve | Z59_S3 | -0.16411 | 0.048103 | 0.000647 | 3.189244 |
| heart failur | Z59_S3 | -0.07296 | 0.031588 | 0.020922 | 1.679406 |

| | | | | | |
|---|---|---|---|---|---|
| hypertensi | Z59_S3 | -0.02405 | 0.010496 | 0.021965 | 1.658268 |
| mitral prob | Z59_S3 | -0.07441 | 0.03793 | 0.04979 | 1.302859 |
| high choles | Z59_S3 | 0.012161 | 0.011533 | 0.291681 | 0.535092 |
| myocardial | Z59_S3 | 0.020113 | 0.024235 | 0.406607 | 0.390825 |
| hypotensio | Z59_S3 | -0.02879 | 0.036077 | 0.424836 | 0.371778 |
| other heart | Z59_S3 | -0.03302 | 0.046251 | 0.475307 | 0.323026 |
| type 2 diab | Z59_S3 | 0.011965 | 0.020722 | 0.56368 | 0.248967 |
| angina pec | Z59_S3 | 0.008874 | 0.022527 | 0.693647 | 0.158861 |
| arrhythmia | Z59_S3 | -0.01016 | 0.041165 | 0.805043 | 0.094181 |
| coronary h | Z59_S3 | 0.004263 | 0.018677 | 0.819453 | 0.086476 |
| conduction | Z59_S3 | -0.00535 | 0.03322 | 0.871965 | 0.059501 |
| type 2 diab | Z1_S5 | -0.14 | 0.026214 | 9.34E-08 | 7.029524 |
| heart failur | Z1_S5 | -0.14222 | 0.040022 | 0.000381 | 3.419119 |
| other heart | Z1_S5 | -0.12341 | 0.058635 | 0.035321 | 1.45197 |
| angina pec | Z1_S5 | -0.0388 | 0.028519 | 0.173706 | 0.760186 |
| coronary h | Z1_S5 | -0.02957 | 0.023633 | 0.210845 | 0.676036 |
| mitral prob | Z1_S5 | -0.05217 | 0.048108 | 0.278223 | 0.555607 |
| conduction | Z1_S5 | 0.044331 | 0.042121 | 0.292589 | 0.533742 |
| atrial fibrill | Z1_S5 | -0.02224 | 0.024048 | 0.355148 | 0.449591 |
| aortic valve | Z1_S5 | 0.048078 | 0.061025 | 0.430795 | 0.365729 |
| hypertensi | Z1_S5 | -0.00993 | 0.013304 | 0.455381 | 0.341625 |
| myocardial | Z1_S5 | -0.00838 | 0.030692 | 0.784779 | 0.105253 |
| arrhythmia | Z1_S5 | -0.01414 | 0.052198 | 0.786514 | 0.104294 |
| hypotensio | Z1_S5 | 0.007171 | 0.045738 | 0.875409 | 0.057789 |
| high choles | Z1_S5 | 0.000592 | 0.014674 | 0.967844 | 0.014195 |
| type 2 diab | Z66_S2 | -0.14829 | 0.027953 | 1.14E-07 | 6.944252 |
| atrial fibrill | Z66_S2 | 0.09631 | 0.02557 | 0.000166 | 3.779987 |
| mitral prob | Z66_S2 | 0.102791 | 0.051222 | 0.044786 | 1.348856 |
| coronary h | Z66_S2 | -0.04797 | 0.0252 | 0.056979 | 1.244287 |
| myocardial | Z66_S2 | -0.05782 | 0.032711 | 0.077117 | 1.11285 |
| angina pec | Z66_S2 | -0.04587 | 0.030365 | 0.130912 | 0.88302 |
| other heart | Z66_S2 | -0.08738 | 0.062462 | 0.161836 | 0.790925 |
| aortic valve | Z66_S2 | 0.087241 | 0.065001 | 0.179556 | 0.745799 |
| conduction | Z66_S2 | 0.056379 | 0.04487 | 0.208957 | 0.679944 |
| high choles | Z66_S2 | -0.01752 | 0.015536 | 0.259525 | 0.585821 |
| arrhythmia | Z66_S2 | -0.04747 | 0.055626 | 0.393475 | 0.405083 |
| hypotensio | Z66_S2 | -0.03991 | 0.048734 | 0.41281 | 0.384249 |
| hypertensi | Z66_S2 | 0.007512 | 0.014117 | 0.594657 | 0.225733 |
| heart failur | Z66_S2 | 7.94E-05 | 0.042674 | 0.998515 | 0.000645 |
| atrial fibrill | Z51_S3 | -0.1443 | 0.027282 | 1.24E-07 | 6.906204 |
| type 2 diab | Z51_S3 | 0.154735 | 0.02981 | 2.11E-07 | 6.675223 |
| mitral prob | Z51_S3 | -0.1967 | 0.054671 | 0.000321 | 3.492825 |
| coronary h | Z51_S3 | 0.079182 | 0.026865 | 0.003208 | 2.493766 |
| angina pec | Z51_S3 | 0.092144 | 0.032397 | 0.004457 | 2.35099 |
| myocardial | Z51_S3 | 0.086509 | 0.034921 | 0.013246 | 1.877926 |

| | | | | | |
|---|---|---|---|---|---|
| heart failur | Z51_S3 | -0.06501 | 0.04554 | 0.153435 | 0.814076 |
| hypertensi | Z51_S3 | -0.0196 | 0.015043 | 0.192632 | 0.715273 |
| arrhythmia | Z51_S3 | 0.040606 | 0.059369 | 0.494012 | 0.306262 |
| high choles | Z51_S3 | 0.01132 | 0.016582 | 0.494836 | 0.305539 |
| hypotensio | Z51_S3 | 0.034807 | 0.052033 | 0.503539 | 0.297967 |
| other heart | Z51_S3 | -0.03083 | 0.066687 | 0.643893 | 0.191187 |
| aortic valve | Z51_S3 | 0.027761 | 0.069403 | 0.689162 | 0.161679 |
| conduction | Z51_S3 | -0.01308 | 0.047911 | 0.784772 | 0.105257 |
| type 2 diab | Z3_S2 | -0.11822 | 0.022878 | 2.39E-07 | 6.621003 |
| angina pec | Z3_S2 | -0.05281 | 0.024899 | 0.033935 | 1.469357 |
| mitral prob | Z3_S2 | 0.074129 | 0.04202 | 0.077723 | 1.109451 |
| atrial fibrill | Z3_S2 | 0.032067 | 0.021021 | 0.127159 | 0.895653 |
| coronary h | Z3_S2 | -0.02762 | 0.020663 | 0.181393 | 0.74138 |
| other heart | Z3_S2 | 0.054576 | 0.051224 | 0.286689 | 0.54259 |
| hypertensi | Z3_S2 | 0.010861 | 0.0116 | 0.349143 | 0.456996 |
| arrhythmia | Z3_S2 | 0.030874 | 0.045583 | 0.498212 | 0.302586 |
| conduction | Z3_S2 | 0.024362 | 0.036761 | 0.507521 | 0.294546 |
| heart failur | Z3_S2 | -0.02205 | 0.034962 | 0.528247 | 0.277163 |
| aortic valve | Z3_S2 | -0.03177 | 0.053297 | 0.551111 | 0.258761 |
| myocardial | Z3_S2 | -0.01588 | 0.026838 | 0.553943 | 0.256535 |
| high choles | Z3_S2 | -0.00302 | 0.012782 | 0.813433 | 0.089678 |
| hypotensio | Z3_S2 | 0.005946 | 0.039946 | 0.881666 | 0.054696 |
| atrial fibrill | Z86_S3 | -0.13092 | 0.025544 | 3.00E-07 | 6.523173 |
| type 2 diab | Z86_S3 | 0.120373 | 0.027872 | 1.58E-05 | 4.802586 |
| conduction | Z86_S3 | -0.13819 | 0.044731 | 0.002008 | 2.697241 |
| mitral prob | Z86_S3 | -0.14707 | 0.05109 | 0.003997 | 2.398258 |
| high choles | Z86_S3 | 0.040772 | 0.015508 | 0.008567 | 2.067173 |
| aortic valve | Z86_S3 | -0.12383 | 0.064821 | 0.056105 | 1.250995 |
| heart failur | Z86_S3 | -0.0755 | 0.042577 | 0.076207 | 1.118008 |
| myocardial | Z86_S3 | -0.04736 | 0.032609 | 0.146438 | 0.834347 |
| hypertensi | Z86_S3 | -0.01486 | 0.014082 | 0.291389 | 0.535527 |
| other heart | Z86_S3 | -0.04588 | 0.062295 | 0.461424 | 0.3359 |
| arrhythmia | Z86_S3 | -0.03864 | 0.055445 | 0.485857 | 0.313491 |
| angina pec | Z86_S3 | 0.020328 | 0.030262 | 0.501773 | 0.299493 |
| hypotensio | Z86_S3 | -0.0228 | 0.048613 | 0.639125 | 0.194414 |
| coronary h | Z86_S3 | 0.005101 | 0.025117 | 0.839072 | 0.076201 |
| myocardial | Z122_S1 | 0.161991 | 0.031845 | 3.67E-07 | 6.435297 |
| coronary h | Z122_S1 | 0.117996 | 0.024509 | 1.49E-06 | 5.828246 |
| angina pec | Z122_S1 | 0.101584 | 0.029567 | 0.000592 | 3.227579 |
| type 2 diab | Z122_S1 | -0.05475 | 0.027262 | 0.044629 | 1.350381 |
| aortic valve | Z122_S1 | 0.122812 | 0.063322 | 0.052453 | 1.280233 |
| arrhythmia | Z122_S1 | 0.079437 | 0.054169 | 0.142538 | 0.846068 |
| hypotensio | Z122_S1 | 0.053593 | 0.047491 | 0.259125 | 0.58649 |
| mitral prob | Z122_S1 | 0.027883 | 0.049935 | 0.576583 | 0.239138 |
| atrial fibrill | Z122_S1 | -0.01127 | 0.025057 | 0.652788 | 0.185228 |

| | | | | | |
|---|---|---|---|---|---|
| other heart | Z122_S1 | -0.01624 | 0.060881 | 0.789644 | 0.102569 |
| conduction | Z122_S1 | 0.004897 | 0.04373 | 0.910833 | 0.040561 |
| high choles | Z122_S1 | 0.001226 | 0.015153 | 0.935507 | 0.028953 |
| hypertensi | Z122_S1 | -0.00068 | 0.013785 | 0.960864 | 0.017338 |
| heart failur | Z122_S1 | -0.00049 | 0.041592 | 0.990537 | 0.004129 |
| type 2 diab | Z56_S5 | 0.111413 | 0.021974 | 4.01E-07 | 6.397337 |
| myocardial | Z56_S5 | -0.07684 | 0.025739 | 0.002836 | 2.547239 |
| aortic valve | Z56_S5 | 0.127249 | 0.051155 | 0.012871 | 1.890402 |
| hypertensi | Z56_S5 | 0.024468 | 0.011149 | 0.028196 | 1.54981 |
| coronary h | Z56_S5 | -0.03243 | 0.019839 | 0.102179 | 0.990638 |
| atrial fibrill | Z56_S5 | 0.030949 | 0.020177 | 0.125064 | 0.902868 |
| angina pec | Z56_S5 | -0.03007 | 0.023914 | 0.208692 | 0.680493 |
| heart failur | Z56_S5 | 0.041661 | 0.033564 | 0.214531 | 0.66851 |
| conduction | Z56_S5 | -0.02971 | 0.035307 | 0.400099 | 0.397833 |
| hypotensio | Z56_S5 | 0.024879 | 0.038311 | 0.516094 | 0.287271 |
| other heart | Z56_S5 | 0.031231 | 0.049141 | 0.525093 | 0.279763 |
| arrhythmia | Z56_S5 | -0.01956 | 0.04374 | 0.654778 | 0.183906 |
| mitral prob | Z56_S5 | -0.00665 | 0.040304 | 0.868862 | 0.061049 |
| high choles | Z56_S5 | -0.00117 | 0.012271 | 0.923794 | 0.034425 |
| atrial fibrill | Z75_S4 | -0.13264 | 0.026527 | 5.77E-07 | 6.238796 |
| type 2 diab | Z75_S4 | 0.13811 | 0.028942 | 1.84E-06 | 5.736023 |
| mitral prob | Z75_S4 | -0.17873 | 0.053134 | 0.00077 | 3.113443 |
| angina pec | Z75_S4 | 0.058561 | 0.03148 | 0.062863 | 1.201605 |
| aortic valve | Z75_S4 | -0.0875 | 0.067417 | 0.19435 | 0.711415 |
| heart failur | Z75_S4 | -0.05486 | 0.044264 | 0.215234 | 0.667088 |
| other heart | Z75_S4 | -0.06011 | 0.064771 | 0.353404 | 0.451729 |
| hypertensi | Z75_S4 | -0.00848 | 0.014619 | 0.561795 | 0.250422 |
| coronary h | Z75_S4 | 0.010436 | 0.026108 | 0.689365 | 0.161551 |
| arrhythmia | Z75_S4 | -0.02168 | 0.057708 | 0.707155 | 0.150485 |
| conduction | Z75_S4 | -0.01713 | 0.046543 | 0.712849 | 0.147003 |
| myocardial | Z75_S4 | -0.00779 | 0.033931 | 0.81837 | 0.08705 |
| high choles | Z75_S4 | 0.002983 | 0.016096 | 0.852979 | 0.069062 |
| hypotensio | Z75_S4 | 0.002736 | 0.05054 | 0.956824 | 0.019168 |
| atrial fibrill | Z62_S2 | 0.101428 | 0.020454 | 7.14E-07 | 6.14636 |
| aortic valve | Z62_S2 | 0.167137 | 0.051764 | 0.001245 | 2.904926 |
| heart failur | Z62_S2 | 0.067324 | 0.033995 | 0.047667 | 1.321778 |
| hypertensi | Z62_S2 | 0.016661 | 0.011288 | 0.139968 | 0.853972 |
| mitral prob | Z62_S2 | 0.057685 | 0.040804 | 0.157459 | 0.802833 |
| type 2 diab | Z62_S2 | 0.026432 | 0.02228 | 0.235488 | 0.628031 |
| hypotensio | Z62_S2 | 0.034033 | 0.038806 | 0.38049 | 0.419657 |
| other heart | Z62_S2 | 0.037645 | 0.049751 | 0.449253 | 0.347509 |
| coronary h | Z62_S2 | 0.010631 | 0.020078 | 0.596468 | 0.224413 |
| myocardial | Z62_S2 | 0.011085 | 0.026057 | 0.670529 | 0.173582 |
| high choles | Z62_S2 | -0.00469 | 0.012409 | 0.705657 | 0.151406 |
| conduction | Z62_S2 | 0.006658 | 0.035746 | 0.852251 | 0.069433 |

| | | | | | |
|---|---|---|---|---|---|
| angina pec | Z62_S2 | -0.00315 | 0.024223 | 0.896695 | 0.047355 |
| arrhythmia | Z62_S2 | 0.000758 | 0.044294 | 0.986338 | 0.005974 |
| heart failur | Z93_S1 | 0.213603 | 0.043259 | 7.96E-07 | 6.099275 |
| coronary h | Z93_S1 | 0.103456 | 0.02552 | 5.05E-05 | 4.296426 |
| myocardial | Z93_S1 | 0.128065 | 0.033157 | 0.000113 | 3.948515 |
| atrial fibrill | Z93_S1 | 0.099354 | 0.025981 | 0.000132 | 3.880741 |
| angina pec | Z93_S1 | 0.105999 | 0.030788 | 0.000577 | 3.239074 |
| mitral prob | Z93_S1 | 0.170799 | 0.051998 | 0.001022 | 2.990366 |
| other heart | Z93_S1 | 0.192477 | 0.06339 | 0.002397 | 2.620337 |
| arrhythmia | Z93_S1 | 0.138435 | 0.0564 | 0.014115 | 1.850326 |
| hypotensio | Z93_S1 | 0.073392 | 0.049425 | 0.137581 | 0.861443 |
| conduction | Z93_S1 | -0.0414 | 0.045516 | 0.363102 | 0.439971 |
| type 2 diab | Z93_S1 | 0.023551 | 0.028323 | 0.405691 | 0.391805 |
| aortic valve | Z93_S1 | 0.011337 | 0.065933 | 0.863475 | 0.06375 |
| high choles | Z93_S1 | -0.00267 | 0.015779 | 0.865848 | 0.062558 |
| hypertensi | Z93_S1 | 0.000599 | 0.014344 | 0.966707 | 0.014705 |
| myocardial | Z74_S4 | -0.15081 | 0.030637 | 8.60E-07 | 6.065493 |
| coronary h | Z74_S4 | -0.09949 | 0.023595 | 2.49E-05 | 4.603691 |
| angina pec | Z74_S4 | -0.08368 | 0.028495 | 0.003319 | 2.479002 |
| heart failur | Z74_S4 | -0.1146 | 0.040025 | 0.004198 | 2.376942 |
| arrhythmia | Z74_S4 | -0.10319 | 0.052166 | 0.047925 | 1.319436 |
| mitral prob | Z74_S4 | -0.06597 | 0.048074 | 0.170013 | 0.769519 |
| aortic valve | Z74_S4 | -0.07039 | 0.061003 | 0.248562 | 0.604565 |
| hypotensio | Z74_S4 | -0.03977 | 0.045711 | 0.384325 | 0.415302 |
| other heart | Z74_S4 | -0.04575 | 0.058627 | 0.435142 | 0.361369 |
| type 2 diab | Z74_S4 | -0.01536 | 0.026216 | 0.558074 | 0.253308 |
| conduction | Z74_S4 | 0.021129 | 0.042103 | 0.615782 | 0.210573 |
| high choles | Z74_S4 | -0.00494 | 0.014595 | 0.735137 | 0.133632 |
| hypertensi | Z74_S4 | -0.0026 | 0.013302 | 0.844891 | 0.073199 |
| atrial fibrill | Z74_S4 | -0.00281 | 0.024092 | 0.907237 | 0.042279 |
| atrial fibrill | Z1_S4 | 0.118062 | 0.024087 | 9.58E-07 | 6.018717 |
| mitral prob | Z1_S4 | 0.143084 | 0.048054 | 0.002908 | 2.536339 |
| heart failur | Z1_S4 | 0.097101 | 0.040021 | 0.015264 | 1.816341 |
| type 2 diab | Z1_S4 | -0.05668 | 0.026211 | 0.030577 | 1.514604 |
| high choles | Z1_S4 | -0.0255 | 0.014599 | 0.080709 | 1.093077 |
| conduction | Z1_S4 | 0.072419 | 0.042095 | 0.085384 | 1.068623 |
| hypertensi | Z1_S4 | -0.0157 | 0.013277 | 0.237022 | 0.625211 |
| aortic valve | Z1_S4 | -0.06536 | 0.060959 | 0.283633 | 0.547243 |
| other heart | Z1_S4 | 0.051027 | 0.058587 | 0.383788 | 0.415909 |
| angina pec | Z1_S4 | -0.00734 | 0.028485 | 0.796653 | 0.098731 |
| myocardial | Z1_S4 | -0.00465 | 0.030669 | 0.879454 | 0.055787 |
| arrhythmia | Z1_S4 | -0.00726 | 0.052143 | 0.88934 | 0.050932 |
| coronary h | Z1_S4 | 0.000809 | 0.023612 | 0.972672 | 0.012033 |
| hypotensio | Z1_S4 | 9.50E-05 | 0.045697 | 0.998342 | 0.000721 |
| atrial fibrill | Z62_S4 | -0.12458 | 0.025787 | 1.37E-06 | 5.864125 |

| | | | | | |
|---|---|---|---|---|---|
| heart failur | Z62_S4 | -0.11861 | 0.04293 | 0.005733 | 2.241632 |
| type 2 diab | Z62_S4 | -0.07588 | 0.028058 | 0.006845 | 2.164613 |
| conduction | Z62_S4 | -0.08405 | 0.045156 | 0.062709 | 1.202671 |
| aortic valve | Z62_S4 | -0.1049 | 0.065425 | 0.108853 | 0.963161 |
| high choles | Z62_S4 | 0.02498 | 0.015639 | 0.110219 | 0.957744 |
| hypertensi | Z62_S4 | -0.02057 | 0.014197 | 0.147283 | 0.831846 |
| hypotensio | Z62_S4 | -0.06077 | 0.049034 | 0.215202 | 0.667153 |
| other heart | Z62_S4 | -0.07712 | 0.062872 | 0.219987 | 0.657603 |
| myocardial | Z62_S4 | 0.039228 | 0.032931 | 0.233585 | 0.631554 |
| arrhythmia | Z62_S4 | -0.03951 | 0.055983 | 0.480379 | 0.318416 |
| mitral prob | Z62_S4 | -0.02158 | 0.051582 | 0.675679 | 0.170259 |
| coronary h | Z62_S4 | 0.010142 | 0.025352 | 0.689139 | 0.161693 |
| angina pec | Z62_S4 | -0.00222 | 0.030562 | 0.941988 | 0.025954 |
| coronary h | Z111_S5 | 0.108317 | 0.022465 | 1.43E-06 | 5.843801 |
| myocardial | Z111_S5 | 0.110196 | 0.029184 | 0.00016 | 3.796399 |
| angina pec | Z111_S5 | 0.091844 | 0.02711 | 0.000706 | 3.151332 |
| aortic valve | Z111_S5 | 0.123359 | 0.057987 | 0.033401 | 1.47624 |
| arrhythmia | Z111_S5 | 0.085681 | 0.049595 | 0.084071 | 1.075353 |
| atrial fibrill | Z111_S5 | 0.02189 | 0.02293 | 0.339773 | 0.468811 |
| conduction | Z111_S5 | -0.03809 | 0.040046 | 0.341555 | 0.46654 |
| mitral prob | Z111_S5 | 0.038476 | 0.045723 | 0.400084 | 0.397848 |
| hypotensio | Z111_S5 | 0.034542 | 0.043477 | 0.426922 | 0.369651 |
| high choles | Z111_S5 | 0.009922 | 0.013879 | 0.474686 | 0.323594 |
| type 2 diab | Z111_S5 | -0.01668 | 0.024961 | 0.504033 | 0.297541 |
| heart failur | Z111_S5 | 0.019916 | 0.038077 | 0.600949 | 0.221162 |
| other heart | Z111_S5 | 0.011016 | 0.055742 | 0.843343 | 0.073996 |
| hypertensi | Z111_S5 | 3.47E-05 | 0.012646 | 0.997809 | 0.000953 |
| atrial fibrill | Z13_S3 | 0.091915 | 0.019077 | 1.46E-06 | 5.8362 |
| aortic valve | Z13_S3 | 0.164731 | 0.048311 | 0.000651 | 3.186215 |
| heart failur | Z13_S3 | 0.10588 | 0.031723 | 0.000846 | 3.072453 |
| mitral prob | Z13_S3 | 0.077237 | 0.038074 | 0.042509 | 1.371524 |
| angina pec | Z13_S3 | 0.044379 | 0.022604 | 0.049618 | 1.304363 |
| coronary h | Z13_S3 | 0.026802 | 0.01876 | 0.153115 | 0.814982 |
| hypertensi | Z13_S3 | 0.013216 | 0.010508 | 0.208534 | 0.680824 |
| hypotensio | Z13_S3 | 0.044964 | 0.036205 | 0.214283 | 0.669012 |
| type 2 diab | Z13_S3 | 0.024804 | 0.020771 | 0.232417 | 0.633732 |
| arrhythmia | Z13_S3 | 0.03841 | 0.041331 | 0.352728 | 0.452559 |
| conduction | Z13_S3 | 0.016922 | 0.033354 | 0.611915 | 0.213309 |
| high choles | Z13_S3 | -0.00568 | 0.011575 | 0.623872 | 0.204904 |
| myocardial | Z13_S3 | 0.006027 | 0.024323 | 0.804282 | 0.094592 |
| other heart | Z13_S3 | 0.006573 | 0.046445 | 0.887459 | 0.051852 |
| atrial fibrill | Z43_S2 | -0.09972 | 0.020717 | 1.49E-06 | 5.82644 |
| aortic valve | Z43_S2 | -0.11836 | 0.052494 | 0.024161 | 1.616881 |
| heart failur | Z43_S2 | -0.06228 | 0.034465 | 0.070765 | 1.150178 |
| mitral prob | Z43_S2 | -0.06003 | 0.041382 | 0.146897 | 0.832986 |

| | | | | | |
|---|---|---|---|---|---|
| hypertensi | Z43_S2 | -0.01616 | 0.01145 | 0.158108 | 0.801047 |
| other heart | Z43_S2 | -0.04243 | 0.050457 | 0.400349 | 0.397561 |
| hypotensio | Z43_S2 | -0.02067 | 0.039336 | 0.599261 | 0.222384 |
| coronary h | Z43_S2 | -0.00732 | 0.020366 | 0.719289 | 0.143096 |
| angina pec | Z43_S2 | -0.00773 | 0.024562 | 0.753027 | 0.123189 |
| conduction | Z43_S2 | 0.010359 | 0.036265 | 0.775143 | 0.110618 |
| high choles | Z43_S2 | 0.002667 | 0.012598 | 0.83234 | 0.079699 |
| type 2 diab | Z43_S2 | 0.003805 | 0.022585 | 0.866212 | 0.062376 |
| myocardial | Z43_S2 | 0.003198 | 0.026429 | 0.903689 | 0.043981 |
| arrhythmia | Z43_S2 | -0.00397 | 0.044914 | 0.929651 | 0.03168 |
| atrial fibrill | Z6_S2 | -0.09042 | 0.018825 | 1.57E-06 | 5.804092 |
| heart failur | Z6_S2 | -0.06195 | 0.031326 | 0.048004 | 1.31872 |
| aortic valve | Z6_S2 | -0.09322 | 0.047723 | 0.050798 | 1.294154 |
| mitral prob | Z6_S2 | -0.05409 | 0.037617 | 0.150508 | 0.822441 |
| hypertensi | Z6_S2 | -0.0107 | 0.010419 | 0.304239 | 0.516785 |
| angina pec | Z6_S2 | -0.01395 | 0.022333 | 0.532357 | 0.273797 |
| other heart | Z6_S2 | -0.02413 | 0.045871 | 0.598874 | 0.222664 |
| coronary h | Z6_S2 | -0.0081 | 0.018522 | 0.662046 | 0.179112 |
| myocardial | Z6_S2 | 0.008968 | 0.024036 | 0.709083 | 0.149303 |
| arrhythmia | Z6_S2 | 0.011154 | 0.040831 | 0.784713 | 0.105289 |
| type 2 diab | Z6_S2 | 0.003509 | 0.020535 | 0.864331 | 0.06332 |
| hypotensio | Z6_S2 | -0.00114 | 0.03577 | 0.974558 | 0.011192 |
| high choles | Z6_S2 | 0.000355 | 0.011472 | 0.975287 | 0.010868 |
| conduction | Z6_S2 | 0.000203 | 0.032963 | 0.99508 | 0.002142 |
| atrial fibrill | Z14_S1 | 0.103231 | 0.021759 | 2.10E-06 | 5.676827 |
| aortic valve | Z14_S1 | 0.156196 | 0.055117 | 0.004602 | 2.337024 |
| heart failur | Z14_S1 | 0.080532 | 0.036164 | 0.025966 | 1.585603 |
| mitral prob | Z14_S1 | 0.082679 | 0.043452 | 0.057083 | 1.243494 |
| hypertensi | Z14_S1 | 0.02206 | 0.012012 | 0.066304 | 1.178462 |
| hypotensio | Z14_S1 | 0.0618 | 0.041307 | 0.134643 | 0.870818 |
| other heart | Z14_S1 | 0.06028 | 0.052962 | 0.255058 | 0.593362 |
| coronary h | Z14_S1 | 0.023398 | 0.02136 | 0.273345 | 0.563289 |
| arrhythmia | Z14_S1 | 0.04125 | 0.047149 | 0.381646 | 0.418339 |
| myocardial | Z14_S1 | 0.022868 | 0.027738 | 0.409693 | 0.387542 |
| type 2 diab | Z14_S1 | 0.003577 | 0.023717 | 0.880127 | 0.055455 |
| conduction | Z14_S1 | -0.0053 | 0.038051 | 0.889233 | 0.050984 |
| high choles | Z14_S1 | -0.00158 | 0.013195 | 0.904857 | 0.04342 |
| angina pec | Z14_S1 | 0.00188 | 0.02578 | 0.941867 | 0.026011 |
| atrial fibrill | Z115_S4 | 0.100299 | 0.021308 | 2.53E-06 | 5.597414 |
| aortic valve | Z115_S4 | 0.159152 | 0.054033 | 0.003228 | 2.49106 |
| myocardial | Z115_S4 | -0.07167 | 0.027209 | 0.008448 | 2.073228 |
| hypertensi | Z115_S4 | 0.029848 | 0.011785 | 0.011326 | 1.945926 |
| angina pec | Z115_S4 | -0.04129 | 0.025278 | 0.102371 | 0.989825 |
| heart failur | Z115_S4 | 0.056947 | 0.035455 | 0.108244 | 0.965596 |
| coronary h | Z115_S4 | -0.02794 | 0.020963 | 0.182553 | 0.738612 |

| | | | | | |
|---|---|---|---|---|---|
| mitral prob | Z115_S4 | 0.052361 | 0.042596 | 0.218998 | 0.65956 |
| type 2 diab | Z115_S4 | 0.025878 | 0.023245 | 0.265595 | 0.57578 |
| hypotensio | Z115_S4 | 0.037831 | 0.040484 | 0.350082 | 0.455831 |
| other heart | Z115_S4 | 0.027037 | 0.051918 | 0.602541 | 0.220013 |
| high choles | Z115_S4 | -0.00538 | 0.012964 | 0.678347 | 0.168548 |
| arrhythmia | Z115_S4 | 0.007493 | 0.046217 | 0.871216 | 0.059874 |
| conduction | Z115_S4 | -0.00017 | 0.037301 | 0.996289 | 0.001615 |
| type 2 diab | Z34_S2 | 0.131673 | 0.027977 | 2.54E-06 | 5.595858 |
| myocardial | Z34_S2 | 0.078173 | 0.032739 | 0.016958 | 1.770617 |
| atrial fibrill | Z34_S2 | -0.06093 | 0.025582 | 0.017236 | 1.763567 |
| coronary h | Z34_S2 | 0.057763 | 0.025213 | 0.021971 | 1.658147 |
| mitral prob | Z34_S2 | -0.106 | 0.05126 | 0.038665 | 1.412682 |
| angina pec | Z34_S2 | 0.060647 | 0.030382 | 0.04593 | 1.337908 |
| hypotensio | Z34_S2 | 0.081459 | 0.048775 | 0.094912 | 1.02268 |
| arrhythmia | Z34_S2 | 0.059874 | 0.055645 | 0.281941 | 0.549841 |
| other heart | Z34_S2 | 0.054275 | 0.062504 | 0.385212 | 0.4143 |
| conduction | Z34_S2 | -0.0287 | 0.044904 | 0.522711 | 0.281738 |
| aortic valve | Z34_S2 | -0.03572 | 0.06503 | 0.58286 | 0.234436 |
| hypertensi | Z34_S2 | -0.00563 | 0.014129 | 0.690253 | 0.160992 |
| high choles | Z34_S2 | 0.004099 | 0.015547 | 0.792024 | 0.101262 |
| heart failur | Z34_S2 | 0.006073 | 0.042697 | 0.886887 | 0.052132 |
| atrial fibrill | Z120_S5 | -0.12736 | 0.027076 | 2.57E-06 | 5.590593 |
| type 2 diab | Z120_S5 | 0.085015 | 0.029541 | 0.004008 | 2.39712 |
| coronary h | Z120_S5 | 0.068807 | 0.02661 | 0.009722 | 2.012262 |
| conduction | Z120_S5 | -0.11662 | 0.047441 | 0.013968 | 1.854854 |
| angina pec | Z120_S5 | 0.065789 | 0.032105 | 0.040452 | 1.393063 |
| mitral prob | Z120_S5 | -0.09393 | 0.054192 | 0.083074 | 1.080537 |
| myocardial | Z120_S5 | 0.048123 | 0.034579 | 0.164032 | 0.785072 |
| heart failur | Z120_S5 | -0.05372 | 0.045115 | 0.233783 | 0.631187 |
| high choles | Z120_S5 | 0.017454 | 0.016446 | 0.288571 | 0.539747 |
| other heart | Z120_S5 | -0.02457 | 0.066087 | 0.710086 | 0.148689 |
| arrhythmia | Z120_S5 | 0.014169 | 0.058833 | 0.809686 | 0.091683 |
| hypotensio | Z120_S5 | 0.009435 | 0.05154 | 0.854749 | 0.068162 |
| hypertensi | Z120_S5 | -0.00167 | 0.014938 | 0.910893 | 0.040533 |
| aortic valve | Z120_S5 | 0.001715 | 0.068761 | 0.980098 | 0.00873 |
| myocardial | Z118_S1 | 0.152323 | 0.032509 | 2.81E-06 | 5.55153 |
| atrial fibrill | Z118_S1 | -0.11728 | 0.025428 | 4.01E-06 | 5.397099 |
| coronary h | Z118_S1 | 0.081049 | 0.025019 | 0.001199 | 2.92108 |
| angina pec | Z118_S1 | 0.077105 | 0.030155 | 0.010566 | 1.976089 |
| heart failur | Z118_S1 | -0.09086 | 0.042401 | 0.032126 | 1.493145 |
| type 2 diab | Z118_S1 | -0.05743 | 0.027778 | 0.038706 | 1.412221 |
| mitral prob | Z118_S1 | -0.08406 | 0.050941 | 0.098938 | 1.004639 |
| other heart | Z118_S1 | -0.08138 | 0.062108 | 0.190132 | 0.720945 |
| high choles | Z118_S1 | 0.016841 | 0.015506 | 0.277421 | 0.55686 |
| arrhythmia | Z118_S1 | 0.054679 | 0.055249 | 0.322339 | 0.491688 |

| | | | | | |
|---|---|---|---|---|---|
| conduction | Z118_S1 | 0.040255 | 0.044637 | 0.367156 | 0.435149 |
| hypotensio | Z118_S1 | 0.037446 | 0.048455 | 0.439655 | 0.356888 |
| hypertensi | Z118_S1 | -0.01016 | 0.014021 | 0.468721 | 0.329086 |
| aortic valve | Z118_S1 | 0.019074 | 0.064617 | 0.767862 | 0.114717 |
| atrial fibrill | Z127_S5 | 0.092068 | 0.019651 | 2.81E-06 | 5.550946 |
| aortic valve | Z127_S5 | 0.172243 | 0.049652 | 0.000523 | 3.281181 |
| type 2 diab | Z127_S5 | -0.06719 | 0.021418 | 0.001708 | 2.76762 |
| coronary h | Z127_S5 | 0.049134 | 0.019276 | 0.010811 | 1.966137 |
| mitral prob | Z127_S5 | 0.091556 | 0.03915 | 0.019367 | 1.712946 |
| myocardial | Z127_S5 | 0.047926 | 0.025017 | 0.055412 | 1.256392 |
| angina pec | Z127_S5 | 0.030526 | 0.023246 | 0.189138 | 0.723222 |
| arrhythmia | Z127_S5 | 0.051443 | 0.042502 | 0.226157 | 0.64559 |
| hypotensio | Z127_S5 | 0.040636 | 0.037258 | 0.275439 | 0.559975 |
| heart failur | Z127_S5 | 0.032405 | 0.032642 | 0.320844 | 0.493707 |
| hypertensi | Z127_S5 | 0.010342 | 0.010834 | 0.339825 | 0.468745 |
| high choles | Z127_S5 | 0.002187 | 0.011894 | 0.854095 | 0.068494 |
| conduction | Z127_S5 | -0.00468 | 0.034309 | 0.891464 | 0.049896 |
| other heart | Z127_S5 | -0.00581 | 0.04775 | 0.903173 | 0.044229 |
| myocardial | Z21_S3 | -0.16654 | 0.035641 | 2.99E-06 | 5.524291 |
| coronary h | Z21_S3 | -0.12554 | 0.027423 | 4.72E-06 | 5.325926 |
| angina pec | Z21_S3 | -0.10151 | 0.033081 | 0.002153 | 2.666866 |
| heart failur | Z21_S3 | -0.10655 | 0.04653 | 0.022036 | 1.656871 |
| other heart | Z21_S3 | -0.14237 | 0.068142 | 0.036689 | 1.435463 |
| atrial fibrill | Z21_S3 | -0.04419 | 0.02793 | 0.113629 | 0.944512 |
| type 2 diab | Z21_S3 | -0.04783 | 0.03048 | 0.116588 | 0.933346 |
| hypertensi | Z21_S3 | 0.01833 | 0.015371 | 0.233074 | 0.632506 |
| conduction | Z21_S3 | -0.04677 | 0.048955 | 0.339402 | 0.469286 |
| arrhythmia | Z21_S3 | -0.05644 | 0.06064 | 0.351978 | 0.453485 |
| hypotensio | Z21_S3 | -0.02556 | 0.053151 | 0.630631 | 0.200225 |
| high choles | Z21_S3 | 0.005835 | 0.016946 | 0.730599 | 0.136321 |
| aortic valve | Z21_S3 | -0.0079 | 0.070894 | 0.911226 | 0.040374 |
| mitral prob | Z21_S3 | -0.00239 | 0.055884 | 0.965889 | 0.015073 |
| atrial fibrill | Z75_S1 | -0.10787 | 0.02337 | 3.94E-06 | 5.404893 |
| myocardial | Z75_S1 | 0.092603 | 0.029923 | 0.001972 | 2.705037 |
| type 2 diab | Z75_S1 | 0.066323 | 0.025572 | 0.009506 | 2.022021 |
| aortic valve | Z75_S1 | -0.1526 | 0.059396 | 0.010199 | 1.991432 |
| angina pec | Z75_S1 | 0.070392 | 0.027765 | 0.011241 | 1.94918 |
| coronary h | Z75_S1 | 0.054189 | 0.023056 | 0.018763 | 1.726692 |
| mitral prob | Z75_S1 | -0.08874 | 0.046817 | 0.058037 | 1.236293 |
| hypertensi | Z75_S1 | -0.01727 | 0.01293 | 0.181732 | 0.740568 |
| conduction | Z75_S1 | -0.05379 | 0.041005 | 0.189573 | 0.722224 |
| arrhythmia | Z75_S1 | 0.05059 | 0.050849 | 0.319797 | 0.495126 |
| other heart | Z75_S1 | 0.034357 | 0.057087 | 0.547286 | 0.261786 |
| hypotensio | Z75_S1 | 0.02008 | 0.04456 | 0.652262 | 0.185578 |
| heart failur | Z75_S1 | -0.01558 | 0.039021 | 0.689681 | 0.161352 |

| | | | | | |
|---|---|---|---|---|---|
| high choles | Z75_S1 | 0.001871 | 0.014212 | 0.895278 | 0.048042 |
| type 2 diab | Z122_S2 | -0.09487 | 0.020682 | 4.52E-06 | 5.344888 |
| aortic valve | Z122_S2 | -0.12857 | 0.048117 | 0.007545 | 2.122345 |
| atrial fibrill | Z122_S2 | -0.03882 | 0.018986 | 0.040903 | 1.388244 |
| myocardial | Z122_S2 | 0.048423 | 0.024226 | 0.045639 | 1.340659 |
| angina pec | Z122_S2 | 0.040132 | 0.022507 | 0.074585 | 1.127348 |
| coronary h | Z122_S2 | 0.023498 | 0.018662 | 0.208011 | 0.681914 |
| other heart | Z122_S2 | 0.056326 | 0.046235 | 0.223141 | 0.65142 |
| conduction | Z122_S2 | 0.039711 | 0.033185 | 0.23146 | 0.635525 |
| arrhythmia | Z122_S2 | 0.048201 | 0.041146 | 0.241422 | 0.617223 |
| heart failur | Z122_S2 | -0.03216 | 0.031565 | 0.308265 | 0.511076 |
| hypotensio | Z122_S2 | -0.02714 | 0.036052 | 0.451562 | 0.345283 |
| hypertensi | Z122_S2 | -0.00682 | 0.01049 | 0.515316 | 0.287926 |
| high choles | Z122_S2 | -0.0058 | 0.011537 | 0.615339 | 0.210886 |
| mitral prob | Z122_S2 | 0.009905 | 0.037911 | 0.793885 | 0.100242 |
| atrial fibrill | Z26_S3 | -0.11553 | 0.025438 | 5.60E-06 | 5.251463 |
| conduction | Z26_S3 | -0.12515 | 0.044533 | 0.004953 | 2.305141 |
| angina pec | Z26_S3 | -0.06482 | 0.030118 | 0.031399 | 1.503079 |
| coronary h | Z26_S3 | -0.04828 | 0.024994 | 0.053421 | 1.272285 |
| myocardial | Z26_S3 | -0.06238 | 0.032451 | 0.054569 | 1.263051 |
| hypertensi | Z26_S3 | 0.026603 | 0.01402 | 0.057772 | 1.238285 |
| type 2 diab | Z26_S3 | 0.04314 | 0.027753 | 0.120099 | 0.92046 |
| aortic valve | Z26_S3 | -0.07626 | 0.064521 | 0.23722 | 0.624848 |
| high choles | Z26_S3 | 0.015767 | 0.015444 | 0.307319 | 0.512411 |
| heart failur | Z26_S3 | -0.0431 | 0.042373 | 0.309106 | 0.509893 |
| other heart | Z26_S3 | 0.034489 | 0.062026 | 0.57819 | 0.23793 |
| hypotensio | Z26_S3 | -0.01149 | 0.048383 | 0.812338 | 0.090263 |
| arrhythmia | Z26_S3 | 0.007904 | 0.055193 | 0.886122 | 0.052506 |
| mitral prob | Z26_S3 | -0.00017 | 0.050866 | 0.997372 | 0.001143 |
| atrial fibrill | Z24_S1 | 0.089851 | 0.019828 | 5.88E-06 | 5.230347 |
| aortic valve | Z24_S1 | 0.164762 | 0.050173 | 0.001026 | 2.989033 |
| mitral prob | Z24_S1 | 0.085277 | 0.03955 | 0.031082 | 1.507495 |
| hypertensi | Z24_S1 | 0.0223 | 0.010961 | 0.041918 | 1.377604 |
| heart failur | Z24_S1 | 0.05702 | 0.032934 | 0.083406 | 1.078805 |
| type 2 diab | Z24_S1 | -0.0267 | 0.021618 | 0.21686 | 0.66382 |
| other heart | Z24_S1 | 0.040914 | 0.048217 | 0.396144 | 0.402147 |
| coronary h | Z24_S1 | 0.01647 | 0.019465 | 0.397506 | 0.400657 |
| hypotensio | Z24_S1 | 0.024781 | 0.037609 | 0.509959 | 0.292465 |
| arrhythmia | Z24_S1 | 0.027433 | 0.042931 | 0.522835 | 0.281636 |
| myocardial | Z24_S1 | 0.005618 | 0.025256 | 0.823966 | 0.084091 |
| conduction | Z24_S1 | 0.004843 | 0.034656 | 0.88887 | 0.051162 |
| angina pec | Z24_S1 | 0.002342 | 0.023483 | 0.920553 | 0.035951 |
| high choles | Z24_S1 | 0.001143 | 0.012029 | 0.924322 | 0.034177 |
| atrial fibrill | Z44_S4 | -0.08865 | 0.019808 | 7.66E-06 | 5.115675 |
| heart failur | Z44_S4 | -0.09646 | 0.032906 | 0.003378 | 2.471324 |

| | | | | | |
|---|---|---|---|---|---|
| aortic valve | Z44_S4 | -0.12232 | 0.050136 | 0.014702 | 1.832612 |
| mitral prob | Z44_S4 | -0.07453 | 0.039509 | 0.059256 | 1.227265 |
| hypertensi | Z44_S4 | -0.01692 | 0.010954 | 0.122447 | 0.912051 |
| myocardial | Z44_S4 | -0.03886 | 0.025229 | 0.123484 | 0.908389 |
| coronary h | Z44_S4 | -0.02706 | 0.01944 | 0.163935 | 0.785327 |
| arrhythmia | Z44_S4 | -0.05332 | 0.042886 | 0.21374 | 0.670114 |
| hypotensio | Z44_S4 | -0.03526 | 0.037573 | 0.348017 | 0.4584 |
| angina pec | Z44_S4 | -0.01836 | 0.02347 | 0.434192 | 0.362318 |
| type 2 diab | Z44_S4 | 0.015736 | 0.021588 | 0.466057 | 0.331561 |
| other heart | Z44_S4 | -0.03142 | 0.048187 | 0.51437 | 0.288724 |
| high choles | Z44_S4 | 0.004283 | 0.012014 | 0.721452 | 0.141793 |
| conduction | Z44_S4 | 0.004435 | 0.034618 | 0.898067 | 0.046691 |
| atrial fibrill | Z4_S4 | -0.09199 | 0.020714 | 8.99E-06 | 5.046047 |
| heart failur | Z4_S4 | -0.11773 | 0.03442 | 0.000626 | 3.203353 |
| myocardial | Z4_S4 | -0.08044 | 0.026374 | 0.002291 | 2.639973 |
| mitral prob | Z4_S4 | -0.12397 | 0.041305 | 0.002692 | 2.569951 |
| type 2 diab | Z4_S4 | 0.061761 | 0.022553 | 0.006177 | 2.209256 |
| coronary h | Z4_S4 | -0.05429 | 0.020326 | 0.007568 | 2.120992 |
| aortic valve | Z4_S4 | -0.13905 | 0.052395 | 0.007963 | 2.098921 |
| arrhythmia | Z4_S4 | -0.10048 | 0.044838 | 0.025037 | 1.601424 |
| hypertensi | Z4_S4 | -0.02262 | 0.011453 | 0.048319 | 1.315878 |
| angina pec | Z4_S4 | -0.03997 | 0.024529 | 0.103232 | 0.986186 |
| other heart | Z4_S4 | -0.06612 | 0.05039 | 0.189447 | 0.722513 |
| hypotensio | Z4_S4 | -0.04338 | 0.039271 | 0.26934 | 0.569698 |
| high choles | Z4_S4 | 0.008712 | 0.012574 | 0.488435 | 0.311193 |
| conduction | Z4_S4 | 0.006316 | 0.036194 | 0.86148 | 0.064755 |
| type 2 diab | Z29_S1 | -0.12069 | 0.027217 | 9.28E-06 | 5.032545 |
| atrial fibrill | Z29_S1 | 0.106993 | 0.024964 | 1.83E-05 | 4.738305 |
| aortic valve | Z29_S1 | 0.223873 | 0.063334 | 0.000409 | 3.388352 |
| mitral prob | Z29_S1 | 0.139783 | 0.049942 | 0.005132 | 2.289735 |
| hypertensi | Z29_S1 | 0.028879 | 0.013758 | 0.035821 | 1.445868 |
| angina pec | Z29_S1 | -0.05699 | 0.029607 | 0.054279 | 1.265366 |
| high choles | Z29_S1 | -0.01932 | 0.015158 | 0.202576 | 0.693412 |
| hypotensio | Z29_S1 | 0.060071 | 0.047498 | 0.205994 | 0.686145 |
| conduction | Z29_S1 | 0.036573 | 0.04373 | 0.402981 | 0.394715 |
| other heart | Z29_S1 | -0.04216 | 0.060883 | 0.488679 | 0.310976 |
| coronary h | Z29_S1 | -0.01011 | 0.024547 | 0.680374 | 0.167253 |
| myocardial | Z29_S1 | -0.01092 | 0.031884 | 0.731957 | 0.135514 |
| heart failur | Z29_S1 | -0.0059 | 0.041609 | 0.887237 | 0.051961 |
| arrhythmia | Z29_S1 | 0.003185 | 0.054233 | 0.953168 | 0.020831 |
| coronary h | Z50_S5 | 0.120397 | 0.027216 | 9.74E-06 | 5.011275 |
| myocardial | Z50_S5 | 0.15152 | 0.03538 | 1.85E-05 | 4.731797 |
| angina pec | Z50_S5 | 0.123883 | 0.032838 | 0.000162 | 3.790481 |
| atrial fibrill | Z50_S5 | -0.08699 | 0.027715 | 0.001698 | 2.770026 |
| mitral prob | Z50_S5 | -0.10484 | 0.055454 | 0.058694 | 1.231407 |

| | | | | | |
|---|---|---|---|---|---|
| arrhythmia | Z50_S5 | 0.106734 | 0.060197 | 0.07623 | 1.117873 |
| hypertensi | Z50_S5 | -0.02694 | 0.015266 | 0.077672 | 1.109733 |
| conduction | Z50_S5 | -0.06464 | 0.048571 | 0.183226 | 0.737014 |
| type 2 diab | Z50_S5 | 0.03543 | 0.030248 | 0.24148 | 0.617118 |
| high choles | Z50_S5 | 0.013167 | 0.016822 | 0.433803 | 0.362707 |
| heart failur | Z50_S5 | -0.03134 | 0.046175 | 0.497257 | 0.303419 |
| hypotensio | Z50_S5 | 0.02446 | 0.052764 | 0.642951 | 0.191822 |
| other heart | Z50_S5 | -0.02814 | 0.067637 | 0.677389 | 0.169162 |
| aortic valve | Z50_S5 | -0.01099 | 0.07037 | 0.87593 | 0.057531 |
| atrial fibrill | Z7_S5 | -0.10965 | 0.02479 | 9.77E-06 | 5.010005 |
| angina pec | Z7_S5 | 0.096467 | 0.029438 | 0.001051 | 2.978486 |
| type 2 diab | Z7_S5 | -0.08771 | 0.027059 | 0.001192 | 2.923825 |
| myocardial | Z7_S5 | 0.09853 | 0.031716 | 0.001894 | 2.722532 |
| hypertensi | Z7_S5 | -0.03666 | 0.013662 | 0.007301 | 2.13662 |
| aortic valve | Z7_S5 | -0.16096 | 0.06302 | 0.010653 | 1.972515 |
| coronary h | Z7_S5 | 0.061289 | 0.024412 | 0.012059 | 1.918684 |
| heart failur | Z7_S5 | -0.07724 | 0.041347 | 0.061756 | 1.209319 |
| hypotensio | Z7_S5 | -0.0691 | 0.047227 | 0.143429 | 0.843363 |
| conduction | Z7_S5 | 0.05117 | 0.043508 | 0.239562 | 0.620581 |
| mitral prob | Z7_S5 | -0.05398 | 0.049674 | 0.277149 | 0.557287 |
| arrhythmia | Z7_S5 | 0.035679 | 0.053917 | 0.508145 | 0.294013 |
| other heart | Z7_S5 | -0.03888 | 0.060559 | 0.52092 | 0.283229 |
| high choles | Z7_S5 | -0.00589 | 0.015057 | 0.695697 | 0.15758 |
| type 2 diab | Z57_S2 | -0.09375 | 0.021205 | 9.86E-06 | 5.005918 |
| atrial fibrill | Z57_S2 | 0.070603 | 0.019461 | 0.000286 | 3.543217 |
| myocardial | Z57_S2 | 0.070365 | 0.024845 | 0.004628 | 2.334594 |
| other heart | Z57_S2 | 0.127459 | 0.047406 | 0.007178 | 2.14397 |
| mitral prob | Z57_S2 | 0.098183 | 0.038861 | 0.011527 | 1.938289 |
| coronary h | Z57_S2 | 0.045493 | 0.019152 | 0.01754 | 1.755959 |
| high choles | Z57_S2 | -0.02424 | 0.011814 | 0.040194 | 1.395837 |
| conduction | Z57_S2 | 0.067706 | 0.034025 | 0.046614 | 1.331484 |
| arrhythmia | Z57_S2 | 0.080946 | 0.042193 | 0.055065 | 1.259125 |
| angina pec | Z57_S2 | 0.039483 | 0.023059 | 0.086868 | 1.061138 |
| aortic valve | Z57_S2 | -0.05582 | 0.049326 | 0.257829 | 0.588668 |
| hypotensio | Z57_S2 | 0.026171 | 0.036965 | 0.478955 | 0.319705 |
| heart failur | Z57_S2 | 0.017703 | 0.032356 | 0.584301 | 0.233364 |
| hypertensi | Z57_S2 | 0.000573 | 0.010749 | 0.957518 | 0.018853 |
| atrial fibrill | Z73_S1 | -0.09661 | 0.021882 | 1.01E-05 | 4.993551 |
| type 2 diab | Z73_S1 | -0.0807 | 0.023872 | 0.000725 | 3.139663 |
| angina pec | Z73_S1 | 0.065516 | 0.025955 | 0.011603 | 1.935437 |
| aortic valve | Z73_S1 | -0.13858 | 0.055531 | 0.012583 | 1.900211 |
| heart failur | Z73_S1 | -0.09052 | 0.036414 | 0.012934 | 1.888279 |
| hypertensi | Z73_S1 | -0.02936 | 0.012099 | 0.015242 | 1.816972 |
| myocardial | Z73_S1 | 0.062146 | 0.027936 | 0.02612 | 1.583034 |
| coronary h | Z73_S1 | 0.044576 | 0.021524 | 0.038367 | 1.416043 |

| | | | | | |
|---|---|---|---|---|---|
| other heart | Z73_S1 | -0.08125 | 0.053344 | 0.127719 | 0.893743 |
| hypotensio | Z73_S1 | -0.05613 | 0.041586 | 0.177115 | 0.751745 |
| conduction | Z73_S1 | 0.039978 | 0.038325 | 0.296892 | 0.527401 |
| mitral prob | Z73_S1 | -0.03528 | 0.043774 | 0.420255 | 0.376487 |
| high choles | Z73_S1 | -0.00554 | 0.013309 | 0.67743 | 0.169136 |
| arrhythmia | Z73_S1 | 0.003316 | 0.047506 | 0.944346 | 0.024869 |
| atrial fibrill | Z51_S4 | -0.08825 | 0.020032 | 1.06E-05 | 4.975147 |
| aortic valve | Z51_S4 | -0.19026 | 0.05066 | 0.000173 | 3.760981 |
| type 2 diab | Z51_S4 | 0.053049 | 0.021843 | 0.015163 | 1.819225 |
| mitral prob | Z51_S4 | -0.09441 | 0.039939 | 0.018101 | 1.7423 |
| hypertensi | Z51_S4 | -0.02441 | 0.01107 | 0.027481 | 1.560973 |
| heart failur | Z51_S4 | -0.06857 | 0.033282 | 0.039385 | 1.40467 |
| coronary h | Z51_S4 | -0.03092 | 0.019655 | 0.115756 | 0.936458 |
| myocardial | Z51_S4 | -0.02601 | 0.025519 | 0.308096 | 0.511314 |
| hypotensio | Z51_S4 | -0.03632 | 0.038 | 0.339147 | 0.469612 |
| arrhythmia | Z51_S4 | -0.04126 | 0.043358 | 0.341355 | 0.466794 |
| high choles | Z51_S4 | 0.009137 | 0.012135 | 0.451497 | 0.345345 |
| angina pec | Z51_S4 | -0.01544 | 0.023726 | 0.515161 | 0.288057 |
| other heart | Z51_S4 | -0.00604 | 0.048716 | 0.901384 | 0.04509 |
| conduction | Z51_S4 | -0.00381 | 0.034987 | 0.91322 | 0.039425 |
| atrial fibrill | Z14_S2 | -0.09815 | 0.022426 | 1.21E-05 | 4.916942 |
| myocardial | Z14_S2 | -0.06146 | 0.028575 | 0.031493 | 1.501789 |
| aortic valve | Z14_S2 | -0.11484 | 0.056782 | 0.043142 | 1.365098 |
| heart failur | Z14_S2 | -0.06679 | 0.037309 | 0.073456 | 1.133972 |
| conduction | Z14_S2 | -0.06498 | 0.039186 | 0.097263 | 1.012051 |
| high choles | Z14_S2 | 0.018717 | 0.013584 | 0.168246 | 0.774056 |
| mitral prob | Z14_S2 | -0.05382 | 0.044751 | 0.229143 | 0.639893 |
| type 2 diab | Z14_S2 | 0.02817 | 0.024439 | 0.249065 | 0.603688 |
| hypotensio | Z14_S2 | -0.02142 | 0.042582 | 0.614975 | 0.211143 |
| coronary h | Z14_S2 | -0.00985 | 0.021998 | 0.654214 | 0.18428 |
| arrhythmia | Z14_S2 | -0.02109 | 0.048572 | 0.66413 | 0.177747 |
| hypertensi | Z14_S2 | -0.00422 | 0.012345 | 0.732681 | 0.135085 |
| other heart | Z14_S2 | -0.01212 | 0.054588 | 0.824339 | 0.083894 |
| angina pec | Z14_S2 | -0.00133 | 0.02649 | 0.959918 | 0.017766 |
| myocardial | Z78_S1 | -0.11795 | 0.027434 | 1.72E-05 | 4.764737 |
| coronary h | Z78_S1 | -0.08035 | 0.021145 | 0.000145 | 3.838055 |
| angina pec | Z78_S1 | -0.08712 | 0.025475 | 0.000627 | 3.202437 |
| atrial fibrill | Z78_S1 | 0.072665 | 0.021474 | 0.000716 | 3.145081 |
| type 2 diab | Z78_S1 | 0.05415 | 0.023454 | 0.020964 | 1.678534 |
| hypertensi | Z78_S1 | 0.026195 | 0.011885 | 0.027526 | 1.56025 |
| aortic valve | Z78_S1 | 0.090006 | 0.054526 | 0.098814 | 1.005179 |
| heart failur | Z78_S1 | 0.0561 | 0.035758 | 0.116684 | 0.932987 |
| conduction | Z78_S1 | -0.05618 | 0.037658 | 0.135763 | 0.867219 |
| mitral prob | Z78_S1 | 0.036219 | 0.042962 | 0.399216 | 0.398792 |
| arrhythmia | Z78_S1 | -0.03677 | 0.046637 | 0.430461 | 0.366066 |

| | | | | | |
|---|---|---|---|---|---|
| other heart | Z78_S1 | 0.033293 | 0.052397 | 0.525175 | 0.279696 |
| hypotensio | Z78_S1 | 0.005121 | 0.040839 | 0.900213 | 0.045655 |
| high choles | Z78_S1 | 0.001546 | 0.013083 | 0.905964 | 0.042889 |
| myocardial | Z3_S4 | 0.152453 | 0.03552 | 1.78E-05 | 4.750258 |
| coronary h | Z3_S4 | 0.115953 | 0.02733 | 2.22E-05 | 4.654329 |
| angina pec | Z3_S4 | 0.129575 | 0.032961 | 8.48E-05 | 4.071585 |
| atrial fibrill | Z3_S4 | -0.07239 | 0.027814 | 0.00926 | 2.033409 |
| type 2 diab | Z3_S4 | 0.054072 | 0.03034 | 0.074735 | 1.126475 |
| arrhythmia | Z3_S4 | 0.092717 | 0.06042 | 0.124914 | 0.903389 |
| mitral prob | Z3_S4 | -0.05279 | 0.055665 | 0.342959 | 0.464758 |
| hypotensio | Z3_S4 | 0.048502 | 0.052947 | 0.359657 | 0.444111 |
| other heart | Z3_S4 | 0.056285 | 0.067878 | 0.406994 | 0.390412 |
| heart failur | Z3_S4 | 0.023532 | 0.046353 | 0.611691 | 0.213468 |
| high choles | Z3_S4 | -0.00506 | 0.016875 | 0.764243 | 0.116768 |
| conduction | Z3_S4 | -0.01386 | 0.04875 | 0.776174 | 0.110041 |
| aortic valve | Z3_S4 | 0.011192 | 0.070624 | 0.87409 | 0.058444 |
| hypertensi | Z3_S4 | 0.001974 | 0.015323 | 0.897483 | 0.046974 |
| myocardial | Z39_S3 | 0.151027 | 0.035474 | 2.08E-05 | 4.682766 |
| coronary h | Z39_S3 | 0.090676 | 0.027297 | 0.000896 | 3.047814 |
| angina pec | Z39_S3 | 0.107668 | 0.03294 | 0.001082 | 2.965708 |
| heart failur | Z39_S3 | 0.136211 | 0.046307 | 0.003269 | 2.485524 |
| atrial fibrill | Z39_S3 | 0.078837 | 0.027814 | 0.004594 | 2.33782 |
| hypertensi | Z39_S3 | -0.03999 | 0.015305 | 0.008989 | 2.04629 |
| type 2 diab | Z39_S3 | 0.061026 | 0.030336 | 0.044264 | 1.353954 |
| arrhythmia | Z39_S3 | 0.109694 | 0.060371 | 0.069232 | 1.159691 |
| other heart | Z39_S3 | 0.078688 | 0.067825 | 0.245995 | 0.609073 |
| hypotensio | Z39_S3 | 0.055773 | 0.052916 | 0.291897 | 0.53477 |
| high choles | Z39_S3 | -0.01442 | 0.016868 | 0.392636 | 0.40601 |
| conduction | Z39_S3 | 0.025246 | 0.048718 | 0.60432 | 0.218733 |
| aortic valve | Z39_S3 | -0.0304 | 0.070562 | 0.666563 | 0.176159 |
| mitral prob | Z39_S3 | -0.00896 | 0.055622 | 0.872008 | 0.05948 |
| atrial fibrill | Z77_S3 | -0.09689 | 0.02298 | 2.49E-05 | 4.603719 |
| coronary h | Z77_S3 | 0.049614 | 0.022602 | 0.028168 | 1.550251 |
| angina pec | Z77_S3 | 0.058687 | 0.027226 | 0.031132 | 1.50679 |
| type 2 diab | Z77_S3 | 0.051614 | 0.025083 | 0.039628 | 1.401995 |
| conduction | Z77_S3 | -0.07794 | 0.040224 | 0.052673 | 1.278409 |
| aortic valve | Z77_S3 | -0.10013 | 0.058312 | 0.085949 | 1.065758 |
| mitral prob | Z77_S3 | -0.06699 | 0.045956 | 0.144949 | 0.838785 |
| high choles | Z77_S3 | 0.017541 | 0.013939 | 0.208251 | 0.681413 |
| heart failur | Z77_S3 | -0.04077 | 0.038304 | 0.287197 | 0.54182 |
| other heart | Z77_S3 | 0.051757 | 0.056052 | 0.355819 | 0.448771 |
| hypertensi | Z77_S3 | -0.01003 | 0.012679 | 0.428989 | 0.367554 |
| myocardial | Z77_S3 | 0.022186 | 0.02937 | 0.450029 | 0.346759 |
| hypotensio | Z77_S3 | -0.02137 | 0.043714 | 0.625021 | 0.204105 |
| arrhythmia | Z77_S3 | 0.016262 | 0.04991 | 0.744561 | 0.1281 |

| | | | | | |
|---|---|---|---|---|---|
| atrial fibrill | Z47_S4 | 0.09454 | 0.023096 | 4.27E-05 | 4.369807 |
| heart failur | Z47_S4 | 0.072193 | 0.038323 | 0.059604 | 1.224728 |
| angina pec | Z47_S4 | -0.05027 | 0.027244 | 0.065016 | 1.186978 |
| other heart | Z47_S4 | 0.103154 | 0.056045 | 0.0657 | 1.182433 |
| mitral prob | Z47_S4 | 0.084104 | 0.045967 | 0.067315 | 1.171889 |
| myocardial | Z47_S4 | -0.05066 | 0.029338 | 0.084239 | 1.074487 |
| coronary h | Z47_S4 | -0.03838 | 0.022593 | 0.089371 | 1.048802 |
| aortic valve | Z47_S4 | -0.06648 | 0.058311 | 0.254291 | 0.594669 |
| hypertensi | Z47_S4 | -0.01219 | 0.012694 | 0.336804 | 0.472623 |
| conduction | Z47_S4 | 0.036466 | 0.040263 | 0.365114 | 0.437571 |
| arrhythmia | Z47_S4 | -0.01566 | 0.049849 | 0.753467 | 0.122936 |
| high choles | Z47_S4 | 0.003937 | 0.013958 | 0.777885 | 0.109085 |
| hypotensio | Z47_S4 | 0.009957 | 0.043732 | 0.819894 | 0.086242 |
| type 2 diab | Z47_S4 | -0.00218 | 0.025085 | 0.930896 | 0.031099 |
| type 2 diab | Z111_S1 | -0.11095 | 0.027212 | 4.57E-05 | 4.340051 |
| conduction | Z111_S1 | 0.092463 | 0.043733 | 0.034505 | 1.462119 |
| heart failur | Z111_S1 | -0.08439 | 0.041559 | 0.042316 | 1.373494 |
| myocardial | Z111_S1 | 0.043104 | 0.031853 | 0.175997 | 0.754495 |
| aortic valve | Z111_S1 | 0.085117 | 0.063321 | 0.17889 | 0.747413 |
| other heart | Z111_S1 | -0.06834 | 0.060878 | 0.261638 | 0.582299 |
| hypertensi | Z111_S1 | 0.011077 | 0.013783 | 0.421589 | 0.375111 |
| atrial fibrill | Z111_S1 | -0.01605 | 0.024961 | 0.520132 | 0.283886 |
| hypotensio | Z111_S1 | 0.020922 | 0.047476 | 0.659437 | 0.180827 |
| coronary h | Z111_S1 | 0.010268 | 0.024519 | 0.675379 | 0.170452 |
| mitral prob | Z111_S1 | -0.0201 | 0.049933 | 0.68728 | 0.162866 |
| arrhythmia | Z111_S1 | -0.01623 | 0.054171 | 0.764455 | 0.116648 |
| high choles | Z111_S1 | 0.001299 | 0.015207 | 0.931949 | 0.030608 |
| angina pec | Z111_S1 | -0.00075 | 0.029577 | 0.979744 | 0.008888 |
| atrial fibrill | Z52_S4 | 0.085085 | 0.020927 | 4.80E-05 | 4.318355 |
| aortic valve | Z52_S4 | 0.173574 | 0.053021 | 0.001063 | 2.973391 |
| hypertensi | Z52_S4 | 0.029268 | 0.011563 | 0.011372 | 1.944157 |
| mitral prob | Z52_S4 | 0.092299 | 0.041802 | 0.027252 | 1.564604 |
| heart failur | Z52_S4 | 0.059205 | 0.034804 | 0.088939 | 1.050907 |
| coronary h | Z52_S4 | 0.023242 | 0.020561 | 0.258328 | 0.587828 |
| other heart | Z52_S4 | 0.043384 | 0.050959 | 0.39458 | 0.403865 |
| hypotensio | Z52_S4 | 0.024678 | 0.03974 | 0.534612 | 0.271962 |
| arrhythmia | Z52_S4 | 0.026505 | 0.045354 | 0.558959 | 0.25262 |
| type 2 diab | Z52_S4 | -0.01077 | 0.022836 | 0.637139 | 0.195766 |
| high choles | Z52_S4 | -0.00508 | 0.012705 | 0.68941 | 0.161523 |
| angina pec | Z52_S4 | 0.009318 | 0.024813 | 0.707261 | 0.15042 |
| myocardial | Z52_S4 | 0.001245 | 0.026704 | 0.962813 | 0.016458 |
| conduction | Z52_S4 | -0.00066 | 0.036616 | 0.985528 | 0.006331 |
| myocardial | Z72_S5 | 0.142132 | 0.035102 | 5.16E-05 | 4.287426 |
| coronary h | Z72_S5 | 0.097318 | 0.027005 | 0.000314 | 3.502647 |
| angina pec | Z72_S5 | 0.104059 | 0.03257 | 0.0014 | 2.853739 |

| | | | | | |
|---|---|---|---|---|---|
| type 2 diab | Z72_S5 | 0.08112 | 0.030018 | 0.00689 | 2.161761 |
| mitral prob | Z72_S5 | -0.10818 | 0.055023 | 0.049299 | 1.307161 |
| hypertensi | Z72_S5 | -0.02973 | 0.015138 | 0.049557 | 1.304893 |
| atrial fibrill | Z72_S5 | -0.03306 | 0.027499 | 0.229323 | 0.639552 |
| arrhythmia | Z72_S5 | 0.053763 | 0.059707 | 0.367887 | 0.434286 |
| high choles | Z72_S5 | 0.012451 | 0.016698 | 0.455884 | 0.341146 |
| aortic valve | Z72_S5 | -0.05106 | 0.069815 | 0.464541 | 0.332976 |
| heart failur | Z72_S5 | 0.019739 | 0.04582 | 0.666622 | 0.176121 |
| hypotensio | Z72_S5 | 0.016616 | 0.05235 | 0.75094 | 0.124395 |
| conduction | Z72_S5 | -0.01108 | 0.048212 | 0.818272 | 0.087102 |
| other heart | Z72_S5 | 0.011072 | 0.067105 | 0.868954 | 0.061003 |
| type 2 diab | Z125_S4 | 0.104395 | 0.025906 | 5.60E-05 | 4.251633 |
| atrial fibrill | Z125_S4 | -0.08264 | 0.023771 | 0.000509 | 3.293512 |
| aortic valve | Z125_S4 | -0.14225 | 0.060295 | 0.018325 | 1.736955 |
| conduction | Z125_S4 | -0.08698 | 0.041613 | 0.036612 | 1.436371 |
| mitral prob | Z125_S4 | -0.08025 | 0.04753 | 0.091336 | 1.039356 |
| other heart | Z125_S4 | 0.055598 | 0.057947 | 0.337338 | 0.471935 |
| myocardial | Z125_S4 | -0.02801 | 0.030349 | 0.355977 | 0.448578 |
| angina pec | Z125_S4 | 0.024317 | 0.028154 | 0.387752 | 0.411446 |
| hypotensio | Z125_S4 | -0.03623 | 0.045204 | 0.422889 | 0.373774 |
| high choles | Z125_S4 | 0.011397 | 0.014446 | 0.430144 | 0.366386 |
| hypertensi | Z125_S4 | -0.00616 | 0.013122 | 0.63901 | 0.194492 |
| heart failur | Z125_S4 | -0.00921 | 0.039603 | 0.816193 | 0.088207 |
| coronary h | Z125_S4 | 0.004601 | 0.023351 | 0.843806 | 0.073757 |
| arrhythmia | Z125_S4 | -0.00818 | 0.05161 | 0.874083 | 0.058447 |
| atrial fibrill | Z39_S1 | 0.08566 | 0.021463 | 6.60E-05 | 4.180624 |
| type 2 diab | Z39_S1 | 0.075868 | 0.023339 | 0.001153 | 2.938281 |
| heart failur | Z39_S1 | 0.097217 | 0.035676 | 0.006435 | 2.19144 |
| aortic valve | Z39_S1 | 0.132259 | 0.054375 | 0.015009 | 1.823655 |
| angina pec | Z39_S1 | 0.034346 | 0.025415 | 0.176578 | 0.753063 |
| coronary h | Z39_S1 | 0.026971 | 0.021076 | 0.200657 | 0.697547 |
| hypertensi | Z39_S1 | 0.014952 | 0.011839 | 0.206606 | 0.684856 |
| conduction | Z39_S1 | 0.044442 | 0.037531 | 0.236371 | 0.626405 |
| high choles | Z39_S1 | -0.01464 | 0.013015 | 0.260784 | 0.583718 |
| mitral prob | Z39_S1 | 0.042979 | 0.042863 | 0.316018 | 0.500288 |
| hypotensio | Z39_S1 | 0.038488 | 0.040743 | 0.344846 | 0.462375 |
| other heart | Z39_S1 | 0.043825 | 0.052246 | 0.401581 | 0.396227 |
| arrhythmia | Z39_S1 | 0.018376 | 0.046512 | 0.692788 | 0.1594 |
| myocardial | Z39_S1 | -0.00901 | 0.02737 | 0.741934 | 0.129635 |
| atrial fibrill | Z80_S5 | 0.105438 | 0.026545 | 7.15E-05 | 4.145775 |
| coronary h | Z80_S5 | -0.07524 | 0.02604 | 0.003863 | 2.413053 |
| mitral prob | Z80_S5 | 0.145833 | 0.053036 | 0.00597 | 2.224044 |
| angina pec | Z80_S5 | -0.08564 | 0.031404 | 0.006394 | 2.19423 |
| myocardial | Z80_S5 | -0.08859 | 0.033849 | 0.008872 | 2.051967 |
| heart failur | Z80_S5 | 0.07746 | 0.044178 | 0.079551 | 1.099356 |

| | | | | | |
|---|---|---|---|---|---|
| type 2 diab | Z80_S5 | -0.04363 | 0.028902 | 0.131195 | 0.882081 |
| high choles | Z80_S5 | -0.02277 | 0.016081 | 0.156784 | 0.804697 |
| arrhythmia | Z80_S5 | -0.06087 | 0.057574 | 0.290408 | 0.536991 |
| conduction | Z80_S5 | 0.031926 | 0.046462 | 0.491995 | 0.30804 |
| hypertensi | Z80_S5 | 0.005607 | 0.014613 | 0.701182 | 0.154169 |
| other heart | Z80_S5 | 0.022608 | 0.064673 | 0.726668 | 0.138664 |
| hypotensio | Z80_S5 | -0.01578 | 0.050445 | 0.754461 | 0.122363 |
| aortic valve | Z80_S5 | -0.00848 | 0.067307 | 0.89977 | 0.045868 |
| type 2 diab | Z5_S5 | -0.10262 | 0.025857 | 7.25E-05 | 4.139475 |
| atrial fibrill | Z5_S5 | 0.054152 | 0.02374 | 0.022557 | 1.646719 |
| aortic valve | Z5_S5 | 0.102859 | 0.060186 | 0.087462 | 1.05818 |
| angina pec | Z5_S5 | -0.04706 | 0.028108 | 0.094122 | 1.02631 |
| high choles | Z5_S5 | 0.017005 | 0.014419 | 0.238267 | 0.622936 |
| myocardial | Z5_S5 | 0.035016 | 0.030297 | 0.247792 | 0.605913 |
| mitral prob | Z5_S5 | 0.05217 | 0.047479 | 0.27187 | 0.565639 |
| heart failur | Z5_S5 | -0.04193 | 0.039523 | 0.28876 | 0.539463 |
| other heart | Z5_S5 | -0.05332 | 0.057848 | 0.356717 | 0.447676 |
| hypotensio | Z5_S5 | 0.02561 | 0.045126 | 0.570367 | 0.243846 |
| arrhythmia | Z5_S5 | 0.004461 | 0.051539 | 0.931025 | 0.031039 |
| conduction | Z5_S5 | 0.003378 | 0.041562 | 0.935231 | 0.029081 |
| coronary h | Z5_S5 | -0.00037 | 0.023316 | 0.987341 | 0.005533 |
| hypertensi | Z5_S5 | -0.00013 | 0.01308 | 0.99227 | 0.00337 |
| atrial fibrill | Z80_S4 | -0.07949 | 0.020273 | 8.85E-05 | 4.053041 |
| aortic valve | Z80_S4 | -0.13443 | 0.051351 | 0.008854 | 2.052846 |
| heart failur | Z80_S4 | -0.07183 | 0.033697 | 0.033048 | 1.48086 |
| hypertensi | Z80_S4 | -0.02238 | 0.011204 | 0.045796 | 1.339172 |
| mitral prob | Z80_S4 | -0.05901 | 0.040474 | 0.14484 | 0.839112 |
| other heart | Z80_S4 | -0.03613 | 0.049342 | 0.464019 | 0.333464 |
| arrhythmia | Z80_S4 | -0.02775 | 0.043917 | 0.527543 | 0.277742 |
| hypotensio | Z80_S4 | -0.02028 | 0.03847 | 0.598055 | 0.223259 |
| coronary h | Z80_S4 | -0.0101 | 0.019913 | 0.611896 | 0.213322 |
| high choles | Z80_S4 | 0.003879 | 0.012317 | 0.752842 | 0.123296 |
| myocardial | Z80_S4 | 0.007318 | 0.025847 | 0.777089 | 0.109529 |
| type 2 diab | Z80_S4 | -0.00523 | 0.022097 | 0.812769 | 0.090033 |
| conduction | Z80_S4 | 0.006854 | 0.035455 | 0.846716 | 0.072262 |
| angina pec | Z80_S4 | -0.00433 | 0.024029 | 0.856869 | 0.067086 |
| type 2 diab | Z105_S5 | -0.09928 | 0.025402 | 9.32E-05 | 4.030466 |
| hypertensi | Z105_S5 | 0.032303 | 0.012833 | 0.011839 | 1.926682 |
| atrial fibrill | Z105_S5 | 0.056295 | 0.023287 | 0.015638 | 1.805818 |
| aortic valve | Z105_S5 | 0.115424 | 0.059016 | 0.050498 | 1.296727 |
| myocardial | Z105_S5 | -0.05511 | 0.029707 | 0.063583 | 1.196658 |
| mitral prob | Z105_S5 | 0.081475 | 0.046515 | 0.079859 | 1.097677 |
| angina pec | Z105_S5 | -0.04749 | 0.027574 | 0.085038 | 1.070387 |
| coronary h | Z105_S5 | -0.02972 | 0.022888 | 0.194144 | 0.711876 |
| heart failur | Z105_S5 | -0.03575 | 0.038763 | 0.356362 | 0.448109 |

| | | | | | |
|---|---|---|---|---|---|
| other heart | Z105_S5 | 0.024243 | 0.056704 | 0.668992 | 0.174579 |
| conduction | Z105_S5 | -0.01419 | 0.040789 | 0.728006 | 0.137865 |
| arrhythmia | Z105_S5 | 0.015647 | 0.050484 | 0.756606 | 0.12113 |
| hypotensio | Z105_S5 | 0.012925 | 0.044225 | 0.7701 | 0.113453 |
| high choles | Z105_S5 | -0.00136 | 0.014127 | 0.923367 | 0.034626 |
| heart failur | Z97_S2 | -0.15518 | 0.039827 | 9.79E-05 | 4.009281 |
| mitral prob | Z97_S2 | -0.1443 | 0.047887 | 0.002586 | 2.587337 |
| atrial fibrill | Z97_S2 | -0.06792 | 0.023941 | 0.004555 | 2.341475 |
| angina pec | Z97_S2 | -0.05549 | 0.028344 | 0.050283 | 1.298576 |
| other heart | Z97_S2 | -0.09193 | 0.058386 | 0.115401 | 0.937791 |
| arrhythmia | Z97_S2 | -0.05846 | 0.051944 | 0.260393 | 0.58437 |
| high choles | Z97_S2 | 0.013533 | 0.014577 | 0.353229 | 0.451944 |
| coronary h | Z97_S2 | -0.02174 | 0.023505 | 0.354945 | 0.449839 |
| aortic valve | Z97_S2 | 0.05293 | 0.060712 | 0.38332 | 0.416438 |
| hypertensi | Z97_S2 | 0.01148 | 0.013225 | 0.385381 | 0.414109 |
| myocardial | Z97_S2 | -0.01644 | 0.030534 | 0.590375 | 0.228872 |
| hypotensio | Z97_S2 | -0.00866 | 0.045534 | 0.849127 | 0.071028 |
| conduction | Z97_S2 | -0.00722 | 0.041934 | 0.863263 | 0.063857 |
| type 2 diab | Z97_S2 | 0.004474 | 0.026104 | 0.863917 | 0.063528 |
| type 2 diab | Z89_S4 | 0.09315 | 0.024192 | 0.000118 | 3.927254 |
| aortic valve | Z89_S4 | -0.18506 | 0.056147 | 0.000982 | 3.007735 |
| coronary h | Z89_S4 | -0.04637 | 0.021769 | 0.033184 | 1.479071 |
| hypertensi | Z89_S4 | -0.02568 | 0.012249 | 0.036029 | 1.443352 |
| myocardial | Z89_S4 | -0.04424 | 0.028274 | 0.117695 | 0.92924 |
| mitral prob | Z89_S4 | -0.0542 | 0.044271 | 0.220835 | 0.655932 |
| angina pec | Z89_S4 | -0.02991 | 0.026266 | 0.254788 | 0.593822 |
| other heart | Z89_S4 | 0.043452 | 0.053987 | 0.420909 | 0.375811 |
| arrhythmia | Z89_S4 | -0.02725 | 0.048042 | 0.570546 | 0.243709 |
| hypotensio | Z89_S4 | -0.0173 | 0.04212 | 0.681307 | 0.166657 |
| heart failur | Z89_S4 | 0.013801 | 0.036902 | 0.708422 | 0.149708 |
| atrial fibrill | Z89_S4 | -0.00544 | 0.022231 | 0.80669 | 0.093293 |
| conduction | Z89_S4 | 0.005806 | 0.038789 | 0.881008 | 0.05502 |
| high choles | Z89_S4 | 0.001134 | 0.013443 | 0.93277 | 0.030226 |
| myocardial | Z113_S2 | -0.13184 | 0.034327 | 0.000123 | 3.910034 |
| angina pec | Z113_S2 | -0.11182 | 0.031854 | 0.000448 | 3.348622 |
| coronary h | Z113_S2 | -0.0895 | 0.02642 | 0.000706 | 3.150985 |
| type 2 diab | Z113_S2 | -0.0737 | 0.029323 | 0.011962 | 1.92221 |
| hypertensi | Z113_S2 | 0.030311 | 0.014786 | 0.040381 | 1.393825 |
| mitral prob | Z113_S2 | 0.098321 | 0.053773 | 0.067496 | 1.170722 |
| aortic valve | Z113_S2 | 0.104851 | 0.068218 | 0.124309 | 0.905497 |
| arrhythmia | Z113_S2 | -0.08145 | 0.058372 | 0.162937 | 0.78798 |
| atrial fibrill | Z113_S2 | 0.023544 | 0.026851 | 0.380595 | 0.419537 |
| hypotensio | Z113_S2 | -0.03811 | 0.051144 | 0.456134 | 0.340908 |
| heart failur | Z113_S2 | -0.02699 | 0.044777 | 0.546722 | 0.262233 |
| other heart | Z113_S2 | -0.03412 | 0.065557 | 0.60271 | 0.219891 |

| | | | | | |
|---|---|---|---|---|---|
| conduction | Z113_S2 | 0.01119 | 0.047099 | 0.812198 | 0.090338 |
| high choles | Z113_S2 | 0.003019 | 0.016297 | 0.853017 | 0.069042 |
| heart failur | Z28_S2 | -0.15723 | 0.041166 | 0.000134 | 3.872621 |
| atrial fibrill | Z28_S2 | -0.0735 | 0.024739 | 0.002971 | 2.527101 |
| angina pec | Z28_S2 | -0.07822 | 0.029327 | 0.007657 | 2.115921 |
| coronary h | Z28_S2 | -0.06425 | 0.024306 | 0.00821 | 2.085636 |
| other heart | Z28_S2 | -0.15279 | 0.060347 | 0.011352 | 1.944945 |
| type 2 diab | Z28_S2 | -0.06446 | 0.026977 | 0.016873 | 1.772799 |
| myocardial | Z28_S2 | -0.06478 | 0.031579 | 0.040255 | 1.395176 |
| mitral prob | Z28_S2 | -0.06432 | 0.049508 | 0.19389 | 0.712445 |
| arrhythmia | Z28_S2 | -0.05941 | 0.053728 | 0.268819 | 0.57054 |
| hypertensi | Z28_S2 | 0.011698 | 0.013691 | 0.392877 | 0.405744 |
| aortic valve | Z28_S2 | 0.046783 | 0.062782 | 0.456182 | 0.340862 |
| hypotensio | Z28_S2 | -0.01887 | 0.04707 | 0.688469 | 0.162116 |
| high choles | Z28_S2 | 0.003211 | 0.015082 | 0.831423 | 0.080178 |
| conduction | Z28_S2 | 0.00344 | 0.043345 | 0.936742 | 0.02838 |
| atrial fibrill | Z120_S2 | 0.076265 | 0.02001 | 0.000139 | 3.858265 |
| aortic valve | Z120_S2 | 0.163981 | 0.050665 | 0.001211 | 2.916698 |
| angina pec | Z120_S2 | -0.05151 | 0.023718 | 0.029882 | 1.524589 |
| myocardial | Z120_S2 | -0.05505 | 0.025519 | 0.030993 | 1.508735 |
| hypertensi | Z120_S2 | 0.023038 | 0.01106 | 0.037265 | 1.428704 |
| coronary h | Z120_S2 | -0.03629 | 0.019671 | 0.065109 | 1.186357 |
| mitral prob | Z120_S2 | 0.045453 | 0.039937 | 0.255082 | 0.59332 |
| conduction | Z120_S2 | -0.03264 | 0.035 | 0.35107 | 0.454606 |
| hypotensio | Z120_S2 | 0.026125 | 0.037988 | 0.491638 | 0.308354 |
| heart failur | Z120_S2 | 0.01859 | 0.033276 | 0.576392 | 0.239282 |
| arrhythmia | Z120_S2 | -0.02073 | 0.043368 | 0.632707 | 0.198797 |
| type 2 diab | Z120_S2 | 0.005766 | 0.021827 | 0.791643 | 0.101471 |
| high choles | Z120_S2 | 0.002982 | 0.012159 | 0.806278 | 0.093515 |
| other heart | Z120_S2 | -1.13E-05 | 0.048697 | 0.999815 | 8.03E-05 |
| atrial fibrill | Z40_S5 | -0.07505 | 0.019716 | 0.000141 | 3.849651 |
| aortic valve | Z40_S5 | -0.16482 | 0.049808 | 0.000937 | 3.028098 |
| type 2 diab | Z40_S5 | 0.04191 | 0.021479 | 0.051042 | 1.292072 |
| coronary h | Z40_S5 | -0.02912 | 0.019331 | 0.131963 | 0.879547 |
| mitral prob | Z40_S5 | -0.05395 | 0.039282 | 0.169634 | 0.770486 |
| hypertensi | Z40_S5 | -0.01328 | 0.010873 | 0.222089 | 0.653473 |
| hypotensio | Z40_S5 | -0.03062 | 0.037384 | 0.412824 | 0.384235 |
| myocardial | Z40_S5 | -0.01784 | 0.025097 | 0.477235 | 0.321268 |
| angina pec | Z40_S5 | -0.0123 | 0.023319 | 0.59779 | 0.223451 |
| arrhythmia | Z40_S5 | -0.02052 | 0.042631 | 0.630312 | 0.200444 |
| high choles | Z40_S5 | -0.00331 | 0.011936 | 0.78178 | 0.106915 |
| heart failur | Z40_S5 | -0.00785 | 0.032742 | 0.810452 | 0.091273 |
| conduction | Z40_S5 | 0.003177 | 0.034414 | 0.926441 | 0.033182 |
| other heart | Z40_S5 | -0.00033 | 0.047896 | 0.994444 | 0.00242 |
| myocardial | Z23_S1 | -0.12512 | 0.033128 | 0.000159 | 3.798096 |

| | | | | | |
|---|---|---|---|---|---|
| type 2 diab | Z23_S1 | 0.097795 | 0.028305 | 0.000551 | 3.258753 |
| coronary h | Z23_S1 | -0.08526 | 0.025495 | 0.000826 | 3.08291 |
| conduction | Z23_S1 | 0.127683 | 0.04547 | 0.004989 | 2.302027 |
| angina pec | Z23_S1 | -0.07684 | 0.030756 | 0.012484 | 1.90366 |
| aortic valve | Z23_S1 | 0.119462 | 0.06587 | 0.069751 | 1.15645 |
| high choles | Z23_S1 | -0.02437 | 0.015775 | 0.122353 | 0.912387 |
| other heart | Z23_S1 | -0.09464 | 0.063332 | 0.13508 | 0.869408 |
| arrhythmia | Z23_S1 | -0.07874 | 0.056349 | 0.162337 | 0.789584 |
| mitral prob | Z23_S1 | -0.06716 | 0.051923 | 0.195837 | 0.708105 |
| hypertensi | Z23_S1 | 0.014069 | 0.014363 | 0.327334 | 0.485009 |
| hypotensio | Z23_S1 | 0.029792 | 0.04938 | 0.546301 | 0.262568 |
| atrial fibrill | Z23_S1 | 0.003167 | 0.025987 | 0.903011 | 0.044307 |
| heart failur | Z23_S1 | -0.00483 | 0.043245 | 0.911146 | 0.040412 |
| type 2 diab | Z85_S5 | -0.08789 | 0.023313 | 0.000164 | 3.785994 |
| coronary h | Z85_S5 | 0.059186 | 0.020966 | 0.004762 | 2.322183 |
| aortic valve | Z85_S5 | 0.149591 | 0.054061 | 0.00566 | 2.247158 |
| myocardial | Z85_S5 | 0.073328 | 0.027219 | 0.007065 | 2.150882 |
| angina pec | Z85_S5 | 0.039687 | 0.02528 | 0.116451 | 0.933856 |
| arrhythmia | Z85_S5 | 0.063591 | 0.046263 | 0.169277 | 0.771402 |
| atrial fibrill | Z85_S5 | 0.026959 | 0.021421 | 0.208212 | 0.681495 |
| mitral prob | Z85_S5 | 0.048064 | 0.042622 | 0.259467 | 0.585918 |
| other heart | Z85_S5 | -0.04647 | 0.051981 | 0.371328 | 0.430242 |
| hypertensi | Z85_S5 | 0.009737 | 0.011781 | 0.408517 | 0.388789 |
| hypotensio | Z85_S5 | 0.029007 | 0.040561 | 0.474535 | 0.323732 |
| high choles | Z85_S5 | 0.008914 | 0.012942 | 0.490989 | 0.308928 |
| heart failur | Z85_S5 | -0.02217 | 0.035541 | 0.532849 | 0.273396 |
| conduction | Z85_S5 | -0.01724 | 0.037351 | 0.644394 | 0.190849 |
| atrial fibrill | Z120_S4 | -0.08472 | 0.022631 | 0.000182 | 3.740417 |
| heart failur | Z120_S4 | -0.12087 | 0.03765 | 0.001327 | 2.87701 |
| aortic valve | Z120_S4 | -0.13949 | 0.057384 | 0.015075 | 1.821744 |
| myocardial | Z120_S4 | -0.0686 | 0.028866 | 0.017492 | 1.757166 |
| mitral prob | Z120_S4 | -0.08894 | 0.045219 | 0.049209 | 1.307958 |
| arrhythmia | Z120_S4 | -0.08486 | 0.04908 | 0.083803 | 1.076742 |
| coronary h | Z120_S4 | -0.03824 | 0.022217 | 0.085203 | 1.069546 |
| hypertensi | Z120_S4 | -0.02111 | 0.012497 | 0.091213 | 1.039944 |
| hypotensio | Z120_S4 | -0.06775 | 0.042978 | 0.114953 | 0.93948 |
| other heart | Z120_S4 | -0.08556 | 0.055153 | 0.120822 | 0.917853 |
| type 2 diab | Z120_S4 | -0.03787 | 0.02467 | 0.124748 | 0.903965 |
| angina pec | Z120_S4 | -0.01705 | 0.026826 | 0.525 | 0.279841 |
| high choles | Z120_S4 | -0.00412 | 0.013724 | 0.764278 | 0.116749 |
| conduction | Z120_S4 | 0.011368 | 0.039602 | 0.774065 | 0.111222 |
| type 2 diab | Z28_S1 | -0.09801 | 0.026546 | 0.000223 | 3.6517 |
| hypertensi | Z28_S1 | -0.0214 | 0.013461 | 0.111973 | 0.950888 |
| mitral prob | Z28_S1 | 0.065232 | 0.04874 | 0.180788 | 0.74283 |
| myocardial | Z28_S1 | 0.041185 | 0.031112 | 0.185589 | 0.731447 |

| | | | | | |
|---|---|---|---|---|---|
| aortic valve | Z28_S1 | -0.07021 | 0.061841 | 0.256276 | 0.591293 |
| atrial fibrill | Z28_S1 | 0.026417 | 0.024393 | 0.278834 | 0.554655 |
| conduction | Z28_S1 | 0.030016 | 0.042697 | 0.482055 | 0.316904 |
| coronary h | Z28_S1 | 0.014886 | 0.023954 | 0.534308 | 0.272208 |
| angina pec | Z28_S1 | 0.011219 | 0.028891 | 0.697791 | 0.156275 |
| other heart | Z28_S1 | -0.02132 | 0.059414 | 0.719712 | 0.142841 |
| arrhythmia | Z28_S1 | 0.01773 | 0.052897 | 0.737492 | 0.132243 |
| high choles | Z28_S1 | -0.00485 | 0.014806 | 0.743223 | 0.128881 |
| hypotensio | Z28_S1 | -0.01188 | 0.046331 | 0.797615 | 0.098207 |
| heart failur | Z28_S1 | -0.0041 | 0.040574 | 0.91947 | 0.036462 |
| atrial fibrill | Z94_S5 | 0.0734 | 0.019946 | 0.000234 | 3.631209 |
| aortic valve | Z94_S5 | 0.165859 | 0.05037 | 0.000994 | 3.002827 |
| type 2 diab | Z94_S5 | -0.06673 | 0.021729 | 0.002134 | 2.670745 |
| coronary h | Z94_S5 | 0.052722 | 0.019551 | 0.00701 | 2.154273 |
| myocardial | Z94_S5 | 0.054866 | 0.025376 | 0.030619 | 1.51401 |
| mitral prob | Z94_S5 | 0.081792 | 0.039724 | 0.039505 | 1.403347 |
| angina pec | Z94_S5 | 0.031794 | 0.023582 | 0.17759 | 0.750582 |
| arrhythmia | Z94_S5 | 0.048788 | 0.043115 | 0.257828 | 0.588671 |
| hypertensi | Z94_S5 | 0.011996 | 0.01099 | 0.275052 | 0.560585 |
| hypotensio | Z94_S5 | 0.033565 | 0.037802 | 0.374589 | 0.426445 |
| heart failur | Z94_S5 | 0.021069 | 0.033115 | 0.52463 | 0.280147 |
| conduction | Z94_S5 | -0.01535 | 0.034806 | 0.659114 | 0.181039 |
| high choles | Z94_S5 | 0.005303 | 0.012069 | 0.660353 | 0.180224 |
| other heart | Z94_S5 | -0.00217 | 0.048443 | 0.964202 | 0.015832 |
| atrial fibrill | Z57_S1 | -0.07788 | 0.021305 | 0.000257 | 3.58937 |
| aortic valve | Z57_S1 | -0.17106 | 0.053973 | 0.00153 | 2.815305 |
| heart failur | Z57_S1 | -0.07667 | 0.035409 | 0.030381 | 1.517399 |
| type 2 diab | Z57_S1 | -0.0469 | 0.023214 | 0.043371 | 1.362796 |
| hypertensi | Z57_S1 | -0.0232 | 0.011759 | 0.048456 | 1.314653 |
| hypotensio | Z57_S1 | -0.07297 | 0.040431 | 0.071112 | 1.148055 |
| other heart | Z57_S1 | -0.04859 | 0.051849 | 0.348654 | 0.457605 |
| angina pec | Z57_S1 | 0.022285 | 0.025238 | 0.377241 | 0.423381 |
| mitral prob | Z57_S1 | -0.03439 | 0.04255 | 0.418908 | 0.377881 |
| arrhythmia | Z57_S1 | -0.02706 | 0.046174 | 0.557824 | 0.253503 |
| high choles | Z57_S1 | -0.00624 | 0.012918 | 0.628949 | 0.201385 |
| myocardial | Z57_S1 | -0.01257 | 0.027152 | 0.643475 | 0.191468 |
| coronary h | Z57_S1 | -0.00619 | 0.020918 | 0.767469 | 0.114939 |
| conduction | Z57_S1 | 0.00791 | 0.03724 | 0.831802 | 0.07998 |
| type 2 diab | Z39_S4 | 0.082745 | 0.022727 | 0.000272 | 3.564936 |
| myocardial | Z39_S4 | -0.08207 | 0.026623 | 0.002055 | 2.68716 |
| coronary h | Z39_S4 | -0.03586 | 0.020517 | 0.080529 | 1.094045 |
| aortic valve | Z39_S4 | 0.084465 | 0.052918 | 0.11047 | 0.956757 |
| other heart | Z39_S4 | -0.07908 | 0.05083 | 0.119783 | 0.921605 |
| atrial fibrill | Z39_S4 | -0.03216 | 0.020872 | 0.123343 | 0.908885 |
| mitral prob | Z39_S4 | -0.05786 | 0.041698 | 0.165288 | 0.781759 |

| | | | | | |
|---|---|---|---|---|---|
| hypertensi | Z39_S4 | 0.011664 | 0.011516 | 0.31117 | 0.507003 |
| angina pec | Z39_S4 | -0.0188 | 0.024722 | 0.446926 | 0.349765 |
| high choles | Z39_S4 | 0.008406 | 0.012664 | 0.506863 | 0.295109 |
| arrhythmia | Z39_S4 | -0.02941 | 0.045241 | 0.51559 | 0.287696 |
| heart failur | Z39_S4 | -0.0085 | 0.034724 | 0.806578 | 0.093353 |
| conduction | Z39_S4 | -0.00813 | 0.036517 | 0.823829 | 0.084163 |
| hypotensio | Z39_S4 | -0.00734 | 0.039625 | 0.853035 | 0.069033 |
| angina pec | Z59_S2 | -0.11096 | 0.030524 | 0.000278 | 3.555274 |
| coronary h | Z59_S2 | -0.08213 | 0.025309 | 0.001176 | 2.92951 |
| myocardial | Z59_S2 | -0.10446 | 0.032886 | 0.001493 | 2.825805 |
| atrial fibrill | Z59_S2 | 0.058504 | 0.025733 | 0.023006 | 1.638168 |
| hypertensi | Z59_S2 | 0.031194 | 0.014176 | 0.027781 | 1.556255 |
| aortic valve | Z59_S2 | 0.098797 | 0.065311 | 0.130368 | 0.884828 |
| type 2 diab | Z59_S2 | -0.03822 | 0.028098 | 0.173749 | 0.760078 |
| mitral prob | Z59_S2 | 0.067033 | 0.051498 | 0.193043 | 0.714346 |
| arrhythmia | Z59_S2 | -0.06625 | 0.055919 | 0.236131 | 0.626847 |
| high choles | Z59_S2 | 0.01657 | 0.015623 | 0.288891 | 0.539266 |
| heart failur | Z59_S2 | -0.03588 | 0.042885 | 0.402755 | 0.39496 |
| other heart | Z59_S2 | -0.04529 | 0.062761 | 0.470571 | 0.327374 |
| conduction | Z59_S2 | -0.03153 | 0.045101 | 0.484543 | 0.314667 |
| hypotensio | Z59_S2 | -0.00808 | 0.048977 | 0.868988 | 0.060986 |
| aortic valve | Z78_S4 | 0.188849 | 0.052118 | 0.000291 | 3.535606 |
| atrial fibrill | Z78_S4 | 0.061001 | 0.020596 | 0.003061 | 2.514096 |
| hypertensi | Z78_S4 | 0.025251 | 0.011374 | 0.026423 | 1.578012 |
| mitral prob | Z78_S4 | 0.072797 | 0.041087 | 0.076442 | 1.116669 |
| coronary h | Z78_S4 | 0.025677 | 0.020216 | 0.204051 | 0.690261 |
| heart failur | Z78_S4 | 0.042089 | 0.034225 | 0.218795 | 0.659962 |
| type 2 diab | Z78_S4 | -0.02664 | 0.022452 | 0.235401 | 0.628193 |
| arrhythmia | Z78_S4 | 0.028543 | 0.044588 | 0.522075 | 0.282267 |
| hypotensio | Z78_S4 | 0.024922 | 0.039076 | 0.523629 | 0.280977 |
| angina pec | Z78_S4 | 0.008241 | 0.024397 | 0.73551 | 0.133411 |
| myocardial | Z78_S4 | 0.007786 | 0.026248 | 0.766765 | 0.115337 |
| conduction | Z78_S4 | -0.00464 | 0.035993 | 0.897483 | 0.046974 |
| other heart | Z78_S4 | 0.004791 | 0.0501 | 0.923817 | 0.034414 |
| high choles | Z78_S4 | -0.00105 | 0.012491 | 0.933296 | 0.029981 |
| aortic valve | Z23_S3 | 0.189882 | 0.052482 | 0.000298 | 3.526494 |
| atrial fibrill | Z23_S3 | 0.057139 | 0.020716 | 0.005815 | 2.235432 |
| hypertensi | Z23_S3 | 0.029612 | 0.011432 | 0.009597 | 2.017878 |
| heart failur | Z23_S3 | 0.064528 | 0.034449 | 0.061058 | 1.214256 |
| type 2 diab | Z23_S3 | 0.040152 | 0.02258 | 0.075383 | 1.122727 |
| myocardial | Z23_S3 | -0.04474 | 0.026429 | 0.090464 | 1.043523 |
| mitral prob | Z23_S3 | 0.061511 | 0.041375 | 0.137115 | 0.862916 |
| conduction | Z23_S3 | -0.03371 | 0.036252 | 0.35239 | 0.452977 |
| hypotensio | Z23_S3 | 0.034445 | 0.039326 | 0.381106 | 0.418954 |
| angina pec | Z23_S3 | -0.01197 | 0.024553 | 0.625895 | 0.203499 |

| | | | | | |
|---|---|---|---|---|---|
| high choles | Z23_S3 | -0.00441 | 0.012587 | 0.726115 | 0.138995 |
| coronary h | Z23_S3 | -0.00583 | 0.020368 | 0.774603 | 0.110921 |
| other heart | Z23_S3 | 0.013953 | 0.050459 | 0.782156 | 0.106707 |
| arrhythmia | Z23_S3 | 0.012037 | 0.044907 | 0.788673 | 0.103103 |
| type 2 diab | Z27_S2 | 0.103865 | 0.028776 | 0.000308 | 3.512028 |
| myocardial | Z27_S2 | 0.109387 | 0.03368 | 0.001165 | 2.933821 |
| angina pec | Z27_S2 | 0.088765 | 0.031284 | 0.004552 | 2.341751 |
| heart failur | Z27_S2 | 0.124183 | 0.043963 | 0.004736 | 2.324565 |
| coronary h | Z27_S2 | 0.070317 | 0.025943 | 0.006725 | 2.172336 |
| other heart | Z27_S2 | 0.136716 | 0.064361 | 0.033665 | 1.472823 |
| arrhythmia | Z27_S2 | 0.092137 | 0.057305 | 0.107888 | 0.967028 |
| conduction | Z27_S2 | -0.07337 | 0.046227 | 0.112507 | 0.948819 |
| atrial fibrill | Z27_S2 | 0.038472 | 0.026392 | 0.144929 | 0.838844 |
| aortic valve | Z27_S2 | -0.08572 | 0.066968 | 0.200538 | 0.697804 |
| hypertensi | Z27_S2 | -0.01677 | 0.014574 | 0.24977 | 0.602459 |
| hypotensio | Z27_S2 | 0.053324 | 0.050206 | 0.288202 | 0.540303 |
| high choles | Z27_S2 | 0.009562 | 0.016028 | 0.550805 | 0.259002 |
| mitral prob | Z27_S2 | 0.017408 | 0.052802 | 0.741637 | 0.129809 |
| atrial fibrill | Z2_S1 | -0.07555 | 0.021253 | 0.000379 | 3.421437 |
| aortic valve | Z2_S1 | -0.1414 | 0.053842 | 0.008641 | 2.063425 |
| heart failur | Z2_S1 | -0.07588 | 0.035323 | 0.031713 | 1.498764 |
| hypertensi | Z2_S1 | -0.02432 | 0.011737 | 0.038286 | 1.416962 |
| mitral prob | Z2_S1 | -0.05494 | 0.042441 | 0.195495 | 0.708863 |
| type 2 diab | Z2_S1 | -0.02952 | 0.02315 | 0.202234 | 0.694146 |
| other heart | Z2_S1 | -0.05344 | 0.051739 | 0.301694 | 0.520433 |
| hypotensio | Z2_S1 | -0.0313 | 0.040333 | 0.43773 | 0.358793 |
| coronary h | Z2_S1 | -0.00906 | 0.020873 | 0.664156 | 0.17773 |
| arrhythmia | Z2_S1 | -0.01852 | 0.046056 | 0.687572 | 0.162682 |
| high choles | Z2_S1 | 0.003872 | 0.012901 | 0.764056 | 0.116875 |
| angina pec | Z2_S1 | -0.00498 | 0.025181 | 0.843138 | 0.074101 |
| myocardial | Z2_S1 | 0.005172 | 0.027099 | 0.848646 | 0.071273 |
| conduction | Z2_S1 | -0.00528 | 0.037176 | 0.8871 | 0.052028 |
| atrial fibrill | Z68_S2 | -0.07412 | 0.020957 | 0.000406 | 3.391847 |
| aortic valve | Z68_S2 | -0.13534 | 0.053023 | 0.010705 | 1.970415 |
| hypertensi | Z68_S2 | -0.01684 | 0.011546 | 0.144711 | 0.839499 |
| myocardial | Z68_S2 | 0.030792 | 0.0267 | 0.24882 | 0.604115 |
| coronary h | Z68_S2 | 0.017488 | 0.02057 | 0.395242 | 0.403137 |
| arrhythmia | Z68_S2 | 0.026569 | 0.045368 | 0.558128 | 0.253266 |
| type 2 diab | Z68_S2 | 0.013311 | 0.022817 | 0.559635 | 0.252095 |
| angina pec | Z68_S2 | 0.01401 | 0.024803 | 0.572178 | 0.242469 |
| other heart | Z68_S2 | 0.028144 | 0.050973 | 0.580868 | 0.235922 |
| hypotensio | Z68_S2 | -0.0209 | 0.039763 | 0.599228 | 0.222408 |
| high choles | Z68_S2 | 0.00355 | 0.01271 | 0.780046 | 0.10788 |
| mitral prob | Z68_S2 | -0.01045 | 0.041797 | 0.802584 | 0.09551 |
| conduction | Z68_S2 | -0.00663 | 0.036619 | 0.856376 | 0.067335 |

| | | | | | |
|---|---|---|---|---|---|
| heart failur | Z68_S2 | -0.00315 | 0.03483 | 0.927927 | 0.032486 |
| aortic valve | Z8_S3 | 0.178685 | 0.050798 | 0.000436 | 3.360073 |
| myocardial | Z8_S3 | -0.07197 | 0.02559 | 0.004919 | 2.30808 |
| atrial fibrill | Z8_S3 | 0.056286 | 0.020046 | 0.004991 | 2.30179 |
| hypertensi | Z8_S3 | 0.030706 | 0.01108 | 0.005587 | 2.252822 |
| coronary h | Z8_S3 | -0.03149 | 0.01972 | 0.110295 | 0.957443 |
| angina pec | Z8_S3 | -0.03526 | 0.023771 | 0.138023 | 0.86005 |
| mitral prob | Z8_S3 | 0.046189 | 0.040052 | 0.248836 | 0.604087 |
| conduction | Z8_S3 | -0.03496 | 0.03509 | 0.319142 | 0.496016 |
| heart failur | Z8_S3 | 0.028999 | 0.033345 | 0.384487 | 0.415118 |
| hypotensio | Z8_S3 | 0.029413 | 0.038077 | 0.439858 | 0.356688 |
| high choles | Z8_S3 | -0.00667 | 0.012182 | 0.583951 | 0.233623 |
| type 2 diab | Z8_S3 | 0.01187 | 0.021872 | 0.58733 | 0.231118 |
| arrhythmia | Z8_S3 | -0.00917 | 0.043463 | 0.832857 | 0.07943 |
| other heart | Z8_S3 | 0.000666 | 0.048843 | 0.989124 | 0.004749 |
| myocardial | Z22_S4 | -0.10674 | 0.030411 | 0.000449 | 3.347716 |
| type 2 diab | Z22_S4 | 0.074535 | 0.025921 | 0.004038 | 2.393839 |
| atrial fibrill | Z22_S4 | 0.060693 | 0.023817 | 0.010831 | 1.965334 |
| aortic valve | Z22_S4 | 0.112072 | 0.060409 | 0.063576 | 1.196707 |
| hypertensi | Z22_S4 | 0.020087 | 0.01313 | 0.126065 | 0.899406 |
| coronary h | Z22_S4 | -0.03157 | 0.02341 | 0.17753 | 0.750729 |
| high choles | Z22_S4 | -0.01804 | 0.014458 | 0.212169 | 0.673318 |
| heart failur | Z22_S4 | 0.027701 | 0.039645 | 0.484729 | 0.314501 |
| hypotensio | Z22_S4 | 0.030012 | 0.045242 | 0.507111 | 0.294897 |
| angina pec | Z22_S4 | -0.01684 | 0.028205 | 0.550391 | 0.259329 |
| other heart | Z22_S4 | -0.01539 | 0.058055 | 0.791001 | 0.101823 |
| conduction | Z22_S4 | 0.008444 | 0.04172 | 0.839611 | 0.075922 |
| arrhythmia | Z22_S4 | -0.01029 | 0.051681 | 0.842168 | 0.074601 |
| mitral prob | Z22_S4 | -0.00715 | 0.047646 | 0.880726 | 0.055159 |
| atrial fibrill | Z65_S5 | -0.09327 | 0.026823 | 0.000507 | 3.294738 |
| type 2 diab | Z65_S5 | 0.092468 | 0.029246 | 0.00157 | 2.80403 |
| conduction | Z65_S5 | -0.14633 | 0.046979 | 0.001843 | 2.734505 |
| hypertensi | Z65_S5 | 0.014604 | 0.0148 | 0.323753 | 0.489786 |
| mitral prob | Z65_S5 | -0.05137 | 0.053667 | 0.338509 | 0.47043 |
| myocardial | Z65_S5 | -0.03202 | 0.034236 | 0.349587 | 0.456445 |
| heart failur | Z65_S5 | -0.03757 | 0.044682 | 0.400427 | 0.397476 |
| high choles | Z65_S5 | 0.013676 | 0.016285 | 0.40102 | 0.396834 |
| hypotensio | Z65_S5 | 0.042554 | 0.051027 | 0.404325 | 0.39327 |
| aortic valve | Z65_S5 | 0.03314 | 0.068076 | 0.626401 | 0.203147 |
| coronary h | Z65_S5 | 0.003432 | 0.026361 | 0.896413 | 0.047492 |
| other heart | Z65_S5 | -0.00822 | 0.065435 | 0.900056 | 0.045731 |
| angina pec | Z65_S5 | 0.003148 | 0.031783 | 0.921111 | 0.035688 |
| arrhythmia | Z65_S5 | 0.002744 | 0.058241 | 0.962429 | 0.016631 |
| type 2 diab | Z94_S2 | -0.09305 | 0.027046 | 0.000582 | 3.235316 |
| angina pec | Z94_S2 | -0.09037 | 0.029388 | 0.002107 | 2.676238 |

| | | | | | |
|---|---|---|---|---|---|
| coronary h | Z94_S2 | -0.05938 | 0.024361 | 0.014802 | 1.829679 |
| hypertensi | Z94_S2 | 0.026558 | 0.013685 | 0.05232 | 1.281332 |
| mitral prob | Z94_S2 | 0.089787 | 0.04962 | 0.070389 | 1.152498 |
| myocardial | Z94_S2 | -0.05193 | 0.031669 | 0.101028 | 0.995556 |
| atrial fibrill | Z94_S2 | 0.037046 | 0.024823 | 0.135612 | 0.867703 |
| other heart | Z94_S2 | 0.085531 | 0.060474 | 0.157276 | 0.803339 |
| arrhythmia | Z94_S2 | 0.037143 | 0.053842 | 0.490291 | 0.309546 |
| hypotensio | Z94_S2 | 0.029685 | 0.047194 | 0.529357 | 0.276251 |
| aortic valve | Z94_S2 | 0.028635 | 0.06294 | 0.64915 | 0.187655 |
| heart failur | Z94_S2 | -0.00944 | 0.041301 | 0.819259 | 0.086579 |
| conduction | Z94_S2 | 0.007515 | 0.043413 | 0.862567 | 0.064207 |
| high choles | Z94_S2 | 0.001452 | 0.015074 | 0.923249 | 0.034681 |
| myocardial | Z24_S5 | -0.10824 | 0.032264 | 0.000796 | 3.09927 |
| coronary h | Z24_S5 | -0.07528 | 0.024842 | 0.002447 | 2.611388 |
| angina pec | Z24_S5 | -0.07658 | 0.029943 | 0.010548 | 1.976809 |
| type 2 diab | Z24_S5 | 0.058013 | 0.027588 | 0.035491 | 1.449887 |
| conduction | Z24_S5 | -0.07852 | 0.044293 | 0.076285 | 1.117561 |
| heart failur | Z24_S5 | 0.073657 | 0.042101 | 0.080212 | 1.095763 |
| hypertensi | Z24_S5 | 0.018559 | 0.01397 | 0.18402 | 0.735135 |
| other heart | Z24_S5 | 0.070259 | 0.061661 | 0.254531 | 0.594259 |
| high choles | Z24_S5 | -0.01614 | 0.015398 | 0.294693 | 0.53063 |
| atrial fibrill | Z24_S5 | 0.025364 | 0.025275 | 0.31562 | 0.500836 |
| mitral prob | Z24_S5 | 0.045532 | 0.050573 | 0.367965 | 0.434194 |
| arrhythmia | Z24_S5 | -0.0183 | 0.054849 | 0.738585 | 0.1316 |
| aortic valve | Z24_S5 | -0.01891 | 0.064152 | 0.768194 | 0.114529 |
| hypotensio | Z24_S5 | -0.00297 | 0.0481 | 0.950698 | 0.021957 |
| myocardial | Z14_S4 | 0.108317 | 0.032622 | 0.0009 | 3.045647 |
| atrial fibrill | Z14_S4 | -0.08152 | 0.025571 | 0.001435 | 2.843115 |
| coronary h | Z14_S4 | 0.07746 | 0.025108 | 0.002037 | 2.690996 |
| angina pec | Z14_S4 | 0.082762 | 0.030271 | 0.006261 | 2.203347 |
| type 2 diab | Z14_S4 | -0.07021 | 0.027897 | 0.011854 | 1.926141 |
| heart failur | Z14_S4 | -0.0921 | 0.042578 | 0.030545 | 1.51506 |
| other heart | Z14_S4 | -0.11391 | 0.062332 | 0.067646 | 1.169759 |
| aortic valve | Z14_S4 | 0.116946 | 0.064851 | 0.071355 | 1.146576 |
| conduction | Z14_S4 | 0.070011 | 0.044795 | 0.118083 | 0.927814 |
| mitral prob | Z14_S4 | -0.04133 | 0.051124 | 0.418877 | 0.377913 |
| hypertensi | Z14_S4 | -0.0041 | 0.014102 | 0.7713 | 0.112777 |
| high choles | Z14_S4 | -0.00281 | 0.015554 | 0.856644 | 0.067199 |
| arrhythmia | Z14_S4 | 0.006025 | 0.055457 | 0.913483 | 0.039299 |
| hypotensio | Z14_S4 | 0.004664 | 0.048636 | 0.923601 | 0.034516 |
| type 2 diab | Z100_S3 | 0.084397 | 0.025575 | 0.000968 | 3.014059 |
| aortic valve | Z100_S3 | 0.1653 | 0.059576 | 0.005531 | 2.257159 |
| myocardial | Z100_S3 | -0.06449 | 0.030005 | 0.031629 | 1.499918 |
| atrial fibrill | Z100_S3 | -0.04939 | 0.023505 | 0.03564 | 1.448067 |
| conduction | Z100_S3 | -0.05892 | 0.041155 | 0.152217 | 0.817538 |

| | | | | | |
|---|---|---|---|---|---|
| heart failur | Z100_S3 | -0.05501 | 0.039117 | 0.159658 | 0.79681 |
| mitral prob | Z100_S3 | -0.06507 | 0.046973 | 0.166016 | 0.779851 |
| hypertensi | Z100_S3 | 0.015222 | 0.01296 | 0.240201 | 0.619424 |
| other heart | Z100_S3 | -0.04571 | 0.057275 | 0.4248 | 0.371815 |
| angina pec | Z100_S3 | 0.01099 | 0.027823 | 0.692842 | 0.159366 |
| hypotensio | Z100_S3 | 0.017213 | 0.044644 | 0.699834 | 0.155005 |
| arrhythmia | Z100_S3 | -0.01731 | 0.050965 | 0.734072 | 0.134261 |
| coronary h | Z100_S3 | 0.004598 | 0.023095 | 0.842194 | 0.074588 |
| high choles | Z100_S3 | 0.002537 | 0.01426 | 0.858818 | 0.066099 |
| type 2 diab | Z53_S2 | -0.09019 | 0.027633 | 0.001101 | 2.95812 |
| atrial fibrill | Z53_S2 | 0.055534 | 0.025388 | 0.028723 | 1.54177 |
| mitral prob | Z53_S2 | 0.092134 | 0.050726 | 0.069335 | 1.159046 |
| conduction | Z53_S2 | 0.07453 | 0.044442 | 0.093557 | 1.028924 |
| heart failur | Z53_S2 | 0.058531 | 0.042233 | 0.165781 | 0.780464 |
| hypertensi | Z53_S2 | -0.01921 | 0.014002 | 0.170146 | 0.769179 |
| myocardial | Z53_S2 | 0.038774 | 0.032384 | 0.231201 | 0.63601 |
| aortic valve | Z53_S2 | -0.05637 | 0.064352 | 0.381063 | 0.419003 |
| high choles | Z53_S2 | -0.0132 | 0.015404 | 0.391646 | 0.407107 |
| coronary h | Z53_S2 | 0.013562 | 0.024924 | 0.586345 | 0.231847 |
| angina pec | Z53_S2 | 0.015493 | 0.030058 | 0.606245 | 0.217352 |
| hypotensio | Z53_S2 | -0.02047 | 0.048218 | 0.67122 | 0.173135 |
| arrhythmia | Z53_S2 | 0.011394 | 0.055049 | 0.836028 | 0.077779 |
| other heart | Z53_S2 | 0.003914 | 0.061847 | 0.949544 | 0.022485 |
| aortic valve | Z98_S3 | -0.16614 | 0.051131 | 0.001158 | 2.936207 |
| atrial fibrill | Z98_S3 | -0.05963 | 0.020227 | 0.003199 | 2.49501 |
| hypertensi | Z98_S3 | -0.02386 | 0.011152 | 0.032372 | 1.489832 |
| myocardial | Z98_S3 | 0.028117 | 0.025757 | 0.275016 | 0.560643 |
| hypotensio | Z98_S3 | -0.03392 | 0.038351 | 0.376408 | 0.424341 |
| angina pec | Z98_S3 | 0.020103 | 0.023935 | 0.400971 | 0.396887 |
| heart failur | Z98_S3 | -0.02817 | 0.033582 | 0.401482 | 0.396333 |
| coronary h | Z98_S3 | 0.013572 | 0.019841 | 0.493957 | 0.306311 |
| mitral prob | Z98_S3 | -0.02756 | 0.040324 | 0.494392 | 0.305929 |
| high choles | Z98_S3 | 0.007159 | 0.012245 | 0.55883 | 0.25272 |
| conduction | Z98_S3 | 0.014513 | 0.035314 | 0.681104 | 0.166787 |
| type 2 diab | Z98_S3 | -0.00531 | 0.022018 | 0.809458 | 0.091806 |
| other heart | Z98_S3 | -0.00822 | 0.049163 | 0.867177 | 0.061892 |
| arrhythmia | Z98_S3 | -0.0051 | 0.043758 | 0.907281 | 0.042258 |
| aortic valve | Z1_S1 | -0.18054 | 0.056469 | 0.00139 | 2.857079 |
| coronary h | Z1_S1 | -0.04904 | 0.021889 | 0.025078 | 1.600712 |
| myocardial | Z1_S1 | -0.05915 | 0.028427 | 0.037484 | 1.426157 |
| hypertensi | Z1_S1 | -0.02047 | 0.012303 | 0.096109 | 1.017235 |
| hypotensio | Z1_S1 | -0.05868 | 0.042336 | 0.165719 | 0.780629 |
| angina pec | Z1_S1 | -0.03225 | 0.026401 | 0.221902 | 0.653838 |
| atrial fibrill | Z1_S1 | -0.02243 | 0.022363 | 0.315787 | 0.500606 |
| arrhythmia | Z1_S1 | -0.03952 | 0.048311 | 0.413308 | 0.383726 |

| | | | | | |
|---|---|---|---|---|---|
| mitral prob | Z1_S1 | -0.02495 | 0.044537 | 0.575413 | 0.24002 |
| high choles | Z1_S1 | -0.00602 | 0.013514 | 0.655753 | 0.18326 |
| heart failur | Z1_S1 | -0.01222 | 0.037093 | 0.741754 | 0.12974 |
| type 2 diab | Z1_S1 | 0.007263 | 0.024301 | 0.76504 | 0.116316 |
| conduction | Z1_S1 | -0.00495 | 0.038991 | 0.899001 | 0.04624 |
| other heart | Z1_S1 | -0.00035 | 0.054281 | 0.994912 | 0.002216 |
| aortic valve | Z52_S3 | 0.162416 | 0.050852 | 0.001406 | 2.852153 |
| atrial fibrill | Z52_S3 | 0.050917 | 0.020075 | 0.011209 | 1.950444 |
| hypertensi | Z52_S3 | 0.022341 | 0.01108 | 0.043781 | 1.358712 |
| myocardial | Z52_S3 | -0.03877 | 0.025613 | 0.130084 | 0.885776 |
| type 2 diab | Z52_S3 | 0.030135 | 0.021874 | 0.16833 | 0.773838 |
| heart failur | Z52_S3 | 0.039573 | 0.033377 | 0.235781 | 0.627492 |
| hypotensio | Z52_S3 | 0.032642 | 0.038115 | 0.391779 | 0.406959 |
| mitral prob | Z52_S3 | 0.026874 | 0.040098 | 0.50274 | 0.298656 |
| conduction | Z52_S3 | -0.02081 | 0.035118 | 0.553505 | 0.256879 |
| high choles | Z52_S3 | -0.00563 | 0.012186 | 0.644191 | 0.190986 |
| coronary h | Z52_S3 | -0.00423 | 0.019733 | 0.830353 | 0.080737 |
| other heart | Z52_S3 | 0.006285 | 0.048892 | 0.897716 | 0.046861 |
| angina pec | Z52_S3 | -0.00174 | 0.023792 | 0.941738 | 0.02607 |
| arrhythmia | Z52_S3 | 0.001159 | 0.043503 | 0.978748 | 0.009329 |
| atrial fibrill | Z60_S2 | -0.06422 | 0.020531 | 0.001764 | 2.753538 |
| aortic valve | Z60_S2 | -0.12075 | 0.051999 | 0.02024 | 1.693799 |
| hypertensi | Z60_S2 | -0.01615 | 0.011333 | 0.154049 | 0.812341 |
| heart failur | Z60_S2 | -0.04807 | 0.034135 | 0.159088 | 0.798362 |
| type 2 diab | Z60_S2 | -0.02801 | 0.022358 | 0.210215 | 0.677336 |
| mitral prob | Z60_S2 | -0.02647 | 0.04099 | 0.518476 | 0.285271 |
| other heart | Z60_S2 | -0.02722 | 0.049972 | 0.58598 | 0.232117 |
| coronary h | Z60_S2 | -0.0103 | 0.020169 | 0.609575 | 0.214973 |
| hypotensio | Z60_S2 | -0.01561 | 0.038964 | 0.688773 | 0.161924 |
| angina pec | Z60_S2 | -0.0094 | 0.024327 | 0.699199 | 0.155399 |
| conduction | Z60_S2 | 0.005256 | 0.035911 | 0.883646 | 0.053722 |
| myocardial | Z60_S2 | 0.003229 | 0.026178 | 0.901822 | 0.044879 |
| arrhythmia | Z60_S2 | 0.004883 | 0.044486 | 0.912597 | 0.039721 |
| high choles | Z60_S2 | -0.00045 | 0.012477 | 0.971076 | 0.012747 |
| atrial fibrill | Z37_S1 | -0.07905 | 0.025572 | 0.001995 | 2.699974 |
| type 2 diab | Z37_S1 | 0.085677 | 0.027829 | 0.002081 | 2.681676 |
| hypertensi | Z37_S1 | 0.03764 | 0.014092 | 0.007569 | 2.120975 |
| myocardial | Z37_S1 | -0.0764 | 0.032635 | 0.019234 | 1.715923 |
| aortic valve | Z37_S1 | 0.136983 | 0.064842 | 0.03465 | 1.460301 |
| mitral prob | Z37_S1 | -0.06949 | 0.051117 | 0.174025 | 0.759387 |
| heart failur | Z37_S1 | -0.05631 | 0.042555 | 0.185774 | 0.731015 |
| arrhythmia | Z37_S1 | -0.06759 | 0.055466 | 0.222986 | 0.651723 |
| other heart | Z37_S1 | -0.04144 | 0.06232 | 0.506105 | 0.295759 |
| high choles | Z37_S1 | -0.00781 | 0.015501 | 0.614221 | 0.211675 |
| coronary h | Z37_S1 | -0.01191 | 0.025113 | 0.635345 | 0.19699 |

| | | | | | |
|---|---|---|---|---|---|
| angina pec | Z37_S1 | -0.01293 | 0.030276 | 0.669351 | 0.174346 |
| conduction | Z37_S1 | 0.005948 | 0.044776 | 0.894315 | 0.04851 |
| hypotensio | Z37_S1 | -0.00429 | 0.048595 | 0.929683 | 0.031665 |
| aortic valve | Z114_S4 | 0.173273 | 0.056687 | 0.002241 | 2.649592 |
| myocardial | Z114_S4 | -0.08137 | 0.028539 | 0.00436 | 2.360519 |
| hypertensi | Z114_S4 | 0.028582 | 0.012361 | 0.020776 | 1.682431 |
| type 2 diab | Z114_S4 | 0.038495 | 0.024381 | 0.114368 | 0.941694 |
| angina pec | Z114_S4 | -0.03542 | 0.026507 | 0.181457 | 0.741226 |
| heart failur | Z114_S4 | -0.04964 | 0.037212 | 0.182269 | 0.739288 |
| atrial fibrill | Z114_S4 | -0.02824 | 0.022381 | 0.207083 | 0.683855 |
| coronary h | Z114_S4 | -0.02674 | 0.021982 | 0.223757 | 0.650224 |
| mitral prob | Z114_S4 | -0.05166 | 0.044694 | 0.247725 | 0.606029 |
| other heart | Z114_S4 | -0.05257 | 0.054484 | 0.334652 | 0.475407 |
| arrhythmia | Z114_S4 | -0.03874 | 0.048488 | 0.424331 | 0.372295 |
| conduction | Z114_S4 | -0.02329 | 0.039139 | 0.551862 | 0.258169 |
| high choles | Z114_S4 | 0.004147 | 0.013598 | 0.760404 | 0.118955 |
| hypotensio | Z114_S4 | 0.008911 | 0.042492 | 0.833893 | 0.07889 |
| type 2 diab | Z62_S3 | 0.078179 | 0.025778 | 0.002426 | 2.615194 |
| aortic valve | Z62_S3 | 0.122841 | 0.060042 | 0.040777 | 1.389585 |
| angina pec | Z62_S3 | 0.042315 | 0.028048 | 0.131401 | 0.8814 |
| mitral prob | Z62_S3 | -0.06128 | 0.047343 | 0.195551 | 0.708741 |
| hypotensio | Z62_S3 | 0.051637 | 0.044992 | 0.251105 | 0.600145 |
| coronary h | Z62_S3 | 0.019072 | 0.023267 | 0.412394 | 0.384687 |
| arrhythmia | Z62_S3 | 0.039421 | 0.051365 | 0.442812 | 0.353781 |
| conduction | Z62_S3 | -0.02096 | 0.04145 | 0.613031 | 0.212517 |
| atrial fibrill | Z62_S3 | -0.00894 | 0.023693 | 0.705779 | 0.151331 |
| hypertensi | Z62_S3 | -0.00442 | 0.013052 | 0.734663 | 0.133912 |
| myocardial | Z62_S3 | 0.008731 | 0.030229 | 0.772704 | 0.111987 |
| other heart | Z62_S3 | -0.01635 | 0.05772 | 0.776998 | 0.10958 |
| heart failur | Z62_S3 | 0.010763 | 0.039414 | 0.784793 | 0.105245 |
| high choles | Z62_S3 | -0.0015 | 0.014349 | 0.916821 | 0.037715 |
| myocardial | Z63_S1 | -0.10276 | 0.034274 | 0.00272 | 2.565465 |
| coronary h | Z63_S1 | -0.05704 | 0.026374 | 0.030563 | 1.514804 |
| conduction | Z63_S1 | 0.082577 | 0.047065 | 0.07935 | 1.100453 |
| angina pec | Z63_S1 | -0.03896 | 0.031825 | 0.220893 | 0.655818 |
| atrial fibrill | Z63_S1 | 0.027906 | 0.026919 | 0.299898 | 0.523026 |
| heart failur | Z63_S1 | 0.044185 | 0.04475 | 0.323474 | 0.490161 |
| type 2 diab | Z63_S1 | 0.027498 | 0.029327 | 0.348447 | 0.457863 |
| arrhythmia | Z63_S1 | -0.0534 | 0.058307 | 0.359742 | 0.444009 |
| hypotensio | Z63_S1 | -0.04514 | 0.051103 | 0.37706 | 0.42359 |
| mitral prob | Z63_S1 | 0.040951 | 0.053746 | 0.44611 | 0.350558 |
| aortic valve | Z63_S1 | -0.03776 | 0.068162 | 0.57963 | 0.236849 |
| high choles | Z63_S1 | -0.00556 | 0.016316 | 0.733478 | 0.134613 |
| hypertensi | Z63_S1 | 0.001945 | 0.014821 | 0.895569 | 0.047901 |
| other heart | Z63_S1 | 0.003646 | 0.065532 | 0.955636 | 0.019708 |

| | | | | | |
|---|---|---|---|---|---|
| atrial fibrill | Z56_S2 | -0.06082 | 0.020326 | 0.002773 | 2.55702 |
| aortic valve | Z56_S2 | -0.09061 | 0.051491 | 0.078459 | 1.105356 |
| type 2 diab | Z56_S2 | -0.03025 | 0.022141 | 0.17186 | 0.764826 |
| other heart | Z56_S2 | 0.059194 | 0.049474 | 0.23152 | 0.635411 |
| high choles | Z56_S2 | 0.01363 | 0.012355 | 0.269947 | 0.568722 |
| angina pec | Z56_S2 | -0.02454 | 0.024052 | 0.307544 | 0.512093 |
| arrhythmia | Z56_S2 | 0.044492 | 0.044017 | 0.312135 | 0.505658 |
| heart failur | Z56_S2 | -0.03338 | 0.033785 | 0.32317 | 0.490569 |
| coronary h | Z56_S2 | -0.01841 | 0.01996 | 0.356229 | 0.448271 |
| myocardial | Z56_S2 | 0.00917 | 0.025921 | 0.723522 | 0.140548 |
| conduction | Z56_S2 | -0.00883 | 0.035508 | 0.803636 | 0.09494 |
| hypertensi | Z56_S2 | 0.002224 | 0.011211 | 0.842747 | 0.074303 |
| mitral prob | Z56_S2 | -0.00485 | 0.040594 | 0.904879 | 0.043409 |
| hypotensio | Z56_S2 | 0.001134 | 0.038591 | 0.976565 | 0.010299 |
| atrial fibrill | Z91_S1 | -0.06633 | 0.022309 | 0.002949 | 2.530385 |
| heart failur | Z91_S1 | -0.0774 | 0.037027 | 0.036586 | 1.43669 |
| aortic valve | Z91_S1 | 0.098288 | 0.056361 | 0.081188 | 1.090509 |
| mitral prob | Z91_S1 | -0.07611 | 0.044433 | 0.086758 | 1.06169 |
| other heart | Z91_S1 | -0.08118 | 0.054164 | 0.133941 | 0.873086 |
| hypertensi | Z91_S1 | 0.015914 | 0.012296 | 0.195592 | 0.708649 |
| coronary h | Z91_S1 | -0.02442 | 0.021851 | 0.263812 | 0.578706 |
| myocardial | Z91_S1 | -0.03131 | 0.028366 | 0.269748 | 0.569041 |
| angina pec | Z91_S1 | -0.02815 | 0.026338 | 0.285168 | 0.544899 |
| high choles | Z91_S1 | 0.010869 | 0.013511 | 0.421154 | 0.375559 |
| arrhythmia | Z91_S1 | -0.03873 | 0.048217 | 0.421801 | 0.374892 |
| hypotensio | Z91_S1 | -0.03068 | 0.042254 | 0.467799 | 0.32994 |
| conduction | Z91_S1 | -0.01301 | 0.038918 | 0.738107 | 0.131881 |
| type 2 diab | Z91_S1 | 0.002127 | 0.024263 | 0.930128 | 0.031457 |
| aortic valve | Z58_S2 | 0.164007 | 0.05517 | 0.002955 | 2.529508 |
| atrial fibrill | Z58_S2 | 0.056858 | 0.021777 | 0.009035 | 2.044066 |
| hypertensi | Z58_S2 | 0.025786 | 0.01202 | 0.031951 | 1.495515 |
| myocardial | Z58_S2 | -0.04382 | 0.027781 | 0.114716 | 0.940374 |
| type 2 diab | Z58_S2 | 0.036557 | 0.023734 | 0.123507 | 0.908308 |
| angina pec | Z58_S2 | -0.02895 | 0.025803 | 0.261832 | 0.581977 |
| conduction | Z58_S2 | -0.03762 | 0.038112 | 0.32359 | 0.490004 |
| coronary h | Z58_S2 | -0.01973 | 0.021401 | 0.356523 | 0.447912 |
| mitral prob | Z58_S2 | 0.033724 | 0.043495 | 0.438143 | 0.358384 |
| hypotensio | Z58_S2 | 0.030087 | 0.041347 | 0.466831 | 0.33084 |
| arrhythmia | Z58_S2 | -0.01718 | 0.047208 | 0.715937 | 0.145125 |
| other heart | Z58_S2 | 0.018638 | 0.053034 | 0.725264 | 0.139504 |
| high choles | Z58_S2 | -0.0038 | 0.013229 | 0.774204 | 0.111145 |
| heart failur | Z58_S2 | 0.008625 | 0.036231 | 0.811846 | 0.090527 |
| coronary h | Z105_S4 | -0.06136 | 0.020704 | 0.003045 | 2.516449 |
| aortic valve | Z105_S4 | -0.15607 | 0.053418 | 0.003484 | 2.457874 |
| myocardial | Z105_S4 | -0.06943 | 0.026887 | 0.009817 | 2.008015 |

| | | | | | |
|---|---|---|---|---|---|
| angina pec | Z105_S4 | -0.05215 | 0.024996 | 0.036942 | 1.432478 |
| atrial fibrill | Z105_S4 | -0.04221 | 0.02113 | 0.045776 | 1.339358 |
| mitral prob | Z105_S4 | -0.06479 | 0.042107 | 0.123886 | 0.906979 |
| type 2 diab | Z105_S4 | 0.034147 | 0.023008 | 0.137784 | 0.8608 |
| hypertensi | Z105_S4 | -0.01689 | 0.011658 | 0.147376 | 0.831572 |
| heart failur | Z105_S4 | -0.04689 | 0.03508 | 0.181355 | 0.74147 |
| arrhythmia | Z105_S4 | -0.05635 | 0.045701 | 0.217602 | 0.662338 |
| other heart | Z105_S4 | -0.02913 | 0.05135 | 0.570493 | 0.24375 |
| hypotensio | Z105_S4 | -0.01879 | 0.040045 | 0.638867 | 0.19459 |
| conduction | Z105_S4 | 0.012383 | 0.036896 | 0.737155 | 0.132441 |
| high choles | Z105_S4 | 0.001398 | 0.012799 | 0.91303 | 0.039515 |
| type 2 diab | Z44_S1 | 0.074861 | 0.025335 | 0.003132 | 2.504245 |
| myocardial | Z44_S1 | -0.08067 | 0.029688 | 0.006586 | 2.181361 |
| hypertensi | Z44_S1 | 0.03033 | 0.012847 | 0.018239 | 1.73901 |
| atrial fibrill | Z44_S1 | -0.04063 | 0.023269 | 0.080819 | 1.092485 |
| coronary h | Z44_S1 | -0.03023 | 0.02287 | 0.186224 | 0.729965 |
| aortic valve | Z44_S1 | 0.065323 | 0.05901 | 0.268315 | 0.571355 |
| mitral prob | Z44_S1 | -0.04561 | 0.046504 | 0.32669 | 0.485864 |
| conduction | Z44_S1 | -0.02946 | 0.040771 | 0.470018 | 0.327885 |
| arrhythmia | Z44_S1 | -0.03616 | 0.050476 | 0.473725 | 0.324474 |
| high choles | Z44_S1 | 0.009288 | 0.014138 | 0.511225 | 0.291388 |
| hypotensio | Z44_S1 | -0.01964 | 0.044192 | 0.656758 | 0.182595 |
| angina pec | Z44_S1 | -0.01197 | 0.027566 | 0.664156 | 0.17773 |
| other heart | Z44_S1 | 0.02104 | 0.0567 | 0.710583 | 0.148385 |
| heart failur | Z44_S1 | -0.00125 | 0.03872 | 0.974345 | 0.011287 |
| type 2 diab | Z96_S3 | 0.071036 | 0.024269 | 0.003425 | 2.465316 |
| aortic valve | Z96_S3 | 0.153192 | 0.056519 | 0.006724 | 2.172354 |
| angina pec | Z96_S3 | 0.033837 | 0.026416 | 0.200231 | 0.698468 |
| hypotensio | Z96_S3 | 0.040366 | 0.042357 | 0.340605 | 0.467749 |
| conduction | Z96_S3 | -0.03668 | 0.039031 | 0.347409 | 0.459158 |
| coronary h | Z96_S3 | 0.019414 | 0.021916 | 0.375712 | 0.425145 |
| hypertensi | Z96_S3 | 0.010103 | 0.012292 | 0.411149 | 0.386001 |
| mitral prob | Z96_S3 | -0.03584 | 0.044572 | 0.421331 | 0.375377 |
| arrhythmia | Z96_S3 | 0.02305 | 0.048354 | 0.633591 | 0.198191 |
| atrial fibrill | Z96_S3 | -0.00798 | 0.022315 | 0.720498 | 0.142367 |
| other heart | Z96_S3 | -0.01333 | 0.054337 | 0.806245 | 0.093533 |
| myocardial | Z96_S3 | -0.0059 | 0.028464 | 0.835867 | 0.077863 |
| heart failur | Z96_S3 | 0.006353 | 0.037101 | 0.864034 | 0.063469 |
| high choles | Z96_S3 | 0.001222 | 0.013514 | 0.927936 | 0.032482 |
| aortic valve | Z31_S2 | 0.17094 | 0.058773 | 0.003636 | 2.439411 |
| atrial fibrill | Z31_S2 | 0.063347 | 0.023187 | 0.006301 | 2.200603 |
| hypertensi | Z31_S2 | 0.029761 | 0.012802 | 0.020101 | 1.696782 |
| type 2 diab | Z31_S2 | 0.049221 | 0.025286 | 0.051595 | 1.287391 |
| angina pec | Z31_S2 | -0.04767 | 0.027482 | 0.082811 | 1.081909 |
| myocardial | Z31_S2 | -0.04858 | 0.02959 | 0.100613 | 0.997345 |

| | | | | | |
|---|---|---|---|---|---|
| coronary h | Z31_S2 | -0.02779 | 0.022792 | 0.22276 | 0.652162 |
| conduction | Z31_S2 | -0.04728 | 0.040595 | 0.244129 | 0.61238 |
| mitral prob | Z31_S2 | 0.047684 | 0.046336 | 0.303445 | 0.51792 |
| hypotensio | Z31_S2 | 0.034303 | 0.044045 | 0.436089 | 0.360425 |
| other heart | Z31_S2 | 0.035299 | 0.056492 | 0.532079 | 0.274024 |
| heart failur | Z31_S2 | 0.021107 | 0.038592 | 0.584437 | 0.233262 |
| arrhythmia | Z31_S2 | -0.01844 | 0.050294 | 0.713895 | 0.146366 |
| high choles | Z31_S2 | -0.00262 | 0.014085 | 0.852466 | 0.069323 |
| type 2 diab | Z76_S3 | -0.0753 | 0.026103 | 0.003921 | 2.406623 |
| aortic valve | Z76_S3 | -0.11328 | 0.060774 | 0.062348 | 1.205175 |
| mitral prob | Z76_S3 | 0.06074 | 0.047914 | 0.204928 | 0.688399 |
| coronary h | Z76_S3 | -0.02551 | 0.023538 | 0.27845 | 0.555253 |
| angina pec | Z76_S3 | -0.03053 | 0.028391 | 0.282177 | 0.549479 |
| hypotensio | Z76_S3 | -0.041 | 0.045557 | 0.368156 | 0.433969 |
| atrial fibrill | Z76_S3 | 0.017553 | 0.024005 | 0.464667 | 0.332858 |
| conduction | Z76_S3 | 0.027366 | 0.041955 | 0.514239 | 0.288835 |
| myocardial | Z76_S3 | -0.01994 | 0.030581 | 0.514462 | 0.288647 |
| hypertensi | Z76_S3 | -0.00721 | 0.013214 | 0.58553 | 0.232451 |
| arrhythmia | Z76_S3 | -0.02313 | 0.051987 | 0.656431 | 0.182811 |
| high choles | Z76_S3 | -0.00331 | 0.014533 | 0.819823 | 0.08628 |
| heart failur | Z76_S3 | -0.00286 | 0.039892 | 0.942917 | 0.025527 |
| other heart | Z76_S3 | 0.002384 | 0.058411 | 0.96744 | 0.014376 |
| coronary h | Z123_S4 | 0.066919 | 0.023728 | 0.004803 | 2.318475 |
| angina pec | Z123_S4 | 0.07219 | 0.028621 | 0.011669 | 1.932984 |
| type 2 diab | Z123_S4 | -0.0612 | 0.026359 | 0.020264 | 1.693267 |
| myocardial | Z123_S4 | 0.056759 | 0.03083 | 0.065634 | 1.182869 |
| aortic valve | Z123_S4 | 0.103204 | 0.06124 | 0.091959 | 1.036404 |
| hypertensi | Z123_S4 | 0.015903 | 0.013334 | 0.233 | 0.632645 |
| high choles | Z123_S4 | -0.01001 | 0.014657 | 0.494793 | 0.305576 |
| mitral prob | Z123_S4 | 0.029326 | 0.048295 | 0.54371 | 0.264633 |
| arrhythmia | Z123_S4 | 0.027411 | 0.052395 | 0.600872 | 0.221218 |
| heart failur | Z123_S4 | -0.0191 | 0.040225 | 0.634946 | 0.197263 |
| hypotensio | Z123_S4 | -0.016 | 0.045913 | 0.727446 | 0.138199 |
| atrial fibrill | Z123_S4 | 0.00585 | 0.02421 | 0.809076 | 0.092011 |
| conduction | Z123_S4 | -0.00352 | 0.042303 | 0.933723 | 0.029782 |
| other heart | Z123_S4 | -0.0011 | 0.05888 | 0.985064 | 0.006536 |
| aortic valve | Z69_S5 | -0.14066 | 0.050356 | 0.005221 | 2.282264 |
| atrial fibrill | Z69_S5 | -0.04065 | 0.019919 | 0.041258 | 1.38449 |
| hypertensi | Z69_S5 | -0.01739 | 0.010992 | 0.113711 | 0.944197 |
| myocardial | Z69_S5 | 0.023395 | 0.025367 | 0.3564 | 0.448063 |
| conduction | Z69_S5 | 0.030318 | 0.034792 | 0.383546 | 0.416183 |
| hypotensio | Z69_S5 | -0.01602 | 0.037771 | 0.671512 | 0.172946 |
| heart failur | Z69_S5 | 0.012632 | 0.033085 | 0.702602 | 0.153291 |
| high choles | Z69_S5 | -0.00451 | 0.012076 | 0.708629 | 0.149581 |
| type 2 diab | Z69_S5 | 0.006722 | 0.021698 | 0.756726 | 0.121062 |

| | | | | | |
|---|---|---|---|---|---|
| mitral prob | Z69_S5 | -0.01107 | 0.039708 | 0.780372 | 0.107699 |
| arrhythmia | Z69_S5 | 0.008575 | 0.043084 | 0.842239 | 0.074565 |
| angina pec | Z69_S5 | 0.004615 | 0.023569 | 0.844775 | 0.073259 |
| other heart | Z69_S5 | 0.009018 | 0.048413 | 0.852242 | 0.069437 |
| coronary h | Z69_S5 | -0.00339 | 0.019542 | 0.862084 | 0.064451 |
| aortic valve | Z122_S5 | 0.143139 | 0.053378 | 0.007332 | 2.134775 |
| myocardial | Z122_S5 | -0.07136 | 0.026874 | 0.007924 | 2.101044 |
| type 2 diab | Z122_S5 | 0.059754 | 0.022954 | 0.009241 | 2.034286 |
| hypertensi | Z122_S5 | 0.022127 | 0.01163 | 0.057098 | 1.243378 |
| angina pec | Z122_S5 | -0.03445 | 0.024957 | 0.167445 | 0.776128 |
| coronary h | Z122_S5 | -0.02638 | 0.020702 | 0.202503 | 0.693568 |
| conduction | Z122_S5 | -0.04212 | 0.036864 | 0.253212 | 0.596517 |
| arrhythmia | Z122_S5 | -0.04581 | 0.045651 | 0.315618 | 0.500838 |
| heart failur | Z122_S5 | -0.03227 | 0.035044 | 0.357217 | 0.447068 |
| mitral prob | Z122_S5 | -0.03843 | 0.042077 | 0.361065 | 0.442414 |
| high choles | Z122_S5 | 0.010615 | 0.012793 | 0.406667 | 0.390762 |
| hypotensio | Z122_S5 | 0.011361 | 0.040008 | 0.776431 | 0.109897 |
| atrial fibrill | Z122_S5 | -0.00571 | 0.021071 | 0.786408 | 0.104352 |
| other heart | Z122_S5 | -0.00796 | 0.051292 | 0.876607 | 0.057195 |
| type 2 diab | Z87_S5 | -0.06689 | 0.024955 | 0.007364 | 2.132914 |
| aortic valve | Z87_S5 | -0.09477 | 0.058128 | 0.103036 | 0.98701 |
| hypertensi | Z87_S5 | -0.017 | 0.012655 | 0.17908 | 0.746954 |
| conduction | Z87_S5 | 0.036206 | 0.040137 | 0.367035 | 0.435293 |
| other heart | Z87_S5 | -0.04542 | 0.055846 | 0.416071 | 0.380832 |
| heart failur | Z87_S5 | -0.02622 | 0.038136 | 0.491811 | 0.308202 |
| coronary h | Z87_S5 | -0.01514 | 0.022523 | 0.501597 | 0.299645 |
| angina pec | Z87_S5 | -0.0175 | 0.027165 | 0.519459 | 0.284449 |
| myocardial | Z87_S5 | 0.017525 | 0.029249 | 0.549076 | 0.260368 |
| hypotensio | Z87_S5 | -0.0214 | 0.043545 | 0.62307 | 0.205463 |
| atrial fibrill | Z87_S5 | -0.01041 | 0.022929 | 0.649844 | 0.187191 |
| arrhythmia | Z87_S5 | -0.02217 | 0.049717 | 0.655619 | 0.183348 |
| high choles | Z87_S5 | -0.0049 | 0.013921 | 0.72497 | 0.13968 |
| mitral prob | Z87_S5 | 0.014988 | 0.045821 | 0.743594 | 0.128664 |
| coronary h | Z5_S1 | -0.05973 | 0.022354 | 0.007543 | 2.122459 |
| angina pec | Z5_S1 | -0.05822 | 0.026973 | 0.030898 | 1.510065 |
| aortic valve | Z5_S1 | -0.12194 | 0.05771 | 0.034618 | 1.460694 |
| myocardial | Z5_S1 | -0.06121 | 0.029035 | 0.03503 | 1.455563 |
| heart failur | Z5_S1 | -0.0378 | 0.03788 | 0.318336 | 0.497114 |
| hypertensi | Z5_S1 | -0.01227 | 0.012575 | 0.329001 | 0.482803 |
| arrhythmia | Z5_S1 | -0.03841 | 0.049366 | 0.436596 | 0.35992 |
| hypotensio | Z5_S1 | -0.0316 | 0.043255 | 0.465117 | 0.332438 |
| type 2 diab | Z5_S1 | -0.0153 | 0.024815 | 0.537515 | 0.26961 |
| atrial fibrill | Z5_S1 | -0.00721 | 0.022817 | 0.752009 | 0.123777 |
| mitral prob | Z5_S1 | -0.01285 | 0.045493 | 0.777643 | 0.10922 |
| other heart | Z5_S1 | -0.01526 | 0.05546 | 0.78319 | 0.106133 |

| | | | | | |
|---|---|---|---|---|---|
| conduction | Z5_S1 | -0.01038 | 0.039846 | 0.794538 | 0.099885 |
| high choles | Z5_S1 | -0.00149 | 0.013815 | 0.914024 | 0.039042 |
| aortic valve | Z17_S1 | 0.145987 | 0.054819 | 0.007748 | 2.110785 |
| hypertensi | Z17_S1 | 0.03093 | 0.01194 | 0.009595 | 2.017938 |
| atrial fibrill | Z17_S1 | 0.049288 | 0.021639 | 0.022753 | 1.642961 |
| type 2 diab | Z17_S1 | 0.048082 | 0.02356 | 0.041281 | 1.384254 |
| myocardial | Z17_S1 | -0.04094 | 0.027579 | 0.137663 | 0.861183 |
| heart failur | Z17_S1 | 0.047295 | 0.035962 | 0.188478 | 0.724739 |
| angina pec | Z17_S1 | -0.02422 | 0.025624 | 0.344477 | 0.46284 |
| coronary h | Z17_S1 | -0.01678 | 0.021244 | 0.429737 | 0.366797 |
| hypotensio | Z17_S1 | 0.028023 | 0.041067 | 0.495013 | 0.305383 |
| mitral prob | Z17_S1 | 0.027546 | 0.043209 | 0.523799 | 0.280835 |
| other heart | Z17_S1 | 0.027072 | 0.052666 | 0.607244 | 0.216637 |
| conduction | Z17_S1 | -0.00919 | 0.037844 | 0.808109 | 0.09253 |
| high choles | Z17_S1 | 0.001586 | 0.013121 | 0.903791 | 0.043932 |
| arrhythmia | Z17_S1 | -0.00406 | 0.046887 | 0.930976 | 0.031062 |
| aortic valve | Z62_S1 | 0.146134 | 0.054891 | 0.007767 | 2.109728 |
| atrial fibrill | Z62_S1 | 0.047913 | 0.021699 | 0.02725 | 1.564636 |
| hypertensi | Z62_S1 | 0.020146 | 0.011965 | 0.092236 | 1.035098 |
| coronary h | Z62_S1 | 0.029884 | 0.021275 | 0.160136 | 0.795511 |
| heart failur | Z62_S1 | 0.040481 | 0.036029 | 0.261204 | 0.58302 |
| mitral prob | Z62_S1 | 0.037914 | 0.043274 | 0.380971 | 0.419109 |
| hypotensio | Z62_S1 | 0.035703 | 0.041137 | 0.385454 | 0.414028 |
| angina pec | Z62_S1 | 0.021035 | 0.025666 | 0.412478 | 0.384599 |
| myocardial | Z62_S1 | 0.020607 | 0.027627 | 0.455743 | 0.34128 |
| arrhythmia | Z62_S1 | 0.024499 | 0.046953 | 0.601832 | 0.220525 |
| type 2 diab | Z62_S1 | 0.010487 | 0.02361 | 0.656913 | 0.182492 |
| other heart | Z62_S1 | 0.022991 | 0.05275 | 0.662949 | 0.17852 |
| conduction | Z62_S1 | 0.006496 | 0.037901 | 0.863921 | 0.063526 |
| high choles | Z62_S1 | -0.00142 | 0.013141 | 0.913896 | 0.039103 |
| atrial fibrill | Z19_S2 | 0.060132 | 0.022625 | 0.007871 | 2.103969 |
| aortic valve | Z19_S2 | 0.112131 | 0.057247 | 0.050157 | 1.299667 |
| myocardial | Z19_S2 | 0.053265 | 0.028805 | 0.064447 | 1.190794 |
| coronary h | Z19_S2 | 0.038152 | 0.022181 | 0.085442 | 1.068328 |
| angina pec | Z19_S2 | 0.041346 | 0.026758 | 0.122322 | 0.912494 |
| type 2 diab | Z19_S2 | 0.026573 | 0.024619 | 0.280442 | 0.552157 |
| heart failur | Z19_S2 | 0.032099 | 0.037598 | 0.393253 | 0.405328 |
| hypertensi | Z19_S2 | 0.008136 | 0.012465 | 0.513945 | 0.289084 |
| hypotensio | Z19_S2 | 0.024032 | 0.042926 | 0.575585 | 0.239891 |
| arrhythmia | Z19_S2 | 0.010078 | 0.048975 | 0.836963 | 0.077294 |
| mitral prob | Z19_S2 | -0.00841 | 0.045124 | 0.852225 | 0.069446 |
| conduction | Z19_S2 | 0.006799 | 0.039524 | 0.863412 | 0.063782 |
| high choles | Z19_S2 | -0.00117 | 0.013708 | 0.931966 | 0.0306 |
| other heart | Z19_S2 | 0.001753 | 0.055023 | 0.974585 | 0.01118 |
| aortic valve | Z7_S1 | -0.14277 | 0.05393 | 0.008117 | 2.09059 |

| | | | | | |
|---|---|---|---|---|---|
| coronary h | Z7_S1 | -0.02765 | 0.020914 | 0.186109 | 0.730232 |
| hypertensi | Z7_S1 | -0.01524 | 0.011781 | 0.195693 | 0.708425 |
| atrial fibrill | Z7_S1 | -0.02659 | 0.021344 | 0.212813 | 0.672002 |
| type 2 diab | Z7_S1 | 0.027366 | 0.023238 | 0.238964 | 0.621668 |
| myocardial | Z7_S1 | -0.02103 | 0.027162 | 0.438883 | 0.357652 |
| angina pec | Z7_S1 | -0.01523 | 0.025234 | 0.546156 | 0.262684 |
| mitral prob | Z7_S1 | -0.02357 | 0.042527 | 0.579407 | 0.237016 |
| hypotensio | Z7_S1 | -0.02112 | 0.04044 | 0.601528 | 0.220744 |
| conduction | Z7_S1 | 0.013544 | 0.037255 | 0.716191 | 0.144971 |
| arrhythmia | Z7_S1 | -0.01169 | 0.046143 | 0.800008 | 0.096906 |
| other heart | Z7_S1 | 0.013068 | 0.051849 | 0.801009 | 0.096363 |
| heart failur | Z7_S1 | 0.007731 | 0.035421 | 0.827216 | 0.082381 |
| high choles | Z7_S1 | -0.00065 | 0.012926 | 0.95999 | 0.017733 |
| aortic valve | Z55_S1 | -0.14405 | 0.054507 | 0.008229 | 2.084668 |
| type 2 diab | Z55_S1 | -0.04214 | 0.023447 | 0.072295 | 1.140894 |
| hypertensi | Z55_S1 | -0.0199 | 0.011883 | 0.093996 | 1.02689 |
| atrial fibrill | Z55_S1 | -0.03563 | 0.021533 | 0.098041 | 1.008591 |
| hypotensio | Z55_S1 | -0.0455 | 0.040835 | 0.265149 | 0.576509 |
| conduction | Z55_S1 | 0.033291 | 0.037628 | 0.376309 | 0.424456 |
| heart failur | Z55_S1 | -0.03038 | 0.03577 | 0.395707 | 0.402626 |
| angina pec | Z55_S1 | 0.010218 | 0.025488 | 0.688499 | 0.162096 |
| high choles | Z55_S1 | -0.00451 | 0.013054 | 0.729877 | 0.13675 |
| other heart | Z55_S1 | -0.01366 | 0.052383 | 0.794218 | 0.10006 |
| mitral prob | Z55_S1 | -0.01001 | 0.042977 | 0.815911 | 0.088357 |
| coronary h | Z55_S1 | -0.00239 | 0.021126 | 0.910047 | 0.040936 |
| myocardial | Z55_S1 | 0.002593 | 0.02744 | 0.924714 | 0.033993 |
| arrhythmia | Z55_S1 | -0.00355 | 0.046621 | 0.939349 | 0.027173 |
| myocardial | Z121_S4 | -0.08602 | 0.032632 | 0.008395 | 2.075992 |
| hypertensi | Z121_S4 | 0.029066 | 0.014104 | 0.039339 | 1.405182 |
| coronary h | Z121_S4 | -0.04712 | 0.025123 | 0.060705 | 1.216776 |
| aortic valve | Z121_S4 | 0.109187 | 0.064856 | 0.09229 | 1.034843 |
| angina pec | Z121_S4 | -0.03536 | 0.030301 | 0.243172 | 0.614087 |
| high choles | Z121_S4 | 0.016914 | 0.015516 | 0.275671 | 0.559609 |
| heart failur | Z121_S4 | -0.04525 | 0.04257 | 0.287845 | 0.540841 |
| mitral prob | Z121_S4 | -0.04417 | 0.051115 | 0.387535 | 0.411689 |
| atrial fibrill | Z121_S4 | -0.02132 | 0.025594 | 0.404952 | 0.392596 |
| other heart | Z121_S4 | -0.02842 | 0.062311 | 0.648326 | 0.188207 |
| arrhythmia | Z121_S4 | -0.01922 | 0.055485 | 0.729078 | 0.137226 |
| conduction | Z121_S4 | 0.013722 | 0.044787 | 0.759321 | 0.119575 |
| type 2 diab | Z121_S4 | 0.006406 | 0.027878 | 0.818266 | 0.087105 |
| hypotensio | Z121_S4 | -0.00902 | 0.048614 | 0.852866 | 0.069119 |
| aortic valve | Z80_S2 | 0.140222 | 0.053339 | 0.008572 | 2.066893 |
| myocardial | Z80_S2 | -0.05819 | 0.02686 | 0.03028 | 1.518848 |
| type 2 diab | Z80_S2 | 0.049476 | 0.022941 | 0.031045 | 1.508007 |
| atrial fibrill | Z80_S2 | 0.044917 | 0.021039 | 0.032774 | 1.484468 |

| | | | | | |
|---|---|---|---|---|---|
| hypertensi | Z80_S2 | 0.021914 | 0.011627 | 0.059478 | 1.225646 |
| angina pec | Z80_S2 | -0.03356 | 0.024951 | 0.178603 | 0.748112 |
| coronary h | Z80_S2 | -0.02695 | 0.020697 | 0.192862 | 0.714753 |
| conduction | Z80_S2 | -0.04354 | 0.03684 | 0.237241 | 0.624809 |
| arrhythmia | Z80_S2 | -0.02954 | 0.045623 | 0.517303 | 0.286255 |
| mitral prob | Z80_S2 | 0.026798 | 0.042041 | 0.52386 | 0.280785 |
| hypotensio | Z80_S2 | 0.024845 | 0.03996 | 0.534123 | 0.272359 |
| heart failur | Z80_S2 | 0.019261 | 0.035014 | 0.582273 | 0.234873 |
| high choles | Z80_S2 | 0.002359 | 0.0128 | 0.853788 | 0.06865 |
| other heart | Z80_S2 | 0.001525 | 0.051265 | 0.976265 | 0.010432 |
| aortic valve | Z50_S3 | -0.14387 | 0.054814 | 0.008676 | 2.061666 |
| hypertensi | Z50_S3 | -0.01652 | 0.011942 | 0.166466 | 0.778675 |
| type 2 diab | Z50_S3 | -0.02979 | 0.023577 | 0.20636 | 0.685374 |
| atrial fibrill | Z50_S3 | -0.02021 | 0.02167 | 0.35108 | 0.454594 |
| hypotensio | Z50_S3 | -0.02897 | 0.041092 | 0.480886 | 0.317958 |
| conduction | Z50_S3 | 0.024898 | 0.037848 | 0.510647 | 0.291879 |
| heart failur | Z50_S3 | -0.0169 | 0.035988 | 0.638631 | 0.19475 |
| coronary h | Z50_S3 | -0.00725 | 0.021258 | 0.733063 | 0.134859 |
| high choles | Z50_S3 | 0.004389 | 0.013127 | 0.738126 | 0.13187 |
| angina pec | Z50_S3 | -0.00757 | 0.025638 | 0.767833 | 0.114733 |
| arrhythmia | Z50_S3 | -0.01325 | 0.046892 | 0.77748 | 0.109311 |
| myocardial | Z50_S3 | 0.007713 | 0.027602 | 0.779925 | 0.107947 |
| mitral prob | Z50_S3 | 0.005388 | 0.043227 | 0.900811 | 0.045366 |
| other heart | Z50_S3 | -0.00523 | 0.052695 | 0.920883 | 0.035796 |
| aortic valve | Z74_S2 | 0.148285 | 0.057089 | 0.009398 | 2.026964 |
| atrial fibrill | Z74_S2 | 0.048832 | 0.022553 | 0.03038 | 1.517413 |
| type 2 diab | Z74_S2 | 0.046585 | 0.024545 | 0.057718 | 1.238687 |
| hypertensi | Z74_S2 | 0.021297 | 0.012426 | 0.086548 | 1.062745 |
| hypotensio | Z74_S2 | 0.035894 | 0.042788 | 0.401549 | 0.396262 |
| angina pec | Z74_S2 | -0.01668 | 0.026693 | 0.532137 | 0.273976 |
| conduction | Z74_S2 | -0.0216 | 0.039421 | 0.583815 | 0.233725 |
| heart failur | Z74_S2 | 0.01632 | 0.037486 | 0.66331 | 0.178283 |
| coronary h | Z74_S2 | -0.00872 | 0.022129 | 0.693658 | 0.158855 |
| myocardial | Z74_S2 | -0.01092 | 0.028732 | 0.70386 | 0.152514 |
| high choles | Z74_S2 | 0.003827 | 0.013666 | 0.779445 | 0.108214 |
| arrhythmia | Z74_S2 | -0.0117 | 0.048846 | 0.81071 | 0.091135 |
| other heart | Z74_S2 | 0.01105 | 0.054869 | 0.840391 | 0.075519 |
| mitral prob | Z74_S2 | 0.004703 | 0.045003 | 0.916776 | 0.037737 |
| atrial fibrill | Z65_S2 | 0.059358 | 0.023199 | 0.010514 | 1.978213 |
| aortic valve | Z65_S2 | 0.113526 | 0.058722 | 0.053215 | 1.273967 |
| type 2 diab | Z65_S2 | -0.02909 | 0.025263 | 0.249531 | 0.602875 |
| conduction | Z65_S2 | 0.041881 | 0.040531 | 0.301468 | 0.520759 |
| other heart | Z65_S2 | -0.05452 | 0.056451 | 0.334134 | 0.47608 |
| myocardial | Z65_S2 | 0.025773 | 0.029546 | 0.383056 | 0.416737 |
| angina pec | Z65_S2 | -0.02108 | 0.02741 | 0.441936 | 0.35464 |

| | | | | | |
|---|---|---|---|---|---|
| hypertensi | Z65_S2 | 0.008755 | 0.012783 | 0.493428 | 0.306776 |
| hypotensio | Z65_S2 | 0.017439 | 0.044045 | 0.692159 | 0.159794 |
| high choles | Z65_S2 | -0.00529 | 0.01407 | 0.706998 | 0.150582 |
| heart failur | Z65_S2 | -0.01205 | 0.038569 | 0.754742 | 0.122201 |
| coronary h | Z65_S2 | -0.00682 | 0.022737 | 0.764121 | 0.116838 |
| arrhythmia | Z65_S2 | -0.01328 | 0.050243 | 0.791485 | 0.101557 |
| mitral prob | Z65_S2 | -0.00993 | 0.046293 | 0.830109 | 0.080865 |
| type 2 diab | Z115_S3 | -0.06378 | 0.026676 | 0.016813 | 1.774348 |
| heart failur | Z115_S3 | -0.07471 | 0.040723 | 0.06659 | 1.17659 |
| myocardial | Z115_S3 | -0.05619 | 0.031222 | 0.071924 | 1.143127 |
| coronary h | Z115_S3 | -0.03954 | 0.024032 | 0.099938 | 1.000269 |
| other heart | Z115_S3 | -0.09305 | 0.059655 | 0.118803 | 0.925172 |
| aortic valve | Z115_S3 | -0.08105 | 0.062083 | 0.19172 | 0.717333 |
| angina pec | Z115_S3 | -0.03607 | 0.029005 | 0.213678 | 0.67024 |
| atrial fibrill | Z115_S3 | -0.03021 | 0.024493 | 0.217425 | 0.662692 |
| hypotensio | Z115_S3 | -0.05102 | 0.046522 | 0.272785 | 0.56418 |
| arrhythmia | Z115_S3 | -0.05561 | 0.053111 | 0.29505 | 0.530104 |
| hypertensi | Z115_S3 | -0.00832 | 0.013515 | 0.538109 | 0.26913 |
| conduction | Z115_S3 | 0.017715 | 0.042855 | 0.679343 | 0.167911 |
| mitral prob | Z115_S3 | -0.00866 | 0.048944 | 0.859527 | 0.06574 |
| high choles | Z115_S3 | 0.000446 | 0.014866 | 0.976073 | 0.010518 |
| type 2 diab | Z92_S1 | -0.05839 | 0.024893 | 0.019006 | 1.721116 |
| aortic valve | Z92_S1 | -0.11325 | 0.057929 | 0.050591 | 1.29593 |
| hypertensi | Z92_S1 | -0.02348 | 0.012617 | 0.062805 | 1.202008 |
| atrial fibrill | Z92_S1 | 0.025285 | 0.022897 | 0.269488 | 0.56946 |
| myocardial | Z92_S1 | 0.028542 | 0.029152 | 0.327563 | 0.484706 |
| mitral prob | Z92_S1 | 0.035779 | 0.045662 | 0.433308 | 0.363203 |
| other heart | Z92_S1 | 0.027242 | 0.055669 | 0.624592 | 0.204403 |
| arrhythmia | Z92_S1 | 0.023086 | 0.049548 | 0.641266 | 0.192962 |
| conduction | Z92_S1 | 0.016962 | 0.039995 | 0.671492 | 0.172959 |
| hypotensio | Z92_S1 | -0.01702 | 0.043408 | 0.694954 | 0.158044 |
| coronary h | Z92_S1 | 0.007836 | 0.022448 | 0.727026 | 0.13845 |
| heart failur | Z92_S1 | 0.012882 | 0.038023 | 0.734765 | 0.133852 |
| high choles | Z92_S1 | -0.00431 | 0.013862 | 0.755836 | 0.121573 |
| angina pec | Z92_S1 | 0.00511 | 0.027074 | 0.850286 | 0.070435 |
| type 2 diab | Z81_S1 | -0.05647 | 0.025497 | 0.026781 | 1.572171 |
| aortic valve | Z81_S1 | -0.11427 | 0.059315 | 0.054046 | 1.267236 |
| hypertensi | Z81_S1 | -0.01687 | 0.012925 | 0.191814 | 0.717121 |
| hypotensio | Z81_S1 | -0.05559 | 0.044442 | 0.210999 | 0.67572 |
| conduction | Z81_S1 | 0.050055 | 0.040949 | 0.221576 | 0.654477 |
| mitral prob | Z81_S1 | 0.031063 | 0.046769 | 0.506582 | 0.29535 |
| coronary h | Z81_S1 | -0.01382 | 0.022977 | 0.547586 | 0.261548 |
| myocardial | Z81_S1 | -0.01281 | 0.029851 | 0.667856 | 0.175317 |
| high choles | Z81_S1 | -0.00442 | 0.0142 | 0.755592 | 0.121713 |
| heart failur | Z81_S1 | -0.01151 | 0.038928 | 0.767411 | 0.114972 |

| | | | | | |
|---|---|---|---|---|---|
| arrhythmia | Z81_S1 | -0.01154 | 0.050734 | 0.820059 | 0.086155 |
| angina pec | Z81_S1 | -0.00488 | 0.027722 | 0.860207 | 0.065397 |
| other heart | Z81_S1 | -0.00557 | 0.057008 | 0.922111 | 0.035217 |
| atrial fibrill | Z81_S1 | -4.93E-05 | 0.023435 | 0.998321 | 0.00073 |
| aortic valve | Z70_S5 | 0.121829 | 0.056247 | 0.030324 | 1.518218 |
| hypertensi | Z70_S5 | 0.018478 | 0.012249 | 0.131418 | 0.881344 |
| myocardial | Z70_S5 | -0.02571 | 0.028313 | 0.363794 | 0.439145 |
| conduction | Z70_S5 | -0.03342 | 0.038848 | 0.389705 | 0.409265 |
| type 2 diab | Z70_S5 | 0.01965 | 0.024179 | 0.416411 | 0.380478 |
| coronary h | Z70_S5 | 0.011812 | 0.02181 | 0.588116 | 0.230537 |
| arrhythmia | Z70_S5 | 0.021309 | 0.048104 | 0.657793 | 0.181911 |
| hypotensio | Z70_S5 | 0.01705 | 0.042147 | 0.685829 | 0.163784 |
| angina pec | Z70_S5 | 0.01031 | 0.026293 | 0.694988 | 0.158023 |
| heart failur | Z70_S5 | -0.01153 | 0.036927 | 0.754938 | 0.122089 |
| other heart | Z70_S5 | -0.01293 | 0.054057 | 0.810901 | 0.091032 |
| atrial fibrill | Z70_S5 | 0.002034 | 0.02222 | 0.927063 | 0.032891 |
| high choles | Z70_S5 | 0.001081 | 0.013469 | 0.936016 | 0.028717 |
| mitral prob | Z70_S5 | 0.001765 | 0.044338 | 0.968248 | 0.014013 |
| type 2 diab | Z11_S5 | -0.05235 | 0.024396 | 0.031907 | 1.496111 |
| aortic valve | Z11_S5 | -0.09953 | 0.056775 | 0.079599 | 1.09909 |
| atrial fibrill | Z11_S5 | -0.03451 | 0.02241 | 0.12356 | 0.908121 |
| hypertensi | Z11_S5 | -0.01637 | 0.012372 | 0.185705 | 0.731176 |
| conduction | Z11_S5 | 0.036698 | 0.039208 | 0.3493 | 0.456802 |
| other heart | Z11_S5 | -0.04691 | 0.054552 | 0.389849 | 0.409104 |
| heart failur | Z11_S5 | -0.02611 | 0.037256 | 0.483442 | 0.315655 |
| coronary h | Z11_S5 | -0.0154 | 0.022004 | 0.483965 | 0.315186 |
| hypotensio | Z11_S5 | -0.02541 | 0.04254 | 0.550279 | 0.259417 |
| arrhythmia | Z11_S5 | -0.02095 | 0.048562 | 0.666175 | 0.176412 |
| myocardial | Z11_S5 | 0.011486 | 0.028574 | 0.687717 | 0.16259 |
| angina pec | Z11_S5 | -0.01021 | 0.026545 | 0.700606 | 0.154526 |
| high choles | Z11_S5 | -0.00408 | 0.013601 | 0.764085 | 0.116859 |
| mitral prob | Z11_S5 | -0.00238 | 0.044757 | 0.957506 | 0.018858 |
| atrial fibrill | Z41_S1 | -0.05144 | 0.024694 | 0.037243 | 1.428956 |
| angina pec | Z41_S1 | 0.045113 | 0.029191 | 0.122255 | 0.912734 |
| type 2 diab | Z41_S1 | 0.041074 | 0.026849 | 0.126084 | 0.899341 |
| aortic valve | Z41_S1 | 0.072382 | 0.062512 | 0.246924 | 0.607436 |
| coronary h | Z41_S1 | 0.024673 | 0.024211 | 0.308188 | 0.511185 |
| hypertensi | Z41_S1 | 0.013289 | 0.013596 | 0.328379 | 0.483625 |
| mitral prob | Z41_S1 | -0.0441 | 0.049271 | 0.370801 | 0.430859 |
| other heart | Z41_S1 | -0.05017 | 0.060083 | 0.403678 | 0.393965 |
| heart failur | Z41_S1 | -0.02201 | 0.041038 | 0.591696 | 0.227901 |
| high choles | Z41_S1 | -0.0078 | 0.014938 | 0.601661 | 0.220648 |
| myocardial | Z41_S1 | -0.00977 | 0.031458 | 0.756236 | 0.121343 |
| conduction | Z41_S1 | 0.005445 | 0.043168 | 0.899627 | 0.045937 |
| arrhythmia | Z41_S1 | -0.005 | 0.053479 | 0.92554 | 0.033605 |

| | | | | | |
|---|---|---|---|---|---|
| hypotensio | Z41_S1 | -0.00238 | 0.04685 | 0.959497 | 0.017957 |
| aortic valve | Z109_S1 | -0.1194 | 0.058372 | 0.040821 | 1.389119 |
| coronary h | Z109_S1 | -0.0462 | 0.022611 | 0.041055 | 1.386638 |
| myocardial | Z109_S1 | -0.05263 | 0.029373 | 0.07319 | 1.13555 |
| angina pec | Z109_S1 | -0.04144 | 0.027285 | 0.128846 | 0.889931 |
| atrial fibrill | Z109_S1 | -0.02864 | 0.023077 | 0.214555 | 0.66846 |
| hypertensi | Z109_S1 | -0.01422 | 0.01272 | 0.263696 | 0.578897 |
| heart failur | Z109_S1 | -0.03623 | 0.038317 | 0.344457 | 0.462865 |
| hypotensio | Z109_S1 | -0.03451 | 0.043754 | 0.430329 | 0.366199 |
| arrhythmia | Z109_S1 | -0.03009 | 0.049938 | 0.546798 | 0.262173 |
| other hear | Z109_S1 | -0.02796 | 0.056104 | 0.61821 | 0.208864 |
| mitral prob | Z109_S1 | -0.01789 | 0.046022 | 0.697445 | 0.15649 |
| type 2 diab | Z109_S1 | -0.0032 | 0.025112 | 0.898515 | 0.046475 |
| conduction | Z109_S1 | -0.0048 | 0.040309 | 0.905126 | 0.043291 |
| high choles | Z109_S1 | -0.00124 | 0.013972 | 0.929116 | 0.03193 |
| aortic valve | Z99_S5 | 0.114855 | 0.056434 | 0.041841 | 1.378403 |
| hypertensi | Z99_S5 | 0.019983 | 0.012297 | 0.104168 | 0.982266 |
| conduction | Z99_S5 | -0.04998 | 0.038976 | 0.199755 | 0.699501 |
| type 2 diab | Z99_S5 | 0.02809 | 0.024266 | 0.247039 | 0.607235 |
| myocardial | Z99_S5 | -0.03078 | 0.028409 | 0.278569 | 0.555068 |
| high choles | Z99_S5 | 0.006921 | 0.013519 | 0.608663 | 0.215623 |
| hypotensio | Z99_S5 | 0.018329 | 0.042284 | 0.664682 | 0.177386 |
| heart failur | Z99_S5 | -0.01595 | 0.037048 | 0.666916 | 0.175929 |
| angina pec | Z99_S5 | -0.00895 | 0.026384 | 0.734497 | 0.13401 |
| arrhythmia | Z99_S5 | 0.015591 | 0.048265 | 0.746669 | 0.126872 |
| atrial fibrill | Z99_S5 | 0.004533 | 0.022297 | 0.838906 | 0.076287 |
| mitral prob | Z99_S5 | -0.00718 | 0.044486 | 0.871763 | 0.059601 |
| coronary h | Z99_S5 | 0.000757 | 0.021879 | 0.972394 | 0.012158 |
| other hear | Z99_S5 | 0.000977 | 0.054227 | 0.985629 | 0.006286 |
