## Supplementary Tables for "Hundreds of cardiac MRI traits derived using 3D diffusion autoencoders share a common genetic architecture"

### Supplementary methods

**Algorithm 1** 3D Convolutional Encoder: Half UNet model with attention

**Input:** Input image $x$, number of latent factors $latent\_dim$, number of channels $n\_ch$, number of initial features $n\_features$, number of residual blocks $n\_res\_blocks$, multiplication factors of the features $feat\_multipliers$, maximum size of a group during group normalisation $group\_limit$
**Output:** Latent embeddings $emb$

**procedure** ENCODER(x)
  **function** RESIDUALBLOCK(x, in_ch, out_ch, group_limit)
    $x \leftarrow \text{GroupNormalisation}(x, \text{num\_groups} = min(\text{group\_limit}, \text{in\_ch}), \text{num\_channels} = \text{in\_ch})$
    $x \leftarrow \text{SiLU}(x)$
    **return** $\text{Conv3D}(x, \text{in\_channels} = \text{in\_ch}, \text{out\_channels} = \text{out\_ch}, \text{kernel\_size=3}, \text{stride=1}, \text{padding=1})$
  **end function**
  **function** ATTENTIONBLOCK(x, n_ch, group_limit)
    $x \leftarrow \text{GroupNormalisation}(x, \text{num\_groups} = min(\text{group\_limit}, \text{n\_ch}), \text{num\_channels} = \text{n\_ch})$

$q, k, v \leftarrow \text{Conv3D}(x, \text{in\_channels} = \text{n\_ch}, \text{out\_channels} = \text{n\_ch} \times 3, \text{kernel\_size} = 1, \text{stride} = 1, \text{padding} = 0)$
    $\text{scale} \leftarrow \frac{1}{\sqrt[4]{\text{n\_ch}}}$
    $\text{weight} \leftarrow\leftarrow (q \times \text{scale})(k \times \text{scale})^T$
    $\text{weight} \leftarrow \text{Softmax}(\text{weight})$
    **return** $weight \cdot v$
  **end function**
  **function** INPUTBLOCKS(x, n_ch, n_features, feat_multipliers, n_res_blocks, group_limit)
    $ch \leftarrow$ n_features × first value of feat_multipliers
    $x \leftarrow \text{Conv3D}(x, \text{in\_channels} = \text{n\_ch}, \text{out\_channels} = \text{ch}, \text{kernel\_size} = 3, \text{stride} = 1, \text{padding} = 1)$
    **for** each multiplier_value in feat_multipliers **do**
      **for** each n_res_blocks **do**
        $x \leftarrow \text{ResidualBlock}(x, ch, \text{multiplier\_value} \times ch, group\_limit)$
        $ch \leftarrow \text{multiplier\_value} \times ch$
      **end for**
      **if** not the final multiplier_value **then**
        $x \leftarrow \text{ResidualBlock}(x, ch, ch, group\_limit)$
        $x \leftarrow \text{AveragePool}(x, \text{stride} = 2)$
      **end if**
    **end for**
    **return** processed $x$ and final value of $ch$
  **end function**
  **function** MIDDLEBLOCK(x, ch, group_limit)
    $x \leftarrow \text{ResidualBlock}(x, ch, ch, group\_limit)$
    $x \leftarrow \text{AttentionBlock}(x, ch, group\_limit)$
    **return** $\text{ResidualBlock}(x, ch, ch, group\_limit)$
  **end function**
  **function** OUTBLOCK(x, ch, group_limit)
    $x \leftarrow \text{GroupNormalisation}(x, \text{num\_groups} = min(\text{group\_limit}, \text{ch}), \text{num\_channels} = \text{ch})$
    $x \leftarrow \text{SiLU}(x)$
    $x \leftarrow \text{AdaptiveAveragePool}(x, \text{output\_size} = 1 \times 1 \times 1)$
    **return** $\text{Conv3D}(x, \text{in\_channels} = \text{ch}, \text{out\_channels} = \text{latent\_dim}, \text{kernel\_size=1}, \text{stride=1}, \text{padding=0})$
  **end function**
  $x \leftarrow \text{InputBlocks}(x, n\_ch, n\_features, feat\_multipliers, n\_res\_blocks, group\_limit)$
  $x \leftarrow \text{MiddleBlock}(x, ch, group\_limit)$
  **return** $\text{OutBlock}(x, ch, group\_limit)$
**end procedure**

**Algorithm 2** Finding Best Matching Latent Factors

**Input:** A collection of latent factors obtained from multiple model runs.
**Output:** A list of the best-matched latent factor pairs across different model runs.

```
for each model run do
    for each latent factor in this reference model run do
        for each model run, excluding the current one do
            for each latent factor in this current model run do
                Compute the absolute Pearson correlation coefficient between the current pair of latent factors.
                if this similarity exceeds the current highest then
                    Update the highest similarity and the details of the pair that is the best matched.
                end if
            end for
        end for
        if a best match has been found then
            Record the reference and best match details, including the similarity score.
        end if
    end for
end for
```
