## Supplementary Figures for "Hundreds of cardiac MRI traits derived using 3D diffusion autoencoders share a common genetic architecture"

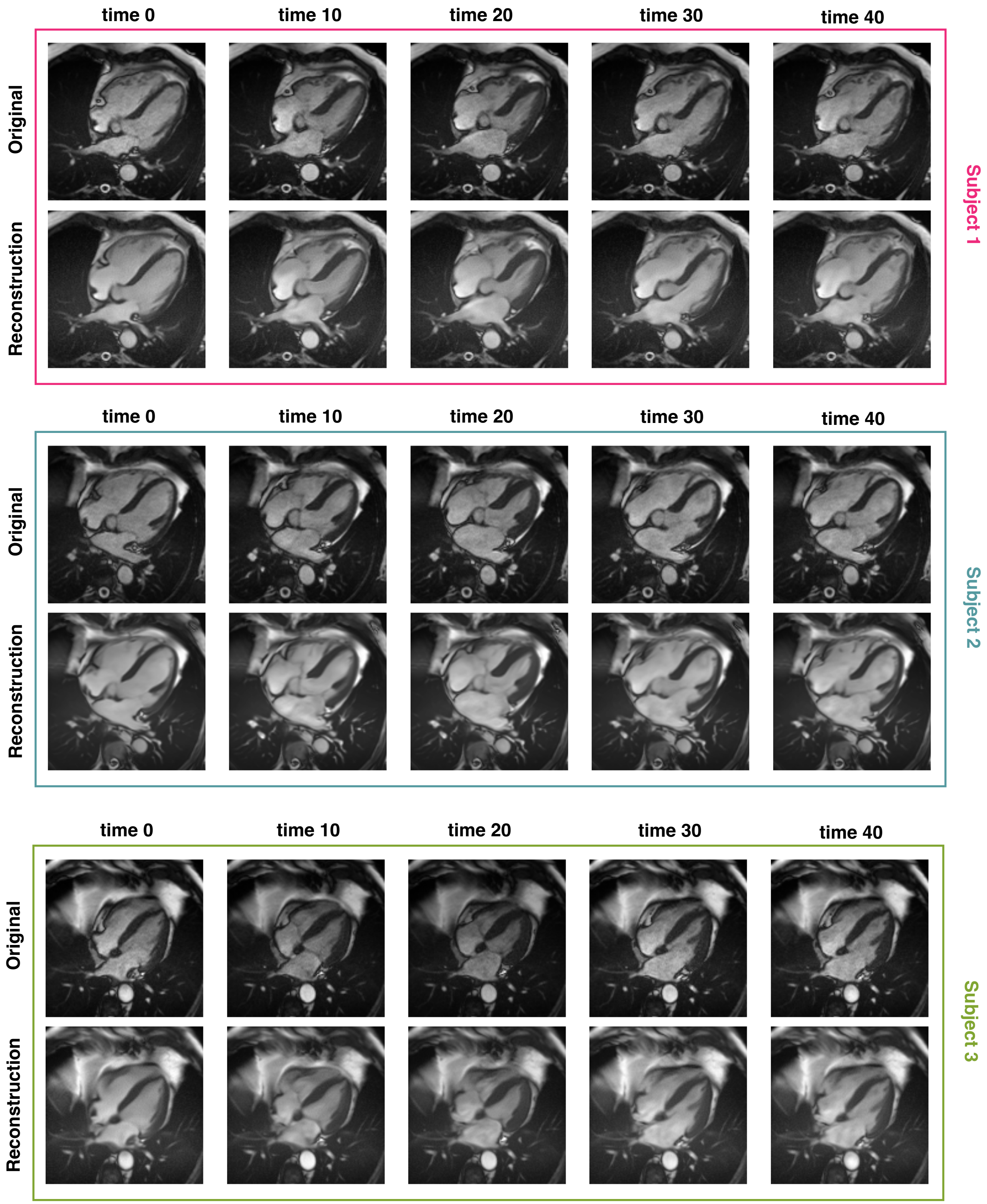


**Supplementary figure 1:** Examples of three 3D DiffAE reconstructions at different time points.

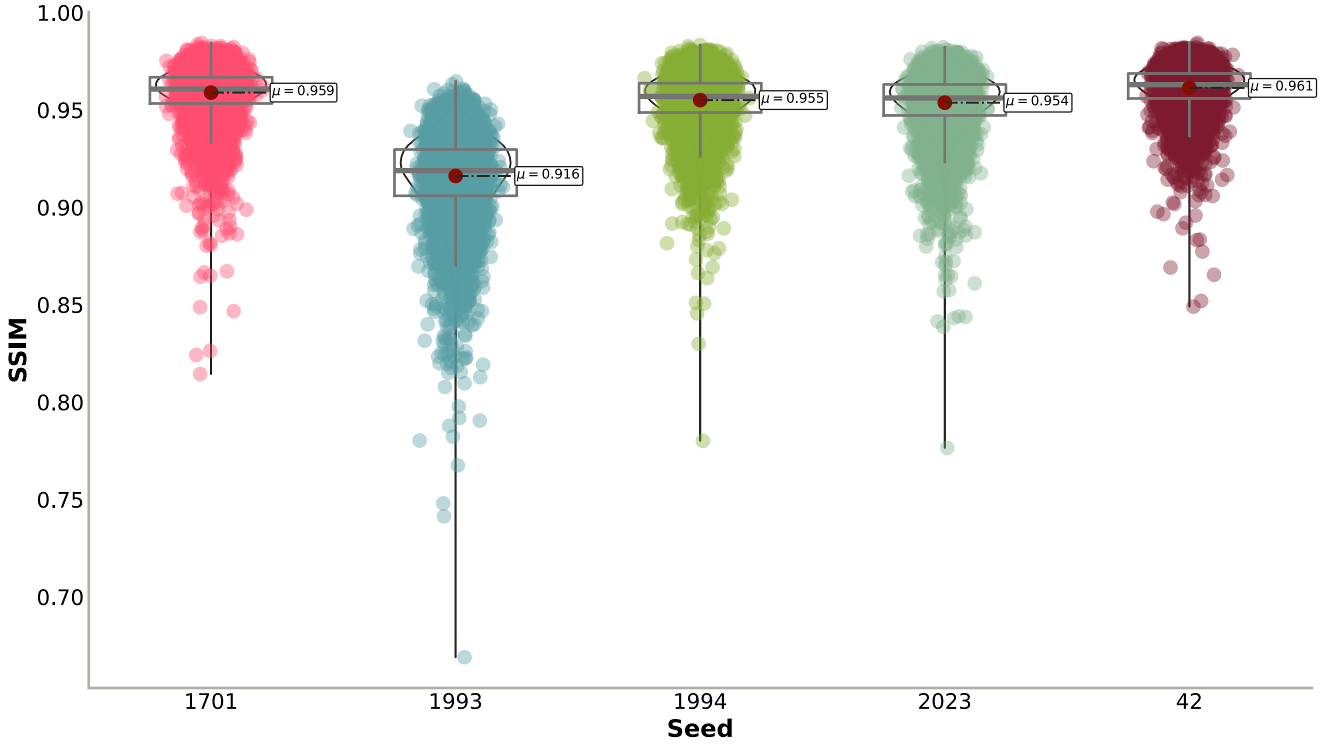


**(a)** Structural Similarity Index (SSIM)

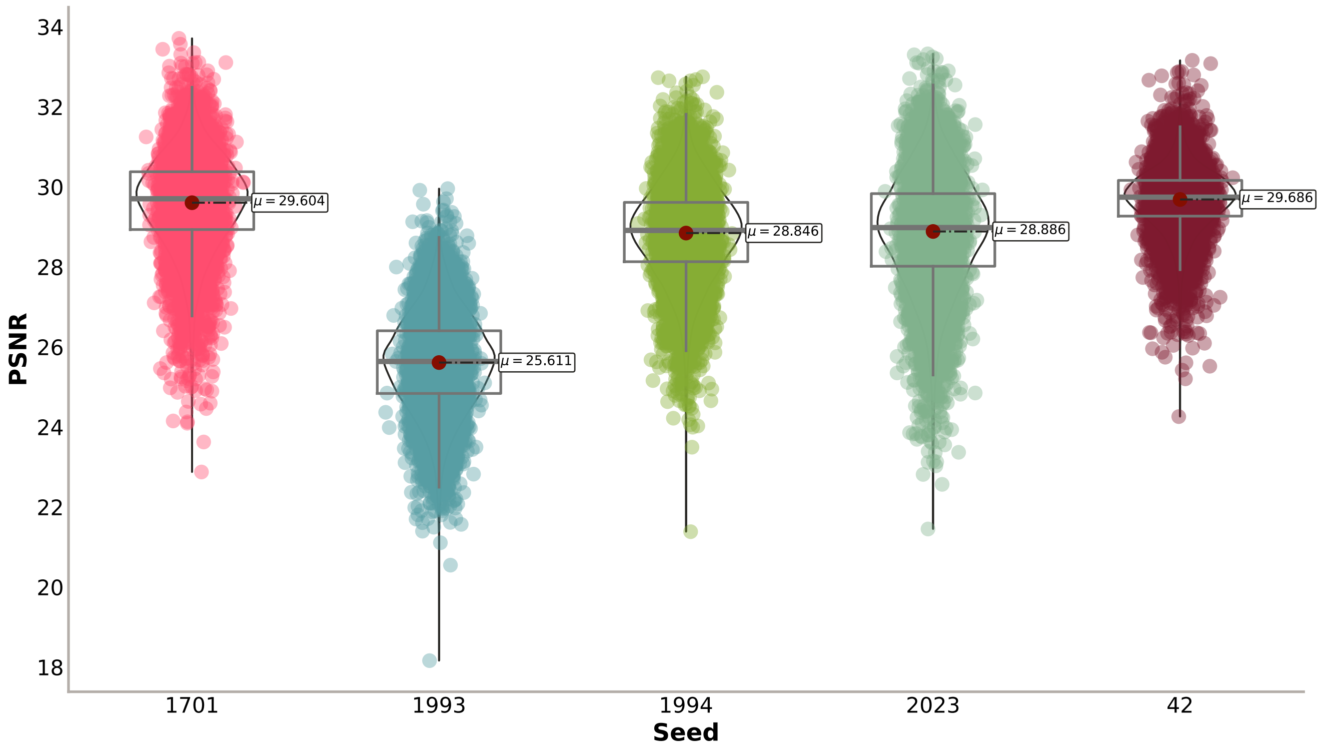


**(b)** Peak Signal-to-noise Ratio (PSNR)

**Supplementary figure 2:** The reconstruction performance of the trained 3D DiffAE models at $t\_step = 20$ across the five different seeds for 10,000 randomly chosen subjects from the test set.

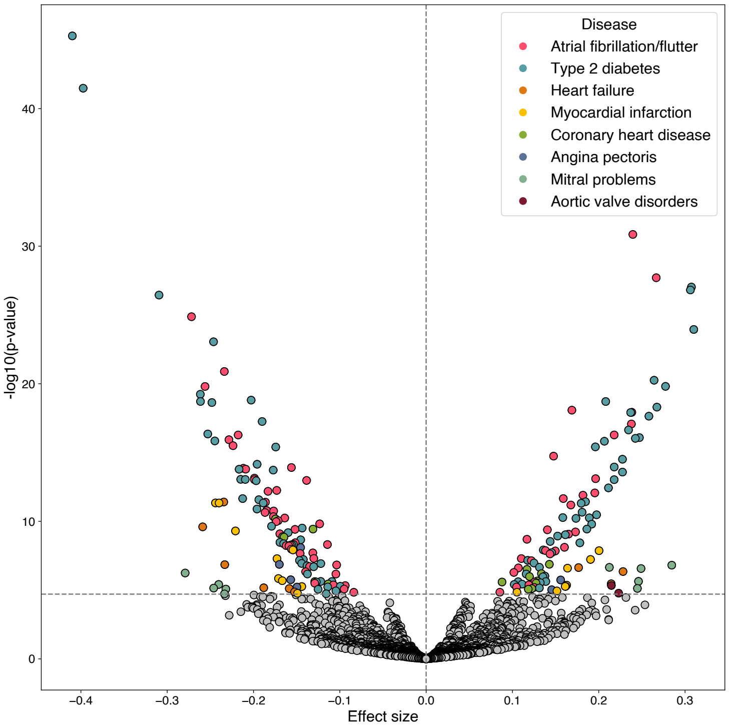


**(a)** Association with diseases

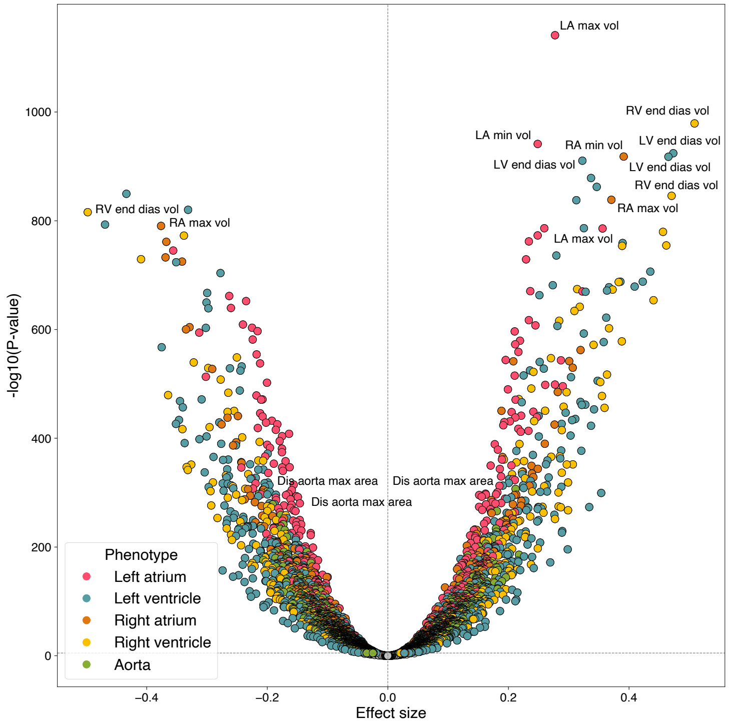


**(b)** Association with continuous traits

**Supplementary figure 3:** Volcano plots of associations between latent phenotype and diseases (a) and MRI-derived continuous traits (b), only including BSA as covariate. Bonferroni adjusted P-value threshold: $p < 1.96 \times 10^{-05}$ for diseases, $p < 9.8 \times 10^{-06}$ for continuous traits. For each category of continuous traits, the top three associations are labelled. LV: left ventricle, RV: right ventricle, LA: left atrium, RA: right atrium.

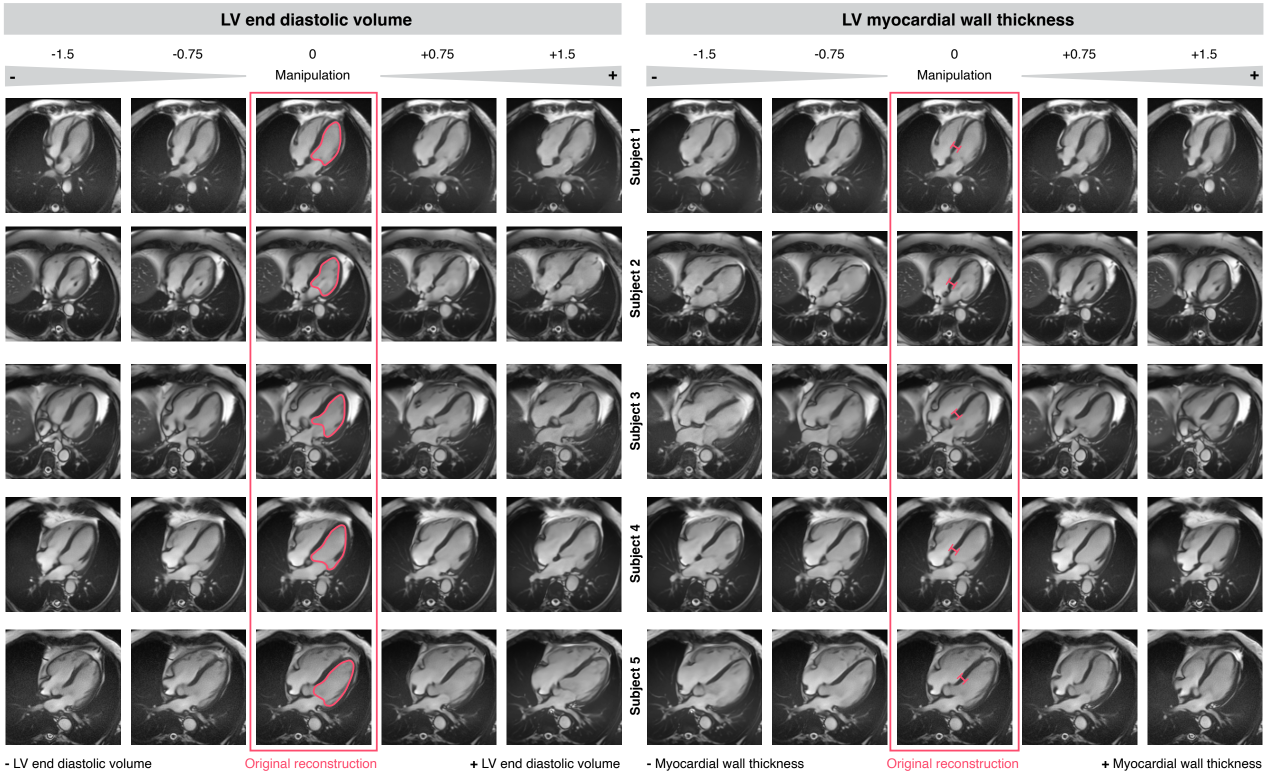


**Supplementary figure 4:** Example reconstructions following latent manipulation for two distinct phenotypes — left ventricular (LV) end-diastolic volume and LV myocardial wall thickness — demonstrate variations in the reconstructed images corresponding to changes in the values of these latent phenotypic traits.

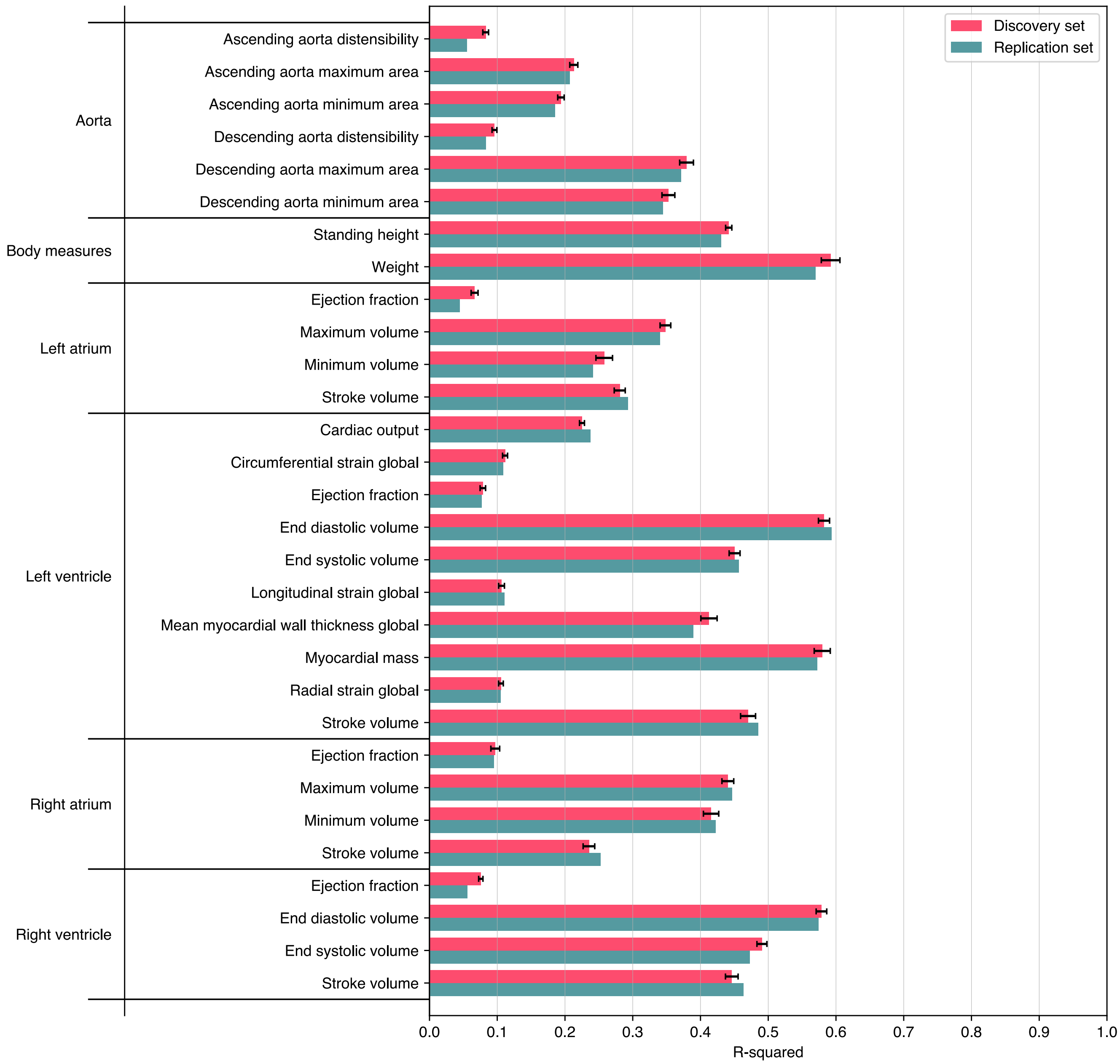


**Supplementary figure 5:** Lasso predictions of continuous traits from the latent phenotypes: Regression R-squared results, in the discovery (5-fold cross-validation mean plus/minus standard deviation) and replication cohorts.

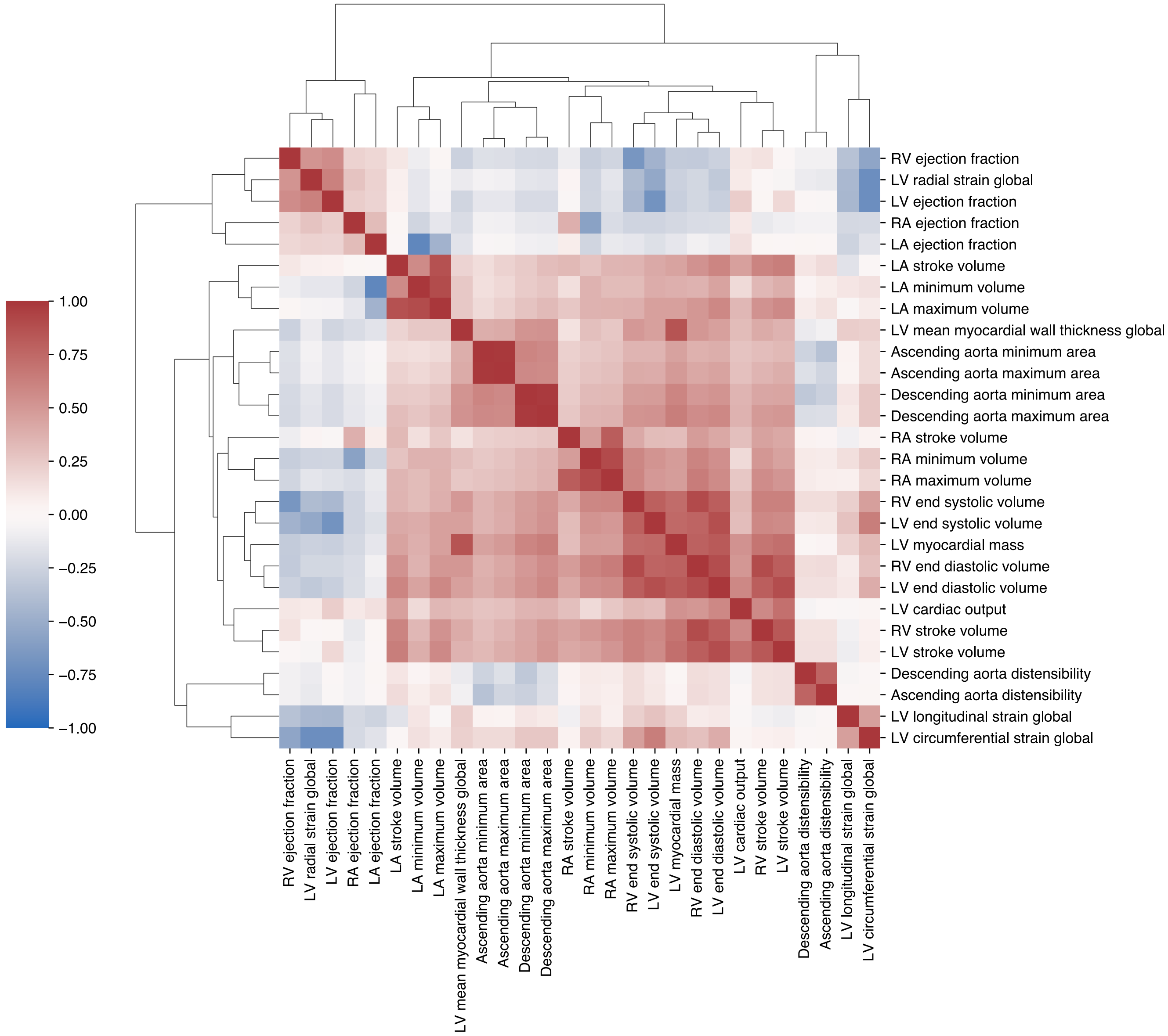


**Supplementary figure 6:** Heatmap of correlations between the MRI-derived cardiac measures, ordered with hierarchical clustering. LV: left ventricle, RV: right ventricle, LA: left atrium, RA: right atrium.

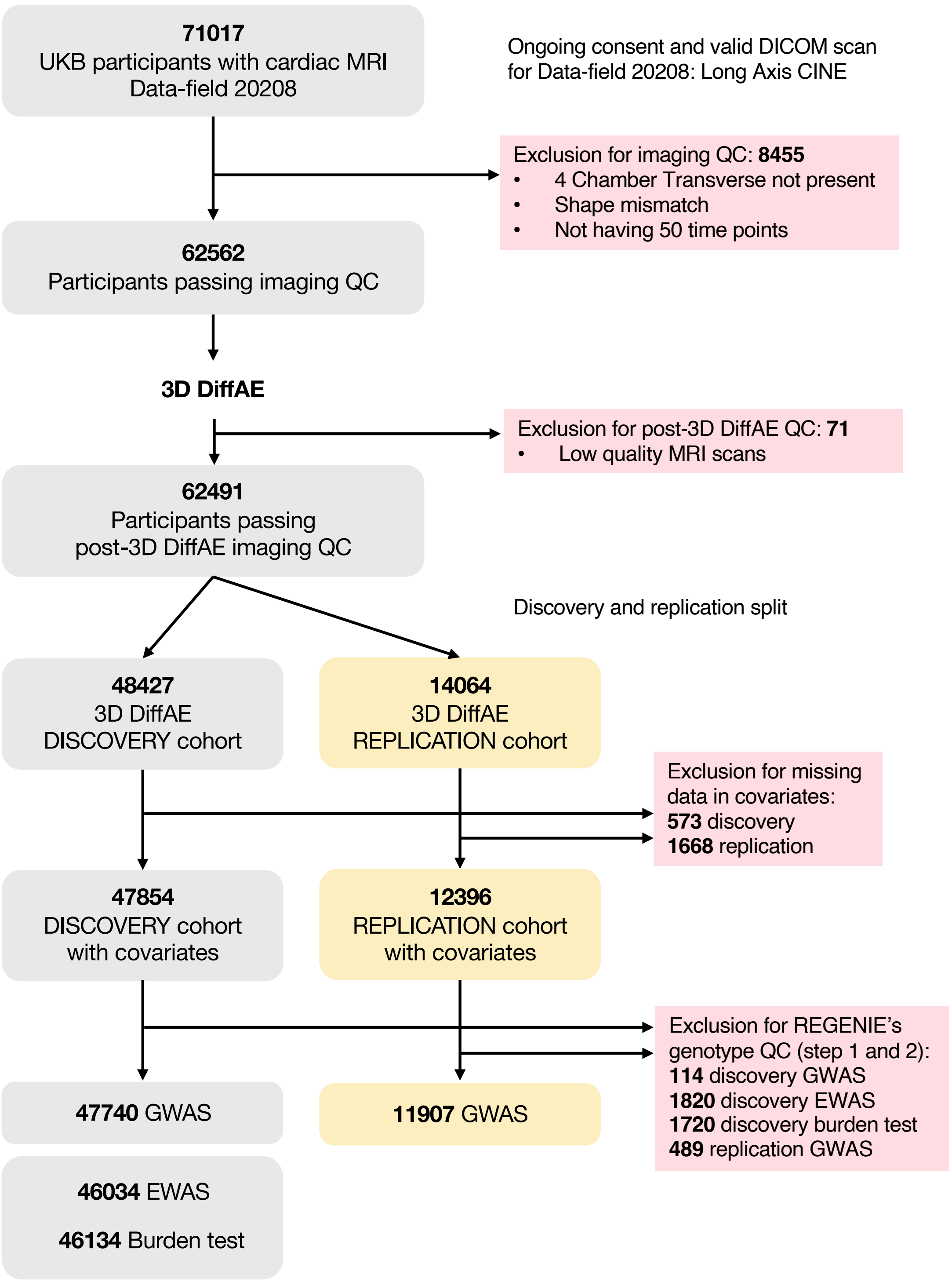


**Supplementary figure 7:** The diagram illustrates the sample filtering process that led to the specific individuals chosen for discovery and replication cohorts to perform GWAS and rare variant analyses.

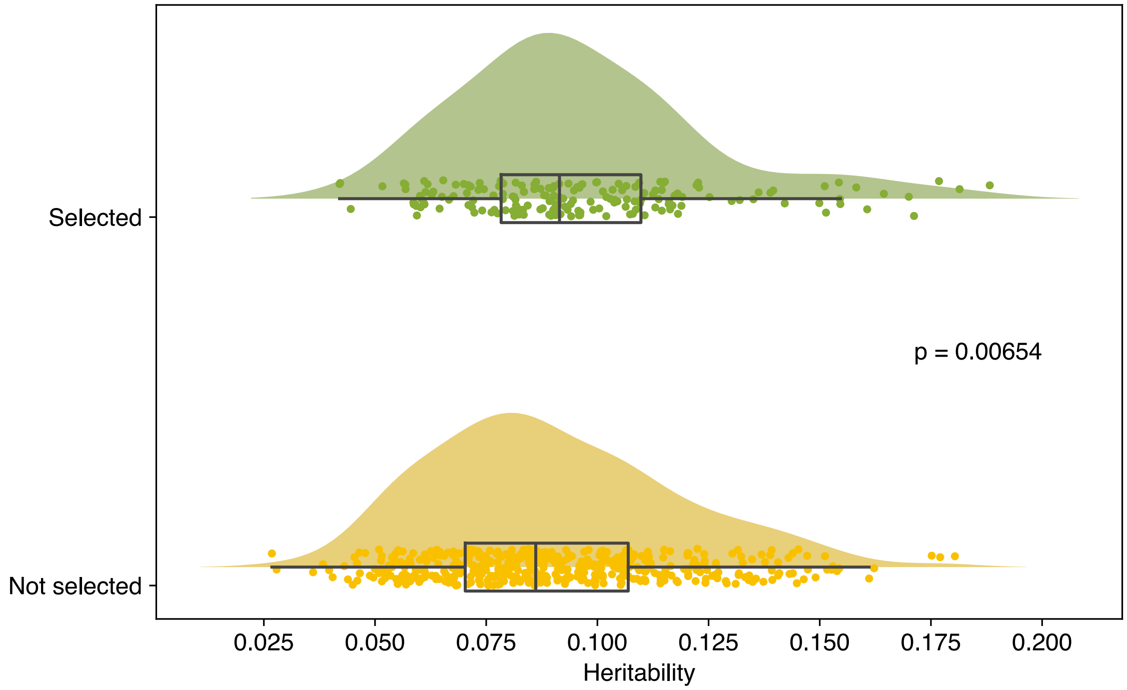


**Supplementary figure 8:** Distribution of latent phenotypes' heritability, comparing the 182 selected traits with the non-selected. P-value from one-sided Mann-Whitney U test confirms that selected latents on average have a higher heritability.

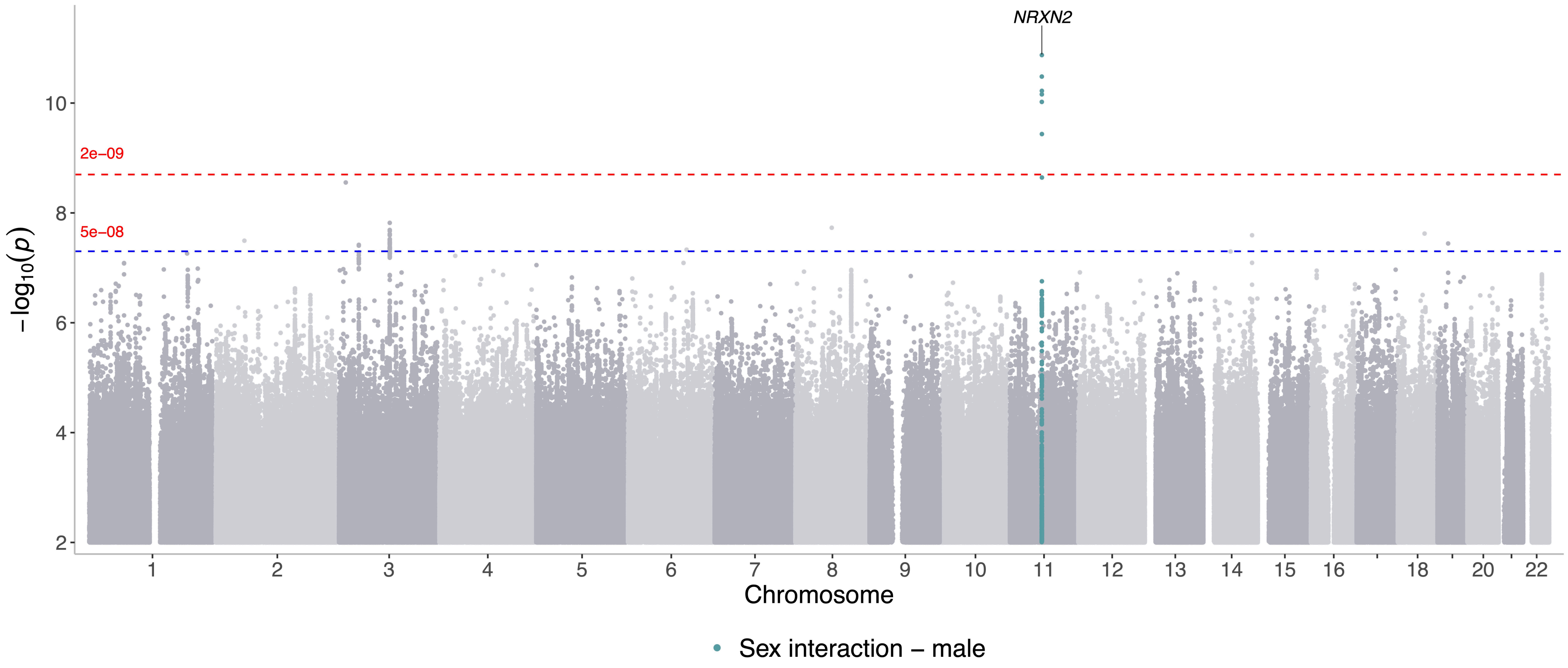


**Supplementary figure 9:** Manhattan plot of gene-by-sex interaction GWAS of all the 182 latent phenotypes. The thresholds are the same as the discovery GWAS, blue line: genome-wide significance ($5 \times 10^{-8}$), red line: study-wide significance ($2.27 \times 10^{-9}$).

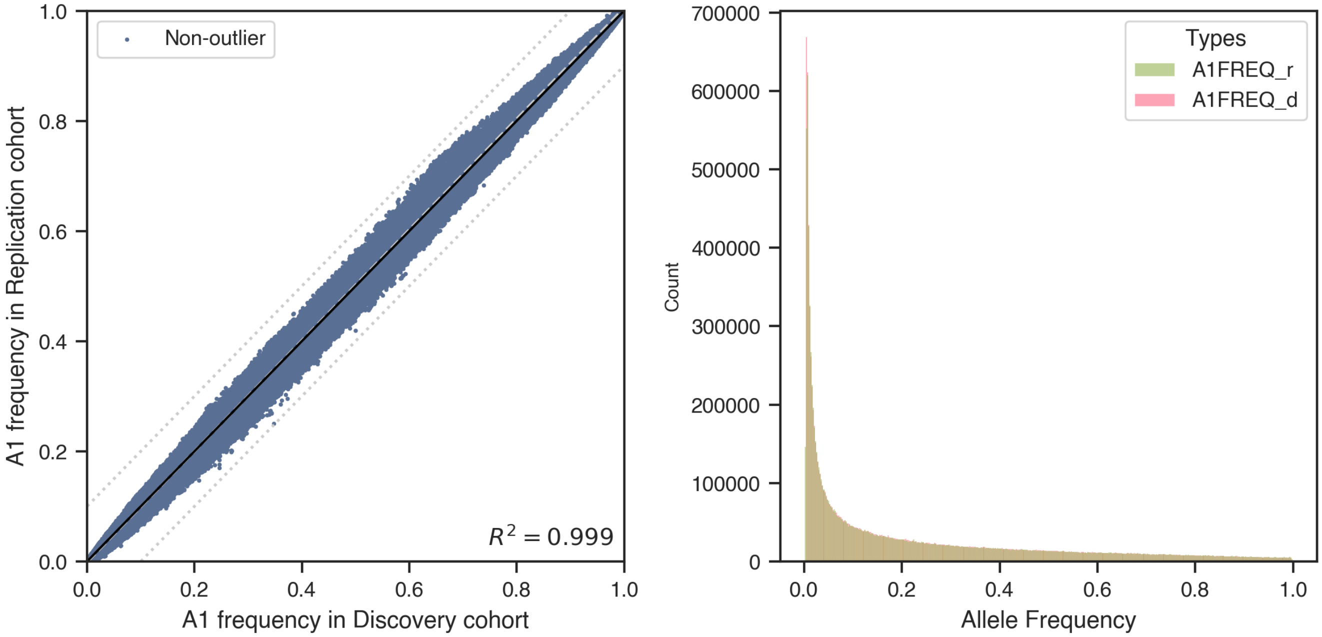


**Supplementary figure 10:** Comparison of allele frequencies between discovery (d) and replication (r) GWAS cohorts. No outliers detected with a 0.1 difference threshold.

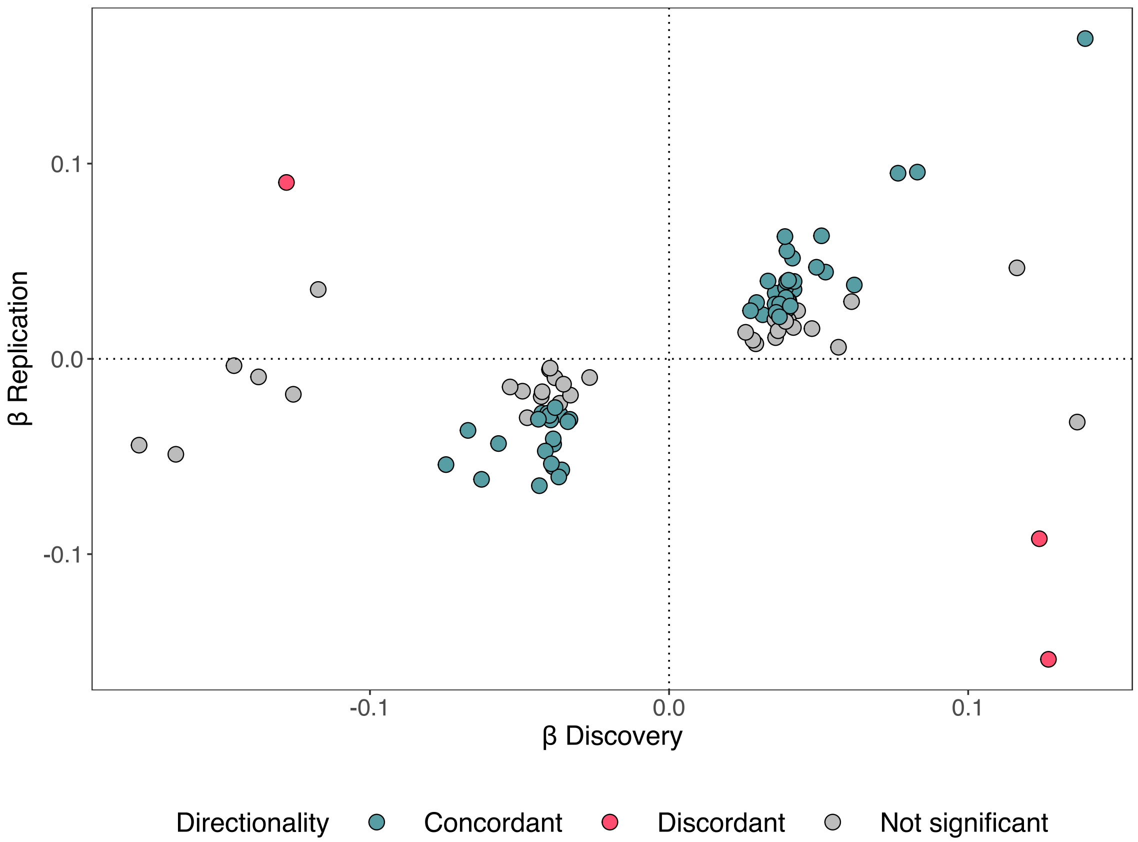


**Supplementary figure 11:** Comparison of effect sizes between discovery and replication for the 89 conditionally independent significant SNPs. In blue the variants with concordant directions of effect, in red those with opposite directions. 51 SNPs with concordant effect were significant at a nominal threshold of $p < 0.05$, those with a higher P-value are coloured in grey.

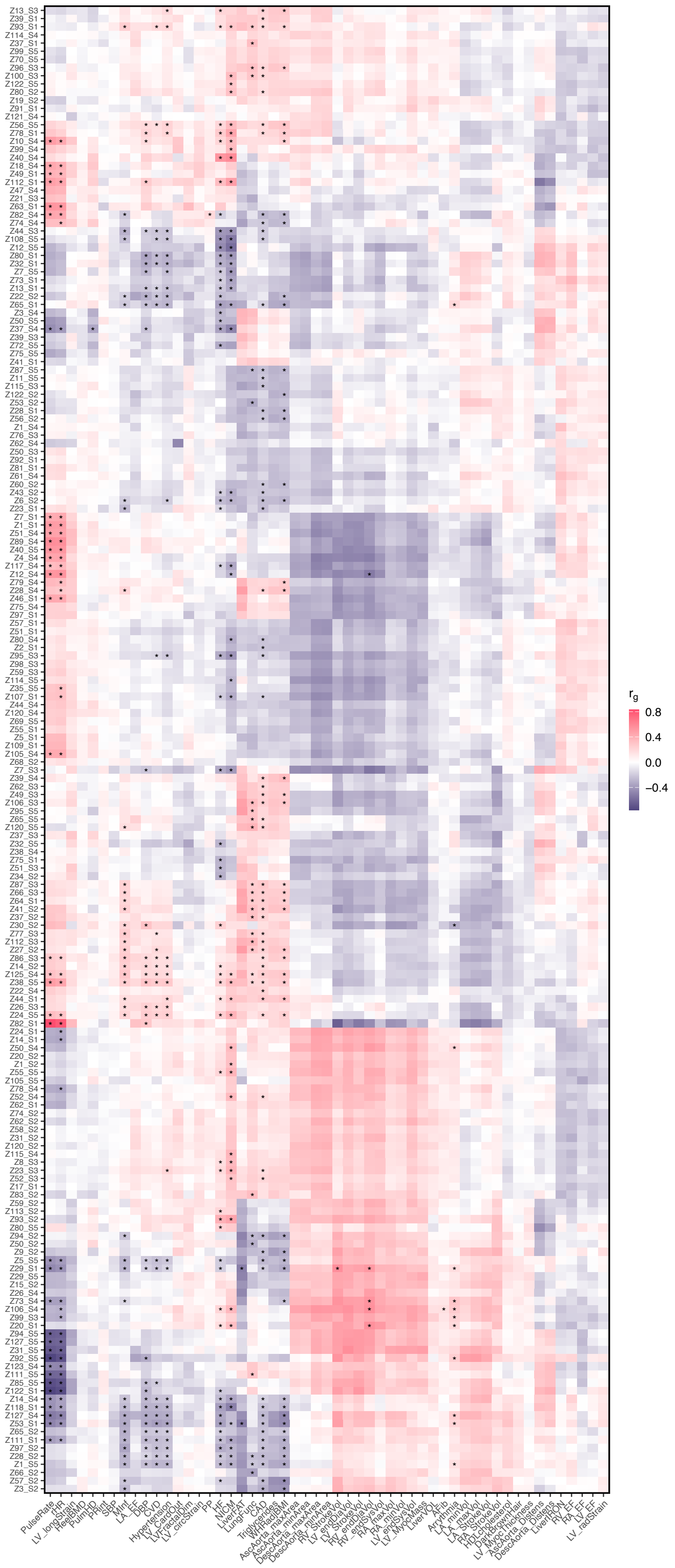


**Supplementary figure 12:** Heatmap of genetic correlations between latent phenotypes (y axis) and relevant cardiovascular traits (x axis). Asterisks are marking significance at Bonferroni-adjusted threshold of P-value $= 4.21 \times 10^{-5}$ (see methods).

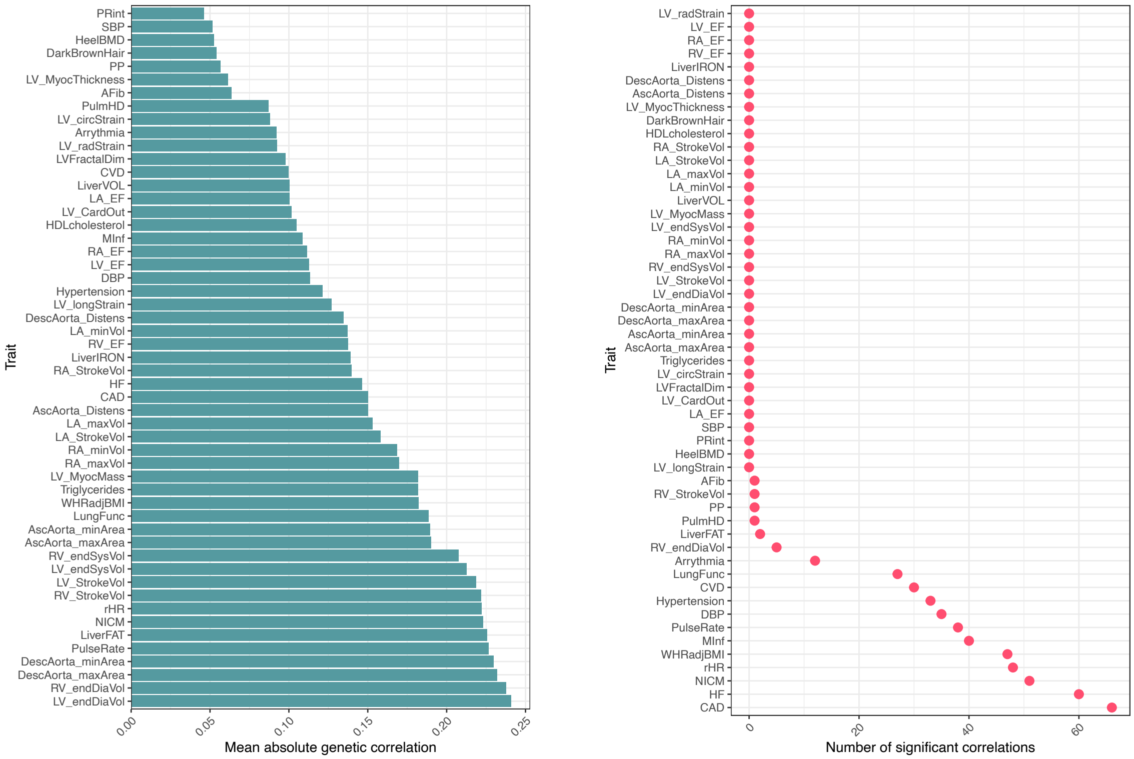


**(a)** Mean absolute correlation **(b)** Number of significant correlations

**Supplementary figure 13:** Summary of the genetic correlation results between latent phenotypes and relevant traits. For each trait, the mean absolute correlation (a) and number of significant correlations (b) across all 182 latent phenotypes is shown, considering the Bonferroni-adjusted threshold of P-value $= 4.21 \times 10^{-05}$ (see methods).

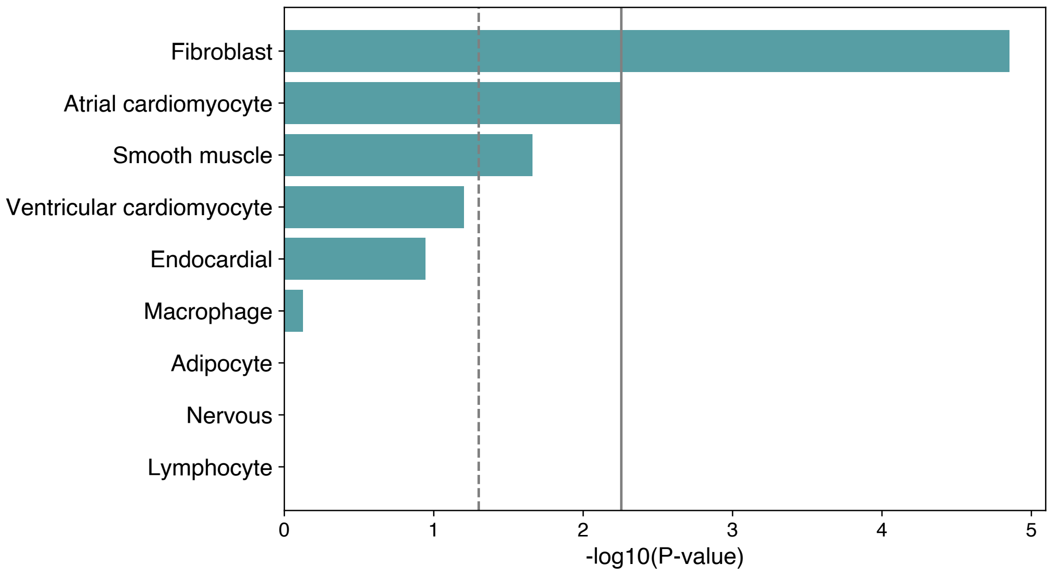


**Supplementary figure 14:** Latent phenotypes' GWAS lead SNPs enrichment in cardiac cell-type specific snATAC-seq peaks using CHEERS method. The solid line represents the Bonferroni-corrected threshold ($p < 0.05/9$ cell types), while the dotted line marks the nominal P-value significance of 0.05.

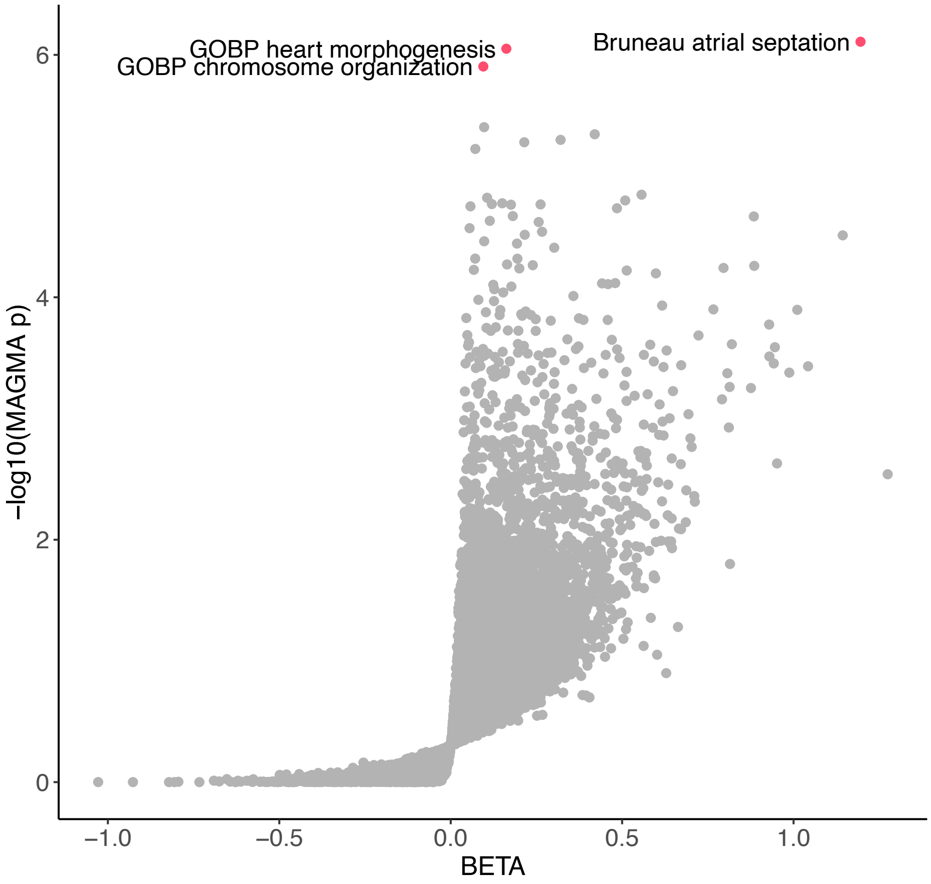


**Supplementary figure 15:** MAGMA gene set analysis on curated gene sets and GO terms. Significant gene sets with Bonferroni-adjusted P-value $p < 0.05$ are annotated.

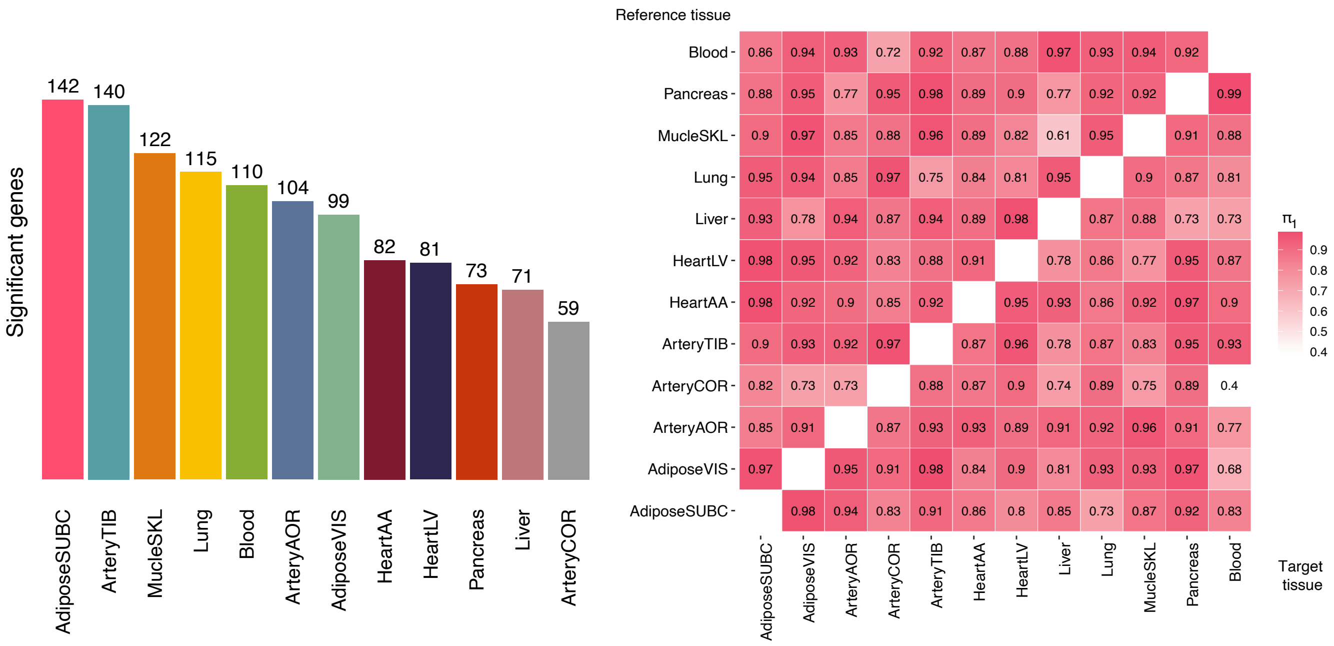


**(a)** Significant TWAS genes per tissue

**(b)** TWAS $\pi_1$ statistic

**Supplementary figure 16:** Tissue distribution of the significant TWAS protein coding genes (a), $\pi_1$ statistic (see methods) to quantify the extent of TWAS significant genes sharing between each tissue pair (b).

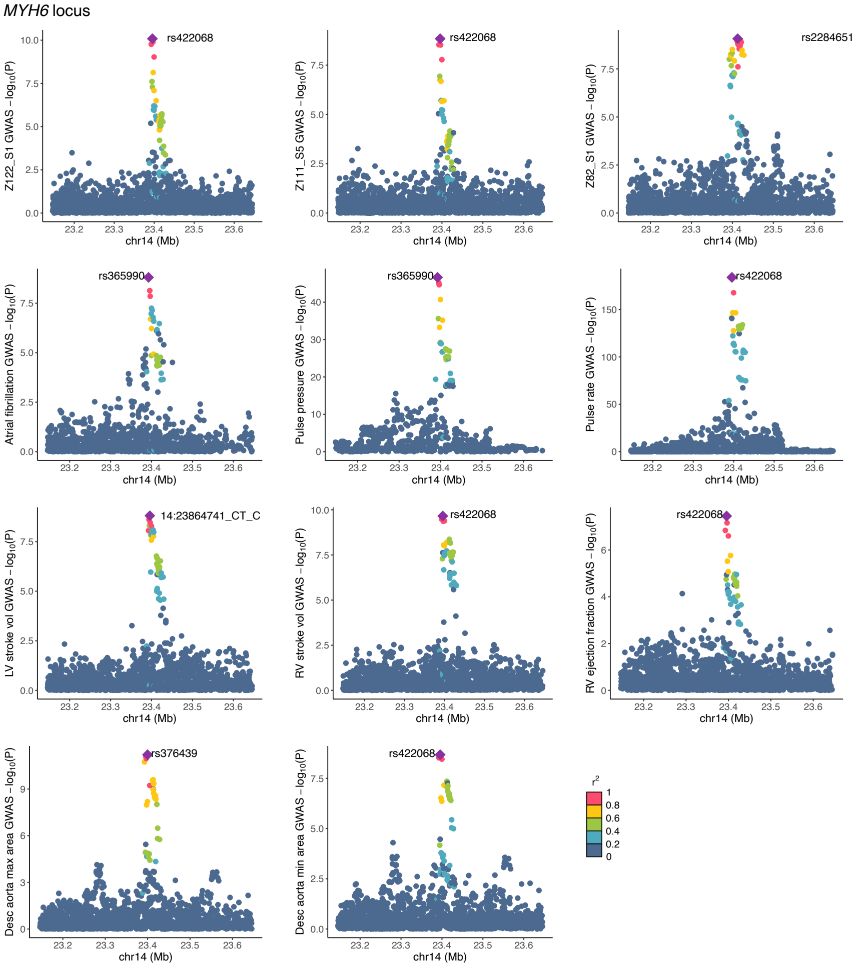


**Supplementary figure 17:** Locuszoom plots showing the colocalising signals in the *MYH6* locus associated with latent phenotypes. For each colocalising trait, the plot shows the -log10(P) of the variants across the genomic region. The lead SNP is annotated and the other variants are coloured according to the LD.

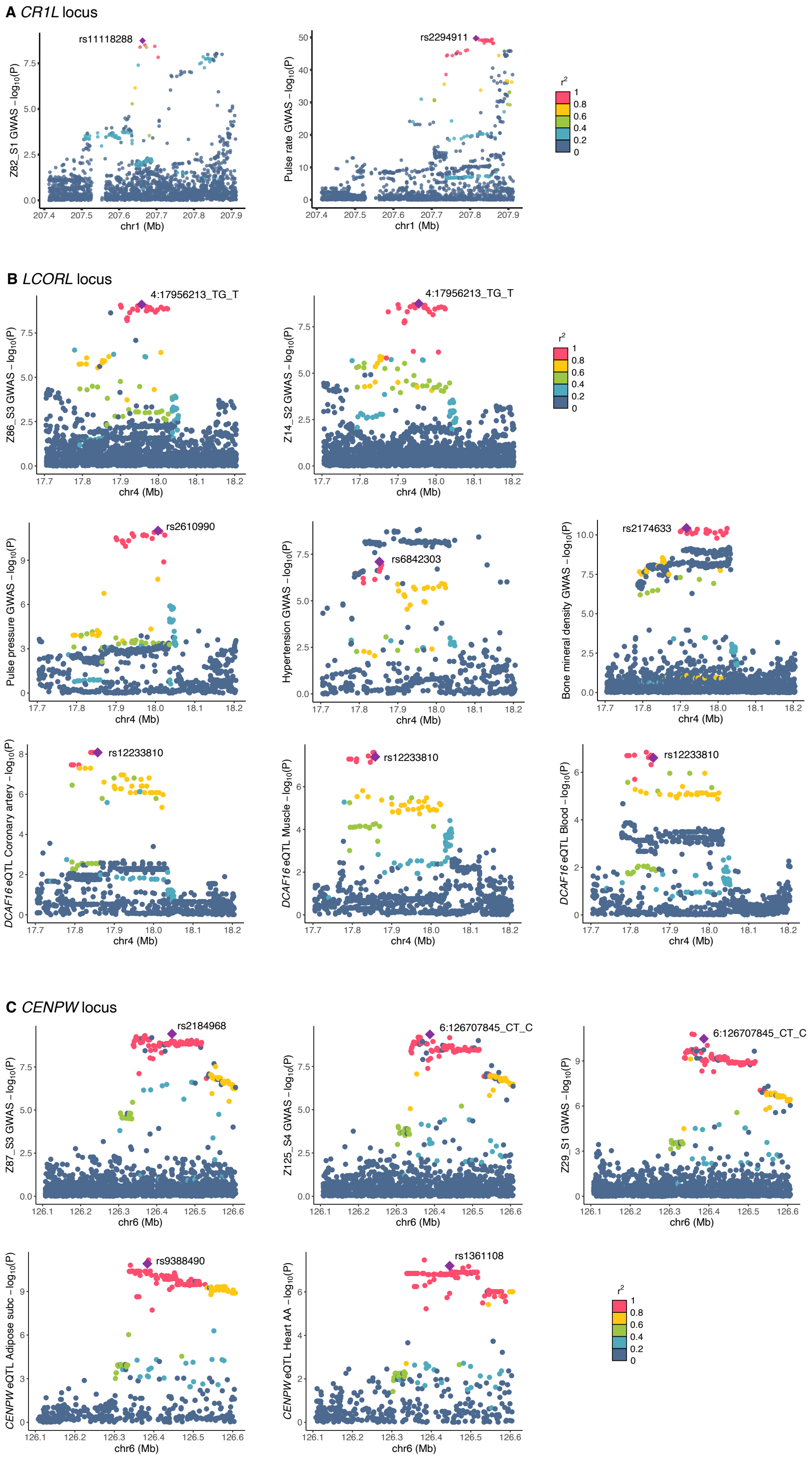


**Supplementary figure 18:** Locuszoom plots showing the colocalising signals of three novel loci associated with latent phenotypes: *CR1L* (**A**), *LCORL* (**B**) and *CENPW* (**C**). For each colocalising trait, the plot shows the -log10(P) of the variants across the genomic region. The lead SNP is annotated and the other variants are coloured according to the LD.

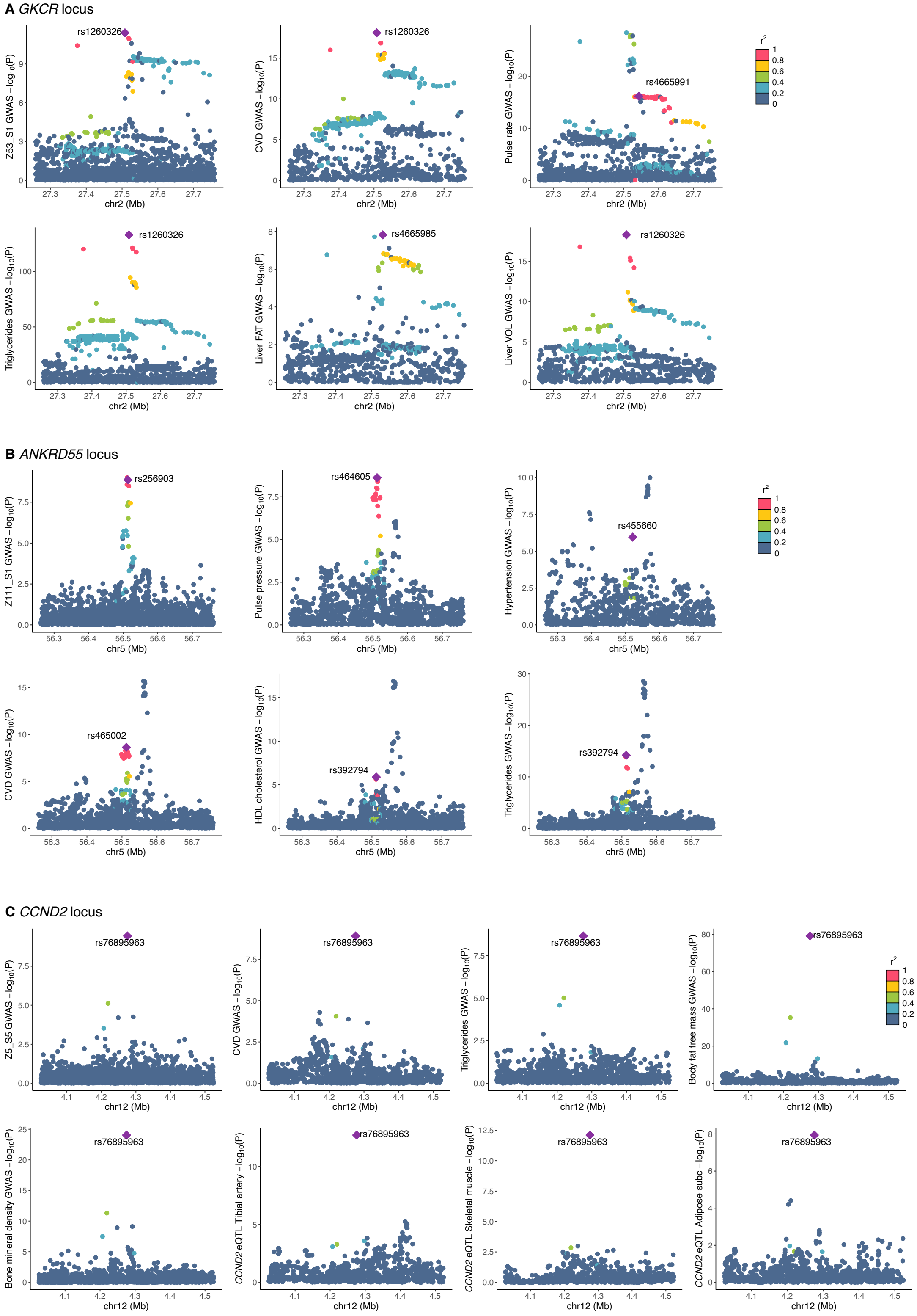


**Supplementary figure 19:** Locuszoom plots showing the colocalising signals of three loci associated with latent phenotypes multiple trait categories: *GKCR* (**A**), *ANKRD55* (**B**) and *CCND2* (**C**). For each colocalising trait, the plot shows the -log10(P) of the variants across the genomic region. The lead SNP is annotated and the other variants are coloured according to the LD.

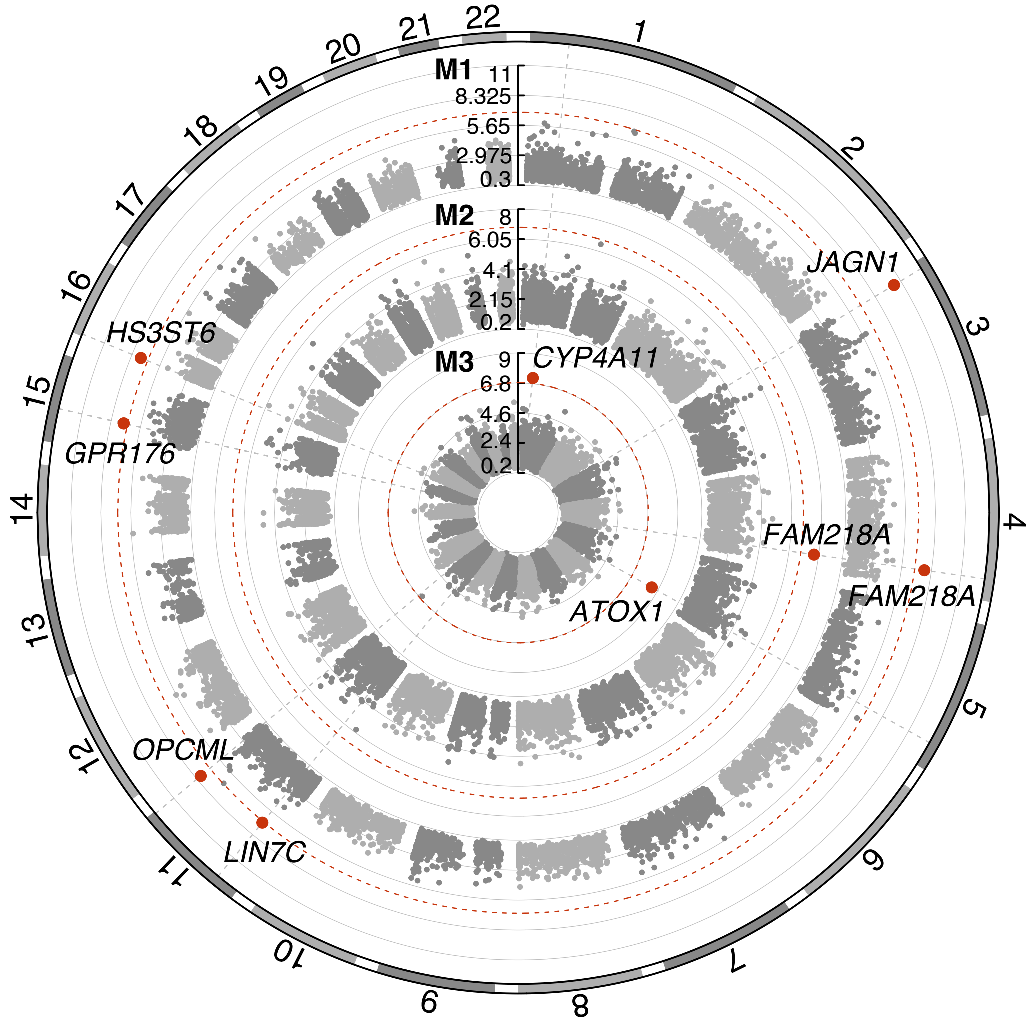


**Supplementary figure 20:** Burden test circular Manhattan plot representing three different masks: M1 LoF, M2 LoF and missense (5/5), M3 LoF and missense (>=1/5). On the y axis -log10(P-value), with significant genes annotated and marked in red according to the P-value threshold of $1.5 \times 10^{-07}$ (dashed red line).

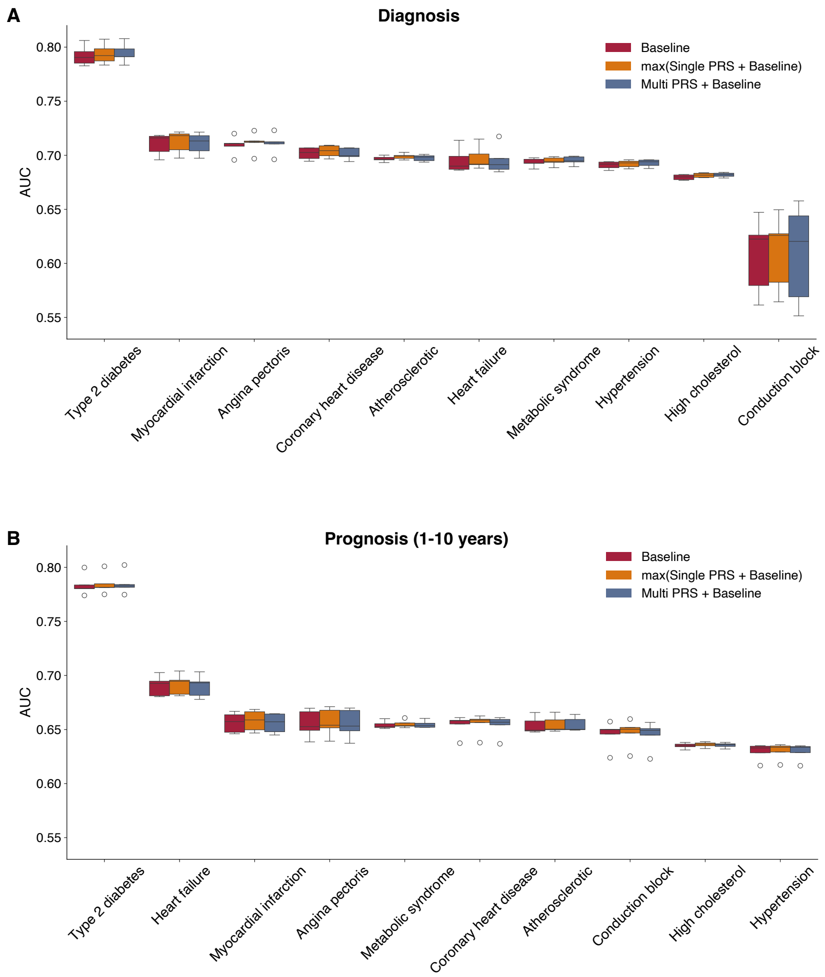


**Supplementary figure 21:** AUC comparison of 3 PRS models for diagnosis **A** and prognosis from 1 to 10 years **B** for 10 diseases.

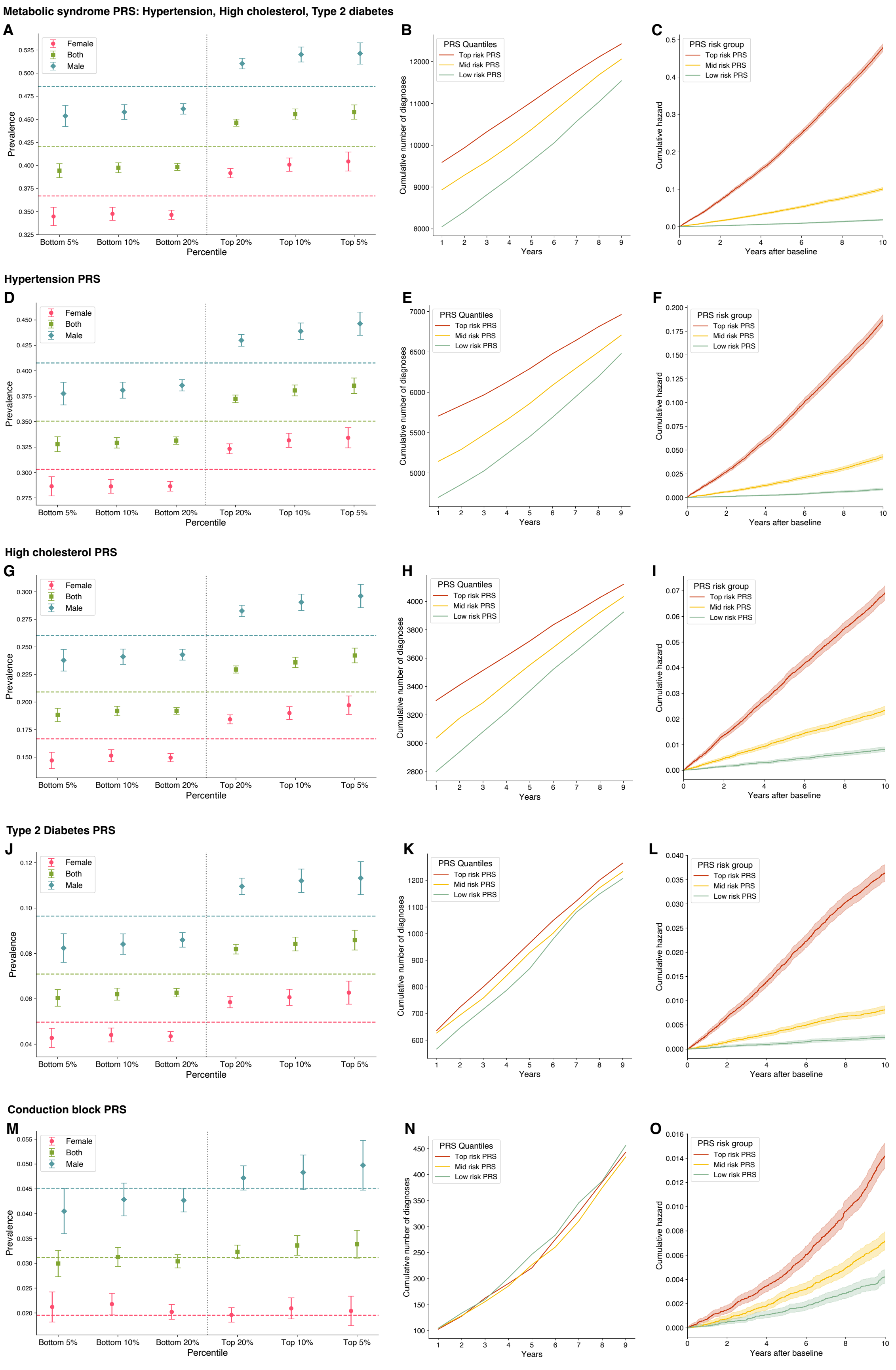


**Supplementary figure 22: A,D,G,J,M**. PRS prevalence plot stratified by sex, **B,E,H,K,N**. Cumulative disease burden, and **C,F,I,L,O** cumulative hazard by PRS risk quantiles, for metabolic syndrome (using its sub-diseases: hypertension, high cholesterol, type 2 diabetes) and conduction block.

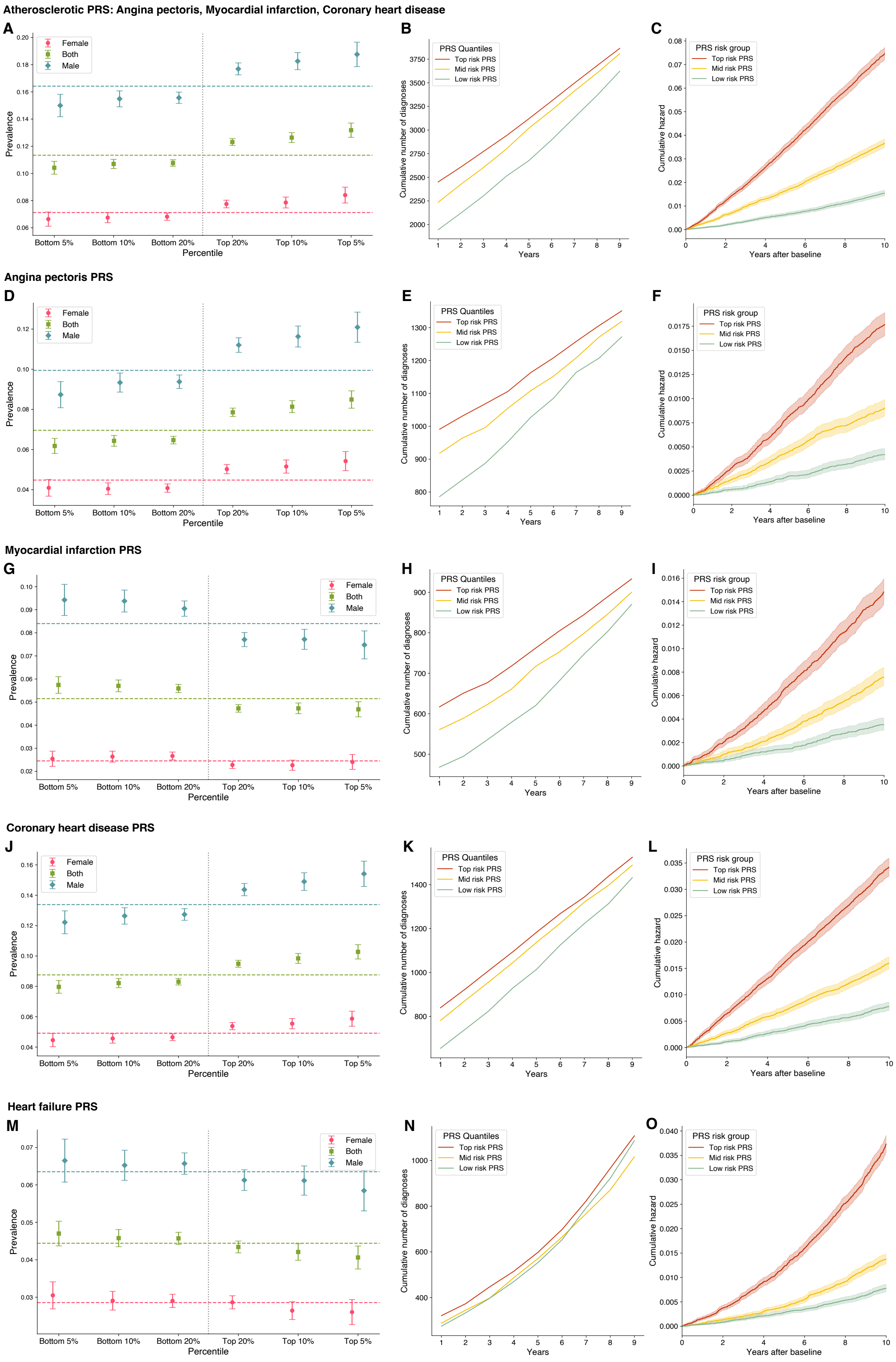


**Supplementary figure 23: A,D,G,J,M**. PRS prevalence plot stratified by sex, **B,E,H,K,N**. Cumulative disease burden, and **C,F,I,L,O** cumulative hazard by PRS risk quantiles, for atherosclerotic diseases (using its sub-diseases: angine pectoris, myocardial infraction, coronary hearth disease) and heart failure.
